## Appendix 1 for "Early and ongoing importations of SARS-CoV-2 in Canada"

| Accession | strain.x | Collection Date | Location | Originating Laboratory | Submitting Laboratory | Authors |
|---|---|---|---|---|---|---|
| EPI_ISL_402119 | Wuhan/IVDC-HB-01/2019 | 2019-12-30 | Asia/China/Hubei/Wuhan | National Institute for Viral Disease Control and Prevention, China CDC | National Institute for Viral Disease Control and Prevention, China CDC | Wenjie Tan et al |
| EPI_ISL_402120 | Wuhan/IVDC-HB-04/2020 | 2020-01-01 | Asia/China/Hubei/Wuhan | National Institute for Viral Disease Control and Prevention, China CDC | National Institute for Viral Disease Control and Prevention, China CDC | Wenjie Tan et al |
| EPI_ISL_402121 | Wuhan/IVDC-HB-05/2019 | 2019-12-30 | Asia/China/Hubei/Wuhan | National Institute for Viral Disease Control and Prevention, China CDC | National Institute for Viral Disease Control and Prevention, China CDC | Wenjie Tan et al |
| EPI_ISL_402123 | Wuhan/IPBCAMS-WH-01/2019 | 2019-12-24 | Asia/China/Hubei/Wuhan | Institute of Pathogen Biology, Chinese Academy of Medical Sciences & Peking Union Medical College | Institute of Pathogen Biology, Chinese Academy of Medical Sciences & Peking Union Medical College | Lili Ren et al |
| EPI_ISL_402124 | Wuhan/WIV04/2019 | 2019-12-30 | Asia/China/Hubei/Wuhan | Wuhan Jinyintan Hospital | Wuhan Institute of Virology, Chinese Academy of Sciences | Peng Zhou et al |
| EPI_ISL_402125 | Wuhan/Hu-1/2019 | 2019-12-26 | Asia/China/Hubei/Wuhan | National Institute for Communicable Disease Control and Prevention (ICDC) Chinese Center for Disease Control and Prevention (China CDC) | National Institute for Communicable Disease Control and Prevention (ICDC) Chinese Center for Disease Control and Prevention (China CDC) | Zhang et al |
| EPI_ISL_402127 | Wuhan/WIV02/2019 | 2019-12-30 | Asia/China/Hubei/Wuhan | Wuhan Jinyintan Hospital | Wuhan Institute of Virology, Chinese Academy of Sciences | Peng Zhou et al |

| | | | | | | |
|---|---|---|---|---|---|---|
| EPI_ISL_402128 | Wuhan/WIV05/2019 | 2019-12-30 | Asia/China/Hubei/Wuhan | Wuhan Jinyintan Hospital | Wuhan Institute of Virology, Chinese Academy of Sciences | Peng Zhou et al |
| EPI_ISL_402129 | Wuhan/WIV06/2019 | 2019-12-30 | Asia/China/Hubei/Wuhan | Wuhan Jinyintan Hospital | Wuhan Institute of Virology, Chinese Academy of Sciences | Peng Zhou et al |
| EPI_ISL_402130 | Wuhan/WIV07/2019 | 2019-12-30 | Asia/China/Hubei/Wuhan | Wuhan Jinyintan Hospital | Wuhan Institute of Virology, Chinese Academy of Sciences | Peng Zhou et al |
| EPI_ISL_402132 | Wuhan/HBCDC-HB-01/2019 | 2019-12-30 | Asia/China/Hubei/Wuhan | Wuhan Jinyintan Hospital | Hubei Provincial Center for Disease Control and Prevention | Bin Fang et al |
| EPI_ISL_403929 | Wuhan/IPBCAMS-WH-04/2019 | 2019-12-30 | Asia/China/Hubei/Wuhan | Institute of Pathogen Biology, Chinese Academy of Medical Sciences & Peking Union Medical College | Institute of Pathogen Biology, Chinese Academy of Medical Sciences & Peking Union Medical College | Lili Ren et al |
| EPI_ISL_403930 | Wuhan/IPBCAMS-WH-03/2019 | 2019-12-30 | Asia/China/Hubei/Wuhan | Institute of Pathogen Biology, Chinese Academy of Medical Sciences & Peking Union Medical College | Institute of Pathogen Biology, Chinese Academy of Medical Sciences & Peking Union Medical College | Lili Ren et al |
| EPI_ISL_403931 | Wuhan/IPBCAMS-WH-02/2019 | 2019-12-30 | Asia/China/Hubei/Wuhan | Institute of Pathogen Biology, Chinese Academy of Medical Sciences & Peking Union Medical College | Institute of Pathogen Biology, Chinese Academy of Medical Sciences & Peking Union Medical College | Lili Ren et al |

| | | | | | | |
|---|---|---|---|---|---|---|
| EPI_ISL_403932 | Guangdong/20SF012/2020 | 2020-01-14 | Asia/China/Guangdong/Shenzhen | Guangdong Provincial Center for Diseases Control and Prevention; Guangdong Provincial Public Health | Department of Microbiology, Guangdong Provincial Center for Diseases Control and Prevention | Min Kang et al |
| EPI_ISL_403933 | Guangdong/20SF013/2020 | 2020-01-15 | Asia/China/Guangdong/Shenzhen | Guangdong Provincial Center for Diseases Control and Prevention; Guangdong Provincial Public Health | Department of Microbiology, Guangdong Provincial Center for Diseases Control and Prevention | Min Kang et al |
| EPI_ISL_403934 | Guangdong/20SF014/2020 | 2020-01-15 | Asia/China/Guangdong/Shenzhen | Guangdong Provincial Center for Diseases Control and Prevention; Guangdong Provincial Public Health | Department of Microbiology, Guangdong Provincial Center for Diseases Control and Prevention | Min Kang et al |
| EPI_ISL_403935 | Guangdong/20SF025/2020 | 2020-01-15 | Asia/China/Guangdong/Shenzhen | Guangdong Provincial Center for Diseases Control and Prevention; Guangdong Provincial Public Health | Department of Microbiology, Guangdong Provincial Center for Diseases Control and Prevention | Min Kang et al |
| EPI_ISL_403936 | Guangdong/20SF028/2020 | 2020-01-17 | Asia/China/Guangdong/Zhuhai | Guangdong Provincial Center for Diseases Control and Prevention; Guangdong Provincial Public Health | Department of Microbiology, Guangdong Provincial Center for Diseases Control and Prevention | Min Kang et al |

| | | | | | | |
|---|---|---|---|---|---|---|
| EPI_ISL_403937 | Guangdong/20SF040/2020 | 2020-01-18 | Asia/China/Guangdong/Zhuhai | Guangdong Provincial Center for Diseases Control and Prevention; Guangdong Provincial Public Health | Department of Microbiology, Guangdong Provincial Center for Diseases Control and Prevention | Min Kang et al |
| EPI_ISL_403962 | Thailand/61/2020 | 2020-01-08 | Asia/Thailand/Nonthaburi/ | Bamrasnaradura Hospital | 1. Department of Medical Sciences, Ministry of Public Health, Thailand 2. Thai Red Cross Emerging Infectious Diseases - Health Science Centre 3. Department of Disease Control, Ministry of Public Health, Thailand | Pilailuk et al |
| EPI_ISL_403963 | Thailand/74/2020 | 2020-01-13 | Asia/Thailand/Nonthaburi/ | Bamrasnaradura Hospital | 1. Department of Medical Sciences, Ministry of Public Health, Thailand 2. Thai Red Cross Emerging Infectious Diseases - Health Science Centre 3. Department of Disease Control, Ministry of Public Health, Thailand | Pilailuk et al |
| EPI_ISL_404227 | Zhejiang/WZ-01/2020 | 2020-01-16 | Asia/China/Zhejiang/Hangzhou | Zhejiang Provincial Center for Disease Control and Prevention | Department of Microbiology, Zhejiang Provincial Center for Disease Control and Prevention | Yin Chen et al |

| | | | | | | |
|---|---|---|---|---|---|---|
| EPI_ISL_404228 | Zhejiang/WZ-02/2020 | 2020-01-17 | Asia/China/Zhejiang/Hangzhou | Zhejiang Provincial Center for Disease Control and Prevention | Department of Microbiology, Zhejiang Provincial Center for Disease Control and Prevention | Yanjun Zhang et al |
| EPI_ISL_404253 | USA/IL1/2020 | 2020-01-21 | North America/USA/Illinois/Chicago | IL Department of Public Health Chicago Laboratory | Pathogen Discovery, Respiratory Viruses Branch, Division of Viral Diseases, Centers for Disease Control and Prevention | Ying Tao et al |
| EPI_ISL_404895 | USA/WA1/2020 | 2020-01-19 | North America/USA/Washington/Snoho | Providence Regional Medical Center | Division of Viral Diseases, Centers for Disease Control and Prevention | Queen et al |
| EPI_ISL_405839 | Shenzhen/HKU-SZ-005/2020 | 2020-01-11 | Asia/China/Guangdong/Shenzhen | The University of Hong Kong - Shenzhen Hospital | Li Ka Shing Faculty of Medicine, The University of Hong Kong | Chan et al |
| EPI_ISL_406030 | Guangdong/HKU-SZ-002/2020 | 2020-01-10 | Asia/China/Guangdong/Shenzhen | The University of Hong Kong - Shenzhen Hospital | Li Ka Shing Faculty of Medicine, The University of Hong Kong | Chan et al |
| EPI_ISL_406031 | Taiwan/2/2020 | 2020-01-23 | Asia/Taiwan/Taiwan/Kaohsiung | Centers for Disease Control, R.O.C. (Taiwan) | Centers for Disease Control, R.O.C. (Taiwan) | Ji-Rong Yang et al |

| | | | | | | |
|---|---|---|---|---|---|---|
| EPI_ISL_406223 | USA/AZ1/2020 | 2020-01-22 | North America/USA/Arizona/Maricopa [illegible] | Arizona Department of Health Services | Pathogen Discovery, Respiratory Viruses Branch, Division of Viral Diseases, Centers for Disease Control and Prevention | Ying Tao et al |
| EPI_ISL_406531 | Guangdong/20SF174/2020 | 2020-01-22 | Asia/China/Guangdong/Zhuhai | Guangdong Provincial Center for Diseases Control and Prevention; Guangdong Provinical Public Health | Guangdong Provincial Center for Disease Control and Prevention | Min Kang et al |
| EPI_ISL_406533 | Guangzhou/20SF206/2020 | 2020-01-22 | Asia/China/Guangdong/Guangzhou | Guangdong Provincial Center for Diseases Control and Prevention; Guangdong Provinical Public Health | Guangdong Provincial Center for Diseases Control and Prevention | Min Kang et al |
| EPI_ISL_406534 | Foshan/20SF207/2020 | 2020-01-22 | Asia/China/Guangdong/Foshan | Guangdong Provincial Center for Diseases Control and Prevention; Guangdong Provincial Public Health | Guangdong Provincial Center for Diseases Control and Prevention | Min Kang et al |
| EPI_ISL_406535 | Foshan/20SF210/2020 | 2020-01-22 | Asia/China/Guangdong/Foshan | Guangdong Provincial Center for Diseases Control and Prevention; Guangdong Provincial Public Health | Guangdong Provincial Center for Diseases Control and Prevention | Min Kang et al |
| EPI_ISL_406536 | Foshan/20SF211/2020 | 2020-01-22 | Asia/China/Guangdong/Foshan | Guangdong Provincial Center for Diseases Control and Prevention; Guangdong Provincial Public Health | Guangdong Provincial Center for Diseases Control and Prevention | Min Kang et al |
| EPI_ISL_406538 | Guangdong/20SF201/2020 | 2020-01-23 | Asia/China/Guangdong/ | Guangdong Provincial Center for Diseases Control and Prevention;Guangdong Provincial Institute of Public Health | Guangdong Provincial Center for Diseases Control and Prevention | Min Kang et al |

| | | | | | | |
|---|---|---|---|---|---|---|
| EPI_ISL_406593 | Shenzhen/SZTH-002/2020 | 2020-01-13 | Asia/China/Guangdong/Shenzhen | Shenzhen Key Laboratory of Pathogen and Immunity, National Clinical Research Center for Infectious Disease, Shenzhen Third People's Hospital | Shenzhen Key Laboratory of Pathogen and Immunity, National Clinical Research Center for Infectious Disease, Shenzhen Third People's Hospital | Yang Yang et al |
| EPI_ISL_406594 | Shenzhen/SZTH-003/2020 | 2020-01-16 | Asia/China/Guangdong/Shenzhen | Shenzhen Key Laboratory of Pathogen and Immunity, National Clinical Research Center for Infectious Disease, Shenzhen Third People's Hospital | Shenzhen Key Laboratory of Pathogen and Immunity, National Clinical Research Center for Infectious Disease, Shenzhen Third People's Hospital | Yang Yang et al |
| EPI_ISL_406596 | France/IDF-0372/2020 | 2020-01-23 | Europe/France/Ile de France/Paris | Department of Infectious and Tropical Diseases, Bichat Claude Bernard Hospital, Paris | National Reference Center for Viruses of Respiratory Infections, Institut Pasteur, Paris | M√©lanie Albert et al |
| EPI_ISL_406597 | France/IDF-0373/2020 | 2020-01-23 | Europe/France/Ile de France/Paris | Department of Infectious and Tropical Diseases, Bichat Claude Bernard Hospital, Paris | National Reference Center for Viruses of Respiratory Infections, Institut Pasteur, Paris | M√©lanie Albert et al |
| EPI_ISL_406716 | Wuhan/WHU01/2020 | 2020-01-02 | Asia/China/Hubei/Wuhan | State Key Laboratory of Virology, Wuhan University | State Key Laboratory of Virology, Wuhan University | Chen et al |
| EPI_ISL_406717 | Wuhan/WHU02/2020 | 2020-01-02 | Asia/China/Hubei/Wuhan | State Key Laboratory of Virology, Wuhan University | State Key Laboratory of Virology, Wuhan University | Chen et al |

| | | | | | | |
|---|---|---|---|---|---|---|
| EPI_ISL_406798 | Wuhan/WH01/2019 | 2019-12-26 | Asia/China/Hubei/Wuhan | General Hospital of Central Theater Command of People's Liberation Army of China | BGI & Institute of Microbiology, Chinese Academy of Sciences & Shandong First Medical University & Shandong Academy of Medical Sciences & General Hospital of Central Theater Command of People's Liberation Army of China | Weijun Chen et al |
| EPI_ISL_406800 | Wuhan/WH03/2020 | 2020-01-01 | Asia/China/Hubei/Wuhan | General Hospital of Central Theater Command of People's Liberation Army of China | BGI & Institute of Microbiology, Chinese Academy of Sciences & Shandong First Medical University & Shandong Academy of Medical Sciences & General Hospital of Central Theater Command of People's Liberation Army of China | Weijun Chen et al |

| | | | | | | |
|---|---|---|---|---|---|---|
| EPI_ISL_406801 | Wuhan/WH04/2020 | 2020-01-05 | Asia/China/Hubei/Wuhan | General Hospital of Central Theater Command of People's Liberation Army of China | BGI & Institute of Microbiology, Chinese Academy of Sciences & Shandong First Medical University & Shandong Academy of Medical Sciences & General Hospital of Central Theater Command of People's Liberation Army of China | Weijun Chen et al |
| EPI_ISL_406844 | Australia/VIC01/2020 | 2020-01-25 | Oceania/Australia/Victoria/Clayton AU | Monash Medical Centre | Collaboration between the University of Melbourne at The Peter Doherty Institute for Infection and Immunity, and the Victorian Infectious Disease Reference Laboratory | Caly et al |
| EPI_ISL_406862 | Germany/BY-ChVir-929/2020 | 2020-01-28 | Europe/Germany/Bavaria/Starnberg | Charit√© Universit√§tsmedizin Berlin, Institute of Virology; Institut f√ºr Mikrobiologie der Bundeswehr, Munich | Charit√© Universit√§tsmedizin Berlin, Institute of Virology | Victor M Corman et al |
| EPI_ISL_406970 | Hangzhou/HZ-1/2020 | 2020-01-20 | Asia/China/Zhejiang/Hangzhou | Hangzhou Center for Disease and Control Microbiology Lab | Hangzhou Center for Disease and Control Microbiology Lab | Yu Hua et al |
| EPI_ISL_406973 | Singapore/1/2020 | 2020-01-23 | Asia/Singapore/Singapore/Singapore | Singapore General Hospital | National Public Health Laboratory | Mak et al |

| | | | | | | |
|---|---|---|---|---|---|---|
| EPI_ISL_407071 | England/01/2020 | 2020-01-29 | Europe/United Kingdom/England/ | Respiratory Virus Unit, Microbiology Services Colindale, Public Health England | Respiratory Virus Unit, Microbiology Services Colindale, Public Health England | Monica Galiano et al |
| EPI_ISL_407073 | England/02/2020 | 2020-01-29 | Europe/United Kingdom/England/ | Respiratory Virus Unit, Microbiology Services Colindale, Public Health England | Respiratory Virus Unit, Microbiology Services Colindale, Public Health England | Monica Galiano et al |
| EPI_ISL_407079 | Finland/1/2020 | 2020-01-29 | Europe/Finland/Finland/Rovaniemi | Lapland Central Hospital | Department of Virology, University of Helsinki and Helsinki University Hospital, Helsinki, Finland | Teemu Smura et al |
| EPI_ISL_407084 | Japan/AI-I-004/2020 | 2020-01-25 | Asia/Japan/Aichi/Aichi | Department of Virology III, National Institute of Infectious Diseases | Pathogen Genomics Center, National Institute of Infectious Diseases | Tsuyoshi Sekizuka et al |
| EPI_ISL_407193 | SouthKorea/KCDC03/2020 | 2020-01-25 | Asia/South Korea/Gyeonggi Province/G | Korea Centers for Disease Control & Prevention (KCDC) Center for Laboratory Control of Infectious Diseases Division of Viral Diseases | Korea Centers for Disease Control & Prevention (KCDC) Center for Laboratory Control of Infectious Diseases Division of Viral Diseases | Jeong-Min Kim et al |
| EPI_ISL_407313 | Hangzhou/HZCDC0001/2020 | 2020-01-19 | Asia/China/Zhejiang/Hangzhou | Hangzhou Center for Disease Control and Prevention | Hangzhou Center for Disease Control and Prevention | Jun Li et al |

| | | | | | | |
|---|---|---|---|---|---|---|
| EPI_ISL_407893 | Australia/NSW01/2020 | 2020-01-24 | Oceania/Australia/New South Wales/Sy | Centre for Infectious Diseases and Microbiology Laboratory Services | NSW Health Pathology - Institute of Clinical Pathology and Medical Research; Westmead Hospital; University of Sydney | Eden J-S et al |
| EPI_ISL_407894 | Australia/QLD01/2020 | 2020-01-28 | Oceania/Australia/Queensland/Gold Coa | Pathology Queensland | Public Health Virology Laboratory | Ben Huang et al |
| EPI_ISL_407896 | Australia/QLD02/2020 | 2020-01-30 | Oceania/Australia/Queensland/Gold Coa | Pathology Queensland | Public Health Virology Laboratory | Ben Huang et al |
| EPI_ISL_407976 | Belgium/GHB-03021/2020 | 2020-02-03 | Europe/Belgium/Leuven/ | KU Leuven, Clinical and Epidemiological Virology | KU Leuven, Clinical and Epidemiological Virology | Bert Vanmechelen et al |
| EPI_ISL_407987 | Singapore/2/2020 | 2020-01-25 | Asia/Singapore/Singapore/Singapore | Singapore General Hospital | Programme in Emerging Infectious Diseases, Duke-NUS Medical School | Danielle E Anderson et al |
| EPI_ISL_407988 | Singapore/3/2020 | 2020-02-01 | Asia/Singapore/Singapore/Singapore | National Centre for Infectious Diseases | Programme in Emerging Infectious Diseases, Duke-NUS Medical School | Danielle E Anderson et al |
| EPI_ISL_408008 | USA/CA-CDC-3/2020 | 2020-01-29 | North America/USA/California/ | California Department of Health | Pathogen Discovery, Respiratory Viruses Branch, Division of Viral Diseases, Centers for Disease Control and Prevention | Krista Queen et al |

| | | | | | | |
|---|---|---|---|---|---|---|
| EPI_ISL_408009 | USA/CA-CDC-4/2020 | 2020-01-29 | North America/USA/California/ | California Department of Health | Pathogen Discovery, Respiratory Viruses Branch, Division of Viral Diseases, Centers for Disease Control and Prevention | Krista Queen et al |
| EPI_ISL_408010 | USA/CA-CDC-5/2020 | 2020-01-29 | North America/USA/California/ | California Department of Health | Pathogen Discovery, Respiratory Viruses Branch, Division of Viral Diseases, Centers for Disease Control and Prevention | Ying Tao et al |
| EPI_ISL_408430 | France/IDF-0515/2020 | 2020-01-29 | Europe/France/Ile de France/Paris | Department of Infectious and Tropical Diseases, Bichat Claude Bernard Hospital, Paris | National Reference Center for Viruses of Respiratory Infections, Institut Pasteur, Paris | M√©lanie Albert et al |
| EPI_ISL_408431 | France/IDF-0626/2020 | 2020-01-29 | Europe/France/Ile de France/Paris | Sorbonne Universit√©, Inserm et Assistance Publique-H√¥pitaux de Paris (Piti√© Salp√©tri√®re) | National Reference Center for Viruses of Respiratory Infections, Institut Pasteur, Paris | M√©lanie Albert et al |
| EPI_ISL_408478 | Chongqing/YC01/2020 | 2020-01-21 | Asia/China/Chongqing/Yongchuan | Yongchuan District Center for Disease Control and Prevention | Chongqing Municipal Center for Disease Control and Prevention | Ye Sheng et al |
| EPI_ISL_408479 | Chongqing/ZX01/2020 | 2020-01-23 | Asia/China/Chongqing/Zhongxian | Zhongxian Center for Disease Control and Prevention | Chongqing Municipal Center for Disease Control and Prevention | Ye Sheng et al |

| | | | | | | |
|---|---|---|---|---|---|---|
| EPI_ISL_408480 | Yunnan/IVDC-YN-003/2020 | 2020-01-17 | Asia/China/Yunnan/Kunming | National Institute for Viral Disease Control and Prevention, China CDC | National Institute for Viral Disease Control & Prevention, CCDC | Wenjie Tan et al |
| EPI_ISL_408481 | Chongqing/IVDC-CQ-001/2020 | 2020-01-18 | Asia/China/Chongqing/ | National Institute for Viral Disease Control and Prevention, China CDC | National Institute for Viral Disease Control & Prevention, CCDC | Wenjie Tan et al |
| EPI_ISL_408482 | Shandong/IVDC-SD-001/2020 | 2020-01-19 | Asia/China/Shandong/Qingdao | National Institute for Viral Disease Control and Prevention, China CDC | National Institute for Viral Disease Control & Prevention, CCDC | Wenjie Tan et al |
| EPI_ISL_408484 | Sichuan/IVDC-SC-001/2020 | 2020-01-15 | Asia/China/Sichuan/Chengdu | National Institute for Viral Disease Control and Prevention, China CDC | National Institute for Viral Disease Control & Prevention, CCDC | Wenjie Tan et al |
| EPI_ISL_408486 | Jiangxi/IVDC-JX-002/2020 | 2020-01-11 | Asia/China/Jiangxi/Pingxiang | National Institute for Viral Disease Control and Prevention, China CDC | National Institute for Viral Disease Control & Prevention, CCDC | Wenjie Tan et al |
| EPI_ISL_408488 | Jiangsu/IVDC-JS-001/2020 | 2020-01-19 | Asia/China/Jiangsu/Huaian | National Institute for Viral Disease Control and Prevention, China CDC | National Institute for Viral Disease Control & Prevention, CCDC | Wenjie Tan et al |
| EPI_ISL_408489 | Taiwan/NTU01/2020 | 2020-01-31 | Asia/Taiwan/Taiwan/Taipei | Department of Laboratory Medicine, National Taiwan University Hospital | Microbial Genomics Core Lab, National Taiwan University Centers of Genomic and Precision Medicine | Shiou-Hwei Yeh et al |
| EPI_ISL_408665 | Japan/TY-WK-012/2020 | 2020-01-29 | Asia/Japan/Tokyo/Tokyo | Dept. of Virology III, National Institute of Infectious Diseases | Pathogen Genomics Center, National Institute of Infectious Diseases | Tsuyoshi Sekizuka et al |

| | | | | | | |
|---|---|---|---|---|---|---|
| EPI_ISL_408666 | Japan/TY-WK-501/2020 | 2020-01-31 | Asia/Japan/Tokyo/Tokyo | Dept. of Virology III, National Institute of Infectious Diseases | Pathogen Genomics Center, National Institute of Infectious Diseases | Tsuyoshi Sekizuka et al |
| EPI_ISL_408667 | Japan/TY-WK-521/2020 | 2020-01-31 | Asia/Japan/Tokyo/Tokyo | Dept. of Virology III, National Institute of Infectious Diseases | Pathogen Genomics Center, National Institute of Infectious Diseases | Tsuyoshi Sekizuka et al |
| EPI_ISL_408668 | Vietnam/VR03-38142/2020 | 2020-01-24 | Asia/Vietnam/Thanh Hoa/ | National Influenza Center - National Institute of Hygiene and Epidemiology (NIHE) | National Influenza Center - National Institute of Hygiene and Epidemiology (NIHE) | Ung Thi Hong Trang et al |
| EPI_ISL_408669 | Japan/KY-V-029/2020 | 2020-01-29 | Asia/Japan/Kyoto/Kyoto | Dept. of Virology III, National Institute of Infectious Diseases | Pathogen Genomics Center, National Institute of Infectious Diseases | Tsuyoshi Sekizuka et al |
| EPI_ISL_408670 | USA/WI1/2020 | 2020-01-31 | North America/USA/Wisconsin/ | Wisconsin Department of Health Services | Pathogen Discovery, Respiratory Viruses Branch, Division of Viral Diseases, Centers for Disease Control and Prevention | Jing Zhang et al |
| EPI_ISL_408976 | Australia/NSW02/2020 | 2020-01-22 | Oceania/Australia/New South Wales/Sy | Centre for Infectious Diseases and Microbiology Laboratory Services | NSW Health Pathology - Institute of Clinical Pathology and Medical Research; Westmead Hospital; University of Sydney | Rockett R et al |

| | | | | | | |
|---|---|---|---|---|---|---|
| EPI_ISL_408977 | Australia/NSW03/2020 | 2020-01-25 | Oceania/Australia/New South Wales/Sy | Serology, Virology and OTDS Laboratories (SAViD), NSW Health Pathology Randwick | NSW Health Pathology - Institute of Clinical Pathology and Medical Research; Centre for Infectious Diseases and Microbiology Laboratory Services; Westmead Hospital; University of Sydney | Eden J-S et al |
| EPI_ISL_408978 | Wuhan/WH05/2020 | 2020-02-07 | Asia/China/Hubei/Wuhan | Wuhan Fourth Hospital | Beijing Genomics Institute (BGI) | Weijun Chen et al |
| EPI_ISL_409067 | USA/MA1/2020 | 2020-01-29 | North America/USA/Massachusetts/Bos | Massachusetts Department of Public Health | Pathogen Discovery, Respiratory Viruses Branch, Division of Viral Diseases, Centers for Disease Control and Prevention | Clinton R. Paden et al |
| EPI_ISL_410044 | USA/CA-CDC-6/2020 | 2020-01-27 | North America/USA/California/ | California Department of Public Health | Pathogen Discovery, Respiratory Viruses Branch, Division of Viral Diseases, Centers for Disease Control and Prevention | Jing Zhang et al |

| | | | | | | |
|---|---|---|---|---|---|---|
| EPI_ISL_410045 | USA/IL2/2020 | 2020-01-28 | North America/USA/Illinois/ | IL Department of Public Health Chicago Laboratory | Pathogen Discovery, Respiratory Viruses Branch, Division of Viral Diseases, Centers for Disease Control and Prevention | Yan Li et al |
| EPI_ISL_410218 | Taiwan/NTU02/2020 | 2020-02-05 | Asia/Taiwan/Taiwan/Taipei | Department of Laboratory Medicine, National Taiwan University Hospital | Microbial Genomics Core Lab, National Taiwan University Centers of Genomic and Precision Medicine | Shiou-Hwei Yeh et al |
| EPI_ISL_410301 | Nepal/61/2020 | 2020-01-13 | Asia/Nepal/Kathmandu/Kathmandu | National Influenza Centre, National Public Health Laboratory, Kathmandu, Nepal | The University of Hong Kong | Ranjit Sah et al |
| EPI_ISL_410486 | France/ARA-739/2020 | 2020-02-08 | Europe/France/Auvergne-Rh√¥ne-Alpes/ | CNR Virus des Infections Respiratoires - France SUD | CNR Virus des Infections Respiratoires - France SUD | Bal et al |
| EPI_ISL_410531 | Japan/NA-20-05-1/2020 | 2020-01-25 | Asia/Japan/Nara/ | Dept. of Pathology, National Institute of Infectious Diseases | Pathogen Genomics Center, National Institute of Infectious Diseases | Tsuyoshi Sekizuka et al |
| EPI_ISL_410532 | Japan/OS-20-07-1/2020 | 2020-01-23 | Asia/Japan/Osaka/ | Dept. of Pathology, National Institute of Infectious Diseases | Pathogen Genomics Center, National Institute of Infectious Diseases | Tsuyoshi Sekizuka et al |
| EPI_ISL_410535 | Singapore/4/2020 | 2020-02-03 | Asia/Singapore/Singapore/Singapore | National Centre for Infectious Diseases | Programme in Emerging Infectious Diseases, Duke-NUS Medical School | Danielle E Anderson et al |

| | | | | | | |
|---|---|---|---|---|---|---|
| EPI_ISL_410536 | Singapore/5/2020 | 2020-02-06 | Asia/Singapore/Singapore/Singapore | Singapore General Hospital, Molecular Laboratory, Division of Pathology | Programme in Emerging Infectious Diseases, Duke-NUS Medical School | Danielle E Anderson et al |
| EPI_ISL_410537 | Singapore/6/2020 | 2020-02-09 | Asia/Singapore/Singapore/Singapore | Singapore General Hospital, Molecular Laboratory, Division of Pathology | Programme in Emerging Infectious Diseases, Duke-NUS Medical School | Danielle E Anderson et al |
| EPI_ISL_410545 | Italy/LAZ-INMI1-isl/2020 | 2020-01-29 | Europe/Italy/Lazio/Rome | INMI Lazzaro Spallanzani IRCCS | Laboratory of Virology, INMI Lazzaro Spallanzani IRCCS | Maria R. Capobianchi et al |
| EPI_ISL_410546 | Italy/LAZ-INMI1-cs/2020 | 2020-01-29 | Europe/Italy/Lazio/Rome | INMI Lazzaro Spallanzani IRCCS | Laboratory of Virology, INMI Lazzaro Spallanzani IRCCS | Maria R. Capobianchi et al |
| EPI_ISL_410713 | Singapore/7/2020 | 2020-01-27 | Asia/Singapore/Singapore/Singapore | National Public Health Laboratory, National Centre for Infectious Diseases | National Public Health Laboratory, National Centre for Infectious Diseases | Octavia S et al |
| EPI_ISL_410714 | Singapore/8/2020 | 2020-02-03 | Asia/Singapore/Singapore/Singapore | National Public Health Laboratory, National Centre for Infectious Diseases | National Public Health Laboratory, National Centre for Infectious Diseases | Octavia S et al |
| EPI_ISL_410715 | Singapore/9/2020 | 2020-02-04 | Asia/Singapore/Singapore/Singapore | National Public Health Laboratory, National Centre for Infectious Diseases | National Public Health Laboratory, National Centre for Infectious Diseases | Octavia S et al |

| | | | | | | |
|---|---|---|---|---|---|---|
| EPI_ISL_410716 | Singapore/10/2020 | 2020-02-04 | Asia/Singapore/Singapore/Singapore | National Public Health Laboratory, National Centre for Infectious Diseases | National Centre for Infectious Diseases, National Centre for Infectious Diseases | Octavia S et al |
| EPI_ISL_410717 | Australia/QLD03/2020 | 2020-02-05 | Oceania/Australia/Queensland/Gold Coa | Pathology Queensland | Public Health Virology Laboratory | Ben Huang et al |
| EPI_ISL_410718 | Australia/QLD04/2020 | 2020-02-05 | Oceania/Australia/Queensland/Gold Coa | Pathology Queensland | Public Health Virology Laboratory | Ben Huang et al |
| EPI_ISL_410719 | Singapore/11/2020 | 2020-02-02 | Asia/Singapore/Singapore/Singapore | National Public Health Laboratory | National Public Health Laboratory | Octavia S et al |
| EPI_ISL_410720 | France/IDF-0372-isl/2020 | 2020-01-23 | Europe/France/Ile de France/Paris | Department of Infectious and Tropical Diseases, Bichat Claude Bernard Hospital, Paris | National Reference Center for Viruses of Respiratory Infections, Institut Pasteur, Paris | M√©lanie Albert et al |
| EPI_ISL_410984 | France/IDF-0515-isl/2020 | 2020-01-29 | Europe/France/Ile de France/Paris | Department of Infectious and Tropical Diseases, Bichat Claude Bernard Hospital, Paris | National Reference Center for Viruses of Respiratory Infections, Institut Pasteur, Paris | M√©lanie Albert et al |
| EPI_ISL_411060 | Fujian/8/2020 | 2020-01-21 | Asia/China/Fujian/ | Fujian Center for Disease Control and Prevention | Fujian Center for Disease Control and Prevention | Chen Wei et al |
| EPI_ISL_411066 | Fujian/13/2020 | 2020-01-22 | Asia/China/Fujian/ | Fujian Center for Disease Control and Prevention | Fujian Center for Disease Control and Prevention | Chen Wei et al |
| EPI_ISL_411218 | France/IDF-0571/2020 | 2020-02-02 | Europe/France/Ile de France/Paris | Department of Infectious and Tropical Diseases, Bichat Claude Bernard Hospital, Paris | Laboratoire Virpath, CIRI U111, UCBL1, INSERM, CNRS, ENS Lyon | Olivier Terrier et al |

| | | | | | | |
|---|---|---|---|---|---|---|
| EPI_ISL_411219 | France/IDF-0386-islP1/2020 | 2020-01-28 | Europe/France/Ile de France/Paris | Department of Infectious and Tropical Diseases, Bichat Claude Bernard Hospital, Paris | Laboratoire Virpath, CIRI U111, UCBL1, INSERM, CNRS, ENS Lyon | Olivier Terrier et al |
| EPI_ISL_411220 | France/IDF-0386-islP3/2020 | 2020-01-28 | Europe/France/Ile de France/Paris | Department of Infectious and Tropical Diseases, Bichat Claude Bernard Hospital, Paris | Laboratoire Virpath, CIRI U111, UCBL1, INSERM, CNRS, ENS Lyon | Olivier Terrier et al |
| EPI_ISL_411902 | Cambodia/0012/2020 | 2020-01-27 | Asia/Cambodia/Sihanoukville/ | Virology Unit, Institut Pasteur du Cambodge. | Virology Unit, Institut Pasteur du Cambodge (Sequencing done by: Jessica E Manning/Jennifer A Bohl at Malaria and Vector Research Research Laboratory, National Institute of Allergy and Infectious Diseases and Vida Ahyong from Chan-Zuckerberg Biohub) | Erik A Karlsson et al |
| EPI_ISL_411915 | Taiwan/CGMH-CGU-01/2020 | 2020-01-25 | Asia/Taiwan/Taiwan/Taoyuan | Laboratory Medicine | Department of Laboratory Medicine, Lin-Kou Chang Gung Memorial Hospital, Taoyuan, Taiwan. | Kuo-Chien Tsao et al |
| EPI_ISL_411926 | Taiwan/3/2020 | 2020-01-24 | Asia/Taiwan/Taiwan/Taipei | Taiwan Centers for Disease Control | Taiwan Centers for Disease Control | Ji-Rong Yang et al |
| EPI_ISL_411927 | Taiwan/4/2020 | 2020-01-28 | Asia/Taiwan/Taiwan/Taipei | Taiwan Centers for Disease Control | Taiwan Centers for Disease Control | Ji-Rong Yang et al |

| | | | | | | |
|---|---|---|---|---|---|---|
| EPI_ISL_411950 | Jiangsu/JS01/2020 | 2020-01-23 | Asia/China/Jiangsu/ | NHC Key laboratory of Enteric Pathogenic Microbiology, Institute of Pathogenic Microbiology | Jiangsu Provincial Center for Disease Control & Prevention | Lunbiao Cui et al |
| EPI_ISL_411951 | Sweden/01/2020 | 2020-02-07 | Europe/Sweden/Sm√•land/J√∂nk√∂ping | Unit for Laboratory Development and Technology Transfer, Public Health Agency of Sweden | Unit for Laboratory Development and Technology Transfer, Public Health Agency of Sweden | Bengner et al |
| EPI_ISL_411952 | Jiangsu/JS02/2020 | 2020-01-24 | Asia/China/Jiangsu/ | NHC Key laboratory of Enteric Pathogenic Microbiology, Institute of Pathogenic Microbiology | Jiangsu Provincial Center for Disease Control & Prevention | Kangchen Zhao et al |
| EPI_ISL_411953 | Jiangsu/JS03/2020 | 2020-01-24 | Asia/China/Jiangsu/ | NHC Key laboratory of Enteric Pathogenic Microbiology, Institute of Pathogenic Microbiology | Jiangsu Provincial Center for Disease Control & Prevention | Kangchen Zhao et al |
| EPI_ISL_411954 | USA/CA-CDC-7/2020 | 2020-02-06 | North America/USA/California/ | California Department of Public Health | Pathogen Discovery, Respiratory Viruses Branch, Division of Viral Diseases, Centers for Disease Control and Prevention | Krista Queen et al |
| EPI_ISL_411955 | USA/CA-CDC-8/2020 | 2020-02-10 | North America/USA/California/ | California Department of Public Health | Pathogen Discovery, Respiratory Viruses Branch, Division of Viral Diseases, Centers for Disease Control and Prevention | Krista Queen et al |

| | | | | | | |
|---|---|---|---|---|---|---|
| EPI_ISL_411956 | USA/TX1/2020 | 2020-02-11 | North America/USA/Texas/ | Texas Department of State Health Services | Pathogen Discovery, Respiratory Viruses Branch, Division of Viral Diseases, Centers for Disease Control and Prevention | Krista Queen et al |
| EPI_ISL_411957 | Beijing/WH-09/2020 | 2020-01-08 | Asia/China/Hubei/Wuhan | Key Laboratory of Human Diseases, Comparative Medicine, Institute of Laboratory Animal Science | Key Laboratory of Human Diseases, Comparative Medicine, Institute of Laboratory Animal Science | Linlin et al |
| EPI_ISL_412028 | HongKong/VM20001061-2/2020 | 2020-01-22 | Asia/Hong Kong/Hong Kong/Hong Kong | Hong Kong Department of Health | School of Public Health, The University of Hon g Kong | Dominic N.C. Tsang et al |
| EPI_ISL_412029 | HongKong/VB20024950-2/2020 | 2020-01-30 | Asia/Hong Kong/Hong Kong/ | Hong Kong Department of Health | The University of Hong Kong | Dominic N.C. Tsang et al |
| EPI_ISL_412030 | HongKong/VB20026565-2/2020 | 2020-02-01 | Asia/Hong Kong/Hong Kong/ | Hong Kong Department of Health | School of Public Health, The University of Hon g Kong | Dominic N.C. Tsang et al |
| EPI_ISL_412116 | England/09c/2020 | 2020-02-09 | Europe/United Kingdom/England/ | Respiratory Virus Unit, Microbiology Services Colindale, Public Health England | Respiratory Virus Unit, Microbiology Services Colindale, Public Health England | Monica Galiano et al |
| EPI_ISL_412386 | Beijing/ICDC-DT005/2020 | 2020-02-05 | Asia/China/Beijing/ | Beijing Ditan Hospital, Capital Medical University | National Institute for Communicable Disease Control and Prevention, Chinese Center for Disease Control and Prevention | Xinmin Xu et al |

| | | | | | | |
|---|---|---|---|---|---|---|
| EPI_ISL_412862 | USA/CA-CDC-9/2020 | 2020-02-23 | North America/USA/California/Solano C | California Department of Public Health | Pathogen Discovery, Respiratory Viruses Branch, Division of Viral Diseases, Centers for Disease Control and Prevention | Krista Queen et al |
| EPI_ISL_412869 | SouthKorea/KCDC05/2020 | 2020-01-30 | Asia/South Korea/Seoul/Seoul | Division of Viral Diseases, Center for Laboratory Control of Infectious Diseases, Korea Centers for Diseases Control and Prevention | Division of Viral Diseases, Center for Laboratory Control of Infectious Diseases, Korea Centers for Diseases Control and Prevention | Jeong-Min Kim et al |
| EPI_ISL_412870 | SouthKorea/KCDC06/2020 | 2020-01-30 | Asia/South Korea/Seoul/Seoul | Division of Viral Diseases, Center for Laboratory Control of Infectious Diseases, Korea Centers for Diseases Control and Prevention | Division of Viral Diseases, Center for Laboratory Control of Infectious Diseases, Korea Centers for Diseases Control and Prevention | Jeong-Min Kim et al |
| EPI_ISL_412871 | SouthKorea/KCDC07/2020 | 2020-01-31 | Asia/South Korea/Seoul/Seoul | Division of Viral Diseases, Center for Laboratory Control of Infectious Diseases, Korea Centers for Diseases Control and Prevention | Division of Viral Diseases, Center for Laboratory Control of Infectious Diseases, Korea Centers for Diseases Control and Prevention | Jeong-Min Kim et al |

| | | | | | | |
|---|---|---|---|---|---|---|
| EPI_ISL_412872 | SouthKorea/KCDC12/2020 | 2020-02-01 | Asia/South Korea/Gyeonggi Province/G\ | Division of Viral Diseases, Center for Laboratory Control of Infectious Diseases, Korea Centers for Diseases Control and Prevention | Division of Viral Diseases, Center for Laboratory Control of Infectious Diseases, Korea Centers for Diseases Control and Prevention | Jeong-Min Kim et al |
| EPI_ISL_412873 | SouthKorea/KCDC24/2020 | 2020-02-06 | Asia/South Korea/Chungcheongnam/Chu | Division of Viral Diseases, Center for Laboratory Control of Infectious Diseases, Korea Centers for Diseases Control and Prevention | Division of Viral Diseases, Center for Laboratory Control of Infectious Diseases, Korea Centers for Diseases Control and Prevention | Jeong-Min Kim et al |
| EPI_ISL_412898 | Wuhan/HBCDC-HB-02/2019 | 2019-12-30 | Asia/China/Hubei/Wuhan | Wuhan Jinyintan Hospital | Hubei Provincial Center for Disease Control and Prevention | Bin Fang et al |
| EPI_ISL_412899 | Wuhan/HBCDC-HB-03/2019 | 2019-12-30 | Asia/China/Hubei/Wuhan | Wuhan Jinyintan Hospital | Hubei Provincial Center for Disease Control and Prevention | Bin Fang et al |
| EPI_ISL_412912 | Germany/BW-ChVir-1577/2020 | 2020-02-25 | Europe/Italy/Baden-Wuerttemberg/ | State Health Office Baden-W√ºrttemberg | Charit√© Universit√§tsmedizin Berlin, Institute of Virology | Victor M Corman et al |
| EPI_ISL_412964 | Brazil/SP-01/2020 | 2020-02-25 | South America/Italy/S√£o Paulo/S√£o P | Hospital Israelita Albert Einstein | Instituto Adolfo Lutz Interdisciplinary Procedures Center Strategic Laboratory | Jaqueline Goes de Jesus et al |
| EPI_ISL_412965 | Canada/BC_37_0-2/2020 | 2020-02-16 | North America/Iran/British Columbia/ | BCCDC Public Health Laboratory | BCCDC Public Health Laboratory | Harrigan et al |

| | | | | | | |
|---|---|---|---|---|---|---|
| EPI_ISL_412966 | Guangzhou/IQTC01/2020 | 2020-02-05 | Asia/China/Guangdong/Guangzhou | Technology Centre, Guangzhou Customs | Technology Centre, Guangzhou Customs | Shi et al |
| EPI_ISL_412967 | Guangzhou/IQTC02/2020 | 2020-01-29 | Asia/China/Guangdong/Guangzhou | Technology Centre, Guangzhou Customs | Technology Centre, Guangzhou Customs | Shi et al |
| EPI_ISL_412968 | Japan/Hu_DP_Kng_19-020/2020 | 2020-02-10 | Asia/Japan/Kanagawa/ | Takayuki Hishiki Kanagawa Prefectural Institute of Public Health | Takayuki Hishiki Kanagawa Prefectural Institute of Public Health | Hishiki et al |
| EPI_ISL_412969 | Japan/Hu_DP_Kng_19-027/2020 | 2020-02-10 | Asia/Japan/Kanagawa/ | Takayuki Hishiki Kanagawa Prefectural Institute of Public Health | Takayuki Hishiki Kanagawa Prefectural Institute of Public Health | Hishiki et al |
| EPI_ISL_412970 | USA/WA2/2020 | 2020-02-24 | North America/USA/Washington/Snoho | Washington State Department of Health | Seattle Flu Study | Helen Chu et al |
| EPI_ISL_412971 | Finland/FIN-25/2020 | 2020-02-25 | Europe/Italy/Helsinki/ | HUS Diagnostiikkakeskus, Hallinto | Department of Virology Faculty of Medicine, Medicum University of Helsinki | Teemu Smura et al |
| EPI_ISL_412972 | Mexico/CMX-InDRE-01/2020 | 2020-02-27 | North America/Italy/Mexico City/ | Instituto Nacional de Enfermedades Respiratorias | Instituto de Diagnostico y Referencia Epidemiologicos (INDRE) | Ramirez-Gonzalez Ernesto et al |
| EPI_ISL_412973 | Italy/LOM-ASST-CDG1/2020 | 2020-02-20 | Europe/Italy/Lombardy/ | Department of Infectious Diseases, Istituto Superiore di Sanit√†, Roma , Italy | Virology Laboratory, Scientific Department, Army Medical Center | Paola Stefanelli et al |

| | | | | | | |
|---|---|---|---|---|---|---|
| EPI_ISL_412974 | Italy/LAZ-INMI-SPL1/2020 | 2020-01-29 | Europe/Italy/Lazio/Rome | Department of Infectious Diseases, Istituto Superiore di Sanitv†, Rome, Italy | Virology Laboratory, Scientific Department, Army Medical Center | Paola Stefanelli et al |
| EPI_ISL_412975 | Australia/NSW05/2020 | 2020-02-28 | Oceania/Iran/New South Wales/Sydney | Centre for Infectious Diseases and Microbiology Laboratory Services | NSW Health Pathology - Institute of Clinical Pathology and Medical Research; Westmead Hospital; University of Sydney | Eden J-S et al |
| EPI_ISL_412978 | Wuhan/HBCDC-HB-02/2020 | 2020-01-17 | Asia/China/Hubei/Wuhan | The Central Hospital Of Wuhan | Hubei Provincial Center for Disease Control and Prevention | Bin Fang et al |
| EPI_ISL_412979 | Wuhan/HBCDC-HB-03/2020 | 2020-01-18 | Asia/China/Hubei/Wuhan | Union Hospital of Tongji Medical College, Huazhong University of Science and Technology | Hubei Provincial Center for Disease Control and Prevention | Bin Fang et al |
| EPI_ISL_412980 | Wuhan/HBCDC-HB-04/2020 | 2020-01-18 | Asia/China/Hubei/Wuhan | Union Hospital of Tongji Medical College, Huazhong University of Science and Technology | Hubei Provincial Center for Disease Control and Prevention | Bin Fang et al |
| EPI_ISL_412981 | Wuhan/HBCDC-HB-05/2020 | 2020-01-18 | Asia/China/Hubei/Wuhan | CR&WISCO GENERAL HOSPITAL | Hubei Provincial Center for Disease Control and Prevention | Bin Fang et al |
| EPI_ISL_412982 | Wuhan/HBCDC-HB-06/2020 | 2020-02-07 | Asia/China/Hubei/Wuhan | Wuhan Lung Hospital | Hubei Provincial Center for Disease Control and Prevention | Bin Fang et al |
| EPI_ISL_412983 | Tianmen/HBCDC-HB-07/2020 | 2020-02-08 | Asia/China/Hubei/Tianmen | Tianmen Center for Disease Control and Prevention | Hubei Provincial Center for Disease Control and Prevention | Bin Fang et al |

| | | | | | | |
|---|---|---|---|---|---|---|
| EPI_ISL_413014 | Canada/ON-PHL-2445/2020 | 20-Jan | North America/Canada/Ontario/ | Public Health Ontario Laboratory | Ontario Agency for Health Protection and Promotion (OAHPP) | Alireza Eshaghi et al |
| EPI_ISL_413015 | Canada/ON_VIDO-01/2020 | 2020-01-23 | North America/Canada/Ontario/ | Public Health Ontario Laboratory | National Microbiology Laboratory | Shari Tyson et al |
| EPI_ISL_413016 | Brazil/SP-02/2020 | 2020-02-28 | South America/Italy/S√£o Paulo/S√£o P | Hospital Israelita Albert Einstein | Instituto Adolfo Lutz, Interdisciplinary Procedures Center, Strategic Laboratory | Jaqueline Goes de Jesus et al |
| EPI_ISL_413017 | SouthKorea/KUMC01/2020 | 2020-02-06 | Asia/South Korea/South Korea/ | Department of Microbiology, Institute for Viral Diseases, College of Medicine, Korea University | Department of Microbiology, Institute for Viral Diseases, College of Medicine, Korea University | Changmin Kang et al |
| EPI_ISL_413018 | SouthKorea/KUMC02/2020 | 2020-02-06 | Asia/South Korea/South Korea/ | Department of Microbiology, Institute for Viral Diseases, College of Medicine, Korea University | Department of Microbiology, Institute for Viral Diseases, College of Medicine, Korea University | Changmin Kang et al |
| EPI_ISL_413019 | Switzerland/ZH-UZH-1000477102/20 | 2020-02-26 | Europe/Switzerland/Z√ºrich/ | Department of Internal Medicine, Triemli Hospital | Institute of Medical Virology, University of Zurich | Stefan Schmutz et al |
| EPI_ISL_413020 | Switzerland/ZH-UZH-1000477377/20 | 2020-02-27 | Europe/Switzerland/Z√ºrich/ | Department of Internal Medicine, Triemli Hospital | Institute of Medical Virology, University of Zurich | Stefan Schmutz et al |
| EPI_ISL_413021 | Switzerland/ZH-UZH-1000477757/20 | 2020-02-29 | Europe/Switzerland/Z√ºrich/ | Klinik Hirslanden Zurich | Institute of Medical Virology, University of Zurich | Stefan Schmutz et al |

| | | | | | | |
|---|---|---|---|---|---|---|
| EPI_ISL_413022 | Switzerland/ZH-UZH-1000477796/20 | 2020-02-29 | Europe/Italy/Zv∫rich/ | Division of Infectious Diseases, University Hospital Zurich | Institute of Medical Virology, University of Zurich | Stefan Schmutz et al |
| EPI_ISL_413023 | Switzerland/ZH-UZH-1000477797/20 | 2020-02-29 | Europe/Italy/Zv∫rich/ | Division of Infectious Diseases, University Hospital Zurich | Institute of Medical Virology, University of Zurich | Stefan Schmutz et al |
| EPI_ISL_413024 | Switzerland/ZH-UZH-1000477806/20 | 2020-02-29 | Europe/Switzerland/Zv∫rich/ | Division of Infectious Diseases, University Hospital Zurich | Institute of Medical Virology, University of Zurich | Stefan Schmutz et al |
| EPI_ISL_413025 | USA/WA3-UW1/2020 | 2020-02-27 | North America/USA/Washington/ | Harborview Medical Center | UW Virology Lab | Pavitra Roychoudhury et al |
| EPI_ISL_413213 | Australia/NSW06/2020 | 2020-02-29 | Oceania/Iran/New South Wales/Sydney | Centre for Infectious Diseases and Microbiology Laboratory Services | NSW Health Pathology - Institute of Clinical Pathology and Medical Research; Westmead Hospital; University of Sydney | Eden J-S et al |
| EPI_ISL_413214 | Australia/NSW07/2020 | 2020-02-29 | Oceania/Australia/New South Wales/Sy | Centre for Infectious Diseases and Microbiology Laboratory Services | NSW Health Pathology - Institute of Clinical Pathology and Medical Research; Westmead Hospital; University of Sydney | Eden J-S et al |
| EPI_ISL_413221 | Scotland/CVR01/2020 | 2020-02-28 | Europe/Italy/Scotland/ | West of Scotland Specialist Virology Centre, NHSGGC | MRC-University of Glasgow Centre for Virus Research | Emma Thomson et al |
| EPI_ISL_413455 | USA/WA4-UW2/2020 | 2020-02-28 | North America/USA/Washington/ | Washington State Public Health Lab | University of Washington Virology Lab | Pavitra Roychoudhury et al |

| | | | | | | |
|---|---|---|---|---|---|---|
| EPI_ISL_413456 | USA/WA-S2/2020 | 2020-02-21 | North America/USA/Washington/King C | Seattle Flu Study, University of Washington Medical Center | Seattle Flu Study, University of Washington Medical Center | Chu et al et al |
| EPI_ISL_413457 | USA/WA6-UW3/2020 | 2020-02-29 | North America/USA/Washington/ | Washington State Public Health Lab | UW Virology Lab | Pavitra Roychoudhury et al |
| EPI_ISL_413459 | Japan/TK-20-31-3/2020 | 2020-02-20 | Asia/Japan/Tokyo/ | Department of Pathology, Toshima Hospital | Pathogen Genomics Center, National Institute of Infectious Diseases | Tsuyoshi Sekizuka et al |
| EPI_ISL_413485 | Anhui/SZ005/2020 | 2020-01-24 | Asia/China/Anhui/Suzhou | Department of microbiology laboratory,Anhui Provincial Center for Disease Control and Prevention | Department of microbiology laboratory,Anhui Provincial Center for Disease Control and Prevention | Weiwei Li et al |
| EPI_ISL_413488 | Germany/NW-HHU-01/2020 | 2020-02-28 | Europe/Germany/North Rhine Westphal | Center of Medical Microbiology, Virology, and Hospital Hygiene, University of Duesseldorf | Center of Medical Microbiology, Virology, and Hospital Hygiene, University of Duesseldorf | Ortwin Adams et al |
| EPI_ISL_413489 | Italy/LOM-UniSR-1/2020 | 2020-03-03 | Europe/Italy/Lombardy/Milan | Laboratorio di Microbiologia e Virologia, Universit√† Vita-Salute San Raffaele, Milano | Laboratorio di Microbiologia e Virologia, Universit√† Vita-Salute San Raffaele, Milano | R.A Diotti et al |
| EPI_ISL_413490 | NewZealand/20VR0174/2020 | 2020-02-27 | Oceania/Iran/Auckland/Auckland | Auckland Hospital | Institute of Environmental Science and Research (ESR) | Matt Storey et al |
| EPI_ISL_413513 | SouthKorea/KUMC03/2020 | 2020-02-27 | Asia/South Korea/South Korea/ | Division of Infectious Diseases, Department of Internal Medicine, Korea University College of Medicine | Department of Microbiology, Institute for Viral Diseases, College of Medicine, Korea University | Changmin Kang et al |

| | | | | | | |
|---|---|---|---|---|---|---|
| EPI_ISL_413515 | SouthKorea/KUMC05/2020 | 2020-02-27 | Asia/South Korea/South Korea/ | Division of Infectious Diseases, Department of Internal Medicine, Korea University College of Medicine | Department of Microbiology, Institute for Viral Diseases, College of Medicine, Korea University | Changmin Kang et al |
| EPI_ISL_413516 | SouthKorea/KUMC06/2020 | 2020-02-27 | Asia/South Korea/South Korea/ | Department of Microbiology, Institute for Viral Diseases, College of Medicine, Korea University | Department of Microbiology, Institute for Viral Diseases, College of Medicine, Korea University | Changmin Kang et al |
| EPI_ISL_413518 | Beijing/105/2020 | 2020-01-26 | Asia/China/Beijing/Beijing | Infectious Disease Control Center, Center for Disease Control and Prevention of PLA | Infectious Disease Control Center, Center for Disease Control and Prevention of PLA | Li et al |
| EPI_ISL_413519 | Beijing/231/2020 | 2020-01-28 | Asia/China/Beijing/ | Infectious Disease Control Center, Center for Disease Control and Prevention of PLA | Infectious Disease Control Center, Center for Disease Control and Prevention of PLA | Li et al |
| EPI_ISL_413520 | Beijing/233/2020 | 2020-01-28 | Asia/China/Beijing/ | Infectious Disease Control Center, Center for Disease Control and Prevention of PLA | Infectious Disease Control Center, Center for Disease Control and Prevention of PLA | Li et al |
| EPI_ISL_413521 | Beijing/235/2020 | 2020-01-28 | Asia/China/Beijing/ | Infectious Disease Control Center, Center for Disease Control and Prevention of PLA | Infectious Disease Control Center, Center for Disease Control and Prevention of PLA | Li et al |
| EPI_ISL_413522 | India/MH-1-27/2020 | 2020-01-27 | Asia/India/Maharashtra/ | Indian Council of Medical Research - National Institute of Virology | National Influenza Center, Indian Council of Medical Research - National Institute of Virology | Potdar V et al |

| | | | | | | |
|---|---|---|---|---|---|---|
| EPI_ISL_413523 | India/MH-1-31/2020 | 2020-01-31 | Asia/India/Maharashtra/ | Indian Council of Medical Research-National Institute of Virology | National Influenza Center, Indian Council of Medical Research-National Institute of Virology | Potdar V et al |
| EPI_ISL_413550 | Nigeria/Lagos01/2020 | 2020-02-27 | Africa/Italy/Lagos/ | Centre for Human and Zoonotic Virology (CHAZVY), College of Medicine University of Lagos/Lagos University Teaching Hospital (LUTH), part of the Laboratory Network of the Nigeria Centre for Disease Control (NCDC) | African Centre of Excellence for Genomics of Infectious Diseases (ACEGID), Redeemer's University, Ede, Osun State, Nigeria | Oluniyi P.E. et al |
| EPI_ISL_413555 | Wales/PHW1/2020 | 2020-02-27 | Europe/Italy/Wales/ | Wales Specialist Virology Centre | Public Health Wales Microbiology Cardiff | Catherine Moore et al |
| EPI_ISL_413557 | USA/CA-CDPH-UC1/2020 | 2020-02-26 | North America/USA/Grand Princess/ | California Department of Public Health | Chiu Laboratory, University of California, San Francisco | Xianding Deng et al |
| EPI_ISL_413558 | USA/CA-CDPH-UC2/2020 | 2020-02-27 | North America/USA/California/Solano C | California Department of Public Health | Chiu Laboratory, University of California, San Francisco | Xianding Deng et al |
| EPI_ISL_413559 | USA/CA-CDPH-UC3/2020 | 2020-02-27 | North America/USA/California/Solano C | California Department of Public Health | Chiu Laboratory, University of California, San Francisco | Xianding Deng et al |
| EPI_ISL_413560 | USA/WA-S3/2020 | 2020-02-28 | North America/USA/Washington/ | Seattle Flu Study | Seattle Flu Study | Chu et al et al |
| EPI_ISL_413561 | USA/CA-CDPH-UC4/2020 | 2020-02-27 | North America/USA/California/Solano C | California Department of Public Health | Chiu Laboratory, University of California, San Francisco | Xianding Deng et al |
| EPI_ISL_413565 | Netherlands/Berlicum_1363564/2020 | 2020-02-24 | Europe/Italy/North Brabant/Berlicum | Foundation Pamm | Erasmus Medical Center | David Nieuwenhuijse et al |

| | | | | | | |
|---|---|---|---|---|---|---|
| EPI_ISL_413569 | Netherlands/Delft_1363424/2020 | 2020-02-28 | Europe/Italy/South Holland/Delft | RIVM | Erasmus Medical Center | David Nieuwenhuijse et al |
| EPI_ISL_413570 | Netherlands/Diemen_1363454/2020 | 2020-02-28 | Europe/Netherlands/North Holland/Die | RIVM | Erasmus Medical Center | David Nieuwenhuijse et al |
| EPI_ISL_413574 | Netherlands/Helmond_1363548/202 | 2020-02-29 | Europe/Netherlands/North Brabant/Hel | MHC West-Brabant | Erasmus Medical Center | David Nieuwenhuijse et al |
| EPI_ISL_413575 | Netherlands/Houten_1363498/2020 | 2020-02-29 | Europe/Netherlands/Utrecht/Houten | RIVM | Erasmus Medical Center | David Nieuwenhuijse et al |
| EPI_ISL_413576 | Netherlands/Loon_op_zand_1363512 | 2020-02-29 | Europe/Netherlands/North Brabant/Loo | RIVM | Erasmus Medical Center | David Nieuwenhuijse et al |
| EPI_ISL_413581 | Netherlands/Oss_1363500/2020 | 2020-02-29 | Europe/Netherlands/North Brabant/Oss | RIVM | Erasmus Medical Center | David Nieuwenhuijse et al |
| EPI_ISL_413586 | Netherlands/Tilburg_1363354/2020 | 2020-02-27 | Europe/Netherlands/North Brabant/Tilb | Foundation Elisabeth-Tweesteden Ziekenhuis | Erasmus Medical Center | David Nieuwenhuijse et al |
| EPI_ISL_413592 | Taiwan/NTU03/2020 | 2020-03-02 | Asia/Italy/Taiwan/Taipei | Department of Laboratory Medicine, National Taiwan University Hospital | Microbial Genomics Core Lab, National Taiwan University Centers of Genomic and Precision Medicine | Shiou-Hwei Yeh et al |
| EPI_ISL_413593 | Luxembourg/Lux1/2020 | 2020-02-29 | Europe/Italy/Luxembourg/ | Laboratoire National de Santé | Erasmus Medical Center | David Nieuwenhuijse et al |
| EPI_ISL_413594 | Australia/NSW08/2020 | 2020-02-28 | Oceania/Australia/New South Wales/Sy | Centre for Infectious Diseases and Microbiology Laboratory Services | NSW Health Pathology - Institute of Clinical Pathology and Medical Research; Westmead Hospital; University of Sydney | Rockett R et al |

| | | | | | | |
|---|---|---|---|---|---|---|
| EPI_ISL_413595 | Australia/NSW09/2020 | 2020-02-28 | Oceania/Australia/New South Wales/Sy | Centre for Infectious Diseases and Microbiology Laboratory Services | NSW Health Pathology - Institute of Clinical Pathology and Medical Research; Westmead Hospital; University of Sydney | Rockett R et al |
| EPI_ISL_413596 | Australia/NSW10/2020 | 2020-02-28 | Oceania/Australia/New South Wales/Sy | Centre for Infectious Diseases and Microbiology - Public Health | NSW Health Pathology - Institute of Clinical Pathology and Medical Research; Westmead Hospital; University of Sydney | Rockett R et al |
| EPI_ISL_413597 | Australia/NSW11/2020 | 2020-03-02 | Oceania/Iran/New South Wales/Sydney | Centre for Infectious Diseases and Microbiology- Public Health | NSW Health Pathology - Institute of Clinical Pathology and Medical Research; Westmead Hospital; University of Sydney | Lam C et al |
| EPI_ISL_413598 | Australia/NSW12/2020 | 2020-03-04 | Oceania/Iran/New South Wales/Sydney | Centre for Infectious Diseases and Microbiology - Public Health | NSW Health Pathology - Institute of Clinical Pathology and Medical Research; Westmead Hospital; University of Sydney | Gray K et al |

| | | | | | | |
|---|---|---|---|---|---|---|
| EPI_ISL_413599 | Australia/NSW13/2020 | 2020-03-04 | Oceania/Iran/New South Wales/Sydney | Centre for Infectious Diseases and Microbiology - Public Health | NSW Health Pathology - Institute of Clinical Pathology and Medical Research; Westmead Hospital; University of Sydney | Timms et al |
| EPI_ISL_413604 | Finland/FIN03032020C/2020 | 2020-03-03 | Europe/Italy/Helsinki/ | Department of Virology and Immunology, University of Helsinki and Helsinki University Hospital, Huslab Finland | Department of Virology, Faculty of Medicine, University of Helsinki, Helsinki, Finland | Teemu Smura et al |
| EPI_ISL_413650 | USA/WA15-UW11/2020 | 2020-03-05 | North America/USA/Washington/ | UW Virology Lab | UW Virology Lab | Pavitra Roychoudhury et al |
| EPI_ISL_413850 | Guangdong/GDFS2020054-P0005/20 | 2020-02-10 | Asia/China/Guangdong/ | Guangdong Provincial Institution of Public Health, Guangdong Provinical Center for Disease Control and Prevention | Guangdong Provincial Institution of Public Health | Jing Lu et al |
| EPI_ISL_413851 | Guangdong/2020XN4373-P0039/2020 | 2020-01-30 | Asia/China/Guangdong/ | Guangdong Provincial Institution of Public Health, Guangdong Provinical Center for Disease Control and Prevention | Guangdong Provincial Institution of Public Health | Jing Lu et al |
| EPI_ISL_413852 | Guangdong/2020XN4433-P0040/2020 | 2020-01-30 | Asia/China/Guangdong/ | Guangdong Provincial Institution of Public Health, Guangdong Provinical Center for Disease Control and Prevention | Guangdong Provincial Institution of Public Health | Jing Lu et al |
| EPI_ISL_413853 | Guangdong/2020XN4243-P0035/2020 | 2020-01-30 | Asia/China/Guangdong/ | Guangdong Provincial Institution of Public Health, Guangdong Provinical Center for Disease Control and Prevention | Guangdong Provincial Institution of Public Health | Jing Lu et al |
| EPI_ISL_413854 | Guangdong/2020XN4475-P0042/2020 | 2020-01-30 | Asia/China/Guangdong/ | Guangdong Provincial Institution of Public Health, Guangdong Provinical Center for Disease Control and Prevention | Guangdong Provincial Institution of Public Health | Jing Lu et al |

| | | | | | | |
|---|---|---|---|---|---|---|
| EPI_ISL_413856 | Guangdong/GD2020233-P0027/2020 | 2020-02-07 | Asia/China/Guangdong/ | Guangdong Provincial Institution of Public Health, Guangdong Provinical Center for Disease Control and Prevention | Guangdong Provincial Institution of Public Health | Jing Lu et al |
| EPI_ISL_413857 | Guangdong/2020XN4448-P0002/2020 | 2020-01-31 | Asia/China/Guangdong/ | Guangdong Provincial Institution of Public Health, Guangdong Provinical Center for Disease Control and Prevention | Guangdong Provincial Institution of Public Health | Jing Lu et al |
| EPI_ISL_413858 | Guangdong/2020XN4459-P0041/2020 | 2020-01-30 | Asia/China/Guangdong/ | Guangdong Provincial Institution of Public Health, Guangdong Provinical Center for Disease Control and Prevention | Guangdong Provincial Institution of Public Health | Jing Lu et al |
| EPI_ISL_413859 | Guangdong/2020XN4276-P0037/2020 | 2020-01-31 | Asia/China/Guangdong/ | Guangdong Provincial Institution of Public Health, Guangdong Provinical Center for Disease Control and Prevention | Guangdong Provincial Institution of Public Health | Jing Lu et al |
| EPI_ISL_413860 | Guangdong/2020XN4273-P0036/2020 | 2020-01-30 | Asia/China/Guangdong/ | Guangdong Provincial Institution of Public Health, Guangdong Provinical Center for Disease Control and Prevention | Guangdong Provincial Institution of Public Health | Jing Lu et al |
| EPI_ISL_413861 | Guangdong/GD2020080-P0010/2020 | 2020-02-01 | Asia/China/Guangdong/ | Guangdong Provincial Institution of Public Health, Guangdong Provinical Center for Disease Control and Prevention | Guangdong Provincial Institution of Public Health | Jing Lu et al |
| EPI_ISL_413862 | Guangdong/GD2020086-P0021/2020 | 2020-02-01 | Asia/China/Guangdong/ | Guangdong Provincial Institution of Public Health, Guangdong Provinical Center for Disease Control and Prevention | Guangdong Provincial Institution of Public Health | Jing Lu et al |
| EPI_ISL_413863 | Guangdong/GD2020087-P0008/2020 | 2020-02-01 | Asia/China/Guangdong/ | Guangdong Provincial Institution of Public Health, Guangdong Provinical Center for Disease Control and Prevention | Guangdong Provincial Institution of Public Health | Jing Lu et al |
| EPI_ISL_413864 | Guangdong/GD2020246-P0028/2020 | 2020-02-09 | Asia/China/Guangdong/ | Guangdong Provincial Institution of Public Health, Guangdong Provinical Center for Disease Control and Prevention | Guangdong Provincial Institution of Public Health | Jing Lu et al |
| EPI_ISL_413866 | Guangdong/GDSZ202008-P0020/2020 | 2020-02-05 | Asia/China/Guangdong/ | Guangdong Provincial Institution of Public Health, Guangdong Provinical Center for Disease Control and Prevention | Guangdong Provincial Institution of Public Health | Jing Lu et al |

| | | | | | | |
|---|---|---|---|---|---|---|
| EPI_ISL_413867 | Guangdong/GDSZ202015-P0019/202[illegible] | 2020-02-05 | Asia/China/Guangdong/ | Guangdong Provincial Institution of Public Health, Guangdong Provinical Center for Disease Control and Prevention | Guangdong Provincial Institution of Public Health | Jing Lu et al |
| EPI_ISL_413868 | Guangdong/DG-S9-P0045/2020 | 2020-02-28 | Asia/China/Guangdong/ | Guangdong Provincial Institution of Public Health, Guangdong Provinical Center for Disease Control and Prevention | Guangdong Provincial Institution of Public Health | Jing Lu et al |
| EPI_ISL_413869 | Guangdong/FS-S42-P0046/2020 | 2020-02-28 | Asia/China/Guangdong/ | Guangdong Provincial Institution of Public Health, Guangdong Provinical Center for Disease Control and Prevention | Guangdong Provincial Institution of Public Health | ‚ÄãJing Lu et al |
| EPI_ISL_413870 | Guangdong/FS-S48-P0047/2020 | 2020-02-28 | Asia/China/Guangdong/ | Guangdong Provincial Institution of Public Health, Guangdong Provinical Center for Disease Control and Prevention | Guangdong Provincial Institution of Public Health | Jing Lu et al |
| EPI_ISL_413871 | Guangdong/MM-S1-P0048/2020 | 2020-02-28 | Asia/China/Guangdong/ | Guangdong Provincial Institution of Public Health, Guangdong Provinical Center for Disease Control and Prevention | Guangdong Provincial Institution of Public Health | Jing Lu et al |
| EPI_ISL_413872 | Guangdong/SZ-N59-P0049/2020 | 2020-02-28 | Asia/China/Guangdong/ | Guangdong Provincial Institution of Public Health, Guangdong Provinical Center for Disease Control and Prevention | Guangdong Provincial Institution of Public Health | Jing Lu et al |
| EPI_ISL_413873 | Guangdong/GZ-S6-P0050/2020 | 2020-02-28 | Asia/China/Guangdong/ | Guangdong Provincial Institution of Public Health, Guangdong Provinical Center for Disease Control and Prevention | Guangdong Provincial Institution of Public Health | Jing Lu et al |
| EPI_ISL_413874 | Guangdong/GDFS2020127-P0026/20[illegible] | 2020-02-12 | Asia/China/Guangdong/ | Guangdong Provincial Institution of Public Health, Guangdong Provinical Center for Disease Control and Prevention | Guangdong Provincial Institution of Public Health | Jing Lu et al |
| EPI_ISL_413875 | Guangdong/GDFS2020052-P0025/20[illegible] | 2020-02-10 | Asia/China/Guangdong/ | Guangdong Provincial Institution of Public Health, Guangdong Provinical Center for Disease Control and Prevention | Guangdong Provincial Institution of Public Health | Jing Lu et al |
| EPI_ISL_413882 | Guangdong/GD2020139-P0007/2020 | 2020-02-02 | Asia/China/Guangdong/ | Guangdong Provincial Institution of Public Health, Guangdong Provinical Center for Disease Control and Prevention | Guangdong Provincial Institution of Public Health | Jing Lu et al |

| | | | | | | |
|---|---|---|---|---|---|---|
| EPI_ISL_413883 | Guangdong/GD2020134-P0031/2020 | 2020-02-02 | Asia/China/Guangdong/ | Guangdong Provincial Institution of Public Health, Guangdong Provinical Center for Disease Control and Prevention | Guangdong Provincial Institution of Public Health | Jing Lu et al |
| EPI_ISL_413884 | Guangdong/GD2020115-P0009/2020 | 2020-02-01 | Asia/China/Guangdong/ | Guangdong Provincial Institution of Public Health, Guangdong Provinical Center for Disease Control and Prevention | Guangdong Provincial Institution of Public Health | Jing Lu et al |
| EPI_ISL_413888 | Guangdong/2020XN4239-P0034/2020 | 2020-01-30 | Asia/China/Guangdong/ | Guangdong Provincial Institution of Public Health, Guangdong Provinical Center for Disease Control and Prevention | Guangdong Provincial Institution of Public Health | Jing Lu et al |
| EPI_ISL_413889 | Guangdong/2020XN4291-P0038/2020 | 2020-01-30 | Asia/China/Guangdong/ | Guangdong Provincial Institution of Public Health, Guangdong Provinical Center for Disease Control and Prevention | Guangdong Provincial Institution of Public Health | Jing Lu et al |
| EPI_ISL_413897 | Guangdong/SZ-N128-P0057/2020 | 2020-02-28 | Asia/China/Guangdong/ | Guangdong Provincial Institution of Public Health, Guangdong Provinical Center for Disease Control and Prevention | Guangdong Provincial Institution of Public Health | Jing Lu et al |
| EPI_ISL_413926 | USA/CA-CDPH-UC8/2020 | 2020-03-05 | North America/USA/Grand Princess/ | California Department of Public Health | Chiu Laboratory, University of California, San Francisco | Xianding Deng et al |
| EPI_ISL_413930 | USA/CA-CDPH-UC10/2020 | 2020-03-05 | North America/USA/California/San Fran | California Department of Public Health | Chiu Laboratory UCSF-Abbott Viral Diagnostics and Discovery Center University of California, San Francisco | Xianding Deng et al |
| EPI_ISL_413996 | Switzerland/TI-SNRCI-29919486/2020 | 2020-02-24 | Europe/Switzerland/Ticino/ | Laboratoire de Virologie, HUG | Swiss National Reference Centre for Influenza | LAUBSCHER Florian et al. et al |
| EPI_ISL_413997 | Switzerland/GE-SNRCI-29933895/202 | 2020-02-26 | Europe/Italy/Geneva/ | Laboratoire de Virologie, HUG | Swiss National Reference Centre for Influenza | LAUBSCHER Florian et al. et al |
| EPI_ISL_413999 | Switzerland/AG-SNRCI-29940361/202 | 2020-02-27 | Europe/Switzerland/Aargau/ | Laboratoire de Virologie, HUG | Swiss National Reference Centre for Influenza | LAUBSCHER Florian et al. et al |

| | | | | | | |
|---|---|---|---|---|---|---|
| EPI_ISL_414005 | England/200940527/2020 | 2020-02-25 | Europe/United Kingdom/England/ | Respiratory Virus Unit, Microbiology Services Colindale, Public Health England | Respiratory Virus Unit, Microbiology Services Colindale, Public Health England | Monica Galiano et al |
| EPI_ISL_414006 | England/200990724/2020 | 2020-02-28 | Europe/United Kingdom/England/ | Respiratory Virus Unit, Microbiology Services Colindale, Public Health England | Respiratory Virus Unit, Microbiology Services Colindale, Public Health England | Monica Galiano et al |
| EPI_ISL_414007 | England/200990725/2020 | 2020-02-28 | Europe/United Kingdom/England/ | Respiratory Virus Unit, Microbiology Services Colindale, Public Health England | Respiratory Virus Unit, Microbiology Services Colindale, Public Health England | Monica Galiano et al |
| EPI_ISL_414008 | England/200960041/2020 | 2020-02-27 | Europe/United Kingdom/England/ | Respiratory Virus Unit, Microbiology Services Colindale, Public Health England | Respiratory Virus Unit, Microbiology Services Colindale, Public Health England | Monica Galiano et al |
| EPI_ISL_414009 | England/200960515/2020 | 2020-02-25 | Europe/United Kingdom/England/ | Respiratory Virus Unit, Microbiology Services Colindale, Public Health England | Respiratory Virus Unit, Microbiology Services Colindale, Public Health England | Monica Galiano et al |
| EPI_ISL_414010 | England/200981386/2020 | 2020-02-26 | Europe/United Kingdom/England/ | Respiratory Virus Unit, Microbiology Services Colindale, Public Health England | Respiratory Virus Unit, Microbiology Services Colindale, Public Health England | Monica Galiano et al |

| | | | | | | |
|---|---|---|---|---|---|---|
| EPI_ISL_414011 | England/200990006/2020 | 2020-02-26 | Europe/United Kingdom/England/ | Respiratory Virus Unit, Microbiology Services Colindale, Public Health England | Respiratory Virus Unit, Microbiology Services Colindale, Public Health England | Monica Galiano et al |
| EPI_ISL_414012 | England/200990723/2020 | 2020-02-27 | Europe/United Kingdom/England/ | Respiratory Virus Unit, Microbiology Services Colindale, Public Health England | Respiratory Virus Unit, Microbiology Services Colindale, Public Health England | Monica Galiano et al |
| EPI_ISL_414015 | Brazil/SP-06/2020 | 2020-02-29 | South America/Brazil/S√£o Paulo/S√£o | Hospital S√£o Joaquim Beneficencia Portuguesa | Instituto Adolfo Lutz, Interdiciplinary Procedures Center, Strategic Laboratory | Claudio Tavares Sacchi et al |
| EPI_ISL_414016 | Brazil/SP-05/2020 | 2020-02-29 | South America/Brazil/S√£o Paulo/S√£o | Hospital S√£o Joaquim Beneficencia Portuguesa | Instituto Adolfo Lutz, Interdiciplinary Procedures Center, Strategic Laboratory | Claudio Tavares Sacchi et al |
| EPI_ISL_414019 | Switzerland/GE-SNRCI-29943121/202 | 2020-02-27 | Europe/Switzerland/Geneva/ | Laboratoire de Virologie, HUG | Swiss National Reference Centre for Influenza | LAUBSCHER Florian et al. et al |
| EPI_ISL_414020 | Switzerland/GE-SNRCI-29945373/202 | 2020-02-27 | Europe/Switzerland/Geneva/ | Laboratoire de Virologie, HUG | Swiss National Reference Centre for Influenza | LAUBSCHER Florian et al. et al |
| EPI_ISL_414021 | Switzerland/BL-SNRCI-29950902/202 | 2020-02-27 | Europe/Switzerland/Basel-Land/ | Laboratoire de Virologie, HUG | Swiss National Reference Centre for Influenza | LAUBSCHER Florian et al. et al |
| EPI_ISL_414022 | Switzerland/GE-SNRCI-29949586/202 | 2020-02-27 | Europe/Italy/Geneva/ | Laboratoire de Virologie, HUG | Swiss National Reference Centre for Influenza | LAUBSCHER Florian et al. et al |

| | | | | | | |
|---|---|---|---|---|---|---|
| EPI_ISL_414040 | England/200641094/2020 | 2020-02-05 | Europe/United Kingdom/England/ | Respiratory Virus Unit, Microbiology Services Colindale, Public Health England | Respiratory Virus Unit, Microbiology Services Colindale, Public Health England | Monica Galiano et al |
| EPI_ISL_414041 | England/200690245/2020 | 2020-02-08 | Europe/United Kingdom/England/ | Respiratory Virus Unit, Microbiology Services Colindale, Public Health England | Respiratory Virus Unit, Microbiology Services Colindale, Public Health England | Monica Galiano et al |
| EPI_ISL_414042 | England/200690300/2020 | 2020-02-08 | Europe/United Kingdom/England/ | Respiratory Virus Unit, Microbiology Services Colindale, Public Health England | Respiratory Virus Unit, Microbiology Services Colindale, Public Health England | Monica Galiano et al |
| EPI_ISL_414043 | England/200690306/2020 | 2020-02-07 | Europe/United Kingdom/England/ | Respiratory Virus Unit, Microbiology Services Colindale, Public Health England | Respiratory Virus Unit, Microbiology Services Colindale, Public Health England | Monica Galiano et al |
| EPI_ISL_414044 | England/200690756/2020 | 2020-02-08 | Europe/United Kingdom/England/ | Respiratory Virus Unit, Microbiology Services Colindale, Public Health England | Respiratory Virus Unit, Microbiology Services Colindale, Public Health England | Monica Galiano et al |
| EPI_ISL_414378 | Singapore/12/2020 | 2020-02-17 | Asia/Singapore/Singapore/Singapore | National Centre for Infectious Diseases | Programme in Emerging Infectious Diseases, Duke-NUS Medical School | Danielle E Anderson et al |

| | | | | | | |
|---|---|---|---|---|---|---|
| EPI_ISL_414379 | Singapore/13/2020 | 2020-02-18 | Asia/Singapore/Singapore/Singapore | National Centre for Infectious Diseases | Programme in Emerging Infectious Diseases, Duke-NUS Medical School | Danielle E Anderson et al |
| EPI_ISL_414380 | Singapore/14/2020 | 2020-02-13 | Asia/Singapore/Singapore/Singapore | National Centre for Infectious Diseases | Programme in Emerging Infectious Diseases, Duke-NUS Medical School | Danielle E Anderson et al |
| EPI_ISL_414414 | Australia/QLD09/2020 | 2020-02-29 | Oceania/Australia/Queensland/Gold Coa | Pathology Queensland | Public Health Virology Laboratory | Bixing Huang et al |
| EPI_ISL_414468 | Netherlands/ZuidHolland_8/2020 | 2020-03-06 | Europe/Netherlands/South Holland/ | Dutch COVID-19 response team | Erasmus Medical Center | David Nieuwenhuijse et al |
| EPI_ISL_414476 | USA/NY1-PV08001/2020 | 2020-02-29 | North America/Iran/New York/ | MSHS Clinical Microbiology Laboratories | MSHS Pathogen Surveillance Program | Gopi Patel et al |
| EPI_ISL_414477 | CzechRepublic/951/2020 | 2020-03-01 | Europe/Italy/Usti nad Labem/ | The National Institute of Public Health Center for Epidemiology and Microbiology | State Veterinary Institute Prague | Alexander Nagy et al |
| EPI_ISL_414495 | Spain/VC-FISABIO-1/2020 | 2020-03-08 | Europe/Spain/Comunitat Valenciana/Va | Servicio Microbiolog√≠a. Hospital Cl√≠nico Universitario. Valencia. | Sequencing and Bioinformatics Service. Molecular Epidemiology Laboratory. FISABIO-Public Health | David Navarro et al |
| EPI_ISL_414497 | Germany/NW-HHU-02-1/2020 | 2020-02-25 | Europe/Germany/North Rhine Westphal | Center of Medical Microbiology, Virology, and Hospital Hygiene, University of Duesseldorf | Center of Medical Microbiology, Virology, and Hospital Hygiene, University of Duesseldorf | Ortwin Adams et al |

| | | | | | | |
|---|---|---|---|---|---|---|
| EPI_ISL_414498 | Germany/NW-HHU-03/2020 | 2020-02-26 | Europe/Germany/North Rhine Westphal | Center of Medical Microbiology, Virology, and Hospital Hygiene, University of Duesseldorf | Center of Medical Microbiology, Virology, and Hospital Hygiene, University of Duesseldorf | Ortwin Adams et al |
| EPI_ISL_414499 | Germany/NW-HHU-04/2020 | 2020-02-26 | Europe/Germany/North Rhine Westphal | Center of Medical Microbiology, Virology, and Hospital Hygiene, University of Duesseldorf | Center of Medical Microbiology, Virology, and Hospital Hygiene, University of Duesseldorf | Ortwin Adams et al |
| EPI_ISL_414504 | Germany/NW-HHU-05/2020 | 2020-02-26 | Europe/Germany/North Rhine Westphal | Center of Medical Microbiology, Virology, and Hospital Hygiene, University of Duesseldorf | Center of Medical Microbiology, Virology, and Hospital Hygiene, University of Duesseldorf | Ortwin Adams et al |
| EPI_ISL_414505 | Germany/NW-HHU-06/2020 | 2020-02-27 | Europe/Germany/North Rhine Westphal | Center of Medical Microbiology, Virology, and Hospital Hygiene, University of Duesseldorf | Center of Medical Microbiology, Virology, and Hospital Hygiene, University of Duesseldorf | Ortwin Adams et al |
| EPI_ISL_414506 | Germany/NW-HHU-07/2020 | 2020-02-27 | Europe/Germany/North Rhine Westphal | Center of Medical Microbiology, Virology, and Hospital Hygiene, University of Duesseldorf | Center of Medical Microbiology, Virology, and Hospital Hygiene, University of Duesseldorf | Ortwin Adams et al |
| EPI_ISL_414508 | Germany/NW-HHU-08/2020 | 2020-02-27 | Europe/Germany/North Rhine Westphal | Center of Medical Microbiology, Virology, and Hospital Hygiene, University of Duesseldorf | Center of Medical Microbiology, Virology, and Hospital Hygiene, University of Duesseldorf | Ortwin Adams et al |

| | | | | | | |
|---|---|---|---|---|---|---|
| EPI_ISL_414509 | Germany/NW-HHU-09/2020 | 2020-02-28 | Europe/Germany/North Rhine Westphal | Center of Medical Microbiology, Virology, and Hospital Hygiene, University of Duesseldorf | Center of Medical Microbiology, Virology, and Hospital Hygiene, University of Duesseldorf | Ortwin Adams et al |
| EPI_ISL_414510 | Shanghai/SH01/2020 | 2020-02-02 | Asia/China/Shanghai/ | Key Laboratory of Medical Molecular Virology (MOE/NHC/CAMS), School of Basic Medicine, Shanghai Medical College, Fudan University | Key Laboratory of Medical Molecular Virology (MOE/NHC/CAMS), School of Basic Medicine, Shanghai Medical College, Fudan University | Zhang et al |
| EPI_ISL_414517 | HongKong/VM20002907/2020 | 2020-02-25 | Asia/Hong Kong/Hong Kong/Hong Kong | Hong Kong Department of Health | School of Public Health, The University of Hong Kong | Dominic N.C. Tsang et al |
| EPI_ISL_414519 | HongKong/VM20002868/2020 | 2020-02-24 | Asia/Hong Kong/Hong Kong/Hong Kong | Hong Kong Department of Health | School of Public Health, The University of Hong Kong | Dominic N.C. Tsang et al |
| EPI_ISL_414520 | Germany/BY-BIM-Pat2/2020 | 2020-03-02 | Europe/Iran/Bavaria/Munich | Bundeswehr Institute of Microbiology | Bundeswehr Institute of Microbiology | Mathias C Walter et al |
| EPI_ISL_414521 | Germany/BY-BIM-Pat3/2020 | 2020-03-02 | Europe/Germany/Bavaria/Munich | Bundeswehr Institute of Microbiology | Bundeswehr Institute of Microbiology | Mathias C Walter et al |
| EPI_ISL_414522 | England/200990002/2020 | 2020-02-28 | Europe/United Kingdom/England/ | Respiratory Virus Unit, Microbiology Services Colindale, Public Health England | Respiratory Virus Unit, Microbiology Services Colindale, Public Health England | Monica Galiano et al |

| | | | | | | |
|---|---|---|---|---|---|---|
| EPI_ISL_414523 | England/200990660/2020 | 2020-02-27 | Europe/United Kingdom/England/ | Respiratory Virus Unit, Microbiology Services Colindale, Public Health England | Respiratory Virus Unit, Microbiology Services Colindale, Public Health England | Monica Galiano et al |
| EPI_ISL_414527 | HongKong/VM20002493-2/2020 | 2020-02-09 | Asia/Hong Kong/Hong Kong/Hong Kong | Hong Kong Department of Health | School of Public Health, The University of Hong Kong | Dominic N.C. Tsang et al |
| EPI_ISL_414528 | HongKong/VM20002507-2/2020 | 2020-02-10 | Asia/Hong Kong/Hong Kong/Hong Kong | Hong Kong Department of Health | School of Public Health, The University of Hong Kong | Dominic N.C. Tsang et al |
| EPI_ISL_414535 | Netherlands/NoordBrabant_26/2020 | 2020-03-06 | Europe/Netherlands/North Brabant/ | Dutch COVID-19 response team | Erasmus Medical Center | David Nieuwenhuijse et al |
| EPI_ISL_414553 | Netherlands/Utrecht_14-2/2020 | 2020-03-09 | Europe/Netherlands/Utrecht/ | Dutch COVID-19 response team | Erasmus Medical Center | David Nieuwenhuijse et al |
| EPI_ISL_414563 | Netherlands/ZuidHolland_21/2020 | 2020-03-03 | Europe/Netherlands/South Holland/ | Dutch COVID-19 response team | Erasmus Medical Center | David Nieuwenhuijse et al |
| EPI_ISL_414569 | HongKong/VM20002582-2/2020 | 2020-02-12 | Asia/Hong Kong/Hong Kong/Hong Kong | Hong Kong Department of Health | School of Public Health, The University of Hong Kong | Dominic N.C. Tsang et al |
| EPI_ISL_414571 | HongKong/VM20002849/2020 | 2020-02-22 | Asia/Hong Kong/Hong Kong/Hong Kong | Hong Kong Department of Health | School of Public Health, The University of Hong Kong | Dominic N.C. Tsang et al |
| EPI_ISL_414574 | Germany/NW-HHU-10/2020 | 2020-02-28 | Europe/Germany/North Rhine Westphal | Center of Medical Microbiology, Virology, and Hospital Hygiene, University of Duesseldorf | Center of Medical Microbiology, Virology, and Hospital Hygiene, University of Duesseldorf | Ortwin Adams et al |
| EPI_ISL_414600 | France/GES-1583/2020 | 2020-02-26 | Europe/France/Grand Est/Strasbourg | Laboratoire de Virologie Institut de Virologie - INSERM U 1109 H√¥pitaux Universitaires de Strasbourg | National Reference Center for Viruses of Respiratory Infections, Institut Pasteur, Paris | M√©lnie Albert et al |

| | | | | | | |
|---|---|---|---|---|---|---|
| EPI_ISL_414623 | France/GES-1583-2/2020 | 2020-02-25 | Europe/France/Grand Est/Strasbourg | Laboratoire de Virologie Institut de Virologie - INSERM U 1109 H√¥pitaux Universitaires de Strasbourg | National Reference Center for Viruses of Respiratory Infections, Institut Pasteur, Paris | M√©lnie Albert et al |
| EPI_ISL_414624 | France/NOR-N1620/2020 | 2020-02-26 | Europe/France/Normandie/Rouen | Centre Hositalier Universitaire de Rouen Laboratoire de Virologie | National Reference Center for Viruses of Respiratory Infections, Institut Pasteur, Paris | M√©lnie Albert et al |
| EPI_ISL_414625 | France/PDL-1643/2020 | 2020-02-26 | Europe/France/Pays de la Loire/Nantes | Centre Hospitalier R√©gional Universitaire de Nantes Laboratoire de Virologie | National Reference Center for Viruses of Respiratory Infections, Institut Pasteur, Paris | M√©lnie Albert et al |
| EPI_ISL_414626 | France/HDF-1684/2020 | 2020-02-29 | Europe/France/Hauts de France/Cr√©p | unknown | National Reference Center for Viruses of Respiratory Infections, Institut Pasteur, Paris | M√©lnie Albert et al |
| EPI_ISL_414632 | France/GES-1977/2020 | 2020-03-04 | Europe/France/Grand Est/Reims | H√¥pital Robert Debr√© Laboratoire de Virologie | National Reference Center for Viruses of Respiratory Infections, Institut Pasteur, Paris | M√©lnie Albert et al |
| EPI_ISL_414635 | France/HDF-1988/2020 | 2020-03-04 | Europe/France/Hauts de France/Compi√ | Centre Hospitalier Compi√®gne Laboratoire de Biologie | National Reference Center for Viruses of Respiratory Infections, Institut Pasteur, Paris | M√©lnie Albert et al |

| | | | | | | |
|---|---|---|---|---|---|---|
| EPI_ISL_414640 | Finland/FIN-114/2020 | 2020-03-01 | Europe/Italy/Finland/ | Department of Virology and Immunology, University of Helsinki and Helsinki University Hospital, Huslab Finland | Department of Virology, Faculty of Medicine, University of Helsinki, Helsinki, Finland | Teemu Smura et al |
| EPI_ISL_414643 | Finland/FIN-508/2020 | 2020-03-07 | Europe/Finland/Finland/ | Department of Virology and Immunology, University of Helsinki and Helsinki University Hospital, Huslab Finland | Department of Virology, Faculty of Medicine, University of Helsinki, Helsinki, Finland | Teemu Smura et al |
| EPI_ISL_414646 | Finland/FIN-266/2020 | 2020-03-04 | Europe/Iran/Finland/ | Department of Virology and Immunology, University of Helsinki and Helsinki University Hospital, Huslab Finland | Department of Virology, Faculty of Medicine, University of Helsinki, Helsinki, Finland | Teemu Smura et al |
| EPI_ISL_414686 | Guangzhou/GZMU0030/2020 | 2020-02-27 | Asia/China/Guangdong/Guangzhou | State Key Laboratory of Respiratory Disease, National Clinical Research Center for Respiratory Disease, Guangzhou Institute of Respiratory Health, the First Affiliated Hospital of Guangzhou Medical University | The First Affiliated Hospital of Guangzhou Medical University & BGI-Shenzhen | Zhao et al et al |
| EPI_ISL_414688 | Guangzhou/GZMU0042/2020 | 2020-02-25 | Asia/China/Guangdong/Guangzhou | State Key Laboratory of Respiratory Disease, National Clinical Research Center for Respiratory Disease, Guangzhou Institute of Respiratory Health, the First Affiliated Hospital of Guangzhou Medical University | The First Affiliated Hospital of Guangzhou Medical University & BGI-Shenzhen | Zhao et al et al |
| EPI_ISL_414689 | Guangzhou/GZMU0044/2020 | 2020-02-25 | Asia/China/Guangdong/Guangzhou | State Key Laboratory of Respiratory Disease, National Clinical Research Center for Respiratory Disease, Guangzhou Institute of Respiratory Health, the First Affiliated Hospital of Guangzhou Medical University | The First Affiliated Hospital of Guangzhou Medical University & BGI-Shenzhen | Zhao et al et al |

| | | | | | | |
|---|---|---|---|---|---|---|
| EPI_ISL_414691 | Guangzhou/GZMU0048/2020 | 2020-02-25 | Asia/China/Guangdong/Guangzhou | State Key Laboratory of Respiratory Disease, National Clinical Research Center for Respiratory Disease, Guangzhou Institute of Respiratory Health, the First Affiliated Hospital of Guangzhou Medical University | The First Affiliated Hospital of Guangzhou Medical University & BGI-Shenzhen | Zhao et al et al |
| EPI_ISL_414692 | Guangzhou/GZMU0014/2020 | 2020-02-25 | Asia/China/Guangdong/Guangzhou | State Key Laboratory of Respiratory Disease, National Clinical Research Center for Respiratory Disease, Guangzhou Institute of Respiratory Health, the First Affiliated Hospital of Guangzhou Medical University | The First Affiliated Hospital of Guangzhou Medical University & BGI-Shenzhen | Zhao et al et al |
| EPI_ISL_414935 | Shandong/LY002-2/2020 | 2020-01-21 | Asia/China/Shandong/ | Shandong Provincial Center for Disease Control and Prevention | Beijing Institute of Microbiology and Epidemiology | Xiao-Lin Jiang et al |
| EPI_ISL_414936 | Shandong/LY003-2/2020 | 2020-01-23 | Asia/China/Shandong/ | Shandong Provincial Center for Disease Control and Prevention | Beijing Institute of Microbiology and Epidemiology | Xiao-Lin Jiang et al |
| EPI_ISL_414937 | Shandong/LY004-2/2020 | 2020-01-26 | Asia/China/Shandong/ | Shandong Provincial Center for Disease Control and Prevention | Beijing Institute of Microbiology and Epidemiology | Xiao-Lin Jiang et al |
| EPI_ISL_414938 | Shandong/LY005-2/2020 | 2020-01-24 | Asia/China/Shandong/ | Shandong Provincial Center for Disease Control and Prevention | Beijing Institute of Microbiology and Epidemiology | Xiao-Lin Jiang et al |
| EPI_ISL_414939 | Shandong/LY006-2/2020 | 2020-01-25 | Asia/China/Shandong/ | Shandong Provincial Center for Disease Control and Prevention | Beijing Institute of Microbiology and Epidemiology | Xiao-Lin Jiang et al |
| EPI_ISL_414940 | Shandong/LY007-2/2020 | 2020-01-25 | Asia/China/Shandong/ | Shandong Provincial Center for Disease Control and Prevention | Beijing Institute of Microbiology and Epidemiology | Xiao-Lin Jiang et al |
| EPI_ISL_414941 | Shandong/LY008-2/2020 | 2020-01-30 | Asia/China/Shandong/ | Shandong Provincial Center for Disease Control and Prevention | Beijing Institute of Microbiology and Epidemiology | Xiao-Lin Jiang et al |
| EPI_ISL_414949 | NorthernIreland/HSCNI01/2020 | 2020-02-26 | Europe/United Kingdom/Northern Irelar | Regional Virus Laboratory, Belfast | Public Health Wales Microbiology Cardiff | Tanya Curran et al |

| | | | | | | |
|---|---|---|---|---|---|---|
| EPI_ISL_415128 | Brazil/ES-225/2020 | 2020-02-29 | South America/Brazil/Espirito Santo/Vil | LACEN/ES - Laboratv≥rio Central de Sav∫de Pv∫blica do Espv≠rito Santo | Instituto Oswaldo Cruz FIOCRUZ - Laboratory of Respiratory Viruses and Measles (LVRS) | Paola Resende et al |
| EPI_ISL_415129 | England/20099038206/2020 | 2020-02-29 | Europe/United Kingdom/England/ | Respiratory Virus Unit, Microbiology Services Colindale, Public Health England | Respiratory Virus Unit, Microbiology Services Colindale, Public Health England | Monica Galiano et al |
| EPI_ISL_415131 | England/20099107406/2020 | 2020-02-29 | Europe/United Kingdom/England/ | Respiratory Virus Unit, Microbiology Services Colindale, Public Health England | Respiratory Virus Unit, Microbiology Services Colindale, Public Health England | Monica Galiano et al |
| EPI_ISL_415132 | England/20100001406/2020 | 2020-02-28 | Europe/United Kingdom/England/ | Respiratory Virus Unit, Microbiology Services Colindale, Public Health England | Respiratory Virus Unit, Microbiology Services Colindale, Public Health England | Monica Galiano et al |
| EPI_ISL_415133 | England/20100004706/2020 | 2020-02-29 | Europe/United Kingdom/England/ | Respiratory Virus Unit, Microbiology Services Colindale, Public Health England | Respiratory Virus Unit, Microbiology Services Colindale, Public Health England | Monica Galiano et al |
| EPI_ISL_415134 | England/20100004806/2020 | 2020-02-29 | Europe/United Kingdom/England/ | Respiratory Virus Unit, Microbiology Services Colindale, Public Health England | Respiratory Virus Unit, Microbiology Services Colindale, Public Health England | Monica Galiano et al |

| | | | | | | |
|---|---|---|---|---|---|---|
| EPI_ISL_415135 | England/20100005406/2020 | 2020-02-28 | Europe/United Kingdom/England/ | Respiratory Virus Unit, Microbiology Services Colindale, Public Health England | Respiratory Virus Unit, Microbiology Services Colindale, Public Health England | Monica Galiano et al |
| EPI_ISL_415136 | England/20100022706/2020 | 2020-02-29 | Europe/United Kingdom/England/ | Respiratory Virus Unit, Microbiology Services Colindale, Public Health England | Respiratory Virus Unit, Microbiology Services Colindale, Public Health England | Monica Galiano et al |
| EPI_ISL_415140 | England/20100121006/2020 | 2020-02-29 | Europe/United Kingdom/England/ | Respiratory Virus Unit, Microbiology Services Colindale, Public Health England | Respiratory Virus Unit, Microbiology Services Colindale, Public Health England | Monica Galiano et al |
| EPI_ISL_415141 | England/20100121007/2020 | 2020-02-29 | Europe/United Kingdom/England/ | Respiratory Virus Unit, Microbiology Services Colindale, Public Health England | Respiratory Virus Unit, Microbiology Services Colindale, Public Health England | Monica Galiano et al |
| EPI_ISL_415144 | England/20102000106/2020 | 2020-02-27 | Europe/United Kingdom/England/ | Respiratory Virus Unit, Microbiology Services Colindale, Public Health England | Respiratory Virus Unit, Microbiology Services Colindale, Public Health England | Monica Galiano et al |
| EPI_ISL_415154 | Belgium/BM-03012/2020 | 2020-03-01 | Europe/Italy/Halle-Vilvoorde/Kraainem | KU Leuven, Clincal and Epidemiological Virology | KU Leuven, Clincal and Epidemiological Virology | Bert Vanmechelen et al |
| EPI_ISL_415158 | Belgium/QKJ-03015/2020 | 2020-03-01 | Europe/Belgium/Brussels/ | KU Leuven, Clinical and Epidemiological Virology | KU Leuven, Clinical and Epidemiological Virology | Bert Vanmechelen et al |

| | | | | | | |
|---|---|---|---|---|---|---|
| EPI_ISL_415159 | Belgium/BA-02291/2020 | 2020-02-29 | Europe/Belgium/Leuven/ | KU Leuven, Clinical and Epidemiological Virology | KU Leuven, Clinical and Epidemiological Virology | Bert Vanmechelen et al |
| EPI_ISL_415454 | Switzerland/GE-HUG-1422/2020 | 2020-02-28 | Europe/Switzerland/Geneva/ | University Hospitals of Geneva Laboratory of Virology | University Hospitals of Geneva Laboratory of Virology | Laubscher F. et al |
| EPI_ISL_415455 | Switzerland/GE-HUG-0199/2020 | 2020-02-28 | Europe/Switzerland/Geneva/ | University Hospitals of Geneva Laboratory of Virology | University Hospitals of Geneva Laboratory of Virology | Laubscher F. et al |
| EPI_ISL_415456 | Switzerland/BE-HUG-6651/2020 | 2020-02-29 | Europe/Switzerland/Bern/ | University Hospitals of Geneva Laboratory of Virology | University Hospitals of Geneva Laboratory of Virology | Laubscher F. et al |
| EPI_ISL_415457 | Switzerland/AG-HUG-7120/2020 | 2020-02-29 | Europe/Switzerland/Aargau/ | University Hospitals of Geneva Laboratory of Virology | University Hospitals of Geneva Laboratory of Virology | Laubscher F. et al |
| EPI_ISL_415459 | Switzerland/GE-HUG-VD0503/2020 | 2020-02-29 | Europe/Switzerland/Vaud/ | University Hospitals of Geneva Laboratory of Virology | University Hospitals of Geneva Laboratory of Virology | Laubscher F. et al |
| EPI_ISL_415497 | Netherlands/NA_8/2020 | 2020-03-09 | Europe/Netherlands/Netherlands/ | Dutch COVID-19 response team | Erasmus Medical Center | David Nieuwenhuijse et al |
| EPI_ISL_415513 | Netherlands/NoordBrabant_57/2020 | 2020-03-11 | Europe/Netherlands/North Brabant/ | Dutch COVID-19 response team | Erasmus Medical Center | David Nieuwenhuijse et al |
| EPI_ISL_415526 | Netherlands/Utrecht_17/2020 | 2020-03-10 | Europe/Netherlands/Utrecht/ | Dutch COVID-19 response team | Erasmus Medical Center | David Nieuwenhuijse et al |
| EPI_ISL_415531 | Netherlands/ZuidHolland_27/2020 | 2020-03-09 | Europe/Netherlands/South Holland/ | Dutch COVID-19 response team | Erasmus Medical Center | David Nieuwenhuijse et al |
| EPI_ISL_415543 | USA/UT-05/2020 | 2020-03-13 | North America/USA/Utah/ | Utah Public Health Laboratory | Utah Public Health Laboratory | Erin Young et al |

| | | | | | | |
|---|---|---|---|---|---|---|
| EPI_ISL_415577 | Canada/BC_69243/2020 | 2020-02-20 | North America/Iran/British Columbia/ | BCCDC Public Health Laboratory | BCCDC Public Health Laboratory | Harrigan et al |
| EPI_ISL_415578 | Canada/BC_13297/2020 | 2020-02-27 | North America/Iran/British Columbia/ | BCCDC Public Health Laboratory | BCCDC Public Health Laboratory | Harrigan et al |
| EPI_ISL_415579 | Canada/BC_25211/2020 | 2020-03-02 | North America/Iran/British Columbia/ | BCCDC Public Health Laboratory | BCCDC Public Health Laboratory | Harrigan et al |
| EPI_ISL_415580 | Canada/BC_17397/2020 | 2020-02-28 | North America/Iran/British Columbia/ | BCCDC Public Health Laboratory | BCCDC Public Health Laboratory | Harrigan et al |
| EPI_ISL_415581 | Canada/BC_02421/2020 | 2020-03-01 | North America/Iran/British Columbia/ | BCCDC Public Health Laboratory | BCCDC Public Health Laboratory | Harrigan et al |
| EPI_ISL_415582 | Canada/BC_35720/2020 | 2020-03-04 | North America/Canada/British Columbia | BCCDC Public Health Laboratory | BCCDC Public Health Laboratory | Harrigan et al |
| EPI_ISL_415583 | Canada/BC_40860/2020 | 2020-03-03 | North America/Iran/British Columbia/ | BCCDC Public Health Laboratory | BCCDC Public Health Laboratory | Harrigan et al |
| EPI_ISL_415584 | Canada/BC_41851/2020 | 2020-03-02 | North America/Canada/British Columbia | BCCDC Public Health Laboratory | BCCDC Public Health Laboratory | Harrigan et al |
| EPI_ISL_415585 | Canada/BC_65034/2020 | 2020-03-04 | North America/Iran/British Columbia/ | BCCDC Public Health Laboratory | BCCDC Public Health Laboratory | Harrigan et al |
| EPI_ISL_415586 | Canada/BC_64686/2020 | 2020-03-05 | North America/Canada/British Columbia | BCCDC Public Health Laboratory | BCCDC Public Health Laboratory | Harrigan et al |
| EPI_ISL_415587 | Canada/BC_66353/2020 | 2020-03-04 | North America/Iran/British Columbia/ | BCCDC Public Health Laboratory | BCCDC Public Health Laboratory | Harrigan et al |
| EPI_ISL_415588 | Canada/BC_83109/2020 | 2020-03-05 | North America/Canada/British Columbia | BCCDC Public Health Laboratory | BCCDC Public Health Laboratory | Harrigan et al |
| EPI_ISL_415589 | Canada/BC_83163/2020 | 2020-03-05 | North America/Canada/British Columbia | BCCDC Public Health Laboratory | BCCDC Public Health Laboratory | Harrigan et al |
| EPI_ISL_415590 | Canada/BC_78548/2020 | 2020-03-05 | North America/Canada/British Columbia | BCCDC Public Health Laboratory | BCCDC Public Health Laboratory | Harrigan et al |
| EPI_ISL_415597 | USA/WA-UW69/2020 | 2020-03-10 | North America/USA/Washington/King C | UW Virology Lab | UW Virology Lab | Pavitra Roychoudhury et al |
| EPI_ISL_415641 | Georgia/Tb-54/2020 | 2020-02-27 | Asia/Georgia/Tbilisi/ | R. G. Lugar Center for Public Health Research, National Center for Disease Control and Public Health (NCDC) of Georgia. | R. G. Lugar Center for Public Health Research, National Center for Disease Control and Public Health (NCDC) of Georgia. | Nato Kotaria et al |

| | | | | | | |
|---|---|---|---|---|---|---|
| EPI_ISL_415644 | Georgia/Tb-82/2020 | 2020-02-28 | Asia/Georgia/Tbilisi/ | R. G. Lugar Center for Public Health Research, National Center for Disease Control and Public Health (NCDC) of Georgia. | R. G. Lugar Center for Public Health Research, National Center for Disease Control and Public Health (NCDC) of Georgia. | Nato Kotaria et al |
| EPI_ISL_415651 | France/BFC-2094/2020 | 2020-03-05 | Europe/France/Bourgogne-France-Comt | unknown | National Reference Center for Viruses of Respiratory Infections, Institut Pasteur, Paris | M√©lnie Albert et al |
| EPI_ISL_415698 | Switzerland/GR-HUG-2988/2020 | 2020-02-27 | Europe/Switzerland/Graub√ºnden/ | University Hospitals of Geneva Laboratory of Virology | University Hospitals of Geneva Laboratory of Virology | Laubscher F. et al |
| EPI_ISL_415699 | Switzerland/GR-HUG-3043/2020 | 2020-02-27 | Europe/Switzerland/Graub√ºnden/ | University Hospitals of Geneva Laboratory of Virology | University Hospitals of Geneva Laboratory of Virology | Laubscher F. et al |
| EPI_ISL_415700 | Switzerland/GE-HUG-1402/2020 | 2020-02-28 | Europe/Switzerland/Geneva/ | University Hospitals of Geneva Laboratory of Virology | University Hospitals of Geneva Laboratory of Virology | Laubscher F. et al |
| EPI_ISL_415703 | Switzerland/TI-HUG-2045/2020 | 2020-03-01 | Europe/Switzerland/Ticino/ | University Hospitals of Geneva Laboratory of Virology | University Hospitals of Geneva Laboratory of Virology | Laubscher F. et al |
| EPI_ISL_415704 | Switzerland/BE-HUG-2536/2020 | 2020-03-04 | Europe/Switzerland/Bern/ | University Hospitals of Geneva Laboratory of Virology | University Hospitals of Geneva Laboratory of Virology | Laubscher F. et al |

| | | | | | | |
|---|---|---|---|---|---|---|
| EPI_ISL_415709 | Hangzhou/ZJU-01/2020 | 2020-01-25 | Asia/China/Zhejiang/Hangzhou | State Key Laboratory for Diagnosis and Treatment of Infectious Diseases, National Clinical Research Center for Infectious Diseases, First Affiliated Hospital, Zhejiang University School of Medicine, Hangzhou, China. 310003 | State Key Laboratory for Diagnosis and Treatment of Infectious Diseases, National Clinical Research Center for Infectious Diseases, First Affiliated Hospital, Zhejiang University School of Medicine, Hangzhou, China. 310003 | Hangping Yao et al |
| EPI_ISL_415710 | Russia/StPetersburg-3524/2020 | 2020-03-15 | Europe/Russia/Saint-Petersburg/ | WHO National Influenza Centre Russian Federation | WHO National Influenza Centre Russian Federation | Andrey Komissarov et al |
| EPI_ISL_415711 | Hangzhou/ZJU-05/2020 | 2020-01-22 | Asia/China/Zhejiang/Hangzhou | State Key Laboratory for Diagnosis and Treatment of Infectious Diseases, National Clinical Research Center for Infectious Diseases, First Affiliated Hospital, Zhejiang University School of Medicine, Hangzhou, China. 310003 | State Key Laboratory for Diagnosis and Treatment of Infectious Diseases, National Clinical Research Center for Infectious Diseases, First Affiliated Hospital, Zhejiang University School of Medicine, Hangzhou, China. 310003 | Hangping Yao et al |

| | | | | | | |
|---|---|---|---|---|---|---|
| EPI_ISL_415741 | Taiwan/CGMH-CGU-03/2020 | 2020-02-26 | Asia/Taiwan/Taiwan/Taoyuan | Laboratory Medicine | Department of Laboratory Medicine, Lin-Kou Chang Gung Memorial Hospital, Taoyuan, Taiwan | Kuo-Chien Tsao et al |
| EPI_ISL_415742 | Taiwan/CGMH-CGU-04/2020 | 2020-02-27 | Asia/Taiwan/Taiwan/Taoyuan | Laboratory Medicine | Department of Laboratory Medicine, Lin-Kou Chang Gung Memorial Hospital, Taoyuan, Taiwan | Kuo-Chien Tsao et al |
| EPI_ISL_415743 | Taiwan/CGMH-CGU-05/2020 | 2020-02-27 | Asia/Taiwan/Taiwan/Taoyuan | Laboratory Medicine | Department of Laboratory Medicine, Lin-Kou Chang Gung Memorial Hospital, Taoyuan, Taiwan | Kuo-Chien Tsao et al |
| EPI_ISL_416036 | Brazil/SP-14/2020 | 2020-03-05 | South America/Italy/S√£o Paulo/S√£o P | National Influenza Center - Instituto Adolfo Lutz | Instituto Adolfo Lutz, Interdiciplinary Procedures Center, Strategic Laboratory | Claudio Tavares Sacchi et al |

| | | | | | | |
|---|---|---|---|---|---|---|
| EPI_ISL_416042 | Hangzhou/ZJU-02/2020 | 2020-01-26 | Asia/China/Zhejiang/Hangzhou | State Key Laboratory for Diagnosis and Treatment of Infectious Diseases, National Clinical Research Center for Infectious Diseases, First Affiliated Hospital, Zhejiang University School of Medicine, Hangzhou, China. 310003 | State Key Laboratory for Diagnosis and Treatment of Infectious Diseases, National Clinical Research Center for Infectious Diseases, First Affiliated Hospital, Zhejiang University School of Medicine, Hangzhou, China. 310003 | Hangping Yao et al |
| EPI_ISL_416044 | Hangzhou/ZJU-03/2020 | 2020-01-25 | Asia/China/Zhejiang/Hangzhou | State Key Laboratory for Diagnosis and Treatment of Infectious Diseases, National Clinical Research Center for Infectious Diseases, First Affiliated Hospital, Zhejiang University School of Medicine, Hangzhou, China 310003 | State Key Laboratory for Diagnosis and Treatment of Infectious Diseases, National Clinical Research Center for Infectious Diseases, First Affiliated Hospital, Zhejiang University School of Medicine, Hangzhou, China 310003 | Hangping Yao et al |

| | | | | | | |
|---|---|---|---|---|---|---|
| EPI_ISL_416046 | Hangzhou/ZJU-04/2020 | 2020-01-24 | Asia/China/Zhejiang/Hangzhou | State Key Laboratory for Diagnosis and Treatment of Infectious Diseases, National Clinical Research Center for Infectious Diseases, First Affiliated Hospital, Zhejiang University School of Medicine, Hangzhou, China 310003 | State Key Laboratory for Diagnosis and Treatment of Infectious Diseases, National Clinical Research Center for Infectious Diseases, First Affiliated Hospital, Zhejiang University School of Medicine, Hangzhou, China 310003 | Hangping Yao et al |
| EPI_ISL_416047 | Hangzhou/ZJU-06/2020 | 2020-02-02 | Asia/China/Zhejiang/Hangzhou | State Key Laboratory for Diagnosis and Treatment of Infectious Diseases, National Clinical Research Center for Infectious Diseases, First Affiliated Hospital, Zhejiang University School of Medicine, Hangzhou, China 310003 | State Key Laboratory for Diagnosis and Treatment of Infectious Diseases, National Clinical Research Center for Infectious Diseases, First Affiliated Hospital, Zhejiang University School of Medicine, Hangzhou, China 310003 | Hangping Yao et al |
| EPI_ISL_416142 | Denmark/SSI-01/2020 | 2020-02-26 | Europe/Denmark/Copenhagen/ | Department of Virus and Microbiological Special diagnostics, Statens Serum Institut, Copenhagen, Denmark. | Statens Serum Institute | Morten Rasmussen et al |
| EPI_ISL_416143 | Denmark/SSI-02/2020 | 2020-02-28 | Europe/Denmark/Copenhagen/ | Department of Virus and Microbiological Special diagnostics, Statens Serum Institut, Copenhagen, Denmark. | ViFU | Morten Rasmussen et al |

| | | | | | | |
|---|---|---|---|---|---|---|
| EPI_ISL_416314 | HongKong/CUHK1/2020 | 2020-02-07 | Asia/Hong Kong/Hong Kong/ | Department of Microbiology, Faculty of Medicine, The Chinese University of Hong Kong, Hong Kong SAR, China | Department of Microbiology, Faculty of Medicine, Chinese University of Hong Kong, Hong Kong SAR, China | Zigui Chen et al |
| EPI_ISL_416316 | Shanghai/SH0002/2020 | 2020-01-25 | Asia/China/Shanghai/ | Shanghai Public Health Clinical Center, Shanghai Medical College, Fudan University | National Research Center for Translational Medicine (Shanghai), Ruijin Hospital affiliated to Shanghai Jiao Tong University School of Medicine & Shanghai Public Health Clinical Center | Shengyue Wang et al |
| EPI_ISL_416317 | Shanghai/SH0003/2020 | 2020-01-25 | Asia/China/Shanghai/ | Shanghai Public Health Clinical Center, Shanghai Medical College, Fudan University | National Research Center for Translational Medicine (Shanghai), Ruijin Hospital affiliated to Shanghai Jiao Tong University School of Medicine & Shanghai Public Health Clinical Center | Shengyue Wang et al |

| | | | | | | |
|---|---|---|---|---|---|---|
| EPI_ISL_416318 | Shanghai/SH0004/2020 | 2020-01-28 | Asia/China/Shanghai/ | Shanghai Public Health Clinical Center, Shanghai Medical College, Fudan University | National Research Center for Translational Medicine (Shanghai), Ruijin Hospital affiliated to Shanghai Jiao Tong University School of Medicine & Shanghai Public Health Clinical Center | Shengyue Wang et al |
| EPI_ISL_416319 | Shanghai/SH0005/2020 | 2020-01-28 | Asia/China/Shanghai/ | Shanghai Public Health Clinical Center, Shanghai Medical College, Fudan University | National Research Center for Translational Medicine (Shanghai), Ruijin Hospital affiliated to Shanghai Jiao Tong University School of Medicine & Shanghai Public Health Clinical Center | Shengyue Wang et al |
| EPI_ISL_416320 | Shanghai/SH0007/2020 | 2020-01-28 | Asia/China/Shanghai/ | Shanghai Public Health Clinical Center, Shanghai Medical College, Fudan University | National Research Center for Translational Medicine (Shanghai), Ruijin Hospital affiliated to Shanghai Jiao Tong University School of Medicine & Shanghai Public Health Clinical Center | Shengyue Wang et al |

| | | | | | | |
|---|---|---|---|---|---|---|
| EPI_ISL_416321 | Shanghai/SH0008/2020 | 2020-01-28 | Asia/China/Shanghai/ | Shanghai Public Health Clinical Center, Shanghai Medical College, Fudan University | National Research Center for Translational Medicine (Shanghai), Ruijin Hospital affiliated to Shanghai Jiao Tong University School of Medicine & Shanghai Public Health Clinical Center | Shengyue Wang et al |
| EPI_ISL_416322 | Shanghai/SH0009/2020 | 2020-01-29 | Asia/China/Shanghai/ | Shanghai Public Health Clinical Center, Shanghai Medical College, Fudan University | National Research Center for Translational Medicine (Shanghai), Ruijin Hospital affiliated to Shanghai Jiao Tong University School of Medicine & Shanghai Public Health Clinical Center | Shengyue Wang et al |
| EPI_ISL_416323 | Shanghai/SH0010/2020 | 2020-01-29 | Asia/China/Shanghai/ | Shanghai Public Health Clinical Center, Shanghai Medical College, Fudan University | National Research Center for Translational Medicine (Shanghai), Ruijin Hospital affiliated to Shanghai Jiao Tong University School of Medicine & Shanghai Public Health Clinical Center | Shengyue Wang et al |

| | | | | | | |
|---|---|---|---|---|---|---|
| EPI_ISL_416324 | Shanghai/SH0011/2020 | 2020-01-29 | Asia/China/Shanghai/ | Shanghai Public Health Clinical Center, Shanghai Medical College, Fudan University | National Research Center for Translational Medicine (Shanghai), Ruijin Hospital affiliated to Shanghai Jiao Tong University School of Medicine & Shanghai Public Health Clinical Center | Shengyue Wang et al |
| EPI_ISL_416326 | Shanghai/SH0013/2020 | 2020-01-30 | Asia/China/Shanghai/ | Shanghai Public Health Clinical Center, Shanghai Medical College, Fudan University | National Research Center for Translational Medicine (Shanghai), Ruijin Hospital affiliated to Shanghai Jiao Tong University School of Medicine & Shanghai Public Health Clinical Center | Shengyue Wang et al |
| EPI_ISL_416327 | Shanghai/SH0014/2020 | 2020-01-28 | Asia/China/Shanghai/ | Shanghai Public Health Clinical Center, Shanghai Medical College, Fudan University | National Research Center for Translational Medicine (Shanghai), Ruijin Hospital affiliated to Shanghai Jiao Tong University School of Medicine & Shanghai Public Health Clinical Center | Shengyue Wang et al |

| | | | | | | |
|---|---|---|---|---|---|---|
| EPI_ISL_416328 | Shanghai/SH0017/2020 | 2020-02-01 | Asia/China/Shanghai/ | Shanghai Public Health Clinical Center, Shanghai Medical College, Fudan University | National Research Center for Translational Medicine (Shanghai), Ruijin Hospital affiliated to Shanghai Jiao Tong University School of Medicine & Shanghai Public Health Clinical Center | Shengyue Wang et al |
| EPI_ISL_416329 | Shanghai/SH0020/2020 | 2020-02-01 | Asia/China/Shanghai/ | Shanghai Public Health Clinical Center, Shanghai Medical College, Fudan University | National Research Center for Translational Medicine (Shanghai), Ruijin Hospital affiliated to Shanghai Jiao Tong University School of Medicine & Shanghai Public Health Clinical Center | Shengyue Wang et al |
| EPI_ISL_416330 | Shanghai/SH0021/2020 | 2020-02-02 | Asia/China/Shanghai/ | Shanghai Public Health Clinical Center, Shanghai Medical College, Fudan University | National Research Center for Translational Medicine (Shanghai), Ruijin Hospital affiliated to Shanghai Jiao Tong University School of Medicine & Shanghai Public Health Clinical Center | Shengyue Wang et al |

| | | | | | | |
|---|---|---|---|---|---|---|
| EPI_ISL_416331 | Shanghai/SH0022/2020 | 2020-01-30 | Asia/China/Shanghai/ | Shanghai Public Health Clinical Center, Shanghai Medical College, Fudan University | National Research Center for Translational Medicine (Shanghai), Ruijin Hospital affiliated to Shanghai Jiao Tong University School of Medicine & Shanghai Public Health Clinical Center | Shengyue Wang et al |
| EPI_ISL_416332 | Shanghai/SH0023/2020 | 2020-01-30 | Asia/China/Shanghai/ | Shanghai Public Health Clinical Center, Shanghai Medical College, Fudan University | National Research Center for Translational Medicine (Shanghai), Ruijin Hospital affiliated to Shanghai Jiao Tong University School of Medicine & Shanghai Public Health Clinical Center | Shengyue Wang et al |
| EPI_ISL_416333 | Shanghai/SH0024/2020 | 2020-01-30 | Asia/China/Shanghai/ | Shanghai Public Health Clinical Center, Shanghai Medical College, Fudan University | National Research Center for Translational Medicine (Shanghai), Ruijin Hospital affiliated to Shanghai Jiao Tong University School of Medicine & Shanghai Public Health Clinical Center | Shengyue Wang et al |

| | | | | | | |
|---|---|---|---|---|---|---|
| EPI_ISL_416334 | Shanghai/SH0025/2020 | 2020-02-06 | Asia/China/Shanghai/ | Shanghai Public Health Clinical Center, Shanghai Medical College, Fudan University | National Research Center for Translational Medicine (Shanghai), Ruijin Hospital affiliated to Shanghai Jiao Tong University School of Medicine & Shanghai Public Health Clinical Center | Shengyue Wang et al |
| EPI_ISL_416335 | Shanghai/SH0026/2020 | 2020-02-02 | Asia/China/Shanghai/ | Shanghai Public Health Clinical Center, Shanghai Medical College, Fudan University | National Research Center for Translational Medicine (Shanghai), Ruijin Hospital affiliated to Shanghai Jiao Tong University School of Medicine & Shanghai Public Health Clinical Center | Shengyue Wang et al |
| EPI_ISL_416336 | Shanghai/SH0027/2020 | 2020-02-01 | Asia/China/Shanghai/ | Shanghai Public Health Clinical Center, Shanghai Medical College, Fudan University | National Research Center for Translational Medicine (Shanghai), Ruijin Hospital affiliated to Shanghai Jiao Tong University School of Medicine & Shanghai Public Health Clinical Center | Shengyue Wang et al |

| | | | | | | |
|---|---|---|---|---|---|---|
| EPI_ISL_416337 | Shanghai/SH0028/2020 | 2020-02-04 | Asia/China/Shanghai/ | Shanghai Public Health Clinical Center, Shanghai Medical College, Fudan University | National Research Center for Translational Medicine (Shanghai), Ruijin Hospital affiliated to Shanghai Jiao Tong University School of Medicine & Shanghai Public Health Clinical Center | Shengyue Wang et al |
| EPI_ISL_416338 | Shanghai/SH0029/2020 | 2020-02-01 | Asia/China/Shanghai/ | Shanghai Public Health Clinical Center, Shanghai Medical College, Fudan University | National Research Center for Translational Medicine (Shanghai), Ruijin Hospital affiliated to Shanghai Jiao Tong University School of Medicine & Shanghai Public Health Clinical Center | Shengyue Wang et al |
| EPI_ISL_416339 | Shanghai/SH0030/2020 | 2020-02-03 | Asia/China/Shanghai/ | Shanghai Public Health Clinical Center, Shanghai Medical College, Fudan University | National Research Center for Translational Medicine (Shanghai), Ruijin Hospital affiliated to Shanghai Jiao Tong University School of Medicine & Shanghai Public Health Clinical Center | Shengyue Wang et al |

| | | | | | | |
|---|---|---|---|---|---|---|
| EPI_ISL_416340 | Shanghai/SH0031/2020 | 2020-02-02 | Asia/China/Shanghai/ | Shanghai Public Health Clinical Center, Shanghai Medical College, Fudan University | National Research Center for Translational Medicine (Shanghai), Ruijin Hospital affiliated to Shanghai Jiao Tong University School of Medicine & Shanghai Public Health Clinical Center | Shengyue Wang et al |
| EPI_ISL_416341 | Shanghai/SH0032/2020 | 2020-02-01 | Asia/China/Shanghai/ | Shanghai Public Health Clinical Center, Shanghai Medical College, Fudan University | National Research Center for Translational Medicine (Shanghai), Ruijin Hospital affiliated to Shanghai Jiao Tong University School of Medicine & Shanghai Public Health Clinical Center | Shengyue Wang et al |
| EPI_ISL_416342 | Shanghai/SH0033/2020 | 2020-02-04 | Asia/China/Shanghai/ | Shanghai Public Health Clinical Center, Shanghai Medical College, Fudan University | National Research Center for Translational Medicine (Shanghai), Ruijin Hospital affiliated to Shanghai Jiao Tong University School of Medicine & Shanghai Public Health Clinical Center | Shengyue Wang et al |

| | | | | | | |
|---|---|---|---|---|---|---|
| EPI_ISL_416343 | Shanghai/SH0034/2020 | 2020-02-04 | Asia/China/Shanghai/ | Shanghai Public Health Clinical Center, Shanghai Medical College, Fudan University | National Research Center for Translational Medicine (Shanghai), Ruijin Hospital affiliated to Shanghai Jiao Tong University School of Medicine & Shanghai Public Health Clinical Center | Shengyue Wang et al |
| EPI_ISL_416344 | Shanghai/SH0035/2020 | 2020-01-30 | Asia/China/Shanghai/ | Shanghai Public Health Clinical Center, Shanghai Medical College, Fudan University | National Research Center for Translational Medicine (Shanghai), Ruijin Hospital affiliated to Shanghai Jiao Tong University School of Medicine & Shanghai Public Health Clinical Center | Shengyue Wang et al |
| EPI_ISL_416345 | Shanghai/SH0036/2020 | 2020-02-04 | Asia/China/Shanghai/ | Shanghai Public Health Clinical Center, Shanghai Medical College, Fudan University | National Research Center for Translational Medicine (Shanghai), Ruijin Hospital affiliated to Shanghai Jiao Tong University School of Medicine & Shanghai Public Health Clinical Center | Shengyue Wang et al |

| | | | | | | |
|---|---|---|---|---|---|---|
| EPI_ISL_416347 | Shanghai/SH0038/2020 | 2020-02-06 | Asia/China/Shanghai/ | Shanghai Public Health Clinical Center, Shanghai Medical College, Fudan University | National Research Center for Translational Medicine (Shanghai), Ruijin Hospital affiliated to Shanghai Jiao Tong University School of Medicine & Shanghai Public Health Clinical Center | Shengyue Wang et al |
| EPI_ISL_416348 | Shanghai/SH0039/2020 | 2020-02-06 | Asia/China/Shanghai/ | Shanghai Public Health Clinical Center, Shanghai Medical College, Fudan University | National Research Center for Translational Medicine (Shanghai), Ruijin Hospital affiliated to Shanghai Jiao Tong University School of Medicine & Shanghai Public Health Clinical Center | Shengyue Wang et al |
| EPI_ISL_416349 | Shanghai/SH0040/2020 | 2020-01-30 | Asia/China/Shanghai/ | Shanghai Public Health Clinical Center, Shanghai Medical College, Fudan University | National Research Center for Translational Medicine (Shanghai), Ruijin Hospital affiliated to Shanghai Jiao Tong University School of Medicine & Shanghai Public Health Clinical Center | Shengyue Wang et al |

| | | | | | | |
|---|---|---|---|---|---|---|
| EPI_ISL_416350 | Shanghai/SH0041/2020 | 2020-01-29 | Asia/China/Shanghai/ | Shanghai Public Health Clinical Center, Shanghai Medical College, Fudan University | National Research Center for Translational Medicine (Shanghai), Ruijin Hospital affiliated to Shanghai Jiao Tong University School of Medicine & Shanghai Public Health Clinical Center | Shengyue Wang et al |
| EPI_ISL_416351 | Shanghai/SH0042/2020 | 2020-02-06 | Asia/China/Shanghai/ | Shanghai Public Health Clinical Center, Shanghai Medical College, Fudan University | National Research Center for Translational Medicine (Shanghai), Ruijin Hospital affiliated to Shanghai Jiao Tong University School of Medicine & Shanghai Public Health Clinical Center | Shengyue Wang et al |
| EPI_ISL_416352 | Shanghai/SH0043/2020 | 2020-01-30 | Asia/China/Shanghai/ | Shanghai Public Health Clinical Center, Shanghai Medical College, Fudan University | National Research Center for Translational Medicine (Shanghai), Ruijin Hospital affiliated to Shanghai Jiao Tong University School of Medicine & Shanghai Public Health Clinical Center | Shengyue Wang et al |

| | | | | | | |
|---|---|---|---|---|---|---|
| EPI_ISL_416353 | Shanghai/SH0044/2020 | 2020-02-04 | Asia/China/Shanghai/ | Shanghai Public Health Clinical Center, Shanghai Medical College, Fudan University | National Research Center for Translational Medicine (Shanghai), Ruijin Hospital affiliated to Shanghai Jiao Tong University School of Medicine & Shanghai Public Health Clinical Center | Shengyue Wang et al |
| EPI_ISL_416354 | Shanghai/SH0045/2020 | 2020-02-04 | Asia/China/Shanghai/ | Shanghai Public Health Clinical Center, Shanghai Medical College, Fudan University | National Research Center for Translational Medicine (Shanghai), Ruijin Hospital affiliated to Shanghai Jiao Tong University School of Medicine & Shanghai Public Health Clinical Center | Shengyue Wang et al |
| EPI_ISL_416355 | Shanghai/SH0047/2020 | 2020-02-04 | Asia/China/Shanghai/ | Shanghai Public Health Clinical Center, Shanghai Medical College, Fudan University | National Research Center for Translational Medicine (Shanghai), Ruijin Hospital affiliated to Shanghai Jiao Tong University School of Medicine & Shanghai Public Health Clinical Center | Shengyue Wang et al |

| | | | | | | |
|---|---|---|---|---|---|---|
| EPI_ISL_416356 | Shanghai/SH0048/2020 | 2020-02-01 | Asia/China/Shanghai/ | Shanghai Public Health Clinical Center, Shanghai Medical College, Fudan University | National Research Center for Translational Medicine (Shanghai), Ruijin Hospital affiliated to Shanghai Jiao Tong University School of Medicine & Shanghai Public Health Clinical Center | Shengyue Wang et al |
| EPI_ISL_416357 | Shanghai/SH0049/2020 | 2020-02-01 | Asia/China/Shanghai/ | Shanghai Public Health Clinical Center, Shanghai Medical College, Fudan University | National Research Center for Translational Medicine (Shanghai), Ruijin Hospital affiliated to Shanghai Jiao Tong University School of Medicine & Shanghai Public Health Clinical Center | Shengyue Wang et al |
| EPI_ISL_416358 | Shanghai/SH0050/2020 | 2020-02-05 | Asia/China/Shanghai/ | Shanghai Public Health Clinical Center, Shanghai Medical College, Fudan University | National Research Center for Translational Medicine (Shanghai), Ruijin Hospital affiliated to Shanghai Jiao Tong University School of Medicine & Shanghai Public Health Clinical Center | Shengyue Wang et al |

| | | | | | | |
|---|---|---|---|---|---|---|
| EPI_ISL_416359 | Shanghai/SH0051/2020 | 2020-02-06 | Asia/China/Shanghai/ | Shanghai Public Health Clinical Center, Shanghai Medical College, Fudan University | National Research Center for Translational Medicine (Shanghai), Ruijin Hospital affiliated to Shanghai Jiao Tong University School of Medicine & Shanghai Public Health Clinical Center | Shengyue Wang et al |
| EPI_ISL_416360 | Shanghai/SH0052/2020 | 2020-02-01 | Asia/China/Shanghai/ | Shanghai Public Health Clinical Center, Shanghai Medical College, Fudan University | National Research Center for Translational Medicine (Shanghai), Ruijin Hospital affiliated to Shanghai Jiao Tong University School of Medicine & Shanghai Public Health Clinical Center | Shengyue Wang et al |
| EPI_ISL_416361 | Shanghai/SH0053/2020 | 2020-02-01 | Asia/China/Shanghai/ | Shanghai Public Health Clinical Center, Shanghai Medical College, Fudan University | National Research Center for Translational Medicine (Shanghai), Ruijin Hospital affiliated to Shanghai Jiao Tong University School of Medicine & Shanghai Public Health Clinical Center | Shengyue Wang et al |

| | | | | | | |
|---|---|---|---|---|---|---|
| EPI_ISL_416362 | Shanghai/SH0054/2020 | 2020-02-02 | Asia/China/Shanghai/ | Shanghai Public Health Clinical Center, Shanghai Medical College, Fudan University | National Research Center for Translational Medicine (Shanghai), Ruijin Hospital affiliated to Shanghai Jiao Tong University School of Medicine & Shanghai Public Health Clinical Center | Shengyue Wang et al |
| EPI_ISL_416363 | Shanghai/SH0055/2020 | 2020-02-08 | Asia/China/Shanghai/ | Shanghai Public Health Clinical Center, Shanghai Medical College, Fudan University | National Research Center for Translational Medicine (Shanghai), Ruijin Hospital affiliated to Shanghai Jiao Tong University School of Medicine & Shanghai Public Health Clinical Center | Shengyue Wang et al |
| EPI_ISL_416364 | Shanghai/SH0056/2020 | 2020-02-08 | Asia/China/Shanghai/ | Shanghai Public Health Clinical Center, Shanghai Medical College, Fudan University | National Research Center for Translational Medicine (Shanghai), Ruijin Hospital affiliated to Shanghai Jiao Tong University School of Medicine & Shanghai Public Health Clinical Center | Shengyue Wang et al |

| | | | | | | |
|---|---|---|---|---|---|---|
| EPI_ISL_416365 | Shanghai/SH0058/2020 | 2020-01-30 | Asia/China/Shanghai/ | Shanghai Public Health Clinical Center, Shanghai Medical College, Fudan University | National Research Center for Translational Medicine (Shanghai), Ruijin Hospital affiliated to Shanghai Jiao Tong University School of Medicine & Shanghai Public Health Clinical Center | Shengyue Wang et al |
| EPI_ISL_416366 | Shanghai/SH0059/2020 | 2020-01-30 | Asia/China/Shanghai/ | Shanghai Public Health Clinical Center, Shanghai Medical College, Fudan University | National Research Center for Translational Medicine (Shanghai), Ruijin Hospital affiliated to Shanghai Jiao Tong University School of Medicine & Shanghai Public Health Clinical Center | Shengyue Wang et al |
| EPI_ISL_416367 | Shanghai/SH0060/2020 | 2020-02-01 | Asia/China/Shanghai/ | Shanghai Public Health Clinical Center, Shanghai Medical College, Fudan University | National Research Center for Translational Medicine (Shanghai), Ruijin Hospital affiliated to Shanghai Jiao Tong University School of Medicine & Shanghai Public Health Clinical Center | Shengyue Wang et al |

| | | | | | | |
|---|---|---|---|---|---|---|
| EPI_ISL_416368 | Shanghai/SH0065/2020 | 2020-02-05 | Asia/China/Shanghai/ | Shanghai Public Health Clinical Center, Shanghai Medical College, Fudan University | National Research Center for Translational Medicine (Shanghai), Ruijin Hospital affiliated to Shanghai Jiao Tong University School of Medicine & Shanghai Public Health Clinical Center | Shengyue Wang et al |
| EPI_ISL_416369 | Shanghai/SH0066/2020 | 2020-02-05 | Asia/China/Shanghai/ | Shanghai Public Health Clinical Center, Shanghai Medical College, Fudan University | National Research Center for Translational Medicine (Shanghai), Ruijin Hospital affiliated to Shanghai Jiao Tong University School of Medicine & Shanghai Public Health Clinical Center | Shengyue Wang et al |
| EPI_ISL_416370 | Shanghai/SH0067/2020 | 2020-02-01 | Asia/China/Shanghai/ | Shanghai Public Health Clinical Center, Shanghai Medical College, Fudan University | National Research Center for Translational Medicine (Shanghai), Ruijin Hospital affiliated to Shanghai Jiao Tong University School of Medicine & Shanghai Public Health Clinical Center | Shengyue Wang et al |

| | | | | | | |
|---|---|---|---|---|---|---|
| EPI_ISL_416372 | Shanghai/SH0069/2020 | 2020-02-01 | Asia/China/Shanghai/ | Shanghai Public Health Clinical Center, Shanghai Medical College, Fudan University | National Research Center for Translational Medicine (Shanghai), Ruijin Hospital affiliated to Shanghai Jiao Tong University School of Medicine & Shanghai Public Health Clinical Center | Shengyue Wang et al |
| EPI_ISL_416373 | Shanghai/SH0070/2020 | 2020-02-07 | Asia/China/Shanghai/ | Shanghai Public Health Clinical Center, Shanghai Medical College, Fudan University | National Research Center for Translational Medicine (Shanghai), Ruijin Hospital affiliated to Shanghai Jiao Tong University School of Medicine & Shanghai Public Health Clinical Center | Shengyue Wang et al |
| EPI_ISL_416375 | Shanghai/SH0072/2020 | 2020-02-01 | Asia/China/Shanghai/ | Shanghai Public Health Clinical Center, Shanghai Medical College, Fudan University | National Research Center for Translational Medicine (Shanghai), Ruijin Hospital affiliated to Shanghai Jiao Tong University School of Medicine & Shanghai Public Health Clinical Center | Shengyue Wang et al |

| | | | | | | |
|---|---|---|---|---|---|---|
| EPI_ISL_416376 | Shanghai/SH0073/2020 | 2020-02-05 | Asia/China/Shanghai/ | Shanghai Public Health Clinical Center, Shanghai Medical College, Fudan University | National Research Center for Translational Medicine (Shanghai), Ruijin Hospital affiliated to Shanghai Jiao Tong University School of Medicine & Shanghai Public Health Clinical Center | Shengyue Wang et al |
| EPI_ISL_416377 | Shanghai/SH0074/2020 | 2020-02-07 | Asia/China/Shanghai/ | Shanghai Public Health Clinical Center, Shanghai Medical College, Fudan University | National Research Center for Translational Medicine (Shanghai), Ruijin Hospital affiliated to Shanghai Jiao Tong University School of Medicine & Shanghai Public Health Clinical Center | Shengyue Wang et al |
| EPI_ISL_416378 | Shanghai/SH0075/2020 | 2020-01-30 | Asia/China/Shanghai/ | Shanghai Public Health Clinical Center, Shanghai Medical College, Fudan University | National Research Center for Translational Medicine (Shanghai), Ruijin Hospital affiliated to Shanghai Jiao Tong University School of Medicine & Shanghai Public Health Clinical Center | Shengyue Wang et al |

| | | | | | | |
|---|---|---|---|---|---|---|
| EPI_ISL_416379 | Shanghai/SH0076/2020 | 2020-02-01 | Asia/China/Shanghai/ | Shanghai Public Health Clinical Center, Shanghai Medical College, Fudan University | National Research Center for Translational Medicine (Shanghai), Ruijin Hospital affiliated to Shanghai Jiao Tong University School of Medicine & Shanghai Public Health Clinical Center | Shengyue Wang et al |
| EPI_ISL_416380 | Shanghai/SH0077/2020 | 2020-02-09 | Asia/China/Shanghai/ | Shanghai Public Health Clinical Center, Shanghai Medical College, Fudan University | National Research Center for Translational Medicine (Shanghai), Ruijin Hospital affiliated to Shanghai Jiao Tong University School of Medicine & Shanghai Public Health Clinical Center | Shengyue Wang et al |
| EPI_ISL_416381 | Shanghai/SH0079/2020 | 2020-02-01 | Asia/China/Shanghai/ | Shanghai Public Health Clinical Center, Shanghai Medical College, Fudan University | National Research Center for Translational Medicine (Shanghai), Ruijin Hospital affiliated to Shanghai Jiao Tong University School of Medicine & Shanghai Public Health Clinical Center | Shengyue Wang et al |

| | | | | | | |
|---|---|---|---|---|---|---|
| EPI_ISL_416382 | Shanghai/SH0080/2020 | 2020-02-08 | Asia/China/Shanghai/ | Shanghai Public Health Clinical Center, Shanghai Medical College, Fudan University | National Research Center for Translational Medicine (Shanghai), Ruijin Hospital affiliated to Shanghai Jiao Tong University School of Medicine & Shanghai Public Health Clinical Center | Shengyue Wang et al |
| EPI_ISL_416384 | Shanghai/SH0083/2020 | 2020-02-04 | Asia/China/Shanghai/ | Shanghai Public Health Clinical Center, Shanghai Medical College, Fudan University | National Research Center for Translational Medicine (Shanghai), Ruijin Hospital affiliated to Shanghai Jiao Tong University School of Medicine & Shanghai Public Health Clinical Center | Shengyue Wang et al |
| EPI_ISL_416386 | Shanghai/SH0086/2020 | 2020-01-31 | Asia/China/Shanghai/ | Shanghai Public Health Clinical Center, Shanghai Medical College, Fudan University | National Research Center for Translational Medicine (Shanghai), Ruijin Hospital affiliated to Shanghai Jiao Tong University School of Medicine & Shanghai Public Health Clinical Center | Shengyue Wang et al |

| | | | | | | |
|---|---|---|---|---|---|---|
| EPI_ISL_416387 | Shanghai/SH0088/2020 | 2020-02-01 | Asia/China/Shanghai/ | Shanghai Public Health Clinical Center, Shanghai Medical College, Fudan University | National Research Center for Translational Medicine (Shanghai), Ruijin Hospital affiliated to Shanghai Jiao Tong University School of Medicine & Shanghai Public Health Clinical Center | Shengyue Wang et al |
| EPI_ISL_416389 | Shanghai/SH0093/2020 | 2020-01-21 | Asia/China/Shanghai/ | Shanghai Public Health Clinical Center, Shanghai Medical College, Fudan University | National Research Center for Translational Medicine (Shanghai), Ruijin Hospital affiliated to Shanghai Jiao Tong University School of Medicine & Shanghai Public Health Clinical Center | Shengyue Wang et al |
| EPI_ISL_416390 | Shanghai/SH0094/2020 | 2020-01-31 | Asia/China/Shanghai/ | Shanghai Public Health Clinical Center, Shanghai Medical College, Fudan University | National Research Center for Translational Medicine (Shanghai), Ruijin Hospital affiliated to Shanghai Jiao Tong University School of Medicine & Shanghai Public Health Clinical Center | Shengyue Wang et al |

| | | | | | | |
|---|---|---|---|---|---|---|
| EPI_ISL_416391 | Shanghai/SH0098/2020 | 2020-02-02 | Asia/China/Shanghai/ | Shanghai Public Health Clinical Center, Shanghai Medical College, Fudan University | National Research Center for Translational Medicine (Shanghai), Ruijin Hospital affiliated to Shanghai Jiao Tong University School of Medicine & Shanghai Public Health Clinical Center | Shengyue Wang et al |
| EPI_ISL_416392 | Shanghai/SH0100/2020 | 2020-02-02 | Asia/China/Shanghai/ | Shanghai Public Health Clinical Center, Shanghai Medical College, Fudan University | National Research Center for Translational Medicine (Shanghai), Ruijin Hospital affiliated to Shanghai Jiao Tong University School of Medicine & Shanghai Public Health Clinical Center | Shengyue Wang et al |
| EPI_ISL_416393 | Shanghai/SH0101/2020 | 2020-02-09 | Asia/China/Shanghai/ | Shanghai Public Health Clinical Center, Shanghai Medical College, Fudan University | National Research Center for Translational Medicine (Shanghai), Ruijin Hospital affiliated to Shanghai Jiao Tong University School of Medicine & Shanghai Public Health Clinical Center | Shengyue Wang et al |

| | | | | | | |
|---|---|---|---|---|---|---|
| EPI_ISL_416394 | Shanghai/SH0102/2020 | 2020-02-14 | Asia/China/Shanghai/ | Shanghai Public Health Clinical Center, Shanghai Medical College, Fudan University | National Research Center for Translational Medicine (Shanghai), Ruijin Hospital affiliated to Shanghai Jiao Tong University School of Medicine & Shanghai Public Health Clinical Center | Shengyue Wang et al |
| EPI_ISL_416396 | Shanghai/SH0106/2020 | 2020-02-10 | Asia/China/Shanghai/ | Shanghai Public Health Clinical Center, Shanghai Medical College, Fudan University | National Research Center for Translational Medicine (Shanghai), Ruijin Hospital affiliated to Shanghai Jiao Tong University School of Medicine & Shanghai Public Health Clinical Center | Shengyue Wang et al |
| EPI_ISL_416397 | Shanghai/SH0107/2020 | 2020-02-02 | Asia/China/Shanghai/ | Shanghai Public Health Clinical Center, Shanghai Medical College, Fudan University | National Research Center for Translational Medicine (Shanghai), Ruijin Hospital affiliated to Shanghai Jiao Tong University School of Medicine & Shanghai Public Health Clinical Center | Shengyue Wang et al |

| | | | | | | |
|---|---|---|---|---|---|---|
| EPI_ISL_416398 | Shanghai/SH0109/2020 | 2020-02-02 | Asia/China/Shanghai/ | Shanghai Public Health Clinical Center, Shanghai Medical College, Fudan University | National Research Center for Translational Medicine (Shanghai), Ruijin Hospital affiliated to Shanghai Jiao Tong University School of Medicine & Shanghai Public Health Clinical Center | Shengyue Wang et al |
| EPI_ISL_416399 | Shanghai/SH0110/2020 | 2020-02-13 | Asia/China/Shanghai/ | Shanghai Public Health Clinical Center, Shanghai Medical College, Fudan University | National Research Center for Translational Medicine (Shanghai), Ruijin Hospital affiliated to Shanghai Jiao Tong University School of Medicine & Shanghai Public Health Clinical Center | Shengyue Wang et al |
| EPI_ISL_416400 | Shanghai/SH0112/2020 | 2020-02-02 | Asia/China/Shanghai/ | Shanghai Public Health Clinical Center, Shanghai Medical College, Fudan University | National Research Center for Translational Medicine (Shanghai), Ruijin Hospital affiliated to Shanghai Jiao Tong University School of Medicine & Shanghai Public Health Clinical Center | Shengyue Wang et al |

| | | | | | | |
|---|---|---|---|---|---|---|
| EPI_ISL_416401 | Shanghai/SH0114/2020 | 2020-02-02 | Asia/China/Shanghai/ | Shanghai Public Health Clinical Center, Shanghai Medical College, Fudan University | National Research Center for Translational Medicine (Shanghai), Ruijin Hospital affiliated to Shanghai Jiao Tong University School of Medicine & Shanghai Public Health Clinical Center | Shengyue Wang et al |
| EPI_ISL_416402 | Shanghai/SH0115/2020 | 2020-02-11 | Asia/China/Shanghai/ | Shanghai Public Health Clinical Center, Shanghai Medical College, Fudan University | National Research Center for Translational Medicine (Shanghai), Ruijin Hospital affiliated to Shanghai Jiao Tong University School of Medicine & Shanghai Public Health Clinical Center | Shengyue Wang et al |
| EPI_ISL_416403 | Shanghai/SH0117/2020 | 2020-02-02 | Asia/China/Shanghai/ | Shanghai Public Health Clinical Center, Shanghai Medical College, Fudan University | National Research Center for Translational Medicine (Shanghai), Ruijin Hospital affiliated to Shanghai Jiao Tong University School of Medicine & Shanghai Public Health Clinical Center | Shengyue Wang et al |

| | | | | | | |
|---|---|---|---|---|---|---|
| EPI_ISL_416404 | Shanghai/SH0119/2020 | 2020-02-09 | Asia/China/Shanghai/ | Shanghai Public Health Clinical Center, Shanghai Medical College, Fudan University | National Research Center for Translational Medicine (Shanghai), Ruijin Hospital affiliated to Shanghai Jiao Tong University School of Medicine & Shanghai Public Health Clinical Center | Shengyue Wang et al |
| EPI_ISL_416405 | Shanghai/SH0121/2020 | 2020-02-02 | Asia/China/Shanghai/ | Shanghai Public Health Clinical Center, Shanghai Medical College, Fudan University | National Research Center for Translational Medicine (Shanghai), Ruijin Hospital affiliated to Shanghai Jiao Tong University School of Medicine & Shanghai Public Health Clinical Center | Shengyue Wang et al |
| EPI_ISL_416406 | Shanghai/SH0125/2020 | 2020-02-15 | Asia/China/Shanghai/ | Shanghai Public Health Clinical Center, Shanghai Medical College, Fudan University | National Research Center for Translational Medicine (Shanghai), Ruijin Hospital affiliated to Shanghai Jiao Tong University School of Medicine & Shanghai Public Health Clinical Center | Shengyue Wang et al |

| | | | | | | |
|---|---|---|---|---|---|---|
| EPI_ISL_416407 | Shanghai/SH0126/2020 | 2020-02-15 | Asia/China/Shanghai/ | Shanghai Public Health Clinical Center, Shanghai Medical College, Fudan University | National Research Center for Translational Medicine (Shanghai), Ruijin Hospital affiliated to Shanghai Jiao Tong University School of Medicine & Shanghai Public Health Clinical Center | Shengyue Wang et al |
| EPI_ISL_416408 | Shanghai/SH0127/2020 | 2020-02-12 | Asia/China/Shanghai/ | Shanghai Public Health Clinical Center, Shanghai Medical College, Fudan University | National Research Center for Translational Medicine (Shanghai), Ruijin Hospital affiliated to Shanghai Jiao Tong University School of Medicine & Shanghai Public Health Clinical Center | Shengyue Wang et al |
| EPI_ISL_416409 | Shanghai/SH0128/2020 | 2020-02-02 | Asia/China/Shanghai/ | Shanghai Public Health Clinical Center, Shanghai Medical College, Fudan University | National Research Center for Translational Medicine (Shanghai), Ruijin Hospital affiliated to Shanghai Jiao Tong University School of Medicine & Shanghai Public Health Clinical Center | Shengyue Wang et al |

| | | | | | | |
|---|---|---|---|---|---|---|
| EPI_ISL_416410 | Australia/VIC02/2020 | 2020-01-24 | Oceania/Australia/Victoria/Melbourne | Victorian Infectious Diseases Reference Laboratory (VIDRL) | Victorian Infectious Diseases Reference Laboratory and Microbiological Diagnostic Unit Public Health Laboratory, Doherty Institute | Caly L. et al |
| EPI_ISL_416411 | Australia/VIC03/2020 | 2020-01-25 | Oceania/Australia/Victoria/Melbourne | Victorian Infectious Diseases Reference Laboratory (VIDRL) | Victorian Infectious Diseases Reference Laboratory and Microbiological Diagnostic Unit Public Health Laboratory, Doherty Institute | Caly L. et al |
| EPI_ISL_416415 | Australia/VIC07/2020 | 2020-02-08 | Oceania/Australia/Victoria/Melbourne | Victorian Infectious Diseases Reference Laboratory (VIDRL) | Victorian Infectious Diseases Reference Laboratory and Microbiological Diagnostic Unit Public Health Laboratory, Doherty Institute | Caly L. et al |

| | | | | | | |
|---|---|---|---|---|---|---|
| EPI_ISL_416425 | Hangzhou/ZJU-07/2020 | 2020-02-03 | Asia/China/Zhejiang/Hangzhou | State Key Laboratory for Diagnosis and Treatment of Infectious Diseases, National Clinical Research Center for Infectious Diseases, First Affiliated Hospital, Zhejiang University School of Medicine, Hangzhou, China 310003 | State Key Laboratory for Diagnosis and Treatment of Infectious Diseases, National Clinical Research Center for Infectious Diseases, First Affiliated Hospital, Zhejiang University School of Medicine, Hangzhou, China 310003 | Hangping Yao et al |
| EPI_ISL_416427 | Vietnam/38142/2020 | 2020-01-24 | Asia/Vietnam/Thanh Hoa/ | National Influenza Center, National Institute of Hygiene and Epidemiology (NIHE) | National Influenza Center, National Institute of Hygiene and Epidemiology (NIHE) | Le Quynh Mai et al |
| EPI_ISL_416429 | Vietnam/CM99/2020 | 2020-02-11 | Asia/Vietnam/Vinhphuc/ | National Influenza Center, National Institute of Hygiene and Epidemiology (NIHE) | National Influenza Center, National Institute of Hygiene and Epidemiology (NIHE) | Le Quynh Mai et al |
| EPI_ISL_416455 | USA/WA-UW38/2020 | 2020-03-07 | North America/USA/Washington/ | UW Virology Lab | UW Virology Lab | Pavitra Roychoudhury et al |
| EPI_ISL_416456 | USA/WA-UW39/2020 | 2020-03-06 | North America/USA/Washington/ | UW Virology Lab | UW Virology Lab | Pavitra Roychoudhury et al |
| EPI_ISL_416458 | Kuwait/KU12/2020 | 2020-03-02 | Asia/Kuwait/Kuwait/Hawali | Virology laboratory Ministry of Health Kuwait sequenced at Dasman Diabetes Institute | Dasman Diabetes Institute | Fahd Al-Mulla et al |
| EPI_ISL_416460 | USA/WA-S5/2020 | 2020-02-29 | North America/USA/Washington/King C | Seattle Flu Study | Seattle Flu Study | Chu et al et al |
| EPI_ISL_416461 | USA/WA-S6/2020 | 2020-02-29 | North America/USA/Washington/King C | Seattle Flu Study | Seattle Flu Study | Chu et al et al |
| EPI_ISL_416462 | USA/WA-S7/2020 | 2020-02-24 | North America/USA/Washington/ | Seattle Flu Study | Seattle Flu Study | Chu et al et al |
| EPI_ISL_416463 | USA/WA-S8/2020 | 2020-02-24 | North America/USA/Washington/King C | Seattle Flu Study | Seattle Flu Study | Chu et al et al |
| EPI_ISL_416465 | USA/WA-S10/2020 | 2020-02-29 | North America/USA/Washington/King C | Seattle Flu Study | Seattle Flu Study | Chu et al et al |

| | | | | | | |
|---|---|---|---|---|---|---|
| EPI_ISL_416472 | Belgium/UMF-03025/2020 | 2020-03-02 | Europe/Belgium/Leuven/Kessel-Lo | KU Leuven, Clinical and Epidemiological Virology | KU Leuven, Clinical and Epidemiological Virology | Bert Vanmechelen et al |
| EPI_ISL_416473 | Hangzhou/ZJU-08/2020 | 2020-01-26 | Asia/China/Zhejiang/Hangzhou | State Key Laboratory for Diagnosis and Treatment of Infectious Diseases, National Clinical Research Center for Infectious Diseases, First Affiliated Hospital, Zhejiang University School of Medicine, Hangzhou, China 310003 | State Key Laboratory for Diagnosis and Treatment of Infectious Diseases, National Clinical Research Center for Infectious Diseases, First Affiliated Hospital, Zhejiang University School of Medicine, Hangzhou, China 310003 | Hangping Yao et al |
| EPI_ISL_416474 | Hangzhou/ZJU-09/2020 | 2020-01-28 | Asia/China/Zhejiang/Hangzhou | State Key Laboratory for Diagnosis and Treatment of Infectious Diseases, National Clinical Research Center for Infectious Diseases, First Affiliated Hospital, Zhejiang University School of Medicine, Hangzhou, China 310003 | State Key Laboratory for Diagnosis and Treatment of Infectious Diseases, National Clinical Research Center for Infectious Diseases, First Affiliated Hospital, Zhejiang University School of Medicine, Hangzhou, China 310003 | Hangping Yao et al |

| | | | | | | |
|---|---|---|---|---|---|---|
| EPI_ISL_416483 | Spain/VC-FISABIO-4/2020 | 2020-02-26 | Europe/Italy/Comunitat Valenciana/Val | Servicio de Microbiolog√≠a. Consorcio Hospital General Universitario de Valencia | Sequencing and Bioinformatics Service and Molecular Epidemiology Research Group. FISABIO-Public Health | Maria Alma Bracho et al |
| EPI_ISL_416484 | Spain/VC-FISABIO-5/2020 | 2020-02-27 | Europe/Spain/Comunitat Valenciana/Va | Servicio de Microbiolog√≠a. Consorcio Hospital General Universitario de Valencia | Sequencing and Bioinformatics Service and Molecular Epidemiology Research Group. FISABIO-Public Health | Maria Dolores Ocete et al |
| EPI_ISL_416485 | Spain/VC-FISABIO-6/2020 | 2020-02-27 | Europe/Spain/Comunitat Valenciana/Va | Servicio de Microbiologia. Consorcio Hospital General Universitario de Valencia | Sequencing and Bioinformatics Service and Molecular Epidemiology Research Group. FISABIO-Public Health | Griselda De Marco et al |
| EPI_ISL_416496 | France/HDF-2237/2020 | 2020-03-10 | Europe/France/Hauts de France/Compiv | Centre Hospitalier Compi√®gne Laboratoire de Biologie | National Reference Center for Viruses of Respiratory Infections, Institut Pasteur, Paris | M√©lnie Albert et al |
| EPI_ISL_416497 | France/HDF-2239/2020 | 2020-03-10 | Europe/France/Hauts de France/Compiv | Centre Hospitalier Compi√®gne Laboratoire de Biologie | National Reference Center for Viruses of Respiratory Infections, Institut Pasteur, Paris | M√©lnie Albert et al |

| | | | | | | |
|---|---|---|---|---|---|---|
| EPI_ISL_416500 | France/IDF-2279/2020 | 2020-03-11 | Europe/France/Ile de France/Longjumea | LABM GH nord Essonne | National Reference Center for Viruses of Respiratory Infections, Institut Pasteur, Paris | M√©lnie Albert et al |
| EPI_ISL_416501 | France/IDF-2284/2020 | 2020-03-10 | Europe/France/Ile de France/Levallois-P | Hopital franco britannique - Service des Urgences | National Reference Center for Viruses of Respiratory Infections, Institut Pasteur, Paris | M√©lnie Albert et al |
| EPI_ISL_416502 | France/BRE-2330/2020 | 2020-02-26 | Europe/France/Bretagne/Rennes | CHRU Pontchaillou - Laboratoire de Virologie | National Reference Center for Viruses of Respiratory Infections, Institut Pasteur, Paris | M√©lnie Albert et al |
| EPI_ISL_416565 | Japan/DP0005/2020 | 2020-02-15 | Asia/Japan/Japan/ | Japanese Quarantine Stations | Pathogen Genomics Center, National Institute of Infectious Diseases | Tsuyoshi Sekizuka et al |
| EPI_ISL_416566 | Japan/DP0027/2020 | 2020-02-15 | Asia/Japan/Japan/ | Japanese Quarantine Stations | Pathogen Genomics Center, National Institute of Infectious Diseases | Tsuyoshi Sekizuka et al |
| EPI_ISL_416567 | Japan/DP0037/2020 | 2020-02-15 | Asia/Japan/Japan/ | Japanese Quarantine Stations | Pathogen Genomics Center, National Institute of Infectious Diseases | Tsuyoshi Sekizuka et al |
| EPI_ISL_416568 | Japan/DP0058/2020 | 2020-02-15 | Asia/Japan/Japan/ | Japanese Quarantine Stations | Pathogen Genomics Center, National Institute of Infectious Diseases | Tsuyoshi Sekizuka et al |

| | | | | | | |
|---|---|---|---|---|---|---|
| EPI_ISL_416569 | Japan/DP0059/2020 | 2020-02-15 | Asia/Japan/Japan/ | Japanese Quarantine Stations | Pathogen Genomics Center, National Institute of Infectious Diseases | Tsuyoshi Sekizuka et al |
| EPI_ISL_416570 | Japan/DP0065/2020 | 2020-02-15 | Asia/Japan/Japan/ | Japanese Quarantine Stations | Pathogen Genomics Center, National Institute of Infectious Diseases | Tsuyoshi Sekizuka et al |
| EPI_ISL_416571 | Japan/DP0077/2020 | 2020-02-15 | Asia/Japan/Japan/ | Japanese Quarantine Stations | Pathogen Genomics Center, National Institute of Infectious Diseases | Tsuyoshi Sekizuka et al |
| EPI_ISL_416572 | Japan/DP0078/2020 | 2020-02-15 | Asia/Japan/Japan/ | Japanese Quarantine Stations | Pathogen Genomics Center, National Institute of Infectious Diseases | Tsuyoshi Sekizuka et al |
| EPI_ISL_416573 | Japan/DP0104/2020 | 2020-02-15 | Asia/Japan/Japan/ | Japanese Quarantine Stations | Pathogen Genomics Center, National Institute of Infectious Diseases | Tsuyoshi Sekizuka et al |
| EPI_ISL_416574 | Japan/DP0107/2020 | 2020-02-15 | Asia/Japan/Japan/ | Japanese Quarantine Stations | Pathogen Genomics Center, National Institute of Infectious Diseases | Tsuyoshi Sekizuka et al |
| EPI_ISL_416575 | Japan/DP0121/2020 | 2020-02-15 | Asia/Japan/Japan/ | Japanese Quarantine Stations | Pathogen Genomics Center, National Institute of Infectious Diseases | Tsuyoshi Sekizuka et al |
| EPI_ISL_416576 | Japan/DP0133/2020 | 2020-02-15 | Asia/Japan/Japan/ | Japanese Quarantine Stations | Pathogen Genomics Center, National Institute of Infectious Diseases | Tsuyoshi Sekizuka et al |

| | | | | | | |
|---|---|---|---|---|---|---|
| EPI_ISL_416577 | Japan/DP0134/2020 | 2020-02-15 | Asia/Japan/Japan/ | Japanese Quarantine Stations | Pathogen Genomics Center, National Institute of Infectious Diseases | Tsuyoshi Sekizuka et al |
| EPI_ISL_416578 | Japan/DP0152/2020 | 2020-02-15 | Asia/Japan/Japan/ | Japanese Quarantine Stations | Pathogen Genomics Center, National Institute of Infectious Diseases | Tsuyoshi Sekizuka et al |
| EPI_ISL_416579 | Japan/DP0158/2020 | 2020-02-15 | Asia/Japan/Japan/ | Japanese Quarantine Stations | Pathogen Genomics Center, National Institute of Infectious Diseases | Tsuyoshi Sekizuka et al |
| EPI_ISL_416580 | Japan/DP0184/2020 | 2020-02-15 | Asia/Japan/Japan/ | Japanese Quarantine Stations | Pathogen Genomics Center, National Institute of Infectious Diseases | Tsuyoshi Sekizuka et al |
| EPI_ISL_416581 | Japan/DP0190/2020 | 2020-02-15 | Asia/Japan/Japan/ | Japanese Quarantine Stations | Pathogen Genomics Center, National Institute of Infectious Diseases | Tsuyoshi Sekizuka et al |
| EPI_ISL_416582 | Japan/DP0191/2020 | 2020-02-15 | Asia/Japan/Japan/ | Japanese Quarantine Stations | Pathogen Genomics Center, National Institute of Infectious Diseases | Tsuyoshi Sekizuka et al |
| EPI_ISL_416583 | Japan/DP0196/2020 | 2020-02-15 | Asia/Japan/Japan/ | Japanese Quarantine Stations | Pathogen Genomics Center, National Institute of Infectious Diseases | Tsuyoshi Sekizuka et al |
| EPI_ISL_416584 | Japan/DP0200/2020 | 2020-02-15 | Asia/Japan/Japan/ | Japanese Quarantine Stations | Pathogen Genomics Center, National Institute of Infectious Diseases | Tsuyoshi Sekizuka et al |

| | | | | | | |
|---|---|---|---|---|---|---|
| EPI_ISL_416585 | Japan/DP0236/2020 | 2020-02-16 | Asia/Japan/Japan/ | Japanese Quarantine Stations | Pathogen Genomics Center, National Institute of Infectious Diseases | Tsuyoshi Sekizuka et al |
| EPI_ISL_416586 | Japan/DP0274/2020 | 2020-02-16 | Asia/Japan/Japan/ | Japanese Quarantine Stations | Pathogen Genomics Center, National Institute of Infectious Diseases | Tsuyoshi Sekizuka et al |
| EPI_ISL_416587 | Japan/DP0278/2020 | 2020-02-16 | Asia/Japan/Japan/ | Japanese Quarantine Stations | Pathogen Genomics Center, National Institute of Infectious Diseases | Tsuyoshi Sekizuka et al |
| EPI_ISL_416588 | Japan/DP0286/2020 | 2020-02-16 | Asia/Japan/Japan/ | Japanese Quarantine Stations | Pathogen Genomics Center, National Institute of Infectious Diseases | Tsuyoshi Sekizuka et al |
| EPI_ISL_416589 | Japan/DP0287/2020 | 2020-02-16 | Asia/Japan/Japan/ | Japanese Quarantine Stations | Pathogen Genomics Center, National Institute of Infectious Diseases | Tsuyoshi Sekizuka et al |
| EPI_ISL_416590 | Japan/DP0289/2020 | 2020-02-16 | Asia/Japan/Japan/ | Japanese Quarantine Stations | Pathogen Genomics Center, National Institute of Infectious Diseases | Tsuyoshi Sekizuka et al |
| EPI_ISL_416591 | Japan/DP0290/2020 | 2020-02-16 | Asia/Japan/Japan/ | Japanese Quarantine Stations | Pathogen Genomics Center, National Institute of Infectious Diseases | Tsuyoshi Sekizuka et al |
| EPI_ISL_416592 | Japan/DP0294/2020 | 2020-02-16 | Asia/Japan/Japan/ | Japanese Quarantine Stations | Pathogen Genomics Center, National Institute of Infectious Diseases | Tsuyoshi Sekizuka et al |

| | | | | | | |
|---|---|---|---|---|---|---|
| EPI_ISL_416593 | Japan/DP0311/2020 | 2020-02-16 | Asia/Japan/Japan/ | Japanese Quarantine Stations | Pathogen Genomics Center, National Institute of Infectious Diseases | Tsuyoshi Sekizuka et al |
| EPI_ISL_416594 | Japan/DP0319/2020 | 2020-02-16 | Asia/Japan/Japan/ | Japanese Quarantine Stations | Pathogen Genomics Center, National Institute of Infectious Diseases | Tsuyoshi Sekizuka et al |
| EPI_ISL_416595 | Japan/DP0328/2020 | 2020-02-16 | Asia/Japan/Japan/ | Japanese Quarantine Stations | Pathogen Genomics Center, National Institute of Infectious Diseases | Tsuyoshi Sekizuka et al |
| EPI_ISL_416596 | Japan/DP0344/2020 | 2020-02-16 | Asia/Japan/Japan/ | Japanese Quarantine Stations | Pathogen Genomics Center, National Institute of Infectious Diseases | Tsuyoshi Sekizuka et al |
| EPI_ISL_416597 | Japan/DP0346/2020 | 2020-02-16 | Asia/Japan/Japan/ | Japanese Quarantine Stations | Pathogen Genomics Center, National Institute of Infectious Diseases | Tsuyoshi Sekizuka et al |
| EPI_ISL_416598 | Japan/DP0357/2020 | 2020-02-16 | Asia/Japan/Japan/ | Japanese Quarantine Stations | Pathogen Genomics Center, National Institute of Infectious Diseases | Tsuyoshi Sekizuka et al |
| EPI_ISL_416599 | Japan/DP0361/2020 | 2020-02-16 | Asia/Japan/Japan/ | Japanese Quarantine Stations | Pathogen Genomics Center, National Institute of Infectious Diseases | Tsuyoshi Sekizuka et al |
| EPI_ISL_416600 | Japan/DP0438/2020 | 2020-02-16 | Asia/Japan/Japan/ | Japanese Quarantine Stations | Pathogen Genomics Center, National Institute of Infectious Diseases | Tsuyoshi Sekizuka et al |

| | | | | | | |
|---|---|---|---|---|---|---|
| EPI_ISL_416601 | Japan/DP0457/2020 | 2020-02-16 | Asia/Japan/Japan/ | Japanese Quarantine Stations | Pathogen Genomics Center, National Institute of Infectious Diseases | Tsuyoshi Sekizuka et al |
| EPI_ISL_416602 | Japan/DP0462/2020 | 2020-02-16 | Asia/Japan/Japan/ | Japanese Quarantine Stations | Pathogen Genomics Center, National Institute of Infectious Diseases | Tsuyoshi Sekizuka et al |
| EPI_ISL_416603 | Japan/DP0464/2020 | 2020-02-16 | Asia/Japan/Japan/ | Japanese Quarantine Stations | Pathogen Genomics Center, National Institute of Infectious Diseases | Tsuyoshi Sekizuka et al |
| EPI_ISL_416604 | Japan/DP0476/2020 | 2020-02-16 | Asia/Japan/Japan/ | Japanese Quarantine Stations | Pathogen Genomics Center, National Institute of Infectious Diseases | Tsuyoshi Sekizuka et al |
| EPI_ISL_416605 | Japan/DP0481/2020 | 2020-02-16 | Asia/Japan/Japan/ | Japanese Quarantine Stations | Pathogen Genomics Center, National Institute of Infectious Diseases | Tsuyoshi Sekizuka et al |
| EPI_ISL_416606 | Japan/DP0482/2020 | 2020-02-16 | Asia/Japan/Japan/ | Japanese Quarantine Stations | Pathogen Genomics Center, National Institute of Infectious Diseases | Tsuyoshi Sekizuka et al |
| EPI_ISL_416607 | Japan/DP0543/2020 | 2020-02-17 | Asia/Japan/Japan/ | Japanese Quarantine Stations | Pathogen Genomics Center, National Institute of Infectious Diseases | Tsuyoshi Sekizuka et al |
| EPI_ISL_416608 | Japan/DP0544/2020 | 2020-02-17 | Asia/Japan/Japan/ | Japanese Quarantine Stations | Pathogen Genomics Center, National Institute of Infectious Diseases | Tsuyoshi Sekizuka et al |

| | | | | | | |
|---|---|---|---|---|---|---|
| EPI_ISL_416609 | Japan/DP0568/2020 | 2020-02-17 | Asia/Japan/Japan/ | Japanese Quarantine Stations | Pathogen Genomics Center, National Institute of Infectious Diseases | Tsuyoshi Sekizuka et al |
| EPI_ISL_416610 | Japan/DP0588/2020 | 2020-02-17 | Asia/Japan/Japan/ | Japanese Quarantine Stations | Pathogen Genomics Center, National Institute of Infectious Diseases | Tsuyoshi Sekizuka et al |
| EPI_ISL_416611 | Japan/DP0644/2020 | 2020-02-17 | Asia/Japan/Japan/ | Japanese Quarantine Stations | Pathogen Genomics Center, National Institute of Infectious Diseases | Tsuyoshi Sekizuka et al |
| EPI_ISL_416612 | Japan/DP0645/2020 | 2020-02-17 | Asia/Japan/Japan/ | Japanese Quarantine Stations | Pathogen Genomics Center, National Institute of Infectious Diseases | Tsuyoshi Sekizuka et al |
| EPI_ISL_416613 | Japan/DP0654/2020 | 2020-02-17 | Asia/Japan/Japan/ | Japanese Quarantine Stations | Pathogen Genomics Center, National Institute of Infectious Diseases | Tsuyoshi Sekizuka et al |
| EPI_ISL_416614 | Japan/DP0687/2020 | 2020-02-17 | Asia/Japan/Japan/ | Japanese Quarantine Stations | Pathogen Genomics Center, National Institute of Infectious Diseases | Tsuyoshi Sekizuka et al |
| EPI_ISL_416615 | Japan/DP0690/2020 | 2020-02-17 | Asia/Japan/Japan/ | Japanese Quarantine Stations | Pathogen Genomics Center, National Institute of Infectious Diseases | Tsuyoshi Sekizuka et al |
| EPI_ISL_416616 | Japan/DP0697/2020 | 2020-02-17 | Asia/Japan/Japan/ | Japanese Quarantine Stations | Pathogen Genomics Center, National Institute of Infectious Diseases | Tsuyoshi Sekizuka et al |

| | | | | | | |
|---|---|---|---|---|---|---|
| EPI_ISL_416617 | Japan/DP0699/2020 | 2020-02-17 | Asia/Japan/Japan/ | Japanese Quarantine Stations | Pathogen Genomics Center, National Institute of Infectious Diseases | Tsuyoshi Sekizuka et al |
| EPI_ISL_416618 | Japan/DP0700/2020 | 2020-02-17 | Asia/Japan/Japan/ | Japanese Quarantine Stations | Pathogen Genomics Center, National Institute of Infectious Diseases | Tsuyoshi Sekizuka et al |
| EPI_ISL_416619 | Japan/DP0703/2020 | 2020-02-17 | Asia/Japan/Japan/ | Japanese Quarantine Stations | Pathogen Genomics Center, National Institute of Infectious Diseases | Tsuyoshi Sekizuka et al |
| EPI_ISL_416620 | Japan/DP0724/2020 | 2020-02-17 | Asia/Japan/Japan/ | Japanese Quarantine Stations | Pathogen Genomics Center, National Institute of Infectious Diseases | Tsuyoshi Sekizuka et al |
| EPI_ISL_416621 | Japan/DP0743/2020 | 2020-02-17 | Asia/Japan/Japan/ | Japanese Quarantine Stations | Pathogen Genomics Center, National Institute of Infectious Diseases | Tsuyoshi Sekizuka et al |
| EPI_ISL_416622 | Japan/DP0752/2020 | 2020-02-17 | Asia/Japan/Japan/ | Japanese Quarantine Stations | Pathogen Genomics Center, National Institute of Infectious Diseases | Tsuyoshi Sekizuka et al |
| EPI_ISL_416623 | Japan/DP0763/2020 | 2020-02-17 | Asia/Japan/Japan/ | Japanese Quarantine Stations | Pathogen Genomics Center, National Institute of Infectious Diseases | Tsuyoshi Sekizuka et al |
| EPI_ISL_416624 | Japan/DP0764/2020 | 2020-02-17 | Asia/Japan/Japan/ | Japanese Quarantine Stations | Pathogen Genomics Center, National Institute of Infectious Diseases | Tsuyoshi Sekizuka et al |

| | | | | | | |
|---|---|---|---|---|---|---|
| EPI_ISL_416625 | Japan/DP0765/2020 | 2020-02-17 | Asia/Japan/Japan/ | Japanese Quarantine Stations | Pathogen Genomics Center, National Institute of Infectious Diseases | Tsuyoshi Sekizuka et al |
| EPI_ISL_416626 | Japan/DP0779/2020 | 2020-02-17 | Asia/Japan/Japan/ | Japanese Quarantine Stations | Pathogen Genomics Center, National Institute of Infectious Diseases | Tsuyoshi Sekizuka et al |
| EPI_ISL_416627 | Japan/DP0785/2020 | 2020-02-17 | Asia/Japan/Japan/ | Japanese Quarantine Stations | Pathogen Genomics Center, National Institute of Infectious Diseases | Tsuyoshi Sekizuka et al |
| EPI_ISL_416628 | Japan/DP0786/2020 | 2020-02-17 | Asia/Japan/Japan/ | Japanese Quarantine Stations | Pathogen Genomics Center, National Institute of Infectious Diseases | Tsuyoshi Sekizuka et al |
| EPI_ISL_416629 | Japan/DP0802/2020 | 2020-02-17 | Asia/Japan/Japan/ | Japanese Quarantine Stations | Pathogen Genomics Center, National Institute of Infectious Diseases | Tsuyoshi Sekizuka et al |
| EPI_ISL_416630 | Japan/DP0803/2020 | 2020-02-17 | Asia/Japan/Japan/ | Japanese Quarantine Stations | Pathogen Genomics Center, National Institute of Infectious Diseases | Tsuyoshi Sekizuka et al |
| EPI_ISL_416631 | Japan/DP0804/2020 | 2020-02-17 | Asia/Japan/Japan/ | Japanese Quarantine Stations | Pathogen Genomics Center, National Institute of Infectious Diseases | Tsuyoshi Sekizuka et al |
| EPI_ISL_416632 | Japan/DP0827/2020 | 2020-02-17 | Asia/Japan/Japan/ | Japanese Quarantine Stations | Pathogen Genomics Center, National Institute of Infectious Diseases | Tsuyoshi Sekizuka et al |

| | | | | | | |
|---|---|---|---|---|---|---|
| EPI_ISL_416633 | Japan/DP0880/2020 | 2020-02-17 | Asia/Japan/Japan/ | Japanese Quarantine Stations | Pathogen Genomics Center, National Institute of Infectious Diseases | Tsuyoshi Sekizuka et al |
| EPI_ISL_416634 | Japan/DP0890/2020 | 2020-02-17 | Asia/Japan/Japan/ | Japanese Quarantine Stations | Pathogen Genomics Center, National Institute of Infectious Diseases | Tsuyoshi Sekizuka et al |
| EPI_ISL_416635 | USA/WA-UW97/2020 | 2020-03-12 | North America/USA/Washington/ | UW Virology Lab | UW Virology Lab | Pavitra Roychoudhury et al |
| EPI_ISL_416636 | USA/WA-UW98/2020 | 2020-03-12 | North America/USA/Washington/ | UW Virology Lab | UW Virology Lab | Pavitra Roychoudhury et al |
| EPI_ISL_416638 | USA/WA-UW100/2020 | 2020-03-12 | North America/USA/Washington/ | UW Virology Lab | UW Virology Lab | Pavitra Roychoudhury et al |
| EPI_ISL_416647 | USA/WA-UW109/2020 | 2020-03-11 | North America/USA/Washington/ | UW Virology Lab | UW Virology Lab | Pavitra Roychoudhury et al |
| EPI_ISL_416652 | USA/WA-UW114/2020 | 2020-03-11 | North America/USA/Washington/ | UW Virology Lab | UW Virology Lab | Pavitra Roychoudhury et al |
| EPI_ISL_416654 | USA/WA-UW116/2020 | 2020-03-11 | North America/USA/Washington/ | UW Virology Lab | UW Virology Lab | Pavitra Roychoudhury et al |
| EPI_ISL_416660 | USA/WA-UW122/2020 | 2020-03-10 | North America/USA/Washington/ | UW Virology Lab | UW Virology Lab | Pavitra Roychoudhury et al |
| EPI_ISL_416685 | USA/WA-UW147/2020 | 2020-03-15 | North America/USA/Washington/ | UW Virology Lab | UW Virology Lab | Pavitra Roychoudhury et al |
| EPI_ISL_416705 | USA/WA-UW167/2020 | 2020-03-13 | North America/USA/Washington/ | UW Virology Lab | UW Virology Lab | Pavitra Roychoudhury et al |
| EPI_ISL_416709 | USA/WA-UW171/2020 | 2020-03-13 | North America/USA/Washington/ | UW Virology Lab | UW Virology Lab | Pavitra Roychoudhury et al |
| EPI_ISL_416719 | USA/WA-UW181/2020 | 2020-03-14 | North America/USA/Washington/ | UW Virology Lab | UW Virology Lab | Pavitra Roychoudhury et al |
| EPI_ISL_416742 | CzechRepublic/ChVir1630/2020 | 2020-02-29 | Europe/Czech Republic/Prague/ | NRL for Influenza, Centrum Epidemiology and Microbiology of National Institute of Public Health, Czech Republic | Charite Universitaetsmedizin Berlin, Institute of Virology | Victor M Corman et al |
| EPI_ISL_416745 | France/ARA-1733/2020 | 2020-03-10 | Europe/France/Auvergne-Rh√¥ne-Alpes/ | CNR Virus des Infections Respiratoires - France SUD | CNR Virus des Infections Respiratoires - France SUD | Bal et al |
| EPI_ISL_416748 | France/ARA-508/2020 | 2020-03-04 | Europe/France/Auvergne-Rh√¥ne-Alpes/ | Institut des Agents Infectieux (IAI) Hospices Civils de Lyon | CNR Virus des Infections Respiratoires - France SUD | Bal et al |

| | | | | | | |
|---|---|---|---|---|---|---|
| EPI_ISL_416754 | France/ARA-06487/2020 | 2020-03-06 | Europe/France/Auvergne-Rhv¥ne-Alpes/ | Institut des Agents Infectieux (IAI) Hospices Civils de Lyon | CNR Virus des Infections Respiratoires - France SUD | Bal et al |
| EPI_ISL_416829 | Malaysia/MKAK-CL-2020-5045/2020 | 2020-01-24 | Asia/Malaysia/Selangor/ | National Public Health Laboratory | Malaysia Genome Institute | Mohd Noor Mat Isa et al |
| EPI_ISL_416866 | Malaysia/MKAK-CL-2020-5047/2020 | 2020-01-24 | Asia/Malaysia/Selangor/ | National Public Health Laboratory | Malaysia Genome Institute | Mohd Noor Mat Isa et al |
| EPI_ISL_416884 | Malaysia/MKAK-CL-2020-5049/2020 | 2020-01-24 | Asia/Malaysia/Selangor/ | National Public Health Laboratory | Malaysia Genome Institute | Mohd Noor Mat Isa et al |
| EPI_ISL_416885 | Malaysia/MKAK-CL-2020-5096/2020 | 2020-01-30 | Asia/Malaysia/Selangor/ | National Public Health Laboratory | Malaysia Genome Institute | Mohd Noor Mat Isa et al |
| EPI_ISL_416886 | Malaysia/MKAK-CL-2020-6430/2020 | 2020-02-04 | Asia/Malaysia/Selangor/ | National Public Health Laboratory | Malaysia Genome Institute | Mohd Noor Mat Isa et al |
| EPI_ISL_416907 | Malaysia/MKAK-CL-2020-7554/2020 | 2020-02-06 | Asia/Malaysia/Selangor/ | National Public Health Laboratory | Malaysia Genome Institute | Mohd Noor Mat Isa et al |
| EPI_ISL_417010 | Spain/MD-ISCIII-201442/2020 | 2020-03-04 | Europe/Spain/Madrid/ | FUNDACION JIMENEZ DIAZ | Instituto de Salud Carlos III | Iglesias-Caballero et al |
| EPI_ISL_417016 | Belgium/ULG-6503/2020 | 2020-03-13 | Europe/Belgium/Liv®ge/ | Department of Clinical Microbiology | GIGA Medical Genomics | Durkin Keith et al |
| EPI_ISL_417030 | Australia/NSW04/2020 | 2020-01-24 | Oceania/Australia/New South Wales/Sy | Centre for Infectious Diseases and Microbiology Laboratory Services | NSW Health Pathology - Institute of Clinical Pathology and Medical Research; Westmead Hospital; University of Sydney | Eden J-S et al |
| EPI_ISL_417034 | Brazil/AM-02/2020 | 2020-03-16 | South America/Brazil/Amazonas BR/Ma | Laboratorio de Ecologia de Doencas Transmissiveis na Amazonia, Instituto Leonidas e Maria Deane - Fiocruz Amazonia | Laboratorio de Ecologia de Doencas Transmissiveis na Amazonia, Instituto Leonidas e Maria Deane - Fiocruz Amazonia | Valdinete Nascimento et al |

| | | | | | | |
|---|---|---|---|---|---|---|
| EPI_ISL_417064 | HongKong/VB20017970-2/2020 | 2020-01-21 | Asia/Hong Kong/Hong Kong/Hong Kong | Prince of Wales Hospital | Hong Kong Department of Health | Alan K.L. Tsang et al |
| EPI_ISL_417074 | USA/WA-S21/2020 | 2020-03-02 | North America/USA/Washington/King C | Washington State Department of Health | Seattle Flu Study | Chu etl al et al |
| EPI_ISL_417078 | USA/WA-S25/2020 | 2020-03-02 | North America/USA/Washington/Snoho | Washington State Department of Health | Seattle Flu Study | Chu etl al et al |
| EPI_ISL_417083 | USA/WA-S30/2020 | 2020-02-28 | North America/USA/Washington/King C | Washington State Department of Health | Seattle Flu Study | Chu etl al et al |
| EPI_ISL_417093 | USA/WA-S40/2020 | 2020-02-28 | North America/USA/Washington/Snoho | Washington State Department of Health | Seattle Flu Study | Chu etl al et al |
| EPI_ISL_417094 | USA/WA-S41/2020 | 2020-02-28 | North America/USA/Washington/King C | Washington State Department of Health | Seattle Flu Study | Chu etl al et al |
| EPI_ISL_417095 | USA/WA-S42/2020 | 2020-02-28 | North America/USA/Washington/King C | Washington State Department of Health | Seattle Flu Study | Chu etl al et al |
| EPI_ISL_417096 | USA/WA-S43/2020 | 2020-02-27 | North America/USA/Washington/ | Washington State Department of Health | Seattle Flu Study | Chu etl al et al |
| EPI_ISL_417098 | USA/WA-S45/2020 | 2020-02-29 | North America/USA/Washington/ | Washington State Department of Health | Seattle Flu Study | Chu etl al et al |
| EPI_ISL_417099 | USA/WA-S46/2020 | 2020-02-29 | North America/USA/Washington/ | Washington State Department of Health | Seattle Flu Study | Chu etl al et al |
| EPI_ISL_417100 | USA/WA-S47/2020 | 2020-02-29 | North America/USA/Washington/Snoho | Washington State Department of Health | Seattle Flu Study | Chu etl al et al |
| EPI_ISL_417101 | USA/WA-S48/2020 | 2020-02-29 | North America/USA/Washington/Snoho | Washington State Department of Health | Seattle Flu Study | Chu etl al et al |
| EPI_ISL_417103 | USA/WA-S50/2020 | 2020-03-05 | North America/USA/Washington/King C | Washington State Department of Health | Seattle Flu Study | Chu etl al et al |
| EPI_ISL_417104 | USA/WA-S51/2020 | 2020-03-03 | North America/USA/Washington/ | Washington State Department of Health | Seattle Flu Study | Chu etl al et al |
| EPI_ISL_417108 | USA/WA-S55/2020 | 2020-02-29 | North America/USA/Washington/King C | Washington State Department of Health | Seattle Flu Study | Chu etl al et al |
| EPI_ISL_417121 | USA/WA-S68/2020 | 2020-03-04 | North America/USA/Washington/King C | Washington State Department of Health | Seattle Flu Study | Chu etl al et al |
| EPI_ISL_417134 | USA/WA-S81/2020 | 2020-02-26 | North America/USA/Washington/King C | Washington State Department of Health | Seattle Flu Study | Chu etl al et al |
| EPI_ISL_417135 | USA/WA-S82/2020 | 2020-02-22 | North America/USA/Washington/King C | Washington State Department of Health | Seattle Flu Study | Chu etl al et al |
| EPI_ISL_417137 | USA/WA-S84/2020 | 2020-02-21 | North America/USA/Washington/King C | Washington State Department of Health | Seattle Flu Study | Chu etl al et al |
| EPI_ISL_417142 | USA/WA-S89/2020 | 2020-02-29 | North America/USA/Oregon/Umatilla C | Washington State Department of Health | Seattle Flu Study | Chu etl al et al |
| EPI_ISL_417143 | USA/WA-S90/2020 | 2020-02-29 | North America/USA/Oregon/Umatilla C | Washington State Department of Health | Seattle Flu Study | Chu etl al et al |
| EPI_ISL_417145 | USA/WA-S92/2020 | 2020-02-29 | North America/USA/Washington/King C | Washington State Department of Health | Seattle Flu Study | Chu etl al et al |
| EPI_ISL_417146 | USA/WA-S93/2020 | 2020-02-29 | North America/USA/Washington/King C | Washington State Department of Health | Seattle Flu Study | Chu etl al et al |
| EPI_ISL_417147 | USA/WA-S94/2020 | 2020-02-28 | North America/USA/Washington/Snoho | Washington State Department of Health | Seattle Flu Study | Chu etl al et al |
| EPI_ISL_417148 | USA/WA-S95/2020 | 2020-02-28 | North America/USA/Washington/King C | Washington State Department of Health | Seattle Flu Study | Chu etl al et al |
| EPI_ISL_417149 | USA/WA-S96/2020 | 2020-02-28 | North America/USA/Washington/King C | Washington State Department of Health | Seattle Flu Study | Chu etl al et al |
| EPI_ISL_417150 | USA/WA-S97/2020 | 2020-02-28 | North America/USA/Washington/King C | Washington State Department of Health | Seattle Flu Study | Chu etl al et al |
| EPI_ISL_417151 | USA/WA-S98/2020 | 2020-02-29 | North America/USA/Washington/King C | Washington State Department of Health | Seattle Flu Study | Chu etl al et al |
| EPI_ISL_417152 | USA/WA-S99/2020 | 2020-02-28 | North America/USA/Washington/King C | Washington State Department of Health | Seattle Flu Study | Chu etl al et al |

| | | | | | | |
|---|---|---|---|---|---|---|
| EPI_ISL_417153 | USA/WA-S100/2020 | 2020-02-29 | North America/USA/Washington/King C | Washington State Department of Health | Seattle Flu Study | Chu etl al et al |
| EPI_ISL_417154 | USA/WA-S101/2020 | 2020-02-28 | North America/USA/Washington/King C | Washington State Department of Health | Seattle Flu Study | Chu etl al et al |
| EPI_ISL_417155 | USA/WA-S102/2020 | 2020-02-28 | North America/USA/Washington/King C | Washington State Department of Health | Seattle Flu Study | Chu etl al et al |
| EPI_ISL_417156 | USA/WA-S103/2020 | 2020-02-28 | North America/USA/Washington/King C | Washington State Department of Health | Seattle Flu Study | Chu etl al et al |
| EPI_ISL_417158 | USA/WA-S105/2020 | 2020-02-28 | North America/USA/Washington/King C | Washington State Department of Health | Seattle Flu Study | Chu etl al et al |
| EPI_ISL_417159 | USA/WA-S106/2020 | 2020-02-29 | North America/USA/Washington/King C | Washington State Department of Health | Seattle Flu Study | Chu etl al et al |
| EPI_ISL_417160 | USA/WA-S107/2020 | 2020-02-29 | North America/USA/Washington/King C | Washington State Department of Health | Seattle Flu Study | Chu etl al et al |
| EPI_ISL_417166 | USA/WA-S113/2020 | 2020-02-29 | North America/USA/Washington/King C | Washington State Department of Health | Seattle Flu Study | Chu etl al et al |
| EPI_ISL_417168 | USA/WA-S115/2020 | 2020-02-29 | North America/USA/Washington/King C | Washington State Department of Health | Seattle Flu Study | Chu etl al et al |
| EPI_ISL_417172 | USA/WA-S119/2020 | 2020-02-29 | North America/USA/Washington/King C | Washington State Department of Health | Seattle Flu Study | Chu etl al et al |
| EPI_ISL_417177 | HongKong/HKPU2_1801/2020 | 2020-01-23 | Asia/Hong Kong/Hong Kong/Hong Kong | Department of Pathology, Princess Margaret Hospital | Department of Health Technology and Informatics, Faculty of Health and Social Science, The Hong Kong Polytechnic University | Kenneth Siu-Sing LEUNG et al |
| EPI_ISL_417178 | HongKong/HKPU6_2101/2020 | 2020-01-25 | Asia/Hong Kong/Hong Kong/Hong Kong | Department of Pathology, Princess Margaret Hospital | Department of Health Technology and Informatics, Faculty of Health and Social Science, The Hong Kong Polytechnic University | Kenneth Siu-Sing LEUNG et al |
| EPI_ISL_417179 | HongKong/HKPU12_2201/2020 | 2020-01-30 | Asia/Hong Kong/Hong Kong/Hong Kong | Department of Pathology, Princess Margaret Hospital | Department of Health Technology and Informatics, Faculty of Health and Social Science, The Hong Kong Polytechnic University | Kenneth Siu-Sing LEUNG et al |

| | | | | | | |
|---|---|---|---|---|---|---|
| EPI_ISL_417180 | HongKong/HKPU17_2201/2020 | 2020-02-03 | Asia/Hong Kong/Hong Kong/Hong Kong | Department of Pathology, United Christian Hospital | Department of Health Technology and Informatics, Faculty of Health and Social Science, The Hong Kong Polytechnic University | Kenneth Siu-Sing LEUNG et al |
| EPI_ISL_417181 | HongKong/HKPU19_0402/2020 | 2020-02-05 | Asia/Hong Kong/Hong Kong/Hong Kong | Department of Pathology, United Christian Hospital | Department of Health Technology and Informatics, Faculty of Health and Social Science, The Hong Kong Polytechnic University | Kenneth Siu-Sing LEUNG et al |
| EPI_ISL_417182 | HongKong/HKPU20_3001/2020 | 2020-02-05 | Asia/Hong Kong/Hong Kong/Hong Kong | Department of Pathology, United Christian Hospital | Department of Health Technology and Informatics, Faculty of Health and Social Science, The Hong Kong Polytechnic University | Kenneth Siu-Sing LEUNG et al |
| EPI_ISL_417183 | HongKong/HKPU23_2601/2020 | 2020-01-30 | Asia/Hong Kong/Hong Kong/Hong Kong | Department of Clinical Pathology, Pamela Youde Nethersole Eastern Hospital | Department of Health Technology and Informatics, Faculty of Health and Social Science, The Hong Kong Polytechnic University | Kenneth Siu-Sing LEUNG et al |

| | | | | | | |
|---|---|---|---|---|---|---|
| EPI_ISL_417184 | HongKong/HKPU27_3001/2020 | 2020-02-08 | Asia/Hong Kong/Hong Kong/Hong Kong | Department of Clinical Pathology, Pamela Youde Nethersole Eastern Hospital | Department of Health Technology and Informatics, Faculty of Health and Social Science, The Hong Kong Polytechnic University | Kenneth Siu-Sing LEUNG et al |
| EPI_ISL_417185 | HongKong/HKPU28_3001/2020 | 2020-02-09 | Asia/Hong Kong/Hong Kong/Hong Kong | Department of Pathology, United Christian Hospital | Department of Health Technology and Informatics, Faculty of Health and Social Science, The Hong Kong Polytechnic University | Kenneth Siu-Sing LEUNG et al |
| EPI_ISL_417187 | HongKong/HKPU29_0102/2020 | 2020-02-08 | Asia/Hong Kong/Hong Kong/Hong Kong | Department of Clinical Pathology, Pamela Youde Nethersole Eastern Hospital | Department of Health Technology and Informatics, Faculty of Health and Social Science, The Hong Kong Polytechnic University | Kenneth Siu-Sing LEUNG et al |
| EPI_ISL_417188 | HongKong/HKPU30_2901/2020 | 2020-02-08 | Asia/Hong Kong/Hong Kong/Hong Kong | Department of Clinical Pathology, Pamela Youde Nethersole Eastern Hospital | Department of Health Technology and Informatics, Faculty of Health and Social Science, The Hong Kong Polytechnic University | Kenneth Siu-Sing LEUNG et al |

| | | | | | | |
|---|---|---|---|---|---|---|
| EPI_ISL_417190 | HongKong/HKPU_2801/2020 | 2020-02-08 | Asia/Hong Kong/Hong Kong/Hong Kong | Department of Clinical Pathology, Pamela Youde Nethersole Eastern Hospital | Department of Health Technology and Informatics, Faculty of Health and Social Science, The Hong Kong Polytechnic University | Kenneth Siu-Sing LEUNG et al |
| EPI_ISL_417193 | HongKong/HKPU32_0402/2020 | 2020-02-09 | Asia/Hong Kong/Hong Kong/Hong Kong | Department of Clinical Pathology, Pamela Youde Nethersole Eastern Hospital | Department of Health Technology and Informatics, Faculty of Health and Social Science, The Hong Kong Polytechnic University | Kenneth Siu-Sing LEUNG et al |
| EPI_ISL_417195 | HongKong/HKPU33_0202/2020 | 2020-02-09 | Asia/Hong Kong/Hong Kong/Hong Kong | Department of Clinical Pathology, Pamela Youde Nethersole Eastern Hospital | Department of Health Technology and Informatics, Faculty of Health and Social Science, The Hong Kong Polytechnic University | Kenneth Siu-Sing LEUNG et al |
| EPI_ISL_417197 | HongKong/HKPU34_3001/2020 | 2020-02-09 | Asia/Hong Kong/Hong Kong/Hong Kong | Department of Clinical Pathology, Pamela Youde Nethersole Eastern Hospital | Department of Health Technology and Informatics, Faculty of Health and Social Science, The Hong Kong Polytechnic University | Kenneth Siu-Sing LEUNG et al |
| EPI_ISL_417198 | USA/MN30-MDH30/2020 | 2020-03-12 | North America/USA/Minnesota/ | Minnesota Department of Health, Public Health Laboratory | Minnesota Department of Health, Public Health Laboratory | Matt Plumb et al |

| | | | | | | |
|---|---|---|---|---|---|---|
| EPI_ISL_417199 | HongKong/HKPU35_0402/2020 | 2020-02-09 | Asia/Hong Kong/Hong Kong/Hong Kong | Department of Clinical Pathology, Pamela Youde Nethersole Eastern Hospital | Department of Health Technology and Informatics, Faculty of Health and Social Science, The Hong Kong Polytechnic University | Chong-Yee YAU et al |
| EPI_ISL_417215 | England/20102087902/2020 | 2020-02-25 | Europe/United Kingdom/England/ | Respiratory Virus Unit, Microbiology Services Colindale, Public Health England | Respiratory Virus Unit, Microbiology Services Colindale, Public Health England | Monica Galiano et al |
| EPI_ISL_417216 | England/20102088002/2020 | 2020-02-25 | Europe/United Kingdom/England/ | Respiratory Virus Unit, Microbiology Services Colindale, Public Health England | Respiratory Virus Unit, Microbiology Services Colindale, Public Health England | Monica Galiano et al |
| EPI_ISL_417318 | USA/CA-SCCPHD-UC14/2020 | 2020-02-29 | North America/USA/California/Santa Cl | Santa Clara County Public Health Department | Chiu Laboratory, University of California, San Francisco | Xianding Deng et al |
| EPI_ISL_417333 | France/ARA-06042/2020 | 2020-03-04 | Europe/France/Auvergne-Rhv¥ne-Alpes/ | Institut des Agents Infectieux (IAI), Hospices Civils de Lyon | CNR Virus des Infections Respiratoires - France SUD | Antonin Bal et al |
| EPI_ISL_417334 | France/ARA-06056/2020 | 2020-03-04 | Europe/France/Auvergne-Rhv¥ne-Alpes/ | Institut des Agents Infectieux (IAI), Hospices Civils de Lyon | CNR Virus des Infections Respiratoires - France SUD | Antonin Bal et al |
| EPI_ISL_417335 | France/ARA-06573/2020 | 2020-03-06 | Europe/France/Auvergne-Rhv¥ne-Alpes/ | Institut des Agents Infectieux (IAI), Hospices Civils de Lyon | CNR Virus des Infections Respiratoires - France SUD | Antonin Bal et al |
| EPI_ISL_417345 | USA/WA-UW196/2020 | 2020-03-13 | North America/USA/Washington/ | UW Virology Lab | UW Virology Lab | Pavitra Roychoudhury et al |
| EPI_ISL_417346 | USA/WA-UW197/2020 | 2020-03-12 | North America/USA/Washington/ | UW Virology Lab | UW Virology Lab | Pavitra Roychoudhury et al |
| EPI_ISL_417353 | USA/WA-UW204/2020 | 2020-03-13 | North America/USA/Washington/ | UW Virology Lab | UW Virology Lab | Pavitra Roychoudhury et al |

| | | | | | | |
|---|---|---|---|---|---|---|
| EPI_ISL_417362 | USA/CT-UW213/2020 | 2020-03-13 | North America/USA/Connecticut/ | UW Virology Lab | UW Virology Lab | Pavitra Roychoudhury et al |
| EPI_ISL_417365 | USA/WA-UW216/2020 | 2020-03-14 | North America/USA/Washington/ | UW Virology Lab | UW Virology Lab | Pavitra Roychoudhury et al |
| EPI_ISL_417370 | USA/WA-UW221/2020 | 2020-03-14 | North America/USA/Washington/ | UW Virology Lab | UW Virology Lab | Pavitra Roychoudhury et al |
| EPI_ISL_417419 | Italy/FVG-ICGEB-S5/2020 | 2020-03-01 | Europe/Italy/Friuli Venezia Giulia/ | Laboratory of Molecular Virology International Center for Genetic Engineering and Biotechnology (ICGEB) | ARGO Open Lab Platform for Genome sequencing | Licastro D et al |
| EPI_ISL_417423 | Italy/FVG-ICGEB-S9/2020 | 2020-03-01 | Europe/Italy/Friuli Venezia Giulia/ | Laboratory of Molecular Virology International Center for Genetic Engineering and Biotechnology (ICGEB) | ARGO Open Lab Platform for Genome sequencing | Licastro D et al |
| EPI_ISL_417429 | Belgium/MMJ-03034/2020 | 2020-03-03 | Europe/Belgium/Leuven/Holsbeek | KU Leuven, Clinical and Epidemiological Virology | KU Leuven, Clinical and Epidemiological Virology | Joan Marti-Carerras et al |
| EPI_ISL_417444 | Pakistan/Gilgit1/2020 | 2020-03-04 | Asia/Iran/Gilgit Baltistan/Gilgit | Department of Healthcare Biotechnology, National University of Sciences and Technology (NUST) | Department of Healthcare Biotechnology, National University of Sciences and Technology (NUST) | Javed et al |
| EPI_ISL_417445 | Italy/LOM-UniMI01/2020 | 2020-02-24 | Europe/Italy/Lombardy/Milan | Laboratory of Infectious Diseases, Department of Biomedical and Clinical Sciences L. Sacco, University of Milan | Laboratory of Infectious Diseases, Department of Biomedical and Clinical Sciences L. Sacco, University of Milan | Gianguglielmo Zehender et al |
| EPI_ISL_417446 | Italy/LOM-UniMI02/2020 | 2020-02-24 | Europe/Italy/Lombardy/Milan | Laboratory of Infectious Diseases, Department of Biomedical and Clinical Sciences L. Sacco, University of Milan | Laboratory of Infectious Diseases, Department of Biomedical and Clinical Sciences L. Sacco, University of Milan | Gianguglielmo Zehender et al |

| | | | | | | |
|---|---|---|---|---|---|---|
| EPI_ISL_417447 | Italy/LOM-UniMI03/2020 | 2020-02-24 | Europe/Italy/Lombardy/Milan | Laboratory of Infectious Diseases, Department of Biomedical and Clinical Sciences L. Sacco, University of Milan | Laboratory of Infectious Diseases, Department of Biomedical and Clinical Sciences L. Sacco, University of Milan | Gianguglielmo Zehender et al |
| EPI_ISL_417448 | USA/WA-UW234/2020 | 2020-03-14 | North America/USA/Washington/ | UW Virology Lab | UW Virology Lab | Pavitra Roychoudhury et al |
| EPI_ISL_417450 | USA/WA-UW236/2020 | 2020-03-14 | North America/USA/Washington/ | UW Virology Lab | UW Virology Lab | Pavitra Roychoudhury et al |
| EPI_ISL_417462 | Germany/NW-HHU-17/2020 | 2020-03-11 | Europe/Germany/North Rhine Westphal | Center of Medical Microbiology, Virology, and Hospital Hygiene, University of Duesseldorf | Center of Medical Microbiology, Virology, and Hospital Hygiene, University of Duesseldorf | Ortwin Adams et al |
| EPI_ISL_417464 | Germany/NW-HHU-19/2020 | 2020-03-14 | Europe/Germany/North Rhine Westphal | Center of Medical Microbiology, Virology, and Hospital Hygiene, University of Duesseldorf | Center of Medical Microbiology, Virology, and Hospital Hygiene, University of Duesseldorf | Ortwin Adams et al |
| EPI_ISL_417466 | Germany/NW-HHU-21/2020 | 2020-03-14 | Europe/Germany/North Rhine Westphal | Center of Medical Microbiology, Virology, and Hospital Hygiene, University of Duesseldorf | Center of Medical Microbiology, Virology, and Hospital Hygiene, University of Duesseldorf | Ortwin Adams et al |
| EPI_ISL_417482 | Ecuador/HEE-01/2020 | 2020-03-09 | South America/Ecuador/Pichincha/Quito | Institute of Microbiology, Universidad San Francisco de Quito | Institute of Microbiology, Universidad San Francisco de Quito | Sully Mv°rquez et al |
| EPI_ISL_417483 | Norway/1379/2020 | 2020-02-27 | Europe/Italy/Oslo/ | Oslo University Hospital, Department of Medical Microbiology | Norwegian Institute of Public Health | Kathrine Stene-Johansen et al |
| EPI_ISL_417484 | Norway/1380/2020 | 2020-02-26 | Europe/Iran/Oslo/ | Oslo University Hospital, Department of Medical Microbiology | Norwegian Institute of Public Health, Department of Virology | Kathrine Stene-Johansen et al |

| | | | | | | |
|---|---|---|---|---|---|---|
| EPI_ISL_417485 | Norway/1526/2020 | 2020-03-02 | Europe/Italy/Oslo/ | University Hospital of Northern Norway, Department for Microbiology and Infectious Disease Control | Norwegian Institute of Public Health, Department of Virology | Kathrine Stene-Johansen et al |
| EPI_ISL_417488 | Norway/1493/2020 | 2020-02-29 | Europe/Norway/Oslo/ | Oslo University Hospital, Department of Medical Microbiology | Norwegian Institute of Public Health, Department of Virology | Kathrine Stene-Johansen et al |
| EPI_ISL_417489 | Norway/1494/2020 | 2020-02-29 | Europe/Norway/Oslo/ | Oslo University Hospital, Department of Medical Microbiology | Norwegian Institute of Public Health, Department of Virology | Kathrine Stene-Johansen et al |
| EPI_ISL_417490 | Norway/1443/2020 | 2020-02-27 | Europe/Italy/Oslo/ | Oslo University Hospital, Department of Medical Microbiology | Norwegian Institute of Public Health, Department of Virology | Kathrine Stene-Johansen et al |
| EPI_ISL_417498 | USA/MN53-MDH53/2020 | 2020-03-13 | North America/USA/Minnesota/ | Minnesota Department of Health, Public Health Laboratory | Minnesota Department of Health, Public Health Laboratory | Matt Plumb et al |
| EPI_ISL_417518 | Taiwan/CGMH-CGU-02/2020 | 2020-02-04 | Asia/Taiwan/Taiwan/Taoyuan | Laboratory Medicine | Department of Laboratory Medicine, Lin-Kou Chang Gung Memorial Hospital, Taoyuan, Taiwan | Kuo-Chien Tsao et al |
| EPI_ISL_417526 | Luxembourg/LNS9371718/2020 | 2020-03-18 | Europe/Luxembourg/Luxembourg/ | Laboratoire Nationale de Santv©, Microbiology, Virology | Laboratoire Nationale de Santv©, Microbiology, Epidemiology and Microbial Genomics | Anke Wienecke-Baldacchino et al |
| EPI_ISL_417557 | Iceland/229/2020 | 2020-03-17 | Europe/Iceland/Reykjavik/ | The National University Hospital of Iceland | deCODE genetics | Daniel F Gudbjartsson et al |
| EPI_ISL_417649 | Iceland/320/2020 | 2020-03-18 | Europe/Iceland/Reykjavik/ | The National University Hospital of Iceland | deCODE genetics | Daniel F Gudbjartsson et al |

| | | | | | | |
|---|---|---|---|---|---|---|
| EPI_ISL_417675 | Iceland/88/2020 | 2020-03-10 | Europe/Iceland/Reykjavik/ | The National University Hospital of Iceland | deCODE genetics | Daniel F Gudbjartsson et al |
| EPI_ISL_417681 | Iceland/37/2020 | 2020-03-04 | Europe/Italy/Reykjavik/ | The National University Hospital of Iceland | deCODE genetics | Daniel F Gudbjartsson et al |
| EPI_ISL_417688 | Iceland/12/2020 | 2020-03-01 | Europe/Italy/Reykjavik/ | The National University Hospital of Iceland | deCODE genetics | Daniel F Gudbjartsson et al |
| EPI_ISL_417691 | Iceland/111/2020 | 2020-03-11 | Europe/Italy/Reykjavik/ | The National University Hospital of Iceland | deCODE genetics | Daniel F Gudbjartsson et al |
| EPI_ISL_417698 | Iceland/103/2020 | 2020-03-10 | Europe/Italy/Reykjavik/ | The National University Hospital of Iceland | deCODE genetics | Daniel F Gudbjartsson et al |
| EPI_ISL_417700 | Iceland/173/2020 | 2020-03-15 | Europe/Italy/Reykjavik/ | The National University Hospital of Iceland | deCODE genetics | Daniel F Gudbjartsson et al |
| EPI_ISL_417702 | Iceland/33/2020 | 2020-03-03 | Europe/Italy/Reykjavik/ | The National University Hospital of Iceland | deCODE genetics | Daniel F Gudbjartsson et al |
| EPI_ISL_417718 | Iceland/49/2020 | 2020-03-05 | Europe/Italy/Reykjavik/ | The National University Hospital of Iceland | deCODE genetics | Daniel F Gudbjartsson et al |
| EPI_ISL_417728 | Iceland/70/2020 | 2020-03-08 | Europe/Italy/Reykjavik/ | The National University Hospital of Iceland | deCODE genetics | Daniel F Gudbjartsson et al |
| EPI_ISL_417730 | Iceland/93/2020 | 2020-03-10 | Europe/Italy/Reykjavik/ | The National University Hospital of Iceland | deCODE genetics | Daniel F Gudbjartsson et al |
| EPI_ISL_417736 | Iceland/14/2020 | 2020-03-01 | Europe/Italy/Reykjavik/ | The National University Hospital of Iceland | deCODE genetics | Daniel F Gudbjartsson et al |
| EPI_ISL_417749 | Iceland/69/2020 | 2020-03-08 | Europe/Italy/Reykjavik/ | The National University Hospital of Iceland | deCODE genetics | Daniel F Gudbjartsson et al |
| EPI_ISL_417751 | Iceland/46/2020 | 2020-03-04 | Europe/Italy/Reykjavik/ | The National University Hospital of Iceland | deCODE genetics | Daniel F Gudbjartsson et al |
| EPI_ISL_417758 | Iceland/164/2020 | 2020-03-14 | Europe/Italy/Reykjavik/ | The National University Hospital of Iceland | deCODE genetics | Daniel F Gudbjartsson et al |
| EPI_ISL_417765 | Iceland/13/2020 | 2020-02-27 | Europe/Italy/Reykjavik/ | The National University Hospital of Iceland | deCODE genetics | Daniel F Gudbjartsson et al |
| EPI_ISL_417775 | Iceland/163/2020 | 2020-03-14 | Europe/Italy/Reykjavik/ | The National University Hospital of Iceland | deCODE genetics | Daniel F Gudbjartsson et al |
| EPI_ISL_417781 | Iceland/113/2020 | 2020-03-11 | Europe/Italy/Reykjavik/ | The National University Hospital of Iceland | deCODE genetics | Daniel F Gudbjartsson et al |
| EPI_ISL_417800 | Iceland/15/2020 | 2020-03-02 | Europe/Italy/Reykjavik/ | The National University Hospital of Iceland | deCODE genetics | Daniel F Gudbjartsson et al |
| EPI_ISL_417807 | Iceland/16/2020 | 2020-03-02 | Europe/Italy/Reykjavik/ | The National University Hospital of Iceland | deCODE genetics | Daniel F Gudbjartsson et al |
| EPI_ISL_417811 | Iceland/17/2020 | 2020-03-02 | Europe/Italy/Reykjavik/ | The National University Hospital of Iceland | deCODE genetics | Daniel F Gudbjartsson et al |
| EPI_ISL_417817 | Iceland/19/2020 | 2020-03-02 | Europe/Italy/Reykjavik/ | The National University Hospital of Iceland | deCODE genetics | Daniel F Gudbjartsson et al |
| EPI_ISL_417840 | Iceland/23/2020 | 2020-03-02 | Europe/Italy/Reykjavik/ | The National University Hospital of Iceland | deCODE genetics | Daniel F Gudbjartsson et al |
| EPI_ISL_417860 | Iceland/72/2020 | 2020-03-08 | Europe/Italy/Reykjavik/ | The National University Hospital of Iceland | deCODE genetics | Daniel F Gudbjartsson et al |
| EPI_ISL_417861 | Iceland/73/2020 | 2020-03-09 | Europe/Italy/Reykjavik/ | The National University Hospital of Iceland | deCODE genetics | Daniel F Gudbjartsson et al |

| | | | | | | |
|---|---|---|---|---|---|---|
| EPI_ISL_417879 | Slovakia/SK-BMC5/2020 | 2020-03-06 | Europe/Slovakia/Kosice/ | Institute of Virology, Biomedical Research Center of the Slovak Academy of Sciences, Bratislava; Public Health Authority of the Slovak Republic, Bratislava | Institute of Virology, Biomedical Research Center of the Slovak Academy of Sciences, Bratislava; Comenius University Science Park, Bratislava | Monika Slv°vikova et al |
| EPI_ISL_417880 | Slovakia/SK-BMC6/2020 | 2020-03-08 | Europe/Slovakia/Martin/ | Institute of Virology, Biomedical Research Center of the Slovak Academy of Sciences, Bratislava; Public Health Authority of the Slovak Republic, Bratislava | Institute of Virology, Biomedical Research Center of the Slovak Academy of Sciences, Bratislava; Comenius University Science Park, Bratislava | Monika Slv°vikova et al |
| EPI_ISL_417921 | Italy/LAZ-INMI3/2020 | 2020-03-01 | Europe/Italy/Lazio/Rome | INMI Lazzaro Spallanzani IRCCS | Laboratory of Virology, INMI Lazzaro Spallanzani IRCCS | Martina Rueca et al |
| EPI_ISL_417922 | Italy/LAZ-INMI4/2020 | 2020-02-28 | Europe/Italy/Lazio/Rome | INMI Lazzaro Spallanzani IRCCS | Laboratory of Virology, INMI Lazzaro Spallanzani IRCCS | Cesare E. M. Gruber et al |
| EPI_ISL_417923 | Italy/LAZ-INMI5/2020 | 2020-03-04 | Europe/Italy/Lazio/Rome | INMI Lazzaro Spallanzani IRCCS | Laboratory of Virology, INMI Lazzaro Spallanzani IRCCS | Francesco Messina et al |
| EPI_ISL_417937 | USA/CA-CZB013/2020 | 2020-03-18 | North America/USA/California/San Fran | UCSF Clinical Microbiology Laboratory | Chan-Zuckerberg Biohub | Shaun Arevalo et al |
| EPI_ISL_417969 | Spain/MD-HLP-LP10-12/2020 | 2020-03-09 | Europe/Spain/Madrid/ | Hospital Universitario La Paz | Hospital Universitario La Paz | Elias Dahdouh et al |
| EPI_ISL_418031 | USA/WA-UW246/2020 | 2020-03-16 | North America/USA/Washington/ | UW Virology Lab | UW Virology Lab | Pavitra Roychoudhury et al |

| | | | | | | |
|---|---|---|---|---|---|---|
| EPI_ISL_418035 | USA/CT-UW250/2020 | 2020-03-14 | North America/USA/Connecticut/ | UW Virology Lab | UW Virology Lab | Pavitra Roychoudhury et al |
| EPI_ISL_418044 | USA/CT-UW259/2020 | 2020-03-14 | North America/USA/Connecticut/ | UW Virology Lab | UW Virology Lab | Pavitra Roychoudhury et al |
| EPI_ISL_418045 | USA/CT-UW260/2020 | 2020-03-14 | North America/USA/Connecticut/ | UW Virology Lab | UW Virology Lab | Pavitra Roychoudhury et al |
| EPI_ISL_418049 | USA/MN-UW264/2020 | 2020-03-13 | North America/USA/Minnesota/ | UW Virology Lab | UW Virology Lab | Pavitra Roychoudhury et al |
| EPI_ISL_418058 | USA/CT-UW273/2020 | 2020-03-13 | North America/USA/Connecticut/ | UW Virology Lab | UW Virology Lab | Pavitra Roychoudhury et al |
| EPI_ISL_418074 | USA/WA-UW289/2020 | 2020-03-15 | North America/USA/Washington/ | UW Virology Lab | UW Virology Lab | Pavitra Roychoudhury et al |
| EPI_ISL_418077 | USA/WA-UW292/2020 | 2020-03-15 | North America/USA/Washington/ | UW Virology Lab | UW Virology Lab | Pavitra Roychoudhury et al |
| EPI_ISL_418079 | USA/WA-UW294/2020 | 2020-03-15 | North America/USA/Washington/ | UW Virology Lab | UW Virology Lab | Pavitra Roychoudhury et al |
| EPI_ISL_418184 | USA/WI-GMF-00018/2020 | 2020-03-18 | North America/USA/Wisconsin/La Crosse | Gundersen Molecular Diagnostics Laboratory | Kabara Cancer Research Institute | Craig S. Richmond et al |
| EPI_ISL_418206 | Senegal/003/2020 | 2020-02-28 | Africa/Senegal/Dakar/ | Institut Pasteur Dakar | Institut Pasteur de Dakar | Ndongo Dia et al |
| EPI_ISL_418216 | Senegal/136/2020 | 2020-03-13 | Africa/Senegal/Dakar/ | Institut Pasteur Dakar | Institut Pasteur de Dakar | Ndongo Dia et al |
| EPI_ISL_418218 | France/HDF-1465/2020 | 2020-02-21 | Europe/France/Hauts de France/Compiv | Centre Hospitalier Compi√®gne Laboratoire de Biologie | National Reference Center for Viruses of Respiratory Infections, Institut Pasteur, Paris | M√©lanie Albert et al |
| EPI_ISL_418219 | France/BRE-1623/2020 | 2020-02-26 | Europe/France/Bretagne/Brest | CHU - H√¥pital Cavale Blanche - Labo. de Virologie | National Reference Center for Viruses of Respiratory Infections, Institut Pasteur, Paris | M√©lanie Albert et al |
| EPI_ISL_418220 | France/HDF-1645/2020 | 2020-02-28 | Europe/France/Hauts de France/Compiv | Centre Hospitalier Compi√®gne Laboratoire de Biologie | National Reference Center for Viruses of Respiratory Infections, Institut Pasteur, Paris | M√©lanie Albert et al |
| EPI_ISL_418222 | France/CVL-2000/2020 | 2020-03-04 | Europe/France/Centre-Val de Loire/Tour | CHRU Bretonneau - Serv. Bacterio-Virol. | National Reference Center for Viruses of Respiratory Infections, Institut Pasteur, Paris | M√©lanie Albert et al |

| | | | | | | |
|---|---|---|---|---|---|---|
| EPI_ISL_418224 | France/HDF-2150/2020 | 2020-03-08 | Europe/France/Hauts de France/Compiv | Centre Hospitalier Compiv®gne Laboratoire de Biologie | National Reference Center for Viruses of Respiratory Infections, Institut Pasteur, Paris | Mv©lanie Albert et al |
| EPI_ISL_418228 | France/HDF-2405/2020 | 2020-03-12 | Europe/France/Hauts de France/Compiv | Centre Hospitalier Compiv®gne Laboratoire de Biologie | National Reference Center for Viruses of Respiratory Infections, Institut Pasteur, Paris | Mv©lanie Albert et al |
| EPI_ISL_418235 | France/IDF-2561/2020 | 2020-03-16 | Europe/France/Ile de France/Vanves | Cabinet mv©dical | National Reference Center for Viruses of Respiratory Infections, Institut Pasteur, Paris | Mv©lanie Albert et al |
| EPI_ISL_418238 | France/HDF-2597/2020 | 2020-03-16 | Europe/France/Hauts de France/Compiv | Centre Hospitalier Compiv®gne Laboratoire de Biologie | National Reference Center for Viruses of Respiratory Infections, Institut Pasteur, Paris | Mv©lanie Albert et al |
| EPI_ISL_418239 | France/HDF-2601/2020 | 2020-03-16 | Europe/France/Hauts de France/Compiv | Centre Hospitalier Compiv®gne Laboratoire de Biologie | National Reference Center for Viruses of Respiratory Infections, Institut Pasteur, Paris | Mv©lanie Albert et al |
| EPI_ISL_418240 | France/IDF-2684/2020 | 2020-03-16 | Europe/France/Ile de France/Longjumea | LABM GH nord Essonne | National Reference Center for Viruses of Respiratory Infections, Institut Pasteur, Paris | Mv©lanie Albert et al |

| | | | | | | |
|---|---|---|---|---|---|---|
| EPI_ISL_418241 | Algeria/G0638_2264/2020 | 2020-03-02 | Africa/Algeria/Boufarik/ | NIC Viral Respiratory Unit - Institut Pasteur of Algeria | National Reference Center for Viruses of Respiratory Infections, Institut Pasteur, Paris | M√©lanie Albert et al |
| EPI_ISL_418242 | Algeria/G0640_2265/2020 | 2020-03-08 | Africa/Algeria/Blida/ | NIC Viral Respiratory Unit - Institut Pasteur of Algeria | National Reference Center for Viruses of Respiratory Infections, Institut Pasteur, Paris | M√©lanie Albert et al |
| EPI_ISL_418243 | Spain/AN-ISCIII-201272/2020 | 2020-02-28 | Europe/Spain/Andalusia/ | HOSPITAL UNIVERSITARIO VIRGEN DE LAS NIEVES | Instituto de Salud Carlos III | Iglesias-Caballero et al |
| EPI_ISL_418244 | Spain/AN-ISCIII-201373/2020 | 2020-03-02 | Europe/Spain/Andalusia/ | HOSPITAL UNIVERSITARIO VIRGEN DE LAS NIEVES | Instituto de Salud Carlos III | Iglesias-Caballero et al |
| EPI_ISL_418247 | Spain/CL-ISCIII-201061/2020 | 2020-02-26 | Europe/Spain/Castilla y Leon/ | HOSPITAL GENERAL DE SEGOVIA | Instituto de Salud Carlos III | Iglesias-Caballero et al |
| EPI_ISL_418251 | Spain/MD-ISCIII-201105/2020 | 2020-02-25 | Europe/Spain/Madrid/ | HOSPITAL UNIVERSITARIO LA PAZ | Instituto de Salud Carlos III | Iglesias-Caballero et al |
| EPI_ISL_418255 | Italy/ABR-IZSGC-TE4925/2020 | 2020-03-14 | Europe/Italy/Abruzzo/ | "Presidio Ospedaliero ""S. Spirito"" - PESCARA" | "Istituto Zooprofilattico Sperimentale dell'Abruzzo e Molise ""G. Caporale""" | Lorusso A et al |
| EPI_ISL_418256 | Italy/ABR-IZSGC-TE4880/2020 | 2020-03-14 | Europe/Italy/Abruzzo/ | Ospedale ‚ÄúSan Liberatore‚Äù di Atri | "Istituto Zooprofilattico Sperimentale dell'Abruzzo e Molise ""G. Caporale""" | Lorusso A et al |
| EPI_ISL_418259 | Italy/ABR-IZSGC-TE4959/2020 | 2020-03-14 | Europe/Italy/Abruzzo/ | "Presidio ospedaliero ""Santo Spirito""" | "Istituto Zooprofilattico Sperimentale dell'Abruzzo e Molise ""G. Caporale""" | Lorusso A et al |

| | | | | | | |
|---|---|---|---|---|---|---|
| EPI_ISL_418260 | Italy/ABR-IZSGC-TE4836/2020 | 2020-03-16 | Europe/Italy/Abruzzo/ | Ospedale Civile Giuseppe Mazzini | "Istituto Zooprofilattico Sperimentale dell'Abruzzo e Molise ""G. Caporale""" | Lorusso A et al |
| EPI_ISL_418267 | Vietnam/19-02S/2020 | 2020-01-22 | Asia/Vietnam/Ho Chi Minh City/ | Microbiology and Immunology department, Pasteur institute in Ho Chi Minh city | Microbiology and Immunology department, Pasteur institute in Ho Chi Minh city | Nguyen et al |
| EPI_ISL_418269 | Vietnam/19-01S/2020 | 2020-01-22 | Asia/Vietnam/Ho Chi Minh City/ | Microbiology and Immunology department, Pasteur institute in Ho Chi Minh city | Microbiology and Immunology department, Pasteur institute in Ho Chi Minh city | Cao et al |
| EPI_ISL_418271 | Switzerland/BS-UHB-42169171/2020 | 2020-02-27 | Europe/Switzerland/Basel-Stadt/ | University Hospital Basel, Clinical Virology | University Hospital Basel, Labormedizin | Hirsch et al |
| EPI_ISL_418273 | Switzerland/BL-UHB-BS-42169471/20 | 2020-02-28 | Europe/Switzerland/Basel-Stadt/ | University Hospital Basel, Clinical Virology | University Hospital Basel, Labormedizin | Hirsch et al |
| EPI_ISL_418280 | Switzerland/BL-UHB-42175220/2020 | 2020-03-03 | Europe/Switzerland/Basel-Stadt/ | University Hospital Basel, Clinical Virology | University Hospital Basel, Clinical Bacteriology | Hirsch et al |
| EPI_ISL_418281 | Switzerland/BL-UHB-42176216/2020 | 2020-03-04 | Europe/Switzerland/Basel-Stadt/ | University Hospital Basel, Clinical Virology | University Hospital Basel, Clinical Bacteriology | Hirsch et al |
| EPI_ISL_418322 | Canada/ON-PHL-3917/2020 | 20-Mar | North America/Canada/Ontario/ | Public Health Ontario Laboratories | Public Health Ontario Laboratories | Alireza Eshaghi et al |
| EPI_ISL_418323 | Canada/ON-PHL-3919/2020 | 20-Mar | North America/Canada/Ontario/ | Public Health Ontario Laboratories | Public Health Ontario Laboratories | Alireza Eshaghi et al |
| EPI_ISL_418324 | Canada/ON-PHL-3318/2020 | 20-Mar | North America/Canada/Ontario/ | Public Health Ontario Laboratories | Public Health Ontario Laboratories | Alireza Eshaghi et al |

| | | | | | | |
|---|---|---|---|---|---|---|
| EPI_ISL_418325 | Canada/ON-PHL-5472/2020 | 20-Feb | North America/Canada/Ontario/ | Public Health Ontario Laboratories | Public Health Ontario Laboratories | Alireza Eshaghi et al |
| EPI_ISL_418326 | Canada/ON-PHL-7972/2020 | 20-Mar | North America/Canada/Ontario/ | Public Health Ontario Laboratories | Public Health Ontario Laboratories | Alireza Eshaghi et al |
| EPI_ISL_418327 | Canada/ON-PHL-4181/2020 | 20-Jan | North America/Canada/Ontario/ | Public Health Ontario Laboratories | Public Health Ontario Laboratories | Alireza Eshaghi et al |
| EPI_ISL_418328 | Canada/ON-PHL-3670/2020 | 20-Mar | North America/Canada/Ontario/ | Public Health Ontario Laboratories | Public Health Ontario Laboratories | Alireza Eshaghi et al |
| EPI_ISL_418329 | Canada/ON-PHL-5694/2020 | 20-Mar | North America/Canada/Ontario/ | Public Health Ontario Laboratories | Public Health Ontario Laboratories | Alireza Eshaghi et al |
| EPI_ISL_418330 | Canada/ON-PHL-8580/2020 | 20-Mar | North America/Canada/Ontario/ | Public Health Ontario Laboratories | Public Health Ontario Laboratories | Alireza Eshaghi et al |
| EPI_ISL_418332 | Canada/ON-PHL-0052/2020 | 20-Mar | North America/Canada/Ontario/ | Public Health Ontario Laboratories | Public Health Ontario Laboratories | Alireza Eshaghi et al |
| EPI_ISL_418335 | Canada/ON-PHL-4464/2020 | 20-Mar | North America/Canada/Ontario/ | Public Health Ontario Laboratories | Public Health Ontario Laboratories | Alireza Eshaghi et al |
| EPI_ISL_418336 | Canada/ON-PHL-5705/2020 | 20-Mar | North America/Canada/Ontario/ | Public Health Ontario Laboratories | Public Health Ontario Laboratories | Alireza Eshaghi et al |
| EPI_ISL_418337 | Canada/ON-PHL-3802/2020 | 20-Mar | North America/Canada/Ontario/ | Public Health Ontario Laboratories | Public Health Ontario Laboratories | Alireza Eshaghi et al |
| EPI_ISL_418338 | Canada/ON-PHL-5710/2020 | 20-Mar | North America/Canada/Ontario/ | Public Health Ontario Laboratories | Public Health Ontario Laboratories | Alireza Eshaghi et al |
| EPI_ISL_418339 | Canada/ON-PHL-3877/2020 | 20-Mar | North America/Canada/Ontario/ | Public Health Ontario Laboratories | Public Health Ontario Laboratories | Alireza Eshaghi et al |
| EPI_ISL_418340 | Canada/ON-PHL-1083/2020 | 20-Mar | North America/Canada/Ontario/ | Public Health Ontario Laboratories | Public Health Ontario Laboratories | Alireza Eshaghi et al |

| | | | | | | |
|---|---|---|---|---|---|---|
| EPI_ISL_418341 | Canada/ON-PHL-0142/2020 | 20-Mar | North America/Canada/Ontario/ | Public Health Ontario Laboratories | Public Health Ontario Laboratories | Alireza Eshaghi et al |
| EPI_ISL_418342 | Canada/ON-PHL-0178/2020 | 20-Mar | North America/Canada/Ontario/ | Public Health Ontario Laboratories | Public Health Ontario Laboratories | Alireza Eshaghi et al |
| EPI_ISL_418343 | Canada/ON-PHL-6884/2020 | 20-Mar | North America/Canada/Ontario/ | Public Health Ontario Laboratories | Public Health Ontario Laboratories | Alireza Eshaghi et al |
| EPI_ISL_418344 | Canada/ON-PHL-2259/2020 | 20-Feb | North America/Canada/Ontario/ | Public Health Ontario Laboratories | Public Health Ontario Laboratories | Alireza Eshaghi et al |
| EPI_ISL_418345 | Canada/ON-PHL-8751/2020 | 20-Feb | North America/Canada/Ontario/ | Public Health Ontario Laboratories | Public Health Ontario Laboratories | Alireza Eshaghi et al |
| EPI_ISL_418346 | Canada/ON-PHL-0743/2020 | 20-Mar | North America/Canada/Ontario/ | Public Health Ontario Laboratories | Public Health Ontario Laboratories | Alireza Eshaghi et al |
| EPI_ISL_418347 | Canada/ON-PHL-3741/2020 | 20-Mar | North America/Canada/Ontario/ | Public Health Ontario Laboratories | Public Health Ontario Laboratories | Alireza Eshaghi et al |
| EPI_ISL_418348 | Canada/ON-PHL-3680/2020 | 20-Mar | North America/Canada/Ontario/ | Public Health Ontario Laboratories | Public Health Ontario Laboratories | Alireza Eshaghi et al |
| EPI_ISL_418349 | Canada/ON-PHL-3650/2020 | 20-Mar | North America/Canada/Ontario/ | Public Health Ontario Laboratories | Public Health Ontario Laboratories | Alireza Eshaghi et al |
| EPI_ISL_418350 | Canada/ON-PHL-3380/2020 | 20-Mar | North America/Canada/Ontario/ | Public Health Ontario Laboratories | Public Health Ontario Laboratories | Alireza Eshaghi et al |
| EPI_ISL_418351 | Canada/ON-PHL-3575/2020 | 20-Mar | North America/Canada/Ontario/ | Public Health Ontario Laboratories | Public Health Ontario Laboratories | Alireza Eshaghi et al |
| EPI_ISL_418352 | Canada/ON-PHL-5672/2020 | 20-Mar | North America/Canada/Ontario/ | Public Health Ontario Laboratories | Public Health Ontario Laboratories | Alireza Eshaghi et al |
| EPI_ISL_418353 | Canada/ON-PHL-0539/2020 | 20-Mar | North America/Canada/Ontario/ | Public Health Ontario Laboratories | Public Health Ontario Laboratories | Alireza Eshaghi et al |

| | | | | | | |
|---|---|---|---|---|---|---|
| EPI_ISL_418354 | Canada/ON-PHL-4088/2020 | 20-Mar | North America/Canada/Ontario/ | Public Health Ontario Laboratories | Public Health Ontario Laboratories | Alireza Eshaghi et al |
| EPI_ISL_418355 | Canada/ON-PHL-U8150/2020 | 20-Mar | North America/Canada/Ontario/ | Public Health Ontario Laboratories | Public Health Ontario Laboratories | Alireza Eshaghi et al |
| EPI_ISL_418356 | Canada/ON-PHL-6980/2020 | 20-Mar | North America/Canada/Ontario/ | Public Health Ontario Laboratories | Public Health Ontario Laboratories | Alireza Eshaghi et al |
| EPI_ISL_418357 | Canada/ON-PHL-H6415/2020 | 20-Mar | North America/Canada/Ontario/ | Public Health Ontario Laboratories | Public Health Ontario Laboratories | Alireza Eshaghi et al |
| EPI_ISL_418358 | Canada/ON-PHL-5756/2020 | 20-Mar | North America/Canada/Ontario/ | Public Health Ontario Laboratories | Public Health Ontario Laboratories | Alireza Eshaghi et al |
| EPI_ISL_418359 | Canada/ON-PHL-5757/2020 | 20-Mar | North America/Canada/Ontario/ | Public Health Ontario Laboratories | Public Health Ontario Laboratories | Alireza Eshaghi et al |
| EPI_ISL_418360 | Canada/ON-PHL-3692/2020 | 20-Mar | North America/Canada/Ontario/ | Public Health Ontario Laboratories | Public Health Ontario Laboratories | Alireza Eshaghi et al |
| EPI_ISL_418361 | Canada/ON-PHL-1095/2020 | 20-Mar | North America/Canada/Ontario/ | Public Health Ontario Laboratories | Public Health Ontario Laboratories | Alireza Eshaghi et al |
| EPI_ISL_418362 | Canada/ON-PHL-3536/2020 | 20-Mar | North America/Canada/Ontario/ | Public Health Ontario Laboratories | Public Health Ontario Laboratories | Alireza Eshaghi et al |
| EPI_ISL_418363 | Canada/ON-PHL-3695/2020 | 20-Mar | North America/Canada/Ontario/ | Public Health Ontario Laboratories | Public Health Ontario Laboratories | Alireza Eshaghi et al |
| EPI_ISL_418364 | Canada/ON-PHL-6922/2020 | 20-Mar | North America/Canada/Ontario/ | Public Health Ontario Laboratories | Public Health Ontario Laboratories | Alireza Eshaghi et al |
| EPI_ISL_418365 | Canada/ON-PHL-0977/2020 | 20-Mar | North America/Canada/Ontario/ | Public Health Ontario Laboratories | Public Health Ontario Laboratories | Alireza Eshaghi et al |
| EPI_ISL_418366 | Canada/ON-PHL-3350/2020 | 20-Mar | North America/Canada/Ontario/ | Public Health Ontario Laboratories | Public Health Ontario Laboratories | Alireza Eshaghi et al |

| | | | | | | |
|---|---|---|---|---|---|---|
| EPI_ISL_418367 | Canada/ON-PHL-7513/2020 | 20-Mar | North America/Canada/Ontario/ | Public Health Ontario Laboratories | Public Health Ontario Laboratories | Alireza Eshaghi et al |
| EPI_ISL_418368 | Canada/ON-PHL-3458/2020 | 20-Mar | North America/Canada/Ontario/ | Public Health Ontario Laboratories | Public Health Ontario Laboratories | Alireza Eshaghi et al |
| EPI_ISL_418369 | Canada/ON-PHL-8539/2020 | 20-Mar | North America/Canada/Ontario/ | Public Health Ontario Laboratories | Public Health Ontario Laboratories | Alireza Eshaghi et al |
| EPI_ISL_418370 | Canada/ON-PHL-0654/2020 | 20-Mar | North America/Canada/Ontario/ | Public Health Ontario Laboratories | Public Health Ontario Laboratories | Alireza Eshaghi et al |
| EPI_ISL_418371 | Canada/ON-PHL-2653/2020 | 20-Mar | North America/Canada/Ontario/ | Public Health Ontario Laboratories | Public Health Ontario Laboratories | Alireza Eshaghi et al |
| EPI_ISL_418372 | Canada/ON-PHL-8458/2020 | 20-Mar | North America/Canada/Ontario/ | Public Health Ontario Laboratories | Public Health Ontario Laboratories | Alireza Eshaghi et al |
| EPI_ISL_418373 | Canada/ON-PHL-7590/2020 | 20-Mar | North America/Canada/Ontario/ | Public Health Ontario Laboratories | Public Health Ontario Laboratories | Alireza Eshaghi et al |
| EPI_ISL_418374 | Canada/ON-PHL-4232/2020 | 20-Mar | North America/Canada/Ontario/ | Public Health Ontario Laboratories | Public Health Ontario Laboratories | Alireza Eshaghi et al |
| EPI_ISL_418375 | Canada/ON-PHL-0141/2020 | 20-Mar | North America/Canada/Ontario/ | Public Health Ontario Laboratories | Public Health Ontario Laboratories | Alireza Eshaghi et al |
| EPI_ISL_418376 | Canada/ON-PHL-3459/2020 | 20-Mar | North America/Canada/Ontario/ | Public Health Ontario Laboratories | Public Health Ontario Laboratories | Alireza Eshaghi et al |
| EPI_ISL_418377 | Canada/ON-PHL-1898/2020 | 20-Mar | North America/Canada/Ontario/ | Public Health Ontario Laboratories | Public Health Ontario Laboratories | Alireza Eshaghi et al |
| EPI_ISL_418378 | Canada/ON-PHL-0976/2020 | 20-Mar | North America/Canada/Ontario/ | Public Health Ontario Laboratories | Public Health Ontario Laboratories | Alireza Eshaghi et al |
| EPI_ISL_418379 | Canada/ON-PHL-7512/2020 | 20-Mar | North America/Canada/Ontario/ | Public Health Ontario Laboratories | Public Health Ontario Laboratories | Alireza Eshaghi et al |

| | | | | | | |
|---|---|---|---|---|---|---|
| EPI_ISL_418380 | Canada/ON-PHL-3476/2020 | 20-Mar | North America/Canada/Ontario/ | Public Health Ontario Laboratories | Public Health Ontario Laboratories | Alireza Eshaghi et al |
| EPI_ISL_418381 | Canada/ON-PHL-2223/2020 | 20-Mar | North America/Canada/Ontario/ | Public Health Ontario Laboratories | Public Health Ontario Laboratories | Alireza Eshaghi et al |
| EPI_ISL_418382 | Canada/ON-PHL-5930/2020 | 20-Feb | North America/Canada/Ontario/ | Public Health Ontario Laboratories | Public Health Ontario Laboratories | Alireza Eshaghi et al |
| EPI_ISL_418383 | Canada/ON-PHL-2273/2020 | 20-Feb | North America/Canada/Ontario/ | Public Health Ontario Laboratories | Public Health Ontario Laboratories | Alireza Eshaghi et al |
| EPI_ISL_418395 | Finland/13M65/2020 | 2020-03-13 | Europe/Finland/Finland/ | Department of Virology and Immunology, University of Helsinki and Helsinki University Hospital, Huslab Finland | Department of Virology, Faculty of Medicine, University of Helsinki, Helsinki, Finland | Teemu Smura et al |
| EPI_ISL_418402 | Finland/14M13/2020 | 2020-03-14 | Europe/Finland/Finland/ | Department of Virology and Immunology, University of Helsinki and Helsinki University Hospital, Huslab Finland | Department of Virology, Faculty of Medicine, University of Helsinki, Helsinki, Finland | Teemu Smura et al |
| EPI_ISL_418404 | Finland/14M16/2020 | 2020-03-14 | Europe/Finland/Finland/ | Department of Virology and Immunology, University of Helsinki and Helsinki University Hospital, Huslab Finland | Department of Virology, Faculty of Medicine, University of Helsinki, Helsinki, Finland | Teemu Smura et al |
| EPI_ISL_418411 | Finland/14M82/2020 | 2020-03-14 | Europe/Finland/Helsinki/ | Department of Virology and Immunology, University of Helsinki and Helsinki University Hospital, Huslab Finland | Department of Virology, Faculty of Medicine, University of Helsinki, Helsinki, Finland | Teemu Smura et al |
| EPI_ISL_418412 | France/ARA-094100/2020 | 2020-03-15 | Europe/France/Auvergne-Rhv¥ne-Alpes/ | Centre Hospitalier des Vals d'Ardeche | CNR Virus des Infections Respiratoires - France SUD | Antonin Bal et al |

| | | | | | | |
|---|---|---|---|---|---|---|
| EPI_ISL_418418 | France/ARA-10163/2020 | 2020-03-16 | Europe/France/Auvergne-Rhv¥ne-Alpes/ | Centre Hospitalier Saint Joseph Saint Luc | CNR Virus des Infections Respiratoires - France SUD | Antonin Bal et al |
| EPI_ISL_418419 | France/ARA-10165/2020 | 2020-03-16 | Europe/France/Auvergne-Rhv¥ne-Alpes/ | Centre Hospitalier Saint Joseph Saint Luc | CNR Virus des Infections Respiratoires - France SUD | Antonin Bal et al |
| EPI_ISL_418422 | France/ARA-10184/2020 | 2020-03-17 | Europe/France/Auvergne-Rhv¥ne-Alpes/ | Institut des Agents Infectieux (IAI), Hospices Civils de Lyon | CNR Virus des Infections Respiratoires - France SUD | Antonin Bal et al |
| EPI_ISL_418424 | France/ARA-10189/2020 | 2020-03-17 | Europe/France/Auvergne-Rhv¥ne-Alpes/ | Institut des Agents Infectieux (IAI), Hospices Civils de Lyon | CNR Virus des Infections Respiratoires - France SUD | Antonin Bal et al |
| EPI_ISL_418432 | France/ARA-11036/2020 | 2020-03-18 | Europe/France/Auvergne-Rhv¥ne-Alpes/ | Institut des Agents Infectieux (IAI), Hospices Civils de Lyon | CNR Virus des Infections Respiratoires - France SUD | Antonin Bal et al |
| EPI_ISL_418441 | Hangzhou/HZ48/2020 | 2020-01-21 | Asia/China/Zhejiang/Hangzhou | Hangzhou Center for Disease Control and Prevention | Inspection Center of Hangzhou Center for Disease Control and Prevention | Yu hua et al |
| EPI_ISL_418442 | Hangzhou/HZ49/2020 | 2020-01-21 | Asia/China/Zhejiang/Hangzhou | Hangzhou Center for Disease Control and Prevention | Inspection Center of Hangzhou Center for Disease Control and Prevention | Yu hua et al |
| EPI_ISL_418502 | Hangzhou/HZ60/2020 | 2020-01-22 | Asia/China/Zhejiang/Hangzhou | Hangzhou Center for Disease Control and Prevention | Inspection Center of Hangzhou Center for Disease Control and Prevention | Yu hua et al |
| EPI_ISL_418503 | Hangzhou/HZ62/2020 | 2020-01-22 | Asia/China/Zhejiang/Hangzhou | Hangzhou Center for Disease Control and Prevention | Inspection Center of Hangzhou Center for Disease Control and Prevention | Yu hua et al |

| | | | | | | |
|---|---|---|---|---|---|---|
| EPI_ISL_418504 | Hangzhou/HZ79/2020 | 2020-01-21 | Asia/China/Zhejiang/Hangzhou | Hangzhou Center for Disease Control and Prevention | Inspection Center of Hangzhou Center for Disease Control and Prevention | Yu hua et al |
| EPI_ISL_418506 | Hangzhou/HZ90/2020 | 2020-01-21 | Asia/China/Zhejiang/Hangzhou | Hangzhou Center for Disease Control and Prevention | Inspection Center of Hanghzou Center for Disease Control and Prevention | Yu hua et al |
| EPI_ISL_418507 | Hangzhou/HZ91/2020 | 2020-01-21 | Asia/China/Zhejiang/Hangzhou | Hangzhou Center for Disease Control and Prevention | Inspection Center of Hangzhou Center for Disease Control and Prevention | Yu hua et al |
| EPI_ISL_418508 | Hangzhou/HZ162/2020 | 2020-01-23 | Asia/China/Zhejiang/Hangzhou | Hangzhou Center for Disease Control and Prevention | Inspection Center of Hangzhou Center for Disease Control and Prevention | Yu hua et al |
| EPI_ISL_418509 | Hangzhou/HZ178/2020 | 2020-01-23 | Asia/China/Zhejiang/Hangzhou | Hangzhou Center for Disease Control and Prevention | Inspection Center of Hangzhou Center for Disease Control and Prevention | Yu hua et al |
| EPI_ISL_418510 | Hangzhou/HZ185/2020 | 2020-01-23 | Asia/China/Zhejiang/Hangzhou | Hangzhou Center for Disease Control and Prevention | Insepction Center of Hangzhou Center for Disease Control and Prevention | Yu hua et al |
| EPI_ISL_418511 | Hangzhou/HZ477/2020 | 2020-01-24 | Asia/China/Zhejiang/Hangzhou | Hangzhou Center for Disease Control and Prevention | Inspection Center of Hangzhou Center for Disease Control and Prevention | Yu hua et al |
| EPI_ISL_418513 | Hangzhou/HZ551/2020 | 2020-01-25 | Asia/China/Zhejiang/Hangzhou | Hangzhou Center for Disease Control and Prevention | Inspection Center of Hangzhou Center for Disease Control and Prevention | Yu hua et al |

| | | | | | | |
|---|---|---|---|---|---|---|
| EPI_ISL_418514 | Hangzhou/HZ576/2020 | 2020-01-25 | Asia/China/Zhejiang/Hangzhou | Hangzhou Center for Disease Control and Prevention | Inspection Center of Hangzhou Center for Disease Control and Prevention | Yu hua et al |
| EPI_ISL_418515 | Hangzhou/HZ638/2020 | 2020-01-25 | Asia/China/Zhejiang/Hangzhou | Hangzhou Center for Disease Control and Prevention | Inspection Center of Hangzhou Center for Disease Control and Prevention | Yu hua et al |
| EPI_ISL_418640 | Belgium/ULG-8634/2020 | 2020-03-19 | Europe/Belgium/Li√®ge/ | Department of Clinical Microbiology | GIGA Medical Genomics | Keith Durkin et al |
| EPI_ISL_418643 | Belgium/ULG-8715/2020 | 2020-03-19 | Europe/Belgium/Li√®ge/ | Department of Clinical Microbiology | GIGA Medical Genomics | Keith Durkin et al |
| EPI_ISL_418736 | England/20124099602/2020 | 2020-03-18 | Europe/United Kingdom/England/ | Respiratory Virus Unit, Microbiology Services Colindale, Public Health England | Respiratory Virus Unit, Microbiology Services Colindale, Public Health England | Monica Galiano et al |
| EPI_ISL_418779 | USA/WA-NH10/2020 | 2020-03-13 | North America/USA/Washington/ | WA State Department of Health | Pathogen Discovery, Respiratory Viruses Branch, Division of Viral Diseases, Centers for Disease Control and Prevention | Ying Tao et al |
| EPI_ISL_418788 | USA/WA-NH21/2020 | 2020-03-13 | North America/USA/Washington/ | WA State Department of Health | Pathogen Discovery, Respiratory Viruses Branch, Division of Viral Diseases, Centers for Disease Control and Prevention | Ying Tao et al |
| EPI_ISL_418799 | Australia/QLDID922/2020 | 2020-02-28 | Oceania/Australia/Queensland/Brisbane | Mater Pathology | Public Health Virology Laboratory | Bixing Huang et al |

| | | | | | | |
|---|---|---|---|---|---|---|
| EPI_ISL_418800 | Belgium/CS-031052/2020 | 2020-03-10 | Europe/Belgium/Turnhout/Gierle | KU Leuven, Clinical and Epidemiological Virology | KU Leuven, Clinical and Epidemiological Virology | Bert Vanmechelen et al |
| EPI_ISL_418809 | Japan/UT-NCGM02/2020 | 2020-02-01 | Asia/Japan/Tokyo/ | University of Wisconsin - Madison: Influenza Research Institute | University of Wisconsin Madison, AIDS Vaccine Research Laboratories | Katarina Braun et al |
| EPI_ISL_418811 | Canada/NB_6/2020 | 2020-03-12 | North America/Canada/New Brunswick/ | Dr. Georges-L.-Dumont University Hospital Centre | National Microbiology Laboratory | Anna Majer et al |
| EPI_ISL_418812 | Canada/MB_8/2020 | 2020-03-12 | North America/Canada/Manitoba/ | Cadham Provincial Laboratory | National Microbiology Laboratory | Anna Majer et al |
| EPI_ISL_418813 | Canada/MB_10/2020 | 2020-03-13 | North America/Canada/Manitoba/ | Cadham Provincial Laboratory | National Microbiology Laboratory | Anna Majer et al |
| EPI_ISL_418814 | Canada/NS_13/2020 | 2020-03-13 | North America/Canada/Nova Scotia/ | Queen Elizabeth II Health Science Centre | National Microbiology Laboratory | Anna Majer et al |
| EPI_ISL_418815 | HongKong/HKPU36-0702/2020 | 2020-02-09 | Asia/Hong Kong/Hong Kong/Hong Kong | Department of Clinical Pathology, Pamela Youde Nethersole Eastern Hospital | Department of Health Technology and Informatics, Faculty of Health and Social Science, The Hong Kong Polytechnic University | Kenneth Siu-Sing LEUNG et al |
| EPI_ISL_418816 | Canada/BC_0443574/2020 | 2020-03-11 | North America/Canada/British Columbia | BCCDC Public Health Laboratory | BCCDC Public Health Laboratory | Harrigan et al |
| EPI_ISL_418817 | Canada/BC_0554880/2020 | 2020-03-13 | North America/Canada/British Columbia | BCCDC Public Health Laboratory | BCCDC Public Health Laboratory | Harrigan et al |
| EPI_ISL_418818 | Canada/BC_1318414/2020 | 2020-03-06 | North America/Canada/British Columbia | BCCDC Public Health Laboratory | BCCDC Public Health Laboratory | Harrigan et al |
| EPI_ISL_418819 | Canada/BC_3808524/2020 | 2020-03-09 | North America/Canada/British Columbia | BCCDC Public Health Laboratory | BCCDC Public Health Laboratory | Harrigan et al |
| EPI_ISL_418820 | Canada/BC_3842755/2020 | 2020-03-09 | North America/Canada/British Columbia | BCCDC Public Health Laboratory | BCCDC Public Health Laboratory | Harrigan et al |

| | | | | | | |
|---|---|---|---|---|---|---|
| EPI_ISL_418821 | Canada/BC_3968175/2020 | 2020-03-10 | North America/Canada/British Columbia | BCCDC Public Health Laboratory | BCCDC Public Health Laboratory | Harrigan et al |
| EPI_ISL_418822 | Canada/BC_3972884/2020 | 2020-03-10 | North America/Canada/British Columbia | BCCDC Public Health Laboratory | BCCDC Public Health Laboratory | Harrigan et al |
| EPI_ISL_418823 | Canada/BC_3989992/2020 | 2020-03-09 | North America/Canada/British Columbia | BCCDC Public Health Laboratory | BCCDC Public Health Laboratory | Harrigan et al |
| EPI_ISL_418824 | Canada/BC_4078583/2020 | 2020-03-03 | North America/Canada/British Columbia | BCCDC Public Health Laboratory | BCCDC Public Health Laboratory | Harrigan et al |
| EPI_ISL_418825 | Canada/BC_4118226/2020 | 2020-03-10 | North America/Canada/British Columbia | BCCDC Public Health Laboratory | BCCDC Public Health Laboratory | Harrigan et al |
| EPI_ISL_418826 | Canada/BC_4122951/2020 | 2020-03-10 | North America/Canada/British Columbia | BCCDC Public Health Laboratory | BCCDC Public Health Laboratory | Harrigan et al |
| EPI_ISL_418827 | Canada/BC_4143842/2020 | 2020-03-04 | North America/Canada/British Columbia | BCCDC Public Health Laboratory | BCCDC Public Health Laboratory | Harrigan et al |
| EPI_ISL_418828 | Canada/BC_4143868/2020 | 2020-03-04 | North America/Canada/British Columbia | BCCDC Public Health Laboratory | BCCDC Public Health Laboratory | Harrigan et al |
| EPI_ISL_418829 | Canada/BC_4540462/2020 | 2020-03-10 | North America/Canada/British Columbia | BCCDC Public Health Laboratory | BCCDC Public Health Laboratory | Harrigan et al |
| EPI_ISL_418830 | Canada/BC_4799711/2020 | 2020-03-09 | North America/Canada/British Columbia | BCCDC Public Health Laboratory | BCCDC Public Health Laboratory | Harrigan et al |
| EPI_ISL_418831 | Canada/BC_5275718/2020 | 2020-03-11 | North America/Canada/British Columbia | BCCDC Public Health Laboratory | BCCDC Public Health Laboratory | Harrigan et al |
| EPI_ISL_418832 | Canada/BC_5282984/2020 | 2020-03-11 | North America/Canada/British Columbia | BCCDC Public Health Laboratory | BCCDC Public Health Laboratory | Harrigan et al |
| EPI_ISL_418833 | Canada/BC_5306970/2020 | 2020-03-11 | North America/Canada/British Columbia | BCCDC Public Health Laboratory | BCCDC Public Health Laboratory | Harrigan et al |
| EPI_ISL_418834 | Canada/BC_5522039/2020 | 2020-03-11 | North America/Canada/British Columbia | BCCDC Public Health Laboratory | BCCDC Public Health Laboratory | Harrigan et al |
| EPI_ISL_418835 | Canada/BC_5979789/2020 | 2020-03-11 | North America/Canada/British Columbia | BCCDC Public Health Laboratory | BCCDC Public Health Laboratory | Harrigan et al |
| EPI_ISL_418836 | Canada/BC_6004567/2020 | 2020-03-10 | North America/Canada/British Columbia | BCCDC Public Health Laboratory | BCCDC Public Health Laboratory | Harrigan et al |
| EPI_ISL_418837 | Canada/BC_6129127/2020 | 2020-03-10 | North America/Canada/British Columbia | BCCDC Public Health Laboratory | BCCDC Public Health Laboratory | Harrigan et al |
| EPI_ISL_418838 | Canada/BC_6404889/2020 | 2020-03-12 | North America/Canada/British Columbia | BCCDC Public Health Laboratory | BCCDC Public Health Laboratory | Harrigan et al |
| EPI_ISL_418839 | Canada/BC_6489864/2020 | 2020-03-10 | North America/Canada/British Columbia | BCCDC Public Health Laboratory | BCCDC Public Health Laboratory | Harrigan et al |
| EPI_ISL_418840 | Canada/BC_6502001/2020 | 2020-03-12 | North America/Canada/British Columbia | BCCDC Public Health Laboratory | BCCDC Public Health Laboratory | Harrigan et al |

| | | | | | | |
|---|---|---|---|---|---|---|
| EPI_ISL_418841 | Canada/BC_6981299/2020 | 2020-03-11 | North America/Canada/British Columbia | BCCDC Public Health Laboratory | BCCDC Public Health Laboratory | Harrigan et al |
| EPI_ISL_418842 | Canada/BC_6997898/2020 | 2020-03-11 | North America/Canada/British Columbia | BCCDC Public Health Laboratory | BCCDC Public Health Laboratory | Harrigan et al |
| EPI_ISL_418843 | Canada/BC_7277381/2020 | 2020-03-12 | North America/Canada/British Columbia | BCCDC Public Health Laboratory | BCCDC Public Health Laboratory | Harrigan et al |
| EPI_ISL_418844 | Canada/BC_7493713/2020 | 2020-03-12 | North America/Canada/British Columbia | BCCDC Public Health Laboratory | BCCDC Public Health Laboratory | Harrigan et al |
| EPI_ISL_418845 | Canada/BC_7515663/2020 | 2020-03-11 | North America/Canada/British Columbia | BCCDC Public Health Laboratory | BCCDC Public Health Laboratory | Harrigan et al |
| EPI_ISL_418846 | Canada/BC_7553799/2020 | 2020-03-12 | North America/Canada/British Columbia | BCCDC Public Health Laboratory | BCCDC Public Health Laboratory | Harrigan et al |
| EPI_ISL_418847 | Canada/BC_8150537/2020 | 2020-03-12 | North America/Canada/British Columbia | BCCDC Public Health Laboratory | BCCDC Public Health Laboratory | Harrigan et al |
| EPI_ISL_418848 | Canada/BC_8159203/2020 | 2020-03-05 | North America/Canada/British Columbia | BCCDC Public Health Laboratory | BCCDC Public Health Laboratory | Harrigan et al |
| EPI_ISL_418849 | Canada/BC_8486790/2020 | 2020-03-13 | North America/Canada/British Columbia | BCCDC Public Health Laboratory | BCCDC Public Health Laboratory | Harrigan et al |
| EPI_ISL_418850 | Canada/BC_8606204/2020 | 2020-03-13 | North America/Canada/British Columbia | BCCDC Public Health Laboratory | BCCDC Public Health Laboratory | Harrigan et al |
| EPI_ISL_418851 | Canada/BC_8622445/2020 | 2020-03-13 | North America/Canada/British Columbia | BCCDC Public Health Laboratory | BCCDC Public Health Laboratory | Harrigan et al |
| EPI_ISL_418852 | Canada/BC_8718874/2020 | 2020-03-07 | North America/Canada/British Columbia | BCCDC Public Health Laboratory | BCCDC Public Health Laboratory | Harrigan et al |
| EPI_ISL_418853 | Canada/BC_8894200/2020 | 2020-03-11 | North America/Canada/British Columbia | BCCDC Public Health Laboratory | BCCDC Public Health Laboratory | Harrigan et al |
| EPI_ISL_418854 | Canada/BC_8896915/2020 | 2020-03-11 | North America/Canada/British Columbia | BCCDC Public Health Laboratory | BCCDC Public Health Laboratory | Harrigan et al |
| EPI_ISL_418855 | Canada/BC_8897642/2020 | 2020-03-11 | North America/Canada/British Columbia | BCCDC Public Health Laboratory | BCCDC Public Health Laboratory | Harrigan et al |
| EPI_ISL_418856 | Canada/BC_9345042/2020 | 2020-03-13 | North America/Canada/British Columbia | BCCDC Public Health Laboratory | BCCDC Public Health Laboratory | Harrigan et al |
| EPI_ISL_418857 | Canada/BC_9345715/2020 | 2020-03-13 | North America/Canada/British Columbia | BCCDC Public Health Laboratory | BCCDC Public Health Laboratory | Harrigan et al |
| EPI_ISL_418858 | Canada/BC_9446031/2020 | 2020-03-13 | North America/Canada/British Columbia | BCCDC Public Health Laboratory | BCCDC Public Health Laboratory | Harrigan et al |
| EPI_ISL_418859 | Canada/BC_9574898/2020 | 2020-03-13 | North America/Canada/British Columbia | BCCDC Public Health Laboratory | BCCDC Public Health Laboratory | Harrigan et al |
| EPI_ISL_418870 | USA/WA-UW302/2020 | 2020-03-16 | North America/USA/Washington/ | UW Virology Lab | UW Virology Lab | Pavitra Roychoudhury et al |
| EPI_ISL_418896 | USA/CT-UW329/2020 | 2020-03-13 | North America/USA/Connecticut/ | UW Virology Lab | UW Virology Lab | Pavitra Roychoudhury et al |

| | | | | | | |
|---|---|---|---|---|---|---|
| EPI_ISL_418907 | USA/WA-UW340/2020 | 2020-03-16 | North America/USA/Washington/ | UW Virology Lab | UW Virology Lab | Pavitra Roychoudhury et al |
| EPI_ISL_418908 | USA/WA-UW341/2020 | 2020-03-16 | North America/USA/Washington/ | UW Virology Lab | UW Virology Lab | Pavitra Roychoudhury et al |
| EPI_ISL_418914 | USA/WA-UW347/2020 | 2020-03-16 | North America/USA/Washington/ | UW Virology Lab | UW Virology Lab | Pavitra Roychoudhury et al |
| EPI_ISL_418940 | USA/WA-UW373/2020 | 2020-03-17 | North America/USA/Washington/ | UW Virology Lab | UW Virology Lab | Pavitra Roychoudhury et al |
| EPI_ISL_418948 | USA/WA-UW383/2020 | 2020-03-16 | North America/USA/Washington/ | UW Virology Lab | UW Virology Lab | Pavitra Roychoudhury et al |
| EPI_ISL_418990 | Hangzhou/ZJU-010/2020 | 2020-02-03 | Asia/China/Zhejiang/Hangzhou | State Key Laboratory for Diagnosis and Treatment of Infectious Diseases, National Clinical Research Center for Infectious Diseases, First Affiliated Hospital, Zhejiang University School of Medicine, Hangzhou, China 310003 | State Key Laboratory for Diagnosis and Treatment of Infectious Diseases, National Clinical Research Center for Infectious Diseases, First Affiliated Hospital, Zhejiang University School of Medicine, Hangzhou, China 310003 | Hangping Yao et al |
| EPI_ISL_418991 | Hangzhou/ZJU-011/2020 | 2020-02-04 | Asia/China/Zhejiang/Hangzhou | State Key Laboratory for Diagnosis and Treatment of Infectious Diseases, National Clinical Research Center for Infectious Diseases, First Affiliated Hospital, Zhejiang University School of Medicine, Hangzhou, China 310003 | State Key Laboratory for Diagnosis and Treatment of Infectious Diseases, National Clinical Research Center for Infectious Diseases, First Affiliated Hospital, Zhejiang University School of Medicine, Hangzhou, China 310003 | Hangping Yao et al |

| | | | | | | |
|---|---|---|---|---|---|---|
| EPI_ISL_418992 | Singapore/17/2020 | 2020-02-10 | Asia/Singapore/Singapore/Singapore | National Public Health Laboratory, National Centre for Infectious Diseases | National Public Health Laboratory, National Centre for Infectious Diseases | Mak TM et al |
| EPI_ISL_418993 | Singapore/20/2020 | 2020-01-29 | Asia/Singapore/Singapore/Singapore | National Public Health Laboratory, National Centre for Infectious Diseases | National Public Health Laboratory, National Centre for Infectious Diseases | Mak TM et al |
| EPI_ISL_418994 | Singapore/3Clin/2020 | 2020-01-29 | Asia/Singapore/Singapore/Singapore | National Public Health Laboratory, National Centre for Infectious Diseases | National Public Health Laboratory, National Centre for Infectious Diseases | Mak TM et al |
| EPI_ISL_418995 | Singapore/12Clin/2020 | 2020-02-28 | Asia/Singapore/Singapore/Singapore | National Public Health Laboratory, National Centre for Infectious Diseases | National Public Health Laboratory, National Centre for Infectious Diseases | Mak TM et al |
| EPI_ISL_418996 | Singapore/15/2020 | 2020-01-27 | Asia/Singapore/Singapore/Singapore | National Public Health Laboratory, National Centre for Infectious Diseases | National Public Health Laboratory, National Centre for Infectious Diseases | Mak TM et al |
| EPI_ISL_418997 | Singapore/16/2020 | 2020-02-06 | Asia/Singapore/Singapore/Singapore | National Public Health Laboratory, National Centre for Infectious Diseases | National Public Health Laboratory, National Centre for Infectious Diseases | Mak TM et al |

| | | | | | | |
|---|---|---|---|---|---|---|
| EPI_ISL_418998 | Singapore/14Clin/2020 | 2020-02-14 | Asia/Singapore/Singapore/Singapore | National Public Health Laboratory, National Centre for Infectious Diseases | National Public Health Laboratory, National Centre for Infectious Diseases | Mak TM et al |
| EPI_ISL_419000 | Singapore/21/2020 | 2020-02-13 | Asia/Singapore/Singapore/Singapore | National Public Health Laboratory, National Centre for Infectious Diseases | National Public Health Laboratory, National Centre for Infectious Diseases | Mak TM et al |
| EPI_ISL_419170 | France/ARA-11943/2020 | 2020-03-21 | Europe/France/Auvergne-Rhv¥ne-Alpes/ | Institut des Agents Infectieux (IAI), Hospices Civils de Lyon | CNR Virus des Infections Respiratoires - France SUD | Antonin Bal et al |
| EPI_ISL_419172 | France/ARA-11950/2020 | 2020-03-21 | Europe/France/Auvergne-Rhv¥ne-Alpes/ | Institut des Agents Infectieux (IAI), Hospices Civils de Lyon | CNR Virus des Infections Respiratoires - France SUD | Antonin Bal et al |
| EPI_ISL_419179 | France/ARA-12222/2020 | 2020-03-22 | Europe/France/Auvergne-Rhv¥ne-Alpes/ | Institut des Agents Infectieux (IAI), Hospices Civils de Lyon | CNR Virus des Infections Respiratoires - France SUD | Antonin Bal et al |
| EPI_ISL_419181 | France/ARA-12249/2020 | 2020-03-22 | Europe/France/Auvergne-Rhv¥ne-Alpes/ | Institut des Agents Infectieux (IAI), Hospices Civils de Lyon | CNR Virus des Infections Respiratoires - France SUD | Antonin Bal et al |
| EPI_ISL_419211 | Israel/ISR_JP0320/2020 | 2020-02-23 | Asia/Israel/Tel Aviv District/Ramat Gan | Central Virology Laboratory | Israel Institute for Biological Research | Inbar Cohen-Gihon et al |
| EPI_ISL_419213 | HongKong/HKPU38-3001/2020 | 2020-02-09 | Asia/Hong Kong/Hong Kong/Hong Kong | Department of Clinical Pathology, Pamela Youde Nethersole Eastern Hospital | Department of Health Technology and Informatics, Faculty of Health and Social Science, The Hong Kong Polytechnic University | Kenneth Siu-Sing LEUNG et al |

| | | | | | | |
|---|---|---|---|---|---|---|
| EPI_ISL_419214 | HongKong/HKPU39-3001/2020 | 2020-02-10 | Asia/Hong Kong/Hong Kong/Hong Kong | Department of Clinical Pathology, Pamela Youde Nethersole Eastern Hospital | Department of Health Technology and Informatics, Faculty of Health and Social Science, The Hong Kong Polytechnic University | Kenneth Siu-Sing LEUNG et al |
| EPI_ISL_419215 | HongKong/HKPU40-2801/2020 | 2020-02-10 | Asia/Hong Kong/Hong Kong/Hong Kong | Department of Clinical Pathology, Pamela Youde Nethersole Eastern Hospital | Department of Health Technology and Informatics, Faculty of Health and Social Science, The Hong Kong Polytechnic University | Kenneth Siu-Sing LEUNG et al |
| EPI_ISL_419216 | HongKong/HKPU41-0802/2020 | 2020-02-10 | Asia/Hong Kong/Hong Kong/Hong Kong | Department of Clinical Pathology, Pamela Youde Nethersole Eastern Hospital | Department of Health Technology and Informatics, Faculty of Health and Social Science, The Hong Kong Polytechnic University | Kenneth Siu-Sing LEUNG et al |
| EPI_ISL_419217 | HongKong/HKPU42-0302/2020 | 2020-02-10 | Asia/Hong Kong/Hong Kong/Hong Kong | Department of Pathology, Princess Margaret Hospital | Department of Health Technology and Informatics, Faculty of Health and Social Science, The Hong Kong Polytechnic University | Kenneth Siu-Sing LEUNG et al |

| | | | | | | |
|---|---|---|---|---|---|---|
| EPI_ISL_419218 | HongKong/HKPU43-0202/2020 | 2020-02-10 | Asia/Hong Kong/Hong Kong/Hong Kong | Department of Clinical Pathology, Pamela Youde Nethersole Eastern Hospital | Department of Health Technology and Informatics, Faculty of Health and Social Science, The Hong Kong Polytechnic University | Kenneth Siu-Sing LEUNG et al |
| EPI_ISL_419219 | HongKong/HKPU44-0102/2020 | 2020-02-10 | Asia/Hong Kong/Hong Kong/Hong Kong | Department of Clinical Pathology, Pamela Youde Nethersole Eastern Hospital | Department of Health Technology and Informatics, Faculty of Health and Social Science, The Hong Kong Polytechnic University | Kenneth Siu-Sing LEUNG et al |
| EPI_ISL_419221 | HongKong/HKPU45-0502/2020 | 2020-02-11 | Asia/Hong Kong/Hong Kong/Hong Kong | Department of Pathology, United Christian Hospital | Department of Health Technology and Informatics, Faculty of Health and Social Science, The Hong Kong Polytechnic University | Kenneth Siu-Sing LEUNG et al |
| EPI_ISL_419222 | HongKong/HKPU48-0202/2020 | 2020-02-11 | Asia/Hong Kong/Hong Kong/Hong Kong | Department of Pathology, Princess Margaret Hospital | Department of Health Technology and Informatics, Faculty of Health and Social Science, The Hong Kong Polytechnic University | Kenneth Siu-Sing LEUNG et al |

| | | | | | | |
|---|---|---|---|---|---|---|
| EPI_ISL_419223 | HongKong/HKPU49-3001/2020 | 2020-02-11 | Asia/Hong Kong/Hong Kong/Hong Kong | Department of Pathology, Princess Margaret Hospital | Department of Health Technology and Informatics, Faculty of Health and Social Science, The Hong Kong Polytechnic University | Kenneth Siu-Sing LEUNG et al |
| EPI_ISL_419224 | HongKong/HKPU52-3101/2020 | 2020-02-13 | Asia/Hong Kong/Hong Kong/Hong Kong | Department of Clinical Pathology, Pamela Youde Nethersole Eastern Hospital | Department of Health Technology and Informatics, Faculty of Health and Social Science, The Hong Kong Polytechnic University | Kenneth Siu-Sing LEUNG et al |
| EPI_ISL_419225 | HongKong/HKPU53-0802/2020 | 2020-02-13 | Asia/Hong Kong/Hong Kong/Hong Kong | Department of Clinical Pathology, Pamela Youde Nethersole Eastern Hospital | Department of Health Technology and Informatics, Faculty of Health and Social Science, The Hong Kong Polytechnic University | Kenneth Siu-Sing LEUNG et al |
| EPI_ISL_419226 | HongKong/HKPU54-0302/2020 | 2020-02-13 | Asia/Hong Kong/Hong Kong/Hong Kong | Department of Clinical Pathology, Pamela Youde Nethersole Eastern Hospital | Department of Health Technology and Informatics, Faculty of Health and Social Science, The Hong Kong Polytechnic University | Kenneth Siu-Sing LEUNG et al |

| | | | | | | |
|---|---|---|---|---|---|---|
| EPI_ISL_419227 | HongKong/HKPU57-0702/2020 | 2020-02-15 | Asia/Hong Kong/Hong Kong/Hong Kong | Department of Clinical Pathology, Pamela Youde Nethersole Eastern Hospital | Department of Health Technology and Informatics, Faculty of Health and Social Science, The Hong Kong Polytechnic University | Kenneth Siu-Sing LEUNG et al |
| EPI_ISL_419228 | HongKong/HKPU60-0802/2020 | 2020-02-17 | Asia/Hong Kong/Hong Kong/Hong Kong | Department of Clinical Pathology, Pamela Youde Nethersole Eastern Hospital | Department of Health Technology and Informatics, Faculty of Health and Social Science, The Hong Kong Polytechnic University | Kenneth Siu-Sing LEUNG et al |
| EPI_ISL_419229 | HongKong/HKPU61-0202/2020 | 2020-02-17 | Asia/Hong Kong/Hong Kong/Hong Kong | Department of Clinical Pathology, Pamela Youde Nethersole Eastern Hospital | Department of Health Technology and Informatics, Faculty of Health and Social Science, The Hong Kong Polytechnic University | Kenneth Siu-Sing LEUNG et al |
| EPI_ISL_419231 | HongKong/HKPU63-1402/2020 | 2020-02-19 | Asia/Hong Kong/Hong Kong/Hong Kong | Department of Clinical Pathology, Tuen Mun Hospital, 23 Tsing Chung Koon Road, Tuen Mun, N.T. | Department of Health Technology and Informatics, Faculty of Health and Social Science, The Hong Kong Polytechnic University | Kenneth Siu-Sing LEUNG et al |

| | | | | | | |
|---|---|---|---|---|---|---|
| EPI_ISL_419232 | HongKong/HKPU64-1202/2020 | 2020-02-19 | Asia/Hong Kong/Hong Kong/Hong Kong | Department of Clinical Pathology, Pamela Youde Nethersole Eastern Hospital | Department of Health Technology and Informatics, Faculty of Health and Social Science, The Hong Kong Polytechnic University | Kenneth Siu-Sing LEUNG et al |
| EPI_ISL_419237 | Spain/MD-ISCIII-201738/2020 | 2020-03-07 | Europe/Spain/Madrid/ | Fundacion Jimenez Diaz | Instituto de Salud Carlos III | Iglesias-Caballero et al |
| EPI_ISL_419240 | Spain/PV-ISCIII-201607/2020 | 2020-03-03 | Europe/Spain/Basque Country/ | HOSPITAL TXAGORRITXU | Instituto de Salud Carlos III | Iglesias-Caballero et al |
| EPI_ISL_419241 | HongKong/HKPU65-1202/2020 | 2020-02-19 | Asia/Hong Kong/Hong Kong/Hong Kong | Department of Clinical Pathology, Pamela Youde Nethersole Eastern Hospital | Department of Health Technology and Informatics, Faculty of Health and Social Science, The Hong Kong Polytechnic University | Kenneth Siu-Sing LEUNG et al |
| EPI_ISL_419242 | HongKong/HKPU66-2501/2020 | 2020-02-23 | Asia/Hong Kong/Hong Kong/Hong Kong | Department of Clinical Pathology, Tuen Mun Hospital | Department of Health Technology and Informatics, Faculty of Health and Social Science, The Hong Kong Polytechnic University | Kenneth Siu-Sing LEUNG et al |
| EPI_ISL_419243 | HongKong/HKPU70-1302/2020 | 2020-02-22 | Asia/Hong Kong/Hong Kong/Hong Kong | Department of Clinical Pathology, Pamela Youde Nethersole Eastern Hospital | Department of Health Technology and Informatics, Faculty of Health and Social Science, The Hong Kong Polytechnic University | Kenneth Siu-Sing LEUNG et al |

| | | | | | | |
|---|---|---|---|---|---|---|
| EPI_ISL_419244 | HongKong/HKPU73-0802/2020 | 2020-02-22 | Asia/Hong Kong/Hong Kong/Hong Kong | Department of Clinical Pathology, Pamela Youde Nethersole Eastern Hospital | Department of Health Technology and Informatics, Faculty of Health and Social Science, The Hong Kong Polytechnic University | Kenneth Siu-Sing LEUNG et al |
| EPI_ISL_419245 | HongKong/HKPU74-1302/2020 | 2020-02-22 | Asia/Hong Kong/Hong Kong/Hong Kong | Department of Clinical Pathology, Pamela Youde Nethersole Eastern Hospital | Department of Health Technology and Informatics, Faculty of Health and Social Science, The Hong Kong Polytechnic University | Kenneth Siu-Sing LEUNG et al |
| EPI_ISL_419246 | HongKong/HKPU76-0802/2020 | 2020-02-23 | Asia/Hong Kong/Hong Kong/Hong Kong | Department of Clinical Pathology, Pamela Youde Nethersole Eastern Hospital | Department of Health Technology and Informatics, Faculty of Health and Social Science, The Hong Kong Polytechnic University | Kenneth Siu-Sing LEUNG et al |
| EPI_ISL_419247 | HongKong/HKPU83-1702/2020 | 2020-02-24 | Asia/Hong Kong/Hong Kong/Hong Kong | Department of Clinical Pathology, Pamela Youde Nethersole Eastern Hospital | Department of Health Technology and Informatics, Faculty of Health and Social Science, The Hong Kong Polytechnic University | Kenneth Siu-Sing LEUNG et al |

| | | | | | | |
|---|---|---|---|---|---|---|
| EPI_ISL_419248 | HongKong/HKPU84-2502/2020 | 2020-02-24 | Asia/Hong Kong/Hong Kong/Hong Kong | Department of Clinical Pathology, Pamela Youde Nethersole Eastern Hospital | Department of Health Technology and Informatics, Faculty of Health and Social Science, The Hong Kong Polytechnic University | Kenneth Siu-Sing LEUNG et al |
| EPI_ISL_419249 | HongKong/HKPU86-1802/2020 | 2020-02-25 | Asia/Hong Kong/Hong Kong/Hong Kong | Department of Clinical Pathology, Pamela Youde Nethersole Eastern Hospital | Department of Health Technology and Informatics, Faculty of Health and Social Science, The Hong Kong Polytechnic University | Kenneth Siu-Sing LEUNG et al |
| EPI_ISL_419250 | HongKong/HKPU89-2502/2020 | 2020-02-26 | Asia/Hong Kong/Hong Kong/Hong Kong | Department of Clinical Pathology, Pamela Youde Nethersole Eastern Hospital | Department of Health Technology and Informatics, Faculty of Health and Social Science, The Hong Kong Polytechnic University | Kenneth Siu-Sing LEUNG et al |
| EPI_ISL_419251 | HongKong/HKPU90-1602/2020 | 2020-02-26 | Asia/Hong Kong/Hong Kong/Hong Kong | Department of Clinical Pathology, Pamela Youde Nethersole Eastern Hospital | Department of Health Technology and Informatics, Faculty of Health and Social Science, The Hong Kong Polytechnic University | Kenneth Siu-Sing LEUNG et al |

| | | | | | | |
|---|---|---|---|---|---|---|
| EPI_ISL_419252 | HongKong/HKPU91-2002/2020 | 2020-02-26 | Asia/Hong Kong/Hong Kong/Hong Kong | Department of Clinical Pathology, Pamela Youde Nethersole Eastern Hospital | Department of Health Technology and Informatics, Faculty of Health and Social Science, The Hong Kong Polytechnic University | Kenneth Siu-Sing LEUNG et al |
| EPI_ISL_419253 | HongKong/HKPU92-1302/2020 | 2020-02-26 | Asia/Hong Kong/Hong Kong/Hong Kong | Department of Pathology, United Christian Hospital | Department of Health Technology and Informatics, Faculty of Health and Social Science, The Hong Kong Polytechnic University | Kenneth Siu-Sing LEUNG et al |
| EPI_ISL_419262 | USA/VA-DCLS-00010/2020 | 2020-03-09 | North America/USA/Virginia/ | Virginia DCLS | Virginia DCLS | Virginia DCLS et al |
| EPI_ISL_419264 | Belgium/UGent-12/2020 | 2020-03-20 | Europe/Belgium/Gent/ | Lab voor klinische biologie | Onderzoeksgroep Virologie | Nick Vereecke et al |
| EPI_ISL_419303 | Japan/P4-3/2020 | 2020-03-12 | Asia/Japan/Saitama/ | Saitama Prefectural Institute of Public Health | Pathogen Genomics Center, National Institute of Infectious Diseases | Tsuyoshi Sekizuka et al |
| EPI_ISL_419413 | Wales/PHWC-24464/2020 | 2020-03-17 | Europe/United Kingdom/Wales/ | Wales Specialist Virology Centre | Public Health Wales Microbiology Cardiff | Catherine Moore et al |
| EPI_ISL_419421 | Wales/PHWC-244FB/2020 | 2020-03-18 | Europe/United Kingdom/Wales/ | Wales Specialist Virology Centre | Public Health Wales Microbiology Cardiff | Catherine Moore et al |
| EPI_ISL_419522 | USA/CT-Yale-029/2020 | 2020-03-18 | North America/USA/Connecticut/ | Yale Clinical Virology Laboratory | Grubaugh Lab - Yale School of Public Health | Joseph Fauver et al |

| | | | | | | |
|---|---|---|---|---|---|---|
| EPI_ISL_419544 | Germany/NW-HHU-27/2020 | 2020-03-15 | Europe/Germany/North Rhine Westphal | Center of Medical Microbiology, Virology, and Hospital Hygiene, University of Duesseldorf | Center of Medical Microbiology, Virology, and Hospital Hygiene, University of Duesseldorf | Ortwin Adams et al |
| EPI_ISL_419546 | Germany/NW-HHU-29/2020 | 2020-03-15 | Europe/Germany/North Rhine Westphal | Center of Medical Microbiology, Virology, and Hospital Hygiene, University of Duesseldorf | Center of Medical Microbiology, Virology, and Hospital Hygiene, University of Duesseldorf | Ortwin Adams et al |
| EPI_ISL_419549 | Germany/NW-HHU-32/2020 | 2020-03-15 | Europe/Germany/North Rhine Westphal | Center of Medical Microbiology, Virology, and Hospital Hygiene, University of Duesseldorf | Center of Medical Microbiology, Virology, and Hospital Hygiene, University of Duesseldorf | Ortwin Adams et al |
| EPI_ISL_419553 | USA/RI_0520/2020 | 2020-02-28 | North America/USA/Rhode Island/ | RI State Health Laboratories | Pathogen Discovery, Respiratory Viruses Branch, Division of Viral Diseases, Centers for Disease Control and Prevention | Ying Tao et al |
| EPI_ISL_419554 | USA/CA-CDC-2602/2020 | 2020-02-26 | North America/USA/California/ | California Department of Public Health | Pathogen Discovery, Respiratory Viruses Branch, Division of Viral Diseases, Centers for Disease Control and Prevention | Ying Tao et al |

| | | | | | | |
|---|---|---|---|---|---|---|
| EPI_ISL_419555 | USA/WA_5030/2020 | 2020-02-27 | North America/USA/Washington/ | WA State Department of Health | Pathogen Discovery, Respiratory Viruses Branch, Division of Viral Diseases, Centers for Disease Control and Prevention | Ying Tao et al |
| EPI_ISL_419556 | USA/GA_2741/2020 | 2020-02-29 | North America/USA/Georgia/ | GA Department of Public Health Laboratory | Pathogen Discovery, Respiratory Viruses Branch, Division of Viral Diseases, Centers for Disease Control and Prevention | Ying Tao et al |
| EPI_ISL_419557 | USA/GA_2742/2020 | 2020-02-29 | North America/USA/Georgia/ | GA Department of Public Health Laboratory | Pathogen Discovery, Respiratory Viruses Branch, Division of Viral Diseases, Centers for Disease Control and Prevention | Ying Tao et al |
| EPI_ISL_419558 | USA/OR_2656/2020 | 2020-02-27 | North America/USA/Oregon/ | OR State PHL-Virology/Immunology Section | Pathogen Discovery, Respiratory Viruses Branch, Division of Viral Diseases, Centers for Disease Control and Prevention | Ying Tao et al |

| | | | | | | |
|---|---|---|---|---|---|---|
| EPI_ISL_419559 | USA/FL_5125/2020 | 2020-02-28 | North America/USA/Florida/ | FL Bureau of Public Health Laboratories-Tampa | Pathogen Discovery, Respiratory Viruses Branch, Division of Viral Diseases, Centers for Disease Control and Prevention | Anna Uehara et al |
| EPI_ISL_419560 | USA/FL_5091/2020 | 2020-02-28 | North America/USA/Florida/ | FL Bureau of Public Health Laboratories-Tampa | Pathogen Discovery, Respiratory Viruses Branch, Division of Viral Diseases, Centers for Disease Control and Prevention | Anna Uehara et al |
| EPI_ISL_419562 | Luxembourg/LNS0000001/2020 | 2020-02-29 | Europe/Luxembourg/Luxembourg/Luxem | Laboratoire National de Sant√©, Microbiology, Virology | Laboratoire National de Sant√©, Microbiology, Epidemiology and Microbial Genomics | Anke Wienecke-Baldacchino et al |
| EPI_ISL_419583 | Luxembourg/LNS3588186/2020 | 2020-03-15 | Europe/Luxembourg/Luxembourg/Luxem | Laboratoire National de Sant√©, Microbiology, Virology | Laboratoire National de Sant√©, Microbiology, Epidemiology and Microbial Genomics | Anke Wienecke-Baldacchino et al |
| EPI_ISL_419592 | Luxembourg/LNS5440244/2020 | 2020-03-15 | Europe/Luxembourg/Luxembourg/Luxem | Laboratoire National de Sant√©, Microbiology, Virology | Laboratoire National de Sant√©, Microbiology, Epidemiology and Microbial Genomics | Anke Wienecke-Baldacchino et al |

| | | | | | | |
|---|---|---|---|---|---|---|
| EPI_ISL_419596 | Luxembourg/LNS7866283/2020 | 2020-03-12 | Europe/Luxembourg/Luxembourg/Luxem | Laboratoire National de Sant√©, Microbiology, Virology | Laboratoire National de Sant√©, Microbiology, Epidemiology and Microbial Genomics | Anke Wienecke-Baldacchino et al |
| EPI_ISL_419655 | Austria/CeMM0002/2020 | 2020-02-26 | Europe/Austria/Vienna/ | Center for Virology, Medical University of Vienna | Bergthaler laboratory, CeMM Research Center for Molecular Medicine of the Austrian Academy of Sciences | Alexandra Popa et al |
| EPI_ISL_419656 | Austria/CeMM0003/2020 | 2020-02-26 | Europe/Italy/Austria/ | Center for Virology, Medical University of Vienna | Bergthaler laboratory, CeMM Research Center for Molecular Medicine of the Austrian Academy of Sciences | Alexandra Popa et al |
| EPI_ISL_419657 | Austria/CeMM0004/2020 | 2020-03-03 | Europe/Iran/Austria/ | Center for Virology, Medical University of Vienna | Bergthaler laboratory, CeMM Research Center for Molecular Medicine of the Austrian Academy of Sciences | Alexandra Popa et al |
| EPI_ISL_419665 | Austria/CeMM0012/2020 | 2020-03-10 | Europe/Austria/Austria/ | Center for Virology, Medical University of Vienna | Bergthaler laboratory, CeMM Research Center for Molecular Medicine of the Austrian Academy of Sciences | Alexandra Popa et al |

| | | | | | | |
|---|---|---|---|---|---|---|
| EPI_ISL_419669 | Austria/CeMM0016/2020 | 2020-03-16 | Europe/Austria/Austria/ | Center for Virology, Medical University of Vienna | Bergthaler laboratory, CeMM Research Center for Molecular Medicine of the Austrian Academy of Sciences | Alexandra Popa et al |
| EPI_ISL_419672 | Austria/CeMM0019/2020 | 2020-03-19 | Europe/Austria/Austria/ | Center for Virology, Medical University of Vienna | Bergthaler laboratory, CeMM Research Center for Molecular Medicine of the Austrian Academy of Sciences | Alexandra Popa et al |
| EPI_ISL_419674 | Austria/CeMM0021/2020 | 2020-03-24 | Europe/Austria/Austria/ | Center for Virology, Medical University of Vienna | Bergthaler laboratory, CeMM Research Center for Molecular Medicine of the Austrian Academy of Sciences | Alexandra Popa et al |
| EPI_ISL_419682 | Spain/VC-FISABIO-18/2020 | 2020-03-10 | Europe/Spain/Comunitat Valenciana/Va | Servicio de Microbiolog√≠a. Consorcio Hospital General Universitario de Valencia | Sequencing and Bioinformatics Service and Molecular Epidemiology Research Group. FISABIO-Public Health | Giuseppe D'Auria et al |
| EPI_ISL_419686 | Spain/VC-FISABIO-22/2020 | 2020-03-09 | Europe/Spain/Comunitat Valenciana/Va | Servicio de Microbiolog√≠a. Consorcio Hospital General Universitario de Valencia | Sequencing and Bioinformatics Service and Molecular Epidemiology Research Group. FISABIO-Public Health | Maria Dolores Ocete et al |

| | | | | | | |
|---|---|---|---|---|---|---|
| EPI_ISL_419687 | Spain/VC-FISABIO-23/2020 | 2020-02-27 | Europe/Spain/Comunitat Valenciana/Va | Servicio de Microbiolog√≠a. Consorcio Hospital General Universitario de Valencia | Sequencing and Bioinformatics Service and Molecular Epidemiology Research Group. FISABIO-Public Health | Giuseppe D'Auria et al |
| EPI_ISL_419688 | Spain/VC-FISABIO-24/2020 | 2020-03-09 | Europe/Spain/Comunitat Valenciana/Va | Servicio de Microbiolog√≠a. Consorcio Hospital General Universitario de Valencia | Sequencing and Bioinformatics Service and Molecular Epidemiology Research Group. FISABIO-Public Health | Griselda De Marco et al |
| EPI_ISL_419689 | Spain/VC-FISABIO-25/2020 | 2020-02-27 | Europe/Spain/Comunitat Valenciana/Va | Servicio de Microbiolog√≠a. Consorcio Hospital General Universitario de Valencia | Sequencing and Bioinformatics Service and Molecular Epidemiology Research Group. FISABIO-Public Health | Neris Garcia-Gonzalez et al |
| EPI_ISL_419724 | Australia/VIC122/2020 | 2020-03-18 | Oceania/Australia/Victoria/ | Microbiological Diagnostic Unit Public Health Laboratory | Microbiological Diagnostic Unit Public Health Laboratory | Seemann T. et al |
| EPI_ISL_419734 | Australia/VIC14/2020 | 2020-02-02 | Oceania/Australia/Victoria/ | Victorian Infectious Diseases Reference Laboratory (VIDRL) | Victorian Infectious Diseases Reference Laboratory and Microbiological Diagnostic Unit Public Health Laboratory, Doherty Institute | Caly L. et al |

| | | | | | | |
|---|---|---|---|---|---|---|
| EPI_ISL_419735 | Australia/VIC15/2020 | 2020-02-24 | Oceania/Australia/Victoria/ | Victorian Infectious Diseases Reference Laboratory (VIDRL) | Victorian Infectious Diseases Reference Laboratory and Microbiological Diagnostic Unit Public Health Laboratory, Doherty Institute | Caly L. et al |
| EPI_ISL_419822 | Australia/VIC113/2020 | 2020-03-18 | Oceania/Australia/Victoria/ | Victorian Infectious Diseases Reference Laboratory (VIDRL) | Victorian Infectious Diseases Reference Laboratory and Microbiological Diagnostic Unit Public Health Laboratory, Doherty Institute | Caly L. et al |
| EPI_ISL_419830 | Australia/VIC137/2020 | 2020-03-22 | Oceania/Australia/Victoria/ | Microbiological Diagnostic Unit Public Health Laboratory | Microbiological Diagnostic Unit Public Health Laboratory | Seemann T. et al |
| EPI_ISL_419932 | Australia/VIC237/2020 | 2020-03-21 | Oceania/Australia/Victoria/ | Victorian Infectious Diseases Reference Laboratory (VIDRL) | Victorian Infectious Diseases Reference Laboratory and Microbiological Diagnostic Unit Public Health Laboratory, Doherty Institute | Caly L. et al |

| | | | | | | |
|---|---|---|---|---|---|---|
| EPI_ISL_419943 | Australia/VIC248/2020 | 2020-03-21 | Oceania/Australia/Victoria/ | Victorian Infectious Diseases Reference Laboratory (VIDRL) | Victorian Infectious Diseases Reference Laboratory and Microbiological Diagnostic Unit Public Health Laboratory, Doherty Institute | Caly L. et al |
| EPI_ISL_419951 | Australia/VIC258/2020 | 2020-03-21 | Oceania/Australia/Victoria/ | Victorian Infectious Diseases Reference Laboratory (VIDRL) | Victorian Infectious Diseases Reference Laboratory and Microbiological Diagnostic Unit Public Health Laboratory, Doherty Institute | Caly L. et al |
| EPI_ISL_420019 | USA/VA-DCLS-0033/2020 | 2020-03-19 | North America/USA/Virginia/ | Virginia DCLS | Virginia DCLS | Virginia DCLS et al |
| EPI_ISL_420025 | USA/VA-DCLS-0044/2020 | 2020-03-27 | North America/USA/Virginia/ | Virginia DCLS | Virginia DCLS | Virginia DCLS et al |
| EPI_ISL_420043 | France/IDF-2792/2020 | 2020-03-18 | Europe/France/Ile de France/Paris | CMIP | National Reference Center for Viruses of Respiratory Infections, Institut Pasteur, Paris | M√©lanie Albert et al |
| EPI_ISL_420048 | France/IDF-2936/2020 | 2020-03-18 | Europe/France/Ile de France/Longjumea | Service de Biologie M√©dicale - BP 125 | National Reference Center for Viruses of Respiratory Infections, Institut Pasteur, Paris | M√©lanie Albert et al |
| EPI_ISL_420050 | France/HDF-2948/2020 | 2020-03-19 | Europe/France/Hauts de France/Compi√ | CH Compi√®gne Laboratoire de Biologie | National Reference Center for Viruses of Respiratory Infections, Institut Pasteur, Paris | M√©lanie Albert et al |

| | | | | | | |
|---|---|---|---|---|---|---|
| EPI_ISL_420056 | France/HDF-3138/2020 | 2020-03-22 | Europe/France/Hauts de France/Compiv | CH Compiv®gne Laboratoire de Biologie | National Reference Center for Viruses of Respiratory Infections, Institut Pasteur, Paris | Mv©lanie Albert et al |
| EPI_ISL_420063 | France/IDF-3230/2020 | 2020-03-22 | Europe/France/Ile de France/Juvisy-sur- | Labo BM - Site de Juvisy - Hopital Gv©nv©ral | National Reference Center for Viruses of Respiratory Infections, Institut Pasteur, Paris | Mv©lanie Albert et al |
| EPI_ISL_420112 | Spain/VC-FISABIO-27/2020 | 2020-03-09 | Europe/Spain/Comunitat Valenciana/Va | Servicio de Microbiologv≠a. Consorcio Hospital General Universitario de Valencia | Sequencing and Bioinformatics Service and Molecular Epidemiology Research Group. FISABIO-Public Health | Lidia Ruiz Roldan et al |
| EPI_ISL_420114 | Spain/VC-FISABIO-29/2020 | 2020-03-09 | Europe/Spain/Comunitat Valenciana/Va | Servicio de Microbiologv≠a. Consorcio Hospital General Universitario de Valencia | Sequencing and Bioinformatics Service and Molecular Epidemiology Research Group. FISABIO-Public Health | Griselda De Marco et al |
| EPI_ISL_420115 | Spain/VC-FISABIO-30/2020 | 2020-03-09 | Europe/Spain/Comunitat Valenciana/Va | Servicio de Microbiologv≠a. Consorcio Hospital General Universitario de Valencia | Sequencing and Bioinformatics Service and Molecular Epidemiology Research Group. FISABIO-Public Health | Marta Pla Diaz et al |

| | | | | | | |
|---|---|---|---|---|---|---|
| EPI_ISL_420116 | Spain/VC-FISABIO-31/2020 | 2020-03-09 | Europe/Spain/Comunitat Valenciana/Va | Servicio de Microbiolog√≠a. Consorcio Hospital General Universitario de Valencia | Sequencing and Bioinformatics Service and Molecular Epidemiology Research Group. FISABIO-Public Health | Neris Garcia-Gonzalez et al |
| EPI_ISL_420118 | Spain/VC-FISABIO-33/2020 | 2020-03-09 | Europe/Spain/Comunitat Valenciana/Va | Servicio de Microbiolog√≠a. Consorcio Hospital General Universitario de Valencia | Sequencing and Bioinformatics Service and Molecular Epidemiology Research Group. FISABIO-Public Health | Inma Galv°n Vendrell et al |
| EPI_ISL_420119 | Spain/VC-FISABIO-34/2020 | 2020-03-09 | Europe/Spain/Comunitat Valenciana/Va | Servicio de Microbiolog√≠a. Consorcio Hospital General Universitario de Valencia | Sequencing and Bioinformatics Service and Molecular Epidemiology Research Group. FISABIO-Public Health | Paula Ruiz-Hueso et al |
| EPI_ISL_420127 | Spain/VC-FISABIO-43/2020 | 2020-03-12 | Europe/Spain/Comunitat Valenciana/Va | Servicio de Microbiologia. Hospital Clinico Universitario de Valencia | Sequencing and Bioinformatics Service and Molecular Epidemiology Research Group. FISABIO-Public Health | Maria Alma Bracho et al |
| EPI_ISL_420128 | Spain/VC-FISABIO-44/2020 | 2020-03-12 | Europe/Spain/Comunitat Valenciana/Va | Servicio de Microbiologia. Hospital Clinico Universitario de Valencia | Sequencing and Bioinformatics Service and Molecular Epidemiology Research Group. FISABIO-Public Health | Griselda De Marco et al |

| | | | | | | |
|---|---|---|---|---|---|---|
| EPI_ISL_420134 | Norway/1495/2020 | 2020-02-28 | Europe/Italy/Norway/ | Akershus University Hospital, Department for Microbiology and Infectious Disease Control | Norwegian Institute of Public Health, Department of Virology | Kathrine Stene-Johansen et al |
| EPI_ISL_420135 | Norway/1455/2020 | 2020-02-28 | Europe/Norway/Norway/ | Oslo University Hospital, Department of Medical Microbiology | Norwegian Institute of Public Health, Department of Virology | Kathrine Stene-Johansen et al |
| EPI_ISL_420136 | Norway/1694/2020 | 2020-03-04 | Europe/Norway/Norway/ | Akershus University Hospital, Department for Microbiology and Infectious Disease Control | Norwegian Institute of Public Health, Department of Virology | Kathrine Stene-Johansen et al |
| EPI_ISL_420137 | Norway/1772/2020 | 2020-03-08 | Europe/Norway/Norway/ | Vestfold Hospital, Tonsberg Department of Microbiology | Norwegian Institute of Public Health, Department of Virology | Kathrine Stene-Johansen et al |
| EPI_ISL_420138 | Norway/1781/2020 | 2020-03-05 | Europe/Norway/Norway/ | University Hospital of Northern Norway, Department for Microbiology and Infectious Disease Control | Norwegian Institute of Public Health, Department of Virology | Kathrine Stene-Johansen et al |
| EPI_ISL_420143 | Norway/1917/2020 | 2020-03-10 | Europe/Norway/Norway/ | Unilabs Laboratory Medicine | Norwegian Institute of Public Health, Department of Virology | Kathrine Stene-Johansen et al |
| EPI_ISL_420146 | Norway/1955/2020 | 2020-03-11 | Europe/Norway/Norway/ | Furst Medical Laboratory | Norwegian Institute of Public Health, Department of Virology | Kathrine Stene-Johansen et al |
| EPI_ISL_420151 | Norway/2087/2020 | 2020-03-17 | Europe/Norway/Norway/ | Nordland Hospital - Bodo, Laboratory Department, Molecular Biology Unit | Norwegian Institute of Public Health, Department of Virology | Kathrine Stene-Johansen et al |

| | | | | | | |
|---|---|---|---|---|---|---|
| EPI_ISL_420152 | Norway/2088/2020 | 2020-03-17 | Europe/Norway/Norway/ | University Hospital of Northern Norway, Department for Microbiology and Infectious Disease Control | Norwegian Institute of Public Health, Department of Virology | Kathrine Stene-Johansen et al |
| EPI_ISL_420153 | Norway/2090/2020 | 2020-03-16 | Europe/Norway/Norway/ | University Hospital of Northern Norway, Department for Microbiology and Infectious Disease Control | Norwegian Institute of Public Health, Department of Virology | Kathrine Stene-Johansen et al |
| EPI_ISL_420167 | England/SHEF-C0804/2020 | 2020-03-25 | Europe/United Kingdom/England/South | Virology Department, Sheffield Teaching Hospitals NHS Foundation Trust | Department of Infection, Immunity and Cardiovascular Disease, The Florey Institute, The Medical School, University of Sheffield | Thushan de Silva et al |
| EPI_ISL_420169 | England/SHEF-C0822/2020 | 2020-03-25 | Europe/United Kingdom/England/South | Virology Department, Sheffield Teaching Hospitals NHS Foundation Trust | Department of Infection, Immunity and Cardiovascular Disease, The Florey Institute, The Medical School, University of Sheffield | Thushan de Silva et al |
| EPI_ISL_420186 | England/SHEF-C0479/2020 | 2020-03-27 | Europe/United Kingdom/England/South | Virology Department, Sheffield Teaching Hospitals NHS Foundation Trust | Department of Infection, Immunity and Cardiovascular Disease, The Florey Institute, The Medical School, University of Sheffield | Thushan de Silva et al |

| | | | | | | |
|---|---|---|---|---|---|---|
| EPI_ISL_420296 | USA/NY-NYUMC45/2020 | 2020-03-18 | North America/USA/New York/Nassau C | NYU Langone Health | Departments of Pathology and Medicine, New York University School of Medicine | Maria Aguero-Rosenfeld et al |
| EPI_ISL_420306 | USA/AK-PHL015/2020 | 2020-03-16 | North America/USA/Alaska/Southeast A | Alaska State Virology Laboratory | Alaska State Virology Laboratory | Chen et al |
| EPI_ISL_420309 | USA/NY-NYUMC54/2020 | 2020-03-19 | North America/USA/New York/Manhatt | NYU Langone Health | Departments of Pathology and Medicine, New York University School of Medicine | Maria Aguero-Rosenfeld et al |
| EPI_ISL_420321 | Belgium/WWM-030665/2020 | 2020-03-06 | Europe/Belgium/Nivelles/Rixensart | KU Leuven, Clinical and Epidemiological Virology | KU Leuven, Clinical and Epidemiological Virology | Joan Marti-Carreras et al |
| EPI_ISL_420323 | Belgium/BJ-030767/2020 | 2020-03-07 | Europe/Belgium/Nivelles/Waterloo | KU Leuven, Clinical and Epidemiological Virology | KU Leuven, Clinical and Epidemiological Virology | Joan Marti-Carreras et al |
| EPI_ISL_420332 | Belgium/WAM-030676/2020 | 2020-03-06 | Europe/Belgium/Li√®ge/Herstal | KU Leuven, Clinical and Epidemiological Virology | KU Leuven, Clinical and Epidemiological Virology | Joan Marti-Carreras et al |
| EPI_ISL_420343 | Belgium/BM-030687/2020 | 2020-03-06 | Europe/Belgium/Namur/Andenne | KU Leuven, Clinical and Epidemiological Virology | KU Leuven, Clinical and Epidemiological Virology | Joan Marti-Carreras et al |
| EPI_ISL_420358 | Belgium/VD-0318102/2020 | 2020-03-18 | Europe/Belgium/Mons/Honnelles | KU Leuven, Clinical and Epidemiological Virology | KU Leuven, Clinical and Epidemiological Virology | Joan Marti-Carreras et al |
| EPI_ISL_420363 | Belgium/VNP-0320107/2020 | 2020-03-20 | Europe/Belgium/Brussels/Watermael-Bo | KU Leuven, Clinical and Epidemiological Virology | KU Leuven, Clinical and Epidemiological Virology | Joan Marti-Carreras et al |

| | | | | | | |
|---|---|---|---|---|---|---|
| EPI_ISL_420364 | Belgium/HP-0319108/2020 | 2020-03-19 | Europe/Belgium/Brussels/Anderlecht | KU Leuven, Clinical and Epidemiological Virology | KU Leuven, Clinical and Epidemiological Virology | Joan Marti-Carreras et al |
| EPI_ISL_420370 | Belgium/SC-0319113/2020 | 2020-03-19 | Europe/Belgium/Namur/ | KU Leuven, Clinical and Epidemiological Virology | KU Leuven, Clinical and Epidemiological Virology | Joan Marti-Carreras et al |
| EPI_ISL_420372 | Belgium/LPDC-0323115/2020 | 2020-03-23 | Europe/Belgium/Brussels/Forest | KU Leuven, Clinical and Epidemiological Virology | KU Leuven, Clinical and Epidemiological Virology | Joan Marti-Carreras et al |
| EPI_ISL_420378 | Belgium/MSE-0324121/2020 | 2020-03-24 | Europe/Belgium/Mons/Frameries | KU Leuven, Clinical and Epidemiological Virology | KU Leuven, Clinical and Epidemiological Virology | Joan Marti-Carreras et al |
| EPI_ISL_420402 | Belgium/GJJ-0325145/2020 | 2020-03-25 | Europe/Belgium/Hasselt/Zolder | KU Leuven, Clinical and Epidemiological Virology | KU Leuven, Clinical and Epidemiological Virology | Joan Marti-Carreras et al |
| EPI_ISL_420424 | Belgium/CCM-0324167/2020 | 2020-03-24 | Europe/Belgium/Mons/ | KU Leuven, Clinical and Epidemiological Virology | KU Leuven, Clinical and Epidemiological Virology | Joan Marti-Carreras et al |
| EPI_ISL_420439 | Belgium/PC-0325182/2020 | 2020-03-25 | Europe/Belgium/Mons/Quaregnon | KU Leuven, Clinical and Epidemiological Virology | KU Leuven, Clinical and Epidemiological Virology | Joan Marti-Carreras et al |
| EPI_ISL_420447 | Belgium/DJ-030452/2020 | 2020-03-04 | Europe/Belgium/Dendermonde/ | KU Leuven, Clinical and Epidemiological Virology | KU Leuven, Clinical and Epidemiological Virology | Joan Marti-Carreras et al |
| EPI_ISL_420448 | Belgium/GSA-030454/2020 | 2020-03-04 | Europe/Belgium/Leuven/Lubbeek | KU Leuven, Clinical and Epidemiological Virology | KU Leuven, Clinical and Epidemiological Virology | Joan Marti-Carreras et al |
| EPI_ISL_420456 | Australia/WA11/2020 | 2020-02-22 | Oceania/Australia/Western Australia/P | PathWest Laboratory Medicine WA | PathWest Laboratory Medicine WA | Chisha Sikazwe et al |

| | | | | | | |
|---|---|---|---|---|---|---|
| EPI_ISL_420547 | India/UN-772/2020 | 2020-03-03 | Asia/Italy/India/ | National Influenza Center, Indian Council of Medical Research - National Institute of Virology | Indian Council of Medical Research-National Institute of Virology, Microbial Containment Complex | Pragya D. Yadav. Savita Patil et al |
| EPI_ISL_420553 | India/UN-781/2020 | 2020-03-03 | Asia/Italy/India/ | National Influenza Center, Indian Council of Medical Research - National Institute of Virology | Indian Council of Medical Research-National Institute of Virology, Microbial Containment Complex | Pragya D. Yadav. Savita Patil et al |
| EPI_ISL_420563 | Italy/ABR-IZSGC-TE5166/2020 | 2020-03-18 | Europe/Italy/Abruzzo/Teramo | Ospedale Civile Giuseppe Mazzini | "Istituto Zooprofilattico Sperimentale dell'Abruzzo e Molise ""G. Caporale""" | Lorusso A et al |
| EPI_ISL_420564 | Italy/ABR-IZSGC-TE5472/2020 | 2020-03-19 | Europe/Italy/Abruzzo/L'Aquila | Ospedale Civile Castel Di Sangro | "Istituto Zooprofilattico Sperimentale dell'Abruzzo e Molise ""G.Caporale""" | Lorusso A et al |
| EPI_ISL_420566 | Italy/ABR-IZSGC-TE5512/2020 | 2020-03-19 | Europe/Italy/Abruzzo/ | Ospedale Regionale San Salvatore | "Istituto Zooprofilattico Sperimentale dell'Abruzzo e Molise ""G.Caporale""" | Lorusso A et al |
| EPI_ISL_420569 | Italy/ABR-IZSGC-TE6195/2020 | 2020-03-23 | Europe/Italy/Abruzzo/ | Ospedale Civile Giuseppe Mazzini | "Istituto Zooprofilattico Sperimentale dell'Abruzzo e Molise ""G.Caporale""" | Lorusso A et al |

| | | | | | | |
|---|---|---|---|---|---|---|
| EPI_ISL_420573 | USA/NY-NYUMC58/2020 | 2020-03-18 | North America/USA/New York/Manhatt | NYU Langone Health | Departments of Pathology and Medicine, New York University School of Medicine | Maria Aguero-Rosenfeld et al |
| EPI_ISL_420575 | USA/NY-NYUMC60/2020 | 2020-03-18 | North America/USA/New York/Nassau ( | NYU Langone Health | Departments of Pathology and Medicine, New York University School of Medicine | Maria Aguero-Rosenfeld et al |
| EPI_ISL_420576 | USA/NY-NYUMC61/2020 | 2020-03-18 | North America/USA/New York/ | NYU Langone Health | Departments of Pathology and Medicine, New York University School of Medicine | Maria Aguero-Rosenfeld et al |
| EPI_ISL_420581 | USA/NY-NYUMC66/2020 | 2020-03-18 | North America/USA/New York/Manhatt | NYU Langone Health | Departments of Pathology and Medicine, New York University School of Medicine | Maria Aguero-Rosenfeld et al |
| EPI_ISL_420583 | Italy/ABR-IZSGC-TE6222/2020 | 2020-03-23 | Europe/Italy/Abruzzo/ | Ospedale Civile Giuseppe Mazzini | "Istituto Zooprofilattico Sperimentale dell'Abruzzo e Molise ""G.Caporale""" | Lorusso A et al |
| EPI_ISL_420585 | USA/NY-NYUMC69/2020 | 2020-03-18 | North America/USA/New York/Manhatt | NYU Langone Health | Departments of Pathology and Medicine, New York University School of Medicine | Maria Aguero-Rosenfeld et al |

| | | | | | | |
|---|---|---|---|---|---|---|
| EPI_ISL_420590 | USA/NY-NYUMC74/2020 | 2020-03-18 | North America/USA/New York/Nassau ( | NYU Langone Health | Departments of Pathology and Medicine, New York University School of Medicine | Maria Aguero-Rosenfeld et al |
| EPI_ISL_420604 | France/ARA-12371/2020 | 2020-03-23 | Europe/France/Auvergne-Rh√¥ne-Alpes/ | Institut des Agents Infectieux (IAI), Hospices Civils de Lyon | CNR Virus des Infections Respiratoires - France SUD | Antonin Bal et al |
| EPI_ISL_420615 | France/ARA-12759/2020 | 2020-03-23 | Europe/France/Auvergne-Rh√¥ne-Alpes/ | Institut des Agents Infectieux (IAI), Hospices Civils de Lyon | CNR Virus des Infections Respiratoires - France SUD | Antonin Bal et al |
| EPI_ISL_420617 | France/ARA-12877/2020 | 2020-03-23 | Europe/France/Auvergne-Rh√¥ne-Alpes/ | Centre Hospitalier Saint Joseph Saint Luc | CNR Virus des Infections Respiratoires - France SUD | Antonin Bal et al |
| EPI_ISL_420682 | England/20139044404/2020 | 2020-03-29 | Europe/United Kingdom/England/ | Respiratory Virus Unit, Microbiology Services Colindale, Public Health England | Respiratory Virus Unit, Microbiology Services Colindale, Public Health England | Monica Galiano et al |
| EPI_ISL_420724 | England/20139058304/2020 | 2020-03-27 | Europe/United Kingdom/England/ | Respiratory Virus Unit, Microbiology Services Colindale, Public Health England | Respiratory Virus Unit, Microbiology Services Colindale, Public Health England | Monica Galiano et al |
| EPI_ISL_420752 | England/20140039004/2020 | 2020-03-26 | Europe/United Kingdom/England/ | Respiratory Virus Unit, Microbiology Services Colindale, Public Health England | Respiratory Virus Unit, Microbiology Services Colindale, Public Health England | Monica Galiano et al |

| | | | | | | |
|---|---|---|---|---|---|---|
| EPI_ISL_420785 | USA/FL_6318/2020 | 2020-03-02 | North America/USA/Florida/ | FL Bureau of Health Laboratories Tampa | Pathogen Discovery, Respiratory Viruses Branch, Division of Viral Diseases, Centers for Disease Control and Prevention | Krista Queen et al |
| EPI_ISL_420789 | USA/IL_1375/2020 | 2020-03-01 | North America/USA/Illinois/ | Illinois Department of Public Health Chicago Laboratory | Pathogen Discovery, Respiratory Viruses Branch, Division of Viral Diseases, Centers for Disease Control and Prevention | Krista Queen et al |
| EPI_ISL_420791 | USA/NH_0004/2020 | 2020-02-29 | North America/USA/New Hampshire/ | NH Department of Health and Human Services Public Health Labs | Pathogen Discovery, Respiratory Viruses Branch, Division of Viral Diseases, Centers for Disease Control and Prevention | Krista Queen et al |
| EPI_ISL_420796 | USA/TX_2039/2020 | 2020-02-29 | North America/USA/Texas/ | Texas DSHS Lab Services | Pathogen Discovery, Respiratory Viruses Branch, Division of Viral Diseases, Centers for Disease Control and Prevention | Krista Queen et al |

| | | | | | | |
|---|---|---|---|---|---|---|
| EPI_ISL_420799 | SouthKorea/BA-ACH_2604/2020 | 2020-02-27 | Asia/South Korea/South Korea/ | Brian D. Allgood Army Community Hospital | Pathogen Discovery, Respiratory Viruses Branch, Division of Viral Diseases, Centers for Disease Control and Prevention | Krista Queen et al |
| EPI_ISL_420800 | SouthKorea/BA-ACH_2718/2020 | 2020-02-29 | Asia/South Korea/South Korea/ | Brian D. Allgood Army Community Hospital | Pathogen Discovery, Respiratory Viruses Branch, Division of Viral Diseases, Centers for Disease Control and Prevention | Krista Queen et al |
| EPI_ISL_420801 | SouthKorea/BA-ACH_2719/2020 | 2020-02-29 | Asia/South Korea/South Korea/ | Brian D. Allgood Army Community Hospital | Pathogen Discovery, Respiratory Viruses Branch, Division of Viral Diseases, Centers for Disease Control and Prevention | Krista Queen et al |
| EPI_ISL_420806 | USA/UT-0284/2020 | 2020-03-14 | North America/USA/Utah/ | Utah Public Health Laboratory | Utah Public Health Laboratory | Erin Young et al |
| EPI_ISL_420817 | USA/UT-0344/2020 | 2020-03-24 | North America/USA/Utah/ | Utah Public Health Laboratory | Utah Public Health Laboratory | Erin Young et al |
| EPI_ISL_420889 | Japan/Hu_DP_Kng_19-031/2020 | 2020-02-14 | Asia/Japan/Japan/ | Takayuki Hishiki Kanagawa Prefectural Institute of Public Health | Takayuki Hishiki Kanagawa Prefectural Institute of Public Health | Hishiki et al |

| | | | | | | |
|---|---|---|---|---|---|---|
| EPI_ISL_420901 | Germany/BY-MVP-V2010840/2020 | 2020-03-12 | Europe/Germany/Bavaria/Munich | Max von Pettenkofer Institute, Virology, National Reference Center for Retroviruses, LMU Munich | Laboratory for Functional Genome Analysis, Dept. Genomics, Gene Center of the LMU Munich | Max Muenchhoff et al |
| EPI_ISL_420903 | Germany/BY-MVP-V2011395/2020 | 2020-03-14 | Europe/Germany/Bavaria/Munich | Max von Pettenkofer Institute, Virology, National Reference Center for Retroviruses, LMU Munich | Laboratory for Functional Genome Analysis, Dept. Genomics, Gene Center of the LMU Munich | Max Muenchhoff et al |
| EPI_ISL_420905 | Germany/BY-MVP-V2011672/2020 | 2020-03-16 | Europe/Germany/Bavaria/Munich | Max von Pettenkofer Institute, Virology, National Reference Center for Retroviruses, LMU Munich | Laboratory for Functional Genome Analysis, Dept. Genomics, Gene Center of the LMU Munich | Max Muenchhoff et al |
| EPI_ISL_420950 | Wales/PHWC-24CE4/2020 | 2020-03-22 | Europe/United Kingdom/Wales/ | Wales Specialist Virology Centre | Public Health Wales Microbiology Cardiff | Catherine Moore et al |
| EPI_ISL_421011 | Wales/PHWC-255F7/2020 | 2020-03-24 | Europe/United Kingdom/Wales/ | Wales Specialist Virology Centre | Public Health Wales Microbiology Cardiff | Catherine Moore et al |
| EPI_ISL_421171 | Spain/MD-H12-1301/2020 | 2020-03-05 | Europe/Spain/Madrid/ | Hospital Universitario 12 de Octubre | Hospital Universitario 12 de Octubre | Esther Viedma et al |
| EPI_ISL_421172 | Spain/MD-H12-H12-1502/2020 | 2020-03-09 | Europe/Spain/Madrid/ | Hospital Universitario 12 de Octubre | Hospital Universitario 12 de Octubre | Esther Viedma et al |
| EPI_ISL_421184 | Belgium/ULG-10004/2020 | 2020-03-31 | Europe/Belgium/Liv®ge/ | Department of Clinical Microbiology | GIGA Medical Genomics | Keith Durkin et al |
| EPI_ISL_421186 | Belgium/ULG-10007/2020 | 2020-03-30 | Europe/Belgium/Liv®ge/ | Department of Clinical Microbiology | GIGA Medical Genomics | Keith Durkin et al |
| EPI_ISL_421194 | Belgium/ULG-10016/2020 | 2020-03-30 | Europe/Belgium/Liv®ge/ | Department of Clinical Microbiology | GIGA Medical Genomics | Keith Durkin et al |

| | | | | | | |
|---|---|---|---|---|---|---|
| EPI_ISL_421223 | Hangzhou/HZCDC0167/2020 | 2020-01-23 | Asia/China/Zhejiang/Hangzhou | Hangzhou Center for Diseases Control and Prevention | Hangzhou Center for Diseases Control and Prevention | Jun Li et al |
| EPI_ISL_421224 | Hangzhou/HZCDC0162/2020 | 2020-01-23 | Asia/China/Zhejiang/Hangzhou | Hangzhou Center for Diseases Control and Prevention | Hangzhou Center for Diseases Control and Prevention | Jun Li et al |
| EPI_ISL_421225 | Hangzhou/HZCDC0135/2020 | 2020-01-22 | Asia/China/Zhejiang/Hangzhou | Hangzhou Center for Diseases Control and Prevention | Hangzhou Center for Diseases Control and Prevention | Jun Li et al |
| EPI_ISL_421226 | Hangzhou/HZCDC0119/2020 | 2020-01-22 | Asia/China/Zhejiang/Hangzhou | Hangzhou Center for Diseases Control and Prevention | Hangzhou Center for Diseases Control and Prevention | Jun Li et al |
| EPI_ISL_421227 | Hangzhou/HZCDC0091L/2020 | 2020-01-21 | Asia/China/Zhejiang/Hangzhou | Hangzhou Center for Diseases Control and Prevention | Hangzhou Center for Diseases Control and Prevention | Jun Li et al |
| EPI_ISL_421228 | Hangzhou/HZCDC0091/2020 | 2020-01-21 | Asia/China/Zhejiang/Hangzhou | Hangzhou Center for Diseases Control and Prevention | Hangzhou Center for Diseases Control and Prevention | Jun Li et al |
| EPI_ISL_421229 | Hangzhou/HZCDC0090L/2020 | 2020-01-21 | Asia/China/Zhejiang/Hangzhou | Hangzhou Center for Diseases Control and Prevention | Hangzhou Center for Diseases Control and Prevention | Jun Li et al |
| EPI_ISL_421230 | Hangzhou/HZCDC0090/2020 | 2020-01-21 | Asia/China/Zhejiang/Hangzhou | Hangzhou Center for Diseases Control and Prevention | Hangzhou Center for Diseases Control and Prevention | Jun Li et al |
| EPI_ISL_421231 | Hangzhou/HZCDC0049L/2020 | 2020-01-21 | Asia/China/Zhejiang/Hangzhou | Hangzhou Center for Diseases Control and Prevention | Hangzhou Center for Diseases Control and Prevention | Jun Li et al |
| EPI_ISL_421232 | Hangzhou/HZCDC0048L/2020 | 2020-01-21 | Asia/China/Zhejiang/Hangzhou | Hangzhou Center for Diseases Control and Prevention | Hangzhou Center for Diseases Control and Prevention | Jun Li et al |

| | | | | | | |
|---|---|---|---|---|---|---|
| EPI_ISL_421233 | Hangzhou/HZCDC0048/2020 | 2020-01-21 | Asia/China/Zhejiang/Hangzhou | Hangzhou Center for Diseases Control and Prevention | Hangzhou Center for Diseases Control and Prevention | Jun Li et al |
| EPI_ISL_421234 | Hangzhou/HZCDC0025/2020 | 2020-01-21 | Asia/China/Zhejiang/Hangzhou | Hangzhou Center for Diseases Control and Prevention | Hangzhou Center for Diseases Control and Prevention | Jun Li et al |
| EPI_ISL_421235 | Hangzhou/HZCDC0013/2020 | 2020-01-20 | Asia/China/Zhejiang/Hangzhou | Hangzhou Center for Diseases Control and Prevention | Hangzhou Center for Diseases Control and Prevention | Jun Li et al |
| EPI_ISL_421236 | Hangzhou/HZCDC0012/2020 | 2020-01-20 | Asia/China/Zhejiang/Hangzhou | Hangzhou Center for Diseases Control and Prevention | Hangzhou Center for Diseases Control and Prevention | Jun Li et al |
| EPI_ISL_421237 | Jiujiang/JX22/2020 | 2020-01-22 | Asia/China/Jiangxi/Jiujiang | Jiangxi Province Center for Disease Control and Prevention | Jiangxi Province Center for Disease Control and Prevention | JianXiong Li et al |
| EPI_ISL_421238 | Nanchang/JX174/2020 | 2020-01-29 | Asia/China/Jiangxi/Nanchang | Jiangxi Province Center for Disease Control and Prevention | Jiangxi Province Center for Disease Control and Prevention | JianXiong Li et al |
| EPI_ISL_421239 | Shangrao/JX105/2020 | 2020-01-26 | Asia/China/Jiangxi/Shangrao | Jiangxi Province Center for Disease Control and Prevention | Jiangxi Province Center for Disease Control and Prevention | JianXiong Li et al |
| EPI_ISL_421240 | Shangrao/JX1948/2020 | 2020-02-05 | Asia/China/Jiangxi/Shangrao | Jiangxi Province Center for Disease Control and Prevention | Jiangxi Province Center for Disease Control and Prevention | JianXiong Li et al |
| EPI_ISL_421241 | Nanchang/JX177/2020 | 2020-01-30 | Asia/China/Jiangxi/Nanchang | Jiangxi Province Center for Disease Control and Prevention | Jiangxi Province Center for Disease Control and Prevention | JianXiong Li et al |
| EPI_ISL_421242 | Ganzhou/JX81/2020 | 2020-01-25 | Asia/China/Jiangxi/Ganzhou | Jiangxi Province Center for Disease Control and Prevention | Jiangxi Province Center for Disease Control and Prevention | JianXiong Li et al |

| | | | | | | |
|---|---|---|---|---|---|---|
| EPI_ISL_421243 | Nanchang/JX14/2020 | 2020-01-21 | Asia/China/Jiangxi/Nanchang | Jiangxi Province Center for Disease Control and Prevention | Jiangxi Province Center for Disease Control and Prevention | JianXiong Li et al |
| EPI_ISL_421244 | Shangrao/JX29/2020 | 2020-01-22 | Asia/China/Jiangxi/Shangrao | Jiangxi Province Center for Disease Control and Prevention | Jiangxi Province Center for Disease Control and Prevention | JianXiong Li et al |
| EPI_ISL_421245 | Nanchang/JX90/2020 | 2020-01-25 | Asia/China/Jiangxi/Nanchang | Jiangxi Province Center for Disease Control and Prevention | Jiangxi Province Center for Disease Control and Prevention | JianXiong Li et al |
| EPI_ISL_421246 | Nanchang/JXN3T4/2020 | 2020-02-26 | Asia/China/Jiangxi/Nanchang | Jiangxi Province Center for Disease Control and Prevention | Jiangxi Province Center for Disease Control and Prevention | JianXiong Li et al |
| EPI_ISL_421247 | Nanchang/JX149/2020 | 2020-01-27 | Asia/China/Jiangxi/Nanchang | Jiangxi Province Center for Disease Control and Prevention | Jiangxi Province Center for Disease Control and Prevention | JianXiong Li et al |
| EPI_ISL_421248 | Nanchang/JX39/2020 | 2020-01-23 | Asia/China/Jiangxi/Nanchang | Jiangxi Province Center for Disease Control and Prevention | Jiangxi Province Center for Disease Control and Prevention | JianXiong Li et al |
| EPI_ISL_421249 | Xinyu/JX122/2020 | 2020-01-25 | Asia/China/Jiangxi/Xinyu | Jiangxi Province Center for Disease Control and Prevention | Jiangxi Province Center for Disease Control and Prevention | JianXiong Li et al |
| EPI_ISL_421250 | Shangrao/JX1177/2020 | 2020-02-03 | Asia/China/Jiangxi/Shangrao | Jiangxi Province Center for Disease Control and Prevention | Jiangxi Province Center for Disease Control and Prevention | JianXiong Li et al |
| EPI_ISL_421251 | Shangrao/JX1176/2020 | 2020-02-03 | Asia/China/Jiangxi/Shangrao | Jiangxi Province Center for Disease Control and Prevention | Jiangxi Province Center for Disease Control and Prevention | JianXiong Li et al |
| EPI_ISL_421252 | Pingxiang/JX5/2020 | 2020-01-11 | Asia/China/Jiangxi/Pingxiang | Jiangxi Province Center for Disease Control and Prevention | Jiangxi Province Center for Disease Control and Prevention | JianXiong Li et al |

| | | | | | | |
|---|---|---|---|---|---|---|
| EPI_ISL_421253 | Jian/JX129/2020 | 2020-01-26 | Asia/China/Jiangxi/Jian | Jiangxi Province Center for Disease Control and Prevention | Jiangxi Province Center for Disease Control and Prevention | JianXiong Li et al |
| EPI_ISL_421254 | Shangrao/JX1178/2020 | 2020-02-03 | Asia/China/Jiangxi/Shangrao | Jiangxi Province Center for Disease Control and Prevention | Jiangxi Province Center for Disease Control and Prevention | JianXiong Li et al |
| EPI_ISL_421256 | Jian/JX169/2020 | 2020-01-27 | Asia/China/Jiangxi/Jian | Jiangxi Province Center for Disease Control and Prevention | Jiangxi Province Center for Disease Control and Prevention | JianXiong Li et al |
| EPI_ISL_421257 | Shangrao/JX1215/2020 | 2020-02-02 | Asia/China/Jiangxi/Shangrao | Jiangxi Province Center for Disease Control and Prevention | Jiangxi Province Center for Disease Control and Prevention | JianXiong Li et al |
| EPI_ISL_421258 | Shangrao/JX1974/2020 | 2020-02-08 | Asia/China/Jiangxi/Shangrao | Jiangxi Province Center for Disease Control and Prevention | Jiangxi Province Center for Disease Control and Prevention | JianXiong Li et al |
| EPI_ISL_421259 | Pingxiang/JX151/2020 | 2020-01-27 | Asia/China/Jiangxi/Pingxiang | Jiangxi Province Center for Disease Control and Prevention | Jiangxi Province Center for Disease Control and Prevention | JianXiong Li et al |
| EPI_ISL_421260 | Xinyu/JX124/2020 | 2020-01-26 | Asia/China/Jiangxi/Xinyu | Jiangxi Province Center for Disease Control and Prevention | Jiangxi Province Center for Disease Control and Prevention | JianXiong Li et al |
| EPI_ISL_421261 | Nanchang/JX176/2020 | 2020-01-29 | Asia/China/Jiangxi/Nanchang | Jiangxi Province Center for Disease Control and Prevention | Jiangxi Province Center for Disease Control and Prevention | JianXiong Li et al |
| EPI_ISL_421262 | Nanchang/JX155/2020 | 2020-01-29 | Asia/China/Jiangxi/Nanchang | Jiangxi Province Center for Disease Control and Prevention | Jiangxi Province Center for Disease Control and Prevention | JianXiong Li et al |

| | | | | | | |
|---|---|---|---|---|---|---|
| EPI_ISL_421281 | USA/IN-Lilly-IPB0170-4bc21/2020 | 2020-03-19 | North America/USA/Indiana/Marion Co | Clinical Diagnostics Laboratory, Diagnostic & Experimental Pathology, Lilly Research Laboratories | Clinical Diagnostics Laboratory, Diagnostic & Experimental Pathology, Lilly Research Laboratories | Tim Holzer et al |
| EPI_ISL_421283 | USA/WI-UW-25/2020 | 2020-03-23 | North America/USA/Wisconsin/Dane Co | University of Wisconsin-Madison AIDS Vaccine Research Laboratories | University of Wisconsin-Madison AIDS Vaccine Research Laboratories | Gage Moreno et al |
| EPI_ISL_421310 | USA/WI-UW-52/2020 | 2020-03-18 | North America/USA/Wisconsin/Dane Co | University of Wisconsin-Madison AIDS Vaccine Research Laboratories | University of Wisconsin-Madison AIDS Vaccine Research Laboratories | Gage Moreno et al |
| EPI_ISL_421315 | USA/WI-UW-57/2020 | 2020-03-19 | North America/USA/Wisconsin/Lafayett | University of Wisconsin-Madison AIDS Vaccine Research Laboratories | University of Wisconsin-Madison AIDS Vaccine Research Laboratories | Gage Moreno et al |
| EPI_ISL_421320 | USA/WI-UW-62/2020 | 2020-03-25 | North America/USA/Wisconsin/Dane Co | University of Wisconsin-Madison AIDS Vaccine Research Laboratories | University of Wisconsin-Madison AIDS Vaccine Research Laboratories | Gage Moreno et al |
| EPI_ISL_421323 | USA/WI-UW-65/2020 | 2020-03-22 | North America/USA/Wisconsin/Dane Co | University of Wisconsin-Madison AIDS Vaccine Research Laboratories | University of Wisconsin-Madison AIDS Vaccine Research Laboratories | Gage Moreno et al |
| EPI_ISL_421363 | USA/NY-PV08143/2020 | 2020-03-17 | North America/USA/New York/Westche | MSHS Clinical Microbiology Laboratories | MSHS Pathogen Surveillance Program | Ana S. Gonzalez-Reiche et al |
| EPI_ISL_421387 | USA/NY-PV08429/2020 | 2020-03-19 | North America/USA/New York/Manhat | MSHS Clinical Microbiology Laboratories | MSHS Pathogen Surveillance Program | Ana S. Gonzalez-Reiche et al |

| | | | | | | |
|---|---|---|---|---|---|---|
| EPI_ISL_421390 | USA/NY-PV08435/2020 | 2020-03-18 | North America/USA/New York/Manhatt | MSHS Clinical Microbiology Laboratories | MSHS Pathogen Surveillance Program | Ana S. Gonzalez-Reiche et al |
| EPI_ISL_421394 | USA/NY-PV08443/2020 | 2020-03-17 | North America/USA/New York/Manhatt | MSHS Clinical Microbiology Laboratories | MSHS Pathogen Surveillance Program | Ana S. Gonzalez-Reiche et al |
| EPI_ISL_421395 | USA/NY-PV08444/2020 | 2020-03-17 | North America/USA/New York/Manhatt | MSHS Clinical Microbiology Laboratories | MSHS Pathogen Surveillance Program | Ana S. Gonzalez-Reiche et al |
| EPI_ISL_421397 | USA/NY-PV08446/2020 | 2020-03-17 | North America/USA/New York/Manhatt | MSHS Clinical Microbiology Laboratories | MSHS Pathogen Surveillance Program | Ana S. Gonzalez-Reiche et al |
| EPI_ISL_421402 | USA/NY-PV08452/2020 | 2020-03-18 | North America/USA/New York/Manhatt | MSHS Clinical Microbiology Laboratories | MSHS Pathogen Surveillance Program | Ana S. Gonzalez-Reiche et al |
| EPI_ISL_421409 | USA/NY-PV08463/2020 | 2020-03-18 | North America/USA/New York/Manhatt | MSHS Clinical Microbiology Laboratories | MSHS Pathogen Surveillance Program | Ana S. Gonzalez-Reiche et al |
| EPI_ISL_421412 | USA/NY-PV08468/2020 | 2020-03-18 | North America/USA/New York/Manhatt | MSHS Clinical Microbiology Laboratories | MSHS Pathogen Surveillance Program | Ana S. Gonzalez-Reiche et al |
| EPI_ISL_421416 | USA/NY-PV08473/2020 | 2020-03-18 | North America/USA/New York/Manhatt | MSHS Clinical Microbiology Laboratories | MSHS Pathogen Surveillance Program | Ana S. Gonzalez-Reiche et al |
| EPI_ISL_421457 | Portugal/PT0054/2020 | 2020-03-18 | Europe/Portugal/Portugal/ | H Dr Nelio Mendonca - Funchal | Instituto Nacional de Saude (INSA) | Guiomar et al et al |
| EPI_ISL_421460 | Portugal/PT0057/2020 | 2020-03-20 | Europe/Portugal/Portugal/ | H Dr. Nelio Mendonca - Funchal | Instituto Nacional de Saude (INSA) | Guiomar et al et al |
| EPI_ISL_421469 | Portugal/PT0066/2020 | 2020-03-21 | Europe/Portugal/Portugal/ | H Santarem | Instituto Nacional de Saude (INSA) | Guiomar et al et al |
| EPI_ISL_421493 | Portugal/PT0090/2020 | 2020-03-21 | Europe/Portugal/Portugal/ | HSE Ilha Terceira - Angra do Heroismo | Instituto Nacional de Saude (INSA) | Guiomar et al et al |
| EPI_ISL_421501 | France/IDF-2359/2020 | 2020-03-12 | Europe/France/Ile de France/Longjumea | Service de Biologie M√©dicale - BP 125 | National Reference Center for Viruses of Respiratory Infections, Institut Pasteur, Paris | M√©lanie Albert et al |

| | | | | | | |
|---|---|---|---|---|---|---|
| EPI_ISL_421503 | France/IDF-2414/2020 | 2020-03-12 | Europe/France/Ile de France/Saint Gern | Parc des Dames | National Reference Center for Viruses of Respiratory Infections, Institut Pasteur, Paris | M√©lanie Albert et al |
| EPI_ISL_421513 | France/IDF-3345/2020 | 2020-03-23 | Europe/France/Ile de France/Orsay | Service de Biologie clinique | National Reference Center for Viruses of Respiratory Infections, Institut Pasteur, Paris | M√©lanie Albert et al |
| EPI_ISL_421518 | Spain/VC-FISABIO-48/2020 | 2020-03-17 | Europe/Spain/Comunitat Valenciana/Va | Servicio de Microbiologia. Hospital Clinico Universitario de Valencia | Sequencing and Bioinformatics Service and Molecular Epidemiology Research Group. FISABIO-Public Health | Inma Galv°n Vendrell et al |
| EPI_ISL_421519 | Spain/VC-FISABIO-52/2020 | 2020-03-21 | Europe/Spain/Comunitat Valenciana/Va | Servicio de Microbiologia. Hospital Clinico Universitario de Valencia | Sequencing and Bioinformatics Service and Molecular Epidemiology Research Group. FISABIO-Public Health | David Navarro et al |
| EPI_ISL_421548 | USA/WY-WYPHL011/2020 | 2020-03-14 | North America/USA/Wyoming/ | Wyoming Public Health Laboratory | Center for Global Health, University of New Mexico Health Sciences Center | Daryl Domman et al |
| EPI_ISL_421553 | USA/WY-WYPHL019/2020 | 2020-03-19 | North America/USA/Wyoming/ | Wyoming Public Health Laboratory | Center for Global Health, University of New Mexico Health Sciences Center | Daryl Domman et al |
| EPI_ISL_421561 | USA/UT-055/2020 | 2020-03-17 | North America/USA/Utah/ | Utah Public Health Laboratory | Utah Public Health Laboratory | Erin Young et al |

| | | | | | | |
|---|---|---|---|---|---|---|
| EPI_ISL_421575 | SouthAfrica/KRISP-011/2020 | 2020-04-01 | Africa/South Africa/KwaZulu-Natal/ | Molecular Diagnostic Services | KRISP, KZN Research Innovation and Sequencing Platform | Giandhari J et al |
| EPI_ISL_421590 | USA/NY-NYUMC89/2020 | 2020-03-18 | North America/USA/New York/Queens | NYU Langone Health | Departments of Pathology and Medicine, New York University School of Medicine | Maria Aguero-Rosenfeld et al |
| EPI_ISL_421598 | USA/NY-PV09025/2020 | 2020-03-21 | North America/USA/New York/Manhatt | MSHS Clinical Microbiology Laboratories | MSHS Pathogen Surveillance Program | Ana S. Gonzalez-Reiche et al |
| EPI_ISL_421604 | USA/NY-PV09039/2020 | 2020-03-21 | North America/USA/New York/Bronx | MSHS Clinical Microbiology Laboratories | MSHS Pathogen Surveillance Program | Ana S. Gonzalez-Reiche et al |
| EPI_ISL_421608 | USA/NY-PV09046/2020 | 2020-03-21 | North America/USA/New York/Nassau C | MSHS Clinical Microbiology Laboratories | MSHS Pathogen Surveillance Program | Ana S. Gonzalez-Reiche et al |
| EPI_ISL_421613 | USA/NY-PV09061/2020 | 2020-03-20 | North America/USA/New York/Bronx | MSHS Clinical Microbiology Laboratories | MSHS Pathogen Surveillance Program | Ana S. Gonzalez-Reiche et al |
| EPI_ISL_421662 | India/UN-1073/2020 | 2020-03-10 | Asia/Iran/India/ | National Influenza Center, Indian Council of Medical Research - National Institute of Virology | Indian Council of Medical Research-National Institute of Virology, Microbial Containment Complex | Pragya D. Yadav et al |
| EPI_ISL_421663 | India/UN-1093/2020 | 2020-03-10 | Asia/Iran/India/ | National Influenza Center, Indian Council of Medical Research - National Institute of Virology | Indian Council of Medical Research-National Institute of Virology, Microbial Containment Complex | Pragya D. Yadav et al |

| | | | | | | |
|---|---|---|---|---|---|---|
| EPI_ISL_421664 | India/UN-1100/2020 | 2020-03-10 | Asia/Iran/India/ | National Influenza Center, Indian Council of Medical Research - National Institute of Virology | Indian Council of Medical Research-National Institute of Virology, Microbial Containment Complex | Pragya D. Yadav et al |
| EPI_ISL_421665 | India/UN-1104/2020 | 2020-03-10 | Asia/Iran/India/ | National Influenza Center, Indian Council of Medical Research - National Institute of Virology | Indian Council of Medical Research-National Institute of Virology, Microbial Containment Complex | Pragya D. Yadav et al |
| EPI_ISL_421666 | India/UN-1111/2020 | 2020-03-10 | Asia/Iran/India/ | National Influenza Center, Indian Council of Medical Research - National Institute of Virology | Indian Council of Medical Research-National Institute of Virology, Microbial Containment Complex | Pragya D. Yadav et al |
| EPI_ISL_421667 | India/UN-1115/2020 | 2020-03-10 | Asia/Iran/India/ | National Influenza Center, Indian Council of Medical Research - National Institute of Virology | Indian Council of Medical Research-National Institute of Virology, Microbial Containment Complex | Pragya D. Yadav et al |
| EPI_ISL_421668 | India/UN-1125/2020 | 2020-03-10 | Asia/Iran/India/ | National Influenza Center, Indian Council of Medical Research - National Institute of Virology | Indian Council of Medical Research-National Institute of Virology, Microbial Containment Complex | Pragya D. Yadav et al |

| | | | | | | |
|---|---|---|---|---|---|---|
| EPI_ISL_421669 | India/UN-1616/2020 | 2020-03-12 | Asia/Iran/India/ | National Influenza Center, Indian Council of Medical Research - National Institute of Virology | Indian Council of Medical Research-National Institute of Virology, Microbial Containment Complex | Pragya D. Yadav et al |
| EPI_ISL_421670 | India/UN-1617/2020 | 2020-03-12 | Asia/Iran/India/ | National Influenza Center, Indian Council of Medical Research - National Institute of Virology | Indian Council of Medical Research-National Institute of Virology, Microbial Containment Complex | Pragya D. Yadav et al |
| EPI_ISL_421671 | India/UN-1621/2020 | 2020-03-12 | Asia/Iran/India/ | National Influenza Center, Indian Council of Medical Research - National Institute of Virology | Indian Council of Medical Research-National Institute of Virology, Microbial Containment Complex | Pragya D. Yadav et al |
| EPI_ISL_421672 | India/UN-1644/2020 | 2020-03-12 | Asia/Iran/India/ | National Influenza Center, Indian Council of Medical Research - National Institute of Virology | Indian Council of Medical Research-National Institute of Virology, Microbial Containment Complex | Pragya D. Yadav et al |
| EPI_ISL_421706 | USA/NY-NYUMC94/2020 | 2020-03-18 | North America/USA/New York/Queens | NYU Langone Health | Departments of Pathology and Medicine, New York University School of Medicine | Maria Aguero-Rosenfeld et al |
| EPI_ISL_421731 | USA/NY-NYUMC119/2020 | 2020-03-18 | North America/USA/New York/Brooklyn | NYU Langone Health | Departments of Pathology and Medicine, New York University School of Medicine | Maria Aguero-Rosenfeld et al |

| | | | | | | |
|---|---|---|---|---|---|---|
| EPI_ISL_421738 | Luxembourg/LNS3404815/2020 | 2020-03-17 | Europe/Luxembourg/Luxembourg/ | Laboratoire National de Sante, Microbiology, Virology | Laboratoire National de Sante, Microbiology, Epidemiology and Microbial Genomics | Anke Wienecke-Baldacchino et al |
| EPI_ISL_421743 | Luxembourg/LNS0272441/2020 | 2020-03-17 | Europe/Luxembourg/Luxembourg/ | Laboratoire National de Sante, Microbiology, Virology | Laboratoire National de Sante, Microbiology, Epidemiology and Microbial Genomics | Anke Wienecke-Baldacchino et al |
| EPI_ISL_421748 | Luxembourg/LNS3052946/2020 | 2020-03-17 | Europe/Luxembourg/Luxembourg/ | Laboratoire National de Sante, Microbiology, Virology | Laboratoire National de Sante, Microbiology, Epidemiology and Microbial Genomics | Anke Wienecke-Baldacchino et al |
| EPI_ISL_421799 | England/201380035/2020 | 2020-03-26 | Europe/United Kingdom/England/ | Respiratory Virus Unit, Microbiology Services Colindale, Public Health England | Respiratory Virus Unit, Microbiology Services Colindale, Public Health England | Monica Galiano et al |
| EPI_ISL_421870 | England/20139009704/2020 | 2020-03-27 | Europe/United Kingdom/England/ | Respiratory Virus Unit, Microbiology Services Colindale, Public Health England | Respiratory Virus Unit, Microbiology Services Colindale, Public Health England | Monica Galiano et al |
| EPI_ISL_421904 | England/20139027404/2020 | 2020-03-26 | Europe/United Kingdom/England/ | Respiratory Virus Unit, Microbiology Services Colindale, Public Health England | Respiratory Virus Unit, Microbiology Services Colindale, Public Health England | Monica Galiano et al |

| | | | | | | |
|---|---|---|---|---|---|---|
| EPI_ISL_421942 | England/20144020004/2020 | 2020-03-31 | Europe/United Kingdom/England/ | Respiratory Virus Unit, Microbiology Services Colindale, Public Health England | Respiratory Virus Unit, Microbiology Services Colindale, Public Health England | Monica Galiano et al |
| EPI_ISL_421971 | England/20146006704/2020 | 2020-04-01 | Europe/United Kingdom/England/ | Respiratory Virus Unit, Microbiology Services Colindale, Public Health England | Respiratory Virus Unit, Microbiology Services Colindale, Public Health England | Monica Galiano et al |
| EPI_ISL_421993 | England/20146064104/2020 | 2020-03-31 | Europe/United Kingdom/England/ | Respiratory Virus Unit, Microbiology Services Colindale, Public Health England | Respiratory Virus Unit, Microbiology Services Colindale, Public Health England | Monica Galiano et al |
| EPI_ISL_422085 | Wales/PHWC-25FAE/2020 | 2020-03-25 | Europe/United Kingdom/Wales/ | Wales Specialist Virology Centre | Public Health Wales Microbiology Cardiff | Catherine Moore et al |
| EPI_ISL_422103 | Wales/PHWC-26C00/2020 | 2020-03-29 | Europe/United Kingdom/Wales/ | Wales Specialist Virology Centre | Public Health Wales Microbiology Cardiff | Catherine Moore et al |
| EPI_ISL_422130 | Wales/PHWC-26A9D/2020 | 2020-03-30 | Europe/United Kingdom/Wales/ | Wales Specialist Virology Centre | Public Health Wales Microbiology Cardiff | Catherine Moore et al |
| EPI_ISL_422191 | Wales/PHWC-26480/2020 | 2020-03-27 | Europe/United Kingdom/Wales/ | Wales Specialist Virology Centre | Public Health Wales Microbiology Cardiff | Catherine Moore et al |
| EPI_ISL_422226 | Wales/PHWC-25F80/2020 | 2020-03-25 | Europe/United Kingdom/Wales/ | Wales Specialist Virology Centre | Public Health Wales Microbiology Cardiff | Catherine Moore et al |

| | | | | | | |
|---|---|---|---|---|---|---|
| EPI_ISL_422293 | Wales/PHWC-26990/2020 | 2020-03-29 | Europe/United Kingdom/Wales/ | Wales Specialist Virology Centre | Public Health Wales Microbiology Cardiff | Catherine Moore et al |
| EPI_ISL_422314 | Wales/PHWC-25858/2020 | 2020-03-25 | Europe/United Kingdom/Wales/ | Wales Specialist Virology Centre | Public Health Wales Microbiology Cardiff | Catherine Moore et al |
| EPI_ISL_422319 | Wales/PHWC-264CC/2020 | 2020-03-28 | Europe/United Kingdom/Wales/ | Wales Specialist Virology Centre | Public Health Wales Microbiology Cardiff | Catherine Moore et al |
| EPI_ISL_422424 | Kuwait/KU006/2020 | 2020-03-15 | Asia/Italy/Kuwait/Dasman | Jaber Al Ahmad Al Sabah Hospital | Dasman diabetes Institute | Fahd Al-Mulla et al |
| EPI_ISL_422425 | Zhejiang/HZ103/2020 | 2020-01-24 | Asia/China/Zhejiang/ | Zhejiang Provincial Center for Disease Control and Prevention | Zhejiang Provincial Center for Disease Control and Prevention | Yanjun Zhang et al |
| EPI_ISL_422428 | Singapore/35/2020 | 2020-02-06 | Asia/Singapore/Singapore/ | National Public Health Laboratory, National Centre for Infectious Diseases | National Public Health Laboratory, National Centre for Infectious Diseases | Mak TM et al |
| EPI_ISL_422429 | Singapore/36/2020 | 2020-02-16 | Asia/Singapore/Singapore/ | National Public Health Laboratory, National Centre for Infectious Diseases | National Public Health Laboratory, National Centre for Infectious Diseases | Mak TM et al |
| EPI_ISL_422437 | Italy/VEN-IZSVe-21-26/2020 | 2020-03-25 | Europe/Italy/Veneto/Verona IT | ULSS9 Distretto di Bussolengo | Istituto Zooprofilattico Sperimentale delle Venezie | Adelaide Milani et al |
| EPI_ISL_422516 | USA/NY-PV09145/2020 | 2020-03-20 | North America/USA/New York/Queens | MSHS Clinical Microbiology Laboratories | MSHS Pathogen Surveillance Program | Ana S. Gonzalez-Reiche et al |

| | | | | | | |
|---|---|---|---|---|---|---|
| EPI_ISL_422530 | USA/NY-PV09168/2020 | 2020-03-22 | North America/USA/New York/Manhatt | MSHS Clinical Microbiology Laboratories | MSHS Pathogen Surveillance Program | Ana S. Gonzalez-Reiche et al |
| EPI_ISL_422543 | USA/NY-PV09186/2020 | 2020-03-20 | North America/USA/New York/Bronx | MSHS Clinical Microbiology Laboratories | MSHS Pathogen Surveillance Program | Ana S. Gonzalez-Reiche et al |
| EPI_ISL_422572 | Netherlands/Friesland_7/2020 | 2020-03-28 | Europe/Netherlands/Friesland/ | Dutch COVID-19 response team | Erasmus Medical Center | Bas Oude Munnink et al |
| EPI_ISL_422578 | Netherlands/NA_267/2020 | 2020-03-30 | Europe/Netherlands/Netherlands/ | Dutch COVID-19 response team | Erasmus Medical Center | Bas Oude Munnink et al |
| EPI_ISL_422582 | Netherlands/NA_271/2020 | 2020-03-30 | Europe/Netherlands/Netherlands/ | Dutch COVID-19 response team | Erasmus Medical Center | Bas Oude Munnink et al |
| EPI_ISL_422588 | Netherlands/NA_277/2020 | 2020-03-31 | Europe/Netherlands/Netherlands/ | Dutch COVID-19 response team | Erasmus Medical Center | Bas Oude Munnink et al |
| EPI_ISL_422632 | Netherlands/ZuidHolland_96/2020 | 2020-03-30 | Europe/Netherlands/South Holland/ | Dutch COVID-19 response team | Erasmus Medical Center | Bas Oude Munnink et al |
| EPI_ISL_422638 | Netherlands/Gelderland_10/2020 | 2020-03-20 | Europe/Netherlands/Gelderland/ | Dutch COVID-19 response team | Erasmus Medical Center | Bas Oude Munnink et al |
| EPI_ISL_422643 | Netherlands/Gelderland_7/2020 | 2020-03-11 | Europe/Netherlands/Gelderland/ | Dutch COVID-19 response team | Erasmus Medical Center | Bas Oude Munnink et al |
| EPI_ISL_422665 | Netherlands/NA_118/2020 | 2020-03-16 | Europe/Netherlands/Netherlands/ | Dutch COVID-19 response team | Erasmus Medical Center | Bas Oude Munnink et al |
| EPI_ISL_422671 | Netherlands/NA_127/2020 | 2020-03-17 | Europe/Netherlands/Netherlands/ | Dutch COVID-19 response team | Erasmus Medical Center | Bas Oude Munnink et al |
| EPI_ISL_422695 | Netherlands/NA_160/2020 | 2020-03-21 | Europe/Netherlands/Netherlands/ | Dutch COVID-19 response team | Erasmus Medical Center | Bas Oude Munnink et al |
| EPI_ISL_422710 | Netherlands/NA_175/2020 | 2020-03-22 | Europe/Netherlands/Netherlands/ | Dutch COVID-19 response team | Erasmus Medical Center | Bas Oude Munnink et al |
| EPI_ISL_422712 | Netherlands/NA_177/2020 | 2020-03-22 | Europe/Netherlands/Netherlands/ | Dutch COVID-19 response team | Erasmus Medical Center | Bas Oude Munnink et al |
| EPI_ISL_422723 | Netherlands/NA_188/2020 | 2020-03-23 | Europe/Netherlands/Netherlands/ | Dutch COVID-19 response team | Erasmus Medical Center | Bas Oude Munnink et al |
| EPI_ISL_422730 | Netherlands/NA_195/2020 | 2020-03-24 | Europe/Netherlands/Netherlands/ | Dutch COVID-19 response team | Erasmus Medical Center | Bas Oude Munnink et al |
| EPI_ISL_422739 | Netherlands/NA_204/2020 | 2020-03-25 | Europe/Netherlands/Netherlands/ | Dutch COVID-19 response team | Erasmus Medical Center | Bas Oude Munnink et al |
| EPI_ISL_422755 | Netherlands/NA_220/2020 | 2020-03-26 | Europe/Netherlands/Netherlands/ | Dutch COVID-19 response team | Erasmus Medical Center | Bas Oude Munnink et al |
| EPI_ISL_422771 | Netherlands/NA_237/2020 | 2020-03-27 | Europe/Netherlands/Netherlands/ | Dutch COVID-19 response team | Erasmus Medical Center | Bas Oude Munnink et al |

| | | | | | | |
|---|---|---|---|---|---|---|
| EPI_ISL_422776 | Netherlands/NA_242/2020 | 2020-03-28 | Europe/Netherlands/Netherlands/ | Dutch COVID-19 response team | Erasmus Medical Center | Bas Oude Munnink et al |
| EPI_ISL_422778 | Netherlands/NA_244/2020 | 2020-03-29 | Europe/Netherlands/Netherlands/ | Dutch COVID-19 response team | Erasmus Medical Center | Bas Oude Munnink et al |
| EPI_ISL_422791 | Netherlands/NA_257/2020 | 2020-03-30 | Europe/Netherlands/Netherlands/ | Dutch COVID-19 response team | Erasmus Medical Center | Bas Oude Munnink et al |
| EPI_ISL_422799 | Netherlands/NA_38/2020 | 2020-03-13 | Europe/Netherlands/Netherlands/ | Dutch COVID-19 response team | Erasmus Medical Center | Bas Oude Munnink et al |
| EPI_ISL_422802 | Netherlands/NA_41/2020 | 2020-03-13 | Europe/Netherlands/Netherlands/ | Dutch COVID-19 response team | Erasmus Medical Center | Bas Oude Munnink et al |
| EPI_ISL_422812 | Netherlands/NA_51/2020 | 2020-03-14 | Europe/Netherlands/Netherlands/ | Dutch COVID-19 response team | Erasmus Medical Center | Bas Oude Munnink et al |
| EPI_ISL_422821 | Netherlands/NA_60/2020 | 2020-03-09 | Europe/Netherlands/Netherlands/ | Dutch COVID-19 response team | Erasmus Medical Center | Bas Oude Munnink et al |
| EPI_ISL_422855 | Netherlands/NA_95/2020 | 2020-03-17 | Europe/Netherlands/Netherlands/ | Dutch COVID-19 response team | Erasmus Medical Center | Bas Oude Munnink et al |
| EPI_ISL_422861 | Netherlands/NoordBrabant_102/202 | 2020-03-21 | Europe/Netherlands/North Brabant/ | Dutch COVID-19 response team | Erasmus Medical Center | Bas Oude Munnink et al |
| EPI_ISL_422876 | Netherlands/NoordBrabant_81/2020 | 2020-03-11 | Europe/Netherlands/North Brabant/ | Dutch COVID-19 response team | Erasmus Medical Center | Bas Oude Munnink et al |
| EPI_ISL_422877 | Netherlands/NoordBrabant_82/2020 | 2020-03-12 | Europe/Netherlands/North Brabant/ | Dutch COVID-19 response team | Erasmus Medical Center | Bas Oude Munnink et al |
| EPI_ISL_422890 | Netherlands/NoordBrabant_96/2020 | 2020-03-18 | Europe/Netherlands/North Brabant/ | Dutch COVID-19 response team | Erasmus Medical Center | Bas Oude Munnink et al |
| EPI_ISL_422891 | Netherlands/NoordBrabant_97/2020 | 2020-03-19 | Europe/Netherlands/North Brabant/ | Dutch COVID-19 response team | Erasmus Medical Center | Bas Oude Munnink et al |
| EPI_ISL_422892 | Netherlands/NoordBrabant_98/2020 | 2020-03-19 | Europe/Netherlands/North Brabant/ | Dutch COVID-19 response team | Erasmus Medical Center | Bas Oude Munnink et al |
| EPI_ISL_422898 | Netherlands/Utrecht_21/2020 | 2020-03-03 | Europe/Netherlands/Utrecht/ | Dutch COVID-19 response team | Erasmus Medical Center | Bas Oude Munnink et al |
| EPI_ISL_422906 | Netherlands/ZuidHolland_34/2020 | 2020-03-16 | Europe/Netherlands/South Holland/ | Dutch COVID-19 response team | Erasmus Medical Center | Bas Oude Munnink et al |
| EPI_ISL_422909 | Netherlands/ZuidHolland_37/2020 | 2020-03-17 | Europe/Netherlands/South Holland/ | Dutch COVID-19 response team | Erasmus Medical Center | Bas Oude Munnink et al |
| EPI_ISL_422910 | Netherlands/ZuidHolland_38/2020 | 2020-03-17 | Europe/Netherlands/South Holland/ | Dutch COVID-19 response team | Erasmus Medical Center | Bas Oude Munnink et al |
| EPI_ISL_422918 | Netherlands/ZuidHolland_47/2020 | 2020-03-19 | Europe/Netherlands/South Holland/ | Dutch COVID-19 response team | Erasmus Medical Center | Bas Oude Munnink et al |
| EPI_ISL_422930 | Netherlands/ZuidHolland_62/2020 | 2020-03-23 | Europe/Netherlands/South Holland/ | Dutch COVID-19 response team | Erasmus Medical Center | Bas Oude Munnink et al |

| | | | | | | |
|---|---|---|---|---|---|---|
| EPI_ISL_422931 | Netherlands/ZuidHolland_63/2020 | 2020-03-23 | Europe/Netherlands/South Holland/ | Dutch COVID-19 response team | Erasmus Medical Center | Bas Oude Munnink et al |
| EPI_ISL_422952 | Netherlands/ZuidHolland_84/2020 | 2020-03-27 | Europe/Netherlands/South Holland/ | Dutch COVID-19 response team | Erasmus Medical Center | Bas Oude Munnink et al |
| EPI_ISL_422994 | USA/WA-UW-1322/2020 | 2020-03-18 | North America/USA/Washington/ | UW Virology Lab | UW Virology Lab | Pavitra Roychoudhury et al |
| EPI_ISL_422995 | USA/WA-UW-1334/2020 | 2020-03-17 | North America/USA/Washington/ | UW Virology Lab | UW Virology Lab | Pavitra Roychoudhury et al |
| EPI_ISL_423001 | USA/WA-UW-1567/2020 | 2020-03-19 | North America/USA/Washington/ | UW Virology Lab | UW Virology Lab | Pavitra Roychoudhury et al |
| EPI_ISL_423008 | USA/WA-UW-1426/2020 | 2020-03-18 | North America/USA/Washington/ | UW Virology Lab | UW Virology Lab | Pavitra Roychoudhury et al |
| EPI_ISL_423020 | USA/WA-UW-1570/2020 | 2020-03-19 | North America/USA/Washington/ | UW Virology Lab | UW Virology Lab | Pavitra Roychoudhury et al |
| EPI_ISL_423024 | USA/WA-UW-1435/2020 | 2020-03-17 | North America/USA/Washington/ | UW Virology Lab | UW Virology Lab | Pavitra Roychoudhury et al |
| EPI_ISL_423170 | England/20134077504/2020 | 2020-03-24 | Europe/United Kingdom/England/ | Respiratory Virus Unit, Microbiology Services Colindale, Public Health England | Respiratory Virus Unit, Microbiology Services Colindale, Public Health England | Monica Galiano et al |
| EPI_ISL_423230 | England/201360176/2020 | 2020-03-24 | Europe/United Kingdom/England/ | Respiratory Virus Unit, Microbiology Services Colindale, Public Health England | Respiratory Virus Unit, Microbiology Services Colindale, Public Health England | Monica Galiano et al |
| EPI_ISL_423399 | England/20140066204/2020 | 2020-03-29 | Europe/United Kingdom/England/ | Respiratory Virus Unit, Microbiology Services Colindale, Public Health England | Respiratory Virus Unit, Microbiology Services Colindale, Public Health England | Monica Galiano et al |
| EPI_ISL_423438 | England/20142018004/2020 | 2020-03-30 | Europe/United Kingdom/England/ | Respiratory Virus Unit, Microbiology Services Colindale, Public Health England | Respiratory Virus Unit, Microbiology Services Colindale, Public Health England | Monica Galiano et al |

| | | | | | | |
|---|---|---|---|---|---|---|
| EPI_ISL_423453 | England/20142026304/2020 | 2020-03-29 | Europe/United Kingdom/England/ | Respiratory Virus Unit, Microbiology Services Colindale, Public Health England | Respiratory Virus Unit, Microbiology Services Colindale, Public Health England | Monica Galiano et al |
| EPI_ISL_423576 | England/20146008504/2020 | 2020-03-31 | Europe/United Kingdom/England/ | Respiratory Virus Unit, Microbiology Services Colindale, Public Health England | Respiratory Virus Unit, Microbiology Services Colindale, Public Health England | Monica Galiano et al |
| EPI_ISL_423632 | England/20130052804/2020 | 2020-03-21 | Europe/United Kingdom/England/ | Respiratory Virus Unit, Microbiology Services Colindale, Public Health England | Respiratory Virus Unit, Microbiology Services Colindale, Public Health England | Monica Galiano et al |
| EPI_ISL_423735 | England/20132018904/2020 | 2020-03-22 | Europe/United Kingdom/England/ | Respiratory Virus Unit, Microbiology Services Colindale, Public Health England | Respiratory Virus Unit, Microbiology Services Colindale, Public Health England | Monica Galiano et al |
| EPI_ISL_423786 | England/20132054604/2020 | 2020-03-23 | Europe/United Kingdom/England/ | Respiratory Virus Unit, Microbiology Services Colindale, Public Health England | Respiratory Virus Unit, Microbiology Services Colindale, Public Health England | Monica Galiano et al |
| EPI_ISL_423868 | England/20126002004/2020 | 2020-03-18 | Europe/United Kingdom/England/ | Respiratory Virus Unit, Microbiology Services Colindale, Public Health England | Respiratory Virus Unit, Microbiology Services Colindale, Public Health England | Monica Galiano et al |

| | | | | | | |
|---|---|---|---|---|---|---|
| EPI_ISL_423952 | England/20128036004/2020 | 2020-03-19 | Europe/United Kingdom/England/ | Respiratory Virus Unit, Microbiology Services Colindale, Public Health England | Respiratory Virus Unit, Microbiology Services Colindale, Public Health England | Monica Galiano et al |
| EPI_ISL_424017 | England/20129025404/2020 | 2020-03-12 | Europe/United Kingdom/England/ | Respiratory Virus Unit, Microbiology Services Colindale, Public Health England | Respiratory Virus Unit, Microbiology Services Colindale, Public Health England | Monica Galiano et al |
| EPI_ISL_424068 | England/20129078704/2020 | 2020-03-13 | Europe/United Kingdom/England/ | Respiratory Virus Unit, Microbiology Services Colindale, Public Health England | Respiratory Virus Unit, Microbiology Services Colindale, Public Health England | Monica Galiano et al |
| EPI_ISL_424218 | USA/WA-UW-1682/2020 | 2020-03-20 | North America/USA/Washington/ | UW Virology Lab | UW Virology Lab | Pavitra Roychoudhury et al |
| EPI_ISL_424229 | USA/WA-UW-1724/2020 | 2020-03-21 | North America/USA/Washington/ | UW Virology Lab | UW Virology Lab | Pavitra Roychoudhury et al |
| EPI_ISL_424234 | USA/WA-UW-1722/2020 | 2020-03-21 | North America/USA/Washington/ | UW Virology Lab | UW Virology Lab | Pavitra Roychoudhury et al |
| EPI_ISL_424255 | USA/UN-UW-1832/2020 | 2020-03-16 | North America/USA/USA/ | UW Virology Lab | UW Virology Lab | Pavitra Roychoudhury et al |
| EPI_ISL_424286 | USA/OR-UW-1849/2020 | 2020-03-20 | North America/USA/Oregon/ | UW Virology Lab | UW Virology Lab | Pavitra Roychoudhury et al |
| EPI_ISL_424314 | USA/WA-UW-1921/2020 | 2020-03-24 | North America/USA/Washington/ | UW Virology Lab | UW Virology Lab | Pavitra Roychoudhury et al |
| EPI_ISL_424322 | USA/ID-UW-1934/2020 | 2020-03-20 | North America/USA/Idaho/ | UW Virology Lab | UW Virology Lab | Pavitra Roychoudhury et al |
| EPI_ISL_424342 | Italy/LAZ-INMI-8/2020 | 2020-03-07 | Europe/Italy/Lazio/Rome | INMI Lazzaro Spallanzani IRCCS | Laboratory of Virology, INMI Lazzaro Spallanzani IRCCS | Concetta Castilletti et al |
| EPI_ISL_424344 | Italy/LAZ-INMI10/2020 | 2020-03-04 | Europe/Italy/Lazio/Rome | INMI Lazzaro Spallanzani IRCCS | Laboratory of Virology, INMI Lazzaro Spallanzani IRCCS | Eleonora Lalle et al |
| EPI_ISL_424355 | Beijing/Wuhan_IME-BJ01/2020 | 2020-01-24 | Asia/China/Beijing/ | Beijing Institute of Microbiology and Epidemiology | Beijing Institute of Microbiology and Epidemiology | Fan et al |

| | | | | | | |
|---|---|---|---|---|---|---|
| EPI_ISL_424356 | Beijing/Wuhan_IME-BJ02/2020 | 2020-01-25 | Asia/China/Beijing/ | Beijing Institute of Microbiology and Epidemiology | Beijing Institute of Microbiology and Epidemiology | Fan et al |
| EPI_ISL_424357 | Beijing/Wuhan_IME-BJ03/2020 | 2020-01-28 | Asia/China/Beijing/ | Beijing Institute of Microbiology and Epidemiology | Beijing Institute of Microbiology and Epidemiology | Fan et al |
| EPI_ISL_424358 | Beijing/Wuhan_IME-BJ04/2020 | 2020-01-28 | Asia/China/Beijing/ | Beijing Institute of Microbiology and Epidemiology | Beijing Institute of Microbiology and Epidemiology | Fan et al |
| EPI_ISL_424359 | Beijing/Wuhan_IME-BJ05/2020 | 2020-01-27 | Asia/China/Beijing/ | Beijing Institute of Microbiology and Epidemiology | Beijing Institute of Microbiology and Epidemiology | Fan et al |
| EPI_ISL_424360 | Beijing/Wuhan_IME-BJ07/2020 | 2020-01-29 | Asia/China/Beijing/ | Beijing Institute of Microbiology and Epidemiology | Beijing Institute of Microbiology and Epidemiology | Fan et al |
| EPI_ISL_424361 | India/UN-1063/2020 | 2020-03-10 | Asia/Iran/India/ | National Influenza Center, Indian Council of Medical Research - National Institute of Virology | Indian Council of Medical Research-National Institute of Virology, Microbial Containment Complex | Pragya D. Yadav et al |
| EPI_ISL_424362 | India/UN-1135/2020 | 2020-03-10 | Asia/Iran/India/ | National Influenza Center, Indian Council of Medical Research - National Institute of Virology | Indian Council of Medical Research-National Institute of Virology, Microbial Containment Complex | Pragya D. Yadav et al |
| EPI_ISL_424363 | India/UN-1652/2020 | 2020-03-12 | Asia/Iran/India/ | National Influenza Center, Indian Council of Medical Research - National Institute of Virology | Indian Council of Medical Research-National Institute of Virology, Microbial Containment Complex | Pragya D. Yadav et al |
| EPI_ISL_424451 | Iceland/427/2020 | 2020-03-20 | Europe/Italy/Reykjavik/ | The National University Hospital of Iceland | deCODE genetics | Daniel F Gudbjartsson et al |
| EPI_ISL_424479 | Iceland/456/2020 | 2020-03-20 | Europe/Iceland/Reykjavik/ | The National University Hospital of Iceland | deCODE genetics | Daniel F Gudbjartsson et al |
| EPI_ISL_424628 | Belgium/ULG-7821/2020 | 2020-03-17 | Europe/Belgium/Liv®ge/ | Department of Clinical Microbiology | GIGA Medical Genomics | Keith Durkin et al |

| | | | | | | |
|---|---|---|---|---|---|---|
| EPI_ISL_424632 | Belgium/ULG-10071/2020 | 2020-04-03 | Europe/Belgium/Liv®ge/ | Department of Clinical Microbiology | GIGA Medical Genomics | Keith Durkin et al |
| EPI_ISL_424635 | Belgium/ULG-10065/2020 | 2020-04-03 | Europe/Belgium/Liv®ge/ | Department of Clinical Microbiology | GIGA Medical Genomics | Keith Durkin et al |
| EPI_ISL_424666 | Mexico/CMX-InDRE_02/2020 | 2020-02-29 | North America/Mexico/Chiapas/ | Instituto de Diagnv≥stico y Referencia Epidemiolv≥gicos | Instituto de Diagnv≥stico y Referencia Epidemiolv≥gicos | Josv© Ernesto Ramv≠rez Gonzv°lez et al |
| EPI_ISL_424670 | Mexico/QUE-InDRE-04/2020 | 2020-03-10 | North America/Mexico/Queretaro/ | Laboratorio Estatal de Salud Publica del Estado de Queretaro | Instituto de Diagnv≥stico y Referencia Epidemiologicos | Gisela Barrera Badillo et al |
| EPI_ISL_424865 | USA/IA_6394/2020 | 2020-03-08 | North America/USA/Iowa/ | IA State Hygienic Laboratory | Pathogen Discovery, Respiratory Viruses Branch, Division of Viral Diseases, Centers for Disease Control and Prevention | Yan Li et al |
| EPI_ISL_424868 | USA/LA_0842/2020 | 2020-03-09 | North America/USA/Louisiana/ | LA Office of Public Health Laboratories | Pathogen Discovery, Respiratory Viruses Branch, Division of Viral Diseases, Centers for Disease Control and Prevention | Yan Li et al |
| EPI_ISL_424878 | USA/NJ_3592/2020 | 2020-03-04 | North America/USA/New Jersey/ | NJ Public Health and Environmental Laboratories | Pathogen Discovery, Respiratory Viruses Branch, Division of Viral Diseases, Centers for Disease Control and Prevention | Yan Li et al |

| | | | | | | |
|---|---|---|---|---|---|---|
| EPI_ISL_424888 | USA/SC_6370/2020 | 2020-03-08 | North America/USA/South Carolina/ | SC Dept of Health and Env. Control-Bureau of Laboratories | Pathogen Discovery, Respiratory Viruses Branch, Division of Viral Diseases, Centers for Disease Control and Prevention | Yan Li et al |
| EPI_ISL_424933 | USA/NY-NYUMC126/2020 | 2020-04-01 | North America/USA/New York/Brooklyn | NYU Langone Health | Departments of Pathology and Medicine, New York University School of Medicine | Maria Aguero-Rosenfeld et al |
| EPI_ISL_424942 | USA/NY-NYUMC135/2020 | 2020-04-01 | North America/USA/New York/Brooklyn | NYU Langone Health | Departments of Pathology and Medicine, New York University School of Medicine | Maria Aguero-Rosenfeld et al |
| EPI_ISL_424949 | USA/NY-NYUMC142/2020 | 2020-04-01 | North America/USA/New York/Manhatt | NYU Langone Health | Departments of Pathology and Medicine, New York University School of Medicine | Maria Aguero-Rosenfeld et al |
| EPI_ISL_424950 | USA/NY-NYUMC143/2020 | 2020-04-01 | North America/USA/New York/Brooklyn | NYU Langone Health | Departments of Pathology and Medicine, New York University School of Medicine | Maria Aguero-Rosenfeld et al |
| EPI_ISL_424951 | USA/NY-NYUMC144/2020 | 2020-04-01 | North America/USA/New York/Brooklyn | NYU Langone Health | Departments of Pathology and Medicine, New York University School of Medicine | Maria Aguero-Rosenfeld et al |

| | | | | | | |
|---|---|---|---|---|---|---|
| EPI_ISL_424959 | USA/NY-NYUMC152/2020 | 2020-04-01 | North America/USA/New York/Brooklyn | NYU Langone Health | Departments of Pathology and Medicine, New York University School of Medicine | Maria Aguero-Rosenfeld et al |
| EPI_ISL_424963 | USA/NY-NYUMC156/2020 | 2020-04-01 | North America/USA/New York/Bronx | NYU Langone Health | Departments of Pathology and Medicine, New York University School of Medicine | Maria Aguero-Rosenfeld et al |
| EPI_ISL_424964 | USA/NY-NYUMC157/2020 | 2020-04-01 | North America/USA/New York/Queens | NYU Langone Health | Departments of Pathology and Medicine, New York University School of Medicine | Maria Aguero-Rosenfeld et al |
| EPI_ISL_424982 | USA/MI-MDHHS-SC20004/2020 | 2020-03-10 | North America/USA/Michigan/ | Bureau of Laboratories, Michigan Department of Health and Human Services | Michigan Department of Health and Human Services | Blankenship HM et al |
| EPI_ISL_424989 | USA/MI-SC2-0007/2020 | 2020-03-12 | North America/USA/Michigan/ | Bureau of Laboratories, Michigan Department of Health and Human Services | Michigan Department of Health and Human Services | Blankenship HM et al |
| EPI_ISL_425054 | Belgium/UGent-10/2020 | 2020-03-25 | Europe/Belgium/Gent/ | Lab voor klinische biologie | Onderzoeksgroep Virologie | Nick Vereecke et al |
| EPI_ISL_425117 | SouthKorea/KCDC2001/2020 | 2020-02-23 | Asia/South Korea/South Korea/ | Division of Viral Diseases, Center for Laboratory Control of Infectious Diseases, Korea Centers for Diseases Control and Prevention | Division of Viral Diseases, Center for Laboratory Control of Infectious Diseases, Korea Centers for Diseases Control and Prevention | Jeong-Min Kim et al |

| | | | | | | |
|---|---|---|---|---|---|---|
| EPI_ISL_425118 | SouthKorea/KCDC2002/2020 | 2020-01-30 | Asia/South Korea/South Korea/ | Division of Viral Diseases, Center for Laboratory Control of Infectious Diseases, Korea Centers for Diseases Control and Prevention | Division of Viral Diseases, Center for Laboratory Control of Infectious Diseases, Korea Centers for Diseases Control and Prevention | Jeong-Min Kim et al |
| EPI_ISL_425123 | Germany/NW-HHU-39/2020 | 2020-03-17 | Europe/Germany/North Rhine Westphal | Center of Medical Microbiology, Virology, and Hospital Hygiene, University of Duesseldorf | Center of Medical Microbiology, Virology, and Hospital Hygiene, University of Duesseldorf | Ortwin Adams et al |
| EPI_ISL_425133 | Germany/NW-HHU-49/2020 | 2020-03-10 | Europe/Germany/North Rhine Westphal | Center of Medical Microbiology, Virology, and Hospital Hygiene, University of Duesseldorf | Center of Medical Microbiology, Virology, and Hospital Hygiene, University of Duesseldorf | Ortwin Adams et al |
| EPI_ISL_425138 | Germany/NW-HHU-43/2020 | 2020-03-19 | Europe/Germany/North Rhine Westphal | Center of Medical Microbiology, Virology, and Hospital Hygiene, University of Duesseldorf | Center of Medical Microbiology, Virology, and Hospital Hygiene, University of Duesseldorf | Ortwin Adams et al |
| EPI_ISL_425139 | Germany/NW-HHU-44/2020 | 2020-03-19 | Europe/Germany/North Rhine Westphal | Center of Medical Microbiology, Virology, and Hospital Hygiene, University of Duesseldorf | Center of Medical Microbiology, Virology, and Hospital Hygiene, University of Duesseldorf | Ortwin Adams et al |
| EPI_ISL_425142 | USA/WI-UW-85/2020 | 2020-04-02 | North America/USA/Wisconsin/Dane Co | University of Wisconsin-Madison AIDS Vaccine Research Laboratories | University of Wisconsin-Madison AIDS Vaccine Research Laboratories | Gage Moreno et al |

| | | | | | | |
|---|---|---|---|---|---|---|
| EPI_ISL_425144 | USA/WI-UW-87/2020 | 2020-04-02 | North America/USA/Wisconsin/Dane Co | University of Wisconsin-Madison AIDS Vaccine Research Laboratories | University of Wisconsin-Madison AIDS Vaccine Research Laboratories | Gage Moreno et al |
| EPI_ISL_425150 | USA/WI-UW-93/2020 | 2020-04-04 | North America/USA/Wisconsin/Dane Co | University of Wisconsin-Madison AIDS Vaccine Research Laboratories | University of Wisconsin-Madison AIDS Vaccine Research Laboratories | Gage Moreno et al |
| EPI_ISL_425154 | USA/WI-UW-97/2020 | 2020-03-30 | North America/USA/Wisconsin/Dane Co | University of Wisconsin-Madison AIDS Vaccine Research Laboratories | University of Wisconsin-Madison AIDS Vaccine Research Laboratories | Gage Moreno et al |
| EPI_ISL_425163 | USA/WI-UW-106/2020 | 2020-03-30 | North America/USA/Wisconsin/Dane Co | University of Wisconsin-Madison AIDS Vaccine Research Laboratories | University of Wisconsin-Madison AIDS Vaccine Research Laboratories | Gage Moreno et al |
| EPI_ISL_425168 | USA/WI-UW-111/2020 | 2020-03-31 | North America/USA/Wisconsin/Dane Co | University of Wisconsin-Madison AIDS Vaccine Research Laboratories | University of Wisconsin-Madison AIDS Vaccine Research Laboratories | Gage Moreno et al |
| EPI_ISL_425177 | Canada/ON_ON-VIDO-01-2/2020 | 2020-01-23 | North America/Canada/Ontario/ | Public Health Ontario | Public Health Agency of Canada - National Microbiology Laboratory | Amrit S. Boese et al |
| EPI_ISL_425178 | Spain/VC-FISABIO-54/2020 | 2020-02-28 | Europe/Spain/Comunitat Valenciana/Va | Servicio de Microbiolog√≠a. Consorcio Hospital General Universitario de Valencia | Sequencing and Bioinformatics Service and Molecular Epidemiology Research Group. FISABIO-Public Health | David Navarro et al |

| | | | | | | |
|---|---|---|---|---|---|---|
| EPI_ISL_425180 | Spain/VC-FISABIO-56/2020 | 2020-03-10 | Europe/Spain/Comunitat Valenciana/Va | Servicio de Microbiolog√≠a. Consorcio Hospital General Universitario de Valencia | Sequencing and Bioinformatics Service and Molecular Epidemiology Research Group. FISABIO-Public Health | Griselda De Marco et al |
| EPI_ISL_425182 | Spain/VC-FISABIO-58/2020 | 2020-03-10 | Europe/Spain/Comunitat Valenciana/Va | Servicio de Microbiolog√≠a. Consorcio Hospital General Universitario de Valencia | Sequencing and Bioinformatics Service and Molecular Epidemiology Research Group. FISABIO-Public Health | Lidia Ruiz Roldan et al |
| EPI_ISL_425191 | Spain/VC-FISABIO-67/2020 | 2020-03-16 | Europe/Spain/Comunitat Valenciana/Va | Servicio de Microbiolog√≠a. Consorcio Hospital General Universitario de Valencia | Sequencing and Bioinformatics Service and Molecular Epidemiology Research Group. FISABIO-Public Health | Ivan Ansari et al |
| EPI_ISL_425193 | Spain/VC-FISABIO-69/2020 | 2020-03-17 | Europe/Spain/Comunitat Valenciana/Va | Servicio de Microbiolog√≠a. Consorcio Hospital General Universitario de Valencia | Sequencing and Bioinformatics Service and Molecular Epidemiology Research Group. FISABIO-Public Health | Maria Alma Bracho et al |
| EPI_ISL_425196 | Spain/VC-FISABIO-72/2020 | 2020-03-17 | Europe/Spain/Comunitat Valenciana/Va | Servicio de Microbiolog√≠a. Consorcio Hospital General Universitario de Valencia | Sequencing and Bioinformatics Service and Molecular Epidemiology Research Group. FISABIO-Public Health | Lidia Ruiz Roldan et al |

| | | | | | | |
|---|---|---|---|---|---|---|
| EPI_ISL_425201 | Spain/VC-FISABIO-77/2020 | 2020-03-19 | Europe/Spain/Comunitat Valenciana/Va | Servicio de Microbiologia. Hospital Clinico Universitario de Valencia | Sequencing and Bioinformatics Service and Molecular Epidemiology Research Group. FISABIO-Public Health | Mariana Reyes-Prieto et al |
| EPI_ISL_425202 | Spain/VC-FISABIO-78/2020 | 2020-03-20 | Europe/Spain/Comunitat Valenciana/Va | Servicio de Microbiologia. Hospital Clinico Universitario de Valencia | Sequencing and Bioinformatics Service and Molecular Epidemiology Research Group. FISABIO-Public Health | Vicente Soriano Chirona et al |
| EPI_ISL_425205 | Spain/VC-FISABIO-81/2020 | 2020-03-20 | Europe/Spain/Comunitat Valenciana/Va | Servicio de Microbiologia. Hospital Clinico Universitario de Valencia | Sequencing and Bioinformatics Service and Molecular Epidemiology Research Group. FISABIO-Public Health | Griselda De Marco et al |
| EPI_ISL_425207 | Spain/VC-FISABIO-83/2020 | 2020-03-21 | Europe/Spain/Comunitat Valenciana/Va | Servicio de Microbiologia. Hospital Clinico Universitario de Valencia | Sequencing and Bioinformatics Service and Molecular Epidemiology Research Group. FISABIO-Public Health | Lidia Ruiz Roldan et al |
| EPI_ISL_425212 | Spain/VC-FISABIO-88/2020 | 2020-03-11 | Europe/Spain/Comunitat Valenciana/Va | Servicio de Microbiolog√≠a. Consorcio Hospital General Universitario de Valencia | Sequencing and Bioinformatics Service and Molecular Epidemiology Research Group. FISABIO-Public Health | David Navarro et al |

| | | | | | | |
|---|---|---|---|---|---|---|
| EPI_ISL_425218 | Spain/VC-FISABIO-94/2020 | 2020-03-17 | Europe/Spain/Comunitat Valenciana/Va | Servicio de Microbiolog√≠a. Consorcio Hospital General Universitario de Valencia | Sequencing and Bioinformatics Service and Molecular Epidemiology Research Group. FISABIO-Public Health | Neris Garcia-Gonzalez et al |
| EPI_ISL_425219 | Spain/VC-FISABIO-95/2020 | 2020-03-17 | Europe/Spain/Comunitat Valenciana/Va | Servicio de Microbiolog√≠a. Consorcio Hospital General Universitario de Valencia | Sequencing and Bioinformatics Service and Molecular Epidemiology Research Group. FISABIO-Public Health | Neris Garcia-Gonzalez et al |
| EPI_ISL_425221 | Spain/VC-FISABIO-97/2020 | 2020-03-17 | Europe/Spain/Comunitat Valenciana/Va | Servicio de Microbiolog√≠a. Consorcio Hospital General Universitario de Valencia | Sequencing and Bioinformatics Service and Molecular Epidemiology Research Group. FISABIO-Public Health | Sandra Carbo et al |
| EPI_ISL_425222 | Spain/VC-FISABIO-98/2020 | 2020-03-17 | Europe/Spain/Comunitat Valenciana/Va | Servicio de Microbiolog√≠a. Consorcio Hospital General Universitario de Valencia | Sequencing and Bioinformatics Service and Molecular Epidemiology Research Group. FISABIO-Public Health | Loreto Ferr√∫s Abad et al |
| EPI_ISL_425473 | England/NOTT-10DF50/2020 | 2020-03-14 | Europe/United Kingdom/England/ | Queens Medical Centre, Clinical Microbiology Department / DeepSeq Nottingham | COVID-19 Genomics UK (COG-UK) Consortium | Gemma Clark et al |
| EPI_ISL_425493 | England/NOTT-10E0A7/2020 | 2020-03-18 | Europe/United Kingdom/England/ | Queens Medical Centre, Clinical Microbiology Department / DeepSeq Nottingham | COVID-19 Genomics UK (COG-UK) Consortium | Gemma Clark et al |
| EPI_ISL_425494 | England/NOTT-10E0B6/2020 | 2020-03-18 | Europe/United Kingdom/England/ | Queens Medical Centre, Clinical Microbiology Department / DeepSeq Nottingham | COVID-19 Genomics UK (COG-UK) Consortium | Gemma Clark et al |

| | | | | | | |
|---|---|---|---|---|---|---|
| EPI_ISL_425516 | England/NOTT-10E247/2020 | 2020-03-19 | Europe/United Kingdom/England/ | Queens Medical Centre, Clinical Microbiology Department / DeepSeq Nottingham | COVID-19 Genomics UK (COG-UK) Consortium | Gemma Clark et al |
| EPI_ISL_425713 | Scotland/CVR173/2020 | 2020-03-17 | Europe/United Kingdom/Scotland/ | West of Scotland Specialist Virology Centre, NHSGGC / MRC-University of Glasgow Centre for Virus Research | COVID-19 Genomics UK (COG-UK) Consortium | Ana da Silva Filipe et al |
| EPI_ISL_425743 | Scotland/CVR215/2020 | 2020-03-18 | Europe/United Kingdom/Scotland/ | West of Scotland Specialist Virology Centre, NHSGGC / MRC-University of Glasgow Centre for Virus Research | COVID-19 Genomics UK (COG-UK) Consortium | Ana da Silva Filipe et al |
| EPI_ISL_425793 | Scotland/CVR634/2020 | 2020-03-25 | Europe/United Kingdom/Scotland/ | West of Scotland Specialist Virology Centre, NHSGGC / MRC-University of Glasgow Centre for Virus Research | COVID-19 Genomics UK (COG-UK) Consortium | Ana da Silva Filipe et al |
| EPI_ISL_425808 | Scotland/CVR78/2020 | 2020-03-06 | Europe/United Kingdom/Scotland/ | West of Scotland Specialist Virology Centre, NHSGGC / MRC-University of Glasgow Centre for Virus Research | COVID-19 Genomics UK (COG-UK) Consortium | Ana da Silva Filipe et al |
| EPI_ISL_425843 | Scotland/EDB037/2020 | 2020-03-12 | Europe/United Kingdom/Scotland/ | Virology Department, Royal Infirmary of Edinburgh, NHS Lothian / School of Biological Sciences, University of Edinburgh / Institute of Genetics and Molecular Medicine, University of Edinburgh | COVID-19 Genomics UK (COG-UK) Consortium | McHugh M et al |
| EPI_ISL_425891 | Scotland/EDB101/2020 | 2020-03-21 | Europe/United Kingdom/Scotland/ | Virology Department, Royal Infirmary of Edinburgh, NHS Lothian / School of Biological Sciences, University of Edinburgh / Institute of Genetics and Molecular Medicine, University of Edinburgh | COVID-19 Genomics UK (COG-UK) Consortium | McHugh M et al |
| EPI_ISL_426063 | USA/ID-UW-2255/2020 | 2020-03-23 | North America/USA/Idaho/ | UW Virology Lab | UW Virology Lab | Pavitra Roychoudhury et al |
| EPI_ISL_426064 | USA/ID-UW-4355/2020 | 2020-03-27 | North America/USA/Idaho/ | UW Virology Lab | UW Virology Lab | Pavitra Roychoudhury et al |
| EPI_ISL_426069 | USA/ID-UW-4396/2020 | 2020-03-28 | North America/USA/Idaho/ | UW Virology Lab | UW Virology Lab | Pavitra Roychoudhury et al |
| EPI_ISL_426070 | USA/ID-UW-4400/2020 | 2020-03-28 | North America/USA/Idaho/ | UW Virology Lab | UW Virology Lab | Pavitra Roychoudhury et al |
| EPI_ISL_426096 | USA/WA-UW-2141/2020 | 2020-03-25 | North America/USA/Washington/ | UW Virology Lab | UW Virology Lab | Pavitra Roychoudhury et al |
| EPI_ISL_426110 | USA/WA-UW-2193/2020 | 2020-03-24 | North America/USA/Washington/ | UW Virology Lab | UW Virology Lab | Pavitra Roychoudhury et al |
| EPI_ISL_426119 | USA/WA-UW-2228/2020 | 2020-03-23 | North America/USA/Washington/ | UW Virology Lab | UW Virology Lab | Pavitra Roychoudhury et al |
| EPI_ISL_426121 | USA/WA-UW-2232/2020 | 2020-03-25 | North America/USA/Washington/ | UW Virology Lab | UW Virology Lab | Pavitra Roychoudhury et al |
| EPI_ISL_426159 | USA/WI-GMF-00986/2020 | 2020-04-06 | North America/USA/Minnesota/Winona | Gundersen Molecular Diagnostics Laboratory | Kabara Cancer Research Institute | Craig S. Richmond et al |
| EPI_ISL_426161 | USA/WI-GMF-00928/2020 | 2020-04-06 | North America/USA/Iowa/Allamakee Co | Gundersen Molecular Diagnostics Laboratory | Kabara Cancer Research Institute | Craig S. Richmond et al |

| | | | | | | |
|---|---|---|---|---|---|---|
| EPI_ISL_426163 | SouthKorea/KCDC2003/2020 | 2020-02-23 | Asia/South Korea/South Korea/ | Division of Viral Diseases, Center for Laboratory Control of Infectious Diseases, Korea Centers for Diseases Control and Prevention | Division of Viral Diseases, Center for Laboratory Control of Infectious Diseases, Korea Centers for Diseases Control and Prevention | Jeong-Min Kim et al |
| EPI_ISL_426164 | SouthKorea/KCDC2004/2020 | 2020-02-02 | Asia/South Korea/South Korea/ | Division of Viral Diseases, Center for Laboratory Control of Infectious Diseases, Korea Centers for Diseases Control and Prevention | Division of Viral Diseases, Center for Laboratory Control of Infectious Diseases, Korea Centers for Diseases Control and Prevention | Jeong-Min Kim et al |
| EPI_ISL_426166 | SouthKorea/KCDC2005/2020 | 2020-02-04 | Asia/South Korea/South Korea/ | Division of Viral Diseases, Center for Laboratory Control of Infectious Diseases, Korea Centers for Diseases Control and Prevention | Division of Viral Diseases, Center for Laboratory Control of Infectious Diseases, Korea Centers for Diseases Control and Prevention | Jeong-Min Kim et al |
| EPI_ISL_426168 | SouthKorea/KCDC2006/2020 | 2020-02-02 | Asia/South Korea/South Korea/ | Division of Viral Diseases, Center for Laboratory Control of Infectious Diseases, Korea Centers for Diseases Control and Prevention | Division of Viral Diseases, Center for Laboratory Control of Infectious Diseases, Korea Centers for Diseases Control and Prevention | Jeong-Min Kim et al |

| | | | | | | |
|---|---|---|---|---|---|---|
| EPI_ISL_426169 | SouthKorea/KCDC2007/2020 | 2020-02-05 | Asia/South Korea/South Korea/ | Division of Viral Diseases, Center for Laboratory Control of Infectious Diseases, Korea Centers for Diseases Control and Prevention | Division of Viral Diseases, Center for Laboratory Control of Infectious Diseases, Korea Centers for Diseases Control and Prevention | Jeong-Min Kim et al |
| EPI_ISL_426171 | SouthKorea/KCDC2008/2020 | 2020-02-05 | Asia/South Korea/South Korea/ | Division of Viral Diseases, Center for Laboratory Control of Infectious Diseases, Korea Centers for Diseases Control and Prevention | Division of Viral Diseases, Center for Laboratory Control of Infectious Diseases, Korea Centers for Diseases Control and Prevention | Jeong-Min Kim et al |
| EPI_ISL_426173 | SouthKorea/KCDC2009/2020 | 2020-02-07 | Asia/South Korea/South Korea/ | Division of Viral Diseases, Center for Laboratory Control of Infectious Diseases, Korea Centers for Diseases Control and Prevention | Division of Viral Diseases, Center for Laboratory Control of Infectious Diseases, Korea Centers for Diseases Control and Prevention | Jeong-Min Kim et al |
| EPI_ISL_426180 | SouthKorea/KCDC2010/2020 | 2020-02-05 | Asia/South Korea/South Korea/ | Division of Viral Diseases, Center for Laboratory Control of Infectious Diseases, Korea Centers for Diseases Control and Prevention | Division of Viral Diseases, Center for Laboratory Control of Infectious Diseases, Korea Centers for Diseases Control and Prevention | Jeong-Min Kim et al |

| | | | | | | |
|---|---|---|---|---|---|---|
| EPI_ISL_426182 | SouthKorea/KCDC2012/2020 | 2020-02-09 | Asia/South Korea/South Korea/ | Division of Viral Diseases, Center for Laboratory Control of Infectious Diseases, Korea Centers for Diseases Control and Prevention | Division of Viral Diseases, Center for Laboratory Control of Infectious Diseases, Korea Centers for Diseases Control and Prevention | Jeong-Min Kim et al |
| EPI_ISL_426183 | SouthKorea/KCDC2013/2020 | 2020-02-23 | Asia/South Korea/South Korea/ | Division of Viral Diseases, Center for Laboratory Control of Infectious Diseases, Korea Centers for Diseases Control and Prevention | Division of Viral Diseases, Center for Laboratory Control of Infectious Diseases, Korea Centers for Diseases Control and Prevention | Jeong-Min Kim et al |
| EPI_ISL_426187 | SouthKorea/KCDC2014/2020 | 2020-02-12 | Asia/South Korea/South Korea/ | Division of Viral Diseases, Center for Laboratory Control of Infectious Diseases, Korea Centers for Diseases Control and Prevention | Division of Viral Diseases, Center for Laboratory Control of Infectious Diseases, Korea Centers for Diseases Control and Prevention | Jeong-Min Kim et al |
| EPI_ISL_426289 | Latvia/09/2020 | 2020-03-30 | Europe/Latvia/Latvia/ | E. Gulbja Laboratorija | Latvian Biomedical Research and Study Centre | Ivars Silamiƒ∑elis et al |
| EPI_ISL_426293 | USA/NY-Wadsworth-12883-02/2020 | 2020-03-10 | North America/USA/New York/Nassau C | Wadsworth Center, New York State Department.of Health | Wadsworth Center, New York State Department.of Health | Kirsten St. George et al |
| EPI_ISL_426320 | USA/NY-Wadsworth-25860-01/2020 | 2020-03-20 | North America/USA/New York/Broome | Wadsworth Center, New York State Department.of Health | Wadsworth Center, New York State Department.of Health | Kirsten St. George et al |

| | | | | | | |
|---|---|---|---|---|---|---|
| EPI_ISL_426322 | USA/NY-Wadsworth-14726-02/2020 | 2020-03-12 | North America/USA/New York/Nassau ( | Wadsworth Center, New York State Department.of Health | Wadsworth Center, New York State Department.of Health | Kirsten St. George et al |
| EPI_ISL_426362 | Mexico/CMX-INCMNSZ-02/2020 | 2020-03-10 | North America/Mexico/Mexico City/ | Instituto Nacional de Ciencias Medicas y Nutricion Salvador Zubiran | Instituto Nacional de Ciencias Medicas y Nutricion Salvador Zubiran | Guillermo M. Ruiz-Palacios et al |
| EPI_ISL_426425 | USA/MD_0027/2020 | 2020-03-04 | North America/USA/Maryland/ | MD DOH Laboratories Administration | Pathogen Discovery, Respiratory Viruses Branch, Division of Viral Diseases, Centers for Disease Control and Prevention | Krista Queen et al |
| EPI_ISL_426435 | USA/RI_0882/2020 | 2020-03-09 | North America/USA/Rhode Island/ | RI State Health Laboratories | Pathogen Discovery, Respiratory Viruses Branch, Division of Viral Diseases, Centers for Disease Control and Prevention | Krista Queen et al |
| EPI_ISL_426439 | USA/WA_0428/2020 | 2020-03-19 | North America/USA/Washington/ | WA State Department of Health | Pathogen Discovery, Respiratory Viruses Branch, Division of Viral Diseases, Centers for Disease Control and Prevention | Jing Zhang et al |

| | | | | | | |
|---|---|---|---|---|---|---|
| EPI_ISL_426440 | USA/WA_0429/2020 | 2020-03-19 | North America/USA/Washington/ | WA State Department of Health | Pathogen Discovery, Respiratory Viruses Branch, Division of Viral Diseases, Centers for Disease Control and Prevention | Ying Tao et al |
| EPI_ISL_426447 | USA/WA_0448/2020 | 2020-03-19 | North America/USA/Washington/ | WA State Department of Health | Pathogen Discovery, Respiratory Viruses Branch, Division of Viral Diseases, Centers for Disease Control and Prevention | Ying Tao et al |
| EPI_ISL_426511 | USA/AZ-TG268770/2020 | 2020-03-19 | North America/USA/Arizona/ | TGen North | TGen North | Jolene Bowers et al |
| EPI_ISL_426515 | USA/AZ-TG268909/2020 | 2020-03-13 | North America/USA/Arizona/ | AZ SPHL, Arizona Department of Health Services | TGen North | Jolene Bowers et al |
| EPI_ISL_426519 | USA/AZ-TG268917/2020 | 2020-03-14 | North America/USA/Arizona/ | AZ SPHL, Arizona Department of Health Services | TGen North | Jolene Bowers et al |
| EPI_ISL_426545 | USA/AZ-TG271884/2020 | 2020-03-25 | North America/USA/Arizona/ | AZ SPHL, Arizona Department of Health Services | TGen North | Jolene Bowers et al |
| EPI_ISL_426552 | USA/AZ-TG271898/2020 | 2020-03-27 | North America/USA/Arizona/ | AZ SPHL, Arizona Department of Health Services | TGen North | Jolene Bowers et al |
| EPI_ISL_426558 | USA/AZ-TG273280/2020 | 2020-03-25 | North America/USA/Arizona/ | AZ SPHL, Arizona Department of Health Services | TGen North | Jolene Bowers et al |
| EPI_ISL_426629 | Taiwan/TSGH-01/2020 | 2020-02-07 | Asia/Taiwan/Taiwan/Taipei | TSGH-CP molecular lab | TSGH-CP molecular lab | Cherng-Lih Perng et al |

| | | | | | | |
|---|---|---|---|---|---|---|
| EPI_ISL_426656 | Australia/VIC347/2020 | 2020-03-20 | Oceania/Australia/Victoria/ | Victorian Infectious Diseases Reference Laboratory (VIDRL) | Microbiological Diagnostic Unit Public Health Laboratory and Victorian Infectious Diseases Reference Laboratory, Doherty Institute | Caly L. et al |
| EPI_ISL_426882 | Australia/VIC913/2020 | 2020-03-24 | Oceania/Australia/Victoria/ | Victorian Infectious Diseases Reference Laboratory (VIDRL) | Microbiological Diagnostic Unit Public Health Laboratory and Victorian Infectious Diseases Reference Laboratory, Doherty Institute | Caly L. et al |
| EPI_ISL_426894 | CzechRepublic/IAB_20/2020 | 2020-03-27 | Europe/Czech Republic/Czech Republic/ | Motol University Hospital | Institute of Applied Biotechnologies a.s. | Petr Bro≈æ et al |
| EPI_ISL_426897 | CzechRepublic/IAB_23/2020 | 2020-03-29 | Europe/Czech Republic/Czech Republic/ | Motol University Hospital | Institute of Applied Biotechnologies a.s. | Petr Bro≈æ et al |
| EPI_ISL_426926 | Australia/VIC622/2020 | 2020-03-29 | Oceania/Australia/Victoria/ | Victorian Infectious Diseases Reference Laboratory (VIDRL) | Microbiological Diagnostic Unit Public Health Laboratory and Victorian Infectious Diseases Reference Laboratory, Doherty Institute | Caly L. et al |

| | | | | | | |
|---|---|---|---|---|---|---|
| EPI_ISL_427041 | Australia/VIC765/2020 | 2020-03-29 | Oceania/Australia/Victoria/ | Victorian Infectious Diseases Reference Laboratory (VIDRL) | Microbiological Diagnostic Unit Public Health Laboratory and Victorian Infectious Diseases Reference Laboratory, Doherty Institute | Caly L. et al |
| EPI_ISL_427103 | Australia/VIC843/2020 | 2020-04-04 | Oceania/Australia/Victoria/ | Victorian Infectious Diseases Reference Laboratory (VIDRL) | Microbiological Diagnostic Unit Public Health Laboratory and Victorian Infectious Diseases Reference Laboratory, Doherty Institute | Caly L. et al |
| EPI_ISL_427161 | USA/CT-UW-3845/2020 | 2020-03-25 | North America/USA/Connecticut/ | UW Virology Lab | UW Virology Lab | Pavitra Roychoudhury et al |
| EPI_ISL_427186 | USA/WA-UW-3833/2020 | 2020-03-26 | North America/USA/Washington/ | UW Virology Lab | UW Virology Lab | Pavitra Roychoudhury et al |
| EPI_ISL_427198 | USA/WA-UW-3871/2020 | 2020-03-26 | North America/USA/Washington/ | UW Virology Lab | UW Virology Lab | Pavitra Roychoudhury et al |
| EPI_ISL_427199 | USA/WA-UW-3880/2020 | 2020-03-26 | North America/USA/Washington/ | UW Virology Lab | UW Virology Lab | Pavitra Roychoudhury et al |
| EPI_ISL_427217 | USA/WA-UW-3946/2020 | 2020-03-27 | North America/USA/Washington/ | UW Virology Lab | UW Virology Lab | Pavitra Roychoudhury et al |
| EPI_ISL_427221 | USA/WA-UW-3963/2020 | 2020-03-30 | North America/USA/Washington/ | UW Virology Lab | UW Virology Lab | Pavitra Roychoudhury et al |
| EPI_ISL_427231 | USA/WA-UW-3992/2020 | 2020-03-29 | North America/USA/Washington/ | UW Virology Lab | UW Virology Lab | Pavitra Roychoudhury et al |
| EPI_ISL_427286 | USA/MN-MDH-147/2020 | 2020-03-23 | North America/USA/Minnesota/ | Minnesota Department of Health, Public Health Laboratory | Minnesota Department of Health, Public Health Laboratory | Matt Plumb et al |
| EPI_ISL_427295 | Brazil/DF-619i/2020 | 2020-03-13 | South America/Brazil/Distrito Federal/B | Instituto Oswaldo Cruz FIOCRUZ - Laboratory of Respiratory Viruses and Measles (LVRS) | Instituto Oswaldo Cruz FIOCRUZ - Laboratory of Respiratory Viruses and Measles (LVRS) | Paola Resende et al |

| | | | | | | |
|---|---|---|---|---|---|---|
| EPI_ISL_427306 | Brazil/SC-769/2020 | 2020-03-10 | South America/Brazil/Santa Catarina/Fl | LACEN-SC - Laboratorio Central de Santa Catarina | Instituto Oswaldo Cruz FIOCRUZ - Laboratory of Respiratory Viruses and Measles (LVRS) | Paola Resende et al |
| EPI_ISL_427310 | Russia/StPetersburg-RII4382V/2020 | 2020-03-29 | Europe/Russia/Saint-Petersburg/ | WHO National Influenza Centre Russian Federation | WHO National Influenza Centre Russian Federation | Andrey Komissarov et al |
| EPI_ISL_427314 | Russia/StPetersburg-RII4546V/2020 | 2020-04-02 | Europe/Russia/Saint-Petersburg/ | WHO National Influenza Centre Russian Federation | WHO National Influenza Centre Russian Federation | Andrey Komissarov et al |
| EPI_ISL_427316 | Russia/StPetersburg-RII4584S/2020 | 2020-04-03 | Europe/Russia/Saint-Petersburg/ | WHO National Influenza Centre Russian Federation | WHO National Influenza Centre Russian Federation | Andrey Komissarov et al |
| EPI_ISL_427317 | Russia/StPetersburg-RII4585S/2020 | 2020-04-03 | Europe/Russia/Saint-Petersburg/ | WHO National Influenza Centre Russian Federation | WHO National Influenza Centre Russian Federation | Andrey Komissarov et al |
| EPI_ISL_427318 | Russia/StPetersburg-RII4591S/2020 | 2020-04-03 | Europe/Russia/Saint-Petersburg/ | WHO National Influenza Centre Russian Federation | WHO National Influenza Centre Russian Federation | Andrey Komissarov et al |
| EPI_ISL_427319 | Russia/StPetersburg-RII4597S/2020 | 2020-04-04 | Europe/Russia/Saint-Petersburg/ | WHO National Influenza Centre Russian Federation | WHO National Influenza Centre Russian Federation | Andrey Komissarov et al |
| EPI_ISL_427323 | Russia/StPetersburg-RII4678S/2020 | 2020-04-06 | Europe/Russia/Saint-Petersburg/ | WHO National Influenza Centre Russian Federation | WHO National Influenza Centre Russian Federation | Andrey Komissarov et al |
| EPI_ISL_427324 | Russia/StPetersburg-RII4686S/2020 | 2020-04-07 | Europe/Russia/Saint-Petersburg/ | WHO National Influenza Centre Russian Federation | WHO National Influenza Centre Russian Federation | Andrey Komissarov et al |

| | | | | | | |
|---|---|---|---|---|---|---|
| EPI_ISL_427326 | Russia/StPetersburg-RII4694S/2020 | 2020-04-07 | Europe/Russia/Saint-Petersburg/ | WHO National Influenza Centre Russian Federation | WHO National Influenza Centre Russian Federation | Andrey Komissarov et al |
| EPI_ISL_427327 | Russia/StPetersburg-RII4696S/2020 | 2020-04-07 | Europe/Russia/Saint-Petersburg/ | WHO National Influenza Centre Russian Federation | WHO National Influenza Centre Russian Federation | Andrey Komissarov et al |
| EPI_ISL_427329 | Russia/StPetersburg-RII4707S/2020 | 2020-04-07 | Europe/Russia/Saint-Petersburg/ | WHO National Influenza Centre Russian Federation | WHO National Influenza Centre Russian Federation | Andrey Komissarov et al |
| EPI_ISL_427335 | Russia/StPetersburg-RII4728S/2020 | 2020-04-07 | Europe/Russia/Saint-Petersburg/ | WHO National Influenza Centre Russian Federation | WHO National Influenza Centre Russian Federation | Andrey Komissarov et al |
| EPI_ISL_427336 | Russia/StPetersburg-RII4781S/2020 | 2020-04-07 | Europe/Russia/Saint-Petersburg/ | WHO National Influenza Centre Russian Federation | WHO National Influenza Centre Russian Federation | Andrey Komissarov et al |
| EPI_ISL_427346 | Belgium/ULG-10038/2020 | 2020-03-30 | Europe/Belgium/Li√®ge/ | Department of Clinical Microbiology | GIGA Medical Genomics | Keith Durkin et al |
| EPI_ISL_427363 | Belgium/ULG-10114/2020 | 2020-04-07 | Europe/Belgium/Li√®ge/ | Department of Clinical Microbiology | GIGA Medical Genomics | Keith Durkin et al |
| EPI_ISL_427365 | Belgium/ULG-10117/2020 | 2020-04-07 | Europe/Belgium/Li√®ge/ | Department of Clinical Microbiology | GIGA Medical Genomics | Keith Durkin et al |
| EPI_ISL_427380 | Belgium/ULG-10135/2020 | 2020-04-08 | Europe/Belgium/Li√®ge/ | Department of Clinical Microbiology | GIGA Medical Genomics | Keith Durkin et al |
| EPI_ISL_427386 | Belgium/ULG-10141/2020 | 2020-04-08 | Europe/Belgium/Li√®ge/ | Department of Clinical Microbiology | GIGA Medical Genomics | Keith Durkin et al |
| EPI_ISL_427391 | Turkey/IST-GLAB-008/2020 | 2020-04-13 | Europe/Turkey/Istanbul/ | Genomic Laboratory (GLAB) (Conjoint lab of Health Directorate of Istanbul and Istanbul Technical University) | Genomic Laboratory (GLAB), Istanbul Technical University | Ilker Karacan et al |
| EPI_ISL_427434 | USA/WI-UW-127/2020 | 2020-04-07 | North America/USA/Wisconsin/Dane Co | University of Wisconsin-Madison AIDS Vaccine Research Laboratories | University of Wisconsin-Madison AIDS Vaccine Research Laboratories | Gage Moreno et al |

| | | | | | | |
|---|---|---|---|---|---|---|
| EPI_ISL_427475 | USA/NY-NYUMC179/2020 | 2020-04-01 | North America/USA/New York/Staten Is | NYU Langone Health | Departments of Pathology and Medicine, New York University School of Medicine | Maria Aguero-Rosenfeld et al |
| EPI_ISL_427480 | USA/NY-NYUMC184/2020 | 2020-04-01 | North America/USA/New York/Queens | NYU Langone Health | Departments of Pathology and Medicine, New York University School of Medicine | Maria Aguero-Rosenfeld et al |
| EPI_ISL_427495 | USA/NY-NYUMC200/2020 | 2020-04-01 | North America/USA/New York/Brooklyr | NYU Langone Health | Departments of Pathology and Medicine, New York University School of Medicine | Maria Aguero-Rosenfeld et al |
| EPI_ISL_427501 | USA/NY-NYUMC206/2020 | 2020-04-01 | North America/USA/New York/Brooklyr | NYU Langone Health | Departments of Pathology and Medicine, New York University School of Medicine | Maria Aguero-Rosenfeld et al |
| EPI_ISL_427504 | USA/NY-NYUMC209/2020 | 2020-04-03 | North America/USA/New York/Brooklyr | NYU Langone Health | Departments of Pathology and Medicine, New York University School of Medicine | Maria Aguero-Rosenfeld et al |
| EPI_ISL_427505 | USA/NY-NYUMC210/2020 | 2020-04-03 | North America/USA/New York/Brooklyr | NYU Langone Health | Departments of Pathology and Medicine, New York University School of Medicine | Maria Aguero-Rosenfeld et al |

| | | | | | | |
|---|---|---|---|---|---|---|
| EPI_ISL_427514 | USA/NY-NYUMC219/2020 | 2020-04-03 | North America/USA/New York/Brooklyn | NYU Langone Health | Departments of Pathology and Medicine, New York University School of Medicine | Maria Aguero-Rosenfeld et al |
| EPI_ISL_427516 | USA/NY-NYUMC221/2020 | 2020-03-18 | North America/USA/New York/Manhatt | NYU Langone Health | Departments of Pathology and Medicine, New York University School of Medicine | Maria Aguero-Rosenfeld et al |
| EPI_ISL_427521 | USA/NY-NYUMC226/2020 | 2020-03-18 | North America/USA/New York/Nassau C | NYU Langone Health | Departments of Pathology and Medicine, New York University School of Medicine | Maria Aguero-Rosenfeld et al |
| EPI_ISL_427544 | USA/NY-WCMP9A05P/2020 | 2020-03-14 | North America/USA/New York/Manhatt | NewYork-Presbyterian & Mason Lab | Mason Lab | Daniel J. Butler et al |
| EPI_ISL_427562 | USA/NY-WCMP10C03P/2020 | 2020-03-15 | North America/USA/New York/Manhatt | NewYork-Presbyterian & Mason Lab | Mason Lab | Daniel J. Butler et al |
| EPI_ISL_427583 | USA/NY-WCMP11A03P/2020 | 2020-03-16 | North America/USA/New York/Manhatt | NewYork-Presbyterian & Mason Lab | Mason Lab | Daniel J. Butler et al |
| EPI_ISL_427585 | USA/NY-WCMP11B03P/2020 | 2020-03-16 | North America/USA/New York/Manhatt | NewYork-Presbyterian & Mason Lab | Mason Lab | Daniel J. Butler et al |
| EPI_ISL_427602 | USA/NY-WCMP12C09P/2020 | 2020-03-16 | North America/USA/New York/Manhatt | NewYork-Presbyterian & Mason Lab | Mason Lab | Daniel J. Butler et al |
| EPI_ISL_427619 | USA/AK-PHL092/2020 | 2020-04-02 | North America/USA/Alaska/Interior Ala | Alaska State Virology Laboratory | Alaska State Virology Laboratory | Chen et al |
| EPI_ISL_427620 | USA/AK-PHL101/2020 | 2020-04-04 | North America/USA/Alaska/Interior Ala | Alaska State Virology Laboratory | Alaska State Virology Laboratory | Chen et al |
| EPI_ISL_427643 | Australia/NSW219/2020 | 2020-02-28 | Oceania/Australia/New South Wales/Sy | Centre for Infectious Diseases and Microbiology Public Health | NSW Health Pathology - Institute of Clinical Pathology and Medical Research; Westmead Hospital; University of Sydney | Timms V et al |

| | | | | | | |
|---|---|---|---|---|---|---|
| EPI_ISL_427664 | Australia/NSW63/2020 | 2020-03-10 | Oceania/Australia/New South Wales/Sy | Centre for Infectious Diseases and Microbiology Public Health | NSW Health Pathology - Institute of Clinical Pathology and Medical Research; Westmead Hospital; University of Sydney | Timms V et al |
| EPI_ISL_427697 | Australia/NSW167/2020 | 2020-03-22 | Oceania/Australia/New South Wales/Sy | Centre for Infectious Diseases and Microbiology Public Health | NSW Health Pathology - Institute of Clinical Pathology and Medical Research; Westmead Hospital; University of Sydney | Gall M et al |
| EPI_ISL_427809 | SouthKorea/KCDC2015/2020 | 2020-02-26 | Asia/South Korea/South Korea/ | Division of Viral Diseases, Center for Laboratory Control of Infectious Diseases, Korea Centers for Diseases Control and Prevention | Division of Viral Diseases, Center for Laboratory Control of Infectious Diseases, Korea Centers for Diseases Control and Prevention | Jeong-Min Kim et al |
| EPI_ISL_427810 | SouthKorea/KCDC2016/2020 | 2020-02-18 | Asia/South Korea/South Korea/ | Division of Viral Diseases, Center for Laboratory Control of Infectious Diseases, Korea Centers for Diseases Control and Prevention | Division of Viral Diseases, Center for Laboratory Control of Infectious Diseases, Korea Centers for Diseases Control and Prevention | Jeong-Min Kim et al |

| | | | | | | |
|---|---|---|---|---|---|---|
| EPI_ISL_427811 | SouthKorea/KCDC2017/2020 | 2020-02-18 | Asia/South Korea/South Korea/ | Division of Viral Diseases, Center for Laboratory Control of Infectious Diseases, Korea Centers for Diseases Control and Prevention | Division of Viral Diseases, Center for Laboratory Control of Infectious Diseases, Korea Centers for Diseases Control and Prevention | Jeong-Min Kim et al |
| EPI_ISL_427812 | SouthKorea/KCDC2018/2020 | 2020-02-19 | Asia/South Korea/South Korea/ | Division of Viral Diseases, Center for Laboratory Control of Infectious Diseases, Korea Centers for Diseases Control and Prevention | Division of Viral Diseases, Center for Laboratory Control of Infectious Diseases, Korea Centers for Diseases Control and Prevention | Jeong-Min Kim et al |
| EPI_ISL_427813 | SouthKorea/KCDC2019/2020 | 2020-02-18 | Asia/South Korea/South Korea/ | Division of Viral Diseases, Center for Laboratory Control of Infectious Diseases, Korea Centers for Diseases Control and Prevention | Division of Viral Diseases, Center for Laboratory Control of Infectious Diseases, Korea Centers for Diseases Control and Prevention | Jeong-Min Kim et al |
| EPI_ISL_427815 | Russia/StPetersburg-RII4706S/2020 | 2020-04-07 | Europe/Russia/Saint-Petersburg/ | WHO National Influenza Centre Russian Federation | WHO National Influenza Centre Russian Federation | Andrey Komissarov et al |
| EPI_ISL_428202 | USA/NE-875/2020 | 2020-03-25 | North America/USA/Nebraska/ | Nebraska Public Health Laboratory | UNMC COVID-19 Response Team | UNMC COVID-19 Response Team et al |
| EPI_ISL_428206 | USA/NE-912/2020 | 2020-03-25 | North America/USA/Nebraska/ | Nebraska Public Health Laboratory | UNMC COVID-19 Response Team | UNMC COVID-19 Response Team et al |
| EPI_ISL_428209 | Poland/Pom1/2020 | 2020-04-06 | Europe/Poland/Pomorskie/ | Laboratory of Molecular Biology, Diagnostyka sp. z o.o. | Laboratory of Recombinant Vaccines | Lukasz Rabalski et al |

| | | | | | | |
|---|---|---|---|---|---|---|
| EPI_ISL_428236 | Poland/1109171/2020 | 2020-03-27 | Europe/Poland/Pomorskie/ | Hematology Laboratory, Section of Molecular Diagnostics, University Clinical Centre, Medical University of Gdansk | Department of Virology, Faculty of Medicine, University of Helsinki, Helsinki, Finland | Marlena Robakowska et al |
| EPI_ISL_428260 | USA/WI-UW-164/2020 | 2020-03-16 | North America/USA/Wisconsin/Waukesl | University of Wisconsin-Madison AIDS Vaccine Research Laboratories | University of Wisconsin-Madison AIDS Vaccine Research Laboratories | Gage Moreno et al |
| EPI_ISL_428263 | USA/WI-UW-167/2020 | 2020-03-18 | North America/USA/Wisconsin/Waukesl | University of Wisconsin-Madison AIDS Vaccine Research Laboratories | University of Wisconsin-Madison AIDS Vaccine Research Laboratories | Gage Moreno et al |
| EPI_ISL_428275 | USA/WI-UW-179/2020 | 2020-03-21 | North America/USA/Wisconsin/Milwauk | University of Wisconsin-Madison AIDS Vaccine Research Laboratories | University of Wisconsin-Madison AIDS Vaccine Research Laboratories | Gage Moreno et al |
| EPI_ISL_428295 | USA/WI-UW-199/2020 | 2020-03-24 | North America/USA/Wisconsin/Waukesl | University of Wisconsin-Madison AIDS Vaccine Research Laboratories | University of Wisconsin-Madison AIDS Vaccine Research Laboratories | Gage Moreno et al |
| EPI_ISL_428311 | USA/WI-UW-215/2020 | 2020-03-25 | North America/USA/Wisconsin/Milwauk | University of Wisconsin-Madison AIDS Vaccine Research Laboratories | University of Wisconsin-Madison AIDS Vaccine Research Laboratories | Gage Moreno et al |
| EPI_ISL_428313 | USA/WI-UW-217/2020 | 2020-03-25 | North America/USA/Wisconsin/Milwauk | University of Wisconsin-Madison AIDS Vaccine Research Laboratories | University of Wisconsin-Madison AIDS Vaccine Research Laboratories | Gage Moreno et al |
| EPI_ISL_428318 | USA/WI-UW-222/2020 | 2020-03-25 | North America/USA/Wisconsin/Milwauk | University of Wisconsin-Madison AIDS Vaccine Research Laboratories | University of Wisconsin-Madison AIDS Vaccine Research Laboratories | Gage Moreno et al |

| | | | | | | |
|---|---|---|---|---|---|---|
| EPI_ISL_428360 | France/HDF-3678/2020 | 2020-03-25 | Europe/France/Hauts de France/Compiv | CH Compiv®gne Laboratoire de Biologie | National Reference Center for Viruses of Respiratory Infections, Institut Pasteur, Paris | Mv©lanie Albert et al |
| EPI_ISL_428361 | France/IDF-3703/2020 | 2020-03-25 | Europe/France/Ile de France/Longjumea | LABM GH nord Essonne de Longjumeau - BP 125 | National Reference Center for Viruses of Respiratory Infections, Institut Pasteur, Paris | Mv©lanie Albert et al |
| EPI_ISL_428368 | Turkey/IST-GLAB-013/2020 | 2020-04-16 | Europe/Turkey/Istanbul/ | Genomic Laboratory (GLAB) (Conjoint lab of Health Directorate of Istanbul and Istanbul Technical University) | Genomic Laboratory (GLAB), Istanbul Technical University | Ilker Karacan et al |
| EPI_ISL_428379 | USA/CT-Yale-059/2020 | 2020-03-28 | North America/USA/Connecticut/ | Yale COVID-19 Biorepository | Grubaugh Lab - Yale School of Public Health | Joseph Fauver et al |
| EPI_ISL_428384 | USA/CT-Yale-076/2020 | 2020-04-04 | North America/USA/Connecticut/ | Yale COVID-19 Biorepository | Grubaugh Lab - Yale School of Public Health | Joseph Fauver et al |
| EPI_ISL_428386 | USA/CT-Yale-068/2020 | 2020-04-03 | North America/USA/Connecticut/ | Yale COVID-19 Biorepository | Grubaugh Lab - Yale School of Public Health | Joseph Fauver et al |
| EPI_ISL_428388 | USA/CT-Yale-058/2020 | 2020-03-28 | North America/USA/Connecticut/ | Yale COVID-19 Biorepository | Grubaugh Lab - Yale School of Public Health | Joseph Fauver et al |
| EPI_ISL_428394 | USA/CT-Yale-086/2020 | 2020-04-07 | North America/USA/Connecticut/ | Yale COVID-19 Biorepository | Grubaugh Lab - Yale School of Public Health | Joseph Fauver et al |
| EPI_ISL_428395 | USA/CT-Yale-091/2020 | 2020-04-07 | North America/USA/Connecticut/ | Yale COVID-19 Biorepository | Grubaugh Lab - Yale School of Public Health | Joseph Fauver et al |
| EPI_ISL_428397 | USA/CT-Yale-094/2020 | 2020-04-07 | North America/USA/Connecticut/ | Yale COVID-19 Biorepository | Grubaugh Lab - Yale School of Public Health | Joseph Fauver et al |

| | | | | | | |
|---|---|---|---|---|---|---|
| EPI_ISL_428441 | Guangdong/20SF1107/2020 | 2020-01-29 | Asia/China/Guangdong/Huizhou | Guangdong Provincial Center for Diseases Control and Prevention;Guangdong Provincial Institute of Public Health | School of Public Health, The University of Hong Kong | Bosheng Li et al |
| EPI_ISL_428442 | Guangdong/20SF1136/2020 | 2020-01-30 | Asia/China/Guangdong/Shanwei | Guangdong Provincial Center for Diseases Control and Prevention;Guangdong Provincial Institute of Public Health | School of Public Health, The University of Hong Kong | Bosheng Li et al |
| EPI_ISL_428443 | Guangdong/20SF115/2020 | 2020-01-21 | Asia/China/Guangdong/Guangzhou | Guangdong Provincial Center for Diseases Control and Prevention;Guangdong Provincial Institute of Public Health | School of Public Health, The University of Hong Kong | Bosheng Li et al |
| EPI_ISL_428444 | Guangdong/20SF1152/2020 | 2020-01-30 | Asia/China/Guangdong/Huizhou | Guangdong Provincial Center for Diseases Control and Prevention;Guangdong Provincial Institute of Public Health | School of Public Health, The University of Hong Kong | Bosheng Li et al |
| EPI_ISL_428445 | Guangdong/20SF1153/2020 | 2020-01-30 | Asia/China/Guangdong/Huizhou | Guangdong Provincial Center for Diseases Control and Prevention;Guangdong Provincial Institute of Public Health | School of Public Health, The University of Hong Kong | Bosheng Li et al |
| EPI_ISL_428446 | Guangdong/20SF1159/2020 | 2020-01-31 | Asia/China/Guangdong/Zhuhai | Guangdong Provincial Center for Diseases Control and Prevention;Guangdong Provincial Institute of Public Health | School of Public Health, The University of Hong Kong | Bosheng Li et al |
| EPI_ISL_428447 | Guangdong/20SF117/2020 | 2020-01-21 | Asia/China/Guangdong/Shenzhen | Guangdong Provincial Center for Diseases Control and Prevention;Guangdong Provincial Institute of Public Health | School of Public Health, The University of Hong Kong | Bosheng Li et al |
| EPI_ISL_428448 | Guangdong/20SF118/2020 | 2020-01-21 | Asia/China/Guangdong/Shenzhen | Guangdong Provincial Center for Diseases Control and Prevention;Guangdong Provincial Institute of Public Health | School of Public Health, The University of Hong Kong | Bosheng Li et al |
| EPI_ISL_428449 | Guangdong/20SF123/2020 | 2020-01-20 | Asia/China/Guangdong/Zhanjiang | Guangdong Provincial Center for Diseases Control and Prevention;Guangdong Provincial Institute of Public Health | School of Public Health, The University of Hong Kong | Bosheng Li et al |
| EPI_ISL_428451 | Guangdong/20SF179/2020 | 2020-01-22 | Asia/China/Guangdong/Zhuhai | Guangdong Provincial Center for Diseases Control and Prevention;Guangdong Provincial Institute of Public Health | School of Public Health, The University of Hong Kong | Bosheng Li et al |

| | | | | | | |
|---|---|---|---|---|---|---|
| EPI_ISL_428452 | Guangdong/20SF190/2020 | 2020-01-23 | Asia/China/Guangdong/Shaoguan | Guangdong Provincial Center for Diseases Control and Prevention;Guangdong Provincial Institute of Public Health | School of Public Health, The University of Hong Kong | Bosheng Li et al |
| EPI_ISL_428453 | Guangdong/20SF195/2020 | 2020-01-22 | Asia/China/Guangdong/Huizhou | Guangdong Provincial Center for Diseases Control and Prevention;Guangdong Provincial Institute of Public Health | School of Public Health, The University of Hong Kong | Bosheng Li et al |
| EPI_ISL_428454 | Guangdong/20SF198/2020 | 2020-01-22 | Asia/China/Guangdong/Huizhou | Guangdong Provincial Center for Diseases Control and Prevention;Guangdong Provincial Institute of Public Health | School of Public Health, The University of Hong Kong | Bosheng Li et al |
| EPI_ISL_428455 | Guangdong/20SF200/2020 | 2020-01-23 | Asia/China/Guangdong/Yangjiang | Guangdong Provincial Center for Diseases Control and Prevention;Guangdong Provincial Institute of Public Health | School of Public Health, The University of Hong Kong | Bosheng Li et al |
| EPI_ISL_428456 | Guangdong/20SF243/2020 | 2020-01-22 | Asia/China/Guangdong/Shenzhen | Guangdong Provincial Center for Diseases Control and Prevention;Guangdong Provincial Institute of Public Health | School of Public Health, The University of Hong Kong | Bosheng Li et al |
| EPI_ISL_428457 | Guangdong/20SF254/2020 | 2020-01-23 | Asia/China/Guangdong/Huizhou | Guangdong Provincial Center for Diseases Control and Prevention;Guangdong Provincial Institute of Public Health | School of Public Health, The University of Hong Kong | Bosheng Li et al |
| EPI_ISL_428458 | Guangdong/20SF2546/2020 | 2020-02-06 | Asia/China/Guangdong/Guangzhou | Guangdong Provincial Center for Diseases Control and Prevention;Guangdong Provincial Institute of Public Health | School of Public Health, The University of Hong Kong | Bosheng Li et al |
| EPI_ISL_428460 | Guangdong/20SF273/2020 | 2020-01-23 | Asia/China/Guangdong/Guangzhou | Guangdong Provincial Center for Diseases Control and Prevention;Guangdong Provincial Institute of Public Health | School of Public Health, The University of Hong Kong | Bosheng Li et al |
| EPI_ISL_428461 | Guangdong/20SF316/2020 | 2020-01-24 | Asia/China/Guangdong/Huizhou | Guangdong Provincial Center for Diseases Control and Prevention;Guangdong Provincial Institute of Public Health | School of Public Health, The University of Hong Kong | Bosheng Li et al |
| EPI_ISL_428462 | Guangdong/20SF4047/2020 | 2020-02-17 | Asia/China/Guangdong/Guangzhou | Guangdong Provincial Center for Diseases Control and Prevention;Guangdong Provincial Institute of Public Health | School of Public Health, The University of Hong Kong | Bosheng Li et al |

| | | | | | | |
|---|---|---|---|---|---|---|
| EPI_ISL_428463 | Guangdong/20SF4051/2020 | 2020-02-17 | Asia/China/Guangdong/Guangzhou | Guangdong Provincial Center for Diseases Control and Prevention;Guangdong Provincial Institute of Public Health | School of Public Health, The University of Hong Kong | Bosheng Li et al |
| EPI_ISL_428464 | Guangdong/20SF602/2020 | 2020-01-25 | Asia/China/Guangdong/Zhanjiang | Guangdong Provincial Center for Diseases Control and Prevention;Guangdong Provincial Institute of Public Health | School of Public Health, The University of Hong Kong | Bosheng Li et al |
| EPI_ISL_428465 | Guangdong/20SF616/2020 | 2020-01-24 | Asia/China/Guangdong/Shenzhen | Guangdong Provincial Center for Diseases Control and Prevention;Guangdong Provincial Institute of Public Health | School of Public Health, The University of Hong Kong | Bosheng Li et al |
| EPI_ISL_428466 | Guangdong/20SF629/2020 | 2020-01-26 | Asia/China/Guangdong/Dongguan | Guangdong Provincial Center for Diseases Control and Prevention;Guangdong Provincial Institute of Public Health | School of Public Health, The University of Hong Kong | Bosheng Li et al |
| EPI_ISL_428467 | Guangdong/20SF630/2020 | 2020-01-26 | Asia/China/Guangdong/Dongguan | Guangdong Provincial Center for Diseases Control and Prevention;Guangdong Provincial Institute of Public Health | School of Public Health, The University of Hong Kong | Bosheng Li et al |
| EPI_ISL_428468 | Guangdong/20SF632/2020 | 2020-01-26 | Asia/China/Guangdong/Dongguan | Guangdong Provincial Center for Diseases Control and Prevention;Guangdong Provincial Institute of Public Health | School of Public Health, The University of Hong Kong | Bosheng Li et al |
| EPI_ISL_428469 | Guangdong/20SF665/2020 | 2020-01-27 | Asia/China/Guangdong/Zhongshan | Guangdong Provincial Center for Diseases Control and Prevention;Guangdong Provincial Institute of Public Health | School of Public Health, The University of Hong Kong | Bosheng Li et al |
| EPI_ISL_428470 | Guangdong/20SF684/2020 | 2020-01-27 | Asia/China/Guangdong/Shantou | Guangdong Provincial Center for Diseases Control and Prevention;Guangdong Provincial Institute of Public Health | School of Public Health, The University of Hong Kong | Bosheng Li et al |
| EPI_ISL_428471 | Guangdong/20SF685/2020 | 2020-01-27 | Asia/China/Guangdong/Shantou | Guangdong Provincial Center for Diseases Control and Prevention;Guangdong Provincial Institute of Public Health | School of Public Health, The University of Hong Kong | Bosheng Li et al |
| EPI_ISL_428472 | Guangdong/20SF753/2020 | 2020-01-28 | Asia/China/Guangdong/Zhuhai | Guangdong Provincial Center for Diseases Control and Prevention;Guangdong Provincial Institute of Public Health | School of Public Health, The University of Hong Kong | Bosheng Li et al |

| | | | | | | |
|---|---|---|---|---|---|---|
| EPI_ISL_428473 | Guangdong/20SF758/2020 | 2020-01-27 | Asia/China/Guangdong/Zhuhai | Guangdong Provincial Center for Diseases Control and Prevention;Guangdong Provincial Institute of Public Health | School of Public Health, The University of Hong Kong | Bosheng Li et al |
| EPI_ISL_428474 | Guangdong/20SF808/2020 | 2020-01-28 | Asia/China/Guangdong/Huizhou | Guangdong Provincial Center for Diseases Control and Prevention;Guangdong Provincial Institute of Public Health | School of Public Health, The University of Hong Kong | Bosheng Li et al |
| EPI_ISL_428475 | Guangdong/20SF812/2020 | 2020-01-28 | Asia/China/Guangdong/Huizhou | Guangdong Provincial Center for Diseases Control and Prevention;Guangdong Provincial Institute of Public Health | School of Public Health, The University of Hong Kong | Bosheng Li et al |
| EPI_ISL_428476 | Guangdong/20SF813/2020 | 2020-01-28 | Asia/China/Guangdong/Huizhou | Guangdong Provincial Center for Diseases Control and Prevention;Guangdong Provincial Institute of Public Health | School of Public Health, The University of Hong Kong | Bosheng Li et al |
| EPI_ISL_428477 | Guangdong/20SF822/2020 | 2020-01-28 | Asia/China/Guangdong/Foshan | Guangdong Provincial Center for Diseases Control and Prevention;Guangdong Provincial Institute of Public Health | School of Public Health, The University of Hong Kong | Bosheng Li et al |
| EPI_ISL_428478 | Guangdong/20SF840/2020 | 2020-01-29 | Asia/China/Guangdong/Dongguan | Guangdong Provincial Center for Diseases Control and Prevention;Guangdong Provincial Institute of Public Health | School of Public Health, The University of Hong Kong | Bosheng Li et al |
| EPI_ISL_428480 | India/KA-nimh-0116/2020 | 2020-04-06 | Asia/India/Karnataka/ | District Surveillence Unit | Department of Neurovirology, National Institute of Mental Health and Neuroscience (NIMHANS) | Chitra Pattabiraman et al |
| EPI_ISL_428481 | India/KA-nimh-0130/2020 | 2020-04-06 | Asia/India/Karnataka/ | District Surveillence Unit | Department of Neurovirology, National Institute of Mental Health and Neuroscience (NIMHANS) | Chitra Pattabiraman et al |
| EPI_ISL_428488 | Taiwan/8/2020 | 2020-01-31 | Asia/Taiwan/Taiwan/ | Centers for Disease Control, R.O.C. (Taiwan) | Centers for Disease Control, R.O.C. (Taiwan) | Ji-Rong Yang et al |

| | | | | | | |
|---|---|---|---|---|---|---|
| EPI_ISL_428489 | Taiwan/5/2020 | 2020-01-31 | Asia/Taiwan/Taiwan/ | Centers for Disease Control, R.O.C. (Taiwan) | Centers for Disease Control, R.O.C. (Taiwan) | Ji-Rong Yang et al |
| EPI_ISL_428675 | Spain/MD-H12-LP16-6193/2020 | 2020-03-09 | Europe/Spain/Madrid/ | Hospital Universitario La Paz | Hospital Universitario 12 de Octubre | Elias Dahdouh et al |
| EPI_ISL_428681 | Spain/MD-H12-LP27-2548/2020 | 2020-03-11 | Europe/Spain/Madrid/ | Hospital Universitario La Paz | Hospital Universitario 12 de Octubre | Elias Dahdouh et al |
| EPI_ISL_428682 | Spain/MD-H12-LP30-2226/2020 | 2020-03-11 | Europe/Spain/Madrid/ | Hospital Universitario La Paz | Hospital Universitario 12 de Octubre | Elias Dahdouh et al |
| EPI_ISL_428683 | Spain/MD-H12-13/2020 | 2020-03-03 | Europe/Spain/Madrid/ | Hospital Universitario 12 de Octubre | Hospital Universitario 12 de Octubre | Sara Gonzv°lez et al |
| EPI_ISL_428694 | Spain/MD-H12-31/2020 | 2020-03-12 | Europe/Spain/Madrid/ | Hospital Universitario 12 de Octubre | Hospital Universitario 12 de Octubre | Sara Gonzv°lez et al |
| EPI_ISL_428701 | Spain/MD-H12-2902/2020 | 2020-03-27 | Europe/Spain/Madrid/ | Hospital Universitario 12 de Octubre | Hospital Universitario 12 de Octubre | Esther Viedma et al |
| EPI_ISL_428702 | Spain/MD-H12-3203/2020 | 2020-03-11 | Europe/Spain/Madrid/ | Hospital Universitario 12 de Octubre | Hospital Universitario 12 de Octubre | Esther Viedma et al |
| EPI_ISL_428704 | Spain/MD-H12-3405/2020 | 2020-03-11 | Europe/Spain/Madrid/ | Hospital Universitario 12 de Octubre | Hospital Universitario 12 de Octubre | Esther Viedma et al |
| EPI_ISL_428706 | Spain/MD-H12-3607/2020 | 2020-03-21 | Europe/Spain/Madrid/ | Hospital Universitario 12 de Octubre | Hospital Universitario 12 de Octubre | Esther Viedma et al |
| EPI_ISL_428709 | Spain/MD-H12-3910/2020 | 2020-03-21 | Europe/Spain/Madrid/ | Hospital Universitario 12 de Octubre | Hospital Universitario 12 de Octubre | Esther Viedma et al |
| EPI_ISL_428713 | Turkey/HSGM-5516/2020 | 2020-03-18 | Europe/Turkey/Ankara/ | Ministry of Health Turkey | Ministry of Health Turkey | Fatma Bayrakdar et al |
| EPI_ISL_428716 | Turkey/HSGM-5711/2020 | 2020-03-18 | Europe/Turkey/Ankara/ | Ministry of Health Turkey | Ministry of Health Turkey | Fatma Bayrakdar et al |
| EPI_ISL_428719 | Turkey/HSGM-7718/2020 | 2020-03-21 | Europe/Turkey/Siirt/ | Ministry of Health Turkey | Ministry of Health Turkey | Fatma Bayrakdar et al |
| EPI_ISL_428721 | Turkey/HSGM-8003/2020 | 2020-03-21 | Europe/Turkey/Ankara/ | Ministry of Health Turkey | Ministry of Health Turkey | Fatma Bayrakdar et al |

| | | | | | | |
|---|---|---|---|---|---|---|
| EPI_ISL_428722 | Turkey/HSGM-8010/2020 | 2020-03-22 | Europe/Turkey/Balikesir/ | Ministry of Health Turkey | Ministry of Health Turkey | Fatma Bayrakdar et al |
| EPI_ISL_428723 | Turkey/HSGM-8964/2020 | 2020-03-22 | Europe/Turkey/Aksaray/ | Ministry of Health Turkey | Ministry of Health Turkey | Fatma Bayrakdar et al |
| EPI_ISL_428724 | USA/WI-UW-250/2020 | 2020-03-13 | North America/USA/Wisconsin/ | University of Wisconsin-Madison AIDS Vaccine Research Laboratories | University of Wisconsin-Madison AIDS Vaccine Research Laboratories | Gage Moreno et al |
| EPI_ISL_428728 | USA/WI-UW-254/2020 | 2020-04-01 | North America/USA/Wisconsin/ | University of Wisconsin-Madison AIDS Vaccine Research Laboratories | University of Wisconsin-Madison AIDS Vaccine Research Laboratories | Gage Moreno et al |
| EPI_ISL_428731 | USA/WI-UW-257/2020 | 2020-04-06 | North America/USA/Wisconsin/ | University of Wisconsin-Madison AIDS Vaccine Research Laboratories | University of Wisconsin-Madison AIDS Vaccine Research Laboratories | Gage Moreno et al |
| EPI_ISL_428746 | USA/CT-Yale-082/2020 | 2020-04-07 | North America/USA/Connecticut/ | Yale COVID-19 Biorepository | Grubaugh Lab - Yale School of Public Health | Joseph Fauver et al |
| EPI_ISL_428749 | USA/CT-Yale-098/2020 | 2020-04-02 | North America/USA/Connecticut/ | Yale COVID-19 Biorepository | Grubaugh Lab - Yale School of Public Health | Joseph Fauver et al |
| EPI_ISL_428750 | USA/CT-Yale-099/2020 | 2020-04-07 | North America/USA/Connecticut/ | Yale COVID-19 Biorepository | Grubaugh Lab - Yale School of Public Health | Joseph Fauver et al |
| EPI_ISL_428781 | USA/NY-NYUMC267/2020 | 2020-04-06 | North America/USA/New York/Brooklyn | NYU Langone Health | Departments of Pathology and Medicine, New York University School of Medicine | Maria Aguero-Rosenfeld et al |
| EPI_ISL_428783 | USA/NY-NYUMC269/2020 | 2020-04-06 | North America/USA/New York/Brooklyn | NYU Langone Health | Departments of Pathology and Medicine, New York University School of Medicine | Maria Aguero-Rosenfeld et al |

| | | | | | | |
|---|---|---|---|---|---|---|
| EPI_ISL_428786 | USA/NY-NYUMC272/2020 | 2020-04-06 | North America/USA/New York/Brooklyn | NYU Langone Health | Departments of Pathology and Medicine, New York University School of Medicine | Maria Aguero-Rosenfeld et al |
| EPI_ISL_428792 | USA/NY-NYUMC278/2020 | 2020-04-04 | North America/USA/New York/Brooklyn | NYU Langone Health | Departments of Pathology and Medicine, New York University School of Medicine | Maria Aguero-Rosenfeld et al |
| EPI_ISL_428822 | Singapore/43/2020 | 2020-02-16 | Asia/Singapore/Singapore/ | National Public Health Laboratory, National Centre for Infectious Diseases | National Public Health Laboratory, National Centre for Infectious Diseases | Mak TM et al |
| EPI_ISL_428830 | Singapore/51/2020 | 2020-02-27 | Asia/Singapore/Singapore/ | National Public Health Laboratory, National Centre for Infectious Diseases | National Public Health Laboratory, National Centre for Infectious Diseases | Mak TM et al |
| EPI_ISL_428854 | Italy/FVG-ICGEB-S18/2020 | 2020-03-30 | Europe/Italy/Friuli Venezia Giulia/ | Laboratory of Molecular Virology International Center for Genetic Engineering and Biotechnology (ICGEB) | ARGO Open Lab Platform for Genome sequencing | Licastro D et al |
| EPI_ISL_428864 | Russia/Moscow-62502/2020 | 2020-03-11 | Europe/Russia/Moscow Oblast/ | State Research Center of Virology and Biotechnology VECTOR, Department of Collection of Microorganisms | State Research Center of Virology and Biotechnology VECTOR, Department of Collection of Microorganisms | Oleg V. Pyankov et al |

| | | | | | | |
|---|---|---|---|---|---|---|
| EPI_ISL_428866 | Russia/Lipetsk-62707/2020 | 2020-03-11 | Europe/Russia/Lipetsk/ | State Research Center of Virology and Biotechnology VECTOR, Department of Collection of Microorganisms | State Research Center of Virology and Biotechnology VECTOR, Department of Collection of Microorganisms | Oleg V. Pyankov et al |
| EPI_ISL_428871 | Russia/Krasnoyarsk-66501/2020 | 2020-03-13 | Europe/Russia/Krasnoyarsk/ | State Research Center of Virology and Biotechnology VECTOR, Department of Collection of Microorganisms | State Research Center of Virology and Biotechnology VECTOR, Department of Collection of Microorganisms | Oleg V. Pyankov et al |
| EPI_ISL_428872 | Russia/Moscow-71605/2020 | 2020-03-19 | Europe/Russia/Moscow Oblast/ | State Research Center of Virology and Biotechnology VECTOR, Department of Collection of Microorganisms | State Research Center of Virology and Biotechnology VECTOR, Department of Collection of Microorganisms | Sergey A. Bodnev et al |
| EPI_ISL_428878 | Russia/Omsk-89001/2020 | 2020-04-01 | Europe/Russia/Omsk/ | State Research Center of Virology and Biotechnology VECTOR, Department of Collection of Microorganisms | State Research Center of Virology and Biotechnology VECTOR, Department of Collection of Microorganisms | Sergey A. Bodnev et al |
| EPI_ISL_428901 | Crimea/SRC-80603/2020 | 2020-03-23 | Europe/Ukraine/Crimea/Crimea | State Research Center of Virology and Biotechnology VECTOR, Department of Collection of Microorganisms | State Research Center of Virology and Biotechnology VECTOR, Department of Collection of Microorganisms | Sergey A. Bodnev et al |

| | | | | | | |
|---|---|---|---|---|---|---|
| EPI_ISL_428907 | Russia/Sakha-89702/2020 | 2020-04-01 | Europe/Russia/Republic of Sakha/ | State Research Center of Virology and Biotechnology VECTOR, Department of Collection of Microorganisms | State Research Center of Virology and Biotechnology VECTOR, Department of Collection of Microorganisms | Oleg V. Pyankov et al |
| EPI_ISL_428910 | Russia/Khabarovsk-88201/2020 | 2020-03-31 | Europe/Russia/Khabarovsk/ | State Research Center of Virology and Biotechnology VECTOR, Department of Collection of Microorganisms | State Research Center of Virology and Biotechnology VECTOR, Department of Collection of Microorganisms | Oleg V. Pyankov et al |
| EPI_ISL_428911 | Russia/Buryatia-88903/2020 | 2020-04-01 | Europe/Russia/Buryatia/ | State Research Center of Virology and Biotechnology VECTOR, Department of Collection of Microorganisms | State Research Center of Virology and Biotechnology VECTOR, Department of Collection of Microorganisms | Oleg V. Pyankov et al |
| EPI_ISL_428917 | Russia/Tatarstan-82208/2020 | 2020-03-24 | Europe/Russia/Tatarstan/ | State Research Center of Virology and Biotechnology VECTOR, Department of Collection of Microorganisms | State Research Center of Virology and Biotechnology VECTOR, Department of Collection of Microorganisms | Sergey A. Bodnev et al |
| EPI_ISL_428929 | Poland/PL_P6/2020 | 2020-03-29 | Europe/Poland/Malopolskie/ | ViroGenetics - BSL3 Laboratory of Virology; Human Genome Variation Research Group & Genomics Centre MCB; Bioinformatics Research Group; Wojewv≥dzka Stacja Sanitarno-Epidemiologiczna w Krakowie | ViroGenetics - BSL3 Laboratory of Virology; Human Genome Variation Research Group & Genomics Centre MCB; Bioinformatics Research Group | Wojciech Branicki et al |

| | | | | | | |
|---|---|---|---|---|---|---|
| EPI_ISL_428935 | USA/WI-UW-259/2020 | 2020-04-17 | North America/USA/Wisconsin/Dane Co | University of Wisconsin-Madison AIDS Vaccine Research Laboratories | University of Wisconsin-Madison AIDS Vaccine Research Laboratories | Gage Moreno et al |
| EPI_ISL_428945 | Luxembourg/LNS3702558/2020 | 2020-04-03 | Europe/Luxembourg/Luxembourg/ | Laboratoire National de Sante, Microbiology, Virology | Laboratoire National de Sante, Microbiology, Epidemiology and Microbial Genomics | Anke Wienecke-Baldacchino et al |
| EPI_ISL_428992 | USA/CA-CZB0138/2020 | 2020-04-08 | North America/USA/California/San Fran | UCSF Clinical Microbiology Laboratory | Chan-Zuckerberg Biohub | CZB Cliahub Consortium et al |
| EPI_ISL_429007 | USA/CA-CZB0458/2020 | 2020-03-18 | North America/USA/California/San Fran | UCSF Clinical Microbiology Laboratory | Chan-Zuckerberg Biohub | CZB Cliahub Consortium et al |
| EPI_ISL_429013 | USA/CA-CZB080/2020 | 2020-03-28 | North America/USA/California/San Fran | UCSF Clinical Microbiology Laboratory | Chan-Zuckerberg Biohub | CZB Cliahub Consortium et al |
| EPI_ISL_429037 | USA/CA-CZB09/2020 | 2020-03-25 | North America/USA/California/San Fran | UCSF Clinical Microbiology Laboratory | Chan-Zuckerberg Biohub | CZB Cliahub Consortium et al |
| EPI_ISL_429040 | USA/CA-CZB0701/2020 | 2020-03-23 | North America/USA/California/San Fran | UCSF Clinical Microbiology Laboratory | Chan-Zuckerberg Biohub | CZB Cliahub Consortium et al |
| EPI_ISL_429043 | USA/CA-CZB0156/2020 | 2020-04-08 | North America/USA/California/San Fran | UCSF Clinical Microbiology Laboratory | Chan-Zuckerberg Biohub | CZB Cliahub Consortium et al |
| EPI_ISL_429044 | USA/CA-CZB0158/2020 | 2020-04-08 | North America/USA/California/San Fran | UCSF Clinical Microbiology Laboratory | Chan-Zuckerberg Biohub | CZB Cliahub Consortium et al |
| EPI_ISL_429049 | USA/CA-CZB090/2020 | 2020-03-28 | North America/USA/California/San Fran | UCSF Clinical Microbiology Laboratory | Chan-Zuckerberg Biohub | CZB Cliahub Consortium et al |
| EPI_ISL_429051 | USA/CA-CZB0741/2020 | 2020-04-03 | North America/USA/California/San Fran | UCSF Clinical Microbiology Laboratory | Chan-Zuckerberg Biohub | CZB Cliahub Consortium et al |
| EPI_ISL_429061 | USA/CA-CZB0151/2020 | 2020-04-08 | North America/USA/California/San Fran | UCSF Clinical Microbiology Laboratory | Chan-Zuckerberg Biohub | CZB Cliahub Consortium et al |
| EPI_ISL_429062 | USA/CA-CZB0145/2020 | 2020-04-08 | North America/USA/California/San Fran | UCSF Clinical Microbiology Laboratory | Chan-Zuckerberg Biohub | CZB Cliahub Consortium et al |
| EPI_ISL_429074 | Guangzhou/GZMU0003/2020 | 2020-02-07 | Asia/China/Guangdong/Guangzhou | The First Affiliated Hospital of Guangzhou Medical University | BGI-shenzhen & The First Affiliated Hospital of Guangzhou Medical University | Yanqun Wang et al |

| | | | | | | |
|---|---|---|---|---|---|---|
| EPI_ISL_429075 | Guangzhou/GZMU0008/2020 | 2020-02-07 | Asia/China/Guangdong/Guangzhou | The First Affiliated Hospital of Guangzhou Medical University | BGI-shenzhen & The First Affiliated Hospital of Guangzhou Medical University | Yanqun Wang et al |
| EPI_ISL_429076 | Guangzhou/GZMU0009/2020 | 2020-02-07 | Asia/China/Guangdong/Guangzhou | The First Affiliated Hospital of Guangzhou Medical University | BGI-shenzhen & The First Affiliated Hospital of Guangzhou Medical University | et al |
| EPI_ISL_429077 | Guangzhou/GZMU0010/2020 | 2020-02-07 | Asia/China/Guangdong/Guangzhou | The First Affiliated Hospital of Guangzhou Medical University | BGI-shenzhen & The First Affiliated Hospital of Guangzhou Medical University | Yanqun Wang et al |
| EPI_ISL_429078 | Guangzhou/GZMU0013/2020 | 2020-02-07 | Asia/China/Guangdong/Guangzhou | The First Affiliated Hospital of Guangzhou Medical University | BGI-shenzhen & The First Affiliated Hospital of Guangzhou Medical University | et al |
| EPI_ISL_429079 | Guangzhou/GZMU0017/2020 | 2020-02-05 | Asia/China/Guangdong/Guangzhou | The First Affiliated Hospital of Guangzhou Medical University | BGI-shenzhen & The First Affiliated Hospital of Guangzhou Medical University | et al |
| EPI_ISL_429080 | Guangzhou/GZMU0019/2020 | 2020-02-05 | Asia/China/Guangdong/Guangzhou | The First Affiliated Hospital of Guangzhou Medical University | BGI-shenzhen & The First Affiliated Hospital of Guangzhou Medical University | et al |

| | | | | | | |
|---|---|---|---|---|---|---|
| EPI_ISL_429081 | Guangzhou/GZMU0020/2020 | 2020-02-05 | Asia/China/Guangdong/Guangzhou | The First Affiliated Hospital of Guangzhou Medical University | BGI-shenzhen & The First Affiliated Hospital of Guangzhou Medical University | et al |
| EPI_ISL_429082 | Guangzhou/GZMU0023/2020 | 2020-02-05 | Asia/China/Guangdong/Guangzhou | The First Affiliated Hospital of Guangzhou Medical University | BGI-shenzhen & The First Affiliated Hospital of Guangzhou Medical University | Yanqun Wang et al |
| EPI_ISL_429083 | Guangzhou/GZMU0024/2020 | 2020-02-05 | Asia/China/Guangdong/Guangzhou | The First Affiliated Hospital of Guangzhou Medical University | BGI-shenzhen & The First Affiliated Hospital of Guangzhou Medical University | Yanqun Wang et al |
| EPI_ISL_429084 | Guangzhou/GZMU0025/2020 | 2020-02-05 | Asia/China/Guangdong/Guangzhou | The First Affiliated Hospital of Guangzhou Medical University | BGI-shenzhen & The First Affiliated Hospital of Guangzhou Medical University | et al |
| EPI_ISL_429085 | Guangzhou/GZMU0033/2020 | 2020-02-01 | Asia/China/Guangdong/Guangzhou | The First Affiliated Hospital of Guangzhou Medical University | BGI-shenzhen & The First Affiliated Hospital of Guangzhou Medical University | Yanqun Wang et al |
| EPI_ISL_429086 | Guangzhou/GZMU0034/2020 | 2020-02-01 | Asia/China/Guangdong/Guangzhou | The First Affiliated Hospital of Guangzhou Medical University | BGI-shenzhen & The First Affiliated Hospital of Guangzhou Medical University | et al |

| | | | | | | |
|---|---|---|---|---|---|---|
| EPI_ISL_429088 | Guangzhou/GZMU0035/2020 | 2020-02-01 | Asia/China/Guangdong/Guangzhou | The First Affiliated Hospital of Guangzhou Medical University | BGI-shenzhen & The First Affiliated Hospital of Guangzhou Medical University | Yanqun Wang et al |
| EPI_ISL_429089 | Guangzhou/GZMU0037/2020 | 2020-02-08 | Asia/China/Guangdong/Guangzhou | The First Affiliated Hospital of Guangzhou Medical University | BGI-shenzhen & The First Affiliated Hospital of Guangzhou Medical University | Yanqun Wang et al |
| EPI_ISL_429090 | Guangzhou/GZMU0039/2020 | 2020-02-08 | Asia/China/Guangdong/Guangzhou | The First Affiliated Hospital of Guangzhou Medical University | BGI-shenzhen & The First Affiliated Hospital of Guangzhou Medical University | Yanqun Wang et al |
| EPI_ISL_429091 | Guangzhou/GZMU0041/2020 | 2020-02-08 | Asia/China/Guangdong/Guangzhou | The First Affiliated Hospital of Guangzhou Medical University | BGI-shenzhen & The First Affiliated Hospital of Guangzhou Medical University | Yanqun Wang et al |
| EPI_ISL_429092 | Guangzhou/GZMU0045/2020 | 2020-02-08 | Asia/China/Guangdong/Guangzhou | The First Affiliated Hospital of Guangzhou Medical University | BGI-shenzhen & The First Affiliated Hospital of Guangzhou Medical University | Yanqun Wang et al |
| EPI_ISL_429093 | Guangzhou/GZMU0046/2020 | 2020-02-08 | Asia/China/Guangdong/Guangzhou | The First Affiliated Hospital of Guangzhou Medical University | BGI-shenzhen & The First Affiliated Hospital of Guangzhou Medical University | Yanqun Wang et al |

| | | | | | | |
|---|---|---|---|---|---|---|
| EPI_ISL_429094 | Guangzhou/GZMU0049/2020 | 2020-02-08 | Asia/China/Guangdong/Guangzhou | The First Affiliated Hospital of Guangzhou Medical University | BGI-shenzhen & The First Affiliated Hospital of Guangzhou Medical University | et al |
| EPI_ISL_429095 | Guangzhou/GZMU0050/2020 | 2020-02-08 | Asia/China/Guangdong/Guangzhou | The First Affiliated Hospital of Guangzhou Medical University | BGI-shenzhen & The First Affiliated Hospital of Guangzhou Medical University | et al |
| EPI_ISL_429096 | Guangzhou/GZMU0051/2020 | 2020-01-29 | Asia/China/Guangdong/Guangzhou | The First Affiliated Hospital of Guangzhou Medical University | BGI-shenzhen & The First Affiliated Hospital of Guangzhou Medical University | Yanqun Wang et al |
| EPI_ISL_429097 | Guangzhou/GZMU0052/2020 | 2020-01-29 | Asia/China/Guangdong/Guangzhou | The First Affiliated Hospital of Guangzhou Medical University | BGI-shenzhen & The First Affiliated Hospital of Guangzhou Medical University | Yanqun Wang et al |
| EPI_ISL_429098 | Guangzhou/GZMU0054/2020 | 2020-01-29 | Asia/China/Guangdong/Guangzhou | The First Affiliated Hospital of Guangzhou Medical University | BGI-shenzhen & The First Affiliated Hospital of Guangzhou Medical University | Yanqun Wang et al |
| EPI_ISL_429099 | Guangzhou/GZMU0056/2020 | 2020-01-29 | Asia/China/Guangdong/Guangzhou | The First Affiliated Hospital of Guangzhou Medical University | BGI-shenzhen & The First Affiliated Hospital of Guangzhou Medical University | et al |

| | | | | | | |
|---|---|---|---|---|---|---|
| EPI_ISL_429100 | Guangzhou/GZMU0057/2020 | 2020-01-27 | Asia/China/Guangdong/Guangzhou | The First Affiliated Hospital of Guangzhou Medical University | BGI-shenzhen & The First Affiliated Hospital of Guangzhou Medical University | et al |
| EPI_ISL_429101 | Guangzhou/GZMU0058/2020 | 2020-01-27 | Asia/China/Guangdong/Guangzhou | The First Affiliated Hospital of Guangzhou Medical University | BGI-shenzhen & The First Affiliated Hospital of Guangzhou Medical University | et al |
| EPI_ISL_429102 | Guangzhou/GZMU0059/2020 | 2020-01-27 | Asia/China/Guangdong/Guangzhou | The First Affiliated Hospital of Guangzhou Medical University | BGI-shenzhen & The First Affiliated Hospital of Guangzhou Medical University | et al |
| EPI_ISL_429103 | Guangzhou/GZMU0060/2020 | 2020-02-09 | Asia/China/Guangdong/Guangzhou | The First Affiliated Hospital of Guangzhou Medical University | BGI-shenzhen & The First Affiliated Hospital of Guangzhou Medical University | et al |
| EPI_ISL_429104 | Guangzhou/GZMU0036/2020 | 2020-02-01 | Asia/China/Guangdong/Guangzhou | The First Affiliated Hospital of Guangzhou Medical University | BGI-shenzhen & The First Affiliated Hospital of Guangzhou Medical University | et al |
| EPI_ISL_429105 | Guangzhou/GZMU0055/2020 | 2020-01-29 | Asia/China/Guangdong/Guangzhou | The First Affiliated Hospital of Guangzhou Medical University | BGI-shenzhen & The First Affiliated Hospital of Guangzhou Medical University | et al |
| EPI_ISL_429208 | Switzerland/GE-HUG-2164/2020 | 2020-03-17 | Europe/Switzerland/Geneva/ | University Hospitals of Geneva Laboratory of Virology | University Hospitals of Geneva Laboratory of Virology | Laubscher F. et al |

| | | | | | | |
|---|---|---|---|---|---|---|
| EPI_ISL_429213 | Switzerland/GE-HUG-8086/2020 | 2020-03-16 | Europe/Switzerland/Geneva/ | University Hospitals of Geneva Laboratory of Virology | University Hospitals of Geneva Laboratory of Virology | Laubscher F. et al |
| EPI_ISL_429218 | Switzerland/GE-HUG-0304/2020 | 2020-04-02 | Europe/Switzerland/Geneva/ | University Hospitals of Geneva Laboratory of Virology | University Hospitals of Geneva Laboratory of Virology | Laubscher F. et al |
| EPI_ISL_429227 | Italy/ABR-IZSGC-TE5553/2020 | 2020-03-17 | Europe/Italy/Abruzzo/ | Presidio Ospedaliero Santo Spirito | "Istituto Zooprofilattico Sperimentale dell'Abruzzo e Molise ""G. Caporale""" | Lorusso A et al |
| EPI_ISL_429228 | Italy/ABR-IZSGC-TE5674/2020 | 2020-03-20 | Europe/Italy/Abruzzo/ | Ospedale Civile Giuseppe Mazzini | "Istituto Zooprofilattico Sperimentale dell'Abruzzo e Molise ""G. Caporale""" | Lorusso A et al |
| EPI_ISL_429231 | Italy/ABR-IZSGC-TE6571/2020 | 2020-03-24 | Europe/Italy/Abruzzo/ | Ospedale Civile Giuseppe Mazzini | "Istituto Zooprofilattico Sperimentale dell'Abruzzo e Molise ""G. Caporale""" | Lorusso A et al |
| EPI_ISL_429233 | Italy/ABR-IZSGC-TE6575/2020 | 2020-03-24 | Europe/Italy/Abruzzo/ | Ospedale Civile Giuseppe Mazzini | "Istituto Zooprofilattico Sperimentale dell'Abruzzo e Molise ""G. Caporale""" | Lorusso A et al |
| EPI_ISL_429234 | Italy/ABR-IZSGC-TE6577/2020 | 2020-03-24 | Europe/Italy/Abruzzo/ | Ospedale Civile Giuseppe Mazzini | "Istituto Zooprofilattico Sperimentale dell'Abruzzo e Molise ""G. Caporale""" | Lorusso A et al |

| | | | | | | |
|---|---|---|---|---|---|---|
| EPI_ISL_429577 | Denmark/ALAB-SSI509/2020 | 2020-03-25 | Europe/Denmark/Denmark/ | Department of Virus and Microbiological Special Diagnostics, Statens Serum Institut, Copenhagen, Denmark, Artillerivej 5, 2300 Copenahgen S | Albertsen lab, Department of Chemistry and Bioscience, Aalborg University, Denmark | Rasmus Kirkegaard et al |
| EPI_ISL_429585 | Denmark/ALAB-SSI520/2020 | 2020-03-26 | Europe/Denmark/Denmark/ | Department of Virus and Microbiological Special Diagnostics, Statens Serum Institut, Copenhagen, Denmark, Artillerivej 5, 2300 Copenahgen S | Albertsen lab, Department of Chemistry and Bioscience, Aalborg University, Denmark | Rasmus Kirkegaard et al |
| EPI_ISL_429598 | USA/WA-UW-4546/2020 | 2020-03-31 | North America/USA/Washington/ | UW Virology Lab | UW Virology Lab | Pavitra Roychoudhury et al |
| EPI_ISL_429611 | USA/WA-UW-4602/2020 | 2020-03-31 | North America/USA/Washington/ | UW Virology Lab | UW Virology Lab | Pavitra Roychoudhury et al |
| EPI_ISL_429618 | USA/WA-UW-4741/2020 | 2020-03-25 | North America/USA/Washington/ | UW Virology Lab | UW Virology Lab | Pavitra Roychoudhury et al |
| EPI_ISL_429629 | USA/WA-UW-4649/2020 | 2020-04-06 | North America/USA/Washington/ | UW Virology Lab | UW Virology Lab | Pavitra Roychoudhury et al |
| EPI_ISL_429633 | USA/WA-UW-4707/2020 | 2020-04-06 | North America/USA/Washington/ | UW Virology Lab | UW Virology Lab | Pavitra Roychoudhury et al |
| EPI_ISL_429646 | USA/CT-UW-5086/2020 | 2020-04-06 | North America/USA/Connecticut/ | UW Virology Lab | UW Virology Lab | Pavitra Roychoudhury et al |
| EPI_ISL_429647 | USA/WA-UW-5095/2020 | 2020-04-08 | North America/USA/Washington/ | UW Virology Lab | UW Virology Lab | Pavitra Roychoudhury et al |
| EPI_ISL_429651 | USA/WA-UW-5158/2020 | 2020-04-02 | North America/USA/Washington/ | UW Virology Lab | UW Virology Lab | Pavitra Roychoudhury et al |
| EPI_ISL_429674 | Brazil/MG-CV12/2020 | 2020-03-11 | South America/Brazil/Minas Gerais/ | Central Public Health Laboratory/Oct√°vio Magalh√£es Institute (IOM) from the Ezequiel Dias Foundation (FUNED) | Instituto Oct√°vio Magalh√£es / Funda√ß√£o Ezequiel Dias (IOM/Funed) | Talita Adelino et al |
| EPI_ISL_429696 | Brazil/MG-CV43/2020 | 2020-03-17 | South America/Brazil/Minas Gerais/ | Central Public Health Laboratory/Oct√°vio Magalh√£es Institute (IOM) from the Ezequiel Dias Foundation (FUNED) | Instituto Oct√°vio Magalh√£es / Funda√ß√£o Ezequiel Dias (IOM/Funed) | Talita Adelino et al |

| | | | | | | |
|---|---|---|---|---|---|---|
| EPI_ISL_429742 | Luxembourg/LNS1658150/2020 | 2020-04-10 | Europe/Luxembourg/Luxembourg/ | Laboratoire National de Sante, Microbiology, Virology | Laboratoire National de Sante, Microbiology, Epidemiology and Microbial Genomics | Anke Wienecke-Baldacchino et al |
| EPI_ISL_429750 | Luxembourg/LNS9598665/2020 | 2020-03-18 | Europe/Luxembourg/Luxembourg/ | Laboratoire National de Sante, Microbiology, Virology | Laboratoire National de Sante, Microbiology, Epidemiology and Microbial Genomics | Anke Wienecke-Baldacchino et al |
| EPI_ISL_429751 | Luxembourg/LNS4576534/2020 | 2020-03-28 | Europe/Luxembourg/Luxembourg/ | Laboratoire National de Sante, Microbiology, Virology | Laboratoire National de Sante, Microbiology, Epidemiology and Microbial Genomics | Anke Wienecke-Baldacchino et al |
| EPI_ISL_429777 | Luxembourg/LNS0994857/2020 | 2020-03-24 | Europe/Luxembourg/Luxembourg/ | Laboratoire National de Sante, Microbiology, Virology | Laboratoire National de Sante, Microbiology, Epidemiology and Microbial Genomics | Anke Wienecke-Baldacchino et al |
| EPI_ISL_429799 | Luxembourg/LNS4284220/2020 | 2020-03-30 | Europe/Luxembourg/Luxembourg/ | Laboratoire National de Sante, Microbiology, Virology | Laboratoire National de Sante, Microbiology, Epidemiology and Microbial Genomics | Anke Wienecke-Baldacchino et al |
| EPI_ISL_429806 | Canada/NB_1/2020 | 2020-03-09 | North America/Canada/New Brunswick/ | Dr. Georges-L.-Dumont University Hospital Centre | National Microbiology Laboratory | Anna Majer et al |
| EPI_ISL_429807 | Canada/MB_2/2020 | 2020-03-10 | North America/Canada/Manitoba/ | Cadham Provincial Laboratory | National Microbiology Laboratory | Anna Majer et al |

| | | | | | | |
|---|---|---|---|---|---|---|
| EPI_ISL_429811 | Canada/NB_21/2020 | 2020-03-12 | North America/Canada/New Brunswick/ | Dr. Georges-L.-Dumont University Hospital Centre | National Microbiology Laboratory | Anna Majer et al |
| EPI_ISL_429812 | Canada/NS_30/2020 | 2020-03-14 | North America/Canada/Nova Scotia/ | Queen Elizabeth II Health Science Centre | National Microbiology Laboratory | Anna Majer et al |
| EPI_ISL_429813 | Canada/NS_31/2020 | 2020-03-15 | North America/Canada/Nova Scotia/ | Queen Elizabeth II Health Science Centre | National Microbiology Laboratory | Anna Majer et al |
| EPI_ISL_429814 | Canada/NS_32/2020 | 2020-03-15 | North America/Canada/Nova Scotia/ | Queen Elizabeth II Health Science Centre | National Microbiology Laboratory | Anna Majer et al |
| EPI_ISL_429815 | Canada/NL_34/2020 | 2020-03-15 | North America/Canada/Newfoundland a | Public Health Laboratory | National Microbiology Laboratory | Anna Majer et al |
| EPI_ISL_429816 | Canada/NB_35/2020 | 2020-03-14 | North America/Canada/New Brunswick/ | Dr. Georges-L.-Dumont University Hospital Centre | National Microbiology Laboratory | Anna Majer et al |
| EPI_ISL_429817 | Canada/MB_36/2020 | 2020-03-14 | North America/Canada/Manitoba/ | Cadham Provincial Laboratory | National Microbiology Laboratory | Anna Majer et al |
| EPI_ISL_429818 | Canada/MB_37/2020 | 2020-03-18 | North America/Canada/Manitoba/ | Cadham Provincial Laboratory | National Microbiology Laboratory | Anna Majer et al |
| EPI_ISL_429819 | Canada/MB_38/2020 | 2020-03-18 | North America/Canada/Manitoba/ | Cadham Provincial Laboratory | National Microbiology Laboratory | Anna Majer et al |
| EPI_ISL_429820 | Canada/MB_39/2020 | 2020-03-14 | North America/Canada/Manitoba/ | Cadham Provincial Laboratory | National Microbiology Laboratory | Anna Majer et al |
| EPI_ISL_429852 | Lishui/LS111/2020 | 2020-01-25 | Asia/China/Zhejiang/Lishui | Centers for Disease Control and Prevention of Lishui | Department of InspectionÔºåCenters for Disease Control and Prevention of Lishui | Wang Xiaoguang et al |

| | | | | | | |
|---|---|---|---|---|---|---|
| EPI_ISL_429853 | Lishui/LS556/2020 | 2020-01-28 | Asia/China/Zhejiang/Lishui | Centers for Disease Control and Prevention of Lishui | Department of InspectionÔºåCenters for Disease Control and Prevention of Lishui | Wang Xiaoguang et al |
| EPI_ISL_429854 | Lishui/LS557/2020 | 2020-02-01 | Asia/China/Zhejiang/Lishui | Centers for Disease Control and Prevention of Lishui | Department of InspectionÔºåCenters for Disease Control and Prevention of Lishui | Wang Xiaoguang et al |
| EPI_ISL_429861 | Turkey/HSGM-8968/2020 | 2020-03-22 | Europe/Turkey/Ankara/ | Ministry of Health Turkey | Ministry of Health Turkey | Fatma Bayrakdar et al |
| EPI_ISL_429862 | Turkey/HSGM-8970/2020 | 2020-03-22 | Europe/Turkey/Ankara/ | Ministry of Health Turkey | Ministry of Health Turkey | Fatma Bayrakdar et al |
| EPI_ISL_429863 | Turkey/HSGM-8975/2020 | 2020-03-22 | Europe/Turkey/Sakarya/ | Ministry of Health Turkey | Ministry of Health Turkey | Fatma Bayrakdar et al |
| EPI_ISL_429864 | Turkey/HSGM-8979/2020 | 2020-03-22 | Europe/Turkey/Sakarya/ | Ministry of Health Turkey | Ministry of Health Turkey | Fatma Bayrakdar et al |
| EPI_ISL_429865 | Turkey/HSGM-8992/2020 | 2020-03-18 | Europe/Turkey/Canakkale/ | Ministry of Health Turkey | Ministry of Health Turkey | Fatma Bayrakdar et al |
| EPI_ISL_429866 | Turkey/HSGM-4236/2020 | 2020-03-16 | Europe/Turkey/Afyon/ | Ministry of Health Turkey | Ministry of Health Turkey | Fatma Bayrakdar et al |
| EPI_ISL_429867 | Turkey/HSGM-4505/2020 | 2020-03-17 | Europe/Turkey/Balikesir/ | Ministry of Health Turkey | Ministry of Health Turkey | Fatma Bayrakdar et al |
| EPI_ISL_429869 | Turkey/HSGM-4701/2020 | 2020-03-17 | Europe/Turkey/Konya/ | Ministry of Health Turkey | Ministry of Health Turkey | Fatma Bayrakdar et al |
| EPI_ISL_429870 | Turkey/HSGM-8990/2020 | 2020-03-22 | Europe/Turkey/Sakarya/ | Ministry of Health Turkey | Ministry of Health Turkey | Fatma Bayrakdar et al |
| EPI_ISL_429872 | Turkey/HSGM-11518/2020 | 2020-03-25 | Europe/Turkey/Kocaeli/ | Ministry of Health Turkey | Ministry of Health Turkey | Fatma Bayrakdar et al |
| EPI_ISL_429873 | Turkey/HSGM-11520/2020 | 2020-03-23 | Europe/Turkey/Kocaeli/ | Ministry of Health Turkey | Ministry of Health Turkey | Fatma Bayrakdar et al |
| EPI_ISL_429884 | Taiwan/284/2020 | 2020-03-30 | Asia/Taiwan/Taiwan/ | Centers for Disease Control, R.O.C. (Taiwan) | Centers for Disease Control, R.O.C. (Taiwan) | Ji-Rong Yang et al |

| | | | | | | |
|---|---|---|---|---|---|---|
| EPI_ISL_429968 | France/HDF-1463/2020 | 2020-02-21 | Europe/France/Hauts de France/Compiv | Centre Hospitalier Compiv®gne Laboratoire de Biologie | National Reference Center for Viruses of Respiratory Infections, Institut Pasteur, Paris | Mv©lanie Albert et al |
| EPI_ISL_429971 | USA/VA-DCLS-0071/2020 | 2020-03-30 | North America/USA/Virginia/ | Virginia DCLS | Virginia DCLS | Virginia DCLS et al |
| EPI_ISL_430014 | Jordan/SR-056/2020 | 2020-03-17 | Asia/Jordan/Amman/ | Biolab Diagnostic Laboratories | Andersen lab at Scripps Research | Issa Abu-Dayyeh et al |
| EPI_ISL_430019 | USA/UT-066/2020 | 2020-03-16 | North America/USA/Utah/ | Utah Public Health Laboratory | Utah Public Health Laboratory | Erin Young et al |
| EPI_ISL_430021 | USA/UT-069/2020 | 2020-03-16 | North America/USA/Utah/ | Utah Public Health Laboratory | Utah Public Health Laboratory | Erin Young et al |
| EPI_ISL_430031 | USA/UT-080/2020 | 2020-03-15 | North America/USA/Utah/ | Utah Public Health Laboratory | Utah Public Health Laboratory | Erin Young et al |
| EPI_ISL_430044 | USA/UT-0446/2020 | 2020-03-30 | North America/USA/Utah/ | Utah Public Health Laboratory | Utah Public Health Laboratory | Erin Young et al |
| EPI_ISL_430048 | USA/UT-0495/2020 | 2020-04-02 | North America/USA/Utah/ | Utah Public Health Laboratory | Utah Public Health Laboratory | Erin Young et al |
| EPI_ISL_430055 | USA/UT-0507/2020 | 2020-04-02 | North America/USA/Utah/ | Utah Public Health Laboratory | Utah Public Health Laboratory | Erin Young et al |
| EPI_ISL_430056 | USA/UT-0508/2020 | 2020-04-02 | North America/USA/Utah/ | Utah Public Health Laboratory | Utah Public Health Laboratory | Erin Young et al |
| EPI_ISL_430067 | Russia/StPetersburg-RII4705S/2020 | 2020-04-07 | Europe/Russia/Saint-Petersburg/ | WHO National Influenza Centre Russian Federation | WHO National Influenza Centre Russian Federation | Andrey Komissarov et al |
| EPI_ISL_430068 | Russia/StPetersburg-RII4712V/2020 | 2020-04-07 | Europe/Russia/Saint-Petersburg/ | WHO National Influenza Centre Russian Federation | WHO National Influenza Centre Russian Federation | Andrey Komissarov et al |
| EPI_ISL_430069 | Russia/StPetersburg-RII4780S/2020 | 2020-04-07 | Europe/Russia/Saint-Petersburg/ | WHO National Influenza Centre Russian Federation | WHO National Influenza Centre Russian Federation | Andrey Komissarov et al |
| EPI_ISL_430070 | Russia/StPetersburg-RII4917S/2020 | 2020-04-07 | Europe/Russia/Saint-Petersburg/ | WHO National Influenza Centre Russian Federation | WHO National Influenza Centre Russian Federation | Andrey Komissarov et al |

| | | | | | | |
|---|---|---|---|---|---|---|
| EPI_ISL_430071 | Russia/StPetersburg-RII4936S/2020 | 2020-04-08 | Europe/Russia/Saint-Petersburg/ | WHO National Influenza Centre Russian Federation | WHO National Influenza Centre Russian Federation | Andrey Komissarov et al |
| EPI_ISL_430073 | Russia/StPetersburg-RII4938S/2020 | 2020-04-08 | Europe/Russia/Saint-Petersburg/ | WHO National Influenza Centre Russian Federation | WHO National Influenza Centre Russian Federation | Andrey Komissarov et al |
| EPI_ISL_430074 | Russia/StPetersburg-RII4944S/2020 | 2020-04-08 | Europe/Russia/Saint-Petersburg/ | WHO National Influenza Centre Russian Federation | WHO National Influenza Centre Russian Federation | Andrey Komissarov et al |
| EPI_ISL_430076 | Russia/StPetersburg-RII4969S/2020 | 2020-04-10 | Europe/Russia/Saint-Petersburg/ | WHO National Influenza Centre Russian Federation | WHO National Influenza Centre Russian Federation | Andrey Komissarov et al |
| EPI_ISL_430079 | Russia/StPetersburg-RII4980S/2020 | 2020-04-10 | Europe/Russia/Saint-Petersburg/ | WHO National Influenza Centre Russian Federation | WHO National Influenza Centre Russian Federation | Andrey Komissarov et al |
| EPI_ISL_430080 | Russia/StPetersburg-RII4983S/2020 | 2020-04-10 | Europe/Russia/Saint-Petersburg/ | WHO National Influenza Centre Russian Federation | WHO National Influenza Centre Russian Federation | Andrey Komissarov et al |
| EPI_ISL_430082 | Russia/StPetersburg-RII4985S/2020 | 2020-04-10 | Europe/Russia/Saint-Petersburg/ | WHO National Influenza Centre Russian Federation | WHO National Influenza Centre Russian Federation | Andrey Komissarov et al |
| EPI_ISL_430083 | Russia/StPetersburg-RII4988S/2020 | 2020-04-10 | Europe/Russia/Saint-Petersburg/ | WHO National Influenza Centre Russian Federation | WHO National Influenza Centre Russian Federation | Andrey Komissarov et al |
| EPI_ISL_430084 | Russia/StPetersburg-RII5017V/2020 | 2020-04-09 | Europe/Russia/Saint-Petersburg/ | WHO National Influenza Centre Russian Federation | WHO National Influenza Centre Russian Federation | Andrey Komissarov et al |
| EPI_ISL_430085 | Russia/StPetersburg-RII5029S/2020 | 2020-04-10 | Europe/Russia/Saint-Petersburg/ | WHO National Influenza Centre Russian Federation | WHO National Influenza Centre Russian Federation | Andrey Komissarov et al |

| | | | | | | |
|---|---|---|---|---|---|---|
| EPI_ISL_430086 | Russia/StPetersburg-RII5033S/2020 | 2020-04-10 | Europe/Russia/Saint-Petersburg/ | WHO National Influenza Centre Russian Federation | WHO National Influenza Centre Russian Federation | Andrey Komissarov et al |
| EPI_ISL_430089 | Russia/StPetersburg-RII5170S/2020 | 2020-04-09 | Europe/Russia/Saint-Petersburg/ | WHO National Influenza Centre Russian Federation | WHO National Influenza Centre Russian Federation | Andrey Komissarov et al |
| EPI_ISL_430090 | Russia/StPetersburg-RII5244S/2020 | 2020-04-10 | Europe/Russia/Saint-Petersburg/ | WHO National Influenza Centre Russian Federation | WHO National Influenza Centre Russian Federation | Andrey Komissarov et al |
| EPI_ISL_430091 | Russia/StPetersburg-RII5247S/2020 | 2020-04-10 | Europe/Russia/Saint-Petersburg/ | WHO National Influenza Centre Russian Federation | WHO National Influenza Centre Russian Federation | Andrey Komissarov et al |
| EPI_ISL_430092 | Russia/StPetersburg-RII5633S/2020 | 2020-04-14 | Europe/Russia/Saint-Petersburg/ | WHO National Influenza Centre Russian Federation | WHO National Influenza Centre Russian Federation | Andrey Komissarov et al |
| EPI_ISL_430093 | Russia/StPetersburg-RII5634S/2020 | 2020-04-14 | Europe/Russia/Saint-Petersburg/ | WHO National Influenza Centre Russian Federation | WHO National Influenza Centre Russian Federation | Andrey Komissarov et al |
| EPI_ISL_430094 | Russia/StPetersburg-RII5635S/2020 | 2020-04-14 | Europe/Russia/Saint-Petersburg/ | WHO National Influenza Centre Russian Federation | WHO National Influenza Centre Russian Federation | Andrey Komissarov et al |
| EPI_ISL_430095 | Russia/StPetersburg-RII5642S/2020 | 2020-04-14 | Europe/Russia/Saint-Petersburg/ | WHO National Influenza Centre Russian Federation | WHO National Influenza Centre Russian Federation | Andrey Komissarov et al |
| EPI_ISL_430096 | Russia/StPetersburg-RII5643S/2020 | 2020-04-14 | Europe/Russia/Saint-Petersburg/ | WHO National Influenza Centre Russian Federation | WHO National Influenza Centre Russian Federation | Andrey Komissarov et al |
| EPI_ISL_430097 | Russia/StPetersburg-RII5644S/2020 | 2020-04-14 | Europe/Russia/Saint-Petersburg/ | WHO National Influenza Centre Russian Federation | WHO National Influenza Centre Russian Federation | Andrey Komissarov et al |

| | | | | | | |
|---|---|---|---|---|---|---|
| EPI_ISL_430098 | Russia/StPetersburg-RII5654S/2020 | 2020-04-14 | Europe/Russia/Saint-Petersburg/ | WHO National Influenza Centre Russian Federation | WHO National Influenza Centre Russian Federation | Andrey Komissarov et al |
| EPI_ISL_430099 | Russia/StPetersburg-RII5942S/2020 | 2020-04-13 | Europe/Russia/Saint-Petersburg/ | WHO National Influenza Centre Russian Federation | WHO National Influenza Centre Russian Federation | Andrey Komissarov et al |
| EPI_ISL_430100 | Russia/StPetersburg-RII6027S/2020 | 2020-04-14 | Europe/Russia/Saint-Petersburg/ | WHO National Influenza Centre Russian Federation | WHO National Influenza Centre Russian Federation | Andrey Komissarov et al |
| EPI_ISL_430101 | Russia/StPetersburg-RII6031S/2020 | 2020-04-14 | Europe/Russia/Saint-Petersburg/ | WHO National Influenza Centre Russian Federation | WHO National Influenza Centre Russian Federation | Andrey Komissarov et al |
| EPI_ISL_430102 | Russia/StPetersburg-RII6039S/2020 | 2020-04-14 | Europe/Russia/Saint-Petersburg/ | WHO National Influenza Centre Russian Federation | WHO National Influenza Centre Russian Federation | Andrey Komissarov et al |
| EPI_ISL_430103 | Russia/StPetersburg-RII6043S/2020 | 2020-04-14 | Europe/Russia/Saint-Petersburg/ | WHO National Influenza Centre Russian Federation | WHO National Influenza Centre Russian Federation | Andrey Komissarov et al |
| EPI_ISL_430104 | Russia/StPetersburg-RII6053S/2020 | 2020-04-15 | Europe/Russia/Saint-Petersburg/ | WHO National Influenza Centre Russian Federation | WHO National Influenza Centre Russian Federation | Andrey Komissarov et al |
| EPI_ISL_430107 | Russia/StPetersburg-RII6060S/2020 | 2020-04-15 | Europe/Russia/Saint-Petersburg/ | WHO National Influenza Centre Russian Federation | WHO National Influenza Centre Russian Federation | Andrey Komissarov et al |
| EPI_ISL_430108 | Russia/StPetersburg-RII6061S/2020 | 2020-04-15 | Europe/Russia/Saint-Petersburg/ | WHO National Influenza Centre Russian Federation | WHO National Influenza Centre Russian Federation | Andrey Komissarov et al |
| EPI_ISL_430110 | Russia/StPetersburg-RII6065S/2020 | 2020-04-15 | Europe/Russia/Saint-Petersburg/ | WHO National Influenza Centre Russian Federation | WHO National Influenza Centre Russian Federation | Andrey Komissarov et al |

| | | | | | | |
|---|---|---|---|---|---|---|
| EPI_ISL_430120 | USA/WA-S266/2020 | 2020-03-23 | North America/USA/Washington/King C | Seattle Flu Study | Seattle Flu Study | Chu et al et al |
| EPI_ISL_430126 | USA/WA-S272/2020 | 2020-02-26 | North America/USA/Washington/King C | Seattle Flu Study | Seattle Flu Study | Chu et al et al |
| EPI_ISL_430131 | USA/WA-S277/2020 | 2020-02-22 | North America/USA/Washington/King C | Seattle Flu Study | Seattle Flu Study | Chu et al et al |
| EPI_ISL_430135 | USA/WA-S281/2020 | 2020-02-29 | North America/USA/Washington/King C | Seattle Flu Study | Seattle Flu Study | Chu et al et al |
| EPI_ISL_430136 | USA/WA-S282/2020 | 2020-03-10 | North America/USA/Washington/King C | Seattle Flu Study | Seattle Flu Study | Chu et al et al |
| EPI_ISL_430160 | USA/WA-S123/2020 | 2020-02-29 | North America/USA/Washington/ | Washington State Department of Health | Seattle Flu Study | Chu et al et al |
| EPI_ISL_430167 | USA/WA-S130/2020 | 2020-03-13 | North America/USA/Washington/King C | Washington State Department of Health | Seattle Flu Study | Chu et al et al |
| EPI_ISL_430215 | USA/WA-S177/2020 | 2020-03-19 | North America/USA/Washington/King C | Washington State Department of Health | Seattle Flu Study | Chu et al et al |
| EPI_ISL_430226 | USA/WA-S188/2020 | 2020-03-20 | North America/USA/Washington/Whatc | Washington State Department of Health | Seattle Flu Study | Chu et al et al |
| EPI_ISL_430231 | USA/WA-S193/2020 | 2020-03-19 | North America/USA/Washington/Grant | Washington State Department of Health | Seattle Flu Study | Chu et al et al |
| EPI_ISL_430243 | USA/WA-S205/2020 | 2020-03-20 | North America/USA/Washington/Whatc | Washington State Department of Health | Seattle Flu Study | Chu et al et al |
| EPI_ISL_430249 | USA/WA-S211/2020 | 2020-03-20 | North America/USA/Washington/King C | Washington State Department of Health | Seattle Flu Study | Chu et al et al |
| EPI_ISL_430267 | USA/WA-S229/2020 | 2020-03-19 | North America/USA/Washington/King C | Washington State Department of Health | Seattle Flu Study | Chu et al et al |
| EPI_ISL_430272 | USA/WA-S234/2020 | 2020-02-28 | North America/USA/Washington/King C | Washington State Department of Health | Seattle Flu Study | Chu et al et al |
| EPI_ISL_430273 | USA/WA-S235/2020 | 2020-02-29 | North America/USA/Washington/King C | Washington State Department of Health | Seattle Flu Study | Chu et al et al |
| EPI_ISL_430276 | USA/WA-S238/2020 | 2020-03-02 | North America/USA/Washington/King C | Washington State Department of Health | Seattle Flu Study | Chu et al et al |
| EPI_ISL_430280 | USA/WA-S242/2020 | 2020-03-01 | North America/USA/Washington/King C | Washington State Department of Health | Seattle Flu Study | Chu et al et al |
| EPI_ISL_430281 | USA/WA-S243/2020 | 2020-02-29 | North America/USA/Washington/King C | Washington State Department of Health | Seattle Flu Study | Chu et al et al |
| EPI_ISL_430289 | USA/WA-S251/2020 | 2020-03-11 | North America/USA/Washington/Skagit | Washington State Department of Health | Seattle Flu Study | Chu et al et al |
| EPI_ISL_430292 | USA/WA-S254/2020 | 2020-02-24 | North America/USA/Washington/Snoho | Washington State Department of Health | Seattle Flu Study | Chu et al et al |
| EPI_ISL_430294 | USA/WA-S256/2020 | 2020-03-02 | North America/USA/Washington/King C | Washington State Department of Health | Seattle Flu Study | Chu et al et al |
| EPI_ISL_430295 | USA/WA-S257/2020 | 2020-02-25 | North America/USA/Washington/King C | Washington State Department of Health | Seattle Flu Study | Chu et al et al |
| EPI_ISL_430329 | USA/NY-NYUMC302/2020 | 2020-03-30 | North America/USA/New York/Staten Is | NYU Langone Health | Departments of Pathology and Medicine, New York University School of Medicine | Maria Aguero-Rosenfeld et al |
| EPI_ISL_430332 | USA/NY-NYUMC305/2020 | 2020-03-30 | North America/USA/New York/Brooklyn | NYU Langone Health | Departments of Pathology and Medicine, New York University School of Medicine | Maria Aguero-Rosenfeld et al |

| | | | | | | |
|---|---|---|---|---|---|---|
| EPI_ISL_430346 | USA/NY-NYUMC319/2020 | 2020-03-30 | North America/USA/New York/Brooklyn | NYU Langone Health | Departments of Pathology and Medicine, New York University School of Medicine | Maria Aguero-Rosenfeld et al |
| EPI_ISL_430355 | USA/NY-NYUMC328/2020 | 2020-04-02 | North America/USA/New York/Nassau C | NYU Langone Health | Departments of Pathology and Medicine, New York University School of Medicine | Maria Aguero-Rosenfeld et al |
| EPI_ISL_430366 | USA/NY-NYUMC339/2020 | 2020-04-08 | North America/USA/New York/Brooklyn | NYU Langone Health | Departments of Pathology and Medicine, New York University School of Medicine | Maria Aguero-Rosenfeld et al |
| EPI_ISL_430395 | USA/NY-NYUMC368/2020 | 2020-04-09 | North America/USA/New York/Brooklyn | NYU Langone Health | Departments of Pathology and Medicine, New York University School of Medicine | Maria Aguero-Rosenfeld et al |
| EPI_ISL_430402 | USA/NY-NYUMC375/2020 | 2020-04-10 | North America/USA/New York/Brooklyn | NYU Langone Health | Departments of Pathology and Medicine, New York University School of Medicine | Maria Aguero-Rosenfeld et al |
| EPI_ISL_430413 | USA/NY-NYUMC386/2020 | 2020-04-09 | North America/USA/New York/Nassau C | NYU Langone Health | Departments of Pathology and Medicine, New York University School of Medicine | Maria Aguero-Rosenfeld et al |

| | | | | | | |
|---|---|---|---|---|---|---|
| EPI_ISL_430424 | USA/NY-NYUMC397/2020 | 2020-04-13 | North America/USA/New York/Brooklyn | NYU Langone Health | Departments of Pathology and Medicine, New York University School of Medicine | Maria Aguero-Rosenfeld et al |
| EPI_ISL_430430 | USA/NY-NYUMC403/2020 | 2020-04-13 | North America/USA/New York/Bronx | NYU Langone Health | Departments of Pathology and Medicine, New York University School of Medicine | Maria Aguero-Rosenfeld et al |
| EPI_ISL_430441 | Malaysia/IMR_WC1097/2020 | 2020-02-29 | Asia/Malaysia/Malaysia/ | Institute for Medical Research, Infectious Disease Research Centre, National Institutes of Health, Ministry of Health Malaysia | Institute for Medical Research, Infectious Disease Research Centre, National Institutes of Health, Ministry of Health Malaysia | Suppiah.J et al |
| EPI_ISL_430442 | Malaysia/IMR_WC1098/2020 | 2020-02-29 | Asia/Malaysia/Malaysia/ | Institute for Medical Research, Infectious Disease Research Centre, National Institutes of Health, Ministry of Health Malaysia | Institute for Medical Research, Infectious Disease Research Centre, National Institutes of Health, Ministry of Health Malaysia | Suppiah.J et al |
| EPI_ISL_430443 | Malaysia/IMR_WC085/2020 | 2020-01-28 | Asia/Malaysia/Malaysia/ | Institute for Medical Research, Infectious Disease Research Centre, National Institutes of Health, Ministry of Health Malaysia | Institute for Medical Research, Infectious Disease Research Centre, National Institutes of Health, Ministry of Health Malaysia | Suppiah.J et al |

| | | | | | | |
|---|---|---|---|---|---|---|
| EPI_ISL_430444 | Malaysia/IMR_WC627/2020 | 2020-02-12 | Asia/Malaysia/Malaysia/ | Institute for Medical Research, Infectious Disease Research Centre, National Institutes of Health, Ministry of Health Malaysia | Institute for Medical Research, Infectious Disease Research Centre, National Institutes of Health, Ministry of Health Malaysia | Suppiah.J et al |
| EPI_ISL_430469 | Greece/127_HPI/2020 | 2020-02-29 | Europe/Greece/Athens/ | Hellenic Pasteur Institute, Public Health Laboratories | Hellenic Pasteur Institute, Public Health Laboratories, Unit of Bioinformatics and Applied Genomics | Vasiliki Pogka et al |
| EPI_ISL_430719 | Spain/MD-H12-LP29/2020 | 2020-03-11 | Europe/Spain/Madrid/ | Hospital Universitario La Paz | Hospital Universitario 12 de Octubre | Elias Dahdouh et al |
| EPI_ISL_430722 | Beijing/BJ287/2020 | 2020-01-29 | Asia/China/Beijing/ | Chinese PLA Institute for Disease Control and Prevention | Chinese PLA Institute for Disease Control and Prevention | Peng Li et al |
| EPI_ISL_430724 | Beijing/BJ752/2020 | 2020-02-08 | Asia/China/Beijing/ | Chinese PLA Institute for Disease Control and Prevention | Chinese PLA Institute for Disease Control and Prevention | Peng Li et al |
| EPI_ISL_430725 | Beijing/BJ288/2020 | 2020-01-29 | Asia/China/Beijing/ | Chinese PLA Institute for Disease Control and Prevention | Chinese PLA Institute for Disease Control and Prevention | Peng Li et al |
| EPI_ISL_430728 | Beijing/BJ492/2020 | 2020-02-02 | Asia/China/Beijing/ | Chinese PLA Institute for Disease Control and Prevention | Chinese PLA Institute for Disease Control and Prevention | Peng Li et al |
| EPI_ISL_430729 | Beijing/BJ709/2020 | 2020-02-07 | Asia/China/Beijing/ | Chinese PLA Institute for Disease Control and Prevention | Chinese PLA Institute for Disease Control and Prevention | Peng Li et al |

| | | | | | | |
|---|---|---|---|---|---|---|
| EPI_ISL_430730 | Beijing/BJ390/2020 | 2020-01-31 | Asia/China/Beijing/ | Chinese PLA Institute for Disease Control and Prevention | Chinese PLA Institute for Disease Control and Prevention | Peng Li et al |
| EPI_ISL_430731 | Beijing/BJ561/2020 | 2020-02-04 | Asia/China/Beijing/ | Chinese PLA Institute for Disease Control and Prevention | Chinese PLA Institute for Disease Control and Prevention | Peng Li et al |
| EPI_ISL_430733 | Beijing/BJ556/2020 | 2020-02-04 | Asia/China/Beijing/ | Chinese PLA Institute for Disease Control and Prevention | Chinese PLA Institute for Disease Control and Prevention | Peng Li et al |
| EPI_ISL_430734 | Beijing/BJ53/2020 | 2020-01-24 | Asia/China/Beijing/ | Chinese PLA Institute for Disease Control and Prevention | Chinese PLA Institute for Disease Control and Prevention | Peng Li et al |
| EPI_ISL_430735 | Beijing/BJ54/2020 | 2020-01-24 | Asia/China/Beijing/ | Chinese PLA Institute for Disease Control and Prevention | Chinese PLA Institute for Disease Control and Prevention | Peng Li et al |
| EPI_ISL_430736 | Beijing/BJ252/2020 | 2020-01-29 | Asia/China/Beijing/ | Chinese PLA Institute for Disease Control and Prevention | Chinese PLA Institute for Disease Control and Prevention | Peng Li et al |
| EPI_ISL_430737 | Beijing/BJ1112/2020 | 2020-02-25 | Asia/China/Beijing/ | Chinese PLA Institute for Disease Control and Prevention | Chinese PLA Institute for Disease Control and Prevention | Peng Li et al |
| EPI_ISL_430738 | Beijing/BJ589/2020 | 2020-02-05 | Asia/China/Beijing/ | Chinese PLA Institute for Disease Control and Prevention | Chinese PLA Institute for Disease Control and Prevention | Peng Li et al |
| EPI_ISL_430739 | Beijing/BJ625/2020 | 2020-02-06 | Asia/China/Beijing/ | Chinese PLA Institute for Disease Control and Prevention | Chinese PLA Institute for Disease Control and Prevention | Peng Li et al |
| EPI_ISL_430740 | Beijing/BJ710/2020 | 2020-02-07 | Asia/China/Beijing/ | Chinese PLA Institute for Disease Control and Prevention | Chinese PLA Institute for Disease Control and Prevention | Peng Li et al |

| | | | | | | |
|---|---|---|---|---|---|---|
| EPI_ISL_430741 | Beijing/BJ782/2020 | 2020-02-10 | Asia/China/Beijing/ | Chinese PLA Institute for Disease Control and Prevention | Chinese PLA Institute for Disease Control and Prevention | Peng Li et al |
| EPI_ISL_430742 | Beijing/BJ251/2020 | 2020-01-29 | Asia/China/Beijing/ | Chinese PLA Institute for Disease Control and Prevention | Chinese PLA Institute for Disease Control and Prevention | Peng Li et al |
| EPI_ISL_430820 | Egypt/NRC-01/2020 | 2020-03-18 | Africa/Egypt/Egypt/ | Center of Scientific Excellence for Influenza Viruses, National Research Centre (NRC), Egypt. | Center of Scientific Excellence for Influenza Viruses, National Research Centre (NRC), Egypt. | Mohamed Ahmed Ali et al |
| EPI_ISL_430837 | Thailand/TNIC-1889/2020 | 2020-03-03 | Asia/Italy/Nonthaburi/ | n/a | Thai National Influenza Center, Department of medical Science, Ministry of Public Health, Thailand | Pilailuk et al |
| EPI_ISL_430843 | Philippines/RITM-05/2020 | 2020-03-22 | Asia/Philippines/Philippines/ | Bethany Hospital | Research Institute for Tropical Medicine | Medado et al |
| EPI_ISL_430847 | Sweden/20-04631/2020 | 2020-02-27 | Europe/Sweden/Stockholm/ | HS mikrobiologi virus | The Public Health Agency of Sweden | Zhibing Yun et al |
| EPI_ISL_430850 | Sweden/20-08150/2020 | 2020-03-11 | Europe/Sweden/Halland/ | Klinisk mikrobiologi och vardhygien Halmstad | The Public Health Agency of Sweden | Arne Kotz et al |
| EPI_ISL_430874 | USA/WA-UW-5190/2020 | 2020-04-01 | North America/USA/Washington/ | UW Virology Lab | UW Virology Lab | Pavitra Roychoudhury et al |
| EPI_ISL_430879 | USA/UN-UW-526/2020 | 2020-03-11 | North America/USA/USA/ | UW Virology Lab | UW Virology Lab | Pavitra Roychoudhury et al |
| EPI_ISL_430884 | USA/WA-UW-543/2020 | 2020-03-12 | North America/USA/Washington/ | UW Virology Lab | UW Virology Lab | Pavitra Roychoudhury et al |
| EPI_ISL_430893 | USA/WA-UW-600/2020 | 2020-03-12 | North America/USA/Washington/ | UW Virology Lab | UW Virology Lab | Pavitra Roychoudhury et al |
| EPI_ISL_430918 | USA/WA-UW-5750/2020 | 2020-04-06 | North America/USA/Washington/ | UW Virology Lab | UW Virology Lab | Pavitra Roychoudhury et al |
| EPI_ISL_430920 | USA/WA-UW-5756/2020 | 2020-04-07 | North America/USA/Washington/ | UW Virology Lab | UW Virology Lab | Pavitra Roychoudhury et al |
| EPI_ISL_430923 | USA/CT-UW-5764/2020 | 2020-04-05 | North America/USA/Connecticut/ | UW Virology Lab | UW Virology Lab | Pavitra Roychoudhury et al |
| EPI_ISL_430924 | USA/UN-UW-5769/2020 | 2020-04-07 | North America/USA/USA/ | UW Virology Lab | UW Virology Lab | Pavitra Roychoudhury et al |
| EPI_ISL_430937 | USA/WA-UW-6178/2020 | 2020-04-01 | North America/USA/Washington/ | UW Virology Lab | UW Virology Lab | Pavitra Roychoudhury et al |

| | | | | | | |
|---|---|---|---|---|---|---|
| EPI_ISL_430939 | USA/WA-UW-4243/2020 | 2020-04-01 | North America/USA/Washington/ | UW Virology Lab | UW Virology Lab | Pavitra Roychoudhury et al |
| EPI_ISL_430945 | USA/CT-UW-4345/2020 | 2020-03-30 | North America/USA/Connecticut/ | UW Virology Lab | UW Virology Lab | Pavitra Roychoudhury et al |
| EPI_ISL_430947 | USA/ID-UW-4354/2020 | 2020-03-27 | North America/USA/Idaho/ | UW Virology Lab | UW Virology Lab | Pavitra Roychoudhury et al |
| EPI_ISL_430953 | USA/WA-UW-6185/2020 | 2020-04-01 | North America/USA/Washington/ | UW Virology Lab | UW Virology Lab | Pavitra Roychoudhury et al |
| EPI_ISL_430961 | USA/CT-UW-6270/2020 | 2020-04-01 | North America/USA/Connecticut/ | UW Virology Lab | UW Virology Lab | Pavitra Roychoudhury et al |
| EPI_ISL_430967 | USA/WA-UW-6327/2020 | 2020-04-02 | North America/USA/Washington/ | UW Virology Lab | UW Virology Lab | Pavitra Roychoudhury et al |
| EPI_ISL_430970 | USA/ID-UW-6349/2020 | 2020-04-01 | North America/USA/Idaho/ | UW Virology Lab | UW Virology Lab | Pavitra Roychoudhury et al |
| EPI_ISL_431013 | USA/AK-PHL010/2020 | 2020-03-17 | North America/USA/Alaska/Interior Ala | Alaska State Virology Laboratory | Alaska State Virology Laboratory | Jack Chen et al |
| EPI_ISL_431080 | USA/CT-Yale-079/2020 | 2020-03-02 | North America/USA/Connecticut/ | Yale COVID-19 Biorepository | Grubaugh Lab - Yale School of Public Health | Joseph Fauver et al |
| EPI_ISL_431102 | India/TG-GMC-KN318/2020 | 2020-03-11 | Asia/Italy/Telangana/Hyderabad | Department of MicroBiology,Gandhi Medical College and Hospital,Secendrabad,Hyderabad,India | Department of Microbiology, Gandhi Medical College and Hospital, Secendrabad, Hyderabad | Nagamani K et al |
| EPI_ISL_431784 | Fujian/IM3520017T/2020 | 2020-03-18 | Asia/China/Fujian/ | Fujian Center for Disease Control and Prevention | Fujian Center for Disease Control and Prevention | Lin Qi et al |
| EPI_ISL_431785 | Fujian/3520022Y/2020 | 2020-01-23 | Asia/China/Fujian/ | Fujian Center for Disease Control and Prevention | Fujian Center for Disease Control and Prevention | Lin Qi et al |
| EPI_ISL_431833 | Philippines/PGC01/2020 | 2020-03-22 | Asia/Philippines/Manila/ | National Institutes of Health, University of the Philippines Manila | Philippine Genome Center, University of the Philippines System | Carlo M. Lapid et al |
| EPI_ISL_432075 | Wales/PHWC-28907/2020 | 2020-03-31 | Europe/United Kingdom/Wales/ | Wales Specialist Virology Centre | Public Health Wales Microbiology Cardiff | Catherine Moore et al |

| | | | | | | |
|---|---|---|---|---|---|---|
| EPI_ISL_432115 | Wales/PHWC-2B1DF/2020 | 2020-04-07 | Europe/United Kingdom/Wales/ | Wales Specialist Virology Centre | Public Health Wales Microbiology Cardiff | Catherine Moore et al |
| EPI_ISL_432268 | Wales/PHWC-2B0D2/2020 | 2020-04-08 | Europe/United Kingdom/Wales/ | Wales Specialist Virology Centre | Public Health Wales Microbiology Cardiff | Catherine Moore et al |
| EPI_ISL_432358 | Wales/PHWC-2B263/2020 | 2020-04-08 | Europe/United Kingdom/Wales/ | Wales Specialist Virology Centre | Public Health Wales Microbiology Cardiff | Catherine Moore et al |
| EPI_ISL_432418 | Wales/PHWC-2B1B1/2020 | 2020-04-07 | Europe/United Kingdom/Wales/ | Wales Specialist Virology Centre | Public Health Wales Microbiology Cardiff | Catherine Moore et al |
| EPI_ISL_432477 | England/SHEF-C9DB1/2020 | 2020-04-02 | Europe/United Kingdom/England/ | Virology Department, Sheffield Teaching Hospitals NHS Foundation Trust / Virology Department, Sheffield Teaching Hospitals NHS Foundation Trust | COVID-19 Genomics UK (COG-UK) Consortium | Thushan de Silva et al |
| EPI_ISL_432503 | England/SHEF-CB1A4/2020 | 2020-04-05 | Europe/United Kingdom/England/ | Virology Department, Sheffield Teaching Hospitals NHS Foundation Trust / Virology Department, Sheffield Teaching Hospitals NHS Foundation Trust | COVID-19 Genomics UK (COG-UK) Consortium | Thushan de Silva et al |
| EPI_ISL_432529 | England/SHEF-CC905/2020 | 2020-03-31 | Europe/United Kingdom/England/ | Virology Department, Sheffield Teaching Hospitals NHS Foundation Trust / Virology Department, Sheffield Teaching Hospitals NHS Foundation Trust | COVID-19 Genomics UK (COG-UK) Consortium | Thushan de Silva et al |
| EPI_ISL_432534 | England/SHEF-CCDCD/2020 | 2020-03-30 | Europe/United Kingdom/England/ | Virology Department, Sheffield Teaching Hospitals NHS Foundation Trust / Virology Department, Sheffield Teaching Hospitals NHS Foundation Trust | COVID-19 Genomics UK (COG-UK) Consortium | Thushan de Silva et al |
| EPI_ISL_432597 | England/SHEF-CFB57/2020 | 2020-03-21 | Europe/United Kingdom/England/ | Virology Department, Sheffield Teaching Hospitals NHS Foundation Trust / Virology Department, Sheffield Teaching Hospitals NHS Foundation Trust | COVID-19 Genomics UK (COG-UK) Consortium | Thushan de Silva et al |
| EPI_ISL_432616 | England/SHEF-D093B/2020 | 2020-04-06 | Europe/United Kingdom/England/ | Virology Department, Sheffield Teaching Hospitals NHS Foundation Trust / Virology Department, Sheffield Teaching Hospitals NHS Foundation Trust | COVID-19 Genomics UK (COG-UK) Consortium | Thushan de Silva et al |

| | | | | | | |
|---|---|---|---|---|---|---|
| EPI_ISL_432704 | England/NOTT-10FC57/2020 | 2020-04-07 | Europe/United Kingdom/England/ | Queens Medical Centre, Clinical Microbiology Department / DeepSeq Nottingham | COVID-19 Genomics UK (COG-UK) Consortium | Gemma Clark et al |
| EPI_ISL_432709 | England/NOTT-10FCC0/2020 | 2020-04-08 | Europe/United Kingdom/England/ | Queens Medical Centre, Clinical Microbiology Department / DeepSeq Nottingham | COVID-19 Genomics UK (COG-UK) Consortium | Gemma Clark et al |
| EPI_ISL_432711 | England/SHEF-BFE7F/2020 | 2020-03-12 | Europe/United Kingdom/England/ | Virology Department, Sheffield Teaching Hospitals NHS Foundation Trust / Virology Department, Sheffield Teaching Hospitals NHS Foundation Trust | COVID-19 Genomics UK (COG-UK) Consortium | Thushan de Silva et al |
| EPI_ISL_432953 | England/NOTT-10EFD7/2020 | 2020-03-30 | Europe/United Kingdom/England/ | Queens Medical Centre, Clinical Microbiology Department / DeepSeq Nottingham | COVID-19 Genomics UK (COG-UK) Consortium | Gemma Clark et al |
| EPI_ISL_432955 | England/NOTT-10F000/2020 | 2020-03-30 | Europe/United Kingdom/England/ | Queens Medical Centre, Clinical Microbiology Department / DeepSeq Nottingham | COVID-19 Genomics UK (COG-UK) Consortium | Gemma Clark et al |
| EPI_ISL_432996 | England/NOTT-10F39E/2020 | 2020-04-02 | Europe/United Kingdom/England/ | Queens Medical Centre, Clinical Microbiology Department / DeepSeq Nottingham | COVID-19 Genomics UK (COG-UK) Consortium | Gemma Clark et al |
| EPI_ISL_433014 | England/NOTT-10F510/2020 | 2020-04-03 | Europe/United Kingdom/England/ | Queens Medical Centre, Clinical Microbiology Department / DeepSeq Nottingham | COVID-19 Genomics UK (COG-UK) Consortium | Gemma Clark et al |
| EPI_ISL_433151 | Scotland/EDB1831/2020 | 2020-04-16 | Europe/United Kingdom/Scotland/ | Virology Department, Royal Infirmary of Edinburgh, NHS Lothian / School of Biological Sciences, University of Edinburgh / Institute of Genetics and Molecular Medicine, University of Edinburgh | COVID-19 Genomics UK (COG-UK) Consortium | McHugh M et al |
| EPI_ISL_433235 | Scotland/EDB736/2020 | 2020-04-02 | Europe/United Kingdom/Scotland/ | Virology Department, Royal Infirmary of Edinburgh, NHS Lothian / School of Biological Sciences, University of Edinburgh / Institute of Genetics and Molecular Medicine, University of Edinburgh | COVID-19 Genomics UK (COG-UK) Consortium | McHugh M et al |
| EPI_ISL_433246 | Scotland/EDB795/2020 | 2020-04-03 | Europe/United Kingdom/Scotland/ | Virology Department, Royal Infirmary of Edinburgh, NHS Lothian / School of Biological Sciences, University of Edinburgh / Institute of Genetics and Molecular Medicine, University of Edinburgh | COVID-19 Genomics UK (COG-UK) Consortium | McHugh M et al |

| | | | | | | |
|---|---|---|---|---|---|---|
| EPI_ISL_433386 | Scotland/EDB1078/2020 | 2020-04-07 | Europe/United Kingdom/Scotland/ | Virology Department, Royal Infirmary of Edinburgh, NHS Lothian / School of Biological Sciences, University of Edinburgh / Institute of Genetics and Molecular Medicine, University of Edinburgh | COVID-19 Genomics UK (COG-UK) Consortium | McHugh M et al |
| EPI_ISL_433420 | Scotland/EDB1170/2020 | 2020-04-09 | Europe/United Kingdom/Scotland/ | Virology Department, Royal Infirmary of Edinburgh, NHS Lothian / School of Biological Sciences, University of Edinburgh / Institute of Genetics and Molecular Medicine, University of Edinburgh | COVID-19 Genomics UK (COG-UK) Consortium | McHugh M et al |
| EPI_ISL_433439 | Scotland/EDB1404/2020 | 2020-04-11 | Europe/United Kingdom/Scotland/ | Virology Department, Royal Infirmary of Edinburgh, NHS Lothian / School of Biological Sciences, University of Edinburgh / Institute of Genetics and Molecular Medicine, University of Edinburgh | COVID-19 Genomics UK (COG-UK) Consortium | McHugh M et al |
| EPI_ISL_433501 | Scotland/CVR1053/2020 | 2020-03-28 | Europe/United Kingdom/Scotland/ | West of Scotland Specialist Virology Centre, NHSGGC / MRC-University of Glasgow Centre for Virus Research | COVID-19 Genomics UK (COG-UK) Consortium | Ana da Silva Filipe et al |
| EPI_ISL_433673 | England/CAMB-73BCB/2020 | 2020-04-03 | Europe/United Kingdom/England/ | Department of Pathology, University of Cambridge | COVID-19 Genomics UK (COG-UK) Consortium | Luke W Meredith et al |
| EPI_ISL_433717 | England/CAMB-78319/2020 | 2020-04-04 | Europe/United Kingdom/England/ | Department of Pathology, University of Cambridge | COVID-19 Genomics UK (COG-UK) Consortium | Luke W Meredith et al |
| EPI_ISL_433724 | England/CAMB-78382/2020 | 2020-04-03 | Europe/United Kingdom/England/ | Department of Pathology, University of Cambridge | COVID-19 Genomics UK (COG-UK) Consortium | Luke W Meredith et al |
| EPI_ISL_433796 | England/CAMB-78962/2020 | 2020-04-07 | Europe/United Kingdom/England/ | Department of Pathology, University of Cambridge | COVID-19 Genomics UK (COG-UK) Consortium | Luke W Meredith et al |
| EPI_ISL_433819 | England/CAMB-79AB9/2020 | 2020-03-27 | Europe/United Kingdom/England/ | Department of Pathology, University of Cambridge | COVID-19 Genomics UK (COG-UK) Consortium | Luke W Meredith et al |
| EPI_ISL_433850 | England/CAMB-7ABE2/2020 | 2020-04-06 | Europe/United Kingdom/England/ | Department of Pathology, University of Cambridge | COVID-19 Genomics UK (COG-UK) Consortium | Luke W Meredith et al |
| EPI_ISL_434031 | England/CAMB-7C351/2020 | 2020-04-14 | Europe/United Kingdom/England/ | Department of Pathology, University of Cambridge | COVID-19 Genomics UK (COG-UK) Consortium | Luke W Meredith et al |
| EPI_ISL_434065 | USA/WA-S305/2020 | 2020-04-01 | North America/USA/Washington/Pierce | Washington State Department of Health | Seattle Flu Study | Chu et al et al |

| | | | | | | |
|---|---|---|---|---|---|---|
| EPI_ISL_434087 | USA/WA-S327/2020 | 2020-04-05 | North America/USA/Washington/Yakim | Washington State Department of Health | Seattle Flu Study | Chu et al et al |
| EPI_ISL_434090 | USA/WA-S330/2020 | 2020-04-02 | North America/USA/Washington/Yakim | Washington State Department of Health | Seattle Flu Study | Chu et al et al |
| EPI_ISL_434097 | USA/WA-S337/2020 | 2020-04-01 | North America/USA/Washington/Pierce | Washington State Department of Health | Seattle Flu Study | Chu et al et al |
| EPI_ISL_434109 | USA/WA-S349/2020 | 2020-04-02 | North America/USA/Washington/Masor | Washington State Department of Health | Seattle Flu Study | Chu et al et al |
| EPI_ISL_434117 | USA/WA-S357/2020 | 2020-03-25 | North America/USA/Washington/Yakim | Washington State Department of Health | Seattle Flu Study | Chu et al et al |
| EPI_ISL_434128 | USA/WA-S368/2020 | 2020-03-26 | North America/USA/Washington/Yakim | Washington State Department of Health | Seattle Flu Study | Chu et al et al |
| EPI_ISL_434131 | USA/WA-S371/2020 | 2020-03-27 | North America/USA/Washington/Skagit | Washington State Department of Health | Seattle Flu Study | Chu et al et al |
| EPI_ISL_434139 | USA/WA-S379/2020 | 2020-03-27 | North America/USA/Washington/Skagit | Washington State Department of Health | Seattle Flu Study | Chu et al et al |
| EPI_ISL_434159 | USA/WA-S399/2020 | 2020-03-28 | North America/USA/Washington/Yakim | Washington State Department of Health | Seattle Flu Study | Chu et al et al |
| EPI_ISL_434166 | USA/WA-S406/2020 | 2020-03-26 | North America/USA/Washington/Yakim | Washington State Department of Health | Seattle Flu Study | Chu et al et al |
| EPI_ISL_434167 | USA/WA-S407/2020 | 2020-03-28 | North America/USA/Washington/Yakim | Washington State Department of Health | Seattle Flu Study | Chu et al et al |
| EPI_ISL_434185 | USA/WA-S425/2020 | 2020-03-28 | North America/USA/Washington/Yakim | Washington State Department of Health | Seattle Flu Study | Chu et al et al |
| EPI_ISL_434192 | USA/WA-S432/2020 | 2020-03-28 | North America/USA/Washington/Yakim | Washington State Department of Health | Seattle Flu Study | Chu et al et al |
| EPI_ISL_434220 | USA/WA-S460/2020 | 2020-04-05 | North America/USA/Washington/King C | Washington State Department of Health | Seattle Flu Study | Chu et al et al |
| EPI_ISL_434222 | USA/WA-S462/2020 | 2020-04-02 | North America/USA/Washington/Yakim | Washington State Department of Health | Seattle Flu Study | Chu et al et al |
| EPI_ISL_434225 | USA/WA-S465/2020 | 2020-04-01 | North America/USA/Washington/Pierce | Washington State Department of Health | Seattle Flu Study | Chu et al et al |
| EPI_ISL_434230 | USA/WA-S470/2020 | 2020-04-05 | North America/USA/Washington/King C | Washington State Department of Health | Seattle Flu Study | Chu et al et al |
| EPI_ISL_434231 | USA/WA-S471/2020 | 2020-04-06 | North America/USA/Washington/Snoho | Washington State Department of Health | Seattle Flu Study | Chu et al et al |
| EPI_ISL_434243 | USA/WA-S483/2020 | 2020-04-05 | North America/USA/Washington/King C | Washington State Department of Health | Seattle Flu Study | Chu et al et al |
| EPI_ISL_434244 | USA/WA-S484/2020 | 2020-04-02 | North America/USA/Washington/Pierce | Washington State Department of Health | Seattle Flu Study | Chu et al et al |
| EPI_ISL_434251 | USA/WA-S491/2020 | 2020-04-02 | North America/USA/Washington/Pierce | Washington State Department of Health | Seattle Flu Study | Chu et al et al |
| EPI_ISL_434253 | USA/WA-S493/2020 | 2020-04-01 | North America/USA/Washington/Yakim | Washington State Department of Health | Seattle Flu Study | Chu et al et al |
| EPI_ISL_434272 | USA/WA-S512/2020 | 2020-04-01 | North America/USA/Washington/Yakim | Washington State Department of Health | Seattle Flu Study | Chu et al et al |
| EPI_ISL_434274 | USA/WA-S514/2020 | 2020-03-29 | North America/USA/Washington/Snoho | Washington State Department of Health | Seattle Flu Study | Chu et al et al |
| EPI_ISL_434290 | USA/WA-S530/2020 | 2020-03-24 | North America/USA/Washington/Yakim | Washington State Department of Health | Seattle Flu Study | Chu et al et al |
| EPI_ISL_434301 | USA/WA-S541/2020 | 2020-03-24 | North America/USA/Washington/Whatc | Washington State Department of Health | Seattle Flu Study | Chu et al et al |
| EPI_ISL_434317 | USA/WA-S557/2020 | 2020-04-01 | North America/USA/Washington/Yakim | Washington State Department of Health | Seattle Flu Study | Chu et al et al |
| EPI_ISL_434323 | USA/WA-S563/2020 | 2020-03-31 | North America/USA/Washington/Pierce | Washington State Department of Health | Seattle Flu Study | Chu et al et al |
| EPI_ISL_434346 | USA/WA-S586/2020 | 2020-03-31 | North America/USA/Washington/Pierce | Washington State Department of Health | Seattle Flu Study | Chu et al et al |
| EPI_ISL_434347 | Belgium/UGent-28/2020 | 2020-03-26 | Europe/Belgium/Gent/ | Lab voor klinische biologie | Onderzoeksgroep Virologie | Laurens Lambrechts et al |

| | | | | | | |
|---|---|---|---|---|---|---|
| EPI_ISL_434356 | Belgium/UGent-39/2020 | 2020-03-27 | Europe/Belgium/Gent/ | Lab voor klinische biologie | Onderzoeksgroep Virologie | Nick Vereecke et al |
| EPI_ISL_434366 | Belgium/ITM_C189/2020 | 2020-03-30 | Europe/Belgium/Antwerpen/ | Hospital AZ Rivierenland | Institute of Tropical Medicine | Philippe Selhorst et al |
| EPI_ISL_434371 | Belgium/ITM_C233/2020 | 2020-04-01 | Europe/Belgium/Antwerpen/ | Hospital AZ Rivierenland | Institute of Tropical Medicine | Philippe Selhorst et al |
| EPI_ISL_434464 | Greece/145/2020 | 2020-03-16 | Europe/Greece/Athens/ | Laboratory of Microbiology, Medical School, National and Kapodistrian University of Athens | Laboratory of Biology, Department of Medicine, Democritus University of Thrace | Kassela K. et al |
| EPI_ISL_434522 | USA/LA-ALSR-0065-NBG/2020 | 2020-03-31 | North America/USA/Louisiana/New Orle | Robert Garry lab | Andersen lab at Scripps Research | Allison Smither et al |
| EPI_ISL_434534 | Wuhan/IVDC-HB-GX02/2019 | 2019-12-30 | Asia/China/Hubei/Wuhan | National Institute for Viral Disease Control and Prevention, China CDC | National Institute for Viral Disease Control and Prevention, China CDC, Yunnan Provincial CDC | Wenjie Tan et al |
| EPI_ISL_434542 | PuertoRico/CDC-S2/2020 | 2020-03-23 | North America/USA/Puerto Rico/ | Puerto Rico Department of Health | Centers for Disease Control and Prevention, Dengue Branch | Gilberto A. Santiago et al |
| EPI_ISL_434551 | PuertoRico/CDC-S11/2020 | 2020-03-24 | North America/USA/Puerto Rico/ | Puerto Rico Department of Health | Centers for Disease Control and Prevention, Dengue Branch | Gilberto A. Santiago et al |
| EPI_ISL_434554 | Philippines/PGC02/2020 | 2020-03-26 | Asia/Philippines/Manila/ | National Institutes of Health, University of the Philippines Manila | Philippine Genome Center | Carlo M. Lapid et al |
| EPI_ISL_434555 | Philippines/PGC03/2020 | 2020-03-26 | Asia/Philippines/Manila/ | National Institutes of Health, University of the Philippines Manila | Philippine Genome Center | Carlo M. Lapid et al |
| EPI_ISL_434556 | Philippines/PGC04/2020 | 2020-03-26 | Asia/Philippines/Manila/ | National Institutes of Health, University of the Philippines Manila | Philippine Genome Center | Carlo M. Lapid et al |
| EPI_ISL_434557 | Philippines/PGC05/2020 | 2020-03-27 | Asia/Philippines/Manila/ | National Institutes of Health, University of the Philippines Manila | Philippine Genome Center | Carlo M. Lapid et al |
| EPI_ISL_434558 | Philippines/PGC06/2020 | 2020-03-28 | Asia/Philippines/Manila/ | National Institutes of Health, University of the Philippines Manila | Philippine Genome Center | Carlo M. Lapid et al |

| | | | | | | |
|---|---|---|---|---|---|---|
| EPI_ISL_434563 | HongKong/HKU-902a/2020 | 2020-01-23 | Asia/Hong Kong/Hong Kong/ | unknown | Microbiology, The University of Hong Kong | To et al |
| EPI_ISL_434564 | HongKong/HKU-902b/2020 | 2020-01-24 | Asia/Hong Kong/Hong Kong/ | unknown | Microbiology, The University of Hong Kong | To et al |
| EPI_ISL_434565 | HongKong/HKU-903a/2020 | 2020-01-24 | Asia/Hong Kong/Hong Kong/ | unknown | Microbiology, The University of Hong Kong | To et al |
| EPI_ISL_434566 | HongKong/HKU-903b/2020 | 2020-01-27 | Asia/Hong Kong/Hong Kong/ | unknown | Microbiology, The University of Hong Kong | To et al |
| EPI_ISL_434567 | HongKong/HKU-907a/2020 | 2020-01-27 | Asia/Hong Kong/Hong Kong/ | unknown | Microbiology, The University of Hong Kong | To et al |
| EPI_ISL_434568 | HongKong/HKU-907b/2020 | 2020-01-29 | Asia/Hong Kong/Hong Kong/ | unknown | Microbiology, The University of Hong Kong | To et al |
| EPI_ISL_434569 | HongKong/HKU-908a/2020 | 2020-01-27 | Asia/Hong Kong/Hong Kong/ | unknown | Microbiology, The University of Hong Kong | To et al |
| EPI_ISL_434570 | HongKong/HKU-908b/2020 | 2020-01-29 | Asia/Hong Kong/Hong Kong/ | unknown | Microbiology | To et al |
| EPI_ISL_434571 | HongKong/HKU-001a/2020 | 2020-01-22 | Asia/Hong Kong/Hong Kong/ | Microbiology, The University of Hong Kong | Microbiology, The University of Hong Kong | Chan et al |
| EPI_ISL_434596 | USA/VA-DCLS-0104/2020 | 2020-04-08 | North America/USA/Virginia/ | Virginia DCLS | Virginia DCLS | Virginia DCLS et al |
| EPI_ISL_434605 | USA/VA-DCLS-0114/2020 | 2020-04-09 | North America/USA/Virginia/ | Virginia DCLS | Virginia DCLS | Virginia DCLS et al |
| EPI_ISL_434615 | USA/WI-UW-269/2020 | 2020-04-06 | North America/USA/Wisconsin/ | University of Wisconsin-Madison AIDS Vaccine Research Laboratories | University of Wisconsin-Madison AIDS Vaccine Research Laboratories | Gage Moreno et al |
| EPI_ISL_434627 | France/OCC-12/2020 | 2020-03-26 | Europe/France/Occitanie/ | CHU Purpan - Laboratoire de Virologie - Institut F√©d√©ratif de Biologie | Laboratoire de virologie - √âcole Nationale V√©t√©rinaire de Toulouse | Guillaume Croville et al |

| | | | | | | |
|---|---|---|---|---|---|---|
| EPI_ISL_434633 | France/OCC-18/2020 | 2020-04-09 | Europe/France/Occitanie/ | CHU Purpan - Laboratoire de Virologie - Institut F√©d√©ratif de Biologie | Laboratoire de virologie - √âcole Nationale V√©t√©rinaire de Toulouse | Guillaume Croville et al |
| EPI_ISL_434641 | Sweden/20-08682/2020 | 2020-03-25 | Europe/Sweden/Norrbotten/ | Laboratoriemedicin | The Public Health Agency of Sweden | Oskar Karlsson Lindsjo et al |
| EPI_ISL_434652 | Sweden/20-06308/2020 | 2020-03-20 | Europe/Sweden/Stockholm/ | Ektorps Vardcentral | The Public Health Agency of Sweden | Eva Espmark et al |
| EPI_ISL_434663 | Sweden/20-07025/2020 | 2020-04-06 | Europe/Sweden/Stockholm/ | Surbrunns VC | The Public Health Agency of Sweden | Erik Embring et al |
| EPI_ISL_434665 | Sweden/20-07042/2020 | 2020-04-04 | Europe/Sweden/Uppsala/ | Uppsala Narakut Aleris | The Public Health Agency of Sweden | Annika Nilsson et al |
| EPI_ISL_434666 | Sweden/20-07171/2020 | 2020-04-08 | Europe/Sweden/Vasterbotten/ | Hornefors Halsocentral | The Public Health Agency of Sweden | Camilla Eiback et al |
| EPI_ISL_434667 | Sweden/20-07298/2020 | 2020-04-14 | Europe/Sweden/Uppsala/ | Ulltuna Vardcentral | The Public Health Agency of Sweden | Heidi Lindback et al |
| EPI_ISL_434668 | Sweden/20-07389/2020 | 2020-04-15 | Europe/Sweden/Vastmanland/ | Kungsors VC | The Public Health Agency of Sweden | Jessica Karlsson et al |
| EPI_ISL_434672 | Sweden/20-07459/2020 | 2020-04-16 | Europe/Sweden/Stockholm/ | Surbrunns VC | The Public Health Agency of Sweden | Erik Embring et al |
| EPI_ISL_434674 | Sweden/20-07572/2020 | 2020-04-21 | Europe/Sweden/Vastra Gotaland/ | Narhalsan Backa vardcentral | The Public Health Agency of Sweden | Mats Olsson et al |
| EPI_ISL_434676 | Sweden/20-07616/2020 | 2020-04-20 | Europe/Sweden/Stockholm/ | Surbrunns VC | The Public Health Agency of Sweden | Erik Embring et al |
| EPI_ISL_434692 | Thailand/NIH-15/2020 | 2020-01-05 | Asia/Thailand/Nonthaburi/ | Bamrasnaradura hospital | National Institute of Health. Department of medical Sciences, Ministry of Public Health, Thailand | Pilailuk et al |
| EPI_ISL_434693 | Thailand/NIH-59/2020 | 2020-01-16 | Asia/Thailand/Nonthaburi/ | Bamrasnaradura hospital | National Institute of Health. Department of medical Sciences, Ministry of Public Health, Thailand | Pilailuk et al |

| | | | | | | |
|---|---|---|---|---|---|---|
| EPI_ISL_434694 | Thailand/NIH-162/2020 | 2020-01-22 | Asia/Thailand/Samut Prakarn/ | Bamrasnaradura hospital | National Institute of Health. Department of medical Sciences, Ministry of Public Health, Thailand | Pilailuk et al |
| EPI_ISL_434695 | Thailand/NIH-190/2020 | 2020-01-22 | Asia/Thailand/Nonthaburi/ | unknown | National Institute of Health. Department of medical Sciences, Ministry of Public Health, Thailand | Pilailuk et al |
| EPI_ISL_434722 | USA/TX-HMH011/2020 | 2020-03-13 | North America/USA/Texas/Greater Hou | Houston Methodist Hospital | Houston Methodist Hospital | S. Wesley Long et al |
| EPI_ISL_434724 | USA/TX-HMH014/2020 | 2020-03-14 | North America/USA/Texas/Greater Hou | Houston Methodist Hospital | Houston Methodist Hospital | S. Wesley Long et al |
| EPI_ISL_434730 | USA/TX-HMH020/2020 | 2020-03-13 | North America/USA/Texas/Greater Hou | Houston Methodist Hospital | Houston Methodist Hospital | S. Wesley Long et al |
| EPI_ISL_434774 | USA/TX-HMH065/2020 | 2020-03-18 | North America/USA/Texas/Greater Hou | Houston Methodist Hospital | Houston Methodist Hospital | S. Wesley Long et al |
| EPI_ISL_434781 | USA/TX-HMH072/2020 | 2020-03-18 | North America/USA/Texas/Greater Hou | Houston Methodist Hospital | Houston Methodist Hospital | S. Wesley Long et al |
| EPI_ISL_434786 | USA/TX-HMH077/2020 | 2020-03-18 | North America/USA/Texas/Greater Hou | Houston Methodist Hospital | Houston Methodist Hospital | S. Wesley Long et al |
| EPI_ISL_434821 | USA/TX-HMH0121/2020 | 2020-03-20 | North America/USA/Texas/Greater Hou | Houston Methodist Hospital | Houston Methodist Hospital | S. Wesley Long et al |
| EPI_ISL_434827 | USA/TX-HMH0127/2020 | 2020-03-20 | North America/USA/Texas/Greater Hou | Houston Methodist Hospital | Houston Methodist Hospital | S. Wesley Long et al |
| EPI_ISL_434835 | USA/TX-HMH0135/2020 | 2020-03-21 | North America/USA/Texas/Greater Hou | Houston Methodist Hospital | Houston Methodist Hospital | S. Wesley Long et al |

| | | | | | | |
|---|---|---|---|---|---|---|
| EPI_ISL_434837 | USA/TX-HMH0137/2020 | 2020-03-21 | North America/USA/Texas/Greater Hou | Houston Methodist Hospital | Houston Methodist Hospital | S. Wesley Long et al |
| EPI_ISL_434838 | USA/TX-HMH0138/2020 | 2020-03-21 | North America/USA/Texas/Greater Hou | Houston Methodist Hospital | Houston Methodist Hospital | S. Wesley Long et al |
| EPI_ISL_434853 | USA/TX-HMH0154/2020 | 2020-03-22 | North America/USA/Texas/Greater Hou | Houston Methodist Hospital | Houston Methodist Hospital | S. Wesley Long et al |
| EPI_ISL_434889 | USA/TX-HMH0214/2020 | 2020-03-24 | North America/USA/Texas/Greater Hou | Houston Methodist Hospital | Houston Methodist Hospital | S. Wesley Long et al |
| EPI_ISL_434890 | USA/TX-HMH0230/2020 | 2020-03-25 | North America/USA/Texas/Greater Hou | Houston Methodist Hospital | Houston Methodist Hospital | S. Wesley Long et al |
| EPI_ISL_434903 | USA/TX-HMH0275/2020 | 2020-03-27 | North America/USA/Texas/Greater Hou | Houston Methodist Hospital | Houston Methodist Hospital | S. Wesley Long et al |
| EPI_ISL_434920 | USA/TX-HMH0295/2020 | 2020-03-27 | North America/USA/Texas/Greater Hou | Houston Methodist Hospital | Houston Methodist Hospital | S. Wesley Long et al |
| EPI_ISL_434942 | USA/TX-HMH0348/2020 | 2020-04-02 | North America/USA/Texas/Greater Hou | Houston Methodist Hospital | Houston Methodist Hospital | S. Wesley Long et al |
| EPI_ISL_434943 | USA/TX-HMH0349/2020 | 2020-04-01 | North America/USA/Texas/Greater Hou | Houston Methodist Hospital | Houston Methodist Hospital | S. Wesley Long et al |
| EPI_ISL_434958 | USA/TX-HMH0366/2020 | 2020-03-25 | North America/USA/Texas/Greater Hou | Houston Methodist Hospital | Houston Methodist Hospital | S. Wesley Long et al |
| EPI_ISL_434976 | USA/TX-HMH0389/2020 | 2020-04-01 | North America/USA/Texas/Greater Hou | Houston Methodist Hospital | Houston Methodist Hospital | S. Wesley Long et al |
| EPI_ISL_434986 | USA/TX-HMH0399/2020 | 2020-03-31 | North America/USA/Texas/Greater Hou | Houston Methodist Hospital | Houston Methodist Hospital | S. Wesley Long et al |
| EPI_ISL_434987 | USA/TX-HMH0400/2020 | 2020-04-01 | North America/USA/Texas/Greater Hou | Houston Methodist Hospital | Houston Methodist Hospital | S. Wesley Long et al |

| | | | | | | |
|---|---|---|---|---|---|---|
| EPI_ISL_434998 | USA/TX-HMH0413/2020 | 2020-04-01 | North America/USA/Texas/Greater Hou | Houston Methodist Hospital | Houston Methodist Hospital | S. Wesley Long et al |
| EPI_ISL_435031 | USA/TX-HMH0452/2020 | 2020-04-03 | North America/USA/Texas/Greater Hou | Houston Methodist Hospital | Houston Methodist Hospital | S. Wesley Long et al |
| EPI_ISL_435034 | USA/LA-EVTL01/2020 | 2020-04-09 | North America/USA/Louisiana/East Feli | LSUHS Emerging Viral Threat Laboratory | Microbial Genome Sequencing Center | Jeremy P. Kamil et al |
| EPI_ISL_435039 | USA/LA-EVTL06/2020 | 2020-04-15 | North America/USA/Louisiana/East Bato | LSUHS Emerging Viral Threat Laboratory | Microbial Genome Sequencing Center | Jeremy P. Kamil et al |
| EPI_ISL_435045 | Kazakhstan/NCB-1/2020 | 2020-03-22 | Asia/Kazakhstan/Nur-Sultan/ | Laboratory of Applied Genetics | "RSE ""National Center for Biotechnology""" | Alexandr Shevtsov et al |
| EPI_ISL_435046 | Kazakhstan/NCB-2/2020 | 2020-03-25 | Asia/Kazakhstan/Nur-Sultan/ | Laboratory of Applied Genetics | "RSE ""National Center for Biotechnology""" | Alexandr Shevtsov et al |
| EPI_ISL_435054 | India/GJ-GBRC7/2020 | 2020-04-14 | Asia/India/Gujarat/Ahmedabad | B.J. Medical College and Civil hospital | Gujarat Biotechnology Research Centre | Monika Gandhi et al |
| EPI_ISL_435057 | Turkey/VETAL1902_1/2020 | 2020-04-09 | Europe/Turkey/Adƒ±yaman/ | T.C. Saƒülƒ±k Bakanlƒ±ƒüƒ± Adƒ±yaman ƒ∞l Saƒülƒ±k Müdürlüğü Adƒ±yaman Eƒüitim Ve Ara≈üƒ±rma Hastanesi | VETAL Animal Health Products Company, BSL3+ Production Laboratuary, Turkey | Fatma Nilay Tutak et al |
| EPI_ISL_435066 | India/DL-NCDC-01711/2020 | 2020-03-18 | Asia/India/Delhi/ | National Centre for Disease control (NCDC), CSIR-Institute of Genomics and Integrative Biology (CSIR-IGIB) | NCDC/CSIR-IGIB | Pramod Kumar et al |
| EPI_ISL_435105 | India/LA-NCDC-01616/2020 | 2020-03-17 | Asia/India/Ladakh/ | National Centre for Disease control (NCDC), CSIR-Institute of Genomics and Integrative Biology (CSIR-IGIB) | NCDC/CSIR-IGIB | Pramod Kumar et al |
| EPI_ISL_435121 | UnitedArabEmirates/L0184/2020 | 2020-02-25 | Asia/United Arab Emirates/United Arab | Mohammed Bin Rashid University of Medicine and Health Sciences | Al Jalila Genomics Center | Ahmad Abou Tayoun et al |
| EPI_ISL_435126 | UnitedArabEmirates/L0904/2020 | 2020-02-25 | Asia/United Arab Emirates/United Arab | Mohammed Bin Rashid University of Medicine and Health Sciences | Al Jalila Genomics Center | Ahmad Abou Tayoun et al |
| EPI_ISL_435131 | UnitedArabEmirates/L2409/2020 | 2020-02-25 | Asia/United Arab Emirates/United Arab | Mohammed Bin Rashid University of Medicine and Health Sciences | Al Jalila Genomics Center | Ahmad Abou Tayoun et al |
| EPI_ISL_435134 | UnitedArabEmirates/L4280/2020 | 2020-02-08 | Asia/United Arab Emirates/United Arab | Mohammed Bin Rashid University of Medicine and Health Sciences | Al Jalila Genomics Center | Ahmad Abou Tayoun et al |

| | | | | | | |
|---|---|---|---|---|---|---|
| EPI_ISL_435135 | UnitedArabEmirates/L4682/2020 | 2020-02-25 | Asia/United Arab Emirates/United Arab | Mohammed Bin Rashid University of Medicine and Health Sciences | Al Jalila Genomics Center | Ahmad Abou Tayoun et al |
| EPI_ISL_435137 | UnitedArabEmirates/L5630/2020 | 2020-01-29 | Asia/United Arab Emirates/United Arab | Mohammed Bin Rashid University of Medicine and Health Sciences | Al Jalila Genomics Center | Ahmad Abou Tayoun et al |
| EPI_ISL_435138 | UnitedArabEmirates/L6599/2020 | 2020-02-25 | Asia/United Arab Emirates/United Arab | Mohammed Bin Rashid University of Medicine and Health Sciences | Al Jalila Genomics Center | Ahmad Abou Tayoun et al |
| EPI_ISL_435139 | UnitedArabEmirates/L6627/2020 | 2020-02-25 | Asia/United Arab Emirates/United Arab | Mohammed Bin Rashid University of Medicine and Health Sciences | Al Jalila Genomics Center | Ahmad Abou Tayoun et al |
| EPI_ISL_435145 | Italy/ABR-IZSGC-TE6644/2020 | 2020-03-24 | Europe/Italy/Abruzzo/ | Ospedale Civile Giuseppe Mazzini | "Istituto Zooprofilattico Sperimentale dell'Abruzzo e Molise ""G.Caporale""" | Lorusso A et al |
| EPI_ISL_435146 | Italy/ABR-IZSGC-TE12337/2020 | 2020-04-07 | Europe/Italy/Abruzzo/ | Villa Serena del Dr. Leonardo Petruzzi | "Istituto Zooprofilattico Sperimentale dell'Abruzzo e Molise ""G.Caporale""" | Lorusso A et al |
| EPI_ISL_435150 | Italy/ABR-IZSGC-TE12774/2020 | 2020-04-08 | Europe/Italy/Abruzzo/ | Ospedale SS Annunziata | "Istituto Zooprofilattico Sperimentale dell'Abruzzo e Molise ""G.Caporale""" | Lorusso A et al |
| EPI_ISL_435151 | Italy/ABR-IZSGC-TE13484/2020 | 2020-04-08 | Europe/Italy/Abruzzo/ | Ospedale SS Annunziata | "Istituto Zooprofilattico Sperimentale dell'Abruzzo e Molise ""G.Caporale""" | Lorusso A et al |
| EPI_ISL_435154 | Italy/ABR-IZSGC-TE14162/2020 | 2020-04-09 | Europe/Italy/Abruzzo/ | SERVIZIO DI IGIENE E SANITVÄ PUBBLICA ASL Teramo | "Istituto Zooprofilattico Sperimentale dell'Abruzzo e Molise ""G.Caporale""" | Lorusso A et al |

| | | | | | | |
|---|---|---|---|---|---|---|
| EPI_ISL_435282 | Indonesia/JK-EIJK-0317/2020 | 2020-03-19 | Asia/Indonesia/Jakarta/ | RS Pondok Indah Hospital ,Äì Pondok Indah | Eijkman Institute for Molecular Biology, Ministry of Research and Technology/National Agency for Research and Innovation | Edison Johar et al |
| EPI_ISL_435308 | Vietnam/VNHN_0299/2020 | 2020-03-09 | Asia/Vietnam/Vietnam/ | National Hospital of Tropical Diseases | Oxford University Clinical Research Unit, Hanoi, Vietnam | Nguyen Thi Tam et al |
| EPI_ISL_435343 | Canada/QC-AG6/2020 | 20-Apr | North America/Canada/Quebec/ | Laboratoire de microbiologie, Hopital de Verdun | Smith Laboratory, Centre de Recherche CHU Sainte-Justine | Martin Smith et al |
| EPI_ISL_435344 | Canada/QC-AG1/2020 | 20-Apr | North America/Canada/Quebec/ | Laboratoire de microbiologie, Hopital de Verdun | Smith Laboratory, Centre de Recherche CHU Sainte-Justine | Martin Smith et al |
| EPI_ISL_435346 | Canada/QC-AI8/2020 | 20-Apr | North America/Canada/Quebec/ | Laboratoire de microbiologie, Hopital de Verdun | Smith Laboratory, Centre de Recherche CHU Sainte-Justine | Martin Smith et al |
| EPI_ISL_435347 | Canada/QC-AB5/2020 | 20-Apr | North America/Canada/Quebec/ | Laboratoire de microbiologie, Hopital de Verdun | Smith Laboratory, Centre de Recherche CHU Sainte-Justine | Martin Smith et al |
| EPI_ISL_435355 | USA/UT-0367/2020 | 2020-03-28 | North America/USA/Utah/ | Utah Public Health Laboratory | Utah Public Health Laboratory | Erin Young et al |
| EPI_ISL_435370 | USA/UT-0536/2020 | 2020-04-04 | North America/USA/Utah/ | Utah Public Health Laboratory | Utah Public Health Laboratory | Erin Young et al |
| EPI_ISL_435371 | USA/UT-0537/2020 | 2020-04-04 | North America/USA/Utah/ | Utah Public Health Laboratory | Utah Public Health Laboratory | Erin Young et al |
| EPI_ISL_435382 | USA/UT-0681/2020 | 2020-04-06 | North America/USA/Utah/ | Utah Public Health Laboratory | Utah Public Health Laboratory | Erin Young et al |
| EPI_ISL_435387 | USA/UT-0886/2020 | 2020-04-06 | North America/USA/Utah/ | Utah Public Health Laboratory | Utah Public Health Laboratory | Erin Young et al |
| EPI_ISL_435391 | USA/UT-0894/2020 | 2020-04-05 | North America/USA/Utah/ | Utah Public Health Laboratory | Utah Public Health Laboratory | Erin Young et al |

| | | | | | | |
|---|---|---|---|---|---|---|
| EPI_ISL_435396 | USA/WI-GMF-01534/2020 | 2020-04-22 | North America/USA/Iowa/Allamakee Co | Gundersen Molecular Diagnostics Laboratory | Kabara Cancer Research Institute | Craig S. Richmond et al |
| EPI_ISL_435397 | USA/WI-GMF-01535/2020 | 2020-04-21 | North America/USA/Iowa/Allamakee Co | Gundersen Molecular Diagnostics Laboratory | Kabara Cancer Research Institute | Craig S. Richmond et al |
| EPI_ISL_435400 | USA/WI-GMF-01602/2020 | 2020-04-23 | North America/USA/Wisconsin/La Cross | Gundersen Molecular Diagnostics Laboratory | Kabara Cancer Research Institute | Craig S. Richmond et al |
| EPI_ISL_435406 | Hungary/SRC-00175/2020 | 2020-03-22 | Europe/Hungary/Baranya County/ | Virological Research Group, Szentv°gothai Research Centre | Bioinformatics Research Group, Szentv°gothai Research Centre | Pv©ter Urbv°n et al |
| EPI_ISL_435415 | Hungary/SRC-00836/2020 | 2020-03-30 | Europe/Hungary/Baranya County/ | Virological Research Group, Szentv°gothai Research Centre | Bioinformatics Research Group, Szentv°gothai Research Centre | Pv©ter Urbv°n et al |
| EPI_ISL_435441 | USA/AK-PHL151/2020 | 2020-04-07 | North America/USA/Alaska/Anchorage | Alaska State Virology Laboratory | Alaska State Virology Laboratory | Jack Chen et al |
| EPI_ISL_435453 | USA/LA-SR0150/2020 | 2020-03-30 | North America/USA/Louisiana/New Orle | Robert Garry lab | Andersen lab at Scripps Research | Allison Smither et al |
| EPI_ISL_435462 | USA/LA-SR0159/2020 | 2020-04-02 | North America/USA/Louisiana/New Orle | Robert Garry lab | Andersen lab at Scripps Research | Allison Smither et al |
| EPI_ISL_435471 | USA/LA-SR0170/2020 | 2020-04-03 | North America/USA/Louisiana/New Orle | Robert Garry lab | Andersen lab at Scripps Research | Allison Smither et al |
| EPI_ISL_435478 | USA/NY-NYUMC411/2020 | 2020-03-30 | North America/USA/New York/Suffolk C | NYU Langone Health | Departments of Pathology and Medicine, New York University School of Medicine | Maria Aguero-Rosenfeld et al |
| EPI_ISL_435487 | USA/NY-NYUMC420/2020 | 2020-04-22 | North America/USA/New York/Nassau C | NYU Langone Health | Departments of Pathology and Medicine, New York University School of Medicine | Maria Aguero-Rosenfeld et al |

| | | | | | | |
|---|---|---|---|---|---|---|
| EPI_ISL_435490 | USA/NY-NYUMC423/2020 | 2020-04-22 | North America/USA/New York/Manhatt | NYU Langone Health | Departments of Pathology and Medicine, New York University School of Medicine | Maria Aguero-Rosenfeld et al |
| EPI_ISL_435491 | USA/NY-NYUMC424/2020 | 2020-04-22 | North America/USA/New York/Nassau ( | NYU Langone Health | Departments of Pathology and Medicine, New York University School of Medicine | Maria Aguero-Rosenfeld et al |
| EPI_ISL_435492 | USA/NY-NYUMC425/2020 | 2020-04-20 | North America/USA/New York/Queens | NYU Langone Health | Departments of Pathology and Medicine, New York University School of Medicine | Maria Aguero-Rosenfeld et al |
| EPI_ISL_435498 | USA/NY-NYUMC431/2020 | 2020-04-07 | North America/USA/New York/Nassau ( | NYU Langone Health | Departments of Pathology and Medicine, New York University School of Medicine | Maria Aguero-Rosenfeld et al |
| EPI_ISL_435499 | USA/NY-NYUMC432/2020 | 2020-04-20 | North America/USA/New York/Queens | NYU Langone Health | Departments of Pathology and Medicine, New York University School of Medicine | Maria Aguero-Rosenfeld et al |
| EPI_ISL_435502 | USA/NY-NYUMC435/2020 | 2020-04-24 | North America/USA/New York/Brooklyr | NYU Langone Health | Departments of Pathology and Medicine, New York University School of Medicine | Maria Aguero-Rosenfeld et al |

| | | | | | | |
|---|---|---|---|---|---|---|
| EPI_ISL_435512 | USA/NY-NYUMC445/2020 | 2020-04-15 | North America/USA/New York/Queens | NYU Langone Health | Departments of Pathology and Medicine, New York University School of Medicine | Maria Aguero-Rosenfeld et al |
| EPI_ISL_435528 | USA/NY-NYUMC462/2020 | 2020-04-20 | North America/USA/New York/Queens | NYU Langone Health | Departments of Pathology and Medicine, New York University School of Medicine | Maria Aguero-Rosenfeld et al |
| EPI_ISL_435532 | USA/NY-NYUMC467/2020 | 2020-04-16 | North America/USA/New York/Brooklyn | NYU Langone Health | Departments of Pathology and Medicine, New York University School of Medicine | Maria Aguero-Rosenfeld et al |
| EPI_ISL_435534 | USA/NY-NYUMC469/2020 | 2020-04-20 | North America/USA/New York/Queens | NYU Langone Health | Departments of Pathology and Medicine, New York University School of Medicine | Maria Aguero-Rosenfeld et al |
| EPI_ISL_435547 | USA/NY-NYUMC482/2020 | 2020-04-24 | North America/USA/New York/ | NYU Langone Health | Departments of Pathology and Medicine, New York University School of Medicine | Maria Aguero-Rosenfeld et al |
| EPI_ISL_435552 | USA/LA-EVTL014/2020 | 2020-04-25 | North America/USA/Louisiana/Caddo Pa | LSUHS Emerging Viral Threat Laboratory | Microbial Genome Sequencing Center | Rona S. Scott et al |
| EPI_ISL_435560 | USA/LA-EVTL022/2020 | 2020-04-24 | North America/USA/Louisiana/Caddo Pa | LSUHS Emerging Viral Threat Laboratory | Microbial Genome Sequencing Center | John A. Vanchiere et al |
| EPI_ISL_435566 | USA/LA-EVTL028/2020 | 2020-04-24 | North America/USA/Louisiana/Caddo Pa | LSUHS Emerging Viral Threat Laboratory | Microbial Genome Sequencing Center | John A. Vanchiere et al |

| | | | | | | |
|---|---|---|---|---|---|---|
| EPI_ISL_435568 | USA/LA-EVTL030/2020 | 2020-04-24 | North America/USA/Louisiana/Caddo Pa | LSUHS Emerging Viral Threat Laboratory | Microbial Genome Sequencing Center | John A. Vanchiere et al |
| EPI_ISL_435570 | USA/LA-EVTL032/2020 | 2020-04-24 | North America/USA/Louisiana/Caddo Pa | LSUHS Emerging Viral Threat Laboratory | Microbial Genome Sequencing Center | Jeremy P. Kamil et al |
| EPI_ISL_435579 | USA/LA-EVTL041/2020 | 2020-04-24 | North America/USA/Louisiana/Webster | LSUHS Emerging Viral Threat Laboratory | Microbial Genome Sequencing Center | Jeremy P. Kamil et al |
| EPI_ISL_435580 | USA/CA-SCCPHD-UC101/2020 | 2020-02-26 | North America/USA/California/Santa Cl | Santa Clara County Public Health Department | Chiu Laboratory, University of California, San Francisco | Xianding Deng et al |
| EPI_ISL_435581 | USA/CA-SCCPHD-UC102/2020 | 2020-02-28 | North America/USA/California/Santa Cl | Santa Clara County Public Health Department | Chiu Laboratory, University of California, San Francisco | Xianding Deng et al |
| EPI_ISL_435582 | USA/CA-SCCPHD-UC103/2020 | 2020-02-28 | North America/USA/California/Santa Cl | Santa Clara County Public Health Department | Chiu Laboratory, University of California, San Francisco | Xianding Deng et al |
| EPI_ISL_435583 | USA/CA-SCCPHD-UC104/2020 | 2020-02-29 | North America/USA/California/Santa Cl | Santa Clara County Public Health Department | Chiu Laboratory, University of California, San Francisco | Xianding Deng et al |
| EPI_ISL_435584 | USA/CA-SCCPHD-UC105/2020 | 2020-02-29 | North America/USA/California/Santa Cl | Santa Clara County Public Health Department | Chiu Laboratory, University of California, San Francisco | Xianding Deng et al |
| EPI_ISL_435585 | USA/CA-SCCPHD-UC106/2020 | 2020-02-29 | North America/USA/California/Santa Cl | Santa Clara County Public Health Department | Chiu Laboratory, University of California, San Francisco | Xianding Deng et al |
| EPI_ISL_435587 | USA/CA-SCCPHD-UC108/2020 | 2020-02-29 | North America/USA/California/Santa Cl | Santa Clara County Public Health Department | Chiu Laboratory, University of California, San Francisco | Xianding Deng et al |

| | | | | | | |
|---|---|---|---|---|---|---|
| EPI_ISL_435631 | USA/CA-SCCPHD-UC152/2020 | 2020-03-11 | North America/USA/California/Santa Cl | Santa Clara County Public Health Department | Chiu Laboratory, University of California, San Francisco | Xianding Deng et al |
| EPI_ISL_435632 | USA/CA-SCCPHD-UC153/2020 | 2020-03-11 | North America/USA/California/Santa Cl | Santa Clara County Public Health Department | Chiu Laboratory, University of California, San Francisco | Xianding Deng et al |
| EPI_ISL_435650 | USA/CA-SCCPHD-UC171/2020 | 2020-03-13 | North America/USA/California/Santa Cl | Santa Clara County Public Health Department | Chiu Laboratory, University of California, San Francisco | Xianding Deng et al |
| EPI_ISL_435653 | USA/CA-SCCPHD-UC174/2020 | 2020-03-13 | North America/USA/California/Santa Cl | Santa Clara County Public Health Department | Chiu Laboratory, University of California, San Francisco | Xianding Deng et al |
| EPI_ISL_435665 | USA/CA-SCCPHD-UC186/2020 | 2020-03-17 | North America/USA/California/Santa Cl | Santa Clara County Public Health Department | Chiu Laboratory, University of California, San Francisco | Xianding Deng et al |
| EPI_ISL_435696 | Singapore/91/2020 | 2020-04-20 | Asia/Singapore/Singapore/ | National Public Health Laboratory, National Centre for Infectious Diseases | National Public Health Laboratory, National Centre for Infectious Diseases | Mak Tze Minn et al |
| EPI_ISL_435706 | USA/CT-Yale-109/2020 | 2020-04-08 | North America/USA/Connecticut/ | Yale COVID-19 Biorepository | Grubaugh Lab - Yale School of Public Health | Joseph Fauver et al |
| EPI_ISL_435719 | USA/CT-Yale-149/2020 | 2020-03-26 | North America/USA/Connecticut/ | Connecticut State Department of Public Health | Grubaugh Lab - Yale School of Public Health | Joseph Fauver et al |
| EPI_ISL_435720 | USA/CT-Yale-151/2020 | 2020-03-13 | North America/USA/Connecticut/ | Yale Clinical Virology Laboratory | Grubaugh Lab - Yale School of Public Health | Joseph Fauver et al |

| | | | | | | |
|---|---|---|---|---|---|---|
| EPI_ISL_436046 | USA/VI-CDC-1993/2020 | 2020-04-01 | North America/USA/Virgin Islands/ | US VI Department of Health | Pathogen Discovery, Respiratory Viruses Branch, Division of Viral Diseases, Centers for Disease Control and Prevention | Ying Tao et al |
| EPI_ISL_436049 | USA/NY-SURV097/2020 | 2020-03-09 | North America/USA/New York/New Yor | NYC Department of Health and Mental Hygiene | Pathogen Discovery, Respiratory Viruses Branch, Division of Viral Diseases, Centers for Disease Control and Prevention | Ying Tao et al |
| EPI_ISL_436058 | USA/NY-SURV0186/2020 | 2020-03-17 | North America/USA/New York/New Yor | NYC Department of Health and Mental Hygiene | Pathogen Discovery, Respiratory Viruses Branch, Division of Viral Diseases, Centers for Disease Control and Prevention | Ying Tao et al |
| EPI_ISL_436080 | USA/NY-SURV0988/2020 | 2020-03-18 | North America/USA/New York/New Yor | NYC Department of Health and Mental Hygiene | Pathogen Discovery, Respiratory Viruses Branch, Division of Viral Diseases, Centers for Disease Control and Prevention | Ying Tao et al |
| EPI_ISL_436108 | Taiwan/TSGH-23/2020 | 2020-04-02 | Asia/Taiwan/Taiwan/Taipei | TSGH-CP molecular lab | TSGH-CP molecular lab | Cherng-Lih Perng et al |

| | | | | | | |
|---|---|---|---|---|---|---|
| EPI_ISL_436220 | Spain/VC-FISABIO-125/2020 | 2020-03-14 | Europe/Spain/Comunitat Valenciana/Va | Servicio de Microbiolog√≠a. Consorcio Hospital General Universitario de Valencia | Sequencing and Bioinformatics Service and Molecular Epidemiology Research Group. FISABIO-Public Health | Griselda De Marco et al |
| EPI_ISL_436221 | Spain/VC-FISABIO-126/2020 | 2020-03-14 | Europe/Spain/Comunitat Valenciana/Va | Servicio de Microbiolog√≠a. Consorcio Hospital General Universitario de Valencia | Sequencing and Bioinformatics Service and Molecular Epidemiology Research Group. FISABIO-Public Health | Beatriz Beamud et al |
| EPI_ISL_436226 | Spain/VC-FISABIO-131/2020 | 2020-03-17 | Europe/Spain/Comunitat Valenciana/Va | Servicio de Microbiolog√≠a. Consorcio Hospital General Universitario de Valencia | Sequencing and Bioinformatics Service and Molecular Epidemiology Research Group. FISABIO-Public Health | Maria Dolores Ocete et al |
| EPI_ISL_436230 | Spain/VC-FISABIO-135/2020 | 2020-03-18 | Europe/Spain/Comunitat Valenciana/Va | Servicio de Microbiolog√≠a. Consorcio Hospital General Universitario de Valencia | Sequencing and Bioinformatics Service and Molecular Epidemiology Research Group. FISABIO-Public Health | Marta Pla Diaz et al |
| EPI_ISL_436232 | Spain/VC-FISABIO-137/2020 | 2020-03-18 | Europe/Spain/Comunitat Valenciana/Va | Servicio de Microbiolog√≠a. Consorcio Hospital General Universitario de Valencia | Sequencing and Bioinformatics Service and Molecular Epidemiology Research Group. FISABIO-Public Health | Loreto Ferr√∫s Abad et al |

| | | | | | | |
|---|---|---|---|---|---|---|
| EPI_ISL_436239 | Spain/VC-FISABIO-144/2020 | 2020-03-18 | Europe/Spain/Comunitat Valenciana/Va | Servicio de Microbiologia. Hospital Universitario Doctor Peset | Sequencing and Bioinformatics Service and Molecular Epidemiology Research Group. FISABIO-Public Health | Juan Alberola Engu√≠danos et al |
| EPI_ISL_436253 | Spain/VC-FISABIO-158/2020 | 2020-03-19 | Europe/Spain/Comunitat Valenciana/Va | Servicio de Microbiologia. Hospital Universitario Doctor Peset | Sequencing and Bioinformatics Service and Molecular Epidemiology Research Group. FISABIO-Public Health | Juan Alberola Engu√≠danos et al |
| EPI_ISL_436256 | Spain/VC-FISABIO-161/2020 | 2020-03-19 | Europe/Spain/Comunitat Valenciana/Va | Servicio de Microbiologia. Hospital Universitario Doctor Peset | Sequencing and Bioinformatics Service and Molecular Epidemiology Research Group. FISABIO-Public Health | Juan Alberola Engu√≠danos et al |
| EPI_ISL_436263 | Spain/VC-FISABIO-168/2020 | 2020-03-20 | Europe/Spain/Comunitat Valenciana/Va | Servicio de Microbiologia. Hospital Universitario Doctor Peset | Sequencing and Bioinformatics Service and Molecular Epidemiology Research Group. FISABIO-Public Health | Juan Alberola Engu√≠danos et al |
| EPI_ISL_436264 | Spain/VC-FISABIO-169/2020 | 2020-03-20 | Europe/Spain/Comunitat Valenciana/Va | Servicio de Microbiologia. Hospital Universitario Doctor Peset | Sequencing and Bioinformatics Service and Molecular Epidemiology Research Group. FISABIO-Public Health | Juan Alberola Engu√≠danos et al |

| | | | | | | |
|---|---|---|---|---|---|---|
| EPI_ISL_436280 | Spain/VC-FISABIO-185/2020 | 2020-03-17 | Europe/Spain/Comunitat Valenciana/Va | Servicio de Microbiologia. Hospital Clinico Universitario de Valencia | Sequencing and Bioinformatics Service and Molecular Epidemiology Research Group. FISABIO-Public Health | Beatriz Beamud et al |
| EPI_ISL_436283 | Spain/VC-FISABIO-188/2020 | 2020-03-17 | Europe/Spain/Comunitat Valenciana/Va | Servicio de Microbiologia. Hospital Clinico Universitario de Valencia | Sequencing and Bioinformatics Service and Molecular Epidemiology Research Group. FISABIO-Public Health | Neris Garcia-Gonzalez et al |
| EPI_ISL_436285 | Spain/VC-FISABIO-190/2020 | 2020-03-24 | Europe/Spain/Comunitat Valenciana/Va | Servicio de Microbiologia. Hospital Clinico Universitario de Valencia | Sequencing and Bioinformatics Service and Molecular Epidemiology Research Group. FISABIO-Public Health | Sandra Carbo et al |
| EPI_ISL_436317 | Spain/VC-FISABIO-222/2020 | 2020-03-26 | Europe/Spain/Comunitat Valenciana/Va | Servicio de Microbiologia. Hospital Clinico Universitario de Valencia | Sequencing and Bioinformatics Service and Molecular Epidemiology Research Group. FISABIO-Public Health | Neris Garcia-Gonzalez et al |
| EPI_ISL_436320 | Spain/VC-FISABIO-225/2020 | 2020-03-19 | Europe/Spain/Comunitat Valenciana/Va | Servicio de Microbiologia. Hospital Clinico Universitario de Valencia | Sequencing and Bioinformatics Service and Molecular Epidemiology Research Group. FISABIO-Public Health | Beatriz Beamud et al |

| | | | | | | |
|---|---|---|---|---|---|---|
| EPI_ISL_436328 | Spain/VC-FISABIO-233/2020 | 2020-03-26 | Europe/Spain/Comunitat Valenciana/Va | Servicio de Microbiologia. Hospital Clinico Universitario de Valencia | Sequencing and Bioinformatics Service and Molecular Epidemiology Research Group. FISABIO-Public Health | Mariana Reyes-Prieto et al |
| EPI_ISL_436336 | Spain/VC-FISABIO-241/2020 | 2020-03-26 | Europe/Spain/Comunitat Valenciana/Va | Servicio de Microbiologia. Hospital Clinico Universitario de Valencia | Sequencing and Bioinformatics Service and Molecular Epidemiology Research Group. FISABIO-Public Health | Marta Pla Diaz et al |
| EPI_ISL_436337 | Spain/VC-FISABIO-242/2020 | 2020-03-26 | Europe/Spain/Comunitat Valenciana/Va | Servicio de Microbiologia. Hospital Clinico Universitario de Valencia | Sequencing and Bioinformatics Service and Molecular Epidemiology Research Group. FISABIO-Public Health | Neris Garcia-Gonzalez et al |
| EPI_ISL_436342 | Spain/VC-FISABIO-247/2020 | 2020-03-25 | Europe/Spain/Comunitat Valenciana/Va | Servicio de Microbiologia. Hospital Clinico Universitario de Valencia | Sequencing and Bioinformatics Service and Molecular Epidemiology Research Group. FISABIO-Public Health | Marta Pla Diaz et al |
| EPI_ISL_436347 | Spain/VC-FISABIO-252/2020 | 2020-03-29 | Europe/Spain/Comunitat Valenciana/Va | Servicio de Microbiologia. Hospital Clinico Universitario de Valencia | Sequencing and Bioinformatics Service and Molecular Epidemiology Research Group. FISABIO-Public Health | Paula Ruiz-Hueso et al |

| | | | | | | |
|---|---|---|---|---|---|---|
| EPI_ISL_436350 | Spain/VC-FISABIO-255/2020 | 2020-03-26 | Europe/Spain/Comunitat Valenciana/Va | Servicio de Microbiologia. Hospital Clinico Universitario de Valencia | Sequencing and Bioinformatics Service and Molecular Epidemiology Research Group. FISABIO-Public Health | Ivan Ansari et al |
| EPI_ISL_436351 | Spain/VC-FISABIO-256/2020 | 2020-03-26 | Europe/Spain/Comunitat Valenciana/Va | Servicio de Microbiologia. Hospital Clinico Universitario de Valencia | Sequencing and Bioinformatics Service and Molecular Epidemiology Research Group. FISABIO-Public Health | David Navarro et al |
| EPI_ISL_436360 | Spain/VC-FISABIO-265/2020 | 2020-03-27 | Europe/Spain/Comunitat Valenciana/Va | Servicio de Microbiologia. Hospital Clinico Universitario de Valencia | Sequencing and Bioinformatics Service and Molecular Epidemiology Research Group. FISABIO-Public Health | Neris Garcia-Gonzalez et al |
| EPI_ISL_436361 | Spain/VC-FISABIO-266/2020 | 2020-02-29 | Europe/Spain/Comunitat Valenciana/Va | Servicio de Microbiolog√≠a. Consorcio Hospital General Universitario de Valencia | Sequencing and Bioinformatics Service and Molecular Epidemiology Research Group. FISABIO-Public Health | Neris Garcia-Gonzalez et al |
| EPI_ISL_436362 | Spain/VC-FISABIO-267/2020 | 2020-03-17 | Europe/Spain/Comunitat Valenciana/Va | Servicio de Microbiologia. Hospital Clinico Universitario de Valencia | Sequencing and Bioinformatics Service and Molecular Epidemiology Research Group. FISABIO-Public Health | David Navarro et al |

| | | | | | | |
|---|---|---|---|---|---|---|
| EPI_ISL_436367 | Spain/VC-FISABIO-272/2020 | 2020-03-25 | Europe/Spain/Comunitat Valenciana/Va | Servicio de Microbiologia. Hospital Clinico Universitario de Valencia | Sequencing and Bioinformatics Service and Molecular Epidemiology Research Group. FISABIO-Public Health | Neris Garcia-Gonzalez et al |
| EPI_ISL_436374 | Spain/VC-FISABIO-279/2020 | 2020-03-16 | Europe/Spain/Comunitat Valenciana/Va | Servicio de Microbiologia. Hospital Universitario Doctor Peset | Sequencing and Bioinformatics Service and Molecular Epidemiology Research Group. FISABIO-Public Health | Juan Alberola Enguv≠danos et al |
| EPI_ISL_436375 | Spain/VC-FISABIO-280/2020 | 2020-03-16 | Europe/Spain/Comunitat Valenciana/Va | Servicio de Microbiologia. Hospital Universitario Doctor Peset | Sequencing and Bioinformatics Service and Molecular Epidemiology Research Group. FISABIO-Public Health | Juan Alberola Enguv≠danos et al |
| EPI_ISL_436376 | Spain/VC-FISABIO-281/2020 | 2020-03-16 | Europe/Spain/Comunitat Valenciana/Va | Servicio de Microbiologia. Hospital Universitario Doctor Peset | Sequencing and Bioinformatics Service and Molecular Epidemiology Research Group. FISABIO-Public Health | Juan Alberola Enguv≠danos et al |
| EPI_ISL_436387 | Spain/VC-FISABIO-292/2020 | 2020-03-17 | Europe/Spain/Comunitat Valenciana/Va | Servicio de Microbiologv≠a. Consorcio Hospital General Universitario de Valencia | Sequencing and Bioinformatics Service and Molecular Epidemiology Research Group. FISABIO-Public Health | Marta Pla Diaz et al |

| | | | | | | |
|---|---|---|---|---|---|---|
| EPI_ISL_436390 | Spain/VC-FISABIO-295/2020 | 2020-03-17 | Europe/Spain/Comunitat Valenciana/Va | Servicio de Microbiolog√≠a. Consorcio Hospital General Universitario de Valencia | Sequencing and Bioinformatics Service and Molecular Epidemiology Research Group. FISABIO-Public Health | Maria Dolores Ocete et al |
| EPI_ISL_436395 | Spain/VC-FISABIO-300/2020 | 2020-03-17 | Europe/Spain/Comunitat Valenciana/Va | Servicio de Microbiolog√≠a. Consorcio Hospital General Universitario de Valencia | Sequencing and Bioinformatics Service and Molecular Epidemiology Research Group. FISABIO-Public Health | Neris Garcia-Gonzalez et al |
| EPI_ISL_436411 | Spain/VC-FISABIO-316/2020 | 2020-03-26 | Europe/Spain/Comunitat Valenciana/Va | Servicio de Microbiologia. Hospital Clinico Universitario de Valencia | Sequencing and Bioinformatics Service and Molecular Epidemiology Research Group. FISABIO-Public Health | Neris Garcia-Gonzalez et al |
| EPI_ISL_436423 | India/DL-NCDC-3162/2020 | 2020-04-05 | Asia/India/Delhi/ | National Centre for Disease control (NCDC) | NCDC/CSIR-IGIB | Pramod Kumar |
| EPI_ISL_436424 | India/DL-NCDC-3163/2020 | 2020-04-05 | Asia/India/Delhi/ | National Centre for Disease control (NCDC) | NCDC/CSIR-IGIB | Pramod Kumar |
| EPI_ISL_436430 | India/DL-NCDC-3195/2020 | 2020-04-05 | Asia/India/Delhi/ | National Centre for Disease control (NCDC) | NCDC/CSIR-IGIB | Pramod Kumar |
| EPI_ISL_436444 | India/MH-NCDC-3965/2020 | 2020-04-09 | Asia/India/Maharashtra/ | National Centre for Disease control (NCDC) | NCDC/CSIR-IGIB | Pramod Kumar |
| EPI_ISL_436446 | India/MH-NCDC-3985/2020 | 2020-04-09 | Asia/India/Maharashtra/ | National Centre for Disease control (NCDC) | NCDC/CSIR-IGIB | Pramod Kumar |
| EPI_ISL_436452 | India/DL-NCDC-4208/2020 | 2020-04-14 | Asia/India/Delhi/ | National Centre for Disease control (NCDC) | NCDC/CSIR-IGIB | Pramod Kumar |
| EPI_ISL_436455 | India/DL-NCDC-4476/2020 | 2020-04-13 | Asia/India/Delhi/ | National Centre for Disease control (NCDC) | NCDC/CSIR-IGIB | Pramod Kumar |
| EPI_ISL_436489 | USA/PA-MGSC41-05/2020 | 2020-03-26 | North America/USA/Pennsylvania/Pittsl | UPMC Clinical Laboratory | Microbial Genome Sequencing Center, Microbial Genomic Epidemiological Laboratory | Dan Snyder et al |

| | | | | | | |
|---|---|---|---|---|---|---|
| EPI_ISL_436490 | USA/PA-MGSC42-05/2020 | 2020-03-26 | North America/USA/Pennsylvania/Pitts | UPMC Clinical Laboratory | Microbial Genome Sequencing Center, Microbial Genomic Epidemiological Laboratory | Dan Snyder et al |
| EPI_ISL_436492 | USA/PA-MGSC44-05/2020 | 2020-03-26 | North America/USA/Pennsylvania/Pitts | UPMC Clinical Laboratory | Microbial Genome Sequencing Center, Microbial Genomic Epidemiological Laboratory | Dan Snyder et al |
| EPI_ISL_436502 | USA/PA-MGSC55-05/2020 | 2020-03-23 | North America/USA/Pennsylvania/Pitts | UPMC Clinical Laboratory | Microbial Genome Sequencing Center, Microbial Genomic Epidemiological Laboratory | Dan Snyder et al |
| EPI_ISL_436512 | USA/FL-BPHL-08/2020 | 2020-04-03 | North America/USA/Florida/ | Florida Bureau of Public Health Laboratories | Florida Bureau of Public Health Laboratories | Sarah Schmedes et al |
| EPI_ISL_436521 | USA/FL-BPHL-017/2020 | 2020-04-08 | North America/USA/Florida/ | Florida Bureau of Public Health Laboratories | Florida Bureau of Public Health Laboratories | Sarah Schmedes et al |
| EPI_ISL_436526 | USA/FL-BPHL-022/2020 | 2020-04-14 | North America/USA/Florida/ | Florida Bureau of Public Health Laboratories | Florida Bureau of Public Health Laboratories | Sarah Schmedes et al |
| EPI_ISL_436545 | USA/FL-BPHL-041/2020 | 2020-04-16 | North America/USA/Florida/ | Florida Bureau of Public Health Laboratories | Florida Bureau of Public Health Laboratories | Sarah Schmedes et al |
| EPI_ISL_436554 | USA/FL-BPHL-050/2020 | 2020-04-15 | North America/USA/Florida/ | Florida Bureau of Public Health Laboratories | Florida Bureau of Public Health Laboratories | Sarah Schmedes et al |
| EPI_ISL_436588 | USA/WI-UW-294/2020 | 2020-04-09 | North America/USA/Wisconsin/Milwauk | University of Wisconsin-Madison AIDS Vaccine Research Laboratories | University of Wisconsin-Madison AIDS Vaccine Research Laboratories | Gage Moreno et al |

| | | | | | | |
|---|---|---|---|---|---|---|
| EPI_ISL_436590 | USA/WI-UW-296/2020 | 2020-04-09 | North America/USA/Wisconsin/Milwauk | University of Wisconsin-Madison AIDS Vaccine Research Laboratories | University of Wisconsin-Madison AIDS Vaccine Research Laboratories | Gage Moreno et al |
| EPI_ISL_436592 | USA/WI-UW-298/2020 | 2020-04-13 | North America/USA/Wisconsin/Milwauk | University of Wisconsin-Madison AIDS Vaccine Research Laboratories | University of Wisconsin-Madison AIDS Vaccine Research Laboratories | Gage Moreno et al |
| EPI_ISL_436599 | USA/WI-UW-305/2020 | 2020-04-14 | North America/USA/Wisconsin/Milwauk | University of Wisconsin-Madison AIDS Vaccine Research Laboratories | University of Wisconsin-Madison AIDS Vaccine Research Laboratories | Gage Moreno et al |
| EPI_ISL_436604 | USA/WI-UW-310/2020 | 2020-04-15 | North America/USA/Wisconsin/Milwauk | University of Wisconsin-Madison AIDS Vaccine Research Laboratories | University of Wisconsin-Madison AIDS Vaccine Research Laboratories | Gage Moreno et al |
| EPI_ISL_436615 | USA/WI-UW-321/2020 | 2020-04-23 | North America/USA/Wisconsin/Milwauk | University of Wisconsin-Madison AIDS Vaccine Research Laboratories | University of Wisconsin-Madison AIDS Vaccine Research Laboratories | Gage Moreno et al |
| EPI_ISL_436624 | USA/WI-UW-330/2020 | 2020-04-26 | North America/USA/Wisconsin/Milwauk | University of Wisconsin-Madison AIDS Vaccine Research Laboratories | University of Wisconsin-Madison AIDS Vaccine Research Laboratories | Gage Moreno et al |
| EPI_ISL_436640 | USA/WI-UW-346/2020 | 2020-04-21 | North America/USA/Wisconsin/Milwauk | University of Wisconsin-Madison AIDS Vaccine Research Laboratories | University of Wisconsin-Madison AIDS Vaccine Research Laboratories | Gage Moreno et al |
| EPI_ISL_436656 | USA/CA-CZB-1015/2020 | 2020-04-09 | North America/USA/California/Santa Cl | County of Santa Clara Public Health Department | Chan-Zuckerberg Biohub | CZB Cliahub Consortium et al |
| EPI_ISL_436659 | USA/CA-CZB-1018/2020 | 2020-04-10 | North America/USA/California/Santa Cl | County of Santa Clara Public Health Department | Chan-Zuckerberg Biohub | CZB Cliahub Consortium et al |

| | | | | | | |
|---|---|---|---|---|---|---|
| EPI_ISL_436660 | USA/CA-CZB-1019/2020 | 2020-04-10 | North America/USA/California/Santa Cl | County of Santa Clara Public Health Department | Chan-Zuckerberg Biohub | CZB Cliahub Consortium et al |
| EPI_ISL_436669 | USA/CA-CZB-1028/2020 | 2020-04-13 | North America/USA/California/Santa Cl | County of Santa Clara Public Health Department | Chan-Zuckerberg Biohub | CZB Cliahub Consortium et al |
| EPI_ISL_436686 | SouthAfrica/KRISP-045/2020 | 2020-03-27 | Africa/South Africa/KwaZulu-Natal/ | KRISP, KZN Research Innovation and Sequencing Platform | KRISP, KZN Research Innovation and Sequencing Platform | Giandhari J et al |
| EPI_ISL_436717 | Russia/Moscow-GCBL3/2020 | 2020-04-14 | Europe/Russia/Moscow Oblast/ | Genomics and Computational Biology Lab, Scientific Research Institute of Physical-Chemical Medicine, FMBA of Russia | Genomics and Computational Biology Lab, Scientific Research Institute of Physical-Chemical Medicine, FMBA of Russia | A. Pavlenko et al |
| EPI_ISL_436720 | Italy/ABR-IZSGC-TE5543/2020 | 2020-03-20 | Europe/Italy/Abruzzo/ | Ospedale Civile S. Liberatore di Atri | "Istituto Zooprofilattico Sperimentale dell'Abruzzo e Molise ""G.Caporale""" | Lorusso A et al |
| EPI_ISL_436722 | Italy/ABR-IZSGC-TE5547/2020 | 2020-03-20 | Europe/Italy/Abruzzo/ | Ospedale Civile S. Liberatore di Atri | "Istituto Zooprofilattico Sperimentale dell'Abruzzo e Molise ""G.Caporale""" | Lorusso A et al |
| EPI_ISL_436732 | Italy/ABR-IZSGC-TE26425/2020 | 2020-04-27 | Europe/Italy/Abruzzo/ | Ospedale Civile S. Liberatore di Atri | "Istituto Zooprofilattico Sperimentale dell'Abruzzo e Molise ""G.Caporale""" | Lorusso A et al |
| EPI_ISL_436801 | USA/MI-MDHHS-SC20122/2020 | 2020-03-10 | North America/USA/Michigan/ | Michigan Department of Health and Human Services, Bureau of Laboratories | Michigan Department of Health and Human Services, Bureau of Laboratories | Blankenship HM et al |

| | | | | | | |
|---|---|---|---|---|---|---|
| EPI_ISL_436807 | USA/MI-MDHHS-SC20169/2020 | 2020-03-12 | North America/USA/Michigan/ | Michigan Department of Health and Human Services, Bureau of Laboratories | Michigan Department of Health and Human Services, Bureau of Laboratories | Blankenship HM et al |
| EPI_ISL_436812 | USA/MI-MDHHS-SC20136/2020 | 2020-03-13 | North America/USA/Michigan/ | Michigan Department of Health and Human Services, Bureau of Laboratories | Michigan Department of Health and Human Services, Bureau of Laboratories | Blankenship HM et al |
| EPI_ISL_436829 | USA/MI-MDHHS-SC20137/2020 | 2020-03-20 | North America/USA/Michigan/ | Michigan Department of Health and Human Services, Bureau of Laboratories | Michigan Department of Health and Human Services, Bureau of Laboratories | Blankenship HM et al |
| EPI_ISL_436842 | USA/MI-MDHHS-SC20152/2020 | 2020-03-23 | North America/USA/Michigan/ | Michigan Department of Health and Human Services, Bureau of Laboratories | Michigan Department of Health and Human Services, Bureau of Laboratories | Blankenship HM et al |
| EPI_ISL_436852 | USA/MI-MDHHS-SC20134/2020 | 2020-03-24 | North America/USA/Michigan/ | Michigan Department of Health and Human Services, Bureau of Laboratories | Michigan Department of Health and Human Services, Bureau of Laboratories | Blankenship HM et al |
| EPI_ISL_436858 | USA/MI-MDHHS-SC20177/2020 | 2020-03-24 | North America/USA/Michigan/ | Michigan Department of Health and Human Services, Bureau of Laboratories | Michigan Department of Health and Human Services, Bureau of Laboratories | Blankenship HM et al |

| | | | | | | |
|---|---|---|---|---|---|---|
| EPI_ISL_436875 | USA/MI-MDHHS-SC20146/2020 | 2020-03-27 | North America/USA/Michigan/ | Michigan Department of Health and Human Services, Bureau of Laboratories | Michigan Department of Health and Human Services, Bureau of Laboratories | Blankenship HM et al |
| EPI_ISL_436888 | USA/MI-MDHHS-SC20172/2020 | 2020-03-26 | North America/USA/Michigan/ | Michigan Department of Health and Human Services, Bureau of Laboratories | Michigan Department of Health and Human Services, Bureau of Laboratories | Blankenship HM et al |
| EPI_ISL_436897 | USA/WI-GMF-02274/2020 | 2020-05-03 | North America/USA/Wisconsin/Vernon ( | Gundersen Molecular Diagnostics Laboratory | Kabara Cancer Research Institute | Craig S. Richmond et al |
| EPI_ISL_436904 | USA/UT-01217/2020 | 2020-04-06 | North America/USA/Utah/ | Utah Public Health Laboratory | Utah Public Health Laboratory | Erin Young et al |
| EPI_ISL_436907 | USA/UT-01222/2020 | 2020-04-08 | North America/USA/Utah/ | Utah Public Health Laboratory | Utah Public Health Laboratory | Erin Young et al |
| EPI_ISL_436912 | USA/UT-01229/2020 | 2020-04-08 | North America/USA/Utah/ | Utah Public Health Laboratory | Utah Public Health Laboratory | Erin Young et al |
| EPI_ISL_436923 | USA/UT-02223/2020 | 2020-04-15 | North America/USA/Utah/ | Utah Public Health Laboratory | Utah Public Health Laboratory | Erin Young et al |
| EPI_ISL_436925 | USA/UT-02227/2020 | 2020-04-14 | North America/USA/Utah/ | Utah Public Health Laboratory | Utah Public Health Laboratory | Erin Young et al |
| EPI_ISL_436927 | USA/UT-02229/2020 | 2020-04-15 | North America/USA/Utah/ | Utah Public Health Laboratory | Utah Public Health Laboratory | Erin Young et al |
| EPI_ISL_436939 | USA/LA-BIE-071/2020 | 2020-04-06 | North America/USA/Louisiana/ | Ochsner Health | Bioinfoexperts, LLC | Amy Feehan et al |
| EPI_ISL_436940 | USA/LA-BIE-072/2020 | 2020-04-08 | North America/USA/Louisiana/ | Ochsner Health | Bioinfoexperts, LLC | Amy Feehan et al |
| EPI_ISL_436943 | USA/LA-BIE-075/2020 | 2020-04-04 | North America/USA/Louisiana/ | Ochsner Health | Bioinfoexperts, LLC | Amy Feehan et al |
| EPI_ISL_436961 | USA/LA-BIE-094/2020 | 2020-04-06 | North America/USA/Louisiana/ | Ochsner Health | Bioinfoexperts, LLC | Amy Feehan et al |
| EPI_ISL_437033 | Denmark/ALAB-SSI-673/2020 | 2020-03-26 | Europe/Denmark/Denmark/ | Department of Virus and Microbiological Special Diagnostics, Statens Serum Institut, Copenhagen, Denmark, Artillerivej 5, 2300 Copenahgen S | Albertsen lab, Department of Chemistry and Bioscience, Aalborg University, Denmark | Rasmus Kirkegaard et al |

| | | | | | | |
|---|---|---|---|---|---|---|
| EPI_ISL_437041 | Denmark/ALAB-SSI-691/2020 | 2020-03-27 | Europe/Denmark/Denmark/ | Department of Virus and Microbiological Special Diagnostics, Statens Serum Institut, Copenhagen, Denmark, Artillerivej 5, 2300 Copenahgen S | Albertsen lab, Department of Chemistry and Bioscience, Aalborg University, Denmark | Rasmus Kirkegaard et al |
| EPI_ISL_437056 | USA/CA-CZB-1056/2020 | 2020-04-07 | North America/USA/California/Santa Cla | County of Santa Clara Public Health | Chan-Zuckerberg Biohub | CZB Cliahub Consortium et al |
| EPI_ISL_437058 | USA/CA-CZB-1058/2020 | 2020-04-06 | North America/USA/California/Santa Cla | County of Santa Clara Public Health | Chan-Zuckerberg Biohub | CZB Cliahub Consortium et al |
| EPI_ISL_437062 | USA/CA-CZB-1062/2020 | 2020-04-10 | North America/USA/California/Santa Cla | County of Santa Clara Public Health | Chan-Zuckerberg Biohub | CZB Cliahub Consortium et al |
| EPI_ISL_437072 | USA/CA-CZB-1072/2020 | 2020-04-21 | North America/USA/California/Santa Cla | County of Santa Clara Public Health | Chan-Zuckerberg Biohub | CZB Cliahub Consortium et al |
| EPI_ISL_437076 | USA/CA-CZB-1076/2020 | 2020-04-07 | North America/USA/California/Santa Cla | County of Santa Clara Public Health | Chan-Zuckerberg Biohub | CZB Cliahub Consortium et al |
| EPI_ISL_437077 | USA/CA-CZB-1077/2020 | 2020-04-07 | North America/USA/California/Santa Cla | County of Santa Clara Public Health | Chan-Zuckerberg Biohub | CZB Cliahub Consortium et al |
| EPI_ISL_437083 | USA/CA-CZB-1083/2020 | 2020-04-09 | North America/USA/California/Santa Cla | County of Santa Clara Public Health | Chan-Zuckerberg Biohub | CZB Cliahub Consortium et al |
| EPI_ISL_437084 | USA/CA-CZB-1084/2020 | 2020-04-07 | North America/USA/California/Santa Cla | County of Santa Clara Public Health | Chan-Zuckerberg Biohub | CZB Cliahub Consortium et al |
| EPI_ISL_437090 | Latvia/011/2020 | 2020-03-24 | Europe/Latvia/Latvia/ | Latvijas Infektoloƒ£ijas centrs | Latvian Biomedical Research and Study Centre | Ivars Silamiƒ∑elis et al |
| EPI_ISL_437104 | USA/MI-MDHHS-SC20017/2020 | 2020-03-13 | North America/USA/Michigan/ | Michigan Department of Health and Human Services, Bureau of Laboratories | Michigan Department of Health and Human Services, Bureau of Laboratories | Blankenship HM et al |
| EPI_ISL_437117 | USA/MI-MDHHS-SC20030/2020 | 2020-03-17 | North America/USA/Michigan/ | Michigan Department of Health and Human Services, Bureau of Laboratories | Michigan Department of Health and Human Services, Bureau of Laboratories | Blankenship HM et al |

| | | | | | | |
|---|---|---|---|---|---|---|
| EPI_ISL_437135 | USA/MI-MDHHS-SC20048/2020 | 2020-03-20 | North America/USA/Michigan/ | Michigan Department of Health and Human Services, Bureau of Laboratories | Michigan Department of Health and Human Services, Bureau of Laboratories | Blankenship HM et al |
| EPI_ISL_437145 | USA/MI-MDHHS-SC20058/2020 | 2020-03-21 | North America/USA/Michigan/ | Michigan Department of Health and Human Services, Bureau of Laboratories | Michigan Department of Health and Human Services, Bureau of Laboratories | Blankenship HM et al |
| EPI_ISL_437183 | USA/MI-MDHHS-SC20096/2020 | 2020-03-30 | North America/USA/Michigan/ | Michigan Department of Health and Human Services, Bureau of Laboratories | Michigan Department of Health and Human Services, Bureau of Laboratories | Blankenship HM et al |
| EPI_ISL_437191 | Indonesia/JK-EIJK-03/2020 | 2020-03-27 | Asia/Indonesia/Jakarta/ | RS Pondok Indah Hospital ‚Äì Pondok Indah | Eijkman Institute for Molecular Biology, Ministry of Research and Technology/National Agency for Research and Innovation | Edison Johar et al |
| EPI_ISL_437203 | Austria/Graz-MUG7/2020 | 2020-04-06 | Europe/Austria/Styria/ | Diagnostic- and Research Institute of Pathology, Medical University of Graz | Diagnostic- and Research Institute of Pathology, Medical University of Graz | Karl Kashofer et al |
| EPI_ISL_437204 | Germany/BY-MVP-0002/2020 | 2020-03-05 | Europe/Germany/Bavaria/Munich | Max von Pettenkofer Institute, Virology, National Reference Center for Retroviruses, LMU M√ºnchen | Laboratory for Functional Genome Analysis, Dept. Genomics, Gene Center of the LMU Munich | Max Muenchhoff et al |

| | | | | | | |
|---|---|---|---|---|---|---|
| EPI_ISL_437215 | Germany/BY-MVP-0014/2020 | 2020-04-08 | Europe/Germany/Bavaria/Munich | Max von Pettenkofer Institute, Virology, National Reference Center for Retroviruses, LMU M√ºnchen | Laboratory for Functional Genome Analysis, Dept. Genomics, Gene Center of the LMU Munich | Max Muenchhoff et al |
| EPI_ISL_437217 | Germany/BY-MVP-0016/2020 | 2020-04-11 | Europe/Germany/Bavaria/Munich | Max von Pettenkofer Institute, Virology, National Reference Center for Retroviruses, LMU M√ºnchen | Laboratory for Functional Genome Analysis, Dept. Genomics, Gene Center of the LMU Munich | Max Muenchhoff et al |
| EPI_ISL_437220 | Germany/BY-MVP-0020/2020 | 2020-04-06 | Europe/Germany/Bavaria/Munich | Max von Pettenkofer Institute, Virology, National Reference Center for Retroviruses, LMU M√ºnchen | Laboratory for Functional Genome Analysis, Dept. Genomics, Gene Center of the LMU Munich | Max Muenchhoff et al |
| EPI_ISL_437222 | Germany/BY-MVP-0022/2020 | 2020-04-11 | Europe/Germany/Bavaria/Munich | Max von Pettenkofer Institute, Virology, National Reference Center for Retroviruses, LMU M√ºnchen | Laboratory for Functional Genome Analysis, Dept. Genomics, Gene Center of the LMU Munich | Max Muenchhoff et al |
| EPI_ISL_437223 | Germany/BY-MVP-0023/2020 | 2020-03-12 | Europe/Germany/Bavaria/Munich | Max von Pettenkofer Institute, Virology, National Reference Center for Retroviruses, LMU M√ºnchen | Laboratory for Functional Genome Analysis, Dept. Genomics, Gene Center of the LMU Munich | Max Muenchhoff et al |
| EPI_ISL_437226 | Germany/BY-MVP-0027/2020 | 2020-04-10 | Europe/Germany/Bavaria/Munich | Max von Pettenkofer Institute, Virology, National Reference Center for Retroviruses, LMU M√ºnchen | Laboratory for Functional Genome Analysis, Dept. Genomics, Gene Center of the LMU Munich | Max Muenchhoff et al |

| | | | | | | |
|---|---|---|---|---|---|---|
| EPI_ISL_437227 | Germany/BY-MVP-0028/2020 | 2020-04-13 | Europe/Germany/Bavaria/Munich | Max von Pettenkofer Institute, Virology, National Reference Center for Retroviruses, LMU M√ºnchen | Laboratory for Functional Genome Analysis, Dept. Genomics, Gene Center of the LMU Munich | Max Muenchhoff et al |
| EPI_ISL_437230 | Germany/BY-MVP-0031/2020 | 2020-03-22 | Europe/Germany/Bavaria/Munich | Max von Pettenkofer Institute, Virology, National Reference Center for Retroviruses, LMU M√ºnchen | Laboratory for Functional Genome Analysis, Dept. Genomics, Gene Center of the LMU Munich | Max Muenchhoff et al |
| EPI_ISL_437231 | Germany/BY-MVP-0032/2020 | 2020-03-25 | Europe/Germany/Bavaria/Munich | Max von Pettenkofer Institute, Virology, National Reference Center for Retroviruses, LMU M√ºnchen | Laboratory for Functional Genome Analysis, Dept. Genomics, Gene Center of the LMU Munich | Max Muenchhoff et al |
| EPI_ISL_437234 | Germany/BY-MVP-0035/2020 | 2020-03-15 | Europe/Germany/Bavaria/Munich | Max von Pettenkofer Institute, Virology, National Reference Center for Retroviruses, LMU M√ºnchen | Laboratory for Functional Genome Analysis, Dept. Genomics, Gene Center of the LMU Munich | Max Muenchhoff et al |
| EPI_ISL_437236 | Germany/BY-MVP-0037/2020 | 2020-03-22 | Europe/Germany/Bavaria/Munich | Max von Pettenkofer Institute, Virology, National Reference Center for Retroviruses, LMU M√ºnchen | Laboratory for Functional Genome Analysis, Dept. Genomics, Gene Center of the LMU Munich | Max Muenchhoff et al |
| EPI_ISL_437241 | Germany/BY-MVP-0042/2020 | 2020-04-07 | Europe/Germany/Bavaria/Munich | Max von Pettenkofer Institute, Virology, National Reference Center for Retroviruses, LMU M√ºnchen | Laboratory for Functional Genome Analysis, Dept. Genomics, Gene Center of the LMU Munich | Max Muenchhoff et al |

| | | | | | | |
|---|---|---|---|---|---|---|
| EPI_ISL_437243 | Germany/BY-MVP-0044/2020 | 2020-04-11 | Europe/Germany/Bavaria/Munich | Max von Pettenkofer Institute, Virology, National Reference Center for Retroviruses, LMU M√ºnchen | Laboratory for Functional Genome Analysis, Dept. Genomics, Gene Center of the LMU Munich | Max Muenchhoff et al |
| EPI_ISL_437244 | Germany/BY-MVP-0045/2020 | 2020-03-08 | Europe/Germany/Bavaria/Munich | Max von Pettenkofer Institute, Virology, National Reference Center for Retroviruses, LMU M√ºnchen | Laboratory for Functional Genome Analysis, Dept. Genomics, Gene Center of the LMU Munich | Max Muenchhoff et al |
| EPI_ISL_437252 | Germany/BY-MVP-0053/2020 | 2020-03-16 | Europe/Germany/Bavaria/Munich | Max von Pettenkofer Institute, Virology, National Reference Center for Retroviruses, LMU M√ºnchen | Laboratory for Functional Genome Analysis, Dept. Genomics, Gene Center of the LMU Munich | Max Muenchhoff et al |
| EPI_ISL_437255 | Germany/BY-MVP-0056/2020 | 2020-03-19 | Europe/Germany/Bavaria/Munich | Max von Pettenkofer Institute, Virology, National Reference Center for Retroviruses, LMU M√ºnchen | Laboratory for Functional Genome Analysis, Dept. Genomics, Gene Center of the LMU Munich | Max Muenchhoff et al |
| EPI_ISL_437257 | Germany/BY-MVP-0058/2020 | 2020-03-20 | Europe/Germany/Bavaria/Munich | Max von Pettenkofer Institute, Virology, National Reference Center for Retroviruses, LMU M√ºnchen | Laboratory for Functional Genome Analysis, Dept. Genomics, Gene Center of the LMU Munich | Max Muenchhoff et al |
| EPI_ISL_437260 | Germany/BY-MVP-0061/2020 | 2020-03-22 | Europe/Germany/Bavaria/Munich | Max von Pettenkofer Institute, Virology, National Reference Center for Retroviruses, LMU M√ºnchen | Laboratory for Functional Genome Analysis, Dept. Genomics, Gene Center of the LMU Munich | Max Muenchhoff et al |

| | | | | | | |
|---|---|---|---|---|---|---|
| EPI_ISL_437264 | Germany/BY-MVP-0067/2020 | 2020-03-25 | Europe/Germany/Bavaria/Munich | Max von Pettenkofer Institute, Virology, National Reference Center for Retroviruses, LMU M√ºnchen | Laboratory for Functional Genome Analysis, Dept. Genomics, Gene Center of the LMU Munich | Max Muenchhoff et al |
| EPI_ISL_437265 | Germany/BY-MVP-0069/2020 | 2020-03-25 | Europe/Germany/Bavaria/Munich | Max von Pettenkofer Institute, Virology, National Reference Center for Retroviruses, LMU M√ºnchen | Laboratory for Functional Genome Analysis, Dept. Genomics, Gene Center of the LMU Munich | Max Muenchhoff et al |
| EPI_ISL_437267 | Germany/BY-MVP-0071/2020 | 2020-03-27 | Europe/Germany/Bavaria/Munich | Max von Pettenkofer Institute, Virology, National Reference Center for Retroviruses, LMU M√ºnchen | Laboratory for Functional Genome Analysis, Dept. Genomics, Gene Center of the LMU Munich | Max Muenchhoff et al |
| EPI_ISL_437273 | Germany/BY-MVP-0079/2020 | 2020-03-30 | Europe/Germany/Bavaria/Munich | Max von Pettenkofer Institute, Virology, National Reference Center for Retroviruses, LMU M√ºnchen | Laboratory for Functional Genome Analysis, Dept. Genomics, Gene Center of the LMU Munich | Max Muenchhoff et al |
| EPI_ISL_437275 | Germany/BY-MVP-0081/2020 | 2020-03-30 | Europe/Germany/Bavaria/Munich | Max von Pettenkofer Institute, Virology, National Reference Center for Retroviruses, LMU M√ºnchen | Laboratory for Functional Genome Analysis, Dept. Genomics, Gene Center of the LMU Munich | Max Muenchhoff et al |
| EPI_ISL_437286 | Germany/BY-MVP-0092/2020 | 2020-04-02 | Europe/Germany/Bavaria/Munich | Max von Pettenkofer Institute, Virology, National Reference Center for Retroviruses, LMU M√ºnchen | Laboratory for Functional Genome Analysis, Dept. Genomics, Gene Center of the LMU Munich | Max Muenchhoff et al |

| | | | | | | |
|---|---|---|---|---|---|---|
| EPI_ISL_437288 | Germany/BY-MVP-0096/2020 | 2020-04-03 | Europe/Germany/Bavaria/Munich | Max von Pettenkofer Institute, Virology, National Reference Center for Retroviruses, LMU M√ºnchen | Laboratory for Functional Genome Analysis, Dept. Genomics, Gene Center of the LMU Munich | Max Muenchhoff et al |
| EPI_ISL_437292 | Germany/BY-MVP-0101/2020 | 2020-04-07 | Europe/Germany/Bavaria/Munich | Max von Pettenkofer Institute, Virology, National Reference Center for Retroviruses, LMU M√ºnchen | Laboratory for Functional Genome Analysis, Dept. Genomics, Gene Center of the LMU Munich | Max Muenchhoff et al |
| EPI_ISL_437293 | Germany/BY-MVP-0103/2020 | 2020-04-09 | Europe/Germany/Bavaria/Munich | Max von Pettenkofer Institute, Virology, National Reference Center for Retroviruses, LMU M√ºnchen | Laboratory for Functional Genome Analysis, Dept. Genomics, Gene Center of the LMU Munich | Max Muenchhoff et al |
| EPI_ISL_437294 | Germany/BY-MVP-0104/2020 | 2020-04-10 | Europe/Germany/Bavaria/Munich | Max von Pettenkofer Institute, Virology, National Reference Center for Retroviruses, LMU M√ºnchen | Laboratory for Functional Genome Analysis, Dept. Genomics, Gene Center of the LMU Munich | Max Muenchhoff et al |
| EPI_ISL_437304 | Turkey/HSGM-674/2020 | 2020-03-26 | Europe/Turkey/Nevsehir/ | Ministry of Health Turkey | Ministry of Health Turkey | Fatma Bayrakdar et al |
| EPI_ISL_437305 | Turkey/HSGM-1418/2020 | 2020-03-27 | Europe/Turkey/Kocaeli/ | Ministry of Health Turkey | Ministry of Health Turkey | Fatma Bayrakdar et al |
| EPI_ISL_437306 | Turkey/HSGM-1450/2020 | 2020-03-27 | Europe/Turkey/Kocaeli/ | Ministry of Health Turkey | Ministry of Health Turkey | Fatma Bayrakdar et al |
| EPI_ISL_437307 | Turkey/HSGM-439/2020 | 2020-03-25 | Europe/Turkey/Mardin/ | Ministry of Health Turkey | Ministry of Health Turkey | Fatma Bayrakdar et al |
| EPI_ISL_437309 | Turkey/HSGM-1049/2020 | 2020-03-26 | Europe/Turkey/Ankara/ | Ministry of Health Turkey | Ministry of Health Turkey | Fatma Bayrakdar et al |
| EPI_ISL_437310 | Turkey/HSGM-1081/2020 | 2020-03-27 | Europe/Turkey/Ankara/ | Ministry of Health Turkey | Ministry of Health Turkey | Fatma Bayrakdar et al |
| EPI_ISL_437311 | Turkey/HSGM-1428/2020 | 2020-03-27 | Europe/Turkey/Ankara/ | Ministry of Health Turkey | Ministry of Health Turkey | Fatma Bayrakdar et al |
| EPI_ISL_437312 | Turkey/HSGM-1468/2020 | 2020-03-25 | Europe/Turkey/Kocaeli/ | Ministry of Health Turkey | Ministry of Health Turkey | Fatma Bayrakdar et al |

| | | | | | | |
|---|---|---|---|---|---|---|
| EPI_ISL_437313 | Turkey/HSGM-302/2020 | 2020-03-27 | Europe/Turkey/Kocaeli/ | Ministry of Health Turkey | Ministry of Health Turkey | Fatma Bayrakdar et al |
| EPI_ISL_437314 | Turkey/HSGM-510/2020 | 2020-03-26 | Europe/Turkey/Ankara/ | Ministry of Health Turkey | Ministry of Health Turkey | Fatma Bayrakdar et al |
| EPI_ISL_437315 | Turkey/HSGM-942/2020 | 2020-03-26 | Europe/Turkey/Ankara/ | Ministry of Health Turkey | Ministry of Health Turkey | Fatma Bayrakdar et al |
| EPI_ISL_437316 | Turkey/HSGM-1004/2020 | 2020-03-25 | Europe/Turkey/Denizli/ | Ministry of Health Turkey | Ministry of Health Turkey | Fatma Bayrakdar et al |
| EPI_ISL_437317 | Turkey/HSGM-1027/2020 | 2020-03-27 | Europe/Turkey/Ankara/ | Ministry of Health Turkey | Ministry of Health Turkey | Fatma Bayrakdar et al |
| EPI_ISL_437318 | Turkey/HSGM-1444/2020 | 2020-03-19 | Europe/Turkey/Ankara/ | Ministry of Health Turkey | Ministry of Health Turkey | Fatma Bayrakdar et al |
| EPI_ISL_437319 | Turkey/HSGM-1490/2020 | 2020-03-19 | Europe/Iran/Kocaeli/ | Ministry of Health Turkey | Ministry of Health Turkey | Fatma Bayrakdar et al |
| EPI_ISL_437320 | Turkey/HSGM-1492/2020 | 2020-03-19 | Europe/Turkey/Istanbul/ | Ministry of Health Turkey | Ministry of Health Turkey | Fatma Bayrakdar et al |
| EPI_ISL_437324 | Turkey/HSGM-1476/2020 | 2020-03-19 | Europe/Iran/Istanbul/ | Ministry of Health Turkey | Ministry of Health Turkey | Fatma Bayrakdar et al |
| EPI_ISL_437325 | Turkey/HSGM-1495/2020 | 2020-03-19 | Europe/Iran/Istanbul/ | Ministry of Health Turkey | Ministry of Health Turkey | Fatma Bayrakdar et al |
| EPI_ISL_437326 | Turkey/HSGM-1432/2020 | 2020-03-19 | Europe/Iran/Istanbul/ | Ministry of Health Turkey | Ministry of Health Turkey | Fatma Bayrakdar et al |
| EPI_ISL_437327 | Turkey/HSGM-1458/2020 | 2020-03-19 | Europe/Iran/Agri/ | Ministry of Health Turkey | Ministry of Health Turkey | Fatma Bayrakdar et al |
| EPI_ISL_437329 | Turkey/HSGM-6204/2020 | 2020-03-19 | Europe/Turkey/Ankara/ | Ministry of Health Turkey | Ministry of Health Turkey | Fatma Bayrakdar et al |
| EPI_ISL_437331 | Turkey/HSGM-1014/2020 | 2020-03-25 | Europe/Turkey/Ankara/ | Ministry of Health Turkey | Ministry of Health Turkey | Fatma Bayrakdar et al |
| EPI_ISL_437332 | Turkey/HSGM-4698/2020 | 2020-03-18 | Europe/Iran/Istanbul/ | Ministry of Health Turkey | Ministry of Health Turkey | Fatma Bayrakdar et al |
| EPI_ISL_437333 | Turkey/HSGM-11640/2020 | 2020-03-25 | Europe/Turkey/Ankara/ | Ministry of Health Turkey | Ministry of Health Turkey | Fatma Bayrakdar et al |
| EPI_ISL_437334 | Turkey/HSGM-10232/2020 | 2020-03-24 | Europe/Turkey/Ankara/ | Ministry of Health Turkey | Ministry of Health Turkey | Fatma Bayrakdar et al |
| EPI_ISL_437335 | Turkey/HSGM-12059/2020 | 2020-03-25 | Europe/Turkey/Denizli/ | Ministry of Health Turkey | Ministry of Health Turkey | Fatma Bayrakdar et al |
| EPI_ISL_437361 | USA/MN-MDH-284/2020 | 2020-03-29 | North America/USA/Minnesota/ | Minnesota Department of Health, Public Health Laboratory | Minnesota Department of Health, Public Health Laboratory | Matt Plumb et al |

| | | | | | | |
|---|---|---|---|---|---|---|
| EPI_ISL_437365 | USA/MN-MDH-372/2020 | 2020-04-05 | North America/USA/Minnesota/ | Minnesota Department of Health, Public Health Laboratory | Minnesota Department of Health, Public Health Laboratory | Matt Plumb et al |
| EPI_ISL_437366 | USA/MN-MDH-374/2020 | 2020-04-06 | North America/USA/Minnesota/ | Minnesota Department of Health, Public Health Laboratory | Minnesota Department of Health, Public Health Laboratory | Matt Plumb et al |
| EPI_ISL_437380 | USA/MN-MDH-564/2020 | 2020-04-14 | North America/USA/Minnesota/ | Minnesota Department of Health, Public Health Laboratory | Minnesota Department of Health, Public Health Laboratory | Matt Plumb et al |
| EPI_ISL_437382 | USA/MN-MDH-585/2020 | 2020-04-14 | North America/USA/Minnesota/ | Minnesota Department of Health, Public Health Laboratory | Minnesota Department of Health, Public Health Laboratory | Matt Plumb et al |
| EPI_ISL_437386 | USA/MN-MDH-666/2020 | 2020-04-17 | North America/USA/Minnesota/ | Minnesota Department of Health, Public Health Laboratory | Minnesota Department of Health, Public Health Laboratory | Matt Plumb et al |
| EPI_ISL_437388 | USA/VA-DCLS-0172/2020 | 2020-04-13 | North America/USA/Virginia/ | Virginia DCLS | Virginia DCLS | Virginia DCLS et al |
| EPI_ISL_437397 | USA/VA-DCLS-0181/2020 | 2020-04-06 | North America/USA/Virginia/ | Virginia DCLS | Virginia DCLS | Virginia DCLS et al |
| EPI_ISL_437402 | USA/VA-DCLS-0186/2020 | 2020-04-07 | North America/USA/Virginia/ | Virginia DCLS | Virginia DCLS | Virginia DCLS et al |
| EPI_ISL_437414 | USA/VA-DCLS-0199/2020 | 2020-04-11 | North America/USA/Virginia/ | Virginia DCLS | Virginia DCLS | Virginia DCLS et al |
| EPI_ISL_437417 | USA/VA-DCLS-0202/2020 | 2020-04-14 | North America/USA/Virginia/ | Virginia DCLS | Virginia DCLS | Virginia DCLS et al |
| EPI_ISL_437430 | USA/VA-DCLS-0215/2020 | 2020-04-15 | North America/USA/Virginia/ | Virginia DCLS | Virginia DCLS | Virginia DCLS et al |
| EPI_ISL_437431 | USA/VA-DCLS-0216/2020 | 2020-04-06 | North America/USA/Virginia/ | Virginia DCLS | Virginia DCLS | Virginia DCLS et al |
| EPI_ISL_437444 | India/GJ-GBRC16/2020 | 2020-04-26 | Asia/India/Gujarat/Surat | Department of MicroBiology, Government Medical College, Surat | Gujarat Biotechnology Research Centre | Dipa Kinariwala et al |
| EPI_ISL_437457 | USA/IN-Lilly-IPB0170-16bc18/2020 | 2020-04-06 | North America/USA/Indiana/Marion Co | Clinical Diagnostics Laboratory, Diagnostic & Experimental Pathology, Lilly Research Laboratories | Clinical Diagnostics Laboratory, Diagnostic & Experimental Pathology, Lilly Research Laboratories | Tim Holzer et al |

| | | | | | | |
|---|---|---|---|---|---|---|
| EPI_ISL_437464 | SaudiArabia/KAUST-Makkah10/2020 | 2020-03-23 | Asia/Saudi Arabia/Makkah/ | Pathogen Genomics Lab King Abdullah University of Science and Technology(KAUST) | Pathogen Genomics Lab King Abdullah University of Science and Technology(KAUST) | Sharif Hala et al |
| EPI_ISL_437482 | SaudiArabia/KAUST-Madinah23/2020 | 2020-03-20 | Asia/Saudi Arabia/Madinah/ | Pathogen Genomics Lab King Abdullah University of Science and Technology(KAUST) | Pathogen Genomics Lab King Abdullah University of Science and Technology(KAUST) | Sara Mfarrej et al |
| EPI_ISL_437518 | USA/AK-PHL283/2020 | 2020-04-26 | North America/USA/Alaska/Interior Ala | Alaska State Virology Laboratory | Alaska State Virology Laboratory | Jack Chen et al |
| EPI_ISL_437520 | USA/OR-OHSU-0010/2020 | 2020-04-08 | North America/USA/Oregon/Columbia C | OHSU Lab Services Molecular Microbiology Lab | Oregon SARS-CoV-2 Genome Sequencing Center | Brendan L. O'Connell et al |
| EPI_ISL_437522 | USA/OR-OHSU-0022/2020 | 2020-04-07 | North America/USA/Oregon/Washingto | OHSU Lab Services Molecular Microbiology Lab | Oregon SARS-CoV-2 Genome Sequencing Center | Brendan L. O'Connell et al |
| EPI_ISL_437525 | USA/OR-OHSU-0043/2020 | 2020-04-20 | North America/USA/Oregon/Multnomal | OHSU Lab Services Molecular Microbiology Lab | Oregon SARS-CoV-2 Genome Sequencing Center | Brendan L. O'Connell et al |
| EPI_ISL_437527 | USA/OR-OHSU-0047/2020 | 2020-04-20 | North America/USA/Oregon/Washingto | OHSU Lab Services Molecular Microbiology Lab | Oregon SARS-CoV-2 Genome Sequencing Center | Brendan L. O'Connell et al |
| EPI_ISL_437530 | USA/OR-OHSU-0050/2020 | 2020-04-16 | North America/USA/Oregon/Washingto | OHSU Lab Services Molecular Microbiology Lab | Oregon SARS-CoV-2 Genome Sequencing Center | Brendan L. O'Connell et al |
| EPI_ISL_437550 | USA/CA-SR0262/2020 | 2020-04-27 | North America/USA/California/San Dieg | Scripps Medical Laboratory | Andersen lab at Scripps Research | SEARCH Alliance San Diego with Michael Quigley et al |

| | | | | | | |
|---|---|---|---|---|---|---|
| EPI_ISL_437558 | USA/CA-SR0253/2020 | 2020-03-27 | North America/USA/California/San Dieg | Scripps Medical Laboratory | Andersen lab at Scripps Research | SEARCH Alliance San Diego with Michael Quigley et al |
| EPI_ISL_437577 | USA/CA-SR0223/2020 | 2020-04-22 | North America/USA/California/San Dieg | Scripps Medical Laboratory | Andersen lab at Scripps Research | SEARCH Alliance San Diego with Michael Quigley et al |
| EPI_ISL_437612 | Thailand/SI203285-NT/2020 | 2020-01-23 | Asia/Thailand/Thailand/ | unknown | Faculty of Medicine | Rodpan et al |
| EPI_ISL_437613 | Thailand/SI202872-NT/2020 | 2020-02-25 | Asia/Thailand/Thailand/ | unknown | Faculty of Medicine | Rodpan et al |
| EPI_ISL_437614 | Thailand/SI202769-NT/2020 | 2020-02-25 | Asia/Thailand/Thailand/ | unknown | Faculty of Medicine | Rodpan et al |
| EPI_ISL_437615 | Thailand/SI202620-NT/2020 | 2020-02-22 | Asia/Thailand/Thailand/ | unknown | Faculty of Medicine | Rodpan et al |
| EPI_ISL_437616 | Thailand/SI202205-NT/2020 | 2020-02-07 | Asia/Thailand/Thailand/ | unknown | Faculty of Medicine | Rodpan et al |
| EPI_ISL_437617 | Thailand/SI201712-NT/2020 | 2020-02-03 | Asia/Thailand/Thailand/ | unknown | Faculty of Medicine | Rodpan et al |
| EPI_ISL_437618 | Thailand/SI200615-NT/2020 | 2020-01-29 | Asia/Thailand/Thailand/ | unknown | Faculty of Medicine | Rodpan et al |
| EPI_ISL_437619 | Thailand/SI200478-NST/2020 | 2020-01-26 | Asia/Thailand/Thailand/ | unknown | Faculty of Medicine | Rodpan et al |
| EPI_ISL_437620 | Thailand/SI200468-NT/2020 | 2020-01-25 | Asia/Thailand/Thailand/ | unknown | Faculty of Medicine | Rodpan et al |
| EPI_ISL_437621 | Thailand/SI200433-NT/2020 | 2020-01-25 | Asia/Thailand/Thailand/ | unknown | Faculty of Medicine | Rodpan et al |
| EPI_ISL_437622 | Thailand/SI200383-NT/2020 | 2020-01-23 | Asia/Thailand/Thailand/ | unknown | Faculty of Medicine | Rodpan et al |
| EPI_ISL_437623 | Thailand/SI200040-NT/2020 | 2020-01-08 | Asia/Thailand/Thailand/ | unknown | Faculty of Medicine | Rodpan et al |
| EPI_ISL_437624 | Thailand/SI200893-NT/2020 | 2020-02-01 | Asia/Thailand/Thailand/ | unknown | Faculty of Medicine | Rodpan et al |
| EPI_ISL_437689 | France/IDF-CIBU200107/2020 | 2020-02-27 | Europe/France/Ile de France/ | Laboratory for Urgent Response to Biological Threats | Institut Pasteur CIBU / ERI | V. Caro et al |
| EPI_ISL_437690 | France/HDF-CIBU200132/2020 | 2020-02-29 | Europe/France/Hauts de France/ | Laboratory for Urgent Response to Biological Threats | Institut Pasteur CIBU /ERI | V. Caro et al |

| | | | | | | |
|---|---|---|---|---|---|---|
| EPI_ISL_437693 | SaudiArabia/KAUST-Makkah138/202 | 2020-03-24 | Asia/Saudi Arabia/Makkah/ | Pathogen Genomics Lab King Abdullah University of Science and Technology(KAUST) | Pathogen Genomics Lab King Abdullah University of Science and Technology(KAUST) | Sharif Hala et al |
| EPI_ISL_437725 | SaudiArabia/KAUST-Makkah229/202 | 2020-04-06 | Asia/Saudi Arabia/Makkah/ | Pathogen Genomics Lab King Abdullah University of Science and Technology(KAUST) | Pathogen Genomics Lab King Abdullah University of Science and Technology(KAUST) | Sharif Hala et al |
| EPI_ISL_437752 | SaudiArabia/KAUST-Madinah270/20 | 2020-03-30 | Asia/Saudi Arabia/Madinah/ | Pathogen Genomics Lab King Abdullah University of Science and Technology(KAUST) | Pathogen Genomics Lab King Abdullah University of Science and Technology(KAUST) | Sharif Hala et al |
| EPI_ISL_437770 | USA/VA-DCLS-0131/2020 | 2020-04-14 | North America/USA/Virginia/ | Virginia DCLS | Virginia DCLS | Virginia DCLS et al |
| EPI_ISL_437778 | USA/VA-DCLS-0139/2020 | 2020-04-16 | North America/USA/Virginia/ | Virginia DCLS | Virginia DCLS | Virginia DCLS et al |
| EPI_ISL_437790 | USA/VA-DCLS-0157/2020 | 2020-04-14 | North America/USA/Virginia/ | Virginia DCLS | Virginia DCLS | Virginia DCLS et al |
| EPI_ISL_437809 | USA/WA-UW-4026/2020 | 2020-03-29 | North America/USA/Washington/ | UW Virology Lab | UW Virology Lab | Pavitra Roychoudhury et al |
| EPI_ISL_437817 | USA/WA-UW-4067/2020 | 2020-04-01 | North America/USA/Washington/ | UW Virology Lab | UW Virology Lab | Pavitra Roychoudhury et al |
| EPI_ISL_437829 | USA/WA-UW-4317/2020 | 2020-03-31 | North America/USA/Washington/ | UW Virology Lab | UW Virology Lab | Pavitra Roychoudhury et al |
| EPI_ISL_437830 | USA/ID-UW-6446/2020 | 2020-04-02 | North America/USA/Idaho/ | UW Virology Lab | UW Virology Lab | Pavitra Roychoudhury et al |
| EPI_ISL_437836 | USA/ID-UW-6467/2020 | 2020-04-02 | North America/USA/Idaho/ | UW Virology Lab | UW Virology Lab | Pavitra Roychoudhury et al |
| EPI_ISL_437841 | USA/WA-UW-6481/2020 | 2020-04-03 | North America/USA/Washington/ | UW Virology Lab | UW Virology Lab | Pavitra Roychoudhury et al |
| EPI_ISL_437844 | USA/WA-UW-6485/2020 | 2020-04-05 | North America/USA/Washington/ | UW Virology Lab | UW Virology Lab | Pavitra Roychoudhury et al |
| EPI_ISL_437850 | USA/WA-UW-6497/2020 | 2020-04-03 | North America/USA/Washington/ | UW Virology Lab | UW Virology Lab | Pavitra Roychoudhury et al |
| EPI_ISL_437857 | USA/WA-UW-6527/2020 | 2020-04-05 | North America/USA/Washington/ | UW Virology Lab | UW Virology Lab | Pavitra Roychoudhury et al |
| EPI_ISL_437859 | USA/WA-UW-6535/2020 | 2020-04-04 | North America/USA/Washington/ | UW Virology Lab | UW Virology Lab | Pavitra Roychoudhury et al |
| EPI_ISL_437860 | USA/WA-UW-6546/2020 | 2020-04-04 | North America/USA/Washington/ | UW Virology Lab | UW Virology Lab | Pavitra Roychoudhury et al |
| EPI_ISL_437866 | USA/CT-UW-6565/2020 | 2020-04-02 | North America/USA/Connecticut/ | UW Virology Lab | UW Virology Lab | Pavitra Roychoudhury et al |

| | | | | | | |
|---|---|---|---|---|---|---|
| EPI_ISL_437872 | USA/CT-UW-6576/2020 | 2020-04-02 | North America/USA/Connecticut/ | UW Virology Lab | UW Virology Lab | Pavitra Roychoudhury et al |
| EPI_ISL_437877 | Greece/204_33112/2020 | 2020-03-12 | Europe/Greece/Athens/ | Laboratory of Microbiology, Medical School, National and Kapodistrian University of Athens | Laboratory of Biology, Department of Medicine, Democritus University of Thrace | Kassela K. et al |
| EPI_ISL_437888 | Greece/221_33038/2020 | 2020-03-12 | Europe/Greece/Athens/ | Laboratory of Microbiology, Medical School, National and Kapodistrian University of Athens | Laboratory of Biology, Department of Medicine, Democritus University of Thrace | Kassela K. et al |
| EPI_ISL_437900 | Greece/248_32261/2020 | 2020-03-10 | Europe/Greece/Athens/ | Laboratory of Microbiology, Medical School, National and Kapodistrian University of Athens | Laboratory of Biology, Department of Medicine, Democritus University of Thrace | Kassela K. et al |
| EPI_ISL_437918 | Austria/CeMM0029/2020 | 2020-03-09 | Europe/Austria/Tyrol/Ischgl | Institut f√ºr Virologie am Department f√ºr Hygiene, Mikrobiologie und Public Health | Bergthaler laboratory, CeMM Research Center for Molecular Medicine of the Austrian Academy of Sciences | Alexandra Popa et al |
| EPI_ISL_437923 | Austria/CeMM0034/2020 | 2020-03-10 | Europe/Austria/Tyrol/Ischgl | Institut f√ºr Virologie am Department f√ºr Hygiene, Mikrobiologie und Public Health | Bergthaler laboratory, CeMM Research Center for Molecular Medicine of the Austrian Academy of Sciences | Alexandra Popa et al |

| | | | | | | |
|---|---|---|---|---|---|---|
| EPI_ISL_437929 | Austria/CeMM0041/2020 | 2020-03-05 | Europe/Austria/Tyrol/Ischgl | Institut f√ºr Virologie am Department f√ºr Hygiene, Mikrobiologie und Public Health | Bergthaler laboratory, CeMM Research Center for Molecular Medicine of the Austrian Academy of Sciences | Alexandra Popa et al |
| EPI_ISL_437932 | Austria/CeMM0045/2020 | 2020-02-24 | Europe/Austria/Tyrol/Ischgl | Institut f√ºr Virologie am Department f√ºr Hygiene, Mikrobiologie und Public Health | Bergthaler laboratory, CeMM Research Center for Molecular Medicine of the Austrian Academy of Sciences | Alexandra Popa et al |
| EPI_ISL_437940 | Austria/CeMM0068/2020 | 2020-03-12 | Europe/Austria/Tyrol/Innsbruck | Universitaetsklinik f√ºr Innere Medizin II Innsbruck | Bergthaler laboratory, CeMM Research Center for Molecular Medicine of the Austrian Academy of Sciences | Alexandra Popa et al |
| EPI_ISL_437951 | Austria/CeMM0088/2020 | 2020-03-24 | Europe/Austria/Tyrol/Innsbruck | Universitaetsklinik f√ºr Innere Medizin II Innsbruck | Bergthaler laboratory, CeMM Research Center for Molecular Medicine of the Austrian Academy of Sciences | Alexandra Popa et al |
| EPI_ISL_437952 | Austria/CeMM0089/2020 | 2020-03-24 | Europe/Austria/Tyrol/Innsbruck | Universitaetsklinik f√ºr Innere Medizin II Innsbruck | Bergthaler laboratory, CeMM Research Center for Molecular Medicine of the Austrian Academy of Sciences | Alexandra Popa et al |

| | | | | | | |
|---|---|---|---|---|---|---|
| EPI_ISL_437960 | Austria/CeMM0099/2020 | 2020-03-20 | Europe/Austria/Tyrol/Innsbruck | Universitaetsklinik f√ºr Innere Medizin II Innsbruck | Bergthaler laboratory, CeMM Research Center for Molecular Medicine of the Austrian Academy of Sciences | Alexandra Popa et al |
| EPI_ISL_437963 | Austria/CeMM0102/2020 | 2020-03-19 | Europe/Austria/Tyrol/Innsbruck | Universitaetsklinik f√ºr Innere Medizin II Innsbruck | Bergthaler laboratory, CeMM Research Center for Molecular Medicine of the Austrian Academy of Sciences | Alexandra Popa et al |
| EPI_ISL_437966 | Austria/CeMM0106/2020 | 2020-03-18 | Europe/Austria/Tyrol/Laengenfeld | Universitaetsklinik f√ºr Innere Medizin II Innsbruck | Bergthaler laboratory, CeMM Research Center for Molecular Medicine of the Austrian Academy of Sciences | Alexandra Popa et al |
| EPI_ISL_437973 | Austria/CeMM0115/2020 | 2020-03-18 | Europe/Austria/Tyrol/Innsbruck | Universitaetsklinik f√ºr Innere Medizin II Innsbruck | Bergthaler laboratory, CeMM Research Center for Molecular Medicine of the Austrian Academy of Sciences | Alexandra Popa et al |
| EPI_ISL_437991 | Austria/CeMM0139/2020 | 2020-03-13 | Europe/Austria/Tyrol/Ischgl | Institut f√ºr Virologie am Department f√ºr Hygiene, Mikrobiologie und Public Health | Bergthaler laboratory, CeMM Research Center for Molecular Medicine of the Austrian Academy of Sciences | Alexandra Popa et al |

| | | | | | | |
|---|---|---|---|---|---|---|
| EPI_ISL_437993 | Austria/CeMM0143/2020 | 2020-02-27 | Europe/Austria/Vienna/ | Center for Virology, Medical University of Vienna | Bergthaler laboratory, CeMM Research Center for Molecular Medicine of the Austrian Academy of Sciences | Alexandra Popa et al |
| EPI_ISL_437994 | Austria/CeMM0146/2020 | 2020-02-28 | Europe/Austria/Vienna/ | Center for Virology, Medical University of Vienna | Bergthaler laboratory, CeMM Research Center for Molecular Medicine of the Austrian Academy of Sciences | Alexandra Popa et al |
| EPI_ISL_437995 | Austria/CeMM0147/2020 | 2020-02-28 | Europe/Austria/Vienna/ | Center for Virology, Medical University of Vienna | Bergthaler laboratory, CeMM Research Center for Molecular Medicine of the Austrian Academy of Sciences | Alexandra Popa et al |
| EPI_ISL_438000 | Austria/CeMM0154/2020 | 2020-03-03 | Europe/Austria/Vienna/ | Center for Virology, Medical University of Vienna | Bergthaler laboratory, CeMM Research Center for Molecular Medicine of the Austrian Academy of Sciences | Alexandra Popa et al |
| EPI_ISL_438001 | Austria/CeMM0155/2020 | 2020-03-03 | Europe/Italy/Italian cruise ship/ | Center for Virology, Medical University of Vienna | Bergthaler laboratory, CeMM Research Center for Molecular Medicine of the Austrian Academy of Sciences | Alexandra Popa et al |

| | | | | | | |
|---|---|---|---|---|---|---|
| EPI_ISL_438002 | Austria/CeMM0156/2020 | 2020-03-03 | Europe/Italy/Italian cruise ship/ | Center for Virology, Medical University of Vienna | Bergthaler laboratory, CeMM Research Center for Molecular Medicine of the Austrian Academy of Sciences | Alexandra Popa et al |
| EPI_ISL_438004 | Austria/CeMM0158/2020 | 2020-03-04 | Europe/Austria/Vienna/ | Center for Virology, Medical University of Vienna | Bergthaler laboratory, CeMM Research Center for Molecular Medicine of the Austrian Academy of Sciences | Alexandra Popa et al |
| EPI_ISL_438026 | Austria/CeMM0184/2020 | 2020-03-11 | Europe/Austria/Tyrol/ | Center for Virology, Medical University of Vienna | Bergthaler laboratory, CeMM Research Center for Molecular Medicine of the Austrian Academy of Sciences | Alexandra Popa et al |
| EPI_ISL_438028 | Austria/CeMM0186/2020 | 2020-03-11 | Europe/Italy/Italian cruise ship/ | Center for Virology, Medical University of Vienna | Bergthaler laboratory, CeMM Research Center for Molecular Medicine of the Austrian Academy of Sciences | Alexandra Popa et al |
| EPI_ISL_438030 | Austria/CeMM0189/2020 | 2020-03-13 | Europe/Italy/Austria/ | Center for Virology, Medical University of Vienna | Bergthaler laboratory, CeMM Research Center for Molecular Medicine of the Austrian Academy of Sciences | Alexandra Popa et al |

| | | | | | | |
|---|---|---|---|---|---|---|
| EPI_ISL_438041 | Austria/CeMM0200/2020 | 2020-03-15 | Europe/Austria/Vienna/ | Center for Virology, Medical University of Vienna | Bergthaler laboratory, CeMM Research Center for Molecular Medicine of the Austrian Academy of Sciences | Alexandra Popa et al |
| EPI_ISL_438046 | Austria/CeMM0209/2020 | 2020-03-16 | Europe/Austria/Vienna/ | Center for Virology, Medical University of Vienna | Bergthaler laboratory, CeMM Research Center for Molecular Medicine of the Austrian Academy of Sciences | Alexandra Popa et al |
| EPI_ISL_438050 | Austria/CeMM0216/2020 | 2020-03-16 | Europe/Austria/Vienna/ | Center for Virology, Medical University of Vienna | Bergthaler laboratory, CeMM Research Center for Molecular Medicine of the Austrian Academy of Sciences | Alexandra Popa et al |
| EPI_ISL_438052 | Austria/CeMM0218/2020 | 2020-03-16 | Europe/Austria/Vienna/ | Center for Virology, Medical University of Vienna | Bergthaler laboratory, CeMM Research Center for Molecular Medicine of the Austrian Academy of Sciences | Alexandra Popa et al |
| EPI_ISL_438058 | Austria/CeMM0224/2020 | 2020-03-17 | Europe/Austria/Vienna/ | Center for Virology, Medical University of Vienna | Bergthaler laboratory, CeMM Research Center for Molecular Medicine of the Austrian Academy of Sciences | Alexandra Popa et al |

| | | | | | | |
|---|---|---|---|---|---|---|
| EPI_ISL_438060 | Austria/CeMM0226/2020 | 2020-03-17 | Europe/Austria/Vienna/ | Center for Virology, Medical University of Vienna | Bergthaler laboratory, CeMM Research Center for Molecular Medicine of the Austrian Academy of Sciences | Alexandra Popa et al |
| EPI_ISL_438069 | Austria/CeMM0239/2020 | 2020-03-19 | Europe/Austria/Vienna/ | Center for Virology, Medical University of Vienna | Bergthaler laboratory, CeMM Research Center for Molecular Medicine of the Austrian Academy of Sciences | Alexandra Popa et al |
| EPI_ISL_438071 | Austria/CeMM0241/2020 | 2020-03-20 | Europe/Austria/Vienna/ | Center for Virology, Medical University of Vienna | Bergthaler laboratory, CeMM Research Center for Molecular Medicine of the Austrian Academy of Sciences | Alexandra Popa et al |
| EPI_ISL_438072 | Austria/CeMM0242/2020 | 2020-03-20 | Europe/Austria/Vienna/ | Center for Virology, Medical University of Vienna | Bergthaler laboratory, CeMM Research Center for Molecular Medicine of the Austrian Academy of Sciences | Alexandra Popa et al |
| EPI_ISL_438075 | Austria/CeMM0245/2020 | 2020-03-20 | Europe/Austria/Vienna/ | Center for Virology, Medical University of Vienna | Bergthaler laboratory, CeMM Research Center for Molecular Medicine of the Austrian Academy of Sciences | Alexandra Popa et al |

| | | | | | | |
|---|---|---|---|---|---|---|
| EPI_ISL_438078 | Austria/CeMM0248/2020 | 2020-03-21 | Europe/Austria/Vienna/ | Center for Virology, Medical University of Vienna | Bergthaler laboratory, CeMM Research Center for Molecular Medicine of the Austrian Academy of Sciences | Alexandra Popa et al |
| EPI_ISL_438094 | Austria/CeMM0265/2020 | 2020-03-23 | Europe/Austria/Vienna/ | Center for Virology, Medical University of Vienna | Bergthaler laboratory, CeMM Research Center for Molecular Medicine of the Austrian Academy of Sciences | Alexandra Popa et al |
| EPI_ISL_438095 | Austria/CeMM0266/2020 | 2020-03-24 | Europe/Austria/Tyrol/ | Center for Virology, Medical University of Vienna | Bergthaler laboratory, CeMM Research Center for Molecular Medicine of the Austrian Academy of Sciences | Alexandra Popa et al |
| EPI_ISL_438096 | Austria/CeMM0267/2020 | 2020-03-24 | Europe/Austria/Styria/ | Center for Virology, Medical University of Vienna | Bergthaler laboratory, CeMM Research Center for Molecular Medicine of the Austrian Academy of Sciences | Alexandra Popa et al |
| EPI_ISL_438104 | Austria/CeMM0277/2020 | 2020-03-25 | Europe/Austria/Vienna/ | Center for Virology, Medical University of Vienna | Bergthaler laboratory, CeMM Research Center for Molecular Medicine of the Austrian Academy of Sciences | Alexandra Popa et al |

| | | | | | | |
|---|---|---|---|---|---|---|
| EPI_ISL_438105 | Austria/CeMM0278/2020 | 2020-03-25 | Europe/Austria/Vienna/ | Center for Virology, Medical University of Vienna | Bergthaler laboratory, CeMM Research Center for Molecular Medicine of the Austrian Academy of Sciences | Alexandra Popa et al |
| EPI_ISL_438119 | Austria/CeMM0296/2020 | 2020-03-27 | Europe/Austria/Vienna/ | Center for Virology, Medical University of Vienna | Bergthaler laboratory, CeMM Research Center for Molecular Medicine of the Austrian Academy of Sciences | Alexandra Popa et al |
| EPI_ISL_438122 | Austria/CeMM0359/2020 | 2020-04-02 | Europe/Austria/Vienna/ | Center for Virology, Medical University of Vienna | Bergthaler laboratory, CeMM Research Center for Molecular Medicine of the Austrian Academy of Sciences | Alexandra Popa et al |
| EPI_ISL_438141 | USA/WA-S588/2020 | 2020-04-02 | North America/USA/Washington/King C | Seattle Flu Study | Seattle Flu Study | Chu et al et al |
| EPI_ISL_438142 | USA/WA-S589/2020 | 2020-04-04 | North America/USA/Washington/King C | Seattle Flu Study | Seattle Flu Study | Chu et al et al |
| EPI_ISL_438146 | USA/WA-S593/2020 | 2020-04-04 | North America/USA/Washington/King C | Seattle Flu Study | Seattle Flu Study | Chu et al et al |
| EPI_ISL_438147 | USA/WA-S594/2020 | 2020-04-12 | North America/USA/Washington/King C | Seattle Flu Study | Seattle Flu Study | Chu et al et al |
| EPI_ISL_438160 | USA/WA-S607/2020 | 2020-04-20 | North America/USA/Washington/King C | Seattle Flu Study | Seattle Flu Study | Chu et al et al |
| EPI_ISL_438166 | USA/WA-S613/2020 | 2020-04-23 | North America/USA/Washington/ | Seattle Flu Study | Seattle Flu Study | Chu et al et al |
| EPI_ISL_438170 | USA/WA-S617/2020 | 2020-04-25 | North America/USA/Washington/King C | Seattle Flu Study | Seattle Flu Study | Chu et al et al |
| EPI_ISL_438177 | USA/WA-S624/2020 | 2020-04-18 | North America/USA/Washington/Yakim | Washington State Department of Health | Seattle Flu Study | Chu et al et al |
| EPI_ISL_438181 | USA/WA-S628/2020 | 2020-04-18 | North America/USA/Washington/Pierce | Washington State Department of Health | Seattle Flu Study | Chu et al et al |
| EPI_ISL_438190 | USA/WA-S637/2020 | 2020-04-17 | North America/USA/Washington/Snoho | Washington State Department of Health | Seattle Flu Study | Chu et al et al |
| EPI_ISL_438195 | USA/WA-S642/2020 | 2020-04-19 | North America/USA/Washington/Pierce | Washington State Department of Health | Seattle Flu Study | Chu et al et al |
| EPI_ISL_438201 | USA/WA-S648/2020 | 2020-04-19 | North America/USA/Washington/Yakim | Washington State Department of Health | Seattle Flu Study | Chu et al et al |

| | | | | | | |
|---|---|---|---|---|---|---|
| EPI_ISL_438202 | USA/WA-S649/2020 | 2020-04-19 | North America/USA/Washington/Yakim | Washington State Department of Health | Seattle Flu Study | Chu et al et al |
| EPI_ISL_438205 | USA/WA-S652/2020 | 2020-04-20 | North America/USA/Washington/Pierce | Washington State Department of Health | Seattle Flu Study | Chu et al et al |
| EPI_ISL_438208 | USA/WA-S655/2020 | 2020-04-16 | North America/USA/Washington/ | Washington State Department of Health | Seattle Flu Study | Chu et al et al |
| EPI_ISL_438210 | USA/WA-S657/2020 | 2020-04-21 | North America/USA/Washington/Pierce | Washington State Department of Health | Seattle Flu Study | Chu et al et al |
| EPI_ISL_438225 | USA/MD-HP00032/2020 | 2020-03-21 | North America/USA/Maryland/ | Johns Hopkins Hospital Department of Pathology | Johns Hopkins Hospital Department of Pathology | Peter M. Thielen et al |
| EPI_ISL_438230 | USA/DC-HP00072/2020 | 2020-03-11 | North America/USA/Washington DC/ | Johns Hopkins Hospital Department of Pathology | Johns Hopkins Hospital Department of Pathology | Peter M. Thielen et al |
| EPI_ISL_438235 | USA/MD-HP00078/2020 | 2020-03-23 | North America/USA/Maryland/ | Johns Hopkins Hospital Department of Pathology | Johns Hopkins Hospital Department of Pathology | Peter M. Thielen et al |
| EPI_ISL_438238 | USA/MD-HP00082/2020 | 2020-03-08 | North America/USA/Maryland/ | Johns Hopkins Hospital Department of Pathology | Johns Hopkins Hospital Department of Pathology | Peter M. Thielen et al |
| EPI_ISL_438245 | USA/DC-HP00100/2020 | 2020-03-14 | North America/USA/Washington DC/ | Johns Hopkins Hospital Department of Pathology | Johns Hopkins Hospital Department of Pathology | Peter M. Thielen et al |
| EPI_ISL_438256 | England/CAMB-76928/2020 | 2020-03-29 | Europe/United Kingdom/England/ | Department of Pathology, University of Cambridge | Wellcome Sanger Institute for the COVID-19 Genomics UK (COG-UK) consortium | Luke W Meredith et al |
| EPI_ISL_438259 | England/CAMB-7674C/2020 | 2020-03-30 | Europe/United Kingdom/England/ | Department of Pathology, University of Cambridge | Wellcome Sanger Institute for the COVID-19 Genomics UK (COG-UK) consortium | Luke W Meredith et al |
| EPI_ISL_438268 | England/CAMB-760F6/2020 | 2020-03-30 | Europe/United Kingdom/England/ | Department of Pathology, University of Cambridge | Wellcome Sanger Institute for the COVID-19 Genomics UK (COG-UK) consortium | Luke W Meredith et al |

| | | | | | | |
|---|---|---|---|---|---|---|
| EPI_ISL_438285 | England/CAMB-76C3E/2020 | 2020-03-29 | Europe/United Kingdom/England/ | Department of Pathology, University of Cambridge | Wellcome Sanger Institute for the COVID-19 Genomics UK (COG-UK) consortium | Luke W Meredith et al |
| EPI_ISL_438368 | England/CAMB-76CD4/2020 | 2020-03-30 | Europe/United Kingdom/England/ | Department of Pathology, University of Cambridge | Wellcome Sanger Institute for the COVID-19 Genomics UK (COG-UK) consortium | Luke W Meredith et al |
| EPI_ISL_438422 | England/CAMB-76B6E/2020 | 2020-03-29 | Europe/United Kingdom/England/ | Department of Pathology, University of Cambridge | Wellcome Sanger Institute for the COVID-19 Genomics UK (COG-UK) consortium | Luke W Meredith et al |
| EPI_ISL_438476 | England/CAMB-759ED/2020 | 2020-03-31 | Europe/United Kingdom/England/ | Department of Pathology, University of Cambridge | Wellcome Sanger Institute for the COVID-19 Genomics UK (COG-UK) consortium | Luke W Meredith et al |
| EPI_ISL_438595 | England/CAMB-81F28/2020 | 2020-04-23 | Europe/United Kingdom/England/ | Department of Pathology, University of Cambridge | COVID-19 Genomics UK (COG-UK) Consortium | Luke W Meredith et al |
| EPI_ISL_438679 | England/CAMB-82CC6/2020 | 2020-04-22 | Europe/United Kingdom/England/ | Department of Pathology, University of Cambridge | COVID-19 Genomics UK (COG-UK) Consortium | Luke W Meredith et al |
| EPI_ISL_438743 | England/CAMB-8463F/2020 | 2020-04-27 | Europe/United Kingdom/England/ | Department of Pathology, University of Cambridge | COVID-19 Genomics UK (COG-UK) Consortium | Luke W Meredith et al |
| EPI_ISL_438913 | Scotland/CVR247/2020 | 2020-03-19 | Europe/United Kingdom/Scotland/ | West of Scotland Specialist Virology Centre, NHSGGC / MRC-University of Glasgow Centre for Virus Research | COVID-19 Genomics UK (COG-UK) Consortium | Ana da Silva Filipe et al |
| EPI_ISL_439063 | Scotland/CVR355/2020 | 2020-03-21 | Europe/United Kingdom/Scotland/ | West of Scotland Specialist Virology Centre, NHSGGC / MRC-University of Glasgow Centre for Virus Research | COVID-19 Genomics UK (COG-UK) Consortium | Ana da Silva Filipe et al |
| EPI_ISL_439122 | Scotland/CVR861/2020 | 2020-03-27 | Europe/United Kingdom/Scotland/ | West of Scotland Specialist Virology Centre, NHSGGC / MRC-University of Glasgow Centre for Virus Research | COVID-19 Genomics UK (COG-UK) Consortium | Ana da Silva Filipe et al |
| EPI_ISL_439127 | Scotland/CVR878/2020 | 2020-03-27 | Europe/United Kingdom/Scotland/ | West of Scotland Specialist Virology Centre, NHSGGC / MRC-University of Glasgow Centre for Virus Research | COVID-19 Genomics UK (COG-UK) Consortium | Ana da Silva Filipe et al |

| | | | | | | |
|---|---|---|---|---|---|---|
| EPI_ISL_439182 | Scotland/EDB2335/2020 | 2020-04-21 | Europe/United Kingdom/Scotland/ | Virology Department, Royal Infirmary of Edinburgh, NHS Lothian / School of Biological Sciences, University of Edinburgh / Institute of Genetics and Molecular Medicine, University of Edinburgh | COVID-19 Genomics UK (COG-UK) Consortium | McHugh M et al |
| EPI_ISL_439276 | Scotland/EDB3503/2020 | 2020-04-13 | Europe/United Kingdom/Scotland/ | Virology Department, Royal Infirmary of Edinburgh, NHS Lothian / School of Biological Sciences, University of Edinburgh / Institute of Genetics and Molecular Medicine, University of Edinburgh | COVID-19 Genomics UK (COG-UK) Consortium | McHugh M et al |
| EPI_ISL_439304 | Scotland/EDB3652/2020 | 2020-04-27 | Europe/United Kingdom/Scotland/ | Virology Department, Royal Infirmary of Edinburgh, NHS Lothian / School of Biological Sciences, University of Edinburgh / Institute of Genetics and Molecular Medicine, University of Edinburgh | COVID-19 Genomics UK (COG-UK) Consortium | McHugh M et al |
| EPI_ISL_439365 | Scotland/EDB3914/2020 | 2020-05-01 | Europe/United Kingdom/Scotland/ | Virology Department, Royal Infirmary of Edinburgh, NHS Lothian / School of Biological Sciences, University of Edinburgh / Institute of Genetics and Molecular Medicine, University of Edinburgh | COVID-19 Genomics UK (COG-UK) Consortium | McHugh M et al |
| EPI_ISL_439377 | England/CAMB-77268/2020 | 2020-04-01 | Europe/United Kingdom/England/ | Department of Pathology, University of Cambridge | Wellcome Sanger Institute for the COVID-19 Genomics UK (COG-UK) consortium | Luke W Meredith et al |
| EPI_ISL_439459 | England/CAMB-7717A/2020 | 2020-04-01 | Europe/United Kingdom/England/ | Department of Pathology, University of Cambridge | Wellcome Sanger Institute for the COVID-19 Genomics UK (COG-UK) consortium | Luke W Meredith et al |
| EPI_ISL_439539 | England/CAMB-71E5B/2020 | 2020-03-28 | Europe/United Kingdom/England/ | Department of Pathology, University of Cambridge | Wellcome Sanger Institute for the COVID-19 Genomics UK (COG-UK) consortium | Luke W Meredith et al |
| EPI_ISL_439601 | England/CAMB-71BFA/2020 | 2020-03-31 | Europe/United Kingdom/England/ | Department of Pathology, University of Cambridge | Wellcome Sanger Institute for the COVID-19 Genomics UK (COG-UK) consortium | Luke W Meredith et al |

| | | | | | | |
|---|---|---|---|---|---|---|
| EPI_ISL_439619 | England/CAMB-71B18/2020 | 2020-03-31 | Europe/United Kingdom/England/ | Department of Pathology, University of Cambridge | Wellcome Sanger Institute for the COVID-19 Genomics UK (COG-UK) consortium | Luke W Meredith et al |
| EPI_ISL_439623 | England/CAMB-71CF7/2020 | 2020-03-30 | Europe/United Kingdom/England/ | Department of Pathology, University of Cambridge | Wellcome Sanger Institute for the COVID-19 Genomics UK (COG-UK) consortium | Luke W Meredith et al |
| EPI_ISL_439702 | England/LIVE-99251/2020 | 2020-04-07 | Europe/United Kingdom/England/ | Liverpool Clinical Laboratories | COVID-19 Genomics UK (COG-UK) Consortium | Sam Haldenby et al |
| EPI_ISL_439704 | England/LIVE-9927F/2020 | 2020-04-07 | Europe/United Kingdom/England/ | Liverpool Clinical Laboratories | COVID-19 Genomics UK (COG-UK) Consortium | Sam Haldenby et al |
| EPI_ISL_439901 | England/CAMB-71C33/2020 | 2020-03-31 | Europe/United Kingdom/England/ | Department of Pathology, University of Cambridge | Wellcome Sanger Institute for the COVID-19 Genomics UK (COG-UK) consortium | Luke W Meredith et al |
| EPI_ISL_439954 | England/CAMB-72F0C/2020 | 2020-03-23 | Europe/United Kingdom/England/ | Department of Pathology, University of Cambridge | Wellcome Sanger Institute for the COVID-19 Genomics UK (COG-UK) consortium | Luke W Meredith et al |
| EPI_ISL_440027 | England/BRIS-121F56/2020 | 2020-03-19 | Europe/United Kingdom/England/ | PHE South West Regional Laboratory, National Infection Service | Wellcome Sanger Institute for the COVID-19 Genomics UK (COG-UK) consortium | Stephanie Hutchings et al |
| EPI_ISL_440071 | England/BRIS-121E59/2020 | 2020-03-19 | Europe/United Kingdom/England/ | PHE South West Regional Laboratory, National Infection Service | Wellcome Sanger Institute for the COVID-19 Genomics UK (COG-UK) consortium | Stephanie Hutchings et al |
| EPI_ISL_440246 | England/CAMB-730F9/2020 | 2020-03-21 | Europe/United Kingdom/England/ | Department of Pathology, University of Cambridge | Wellcome Sanger Institute for the COVID-19 Genomics UK (COG-UK) consortium | Luke W Meredith et al |

| | | | | | | |
|---|---|---|---|---|---|---|
| EPI_ISL_440346 | England/CAMB-73F74/2020 | 2020-03-20 | Europe/United Kingdom/England/ | Department of Pathology, University of Cambridge | Wellcome Sanger Institute for the COVID-19 Genomics UK (COG-UK) consortium | Luke W Meredith et al |
| EPI_ISL_440386 | England/CAMB-7508E/2020 | 2020-03-16 | Europe/United Kingdom/England/ | Department of Pathology, University of Cambridge | Wellcome Sanger Institute for the COVID-19 Genomics UK (COG-UK) consortium | Luke W Meredith et al |
| EPI_ISL_440441 | England/CAMB-756D7/2020 | 2020-03-15 | Europe/United Kingdom/England/ | Department of Pathology, University of Cambridge | Wellcome Sanger Institute for the COVID-19 Genomics UK (COG-UK) consortium | Luke W Meredith et al |
| EPI_ISL_440442 | England/CAMB-75798/2020 | 2020-03-14 | Europe/United Kingdom/England/ | Department of Pathology, University of Cambridge | Wellcome Sanger Institute for the COVID-19 Genomics UK (COG-UK) consortium | Luke W Meredith et al |
| EPI_ISL_440456 | England/CAMB-84FCB/2020 | 2020-03-12 | Europe/United Kingdom/England/ | Department of Pathology, University of Cambridge | Wellcome Sanger Institute for the COVID-19 Genomics UK (COG-UK) consortium | Luke W Meredith et al |
| EPI_ISL_440483 | England/CAMB-84769/2020 | 2020-02-28 | Europe/United Kingdom/England/ | Department of Pathology, University of Cambridge | Wellcome Sanger Institute for the COVID-19 Genomics UK (COG-UK) consortium | Luke W Meredith et al |
| EPI_ISL_440486 | England/CAMB-8475A/2020 | 2020-02-27 | Europe/United Kingdom/England/ | Department of Pathology, University of Cambridge | Wellcome Sanger Institute for the COVID-19 Genomics UK (COG-UK) consortium | Luke W Meredith et al |
| EPI_ISL_440489 | England/CAMB-84BA9/2020 | 2020-03-11 | Europe/United Kingdom/England/ | Department of Pathology, University of Cambridge | Wellcome Sanger Institute for the COVID-19 Genomics UK (COG-UK) consortium | Luke W Meredith et al |

| | | | | | | |
|---|---|---|---|---|---|---|
| EPI_ISL_440499 | England/CAMB-8473C/2020 | 2020-02-26 | Europe/United Kingdom/England/ | Department of Pathology, University of Cambridge | Wellcome Sanger Institute for the COVID-19 Genomics UK (COG-UK) consortium | Luke W Meredith et al |
| EPI_ISL_440514 | England/CAMB-84778/2020 | 2020-02-28 | Europe/United Kingdom/England/ | Department of Pathology, University of Cambridge | Wellcome Sanger Institute for the COVID-19 Genomics UK (COG-UK) consortium | Luke W Meredith et al |
| EPI_ISL_440585 | England/CAMB-7906C/2020 | 2020-04-03 | Europe/United Kingdom/England/ | Department of Pathology, University of Cambridge | Wellcome Sanger Institute for the COVID-19 Genomics UK (COG-UK) consortium | Luke W Meredith et al |
| EPI_ISL_440597 | England/CAMB-79E44/2020 | 2020-04-03 | Europe/United Kingdom/England/ | Department of Pathology, University of Cambridge | Wellcome Sanger Institute for the COVID-19 Genomics UK (COG-UK) consortium | Luke W Meredith et al |
| EPI_ISL_440700 | England/BRIS-122FEC/2020 | 2020-04-01 | Europe/United Kingdom/England/ | PHE South West Regional Laboratory, National Infection Service | Wellcome Sanger Institute for the COVID-19 Genomics UK (COG-UK) consortium | Stephanie Hutchings et al |
| EPI_ISL_440788 | England/BRIS-122605/2020 | 2020-04-04 | Europe/United Kingdom/England/ | PHE South West Regional Laboratory, National Infection Service | Wellcome Sanger Institute for the COVID-19 Genomics UK (COG-UK) consortium | Stephanie Hutchings et al |
| EPI_ISL_440833 | England/CAMB-77BD6/2020 | 2020-04-03 | Europe/United Kingdom/England/ | Department of Pathology, University of Cambridge | Wellcome Sanger Institute for the COVID-19 Genomics UK (COG-UK) consortium | Luke W Meredith et al |
| EPI_ISL_440835 | England/CAMB-77E37/2020 | 2020-04-03 | Europe/United Kingdom/England/ | Department of Pathology, University of Cambridge | Wellcome Sanger Institute for the COVID-19 Genomics UK (COG-UK) consortium | Luke W Meredith et al |

| | | | | | | |
|---|---|---|---|---|---|---|
| EPI_ISL_440838 | England/CAMB-77C00/2020 | 2020-04-01 | Europe/United Kingdom/England/ | Department of Pathology, University of Cambridge | Wellcome Sanger Institute for the COVID-19 Genomics UK (COG-UK) consortium | Luke W Meredith et al |
| EPI_ISL_440877 | England/LIVE-9B7AB/2020 | 2020-04-22 | Europe/United Kingdom/England/ | Liverpool Clinical Laboratories | COVID-19 Genomics UK (COG-UK) Consortium | Sam Haldenby et al |
| EPI_ISL_441034 | England/LOND-D3ACC/2020 | 2020-04-06 | Europe/United Kingdom/England/ | University College London, Great Ormond Street Hospital for Children NHS Foundation Trust, Imperial College Healthcare NHS Trust | COVID-19 Genomics UK (COG-UK) Consortium | Sergi Castellano et al |
| EPI_ISL_441173 | England/CAMB-798A0/2020 | 2020-04-01 | Europe/United Kingdom/England/ | Department of Pathology, University of Cambridge | Wellcome Sanger Institute for the COVID-19 Genomics UK (COG-UK) consortium | Luke W Meredith et al |
| EPI_ISL_441185 | England/CAMB-77EA0/2020 | 2020-04-01 | Europe/United Kingdom/England/ | Department of Pathology, University of Cambridge | Wellcome Sanger Institute for the COVID-19 Genomics UK (COG-UK) consortium | Luke W Meredith et al |
| EPI_ISL_441201 | England/CAMB-795D6/2020 | 2020-04-03 | Europe/United Kingdom/England/ | Department of Pathology, University of Cambridge | Wellcome Sanger Institute for the COVID-19 Genomics UK (COG-UK) consortium | Luke W Meredith et al |
| EPI_ISL_441244 | England/CAMB-7A12C/2020 | 2020-04-03 | Europe/United Kingdom/England/ | Department of Pathology, University of Cambridge | Wellcome Sanger Institute for the COVID-19 Genomics UK (COG-UK) consortium | Luke W Meredith et al |
| EPI_ISL_441340 | England/CAMB-79F7E/2020 | 2020-04-02 | Europe/United Kingdom/England/ | Department of Pathology, University of Cambridge | Wellcome Sanger Institute for the COVID-19 Genomics UK (COG-UK) consortium | Luke W Meredith et al |

| | | | | | | |
|---|---|---|---|---|---|---|
| EPI_ISL_441343 | England/CAMB-7A344/2020 | 2020-04-02 | Europe/United Kingdom/England/ | Department of Pathology, University of Cambridge | Wellcome Sanger Institute for the COVID-19 Genomics UK (COG-UK) consortium | Luke W Meredith et al |
| EPI_ISL_441427 | NorthernIreland/NIRE-FB475/2020 | 2020-03-24 | Europe/United Kingdom/Northern Irelar | Regional Virus Laboratory, Belfast Health and Social Care Trust | COVID-19 Genomics UK (COG-UK) Consortium | Conall McCaughey et al |
| EPI_ISL_441433 | NorthernIreland/NIRE-FB4DF/2020 | 2020-03-18 | Europe/United Kingdom/Northern Irelar | Regional Virus Laboratory, Belfast Health and Social Care Trust | COVID-19 Genomics UK (COG-UK) Consortium | Conall McCaughey et al |
| EPI_ISL_441569 | England/CAMB-7B97E/2020 | 2020-04-05 | Europe/United Kingdom/England/ | Department of Pathology, University of Cambridge | Wellcome Sanger Institute for the COVID-19 Genomics UK (COG-UK) consortium | Luke W Meredith et al |
| EPI_ISL_441590 | England/CAMB-7D2EA/2020 | 2020-04-06 | Europe/United Kingdom/England/ | Department of Pathology, University of Cambridge | Wellcome Sanger Institute for the COVID-19 Genomics UK (COG-UK) consortium | Luke W Meredith et al |
| EPI_ISL_441601 | England/CAMB-7D402/2020 | 2020-04-06 | Europe/United Kingdom/England/ | Department of Pathology, University of Cambridge | Wellcome Sanger Institute for the COVID-19 Genomics UK (COG-UK) consortium | Luke W Meredith et al |
| EPI_ISL_441607 | England/CAMB-7B941/2020 | 2020-04-04 | Europe/United Kingdom/England/ | Department of Pathology, University of Cambridge | Wellcome Sanger Institute for the COVID-19 Genomics UK (COG-UK) consortium | Luke W Meredith et al |
| EPI_ISL_441936 | England/SHEF-C2E4C/2020 | 2020-04-08 | Europe/United Kingdom/England/ | Virology Department, Sheffield Teaching Hospitals NHS Foundation Trust/Department of Infection, Immunity and Cardiovascular Disease, The Medical School, University of Sheffield | COVID-19 Genomics UK (COG-UK) Consortium | Thushan de Silva et al |
| EPI_ISL_441959 | England/SHEF-C4132/2020 | 2020-03-25 | Europe/United Kingdom/England/ | Virology Department, Sheffield Teaching Hospitals NHS Foundation Trust/Department of Infection, Immunity and Cardiovascular Disease, The Medical School, University of Sheffield | COVID-19 Genomics UK (COG-UK) Consortium | Thushan de Silva et al |

| | | | | | | |
|---|---|---|---|---|---|---|
| EPI_ISL_442012 | England/SHEF-C78EF/2020 | 2020-03-24 | Europe/United Kingdom/England/ | Virology Department, Sheffield Teaching Hospitals NHS Foundation Trust/Department of Infection, Immunity and Cardiovascular Disease, The Medical School, University of Sheffield | COVID-19 Genomics UK (COG-UK) Consortium | Thushan de Silva et al |
| EPI_ISL_442029 | England/SHEF-C84CC/2020 | 2020-03-25 | Europe/United Kingdom/England/ | Virology Department, Sheffield Teaching Hospitals NHS Foundation Trust/Department of Infection, Immunity and Cardiovascular Disease, The Medical School, University of Sheffield | COVID-19 Genomics UK (COG-UK) Consortium | Thushan de Silva et al |
| EPI_ISL_442044 | Iran/KHGRC-2-2162/2020 | 2020-03-26 | Asia/Iran/Tehran/ | Kawsar Human Genetic Research Center | Kawsar Human Genetic Research Center | Mohammad Ali Khosravi et al |
| EPI_ISL_442074 | England/CAMB-7C9E6/2020 | 2020-04-06 | Europe/United Kingdom/England/ | Department of Pathology, University of Cambridge | Wellcome Sanger Institute for the COVID-19 Genomics UK (COG-UK) consortium | Luke W Meredith et al |
| EPI_ISL_442143 | England/CAMB-7CBFF/2020 | 2020-04-05 | Europe/United Kingdom/England/ | Department of Pathology, University of Cambridge | Wellcome Sanger Institute for the COVID-19 Genomics UK (COG-UK) consortium | Luke W Meredith et al |
| EPI_ISL_442153 | England/CAMB-7C8BC/2020 | 2020-04-06 | Europe/United Kingdom/England/ | Department of Pathology, University of Cambridge | Wellcome Sanger Institute for the COVID-19 Genomics UK (COG-UK) consortium | Luke W Meredith et al |
| EPI_ISL_442158 | England/CAMB-7E085/2020 | 2020-04-06 | Europe/United Kingdom/England/ | Department of Pathology, University of Cambridge | Wellcome Sanger Institute for the COVID-19 Genomics UK (COG-UK) consortium | Luke W Meredith et al |
| EPI_ISL_442173 | England/CAMB-7C764/2020 | 2020-04-05 | Europe/United Kingdom/England/ | Department of Pathology, University of Cambridge | Wellcome Sanger Institute for the COVID-19 Genomics UK (COG-UK) consortium | Luke W Meredith et al |

| | | | | | | |
|---|---|---|---|---|---|---|
| EPI_ISL_442191 | England/CAMB-7BD72/2020 | 2020-04-04 | Europe/United Kingdom/England/ | Department of Pathology, University of Cambridge | Wellcome Sanger Institute for the COVID-19 Genomics UK (COG-UK) consortium | Luke W Meredith et al |
| EPI_ISL_442244 | England/CAMB-7C54C/2020 | 2020-04-05 | Europe/United Kingdom/England/ | Department of Pathology, University of Cambridge | Wellcome Sanger Institute for the COVID-19 Genomics UK (COG-UK) consortium | Luke W Meredith et al |
| EPI_ISL_442523 | Iran/KHGRC-1.1-IPI-8206/2020 | 2020-03-09 | Asia/Iran/Semnan/ | Pasteur Institute of Iran | Kawsar Human Genetic Research Company | Sirous Zeinali et al |
| EPI_ISL_442548 | England/CAMB-8174E/2020 | 2020-04-12 | Europe/United Kingdom/England/ | Department of Pathology, University of Cambridge | Wellcome Sanger Institute for the COVID-19 Genomics UK (COG-UK) consortium | Luke W Meredith et al |
| EPI_ISL_442565 | England/CAMB-81CA9/2020 | 2020-04-12 | Europe/United Kingdom/England/ | Department of Pathology, University of Cambridge | Wellcome Sanger Institute for the COVID-19 Genomics UK (COG-UK) consortium | Luke W Meredith et al |
| EPI_ISL_442605 | England/CAMB-81B7F/2020 | 2020-04-11 | Europe/United Kingdom/England/ | Department of Pathology, University of Cambridge | Wellcome Sanger Institute for the COVID-19 Genomics UK (COG-UK) consortium | Luke W Meredith et al |
| EPI_ISL_442624 | England/BRIS-1238FF/2020 | 2020-04-06 | Europe/United Kingdom/England/ | PHE South West Regional Laboratory, National Infection Service | Wellcome Sanger Institute for the COVID-19 Genomics UK (COG-UK) consortium | Stephanie Hutchings et al |
| EPI_ISL_442627 | England/BRIS-123789/2020 | 2020-04-03 | Europe/United Kingdom/England/ | PHE South West Regional Laboratory, National Infection Service | Wellcome Sanger Institute for the COVID-19 Genomics UK (COG-UK) consortium | Stephanie Hutchings et al |

| | | | | | | |
|---|---|---|---|---|---|---|
| EPI_ISL_442633 | England/BRIS-123ADB/2020 | 2020-04-07 | Europe/United Kingdom/England/ | PHE South West Regional Laboratory, National Infection Service | Wellcome Sanger Institute for the COVID-19 Genomics UK (COG-UK) consortium | Stephanie Hutchings et al |
| EPI_ISL_442740 | England/BRIS-1235CB/2020 | 2020-04-05 | Europe/United Kingdom/England/ | PHE South West Regional Laboratory, National Infection Service | Wellcome Sanger Institute for the COVID-19 Genomics UK (COG-UK) consortium | Stephanie Hutchings et al |
| EPI_ISL_442759 | England/BRIS-1234DD/2020 | 2020-04-06 | Europe/United Kingdom/England/ | PHE South West Regional Laboratory, National Infection Service | Wellcome Sanger Institute for the COVID-19 Genomics UK (COG-UK) consortium | Stephanie Hutchings et al |
| EPI_ISL_442770 | England/BRIS-123200/2020 | 2020-04-05 | Europe/United Kingdom/England/ | PHE South West Regional Laboratory, National Infection Service | Wellcome Sanger Institute for the COVID-19 Genomics UK (COG-UK) consortium | Stephanie Hutchings et al |
| EPI_ISL_442841 | England/CAMB-7F260/2020 | 2020-04-08 | Europe/United Kingdom/England/ | Department of Pathology, University of Cambridge | Wellcome Sanger Institute for the COVID-19 Genomics UK (COG-UK) consortium | Luke W Meredith et al |
| EPI_ISL_443038 | England/CAMB-8124D/2020 | 2020-04-10 | Europe/United Kingdom/England/ | Department of Pathology, University of Cambridge | Wellcome Sanger Institute for the COVID-19 Genomics UK (COG-UK) consortium | Luke W Meredith et al |
| EPI_ISL_443213 | Singapore/121/2020 | 2020-04-08 | Asia/Singapore/Singapore/ | National Public Health Laboratory, National Centre for Infectious Diseases | National Public Health Laboratory, National Centre for Infectious Diseases | Mak Tze Minn et al |

| | | | | | | |
|---|---|---|---|---|---|---|
| EPI_ISL_443261 | France/BFC-5011/2020 | 2020-04-03 | Europe/France/Bourgogne-France-Comt | CHU de Dijon - Laboratoire de Virologie | National Reference Center for Viruses of Respiratory Infections, Institut Pasteur, Paris | M√©lanie Albert et al |
| EPI_ISL_443263 | France/BFC-5017/2020 | 2020-04-02 | Europe/France/Bourgogne-France-Comt | CHU de Dijon - Laboratoire de Virologie | National Reference Center for Viruses of Respiratory Infections, Institut Pasteur, Paris | M√©lanie Albert et al |
| EPI_ISL_443265 | France/BRE-5310/2020 | 2020-04-03 | Europe/France/Bretagne/Brest | CHU - H√¥pital Cavale Blanche - Labo. de Virologie | National Reference Center for Viruses of Respiratory Infections, Institut Pasteur, Paris | M√©lanie Albert et al |
| EPI_ISL_443267 | France/BRE-5325/2020 | 2020-03-18 | Europe/France/Bretagne/Brest | CHU - H√¥pital Cavale Blanche - Labo. de Virologie | National Reference Center for Viruses of Respiratory Infections, Institut Pasteur, Paris | M√©lanie Albert et al |
| EPI_ISL_443272 | France/BRE-5359/2020 | 2020-03-24 | Europe/France/Bretagne/Brest | CHU - H√¥pital Cavale Blanche - Labo. de Virologie | National Reference Center for Viruses of Respiratory Infections, Institut Pasteur, Paris | M√©lanie Albert et al |
| EPI_ISL_443273 | France/BRE-5363/2020 | 2020-03-24 | Europe/France/Bretagne/Brest | CHU - H√¥pital Cavale Blanche - Labo. de Virologie | National Reference Center for Viruses of Respiratory Infections, Institut Pasteur, Paris | M√©lanie Albert et al |

| | | | | | | |
|---|---|---|---|---|---|---|
| EPI_ISL_443275 | France/BRE-5397/2020 | 2020-03-27 | Europe/France/Bretagne/Brest | CHU - H√¥pital Cavale Blanche - Labo. de Virologie | National Reference Center for Viruses of Respiratory Infections, Institut Pasteur, Paris | M√©lanie Albert et al |
| EPI_ISL_443277 | France/BRE-5413/2020 | 2020-03-28 | Europe/France/Bretagne/Brest | CHU - H√¥pital Cavale Blanche - Labo. de Virologie | National Reference Center for Viruses of Respiratory Infections, Institut Pasteur, Paris | M√©lanie Albert et al |
| EPI_ISL_443283 | France/BRE-5465/2020 | 2020-03-23 | Europe/France/Bretagne/Brest | CHU - H√¥pital Cavale Blanche - Labo. de Virologie | National Reference Center for Viruses of Respiratory Infections, Institut Pasteur, Paris | M√©lanie Albert et al |
| EPI_ISL_443287 | France/HDF-5662/2020 | 2020-04-03 | Europe/France/Ile de France/Pontoise | Laboratoire de Microbiologie - B√¢t A - CH Ren√© Dubois | National Reference Center for Viruses of Respiratory Infections, Institut Pasteur, Paris | M√©lanie Albert et al |
| EPI_ISL_443293 | France/BRE-5688/2020 | 2020-04-03 | Europe/France/Bretagne/Rennes | CHRU Pontchaillou - Laboratoire de Virologie | National Reference Center for Viruses of Respiratory Infections, Institut Pasteur, Paris | M√©lanie Albert et al |
| EPI_ISL_443295 | France/IDF-5847/2020 | 2020-03-19 | Europe/France/Ile de France/Paris | H√¥pital Necker - Enfants - Malades Laboratoire de Virologie | National Reference Center for Viruses of Respiratory Infections, Institut Pasteur, Paris | M√©lanie Albert et al |

| | | | | | | |
|---|---|---|---|---|---|---|
| EPI_ISL_443299 | France/IDF-5853/2020 | 2020-03-25 | Europe/France/Ile de France/Paris | H√¥pital Necker - Enfants - Malades Laboratoire de Virologie | National Reference Center for Viruses of Respiratory Infections, Institut Pasteur, Paris | M√©lanie Albert et al |
| EPI_ISL_443302 | France/IDF-3368/2020 | 2020-03-23 | Europe/France/Ile de France/Cergy | Cabinet M√©dical | National Reference Center for Viruses of Respiratory Infections, Institut Pasteur, Paris | M√©lanie Albert et al |
| EPI_ISL_443306 | France/IDF-6037/2020 | 2020-04-02 | Europe/France/Ile de France/Moussy Le | Cabinet M√©dical | National Reference Center for Viruses of Respiratory Infections, Institut Pasteur, Paris | M√©lanie Albert et al |
| EPI_ISL_443307 | France/BFC-6053/2020 | 2020-04-09 | Europe/France/Bourgogne-France-Comt | La Villa Papyri | National Reference Center for Viruses of Respiratory Infections, Institut Pasteur, Paris | M√©lanie Albert et al |
| EPI_ISL_443309 | France/HDF-3669/2020 | 2020-03-25 | Europe/France/Hauts de France/Compiv | CH Compi√®gne Laboratoire de Biologie | National Reference Center for Viruses of Respiratory Infections, Institut Pasteur, Paris | M√©lanie Albert et al |
| EPI_ISL_443315 | France/HDF-6466/2020 | 2020-04-02 | Europe/France/Hauts de France/Nogent | Ch√¢teau de la Source | National Reference Center for Viruses of Respiratory Infections, Institut Pasteur, Paris | M√©lanie Albert et al |

| | | | | | | |
|---|---|---|---|---|---|---|
| EPI_ISL_443317 | France/IDF-4432/2020 | 2020-03-23 | Europe/France/Ile de France/Cergy | Cabinet M√©dical | National Reference Center for Viruses of Respiratory Infections, Institut Pasteur, Paris | M√©lanie Albert et al |
| EPI_ISL_443376 | England/CAMB-83C3E/2020 | 2020-04-16 | Europe/United Kingdom/England/ | Department of Pathology, University of Cambridge | Wellcome Sanger Institute for the COVID-19 Genomics UK (COG-UK) consortium | Luke W Meredith et al |
| EPI_ISL_443378 | England/CAMB-83991/2020 | 2020-04-16 | Europe/United Kingdom/England/ | Department of Pathology, University of Cambridge | Wellcome Sanger Institute for the COVID-19 Genomics UK (COG-UK) consortium | Luke W Meredith et al |
| EPI_ISL_443408 | England/CAMB-1AA2D4/2020 | 2020-04-16 | Europe/United Kingdom/England/ | Department of Pathology, University of Cambridge | Wellcome Sanger Institute for the COVID-19 Genomics UK (COG-UK) consortium | Luke W Meredith et al |
| EPI_ISL_443413 | England/CAMB-1AA438/2020 | 2020-04-17 | Europe/United Kingdom/England/ | Department of Pathology, University of Cambridge | Wellcome Sanger Institute for the COVID-19 Genomics UK (COG-UK) consortium | Luke W Meredith et al |
| EPI_ISL_443425 | England/CAMB-82F72/2020 | 2020-04-12 | Europe/United Kingdom/England/ | Department of Pathology, University of Cambridge | Wellcome Sanger Institute for the COVID-19 Genomics UK (COG-UK) consortium | Luke W Meredith et al |
| EPI_ISL_443435 | England/CAMB-836D6/2020 | 2020-04-15 | Europe/United Kingdom/England/ | Department of Pathology, University of Cambridge | Wellcome Sanger Institute for the COVID-19 Genomics UK (COG-UK) consortium | Luke W Meredith et al |
| EPI_ISL_443447 | England/CAMB-834DC/2020 | 2020-04-15 | Europe/United Kingdom/England/ | Department of Pathology, University of Cambridge | Wellcome Sanger Institute for the COVID-19 Genomics UK (COG-UK) consortium | Luke W Meredith et al |

| | | | | | | |
|---|---|---|---|---|---|---|
| EPI_ISL_443465 | England/CAMB-823EF/2020 | 2020-04-12 | Europe/United Kingdom/England/ | Department of Pathology, University of Cambridge | Wellcome Sanger Institute for the COVID-19 Genomics UK (COG-UK) consortium | Luke W Meredith et al |
| EPI_ISL_443504 | England/CAMB-8267D/2020 | 2020-04-12 | Europe/United Kingdom/England/ | Department of Pathology, University of Cambridge | Wellcome Sanger Institute for the COVID-19 Genomics UK (COG-UK) consortium | Luke W Meredith et al |
| EPI_ISL_443534 | England/CAMB-844CC/2020 | 2020-04-16 | Europe/United Kingdom/England/ | Department of Pathology, University of Cambridge | Wellcome Sanger Institute for the COVID-19 Genomics UK (COG-UK) consortium | Luke W Meredith et al |
| EPI_ISL_443564 | England/CAMB-8424A/2020 | 2020-04-16 | Europe/United Kingdom/England/ | Department of Pathology, University of Cambridge | Wellcome Sanger Institute for the COVID-19 Genomics UK (COG-UK) consortium | Luke W Meredith et al |
| EPI_ISL_443677 | England/CAMB-82E1B/2020 | 2020-04-14 | Europe/United Kingdom/England/ | Department of Pathology, University of Cambridge | Wellcome Sanger Institute for the COVID-19 Genomics UK (COG-UK) consortium | Luke W Meredith et al |
| EPI_ISL_443815 | England/BRIS-124ADA/2020 | 2020-03-26 | Europe/United Kingdom/England/ | PHE South West Regional Laboratory, National Infection Service | Wellcome Sanger Institute for the COVID-19 Genomics UK (COG-UK) consortium | Stephanie Hutchings et al |
| EPI_ISL_443826 | England/BRIS-125BA9/2020 | 2020-03-30 | Europe/United Kingdom/England/ | PHE South West Regional Laboratory, National Infection Service | Wellcome Sanger Institute for the COVID-19 Genomics UK (COG-UK) consortium | Stephanie Hutchings et al |
| EPI_ISL_443831 | England/BRIS-125ECE/2020 | 2020-03-30 | Europe/United Kingdom/England/ | PHE South West Regional Laboratory, National Infection Service | Wellcome Sanger Institute for the COVID-19 Genomics UK (COG-UK) consortium | Stephanie Hutchings et al |

| | | | | | | |
|---|---|---|---|---|---|---|
| EPI_ISL_443832 | England/BRIS-124F44/2020 | 2020-03-26 | Europe/United Kingdom/England/ | PHE South West Regional Laboratory, National Infection Service | Wellcome Sanger Institute for the COVID-19 Genomics UK (COG-UK) consortium | Stephanie Hutchings et al |
| EPI_ISL_443918 | England/BRIS-124982/2020 | 2020-03-25 | Europe/United Kingdom/England/ | PHE South West Regional Laboratory, National Infection Service | Wellcome Sanger Institute for the COVID-19 Genomics UK (COG-UK) consortium | Stephanie Hutchings et al |
| EPI_ISL_443995 | England/BRIS-1249FB/2020 | 2020-03-25 | Europe/United Kingdom/England/ | PHE South West Regional Laboratory, National Infection Service | Wellcome Sanger Institute for the COVID-19 Genomics UK (COG-UK) consortium | Stephanie Hutchings et al |
| EPI_ISL_444000 | England/BRIS-124FAE/2020 | 2020-03-27 | Europe/United Kingdom/England/ | PHE South West Regional Laboratory, National Infection Service | Wellcome Sanger Institute for the COVID-19 Genomics UK (COG-UK) consortium | Stephanie Hutchings et al |
| EPI_ISL_444011 | England/BRIS-1256B7/2020 | 2020-03-28 | Europe/United Kingdom/England/ | PHE South West Regional Laboratory, National Infection Service | Wellcome Sanger Institute for the COVID-19 Genomics UK (COG-UK) consortium | Stephanie Hutchings et al |
| EPI_ISL_444058 | USA/CA-CZB-1100/2020 | 2020-04-28 | North America/USA/California/San Fran | UCSF Clinical Microbiology Laboratory | Chan-Zuckerberg Biohub | CZB Cliahub Consortium et al |
| EPI_ISL_444074 | USA/CA-CZB-1116/2020 | 2020-04-30 | North America/USA/California/San Fran | UCSF Clinical Microbiology Laboratory | Chan-Zuckerberg Biohub | CZB Cliahub Consortium et al |
| EPI_ISL_444166 | England/LOND-D49EC/2020 | 2020-04-10 | Europe/United Kingdom/England/ | University College London, Great Ormond Street Hospital for Children NHS Foundation Trust, Imperial College Healthcare NHS Trust | COVID-19 Genomics UK (COG-UK) Consortium | Sergi Castellano et al |
| EPI_ISL_444261 | England/LOND-D5F07/2020 | 2020-04-18 | Europe/United Kingdom/England/ | University College London, Great Ormond Street Hospital for Children NHS Foundation Trust, Imperial College Healthcare NHS Trust | COVID-19 Genomics UK (COG-UK) Consortium | Sergi Castellano et al |

| | | | | | | |
|---|---|---|---|---|---|---|
| EPI_ISL_444273 | Guangzhou/IQTC05/2020 | 2020-02-05 | Asia/China/Guangdong/Guangzhou | State Key Laboratory of Respiratory Disease, National Clinical Research Center for Respiratory Disease, Guangzhou Institute of Respiratory Health, the First Affiliated Hospital of Guangzhou Medical University | State Key Laboratory of Respiratory Disease, National Clinical Research Center for Respiratory Disease, Guangzhou Institute of Respiratory Health, the First Affiliated Hospital of Guangzhou Medical University | Sun et al |
| EPI_ISL_444274 | Taiwan/CGMH-CGU-21/2020 | 2020-02-29 | Asia/Taiwan/Taiwan/Taoyuan | Laboratory Medicine | Department of Laboratory Medicine, Lin-Kou Chang Gung Memorial Hospital, Taoyuan, Taiwan | Kuo-Chien Tsao et al |
| EPI_ISL_444478 | India/GJ-GBRC47/2020 | 2020-04-30 | Asia/India/Gujarat/Ahmedabad | B.J. Medical College and Civil hospital | Gujarat Biotechnology Research Centre | Geeta Vaghela et al |
| EPI_ISL_444485 | India/GJ-GBRC54/2020 | 2020-05-04 | Asia/India/Gujarat/Modasa | Gujarat Biotechnology Research Centre | Gujarat Biotechnology Research Centre | R D Dixit et al |
| EPI_ISL_444494 | Canada/QC_AA2/2020 | 2020-04-11 | North America/Canada/Quebec/ | Laboratoire de microbiologie, Hopital de Verdun | Smith Laboratory, Centre de Recherche CHU Sainte-Justine | Martin Smith et al |
| EPI_ISL_444495 | Canada/QC_AA3/2020 | 2020-04-05 | North America/Canada/Quebec/ | Laboratoire de microbiologie, Hopital de Verdun | Smith Laboratory, Centre de Recherche CHU Sainte-Justine | Martin Smith et al |
| EPI_ISL_444496 | Canada/QC_AA6/2020 | 2020-04-07 | North America/Canada/Quebec/ | Laboratoire de microbiologie, Hopital de Verdun | Smith Laboratory, Centre de Recherche CHU Sainte-Justine | Martin Smith et al |

| | | | | | | |
|---|---|---|---|---|---|---|
| EPI_ISL_444497 | Canada/QC_AA8/2020 | 2020-04-07 | North America/Canada/Quebec/ | Laboratoire de microbiologie, Hopital de Verdun | Smith Laboratory, Centre de Recherche CHU Sainte-Justine | Martin Smith et al |
| EPI_ISL_444498 | Canada/QC_AA9/2020 | 2020-04-06 | North America/Canada/Quebec/ | Laboratoire de microbiologie, Hopital de Verdun | Smith Laboratory, Centre de Recherche CHU Sainte-Justine | Martin Smith et al |
| EPI_ISL_444499 | Canada/QC_AB1/2020 | 2020-04-05 | North America/Canada/Quebec/ | Laboratoire de microbiologie, Hopital de Verdun | Smith Laboratory, Centre de Recherche CHU Sainte-Justine | Martin Smith et al |
| EPI_ISL_444500 | Canada/QC_AB2/2020 | 2020-04-07 | North America/Canada/Quebec/ | Laboratoire de microbiologie, Hopital de Verdun | Smith Laboratory, Centre de Recherche CHU Sainte-Justine | Martin Smith et al |
| EPI_ISL_444501 | Canada/QC_AB3/2020 | 2020-04-06 | North America/Canada/Quebec/ | Laboratoire de microbiologie, Hopital de Verdun | Smith Laboratory, Centre de Recherche CHU Sainte-Justine | Martin Smith et al |
| EPI_ISL_444502 | Canada/QC_AB4/2020 | 2020-04-06 | North America/Canada/Quebec/ | Laboratoire de microbiologie, Hopital de Verdun | Smith Laboratory, Centre de Recherche CHU Sainte-Justine | Martin Smith et al |
| EPI_ISL_444504 | Canada/QC_AB7/2020 | 2020-04-05 | North America/Canada/Quebec/ | Laboratoire de microbiologie, Hopital de Verdun | Smith Laboratory, Centre de Recherche CHU Sainte-Justine | Martin Smith et al |
| EPI_ISL_444505 | Canada/QC_AC1/2020 | 2020-04-05 | North America/Canada/Quebec/ | Laboratoire de microbiologie, Hopital de Verdun | Smith Laboratory, Centre de Recherche CHU Sainte-Justine | Martin Smith et al |
| EPI_ISL_444507 | Canada/QC_AC3/2020 | 2020-04-06 | North America/Canada/Quebec/ | Laboratoire de microbiologie, Hopital de Verdun | Smith Laboratory, Centre de Recherche CHU Sainte-Justine | Martin Smith et al |
| EPI_ISL_444509 | Canada/QC_AC6/2020 | 2020-04-05 | North America/Canada/Quebec/ | Laboratoire de microbiologie, Hopital de Verdun | Smith Laboratory, Centre de Recherche CHU Sainte-Justine | Martin Smith et al |

| | | | | | | |
|---|---|---|---|---|---|---|
| EPI_ISL_444510 | Canada/QC_AC7/2020 | 2020-04-06 | North America/Canada/Quebec/ | Laboratoire de microbiologie, Hopital de Verdun | Smith Laboratory, Centre de Recherche CHU Sainte-Justine | Martin Smith et al |
| EPI_ISL_444515 | Canada/QC_AD5/2020 | 2020-04-06 | North America/Canada/Quebec/ | Laboratoire de microbiologie, Hopital de Verdun | Smith Laboratory, Centre de Recherche CHU Sainte-Justine | Martin Smith et al |
| EPI_ISL_444521 | USA/IL-NM08/2020 | 2020-03-17 | North America/USA/Illinois/Chicago | Northwestern Memorial Hospital | Ozer Lab | Ramon Lorenzo-Redondo et al |
| EPI_ISL_444523 | USA/IL-NM010/2020 | 2020-03-17 | North America/USA/Illinois/Chicago | Northwestern Memorial Hospital | Ozer Lab | Ramon Lorenzo-Redondo et al |
| EPI_ISL_444548 | USA/IL-NM051/2020 | 2020-03-14 | North America/USA/Illinois/Chicago | Northwestern Memorial Hospital | Ozer Lab | Ramon Lorenzo-Redondo et al |
| EPI_ISL_444553 | USA/IL-NM057/2020 | 2020-03-18 | North America/USA/Illinois/Chicago | Northwestern Memorial Hospital | Ozer Lab | Ramon Lorenzo-Redondo et al |
| EPI_ISL_444573 | USA/IL-NM082/2020 | 2020-03-19 | North America/USA/Illinois/Chicago | Northwestern Memorial Hospital | Ozer Lab | Ramon Lorenzo-Redondo et al |
| EPI_ISL_444582 | USA/IL-NM093/2020 | 2020-03-19 | North America/USA/Illinois/Chicago | Northwestern Memorial Hospital | Ozer Lab | Ramon Lorenzo-Redondo et al |
| EPI_ISL_444594 | USA/IL-NM0109/2020 | 2020-03-20 | North America/USA/Illinois/Chicago | Northwestern Memorial Hospital | Ozer Lab | Ramon Lorenzo-Redondo et al |
| EPI_ISL_444613 | USA/NY-NYUMC496/2020 | 2020-04-03 | North America/USA/New York/Queens | NYU Langone Health | Departments of Pathology and Medicine, New York University School of Medicine | Maria Aguero-Rosenfeld et al |
| EPI_ISL_444619 | USA/NY-NYUMC502/2020 | 2020-03-30 | North America/USA/New York/Nassau ( | NYU Langone Health | Departments of Pathology and Medicine, New York University School of Medicine | Maria Aguero-Rosenfeld et al |
| EPI_ISL_444647 | USA/NY-NYUMC530/2020 | 2020-03-29 | North America/USA/New York/ | NYU Langone Health | Departments of Pathology and Medicine, New York University School of Medicine | Maria Aguero-Rosenfeld et al |

| | | | | | | |
|---|---|---|---|---|---|---|
| EPI_ISL_444655 | USA/NJ-NYUMC538/2020 | 2020-03-30 | North America/USA/New Jersey/ | NYU Langone Health | Departments of Pathology and Medicine, New York University School of Medicine | Maria Aguero-Rosenfeld et al |
| EPI_ISL_444679 | USA/NY-NYUMC562/2020 | 2020-03-30 | North America/USA/New York/Manhatt | NYU Langone Health | Departments of Pathology and Medicine, New York University School of Medicine | Maria Aguero-Rosenfeld et al |
| EPI_ISL_444680 | USA/NY-NYUMC563/2020 | 2020-03-30 | North America/USA/New York/Nassau ( | NYU Langone Health | Departments of Pathology and Medicine, New York University School of Medicine | Maria Aguero-Rosenfeld et al |
| EPI_ISL_444707 | USA/NY-NYUMC590/2020 | 2020-04-03 | North America/USA/New York/Queens | NYU Langone Health | Departments of Pathology and Medicine, New York University School of Medicine | Maria Aguero-Rosenfeld et al |
| EPI_ISL_444714 | USA/NY-NYUMC597/2020 | 2020-04-03 | North America/USA/New York/Queens | NYU Langone Health | Departments of Pathology and Medicine, New York University School of Medicine | Maria Aguero-Rosenfeld et al |
| EPI_ISL_444715 | USA/NY-NYUMC598/2020 | 2020-04-03 | North America/USA/New York/Queens | NYU Langone Health | Departments of Pathology and Medicine, New York University School of Medicine | Maria Aguero-Rosenfeld et al |

| | | | | | | |
|---|---|---|---|---|---|---|
| EPI_ISL_444716 | USA/NY-NYUMC599/2020 | 2020-04-03 | North America/USA/New York/Manhatt | NYU Langone Health | Departments of Pathology and Medicine, New York University School of Medicine | Maria Aguero-Rosenfeld et al |
| EPI_ISL_444722 | USA/NY-NYUMC605/2020 | 2020-04-03 | North America/USA/New York/Brooklyn | NYU Langone Health | Departments of Pathology and Medicine, New York University School of Medicine | Maria Aguero-Rosenfeld et al |
| EPI_ISL_444723 | USA/NY-NYUMC606/2020 | 2020-04-03 | North America/USA/New York/Manhatt | NYU Langone Health | Departments of Pathology and Medicine, New York University School of Medicine | Maria Aguero-Rosenfeld et al |
| EPI_ISL_444726 | USA/NY-NYUMC609/2020 | 2020-04-02 | North America/USA/New York/Manhatt | NYU Langone Health | Departments of Pathology and Medicine, New York University School of Medicine | Maria Aguero-Rosenfeld et al |
| EPI_ISL_444729 | USA/NY-NYUMC612/2020 | 2020-04-01 | North America/USA/New York/Nassau C | NYU Langone Health | Departments of Pathology and Medicine, New York University School of Medicine | Maria Aguero-Rosenfeld et al |
| EPI_ISL_444732 | USA/NY-NYUMC615/2020 | 2020-04-02 | North America/USA/New York/Nassau C | NYU Langone Health | Departments of Pathology and Medicine, New York University School of Medicine | Maria Aguero-Rosenfeld et al |

| | | | | | | |
|---|---|---|---|---|---|---|
| EPI_ISL_444740 | USA/NY-NYUMC623/2020 | 2020-04-02 | North America/USA/New York/Bronx | NYU Langone Health | Departments of Pathology and Medicine, New York University School of Medicine | Maria Aguero-Rosenfeld et al |
| EPI_ISL_444749 | USA/NY-NYUMC632/2020 | 2020-04-02 | North America/USA/New York/Manhatt | NYU Langone Health | Departments of Pathology and Medicine, New York University School of Medicine | Maria Aguero-Rosenfeld et al |
| EPI_ISL_444759 | USA/NY-NYUMC642/2020 | 2020-04-03 | North America/USA/New York/Brooklyı | NYU Langone Health | Departments of Pathology and Medicine, New York University School of Medicine | Maria Aguero-Rosenfeld et al |
| EPI_ISL_444764 | USA/NY-NYUMC647/2020 | 2020-04-02 | North America/USA/New York/Queens | NYU Langone Health | Departments of Pathology and Medicine, New York University School of Medicine | Maria Aguero-Rosenfeld et al |
| EPI_ISL_444766 | USA/NY-NYUMC649/2020 | 2020-04-03 | North America/USA/New York/Nassau ( | NYU Langone Health | Departments of Pathology and Medicine, New York University School of Medicine | Maria Aguero-Rosenfeld et al |
| EPI_ISL_444769 | USA/NY-NYUMC652/2020 | 2020-04-06 | North America/USA/New York/Suffolk ( | NYU Langone Health | Departments of Pathology and Medicine, New York University School of Medicine | Maria Aguero-Rosenfeld et al |

| | | | | | | |
|---|---|---|---|---|---|---|
| EPI_ISL_444772 | USA/NY-NYUMC655/2020 | 2020-04-10 | North America/USA/New York/Brooklyn | NYU Langone Health | Departments of Pathology and Medicine, New York University School of Medicine | Maria Aguero-Rosenfeld et al |
| EPI_ISL_444782 | USA/NY-NYUMC665/2020 | 2020-03-18 | North America/USA/New York/Nassau ( | NYU Langone Health | Departments of Pathology and Medicine, New York University School of Medicine | Maria Aguero-Rosenfeld et al |
| EPI_ISL_444865 | Denmark/ALAB-SSI-702/2020 | 2020-03-27 | Europe/Denmark/Denmark/ | Department of Virus and Microbiological Special Diagnostics, Statens Serum Institut, Copenhagen, Denmark, Artillerivej 5, 2300 Copenahgen S | Albertsen lab, Department of Chemistry and Bioscience, Aalborg University, Denmark | Rasmus Kirkegaard et al |
| EPI_ISL_444885 | Denmark/ALAB-SSI-743/2020 | 2020-03-28 | Europe/Denmark/Denmark/ | Department of Virus and Microbiological Special Diagnostics, Statens Serum Institut, Copenhagen, Denmark, Artillerivej 5, 2300 Copenahgen S | Albertsen lab, Department of Chemistry and Bioscience, Aalborg University, Denmark | Rasmus Kirkegaard et al |
| EPI_ISL_444911 | Denmark/ALAB-SSI-797/2020 | 2020-03-30 | Europe/Denmark/Denmark/ | Department of Virus and Microbiological Special Diagnostics, Statens Serum Institut, Copenhagen, Denmark, Artillerivej 5, 2300 Copenahgen S | Albertsen lab, Department of Chemistry and Bioscience, Aalborg University, Denmark | Rasmus Kirkegaard et al |
| EPI_ISL_444969 | Guangdong/SYSU-IHV/2020 | 2020-04-16 | Asia/China/Guangdong/Guangzhou | Guangzhou Eighth People's Hospital (Jiahe Sector) | Institute of Human Virology, Zhongshan School of Medicine, Sun Yat-sen University | Junsong Zhang et al |
| EPI_ISL_444971 | Spain/CT-HUVH-VH6372/2020 | 2020-04-04 | Europe/Spain/Catalunya/ | Hospital Universitari Vall d'Hebron - Vall d'Hebron Institut de Recerca | Hospital Universitari Vall d'Hebron | Cristina Andr√©s et al |

| | | | | | | |
|---|---|---|---|---|---|---|
| EPI_ISL_444978 | Spain/CT-HUVH-VH4141/2020 | 2020-03-20 | Europe/Spain/Catalunya/ | Hospital Universitari Vall d'Hebron - Vall d'Hebron Institut de Recerca | Hospital Universitari Vall d'Hebron | Cristina Andr√©s et al |
| EPI_ISL_444981 | Spain/CT-HUVH-VH5081/2020 | 2020-03-21 | Europe/Spain/Catalunya/ | "Hospital Universitari Vall d'Hebron - Vall d""hebron Institut de Recerca" | Hospital Universitari Vall d'Hebron | Cristina Andr√©s et al |
| EPI_ISL_445008 | USA/FL-BPHL-0065/2020 | 2020-04-15 | North America/USA/Florida/ | Florida Bureau of Public Health Laboratories | Florida Bureau of Public Health Laboratories | Sarah Schmedes et al |
| EPI_ISL_445009 | USA/FL-BPHL-0066/2020 | 2020-04-15 | North America/USA/Florida/ | Florida Bureau of Public Health Laboratories | Florida Bureau of Public Health Laboratories | Sarah Schmedes et al |
| EPI_ISL_445015 | USA/FL-BPHL-0072/2020 | 2020-04-20 | North America/USA/Florida/ | Florida Bureau of Public Health Laboratories | Florida Bureau of Public Health Laboratories | Sarah Schmedes et al |
| EPI_ISL_445040 | USA/FL-BPHL-0097/2020 | 2020-04-27 | North America/USA/Florida/ | Florida Bureau of Public Health Laboratories | Florida Bureau of Public Health Laboratories | Sarah Schmedes et al |
| EPI_ISL_445057 | Luxembourg/LNS8705424/2020 | 2020-04-23 | Europe/Luxembourg/Luxembourg/ | Laboratoire National de Sante, Microbiology, Virology | Laboratoire National de Sante, Microbiology, Epidemiology and Microbial Genomics | Anke Wienecke-Baldacchino et al |
| EPI_ISL_445062 | Luxembourg/LNS7543365/2020 | 2020-04-29 | Europe/Luxembourg/Luxembourg/ | Laboratoire National de Sante, Microbiology, Virology | Laboratoire National de Sante, Microbiology, Epidemiology and Microbial Genomics | Anke Wienecke-Baldacchino et al |
| EPI_ISL_445104 | USA/CA-SR0108/2020 | 2020-03-20 | North America/USA/California/San Dieg | UC San Diego Center for Advanced Laboratory Medicine | Andersen lab at Scripps Research | SEARCH Alliance San Diego with David Pride et al |
| EPI_ISL_445105 | USA/CA-SR0111/2020 | 2020-03-24 | North America/USA/California/San Dieg | UC San Diego Center for Advanced Laboratory Medicine | Andersen lab at Scripps Research | SEARCH Alliance San Diego with David Pride et al |
| EPI_ISL_445109 | USA/CA-SR0118/2020 | 2020-03-12 | North America/USA/California/San Dieg | UC San Diego Center for Advanced Laboratory Medicine | Andersen lab at Scripps Research | SEARCH Alliance San Diego with David Pride et al |
| EPI_ISL_445111 | USA/CA-SR0120/2020 | 2020-03-26 | North America/USA/California/San Dieg | UC San Diego Center for Advanced Laboratory Medicine | Andersen lab at Scripps Research | SEARCH Alliance San Diego with David Pride et al |

| | | | | | | |
|---|---|---|---|---|---|---|
| EPI_ISL_445118 | USA/CA-SR0217/2020 | 2020-05-04 | North America/USA/California/San Dieg | Rady's Childrens Hospital | Andersen lab at Scripps Research | SEARCH Alliance San Diego et al |
| EPI_ISL_445124 | USA/LA-SR0269/2020 | 2020-04-02 | North America/USA/Louisiana/New Orle | Robert Garry lab | Andersen lab at Scripps Research | Allison Smither et al |
| EPI_ISL_445134 | USA/LA-SR0279/2020 | 2020-03-31 | North America/USA/Louisiana/New Orle | Robert Garry lab | Andersen lab at Scripps Research | Allison Smither et al |
| EPI_ISL_445136 | USA/LA-SR0281/2020 | 2020-03-31 | North America/USA/Louisiana/New Orle | Robert Garry lab | Andersen lab at Scripps Research | Allison Smither et al |
| EPI_ISL_445142 | USA/LA-SR0287/2020 | 2020-04-01 | North America/USA/Louisiana/New Orle | Robert Garry lab | Andersen lab at Scripps Research | Allison Smither et al |
| EPI_ISL_445148 | USA/LA-SR0293/2020 | 2020-04-07 | North America/USA/Louisiana/New Orle | Robert Garry lab | Andersen lab at Scripps Research | Allison Smither et al |
| EPI_ISL_445149 | USA/LA-SR0294/2020 | 2020-04-08 | North America/USA/Louisiana/New Orle | Robert Garry lab | Andersen lab at Scripps Research | Allison Smither et al |
| EPI_ISL_445154 | USA/LA-SR0299/2020 | 2020-04-07 | North America/USA/Louisiana/New Orle | Robert Garry lab | Andersen lab at Scripps Research | Allison Smither et al |
| EPI_ISL_445155 | USA/LA-SR0300/2020 | 2020-04-09 | North America/USA/Louisiana/New Orle | Robert Garry lab | Andersen lab at Scripps Research | Allison Smither et al |
| EPI_ISL_445161 | USA/LA-SR0306/2020 | 2020-04-07 | North America/USA/Louisiana/New Orle | Robert Garry lab | Andersen lab at Scripps Research | Allison Smither et al |
| EPI_ISL_445163 | USA/LA-SR0309/2020 | 2020-04-08 | North America/USA/Louisiana/New Orle | Robert Garry lab | Andersen lab at Scripps Research | Allison Smither et al |
| EPI_ISL_445175 | USA/CA-CZB-1127/2020 | 2020-04-05 | North America/USA/California/San Fran | UCSF Clinical Microbiology Laboratory | Chan-Zuckerberg Biohub | CZB Cliahub Consortium et al |
| EPI_ISL_445240 | Sweden/20-07691/2020 | 2020-04-22 | Europe/Sweden/Uppsala/ | Ulltuna Vardcentral | The Public Health Agency of Sweden | Heidi Lindback et al |
| EPI_ISL_445242 | Sweden/20-07772/2020 | 2020-04-22 | Europe/Sweden/Norrbotten/ | Jokkmokks Halsocentral | The Public Health Agency of Sweden | Markus Beland et al |
| EPI_ISL_445243 | Romania/279067/2020 | 2020-04-30 | Europe/Romania/Constanta/ | Laboratory for Respiratory Viruses, Cantacuzino National Military-Medical Institute for Research and Development | Cantacuzino Institute | M.Lazar et al |
| EPI_ISL_445286 | Chile/VS-ISPCH-3/2020 | 2020-03-19 | South America/Chile/Valparaiso/ | HOSPITAL HANGA ROA | Instituto de Salud Publica de Chile | Andr√©s E Castillo et al |
| EPI_ISL_445323 | Chile/RM-ISPCH-44/2020 | 2020-03-16 | South America/Chile/Region Metropolit | HOSP.ENFERMEDADES INFECCIOSAS | Instituto de Salud Publica de Chile | Andr√©s E Castillo et al |
| EPI_ISL_445330 | Chile/RM-ISPCH-51/2020 | 2020-03-23 | South America/Chile/Region Metropolit | CLINICA VESPUCIO S. A. | Instituto de Salud Publica de Chile | Andr√©s E Castillo et al |
| EPI_ISL_445349 | Chile/RM-ISPCH-52/2020 | 2020-04-01 | South America/Chile/Region Metropolit | HOSPITAL SAN JUAN DE DIOS | Instituto de Salud Publica de Chile | Andr√©s E Castillo et al |

| | | | | | | |
|---|---|---|---|---|---|---|
| EPI_ISL_445350 | Chile/RM-ISPCH-54/2020 | 2020-04-01 | South America/Chile/Region Metropolit | HOSPITAL SAN JUAN DE DIOS | Instituto de Salud Publica de Chile | Andr√©s E Castillo et al |
| EPI_ISL_445352 | Chile/RM-ISPCH-56/2020 | 2020-04-01 | South America/Chile/Region Metropolit | HOSPITAL DEL PROFESOR | Instituto de Salud Publica de Chile | Andr√©s E Castillo et al |
| EPI_ISL_445367 | Chile/RM-ISPCH-71/2020 | 2020-03-21 | South America/Chile/Region Metropolit | ASISTENCIA PUBLICA DR.ALEJANDRO DEL RIO | Instituto de Salud Publica de Chile | Andr√©s E Castillo et al |
| EPI_ISL_445369 | Chile/RM-ISPCH-73/2020 | 2020-04-06 | South America/Chile/Region Metropolit | HOSPITAL DE CARABINEROS | Instituto de Salud Publica de Chile | Andr√©s E Castillo et al |
| EPI_ISL_445382 | Wales/PHWC-2899E/2020 | 2020-04-02 | Europe/United Kingdom/Wales/ | Wales Specialist Virology Centre | Public Health Wales Microbiology Cardiff | Catherine Moore et al |
| EPI_ISL_445411 | Wales/PHWC-28B98/2020 | 2020-04-03 | Europe/United Kingdom/Wales/ | Wales Specialist Virology Centre | Public Health Wales Microbiology Cardiff | Catherine Moore et al |
| EPI_ISL_445470 | Wales/PHWC-2902F/2020 | 2020-04-03 | Europe/United Kingdom/Wales/ | Wales Specialist Virology Centre | Public Health Wales Microbiology Cardiff | Catherine Moore et al |
| EPI_ISL_445489 | Wales/PHWC-29177/2020 | 2020-04-04 | Europe/United Kingdom/Wales/ | Wales Specialist Virology Centre | Public Health Wales Microbiology Cardiff | Catherine Moore et al |
| EPI_ISL_445586 | Wales/PHWC-2A589/2020 | 2020-04-07 | Europe/United Kingdom/Wales/ | Wales Specialist Virology Centre | Public Health Wales Microbiology Cardiff | Catherine Moore et al |
| EPI_ISL_445604 | Wales/PHWC-2A6A4/2020 | 2020-04-07 | Europe/United Kingdom/Wales/ | Wales Specialist Virology Centre | Public Health Wales Microbiology Cardiff | Catherine Moore et al |
| EPI_ISL_445635 | Wales/PHWC-2A905/2020 | 2020-04-08 | Europe/United Kingdom/Wales/ | Wales Specialist Virology Centre | Public Health Wales Microbiology Cardiff | Catherine Moore et al |
| EPI_ISL_445737 | Wales/PHWC-2B62B/2020 | 2020-04-10 | Europe/United Kingdom/Wales/ | Wales Specialist Virology Centre | Public Health Wales Microbiology Cardiff | Catherine Moore et al |

| | | | | | | |
|---|---|---|---|---|---|---|
| EPI_ISL_445801 | Wales/PHWC-2BA89/2020 | 2020-03-28 | Europe/United Kingdom/Wales/ | Wales Specialist Virology Centre | Public Health Wales Microbiology Cardiff | Catherine Moore et al |
| EPI_ISL_445912 | Wales/PHWC-2C7DC/2020 | 2020-03-31 | Europe/United Kingdom/Wales/ | Wales Specialist Virology Centre | Public Health Wales Microbiology Cardiff | Catherine Moore et al |
| EPI_ISL_445927 | Wales/PHWC-2C97C/2020 | 2020-03-31 | Europe/United Kingdom/Wales/ | Wales Specialist Virology Centre | Public Health Wales Microbiology Cardiff | Catherine Moore et al |
| EPI_ISL_445936 | Wales/PHWC-2CA2E/2020 | 2020-04-01 | Europe/United Kingdom/Wales/ | Wales Specialist Virology Centre | Public Health Wales Microbiology Cardiff | Catherine Moore et al |
| EPI_ISL_445951 | Wales/PHWC-2CB2B/2020 | 2020-04-01 | Europe/United Kingdom/Wales/ | Wales Specialist Virology Centre | Public Health Wales Microbiology Cardiff | Catherine Moore et al |
| EPI_ISL_445986 | Wales/PHWC-2D44D/2020 | 2020-04-04 | Europe/United Kingdom/Wales/ | Wales Specialist Virology Centre | Public Health Wales Microbiology Cardiff | Catherine Moore et al |
| EPI_ISL_446016 | Wales/PHWC-2D708/2020 | 2020-04-05 | Europe/United Kingdom/Wales/ | Wales Specialist Virology Centre | Public Health Wales Microbiology Cardiff | Catherine Moore et al |
| EPI_ISL_446027 | Wales/PHWC-2DE21/2020 | 2020-04-14 | Europe/United Kingdom/Wales/ | Wales Specialist Virology Centre | Public Health Wales Microbiology Cardiff | Catherine Moore et al |
| EPI_ISL_446068 | Wales/PHWC-2E1BE/2020 | 2020-04-07 | Europe/United Kingdom/Wales/ | Wales Specialist Virology Centre | Public Health Wales Microbiology Cardiff | Catherine Moore et al |
| EPI_ISL_446121 | Wales/PHWC-2EB92/2020 | 2020-04-07 | Europe/United Kingdom/Wales/ | Wales Specialist Virology Centre | Public Health Wales Microbiology Cardiff | Catherine Moore et al |

| | | | | | | |
|---|---|---|---|---|---|---|
| EPI_ISL_446140 | Wales/PHWC-2ECEA/2020 | 2020-04-07 | Europe/United Kingdom/Wales/ | Wales Specialist Virology Centre | Public Health Wales Microbiology Cardiff | Catherine Moore et al |
| EPI_ISL_446192 | Wales/PHWC-2F056/2020 | 2020-04-05 | Europe/United Kingdom/Wales/ | Wales Specialist Virology Centre | Public Health Wales Microbiology Cardiff | Catherine Moore et al |
| EPI_ISL_446343 | Wales/PHWC-30C7C/2020 | 2020-04-11 | Europe/United Kingdom/Wales/ | Wales Specialist Virology Centre | Public Health Wales Microbiology Cardiff | Catherine Moore et al |
| EPI_ISL_446429 | Wales/PHWC-31297/2020 | 2020-04-10 | Europe/United Kingdom/Wales/ | Wales Specialist Virology Centre | Public Health Wales Microbiology Cardiff | Catherine Moore et al |
| EPI_ISL_446441 | Wales/PHWC-313EF/2020 | 2020-04-13 | Europe/United Kingdom/Wales/ | Wales Specialist Virology Centre | Public Health Wales Microbiology Cardiff | Catherine Moore et al |
| EPI_ISL_446597 | Wales/PHWC-31FFA/2020 | 2020-04-13 | Europe/United Kingdom/Wales/ | Wales Specialist Virology Centre | Public Health Wales Microbiology Cardiff | Catherine Moore et al |
| EPI_ISL_446601 | Wales/PHWC-32050/2020 | 2020-04-13 | Europe/United Kingdom/Wales/ | Wales Specialist Virology Centre | Public Health Wales Microbiology Cardiff | Catherine Moore et al |
| EPI_ISL_446607 | Wales/PHWC-32111/2020 | 2020-04-14 | Europe/United Kingdom/Wales/ | Wales Specialist Virology Centre | Public Health Wales Microbiology Cardiff | Catherine Moore et al |
| EPI_ISL_446635 | Wales/PHWC-32339/2020 | 2020-04-08 | Europe/United Kingdom/Wales/ | Wales Specialist Virology Centre | Public Health Wales Microbiology Cardiff | Catherine Moore et al |
| EPI_ISL_446715 | Wales/PHWC-32F71/2020 | 2020-04-13 | Europe/United Kingdom/Wales/ | Wales Specialist Virology Centre | Public Health Wales Microbiology Cardiff | Catherine Moore et al |

| | | | | | | |
|---|---|---|---|---|---|---|
| EPI_ISL_447025 | Thailand/Bangkok-0084/2020 | 2020-04-02 | Asia/Thailand/Bangkok/ | Ramathibodi Hospital | COVID-19 Network Investigations (CONI) Alliance | Elizabeth Batty et al |
| EPI_ISL_447033 | India/GJ-GBRC58/2020 | 2020-05-03 | Asia/India/Gujarat/Ahmedabad | B.J. Medical College and Civil hospital | Gujarat Biotechnology Research Centre | Ankit Hinsu et al |
| EPI_ISL_447043 | India/GJ-GBRC67a/2020 | 2020-05-03 | Asia/India/Gujarat/Ahmedabad | B.J. Medical College and Civil hospital | Gujarat Biotechnology Research Centre | Raghawendra Kumar et al |
| EPI_ISL_447044 | India/GJ-GBRC67b/2020 | 2020-05-03 | Asia/India/Gujarat/Ahmedabad | B.J. Medical College and Civil hospital | Gujarat Biotechnology Research Centre | Dinesh Kumar et al |
| EPI_ISL_447055 | Georgia/Tb-6598/2020 | 2020-04-29 | Asia/Georgia/Tbilisi/ | Department for Virology, Molecular Biology and Genome Research, R. G. Lugar Center for Public Health Research, National Center for Disease Control and Public Health (NCDC) of Georgia. | Department for Virology, Molecular Biology and Genome Research, R. G. Lugar Center for Public Health Research, National Center for Disease Control and Public Health (NCDC) of Georgia. | Meri Pantsulaia et al |
| EPI_ISL_447093 | USA/MI-MDHHS-SC20296/2020 | 2020-03-14 | North America/USA/Michigan/ | Michigan Department of Health and Human Services, Bureau of Laboratories | Michigan Department of Health and Human Services, Bureau of Laboratories | Blankenship HM et al |
| EPI_ISL_447102 | USA/MI-MDHHS-SC20305/2020 | 2020-04-02 | North America/USA/Michigan/ | Michigan Department of Health and Human Services, Bureau of Laboratories | Michigan Department of Health and Human Services, Bureau of Laboratories | Blankenship HM et al |

| | | | | | | |
|---|---|---|---|---|---|---|
| EPI_ISL_447104 | USA/MI-MDHHS-SC20307/2020 | 2020-04-03 | North America/USA/Michigan/ | Michigan Department of Health and Human Services, Bureau of Laboratories | Michigan Department of Health and Human Services, Bureau of Laboratories | Blankenship HM et al |
| EPI_ISL_447105 | USA/MI-MDHHS-SC20308/2020 | 2020-04-02 | North America/USA/Michigan/ | Michigan Department of Health and Human Services, Bureau of Laboratories | Michigan Department of Health and Human Services, Bureau of Laboratories | Blankenship HM et al |
| EPI_ISL_447121 | Belgium/ULG-6842/2020 | 2020-03-14 | Europe/Belgium/Liv®ge/ | Department of Clinical Microbiology | GIGA Medical Genomics | Keith Durkin et al |
| EPI_ISL_447147 | Belgium/ULG-10150/2020 | 2020-04-18 | Europe/Belgium/Liv®ge/ | Department of Clinical Microbiology | GIGA Medical Genomics | Keith Durkin et al |
| EPI_ISL_447151 | Belgium/ULG-10155/2020 | 2020-04-18 | Europe/Belgium/Liv®ge/ | Department of Clinical Microbiology | GIGA Medical Genomics | Keith Durkin et al |
| EPI_ISL_447154 | Belgium/ULG-10159/2020 | 2020-04-18 | Europe/Belgium/Liv®ge/ | Department of Clinical Microbiology | GIGA Medical Genomics | Keith Durkin et al |
| EPI_ISL_447198 | USA/MI-MDHHS-SC20228/2020 | 2020-03-19 | North America/USA/Michigan/ | Michigan Department of Health and Human Services, Bureau of Laboratories | Michigan Department of Health and Human Services, Bureau of Laboratories | Blankenship HM et al |
| EPI_ISL_447199 | USA/MI-MDHHS-SC20229/2020 | 2020-03-22 | North America/USA/Michigan/ | Michigan Department of Health and Human Services, Bureau of Laboratories | Michigan Department of Health and Human Services, Bureau of Laboratories | Blankenship HM et al |
| EPI_ISL_447205 | USA/MI-MDHHS-SC20235/2020 | 2020-04-22 | North America/USA/Michigan/ | Michigan Department of Health and Human Services, Bureau of Laboratories | Michigan Department of Health and Human Services, Bureau of Laboratories | Blankenship HM et al |
| EPI_ISL_447252 | Taiwan/TSGH-26/2020 | 2020-02-25 | Asia/Taiwan/New Taipei City/ | TSGH-CP molecular lab | TSGH-CP molecular lab | Cherng-Lih Perng et al |

| | | | | | | |
|---|---|---|---|---|---|---|
| EPI_ISL_447255 | Taiwan/TSGH-29/2020 | 2020-03-17 | Asia/Taiwan/New Taipei City/ | TSGH-CP molecular lab | TSGH-CP molecular lab | Cherng-Lih Perng et al |
| EPI_ISL_447259 | Israel/990430264/2020 | 2020-03-29 | Asia/Israel/South District/South Coast | Microbiology laboratory, Assuta Ashdod University-Affiliated Hospital | Stern Lab | Stern Lab et al |
| EPI_ISL_447267 | Israel/990300681/2020 | 2020-03-30 | Asia/Israel/South District/South Coast | Microbiology laboratory, Assuta Ashdod University-Affiliated Hospital | Stern Lab | Stern Lab et al |
| EPI_ISL_447269 | Israel/990059251/2020 | 2020-03-25 | Asia/Israel/South District/South Coast | Microbiology laboratory, Assuta Ashdod University-Affiliated Hospital | Stern Lab | Stern Lab et al |
| EPI_ISL_447270 | Israel/990059244/2020 | 2020-03-25 | Asia/Israel/South District/South Coast | Microbiology laboratory, Assuta Ashdod University-Affiliated Hospital | Stern Lab | Stern Lab et al |
| EPI_ISL_447273 | Israel/990059233/2020 | 2020-03-25 | Asia/Israel/South District/South Coast | Microbiology laboratory, Assuta Ashdod University-Affiliated Hospital | Stern Lab | Stern Lab et al |
| EPI_ISL_447275 | Israel/990059231/2020 | 2020-03-25 | Asia/Israel/South District/South Coast | Microbiology laboratory, Assuta Ashdod University-Affiliated Hospital | Stern Lab | Stern Lab et al |
| EPI_ISL_447292 | Israel/701002540/2020 | 2020-03-25 | Asia/Israel/South District/South Coast | Microbiology Division, Barzilai University Medical Center | Stern Lab | Stern Lab et al |
| EPI_ISL_447301 | Israel/701002440/2020 | 2020-03-24 | Asia/Israel/South District/South Coast | Microbiology Division, Barzilai University Medical Center | Stern Lab | Stern Lab et al |
| EPI_ISL_447305 | Israel/701002403/2020 | 2020-03-23 | Asia/Israel/South District/South Coast | Microbiology Division, Barzilai University Medical Center | Stern Lab | Stern Lab et al |
| EPI_ISL_447307 | Israel/701002327/2020 | 2020-03-22 | Asia/Israel/South District/South Coast | Microbiology Division, Barzilai University Medical Center | Stern Lab | Stern Lab et al |
| EPI_ISL_447346 | Israel/2113174/2020 | 2020-04-15 | Asia/Israel/Tel Aviv District/ | Clinical Virology Unit, Hadassah Hebrew University Medical Center | Stern Lab | Stern Lab et al |
| EPI_ISL_447358 | Israel/2099018/2020 | 2020-04-02 | Asia/Israel/Jerusalem District/ | Clinical Virology Unit, Hadassah Hebrew University Medical Center | Stern Lab | Stern Lab et al |
| EPI_ISL_447360 | Israel/2089863/2020 | 2020-04-01 | Asia/Israel/Jerusalem District/ | Clinical Virology Unit, Hadassah Hebrew University Medical Center | Stern Lab | Stern Lab et al |
| EPI_ISL_447363 | Israel/2089839/2020 | 2020-04-01 | Asia/Israel/Jerusalem District/ | Clinical Virology Unit, Hadassah Hebrew University Medical Center | Stern Lab | Stern Lab et al |
| EPI_ISL_447366 | Israel/2089718/2020 | 2020-04-01 | Asia/Israel/Jerusalem District/ | Clinical Virology Unit, Hadassah Hebrew University Medical Center | Stern Lab | Stern Lab et al |
| EPI_ISL_447374 | Israel/2089366/2020 | 2020-04-01 | Asia/Israel/Jerusalem District/ | Clinical Virology Unit, Hadassah Hebrew University Medical Center | Stern Lab | Stern Lab et al |
| EPI_ISL_447376 | Israel/2086034/2020 | 2020-04-01 | Asia/Israel/Jerusalem District/ | Clinical Virology Unit, Hadassah Hebrew University Medical Center | Stern Lab | Stern Lab et al |
| EPI_ISL_447378 | Israel/2086022/2020 | 2020-04-01 | Asia/Israel/Jerusalem District/ | Clinical Virology Unit, Hadassah Hebrew University Medical Center | Stern Lab | Stern Lab et al |
| EPI_ISL_447382 | Israel/2086001/2020 | 2020-04-01 | Asia/Israel/Jerusalem District/ | Clinical Virology Unit, Hadassah Hebrew University Medical Center | Stern Lab | Stern Lab et al |

| | | | | | | |
|---|---|---|---|---|---|---|
| EPI_ISL_447384 | Israel/2047883/2020 | 2020-03-31 | Asia/Israel/North District/ | Clinical Microbiology Laboratory, The Baruch Padeh Medical Center, Poriya | Stern Lab | Stern Lab et al |
| EPI_ISL_447392 | Israel/2047392/2020 | 2020-03-29 | Asia/Israel/North District/ | Clinical Microbiology Laboratory, The Baruch Padeh Medical Center, Poriya | Stern Lab | Stern Lab et al |
| EPI_ISL_447406 | Israel/2046129/2020 | 2020-03-31 | Asia/Israel/North District/ | Clinical Microbiology Laboratory, The Baruch Padeh Medical Center, Poriya | Stern Lab | Stern Lab et al |
| EPI_ISL_447417 | Israel/1639996/2020 | 2020-03-26 | Asia/Israel/North District/ | Clinical Microbiology Laboratory, The Baruch Padeh Medical Center, Poriya | Stern Lab | Stern Lab et al |
| EPI_ISL_447424 | Israel/13077823/2020 | 2020-04-04 | Asia/Israel/South District/South Coast D | Clinical Microbiology Laboratory, Sheba Medical Center | Stern Lab | Stern Lab et al |
| EPI_ISL_447438 | Israel/13077413/2020 | 2020-04-03 | Asia/Israel/Tel Aviv District/ | Clinical Microbiology Laboratory, Sheba Medical Center | Stern Lab | Stern Lab et al |
| EPI_ISL_447439 | Israel/13077383/2020 | 2020-04-02 | Asia/Israel/Jerusalem District/ | Clinical Microbiology Laboratory, Sheba Medical Center | Stern Lab | Stern Lab et al |
| EPI_ISL_447449 | Israel/13075719/2020 | 2020-03-29 | Asia/Israel/Tel Aviv District/ | Clinical Microbiology Laboratory, Sheba Medical Center | Stern Lab | Stern Lab et al |
| EPI_ISL_447453 | Israel/130711116/2020 | 2020-04-21 | Asia/Israel/Tel Aviv District/ | Clinical Microbiology Laboratory, Sheba Medical Center | Stern Lab | Stern Lab et al |
| EPI_ISL_447460 | Israel/130710414/2020 | 2020-04-17 | Asia/Israel/Tel Aviv District/ | Clinical Microbiology Laboratory, Sheba Medical Center | Stern Lab | Stern Lab et al |
| EPI_ISL_447463 | Israel/130710211/2020 | 2020-04-16 | Asia/Israel/Tel Aviv District/ | Clinical Microbiology Laboratory, Sheba Medical Center | Stern Lab | Stern Lab et al |
| EPI_ISL_447470 | Spain/VC-FISABIO-470/2020 | 2020-03-26 | Europe/Spain/Comunitat Valenciana/Ca | Servicio de Microbiologia. Hospital Clinico Universitario de Valencia | Sequencing and Bioinformatics Service and Molecular Epidemiology Research Group. FISABIO-Public Health | David Navarro et al |
| EPI_ISL_447473 | Spain/VC-FISABIO-522/2020 | 2020-03-26 | Europe/Spain/Comunitat Valenciana/Sa | Servicio de Microbiologia. Hospital Clinico Universitario de Valencia | Sequencing and Bioinformatics Service and Molecular Epidemiology Research Group. FISABIO-Public Health | Griselda De Marco et al |

| | | | | | | |
|---|---|---|---|---|---|---|
| EPI_ISL_447477 | Spain/VC-FISABIO-526/2020 | 2020-03-29 | Europe/Spain/Comunitat Valenciana/Va | Servicio de Microbiologia. Hospital Clinico Universitario de Valencia | Sequencing and Bioinformatics Service and Molecular Epidemiology Research Group. FISABIO-Public Health | Sandra Carbo et al |
| EPI_ISL_447486 | Spain/VC-FISABIO-541/2020 | 2020-03-29 | Europe/Spain/Comunitat Valenciana/Va | Servicio de Microbiologia. Hospital Clinico Universitario de Valencia | Sequencing and Bioinformatics Service and Molecular Epidemiology Research Group. FISABIO-Public Health | Maria Alma Bracho et al |
| EPI_ISL_447490 | Spain/VC-FISABIO-548/2020 | 2020-03-26 | Europe/Spain/Comunitat Valenciana/Be | Servicio de Microbiologia. Hospital Clinico Universitario de Valencia | Sequencing and Bioinformatics Service and Molecular Epidemiology Research Group. FISABIO-Public Health | Inma GalV°n Vendrell et al |
| EPI_ISL_447495 | Spain/VC-FISABIO-555/2020 | 2020-03-27 | Europe/Spain/Comunitat Valenciana/Oli | Servicio de Microbiologia. Hospital Clinico Universitario de Valencia | Sequencing and Bioinformatics Service and Molecular Epidemiology Research Group. FISABIO-Public Health | Vicente Soriano Chirona et al |
| EPI_ISL_447523 | Spain/VC-FISABIO-604/2020 | 2020-04-01 | Europe/Spain/Comunitat Valenciana/Gi | Servicio de Microbiologia. Hospital Clinico Universitario de Valencia | Sequencing and Bioinformatics Service and Molecular Epidemiology Research Group. FISABIO-Public Health | Giuseppe 'Auria et al |

| | | | | | | |
|---|---|---|---|---|---|---|
| EPI_ISL_447538 | India/GJ-GBRC79a/2020 | 2020-05-05 | Asia/India/Gujarat/Modasa | Gujarat Biotechnology Research Centre | Gujarat Biotechnology Research Centre | Snehal Bagatharia et al |
| EPI_ISL_447544 | India/GJ-GBRC83/2020 | 2020-05-05 | Asia/India/Gujarat/Modasa | Gujarat Biotechnology Research Centre | Gujarat Biotechnology Research Centre | Apurvasinh Puvar et al |
| EPI_ISL_447551 | India/GJ-GBRC89/2020 | 2020-04-25 | Asia/India/Gujarat/Gandhinagar | GMERS Medical College and Hospital, Gandhinagar | Gujarat Biotechnology Research Centre | Nitin Savaliya et al |
| EPI_ISL_447553 | India/GJ-GBRC91/2020 | 2020-04-27 | Asia/India/Gujarat/Gandhinagar | GMERS Medical College and Hospital, Gandhinagar | Gujarat Biotechnology Research Centre | Dinesh Kumar et al |
| EPI_ISL_447561 | India/TG-CCMB-J067/2020 | 2020-03-31 | Asia/India/Telangana/Gadwal | CSIR-Centre for Cellular and Molecular Biology | CSIR-Centre for Cellular and Molecular Biology | Shagufta Khan et al |
| EPI_ISL_447571 | India/TG-CCMB-J224/2020 | 2020-04-01 | Asia/India/Telangana/Nizamabad | CSIR-Centre for Cellular and Molecular Biology | CSIR-Centre for Cellular and Molecular Biology | Lamuk Zaveri et al |
| EPI_ISL_447575 | India/TG-CCMB-J458/2020 | 2020-04-02 | Asia/India/Telangana/Nirmal | CSIR-Centre for Cellular and Molecular Biology | CSIR-Centre for Cellular and Molecular Biology | Sofia Banu et al |
| EPI_ISL_447741 | Colombia/BOY-GUR-0210/2020 | 2020-03-28 | South America/Colombia/Boyaca/Togui | Grupo de Investigaciones Microbiol√≥gicas-UR (GIMUR), Departamento de Biolog√≠a, Facultad de Ciencias Naturales, Universidad del Rosario, Bogot√°, Colombia | Grupo de Investigaciones Microbiol√≥gicas-UR (GIMUR), Departamento de Biolog√≠a, Facultad de Ciencias Naturales, Universidad del Rosario, Bogot√°, Colombia Instituto Nacional de Salud, Bogot√°, Colombia Icahn School of Medicine at Mount Sinai, New York, USA | Juan David Ram√≠rez et al |

| | | | | | | |
|---|---|---|---|---|---|---|
| EPI_ISL_447780 | Colombia/VAC-GVI-93470/2020 | 2020-04-01 | South America/Colombia/Valle del Cauc | Instituto Nacional de Salud, Bogotv°, Colombia | Grupo de Investigaciones Microbiolv≥gicas-UR (GIMUR), Departamento de Biologv≠a, Facultad de Ciencias Naturales, Universidad del Rosario, Bogotv°, Colombia Instituto Nacional de Salud, Bogotv°, Colombia Icahn School of Medicine at Mount Sinai, New York, USA | Juan David Ramv≠rez et al |
| EPI_ISL_447781 | Colombia/VAC-GVI-93473/2020 | 2020-04-01 | South America/Colombia/Valle del Cauc | Instituto Nacional de Salud, Bogotv°, Colombia | Grupo de Investigaciones Microbiolv≥gicas-UR (GIMUR), Departamento de Biologv≠a, Facultad de Ciencias Naturales, Universidad del Rosario, Bogotv°, Colombia Instituto Nacional de Salud, Bogotv°, Colombia Icahn School of Medicine at Mount Sinai, New York, USA | Juan David Ramv≠rez et al |

| | | | | | | |
|---|---|---|---|---|---|---|
| EPI_ISL_447841 | USA/FL-CDC-7619/2020 | 2020-03-06 | North America/USA/Florida/ | FL Bureau of Public Health Laboratories-Tampa | Pathogen Discovery, Respiratory Viruses Branch, Division of Viral Diseases, Centers for Disease Control and Prevention | Krista Queen et al |
| EPI_ISL_447843 | USA/MD-CDC-0025/2020 | 2020-03-04 | North America/USA/Maryland/ | MD DOH Laboratories Administration | Pathogen Discovery, Respiratory Viruses Branch, Division of Viral Diseases, Centers for Disease Control and Prevention | Krista Queen et al |
| EPI_ISL_447852 | India/TG-CCMB-J235/2020 | 2020-04-01 | Asia/India/Telangana/Nizamabad | CSIR-Centre for Cellular and Molecular Biology | CSIR-Centre for Cellular and Molecular Biology | Lamuk Zaveri et al |
| EPI_ISL_447860 | India/TG-CCMB-K849/2020 | 2020-04-16 | Asia/India/Telangana/Rangareddy | CSIR-Centre for Cellular and Molecular Biology | CSIR-Centre for Cellular and Molecular Biology | Tulasi Nagabandi et al |
| EPI_ISL_447862 | India/TG-CCMB-J047/2020 | 2020-03-31 | Asia/India/Telangana/Kamareddy | CSIR-Centre for Cellular and Molecular Biology | CSIR-Centre for Cellular and Molecular Biology | Payel Mukherjee et al |
| EPI_ISL_447863 | India/TG-CCMB-K599/2020 | 2020-04-14 | Asia/India/Telangana/Nizamabad | CSIR-Centre for Cellular and Molecular Biology | CSIR-Centre for Cellular and Molecular Biology | Payel Mukherjee et al |
| EPI_ISL_447887 | USA/CA-CZB-1136/2020 | 2020-03-27 | North America/USA/California/Yolo Cou | University of California, Davis | Chan-Zuckerberg Biohub | CZB Cliahub Consortium et al |
| EPI_ISL_447904 | Bangladesh/NIB-01/2020 | 2020-05-11 | Asia/Bangladesh/Bangladesh/ | National Institute of Biotechnology | National Institute of Biotechnology | Md. Moniruzzaman et al |
| EPI_ISL_447909 | Thailand/Nonthaburi_59/2020 | 2020-01-13 | Asia/Thailand/Nonthaburi/ | n/a | National Institute of Health. Department of medical Sciences, Ministry of Public Health, Thailand | Pilailuk et al |

| | | | | | | |
|---|---|---|---|---|---|---|
| EPI_ISL_447910 | Thailand/Nonthaburi_68/2020 | 2020-01-15 | Asia/Thailand/Nonthaburi/ | n/a | National Institute of Health. Department of medical Sciences, Ministry of Public Health, Thailand | Pilailuk et al |
| EPI_ISL_447911 | Thailand/Nonthaburi_193/2020 | 2020-01-24 | Asia/Thailand/Nonthaburi/ | n/a | National Institute of Health. Department of medical Sciences, Ministry of Public Health, Thailand | Pilailuk et al |
| EPI_ISL_447912 | Thailand/Bangkok_237/2020 | 2020-01-25 | Asia/Thailand/Bangkok/ | n/a | National Institute of Health. Department of medical Sciences, Ministry of Public Health, Thailand | Pilailuk et al |
| EPI_ISL_447913 | Thailand/Bangkok_238/2020 | 2020-01-31 | Asia/Thailand/Bangkok/ | n/a | National Institute of Health. Department of medical Sciences, Ministry of Public Health, Thailand | Pilailuk et al |
| EPI_ISL_447914 | Thailand/Phuket_247/2020 | 2020-01-25 | Asia/Thailand/Phuket/ | n/a | National Institute of Health. Department of medical Sciences, Ministry of Public Health, Thailand | Pilailuk et al |
| EPI_ISL_447915 | Thailand/Bangkok_269/2020 | 2020-01-26 | Asia/Thailand/Bangkok/ | n/a | National Institute of Health. Department of medical Sciences, Ministry of Public Health, Thailand | Pilailuk et al |

| | | | | | | |
|---|---|---|---|---|---|---|
| EPI_ISL_447916 | Thailand/Bangkok_323/2020 | 2020-01-29 | Asia/Thailand/Bangkok/ | n/a | National Institute of Health. Department of medical Sciences, Ministry of Public Health, Thailand | Pilailuk et al |
| EPI_ISL_447917 | Thailand/Nonthaburi_363/2020 | 2020-01-29 | Asia/Thailand/Nonthaburi/ | n/a | National Institute of Health. Department of medical Sciences, Ministry of Public Health, Thailand | Pilailuk et al |
| EPI_ISL_447918 | Thailand/Bangkok_580/2020 | 2020-02-01 | Asia/Thailand/Bangkok/ | n/a | National Institute of Health. Department of medical Sciences, Ministry of Public Health, Thailand | Pilailuk et al |
| EPI_ISL_447919 | Thailand/Samutprakarn_840/2020 | 2020-02-04 | Asia/Thailand/Samut Prakarn/ | n/a | National Institute of Health. Department of medical Sciences, Ministry of Public Health, Thailand | Pilailuk et al |
| EPI_ISL_447920 | Thailand/Bangkok_1716/2020 | 2020-02-25 | Asia/Thailand/Bangkok/ | n/a | National Institute of Health. Department of medical Sciences, Ministry of Public Health, Thailand | Pilailuk et al |
| EPI_ISL_447935 | England/BIRM-5EC3C/2020 | 2020-04-28 | Europe/United Kingdom/England/ | University of Birmingham | COVID-19 Genomics UK (COG-UK) Consortium | Claire McMurray et al |
| EPI_ISL_447954 | England/CAMB-1ACFEA/2020 | 2020-04-29 | Europe/United Kingdom/England/ | Department of Pathology, University of Cambridge | COVID-19 Genomics UK (COG-UK) Consortium | Luke W Meredith et al |
| EPI_ISL_447958 | England/CAMB-1AD031/2020 | 2020-04-29 | Europe/United Kingdom/England/ | Department of Pathology, University of Cambridge | COVID-19 Genomics UK (COG-UK) Consortium | Luke W Meredith et al |

| | | | | | | |
|---|---|---|---|---|---|---|
| EPI_ISL_447990 | England/CAMB-1AD602/2020 | 2020-04-30 | Europe/United Kingdom/England/ | Department of Pathology, University of Cambridge | COVID-19 Genomics UK (COG-UK) Consortium | Luke W Meredith et al |
| EPI_ISL_448100 | England/CAMB-1B003B/2020 | 2020-05-08 | Europe/United Kingdom/England/ | Department of Pathology, University of Cambridge | COVID-19 Genomics UK (COG-UK) Consortium | Luke W Meredith et al |
| EPI_ISL_448205 | Scotland/CVR2893/2020 | 2020-04-21 | Europe/United Kingdom/Scotland/ | West of Scotland Specialist Virology Centre, NHSGGC / MRC-University of Glasgow Centre for Virus Research | COVID-19 Genomics UK (COG-UK) Consortium | Ana da Silva Filipe et al |
| EPI_ISL_448352 | England/NORW-E9B48/2020 | 2020-04-30 | Europe/United Kingdom/England/ | Quadram Institute Bioscience | COVID-19 Genomics UK (COG-UK) Consortium | Dave J. Baker et al |
| EPI_ISL_448365 | England/NORW-E9D8E/2020 | 2020-05-02 | Europe/United Kingdom/England/ | Quadram Institute Bioscience | COVID-19 Genomics UK (COG-UK) Consortium | Dave J. Baker et al |
| EPI_ISL_448401 | England/NOTT-110C9F/2020 | 2020-05-01 | Europe/United Kingdom/England/ | Queens Medical Centre, Clinical Microbiology Department / DeepSeq Nottingham | COVID-19 Genomics UK (COG-UK) Consortium | Gemma Clark et al |
| EPI_ISL_448422 | England/NOTT-110E02/2020 | 2020-04-29 | Europe/United Kingdom/England/ | Queens Medical Centre, Clinical Microbiology Department / DeepSeq Nottingham | COVID-19 Genomics UK (COG-UK) Consortium | Gemma Clark et al |
| EPI_ISL_448507 | England/OXON-ACADB/2020 | 2020-04-05 | Europe/United Kingdom/England/ | Oxford Viromics, NDM, University of Oxford; Oxford University Hospitals; Basingstoke and North Hampshire Hospital | COVID-19 Genomics UK (COG-UK) Consortium | Tanya Golubchik et al |
| EPI_ISL_448537 | England/OXON-ACDA5/2020 | 2020-04-06 | Europe/United Kingdom/England/ | Oxford Viromics, NDM, University of Oxford; Oxford University Hospitals; Basingstoke and North Hampshire Hospital | COVID-19 Genomics UK (COG-UK) Consortium | Tanya Golubchik et al |
| EPI_ISL_448778 | England/OXON-AECD3/2020 | 2020-04-08 | Europe/United Kingdom/England/ | Oxford Viromics, NDM, University of Oxford; Oxford University Hospitals; Basingstoke and North Hampshire Hospital | COVID-19 Genomics UK (COG-UK) Consortium | Tanya Golubchik et al |
| EPI_ISL_448820 | England/OXON-AF249/2020 | 2020-03-24 | Europe/United Kingdom/England/ | Oxford Viromics, NDM, University of Oxford; Oxford University Hospitals; Basingstoke and North Hampshire Hospital | COVID-19 Genomics UK (COG-UK) Consortium | Tanya Golubchik et al |
| EPI_ISL_448823 | England/OXON-AF2C1/2020 | 2020-03-24 | Europe/United Kingdom/England/ | Oxford Viromics, NDM, University of Oxford; Oxford University Hospitals; Basingstoke and North Hampshire Hospital | COVID-19 Genomics UK (COG-UK) Consortium | Tanya Golubchik et al |
| EPI_ISL_448892 | England/SHEF-CECFB/2020 | 2020-04-16 | Europe/United Kingdom/England/ | Virology Department, Sheffield Teaching Hospitals NHS Foundation Trust/Department of Infection, Immunity and Cardiovascular Disease, The Medical School, University of Sheffield | COVID-19 Genomics UK (COG-UK) Consortium | Thushan de Silva et al |

| | | | | | | |
|---|---|---|---|---|---|---|
| EPI_ISL_449036 | England/NORW-E744F/2020 | 2020-05-03 | Europe/United Kingdom/England/ | Quadram Institute Bioscience | COVID-19 Genomics UK (COG-UK) Consortium | Dave J. Baker et al |
| EPI_ISL_449046 | England/NORW-E753D/2020 | 2020-05-04 | Europe/United Kingdom/England/ | Quadram Institute Bioscience | COVID-19 Genomics UK (COG-UK) Consortium | Dave J. Baker et al |
| EPI_ISL_449166 | England/NORW-E85F0/2020 | 2020-04-18 | Europe/United Kingdom/England/ | Quadram Institute Bioscience | COVID-19 Genomics UK (COG-UK) Consortium | Dave J. Baker et al |
| EPI_ISL_449191 | Scotland/CVR3154/2020 | 2020-04-22 | Europe/United Kingdom/Scotland/ | West of Scotland Specialist Virology Centre, NHSGGC / MRC-University of Glasgow Centre for Virus Research | COVID-19 Genomics UK (COG-UK) Consortium | Ana da Silva Filipe et al |
| EPI_ISL_449274 | Scotland/EDB4098/2020 | 2020-04-16 | Europe/United Kingdom/Scotland/ | Virology Department, Royal Infirmary of Edinburgh, NHS Lothian / School of Biological Sciences, University of Edinburgh / Institute of Genetics and Molecular Medicine, University of Edinburgh | COVID-19 Genomics UK (COG-UK) Consortium | McHugh M et al |
| EPI_ISL_449289 | Scotland/EDB4279/2020 | 2020-04-20 | Europe/United Kingdom/Scotland/ | Virology Department, Royal Infirmary of Edinburgh, NHS Lothian / School of Biological Sciences, University of Edinburgh / Institute of Genetics and Molecular Medicine, University of Edinburgh | COVID-19 Genomics UK (COG-UK) Consortium | McHugh M et al |
| EPI_ISL_449327 | Scotland/EDB4711/2020 | 2020-04-27 | Europe/United Kingdom/Scotland/ | Virology Department, Royal Infirmary of Edinburgh, NHS Lothian / School of Biological Sciences, University of Edinburgh / Institute of Genetics and Molecular Medicine, University of Edinburgh | COVID-19 Genomics UK (COG-UK) Consortium | McHugh M et al |
| EPI_ISL_449358 | England/LIVE-9A672/2020 | 2020-03-28 | Europe/United Kingdom/England/ | Liverpool Clinical Laboratories | COVID-19 Genomics UK (COG-UK) Consortium | Sam Haldenby et al |
| EPI_ISL_449411 | England/LIVE-9B107/2020 | 2020-03-25 | Europe/United Kingdom/England/ | Liverpool Clinical Laboratories | COVID-19 Genomics UK (COG-UK) Consortium | Sam Haldenby et al |
| EPI_ISL_449476 | Wuhan/WHUH001/2020 | 2020-01-22 | Asia/China/Hubei/Wuhan | unknown | Department of Respiratory and Critical Care | Wang et al |
| EPI_ISL_449477 | Wuhan/WHUH002/2020 | 2020-01-22 | Asia/China/Hubei/Wuhan | unknown | Department of Respiratory and Critical Care | Wang et al |

| | | | | | | |
|---|---|---|---|---|---|---|
| EPI_ISL_449482 | Wuhan/WHUH007/2020 | 2020-01-22 | Asia/China/Hubei/Wuhan | unknown | Department of Respiratory and Critical Care | Wang et al |
| EPI_ISL_449484 | Wuhan/WHUH011/2020 | 2020-01-22 | Asia/China/Hubei/Wuhan | unknown | Department of Respiratory and Critical Care | Wang et al |
| EPI_ISL_449485 | Wuhan/WHUH012/2020 | 2020-01-22 | Asia/China/Hubei/Wuhan | unknown | Department of Respiratory and Critical Care | Wang et al |
| EPI_ISL_449486 | Wuhan/WHUH020/2020 | 2020-01-22 | Asia/China/Hubei/Wuhan | unknown | Department of Respiratory and Critical Care | Wang et al |
| EPI_ISL_449487 | Wuhan/WHUH021/2020 | 2020-01-22 | Asia/China/Hubei/Wuhan | unknown | Department of Respiratory and Critical Care | Wang et al |
| EPI_ISL_449495 | England/LIVE-9BFA3/2020 | 2020-03-23 | Europe/United Kingdom/England/ | Liverpool Clinical Laboratories | COVID-19 Genomics UK (COG-UK) Consortium | Sam Haldenby et al |
| EPI_ISL_449676 | England/LOND-D4CD4/2020 | 2020-04-15 | Europe/United Kingdom/England/ | University College London, Great Ormond Street Hospital for Children NHS Foundation Trust, Imperial College Healthcare NHS Trust | COVID-19 Genomics UK (COG-UK) Consortium | Sergi Castellano et al |
| EPI_ISL_449720 | England/LOND-D5796/2020 | 2020-04-17 | Europe/United Kingdom/England/ | University College London, Great Ormond Street Hospital for Children NHS Foundation Trust, Imperial College Healthcare NHS Trust | COVID-19 Genomics UK (COG-UK) Consortium | Sergi Castellano et al |
| EPI_ISL_449721 | England/LOND-D57A5/2020 | 2020-04-17 | Europe/United Kingdom/England/ | University College London, Great Ormond Street Hospital for Children NHS Foundation Trust, Imperial College Healthcare NHS Trust | COVID-19 Genomics UK (COG-UK) Consortium | Sergi Castellano et al |
| EPI_ISL_449818 | USA/UT-01566/2020 | 2020-04-07 | North America/USA/Utah/ | Utah Public Health Laboratory | Utah Public Health Laboratory | Erin Young et al |
| EPI_ISL_449821 | USA/UT-01572/2020 | 2020-04-07 | North America/USA/Utah/ | Utah Public Health Laboratory | Utah Public Health Laboratory | Erin Young et al |
| EPI_ISL_449828 | USA/UT-01583/2020 | 2020-04-07 | North America/USA/Utah/ | Utah Public Health Laboratory | Utah Public Health Laboratory | Erin Young et al |
| EPI_ISL_449829 | USA/UT-01585/2020 | 2020-04-07 | North America/USA/Utah/ | Utah Public Health Laboratory | Utah Public Health Laboratory | Erin Young et al |
| EPI_ISL_449837 | USA/UT-01607/2020 | 2020-04-06 | North America/USA/Utah/ | Utah Public Health Laboratory | Utah Public Health Laboratory | Erin Young et al |

| | | | | | | |
|---|---|---|---|---|---|---|
| EPI_ISL_449839 | USA/WA-S669/2020 | 2020-04-08 | North America/USA/Washington/Yakim | Washington State Department of Health | Seattle Flu Study | Chu et al et al |
| EPI_ISL_449844 | USA/WA-S674/2020 | 2020-04-08 | North America/USA/Washington/Yakim | Washington State Department of Health | Seattle Flu Study | Chu et al et al |
| EPI_ISL_449855 | USA/WA-S685/2020 | 2020-04-11 | North America/USA/Washington/Yakim | Washington State Department of Health | Seattle Flu Study | Chu et al et al |
| EPI_ISL_449856 | USA/WA-S686/2020 | 2020-04-08 | North America/USA/Washington/Yakim | Washington State Department of Health | Seattle Flu Study | Chu et al et al |
| EPI_ISL_449857 | USA/WA-S687/2020 | 2020-04-08 | North America/USA/Washington/Pierce | Washington State Department of Health | Seattle Flu Study | Chu et al et al |
| EPI_ISL_449858 | USA/WA-S688/2020 | 2020-04-08 | North America/USA/Washington/Pierce | Washington State Department of Health | Seattle Flu Study | Chu et al et al |
| EPI_ISL_449863 | USA/WA-S693/2020 | 2020-04-08 | North America/USA/Washington/Yakim | Washington State Department of Health | Seattle Flu Study | Chu et al et al |
| EPI_ISL_449867 | USA/WA-S697/2020 | 2020-04-08 | North America/USA/Washington/Yakim | Washington State Department of Health | Seattle Flu Study | Chu et al et al |
| EPI_ISL_449879 | USA/WA-S709/2020 | 2020-04-09 | North America/USA/Washington/Yakim | Washington State Department of Health | Seattle Flu Study | Chu et al et al |
| EPI_ISL_449886 | USA/WA-S716/2020 | 2020-04-11 | North America/USA/Washington/Pierce | Washington State Department of Health | Seattle Flu Study | Chu et al et al |
| EPI_ISL_449899 | USA/WA-S729/2020 | 2020-04-11 | North America/USA/Washington/King C | Washington State Department of Health | Seattle Flu Study | Chu et al et al |
| EPI_ISL_449902 | USA/WA-S732/2020 | 2020-04-13 | North America/USA/Washington/Yakim | Washington State Department of Health | Seattle Flu Study | Chu et al et al |
| EPI_ISL_449906 | USA/WA-S736/2020 | 2020-04-13 | North America/USA/Washington/Yakim | Washington State Department of Health | Seattle Flu Study | Chu et al et al |
| EPI_ISL_449920 | USA/WA-S750/2020 | 2020-04-13 | North America/USA/Washington/Snoho | Washington State Department of Health | Seattle Flu Study | Chu et al et al |
| EPI_ISL_449926 | USA/WA-S756/2020 | 2020-04-14 | North America/USA/Washington/Yakim | Washington State Department of Health | Seattle Flu Study | Chu et al et al |
| EPI_ISL_449927 | USA/WA-S757/2020 | 2020-04-13 | North America/USA/Washington/Yakim | Washington State Department of Health | Seattle Flu Study | Chu et al et al |
| EPI_ISL_449939 | USA/WA-S769/2020 | 2020-04-14 | North America/USA/Washington/Snoho | Washington State Department of Health | Seattle Flu Study | Chu et al et al |
| EPI_ISL_449946 | USA/WA-S776/2020 | 2020-04-14 | North America/USA/Washington/King C | Washington State Department of Health | Seattle Flu Study | Chu et al et al |
| EPI_ISL_449947 | USA/WA-S777/2020 | 2020-04-14 | North America/USA/Washington/King C | Washington State Department of Health | Seattle Flu Study | Chu et al et al |
| EPI_ISL_449948 | USA/WA-S778/2020 | 2020-04-14 | North America/USA/Washington/King C | Washington State Department of Health | Seattle Flu Study | Chu et al et al |
| EPI_ISL_449957 | USA/WA-S787/2020 | 2020-04-14 | North America/USA/Washington/Yakim | Washington State Department of Health | Seattle Flu Study | Chu et al et al |
| EPI_ISL_449958 | USA/WA-S788/2020 | 2020-04-14 | North America/USA/Washington/Yakim | Washington State Department of Health | Seattle Flu Study | Chu et al et al |
| EPI_ISL_449978 | USA/WA-S808/2020 | 2020-04-15 | North America/USA/Washington/Yakim | Washington State Department of Health | Seattle Flu Study | Chu et al et al |
| EPI_ISL_449980 | USA/WA-S810/2020 | 2020-04-15 | North America/USA/Washington/Yakim | Washington State Department of Health | Seattle Flu Study | Chu et al et al |
| EPI_ISL_449982 | USA/WA-S812/2020 | 2020-04-15 | North America/USA/Washington/Yakim | Washington State Department of Health | Seattle Flu Study | Chu et al et al |
| EPI_ISL_449984 | USA/WA-S814/2020 | 2020-04-17 | North America/USA/Washington/Yakim | Washington State Department of Health | Seattle Flu Study | Chu et al et al |
| EPI_ISL_449989 | USA/WA-S819/2020 | 2020-04-15 | North America/USA/Washington/Asotir | Washington State Department of Health | Seattle Flu Study | Chu et al et al |
| EPI_ISL_449991 | USA/WA-S821/2020 | 2020-04-16 | North America/USA/Washington/Yakim | Washington State Department of Health | Seattle Flu Study | Chu et al et al |
| EPI_ISL_449994 | USA/WA-S824/2020 | 2020-04-17 | North America/USA/Washington/Yakim | Washington State Department of Health | Seattle Flu Study | Chu et al et al |
| EPI_ISL_450000 | USA/WA-S830/2020 | 2020-04-16 | North America/USA/Washington/Snoho | Washington State Department of Health | Seattle Flu Study | Chu et al et al |
| EPI_ISL_450001 | USA/WA-S831/2020 | 2020-04-16 | North America/USA/Washington/Whatc | Washington State Department of Health | Seattle Flu Study | Chu et al et al |

| | | | | | | |
|---|---|---|---|---|---|---|
| EPI_ISL_450003 | USA/WA-S833/2020 | 2020-04-16 | North America/USA/Washington/King C | Washington State Department of Health | Seattle Flu Study | Chu et al et al |
| EPI_ISL_450021 | USA/NY-PV09333/2020 | 2020-03-18 | North America/USA/New York/Brooklyn | MSHS Clinical Microbiology Laboratories | MSHS Pathogen Surveillance Program | Ana S. Gonzalez-Reiche et al |
| EPI_ISL_450026 | USA/NY-PV09338/2020 | 2020-03-24 | North America/USA/New York/Nassau C | MSHS Clinical Microbiology Laboratories | MSHS Pathogen Surveillance Program | Ana S. Gonzalez-Reiche et al |
| EPI_ISL_450042 | USA/NY-PV09354/2020 | 2020-03-24 | North America/USA/New York/Queens | MSHS Clinical Microbiology Laboratories | MSHS Pathogen Surveillance Program | Ana S. Gonzalez-Reiche et al |
| EPI_ISL_450058 | USA/NY-PV09374/2020 | 2020-03-19 | North America/USA/New York/Manhatt | MSHS Clinical Microbiology Laboratories | MSHS Pathogen Surveillance Program | Ana S. Gonzalez-Reiche et al |
| EPI_ISL_450060 | USA/NY-PV09377/2020 | 2020-03-19 | North America/USA/New York/Manhatt | MSHS Clinical Microbiology Laboratories | MSHS Pathogen Surveillance Program | Ana S. Gonzalez-Reiche et al |
| EPI_ISL_450061 | USA/NY-PV09378/2020 | 2020-03-19 | North America/USA/New York/Queens | MSHS Clinical Microbiology Laboratories | MSHS Pathogen Surveillance Program | Ana S. Gonzalez-Reiche et al |
| EPI_ISL_450067 | USA/NY-PV09384/2020 | 2020-03-19 | North America/USA/New York/Bronx | MSHS Clinical Microbiology Laboratories | MSHS Pathogen Surveillance Program | Ana S. Gonzalez-Reiche et al |
| EPI_ISL_450076 | USA/NY-PV09395/2020 | 2020-03-19 | North America/USA/New York/Bronx | MSHS Clinical Microbiology Laboratories | MSHS Pathogen Surveillance Program | Ana S. Gonzalez-Reiche et al |
| EPI_ISL_450079 | USA/NY-PV09398/2020 | 2020-03-19 | North America/USA/New York/Manhatt | MSHS Clinical Microbiology Laboratories | MSHS Pathogen Surveillance Program | Ana S. Gonzalez-Reiche et al |
| EPI_ISL_450081 | USA/NY-PV09400/2020 | 2020-03-19 | North America/USA/New York/Manhatt | MSHS Clinical Microbiology Laboratories | MSHS Pathogen Surveillance Program | Ana S. Gonzalez-Reiche et al |
| EPI_ISL_450089 | USA/NY-PV09409/2020 | 2020-03-11 | North America/USA/New York/Manhatt | MSHS Clinical Microbiology Laboratories | MSHS Pathogen Surveillance Program | Ana S. Gonzalez-Reiche et al |
| EPI_ISL_450100 | USA/NY-PV09421/2020 | 2020-03-13 | North America/USA/New York/Manhatt | MSHS Clinical Microbiology Laboratories | MSHS Pathogen Surveillance Program | Ana S. Gonzalez-Reiche et al |
| EPI_ISL_450101 | USA/NY-PV09422/2020 | 2020-03-13 | North America/USA/New York/Brooklyn | MSHS Clinical Microbiology Laboratories | MSHS Pathogen Surveillance Program | Ana S. Gonzalez-Reiche et al |

| | | | | | | |
|---|---|---|---|---|---|---|
| EPI_ISL_450110 | USA/NY-PV09433/2020 | 2020-03-11 | North America/USA/New York/Manhatt | MSHS Clinical Microbiology Laboratories | MSHS Pathogen Surveillance Program | Ana S. Gonzalez-Reiche et al |
| EPI_ISL_450112 | USA/NY-PV09435/2020 | 2020-03-13 | North America/USA/New York/Manhatt | MSHS Clinical Microbiology Laboratories | MSHS Pathogen Surveillance Program | Ana S. Gonzalez-Reiche et al |
| EPI_ISL_450124 | USA/NY-PV09450/2020 | 2020-03-19 | North America/USA/New York/Manhatt | MSHS Clinical Microbiology Laboratories | MSHS Pathogen Surveillance Program | Ana S. Gonzalez-Reiche et al |
| EPI_ISL_450196 | Thailand/Bangkok_168/2020 | 2020-01-23 | Asia/Thailand/Bangkok/ | bumrungrad international hospital | National Institute of Health. Department of medical Sciences, Ministry of Public Health, Thailand | Pilailuk et al |
| EPI_ISL_450198 | Germany/BY-ChVir-929-2/2020 | 2020-01-28 | Europe/Germany/Bavaria/ | Department of Virology | Department of Virology | Boehmer et al |
| EPI_ISL_450199 | Germany/BY-ChVir-1019/2020 | 2020-01-29 | Europe/Germany/Bavaria/ | Department of Virology | Department of Virology | Boehmer et al |
| EPI_ISL_450200 | Germany/BY-ChVir-1020/2020 | 2020-01-28 | Europe/Germany/Bavaria/ | Department of Virology | Department of Virology | Boehmer et al |
| EPI_ISL_450203 | Germany/BY-ChVir-1289/2020 | 2020-02-01 | Europe/Germany/Bavaria/ | Department of Virology | Department of Virology | Boehmer et al |
| EPI_ISL_450204 | Germany/BY-ChVir-1356/2020 | 2020-02-03 | Europe/Germany/Bavaria/ | Department of Virology | Department of Virology | Boehmer et al |
| EPI_ISL_450207 | Germany/BY-ChVir-1482/2020 | 2020-02-07 | Europe/Germany/Bavaria/ | Department of Virology | Department of Virology | Boehmer et al |
| EPI_ISL_450208 | Germany/BY-ChVir-1483/2020 | 2020-02-08 | Europe/Germany/Bavaria/ | Department of Virology | Department of Virology | Boehmer et al |
| EPI_ISL_450209 | Germany/BY-ChVir-1017/2020 | 2020-01-30 | Europe/Germany/Bavaria/ | Department of Virology | Department of Virology | Boehmer et al |
| EPI_ISL_450241 | Russia/StPetersburg-RII6251S/2020 | 2020-04-15 | Europe/Russia/Saint-Petersburg/ | WHO National Influenza Centre Russian Federation | WHO National Influenza Centre Russian Federation | Andrey Komissarov et al |
| EPI_ISL_450242 | Russia/StPetersburg-RII6704V/2020 | 2020-04-16 | Europe/Russia/Saint-Petersburg/ | WHO National Influenza Centre Russian Federation | WHO National Influenza Centre Russian Federation | Andrey Komissarov et al |

| | | | | | | |
|---|---|---|---|---|---|---|
| EPI_ISL_450244 | Russia/StPetersburg-RII6840S/2020 | 2020-04-15 | Europe/Russia/Saint-Petersburg/ | WHO National Influenza Centre Russian Federation | WHO National Influenza Centre Russian Federation | Andrey Komissarov et al |
| EPI_ISL_450246 | Russia/Kipen-RII6907V/2020 | 2020-04-19 | Europe/Russia/Leningrad Oblast/Kipen | WHO National Influenza Centre Russian Federation | WHO National Influenza Centre Russian Federation | Andrey Komissarov et al |
| EPI_ISL_450248 | Russia/StPetersburg-RII7318V/2020 | 2020-04-21 | Europe/Russia/Saint-Petersburg/ | WHO National Influenza Centre Russian Federation | WHO National Influenza Centre Russian Federation | Andrey Komissarov et al |
| EPI_ISL_450249 | Russia/StPetersburg-RII7319V/2020 | 2020-04-21 | Europe/Russia/Saint-Petersburg/ | WHO National Influenza Centre Russian Federation | WHO National Influenza Centre Russian Federation | Andrey Komissarov et al |
| EPI_ISL_450251 | Russia/StPetersburg-RII7340V/2020 | 2020-04-20 | Europe/Russia/Saint-Petersburg/ | WHO National Influenza Centre Russian Federation | WHO National Influenza Centre Russian Federation | Andrey Komissarov et al |
| EPI_ISL_450252 | Russia/StPetersburg-RII7464S/2020 | 2020-04-20 | Europe/Russia/Saint-Petersburg/ | WHO National Influenza Centre Russian Federation | WHO National Influenza Centre Russian Federation | Andrey Komissarov et al |
| EPI_ISL_450253 | Russia/StPetersburg-RII7488S/2020 | 2020-04-20 | Europe/Russia/Saint-Petersburg/ | WHO National Influenza Centre Russian Federation | WHO National Influenza Centre Russian Federation | Andrey Komissarov et al |
| EPI_ISL_450254 | Russia/StPetersburg-RII7501S/2020 | 2020-04-20 | Europe/Russia/Saint-Petersburg/ | WHO National Influenza Centre Russian Federation | WHO National Influenza Centre Russian Federation | Andrey Komissarov et al |
| EPI_ISL_450255 | Russia/StPetersburg-RII7505S/2020 | 2020-04-20 | Europe/Russia/Saint-Petersburg/ | WHO National Influenza Centre Russian Federation | WHO National Influenza Centre Russian Federation | Andrey Komissarov et al |
| EPI_ISL_450256 | Russia/StPetersburg-RII7520S/2020 | 2020-04-20 | Europe/Russia/Saint-Petersburg/ | WHO National Influenza Centre Russian Federation | WHO National Influenza Centre Russian Federation | Andrey Komissarov et al |

| | | | | | | |
|---|---|---|---|---|---|---|
| EPI_ISL_450257 | Russia/StPetersburg-RII7545S/2020 | 2020-04-20 | Europe/Russia/Saint-Petersburg/ | WHO National Influenza Centre Russian Federation | WHO National Influenza Centre Russian Federation | Andrey Komissarov et al |
| EPI_ISL_450259 | Russia/StPetersburg-RII7555S/2020 | 2020-04-21 | Europe/Russia/Saint-Petersburg/ | WHO National Influenza Centre Russian Federation | WHO National Influenza Centre Russian Federation | Andrey Komissarov et al |
| EPI_ISL_450260 | Russia/StPetersburg-RII7557S/2020 | 2020-04-21 | Europe/Russia/Saint-Petersburg/ | WHO National Influenza Centre Russian Federation | WHO National Influenza Centre Russian Federation | Andrey Komissarov et al |
| EPI_ISL_450261 | Russia/StPetersburg-RII7581S/2020 | 2020-04-21 | Europe/Russia/Saint-Petersburg/ | WHO National Influenza Centre Russian Federation | WHO National Influenza Centre Russian Federation | Andrey Komissarov et al |
| EPI_ISL_450263 | Russia/StPetersburg-RII7603S/2020 | 2020-04-21 | Europe/Russia/Saint-Petersburg/ | WHO National Influenza Centre Russian Federation | WHO National Influenza Centre Russian Federation | Andrey Komissarov et al |
| EPI_ISL_450265 | Russia/StPetersburg-RII7658S/2020 | 2020-04-21 | Europe/Russia/Saint-Petersburg/ | WHO National Influenza Centre Russian Federation | WHO National Influenza Centre Russian Federation | Andrey Komissarov et al |
| EPI_ISL_450266 | Russia/StPetersburg-RII7880S/2020 | 2020-04-22 | Europe/Russia/Saint-Petersburg/ | WHO National Influenza Centre Russian Federation | WHO National Influenza Centre Russian Federation | Andrey Komissarov et al |
| EPI_ISL_450267 | Russia/StPetersburg-RII7896S/2020 | 2020-04-21 | Europe/Russia/Saint-Petersburg/ | WHO National Influenza Centre Russian Federation | WHO National Influenza Centre Russian Federation | Andrey Komissarov et al |
| EPI_ISL_450269 | Russia/StPetersburg-RII8289S/2020 | 2020-04-21 | Europe/Russia/Saint-Petersburg/ | WHO National Influenza Centre Russian Federation | WHO National Influenza Centre Russian Federation | Andrey Komissarov et al |
| EPI_ISL_450272 | Russia/StPetersburg-RII8319S/2020 | 2020-04-21 | Europe/Russia/Saint-Petersburg/ | WHO National Influenza Centre Russian Federation | WHO National Influenza Centre Russian Federation | Andrey Komissarov et al |

| | | | | | | |
|---|---|---|---|---|---|---|
| EPI_ISL_450273 | Russia/StPetersburg-RII8350S/2020 | 2020-04-22 | Europe/Russia/Saint-Petersburg/ | WHO National Influenza Centre Russian Federation | WHO National Influenza Centre Russian Federation | Andrey Komissarov et al |
| EPI_ISL_450274 | Russia/StPetersburg-RII8371S/2020 | 2020-04-22 | Europe/Russia/Saint-Petersburg/ | WHO National Influenza Centre Russian Federation | WHO National Influenza Centre Russian Federation | Andrey Komissarov et al |
| EPI_ISL_450277 | Russia/StPetersburg-RII8372S/2020 | 2020-04-22 | Europe/Russia/Saint-Petersburg/ | WHO National Influenza Centre Russian Federation | WHO National Influenza Centre Russian Federation | Andrey Komissarov et al |
| EPI_ISL_450279 | Russia/StPetersburg-RII8393S/2020 | 2020-04-22 | Europe/Russia/Saint-Petersburg/ | WHO National Influenza Centre Russian Federation | WHO National Influenza Centre Russian Federation | Andrey Komissarov et al |
| EPI_ISL_450280 | Russia/StPetersburg-RII8397S/2020 | 2020-04-22 | Europe/Russia/Saint-Petersburg/ | WHO National Influenza Centre Russian Federation | WHO National Influenza Centre Russian Federation | Andrey Komissarov et al |
| EPI_ISL_450281 | Russia/StPetersburg-RII8402S/2020 | 2020-04-22 | Europe/Russia/Saint-Petersburg/ | WHO National Influenza Centre Russian Federation | WHO National Influenza Centre Russian Federation | Andrey Komissarov et al |
| EPI_ISL_450282 | Russia/StPetersburg-RII8409S/2020 | 2020-04-22 | Europe/Russia/Saint-Petersburg/ | WHO National Influenza Centre Russian Federation | WHO National Influenza Centre Russian Federation | Andrey Komissarov et al |
| EPI_ISL_450284 | Russia/StPetersburg-RII8418S/2020 | 2020-04-22 | Europe/Russia/Saint-Petersburg/ | WHO National Influenza Centre Russian Federation | WHO National Influenza Centre Russian Federation | Andrey Komissarov et al |
| EPI_ISL_450285 | Russia/StPetersburg-RII8425S/2020 | 2020-04-22 | Europe/Russia/Saint-Petersburg/ | WHO National Influenza Centre Russian Federation | WHO National Influenza Centre Russian Federation | Andrey Komissarov et al |
| EPI_ISL_450287 | Russia/StPetersburg-RII8429S/2020 | 2020-04-22 | Europe/Russia/Saint-Petersburg/ | WHO National Influenza Centre Russian Federation | WHO National Influenza Centre Russian Federation | Andrey Komissarov et al |

| | | | | | | |
|---|---|---|---|---|---|---|
| EPI_ISL_450288 | Russia/StPetersburg-RII8876S/2020 | 2020-04-23 | Europe/Russia/Saint-Petersburg/ | WHO National Influenza Centre Russian Federation | WHO National Influenza Centre Russian Federation | Andrey Komissarov et al |
| EPI_ISL_450289 | Russia/StPetersburg-RII8890S/2020 | 2020-04-23 | Europe/Russia/Saint-Petersburg/ | WHO National Influenza Centre Russian Federation | WHO National Influenza Centre Russian Federation | Andrey Komissarov et al |
| EPI_ISL_450290 | Russia/StPetersburg-RII8913S/2020 | 2020-04-23 | Europe/Russia/Saint-Petersburg/ | WHO National Influenza Centre Russian Federation | WHO National Influenza Centre Russian Federation | Andrey Komissarov et al |
| EPI_ISL_450292 | Russia/StPetersburg-RII8935S/2020 | 2020-04-21 | Europe/Russia/Saint-Petersburg/ | WHO National Influenza Centre Russian Federation | WHO National Influenza Centre Russian Federation | Andrey Komissarov et al |
| EPI_ISL_450293 | Russia/StPetersburg-RII8955S/2020 | 2020-04-22 | Europe/Russia/Saint-Petersburg/ | WHO National Influenza Centre Russian Federation | WHO National Influenza Centre Russian Federation | Andrey Komissarov et al |
| EPI_ISL_450301 | SouthAfrica/R07719/2020 | 2020-04-01 | Africa/South Africa/South Africa/ | National Institute for Communicable Diseases of the National Health Laboratory Service | National Institute for Communicable Diseases of the National Health Laboratory Service | Allam M et al |
| EPI_ISL_450302 | Canada/Qc-L00240556/2020 | 2020-03-27 | North America/Canada/Quebec/ | Centre hospitalier Anna-Laberge | Laboratoire de santv© publique du Quv©bec | Sandrine Moreira et al |
| EPI_ISL_450303 | Canada/Qc-L00240587/2020 | 2020-03-27 | North America/Canada/Quebec/Montv© | Centre hospitalier Anna-Laberge | Laboratoire de santv© publique du Quv©bec | Sandrine Moreira et al |
| EPI_ISL_450304 | Canada/Qc-L00240652/2020 | 2020-03-27 | North America/Canada/Quebec/Montv© | Hv¥pital Charles-LeMoyne | Laboratoire de santv© publique du Quv©bec | Sandrine Moreira et al |
| EPI_ISL_450305 | Canada/Qc-L00240685/2020 | 2020-03-27 | North America/Canada/Quebec/Montv© | Hv¥pital Charles-LeMoyne | Laboratoire de santv© publique du Quv©bec | Sandrine Moreira et al |

| | | | | | | |
|---|---|---|---|---|---|---|
| EPI_ISL_450306 | Canada/Qc-L00240857/2020 | 2020-03-27 | North America/Canada/Quebec/Montv© | CSSS Haut-Richelieu/Rouville (Hv¥pital) | Laboratoire de santv© publique du Quv©bec | Sandrine Moreira et al |
| EPI_ISL_450307 | Canada/Qc-L00240954/2020 | 2020-03-27 | North America/Canada/Quebec/Montv© | Hv¥pital du SurovÆt | Laboratoire de santv© publique du Quv©bec | Sandrine Moreira et al |
| EPI_ISL_450308 | Canada/Qc-L00240979/2020 | 2020-03-27 | North America/Canada/Quebec/Montv© | Hv¥pital Charles-LeMoyne | Laboratoire de santv© publique du Quv©bec | Sandrine Moreira et al |
| EPI_ISL_450309 | Canada/Qc-L00241026/2020 | 2020-03-27 | North America/Canada/Quebec/Montv© | Hv¥pital Pierre-Boucher | Laboratoire de santv© publique du Quv©bec | Sandrine Moreira et al |
| EPI_ISL_450310 | Canada/Qc-L00241042/2020 | 2020-03-27 | North America/Canada/Quebec/Montv© | Hv¥pital Charles-LeMoyne | Laboratoire de santv© publique du Quv©bec | Sandrine Moreira et al |
| EPI_ISL_450311 | Canada/Qc-L00241168/2020 | 2020-03-27 | North America/Canada/Quebec/Montv© | Hv¥pital du SurovÆt | Laboratoire de santv© publique du Quv©bec | Sandrine Moreira et al |
| EPI_ISL_450312 | Canada/Qc-L00241192/2020 | 2020-03-27 | North America/Canada/Quebec/Montv© | Hv¥pital du SurovÆt | Laboratoire de santv© publique du Quv©bec | Sandrine Moreira et al |
| EPI_ISL_450313 | Canada/Qc-L00241205/2020 | 2020-03-27 | North America/Canada/Quebec/Montv© | Hv¥pital du SurovÆt | Laboratoire de santv© publique du Quv©bec | Sandrine Moreira et al |
| EPI_ISL_450314 | Canada/Qc-L00241208/2020 | 2020-03-27 | North America/Canada/Quebec/Montv© | Hv¥pital Pierre-Boucher | Laboratoire de santv© publique du Quv©bec | Sandrine Moreira et al |
| EPI_ISL_450315 | Canada/Qc-L00241245/2020 | 2020-03-27 | North America/Canada/Quebec/Montv© | Hv¥pital Pierre-Boucher | Laboratoire de santv© publique du Quv©bec | Sandrine Moreira et al |
| EPI_ISL_450316 | Canada/Qc-L00241292/2020 | 2020-03-27 | North America/Canada/Quebec/Lanaudi | Hv¥pital Charles-LeMoyne | Laboratoire de santv© publique du Quv©bec | Sandrine Moreira et al |
| EPI_ISL_450317 | Canada/Qc-L00241344/2020 | 2020-03-27 | North America/Canada/Quebec/Montv© | Hv¥pital Pierre-Boucher | Laboratoire de santv© publique du Quv©bec | Sandrine Moreira et al |
| EPI_ISL_450318 | Canada/Qc-L00241355/2020 | 2020-03-27 | North America/Canada/Quebec/Montv© | Hv¥pital Pierre-Boucher | Laboratoire de santv© publique du Quv©bec | Sandrine Moreira et al |

| | | | | | | |
|---|---|---|---|---|---|---|
| EPI_ISL_450338 | Poland/IHG_PAS_1_77/2020 | 2020-04-11 | Europe/Poland/Wielkopolskie/ | Institute of Human Genetics, Polish Academy of Sciences; Sanitary and Epidemiological Station in Pozna≈Ñ | Institute of Human Genetics, Polish Academy of Sciences | Szymon Hryhorowicz et al |
| EPI_ISL_450339 | Bangladesh/BARJ-CVASU-CTG-501/20 | 2020-05-10 | Asia/Bangladesh/Chattogram/ | Bangladesh Institute of Tropical & Infectious Diseases, COVID-19 Testing Laboratory | Basic and Applied Research on Jute Project | Rasel Ahmed et al |
| EPI_ISL_450340 | Bangladesh/BARJ-CVASU-CTG-502/20 | 2020-05-13 | Asia/Bangladesh/Chattogram/ | Bangladesh Institute of Tropical & Infectious Diseases, COVID-19 Testing Laboratory | Basic and Applied Research on Jute Project | Rasel Ahmed et al |
| EPI_ISL_450341 | Bangladesh/BARJ-CVASU-CTG-503/20 | 2020-05-13 | Asia/Bangladesh/Chattogram/ | Bangladesh Institute of Tropical & Infectious Diseases, COVID-19 Testing Laboratory | Basic and Applied Research on Jute Project | Md. Sabbir Hossain et al |
| EPI_ISL_450344 | Bangladesh/BARJ-CVASU-CTG-517/20 | 2020-05-03 | Asia/Bangladesh/Chattogram/ | Bangladesh Institute of Tropical & Infectious Diseases, COVID-19 Testing Laboratory | Basic and Applied Research on Jute Project | Rasel Ahmed et al |
| EPI_ISL_450345 | Bangladesh/BARJ-CVASU-CTG-518/20 | 2020-05-10 | Asia/Bangladesh/Chattogram/ | Bangladesh Institute of Tropical & Infectious Diseases, COVID-19 Testing Laboratory | Basic and Applied Research on Jute Project | Md. Sabbir Hossain et al |
| EPI_ISL_450346 | Norway/Trondheim-S4/2020 | 2020-03-25 | Europe/Norway/Trondheim/ | St.Olavs hospital/NTNU | Institute of Genomics Core Facility, University of Tartu | Aleksandr Ianevski et al |
| EPI_ISL_450347 | Norway/Trondheim-S5/2020 | 2020-03-25 | Europe/Norway/Trondheim/ | St.Olavs hospital/NTNU | Institute of Genomics Core Facility, University of Tartu | Aleksandr Ianevski et al |
| EPI_ISL_450352 | Norway/Trondheim-E10/2020 | 2020-03-11 | Europe/Norway/Trondheim/ | St.Olavs hospital/NTNU | Institute of Genomics Core Facility, University of Tartu | Aleksandr Ianevski et al |
| EPI_ISL_450401 | USA/NY-NYUMC684/2020 | 2020-04-01 | North America/USA/New York/Queens | NYU Langone Health | Departments of Pathology and Medicine, New York University School of Medicine | Maria Aguero-Rosenfeld et al |

| | | | | | | |
|---|---|---|---|---|---|---|
| EPI_ISL_450408 | HongKong/HKU-905a/2020 | 2020-01-24 | Asia/Hong Kong/Hong Kong/ | unknown | Microbiology | To et al |
| EPI_ISL_450409 | HongKong/HKU-911a/2020 | 2020-01-30 | Asia/Hong Kong/Hong Kong/ | unknown | Microbiology | To et al |
| EPI_ISL_450410 | HongKong/HKU-913a/2020 | 2020-01-31 | Asia/Hong Kong/Hong Kong/ | unknown | Microbiology | To et al |
| EPI_ISL_450411 | HongKong/HKU-915a/2020 | 2020-02-02 | Asia/Hong Kong/Hong Kong/ | unknown | Microbiology | To et al |
| EPI_ISL_450412 | HongKong/HKU-904a/2020 | 2020-01-30 | Asia/Hong Kong/Hong Kong/ | unknown | Microbiology | To et al |
| EPI_ISL_450413 | Germany/HH-UMC-01/2020 | 2020-03-19 | Europe/Germany/Hamburg/ | Center for Diagnostics, Institute of Medical Microbiology, Virology and Hygiene | University Medical Center Hamburg-Eppendorf | Huang et al |
| EPI_ISL_450442 | Henan/HN03/2020 | 2020-02-24 | Asia/China/Henan/ | The Department of Infectious Disease Prevention and Control, Henan Provincial Center for Disease Control and Prevention | The Department of Infectious Disease Prevention and Control, Henan Provincial Center for Disease Control and Prevention | Li et al |
| EPI_ISL_450444 | Beijing/IME-HZ01/2020 | 2020-01-22 | Asia/China/Beijing/ | 20 Dongda Street, Fengtai District, Beijing, Beijing 100071, China | Dept. OPA, Beijing Institute of Microbiology and Epidemiology | Zhang et al |
| EPI_ISL_450460 | USA/CA-CZB-1211/2020 | 2020-03-24 | North America/USA/California/Santa Cl | Stanford clinical virology lab | Chan-Zuckerberg Biohub | Benjamin Pinksy et al |
| EPI_ISL_450464 | USA/CA-CZB-1215/2020 | 2020-03-23 | North America/USA/California/Santa Cl | Stanford clinical virology lab | Chan-Zuckerberg Biohub | Benjamin Pinksy et al |
| EPI_ISL_450465 | USA/CA-CZB-1216/2020 | 2020-03-23 | North America/USA/California/Santa Cl | Stanford clinical virology lab | Chan-Zuckerberg Biohub | Benjamin Pinksy et al |
| EPI_ISL_450475 | USA/CA-CZB-1226/2020 | 2020-03-19 | North America/USA/California/Santa Cl | Stanford clinical virology lab | Chan-Zuckerberg Biohub | Benjamin Pinksy et al |
| EPI_ISL_450485 | USA/CA-CZB-59X001/2020 | 2020-03-24 | North America/USA/California/ | unknown | Data Science | Carroll et al |
| EPI_ISL_450489 | Wuhan/YB012504/2020 | 2020-01-25 | Asia/China/Hubei/Wuhan | Wuhan Institute of Virology, Chinese Academy of Sciences | Wuhan Institute of Virology, Chinese Academy of Sciences | Si et al |

| | | | | | | |
|---|---|---|---|---|---|---|
| EPI_ISL_450500 | Wuhan/OS52/2020 | 2020-01-10 | Asia/China/Hubei/Wuhan | unknown | CAS Key Laboratory of Special Pathogens and Biosafety and Center for Emerging Infectious Diseases | Si et al |
| EPI_ISL_450501 | Wuhan/YB012611/2020 | 2020-01-26 | Asia/China/Hubei/Wuhan | unknown | CAS Key Laboratory of Special Pathogens and Biosafety and Center for Emerging Infectious Diseases | Si et al |
| EPI_ISL_450502 | Wuhan/YB012605/2020 | 2020-01-26 | Asia/China/Hubei/Wuhan | unknown | CAS Key Laboratory of Special Pathogens and Biosafety and Center for Emerging Infectious Diseases | Si et al |
| EPI_ISL_450503 | Wuhan/YB012602/2020 | 2020-01-26 | Asia/China/Hubei/Wuhan | unknown | CAS Key Laboratory of Special Pathogens and Biosafety and Center for Emerging Infectious Diseases | Si et al |
| EPI_ISL_450504 | Wuhan/YB012506/2020 | 2020-01-25 | Asia/China/Hubei/Wuhan | unknown | CAS Key Laboratory of Special Pathogens and Biosafety and Center for Emerging Infectious Diseases | Si et al |

| | | | | | | |
|---|---|---|---|---|---|---|
| EPI_ISL_450511 | Lebanon/S4_761/2020 | 2020-03-04 | Asia/Lebanon/Beirut/ | Rafik Hariri University Hospital | Rafik Hariri University Hospital | Rita Feghali et al |
| EPI_ISL_450512 | Lebanon/S5_762/2020 | 2020-02-27 | Asia/Iran/Beirut/ | Rafik Hariri University Hospital | Rafik Hariri University Hospital | Rita Feghali et al |
| EPI_ISL_450530 | Poland/1111628/2020 | 2020-04-01 | Europe/Poland/Pomorskie/ | Hematology Laboratory, Section of Molecular Diagnostics, University Clinical Centre, Medical University of Gdansk | Department of Virology, Faculty of Medicine, University of Helsinki, Helsinki, Finland | Maciej Grzybek et al |
| EPI_ISL_450542 | USA/UT-03475/2020 | 2020-03-19 | North America/USA/Utah/ | Utah Public Health Laboratory | Utah Public Health Laboratory | Erin Young et al |
| EPI_ISL_450551 | USA/UT-03696/2020 | 2020-04-07 | North America/USA/Utah/ | Utah Public Health Laboratory | Utah Public Health Laboratory | Erin Young et al |
| EPI_ISL_450564 | USA/UT-03879/2020 | 2020-04-13 | North America/USA/Utah/ | Utah Public Health Laboratory | Utah Public Health Laboratory | Erin Young et al |
| EPI_ISL_450568 | USA/UT-03911/2020 | 2020-04-13 | North America/USA/Utah/ | Utah Public Health Laboratory | Utah Public Health Laboratory | Erin Young et al |
| EPI_ISL_450571 | USA/MI-MDHHS-SC20360/2020 | 2020-03-13 | North America/USA/Michigan/ | Michigan Department of Health and Human Services, Bureau of Laboratories | Michigan Department of Health and Human Services, Bureau of Laboratories | Blankenship HM et al |
| EPI_ISL_450579 | USA/MI-MDHHS-SC20368/2020 | 2020-03-13 | North America/USA/Michigan/ | Michigan Department of Health and Human Services, Bureau of Laboratories | Michigan Department of Health and Human Services, Bureau of Laboratories | Blankenship HM et al |
| EPI_ISL_450597 | USA/MI-MDHHS-SC20386/2020 | 2020-03-25 | North America/USA/Michigan/ | Michigan Department of Health and Human Services, Bureau of Laboratories | Michigan Department of Health and Human Services, Bureau of Laboratories | Blankenship HM et al |

| | | | | | | |
|---|---|---|---|---|---|---|
| EPI_ISL_450601 | USA/MI-MDHHS-SC20322/2020 | 2020-03-11 | North America/USA/Michigan/ | Michigan Department of Health and Human Services, Bureau of Laboratories | Michigan Department of Health and Human Services, Bureau of Laboratories | Blankenship HM et al |
| EPI_ISL_450606 | USA/MI-MDHHS-SC20327/2020 | 2020-03-15 | North America/USA/Michigan/ | Michigan Department of Health and Human Services, Bureau of Laboratories | Michigan Department of Health and Human Services, Bureau of Laboratories | Blankenship HM et al |
| EPI_ISL_450638 | Canada/QC-AJ2/2020 | 2020-04-08 | North America/Canada/Quebec/ | Laboratoire de microbiologie, Hopital de Verdun | Smith Laboratory, Centre de Recherche CHU Sainte-Justine | Martin Smith et al |
| EPI_ISL_450639 | Canada/QC-AJ7/2020 | 2020-04-11 | North America/Canada/Quebec/ | Laboratoire de microbiologie, Hopital de Verdun | Smith Laboratory, Centre de Recherche CHU Sainte-Justine | Martin Smith et al |
| EPI_ISL_450640 | Canada/QC-AJ9/2020 | 2020-04-08 | North America/Canada/Quebec/ | Laboratoire de microbiologie, Hopital de Verdun | Smith Laboratory, Centre de Recherche CHU Sainte-Justine | Martin Smith et al |
| EPI_ISL_450641 | Canada/QC-AK2/2020 | 2020-04-07 | North America/Canada/Quebec/ | Laboratoire de microbiologie, Hopital de Verdun | Smith Laboratory, Centre de Recherche CHU Sainte-Justine | Martin Smith et al |
| EPI_ISL_450642 | Canada/QC-AK3/2020 | 2020-04-08 | North America/Canada/Quebec/ | Laboratoire de microbiologie, Hopital de Verdun | Smith Laboratory, Centre de Recherche CHU Sainte-Justine | Martin Smith et al |
| EPI_ISL_450643 | Canada/QC-AK4/2020 | 2020-04-11 | North America/Canada/Quebec/ | Laboratoire de microbiologie, Hopital de Verdun | Smith Laboratory, Centre de Recherche CHU Sainte-Justine | Martin Smith et al |
| EPI_ISL_450644 | Canada/QC-AK7/2020 | 2020-04-08 | North America/Canada/Quebec/ | Laboratoire de microbiologie, Hopital de Verdun | Smith Laboratory, Centre de Recherche CHU Sainte-Justine | Martin Smith et al |

| | | | | | | |
|---|---|---|---|---|---|---|
| EPI_ISL_450645 | Canada/QC-AK8/2020 | 2020-04-08 | North America/Canada/Quebec/ | Laboratoire de microbiologie, Hopital de Verdun | Smith Laboratory, Centre de Recherche CHU Sainte-Justine | Martin Smith et al |
| EPI_ISL_450646 | Canada/QC-AL1/2020 | 2020-04-11 | North America/Canada/Quebec/ | Laboratoire de microbiologie, Hopital de Verdun | Smith Laboratory, Centre de Recherche CHU Sainte-Justine | Martin Smith et al |
| EPI_ISL_450647 | Canada/QC-AL2/2020 | 2020-04-08 | North America/Canada/Quebec/ | Laboratoire de microbiologie, Hopital de Verdun | Smith Laboratory, Centre de Recherche CHU Sainte-Justine | Martin Smith et al |
| EPI_ISL_450648 | Canada/QC-AL5/2020 | 2020-04-13 | North America/Canada/Quebec/ | Laboratoire de microbiologie, Hopital de Verdun | Smith Laboratory, Centre de Recherche CHU Sainte-Justine | Martin Smith et al |
| EPI_ISL_450649 | Canada/QC-AL6/2020 | 2020-04-06 | North America/Canada/Quebec/ | Laboratoire de microbiologie, Hopital de Verdun | Smith Laboratory, Centre de Recherche CHU Sainte-Justine | Martin Smith et al |
| EPI_ISL_450650 | Canada/QC-AL7/2020 | 2020-04-08 | North America/Canada/Quebec/ | Laboratoire de microbiologie, Hopital de Verdun | Smith Laboratory, Centre de Recherche CHU Sainte-Justine | Martin Smith et al |
| EPI_ISL_450651 | Canada/QC-AL8/2020 | 2020-04-07 | North America/Canada/Quebec/ | Laboratoire de microbiologie, Hopital de Verdun | Smith Laboratory, Centre de Recherche CHU Sainte-Justine | Martin Smith et al |
| EPI_ISL_450652 | Canada/QC-AL9/2020 | 2020-04-10 | North America/Canada/Quebec/ | Laboratoire de microbiologie, Hopital de Verdun | Smith Laboratory, Centre de Recherche CHU Sainte-Justine | Martin Smith et al |
| EPI_ISL_450653 | Canada/QC-AM3/2020 | 2020-04-18 | North America/Canada/Quebec/ | Laboratoire de microbiologie, Hopital de Verdun | Smith Laboratory, Centre de Recherche CHU Sainte-Justine | Martin Smith et al |

| | | | | | | |
|---|---|---|---|---|---|---|
| EPI_ISL_450701 | USA/IL-UW-347/2020 | 2020-04-18 | North America/USA/California/San Dieg | University of Wisconsin-Madison AIDS Vaccine Research Laboratories | University of Wisconsin-Madison AIDS Vaccine Research Laboratories | Gage Moreno et al |
| EPI_ISL_450706 | USA/IL-UW-352/2020 | 2020-04-18 | North America/USA/Illinois/Winnebago | University of Wisconsin-Madison AIDS Vaccine Research Laboratories | University of Wisconsin-Madison AIDS Vaccine Research Laboratories | Gage Moreno et al |
| EPI_ISL_450713 | USA/WI-UW-359/2020 | 2020-04-18 | North America/USA/Wisconsin/Verona | University of Wisconsin-Madison AIDS Vaccine Research Laboratories | University of Wisconsin-Madison AIDS Vaccine Research Laboratories | Gage Moreno et al |
| EPI_ISL_450715 | USA/IL-UW-361/2020 | 2020-04-18 | North America/USA/Illinois/Winnebago | University of Wisconsin-Madison AIDS Vaccine Research Laboratories | University of Wisconsin-Madison AIDS Vaccine Research Laboratories | Gage Moreno et al |
| EPI_ISL_450718 | USA/IL-UW-364/2020 | 2020-04-16 | North America/USA/Illinois/Winnebago | University of Wisconsin-Madison AIDS Vaccine Research Laboratories | University of Wisconsin-Madison AIDS Vaccine Research Laboratories | Gage Moreno et al |
| EPI_ISL_450729 | Belgium/ITM_C169/2020 | 2020-03-29 | Europe/Belgium/Antwerpen/ | Hospital AZ Rivierenland | Institute of Tropical Medicine | Philippe Selhorst et al |
| EPI_ISL_450731 | Belgium/ITM_C193/2020 | 2020-03-31 | Europe/Belgium/Antwerpen/ | Hospital AZ Rivierenland | Institute of Tropical Medicine | Philippe Selhorst et al |
| EPI_ISL_450741 | Vietnam/HCMC-35016/2020 | 2020-03-20 | Asia/Vietnam/Ho Chi Minh City/ | OUCRU/HTD | OUCRU/HTD | Nguyen Van Vinh Chau et al |
| EPI_ISL_450747 | Canada/ON_SHSC-01/2020 | 2020-02-26 | North America/Iran/Ontario/ | Sunnybrook Health Sciences Centre | Department of Laboratory Medicine and Molecular Diagnostics, Sunnybrook Health Sciences Centre | Jalees A. Nasir et al |

| | | | | | | |
|---|---|---|---|---|---|---|
| EPI_ISL_450752 | USA/MN-MDH-278/2020 | 2020-03-29 | North America/USA/Minnesota/ | Minnesota Department of Health, Public Health Laboratory | Minnesota Department of Health, Public Health Laboratory | Matt Plumb et al |
| EPI_ISL_450777 | USA/MN-MDH-319/2020 | 2020-03-30 | North America/USA/Minnesota/ | Minnesota Department of Health, Public Health Laboratory | Minnesota Department of Health, Public Health Laboratory | Matt Plumb et al |
| EPI_ISL_450789 | India/GJ-GBRC100/2020 | 2020-05-10 | Asia/India/Gujarat/Kodinar | Pandit Deendayal Upadhyay Government Medical College, Rajkot | Gujarat Biotechnology Research Centre | Ankit Hinsu et al |
| EPI_ISL_450791 | India/GJ-GBRC102/2020 | 2020-04-27 | Asia/India/Gujarat/Rajkot | Pandit Deendayal Upadhyay Government Medical College, Rajkot | Gujarat Biotechnology Research Centre | Zarna Patel et al |
| EPI_ISL_450801 | USA/GA-CDC-4518/2020 | 2020-04-02 | North America/USA/Georgia/ | Georgia Department of Health | Pathogen Discovery, Respiratory Viruses Branch, Division of Viral Diseases, Centers for Disease Control and Prevention | Yan Li et al |
| EPI_ISL_450819 | Sweden/20-07833/2020 | 2020-04-24 | Europe/Sweden/Vastra Gotaland/ | Hovas Askim Familjelakare och BVC | The Public Health Agency of Sweden | Anna Wendel et al |
| EPI_ISL_450821 | Sweden/20-07860/2020 | 2020-04-23 | Europe/Sweden/Jamtland/ | Jarpens HC | The Public Health Agency of Sweden | Gunilla Johansson et al |
| EPI_ISL_450822 | Sweden/20-07861/2020 | 2020-04-23 | Europe/Sweden/Jamtland/ | Jarpens HC | The Public Health Agency of Sweden | Gunilla Johansson et al |
| EPI_ISL_450825 | Sweden/20-14084/2020 | 2020-04-27 | Europe/Sweden/Vastra Gotaland/ | Narhalsan Backa vardcentral | The Public Health Agency of Sweden | Mats Olsson et al |
| EPI_ISL_450827 | Sweden/20-14089/2020 | 2020-04-27 | Europe/Sweden/Uppsala/ | Uppsala Narakut Aleris | The Public Health Agency of Sweden | Annika Nilsson et al |
| EPI_ISL_450829 | Sweden/20-14094/2020 | 2020-04-28 | Europe/Sweden/Vastra Gotaland/ | Narhalsan Sjobo vardcentral | The Public Health Agency of Sweden | Lovisa Hjerten et al |
| EPI_ISL_450833 | Sweden/20-51311/2020 | 2020-05-07 | Europe/Sweden/Jonkoping/ | Wetterhalsan | The Public Health Agency of Sweden | Anders Tengblad et al |
| EPI_ISL_450839 | Bangladesh/DU-50740/2020 | 2020-05-06 | Asia/Bangladesh/Dhaka/ | COVID-19 Laboratory Centre for Advanced Research in Sciences (CARS), University of Dhaka, Dhaka-1000, Bangladesh | DNA Solution Ltd | Sharif Akhteruzzaman et al |

| | | | | | | |
|---|---|---|---|---|---|---|
| EPI_ISL_450840 | Bangladesh/DU-50758/2020 | 2020-05-06 | Asia/Bangladesh/Dhaka/ | COVID-19 Laboratory | DNA Solution Ltd. L-5 | Sharif Akhteruzzaman et al |
| EPI_ISL_450842 | Bangladesh/DU-50761/2020 | 2020-05-06 | Asia/Bangladesh/Dhaka/ | COVID-19 Laboratory | DNA Solution Ltd. | Sharif Akhteruzzaman et al |
| EPI_ISL_450843 | Bangladesh/DU-50768/2020 | 2020-05-06 | Asia/Bangladesh/Dhaka/ | COVID-19 Laboratory | DNA Solution Ltd. | Sharif Akhteruzzaman et al |
| EPI_ISL_450849 | USA/FL-BPHL-0115/2020 | 2020-04-07 | North America/USA/Florida/ | Florida Bureau of Public Health Laboratories | Florida Bureau of Public Health Laboratories | Sarah Schmedes et al |
| EPI_ISL_450867 | USA/FL-BPHL-0133/2020 | 2020-04-09 | North America/USA/Florida/ | Florida Bureau of Public Health Laboratories | Florida Bureau of Public Health Laboratories | Sarah Schmedes et al |
| EPI_ISL_450870 | USA/FL-BPHL-0136/2020 | 2020-04-10 | North America/USA/Florida/ | Florida Bureau of Public Health Laboratories | Florida Bureau of Public Health Laboratories | Sarah Schmedes et al |
| EPI_ISL_451076 | Sichuan/SC-WCH2-006/2020 | 2020-02-08 | Asia/China/Sichuan/ | West China Hospital of Sichuan University | State Key Laboratory of Biotherapy of Sichuan University | Baowen Du et al |
| EPI_ISL_451150 | India/GJ-GBRC105/2020 | 2020-05-06 | Asia/India/Gujarat/Jamnagar | M.P Shah Government Medocal college Jamnagar | Gujarat Biotechnology Research Centre | Zarna Patel et al |
| EPI_ISL_451155 | India/GJ-GBRC110/2020 | 2020-05-03 | Asia/India/Gujarat/Vadodara | Government Medical College, Vadodara | Gujarat Biotechnology Research Centre | Tanuja Javadekar et al |
| EPI_ISL_451158 | India/GJ-GBRC113/2020 | 2020-05-03 | Asia/India/Gujarat/Vadodara | Government Medical College, Vadodara | Gujarat Biotechnology Research Centre | Nitin Savaliya et al |
| EPI_ISL_451163 | India/GJ-GBRC118/2020 | 2020-05-02 | Asia/India/Gujarat/Vadodara | Government Medical College, Vadodara | Gujarat Biotechnology Research Centre | Labdhi Pandya et al |
| EPI_ISL_451168 | Belgium/UGent-57/2020 | 2020-04-03 | Europe/Belgium/Gent/ | Lab voor klinische biologie | Onderzoeksgroep Virologie | Laurens Lambrechts et al |
| EPI_ISL_451175 | Belgium/UGent-64/2020 | 2020-04-07 | Europe/Belgium/Gent/ | Lab voor klinische biologie | Onderzoeksgroep Virologie | Nick Vereecke et al |
| EPI_ISL_451205 | USA/LA-EVTL044/2020 | 2020-05-01 | North America/USA/Louisiana/Caddo Pa | LSUHS Emerging Viral Threat Laboratory | Microbial Genome Sequencing Center | Jeremy P. Kamil et al |
| EPI_ISL_451211 | USA/LA-EVTL050/2020 | 2020-05-07 | North America/USA/Louisiana/Webster | LSUHS Emerging Viral Threat Laboratory | Microbial Genome Sequencing Center | Jeremy P. Kamil et al |

| | | | | | | |
|---|---|---|---|---|---|---|
| EPI_ISL_451214 | USA/LA-EVTL053/2020 | 2020-04-27 | North America/USA/Louisiana/Caddo Pa | LSUHS Emerging Viral Threat Laboratory | Microbial Genome Sequencing Center | Jeremy P. Kamil et al |
| EPI_ISL_451217 | USA/LA-EVTL056/2020 | 2020-05-02 | North America/USA/Louisiana/Caddo Pa | LSUHS Emerging Viral Threat Laboratory | Microbial Genome Sequencing Center | Jeremy P. Kamil et al |
| EPI_ISL_451222 | USA/LA-EVTL061/2020 | 2020-04-28 | North America/USA/Louisiana/Caddo Pa | LSUHS Emerging Viral Threat Laboratory | Microbial Genome Sequencing Center | Rona S. Scott et al |
| EPI_ISL_451226 | USA/LA-EVTL065/2020 | 2020-05-02 | North America/USA/Louisiana/Caddo Pa | LSUHS Emerging Viral Threat Laboratory | Microbial Genome Sequencing Center | Rona S. Scott et al |
| EPI_ISL_451232 | USA/LA-EVTL071/2020 | 2020-05-04 | North America/USA/Louisiana/Caddo Pa | LSUHS Emerging Viral Threat Laboratory | Microbial Genome Sequencing Center | John A. Vanchiere et al |
| EPI_ISL_451233 | USA/LA-EVTL072/2020 | 2020-05-04 | North America/USA/Louisiana/Caddo Pa | LSUHS Emerging Viral Threat Laboratory | Microbial Genome Sequencing Center | John A. Vanchiere et al |
| EPI_ISL_451238 | USA/LA-EVTL077/2020 | 2020-05-08 | North America/USA/Louisiana/Caddo Pa | LSUHS Emerging Viral Threat Laboratory | Microbial Genome Sequencing Center | John A. Vanchiere et al |
| EPI_ISL_451239 | USA/LA-EVTL078/2020 | 2020-05-04 | North America/USA/Louisiana/Caddo Pa | LSUHS Emerging Viral Threat Laboratory | Microbial Genome Sequencing Center | John A. Vanchiere et al |
| EPI_ISL_451247 | USA/LA-EVTL086/2020 | 2020-05-04 | North America/USA/Louisiana/Caddo Pa | LSUHS Emerging Viral Threat Laboratory | Microbial Genome Sequencing Center | John A. Vanchiere et al |
| EPI_ISL_451264 | USA/LA-EVTL103/2020 | 2020-05-06 | North America/USA/Louisiana/Caddo Pa | LSUHS Emerging Viral Threat Laboratory | Microbial Genome Sequencing Center | Jeremy P. Kamil et al |
| EPI_ISL_451265 | USA/LA-EVTL104/2020 | 2020-05-06 | North America/USA/Louisiana/Caddo Pa | LSUHS Emerging Viral Threat Laboratory | Microbial Genome Sequencing Center | Jeremy P. Kamil et al |
| EPI_ISL_451268 | USA/LA-EVTL107/2020 | 2020-05-07 | North America/USA/Louisiana/Rapides P | LSUHS Emerging Viral Threat Laboratory | Microbial Genome Sequencing Center | Jeremy P. Kamil et al |
| EPI_ISL_451272 | USA/LA-EVTL111/2020 | 2020-05-06 | North America/USA/Louisiana/Caddo Pa | LSUHS Emerging Viral Threat Laboratory | Microbial Genome Sequencing Center | Jeremy P. Kamil et al |

| | | | | | | |
|---|---|---|---|---|---|---|
| EPI_ISL_451281 | USA/LA-EVTL120/2020 | 2020-05-07 | North America/USA/Louisiana/Caddo Pa | LSUHS Emerging Viral Threat Laboratory | Microbial Genome Sequencing Center | Jeremy P. Kamil et al |
| EPI_ISL_451286 | USA/LA-EVTL125/2020 | 2020-05-06 | North America/USA/Louisiana/Caddo Pa | LSUHS Emerging Viral Threat Laboratory | Microbial Genome Sequencing Center | Jeremy P. Kamil et al |
| EPI_ISL_451296 | USA/LA-EVTL135/2020 | 2020-05-09 | North America/USA/Louisiana/Rapides P | LSUHS Emerging Viral Threat Laboratory | Microbial Genome Sequencing Center | Jeremy P. Kamil et al |
| EPI_ISL_451298 | Italy/LAZ-INMI1-B1/2020 | 2020-02-12 | Europe/Italy/Lazio/ | Laboratory of Virology, INMI Lazzaro Spallanzani IRCCS | Laboratory of Virology, INMI Lazzaro Spallanzani IRCCS | Cesare E.M. Gruber et al |
| EPI_ISL_451299 | Italy/LAZ-INMI1-B2/2020 | 2020-02-04 | Europe/Italy/Lazio/ | Laboratory of Virology, INMI Lazzaro Spallanzani IRCCS | Laboratory of Virology, INMI Lazzaro Spallanzani IRCCS | Martina Rueca et al |
| EPI_ISL_451300 | Italy/LAZ-INMI1-N/2020 | 2020-02-03 | Europe/Italy/Lazio/ | Laboratory of Virology, INMI Lazzaro Spallanzani IRCCS | Laboratory of Virology, INMI Lazzaro Spallanzani IRCCS | Cesare E.M. Gruber et al |
| EPI_ISL_451301 | Italy/LAZ-INMI2-B/2020 | 2020-02-03 | Europe/Italy/Lazio/ | Laboratory of Virology, INMI Lazzaro Spallanzani IRCCS | Laboratory of Virology, INMI Lazzaro Spallanzani IRCCS | Martina Rueca et al |
| EPI_ISL_451302 | Italy/LAZ-INMI2-N/2020 | 2020-01-30 | Europe/Italy/Lazio/ | Laboratory of Virology, INMI Lazzaro Spallanzani IRCCS | Laboratory of Virology, INMI Lazzaro Spallanzani IRCCS | Cesare E.M. Gruber et al |
| EPI_ISL_451303 | Italy/LAZ-INMI5-N/2020 | 2020-03-04 | Europe/Italy/Lazio/ | Laboratory of Virology, INMI Lazzaro Spallanzani IRCCS | Laboratory of Virology, INMI Lazzaro Spallanzani IRCCS | Martina Rueca et al |
| EPI_ISL_451304 | Italy/LAZ-INMI11-B/2020 | 2020-03-23 | Europe/Italy/Lazio/ | Laboratory of Virology, INMI Lazzaro Spallanzani IRCCS | Laboratory of Virology, INMI Lazzaro Spallanzani IRCCS | Cesare E.M. Gruber et al |

| | | | | | | |
|---|---|---|---|---|---|---|
| EPI_ISL_451306 | Italy/LOM-INMI-5314-B/2020 | 2020-02-21 | Europe/Italy/Lombardy/ | Molecular Virology Unit, Fondazione IRCCS Policlinico San Matteo , Pavia | Laboratory of Virology, INMI Lazzaro Spallanzani IRCCS | Antonio Piralla et al |
| EPI_ISL_451307 | Italy/LOM-INMI-5314-N/2020 | 2020-02-21 | Europe/Italy/Lombardy/ | Molecular Virology Unit, Fondazione IRCCS Policlinico San Matteo , Pavia | Laboratory of Virology, INMI Lazzaro Spallanzani IRCCS | Fausto Baldanti et al |
| EPI_ISL_451309 | Italy/LOM-INMI-13075-N/2020 | 2020-03-01 | Europe/Italy/Lombardy/ | Molecular Virology Unit, Fondazione IRCCS Policlinico San Matteo , Pavia | Laboratory of Virology, INMI Lazzaro Spallanzani IRCCS | Fausto Baldanti et al |
| EPI_ISL_451310 | Greece/HPI-6478/2020 | 2020-03-28 | Europe/Greece/Athens/ | Hellenic Pasteur Institute, National Influenza Reference laboratory of Southern Greece & Unit of Bioinformatics and Applied Genomics | Hellenic Pasteur Institute, National Influenza Reference laboratory of Southern Greece & Unit of Bioinformatics and Applied Genomics | Vasiliki Pogka et al |
| EPI_ISL_451312 | Sichuan/SC-GA-061/2020 | 2020-01-26 | Asia/China/Sichuan/ | West China Hospital of Sichuan University | State Key Laboratory of Biotherapy of Sichuan University | Baowen Du et al |
| EPI_ISL_451313 | Sichuan/SC-GA-062/2020 | 2020-01-26 | Asia/China/Sichuan/ | West China Hospital of Sichuan University | State Key Laboratory of Biotherapy of Sichuan University | Baowen Du et al |
| EPI_ISL_451314 | Sichuan/SC-GA-063/2020 | 2020-01-30 | Asia/China/Sichuan/ | West China Hospital of Sichuan University | State Key Laboratory of Biotherapy of Sichuan University | Baowen Du et al |

| | | | | | | |
|---|---|---|---|---|---|---|
| EPI_ISL_451315 | Sichuan/SC-GA-064/2020 | 2020-01-30 | Asia/China/Sichuan/ | West China Hospital of Sichuan University | State Key Laboratory of Biotherapy of Sichuan University | Baowen Du et al |
| EPI_ISL_451316 | Sichuan/SC-GA-065/2020 | 2020-02-01 | Asia/China/Sichuan/ | West China Hospital of Sichuan University | State Key Laboratory of Biotherapy of Sichuan University | Baowen Du et al |
| EPI_ISL_451317 | Sichuan/SC-GA-066/2020 | 2020-02-03 | Asia/China/Sichuan/ | West China Hospital of Sichuan University | State Key Laboratory of Biotherapy of Sichuan University | Baowen Du et al |
| EPI_ISL_451318 | Sichuan/SC-LZ-079/2020 | 2020-02-19 | Asia/China/Sichuan/ | West China Hospital of Sichuan University | State Key Laboratory of Biotherapy of Sichuan University | Baowen Du et al |
| EPI_ISL_451319 | Sichuan/SC-MY-053/2020 | 2020-01-27 | Asia/China/Sichuan/ | West China Hospital of Sichuan University | State Key Laboratory of Biotherapy of Sichuan University | Baowen Du et al |
| EPI_ISL_451320 | Sichuan/SC-MY-054/2020 | 2020-01-27 | Asia/China/Sichuan/ | West China Hospital of Sichuan University | State Key Laboratory of Biotherapy of Sichuan University | Baowen Du et al |
| EPI_ISL_451321 | Sichuan/SC-MY-055/2020 | 2020-02-02 | Asia/China/Sichuan/ | West China Hospital of Sichuan University | State Key Laboratory of Biotherapy of Sichuan University | Baowen Du et al |
| EPI_ISL_451322 | Sichuan/SC-MY-056/2020 | 2020-02-01 | Asia/China/Sichuan/ | West China Hospital of Sichuan University | State Key Laboratory of Biotherapy of Sichuan University | Baowen Du et al |

| | | | | | | |
|---|---|---|---|---|---|---|
| EPI_ISL_451323 | Sichuan/SC-MY-057/2020 | 2020-01-22 | Asia/China/Sichuan/ | West China Hospital of Sichuan University | State Key Laboratory of Biotherapy of Sichuan University | Baowen Du et al |
| EPI_ISL_451324 | Sichuan/SC-MY-058/2020 | 2020-02-02 | Asia/China/Sichuan/ | West China Hospital of Sichuan University | State Key Laboratory of Biotherapy of Sichuan University | Baowen Du et al |
| EPI_ISL_451325 | Sichuan/SC-MY-059/2020 | 2020-01-31 | Asia/China/Sichuan/ | West China Hospital of Sichuan University | State Key Laboratory of Biotherapy of Sichuan University | Baowen Du et al |
| EPI_ISL_451326 | Sichuan/SC-MY-060/2020 | 2020-02-02 | Asia/China/Sichuan/ | West China Hospital of Sichuan University | State Key Laboratory of Biotherapy of Sichuan University | Baowen Du et al |
| EPI_ISL_451327 | Sichuan/SC-NC-067/2020 | 2020-02-01 | Asia/China/Sichuan/ | West China Hospital of Sichuan University | State Key Laboratory of Biotherapy of Sichuan University | Baowen Du et al |
| EPI_ISL_451328 | Sichuan/SC-NC-068/2020 | 2020-02-02 | Asia/China/Sichuan/ | West China Hospital of Sichuan University | State Key Laboratory of Biotherapy of Sichuan University | Baowen Du et al |
| EPI_ISL_451329 | Sichuan/SC-NC-069/2020 | 2020-02-05 | Asia/China/Sichuan/ | West China Hospital of Sichuan University | State Key Laboratory of Biotherapy of Sichuan University | Baowen Du et al |
| EPI_ISL_451330 | Sichuan/SC-NC-071/2020 | 2020-01-29 | Asia/China/Sichuan/ | West China Hospital of Sichuan University | State Key Laboratory of Biotherapy of Sichuan University | Baowen Du et al |

| | | | | | | |
|---|---|---|---|---|---|---|
| EPI_ISL_451331 | Sichuan/SC-NC-072/2020 | 2020-02-02 | Asia/China/Sichuan/ | West China Hospital of Sichuan University | State Key Laboratory of Biotherapy of Sichuan University | Baowen Du et al |
| EPI_ISL_451332 | Sichuan/SC-NC-073/2020 | 2020-01-30 | Asia/China/Sichuan/ | West China Hospital of Sichuan University | State Key Laboratory of Biotherapy of Sichuan University | Baowen Du et al |
| EPI_ISL_451333 | Sichuan/SC-NC-074/2020 | 2020-02-01 | Asia/China/Sichuan/ | West China Hospital of Sichuan University | State Key Laboratory of Biotherapy of Sichuan University | Baowen Du et al |
| EPI_ISL_451334 | Sichuan/SC-NC-075/2020 | 2020-01-30 | Asia/China/Sichuan/ | West China Hospital of Sichuan University | State Key Laboratory of Biotherapy of Sichuan University | Baowen Du et al |
| EPI_ISL_451335 | Sichuan/SC-NC-076/2020 | 2020-01-29 | Asia/China/Sichuan/ | West China Hospital of Sichuan University | State Key Laboratory of Biotherapy of Sichuan University | Baowen Du et al |
| EPI_ISL_451336 | Sichuan/SC-NC-077/2020 | 2020-01-29 | Asia/China/Sichuan/ | West China Hospital of Sichuan University | State Key Laboratory of Biotherapy of Sichuan University | Baowen Du et al |
| EPI_ISL_451337 | Sichuan/SC-NC-078/2020 | 2020-01-30 | Asia/China/Sichuan/ | West China Hospital of Sichuan University | State Key Laboratory of Biotherapy of Sichuan University | Baowen Du et al |
| EPI_ISL_451338 | Sichuan/SC-NJ-089/2020 | 2020-02-05 | Asia/China/Sichuan/ | West China Hospital of Sichuan University | State Key Laboratory of Biotherapy of Sichuan University | Baowen Du et al |

| | | | | | | |
|---|---|---|---|---|---|---|
| EPI_ISL_451339 | Sichuan/SC-NJ-090/2020 | 2020-01-27 | Asia/China/Sichuan/ | West China Hospital of Sichuan University | State Key Laboratory of Biotherapy of Sichuan University | Baowen Du et al |
| EPI_ISL_451340 | Sichuan/SC-NJ-091/2020 | 2020-02-06 | Asia/China/Sichuan/ | West China Hospital of Sichuan University | State Key Laboratory of Biotherapy of Sichuan University | Baowen Du et al |
| EPI_ISL_451341 | Sichuan/SC-NJ-092/2020 | 2020-02-03 | Asia/China/Sichuan/ | West China Hospital of Sichuan University | State Key Laboratory of Biotherapy of Sichuan University | Baowen Du et al |
| EPI_ISL_451342 | Sichuan/SC-NJ-093/2020 | 2020-02-11 | Asia/China/Sichuan/ | West China Hospital of Sichuan University | State Key Laboratory of Biotherapy of Sichuan University | Baowen Du et al |
| EPI_ISL_451343 | Sichuan/SC-NJ-094/2020 | 2020-01-30 | Asia/China/Sichuan/ | West China Hospital of Sichuan University | State Key Laboratory of Biotherapy of Sichuan University | Baowen Du et al |
| EPI_ISL_451344 | Sichuan/SC-NJ-095/2020 | 2020-01-31 | Asia/China/Sichuan/ | West China Hospital of Sichuan University | State Key Laboratory of Biotherapy of Sichuan University | Baowen Du et al |
| EPI_ISL_451345 | Sichuan/SC-PHCC1-022/2020 | 2020-01-24 | Asia/China/Sichuan/ | West China Hospital of Sichuan University | State Key Laboratory of Biotherapy of Sichuan University | Baowen Du et al |
| EPI_ISL_451346 | Sichuan/SC-PHCC1-025/2020 | 2020-02-04 | Asia/China/Sichuan/ | West China Hospital of Sichuan University | State Key Laboratory of Biotherapy of Sichuan University | Baowen Du et al |

| | | | | | | |
|---|---|---|---|---|---|---|
| EPI_ISL_451347 | Sichuan/SC-PHCC1-026/2020 | 2020-02-20 | Asia/China/Sichuan/ | West China Hospital of Sichuan University | State Key Laboratory of Biotherapy of Sichuan University | Baowen Du et al |
| EPI_ISL_451348 | Sichuan/SC-PHCC1-027/2020 | 2020-02-09 | Asia/China/Sichuan/ | West China Hospital of Sichuan University | State Key Laboratory of Biotherapy of Sichuan University | Baowen Du et al |
| EPI_ISL_451349 | Sichuan/SC-PHCC1-028/2020 | 2020-02-08 | Asia/China/Sichuan/ | West China Hospital of Sichuan University | State Key Laboratory of Biotherapy of Sichuan University | Baowen Du et al |
| EPI_ISL_451350 | Sichuan/SC-PHCC1-029/2020 | 2020-02-08 | Asia/China/Sichuan/ | West China Hospital of Sichuan University | State Key Laboratory of Biotherapy of Sichuan University | Baowen Du et al |
| EPI_ISL_451351 | Sichuan/SC-PHCC1-030/2020 | 2020-01-27 | Asia/China/Sichuan/ | West China Hospital of Sichuan University | State Key Laboratory of Biotherapy of Sichuan University | Baowen Du et al |
| EPI_ISL_451352 | Sichuan/SC-PHCC1-031/2020 | 2020-02-08 | Asia/China/Sichuan/ | West China Hospital of Sichuan University | State Key Laboratory of Biotherapy of Sichuan University | Baowen Du et al |
| EPI_ISL_451353 | Sichuan/SC-PHCC1-032/2020 | 2020-01-27 | Asia/China/Sichuan/ | West China Hospital of Sichuan University | State Key Laboratory of Biotherapy of Sichuan University | Baowen Du et al |
| EPI_ISL_451354 | Sichuan/SC-PHCC1-033/2020 | 2020-02-09 | Asia/China/Sichuan/ | West China Hospital of Sichuan University | State Key Laboratory of Biotherapy of Sichuan University | Baowen Du et al |

| | | | | | | |
|---|---|---|---|---|---|---|
| EPI_ISL_451355 | Sichuan/SC-PHCC1-034/2020 | 2020-02-12 | Asia/China/Sichuan/ | West China Hospital of Sichuan University | State Key Laboratory of Biotherapy of Sichuan University | Baowen Du et al |
| EPI_ISL_451356 | Sichuan/SC-PHCC1-035/2020 | 2020-02-07 | Asia/China/Sichuan/ | West China Hospital of Sichuan University | State Key Laboratory of Biotherapy of Sichuan University | Baowen Du et al |
| EPI_ISL_451357 | Sichuan/SC-PHCC1-036/2020 | 2020-02-03 | Asia/China/Sichuan/ | West China Hospital of Sichuan University | State Key Laboratory of Biotherapy of Sichuan University | Baowen Du et al |
| EPI_ISL_451358 | Sichuan/SC-PHCC2-038/2020 | 2020-02-03 | Asia/China/Sichuan/ | West China Hospital of Sichuan University | State Key Laboratory of Biotherapy of Sichuan University | Baowen Du et al |
| EPI_ISL_451359 | Sichuan/SC-PHCC2-039/2020 | 2020-01-27 | Asia/China/Sichuan/ | West China Hospital of Sichuan University | State Key Laboratory of Biotherapy of Sichuan University | Baowen Du et al |
| EPI_ISL_451360 | Sichuan/SC-PHCC2-040/2020 | 2020-01-24 | Asia/China/Sichuan/ | West China Hospital of Sichuan University | State Key Laboratory of Biotherapy of Sichuan University | Baowen Du et al |
| EPI_ISL_451361 | Sichuan/SC-PHCC2-041/2020 | 2020-01-24 | Asia/China/Sichuan/ | West China Hospital of Sichuan University | State Key Laboratory of Biotherapy of Sichuan University | Baowen Du et al |
| EPI_ISL_451362 | Sichuan/SC-PHCC2-042/2020 | 2020-02-13 | Asia/China/Sichuan/ | West China Hospital of Sichuan University | State Key Laboratory of Biotherapy of Sichuan University | Baowen Du et al |

| | | | | | | |
|---|---|---|---|---|---|---|
| EPI_ISL_451363 | Sichuan/SC-PHCC2-043/2020 | 2020-01-29 | Asia/China/Sichuan/ | West China Hospital of Sichuan University | State Key Laboratory of Biotherapy of Sichuan University | Baowen Du et al |
| EPI_ISL_451364 | Sichuan/SC-PHCC2-044/2020 | 2020-02-12 | Asia/China/Sichuan/ | West China Hospital of Sichuan University | State Key Laboratory of Biotherapy of Sichuan University | Baowen Du et al |
| EPI_ISL_451365 | Sichuan/SC-PHCC2-045/2020 | 2020-01-26 | Asia/China/Sichuan/ | West China Hospital of Sichuan University | State Key Laboratory of Biotherapy of Sichuan University | Baowen Du et al |
| EPI_ISL_451366 | Sichuan/SC-PHCC2-046/2020 | 2020-01-29 | Asia/China/Sichuan/ | West China Hospital of Sichuan University | State Key Laboratory of Biotherapy of Sichuan University | Baowen Du et al |
| EPI_ISL_451367 | Sichuan/SC-PHCC2-047/2020 | 2020-02-08 | Asia/China/Sichuan/ | West China Hospital of Sichuan University | State Key Laboratory of Biotherapy of Sichuan University | Baowen Du et al |
| EPI_ISL_451368 | Sichuan/SC-PHCC2-048/2020 | 2020-02-14 | Asia/China/Sichuan/ | West China Hospital of Sichuan University | State Key Laboratory of Biotherapy of Sichuan University | Baowen Du et al |
| EPI_ISL_451369 | Sichuan/SC-PHCC2-049/2020 | 2020-02-08 | Asia/China/Sichuan/ | West China Hospital of Sichuan University | State Key Laboratory of Biotherapy of Sichuan University | Baowen Du et al |
| EPI_ISL_451370 | Sichuan/SC-PHCC2-050/2020 | 2020-02-04 | Asia/China/Sichuan/ | West China Hospital of Sichuan University | State Key Laboratory of Biotherapy of Sichuan University | Baowen Du et al |

| | | | | | | |
|---|---|---|---|---|---|---|
| EPI_ISL_451371 | Sichuan/SC-PHCC2-051/2020 | 2020-02-09 | Asia/China/Sichuan/ | West China Hospital of Sichuan University | State Key Laboratory of Biotherapy of Sichuan University | Baowen Du et al |
| EPI_ISL_451372 | Sichuan/SC-PHCC2-052/2020 | 2020-02-03 | Asia/China/Sichuan/ | West China Hospital of Sichuan University | State Key Laboratory of Biotherapy of Sichuan University | Baowen Du et al |
| EPI_ISL_451373 | Sichuan/SC-WCH-081/2020 | 2020-02-03 | Asia/China/Sichuan/ | West China Hospital of Sichuan University | State Key Laboratory of Biotherapy of Sichuan University | Baowen Du et al |
| EPI_ISL_451374 | Sichuan/SC-WCH-082/2020 | 2020-01-31 | Asia/China/Sichuan/ | West China Hospital of Sichuan University | State Key Laboratory of Biotherapy of Sichuan University | Baowen Du et al |
| EPI_ISL_451375 | Sichuan/SC-WCH1-002/2020 | 2020-02-08 | Asia/China/Sichuan/ | West China Hospital of Sichuan University | State Key Laboratory of Biotherapy of Sichuan University | Baowen Du et al |
| EPI_ISL_451376 | Sichuan/SC-WCH1-005/2020 | 2020-01-28 | Asia/China/Sichuan/ | West China Hospital of Sichuan University | State Key Laboratory of Biotherapy of Sichuan University | Baowen Du et al |
| EPI_ISL_451378 | Sichuan/SC-WCH2-017/2020 | 2020-01-28 | Asia/China/Sichuan/ | West China Hospital of Sichuan University | State Key Laboratory of Biotherapy of Sichuan University | Baowen Du et al |
| EPI_ISL_451379 | Sichuan/SC-WCH-308/2020 | 2020-01-23 | Asia/China/Sichuan/ | West China Hospital of Sichuan University | State Key Laboratory of Biotherapy of Sichuan University | Baowen Du et al |

| | | | | | | |
|---|---|---|---|---|---|---|
| EPI_ISL_451380 | Sichuan/SC-WCH-310/2020 | 2020-01-24 | Asia/China/Sichuan/ | West China Hospital of Sichuan University | State Key Laboratory of Biotherapy of Sichuan University | Baowen Du et al |
| EPI_ISL_451381 | Sichuan/SC-WCH-311/2020 | 2020-01-27 | Asia/China/Sichuan/ | West China Hospital of Sichuan University | State Key Laboratory of Biotherapy of Sichuan University | Baowen Du et al |
| EPI_ISL_451382 | Sichuan/SC-WCH-315/2020 | 2020-01-25 | Asia/China/Sichuan/ | West China Hospital of Sichuan University | State Key Laboratory of Biotherapy of Sichuan University | Baowen Du et al |
| EPI_ISL_451384 | Sichuan/SC-WCH3-255/2020 | 2020-01-27 | Asia/China/Sichuan/ | West China Hospital of Sichuan University | State Key Laboratory of Biotherapy of Sichuan University | Baowen Du et al |
| EPI_ISL_451385 | Sichuan/SC-WCH3-259/2020 | 2020-02-03 | Asia/China/Sichuan/ | West China Hospital of Sichuan University | State Key Laboratory of Biotherapy of Sichuan University | Baowen Du et al |
| EPI_ISL_451386 | Sichuan/SC-WCH4-284/2020 | 2020-02-02 | Asia/China/Sichuan/ | West China Hospital of Sichuan University | State Key Laboratory of Biotherapy of Sichuan University | Baowen Du et al |
| EPI_ISL_451387 | Sichuan/SC-WCH4-285/2020 | 2020-01-27 | Asia/China/Sichuan/ | West China Hospital of Sichuan University | State Key Laboratory of Biotherapy of Sichuan University | Baowen Du et al |
| EPI_ISL_451388 | Sichuan/SC-WCH4-286/2020 | 2020-02-10 | Asia/China/Sichuan/ | West China Hospital of Sichuan University | State Key Laboratory of Biotherapy of Sichuan University | Baowen Du et al |

| | | | | | | |
|---|---|---|---|---|---|---|
| EPI_ISL_451389 | Sichuan/SC-WCH4-287/2020 | 2020-01-30 | Asia/China/Sichuan/ | West China Hospital of Sichuan University | State Key Laboratory of Biotherapy of Sichuan University | Baowen Du et al |
| EPI_ISL_451390 | Sichuan/SC-WCH4-288/2020 | 2020-01-23 | Asia/China/Sichuan/ | West China Hospital of Sichuan University | State Key Laboratory of Biotherapy of Sichuan University | Baowen Du et al |
| EPI_ISL_451391 | Sichuan/SC-WCH4-289/2020 | 2020-02-13 | Asia/China/Sichuan/ | West China Hospital of Sichuan University | State Key Laboratory of Biotherapy of Sichuan University | Baowen Du et al |
| EPI_ISL_451392 | Sichuan/SC-WCH4-290/2020 | 2020-01-24 | Asia/China/Sichuan/ | West China Hospital of Sichuan University | State Key Laboratory of Biotherapy of Sichuan University | Baowen Du et al |
| EPI_ISL_451393 | Sichuan/SC-WCH4-291/2020 | 2020-02-15 | Asia/China/Sichuan/ | West China Hospital of Sichuan University | State Key Laboratory of Biotherapy of Sichuan University | Baowen Du et al |
| EPI_ISL_451394 | Sichuan/SC-WCH4-293/2020 | 2020-02-03 | Asia/China/Sichuan/ | West China Hospital of Sichuan University | State Key Laboratory of Biotherapy of Sichuan University | Baowen Du et al |
| EPI_ISL_451395 | Sichuan/SC-WCH4-301/2020 | 2020-01-30 | Asia/China/Sichuan/ | West China Hospital of Sichuan University | State Key Laboratory of Biotherapy of Sichuan University | Baowen Du et al |
| EPI_ISL_451396 | Sichuan/SC-YA-087/2020 | 2020-01-30 | Asia/China/Sichuan/ | West China Hospital of Sichuan University | State Key Laboratory of Biotherapy of Sichuan University | Baowen Du et al |

| | | | | | | |
|---|---|---|---|---|---|---|
| EPI_ISL_451397 | Sichuan/SC-YA-088/2020 | 2020-01-31 | Asia/China/Sichuan/ | West China Hospital of Sichuan University | State Key Laboratory of Biotherapy of Sichuan University | Baowen Du et al |
| EPI_ISL_451398 | Sichuan/SC-YB-085/2020 | 2020-01-31 | Asia/China/Sichuan/ | West China Hospital of Sichuan University | State Key Laboratory of Biotherapy of Sichuan University | Baowen Du et al |
| EPI_ISL_451399 | Sichuan/SC-YB-086/2020 | 2020-02-04 | Asia/China/Sichuan/ | West China Hospital of Sichuan University | State Key Laboratory of Biotherapy of Sichuan University | Baowen Du et al |
| EPI_ISL_451424 | USA/NY-NYUMC709/2020 | 2020-03-20 | North America/USA/New York/Manhat | NYU Langone Health | Departments of Pathology and Medicine, New York University School of Medicine | Maria Aguero-Rosenfeld et al |
| EPI_ISL_451432 | USA/NY-NYUMC717/2020 | 2020-03-26 | North America/USA/New York/Nassau ( | NYU Langone Health | Departments of Pathology and Medicine, New York University School of Medicine | Maria Aguero-Rosenfeld et al |
| EPI_ISL_451645 | Poland/Pom4/2020 | 2020-05-02 | Europe/Poland/Pomorskie/ | Laboratory of Molecular Biology, Diagnostyka sp. z o.o. | Laboratory of Recombinant Vaccines | Lukasz Rabalski et al |
| EPI_ISL_451656 | Poland/Sla2/2020 | 2020-05-05 | Europe/Poland/Slask/ | State Sanitary Inspectorate | Laboratory of Recombinant Vaccines | Lukasz Rabalski et al |
| EPI_ISL_451658 | Poland/Sla4/2020 | 2020-05-05 | Europe/Poland/Slask/ | State Sanitary Inspectorate | Laboratory of Recombinant Vaccines | Lukasz Rabalski et al |
| EPI_ISL_451666 | India/GJ-GBRC103/2020 | 2020-05-04 | Asia/India/Gujarat/Jamnagar | M.P Shah Government Medocal college Jamnagar | Gujarat Biotechnology Research Centre | Binita Aring et al |

| | | | | | | |
|---|---|---|---|---|---|---|
| EPI_ISL_451693 | Switzerland/BL-ETHZ-100109/2020 | 2020-03-14 | Europe/Switzerland/Basel-Land/ | Viollier AG | Department of Biosystems Science and Engineering, ETH Zv∫rich | Christian Beisel et al |
| EPI_ISL_451695 | Switzerland/BS-ETHZ-100111/2020 | 2020-03-16 | Europe/Switzerland/Basel-Stadt/ | Viollier AG | Department of Biosystems Science and Engineering, ETH Zv∫rich | Christian Beisel et al |
| EPI_ISL_451709 | Switzerland/AG-ETHZ-100125/2020 | 2020-03-16 | Europe/Switzerland/Aargau/ | Viollier AG | Department of Biosystems Science and Engineering, ETH Zv∫rich | Christian Beisel et al |
| EPI_ISL_451710 | Switzerland/ZH-ETHZ-100126/2020 | 2020-03-16 | Europe/Switzerland/Zv∫rich/ | Viollier AG | Department of Biosystems Science and Engineering, ETH Zv∫rich | Christian Beisel et al |
| EPI_ISL_451711 | Switzerland/TI-ETHZ-100127/2020 | 2020-03-16 | Europe/Switzerland/Ticino/ | Viollier AG | Department of Biosystems Science and Engineering, ETH Zv∫rich | Christian Beisel et al |
| EPI_ISL_451717 | Switzerland/AG-ETHZ-100133/2020 | 2020-03-16 | Europe/Switzerland/Aargau/ | Viollier AG | Department of Biosystems Science and Engineering, ETH Zv∫rich | Christian Beisel et al |
| EPI_ISL_451719 | Switzerland/BL-ETHZ-100135/2020 | 2020-03-16 | Europe/Switzerland/Basel-Land/ | Viollier AG | Department of Biosystems Science and Engineering, ETH Zv∫rich | Christian Beisel et al |
| EPI_ISL_451730 | Switzerland/TI-ETHZ-100146/2020 | 2020-03-16 | Europe/Switzerland/Ticino/ | Viollier AG | Department of Biosystems Science and Engineering, ETH Zv∫rich | Christian Beisel et al |

| | | | | | | |
|---|---|---|---|---|---|---|
| EPI_ISL_451753 | Switzerland/ZH-ETHZ-100803/2020 | 2020-03-25 | Europe/Switzerland/Z√ºrich/ | Viollier AG | Department of Biosystems Science and Engineering, ETH Z√ºrich | Christian Beisel et al |
| EPI_ISL_451770 | Switzerland/VD-ETHZ-100821/2020 | 2020-03-26 | Europe/Switzerland/Vaud/ | Viollier AG | Department of Biosystems Science and Engineering, ETH Z√ºrich | Christian Beisel et al |
| EPI_ISL_451780 | Switzerland/UR-ETHZ-100832/2020 | 2020-03-25 | Europe/Switzerland/Uri/ | Viollier AG | Department of Biosystems Science and Engineering, ETH Z√ºrich | Christian Beisel et al |
| EPI_ISL_451782 | Switzerland/BL-ETHZ-100834/2020 | 2020-03-25 | Europe/Switzerland/Basel-Land/ | Viollier AG | Department of Biosystems Science and Engineering, ETH Z√ºrich | Christian Beisel et al |
| EPI_ISL_451827 | Switzerland/LU-ETHZ-101161/2020 | 2020-03-31 | Europe/Switzerland/Lucerne/ | Viollier AG | Department of Biosystems Science and Engineering, ETH Z√ºrich | Christian Beisel et al |
| EPI_ISL_451874 | Switzerland/BS-ETHZ-110454/2020 | 2020-04-07 | Europe/Switzerland/Basel-Stadt/ | Viollier AG | Department of Biosystems Science and Engineering, ETH Z√ºrich | Christian Beisel et al |
| EPI_ISL_451885 | Switzerland/BS-ETHZ-110467/2020 | 2020-04-06 | Europe/Switzerland/Basel-Stadt/ | Viollier AG | Department of Biosystems Science and Engineering, ETH Z√ºrich | Christian Beisel et al |
| EPI_ISL_451893 | Switzerland/BE-ETHZ-110481/2020 | 2020-04-07 | Europe/Switzerland/Bern/ | Viollier AG | Department of Biosystems Science and Engineering, ETH Z√ºrich | Christian Beisel et al |

| | | | | | | |
|---|---|---|---|---|---|---|
| EPI_ISL_451900 | Switzerland/BL-ETHZ-110489/2020 | 2020-04-07 | Europe/Switzerland/Basel-Land/ | Viollier AG | Department of Biosystems Science and Engineering, ETH Z√ºrich | Christian Beisel et al |
| EPI_ISL_451937 | Germany/BY-MVP-0113/2020 | 2020-03-17 | Europe/Germany/Bavaria/Munich | Max von Pettenkofer Institute, Virology, National Reference Center for Retroviruses, LMU M√ºnchen | Laboratory for Functional Genome Analysis, Dept. Genomics, Gene Center of the LMU Munich | Max Muenchhoff et al |
| EPI_ISL_451940 | Germany/BY-MVP-0123/2020 | 2020-03-23 | Europe/Germany/Bavaria/Munich | Max von Pettenkofer Institute, Virology, National Reference Center for Retroviruses, LMU M√ºnchen | Laboratory for Functional Genome Analysis, Dept. Genomics, Gene Center of the LMU Munich | Max Muenchhoff et al |
| EPI_ISL_451941 | Germany/BY-MVP-0124/2020 | 2020-03-23 | Europe/Germany/Bavaria/Munich | Max von Pettenkofer Institute, Virology, National Reference Center for Retroviruses, LMU M√ºnchen | Laboratory for Functional Genome Analysis, Dept. Genomics, Gene Center of the LMU Munich | Max Muenchhoff et al |
| EPI_ISL_451943 | Germany/BY-MVP-0151/2020 | 2020-03-30 | Europe/Germany/Bavaria/Munich | Max von Pettenkofer Institute, Virology, National Reference Center for Retroviruses, LMU M√ºnchen | Laboratory for Functional Genome Analysis, Dept. Genomics, Gene Center of the LMU Munich | Max Muenchhoff et al |
| EPI_ISL_451944 | Germany/BY-MVP-0153/2020 | 2020-03-31 | Europe/Germany/Bavaria/Munich | Max von Pettenkofer Institute, Virology, National Reference Center for Retroviruses, LMU M√ºnchen | Laboratory for Functional Genome Analysis, Dept. Genomics, Gene Center of the LMU Munich | Max Muenchhoff et al |
| EPI_ISL_451946 | Germany/BY-MVP-0157/2020 | 2020-04-02 | Europe/Germany/Bavaria/Munich | Max von Pettenkofer Institute, Virology, National Reference Center for Retroviruses, LMU M√ºnchen | Laboratory for Functional Genome Analysis, Dept. Genomics, Gene Center of the LMU Munich | Max Muenchhoff et al |

| | | | | | | |
|---|---|---|---|---|---|---|
| EPI_ISL_451947 | Germany/BY-MVP-0173/2020 | 2020-04-08 | Europe/Germany/Bavaria/Munich | Max von Pettenkofer Institute, Virology, National Reference Center for Retroviruses, LMU M√ºnchen | Laboratory for Functional Genome Analysis, Dept. Genomics, Gene Center of the LMU Munich | Max Muenchhoff et al |
| EPI_ISL_451958 | Pakistan/KHI1/2020 | 2020-03-16 | Asia/Pakistan/Karachi/ | Jamil-ur-Rahman Center for Genome Research, Dr. Panjwani Center for Molecular Medicine and Drug Research, International Center for Chemical and Biological Sciences, University of Karachi | Jamil-ur-Rahman Center for Genome Research, Dr. Panjwani Center for Molecular Medicine and Drug Research, International Center for Chemical and Biological Sciences, University of Karachi | Shakeel et al |
| EPI_ISL_451963 | Russia/SCPM-O-01/2020 | 2020-03-20 | Europe/Russia/Moscow Oblast/ | Federal Budget Institution of Science, State Research Center for Applied Microbiology & Biotechnology | Federal Budget Institution of Science, State Research Center for Applied Microbiology & Biotechnology | Dyatlov I et al |

| | | | | | | |
|---|---|---|---|---|---|---|
| EPI_ISL_451974 | Poland/PL_P13/2020 | 2020-04-03 | Europe/Poland/Malopolskie/Krakv≥w | 1. ViroGenetics - BSL3 Laboratory of Virology, Ma≈Çopolska Centre of Biotechnology, Jagiellonian University; 2. II Department of Internal Medicine, Faculty of Medicine, Jagiellonian University Medical College; 3. DIAGNOSTYKA Ltd. | 1. ViroGenetics - BSL3 Laboratory of Virology, Ma≈Çopolska Centre of Biotechnology, Jagiellonian University; 2. II Department of Internal Medicine, Faculty of Medicine, Jagiellonian University Medical College. | Marek Sanak et al |
| EPI_ISL_451979 | Poland/PL_P18/2020 | 2020-03-31 | Europe/Poland/Malopolskie/Krakv≥w | 1. ViroGenetics - BSL3 Laboratory of Virology, Ma≈Çopolska Centre of Biotechnology, Jagiellonian University; 2. II Department of Internal Medicine, Faculty of Medicine, Jagiellonian University Medical College; 3. DIAGNOSTYKA Ltd. | 1. ViroGenetics - BSL3 Laboratory of Virology, Ma≈Çopolska Centre of Biotechnology, Jagiellonian University; 2. II Department of Internal Medicine, Faculty of Medicine, Jagiellonian University Medical College. | Marek Sanak et al |

| | | | | | | |
|---|---|---|---|---|---|---|
| EPI_ISL_451983 | Poland/PL_P22/2020 | 2020-04-02 | Europe/Poland/Malopolskie/ | 1. ViroGenetics - BSL3 Laboratory of Virology, Ma≈Çopolska Centre of Biotechnology, Jagiellonian University; 2. II Department of Internal Medicine, Faculty of Medicine, Jagiellonian University Medical College; 3. DIAGNOSTYKA Ltd. | 1. ViroGenetics - BSL3 Laboratory of Virology, Ma≈Çopolska Centre of Biotechnology, Jagiellonian University; 2. II Department of Internal Medicine, Faculty of Medicine, Jagiellonian University Medical College. | Marek Sanak et al |
| EPI_ISL_451984 | Poland/PL_P23/2020 | 2020-03-29 | Europe/Poland/Malopolskie/Krakv≥w | 1. ViroGenetics - BSL3 Laboratory of Virology, Ma≈Çopolska Centre of Biotechnology, Jagiellonian University; 2. II Department of Internal Medicine, Faculty of Medicine, Jagiellonian University Medical College; 3. DIAGNOSTYKA Ltd. | 1. ViroGenetics - BSL3 Laboratory of Virology, Ma≈Çopolska Centre of Biotechnology, Jagiellonian University; 2. II Department of Internal Medicine, Faculty of Medicine, Jagiellonian University Medical College. | Marek Sanak et al |
| EPI_ISL_452114 | USA/KS-CDC-4589/2020 | 2020-03-10 | North America/USA/Kansas/ | KS Health and Environmental Laboratories | Pathogen Discovery, Respiratory Viruses Branch, Division of Viral Diseases, Centers for Disease Control and Prevention | Yan Li et al |

| | | | | | | |
|---|---|---|---|---|---|---|
| EPI_ISL_452132 | USA/FL-CDC-2316/2020 | 2020-03-09 | North America/USA/Florida/ | FL Bureau of Public Health Laboratories | Pathogen Discovery, Respiratory Viruses Branch, Division of Viral Diseases, Centers for Disease Control and Prevention | Krista Queen et al |
| EPI_ISL_452139 | Mexico/CMX-InDRE-07/2020 | 2020-02-28 | North America/Italy/Estado de Mexico/ | Instituto de Diagnostico y Referencia Epidemiologicos (INDRE) | Instituto de diagn√≥stico y Referencia Epidemiologicos (INDRE) | Ramirez-Gonzalez Ernesto et al |
| EPI_ISL_452141 | Mexico/CMX-InDRE-08/2020 | 2020-03-11 | North America/Italy/Estado de Mexico/ | Instituto de Diagnostico y Referencia Epidemiologicos (INDRE) | Instituto de diagn√≥stico y Referencia Epidemiologicos (INDRE) | Ramirez-Gonzalez Ernesto et al |
| EPI_ISL_452153 | USA/UT-01440/2020 | 2020-04-10 | North America/USA/Utah/ | Utah Public Health Laboratory | Utah Public Health Laboratory | Erin Young et al |
| EPI_ISL_452166 | USA/UT-01979/2020 | 2020-04-16 | North America/USA/Utah/ | Utah Public Health Laboratory | Utah Public Health Laboratory | Erin Young et al |
| EPI_ISL_452180 | Germany/NW-HHU-54/2020 | 2020-03-20 | Europe/Germany/North Rhine Westphal | Center of Medical Microbiology, Virology, and Hospital Hygiene, University of Duesseldorf | Center of Medical Microbiology, Virology, and Hospital Hygiene, University of Duesseldorf | Ortwin Adams et al |
| EPI_ISL_452182 | Italy/VEN-IZSVe-11-181/2020 | 2020-03-23 | Europe/Italy/Veneto/Verona IT | ULSS9 Distretto di Bussolengo | Istituto Zooprofilattico Sperimentale delle Venezie | Adelaide Milani et al |
| EPI_ISL_452184 | Italy/VEN-IZSVe-18-48/2020 | 2020-03-24 | Europe/Italy/Veneto/Verona IT | ULSS9 Distretto di Bussolengo | Istituto Zooprofilattico Sperimentale delle Venezie | Adelaide Milani et al |

| | | | | | | |
|---|---|---|---|---|---|---|
| EPI_ISL_452188 | Italy/VEN-IZSVe-31-19/2020 | 2020-03-31 | Europe/Italy/Veneto/Verona IT | ULSS9 Distretto di Bussolengo | Istituto Zooprofilattico Sperimentale delle Venezie | Adelaide Milani et al |
| EPI_ISL_452189 | Italy/VEN-IZSVe-33-126/2020 | 2020-04-01 | Europe/Italy/Veneto/Verona IT | ULSS9 Distretto di Bussolengo | Istituto Zooprofilattico Sperimentale delle Venezie | Adelaide Milani et al |
| EPI_ISL_452193 | India/MH-NIV-10419/2020 | 2020-04-16 | Asia/India/Maharashtra/ | NIV Influenza | NIV Influenza | Potdar V et al |
| EPI_ISL_452198 | India/MH-NIV-11855/2020 | 2020-04-20 | Asia/India/Maharashtra/ | NIV Influenza | NIV Influenza | Potdar V et al |
| EPI_ISL_452212 | India/MH-NIV-9768/2020 | 2020-04-15 | Asia/India/Maharashtra/ | NIV Influenza | NIV Influenza | Potdar V et al |
| EPI_ISL_452214 | India/MH-NIV-QA-709/2020 | 2020-04-07 | Asia/India/Maharashtra/ | NIV Influenza | NIV Influenza | Potdar V et al |
| EPI_ISL_452220 | Germany/HE-FFM4/2020 | 2020-03-02 | Europe/Italy/Hesse/Frankfurt | Goethe University Hospital Frankfurt | Institute for Medical Virology, Goethe University Hospital Frankfurt | Tuna Toptan et al |
| EPI_ISL_452242 | Sweden/20-51160/2020 | 2020-05-04 | Europe/Sweden/Vastra Gotaland/ | Wernstedt Medical AB | The Public Health Agency of Sweden | Eva Sandberg et al |
| EPI_ISL_452265 | USA/MI-MDHHS-SC20391/2020 | 2020-03-23 | North America/USA/Michigan/ | Michigan Department of Health and Human Services, Bureau of Laboratories | Michigan Department of Health and Human Services, Bureau of Laboratories | Blankenship HM et al |
| EPI_ISL_452275 | USA/MI-MDHHS-SC20401/2020 | 2020-03-26 | North America/USA/Michigan/ | Michigan Department of Health and Human Services, Bureau of Laboratories | Michigan Department of Health and Human Services, Bureau of Laboratories | Blankenship HM et al |
| EPI_ISL_452288 | USA/MI-MDHHS-SC20414/2020 | 2020-03-15 | North America/USA/Michigan/ | Michigan Department of Health and Human Services, Bureau of Laboratories | Michigan Department of Health and Human Services, Bureau of Laboratories | Blankenship HM et al |

| | | | | | | |
|---|---|---|---|---|---|---|
| EPI_ISL_452300 | USA/MI-MDHHS-SC20426/2020 | 2020-03-18 | North America/USA/Michigan/ | Michigan Department of Health and Human Services, Bureau of Laboratories | Michigan Department of Health and Human Services, Bureau of Laboratories | Blankenship HM et al |
| EPI_ISL_452307 | USA/MI-MDHHS-SC20433/2020 | 2020-03-30 | North America/USA/Michigan/ | Michigan Department of Health and Human Services, Bureau of Laboratories | Michigan Department of Health and Human Services, Bureau of Laboratories | Blankenship HM et al |
| EPI_ISL_452322 | USA/MI-MDHHS-SC20448/2020 | 2020-03-16 | North America/USA/Michigan/ | Michigan Department of Health and Human Services, Bureau of Laboratories | Michigan Department of Health and Human Services, Bureau of Laboratories | Blankenship HM et al |
| EPI_ISL_452332 | Beijing/DT-travelUSA01/2020 | 2020-02-29 | Asia/Iran/Beijing/ | Laboratory of Infectious Diseases Center of Beijing Ditan Hospital | Laboratory of Infectious Diseases Center of Beijing Ditan Hospital | Siyuan Yang et al |
| EPI_ISL_452333 | Beijing/DT-travelIRI01/2020 | 2020-03-12 | Asia/Italy/Beijing/ | Laboratory of Infectious Diseases Center of Beijing Ditan Hospital | Laboratory of Infectious Diseases Center of Beijing Ditan Hospital | Siyuan Yang et al |
| EPI_ISL_452336 | Beijing/DT-travelUK04/2020 | 2020-03-20 | Asia/China/Beijing/ | Laboratory of Infectious Diseases Center of Beijing Ditan Hospital | Laboratory of Infectious Diseases Center of Beijing Ditan Hospital | Siyuan Yang et al |
| EPI_ISL_452340 | Beijing/DT-travelUSA03/2020 | 2020-03-19 | Asia/China/Beijing/ | Laboratory of Infectious Diseases Center of Beijing Ditan Hospital | Laboratory of Infectious Diseases Center of Beijing Ditan Hospital | Siyuan Yang et al |

| | | | | | | |
|---|---|---|---|---|---|---|
| EPI_ISL_452357 | Beijing/DT-WH02/2020 | 2020-02-04 | Asia/China/Beijing/ | Laboratory of Infectious Diseases Center of Beijing Ditan Hospital | Laboratory of Infectious Diseases Center of Beijing Ditan Hospital | Siyuan Yang et al |
| EPI_ISL_452358 | Beijing/DT-BJ01/2020 | 2020-02-11 | Asia/China/Beijing/ | Laboratory of Infectious Diseases Center of Beijing Ditan Hospital | Laboratory of Infectious Diseases Center of Beijing Ditan Hospital | Siyuan Yang et al |
| EPI_ISL_452359 | Beijing/DT-BJ02/2020 | 2020-02-08 | Asia/China/Beijing/ | Laboratory of Infectious Diseases Center of Beijing Ditan Hospital | Laboratory of Infectious Diseases Center of Beijing Ditan Hospital | Siyuan Yang et al |
| EPI_ISL_452360 | Beijing/DT-WH01/2020 | 2020-02-01 | Asia/China/Beijing/ | Laboratory of Infectious Diseases Center of Beijing Ditan Hospital | Laboratory of Infectious Diseases Center of Beijing Ditan Hospital | Siyuan Yang et al |
| EPI_ISL_452361 | Beijing/DT-BJ03/2020 | 2020-02-11 | Asia/China/Beijing/ | Laboratory of Infectious Diseases Center of Beijing Ditan Hospital | Laboratory of Infectious Diseases Center of Beijing Ditan Hospital | Siyuan Yang et al |
| EPI_ISL_452362 | Beijing/DT-WH03/2020 | 2020-02-01 | Asia/China/Beijing/ | Laboratory of Infectious Diseases Center of Beijing Ditan Hospital | Laboratory of Infectious Diseases Center of Beijing Ditan Hospital | Siyuan Yang et al |
| EPI_ISL_452363 | Beijing/DT-WH04/2020 | 2020-01-31 | Asia/China/Beijing/ | Laboratory of Infectious Diseases Center of Beijing Ditan Hospital | Laboratory of Infectious Diseases Center of Beijing Ditan Hospital | Siyuan Yang et al |
| EPI_ISL_452364 | Beijing/DT-BJ04/2020 | 2020-02-05 | Asia/China/Beijing/ | Laboratory of Infectious Diseases Center of Beijing Ditan Hospital | Laboratory of Infectious Diseases Center of Beijing Ditan Hospital | Siyuan Yang et al |

| | | | | | | |
|---|---|---|---|---|---|---|
| EPI_ISL_452370 | Spain/AN-IBV-002059/2020 | 2020-04-05 | Europe/Spain/Andalusia/Velez Malaga | Servicio de Microbiolog√≠a. HRU de M√°laga. Servicio Andaluz de Salud | SeqCOVID-SPAIN consortium/IBV(CSIC) | Inmaculada de Toro Peinado et al |
| EPI_ISL_452375 | Spain/AN-IBV-002054/2020 | 2020-03-15 | Europe/Spain/Andalusia/Velez Malaga | Servicio de Microbiolog√≠a. HRU de M√°laga. Servicio Andaluz de Salud | SeqCOVID-SPAIN consortium/IBV(CSIC) | Inmaculada de Toro Peinado et al |
| EPI_ISL_452380 | Spain/AN-IBV-002047/2020 | 2020-03-16 | Europe/Spain/Andalusia/Malaga | Servicio de Microbiolog√≠a. HRU de M√°laga. Servicio Andaluz de Salud | SeqCOVID-SPAIN consortium/IBV(CSIC) | Inmaculada de Toro Peinado et al |
| EPI_ISL_452385 | Spain/AN-IBV-002041/2020 | 2020-03-16 | Europe/Spain/Andalusia/Malaga | Servicio de Microbiolog√≠a. HRU de M√°laga. Servicio Andaluz de Salud | SeqCOVID-SPAIN consortium/IBV(CSIC) | Inmaculada de Toro Peinado et al |
| EPI_ISL_452391 | Spain/AN-IBV-002028/2020 | 2020-03-17 | Europe/Spain/Andalusia/Velez Malaga | Servicio de Microbiolog√≠a. HRU de M√°laga. Servicio Andaluz de Salud | SeqCOVID-SPAIN consortium/IBV(CSIC) | Inmaculada de Toro Peinado et al |
| EPI_ISL_452397 | Spain/AN-IBV-002001/2020 | 2020-03-16 | Europe/Spain/Andalusia/Malaga | Servicio de Microbiolog√≠a. HRU de M√°laga. Servicio Andaluz de Salud | SeqCOVID-SPAIN consortium/IBV(CSIC) | Inmaculada de Toro Peinado et al |
| EPI_ISL_452413 | Spain/AN-IBV-001969/2020 | 2020-03-14 | Europe/Spain/Andalusia/Almeria | Servicio de Microbiolog√≠a. HRU de M√°laga. Servicio Andaluz de Salud | SeqCOVID-SPAIN consortium/IBV(CSIC) | Inmaculada de Toro Peinado et al |
| EPI_ISL_452421 | Spain/AN-IBV-001960/2020 | 2020-03-14 | Europe/Spain/Andalusia/Malaga | Servicio de Microbiolog√≠a. HRU de M√°laga. Servicio Andaluz de Salud | SeqCOVID-SPAIN consortium/IBV(CSIC) | Inmaculada de Toro Peinado et al |
| EPI_ISL_452436 | Spain/AN-IBV-001941/2020 | 2020-03-12 | Europe/Spain/Andalusia/Cadiz | Servicio de Microbiolog√≠a. HRU de M√°laga. Servicio Andaluz de Salud | SeqCOVID-SPAIN consortium/IBV(CSIC) | Inmaculada de Toro Peinado et al |
| EPI_ISL_452440 | Spain/AN-IBV-001936/2020 | 2020-03-12 | Europe/Spain/Andalusia/Malaga | Servicio de Microbiolog√≠a. HRU de M√°laga. Servicio Andaluz de Salud | SeqCOVID-SPAIN consortium/IBV(CSIC) | Inmaculada de Toro Peinado et al |
| EPI_ISL_452441 | Spain/AN-IBV-001935/2020 | 2020-03-12 | Europe/Spain/Andalusia/Malaga | Servicio de Microbiolog√≠a. HRU de M√°laga. Servicio Andaluz de Salud | SeqCOVID-SPAIN consortium/IBV(CSIC) | Inmaculada de Toro Peinado et al |
| EPI_ISL_452444 | Spain/VC-IBV-001930/2020 | 2020-03-09 | Europe/Spain/Comunitat Valenciana/Ch | Servicio de Microbiolog√≠a. HRU de M√°laga. Servicio Andaluz de Salud | SeqCOVID-SPAIN consortium/IBV(CSIC) | Inmaculada de Toro Peinado et al |
| EPI_ISL_452452 | Spain/AN-IBV-001922/2020 | 2020-03-07 | Europe/Spain/Andalusia/Malaga | Servicio de Microbiolog√≠a. HRU de M√°laga. Servicio Andaluz de Salud | SeqCOVID-SPAIN consortium/IBV(CSIC) | Inmaculada de Toro Peinado et al |

| | | | | | | |
|---|---|---|---|---|---|---|
| EPI_ISL_452466 | Spain/AN-IBV-001880/2020 | 2020-03-11 | Europe/Spain/Andalusia/Mairena Aljara | Hospital Universitario Puerta del Mar de CV°diz - INIBICA | SeqCOVID-SPAIN consortium/IBV(CSIC) | Salud Rodrv≠guez-Pallares et al |
| EPI_ISL_452469 | Spain/AN-IBV-001876/2020 | 2020-03-09 | Europe/Spain/Andalusia/Puerto Santa M | Hospital Universitario Puerta del Mar de CV°diz - INIBICA | SeqCOVID-SPAIN consortium/IBV(CSIC) | Salud Rodrv≠guez-Pallares et al |
| EPI_ISL_452475 | Spain/NC-IBV-001426/2020 | 2020-03-23 | Europe/Spain/Navarra/Pamplona | ClV≠nica Universidad de Navarra. Servicio de Enfermedades Infecciosas y MicrobiologV≠a clV≠nica | SeqCOVID-SPAIN consortium/IBV(CSIC) | Mirian FernV°ndez-Alonso et al |
| EPI_ISL_452477 | Spain/NC-IBV-001424/2020 | 2020-03-22 | Europe/Spain/Navarra/Pamplona | ClV≠nica Universidad de Navarra. Servicio de Enfermedades Infecciosas y MicrobiologV≠a clV≠nica | SeqCOVID-SPAIN consortium/IBV(CSIC) | Mirian FernV°ndez-Alonso et al |
| EPI_ISL_452484 | Spain/NC-IBV-001417/2020 | 2020-03-22 | Europe/Spain/Navarra/Pamplona | ClV≠nica Universidad de Navarra. Servicio de Enfermedades Infecciosas y MicrobiologV≠a clV≠nica | SeqCOVID-SPAIN consortium/IBV(CSIC) | Mirian FernV°ndez-Alonso et al |
| EPI_ISL_452488 | Spain/NC-IBV-001413/2020 | 2020-03-20 | Europe/Spain/Navarra/Pamplona | ClV≠nica Universidad de Navarra. Servicio de Enfermedades Infecciosas y MicrobiologV≠a clV≠nica | SeqCOVID-SPAIN consortium/IBV(CSIC) | Mirian FernV°ndez-Alonso et al |
| EPI_ISL_452489 | Spain/NC-IBV-001412/2020 | 2020-03-20 | Europe/Spain/Navarra/Pamplona | ClV≠nica Universidad de Navarra. Servicio de Enfermedades Infecciosas y MicrobiologV≠a clV≠nica | SeqCOVID-SPAIN consortium/IBV(CSIC) | Mirian FernV°ndez-Alonso et al |
| EPI_ISL_452514 | Spain/NC-IBV-001387/2020 | 2020-03-16 | Europe/Spain/Navarra/Pamplona | ClV≠nica Universidad de Navarra. Servicio de Enfermedades Infecciosas y MicrobiologV≠a clV≠nica | SeqCOVID-SPAIN consortium/IBV(CSIC) | Mirian FernV°ndez-Alonso et al |
| EPI_ISL_452515 | Spain/NC-IBV-001386/2020 | 2020-03-15 | Europe/Spain/Navarra/Pamplona | ClV≠nica Universidad de Navarra. Servicio de Enfermedades Infecciosas y MicrobiologV≠a clV≠nica | SeqCOVID-SPAIN consortium/IBV(CSIC) | Mirian FernV°ndez-Alonso et al |
| EPI_ISL_452521 | Spain/NC-IBV-001380/2020 | 2020-03-14 | Europe/Spain/Navarra/Pamplona | ClV≠nica Universidad de Navarra. Servicio de Enfermedades Infecciosas y MicrobiologV≠a clV≠nica | SeqCOVID-SPAIN consortium/IBV(CSIC) | Mirian FernV°ndez-Alonso et al |
| EPI_ISL_452522 | Spain/NC-IBV-001379/2020 | 2020-03-14 | Europe/Spain/Navarra/Pamplona | ClV≠nica Universidad de Navarra. Servicio de Enfermedades Infecciosas y MicrobiologV≠a clV≠nica | SeqCOVID-SPAIN consortium/IBV(CSIC) | Mirian FernV°ndez-Alonso et al |
| EPI_ISL_452523 | Spain/NC-IBV-001378/2020 | 2020-03-14 | Europe/Spain/Navarra/Pamplona | ClV≠nica Universidad de Navarra. Servicio de Enfermedades Infecciosas y MicrobiologV≠a clV≠nica | SeqCOVID-SPAIN consortium/IBV(CSIC) | Mirian FernV°ndez-Alonso et al |
| EPI_ISL_452527 | Spain/NC-IBV-001374/2020 | 2020-03-13 | Europe/Spain/Navarra/Cizur Menor | ClV≠nica Universidad de Navarra. Servicio de Enfermedades Infecciosas y MicrobiologV≠a clV≠nica | SeqCOVID-SPAIN consortium/IBV(CSIC) | Mirian FernV°ndez-Alonso et al |

| | | | | | | |
|---|---|---|---|---|---|---|
| EPI_ISL_452535 | Spain/MD-IBV-001366/2020 | 2020-03-12 | Europe/Spain/Madrid/Alcobendas | Cl√≠nica Universidad de Navarra. Servicio de Enfermedades Infecciosas y Microbiolog√≠a cl√≠nica | SeqCOVID-SPAIN consortium/IBV(CSIC) | Mirian Fern√°ndez-Alonso et al |
| EPI_ISL_452541 | Spain/NC-IBV-001360/2020 | 2020-03-10 | Europe/Spain/Navarra/Baranain | Cl√≠nica Universidad de Navarra. Servicio de Enfermedades Infecciosas y Microbiolog√≠a cl√≠nica | SeqCOVID-SPAIN consortium/IBV(CSIC) | Mirian Fern√°ndez-Alonso et al |
| EPI_ISL_452542 | Spain/NC-IBV-001359/2020 | 2020-03-09 | Europe/Spain/Navarra/Pamplona | Cl√≠nica Universidad de Navarra. Servicio de Enfermedades Infecciosas y Microbiolog√≠a cl√≠nica | SeqCOVID-SPAIN consortium/IBV(CSIC) | Mirian Fern√°ndez-Alonso et al |
| EPI_ISL_452546 | Spain/AN-IBV-001327/2020 | 2020-03-23 | Europe/Spain/Andalusia/Sevilla | Servicio de Microbiolog√≠a y Parasitolog√≠a cl√≠nica. UCEIMP. Hospital Universitario Virgen del Roc√≠o/IBIS/CSIC/US. | SeqCOVID-SPAIN consortium/IBV(CSIC) | Guillermo Mart√≠¬≠n Guti√©rrez et al |
| EPI_ISL_452547 | Spain/AN-IBV-001326/2020 | 2020-03-23 | Europe/Spain/Andalusia/Sevilla | Servicio de Microbiolog√≠a y Parasitolog√≠a cl√≠nica. UCEIMP. Hospital Universitario Virgen del Roc√≠o/IBIS/CSIC/US. | SeqCOVID-SPAIN consortium/IBV(CSIC) | Guillermo Mart√≠¬≠n Guti√©rrez et al |
| EPI_ISL_452570 | Spain/AN-IBV-001169/2020 | 2020-03-26 | Europe/Spain/Andalusia/Sevilla | Servicio de Microbiolog√≠a y Parasitolog√≠a cl√≠nica. UCEIMP. Hospital Universitario Virgen del Roc√≠o/IBIS/CSIC/US. | SeqCOVID-SPAIN consortium/IBV(CSIC) | Guillermo Mart√≠¬≠n Guti√©rrez et al |
| EPI_ISL_452574 | Spain/AN-IBV-001162/2020 | 2020-03-25 | Europe/Spain/Andalusia/Sevilla | Servicio de Microbiolog√≠a y Parasitolog√≠a cl√≠nica. UCEIMP. Hospital Universitario Virgen del Roc√≠o/IBIS/CSIC/US. | SeqCOVID-SPAIN consortium/IBV(CSIC) | Guillermo Mart√≠¬≠n Guti√©rrez et al |
| EPI_ISL_452575 | Spain/AN-IBV-001161/2020 | 2020-03-25 | Europe/Spain/Andalusia/Sevilla | Servicio de Microbiolog√≠a y Parasitolog√≠a cl√≠nica. UCEIMP. Hospital Universitario Virgen del Roc√≠o/IBIS/CSIC/US. | SeqCOVID-SPAIN consortium/IBV(CSIC) | Guillermo Mart√≠¬≠n Guti√©rrez et al |
| EPI_ISL_452577 | Spain/AN-IBV-001155/2020 | 2020-03-25 | Europe/Spain/Andalusia/Sevilla | Servicio de Microbiolog√≠a y Parasitolog√≠a cl√≠nica. UCEIMP. Hospital Universitario Virgen del Roc√≠o/IBIS/CSIC/US. | SeqCOVID-SPAIN consortium/IBV(CSIC) | Guillermo Mart√≠¬≠n Guti√©rrez et al |
| EPI_ISL_452581 | Spain/AN-IBV-001120/2020 | 2020-03-28 | Europe/Spain/Andalusia/Sevilla | Servicio de Microbiolog√≠a y Parasitolog√≠a cl√≠nica. UCEIMP. Hospital Universitario Virgen del Roc√≠o/IBIS/CSIC/US. | SeqCOVID-SPAIN consortium/IBV(CSIC) | Guillermo Mart√≠¬≠n Guti√©rrez et al |

| | | | | | | |
|---|---|---|---|---|---|---|
| EPI_ISL_452585 | Spain/AN-IBV-001106/2020 | 2020-03-29 | Europe/Spain/Andalusia/Sevilla | Servicio de Microbiolog√≠a y Parasitolog√≠a cl√≠nica. UCEIMP. Hospital Universitario Virgen del Roc√≠o/IBIS/CSIC/US. | SeqCOVID-SPAIN consortium/IBV(CSIC) | Guillermo Mart√≠¬≠n Guti√©rrez et al |
| EPI_ISL_452587 | Spain/AN-IBV-001101/2020 | 2020-03-29 | Europe/Spain/Andalusia/Sevilla | Servicio de Microbiolog√≠a y Parasitolog√≠a cl√≠nica. UCEIMP. Hospital Universitario Virgen del Roc√≠o/IBIS/CSIC/US. | SeqCOVID-SPAIN consortium/IBV(CSIC) | Guillermo Mart√≠¬≠n Guti√©rrez et al |
| EPI_ISL_452592 | Spain/AN-IBV-001094/2020 | 2020-03-27 | Europe/Spain/Andalusia/Sevilla | Servicio de Microbiolog√≠a y Parasitolog√≠a cl√≠nica. UCEIMP. Hospital Universitario Virgen del Roc√≠o/IBIS/CSIC/US. | SeqCOVID-SPAIN consortium/IBV(CSIC) | Guillermo Mart√≠¬≠n Guti√©rrez et al |
| EPI_ISL_452597 | Spain/AN-IBV-001089/2020 | 2020-03-26 | Europe/Spain/Andalusia/Sevilla | Servicio de Microbiolog√≠a y Parasitolog√≠a cl√≠nica. UCEIMP. Hospital Universitario Virgen del Roc√≠o/IBIS/CSIC/US. | SeqCOVID-SPAIN consortium/IBV(CSIC) | Guillermo Mart√≠¬≠n Guti√©rrez et al |
| EPI_ISL_452619 | Spain/PV-IBV-000956/2020 | 2020-03-28 | Europe/Spain/Basque Country/Donostia | Servicio de Microbiolog√≠a. Hospital Universitario Donostia. OSI Donostialdea. √Årea de Enfermedades Infecciosas, Grupo de Infecci√≥n Respiratoria y Resistencia Antimicrobiana. Instituto de Investigaci√≥n Sanitaria Biodonostia. | SeqCOVID-SPAIN consortium/IBV(CSIC) | Gustavo Cilla et al |
| EPI_ISL_452627 | Spain/PV-IBV-000946/2020 | 2020-03-28 | Europe/Spain/Basque Country/Donostia | Servicio de Microbiolog√≠a. Hospital Universitario Donostia. OSI Donostialdea. √Årea de Enfermedades Infecciosas, Grupo de Infecci√≥n Respiratoria y Resistencia Antimicrobiana. Instituto de Investigaci√≥n Sanitaria Biodonostia. | SeqCOVID-SPAIN consortium/IBV(CSIC) | Gustavo Cilla et al |
| EPI_ISL_452639 | Spain/PV-IBV-000933/2020 | 2020-03-27 | Europe/Spain/Basque Country/Donostia | Servicio de Microbiolog√≠a. Hospital Universitario Donostia. OSI Donostialdea. √Årea de Enfermedades Infecciosas, Grupo de Infecci√≥n Respiratoria y Resistencia Antimicrobiana. Instituto de Investigaci√≥n Sanitaria Biodonostia. | SeqCOVID-SPAIN consortium/IBV(CSIC) | Gustavo Cilla et al |
| EPI_ISL_452642 | Spain/PV-IBV-000929/2020 | 2020-03-27 | Europe/Spain/Basque Country/Donostia | Servicio de Microbiolog√≠a. Hospital Universitario Donostia. OSI Donostialdea. √Årea de Enfermedades Infecciosas, Grupo de Infecci√≥n Respiratoria y Resistencia Antimicrobiana. Instituto de Investigaci√≥n Sanitaria Biodonostia. | SeqCOVID-SPAIN consortium/IBV(CSIC) | Gustavo Cilla et al |

| | | | | | | |
|---|---|---|---|---|---|---|
| EPI_ISL_452644 | Spain/PV-IBV-000927/2020 | 2020-03-27 | Europe/Spain/Basque Country/Donostia | Servicio de Microbiolog√≠a. Hospital Universitario Donostia. OSI Donostialdea. √Årea de Enfermedades Infecciosas, Grupo de Infecci√≥n Respiratoria y Resistencia Antimicrobiana. Instituto de Investigaci√≥n Sanitaria Biodonostia. | SeqCOVID-SPAIN consortium/IBV(CSIC) | Gustavo Cilla et al |
| EPI_ISL_452647 | Spain/PV-IBV-000924/2020 | 2020-03-27 | Europe/Spain/Basque Country/Donostia | Servicio de Microbiolog√≠a. Hospital Universitario Donostia. OSI Donostialdea. √Årea de Enfermedades Infecciosas, Grupo de Infecci√≥n Respiratoria y Resistencia Antimicrobiana. Instituto de Investigaci√≥n Sanitaria Biodonostia. | SeqCOVID-SPAIN consortium/IBV(CSIC) | Gustavo Cilla et al |
| EPI_ISL_452653 | Spain/PV-IBV-000915/2020 | 2020-03-27 | Europe/Spain/Basque Country/Donostia | Servicio de Microbiolog√≠a. Hospital Universitario Donostia. OSI Donostialdea. √Årea de Enfermedades Infecciosas, Grupo de Infecci√≥n Respiratoria y Resistencia Antimicrobiana. Instituto de Investigaci√≥n Sanitaria Biodonostia. | SeqCOVID-SPAIN consortium/IBV(CSIC) | Gustavo Cilla et al |
| EPI_ISL_452655 | Spain/PV-IBV-000912/2020 | 2020-03-27 | Europe/Spain/Basque Country/Donostia | Servicio de Microbiolog√≠a. Hospital Universitario Donostia. OSI Donostialdea. √Årea de Enfermedades Infecciosas, Grupo de Infecci√≥n Respiratoria y Resistencia Antimicrobiana. Instituto de Investigaci√≥n Sanitaria Biodonostia. | SeqCOVID-SPAIN consortium/IBV(CSIC) | Gustavo Cilla et al |
| EPI_ISL_452656 | Spain/PV-IBV-000911/2020 | 2020-03-26 | Europe/Spain/Basque Country/Donostia | Servicio de Microbiolog√≠a. Hospital Universitario Donostia. OSI Donostialdea. √Årea de Enfermedades Infecciosas, Grupo de Infecci√≥n Respiratoria y Resistencia Antimicrobiana. Instituto de Investigaci√≥n Sanitaria Biodonostia. | SeqCOVID-SPAIN consortium/IBV(CSIC) | Gustavo Cilla et al |
| EPI_ISL_452658 | Spain/PV-IBV-000909/2020 | 2020-03-26 | Europe/Spain/Basque Country/Donostia | Servicio de Microbiolog√≠a. Hospital Universitario Donostia. OSI Donostialdea. √Årea de Enfermedades Infecciosas, Grupo de Infecci√≥n Respiratoria y Resistencia Antimicrobiana. Instituto de Investigaci√≥n Sanitaria Biodonostia. | SeqCOVID-SPAIN consortium/IBV(CSIC) | Gustavo Cilla et al |

| | | | | | | |
|---|---|---|---|---|---|---|
| EPI_ISL_452667 | Spain/PV-IBV-000896/2020 | 2020-03-26 | Europe/Spain/Basque Country/Donostia | Servicio de Microbiolog√≠a. Hospital Universitario Donostia. OSI Donostialdea. √Årea de Enfermedades Infecciosas, Grupo de Infecci√≥n Respiratoria y Resistencia Antimicrobiana. Instituto de Investigaci√≥n Sanitaria Biodonostia. | SeqCOVID-SPAIN consortium/IBV(CSIC) | Gustavo Cilla et al |
| EPI_ISL_452673 | Spain/PV-IBV-000890/2020 | 2020-03-26 | Europe/Spain/Basque Country/Donostia | Servicio de Microbiolog√≠a. Hospital Universitario Donostia. OSI Donostialdea. √Årea de Enfermedades Infecciosas, Grupo de Infecci√≥n Respiratoria y Resistencia Antimicrobiana. Instituto de Investigaci√≥n Sanitaria Biodonostia. | SeqCOVID-SPAIN consortium/IBV(CSIC) | Gustavo Cilla et al |
| EPI_ISL_452692 | Spain/PV-IBV-000860/2020 | 2020-02-29 | Europe/Spain/Basque Country/Vitoria-G | Hospital Universitario Araba. Vitoria-Gasteiz, | SeqCOVID-SPAIN consortium/IBV(CSIC) | Silvia Hern√°ez Crespo et al |
| EPI_ISL_452695 | Spain/PV-IBV-000857/2020 | 2020-03-12 | Europe/Spain/Basque Country/Vitoria-G | Hospital Universitario Araba. Vitoria-Gasteiz, | SeqCOVID-SPAIN consortium/IBV(CSIC) | Silvia Hern√°ez Crespo et al |
| EPI_ISL_452703 | Spain/PV-IBV-000843/2020 | 2020-03-11 | Europe/Spain/Basque Country/Vitoria-G | Hospital Universitario Araba. Vitoria-Gasteiz, | SeqCOVID-SPAIN consortium/IBV(CSIC) | Silvia Hern√°ez Crespo et al |
| EPI_ISL_452705 | Spain/PV-IBV-000840/2020 | 2020-03-11 | Europe/Spain/Basque Country/Vitoria-G | Hospital Universitario Araba. Vitoria-Gasteiz, | SeqCOVID-SPAIN consortium/IBV(CSIC) | Silvia Hern√°ez Crespo et al |
| EPI_ISL_452712 | Spain/PV-IBV-000816/2020 | 2020-03-10 | Europe/Spain/Basque Country/Vitoria-G | Hospital Universitario Araba. Vitoria-Gasteiz, | SeqCOVID-SPAIN consortium/IBV(CSIC) | Silvia Hern√°ez Crespo et al |
| EPI_ISL_452717 | Spain/PV-IBV-000805/2020 | 2020-03-08 | Europe/Spain/Basque Country/Vitoria-G | Hospital Universitario Araba. Vitoria-Gasteiz, | SeqCOVID-SPAIN consortium/IBV(CSIC) | Silvia Hern√°ez Crespo et al |
| EPI_ISL_452718 | Spain/PV-IBV-000804/2020 | 2020-03-08 | Europe/Spain/Basque Country/Vitoria-G | Hospital Universitario Araba. Vitoria-Gasteiz, | SeqCOVID-SPAIN consortium/IBV(CSIC) | Silvia Hern√°ez Crespo et al |
| EPI_ISL_452721 | Spain/PV-IBV-000793/2020 | 2020-03-09 | Europe/Spain/Basque Country/Vitoria-G | Hospital Universitario Araba. Vitoria-Gasteiz, | SeqCOVID-SPAIN consortium/IBV(CSIC) | Silvia Hern√°ez Crespo et al |
| EPI_ISL_452723 | Spain/PV-IBV-000786/2020 | 2020-03-09 | Europe/Spain/Basque Country/Vitoria-G | Hospital Universitario Araba. Vitoria-Gasteiz, | SeqCOVID-SPAIN consortium/IBV(CSIC) | Silvia Hern√°ez Crespo et al |

| | | | | | | |
|---|---|---|---|---|---|---|
| EPI_ISL_452728 | Spain/PV-IBV-000775/2020 | 2020-03-09 | Europe/Spain/Basque Country/Vitoria-G | Hospital Universitario Araba. Vitoria-Gasteiz, | SeqCOVID-SPAIN consortium/IBV(CSIC) | Silvia Hernv°ez Crespo et al |
| EPI_ISL_452730 | Spain/PV-IBV-000766/2020 | 2020-03-07 | Europe/Spain/Basque Country/Vitoria-G | Hospital Universitario Araba. Vitoria-Gasteiz, | SeqCOVID-SPAIN consortium/IBV(CSIC) | Silvia Hernv°ez Crespo et al |
| EPI_ISL_452732 | Spain/PV-IBV-000759/2020 | 2020-03-06 | Europe/Spain/Basque Country/Vitoria-G | Hospital Universitario Araba. Vitoria-Gasteiz, | SeqCOVID-SPAIN consortium/IBV(CSIC) | Silvia Hernv°ez Crespo et al |
| EPI_ISL_452744 | Spain/PV-IBV-000736/2020 | 2020-03-07 | Europe/Spain/Basque Country/Vitoria-G | Hospital Universitario Araba. Vitoria-Gasteiz, | SeqCOVID-SPAIN consortium/IBV(CSIC) | Silvia Hernv°ez Crespo et al |
| EPI_ISL_452746 | Spain/PV-IBV-000729/2020 | 2020-03-07 | Europe/Spain/Basque Country/Vitoria-G | Hospital Universitario Araba. Vitoria-Gasteiz, | SeqCOVID-SPAIN consortium/IBV(CSIC) | Silvia Hernv°ez Crespo et al |
| EPI_ISL_452750 | Spain/PV-IBV-000718/2020 | 2020-03-02 | Europe/Spain/Basque Country/Vitoria-G | Hospital Universitario Araba. Vitoria-Gasteiz, | SeqCOVID-SPAIN consortium/IBV(CSIC) | Silvia Hernv°ez Crespo et al |
| EPI_ISL_452752 | Spain/PV-IBV-000716/2020 | 2020-03-05 | Europe/Spain/Basque Country/Vitoria-G | Hospital Universitario Araba. Vitoria-Gasteiz, | SeqCOVID-SPAIN consortium/IBV(CSIC) | Silvia Hernv°ez Crespo et al |
| EPI_ISL_452753 | Spain/PV-IBV-000715/2020 | 2020-03-08 | Europe/Spain/Basque Country/Vitoria-G | Hospital Universitario Araba. Vitoria-Gasteiz, | SeqCOVID-SPAIN consortium/IBV(CSIC) | Silvia Hernv°ez Crespo et al |
| EPI_ISL_452757 | Spain/PV-IBV-000709/2020 | 2020-03-04 | Europe/Spain/Basque Country/Vitoria-G | Hospital Universitario Araba. Vitoria-Gasteiz, | SeqCOVID-SPAIN consortium/IBV(CSIC) | Silvia Hernv°ez Crespo et al |
| EPI_ISL_452763 | Spain/PV-IBV-000703/2020 | 2020-03-06 | Europe/Spain/Basque Country/Vitoria-G | Hospital Universitario Araba. Vitoria-Gasteiz, | SeqCOVID-SPAIN consortium/IBV(CSIC) | Silvia Hernv°ez Crespo et al |
| EPI_ISL_452764 | Spain/PV-IBV-000702/2020 | 2020-03-04 | Europe/Spain/Basque Country/Vitoria-G | Hospital Universitario Araba. Vitoria-Gasteiz, | SeqCOVID-SPAIN consortium/IBV(CSIC) | Silvia Hernv°ez Crespo et al |
| EPI_ISL_452772 | Spain/PV-IBV-000692/2020 | 2020-02-29 | Europe/Spain/Basque Country/Vitoria-G | Hospital Universitario Araba. Vitoria-Gasteiz, | SeqCOVID-SPAIN consortium/IBV(CSIC) | Silvia Hernv°ez Crespo et al |
| EPI_ISL_452776 | Spain/PV-IBV-000686/2020 | 2020-03-04 | Europe/Spain/Basque Country/Vitoria-G | Hospital Universitario Araba. Vitoria-Gasteiz, | SeqCOVID-SPAIN consortium/IBV(CSIC) | Silvia Hernv°ez Crespo et al |

| | | | | | | |
|---|---|---|---|---|---|---|
| EPI_ISL_452782 | Spain/PV-IBV-000670/2020 | 2020-03-06 | Europe/Spain/Basque Country/Vitoria-G | Hospital Universitario Araba. Vitoria-Gasteiz, | SeqCOVID-SPAIN consortium/IBV(CSIC) | Silvia Hernv°ez Crespo et al |
| EPI_ISL_452783 | Spain/PV-IBV-000669/2020 | 2020-03-06 | Europe/Spain/Basque Country/Vitoria-G | Hospital Universitario Araba. Vitoria-Gasteiz, | SeqCOVID-SPAIN consortium/IBV(CSIC) | Silvia Hernv°ez Crespo et al |
| EPI_ISL_452785 | Spain/PV-IBV-000665/2020 | 2020-03-06 | Europe/Spain/Basque Country/Vitoria-G | Hospital Universitario Araba. Vitoria-Gasteiz, | SeqCOVID-SPAIN consortium/IBV(CSIC) | Silvia Hernv°ez Crespo et al |
| EPI_ISL_452794 | India/MP-NIHSAD-0905-216/2020 | 2020-05-08 | Asia/India/Madhya Pradesh/Neemach | ICAR-National Institute of High Security Animal Diseases | ICAR-National Institute of High Security Animal Diseases | Anamika Mishra et al |
| EPI_ISL_452798 | USA/VA-DCLS-0220/2020 | 2020-04-03 | North America/USA/Virginia/ | Virginia DCLS | Virginia DCLS | Virginia DCLS et al |
| EPI_ISL_452803 | USA/VA-DCLS-0226/2020 | 2020-04-03 | North America/USA/Virginia/ | Virginia DCLS | Virginia DCLS | Virginia DCLS et al |
| EPI_ISL_452807 | USA/VA-DCLS-0230/2020 | 2020-04-05 | North America/USA/Virginia/ | Virginia DCLS | Virginia DCLS | Virginia DCLS et al |
| EPI_ISL_452813 | USA/VA-DCLS-0238/2020 | 2020-04-15 | North America/USA/Virginia/ | Virginia DCLS | Virginia DCLS | Virginia DCLS et al |
| EPI_ISL_452818 | USA/VA-DCLS-0244/2020 | 2020-04-16 | North America/USA/Virginia/ | Virginia DCLS | Virginia DCLS | Virginia DCLS et al |
| EPI_ISL_452825 | USA/VA-DCLS-0251/2020 | 2020-04-20 | North America/USA/Virginia/ | Virginia DCLS | Virginia DCLS | Virginia DCLS et al |
| EPI_ISL_452826 | USA/VA-DCLS-0252/2020 | 2020-04-20 | North America/USA/Virginia/ | Virginia DCLS | Virginia DCLS | Virginia DCLS et al |
| EPI_ISL_452827 | USA/VA-DCLS-0253/2020 | 2020-04-20 | North America/USA/Virginia/ | Virginia DCLS | Virginia DCLS | Virginia DCLS et al |
| EPI_ISL_452829 | USA/VA-DCLS-0255/2020 | 2020-04-20 | North America/USA/Virginia/ | Virginia DCLS | Virginia DCLS | Virginia DCLS et al |
| EPI_ISL_452839 | USA/VA-DCLS-0266/2020 | 2020-04-22 | North America/USA/Virginia/ | Virginia DCLS | Virginia DCLS | Virginia DCLS et al |
| EPI_ISL_452840 | USA/VA-DCLS-0271/2020 | 2020-04-22 | North America/USA/Virginia/ | Virginia DCLS | Virginia DCLS | Virginia DCLS et al |
| EPI_ISL_452844 | USA/VA-DCLS-0275/2020 | 2020-04-22 | North America/USA/Virginia/ | Virginia DCLS | Virginia DCLS | Virginia DCLS et al |
| EPI_ISL_452856 | USA/VA-DCLS-0287/2020 | 2020-04-21 | North America/USA/Virginia/ | Virginia DCLS | Virginia DCLS | Virginia DCLS et al |
| EPI_ISL_452859 | England/CAMB-1AED75/2020 | 2020-05-07 | Europe/United Kingdom/England/ | Department of Pathology, University of Cambridge | COVID-19 Genomics UK (COG-UK) Consortium | Luke W Meredith et al |
| EPI_ISL_452896 | England/CAMB-1B05B4/2020 | 2020-05-11 | Europe/United Kingdom/England/ | Department of Pathology, University of Cambridge | COVID-19 Genomics UK (COG-UK) Consortium | Luke W Meredith et al |
| EPI_ISL_452946 | England/CAMB-1B0B76/2020 | 2020-05-13 | Europe/United Kingdom/England/ | Department of Pathology, University of Cambridge | COVID-19 Genomics UK (COG-UK) Consortium | Luke W Meredith et al |

| | | | | | | |
|---|---|---|---|---|---|---|
| EPI_ISL_452978 | England/CAMB-1B179F/2020 | 2020-05-13 | Europe/United Kingdom/England/ | Department of Pathology, University of Cambridge | COVID-19 Genomics UK (COG-UK) Consortium | Luke W Meredith et al |
| EPI_ISL_453014 | Scotland/CVR2013/2020 | 2020-04-09 | Europe/United Kingdom/Scotland/ | West of Scotland Specialist Virology Centre, NHSGGC / MRC-University of Glasgow Centre for Virus Research | COVID-19 Genomics UK (COG-UK) Consortium | Ana da Silva Filipe et al |
| EPI_ISL_453192 | Scotland/EDB5113/2020 | 2020-04-24 | Europe/United Kingdom/Scotland/ | Virology Department, Royal Infirmary of Edinburgh, NHS Lothian / School of Biological Sciences, University of Edinburgh / Institute of Genetics and Molecular Medicine, University of Edinburgh | COVID-19 Genomics UK (COG-UK) Consortium | McHugh M et al |
| EPI_ISL_453272 | England/LIVE-9E17D/2020 | 2020-04-03 | Europe/United Kingdom/England/ | Liverpool Clinical Laboratories | COVID-19 Genomics UK (COG-UK) Consortium | Sam Haldenby et al |
| EPI_ISL_453362 | England/LIVE-9EABE/2020 | 2020-04-04 | Europe/United Kingdom/England/ | Liverpool Clinical Laboratories | COVID-19 Genomics UK (COG-UK) Consortium | Sam Haldenby et al |
| EPI_ISL_453408 | England/LIVE-9EEEF/2020 | 2020-04-09 | Europe/United Kingdom/England/ | Liverpool Clinical Laboratories | COVID-19 Genomics UK (COG-UK) Consortium | Sam Haldenby et al |
| EPI_ISL_453433 | England/LIVE-9F19A/2020 | 2020-05-03 | Europe/United Kingdom/England/ | Liverpool Clinical Laboratories | COVID-19 Genomics UK (COG-UK) Consortium | Sam Haldenby et al |
| EPI_ISL_453563 | England/NORW-E8EF5/2020 | 2020-04-24 | Europe/United Kingdom/England/ | Quadram Institute Bioscience | COVID-19 Genomics UK (COG-UK) Consortium | Dave J. Baker et al |
| EPI_ISL_453565 | England/NORW-E8F3E/2020 | 2020-04-26 | Europe/United Kingdom/England/ | Quadram Institute Bioscience | COVID-19 Genomics UK (COG-UK) Consortium | Dave J. Baker et al |
| EPI_ISL_453638 | England/NOTT-11127E/2020 | 2020-04-23 | Europe/United Kingdom/England/ | Queens Medical Centre, Clinical Microbiology Department / DeepSeq Nottingham | COVID-19 Genomics UK (COG-UK) Consortium | Gemma Clark et al |
| EPI_ISL_453684 | England/PORT-2CF496/2020 | 2020-04-27 | Europe/United Kingdom/England/ | Centre for Enzyme Innovation, University of Portsmouth / Translational Research Laboratory, Portsmouth Hospitals NHS Trust | COVID-19 Genomics UK (COG-UK) Consortium | Angela Beckett et al |
| EPI_ISL_453716 | England/SHEF-C576C/2020 | 2020-04-22 | Europe/United Kingdom/England/ | Virology Department, Sheffield Teaching Hospitals NHS Foundation Trust/Department of Infection, Immunity and Cardiovascular Disease, The Medical School, University of Sheffield | COVID-19 Genomics UK (COG-UK) Consortium | Thushan de Silva et al |

| | | | | | | |
|---|---|---|---|---|---|---|
| EPI_ISL_453863 | Portugal/PT0147/2020 | 2020-03-27 | Europe/Portugal/Portugal/ | unknown | Instituto Nacional de Saude (INSA) | Borges et al et al |
| EPI_ISL_453922 | Portugal/PT0208/2020 | 2020-03-30 | Europe/Portugal/Portugal/ | unknown | Instituto Nacional de Saude (INSA) | Borges et al et al |
| EPI_ISL_453953 | Portugal/PT0239/2020 | 2020-03-31 | Europe/Portugal/Portugal/ | unknown | Instituto Nacional de Saude (INSA) | Borges et al et al |
| EPI_ISL_453962 | Portugal/PT0248/2020 | 2020-04-02 | Europe/Portugal/Portugal/ | unknown | Instituto Nacional de Saude (INSA) | Borges et al et al |
| EPI_ISL_453975 | Portugal/PT0261/2020 | 2020-04-03 | Europe/Portugal/Portugal/ | unknown | Instituto Nacional de Saude (INSA) | Borges et al et al |
| EPI_ISL_454017 | Portugal/PT0301/2020 | 2020-03-14 | Europe/Portugal/Portugal/ | unknown | Instituto Nacional de Saude (INSA) | Borges et al et al |
| EPI_ISL_454021 | Portugal/PT0305/2020 | 2020-03-14 | Europe/Portugal/Portugal/ | unknown | Instituto Nacional de Saude (INSA) | Borges et al et al |
| EPI_ISL_454044 | Portugal/PT0328/2020 | 2020-03-16 | Europe/Portugal/Portugal/ | unknown | Instituto Nacional de Saude (INSA) | Borges et al et al |
| EPI_ISL_454101 | Portugal/PT0385/2020 | 2020-03-20 | Europe/Portugal/Portugal/ | unknown | Instituto Nacional de Saude (INSA) | Borges et al et al |
| EPI_ISL_454107 | Portugal/PT0389c/2020 | 2020-04-23 | Europe/Portugal/Portugal/ | unknown | Instituto Nacional de Saude (INSA) | Borges et al et al |
| EPI_ISL_454119 | Portugal/PT0397a/2020 | 2020-04-19 | Europe/Portugal/Portugal/ | unknown | Instituto Nacional de Saude (INSA) | Borges et al et al |
| EPI_ISL_454135 | Portugal/PT0411/2020 | 2020-03-21 | Europe/Portugal/Portugal/ | unknown | Instituto Nacional de Saude (INSA) | Borges et al et al |
| EPI_ISL_454155 | Portugal/PT0431/2020 | 2020-04-01 | Europe/Portugal/Portugal/ | unknown | Instituto Nacional de Saude (INSA) | Borges et al et al |
| EPI_ISL_454182 | Portugal/PT0458/2020 | 2020-04-19 | Europe/Portugal/Portugal/ | unknown | Instituto Nacional de Saude (INSA) | Borges et al et al |
| EPI_ISL_454185 | Portugal/PT0461/2020 | 2020-04-17 | Europe/Portugal/Portugal/ | unknown | Instituto Nacional de Saude (INSA) | Borges et al et al |
| EPI_ISL_454191 | Portugal/PT0467/2020 | 2020-04-18 | Europe/Portugal/Portugal/ | unknown | Instituto Nacional de Saude (INSA) | Borges et al et al |
| EPI_ISL_454192 | Portugal/PT0468/2020 | 2020-04-18 | Europe/Portugal/Portugal/ | unknown | Instituto Nacional de Saude (INSA) | Borges et al et al |
| EPI_ISL_454194 | Portugal/PT0470a/2020 | 2020-04-18 | Europe/Portugal/Portugal/ | unknown | Instituto Nacional de Saude (INSA) | Borges et al et al |
| EPI_ISL_454201 | Portugal/PT0477/2020 | 2020-04-26 | Europe/Portugal/Portugal/ | unknown | Instituto Nacional de Saude (INSA) | Borges et al et al |
| EPI_ISL_454211 | Portugal/PT0487/2020 | 2020-04-01 | Europe/Portugal/Portugal/ | unknown | Instituto Nacional de Saude (INSA) | Borges et al et al |

| | | | | | | |
|---|---|---|---|---|---|---|
| EPI_ISL_454231 | Portugal/PT0507/2020 | 2020-03-19 | Europe/Portugal/Portugal/ | unknown | Instituto Nacional de Saude (INSA) | Borges et al et al |
| EPI_ISL_454238 | Portugal/PT0514/2020 | 2020-03-20 | Europe/Portugal/Portugal/ | unknown | Instituto Nacional de Saude (INSA) | Borges et al et al |
| EPI_ISL_454254 | Portugal/PT0530/2020 | 2020-04-06 | Europe/Portugal/Portugal/ | unknown | Instituto Nacional de Saude (INSA) | Borges et al et al |
| EPI_ISL_454257 | Portugal/PT0533/2020 | 2020-04-07 | Europe/Portugal/Portugal/ | unknown | Instituto Nacional de Saude (INSA) | Borges et al et al |
| EPI_ISL_454262 | Portugal/PT0538/2020 | 2020-04-07 | Europe/Portugal/Portugal/ | unknown | Instituto Nacional de Saude (INSA) | Borges et al et al |
| EPI_ISL_454266 | Portugal/PT0542/2020 | 2020-04-08 | Europe/Portugal/Portugal/ | unknown | Instituto Nacional de Saude (INSA) | Borges et al et al |
| EPI_ISL_454267 | Portugal/PT0543/2020 | 2020-04-08 | Europe/Portugal/Portugal/ | unknown | Instituto Nacional de Saude (INSA) | Borges et al et al |
| EPI_ISL_454270 | Portugal/PT0546/2020 | 2020-04-14 | Europe/Portugal/Portugal/ | unknown | Instituto Nacional de Saude (INSA) | Borges et al et al |
| EPI_ISL_454271 | Portugal/PT0547/2020 | 2020-04-03 | Europe/Portugal/Portugal/ | unknown | Instituto Nacional de Saude (INSA) | Borges et al et al |
| EPI_ISL_454276 | Portugal/PT0553/2020 | 2020-04-07 | Europe/Portugal/Portugal/ | unknown | Instituto Nacional de Saude (INSA) | Borges et al et al |
| EPI_ISL_454317 | Portugal/PT0594/2020 | 2020-04-03 | Europe/Portugal/Portugal/ | unknown | Instituto Nacional de Saude (INSA) | Borges et al et al |
| EPI_ISL_454324 | Portugal/PT0601/2020 | 2020-04-06 | Europe/Portugal/Portugal/ | unknown | Instituto Nacional de Saude (INSA) | Borges et al et al |
| EPI_ISL_454341 | Portugal/PT0618/2020 | 2020-03-17 | Europe/Portugal/Portugal/ | unknown | Instituto Nacional de Saude (INSA) | Borges et al et al |
| EPI_ISL_454353 | USA/PA-MGEL62/2020 | 2020-03-30 | North America/USA/Pennsylvania/Alleg | Microbial Genome Sequencing Center; Microbial Genomic Epidemiology Laboratory | Microbial Genomic Epidemiology Laboratory, University of Pittsburgh | Mustapha M. Mustapha et al |
| EPI_ISL_454356 | USA/PA-MGEL66/2020 | 2020-03-31 | North America/USA/Pennsylvania/Alleg | UPMC Clinical Microbiology Laboratory | Microbial Genome Sequencing Center, Microbial Genomic Epidemiological Laboratory | Mustapha M. Mustapha et al |

| | | | | | | |
|---|---|---|---|---|---|---|
| EPI_ISL_454358 | USA/PA-MGEL70/2020 | 2020-03-30 | North America/USA/Pennsylvania/Alleg | UPMC Clinical Microbiology Laboratory | Microbial Genome Sequencing Center, Microbial Genomic Epidemiological Laboratory | Mustapha M. Mustapha et al |
| EPI_ISL_454370 | USA/PA-MGEL86/2020 | 2020-04-01 | North America/USA/Pennsylvania/Alleg | UPMC Clinical Microbiology Laboratory | Microbial Genome Sequencing Center, Microbial Genomic Epidemiological Laboratory | Mustapha M. Mustapha et al |
| EPI_ISL_454407 | USA/PA-MGEL149/2020 | 2020-04-10 | North America/USA/Pennsylvania/Alleg | UPMC Clinical Microbiology Laboratory | Microbial Genome Sequencing Center, Microbial Genomic Epidemiological Laboratory | Mustapha M. Mustapha et al |
| EPI_ISL_454411 | USA/PA-MGEL155/2020 | 2020-04-08 | North America/USA/Pennsylvania/Alleg | UPMC Clinical Microbiology Laboratory | Microbial Genome Sequencing Center, Microbial Genomic Epidemiological Laboratory | Mustapha M. Mustapha et al |
| EPI_ISL_454417 | Guangdong/MZ01/2020 | 2020-02-13 | Asia/China/Guangdong/Meizhou | Research and Experiment Center, Meizhou People Hospital | Research and Experiment Center, Meizhou People Hospital | Guo et al |
| EPI_ISL_454418 | Guangdong/MZ02/2020 | 2020-02-15 | Asia/China/Guangdong/Meizhou | Research and Experiment Center, Meizhou People Hospital | Research and Experiment Center, Meizhou People Hospital | Guo et al |
| EPI_ISL_454420 | Lebanon/S7_763/2020 | 2020-02-21 | Asia/Iran/Beirut/ | Rafik Hariri University Hospital | Rafik Hariri University Hospital | Rita Feghali et al |
| EPI_ISL_454444 | Sweden/20-08129/2020 | 2020-03-10 | Europe/Sweden/Halland/ | Halmstad klinisk mikrobiologi | The Public Health Agency of Sweden | Anna-Malin Linde et al |
| EPI_ISL_454454 | Sweden/20-08079/2020 | 2020-03-05 | Europe/Sweden/Stockholm/ | Karolinska Universitetslaboratoriet | The Public Health Agency of Sweden | Anna-Malin Linde et al |
| EPI_ISL_454464 | Sweden/20-08092/2020 | 2020-03-06 | Europe/Sweden/Stockholm/ | Karolinska Universitetslaboratoriet | The Public Health Agency of Sweden | Anna-Malin Linde et al |

| | | | | | | |
|---|---|---|---|---|---|---|
| EPI_ISL_454479 | Sweden/20-08108/2020 | 2020-03-09 | Europe/Sweden/Stockholm/ | Karolinska Universitetslaboratoriet | The Public Health Agency of Sweden | Anna-Malin Linde et al |
| EPI_ISL_454483 | Sweden/20-08112/2020 | 2020-03-09 | Europe/Sweden/Stockholm/ | Karolinska Universitetslaboratoriet | The Public Health Agency of Sweden | Anna-Malin Linde et al |
| EPI_ISL_454497 | Kazakhstan/16173/2020 | 2020-05-14 | Asia/Kazakhstan/Nur-Sultan/ | "RSE ""National Center for Biotechnology""" | "RSE ""National Center for Biotechnology""" | Alexandr Shevtsov et al |
| EPI_ISL_454499 | Kazakhstan/16537/2020 | 2020-04-16 | Asia/Kazakhstan/Nur-Sultan/ | "RSE ""National Center for Biotechnology""" | "RSE ""National Center for Biotechnology""" | Alexandr Shevtsov et al |
| EPI_ISL_454501 | Kazakhstan/20679/2020 | 2020-04-20 | Asia/Kazakhstan/Nur-Sultan/ | "RSE ""National Center for Biotechnology""" | "RSE ""National Center for Biotechnology""" | Alexandr Shevtsov et al |
| EPI_ISL_454504 | Kazakhstan/26473/2020 | 2020-04-26 | Asia/Kazakhstan/Nur-Sultan/ | "RSE ""National Center for Biotechnology""" | "RSE ""National Center for Biotechnology""" | Alexandr Shevtsov et al |
| EPI_ISL_454549 | India/MH-NIV-7558/2020 | 2020-04-05 | Asia/India/Maharashtra/ | NIV Influenza | NIV Influenza | Potdar V et al |
| EPI_ISL_454552 | India/MH-NIV-7966/2020 | 2020-04-06 | Asia/India/Maharashtra/ | NIV Influenza | NIV Influenza | Potdar V et al |
| EPI_ISL_454555 | India/MH-NIV-8404/2020 | 2020-04-07 | Asia/India/Maharashtra/ | NIV Influenza | NIV Influenza | Potdar V et al |
| EPI_ISL_454574 | Croatia/OY-S1new/2020 | 2020-04-09 | Europe/Croatia/Istria/ | nstitute for Public Health | Laboratory for advanced genomics | Filip Rokifá et al |
| EPI_ISL_454575 | Kazakhstan/38716/2020 | 2020-05-09 | Asia/Kazakhstan/Nur-Sultan/ | Laboratory of virology, National Center of Expertise | Laboratory of molecular-genetic research, National Center for Expertise, Kazakhstan National Center for Biotechnology, Kazakhstan | Abdaliyev Askar et al |
| EPI_ISL_454581 | Croatia/1146_Split/2020 | 2020-03-18 | Europe/Croatia/Split/ | "University Hospital for Infectious Diseases ""Dr. Fran Mihaljevifá"", Research Unit" | University of Zagreb, Centre for research and knowledge transfer in biotechnology | Ivan-Christian Kurolt et al |

| | | | | | | |
|---|---|---|---|---|---|---|
| EPI_ISL_454585 | Kazakhstan/33496/2020 | 2020-05-04 | Asia/Kazakhstan/Nur-Sultan/ | Laboratory of virology, National Center of Expertise | Laboratory of molecular-genetic research, National Center for Expertise, Kazakhstan National Center for Biotechnology, Kazakhstan | Abdaliyev Askar et al |
| EPI_ISL_454592 | Croatia/297_Varazdin/2020 | 2020-03-05 | Europe/Croatia/Varazdin/ | "University Hospital for Infectious Diseases ""Dr. Fran Mihaljevifá"", Research Unit" | University of Zagreb, Centre for research and knowledge transfer in biotechnology | Ivan-Christian Kurolt et al |
| EPI_ISL_454594 | Kazakhstan/26576/2020 | 2020-04-26 | Asia/Kazakhstan/Nur-Sultan/ | Laboratory of virology, National Center of Expertise | Laboratory of molecular-genetic research, National Center of Expertise, Kazakhstan National Center for Biotechnology, Kazakhstan | Abdaliyev Askar et al |
| EPI_ISL_454599 | Kazakhstan/26562/2020 | 2020-04-26 | Asia/Kazakhstan/Nur-Sultan/ | Laboratory of virology, National Center of Expertise | Laboratory of molecular-genetic research, National Center for Expertise, Kazakhstan National Center for Biotechnology, Kazakhstan | Abdaliyev Askar et al |

| | | | | | | |
|---|---|---|---|---|---|---|
| EPI_ISL_454600 | Kazakhstan/26565/2020 | 2020-04-26 | Asia/Kazakhstan/Nur-Sultan/ | Laboratory of virology, National Center of Expertise | Laboratory of molecular-genetic research, National Center for Expertise, Kazakhstan National Center for Biotechnology, Kazakhstan | Abdaliyev Askar et al |
| EPI_ISL_454602 | Croatia/14412_Koprivnica/2020 | 2020-04-21 | Europe/Croatia/Koprivnica/ | Croatian Institute of Public Health | University of Zagreb, Centre for research and knowledge transfer in biotechnology | Irena Tabain et al |
| EPI_ISL_454604 | Kazakhstan/26580/2020 | 2020-04-26 | Asia/Kazakhstan/Nur-Sultan/ | Laboratory of virology, National Center of Expertise | Laboratory of molecular-genetic research, National Center for Expertise, Kazakhstan National Center for Biotechnology, Kazakhstan | Abdaliyev Askar et al |
| EPI_ISL_454605 | Croatia/8U-S17new/2020 | 2020-03-31 | Europe/Croatia/Istria/ | Institute for Public Health | Laboratory for advanced genomics | Filip Rokifá et al |
| EPI_ISL_454606 | Croatia/I7-S21new/2020 | 2020-03-30 | Europe/Croatia/Istria/ | Institute for Public Health | Laboratory for advanced genomics | Filip Rokifá et al |
| EPI_ISL_454607 | USA/CA-CZB-1270/2020 | 2020-03-19 | North America/USA/California/Alameda | Alameda County Public Health Lab | Chan-Zuckerberg Biohub | CZB Cliahub Consortium et al |
| EPI_ISL_454615 | USA/CA-CZB-1150/2020 | 2020-04-26 | North America/USA/California/San Fran | UCSF Clinical Microbiology Laboratory | Chan-Zuckerberg Biohub | CZB Cliahub Consortium et al |
| EPI_ISL_454621 | USA/CA-CZB-1164/2020 | 2020-04-27 | North America/USA/California/San Fran | UCSF Clinical Microbiology Laboratory | Chan-Zuckerberg Biohub | CZB Cliahub Consortium et al |
| EPI_ISL_454625 | USA/CA-CZB-1174/2020 | 2020-04-30 | North America/USA/California/San Fran | UCSF Clinical Microbiology Laboratory | Chan-Zuckerberg Biohub | CZB Cliahub Consortium et al |
| EPI_ISL_454628 | USA/CA-CZB-1178/2020 | 2020-04-30 | North America/USA/California/San Fran | UCSF Clinical Microbiology Laboratory | Chan-Zuckerberg Biohub | CZB Cliahub Consortium et al |

| | | | | | | |
|---|---|---|---|---|---|---|
| EPI_ISL_454638 | USA/CA-CZB-1280/2020 | 2020-03-31 | North America/USA/California/Humbold | Humboldt County Public Health Laboratory | Chan-Zuckerberg Biohub | CZB Cliahub Consortium et al |
| EPI_ISL_454642 | USA/CT-CDC-6001/2020 | 2020-04-07 | North America/USA/Connecticut/ | CT-Dr. Katherine A. Kelley State Public Health Lab | Pathogen Discovery, Respiratory Viruses Branch, Division of Viral Diseases, Centers for Disease Control and Prevention | Jing Zhang et al |
| EPI_ISL_454644 | USA/VI-CDC-4411/2020 | 2020-04-27 | North America/USA/Virgin Islands/ | VI-US Virgin Islands Department of Health | Pathogen Discovery, Respiratory Viruses Branch, Division of Viral Diseases, Centers for Disease Control and Prevention | Jing Zhang et al |
| EPI_ISL_454649 | USA/VI-CDC-4371/2020 | 2020-04-26 | North America/USA/Virgin Islands/ | VI-US Virgin Islands Department of Health | Pathogen Discovery, Respiratory Viruses Branch, Division of Viral Diseases, Centers for Disease Control and Prevention | Ying Tao et al |
| EPI_ISL_454651 | USA/VI-CDC-4408/2020 | 2020-04-27 | North America/USA/Virgin Islands/ | VI-US Virgin Islands Department of Health | Pathogen Discovery, Respiratory Viruses Branch, Division of Viral Diseases, Centers for Disease Control and Prevention | Ying Tao et al |
| EPI_ISL_454655 | USA/CA-CZB-1235/2020 | 2020-04-11 | North America/USA/California/Santa Cl | County of Santa Clara Public Health Department | Chan-Zuckerberg Biohub | CZB Cliahub Consortium et al |

| | | | | | | |
|---|---|---|---|---|---|---|
| EPI_ISL_454665 | USA/CA-CZB-1245/2020 | 2020-04-14 | North America/USA/California/Santa Cl | County of Santa Clara Public Health Department | Chan-Zuckerberg Biohub | CZB Cliahub Consortium et al |
| EPI_ISL_454670 | USA/CA-CZB-1250/2020 | 2020-04-12 | North America/USA/California/Santa Cl | County of Santa Clara Public Health Department | Chan-Zuckerberg Biohub | CZB Cliahub Consortium et al |
| EPI_ISL_454672 | USA/CA-CZB-1252/2020 | 2020-05-12 | North America/USA/California/Santa Cl | County of Santa Clara Public Health Department | Chan-Zuckerberg Biohub | CZB Cliahub Consortium et al |
| EPI_ISL_454674 | USA/CA-CZB-1254/2020 | 2020-04-12 | North America/USA/California/Santa Cl | County of Santa Clara Public Health Department | Chan-Zuckerberg Biohub | CZB Cliahub Consortium et al |
| EPI_ISL_454681 | USA/CA-CZB-1261/2020 | 2020-05-12 | North America/USA/California/Santa Cl | County of Santa Clara Public Health Department | Chan-Zuckerberg Biohub | CZB Cliahub Consortium et al |
| EPI_ISL_454690 | USA/GA-EHC-083E/2020 | 2020-03-09 | North America/USA/Georgia/ | Emory Molecular Diagnostics Laboratory, Emory Healthcare | Piantadosi Lab, Emory Department of Pathology | Ahmed Babiker et al |
| EPI_ISL_454711 | USA/UT-01687/2020 | 2020-04-13 | North America/USA/Utah/ | Utah Public Health Laboratory | Utah Public Health Laboratory | Erin Young et al |
| EPI_ISL_454716 | USA/UT-01696/2020 | 2020-04-13 | North America/USA/Utah/ | Utah Public Health Laboratory | Utah Public Health Laboratory | Erin Young et al |
| EPI_ISL_454718 | USA/UT-01893/2020 | 2020-04-13 | North America/USA/Utah/ | Utah Public Health Laboratory | Utah Public Health Laboratory | Erin Young et al |
| EPI_ISL_454720 | USA/UT-01895/2020 | 2020-04-13 | North America/USA/Utah/ | Utah Public Health Laboratory | Utah Public Health Laboratory | Erin Young et al |
| EPI_ISL_454726 | USA/UT-02008/2020 | 2020-04-16 | North America/USA/Utah/ | Utah Public Health Laboratory | Utah Public Health Laboratory | Erin Young et al |
| EPI_ISL_454729 | USA/UT-02016/2020 | 2020-04-16 | North America/USA/Utah/ | Utah Public Health Laboratory | Utah Public Health Laboratory | Erin Young et al |
| EPI_ISL_454730 | USA/UT-02024/2020 | 2020-04-17 | North America/USA/Utah/ | Utah Public Health Laboratory | Utah Public Health Laboratory | Erin Young et al |
| EPI_ISL_454732 | Russia/Moscow_PMVL-6/2020 | 2020-04-02 | Europe/Russia/Moscow Oblast/ | Russian State Collection of Viruses | Pathogenic Microorganisms Variability Laboratory | Denis Protsenko et al |
| EPI_ISL_454733 | Italy/TUS-UniSI-1/2020 | 2020-03-01 | Europe/Italy/Tuscany/Siena | Department of Medical, Biotechnologies University of Siena | Department of Medical, Biotechnologies University of Siena | Cusi et al |

| | | | | | | |
|---|---|---|---|---|---|---|
| EPI_ISL_454749 | Japan/DP0321/2020 | 2020-02-16 | Asia/Japan/Japan/ | Japanese Quarantine Stations | Pathogen Genomics Center, National Institute of Infectious Diseases | Tsuyoshi Sekizuka et al |
| EPI_ISL_454750 | Netherlands/NoordHolland_10001/2( | 2020-02-27 | Europe/Netherlands/Utrecht/ | Dutch COVID-19 response team | National Institute for Public Health and the Environment (RIVM) | Adam Meijer et al |
| EPI_ISL_454751 | Netherlands/NoordHolland_10002/2( | 2020-02-28 | Europe/Netherlands/Utrecht/ | Dutch COVID-19 response team | National Institute for Public Health and the Environment (RIVM) | Adam Meijer et al |
| EPI_ISL_454752 | Netherlands/NoordBrabant_10003/2 | 2020-02-28 | Europe/Netherlands/Utrecht/ | Dutch COVID-19 response team | National Institute for Public Health and the Environment (RIVM) | Adam Meijer et al |
| EPI_ISL_454753 | Netherlands/ZuidHolland_10004/202 | 2020-02-28 | Europe/Netherlands/Utrecht/ | Dutch COVID-19 response team | National Institute for Public Health and the Environment (RIVM) | Adam Meijer et al |
| EPI_ISL_454754 | Netherlands/ZuidHolland_10005/202 | 2020-02-28 | Europe/Netherlands/Utrecht/ | Dutch COVID-19 response team | National Institute for Public Health and the Environment (RIVM) | Adam Meijer et al |
| EPI_ISL_454778 | Netherlands/Utrecht_10029/2020 | 2020-04-03 | Europe/Netherlands/Utrecht/ | Dutch COVID-19 response team | National Institute for Public Health and the Environment (RIVM) | Adam Meijer et al |
| EPI_ISL_454791 | Netherlands/NoordBrabant_10042/2 | 2020-03-25 | Europe/Netherlands/Utrecht/ | Dutch COVID-19 response team | National Institute for Public Health and the Environment (RIVM) | Adam Meijer et al |

| | | | | | | |
|---|---|---|---|---|---|---|
| EPI_ISL_454830 | India/RJ-SMSCOV109/2020 | 2020-04-23 | Asia/India/Rajasthan/Jaipur | SMS Medical College, Jaipur | CSIR Institute of Genomics and Integrative Biology | Sudhir Bhandari et al |
| EPI_ISL_454875 | Sweden/20-50093/2020 | 2020-03-04 | Europe/Sweden/Stockholm/ | Karolinska Universitetslaboratoriet | The Public Health Agency of Sweden | Anna-Malin Linde et al |
| EPI_ISL_454889 | Sweden/20-50208/2020 | 2020-03-06 | Europe/Sweden/Stockholm/ | Karolinska Universitetslaboratoriet | The Public Health Agency of Sweden | Anna-Malin Linde et al |
| EPI_ISL_454906 | Wuhan/HB-WH1-125/2020 | 2020-03-02 | Asia/China/Hubei/Wuhan | Wuhan Chain Medical Labs (CMLabs) | State Key Laboratory of Biotherapy of Sichuan University | Baowen Du et al |
| EPI_ISL_454911 | Wuhan/HB-WH1-134/2020 | 2020-02-22 | Asia/China/Hubei/Wuhan | Wuhan Chain Medical Labs (CMLabs) | State Key Laboratory of Biotherapy of Sichuan University | Baowen Du et al |
| EPI_ISL_454912 | Wuhan/HB-WH1-135/2020 | 2020-02-25 | Asia/China/Hubei/Wuhan | Wuhan Chain Medical Labs (CMLabs) | State Key Laboratory of Biotherapy of Sichuan University | Baowen Du et al |
| EPI_ISL_454915 | Wuhan/HB-WH1-138/2020 | 2020-02-24 | Asia/China/Hubei/Wuhan | Wuhan Chain Medical Labs (CMLabs) | State Key Laboratory of Biotherapy of Sichuan University | Baowen Du et al |
| EPI_ISL_454916 | Wuhan/HB-WH1-140/2020 | 2020-02-27 | Asia/China/Hubei/Wuhan | Wuhan Chain Medical Labs (CMLabs) | State Key Laboratory of Biotherapy of Sichuan University | Baowen Du et al |
| EPI_ISL_454919 | Wuhan/HB-WH1-143/2020 | 2020-02-03 | Asia/China/Hubei/Wuhan | Wuhan Chain Medical Labs (CMLabs) | State Key Laboratory of Biotherapy of Sichuan University | Baowen Du et al |

| | | | | | | |
|---|---|---|---|---|---|---|
| EPI_ISL_454920 | Wuhan/HB-WH2-148/2020 | 2020-02-21 | Asia/China/Hubei/Wuhan | Wuhan Chain Medical Labs (CMLabs) | State Key Laboratory of Biotherapy of Sichuan University | Baowen Du et al |
| EPI_ISL_454921 | Wuhan/HB-WH2-149/2020 | 2020-02-21 | Asia/China/Hubei/Wuhan | Wuhan Chain Medical Labs (CMLabs) | State Key Laboratory of Biotherapy of Sichuan University | Baowen Du et al |
| EPI_ISL_454922 | Wuhan/HB-WH2-152/2020 | 2020-02-28 | Asia/China/Hubei/Wuhan | Wuhan Chain Medical Labs (CMLabs) | State Key Laboratory of Biotherapy of Sichuan University | Baowen Du et al |
| EPI_ISL_454923 | Wuhan/HB-WH2-155/2020 | 2020-02-28 | Asia/China/Hubei/Wuhan | Wuhan Chain Medical Labs (CMLabs) | State Key Laboratory of Biotherapy of Sichuan University | Baowen Du et al |
| EPI_ISL_454924 | Wuhan/HB-WH2-156/2020 | 2020-02-28 | Asia/China/Hubei/Wuhan | Wuhan Chain Medical Labs (CMLabs) | State Key Laboratory of Biotherapy of Sichuan University | Baowen Du et al |
| EPI_ISL_454925 | Wuhan/HB-WH2-157/2020 | 2020-02-28 | Asia/China/Hubei/Wuhan | Wuhan Chain Medical Labs (CMLabs) | State Key Laboratory of Biotherapy of Sichuan University | Baowen Du et al |
| EPI_ISL_454926 | Wuhan/HB-WH2-158/2020 | 2020-02-28 | Asia/China/Hubei/Wuhan | Wuhan Chain Medical Labs (CMLabs) | State Key Laboratory of Biotherapy of Sichuan University | Baowen Du et al |
| EPI_ISL_454927 | Wuhan/HB-WH2-159/2020 | 2020-02-28 | Asia/China/Hubei/Wuhan | Wuhan Chain Medical Labs (CMLabs) | State Key Laboratory of Biotherapy of Sichuan University | Baowen Du et al |

| | | | | | | |
|---|---|---|---|---|---|---|
| EPI_ISL_454928 | Wuhan/HB-WH2-160/2020 | 2020-02-28 | Asia/China/Hubei/Wuhan | Wuhan Chain Medical Labs (CMLabs) | State Key Laboratory of Biotherapy of Sichuan University | Baowen Du et al |
| EPI_ISL_454929 | Wuhan/HB-WH2-161/2020 | 2020-02-28 | Asia/China/Hubei/Wuhan | Wuhan Chain Medical Labs (CMLabs) | State Key Laboratory of Biotherapy of Sichuan University | Baowen Du et al |
| EPI_ISL_454930 | Wuhan/HB-WH2-163/2020 | 2020-02-10 | Asia/China/Hubei/Wuhan | Wuhan Chain Medical Labs (CMLabs) | State Key Laboratory of Biotherapy of Sichuan University | Baowen Du et al |
| EPI_ISL_454931 | Wuhan/HB-WH2-164/2020 | 2020-02-10 | Asia/China/Hubei/Wuhan | Wuhan Chain Medical Labs (CMLabs) | State Key Laboratory of Biotherapy of Sichuan University | Baowen Du et al |
| EPI_ISL_454932 | Wuhan/HB-WH2-166/2020 | 2020-02-11 | Asia/China/Hubei/Wuhan | Wuhan Chain Medical Labs (CMLabs) | State Key Laboratory of Biotherapy of Sichuan University | Baowen Du et al |
| EPI_ISL_454933 | Wuhan/HB-WH2-167/2020 | 2020-02-11 | Asia/China/Hubei/Wuhan | Wuhan Chain Medical Labs (CMLabs) | State Key Laboratory of Biotherapy of Sichuan University | Baowen Du et al |
| EPI_ISL_454934 | Wuhan/HB-WH2-168/2020 | 2020-02-11 | Asia/China/Hubei/Wuhan | Wuhan Chain Medical Labs (CMLabs) | State Key Laboratory of Biotherapy of Sichuan University | Baowen Du et al |
| EPI_ISL_454935 | Wuhan/HB-WH2-169/2020 | 2020-02-11 | Asia/China/Hubei/Wuhan | Wuhan Chain Medical Labs (CMLabs) | State Key Laboratory of Biotherapy of Sichuan University | Baowen Du et al |

| | | | | | | |
|---|---|---|---|---|---|---|
| EPI_ISL_454936 | Wuhan/HB-WH3-170/2020 | 2020-02-11 | Asia/China/Hubei/Wuhan | Wuhan Chain Medical Labs (CMLabs) | State Key Laboratory of Biotherapy of Sichuan University | Baowen Du et al |
| EPI_ISL_454937 | Wuhan/HB-WH3-171/2020 | 2020-02-11 | Asia/China/Hubei/Wuhan | Wuhan Chain Medical Labs (CMLabs) | State Key Laboratory of Biotherapy of Sichuan University | Baowen Du et al |
| EPI_ISL_454939 | Wuhan/HB-WH3-173/2020 | 2020-02-28 | Asia/China/Hubei/Wuhan | Wuhan Chain Medical Labs (CMLabs) | State Key Laboratory of Biotherapy of Sichuan University | Baowen Du et al |
| EPI_ISL_454940 | Wuhan/HB-WH3-176/2020 | 2020-02-28 | Asia/China/Hubei/Wuhan | Wuhan Chain Medical Labs (CMLabs) | State Key Laboratory of Biotherapy of Sichuan University | Baowen Du et al |
| EPI_ISL_454952 | Wuhan/HB-WH4-200/2020 | 2020-02-19 | Asia/China/Hubei/Wuhan | Wuhan Chain Medical Labs (CMLabs) | State Key Laboratory of Biotherapy of Sichuan University | Baowen Du et al |
| EPI_ISL_454953 | Wuhan/HB-WH4-201/2020 | 2020-03-18 | Asia/China/Hubei/Wuhan | Wuhan Chain Medical Labs (CMLabs) | State Key Laboratory of Biotherapy of Sichuan University | Baowen Du et al |
| EPI_ISL_454955 | Wuhan/HB-WH4-205/2020 | 2020-03-01 | Asia/China/Hubei/Wuhan | Wuhan Chain Medical Labs (CMLabs) | State Key Laboratory of Biotherapy of Sichuan University | Baowen Du et al |
| EPI_ISL_454964 | Wuhan/HB-WH4-216/2020 | 2020-02-06 | Asia/China/Hubei/Wuhan | Wuhan Chain Medical Labs (CMLabs) | State Key Laboratory of Biotherapy of Sichuan University | Baowen Du et al |

| | | | | | | |
|---|---|---|---|---|---|---|
| EPI_ISL_454965 | Wuhan/HB-WH5-220/2020 | 2020-02-20 | Asia/China/Hubei/Wuhan | Wuhan Chain Medical Labs (CMLabs) | State Key Laboratory of Biotherapy of Sichuan University | Baowen Du et al |
| EPI_ISL_454973 | Wuhan/HB-WH5-229/2020 | 2020-02-22 | Asia/China/Hubei/Wuhan | Wuhan Chain Medical Labs (CMLabs) | State Key Laboratory of Biotherapy of Sichuan University | Baowen Du et al |
| EPI_ISL_454974 | Wuhan/HB-WH5-231/2020 | 2020-02-17 | Asia/China/Hubei/Wuhan | Wuhan Chain Medical Labs (CMLabs) | State Key Laboratory of Biotherapy of Sichuan University | Baowen Du et al |
| EPI_ISL_454976 | Wuhan/HB-WH5-233/2020 | 2020-03-04 | Asia/China/Hubei/Wuhan | Wuhan Chain Medical Labs (CMLabs) | State Key Laboratory of Biotherapy of Sichuan University | Baowen Du et al |
| EPI_ISL_454980 | Wuhan/HB-WH5-239/2020 | 2020-02-09 | Asia/China/Hubei/Wuhan | Wuhan Chain Medical Labs (CMLabs) | State Key Laboratory of Biotherapy of Sichuan University | Baowen Du et al |
| EPI_ISL_454981 | Wuhan/HB-WH5-240/2020 | 2020-02-04 | Asia/China/Hubei/Wuhan | Wuhan Chain Medical Labs (CMLabs) | State Key Laboratory of Biotherapy of Sichuan University | Baowen Du et al |
| EPI_ISL_454982 | Wuhan/HB-WH6-242/2020 | 2020-03-14 | Asia/China/Hubei/Wuhan | Wuhan Chain Medical Labs (CMLabs) | State Key Laboratory of Biotherapy of Sichuan University | Baowen Du et al |
| EPI_ISL_454983 | Wuhan/HB-WH6-246/2020 | 2020-03-12 | Asia/China/Hubei/Wuhan | Wuhan Chain Medical Labs (CMLabs) | State Key Laboratory of Biotherapy of Sichuan University | Baowen Du et al |

| | | | | | | |
|---|---|---|---|---|---|---|
| EPI_ISL_454990 | Wuhan/HB-WHCM-107/2020 | 2020-03-02 | Asia/China/Hubei/Wuhan | Wuhan Chain Medical Labs (CMLabs) | State Key Laboratory of Biotherapy of Sichuan University | Baowen Du et al |
| EPI_ISL_454992 | Wuhan/HB-WHCM-109/2020 | 2020-02-27 | Asia/China/Hubei/Wuhan | Wuhan Chain Medical Labs (CMLabs) | State Key Laboratory of Biotherapy of Sichuan University | Baowen Du et al |
| EPI_ISL_454993 | Wuhan/HB-WHCM-113/2020 | 2020-02-28 | Asia/China/Hubei/Wuhan | Wuhan Chain Medical Labs (CMLabs) | State Key Laboratory of Biotherapy of Sichuan University | Baowen Du et al |
| EPI_ISL_454995 | Wuhan/HB-WHCM-115/2020 | 2020-02-21 | Asia/China/Hubei/Wuhan | Wuhan Chain Medical Labs (CMLabs) | State Key Laboratory of Biotherapy of Sichuan University | Baowen Du et al |
| EPI_ISL_454996 | Wuhan/HB-WHCM-116/2020 | 2020-02-26 | Asia/China/Hubei/Wuhan | Wuhan Chain Medical Labs (CMLabs) | State Key Laboratory of Biotherapy of Sichuan University | Baowen Du et al |
| EPI_ISL_454998 | Wuhan/HB-WHCM-120/2020 | 2020-02-24 | Asia/China/Hubei/Wuhan | Wuhan Chain Medical Labs (CMLabs) | State Key Laboratory of Biotherapy of Sichuan University | Baowen Du et al |
| EPI_ISL_454999 | Wuhan/HB-WHCM2-264/2020 | 2020-02-13 | Asia/China/Hubei/Wuhan | Wuhan Chain Medical Labs (CMLabs) | State Key Laboratory of Biotherapy of Sichuan University | Baowen Du et al |
| EPI_ISL_455000 | Wuhan/HB-WHCM2-265/2020 | 2020-02-14 | Asia/China/Hubei/Wuhan | Wuhan Chain Medical Labs (CMLabs) | State Key Laboratory of Biotherapy of Sichuan University | Baowen Du et al |

| | | | | | | |
|---|---|---|---|---|---|---|
| EPI_ISL_455002 | Wuhan/HB-WHCM2-267/2020 | 2020-02-17 | Asia/China/Hubei/Wuhan | Wuhan Chain Medical Labs (CMLabs) | State Key Laboratory of Biotherapy of Sichuan University | Baowen Du et al |
| EPI_ISL_455003 | Wuhan/HB-WHCM2-268/2020 | 2020-02-15 | Asia/China/Hubei/Wuhan | Wuhan Chain Medical Labs (CMLabs) | State Key Laboratory of Biotherapy of Sichuan University | Baowen Du et al |
| EPI_ISL_455004 | Wuhan/HB-WHCM2-269/2020 | 2020-02-24 | Asia/China/Hubei/Wuhan | Wuhan Chain Medical Labs (CMLabs) | State Key Laboratory of Biotherapy of Sichuan University | Baowen Du et al |
| EPI_ISL_455006 | Wuhan/HB-WHCM2-271/2020 | 2020-02-17 | Asia/China/Hubei/Wuhan | Wuhan Chain Medical Labs (CMLabs) | State Key Laboratory of Biotherapy of Sichuan University | Baowen Du et al |
| EPI_ISL_455007 | Wuhan/HB-WHCM2-272/2020 | 2020-02-26 | Asia/China/Hubei/Wuhan | Wuhan Chain Medical Labs (CMLabs) | State Key Laboratory of Biotherapy of Sichuan University | Baowen Du et al |
| EPI_ISL_455008 | Wuhan/HB-WHCM2-274/2020 | 2020-02-23 | Asia/China/Hubei/Wuhan | Wuhan Chain Medical Labs (CMLabs) | State Key Laboratory of Biotherapy of Sichuan University | Baowen Du et al |
| EPI_ISL_455009 | Wuhan/HB-WHCM2-276/2020 | 2020-02-24 | Asia/China/Hubei/Wuhan | Wuhan Chain Medical Labs (CMLabs) | State Key Laboratory of Biotherapy of Sichuan University | Baowen Du et al |
| EPI_ISL_455010 | Wuhan/HB-WHCM2-278/2020 | 2020-02-22 | Asia/China/Hubei/Wuhan | Wuhan Chain Medical Labs (CMLabs) | State Key Laboratory of Biotherapy of Sichuan University | Baowen Du et al |

| | | | | | | |
|---|---|---|---|---|---|---|
| EPI_ISL_455011 | Wuhan/HB-WHCM2-279/2020 | 2020-02-11 | Asia/China/Hubei/Wuhan | Wuhan Chain Medical Labs (CMLabs) | State Key Laboratory of Biotherapy of Sichuan University | Baowen Du et al |
| EPI_ISL_455012 | Wuhan/HB-WHCM2-280/2020 | 2020-02-25 | Asia/China/Hubei/Wuhan | Wuhan Chain Medical Labs (CMLabs) | State Key Laboratory of Biotherapy of Sichuan University | Baowen Du et al |
| EPI_ISL_455013 | Wuhan/HB-WHCM2-282/2020 | 2020-02-24 | Asia/China/Hubei/Wuhan | Wuhan Chain Medical Labs (CMLabs) | State Key Laboratory of Biotherapy of Sichuan University | Baowen Du et al |
| EPI_ISL_455017 | India/GJ-GBRC120/2020 | 2020-05-02 | Asia/India/Gujarat/Vadodara | Government Medical College, Vadodara | Gujarat Biotechnology Research Centre | Tanuja Javadekar et al |
| EPI_ISL_455019 | India/GJ-GBRC122/2020 | 2020-05-02 | Asia/India/Gujarat/Vadodara | Government Medical College, Vadodara | Gujarat Biotechnology Research Centre | Ramesh Pandit et al |
| EPI_ISL_455020 | India/GJ-GBRC123/2020 | 2020-05-02 | Asia/India/Gujarat/Vadodara | Government Medical College, Vadodara | Gujarat Biotechnology Research Centre | Tejas Shah et al |
| EPI_ISL_455026 | India/GJ-GBRC129/2020 | 2020-05-02 | Asia/India/Gujarat/Vadodara | Government Medical College, Vadodara | Gujarat Biotechnology Research Centre | Monika Gandhi et al |
| EPI_ISL_455027 | India/GJ-GBRC130/2020 | 2020-05-02 | Asia/India/Gujarat/Vadodara | Government Medical College, Vadodara | Gujarat Biotechnology Research Centre | Pinal Trivedi et al |
| EPI_ISL_455104 | Sweden/20-51264/2020 | 2020-05-06 | Europe/Sweden/Uppsala/ | Ulltuna Vardcentral | The Public Health Agency of Sweden | Heidi Lindback et al |
| EPI_ISL_455108 | Sweden/20-51391/2020 | 2020-05-13 | Europe/Sweden/Kalmar/ | Smedby HC | The Public Health Agency of Sweden | Susanne Brunby et al |
| EPI_ISL_455143 | Netherlands/NA_338/2020 | 2020-04-04 | Europe/Netherlands/Netherlands/ | Dutch COVID-19 response team | Erasmus Medical Center | Bas Oude Munnink et al |
| EPI_ISL_455151 | Netherlands/NA_346/2020 | 2020-04-05 | Europe/Netherlands/Netherlands/ | Dutch COVID-19 response team | Erasmus Medical Center | Bas Oude Munnink et al |
| EPI_ISL_455160 | Netherlands/NA_356/2020 | 2020-04-07 | Europe/Netherlands/Netherlands/ | Dutch COVID-19 response team | Erasmus Medical Center | Bas Oude Munnink et al |

| | | | | | | |
|---|---|---|---|---|---|---|
| EPI_ISL_455165 | Netherlands/NA_361/2020 | 2020-03-30 | Europe/Netherlands/Netherlands/ | Dutch COVID-19 response team | Erasmus Medical Center | Bas Oude Munnink et al |
| EPI_ISL_455172 | Netherlands/NA_368/2020 | 2020-04-08 | Europe/Netherlands/Netherlands/ | Dutch COVID-19 response team | Erasmus Medical Center | Bas Oude Munnink et al |
| EPI_ISL_455235 | Netherlands/NA_433/2020 | 2020-04-06 | Europe/Netherlands/Netherlands/ | Dutch COVID-19 response team | Erasmus Medical Center | Bas Oude Munnink et al |
| EPI_ISL_455236 | Netherlands/NA_434/2020 | 2020-04-07 | Europe/Netherlands/Netherlands/ | Dutch COVID-19 response team | Erasmus Medical Center | Bas Oude Munnink et al |
| EPI_ISL_455240 | Netherlands/NA_438/2020 | 2020-04-09 | Europe/Netherlands/Netherlands/ | Dutch COVID-19 response team | Erasmus Medical Center | Bas Oude Munnink et al |
| EPI_ISL_455242 | Netherlands/NA_440/2020 | 2020-04-10 | Europe/Netherlands/Netherlands/ | Dutch COVID-19 response team | Erasmus Medical Center | Bas Oude Munnink et al |
| EPI_ISL_455257 | Netherlands/NA_455/2020 | 2020-03-24 | Europe/Netherlands/Netherlands/ | Dutch COVID-19 response team | Erasmus Medical Center | Bas Oude Munnink et al |
| EPI_ISL_455277 | Netherlands/NA_475/2020 | 2020-04-14 | Europe/Netherlands/Netherlands/ | Dutch COVID-19 response team | Erasmus Medical Center | Bas Oude Munnink et al |
| EPI_ISL_455297 | Netherlands/ZuidHolland_108/2020 | 2020-04-07 | Europe/Netherlands/South Holland/ | Dutch COVID-19 response team | Erasmus Medical Center | Bas Oude Munnink et al |
| EPI_ISL_455308 | India/OR-ILC-RMRC25/2020 | 2020-04-04 | Asia/India/Odisha/ | REGIONAL VRDL,ICMR-RMRC BBSR | Immunogenomics group, Institute of Life Sciences, Bhubaneswar | Sunil Raghav et al |
| EPI_ISL_455314 | Spain/AN-ISCIII-201257/2020 | 2020-02-28 | Europe/Spain/Andalusia/ | Hospital Virgen del Rocio | Instituto de Salud Carlos III | Iglesias-Caballero et al |
| EPI_ISL_455315 | Spain/AN-ISCIII-201488/2020 | 2020-03-04 | Europe/Spain/Andalusia/ | Hospital Virgen de las Nieves | Instituto de Salud Carlos III | Iglesias-Caballero et al |
| EPI_ISL_455317 | Spain/AN-ISCIII-201614/2020 | 2020-03-05 | Europe/Spain/Andalusia/ | Hospital Virgen de las Nieves | Instituto de Salud Carlos III | Iglesias-Caballero et al |
| EPI_ISL_455319 | Spain/AN-ISCIII-201616/2020 | 2020-03-05 | Europe/Spain/Andalusia/ | Hospital Virgen de las Nieves | Instituto de Salud Carlos III | Iglesias-Caballero et al |
| EPI_ISL_455322 | Spain/AN-ISCIII-201623/2020 | 2020-03-05 | Europe/Spain/Andalusia/ | Hospital Virgen de las Nieves | Instituto de Salud Carlos III | Iglesias-Caballero et al |
| EPI_ISL_455326 | Spain/CN-ISCIII-201939/2020 | 2020-02-29 | Europe/Spain/Canary Islands/ | Hospital Universitario Insular de Gran Canaria | Instituto de Salud Carlos III | Iglesias-Caballero et al |
| EPI_ISL_455328 | Spain/GA-ISCIII-201792/2020 | 2020-03-09 | Europe/Spain/Galicia/ | Complejo Hospitalario Universitario La Coruna | Instituto de Salud Carlos III | Iglesias-Caballero et al |
| EPI_ISL_455331 | Spain/GA-ISCIII-201863/2020 | 2020-03-10 | Europe/Spain/Galicia/ | Complejo Hospitalario Universitario La Coruna | Instituto de Salud Carlos III | Iglesias-Caballero et al |
| EPI_ISL_455333 | Spain/GA-ISCIII-202230/2020 | 2020-03-11 | Europe/Spain/Galicia/ | Complejo Hospitalario Universitario de Santiago | Instituto de Salud Carlos III | Iglesias-Caballero et al |

| | | | | | | |
|---|---|---|---|---|---|---|
| EPI_ISL_455335 | Spain/GA-ISCIII-202297/2020 | 2020-03-11 | Europe/Spain/Galicia/ | Complejo Hospitalario de Orense | Instituto de Salud Carlos III | Iglesias-Caballero et al |
| EPI_ISL_455336 | Spain/RI-ISCIII-201277/2020 | 2020-02-29 | Europe/Spain/La Rioja/ | Hospital San Pedro | Instituto de Salud Carlos III | Iglesias-Caballero et al |
| EPI_ISL_455340 | Spain/RI-ISCIII-201568/2020 | 2020-03-05 | Europe/Spain/La Rioja/ | Hospital San Pedro | Instituto de Salud Carlos III | Iglesias-Caballero et al |
| EPI_ISL_455344 | Spain/ML-ISCIII-202020/2020 | 2020-03-11 | Europe/Spain/Melilla/ | Hospital Comarcal de Melilla | Instituto de Salud Carlos III | Iglesias-Caballero et al |
| EPI_ISL_455350 | Spain/PV-ISCIII-201385/2020 | 2020-02-29 | Europe/Spain/Basque Country/ | Hospital Txagorritxu | Instituto de Salud Carlos III | Iglesias-Caballero et al |
| EPI_ISL_455351 | Spain/PV-ISCIII-201388/2020 | 2020-02-29 | Europe/Spain/Basque Country/ | Hospital Txagorritxu | Instituto de Salud Carlos III | Iglesias-Caballero et al |
| EPI_ISL_455353 | Spain/PV-ISCIII-201492/2020 | 2020-03-04 | Europe/Spain/Basque Country/ | Hospital de Cruces | Instituto de Salud Carlos III | Iglesias-Caballero et al |
| EPI_ISL_455365 | Wuhan/HB-WH1-132/2020 | 2020-02-23 | Asia/China/Hubei/Wuhan | Wuhan Chain Medical Labs (CMLabs) | State Key Laboratory of Biotherapy of Sichuan University | Baowen Du et al |
| EPI_ISL_455366 | Wuhan/HB-WH1-133/2020 | 2020-02-23 | Asia/China/Hubei/Wuhan | Wuhan Chain Medical Labs (CMLabs) | State Key Laboratory of Biotherapy of Sichuan University | Baowen Du et al |
| EPI_ISL_455367 | Wuhan/HB-WH1-139/2020 | 2020-02-23 | Asia/China/Hubei/Wuhan | Wuhan Chain Medical Labs (CMLabs) | State Key Laboratory of Biotherapy of Sichuan University | Baowen Du et al |
| EPI_ISL_455368 | Wuhan/HB-WH1-144/2020 | 2020-02-23 | Asia/China/Hubei/Wuhan | Wuhan Chain Medical Labs (CMLabs) | State Key Laboratory of Biotherapy of Sichuan University | Baowen Du et al |
| EPI_ISL_455370 | Wuhan/HB-WH2-146/2020 | 2020-02-24 | Asia/China/Hubei/Wuhan | Wuhan Chain Medical Labs (CMLabs) | State Key Laboratory of Biotherapy of Sichuan University | Baowen Du et al |

| | | | | | | |
|---|---|---|---|---|---|---|
| EPI_ISL_455373 | Wuhan/HB-WH2-154/2020 | 2020-02-28 | Asia/China/Hubei/Wuhan | Wuhan Chain Medical Labs (CMLabs) | State Key Laboratory of Biotherapy of Sichuan University | Baowen Du et al |
| EPI_ISL_455374 | Wuhan/HB-WH2-162/2020 | 2020-02-10 | Asia/China/Hubei/Wuhan | Wuhan Chain Medical Labs (CMLabs) | State Key Laboratory of Biotherapy of Sichuan University | Baowen Du et al |
| EPI_ISL_455375 | Wuhan/HB-WH2-165/2020 | 2020-02-11 | Asia/China/Hubei/Wuhan | Wuhan Chain Medical Labs (CMLabs) | State Key Laboratory of Biotherapy of Sichuan University | Baowen Du et al |
| EPI_ISL_455376 | Wuhan/HB-WH3-174/2020 | 2020-02-28 | Asia/China/Hubei/Wuhan | Wuhan Chain Medical Labs (CMLabs) | State Key Laboratory of Biotherapy of Sichuan University | Baowen Du et al |
| EPI_ISL_455380 | Wuhan/HB-WH3-184/2020 | 2020-03-12 | Asia/China/Hubei/Wuhan | Wuhan Chain Medical Labs (CMLabs) | State Key Laboratory of Biotherapy of Sichuan University | Baowen Du et al |
| EPI_ISL_455385 | Wuhan/HB-WH4-193/2020 | 2020-02-07 | Asia/China/Hubei/Wuhan | Wuhan Chain Medical Labs (CMLabs) | State Key Laboratory of Biotherapy of Sichuan University | Baowen Du et al |
| EPI_ISL_455386 | Wuhan/HB-WH4-198/2020 | 2020-02-12 | Asia/China/Hubei/Wuhan | Wuhan Chain Medical Labs (CMLabs) | State Key Laboratory of Biotherapy of Sichuan University | Baowen Du et al |
| EPI_ISL_455391 | Wuhan/HB-WH5-228/2020 | 2020-02-04 | Asia/China/Hubei/Wuhan | Wuhan Chain Medical Labs (CMLabs) | State Key Laboratory of Biotherapy of Sichuan University | Baowen Du et al |

| | | | | | | |
|---|---|---|---|---|---|---|
| EPI_ISL_455392 | Wuhan/HB-WH5-230/2020 | 2020-03-17 | Asia/China/Hubei/Wuhan | Wuhan Chain Medical Labs (CMLabs) | State Key Laboratory of Biotherapy of Sichuan University | Baowen Du et al |
| EPI_ISL_455399 | Wuhan/HB-WHCM-101/2020 | 2020-03-02 | Asia/China/Hubei/Wuhan | Wuhan Chain Medical Labs (CMLabs) | State Key Laboratory of Biotherapy of Sichuan University | Baowen Du et al |
| EPI_ISL_455420 | Bangladesh/BCSIR-NILMRC-007/2020 | 2020-05-21 | Asia/Bangladesh/Dhaka/Mirpur | National Institute of Laboratory Medicine and Referral Center | Genomic Research Lab, BCSIR | Abu Sayeed Mohammad Mahmud et al |
| EPI_ISL_455422 | Nigeria/OS070-CV11/2020 | 2020-03-29 | Africa/Nigeria/Osun State/ | Nigeria Centre for Disease Control | African Centre of Excellence for Genomics of Infectious Diseases (ACEGID), Redeemer's University, Ede, Osun State, Nigeria | Oluniyi P.E. et al |

| | | | | | | |
|---|---|---|---|---|---|---|
| EPI_ISL_455440 | Poland/PL_P27/2020 | 2020-03-27 | Europe/Poland/Mazowieckie/Plock | 1. ViroGenetics - BSL3 Laboratory of Virology, Ma≈Çopolska Centre of Biotechnology, Jagiellonian University; 2. II Department of Internal Medicine, Faculty of Medicine, Jagiellonian University Medical College; 3. Narodowy Instytut Zdrowia Publicznego ‚Äì Pa≈Ñstwowy Zak≈Çad Higieny (NIZP-PZH) | 1. ViroGenetics - BSL3 Laboratory of Virology, Ma≈Çopolska Centre of Biotechnology, Jagiellonian University; 2. II Department of Internal Medicine, Faculty of Medicine, Jagiellonian University Medical College; 3. Narodowy Instytut Zdrowia Publicznego ‚Äì Pa≈Ñstwowy Zak≈Çad Higieny (NIZP-PZH). | Katarzyna Pancer et al |

| | | | | | | |
|---|---|---|---|---|---|---|
| EPI_ISL_455451 | Poland/PL_P38/2020 | 2020-03-17 | Europe/Poland/Mazowieckie/Warszawa | 1. ViroGenetics - BSL3 Laboratory of Virology, Ma≈Çopolska Centre of Biotechnology, Jagiellonian University; 2. II Department of Internal Medicine, Faculty of Medicine, Jagiellonian University Medical College; 3. Narodowy Instytut Zdrowia Publicznego ‚Äì Pa≈Ñstwowy Zak≈Çad Higieny (NIZP-PZH) | 1. ViroGenetics - BSL3 Laboratory of Virology, Ma≈Çopolska Centre of Biotechnology, Jagiellonian University; 2. II Department of Internal Medicine, Faculty of Medicine, Jagiellonian University Medical College; 3. Narodowy Instytut Zdrowia Publicznego ‚Äì Pa≈Ñstwowy Zak≈Çad Higieny (NIZP-PZH). | Katarzyna Pancer et al |
| EPI_ISL_455456 | Mexico/QUE-InDRE-18/2020 | 2020-03-12 | North America/Mexico/Queretaro/ | Instituto de Diagnostico y Referencia Epidemiologicos (INDRE) | Instituto de Diagnostico y Referencia Epidemiologicos (INDRE) | Rodriguez-Maldonado Abril et al |
| EPI_ISL_455459 | Bangladesh/BCSIR-NILMRC-008/2020 | 2020-05-21 | Asia/Bangladesh/Dhaka/Mirpur | National Institute of Laboratory Medicine and Referral Center | Genomic Research Lab, BCSIR | Abu Sayeed Mohammad Mahmud et al |
| EPI_ISL_455460 | Yichun/JX2325/2020 | 2020-02-09 | Asia/China/Jiangxi/Yichun | Jiangxi Province Center for Disease Control and Prevention | Jiangxi Province Center for Disease Control and Prevention | JianXiong Li et al |
| EPI_ISL_455461 | Yichun/JX2391/2020 | 2020-02-06 | Asia/China/Jiangxi/Yichun | Jiangxi Province Center for Disease Control and Prevention | Jiangxi Province Center for Disease Control and Prevention | JianXiong Li et al |
| EPI_ISL_455462 | Fuzhou/JX2012/2020 | 2020-02-05 | Asia/China/Jiangxi/Fuzhou | Jiangxi Province Center for Disease Control and Prevention | Jiangxi Province Center for Disease Control and Prevention | JianXiong Li et al |

| | | | | | | |
|---|---|---|---|---|---|---|
| EPI_ISL_455463 | Jiujiang/JX490/2020 | 2020-02-03 | Asia/China/Jiangxi/Jiujiang | Jiangxi Province Center for Disease Control and Prevention | Jiangxi Province Center for Disease Control and Prevention | JianXiong Li et al |
| EPI_ISL_455464 | Fuzhou/JX2009/2020 | 2020-02-05 | Asia/China/Jiangxi/Fuzhou | Jiangxi Province Center for Disease Control and Prevention | Jiangxi Province Center for Disease Control and Prevention | JianXiong Li et al |
| EPI_ISL_455465 | Fuzhou/JX2019/2020 | 2020-02-10 | Asia/China/Jiangxi/Fuzhou | Jiangxi Province Center for Disease Control and Prevention | Jiangxi Province Center for Disease Control and Prevention | JianXiong Li et al |
| EPI_ISL_455466 | Yingtan/JX2480/2020 | 2020-02-01 | Asia/China/Jiangxi/Yingtan | Jiangxi Province Center for Disease Control and Prevention | Jiangxi Province Center for Disease Control and Prevention | JianXiong Li et al |
| EPI_ISL_455467 | Yichun/JX2424/2020 | 2020-02-12 | Asia/China/Jiangxi/Yichun | Jiangxi Province Center for Disease Control and Prevention | Jiangxi Province Center for Disease Control and Prevention | JianXiong Li et al |
| EPI_ISL_455468 | Romania/284213/2020 | 2020-05-13 | Europe/Romania/Bucharest/ | Laboratory for Respiratory Viruses, Cantacuzino National Military-Medical Institute for Research and Development | Cantacuzino Institute | M.Lazar et al |
| EPI_ISL_455469 | Romania/283584/2020 | 2020-05-12 | Europe/Romania/Bucharest/ | Laboratory for Respiratory Viruses, Cantacuzino National Military-Medical Institute for Research and Development | Cantacuzino Institute | M.Lazar et al |
| EPI_ISL_455470 | Romania/284489/2020 | 2020-05-14 | Europe/Romania/Bucharest/ | Laboratory for Respiratory Viruses, Cantacuzino National Military-Medical Institute for Research and Development | Cantacuzino Institute | M.Lazar et al |
| EPI_ISL_455471 | Romania/284508/2020 | 2020-05-14 | Europe/Romania/Bucharest/ | Laboratory for Respiratory Viruses, Cantacuzino National Military-Medical Institute for Research and Development | Cantacuzino Institute | M.Lazar et al |
| EPI_ISL_455474 | Romania/284783/2020 | 2020-05-14 | Europe/Romania/Arges/ | Laboratory for Respiratory Viruses, Cantacuzino National Military-Medical Institute for Research and Development | Cantacuzino Institute | M.Lazar et al |
| EPI_ISL_455475 | Romania/284762/2020 | 2020-05-14 | Europe/Romania/Arges/ | Laboratory for Respiratory Viruses, Cantacuzino National Military-Medical Institute for Research and Development | Cantacuzino Institute | M.Lazar et al |

| | | | | | | |
|---|---|---|---|---|---|---|
| EPI_ISL_455476 | Romania/285388/2020 | 2020-05-17 | Europe/Romania/Bucharest/ | Laboratory for Respiratory Viruses, Cantacuzino National Military-Medical Institute for Research and Development | Cantacuzino Institute | M.Lazar et al |
| EPI_ISL_455479 | Romania/284468/2020 | 2020-05-14 | Europe/Romania/Bucharest/ | Laboratory for Respiratory Viruses, Cantacuzino National Military-Medical Institute for Research and Development | Cantacuzino Institute | M.Lazar et al |
| EPI_ISL_455566 | Croatia/17-S7new/2020 | 2020-03-30 | Europe/Croatia/Istria/ | Institute for Public Health | Laboratory for advanced genomics | Filip Rokifá et al |
| EPI_ISL_455568 | USA/IA-GMF-02699/2020 | 2020-05-08 | North America/USA/Iowa/Allamakee Co | Gundersen Molecular Diagnostics Laboratory | Kabara Cancer Research Institute | Craig S. Richmond et al |
| EPI_ISL_455569 | USA/WI-GMF-02728/2020 | 2020-05-08 | North America/USA/Wisconsin/Grant Co | Gundersen Molecular Diagnostics Laboratory | Kabara Cancer Research Institute | Craig S. Richmond et al |
| EPI_ISL_455572 | USA/WI-GMF-03156/2020 | 2020-05-12 | North America/USA/Wisconsin/La Crosse | Gundersen Molecular Diagnostics Laboratory | Kabara Cancer Research Institute | Craig S. Richmond et al |
| EPI_ISL_455573 | USA/WI-GMF-M00001/2020 | 2020-05-10 | North America/USA/Wisconsin/La Crosse | Gundersen Clinical Microbiology Laboratory | Kabara Cancer Research Institute | Craig S. Richmond et al |
| EPI_ISL_455574 | USA/WI-GMF-M00002/2020 | 2020-05-10 | North America/USA/Wisconsin/Trempea | Gundersen Clinical Microbiology Laboratory | Kabara Cancer Research Institute | Craig S. Richmond et al |
| EPI_ISL_455576 | USA/WI-GMF-03823/2020 | 2020-05-19 | North America/USA/Wisconsin/Trempea | Gundersen Molecular Diagnostics Laboratory | Kabara Cancer Research Institute | Craig S. Richmond et al |
| EPI_ISL_455580 | USA/WI-GMF-M00003/2020 | 2020-05-12 | North America/USA/Wisconsin/La Crosse | Gundersen Clinical Microbiology Laboratory | Kabara Cancer Research Institute | Craig S. Richmond et al |
| EPI_ISL_455603 | Australia/VPRL014/2020 | 2020-01-30 | Oceania/Australia/South Australia/ | SA Pathology | VPRL | Beard et al |
| EPI_ISL_455608 | Thailand/Bangkok_2504/2020 | 2020-03-14 | Asia/Thailand/Bangkok/ | Phramongkutklao Hospital | National Institute of Health. Department of medical Sciences, Ministry of Public Health, Thailand | Pilailuk et al |
| EPI_ISL_455619 | USA/LA-BIE-065/2020 | 2020-04-06 | North America/USA/Louisiana/New Orle | Ochsner Health | Bioinfoexperts, LLC | Susanna L. Lamers et al |
| EPI_ISL_455620 | USA/LA-BIE-066/2020 | 2020-04-06 | North America/USA/Louisiana/New Orle | Ochsner Health | Bioinfoexperts, LLC | Susanna L. Lamers et al |
| EPI_ISL_455623 | USA/LA-BIE-070/2020 | 2020-04-06 | North America/USA/Louisiana/Jefferson | Ochsner Health | Bioinfoexperts, LLC | Susanna L. Lamers et al |
| EPI_ISL_455637 | SouthAfrica/KRISP-107/2020 | 2020-05-04 | Africa/South Africa/KwaZulu-Natal/ | KRISP, KZN Research Innovation and Sequencing Platform | KRISP, KZN Research Innovation and Sequencing Platform | Giandhari J et al |

| | | | | | | |
|---|---|---|---|---|---|---|
| EPI_ISL_455649 | India/WB-S19/2020 | 2020-04-20 | Asia/India/West Bengal/Kolkata | ICMR-National Institute of Cholera and Enteric Diseases | National Institute of Biomedical Genomics | Arindam Maitra et al |
| EPI_ISL_455653 | India/WB-S23/2020 | 2020-04-21 | Asia/India/West Bengal/South 24 Parga | ICMR-National Institute of Cholera and Enteric Diseases | National Institute of Biomedical Genomics | Arindam Maitra et al |
| EPI_ISL_455655 | India/RJ-S25/2020 | 2020-04-30 | Asia/India/Rajasthan/Bikaner | ICMR-National Institute of Cholera and Enteric Diseases | National Institute of Biomedical Genomics | Arindam Maitra et al |
| EPI_ISL_455666 | India/WB-S45/2020 | 2020-05-01 | Asia/India/West Bengal/Kolkata | ICMR-National Institute of Cholera and Enteric Diseases | National Institute of Biomedical Genomics | Arindam Maitra et al |
| EPI_ISL_455671 | India/WB-S51/2020 | 2020-05-03 | Asia/India/West Bengal/Kolkata | ICMR-National Institute of Cholera and Enteric Diseases | National Institute of Biomedical Genomics | Arindam Maitra et al |
| EPI_ISL_455672 | India/WB-S52/2020 | 2020-05-03 | Asia/India/West Bengal/North 24 Parga | ICMR-National Institute of Cholera and Enteric Diseases | National Institute of Biomedical Genomics | Arindam Maitra et al |
| EPI_ISL_455674 | India/WB-S55/2020 | 2020-05-03 | Asia/India/West Bengal/Kolkata | ICMR-National Institute of Cholera and Enteric Diseases | National Institute of Biomedical Genomics | Arindam Maitra et al |
| EPI_ISL_455680 | Changzhou/JS27/2020 | 2020-01-26 | Asia/China/Jiangsu/Changzhou | Institute of pathogenic microbiology, Jiangsu Provincial Center for Disease Control and Prevention | Institute of pathogenic microbiology, Jiangsu Provincial Center for Disease Control and Prevention | Cui et al |
| EPI_ISL_455683 | Zhejiang/CN1/2020 | 2020-01-22 | Asia/China/Zhejiang/ | unknown | Department of Microbiology | Gao et al |
| EPI_ISL_455684 | Zhejiang/CN2/2020 | 2020-01-22 | Asia/China/Zhejiang/ | unknown | Department of Microbiology | Gao et al |
| EPI_ISL_455685 | Zhejiang/CN5/2020 | 2020-01-22 | Asia/China/Zhejiang/ | unknown | Department of Microbiology | Gao et al |
| EPI_ISL_455686 | Zhejiang/CN3/2020 | 2020-01-26 | Asia/China/Zhejiang/ | unknown | Department of Microbiology | Gao et al |
| EPI_ISL_455687 | Zhejiang/CN4/2020 | 2020-01-26 | Asia/China/Zhejiang/ | unknown | Department of Microbiology | Gao et al |
| EPI_ISL_455688 | Zhejiang/OS1/2020 | 2020-03-24 | Asia/China/Zhejiang/ | unknown | Department of Microbiology | Gao et al |

| | | | | | | |
|---|---|---|---|---|---|---|
| EPI_ISL_455692 | Zhejiang/OS2/2020 | 2020-03-24 | Asia/China/Zhejiang/ | unknown | Department of Microbiology | Gao et al |
| EPI_ISL_455719 | Turkey/VETAL1902_2/2020 | 2020-04-09 | Europe/Turkey/Mardin/ | T.C. Saƒülƒ±k Bakanlƒ±ƒüƒ± Adƒ±yaman ƒ∞l Saƒülƒ±k M√ºd√ºrl√ºƒü√º Adƒ±yaman Eƒüitim Ve Ara≈üƒ±rma Hastanesi | VETAL Animal Health Products Company, BSL3+ Production Laboratuary /Turkey | Mehmet Turgut et al |
| EPI_ISL_455721 | Spain/VC-FISABIO-648/2020 | 2020-03-31 | Europe/Spain/Comunitat Valenciana/Al | Servicio de Microbiologia. Hospital Clinico Universitario de Valencia | Sequencing and Bioinformatics Service and Molecular Epidemiology Research Group. FISABIO-Public Health, and SeqCOVID-Spain Consortium | Sandra Carbo et al |
| EPI_ISL_455724 | Spain/VC-FISABIO-618/2020 | 2020-03-31 | Europe/Spain/Comunitat Valenciana/Ga | Servicio de Microbiologia. Hospital Clinico Universitario de Valencia | Sequencing and Bioinformatics Service and Molecular Epidemiology Research Group. FISABIO-Public Health, and SeqCOVID-Spain Consortium | Loreto Ferr√∫s Abad et al |
| EPI_ISL_455728 | Spain/VC-FISABIO-617/2020 | 2020-03-31 | Europe/Spain/Comunitat Valenciana/Mo | Servicio de Microbiologia. Hospital Clinico Universitario de Valencia | Sequencing and Bioinformatics Service and Molecular Epidemiology Research Group. FISABIO-Public Health, and SeqCOVID-Spain Consortium | Maria Alma Bracho et al |

| | | | | | | |
|---|---|---|---|---|---|---|
| EPI_ISL_455742 | Spain/VC-FISABIO-652/2020 | 2020-03-20 | Europe/Spain/Comunitat Valenciana/Va | Servicio de Microbiologia. Hospital Clinico Universitario de Valencia | Sequencing and Bioinformatics Service and Molecular Epidemiology Research Group. FISABIO-Public Health, and SeqCOVID-Spain Consortium | Neris Garcia-Gonzalez et al |
| EPI_ISL_455743 | Spain/VC-FISABIO-654/2020 | 2020-03-30 | Europe/Spain/Comunitat Valenciana/Va | Servicio de Microbiologia. Hospital Clinico Universitario de Valencia | Sequencing and Bioinformatics Service and Molecular Epidemiology Research Group. FISABIO-Public Health, and SeqCOVID-Spain Consortium | Eliseo Albert et al |
| EPI_ISL_455753 | India/OR-RMRC110/2020 | 2020-05-06 | Asia/India/Odisha/ | REGIONAL VRDL,ICMR-RMRC BBSR | Immunogenomics lab, Institute of Life Sciences, Bhubaneswar | Sunil Raghav et al |
| EPI_ISL_455759 | India/OR-RMRC158/2020 | 2020-05-07 | Asia/India/Odisha/ | REGIONAL VRDL,ICMR-RMRC BBSR | Immunogenomics lab, Institute of Life Sciences, Bhubaneswar | Sunil Raghav et al |
| EPI_ISL_455763 | India/OR-RMRC163/2020 | 2020-05-07 | Asia/India/Odisha/ | REGIONAL VRDL,ICMR-RMRC BBSR | Immunogenomics lab, Institute of Life Sciences, Bhubaneswar | Sunil Raghav et al |
| EPI_ISL_455764 | India/OR-RMRC164/2020 | 2020-05-07 | Asia/India/Odisha/ | REGIONAL VRDL,ICMR-RMRC BBSR | Immunogenomics lab, Institute of Life Sciences, Bhubaneswar | Sunil Raghav et al |
| EPI_ISL_455769 | India/OR-RMRC170/2020 | 2020-05-07 | Asia/India/Odisha/ | REGIONAL VRDL,ICMR-RMRC BBSR | Immunogenomics lab, Institute of Life Sciences, Bhubaneswar | Sunil Raghav et al |

| | | | | | | |
|---|---|---|---|---|---|---|
| EPI_ISL_455770 | India/OR-RMRC171/2020 | 2020-05-07 | Asia/India/Odisha/ | REGIONAL VRDL,ICMR-RMRC BBSR | Immunogenomics lab, Institute of Life Sciences, Bhubaneswar | Sunil Raghav et al |
| EPI_ISL_455775 | India/OR-RMRC24/2020 | 2020-04-04 | Asia/India/Odisha/ | REGIONAL VRDL,ICMR-RMRC BBSR | Immunogenomics lab, Institute of Life Sciences, Bhubaneswar | Sunil Raghav et al |
| EPI_ISL_455780 | India/OR-RMRC46/2020 | 2020-04-08 | Asia/India/Odisha/ | REGIONAL VRDL,ICMR-RMRC BBSR | Immunogenomics lab, Institute of Life Sciences, Bhubaneswar | Sunil Raghav et al |
| EPI_ISL_455786 | India/OR-RMRC90/2020 | 2020-05-04 | Asia/India/Odisha/ | REGIONAL VRDL,ICMR-RMRC BBSR | Immunogenomics lab, Institute of Life Sciences, Bhubaneswar | Sunil Raghav et al |
| EPI_ISL_455787 | India/OR-RMRC92/2020 | 2020-05-04 | Asia/India/Odisha/ | REGIONAL VRDL,ICMR-RMRC BBSR | Immunogenomics lab, Institute of Life Sciences, Bhubaneswar | Sunil Raghav et al |
| EPI_ISL_455790 | Malaysia/IMR_WC119/2020 | 2020-01-30 | Asia/Malaysia/Selangor/ | Institute for Medical Research, Infectious Disease Research Centre, National Institutes of Health, Ministry of Health Malaysia | Malaysia Genome Institute | Mohd Noor Mat Isa et al |
| EPI_ISL_455791 | Malaysia/IMR_WC413/2020 | 2020-02-08 | Asia/Malaysia/Selangor/ | Institute for Medical Research, Infectious Disease Research Centre, National Institutes of Health, Ministry of Health Malaysia | Malaysia Genome Institute | Mohd Noor Mat Isa et al |
| EPI_ISL_455792 | Malaysia/IMR_WC458/2020 | 2020-02-09 | Asia/Malaysia/Selangor/ | Institute for Medical Research, Infectious Disease Research Centre, National Institutes of Health, Ministry of Health Malaysia | Malaysia Genome Institute | Mohd Noor Mat Isa et al |
| EPI_ISL_455793 | Malaysia/IMR_WC1114/2020 | 2020-02-29 | Asia/Malaysia/Selangor/ | Institute for Medical Research, Infectious Disease Research Centre, National Institutes of Health, Ministry of Health Malaysia | Malaysia Genome Institute | Mohd Noor Mat Isa et al |
| EPI_ISL_455872 | Sweden/20-50213/2020 | 2020-03-06 | Europe/Sweden/Stockholm/ | Karolinska Universitetslaboratoriet | The Public Health Agency of Sweden | Anna-Malin Linde et al |
| EPI_ISL_455879 | Sweden/20-50229/2020 | 2020-03-07 | Europe/Sweden/Stockholm/ | Karolinska Universitetslaboratoriet | The Public Health Agency of Sweden | Anna-Malin Linde et al |

| | | | | | | |
|---|---|---|---|---|---|---|
| EPI_ISL_455898 | Sweden/20-50284/2020 | 2020-03-10 | Europe/Sweden/Stockholm/ | Karolinska Universitetslaboratoriet | The Public Health Agency of Sweden | Anna-Malin Linde et al |
| EPI_ISL_455908 | Thailand/Bangkok-CONI-0105/2020 | 2020-03-09 | Asia/Thailand/Bangkok/ | Ramathibodi Hospital | COVID-19 Network Investigations (CONI) Alliance | Elizabeth Batty et al |
| EPI_ISL_455934 | Thailand/Bangkok-CONI-0140/2020 | 2020-03-14 | Asia/Thailand/Bangkok/ | Ramathibodi Hospital | COVID-19 Network Investigations (CONI) Alliance | Elizabeth Batty et al |
| EPI_ISL_455968 | Belgium/ULG-10180/2020 | 2020-05-09 | Europe/Belgium/Liv®ge/ | Department of Clinical Microbiology | GIGA Medical Genomics | Keith Durkin et al |
| EPI_ISL_455981 | USA/LA-EVTL138/2020 | 2020-03-28 | North America/USA/Louisiana/Caddo Pa | LSUHS Emerging Viral Threat Laboratory | Microbial Genome Sequencing Center | Jeremy P. Kamil et al |
| EPI_ISL_455982 | USA/LA-EVTL139/2020 | 2020-03-28 | North America/USA/Louisiana/Caddo Pa | LSUHS Emerging Viral Threat Laboratory | Microbial Genome Sequencing Center | Jeremy P. Kamil et al |
| EPI_ISL_455996 | USA/LA-EVTL153/2020 | 2020-04-01 | North America/USA/Louisiana/Caddo Pa | LSUHS Emerging Viral Threat Laboratory | Microbial Genome Sequencing Center | John A. Vanchiere et al |
| EPI_ISL_455998 | USA/LA-EVTL155/2020 | 2020-04-01 | North America/USA/Louisiana/Caddo Pa | LSUHS Emerging Viral Threat Laboratory | Microbial Genome Sequencing Center | John A. Vanchiere et al |
| EPI_ISL_456013 | USA/NY-NYUMC776/2020 | 2020-03-27 | North America/USA/New York/Nassau ( | NYU Langone Health | Departments of Pathology and Medicine, New York University School of Medicine | Maria Aguero-Rosenfeld et al |
| EPI_ISL_456016 | USA/NY-NYUMC779/2020 | 2020-03-27 | North America/USA/New York/Brooklyn | NYU Langone Health | Departments of Pathology and Medicine, New York University School of Medicine | Maria Aguero-Rosenfeld et al |

| | | | | | | |
|---|---|---|---|---|---|---|
| EPI_ISL_456018 | USA/NY-NYUMC781/2020 | 2020-03-27 | North America/USA/New York/Manhatt | NYU Langone Health | Departments of Pathology and Medicine, New York University School of Medicine | Maria Aguero-Rosenfeld et al |
| EPI_ISL_456027 | USA/NY-NYUMC790/2020 | 2020-03-25 | North America/USA/New York/Brooklyn | NYU Langone Health | Departments of Pathology and Medicine, New York University School of Medicine | Maria Aguero-Rosenfeld et al |
| EPI_ISL_456072 | Brazil/RJ-1056/2020 | 2020-04-01 | South America/Brazil/Rio de Janeiro/ | Laboratory of Respiratory Viruses and Measles, Oswaldo Cruz Institute, FIOCRUZ | Laboratory of Respiratory Viruses and Measles, Oswaldo Cruz Institute, FIOCRUZ | Paola Resende et al |
| EPI_ISL_456073 | Brazil/RJ-1058/2020 | 2020-04-01 | South America/Brazil/Rio de Janeiro/ | Laboratory of Respiratory Viruses and Measles, Oswaldo Cruz Institute, FIOCRUZ | Laboratory of Respiratory Viruses and Measles, Oswaldo Cruz Institute, FIOCRUZ | Paola Resende et al |
| EPI_ISL_456074 | Brazil/RJ-1065/2020 | 2020-04-01 | South America/Brazil/Rio de Janeiro/ | Laboratory of Respiratory Viruses and Measles, Oswaldo Cruz Institute, FIOCRUZ | Laboratory of Respiratory Viruses and Measles, Oswaldo Cruz Institute, FIOCRUZ | Paola Resende et al |
| EPI_ISL_456075 | Brazil/RJ-1100/2020 | 2020-04-02 | South America/Brazil/Rio de Janeiro/Be | Laboratory of Respiratory Viruses and Measles, Oswaldo Cruz Institute, FIOCRUZ | Laboratory of Respiratory Viruses and Measles, Oswaldo Cruz Institute, FIOCRUZ | Paola Resende et al |

| | | | | | | |
|---|---|---|---|---|---|---|
| EPI_ISL_456097 | Brazil/RJ-1966/2020 | 2020-04-13 | South America/Brazil/Rio de Janeiro/ | Laboratory of Respiratory Viruses and Measles, Oswaldo Cruz Institute, FIOCRUZ | Laboratory of Respiratory Viruses and Measles, Oswaldo Cruz Institute, FIOCRUZ | Paola Resende et al |
| EPI_ISL_456102 | Brazil/RJ-2057/2020 | 2020-04-16 | South America/Brazil/Rio de Janeiro/ | Laboratory of Respiratory Viruses and Measles, Oswaldo Cruz Institute, FIOCRUZ | Laboratory of Respiratory Viruses and Measles, Oswaldo Cruz Institute, FIOCRUZ | Paola Resende et al |
| EPI_ISL_456103 | Brazil/RJ-2062/2020 | 2020-04-16 | South America/Brazil/Rio de Janeiro/ | Laboratory of Respiratory Viruses and Measles, Oswaldo Cruz Institute, FIOCRUZ | Laboratory of Respiratory Viruses and Measles, Oswaldo Cruz Institute, FIOCRUZ | Paola Resende et al |
| EPI_ISL_456104 | Brazil/RJ-2072/2020 | 2020-04-16 | South America/Brazil/Rio de Janeiro/ | Laboratory of Respiratory Viruses and Measles, Oswaldo Cruz Institute, FIOCRUZ | Laboratory of Respiratory Viruses and Measles, Oswaldo Cruz Institute, FIOCRUZ | Paola Resende et al |
| EPI_ISL_456111 | USA/NY-NYUMC839/2020 | 2020-04-24 | North America/USA/New York/Brooklyn | NYU Langone Health | Departments of Pathology and Medicine, New York University School of Medicine | Maria Aguero-Rosenfeld et al |

| | | | | | | |
|---|---|---|---|---|---|---|
| EPI_ISL_456126 | Colombia/NSA-INS-80656/2020 | 2020-03-18 | South America/Colombia/Norte de Sant | Instituto Nacional de Salud - Unidad de Secuenciaci√≥n y An√°lisis Gen√≥mico | Instituto Nacional de Salud, Universidad Cooperativa de Colombia, Instituto Alexander von Humboldt, Imperial College-London, London School of Hygiene & Tropical Medicine | Katherine Laiton-Donato et al |
| EPI_ISL_456131 | Colombia/SAN-INS-79752/2020 | 2020-03-14 | South America/Colombia/Santander/Bu | Instituto Nacional de Salud - Unidad de Secuenciaci√≥n y An√°lisis Gen√≥mico | Instituto Nacional de Salud, Universidad Cooperativa de Colombia, Instituto Alexander von Humboldt, Imperial College-London, London School of Hygiene & Tropical Medicine | Katherine Laiton-Donato et al |
| EPI_ISL_456313 | NewZealand/20VR1929/2020 | 2020-04-03 | Oceania/New Zealand/Southern/ | Southern Community Labs Dunedin | Institute of Environmental Science and Research (ESR) | Matt Storey et al |
| EPI_ISL_456318 | NewZealand/20VR1947/2020 | 2020-04-03 | Oceania/New Zealand/Southern/ | Southern Community Labs Dunedin | Institute of Environmental Science and Research (ESR) | Matt Storey et al |
| EPI_ISL_456321 | NewZealand/20VR1984/2020 | 2020-03-25 | Oceania/New Zealand/Canterbury/ | Canterbury Health Laboratories | Institute of Environmental Science and Research (ESR) | Matt Storey et al |

| | | | | | | |
|---|---|---|---|---|---|---|
| EPI_ISL_456330 | NewZealand/20VR2004/2020 | 2020-04-01 | Oceania/New Zealand/Wellington/ | Wellington SCL | Institute of Environmental Science and Research (ESR) | Matt Storey et al |
| EPI_ISL_456373 | NewZealand/20VR2092/2020 | 2020-04-05 | Oceania/New Zealand/Canterbury/ | Canterbury Health Laboratories | Institute of Environmental Science and Research (ESR) | Matt Storey et al |
| EPI_ISL_456673 | England/BIRM-5F372/2020 | 2020-05-04 | Europe/United Kingdom/England/ | University of Birmingham | COVID-19 Genomics UK (COG-UK) Consortium | Loman Lab: Claire McMurray et al |
| EPI_ISL_456767 | Scotland/CVR3167/2020 | 2020-04-22 | Europe/United Kingdom/Scotland/ | West of Scotland Specialist Virology Centre, NHSGGC / MRC-University of Glasgow Centre for Virus Research | COVID-19 Genomics UK (COG-UK) Consortium | Ana da Silva Filipe et al |
| EPI_ISL_456822 | Scotland/CVR3395/2020 | 2020-05-09 | Europe/United Kingdom/Scotland/ | West of Scotland Specialist Virology Centre, NHSGGC / MRC-University of Glasgow Centre for Virus Research | COVID-19 Genomics UK (COG-UK) Consortium | Ana da Silva Filipe et al |
| EPI_ISL_456848 | Scotland/CVR3427/2020 | 2020-05-07 | Europe/United Kingdom/Scotland/ | West of Scotland Specialist Virology Centre, NHSGGC / MRC-University of Glasgow Centre for Virus Research | COVID-19 Genomics UK (COG-UK) Consortium | Ana da Silva Filipe et al |
| EPI_ISL_456914 | Scotland/EDB3295/2020 | 2020-04-08 | Europe/United Kingdom/Scotland/ | Virology Department, Royal Infirmary of Edinburgh, NHS Lothian / School of Biological Sciences, University of Edinburgh / Institute of Genetics and Molecular Medicine, University of Edinburgh | COVID-19 Genomics UK (COG-UK) Consortium | McHugh M et al |
| EPI_ISL_457007 | Scotland/EDB5358/2020 | 2020-05-24 | Europe/United Kingdom/Scotland/ | Virology Department, Royal Infirmary of Edinburgh, NHS Lothian / School of Biological Sciences, University of Edinburgh / Institute of Genetics and Molecular Medicine, University of Edinburgh | COVID-19 Genomics UK (COG-UK) Consortium | McHugh M et al |
| EPI_ISL_457013 | Scotland/EDB554/2020 | 2020-03-31 | Europe/United Kingdom/Scotland/ | Virology Department, Royal Infirmary of Edinburgh, NHS Lothian / School of Biological Sciences, University of Edinburgh / Institute of Genetics and Molecular Medicine, University of Edinburgh | COVID-19 Genomics UK (COG-UK) Consortium | McHugh M et al |
| EPI_ISL_457054 | England/EXET-1353DB/2020 | 2020-04-03 | Europe/United Kingdom/England/ | University of Exeter | COVID-19 Genomics UK (COG-UK) Consortium | Ben Temperton et al |
| EPI_ISL_457205 | England/EXET-136185/2020 | 2020-04-14 | Europe/United Kingdom/England/ | University of Exeter | COVID-19 Genomics UK (COG-UK) Consortium | Ben Temperton et al |

| | | | | | | |
|---|---|---|---|---|---|---|
| EPI_ISL_457235 | England/EXET-1367DE/2020 | 2020-05-06 | Europe/United Kingdom/England/ | University of Exeter | COVID-19 Genomics UK (COG-UK) Consortium | Ben Temperton et al |
| EPI_ISL_457251 | England/EXET-136B78/2020 | 2020-05-01 | Europe/United Kingdom/England/ | University of Exeter | COVID-19 Genomics UK (COG-UK) Consortium | Ben Temperton et al |
| EPI_ISL_457253 | England/EXET-136B96/2020 | 2020-05-03 | Europe/United Kingdom/England/ | University of Exeter | COVID-19 Genomics UK (COG-UK) Consortium | Ben Temperton et al |
| EPI_ISL_457314 | England/NORT-28BCDA/2020 | 2020-05-08 | Europe/United Kingdom/England/ | Northumbria University / South Tees Hospitals NHS Foundation Trust / North Cumbria Integrated Care NHS Foundation Trust / North Tees and Hartlepool NHS Foundation Trust / Newcastle Hospitals NHS Foundation Trust | COVID-19 Genomics UK (COG-UK) Consortium | Darren L Smith et al |
| EPI_ISL_457323 | England/NORT-292450/2020 | 2020-05-04 | Europe/United Kingdom/England/ | Northumbria University / South Tees Hospitals NHS Foundation Trust / North Cumbria Integrated Care NHS Foundation Trust / North Tees and Hartlepool NHS Foundation Trust / Newcastle Hospitals NHS Foundation Trust | COVID-19 Genomics UK (COG-UK) Consortium | Darren L Smith et al |
| EPI_ISL_457432 | England/NORW-EB44B/2020 | 2020-05-02 | Europe/United Kingdom/England/ | Quadram Institute Bioscience | COVID-19 Genomics UK (COG-UK) Consortium | Dave J. Baker et al |
| EPI_ISL_457461 | England/NORW-EB997/2020 | 2020-05-07 | Europe/United Kingdom/England/ | Quadram Institute Bioscience | COVID-19 Genomics UK (COG-UK) Consortium | Dave J. Baker et al |
| EPI_ISL_457476 | England/NORW-EBAEF/2020 | 2020-05-05 | Europe/United Kingdom/England/ | Quadram Institute Bioscience | COVID-19 Genomics UK (COG-UK) Consortium | Dave J. Baker et al |
| EPI_ISL_457490 | England/NORW-EBC07/2020 | 2020-05-06 | Europe/United Kingdom/England/ | Quadram Institute Bioscience | COVID-19 Genomics UK (COG-UK) Consortium | Dave J. Baker et al |
| EPI_ISL_457495 | England/NORW-EBC70/2020 | 2020-05-05 | Europe/United Kingdom/England/ | Quadram Institute Bioscience | COVID-19 Genomics UK (COG-UK) Consortium | Dave J. Baker et al |
| EPI_ISL_457502 | England/NORW-EBD13/2020 | 2020-05-01 | Europe/United Kingdom/England/ | Quadram Institute Bioscience | COVID-19 Genomics UK (COG-UK) Consortium | Dave J. Baker et al |

| | | | | | | |
|---|---|---|---|---|---|---|
| EPI_ISL_457526 | England/NORW-EC44A/2020 | 2020-05-11 | Europe/United Kingdom/England/ | Quadram Institute Bioscience | COVID-19 Genomics UK (COG-UK) Consortium | Dave J. Baker et al |
| EPI_ISL_457532 | England/NORW-EC50B/2020 | 2020-05-11 | Europe/United Kingdom/England/ | Quadram Institute Bioscience | COVID-19 Genomics UK (COG-UK) Consortium | Dave J. Baker et al |
| EPI_ISL_457581 | England/SHEF-C0989/2020 | 2020-05-02 | Europe/United Kingdom/England/ | Virology Department, Sheffield Teaching Hospitals NHS Foundation Trust/Department of Infection, Immunity and Cardiovascular Disease, The Medical School, University of Sheffield | COVID-19 Genomics UK (COG-UK) Consortium | Thushan de Silva et al |
| EPI_ISL_457687 | Guangzhou/GZMU0074/2020 | 2020-02-11 | Asia/China/Guangdong/Guangzhou | The First Affiliated Hospital of Guangzhou Medical University, Guangzhou, China | BGI-shenzhen & The First Affiliated Hospital of Guangzhou Medical University | Yanqun Wang et al |
| EPI_ISL_457688 | Guangzhou/GZMU0075/2020 | 2020-02-11 | Asia/China/Guangdong/Guangzhou | The First Affiliated Hospital of Guangzhou Medical University, Guangzhou, China | BGI-shenzhen & The First Affiliated Hospital of Guangzhou Medical University | Yanqun Wang et al |
| EPI_ISL_457689 | Guangzhou/GZMU0077/2020 | 2020-02-11 | Asia/China/Guangdong/Guangzhou | The First Affiliated Hospital of Guangzhou Medical University, Guangzhou, China | BGI-shenzhen & The First Affiliated Hospital of Guangzhou Medical University | Yanqun Wang et al |
| EPI_ISL_457690 | Guangzhou/GZMU0078/2020 | 2020-02-11 | Asia/China/Guangdong/Guangzhou | The First Affiliated Hospital of Guangzhou Medical University, Guangzhou, China | BGI-shenzhen & The First Affiliated Hospital of Guangzhou Medical University | Yanqun Wang et al |
| EPI_ISL_457691 | Guangzhou/GZMU0079/2020 | 2020-02-11 | Asia/China/Guangdong/Guangzhou | The First Affiliated Hospital of Guangzhou Medical University, Guangzhou, China | BGI-shenzhen & The First Affiliated Hospital of Guangzhou Medical University | Yanqun Wang et al |

| | | | | | | |
|---|---|---|---|---|---|---|
| EPI_ISL_457692 | Guangzhou/GZMU0081/2020 | 2020-02-14 | Asia/China/Guangdong/Guangzhou | The First Affiliated Hospital of Guangzhou Medical University, Guangzhou, China | BGI-shenzhen & The First Affiliated Hospital of Guangzhou Medical University | Yanqun Wang et al |
| EPI_ISL_457693 | Guangzhou/GZMU0083/2020 | 2020-02-14 | Asia/China/Guangdong/Guangzhou | The First Affiliated Hospital of Guangzhou Medical University, Guangzhou, China | BGI-shenzhen & The First Affiliated Hospital of Guangzhou Medical University | Yanqun Wang et al |
| EPI_ISL_457694 | Guangzhou/GZMU0092/2020 | 2020-02-17 | Asia/China/Guangdong/Guangzhou | The First Affiliated Hospital of Guangzhou Medical University, Guangzhou, China | BGI-shenzhen & The First Affiliated Hospital of Guangzhou Medical University | Yanqun Wang et al |
| EPI_ISL_457695 | Guangzhou/GZMU0095/2020 | 2020-02-17 | Asia/China/Guangdong/Guangzhou | The First Affiliated Hospital of Guangzhou Medical University, Guangzhou, China | BGI-shenzhen & The First Affiliated Hospital of Guangzhou Medical University | Yanqun Wang et al |
| EPI_ISL_457696 | Guangzhou/GZMU0099/2020 | 2020-02-20 | Asia/China/Guangdong/Guangzhou | The First Affiliated Hospital of Guangzhou Medical University, Guangzhou, China | BGI-shenzhen & The First Affiliated Hospital of Guangzhou Medical University | Yanqun Wang et al |
| EPI_ISL_457697 | Guangzhou/GZMU0101/2020 | 2020-02-20 | Asia/China/Guangdong/Guangzhou | The First Affiliated Hospital of Guangzhou Medical University, Guangzhou, China | BGI-shenzhen & The First Affiliated Hospital of Guangzhou Medical University | Yanqun Wang et al |

| | | | | | | |
|---|---|---|---|---|---|---|
| EPI_ISL_457698 | Guangzhou/GZMU0117/2020 | 2020-02-26 | Asia/China/Guangdong/Guangzhou | The First Affiliated Hospital of Guangzhou Medical University, Guangzhou, China | BGI-shenzhen & The First Affiliated Hospital of Guangzhou Medical University | Yanqun Wang et al |
| EPI_ISL_457699 | Italy/EMR-AMVRC-206/2020 | 2020-02-22 | Europe/Italy/Emilia-Romagna/ | Department of Infectious Diseases, Istituto Superiore di Sanitv†, Roma , Italy | Army Medical and Veterinary Research Center | Paola Stefanelli et al |
| EPI_ISL_457700 | Italy/TAA-AMVRC-2629/2020 | 2020-03-06 | Europe/Italy/Trentino-Alto Adige/ | Department of Infectious Diseases, Istituto Superiore di Sanitv†, Roma , Italy | Army Medical and Veterinary Research Center | Paola Stefanelli et al |
| EPI_ISL_457701 | Oman/RESP-20-797/2020 | 2020-02-23 | Asia/Oman/Muscat/ | Oman-NIC | Oman-NIC | Samira Al-Maruqi et al |
| EPI_ISL_457704 | Oman/RESP-20-837/2020 | 2020-02-24 | Asia/Oman/Muscat/ | Oman-NIC | Oman-NIC | Samira Al-Maruqi et al |
| EPI_ISL_457721 | Italy/SIC-AMVRC-4411/2020 | 2020-03-12 | Europe/Italy/Sicily/ | Department of Infectious Diseases, Istituto Superiore di Sanitv†, Roma , Italy | Army Medical and Veterinary Research Center | Paola Stefanelli et al |
| EPI_ISL_457724 | Italy/AVA-AMVRC-4875/2020 | 2020-03-15 | Europe/Italy/Valle D'Aosta/ | Department of Infectious Diseases, Istituto Superiore di Sanitv†, Roma , Italy | Army Medical and Veterinary Research Center | Paola Stefanelli et al |
| EPI_ISL_457726 | Taiwan/TSGH-35/2020 | 2020-02-06 | Asia/Taiwan/Taiwan/Taipei | TSGH-CP molecular lab | TSGH-CP molecular lab | Cherng-Lih Perng et al |
| EPI_ISL_457728 | Italy/MOL-AMVRC-2389/2020 | 2020-03-04 | Europe/Italy/Molise/ | Department of Infectious Diseases, Istituto Superiore di Sanitv†, Roma , Italy | Army Medical and Veterinary Research Center | Paola Stefanelli et al |
| EPI_ISL_457730 | Taiwan/TSGH-36/2020 | 2020-02-06 | Asia/Taiwan/Taiwan/Taipei | TSGH-CP molecular lab | TSGH-CP molecular lab | Cherng-Lih Perng et al |
| EPI_ISL_457732 | Italy/AVA-AMVRC-4870/2020 | 2020-03-14 | Europe/Italy/Valle D'Aosta/ | Department of Infectious Diseases, Istituto Superiore di Sanitv†, Roma , Italy | Army Medical and Veterinary Research Center | Paola Stefanelli et al |
| EPI_ISL_457733 | Taiwan/TSGH-37/2020 | 2020-02-08 | Asia/Taiwan/Taiwan/Taipei | TSGH-CP molecular lab | TSGH-CP molecular lab | Cherng-Lih Perng et al |
| EPI_ISL_457749 | Italy/ABR-IZSGC-484/2020 | 2020-02-27 | Europe/Italy/Abruzzo/ | Department of Infectious Diseases, Istituto Superiore di Sanitv†, Roma , Italy | Army Medical and Veterinary Research Center | Paola Stefanelli et al |
| EPI_ISL_457761 | USA/MD-HP00097/2020 | 2020-03-24 | North America/USA/Maryland/ | Johns Hopkins Hospital Department of Pathology | Johns Hopkins Hospital Department of Pathology | Peter M. Thielen et al |

| | | | | | | |
|---|---|---|---|---|---|---|
| EPI_ISL_457775 | USA/DC-HP00145/2020 | 2020-03-26 | North America/USA/Washington DC/ | Johns Hopkins Hospital Department of Pathology | Johns Hopkins Hospital Department of Pathology | Peter M. Thielen et al |
| EPI_ISL_457781 | USA/DC-HP00152/2020 | 2020-03-27 | North America/USA/Washington DC/ | Johns Hopkins Hospital Department of Pathology | Johns Hopkins Hospital Department of Pathology | Peter M. Thielen et al |
| EPI_ISL_457794 | USA/MD-HP00050/2020 | 2020-03-19 | North America/USA/Maryland/ | Johns Hopkins Hospital Department of Pathology | Johns Hopkins Hospital Department of Pathology | Peter M. Thielen et al |
| EPI_ISL_457795 | USA/MD-HP00051/2020 | 2020-03-19 | North America/USA/Maryland/ | Johns Hopkins Hospital Department of Pathology | Johns Hopkins Hospital Department of Pathology | Peter M. Thielen et al |
| EPI_ISL_457797 | USA/DC-HP00054/2020 | 2020-03-07 | North America/USA/Washington DC/ | Johns Hopkins Hospital Department of Pathology | Johns Hopkins Hospital Department of Pathology | Peter M. Thielen et al |
| EPI_ISL_457812 | USA/MD-HP00119/2020 | 2020-03-18 | North America/USA/Maryland/ | Johns Hopkins Hospital Department of Pathology | Johns Hopkins Hospital Department of Pathology | Peter M. Thielen et al |
| EPI_ISL_457817 | USA/MD-HP00128/2020 | 2020-03-24 | North America/USA/Maryland/ | Johns Hopkins Hospital Department of Pathology | Johns Hopkins Hospital Department of Pathology | Peter M. Thielen et al |
| EPI_ISL_457824 | Turkey/Istanbul-BezmCoV1/2020 | 2020-03-24 | Europe/Turkey/Istanbul/ | Bezmialem Vakif University, Dept Microbiology, Medical School, Fatih, Istanbul, Turkey | Bezmialem Vakif University, Medical School & Beykoz Institute of Life Sciences & Biotechnology | Mehmet Z. Doymaz et al |
| EPI_ISL_457943 | Uruguay/UY-NYUMC847/2020 | 2020-03-17 | South America/Uruguay/Montevideo/ | Laboratorio de Biolog√≠a Molecular Asociaci√≥n Espa√±ola Primera en Salud | Departments of Pathology and Medicine, New York University School of Medicine | Maria Victoria Elizondo et al |

| | | | | | | |
|---|---|---|---|---|---|---|
| EPI_ISL_458028 | Thailand/Bangkok-CONI-0099/2020 | 2020-04-04 | Asia/Thailand/Bangkok/ | Hospital for Tropical Diseases | COVID-19 Network Investigations (CONI) Alliance | Elizabeth Batty et al |
| EPI_ISL_458033 | India/TN-CCMB-C5/2020 | 2020-04-29 | Asia/India/Tamil Nadu/Chennai | King Institute of Preventive Medicine & Research | CSIR-Centre for Cellular and Molecular Biology | K.Kaveri et al |
| EPI_ISL_458052 | India/TG-CCMB-L1063/2020 | 2020-05-12 | Asia/India/Telangana/Mancherial | CSIR-Centre for Cellular and Molecular Biology | CSIR-Centre for Cellular and Molecular Biology | Sofia Banu et al |
| EPI_ISL_458056 | India/TG-CCMB-L1085/2020 | 2020-05-13 | Asia/India/Telangana/Hyderabad | CSIR-Centre for Cellular and Molecular Biology | CSIR-Centre for Cellular and Molecular Biology | Tulasi Nagabandi et al |
| EPI_ISL_458059 | India/TG-CCMB-L1100/2020 | 2020-05-13 | Asia/India/Telangana/Hyderabad | CSIR-Centre for Cellular and Molecular Biology | CSIR-Centre for Cellular and Molecular Biology | Shagufta Khan et al |
| EPI_ISL_458064 | India/TG-CCMB-L988/2020 | 2020-05-11 | Asia/India/Telangana/Hyderabad | CSIR-Centre for Cellular and Molecular Biology | CSIR-Centre for Cellular and Molecular Biology | Shagufta Khan et al |
| EPI_ISL_458065 | India/TG-CCMB-L989/2020 | 2020-05-11 | Asia/India/Telangana/Hyderabad | CSIR-Centre for Cellular and Molecular Biology | CSIR-Centre for Cellular and Molecular Biology | Lamuk Zaveri et al |
| EPI_ISL_458066 | India/TG-CCMB-O1/2020 | 2020-04-13 | Asia/India/Telangana/Hyderabad | Osmania Medical College | CSIR-Centre for Cellular and Molecular Biology | Shashikala Reddy et al |
| EPI_ISL_458074 | India/TG-CCMB-L301-P1/2020 | 2020-04-20 | Asia/India/Telangana/Suryapet | CSIR-Centre for Cellular and Molecular Biology | CSIR-Centre for Cellular and Molecular Biology | Dhiviya Vedagiri et al |
| EPI_ISL_458085 | Italy/SAR-AMVRC-28/2020 | 2020-04-12 | Europe/Italy/Sardinia/ | "Laboratorio Biologia Molecolare Sars Cov2 - UOC Laboratorio Analisi - Servizio Medicina di Laboratorio , Ospedale ""San Francesco"" - ATS- ASSL Nuoro" | "Laboratorio specialistico UOC Ematologia - Ospedale ""San Francesco"" - ATS- ASSL Nuoro" | Piras Giovanna et al |
| EPI_ISL_458089 | India/GJ-GBRC134/2020 | 2020-05-24 | Asia/India/Gujarat/Ahmedabad | B.J. Medical College and Civil hospital | Gujarat Biotechnology Research Centre | Pranay Shah et al |
| EPI_ISL_458092 | India/GJ-GBRC136a/2020 | 2020-05-24 | Asia/India/Gujarat/Ahmedabad | B.J. Medical College and Civil hospital | Gujarat Biotechnology Research Centre | Amit Kanani et al |

| | | | | | | |
|---|---|---|---|---|---|---|
| EPI_ISL_458094 | India/GJ-GBRC137a/2020 | 2020-05-24 | Asia/India/Gujarat/Ahmedabad | B.J. Medical College and Civil hospital | Gujarat Biotechnology Research Centre | Nitin Savaliya et al |
| EPI_ISL_458095 | India/GJ-GBRC137b/2020 | 2020-05-24 | Asia/India/Gujarat/Ahmedabad | B.J. Medical College and Civil hospital | Gujarat Biotechnology Research Centre | Raghawendra Kumar et al |
| EPI_ISL_458099 | India/GJ-GBRC140/2020 | 2020-05-24 | Asia/India/Gujarat/Ahmedabad | B.J. Medical College and Civil hospital | Gujarat Biotechnology Research Centre | Labdhi Pandya et al |
| EPI_ISL_458107 | India/GJ-GBRC146b/2020 | 2020-05-16 | Asia/India/Gujarat/Himatnagar | Gujarat Biotechnology Research Centre | Gujarat Biotechnology Research Centre | Apurvasinh Puvar et al |
| EPI_ISL_458108 | India/GJ-GBRC147/2020 | 2020-05-16 | Asia/India/Gujarat/Khedbrahma | Gujarat Biotechnology Research Centre | Gujarat Biotechnology Research Centre | Janvi Raval et al |
| EPI_ISL_458110 | India/GJ-GBRC148b/2020 | 2020-05-17 | Asia/India/Gujarat/Khedbrahma | Gujarat Biotechnology Research Centre | Gujarat Biotechnology Research Centre | Monika Gandhi et al |
| EPI_ISL_458118 | Oman/205033010/2020 | 2020-05-01 | Asia/Oman/Muscat/ | Oman National Influenza Centre | Department of Microbiology and Immunology-SQUH | Fahad Zadjali et al |
| EPI_ISL_458150 | Morocco/15N/2020 | 2020-05-15 | Africa/Morocco/Casablanca/ | ANOUAL | ANOUAL | Jouali Farah et al |
| EPI_ISL_458156 | Belgium/Rega-0328186/2020 | 2020-03-28 | Europe/Belgium/Leuven/Kaggevinne | KU Leuven, Rega Institute, Clinical and Epidemiological Virology | KU Leuven, Rega Institute, Clinical and Epidemiological Virology | Tony Wawina-Bokalanga et al |
| EPI_ISL_458158 | Belgium/Rega-0328188/2020 | 2020-03-28 | Europe/Belgium/Leuven/Geetbets | KU Leuven, Rega Institute, Clinical and Epidemiological Virology | KU Leuven, Rega Institute, Clinical and Epidemiological Virology | Tony Wawina-Bokalanga et al |
| EPI_ISL_458160 | Belgium/Rega-0328190/2020 | 2020-03-28 | Europe/Belgium/Mons/Colfontaine | KU Leuven, Rega Institute, Clinical and Epidemiological Virology | KU Leuven, Rega Institute, Clinical and Epidemiological Virology | Tony Wawina-Bokalanga et al |

| | | | | | | |
|---|---|---|---|---|---|---|
| EPI_ISL_458162 | Belgium/Rega-0328192/2020 | 2020-03-28 | Europe/Belgium/La LouviV®re/ | KU Leuven, Rega Institute, Clinical and Epidemiological Virology | KU Leuven, Rega Institute, Clinical and Epidemiological Virology | Tony Wawina-Bokalanga et al |
| EPI_ISL_458181 | Belgium/Rega-0327211/2020 | 2020-03-27 | Europe/Belgium/Leuven/Molenstede | KU Leuven, Rega Institute, Clinical and Epidemiological Virology | KU Leuven, Rega Institute, Clinical and Epidemiological Virology | Tony Wawina-Bokalanga et al |
| EPI_ISL_458194 | Belgium/Rega-0329224/2020 | 2020-03-29 | Europe/Belgium/Sint-Niklaas/Temse | KU Leuven, Rega Institute, Clinical and Epidemiological Virology | KU Leuven, Rega Institute, Clinical and Epidemiological Virology | Tony Wawina-Bokalanga et al |
| EPI_ISL_458204 | Belgium/Rega-0329234/2020 | 2020-03-29 | Europe/Belgium/Hasselt/Paal | KU Leuven, Rega Institute, Clinical and Epidemiological Virology | KU Leuven, Rega Institute, Clinical and Epidemiological Virology | Tony Wawina-Bokalanga et al |
| EPI_ISL_458208 | Belgium/Rega-0329238/2020 | 2020-03-29 | Europe/Belgium/Brussels/Ixelles | KU Leuven, Rega Institute, Clinical and Epidemiological Virology | KU Leuven, Rega Institute, Clinical and Epidemiological Virology | Tony Wawina-Bokalanga et al |
| EPI_ISL_458211 | Belgium/Rega-0329241/2020 | 2020-03-29 | Europe/Belgium/Hasselt/Zolder | KU Leuven, Rega Institute, Clinical and Epidemiological Virology | KU Leuven, Rega Institute, Clinical and Epidemiological Virology | Tony Wawina-Bokalanga et al |
| EPI_ISL_458221 | Belgium/Rega-0330251/2020 | 2020-03-30 | Europe/Belgium/Antwerpen/Malle | KU Leuven, Rega Institute, Clinical and Epidemiological Virology | KU Leuven, Rega Institute, Clinical and Epidemiological Virology | Tony Wawina-Bokalanga et al |
| EPI_ISL_458231 | Belgium/Rega-0326261/2020 | 2020-03-26 | Europe/Belgium/Leuven/Wespelaar | KU Leuven, Rega Institute, Clinical and Epidemiological Virology | KU Leuven, Rega Institute, Clinical and Epidemiological Virology | Tony Wawina-Bokalanga et al |

| | | | | | | |
|---|---|---|---|---|---|---|
| EPI_ISL_458242 | USA/CA-SR0358/2020 | 2020-04-20 | North America/USA/California/San Dieg | Scripps Medical Laboratory | Andersen lab at Scripps Research | SEARCH Alliance San Diego with Michael Quigley et al |
| EPI_ISL_458243 | USA/CA-SR0359/2020 | 2020-04-12 | North America/USA/California/San Dieg | Scripps Medical Laboratory | Andersen lab at Scripps Research | SEARCH Alliance San Diego with Michael Quigley et al |
| EPI_ISL_458247 | USA/CA-SR0364/2020 | 2020-04-14 | North America/USA/California/San Dieg | Scripps Medical Laboratory | Andersen lab at Scripps Research | SEARCH Alliance San Diego with Michael Quigley et al |
| EPI_ISL_458256 | USA/CA-SR0373/2020 | 2020-04-07 | North America/USA/California/San Dieg | Scripps Medical Laboratory | Andersen lab at Scripps Research | SEARCH Alliance San Diego with Michael Quigley et al |
| EPI_ISL_458270 | USA/CA-SR0395/2020 | 2020-04-04 | North America/USA/California/San Dieg | Scripps Medical Laboratory | Andersen lab at Scripps Research | SEARCH Alliance San Diego with Michael Quigley et al |
| EPI_ISL_458277 | USA/CA-SR0407/2020 | 2020-04-20 | North America/USA/California/San Dieg | Scripps Medical Laboratory | Andersen lab at Scripps Research | SEARCH Alliance San Diego with Michael Quigley et al |
| EPI_ISL_458325 | England/BRIS-12AAC5/2020 | 2020-04-30 | Europe/United Kingdom/England/ | PHE South West Regional Laboratory, National Infection Service | Wellcome Sanger Institute for the COVID-19 Genomics UK (COG-UK) consortium | Stephanie Hutchings et al |
| EPI_ISL_458367 | England/BRIS-12ACFC/2020 | 2020-05-01 | Europe/United Kingdom/England/ | PHE South West Regional Laboratory, National Infection Service | Wellcome Sanger Institute for the COVID-19 Genomics UK (COG-UK) consortium | Stephanie Hutchings et al |
| EPI_ISL_458427 | England/BRIS-12ADBD/2020 | 2020-05-02 | Europe/United Kingdom/England/ | PHE South West Regional Laboratory, National Infection Service | Wellcome Sanger Institute for the COVID-19 Genomics UK (COG-UK) consortium | Stephanie Hutchings et al |
| EPI_ISL_458474 | England/BRIS-12A834/2020 | 2020-04-29 | Europe/United Kingdom/England/ | PHE South West Regional Laboratory, National Infection Service | Wellcome Sanger Institute for the COVID-19 Genomics UK (COG-UK) consortium | Stephanie Hutchings et al |
| EPI_ISL_458578 | England/NORT-28995D/2020 | 2020-04-02 | Europe/United Kingdom/England/ | NU-OMICS DNA Sequencing research facility, Northumbria University | Wellcome Sanger Institute for the COVID-19 Genomics UK (COG-UK) consortium | Chris Duncan et al |

| | | | | | | |
|---|---|---|---|---|---|---|
| EPI_ISL_458604 | England/NORT-28901C/2020 | 2020-03-26 | Europe/United Kingdom/England/ | NU-OMICS DNA Sequencing research facility, Northumbria University | Wellcome Sanger Institute for the COVID-19 Genomics UK (COG-UK) consortium | Chris Duncan et al |
| EPI_ISL_458682 | England/NORT-289498/2020 | 2020-03-29 | Europe/United Kingdom/England/ | NU-OMICS DNA Sequencing research facility, Northumbria University | Wellcome Sanger Institute for the COVID-19 Genomics UK (COG-UK) consortium | Chris Duncan et al |
| EPI_ISL_458725 | England/BRIS-128AB8/2020 | 2020-04-14 | Europe/United Kingdom/England/ | PHE South West Regional Laboratory, National Infection Service | Wellcome Sanger Institute for the COVID-19 Genomics UK (COG-UK) consortium | Stephanie Hutchings et al |
| EPI_ISL_458766 | England/BRIS-1294E6/2020 | 2020-04-20 | Europe/United Kingdom/England/ | PHE South West Regional Laboratory, National Infection Service | Wellcome Sanger Institute for the COVID-19 Genomics UK (COG-UK) consortium | Stephanie Hutchings et al |
| EPI_ISL_458883 | England/BRIS-128BB5/2020 | 2020-04-14 | Europe/United Kingdom/England/ | PHE South West Regional Laboratory, National Infection Service | Wellcome Sanger Institute for the COVID-19 Genomics UK (COG-UK) consortium | Stephanie Hutchings et al |
| EPI_ISL_458991 | England/BRIS-126BF3/2020 | 2020-04-10 | Europe/United Kingdom/England/ | PHE South West Regional Laboratory, National Infection Service | Wellcome Sanger Institute for the COVID-19 Genomics UK (COG-UK) consortium | Stephanie Hutchings et al |
| EPI_ISL_459154 | Scotland/GCVR-172BAD/2020 | 2020-04-06 | Europe/United Kingdom/Scotland/ | NHSGGC West of Scotland Specialist Virology Centre / MRC-University of Glasgow Centre for Virus Research | Wellcome Sanger Institute for the COVID-19 Genomics UK (COG-UK) consortium | Ana da Silva Filipe et al |
| EPI_ISL_459196 | England/CAMB-1AFF6F/2020 | 2020-05-04 | Europe/United Kingdom/England/ | Department of Pathology, University of Cambridge | Wellcome Sanger Institute for the COVID-19 Genomics UK (COG-UK) consortium | Luke W Meredith et al |

| | | | | | | |
|---|---|---|---|---|---|---|
| EPI_ISL_459235 | England/CAMB-1B095E/2020 | 2020-05-04 | Europe/United Kingdom/England/ | Department of Pathology, University of Cambridge | Wellcome Sanger Institute for the COVID-19 Genomics UK (COG-UK) consortium | Luke W Meredith et al |
| EPI_ISL_459294 | England/CAMB-1AFB01/2020 | 2020-05-03 | Europe/United Kingdom/England/ | Department of Pathology, University of Cambridge | Wellcome Sanger Institute for the COVID-19 Genomics UK (COG-UK) consortium | Luke W Meredith et al |
| EPI_ISL_459346 | NorthernIreland/NIRE-FAB35/2020 | 2020-04-15 | Europe/United Kingdom/Northern Irelar | Regional Virus Laboratory, Belfast Health and Social Care Trust | Wellcome Sanger Institute for the COVID-19 Genomics UK (COG-UK) consortium | Conall McCaughey et al |
| EPI_ISL_459460 | England/CAMB-1AE531/2020 | 2020-04-28 | Europe/United Kingdom/England/ | Department of Pathology, University of Cambridge | Wellcome Sanger Institute for the COVID-19 Genomics UK (COG-UK) consortium | Luke W Meredith et al |
| EPI_ISL_459471 | England/CAMB-1ADD67/2020 | 2020-04-26 | Europe/United Kingdom/England/ | Department of Pathology, University of Cambridge | Wellcome Sanger Institute for the COVID-19 Genomics UK (COG-UK) consortium | Luke W Meredith et al |
| EPI_ISL_459491 | England/CAMB-1AE67A/2020 | 2020-04-27 | Europe/United Kingdom/England/ | Department of Pathology, University of Cambridge | Wellcome Sanger Institute for the COVID-19 Genomics UK (COG-UK) consortium | Luke W Meredith et al |
| EPI_ISL_459568 | Scotland/GCVR-1712D6/2020 | 2020-03-26 | Europe/United Kingdom/Scotland/ | NHSGGC West of Scotland Specialist Virology Centre / MRC-University of Glasgow Centre for Virus Research | Wellcome Sanger Institute for the COVID-19 Genomics UK (COG-UK) consortium | Ana da Silva Filipe et al |
| EPI_ISL_459598 | Scotland/GCVR-170529/2020 | 2020-03-23 | Europe/United Kingdom/Scotland/ | NHSGGC West of Scotland Specialist Virology Centre / MRC-University of Glasgow Centre for Virus Research | Wellcome Sanger Institute for the COVID-19 Genomics UK (COG-UK) consortium | Ana da Silva Filipe et al |

| | | | | | | |
|---|---|---|---|---|---|---|
| EPI_ISL_459603 | Scotland/GCVR-1708D5/2020 | 2020-03-24 | Europe/United Kingdom/Scotland/ | NHSGGC West of Scotland Specialist Virology Centre / MRC-University of Glasgow Centre for Virus Research | Wellcome Sanger Institute for the COVID-19 Genomics UK (COG-UK) consortium | Ana da Silva Filipe et al |
| EPI_ISL_459866 | Canada/ON_QGLO-01/2020 | 2020-03-18 | North America/Canada/Ontario/ | Kingston Health Sciences Center | Queen's Genomics Lab at Ongwanada (Q-GLO) | Sjaarda CP et al |
| EPI_ISL_459867 | Canada/ON_QGLO-02/2020 | 2020-03-22 | North America/Canada/Ontario/ | Kingston Health Sciences Center | Queen's Genomics Lab at Ongwanada (Q-GLO) | Sjaarda CP et al |
| EPI_ISL_459868 | Canada/ON_QGLO-04/2020 | 2020-03-24 | North America/Canada/Ontario/ | Kingston Health Sciences Center | Queen's Genomics Lab at Ongwanada (Q-GLO) | Sjaarda CP et al |
| EPI_ISL_459869 | Canada/ON_QGLO-010/2020 | 2020-03-26 | North America/Canada/Ontario/ | Kingston Health Sciences Center | Queen's Genomics Lab at Ongwanada (Q-GLO) | Sjaarda CP et al |
| EPI_ISL_459871 | Canada/ON_QGLO-011/2020 | 2020-03-26 | North America/Canada/Ontario/ | Kingston Health Sciences Center | Queen's Genomics Lab at Ongwanada (Q-GLO) | Sjaarda CP et al |
| EPI_ISL_459872 | Canada/ON_QGLO-012/2020 | 2020-03-26 | North America/Canada/Ontario/ | Kingston Health Sciences Center | Queen's Genomics Lab at Ongwanada (Q-GLO) | Sjaarda CP et al |
| EPI_ISL_459873 | Canada/ON_QGLO-016/2020 | 2020-03-26 | North America/Canada/Ontario/ | Kingston Health Sciences Center | Queen's Genomics Lab at Ongwanada (Q-GLO) | Sjaarda CP et al |
| EPI_ISL_459874 | Canada/ON_QGLO-017/2020 | 2020-03-27 | North America/Canada/Ontario/ | Kingston Health Sciences Center | Queen's Genomics Lab at Ongwanada (Q-GLO) | Sjaarda CP et al |

| | | | | | | |
|---|---|---|---|---|---|---|
| EPI_ISL_459875 | Canada/ON_QGLO-018/2020 | 2020-03-27 | North America/Canada/Ontario/ | Kingston Health Sciences Center | Queen's Genomics Lab at Ongwanada (Q-GLO) | Sjaarda CP et al |
| EPI_ISL_459877 | Canada/ON-QGLO-019/2020 | 2020-03-27 | North America/Canada/Ontario/ | Kingston Health Sciences Center | Queen's Genomics Lab at Ongwanada (Q-GLO) | Sjaarda CP et al |
| EPI_ISL_459878 | Canada/ON-QGLO-021/2020 | 2020-03-27 | North America/Canada/Ontario/ | Kingston Health Sciences Center | Queen's Genomics Lab at Ongwanada (Q-GLO) | Sjaarda CP et al |
| EPI_ISL_459879 | Canada/ON_QGLO-023/2020 | 2020-03-28 | North America/Canada/Ontario/ | Kingston Health Sciences Center | Queen's Genomics Lab at Ongwanada (Q-GLO) | Sjaarda CP et al |
| EPI_ISL_459880 | Canada/ON_QGLO-024/2020 | 2020-03-28 | North America/Canada/Ontario/ | Kingston Health Sciences Center | Queen's Genomics Lab at Ongwanada (Q-GLO) | Sjaarda CP et al |
| EPI_ISL_459881 | Canada/ON_QGLO-025/2020 | 2020-03-28 | North America/Canada/Ontario/ | Kingston Health Sciences Center | Queen's Genomics Lab at Ongwanada (Q-GLO) | Sjaarda CP et al |
| EPI_ISL_459882 | Canada/ON_QGLO-026/2020 | 2020-03-28 | North America/Canada/Ontario/ | Kingston Health Sciences Center | Queen's Genomics Lab at Ongwanada (Q-GLO) | Sjaarda CP et al |
| EPI_ISL_459883 | Canada/ON_QGLO-029/2020 | 2020-03-28 | North America/Canada/Ontario/ | Kingston Health Sciences Center | Queen's Genomics Lab at Ongwanada (Q-GLO) | Sjaarda CP et al |
| EPI_ISL_459884 | Canada/ON-QGLO-030/2020 | 2020-03-28 | North America/Canada/Ontario/ | Kingston Health Sciences Center | Queen's Genomics Lab at Ongwanada (Q-GLO) | Sjaarda CP et al |
| EPI_ISL_459885 | Canada/ON_QGLO-034/2020 | 2020-03-29 | North America/Canada/Ontario/ | Kingston Health Sciences Center | Queen's Genomics Lab at Ongwanada (Q-GLO) | Sjaarda CP et al |

| | | | | | | |
|---|---|---|---|---|---|---|
| EPI_ISL_459886 | Canada/ON_QGLO-035/2020 | 2020-03-30 | North America/Canada/Ontario/ | Kingston Health Sciences Center | Queen's Genomics Lab at Ongwanada (Q-GLO) | Sjaarda CP et al |
| EPI_ISL_459887 | Canada/ON_QGLO-036/2020 | 2020-03-30 | North America/Canada/Ontario/ | Kingston Health Sciences Center | Queen's Genomics Lab at Ongwanada (Q-GLO) | Sjaarda CP et al |
| EPI_ISL_459888 | Canada/ON_QGLO-037/2020 | 2020-03-30 | North America/Canada/Ontario/ | Kingston Health Sciences Center | Queen's Genomics Lab at Ongwanada (Q-GLO) | Sjaarda CP et al |
| EPI_ISL_459889 | Canada/ON_QGLO-038/2020 | 2020-03-30 | North America/Canada/Ontario/ | Kingston Health Sciences Center | Queen's Genomics Lab at Ongwanada (Q-GLO) | Sjaarda CP et al |
| EPI_ISL_459890 | Canada/ON_QGLO-039/2020 | 2020-03-29 | North America/Canada/Ontario/ | Kingston Health Sciences Center | Queen's Genomics Lab at Ongwanada (Q-GLO) | Sjaarda CP et al |
| EPI_ISL_459891 | Canada/ON_QGLO-041/2020 | 2020-03-30 | North America/Canada/Ontario/ | Kingston Health Sciences Center | Queen's Genomics Lab at Ongwanada (Q-GLO) | Sjaarda CP et al |
| EPI_ISL_459892 | Canada/ON_QGLO-042/2020 | 2020-03-30 | North America/Canada/Ontario/ | Kingston Health Sciences Center | Queen's Genomics Lab at Ongwanada (Q-GLO) | Sjaarda CP et al |
| EPI_ISL_459901 | Luxembourg/LNS6469209/2020 | 2020-05-19 | Europe/Luxembourg/Luxembourg/ | Laboratoire National de Sante, Microbiology, Virology | Laboratoire National de Sante, Microbiology, Epidemiology and Microbial Genomics | Anke Wienecke-Baldacchino et al |
| EPI_ISL_459922 | India/DL-MaxCov0020-CSIR-IGIB/2020 | 2020-05-10 | Asia/India/Delhi/ | Devki Devi Foundation, a unit of Max Healthcare | CSIR-IGIB/Max | Rajesh Pandey |
| EPI_ISL_459933 | India/DL-MaxCov0031-CSIR-IGIB/2020 | 2020-05-05 | Asia/India/Delhi/ | Devki Devi Foundation, a unit of Max Healthcare | CSIR-IGIB/Max | Rajesh Pandey |

| | | | | | | |
|---|---|---|---|---|---|---|
| EPI_ISL_459947 | India/DL-MaxCov0045-CSIR-IGIB/2020 | 2020-04-28 | Asia/India/Delhi/ | Devki Devi Foundation, a unit of Max Healthcare | CSIR-IGIB/Max | Rajesh Pandey |
| EPI_ISL_459950 | India/DL-MaxCov0048-CSIR-IGIB/2020 | 2020-05-01 | Asia/India/Delhi/ | Devki Devi Foundation, a unit of Max Healthcare | CSIR-IGIB/Max | Rajesh Pandey |
| EPI_ISL_459957 | Malaysia/IMR_WC10180/2020 | 2020-04-01 | Asia/Malaysia/Malaysia/ | Institute for Medical Research, Infectious Disease Research Centre, National Institutes of Health, Minis | Institute for Medical Research, Infectious Disease Research Centre, National Institutes of Health, Minis | Suppiah J et al |
| EPI_ISL_459968 | Morocco/6890/2020 | 2020-03-17 | Africa/Morocco/Morocco/ | Institut Pasteur du Maroc | Institut Pasteur du Maroc | Marion Barbet et al |
| EPI_ISL_459969 | Morocco/6891/2020 | 2020-03-20 | Africa/Morocco/Morocco/ | Institut Pasteur du Maroc | Institut Pasteur du Maroc | Marion Barbet et al |
| EPI_ISL_459970 | Morocco/6892/2020 | 2020-03-17 | Africa/Morocco/Morocco/ | Institut Pasteur du Maroc | Institut Pasteur du Maroc | Marion Barbet et al |
| EPI_ISL_459973 | Morocco/6895/2020 | 2020-03-20 | Africa/Morocco/Morocco/ | Institut Pasteur du Maroc | Institut Pasteur du Maroc | Marion Barbet et al |
| EPI_ISL_459974 | Morocco/6896/2020 | 2020-03-20 | Africa/Morocco/Morocco/ | Institut Pasteur du Maroc | Institut Pasteur du Maroc | Marion Barbet et al |
| EPI_ISL_459977 | Morocco/6899/2020 | 2020-04-21 | Africa/Morocco/Morocco/ | Institut Pasteur du Maroc | Institut Pasteur du Maroc | Marion Barbet et al |
| EPI_ISL_459984 | Morocco/6906/2020 | 2020-04-06 | Africa/Morocco/Morocco/ | Institut Pasteur du Maroc | Institut Pasteur du Maroc | Marion Barbet et al |
| EPI_ISL_459998 | USA/MI-MDHHS-SC20459/2020 | 2020-05-06 | North America/USA/Michigan/ | Michigan Department of Health and Human Services, Bureau of Laboratories | Michigan Department of Health and Human Services, Bureau of Laboratories | Blankenship HM et al |
| EPI_ISL_460002 | USA/MI-MDHHS-SC20463/2020 | 2020-03-30 | North America/USA/Michigan/ | Michigan Department of Health and Human Services, Bureau of Laboratories | Michigan Department of Health and Human Services, Bureau of Laboratories | Blankenship HM et al |

| | | | | | | |
|---|---|---|---|---|---|---|
| EPI_ISL_460006 | USA/MI-MDHHS-SC20467/2020 | 2020-03-30 | North America/USA/Michigan/ | Michigan Department of Health and Human Services, Bureau of Laboratories | Michigan Department of Health and Human Services, Bureau of Laboratories | Blankenship HM et al |
| EPI_ISL_460011 | USA/MI-MDHHS-SC20472/2020 | 2020-03-31 | North America/USA/Michigan/ | Michigan Department of Health and Human Services, Bureau of Laboratories | Michigan Department of Health and Human Services, Bureau of Laboratories | Blankenship HM et al |
| EPI_ISL_460020 | USA/MI-MDHHS-SC20481/2020 | 2020-03-31 | North America/USA/Michigan/ | Michigan Department of Health and Human Services, Bureau of Laboratories | Michigan Department of Health and Human Services, Bureau of Laboratories | Blankenship HM et al |
| EPI_ISL_460022 | USA/MI-MDHHS-SC20483/2020 | 2020-04-01 | North America/USA/Michigan/ | Michigan Department of Health and Human Services, Bureau of Laboratories | Michigan Department of Health and Human Services, Bureau of Laboratories | Blankenship HM et al |
| EPI_ISL_460023 | USA/MI-MDHHS-SC20484/2020 | 2020-04-01 | North America/USA/Michigan/ | Michigan Department of Health and Human Services, Bureau of Laboratories | Michigan Department of Health and Human Services, Bureau of Laboratories | Blankenship HM et al |
| EPI_ISL_460027 | USA/MI-MDHHS-SC20488/2020 | 2020-04-01 | North America/USA/Michigan/ | Michigan Department of Health and Human Services, Bureau of Laboratories | Michigan Department of Health and Human Services, Bureau of Laboratories | Blankenship HM et al |

| | | | | | | |
|---|---|---|---|---|---|---|
| EPI_ISL_460029 | USA/MI-MDHHS-SC20490/2020 | 2020-03-30 | North America/USA/Michigan/ | Michigan Department of Health and Human Services, Bureau of Laboratories | Michigan Department of Health and Human Services, Bureau of Laboratories | Blankenship HM et al |
| EPI_ISL_460030 | USA/MI-MDHHS-SC20491/2020 | 2020-04-01 | North America/USA/Michigan/ | Michigan Department of Health and Human Services, Bureau of Laboratories | Michigan Department of Health and Human Services, Bureau of Laboratories | Blankenship HM et al |
| EPI_ISL_460034 | USA/MI-MDHHS-SC20496/2020 | 2020-05-06 | North America/USA/Michigan/ | Michigan Department of Health and Human Services, Bureau of Laboratories | Michigan Department of Health and Human Services, Bureau of Laboratories | Blankenship HM et al |
| EPI_ISL_460076 | USA/MN-MDH-95/2020 | 2020-03-15 | North America/USA/Minnesota/ | Minnesota Department of Health, Public Health Laboratory | Minnesota Department of Health, Public Health Laboratory | Matt Plumb et al |
| EPI_ISL_460079 | Italy/LOM-INMI-BG-5825/2020 | 2020-02-22 | Europe/Italy/Lombardy/ | Molecular Virology Unit, Fondazione IRCCS Policlinico San Matteo , Pavia | Laboratory of Virology, INMI Lazzaro Spallanzani IRCCS | Barbara Bartolini et al |
| EPI_ISL_460080 | Italy/LOM-INMI-BG-11639/2020 | 2020-02-28 | Europe/Italy/Lombardy/ | Molecular Virology Unit, Fondazione IRCCS Policlinico San Matteo , Pavia | Laboratory of Virology, INMI Lazzaro Spallanzani IRCCS | Antonio Piralla et al |
| EPI_ISL_460081 | Italy/LOM-INMI-BG-7158/2020 | 2020-02-24 | Europe/Italy/Lombardy/ | Molecular Virology Unit, Fondazione IRCCS Policlinico San Matteo , Pavia | Laboratory of Virology, INMI Lazzaro Spallanzani IRCCS | Fausto Baldanti et al |
| EPI_ISL_460082 | Italy/LOM-INMI-BG-9209/2020 | 2020-02-26 | Europe/Italy/Lombardy/ | Molecular Virology Unit, Fondazione IRCCS Policlinico San Matteo , Pavia | Laboratory of Virology, INMI Lazzaro Spallanzani IRCCS | Martina Rueca et al |

| | | | | | | |
|---|---|---|---|---|---|---|
| EPI_ISL_460083 | Italy/LOM-INMI-BG-9225/2020 | 2020-02-26 | Europe/Italy/Lombardy/ | Molecular Virology Unit, Fondazione IRCCS Policlinico San Matteo , Pavia | Laboratory of Virology, INMI Lazzaro Spallanzani IRCCS | Martina Rueca et al |
| EPI_ISL_460084 | Italy/LOM-INMI-BS-12256/2020 | 2020-02-29 | Europe/Italy/Lombardy/ | Molecular Virology Unit, Fondazione IRCCS Policlinico San Matteo , Pavia | Laboratory of Virology, INMI Lazzaro Spallanzani IRCCS | Fausto Baldanti et al |
| EPI_ISL_460086 | Italy/LOM-INMI-7070/2020 | 2020-02-24 | Europe/Italy/Lombardy/ | Molecular Virology Unit, Fondazione IRCCS Policlinico San Matteo , Pavia | Laboratory of Virology, INMI Lazzaro Spallanzani IRCCS | Maria R. Capobianchi et al |
| EPI_ISL_460087 | Italy/LOM-INMI-9206/2020 | 2020-02-26 | Europe/Italy/Lombardy/ | Molecular Virology Unit, Fondazione IRCCS Policlinico San Matteo , Pavia | Laboratory of Virology, INMI Lazzaro Spallanzani IRCCS | Cesare E.M. Gruber et al |
| EPI_ISL_460088 | Italy/LOM-INMI-9214/2020 | 2020-02-26 | Europe/Italy/Lombardy/ | Molecular Virology Unit, Fondazione IRCCS Policlinico San Matteo , Pavia | Laboratory of Virology, INMI Lazzaro Spallanzani IRCCS | Martina Rueca et al |
| EPI_ISL_460089 | Italy/LOM-INMI-9491/2020 | 2020-02-26 | Europe/Italy/Lombardy/ | Molecular Virology Unit, Fondazione IRCCS Policlinico San Matteo , Pavia | Laboratory of Virology, INMI Lazzaro Spallanzani IRCCS | Antonino Di Caro et al |
| EPI_ISL_460090 | Italy/LOM-INMI-11550/2020 | 2020-02-28 | Europe/Italy/Lombardy/ | Molecular Virology Unit, Fondazione IRCCS Policlinico San Matteo , Pavia | Laboratory of Virology, INMI Lazzaro Spallanzani IRCCS | Antonio Piralla et al |
| EPI_ISL_460091 | Italy/LOM-INMI-5925/2020 | 2020-02-22 | Europe/Italy/Lombardy/ | Molecular Virology Unit, Fondazione IRCCS Policlinico San Matteo , Pavia | Laboratory of Virology, INMI Lazzaro Spallanzani IRCCS | Antonino Di Caro et al |
| EPI_ISL_460092 | Italy/LOM-INMI-11951/2020 | 2020-02-28 | Europe/Italy/Lombardy/ | Molecular Virology Unit, Fondazione IRCCS Policlinico San Matteo , Pavia | Laboratory of Virology, INMI Lazzaro Spallanzani IRCCS | Cesare E.M. Gruber et al |
| EPI_ISL_460093 | Italy/LOM-INMI-12578/2020 | 2020-02-29 | Europe/Italy/Lombardy/ | Molecular Virology Unit, Fondazione IRCCS Policlinico San Matteo , Pavia | Laboratory of Virology, INMI Lazzaro Spallanzani IRCCS | Maria R. Capobianchi et al |

| | | | | | | |
|---|---|---|---|---|---|---|
| EPI_ISL_460094 | Italy/LOM-INMI-8058/2020 | 2020-02-25 | Europe/Italy/Lombardy/ | Molecular Virology Unit, Fondazione IRCCS Policlinico San Matteo , Pavia | Laboratory of Virology, INMI Lazzaro Spallanzani IRCCS | Barbara Bartolini et al |
| EPI_ISL_460095 | Italy/LOM-INMI-9675/2020 | 2020-02-26 | Europe/Italy/Lombardy/ | Molecular Virology Unit, Fondazione IRCCS Policlinico San Matteo , Pavia | Laboratory of Virology, INMI Lazzaro Spallanzani IRCCS | Barbara Bartolini et al |
| EPI_ISL_460097 | USA/MA-MGH-00568/2020 | 2020-03-22 | North America/USA/Massachusetts/ | Massachusetts General Hospital | Infectious Disease Program, Broad Institute of Harvard and MIT | Lemieux et al |
| EPI_ISL_460107 | USA/MA-MGH-00157/2020 | 2020-03-26 | North America/USA/Massachusetts/ | Massachusetts General Hospital | Infectious Disease Program, Broad Institute of Harvard and MIT | Lemieux et al |
| EPI_ISL_460115 | USA/MA-MGH-00588/2020 | 2020-03-28 | North America/USA/Massachusetts/ | Massachusetts General Hospital | Infectious Disease Program, Broad Institute of Harvard and MIT | Lemieux et al |
| EPI_ISL_460120 | USA/MA-MGH-00656/2020 | 2020-04-01 | North America/USA/Massachusetts/ | Massachusetts General Hospital | Infectious Disease Program, Broad Institute of Harvard and MIT | Lemieux et al |
| EPI_ISL_460123 | USA/MA-MGH-00711/2020 | 2020-04-08 | North America/USA/Massachusetts/ | Massachusetts General Hospital | Infectious Disease Program, Broad Institute of Harvard and MIT | Lemieux et al |
| EPI_ISL_460130 | USA/MA-MGH-00706/2020 | 2020-04-07 | North America/USA/Massachusetts/ | Massachusetts General Hospital | Infectious Disease Program, Broad Institute of Harvard and MIT | Lemieux et al |
| EPI_ISL_460132 | USA/MA-MGH-00687/2020 | 2020-04-06 | North America/USA/Massachusetts/ | Massachusetts General Hospital | Infectious Disease Program, Broad Institute of Harvard and MIT | Lemieux et al |
| EPI_ISL_460138 | USA/MA-MGH-00544/2020 | 2020-03-20 | North America/USA/Massachusetts/ | Massachusetts General Hospital | Infectious Disease Program, Broad Institute of Harvard and MIT | Lemieux et al |

| | | | | | | |
|---|---|---|---|---|---|---|
| EPI_ISL_460146 | USA/MA-MGH-00683/2020 | 2020-03-21 | North America/USA/Massachusetts/ | Massachusetts General Hospital | Infectious Disease Program, Broad Institute of Harvard and MIT | Lemieux et al |
| EPI_ISL_460149 | USA/MA-MGH-00023/2020 | 2020-03-24 | North America/USA/Massachusetts/ | Massachusetts General Hospital | Infectious Disease Program, Broad Institute of Harvard and MIT | Lemieux et al |
| EPI_ISL_460159 | USA/MA-MGH-00467/2020 | 2020-04-04 | North America/USA/Massachusetts/ | Massachusetts General Hospital | Infectious Disease Program, Broad Institute of Harvard and MIT | Lemieux et al |
| EPI_ISL_460160 | USA/MA-MGH-00264/2020 | 2020-04-01 | North America/USA/Massachusetts/ | Massachusetts General Hospital | Infectious Disease Program, Broad Institute of Harvard and MIT | Lemieux et al |
| EPI_ISL_460171 | USA/MA-MGH-00291/2020 | 2020-04-01 | North America/USA/Massachusetts/ | Massachusetts General Hospital | Infectious Disease Program, Broad Institute of Harvard and MIT | Lemieux et al |
| EPI_ISL_460172 | USA/MA-MGH-00276/2020 | 2020-04-01 | North America/USA/Massachusetts/ | Massachusetts General Hospital | Infectious Disease Program, Broad Institute of Harvard and MIT | Lemieux et al |
| EPI_ISL_460174 | USA/MA-MGH-00059/2020 | 2020-03-23 | North America/USA/Massachusetts/ | Massachusetts General Hospital | Infectious Disease Program, Broad Institute of Harvard and MIT | Lemieux et al |
| EPI_ISL_460187 | USA/MA-MGH-00690/2020 | 2020-04-06 | North America/USA/Massachusetts/ | Massachusetts General Hospital | Infectious Disease Program, Broad Institute of Harvard and MIT | Lemieux et al |
| EPI_ISL_460189 | USA/MA-MGH-00633/2020 | 2020-04-02 | North America/USA/Massachusetts/ | Massachusetts General Hospital | Infectious Disease Program, Broad Institute of Harvard and MIT | Lemieux et al |
| EPI_ISL_460190 | USA/MA-MGH-00669/2020 | 2020-04-04 | North America/USA/Massachusetts/ | Massachusetts General Hospital | Infectious Disease Program, Broad Institute of Harvard and MIT | Lemieux et al |

| | | | | | | |
|---|---|---|---|---|---|---|
| EPI_ISL_460193 | USA/MA-MGH-00286/2020 | 2020-04-01 | North America/USA/Massachusetts/ | Massachusetts General Hospital | Infectious Disease Program, Broad Institute of Harvard and MIT | Lemieux et al |
| EPI_ISL_460196 | USA/MA-MGH-00290/2020 | 2020-04-01 | North America/USA/Massachusetts/ | Massachusetts General Hospital | Infectious Disease Program, Broad Institute of Harvard and MIT | Lemieux et al |
| EPI_ISL_460208 | USA/MA-MGH-00477/2020 | 2020-04-04 | North America/USA/Massachusetts/ | Massachusetts General Hospital | Infectious Disease Program, Broad Institute of Harvard and MIT | Lemieux et al |
| EPI_ISL_460226 | USA/MA-MGH-00469/2020 | 2020-04-04 | North America/USA/Massachusetts/ | Massachusetts General Hospital | Infectious Disease Program, Broad Institute of Harvard and MIT | Lemieux et al |
| EPI_ISL_460240 | USA/MA-MGH-00037/2020 | 2020-03-24 | North America/USA/Massachusetts/ | Massachusetts General Hospital | Infectious Disease Program, Broad Institute of Harvard and MIT | Lemieux et al |
| EPI_ISL_460265 | USA/MA-MGH-00298/2020 | 2020-04-01 | North America/USA/Massachusetts/ | Massachusetts General Hospital | Infectious Disease Program, Broad Institute of Harvard and MIT | Lemieux et al |
| EPI_ISL_460272 | USA/MA-MGH-00458/2020 | 2020-04-04 | North America/USA/Massachusetts/ | Massachusetts General Hospital | Infectious Disease Program, Broad Institute of Harvard and MIT | Lemieux et al |
| EPI_ISL_460274 | USA/MA-MGH-00294/2020 | 2020-04-01 | North America/USA/Massachusetts/ | Massachusetts General Hospital | Infectious Disease Program, Broad Institute of Harvard and MIT | Lemieux et al |
| EPI_ISL_460280 | USA/MA-MGH-00209/2020 | 2020-03-26 | North America/USA/Massachusetts/ | Massachusetts General Hospital | Infectious Disease Program, Broad Institute of Harvard and MIT | Lemieux et al |
| EPI_ISL_460283 | USA/MA-MGH-00170/2020 | 2020-03-27 | North America/USA/Massachusetts/ | Massachusetts General Hospital | Infectious Disease Program, Broad Institute of Harvard and MIT | Lemieux et al |

| | | | | | | |
|---|---|---|---|---|---|---|
| EPI_ISL_460287 | USA/MA-MGH-00644/2020 | 2020-04-01 | North America/USA/Massachusetts/ | Massachusetts General Hospital | Infectious Disease Program, Broad Institute of Harvard and MIT | Lemieux et al |
| EPI_ISL_460300 | USA/MA-MGH-00419/2020 | 2020-03-07 | North America/USA/Massachusetts/ | Massachusetts General Hospital | Infectious Disease Program, Broad Institute of Harvard and MIT | Lemieux et al |
| EPI_ISL_460344 | USA/MA-MGH-00528/2020 | 2020-03-18 | North America/USA/Massachusetts/ | Massachusetts General Hospital | Infectious Disease Program, Broad Institute of Harvard and MIT | Lemieux et al |
| EPI_ISL_460351 | USA/MA-MGH-00318/2020 | 2020-04-04 | North America/USA/Massachusetts/ | Massachusetts General Hospital | Infectious Disease Program, Broad Institute of Harvard and MIT | Lemieux et al |
| EPI_ISL_460355 | USA/MA-MGH-00153/2020 | 2020-03-26 | North America/USA/Massachusetts/ | Massachusetts General Hospital | Infectious Disease Program, Broad Institute of Harvard and MIT | Lemieux et al |
| EPI_ISL_460361 | USA/MA-MGH-00472/2020 | 2020-04-04 | North America/USA/Massachusetts/ | Massachusetts General Hospital | Infectious Disease Program, Broad Institute of Harvard and MIT | Lemieux et al |
| EPI_ISL_460377 | USA/MA-MGH-00663/2020 | 2020-04-03 | North America/USA/Massachusetts/ | Massachusetts General Hospital | Infectious Disease Program, Broad Institute of Harvard and MIT | Lemieux et al |
| EPI_ISL_460394 | USA/MA-MGH-00160/2020 | 2020-03-26 | North America/USA/Massachusetts/ | Massachusetts General Hospital | Infectious Disease Program, Broad Institute of Harvard and MIT | Lemieux et al |
| EPI_ISL_460407 | USA/MA-MGH-00244/2020 | 2020-04-06 | North America/USA/Massachusetts/ | Massachusetts General Hospital | Infectious Disease Program, Broad Institute of Harvard and MIT | Lemieux et al |
| EPI_ISL_460409 | USA/MA-MGH-00149/2020 | 2020-03-26 | North America/USA/Massachusetts/ | Massachusetts General Hospital | Infectious Disease Program, Broad Institute of Harvard and MIT | Lemieux et al |

| | | | | | | |
|---|---|---|---|---|---|---|
| EPI_ISL_460415 | USA/MA-MGH-00517/2020 | 2020-03-14 | North America/USA/Massachusetts/ | Massachusetts General Hospital | Infectious Disease Program, Broad Institute of Harvard and MIT | Lemieux et al |
| EPI_ISL_460417 | USA/MA-MGH-00164/2020 | 2020-03-26 | North America/USA/Massachusetts/ | Massachusetts General Hospital | Infectious Disease Program, Broad Institute of Harvard and MIT | Lemieux et al |
| EPI_ISL_460419 | USA/MA-MGH-00646/2020 | 2020-04-02 | North America/USA/Massachusetts/ | Massachusetts General Hospital | Infectious Disease Program, Broad Institute of Harvard and MIT | Lemieux et al |
| EPI_ISL_460439 | USA/MA-MGH-00255/2020 | 2020-04-06 | North America/USA/Massachusetts/ | Massachusetts General Hospital | Infectious Disease Program, Broad Institute of Harvard and MIT | Lemieux et al |
| EPI_ISL_460442 | USA/MA-MGH-00432/2020 | 2020-03-07 | North America/USA/Massachusetts/ | Massachusetts General Hospital | Infectious Disease Program, Broad Institute of Harvard and MIT | Lemieux et al |
| EPI_ISL_460450 | USA/MA-MGH-00233/2020 | 2020-04-06 | North America/USA/Massachusetts/ | Massachusetts General Hospital | Infectious Disease Program, Broad Institute of Harvard and MIT | Lemieux et al |
| EPI_ISL_460454 | USA/MA-MGH-00459/2020 | 2020-04-04 | North America/USA/Massachusetts/ | Massachusetts General Hospital | Infectious Disease Program, Broad Institute of Harvard and MIT | Lemieux et al |
| EPI_ISL_460455 | USA/MA-MGH-00250/2020 | 2020-04-06 | North America/USA/Massachusetts/ | Massachusetts General Hospital | Infectious Disease Program, Broad Institute of Harvard and MIT | Lemieux et al |
| EPI_ISL_460467 | USA/MA-MGH-00004/2020 | 2020-03-11 | North America/USA/Massachusetts/ | Massachusetts General Hospital | Infectious Disease Program, Broad Institute of Harvard and MIT | Lemieux et al |

| | | | | | | |
|---|---|---|---|---|---|---|
| EPI_ISL_460566 | USA/MI-MDHHS-SC20518/2020 | 2020-04-28 | North America/USA/Michigan/ | Michigan Department of Health and Human Services, Bureau of Laboratories | Michigan Department of Health and Human Services, Bureau of Laboratories | Blankenship HM et al |
| EPI_ISL_460571 | USA/MI-MDHHS-SC20523/2020 | 2020-05-14 | North America/USA/Michigan/ | Michigan Department of Health and Human Services, Bureau of Laboratories | Michigan Department of Health and Human Services, Bureau of Laboratories | Blankenship HM et al |
| EPI_ISL_460581 | USA/MI-MDHHS-SC20533/2020 | 2020-05-14 | North America/USA/Michigan/ | Michigan Department of Health and Human Services, Bureau of Laboratories | Michigan Department of Health and Human Services, Bureau of Laboratories | Blankenship HM et al |
| EPI_ISL_460586 | USA/MI-MDHHS-SC20538/2020 | 2020-05-14 | North America/USA/Michigan/ | Michigan Department of Health and Human Services, Bureau of Laboratories | Michigan Department of Health and Human Services, Bureau of Laboratories | Blankenship HM et al |
| EPI_ISL_460590 | USA/MI-MDHHS-SC20542/2020 | 2020-05-04 | North America/USA/Michigan/ | Michigan Department of Health and Human Services, Bureau of Laboratories | Michigan Department of Health and Human Services, Bureau of Laboratories | Blankenship HM et al |
| EPI_ISL_460604 | Russia/CRIE145922/2020 | 2020-03-29 | Europe/Russia/Moscow Oblast/ | "Molecular diagnostic laboratory of Federal Budget Institution of Science ""Central Research Institute of Epidemiology"" of The Federal Service on Customers' Rights Protection and Human Well-being Surveillance" | Group of Genomics and Postgenomic Technologies of Central Research Institute of Epidemilology | Speranskaya AS et al |
| EPI_ISL_460607 | Canada/BC_08719828/2020 | 20-Mar | North America/Canada/British Columbia | BCCDC Public Health Laboratory | BCCDC Public Health Laboratory | Harrigan et al |
| EPI_ISL_460608 | Canada/BC_13871523/2020 | 20-Mar | North America/Canada/British Columbia | BCCDC Public Health Laboratory | BCCDC Public Health Laboratory | Harrigan et al |

| | | | | | | |
|---|---|---|---|---|---|---|
| EPI_ISL_460609 | Canada/BC_15926629/2020 | 20-Mar | North America/Canada/British Columbia | BCCDC Public Health Laboratory | BCCDC Public Health Laboratory | Harrigan et al |
| EPI_ISL_460610 | Canada/BC_16344159/2020 | 20-Mar | North America/Canada/British Columbia | BCCDC Public Health Laboratory | BCCDC Public Health Laboratory | Harrigan et al |
| EPI_ISL_460611 | Canada/BC_17045358/2020 | 20-Mar | North America/Canada/British Columbia | BCCDC Public Health Laboratory | BCCDC Public Health Laboratory | Harrigan et al |
| EPI_ISL_460612 | Canada/BC_17422770/2020 | 20-Mar | North America/Canada/British Columbia | BCCDC Public Health Laboratory | BCCDC Public Health Laboratory | Harrigan et al |
| EPI_ISL_460613 | Canada/BC_17428296/2020 | 20-Mar | North America/Canada/British Columbia | BCCDC Public Health Laboratory | BCCDC Public Health Laboratory | Harrigan et al |
| EPI_ISL_460614 | Canada/BC_17548428/2020 | 20-Mar | North America/Canada/British Columbia | BCCDC Public Health Laboratory | BCCDC Public Health Laboratory | Harrigan et al |
| EPI_ISL_460615 | Canada/BC_18097749/2020 | 20-Mar | North America/Canada/British Columbia | BCCDC Public Health Laboratory | BCCDC Public Health Laboratory | Harrigan et al |
| EPI_ISL_460616 | Canada/BC_18111443/2020 | 20-Mar | North America/Canada/British Columbia | BCCDC Public Health Laboratory | BCCDC Public Health Laboratory | Harrigan et al |
| EPI_ISL_460617 | Turkey/IMU-SP-02/2020 | 2020-04-16 | Europe/Turkey/Turkey/ | unknown | Physiology | Pence et al |
| EPI_ISL_460618 | Turkey/IMU-SP-01/2020 | 2020-04-15 | Europe/Turkey/Turkey/ | unknown | Physiology | Pence et al |
| EPI_ISL_460619 | Turkey/IMU-SP-04/2020 | 2020-04-16 | Europe/Turkey/Turkey/ | unknown | Physiology | Pence et al |
| EPI_ISL_460653 | Netherlands/Gelderland_17/2020 | 2020-04-08 | Europe/Netherlands/Gelderland/ | Dutch COVID-19 response team | Erasmus Medical Center | Bas Oude Munnink et al |
| EPI_ISL_460655 | Netherlands/Gelderland_19/2020 | 2020-03-18 | Europe/Netherlands/Gelderland/ | Dutch COVID-19 response team | Erasmus Medical Center | Bas Oude Munnink et al |
| EPI_ISL_460673 | Netherlands/NA_500/2020 | 2020-03-27 | Europe/Netherlands/Netherlands/ | Dutch COVID-19 response team | Erasmus Medical Center | Bas Oude Munnink et al |
| EPI_ISL_460675 | Netherlands/NA_502/2020 | 2020-03-31 | Europe/Netherlands/Netherlands/ | Dutch COVID-19 response team | Erasmus Medical Center | Bas Oude Munnink et al |
| EPI_ISL_460697 | Netherlands/NA_524/2020 | 2020-03-22 | Europe/Netherlands/Netherlands/ | Dutch COVID-19 response team | Erasmus Medical Center | Bas Oude Munnink et al |
| EPI_ISL_460702 | Netherlands/NA_529/2020 | 2020-04-08 | Europe/Netherlands/Netherlands/ | Dutch COVID-19 response team | Erasmus Medical Center | Bas Oude Munnink et al |
| EPI_ISL_460726 | Netherlands/NA_561/2020 | 2020-04-11 | Europe/Netherlands/Netherlands/ | Dutch COVID-19 response team | Erasmus Medical Center | Bas Oude Munnink et al |
| EPI_ISL_460730 | Netherlands/NA_566/2020 | 2020-04-10 | Europe/Netherlands/Netherlands/ | Dutch COVID-19 response team | Erasmus Medical Center | Bas Oude Munnink et al |
| EPI_ISL_460734 | Netherlands/NA_570/2020 | 2020-04-11 | Europe/Netherlands/Netherlands/ | Dutch COVID-19 response team | Erasmus Medical Center | Bas Oude Munnink et al |
| EPI_ISL_460759 | Netherlands/NA_596/2020 | 2020-04-19 | Europe/Netherlands/Netherlands/ | Dutch COVID-19 response team | Erasmus Medical Center | Bas Oude Munnink et al |

| | | | | | | |
|---|---|---|---|---|---|---|
| EPI_ISL_460795 | Netherlands/NoordHolland_6/2020 | 2020-04-02 | Europe/Netherlands/North Holland/ | Dutch COVID-19 response team | Erasmus Medical Center | Bas Oude Munnink et al |
| EPI_ISL_460835 | Netherlands/Flevoland_12/2020 | 2020-05-01 | Europe/Netherlands/Flevoland/ | Dutch COVID-19 response team | Erasmus Medical Center | Bas Oude Munnink et al |
| EPI_ISL_460853 | Netherlands/Friesland_38/2020 | 2020-04-14 | Europe/Netherlands/Friesland/ | Dutch COVID-19 response team | Erasmus Medical Center | Bas Oude Munnink et al |
| EPI_ISL_460859 | Netherlands/Friesland_44/2020 | 2020-05-04 | Europe/Netherlands/Friesland/ | Dutch COVID-19 response team | Erasmus Medical Center | Bas Oude Munnink et al |
| EPI_ISL_460877 | Netherlands/Gelderland_116/2020 | 2020-05-22 | Europe/Netherlands/Gelderland/ | Dutch COVID-19 response team | Erasmus Medical Center | Bas Oude Munnink et al |
| EPI_ISL_460880 | Netherlands/Gelderland_119/2020 | 2020-05-22 | Europe/Netherlands/Gelderland/ | Dutch COVID-19 response team | Erasmus Medical Center | Bas Oude Munnink et al |
| EPI_ISL_460924 | Netherlands/Gelderland_24/2020 | 2020-04-22 | Europe/Netherlands/Gelderland/ | Dutch COVID-19 response team | Erasmus Medical Center | Bas Oude Munnink et al |
| EPI_ISL_460926 | Netherlands/Gelderland_26/2020 | 2020-04-26 | Europe/Netherlands/Gelderland/ | Dutch COVID-19 response team | Erasmus Medical Center | Bas Oude Munnink et al |
| EPI_ISL_460951 | Netherlands/Gelderland_52/2020 | 2020-05-08 | Europe/Netherlands/Gelderland/ | Dutch COVID-19 response team | Erasmus Medical Center | Bas Oude Munnink et al |
| EPI_ISL_460964 | Netherlands/Gelderland_65/2020 | 2020-05-18 | Europe/Netherlands/Gelderland/ | Dutch COVID-19 response team | Erasmus Medical Center | Bas Oude Munnink et al |
| EPI_ISL_460977 | Netherlands/Gelderland_78/2020 | 2020-05-19 | Europe/Netherlands/Gelderland/ | Dutch COVID-19 response team | Erasmus Medical Center | Bas Oude Munnink et al |
| EPI_ISL_461010 | Netherlands/Limburg_22/2020 | 2020-04-25 | Europe/Netherlands/Limburg/ | Dutch COVID-19 response team | Erasmus Medical Center | Bas Oude Munnink et al |
| EPI_ISL_461020 | Netherlands/Limburg_32/2020 | 2020-04-22 | Europe/Netherlands/Limburg/ | Dutch COVID-19 response team | Erasmus Medical Center | Bas Oude Munnink et al |
| EPI_ISL_461027 | Netherlands/Limburg_39/2020 | 2020-04-19 | Europe/Netherlands/Limburg/ | Dutch COVID-19 response team | Erasmus Medical Center | Bas Oude Munnink et al |
| EPI_ISL_461047 | Netherlands/Limburg_61/2020 | 2020-04-25 | Europe/Netherlands/Limburg/ | Dutch COVID-19 response team | Erasmus Medical Center | Bas Oude Munnink et al |
| EPI_ISL_461101 | Netherlands/NA_666/2020 | 2020-02-29 | Europe/Netherlands/Netherlands/ | Dutch COVID-19 response team | Erasmus Medical Center | Bas Oude Munnink et al |
| EPI_ISL_461105 | Netherlands/NA_670/2020 | 2020-03-09 | Europe/Netherlands/Netherlands/ | Dutch COVID-19 response team | Erasmus Medical Center | Bas Oude Munnink et al |
| EPI_ISL_461151 | Netherlands/NoordBrabant_116/202 | 2020-03-23 | Europe/Netherlands/North Brabant/ | Dutch COVID-19 response team | Erasmus Medical Center | Bas Oude Munnink et al |
| EPI_ISL_461193 | Netherlands/NoordBrabant_158/202 | 2020-04-22 | Europe/Netherlands/North Brabant/ | Dutch COVID-19 response team | Erasmus Medical Center | Bas Oude Munnink et al |
| EPI_ISL_461230 | Netherlands/Utrecht_32/2020 | 2020-04-23 | Europe/Netherlands/Utrecht/ | Dutch COVID-19 response team | Erasmus Medical Center | Bas Oude Munnink et al |

| | | | | | | |
|---|---|---|---|---|---|---|
| EPI_ISL_461247 | Netherlands/Zeeland_1/2020 | 2020-03-21 | Europe/Netherlands/Zeeland/ | Dutch COVID-19 response team | Erasmus Medical Center | Bas Oude Munnink et al |
| EPI_ISL_461256 | Netherlands/Zeeland_18/2020 | 2020-04-03 | Europe/Netherlands/Zeeland/ | Dutch COVID-19 response team | Erasmus Medical Center | Bas Oude Munnink et al |
| EPI_ISL_461268 | Netherlands/Zeeland_3/2020 | 2020-03-29 | Europe/Netherlands/Zeeland/ | Dutch COVID-19 response team | Erasmus Medical Center | Bas Oude Munnink et al |
| EPI_ISL_461278 | Netherlands/Zeeland_39/2020 | 2020-03-22 | Europe/Netherlands/Zeeland/ | Dutch COVID-19 response team | Erasmus Medical Center | Bas Oude Munnink et al |
| EPI_ISL_461284 | Netherlands/Zeeland_45/2020 | 2020-03-24 | Europe/Netherlands/Zeeland/ | Dutch COVID-19 response team | Erasmus Medical Center | Bas Oude Munnink et al |
| EPI_ISL_461285 | Netherlands/Zeeland_46/2020 | 2020-03-22 | Europe/Netherlands/Zeeland/ | Dutch COVID-19 response team | Erasmus Medical Center | Bas Oude Munnink et al |
| EPI_ISL_461293 | Netherlands/ZuidHolland_120/2020 | 2020-04-20 | Europe/Netherlands/South Holland/ | Dutch COVID-19 response team | Erasmus Medical Center | Bas Oude Munnink et al |
| EPI_ISL_461347 | Netherlands/ZuidHolland_175/2020 | 2020-04-17 | Europe/Netherlands/South Holland/ | Dutch COVID-19 response team | Erasmus Medical Center | Bas Oude Munnink et al |
| EPI_ISL_461370 | Netherlands/ZuidHolland_198/2020 | 2020-04-28 | Europe/Netherlands/South Holland/ | Dutch COVID-19 response team | Erasmus Medical Center | Bas Oude Munnink et al |
| EPI_ISL_461419 | USA/WA-UW-6601/2020 | 2020-04-05 | North America/USA/Washington/ | UW Virology Lab | UW Virology Lab | Pavitra Roychoudhury et al |
| EPI_ISL_461421 | USA/WA-UW-6605/2020 | 2020-04-05 | North America/USA/Washington/ | UW Virology Lab | UW Virology Lab | Pavitra Roychoudhury et al |
| EPI_ISL_461453 | USA/WA-UW-10031/2020 | 2020-05-29 | North America/USA/Washington/ | UW Virology Lab | UW Virology Lab | Pavitra Roychoudhury et al |
| EPI_ISL_461458 | USA/WA-UW-10088/2020 | 2020-05-29 | North America/USA/Washington/ | UW Virology Lab | UW Virology Lab | Pavitra Roychoudhury et al |
| EPI_ISL_461459 | USA/WA-UW-10089/2020 | 2020-05-29 | North America/USA/Washington/ | UW Virology Lab | UW Virology Lab | Pavitra Roychoudhury et al |
| EPI_ISL_461469 | USA/WA-UW-10117/2020 | 2020-05-29 | North America/USA/Washington/ | UW Virology Lab | UW Virology Lab | Pavitra Roychoudhury et al |
| EPI_ISL_461492 | India/GJ-GBRC160b/2020 | 2020-05-27 | Asia/India/Gujarat/Ahmedabad | B.J. Medical College and Civil hospital | Gujarat Biotechnology Research Centre | Neelam Nathani et al |
| EPI_ISL_461494 | India/GJ-GBRC161b/2020 | 2020-05-27 | Asia/India/Gujarat/Ahmedabad | B.J. Medical College and Civil hospital | Gujarat Biotechnology Research Centre | Bhavya Jindal et al |
| EPI_ISL_461499 | India/GJ-GBRC164/2020 | 2020-05-27 | Asia/India/Gujarat/Ahmedabad | B.J. Medical College and Civil hospital | Gujarat Biotechnology Research Centre | Akanksha Verma et al |
| EPI_ISL_461501 | India/GJ-GBRC166a/2020 | 2020-05-27 | Asia/India/Gujarat/Ahmedabad | B.J. Medical College and Civil hospital | Gujarat Biotechnology Research Centre | Pragya Sharma et al |
| EPI_ISL_461505 | India/GJ-GBRC168a/2020 | 2020-05-27 | Asia/India/Gujarat/Ahmedabad | B.J. Medical College and Civil hospital | Gujarat Biotechnology Research Centre | Pranay Shah et al |

| | | | | | | |
|---|---|---|---|---|---|---|
| EPI_ISL_461573 | England/CAMB-1B3A94/2020 | 2020-05-27 | Europe/United Kingdom/England/ | Department of Pathology, University of Cambridge | COVID-19 Genomics UK (COG-UK) Consortium | Luke W Meredith et al |
| EPI_ISL_461663 | Scotland/CVR3079/2020 | 2020-04-24 | Europe/United Kingdom/Scotland/ | West of Scotland Specialist Virology Centre, NHSGGC / MRC-University of Glasgow Centre for Virus Research | COVID-19 Genomics UK (COG-UK) Consortium | Ana da Silva Filipe et al |
| EPI_ISL_461842 | England/NORW-E9A69/2020 | 2020-04-30 | Europe/United Kingdom/England/ | Quadram Institute Bioscience | COVID-19 Genomics UK (COG-UK) Consortium | Dave J. Baker et al |
| EPI_ISL_461866 | England/NORW-EA7CB/2020 | 2020-05-17 | Europe/United Kingdom/England/ | Quadram Institute Bioscience | COVID-19 Genomics UK (COG-UK) Consortium | Dave J. Baker et al |
| EPI_ISL_461871 | England/NORW-EA8AA/2020 | 2020-05-20 | Europe/United Kingdom/England/ | Quadram Institute Bioscience | COVID-19 Genomics UK (COG-UK) Consortium | Dave J. Baker et al |
| EPI_ISL_461909 | England/NOTT-1116DC/2020 | 2020-05-22 | Europe/United Kingdom/England/ | Queens Medical Centre, Clinical Microbiology Department / DeepSeq Nottingham | COVID-19 Genomics UK (COG-UK) Consortium | Gemma Clark et al |
| EPI_ISL_461919 | England/NOTT-11177F/2020 | 2020-05-19 | Europe/United Kingdom/England/ | Queens Medical Centre, Clinical Microbiology Department / DeepSeq Nottingham | COVID-19 Genomics UK (COG-UK) Consortium | Gemma Clark et al |
| EPI_ISL_462032 | England/SHEF-C7885/2020 | 2020-04-18 | Europe/United Kingdom/England/ | Virology Department, Sheffield Teaching Hospitals NHS Foundation Trust/Department of Infection, Immunity and Cardiovascular Disease, The Medical School, University of Sheffield | COVID-19 Genomics UK (COG-UK) Consortium | Thushan de Silva et al |
| EPI_ISL_462090 | Bangladesh/BCSIR-NILMRC-004/2020 | 2020-05-23 | Asia/Bangladesh/Dhaka/ | National Institute of Laboratory Medicine and Referral Center | Genomic Research Lab, BCSIR | Barna Goswami et al |
| EPI_ISL_462091 | Bangladesh/BCSIR-NILMRC-002/2020 | 2020-05-23 | Asia/Bangladesh/Dhaka/ | National Institute of Laboratory Medicine and Referral Center | Genomic Research Lab, BCSIR | Iffat Jahan et al |
| EPI_ISL_462092 | Bangladesh/BCSIR-NILMRC-003/2020 | 2020-05-23 | Asia/Bangladesh/Dhaka/ | National Institute of Laboratory Medicine and Referral Center | Genomic Research Lab, BCSIR | Shahina Akter et al |
| EPI_ISL_462093 | Bangladesh/BCSIR-NILMRC-005/2020 | 2020-05-23 | Asia/Bangladesh/Dhaka/Dhaka | National Institute of Laboratory Medicine and Referral Center | Genomic Research Lab, BCSIR | Abu Sayeed Mohammad Mahmud et al |
| EPI_ISL_462095 | Bangladesh/BCSIR-NILMRC-009/2020 | 2020-05-23 | Asia/Bangladesh/Dhaka/Dhaka | National Institute of Laboratory Medicine and Referral Center | Genomic Research Lab, BCSIR | Abu Sayeed Mohammad Mahmud et al |
| EPI_ISL_462096 | Bangladesh/BCSIR-NILMRC-015/2020 | 2020-05-23 | Asia/Bangladesh/Dhaka/Dhaka | National Institute of Laboratory Medicine and Referral Center | Genomic Research Lab, BCSIR | Abu Sayeed Mohammad Mahmud et al |
| EPI_ISL_462097 | Bangladesh/BCSIR-NILMRC-042/2020 | 2020-05-23 | Asia/Bangladesh/Dhaka/Dhaka | National Institute of Laboratory Medicine and Referral Center | Genomic Research Lab, BCSIR | Abu Sayeed Mohammad Mahmud et al |

| | | | | | | |
|---|---|---|---|---|---|---|
| EPI_ISL_462098 | Bangladesh/BCSIR-NILMRC-050/2020 | 2020-05-21 | Asia/Bangladesh/Dhaka/Dhaka | National Institute of Laboratory Medicine and Referral Center | Genomic Research Lab, BCSIR | Abu Sayeed Mohammad Mahmud et al |
| EPI_ISL_462151 | Belgium/rega-0423297/2020 | 2020-04-23 | Europe/Belgium/Verviers/Kelmis | KU Leuven, Rega Institute, Clinical and Epidemiological Virology | KU Leuven, Rega Institute, Clinical and Epidemiological Virology | Tony Wawina-Bokalanga et al |
| EPI_ISL_462155 | Belgium/rega-0423301/2020 | 2020-04-23 | Europe/Belgium/Verviers/Kelmis | KU Leuven, Rega Institute, Clinical and Epidemiological Virology | KU Leuven, Rega Institute, Clinical and Epidemiological Virology | Tony Wawina-Bokalanga et al |
| EPI_ISL_462170 | Belgium/PF-0326316/2020 | 2020-03-26 | Europe/Belgium/Nivelles/Genval | KU Leuven, Rega Institute, Clinical and Epidemiological Virology | KU Leuven, Rega Institute, Clinical and Epidemiological Virology | Tony Wawina-Bokalanga et al |
| EPI_ISL_462176 | Belgium/IL-0326322/2020 | 2020-03-26 | Europe/Belgium/Charleroi/Mellet | KU Leuven, Rega Institute, Clinical and Epidemiological Virology | KU Leuven, Rega Institute, Clinical and Epidemiological Virology | Tony Wawina-Bokalanga et al |
| EPI_ISL_462180 | Belgium/JBE-0317326/2020 | 2020-03-17 | Europe/Belgium/Li√®ge/ | KU Leuven, Rega Institute, Clinical and Epidemiological Virology | KU Leuven, Rega Institute, Clinical and Epidemiological Virology | Tony Wawina-Bokalanga et al |
| EPI_ISL_462181 | Belgium/HMM-0317327/2020 | 2020-03-17 | Europe/Belgium/Li√®ge/Voroux-lez-Liers | KU Leuven, Rega Institute, Clinical and Epidemiological Virology | KU Leuven, Rega Institute, Clinical and Epidemiological Virology | Tony Wawina-Bokalanga et al |
| EPI_ISL_462186 | Belgium/CL-0330332/2020 | 2020-03-30 | Europe/Belgium/Antwerpen/Wijnegem | KU Leuven, Rega Institute, Clinical and Epidemiological Virology | KU Leuven, Rega Institute, Clinical and Epidemiological Virology | Tony Wawina-Bokalanga et al |

| | | | | | | |
|---|---|---|---|---|---|---|
| EPI_ISL_462189 | Belgium/BA-0330335/2020 | 2020-03-30 | Europe/Belgium/Namur/Champion | KU Leuven, Rega Institute, Clinical and Epidemiological Virology | KU Leuven, Rega Institute, Clinical and Epidemiological Virology | Tony Wawina-Bokalanga et al |
| EPI_ISL_462197 | Belgium/NA-0328342/2020 | 2020-03-28 | Europe/Belgium/Maaseik/ | KU Leuven, Rega Institute, Clinical and Epidemiological Virology | KU Leuven, Rega Institute, Clinical and Epidemiological Virology | Tony Wawina-Bokalanga et al |
| EPI_ISL_462201 | Belgium/BN-0330347/2020 | 2020-03-30 | Europe/Belgium/Brugge/Torhout | KU Leuven, Rega Institute, Clinical and Epidemiological Virology | KU Leuven, Rega Institute, Clinical and Epidemiological Virology | Tony Wawina-Bokalanga et al |
| EPI_ISL_462208 | Belgium/GJM-0328354/2020 | 2020-03-28 | Europe/Belgium/Namur/Andenne | KU Leuven, Rega Institute, Clinical and Epidemiological Virology | KU Leuven, Rega Institute, Clinical and Epidemiological Virology | Tony Wawina-Bokalanga et al |
| EPI_ISL_462215 | Belgium/BS-0331361/2020 | 2020-03-31 | Europe/Belgium/Charleroi/Buzet | KU Leuven, Rega Institute, Clinical and Epidemiological Virology | KU Leuven, Rega Institute, Clinical and Epidemiological Virology | Tony Wawina-Bokalanga et al |
| EPI_ISL_462219 | Belgium/PBT-0330365/2020 | 2020-03-30 | Europe/Belgium/Waremme/Berloz | KU Leuven, Rega Institute, Clinical and Epidemiological Virology | KU Leuven, Rega Institute, Clinical and Epidemiological Virology | Tony Wawina-Bokalanga et al |
| EPI_ISL_462224 | Belgium/VAJ-0328370/2020 | 2020-03-28 | Europe/Belgium/Oudenaarde/Bevere | KU Leuven, Rega Institute, Clinical and Epidemiological Virology | KU Leuven, Rega Institute, Clinical and Epidemiological Virology | Tony Wawina-Bokalanga et al |
| EPI_ISL_462235 | Belgium/DEJ-0403381/2020 | 2020-04-03 | Europe/Belgium/Charleroi/Goutroux | KU Leuven, Rega Institute, Clinical and Epidemiological Virology | KU Leuven, Rega Institute, Clinical and Epidemiological Virology | Tony Wawina-Bokalanga et al |

| | | | | | | |
|---|---|---|---|---|---|---|
| EPI_ISL_462238 | Belgium/LMA-0330384/2020 | 2020-03-30 | Europe/Belgium/Gent/Mariakerke | KU Leuven, Rega Institute, Clinical and Epidemiological Virology | KU Leuven, Rega Institute, Clinical and Epidemiological Virology | Tony Wawina-Bokalanga et al |
| EPI_ISL_462240 | Belgium/RMO-0329386/2020 | 2020-03-29 | Europe/Belgium/Dendermonde/Kalken | KU Leuven, Rega Institute, Clinical and Epidemiological Virology | KU Leuven, Rega Institute, Clinical and Epidemiological Virology | Tony Wawina-Bokalanga et al |
| EPI_ISL_462243 | Belgium/LHL-0329389/2020 | 2020-03-29 | Europe/Belgium/Maaseik/Lommel | KU Leuven, Rega Institute, Clinical and Epidemiological Virology | KU Leuven, Rega Institute, Clinical and Epidemiological Virology | Tony Wawina-Bokalanga et al |
| EPI_ISL_462248 | Belgium/SB-0407394/2020 | 2020-04-07 | Europe/Belgium/Halle-Vilvoorde/Berg | KU Leuven, Rega Institute, Clinical and Epidemiological Virology | KU Leuven, Rega Institute, Clinical and Epidemiological Virology | Tony Wawina-Bokalanga et al |
| EPI_ISL_462256 | Belgium/REGA-0325402/2020 | 2020-03-25 | Europe/Belgium/Charleroi/Aiseau | KU Leuven, Rega Institute, Clinical and Epidemiological Virology | KU Leuven, Rega Institute, Clinical and Epidemiological Virology | Tony Wawina-Bokalanga et al |
| EPI_ISL_462258 | Belgium/AO-04074040/2020 | 2020-04-07 | Europe/Belgium/Brussels/Molenbeek-Sa | KU Leuven, Rega Institute, Clinical and Epidemiological Virology | KU Leuven, Rega Institute, Clinical and Epidemiological Virology | Tony Wawina-Bokalanga et al |
| EPI_ISL_462260 | Belgium/BJA-0409406/2020 | 2020-04-09 | Europe/Belgium/Tournai-Mouscron/Ere | KU Leuven, Rega Institute, Clinical and Epidemiological Virology | KU Leuven, Rega Institute, Clinical and Epidemiological Virology | Tony Wawina-Bokalanga et al |
| EPI_ISL_462263 | Belgium/DJ-0323409/2020 | 2020-03-23 | Europe/Belgium/Antwerpen/ | KU Leuven, Rega Institute, Clinical and Epidemiological Virology | KU Leuven, Rega Institute, Clinical and Epidemiological Virology | Tony Wawina-Bokalanga et al |

| | | | | | | |
|---|---|---|---|---|---|---|
| EPI_ISL_462270 | Belgium/VAM-0506416/2020 | 2020-05-06 | Europe/Belgium/Mons/Bauffe | KU Leuven, Rega Institute, Clinical and Epidemiological Virology | KU Leuven, Rega Institute, Clinical and Epidemiological Virology | Tony Wawina-Bokalanga et al |
| EPI_ISL_462283 | Singapore/165/2020 | 2020-01-27 | Asia/Singapore/Singapore/ | National Public Health Laboratory, National Centre for Infectious Diseases | National Public Health Laboratory, National Centre for Infectious Diseases | Mak TM et al |
| EPI_ISL_462289 | Singapore/171/2020 | 2020-04-15 | Asia/Singapore/Singapore/ | National Public Health Laboratory, National Centre for Infectious Diseases | National Public Health Laboratory, National Centre for Infectious Diseases | Mak TM et al |
| EPI_ISL_462293 | Singapore/175/2020 | 2020-02-09 | Asia/Singapore/Singapore/ | National Public Health Laboratory, National Centre for Infectious Diseases | National Public Health Laboratory, National Centre for Infectious Diseases | Mak TM et al |
| EPI_ISL_462304 | Singapore/186/2020 | 2020-02-08 | Asia/Singapore/Singapore/ | National Public Health Laboratory, National Centre for Infectious Diseases | National Public Health Laboratory, National Centre for Infectious Diseases | Mak TM et al |
| EPI_ISL_462305 | Singapore/187/2020 | 2020-02-27 | Asia/Singapore/Singapore/ | National Public Health Laboratory, National Centre for Infectious Diseases | National Public Health Laboratory, National Centre for Infectious Diseases | Mak TM et al |
| EPI_ISL_462306 | Singapore/188/2020 | 2020-02-14 | Asia/Singapore/Singapore/ | National Public Health Laboratory, National Centre for Infectious Diseases | National Public Health Laboratory, National Centre for Infectious Diseases | Mak TM et al |

| | | | | | | |
|---|---|---|---|---|---|---|
| EPI_ISL_462314 | Singapore/196/2020 | 2020-04-20 | Asia/Singapore/Singapore/ | National Public Health Laboratory, National Centre for Infectious Diseases | National Public Health Laboratory, National Centre for Infectious Diseases | Mak TM et al |
| EPI_ISL_462315 | Singapore/197/2020 | 2020-04-22 | Asia/Singapore/Singapore/ | National Public Health Laboratory, National Centre for Infectious Diseases | National Public Health Laboratory, National Centre for Infectious Diseases | Mak TM et al |
| EPI_ISL_462340 | Singapore/222/2020 | 2020-04-07 | Asia/Singapore/Singapore/ | National Public Health Laboratory, National Centre for Infectious Diseases | National Public Health Laboratory, National Centre for Infectious Diseases | Mak TM et al |
| EPI_ISL_462388 | Singapore/270/2020 | 2020-04-14 | Asia/Singapore/Singapore/ | National Public Health Laboratory, National Centre for Infectious Diseases | National Public Health Laboratory, National Centre for Infectious Diseases | Mak TM et al |
| EPI_ISL_462399 | Singapore/281/2020 | 2020-04-14 | Asia/Singapore/Singapore/ | National Public Health Laboratory, National Centre for Infectious Diseases | National Public Health Laboratory, National Centre for Infectious Diseases | Mak TM et al |
| EPI_ISL_462412 | Singapore/294/2020 | 2020-05-03 | Asia/Singapore/Singapore/ | National Public Health Laboratory, National Centre for Infectious Diseases | National Public Health Laboratory, National Centre for Infectious Diseases | Mak TM et al |

| | | | | | | |
|---|---|---|---|---|---|---|
| EPI_ISL_462433 | Singapore/315/2020 | 2020-02-19 | Asia/Singapore/Singapore/ | National Public Health Laboratory, National Centre for Infectious Diseases | National Public Health Laboratory, National Centre for Infectious Diseases | Mak TM et al |
| EPI_ISL_462437 | Serbia/KV27-04/2020 | 2020-04-23 | Europe/Serbia/Ra≈°ka District/Kraljevo | unknown | Laboratory Diagnostic | Vidanovic et al |
| EPI_ISL_462438 | Serbia/KV77-04/2020 | 2020-04-22 | Europe/Serbia/Ra≈°ka District/Kraljevo | unknown | Laboratory Diagnostic | Vidanovic et al |
| EPI_ISL_462470 | BosniaandHerzegovina/ChVir7402/20 | 2020-04-22 | Europe/Bosnia and Herzegovina/Sarajev | Clinical Center, University of Sarajevo | Charite Universitatsmedizin Berlin, Institute of Virology | Victor M Corman et al |
| EPI_ISL_462472 | BosniaandHerzegovina/ChVir7382/20 | 2020-04-01 | Europe/Bosnia and Herzegovina/Vitez/ | Clinical Center, University of Sarajevo | Charite Universitatsmedizin Berlin, Institute of Virology | Victor M Corman et al |
| EPI_ISL_462473 | BosniaandHerzegovina/ChVir7372/20 | 2020-03-29 | Europe/Bosnia and Herzegovina/Zepce/ | Clinical Center, University of Sarajevo | Charite Universitatsmedizin Berlin, Institute of Virology | Victor M Corman et al |
| EPI_ISL_462477 | Spain/AN-ISCIII-202659/2020 | 2020-03-13 | Europe/Spain/Andalusia/ | Hospital Costa del Sol | Instituto de Salud Carlos III | Iglesias-Caballero et al |
| EPI_ISL_462479 | Spain/CT-ISCIII-202052/2020 | 2020-03-02 | Europe/Spain/Catalunya/ | Hospital Clinic | Instituto de Salud Carlos III | Iglesias-Caballero et al |
| EPI_ISL_462480 | Poland/IHG_PAS_3_57/2020 | 2020-05-13 | Europe/Poland/Wielkopolskie/ | Institute of Human Genetics, Polish Academy of Sciences | Institute of Human Genetics, Polish Academy of Sciences | Szymon Hryhorowicz et al |
| EPI_ISL_462639 | USA/MI-MDHHS-SC20557/2020 | 2020-04-30 | North America/USA/Michigan/ | Michigan Department of Health and Human Services, Bureau of Laboratories | Michigan Department of Health and Human Services, Bureau of Laboratories | Blankenship HM et al |

| | | | | | | |
|---|---|---|---|---|---|---|
| EPI_ISL_462642 | USA/MI-MDHHS-SC20560/2020 | 2020-04-29 | North America/USA/Michigan/ | Michigan Department of Health and Human Services, Bureau of Laboratories | Michigan Department of Health and Human Services, Bureau of Laboratories | Blankenship HM et al |
| EPI_ISL_462657 | USA/MI-MDHHS-SC20575/2020 | 2020-04-30 | North America/USA/Michigan/ | Michigan Department of Health and Human Services, Bureau of Laboratories | Michigan Department of Health and Human Services, Bureau of Laboratories | Blankenship HM et al |
| EPI_ISL_462660 | USA/MI-MDHHS-SC20578/2020 | 2020-04-29 | North America/USA/Michigan/ | Michigan Department of Health and Human Services, Bureau of Laboratories | Michigan Department of Health and Human Services, Bureau of Laboratories | Blankenship HM et al |
| EPI_ISL_462662 | USA/MI-MDHHS-SC20580/2020 | 2020-04-30 | North America/USA/Michigan/ | Michigan Department of Health and Human Services, Bureau of Laboratories | Michigan Department of Health and Human Services, Bureau of Laboratories | Blankenship HM et al |
| EPI_ISL_462665 | USA/MI-MDHHS-SC20583/2020 | 2020-04-29 | North America/USA/Michigan/ | Michigan Department of Health and Human Services, Bureau of Laboratories | Michigan Department of Health and Human Services, Bureau of Laboratories | Blankenship HM et al |
| EPI_ISL_462669 | USA/MI-MDHHS-SC20587/2020 | 2020-04-29 | North America/USA/Michigan/ | Michigan Department of Health and Human Services, Bureau of Laboratories | Michigan Department of Health and Human Services, Bureau of Laboratories | Blankenship HM et al |

| | | | | | | |
|---|---|---|---|---|---|---|
| EPI_ISL_462675 | USA/MI-MDHHS-SC20593/2020 | 2020-04-29 | North America/USA/Michigan/ | Michigan Department of Health and Human Services, Bureau of Laboratories | Michigan Department of Health and Human Services, Bureau of Laboratories | Blankenship HM et al |
| EPI_ISL_462687 | USA/MI-MDHHS-SC20605/2020 | 2020-04-29 | North America/USA/Michigan/ | Michigan Department of Health and Human Services, Bureau of Laboratories | Michigan Department of Health and Human Services, Bureau of Laboratories | Blankenship HM et al |
| EPI_ISL_462699 | USA/MI-MDHHS-SC20617/2020 | 2020-04-30 | North America/USA/Michigan/ | Michigan Department of Health and Human Services, Bureau of Laboratories | Michigan Department of Health and Human Services, Bureau of Laboratories | Blankenship HM et al |
| EPI_ISL_462702 | USA/MI-MDHHS-SC20620/2020 | 2020-04-16 | North America/USA/Michigan/ | Michigan Department of Health and Human Services, Bureau of Laboratories | Michigan Department of Health and Human Services, Bureau of Laboratories | Blankenship HM et al |
| EPI_ISL_462706 | USA/MI-MDHHS-SC20624/2020 | 2020-04-17 | North America/USA/Michigan/ | Michigan Department of Health and Human Services, Bureau of Laboratories | Michigan Department of Health and Human Services, Bureau of Laboratories | Blankenship HM et al |
| EPI_ISL_462707 | USA/MI-MDHHS-SC20625/2020 | 2020-04-17 | North America/USA/Michigan/ | Michigan Department of Health and Human Services, Bureau of Laboratories | Michigan Department of Health and Human Services, Bureau of Laboratories | Blankenship HM et al |

| | | | | | | |
|---|---|---|---|---|---|---|
| EPI_ISL_462708 | USA/MI-MDHHS-SC20626/2020 | 2020-04-17 | North America/USA/Michigan/ | Michigan Department of Health and Human Services, Bureau of Laboratories | Michigan Department of Health and Human Services, Bureau of Laboratories | Blankenship HM et al |
| EPI_ISL_462712 | USA/MI-MDHHS-SC20630/2020 | 2020-04-17 | North America/USA/Michigan/ | Michigan Department of Health and Human Services, Bureau of Laboratories | Michigan Department of Health and Human Services, Bureau of Laboratories | Blankenship HM et al |
| EPI_ISL_462717 | USA/MI-MDHHS-SC20635/2020 | 2020-04-18 | North America/USA/Michigan/ | Michigan Department of Health and Human Services, Bureau of Laboratories | Michigan Department of Health and Human Services, Bureau of Laboratories | Blankenship HM et al |
| EPI_ISL_462727 | USA/MI-MDHHS-SC20645/2020 | 2020-04-24 | North America/USA/Michigan/ | Michigan Department of Health and Human Services, Bureau of Laboratories | Michigan Department of Health and Human Services, Bureau of Laboratories | Blankenship HM et al |
| EPI_ISL_462733 | USA/MI-MDHHS-SC20651/2020 | 2020-04-23 | North America/USA/Michigan/ | Michigan Department of Health and Human Services, Bureau of Laboratories | Michigan Department of Health and Human Services, Bureau of Laboratories | Blankenship HM et al |
| EPI_ISL_462735 | USA/MI-MDHHS-SC20653/2020 | 2020-04-27 | North America/USA/Michigan/ | Michigan Department of Health and Human Services, Bureau of Laboratories | Michigan Department of Health and Human Services, Bureau of Laboratories | Blankenship HM et al |
| EPI_ISL_462754 | Canada/BC_18114270/2020 | 20-Mar | North America/Canada/British Columbia | BCCDC Public Health Laboratory | BCCDC Public Health Laboratory | Harrigan et al |
| EPI_ISL_462755 | Canada/BC_18135843/2020 | 20-Mar | North America/Canada/British Columbia | BCCDC Public Health Laboratory | BCCDC Public Health Laboratory | Harrigan et al |

| | | | | | | |
|---|---|---|---|---|---|---|
| EPI_ISL_462756 | Canada/BC_18138681/2020 | 20-Mar | North America/Canada/British Columbia | BCCDC Public Health Laboratory | BCCDC Public Health Laboratory | Harrigan et al |
| EPI_ISL_462757 | Canada/BC_18165111/2020 | 20-Mar | North America/Canada/British Columbia | BCCDC Public Health Laboratory | BCCDC Public Health Laboratory | Harrigan et al |
| EPI_ISL_462758 | Canada/BC_18507914/2020 | 20-Mar | North America/Canada/British Columbia | BCCDC Public Health Laboratory | BCCDC Public Health Laboratory | Harrigan et al |
| EPI_ISL_462759 | Canada/BC_18547866/2020 | 20-Mar | North America/Canada/British Columbia | BCCDC Public Health Laboratory | BCCDC Public Health Laboratory | Harrigan et al |
| EPI_ISL_462760 | Canada/BC_18969070/2020 | 20-Mar | North America/Canada/British Columbia | BCCDC Public Health Laboratory | BCCDC Public Health Laboratory | Harrigan et al |
| EPI_ISL_462761 | Canada/BC_19421714/2020 | 20-Mar | North America/Canada/British Columbia | BCCDC Public Health Laboratory | BCCDC Public Health Laboratory | Harrigan et al |
| EPI_ISL_462762 | Canada/BC_19421715/2020 | 20-Mar | North America/Canada/British Columbia | BCCDC Public Health Laboratory | BCCDC Public Health Laboratory | Harrigan et al |
| EPI_ISL_462763 | Canada/BC_19431931/2020 | 20-Mar | North America/Canada/British Columbia | BCCDC Public Health Laboratory | BCCDC Public Health Laboratory | Harrigan et al |
| EPI_ISL_462764 | Canada/BC_19431939/2020 | 20-Mar | North America/Canada/British Columbia | BCCDC Public Health Laboratory | BCCDC Public Health Laboratory | Harrigan et al |
| EPI_ISL_462765 | Canada/BC_19518330/2020 | 20-Mar | North America/Canada/British Columbia | BCCDC Public Health Laboratory | BCCDC Public Health Laboratory | Harrigan et al |
| EPI_ISL_462766 | Canada/BC_19677629/2020 | 20-Mar | North America/Canada/British Columbia | BCCDC Public Health Laboratory | BCCDC Public Health Laboratory | Harrigan et al |
| EPI_ISL_462767 | Canada/BC_19703911/2020 | 20-Mar | North America/Canada/British Columbia | BCCDC Public Health Laboratory | BCCDC Public Health Laboratory | Harrigan et al |
| EPI_ISL_462768 | Canada/BC_19704814/2020 | 20-Mar | North America/Canada/British Columbia | BCCDC Public Health Laboratory | BCCDC Public Health Laboratory | Harrigan et al |
| EPI_ISL_462769 | Canada/BC_19785113/2020 | 20-Mar | North America/Canada/British Columbia | BCCDC Public Health Laboratory | BCCDC Public Health Laboratory | Harrigan et al |
| EPI_ISL_462771 | Canada/BC_20332712/2020 | 20-Mar | North America/Canada/British Columbia | BCCDC Public Health Laboratory | BCCDC Public Health Laboratory | Harrigan et al |
| EPI_ISL_462772 | Canada/BC_20363111/2020 | 20-Mar | North America/Canada/British Columbia | BCCDC Public Health Laboratory | BCCDC Public Health Laboratory | Harrigan et al |
| EPI_ISL_462773 | Canada/BC_20425011/2020 | 20-Mar | North America/Canada/British Columbia | BCCDC Public Health Laboratory | BCCDC Public Health Laboratory | Harrigan et al |
| EPI_ISL_462774 | Canada/BC_20432317/2020 | 20-Mar | North America/Canada/British Columbia | BCCDC Public Health Laboratory | BCCDC Public Health Laboratory | Harrigan et al |
| EPI_ISL_462775 | Canada/BC_20505516/2020 | 20-Mar | North America/Canada/British Columbia | BCCDC Public Health Laboratory | BCCDC Public Health Laboratory | Harrigan et al |
| EPI_ISL_462776 | Canada/BC_20534610/2020 | 20-Mar | North America/Canada/British Columbia | BCCDC Public Health Laboratory | BCCDC Public Health Laboratory | Harrigan et al |

| | | | | | | |
|---|---|---|---|---|---|---|
| EPI_ISL_462777 | Canada/BC_20639310/2020 | 20-Mar | North America/Canada/British Columbia | BCCDC Public Health Laboratory | BCCDC Public Health Laboratory | Harrigan et al |
| EPI_ISL_462778 | Canada/BC_20693717/2020 | 20-Mar | North America/Canada/British Columbia | BCCDC Public Health Laboratory | BCCDC Public Health Laboratory | Harrigan et al |
| EPI_ISL_462779 | Canada/BC_20722019/2020 | 20-Mar | North America/Canada/British Columbia | BCCDC Public Health Laboratory | BCCDC Public Health Laboratory | Harrigan et al |
| EPI_ISL_462780 | Canada/BC_20730616/2020 | 20-Mar | North America/Canada/British Columbia | BCCDC Public Health Laboratory | BCCDC Public Health Laboratory | Harrigan et al |
| EPI_ISL_462781 | Canada/BC_20734315/2020 | 20-Mar | North America/Canada/British Columbia | BCCDC Public Health Laboratory | BCCDC Public Health Laboratory | Harrigan et al |
| EPI_ISL_462782 | Canada/BC_20847524/2020 | 20-Mar | North America/Canada/British Columbia | BCCDC Public Health Laboratory | BCCDC Public Health Laboratory | Harrigan et al |
| EPI_ISL_462783 | Canada/BC_20855313/2020 | 20-Mar | North America/Canada/British Columbia | BCCDC Public Health Laboratory | BCCDC Public Health Laboratory | Harrigan et al |
| EPI_ISL_462784 | Canada/BC_21008811/2020 | 20-Mar | North America/Canada/British Columbia | BCCDC Public Health Laboratory | BCCDC Public Health Laboratory | Harrigan et al |
| EPI_ISL_462785 | Canada/BC_21008826/2020 | 20-Mar | North America/Canada/British Columbia | BCCDC Public Health Laboratory | BCCDC Public Health Laboratory | Harrigan et al |
| EPI_ISL_462786 | Canada/BC_21016029/2020 | 20-Mar | North America/Canada/British Columbia | BCCDC Public Health Laboratory | BCCDC Public Health Laboratory | Harrigan et al |
| EPI_ISL_462787 | Canada/BC_21022514/2020 | 20-Mar | North America/Canada/British Columbia | BCCDC Public Health Laboratory | BCCDC Public Health Laboratory | Harrigan et al |
| EPI_ISL_462788 | Canada/BC_21084910/2020 | 20-Feb | North America/Canada/British Columbia | BCCDC Public Health Laboratory | BCCDC Public Health Laboratory | Harrigan et al |
| EPI_ISL_462789 | Canada/BC_21085515/2020 | 20-Mar | North America/Canada/British Columbia | BCCDC Public Health Laboratory | BCCDC Public Health Laboratory | Harrigan et al |
| EPI_ISL_462790 | Canada/BC_21105219/2020 | 20-Mar | North America/Canada/British Columbia | BCCDC Public Health Laboratory | BCCDC Public Health Laboratory | Harrigan et al |
| EPI_ISL_462791 | Canada/BC_21110314/2020 | 20-Mar | North America/Canada/British Columbia | BCCDC Public Health Laboratory | BCCDC Public Health Laboratory | Harrigan et al |
| EPI_ISL_462792 | Canada/BC_21127214/2020 | 20-Mar | North America/Canada/British Columbia | BCCDC Public Health Laboratory | BCCDC Public Health Laboratory | Harrigan et al |
| EPI_ISL_462793 | Canada/BC_21127913/2020 | 20-Mar | North America/Canada/British Columbia | BCCDC Public Health Laboratory | BCCDC Public Health Laboratory | Harrigan et al |
| EPI_ISL_462794 | Canada/BC_21146111/2020 | 20-Mar | North America/Canada/British Columbia | BCCDC Public Health Laboratory | BCCDC Public Health Laboratory | Harrigan et al |
| EPI_ISL_462795 | Canada/BC_21154418/2020 | 20-Mar | North America/Canada/British Columbia | BCCDC Public Health Laboratory | BCCDC Public Health Laboratory | Harrigan et al |
| EPI_ISL_462796 | Canada/BC_21174010/2020 | 20-Mar | North America/Canada/British Columbia | BCCDC Public Health Laboratory | BCCDC Public Health Laboratory | Harrigan et al |

| | | | | | | |
|---|---|---|---|---|---|---|
| EPI_ISL_462797 | Canada/BC_21174021/2020 | 20-Mar | North America/Canada/British Columbia | BCCDC Public Health Laboratory | BCCDC Public Health Laboratory | Harrigan et al |
| EPI_ISL_462798 | Canada/BC_21187418/2020 | 20-Mar | North America/Canada/British Columbia | BCCDC Public Health Laboratory | BCCDC Public Health Laboratory | Harrigan et al |
| EPI_ISL_462799 | Canada/BC_21216919/2020 | 20-Mar | North America/Canada/British Columbia | BCCDC Public Health Laboratory | BCCDC Public Health Laboratory | Harrigan et al |
| EPI_ISL_462800 | Canada/BC_21221818/2020 | 20-Mar | North America/Canada/British Columbia | BCCDC Public Health Laboratory | BCCDC Public Health Laboratory | Harrigan et al |
| EPI_ISL_462801 | Canada/BC_21228217/2020 | 20-Mar | North America/Canada/British Columbia | BCCDC Public Health Laboratory | BCCDC Public Health Laboratory | Harrigan et al |
| EPI_ISL_462802 | Canada/BC_21228218/2020 | 20-Mar | North America/Canada/British Columbia | BCCDC Public Health Laboratory | BCCDC Public Health Laboratory | Harrigan et al |
| EPI_ISL_462803 | Canada/BC_21262415/2020 | 20-Mar | North America/Canada/British Columbia | BCCDC Public Health Laboratory | BCCDC Public Health Laboratory | Harrigan et al |
| EPI_ISL_462804 | Canada/BC_21282517/2020 | 20-Mar | North America/Canada/British Columbia | BCCDC Public Health Laboratory | BCCDC Public Health Laboratory | Harrigan et al |
| EPI_ISL_462805 | Canada/BC_21317517/2020 | 20-Mar | North America/Canada/British Columbia | BCCDC Public Health Laboratory | BCCDC Public Health Laboratory | Harrigan et al |
| EPI_ISL_462806 | Canada/BC_21333019/2020 | 20-Mar | North America/Canada/British Columbia | BCCDC Public Health Laboratory | BCCDC Public Health Laboratory | Harrigan et al |
| EPI_ISL_462807 | Canada/BC_21346429/2020 | 20-Mar | North America/Canada/British Columbia | BCCDC Public Health Laboratory | BCCDC Public Health Laboratory | Harrigan et al |
| EPI_ISL_462808 | Canada/BC_21350918/2020 | 20-Mar | North America/Canada/British Columbia | BCCDC Public Health Laboratory | BCCDC Public Health Laboratory | Harrigan et al |
| EPI_ISL_462809 | Canada/BC_21368114/2020 | 20-Mar | North America/Canada/British Columbia | BCCDC Public Health Laboratory | BCCDC Public Health Laboratory | Harrigan et al |
| EPI_ISL_462810 | Canada/BC_21385821/2020 | 20-Mar | North America/Canada/British Columbia | BCCDC Public Health Laboratory | BCCDC Public Health Laboratory | Harrigan et al |
| EPI_ISL_462811 | Canada/BC_21388915/2020 | 20-Mar | North America/Canada/British Columbia | BCCDC Public Health Laboratory | BCCDC Public Health Laboratory | Harrigan et al |
| EPI_ISL_462812 | Canada/BC_21426715/2020 | 20-Mar | North America/Canada/British Columbia | BCCDC Public Health Laboratory | BCCDC Public Health Laboratory | Harrigan et al |
| EPI_ISL_462813 | Canada/BC_21446719/2020 | 20-Mar | North America/Canada/British Columbia | BCCDC Public Health Laboratory | BCCDC Public Health Laboratory | Harrigan et al |
| EPI_ISL_462814 | Canada/BC_21461212/2020 | 20-Mar | North America/Canada/British Columbia | BCCDC Public Health Laboratory | BCCDC Public Health Laboratory | Harrigan et al |
| EPI_ISL_462815 | Canada/BC_21462515/2020 | 20-Mar | North America/Canada/British Columbia | BCCDC Public Health Laboratory | BCCDC Public Health Laboratory | Harrigan et al |
| EPI_ISL_462816 | Canada/BC_21466919/2020 | 20-Mar | North America/Canada/British Columbia | BCCDC Public Health Laboratory | BCCDC Public Health Laboratory | Harrigan et al |

| | | | | | | |
|---|---|---|---|---|---|---|
| EPI_ISL_462817 | Canada/BC_21467217/2020 | 20-Mar | North America/Canada/British Columbia | BCCDC Public Health Laboratory | BCCDC Public Health Laboratory | Harrigan et al |
| EPI_ISL_462818 | Canada/BC_21480617/2020 | 20-Mar | North America/Canada/British Columbia | BCCDC Public Health Laboratory | BCCDC Public Health Laboratory | Harrigan et al |
| EPI_ISL_462819 | Canada/BC_21927217/2020 | 20-Mar | North America/Canada/British Columbia | BCCDC Public Health Laboratory | BCCDC Public Health Laboratory | Harrigan et al |
| EPI_ISL_462820 | Canada/BC_21993212/2020 | 20-Mar | North America/Canada/British Columbia | BCCDC Public Health Laboratory | BCCDC Public Health Laboratory | Harrigan et al |
| EPI_ISL_462821 | Canada/BC_22032711/2020 | 20-Mar | North America/Canada/British Columbia | BCCDC Public Health Laboratory | BCCDC Public Health Laboratory | Harrigan et al |
| EPI_ISL_462822 | Canada/BC_22037912/2020 | 20-Mar | North America/Canada/British Columbia | BCCDC Public Health Laboratory | BCCDC Public Health Laboratory | Harrigan et al |
| EPI_ISL_462824 | Canada/BC_22070527/2020 | 20-Mar | North America/Canada/British Columbia | BCCDC Public Health Laboratory | BCCDC Public Health Laboratory | Harrigan et al |
| EPI_ISL_462825 | Canada/BC_22156718/2020 | 20-Mar | North America/Canada/British Columbia | BCCDC Public Health Laboratory | BCCDC Public Health Laboratory | Harrigan et al |
| EPI_ISL_462826 | Canada/BC_22218811/2020 | 20-Mar | North America/Canada/British Columbia | BCCDC Public Health Laboratory | BCCDC Public Health Laboratory | Harrigan et al |
| EPI_ISL_462827 | Canada/BC_22230811/2020 | 20-Mar | North America/Canada/British Columbia | BCCDC Public Health Laboratory | BCCDC Public Health Laboratory | Harrigan et al |
| EPI_ISL_462828 | Canada/BC_22332113/2020 | 20-Mar | North America/Canada/British Columbia | BCCDC Public Health Laboratory | BCCDC Public Health Laboratory | Harrigan et al |
| EPI_ISL_462829 | Canada/BC_22349316/2020 | 20-Mar | North America/Canada/British Columbia | BCCDC Public Health Laboratory | BCCDC Public Health Laboratory | Harrigan et al |
| EPI_ISL_462830 | Canada/BC_22430814/2020 | 20-Mar | North America/Canada/British Columbia | BCCDC Public Health Laboratory | BCCDC Public Health Laboratory | Harrigan et al |
| EPI_ISL_462831 | Canada/BC_22451513/2020 | 20-Mar | North America/Canada/British Columbia | BCCDC Public Health Laboratory | BCCDC Public Health Laboratory | Harrigan et al |
| EPI_ISL_462832 | Canada/BC_22466713/2020 | 20-Mar | North America/Canada/British Columbia | BCCDC Public Health Laboratory | BCCDC Public Health Laboratory | Harrigan et al |
| EPI_ISL_462833 | Canada/BC_22470913/2020 | 20-Mar | North America/Canada/British Columbia | BCCDC Public Health Laboratory | BCCDC Public Health Laboratory | Harrigan et al |
| EPI_ISL_462834 | Canada/BC_22473210/2020 | 20-Mar | North America/Canada/British Columbia | BCCDC Public Health Laboratory | BCCDC Public Health Laboratory | Harrigan et al |
| EPI_ISL_462835 | Canada/BC_22475116/2020 | 20-Mar | North America/Canada/British Columbia | BCCDC Public Health Laboratory | BCCDC Public Health Laboratory | Harrigan et al |
| EPI_ISL_462836 | Canada/BC_22511314/2020 | 20-Mar | North America/Canada/British Columbia | BCCDC Public Health Laboratory | BCCDC Public Health Laboratory | Harrigan et al |
| EPI_ISL_462837 | Canada/BC_22514413/2020 | 20-Mar | North America/Canada/British Columbia | BCCDC Public Health Laboratory | BCCDC Public Health Laboratory | Harrigan et al |

| | | | | | | |
|---|---|---|---|---|---|---|
| EPI_ISL_462838 | Canada/BC_22946611/2020 | 20-Mar | North America/Canada/British Columbia | BCCDC Public Health Laboratory | BCCDC Public Health Laboratory | Harrigan et al |
| EPI_ISL_462839 | Canada/BC_23052516/2020 | 20-Mar | North America/Canada/British Columbia | BCCDC Public Health Laboratory | BCCDC Public Health Laboratory | Harrigan et al |
| EPI_ISL_462840 | Canada/BC_23064916/2020 | 20-Mar | North America/Canada/British Columbia | BCCDC Public Health Laboratory | BCCDC Public Health Laboratory | Harrigan et al |
| EPI_ISL_462841 | Canada/BC_23094312/2020 | 20-Mar | North America/Canada/British Columbia | BCCDC Public Health Laboratory | BCCDC Public Health Laboratory | Harrigan et al |
| EPI_ISL_462842 | Canada/BC_23098110/2020 | 20-Mar | North America/Canada/British Columbia | BCCDC Public Health Laboratory | BCCDC Public Health Laboratory | Harrigan et al |
| EPI_ISL_462843 | Canada/BC_23108423/2020 | 20-Mar | North America/Canada/British Columbia | BCCDC Public Health Laboratory | BCCDC Public Health Laboratory | Harrigan et al |
| EPI_ISL_462844 | Canada/BC_23348019/2020 | 20-Mar | North America/Canada/British Columbia | BCCDC Public Health Laboratory | BCCDC Public Health Laboratory | Harrigan et al |
| EPI_ISL_462860 | USA/MN-MDH-1015/2020 | 2020-04-30 | North America/USA/Minnesota/ | Minnesota Department of Health, Public Health Laboratory | Minnesota Department of Health, Public Health Laboratory | Matt Plumb et al |
| EPI_ISL_462862 | USA/MN-MDH-1017/2020 | 2020-04-30 | North America/USA/Minnesota/ | Minnesota Department of Health, Public Health Laboratory | Minnesota Department of Health, Public Health Laboratory | Matt Plumb et al |
| EPI_ISL_462863 | USA/MN-MDH-1018/2020 | 2020-04-30 | North America/USA/Minnesota/ | Minnesota Department of Health, Public Health Laboratory | Minnesota Department of Health, Public Health Laboratory | Matt Plumb et al |
| EPI_ISL_462865 | USA/MN-MDH-1020/2020 | 2020-05-07 | North America/USA/Minnesota/ | Minnesota Department of Health, Public Health Laboratory | Minnesota Department of Health, Public Health Laboratory | Matt Plumb et al |
| EPI_ISL_462869 | USA/MN-MDH-1024/2020 | 2020-05-07 | North America/USA/Minnesota/ | Minnesota Department of Health, Public Health Laboratory | Minnesota Department of Health, Public Health Laboratory | Matt Plumb et al |
| EPI_ISL_462875 | USA/MN-MDH-1030/2020 | 2020-05-07 | North America/USA/Minnesota/ | Minnesota Department of Health, Public Health Laboratory | Minnesota Department of Health, Public Health Laboratory | Matt Plumb et al |

| | | | | | | |
|---|---|---|---|---|---|---|
| EPI_ISL_462880 | USA/MN-MDH-1035/2020 | 2020-05-07 | North America/USA/Minnesota/ | Minnesota Department of Health, Public Health Laboratory | Minnesota Department of Health, Public Health Laboratory | Matt Plumb et al |
| EPI_ISL_462894 | USA/MN-MDH-1049/2020 | 2020-05-14 | North America/USA/Minnesota/ | Minnesota Department of Health, Public Health Laboratory | Minnesota Department of Health, Public Health Laboratory | Matt Plumb et al |
| EPI_ISL_462899 | USA/MN-MDH-1054/2020 | 2020-05-14 | North America/USA/Minnesota/ | Minnesota Department of Health, Public Health Laboratory | Minnesota Department of Health, Public Health Laboratory | Matt Plumb et al |
| EPI_ISL_462908 | USA/MN-MDH-1063/2020 | 2020-05-14 | North America/USA/Minnesota/ | Minnesota Department of Health, Public Health Laboratory | Minnesota Department of Health, Public Health Laboratory | Matt Plumb et al |
| EPI_ISL_462922 | USA/WY-WYPHL-00029/2020 | 2020-03-21 | North America/USA/Wyoming/ | Wyoming Public Health Laboratory | Center for Global Health, University of New Mexico Health Sciences Center | Daryl Domman et al |
| EPI_ISL_462941 | USA/WY-WYPHL-00049/2020 | 2020-03-25 | North America/USA/Wyoming/ | Wyoming Public Health Laboratory | Center for Global Health, University of New Mexico Health Sciences Center | Daryl Domman et al |
| EPI_ISL_462954 | USA/WY-WYPHL-00062/2020 | 2020-03-27 | North America/USA/Wyoming/ | Wyoming Public Health Laboratory | Center for Global Health, University of New Mexico Health Sciences Center | Daryl Domman et al |
| EPI_ISL_462958 | USA/WY-WYPHL-00066/2020 | 2020-03-27 | North America/USA/Wyoming/ | Wyoming Public Health Laboratory | Center for Global Health, University of New Mexico Health Sciences Center | Daryl Domman et al |

| | | | | | | |
|---|---|---|---|---|---|---|
| EPI_ISL_462987 | USA/WY-WYPHL-00097/2020 | 2020-03-30 | North America/USA/Wyoming/ | Wyoming Public Health Laboratory | Center for Global Health, University of New Mexico Health Sciences Center | Daryl Domman et al |
| EPI_ISL_462990 | BosniaandHerzegovina/02-BanjaLuka | 2020-04-29 | Europe/Bosnia and Herzegovina/ƒåelina | University Clinical Centre of the Republic of Srpska | University of Sarajevo, Veterinary Faculty | Teufik et al |
| EPI_ISL_462996 | Spain/AS-MG-68-19/2020 | 2020-05-30 | Europe/Spain/Asturias/ | Molecular Genetics | Molecular Genetics | Gomez et al |
| EPI_ISL_462997 | Spain/AS-MG-80-19/2020 | 2020-05-30 | Europe/Spain/Asturias/ | Molecular Genetics | Molecular Genetics | Gomez et al |
| EPI_ISL_463004 | Tunisia/COV1663/2020 | 2020-04-01 | Africa/Tunisia/Tunisia/ | unknown | Clinical virology | Fares et al |
| EPI_ISL_463013 | India/OR-ILSCV12742/2020 | 2020-05-09 | Asia/India/Odisha/ | Institute of Life Sciences, Bhubaneswar | Immunogenomics lab, Institute of Life Sciences, Bhubaneswar | Sunil Raghav et al |
| EPI_ISL_463014 | India/OR-ILSCV12743/2020 | 2020-05-09 | Asia/India/Odisha/ | Institute of Life Sciences, Bhubaneswar | Immunogenomics lab, Institute of Life Sciences, Bhubaneswar | Sunil Raghav et al |
| EPI_ISL_463020 | India/OR-ILSCV13678/2020 | 2020-05-09 | Asia/India/Odisha/ | Institute of Life Sciences, Bhubaneswar | Immunogenomics lab, Institute of Life Sciences, Bhubaneswar | Sunil Raghav et al |
| EPI_ISL_463026 | India/OR-ILSCV13929/2020 | 2020-05-10 | Asia/India/Odisha/ | Institute of Life Sciences, Bhubaneswar | Immunogenomics lab, Institute of Life Sciences, Bhubaneswar | Sunil Raghav et al |
| EPI_ISL_463027 | India/OR-ILSCV14004/2020 | 2020-05-09 | Asia/India/Odisha/ | Institute of Life Sciences, Bhubaneswar | Immunogenomics lab, Institute of Life Sciences, Bhubaneswar | Sunil Raghav et al |
| EPI_ISL_463029 | India/OR-ILSCV14262/2020 | 2020-05-11 | Asia/India/Odisha/ | Institute of Life Sciences, Bhubaneswar | Immunogenomics lab, Institute of Life Sciences, Bhubaneswar | Sunil Raghav et al |

| | | | | | | |
|---|---|---|---|---|---|---|
| EPI_ISL_463039 | India/OR-ILSCV15917/2020 | 2020-05-11 | Asia/India/Odisha/ | Institute of Life Sciences, Bhubaneswar | Immunogenomics lab, Institute of Life Sciences, Bhubaneswar | Sunil Raghav et al |
| EPI_ISL_463041 | India/OR-ILSCV15937/2020 | 2020-05-11 | Asia/India/Odisha/ | Institute of Life Sciences, Bhubaneswar | Immunogenomics lab, Institute of Life Sciences, Bhubaneswar | Sunil Raghav et al |
| EPI_ISL_463052 | India/OR-ILSCV16514/2020 | 2020-05-12 | Asia/India/Odisha/ | Institute of Life Sciences, Bhubaneswar | Immunogenomics lab, Institute of Life Sciences, Bhubaneswar | Sunil Raghav et al |
| EPI_ISL_463054 | India/OR-ILSCV16809/2020 | 2020-05-11 | Asia/India/Odisha/ | Institute of Life Sciences, Bhubaneswar | Immunogenomics lab, Institute of Life Sciences, Bhubaneswar | Sunil Raghav et al |
| EPI_ISL_463064 | India/OR-ILSCV19048/2020 | 2020-05-16 | Asia/India/Odisha/ | Institute of Life Sciences, Bhubaneswar | Immunogenomics lab, Institute of Life Sciences, Bhubaneswar | Sunil Raghav et al |
| EPI_ISL_463076 | India/OR-ILSCV20408/2020 | 2020-05-15 | Asia/India/Odisha/ | Institute of Life Sciences, Bhubaneswar | Immunogenomics lab, Institute of Life Sciences, Bhubaneswar | Sunil Raghav et al |
| EPI_ISL_463081 | India/OR-ILSCV20600/2020 | 2020-05-15 | Asia/India/Odisha/ | Institute of Life Sciences, Bhubaneswar | Immunogenomics lab, Institute of Life Sciences, Bhubaneswar | Sunil Raghav et al |
| EPI_ISL_463085 | India/OR-ILSCV21721/2020 | 2020-05-16 | Asia/India/Odisha/ | Institute of Life Sciences, Bhubaneswar | Immunogenomics lab, Institute of Life Sciences, Bhubaneswar | Sunil Raghav et al |
| EPI_ISL_463091 | India/OR-ILSCV27412/2020 | 2020-05-21 | Asia/India/Odisha/ | Institute of Life Sciences, Bhubaneswar | Immunogenomics lab, Institute of Life Sciences, Bhubaneswar | Sunil Raghav et al |
| EPI_ISL_463113 | USA/VA-DCLS-0310/2020 | 2020-05-18 | North America/USA/Virginia/ | Virginia DCLS | Virginia DCLS | Virginia DCLS et al |
| EPI_ISL_463117 | USA/VA-DCLS-0350/2020 | 2020-05-08 | North America/USA/Virginia/ | Virginia DCLS | Virginia DCLS | Virginia DCLS et al |
| EPI_ISL_463127 | USA/VA-DCLS-0371/2020 | 2020-05-08 | North America/USA/Virginia/ | Virginia DCLS | Virginia DCLS | Virginia DCLS et al |

| | | | | | | |
|---|---|---|---|---|---|---|
| EPI_ISL_463130 | USA/VA-DCLS-0374/2020 | 2020-05-12 | North America/USA/Virginia/ | Virginia DCLS | Virginia DCLS | Virginia DCLS et al |
| EPI_ISL_463137 | USA/VA-DCLS-0381/2020 | 2020-05-13 | North America/USA/Virginia/ | Virginia DCLS | Virginia DCLS | Virginia DCLS et al |
| EPI_ISL_463154 | USA/CT-Yale-218/2020 | 2020-03-31 | North America/USA/Connecticut/New H | Yale Clinical Virology Laboratory | Grubaugh Lab - Yale School of Public Health | Joseph Fauver et al |
| EPI_ISL_463160 | USA/CT-Yale-226/2020 | 2020-03-31 | North America/USA/Connecticut/New H | Yale Clinical Virology Laboratory | Grubaugh Lab - Yale School of Public Health | Joseph Fauver et al |
| EPI_ISL_463167 | USA/CT-Yale-234/2020 | 2020-04-01 | North America/USA/Connecticut/New H | Yale Clinical Virology Laboratory | Grubaugh Lab - Yale School of Public Health | Joseph Fauver et al |
| EPI_ISL_463168 | USA/CT-Yale-236/2020 | 2020-04-01 | North America/USA/Connecticut/West H | Yale Clinical Virology Laboratory | Grubaugh Lab - Yale School of Public Health | Joseph Fauver et al |
| EPI_ISL_463170 | USA/CT-Yale-238/2020 | 2020-04-01 | North America/USA/Connecticut/New H | Yale Clinical Virology Laboratory | Grubaugh Lab - Yale School of Public Health | Joseph Fauver et al |
| EPI_ISL_463186 | Canada/BC_23745918/2020 | 20-Mar | North America/Canada/British Columbia | BCCDC Public Health Laboratory | BCCDC Public Health Laboratory | Richard Harrigan et al |
| EPI_ISL_463187 | Canada/BC_23815516/2020 | 20-Mar | North America/Canada/British Columbia | BCCDC Public Health Laboratory | BCCDC Public Health Laboratory | Richard Harrigan et al |
| EPI_ISL_463188 | Canada/BC_23840285/2020 | 20-Mar | North America/Canada/British Columbia | BCCDC Public Health Laboratory | BCCDC Public Health Laboratory | Richard Harrigan et al |
| EPI_ISL_463189 | Canada/BC_23918026/2020 | 20-Mar | North America/Canada/British Columbia | BCCDC Public Health Laboratory | BCCDC Public Health Laboratory | Richard Harrigan et al |
| EPI_ISL_463191 | Canada/BC_24044059/2020 | 20-Mar | North America/Canada/British Columbia | BCCDC Public Health Laboratory | BCCDC Public Health Laboratory | Richard Harrigan et al |
| EPI_ISL_463193 | Canada/BC_24150410/2020 | 20-Mar | North America/Canada/British Columbia | BCCDC Public Health Laboratory | BCCDC Public Health Laboratory | Richard Harrigan et al |
| EPI_ISL_463194 | Canada/BC_24165067/2020 | 20-Mar | North America/Canada/British Columbia | BCCDC Public Health Laboratory | BCCDC Public Health Laboratory | Richard Harrigan et al |
| EPI_ISL_463195 | Canada/BC_24190425/2020 | 20-Mar | North America/Canada/British Columbia | BCCDC Public Health Laboratory | BCCDC Public Health Laboratory | Richard Harrigan et al |
| EPI_ISL_463196 | Canada/BC_24192775/2020 | 20-Mar | North America/Canada/British Columbia | BCCDC Public Health Laboratory | BCCDC Public Health Laboratory | Richard Harrigan et al |
| EPI_ISL_463198 | Canada/BC_24201275/2020 | 20-Mar | North America/Canada/British Columbia | BCCDC Public Health Laboratory | BCCDC Public Health Laboratory | Richard Harrigan et al |
| EPI_ISL_463199 | Canada/BC_24225776/2020 | 20-Mar | North America/Canada/British Columbia | BCCDC Public Health Laboratory | BCCDC Public Health Laboratory | Richard Harrigan et al |

| | | | | | | |
|---|---|---|---|---|---|---|
| EPI_ISL_463200 | Canada/BC_24228539/2020 | 20-Mar | North America/Canada/British Columbia | BCCDC Public Health Laboratory | BCCDC Public Health Laboratory | Richard Harrigan et al |
| EPI_ISL_463201 | Canada/BC_24232310/2020 | 20-Mar | North America/Canada/British Columbia | BCCDC Public Health Laboratory | BCCDC Public Health Laboratory | Richard Harrigan et al |
| EPI_ISL_463202 | Canada/BC_24234949/2020 | 20-Mar | North America/Canada/British Columbia | BCCDC Public Health Laboratory | BCCDC Public Health Laboratory | Richard Harrigan et al |
| EPI_ISL_463203 | Canada/BC_24235415/2020 | 20-Mar | North America/Canada/British Columbia | BCCDC Public Health Laboratory | BCCDC Public Health Laboratory | Richard Harrigan et al |
| EPI_ISL_463204 | Canada/BC_24239917/2020 | 20-Mar | North America/Canada/British Columbia | BCCDC Public Health Laboratory | BCCDC Public Health Laboratory | Richard Harrigan et al |
| EPI_ISL_463205 | Canada/BC_24245374/2020 | 20-Mar | North America/Canada/British Columbia | BCCDC Public Health Laboratory | BCCDC Public Health Laboratory | Richard Harrigan et al |
| EPI_ISL_463206 | Canada/BC_24252365/2020 | 20-Mar | North America/Canada/British Columbia | BCCDC Public Health Laboratory | BCCDC Public Health Laboratory | Richard Harrigan et al |
| EPI_ISL_463207 | Canada/BC_24258528/2020 | 20-Mar | North America/Canada/British Columbia | BCCDC Public Health Laboratory | BCCDC Public Health Laboratory | Richard Harrigan et al |
| EPI_ISL_463208 | Canada/BC_24261010/2020 | 20-Mar | North America/Canada/British Columbia | BCCDC Public Health Laboratory | BCCDC Public Health Laboratory | Richard Harrigan et al |
| EPI_ISL_463209 | Canada/BC_24271871/2020 | 20-Mar | North America/Canada/British Columbia | BCCDC Public Health Laboratory | BCCDC Public Health Laboratory | Richard Harrigan et al |
| EPI_ISL_463210 | Canada/BC_24302962/2020 | 20-Mar | North America/Canada/British Columbia | BCCDC Public Health Laboratory | BCCDC Public Health Laboratory | Richard Harrigan et al |
| EPI_ISL_463211 | Canada/BC_24319756/2020 | 20-Mar | North America/Canada/British Columbia | BCCDC Public Health Laboratory | BCCDC Public Health Laboratory | Richard Harrigan et al |
| EPI_ISL_463212 | Canada/BC_24340030/2020 | 20-Mar | North America/Canada/British Columbia | BCCDC Public Health Laboratory | BCCDC Public Health Laboratory | Richard Harrigan et al |
| EPI_ISL_463213 | Canada/BC_24340319/2020 | 20-Mar | North America/Canada/British Columbia | BCCDC Public Health Laboratory | BCCDC Public Health Laboratory | Richard Harrigan et al |
| EPI_ISL_463214 | Canada/BC_24360831/2020 | 20-Mar | North America/Canada/British Columbia | BCCDC Public Health Laboratory | BCCDC Public Health Laboratory | Richard Harrigan et al |
| EPI_ISL_463215 | Canada/BC_24380644/2020 | 20-Mar | North America/Canada/British Columbia | BCCDC Public Health Laboratory | BCCDC Public Health Laboratory | Richard Harrigan et al |
| EPI_ISL_463216 | Canada/BC_24825751/2020 | 20-Mar | North America/Canada/British Columbia | BCCDC Public Health Laboratory | BCCDC Public Health Laboratory | Richard Harrigan et al |
| EPI_ISL_463217 | Canada/BC_25143276/2020 | 20-Mar | North America/Canada/British Columbia | BCCDC Public Health Laboratory | BCCDC Public Health Laboratory | Richard Harrigan et al |
| EPI_ISL_463218 | Canada/BC_25156715/2020 | 20-Mar | North America/Canada/British Columbia | BCCDC Public Health Laboratory | BCCDC Public Health Laboratory | Richard Harrigan et al |
| EPI_ISL_463219 | Canada/BC_25336669/2020 | 20-Mar | North America/Canada/British Columbia | BCCDC Public Health Laboratory | BCCDC Public Health Laboratory | Richard Harrigan et al |

| | | | | | | |
|---|---|---|---|---|---|---|
| EPI_ISL_463220 | Canada/BC_25367465/2020 | 20-Feb | North America/Canada/British Columbia | BCCDC Public Health Laboratory | BCCDC Public Health Laboratory | Richard Harrigan et al |
| EPI_ISL_463221 | Canada/BC_25376719/2020 | 20-Mar | North America/Canada/British Columbia | BCCDC Public Health Laboratory | BCCDC Public Health Laboratory | Richard Harrigan et al |
| EPI_ISL_463222 | Canada/BC_25384316/2020 | 20-Mar | North America/Canada/British Columbia | BCCDC Public Health Laboratory | BCCDC Public Health Laboratory | Richard Harrigan et al |
| EPI_ISL_463223 | Canada/BC_25390851/2020 | 20-Mar | North America/Canada/British Columbia | BCCDC Public Health Laboratory | BCCDC Public Health Laboratory | Richard Harrigan et al |
| EPI_ISL_463224 | Canada/BC_25416870/2020 | 20-Mar | North America/Canada/British Columbia | BCCDC Public Health Laboratory | BCCDC Public Health Laboratory | Richard Harrigan et al |
| EPI_ISL_463225 | Canada/BC_25515215/2020 | 20-Mar | North America/Canada/British Columbia | BCCDC Public Health Laboratory | BCCDC Public Health Laboratory | Richard Harrigan et al |
| EPI_ISL_463226 | Canada/BC_25574199/2020 | 20-Mar | North America/Canada/British Columbia | BCCDC Public Health Laboratory | BCCDC Public Health Laboratory | Richard Harrigan et al |
| EPI_ISL_463227 | Canada/BC_25593681/2020 | 20-Mar | North America/Canada/British Columbia | BCCDC Public Health Laboratory | BCCDC Public Health Laboratory | Richard Harrigan et al |
| EPI_ISL_463228 | Canada/BC_25856970/2020 | 20-Mar | North America/Canada/British Columbia | BCCDC Public Health Laboratory | BCCDC Public Health Laboratory | Richard Harrigan et al |
| EPI_ISL_463229 | Canada/BC_25857092/2020 | 20-Mar | North America/Canada/British Columbia | BCCDC Public Health Laboratory | BCCDC Public Health Laboratory | Richard Harrigan et al |
| EPI_ISL_463230 | Canada/BC_26101196/2020 | 20-Mar | North America/Canada/British Columbia | BCCDC Public Health Laboratory | BCCDC Public Health Laboratory | Richard Harrigan et al |
| EPI_ISL_463231 | Canada/BC_26172976/2020 | 20-Mar | North America/Canada/British Columbia | BCCDC Public Health Laboratory | BCCDC Public Health Laboratory | Richard Harrigan et al |
| EPI_ISL_463235 | Canada/BC_26509737/2020 | 20-Mar | North America/Canada/British Columbia | BCCDC Public Health Laboratory | BCCDC Public Health Laboratory | Richard Harrigan et al |
| EPI_ISL_463236 | Canada/BC_26564890/2020 | 20-Mar | North America/Canada/British Columbia | BCCDC Public Health Laboratory | BCCDC Public Health Laboratory | Richard Harrigan et al |
| EPI_ISL_463237 | Canada/BC_26584470/2020 | 20-Mar | North America/Canada/British Columbia | BCCDC Public Health Laboratory | BCCDC Public Health Laboratory | Richard Harrigan et al |
| EPI_ISL_463238 | Canada/BC_26585549/2020 | 20-Mar | North America/Canada/British Columbia | BCCDC Public Health Laboratory | BCCDC Public Health Laboratory | Richard Harrigan et al |
| EPI_ISL_463239 | Canada/BC_26590014/2020 | 20-Mar | North America/Canada/British Columbia | BCCDC Public Health Laboratory | BCCDC Public Health Laboratory | Richard Harrigan et al |
| EPI_ISL_463240 | Canada/BC_26590284/2020 | 20-Mar | North America/Canada/British Columbia | BCCDC Public Health Laboratory | BCCDC Public Health Laboratory | Richard Harrigan et al |
| EPI_ISL_463241 | Canada/BC_26591461/2020 | 20-Mar | North America/Canada/British Columbia | BCCDC Public Health Laboratory | BCCDC Public Health Laboratory | Richard Harrigan et al |
| EPI_ISL_463242 | Canada/BC_26602426/2020 | 20-Mar | North America/Canada/British Columbia | BCCDC Public Health Laboratory | BCCDC Public Health Laboratory | Richard Harrigan et al |

| | | | | | | |
|---|---|---|---|---|---|---|
| EPI_ISL_463243 | Canada/BC_26605771/2020 | 20-Mar | North America/Canada/British Columbia | BCCDC Public Health Laboratory | BCCDC Public Health Laboratory | Richard Harrigan et al |
| EPI_ISL_463244 | Canada/BC_26611727/2020 | 20-Mar | North America/Canada/British Columbia | BCCDC Public Health Laboratory | BCCDC Public Health Laboratory | Richard Harrigan et al |
| EPI_ISL_463245 | Canada/BC_26624482/2020 | 20-Mar | North America/Canada/British Columbia | BCCDC Public Health Laboratory | BCCDC Public Health Laboratory | Richard Harrigan et al |
| EPI_ISL_463246 | Canada/BC_26625053/2020 | 20-Mar | North America/Canada/British Columbia | BCCDC Public Health Laboratory | BCCDC Public Health Laboratory | Richard Harrigan et al |
| EPI_ISL_463247 | Canada/BC_26628654/2020 | 20-Mar | North America/Canada/British Columbia | BCCDC Public Health Laboratory | BCCDC Public Health Laboratory | Richard Harrigan et al |
| EPI_ISL_463248 | Canada/BC_26646523/2020 | 20-Mar | North America/Canada/British Columbia | BCCDC Public Health Laboratory | BCCDC Public Health Laboratory | Richard Harrigan et al |
| EPI_ISL_463249 | Canada/BC_26649386/2020 | 20-Mar | North America/Canada/British Columbia | BCCDC Public Health Laboratory | BCCDC Public Health Laboratory | Richard Harrigan et al |
| EPI_ISL_463250 | Canada/BC_26650347/2020 | 20-Mar | North America/Canada/British Columbia | BCCDC Public Health Laboratory | BCCDC Public Health Laboratory | Richard Harrigan et al |
| EPI_ISL_463251 | Canada/BC_26654155/2020 | 20-Mar | North America/Canada/British Columbia | BCCDC Public Health Laboratory | BCCDC Public Health Laboratory | Richard Harrigan et al |
| EPI_ISL_463252 | Canada/BC_26656159/2020 | 20-Mar | North America/Canada/British Columbia | BCCDC Public Health Laboratory | BCCDC Public Health Laboratory | Richard Harrigan et al |
| EPI_ISL_463253 | Canada/BC_26659043/2020 | 20-Mar | North America/Canada/British Columbia | BCCDC Public Health Laboratory | BCCDC Public Health Laboratory | Richard Harrigan et al |
| EPI_ISL_463254 | Canada/BC_26693628/2020 | 20-Mar | North America/Canada/British Columbia | BCCDC Public Health Laboratory | BCCDC Public Health Laboratory | Richard Harrigan et al |
| EPI_ISL_463255 | Canada/BC_26943931/2020 | 20-Mar | North America/Canada/British Columbia | BCCDC Public Health Laboratory | BCCDC Public Health Laboratory | Richard Harrigan et al |
| EPI_ISL_463256 | Canada/BC_26964094/2020 | 20-Mar | North America/Canada/British Columbia | BCCDC Public Health Laboratory | BCCDC Public Health Laboratory | Richard Harrigan et al |
| EPI_ISL_463257 | Canada/BC_27007571/2020 | 20-Mar | North America/Canada/British Columbia | BCCDC Public Health Laboratory | BCCDC Public Health Laboratory | Richard Harrigan et al |
| EPI_ISL_463258 | Canada/BC_27017413/2020 | 20-Mar | North America/Canada/British Columbia | BCCDC Public Health Laboratory | BCCDC Public Health Laboratory | Richard Harrigan et al |
| EPI_ISL_463259 | Canada/BC_27081621/2020 | 20-Mar | North America/Canada/British Columbia | BCCDC Public Health Laboratory | BCCDC Public Health Laboratory | Richard Harrigan et al |
| EPI_ISL_463260 | Canada/BC_27121841/2020 | 20-Mar | North America/Canada/British Columbia | BCCDC Public Health Laboratory | BCCDC Public Health Laboratory | Richard Harrigan et al |
| EPI_ISL_463261 | Canada/BC_27180562/2020 | 20-Mar | North America/Canada/British Columbia | BCCDC Public Health Laboratory | BCCDC Public Health Laboratory | Richard Harrigan et al |
| EPI_ISL_463262 | Canada/BC_27222254/2020 | 20-Mar | North America/Canada/British Columbia | BCCDC Public Health Laboratory | BCCDC Public Health Laboratory | Richard Harrigan et al |

| | | | | | | |
|---|---|---|---|---|---|---|
| EPI_ISL_463263 | Canada/BC_27279444/2020 | 20-Mar | North America/Canada/British Columbia | BCCDC Public Health Laboratory | BCCDC Public Health Laboratory | Richard Harrigan et al |
| EPI_ISL_463264 | Canada/BC_27280425/2020 | 20-Mar | North America/Canada/British Columbia | BCCDC Public Health Laboratory | BCCDC Public Health Laboratory | Richard Harrigan et al |
| EPI_ISL_463265 | Canada/BC_27280641/2020 | 20-Mar | North America/Canada/British Columbia | BCCDC Public Health Laboratory | BCCDC Public Health Laboratory | Richard Harrigan et al |
| EPI_ISL_463266 | Canada/BC_27280776/2020 | 20-Mar | North America/Canada/British Columbia | BCCDC Public Health Laboratory | BCCDC Public Health Laboratory | Richard Harrigan et al |
| EPI_ISL_463267 | Canada/BC_27283916/2020 | 20-Mar | North America/Canada/British Columbia | BCCDC Public Health Laboratory | BCCDC Public Health Laboratory | Richard Harrigan et al |
| EPI_ISL_463268 | Canada/BC_27285211/2020 | 20-Mar | North America/Canada/British Columbia | BCCDC Public Health Laboratory | BCCDC Public Health Laboratory | Richard Harrigan et al |
| EPI_ISL_463270 | Canada/BC_27298990/2020 | 20-Mar | North America/Canada/British Columbia | BCCDC Public Health Laboratory | BCCDC Public Health Laboratory | Richard Harrigan et al |
| EPI_ISL_463271 | Canada/BC_27302671/2020 | 20-Mar | North America/Canada/British Columbia | BCCDC Public Health Laboratory | BCCDC Public Health Laboratory | Richard Harrigan et al |
| EPI_ISL_463272 | Canada/BC_27303787/2020 | 20-Mar | North America/Canada/British Columbia | BCCDC Public Health Laboratory | BCCDC Public Health Laboratory | Richard Harrigan et al |
| EPI_ISL_463273 | Canada/BC_27303944/2020 | 20-Mar | North America/Canada/British Columbia | BCCDC Public Health Laboratory | BCCDC Public Health Laboratory | Richard Harrigan et al |
| EPI_ISL_463274 | Canada/BC_27304064/2020 | 20-Mar | North America/Canada/British Columbia | BCCDC Public Health Laboratory | BCCDC Public Health Laboratory | Richard Harrigan et al |
| EPI_ISL_463275 | Canada/BC_27304554/2020 | 20-Mar | North America/Canada/British Columbia | BCCDC Public Health Laboratory | BCCDC Public Health Laboratory | Richard Harrigan et al |
| EPI_ISL_463276 | Canada/BC_27305318/2020 | 20-Mar | North America/Canada/British Columbia | BCCDC Public Health Laboratory | BCCDC Public Health Laboratory | Richard Harrigan et al |
| EPI_ISL_463280 | USA/LA-BIE-032/2020 | 2020-04-06 | North America/USA/Louisiana/ | Ochsner Health | Bioinfoexperts, LLC | Susanna L. Lamers et al |
| EPI_ISL_463288 | USA/LA-BIE-040/2020 | 2020-04-03 | North America/USA/Louisiana/ | Ochsner Health | Bioinfoexperts, LLC | Susanna L. Lamers et al |
| EPI_ISL_463289 | USA/LA-BIE-041/2020 | 2020-04-06 | North America/USA/Louisiana/ | Ochsner Health | Bioinfoexperts, LLC | Susanna L. Lamers et al |
| EPI_ISL_463292 | USA/LA-BIE-044/2020 | 2020-04-07 | North America/USA/Louisiana/ | Ochsner Health | Bioinfoexperts, LLC | Susanna L. Lamers et al |
| EPI_ISL_463345 | USA/WA-S855/2020 | 2020-04-24 | North America/USA/Washington/King C | Washington State Department of Health | Seattle Flu Study | Chu et al et al |
| EPI_ISL_463353 | USA/WA-S863/2020 | 2020-04-22 | North America/USA/Washington/Yakim | Washington State Department of Health | Seattle Flu Study | Chu et al et al |
| EPI_ISL_463364 | USA/WA-S874/2020 | 2020-04-16 | North America/USA/Washington/Pierce | Washington State Department of Health | Seattle Flu Study | Chu et al et al |
| EPI_ISL_463380 | USA/WA-S890/2020 | 2020-04-28 | North America/USA/Washington/Yakim | Washington State Department of Health | Seattle Flu Study | Chu et al et al |
| EPI_ISL_463386 | USA/WA-S896/2020 | 2020-04-28 | North America/USA/Washington/Snoho | Washington State Department of Health | Seattle Flu Study | Chu et al et al |
| EPI_ISL_463396 | USA/WA-S906/2020 | 2020-04-27 | North America/USA/Washington/Yakim | Washington State Department of Health | Seattle Flu Study | Chu et al et al |
| EPI_ISL_463405 | USA/WA-S915/2020 | 2020-04-27 | North America/USA/Washington/Snoho | Washington State Department of Health | Seattle Flu Study | Chu et al et al |

| | | | | | | |
|---|---|---|---|---|---|---|
| EPI_ISL_463411 | USA/WA-S921/2020 | 2020-04-25 | North America/USA/Washington/Yakim | Washington State Department of Health | Seattle Flu Study | Chu et al et al |
| EPI_ISL_463414 | USA/WA-S924/2020 | 2020-04-26 | North America/USA/Washington/Yakim | Washington State Department of Health | Seattle Flu Study | Chu et al et al |
| EPI_ISL_463418 | USA/WA-S928/2020 | 2020-04-23 | North America/USA/Washington/Pierce | Washington State Department of Health | Seattle Flu Study | Chu et al et al |
| EPI_ISL_463428 | USA/WA-S938/2020 | 2020-04-23 | North America/USA/Washington/Yakim | Washington State Department of Health | Seattle Flu Study | Chu et al et al |
| EPI_ISL_463433 | USA/WA-S943/2020 | 2020-04-27 | North America/USA/Washington/Snoho | Washington State Department of Health | Seattle Flu Study | Chu et al et al |
| EPI_ISL_463434 | USA/WA-S944/2020 | 2020-04-27 | North America/USA/Washington/Snoho | Washington State Department of Health | Seattle Flu Study | Chu et al et al |
| EPI_ISL_463444 | USA/WA-S954/2020 | 2020-05-01 | North America/USA/Washington/Yakim | Washington State Department of Health | Seattle Flu Study | Chu et al et al |
| EPI_ISL_463445 | USA/WA-S955/2020 | 2020-05-01 | North America/USA/Washington/Yakim | Washington State Department of Health | Seattle Flu Study | Chu et al et al |
| EPI_ISL_463457 | USA/WA-S967/2020 | 2020-05-01 | North America/USA/Washington/Skagit | Washington State Department of Health | Seattle Flu Study | Chu et al et al |
| EPI_ISL_463461 | USA/WA-S971/2020 | 2020-05-01 | North America/USA/Washington/Yakim | Washington State Department of Health | Seattle Flu Study | Chu et al et al |
| EPI_ISL_463467 | USA/WA-S977/2020 | 2020-05-01 | North America/USA/Washington/Yakim | Washington State Department of Health | Seattle Flu Study | Chu et al et al |
| EPI_ISL_463479 | USA/WA-S989/2020 | 2020-04-28 | North America/USA/Washington/Spoka | Washington State Department of Health | Seattle Flu Study | Chu et al et al |
| EPI_ISL_463486 | USA/WA-S996/2020 | 2020-04-28 | North America/USA/Washington/Skagit | Washington State Department of Health | Seattle Flu Study | Chu et al et al |
| EPI_ISL_463493 | USA/WA-S1003/2020 | 2020-04-29 | North America/USA/Washington/Yakim | Washington State Department of Health | Seattle Flu Study | Chu et al et al |
| EPI_ISL_463494 | USA/WA-S1004/2020 | 2020-04-29 | North America/USA/Washington/Yakim | Washington State Department of Health | Seattle Flu Study | Chu et al et al |
| EPI_ISL_463495 | USA/WA-S1005/2020 | 2020-04-29 | North America/USA/Washington/Yakim | Washington State Department of Health | Seattle Flu Study | Chu et al et al |
| EPI_ISL_463500 | USA/WA-S1010/2020 | 2020-04-29 | North America/USA/Washington/Skagit | Washington State Department of Health | Seattle Flu Study | Chu et al et al |
| EPI_ISL_463505 | USA/WA-S1015/2020 | 2020-04-30 | North America/USA/Washington/Yakim | Washington State Department of Health | Seattle Flu Study | Chu et al et al |
| EPI_ISL_463508 | USA/WA-S1018/2020 | 2020-04-30 | North America/USA/Washington/Yakim | Washington State Department of Health | Seattle Flu Study | Chu et al et al |
| EPI_ISL_463514 | USA/WA-S1024/2020 | 2020-05-01 | North America/USA/Washington/Yakim | Washington State Department of Health | Seattle Flu Study | Chu et al et al |
| EPI_ISL_463516 | USA/WA-S1026/2020 | 2020-04-30 | North America/USA/Washington/Yakim | Washington State Department of Health | Seattle Flu Study | Chu et al et al |
| EPI_ISL_463519 | USA/WA-S1029/2020 | 2020-05-01 | North America/USA/Washington/Yakim | Washington State Department of Health | Seattle Flu Study | Chu et al et al |
| EPI_ISL_463529 | USA/WA-S1039/2020 | 2020-05-03 | North America/USA/Washington/Yakim | Washington State Department of Health | Seattle Flu Study | Chu et al et al |
| EPI_ISL_463530 | USA/WA-S1040/2020 | 2020-05-03 | North America/USA/Washington/Yakim | Washington State Department of Health | Seattle Flu Study | Chu et al et al |
| EPI_ISL_463535 | USA/WA-S1045/2020 | 2020-05-03 | North America/USA/Washington/Pierce | Washington State Department of Health | Seattle Flu Study | Chu et al et al |
| EPI_ISL_463536 | USA/WA-S1046/2020 | 2020-05-03 | North America/USA/Washington/Pierce | Washington State Department of Health | Seattle Flu Study | Chu et al et al |
| EPI_ISL_463537 | USA/WA-S1047/2020 | 2020-05-03 | North America/USA/Washington/Yakim | Washington State Department of Health | Seattle Flu Study | Chu et al et al |
| EPI_ISL_463553 | USA/WA-S1063/2020 | 2020-04-30 | North America/USA/Washington/Yakim | Washington State Department of Health | Seattle Flu Study | Chu et al et al |
| EPI_ISL_463555 | USA/WA-S1065/2020 | 2020-04-29 | North America/USA/Washington/Yakim | Washington State Department of Health | Seattle Flu Study | Chu et al et al |
| EPI_ISL_463560 | USA/WA-S1070/2020 | 2020-04-30 | North America/USA/Washington/Yakim | Washington State Department of Health | Seattle Flu Study | Chu et al et al |
| EPI_ISL_463561 | USA/WA-S1071/2020 | 2020-04-30 | North America/USA/Washington/Yakim | Washington State Department of Health | Seattle Flu Study | Chu et al et al |

| | | | | | | |
|---|---|---|---|---|---|---|
| EPI_ISL_463575 | USA/WA-S1085/2020 | 2020-04-29 | North America/USA/Washington/King C | Washington State Department of Health | Seattle Flu Study | Chu et al et al |
| EPI_ISL_463576 | USA/WA-S1086/2020 | 2020-04-29 | North America/USA/Washington/Snoho | Washington State Department of Health | Seattle Flu Study | Chu et al et al |
| EPI_ISL_463580 | USA/WA-S1090/2020 | 2020-04-29 | North America/USA/Washington/Yakim | Washington State Department of Health | Seattle Flu Study | Chu et al et al |
| EPI_ISL_463586 | USA/WA-S1096/2020 | 2020-05-01 | North America/USA/Washington/Yakim | Washington State Department of Health | Seattle Flu Study | Chu et al et al |
| EPI_ISL_463587 | USA/WA-S1097/2020 | 2020-05-01 | North America/USA/Washington/Yakim | Washington State Department of Health | Seattle Flu Study | Chu et al et al |
| EPI_ISL_463588 | USA/WA-S1098/2020 | 2020-05-03 | North America/USA/Washington/Snoho | Washington State Department of Health | Seattle Flu Study | Chu et al et al |
| EPI_ISL_463589 | USA/WA-S1099/2020 | 2020-05-03 | North America/USA/Washington/Yakim | Washington State Department of Health | Seattle Flu Study | Chu et al et al |
| EPI_ISL_463592 | USA/WA-S1102/2020 | 2020-05-04 | North America/USA/Washington/Pierce | Washington State Department of Health | Seattle Flu Study | Chu et al et al |
| EPI_ISL_463610 | USA/WA-S1120/2020 | 2020-05-04 | North America/USA/Washington/Pierce | Washington State Department of Health | Seattle Flu Study | Chu et al et al |
| EPI_ISL_463614 | USA/WA-S1124/2020 | 2020-05-04 | North America/USA/Washington/Yakim | Washington State Department of Health | Seattle Flu Study | Chu et al et al |
| EPI_ISL_463619 | USA/WA-S1129/2020 | 2020-05-04 | North America/USA/Washington/Pierce | Washington State Department of Health | Seattle Flu Study | Chu et al et al |
| EPI_ISL_463625 | USA/WA-S1135/2020 | 2020-05-04 | North America/USA/Washington/Snoho | Washington State Department of Health | Seattle Flu Study | Chu et al et al |
| EPI_ISL_463626 | USA/WA-S1136/2020 | 2020-05-04 | North America/USA/Washington/Pierce | Washington State Department of Health | Seattle Flu Study | Chu et al et al |
| EPI_ISL_463635 | USA/WA-S1145/2020 | 2020-05-04 | North America/USA/Washington/Yakim | Washington State Department of Health | Seattle Flu Study | Chu et al et al |
| EPI_ISL_463637 | USA/WA-S1147/2020 | 2020-05-05 | North America/USA/Washington/Yakim | Washington State Department of Health | Seattle Flu Study | Chu et al et al |
| EPI_ISL_463638 | USA/WA-S1148/2020 | 2020-05-05 | North America/USA/Washington/Snoho | Washington State Department of Health | Seattle Flu Study | Chu et al et al |
| EPI_ISL_463651 | USA/WA-S1161/2020 | 2020-05-06 | North America/USA/Washington/Yakim | Washington State Department of Health | Seattle Flu Study | Chu et al et al |
| EPI_ISL_463657 | USA/WA-S1167/2020 | 2020-05-05 | North America/USA/Washington/Yakim | Washington State Department of Health | Seattle Flu Study | Chu et al et al |
| EPI_ISL_463659 | USA/WA-S1169/2020 | 2020-05-05 | North America/USA/Washington/Yakim | Washington State Department of Health | Seattle Flu Study | Chu et al et al |
| EPI_ISL_463660 | USA/WA-S1170/2020 | 2020-05-04 | North America/USA/Washington/Pierce | Washington State Department of Health | Seattle Flu Study | Chu et al et al |
| EPI_ISL_463672 | USA/WA-S1182/2020 | 2020-05-06 | North America/USA/Washington/Yakim | Washington State Department of Health | Seattle Flu Study | Chu et al et al |
| EPI_ISL_463673 | USA/WA-S1183/2020 | 2020-05-05 | North America/USA/Washington/Snoho | Washington State Department of Health | Seattle Flu Study | Chu et al et al |
| EPI_ISL_463674 | USA/WA-S1184/2020 | 2020-05-06 | North America/USA/Washington/Snoho | Washington State Department of Health | Seattle Flu Study | Chu et al et al |
| EPI_ISL_463679 | USA/WA-S1189/2020 | 2020-05-05 | North America/USA/Washington/King C | Washington State Department of Health | Seattle Flu Study | Chu et al et al |
| EPI_ISL_463696 | USA/WA-S1206/2020 | 2020-05-06 | North America/USA/Washington/Yakim | Washington State Department of Health | Seattle Flu Study | Chu et al et al |
| EPI_ISL_463702 | USA/WA-S1212/2020 | 2020-05-07 | North America/USA/Washington/Yakim | Washington State Department of Health | Seattle Flu Study | Chu et al et al |

| | | | | | | |
|---|---|---|---|---|---|---|
| EPI_ISL_463889 | Shaoxing/01/2020 | 2020-01-27 | Asia/China/Zhejiang/Keqiao | Shaoxing Center for Disease Control and Prevention | Department of Pathology and Laboratory Medicine, University of California Los Angeles | Jinkun Chen et al |
| EPI_ISL_463894 | Shaoxing/02/2020 | 2020-01-30 | Asia/China/Zhejiang/Keqiao | Shaoxing Center for Disease Control and Prevention | Department of Pathology and Laboratory Medicine, University of California Los Angeles | Jinkun Chen et al |
| EPI_ISL_463895 | Shaoxing/03/2020 | 2020-01-30 | Asia/China/Zhejiang/Keqiao | Shaoxing Center for Disease Control and Prevention | Department of Pathology and Laboratory Medicine, University of California Los Angeles | Jinkun Chen et al |
| EPI_ISL_463896 | Shaoxing/04/2020 | 2020-01-31 | Asia/China/Zhejiang/Keqiao | Shaoxing Center for Disease Control and Prevention | Department of Pathology and Laboratory Medicine, University of California Los Angeles | Jinkun Chen et al |
| EPI_ISL_463897 | Shaoxing/05/2020 | 2020-01-31 | Asia/China/Zhejiang/Keqiao | Shaoxing Center for Disease Control and Prevention | Department of Pathology and Laboratory Medicine, University of California Los Angeles | Jinkun Chen et al |

| | | | | | | |
|---|---|---|---|---|---|---|
| EPI_ISL_463898 | Shaoxing/06/2020 | 2020-01-31 | Asia/China/Zhejiang/Yuecheng | Shaoxing Center for Disease Control and Prevention | Department of Pathology and Laboratory Medicine, University of California Los Angeles | Jinkun Chen et al |
| EPI_ISL_463899 | Shaoxing/07/2020 | 2020-01-30 | Asia/China/Zhejiang/Keqiao | Shaoxing Center for Disease Control and Prevention | Department of Pathology and Laboratory Medicine, University of California Los Angeles | Jinkun Chen et al |
| EPI_ISL_463900 | Shaoxing/08/2020 | 2020-02-07 | Asia/China/Zhejiang/Yuecheng | Shaoxing Center for Disease Control and Prevention | Department of Pathology and Laboratory Medicine, University of California Los Angeles | Jinkun Chen et al |
| EPI_ISL_463901 | Shaoxing/09/2020 | 2020-02-05 | Asia/China/Zhejiang/Shangyu | Shaoxing Center for Disease Control and Prevention | Department of Pathology and Laboratory Medicine, University of California Los Angeles | Jinkun Chen et al |
| EPI_ISL_463902 | Shaoxing/10/2020 | 2020-02-06 | Asia/China/Zhejiang/Shangyu | Shaoxing Center for Disease Control and Prevention | Department of Pathology and Laboratory Medicine, University of California Los Angeles | Jinkun Chen et al |
| EPI_ISL_463903 | Canada/QC_AC4/2020 | 2020-04-04 | North America/Canada/Quebec/ | Laboratoire de microbiologie, Hopital de Verdun | Smith Laboratory, Centre de Recherche CHU Sainte-Justine | Martin Smith et al |

| | | | | | | |
|---|---|---|---|---|---|---|
| EPI_ISL_463904 | Canada/QC_AC6.1/2020 | 2020-04-05 | North America/Canada/Quebec/ | Laboratoire de microbiologie, Hopital de Verdun | Smith Laboratory, Centre de Recherche CHU Sainte-Justine | Martin Smith et al |
| EPI_ISL_463905 | Canada/QC_AE4/2020 | 2020-04-08 | North America/Canada/Quebec/ | Laboratoire de microbiologie, Hopital de Verdun | Smith Laboratory, Centre de Recherche CHU Sainte-Justine | Martin Smith et al |
| EPI_ISL_463906 | Canada/QC_AE6/2020 | 2020-04-06 | North America/Canada/Quebec/ | Laboratoire de microbiologie, Hopital de Verdun | Smith Laboratory, Centre de Recherche CHU Sainte-Justine | Martin Smith et al |
| EPI_ISL_463907 | Canada/QC_AE8/2020 | 2020-04-07 | North America/Canada/Quebec/ | Laboratoire de microbiologie, Hopital de Verdun | Smith Laboratory, Centre de Recherche CHU Sainte-Justine | Martin Smith et al |
| EPI_ISL_463908 | Canada/QC_AE9/2020 | 2020-04-06 | North America/Canada/Quebec/ | Laboratoire de microbiologie, Hopital de Verdun | Smith Laboratory, Centre de Recherche CHU Sainte-Justine | Martin Smith et al |
| EPI_ISL_463909 | Canada/QC_AF1/2020 | 2020-04-07 | North America/Canada/Quebec/ | Laboratoire de microbiologie, Hopital de Verdun | Smith Laboratory, Centre de Recherche CHU Sainte-Justine | Martin Smith et al |
| EPI_ISL_463910 | Canada/QC_AF2/2020 | 2020-04-05 | North America/Canada/Quebec/ | Laboratoire de microbiologie, Hopital de Verdun | Smith Laboratory, Centre de Recherche CHU Sainte-Justine | Martin Smith et al |
| EPI_ISL_463911 | Canada/QC_AF4/2020 | 2020-04-04 | North America/Canada/Quebec/ | Laboratoire de microbiologie, Hopital de Verdun | Smith Laboratory, Centre de Recherche CHU Sainte-Justine | Martin Smith et al |
| EPI_ISL_463912 | Canada/QC_AF6/2020 | 2020-04-06 | North America/Canada/Quebec/ | Laboratoire de microbiologie, Hopital de Verdun | Smith Laboratory, Centre de Recherche CHU Sainte-Justine | Martin Smith et al |
| EPI_ISL_463913 | Canada/QC_AG4/2020 | 2020-04-11 | North America/Canada/Quebec/ | Laboratoire de microbiologie, Hopital de Verdun | Smith Laboratory, Centre de Recherche CHU Sainte-Justine | Martin Smith et al |

| | | | | | | |
|---|---|---|---|---|---|---|
| EPI_ISL_463914 | Canada/QC_AH2/2020 | 2020-04-13 | North America/Canada/Quebec/ | Laboratoire de microbiologie, Hopital de Verdun | Smith Laboratory, Centre de Recherche CHU Sainte-Justine | Martin Smith et al |
| EPI_ISL_463915 | Canada/QC_AH4/2020 | 2020-04-08 | North America/Canada/Quebec/ | Laboratoire de microbiologie, Hopital de Verdun | Smith Laboratory, Centre de Recherche CHU Sainte-Justine | Martin Smith et al |
| EPI_ISL_463916 | Canada/QC_AI3/2020 | 2020-04-10 | North America/Canada/Quebec/ | Laboratoire de microbiologie, Hopital de Verdun | Smith Laboratory, Centre de Recherche CHU Sainte-Justine | Martin Smith et al |
| EPI_ISL_463917 | Canada/QC_AI5/2020 | 2020-04-09 | North America/Canada/Quebec/ | Laboratoire de microbiologie, Hopital de Verdun | Smith Laboratory, Centre de Recherche CHU Sainte-Justine | Martin Smith et al |
| EPI_ISL_463918 | Canada/QC_AI9/2020 | 2020-04-07 | North America/Canada/Quebec/ | Laboratoire de microbiologie, Hopital de Verdun | Smith Laboratory, Centre de Recherche CHU Sainte-Justine | Martin Smith et al |
| EPI_ISL_463919 | Canada/QC_AK5/2020 | 2020-04-08 | North America/Canada/Quebec/ | Laboratoire de microbiologie, Hopital de Verdun | Smith Laboratory, Centre de Recherche CHU Sainte-Justine | Martin Smith et al |
| EPI_ISL_463920 | Canada/QC_AO3/2020 | 2020-04-09 | North America/Canada/Quebec/ | Laboratoire de microbiologie, Hopital de Verdun | Smith Laboratory, Centre de Recherche CHU Sainte-Justine | Martin Smith et al |
| EPI_ISL_463921 | Canada/QC_AO6/2020 | 2020-04-08 | North America/Canada/Quebec/ | Laboratoire de microbiologie, Hopital de Verdun | Smith Laboratory, Centre de Recherche CHU Sainte-Justine | Martin Smith et al |
| EPI_ISL_463922 | Canada/QC_AO7/2020 | 2020-04-09 | North America/Canada/Quebec/ | Laboratoire de microbiologie, Hopital de Verdun | Smith Laboratory, Centre de Recherche CHU Sainte-Justine | Martin Smith et al |
| EPI_ISL_463923 | Canada/QC_AO9/2020 | 2020-04-09 | North America/Canada/Quebec/ | Laboratoire de microbiologie, Hopital de Verdun | Smith Laboratory, Centre de Recherche CHU Sainte-Justine | Martin Smith et al |

| | | | | | | |
|---|---|---|---|---|---|---|
| EPI_ISL_463924 | Canada/QC_AP1/2020 | 2020-04-06 | North America/Canada/Quebec/ | Laboratoire de microbiologie, Hopital de Verdun | Smith Laboratory, Centre de Recherche CHU Sainte-Justine | Martin Smith et al |
| EPI_ISL_463925 | Canada/QC_AP2/2020 | 2020-04-07 | North America/Canada/Quebec/ | Laboratoire de microbiologie, Hopital de Verdun | Smith Laboratory, Centre de Recherche CHU Sainte-Justine | Martin Smith et al |
| EPI_ISL_463926 | Canada/QC_AP6/2020 | 2020-04-14 | North America/Canada/Quebec/ | Laboratoire de microbiologie, Hopital de Verdun | Smith Laboratory, Centre de Recherche CHU Sainte-Justine | Martin Smith et al |
| EPI_ISL_463927 | Canada/QC_AQ1/2020 | 2020-04-02 | North America/Canada/Quebec/ | Laboratoire de microbiologie, Hopital de Verdun | Smith Laboratory, Centre de Recherche CHU Sainte-Justine | Martin Smith et al |
| EPI_ISL_463928 | Canada/QC_AQ4/2020 | 2020-04-01 | North America/Canada/Quebec/ | Laboratoire de microbiologie, Hopital de Verdun | Smith Laboratory, Centre de Recherche CHU Sainte-Justine | Martin Smith et al |
| EPI_ISL_463929 | Canada/QC_AQ5/2020 | 2020-04-03 | North America/Canada/Quebec/ | Laboratoire de microbiologie, Hopital de Verdun | Smith Laboratory, Centre de Recherche CHU Sainte-Justine | Martin Smith et al |
| EPI_ISL_463930 | Canada/QC_AQ6/2020 | 2020-04-02 | North America/Canada/Quebec/ | Laboratoire de microbiologie, Hopital de Verdun | Smith Laboratory, Centre de Recherche CHU Sainte-Justine | Martin Smith et al |
| EPI_ISL_463931 | Canada/QC_AR3/2020 | 2020-04-04 | North America/Canada/Quebec/ | Laboratoire de microbiologie, Hopital de Verdun | Smith Laboratory, Centre de Recherche CHU Sainte-Justine | Martin Smith et al |
| EPI_ISL_463932 | Canada/QC_AR5/2020 | 2020-04-04 | North America/Canada/Quebec/ | Laboratoire de microbiologie, Hopital de Verdun | Smith Laboratory, Centre de Recherche CHU Sainte-Justine | Martin Smith et al |
| EPI_ISL_463933 | Canada/QC_AR6/2020 | 2020-04-01 | North America/Canada/Quebec/ | Laboratoire de microbiologie, Hopital de Verdun | Smith Laboratory, Centre de Recherche CHU Sainte-Justine | Martin Smith et al |

| | | | | | | |
|---|---|---|---|---|---|---|
| EPI_ISL_463934 | Canada/QC_AR9/2020 | 2020-04-05 | North America/Canada/Quebec/ | Laboratoire de microbiologie, Hopital de Verdun | Smith Laboratory, Centre de Recherche CHU Sainte-Justine | Martin Smith et al |
| EPI_ISL_463935 | Canada/QC_AS1/2020 | 2020-04-02 | North America/Canada/Quebec/ | Laboratoire de microbiologie, Hopital de Verdun | Smith Laboratory, Centre de Recherche CHU Sainte-Justine | Martin Smith et al |
| EPI_ISL_463936 | Canada/QC_AS4/2020 | 2020-04-02 | North America/Canada/Quebec/ | Laboratoire de microbiologie, Hopital de Verdun | Smith Laboratory, Centre de Recherche CHU Sainte-Justine | Martin Smith et al |
| EPI_ISL_463937 | Canada/QC_AS7/2020 | 2020-04-03 | North America/Canada/Quebec/ | Laboratoire de microbiologie, Hopital de Verdun | Smith Laboratory, Centre de Recherche CHU Sainte-Justine | Martin Smith et al |
| EPI_ISL_463938 | Canada/QC_AS8/2020 | 2020-04-04 | North America/Canada/Quebec/ | Laboratoire de microbiologie, Hopital de Verdun | Smith Laboratory, Centre de Recherche CHU Sainte-Justine | Martin Smith et al |
| EPI_ISL_463939 | Canada/QC_AS9/2020 | 2020-04-03 | North America/Canada/Quebec/ | Laboratoire de microbiologie, Hopital de Verdun | Smith Laboratory, Centre de Recherche CHU Sainte-Justine | Martin Smith et al |
| EPI_ISL_463940 | Canada/QC_AT3/2020 | 2020-03-31 | North America/Canada/Quebec/ | Laboratoire de microbiologie, Hopital de Verdun | Smith Laboratory, Centre de Recherche CHU Sainte-Justine | Martin Smith et al |
| EPI_ISL_463941 | Canada/QC_AT6/2020 | 2020-04-06 | North America/Canada/Quebec/ | Laboratoire de microbiologie, Hopital de Verdun | Smith Laboratory, Centre de Recherche CHU Sainte-Justine | Martin Smith et al |
| EPI_ISL_463942 | Canada/QC_AT7/2020 | 2020-04-03 | North America/Canada/Quebec/ | Laboratoire de microbiologie, Hopital de Verdun | Smith Laboratory, Centre de Recherche CHU Sainte-Justine | Martin Smith et al |
| EPI_ISL_463943 | Canada/QC_AT8/2020 | 2020-04-04 | North America/Canada/Quebec/ | Laboratoire de microbiologie, Hopital de Verdun | Smith Laboratory, Centre de Recherche CHU Sainte-Justine | Martin Smith et al |

| | | | | | | |
|---|---|---|---|---|---|---|
| EPI_ISL_463944 | Canada/QC_AT9/2020 | 2020-04-05 | North America/Canada/Quebec/ | Laboratoire de microbiologie, Hopital de Verdun | Smith Laboratory, Centre de Recherche CHU Sainte-Justine | Martin Smith et al |
| EPI_ISL_463945 | Canada/QC_AU2/2020 | 2020-04-08 | North America/Canada/Quebec/ | Laboratoire de microbiologie, Hopital de Verdun | Smith Laboratory, Centre de Recherche CHU Sainte-Justine | Martin Smith et al |
| EPI_ISL_463946 | Canada/QC_AU7/2020 | 2020-04-03 | North America/Canada/Quebec/ | Laboratoire de microbiologie, Hopital de Verdun | Smith Laboratory, Centre de Recherche CHU Sainte-Justine | Martin Smith et al |
| EPI_ISL_463947 | Canada/QC_AU8/2020 | 2020-04-04 | North America/Canada/Quebec/ | Laboratoire de microbiologie, Hopital de Verdun | Smith Laboratory, Centre de Recherche CHU Sainte-Justine | Martin Smith et al |
| EPI_ISL_463948 | Canada/QC_AU9/2020 | 2020-04-06 | North America/Canada/Quebec/ | Laboratoire de microbiologie, Hopital de Verdun | Smith Laboratory, Centre de Recherche CHU Sainte-Justine | Martin Smith et al |
| EPI_ISL_463949 | Canada/QC_AV4/2020 | 2020-04-04 | North America/Canada/Quebec/ | Laboratoire de microbiologie, Hopital de Verdun | Smith Laboratory, Centre de Recherche CHU Sainte-Justine | Martin Smith et al |
| EPI_ISL_463950 | Canada/QC_AV6/2020 | 2020-04-04 | North America/Canada/Quebec/ | Laboratoire de microbiologie, Hopital de Verdun | Smith Laboratory, Centre de Recherche CHU Sainte-Justine | Martin Smith et al |
| EPI_ISL_463951 | Canada/QC_AV9/2020 | 2020-04-04 | North America/Canada/Quebec/ | Laboratoire de microbiologie, Hopital de Verdun | Smith Laboratory, Centre de Recherche CHU Sainte-Justine | Martin Smith et al |
| EPI_ISL_463952 | Canada/QC_AW2/2020 | 2020-04-01 | North America/Canada/Quebec/ | Laboratoire de microbiologie, Hopital de Verdun | Smith Laboratory, Centre de Recherche CHU Sainte-Justine | Martin Smith et al |
| EPI_ISL_463953 | Canada/QC_AX6/2020 | 2020-04-13 | North America/Canada/Quebec/ | Laboratoire de microbiologie, Hopital de Verdun | Smith Laboratory, Centre de Recherche CHU Sainte-Justine | Martin Smith et al |

| | | | | | | |
|---|---|---|---|---|---|---|
| EPI_ISL_463954 | Canada/QC_AY1/2020 | 2020-04-12 | North America/Canada/Quebec/ | Laboratoire de microbiologie, Hopital de Verdun | Smith Laboratory, Centre de Recherche CHU Sainte-Justine | Martin Smith et al |
| EPI_ISL_463955 | Canada/QC_AY3/2020 | 2020-04-12 | North America/Canada/Quebec/ | Laboratoire de microbiologie, Hopital de Verdun | Smith Laboratory, Centre de Recherche CHU Sainte-Justine | Martin Smith et al |
| EPI_ISL_463956 | Canada/QC_AY8/2020 | 2020-04-12 | North America/Canada/Quebec/ | Laboratoire de microbiologie, Hopital de Verdun | Smith Laboratory, Centre de Recherche CHU Sainte-Justine | Martin Smith et al |
| EPI_ISL_463957 | Canada/QC_AY9/2020 | 2020-04-11 | North America/Canada/Quebec/ | Laboratoire de microbiologie, Hopital de Verdun | Smith Laboratory, Centre de Recherche CHU Sainte-Justine | Martin Smith et al |
| EPI_ISL_463958 | Canada/QC_AZ1/2020 | 2020-04-12 | North America/Canada/Quebec/ | Laboratoire de microbiologie, Hopital de Verdun | Smith Laboratory, Centre de Recherche CHU Sainte-Justine | Martin Smith et al |
| EPI_ISL_463959 | Canada/QC_AZ3/2020 | 2020-04-11 | North America/Canada/Quebec/ | Laboratoire de microbiologie, Hopital de Verdun | Smith Laboratory, Centre de Recherche CHU Sainte-Justine | Martin Smith et al |
| EPI_ISL_463960 | Canada/QC_AZ8/2020 | 2020-04-12 | North America/Canada/Quebec/ | Laboratoire de microbiologie, Hopital de Verdun | Smith Laboratory, Centre de Recherche CHU Sainte-Justine | Martin Smith et al |
| EPI_ISL_463961 | Canada/QC_AZ9/2020 | 2020-04-14 | North America/Canada/Quebec/ | Laboratoire de microbiologie, Hopital de Verdun | Smith Laboratory, Centre de Recherche CHU Sainte-Justine | Martin Smith et al |
| EPI_ISL_463962 | Canada/QC_BA5/2020 | 2020-04-07 | North America/Canada/Quebec/ | Laboratoire de microbiologie, Hopital de Verdun | Smith Laboratory, Centre de Recherche CHU Sainte-Justine | Martin Smith et al |
| EPI_ISL_463963 | Canada/QC_BA6/2020 | 2020-04-13 | North America/Canada/Quebec/ | Laboratoire de microbiologie, Hopital de Verdun | Smith Laboratory, Centre de Recherche CHU Sainte-Justine | Martin Smith et al |

| | | | | | | |
|---|---|---|---|---|---|---|
| EPI_ISL_463964 | Canada/QC_BB1/2020 | 2020-04-14 | North America/Canada/Quebec/ | Laboratoire de microbiologie, Hopital de Verdun | Smith Laboratory, Centre de Recherche CHU Sainte-Justine | Martin Smith et al |
| EPI_ISL_463965 | Canada/QC_BB7/2020 | 2020-04-13 | North America/Canada/Quebec/ | Laboratoire de microbiologie, Hopital de Verdun | Smith Laboratory, Centre de Recherche CHU Sainte-Justine | Martin Smith et al |
| EPI_ISL_463966 | Canada/QC_BC1/2020 | 2020-04-14 | North America/Canada/Quebec/ | Laboratoire de microbiologie, Hopital de Verdun | Smith Laboratory, Centre de Recherche CHU Sainte-Justine | Martin Smith et al |
| EPI_ISL_463967 | Canada/QC_BC3/2020 | 2020-04-09 | North America/Canada/Quebec/ | Laboratoire de microbiologie, Hopital de Verdun | Smith Laboratory, Centre de Recherche CHU Sainte-Justine | Martin Smith et al |
| EPI_ISL_463968 | Canada/QC_BC5/2020 | 2020-04-09 | North America/Canada/Quebec/ | Laboratoire de microbiologie, Hopital de Verdun | Smith Laboratory, Centre de Recherche CHU Sainte-Justine | Martin Smith et al |
| EPI_ISL_463969 | Canada/QC_BD5/2020 | 2020-04-13 | North America/Canada/Quebec/ | Laboratoire de microbiologie, Hopital de Verdun | Smith Laboratory, Centre de Recherche CHU Sainte-Justine | Martin Smith et al |
| EPI_ISL_463970 | Canada/ON_MU-S1/2020 | 2020-03-13 | North America/Canada/Ontario/Toront | Toronto Invasive Bacterial Diseases Network | McMaster University | Allison McGeer et al |
| EPI_ISL_463971 | Canada/ON_MU-S13/2020 | 2020-03-14 | North America/Canada/Ontario/Toront | Toronto Invasive Bacterial Diseases Network | McMaster University | Allison McGeer et al |
| EPI_ISL_463972 | Canada/ON_MU-S181/2020 | 2020-03-24 | North America/Canada/Ontario/Toront | Toronto Invasive Bacterial Diseases Network | McMaster University | Allison McGeer et al |
| EPI_ISL_463973 | Canada/ON_MU-S212/2020 | 2020-03-26 | North America/Canada/Ontario/Toront | Toronto Invasive Bacterial Diseases Network | McMaster University | Allison McGeer et al |
| EPI_ISL_463974 | Canada/ON_MU-S218/2020 | 2020-03-26 | North America/Canada/Ontario/Toront | Toronto Invasive Bacterial Diseases Network | McMaster University | Allison McGeer et al |
| EPI_ISL_463975 | Canada/ON_MU-S26/2020 | 2020-03-16 | North America/Canada/Ontario/Toront | Toronto Invasive Bacterial Diseases Network | McMaster University | Allison McGeer et al |
| EPI_ISL_463976 | Canada/ON_MU-S284/2020 | 2020-03-28 | North America/Canada/Ontario/Toront | Toronto Invasive Bacterial Diseases Network | McMaster University | Allison McGeer et al |
| EPI_ISL_463977 | Canada/ON_MU-S29/2020 | 2020-03-16 | North America/Canada/Ontario/Toront | Toronto Invasive Bacterial Diseases Network | McMaster University | Allison McGeer et al |

| | | | | | | |
|---|---|---|---|---|---|---|
| EPI_ISL_463978 | Canada/ON_MU-S305/2020 | 2020-03-28 | North America/Canada/Ontario/Toronto | Toronto Invasive Bacterial Diseases Network | McMaster University | Allison McGeer et al |
| EPI_ISL_463979 | Canada/ON_MU-S306/2020 | 2020-03-28 | North America/Canada/Ontario/Toronto | Toronto Invasive Bacterial Diseases Network | McMaster University | Allison McGeer et al |
| EPI_ISL_463980 | Canada/ON_MU-S312/2020 | 2020-03-29 | North America/Canada/Ontario/Toronto | Toronto Invasive Bacterial Diseases Network | McMaster University | Allison McGeer et al |
| EPI_ISL_463981 | Canada/ON_MU-S328/2020 | 2020-03-29 | North America/Canada/Ontario/Toronto | Toronto Invasive Bacterial Diseases Network | McMaster University | Allison McGeer et al |
| EPI_ISL_463982 | Canada/ON_MU-S349/2020 | 2020-03-29 | North America/Canada/Ontario/Toronto | Toronto Invasive Bacterial Diseases Network | McMaster University | Allison McGeer et al |
| EPI_ISL_463983 | Canada/ON_MU-S350/2020 | 2020-03-29 | North America/Canada/Ontario/Toronto | Toronto Invasive Bacterial Diseases Network | McMaster University | Allison McGeer et al |
| EPI_ISL_463984 | Canada/ON_MU-S40/2020 | 2020-03-17 | North America/Canada/Ontario/Toronto | Toronto Invasive Bacterial Diseases Network | McMaster University | Allison McGeer et al |
| EPI_ISL_463985 | Canada/ON_MU-S401/2020 | 2020-03-30 | North America/Canada/Ontario/Toronto | Toronto Invasive Bacterial Diseases Network | McMaster University | Allison McGeer et al |
| EPI_ISL_463986 | Canada/ON_MU-S461/2020 | 2020-04-01 | North America/Canada/Ontario/Toronto | Toronto Invasive Bacterial Diseases Network | McMaster University | Allison McGeer et al |
| EPI_ISL_463987 | Canada/ON-MU-S49/2020 | 2020-03-17 | North America/Canada/Ontario/Toronto | Toronto Invasive Bacterial Diseases Network | McMaster University | Allison McGeer et al |
| EPI_ISL_463988 | Canada/ON_MU-S5/2020 | 2020-03-13 | North America/Canada/Ontario/Toronto | Toronto Invasive Bacterial Diseases Network | McMaster University | Allison McGeer et al |
| EPI_ISL_463989 | Canada/ON_MU-S51/2020 | 2020-03-17 | North America/Canada/Ontario/Toronto | Toronto Invasive Bacterial Diseases Network | McMaster University | Allison McGeer et al |
| EPI_ISL_463990 | Canada/ON_MU-S53/2020 | 2020-03-18 | North America/Canada/Ontario/Toronto | Toronto Invasive Bacterial Diseases Network | McMaster University | Allison McGeer et al |
| EPI_ISL_463991 | Canada/ON_MU-S63/2020 | 2020-03-19 | North America/Canada/Ontario/Toronto | Toronto Invasive Bacterial Diseases Network | McMaster University | Allison McGeer et al |
| EPI_ISL_463992 | Canada/ON_MU-S679/2020 | 2020-04-09 | North America/Canada/Ontario/Toronto | Toronto Invasive Bacterial Diseases Network | McMaster University | Allison McGeer et al |
| EPI_ISL_463993 | Canada/ON_MU-S7/2020 | 2020-03-13 | North America/Canada/Ontario/Toronto | Toronto Invasive Bacterial Diseases Network | McMaster University | Allison McGeer et al |
| EPI_ISL_463994 | Canada/ON_MU-S82/2020 | 2020-03-19 | North America/Canada/Ontario/Toronto | Toronto Invasive Bacterial Diseases Network | McMaster University | Allison McGeer et al |
| EPI_ISL_463995 | Canada/ON-UHTC_0078/2020 | 2020-04-17 | North America/Canada/Ontario/ | Unity Health Toronto | Ontario Institute for Cancer Research | Ramzi Fattouh et al |
| EPI_ISL_463996 | Canada/ON-UHTC_0071/2020 | 2020-04-23 | North America/Canada/Ontario/ | Unity Health Toronto | Ontario Institute for Cancer Research | Ramzi Fattouh et al |

| | | | | | | |
|---|---|---|---|---|---|---|
| EPI_ISL_463997 | Canada/ON-UHTC_0067/2020 | 2020-04-10 | North America/Canada/Ontario/ | Unity Health Toronto | Ontario Institute for Cancer Research | Ramzi Fattouh et al |
| EPI_ISL_463998 | Canada/ON-UHTC_0082/2020 | 2020-04-15 | North America/Canada/Ontario/ | Unity Health Toronto | Ontario Institute for Cancer Research | Ramzi Fattouh et al |
| EPI_ISL_463999 | Canada/ON-UHTC_0075/2020 | 2020-04-11 | North America/Canada/Ontario/ | Unity Health Toronto | Ontario Institute for Cancer Research | Ramzi Fattouh et al |
| EPI_ISL_464000 | Canada/ON-UHTC_0023/2020 | 2020-03-31 | North America/Canada/Ontario/ | Unity Health Toronto | Ontario Institute for Cancer Research | Ramzi Fattouh et al |
| EPI_ISL_464001 | Canada/ON-UHTC_0061/2020 | 2020-04-06 | North America/Canada/Ontario/ | Unity Health Toronto | Ontario Institute for Cancer Research | Ramzi Fattouh et al |
| EPI_ISL_464002 | Canada/ON-UHTC_0008/2020 | 2020-03-27 | North America/Canada/Ontario/ | Unity Health Toronto | Ontario Institute for Cancer Research | Ramzi Fattouh et al |
| EPI_ISL_464003 | Canada/ON-UHTC_0019/2020 | 2020-03-30 | North America/Canada/Ontario/ | Unity Health Toronto | Ontario Institute for Cancer Research | Ramzi Fattouh et al |
| EPI_ISL_464004 | Canada/ON-UHTC_0053/2020 | 2020-04-07 | North America/Canada/Ontario/ | Unity Health Toronto | Ontario Institute for Cancer Research | Ramzi Fattouh et al |
| EPI_ISL_464005 | Canada/ON-UHTC_0025/2020 | 2020-04-01 | North America/Canada/Ontario/ | Unity Health Toronto | Ontario Institute for Cancer Research | Ramzi Fattouh et al |
| EPI_ISL_464006 | Canada/ON-UHTC_0016/2020 | 2020-03-29 | North America/Canada/Ontario/ | Unity Health Toronto | Ontario Institute for Cancer Research | Ramzi Fattouh et al |
| EPI_ISL_464007 | Canada/ON-UHTC_0063/2020 | 2020-04-12 | North America/Canada/Ontario/ | Unity Health Toronto | Ontario Institute for Cancer Research | Ramzi Fattouh et al |
| EPI_ISL_464008 | Canada/ON-UHTC_0068/2020 | 2020-04-28 | North America/Canada/Ontario/ | Unity Health Toronto | Ontario Institute for Cancer Research | Ramzi Fattouh et al |
| EPI_ISL_464009 | Canada/ON-UHTC_0003/2020 | 2020-03-18 | North America/Canada/Ontario/ | Unity Health Toronto | Ontario Institute for Cancer Research | Ramzi Fattouh et al |

| | | | | | | |
|---|---|---|---|---|---|---|
| EPI_ISL_464010 | Canada/ON-UHTC_0028/2020 | 2020-04-02 | North America/Canada/Ontario/ | Unity Health Toronto | Ontario Institute for Cancer Research | Ramzi Fattouh et al |
| EPI_ISL_464011 | Canada/ON-UHTC_0091/2020 | 2020-04-11 | North America/Canada/Ontario/ | Unity Health Toronto | Ontario Institute for Cancer Research | Ramzi Fattouh et al |
| EPI_ISL_464012 | Canada/ON-UHTC_0047/2020 | 2020-04-02 | North America/Canada/Ontario/ | Unity Health Toronto | Ontario Institute for Cancer Research | Ramzi Fattouh et al |
| EPI_ISL_464013 | Canada/ON-UHTC_0049/2020 | 2020-04-05 | North America/Canada/Ontario/ | Unity Health Toronto | Ontario Institute for Cancer Research | Ramzi Fattouh et al |
| EPI_ISL_464014 | Canada/ON-UHTC_0048/2020 | 2020-04-04 | North America/Canada/Ontario/ | Unity Health Toronto | Ontario Institute for Cancer Research | Ramzi Fattouh et al |
| EPI_ISL_464015 | Canada/ON-UHTC_0006/2020 | 2020-03-25 | North America/Canada/Ontario/ | Unity Health Toronto | Ontario Institute for Cancer Research | Ramzi Fattouh et al |
| EPI_ISL_464016 | Canada/ON-UHTC_0033/2020 | 2020-04-01 | North America/Canada/Ontario/ | Unity Health Toronto | Ontario Institute for Cancer Research | Ramzi Fattouh et al |
| EPI_ISL_464017 | Canada/ON-UHTC_0011/2020 | 2020-03-27 | North America/Canada/Ontario/ | Unity Health Toronto | Ontario Institute for Cancer Research | Ramzi Fattouh et al |
| EPI_ISL_464018 | Canada/ON-UHTC_0038/2020 | 2020-04-03 | North America/Canada/Ontario/ | Unity Health Toronto | Ontario Institute for Cancer Research | Ramzi Fattouh et al |
| EPI_ISL_464019 | Canada/ON-UHTC_0076/2020 | 2020-04-13 | North America/Canada/Ontario/ | Unity Health Toronto | Ontario Institute for Cancer Research | Ramzi Fattouh et al |
| EPI_ISL_464020 | Canada/ON-UHTC_0043/2020 | 2020-04-02 | North America/Canada/Ontario/ | Unity Health Toronto | Ontario Institute for Cancer Research | Ramzi Fattouh et al |
| EPI_ISL_464021 | Canada/ON-UHTC_0069/2020 | 2020-04-25 | North America/Canada/Ontario/ | Unity Health Toronto | Ontario Institute for Cancer Research | Ramzi Fattouh et al |
| EPI_ISL_464022 | Canada/ON-UHTC_0070/2020 | 2020-04-12 | North America/Canada/Ontario/ | Unity Health Toronto | Ontario Institute for Cancer Research | Ramzi Fattouh et al |

| | | | | | | |
|---|---|---|---|---|---|---|
| EPI_ISL_464023 | Canada/ON-UHTC_0001/2020 | 2020-03-23 | North America/Canada/Ontario/ | Unity Health Toronto | Ontario Institute for Cancer Research | Ramzi Fattouh et al |
| EPI_ISL_464024 | Canada/ON-UHTC_0058/2020 | 2020-04-03 | North America/Canada/Ontario/ | Unity Health Toronto | Ontario Institute for Cancer Research | Ramzi Fattouh et al |
| EPI_ISL_464025 | Canada/ON-UHTC_0013/2020 | 2020-03-28 | North America/Canada/Ontario/ | Unity Health Toronto | Ontario Institute for Cancer Research | Ramzi Fattouh et al |
| EPI_ISL_464026 | Canada/ON-UHTC_0039/2020 | 2020-04-10 | North America/Canada/Ontario/ | Unity Health Toronto | Ontario Institute for Cancer Research | Ramzi Fattouh et al |
| EPI_ISL_464027 | Canada/ON-UHTC_0021/2020 | 2020-03-30 | North America/Canada/Ontario/ | Unity Health Toronto | Ontario Institute for Cancer Research | Ramzi Fattouh et al |
| EPI_ISL_464028 | Canada/ON-UHTC_0037/2020 | 2020-04-03 | North America/Canada/Ontario/ | Unity Health Toronto | Ontario Institute for Cancer Research | Ramzi Fattouh et al |
| EPI_ISL_464029 | Canada/ON-UHTC_0085/2020 | 2020-04-27 | North America/Canada/Ontario/ | Unity Health Toronto | Ontario Institute for Cancer Research | Ramzi Fattouh et al |
| EPI_ISL_464030 | Canada/ON-UHTC_0065/2020 | 2020-04-22 | North America/Canada/Ontario/ | Unity Health Toronto | Ontario Institute for Cancer Research | Ramzi Fattouh et al |
| EPI_ISL_464031 | Canada/ON-UHTC_0002/2020 | 2020-03-19 | North America/Canada/Ontario/ | Unity Health Toronto | Ontario Institute for Cancer Research | Ramzi Fattouh et al |
| EPI_ISL_464033 | Canada/ON-UHTC_0035/2020 | 2020-04-07 | North America/Canada/Ontario/ | Unity Health Toronto | Ontario Institute for Cancer Research | Ramzi Fattouh et al |
| EPI_ISL_464034 | Canada/ON-UHTC_0009/2020 | 2020-03-26 | North America/Canada/Ontario/ | Unity Health Toronto | Ontario Institute for Cancer Research | Ramzi Fattouh et al |
| EPI_ISL_464035 | Canada/ON-UHTC_0024/2020 | 2020-04-01 | North America/Canada/Ontario/ | Unity Health Toronto | Ontario Institute for Cancer Research | Ramzi Fattouh et al |
| EPI_ISL_464036 | Canada/ON-UHTC_0050/2020 | 2020-04-08 | North America/Canada/Ontario/ | Unity Health Toronto | Ontario Institute for Cancer Research | Ramzi Fattouh et al |

| | | | | | | |
|---|---|---|---|---|---|---|
| EPI_ISL_464037 | Canada/ON-UHTC_0041/2020 | 2020-04-03 | North America/Canada/Ontario/ | Unity Health Toronto | Ontario Institute for Cancer Research | Ramzi Fattouh et al |
| EPI_ISL_464038 | Canada/ON-UHTC_0066/2020 | 2020-04-24 | North America/Canada/Ontario/ | Unity Health Toronto | Ontario Institute for Cancer Research | Ramzi Fattouh et al |
| EPI_ISL_464039 | Canada/ON-UHTC_0017/2020 | 2020-03-29 | North America/Canada/Ontario/ | Unity Health Toronto | Ontario Institute for Cancer Research | Ramzi Fattouh et al |
| EPI_ISL_464040 | Canada/ON-UHTC_0080/2020 | 2020-04-10 | North America/Canada/Ontario/ | Unity Health Toronto | Ontario Institute for Cancer Research | Ramzi Fattouh et al |
| EPI_ISL_464041 | Canada/ON-UHTC_0042/2020 | 2020-04-20 | North America/Canada/Ontario/ | Unity Health Toronto | Ontario Institute for Cancer Research | Ramzi Fattouh et al |
| EPI_ISL_464042 | Canada/ON-UHTC_0057/2020 | 2020-04-09 | North America/Canada/Ontario/ | Unity Health Toronto | Ontario Institute for Cancer Research | Ramzi Fattouh et al |
| EPI_ISL_464043 | Canada/ON-UHTC_0005/2020 | 2020-03-24 | North America/Canada/Ontario/ | Unity Health Toronto | Ontario Institute for Cancer Research | Ramzi Fattouh et al |
| EPI_ISL_464045 | Canada/ON-UHTC_0032/2020 | 2020-03-31 | North America/Canada/Ontario/ | Unity Health Toronto | Ontario Institute for Cancer Research | Ramzi Fattouh et al |
| EPI_ISL_464046 | Canada/ON-UHTC_0055/2020 | 2020-04-02 | North America/Canada/Ontario/ | Unity Health Toronto | Ontario Institute for Cancer Research | Ramzi Fattouh et al |
| EPI_ISL_464047 | Canada/ON-UHTC_0036/2020 | 2020-04-06 | North America/Canada/Ontario/ | Unity Health Toronto | Ontario Institute for Cancer Research | Ramzi Fattouh et al |
| EPI_ISL_464048 | Canada/ON-UHTC_0062/2020 | 2020-04-01 | North America/Canada/Ontario/ | Unity Health Toronto | Ontario Institute for Cancer Research | Ramzi Fattouh et al |
| EPI_ISL_464049 | Canada/ON-UHTC_0018/2020 | 2020-03-30 | North America/Canada/Ontario/ | Unity Health Toronto | Ontario Institute for Cancer Research | Ramzi Fattouh et al |
| EPI_ISL_464050 | Canada/ON-UHTC_0054/2020 | 2020-04-10 | North America/Canada/Ontario/ | Unity Health Toronto | Ontario Institute for Cancer Research | Ramzi Fattouh et al |

| | | | | | | |
|---|---|---|---|---|---|---|
| EPI_ISL_464051 | Canada/ON-UHTC_0004/2020 | 2020-03-18 | North America/Canada/Ontario/ | Unity Health Toronto | Ontario Institute for Cancer Research | Ramzi Fattouh et al |
| EPI_ISL_464054 | Canada/ON-UHTC_0088/2020 | 2020-04-24 | North America/Canada/Ontario/ | Unity Health Toronto | Ontario Institute for Cancer Research | Ramzi Fattouh et al |
| EPI_ISL_464055 | Canada/ON-UHTC_0022/2020 | 2020-03-31 | North America/Canada/Ontario/ | Unity Health Toronto | Ontario Institute for Cancer Research | Ramzi Fattouh et al |
| EPI_ISL_464056 | Canada/ON-UHTC_0052/2020 | 2020-04-02 | North America/Canada/Ontario/ | Unity Health Toronto | Ontario Institute for Cancer Research | Ramzi Fattouh et al |
| EPI_ISL_464057 | Canada/ON-UHTC_0089/2020 | 2020-04-19 | North America/Canada/Ontario/ | Unity Health Toronto | Ontario Institute for Cancer Research | Ramzi Fattouh et al |
| EPI_ISL_464058 | Canada/ON-UHTC_0086/2020 | 2020-04-15 | North America/Canada/Ontario/ | Unity Health Toronto | Ontario Institute for Cancer Research | Ramzi Fattouh et al |
| EPI_ISL_464059 | Canada/ON-UHTC_0056/2020 | 2020-04-10 | North America/Canada/Ontario/ | Unity Health Toronto | Ontario Institute for Cancer Research | Ramzi Fattouh et al |
| EPI_ISL_464060 | Canada/ON-UHTC_0029/2020 | 2020-04-01 | North America/Canada/Ontario/ | Unity Health Toronto | Ontario Institute for Cancer Research | Ramzi Fattouh et al |
| EPI_ISL_464061 | Canada/ON-UHTC_0045/2020 | 2020-04-22 | North America/Canada/Ontario/ | Unity Health Toronto | Ontario Institute for Cancer Research | Ramzi Fattouh et al |
| EPI_ISL_464062 | Canada/ON-UHTC_0074/2020 | 2020-04-24 | North America/Canada/Ontario/ | Unity Health Toronto | Ontario Institute for Cancer Research | Ramzi Fattouh et al |
| EPI_ISL_464063 | Canada/ON-UHTC_0051/2020 | 2020-04-03 | North America/Canada/Ontario/ | Unity Health Toronto | Ontario Institute for Cancer Research | Ramzi Fattouh et al |
| EPI_ISL_464064 | Canada/ON-UHTC_0081/2020 | 2020-04-29 | North America/Canada/Ontario/ | Unity Health Toronto | Ontario Institute for Cancer Research | Ramzi Fattouh et al |
| EPI_ISL_464067 | Belgium/MA-0406273/2020 | 2020-04-06 | Europe/Belgium/Brussels/Uccle | KU Leuven, Rega Institute, Clinical and Epidemiological Virology | KU Leuven, Rega Institute, Clinical and Epidemiological Virology | Tony Wawina-Bokalanga et al |

| | | | | | | |
|---|---|---|---|---|---|---|
| EPI_ISL_464086 | Belgium/RD-0408292/2020 | 2020-04-08 | Europe/Belgium/Tournai-Mouscron/Brur | KU Leuven, Rega Institute, Clinical and Epidemiological Virology | KU Leuven, Rega Institute, Clinical and Epidemiological Virology | Tony Wawina-Bokalanga et al |
| EPI_ISL_464159 | Bangladesh/BCSIR-NILMRC-087/2020 | 2020-05-09 | Asia/Bangladesh/Dhaka/Narayanganj | National Institute of Laboratory Medicine and Referral Center | Genomic Research Lab, BCSIR | Shahina Akter et al |
| EPI_ISL_464163 | Bangladesh/BCSIR-NILMRC-064/2020 | 2020-05-31 | Asia/Bangladesh/Dhaka/Narayanganj | National Institute of Laboratory Medicine and Referral Center | Genomic Research Lab, BCSIR | Tanjina Akhter Banu et al |
| EPI_ISL_464164 | Bangladesh/BCSIR-NILMRC-066/2020 | 2020-05-26 | Asia/Bangladesh/Chattogram/ | National Institute of Laboratory Medicine and Referral Center | Genomic Research Lab, BCSIR | Tanjina Akhter Banu et al |
| EPI_ISL_464165 | Bangladesh/BCSIR-NILMRC-060/2020 | 2020-05-10 | Asia/Bangladesh/Dhaka/Narayanganj | National Institute of Laboratory Medicine and Referral Center | Genomic Research Lab, BCSIR | Barna Goswami et al |
| EPI_ISL_464168 | England/20072030304/2020 | 2020-02-09 | Europe/United Kingdom/England/ | Respiratory Virus Unit, Microbiology Services Colindale, Public Health England | Respiratory Virus Unit, Microbiology Services Colindale, Public Health England | PHE Covid Sequencing Team et al |
| EPI_ISL_464169 | England/20078003804/2020 | 2020-02-13 | Europe/United Kingdom/England/ | Respiratory Virus Unit, Microbiology Services Colindale, Public Health England | Respiratory Virus Unit, Microbiology Services Colindale, Public Health England | PHE Covid Sequencing Team et al |
| EPI_ISL_464173 | England/20079022704/2020 | 2020-02-15 | Europe/United Kingdom/England/ | Respiratory Virus Unit, Microbiology Services Colindale, Public Health England | Respiratory Virus Unit, Microbiology Services Colindale, Public Health England | PHE Covid Sequencing Team et al |
| EPI_ISL_464175 | England/20080000104/2020 | 2020-02-16 | Europe/United Kingdom/England/ | Respiratory Virus Unit, Microbiology Services Colindale, Public Health England | Respiratory Virus Unit, Microbiology Services Colindale, Public Health England | PHE Covid Sequencing Team et al |

| | | | | | | |
|---|---|---|---|---|---|---|
| EPI_ISL_464176 | England/20080000204/2020 | 2020-02-16 | Europe/United Kingdom/England/ | Respiratory Virus Unit, Microbiology Services Colindale, Public Health England | Respiratory Virus Unit, Microbiology Services Colindale, Public Health England | PHE Covid Sequencing Team et al |
| EPI_ISL_464177 | England/20080000404/2020 | 2020-02-16 | Europe/United Kingdom/England/ | Respiratory Virus Unit, Microbiology Services Colindale, Public Health England | Respiratory Virus Unit, Microbiology Services Colindale, Public Health England | PHE Covid Sequencing Team et al |
| EPI_ISL_464178 | England/20084031204/2020 | 2020-02-18 | Europe/United Kingdom/England/ | Respiratory Virus Unit, Microbiology Services Colindale, Public Health England | Respiratory Virus Unit, Microbiology Services Colindale, Public Health England | PHE Covid Sequencing Team et al |
| EPI_ISL_464180 | England/20088028304/2020 | 2020-02-20 | Europe/United Kingdom/England/ | Respiratory Virus Unit, Microbiology Services Colindale, Public Health England | Respiratory Virus Unit, Microbiology Services Colindale, Public Health England | PHE Covid Sequencing Team et al |
| EPI_ISL_464181 | England/20092000704/2020 | 2020-02-24 | Europe/United Kingdom/England/ | Respiratory Virus Unit, Microbiology Services Colindale, Public Health England | Respiratory Virus Unit, Microbiology Services Colindale, Public Health England | PHE Covid Sequencing Team et al |
| EPI_ISL_464182 | England/20092000804/2020 | 2020-02-24 | Europe/United Kingdom/England/ | Respiratory Virus Unit, Microbiology Services Colindale, Public Health England | Respiratory Virus Unit, Microbiology Services Colindale, Public Health England | PHE Covid Sequencing Team et al |

| | | | | | | |
|---|---|---|---|---|---|---|
| EPI_ISL_464183 | England/20092096704/2020 | 2020-02-24 | Europe/United Kingdom/England/ | Respiratory Virus Unit, Microbiology Services Colindale, Public Health England | Respiratory Virus Unit, Microbiology Services Colindale, Public Health England | PHE Covid Sequencing Team et al |
| EPI_ISL_464184 | England/20094007004/2020 | 2020-02-25 | Europe/United Kingdom/England/ | Respiratory Virus Unit, Microbiology Services Colindale, Public Health England | Respiratory Virus Unit, Microbiology Services Colindale, Public Health England | PHE Covid Sequencing Team et al |
| EPI_ISL_464185 | England/20094061404/2020 | 2020-02-25 | Europe/United Kingdom/England/ | Respiratory Virus Unit, Microbiology Services Colindale, Public Health England | Respiratory Virus Unit, Microbiology Services Colindale, Public Health England | PHE Covid Sequencing Team et al |
| EPI_ISL_464186 | England/20094061504/2020 | 2020-02-25 | Europe/United Kingdom/England/ | Respiratory Virus Unit, Microbiology Services Colindale, Public Health England | Respiratory Virus Unit, Microbiology Services Colindale, Public Health England | PHE Covid Sequencing Team et al |
| EPI_ISL_464187 | England/20096051004/2020 | 2020-02-26 | Europe/United Kingdom/England/ | Respiratory Virus Unit, Microbiology Services Colindale, Public Health England | Respiratory Virus Unit, Microbiology Services Colindale, Public Health England | PHE Covid Sequencing Team et al |
| EPI_ISL_464188 | England/20096051104/2020 | 2020-02-26 | Europe/United Kingdom/England/ | Respiratory Virus Unit, Microbiology Services Colindale, Public Health England | Respiratory Virus Unit, Microbiology Services Colindale, Public Health England | PHE Covid Sequencing Team et al |

| | | | | | | |
|---|---|---|---|---|---|---|
| EPI_ISL_464189 | England/20096101804/2020 | 2020-02-27 | Europe/United Kingdom/England/ | Respiratory Virus Unit, Microbiology Services Colindale, Public Health England | Respiratory Virus Unit, Microbiology Services Colindale, Public Health England | PHE Covid Sequencing Team et al |
| EPI_ISL_464190 | England/20096115304/2020 | 2020-02-27 | Europe/United Kingdom/England/ | Respiratory Virus Unit, Microbiology Services Colindale, Public Health England | Respiratory Virus Unit, Microbiology Services Colindale, Public Health England | PHE Covid Sequencing Team et al |
| EPI_ISL_464192 | England/20098017104/2020 | 2020-02-27 | Europe/United Kingdom/England/ | Respiratory Virus Unit, Microbiology Services Colindale, Public Health England | Respiratory Virus Unit, Microbiology Services Colindale, Public Health England | PHE Covid Sequencing Team et al |
| EPI_ISL_464193 | England/20098017204/2020 | 2020-02-27 | Europe/United Kingdom/England/ | Respiratory Virus Unit, Microbiology Services Colindale, Public Health England | Respiratory Virus Unit, Microbiology Services Colindale, Public Health England | PHE Covid Sequencing Team et al |
| EPI_ISL_464194 | England/20098017504/2020 | 2020-02-27 | Europe/United Kingdom/England/ | Respiratory Virus Unit, Microbiology Services Colindale, Public Health England | Respiratory Virus Unit, Microbiology Services Colindale, Public Health England | PHE Covid Sequencing Team et al |
| EPI_ISL_464195 | England/20098017604/2020 | 2020-02-26 | Europe/United Kingdom/England/ | Respiratory Virus Unit, Microbiology Services Colindale, Public Health England | Respiratory Virus Unit, Microbiology Services Colindale, Public Health England | PHE Covid Sequencing Team et al |

| | | | | | | |
|---|---|---|---|---|---|---|
| EPI_ISL_464196 | England/20098017704/2020 | 2020-02-27 | Europe/United Kingdom/England/ | Respiratory Virus Unit, Microbiology Services Colindale, Public Health England | Respiratory Virus Unit, Microbiology Services Colindale, Public Health England | PHE Covid Sequencing Team et al |
| EPI_ISL_464197 | England/20098017804/2020 | 2020-02-27 | Europe/United Kingdom/England/ | Respiratory Virus Unit, Microbiology Services Colindale, Public Health England | Respiratory Virus Unit, Microbiology Services Colindale, Public Health England | PHE Covid Sequencing Team et al |
| EPI_ISL_464198 | England/20099000104/2020 | 2020-02-28 | Europe/United Kingdom/England/ | Respiratory Virus Unit, Microbiology Services Colindale, Public Health England | Respiratory Virus Unit, Microbiology Services Colindale, Public Health England | PHE Covid Sequencing Team et al |
| EPI_ISL_464199 | England/20099000304/2020 | 2020-02-28 | Europe/United Kingdom/England/ | Respiratory Virus Unit, Microbiology Services Colindale, Public Health England | Respiratory Virus Unit, Microbiology Services Colindale, Public Health England | PHE Covid Sequencing Team et al |
| EPI_ISL_464200 | England/20099000504/2020 | 2020-02-27 | Europe/United Kingdom/England/ | Respiratory Virus Unit, Microbiology Services Colindale, Public Health England | Respiratory Virus Unit, Microbiology Services Colindale, Public Health England | PHE Covid Sequencing Team et al |
| EPI_ISL_464201 | England/20099000804/2020 | 2020-02-28 | Europe/United Kingdom/England/ | Respiratory Virus Unit, Microbiology Services Colindale, Public Health England | Respiratory Virus Unit, Microbiology Services Colindale, Public Health England | PHE Covid Sequencing Team et al |

| | | | | | | |
|---|---|---|---|---|---|---|
| EPI_ISL_464202 | England/20099015104/2020 | 2020-02-29 | Europe/United Kingdom/England/ | Respiratory Virus Unit, Microbiology Services Colindale, Public Health England | Respiratory Virus Unit, Microbiology Services Colindale, Public Health England | PHE Covid Sequencing Team et al |
| EPI_ISL_464203 | England/20099038304/2020 | 2020-02-29 | Europe/United Kingdom/England/ | Respiratory Virus Unit, Microbiology Services Colindale, Public Health England | Respiratory Virus Unit, Microbiology Services Colindale, Public Health England | PHE Covid Sequencing Team et al |
| EPI_ISL_464204 | England/20099044904/2020 | 2020-02-26 | Europe/United Kingdom/England/ | Respiratory Virus Unit, Microbiology Services Colindale, Public Health England | Respiratory Virus Unit, Microbiology Services Colindale, Public Health England | PHE Covid Sequencing Team et al |
| EPI_ISL_464205 | England/20099048404/2020 | 2020-02-28 | Europe/United Kingdom/England/ | Respiratory Virus Unit, Microbiology Services Colindale, Public Health England | Respiratory Virus Unit, Microbiology Services Colindale, Public Health England | PHE Covid Sequencing Team et al |
| EPI_ISL_464206 | England/20099048504/2020 | 2020-02-28 | Europe/United Kingdom/England/ | Respiratory Virus Unit, Microbiology Services Colindale, Public Health England | Respiratory Virus Unit, Microbiology Services Colindale, Public Health England | PHE Covid Sequencing Team et al |
| EPI_ISL_464207 | England/20099048604/2020 | 2020-02-28 | Europe/United Kingdom/England/ | Respiratory Virus Unit, Microbiology Services Colindale, Public Health England | Respiratory Virus Unit, Microbiology Services Colindale, Public Health England | PHE Covid Sequencing Team et al |

| | | | | | | |
|---|---|---|---|---|---|---|
| EPI_ISL_464208 | England/20099066104/2020 | 2020-02-27 | Europe/United Kingdom/England/ | Respiratory Virus Unit, Microbiology Services Colindale, Public Health England | Respiratory Virus Unit, Microbiology Services Colindale, Public Health England | PHE Covid Sequencing Team et al |
| EPI_ISL_464209 | England/20099066204/2020 | 2020-02-27 | Europe/United Kingdom/England/ | Respiratory Virus Unit, Microbiology Services Colindale, Public Health England | Respiratory Virus Unit, Microbiology Services Colindale, Public Health England | PHE Covid Sequencing Team et al |
| EPI_ISL_464210 | England/20099068104/2020 | 2020-02-29 | Europe/United Kingdom/England/ | Respiratory Virus Unit, Microbiology Services Colindale, Public Health England | Respiratory Virus Unit, Microbiology Services Colindale, Public Health England | PHE Covid Sequencing Team et al |
| EPI_ISL_464211 | England/20099068304/2020 | 2020-02-29 | Europe/United Kingdom/England/ | Respiratory Virus Unit, Microbiology Services Colindale, Public Health England | Respiratory Virus Unit, Microbiology Services Colindale, Public Health England | PHE Covid Sequencing Team et al |
| EPI_ISL_464212 | England/20099068604/2020 | 2020-02-29 | Europe/United Kingdom/England/ | Respiratory Virus Unit, Microbiology Services Colindale, Public Health England | Respiratory Virus Unit, Microbiology Services Colindale, Public Health England | PHE Covid Sequencing Team et al |
| EPI_ISL_464214 | England/20099069004/2020 | 2020-02-29 | Europe/United Kingdom/England/ | Respiratory Virus Unit, Microbiology Services Colindale, Public Health England | Respiratory Virus Unit, Microbiology Services Colindale, Public Health England | PHE Covid Sequencing Team et al |

| | | | | | | |
|---|---|---|---|---|---|---|
| EPI_ISL_464215 | England/20099069504/2020 | 2020-02-29 | Europe/United Kingdom/England/ | Respiratory Virus Unit, Microbiology Services Colindale, Public Health England | Respiratory Virus Unit, Microbiology Services Colindale, Public Health England | PHE Covid Sequencing Team et al |
| EPI_ISL_464216 | England/20099069904/2020 | 2020-02-29 | Europe/United Kingdom/England/ | Respiratory Virus Unit, Microbiology Services Colindale, Public Health England | Respiratory Virus Unit, Microbiology Services Colindale, Public Health England | PHE Covid Sequencing Team et al |
| EPI_ISL_464217 | England/20099070104/2020 | 2020-02-29 | Europe/United Kingdom/England/ | Respiratory Virus Unit, Microbiology Services Colindale, Public Health England | Respiratory Virus Unit, Microbiology Services Colindale, Public Health England | PHE Covid Sequencing Team et al |
| EPI_ISL_464218 | England/20099071404/2020 | 2020-02-29 | Europe/United Kingdom/England/ | Respiratory Virus Unit, Microbiology Services Colindale, Public Health England | Respiratory Virus Unit, Microbiology Services Colindale, Public Health England | PHE Covid Sequencing Team et al |
| EPI_ISL_464219 | England/20099071804/2020 | 2020-02-28 | Europe/United Kingdom/England/ | Respiratory Virus Unit, Microbiology Services Colindale, Public Health England | Respiratory Virus Unit, Microbiology Services Colindale, Public Health England | PHE Covid Sequencing Team et al |
| EPI_ISL_464221 | England/20099072004/2020 | 2020-02-28 | Europe/United Kingdom/England/ | Respiratory Virus Unit, Microbiology Services Colindale, Public Health England | Respiratory Virus Unit, Microbiology Services Colindale, Public Health England | PHE Covid Sequencing Team et al |

| | | | | | | |
|---|---|---|---|---|---|---|
| EPI_ISL_464222 | England/20099072104/2020 | 2020-03-01 | Europe/United Kingdom/England/ | Respiratory Virus Unit, Microbiology Services Colindale, Public Health England | Respiratory Virus Unit, Microbiology Services Colindale, Public Health England | PHE Covid Sequencing Team et al |
| EPI_ISL_464223 | England/20099072204/2020 | 2020-02-28 | Europe/United Kingdom/England/ | Respiratory Virus Unit, Microbiology Services Colindale, Public Health England | Respiratory Virus Unit, Microbiology Services Colindale, Public Health England | PHE Covid Sequencing Team et al |
| EPI_ISL_464224 | England/20099072604/2020 | 2020-02-28 | Europe/United Kingdom/England/ | Respiratory Virus Unit, Microbiology Services Colindale, Public Health England | Respiratory Virus Unit, Microbiology Services Colindale, Public Health England | PHE Covid Sequencing Team et al |
| EPI_ISL_464226 | England/20099073004/2020 | 2020-02-27 | Europe/United Kingdom/England/ | Respiratory Virus Unit, Microbiology Services Colindale, Public Health England | Respiratory Virus Unit, Microbiology Services Colindale, Public Health England | PHE Covid Sequencing Team et al |
| EPI_ISL_464228 | England/20099089504/2020 | 2020-02-28 | Europe/United Kingdom/England/ | Respiratory Virus Unit, Microbiology Services Colindale, Public Health England | Respiratory Virus Unit, Microbiology Services Colindale, Public Health England | PHE Covid Sequencing Team et al |
| EPI_ISL_464229 | England/20099089604/2020 | 2020-02-27 | Europe/United Kingdom/England/ | Respiratory Virus Unit, Microbiology Services Colindale, Public Health England | Respiratory Virus Unit, Microbiology Services Colindale, Public Health England | PHE Covid Sequencing Team et al |

| | | | | | | |
|---|---|---|---|---|---|---|
| EPI_ISL_464247 | England/20100004404/2020 | 2020-02-29 | Europe/United Kingdom/England/ | Respiratory Virus Unit, Microbiology Services Colindale, Public Health England | Respiratory Virus Unit, Microbiology Services Colindale, Public Health England | PHE Covid Sequencing Team et al |
| EPI_ISL_464251 | England/20100005304/2020 | 2020-02-28 | Europe/United Kingdom/England/ | Respiratory Virus Unit, Microbiology Services Colindale, Public Health England | Respiratory Virus Unit, Microbiology Services Colindale, Public Health England | PHE Covid Sequencing Team et al |
| EPI_ISL_464252 | England/20100005504/2020 | 2020-02-28 | Europe/United Kingdom/England/ | Respiratory Virus Unit, Microbiology Services Colindale, Public Health England | Respiratory Virus Unit, Microbiology Services Colindale, Public Health England | PHE Covid Sequencing Team et al |
| EPI_ISL_464253 | England/20100005604/2020 | 2020-02-28 | Europe/United Kingdom/England/ | Respiratory Virus Unit, Microbiology Services Colindale, Public Health England | Respiratory Virus Unit, Microbiology Services Colindale, Public Health England | PHE Covid Sequencing Team et al |
| EPI_ISL_464254 | England/20100007104/2020 | 2020-02-29 | Europe/United Kingdom/England/ | Respiratory Virus Unit, Microbiology Services Colindale, Public Health England | Respiratory Virus Unit, Microbiology Services Colindale, Public Health England | PHE Covid Sequencing Team et al |
| EPI_ISL_464259 | England/20100120904/2020 | 2020-02-29 | Europe/United Kingdom/England/ | Respiratory Virus Unit, Microbiology Services Colindale, Public Health England | Respiratory Virus Unit, Microbiology Services Colindale, Public Health England | PHE Covid Sequencing Team et al |

| | | | | | | |
|---|---|---|---|---|---|---|
| EPI_ISL_464371 | England/201060032/2020 | 2020-03-05 | Europe/United Kingdom/England/ | Respiratory Virus Unit, Microbiology Services Colindale, Public Health England | Respiratory Virus Unit, Microbiology Services Colindale, Public Health England | PHE Covid Sequencing Team et al |
| EPI_ISL_464401 | England/201060300/2020 | 2020-03-03 | Europe/United Kingdom/England/ | Respiratory Virus Unit, Microbiology Services Colindale, Public Health England | Respiratory Virus Unit, Microbiology Services Colindale, Public Health England | PHE Covid Sequencing Team et al |
| EPI_ISL_464411 | England/201061453/2020 | 2020-02-27 | Europe/United Kingdom/England/ | Respiratory Virus Unit, Microbiology Services Colindale, Public Health England | Respiratory Virus Unit, Microbiology Services Colindale, Public Health England | PHE Covid Sequencing Team et al |
| EPI_ISL_464412 | England/201061454/2020 | 2020-02-29 | Europe/United Kingdom/England/ | Respiratory Virus Unit, Microbiology Services Colindale, Public Health England | Respiratory Virus Unit, Microbiology Services Colindale, Public Health England | PHE Covid Sequencing Team et al |
| EPI_ISL_464483 | England/20108134404/2020 | 2020-03-04 | Europe/United Kingdom/England/ | Respiratory Virus Unit, Microbiology Services Colindale, Public Health England | Respiratory Virus Unit, Microbiology Services Colindale, Public Health England | PHE Covid Sequencing Team et al |
| EPI_ISL_464816 | England/201161176/2020 | 2020-03-11 | Europe/United Kingdom/England/ | Respiratory Virus Unit, Microbiology Services Colindale, Public Health England | Respiratory Virus Unit, Microbiology Services Colindale, Public Health England | PHE Covid Sequencing Team et al |

| | | | | | | |
|---|---|---|---|---|---|---|
| EPI_ISL_465030 | England/20119011704/2020 | 2020-03-12 | Europe/United Kingdom/England/ | Respiratory Virus Unit, Microbiology Services Colindale, Public Health England | Respiratory Virus Unit, Microbiology Services Colindale, Public Health England | PHE Covid Sequencing Team et al |
| EPI_ISL_465162 | England/20120037806/2020 | 2020-03-14 | Europe/United Kingdom/England/ | Respiratory Virus Unit, Microbiology Services Colindale, Public Health England | Respiratory Virus Unit, Microbiology Services Colindale, Public Health England | PHE Covid Sequencing Team et al |
| EPI_ISL_465164 | Bangladesh/BCSIR-NILMRC-059/2020 | 2020-05-10 | Asia/Bangladesh/Dhaka/Narayanganj | National Institute of Laboratory Medicine and Referral Center | Genomic Research Lab, BCSIR | Iffat Jahan et al |
| EPI_ISL_465227 | England/20169032504/2020 | 2020-04-17 | Europe/United Kingdom/England/ | Respiratory Virus Unit, Microbiology Services Colindale, Public Health England | Respiratory Virus Unit, Microbiology Services Colindale, Public Health England | PHE Covid Sequencing Team et al |
| EPI_ISL_465270 | England/20170003404/2020 | 2020-04-19 | Europe/United Kingdom/England/ | Respiratory Virus Unit, Microbiology Services Colindale, Public Health England | Respiratory Virus Unit, Microbiology Services Colindale, Public Health England | PHE Covid Sequencing Team et al |
| EPI_ISL_465273 | England/20170030604/2020 | 2020-04-17 | Europe/United Kingdom/England/ | Respiratory Virus Unit, Microbiology Services Colindale, Public Health England | Respiratory Virus Unit, Microbiology Services Colindale, Public Health England | PHE Covid Sequencing Team et al |
| EPI_ISL_465417 | England/20176047004/2020 | 2020-04-22 | Europe/United Kingdom/England/ | Respiratory Virus Unit, Microbiology Services Colindale, Public Health England | Respiratory Virus Unit, Microbiology Services Colindale, Public Health England | PHE Covid Sequencing Team et al |

| | | | | | | |
|---|---|---|---|---|---|---|
| EPI_ISL_465564 | England/20189159601/2020 | 2020-05-02 | Europe/United Kingdom/England/ | Respiratory Virus Unit, Microbiology Services Colindale, Public Health England | Respiratory Virus Unit, Microbiology Services Colindale, Public Health England | PHE Covid Sequencing Team et al |
| EPI_ISL_465660 | England/20206035304/2020 | 2020-05-13 | Europe/United Kingdom/England/ | Respiratory Virus Unit, Microbiology Services Colindale, Public Health England | Respiratory Virus Unit, Microbiology Services Colindale, Public Health England | PHE Covid Sequencing Team et al |
| EPI_ISL_465679 | Canada/Qc-L00240569/2020 | 2020-03-27 | North America/Canada/Quebec/ | H√¥pital du Suro√Æt | Laboratoire de sant√© publique du Qu√©bec | Sandrine Moreira et al |
| EPI_ISL_465680 | Canada/Qc-L00240594/2020 | 2020-03-27 | North America/Canada/Quebec/ | H√¥pital du Suro√Æt | Laboratoire de sant√© publique du Qu√©bec | Sandrine Moreira et al |
| EPI_ISL_465681 | Canada/Qc-L00240642/2020 | 2020-03-27 | North America/Canada/Quebec/ | H√¥pital Charles-LeMoyne | Laboratoire de sant√© publique du Qu√©bec | Sandrine Moreira et al |
| EPI_ISL_465682 | Canada/Qc-L00240666/2020 | 2020-03-27 | North America/Canada/Quebec/ | H√¥pital Charles-LeMoyne | Laboratoire de sant√© publique du Qu√©bec | Sandrine Moreira et al |
| EPI_ISL_465683 | Canada/Qc-L00240746/2020 | 2020-03-26 | North America/Canada/Quebec/ | H√¥pital de Maria | Laboratoire de sant√© publique du Qu√©bec | Sandrine Moreira et al |
| EPI_ISL_465684 | Canada/Qc-L00240750/2020 | 2020-03-26 | North America/Canada/Quebec/ | H√¥pital de Maria | Laboratoire de sant√© publique du Qu√©bec | Sandrine Moreira et al |
| EPI_ISL_465685 | Canada/Qc-L00240788/2020 | 2020-03-27 | North America/Canada/Quebec/ | CSSS Haut-Richelieu/Rouville (H√¥pital) | Laboratoire de sant√© publique du Qu√©bec | Sandrine Moreira et al |
| EPI_ISL_465686 | Canada/Qc-L00240796/2020 | 2020-03-27 | North America/Canada/Quebec/ | H√¥pital de Hull | Laboratoire de sant√© publique du Qu√©bec | Sandrine Moreira et al |
| EPI_ISL_465687 | Canada/Qc-L00240804/2020 | 2020-03-27 | North America/Canada/Quebec/ | CSSS Haut-Richelieu/Rouville (H√¥pital) | Laboratoire de sant√© publique du Qu√©bec | Sandrine Moreira et al |

| | | | | | | |
|---|---|---|---|---|---|---|
| EPI_ISL_465688 | Canada/Qc-L00240846/2020 | 2020-03-27 | North America/Canada/Quebec/ | Centre hospitalier Anna-Laberge | Laboratoire de santv© publique du Quv©bec | Sandrine Moreira et al |
| EPI_ISL_465689 | Canada/Qc-L00240851/2020 | 2020-03-27 | North America/Canada/Quebec/ | Centre hospitalier Anna-Laberge | Laboratoire de santv© publique du Quv©bec | Sandrine Moreira et al |
| EPI_ISL_465690 | Canada/Qc-L00240866/2020 | 2020-03-27 | North America/Canada/Quebec/ | Centre hospitalier Anna-Laberge | Laboratoire de santv© publique du Quv©bec | Sandrine Moreira et al |
| EPI_ISL_465691 | Canada/Qc-L00240891/2020 | 2020-03-27 | North America/Canada/Quebec/ | Hv¥pital de Gatineau | Laboratoire de santv© publique du Quv©bec | Sandrine Moreira et al |
| EPI_ISL_465692 | Canada/Qc-L00240893/2020 | 2020-03-27 | North America/Canada/Quebec/ | CSSS Haut-Richelieu/Rouville (Hv¥pital) | Laboratoire de santv© publique du Quv©bec | Sandrine Moreira et al |
| EPI_ISL_465693 | Canada/Qc-L00240947/2020 | 2020-03-27 | North America/Canada/Quebec/ | Hv¥pital du SurovÆt | Laboratoire de santv© publique du Quv©bec | Sandrine Moreira et al |
| EPI_ISL_465694 | Canada/Qc-L00241007/2020 | 2020-03-27 | North America/Canada/Quebec/ | Hv¥pital Charles-LeMoyne | Laboratoire de santv© publique du Quv©bec | Sandrine Moreira et al |
| EPI_ISL_465695 | Canada/Qc-L00241074/2020 | 2020-03-27 | North America/Canada/Quebec/ | Hv¥pital Pierre-Boucher | Laboratoire de santv© publique du Quv©bec | Sandrine Moreira et al |
| EPI_ISL_465696 | Canada/Qc-L00241102/2020 | 2020-03-27 | North America/Canada/Quebec/ | Hv¥pital Pierre-Boucher | Laboratoire de santv© publique du Quv©bec | Sandrine Moreira et al |
| EPI_ISL_465697 | Canada/Qc-L00241112/2020 | 2020-03-26 | North America/Canada/Quebec/ | Centre de santv© Inuulitsivik | Laboratoire de santv© publique du Quv©bec | Sandrine Moreira et al |
| EPI_ISL_465698 | Canada/Qc-L00241115/2020 | 2020-03-27 | North America/Canada/Quebec/ | Hv¥pital Charles-LeMoyne | Laboratoire de santv© publique du Quv©bec | Sandrine Moreira et al |
| EPI_ISL_465699 | Canada/Qc-L00241158/2020 | 2020-03-27 | North America/Canada/Quebec/ | Hv¥pital Charles-LeMoyne | Laboratoire de santv© publique du Quv©bec | Sandrine Moreira et al |
| EPI_ISL_465700 | Canada/Qc-L00241214/2020 | 2020-03-23 | North America/Canada/Quebec/ | Hv¥pital Pierre-Boucher | Laboratoire de santv© publique du Quv©bec | Sandrine Moreira et al |

| | | | | | | |
|---|---|---|---|---|---|---|
| EPI_ISL_465701 | Canada/Qc-L00241251/2020 | 2020-03-27 | North America/Canada/Quebec/ | H√¥pital du Surov√Æt | Laboratoire de sant√© publique du Qu√©bec | Sandrine Moreira et al |
| EPI_ISL_465702 | Canada/Qc-L00241319/2020 | 2020-03-27 | North America/Canada/Quebec/ | H√¥pital Pierre-Boucher | Laboratoire de sant√© publique du Qu√©bec | Sandrine Moreira et al |
| EPI_ISL_465703 | Canada/Qc-L00241349/2020 | 2020-03-27 | North America/Canada/Quebec/ | H√¥pital Pierre-Boucher | Laboratoire de sant√© publique du Qu√©bec | Sandrine Moreira et al |
| EPI_ISL_465812 | England/20134075204/2020 | 2020-03-24 | Europe/United Kingdom/England/ | Respiratory Virus Unit, Microbiology Services Colindale, Public Health England | Respiratory Virus Unit, Microbiology Services Colindale, Public Health England | PHE Covid Sequencing Team et al |
| EPI_ISL_465930 | England/201420209/2020 | 2020-03-29 | Europe/United Kingdom/England/ | Respiratory Virus Unit, Microbiology Services Colindale, Public Health England | Respiratory Virus Unit, Microbiology Services Colindale, Public Health England | PHE Covid Sequencing Team et al |
| EPI_ISL_465977 | England/20144011404/2020 | 2020-03-30 | Europe/United Kingdom/England/ | Respiratory Virus Unit, Microbiology Services Colindale, Public Health England | Respiratory Virus Unit, Microbiology Services Colindale, Public Health England | PHE Covid Sequencing Team et al |
| EPI_ISL_465999 | England/20144037104/2020 | 2020-03-31 | Europe/United Kingdom/England/ | Respiratory Virus Unit, Microbiology Services Colindale, Public Health England | Respiratory Virus Unit, Microbiology Services Colindale, Public Health England | PHE Covid Sequencing Team et al |
| EPI_ISL_466370 | England/201590076/2020 | 2020-04-09 | Europe/United Kingdom/England/ | Respiratory Virus Unit, Microbiology Services Colindale, Public Health England | Respiratory Virus Unit, Microbiology Services Colindale, Public Health England | PHE Covid Sequencing Team et al |

| | | | | | | |
|---|---|---|---|---|---|---|
| EPI_ISL_466408 | England/20162009704/2020 | 2020-04-12 | Europe/United Kingdom/England/ | Respiratory Virus Unit, Microbiology Services Colindale, Public Health England | Respiratory Virus Unit, Microbiology Services Colindale, Public Health England | PHE Covid Sequencing Team et al |
| EPI_ISL_466409 | England/20162010504/2020 | 2020-04-12 | Europe/United Kingdom/England/ | Respiratory Virus Unit, Microbiology Services Colindale, Public Health England | Respiratory Virus Unit, Microbiology Services Colindale, Public Health England | PHE Covid Sequencing Team et al |
| EPI_ISL_466424 | England/201620530/2020 | 2020-04-09 | Europe/United Kingdom/England/ | Respiratory Virus Unit, Microbiology Services Colindale, Public Health England | Respiratory Virus Unit, Microbiology Services Colindale, Public Health England | PHE Covid Sequencing Team et al |
| EPI_ISL_466627 | Bangladesh/BCSIR-NILMRC-052/2020 | 2020-06-01 | Asia/Bangladesh/Dhaka/Narayanganj | National Institute of Laboratory Medicine and Referral Center | Genomic Research Lab, BCSIR | Abu Sayeed Mohammad Mahmud et al |
| EPI_ISL_466636 | Bangladesh/BCSIR-NILMRC-058/2020 | 2020-05-10 | Asia/Bangladesh/Dhaka/Narayanganj | National Institute of Laboratory Medicine and Referral Center | Genomic Research Lab, BCSIR | Abu Sayeed Mohammad Mahmud et al |
| EPI_ISL_466637 | Bangladesh/BCSIR-NILMRC-067/2020 | 2020-05-26 | Asia/Bangladesh/Chattogram/ | National Institute of Laboratory Medicine and Referral Center | Genomic Research Lab, BCSIR | Abu Sayeed Mohammad Mahmud et al |
| EPI_ISL_466638 | Bangladesh/BCSIR-NILMRC-069/2020 | 2020-05-26 | Asia/Bangladesh/Chattogram/ | National Institute of Laboratory Medicine and Referral Center | Genomic Research Lab, BCSIR | Abu Sayeed Mohammad Mahmud et al |
| EPI_ISL_466649 | Bangladesh/BCSIR-NILMRC-082/2020 | 2020-06-01 | Asia/Bangladesh/Dhaka/Agargaon | National Institute of Laboratory Medicine and Referral Center | Genomic Research Lab, BCSIR | Abu Sayeed Mohammad Mahmud et al |
| EPI_ISL_466650 | Bangladesh/BCSIR-NILMRC-085/2020 | 2020-06-01 | Asia/Bangladesh/Dhaka/ | National Institute of Laboratory Medicine and Referral Center | Genomic Research Lab, BCSIR | Abu Sayeed Mohammad Mahmud et al |
| EPI_ISL_466653 | USA/NE-10746/2020 | 2020-05-05 | North America/USA/Nebraska/ | Nebraska Public Health Laboratory | UNMC COVID-19 Response Team | UNMC COVID-19 Response Team et al |
| EPI_ISL_466655 | USA/NE-10940/2020 | 2020-05-04 | North America/USA/Nebraska/ | Nebraska Public Health Laboratory | UNMC COVID-19 Response Team | UNMC COVID-19 Response Team et al |
| EPI_ISL_466658 | USA/NE-10969/2020 | 2020-05-04 | North America/USA/Nebraska/ | Nebraska Public Health Laboratory | UNMC COVID-19 Response Team | UNMC COVID-19 Response Team et al |
| EPI_ISL_466664 | USA/NE-10732/2020 | 2020-05-05 | North America/USA/Nebraska/ | Nebraska Public Health Laboratory | UNMC COVID-19 Response Team | UNMC COVID-19 Response Team et al |
| EPI_ISL_466668 | USA/NE-10928/2020 | 2020-05-04 | North America/USA/Nebraska/ | Nebraska Public Health Laboratory | UNMC COVID-19 Response Team | UNMC COVID-19 Response Team et al |

| | | | | | | |
|---|---|---|---|---|---|---|
| EPI_ISL_466672 | USA/NE-11120/2020 | 2020-05-04 | North America/USA/Nebraska/ | Nebraska Public Health Laboratory | UNMC COVID-19 Response Team | UNMC COVID-19 Response Team et al |
| EPI_ISL_466679 | USA/NE-11552/2020 | 2020-05-05 | North America/USA/Nebraska/ | Nebraska Public Health Laboratory | UNMC COVID-19 Response Team | UNMC COVID-19 Response Team et al |
| EPI_ISL_466686 | Bangladesh/BCSIR-NILMRC-088/2020 | 2020-05-09 | Asia/Bangladesh/Dhaka/Narayanganj | National Institute of Laboratory Medicine and Referral Center | Genomic Research Lab, BCSIR | Abu Sayeed Mohammad Mahmud et al |
| EPI_ISL_466687 | Bangladesh/BCSIR-NILMRC-072/2020 | 2020-05-26 | Asia/Bangladesh/Chattogram/ | National Institute of Laboratory Medicine and Referral Center | Genomic Research Lab, BCSIR | Abu Sayeed Mohammad Mahmud et al |
| EPI_ISL_466688 | Bangladesh/BCSIR-NILMRC-086/2020 | 2020-06-01 | Asia/Bangladesh/Dhaka/ | National Institute of Laboratory Medicine and Referral Center | Genomic Research Lab, BCSIR | Abu Sayeed Mohammad Mahmud et al |
| EPI_ISL_466691 | Bangladesh/BCSIR-NILMRC-075/2020 | 2020-06-01 | Asia/Bangladesh/Dhaka/Uttara | National Institute of Laboratory Medicine and Referral Center | Genomic Research Lab, BCSIR | Abu Sayeed Mohammad Mahmud et al |
| EPI_ISL_466692 | Bangladesh/BCSIR-NILMRC-071/2020 | 2020-05-26 | Asia/Bangladesh/Chattogram/ | National Institute of Laboratory Medicine and Referral Center | Genomic Research Lab, BCSIR | Abu Sayeed Mohammad Mahmud et al |
| EPI_ISL_466693 | Bangladesh/BCSIR-NILMRC-078/2020 | 2020-06-01 | Asia/Bangladesh/Dhaka/ | National Institute of Laboratory Medicine and Referral Center | Genomic Research Lab, BCSIR | Abu Sayeed Mohammad Mahmud et al |
| EPI_ISL_466694 | Bangladesh/BCSIR-NILMRC-081/2020 | 2020-06-01 | Asia/Bangladesh/Dhaka/Mirpur | National Institute of Laboratory Medicine and Referral Center | Genomic Research Lab, BCSIR | Abu Sayeed Mohammad Mahmud et al |
| EPI_ISL_466696 | Canada/BC_00576317/2020 | 20-Apr | North America/Canada/British Columbia | BCCDC Public Health Laboratory | BCCDC Public Health Laboratory | Richard Harrigan et al |
| EPI_ISL_466697 | Canada/BC_00646519/2020 | 20-Apr | North America/Canada/British Columbia | BCCDC Public Health Laboratory | BCCDC Public Health Laboratory | Richard Harrigan et al |
| EPI_ISL_466698 | Canada/BC_00655815/2020 | 20-Apr | North America/Canada/British Columbia | BCCDC Public Health Laboratory | BCCDC Public Health Laboratory | Richard Harrigan et al |
| EPI_ISL_466699 | Canada/BC_00696199/2020 | 20-Apr | North America/Canada/British Columbia | BCCDC Public Health Laboratory | BCCDC Public Health Laboratory | Richard Harrigan et al |
| EPI_ISL_466700 | Canada/BC_00713032/2020 | 20-Apr | North America/Canada/British Columbia | BCCDC Public Health Laboratory | BCCDC Public Health Laboratory | Richard Harrigan et al |
| EPI_ISL_466701 | Canada/BC_00714140/2020 | 20-Apr | North America/Canada/British Columbia | BCCDC Public Health Laboratory | BCCDC Public Health Laboratory | Richard Harrigan et al |
| EPI_ISL_466703 | Canada/BC_01210684/2020 | 20-Apr | North America/Canada/British Columbia | BCCDC Public Health Laboratory | BCCDC Public Health Laboratory | Richard Harrigan et al |
| EPI_ISL_466704 | Canada/BC_01218976/2020 | 20-Apr | North America/Canada/British Columbia | BCCDC Public Health Laboratory | BCCDC Public Health Laboratory | Richard Harrigan et al |
| EPI_ISL_466705 | Canada/BC_01222955/2020 | 20-Apr | North America/Canada/British Columbia | BCCDC Public Health Laboratory | BCCDC Public Health Laboratory | Richard Harrigan et al |
| EPI_ISL_466706 | Canada/BC_01254071/2020 | 20-Apr | North America/Canada/British Columbia | BCCDC Public Health Laboratory | BCCDC Public Health Laboratory | Richard Harrigan et al |
| EPI_ISL_466707 | Canada/BC_01294861/2020 | 20-Apr | North America/Canada/British Columbia | BCCDC Public Health Laboratory | BCCDC Public Health Laboratory | Richard Harrigan et al |

| | | | | | | |
|---|---|---|---|---|---|---|
| EPI_ISL_466708 | Canada/BC_01296812/2020 | 20-Apr | North America/Canada/British Columbia | BCCDC Public Health Laboratory | BCCDC Public Health Laboratory | Richard Harrigan et al |
| EPI_ISL_466709 | Canada/BC_01468719/2020 | 20-Apr | North America/Canada/British Columbia | BCCDC Public Health Laboratory | BCCDC Public Health Laboratory | Richard Harrigan et al |
| EPI_ISL_466710 | Canada/BC_01560118/2020 | 20-Apr | North America/Canada/British Columbia | BCCDC Public Health Laboratory | BCCDC Public Health Laboratory | Richard Harrigan et al |
| EPI_ISL_466711 | Canada/BC_01594615/2020 | 20-Apr | North America/Canada/British Columbia | BCCDC Public Health Laboratory | BCCDC Public Health Laboratory | Richard Harrigan et al |
| EPI_ISL_466712 | Canada/BC_02045287/2020 | 20-Apr | North America/Canada/British Columbia | BCCDC Public Health Laboratory | BCCDC Public Health Laboratory | Richard Harrigan et al |
| EPI_ISL_466713 | Canada/BC_02051350/2020 | 20-Apr | North America/Canada/British Columbia | BCCDC Public Health Laboratory | BCCDC Public Health Laboratory | Richard Harrigan et al |
| EPI_ISL_466714 | Canada/BC_02077392/2020 | 20-Apr | North America/Canada/British Columbia | BCCDC Public Health Laboratory | BCCDC Public Health Laboratory | Richard Harrigan et al |
| EPI_ISL_466715 | Canada/BC_02083468/2020 | 20-Apr | North America/Canada/British Columbia | BCCDC Public Health Laboratory | BCCDC Public Health Laboratory | Richard Harrigan et al |
| EPI_ISL_466716 | Canada/BC_02091552/2020 | 20-Apr | North America/Canada/British Columbia | BCCDC Public Health Laboratory | BCCDC Public Health Laboratory | Richard Harrigan et al |
| EPI_ISL_466717 | Canada/BC_02168448/2020 | 20-Apr | North America/Canada/British Columbia | BCCDC Public Health Laboratory | BCCDC Public Health Laboratory | Richard Harrigan et al |
| EPI_ISL_466718 | Canada/BC_02169597/2020 | 20-Apr | North America/Canada/British Columbia | BCCDC Public Health Laboratory | BCCDC Public Health Laboratory | Richard Harrigan et al |
| EPI_ISL_466719 | Canada/BC_02185653/2020 | 20-Apr | North America/Canada/British Columbia | BCCDC Public Health Laboratory | BCCDC Public Health Laboratory | Richard Harrigan et al |
| EPI_ISL_466720 | Canada/BC_02246883/2020 | 20-Apr | North America/Canada/British Columbia | BCCDC Public Health Laboratory | BCCDC Public Health Laboratory | Richard Harrigan et al |
| EPI_ISL_466721 | Canada/BC_02415563/2020 | 20-Apr | North America/Canada/British Columbia | BCCDC Public Health Laboratory | BCCDC Public Health Laboratory | Richard Harrigan et al |
| EPI_ISL_466722 | Canada/BC_02472119/2020 | 20-Apr | North America/Canada/British Columbia | BCCDC Public Health Laboratory | BCCDC Public Health Laboratory | Richard Harrigan et al |
| EPI_ISL_466723 | Canada/BC_02502338/2020 | 20-Apr | North America/Canada/British Columbia | BCCDC Public Health Laboratory | BCCDC Public Health Laboratory | Richard Harrigan et al |
| EPI_ISL_466724 | Canada/BC_02511020/2020 | 20-Apr | North America/Canada/British Columbia | BCCDC Public Health Laboratory | BCCDC Public Health Laboratory | Richard Harrigan et al |
| EPI_ISL_466725 | Canada/BC_02585169/2020 | 20-Apr | North America/Canada/British Columbia | BCCDC Public Health Laboratory | BCCDC Public Health Laboratory | Richard Harrigan et al |
| EPI_ISL_466726 | Canada/BC_02594336/2020 | 20-Apr | North America/Canada/British Columbia | BCCDC Public Health Laboratory | BCCDC Public Health Laboratory | Richard Harrigan et al |
| EPI_ISL_466727 | Canada/BC_02803451/2020 | 20-Apr | North America/Canada/British Columbia | BCCDC Public Health Laboratory | BCCDC Public Health Laboratory | Richard Harrigan et al |

| | | | | | | |
|---|---|---|---|---|---|---|
| EPI_ISL_466728 | Canada/BC_02807114/2020 | 20-Apr | North America/Canada/British Columbia | BCCDC Public Health Laboratory | BCCDC Public Health Laboratory | Richard Harrigan et al |
| EPI_ISL_466729 | Canada/BC_02811162/2020 | 20-Apr | North America/Canada/British Columbia | BCCDC Public Health Laboratory | BCCDC Public Health Laboratory | Richard Harrigan et al |
| EPI_ISL_466730 | Canada/BC_02820264/2020 | 20-Apr | North America/Canada/British Columbia | BCCDC Public Health Laboratory | BCCDC Public Health Laboratory | Richard Harrigan et al |
| EPI_ISL_466731 | Canada/BC_02822314/2020 | 20-Apr | North America/Canada/British Columbia | BCCDC Public Health Laboratory | BCCDC Public Health Laboratory | Richard Harrigan et al |
| EPI_ISL_466732 | Canada/BC_02887476/2020 | 20-Apr | North America/Canada/British Columbia | BCCDC Public Health Laboratory | BCCDC Public Health Laboratory | Richard Harrigan et al |
| EPI_ISL_466733 | Canada/BC_02888326/2020 | 20-Apr | North America/Canada/British Columbia | BCCDC Public Health Laboratory | BCCDC Public Health Laboratory | Richard Harrigan et al |
| EPI_ISL_466734 | Canada/BC_03055797/2020 | 20-Apr | North America/Canada/British Columbia | BCCDC Public Health Laboratory | BCCDC Public Health Laboratory | Richard Harrigan et al |
| EPI_ISL_466735 | Canada/BC_03069740/2020 | 20-Apr | North America/Canada/British Columbia | BCCDC Public Health Laboratory | BCCDC Public Health Laboratory | Richard Harrigan et al |
| EPI_ISL_466736 | Canada/BC_03120111/2020 | 20-Apr | North America/Canada/British Columbia | BCCDC Public Health Laboratory | BCCDC Public Health Laboratory | Richard Harrigan et al |
| EPI_ISL_466737 | Canada/BC_03385157/2020 | 20-Apr | North America/Canada/British Columbia | BCCDC Public Health Laboratory | BCCDC Public Health Laboratory | Richard Harrigan et al |
| EPI_ISL_466738 | Canada/BC_03386598/2020 | 20-Apr | North America/Canada/British Columbia | BCCDC Public Health Laboratory | BCCDC Public Health Laboratory | Richard Harrigan et al |
| EPI_ISL_466739 | Canada/BC_03405412/2020 | 20-Apr | North America/Canada/British Columbia | BCCDC Public Health Laboratory | BCCDC Public Health Laboratory | Richard Harrigan et al |
| EPI_ISL_466740 | Canada/BC_03423088/2020 | 20-Apr | North America/Canada/British Columbia | BCCDC Public Health Laboratory | BCCDC Public Health Laboratory | Richard Harrigan et al |
| EPI_ISL_466741 | Canada/BC_03452391/2020 | 20-Apr | North America/Canada/British Columbia | BCCDC Public Health Laboratory | BCCDC Public Health Laboratory | Richard Harrigan et al |
| EPI_ISL_466742 | Canada/BC_03469645/2020 | 20-Apr | North America/Canada/British Columbia | BCCDC Public Health Laboratory | BCCDC Public Health Laboratory | Richard Harrigan et al |
| EPI_ISL_466743 | Canada/BC_03469815/2020 | 20-Apr | North America/Canada/British Columbia | BCCDC Public Health Laboratory | BCCDC Public Health Laboratory | Richard Harrigan et al |
| EPI_ISL_466744 | Canada/BC_03469913/2020 | 20-Apr | North America/Canada/British Columbia | BCCDC Public Health Laboratory | BCCDC Public Health Laboratory | Richard Harrigan et al |
| EPI_ISL_466745 | Canada/BC_03470015/2020 | 20-Apr | North America/Canada/British Columbia | BCCDC Public Health Laboratory | BCCDC Public Health Laboratory | Richard Harrigan et al |
| EPI_ISL_466746 | Canada/BC_03470080/2020 | 20-Apr | North America/Canada/British Columbia | BCCDC Public Health Laboratory | BCCDC Public Health Laboratory | Richard Harrigan et al |
| EPI_ISL_466747 | Canada/BC_03470522/2020 | 20-Apr | North America/Canada/British Columbia | BCCDC Public Health Laboratory | BCCDC Public Health Laboratory | Richard Harrigan et al |

| | | | | | | |
|---|---|---|---|---|---|---|
| EPI_ISL_466748 | Canada/BC_03470614/2020 | 20-Apr | North America/Canada/British Columbia | BCCDC Public Health Laboratory | BCCDC Public Health Laboratory | Richard Harrigan et al |
| EPI_ISL_466749 | Canada/BC_03471049/2020 | 20-Apr | North America/Canada/British Columbia | BCCDC Public Health Laboratory | BCCDC Public Health Laboratory | Richard Harrigan et al |
| EPI_ISL_466750 | Canada/BC_03471188/2020 | 20-Apr | North America/Canada/British Columbia | BCCDC Public Health Laboratory | BCCDC Public Health Laboratory | Richard Harrigan et al |
| EPI_ISL_466751 | Canada/BC_03477384/2020 | 20-Apr | North America/Canada/British Columbia | BCCDC Public Health Laboratory | BCCDC Public Health Laboratory | Richard Harrigan et al |
| EPI_ISL_466752 | Canada/BC_03489456/2020 | 20-Apr | North America/Canada/British Columbia | BCCDC Public Health Laboratory | BCCDC Public Health Laboratory | Richard Harrigan et al |
| EPI_ISL_466753 | Canada/BC_03489776/2020 | 20-Apr | North America/Canada/British Columbia | BCCDC Public Health Laboratory | BCCDC Public Health Laboratory | Richard Harrigan et al |
| EPI_ISL_466754 | Canada/BC_03493786/2020 | 20-Apr | North America/Canada/British Columbia | BCCDC Public Health Laboratory | BCCDC Public Health Laboratory | Richard Harrigan et al |
| EPI_ISL_466755 | Canada/BC_03907778/2020 | 20-Apr | North America/Canada/British Columbia | BCCDC Public Health Laboratory | BCCDC Public Health Laboratory | Richard Harrigan et al |
| EPI_ISL_466756 | Canada/BC_03914944/2020 | 20-Apr | North America/Canada/British Columbia | BCCDC Public Health Laboratory | BCCDC Public Health Laboratory | Richard Harrigan et al |
| EPI_ISL_466757 | Canada/BC_03917951/2020 | 20-Apr | North America/Canada/British Columbia | BCCDC Public Health Laboratory | BCCDC Public Health Laboratory | Richard Harrigan et al |
| EPI_ISL_466758 | Canada/BC_03959087/2020 | 20-Apr | North America/Canada/British Columbia | BCCDC Public Health Laboratory | BCCDC Public Health Laboratory | Richard Harrigan et al |
| EPI_ISL_466759 | Canada/BC_03970582/2020 | 20-Apr | North America/Canada/British Columbia | BCCDC Public Health Laboratory | BCCDC Public Health Laboratory | Richard Harrigan et al |
| EPI_ISL_466760 | Canada/BC_04013424/2020 | 20-Apr | North America/Canada/British Columbia | BCCDC Public Health Laboratory | BCCDC Public Health Laboratory | Richard Harrigan et al |
| EPI_ISL_466761 | Canada/BC_04020783/2020 | 20-Apr | North America/Canada/British Columbia | BCCDC Public Health Laboratory | BCCDC Public Health Laboratory | Richard Harrigan et al |
| EPI_ISL_466762 | Canada/BC_04033837/2020 | 20-Apr | North America/Canada/British Columbia | BCCDC Public Health Laboratory | BCCDC Public Health Laboratory | Richard Harrigan et al |
| EPI_ISL_466763 | Canada/BC_04048819/2020 | 20-Apr | North America/Canada/British Columbia | BCCDC Public Health Laboratory | BCCDC Public Health Laboratory | Richard Harrigan et al |
| EPI_ISL_466764 | Canada/BC_04164497/2020 | 20-Apr | North America/Canada/British Columbia | BCCDC Public Health Laboratory | BCCDC Public Health Laboratory | Richard Harrigan et al |
| EPI_ISL_466765 | Canada/BC_04286257/2020 | 20-Apr | North America/Canada/British Columbia | BCCDC Public Health Laboratory | BCCDC Public Health Laboratory | Richard Harrigan et al |
| EPI_ISL_466766 | Canada/BC_04346268/2020 | 20-Apr | North America/Canada/British Columbia | BCCDC Public Health Laboratory | BCCDC Public Health Laboratory | Richard Harrigan et al |
| EPI_ISL_466767 | Canada/BC_04837165/2020 | 20-Apr | North America/Canada/British Columbia | BCCDC Public Health Laboratory | BCCDC Public Health Laboratory | Richard Harrigan et al |

| | | | | | | |
|---|---|---|---|---|---|---|
| EPI_ISL_466768 | Canada/BC_04837932/2020 | 20-Apr | North America/Canada/British Columbia | BCCDC Public Health Laboratory | BCCDC Public Health Laboratory | Richard Harrigan et al |
| EPI_ISL_466769 | Canada/BC_04869329/2020 | 20-Apr | North America/Canada/British Columbia | BCCDC Public Health Laboratory | BCCDC Public Health Laboratory | Richard Harrigan et al |
| EPI_ISL_466770 | Canada/BC_04882357/2020 | 20-Apr | North America/Canada/British Columbia | BCCDC Public Health Laboratory | BCCDC Public Health Laboratory | Richard Harrigan et al |
| EPI_ISL_466771 | Canada/BC_04903777/2020 | 20-Apr | North America/Canada/British Columbia | BCCDC Public Health Laboratory | BCCDC Public Health Laboratory | Richard Harrigan et al |
| EPI_ISL_466772 | Canada/BC_04930828/2020 | 20-Apr | North America/Canada/British Columbia | BCCDC Public Health Laboratory | BCCDC Public Health Laboratory | Richard Harrigan et al |
| EPI_ISL_466773 | Canada/BC_04935664/2020 | 20-Apr | North America/Canada/British Columbia | BCCDC Public Health Laboratory | BCCDC Public Health Laboratory | Richard Harrigan et al |
| EPI_ISL_466774 | Canada/BC_04975041/2020 | 20-Apr | North America/Canada/British Columbia | BCCDC Public Health Laboratory | BCCDC Public Health Laboratory | Richard Harrigan et al |
| EPI_ISL_466775 | Canada/BC_05017368/2020 | 20-Apr | North America/Canada/British Columbia | BCCDC Public Health Laboratory | BCCDC Public Health Laboratory | Richard Harrigan et al |
| EPI_ISL_466776 | Canada/BC_05023412/2020 | 20-Apr | North America/Canada/British Columbia | BCCDC Public Health Laboratory | BCCDC Public Health Laboratory | Richard Harrigan et al |
| EPI_ISL_466777 | Canada/BC_05075366/2020 | 20-Apr | North America/Canada/British Columbia | BCCDC Public Health Laboratory | BCCDC Public Health Laboratory | Richard Harrigan et al |
| EPI_ISL_466779 | Canada/BC_05075882/2020 | 20-Apr | North America/Canada/British Columbia | BCCDC Public Health Laboratory | BCCDC Public Health Laboratory | Richard Harrigan et al |
| EPI_ISL_466780 | Canada/BC_05075921/2020 | 20-Apr | North America/Canada/British Columbia | BCCDC Public Health Laboratory | BCCDC Public Health Laboratory | Richard Harrigan et al |
| EPI_ISL_466781 | Canada/BC_05076038/2020 | 20-Apr | North America/Canada/British Columbia | BCCDC Public Health Laboratory | BCCDC Public Health Laboratory | Richard Harrigan et al |
| EPI_ISL_466782 | Canada/BC_05132390/2020 | 20-Apr | North America/Canada/British Columbia | BCCDC Public Health Laboratory | BCCDC Public Health Laboratory | Richard Harrigan et al |
| EPI_ISL_466783 | Canada/BC_05248019/2020 | 20-Apr | North America/Canada/British Columbia | BCCDC Public Health Laboratory | BCCDC Public Health Laboratory | Richard Harrigan et al |
| EPI_ISL_466784 | Canada/BC_05256838/2020 | 20-Apr | North America/Canada/British Columbia | BCCDC Public Health Laboratory | BCCDC Public Health Laboratory | Richard Harrigan et al |
| EPI_ISL_466785 | Canada/BC_05294838/2020 | 20-Apr | North America/Canada/British Columbia | BCCDC Public Health Laboratory | BCCDC Public Health Laboratory | Richard Harrigan et al |
| EPI_ISL_466786 | Canada/BC_05295317/2020 | 20-Apr | North America/Canada/British Columbia | BCCDC Public Health Laboratory | BCCDC Public Health Laboratory | Richard Harrigan et al |
| EPI_ISL_466787 | Canada/BC_05300317/2020 | 20-Apr | North America/Canada/British Columbia | BCCDC Public Health Laboratory | BCCDC Public Health Laboratory | Richard Harrigan et al |
| EPI_ISL_466788 | Canada/BC_05333597/2020 | 20-Apr | North America/Canada/British Columbia | BCCDC Public Health Laboratory | BCCDC Public Health Laboratory | Richard Harrigan et al |

| | | | | | | |
|---|---|---|---|---|---|---|
| EPI_ISL_466789 | Canada/BC_05740462/2020 | 20-Apr | North America/Canada/British Columbia | BCCDC Public Health Laboratory | BCCDC Public Health Laboratory | Richard Harrigan et al |
| EPI_ISL_466790 | Canada/BC_05844438/2020 | 20-Apr | North America/Canada/British Columbia | BCCDC Public Health Laboratory | BCCDC Public Health Laboratory | Richard Harrigan et al |
| EPI_ISL_466791 | Canada/BC_05845391/2020 | 20-Apr | North America/Canada/British Columbia | BCCDC Public Health Laboratory | BCCDC Public Health Laboratory | Richard Harrigan et al |
| EPI_ISL_466792 | Canada/BC_05941640/2020 | 20-Apr | North America/Canada/British Columbia | BCCDC Public Health Laboratory | BCCDC Public Health Laboratory | Richard Harrigan et al |
| EPI_ISL_466793 | Canada/BC_05941876/2020 | 20-Apr | North America/Canada/British Columbia | BCCDC Public Health Laboratory | BCCDC Public Health Laboratory | Richard Harrigan et al |
| EPI_ISL_466794 | Canada/BC_05941974/2020 | 20-Apr | North America/Canada/British Columbia | BCCDC Public Health Laboratory | BCCDC Public Health Laboratory | Richard Harrigan et al |
| EPI_ISL_466795 | Canada/BC_05942483/2020 | 20-Apr | North America/Canada/British Columbia | BCCDC Public Health Laboratory | BCCDC Public Health Laboratory | Richard Harrigan et al |
| EPI_ISL_466796 | Canada/BC_05943217/2020 | 20-Apr | North America/Canada/British Columbia | BCCDC Public Health Laboratory | BCCDC Public Health Laboratory | Richard Harrigan et al |
| EPI_ISL_466797 | Canada/BC_06238282/2020 | 20-Apr | North America/Canada/British Columbia | BCCDC Public Health Laboratory | BCCDC Public Health Laboratory | Richard Harrigan et al |
| EPI_ISL_466798 | Canada/BC_06364232/2020 | 20-Apr | North America/Canada/British Columbia | BCCDC Public Health Laboratory | BCCDC Public Health Laboratory | Richard Harrigan et al |
| EPI_ISL_466799 | Canada/BC_06370010/2020 | 20-Apr | North America/Canada/British Columbia | BCCDC Public Health Laboratory | BCCDC Public Health Laboratory | Richard Harrigan et al |
| EPI_ISL_466800 | Canada/BC_06378958/2020 | 20-Apr | North America/Canada/British Columbia | BCCDC Public Health Laboratory | BCCDC Public Health Laboratory | Richard Harrigan et al |
| EPI_ISL_466802 | Canada/BC_06764319/2020 | 20-Apr | North America/Canada/British Columbia | BCCDC Public Health Laboratory | BCCDC Public Health Laboratory | Richard Harrigan et al |
| EPI_ISL_466803 | Canada/BC_06768018/2020 | 20-Apr | North America/Canada/British Columbia | BCCDC Public Health Laboratory | BCCDC Public Health Laboratory | Richard Harrigan et al |
| EPI_ISL_466804 | Canada/BC_06779312/2020 | 20-Apr | North America/Canada/British Columbia | BCCDC Public Health Laboratory | BCCDC Public Health Laboratory | Richard Harrigan et al |
| EPI_ISL_466805 | Canada/BC_06802412/2020 | 20-Apr | North America/Canada/British Columbia | BCCDC Public Health Laboratory | BCCDC Public Health Laboratory | Richard Harrigan et al |
| EPI_ISL_466806 | Canada/BC_06803213/2020 | 20-Apr | North America/Canada/British Columbia | BCCDC Public Health Laboratory | BCCDC Public Health Laboratory | Richard Harrigan et al |
| EPI_ISL_466807 | Canada/BC_06852453/2020 | 20-Apr | North America/Canada/British Columbia | BCCDC Public Health Laboratory | BCCDC Public Health Laboratory | Richard Harrigan et al |
| EPI_ISL_466808 | Canada/BC_06944842/2020 | 20-Apr | North America/Canada/British Columbia | BCCDC Public Health Laboratory | BCCDC Public Health Laboratory | Richard Harrigan et al |
| EPI_ISL_466809 | Canada/BC_06946792/2020 | 20-Apr | North America/Canada/British Columbia | BCCDC Public Health Laboratory | BCCDC Public Health Laboratory | Richard Harrigan et al |

| | | | | | | |
|---|---|---|---|---|---|---|
| EPI_ISL_466810 | Canada/BC_06955362/2020 | 20-Apr | North America/Canada/British Columbia | BCCDC Public Health Laboratory | BCCDC Public Health Laboratory | Richard Harrigan et al |
| EPI_ISL_466811 | Canada/BC_06975953/2020 | 20-Apr | North America/Canada/British Columbia | BCCDC Public Health Laboratory | BCCDC Public Health Laboratory | Richard Harrigan et al |
| EPI_ISL_466812 | Canada/BC_06998971/2020 | 20-Apr | North America/Canada/British Columbia | BCCDC Public Health Laboratory | BCCDC Public Health Laboratory | Richard Harrigan et al |
| EPI_ISL_466813 | Canada/BC_07138890/2020 | 20-Apr | North America/Canada/British Columbia | BCCDC Public Health Laboratory | BCCDC Public Health Laboratory | Richard Harrigan et al |
| EPI_ISL_466814 | Canada/BC_07229190/2020 | 20-Apr | North America/Canada/British Columbia | BCCDC Public Health Laboratory | BCCDC Public Health Laboratory | Richard Harrigan et al |
| EPI_ISL_466815 | Canada/BC_07312838/2020 | 20-Apr | North America/Canada/British Columbia | BCCDC Public Health Laboratory | BCCDC Public Health Laboratory | Richard Harrigan et al |
| EPI_ISL_466816 | Canada/BC_07360020/2020 | 20-Apr | North America/Canada/British Columbia | BCCDC Public Health Laboratory | BCCDC Public Health Laboratory | Richard Harrigan et al |
| EPI_ISL_466818 | Canada/BC_07396328/2020 | 20-Apr | North America/Canada/British Columbia | BCCDC Public Health Laboratory | BCCDC Public Health Laboratory | Richard Harrigan et al |
| EPI_ISL_466819 | Canada/BC_07720149/2020 | 20-Apr | North America/Canada/British Columbia | BCCDC Public Health Laboratory | BCCDC Public Health Laboratory | Richard Harrigan et al |
| EPI_ISL_466820 | Canada/BC_07788742/2020 | 20-Apr | North America/Canada/British Columbia | BCCDC Public Health Laboratory | BCCDC Public Health Laboratory | Richard Harrigan et al |
| EPI_ISL_466821 | Canada/BC_07812071/2020 | 20-Apr | North America/Canada/British Columbia | BCCDC Public Health Laboratory | BCCDC Public Health Laboratory | Richard Harrigan et al |
| EPI_ISL_466822 | Canada/BC_07833956/2020 | 20-Apr | North America/Canada/British Columbia | BCCDC Public Health Laboratory | BCCDC Public Health Laboratory | Richard Harrigan et al |
| EPI_ISL_466823 | Canada/BC_07850299/2020 | 20-Apr | North America/Canada/British Columbia | BCCDC Public Health Laboratory | BCCDC Public Health Laboratory | Richard Harrigan et al |
| EPI_ISL_466824 | Canada/BC_07884511/2020 | 20-Apr | North America/Canada/British Columbia | BCCDC Public Health Laboratory | BCCDC Public Health Laboratory | Richard Harrigan et al |
| EPI_ISL_466825 | Canada/BC_07943734/2020 | 20-Apr | North America/Canada/British Columbia | BCCDC Public Health Laboratory | BCCDC Public Health Laboratory | Richard Harrigan et al |
| EPI_ISL_466826 | Canada/BC_07986535/2020 | 20-Apr | North America/Canada/British Columbia | BCCDC Public Health Laboratory | BCCDC Public Health Laboratory | Richard Harrigan et al |
| EPI_ISL_466827 | Canada/BC_08269684/2020 | 20-Apr | North America/Canada/British Columbia | BCCDC Public Health Laboratory | BCCDC Public Health Laboratory | Richard Harrigan et al |
| EPI_ISL_466828 | Canada/BC_08283773/2020 | 20-Apr | North America/Canada/British Columbia | BCCDC Public Health Laboratory | BCCDC Public Health Laboratory | Richard Harrigan et al |
| EPI_ISL_466829 | Canada/BC_08323846/2020 | 20-Apr | North America/Canada/British Columbia | BCCDC Public Health Laboratory | BCCDC Public Health Laboratory | Richard Harrigan et al |
| EPI_ISL_466830 | Canada/BC_08343934/2020 | 20-Apr | North America/Canada/British Columbia | BCCDC Public Health Laboratory | BCCDC Public Health Laboratory | Richard Harrigan et al |

| | | | | | | |
|---|---|---|---|---|---|---|
| EPI_ISL_466831 | Canada/BC_08347845/2020 | 20-Apr | North America/Canada/British Columbia | BCCDC Public Health Laboratory | BCCDC Public Health Laboratory | Richard Harrigan et al |
| EPI_ISL_466832 | Canada/BC_08419582/2020 | 20-Apr | North America/Canada/British Columbia | BCCDC Public Health Laboratory | BCCDC Public Health Laboratory | Richard Harrigan et al |
| EPI_ISL_466833 | Canada/BC_08797966/2020 | 20-Apr | North America/Canada/British Columbia | BCCDC Public Health Laboratory | BCCDC Public Health Laboratory | Richard Harrigan et al |
| EPI_ISL_466834 | Canada/BC_08847088/2020 | 20-Apr | North America/Canada/British Columbia | BCCDC Public Health Laboratory | BCCDC Public Health Laboratory | Richard Harrigan et al |
| EPI_ISL_466835 | Canada/BC_08847743/2020 | 20-Apr | North America/Canada/British Columbia | BCCDC Public Health Laboratory | BCCDC Public Health Laboratory | Richard Harrigan et al |
| EPI_ISL_466836 | Canada/BC_08848124/2020 | 20-Apr | North America/Canada/British Columbia | BCCDC Public Health Laboratory | BCCDC Public Health Laboratory | Richard Harrigan et al |
| EPI_ISL_466837 | Canada/BC_08851080/2020 | 20-Apr | North America/Canada/British Columbia | BCCDC Public Health Laboratory | BCCDC Public Health Laboratory | Richard Harrigan et al |
| EPI_ISL_466838 | Canada/BC_08940467/2020 | 20-Apr | North America/Canada/British Columbia | BCCDC Public Health Laboratory | BCCDC Public Health Laboratory | Richard Harrigan et al |
| EPI_ISL_466839 | India/TG-NGC-CDFD-01/2020 | 2020-05-10 | Asia/India/Telangana/ | National Genomics Core-Center for DNA Fingerprinting and Diagnostics | National Genomics Core- Center for DNA Fingerprinting and Diagnostics (NGC-CDFD)- DBT's PAN-INDIA-1000 Genome consortium | Bala Pratyusha et al |
| EPI_ISL_466848 | India/TG-NGC-CDFD-10/2020 | 2020-05-13 | Asia/India/Telangana/ | National Genomics Core-Center for DNA Fingerprinting and Diagnostics | National Genomics Core- Center for DNA Fingerprinting and Diagnostics (NGC-CDFD)- DBT's PAN-INDIA-1000 Genome consortium | Bala Pratyusha et al |

| | | | | | | |
|---|---|---|---|---|---|---|
| EPI_ISL_466852 | India/TG-NGC-CDFD-14/2020 | 2020-05-27 | Asia/India/Telangana/ | National Genomics Core-Center for DNA Fingerprinting and Diagnostics | National Genomics Core- Center for DNA Fingerprinting and Diagnostics (NGC-CDFD)- DBT's PAN-INDIA-1000 Genome consortium | Bala Pratyusha et al |
| EPI_ISL_466856 | India/TG-NGC-CDFD-18/2020 | 2020-05-27 | Asia/India/Telangana/ | National Genomics Core-Center for DNA Fingerprinting and Diagnostics | National Genomics Core- Center for DNA Fingerprinting and Diagnostics (NGC-CDFD)- DBT's PAN-INDIA-1000 Genome consortium | Bala Pratyusha et al |
| EPI_ISL_466860 | India/TG-NGC-CDFD-22/2020 | 2020-05-30 | Asia/India/Telangana/ | National Genomics Core-Center for DNA Fingerprinting and Diagnostics | National Genomics Core- Center for DNA Fingerprinting and Diagnostics (NGC-CDFD)- DBT's PAN-INDIA-1000 Genome consortium | Bala Pratyusha et al |
| EPI_ISL_466862 | India/TG-NGC-CDFD-24/2020 | 2020-05-30 | Asia/India/Telangana/ | National Genomics Core-Center for DNA Fingerprinting and Diagnostics | National Genomics Core- Center for DNA Fingerprinting and Diagnostics (NGC-CDFD)- DBT's PAN-INDIA-1000 Genome consortium | Bala Pratyusha et al |

| | | | | | | |
|---|---|---|---|---|---|---|
| EPI_ISL_466869 | India/TG-NGC-CDFD-31/2020 | 2020-05-30 | Asia/India/Telangana/ | National Genomics Core-Center for DNA Fingerprinting and Diagnostics | National Genomics Core- Center for DNA Fingerprinting and Diagnostics (NGC-CDFD)- DBT's PAN-INDIA-1000 Genome consortium | Bala Pratyusha et al |
| EPI_ISL_466870 | India/TG-NGC-CDFD-32/2020 | 2020-05-30 | Asia/India/Telangana/ | National Genomics Core-Center for DNA Fingerprinting and Diagnostics | National Genomics Core- Center for DNA Fingerprinting and Diagnostics (NGC-CDFD)- DBT's PAN-INDIA-1000 Genome consortium | Bala Pratyusha et al |
| EPI_ISL_466872 | India/TG-NGC-CDFD-34/2020 | 2020-05-30 | Asia/India/Telangana/ | National Genomics Core-Center for DNA Fingerprinting and Diagnostics | National Genomics Core- Center for DNA Fingerprinting and Diagnostics (NGC-CDFD)- DBT's PAN-INDIA-1000 Genome consortium | Bala Pratyusha et al |
| EPI_ISL_466875 | Germany/BY-MVP-0182/2020 | 2020-04-13 | Europe/Germany/Bavaria/Munich | Max von Pettenkofer Institute, Virology, National Reference Center for Retroviruses, LMU M√ºnchen | Laboratory for Functional Genome Analysis, Dept. Genomics, Gene Center of the LMU Munich | Max Muenchhoff et al |
| EPI_ISL_466876 | Germany/BY-MVP-0183/2020 | 2020-04-13 | Europe/Germany/Bavaria/Munich | Max von Pettenkofer Institute, Virology, National Reference Center for Retroviruses, LMU M√ºnchen | Laboratory for Functional Genome Analysis, Dept. Genomics, Gene Center of the LMU Munich | Max Muenchhoff et al |

| | | | | | | |
|---|---|---|---|---|---|---|
| EPI_ISL_466879 | Germany/BY-MVP-0186/2020 | 2020-04-13 | Europe/Germany/Bavaria/Munich | Max von Pettenkofer Institute, Virology, National Reference Center for Retroviruses, LMU M√ºnchen | Laboratory for Functional Genome Analysis, Dept. Genomics, Gene Center of the LMU Munich | Max Muenchhoff et al |
| EPI_ISL_466880 | Germany/BY-MVP-0187/2020 | 2020-04-13 | Europe/Germany/Bavaria/Munich | Max von Pettenkofer Institute, Virology, National Reference Center for Retroviruses, LMU M√ºnchen | Laboratory for Functional Genome Analysis, Dept. Genomics, Gene Center of the LMU Munich | Max Muenchhoff et al |
| EPI_ISL_466884 | Germany/BY-MVP-0196/2020 | 2020-04-16 | Europe/Germany/Bavaria/Munich | Max von Pettenkofer Institute, Virology, National Reference Center for Retroviruses, LMU M√ºnchen | Laboratory for Functional Genome Analysis, Dept. Genomics, Gene Center of the LMU Munich | Max Muenchhoff et al |
| EPI_ISL_466885 | Germany/BY-MVP-0200/2020 | 2020-04-16 | Europe/Germany/Bavaria/Munich | Max von Pettenkofer Institute, Virology, National Reference Center for Retroviruses, LMU M√ºnchen | Laboratory for Functional Genome Analysis, Dept. Genomics, Gene Center of the LMU Munich | Max Muenchhoff et al |
| EPI_ISL_466886 | Germany/BY-MVP-0201/2020 | 2020-04-17 | Europe/Germany/Bavaria/Munich | Max von Pettenkofer Institute, Virology, National Reference Center for Retroviruses, LMU M√ºnchen | Laboratory for Functional Genome Analysis, Dept. Genomics, Gene Center of the LMU Munich | Max Muenchhoff et al |
| EPI_ISL_466888 | Germany/BY-MVP-0207/2020 | 2020-04-20 | Europe/Germany/Bavaria/Munich | Max von Pettenkofer Institute, Virology, National Reference Center for Retroviruses, LMU M√ºnchen | Laboratory for Functional Genome Analysis, Dept. Genomics, Gene Center of the LMU Munich | Max Muenchhoff et al |

| | | | | | | |
|---|---|---|---|---|---|---|
| EPI_ISL_466889 | Germany/BY-MVP-0208/2020 | 2020-04-19 | Europe/Germany/Bavaria/Munich | Max von Pettenkofer Institute, Virology, National Reference Center for Retroviruses, LMU M√ºnchen | Laboratory for Functional Genome Analysis, Dept. Genomics, Gene Center of the LMU Munich | Max Muenchhoff et al |
| EPI_ISL_466901 | Germany/BY-MVP-0246/2020 | 2020-04-30 | Europe/Germany/Bavaria/Munich | Max von Pettenkofer Institute, Virology, National Reference Center for Retroviruses, LMU M√ºnchen | Laboratory for Functional Genome Analysis, Dept. Genomics, Gene Center of the LMU Munich | Max Muenchhoff et al |
| EPI_ISL_466907 | Germany/BY-MVP-0254/2020 | 2020-05-04 | Europe/Germany/Bavaria/Munich | Max von Pettenkofer Institute, Virology, National Reference Center for Retroviruses, LMU M√ºnchen | Laboratory for Functional Genome Analysis, Dept. Genomics, Gene Center of the LMU Munich | Max Muenchhoff et al |
| EPI_ISL_466909 | Germany/BY-MVP-0283/2020 | 2020-05-11 | Europe/Germany/Bavaria/Munich | Max von Pettenkofer Institute, Virology, National Reference Center for Retroviruses, LMU M√ºnchen | Laboratory for Functional Genome Analysis, Dept. Genomics, Gene Center of the LMU Munich | Max Muenchhoff et al |
| EPI_ISL_466911 | Germany/BY-MVP-0269/2020 | 2020-05-18 | Europe/Germany/Bavaria/Munich | Max von Pettenkofer Institute, Virology, National Reference Center for Retroviruses, LMU M√ºnchen | Laboratory for Functional Genome Analysis, Dept. Genomics, Gene Center of the LMU Munich | Max Muenchhoff et al |
| EPI_ISL_466912 | Germany/BY-MVP-0270/2020 | 2020-05-19 | Europe/Germany/Bavaria/Munich | Max von Pettenkofer Institute, Virology, National Reference Center for Retroviruses, LMU M√ºnchen | Laboratory for Functional Genome Analysis, Dept. Genomics, Gene Center of the LMU Munich | Max Muenchhoff et al |

| | | | | | | |
|---|---|---|---|---|---|---|
| EPI_ISL_466917 | Germany/BY-MVP-0284/2020 | 2020-05-23 | Europe/Germany/Bavaria/Munich | Max von Pettenkofer Institute, Virology, National Reference Center for Retroviruses, LMU M√ºnchen | Laboratory for Functional Genome Analysis, Dept. Genomics, Gene Center of the LMU Munich | Max Muenchhoff et al |
| EPI_ISL_466922 | Germany/BY-MVP-0289/2020 | 2020-05-25 | Europe/Germany/Bavaria/Munich | Max von Pettenkofer Institute, Virology, National Reference Center for Retroviruses, LMU M√ºnchen | Laboratory for Functional Genome Analysis, Dept. Genomics, Gene Center of the LMU Munich | Max Muenchhoff et al |
| EPI_ISL_466931 | Switzerland/BS-ETHZ-100006/2020 | 2020-03-06 | Europe/Switzerland/Basel-Stadt/ | Viollier AG | Department of Biosystems Science and Engineering, ETH Z√ºrich | Christian Beisel et al |
| EPI_ISL_466937 | Switzerland/TI-ETHZ-100012/2020 | 2020-03-06 | Europe/Switzerland/Ticino/ | Viollier AG | Department of Biosystems Science and Engineering, ETH Z√ºrich | Christian Beisel et al |
| EPI_ISL_466986 | Switzerland/SZ-ETHZ-100064/2020 | 2020-03-12 | Europe/Switzerland/Schwyz/ | Viollier AG | Department of Biosystems Science and Engineering, ETH Z√ºrich | Christian Beisel et al |
| EPI_ISL_466991 | Switzerland/TI-ETHZ-100069/2020 | 2020-03-12 | Europe/Switzerland/Ticino/ | Viollier AG | Department of Biosystems Science and Engineering, ETH Z√ºrich | Christian Beisel et al |
| EPI_ISL_467007 | Switzerland/SG-ETHZ-130013/2020 | 2020-05-07 | Europe/Switzerland/Sankt Gallen/ | Viollier AG | Department of Biosystems Science and Engineering, ETH Z√ºrich | Christian Beisel et al |
| EPI_ISL_467030 | India/GJ-GBRC169b/2020 | 2020-06-05 | Asia/India/Gujarat/Gandhinagar | GMERS Medical College and Hospital, Gandhinagar | Gujarat Biotechnology Research Centre | Gaurishankar Shrimali et al |

| | | | | | | |
|---|---|---|---|---|---|---|
| EPI_ISL_467033 | India/GJ-GBRC171/2020 | 2020-06-05 | Asia/India/Gujarat/Gandhinagar | GMERS Medical College and Hospital, Gandhinagar | Gujarat Biotechnology Research Centre | Tejas Shah et al |
| EPI_ISL_467034 | India/GJ-GBRC172/2020 | 2020-06-05 | Asia/India/Gujarat/Kalol | GMERS Medical College and Hospital, Gandhinagar | Gujarat Biotechnology Research Centre | Ankit Hinsu et al |
| EPI_ISL_467036 | India/GJ-GBRC173b/2020 | 2020-06-05 | Asia/India/Gujarat/Kalol | GMERS Medical College and Hospital, Gandhinagar | Gujarat Biotechnology Research Centre | Apurvasinh Puvar et al |
| EPI_ISL_467042 | India/GJ-GBRC177b/2020 | 2020-06-03 | Asia/India/Gujarat/Ahmedabad | B.J. Medical College and Civil hospital | Gujarat Biotechnology Research Centre | Pinal Trivedi et al |
| EPI_ISL_467045 | India/GJ-GBRC179a/2020 | 2020-06-03 | Asia/India/Gujarat/Ahmedabad | B.J. Medical College and Civil hospital | Gujarat Biotechnology Research Centre | Nitin Savaliya et al |
| EPI_ISL_467047 | India/GJ-GBRC180a/2020 | 2020-06-03 | Asia/India/Gujarat/Ahmedabad | B.J. Medical College and Civil hospital | Gujarat Biotechnology Research Centre | Dinesh Kumar et al |
| EPI_ISL_467048 | India/GJ-GBRC180b/2020 | 2020-06-03 | Asia/India/Gujarat/Ahmedabad | B.J. Medical College and Civil hospital | Gujarat Biotechnology Research Centre | Zuber Saiyed et al |
| EPI_ISL_467049 | India/GJ-GBRC181a/2020 | 2020-06-03 | Asia/India/Gujarat/Ahmedabad | B.J. Medical College and Civil hospital | Gujarat Biotechnology Research Centre | Komal Patel et al |
| EPI_ISL_467051 | India/GJ-GBRC182a/2020 | 2020-06-03 | Asia/India/Gujarat/Ahmedabad | B.J. Medical College and Civil hospital | Gujarat Biotechnology Research Centre | Snehal Bagatharia et al |
| EPI_ISL_467054 | India/GJ-GBRC183b/2020 | 2020-06-03 | Asia/India/Gujarat/Ahmedabad | B.J. Medical College and Civil hospital | Gujarat Biotechnology Research Centre | Nirav Mungalpara et al |
| EPI_ISL_467064 | Spain/AN-IBV-001900/2020 | 2020-03-15 | Europe/Spain/Andalusia/Puerto Santa M | Hospital Universitario Puerta del Mar de CV°diz - INIBICA | SeqCOVID-SPAIN consortium/IBV(CSIC) | Salud Rodrv≠guez-Pallares et al |
| EPI_ISL_467070 | Spain/AN-IBV-001901/2020 | 2020-03-15 | Europe/Spain/Andalusia/Puerto Santa M | Hospital Universitario Puerta del Mar de CV°diz - INIBICA | SeqCOVID-SPAIN consortium/IBV(CSIC) | Salud Rodrv≠guez-Pallares et al |
| EPI_ISL_467072 | Spain/AN-IBV-001907/2020 | 2020-03-16 | Europe/Spain/Andalusia/Algeciras | Hospital Universitario Puerta del Mar de CV°diz - INIBICA | SeqCOVID-SPAIN consortium/IBV(CSIC) | Salud Rodrv≠guez-Pallares et al |

| | | | | | | |
|---|---|---|---|---|---|---|
| EPI_ISL_467073 | Spain/AN-IBV-001904/2020 | 2020-03-16 | Europe/Spain/Andalusia/Tarifa | Hospital Universitario Puerta del Mar de C√°diz - INIBICA | SeqCOVID-SPAIN consortium/IBV(CSIC) | Salud Rodr√≠guez-Pallares et al |
| EPI_ISL_467077 | Spain/AN-IBV-001903/2020 | 2020-03-16 | Europe/Spain/Andalusia/Cadiz | Hospital Universitario Puerta del Mar de C√°diz - INIBICA | SeqCOVID-SPAIN consortium/IBV(CSIC) | Salud Rodr√≠guez-Pallares et al |
| EPI_ISL_467083 | Spain/MD-IBV-001898/2020 | 2020-03-15 | Europe/Spain/Madrid/ | Hospital Universitario Puerta del Mar de C√°diz - INIBICA | SeqCOVID-SPAIN consortium/IBV(CSIC) | Salud Rodr√≠guez-Pallares et al |
| EPI_ISL_467090 | Spain/CN-IBV-002595/2020 | 2020-03-12 | Europe/Spain/Canary Islands/Las Palmas | Hospital Universitario de Gran Canaria Dr. Negr√≠n | SeqCOVID-SPAIN consortium/IBV(CSIC) | M. Carmen P√©rez Gonz√°lez et al |
| EPI_ISL_467091 | Spain/CN-IBV-002592/2020 | 2020-03-10 | Europe/Spain/Canary Islands/Las Palmas | Hospital Universitario de Gran Canaria Dr. Negr√≠n | SeqCOVID-SPAIN consortium/IBV(CSIC) | M. Carmen P√©rez Gonz√°lez et al |
| EPI_ISL_467105 | Spain/PV-IBV-001575/2020 | 2020-03-13 | Europe/Spain/Basque Country/Vitoria-G | Hospital Universitario Araba. Vitoria-Gasteiz | SeqCOVID-SPAIN consortium/IBV(CSIC) | Silvia Hern√°ez Crespo et al |
| EPI_ISL_467108 | Spain/PV-IBV-001438/2020 | 2020-03-17 | Europe/Spain/Basque Country/Vitoria-G | Hospital Universitario Araba. Vitoria-Gasteiz | SeqCOVID-SPAIN consortium/IBV(CSIC) | Silvia Hern√°ez Crespo et al |
| EPI_ISL_467110 | Spain/PV-IBV-001547/2020 | 2020-03-16 | Europe/Spain/Basque Country/Vitoria-G | Hospital Universitario Araba. Vitoria-Gasteiz | SeqCOVID-SPAIN consortium/IBV(CSIC) | Silvia Hern√°ez Crespo et al |
| EPI_ISL_467112 | Spain/PV-IBV-001504/2020 | 2020-03-19 | Europe/Spain/Basque Country/Vitoria-G | Hospital Universitario Araba. Vitoria-Gasteiz | SeqCOVID-SPAIN consortium/IBV(CSIC) | Silvia Hern√°ez Crespo et al |
| EPI_ISL_467114 | Spain/PV-IBV-001553/2020 | 2020-03-12 | Europe/Spain/Basque Country/Vitoria-G | Hospital Universitario Araba. Vitoria-Gasteiz | SeqCOVID-SPAIN consortium/IBV(CSIC) | Silvia Hern√°ez Crespo et al |
| EPI_ISL_467123 | Spain/PV-IBV-001523/2020 | 2020-03-17 | Europe/Spain/Basque Country/Vitoria-G | Hospital Universitario Araba. Vitoria-Gasteiz | SeqCOVID-SPAIN consortium/IBV(CSIC) | Silvia Hern√°ez Crespo et al |
| EPI_ISL_467125 | Spain/PV-IBV-001530/2020 | 2020-03-18 | Europe/Spain/Basque Country/Vitoria-G | Hospital Universitario Araba. Vitoria-Gasteiz | SeqCOVID-SPAIN consortium/IBV(CSIC) | Silvia Hern√°ez Crespo et al |
| EPI_ISL_467147 | Spain/PV-IBV-001576/2020 | 2020-03-14 | Europe/Spain/Basque Country/Vitoria-G | Hospital Universitario Araba. Vitoria-Gasteiz | SeqCOVID-SPAIN consortium/IBV(CSIC) | Silvia Hern√°ez Crespo et al |

| | | | | | | |
|---|---|---|---|---|---|---|
| EPI_ISL_467151 | Spain/PV-IBV-001561/2020 | 2020-03-16 | Europe/Spain/Basque Country/Vitoria-G | Hospital Universitario Araba. Vitoria-Gasteiz | SeqCOVID-SPAIN consortium/IBV(CSIC) | Silvia Hernv°ez Crespo et al |
| EPI_ISL_467153 | Spain/PV-IBV-001612/2020 | 2020-03-12 | Europe/Spain/Basque Country/Vitoria-G | Hospital Universitario Araba. Vitoria-Gasteiz | SeqCOVID-SPAIN consortium/IBV(CSIC) | Silvia Hernv°ez Crespo et al |
| EPI_ISL_467156 | Spain/PV-IBV-001577/2020 | 2020-03-13 | Europe/Spain/Basque Country/Vitoria-G | Hospital Universitario Araba. Vitoria-Gasteiz | SeqCOVID-SPAIN consortium/IBV(CSIC) | Silvia Hernv°ez Crespo et al |
| EPI_ISL_467157 | Spain/PV-IBV-001498/2020 | 2020-03-19 | Europe/Spain/Basque Country/Vitoria-G | Hospital Universitario Araba. Vitoria-Gasteiz | SeqCOVID-SPAIN consortium/IBV(CSIC) | Silvia Hernv°ez Crespo et al |
| EPI_ISL_467175 | Spain/PV-IBV-001604/2020 | 2020-03-13 | Europe/Spain/Basque Country/Vitoria-G | Hospital Universitario Araba. Vitoria-Gasteiz | SeqCOVID-SPAIN consortium/IBV(CSIC) | Silvia Hernv°ez Crespo et al |
| EPI_ISL_467177 | Spain/PV-IBV-001440/2020 | 2020-03-19 | Europe/Spain/Basque Country/Vitoria-G | Hospital Universitario Araba. Vitoria-Gasteiz | SeqCOVID-SPAIN consortium/IBV(CSIC) | Silvia Hernv°ez Crespo et al |
| EPI_ISL_467211 | Spain/MD-IBV-003701/2020 | 2020-04-02 | Europe/Spain/Madrid/ | Hospital General Universitario Gregorio Marav±v≥n | SeqCOVID-SPAIN consortium/IBV(CSIC) | Laura Pv©rez-Lago et al |
| EPI_ISL_467234 | Spain/MD-IBV-003723/2020 | 2020-05-07 | Europe/Spain/Madrid/ | Hospital General Universitario Gregorio Marav±v≥n | SeqCOVID-SPAIN consortium/IBV(CSIC) | Laura Pv©rez-Lago et al |
| EPI_ISL_467236 | Spain/MD-IBV-003720/2020 | 2020-04-03 | Europe/Spain/Madrid/ | Hospital General Universitario Gregorio Marav±v≥n | SeqCOVID-SPAIN consortium/IBV(CSIC) | Laura Pv©rez-Lago et al |
| EPI_ISL_467238 | Spain/MD-IBV-003716/2020 | 2020-04-03 | Europe/Spain/Madrid/ | Hospital General Universitario Gregorio Marav±v≥n | SeqCOVID-SPAIN consortium/IBV(CSIC) | Laura Pv©rez-Lago et al |
| EPI_ISL_467242 | Spain/MD-IBV-003718/2020 | 2020-04-03 | Europe/Spain/Madrid/ | Hospital General Universitario Gregorio Marav±v≥n | SeqCOVID-SPAIN consortium/IBV(CSIC) | Laura Pv©rez-Lago et al |
| EPI_ISL_467253 | Spain/MD-IBV-003672/2020 | 2020-04-01 | Europe/Spain/Madrid/ | Hospital General Universitario Gregorio Marav±v≥n | SeqCOVID-SPAIN consortium/IBV(CSIC) | Laura Pv©rez-Lago et al |
| EPI_ISL_467254 | Spain/MD-IBV-003664/2020 | 2020-04-01 | Europe/Spain/Madrid/ | Hospital General Universitario Gregorio Marav±v≥n | SeqCOVID-SPAIN consortium/IBV(CSIC) | Laura Pv©rez-Lago et al |

| | | | | | | |
|---|---|---|---|---|---|---|
| EPI_ISL_467263 | Spain/GA-IBV-002714/2020 | 2020-04-07 | Europe/Spain/Galicia/Santiago de Comp | Hospital Cl√≠nico Universitario de Santiago de Compostela | SeqCOVID-SPAIN consortium/IBV(CSIC) | Jos√© Javier Costa Alcalde et al |
| EPI_ISL_467268 | Spain/GA-IBV-002743/2020 | 2020-04-13 | Europe/Spain/Galicia/Negreira | Hospital Cl√≠nico Universitario de Santiago de Compostela | SeqCOVID-SPAIN consortium/IBV(CSIC) | Jos√© Javier Costa Alcalde et al |
| EPI_ISL_467269 | Spain/GA-IBV-002748/2020 | 2020-04-13 | Europe/Spain/Galicia/Boiro | Hospital Cl√≠nico Universitario de Santiago de Compostela | SeqCOVID-SPAIN consortium/IBV(CSIC) | Jos√© Javier Costa Alcalde et al |
| EPI_ISL_467272 | Spain/GA-IBV-002730/2020 | 2020-04-13 | Europe/Spain/Galicia/Santiago de Comp | Hospital Cl√≠nico Universitario de Santiago de Compostela | SeqCOVID-SPAIN consortium/IBV(CSIC) | Jos√© Javier Costa Alcalde et al |
| EPI_ISL_467282 | Spain/GA-IBV-002750/2020 | 2020-04-13 | Europe/Spain/Galicia/Santiago de Comp | Hospital Cl√≠nico Universitario de Santiago de Compostela | SeqCOVID-SPAIN consortium/IBV(CSIC) | Jos√© Javier Costa Alcalde et al |
| EPI_ISL_467287 | Spain/GA-IBV-002722/2020 | 2020-04-07 | Europe/Spain/Galicia/Oroso | Hospital Cl√≠nico Universitario de Santiago de Compostela | SeqCOVID-SPAIN consortium/IBV(CSIC) | Jos√© Javier Costa Alcalde et al |
| EPI_ISL_467294 | Spain/GA-IBV-002715/2020 | 2020-04-08 | Europe/Spain/Galicia/Santiago de Comp | Hospital Cl√≠nico Universitario de Santiago de Compostela | SeqCOVID-SPAIN consortium/IBV(CSIC) | Jos√© Javier Costa Alcalde et al |
| EPI_ISL_467300 | BosniaandHerzegovina/04-Sarajevo/ | 2020-04-08 | Europe/Bosnia and Herzegovina/Sarajev | "General Hospital ""Abdulah Nakas""" | Alea Genetic Center | Rijad Konjhodzic et al |
| EPI_ISL_467303 | Canada/BC_09320741/2020 | 20-Apr | North America/Canada/British Columbia | BCCDC Public Health Laboratory | BCCDC Public Health Laboratory | Richard Harrigan et al |
| EPI_ISL_467305 | Canada/BC_09329927/2020 | 20-Apr | North America/Canada/British Columbia | BCCDC Public Health Laboratory | BCCDC Public Health Laboratory | Richard Harrigan et al |
| EPI_ISL_467306 | Canada/BC_09452317/2020 | 20-Apr | North America/Canada/British Columbia | BCCDC Public Health Laboratory | BCCDC Public Health Laboratory | Richard Harrigan et al |
| EPI_ISL_467307 | Canada/BC_09585059/2020 | 20-Apr | North America/Canada/British Columbia | BCCDC Public Health Laboratory | BCCDC Public Health Laboratory | Richard Harrigan et al |
| EPI_ISL_467308 | Canada/BC_09671877/2020 | 20-Apr | North America/Canada/British Columbia | BCCDC Public Health Laboratory | BCCDC Public Health Laboratory | Richard Harrigan et al |
| EPI_ISL_467309 | Canada/BC_09978786/2020 | 20-Apr | North America/Canada/British Columbia | BCCDC Public Health Laboratory | BCCDC Public Health Laboratory | Richard Harrigan et al |
| EPI_ISL_467310 | Canada/BC_10063599/2020 | 20-Apr | North America/Canada/British Columbia | BCCDC Public Health Laboratory | BCCDC Public Health Laboratory | Richard Harrigan et al |
| EPI_ISL_467311 | Canada/BC_10110624/2020 | 20-Apr | North America/Canada/British Columbia | BCCDC Public Health Laboratory | BCCDC Public Health Laboratory | Richard Harrigan et al |

| | | | | | | |
|---|---|---|---|---|---|---|
| EPI_ISL_467312 | Canada/BC_10111761/2020 | 20-Apr | North America/Canada/British Columbia | BCCDC Public Health Laboratory | BCCDC Public Health Laboratory | Richard Harrigan et al |
| EPI_ISL_467313 | Canada/BC_10111910/2020 | 20-Apr | North America/Canada/British Columbia | BCCDC Public Health Laboratory | BCCDC Public Health Laboratory | Richard Harrigan et al |
| EPI_ISL_467314 | Canada/BC_10589793/2020 | 20-Apr | North America/Canada/British Columbia | BCCDC Public Health Laboratory | BCCDC Public Health Laboratory | Richard Harrigan et al |
| EPI_ISL_467315 | Canada/BC_10592956/2020 | 20-Apr | North America/Canada/British Columbia | BCCDC Public Health Laboratory | BCCDC Public Health Laboratory | Richard Harrigan et al |
| EPI_ISL_467316 | Canada/BC_11025311/2020 | 20-Apr | North America/Canada/British Columbia | BCCDC Public Health Laboratory | BCCDC Public Health Laboratory | Richard Harrigan et al |
| EPI_ISL_467317 | Canada/BC_11028219/2020 | 20-Apr | North America/Canada/British Columbia | BCCDC Public Health Laboratory | BCCDC Public Health Laboratory | Richard Harrigan et al |
| EPI_ISL_467318 | Canada/BC_11135150/2020 | 20-Apr | North America/Canada/British Columbia | BCCDC Public Health Laboratory | BCCDC Public Health Laboratory | Richard Harrigan et al |
| EPI_ISL_467319 | Canada/BC_11180253/2020 | 20-Apr | North America/Canada/British Columbia | BCCDC Public Health Laboratory | BCCDC Public Health Laboratory | Richard Harrigan et al |
| EPI_ISL_467320 | Canada/BC_11184415/2020 | 20-Apr | North America/Canada/British Columbia | BCCDC Public Health Laboratory | BCCDC Public Health Laboratory | Richard Harrigan et al |
| EPI_ISL_467321 | Canada/BC_11186723/2020 | 20-Apr | North America/Canada/British Columbia | BCCDC Public Health Laboratory | BCCDC Public Health Laboratory | Richard Harrigan et al |
| EPI_ISL_467322 | Canada/BC_11303557/2020 | 20-Apr | North America/Canada/British Columbia | BCCDC Public Health Laboratory | BCCDC Public Health Laboratory | Richard Harrigan et al |
| EPI_ISL_467323 | Canada/BC_11306197/2020 | 20-Apr | North America/Canada/British Columbia | BCCDC Public Health Laboratory | BCCDC Public Health Laboratory | Richard Harrigan et al |
| EPI_ISL_467324 | Canada/BC_11637543/2020 | 20-Apr | North America/Canada/British Columbia | BCCDC Public Health Laboratory | BCCDC Public Health Laboratory | Richard Harrigan et al |
| EPI_ISL_467325 | Canada/BC_11647011/2020 | 20-Apr | North America/Canada/British Columbia | BCCDC Public Health Laboratory | BCCDC Public Health Laboratory | Richard Harrigan et al |
| EPI_ISL_467326 | Canada/BC_11649571/2020 | 20-Apr | North America/Canada/British Columbia | BCCDC Public Health Laboratory | BCCDC Public Health Laboratory | Richard Harrigan et al |
| EPI_ISL_467327 | Canada/BC_11656063/2020 | 20-Apr | North America/Canada/British Columbia | BCCDC Public Health Laboratory | BCCDC Public Health Laboratory | Richard Harrigan et al |
| EPI_ISL_467328 | Canada/BC_11656112/2020 | 20-Apr | North America/Canada/British Columbia | BCCDC Public Health Laboratory | BCCDC Public Health Laboratory | Richard Harrigan et al |
| EPI_ISL_467329 | Canada/BC_11726488/2020 | 20-Apr | North America/Canada/British Columbia | BCCDC Public Health Laboratory | BCCDC Public Health Laboratory | Richard Harrigan et al |
| EPI_ISL_467330 | Canada/BC_12186719/2020 | 20-Apr | North America/Canada/British Columbia | BCCDC Public Health Laboratory | BCCDC Public Health Laboratory | Richard Harrigan et al |
| EPI_ISL_467331 | Canada/BC_12199086/2020 | 20-Apr | North America/Canada/British Columbia | BCCDC Public Health Laboratory | BCCDC Public Health Laboratory | Richard Harrigan et al |

| | | | | | | |
|---|---|---|---|---|---|---|
| EPI_ISL_467332 | Canada/BC_12210980/2020 | 20-Apr | North America/Canada/British Columbia | BCCDC Public Health Laboratory | BCCDC Public Health Laboratory | Richard Harrigan et al |
| EPI_ISL_467333 | Canada/BC_12246693/2020 | 20-Apr | North America/Canada/British Columbia | BCCDC Public Health Laboratory | BCCDC Public Health Laboratory | Richard Harrigan et al |
| EPI_ISL_467334 | Canada/BC_12251717/2020 | 20-Apr | North America/Canada/British Columbia | BCCDC Public Health Laboratory | BCCDC Public Health Laboratory | Richard Harrigan et al |
| EPI_ISL_467335 | Canada/BC_12477112/2020 | 20-Apr | North America/Canada/British Columbia | BCCDC Public Health Laboratory | BCCDC Public Health Laboratory | Richard Harrigan et al |
| EPI_ISL_467336 | Canada/BC_12661418/2020 | 20-Apr | North America/Canada/British Columbia | BCCDC Public Health Laboratory | BCCDC Public Health Laboratory | Richard Harrigan et al |
| EPI_ISL_467337 | Canada/BC_12775269/2020 | 20-Apr | North America/Canada/British Columbia | BCCDC Public Health Laboratory | BCCDC Public Health Laboratory | Richard Harrigan et al |
| EPI_ISL_467338 | Canada/BC_12793935/2020 | 20-Apr | North America/Canada/British Columbia | BCCDC Public Health Laboratory | BCCDC Public Health Laboratory | Richard Harrigan et al |
| EPI_ISL_467339 | Canada/BC_13086747/2020 | 20-Apr | North America/Canada/British Columbia | BCCDC Public Health Laboratory | BCCDC Public Health Laboratory | Richard Harrigan et al |
| EPI_ISL_467340 | Canada/BC_13088259/2020 | 20-Apr | North America/Canada/British Columbia | BCCDC Public Health Laboratory | BCCDC Public Health Laboratory | Richard Harrigan et al |
| EPI_ISL_467341 | Canada/BC_13610520/2020 | 20-Apr | North America/Canada/British Columbia | BCCDC Public Health Laboratory | BCCDC Public Health Laboratory | Richard Harrigan et al |
| EPI_ISL_467342 | Canada/BC_13662361/2020 | 20-Apr | North America/Canada/British Columbia | BCCDC Public Health Laboratory | BCCDC Public Health Laboratory | Richard Harrigan et al |
| EPI_ISL_467343 | Canada/BC_13675191/2020 | 20-Apr | North America/Canada/British Columbia | BCCDC Public Health Laboratory | BCCDC Public Health Laboratory | Richard Harrigan et al |
| EPI_ISL_467345 | Brazil/RJ-1574/2020 | 2020-04-02 | South America/Brazil/Rio de Janeiro/Rio | Laboratory of Respiratory Viruses and Measles, Oswaldo Cruz Institute, FIOCRUZ | Laboratory of Respiratory Viruses and Measles, Oswaldo Cruz Institute, FIOCRUZ | Paola Resende et al |
| EPI_ISL_467346 | Brazil/RJ-1595/2020 | 2020-04-02 | South America/Brazil/Rio de Janeiro/Rio | Laboratory of Respiratory Viruses and Measles, Oswaldo Cruz Institute, FIOCRUZ | Laboratory of Respiratory Viruses and Measles, Oswaldo Cruz Institute, FIOCRUZ | Paola Resende et al |

| | | | | | | |
|---|---|---|---|---|---|---|
| EPI_ISL_467360 | Brazil/RJ-2733/2020 | 2020-04-25 | South America/Brazil/Rio de Janeiro/Du | Laboratory of Respiratory Viruses and Measles, Oswaldo Cruz Institute, FIOCRUZ | Laboratory of Respiratory Viruses and Measles, Oswaldo Cruz Institute, FIOCRUZ | Paola Resende et al |
| EPI_ISL_467374 | Indonesia/KI-EIJK-05/2020 | 2020-03-18 | Asia/Indonesia/East Kalimantan/Samari | Dinkes Samarinda | Eijkman Institute for Molecular Biology, Ministry of Research and Technology/National Agency for Research and Innovation | Edison Johar et al |
| EPI_ISL_467375 | Indonesia/SA-EIJK-06/2020 | 2020-03-23 | Asia/Indonesia/North Sulawesi/Manado | RSUP Prof. Dr. R. Kandou Manado | Eijkman Institute for Molecular Biology, Ministry of Research and Technology/National Agency for Research and Innovation | Edison Johar et al |
| EPI_ISL_467377 | USA/NY-NYUMC884/2020 | 2020-05-03 | North America/USA/New York/Manhatt | NYU Langone Health | Departments of Pathology and Medicine, New York University School of Medicine | Maria Aguero-Rosenfeld et al |
| EPI_ISL_467393 | USA/NY-NYUMC900/2020 | 2020-04-25 | North America/USA/New York/Brooklyn | NYU Langone Health | Departments of Pathology and Medicine, New York University School of Medicine | Maria Aguero-Rosenfeld et al |
| EPI_ISL_467398 | USA/NY-NYUMC905/2020 | 2020-04-28 | North America/USA/New York/Nassau C | NYU Langone Health | Departments of Pathology and Medicine, New York University School of Medicine | Maria Aguero-Rosenfeld et al |

| | | | | | | |
|---|---|---|---|---|---|---|
| EPI_ISL_467407 | USA/NY-NYUMC914/2020 | 2020-05-08 | North America/USA/New York/Bronx | NYU Langone Health | Departments of Pathology and Medicine, New York University School of Medicine | Maria Aguero-Rosenfeld et al |
| EPI_ISL_467417 | USA/NY-NYUMC924/2020 | 2020-03-18 | North America/USA/New York/Brooklyn | NYU Langone Health | Departments of Pathology and Medicine, New York University School of Medicine | Maria Aguero-Rosenfeld et al |
| EPI_ISL_467423 | Canada/BC_13704688/2020 | 20-Apr | North America/Canada/British Columbia | BCCDC Public Health Laboratory | BCCDC Public Health Laboratory | Richard Harrigan et al |
| EPI_ISL_467424 | Canada/BC_13705728/2020 | 20-Apr | North America/Canada/British Columbia | BCCDC Public Health Laboratory | BCCDC Public Health Laboratory | Richard Harrigan et al |
| EPI_ISL_467425 | Canada/BC_13949840/2020 | 20-Apr | North America/Canada/British Columbia | BCCDC Public Health Laboratory | BCCDC Public Health Laboratory | Richard Harrigan et al |
| EPI_ISL_467426 | Canada/BC_14260476/2020 | 20-Apr | North America/Canada/British Columbia | BCCDC Public Health Laboratory | BCCDC Public Health Laboratory | Richard Harrigan et al |
| EPI_ISL_467427 | Canada/BC_14282914/2020 | 20-Apr | North America/Canada/British Columbia | BCCDC Public Health Laboratory | BCCDC Public Health Laboratory | Richard Harrigan et al |
| EPI_ISL_467428 | Canada/BC_14592296/2020 | 20-Apr | North America/Canada/British Columbia | BCCDC Public Health Laboratory | BCCDC Public Health Laboratory | Richard Harrigan et al |
| EPI_ISL_467429 | Canada/BC_14592943/2020 | 20-Apr | North America/Canada/British Columbia | BCCDC Public Health Laboratory | BCCDC Public Health Laboratory | Richard Harrigan et al |
| EPI_ISL_467438 | SouthAfrica/KRISP-0028/2020 | 2020-04-02 | Africa/South Africa/KwaZulu-Natal/Bere | NHLS-IALCH | KRISP, KZN Research Innovation and Sequencing Platform | Giandhari J et al |
| EPI_ISL_467439 | SouthAfrica/KRISP-0029/2020 | 2020-04-02 | Africa/South Africa/KwaZulu-Natal/Bere | NHLS-IALCH | KRISP, KZN Research Innovation and Sequencing Platform | Giandhari J et al |

| | | | | | | |
|---|---|---|---|---|---|---|
| EPI_ISL_467446 | SouthAfrica/KRISP-0057/2020 | 2020-03-29 | Africa/South Africa/KwaZulu-Natal/Bere | Molecular Diagnostics Services (MDS) | KRISP, KZN Research Innovation and Sequencing Platform | Giandhari J et al |
| EPI_ISL_467458 | SouthAfrica/KRISP-0089/2020 | 2020-04-05 | Africa/South Africa/KwaZulu-Natal/Bere | AMPATH-DBN | KRISP, KZN Research Innovation and Sequencing Platform | Giandhari J et al |
| EPI_ISL_467459 | SouthAfrica/KRISP-0090/2020 | 2020-04-03 | Africa/South Africa/KwaZulu-Natal/Bere | AMPATH-DBN | KRISP, KZN Research Innovation and Sequencing Platform | Giandhari J et al |
| EPI_ISL_467464 | SouthAfrica/KRISP-0100/2020 | 2020-04-01 | Africa/South Africa/KwaZulu-Natal/Bere | AMPATH-DBN | KRISP, KZN Research Innovation and Sequencing Platform | Giandhari J et al |
| EPI_ISL_467467 | SouthAfrica/KRISP-0113/2020 | 2020-04-05 | Africa/South Africa/KwaZulu-Natal/Bere | AMPATH-DBN | KRISP, KZN Research Innovation and Sequencing Platform | Giandhari J et al |
| EPI_ISL_467475 | SouthAfrica/KRISP-0123/2020 | 2020-05-14 | Africa/South Africa/KwaZulu-Natal/ | Molecular Diagnostics Services (MDS) | KRISP, KZN Research Innovation and Sequencing Platform | Giandhari J et al |
| EPI_ISL_467476 | SouthAfrica/KRISP-0124/2020 | 2020-05-13 | Africa/South Africa/KwaZulu-Natal/ | Molecular Diagnostics Services (MDS) | KRISP, KZN Research Innovation and Sequencing Platform | Giandhari J et al |
| EPI_ISL_467477 | SouthAfrica/KRISP-0125/2020 | 2020-05-13 | Africa/South Africa/KwaZulu-Natal/ | Molecular Diagnostics Services (MDS) | KRISP, KZN Research Innovation and Sequencing Platform | Giandhari J et al |

| | | | | | | |
|---|---|---|---|---|---|---|
| EPI_ISL_467478 | SouthAfrica/KRISP-0126/2020 | 2020-05-13 | Africa/South Africa/KwaZulu-Natal/ | Molecular Diagnostics Services (MDS) | KRISP, KZN Research Innovation and Sequencing Platform | Giandhari J et al |
| EPI_ISL_467481 | SouthAfrica/KRISP-0129/2020 | 2020-05-13 | Africa/South Africa/KwaZulu-Natal/ | Molecular Diagnostics Services (MDS) | KRISP, KZN Research Innovation and Sequencing Platform | Giandhari J et al |
| EPI_ISL_467486 | SouthAfrica/KRISP-0134/2020 | 2020-05-14 | Africa/South Africa/KwaZulu-Natal/ | Molecular Diagnostics Services (MDS) | KRISP, KZN Research Innovation and Sequencing Platform | Giandhari J et al |
| EPI_ISL_467487 | SouthAfrica/KRISP-0135/2020 | 2020-05-14 | Africa/South Africa/KwaZulu-Natal/ | Molecular Diagnostics Services (MDS) | KRISP, KZN Research Innovation and Sequencing Platform | Giandhari J et al |
| EPI_ISL_467491 | SouthAfrica/KRISP-0139/2020 | 2020-05-16 | Africa/South Africa/KwaZulu-Natal/ | Molecular Diagnostics Services (MDS) | KRISP, KZN Research Innovation and Sequencing Platform | Giandhari J et al |
| EPI_ISL_467494 | SouthAfrica/KRISP-0144/2020 | 2020-05-30 | Africa/South Africa/KwaZulu-Natal/ | Molecular Diagnostics Services (MDS) | KRISP, KZN Research Innovation and Sequencing Platform | Giandhari J et al |
| EPI_ISL_467501 | SouthAfrica/KRISP-0151/2020 | 2020-06-01 | Africa/South Africa/KwaZulu-Natal/ | Molecular Diagnostics Services (MDS) | KRISP, KZN Research Innovation and Sequencing Platform | Giandhari J et al |
| EPI_ISL_467502 | SouthAfrica/KRISP-0152/2020 | 2020-06-01 | Africa/South Africa/KwaZulu-Natal/ | Molecular Diagnostics Services (MDS) | KRISP, KZN Research Innovation and Sequencing Platform | Giandhari J et al |

| | | | | | | |
|---|---|---|---|---|---|---|
| EPI_ISL_467504 | SouthAfrica/KRISP-0154/2020 | 2020-06-01 | Africa/South Africa/KwaZulu-Natal/ | Molecular Diagnostics Services (MDS) | KRISP, KZN Research Innovation and Sequencing Platform | Giandhari J et al |
| EPI_ISL_467505 | SouthAfrica/KRISP-0155/2020 | 2020-06-01 | Africa/South Africa/KwaZulu-Natal/ | Molecular Diagnostics Services (MDS) | KRISP, KZN Research Innovation and Sequencing Platform | Giandhari J et al |
| EPI_ISL_467506 | SouthAfrica/KRISP-0156/2020 | 2020-06-01 | Africa/South Africa/KwaZulu-Natal/ | Molecular Diagnostics Services (MDS) | KRISP, KZN Research Innovation and Sequencing Platform | Giandhari J et al |
| EPI_ISL_467507 | SouthAfrica/KRISP-0157/2020 | 2020-05-25 | Africa/South Africa/KwaZulu-Natal/Umb | NHLS-IALCH | KRISP, KZN Research Innovation and Sequencing Platform | Giandhari J et al |
| EPI_ISL_467511 | SouthAfrica/KRISP-0161/2020 | 2020-06-01 | Africa/South Africa/KwaZulu-Natal/Umb | NHLS-IALCH | KRISP, KZN Research Innovation and Sequencing Platform | Giandhari J et al |
| EPI_ISL_467512 | SouthAfrica/KRISP-0162/2020 | 2020-06-01 | Africa/South Africa/KwaZulu-Natal/Umb | NHLS-IALCH | KRISP, KZN Research Innovation and Sequencing Platform | Giandhari J et al |
| EPI_ISL_467513 | SouthAfrica/KRISP-0163/2020 | 2020-06-01 | Africa/South Africa/KwaZulu-Natal/Umb | NHLS-IALCH | KRISP, KZN Research Innovation and Sequencing Platform | Giandhari J et al |
| EPI_ISL_467514 | SouthAfrica/KRISP-0164/2020 | 2020-06-01 | Africa/South Africa/KwaZulu-Natal/Umb | NHLS-IALCH | KRISP, KZN Research Innovation and Sequencing Platform | Giandhari J et al |

| | | | | | | |
|---|---|---|---|---|---|---|
| EPI_ISL_467516 | SouthAfrica/KRISP-0166/2020 | 2020-04-14 | Africa/South Africa/KwaZulu-Natal/ | CAPRISA | KRISP, KZN Research Innovation and Sequencing Platform | Giandhari J et al |
| EPI_ISL_467518 | SouthAfrica/KRISP-0168/2020 | 2020-05-28 | Africa/South Africa/KwaZulu-Natal/Umb | NHLS-IALCH | KRISP, KZN Research Innovation and Sequencing Platform | Giandhari J et al |
| EPI_ISL_467519 | SouthAfrica/KRISP-0169/2020 | 2020-05-31 | Africa/South Africa/KwaZulu-Natal/Umb | NHLS-IALCH | KRISP, KZN Research Innovation and Sequencing Platform | Giandhari J et al |
| EPI_ISL_467520 | SouthAfrica/KRISP-0170/2020 | 2020-05-31 | Africa/South Africa/KwaZulu-Natal/Umb | NHLS-IALCH | KRISP, KZN Research Innovation and Sequencing Platform | Giandhari J et al |
| EPI_ISL_467522 | SouthAfrica/KRISP-0172/2020 | 2020-06-02 | Africa/South Africa/KwaZulu-Natal/Umb | NHLS-IALCH | KRISP, KZN Research Innovation and Sequencing Platform | Giandhari J et al |
| EPI_ISL_467523 | SouthAfrica/KRISP-0173/2020 | 2020-06-02 | Africa/South Africa/KwaZulu-Natal/Umb | NHLS-IALCH | KRISP, KZN Research Innovation and Sequencing Platform | Giandhari J et al |
| EPI_ISL_467524 | SouthAfrica/KRISP-0174/2020 | 2020-06-04 | Africa/South Africa/KwaZulu-Natal/Umb | NHLS-IALCH | KRISP, KZN Research Innovation and Sequencing Platform | Giandhari J et al |
| EPI_ISL_467541 | USA/NM-UNM-00090/2020 | 2020-04-01 | North America/USA/New Mexico/ | New Mexico Department of Health Scientific Laboratory Division | Center for Global Health, University of New Mexico Health Sciences Center | Daryl Domman et al |

| | | | | | | |
|---|---|---|---|---|---|---|
| EPI_ISL_467544 | USA/NM-UNM-00093/2020 | 2020-04-01 | North America/USA/New Mexico/ | New Mexico Department of Health Scientific Laboratory Division | Center for Global Health, University of New Mexico Health Sciences Center | Daryl Domman et al |
| EPI_ISL_467547 | USA/NM-UNM-00102/2020 | 2020-04-01 | North America/USA/New Mexico/ | New Mexico Department of Health Scientific Laboratory Division | Center for Global Health, University of New Mexico Health Sciences Center | Daryl Domman et al |
| EPI_ISL_467549 | USA/NM-UNM-00104/2020 | 2020-04-02 | North America/USA/New Mexico/ | New Mexico Department of Health Scientific Laboratory Division | Center for Global Health, University of New Mexico Health Sciences Center | Daryl Domman et al |
| EPI_ISL_467550 | USA/NM-UNM-00105/2020 | 2020-04-02 | North America/USA/New Mexico/ | New Mexico Department of Health Scientific Laboratory Division | Center for Global Health, University of New Mexico Health Sciences Center | Daryl Domman et al |
| EPI_ISL_467576 | USA/NM-UNM-00161/2020 | 2020-04-03 | North America/USA/New Mexico/ | New Mexico Department of Health Scientific Laboratory Division | Center for Global Health, University of New Mexico Health Sciences Center | Daryl Domman et al |
| EPI_ISL_467599 | USA/NM-UNM-00018/2020 | 2020-03-12 | North America/USA/New Mexico/ | New Mexico Department of Health Scientific Laboratory Division | Center for Global Health, University of New Mexico Health Sciences Center | Daryl Domman et al |
| EPI_ISL_467612 | USA/NM-UNM-00031/2020 | 2020-03-17 | North America/USA/New Mexico/ | New Mexico Department of Health Scientific Laboratory Division | Center for Global Health, University of New Mexico Health Sciences Center | Daryl Domman et al |
| EPI_ISL_467643 | USA/NM-UNM-00109/2020 | 2020-04-02 | North America/USA/New Mexico/ | New Mexico Department of Health Scientific Laboratory Division | Center for Global Health, University of New Mexico Health Sciences Center | Daryl Domman et al |

| | | | | | | |
|---|---|---|---|---|---|---|
| EPI_ISL_467656 | USA/NM-UNM-00154/2020 | 2020-04-03 | North America/USA/New Mexico/ | New Mexico Department of Health Scientific Laboratory Division | Center for Global Health, University of New Mexico Health Sciences Center | Daryl Domman et al |
| EPI_ISL_467659 | USA/NM-UNM-00158/2020 | 2020-04-03 | North America/USA/New Mexico/ | New Mexico Department of Health Scientific Laboratory Division | Center for Global Health, University of New Mexico Health Sciences Center | Daryl Domman et al |
| EPI_ISL_467660 | USA/NM-UNM-00159/2020 | 2020-04-03 | North America/USA/New Mexico/ | New Mexico Department of Health Scientific Laboratory Division | Center for Global Health, University of New Mexico Health Sciences Center | Daryl Domman et al |
| EPI_ISL_467774 | Russia/CRIE163844/2020 | 2020-03-31 | Europe/Russia/Moscow Oblast/ | "Molecular diagnostic laboratory of Federal Budget Institution of Science ""Central Research Institute of Epidemiology"" of The Federal Service on Customers' Rights Protection and Human Well-being Surveillance" | Group of Genomics and Postgenomic Technologies of Central Research Institute of Epidemilology | Speranskaya AS et al |
| EPI_ISL_467779 | Bucuresti/ChVir7245/2020 | 2020-04-27 | Europe/Romania/Bucharest/ | National Influenza Centre Romania | Charite Universitatsmedizin Berlin, Institute of Virology | Victor M Corman et al |
| EPI_ISL_467786 | USA/VA-DCLS-0389/2020 | 2020-04-18 | North America/USA/Virginia/ | Virginia DCLS | Virginia DCLS | Virginia DCLS et al |
| EPI_ISL_467787 | USA/VA-DCLS-0390/2020 | 2020-04-18 | North America/USA/Virginia/ | Virginia DCLS | Virginia DCLS | Virginia DCLS et al |
| EPI_ISL_467789 | USA/VA-DCLS-0393/2020 | 2020-04-19 | North America/USA/Virginia/ | Virginia DCLS | Virginia DCLS | Virginia DCLS et al |
| EPI_ISL_467790 | USA/VA-DCLS-0395/2020 | 2020-04-18 | North America/USA/Virginia/ | Virginia DCLS | Virginia DCLS | Virginia DCLS et al |
| EPI_ISL_467794 | USA/VA-DCLS-0399/2020 | 2020-04-21 | North America/USA/Virginia/ | Virginia DCLS | Virginia DCLS | Virginia DCLS et al |
| EPI_ISL_467801 | USA/VA-DCLS-0407/2020 | 2020-04-22 | North America/USA/Virginia/ | Virginia DCLS | Virginia DCLS | Virginia DCLS et al |
| EPI_ISL_467814 | USA/CA-QDX-148/2020 | 2020-03-16 | North America/USA/California/ | Quest Diagnostics | Quest Diagnostics | Anderson et al |
| EPI_ISL_467827 | USA/CA-QDX-33/2020 | 2020-03-15 | North America/USA/California/ | Quest Diagnostics | Quest Diagnostics | Anderson et al |
| EPI_ISL_467828 | USA/CA-QDX-34/2020 | 2020-03-15 | North America/USA/California/ | Quest Diagnostics | Quest Diagnostics | Anderson et al |
| EPI_ISL_467830 | USA/CA-QDX-38/2020 | 2020-03-15 | North America/USA/California/ | Quest Diagnostics | Quest Diagnostics | Anderson et al |
| EPI_ISL_467853 | USA/MA-QDX-145/2020 | 2020-03-14 | North America/USA/Massachusetts/ | Quest Diagnostics | Quest Diagnostics | Anderson et al |

| | | | | | | |
|---|---|---|---|---|---|---|
| EPI_ISL_467863 | USA/MA-QDX-64/2020 | 2020-03-15 | North America/USA/Massachusetts/ | Quest Diagnostics | Quest Diagnostics | Anderson et al |
| EPI_ISL_467878 | USA/NJ-QDX-144/2020 | 2020-03-13 | North America/USA/New Jersey/ | Quest Diagnostics | Quest Diagnostics | Anderson et al |
| EPI_ISL_467886 | USA/NY-QDX-128/2020 | 2020-03-16 | North America/USA/New York/ | Quest Diagnostics | Quest Diagnostics | Anderson et al |
| EPI_ISL_467892 | USA/NY-QDX-140/2020 | 2020-03-15 | North America/USA/New York/ | Quest Diagnostics | Quest Diagnostics | Anderson et al |
| EPI_ISL_467902 | USA/NY-QDX-75/2020 | 2020-03-13 | North America/USA/New York/ | Quest Diagnostics | Quest Diagnostics | Anderson et al |
| EPI_ISL_467916 | USA/UT-QDX-18/2020 | 2020-03-13 | North America/USA/Utah/ | Quest Diagnostics | Quest Diagnostics | Anderson et al |
| EPI_ISL_467931 | USA/VA-DCLS-0320/2020 | 2020-05-04 | North America/USA/Virginia/ | Virginia DCLS | Virginia DCLS | Virginia DCLS et al |
| EPI_ISL_467933 | USA/VA-DCLS-0322/2020 | 2020-05-04 | North America/USA/Virginia/ | Virginia DCLS | Virginia DCLS | Virginia DCLS et al |
| EPI_ISL_467948 | USA/CA-IGI-0031/2020 | 2020-05-15 | North America/USA/California/Alameda | Innovative Genomics Institute, UC Berkeley | Innovative Genomics Institute, UC Berkeley | Stacia Wyman et al |
| EPI_ISL_467958 | USA/CA-SR0428/2020 | 2020-03-10 | North America/USA/California/San Dieg | San Diego County Public Health Laboratory | Andersen lab at Scripps Research | SEARCH Alliance San Diego with Tracy Basler et al |
| EPI_ISL_467980 | USA/CA-SR0458/2020 | 2020-04-02 | North America/USA/California/San Dieg | San Diego County Public Health Laboratory | Andersen lab at Scripps Research | SEARCH Alliance San Diego with Tracy Basler et al |
| EPI_ISL_468015 | Australia/SAP243/2020 | 2020-03-25 | Oceania/Australia/South Australia/ | SA Pathology | SA Pathology | Lex Leong et al |
| EPI_ISL_468032 | Australia/SAP214/2020 | 2020-01-30 | Oceania/Australia/South Australia/ | SA Pathology | SA Pathology | Lex Leong et al |
| EPI_ISL_468037 | Australia/SAP305/2020 | 2020-01-30 | Oceania/Australia/South Australia/ | SA Pathology | SA Pathology | Lex Leong et al |
| EPI_ISL_468039 | Australia/SAP097/2020 | 2020-02-22 | Oceania/Australia/South Australia/ | SA Pathology | SA Pathology | Lex Leong et al |
| EPI_ISL_468045 | Egypt/CUNCI-HGC002/2020 | 2020-05-02 | Africa/Egypt/Egypt/ | Egyptian National Cancer Institute (ENCI) | Egyptian National Cancer Institute (ENCI) | Zekri et al |
| EPI_ISL_468047 | Egypt/CUNCI-HGC013/2020 | 2020-05-02 | Africa/Egypt/Egypt/ | Egyptian National Cancer Institute (ENCI) | Egyptian National Cancer Institute (ENCI) | Zekri et al |
| EPI_ISL_468048 | Egypt/CUNCI-HGC004/2020 | 2020-05-02 | Africa/Egypt/Egypt/ | Egyptian National Cancer Institute (ENCI) | Egyptian National Cancer Institute (ENCI) | Zekri et al |
| EPI_ISL_468049 | Egypt/CUNCI-HGC003/2020 | 2020-05-02 | Africa/Egypt/Egypt/ | Egyptian National Cancer Institute (ENCI) | Egyptian National Cancer Institute (ENCI) | Zekri et al |
| EPI_ISL_468051 | Egypt/CUNCI-HGC012/2020 | 2020-05-02 | Africa/Egypt/Egypt/ | Egyptian National Cancer Institute (ENCI) | Egyptian National Cancer Institute (ENCI) | Zekri et al |

| | | | | | | |
|---|---|---|---|---|---|---|
| EPI_ISL_468052 | Egypt/CUNCI-HGC005/2020 | 2020-05-02 | Africa/Egypt/Egypt/ | Egyptian National Cancer Institute (ENCI) | Egyptian National Cancer Institute (ENCI) | Zekri et al |
| EPI_ISL_468053 | Egypt/CUNCI-HGC009/2020 | 2020-05-02 | Africa/Egypt/Egypt/ | Egyptian National Cancer Institute (ENCI) | Egyptian National Cancer Institute (ENCI) | Zekri et al |
| EPI_ISL_468057 | Egypt/CUNCI-HGC023/2020 | 2020-05-02 | Africa/Egypt/Egypt/ | Egyptian National Cancer Institute (ENCI) | Egyptian National Cancer Institute (ENCI) | Zekri et al |
| EPI_ISL_468058 | Egypt/CUNCI-HGC016/2020 | 2020-05-02 | Africa/Egypt/Egypt/ | Egyptian National Cancer Institute (ENCI) | Egyptian National Cancer Institute (ENCI) | Zekri et al |
| EPI_ISL_468059 | Egypt/CUNCI-HGC011/2020 | 2020-05-02 | Africa/Egypt/Egypt/ | Egyptian National Cancer Institute (ENCI) | Egyptian National Cancer Institute (ENCI) | Zekri et al |
| EPI_ISL_468060 | Egypt/CUNCI-HGC014/2020 | 2020-05-02 | Africa/Egypt/Egypt/ | Egyptian National Cancer Institute (ENCI) | Egyptian National Cancer Institute (ENCI) | Zekri et al |
| EPI_ISL_468061 | Egypt/CUNCI-HGC021/2020 | 2020-05-02 | Africa/Egypt/Egypt/ | Egyptian National Cancer Institute (ENCI) | Egyptian National Cancer Institute (ENCI) | Zekri et al |
| EPI_ISL_468062 | Egypt/CUNCI-HGC028/2020 | 2020-05-02 | Africa/Egypt/Egypt/ | Egyptian National Cancer Institute (ENCI) | Egyptian National Cancer Institute (ENCI) | Zekri et al |
| EPI_ISL_468064 | USA/KY-9A10/2020 | 2020-03-18 | North America/USA/Kentucky/ | unknown | Computer Science and Engineering | Rouchka et al |
| EPI_ISL_468066 | Turkey/IMU-SP-03/2020 | 2020-04-16 | Europe/Turkey/Turkey/ | Physiology, Istanbul Medeniyet University | Physiology, Istanbul Medeniyet University | Pence et al |
| EPI_ISL_468074 | Bangladesh/CHRF-0006/2020 | 2020-05-17 | Asia/Bangladesh/Chattogram/ | Child Health Research Foundation | Child Health Research Foundation | Senjuti Saha et al |
| EPI_ISL_468075 | Bangladesh/CHRF-0007/2020 | 2020-05-14 | Asia/Bangladesh/Dhaka/ | Child Health Research Foundation | Child Health Research Foundation | Senjuti Saha et al |
| EPI_ISL_468078 | Bangladesh/CHRF-0010/2020 | 2020-05-03 | Asia/Bangladesh/Dhaka/ | Child Health Research Foundation | Child Health Research Foundation | Senjuti Saha et al |

| | | | | | | |
|---|---|---|---|---|---|---|
| EPI_ISL_468086 | USA/OR-OHSU-0049/2020 | 2020-04-16 | North America/USA/Oregon/Washingto | OHSU Lab Services Molecular Microbiology Lab | Oregon SARS-CoV-2 Genome Sequencing Center | Brendan L. O'Connell et al |
| EPI_ISL_468095 | USA/OR-OHSU-0075/2020 | 2020-04-23 | North America/USA/Oregon/Washingto | OHSU Lab Services Molecular Microbiology Lab | Oregon SARS-CoV-2 Genome Sequencing Center | Brendan L. O'Connell et al |
| EPI_ISL_468104 | USA/OR-OHSU-0103/2020 | 2020-04-28 | North America/USA/Oregon/Washingto | OHSU Lab Services Molecular Microbiology Lab | Oregon SARS-CoV-2 Genome Sequencing Center | Brendan L. O'Connell et al |
| EPI_ISL_468119 | USA/OR-OHSU-0176/2020 | 2020-05-08 | North America/USA/Oregon/ | OHSU Lab Services Molecular Microbiology Lab | Oregon SARS-CoV-2 Genome Sequencing Center | Brendan L. O'Connell et al |
| EPI_ISL_468121 | USA/OR-OHSU-0178/2020 | 2020-05-06 | North America/USA/Oregon/ | OHSU Lab Services Molecular Microbiology Lab | Oregon SARS-CoV-2 Genome Sequencing Center | Brendan L. O'Connell et al |
| EPI_ISL_468122 | USA/OR-OHSU-0179/2020 | 2020-05-06 | North America/USA/Oregon/ | OHSU Lab Services Molecular Microbiology Lab | Oregon SARS-CoV-2 Genome Sequencing Center | Brendan L. O'Connell et al |
| EPI_ISL_468125 | USA/OR-OHSU-0183/2020 | 2020-05-11 | North America/USA/Oregon/ | OHSU Lab Services Molecular Microbiology Lab | Oregon SARS-CoV-2 Genome Sequencing Center | Brendan L. O'Connell et al |
| EPI_ISL_468134 | Romania/Bucuresti-4105/2020 | 2020-03-21 | Europe/Romania/Bucharest/ | [Romania, Bucharest] National Institute for Infectious Diseases ‚ÄúProf. Dr. Matei Bal»ô‚Äù | [Romania, Bucharest] National Institute for Infectious Diseases ‚ÄúProf. Dr. Matei Bal»ô‚Äù | Leontina Banica et al |

| | | | | | | |
|---|---|---|---|---|---|---|
| EPI_ISL_468135 | Romania/Bucuresti-4284/2020 | 2020-03-22 | Europe/Romania/Bucharest/ | [Romania, Bucharest] National Institute for Infectious Diseases ‚ÄúProf. Dr. Matei Bal»ô‚Äù | [Romania, Bucharest] National Institute for Infectious Diseases ‚ÄúProf. Dr. Matei Bal»ô‚Äù | Leontina Banica et al |
| EPI_ISL_468137 | Romania/Suceava-4682/2020 | 2020-03-22 | Europe/Romania/Suceava/ | [Romania, Bucharest] National Institute for Infectious Diseases ‚ÄúProf. Dr. Matei Bal»ô‚Äù | [Romania, Bucharest] National Institute for Infectious Diseases ‚ÄúProf. Dr. Matei Bal»ô‚Äù | Leontina Banica et al |
| EPI_ISL_468138 | Romania/Suceava-5709/2020 | 2020-03-25 | Europe/Romania/Suceava/ | [Romania, Bucharest] National Institute for Infectious Diseases ‚ÄúProf. Dr. Matei Bal»ô‚Äù | [Romania, Bucharest] National Institute for Infectious Diseases ‚ÄúProf. Dr. Matei Bal»ô‚Äù | Leontina Banica et al |
| EPI_ISL_468139 | Romania/Suceava-5710/2020 | 2020-03-25 | Europe/Romania/Suceava/ | [Romania, Bucharest] National Institute for Infectious Diseases ‚ÄúProf. Dr. Matei Bal»ô‚Äù | [Romania, Bucharest] National Institute for Infectious Diseases ‚ÄúProf. Dr. Matei Bal»ô‚Äù | Leontina Banica et al |
| EPI_ISL_468143 | Romania/Suceava-5893/2020 | 2020-03-25 | Europe/Romania/Suceava/ | [Romania, Bucharest] National Institute for Infectious Diseases ‚ÄúProf. Dr. Matei Bal»ô‚Äù | [Romania, Bucharest] National Institute for Infectious Diseases ‚ÄúProf. Dr. Matei Bal»ô‚Äù | Leontina Banica et al |

| | | | | | | |
|---|---|---|---|---|---|---|
| EPI_ISL_468145 | Romania/Bucuresti-6451/2020 | 2020-03-28 | Europe/Romania/Bucharest/ | [Romania, Bucharest] National Institute for Infectious Diseases ‚ÄúProf. Dr. Matei Bal»ô‚Äù | [Romania, Bucharest] National Institute for Infectious Diseases ‚ÄúProf. Dr. Matei Bal»ô‚Äù | Leontina Banica et al |
| EPI_ISL_468148 | Romania/Bucuresti-12296/2020 | 2020-04-11 | Europe/Romania/Bucharest/ | [Romania, Bucharest] National Institute for Infectious Diseases ‚ÄúProf. Dr. Matei Bal»ô‚Äù | [Romania, Bucharest] National Institute for Infectious Diseases ‚ÄúProf. Dr. Matei Bal»ô‚Äù | Leontina Banica et al |
| EPI_ISL_468150 | Romania/Bucuresti-12547/2020 | 2020-04-12 | Europe/Romania/Bucharest/ | [Romania, Bucharest] National Institute for Infectious Diseases ‚ÄúProf. Dr. Matei Bal»ô‚Äù | [Romania, Bucharest] National Institute for Infectious Diseases ‚ÄúProf. Dr. Matei Bal»ô‚Äù | Leontina Banica et al |
| EPI_ISL_468151 | Romania/Bucuresti-13907/2020 | 2020-04-15 | Europe/Romania/Bucharest/ | [Romania, Bucharest] National Institute for Infectious Diseases ‚ÄúProf. Dr. Matei Bal»ô‚Äù | [Romania, Bucharest] National Institute for Infectious Diseases ‚ÄúProf. Dr. Matei Bal»ô‚Äù | Leontina Banica et al |
| EPI_ISL_468155 | Romania/Bucuresti-15503/2020 | 2020-04-18 | Europe/Romania/Bucharest/ | [Romania, Bucharest] National Institute for Infectious Diseases ‚ÄúProf. Dr. Matei Bal»ô‚Äù | [Romania, Bucharest] National Institute for Infectious Diseases ‚ÄúProf. Dr. Matei Bal»ô‚Äù | Leontina Banica et al |

| | | | | | | |
|---|---|---|---|---|---|---|
| EPI_ISL_468156 | Romania/Mioveni-24095/2020 | 2020-05-08 | Europe/Romania/Mioveni/ | [Romania, Bucharest] National Institute for Infectious Diseases ‚ÄúProf. Dr. Matei Bal»ô‚Äù | [Romania, Bucharest] National Institute for Infectious Diseases ‚ÄúProf. Dr. Matei Bal»ô‚Äù | Leontina Banica et al |
| EPI_ISL_468157 | Romania/Suceava-31645/2020 | 2020-05-29 | Europe/Romania/Suceava/ | [Romania, Bucharest] National Institute for Infectious Diseases ‚ÄúProf. Dr. Matei Bal»ô‚Äù | [Romania, Bucharest] National Institute for Infectious Diseases ‚ÄúProf. Dr. Matei Bal»ô‚Äù | Leontina Banica et al |
| EPI_ISL_468158 | Romania/Suceava-31646/2020 | 2020-05-29 | Europe/Romania/Suceava/ | [Romania, Bucharest] National Institute for Infectious Diseases ‚ÄúProf. Dr. Matei Bal»ô‚Äù | [Romania, Bucharest] National Institute for Infectious Diseases ‚ÄúProf. Dr. Matei Bal»ô‚Äù | Leontina Banica et al |
| EPI_ISL_468159 | Pakistan/NIH-44905/2020 | 2020-06-02 | Asia/Pakistan/Islamabad/ | unknown | Department of Virology, Public Health Laboratories Division, National Institute of Health | Massab Umair et al |
| EPI_ISL_468160 | Pakistan/NIH-45143/2020 | 2020-06-02 | Asia/Pakistan/Rawalpindi/ | unknown | Department of Virology, Public Health Laboratories Division, National Institute of Health | Massab Umair et al |
| EPI_ISL_468161 | Pakistan/NIH-45090/2020 | 2020-06-02 | Asia/Pakistan/Islamabad/ | Department of Virology, Public Health Laboratories Division, National Institute of Health | Department of Virology, Public Health Laboratories Division, National Institute of Health | Massab Umair et al |

| | | | | | | |
|---|---|---|---|---|---|---|
| EPI_ISL_468162 | Pakistan/NIH-45579/2020 | 2020-06-02 | Asia/Pakistan/Islamabad/ | unknown | Department of Virology, Public Health Laboratories Division, National Institute of Health | Massab Umair et al |
| EPI_ISL_468163 | Pakistan/NIH-HAS001/2020 | 2020-06-02 | Asia/Pakistan/Islamabad/ | Department of Virology, Public Health Laboratories Division, National Institute of Health | Department of Virology, Public Health Laboratories Division, National Institute of Health | Massab Umair et al |
| EPI_ISL_468239 | Switzerland/BE-ETHZ-120067/2020 | 2020-04-20 | Europe/Switzerland/Bern/ | Viollier AG | Department of Biosystems Science and Engineering, ETH Zvºrich | Christian Beisel et al |
| EPI_ISL_468242 | Switzerland/ZH-ETHZ-120072/2020 | 2020-04-21 | Europe/Switzerland/Zvºrich/ | Viollier AG | Department of Biosystems Science and Engineering, ETH Zvºrich | Christian Beisel et al |
| EPI_ISL_468250 | Switzerland/AG-ETHZ-120172/2020 | 2020-04-27 | Europe/Switzerland/Aargau/ | Viollier AG | Department of Biosystems Science and Engineering, ETH Zvºrich | Christian Beisel et al |
| EPI_ISL_468255 | Switzerland/VD-ETHZ-120179/2020 | 2020-04-27 | Europe/Switzerland/Vaud/ | Viollier AG | Department of Biosystems Science and Engineering, ETH Zvºrich | Christian Beisel et al |
| EPI_ISL_468270 | Switzerland/BL-ETHZ-120196/2020 | 2020-04-28 | Europe/Switzerland/Basel-Land/ | Viollier AG | Department of Biosystems Science and Engineering, ETH Zvºrich | Christian Beisel et al |

| | | | | | | |
|---|---|---|---|---|---|---|
| EPI_ISL_468287 | Switzerland/BE-ETHZ-120225/2020 | 2020-04-30 | Europe/Switzerland/Bern/ | Viollier AG | Department of Biosystems Science and Engineering, ETH Zvºrich | Christian Beisel et al |
| EPI_ISL_468335 | Spain/GA-USC-005/2020 | 2020-04-29 | Europe/Spain/Galicia/A Coruna | Microbiology Service, University Hospital of A Coruna-Biomedical Research Institute | Genomes & Disease, Center for Research in Molecular Medicine and Chronic Diseases, University of Santiago de Compostela | Kelly Conde et al |
| EPI_ISL_468336 | Spain/GA-USC-006/2020 | 2020-05-10 | Europe/Spain/Galicia/A Coruna | Microbiology Service, University Hospital of A Coruna-Biomedical Research Institute | Genomes & Disease, Center for Research in Molecular Medicine and Chronic Diseases, University of Santiago de Compostela | Kelly Conde et al |
| EPI_ISL_468338 | Spain/GA-USC-008/2020 | 2020-05-10 | Europe/Spain/Galicia/A Coruna | Microbiology Service, University Hospital of A Coruna-Biomedical Research Institute | Genomes & Disease, Center for Research in Molecular Medicine and Chronic Diseases, University of Santiago de Compostela | Kelly Conde et al |
| EPI_ISL_468340 | Spain/GA-USC-010/2020 | 2020-04-22 | Europe/Spain/Galicia/A Coruna | Microbiology Service, University Hospital of A Coruna-Biomedical Research Institute | Genomes & Disease, Center for Research in Molecular Medicine and Chronic Diseases, University of Santiago de Compostela | Kelly Conde et al |

| | | | | | | |
|---|---|---|---|---|---|---|
| EPI_ISL_468343 | Spain/GA-USC-013/2020 | 2020-05-29 | Europe/Spain/Galicia/A Coruna | Microbiology Service, University Hospital of A Coruna-Biomedical Research Institute | Genomes & Disease, Center for Research in Molecular Medicine and Chronic Diseases, University of Santiago de Compostela | Kelly Conde et al |
| EPI_ISL_468348 | USA/CA-CZB-1295/2020 | 2020-04-25 | North America/USA/California/Santa Cla | County of Santa Clara Public Health Department | Chan-Zuckerberg Biohub | CZB Cliahub Consortium et al |
| EPI_ISL_468352 | USA/CA-CZB-1312/2020 | 2020-05-07 | North America/USA/California/Santa Cla | County of Santa Clara Public Health Department | Chan-Zuckerberg Biohub | CZB Cliahub Consortium et al |
| EPI_ISL_468353 | USA/CA-CZB-1317/2020 | 2020-05-16 | North America/USA/California/Santa Cla | County of Santa Clara Public Health Department | Chan-Zuckerberg Biohub | CZB Cliahub Consortium et al |
| EPI_ISL_468355 | USA/CA-CZB-1322/2020 | 2020-05-20 | North America/USA/California/Santa Cla | County of Santa Clara Public Health Department | Chan-Zuckerberg Biohub | CZB Cliahub Consortium et al |
| EPI_ISL_468363 | USA/CA-CZB-1347/2020 | 2020-04-09 | North America/USA/California/Alameda | Alameda County Public Health Lab | Chan-Zuckerberg Biohub | CZB Cliahub Consortium et al |
| EPI_ISL_468375 | USA/CA-CZB-1374/2020 | 2020-04-17 | North America/USA/California/Alameda | Alameda County Public Health Lab | Chan-Zuckerberg Biohub | CZB Cliahub Consortium et al |
| EPI_ISL_468376 | USA/CA-CZB-1391/2020 | 2020-04-18 | North America/USA/California/Alameda | Alameda County Public Health Lab | Chan-Zuckerberg Biohub | CZB Cliahub Consortium et al |
| EPI_ISL_468378 | USA/CA-CZB-1396/2020 | 2020-04-19 | North America/USA/California/Alameda | Alameda County Public Health Lab | Chan-Zuckerberg Biohub | CZB Cliahub Consortium et al |
| EPI_ISL_468392 | USA/CA-CZB-1418/2020 | 2020-03-17 | North America/USA/California/San Luis | County of San Luis Obispo Public Health Laboratory | Chan-Zuckerberg Biohub | CZB Cliahub Consortium et al |
| EPI_ISL_468407 | USA/CA-CZB-1437/2020 | 2020-03-26 | North America/USA/California/San Luis | County of San Luis Obispo Public Health Laboratory | Chan-Zuckerberg Biohub | CZB Cliahub Consortium et al |
| EPI_ISL_468425 | USA/CA-CZB-1466/2020 | 2020-04-27 | North America/USA/California/San Luis | County of San Luis Obispo Public Health Laboratory | Chan-Zuckerberg Biohub | CZB Cliahub Consortium et al |
| EPI_ISL_468436 | USA/CA-CZB-1485/2020 | 2020-05-06 | North America/USA/California/San Luis | County of San Luis Obispo Public Health Laboratory | Chan-Zuckerberg Biohub | CZB Cliahub Consortium et al |
| EPI_ISL_468445 | USA/CA-CZB-1495/2020 | 2020-03-24 | North America/USA/California/Humbold | Humboldt County Public Health Laboratory | Chan-Zuckerberg Biohub | CZB Cliahub Consortium et al |
| EPI_ISL_468463 | USA/CA-CZB-1517/2020 | 2020-05-13 | North America/USA/California/Ventura | Ventura County Public Health Lab | Chan-Zuckerberg Biohub | CZB Cliahub Consortium et al |
| EPI_ISL_468465 | USA/CA-CZB-1519/2020 | 2020-05-11 | North America/USA/California/Ventura | Ventura County Public Health Lab | Chan-Zuckerberg Biohub | CZB Cliahub Consortium et al |

| | | | | | | |
|---|---|---|---|---|---|---|
| EPI_ISL_468497 | USA/CA-CZB-1560/2020 | 2020-03-30 | North America/USA/California/Ventura | Ventura County Public Health Lab | Chan-Zuckerberg Biohub | CZB Cliahub Consortium et al |
| EPI_ISL_468499 | USA/CA-CZB-1562/2020 | 2020-04-02 | North America/USA/California/Ventura | Ventura County Public Health Lab | Chan-Zuckerberg Biohub | CZB Cliahub Consortium et al |
| EPI_ISL_468511 | USA/CA-CZB-1581/2020 | 2020-03-21 | North America/USA/California/San Joaq | San Joaquin County Public Health Lab | Chan-Zuckerberg Biohub | CZB Cliahub Consortium et al |
| EPI_ISL_468524 | USA/CA-CZB-1594/2020 | 2020-03-28 | North America/USA/California/San Joaq | San Joaquin County Public Health Lab | Chan-Zuckerberg Biohub | CZB Cliahub Consortium et al |
| EPI_ISL_468530 | USA/CA-CZB-1601/2020 | 2020-04-02 | North America/USA/California/San Joaq | San Joaquin County Public Health Lab | Chan-Zuckerberg Biohub | CZB Cliahub Consortium et al |
| EPI_ISL_468546 | USA/CA-CZB-1617/2020 | 2020-04-17 | North America/USA/California/San Joaq | San Joaquin County Public Health Lab | Chan-Zuckerberg Biohub | CZB Cliahub Consortium et al |
| EPI_ISL_468549 | USA/CA-CZB-1620/2020 | 2020-04-21 | North America/USA/California/San Joaq | San Joaquin County Public Health Lab | Chan-Zuckerberg Biohub | CZB Cliahub Consortium et al |
| EPI_ISL_468568 | USA/CA-QDX-97/2020 | 2020-03-13 | North America/USA/California/ | Quest Diagnostics | Quest Diagnostics | Anderson et al |
| EPI_ISL_468573 | USA/HI-QDX-163/2020 | 2020-03-12 | North America/USA/Hawaii/ | Quest Diagnostics | Quest Diagnostics | Anderson et al |
| EPI_ISL_468584 | USA/NY-QDX-90/2020 | 2020-03-12 | North America/USA/New York/ | Quest Diagnostics | Quest Diagnostics | Anderson et al |
| EPI_ISL_468594 | USA/CA-CZB-1650/2020 | 2020-04-18 | North America/USA/California/Orange C | Orange County Public Health Lab | Chan-Zuckerberg Biohub | CZB Cliahub Consortium et al |
| EPI_ISL_468614 | USA/CA-CZB-1672/2020 | 2020-05-14 | North America/USA/California/Orange C | Orange County Public Health Lab | Chan-Zuckerberg Biohub | CZB Cliahub Consortium et al |
| EPI_ISL_468616 | USA/CA-CZB-1676/2020 | 2020-03-30 | North America/USA/California/Contra C | Contra Costa Public Health Lab | Chan-Zuckerberg Biohub | CZB Cliahub Consortium et al |
| EPI_ISL_468617 | USA/CA-CZB-1677/2020 | 2020-03-30 | North America/USA/California/Contra C | Contra Costa Public Health Lab | Chan-Zuckerberg Biohub | CZB Cliahub Consortium et al |
| EPI_ISL_468622 | USA/CA-CZB-1684/2020 | 2020-04-01 | North America/USA/California/Contra C | Contra Costa Public Health Lab | Chan-Zuckerberg Biohub | CZB Cliahub Consortium et al |
| EPI_ISL_468629 | USA/CA-CZB-1694/2020 | 2020-04-02 | North America/USA/California/Contra C | Contra Costa Public Health Lab | Chan-Zuckerberg Biohub | CZB Cliahub Consortium et al |
| EPI_ISL_468634 | USA/CA-CZB-1700/2020 | 2020-04-02 | North America/USA/California/Contra C | Contra Costa Public Health Lab | Chan-Zuckerberg Biohub | CZB Cliahub Consortium et al |
| EPI_ISL_468641 | USA/CA-CZB-1710/2020 | 2020-04-02 | North America/USA/California/Contra C | Contra Costa Public Health Lab | Chan-Zuckerberg Biohub | CZB Cliahub Consortium et al |
| EPI_ISL_468643 | USA/CA-CZB-1713/2020 | 2020-04-03 | North America/USA/California/Contra C | Contra Costa Public Health Lab | Chan-Zuckerberg Biohub | CZB Cliahub Consortium et al |
| EPI_ISL_468656 | Croatia/AU-S10new/2020 | 2020-04-09 | Europe/Croatia/Istria/ | Institute for Public Health | Laboratory for advanced genomics | Filip Rokifá et al |

| | | | | | | |
|---|---|---|---|---|---|---|
| EPI_ISL_468657 | Canada/BC_14605710/2020 | 20-Apr | North America/Canada/British Columbia | BCCDC Public Health Laboratory | BCCDC Public Health Laboratory | Richard Harrigan et al |
| EPI_ISL_468658 | Canada/BC_14667611/2020 | 20-Apr | North America/Canada/British Columbia | BCCDC Public Health Laboratory | BCCDC Public Health Laboratory | Richard Harrigan et al |
| EPI_ISL_468659 | Canada/BC_14727596/2020 | 20-Apr | North America/Canada/British Columbia | BCCDC Public Health Laboratory | BCCDC Public Health Laboratory | Richard Harrigan et al |
| EPI_ISL_468660 | Canada/BC_14739124/2020 | 20-Apr | North America/Canada/British Columbia | BCCDC Public Health Laboratory | BCCDC Public Health Laboratory | Richard Harrigan et al |
| EPI_ISL_468661 | Canada/BC_14763711/2020 | 20-Apr | North America/Canada/British Columbia | BCCDC Public Health Laboratory | BCCDC Public Health Laboratory | Richard Harrigan et al |
| EPI_ISL_468662 | Canada/BC_15188754/2020 | 20-Apr | North America/Canada/British Columbia | BCCDC Public Health Laboratory | BCCDC Public Health Laboratory | Richard Harrigan et al |
| EPI_ISL_468663 | Canada/BC_15209047/2020 | 20-Apr | North America/Canada/British Columbia | BCCDC Public Health Laboratory | BCCDC Public Health Laboratory | Richard Harrigan et al |
| EPI_ISL_468664 | Canada/BC_15222281/2020 | 20-Apr | North America/Canada/British Columbia | BCCDC Public Health Laboratory | BCCDC Public Health Laboratory | Richard Harrigan et al |
| EPI_ISL_468665 | Canada/BC_15222496/2020 | 20-Apr | North America/Canada/British Columbia | BCCDC Public Health Laboratory | BCCDC Public Health Laboratory | Richard Harrigan et al |
| EPI_ISL_468666 | Canada/BC_15443411/2020 | 20-Apr | North America/Canada/British Columbia | BCCDC Public Health Laboratory | BCCDC Public Health Laboratory | Richard Harrigan et al |
| EPI_ISL_468667 | Canada/BC_15448387/2020 | 20-Apr | North America/Canada/British Columbia | BCCDC Public Health Laboratory | BCCDC Public Health Laboratory | Richard Harrigan et al |
| EPI_ISL_468668 | Canada/BC_15467213/2020 | 20-Apr | North America/Canada/British Columbia | BCCDC Public Health Laboratory | BCCDC Public Health Laboratory | Richard Harrigan et al |
| EPI_ISL_468669 | Canada/BC_15474127/2020 | 20-Apr | North America/Canada/British Columbia | BCCDC Public Health Laboratory | BCCDC Public Health Laboratory | Richard Harrigan et al |
| EPI_ISL_468670 | Canada/BC_15478567/2020 | 20-Apr | North America/Canada/British Columbia | BCCDC Public Health Laboratory | BCCDC Public Health Laboratory | Richard Harrigan et al |
| EPI_ISL_468671 | Canada/BC_15550440/2020 | 20-Apr | North America/Canada/British Columbia | BCCDC Public Health Laboratory | BCCDC Public Health Laboratory | Richard Harrigan et al |
| EPI_ISL_468672 | Canada/BC_15678040/2020 | 20-Apr | North America/Canada/British Columbia | BCCDC Public Health Laboratory | BCCDC Public Health Laboratory | Richard Harrigan et al |
| EPI_ISL_468673 | Canada/BC_15682745/2020 | 20-Apr | North America/Canada/British Columbia | BCCDC Public Health Laboratory | BCCDC Public Health Laboratory | Richard Harrigan et al |
| EPI_ISL_468674 | Canada/BC_15708725/2020 | 20-Apr | North America/Canada/British Columbia | BCCDC Public Health Laboratory | BCCDC Public Health Laboratory | Richard Harrigan et al |
| EPI_ISL_468675 | Canada/BC_15710616/2020 | 20-Apr | North America/Canada/British Columbia | BCCDC Public Health Laboratory | BCCDC Public Health Laboratory | Richard Harrigan et al |
| EPI_ISL_468676 | Canada/BC_15804748/2020 | 20-Apr | North America/Canada/British Columbia | BCCDC Public Health Laboratory | BCCDC Public Health Laboratory | Richard Harrigan et al |

| | | | | | | |
|---|---|---|---|---|---|---|
| EPI_ISL_468677 | Canada/BC_15820631/2020 | 20-Apr | North America/Canada/British Columbia | BCCDC Public Health Laboratory | BCCDC Public Health Laboratory | Richard Harrigan et al |
| EPI_ISL_468678 | Canada/BC_15918444/2020 | 20-Apr | North America/Canada/British Columbia | BCCDC Public Health Laboratory | BCCDC Public Health Laboratory | Richard Harrigan et al |
| EPI_ISL_468679 | Canada/BC_16223737/2020 | 20-Apr | North America/Canada/British Columbia | BCCDC Public Health Laboratory | BCCDC Public Health Laboratory | Richard Harrigan et al |
| EPI_ISL_468680 | Canada/BC_16537755/2020 | 20-Apr | North America/Canada/British Columbia | BCCDC Public Health Laboratory | BCCDC Public Health Laboratory | Richard Harrigan et al |
| EPI_ISL_468681 | Canada/BC_16567748/2020 | 20-Apr | North America/Canada/British Columbia | BCCDC Public Health Laboratory | BCCDC Public Health Laboratory | Richard Harrigan et al |
| EPI_ISL_468682 | Canada/BC_16569820/2020 | 20-Apr | North America/Canada/British Columbia | BCCDC Public Health Laboratory | BCCDC Public Health Laboratory | Richard Harrigan et al |
| EPI_ISL_468683 | Canada/BC_16588829/2020 | 20-Apr | North America/Canada/British Columbia | BCCDC Public Health Laboratory | BCCDC Public Health Laboratory | Richard Harrigan et al |
| EPI_ISL_468684 | Canada/BC_16839757/2020 | 20-Apr | North America/Canada/British Columbia | BCCDC Public Health Laboratory | BCCDC Public Health Laboratory | Richard Harrigan et al |
| EPI_ISL_468685 | Canada/BC_16917931/2020 | 20-Apr | North America/Canada/British Columbia | BCCDC Public Health Laboratory | BCCDC Public Health Laboratory | Richard Harrigan et al |
| EPI_ISL_468686 | Canada/BC_16943416/2020 | 20-Apr | North America/Canada/British Columbia | BCCDC Public Health Laboratory | BCCDC Public Health Laboratory | Richard Harrigan et al |
| EPI_ISL_468687 | Canada/BC_16976214/2020 | 20-Apr | North America/Canada/British Columbia | BCCDC Public Health Laboratory | BCCDC Public Health Laboratory | Richard Harrigan et al |
| EPI_ISL_468688 | Canada/BC_17080413/2020 | 20-Apr | North America/Canada/British Columbia | BCCDC Public Health Laboratory | BCCDC Public Health Laboratory | Richard Harrigan et al |
| EPI_ISL_468689 | Canada/BC_17607523/2020 | 20-Apr | North America/Canada/British Columbia | BCCDC Public Health Laboratory | BCCDC Public Health Laboratory | Richard Harrigan et al |
| EPI_ISL_468690 | Canada/BC_17670489/2020 | 20-Apr | North America/Canada/British Columbia | BCCDC Public Health Laboratory | BCCDC Public Health Laboratory | Richard Harrigan et al |
| EPI_ISL_468691 | Canada/BC_17754491/2020 | 20-Apr | North America/Canada/British Columbia | BCCDC Public Health Laboratory | BCCDC Public Health Laboratory | Richard Harrigan et al |
| EPI_ISL_468692 | Canada/BC_17847248/2020 | 20-Apr | North America/Canada/British Columbia | BCCDC Public Health Laboratory | BCCDC Public Health Laboratory | Richard Harrigan et al |
| EPI_ISL_468693 | Canada/BC_17906513/2020 | 20-Apr | North America/Canada/British Columbia | BCCDC Public Health Laboratory | BCCDC Public Health Laboratory | Richard Harrigan et al |
| EPI_ISL_468694 | Canada/BC_17943129/2020 | 20-Apr | North America/Canada/British Columbia | BCCDC Public Health Laboratory | BCCDC Public Health Laboratory | Richard Harrigan et al |
| EPI_ISL_468695 | Canada/BC_18101737/2020 | 20-Apr | North America/Canada/British Columbia | BCCDC Public Health Laboratory | BCCDC Public Health Laboratory | Richard Harrigan et al |
| EPI_ISL_468696 | Canada/BC_18115071/2020 | 20-Apr | North America/Canada/British Columbia | BCCDC Public Health Laboratory | BCCDC Public Health Laboratory | Richard Harrigan et al |

| | | | | | | |
|---|---|---|---|---|---|---|
| EPI_ISL_468697 | Canada/BC_18115447/2020 | 20-Apr | North America/Canada/British Columbia | BCCDC Public Health Laboratory | BCCDC Public Health Laboratory | Richard Harrigan et al |
| EPI_ISL_468698 | Canada/BC_18116391/2020 | 20-Apr | North America/Canada/British Columbia | BCCDC Public Health Laboratory | BCCDC Public Health Laboratory | Richard Harrigan et al |
| EPI_ISL_468699 | Canada/BC_18141519/2020 | 20-Apr | North America/Canada/British Columbia | BCCDC Public Health Laboratory | BCCDC Public Health Laboratory | Richard Harrigan et al |
| EPI_ISL_468700 | Canada/BC_18143554/2020 | 20-Apr | North America/Canada/British Columbia | BCCDC Public Health Laboratory | BCCDC Public Health Laboratory | Richard Harrigan et al |
| EPI_ISL_468701 | USA/LA-BIE-001/2020 | 2020-04-06 | North America/USA/Louisiana/New Orle | Ochsner Health | Bioinfoexperts, LLC | Rebecca Rose et al |
| EPI_ISL_468707 | USA/LA-BIE-015/2020 | 2020-04-08 | North America/USA/Louisiana/New Orle | Ochsner Health | Bioinfoexperts, LLC | Rebecca Rose et al |
| EPI_ISL_468733 | Belgium/UGent-85/2020 | 2020-03-11 | Europe/Belgium/Gent/ | Lab voor klinische biologie | Onderzoeksgroep Virologie | Laurens Lambrechts et al |
| EPI_ISL_468737 | Belgium/UGent-89/2020 | 2020-03-17 | Europe/Belgium/Gent/ | Lab voor klinische biologie | Onderzoeksgroep Virologie | Nick Vereecke et al |
| EPI_ISL_468743 | Belgium/UGent-95/2020 | 2020-03-17 | Europe/Belgium/Gent/ | Lab voor klinische biologie | Onderzoeksgroep Virologie | Nick Vereecke et al |
| EPI_ISL_468748 | Chile/RM-CMM-05207/2020 | 2020-04-30 | South America/Chile/Region Metropolit | Facultad de Medicina UC | Center for Mathematical Modeling and Center for Genome Regulation. Santiago, Chile | Gaete A et al |
| EPI_ISL_468758 | Chile/AT-CMM-00063/2020 | 2020-05-01 | South America/Chile/Atacama/Copiapo | Laboratorio de Biologia Molecular, Facultad de Medicina, Universidad de Atacama | Center for Mathematical Modeling and Center for Genome Regulation. Santiago, Chile | Gaete A et al |
| EPI_ISL_468759 | Chile/AT-CMM-00064/2020 | 2020-05-01 | South America/Chile/Atacama/Copiapo | Laboratorio de Biologia Molecular, Facultad de Medicina, Universidad de Atacama | Center for Mathematical Modeling and Center for Genome Regulation. Santiago, Chile | Gaete A et al |
| EPI_ISL_468764 | Spain/RI-IBV-h2285/2020 | 2020-03-01 | Europe/Spain/La Rioja/Logrono | Centro de Investigaci√≥n Biom√©dica de La Rioja - Hospital San Pedro Logro√±o | SeqCOVID-SPAIN consortium/IBV(CSIC) | Mar√≠a de Toro et al |

| | | | | | | |
|---|---|---|---|---|---|---|
| EPI_ISL_468769 | Spain/AR-IBV-2476/2020 | 2020-03-17 | Europe/Spain/Aragon/Zaragoza | Servicio de Microbiolog√≠a, Hospital Miguel Servet, Zaragoza | SeqCOVID-SPAIN consortium/IBV(CSIC) | Antonio Rezusta L√≥pez et al |
| EPI_ISL_468772 | Spain/AR-IBV-2414/2020 | 2020-03-24 | Europe/Spain/Aragon/Teruel | Servicio de Microbiolog√≠a, Hospital Miguel Servet, Zaragoza | SeqCOVID-SPAIN consortium/IBV(CSIC) | Antonio Rezusta L√≥pez et al |
| EPI_ISL_468774 | Spain/AR-IBV-2454/2020 | 2020-03-24 | Europe/Spain/Aragon/Teruel | Servicio de Microbiolog√≠a, Hospital Miguel Servet, Zaragoza | SeqCOVID-SPAIN consortium/IBV(CSIC) | Antonio Rezusta L√≥pez et al |
| EPI_ISL_468785 | Spain/AR-IBV-2483/2020 | 2020-03-24 | Europe/Spain/Aragon/Zaragoza | Servicio de Microbiolog√≠a, Hospital Miguel Servet, Zaragoza | SeqCOVID-SPAIN consortium/IBV(CSIC) | Antonio Rezusta L√≥pez et al |
| EPI_ISL_468791 | Spain/AR-IBV-2458/2020 | 2020-03-26 | Europe/Spain/Aragon/Zaragoza | Servicio de Microbiolog√≠a, Hospital Miguel Servet, Zaragoza | SeqCOVID-SPAIN consortium/IBV(CSIC) | Antonio Rezusta L√≥pez et al |
| EPI_ISL_468796 | Spain/AR-IBV-2403/2020 | 2020-03-23 | Europe/Spain/Aragon/Zaragoza | Servicio de Microbiolog√≠a, Hospital Miguel Servet, Zaragoza | SeqCOVID-SPAIN consortium/IBV(CSIC) | Antonio Rezusta L√≥pez et al |
| EPI_ISL_468800 | Spain/AR-IBV-2390/2020 | 2020-03-27 | Europe/Spain/Aragon/Alcaniz | Servicio de Microbiolog√≠a, Hospital Miguel Servet, Zaragoza | SeqCOVID-SPAIN consortium/IBV(CSIC) | Antonio Rezusta L√≥pez et al |
| EPI_ISL_468808 | Spain/AR-IBV-2445/2020 | 2020-03-23 | Europe/Spain/Aragon/Zaragoza | Servicio de Microbiolog√≠a, Hospital Miguel Servet, Zaragoza | SeqCOVID-SPAIN consortium/IBV(CSIC) | Antonio Rezusta L√≥pez et al |
| EPI_ISL_468818 | Spain/AR-IBV-2437/2020 | 2020-03-18 | Europe/Spain/Aragon/Zaragoza | Servicio de Microbiolog√≠a, Hospital Miguel Servet, Zaragoza | SeqCOVID-SPAIN consortium/IBV(CSIC) | Antonio Rezusta L√≥pez et al |
| EPI_ISL_468829 | Spain/AR-IBV-2434/2020 | 2020-03-24 | Europe/Spain/Aragon/Teruel | Servicio de Microbiolog√≠a, Hospital Miguel Servet, Zaragoza | SeqCOVID-SPAIN consortium/IBV(CSIC) | Antonio Rezusta L√≥pez et al |
| EPI_ISL_468831 | Spain/AR-IBV-2453/2020 | 2020-03-24 | Europe/Spain/Aragon/Zaragoza | Servicio de Microbiolog√≠a, Hospital Miguel Servet, Zaragoza | SeqCOVID-SPAIN consortium/IBV(CSIC) | Antonio Rezusta L√≥pez et al |
| EPI_ISL_468837 | Spain/AR-IBV-2400/2020 | 2020-03-28 | Europe/Spain/Aragon/Zaragoza | Servicio de Microbiolog√≠a, Hospital Miguel Servet, Zaragoza | SeqCOVID-SPAIN consortium/IBV(CSIC) | Antonio Rezusta L√≥pez et al |
| EPI_ISL_468842 | Spain/AR-IBV-2419/2020 | 2020-03-27 | Europe/Spain/Aragon/Teruel | Servicio de Microbiolog√≠a, Hospital Miguel Servet, Zaragoza | SeqCOVID-SPAIN consortium/IBV(CSIC) | Antonio Rezusta L√≥pez et al |

| | | | | | | |
|---|---|---|---|---|---|---|
| EPI_ISL_468857 | Spain/AR-IBV-2482/2020 | 2020-03-23 | Europe/Spain/Aragon/Zaragoza | Servicio de Microbiolog√≠a, Hospital Miguel Servet, Zaragoza | SeqCOVID-SPAIN consortium/IBV(CSIC) | Antonio Rezusta L√≥pez et al |
| EPI_ISL_468887 | Spain/PV-IBV-2119/2020 | 2020-04-11 | Europe/Spain/Basque Country/San Seba | Servicio de Microbiolog√≠a. Hospital Universitario Donostia. OSI Donostialdea. √Årea de Enfermedades Infecciosas, Grupo de Infecci√≥n Respiratoria y Resistencia Antimicrobiana. Instituto de Investigaci√≥n Sanitaria Biodonostia. | SeqCOVID-SPAIN consortium/IBV(CSIC) | Gustavo Cilla et al |
| EPI_ISL_468900 | Spain/PV-IBV-2079/2020 | 2020-04-10 | Europe/Spain/Basque Country/San Seba | Servicio de Microbiolog√≠a. Hospital Universitario Donostia. OSI Donostialdea. √Årea de Enfermedades Infecciosas, Grupo de Infecci√≥n Respiratoria y Resistencia Antimicrobiana. Instituto de Investigaci√≥n Sanitaria Biodonostia. | SeqCOVID-SPAIN consortium/IBV(CSIC) | Gustavo Cilla et al |
| EPI_ISL_468901 | Spain/PV-IBV-2090/2020 | 2020-04-10 | Europe/Spain/Basque Country/San Seba | Servicio de Microbiolog√≠a. Hospital Universitario Donostia. OSI Donostialdea. √Årea de Enfermedades Infecciosas, Grupo de Infecci√≥n Respiratoria y Resistencia Antimicrobiana. Instituto de Investigaci√≥n Sanitaria Biodonostia. | SeqCOVID-SPAIN consortium/IBV(CSIC) | Gustavo Cilla et al |
| EPI_ISL_468907 | Spain/PV-IBV-2109/2020 | 2020-04-11 | Europe/Spain/Basque Country/San Seba | Servicio de Microbiolog√≠a. Hospital Universitario Donostia. OSI Donostialdea. √Årea de Enfermedades Infecciosas, Grupo de Infecci√≥n Respiratoria y Resistencia Antimicrobiana. Instituto de Investigaci√≥n Sanitaria Biodonostia. | SeqCOVID-SPAIN consortium/IBV(CSIC) | Gustavo Cilla et al |
| EPI_ISL_468922 | Spain/PV-IBV-2099/2020 | 2020-04-11 | Europe/Spain/Basque Country/San Seba | Servicio de Microbiolog√≠a. Hospital Universitario Donostia. OSI Donostialdea. √Årea de Enfermedades Infecciosas, Grupo de Infecci√≥n Respiratoria y Resistencia Antimicrobiana. Instituto de Investigaci√≥n Sanitaria Biodonostia. | SeqCOVID-SPAIN consortium/IBV(CSIC) | Gustavo Cilla et al |
| EPI_ISL_468925 | Spain/PV-IBV-2082/2020 | 2020-04-10 | Europe/Spain/Basque Country/San Seba | Servicio de Microbiolog√≠a. Hospital Universitario Donostia. OSI Donostialdea. √Årea de Enfermedades Infecciosas, Grupo de Infecci√≥n Respiratoria y Resistencia Antimicrobiana. Instituto de Investigaci√≥n Sanitaria Biodonostia. | SeqCOVID-SPAIN consortium/IBV(CSIC) | Gustavo Cilla et al |

| | | | | | | |
|---|---|---|---|---|---|---|
| EPI_ISL_468936 | Spain/PV-IBV-2094/2020 | 2020-04-11 | Europe/Spain/Basque Country/San Seba | Servicio de Microbiolog√≠a. Hospital Universitario Donostia. OSI Donostialdea. √Årea de Enfermedades Infecciosas, Grupo de Infecci√≥n Respiratoria y Resistencia Antimicrobiana. Instituto de Investigaci√≥n Sanitaria Biodonostia. | SeqCOVID-SPAIN consortium/IBV(CSIC) | Gustavo Cilla et al |
| EPI_ISL_468945 | Spain/PV-IBV-2105/2020 | 2020-04-11 | Europe/Spain/Basque Country/San Seba | Servicio de Microbiolog√≠a. Hospital Universitario Donostia. OSI Donostialdea. √Årea de Enfermedades Infecciosas, Grupo de Infecci√≥n Respiratoria y Resistencia Antimicrobiana. Instituto de Investigaci√≥n Sanitaria Biodonostia. | SeqCOVID-SPAIN consortium/IBV(CSIC) | Gustavo Cilla et al |
| EPI_ISL_468947 | Spain/PV-IBV-2124/2020 | 2020-04-11 | Europe/Spain/Basque Country/San Seba | Servicio de Microbiolog√≠a. Hospital Universitario Donostia. OSI Donostialdea. √Årea de Enfermedades Infecciosas, Grupo de Infecci√≥n Respiratoria y Resistencia Antimicrobiana. Instituto de Investigaci√≥n Sanitaria Biodonostia. | SeqCOVID-SPAIN consortium/IBV(CSIC) | Gustavo Cilla et al |
| EPI_ISL_468951 | Spain/PV-IBV-2167/2020 | 2020-04-12 | Europe/Spain/Basque Country/San Seba | Servicio de Microbiolog√≠a. Hospital Universitario Donostia. OSI Donostialdea. √Årea de Enfermedades Infecciosas, Grupo de Infecci√≥n Respiratoria y Resistencia Antimicrobiana. Instituto de Investigaci√≥n Sanitaria Biodonostia. | SeqCOVID-SPAIN consortium/IBV(CSIC) | Gustavo Cilla et al |
| EPI_ISL_468969 | Spain/IB-IBV-2540/2020 | 2020-03-28 | Europe/Spain/Balear Islands/Palma | Servicio de Microbiolog√≠a, Hospital Universitario Son Espases | SeqCOVID-SPAIN consortium/IBV(CSIC) | Carla L√≥pez-Causap√© et al |
| EPI_ISL_468970 | Spain/IB-IBV-2509/2020 | 2020-03-17 | Europe/Spain/Balear Islands/Palma | Servicio de Microbiolog√≠a, Hospital Universitario Son Espases | SeqCOVID-SPAIN consortium/IBV(CSIC) | Carla L√≥pez-Causap√© et al |
| EPI_ISL_468973 | Spain/IB-IBV-2555/2020 | 2020-03-25 | Europe/Spain/Balear Islands/Palma | Servicio de Microbiolog√≠a, Hospital Universitario Son Espases | SeqCOVID-SPAIN consortium/IBV(CSIC) | Carla L√≥pez-Causap√© et al |
| EPI_ISL_468978 | Spain/IB-IBV-2499/2020 | 2020-03-20 | Europe/Spain/Balear Islands/Santa Mar | Servicio de Microbiolog√≠a, Hospital Universitario Son Espases | SeqCOVID-SPAIN consortium/IBV(CSIC) | Carla L√≥pez-Causap√© et al |
| EPI_ISL_468980 | Spain/IB-IBV-2571/2020 | 2020-04-07 | Europe/Spain/Balear Islands/Palma | Servicio de Microbiolog√≠a, Hospital Universitario Son Espases | SeqCOVID-SPAIN consortium/IBV(CSIC) | Carla L√≥pez-Causap√© et al |

| | | | | | | |
|---|---|---|---|---|---|---|
| EPI_ISL_468997 | Spain/IB-IBV-2562/2020 | 2020-04-05 | Europe/Spain/Balear Islands/Palma | Servicio de Microbiolog√≠a, Hospital Universitario Son Espases | SeqCOVID-SPAIN consortium/IBV(CSIC) | Carla L√≥pez-Causap√© et al |
| EPI_ISL_468998 | Spain/IB-IBV-2554/2020 | 2020-03-28 | Europe/Spain/Balear Islands/Manacor | Servicio de Microbiolog√≠a, Hospital Universitario Son Espases | SeqCOVID-SPAIN consortium/IBV(CSIC) | Carla L√≥pez-Causap√© et al |
| EPI_ISL_468999 | Spain/IB-IBV-h2551/2020 | 2020-03-20 | Europe/Spain/Balear Islands/Palma de M | Servicio de Microbiolog√≠a, Hospital Universitario Son Espases | SeqCOVID-SPAIN consortium/IBV(CSIC) | Carla L√≥pez-Causap√© et al |
| EPI_ISL_469001 | Spain/IB-IBV-2512/2020 | 2020-03-20 | Europe/Spain/Balear Islands/Palma | Servicio de Microbiolog√≠a, Hospital Universitario Son Espases | SeqCOVID-SPAIN consortium/IBV(CSIC) | Carla L√≥pez-Causap√© et al |
| EPI_ISL_469002 | Spain/IB-IBV-2508/2020 | 2020-03-21 | Europe/Spain/Balear Islands/Palma | Servicio de Microbiolog√≠a, Hospital Universitario Son Espases | SeqCOVID-SPAIN consortium/IBV(CSIC) | Carla L√≥pez-Causap√© et al |
| EPI_ISL_469009 | Spain/IB-IBV-2552/2020 | 2020-03-24 | Europe/Spain/Balear Islands/Son Server | Servicio de Microbiolog√≠a, Hospital Universitario Son Espases | SeqCOVID-SPAIN consortium/IBV(CSIC) | Carla L√≥pez-Causap√© et al |
| EPI_ISL_469018 | Italy/APU-UniMI-802/2020 | 2020-03-19 | Europe/Italy/Apulia/ | Istituto Zooprofilattico Sperimentale Puglia e Basilicata; Dipartimento di Bioscienze, Biotecnologie e Biofarmaceutica dell‚ÄôUniversit√† degli Studi di Bari ‚ÄúA.Moro‚Äù; Istituto di Biomembrane. Bioenergetica e Biotecnologie Molecolari del Consiglio Nazionale delle Ricerche di Bari | Beaconlab (Bioinformatics, Evolution and Comparative Genomics lab), Dept of Biosciences, University on Milan | Parisi A. et al |
| EPI_ISL_469021 | Italy/APU-UniMI-813/2020 | 2020-03-20 | Europe/Italy/Apulia/Lecce | Istituto Zooprofilattico Sperimentale Puglia e Basilicata; Dipartimento di Bioscienze, Biotecnologie e Biofarmaceutica dell‚ÄôUniversit√† degli Studi di Bari ‚ÄúA.Moro‚Äù; Istituto di Biomembrane. Bioenergetica e Biotecnologie Molecolari del Consiglio Nazionale delle Ricerche di Bari | Beaconlab (Bioinformatics, Evolution and Comparative Genomics lab), Dept of Biosciences, University on Milan | Parisi A. et al |

| | | | | | | |
|---|---|---|---|---|---|---|
| EPI_ISL_469022 | Italy/APU-UniMI-847/2020 | 2020-03-20 | Europe/Italy/Apulia/Lecce | Istituto Zooprofilattico Sperimentale Puglia e Basilicata; Dipartimento di Bioscienze, Biotecnologie e Biofarmaceutica dell‚ÄôUniversit√† degli Studi di Bari ‚ÄúA.Moro‚Äù; Istituto di Biomembrane. Bioenergetica e Biotecnologie Molecolari del Consiglio Nazionale delle Ricerche di Bari | Beaconlab (Bioinformatics, Evolution and Comparative Genomics lab), Dept of Biosciences, University on Milan | Parisi A. et al |
| EPI_ISL_469023 | Italy/APU-UniMI-857/2020 | 2020-03-20 | Europe/Italy/Apulia/Brindisi | Istituto Zooprofilattico Sperimentale Puglia e Basilicata; Dipartimento di Bioscienze, Biotecnologie e Biofarmaceutica dell‚ÄôUniversit√† degli Studi di Bari ‚ÄúA.Moro‚Äù; Istituto di Biomembrane. Bioenergetica e Biotecnologie Molecolari del Consiglio Nazionale delle Ricerche di Bari | Beaconlab (Bioinformatics, Evolution and Comparative Genomics lab), Dept of Biosciences, University on Milan | Parisi A. et al |
| EPI_ISL_469028 | India/GJ-GBRC186b/2020 | 2020-06-03 | Asia/India/Gujarat/Ahmedabad | B.J. Medical College and Civil hospital | Gujarat Biotechnology Research Centre | Janvi Raval et al |
| EPI_ISL_469029 | India/GJ-GBRC187a/2020 | 2020-06-05 | Asia/India/Gujarat/Vadodara | Government Medical College, Vadodara | Gujarat Biotechnology Research Centre | Zarna Patel et al |
| EPI_ISL_469031 | India/GJ-GBRC188/2020 | 2020-06-05 | Asia/India/Gujarat/Vadodara | Government Medical College, Vadodara | Gujarat Biotechnology Research Centre | Pinal Trivedi et al |
| EPI_ISL_469033 | India/GJ-GBRC190/2020 | 2020-06-05 | Asia/India/Gujarat/Vadodara | Government Medical College, Vadodara | Gujarat Biotechnology Research Centre | Nidhi Patel et al |
| EPI_ISL_469035 | India/GJ-GBRC191b/2020 | 2020-06-05 | Asia/India/Gujarat/Vadodara | Government Medical College, Vadodara | Gujarat Biotechnology Research Centre | Raghawendra Kumar et al |
| EPI_ISL_469039 | India/GJ-GBRC195a/2020 | 2020-06-08 | Asia/India/Gujarat/Vadodara | GMERS Medical College & Hospital | Gujarat Biotechnology Research Centre | Labdhi Pandya et al |
| EPI_ISL_469040 | India/GJ-GBRC195b/2020 | 2020-06-08 | Asia/India/Gujarat/Vadodara | GMERS Medical College & Hospital | Gujarat Biotechnology Research Centre | Snehal Bagatharia et al |

| | | | | | | |
|---|---|---|---|---|---|---|
| EPI_ISL_469041 | India/GJ-GBRC196/2020 | 2020-06-08 | Asia/India/Gujarat/Vadodara | GMERS Medical College & Hospital | Gujarat Biotechnology Research Centre | Meenakshi Shah et al |
| EPI_ISL_469043 | India/GJ-GBRC198/2020 | 2020-06-08 | Asia/India/Gujarat/Nadiad | Dr. N. D. Desai Medical College & Hospital | Gujarat Biotechnology Research Centre | J G Buch et al |
| EPI_ISL_469044 | India/GJ-GBRC199/2020 | 2020-06-08 | Asia/India/Gujarat/Nadiad | Dr. N. D. Desai Medical College & Hospital | Gujarat Biotechnology Research Centre | Jigar Gusani et al |
| EPI_ISL_469045 | India/GJ-GBRC200/2020 | 2020-06-08 | Asia/India/Gujarat/Kapadvanj | Dr. N. D. Desai Medical College & Hospital | Gujarat Biotechnology Research Centre | Supreet Prabhu et al |
| EPI_ISL_469046 | India/GJ-GBRC201a/2020 | 2020-06-08 | Asia/India/Gujarat/Mahemdavad | Dr. N. D. Desai Medical College & Hospital | Gujarat Biotechnology Research Centre | Tejas Shah et al |
| EPI_ISL_469047 | India/GJ-GBRC201b/2020 | 2020-06-08 | Asia/India/Gujarat/Mahemdavad | Dr. N. D. Desai Medical College & Hospital | Gujarat Biotechnology Research Centre | Ankit Hinsu et al |
| EPI_ISL_469048 | India/GJ-GBRC202/2020 | 2020-06-05 | Asia/India/Gujarat/Dhanera | Banas Medical College and Research Institute | Gujarat Biotechnology Research Centre | Radhika Khara et al |
| EPI_ISL_469052 | Morocco/RMPS-04/2020 | 2020-03-30 | Africa/Morocco/Morocco/ | LNR National Reference Laboratory, Mohammed VI University of Health Sciences | Medical Biotechnology Laboratory, Rabat Medical and Pharmacy School, Mohammed The Vth University in Rabat | Meriem LAAMARTI et al |
| EPI_ISL_469054 | Morocco/RMPS-06/2020 | 2020-04-01 | Africa/Morocco/Morocco/ | LNR National Reference Laboratory, Mohammed VI University of Health Sciences | Medical Biotechnology Laboratory, Rabat Medical and Pharmacy School, Mohammed The Vth University in Rabat | Meriem LAAMARTI et al |
| EPI_ISL_469055 | Sweden/20-14262/2020 | 2020-05-02 | Europe/Sweden/Ostergotland/ | Jourcentralen | The Public Health Agency of Sweden | Oskar Karlsson Lindsjo et al |
| EPI_ISL_469067 | Sweden/20-51598/2020 | 2020-05-19 | Europe/Sweden/Stockholm/ | Kungsholmsdoktorn | The Public Health Agency of Sweden | Oskar Karlsson Lindsjo et al |

| | | | | | | |
|---|---|---|---|---|---|---|
| EPI_ISL_469068 | Sweden/20-51618/2020 | 2020-05-20 | Europe/Sweden/Vastra Gotaland/ | Hovas Askim Familjelakare och BVC | The Public Health Agency of Sweden | Oskar Karlsson Lindsjo et al |
| EPI_ISL_469100 | Singapore/323ilmn/2020 | 2020-03-26 | Asia/Singapore/Singapore/ | National Public Health Laboratory, National Centre for Infectious Diseases | National Public Health Laboratory, National Centre for Infectious Diseases | Mak TM et al |
| EPI_ISL_469105 | Singapore/350/2020 | 2020-04-06 | Asia/Singapore/Singapore/ | National Public Health Laboratory, National Centre for Infectious Diseases | National Public Health Laboratory, National Centre for Infectious Diseases | Mak TM et al |
| EPI_ISL_469108 | Singapore/353/2020 | 2020-05-14 | Asia/Singapore/Singapore/ | National Public Health Laboratory, National Centre for Infectious Diseases | National Public Health Laboratory, National Centre for Infectious Diseases | Mak TM et al |
| EPI_ISL_469113 | Singapore/358/2020 | 2020-04-06 | Asia/Singapore/Singapore/ | National Public Health Laboratory, National Centre for Infectious Diseases | National Public Health Laboratory, National Centre for Infectious Diseases | Mak TM et al |
| EPI_ISL_469120 | Singapore/365/2020 | 2020-04-06 | Asia/Singapore/Singapore/ | National Public Health Laboratory, National Centre for Infectious Diseases | National Public Health Laboratory, National Centre for Infectious Diseases | Mak TM et al |
| EPI_ISL_469132 | Singapore/383/2020 | 2020-02-15 | Asia/Singapore/Singapore/ | National Public Health Laboratory, National Centre for Infectious Diseases | National Public Health Laboratory, National Centre for Infectious Diseases | Mak TM et al |

| | | | | | | |
|---|---|---|---|---|---|---|
| EPI_ISL_469136 | Singapore/387/2020 | 2020-04-17 | Asia/Singapore/Singapore/ | National Public Health Laboratory, National Centre for Infectious Diseases | National Public Health Laboratory, National Centre for Infectious Diseases | Mak TM et al |
| EPI_ISL_469176 | USA/CT-Yale-248/2020 | 2020-04-01 | North America/USA/Connecticut/New H | Yale Clinical Virology Laboratory | Grubaugh Lab - Yale School of Public Health | Joseph Fauver et al |
| EPI_ISL_469184 | USA/CT-Yale-258/2020 | 2020-04-01 | North America/USA/Connecticut/New H | Yale Clinical Virology Laboratory | Grubaugh Lab - Yale School of Public Health | Joseph Fauver et al |
| EPI_ISL_469203 | USA/CT-Yale-280/2020 | 2020-04-02 | North America/USA/Connecticut/Fairfie | Yale Clinical Virology Laboratory | Grubaugh Lab - Yale School of Public Health | Joseph Fauver et al |
| EPI_ISL_469209 | Canada/BC_18173416/2020 | 20-Apr | North America/Canada/British Columbia | BCCDC Public Health Laboratory | BCCDC Public Health Laboratory | Richard Harrigan et al |
| EPI_ISL_469210 | Canada/BC_18604452/2020 | 20-Apr | North America/Canada/British Columbia | BCCDC Public Health Laboratory | BCCDC Public Health Laboratory | Richard Harrigan et al |
| EPI_ISL_469211 | Canada/BC_18717764/2020 | 20-Apr | North America/Canada/British Columbia | BCCDC Public Health Laboratory | BCCDC Public Health Laboratory | Richard Harrigan et al |
| EPI_ISL_469212 | Canada/BC_18839283/2020 | 20-Apr | North America/Canada/British Columbia | BCCDC Public Health Laboratory | BCCDC Public Health Laboratory | Richard Harrigan et al |
| EPI_ISL_469213 | Canada/BC_19071912/2020 | 20-Apr | North America/Canada/British Columbia | BCCDC Public Health Laboratory | BCCDC Public Health Laboratory | Richard Harrigan et al |
| EPI_ISL_469214 | Canada/BC_19072825/2020 | 20-Apr | North America/Canada/British Columbia | BCCDC Public Health Laboratory | BCCDC Public Health Laboratory | Richard Harrigan et al |
| EPI_ISL_469215 | Canada/BC_19072819/2020 | 20-Apr | North America/Canada/British Columbia | BCCDC Public Health Laboratory | BCCDC Public Health Laboratory | Richard Harrigan et al |
| EPI_ISL_469216 | Canada/BC_19100589/2020 | 20-Apr | North America/Canada/British Columbia | BCCDC Public Health Laboratory | BCCDC Public Health Laboratory | Richard Harrigan et al |
| EPI_ISL_469217 | Canada/BC_19131446/2020 | 20-Apr | North America/Canada/British Columbia | BCCDC Public Health Laboratory | BCCDC Public Health Laboratory | Richard Harrigan et al |
| EPI_ISL_469218 | Canada/BC_19133870/2020 | 20-Apr | North America/Canada/British Columbia | BCCDC Public Health Laboratory | BCCDC Public Health Laboratory | Richard Harrigan et al |
| EPI_ISL_469220 | Canada/BC_19196568/2020 | 20-Apr | North America/Canada/British Columbia | BCCDC Public Health Laboratory | BCCDC Public Health Laboratory | Richard Harrigan et al |
| EPI_ISL_469222 | Canada/BC_19677621/2020 | 20-Apr | North America/Canada/British Columbia | BCCDC Public Health Laboratory | BCCDC Public Health Laboratory | Richard Harrigan et al |

| | | | | | | |
|---|---|---|---|---|---|---|
| EPI_ISL_469223 | Canada/BC_19803546/2020 | 20-Apr | North America/Canada/British Columbia | BCCDC Public Health Laboratory | BCCDC Public Health Laboratory | Richard Harrigan et al |
| EPI_ISL_469224 | Canada/NL_82/2020 | 2020-03-22 | North America/Canada/Newfoundland a | Public Health Laboratory | National Microbiology Laboratory | Anna Majer et al |
| EPI_ISL_469225 | Canada/NL_83/2020 | 2020-03-21 | North America/Canada/Newfoundland a | Public Health Laboratory | National Microbiology Laboratory | Anna Majer et al |
| EPI_ISL_469226 | Canada/NL_84/2020 | 2020-03-22 | North America/Canada/Newfoundland a | Public Health Laboratory | National Microbiology Laboratory | Anna Majer et al |
| EPI_ISL_469227 | Canada/NL_85/2020 | 2020-03-21 | North America/Canada/Newfoundland a | Public Health Laboratory | National Microbiology Laboratory | Anna Majer et al |
| EPI_ISL_469228 | Canada/NL_86/2020 | 2020-03-22 | North America/Canada/Newfoundland a | Public Health Laboratory | National Microbiology Laboratory | Anna Majer et al |
| EPI_ISL_469229 | Canada/NL_87/2020 | 2020-03-20 | North America/Canada/Newfoundland a | Public Health Laboratory | National Microbiology Laboratory | Anna Majer et al |
| EPI_ISL_469230 | Canada/NL_88/2020 | 2020-03-21 | North America/Canada/Newfoundland a | Public Health Laboratory | National Microbiology Laboratory | Anna Majer et al |
| EPI_ISL_469231 | Canada/NL_89/2020 | 2020-03-21 | North America/Canada/Newfoundland a | Public Health Laboratory | National Microbiology Laboratory | Anna Majer et al |
| EPI_ISL_469232 | Canada/NL_90/2020 | 2020-03-21 | North America/Canada/Newfoundland a | Public Health Laboratory | National Microbiology Laboratory | Anna Majer et al |
| EPI_ISL_469233 | Canada/NL_93/2020 | 2020-03-22 | North America/Canada/Newfoundland a | Public Health Laboratory | National Microbiology Laboratory | Anna Majer et al |
| EPI_ISL_469234 | Canada/NL_95/2020 | 2020-03-22 | North America/Canada/Newfoundland a | Public Health Laboratory | National Microbiology Laboratory | Anna Majer et al |
| EPI_ISL_469235 | Canada/NL_96/2020 | 2020-03-22 | North America/Canada/Newfoundland a | Public Health Laboratory | National Microbiology Laboratory | Anna Majer et al |
| EPI_ISL_469236 | Canada/NL_97/2020 | 2020-03-22 | North America/Canada/Newfoundland a | Public Health Laboratory | National Microbiology Laboratory | Anna Majer et al |

| | | | | | | |
|---|---|---|---|---|---|---|
| EPI_ISL_469237 | Canada/NL_98/2020 | 2020-03-22 | North America/Canada/Newfoundland a | Public Health Laboratory | National Microbiology Laboratory | Anna Majer et al |
| EPI_ISL_469238 | Canada/NL_99/2020 | 2020-03-22 | North America/Canada/Newfoundland a | Public Health Laboratory | National Microbiology Laboratory | Anna Majer et al |
| EPI_ISL_469239 | Canada/NL_100/2020 | 2020-03-22 | North America/Canada/Newfoundland a | Public Health Laboratory | National Microbiology Laboratory | Anna Majer et al |
| EPI_ISL_469240 | Canada/NL_101/2020 | 2020-03-22 | North America/Canada/Newfoundland a | Public Health Laboratory | National Microbiology Laboratory | Anna Majer et al |
| EPI_ISL_469243 | SaudiArabia/85613/2020 | 2020-03-15 | Asia/Saudi Arabia/Jeddah/ | Special Infectious Agents Unit | Special Infectious Agents Unit | Azhar et al |
| EPI_ISL_469253 | Shanghai/CH-02/2020 | 2020-02-28 | Asia/China/Shanghai/ | Second Military Medical University, Department of Microbiology | Second Military Medical University, Department of Microbiology | Peng et al |
| EPI_ISL_469254 | Beijing/IVDC-01-06/2020 | 2020-06-11 | Asia/China/Beijing/ | National Institute for Viral Disease Control and Prevention, China CDC | Institute of Viral Disease Control and Prevention, China CDC | Wenjie Tan et al |
| EPI_ISL_469255 | Beijing/IVDC-02-06/2020 | 2020-06-11 | Asia/China/Beijing/ | National Institute for Viral Disease Control and Prevention, China CDC | Institute of Viral Disease Control and Prevention, China CDC | Xiang Zhao et al |
| EPI_ISL_469279 | UnitedArabEmirates/L5857/2020 | 2020-03-31 | Asia/United Arab Emirates/United Arab | Mohammed Bin Rashid University of Medicine and Health Sciences | Al Jalila Genomics Center | Ahmad Abou Tayoun et al |
| EPI_ISL_469280 | UnitedArabEmirates/L6841/2020 | 2020-03-27 | Asia/United Arab Emirates/United Arab | Mohammed Bin Rashid University of Medicine and Health Sciences | Al Jalila Genomics Center | Ahmad Abou Tayoun et al |
| EPI_ISL_469297 | Bangladesh/BCSIR-NILMRC-095/2020 | 2020-05-23 | Asia/Bangladesh/Rangpur/ | National Institute of Laboratory Medicine and Referral Center | Genomic Research Lab, BCSIR | Barna Goswami et al |
| EPI_ISL_469298 | Bangladesh/BCSIR-NILMRC-096/2020 | 2020-05-23 | Asia/Bangladesh/Rangpur/ | National Institute of Laboratory Medicine and Referral Center | Genomic Research Lab, BCSIR | Md. Murshed Hasan Sarkar et al |
| EPI_ISL_469417 | England/BRIS-12C38C/2020 | 2020-05-11 | Europe/United Kingdom/England/ | PHE South West Regional Laboratory, National Infection Service | Wellcome Sanger Institute for the COVID-19 Genomics UK (COG-UK) consortium | Stephanie Hutchings et al |

| | | | | | | |
|---|---|---|---|---|---|---|
| EPI_ISL_469418 | England/BRIS-12BAE2/2020 | 2020-05-18 | Europe/United Kingdom/England/ | PHE South West Regional Laboratory, National Infection Service | Wellcome Sanger Institute for the COVID-19 Genomics UK (COG-UK) consortium | Stephanie Hutchings et al |
| EPI_ISL_469450 | England/BRIS-12BE40/2020 | 2020-05-08 | Europe/United Kingdom/England/ | PHE South West Regional Laboratory, National Infection Service | Wellcome Sanger Institute for the COVID-19 Genomics UK (COG-UK) consortium | Stephanie Hutchings et al |
| EPI_ISL_469744 | England/BRIS-1279E9/2020 | 2020-04-17 | Europe/United Kingdom/England/ | PHE South West Regional Laboratory, National Infection Service | Wellcome Sanger Institute for the COVID-19 Genomics UK (COG-UK) consortium | Stephanie Hutchings et al |
| EPI_ISL_469787 | England/BRIS-128362/2020 | 2020-04-16 | Europe/United Kingdom/England/ | PHE South West Regional Laboratory, National Infection Service | Wellcome Sanger Institute for the COVID-19 Genomics UK (COG-UK) consortium | Stephanie Hutchings et al |
| EPI_ISL_469836 | England/BRIS-12B02C/2020 | 2020-05-04 | Europe/United Kingdom/England/ | PHE South West Regional Laboratory, National Infection Service | Wellcome Sanger Institute for the COVID-19 Genomics UK (COG-UK) consortium | Stephanie Hutchings et al |
| EPI_ISL_469874 | NorthernIreland/NIRE-1015A5/2020 | 2020-04-29 | Europe/United Kingdom/Northern Irelar | Regional Virus Laboratory, Belfast Health and Social Care Trust | Wellcome Sanger Institute for the COVID-19 Genomics UK (COG-UK) consortium | Conall McCaughey et al |
| EPI_ISL_469886 | NorthernIreland/NIRE-100B86/2020 | 2020-04-29 | Europe/United Kingdom/Northern Irelar | Regional Virus Laboratory, Belfast Health and Social Care Trust | Wellcome Sanger Institute for the COVID-19 Genomics UK (COG-UK) consortium | Conall McCaughey et al |
| EPI_ISL_469982 | Scotland/GCVR-172396/2020 | 2020-03-30 | Europe/United Kingdom/Scotland/ | NHSGGC West of Scotland Specialist Virology Centre / MRC-University of Glasgow Centre for Virus Research | Wellcome Sanger Institute for the COVID-19 Genomics UK (COG-UK) consortium | Ana da Silva Filipe et al |

| | | | | | | |
|---|---|---|---|---|---|---|
| EPI_ISL_470010 | Scotland/GCVR-171F2A/2020 | 2020-03-31 | Europe/United Kingdom/Scotland/ | NHSGGC West of Scotland Specialist Virology Centre / MRC-University of Glasgow Centre for Virus Research | Wellcome Sanger Institute for the COVID-19 Genomics UK (COG-UK) consortium | Ana da Silva Filipe et al |
| EPI_ISL_470078 | NorthernIreland/NIRE-FCBE8/2020 | 2020-04-22 | Europe/United Kingdom/Northern Irelar | Regional Virus Laboratory, Belfast Health and Social Care Trust | Wellcome Sanger Institute for the COVID-19 Genomics UK (COG-UK) consortium | Conall McCaughey et al |
| EPI_ISL_470114 | England/CAMB-1AA6AB/2020 | 2020-04-17 | Europe/United Kingdom/England/ | Department of Pathology, University of Cambridge | Wellcome Sanger Institute for the COVID-19 Genomics UK (COG-UK) consortium | Luke W Meredith et al |
| EPI_ISL_470125 | England/CAMB-1AA9FD/2020 | 2020-04-17 | Europe/United Kingdom/England/ | Department of Pathology, University of Cambridge | Wellcome Sanger Institute for the COVID-19 Genomics UK (COG-UK) consortium | Luke W Meredith et al |
| EPI_ISL_470132 | England/CAMB-1AB6F5/2020 | 2020-04-21 | Europe/United Kingdom/England/ | Department of Pathology, University of Cambridge | Wellcome Sanger Institute for the COVID-19 Genomics UK (COG-UK) consortium | Luke W Meredith et al |
| EPI_ISL_470237 | England/CAMB-1AA6C9/2020 | 2020-04-18 | Europe/United Kingdom/England/ | Department of Pathology, University of Cambridge | Wellcome Sanger Institute for the COVID-19 Genomics UK (COG-UK) consortium | Luke W Meredith et al |
| EPI_ISL_470290 | England/CAMB-1AB4EC/2020 | 2020-04-22 | Europe/United Kingdom/England/ | Department of Pathology, University of Cambridge | Wellcome Sanger Institute for the COVID-19 Genomics UK (COG-UK) consortium | Luke W Meredith et al |
| EPI_ISL_470332 | England/CAMB-1AB65F/2020 | 2020-04-21 | Europe/United Kingdom/England/ | Department of Pathology, University of Cambridge | Wellcome Sanger Institute for the COVID-19 Genomics UK (COG-UK) consortium | Luke W Meredith et al |

| | | | | | | |
|---|---|---|---|---|---|---|
| EPI_ISL_470388 | England/CAMB-80ACE/2020 | 2020-04-10 | Europe/United Kingdom/England/ | Department of Pathology, University of Cambridge | Wellcome Sanger Institute for the COVID-19 Genomics UK (COG-UK) consortium | Luke W Meredith et al |
| EPI_ISL_470485 | England/CAMB-7DF98/2020 | 2020-04-06 | Europe/United Kingdom/England/ | Department of Pathology, University of Cambridge | Wellcome Sanger Institute for the COVID-19 Genomics UK (COG-UK) consortium | Luke W Meredith et al |
| EPI_ISL_470541 | USA/UT-01909/2020 | 2020-04-13 | North America/USA/Utah/ | Utah Public Health Laboratory | Utah Public Health Laboratory | Erin Young et al |
| EPI_ISL_470551 | USA/UT-02435/2020 | 2020-04-22 | North America/USA/Utah/ | Utah Public Health Laboratory | Utah Public Health Laboratory | Erin Young et al |
| EPI_ISL_470552 | USA/UT-02437/2020 | 2020-04-21 | North America/USA/Utah/ | Utah Public Health Laboratory | Utah Public Health Laboratory | Erin Young et al |
| EPI_ISL_470553 | USA/UT-02438/2020 | 2020-04-21 | North America/USA/Utah/ | Utah Public Health Laboratory | Utah Public Health Laboratory | Erin Young et al |
| EPI_ISL_470555 | USA/UT-03893/2020 | 2020-04-13 | North America/USA/Utah/ | Utah Public Health Laboratory | Utah Public Health Laboratory | Erin Young et al |
| EPI_ISL_470561 | USA/UT-03906/2020 | 2020-04-13 | North America/USA/Utah/ | Utah Public Health Laboratory | Utah Public Health Laboratory | Erin Young et al |
| EPI_ISL_470576 | Brazil/GO-0209/2020 | 2020-04-03 | South America/Brazil/Goiv°s/ | Hermes Pardini | Bioinformatics Laboratory / LNCC | Alexandra Gerber et al |
| EPI_ISL_470585 | Brazil/MG-0219/2020 | 2020-04-03 | South America/Brazil/Minas Gerais/ | Hermes Pardini | Bioinformatics Laboratory / LNCC | Alexandra Gerber et al |
| EPI_ISL_470586 | Brazil/MG-0220/2020 | 2020-04-04 | South America/Brazil/Minas Gerais/ | Hermes Pardini | Bioinformatics Laboratory / LNCC | Alexandra Gerber et al |
| EPI_ISL_470590 | Brazil/MG-0285/2020 | 2020-04-08 | South America/Brazil/Minas Gerais/ | Simile | Bioinformatics Laboratory / LNCC | Alexandra Gerber et al |
| EPI_ISL_470611 | Brazil/PA-0237/2020 | 2020-04-03 | South America/Brazil/Para/ | Hermes Pardini | Bioinformatics Laboratory / LNCC | Alexandra Gerber et al |
| EPI_ISL_470612 | Brazil/PE-0238/2020 | 2020-04-03 | South America/Brazil/Pernambuco/ | Hermes Pardini | Bioinformatics Laboratory / LNCC | Alexandra Gerber et al |
| EPI_ISL_470632 | Brazil/RJ-0266/2020 | 2020-04-14 | South America/Brazil/Rio de Janeiro/ | Laboratorio de Virologia Molecular / UFRJ | Bioinformatics Laboratory / LNCC | Alexandra Gerber et al |
| EPI_ISL_470638 | Brazil/RJ-0272/2020 | 2020-04-17 | South America/Brazil/Rio de Janeiro/ | Laboratorio de Virologia Molecular / UFRJ | Bioinformatics Laboratory / LNCC | Alexandra Gerber et al |
| EPI_ISL_470640 | Brazil/RJ-0274/2020 | 2020-04-20 | South America/Brazil/Rio de Janeiro/ | Laboratorio de Virologia Molecular / UFRJ | Bioinformatics Laboratory / LNCC | Alexandra Gerber et al |

| | | | | | | |
|---|---|---|---|---|---|---|
| EPI_ISL_470642 | Brazil/RJ-0276/2020 | 2020-04-22 | South America/Brazil/Rio de Janeiro/ | Laboratorio de Virologia Molecular / UFRJ | Bioinformatics Laboratory / LNCC | Alexandra Gerber et al |
| EPI_ISL_470661 | USA/UT-02034/2020 | 2020-04-17 | North America/USA/Utah/ | Utah Public Health Laboratory | Utah Public Health Laboratory | Erin Young et al |
| EPI_ISL_470663 | USA/UT-02040/2020 | 2020-04-14 | North America/USA/Utah/ | Utah Public Health Laboratory | Utah Public Health Laboratory | Erin Young et al |
| EPI_ISL_470664 | USA/UT-02041/2020 | 2020-04-14 | North America/USA/Utah/ | Utah Public Health Laboratory | Utah Public Health Laboratory | Erin Young et al |
| EPI_ISL_470667 | USA/UT-02046/2020 | 2020-04-19 | North America/USA/Utah/ | Utah Public Health Laboratory | Utah Public Health Laboratory | Erin Young et al |
| EPI_ISL_470677 | USA/UT-02429/2020 | 2020-04-21 | North America/USA/Utah/ | Utah Public Health Laboratory | Utah Public Health Laboratory | Erin Young et al |
| EPI_ISL_470680 | USA/UT-01837/2020 | 2020-04-13 | North America/USA/Utah/ | Utah Public Health Laboratory | Utah Public Health Laboratory | Erin Young et al |
| EPI_ISL_470681 | USA/UT-01838/2020 | 2020-04-13 | North America/USA/Utah/ | Utah Public Health Laboratory | Utah Public Health Laboratory | Erin Young et al |
| EPI_ISL_470682 | USA/UT-01840/2020 | 2020-04-13 | North America/USA/Utah/ | Utah Public Health Laboratory | Utah Public Health Laboratory | Erin Young et al |
| EPI_ISL_470688 | USA/UT-01846/2020 | 2020-04-13 | North America/USA/Utah/ | Utah Public Health Laboratory | Utah Public Health Laboratory | Erin Young et al |
| EPI_ISL_470694 | USA/UT-01870/2020 | 2020-04-13 | North America/USA/Utah/ | Utah Public Health Laboratory | Utah Public Health Laboratory | Erin Young et al |
| EPI_ISL_470696 | USA/UT-01872/2020 | 2020-04-13 | North America/USA/Utah/ | Utah Public Health Laboratory | Utah Public Health Laboratory | Erin Young et al |
| EPI_ISL_470709 | USA/UT-02137/2020 | 2020-04-14 | North America/USA/Utah/ | Utah Public Health Laboratory | Utah Public Health Laboratory | Erin Young et al |
| EPI_ISL_470712 | USA/UT-03810/2020 | 2020-04-03 | North America/USA/Utah/ | Utah Public Health Laboratory | Utah Public Health Laboratory | Erin Young et al |
| EPI_ISL_470714 | USA/UT-03816/2020 | 2020-04-03 | North America/USA/Utah/ | Utah Public Health Laboratory | Utah Public Health Laboratory | Erin Young et al |
| EPI_ISL_470744 | USA/UT-01887/2020 | 2020-04-13 | North America/USA/Utah/ | Utah Public Health Laboratory | Utah Public Health Laboratory | Heidi Butz et al |
| EPI_ISL_470747 | USA/MN-MDH-1067/2020 | 2020-05-21 | North America/USA/Minnesota/ | Minnesota Department of Health, Public Health Laboratory | Minnesota Department of Health, Public Health Laboratory | Matt Plumb et al |
| EPI_ISL_470748 | USA/MN-MDH-1068/2020 | 2020-05-21 | North America/USA/Minnesota/ | Minnesota Department of Health, Public Health Laboratory | Minnesota Department of Health, Public Health Laboratory | Matt Plumb et al |

| | | | | | | |
|---|---|---|---|---|---|---|
| EPI_ISL_470752 | USA/MN-MDH-1072/2020 | 2020-06-03 | North America/USA/Minnesota/ | Minnesota Department of Health, Public Health Laboratory | Minnesota Department of Health, Public Health Laboratory | Matt Plumb et al |
| EPI_ISL_470756 | USA/MN-MDH-1076/2020 | 2020-06-03 | North America/USA/Minnesota/ | Minnesota Department of Health, Public Health Laboratory | Minnesota Department of Health, Public Health Laboratory | Matt Plumb et al |
| EPI_ISL_470759 | USA/MN-MDH-1079/2020 | 2020-06-03 | North America/USA/Minnesota/ | Minnesota Department of Health, Public Health Laboratory | Minnesota Department of Health, Public Health Laboratory | Matt Plumb et al |
| EPI_ISL_470764 | USA/MN-MDH-1084/2020 | 2020-06-03 | North America/USA/Minnesota/ | Minnesota Department of Health, Public Health Laboratory | Minnesota Department of Health, Public Health Laboratory | Matt Plumb et al |
| EPI_ISL_470773 | USA/MN-MDH-1093/2020 | 2020-06-03 | North America/USA/Minnesota/ | Minnesota Department of Health, Public Health Laboratory | Minnesota Department of Health, Public Health Laboratory | Matt Plumb et al |
| EPI_ISL_470783 | USA/MN-MDH-1103/2020 | 2020-06-03 | North America/USA/Minnesota/ | Minnesota Department of Health, Public Health Laboratory | Minnesota Department of Health, Public Health Laboratory | Matt Plumb et al |
| EPI_ISL_470785 | USA/MN-MDH-1105/2020 | 2020-06-03 | North America/USA/Minnesota/ | Minnesota Department of Health, Public Health Laboratory | Minnesota Department of Health, Public Health Laboratory | Matt Plumb et al |
| EPI_ISL_470789 | USA/MN-MDH-1109/2020 | 2020-06-03 | North America/USA/Minnesota/ | Minnesota Department of Health, Public Health Laboratory | Minnesota Department of Health, Public Health Laboratory | Matt Plumb et al |
| EPI_ISL_470792 | USA/MN-MDH-1112/2020 | 2020-05-03 | North America/USA/Minnesota/ | M Health Fairview | Minnesota Department of Health, Public Health Laboratory | Matt Plumb et al |
| EPI_ISL_470793 | USA/MN-MDH-1113/2020 | 2020-05-03 | North America/USA/Minnesota/ | M Health Fairview | Minnesota Department of Health, Public Health Laboratory | Matt Plumb et al |

| | | | | | | |
|---|---|---|---|---|---|---|
| EPI_ISL_470851 | Australia/WA34/2020 | 2020-02-27 | Oceania/Australia/Western Australia/ | PathWest Laboratory Medicine WA | PathWest Laboratory Medicine WA | Chisha Sikazwe et al |
| EPI_ISL_470868 | Australia/WA51/2020 | 2020-03-20 | Oceania/Australia/Western Australia/ | PathWest Laboratory Medicine WA | PathWest Laboratory Medicine WA | Chisha Sikazwe et al |
| EPI_ISL_470877 | Georgia/Tb-3118/2020 | 2020-04-15 | Asia/Georgia/Tbilisi/ | Department for Virology, Molecular Biology and Genome Research, R. G. Lugar Center for Public Health Research, National Center for Disease Control and Public Health (NCDC) of Georgia. | Department for Virology, Molecular Biology and Genome Research, R. G. Lugar Center for Public Health Research, National Center for Disease Control and Public Health (NCDC) of Georgia. | Gvantsa Brachveli et al |
| EPI_ISL_470897 | Russia/Moscow_PMVL-3/2020 | 2020-05-05 | Europe/Russia/Moscow Oblast/ | Pathogenic Microorganisms Variability Laboratory | Pathogenic Microorganisms Variability Laboratory | Alexey Shchetinin et al |
| EPI_ISL_470899 | Russia/Moscow_PMVL-5/2020 | 2020-05-05 | Europe/Russia/Moscow Oblast/ | Pathogenic Microorganisms Variability Laboratory | Pathogenic Microorganisms Variability Laboratory | Alexey Shchetinin et al |
| EPI_ISL_470902 | Russia/Moscow_PMVL-9/2020 | 2020-04-08 | Europe/Russia/Moscow Oblast/ | Influenza etiology and epidemiology laboratory | Pathogenic Microorganisms Variability Laboratory | Alexey Shchetinin et al |
| EPI_ISL_470903 | Russia/Moscow_PMVL-10/2020 | 2020-04-10 | Europe/Russia/Moscow Oblast/ | Influenza etiology and epidemiology laboratory | Pathogenic Microorganisms Variability Laboratory | Alexey Shchetinin et al |
| EPI_ISL_470904 | Russia/Moscow_PMVL-11/2020 | 2020-04-10 | Europe/Russia/Moscow Oblast/ | Influenza etiology and epidemiology laboratory | Pathogenic Microorganisms Variability Laboratory | Alexey Shchetinin et al |

| | | | | | | |
|---|---|---|---|---|---|---|
| EPI_ISL_471147 | USA/WI-GMF-05641/2020 | 2020-06-08 | North America/USA/Wisconsin/La Crosse | Gundersen Molecular Diagnostics Laboratory | Kabara Cancer Research Institute | Craig S. Richmond et al |
| EPI_ISL_471188 | USA/WI-WSLH-200023/2020 | 2020-04-10 | North America/USA/Wisconsin/ | Wisconsin State Laboratory of Hygiene Communicable Disease Division | Wisconsin State Laboratory of Hygiene Communicable Disease Division | Kelsey R. Florek et al |
| EPI_ISL_471210 | USA/WI-WSLH-200045/2020 | 2020-03-30 | North America/USA/Wisconsin/ | Wisconsin State Laboratory of Hygiene Communicable Disease Division | Wisconsin State Laboratory of Hygiene Communicable Disease Division | Kelsey R. Florek et al |
| EPI_ISL_471218 | USA/WI-WSLH-200053/2020 | 2020-04-20 | North America/USA/Wisconsin/ | Wisconsin State Laboratory of Hygiene Communicable Disease Division | Wisconsin State Laboratory of Hygiene Communicable Disease Division | Kelsey R. Florek et al |
| EPI_ISL_471231 | USA/WI-WSLH-200066/2020 | 2020-04-15 | North America/USA/Wisconsin/ | Wisconsin State Laboratory of Hygiene Communicable Disease Division | Wisconsin State Laboratory of Hygiene Communicable Disease Division | Kelsey R. Florek et al |
| EPI_ISL_471241 | USA/WI-WSLH-200076/2020 | 2020-04-04 | North America/USA/Wisconsin/ | Wisconsin State Laboratory of Hygiene Communicable Disease Division | Wisconsin State Laboratory of Hygiene Communicable Disease Division | Kelsey R. Florek et al |
| EPI_ISL_471247 | USA/WI-WSLH-200083/2020 | 2020-04-13 | North America/USA/Wisconsin/ | Wisconsin State Laboratory of Hygiene Communicable Disease Division | Wisconsin State Laboratory of Hygiene Communicable Disease Division | Kelsey R. Florek et al |
| EPI_ISL_471250 | USA/WI-WSLH-200086/2020 | 2020-03-16 | North America/USA/Wisconsin/ | Wisconsin State Laboratory of Hygiene Communicable Disease Division | Wisconsin State Laboratory of Hygiene Communicable Disease Division | Kelsey R. Florek et al |

| | | | | | | |
|---|---|---|---|---|---|---|
| EPI_ISL_471262 | USA/WI-WSLH-200098/2020 | 2020-03-27 | North America/USA/Wisconsin/ | Wisconsin State Laboratory of Hygiene Communicable Disease Division | Wisconsin State Laboratory of Hygiene Communicable Disease Division | Kelsey R. Florek et al |
| EPI_ISL_471265 | USA/WI-WSLH-200101/2020 | 2020-03-30 | North America/USA/Wisconsin/ | Wisconsin State Laboratory of Hygiene Communicable Disease Division | Wisconsin State Laboratory of Hygiene Communicable Disease Division | Kelsey R. Florek et al |
| EPI_ISL_471267 | Ecuador/USFQ-020/2020 | 2020-04-17 | South America/Ecuador/Los Rios/Babaho | Hospital IESS Babahoyo | Institute of Microbiology, Universidad San Francisco de Quito | Sully M√°rquez et al |
| EPI_ISL_471268 | Ecuador/USFQ-045/2020 | 2020-04-17 | South America/Ecuador/Los Rios/Babaho | Hospital IESS Babahoyo | Institute of Microbiology, Universidad San Francisco de Quito | Bel√©n Prado-Vivar et al |
| EPI_ISL_471270 | Ecuador/USFQ-112/2020 | 2020-05-19 | South America/Ecuador/Pichincha/Quito | Hospital Oncol√≥gico Solca N√∫cleo de Quito | Institute of Microbiology, Universidad San Francisco de Quito | Sully M√°rquez et al |
| EPI_ISL_471271 | Ecuador/USFQ-106/2020 | 2020-05-19 | South America/Ecuador/Pichincha/Quito | Hospital Oncol√≥gico Solca N√∫cleo de Quito | Institute of Microbiology, Universidad San Francisco de Quito | Sully M√°rquez et al |
| EPI_ISL_471417 | Romania/Buzau-292711/2020 | 2020-06-03 | Europe/Romania/Buzau/ | Laboratory for Respiratory Viruses, National Influenza Centre, Cantacuzino National Military-Medical Institute for Research and Development | Cantacuzino Institute | Luiza Ustea et al |
| EPI_ISL_471419 | Romania/Buzau-293197/2020 | 2020-06-03 | Europe/Romania/Buzau/ | Laboratory for Respiratory Viruses, National Influenza Centre, Cantacuzino National Military-Medical Institute for Research and Development | Cantacuzino Institute | Luiza Ustea et al |
| EPI_ISL_471420 | Romania/Bucuresti-293034/2020 | 2020-06-04 | Europe/Romania/Bucharest/ | Laboratory for Respiratory Viruses, National Influenza Centre, Cantacuzino National Military-Medical Institute for Research and Development | Cantacuzino Institute | Luiza Ustea et al |

| | | | | | | |
|---|---|---|---|---|---|---|
| EPI_ISL_471421 | Romania/Iasi-296512/2020 | 2020-06-10 | Europe/Romania/Suceava/Iasi | Laboratory for Respiratory Viruses, National Influenza Centre, Cantacuzino National Military-Medical Institute for Research and Development | Cantacuzino Institute | Luiza Ustea et al |
| EPI_ISL_471423 | Romania/Buzau-292638/2020 | 2020-06-04 | Europe/Romania/Buzau/ | Laboratory for Respiratory Viruses, National Influenza Centre, Cantacuzino National Military-Medical Institute for Research and Development | Cantacuzino Institute | Luiza Ustea et al |
| EPI_ISL_471425 | SouthKorea/KCDC2020/2020 | 2020-02-18 | Asia/South Korea/South Korea/ | Division of Viral Diseases, Center for Laboratory Control of Infectious Diseases, Korea Centers for Diseases Control and Prevention | Division of Viral Diseases, Center for Laboratory Control of Infectious Diseases, Korea Centers for Diseases Control and Prevention | Jeong-Min Kim et al |
| EPI_ISL_471426 | SouthKorea/KCDC2021/2020 | 2020-02-18 | Asia/South Korea/South Korea/ | Division of Viral Diseases, Center for Laboratory Control of Infectious Diseases, Korea Centers for Diseases Control and Prevention | Division of Viral Diseases, Center for Laboratory Control of Infectious Diseases, Korea Centers for Diseases Control and Prevention | Jeong-Min Kim et al |
| EPI_ISL_471429 | Belgium/ULG-10194/2020 | 2020-05-20 | Europe/Belgium/Liv®ge/ | Department of Clinical Microbiology | GIGA Medical Genomics | Keith Durkin et al |
| EPI_ISL_471438 | SouthKorea/KCDC2022/2020 | 2020-02-18 | Asia/South Korea/South Korea/ | Division of Viral Diseases, Center for Laboratory Control of Infectious Diseases, Korea Centers for Diseases Control and Prevention | Division of Viral Diseases, Center for Laboratory Control of Infectious Diseases, Korea Centers for Diseases Control and Prevention | Jeong-Min Kim et al |

| | | | | | | |
|---|---|---|---|---|---|---|
| EPI_ISL_471439 | SouthKorea/KCDC2023/2020 | 2020-02-19 | Asia/South Korea/South Korea/ | Division of Viral Diseases, Center for Laboratory Control of Infectious Diseases, Korea Centers for Diseases Control and Prevention | Division of Viral Diseases, Center for Laboratory Control of Infectious Diseases, Korea Centers for Diseases Control and Prevention | Jeong-Min Kim et al |
| EPI_ISL_471440 | SouthKorea/KCDC2024/2020 | 2020-02-19 | Asia/South Korea/South Korea/ | Division of Viral Diseases, Center for Laboratory Control of Infectious Diseases, Korea Centers for Diseases Control and Prevention | Division of Viral Diseases, Center for Laboratory Control of Infectious Diseases, Korea Centers for Diseases Control and Prevention | Jeong-Min Kim et al |
| EPI_ISL_471441 | SouthKorea/KCDC2025/2020 | 2020-02-19 | Asia/South Korea/South Korea/ | Division of Viral Diseases, Center for Laboratory Control of Infectious Diseases, Korea Centers for Diseases Control and Prevention | Division of Viral Diseases, Center for Laboratory Control of Infectious Diseases, Korea Centers for Diseases Control and Prevention | Jeong-Min Kim et al |
| EPI_ISL_471442 | SouthKorea/KCDC2026/2020 | 2020-02-19 | Asia/South Korea/South Korea/ | Division of Viral Diseases, Center for Laboratory Control of Infectious Diseases, Korea Centers for Diseases Control and Prevention | Division of Viral Diseases, Center for Laboratory Control of Infectious Diseases, Korea Centers for Diseases Control and Prevention | Jeong-Min Kim et al |

| | | | | | | |
|---|---|---|---|---|---|---|
| EPI_ISL_471443 | SouthKorea/KCDC2027/2020 | 2020-02-19 | Asia/South Korea/South Korea/ | Division of Viral Diseases, Center for Laboratory Control of Infectious Diseases, Korea Centers for Diseases Control and Prevention | Division of Viral Diseases, Center for Laboratory Control of Infectious Diseases, Korea Centers for Diseases Control and Prevention | Jeong-Min Kim et al |
| EPI_ISL_471444 | SouthKorea/KCDC2028/2020 | 2020-02-20 | Asia/South Korea/South Korea/ | Division of Viral Diseases, Center for Laboratory Control of Infectious Diseases, Korea Centers for Diseases Control and Prevention | Division of Viral Diseases, Center for Laboratory Control of Infectious Diseases, Korea Centers for Diseases Control and Prevention | Jeong-Min Kim et al |
| EPI_ISL_471445 | SouthKorea/KCDC2029/2020 | 2020-02-20 | Asia/South Korea/South Korea/ | Division of Viral Diseases, Center for Laboratory Control of Infectious Diseases, Korea Centers for Diseases Control and Prevention | Division of Viral Diseases, Center for Laboratory Control of Infectious Diseases, Korea Centers for Diseases Control and Prevention | Jeong-Min Kim et al |
| EPI_ISL_471446 | SouthKorea/KCDC2040/2020 | 2020-02-19 | Asia/South Korea/South Korea/ | Division of Viral Diseases, Center for Laboratory Control of Infectious Diseases, Korea Centers for Diseases Control and Prevention | Division of Viral Diseases, Center for Laboratory Control of Infectious Diseases, Korea Centers for Diseases Control and Prevention | Jeong-Min Kim et al |

| | | | | | | |
|---|---|---|---|---|---|---|
| EPI_ISL_471447 | SouthKorea/KCDC2041/2020 | 2020-02-19 | Asia/South Korea/South Korea/ | Division of Viral Diseases, Center for Laboratory Control of Infectious Diseases, Korea Centers for Diseases Control and Prevention | Division of Viral Diseases, Center for Laboratory Control of Infectious Diseases, Korea Centers for Diseases Control and Prevention | Jeong-Min Kim et al |
| EPI_ISL_471448 | SouthKorea/KCDC2042/2020 | 2020-02-19 | Asia/South Korea/South Korea/ | Division of Viral Diseases, Center for Laboratory Control of Infectious Diseases, Korea Centers for Diseases Control and Prevention | Division of Viral Diseases, Center for Laboratory Control of Infectious Diseases, Korea Centers for Diseases Control and Prevention | Jeong-Min Kim et al |
| EPI_ISL_471449 | SouthKorea/KCDC2043/2020 | 2020-02-19 | Asia/South Korea/South Korea/ | Division of Viral Diseases, Center for Laboratory Control of Infectious Diseases, Korea Centers for Diseases Control and Prevention | Division of Viral Diseases, Center for Laboratory Control of Infectious Diseases, Korea Centers for Diseases Control and Prevention | Jeong-Min Kim et al |
| EPI_ISL_471450 | SouthKorea/KCDC2044/2020 | 2020-02-19 | Asia/South Korea/South Korea/ | Division of Viral Diseases, Center for Laboratory Control of Infectious Diseases, Korea Centers for Diseases Control and Prevention | Division of Viral Diseases, Center for Laboratory Control of Infectious Diseases, Korea Centers for Diseases Control and Prevention | Jeong-Min Kim et al |

| | | | | | | |
|---|---|---|---|---|---|---|
| EPI_ISL_471451 | SouthKorea/KCDC2045/2020 | 2020-02-20 | Asia/South Korea/South Korea/ | Division of Viral Diseases, Center for Laboratory Control of Infectious Diseases, Korea Centers for Diseases Control and Prevention | Division of Viral Diseases, Center for Laboratory Control of Infectious Diseases, Korea Centers for Diseases Control and Prevention | Jeong-Min Kim et al |
| EPI_ISL_471452 | SouthKorea/KCDC2046/2020 | 2020-02-21 | Asia/South Korea/South Korea/ | Division of Viral Diseases, Center for Laboratory Control of Infectious Diseases, Korea Centers for Diseases Control and Prevention | Division of Viral Diseases, Center for Laboratory Control of Infectious Diseases, Korea Centers for Diseases Control and Prevention | Jeong-Min Kim et al |
| EPI_ISL_471453 | SouthKorea/KCDC2047/2020 | 2020-02-20 | Asia/South Korea/South Korea/ | Division of Viral Diseases, Center for Laboratory Control of Infectious Diseases, Korea Centers for Diseases Control and Prevention | Division of Viral Diseases, Center for Laboratory Control of Infectious Diseases, Korea Centers for Diseases Control and Prevention | Jeong-Min Kim et al |
| EPI_ISL_471454 | SouthKorea/KCDC2048/2020 | 2020-02-21 | Asia/South Korea/South Korea/ | Division of Viral Diseases, Center for Laboratory Control of Infectious Diseases, Korea Centers for Diseases Control and Prevention | Division of Viral Diseases, Center for Laboratory Control of Infectious Diseases, Korea Centers for Diseases Control and Prevention | Jeong-Min Kim et al |

| | | | | | | |
|---|---|---|---|---|---|---|
| EPI_ISL_471455 | SouthKorea/KCDC2049/2020 | 2020-02-21 | Asia/South Korea/South Korea/ | Division of Viral Diseases, Center for Laboratory Control of Infectious Diseases, Korea Centers for Diseases Control and Prevention | Division of Viral Diseases, Center for Laboratory Control of Infectious Diseases, Korea Centers for Diseases Control and Prevention | Jeong-Min Kim et al |
| EPI_ISL_471458 | Morocco/HMIMV-Rabat1429-04/202[illegible] | 2020-04-19 | Africa/Morocco/Rabat/ | Centre de Virologie des Maladies Tropicales | Functional Genomic Platform/Service Analyses Biologique/UATRS/ Centre National Pour la Recherche Scientifique Et Technique (CNRST) | Hicham ANNAZ et al |
| EPI_ISL_471528 | CzechRepublic/NRL_1751/2020 | 2020-03-09 | Europe/Italy/Litomerice-Brnay/ | The National Institute of Public Health | State Veterinary Institute Prague and The National Institute of Public Health | Nagy et al |
| EPI_ISL_471530 | CzechRepublic/NRL_1885/2020 | 2020-03-10 | Europe/Italy/Melnik/ | The National Institute of Public Health | State Veterinary Institute Prague and The National Institute of Public Health | Nagy et al |
| EPI_ISL_471554 | Brazil/SP-551/2020 | 2020-03-29 | South America/Brazil/S√£o Paulo/S√£o [illegible] | Hospital Bosque da Sa√∫de | Instituto Adolfo Lutz, Interdiciplinary Procedures Center, Strategic Laboratory | Claudio Tavares Sacchi et al |
| EPI_ISL_471590 | India/TG-CCMB-L1451/2020 | 2020-05-25 | Asia/India/Telangana/Hyderabad | CSIR-Centre for Cellular and Molecular Biology | CSIR-Centre for Cellular and Molecular Biology | Lamuk Zaveri et al |
| EPI_ISL_471591 | India/TG-CCMB-L1452/2020 | 2020-05-25 | Asia/India/Telangana/Hyderabad | CSIR-Centre for Cellular and Molecular Biology | CSIR-Centre for Cellular and Molecular Biology | Namami Gaur et al |

| | | | | | | |
|---|---|---|---|---|---|---|
| EPI_ISL_471592 | India/TG-CCMB-L1453/2020 | 2020-05-25 | Asia/India/Telangana/Hyderabad | CSIR-Centre for Cellular and Molecular Biology | CSIR-Centre for Cellular and Molecular Biology | Namami Gaur et al |
| EPI_ISL_471594 | India/TG-CCMB-L1455/2020 | 2020-05-25 | Asia/India/Telangana/Hyderabad | CSIR-Centre for Cellular and Molecular Biology | CSIR-Centre for Cellular and Molecular Biology | Namami Gaur et al |
| EPI_ISL_471596 | India/TG-CCMB-L1459/2020 | 2020-05-25 | Asia/India/Telangana/Hyderabad | CSIR-Centre for Cellular and Molecular Biology | CSIR-Centre for Cellular and Molecular Biology | Payel Mukherjee et al |
| EPI_ISL_471600 | India/TG-CCMB-L1469/2020 | 2020-05-25 | Asia/India/Telangana/Hyderabad | CSIR-Centre for Cellular and Molecular Biology | CSIR-Centre for Cellular and Molecular Biology | Sakshi Shambhavi et al |
| EPI_ISL_471602 | India/TG-CCMB-L1471/2020 | 2020-05-25 | Asia/India/Telangana/Hyderabad | CSIR-Centre for Cellular and Molecular Biology | CSIR-Centre for Cellular and Molecular Biology | Sakshi Shambhavi et al |
| EPI_ISL_471604 | India/TG-CCMB-L1476/2020 | 2020-05-25 | Asia/India/Telangana/Hyderabad | CSIR-Centre for Cellular and Molecular Biology | CSIR-Centre for Cellular and Molecular Biology | Shagufta Khan et al |
| EPI_ISL_471606 | India/TG-CCMB-L1478/2020 | 2020-05-25 | Asia/India/Telangana/Hyderabad | CSIR-Centre for Cellular and Molecular Biology | CSIR-Centre for Cellular and Molecular Biology | Shagufta Khan et al |
| EPI_ISL_471607 | India/TG-CCMB-L1484/2020 | 2020-05-25 | Asia/India/Telangana/Hyderabad | CSIR-Centre for Cellular and Molecular Biology | CSIR-Centre for Cellular and Molecular Biology | Sofia Banu et al |
| EPI_ISL_471608 | India/TG-CCMB-L1487/2020 | 2020-05-25 | Asia/India/Telangana/Hyderabad | CSIR-Centre for Cellular and Molecular Biology | CSIR-Centre for Cellular and Molecular Biology | Sofia Banu et al |
| EPI_ISL_471611 | India/TG-CCMB-L1495/2020 | 2020-05-25 | Asia/India/Telangana/Hyderabad | CSIR-Centre for Cellular and Molecular Biology | CSIR-Centre for Cellular and Molecular Biology | Tulasi Nagabandi et al |
| EPI_ISL_471614 | India/TG-CCMB-L1499/2020 | 2020-05-25 | Asia/India/Telangana/Hyderabad | CSIR-Centre for Cellular and Molecular Biology | CSIR-Centre for Cellular and Molecular Biology | Tulasi Nagabandi et al |
| EPI_ISL_471622 | India/TG-CCMB-L1684/2020 | 2020-05-29 | Asia/India/Telangana/Hyderabad | CSIR-Centre for Cellular and Molecular Biology | CSIR-Centre for Cellular and Molecular Biology | Namami Gaur et al |
| EPI_ISL_471625 | India/TG-CCMB-L1721/2020 | 2020-05-30 | Asia/India/Telangana/Hyderabad | CSIR-Centre for Cellular and Molecular Biology | CSIR-Centre for Cellular and Molecular Biology | Payel Mukherjee et al |

| | | | | | | |
|---|---|---|---|---|---|---|
| EPI_ISL_471628 | India/TG-CCMB-L1728/2020 | 2020-05-30 | Asia/India/Telangana/Hyderabad | CSIR-Centre for Cellular and Molecular Biology | CSIR-Centre for Cellular and Molecular Biology | Sakshi Shambhavi et al |
| EPI_ISL_471631 | India/TG-CCMB-L1746/2020 | 2020-06-01 | Asia/India/Telangana/Hyderabad | CSIR-Centre for Cellular and Molecular Biology | CSIR-Centre for Cellular and Molecular Biology | Shagufta Khan et al |
| EPI_ISL_471632 | India/TG-CCMB-L1762/2020 | 2020-06-01 | Asia/India/Telangana/Hyderabad | CSIR-Centre for Cellular and Molecular Biology | CSIR-Centre for Cellular and Molecular Biology | Shagufta Khan et al |
| EPI_ISL_471633 | India/TG-CCMB-L1781/2020 | 2020-06-02 | Asia/India/Telangana/Hyderabad | CSIR-Centre for Cellular and Molecular Biology | CSIR-Centre for Cellular and Molecular Biology | Shagufta Khan et al |
| EPI_ISL_471634 | India/TG-CCMB-L1806/2020 | 2020-06-02 | Asia/India/Telangana/Hyderabad | CSIR-Centre for Cellular and Molecular Biology | CSIR-Centre for Cellular and Molecular Biology | Shagufta Khan et al |
| EPI_ISL_471635 | India/TG-CCMB-L1809/2020 | 2020-06-02 | Asia/India/Telangana/Hyderabad | CSIR-Centre for Cellular and Molecular Biology | CSIR-Centre for Cellular and Molecular Biology | Sofia Banu et al |
| EPI_ISL_471636 | India/TG-CCMB-L1810/2020 | 2020-06-02 | Asia/India/Telangana/Hyderabad | CSIR-Centre for Cellular and Molecular Biology | CSIR-Centre for Cellular and Molecular Biology | Sofia Banu et al |
| EPI_ISL_471637 | India/TG-CCMB-L1811/2020 | 2020-06-02 | Asia/India/Telangana/Hyderabad | CSIR-Centre for Cellular and Molecular Biology | CSIR-Centre for Cellular and Molecular Biology | Sofia Banu et al |
| EPI_ISL_471687 | USA/MI-MDHHS-SC20663/2020 | 2020-04-29 | North America/USA/Michigan/ | Michigan Department of Health and Human Services, Bureau of Laboratories | Michigan Department of Health and Human Services, Bureau of Laboratories | Blankenship HM et al |
| EPI_ISL_471695 | USA/MI-MDHHS-SC20671/2020 | 2020-05-14 | North America/USA/Michigan/ | Michigan Department of Health and Human Services, Bureau of Laboratories | Michigan Department of Health and Human Services, Bureau of Laboratories | Blankenship HM et al |

| | | | | | | |
|---|---|---|---|---|---|---|
| EPI_ISL_471705 | USA/MI-MDHHS-SC20681/2020 | 2020-05-14 | North America/USA/Michigan/ | Michigan Department of Health and Human Services, Bureau of Laboratories | Michigan Department of Health and Human Services, Bureau of Laboratories | Blankenship HM et al |
| EPI_ISL_471710 | USA/MI-MDHHS-SC20686/2020 | 2020-05-15 | North America/USA/Michigan/ | Michigan Department of Health and Human Services, Bureau of Laboratories | Michigan Department of Health and Human Services, Bureau of Laboratories | Blankenship HM et al |
| EPI_ISL_471717 | USA/MI-MDHHS-SC20693/2020 | 2020-05-20 | North America/USA/Michigan/ | Michigan Department of Health and Human Services, Bureau of Laboratories | Michigan Department of Health and Human Services, Bureau of Laboratories | Blankenship HM et al |
| EPI_ISL_471720 | USA/MI-MDHHS-SC20696/2020 | 2020-04-27 | North America/USA/Michigan/ | Michigan Department of Health and Human Services, Bureau of Laboratories | Michigan Department of Health and Human Services, Bureau of Laboratories | Blankenship HM et al |
| EPI_ISL_471734 | USA/MI-MDHHS-SC20710/2020 | 2020-04-27 | North America/USA/Michigan/ | Michigan Department of Health and Human Services, Bureau of Laboratories | Michigan Department of Health and Human Services, Bureau of Laboratories | Blankenship HM et al |
| EPI_ISL_471740 | USA/MI-MDHHS-SC20716/2020 | 2020-04-14 | North America/USA/Michigan/ | Michigan Department of Health and Human Services, Bureau of Laboratories | Michigan Department of Health and Human Services, Bureau of Laboratories | Blankenship HM et al |

| | | | | | | |
|---|---|---|---|---|---|---|
| EPI_ISL_471746 | USA/MI-MDHHS-SC20722/2020 | 2020-04-20 | North America/USA/Michigan/ | Michigan Department of Health and Human Services, Bureau of Laboratories | Michigan Department of Health and Human Services, Bureau of Laboratories | Blankenship HM et al |
| EPI_ISL_471749 | USA/MI-MDHHS-SC20725/2020 | 2020-04-17 | North America/USA/Michigan/ | Michigan Department of Health and Human Services, Bureau of Laboratories | Michigan Department of Health and Human Services, Bureau of Laboratories | Blankenship HM et al |
| EPI_ISL_471755 | USA/MI-MDHHS-SC20734/2020 | 2020-04-29 | North America/USA/Michigan/ | Michigan Department of Health and Human Services, Bureau of Laboratories | Michigan Department of Health and Human Services, Bureau of Laboratories | Blankenship HM et al |
| EPI_ISL_471758 | USA/MI-MDHHS-SC20737/2020 | 2020-04-29 | North America/USA/Michigan/ | Michigan Department of Health and Human Services, Bureau of Laboratories | Michigan Department of Health and Human Services, Bureau of Laboratories | Blankenship HM et al |
| EPI_ISL_471772 | USA/MI-MDHHS-SC20751/2020 | 2020-04-29 | North America/USA/Michigan/ | Michigan Department of Health and Human Services, Bureau of Laboratories | Michigan Department of Health and Human Services, Bureau of Laboratories | Blankenship HM et al |
| EPI_ISL_471773 | USA/MI-MDHHS-SC20752/2020 | 2020-04-30 | North America/USA/Michigan/ | Michigan Department of Health and Human Services, Bureau of Laboratories | Michigan Department of Health and Human Services, Bureau of Laboratories | Blankenship HM et al |

| | | | | | | |
|---|---|---|---|---|---|---|
| EPI_ISL_471783 | USA/MI-MDHHS-SC20762/2020 | 2020-04-22 | North America/USA/Michigan/ | Michigan Department of Health and Human Services, Bureau of Laboratories | Michigan Department of Health and Human Services, Bureau of Laboratories | Blankenship HM et al |
| EPI_ISL_471787 | USA/MI-MDHHS-SC20766/2020 | 2020-04-22 | North America/USA/Michigan/ | Michigan Department of Health and Human Services, Bureau of Laboratories | Michigan Department of Health and Human Services, Bureau of Laboratories | Blankenship HM et al |
| EPI_ISL_471795 | USA/MI-MDHHS-SC20774/2020 | 2020-04-30 | North America/USA/Michigan/ | Michigan Department of Health and Human Services, Bureau of Laboratories | Michigan Department of Health and Human Services, Bureau of Laboratories | Blankenship HM et al |
| EPI_ISL_471796 | USA/MI-MDHHS-SC20775/2020 | 2020-04-16 | North America/USA/Michigan/ | Michigan Department of Health and Human Services, Bureau of Laboratories | Michigan Department of Health and Human Services, Bureau of Laboratories | Blankenship HM et al |
| EPI_ISL_471814 | USA/MI-MDHHS-SC20793/2020 | 2020-04-29 | North America/USA/Michigan/ | Michigan Department of Health and Human Services, Bureau of Laboratories | Michigan Department of Health and Human Services, Bureau of Laboratories | Blankenship HM et al |
| EPI_ISL_471821 | USA/MI-MDHHS-SC20800/2020 | 2020-05-19 | North America/USA/Michigan/ | Michigan Department of Health and Human Services, Bureau of Laboratories | Michigan Department of Health and Human Services, Bureau of Laboratories | Blankenship HM et al |

| | | | | | | |
|---|---|---|---|---|---|---|
| EPI_ISL_471825 | USA/MI-MDHHS-SC20804/2020 | 2020-05-18 | North America/USA/Michigan/ | Michigan Department of Health and Human Services, Bureau of Laboratories | Michigan Department of Health and Human Services, Bureau of Laboratories | Blankenship HM et al |
| EPI_ISL_471830 | USA/MI-MDHHS-SC20809/2020 | 2020-05-20 | North America/USA/Michigan/ | Michigan Department of Health and Human Services, Bureau of Laboratories | Michigan Department of Health and Human Services, Bureau of Laboratories | Blankenship HM et al |
| EPI_ISL_471831 | USA/MI-MDHHS-SC20810/2020 | 2020-05-20 | North America/USA/Michigan/ | Michigan Department of Health and Human Services, Bureau of Laboratories | Michigan Department of Health and Human Services, Bureau of Laboratories | Blankenship HM et al |
| EPI_ISL_471839 | USA/MI-MDHHS-SC20818/2020 | 2020-05-13 | North America/USA/Michigan/ | Michigan Department of Health and Human Services, Bureau of Laboratories | Michigan Department of Health and Human Services, Bureau of Laboratories | Blankenship HM et al |
| EPI_ISL_471842 | USA/MI-MDHHS-SC20821/2020 | 2020-05-13 | North America/USA/Michigan/ | Michigan Department of Health and Human Services, Bureau of Laboratories | Michigan Department of Health and Human Services, Bureau of Laboratories | Blankenship HM et al |
| EPI_ISL_471843 | USA/MI-MDHHS-SC20822/2020 | 2020-05-12 | North America/USA/Michigan/ | Michigan Department of Health and Human Services, Bureau of Laboratories | Michigan Department of Health and Human Services, Bureau of Laboratories | Blankenship HM et al |

| | | | | | | |
|---|---|---|---|---|---|---|
| EPI_ISL_471854 | USA/MI-MDHHS-SC20833/2020 | 2020-04-17 | North America/USA/Michigan/ | Michigan Department of Health and Human Services, Bureau of Laboratories | Michigan Department of Health and Human Services, Bureau of Laboratories | Blankenship HM et al |
| EPI_ISL_471856 | USA/MI-MDHHS-SC20835/2020 | 2020-04-17 | North America/USA/Michigan/ | Michigan Department of Health and Human Services, Bureau of Laboratories | Michigan Department of Health and Human Services, Bureau of Laboratories | Blankenship HM et al |
| EPI_ISL_471861 | USA/MI-MDHHS-SC20840/2020 | 2020-04-29 | North America/USA/Michigan/ | Michigan Department of Health and Human Services, Bureau of Laboratories | Michigan Department of Health and Human Services, Bureau of Laboratories | Blankenship HM et al |
| EPI_ISL_471862 | USA/MI-MDHHS-SC20841/2020 | 2020-04-17 | North America/USA/Michigan/ | Michigan Department of Health and Human Services, Bureau of Laboratories | Michigan Department of Health and Human Services, Bureau of Laboratories | Blankenship HM et al |
| EPI_ISL_471866 | USA/MI-MDHHS-SC20845/2020 | 2020-04-29 | North America/USA/Michigan/ | Michigan Department of Health and Human Services, Bureau of Laboratories | Michigan Department of Health and Human Services, Bureau of Laboratories | Blankenship HM et al |
| EPI_ISL_471876 | USA/MI-MDHHS-SC20855/2020 | 2020-05-08 | North America/USA/Michigan/ | Michigan Department of Health and Human Services, Bureau of Laboratories | Michigan Department of Health and Human Services, Bureau of Laboratories | Blankenship HM et al |

| | | | | | | |
|---|---|---|---|---|---|---|
| EPI_ISL_471881 | USA/MI-MDHHS-SC20860/2020 | 2020-04-29 | North America/USA/Michigan/ | Michigan Department of Health and Human Services, Bureau of Laboratories | Michigan Department of Health and Human Services, Bureau of Laboratories | Blankenship HM et al |
| EPI_ISL_471898 | USA/MI-MDHHS-SC20877/2020 | 2020-05-19 | North America/USA/Michigan/ | Michigan Department of Health and Human Services, Bureau of Laboratories | Michigan Department of Health and Human Services, Bureau of Laboratories | Blankenship HM et al |
| EPI_ISL_471904 | USA/MI-MDHHS-SC20883/2020 | 2020-05-17 | North America/USA/Michigan/ | Michigan Department of Health and Human Services, Bureau of Laboratories | Michigan Department of Health and Human Services, Bureau of Laboratories | Blankenship HM et al |
| EPI_ISL_471949 | England/EXET-1370F0/2020 | 2020-04-21 | Europe/United Kingdom/England/ | University of Exeter | COVID-19 Genomics UK (COG-UK) Consortium | Ben Temperton et al |
| EPI_ISL_471998 | England/LIVE-9F69B/2020 | 2020-04-22 | Europe/United Kingdom/England/ | Liverpool Clinical Laboratories | COVID-19 Genomics UK (COG-UK) Consortium | Sam Haldenby et al |
| EPI_ISL_472050 | England/LIVE-9FC3F/2020 | 2020-06-01 | Europe/United Kingdom/England/ | Liverpool Clinical Laboratories | COVID-19 Genomics UK (COG-UK) Consortium | Sam Haldenby et al |
| EPI_ISL_472231 | England/NORT-282D3A/2020 | 2020-03-29 | Europe/United Kingdom/England/ | Northumbria University / South Tees Hospitals NHS Foundation Trust / North Cumbria Integrated Care NHS Foundation Trust / North Tees and Hartlepool NHS Foundation Trust / Newcastle Hospitals NHS Foundation Trust | COVID-19 Genomics UK (COG-UK) Consortium | Darren L Smith et al |
| EPI_ISL_472268 | England/NORT-284169/2020 | 2020-05-06 | Europe/United Kingdom/England/ | Northumbria University / South Tees Hospitals NHS Foundation Trust / North Cumbria Integrated Care NHS Foundation Trust / North Tees and Hartlepool NHS Foundation Trust / Newcastle Hospitals NHS Foundation Trust | COVID-19 Genomics UK (COG-UK) Consortium | Darren L Smith et al |

| | | | | | | |
|---|---|---|---|---|---|---|
| EPI_ISL_472284 | England/NORT-292E70/2020 | 2020-05-15 | Europe/United Kingdom/England/ | Northumbria University / South Tees Hospitals NHS Foundation Trust / North Cumbria Integrated Care NHS Foundation Trust / North Tees and Hartlepool NHS Foundation Trust / Newcastle Hospitals NHS Foundation Trust | COVID-19 Genomics UK (COG-UK) Consortium | Darren L Smith et al |
| EPI_ISL_472357 | England/NORW-ED26D/2020 | 2020-05-25 | Europe/United Kingdom/England/ | Quadram Institute Bioscience | COVID-19 Genomics UK (COG-UK) Consortium | Dave J. Baker et al |
| EPI_ISL_472364 | England/NORW-ED40D/2020 | 2020-05-26 | Europe/United Kingdom/England/ | Quadram Institute Bioscience | COVID-19 Genomics UK (COG-UK) Consortium | Dave J. Baker et al |
| EPI_ISL_472621 | Wales/PHWC-15CDD0/2020 | 2020-05-08 | Europe/United Kingdom/Wales/ | Wales Specialist Virology Centre Sequencing lab: Pathogen Genomics Unit | COVID-19 Genomics UK (COG-UK) Consortium | Catherine Moore et al |
| EPI_ISL_472640 | Wales/PHWC-15CF16/2020 | 2020-05-05 | Europe/United Kingdom/Wales/ | Wales Specialist Virology Centre Sequencing lab: Pathogen Genomics Unit | COVID-19 Genomics UK (COG-UK) Consortium | Catherine Moore et al |
| EPI_ISL_472669 | Wales/PHWC-15D11F/2020 | 2020-05-04 | Europe/United Kingdom/Wales/ | Wales Specialist Virology Centre Sequencing lab: Pathogen Genomics Unit | COVID-19 Genomics UK (COG-UK) Consortium | Catherine Moore et al |
| EPI_ISL_472686 | Wales/PHWC-15D22B/2020 | 2020-04-22 | Europe/United Kingdom/Wales/ | Wales Specialist Virology Centre Sequencing lab: Pathogen Genomics Unit | COVID-19 Genomics UK (COG-UK) Consortium | Catherine Moore et al |
| EPI_ISL_472716 | Wales/PHWC-15D416/2020 | 2020-05-04 | Europe/United Kingdom/Wales/ | Wales Specialist Virology Centre Sequencing lab: Pathogen Genomics Unit | COVID-19 Genomics UK (COG-UK) Consortium | Catherine Moore et al |
| EPI_ISL_472719 | Wales/PHWC-15D443/2020 | 2020-05-02 | Europe/United Kingdom/Wales/ | Wales Specialist Virology Centre Sequencing lab: Pathogen Genomics Unit | COVID-19 Genomics UK (COG-UK) Consortium | Catherine Moore et al |
| EPI_ISL_472771 | Wales/PHWC-15DCF0/2020 | 2020-05-14 | Europe/United Kingdom/Wales/ | Wales Specialist Virology Centre Sequencing lab: Pathogen Genomics Unit | COVID-19 Genomics UK (COG-UK) Consortium | Catherine Moore et al |
| EPI_ISL_472805 | Wales/PHWC-15E327/2020 | 2020-05-15 | Europe/United Kingdom/Wales/ | Wales Specialist Virology Centre Sequencing lab: Pathogen Genomics Unit | COVID-19 Genomics UK (COG-UK) Consortium | Catherine Moore et al |
| EPI_ISL_472823 | Wales/PHWC-15E61F/2020 | 2020-05-15 | Europe/United Kingdom/Wales/ | Wales Specialist Virology Centre Sequencing lab: Pathogen Genomics Unit | COVID-19 Genomics UK (COG-UK) Consortium | Catherine Moore et al |

| | | | | | | |
|---|---|---|---|---|---|---|
| EPI_ISL_472912 | Wales/PHWC-160252/2020 | 2020-05-19 | Europe/United Kingdom/Wales/ | Wales Specialist Virology Centre Sequencing lab: Pathogen Genomics Unit | COVID-19 Genomics UK (COG-UK) Consortium | Catherine Moore et al |
| EPI_ISL_472948 | Wales/PHWC-16054A/2020 | 2020-04-28 | Europe/United Kingdom/Wales/ | Wales Specialist Virology Centre Sequencing lab: Pathogen Genomics Unit | COVID-19 Genomics UK (COG-UK) Consortium | Catherine Moore et al |
| EPI_ISL_473012 | Wales/PHWC-160AE1/2020 | 2020-05-20 | Europe/United Kingdom/Wales/ | Wales Specialist Virology Centre Sequencing lab: Pathogen Genomics Unit | COVID-19 Genomics UK (COG-UK) Consortium | Catherine Moore et al |
| EPI_ISL_473051 | Wales/PHWC-160DBB/2020 | 2020-05-23 | Europe/United Kingdom/Wales/ | Wales Specialist Virology Centre Sequencing lab: Pathogen Genomics Unit | COVID-19 Genomics UK (COG-UK) Consortium | Catherine Moore et al |
| EPI_ISL_473055 | Wales/PHWC-160DF7/2020 | 2020-05-26 | Europe/United Kingdom/Wales/ | Wales Specialist Virology Centre Sequencing lab: Pathogen Genomics Unit | COVID-19 Genomics UK (COG-UK) Consortium | Catherine Moore et al |
| EPI_ISL_473061 | Wales/PHWC-160E8B/2020 | 2020-05-18 | Europe/United Kingdom/Wales/ | Wales Specialist Virology Centre Sequencing lab: Pathogen Genomics Unit | COVID-19 Genomics UK (COG-UK) Consortium | Catherine Moore et al |
| EPI_ISL_473110 | Wales/PHWC-162706/2020 | 2020-05-14 | Europe/United Kingdom/Wales/ | Wales Specialist Virology Centre Sequencing lab: Pathogen Genomics Unit | COVID-19 Genomics UK (COG-UK) Consortium | Catherine Moore et al |
| EPI_ISL_473113 | Wales/PHWC-162733/2020 | 2020-05-28 | Europe/United Kingdom/Wales/ | Wales Specialist Virology Centre Sequencing lab: Pathogen Genomics Unit | COVID-19 Genomics UK (COG-UK) Consortium | Catherine Moore et al |
| EPI_ISL_473125 | Wales/PHWC-162803/2020 | 2020-05-18 | Europe/United Kingdom/Wales/ | Wales Specialist Virology Centre Sequencing lab: Pathogen Genomics Unit | COVID-19 Genomics UK (COG-UK) Consortium | Catherine Moore et al |
| EPI_ISL_473256 | Wales/PHWC-16318F/2020 | 2020-05-20 | Europe/United Kingdom/Wales/ | Wales Specialist Virology Centre Sequencing lab: Pathogen Genomics Unit | COVID-19 Genomics UK (COG-UK) Consortium | Catherine Moore et al |
| EPI_ISL_473260 | Wales/PHWC-1631DA/2020 | 2020-05-21 | Europe/United Kingdom/Wales/ | Wales Specialist Virology Centre Sequencing lab: Pathogen Genomics Unit | COVID-19 Genomics UK (COG-UK) Consortium | Catherine Moore et al |
| EPI_ISL_473276 | Wales/PHWC-1632F5/2020 | 2020-05-14 | Europe/United Kingdom/Wales/ | Wales Specialist Virology Centre Sequencing lab: Pathogen Genomics Unit | COVID-19 Genomics UK (COG-UK) Consortium | Catherine Moore et al |
| EPI_ISL_473277 | Wales/PHWC-163301/2020 | 2020-05-21 | Europe/United Kingdom/Wales/ | Wales Specialist Virology Centre Sequencing lab: Pathogen Genomics Unit | COVID-19 Genomics UK (COG-UK) Consortium | Catherine Moore et al |

| | | | | | | |
|---|---|---|---|---|---|---|
| EPI_ISL_473347 | England/BIRM-5F8CE/2020 | 2020-05-18 | Europe/United Kingdom/England/ | University of Birmingham | COVID-19 Genomics UK (COG-UK) Consortium | Institute of Microbiology et al |
| EPI_ISL_473424 | England/BIRM-60569/2020 | 2020-05-27 | Europe/United Kingdom/England/ | University of Birmingham | COVID-19 Genomics UK (COG-UK) Consortium | Institute of Microbiology et al |
| EPI_ISL_473431 | England/BIRM-61058/2020 | 2020-05-27 | Europe/United Kingdom/England/ | University of Birmingham | COVID-19 Genomics UK (COG-UK) Consortium | Institute of Microbiology et al |
| EPI_ISL_473479 | England/CAMB-1B4671/2020 | 2020-05-27 | Europe/United Kingdom/England/ | Department of Pathology, University of Cambridge | COVID-19 Genomics UK (COG-UK) Consortium | Luke W Meredith et al |
| EPI_ISL_473566 | Scotland/CVR2787/2020 | 2020-04-20 | Europe/United Kingdom/Scotland/ | West of Scotland Specialist Virology Centre, NHSGGC / MRC-University of Glasgow Centre for Virus Research | COVID-19 Genomics UK (COG-UK) Consortium | Ana da Silva Filipe et al |
| EPI_ISL_473653 | Scotland/CVR3513/2020 | 2020-05-12 | Europe/United Kingdom/Scotland/ | West of Scotland Specialist Virology Centre, NHSGGC / MRC-University of Glasgow Centre for Virus Research | COVID-19 Genomics UK (COG-UK) Consortium | Ana da Silva Filipe et al |
| EPI_ISL_473723 | Scotland/CVR3594/2020 | 2020-05-19 | Europe/United Kingdom/Scotland/ | West of Scotland Specialist Virology Centre, NHSGGC / MRC-University of Glasgow Centre for Virus Research | COVID-19 Genomics UK (COG-UK) Consortium | Ana da Silva Filipe et al |
| EPI_ISL_473731 | Scotland/CVR3611/2020 | 2020-05-20 | Europe/United Kingdom/Scotland/ | West of Scotland Specialist Virology Centre, NHSGGC / MRC-University of Glasgow Centre for Virus Research | COVID-19 Genomics UK (COG-UK) Consortium | Ana da Silva Filipe et al |
| EPI_ISL_473734 | Scotland/CVR3614/2020 | 2020-05-20 | Europe/United Kingdom/Scotland/ | West of Scotland Specialist Virology Centre, NHSGGC / MRC-University of Glasgow Centre for Virus Research | COVID-19 Genomics UK (COG-UK) Consortium | Ana da Silva Filipe et al |
| EPI_ISL_473815 | Scotland/EDB5196/2020 | 2020-05-18 | Europe/United Kingdom/Scotland/ | Virology Department, Royal Infirmary of Edinburgh, NHS Lothian / School of Biological Sciences, University of Edinburgh / Institute of Genetics and Molecular Medicine, University of Edinburgh | COVID-19 Genomics UK (COG-UK) Consortium | McHugh M et al |
| EPI_ISL_473909 | Scotland/EDB6193/2020 | 2020-05-18 | Europe/United Kingdom/Scotland/ | Virology Department, Royal Infirmary of Edinburgh, NHS Lothian / School of Biological Sciences, University of Edinburgh / Institute of Genetics and Molecular Medicine, University of Edinburgh | COVID-19 Genomics UK (COG-UK) Consortium | McHugh M et al |

| | | | | | | |
|---|---|---|---|---|---|---|
| EPI_ISL_473930 | Scotland/EDB6248/2020 | 2020-05-21 | Europe/United Kingdom/Scotland/ | Virology Department, Royal Infirmary of Edinburgh, NHS Lothian / School of Biological Sciences, University of Edinburgh / Institute of Genetics and Molecular Medicine, University of Edinburgh | COVID-19 Genomics UK (COG-UK) Consortium | McHugh M et al |
| EPI_ISL_473972 | Wales/PHWC-163617/2020 | 2020-05-06 | Europe/United Kingdom/Wales/ | Wales Specialist Virology Centre Sequencing lab: Pathogen Genomics Unit | COVID-19 Genomics UK (COG-UK) Consortium | Catherine Moore et al |
| EPI_ISL_474019 | Wales/PHWC-16395A/2020 | 2020-05-05 | Europe/United Kingdom/Wales/ | Wales Specialist Virology Centre Sequencing lab: Pathogen Genomics Unit | COVID-19 Genomics UK (COG-UK) Consortium | Catherine Moore et al |
| EPI_ISL_474069 | Wales/PHWC-163CE8/2020 | 2020-04-24 | Europe/United Kingdom/Wales/ | Wales Specialist Virology Centre Sequencing lab: Pathogen Genomics Unit | COVID-19 Genomics UK (COG-UK) Consortium | Catherine Moore et al |
| EPI_ISL_474085 | Wales/PHWC-163DE5/2020 | 2020-05-08 | Europe/United Kingdom/Wales/ | Wales Specialist Virology Centre Sequencing lab: Pathogen Genomics Unit | COVID-19 Genomics UK (COG-UK) Consortium | Catherine Moore et al |
| EPI_ISL_474113 | Wales/PHWC-163FC1/2020 | 2020-05-09 | Europe/United Kingdom/Wales/ | Wales Specialist Virology Centre Sequencing lab: Pathogen Genomics Unit | COVID-19 Genomics UK (COG-UK) Consortium | Catherine Moore et al |
| EPI_ISL_474114 | Wales/PHWC-163FFE/2020 | 2020-05-08 | Europe/United Kingdom/Wales/ | Wales Specialist Virology Centre Sequencing lab: Pathogen Genomics Unit | COVID-19 Genomics UK (COG-UK) Consortium | Catherine Moore et al |
| EPI_ISL_474165 | Wales/PHWC-164379/2020 | 2020-05-09 | Europe/United Kingdom/Wales/ | Wales Specialist Virology Centre Sequencing lab: Pathogen Genomics Unit | COVID-19 Genomics UK (COG-UK) Consortium | Catherine Moore et al |
| EPI_ISL_474183 | Wales/PHWC-1644A3/2020 | 2020-05-09 | Europe/United Kingdom/Wales/ | Wales Specialist Virology Centre Sequencing lab: Pathogen Genomics Unit | COVID-19 Genomics UK (COG-UK) Consortium | Catherine Moore et al |
| EPI_ISL_474281 | Wales/PHWC-2C156/2020 | 2020-04-04 | Europe/United Kingdom/Wales/ | Wales Specialist Virology Centre Sequencing lab: Pathogen Genomics Unit | COVID-19 Genomics UK (COG-UK) Consortium | Catherine Moore et al |
| EPI_ISL_474505 | Wales/PHWC-3599E/2020 | 2020-04-24 | Europe/United Kingdom/Wales/ | Wales Specialist Virology Centre Sequencing lab: Pathogen Genomics Unit | COVID-19 Genomics UK (COG-UK) Consortium | Catherine Moore et al |
| EPI_ISL_474541 | Wales/PHWC-35BE3/2020 | 2020-04-27 | Europe/United Kingdom/Wales/ | Wales Specialist Virology Centre Sequencing lab: Pathogen Genomics Unit | COVID-19 Genomics UK (COG-UK) Consortium | Catherine Moore et al |
| EPI_ISL_474562 | Wales/PHWC-35D47/2020 | 2020-04-27 | Europe/United Kingdom/Wales/ | Wales Specialist Virology Centre Sequencing lab: Pathogen Genomics Unit | COVID-19 Genomics UK (COG-UK) Consortium | Catherine Moore et al |

| | | | | | | |
|---|---|---|---|---|---|---|
| EPI_ISL_474690 | Wales/PHWC-3660F/2020 | 2020-05-01 | Europe/United Kingdom/Wales/ | Wales Specialist Virology Centre Sequencing lab: Pathogen Genomics Unit | COVID-19 Genomics UK (COG-UK) Consortium | Catherine Moore et al |
| EPI_ISL_474721 | Wales/PHWC-36872/2020 | 2020-04-27 | Europe/United Kingdom/Wales/ | Wales Specialist Virology Centre Sequencing lab: Pathogen Genomics Unit | COVID-19 Genomics UK (COG-UK) Consortium | Catherine Moore et al |
| EPI_ISL_474756 | Wales/PHWC-36AB8/2020 | 2020-05-01 | Europe/United Kingdom/Wales/ | Wales Specialist Virology Centre Sequencing lab: Pathogen Genomics Unit | COVID-19 Genomics UK (COG-UK) Consortium | Catherine Moore et al |
| EPI_ISL_474776 | Wales/PHWC-36BF1/2020 | 2020-05-01 | Europe/United Kingdom/Wales/ | Wales Specialist Virology Centre Sequencing lab: Pathogen Genomics Unit | COVID-19 Genomics UK (COG-UK) Consortium | Catherine Moore et al |
| EPI_ISL_474802 | Spain/CM-IBV-003757/2020 | 2020-04-03 | Europe/Spain/Castilla la Mancha/Albace | Complejo Hospitalario Universitario de Albacete | SeqCOVID-SPAIN consortium/IBV(CSIC) | Encarnacion Simarro Cv≥rdoba et al |
| EPI_ISL_474821 | Spain/CM-IBV-003838/2020 | 2020-05-25 | Europe/Spain/Castilla la Mancha/Albace | Complejo Hospitalario Universitario de Albacete | SeqCOVID-SPAIN consortium/IBV(CSIC) | Encarnacion Simarro Cv≥rdoba et al |
| EPI_ISL_474827 | Spain/CM-IBV-003884/2020 | 2020-05-18 | Europe/Spain/Castilla la Mancha/Albace | Complejo Hospitalario Universitario de Albacete | SeqCOVID-SPAIN consortium/IBV(CSIC) | Encarnacion Simarro Cv≥rdoba et al |
| EPI_ISL_474832 | Spain/AN-IBV-003009/2020 | 2020-04-01 | Europe/Spain/Andalusia/Albolote | Hospital Universitario Virgen de las Nieves de Granada-SAS | SeqCOVID-SPAIN consortium/IBV(CSIC) | Mercedes Pv©rez Ruiz et al |
| EPI_ISL_474840 | Spain/CM-IBV-003806/2020 | 2020-05-14 | Europe/Spain/Castilla la Mancha/Alman | Complejo Hospitalario Universitario de Albacete | SeqCOVID-SPAIN consortium/IBV(CSIC) | Encarnacion Simarro Cv≥rdoba et al |
| EPI_ISL_474841 | Spain/AN-IBV-002842/2020 | 2020-03-09 | Europe/Spain/Andalusia/Almeria | Hospital Universitario Virgen de las Nieves de Granada-SAS | SeqCOVID-SPAIN consortium/IBV(CSIC) | Mercedes Pv©rez Ruiz et al |
| EPI_ISL_474853 | Spain/CM-IBV-003771/2020 | 2020-04-02 | Europe/Spain/Castilla la Mancha/Cuenca | Complejo Hospitalario Universitario de Albacete | SeqCOVID-SPAIN consortium/IBV(CSIC) | Encarnacion Simarro Cv≥rdoba et al |
| EPI_ISL_474854 | Spain/GA-IBV-002922/2020 | 2020-03-21 | Europe/Spain/Galicia/El Rosal | Hospital Universitario Virgen de las Nieves de Granada-SAS | SeqCOVID-SPAIN consortium/IBV(CSIC) | Mercedes Pv©rez Ruiz et al |
| EPI_ISL_474858 | Spain/AN-IBV-002859/2020 | 2020-03-13 | Europe/Spain/Andalusia/Granada ES | Hospital Universitario Virgen de las Nieves de Granada-SAS | SeqCOVID-SPAIN consortium/IBV(CSIC) | Mercedes Pv©rez Ruiz et al |

| | | | | | | |
|---|---|---|---|---|---|---|
| EPI_ISL_474859 | Spain/AN-IBV-002867/2020 | 2020-03-14 | Europe/Spain/Andalusia/Granada ES | Hospital Universitario Virgen de las Nieves de Granada-SAS | SeqCOVID-SPAIN consortium/IBV(CSIC) | Mercedes P√©rez Ruiz et al |
| EPI_ISL_474863 | Spain/AN-IBV-002877/2020 | 2020-03-16 | Europe/Spain/Andalusia/Granada ES | Hospital Universitario Virgen de las Nieves de Granada-SAS | SeqCOVID-SPAIN consortium/IBV(CSIC) | Mercedes P√©rez Ruiz et al |
| EPI_ISL_474865 | Spain/AN-IBV-002882/2020 | 2020-03-16 | Europe/Spain/Andalusia/Granada ES | Hospital Universitario Virgen de las Nieves de Granada-SAS | SeqCOVID-SPAIN consortium/IBV(CSIC) | Mercedes P√©rez Ruiz et al |
| EPI_ISL_474866 | Spain/AN-IBV-002888/2020 | 2020-03-17 | Europe/Spain/Andalusia/Granada ES | Hospital Universitario Virgen de las Nieves de Granada-SAS | SeqCOVID-SPAIN consortium/IBV(CSIC) | Mercedes P√©rez Ruiz et al |
| EPI_ISL_474870 | Spain/AN-IBV-002915/2020 | 2020-03-19 | Europe/Spain/Andalusia/Granada ES | Hospital Universitario Virgen de las Nieves de Granada-SAS | SeqCOVID-SPAIN consortium/IBV(CSIC) | Mercedes P√©rez Ruiz et al |
| EPI_ISL_474887 | Spain/AN-IBV-002989/2020 | 2020-04-01 | Europe/Spain/Andalusia/Granada ES | Hospital Universitario Virgen de las Nieves de Granada-SAS | SeqCOVID-SPAIN consortium/IBV(CSIC) | Mercedes P√©rez Ruiz et al |
| EPI_ISL_474897 | Spain/AN-IBV-002887/2020 | 2020-03-16 | Europe/Spain/Andalusia/Guadix | Hospital Universitario Virgen de las Nieves de Granada-SAS | SeqCOVID-SPAIN consortium/IBV(CSIC) | Mercedes P√©rez Ruiz et al |
| EPI_ISL_474901 | Spain/CM-IBV-003753/2020 | 2020-04-03 | Europe/Spain/Castilla la Mancha/Hellin | Complejo Hospitalario Universitario de Albacete | SeqCOVID-SPAIN consortium/IBV(CSIC) | Encarnacion Simarro C√≥rdoba et al |
| EPI_ISL_474902 | Spain/CM-IBV-003761/2020 | 2020-04-03 | Europe/Spain/Castilla la Mancha/Hellin | Complejo Hospitalario Universitario de Albacete | SeqCOVID-SPAIN consortium/IBV(CSIC) | Encarnacion Simarro C√≥rdoba et al |
| EPI_ISL_474912 | Spain/CN-IBV-002596/2020 | 2020-03-15 | Europe/Spain/Canary Islands/Las Palmas | Hospital Universitario de Gran Canaria Dr. Negr√≠n | SeqCOVID-SPAIN consortium/IBV(CSIC) | M. Carmen P√©rez Gonz√°lez et al |
| EPI_ISL_474916 | Spain/CN-IBV-002602/2020 | 2020-03-17 | Europe/Spain/Canary Islands/Las Palmas | Hospital Universitario de Gran Canaria Dr. Negr√≠n | SeqCOVID-SPAIN consortium/IBV(CSIC) | M. Carmen P√©rez Gonz√°lez et al |
| EPI_ISL_474936 | Spain/AN-IBV-002885/2020 | 2020-03-17 | Europe/Spain/Andalusia/Otura | Hospital Universitario Virgen de las Nieves de Granada-SAS | SeqCOVID-SPAIN consortium/IBV(CSIC) | Mercedes P√©rez Ruiz et al |
| EPI_ISL_474940 | Spain/CM-IBV-003780/2020 | 2020-04-03 | Europe/Spain/Castilla la Mancha/Riopar | Complejo Hospitalario Universitario de Albacete | SeqCOVID-SPAIN consortium/IBV(CSIC) | Encarnacion Simarro C√≥rdoba et al |

| | | | | | | |
|---|---|---|---|---|---|---|
| EPI_ISL_474941 | Spain/CM-IBV-003886/2020 | 2020-05-20 | Europe/Spain/Castilla la Mancha/San Cl | Complejo Hospitalario Universitario de Albacete | SeqCOVID-SPAIN consortium/IBV(CSIC) | Encarnacion Simarro CV≥rdoba et al |
| EPI_ISL_474942 | Spain/AN-IBV-002919/2020 | 2020-03-18 | Europe/Spain/Andalusia/Santa Fe ES | Hospital Universitario Virgen de las Nieves de Granada-SAS | SeqCOVID-SPAIN consortium/IBV(CSIC) | Mercedes PV©rez Ruiz et al |
| EPI_ISL_474946 | Spain/CM-IBV-003768/2020 | 2020-04-03 | Europe/Spain/Castilla la Mancha/Tazona | Complejo Hospitalario Universitario de Albacete | SeqCOVID-SPAIN consortium/IBV(CSIC) | Encarnacion Simarro CV≥rdoba et al |
| EPI_ISL_474954 | Spain/CM-IBV-003790/2020 | 2020-04-03 | Europe/Spain/Castilla la Mancha/Villarr | Complejo Hospitalario Universitario de Albacete | SeqCOVID-SPAIN consortium/IBV(CSIC) | Encarnacion Simarro CV≥rdoba et al |
| EPI_ISL_474963 | Israel/CVL-n-7216/2020 | 2020-04-11 | Asia/Israel/Tel Aviv District/Tel-Aviv | Israel Central Virology laboratory | Israel Central Virology laboratory | Neta Zuckerman et al |
| EPI_ISL_474988 | Israel/CVL-n-6118/2020 | 2020-03-15 | Asia/Israel/Central District/Yehud | Israel Central Virology laboratory | Israel Central Virology laboratory | Neta Zuckerman et al |
| EPI_ISL_474989 | Israel/CVL-n-7224/2020 | 2020-04-11 | Asia/Israel/Jerusalem District/Beit Shem | Israel Central Virology laboratory | Israel Central Virology laboratory | Neta Zuckerman et al |
| EPI_ISL_474999 | Israel/CVL-n-6104/2020 | 2020-03-15 | Asia/Israel/Tel Aviv District/Ramat Gan | Israel Central Virology laboratory | Israel Central Virology laboratory | Neta Zuckerman et al |
| EPI_ISL_475000 | Israel/CVL-n-6105/2020 | 2020-03-15 | Asia/Israel/Central District/Gedera | Israel Central Virology laboratory | Israel Central Virology laboratory | Neta Zuckerman et al |
| EPI_ISL_475023 | Israel/CVL-n-7422/2020 | 2020-04-11 | Asia/Israel/Israel/ | Israel Central Virology laboratory | Israel Central Virology laboratory | Neta Zuckerman et al |
| EPI_ISL_475026 | India/GJ-GBRC203a/2020 | 2020-06-07 | Asia/India/Gujarat/Palanpur | Banas Medical College and Research Institute | Gujarat Biotechnology Research Centre | Sunil R Joshi et al |
| EPI_ISL_475027 | India/GJ-GBRC203b/2020 | 2020-06-07 | Asia/India/Gujarat/Palanpur | Banas Medical College and Research Institute | Gujarat Biotechnology Research Centre | Viren s Doshi et al |
| EPI_ISL_475028 | India/GJ-GBRC204a/2020 | 2020-06-07 | Asia/India/Gujarat/Palanpur | Banas Medical College and Research Institute | Gujarat Biotechnology Research Centre | Pritesh Sabara et al |

| | | | | | | |
|---|---|---|---|---|---|---|
| EPI_ISL_475031 | India/GJ-GBRC205b/2020 | 2020-06-11 | Asia/India/Gujarat/Surat | Department of MicroBiology, Government Medical College, Surat | Gujarat Biotechnology Research Centre | Zarna Patel et al |
| EPI_ISL_475032 | India/GJ-GBRC206/2020 | 2020-06-11 | Asia/India/Gujarat/Surat | Department of MicroBiology, Government Medical College, Surat | Gujarat Biotechnology Research Centre | Monika Gandhi et al |
| EPI_ISL_475034 | India/GJ-GBRC207b/2020 | 2020-06-11 | Asia/India/Gujarat/Surat | Department of MicroBiology, Government Medical College, Surat | Gujarat Biotechnology Research Centre | Maharshi Pandya et al |
| EPI_ISL_475036 | India/GJ-GBRC208b/2020 | 2020-06-11 | Asia/India/Gujarat/Surat | Department of MicroBiology, Government Medical College, Surat | Gujarat Biotechnology Research Centre | Nitin Savaliya et al |
| EPI_ISL_475039 | India/GJ-GBRC210a/2020 | 2020-06-11 | Asia/India/Gujarat/Surat | Department of MicroBiology, Government Medical College, Surat | Gujarat Biotechnology Research Centre | Zuber Saiyed et al |
| EPI_ISL_475040 | India/GJ-GBRC210b/2020 | 2020-06-11 | Asia/India/Gujarat/Surat | Department of MicroBiology, Government Medical College, Surat | Gujarat Biotechnology Research Centre | Komal Patel et al |
| EPI_ISL_475043 | India/GJ-GBRC213a/2020 | 2020-06-11 | Asia/India/Gujarat/Surat | Department of MicroBiology, Government Medical College, Surat | Gujarat Biotechnology Research Centre | Naresh Chauhan et al |
| EPI_ISL_475047 | India/GJ-GBRC194a/2020 | 2020-06-05 | Asia/India/Gujarat/Savli | GMERS Medical College & Hospital | Gujarat Biotechnology Research Centre | Apurvasinh Puvar et al |
| EPI_ISL_475048 | India/GJ-GBRC194b/2020 | 2020-06-05 | Asia/India/Gujarat/Savli | GMERS Medical College & Hospital | Gujarat Biotechnology Research Centre | Janvi Raval et al |
| EPI_ISL_475050 | India/GJ-GBRC216a/2020 | 2020-06-08 | Asia/India/Gujarat/Vadodara | GMERS Medical College & Hospital | Gujarat Biotechnology Research Centre | Monika Gandhi et al |
| EPI_ISL_475053 | India/GJ-GBRC217b/2020 | 2020-06-08 | Asia/India/Gujarat/Vadodara | GMERS Medical College & Hospital | Gujarat Biotechnology Research Centre | Nidhi Patel et al |
| EPI_ISL_475055 | India/GJ-GBRC218b/2020 | 2020-06-08 | Asia/India/Gujarat/Vadodara | GMERS Medical College & Hospital | Gujarat Biotechnology Research Centre | Raghawendra Kumar et al |
| EPI_ISL_475066 | Belgium/UGent-106/2020 | 2020-03-18 | Europe/Belgium/Gent/ | Lab voor klinische biologie | Onderzoeksgroep Virologie | Laurens Lambrechts et al |
| EPI_ISL_475068 | Belgium/UGent-108/2020 | 2020-03-18 | Europe/Belgium/Gent/ | Lab voor klinische biologie | Onderzoeksgroep Virologie | Laurens Lambrechts et al |

| | | | | | | |
|---|---|---|---|---|---|---|
| EPI_ISL_475070 | Belgium/UGent-110/2020 | 2020-03-18 | Europe/Belgium/Gent/ | Lab voor klinische biologie | Onderzoeksgroep Virologie | Laurens Lambrechts et al |
| EPI_ISL_475093 | Sweden/20-08210/2020 | 2020-03-16 | Europe/Sweden/Halland/ | Halmstad klinisk mikrobiologi | The Public Health Agency of Sweden | Oskar Karlsson Lindsjo et al |
| EPI_ISL_475140 | Sweden/20-08339/2020 | 2020-03-22 | Europe/Sweden/Vastra Gotaland/ | Skovde/Unilabs | The Public Health Agency of Sweden | Oskar Karlsson Lindsjo et al |
| EPI_ISL_475156 | Sweden/20-08842/2020 | 2020-04-09 | Europe/Sweden/Halland/ | Halmstad klinisk mikrobiologi | The Public Health Agency of Sweden | Oskar Karlsson Lindsjo et al |
| EPI_ISL_475167 | Bangladesh/BCSIR-NILMRC-111/2020 | 2020-06-07 | Asia/Bangladesh/Rangpur/ | National Institute of Laboratory Medicine and Referral Center | Genomic Research Lab, BCSIR | Barna Goswami et al |
| EPI_ISL_475168 | Bangladesh/BCSIR-NILMRC-117/2020 | 2020-06-07 | Asia/Bangladesh/Rangpur/ | National Institute of Laboratory Medicine and Referral Center | Genomic Research Lab, BCSIR | Iffat Jahan et al |
| EPI_ISL_475169 | Bangladesh/BCSIR-NILMRC-018/2020 | 2020-05-24 | Asia/Bangladesh/Dhaka/ | National Institute of Laboratory Medicine and Referral Center | Genomic Research Lab, BCSIR | Md. Saddam Hossain et al |
| EPI_ISL_475170 | Bangladesh/BCSIR-NILMRC-017/2020 | 2020-05-24 | Asia/Bangladesh/Dhaka/ | National Institute of Laboratory Medicine and Referral Center | Genomic Research Lab, BCSIR | Abu Sayeed Mohammad Mahmud et al |
| EPI_ISL_475172 | Bangladesh/BCSIR-NILMRC-109/2020 | 2020-06-07 | Asia/Bangladesh/Rangpur/ | National Institute of Laboratory Medicine and Referral Center | Genomic Research Lab, BCSIR | Abu Sayeed Mohammad Mahmud et al |
| EPI_ISL_475181 | USA/NE-10764/2020 | 2020-05-04 | North America/USA/Nebraska/ | Nebraska Public Health Laboratory | UNMC COVID-19 Response Team | UNMC COVID-19 Response Team et al |
| EPI_ISL_475184 | USA/NE-10832/2020 | 2020-05-04 | North America/USA/Nebraska/ | Nebraska Public Health Laboratory | UNMC COVID-19 Response Team | UNMC COVID-19 Response Team et al |
| EPI_ISL_475192 | USA/NE-10866/2020 | 2020-05-04 | North America/USA/Nebraska/ | Nebraska Public Health Laboratory | UNMC COVID-19 Response Team | UNMC COVID-19 Response Team et al |
| EPI_ISL_475195 | USA/NE-10943/2020 | 2020-05-04 | North America/USA/Nebraska/ | Nebraska Public Health Laboratory | UNMC COVID-19 Response Team | UNMC COVID-19 Response Team et al |
| EPI_ISL_475196 | USA/NE-10964/2020 | 2020-05-04 | North America/USA/Nebraska/ | Nebraska Public Health Laboratory | UNMC COVID-19 Response Team | UNMC COVID-19 Response Team et al |
| EPI_ISL_475199 | USA/NE-11118/2020 | 2020-05-04 | North America/USA/Nebraska/ | Nebraska Public Health Laboratory | UNMC COVID-19 Response Team | UNMC COVID-19 Response Team et al |
| EPI_ISL_475203 | USA/NE-11288/2020 | 2020-05-04 | North America/USA/Nebraska/ | Nebraska Public Health Laboratory | UNMC COVID-19 Response Team | UNMC COVID-19 Response Team et al |
| EPI_ISL_475209 | USA/NE-11343/2020 | 2020-05-03 | North America/USA/Nebraska/ | Nebraska Public Health Laboratory | UNMC COVID-19 Response Team | UNMC COVID-19 Response Team et al |
| EPI_ISL_475213 | USA/NE-11445/2020 | 2020-05-05 | North America/USA/Nebraska/ | Nebraska Public Health Laboratory | UNMC COVID-19 Response Team | UNMC COVID-19 Response Team et al |
| EPI_ISL_475216 | USA/NE-11471/2020 | 2020-05-04 | North America/USA/Nebraska/ | Nebraska Public Health Laboratory | UNMC COVID-19 Response Team | UNMC COVID-19 Response Team et al |
| EPI_ISL_475218 | USA/NE-11483/2020 | 2020-05-05 | North America/USA/Nebraska/ | Nebraska Public Health Laboratory | UNMC COVID-19 Response Team | UNMC COVID-19 Response Team et al |

| | | | | | | |
|---|---|---|---|---|---|---|
| EPI_ISL_475222 | USA/NE-11507/2020 | 2020-05-05 | North America/USA/Nebraska/ | Nebraska Public Health Laboratory | UNMC COVID-19 Response Team | UNMC COVID-19 Response Team et al |
| EPI_ISL_475225 | USA/NE-11518/2020 | 2020-05-05 | North America/USA/Nebraska/ | Nebraska Public Health Laboratory | UNMC COVID-19 Response Team | UNMC COVID-19 Response Team et al |
| EPI_ISL_475228 | USA/NE-11549/2020 | 2020-05-05 | North America/USA/Nebraska/ | Nebraska Public Health Laboratory | UNMC COVID-19 Response Team | UNMC COVID-19 Response Team et al |
| EPI_ISL_475231 | USA/NE-11566/2020 | 2020-05-05 | North America/USA/Nebraska/ | Nebraska Public Health Laboratory | UNMC COVID-19 Response Team | UNMC COVID-19 Response Team et al |
| EPI_ISL_475233 | USA/NE-11644/2020 | 2020-05-05 | North America/USA/Nebraska/ | Nebraska Public Health Laboratory | UNMC COVID-19 Response Team | UNMC COVID-19 Response Team et al |
| EPI_ISL_475234 | USA/NE-11669/2020 | 2020-05-05 | North America/USA/Nebraska/ | Nebraska Public Health Laboratory | UNMC COVID-19 Response Team | UNMC COVID-19 Response Team et al |
| EPI_ISL_475238 | Bangladesh/BCSIR-NILMRC-114/2020 | 2020-06-07 | Asia/Bangladesh/Rangpur/ | National Institute of Laboratory Medicine and Referral Center | Genomic Research Lab, BCSIR | Abu Sayeed Mohammad Mahmud et al |
| EPI_ISL_475297 | England/PORT-2D0E85/2020 | 2020-03-31 | Europe/United Kingdom/England/ | Centre for Enzyme Innovation, University of Portsmouth / Translational Research Laboratory, Portsmouth Hospitals NHS Trust | COVID-19 Genomics UK (COG-UK) Consortium | Angela Beckett et al |
| EPI_ISL_475403 | England/SHEF-C7E29/2020 | 2020-05-24 | Europe/United Kingdom/England/ | Virology Department, Sheffield Teaching Hospitals NHS Foundation Trust/Department of Infection, Immunity and Cardiovascular Disease, The Medical School, University of Sheffield | COVID-19 Genomics UK (COG-UK) Consortium | Thushan de Silva et al |
| EPI_ISL_475523 | Sweden/20-14941/2020 | 2020-05-25 | Europe/Sweden/Stockholm/ | Huddinge VC | The Public Health Agency of Sweden | Oskar Karlsson Lindsjo et al |
| EPI_ISL_475528 | Sweden/20-51755/2020 | 2020-05-27 | Europe/Sweden/Vastra Gotaland/ | Omtanken Grimmered | The Public Health Agency of Sweden | Oskar Karlsson Lindsjo et al |
| EPI_ISL_475532 | Sweden/20-51831/2020 | 2020-06-02 | Europe/Sweden/Vastmanland/ | Kungsors VC | The Public Health Agency of Sweden | Oskar Karlsson Lindsjo et al |
| EPI_ISL_475535 | Sweden/20-51955/2020 | 2020-06-09 | Europe/Sweden/Vastra Gotaland/ | Omtanken Grimmered | The Public Health Agency of Sweden | Oskar Karlsson Lindsjo et al |
| EPI_ISL_475536 | Sweden/20-06070/2020 | 2020-03-20 | Europe/Sweden/Jamtland/ | Follinge Halsocentral | The Public Health Agency of Sweden | Oskar Karlsson Lindsjo et al |
| EPI_ISL_475539 | Sweden/20-07017/2020 | 2020-04-07 | Europe/Sweden/Vastra Gotaland/ | Narhalsan Sjobo vardcentral | The Public Health Agency of Sweden | Oskar Karlsson Lindsjo et al |
| EPI_ISL_475544 | Sweden/20-04736/2020 | 2020-02-26 | Europe/Sweden/Uppsala/ | Karolinska Universitetslaboratoriet | The Public Health Agency of Sweden | Oskar Karlsson Lindsjo et al |
| EPI_ISL_475545 | Sweden/20-04850/2020 | 2020-02-28 | Europe/Sweden/Stockholm/ | Karolinska Universitetslaboratoriet | The Public Health Agency of Sweden | Oskar Karlsson Lindsjo et al |

| | | | | | | |
|---|---|---|---|---|---|---|
| EPI_ISL_475546 | Sweden/20-04851/2020 | 2020-02-28 | Europe/Sweden/Stockholm/ | Karolinska Universitetslaboratoriet | The Public Health Agency of Sweden | Oskar Karlsson Lindsjo et al |
| EPI_ISL_475547 | Sweden/20-04852/2020 | 2020-02-28 | Europe/Sweden/Stockholm/ | Karolinska Universitetslaboratoriet | The Public Health Agency of Sweden | Oskar Karlsson Lindsjo et al |
| EPI_ISL_475565 | Sweden/20-06911/2020 | 2020-04-06 | Europe/Sweden/Vastra Gotaland/ | Narhalsan Backa vardcentral | The Public Health Agency of Sweden | Oskar Karlsson Lindsjo et al |
| EPI_ISL_475566 | Sweden/20-14611/2020 | 2020-05-13 | Europe/Sweden/Ostergotland/ | Vardcentralen Brinken | The Public Health Agency of Sweden | Oskar Karlsson Lindsjo et al |
| EPI_ISL_475568 | Sweden/20-51816/2020 | 2020-06-01 | Europe/Sweden/Vastmanland/ | Kungsors VC | The Public Health Agency of Sweden | Oskar Karlsson Lindsjo et al |
| EPI_ISL_475569 | Sweden/20-51988/2020 | 2020-06-11 | Europe/Sweden/Stockholm/ | Kungsholmsdoktorn | The Public Health Agency of Sweden | Oskar Karlsson Lindsjo et al |
| EPI_ISL_475570 | Bangladesh/JUST-GC15.09/2020 | 2020-05-06 | Asia/Bangladesh/Khulna/Jhenaidah | Genome Center | Genome Center | A. S. M. Rubayet- Ul- Alam et al |
| EPI_ISL_475571 | Bangladesh/JUST-GC40.38/2020 | 2020-06-11 | Asia/Bangladesh/Khulna/Narail | Genome Center | Genome Center | Hassan M. Al-Emran et al |
| EPI_ISL_475573 | Bangladesh/JUST-GC40.86/2020 | 2020-06-11 | Asia/Bangladesh/Khulna/Bagerhat | Genome Center | Genome Center | Md. Shazid Hasan et al |
| EPI_ISL_475576 | USA/CA-CSMC101/2020 | 2020-04-02 | North America/USA/California/Los Ange | Cedars-Sinai Medical Center, Department of Pathology & Laboratory Medicine, Molecular Pathology Laboratory | Cedars-Sinai Medical Center, Molecular Pathology Laboratory of Department of Pathology & Laboratory Medicine and Genomic Core | Wenjuan Zhang et al |
| EPI_ISL_475582 | USA/CA-CSMC107/2020 | 2020-04-02 | North America/USA/California/Los Ange | Cedars-Sinai Medical Center, Department of Pathology & Laboratory Medicine, Molecular Pathology Laboratory | Cedars-Sinai Medical Center, Molecular Pathology Laboratory of Department of Pathology & Laboratory Medicine and Genomic Core | Wenjuan Zhang et al |

| | | | | | | |
|---|---|---|---|---|---|---|
| EPI_ISL_475585 | USA/CA-CSMC11/2020 | 2020-04-01 | North America/USA/California/Los Ange | Cedars-Sinai Medical Center, Department of Pathology & Laboratory Medicine, Molecular Pathology Laboratory | Cedars-Sinai Medical Center, Molecular Pathology Laboratory of Department of Pathology & Laboratory Medicine and Genomic Core | Wenjuan Zhang et al |
| EPI_ISL_475588 | USA/CA-CSMC112/2020 | 2020-04-02 | North America/USA/California/Los Ange | Cedars-Sinai Medical Center, Department of Pathology & Laboratory Medicine, Molecular Pathology Laboratory | Cedars-Sinai Medical Center, Molecular Pathology Laboratory of Department of Pathology & Laboratory Medicine and Genomic Core | Wenjuan Zhang et al |
| EPI_ISL_475597 | USA/CA-CSMC120/2020 | 2020-04-05 | North America/USA/California/Los Ange | Cedars-Sinai Medical Center, Department of Pathology & Laboratory Medicine, Molecular Pathology Laboratory | Cedars-Sinai Medical Center, Molecular Pathology Laboratory of Department of Pathology & Laboratory Medicine and Genomic Core | Wenjuan Zhang et al |
| EPI_ISL_475603 | USA/CA-CSMC126/2020 | 2020-04-09 | North America/USA/California/Los Ange | Cedars-Sinai Medical Center, Department of Pathology & Laboratory Medicine, Molecular Pathology Laboratory | Cedars-Sinai Medical Center, Molecular Pathology Laboratory of Department of Pathology & Laboratory Medicine and Genomic Core | Wenjuan Zhang et al |

| | | | | | | |
|---|---|---|---|---|---|---|
| EPI_ISL_475607 | USA/CA-CSMC13/2020 | 2020-03-28 | North America/USA/California/Los Ange | Cedars-Sinai Medical Center, Department of Pathology & Laboratory Medicine, Molecular Pathology Laboratory | Cedars-Sinai Medical Center, Molecular Pathology Laboratory of Department of Pathology & Laboratory Medicine and Genomic Core | Wenjuan Zhang et al |
| EPI_ISL_475619 | USA/CA-CSMC140/2020 | 2020-04-12 | North America/USA/California/Los Ange | Cedars-Sinai Medical Center, Department of Pathology & Laboratory Medicine, Molecular Pathology Laboratory | Cedars-Sinai Medical Center, Molecular Pathology Laboratory of Department of Pathology & Laboratory Medicine and Genomic Core | Wenjuan Zhang et al |
| EPI_ISL_475620 | USA/CA-CSMC141/2020 | 2020-04-08 | North America/USA/California/Los Ange | Cedars-Sinai Medical Center, Department of Pathology & Laboratory Medicine, Molecular Pathology Laboratory | Cedars-Sinai Medical Center, Molecular Pathology Laboratory of Department of Pathology & Laboratory Medicine and Genomic Core | Wenjuan Zhang et al |
| EPI_ISL_475635 | USA/CA-CSMC25/2020 | 2020-03-25 | North America/USA/California/Los Ange | Cedars-Sinai Medical Center, Department of Pathology & Laboratory Medicine, Molecular Pathology Laboratory | Cedars-Sinai Medical Center, Molecular Pathology Laboratory of Department of Pathology & Laboratory Medicine and Genomic Core | Wenjuan Zhang et al |

| | | | | | | |
|---|---|---|---|---|---|---|
| EPI_ISL_475654 | USA/CA-CSMC42/2020 | 2020-04-01 | North America/USA/California/Los Ange | Cedars-Sinai Medical Center, Department of Pathology & Laboratory Medicine, Molecular Pathology Laboratory | Cedars-Sinai Medical Center, Molecular Pathology Laboratory of Department of Pathology & Laboratory Medicine and Genomic Core | Wenjuan Zhang et al |
| EPI_ISL_475659 | USA/CA-CSMC47/2020 | 2020-04-09 | North America/USA/California/Los Ange | Cedars-Sinai Medical Center, Department of Pathology & Laboratory Medicine, Molecular Pathology Laboratory | Cedars-Sinai Medical Center, Molecular Pathology Laboratory of Department of Pathology & Laboratory Medicine and Genomic Core | Wenjuan Zhang et al |
| EPI_ISL_475661 | USA/CA-CSMC49/2020 | 2020-04-01 | North America/USA/California/Los Ange | Cedars-Sinai Medical Center, Department of Pathology & Laboratory Medicine, Molecular Pathology Laboratory | Cedars-Sinai Medical Center, Molecular Pathology Laboratory of Department of Pathology & Laboratory Medicine and Genomic Core | Wenjuan Zhang et al |
| EPI_ISL_475663 | USA/CA-CSMC50/2020 | 2020-04-06 | North America/USA/California/Los Ange | Cedars-Sinai Medical Center, Department of Pathology & Laboratory Medicine, Molecular Pathology Laboratory | Cedars-Sinai Medical Center, Molecular Pathology Laboratory of Department of Pathology & Laboratory Medicine and Genomic Core | Wenjuan Zhang et al |

| | | | | | | |
|---|---|---|---|---|---|---|
| EPI_ISL_475665 | USA/CA-CSMC52/2020 | 2020-04-06 | North America/USA/California/Los Ange | Cedars-Sinai Medical Center, Department of Pathology & Laboratory Medicine, Molecular Pathology Laboratory | Cedars-Sinai Medical Center, Molecular Pathology Laboratory of Department of Pathology & Laboratory Medicine and Genomic Core | Wenjuan Zhang et al |
| EPI_ISL_475666 | USA/CA-CSMC53/2020 | 2020-04-01 | North America/USA/California/Los Ange | Cedars-Sinai Medical Center, Department of Pathology & Laboratory Medicine, Molecular Pathology Laboratory | Cedars-Sinai Medical Center, Molecular Pathology Laboratory of Department of Pathology & Laboratory Medicine and Genomic Core | Wenjuan Zhang et al |
| EPI_ISL_475671 | USA/CA-CSMC58/2020 | 2020-04-01 | North America/USA/California/Los Ange | Cedars-Sinai Medical Center, Department of Pathology & Laboratory Medicine, Molecular Pathology Laboratory | Cedars-Sinai Medical Center, Molecular Pathology Laboratory of Department of Pathology & Laboratory Medicine and Genomic Core | Wenjuan Zhang et al |
| EPI_ISL_475674 | USA/CA-CSMC60/2020 | 2020-04-01 | North America/USA/California/Los Ange | Cedars-Sinai Medical Center, Department of Pathology & Laboratory Medicine, Molecular Pathology Laboratory | Cedars-Sinai Medical Center, Molecular Pathology Laboratory of Department of Pathology & Laboratory Medicine and Genomic Core | Wenjuan Zhang et al |

| | | | | | | |
|---|---|---|---|---|---|---|
| EPI_ISL_475682 | USA/CA-CSMC68/2020 | 2020-03-28 | North America/USA/California/Los Ange | Cedars-Sinai Medical Center, Department of Pathology & Laboratory Medicine, Molecular Pathology Laboratory | Cedars-Sinai Medical Center, Molecular Pathology Laboratory of Department of Pathology & Laboratory Medicine and Genomic Core | Wenjuan Zhang et al |
| EPI_ISL_475683 | USA/CA-CSMC69/2020 | 2020-03-28 | North America/USA/California/Los Ange | Cedars-Sinai Medical Center, Department of Pathology & Laboratory Medicine, Molecular Pathology Laboratory | Cedars-Sinai Medical Center, Molecular Pathology Laboratory of Department of Pathology & Laboratory Medicine and Genomic Core | Wenjuan Zhang et al |
| EPI_ISL_475696 | USA/CA-CSMC80/2020 | 2020-03-26 | North America/USA/California/Los Ange | Cedars-Sinai Medical Center, Department of Pathology & Laboratory Medicine, Molecular Pathology Laboratory | Cedars-Sinai Medical Center, Molecular Pathology Laboratory of Department of Pathology & Laboratory Medicine and Genomic Core | Wenjuan Zhang et al |
| EPI_ISL_475724 | Egypt/CUNCI-HGC6I031/2020 | 2020-06-02 | Africa/Egypt/Egypt/ | Egyptian National Cancer Institute (ENCI) | Egyptian National Cancer Institute (ENCI) | Zekri et al |
| EPI_ISL_475728 | USA/UT-04034/2020 | 2020-05-12 | North America/USA/Utah/ | Utah Public Health Laboratory | Utah Public Health Laboratory | Erin Young et al |
| EPI_ISL_475729 | USA/UT-04035/2020 | 2020-05-12 | North America/USA/Utah/ | Utah Public Health Laboratory | Utah Public Health Laboratory | Erin Young et al |
| EPI_ISL_475740 | USA/UT-04091/2020 | 2020-05-12 | North America/USA/Utah/ | Utah Public Health Laboratory | Utah Public Health Laboratory | Erin Young et al |
| EPI_ISL_475742 | USA/UT-04102/2020 | 2020-05-12 | North America/USA/Utah/ | Utah Public Health Laboratory | Utah Public Health Laboratory | Erin Young et al |

| | | | | | | |
|---|---|---|---|---|---|---|
| EPI_ISL_475754 | Bangladesh/BCSIR-NILMRC-025/2020 | 2020-05-24 | Asia/Bangladesh/Dhaka/ | National Institute of Laboratory Medicine and Referral Center | Genomic Research Lab, BCSIR | Shahina Akter et al |
| EPI_ISL_475756 | Bangladesh/BCSIR-NILMRC-108/2020 | 2020-06-07 | Asia/Bangladesh/Rangpur/ | National Institute of Laboratory Medicine and Referral Center | Genomic Research Lab, BCSIR | Tanjina Akhter Banu et al |
| EPI_ISL_475757 | Bangladesh/BCSIR-NILMRC-105/2020 | 2020-06-07 | Asia/Bangladesh/Rangpur/ | National Institute of Laboratory Medicine and Referral Center | Genomic Research Lab, BCSIR | Barna Goswami et al |
| EPI_ISL_475761 | Bangladesh/BCSIR-NILMRC-112/2020 | 2020-06-07 | Asia/Bangladesh/Rangpur/ | National Institute of Laboratory Medicine and Referral Center | Genomic Research Lab, BCSIR | Abu Sayeed Mohammad Mahmud et al |
| EPI_ISL_475790 | Austria/CeMM0319/2020 | 2020-03-30 | Europe/Austria/Vienna/ | Center for Virology, Medical University of Vienna | Bergthaler laboratory, CeMM Research Center for Molecular Medicine of the Austrian Academy of Sciences | Alexandra Popa et al |
| EPI_ISL_475801 | Austria/CeMM0340/2020 | 2020-04-03 | Europe/Austria/Vienna/ | Center for Virology, Medical University of Vienna | Bergthaler laboratory, CeMM Research Center for Molecular Medicine of the Austrian Academy of Sciences | Alexandra Popa et al |
| EPI_ISL_475836 | Austria/CeMM0401/2020 | 2020-02-28 | Europe/Austria/Austria/ | Austrian Agency for Health and Food Safety (AGES) | Bergthaler laboratory, CeMM Research Center for Molecular Medicine of the Austrian Academy of Sciences | Alexandra Popa et al |
| EPI_ISL_475841 | Austria/CeMM0409/2020 | 2020-03-26 | Europe/Austria/Austria/ | Austrian Agency for Health and Food Safety (AGES) | Bergthaler laboratory, CeMM Research Center for Molecular Medicine of the Austrian Academy of Sciences | Alexandra Popa et al |

| | | | | | | |
|---|---|---|---|---|---|---|
| EPI_ISL_475845 | Austria/CeMM0417/2020 | 2020-03-22 | Europe/Austria/Austria/ | Austrian Agency for Health and Food Safety (AGES) | Bergthaler laboratory, CeMM Research Center for Molecular Medicine of the Austrian Academy of Sciences | Alexandra Popa et al |
| EPI_ISL_475857 | Austria/CeMM0430/2020 | 2020-03-24 | Europe/Austria/Austria/ | Austrian Agency for Health and Food Safety (AGES) | Bergthaler laboratory, CeMM Research Center for Molecular Medicine of the Austrian Academy of Sciences | Alexandra Popa et al |
| EPI_ISL_475887 | Austria/CeMM0514/2020 | 2020-03-27 | Europe/Austria/Austria/ | Zentralinstitut f√ºr medizinische und chemische Labordiagnostik, Universit√§tskliniken Insbruck | Bergthaler laboratory, CeMM Research Center for Molecular Medicine of the Austrian Academy of Sciences | Alexandra Popa et al |
| EPI_ISL_475888 | Austria/CeMM0515/2020 | 2020-03-28 | Europe/Austria/Austria/ | Zentralinstitut f√ºr medizinische und chemische Labordiagnostik, Universit√§tskliniken Insbruck | Bergthaler laboratory, CeMM Research Center for Molecular Medicine of the Austrian Academy of Sciences | Alexandra Popa et al |
| EPI_ISL_475923 | Austria/CeMM0576/2020 | 2020-04-01 | Europe/Austria/Tyrol/Innsbruck | Institut f√ºr Virologie am Department f√ºr Hygiene, Mikrobiologie und Public Health | Bergthaler laboratory, CeMM Research Center for Molecular Medicine of the Austrian Academy of Sciences | Alexandra Popa et al |

| | | | | | | |
|---|---|---|---|---|---|---|
| EPI_ISL_475925 | Austria/CeMM0578/2020 | 2020-03-30 | Europe/Austria/Tyrol/Innsbruck | Institut f√ºr Virologie am Department f√ºr Hygiene, Mikrobiologie und Public Health | Bergthaler laboratory, CeMM Research Center for Molecular Medicine of the Austrian Academy of Sciences | Alexandra Popa et al |
| EPI_ISL_475937 | Singapore/320/2020 | 2020-05-19 | Asia/Singapore/Singapore/ | National Public Health Laboratory, National Centre for Infectious Diseases | National Public Health Laboratory, National Centre for Infectious Diseases | Mak TM et al |
| EPI_ISL_475945 | Singapore/414/2020 | 2020-04-24 | Asia/Singapore/Singapore/ | National Public Health Laboratory, National Centre for Infectious Diseases | National Public Health Laboratory, National Centre for Infectious Diseases | Mak TM et al |
| EPI_ISL_475951 | Singapore/420/2020 | 2020-04-09 | Asia/Singapore/Singapore/ | National Public Health Laboratory, National Centre for Infectious Diseases | National Public Health Laboratory, National Centre for Infectious Diseases | Mak TM et al |
| EPI_ISL_475974 | Singapore/443/2020 | 2020-04-15 | Asia/Singapore/Singapore/ | National Public Health Laboratory, National Centre for Infectious Diseases | National Public Health Laboratory, National Centre for Infectious Diseases | Mak TM et al |
| EPI_ISL_475979 | Singapore/448/2020 | 2020-04-15 | Asia/Singapore/Singapore/ | National Public Health Laboratory, National Centre for Infectious Diseases | National Public Health Laboratory, National Centre for Infectious Diseases | Mak TM et al |

| | | | | | | |
|---|---|---|---|---|---|---|
| EPI_ISL_475997 | Singapore/466/2020 | 2020-01-31 | Asia/Singapore/Singapore/ | National Public Health Laboratory, National Centre for Infectious Diseases | National Public Health Laboratory, National Centre for Infectious Diseases | Mak TM et al |
| EPI_ISL_475998 | Singapore/467/2020 | 2020-02-28 | Asia/Singapore/Singapore/ | National Public Health Laboratory, National Centre for Infectious Diseases | National Public Health Laboratory, National Centre for Infectious Diseases | Mak TM et al |
| EPI_ISL_475999 | Singapore/468/2020 | 2020-02-14 | Asia/Singapore/Singapore/ | National Public Health Laboratory, National Centre for Infectious Diseases | National Public Health Laboratory, National Centre for Infectious Diseases | Mak TM et al |
| EPI_ISL_476018 | USA/MO-WUSTL070/2020 | 2020-06-19 | North America/USA/Missouri/St. Louis M | Washington University in St. Louis | Washington University in St. Louis | David Wang et al |
| EPI_ISL_476019 | USA/MO-WUSTL066/2020 | 2020-06-19 | North America/USA/Missouri/St. Louis M | Washington University in St. Louis | Washington University in St. Louis | David Wang et al |
| EPI_ISL_476024 | Morocco/moh100/2020 | 2020-05-05 | Africa/Morocco/Morocco/ | Laboratoire de Recherche et d'Analyses M√©dicales de la Gendarmerie Royale | Laboratoire de Recherche et d'Analyses M√©dicales de la Gendarmerie Royale | Sana√¢ Lemriss et al |
| EPI_ISL_476025 | Morocco/CA24/2020 | 2020-05-02 | Africa/Morocco/Morocco/ | Laboratoire de Recherche et d'Analyses M√©dicales de la Gendarmerie Royale | Laboratoire de Recherche et d'Analyses M√©dicales de la Gendarmerie Royale | Sana√¢ LEMRISS et al |

| | | | | | | |
|---|---|---|---|---|---|---|
| EPI_ISL_476027 | USA/MI-MDHHS-SC20890/2020 | 2020-05-21 | North America/USA/Michigan/ | Michigan Department of Health and Human Services, Bureau of Laboratories | Michigan Department of Health and Human Services, Bureau of Laboratories | Blankenship HM et al |
| EPI_ISL_476028 | USA/MI-MDHHS-SC20891/2020 | 2020-05-20 | North America/USA/Michigan/ | Michigan Department of Health and Human Services, Bureau of Laboratories | Michigan Department of Health and Human Services, Bureau of Laboratories | Blankenship HM et al |
| EPI_ISL_476036 | USA/MI-MDHHS-SC20899/2020 | 2020-04-24 | North America/USA/Michigan/ | Michigan Department of Health and Human Services, Bureau of Laboratories | Michigan Department of Health and Human Services, Bureau of Laboratories | Blankenship HM et al |
| EPI_ISL_476040 | USA/MI-MDHHS-SC20903/2020 | 2020-04-24 | North America/USA/Michigan/ | Michigan Department of Health and Human Services, Bureau of Laboratories | Michigan Department of Health and Human Services, Bureau of Laboratories | Blankenship HM et al |
| EPI_ISL_476041 | USA/MI-MDHHS-SC20905/2020 | 2020-04-29 | North America/USA/Michigan/ | Michigan Department of Health and Human Services, Bureau of Laboratories | Michigan Department of Health and Human Services, Bureau of Laboratories | Blankenship HM et al |
| EPI_ISL_476046 | USA/MI-MDHHS-SC20910/2020 | 2020-04-29 | North America/USA/Michigan/ | Michigan Department of Health and Human Services, Bureau of Laboratories | Michigan Department of Health and Human Services, Bureau of Laboratories | Blankenship HM et al |

| | | | | | | |
|---|---|---|---|---|---|---|
| EPI_ISL_476049 | USA/MI-MDHHS-SC20913/2020 | 2020-04-29 | North America/USA/Michigan/ | Michigan Department of Health and Human Services, Bureau of Laboratories | Michigan Department of Health and Human Services, Bureau of Laboratories | Blankenship HM et al |
| EPI_ISL_476057 | USA/MI-MDHHS-SC20921/2020 | 2020-05-26 | North America/USA/Michigan/ | Michigan Department of Health and Human Services, Bureau of Laboratories | Michigan Department of Health and Human Services, Bureau of Laboratories | Blankenship HM et al |
| EPI_ISL_476058 | USA/MI-MDHHS-SC20922/2020 | 2020-05-26 | North America/USA/Michigan/ | Michigan Department of Health and Human Services, Bureau of Laboratories | Michigan Department of Health and Human Services, Bureau of Laboratories | Blankenship HM et al |
| EPI_ISL_476064 | USA/MI-MDHHS-SC20928/2020 | 2020-05-28 | North America/USA/Michigan/ | Michigan Department of Health and Human Services, Bureau of Laboratories | Michigan Department of Health and Human Services, Bureau of Laboratories | Blankenship HM et al |
| EPI_ISL_476071 | Hungary/UD-6858/2020 | 2020-04-08 | Europe/Hungary/Debrecen/ | University of Debrecen, Department of Medical Microbiology | National Laboratory of Virology, Szentv°gothai Research Centre | Endre Gv°bor Tv≥th et al |
| EPI_ISL_476074 | Hungary/UD-7373/2020 | 2020-04-08 | Europe/Hungary/Heves County/ | University of Debrecen, Department of Medical Microbiology | National Laboratory of Virology, Szentv°gothai Research Centre | Endre Gv°bor Tv≥th et al |
| EPI_ISL_476116 | Switzerland/BE-ETHZ-140068/2020 | 2020-04-29 | Europe/Switzerland/Bern/ | Viollier AG | Department of Biosystems Science and Engineering, ETH Zvºrich | Christian Beisel et al |

| | | | | | | |
|---|---|---|---|---|---|---|
| EPI_ISL_476121 | Switzerland/UR-ETHZ-140074/2020 | 2020-04-24 | Europe/Switzerland/Uri/ | Viollier AG | Department of Biosystems Science and Engineering, ETH Z√ºrich | Christian Beisel et al |
| EPI_ISL_476129 | Switzerland/AG-ETHZ-140086/2020 | 2020-04-20 | Europe/Switzerland/Aargau/ | Viollier AG | Department of Biosystems Science and Engineering, ETH Z√ºrich | Christian Beisel et al |
| EPI_ISL_476136 | Sweden/20-51636/2020 | 2020-05-22 | Europe/Sweden/Stockholm/ | Surbrunns VC | The Public Health Agency of Sweden | Oskar Karlsson Lindsjo et al |
| EPI_ISL_476139 | Sweden/20-02114/2020 | 2020-01-31 | Europe/Sweden/Jonkoping/ | Folkhalsomyndigheten | The Public Health Agency of Sweden | Oskar Karlsson Lindsjo et al |
| EPI_ISL_476143 | Sweden/20-08533/2020 | 2020-03-27 | Europe/Sweden/Vastra Gotaland/ | Skovde/Unilabs | The Public Health Agency of Sweden | Oskar Karlsson Lindsjo et al |
| EPI_ISL_476146 | Sweden/20-08866/2020 | 2020-04-09 | Europe/Sweden/Jamtland Harjedalen/ | Ostersund klinisk mikrobiologi | The Public Health Agency of Sweden | Oskar Karlsson Lindsjo et al |
| EPI_ISL_476147 | Sweden/20-08869/2020 | 2020-04-09 | Europe/Sweden/Jamtland Harjedalen/ | Ostersund klinisk mikrobiologi | The Public Health Agency of Sweden | Oskar Karlsson Lindsjo et al |
| EPI_ISL_476163 | Brazil/SP-L1-CAMPI29/2020 | 2020-04-14 | South America/Brazil/S√£o Paulo/ | Laborat√≥rio de Patologia Cl√≠nica - UNICAMP | Laborat√≥rio de Estudos de V√≠rus Emergentes - UNICAMP | Jos√© Luiz Proen√ßa-Modena et al |
| EPI_ISL_476169 | Brazil/SP-L1-CAMPI7/2020 | 2020-04-03 | South America/Brazil/S√£o Paulo/ | Laborat√≥rio de Patologia Cl√≠nica - UNICAMP | Laborat√≥rio de Estudos de V√≠rus Emergentes - UNICAMP | Jos√© Luiz Proen√ßa-Modena et al |
| EPI_ISL_476170 | Brazil/SP-L1-CAMPI8/2020 | 2020-04-03 | South America/Brazil/S√£o Paulo/ | Laborat√≥rio de Patologia Cl√≠nica - UNICAMP | Laborat√≥rio de Estudos de V√≠rus Emergentes - UNICAMP | Jos√© Luiz Proen√ßa-Modena et al |
| EPI_ISL_476180 | Brazil/RJ-L10-CD158/2020 | 2020-03-16 | South America/Brazil/Rio de Janeiro/ | DB Diagn√≥sticos do Brasil | Instituto de Medicina Tropical da Univesidade de S√£o Paulo | Samples: Nelson Gaburo Jr et al |
| EPI_ISL_476214 | Brazil/CE-L12-CD214/2020 | 2020-03-16 | South America/Brazil/Cear√°/ | DB Diagn√≥sticos do Brasil | Instituto de Medicina Tropical da Univesidade de S√£o Paulo | Samples: Nelson Gaburo Jr et al |

| | | | | | | |
|---|---|---|---|---|---|---|
| EPI_ISL_476243 | Brazil/SP-L14-CD239/2020 | 2020-03-26 | South America/Brazil/S√£o Paulo/ | Hospital da Cl√≠nicas da Faculdade de Medicina da Universidade de S√£o Paulo | Instituto de Medicina Tropical da Univesidade de S√£o Paulo | Samples: Ingra Morales Claro et al |
| EPI_ISL_476267 | Brazil/SP-L15-CD274/2020 | 2020-03-30 | South America/Brazil/S√£o Paulo/ | Hospital da Cl√≠nicas da Faculdade de Medicina da Universidade de S√£o Paulo | Instituto de Medicina Tropical da Univesidade de S√£o Paulo | Samples: Ingra Morales Claro et al |
| EPI_ISL_476277 | Brazil/SP-L15-CD286/2020 | 2020-03-30 | South America/Brazil/S√£o Paulo/ | Hospital da Cl√≠nicas da Faculdade de Medicina da Universidade de S√£o Paulo | Instituto de Medicina Tropical da Univesidade de S√£o Paulo | Samples: Ingra Morales Claro et al |
| EPI_ISL_476281 | Brazil/CE-L16-CD332/2020 | 2020-03-28 | South America/Brazil/Cear√°/ | DB Diagn√≥sticos do Brasil | Instituto de Medicina Tropical da Univesidade de S√£o Paulo | Samples: Nelson Gaburo Jr et al |
| EPI_ISL_476320 | Brazil/SP-L19-CD391/2020 | 2020-04-01 | South America/Brazil/S√£o Paulo/ | DB Diagn√≥sticos do Brasil | Instituto de Medicina Tropical da Univesidade de S√£o Paulo | Samples: Nelson Gaburo Jr et al |
| EPI_ISL_476330 | Brazil/MA-L19-CD407/2020 | 2020-04-02 | South America/Brazil/Maranh√£o/ | DB Diagn√≥sticos do Brasil | Instituto de Medicina Tropical da Univesidade de S√£o Paulo | Samples: Nelson Gaburo Jr et al |
| EPI_ISL_476331 | Brazil/CE-L19-CD408/2020 | 2020-04-02 | South America/Brazil/Cear√°/ | DB Diagn√≥sticos do Brasil | Instituto de Medicina Tropical da Univesidade de S√£o Paulo | Samples: Nelson Gaburo Jr et al |
| EPI_ISL_476351 | Brazil/SP-L20-CD416/2020 | 2020-04-02 | South America/Brazil/S√£o Paulo/ | DB Diagn√≥sticos do Brasil | Instituto de Medicina Tropical da Univesidade de S√£o Paulo | Samples: Nelson Gaburo Jr et al |
| EPI_ISL_476360 | Brazil/SP-L20-CD434/2020 | 2020-04-04 | South America/Brazil/S√£o Paulo/ | DB Diagn√≥sticos do Brasil | Instituto de Medicina Tropical da Univesidade de S√£o Paulo | Samples: Nelson Gaburo Jr et al |
| EPI_ISL_476425 | Brazil/SP-L5-CAMPI99/2020 | 2020-04-25 | South America/Brazil/S√£o Paulo/ | Laborat√≥rio de Patologia Cl√≠nica - UNICAMP | Laborat√≥rio de Estudos de V√≠rus Emergentes - UNICAMP | Jos√© Luiz Proen√ßa-Modena et al |

| | | | | | | |
|---|---|---|---|---|---|---|
| EPI_ISL_476435 | Brazil/SP-L7-CD67/2020 | 2020-03-19 | South America/Brazil/S√£o Paulo/ | Hospital da Cl√≠nicas da Faculdade de Medicina da Universidade de S√£o Paulo | Instituto de Medicina Tropical da Univesidade de S√£o Paulo | Samples: Ingra Morales Claro et al |
| EPI_ISL_476452 | Brazil/SP-L8-CD105/2020 | 2020-03-21 | South America/Brazil/S√£o Paulo/ | Hospital da Cl√≠nicas da Faculdade de Medicina da Universidade de S√£o Paulo | Instituto de Medicina Tropical da Univesidade de S√£o Paulo | Samples: Ingra Morales Claro et al |
| EPI_ISL_476498 | Canada/QC_AA4/2020 | 2020-04-05 | North America/Canada/Quebec/ | Laboratoire de microbiologie, Hopital de Verdun | Smith Laboratory, Centre de Recherche CHU Sainte-Justine | Martin Smith et al |
| EPI_ISL_476499 | Canada/QC_AC9/2020 | 2020-04-06 | North America/Canada/Quebec/ | Laboratoire de microbiologie, Hopital de Verdun | Smith Laboratory, Centre de Recherche CHU Sainte-Justine | Martin Smith et al |
| EPI_ISL_476500 | Canada/QC_AD6/2020 | 2020-04-06 | North America/Canada/Quebec/ | Laboratoire de microbiologie, Hopital de Verdun | Smith Laboratory, Centre de Recherche CHU Sainte-Justine | Martin Smith et al |
| EPI_ISL_476501 | Canada/QC_AD7/2020 | 2020-04-04 | North America/Canada/Quebec/ | Laboratoire de microbiologie, Hopital de Verdun | Smith Laboratory, Centre de Recherche CHU Sainte-Justine | Martin Smith et al |
| EPI_ISL_476502 | Canada/QC_AG3/2020 | 2020-04-08 | North America/Canada/Quebec/ | Laboratoire de microbiologie, Hopital de Verdun | Smith Laboratory, Centre de Recherche CHU Sainte-Justine | Martin Smith et al |
| EPI_ISL_476503 | Canada/QC_AG8/2020 | 2020-04-14 | North America/Canada/Quebec/ | Laboratoire de microbiologie, Hopital de Verdun | Smith Laboratory, Centre de Recherche CHU Sainte-Justine | Martin Smith et al |
| EPI_ISL_476504 | Canada/QC_AH6/2020 | 2020-04-07 | North America/Canada/Quebec/ | Laboratoire de microbiologie, Hopital de Verdun | Smith Laboratory, Centre de Recherche CHU Sainte-Justine | Martin Smith et al |
| EPI_ISL_476505 | Canada/QC_AI6/2020 | 2020-04-08 | North America/Canada/Quebec/ | Laboratoire de microbiologie, Hopital de Verdun | Smith Laboratory, Centre de Recherche CHU Sainte-Justine | Martin Smith et al |

| | | | | | | |
|---|---|---|---|---|---|---|
| EPI_ISL_476506 | Canada/QC_AJ8/2020 | 2020-04-08 | North America/Canada/Quebec/ | Laboratoire de microbiologie, Hopital de Verdun | Smith Laboratory, Centre de Recherche CHU Sainte-Justine | Martin Smith et al |
| EPI_ISL_476507 | Canada/QC_AK1/2020 | 2020-04-11 | North America/Canada/Quebec/ | Laboratoire de microbiologie, Hopital de Verdun | Smith Laboratory, Centre de Recherche CHU Sainte-Justine | Martin Smith et al |
| EPI_ISL_476508 | Canada/QC_AM1/2020 | 2020-04-09 | North America/Canada/Quebec/ | Laboratoire de microbiologie, Hopital de Verdun | Smith Laboratory, Centre de Recherche CHU Sainte-Justine | Martin Smith et al |
| EPI_ISL_476509 | Canada/QC_AM6/2020 | 2020-04-18 | North America/Canada/Quebec/ | Laboratoire de microbiologie, Hopital de Verdun | Smith Laboratory, Centre de Recherche CHU Sainte-Justine | Martin Smith et al |
| EPI_ISL_476511 | Canada/QC_AQ3/2020 | 2020-04-03 | North America/Canada/Quebec/ | Laboratoire de microbiologie, Hopital de Verdun | Smith Laboratory, Centre de Recherche CHU Sainte-Justine | Martin Smith et al |
| EPI_ISL_476512 | Canada/QC_AR2/2020 | 2020-04-03 | North America/Canada/Quebec/ | Laboratoire de microbiologie, Hopital de Verdun | Smith Laboratory, Centre de Recherche CHU Sainte-Justine | Martin Smith et al |
| EPI_ISL_476513 | Canada/QC_AS5/2020 | 2020-04-05 | North America/Canada/Quebec/ | Laboratoire de microbiologie, Hopital de Verdun | Smith Laboratory, Centre de Recherche CHU Sainte-Justine | Martin Smith et al |
| EPI_ISL_476519 | USA/CT-Yale-289/2020 | 2020-03-28 | North America/USA/Connecticut/Fairfie | Yale Clinical Virology Laboratory | Grubaugh Lab - Yale School of Public Health | Joseph Fauver et al |
| EPI_ISL_476529 | USA/CT-Yale-300/2020 | 2020-03-28 | North America/USA/Connecticut/Fairfie | Yale Clinical Virology Laboratory | Grubaugh Lab - Yale School of Public Health | Joseph Fauver et al |
| EPI_ISL_476535 | USA/CT-Yale-306/2020 | 2020-03-30 | North America/USA/Connecticut/Hartfo | Yale Clinical Virology Laboratory | Grubaugh Lab - Yale School of Public Health | Joseph Fauver et al |
| EPI_ISL_476543 | USA/CT-Yale-315/2020 | 2020-03-31 | North America/USA/Connecticut/New H | Yale Clinical Virology Laboratory | Grubaugh Lab - Yale School of Public Health | Joseph Fauver et al |

| | | | | | | |
|---|---|---|---|---|---|---|
| EPI_ISL_476559 | Morocco/refstage1/2020 | 2020-02-27 | Africa/Morocco/Morocco/ | unknown | Laboratoire Sciences et Technologies de la Sant√© (STS) Institut Sup√©rieur des Sciences de la Sant√© Universit√© Hassan 1er, Settat, Morocco | Hajar Lemriss et al |
| EPI_ISL_476574 | Senegal/65052/2020 | 2020-06-13 | Africa/Senegal/Dakar/ | Institut Pasteur Dakar | Institut Pasteur de Dakar | Ndongo Dia et al |
| EPI_ISL_476708 | USA/MN-MDH-1123/2020 | 2020-04-23 | North America/USA/Minnesota/ | Minnesota Department of Health, Public Health Laboratory | Minnesota Department of Health, Public Health Laboratory | Matt Plumb et al |
| EPI_ISL_476709 | USA/MN-MDH-1124/2020 | 2020-04-23 | North America/USA/Minnesota/ | Minnesota Department of Health, Public Health Laboratory | Minnesota Department of Health, Public Health Laboratory | Matt Plumb et al |
| EPI_ISL_476716 | USA/MN-MDH-1131/2020 | 2020-04-25 | North America/USA/Minnesota/ | Minnesota Department of Health, Public Health Laboratory | Minnesota Department of Health, Public Health Laboratory | Matt Plumb et al |
| EPI_ISL_476724 | USA/MN-MDH-1139/2020 | 2020-04-28 | North America/USA/Minnesota/ | Minnesota Department of Health, Public Health Laboratory | Minnesota Department of Health, Public Health Laboratory | Matt Plumb et al |
| EPI_ISL_476725 | USA/MN-MDH-1140/2020 | 2020-04-28 | North America/USA/Minnesota/ | Minnesota Department of Health, Public Health Laboratory | Minnesota Department of Health, Public Health Laboratory | Matt Plumb et al |
| EPI_ISL_476726 | USA/MN-MDH-1141/2020 | 2020-04-28 | North America/USA/Minnesota/ | Minnesota Department of Health, Public Health Laboratory | Minnesota Department of Health, Public Health Laboratory | Matt Plumb et al |

| | | | | | | |
|---|---|---|---|---|---|---|
| EPI_ISL_476728 | USA/MN-MDH-1143/2020 | 2020-04-28 | North America/USA/Minnesota/ | Minnesota Department of Health, Public Health Laboratory | Minnesota Department of Health, Public Health Laboratory | Matt Plumb et al |
| EPI_ISL_476731 | USA/MN-MDH-1146/2020 | 2020-04-28 | North America/USA/Minnesota/ | Minnesota Department of Health, Public Health Laboratory | Minnesota Department of Health, Public Health Laboratory | Matt Plumb et al |
| EPI_ISL_476736 | USA/MN-MDH-1151/2020 | 2020-04-30 | North America/USA/Minnesota/ | Minnesota Department of Health, Public Health Laboratory | Minnesota Department of Health, Public Health Laboratory | Matt Plumb et al |
| EPI_ISL_476737 | USA/MN-MDH-1152/2020 | 2020-05-01 | North America/USA/Minnesota/ | Minnesota Department of Health, Public Health Laboratory | Minnesota Department of Health, Public Health Laboratory | Matt Plumb et al |
| EPI_ISL_476742 | USA/MN-MDH-1157/2020 | 2020-05-04 | North America/USA/Minnesota/ | Minnesota Department of Health, Public Health Laboratory | Minnesota Department of Health, Public Health Laboratory | Matt Plumb et al |
| EPI_ISL_476746 | USA/MN-MDH-1161/2020 | 2020-05-04 | North America/USA/Minnesota/ | Minnesota Department of Health, Public Health Laboratory | Minnesota Department of Health, Public Health Laboratory | Matt Plumb et al |
| EPI_ISL_476750 | USA/MN-MDH-1165/2020 | 2020-05-05 | North America/USA/Minnesota/ | Minnesota Department of Health, Public Health Laboratory | Minnesota Department of Health, Public Health Laboratory | Matt Plumb et al |
| EPI_ISL_476752 | USA/MN-MDH-1167/2020 | 2020-05-05 | North America/USA/Minnesota/ | Minnesota Department of Health, Public Health Laboratory | Minnesota Department of Health, Public Health Laboratory | Matt Plumb et al |
| EPI_ISL_476761 | USA/MN-MDH-1176/2020 | 2020-05-21 | North America/USA/Minnesota/ | Minnesota Department of Health, Public Health Laboratory | Minnesota Department of Health, Public Health Laboratory | Matt Plumb et al |

| | | | | | | |
|---|---|---|---|---|---|---|
| EPI_ISL_476795 | Singapore/469/2020 | 2020-02-19 | Asia/Singapore/Singapore/ | Department of Laboratory Medicine, Tan Tock Seng Hospital | Department of Laboratory Medicine, Tan Tock Seng Hospital | Chen YYC et al |
| EPI_ISL_476832 | Turkey/KOU-MG/2020 | 2020-04-27 | Europe/Turkey/Kocaeli/ | Medical Biology Department, Kocaeli University | Medical Genetics Department, Kocaeli University | Savli H et al |
| EPI_ISL_476837 | Greece/AUTH_6/2020 | 2020-04-15 | Europe/Greece/Thessaloniki/ | National Influenza Centre for Northern Greece | National Influenza Centre for Northern Greece | Maria Christoforidi et al |
| EPI_ISL_476842 | India/MP-DRDE4764/2020 | 2020-05-28 | Asia/India/Madhya Pradesh/ | Defence Research & Development Establishment (DRDE) | Defence Research & Development Establishment (DRDE) | Shashi Sharma et al |
| EPI_ISL_476850 | India/MP-DRDE2429/2020 | 2020-05-03 | Asia/India/Madhya Pradesh/ | Defence Research & Development Establishment (DRDE) | Defence Research & Development Establishment (DRDE) | Shashi Sharma et al |
| EPI_ISL_476860 | India/GJ-GBRC224b/2020 | 2020-06-08 | Asia/India/Gujarat/Vadodara | GMERS Medical College & Hospital, Gotri, Vadodara | Gujarat Biotechnology Research Centre | Maharshi Pandya et al |
| EPI_ISL_476867 | India/GJ-GBRC229a/2020 | 2020-06-12 | Asia/India/Gujarat/Palanpur | Banas Medical College and Research Institute | Gujarat Biotechnology Research Centre | Labdhi Pandya et al |
| EPI_ISL_476868 | India/GJ-GBRC229b/2020 | 2020-06-12 | Asia/India/Gujarat/Palanpur | Banas Medical College and Research Institute | Gujarat Biotechnology Research Centre | Afzal Ansari et al |
| EPI_ISL_476871 | India/GJ-GBRC231b/2020 | 2020-06-11 | Asia/India/Gujarat/Surat | Department of MicroBiology, Government Medical College, Surat | Gujarat Biotechnology Research Centre | Summaiya Mullan et al |
| EPI_ISL_476872 | India/GJ-GBRC232a/2020 | 2020-06-11 | Asia/India/Gujarat/Surat | Department of MicroBiology, Government Medical College, Surat | Gujarat Biotechnology Research Centre | Amit gamit et al |
| EPI_ISL_476873 | India/GJ-GBRC232b/2020 | 2020-06-11 | Asia/India/Gujarat/Surat | Department of MicroBiology, Government Medical College, Surat | Gujarat Biotechnology Research Centre | Apurvasinh Puvar et al |
| EPI_ISL_476875 | India/GJ-GBRC233b/2020 | 2020-06-12 | Asia/India/Gujarat/Surat | Department of MicroBiology, Government Medical College, Surat | Gujarat Biotechnology Research Centre | Zarna Patel et al |

| | | | | | | |
|---|---|---|---|---|---|---|
| EPI_ISL_476876 | India/GJ-GBRC235a/2020 | 2020-06-12 | Asia/India/Gujarat/Surat | Department of MicroBiology, Government Medical College, Surat | Gujarat Biotechnology Research Centre | Pinal Trivedi et al |
| EPI_ISL_476878 | India/GJ-GBRC236/2020 | 2020-06-12 | Asia/India/Gujarat/Surat | Department of MicroBiology, Government Medical College, Surat | Gujarat Biotechnology Research Centre | Nidhi Patel et al |
| EPI_ISL_476880 | India/GJ-GBRC237b/2020 | 2020-06-12 | Asia/India/Gujarat/Bardoli | Department of MicroBiology, Government Medical College, Surat | Gujarat Biotechnology Research Centre | Raghawendra Kumar et al |
| EPI_ISL_476881 | India/GJ-GBRC238a/2020 | 2020-06-12 | Asia/India/Gujarat/Choryasi | Department of MicroBiology, Government Medical College, Surat | Gujarat Biotechnology Research Centre | Dinesh Kumar et al |
| EPI_ISL_476887 | India/MP-DRDE-4093/2020 | 2020-05-17 | Asia/India/Madhya Pradesh/ | Defence Research & Development Establishment (DRDE) | Defence Research & Development Establishment (DRDE) | Shashi Sharma et al |
| EPI_ISL_476889 | India/MP-DRDE-3987/2020 | 2020-05-16 | Asia/India/Madhya Pradesh/ | Defence Research & Development Establishment (DRDE) | Defence Research & Development Establishment (DRDE) | Shashi Sharma et al |
| EPI_ISL_476892 | India/MP-DRDE-4105/2020 | 2020-05-17 | Asia/India/Madhya Pradesh/ | Defence Research & Development Establishment (DRDE) | Defence Research & Development Establishment (DRDE) | Shashi Sharma et al |
| EPI_ISL_476905 | USA/WA-UW-1274/2020 | 2020-03-17 | North America/USA/Washington/ | UW Virology Lab | UW Virology Lab | Pavitra Roychoudhury et al |
| EPI_ISL_476906 | USA/WA-UW-571/2020 | 2020-03-12 | North America/USA/Washington/ | UW Virology Lab | UW Virology Lab | Pavitra Roychoudhury et al |
| EPI_ISL_476913 | USA/CT-UW-647/2020 | 2020-03-13 | North America/USA/Connecticut/ | UW Virology Lab | UW Virology Lab | Pavitra Roychoudhury et al |
| EPI_ISL_476920 | USA/WA-UW-661/2020 | 2020-03-12 | North America/USA/Washington/ | UW Virology Lab | UW Virology Lab | Pavitra Roychoudhury et al |
| EPI_ISL_476948 | Belgium/rega-0409439/2020 | 2020-04-09 | Europe/Belgium/Leuven/Tienen | KU Leuven, Rega Institute, Clinical and Epidemiological Virology | KU Leuven, Rega Institute, Clinical and Epidemiological Virology | Tony Wawina-Bokalanga et al |
| EPI_ISL_476955 | Belgium/rega-0411445/2020 | 2020-04-11 | Europe/Belgium/Leuven/Tienen | KU Leuven, Rega Institute, Clinical and Epidemiological Virology | KU Leuven, Rega Institute, Clinical and Epidemiological Virology | Tony Wawina-Bokalanga et al |

| | | | | | | |
|---|---|---|---|---|---|---|
| EPI_ISL_476956 | Belgium/rega-0411446/2020 | 2020-04-11 | Europe/Belgium/Tournai-Mouscron/Froy | KU Leuven, Rega Institute, Clinical and Epidemiological Virology | KU Leuven, Rega Institute, Clinical and Epidemiological Virology | Tony Wawina-Bokalanga et al |
| EPI_ISL_476957 | Belgium/rega-0411447/2020 | 2020-04-11 | Europe/Belgium/Leuven/Haasrode | KU Leuven, Rega Institute, Clinical and Epidemiological Virology | KU Leuven, Rega Institute, Clinical and Epidemiological Virology | Tony Wawina-Bokalanga et al |
| EPI_ISL_476960 | Belgium/rega-0411450/2020 | 2020-04-11 | Europe/Belgium/Ath/ | KU Leuven, Rega Institute, Clinical and Epidemiological Virology | KU Leuven, Rega Institute, Clinical and Epidemiological Virology | Tony Wawina-Bokalanga et al |
| EPI_ISL_476962 | Belgium/rega-0411452/2020 | 2020-04-11 | Europe/Belgium/Leuven/Scherpenheuve | KU Leuven, Rega Institute, Clinical and Epidemiological Virology | KU Leuven, Rega Institute, Clinical and Epidemiological Virology | Tony Wawina-Bokalanga et al |
| EPI_ISL_476970 | Belgium/rega-0411460/2020 | 2020-04-11 | Europe/Belgium/Leuven/Diest | KU Leuven, Rega Institute, Clinical and Epidemiological Virology | KU Leuven, Rega Institute, Clinical and Epidemiological Virology | Tony Wawina-Bokalanga et al |
| EPI_ISL_476976 | Belgium/rega-0411466/2020 | 2020-04-11 | Europe/Belgium/La Louvi√®re/Maurage | KU Leuven, Rega Institute, Clinical and Epidemiological Virology | KU Leuven, Rega Institute, Clinical and Epidemiological Virology | Tony Wawina-Bokalanga et al |
| EPI_ISL_476992 | Belgium/rega-0427424/2020 | 2020-04-27 | Europe/Belgium/Leuven/Kersbeek-Misk | KU Leuven, Rega Institute, Clinical and Epidemiological Virology | KU Leuven, Rega Institute, Clinical and Epidemiological Virology | Tony Wawina-Bokalanga et al |
| EPI_ISL_476995 | Belgium/rega-0429426/2020 | 2020-04-29 | Europe/Belgium/Tournai-Mouscron/Luin | KU Leuven, Rega Institute, Clinical and Epidemiological Virology | KU Leuven, Rega Institute, Clinical and Epidemiological Virology | Tony Wawina-Bokalanga et al |

| | | | | | | |
|---|---|---|---|---|---|---|
| EPI_ISL_477004 | Belgium/rega-0506487/2020 | 2020-05-06 | Europe/Belgium/Brussels/Laken | KU Leuven, Rega Institute, Clinical and Epidemiological Virology | KU Leuven, Rega Institute, Clinical and Epidemiological Virology | Tony Wawina-Bokalanga et al |
| EPI_ISL_477012 | Hungary/UD-7364/2020 | 2020-04-08 | Europe/Hungary/Borsod-Aba√∫j-Zempl√ | University of Debrecen, Department of Medical Microbiology | National Laboratory of Virology, Szent√°gothai Research Centre | Endre G√°bor T√≥th et al |
| EPI_ISL_477015 | Ecuador/USFQ-096/2020 | 2020-06-17 | South America/Ecuador/Pichincha/Quito | Institute of Microbiology, Universidad San Francisco de Quito | Institute of Microbiology, Universidad San Francisco de Quito | Sully M√°rquez et al |
| EPI_ISL_477016 | Ecuador/USFQ-097/2020 | 2020-06-17 | South America/Ecuador/Pichincha/Quito | Institute of Microbiology, Universidad San Francisco de Quito | Institute of Microbiology, Universidad San Francisco de Quito | Juan Jos√© Guadalupe et al |
| EPI_ISL_477017 | Canada/BC_19829412/2020 | 20-Apr | North America/Canada/British Columbia | BCCDC Public Health Laboratory | BCCDC Public Health Laboratory | Richard Harrigan et al |
| EPI_ISL_477019 | Canada/BC_19860669/2020 | 20-Apr | North America/Canada/British Columbia | BCCDC Public Health Laboratory | BCCDC Public Health Laboratory | Richard Harrigan et al |
| EPI_ISL_477020 | Canada/BC_19909443/2020 | 20-Apr | North America/Canada/British Columbia | BCCDC Public Health Laboratory | BCCDC Public Health Laboratory | Richard Harrigan et al |
| EPI_ISL_477021 | Canada/BC_19909482/2020 | 20-Apr | North America/Canada/British Columbia | BCCDC Public Health Laboratory | BCCDC Public Health Laboratory | Richard Harrigan et al |
| EPI_ISL_477022 | Canada/BC_20367856/2020 | 20-Apr | North America/Canada/British Columbia | BCCDC Public Health Laboratory | BCCDC Public Health Laboratory | Richard Harrigan et al |
| EPI_ISL_477023 | Canada/BC_21355315/2020 | 20-Apr | North America/Canada/British Columbia | BCCDC Public Health Laboratory | BCCDC Public Health Laboratory | Richard Harrigan et al |
| EPI_ISL_477024 | Canada/BC_21358214/2020 | 20-Apr | North America/Canada/British Columbia | BCCDC Public Health Laboratory | BCCDC Public Health Laboratory | Richard Harrigan et al |
| EPI_ISL_477025 | Canada/BC_21426895/2020 | 20-Apr | North America/Canada/British Columbia | BCCDC Public Health Laboratory | BCCDC Public Health Laboratory | Richard Harrigan et al |
| EPI_ISL_477028 | Canada/BC_21449836/2020 | 20-Apr | North America/Canada/British Columbia | BCCDC Public Health Laboratory | BCCDC Public Health Laboratory | Richard Harrigan et al |
| EPI_ISL_477029 | Canada/BC_21451231/2020 | 20-Apr | North America/Canada/British Columbia | BCCDC Public Health Laboratory | BCCDC Public Health Laboratory | Richard Harrigan et al |
| EPI_ISL_477030 | Canada/BC_21960731/2020 | 20-Apr | North America/Canada/British Columbia | BCCDC Public Health Laboratory | BCCDC Public Health Laboratory | Richard Harrigan et al |

| | | | | | | |
|---|---|---|---|---|---|---|
| EPI_ISL_477031 | Canada/BC_22422949/2020 | 20-Apr | North America/Canada/British Columbia | BCCDC Public Health Laboratory | BCCDC Public Health Laboratory | Richard Harrigan et al |
| EPI_ISL_477032 | Canada/BC_22502164/2020 | 20-Apr | North America/Canada/British Columbia | BCCDC Public Health Laboratory | BCCDC Public Health Laboratory | Richard Harrigan et al |
| EPI_ISL_477033 | Canada/BC_22510360/2020 | 20-Apr | North America/Canada/British Columbia | BCCDC Public Health Laboratory | BCCDC Public Health Laboratory | Richard Harrigan et al |
| EPI_ISL_477034 | Canada/BC_22611474/2020 | 20-Apr | North America/Canada/British Columbia | BCCDC Public Health Laboratory | BCCDC Public Health Laboratory | Richard Harrigan et al |
| EPI_ISL_477035 | Canada/BC_23246810/2020 | 20-Apr | North America/Canada/British Columbia | BCCDC Public Health Laboratory | BCCDC Public Health Laboratory | Richard Harrigan et al |
| EPI_ISL_477036 | Canada/BC_23252816/2020 | 20-Apr | North America/Canada/British Columbia | BCCDC Public Health Laboratory | BCCDC Public Health Laboratory | Richard Harrigan et al |
| EPI_ISL_477037 | Canada/BC_23279341/2020 | 20-Apr | North America/Canada/British Columbia | BCCDC Public Health Laboratory | BCCDC Public Health Laboratory | Richard Harrigan et al |
| EPI_ISL_477038 | Canada/BC_26060645/2020 | 20-Apr | North America/Canada/British Columbia | BCCDC Public Health Laboratory | BCCDC Public Health Laboratory | Richard Harrigan et al |
| EPI_ISL_477039 | Canada/BC_00740314/2020 | 20-May | North America/Canada/British Columbia | BCCDC Public Health Laboratory | BCCDC Public Health Laboratory | Richard Harrigan et al |
| EPI_ISL_477040 | Canada/BC_01775252/2020 | 20-May | North America/Canada/British Columbia | BCCDC Public Health Laboratory | BCCDC Public Health Laboratory | Richard Harrigan et al |
| EPI_ISL_477041 | Canada/BC_02281789/2020 | 20-May | North America/Canada/British Columbia | BCCDC Public Health Laboratory | BCCDC Public Health Laboratory | Richard Harrigan et al |
| EPI_ISL_477043 | Canada/BC_04483654/2020 | 20-May | North America/Canada/British Columbia | BCCDC Public Health Laboratory | BCCDC Public Health Laboratory | Richard Harrigan et al |
| EPI_ISL_477044 | Canada/BC_04528452/2020 | 20-May | North America/Canada/British Columbia | BCCDC Public Health Laboratory | BCCDC Public Health Laboratory | Richard Harrigan et al |
| EPI_ISL_477045 | Canada/BC_04547356/2020 | 20-May | North America/Canada/British Columbia | BCCDC Public Health Laboratory | BCCDC Public Health Laboratory | Richard Harrigan et al |
| EPI_ISL_477046 | Canada/BC_04579295/2020 | 20-May | North America/Canada/British Columbia | BCCDC Public Health Laboratory | BCCDC Public Health Laboratory | Richard Harrigan et al |
| EPI_ISL_477047 | Canada/BC_04583818/2020 | 20-May | North America/Canada/British Columbia | BCCDC Public Health Laboratory | BCCDC Public Health Laboratory | Richard Harrigan et al |
| EPI_ISL_477048 | Canada/BC_04677077/2020 | 20-May | North America/Canada/British Columbia | BCCDC Public Health Laboratory | BCCDC Public Health Laboratory | Richard Harrigan et al |
| EPI_ISL_477049 | Canada/BC_04684452/2020 | 20-May | North America/Canada/British Columbia | BCCDC Public Health Laboratory | BCCDC Public Health Laboratory | Richard Harrigan et al |
| EPI_ISL_477050 | Canada/BC_04701167/2020 | 20-May | North America/Canada/British Columbia | BCCDC Public Health Laboratory | BCCDC Public Health Laboratory | Richard Harrigan et al |
| EPI_ISL_477051 | Canada/BC_04709878/2020 | 20-May | North America/Canada/British Columbia | BCCDC Public Health Laboratory | BCCDC Public Health Laboratory | Richard Harrigan et al |

| | | | | | | |
|---|---|---|---|---|---|---|
| EPI_ISL_477052 | Canada/BC_04710049/2020 | 20-May | North America/Canada/British Columbia | BCCDC Public Health Laboratory | BCCDC Public Health Laboratory | Richard Harrigan et al |
| EPI_ISL_477053 | Canada/BC_04803768/2020 | 20-May | North America/Canada/British Columbia | BCCDC Public Health Laboratory | BCCDC Public Health Laboratory | Richard Harrigan et al |
| EPI_ISL_477054 | Canada/BC_04806249/2020 | 20-May | North America/Canada/British Columbia | BCCDC Public Health Laboratory | BCCDC Public Health Laboratory | Richard Harrigan et al |
| EPI_ISL_477056 | Canada/BC_04806819/2020 | 20-May | North America/Canada/British Columbia | BCCDC Public Health Laboratory | BCCDC Public Health Laboratory | Richard Harrigan et al |
| EPI_ISL_477057 | Canada/BC_04865055/2020 | 20-May | North America/Canada/British Columbia | BCCDC Public Health Laboratory | BCCDC Public Health Laboratory | Richard Harrigan et al |
| EPI_ISL_477058 | Canada/BC_06074915/2020 | 20-May | North America/Canada/British Columbia | BCCDC Public Health Laboratory | BCCDC Public Health Laboratory | Richard Harrigan et al |
| EPI_ISL_477059 | Canada/BC_06514625/2020 | 20-May | North America/Canada/British Columbia | BCCDC Public Health Laboratory | BCCDC Public Health Laboratory | Richard Harrigan et al |
| EPI_ISL_477060 | Canada/BC_07353444/2020 | 20-May | North America/Canada/British Columbia | BCCDC Public Health Laboratory | BCCDC Public Health Laboratory | Richard Harrigan et al |
| EPI_ISL_477061 | Canada/BC_07653696/2020 | 20-May | North America/Canada/British Columbia | BCCDC Public Health Laboratory | BCCDC Public Health Laboratory | Richard Harrigan et al |
| EPI_ISL_477062 | Canada/BC_08164457/2020 | 20-May | North America/Canada/British Columbia | BCCDC Public Health Laboratory | BCCDC Public Health Laboratory | Richard Harrigan et al |
| EPI_ISL_477063 | Canada/BC_08312782/2020 | 20-May | North America/Canada/British Columbia | BCCDC Public Health Laboratory | BCCDC Public Health Laboratory | Richard Harrigan et al |
| EPI_ISL_477064 | Canada/BC_08534834/2020 | 20-Apr | North America/Canada/British Columbia | BCCDC Public Health Laboratory | BCCDC Public Health Laboratory | Richard Harrigan et al |
| EPI_ISL_477065 | Canada/BC_08855516/2020 | 20-May | North America/Canada/British Columbia | BCCDC Public Health Laboratory | BCCDC Public Health Laboratory | Richard Harrigan et al |
| EPI_ISL_477066 | Canada/BC_08859333/2020 | 20-May | North America/Canada/British Columbia | BCCDC Public Health Laboratory | BCCDC Public Health Laboratory | Richard Harrigan et al |
| EPI_ISL_477067 | Canada/BC_08865483/2020 | 20-May | North America/Canada/British Columbia | BCCDC Public Health Laboratory | BCCDC Public Health Laboratory | Richard Harrigan et al |
| EPI_ISL_477068 | Canada/BC_08984631/2020 | 20-May | North America/Canada/British Columbia | BCCDC Public Health Laboratory | BCCDC Public Health Laboratory | Richard Harrigan et al |
| EPI_ISL_477069 | Canada/BC_08987762/2020 | 20-May | North America/Canada/British Columbia | BCCDC Public Health Laboratory | BCCDC Public Health Laboratory | Richard Harrigan et al |
| EPI_ISL_477070 | Canada/BC_09273580/2020 | 20-May | North America/Canada/British Columbia | BCCDC Public Health Laboratory | BCCDC Public Health Laboratory | Richard Harrigan et al |
| EPI_ISL_477071 | Canada/BC_09625012/2020 | 20-May | North America/Canada/British Columbia | BCCDC Public Health Laboratory | BCCDC Public Health Laboratory | Richard Harrigan et al |
| EPI_ISL_477072 | Canada/BC_09819716/2020 | 20-May | North America/Canada/British Columbia | BCCDC Public Health Laboratory | BCCDC Public Health Laboratory | Richard Harrigan et al |

| | | | | | | |
|---|---|---|---|---|---|---|
| EPI_ISL_477073 | Canada/BC_09963377/2020 | 20-May | North America/Canada/British Columbia | BCCDC Public Health Laboratory | BCCDC Public Health Laboratory | Richard Harrigan et al |
| EPI_ISL_477074 | Canada/BC_09965869/2020 | 20-May | North America/Canada/British Columbia | BCCDC Public Health Laboratory | BCCDC Public Health Laboratory | Richard Harrigan et al |
| EPI_ISL_477075 | Canada/BC_09986014/2020 | 20-May | North America/Canada/British Columbia | BCCDC Public Health Laboratory | BCCDC Public Health Laboratory | Richard Harrigan et al |
| EPI_ISL_477076 | Canada/BC_19879538/2020 | 20-Apr | North America/Canada/British Columbia | BCCDC Public Health Laboratory | BCCDC Public Health Laboratory | Richard Harrigan et al |
| EPI_ISL_477078 | Canada/BC_10404289/2020 | 20-Mar | North America/Canada/British Columbia | BCCDC Public Health Laboratory | BCCDC Public Health Laboratory | Richard Harrigan et al |
| EPI_ISL_477079 | Canada/BC_10404688/2020 | 20-Mar | North America/Canada/British Columbia | BCCDC Public Health Laboratory | BCCDC Public Health Laboratory | Richard Harrigan et al |
| EPI_ISL_477080 | Canada/BC_10420892/2020 | 20-Mar | North America/Canada/British Columbia | BCCDC Public Health Laboratory | BCCDC Public Health Laboratory | Richard Harrigan et al |
| EPI_ISL_477081 | Canada/BC_13929277/2020 | 20-Mar | North America/Canada/British Columbia | BCCDC Public Health Laboratory | BCCDC Public Health Laboratory | Richard Harrigan et al |
| EPI_ISL_477082 | Canada/BC_13929332/2020 | 20-Mar | North America/Canada/British Columbia | BCCDC Public Health Laboratory | BCCDC Public Health Laboratory | Richard Harrigan et al |
| EPI_ISL_477083 | Canada/BC_16492879/2020 | 20-Mar | North America/Canada/British Columbia | BCCDC Public Health Laboratory | BCCDC Public Health Laboratory | Richard Harrigan et al |
| EPI_ISL_477084 | Canada/BC_16502858/2020 | 20-Mar | North America/Canada/British Columbia | BCCDC Public Health Laboratory | BCCDC Public Health Laboratory | Richard Harrigan et al |
| EPI_ISL_477085 | Canada/BC_16505592/2020 | 20-Mar | North America/Canada/British Columbia | BCCDC Public Health Laboratory | BCCDC Public Health Laboratory | Richard Harrigan et al |
| EPI_ISL_477086 | Canada/BC_16510082/2020 | 20-Mar | North America/Canada/British Columbia | BCCDC Public Health Laboratory | BCCDC Public Health Laboratory | Richard Harrigan et al |
| EPI_ISL_477087 | Canada/BC_02018462/2020 | 20-Mar | North America/Canada/British Columbia | BCCDC Public Health Laboratory | BCCDC Public Health Laboratory | Richard Harrigan et al |
| EPI_ISL_477088 | Canada/BC_02038211/2020 | 20-Mar | North America/Canada/British Columbia | BCCDC Public Health Laboratory | BCCDC Public Health Laboratory | Richard Harrigan et al |
| EPI_ISL_477089 | Canada/BC_02349958/2020 | 20-Mar | North America/Canada/British Columbia | BCCDC Public Health Laboratory | BCCDC Public Health Laboratory | Richard Harrigan et al |
| EPI_ISL_477090 | Canada/BC_03990815/2020 | 20-Apr | North America/Canada/British Columbia | BCCDC Public Health Laboratory | BCCDC Public Health Laboratory | Richard Harrigan et al |
| EPI_ISL_477091 | Canada/BC_03998412/2020 | 20-Apr | North America/Canada/British Columbia | BCCDC Public Health Laboratory | BCCDC Public Health Laboratory | Richard Harrigan et al |
| EPI_ISL_477092 | Canada/BC_04000917/2020 | 20-Apr | North America/Canada/British Columbia | BCCDC Public Health Laboratory | BCCDC Public Health Laboratory | Richard Harrigan et al |
| EPI_ISL_477093 | Canada/BC_07905258/2020 | 20-Apr | North America/Canada/British Columbia | BCCDC Public Health Laboratory | BCCDC Public Health Laboratory | Richard Harrigan et al |

| | | | | | | |
|---|---|---|---|---|---|---|
| EPI_ISL_477094 | Canada/BC_07905790/2020 | 20-Apr | North America/Canada/British Columbia | BCCDC Public Health Laboratory | BCCDC Public Health Laboratory | Richard Harrigan et al |
| EPI_ISL_477095 | Canada/BC_07911261/2020 | 20-Apr | North America/Canada/British Columbia | BCCDC Public Health Laboratory | BCCDC Public Health Laboratory | Richard Harrigan et al |
| EPI_ISL_477096 | Canada/BC_07912118/2020 | 20-Apr | North America/Canada/British Columbia | BCCDC Public Health Laboratory | BCCDC Public Health Laboratory | Richard Harrigan et al |
| EPI_ISL_477097 | Canada/BC_07912722/2020 | 20-Apr | North America/Canada/British Columbia | BCCDC Public Health Laboratory | BCCDC Public Health Laboratory | Richard Harrigan et al |
| EPI_ISL_477098 | Canada/BC_07916655/2020 | 20-Apr | North America/Canada/British Columbia | BCCDC Public Health Laboratory | BCCDC Public Health Laboratory | Richard Harrigan et al |
| EPI_ISL_477099 | Canada/BC_07917243/2020 | 20-Apr | North America/Canada/British Columbia | BCCDC Public Health Laboratory | BCCDC Public Health Laboratory | Richard Harrigan et al |
| EPI_ISL_477100 | Canada/BC_07917795/2020 | 20-Apr | North America/Canada/British Columbia | BCCDC Public Health Laboratory | BCCDC Public Health Laboratory | Richard Harrigan et al |
| EPI_ISL_477101 | Canada/BC_07920084/2020 | 20-Apr | North America/Canada/British Columbia | BCCDC Public Health Laboratory | BCCDC Public Health Laboratory | Richard Harrigan et al |
| EPI_ISL_477102 | Canada/BC_07924910/2020 | 20-Apr | North America/Canada/British Columbia | BCCDC Public Health Laboratory | BCCDC Public Health Laboratory | Richard Harrigan et al |
| EPI_ISL_477103 | Canada/BC_07925997/2020 | 20-Apr | North America/Canada/British Columbia | BCCDC Public Health Laboratory | BCCDC Public Health Laboratory | Richard Harrigan et al |
| EPI_ISL_477104 | Canada/BC_07926740/2020 | 20-Apr | North America/Canada/British Columbia | BCCDC Public Health Laboratory | BCCDC Public Health Laboratory | Richard Harrigan et al |
| EPI_ISL_477105 | Canada/BC_09009433/2020 | 20-Apr | North America/Canada/British Columbia | BCCDC Public Health Laboratory | BCCDC Public Health Laboratory | Richard Harrigan et al |
| EPI_ISL_477106 | Canada/BC_09014362/2020 | 20-Apr | North America/Canada/British Columbia | BCCDC Public Health Laboratory | BCCDC Public Health Laboratory | Richard Harrigan et al |
| EPI_ISL_477107 | Canada/BC_09021025/2020 | 20-Apr | North America/Canada/British Columbia | BCCDC Public Health Laboratory | BCCDC Public Health Laboratory | Richard Harrigan et al |
| EPI_ISL_477108 | Canada/BC_09023420/2020 | 20-Apr | North America/Canada/British Columbia | BCCDC Public Health Laboratory | BCCDC Public Health Laboratory | Richard Harrigan et al |
| EPI_ISL_477109 | Canada/BC_09025513/2020 | 20-Apr | North America/Canada/British Columbia | BCCDC Public Health Laboratory | BCCDC Public Health Laboratory | Richard Harrigan et al |
| EPI_ISL_477110 | Canada/BC_09026743/2020 | 20-Apr | North America/Canada/British Columbia | BCCDC Public Health Laboratory | BCCDC Public Health Laboratory | Richard Harrigan et al |
| EPI_ISL_477111 | Canada/BC_09029034/2020 | 20-Apr | North America/Canada/British Columbia | BCCDC Public Health Laboratory | BCCDC Public Health Laboratory | Richard Harrigan et al |
| EPI_ISL_477112 | Canada/BC_09035190/2020 | 20-Apr | North America/Canada/British Columbia | BCCDC Public Health Laboratory | BCCDC Public Health Laboratory | Richard Harrigan et al |
| EPI_ISL_477113 | Canada/BC_09040534/2020 | 20-Apr | North America/Canada/British Columbia | BCCDC Public Health Laboratory | BCCDC Public Health Laboratory | Richard Harrigan et al |

| | | | | | | |
|---|---|---|---|---|---|---|
| EPI_ISL_477114 | Canada/BC_10600259/2020 | 20-Apr | North America/Canada/British Columbia | BCCDC Public Health Laboratory | BCCDC Public Health Laboratory | Richard Harrigan et al |
| EPI_ISL_477115 | Canada/BC_10600634/2020 | 20-Apr | North America/Canada/British Columbia | BCCDC Public Health Laboratory | BCCDC Public Health Laboratory | Richard Harrigan et al |
| EPI_ISL_477116 | Canada/BC_10601268/2020 | 20-Apr | North America/Canada/British Columbia | BCCDC Public Health Laboratory | BCCDC Public Health Laboratory | Richard Harrigan et al |
| EPI_ISL_477117 | Canada/BC_10601737/2020 | 20-Apr | North America/Canada/British Columbia | BCCDC Public Health Laboratory | BCCDC Public Health Laboratory | Richard Harrigan et al |
| EPI_ISL_477118 | Canada/BC_10603886/2020 | 20-Apr | North America/Canada/British Columbia | BCCDC Public Health Laboratory | BCCDC Public Health Laboratory | Richard Harrigan et al |
| EPI_ISL_477119 | Canada/BC_10616731/2020 | 20-Apr | North America/Canada/British Columbia | BCCDC Public Health Laboratory | BCCDC Public Health Laboratory | Richard Harrigan et al |
| EPI_ISL_477125 | Bangladesh/CHRF-0012/2020 | 2020-05-05 | Asia/Bangladesh/Bangladesh/ | Child Health Research Foundation | Child Health Research Foundation | Senjuti Saha et al |
| EPI_ISL_477126 | Bangladesh/CHRF-0013/2020 | 2020-05-09 | Asia/Bangladesh/Bangladesh/ | Child Health Research Foundation | Child Health Research Foundation | Senjuti Saha et al |
| EPI_ISL_477128 | Bangladesh/CHRF-0015/2020 | 2020-05-02 | Asia/Bangladesh/Bangladesh/ | Child Health Research Foundation | Child Health Research Foundation | Senjuti Saha et al |
| EPI_ISL_477130 | Bangladesh/CHRF-0017/2020 | 2020-05-30 | Asia/Bangladesh/Bangladesh/ | Child Health Research Foundation | Child Health Research Foundation | Senjuti Saha et al |
| EPI_ISL_477131 | Bangladesh/CHRF-0020/2020 | 2020-05-30 | Asia/Bangladesh/Bangladesh/ | Child Health Research Foundation | Child Health Research Foundation | Senjuti Saha et al |
| EPI_ISL_477132 | Bangladesh/CHRF-0022/2020 | 2020-06-14 | Asia/Bangladesh/Bangladesh/ | Child Health Research Foundation | Child Health Research Foundation | Senjuti Saha et al |
| EPI_ISL_477135 | Bangladesh/CHRF-0025/2020 | 2020-06-14 | Asia/Bangladesh/Bangladesh/ | Child Health Research Foundation | Child Health Research Foundation | Senjuti Saha et al |
| EPI_ISL_477137 | Bangladesh/CHRF-0028/2020 | 2020-06-18 | Asia/Bangladesh/Bangladesh/ | Child Health Research Foundation | Child Health Research Foundation | Senjuti Saha et al |
| EPI_ISL_477139 | Bangladesh/CHRF-0030/2020 | 2020-06-11 | Asia/Bangladesh/Bangladesh/ | Child Health Research Foundation | Child Health Research Foundation | Senjuti Saha et al |

| | | | | | | |
|---|---|---|---|---|---|---|
| EPI_ISL_477140 | Bangladesh/CHRF-0031/2020 | 2020-06-16 | Asia/Bangladesh/Bangladesh/ | Child Health Research Foundation | Child Health Research Foundation | Senjuti Saha et al |
| EPI_ISL_477161 | Egypt/CUNCI-HGC6I029/2020 | 2020-06-02 | Africa/Egypt/Egypt/ | Egyptian National Cancer Institute (ENCI) | Egyptian National Cancer Institute (ENCI) | Zekri et al |
| EPI_ISL_477169 | Georgia/Tb-7851/2020 | 2020-05-01 | Asia/Georgia/Tbilisi/ | Department for Virology, Molecular Biology and Genome Research, R. G. Lugar Center for Public Health Research, National Center for Disease Control and Public Health (NCDC) of Georgia. | Department for Virology, Molecular Biology and Genome Research, R. G. Lugar Center for Public Health Research, National Center for Disease Control and Public Health (NCDC) of Georgia. | Tata Imnadze et al |
| EPI_ISL_477170 | Singapore/470/2020 | 2020-02-16 | Asia/Singapore/Singapore/ | Department of Laboratory, Medicine Tan Tock Seng Hospital | Department of Laboratory Medicine Tan Tock Seng Hospital | Chen YYC et al |
| EPI_ISL_477182 | Singapore/488/2020 | 2020-04-13 | Asia/Singapore/Singapore/ | Department of Laboratory Medicine Tan Tock Seng Hospital | Department of Laboratory Medicine Tan Tock Seng Hospital | Chen YYC et al |
| EPI_ISL_477183 | India/GJ-GBRC234/2020 | 2020-06-12 | Asia/India/Gujarat/Choryasi | Department of MicroBiology, Government Medical College, Surat | Gujarat Biotechnology Research Centre | Monika Gandhi et al |
| EPI_ISL_477194 | Italy/APU-UniMI-39/2020 | 2020-03-16 | Europe/Italy/Apulia/ | Istituto Zooprofilattico Sperimentale Puglia e Basilicata; | Beaconlab (Bioinformatics, Evolution and Comparative Genomics lab), Dept of Biosciences, University on Mila | Parisi A. et al |

| | | | | | | |
|---|---|---|---|---|---|---|
| EPI_ISL_477196 | Italy/BAS-UniMI-4028/2020 | 2020-04-06 | Europe/Italy/Basilicata/ | Istituto Zooprofilattico Sperimentale Puglia e Basilicata; | Beaconlab (Bioinformatics, Evolution and Comparative Genomics lab), Dept of Biosciences, University on Milan | Parisi A. et al |
| EPI_ISL_477197 | Italy/BAS-UniMI-3725/2020 | 2020-04-03 | Europe/Italy/Basilicata/ | Istituto Zooprofilattico Sperimentale Puglia e Basilicata; | Beaconlab (Bioinformatics, Evolution and Comparative Genomics lab), Dept of Biosciences, University on Milan | Parisi A. et al |
| EPI_ISL_477199 | Italy/BAS-UniMI-2167/2020 | 2020-03-23 | Europe/Italy/Basilicata/ | Istituto Zooprofilattico Sperimentale Puglia e Basilicata; | Beaconlab (Bioinformatics, Evolution and Comparative Genomics lab), Dept of Biosciences, University on Milan | Parisi A. et al |
| EPI_ISL_477202 | Italy/APU-UniMI-854/2020 | 2020-03-20 | Europe/Italy/Apulia/ | Istituto Zooprofilattico Sperimentale Puglia e Basilicata; | Beaconlab (Bioinformatics, Evolution and Comparative Genomics lab), Dept of Biosciences, University on Mila | Parisi A. et al |

| | | | | | | |
|---|---|---|---|---|---|---|
| EPI_ISL_477203 | Italy/APU-UniMI-694/2020 | 2020-03-19 | Europe/Italy/Apulia/ | Istituto Zooprofilattico Sperimentale Puglia e Basilicata; | Beaconlab (Bioinformatics, Evolution and Comparative Genomics lab), Dept of Biosciences, University on Mila | Parisi A. et al |
| EPI_ISL_477204 | Italy/CAM-CEINGE-1/2020 | 2020-05-30 | Europe/Italy/Campania/Naples | Prof. Massimo Zollo CEINGE TASK-FORCE COVID19 - Regione Campania | Prof. Massimo Zollo CEINGE TASK-FORCE COVID19 - Regione Campania | Veronica Ferrucci1 et al |
| EPI_ISL_477209 | India/KA-InStem-NCBS-0006/2020 | 2020-06-01 | Asia/India/Karnataka/ | Institute for Stem Cell Science and Regenerative Medicine | National Centre for Biological Sciences | Farhan Ali et al |
| EPI_ISL_477212 | India/KA-InStem-NCBS-0009/2020 | 2020-05-27 | Asia/India/Karnataka/ | Institute for Stem Cell Science and Regenerative Medicine | National Centre for Biological Sciences | Farhan Ali et al |
| EPI_ISL_477213 | India/KA-InStem-NCBS-0010/2020 | 2020-05-27 | Asia/India/Karnataka/ | Institute for Stem Cell Science and Regenerative Medicine | National Centre for Biological Sciences | Farhan Ali et al |
| EPI_ISL_477222 | India/KA-InStem-NCBS-0020/2020 | 2020-06-11 | Asia/India/Karnataka/ | Institute for Stem Cell Science and Regenerative Medicine | National Centre for Biological Sciences | Farhan Ali et al |
| EPI_ISL_477224 | India/KA-InStem-NCBS-0022/2020 | 2020-06-11 | Asia/India/Karnataka/ | Institute for Stem Cell Science and Regenerative Medicine | National Centre for Biological Sciences | Farhan Ali et al |
| EPI_ISL_477229 | India/KA-InStem-NCBS-0028/2020 | 2020-06-15 | Asia/India/Karnataka/ | Institute for Stem Cell Science and Regenerative Medicine | National Centre for Biological Sciences | Farhan Ali et al |
| EPI_ISL_477239 | India/KA-InStem-NCBS-0038/2020 | 2020-06-17 | Asia/India/Karnataka/ | Institute for Stem Cell Science and Regenerative Medicine | National Centre for Biological Sciences | Farhan Ali et al |
| EPI_ISL_477243 | India/KA-InStem-NCBS-0042/2020 | 2020-06-16 | Asia/India/Karnataka/ | Institute for Stem Cell Science and Regenerative Medicine | National Centre for Biological Sciences | Farhan Ali et al |
| EPI_ISL_477248 | India/KA-InStem-NCBS-0048/2020 | 2020-05-07 | Asia/India/Karnataka/ | Institute for Stem Cell Science and Regenerative Medicine | National Centre for Biological Sciences | Farhan Ali et al |

| | | | | | | |
|---|---|---|---|---|---|---|
| EPI_ISL_477255 | India/KA-InStem-NCBS-0055/2020 | 2020-05-16 | Asia/India/Karnataka/ | Institute for Stem Cell Science and Regenerative Medicine | National Centre for Biological Sciences | Farhan Ali et al |
| EPI_ISL_477273 | USA/MN-MDH-1183/2020 | 2020-05-20 | North America/USA/Minnesota/ | Mayo Clinic & Mayo Clinic Laboratories | Minnesota Department of Health, Public Health Laboratory | Matt Plumb et al |
| EPI_ISL_477278 | USA/MN-MDH-1188/2020 | 2020-05-02 | North America/USA/Minnesota/ | M Health Fairview | Minnesota Department of Health, Public Health Laboratory | Matt Plumb et al |
| EPI_ISL_477279 | USA/MN-MDH-1189/2020 | 2020-05-02 | North America/USA/Minnesota/ | M Health Fairview | Minnesota Department of Health, Public Health Laboratory | Matt Plumb et al |
| EPI_ISL_477294 | USA/MN-MDH-1205/2020 | 2020-04-30 | North America/USA/Minnesota/ | Mayo Clinic & Mayo Clinic Laboratories | Minnesota Department of Health, Public Health Laboratory | Matt Plumb et al |
| EPI_ISL_477300 | USA/MN-MDH-1211/2020 | 2020-05-20 | North America/USA/Minnesota/ | Mayo Clinic & Mayo Clinic Laboratories | Minnesota Department of Health, Public Health Laboratory | Matt Plumb et al |
| EPI_ISL_477301 | USA/MN-MDH-1212/2020 | 2020-05-21 | North America/USA/Minnesota/ | Mayo Clinic & Mayo Clinic Laboratories | Minnesota Department of Health, Public Health Laboratory | Matt Plumb et al |
| EPI_ISL_477303 | USA/MN-MDH-1214/2020 | 2020-05-22 | North America/USA/Minnesota/ | Mayo Clinic & Mayo Clinic Laboratories | Minnesota Department of Health, Public Health Laboratory | Matt Plumb et al |
| EPI_ISL_477304 | USA/MN-MDH-1215/2020 | 2020-05-23 | North America/USA/Minnesota/ | Mayo Clinic & Mayo Clinic Laboratories | Minnesota Department of Health, Public Health Laboratory | Matt Plumb et al |
| EPI_ISL_477306 | USA/MN-MDH-1217/2020 | 2020-05-26 | North America/USA/Minnesota/ | Mayo Clinic & Mayo Clinic Laboratories | Minnesota Department of Health, Public Health Laboratory | Matt Plumb et al |

| | | | | | | |
|---|---|---|---|---|---|---|
| EPI_ISL_477308 | USA/MN-MDH-1219/2020 | 2020-05-29 | North America/USA/Minnesota/ | Mayo Clinic & Mayo Clinic Laboratories | Minnesota Department of Health, Public Health Laboratory | Matt Plumb et al |
| EPI_ISL_477311 | USA/MN-MDH-1222/2020 | 2020-06-03 | North America/USA/Minnesota/ | Mayo Clinic & Mayo Clinic Laboratories | Minnesota Department of Health, Public Health Laboratory | Matt Plumb et al |
| EPI_ISL_477641 | USA/VA-DCLS-0425/2020 | 2020-04-29 | North America/USA/Virginia/ | Virginia DCLS | Virginia DCLS | Virginia DCLS et al |
| EPI_ISL_477642 | USA/VA-DCLS-0426/2020 | 2020-04-30 | North America/USA/Virginia/ | Virginia DCLS | Virginia DCLS | Virginia DCLS et al |
| EPI_ISL_477649 | USA/VA-DCLS-0433/2020 | 2020-05-01 | North America/USA/Virginia/ | Virginia DCLS | Virginia DCLS | Virginia DCLS et al |
| EPI_ISL_477655 | USA/VA-DCLS-0439/2020 | 2020-05-02 | North America/USA/Virginia/ | Virginia DCLS | Virginia DCLS | Virginia DCLS et al |
| EPI_ISL_477656 | USA/VA-DCLS-0440/2020 | 2020-05-02 | North America/USA/Virginia/ | Virginia DCLS | Virginia DCLS | Virginia DCLS et al |
| EPI_ISL_477657 | USA/VA-DCLS-0441/2020 | 2020-05-02 | North America/USA/Virginia/ | Virginia DCLS | Virginia DCLS | Virginia DCLS et al |
| EPI_ISL_477660 | USA/VA-DCLS-0444/2020 | 2020-05-04 | North America/USA/Virginia/ | Virginia DCLS | Virginia DCLS | Virginia DCLS et al |
| EPI_ISL_477669 | USA/VA-DCLS-0459/2020 | 2020-05-05 | North America/USA/Virginia/ | Virginia DCLS | Virginia DCLS | Virginia DCLS et al |
| EPI_ISL_477671 | USA/VA-DCLS-0462/2020 | 2020-05-05 | North America/USA/Virginia/ | Virginia DCLS | Virginia DCLS | Virginia DCLS et al |
| EPI_ISL_477672 | USA/VA-DCLS-0466/2020 | 2020-05-06 | North America/USA/Virginia/ | Virginia DCLS | Virginia DCLS | Virginia DCLS et al |
| EPI_ISL_477677 | USA/CT-UW-2468/2020 | 2020-03-30 | North America/USA/Connecticut/ | UW Virology Lab | UW Virology Lab | Pavitra Roychoudhury et al |
| EPI_ISL_477681 | USA/ID-UW-2067/2020 | 2020-03-22 | North America/USA/Idaho/ | UW Virology Lab | UW Virology Lab | Pavitra Roychoudhury et al |
| EPI_ISL_477686 | USA/WA-UW-1412/2020 | 2020-03-18 | North America/USA/Washington/ | UW Virology Lab | UW Virology Lab | Pavitra Roychoudhury et al |
| EPI_ISL_477706 | USA/WA-UW-2270/2020 | 2020-03-31 | North America/USA/Washington/ | UW Virology Lab | UW Virology Lab | Pavitra Roychoudhury et al |
| EPI_ISL_477709 | USA/WA-UW-2281/2020 | 2020-03-30 | North America/USA/Washington/ | UW Virology Lab | UW Virology Lab | Pavitra Roychoudhury et al |
| EPI_ISL_477715 | USA/WA-UW-2376/2020 | 2020-03-30 | North America/USA/Washington/ | UW Virology Lab | UW Virology Lab | Pavitra Roychoudhury et al |
| EPI_ISL_477779 | England/BIRM-61595/2020 | 2020-04-28 | Europe/United Kingdom/England/ | University of Birmingham | COVID-19 Genomics UK (COG-UK) Consortium | Institute of Microbiology et al |
| EPI_ISL_477834 | Scotland/CVR1911/2020 | 2020-04-08 | Europe/United Kingdom/Scotland/ | West of Scotland Specialist Virology Centre, NHSGGC / MRC-University of Glasgow Centre for Virus Research | COVID-19 Genomics UK (COG-UK) Consortium | Ana da Silva Filipe et al |
| EPI_ISL_477961 | Scotland/CVR2520/2020 | 2020-04-15 | Europe/United Kingdom/Scotland/ | West of Scotland Specialist Virology Centre, NHSGGC / MRC-University of Glasgow Centre for Virus Research | COVID-19 Genomics UK (COG-UK) Consortium | Ana da Silva Filipe et al |

| | | | | | | |
|---|---|---|---|---|---|---|
| EPI_ISL_478050 | Scotland/CVR2955/2020 | 2020-04-23 | Europe/United Kingdom/Scotland/ | West of Scotland Specialist Virology Centre, NHSGGC / MRC-University of Glasgow Centre for Virus Research | COVID-19 Genomics UK (COG-UK) Consortium | Ana da Silva Filipe et al |
| EPI_ISL_478060 | Scotland/CVR3450/2020 | 2020-05-10 | Europe/United Kingdom/Scotland/ | West of Scotland Specialist Virology Centre, NHSGGC / MRC-University of Glasgow Centre for Virus Research | COVID-19 Genomics UK (COG-UK) Consortium | Ana da Silva Filipe et al |
| EPI_ISL_478140 | Scotland/CVR3814/2020 | 2020-05-19 | Europe/United Kingdom/Scotland/ | West of Scotland Specialist Virology Centre, NHSGGC / MRC-University of Glasgow Centre for Virus Research | COVID-19 Genomics UK (COG-UK) Consortium | Ana da Silva Filipe et al |
| EPI_ISL_478329 | England/LCST-24D6ACF/2020 | 2020-06-05 | Europe/United Kingdom/England/ | University Hospitals Of Leicester NHS Trust and DeepSeq Nottingham | COVID-19 Genomics UK (COG-UK) Consortium | Christopher Holmes et al |
| EPI_ISL_478565 | England/NORT-284767/2020 | 2020-03-23 | Europe/United Kingdom/England/ | Northumbria University / South Tees Hospitals NHS Foundation Trust / North Cumbria Integrated Care NHS Foundation Trust / North Tees and Hartlepool NHS Foundation Trust / Newcastle Hospitals NHS Foundation Trust | COVID-19 Genomics UK (COG-UK) Consortium | Darren L Smith et al |
| EPI_ISL_478606 | England/NORT-28C8F3/2020 | 2020-05-16 | Europe/United Kingdom/England/ | Northumbria University / South Tees Hospitals NHS Foundation Trust / North Cumbria Integrated Care NHS Foundation Trust / North Tees and Hartlepool NHS Foundation Trust / Newcastle Hospitals NHS Foundation Trust | COVID-19 Genomics UK (COG-UK) Consortium | Darren L Smith et al |
| EPI_ISL_478631 | England/NORT-295129/2020 | 2020-03-30 | Europe/United Kingdom/England/ | Northumbria University / South Tees Hospitals NHS Foundation Trust / North Cumbria Integrated Care NHS Foundation Trust / North Tees and Hartlepool NHS Foundation Trust / Newcastle Hospitals NHS Foundation Trust | COVID-19 Genomics UK (COG-UK) Consortium | Darren L Smith et al |
| EPI_ISL_478662 | England/NORT-295D52/2020 | 2020-05-08 | Europe/United Kingdom/England/ | Northumbria University / South Tees Hospitals NHS Foundation Trust / North Cumbria Integrated Care NHS Foundation Trust / North Tees and Hartlepool NHS Foundation Trust / Newcastle Hospitals NHS Foundation Trust | COVID-19 Genomics UK (COG-UK) Consortium | Darren L Smith et al |
| EPI_ISL_478669 | Turkey/KOC-IST-B303/2020 | 2020-03-27 | Europe/Turkey/Turkey/ | unknown | Microbiology, Koc University | Can et al |
| EPI_ISL_478672 | Egypt/CUNCI-HGC6I010/2020 | 2020-06-02 | Africa/Egypt/Egypt/ | Egyptian National Cancer Institute (ENCI) | Egyptian National Cancer Institute (ENCI) | Zekri et al |

| | | | | | | |
|---|---|---|---|---|---|---|
| EPI_ISL_478873 | England/OXON-B02FA/2020 | 2020-04-19 | Europe/United Kingdom/England/ | Oxford Viromics, NDM, University of Oxford; Oxford University Hospitals; Basingstoke and North Hampshire Hospital | COVID-19 Genomics UK (COG-UK) Consortium | Tanya Golubchik et al |
| EPI_ISL_479099 | England/OXON-B1EAA/2020 | 2020-05-02 | Europe/United Kingdom/England/ | Oxford Viromics, NDM, University of Oxford; Oxford University Hospitals; Basingstoke and North Hampshire Hospital | COVID-19 Genomics UK (COG-UK) Consortium | Tanya Golubchik et al |
| EPI_ISL_479128 | England/OXON-B2207/2020 | 2020-04-20 | Europe/United Kingdom/England/ | Oxford Viromics, NDM, University of Oxford; Oxford University Hospitals; Basingstoke and North Hampshire Hospital | COVID-19 Genomics UK (COG-UK) Consortium | Tanya Golubchik et al |
| EPI_ISL_479176 | England/PORT-2D12C4/2020 | 2020-03-24 | Europe/United Kingdom/England/ | Centre for Enzyme Innovation, University of Portsmouth / Translational Research Laboratory, Portsmouth Hospitals NHS Trust | COVID-19 Genomics UK (COG-UK) Consortium | Angela Beckett et al |
| EPI_ISL_479191 | England/PORT-2D13B2/2020 | 2020-03-25 | Europe/United Kingdom/England/ | Centre for Enzyme Innovation, University of Portsmouth / Translational Research Laboratory, Portsmouth Hospitals NHS Trust | COVID-19 Genomics UK (COG-UK) Consortium | Angela Beckett et al |
| EPI_ISL_479239 | England/SHEF-C9E54/2020 | 2020-05-11 | Europe/United Kingdom/England/ | Virology Department, Sheffield Teaching Hospitals NHS Foundation Trust/Department of Infection, Immunity and Cardiovascular Disease, The Medical School, University of Sheffield | COVID-19 Genomics UK (COG-UK) Consortium | Thushan de Silva et al |
| EPI_ISL_479286 | Wales/PHWC-15FE52/2020 | 2020-05-19 | Europe/United Kingdom/Wales/ | Wales Specialist Virology Centre Sequencing lab: Pathogen Genomics Unit | COVID-19 Genomics UK (COG-UK) Consortium | Catherine Moore et al |
| EPI_ISL_479342 | Wales/PHWC-164A92/2020 | 2020-05-11 | Europe/United Kingdom/Wales/ | Wales Specialist Virology Centre Sequencing lab: Pathogen Genomics Unit | COVID-19 Genomics UK (COG-UK) Consortium | Catherine Moore et al |
| EPI_ISL_479394 | Wales/PHWC-164ED2/2020 | 2020-05-28 | Europe/United Kingdom/Wales/ | Wales Specialist Virology Centre Sequencing lab: Pathogen Genomics Unit | COVID-19 Genomics UK (COG-UK) Consortium | Catherine Moore et al |
| EPI_ISL_479472 | Wales/PHWC-1654DF/2020 | 2020-05-15 | Europe/United Kingdom/Wales/ | Wales Specialist Virology Centre Sequencing lab: Pathogen Genomics Unit | COVID-19 Genomics UK (COG-UK) Consortium | Catherine Moore et al |
| EPI_ISL_479533 | India/MH-NIV-31191/2020 | 2020-05-15 | Asia/India/Maharashtra/ | NIV Influenza | NIV Influenza | Potdar V et al |
| EPI_ISL_479535 | India/MH-NIV-36405/2020 | 2020-05-18 | Asia/India/Maharashtra/ | NIV Influenza | NIV Influenza | Potdar V et al |
| EPI_ISL_479537 | India/MH-NIV-36461/2020 | 2020-05-20 | Asia/India/Maharashtra/ | NIV Influenza | NIV Influenza | Potdar V et al |
| EPI_ISL_479540 | India/MH-NIV-37681/2020 | 2020-05-21 | Asia/India/Maharashtra/ | NIV Influenza | NIV Influenza | Potdar V et al |
| EPI_ISL_479541 | India/MH-NIV-37985/2020 | 2020-05-21 | Asia/India/Maharashtra/ | NIV Influenza | NIV Influenza | Potdar V et al |

| | | | | | | |
|---|---|---|---|---|---|---|
| EPI_ISL_479542 | India/MH-NIV-38793/2020 | 2020-05-22 | Asia/India/Maharashtra/ | NIV Influenza | NIV Influenza | Potdar V et al |
| EPI_ISL_479544 | India/MH-NIV-42694/2020 | 2020-05-24 | Asia/India/Maharashtra/ | NIV Influenza | NIV Influenza | Potdar V et al |
| EPI_ISL_479545 | India/MH-NIV-46430/2020 | 2020-05-26 | Asia/India/Maharashtra/ | NIV Influenza | NIV Influenza | Potdar V et al |
| EPI_ISL_479548 | India/MH-NIV-45936/2020 | 2020-05-28 | Asia/India/Maharashtra/ | NIV Influenza | NIV Influenza | Potdar V et al |
| EPI_ISL_479551 | India/MH-NIV-46348/2020 | 2020-05-28 | Asia/India/Maharashtra/ | NIV Influenza | NIV Influenza | Potdar V et al |
| EPI_ISL_479555 | India/MH-NIV-64478/2020 | 2020-06-15 | Asia/India/Maharashtra/ | NIV Influenza | NIV Influenza | Potdar V et al |
| EPI_ISL_479556 | India/MH-NIV-64850/2020 | 2020-06-16 | Asia/India/Maharashtra/ | NIV Influenza | NIV Influenza | Potdar V et al |
| EPI_ISL_479557 | India/MH-NIV-64877/2020 | 2020-06-16 | Asia/India/Maharashtra/ | NIV Influenza | NIV Influenza | Potdar V et al |
| EPI_ISL_479558 | India/MH-NIV-64904/2020 | 2020-06-16 | Asia/India/Maharashtra/ | NIV Influenza | NIV Influenza | Potdar V et al |
| EPI_ISL_479559 | India/MH-NIV-64905/2020 | 2020-06-16 | Asia/India/Maharashtra/ | NIV Influenza | NIV Influenza | Potdar V et al |
| EPI_ISL_479560 | India/MH-NIV-64908/2020 | 2020-06-16 | Asia/India/Maharashtra/ | NIV Influenza | NIV Influenza | Potdar V et al |
| EPI_ISL_479562 | India/MH-NIV-65282/2020 | 2020-06-16 | Asia/India/Maharashtra/ | NIV Influenza | NIV Influenza | Potdar V et al |
| EPI_ISL_479563 | India/MH-NIV-65417/2020 | 2020-06-16 | Asia/India/Maharashtra/ | NIV Influenza | NIV Influenza | Potdar V et al |
| EPI_ISL_479564 | India/MH-NIV-65433/2020 | 2020-06-16 | Asia/India/Maharashtra/ | NIV Influenza | NIV Influenza | Potdar V et al |
| EPI_ISL_479565 | India/MH-NIV-65813/2020 | 2020-06-16 | Asia/India/Maharashtra/ | NIV Influenza | NIV Influenza | Potdar V et al |
| EPI_ISL_479569 | India/MH-NIV-65861/2020 | 2020-06-16 | Asia/India/Maharashtra/ | NIV Influenza | NIV Influenza | Potdar V et al |
| EPI_ISL_479570 | India/MH-NIV-65877/2020 | 2020-06-16 | Asia/India/Maharashtra/ | NIV Influenza | NIV Influenza | Potdar V et al |
| EPI_ISL_479574 | Singapore/496/2020 | 2020-04-20 | Asia/Singapore/Singapore/ | National Public Health Laboratory, National Centre for Infectious Diseases | National Public Health Laboratory, National Centre for Infectious Diseases | Mak TM et al |
| EPI_ISL_479577 | Singapore/499/2020 | 2020-04-17 | Asia/Singapore/Singapore/ | National Public Health Laboratory, National Centre for Infectious Diseases | National Public Health Laboratory, National Centre for Infectious Diseases | Mak TM et al |
| EPI_ISL_479585 | Singapore/507/2020 | 2020-05-15 | Asia/Singapore/Singapore/ | National Public Health Laboratory, National Centre for Infectious Diseases | National Public Health Laboratory, National Centre for Infectious Diseases | Mak TM et al |

| | | | | | | |
|---|---|---|---|---|---|---|
| EPI_ISL_479587 | Singapore/509/2020 | 2020-05-20 | Asia/Singapore/Singapore/ | National Public Health Laboratory, National Centre for Infectious Diseases | National Public Health Laboratory, National Centre for Infectious Diseases | Mak TM et al |
| EPI_ISL_479588 | Singapore/510/2020 | 2020-05-22 | Asia/Singapore/Singapore/ | National Public Health Laboratory, National Centre for Infectious Diseases | National Public Health Laboratory, National Centre for Infectious Diseases | Mak TM et al |
| EPI_ISL_479589 | Singapore/511/2020 | 2020-05-22 | Asia/Singapore/Singapore/ | National Public Health Laboratory, National Centre for Infectious Diseases | National Public Health Laboratory, National Centre for Infectious Diseases | Mak TM et al |
| EPI_ISL_479590 | Singapore/512/2020 | 2020-05-23 | Asia/Singapore/Singapore/ | National Public Health Laboratory, National Centre for Infectious Diseases | National Public Health Laboratory, National Centre for Infectious Diseases | Mak TM et al |
| EPI_ISL_479616 | Italy/FVG-ICGEB-bal1A/2020 | 2020-03-27 | Europe/Italy/Friuli Venezia Giulia/ | Laboratory of Molecular Virology of the International Centre for Genetic Engineering and Biotechnology (ICGEB) | ARGO Open Lab Platform for Genome Sequencing | Licastro et al |
| EPI_ISL_479625 | Canada/NB-202/2020 | 2020-03-22 | North America/Canada/New Brunswick/ | Dr. Georges-L.-Dumont University Hospital Centre | National Microbiology Laboratory | Anna Majer et al |
| EPI_ISL_479626 | Canada/NB-204/2020 | 2020-03-29 | North America/Canada/New Brunswick/ | Dr. Georges-L.-Dumont University Hospital Centre | National Microbiology Laboratory | Anna Majer et al |
| EPI_ISL_479628 | Canada/NB-209/2020 | 2020-03-29 | North America/Canada/New Brunswick/ | Dr. Georges-L.-Dumont University Hospital Centre | National Microbiology Laboratory | Anna Majer et al |
| EPI_ISL_479629 | Canada/NB-213/2020 | 2020-03-30 | North America/Canada/New Brunswick/ | Dr. Georges-L.-Dumont University Hospital Centre | National Microbiology Laboratory | Anna Majer et al |

| | | | | | | |
|---|---|---|---|---|---|---|
| EPI_ISL_479630 | Canada/NB-214/2020 | 2020-03-27 | North America/Canada/New Brunswick/ | Dr. Georges-L.-Dumont University Hospital Centre | National Microbiology Laboratory | Anna Majer et al |
| EPI_ISL_479631 | Canada/NB-215/2020 | 2020-03-27 | North America/Canada/New Brunswick/ | Dr. Georges-L.-Dumont University Hospital Centre | National Microbiology Laboratory | Anna Majer et al |
| EPI_ISL_479632 | Canada/NB-216/2020 | 2020-03-30 | North America/Canada/New Brunswick/ | Dr. Georges-L.-Dumont University Hospital Centre | National Microbiology Laboratory | Anna Majer et al |
| EPI_ISL_479633 | Canada/NB-217/2020 | 2020-03-30 | North America/Canada/New Brunswick/ | Dr. Georges-L.-Dumont University Hospital Centre | National Microbiology Laboratory | Anna Majer et al |
| EPI_ISL_479634 | Canada/NB-218/2020 | 2020-03-25 | North America/Canada/New Brunswick/ | Dr. Georges-L.-Dumont University Hospital Centre | National Microbiology Laboratory | Anna Majer et al |
| EPI_ISL_479635 | Canada/NB-219/2020 | 2020-03-25 | North America/Canada/New Brunswick/ | Dr. Georges-L.-Dumont University Hospital Centre | National Microbiology Laboratory | Anna Majer et al |
| EPI_ISL_479636 | Canada/NB-220/2020 | 2020-03-25 | North America/Canada/New Brunswick/ | Dr. Georges-L.-Dumont University Hospital Centre | National Microbiology Laboratory | Anna Majer et al |
| EPI_ISL_479637 | Canada/NB-221/2020 | 2020-03-26 | North America/Canada/New Brunswick/ | Dr. Georges-L.-Dumont University Hospital Centre | National Microbiology Laboratory | Anna Majer et al |
| EPI_ISL_479638 | Canada/NB-222/2020 | 2020-03-25 | North America/Canada/New Brunswick/ | Dr. Georges-L.-Dumont University Hospital Centre | National Microbiology Laboratory | Anna Majer et al |
| EPI_ISL_479639 | Canada/NB-223/2020 | 2020-03-26 | North America/Canada/New Brunswick/ | Dr. Georges-L.-Dumont University Hospital Centre | National Microbiology Laboratory | Anna Majer et al |
| EPI_ISL_479640 | Canada/NB-224/2020 | 2020-03-26 | North America/Canada/New Brunswick/ | Dr. Georges-L.-Dumont University Hospital Centre | National Microbiology Laboratory | Anna Majer et al |
| EPI_ISL_479641 | Canada/NB-225/2020 | 2020-03-26 | North America/Canada/New Brunswick/ | Dr. Georges-L.-Dumont University Hospital Centre | National Microbiology Laboratory | Anna Majer et al |
| EPI_ISL_479642 | Canada/NB-226/2020 | 2020-03-23 | North America/Canada/New Brunswick/ | Dr. Georges-L.-Dumont University Hospital Centre | National Microbiology Laboratory | Anna Majer et al |

| | | | | | | |
|---|---|---|---|---|---|---|
| EPI_ISL_479643 | Canada/NB-228/2020 | 2020-03-30 | North America/Canada/New Brunswick/ | Dr. Georges-L.-Dumont University Hospital Centre | National Microbiology Laboratory | Anna Majer et al |
| EPI_ISL_479644 | Canada/NB-231/2020 | 2020-03-24 | North America/Canada/New Brunswick/ | Dr. Georges-L.-Dumont University Hospital Centre | National Microbiology Laboratory | Anna Majer et al |
| EPI_ISL_479645 | Canada/NB-232/2020 | 2020-03-25 | North America/Canada/New Brunswick/ | Dr. Georges-L.-Dumont University Hospital Centre | National Microbiology Laboratory | Anna Majer et al |
| EPI_ISL_479646 | Canada/NB-233/2020 | 2020-03-24 | North America/Canada/New Brunswick/ | Dr. Georges-L.-Dumont University Hospital Centre | National Microbiology Laboratory | Anna Majer et al |
| EPI_ISL_479647 | Canada/NB-234/2020 | 2020-03-25 | North America/Canada/New Brunswick/ | Dr. Georges-L.-Dumont University Hospital Centre | National Microbiology Laboratory | Anna Majer et al |
| EPI_ISL_479648 | Canada/NB-235/2020 | 2020-03-31 | North America/Canada/New Brunswick/ | Dr. Georges-L.-Dumont University Hospital Centre | National Microbiology Laboratory | Anna Majer et al |
| EPI_ISL_479649 | Canada/NB-236/2020 | 2020-03-31 | North America/Canada/New Brunswick/ | Dr. Georges-L.-Dumont University Hospital Centre | National Microbiology Laboratory | Anna Majer et al |
| EPI_ISL_479650 | Canada/NB-237/2020 | 2020-03-31 | North America/Canada/New Brunswick/ | Dr. Georges-L.-Dumont University Hospital Centre | National Microbiology Laboratory | Anna Majer et al |
| EPI_ISL_479651 | Canada/NB-238/2020 | 2020-03-25 | North America/Canada/New Brunswick/ | Dr. Georges-L.-Dumont University Hospital Centre | National Microbiology Laboratory | Anna Majer et al |
| EPI_ISL_479657 | India/MH-NIV-25949/2020 | 2020-05-09 | Asia/India/Maharashtra/ | NIV Influenza | NIV Influenza | Potdar V et al |
| EPI_ISL_479674 | USA/NY-NYU-VC-022/2020 | 2020-04-08 | North America/USA/New York/ | Center for Genomics and System Biology, New York University | Center for Genomics and System Biology, New York University | Roder et al |
| EPI_ISL_479738 | India/KA-InStem-NCBS-0067/2020 | 2020-06-02 | Asia/India/Karnataka/ | Institute for Stem Cell Science and Regenerative Medicine | National Centre for Biological Sciences | Farhan Ali et al |
| EPI_ISL_479740 | India/KA-InStem-NCBS-0069/2020 | 2020-05-28 | Asia/India/Karnataka/ | Institute for Stem Cell Science and Regenerative Medicine | National Centre for Biological Sciences | Farhan Ali et al |

| | | | | | | |
|---|---|---|---|---|---|---|
| EPI_ISL_479743 | India/KA-InStem-NCBS-0072/2020 | 2020-06-17 | Asia/India/Karnataka/ | Institute for Stem Cell Science and Regenerative Medicine | National Centre for Biological Sciences | Farhan Ali et al |
| EPI_ISL_479747 | India/KA-InStem-NCBS-0076/2020 | 2020-06-17 | Asia/India/Karnataka/ | Institute for Stem Cell Science and Regenerative Medicine | National Centre for Biological Sciences | Farhan Ali et al |
| EPI_ISL_479751 | India/KA-InStem-NCBS-0080/2020 | 2020-06-17 | Asia/India/Karnataka/ | Institute for Stem Cell Science and Regenerative Medicine | National Centre for Biological Sciences | Farhan Ali et al |
| EPI_ISL_479752 | India/KA-InStem-NCBS-0081/2020 | 2020-06-17 | Asia/India/Karnataka/ | Institute for Stem Cell Science and Regenerative Medicine | National Centre for Biological Sciences | Farhan Ali et al |
| EPI_ISL_479753 | India/KA-InStem-NCBS-0082/2020 | 2020-06-17 | Asia/India/Karnataka/ | Institute for Stem Cell Science and Regenerative Medicine | National Centre for Biological Sciences | Farhan Ali et al |
| EPI_ISL_479754 | India/KA-InStem-NCBS-0083/2020 | 2020-06-17 | Asia/India/Karnataka/ | Institute for Stem Cell Science and Regenerative Medicine | National Centre for Biological Sciences | Farhan Ali et al |
| EPI_ISL_479755 | India/KA-InStem-NCBS-0084/2020 | 2020-05-07 | Asia/India/Karnataka/ | Institute for Stem Cell Science and Regenerative Medicine | National Centre for Biological Sciences | Farhan Ali et al |
| EPI_ISL_479764 | USA/FL-05-UMTL-9TXV/2020 | 2020-05-21 | North America/USA/Florida/Miami-Dad | University of Miami Immunology and Histocompatibility Laboratory | University of Miami Immunology and Histocompatibility Laboratory | Emilio Margolles-Clark et al |
| EPI_ISL_479766 | USA/FL-06-UMTL-A3MJ/2020 | 2020-04-09 | North America/USA/Florida/Miami-Dad | University of Miami Immunology and Histocompatibility Laboratory | University of Miami Immunology and Histocompatibility Laboratory | Emilio Margolles-Clark et al |
| EPI_ISL_479769 | USA/FL-06-UMTL-A6U2/2020 | 2020-04-15 | North America/USA/Florida/Miami-Dad | University of Miami Immunology and Histocompatibility Laboratory | University of Miami Immunology and Histocompatibility Laboratory | Emilio Margolles-Clark et al |
| EPI_ISL_479797 | Japan/PG-0010/2020 | 2020-02-18 | Asia/Japan/Kanto/ | Sagamihara City Public Health Research Institute | Pathogen Genomics Center, National Institute of Infectious Diseases | Tsuyoshi Sekizuka et al |

| | | | | | | |
|---|---|---|---|---|---|---|
| EPI_ISL_479798 | Japan/PG-0011/2020 | 2020-02-18 | Asia/Japan/Kanto/ | Sagamihara City Public Health Research Institute | Pathogen Genomics Center, National Institute of Infectious Diseases | Tsuyoshi Sekizuka et al |
| EPI_ISL_479799 | Japan/PG-0015/2020 | 2020-01-20 | Asia/Japan/Hokkaido/ | Sapporo City Institute of Public Health | Pathogen Genomics Center, National Institute of Infectious Diseases | Tsuyoshi Sekizuka et al |
| EPI_ISL_479800 | Japan/PG-0016/2020 | 2020-01-20 | Asia/Japan/Hokkaido/ | Sapporo City Institute of Public Health | Pathogen Genomics Center, National Institute of Infectious Diseases | Tsuyoshi Sekizuka et al |
| EPI_ISL_479802 | Japan/PG-0031/2020 | 2020-02-25 | Asia/Japan/Kanto/ | Sagamihara City Public Health Research Institute | Pathogen Genomics Center, National Institute of Infectious Diseases | Tsuyoshi Sekizuka et al |
| EPI_ISL_479803 | Japan/PG-0033/2020 | 2020-02-28 | Asia/Japan/Kanto/ | Sagamihara City Public Health Research Institute | Pathogen Genomics Center, National Institute of Infectious Diseases | Tsuyoshi Sekizuka et al |
| EPI_ISL_479804 | Japan/PG-0036/2020 | 2020-02-29 | Asia/Japan/Kanto/ | Sagamihara City Public Health Research Institute | Pathogen Genomics Center, National Institute of Infectious Diseases | Tsuyoshi Sekizuka et al |
| EPI_ISL_479821 | Japan/PG-0058/2020 | 2020-03-02 | Asia/Japan/Kansai/ | Department of Infectious Diseases, Kobe Institute of Health | Pathogen Genomics Center, National Institute of Infectious Diseases | Tsuyoshi Sekizuka et al |
| EPI_ISL_479824 | Japan/PG-0066/2020 | 2020-02-15 | Asia/Japan/Kyusyu/ | Kumamoto PrefecturalInstituteof Public-Health and Environmental Science | Pathogen Genomics Center, National Institute of Infectious Diseases | Tsuyoshi Sekizuka et al |

| | | | | | | |
|---|---|---|---|---|---|---|
| EPI_ISL_479826 | Japan/PG-0071/2020 | 2020-02-26 | Asia/Japan/Hokkaido/ | Sapporo City Institute of Public Health | Pathogen Genomics Center, National Institute of Infectious Diseases | Tsuyoshi Sekizuka et al |
| EPI_ISL_479827 | Japan/PG-0072/2020 | 2020-02-27 | Asia/Japan/Hokkaido/ | Sapporo City Institute of Public Health | Pathogen Genomics Center, National Institute of Infectious Diseases | Tsuyoshi Sekizuka et al |
| EPI_ISL_479828 | Japan/PG-0074/2020 | 2020-02-27 | Asia/Japan/Hokkaido/ | Sapporo City Institute of Public Health | Pathogen Genomics Center, National Institute of Infectious Diseases | Tsuyoshi Sekizuka et al |
| EPI_ISL_479829 | Japan/PG-0076/2020 | 2020-02-28 | Asia/Japan/Hokkaido/ | Sapporo City Institute of Public Health | Pathogen Genomics Center, National Institute of Infectious Diseases | Tsuyoshi Sekizuka et al |
| EPI_ISL_479830 | Japan/PG-0077/2020 | 2020-02-28 | Asia/Japan/Hokkaido/ | Sapporo City Institute of Public Health | Pathogen Genomics Center, National Institute of Infectious Diseases | Tsuyoshi Sekizuka et al |
| EPI_ISL_479850 | Japan/PG-0114/2020 | 2020-02-13 | Asia/Japan/Kanto/ | Gunma Prefectural Institute of Public Health and Environmental Sciences | Pathogen Genomics Center, National Institute of Infectious Diseases | Tsuyoshi Sekizuka et al |
| EPI_ISL_479851 | Japan/PG-0115/2020 | 2020-02-17 | Asia/Japan/Kanto/ | Gunma Prefectural Institute of Public Health and Environmental Sciences | Pathogen Genomics Center, National Institute of Infectious Diseases | Tsuyoshi Sekizuka et al |
| EPI_ISL_479852 | Japan/PG-0119/2020 | 2020-02-25 | Asia/Japan/Kanto/ | Gunma Prefectural Institute of Public Health and Environmental Sciences | Pathogen Genomics Center, National Institute of Infectious Diseases | Tsuyoshi Sekizuka et al |

| | | | | | | |
|---|---|---|---|---|---|---|
| EPI_ISL_479863 | Japan/PG-0140/2020 | 2020-02-17 | Asia/Japan/Kansai/ | Wakayama Prefectural Research Center of Environment and Public Health | Pathogen Genomics Center, National Institute of Infectious Diseases | Tsuyoshi Sekizuka et al |
| EPI_ISL_479864 | Japan/PG-0141/2020 | 2020-02-23 | Asia/Japan/Kansai/ | Wakayama Prefectural Research Center of Environment and Public Health | Pathogen Genomics Center, National Institute of Infectious Diseases | Tsuyoshi Sekizuka et al |
| EPI_ISL_479870 | Japan/PG-0159/2020 | 2020-02-21 | Asia/Japan/Kanto/ | Sagamihara City Public Health Research Institute | Pathogen Genomics Center, National Institute of Infectious Diseases | Tsuyoshi Sekizuka et al |
| EPI_ISL_479871 | Japan/PG-0160/2020 | 2020-02-25 | Asia/Japan/Kanto/ | Sagamihara City Public Health Research Institute | Pathogen Genomics Center, National Institute of Infectious Diseases | Tsuyoshi Sekizuka et al |
| EPI_ISL_479881 | Japan/PG-0172/2020 | 2020-03-15 | Asia/Japan/Hokkaido/ | Sapporo City Institute of Public Health | Pathogen Genomics Center, National Institute of Infectious Diseases | Tsuyoshi Sekizuka et al |
| EPI_ISL_479913 | Japan/PG-0213/2020 | 2020-02-28 | Asia/Japan/Kanto/ | Niigata City Public Health Research Institute | Pathogen Genomics Center, National Institute of Infectious Diseases | Tsuyoshi Sekizuka et al |
| EPI_ISL_479916 | Japan/PG-0216/2020 | 2020-03-01 | Asia/Japan/Kanto/ | Niigata City Public Health Research Institute | Pathogen Genomics Center, National Institute of Infectious Diseases | Tsuyoshi Sekizuka et al |
| EPI_ISL_479936 | Japan/PG-0251/2020 | 2020-03-23 | Asia/Japan/Kanto/ | Ibaraki Prefectural Institute of Public Health | Pathogen Genomics Center, National Institute of Infectious Diseases | Tsuyoshi Sekizuka et al |

| | | | | | | |
|---|---|---|---|---|---|---|
| EPI_ISL_479971 | Japan/PG-0294/2020 | 2020-03-26 | Asia/Japan/Hokurikushinsyu/ | Fukui Prefectural Institute of Public Health and Environmental Science | Pathogen Genomics Center, National Institute of Infectious Diseases | Tsuyoshi Sekizuka et al |
| EPI_ISL_479997 | Japan/PG-0322/2020 | 2020-02-24 | Asia/Japan/Hokurikushinsyu/ | Nagano Environmental Conservation Research Institute | Pathogen Genomics Center, National Institute of Infectious Diseases | Tsuyoshi Sekizuka et al |
| EPI_ISL_479998 | Japan/PG-0323/2020 | 2020-02-25 | Asia/Japan/Hokurikushinsyu/ | Nagano Environmental Conservation Research Institute | Pathogen Genomics Center, National Institute of Infectious Diseases | Tsuyoshi Sekizuka et al |
| EPI_ISL_480030 | Japan/PG-0359/2020 | 2020-02-17 | Asia/Japan/Kanto/ | Tochigi Prefectural Institute of Public Health and Environmental Science | Pathogen Genomics Center, National Institute of Infectious Diseases | Tsuyoshi Sekizuka et al |
| EPI_ISL_480031 | Japan/PG-0361/2020 | 2020-02-19 | Asia/Japan/Kanto/ | Tochigi Prefectural Institute of Public Health and Environmental Science | Pathogen Genomics Center, National Institute of Infectious Diseases | Tsuyoshi Sekizuka et al |
| EPI_ISL_480032 | Japan/PG-0362/2020 | 2020-02-21 | Asia/Japan/Kanto/ | Tochigi Prefectural Institute of Public Health and Environmental Science | Pathogen Genomics Center, National Institute of Infectious Diseases | Tsuyoshi Sekizuka et al |
| EPI_ISL_480033 | Japan/PG-0363/2020 | 2020-02-22 | Asia/Japan/Kanto/ | Tochigi Prefectural Institute of Public Health and Environmental Science | Pathogen Genomics Center, National Institute of Infectious Diseases | Tsuyoshi Sekizuka et al |
| EPI_ISL_480042 | Japan/PG-0372/2020 | 2020-02-15 | Asia/Japan/Chubu/ | Nagoya City Public Health Research Institute | Pathogen Genomics Center, National Institute of Infectious Diseases | Tsuyoshi Sekizuka et al |

| | | | | | | |
|---|---|---|---|---|---|---|
| EPI_ISL_480043 | Japan/PG-0373/2020 | 2020-02-21 | Asia/Japan/Chubu/ | Nagoya City Public Health Research Institute | Pathogen Genomics Center, National Institute of Infectious Diseases | Tsuyoshi Sekizuka et al |
| EPI_ISL_480044 | Japan/PG-0374/2020 | 2020-02-22 | Asia/Japan/Chubu/ | Nagoya City Public Health Research Institute | Pathogen Genomics Center, National Institute of Infectious Diseases | Tsuyoshi Sekizuka et al |
| EPI_ISL_480045 | Japan/PG-0375/2020 | 2020-02-22 | Asia/Japan/Chubu/ | Nagoya City Public Health Research Institute | Pathogen Genomics Center, National Institute of Infectious Diseases | Tsuyoshi Sekizuka et al |
| EPI_ISL_480046 | Japan/PG-0376/2020 | 2020-02-22 | Asia/Japan/Chubu/ | Nagoya City Public Health Research Institute | Pathogen Genomics Center, National Institute of Infectious Diseases | Tsuyoshi Sekizuka et al |
| EPI_ISL_480047 | Japan/PG-0377/2020 | 2020-02-25 | Asia/Japan/Chubu/ | Nagoya City Public Health Research Institute | Pathogen Genomics Center, National Institute of Infectious Diseases | Tsuyoshi Sekizuka et al |
| EPI_ISL_480048 | Japan/PG-0378/2020 | 2020-02-26 | Asia/Japan/Chubu/ | Nagoya City Public Health Research Institute | Pathogen Genomics Center, National Institute of Infectious Diseases | Tsuyoshi Sekizuka et al |
| EPI_ISL_480049 | Japan/PG-0379/2020 | 2020-02-26 | Asia/Japan/Chubu/ | Nagoya City Public Health Research Institute | Pathogen Genomics Center, National Institute of Infectious Diseases | Tsuyoshi Sekizuka et al |
| EPI_ISL_480059 | Japan/PG-0390/2020 | 2020-03-05 | Asia/Japan/Chubu/ | Nagoya City Public Health Research Institute | Pathogen Genomics Center, National Institute of Infectious Diseases | Tsuyoshi Sekizuka et al |

| | | | | | | |
|---|---|---|---|---|---|---|
| EPI_ISL_480097 | Japan/PG-0430/2020 | 2020-04-05 | Asia/Japan/Kansai/ | Department of Infectious Diseases, Kobe Institute of Health | Pathogen Genomics Center, National Institute of Infectious Diseases | Tsuyoshi Sekizuka et al |
| EPI_ISL_480117 | Japan/PG-0455/2020 | 2020-03-19 | Asia/Japan/Kyusyu/ | Oita Prefectural Institute of Public Health and Environmental Science | Pathogen Genomics Center, National Institute of Infectious Diseases | Tsuyoshi Sekizuka et al |
| EPI_ISL_480208 | Japan/PG-0557/2020 | 2020-04-07 | Asia/Japan/Kansai/ | Pathogen Genomics Center, National Institute of Infectious Diseases | Pathogen Genomics Center, National Institute of Infectious Diseases | Tsuyoshi Sekizuka et al |
| EPI_ISL_480229 | Turkey/IST-GLAB-004/2020 | 2020-04-13 | Europe/Turkey/Istanbul/ | Genomic Laboratory (GLAB) (Conjoint lab of Health Directorate of Istanbul and Istanbul Technical University) | Genomic Laboratory (GLAB), Istanbul Technical University | Ilker Karacan et al |
| EPI_ISL_480230 | Turkey/IST-GLAB-006/2020 | 2020-04-13 | Europe/Turkey/Istanbul/ | Genomic Laboratory (GLAB) (Conjoint lab of Health Directorate of Istanbul and Istanbul Technical University) | Genomic Laboratory (GLAB), Istanbul Technical University | Ilker Karacan et al |
| EPI_ISL_480231 | Turkey/IST-GLAB-007/2020 | 2020-04-13 | Europe/Turkey/Istanbul/ | Genomic Laboratory (GLAB) (Conjoint lab of Health Directorate of Istanbul and Istanbul Technical University) | Genomic Laboratory (GLAB), Istanbul Technical University | Ilker Karacan et al |
| EPI_ISL_480232 | Turkey/IST-GLAB-014/2020 | 2020-04-17 | Europe/Turkey/Istanbul/ | Genomic Laboratory (GLAB) (Conjoint lab of Health Directorate of Istanbul and Istanbul Technical University) | Genomic Laboratory (GLAB), Istanbul Technical University | Ilker Karacan et al |
| EPI_ISL_480233 | Turkey/IST-GLAB-021/2020 | 2020-04-17 | Europe/Turkey/Istanbul/ | Genomic Laboratory (GLAB) (Conjoint lab of Health Directorate of Istanbul and Istanbul Technical University) | Genomic Laboratory (GLAB), Istanbul Technical University | Ilker Karacan et al |

| | | | | | | |
|---|---|---|---|---|---|---|
| EPI_ISL_480234 | Turkey/IST-GLAB-030/2020 | 2020-04-18 | Europe/Turkey/Istanbul/ | Genomic Laboratory (GLAB) (Conjoint lab of Health Directorate of Istanbul and Istanbul Technical University) | Genomic Laboratory (GLAB), Istanbul Technical University | Ilker Karacan et al |
| EPI_ISL_480235 | Turkey/IST-GLAB-054/2020 | 2020-04-22 | Europe/Turkey/Istanbul/ | Genomic Laboratory (GLAB) (Conjoint lab of Health Directorate of Istanbul and Istanbul Technical University) | Genomic Laboratory (GLAB), Istanbul Technical University | Ilker Karacan et al |
| EPI_ISL_480236 | Turkey/IST-GLAB-055/2020 | 2020-04-22 | Europe/Turkey/Istanbul/ | Genomic Laboratory (GLAB) (Conjoint lab of Health Directorate of Istanbul and Istanbul Technical University) | Genomic Laboratory (GLAB), Istanbul Technical University | Ilker Karacan et al |
| EPI_ISL_480237 | Turkey/IST-GLAB-059/2020 | 2020-04-22 | Europe/Turkey/Istanbul/ | Genomic Laboratory (GLAB) (Conjoint lab of Health Directorate of Istanbul and Istanbul Technical University) | Genomic Laboratory (GLAB), Istanbul Technical University | Ilker Karacan et al |
| EPI_ISL_480238 | Turkey/IST-GLAB-062/2020 | 2020-04-22 | Europe/Turkey/Istanbul/ | Genomic Laboratory (GLAB) (Conjoint lab of Health Directorate of Istanbul and Istanbul Technical University) | Genomic Laboratory (GLAB), Istanbul Technical University | Ilker Karacan et al |
| EPI_ISL_480239 | Turkey/IST-GLAB-063/2020 | 2020-04-22 | Europe/Turkey/Istanbul/ | Genomic Laboratory (GLAB) (Conjoint lab of Health Directorate of Istanbul and Istanbul Technical University) | Genomic Laboratory (GLAB), Istanbul Technical University | Ilker Karacan et al |
| EPI_ISL_480240 | Turkey/IST-GLAB-064/2020 | 2020-04-22 | Europe/Turkey/Istanbul/ | Genomic Laboratory (GLAB) (Conjoint lab of Health Directorate of Istanbul and Istanbul Technical University) | Genomic Laboratory (GLAB), Istanbul Technical University | Ilker Karacan et al |
| EPI_ISL_480241 | Turkey/IST-GLAB-086/2020 | 2020-04-23 | Europe/Turkey/Istanbul/ | Genomic Laboratory (GLAB) (Conjoint lab of Health Directorate of Istanbul and Istanbul Technical University) | Genomic Laboratory (GLAB), Istanbul Technical University | Ilker Karacan et al |

| | | | | | | |
|---|---|---|---|---|---|---|
| EPI_ISL_480242 | Turkey/IST-GLAB-089/2020 | 2020-04-29 | Europe/Turkey/Istanbul/ | Genomic Laboratory (GLAB) (Conjoint lab of Health Directorate of Istanbul and Istanbul Technical University) | Genomic Laboratory (GLAB), Istanbul Technical University | Ilker Karacan et al |
| EPI_ISL_480243 | Turkey/IST-GLAB-091/2020 | 2020-04-23 | Europe/Turkey/Istanbul/ | Genomic Laboratory (GLAB) (Conjoint lab of Health Directorate of Istanbul and Istanbul Technical University) | Genomic Laboratory (GLAB), Istanbul Technical University | Ilker Karacan et al |
| EPI_ISL_480244 | Turkey/IST-GLAB-092/2020 | 2020-04-23 | Europe/Turkey/Istanbul/ | Genomic Laboratory (GLAB) (Conjoint lab of Health Directorate of Istanbul and Istanbul Technical University) | Genomic Laboratory (GLAB), Istanbul Technical University | Ilker Karacan et al |
| EPI_ISL_480245 | Turkey/IST-GLAB-094/2020 | 2020-04-26 | Europe/Turkey/Istanbul/ | Genomic Laboratory (GLAB) (Conjoint lab of Health Directorate of Istanbul and Istanbul Technical University) | Genomic Laboratory (GLAB), Istanbul Technical University | Ilker Karacan et al |
| EPI_ISL_480246 | Turkey/IST-GLAB-114/2020 | 2020-04-26 | Europe/Turkey/Istanbul/ | Genomic Laboratory (GLAB) (Conjoint lab of Health Directorate of Istanbul and Istanbul Technical University) | Genomic Laboratory (GLAB), Istanbul Technical University | Ilker Karacan et al |
| EPI_ISL_480247 | Turkey/IST-GLAB-115/2020 | 2020-04-26 | Europe/Turkey/Istanbul/ | Genomic Laboratory (GLAB) (Conjoint lab of Health Directorate of Istanbul and Istanbul Technical University) | Genomic Laboratory (GLAB), Istanbul Technical University | Ilker Karacan et al |
| EPI_ISL_480249 | Turkey/IST-GLAB-139/2020 | 2020-04-30 | Europe/Turkey/Istanbul/ | Genomic Laboratory (GLAB) (Conjoint lab of Health Directorate of Istanbul and Istanbul Technical University) | Genomic Laboratory (GLAB), Istanbul Technical University | Ilker Karacan et al |
| EPI_ISL_480250 | Turkey/IST-GLAB-149/2020 | 2020-05-02 | Europe/Turkey/Istanbul/ | Genomic Laboratory (GLAB) (Conjoint lab of Health Directorate of Istanbul and Istanbul Technical University) | Genomic Laboratory (GLAB), Istanbul Technical University | Ilker Karacan et al |

| | | | | | | |
|---|---|---|---|---|---|---|
| EPI_ISL_480251 | Turkey/IST-GLAB-152/2020 | 2020-05-02 | Europe/Turkey/Istanbul/ | Genomic Laboratory (GLAB) (Conjoint lab of Health Directorate of Istanbul and Istanbul Technical University) | Genomic Laboratory (GLAB), Istanbul Technical University | Ilker Karacan et al |
| EPI_ISL_480253 | Turkey/IST-GLAB-155/2020 | 2020-05-01 | Europe/Turkey/Istanbul/ | Genomic Laboratory (GLAB) (Conjoint lab of Health Directorate of Istanbul and Istanbul Technical University) | Genomic Laboratory (GLAB), Istanbul Technical University | Ilker Karacan et al |
| EPI_ISL_480257 | Turkey/IST-GLAB-162/2020 | 2020-05-03 | Europe/Turkey/Istanbul/ | Genomic Laboratory (GLAB) (Conjoint lab of Health Directorate of Istanbul and Istanbul Technical University) | Genomic Laboratory (GLAB), Istanbul Technical University | Ilker Karacan et al |
| EPI_ISL_480258 | Turkey/IST-GLAB-163/2020 | 2020-05-03 | Europe/Turkey/Istanbul/ | Genomic Laboratory (GLAB) (Conjoint lab of Health Directorate of Istanbul and Istanbul Technical University) | Genomic Laboratory (GLAB), Istanbul Technical University | Ilker Karacan et al |
| EPI_ISL_480261 | Turkey/IST-GLAB-168/2020 | 2020-05-03 | Europe/Turkey/Istanbul/ | Genomic Laboratory (GLAB) (Conjoint lab of Health Directorate of Istanbul and Istanbul Technical University) | Genomic Laboratory (GLAB), Istanbul Technical University | Ilker Karacan et al |
| EPI_ISL_480262 | Turkey/IST-GLAB-169/2020 | 2020-05-01 | Europe/Turkey/Istanbul/ | Genomic Laboratory (GLAB) (Conjoint lab of Health Directorate of Istanbul and Istanbul Technical University) | Genomic Laboratory (GLAB), Istanbul Technical University | Ilker Karacan et al |
| EPI_ISL_480264 | Turkey/IST-GLAB-171/2020 | 2020-04-20 | Europe/Turkey/Istanbul/ | Genomic Laboratory (GLAB) (Conjoint lab of Health Directorate of Istanbul and Istanbul Technical University) | Genomic Laboratory (GLAB), Istanbul Technical University | Ilker Karacan et al |
| EPI_ISL_480265 | Turkey/IST-GLAB-173/2020 | 2020-05-04 | Europe/Turkey/Istanbul/ | Genomic Laboratory (GLAB) (Conjoint lab of Health Directorate of Istanbul and Istanbul Technical University) | Genomic Laboratory (GLAB), Istanbul Technical University | Ilker Karacan et al |

| | | | | | | |
|---|---|---|---|---|---|---|
| EPI_ISL_480266 | Turkey/IST-GLAB-174/2020 | 2020-05-04 | Europe/Turkey/Istanbul/ | Genomic Laboratory (GLAB) (Conjoint lab of Health Directorate of Istanbul and Istanbul Technical University) | Genomic Laboratory (GLAB), Istanbul Technical University | Ilker Karacan et al |
| EPI_ISL_480267 | Turkey/IST-GLAB-176/2020 | 2020-05-04 | Europe/Turkey/Istanbul/ | Genomic Laboratory (GLAB) (Conjoint lab of Health Directorate of Istanbul and Istanbul Technical University) | Genomic Laboratory (GLAB), Istanbul Technical University | Ilker Karacan et al |
| EPI_ISL_480268 | Turkey/IST-GLAB-178/2020 | 2020-05-04 | Europe/Turkey/Istanbul/ | Genomic Laboratory (GLAB) (Conjoint lab of Health Directorate of Istanbul and Istanbul Technical University) | Genomic Laboratory (GLAB), Istanbul Technical University | Ilker Karacan et al |
| EPI_ISL_480269 | Turkey/IST-GLAB-179/2020 | 2020-05-04 | Europe/Turkey/Istanbul/ | Genomic Laboratory (GLAB) (Conjoint lab of Health Directorate of Istanbul and Istanbul Technical University) | Genomic Laboratory (GLAB), Istanbul Technical University | Ilker Karacan et al |
| EPI_ISL_480271 | Turkey/IST-GLAB-182/2020 | 2020-05-05 | Europe/Turkey/Istanbul/ | Genomic Laboratory (GLAB) (Conjoint lab of Health Directorate of Istanbul and Istanbul Technical University) | Genomic Laboratory (GLAB), Istanbul Technical University | Ilker Karacan et al |
| EPI_ISL_480272 | Turkey/IST-GLAB-183/2020 | 2020-05-07 | Europe/Turkey/Istanbul/ | Genomic Laboratory (GLAB) (Conjoint lab of Health Directorate of Istanbul and Istanbul Technical University) | Genomic Laboratory (GLAB), Istanbul Technical University | Ilker Karacan et al |
| EPI_ISL_480274 | Turkey/IST-GLAB-187/2020 | 2020-05-07 | Europe/Turkey/Istanbul/ | Genomic Laboratory (GLAB) (Conjoint lab of Health Directorate of Istanbul and Istanbul Technical University) | Genomic Laboratory (GLAB), Istanbul Technical University | Ilker Karacan et al |
| EPI_ISL_480275 | Turkey/IST-GLAB-188/2020 | 2020-05-07 | Europe/Turkey/Istanbul/ | Genomic Laboratory (GLAB) (Conjoint lab of Health Directorate of Istanbul and Istanbul Technical University) | Genomic Laboratory (GLAB), Istanbul Technical University | Ilker Karacan et al |

| | | | | | | |
|---|---|---|---|---|---|---|
| EPI_ISL_480276 | Turkey/IST-GLAB-189/2020 | 2020-05-05 | Europe/Turkey/Istanbul/ | Genomic Laboratory (GLAB) (Conjoint lab of Health Directorate of Istanbul and Istanbul Technical University) | Genomic Laboratory (GLAB), Istanbul Technical University | Ilker Karacan et al |
| EPI_ISL_480279 | Turkey/IST-GLAB-195/2020 | 2020-05-07 | Europe/Turkey/Istanbul/ | Genomic Laboratory (GLAB) (Conjoint lab of Health Directorate of Istanbul and Istanbul Technical University) | Genomic Laboratory (GLAB), Istanbul Technical University | Ilker Karacan et al |
| EPI_ISL_480282 | Turkey/IST-GLAB-199/2020 | 2020-05-09 | Europe/Turkey/Istanbul/ | Genomic Laboratory (GLAB) (Conjoint lab of Health Directorate of Istanbul and Istanbul Technical University) | Genomic Laboratory (GLAB), Istanbul Technical University | Ilker Karacan et al |
| EPI_ISL_480283 | Turkey/IST-GLAB-204/2020 | 2020-05-08 | Europe/Turkey/Istanbul/ | Genomic Laboratory (GLAB) (Conjoint lab of Health Directorate of Istanbul and Istanbul Technical University) | Genomic Laboratory (GLAB), Istanbul Technical University | Ilker Karacan et al |
| EPI_ISL_480286 | Turkey/IST-GLAB-207/2020 | 2020-05-10 | Europe/Turkey/Istanbul/ | Genomic Laboratory (GLAB) (Conjoint lab of Health Directorate of Istanbul and Istanbul Technical University) | Genomic Laboratory (GLAB), Istanbul Technical University | Ilker Karacan et al |
| EPI_ISL_480287 | Turkey/IST-GLAB-208/2020 | 2020-05-09 | Europe/Turkey/Istanbul/ | Genomic Laboratory (GLAB) (Conjoint lab of Health Directorate of Istanbul and Istanbul Technical University) | Genomic Laboratory (GLAB), Istanbul Technical University | Ilker Karacan et al |
| EPI_ISL_480288 | Turkey/IST-GLAB-209/2020 | 2020-05-09 | Europe/Turkey/Istanbul/ | Genomic Laboratory (GLAB) (Conjoint lab of Health Directorate of Istanbul and Istanbul Technical University) | Genomic Laboratory (GLAB), Istanbul Technical University | Ilker Karacan et al |
| EPI_ISL_480290 | Turkey/IST-GLAB-214/2020 | 2020-05-09 | Europe/Turkey/Istanbul/ | Genomic Laboratory (GLAB) (Conjoint lab of Health Directorate of Istanbul and Istanbul Technical University) | Genomic Laboratory (GLAB), Istanbul Technical University | Ilker Karacan et al |

| | | | | | | |
|---|---|---|---|---|---|---|
| EPI_ISL_480291 | Turkey/IST-GLAB-216/2020 | 2020-05-09 | Europe/Turkey/Istanbul/ | Genomic Laboratory (GLAB) (Conjoint lab of Health Directorate of Istanbul and Istanbul Technical University) | Genomic Laboratory (GLAB), Istanbul Technical University | Ilker Karacan et al |
| EPI_ISL_480292 | Turkey/IST-GLAB-217/2020 | 2020-05-10 | Europe/Turkey/Istanbul/ | Genomic Laboratory (GLAB) (Conjoint lab of Health Directorate of Istanbul and Istanbul Technical University) | Genomic Laboratory (GLAB), Istanbul Technical University | Ilker Karacan et al |
| EPI_ISL_480293 | India/KA-InStem-NCBS-0015/2020 | 2020-06-11 | Asia/India/Karnataka/ | Institute for Stem Cell Science and Regenerative Medicine | National Centre for Biological Sciences | Farhan Ali et al |
| EPI_ISL_480298 | Bulgaria/15/2020 | 2020-05-13 | Europe/Bulgaria/Blagoevgrad/ | "National Reference Laboratory ""Influenza and acute respiratory diseases""" | NRL-HIV | Ivan Ivanov et al |
| EPI_ISL_480302 | Bulgaria/21/2020 | 2020-05-07 | Europe/Bulgaria/Dupnitza/ | "National Reference Laboratory ""Influenza and acute respiratory diseases""" | NRL-HIV | Ivan Ivanov et al |
| EPI_ISL_480306 | Bulgaria/34/2020 | 2020-04-25 | Europe/Bulgaria/Vidin/ | "National Reference Laboratory ""Influenza and acute respiratory diseases""" | NRL-HIV | Ivan Ivanov et al |
| EPI_ISL_480307 | Bulgaria/36/2020 | 2020-03-30 | Europe/Bulgaria/Sofia/ | "National Reference Laboratory ""Influenza and acute respiratory diseases""" | NRL-HIV | Ivan Ivanov et al |
| EPI_ISL_480308 | Bulgaria/37/2020 | 2020-05-11 | Europe/Bulgaria/Pazardzhik/ | "National Reference Laboratory ""Influenza and acute respiratory diseases""" | NRL-HIV | Ivan Ivanov et al |
| EPI_ISL_480309 | Bulgaria/40/2020 | 2020-04-19 | Europe/Bulgaria/Sofia/ | "National Reference Laboratory ""Influenza and acute respiratory diseases""" | NRL-HIV | Ivan Ivanov et al |
| EPI_ISL_480310 | Bulgaria/41/2020 | 2020-03-10 | Europe/Bulgaria/Sofia/ | "National Reference Laboratory ""Influenza and acute respiratory diseases""" | NRL-HIV | Ivan Ivanov et al |
| EPI_ISL_480323 | CostaRica/CRV-0019/2020 | 2020-03-18 | North America/Costa Rica/San Jose/ | Hospital Nacional de Ni√±os | Charit√© Virology-University of Costa Rica | Andres Moreira-Soto et al |
| EPI_ISL_480325 | CostaRica/CRV-0021/2020 | 2020-03-26 | North America/Costa Rica/San Jose/ | Hospital Nacional de Ni√±os | Charit√© Virology-University of Costa Rica | Andres Moreira-Soto et al |
| EPI_ISL_480353 | USA/WI-UW-371/2020 | 2020-05-20 | North America/USA/Wisconsin/Dane Co | University of Wisconsin-Madison AIDS Vaccine Research Laboratories | University of Wisconsin-Madison AIDS Vaccine Research Laboratories | Gage Moreno et al |

| | | | | | | |
|---|---|---|---|---|---|---|
| EPI_ISL_480354 | USA/WI-UW-372/2020 | 2020-05-20 | North America/USA/Wisconsin/Lafayett | University of Wisconsin-Madison AIDS Vaccine Research Laboratories | University of Wisconsin-Madison AIDS Vaccine Research Laboratories | Gage Moreno et al |
| EPI_ISL_480356 | USA/WI-UW-374/2020 | 2020-05-20 | North America/USA/Wisconsin/Jefferson | University of Wisconsin-Madison AIDS Vaccine Research Laboratories | University of Wisconsin-Madison AIDS Vaccine Research Laboratories | Gage Moreno et al |
| EPI_ISL_480359 | USA/WI-UW-377/2020 | 2020-05-21 | North America/USA/Wisconsin/Dane Co | University of Wisconsin-Madison AIDS Vaccine Research Laboratories | University of Wisconsin-Madison AIDS Vaccine Research Laboratories | Gage Moreno et al |
| EPI_ISL_480361 | USA/IL-UW-379/2020 | 2020-05-22 | North America/USA/Illinois/Winnebago | University of Wisconsin-Madison AIDS Vaccine Research Laboratories | University of Wisconsin-Madison AIDS Vaccine Research Laboratories | Gage Moreno et al |
| EPI_ISL_480367 | USA/IL-UW-385/2020 | 2020-05-23 | North America/USA/Illinois/Winnebago | University of Wisconsin-Madison AIDS Vaccine Research Laboratories | University of Wisconsin-Madison AIDS Vaccine Research Laboratories | Gage Moreno et al |
| EPI_ISL_480371 | USA/WI-UW-389/2020 | 2020-05-26 | North America/USA/Wisconsin/Dane Co | University of Wisconsin-Madison AIDS Vaccine Research Laboratories | University of Wisconsin-Madison AIDS Vaccine Research Laboratories | Gage Moreno et al |
| EPI_ISL_480373 | USA/WI-UW-391/2020 | 2020-05-26 | North America/USA/Wisconsin/Dane Co | University of Wisconsin-Madison AIDS Vaccine Research Laboratories | University of Wisconsin-Madison AIDS Vaccine Research Laboratories | Gage Moreno et al |
| EPI_ISL_480374 | USA/WI-UW-392/2020 | 2020-05-20 | North America/USA/Wisconsin/Dane Co | University of Wisconsin-Madison AIDS Vaccine Research Laboratories | University of Wisconsin-Madison AIDS Vaccine Research Laboratories | Gage Moreno et al |

| | | | | | | |
|---|---|---|---|---|---|---|
| EPI_ISL_480396 | USA/WI-UW-414/2020 | 2020-04-06 | North America/USA/Wisconsin/Milwauk | University of Wisconsin-Madison AIDS Vaccine Research Laboratories | University of Wisconsin-Madison AIDS Vaccine Research Laboratories | Gage Moreno et al |
| EPI_ISL_480402 | USA/WI-UW-420/2020 | 2020-04-21 | North America/USA/Wisconsin/Milwauk | University of Wisconsin-Madison AIDS Vaccine Research Laboratories | University of Wisconsin-Madison AIDS Vaccine Research Laboratories | Gage Moreno et al |
| EPI_ISL_480405 | USA/WI-UW-423/2020 | 2020-04-27 | North America/USA/Wisconsin/Milwauk | University of Wisconsin-Madison AIDS Vaccine Research Laboratories | University of Wisconsin-Madison AIDS Vaccine Research Laboratories | Gage Moreno et al |
| EPI_ISL_480407 | USA/WI-UW-425/2020 | 2020-04-27 | North America/USA/Wisconsin/Milwauk | University of Wisconsin-Madison AIDS Vaccine Research Laboratories | University of Wisconsin-Madison AIDS Vaccine Research Laboratories | Gage Moreno et al |
| EPI_ISL_480414 | Bangladesh/BCSIR-NILMRC-130/2020 | 2020-06-17 | Asia/Bangladesh/Dhaka/Kodomtoly | National Institute of Laboratory Medicine and Referral Center | Bangladesh Council of Scientific and Industrial Research | Md. Saddam Hossain et al |
| EPI_ISL_480416 | Bangladesh/BCSIR-NILMRC-132/2020 | 2020-06-18 | Asia/Bangladesh/Dhaka/Mirpur | National Institute of Laboratory Medicine and Referral Center | Bangladesh Council of Scientific and Industrial Research | Md. Saddam Hossain et al |
| EPI_ISL_480418 | Bangladesh/BCSIR-NILMRC-144/2020 | 2020-06-18 | Asia/Bangladesh/Dhaka/Narayanganj | National Institute of Laboratory Medicine and Referral Center | Bangladesh Council of Scientific and Industrial Research | Md. Saddam Hossain et al |
| EPI_ISL_480419 | Bangladesh/BCSIR-NILMRC-133/2020 | 2020-06-18 | Asia/Bangladesh/Dhaka/Mohammadpur | National Institute of Laboratory Medicine and Referral Center | Bangladesh Council of Scientific and Industrial Research | Md. Murshed Hasan Sarkar et al |

| | | | | | | |
|---|---|---|---|---|---|---|
| EPI_ISL_480420 | Bangladesh/BCSIR-NILMRC-135/2020 | 2020-06-18 | Asia/Bangladesh/Dhaka/Mirpur | National Institute of Laboratory Medicine and Referral Center | Bangladesh Council of Scientific and Industrial Research | Md. Murshed Hasan Sarkar et al |
| EPI_ISL_480427 | Bangladesh/BCSIR-NILMRC-146/2020 | 2020-06-18 | Asia/Bangladesh/Dhaka/Tejgaon | National Institute of Laboratory Medicine and Referral Center | Bangladesh Council of Scientific and Industrial Research | Shahina Akter et al |
| EPI_ISL_480442 | Bangladesh/BCSIR-NILMRC-153/2020 | 2020-06-18 | Asia/Bangladesh/Rajshahi/ | National Institute of Laboratory Medicine and Referral Center | Bangladesh Council of Scientific and Industrial Research | Barna Goswami et al |
| EPI_ISL_480445 | Bangladesh/BCSIR-NILMRC-154/2020 | 2020-06-18 | Asia/Bangladesh/Rajshahi/ | National Institute of Laboratory Medicine and Referral Center | Genomic Research Lab, BCSIR | Md. Ahasan Habib et al |
| EPI_ISL_480449 | Bangladesh/BCSIR-NILMRC-172/2020 | 2020-06-18 | Asia/Bangladesh/Rajshahi/ | National Institute of Laboratory Medicine and Referral Center | Genomic Research Lab, BCSIR | Abu Sayeed Mohammad Mahmud et al |
| EPI_ISL_480778 | Australia/SA01/2020 | 2020-01-30 | Oceania/Australia/South Australia/ | Victorian Infectious Diseases Reference Laboratory (VIDRL) | VIDRL and MDU-PHL | Caly L. et al |
| EPI_ISL_480779 | Australia/SA02/2020 | 2020-01-30 | Oceania/Australia/South Australia/ | Victorian Infectious Diseases Reference Laboratory (VIDRL) | VIDRL and MDU-PHL | Caly L. et al |
| EPI_ISL_480780 | Australia/SA03/2020 | 2020-01-30 | Oceania/Australia/South Australia/ | Victorian Infectious Diseases Reference Laboratory (VIDRL) | VIDRL and MDU-PHL | Caly L. et al |
| EPI_ISL_480781 | Australia/SA04/2020 | 2020-01-30 | Oceania/Australia/South Australia/ | Victorian Infectious Diseases Reference Laboratory (VIDRL) | VIDRL and MDU-PHL | Caly L. et al |
| EPI_ISL_480785 | USA/NY-CDC-6684/2020 | 2020-04-06 | North America/USA/New York/New Yor | NYC Department of Health and Mental Hygiene | Pathogen Discovery, Respiratory Viruses Branch, Division of Viral Diseases, Centers for Disease Control and Prevention | Krista Queen et al |
| EPI_ISL_480814 | USA/FL-BPHL-0163/2020 | 2020-04-01 | North America/USA/Florida/ | Florida Bureau of Public Health Laboratories | Florida Bureau of Public Health Laboratories | Sarah Schmedes et al |

| | | | | | | |
|---|---|---|---|---|---|---|
| EPI_ISL_480817 | USA/FL-BPHL-0166/2020 | 2020-04-02 | North America/USA/Florida/ | Florida Bureau of Public Health Laboratories | Florida Bureau of Public Health Laboratories | Sarah Schmedes et al |
| EPI_ISL_480828 | USA/FL-BPHL-0177/2020 | 2020-04-04 | North America/USA/Florida/ | Florida Bureau of Public Health Laboratories | Florida Bureau of Public Health Laboratories | Sarah Schmedes et al |
| EPI_ISL_480833 | USA/FL-BPHL-0182/2020 | 2020-04-03 | North America/USA/Florida/ | Florida Bureau of Public Health Laboratories | Florida Bureau of Public Health Laboratories | Sarah Schmedes et al |
| EPI_ISL_480835 | USA/FL-BPHL-0184/2020 | 2020-04-03 | North America/USA/Florida/ | Florida Bureau of Public Health Laboratories | Florida Bureau of Public Health Laboratories | Sarah Schmedes et al |
| EPI_ISL_480844 | USA/FL-BPHL-0193/2020 | 2020-04-04 | North America/USA/Florida/ | Florida Bureau of Public Health Laboratories | Florida Bureau of Public Health Laboratories | Sarah Schmedes et al |
| EPI_ISL_480858 | USA/FL-BPHL-0207/2020 | 2020-04-14 | North America/USA/Florida/ | Florida Bureau of Public Health Laboratories | Florida Bureau of Public Health Laboratories | Sarah Schmedes et al |
| EPI_ISL_480879 | USA/FL-BPHL-0228/2020 | 2020-04-20 | North America/USA/Florida/ | Florida Bureau of Public Health Laboratories | Florida Bureau of Public Health Laboratories | Sarah Schmedes et al |
| EPI_ISL_480887 | USA/FL-BPHL-0236/2020 | 2020-04-24 | North America/USA/Florida/ | Florida Bureau of Public Health Laboratories | Florida Bureau of Public Health Laboratories | Sarah Schmedes et al |
| EPI_ISL_480900 | USA/FL-BPHL-0249/2020 | 2020-05-02 | North America/USA/Florida/ | Florida Bureau of Public Health Laboratories | Florida Bureau of Public Health Laboratories | Sarah Schmedes et al |
| EPI_ISL_480901 | USA/FL-BPHL-0250/2020 | 2020-05-02 | North America/USA/Florida/ | Florida Bureau of Public Health Laboratories | Florida Bureau of Public Health Laboratories | Sarah Schmedes et al |
| EPI_ISL_480902 | USA/FL-BPHL-0251/2020 | 2020-05-04 | North America/USA/Florida/ | Florida Bureau of Public Health Laboratories | Florida Bureau of Public Health Laboratories | Sarah Schmedes et al |
| EPI_ISL_480921 | USA/FL-BPHL-0270/2020 | 2020-05-05 | North America/USA/Florida/ | Florida Bureau of Public Health Laboratories | Florida Bureau of Public Health Laboratories | Sarah Schmedes et al |
| EPI_ISL_480923 | USA/FL-BPHL-0272/2020 | 2020-05-05 | North America/USA/Florida/ | Florida Bureau of Public Health Laboratories | Florida Bureau of Public Health Laboratories | Sarah Schmedes et al |

| | | | | | | |
|---|---|---|---|---|---|---|
| EPI_ISL_480928 | USA/FL-BPHL-0277/2020 | 2020-05-07 | North America/USA/Florida/ | Florida Bureau of Public Health Laboratories | Florida Bureau of Public Health Laboratories | Sarah Schmedes et al |
| EPI_ISL_480929 | USA/FL-BPHL-0278/2020 | 2020-05-07 | North America/USA/Florida/ | Florida Bureau of Public Health Laboratories | Florida Bureau of Public Health Laboratories | Sarah Schmedes et al |
| EPI_ISL_480932 | USA/FL-BPHL-0281/2020 | 2020-05-12 | North America/USA/Florida/ | Florida Bureau of Public Health Laboratories | Florida Bureau of Public Health Laboratories | Sarah Schmedes et al |
| EPI_ISL_480944 | USA/FL-BPHL-0293/2020 | 2020-05-15 | North America/USA/Florida/ | Florida Bureau of Public Health Laboratories | Florida Bureau of Public Health Laboratories | Sarah Schmedes et al |
| EPI_ISL_480946 | USA/FL-BPHL-0295/2020 | 2020-05-14 | North America/USA/Florida/ | Florida Bureau of Public Health Laboratories | Florida Bureau of Public Health Laboratories | Sarah Schmedes et al |
| EPI_ISL_480955 | Spain/PV-IBV-003263/2020 | 2020-03-17 | Europe/Spain/Basque Country/San Seba | Servicio de Microbiolog√≠a. Hospital Universitario Donostia. OSI Donostialdea. √Årea de Enfermedades Infecciosas, Grupo de Infecci√≥n Respiratoria y Resistencia Antimicrobiana. Instituto de Investigaci√≥n Sanitaria Biodonostia | SeqCOVID-SPAIN consortium/IBV(CSIC) | Gustavo Cilla et al |
| EPI_ISL_480963 | Spain/PV-IBV-003221/2020 | 2020-03-14 | Europe/Spain/Basque Country/San Seba | Servicio de Microbiolog√≠a. Hospital Universitario Donostia. OSI Donostialdea. √Årea de Enfermedades Infecciosas, Grupo de Infecci√≥n Respiratoria y Resistencia Antimicrobiana. Instituto de Investigaci√≥n Sanitaria Biodonostia | SeqCOVID-SPAIN consortium/IBV(CSIC) | Gustavo Cilla et al |
| EPI_ISL_480966 | Spain/PV-IBV-003251/2020 | 2020-03-17 | Europe/Spain/Basque Country/San Seba | Servicio de Microbiolog√≠a. Hospital Universitario Donostia. OSI Donostialdea. √Årea de Enfermedades Infecciosas, Grupo de Infecci√≥n Respiratoria y Resistencia Antimicrobiana. Instituto de Investigaci√≥n Sanitaria Biodonostia | SeqCOVID-SPAIN consortium/IBV(CSIC) | Gustavo Cilla et al |
| EPI_ISL_480974 | Spain/PV-IBV-003211/2020 | 2020-03-13 | Europe/Spain/Basque Country/San Seba | Servicio de Microbiolog√≠a. Hospital Universitario Donostia. OSI Donostialdea. √Årea de Enfermedades Infecciosas, Grupo de Infecci√≥n Respiratoria y Resistencia Antimicrobiana. Instituto de Investigaci√≥n Sanitaria Biodonostia | SeqCOVID-SPAIN consortium/IBV(CSIC) | Gustavo Cilla et al |

| | | | | | | |
|---|---|---|---|---|---|---|
| EPI_ISL_480977 | Spain/PV-IBV-003266/2020 | 2020-03-18 | Europe/Spain/Basque Country/San Seba | Servicio de Microbiolog√≠a. Hospital Universitario Donostia. OSI Donostialdea. √Årea de Enfermedades Infecciosas, Grupo de Infecci√≥n Respiratoria y Resistencia Antimicrobiana. Instituto de Investigaci√≥n Sanitaria Biodonostia | SeqCOVID-SPAIN consortium/IBV(CSIC) | Gustavo Cilla et al |
| EPI_ISL_480979 | Spain/PV-IBV-003191/2020 | 2020-03-02 | Europe/Spain/Basque Country/San Seba | Servicio de Microbiolog√≠a. Hospital Universitario Donostia. OSI Donostialdea. √Årea de Enfermedades Infecciosas, Grupo de Infecci√≥n Respiratoria y Resistencia Antimicrobiana. Instituto de Investigaci√≥n Sanitaria Biodonostia | SeqCOVID-SPAIN consortium/IBV(CSIC) | Gustavo Cilla et al |
| EPI_ISL_480982 | Spain/PV-IBV-003196/2020 | 2020-03-10 | Europe/Spain/Basque Country/San Seba | Servicio de Microbiolog√≠a. Hospital Universitario Donostia. OSI Donostialdea. √Årea de Enfermedades Infecciosas, Grupo de Infecci√≥n Respiratoria y Resistencia Antimicrobiana. Instituto de Investigaci√≥n Sanitaria Biodonostia | SeqCOVID-SPAIN consortium/IBV(CSIC) | Gustavo Cilla et al |
| EPI_ISL_480988 | Spain/PV-IBV-003230/2020 | 2020-03-15 | Europe/Spain/Basque Country/San Seba | Servicio de Microbiolog√≠a. Hospital Universitario Donostia. OSI Donostialdea. √Årea de Enfermedades Infecciosas, Grupo de Infecci√≥n Respiratoria y Resistencia Antimicrobiana. Instituto de Investigaci√≥n Sanitaria Biodonostia | SeqCOVID-SPAIN consortium/IBV(CSIC) | Gustavo Cilla et al |
| EPI_ISL_480990 | Spain/PV-IBV-003220/2020 | 2020-03-14 | Europe/Spain/Basque Country/San Seba | Servicio de Microbiolog√≠a. Hospital Universitario Donostia. OSI Donostialdea. √Årea de Enfermedades Infecciosas, Grupo de Infecci√≥n Respiratoria y Resistencia Antimicrobiana. Instituto de Investigaci√≥n Sanitaria Biodonostia | SeqCOVID-SPAIN consortium/IBV(CSIC) | Gustavo Cilla et al |
| EPI_ISL_480994 | Spain/PV-IBV-003256/2020 | 2020-03-17 | Europe/Spain/Basque Country/San Seba | Servicio de Microbiolog√≠a. Hospital Universitario Donostia. OSI Donostialdea. √Årea de Enfermedades Infecciosas, Grupo de Infecci√≥n Respiratoria y Resistencia Antimicrobiana. Instituto de Investigaci√≥n Sanitaria Biodonostia | SeqCOVID-SPAIN consortium/IBV(CSIC) | Gustavo Cilla et al |
| EPI_ISL_480995 | Spain/CT-IBV-003733/2020 | 2020-04-04 | Europe/Spain/Catalunya/Barcelona | ISGlobal, Institut de Salut Global de Barcelona | SeqCOVID-SPAIN consortium/IBV(CSIC) | Alfredo Mayor et al |

| | | | | | | |
|---|---|---|---|---|---|---|
| EPI_ISL_481010 | Spain/PV-IBV-003198/2020 | 2020-03-11 | Europe/Spain/Basque Country/San Seba | Servicio de Microbiolog√≠a. Hospital Universitario Donostia. OSI Donostialdea. √Årea de Enfermedades Infecciosas, Grupo de Infecci√≥n Respiratoria y Resistencia Antimicrobiana. Instituto de Investigaci√≥n Sanitaria Biodonostia | SeqCOVID-SPAIN consortium/IBV(CSIC) | Gustavo Cilla et al |
| EPI_ISL_481014 | Spain/PV-IBV-003232/2020 | 2020-03-15 | Europe/Spain/Basque Country/San Seba | Servicio de Microbiolog√≠a. Hospital Universitario Donostia. OSI Donostialdea. √Årea de Enfermedades Infecciosas, Grupo de Infecci√≥n Respiratoria y Resistencia Antimicrobiana. Instituto de Investigaci√≥n Sanitaria Biodonostia | SeqCOVID-SPAIN consortium/IBV(CSIC) | Gustavo Cilla et al |
| EPI_ISL_481015 | Spain/PV-IBV-003208/2020 | 2020-03-12 | Europe/Spain/Basque Country/San Seba | Servicio de Microbiolog√≠a. Hospital Universitario Donostia. OSI Donostialdea. √Årea de Enfermedades Infecciosas, Grupo de Infecci√≥n Respiratoria y Resistencia Antimicrobiana. Instituto de Investigaci√≥n Sanitaria Biodonostia | SeqCOVID-SPAIN consortium/IBV(CSIC) | Gustavo Cilla et al |
| EPI_ISL_481017 | Spain/CT-IBV-003735/2020 | 2020-04-08 | Europe/Spain/Catalunya/Barcelona | ISGlobal, Institut de Salut Global de Barcelona | SeqCOVID-SPAIN consortium/IBV(CSIC) | Alfredo Mayor et al |
| EPI_ISL_481020 | Spain/PV-IBV-003199/2020 | 2020-03-11 | Europe/Spain/Basque Country/San Seba | Servicio de Microbiolog√≠a. Hospital Universitario Donostia. OSI Donostialdea. √Årea de Enfermedades Infecciosas, Grupo de Infecci√≥n Respiratoria y Resistencia Antimicrobiana. Instituto de Investigaci√≥n Sanitaria Biodonostia | SeqCOVID-SPAIN consortium/IBV(CSIC) | Gustavo Cilla et al |
| EPI_ISL_481029 | Spain/CT-IBV-003736/2020 | 2020-04-08 | Europe/Spain/Catalunya/Barcelona | ISGlobal, Institut de Salut Global de Barcelona | SeqCOVID-SPAIN consortium/IBV(CSIC) | Alfredo Mayor et al |
| EPI_ISL_481035 | Spain/CT-IBV-003731/2020 | 2020-04-04 | Europe/Spain/Catalunya/Barcelona | ISGlobal, Institut de Salut Global de Barcelona | SeqCOVID-SPAIN consortium/IBV(CSIC) | Alfredo Mayor et al |
| EPI_ISL_481038 | Spain/PV-IBV-003218/2020 | 2020-03-14 | Europe/Spain/Basque Country/San Seba | Servicio de Microbiolog√≠a. Hospital Universitario Donostia. OSI Donostialdea. √Årea de Enfermedades Infecciosas, Grupo de Infecci√≥n Respiratoria y Resistencia Antimicrobiana. Instituto de Investigaci√≥n Sanitaria Biodonostia | SeqCOVID-SPAIN consortium/IBV(CSIC) | Gustavo Cilla et al |

| | | | | | | |
|---|---|---|---|---|---|---|
| EPI_ISL_481041 | Spain/MD-IBV-004968/2020 | 2020-05-07 | Europe/Spain/Madrid/ | Hospital General Universitario Gregorio Marav√±√≥n | SeqCOVID-SPAIN consortium/IBV(CSIC) | Laura P√©rez-Lago et al |
| EPI_ISL_481042 | Spain/MD-IBV-004960/2020 | 2020-05-03 | Europe/Spain/Madrid/ | Hospital General Universitario Gregorio Marav√±√≥n | SeqCOVID-SPAIN consortium/IBV(CSIC) | Laura P√©rez-Lago et al |
| EPI_ISL_481055 | Spain/MD-IBV-004915/2020 | 2020-03-11 | Europe/Spain/Madrid/ | Hospital General Universitario Gregorio Marav√±√≥n | SeqCOVID-SPAIN consortium/IBV(CSIC) | Laura P√©rez-Lago et al |
| EPI_ISL_481063 | Spain/MD-IBV-004982/2020 | 2020-05-29 | Europe/Spain/Madrid/ | Hospital General Universitario Gregorio Marav√±√≥n | SeqCOVID-SPAIN consortium/IBV(CSIC) | Laura P√©rez-Lago et al |
| EPI_ISL_481081 | Spain/MD-IBV-004939/2020 | 2020-04-10 | Europe/Spain/Madrid/ | Hospital General Universitario Gregorio Marav√±√≥n | SeqCOVID-SPAIN consortium/IBV(CSIC) | Laura P√©rez-Lago et al |
| EPI_ISL_481094 | Spain/MD-IBV-004901/2020 | 2020-03-09 | Europe/Spain/Madrid/ | Hospital General Universitario Gregorio Marav√±√≥n | SeqCOVID-SPAIN consortium/IBV(CSIC) | Laura P√©rez-Lago et al |
| EPI_ISL_481095 | Spain/MD-IBV-004974/2020 | 2020-05-11 | Europe/Spain/Madrid/ | Hospital General Universitario Gregorio Marav√±√≥n | SeqCOVID-SPAIN consortium/IBV(CSIC) | Laura P√©rez-Lago et al |
| EPI_ISL_481099 | Spain/MD-IBV-004967/2020 | 2020-05-06 | Europe/Spain/Madrid/ | Hospital General Universitario Gregorio Marav√±√≥n | SeqCOVID-SPAIN consortium/IBV(CSIC) | Laura P√©rez-Lago et al |
| EPI_ISL_481103 | Spain/MD-IBV-004919/2020 | 2020-03-13 | Europe/Spain/Madrid/ | Hospital General Universitario Gregorio Marav√±√≥n | SeqCOVID-SPAIN consortium/IBV(CSIC) | Laura P√©rez-Lago et al |
| EPI_ISL_481113 | India/OR-ILSCV25524/2020 | 2020-05-19 | Asia/India/Odisha/ | Immunogenomics lab, Institute of Life Sciences, Bhubaneswar | Immunogenomics lab, Institute of Life Sciences, Bhubaneswar | Sunil Raghav et al |
| EPI_ISL_481119 | India/OR-ILSCV27406/2020 | 2020-05-21 | Asia/India/Odisha/ | Immunogenomics lab, Institute of Life Sciences, Bhubaneswar | Immunogenomics lab, Institute of Life Sciences, Bhubaneswar | Sunil Raghav et al |
| EPI_ISL_481125 | India/OR-ILSCV28062/2020 | 2020-05-21 | Asia/India/Odisha/ | Immunogenomics lab, Institute of Life Sciences, Bhubaneswar | Immunogenomics lab, Institute of Life Sciences, Bhubaneswar | Sunil Raghav et al |

| | | | | | | |
|---|---|---|---|---|---|---|
| EPI_ISL_481128 | India/OR-ILSCV29215/2020 | 2020-05-23 | Asia/India/Odisha/ | Immunogenomics lab, Institute of Life Sciences, Bhubaneswar | Immunogenomics lab, Institute of Life Sciences, Bhubaneswar | Sunil Raghav et al |
| EPI_ISL_481133 | India/OR-ILSCV29960/2020 | 2020-05-23 | Asia/India/Odisha/ | Immunogenomics lab, Institute of Life Sciences, Bhubaneswar | Immunogenomics lab, Institute of Life Sciences, Bhubaneswar | Sunil Raghav et al |
| EPI_ISL_481138 | India/OR-ILSCV30115/2020 | 2020-05-23 | Asia/India/Odisha/ | Immunogenomics lab, Institute of Life Sciences, Bhubaneswar | Immunogenomics lab, Institute of Life Sciences, Bhubaneswar | Sunil Raghav et al |
| EPI_ISL_481140 | India/OR-ILSCV30141/2020 | 2020-05-23 | Asia/India/Odisha/ | Immunogenomics lab, Institute of Life Sciences, Bhubaneswar | Immunogenomics lab, Institute of Life Sciences, Bhubaneswar | Sunil Raghav et al |
| EPI_ISL_481143 | India/OR-ILSCV30220/2020 | 2020-05-23 | Asia/India/Odisha/ | Immunogenomics lab, Institute of Life Sciences, Bhubaneswar | Immunogenomics lab, Institute of Life Sciences, Bhubaneswar | Sunil Raghav et al |
| EPI_ISL_481145 | India/OR-ILSCV30273/2020 | 2020-05-23 | Asia/India/Odisha/ | Immunogenomics lab, Institute of Life Sciences, Bhubaneswar | Immunogenomics lab, Institute of Life Sciences, Bhubaneswar | Sunil Raghav et al |
| EPI_ISL_481146 | India/OR-ILSCV30442/2020 | 2020-05-24 | Asia/India/Odisha/ | Immunogenomics lab, Institute of Life Sciences, Bhubaneswar | Immunogenomics lab, Institute of Life Sciences, Bhubaneswar | Sunil Raghav et al |
| EPI_ISL_481152 | India/OR-ILSCV31694/2020 | 2020-05-26 | Asia/India/Odisha/ | Immunogenomics lab, Institute of Life Sciences, Bhubaneswar | Immunogenomics lab, Institute of Life Sciences, Bhubaneswar | Sunil Raghav et al |
| EPI_ISL_481157 | India/OR-ILSCV32485/2020 | 2020-06-02 | Asia/India/Odisha/ | Immunogenomics lab, Institute of Life Sciences, Bhubaneswar | Immunogenomics lab, Institute of Life Sciences, Bhubaneswar | Sunil Raghav et al |
| EPI_ISL_481160 | India/OR-ILSCV32652/2020 | 2020-06-03 | Asia/India/Odisha/ | Immunogenomics lab, Institute of Life Sciences, Bhubaneswar | Immunogenomics lab, Institute of Life Sciences, Bhubaneswar | Sunil Raghav et al |

| | | | | | | |
|---|---|---|---|---|---|---|
| EPI_ISL_481161 | India/OR-ILSCV32656/2020 | 2020-06-03 | Asia/India/Odisha/ | Immunogenomics lab, Institute of Life Sciences, Bhubaneswar | Immunogenomics lab, Institute of Life Sciences, Bhubaneswar | Sunil Raghav et al |
| EPI_ISL_481162 | India/OR-ILSCV32670/2020 | 2020-06-03 | Asia/India/Odisha/ | Immunogenomics lab, Institute of Life Sciences, Bhubaneswar | Immunogenomics lab, Institute of Life Sciences, Bhubaneswar | Sunil Raghav et al |
| EPI_ISL_481166 | India/OR-ILSCV32817/2020 | 2020-06-04 | Asia/India/Odisha/ | Immunogenomics lab, Institute of Life Sciences, Bhubaneswar | Immunogenomics lab, Institute of Life Sciences, Bhubaneswar | Sunil Raghav et al |
| EPI_ISL_481169 | India/OR-ILSCV32872/2020 | 2020-06-04 | Asia/India/Odisha/ | Immunogenomics lab, Institute of Life Sciences, Bhubaneswar | Immunogenomics lab, Institute of Life Sciences, Bhubaneswar | Sunil Raghav et al |
| EPI_ISL_481170 | India/OR-ILSCV32879/2020 | 2020-06-05 | Asia/India/Odisha/ | Immunogenomics lab, Institute of Life Sciences, Bhubaneswar | Immunogenomics lab, Institute of Life Sciences, Bhubaneswar | Sunil Raghav et al |
| EPI_ISL_481171 | India/OR-ILSCV32889/2020 | 2020-06-05 | Asia/India/Odisha/ | Immunogenomics lab, Institute of Life Sciences, Bhubaneswar | Immunogenomics lab, Institute of Life Sciences, Bhubaneswar | Sunil Raghav et al |
| EPI_ISL_481172 | India/OR-ILSCV32892/2020 | 2020-06-05 | Asia/India/Odisha/ | Immunogenomics lab, Institute of Life Sciences, Bhubaneswar | Immunogenomics lab, Institute of Life Sciences, Bhubaneswar | Sunil Raghav et al |
| EPI_ISL_481173 | India/OR-ILSCV32893/2020 | 2020-06-05 | Asia/India/Odisha/ | Immunogenomics lab, Institute of Life Sciences, Bhubaneswar | Immunogenomics lab, Institute of Life Sciences, Bhubaneswar | Sunil Raghav et al |
| EPI_ISL_481174 | India/OR-ILSCV32895/2020 | 2020-06-05 | Asia/India/Odisha/ | Immunogenomics lab, Institute of Life Sciences, Bhubaneswar | Immunogenomics lab, Institute of Life Sciences, Bhubaneswar | Sunil Raghav et al |
| EPI_ISL_481181 | India/OR-ILSCV33983/2020 | 2020-06-08 | Asia/India/Odisha/ | Immunogenomics lab, Institute of Life Sciences, Bhubaneswar | Immunogenomics lab, Institute of Life Sciences, Bhubaneswar | Sunil Raghav et al |

| | | | | | | |
|---|---|---|---|---|---|---|
| EPI_ISL_481183 | India/OR-ILSCV34479/2020 | 2020-06-09 | Asia/India/Odisha/ | Immunogenomics lab, Institute of Life Sciences, Bhubaneswar | Immunogenomics lab, Institute of Life Sciences, Bhubaneswar | Sunil Raghav et al |
| EPI_ISL_481185 | India/OR-ILSCV34933/2020 | 2020-06-10 | Asia/India/Odisha/ | Immunogenomics lab, Institute of Life Sciences, Bhubaneswar | Immunogenomics lab, Institute of Life Sciences, Bhubaneswar | Sunil Raghav et al |
| EPI_ISL_481186 | India/OR-ILSCV34939/2020 | 2020-06-10 | Asia/India/Odisha/ | Immunogenomics lab, Institute of Life Sciences, Bhubaneswar | Immunogenomics lab, Institute of Life Sciences, Bhubaneswar | Sunil Raghav et al |
| EPI_ISL_481187 | India/OR-ILSCV35141/2020 | 2020-06-10 | Asia/India/Odisha/ | Immunogenomics lab, Institute of Life Sciences, Bhubaneswar | Immunogenomics lab, Institute of Life Sciences, Bhubaneswar | Sunil Raghav et al |
| EPI_ISL_481188 | India/OR-ILSCV35308/2020 | 2020-06-10 | Asia/India/Odisha/ | Immunogenomics lab, Institute of Life Sciences, Bhubaneswar | Immunogenomics lab, Institute of Life Sciences, Bhubaneswar | Sunil Raghav et al |
| EPI_ISL_481191 | India/OR-ILSCV35350/2020 | 2020-06-10 | Asia/India/Odisha/ | Immunogenomics lab, Institute of Life Sciences, Bhubaneswar | Immunogenomics lab, Institute of Life Sciences, Bhubaneswar | Sunil Raghav et al |
| EPI_ISL_481192 | India/OR-ILSCV35351/2020 | 2020-06-10 | Asia/India/Odisha/ | Immunogenomics lab, Institute of Life Sciences, Bhubaneswar | Immunogenomics lab, Institute of Life Sciences, Bhubaneswar | Sunil Raghav et al |
| EPI_ISL_481194 | India/OR-ILSCV35366/2020 | 2020-06-10 | Asia/India/Odisha/ | Immunogenomics lab, Institute of Life Sciences, Bhubaneswar | Immunogenomics lab, Institute of Life Sciences, Bhubaneswar | Sunil Raghav et al |
| EPI_ISL_481198 | India/OR-ILSCV35461/2020 | 2020-06-10 | Asia/India/Odisha/ | Immunogenomics lab, Institute of Life Sciences, Bhubaneswar | Immunogenomics lab, Institute of Life Sciences, Bhubaneswar | Sunil Raghav et al |
| EPI_ISL_481200 | India/OR-ILSCV35476/2020 | 2020-06-10 | Asia/India/Odisha/ | Immunogenomics lab, Institute of Life Sciences, Bhubaneswar | Immunogenomics lab, Institute of Life Sciences, Bhubaneswar | Sunil Raghav et al |

| | | | | | | |
|---|---|---|---|---|---|---|
| EPI_ISL_481202 | India/OR-ILSCV35574/2020 | 2020-06-11 | Asia/India/Odisha/ | Immunogenomics lab, Institute of Life Sciences, Bhubaneswar | Immunogenomics lab, Institute of Life Sciences, Bhubaneswar | Sunil Raghav et al |
| EPI_ISL_481204 | India/OR-ILSCV35615/2020 | 2020-06-12 | Asia/India/Odisha/ | Immunogenomics lab, Institute of Life Sciences, Bhubaneswar | Immunogenomics lab, Institute of Life Sciences, Bhubaneswar | Sunil Raghav et al |
| EPI_ISL_481207 | Norway/2596/2020 | 2020-05-12 | Europe/Norway/Norway/ | Ostfold Hospital Trust - Kalnes, Centre for Laboratory Medicine, Section for gene technology and infection serology | Norwegian Institute of Public Health, Department of Virology | Kathrine Stene-Johansen et al |
| EPI_ISL_481208 | Norway/2448/2020 | 2020-04-20 | Europe/Norway/Norway/ | Furst Medical Laboratory | Norwegian Institute of Public Health, Department of Virology | Kathrine Stene-Johansen et al |
| EPI_ISL_481210 | Norway/2498/2020 | 2020-05-04 | Europe/Norway/Norway/ | Ostfold Hospital Trust - Kalnes, Centre for Laboratory Medicine, Section for gene technology and infection serology | Norwegian Institute of Public Health, Department of Virology | Kathrine Stene-Johansen et al |
| EPI_ISL_481218 | Norway/2508/2020 | 2020-04-13 | Europe/Norway/Norway/ | Oslo University Hospital, Department of Medical Microbiology | Norwegian Institute of Public Health, Department of Virology | Kathrine Stene-Johansen et al |
| EPI_ISL_481220 | Senegal/2840/2020 | 2020-04-06 | Africa/Senegal/Dakar/ | Institut Pasteur Dakar | Institut Pasteur de Dakar | Ndongo Dia et al |
| EPI_ISL_481227 | Belgium/UGent-133/2020 | 2020-04-28 | Europe/Belgium/Gent/ | Lab voor klinische biologie | Onderzoeksgroep Virologie | Nick Vereecke et al |
| EPI_ISL_481229 | Belgium/UGent-136/2020 | 2020-04-16 | Europe/Belgium/Gent/ | Lab voor klinische biologie | Onderzoeksgroep Virologie | Nick Vereecke et al |
| EPI_ISL_481230 | Belgium/UGent-138/2020 | 2020-04-27 | Europe/Belgium/Gent/ | Lab voor klinische biologie | Onderzoeksgroep Virologie | Nick Vereecke et al |
| EPI_ISL_481242 | USA/MN-MDH-1224/2020 | 2020-06-06 | North America/USA/Minnesota/ | Mayo Clinic & Mayo Clinic Laboratories | Minnesota Department of Health, Public Health Laboratory | Matt Plumb et al |

| | | | | | | |
|---|---|---|---|---|---|---|
| EPI_ISL_481244 | Ecuador/USFQ-026/2020 | 2020-04-17 | South America/Ecuador/Los Rios/Babaho | Hospital IESS Babahoyo | Institute of Microbiology, Universidad San Francisco de Quito | Bel√©n Prado-Vivar et al |
| EPI_ISL_481245 | Ecuador/USFQ-039/2020 | 2020-04-17 | South America/Ecuador/Los Rios/Babaho | Hospital IESS Babahoyo | Institute of Microbiology, Universidad San Francisco de Quito | Bel√©n Prado-Vivar et al |
| EPI_ISL_481246 | Ecuador/USFQ-054/2020 | 2020-04-17 | South America/Ecuador/Los Rios/Babaho | Hospital IESS Babahoyo | Institute of Microbiology, Universidad San Francisco de Quito | Bel√©n Prado-Vivar et al |
| EPI_ISL_481247 | Ecuador/USFQ-065/2020 | 2020-04-17 | South America/Ecuador/Los Rios/Babaho | Hospital IESS Babahoyo | Institute of Microbiology, Universidad San Francisco de Quito | Bel√©n Prado-Vivar et al |
| EPI_ISL_481248 | Ecuador/USFQ-066/2020 | 2020-04-17 | South America/Ecuador/Los Rios/Babaho | Hospital IESS Babahoyo | Institute of Microbiology, Universidad San Francisco de Quito | Bel√©n Prado-Vivar et al |
| EPI_ISL_481253 | Germany/BW-RKI-N-0001/2020 | 2020-03-12 | Europe/Germany/Baden-Wuerttemberg | Robert Koch Institute, National Reference center for Influenza, Berlin, Germany | Robert Koch Institute, Bioinformatics MF1, Berlin, Germany | Marianne Wedde et al |
| EPI_ISL_481264 | Germany/BE-RKI-N-0025/2020 | 2020-03-27 | Europe/Germany/Berlin/ | Robert Koch Institute, National Reference center for Influenza, Berlin, Germany | Robert Koch Institute, Bioinformatics MF1, Berlin, Germany | Marianne Wedde et al |
| EPI_ISL_481266 | USA/MD-MDH-0017/2020 | 2020-04-01 | North America/USA/Maryland/ | Maryland Department of Health | Maryland Department of Health | Keller et al |
| EPI_ISL_481274 | USA/MD-MDH-0028/2020 | 2020-04-04 | North America/USA/Maryland/ | Maryland Department of Health | Maryland Department of Health | Keller et al |

| | | | | | | |
|---|---|---|---|---|---|---|
| EPI_ISL_481370 | SouthKorea/KCDC2050/2020 | 2020-02-21 | Asia/South Korea/South Korea/ | Division of Viral Diseases, Center for Laboratory Control of Infectious Diseases, Korea Centers for Diseases Control and Prevention | Division of Viral Diseases, Center for Laboratory Control of Infectious Diseases, Korea Centers for Diseases Control and Prevention | Jeong-Min Kim et al |
| EPI_ISL_481371 | SouthKorea/KCDC2051/2020 | 2020-02-21 | Asia/South Korea/South Korea/ | Division of Viral Diseases, Center for Laboratory Control of Infectious Diseases, Korea Centers for Diseases Control and Prevention | Division of Viral Diseases, Center for Laboratory Control of Infectious Diseases, Korea Centers for Diseases Control and Prevention | Jeong-Min Kim et al |
| EPI_ISL_481372 | SouthKorea/KCDC2052/2020 | 2020-02-21 | Asia/South Korea/South Korea/ | Division of Viral Diseases, Center for Laboratory Control of Infectious Diseases, Korea Centers for Diseases Control and Prevention | Division of Viral Diseases, Center for Laboratory Control of Infectious Diseases, Korea Centers for Diseases Control and Prevention | Jeong-Min Kim et al |
| EPI_ISL_481373 | SouthKorea/KCDC2053/2020 | 2020-02-23 | Asia/South Korea/South Korea/ | Division of Viral Diseases, Center for Laboratory Control of Infectious Diseases, Korea Centers for Diseases Control and Prevention | Division of Viral Diseases, Center for Laboratory Control of Infectious Diseases, Korea Centers for Diseases Control and Prevention | Jeong-Min Kim et al |

| | | | | | | |
|---|---|---|---|---|---|---|
| EPI_ISL_481374 | SouthKorea/KCDC2054/2020 | 2020-02-21 | Asia/South Korea/South Korea/ | Division of Viral Diseases, Center for Laboratory Control of Infectious Diseases, Korea Centers for Diseases Control and Prevention | Division of Viral Diseases, Center for Laboratory Control of Infectious Diseases, Korea Centers for Diseases Control and Prevention | Jeong-Min Kim et al |
| EPI_ISL_481375 | SouthKorea/KCDC2055/2020 | 2020-02-22 | Asia/South Korea/South Korea/ | Division of Viral Diseases, Center for Laboratory Control of Infectious Diseases, Korea Centers for Diseases Control and Prevention | Division of Viral Diseases, Center for Laboratory Control of Infectious Diseases, Korea Centers for Diseases Control and Prevention | Jeong-Min Kim et al |
| EPI_ISL_481376 | SouthKorea/KCDC2056/2020 | 2020-02-20 | Asia/South Korea/South Korea/ | Division of Viral Diseases, Center for Laboratory Control of Infectious Diseases, Korea Centers for Diseases Control and Prevention | Division of Viral Diseases, Center for Laboratory Control of Infectious Diseases, Korea Centers for Diseases Control and Prevention | Jeong-Min Kim et al |
| EPI_ISL_481377 | SouthKorea/KCDC2057/2020 | 2020-02-23 | Asia/South Korea/South Korea/ | Division of Viral Diseases, Center for Laboratory Control of Infectious Diseases, Korea Centers for Diseases Control and Prevention | Division of Viral Diseases, Center for Laboratory Control of Infectious Diseases, Korea Centers for Diseases Control and Prevention | Jeong-Min Kim et al |

| | | | | | | |
|---|---|---|---|---|---|---|
| EPI_ISL_481378 | SouthKorea/KCDC2058/2020 | 2020-02-22 | Asia/South Korea/South Korea/ | Division of Viral Diseases, Center for Laboratory Control of Infectious Diseases, Korea Centers for Diseases Control and Prevention | Division of Viral Diseases, Center for Laboratory Control of Infectious Diseases, Korea Centers for Diseases Control and Prevention | Jeong-Min Kim et al |
| EPI_ISL_481379 | SouthKorea/KCDC2059/2020 | 2020-02-24 | Asia/South Korea/South Korea/ | Division of Viral Diseases, Center for Laboratory Control of Infectious Diseases, Korea Centers for Diseases Control and Prevention | Division of Viral Diseases, Center for Laboratory Control of Infectious Diseases, Korea Centers for Diseases Control and Prevention | Jeong-Min Kim et al |
| EPI_ISL_481380 | Georgia/Tb-7856/2020 | 2020-05-01 | Asia/Georgia/Tbilisi/ | Department for Virology, Molecular Biology and Genome Research, R. G. Lugar Center for Public Health Research, National Center for Disease Control and Public Health (NCDC) of Georgia. | Department for Virology, Molecular Biology and Genome Research, R. G. Lugar Center for Public Health Research, National Center for Disease Control and Public Health (NCDC) of Georgia. | Ana Papkiauri et al |
| EPI_ISL_481524 | Finland/10A30S14/2020 | 2020-04-10 | Europe/Finland/Uusimaa/ | Department of Virology and Immunology, University of Helsinki and Helsinki University Hospital, Huslab Finland | Department of Virology, Faculty of Medicine, University of Helsinki, Helsinki, Finland | Teemu Smura et al |

| | | | | | | |
|---|---|---|---|---|---|---|
| EPI_ISL_481528 | Finland/10A57S10/2020 | 2020-04-10 | Europe/Finland/Uusimaa/ | Department of Virology and Immunology, University of Helsinki and Helsinki University Hospital, Huslab Finland | Department of Virology, Faculty of Medicine, University of Helsinki, Helsinki, Finland | Teemu Smura et al |
| EPI_ISL_481537 | Finland/11May91S10/2020 | 2020-05-11 | Europe/Finland/Uusimaa/ | Department of Virology and Immunology, University of Helsinki and Helsinki University Hospital, Huslab Finland | Department of Virology, Faculty of Medicine, University of Helsinki, Helsinki, Finland | Teemu Smura et al |
| EPI_ISL_481571 | Finland/16May68S11/2020 | 2020-05-16 | Europe/Finland/Uusimaa/ | Department of Virology and Immunology, University of Helsinki and Helsinki University Hospital, Huslab Finland | Department of Virology, Faculty of Medicine, University of Helsinki, Helsinki, Finland | Teemu Smura et al |
| EPI_ISL_481601 | Finland/20A51S7/2020 | 2020-04-20 | Europe/Finland/Uusimaa/ | Department of Virology and Immunology, University of Helsinki and Helsinki University Hospital, Huslab Finland | Department of Virology, Faculty of Medicine, University of Helsinki, Helsinki, Finland | Teemu Smura et al |
| EPI_ISL_481624 | Finland/28A30S6/2020 | 2020-04-28 | Europe/Finland/Uusimaa/ | Department of Virology and Immunology, University of Helsinki and Helsinki University Hospital, Huslab Finland | Department of Virology, Faculty of Medicine, University of Helsinki, Helsinki, Finland | Teemu Smura et al |
| EPI_ISL_481694 | Finland/5May13S12/2020 | 2020-05-05 | Europe/Finland/Uusimaa/ | Department of Virology and Immunology, University of Helsinki and Helsinki University Hospital, Huslab Finland | Department of Virology, Faculty of Medicine, University of Helsinki, Helsinki, Finland | Teemu Smura et al |

| | | | | | | |
|---|---|---|---|---|---|---|
| EPI_ISL_481697 | Finland/5May26S2/2020 | 2020-05-05 | Europe/Finland/Uusimaa/ | Department of Virology and Immunology, University of Helsinki and Helsinki University Hospital, Huslab Finland | Department of Virology, Faculty of Medicine, University of Helsinki, Helsinki, Finland | Teemu Smura et al |
| EPI_ISL_481698 | Finland/5May36S12/2020 | 2020-05-05 | Europe/Finland/Uusimaa/ | Department of Virology and Immunology, University of Helsinki and Helsinki University Hospital, Huslab Finland | Department of Virology, Faculty of Medicine, University of Helsinki, Helsinki, Finland | Teemu Smura et al |
| EPI_ISL_481712 | Finland/6A962SIIC/2020 | 2020-04-06 | Europe/Finland/Uusimaa/ | Department of Virology and Immunology, University of Helsinki and Helsinki University Hospital, Huslab Finland | Department of Virology, Faculty of Medicine, University of Helsinki, Helsinki, Finland | Teemu Smura et al |
| EPI_ISL_481715 | Finland/7May68S22/2020 | 2020-05-07 | Europe/Finland/Uusimaa/ | Department of Virology and Immunology, University of Helsinki and Helsinki University Hospital, Huslab Finland | Department of Virology, Faculty of Medicine, University of Helsinki, Helsinki, Finland | Teemu Smura et al |
| EPI_ISL_481717 | Finland/7May75S23/2020 | 2020-05-07 | Europe/Finland/Uusimaa/ | Department of Virology and Immunology, University of Helsinki and Helsinki University Hospital, Huslab Finland | Department of Virology, Faculty of Medicine, University of Helsinki, Helsinki, Finland | Teemu Smura et al |
| EPI_ISL_481731 | Finland/9A29S10/2020 | 2020-04-09 | Europe/Finland/Uusimaa/ | Department of Virology and Immunology, University of Helsinki and Helsinki University Hospital, Huslab Finland | Department of Virology, Faculty of Medicine, University of Helsinki, Helsinki, Finland | Teemu Smura et al |

| | | | | | | |
|---|---|---|---|---|---|---|
| EPI_ISL_481735 | Finland/9A785S11A/2020 | 2020-04-09 | Europe/Finland/Uusimaa/ | Department of Virology and Immunology, University of Helsinki and Helsinki University Hospital, Huslab Finland | Department of Virology, Faculty of Medicine, University of Helsinki, Helsinki, Finland | Teemu Smura et al |
| EPI_ISL_481742 | Canada/NB_243/2020 | 2020-03-23 | North America/Canada/New Brunswick/ | Dr. Georges-L.-Dumont University Hospital Centre | National Microbiology Laboratory | Anna Majer et al |
| EPI_ISL_481743 | Canada/NB_245/2020 | 2020-03-24 | North America/Canada/New Brunswick/ | Dr. Georges-L.-Dumont University Hospital Centre | National Microbiology Laboratory | Anna Majer et al |
| EPI_ISL_481744 | Canada/NB_246/2020 | 2020-03-31 | North America/Canada/New Brunswick/ | Dr. Georges-L.-Dumont University Hospital Centre | National Microbiology Laboratory | Anna Majer et al |
| EPI_ISL_481745 | Canada/NB_247/2020 | 2020-03-31 | North America/Canada/New Brunswick/ | Dr. Georges-L.-Dumont University Hospital Centre | National Microbiology Laboratory | Anna Majer et al |
| EPI_ISL_481746 | Canada/NB_248/2020 | 2020-04-01 | North America/Canada/New Brunswick/ | Dr. Georges-L.-Dumont University Hospital Centre | National Microbiology Laboratory | Anna Majer et al |
| EPI_ISL_481747 | Canada/NB_249/2020 | 2020-03-30 | North America/Canada/New Brunswick/ | Dr. Georges-L.-Dumont University Hospital Centre | National Microbiology Laboratory | Anna Majer et al |
| EPI_ISL_481748 | Canada/NB_250/2020 | 2020-04-02 | North America/Canada/New Brunswick/ | Dr. Georges-L.-Dumont University Hospital Centre | National Microbiology Laboratory | Anna Majer et al |
| EPI_ISL_481749 | Canada/NB_252/2020 | 2020-04-01 | North America/Canada/New Brunswick/ | Dr. Georges-L.-Dumont University Hospital Centre | National Microbiology Laboratory | Anna Majer et al |
| EPI_ISL_481750 | Canada/NB_257/2020 | 2020-04-03 | North America/Canada/New Brunswick/ | Dr. Georges-L.-Dumont University Hospital Centre | National Microbiology Laboratory | Anna Majer et al |
| EPI_ISL_481753 | Canada/NB_263/2020 | 2020-04-06 | North America/Canada/New Brunswick/ | Dr. Georges-L.-Dumont University Hospital Centre | National Microbiology Laboratory | Anna Majer et al |
| EPI_ISL_481754 | Canada/NB_264/2020 | 2020-04-07 | North America/Canada/New Brunswick/ | Dr. Georges-L.-Dumont University Hospital Centre | National Microbiology Laboratory | Anna Majer et al |

| | | | | | | |
|---|---|---|---|---|---|---|
| EPI_ISL_481755 | Canada/NB_270/2020 | 2020-04-10 | North America/Canada/New Brunswick/ | Dr. Georges-L.-Dumont University Hospital Centre | National Microbiology Laboratory | Anna Majer et al |
| EPI_ISL_481756 | Canada/NB_271/2020 | 2020-04-10 | North America/Canada/New Brunswick/ | Dr. Georges-L.-Dumont University Hospital Centre | National Microbiology Laboratory | Anna Majer et al |
| EPI_ISL_481757 | Canada/NB_274/2020 | 2020-04-11 | North America/Canada/New Brunswick/ | Dr. Georges-L.-Dumont University Hospital Centre | National Microbiology Laboratory | Anna Majer et al |
| EPI_ISL_481758 | Canada/NB_277/2020 | 2020-04-13 | North America/Canada/New Brunswick/ | Dr. Georges-L.-Dumont University Hospital Centre | National Microbiology Laboratory | Anna Majer et al |
| EPI_ISL_481769 | England/BRIS-12D6EC/2020 | 2020-05-07 | Europe/United Kingdom/England/ | PHE South West Regional Laboratory, National Infection Service | Wellcome Sanger Institute for the COVID-19 Genomics UK (COG-UK) consortium | Stephanie Hutchings et al |
| EPI_ISL_481780 | England/BRIS-12D084/2020 | 2020-04-25 | Europe/United Kingdom/England/ | PHE South West Regional Laboratory, National Infection Service | Wellcome Sanger Institute for the COVID-19 Genomics UK (COG-UK) consortium | Stephanie Hutchings et al |
| EPI_ISL_481844 | England/BRIS-12D89B/2020 | 2020-05-06 | Europe/United Kingdom/England/ | PHE South West Regional Laboratory, National Infection Service | Wellcome Sanger Institute for the COVID-19 Genomics UK (COG-UK) consortium | Stephanie Hutchings et al |
| EPI_ISL_481882 | England/BRIS-12D1CD/2020 | 2020-05-04 | Europe/United Kingdom/England/ | PHE South West Regional Laboratory, National Infection Service | Wellcome Sanger Institute for the COVID-19 Genomics UK (COG-UK) consortium | Stephanie Hutchings et al |
| EPI_ISL_481928 | England/BRIS-12D9B6/2020 | 2020-05-07 | Europe/United Kingdom/England/ | PHE South West Regional Laboratory, National Infection Service | Wellcome Sanger Institute for the COVID-19 Genomics UK (COG-UK) consortium | Stephanie Hutchings et al |

| | | | | | | |
|---|---|---|---|---|---|---|
| EPI_ISL_481941 | England/BRIS-12D3C7/2020 | 2020-05-04 | Europe/United Kingdom/England/ | PHE South West Regional Laboratory, National Infection Service | Wellcome Sanger Institute for the COVID-19 Genomics UK (COG-UK) consortium | Stephanie Hutchings et al |
| EPI_ISL_481952 | England/BRIS-12D45B/2020 | 2020-05-04 | Europe/United Kingdom/England/ | PHE South West Regional Laboratory, National Infection Service | Wellcome Sanger Institute for the COVID-19 Genomics UK (COG-UK) consortium | Stephanie Hutchings et al |
| EPI_ISL_482016 | England/BRIS-12E029/2020 | 2020-04-20 | Europe/United Kingdom/England/ | PHE South West Regional Laboratory, National Infection Service | Wellcome Sanger Institute for the COVID-19 Genomics UK (COG-UK) consortium | Stephanie Hutchings et al |
| EPI_ISL_482119 | England/HECH-2599EBC/2020 | 2020-05-02 | Europe/United Kingdom/England/ | Microbiology Department, Hereford County Hospital | Wellcome Sanger Institute for the COVID-19 Genomics UK (COG-UK) consortium | Alison Johnson et al |
| EPI_ISL_482140 | NorthernIreland/NIRE-1016FD/2020 | 2020-04-28 | Europe/United Kingdom/Northern Irelar | Regional Virus Laboratory, Belfast Health and Social Care Trust | Wellcome Sanger Institute for the COVID-19 Genomics UK (COG-UK) consortium | Conall McCaughey et al |
| EPI_ISL_482190 | England/LOND-D61C4/2020 | 2020-04-20 | Europe/United Kingdom/England/ | University College London, Great Ormond Street Hospital for Children NHS Foundation Trust, Imperial College Healthcare NHS Trust | Wellcome Sanger Institute for the COVID-19 Genomics UK (COG-UK) consortium | Sergi Castellano et al |
| EPI_ISL_482305 | USA/OR-PROV-018/2020 | 2020-04-06 | North America/USA/Oregon/ | Providence St. Joseph Health Molecular Genomics Laboratory | Providence St. Joseph Health Molecular Genomics Laboratory | Alexa K Dowdell et al |
| EPI_ISL_482319 | USA/OR-PROV-037/2020 | 2020-03-26 | North America/USA/Oregon/ | Providence St. Joseph Health Molecular Genomics Laboratory | Providence St. Joseph Health Molecular Genomics Laboratory | Alexa K Dowdell et al |

| | | | | | | |
|---|---|---|---|---|---|---|
| EPI_ISL_482327 | USA/OR-PROV-045/2020 | 2020-03-21 | North America/USA/Oregon/ | Providence St. Joseph Health Molecular Genomics Laboratory | Providence St. Joseph Health Molecular Genomics Laboratory | Alexa K Dowdell et al |
| EPI_ISL_482332 | USA/OR-PROV-052/2020 | 2020-03-24 | North America/USA/Oregon/ | Providence St. Joseph Health Molecular Genomics Laboratory | Providence St. Joseph Health Molecular Genomics Laboratory | Alexa K Dowdell et al |
| EPI_ISL_482344 | USA/OR-PROV-065/2020 | 2020-04-12 | North America/USA/Oregon/ | Providence St. Joseph Health Molecular Genomics Laboratory | Providence St. Joseph Health Molecular Genomics Laboratory | Alexa K Dowdell et al |
| EPI_ISL_482345 | USA/OR-PROV-066/2020 | 2020-04-12 | North America/USA/Oregon/ | Providence St. Joseph Health Molecular Genomics Laboratory | Providence St. Joseph Health Molecular Genomics Laboratory | Alexa K Dowdell et al |
| EPI_ISL_482351 | USA/OR-PROV-078/2020 | 2020-04-17 | North America/USA/Oregon/ | Providence St. Joseph Health Molecular Genomics Laboratory | Providence St. Joseph Health Molecular Genomics Laboratory | Alexa K Dowdell et al |
| EPI_ISL_482362 | USA/OR-PROV-091/2020 | 2020-04-20 | North America/USA/Oregon/ | Providence St. Joseph Health Molecular Genomics Laboratory | Providence St. Joseph Health Molecular Genomics Laboratory | Alexa K Dowdell et al |
| EPI_ISL_482366 | USA/OR-PROV-095/2020 | 2020-04-21 | North America/USA/Oregon/ | Providence St. Joseph Health Molecular Genomics Laboratory | Providence St. Joseph Health Molecular Genomics Laboratory | Alexa K Dowdell et al |
| EPI_ISL_482388 | USA/OR-PROV-124/2020 | 2020-04-28 | North America/USA/Oregon/ | Providence St. Joseph Health Molecular Genomics Laboratory | Providence St. Joseph Health Molecular Genomics Laboratory | Alexa K Dowdell et al |

| | | | | | | |
|---|---|---|---|---|---|---|
| EPI_ISL_482389 | USA/OR-PROV-125/2020 | 2020-04-28 | North America/USA/Oregon/ | Providence St. Joseph Health Molecular Genomics Laboratory | Providence St. Joseph Health Molecular Genomics Laboratory | Alexa K Dowdell et al |
| EPI_ISL_482392 | USA/OR-PROV-128/2020 | 2020-04-29 | North America/USA/Oregon/ | Providence St. Joseph Health Molecular Genomics Laboratory | Providence St. Joseph Health Molecular Genomics Laboratory | Alexa K Dowdell et al |
| EPI_ISL_482401 | USA/OR-PROV-143/2020 | 2020-05-02 | North America/USA/Oregon/ | Providence St. Joseph Health Molecular Genomics Laboratory | Providence St. Joseph Health Molecular Genomics Laboratory | Alexa K Dowdell et al |
| EPI_ISL_482404 | USA/OR-PROV-146/2020 | 2020-05-02 | North America/USA/Oregon/ | Providence St. Joseph Health Molecular Genomics Laboratory | Providence St. Joseph Health Molecular Genomics Laboratory | Alexa K Dowdell et al |
| EPI_ISL_482405 | USA/OR-PROV-147/2020 | 2020-05-02 | North America/USA/Oregon/ | Providence St. Joseph Health Molecular Genomics Laboratory | Providence St. Joseph Health Molecular Genomics Laboratory | Alexa K Dowdell et al |
| EPI_ISL_482408 | USA/OR-PROV-153/2020 | 2020-05-04 | North America/USA/Oregon/ | Providence St. Joseph Health Molecular Genomics Laboratory | Providence St. Joseph Health Molecular Genomics Laboratory | Alexa K Dowdell et al |
| EPI_ISL_482410 | USA/OR-PROV-155/2020 | 2020-05-05 | North America/USA/Oregon/ | Providence St. Joseph Health Molecular Genomics Laboratory | Providence St. Joseph Health Molecular Genomics Laboratory | Alexa K Dowdell et al |
| EPI_ISL_482413 | USA/OR-PROV-158/2020 | 2020-05-06 | North America/USA/Oregon/ | Providence St. Joseph Health Molecular Genomics Laboratory | Providence St. Joseph Health Molecular Genomics Laboratory | Alexa K Dowdell et al |

| | | | | | | |
|---|---|---|---|---|---|---|
| EPI_ISL_482418 | USA/OR-PROV-167/2020 | 2020-05-08 | North America/USA/Oregon/ | Providence St. Joseph Health Molecular Genomics Laboratory | Providence St. Joseph Health Molecular Genomics Laboratory | Alexa K Dowdell et al |
| EPI_ISL_482419 | USA/OR-PROV-168/2020 | 2020-05-08 | North America/USA/Oregon/ | Providence St. Joseph Health Molecular Genomics Laboratory | Providence St. Joseph Health Molecular Genomics Laboratory | Alexa K Dowdell et al |
| EPI_ISL_482422 | USA/OR-PROV-171/2020 | 2020-05-11 | North America/USA/Oregon/ | Providence St. Joseph Health Molecular Genomics Laboratory | Providence St. Joseph Health Molecular Genomics Laboratory | Alexa K Dowdell et al |
| EPI_ISL_482423 | USA/OR-PROV-173/2020 | 2020-05-11 | North America/USA/Oregon/ | Providence St. Joseph Health Molecular Genomics Laboratory | Providence St. Joseph Health Molecular Genomics Laboratory | Alexa K Dowdell et al |
| EPI_ISL_482425 | USA/OR-PROV-175/2020 | 2020-05-12 | North America/USA/Oregon/ | Providence St. Joseph Health Molecular Genomics Laboratory | Providence St. Joseph Health Molecular Genomics Laboratory | Alexa K Dowdell et al |
| EPI_ISL_482426 | USA/OR-PROV-176/2020 | 2020-05-12 | North America/USA/Oregon/ | Providence St. Joseph Health Molecular Genomics Laboratory | Providence St. Joseph Health Molecular Genomics Laboratory | Alexa K Dowdell et al |
| EPI_ISL_482438 | USA/OR-PROV-193/2020 | 2020-05-17 | North America/USA/Oregon/ | Providence St. Joseph Health Molecular Genomics Laboratory | Providence St. Joseph Health Molecular Genomics Laboratory | Alexa K Dowdell et al |
| EPI_ISL_482443 | USA/OR-PROV-201/2020 | 2020-05-08 | North America/USA/Oregon/ | Providence St. Joseph Health Molecular Genomics Laboratory | Providence St. Joseph Health Molecular Genomics Laboratory | Alexa K Dowdell et al |

| | | | | | | |
|---|---|---|---|---|---|---|
| EPI_ISL_482444 | USA/OR-PROV-202/2020 | 2020-05-10 | North America/USA/Oregon/ | Providence St. Joseph Health Molecular Genomics Laboratory | Providence St. Joseph Health Molecular Genomics Laboratory | Alexa K Dowdell et al |
| EPI_ISL_482452 | USA/OR-PROV-217/2020 | 2020-03-30 | North America/USA/Oregon/ | Providence St. Joseph Health Molecular Genomics Laboratory | Providence St. Joseph Health Molecular Genomics Laboratory | Alexa K Dowdell et al |
| EPI_ISL_482459 | USA/OR-PROV-224/2020 | 2020-03-28 | North America/USA/Oregon/ | Providence St. Joseph Health Molecular Genomics Laboratory | Providence St. Joseph Health Molecular Genomics Laboratory | Alexa K Dowdell et al |
| EPI_ISL_482469 | Canada/NS_40/2020 | 2020-03-05 | North America/Canada/Nova Scotia/ | Queen Elizabeth II Health Science Centre | National Microbiology Laboratory | Anna Majer et al |
| EPI_ISL_482470 | Canada/NS_42/2020 | 2020-03-16 | North America/Canada/Nova Scotia/ | Queen Elizabeth II Health Science Centre | National Microbiology Laboratory | Anna Majer et al |
| EPI_ISL_482471 | Canada/NB_54/2020 | 2020-03-17 | North America/Canada/New Brunswick/ | Dr. Georges-L.-Dumont University Hospital Centre | National Microbiology Laboratory | Anna Majer et al |
| EPI_ISL_482472 | Canada/NB_55/2020 | 2020-03-15 | North America/Canada/New Brunswick/ | Dr. Georges-L.-Dumont University Hospital Centre | National Microbiology Laboratory | Anna Majer et al |
| EPI_ISL_482473 | Canada/NB_56/2020 | 2020-03-15 | North America/Canada/New Brunswick/ | Dr. Georges-L.-Dumont University Hospital Centre | National Microbiology Laboratory | Anna Majer et al |
| EPI_ISL_482474 | Canada/MB_58/2020 | 2020-03-19 | North America/Canada/Manitoba/ | Cadham Provincial Laboratory | National Microbiology Laboratory | Anna Majer et al |
| EPI_ISL_482475 | Canada/MB_59/2020 | 2020-03-15 | North America/Canada/Manitoba/ | Cadham Provincial Laboratory | National Microbiology Laboratory | Anna Majer et al |
| EPI_ISL_482476 | Canada/NS_60/2020 | 2020-03-17 | North America/Canada/Nova Scotia/ | Queen Elizabeth II Health Science Centre | National Microbiology Laboratory | Anna Majer et al |

| | | | | | | |
|---|---|---|---|---|---|---|
| EPI_ISL_482477 | Canada/NS_61/2020 | 2020-03-18 | North America/Canada/Nova Scotia/ | Queen Elizabeth II Health Science Centre | National Microbiology Laboratory | Anna Majer et al |
| EPI_ISL_482478 | Canada/MB_71/2020 | 2020-03-16 | North America/Canada/Manitoba/ | Cadham Provincial Laboratory | National Microbiology Laboratory | Anna Majer et al |
| EPI_ISL_482479 | Canada/NL_72/2020 | 2020-03-19 | North America/Canada/Newfoundland a | Public Health Laboratory | National Microbiology Laboratory | Anna Majer et al |
| EPI_ISL_482480 | Canada/MB_75/2020 | 2020-03-23 | North America/Canada/Manitoba/ | Cadham Provincial Laboratory | National Microbiology Laboratory | Anna Majer et al |
| EPI_ISL_482481 | Canada/MB_76/2020 | 2020-03-16 | North America/Canada/Manitoba/ | Cadham Provincial Laboratory | National Microbiology Laboratory | Anna Majer et al |
| EPI_ISL_482482 | Canada/MB_78/2020 | 2020-03-16 | North America/Canada/Manitoba/ | Cadham Provincial Laboratory | National Microbiology Laboratory | Anna Majer et al |
| EPI_ISL_482483 | Canada/MB_79/2020 | 2020-03-11 | North America/Canada/Manitoba/ | Cadham Provincial Laboratory | National Microbiology Laboratory | Anna Majer et al |
| EPI_ISL_482484 | Canada/MB_80/2020 | 2020-03-13 | North America/Canada/Manitoba/ | Cadham Provincial Laboratory | National Microbiology Laboratory | Anna Majer et al |
| EPI_ISL_482486 | Bangladesh/BCSIR-NILMRC-160/2020 | 2020-06-18 | Asia/Bangladesh/Rajshahi/ | National Institute of Laboratory Medicine and Referral Center | Genomic Research Lab, BCSIR | Abu Sayeed Mohammad Mahmud et al |
| EPI_ISL_482487 | Bangladesh/BCSIR-NILMRC-118a/2020 | 2020-06-18 | Asia/Bangladesh/Dhaka/Dhaka | National Institute of Laboratory Medicine and Referral Center | Genomic Research Lab, BCSIR | Abu Sayeed Mohammad Mahmud et al |
| EPI_ISL_482488 | Bangladesh/BCSIR-NILMRC-161/2020 | 2020-06-18 | Asia/Bangladesh/Rajshahi/ | National Institute of Laboratory Medicine and Referral Center | Genomic Research Lab, BCSIR | Md. Murshed Hasan Sarkar et al |
| EPI_ISL_482492 | India/DL-NCDC1520-CSIR-IGIB/2020 | 2020-05-17 | Asia/India/Delhi/ | National Centre for Disease control (NCDC) | NCDC/CSIR-IGIB | Pramod Kumar |
| EPI_ISL_482494 | India/DL-NCDC1555-CSIR-IGIB/2020 | 2020-05-16 | Asia/India/Delhi/ | National Centre for Disease control (NCDC) | NCDC/CSIR-IGIB | Pramod Kumar |
| EPI_ISL_482495 | India/DL-NCDC1558-CSIR-IGIB/2020 | 2020-05-16 | Asia/India/Delhi/ | National Centre for Disease control (NCDC) | NCDC/CSIR-IGIB | Pramod Kumar |
| EPI_ISL_482502 | India/DL-NCDC1727-CSIR-IGIB/2020 | 2020-05-17 | Asia/India/Delhi/ | National Centre for Disease control (NCDC) | NCDC/CSIR-IGIB | Pramod Kumar |
| EPI_ISL_482506 | India/DL-NCDC1768-CSIR-IGIB/2020 | 2020-05-18 | Asia/India/Delhi/ | National Centre for Disease control (NCDC) | NCDC/CSIR-IGIB | Pramod Kumar |
| EPI_ISL_482509 | India/DL-NCDC1873-CSIR-IGIB/2020 | 2020-05-18 | Asia/India/Delhi/ | National Centre for Disease control (NCDC) | NCDC/CSIR-IGIB | Pramod Kumar |
| EPI_ISL_482512 | India/DL-NCDC1895-CSIR-IGIB/2020 | 2020-05-18 | Asia/India/Delhi/ | National Centre for Disease control (NCDC) | NCDC/CSIR-IGIB | Pramod Kumar |
| EPI_ISL_482513 | India/DL-NCDC1934-CSIR-IGIB/2020 | 2020-05-18 | Asia/India/Delhi/ | National Centre for Disease control (NCDC) | NCDC/CSIR-IGIB | Pramod Kumar |

| | | | | | | |
|---|---|---|---|---|---|---|
| EPI_ISL_482519 | India/DL-NCDC1993-CSIR-IGIB/2020 | 2020-05-18 | Asia/India/Delhi/ | National Centre for Disease control (NCDC) | NCDC/CSIR-IGIB | Pramod Kumar |
| EPI_ISL_482526 | India/DL-NCDC2019-CSIR-IGIB/2020 | 2020-05-18 | Asia/India/Delhi/ | National Centre for Disease control (NCDC) | NCDC/CSIR-IGIB | Pramod Kumar |
| EPI_ISL_482527 | India/DL-NCDC2020-CSIR-IGIB/2020 | 2020-05-18 | Asia/India/Delhi/ | National Centre for Disease control (NCDC) | NCDC/CSIR-IGIB | Pramod Kumar |
| EPI_ISL_482530 | India/DL-NCDC2032-CSIR-IGIB/2020 | 2020-05-18 | Asia/India/Delhi/ | National Centre for Disease control (NCDC) | NCDC/CSIR-IGIB | Pramod Kumar |
| EPI_ISL_482541 | India/DL-NCDC2068-CSIR-IGIB/2020 | 2020-05-19 | Asia/India/Delhi/ | National Centre for Disease control (NCDC) | NCDC/CSIR-IGIB | Pramod Kumar |
| EPI_ISL_482547 | India/DL-NCDC2192-CSIR-IGIB/2020 | 2020-05-18 | Asia/India/Delhi/ | National Centre for Disease control (NCDC) | NCDC/CSIR-IGIB | Pramod Kumar |
| EPI_ISL_482559 | India/DL-NCDC2330-CSIR-IGIB/2020 | 2020-05-19 | Asia/India/Delhi/ | National Centre for Disease control (NCDC) | NCDC/CSIR-IGIB | Pramod Kumar |
| EPI_ISL_482562 | India/DL-NCDC2343-CSIR-IGIB/2020 | 2020-05-19 | Asia/India/Delhi/ | National Centre for Disease control (NCDC) | NCDC/CSIR-IGIB | Pramod Kumar |
| EPI_ISL_482568 | India/DL-NCDC3743-CSIR-IGIB/2020 | 2020-05-20 | Asia/India/Delhi/ | National Centre for Disease control (NCDC) | NCDC/CSIR-IGIB | Pramod Kumar |
| EPI_ISL_482573 | India/DL-NCDC3803-CSIR-IGIB/2020 | 2020-05-20 | Asia/India/Delhi/ | National Centre for Disease control (NCDC) | NCDC/CSIR-IGIB | Pramod Kumar |
| EPI_ISL_482574 | India/DL-NCDC3820-CSIR-IGIB/2020 | 2020-05-20 | Asia/India/Delhi/ | National Centre for Disease control (NCDC) | NCDC/CSIR-IGIB | Pramod Kumar |
| EPI_ISL_482595 | India/DL-NCDC4590-CSIR-IGIB/2020 | 2020-05-23 | Asia/India/Delhi/ | National Centre for Disease control (NCDC) | NCDC/CSIR-IGIB | Pramod Kumar |
| EPI_ISL_482604 | India/DL-NCDC5640-CSIR-IGIB/2020 | 2020-05-25 | Asia/India/Delhi/ | National Centre for Disease control (NCDC) | NCDC/CSIR-IGIB | Pramod Kumar |
| EPI_ISL_482606 | India/DL-NCDC5642-CSIR-IGIB/2020 | 2020-05-25 | Asia/India/Delhi/ | National Centre for Disease control (NCDC) | NCDC/CSIR-IGIB | Pramod Kumar |
| EPI_ISL_482615 | India/DL-NCDC6079-CSIR-IGIB/2020 | 2020-05-25 | Asia/India/Delhi/ | National Centre for Disease control (NCDC) | NCDC/CSIR-IGIB | Pramod Kumar |
| EPI_ISL_482622 | India/DL-NCDC7067-CSIR-IGIB/2020 | 2020-05-27 | Asia/India/Delhi/ | National Centre for Disease control (NCDC) | NCDC/CSIR-IGIB | Pramod Kumar |
| EPI_ISL_482629 | India/DL-NCDC7101-CSIR-IGIB/2020 | 2020-05-27 | Asia/India/Delhi/ | National Centre for Disease control (NCDC) | NCDC/CSIR-IGIB | Pramod Kumar |
| EPI_ISL_482649 | India/DL-NCDC7620-CSIR-IGIB/2020 | 2020-05-28 | Asia/India/Delhi/ | National Centre for Disease control (NCDC) | NCDC/CSIR-IGIB | Pramod Kumar |
| EPI_ISL_482655 | India/DL-NCDC7649-CSIR-IGIB/2020 | 2020-05-28 | Asia/India/Delhi/ | National Centre for Disease control (NCDC) | NCDC/CSIR-IGIB | Pramod Kumar |
| EPI_ISL_482656 | India/DL-NCDC7661-CSIR-IGIB/2020 | 2020-05-28 | Asia/India/Delhi/ | National Centre for Disease control (NCDC) | NCDC/CSIR-IGIB | Pramod Kumar |
| EPI_ISL_482660 | India/DL-NCDC7686-CSIR-IGIB/2020 | 2020-05-28 | Asia/India/Delhi/ | National Centre for Disease control (NCDC) | NCDC/CSIR-IGIB | Pramod Kumar |
| EPI_ISL_482663 | India/DL-NCDC7717-CSIR-IGIB/2020 | 2020-05-28 | Asia/India/Delhi/ | National Centre for Disease control (NCDC) | NCDC/CSIR-IGIB | Pramod Kumar |
| EPI_ISL_482666 | India/DL-NCDC7733-CSIR-IGIB/2020 | 2020-05-28 | Asia/India/Delhi/ | National Centre for Disease control (NCDC) | NCDC/CSIR-IGIB | Pramod Kumar |
| EPI_ISL_482678 | Singapore/1nan/2020 | 2020-01-23 | Asia/Singapore/Singapore/ | Singapore General Hospital | Department of Microbiology | Nurdyana Abdul Rahman et al |
| EPI_ISL_482679 | Singapore/2nan/2020 | 2020-01-27 | Asia/Singapore/Singapore/ | Singapore General Hospital | Department of Microbiology | Nurdyana Abdul Rahman et al |
| EPI_ISL_482680 | Singapore/6nan/2020 | 2020-02-09 | Asia/Singapore/Singapore/ | Singapore General Hospital | Department of Microbiology | Nurdyana Abdul Rahman et al |
| EPI_ISL_482688 | Singapore/540/2020 | 2020-06-02 | Asia/Singapore/Singapore/ | Singapore General Hospital | Department of Microbiology | Nurdyana Abdul Rahman et al |
| EPI_ISL_482689 | Singapore/541/2020 | 2020-06-02 | Asia/Singapore/Singapore/ | Singapore General Hospital | Department of Microbiology | Nurdyana Abdul Rahman et al |

| | | | | | | |
|---|---|---|---|---|---|---|
| EPI_ISL_482691 | Singapore/543/2020 | 2020-06-02 | Asia/Singapore/Singapore/ | Singapore General Hospital | Department of Microbiology | Nurdyana Abdul Rahman et al |
| EPI_ISL_482696 | Singapore/549/2020 | 2020-05-18 | Asia/Singapore/Singapore/ | Singapore General Hospital | Department of Microbiology | Nurdyana Abdul Rahman et al |
| EPI_ISL_482699 | Singapore/552/2020 | 2020-06-06 | Asia/Singapore/Singapore/ | Singapore General Hospital | Department of Microbiology | Nurdyana Abdul Rahman et al |
| EPI_ISL_482700 | Bangladesh/BCSIR-NILMRC-118/2020 | 2020-06-18 | Asia/Bangladesh/Dhaka/Dhaka | National Institute of Laboratory Medicine and Referral Center | Genomic Research Lab, BCSIR | Abu Sayeed Mohammad Mahmud et al |
| EPI_ISL_482702 | SouthAfrica/KRISP-0175/2020 | 2020-06-05 | Africa/South Africa/KwaZulu-Natal/ | Molecular Diagnostics Services (MDS) | KRISP, KZN Research Innovation and Sequencing Platform | Giandhari J et al |
| EPI_ISL_482705 | SouthAfrica/KRISP-0179/2020 | 2020-06-06 | Africa/South Africa/KwaZulu-Natal/ | Molecular Diagnostics Services (MDS) | KRISP, KZN Research Innovation and Sequencing Platform | Giandhari J et al |
| EPI_ISL_482707 | SouthAfrica/KRISP-0184/2020 | 2020-06-07 | Africa/South Africa/KwaZulu-Natal/ | Molecular Diagnostics Services (MDS) | KRISP, KZN Research Innovation and Sequencing Platform | Giandhari J et al |
| EPI_ISL_482708 | SouthAfrica/KRISP-0185/2020 | 2020-06-07 | Africa/South Africa/KwaZulu-Natal/ | Molecular Diagnostics Services (MDS) | KRISP, KZN Research Innovation and Sequencing Platform | Giandhari J et al |
| EPI_ISL_482709 | SouthAfrica/KRISP-0190/2020 | 2020-06-06 | Africa/South Africa/KwaZulu-Natal/ | Molecular Diagnostics Services (MDS) | KRISP, KZN Research Innovation and Sequencing Platform | Giandhari J et al |
| EPI_ISL_482710 | SouthAfrica/KRISP-0191/2020 | 2020-06-04 | Africa/South Africa/KwaZulu-Natal/eThe | NHLS-IALCH | KRISP, KZN Research Innovation and Sequencing Platform | Giandhari J et al |

| | | | | | | |
|---|---|---|---|---|---|---|
| EPI_ISL_482711 | SouthAfrica/KRISP-0192/2020 | 2020-06-04 | Africa/South Africa/KwaZulu-Natal/eThe | NHLS-IALCH | KRISP, KZN Research Innovation and Sequencing Platform | Giandhari J et al |
| EPI_ISL_482714 | SouthAfrica/KRISP-0199/2020 | 2020-06-11 | Africa/South Africa/KwaZulu-Natal/ | Molecular Diagnostics Services (MDS) | KRISP, KZN Research Innovation and Sequencing Platform | Giandhari J et al |
| EPI_ISL_482715 | SouthAfrica/KRISP-0200/2020 | 2020-06-11 | Africa/South Africa/KwaZulu-Natal/ | Molecular Diagnostics Services (MDS) | KRISP, KZN Research Innovation and Sequencing Platform | Giandhari J et al |
| EPI_ISL_482716 | SouthAfrica/KRISP-0201/2020 | 2020-06-11 | Africa/South Africa/KwaZulu-Natal/ | Molecular Diagnostics Services (MDS) | KRISP, KZN Research Innovation and Sequencing Platform | Giandhari J et al |
| EPI_ISL_482717 | SouthAfrica/KRISP-0202/2020 | 2020-06-11 | Africa/South Africa/KwaZulu-Natal/ | Molecular Diagnostics Services (MDS) | KRISP, KZN Research Innovation and Sequencing Platform | Giandhari J et al |
| EPI_ISL_482718 | SouthAfrica/KRISP-0205/2020 | 2020-06-11 | Africa/South Africa/KwaZulu-Natal/ | Molecular Diagnostics Services (MDS) | KRISP, KZN Research Innovation and Sequencing Platform | Giandhari J et al |
| EPI_ISL_482719 | SouthAfrica/KRISP-0206/2020 | 2020-06-11 | Africa/South Africa/KwaZulu-Natal/ | Molecular Diagnostics Services (MDS) | KRISP, KZN Research Innovation and Sequencing Platform | Giandhari J et al |
| EPI_ISL_482721 | SouthAfrica/KRISP-0212/2020 | 2020-06-11 | Africa/South Africa/KwaZulu-Natal/ | Molecular Diagnostics Services (MDS) | KRISP, KZN Research Innovation and Sequencing Platform | Giandhari J et al |

| | | | | | | |
|---|---|---|---|---|---|---|
| EPI_ISL_482722 | SouthAfrica/KRISP-0213/2020 | 2020-06-11 | Africa/South Africa/KwaZulu-Natal/ | Molecular Diagnostics Services (MDS) | KRISP, KZN Research Innovation and Sequencing Platform | Giandhari J et al |
| EPI_ISL_482723 | SouthAfrica/KRISP-0214/2020 | 2020-06-11 | Africa/South Africa/KwaZulu-Natal/ | Molecular Diagnostics Services (MDS) | KRISP, KZN Research Innovation and Sequencing Platform | Giandhari J et al |
| EPI_ISL_482730 | SouthAfrica/KRISP-0260/2020 | 2020-06-15 | Africa/South Africa/KwaZulu-Natal/eThe | NHLS-IALCH | KRISP, KZN Research Innovation and Sequencing Platform | Giandhari J et al |
| EPI_ISL_482731 | SouthAfrica/KRISP-0261/2020 | 2020-06-15 | Africa/South Africa/KwaZulu-Natal/eThe | NHLS-IALCH | KRISP, KZN Research Innovation and Sequencing Platform | Giandhari J et al |
| EPI_ISL_482739 | Morocco/RMPS-14/2020 | 2020-04-22 | Africa/Morocco/Morocco/ | LNR National Reference Laboratory, Mohammed VI University of Health Sciences | Medical Biotechnology Laboratory, Rabat Medical and Pharmacy School, Mohammed The Vth University in Rabat | Meriem LAAMARTI et al |
| EPI_ISL_482740 | Morocco/RMPS-15/2020 | 2020-05-26 | Africa/Morocco/Morocco/ | LNR National Reference Laboratory, Mohammed VI University of Health Sciences | Medical Biotechnology Laboratory, Rabat Medical and Pharmacy School, Mohammed The Vth University in Rabat | Meriem LAAMARTI et al |
| EPI_ISL_482759 | Egypt/MASRI-005/2020 | 2020-04-30 | Africa/Egypt/Egypt/ | Medical Ain Shams Research Institute (MASRI), Ain Shams University | Medical Ain Shams Research Institute (MASRI), Ain Shams University | Hesham Elghazaly et al |

| | | | | | | |
|---|---|---|---|---|---|---|
| EPI_ISL_482760 | Egypt/MASRI-007/2020 | 2020-04-28 | Africa/Egypt/Egypt/ | Medical Ain Shams Research Institute (MASRI), Ain Shams University | Medical Ain Shams Research Institute (MASRI), Ain Shams University | Hesham Elghazaly et al |
| EPI_ISL_482761 | Egypt/MASRI-008/2020 | 2020-04-30 | Africa/Egypt/Egypt/ | Medical Ain Shams Research Institute (MASRI), Ain Shams University | Medical Ain Shams Research Institute (MASRI), Ain Shams University | Hesham Elghazaly et al |
| EPI_ISL_482764 | Egypt/MASRI-013/2020 | 2020-05-05 | Africa/Egypt/Egypt/ | Medical Ain Shams Research Institute (MASRI), Ain Shams University | Medical Ain Shams Research Institute (MASRI), Ain Shams University | Hesham Elghazaly et al |
| EPI_ISL_482766 | Egypt/MASRI-015/2020 | 2020-05-11 | Africa/Egypt/Egypt/ | Medical Ain Shams Research Institute (MASRI), Ain Shams University | Medical Ain Shams Research Institute (MASRI), Ain Shams University | Hesham Elghazaly et al |
| EPI_ISL_482767 | Egypt/MASRI-016/2020 | 2020-05-11 | Africa/Egypt/Egypt/ | Medical Ain Shams Research Institute (MASRI), Ain Shams University | Medical Ain Shams Research Institute (MASRI), Ain Shams University | Hesham Elghazaly et al |
| EPI_ISL_482768 | Egypt/MASRI-022/2020 | 2020-05-13 | Africa/Egypt/Egypt/ | Medical Ain Shams Research Institute (MASRI), Ain Shams University | Medical Ain Shams Research Institute (MASRI), Ain Shams University | Hesham Elghazaly et al |
| EPI_ISL_482769 | Egypt/MASRI-023/2020 | 2020-05-11 | Africa/Egypt/Egypt/ | Medical Ain Shams Research Institute (MASRI), Ain Shams University | Medical Ain Shams Research Institute (MASRI), Ain Shams University | Hesham Elghazaly et al |
| EPI_ISL_482771 | Egypt/MASRI-025/2020 | 2020-05-10 | Africa/Egypt/Egypt/ | Medical Ain Shams Research Institute (MASRI), Ain Shams University | Medical Ain Shams Research Institute (MASRI), Ain Shams University | Hesham Elghazaly et al |

| | | | | | | |
|---|---|---|---|---|---|---|
| EPI_ISL_482772 | Egypt/MASRI-026/2020 | 2020-05-10 | Africa/Egypt/Egypt/ | Medical Ain Shams Research Institute (MASRI), Ain Shams University | Medical Ain Shams Research Institute (MASRI), Ain Shams University | Hesham Elghazaly et al |
| EPI_ISL_482775 | Egypt/MASRI-030/2020 | 2020-04-14 | Africa/Egypt/Egypt/ | Medical Ain Shams Research Institute (MASRI), Ain Shams University | Medical Ain Shams Research Institute (MASRI), Ain Shams University | Hesham Elghazaly et al |
| EPI_ISL_482848 | SouthAfrica/KRISP-0193/2020 | 2020-06-03 | Africa/South Africa/KwaZulu-Natal/Umb | NHLS-IALCH | KRISP, KZN Research Innovation and Sequencing Platform | Giandhari J et al |
| EPI_ISL_482849 | SouthAfrica/KRISP-0194/2020 | 2020-06-06 | Africa/South Africa/KwaZulu-Natal/Umb | NHLS-IALCH | KRISP, KZN Research Innovation and Sequencing Platform | Giandhari J et al |
| EPI_ISL_482851 | SouthAfrica/KRISP-0198/2020 | 2020-06-11 | Africa/South Africa/KwaZulu-Natal/eThe | Molecular Diagnostics Services (MDS) | KRISP, KZN Research Innovation and Sequencing Platform | Giandhari J et al |
| EPI_ISL_482852 | SouthAfrica/KRISP-0203/2020 | 2020-06-11 | Africa/South Africa/KwaZulu-Natal/eThe | Molecular Diagnostics Services (MDS) | KRISP, KZN Research Innovation and Sequencing Platform | Giandhari J et al |
| EPI_ISL_482854 | SouthAfrica/KRISP-0267/2020 | 2020-06-17 | Africa/South Africa/KwaZulu-Natal/eThe | Molecular Diagnostics Services (MDS) | KRISP, KZN Research Innovation and Sequencing Platform | Giandhari J et al |
| EPI_ISL_482856 | SouthAfrica/KRISP-0269/2020 | 2020-06-17 | Africa/South Africa/KwaZulu-Natal/eThe | Molecular Diagnostics Services (MDS) | KRISP, KZN Research Innovation and Sequencing Platform | Giandhari J et al |

| | | | | | | |
|---|---|---|---|---|---|---|
| EPI_ISL_482857 | SouthAfrica/KRISP-0270/2020 | 2020-06-17 | Africa/South Africa/KwaZulu-Natal/eThe | Molecular Diagnostics Services (MDS) | KRISP, KZN Research Innovation and Sequencing Platform | Giandhari J et al |
| EPI_ISL_482860 | SouthAfrica/KRISP-0273/2020 | 2020-06-18 | Africa/South Africa/KwaZulu-Natal/eThe | Molecular Diagnostics Services (MDS) | KRISP, KZN Research Innovation and Sequencing Platform | Giandhari J et al |
| EPI_ISL_482861 | SouthAfrica/KRISP-0274/2020 | 2020-06-18 | Africa/South Africa/KwaZulu-Natal/eThe | Molecular Diagnostics Services (MDS) | KRISP, KZN Research Innovation and Sequencing Platform | Giandhari J et al |
| EPI_ISL_482863 | SouthAfrica/KRISP-0276/2020 | 2020-06-18 | Africa/South Africa/KwaZulu-Natal/eThe | Molecular Diagnostics Services (MDS) | KRISP, KZN Research Innovation and Sequencing Platform | Giandhari J et al |
| EPI_ISL_482864 | SouthAfrica/KRISP-0277/2020 | 2020-06-16 | Africa/South Africa/KwaZulu-Natal/eThe | Molecular Diagnostics Services (MDS) | KRISP, KZN Research Innovation and Sequencing Platform | Giandhari J et al |
| EPI_ISL_482867 | SouthAfrica/KRISP-0280/2020 | 2020-06-16 | Africa/South Africa/KwaZulu-Natal/eThe | Molecular Diagnostics Services (MDS) | KRISP, KZN Research Innovation and Sequencing Platform | Giandhari J et al |
| EPI_ISL_482869 | SouthAfrica/KRISP-0282/2020 | 2020-06-16 | Africa/South Africa/KwaZulu-Natal/eThe | Molecular Diagnostics Services (MDS) | KRISP, KZN Research Innovation and Sequencing Platform | Giandhari J et al |
| EPI_ISL_482871 | SouthAfrica/KRISP-0284/2020 | 2020-06-16 | Africa/South Africa/KwaZulu-Natal/eThe | Molecular Diagnostics Services (MDS) | KRISP, KZN Research Innovation and Sequencing Platform | Giandhari J et al |

| | | | | | | |
|---|---|---|---|---|---|---|
| EPI_ISL_482872 | SouthAfrica/KRISP-0285/2020 | 2020-06-16 | Africa/South Africa/KwaZulu-Natal/eThe | Molecular Diagnostics Services (MDS) | KRISP, KZN Research Innovation and Sequencing Platform | Giandhari J et al |
| EPI_ISL_482880 | France/OCC-22/2020 | 2020-06-08 | Europe/France/Occitanie/ | CHU Purpan - Laboratoire de Virologie - Institut F√©d√©ratif de Biologie | Laboratoire de virologie - √âcole Nationale V√©t√©rinaire de Toulouse | Guillaume Croville et al |
| EPI_ISL_482881 | France/OCC-23/2020 | 2020-06-09 | Europe/France/Occitanie/ | CHU Purpan - Laboratoire de Virologie - Institut F√©d√©ratif de Biologie | Laboratoire de virologie - √âcole Nationale V√©t√©rinaire de Toulouse | Guillaume Croville et al |
| EPI_ISL_482884 | France/OCC-26/2020 | 2020-06-10 | Europe/France/Occitanie/ | CHU Purpan - Laboratoire de Virologie - Institut F√©d√©ratif de Biologie | Laboratoire de virologie - √âcole Nationale V√©t√©rinaire de Toulouse | Guillaume Croville et al |
| EPI_ISL_482886 | France/OCC-28/2020 | 2020-06-12 | Europe/France/Occitanie/ | CHU Purpan - Laboratoire de Virologie - Institut F√©d√©ratif de Biologie | Laboratoire de virologie - √âcole Nationale V√©t√©rinaire de Toulouse | Guillaume Croville et al |
| EPI_ISL_482951 | USA/MN-MDH-1230/2020 | 2020-05-18 | North America/USA/Minnesota/ | Minnesota Department of Health, Public Health Laboratory | Minnesota Department of Health, Public Health Laboratory | Matt Plumb et al |
| EPI_ISL_482955 | USA/MN-MDH-1234/2020 | 2020-05-18 | North America/USA/Minnesota/ | Minnesota Department of Health, Public Health Laboratory | Minnesota Department of Health, Public Health Laboratory | Matt Plumb et al |
| EPI_ISL_482965 | USA/MN-MDH-1244/2020 | 2020-05-27 | North America/USA/Minnesota/ | Minnesota Department of Health, Public Health Laboratory | Minnesota Department of Health, Public Health Laboratory | Matt Plumb et al |

| | | | | | | |
|---|---|---|---|---|---|---|
| EPI_ISL_482968 | USA/MN-MDH-1247/2020 | 2020-04-21 | North America/USA/Minnesota/ | Mayo Clinic & Mayo Clinic Laboratories | Minnesota Department of Health, Public Health Laboratory | Matt Plumb et al |
| EPI_ISL_482973 | USA/MN-MDH-1252/2020 | 2020-06-09 | North America/USA/Minnesota/ | Mayo Clinic & Mayo Clinic Laboratories | Minnesota Department of Health, Public Health Laboratory | Matt Plumb et al |
| EPI_ISL_482975 | USA/MN-MDH-1254/2020 | 2020-06-09 | North America/USA/Minnesota/ | Mayo Clinic & Mayo Clinic Laboratories | Minnesota Department of Health, Public Health Laboratory | Matt Plumb et al |
| EPI_ISL_482976 | USA/MN-MDH-1255/2020 | 2020-06-09 | North America/USA/Minnesota/ | Mayo Clinic & Mayo Clinic Laboratories | Minnesota Department of Health, Public Health Laboratory | Matt Plumb et al |
| EPI_ISL_482981 | USA/MN-MDH-1260/2020 | 2020-06-09 | North America/USA/Minnesota/ | Mayo Clinic & Mayo Clinic Laboratories | Minnesota Department of Health, Public Health Laboratory | Matt Plumb et al |
| EPI_ISL_482985 | USA/MN-MDH-1264/2020 | 2020-06-09 | North America/USA/Minnesota/ | Mayo Clinic & Mayo Clinic Laboratories | Minnesota Department of Health, Public Health Laboratory | Matt Plumb et al |
| EPI_ISL_483016 | USA/MN-MDH-1296/2020 | 2020-03-24 | North America/USA/Minnesota/ | Minnesota Department of Health, Public Health Laboratory | Minnesota Department of Health, Public Health Laboratory | Matt Plumb et al |
| EPI_ISL_483021 | USA/UT-04036/2020 | 2020-05-11 | North America/USA/Utah/ | Utah Public Health Laboratory | Utah Public Health Laboratory | Heidi Butz et al |
| EPI_ISL_483024 | USA/UT-04056/2020 | 2020-05-12 | North America/USA/Utah/ | Utah Public Health Laboratory | Utah Public Health Laboratory | Heidi Butz et al |
| EPI_ISL_483035 | Egypt/MASRI-009/2020 | 2020-04-30 | Africa/Egypt/Cairo/ | Medical Ain Shams Research Institute (MASRI), Ain Shams University | Medical Ain Shams Research Institute (MASRI), Ain Shams University | Hesham Elghazaly et al |

| | | | | | | |
|---|---|---|---|---|---|---|
| EPI_ISL_483037 | Egypt/MASRI-020/2020 | 2020-05-13 | Africa/Egypt/Kalyoubia/ | Medical Ain Shams Research Institute (MASRI), Ain Shams University | Medical Ain Shams Research Institute (MASRI), Ain Shams University | Hesham Elghazaly et al |
| EPI_ISL_483088 | Australia/SAP291/2020 | 2020-03-27 | Oceania/Australia/South Australia/ | SA Pathology | SA Pathology | Lex Leong et al |
| EPI_ISL_483139 | Germany/BE-RKI-Z-0004/2020 | 2020-04-18 | Europe/Germany/Berlin/ | Robert Koch Institute, ZBS1 Highly Pathogenic Viruses, Berlin, Germany | Robert Koch Institute, Bioinformatics MF1, Berlin, Germany | Janine Michel et al |
| EPI_ISL_483140 | Germany/BE-RKI-Z-0005/2020 | 2020-04-18 | Europe/Germany/Berlin/ | Robert Koch Institute, ZBS1 Highly Pathogenic Viruses, Berlin, Germany | Robert Koch Institute, Bioinformatics MF1, Berlin, Germany | Janine Michel et al |
| EPI_ISL_483143 | Germany/BE-RKI-Z-0008/2020 | 2020-04-21 | Europe/Germany/Berlin/ | Robert Koch Institute, ZBS1 Highly Pathogenic Viruses, Berlin, Germany | Robert Koch Institute, Bioinformatics MF1, Berlin, Germany | Janine Michel et al |
| EPI_ISL_483145 | Germany/BE-RKI-Z-0010/2020 | 2020-04-25 | Europe/Germany/Berlin/ | Robert Koch Institute, ZBS1 Highly Pathogenic Viruses, Berlin, Germany | Robert Koch Institute, Bioinformatics MF1, Berlin, Germany | Janine Michel et al |
| EPI_ISL_483151 | Germany/BE-RKI-Z-0017/2020 | 2020-04-28 | Europe/Germany/Berlin/ | Robert Koch Institute, ZBS1 Highly Pathogenic Viruses, Berlin, Germany | Robert Koch Institute, Bioinformatics MF1, Berlin, Germany | Janine Michel et al |
| EPI_ISL_483153 | Germany/BE-RKI-Z-0019/2020 | 2020-04-21 | Europe/Germany/Berlin/ | Robert Koch Institute, ZBS1 Highly Pathogenic Viruses, Berlin, Germany | Robert Koch Institute, Bioinformatics MF1, Berlin, Germany | Janine Michel et al |

| | | | | | | |
|---|---|---|---|---|---|---|
| EPI_ISL_483155 | Germany/BE-RKI-Z-0022/2020 | 2020-04-27 | Europe/Germany/Berlin/ | Robert Koch Institute, ZBS1 Highly Pathogenic Viruses, Berlin, Germany | Robert Koch Institute, Bioinformatics MF1, Berlin, Germany | Janine Michel et al |
| EPI_ISL_483157 | Germany/BE-RKI-Z-0024/2020 | 2020-04-28 | Europe/Germany/Berlin/ | Robert Koch Institute, ZBS1 Highly Pathogenic Viruses, Berlin, Germany | Robert Koch Institute, Bioinformatics MF1, Berlin, Germany | Janine Michel et al |
| EPI_ISL_483160 | USA/CA-ALSR-0469-SAN/2020 | 2020-04-08 | North America/USA/California/San Dieg | San Diego County Public Health Laboratory | Andersen lab at Scripps Research | SEARCH Alliance San Diego with Tracy Basler et al |
| EPI_ISL_483162 | USA/CA-ALSR-0472-SAN/2020 | 2020-04-15 | North America/USA/California/San Dieg | San Diego County Public Health Laboratory | Andersen lab at Scripps Research | SEARCH Alliance San Diego with Tracy Basler et al |
| EPI_ISL_483168 | USA/CA-ALSR-0479-SAN/2020 | 2020-04-10 | North America/USA/California/San Dieg | UC San Diego Center for Advanced Laboratory Medicine | Andersen lab at Scripps Research | SEARCH Alliance San Diego with David Pride et al |
| EPI_ISL_483186 | USA/CA-ALSR-0502-SAN/2020 | 2020-04-15 | North America/USA/California/San Dieg | UC San Diego Center for Advanced Laboratory Medicine | Andersen lab at Scripps Research | SEARCH Alliance San Diego with David Pride et al |
| EPI_ISL_483187 | USA/CA-ALSR-0503-SAN/2020 | 2020-04-02 | North America/USA/California/San Dieg | UC San Diego Center for Advanced Laboratory Medicine | Andersen lab at Scripps Research | SEARCH Alliance San Diego with David Pride et al |
| EPI_ISL_483188 | USA/CA-ALSR-0505-SAN/2020 | 2020-04-01 | North America/USA/California/San Dieg | UC San Diego Center for Advanced Laboratory Medicine | Andersen lab at Scripps Research | SEARCH Alliance San Diego with David Pride et al |
| EPI_ISL_483211 | USA/CA-ALSR-0539-IPL/2020 | 2020-05-05 | North America/USA/California/Imperial | UC San Diego Center for Advanced Laboratory Medicine | Andersen lab at Scripps Research | SEARCH Alliance San Diego with David Pride et al |
| EPI_ISL_483223 | USA/CA-ALSR-0559-SAN/2020 | 2020-05-11 | North America/USA/California/San Dieg | UC San Diego Center for Advanced Laboratory Medicine | Andersen lab at Scripps Research | SEARCH Alliance San Diego with David Pride et al |
| EPI_ISL_483228 | USA/CA-ALSR-0567-IPL/2020 | 2020-05-12 | North America/USA/California/Imperial | UC San Diego Center for Advanced Laboratory Medicine | Andersen lab at Scripps Research | SEARCH Alliance San Diego with David Pride et al |
| EPI_ISL_483250 | USA/CA-ALSR-0602-IPL/2020 | 2020-04-15 | North America/USA/California/Imperial | UC San Diego Center for Advanced Laboratory Medicine | Andersen lab at Scripps Research | SEARCH Alliance San Diego with David Pride et al |
| EPI_ISL_483258 | USA/CA-ALSR-0614-SAN/2020 | 2020-04-04 | North America/USA/California/San Dieg | UC San Diego Center for Advanced Laboratory Medicine | Andersen lab at Scripps Research | SEARCH Alliance San Diego with David Pride et al |
| EPI_ISL_483260 | USA/CA-ALSR-0616-SAN/2020 | 2020-04-07 | North America/USA/California/San Dieg | UC San Diego Center for Advanced Laboratory Medicine | Andersen lab at Scripps Research | SEARCH Alliance San Diego with David Pride et al |
| EPI_ISL_483268 | USA/CA-ALSR-0629-IPL/2020 | 2020-04-23 | North America/USA/California/Imperial | UC San Diego Center for Advanced Laboratory Medicine | Andersen lab at Scripps Research | SEARCH Alliance San Diego with David Pride et al |
| EPI_ISL_483276 | USA/CA-ALSR-0643-IPL/2020 | 2020-04-30 | North America/USA/California/Imperial | UC San Diego Center for Advanced Laboratory Medicine | Andersen lab at Scripps Research | SEARCH Alliance San Diego with David Pride et al |
| EPI_ISL_483279 | USA/CA-ALSR-0649-SAN/2020 | 2020-05-15 | North America/USA/California/San Dieg | UC San Diego Center for Advanced Laboratory Medicine | Andersen lab at Scripps Research | SEARCH Alliance San Diego with David Pride et al |

| | | | | | | |
|---|---|---|---|---|---|---|
| EPI_ISL_483285 | USA/CA-ALSR-0660-IPL/2020 | 2020-04-29 | North America/USA/California/Imperial | UC San Diego Center for Advanced Laboratory Medicine | Andersen lab at Scripps Research | SEARCH Alliance San Diego with David Pride et al |
| EPI_ISL_483303 | USA/CA-ALSR-0687-SAN/2020 | 2020-03-29 | North America/USA/California/San Dieg | UC San Diego Center for Advanced Laboratory Medicine | Andersen lab at Scripps Research | SEARCH Alliance San Diego with David Pride et al |
| EPI_ISL_483340 | USA/CA-ALSR-0761-IPL/2020 | 2020-04-11 | North America/USA/California/Imperial | UC San Diego Center for Advanced Laboratory Medicine | Andersen lab at Scripps Research | SEARCH Alliance San Diego with David Pride et al |
| EPI_ISL_483341 | USA/CA-ALSR-0762-SAN/2020 | 2020-04-11 | North America/USA/California/San Dieg | UC San Diego Center for Advanced Laboratory Medicine | Andersen lab at Scripps Research | SEARCH Alliance San Diego with David Pride et al |
| EPI_ISL_483347 | USA/CA-ALSR-0771-IPL/2020 | 2020-04-10 | North America/USA/California/Imperial | UC San Diego Center for Advanced Laboratory Medicine | Andersen lab at Scripps Research | SEARCH Alliance San Diego with David Pride et al |
| EPI_ISL_483352 | USA/CA-ALSR-0778-IPL/2020 | 2020-04-08 | North America/USA/California/Imperial | UC San Diego Center for Advanced Laboratory Medicine | Andersen lab at Scripps Research | SEARCH Alliance San Diego with David Pride et al |
| EPI_ISL_483371 | USA/CA-ALSR-0805-SAN/2020 | 2020-03-30 | North America/USA/California/San Dieg | UC San Diego Center for Advanced Laboratory Medicine | Andersen lab at Scripps Research | SEARCH Alliance San Diego with David Pride et al |
| EPI_ISL_483401 | USA/CA-ALSR-0854-NBG/2020 | 2020-05-14 | North America/USA/Louisiana/New Orle | UC San Diego Center for Advanced Laboratory Medicine | Andersen lab at Scripps Research | Allison Smither et al |
| EPI_ISL_483405 | USA/CA-ALSR-0902-IPL/2020 | 2020-04-29 | North America/USA/California/Imperial | UC San Diego Center for Advanced Laboratory Medicine | Andersen lab at Scripps Research | SEARCH Alliance San Diego with David Pride et al |
| EPI_ISL_483415 | USA/CA-ALSR-0923-IPL/2020 | 2020-05-03 | North America/USA/California/Imperial | UC San Diego Center for Advanced Laboratory Medicine | Andersen lab at Scripps Research | SEARCH Alliance San Diego with David Pride et al |
| EPI_ISL_483416 | USA/CA-ALSR-0925-IPL/2020 | 2020-05-03 | North America/USA/California/Imperial | UC San Diego Center for Advanced Laboratory Medicine | Andersen lab at Scripps Research | SEARCH Alliance San Diego with David Pride et al |
| EPI_ISL_483437 | USA/CA-ALSR-0978-IPL/2020 | 2020-04-04 | North America/USA/California/Imperial | UC San Diego Center for Advanced Laboratory Medicine | Andersen lab at Scripps Research | SEARCH Alliance San Diego with David Pride et al |
| EPI_ISL_483452 | USA/CA-ALSR-0998-IPL/2020 | 2020-04-23 | North America/USA/California/Imperial | UC San Diego Center for Advanced Laboratory Medicine | Andersen lab at Scripps Research | SEARCH Alliance San Diego with David Pride et al |
| EPI_ISL_483454 | USA/CA-ALSR-1000-SAN/2020 | 2020-04-22 | North America/USA/California/San Dieg | UC San Diego Center for Advanced Laboratory Medicine | Andersen lab at Scripps Research | SEARCH Alliance San Diego with David Pride et al |
| EPI_ISL_483460 | USA/CA-ALSR-1012-SAN/2020 | 2020-04-19 | North America/USA/California/San Dieg | UC San Diego Center for Advanced Laboratory Medicine | Andersen lab at Scripps Research | SEARCH Alliance San Diego with David Pride et al |
| EPI_ISL_483469 | USA/CA-ALSR-1023-SAN/2020 | 2020-04-20 | North America/USA/California/San Dieg | UC San Diego Center for Advanced Laboratory Medicine | Andersen lab at Scripps Research | SEARCH Alliance San Diego with David Pride et al |
| EPI_ISL_483473 | USA/CA-ALSR-1029-SAN/2020 | 2020-04-24 | North America/USA/California/San Dieg | UC San Diego Center for Advanced Laboratory Medicine | Andersen lab at Scripps Research | SEARCH Alliance San Diego with David Pride et al |
| EPI_ISL_483480 | USA/CA-ALSR-1044-SAN/2020 | 2020-06-19 | North America/USA/California/San Dieg | UC San Diego Center for Advanced Laboratory Medicine | Andersen lab at Scripps Research | SEARCH Alliance San Diego with David Pride et al |
| EPI_ISL_483483 | USA/CA-ALSR-1050-SAN/2020 | 2020-06-21 | North America/USA/California/San Dieg | UC San Diego Center for Advanced Laboratory Medicine | Andersen lab at Scripps Research | SEARCH Alliance San Diego with David Pride et al |
| EPI_ISL_483503 | USA/CA-ALSR-1076-SAN/2020 | 2020-06-22 | North America/USA/California/San Dieg | UC San Diego Center for Advanced Laboratory Medicine | Andersen lab at Scripps Research | SEARCH Alliance San Diego with David Pride et al |

| | | | | | | |
|---|---|---|---|---|---|---|
| EPI_ISL_483517 | USA/CA-ALSR-1090-SAN/2020 | 2020-06-15 | North America/USA/California/San Dieg | UC San Diego Center for Advanced Laboratory Medicine | Andersen lab at Scripps Research | SEARCH Alliance San Diego with David Pride et al |
| EPI_ISL_483519 | USA/CA-ALSR-1092-SAN/2020 | 2020-06-13 | North America/USA/California/San Dieg | UC San Diego Center for Advanced Laboratory Medicine | Andersen lab at Scripps Research | SEARCH Alliance San Diego with David Pride et al |
| EPI_ISL_483520 | USA/CA-ALSR-1093-SAN/2020 | 2020-06-18 | North America/USA/California/San Dieg | UC San Diego Center for Advanced Laboratory Medicine | Andersen lab at Scripps Research | SEARCH Alliance San Diego with David Pride et al |
| EPI_ISL_483521 | USA/CA-ALSR-1094-SAN/2020 | 2020-06-19 | North America/USA/California/San Dieg | UC San Diego Center for Advanced Laboratory Medicine | Andersen lab at Scripps Research | SEARCH Alliance San Diego with David Pride et al |
| EPI_ISL_483533 | USA/CA-ALSR-1110-SAN/2020 | 2020-06-18 | North America/USA/California/San Dieg | San Diego County Public Health Laboratory | Andersen lab at Scripps Research | SEARCH Alliance San Diego with Tracy Basler et al |
| EPI_ISL_483536 | USA/CA-ALSR-1113-SAN/2020 | 2020-06-23 | North America/USA/California/San Dieg | San Diego County Public Health Laboratory | Andersen lab at Scripps Research | SEARCH Alliance San Diego with Tracy Basler et al |
| EPI_ISL_483538 | USA/CA-ALSR-1115-SAN/2020 | 2020-06-23 | North America/USA/California/San Dieg | San Diego County Public Health Laboratory | Andersen lab at Scripps Research | SEARCH Alliance San Diego with Tracy Basler et al |
| EPI_ISL_483541 | USA/CA-ALSR-1118-SAN/2020 | 2020-06-21 | North America/USA/California/San Dieg | San Diego County Public Health Laboratory | Andersen lab at Scripps Research | SEARCH Alliance San Diego with Tracy Basler et al |
| EPI_ISL_483546 | Bahrein/BAH-05/2020 | 2020-03-15 | Asia/Bahrain/Bahrain/ | Kingdom of Bahrein Ministry of Health | Erasmus Medical Center | Bas Oude Munnink et al |
| EPI_ISL_483552 | Bahrein/BAH-11/2020 | 2020-03-30 | Asia/Bahrain/Bahrain/ | Kingdom of Bahrein Ministry of Health | Erasmus Medical Center | Bas Oude Munnink et al |
| EPI_ISL_483556 | Bahrein/BAH-15/2020 | 2020-04-04 | Asia/Bahrain/Bahrain/ | Kingdom of Bahrein Ministry of Health | Erasmus Medical Center | Bas Oude Munnink et al |
| EPI_ISL_483577 | Singapore/554/2020 | 2020-06-01 | Asia/Singapore/Singapore/ | National Public Health Laboratory, National Centre for Infectious Diseases | National Public Health Laboratory, National Centre for Infectious Diseases | Mak TM et al |
| EPI_ISL_483578 | Singapore/555/2020 | 2020-05-31 | Asia/Singapore/Singapore/ | National Public Health Laboratory, National Centre for Infectious Diseases | National Public Health Laboratory, National Centre for Infectious Diseases | Mak TM et al |
| EPI_ISL_483580 | Singapore/557/2020 | 2020-05-10 | Asia/Singapore/Singapore/ | National Public Health Laboratory, National Centre for Infectious Diseases | National Public Health Laboratory, National Centre for Infectious Diseases | Mak TM et al |

| | | | | | | |
|---|---|---|---|---|---|---|
| EPI_ISL_483588 | Singapore/565/2020 | 2020-05-10 | Asia/Singapore/Singapore/ | National Public Health Laboratory, National Centre for Infectious Diseases | National Public Health Laboratory, National Centre for Infectious Diseases | Mak TM et al |
| EPI_ISL_483592 | Singapore/569/2020 | 2020-05-29 | Asia/Singapore/Singapore/ | National Public Health Laboratory, National Centre for Infectious Diseases | National Public Health Laboratory, National Centre for Infectious Diseases | Mak TM et al |
| EPI_ISL_483599 | Singapore/576/2020 | 2020-05-20 | Asia/Singapore/Singapore/ | National Public Health Laboratory, National Centre for Infectious Diseases | National Public Health Laboratory, National Centre for Infectious Diseases | Mak TM et al |
| EPI_ISL_483601 | Singapore/578/2020 | 2020-04-21 | Asia/Singapore/Singapore/ | National Public Health Laboratory, National Centre for Infectious Diseases | National Public Health Laboratory, National Centre for Infectious Diseases | Mak TM et al |
| EPI_ISL_483614 | Singapore/591/2020 | 2020-06-26 | Asia/Singapore/Singapore/ | National Public Health Laboratory, National Centre for Infectious Diseases | National Public Health Laboratory, National Centre for Infectious Diseases | Mak TM et al |
| EPI_ISL_483624 | Bangladesh/BCSIR-NILMRC-177/2020 | 2020-06-12 | Asia/Bangladesh/Rajshahi/Pabna | National Institute of Laboratory Medicine and Referral Center | Genomic Research Lab, BCSIR | Md. Maruf Ahmed Molla et al |
| EPI_ISL_483631 | Bangladesh/BCSIR-NILMRC-184/2020 | 2020-06-20 | Asia/Bangladesh/Rajshahi/Pabna | National Institute of Laboratory Medicine and Referral Center | Genomic Research Lab, BCSIR | Md. Ahasan Habib et al |
| EPI_ISL_483632 | Bangladesh/BCSIR-NILMRC-187/2020 | 2020-06-16 | Asia/Bangladesh/Chattogram/Brahmanb | National Institute of Laboratory Medicine and Referral Center | Genomic Research Lab, BCSIR | Md. Ahasan Habib et al |
| EPI_ISL_483633 | Bangladesh/BCSIR-NILMRC-188/2020 | 2020-06-16 | Asia/Bangladesh/Chattogram/Brahmanb | National Institute of Laboratory Medicine and Referral Center | Genomic Research Lab, BCSIR | Shahina Akter et al |
| EPI_ISL_483636 | Bangladesh/BCSIR-NILMRC-196/2020 | 2020-06-20 | Asia/Bangladesh/Chattogram/Chandpur | National Institute of Laboratory Medicine and Referral Center | Genomic Research Lab, BCSIR | Tanjina Akhter Banu et al |

| | | | | | | |
|---|---|---|---|---|---|---|
| EPI_ISL_483639 | Bahrein/BAH-25/2020 | 2020-04-08 | Asia/Bahrain/Bahrain/ | Kingdom of Bahrein Ministry of Health | Erasmus Medical Center | Bas Oude Munnink et al |
| EPI_ISL_483641 | Bangladesh/BCSIR-NILMRC-195/2020 | 2020-06-20 | Asia/Bangladesh/Chattogram/Chandpur | National Institute of Laboratory Medicine and Referral Center | Genomic Research Lab, BCSIR | Barna Goswami et al |
| EPI_ISL_483643 | Bangladesh/BCSIR-NILMRC-199/2020 | 2020-06-20 | Asia/Bangladesh/Chattogram/Chandpur | National Institute of Laboratory Medicine and Referral Center | Genomic Research Lab, BCSIR | Iffat Jahan et al |
| EPI_ISL_483644 | Bangladesh/BCSIR-NILMRC-201/2020 | 2020-06-20 | Asia/Bangladesh/Chattogram/ | National Institute of Laboratory Medicine and Referral Center | Genomic Research Lab, BCSIR | Iffat Jahan et al |
| EPI_ISL_483646 | Bangladesh/BCSIR-NILMRC-212/2020 | 2020-06-14 | Asia/Bangladesh/Sylhet/Habiganj | National Institute of Laboratory Medicine and Referral Center | Genomic Research Lab, BCSIR | Md. Saddam Hossain et al |
| EPI_ISL_483647 | Bangladesh/BCSIR-NILMRC-213/2020 | 2020-06-14 | Asia/Bangladesh/Sylhet/Habiganj | National Institute of Laboratory Medicine and Referral Center | Genomic Research Lab, BCSIR | Md. Saddam Hossain et al |
| EPI_ISL_483687 | Bangladesh/BCSIR-NILMRC-205/2020 | 2020-06-26 | Asia/Bangladesh/Chattogram/Chandpur | National Institute of Laboratory Medicine and Referral Center | Genomic Research Lab, BCSIR | Md. Murshed Hasan Sarkar et al |
| EPI_ISL_483688 | Bangladesh/BCSIR-NILMRC-206/2020 | 2020-06-27 | Asia/Bangladesh/Chattogram/Chandpur | Genomic Research Lab, BCSIR | Genomic Research Lab, BCSIR | Md. Murshed Hasan Sarkar et al |
| EPI_ISL_483690 | Bangladesh/BCSIR-NILMRC-208/2020 | 2020-06-27 | Asia/Bangladesh/Chattogram/Chandpur | National Institute of Laboratory Medicine and Referral Center | Genomic Research Lab, BCSIR | Md. Murshed Hasan Sarkar et al |
| EPI_ISL_483691 | Bangladesh/BCSIR-NILMRC-210/2020 | 2020-06-14 | Asia/Bangladesh/Sylhet/Habiganj | National Institute of Laboratory Medicine and Referral Center | Genomic Research Lab, BCSIR | Md. Murshed Hasan Sarkar et al |
| EPI_ISL_483692 | Bangladesh/BCSIR-NILMRC-217/2020 | 2020-06-24 | Asia/Bangladesh/Sylhet/Moulvibazar | National Institute of Laboratory Medicine and Referral Center | Genomic Research Lab, BCSIR | Md. Murshed Hasan Sarkar et al |
| EPI_ISL_483694 | Bangladesh/BCSIR-NILMRC-186/2020 | 2020-06-16 | Asia/Bangladesh/Chattogram/Brahmanb | National Institute of Laboratory Medicine and Referral Center | Genomic Research Lab, BCSIR | Abu Sayeed Mohammad Mahmud et al |
| EPI_ISL_483695 | Bangladesh/BCSIR-NILMRC-190/2020 | 2020-06-17 | Asia/Bangladesh/Chattogram/Brahmanb | National Institute of Laboratory Medicine and Referral Center | Genomic Research Lab, BCSIR | Abu Sayeed Mohammad Mahmud et al |
| EPI_ISL_483699 | Bangladesh/BCSIR-NILMRC-194/2020 | 2020-06-20 | Asia/Bangladesh/Chattogram/Chandpur | National Institute of Laboratory Medicine and Referral Center | Genomic Research Lab, BCSIR | Abu Sayeed Mohammad Mahmud et al |
| EPI_ISL_483700 | Bangladesh/BCSIR-NILMRC-203/2020 | 2020-06-20 | Asia/Bangladesh/Chattogram/Chandpur | National Institute of Laboratory Medicine and Referral Center | Genomic Research Lab, BCSIR | Abu Sayeed Mohammad Mahmud et al |
| EPI_ISL_483705 | Bangladesh/BCSIR-NILMRC-218/2020 | 2020-06-24 | Asia/Bangladesh/Sylhet/Habiganj | National Institute of Laboratory Medicine and Referral Center | Genomic Research Lab, BCSIR | Abu Sayeed Mohammad Mahmud et al |
| EPI_ISL_483707 | Bangladesh/BCSIR-NILMRC-219/2020 | 2020-06-24 | Asia/Bangladesh/Sylhet/Habiganj | National Institute of Laboratory Medicine and Referral Center | Genomic Research Lab, BCSIR | Abu Sayeed Mohammad Mahmud et al |
| EPI_ISL_483716 | Israel/n14288/2020 | 2020-04-05 | Asia/Israel/Central District/Kfar Saba | Israel Central Virology laboratory | Israel Central Virology laboratory | Neta Zuckerman et al |
| EPI_ISL_483719 | Israel/n14291/2020 | 2020-04-05 | Asia/Israel/Central District/Kfar Saba | Israel Central Virology laboratory | Israel Central Virology laboratory | Neta Zuckerman et al |

| | | | | | | |
|---|---|---|---|---|---|---|
| EPI_ISL_483824 | India/GJ-GBRC243/2020 | 2020-06-14 | Asia/India/Gujarat/Himatnagar | GMERS Medical College Himmatnagar | Gujarat Biotechnology Research Centre | Himanshu Khatri et al |
| EPI_ISL_483826 | India/GJ-GBRC245/2020 | 2020-06-14 | Asia/India/Gujarat/Modasa | GMERS Medical College Himmatnagar | Gujarat Biotechnology Research Centre | Apurvasinh Puvar et al |
| EPI_ISL_483832 | India/GJ-GBRC249b/2020 | 2020-06-12 | Asia/India/Gujarat/Rajkot | Pandit Deendayal Upadhyay Government Medical College, Rajkot | Gujarat Biotechnology Research Centre | Nidhi Patel et al |
| EPI_ISL_483833 | India/GJ-GBRC250a/2020 | 2020-06-12 | Asia/India/Gujarat/Chuda | Pandit Deendayal Upadhyay Government Medical College, Rajkot | Gujarat Biotechnology Research Centre | Nitin Savaliya et al |
| EPI_ISL_483835 | India/GJ-GBRC251a/2020 | 2020-06-12 | Asia/India/Gujarat/Junagadh | Pandit Deendayal Upadhyay Government Medical College, Rajkot | Gujarat Biotechnology Research Centre | Dinesh Kumar et al |
| EPI_ISL_483836 | India/GJ-GBRC251b/2020 | 2020-06-12 | Asia/India/Gujarat/Junagadh | Pandit Deendayal Upadhyay Government Medical College, Rajkot | Gujarat Biotechnology Research Centre | Zuber Saiyed et al |
| EPI_ISL_483837 | India/GJ-GBRC252/2020 | 2020-06-12 | Asia/India/Gujarat/Rajkot | Pandit Deendayal Upadhyay Government Medical College, Rajkot | Gujarat Biotechnology Research Centre | Komal Patel et al |
| EPI_ISL_483839 | India/GJ-GBRC253b/2020 | 2020-06-12 | Asia/India/Gujarat/Rajkot | Pandit Deendayal Upadhyay Government Medical College, Rajkot | Gujarat Biotechnology Research Centre | Afzal Ansari et al |
| EPI_ISL_483842 | India/GJ-GBRC255a/2020 | 2020-06-12 | Asia/India/Gujarat/Rajkot | Pandit Deendayal Upadhyay Government Medical College, Rajkot | Gujarat Biotechnology Research Centre | Arti Trivedi et al |
| EPI_ISL_483843 | India/GJ-GBRC255b/2020 | 2020-06-12 | Asia/India/Gujarat/Rajkot | Pandit Deendayal Upadhyay Government Medical College, Rajkot | Gujarat Biotechnology Research Centre | Apurvasinh Puvar et al |
| EPI_ISL_483844 | India/GJ-GBRC256a/2020 | 2020-06-12 | Asia/India/Gujarat/Jetpur | Pandit Deendayal Upadhyay Government Medical College, Rajkot | Gujarat Biotechnology Research Centre | Janvi Raval et al |
| EPI_ISL_483846 | India/GJ-GBRC257a/2020 | 2020-06-12 | Asia/India/Gujarat/Rajkot | Pandit Deendayal Upadhyay Government Medical College, Rajkot | Gujarat Biotechnology Research Centre | Monika Gandhi et al |
| EPI_ISL_483848 | India/GJ-GBRC258a/2020 | 2020-06-12 | Asia/India/Gujarat/Gondal | Pandit Deendayal Upadhyay Government Medical College, Rajkot | Gujarat Biotechnology Research Centre | Maharshi Pandya et al |

| | | | | | | |
|---|---|---|---|---|---|---|
| EPI_ISL_483849 | India/GJ-GBRC258b/2020 | 2020-06-12 | Asia/India/Gujarat/Gondal | Pandit Deendayal Upadhyay Government Medical College, Rajkot | Gujarat Biotechnology Research Centre | Nidhi Patel et al |
| EPI_ISL_483851 | India/GJ-GBRC260a/2020 | 2020-06-11 | Asia/India/Gujarat/Surat | Department of Microbiology, Government Medical College, Surat | Gujarat Biotechnology Research Centre | Raghawendra Kumar et al |
| EPI_ISL_483854 | India/GJ-GBRC261b/2020 | 2020-06-11 | Asia/India/Gujarat/Surat | Department of Microbiology, Government Medical College, Surat | Gujarat Biotechnology Research Centre | Komal Patel et al |
| EPI_ISL_483855 | India/GJ-GBRC262/2020 | 2020-06-11 | Asia/India/Gujarat/Surat | Department of Microbiology, Government Medical College, Surat | Gujarat Biotechnology Research Centre | Labdhi Pandya et al |
| EPI_ISL_483857 | India/GJ-GBRC263b/2020 | 2020-06-12 | Asia/India/Gujarat/Surat | Department of Microbiology, Government Medical College, Surat | Gujarat Biotechnology Research Centre | Nikha Trivedi et al |
| EPI_ISL_483860 | India/GJ-GBRC265/2020 | 2020-06-13 | Asia/India/Gujarat/Surat | Department of Microbiology, Government Medical College, Surat | Gujarat Biotechnology Research Centre | Amit gamit et al |
| EPI_ISL_483861 | India/GJ-GBRC266/2020 | 2020-06-13 | Asia/India/Gujarat/Surat | Department of Microbiology, Government Medical College, Surat | Gujarat Biotechnology Research Centre | Apurvasinh Puvar et al |
| EPI_ISL_483862 | India/GJ-GBRC267a/2020 | 2020-06-13 | Asia/India/Gujarat/Choryasi | Department of Microbiology, Government Medical College, Surat | Gujarat Biotechnology Research Centre | Janvi Raval et al |
| EPI_ISL_483864 | India/GJ-GBRC268a/2020 | 2020-06-13 | Asia/India/Gujarat/Surat | Department of Microbiology, Government Medical College, Surat | Gujarat Biotechnology Research Centre | Monika Gandhi et al |
| EPI_ISL_483866 | India/GJ-GBRC269/2020 | 2020-06-13 | Asia/India/Gujarat/Choryasi | Department of Microbiology, Government Medical College, Surat | Gujarat Biotechnology Research Centre | Maharshi Pandya et al |
| EPI_ISL_483872 | India/GJ-GBRC273a/2020 | 2020-06-13 | Asia/India/Gujarat/Choryasi | Department of Microbiology, Government Medical College, Surat | Gujarat Biotechnology Research Centre | Komal Patel et al |
| EPI_ISL_483873 | India/GJ-GBRC273b/2020 | 2020-06-13 | Asia/India/Gujarat/Choryasi | Department of Microbiology, Government Medical College, Surat | Gujarat Biotechnology Research Centre | Labdhi Pandya et al |
| EPI_ISL_483874 | India/GJ-GBRC274/2020 | 2020-06-13 | Asia/India/Gujarat/Surat | Department of Microbiology, Government Medical College, Surat | Gujarat Biotechnology Research Centre | Afzal Ansari et al |

| | | | | | | |
|---|---|---|---|---|---|---|
| EPI_ISL_483875 | India/GJ-GBRC275a/2020 | 2020-06-13 | Asia/India/Gujarat/Surat | Department of Microbiology, Government Medical College, Surat | Gujarat Biotechnology Research Centre | Nikha Trivedi et al |
| EPI_ISL_483877 | India/GJ-GBRC276/2020 | 2020-06-13 | Asia/India/Gujarat/Surat | Department of Microbiology, Government Medical College, Surat | Gujarat Biotechnology Research Centre | Summaiya Mullan et al |
| EPI_ISL_483878 | India/GJ-GBRC277a/2020 | 2020-06-13 | Asia/India/Gujarat/Olpad | Department of Microbiology, Government Medical College, Surat | Gujarat Biotechnology Research Centre | Amit gamit et al |
| EPI_ISL_483896 | England/BIRM-6088E/2020 | 2020-06-12 | Europe/United Kingdom/England/ | University of Birmingham | COVID-19 Genomics UK (COG-UK) Consortium | Institute of Microbiology et al |
| EPI_ISL_483943 | England/GSTT-265D33C/2020 | 2020-03-17 | Europe/United Kingdom/England/ | Centre for Clinical Infection and Diagnostics Research and Genomics Innovation Unit, Guy's and St. Thomas' NHS Trust | COVID-19 Genomics UK (COG-UK) Consortium | Chloe Fisher et al |
| EPI_ISL_483972 | England/GSTT-265D518/2020 | 2020-03-23 | Europe/United Kingdom/England/ | Centre for Clinical Infection and Diagnostics Research and Genomics Innovation Unit, Guy's and St. Thomas' NHS Trust | COVID-19 Genomics UK (COG-UK) Consortium | Chloe Fisher et al |
| EPI_ISL_483980 | England/GSTT-265D590/2020 | 2020-03-23 | Europe/United Kingdom/England/ | Centre for Clinical Infection and Diagnostics Research and Genomics Innovation Unit, Guy's and St. Thomas' NHS Trust | COVID-19 Genomics UK (COG-UK) Consortium | Chloe Fisher et al |
| EPI_ISL_484009 | England/GSTT-265D77C/2020 | 2020-03-24 | Europe/United Kingdom/England/ | Centre for Clinical Infection and Diagnostics Research and Genomics Innovation Unit, Guy's and St. Thomas' NHS Trust | COVID-19 Genomics UK (COG-UK) Consortium | Chloe Fisher et al |
| EPI_ISL_484024 | England/GSTT-265D86A/2020 | 2020-03-25 | Europe/United Kingdom/England/ | Centre for Clinical Infection and Diagnostics Research and Genomics Innovation Unit, Guy's and St. Thomas' NHS Trust | COVID-19 Genomics UK (COG-UK) Consortium | Chloe Fisher et al |
| EPI_ISL_484055 | England/GSTT-265DA64/2020 | 2020-03-26 | Europe/United Kingdom/England/ | Centre for Clinical Infection and Diagnostics Research and Genomics Innovation Unit, Guy's and St. Thomas' NHS Trust | COVID-19 Genomics UK (COG-UK) Consortium | Chloe Fisher et al |
| EPI_ISL_484069 | England/GSTT-265DB43/2020 | 2020-03-27 | Europe/United Kingdom/England/ | Centre for Clinical Infection and Diagnostics Research and Genomics Innovation Unit, Guy's and St. Thomas' NHS Trust | COVID-19 Genomics UK (COG-UK) Consortium | Chloe Fisher et al |
| EPI_ISL_484169 | England/GSTT-265E1F5/2020 | 2020-03-31 | Europe/United Kingdom/England/ | Centre for Clinical Infection and Diagnostics Research and Genomics Innovation Unit, Guy's and St. Thomas' NHS Trust | COVID-19 Genomics UK (COG-UK) Consortium | Chloe Fisher et al |
| EPI_ISL_484184 | England/GSTT-265E2F2/2020 | 2020-03-12 | Europe/United Kingdom/England/ | Centre for Clinical Infection and Diagnostics Research and Genomics Innovation Unit, Guy's and St. Thomas' NHS Trust | COVID-19 Genomics UK (COG-UK) Consortium | Chloe Fisher et al |

| | | | | | | |
|---|---|---|---|---|---|---|
| EPI_ISL_484208 | England/GSTT-265E4B0/2020 | 2020-03-31 | Europe/United Kingdom/England/ | Centre for Clinical Infection and Diagnostics Research and Genomics Innovation Unit, Guy's and St. Thomas' NHS Trust | COVID-19 Genomics UK (COG-UK) Consortium | Chloe Fisher et al |
| EPI_ISL_484286 | England/NORT-29C8C3/2020 | 2020-05-03 | Europe/United Kingdom/England/ | Northumbria University / South Tees Hospitals NHS Foundation Trust / North Cumbria Integrated Care NHS Foundation Trust / North Tees and Hartlepool NHS Foundation Trust / Newcastle Hospitals NHS Foundation Trust | COVID-19 Genomics UK (COG-UK) Consortium | Darren L Smith et al |
| EPI_ISL_484315 | England/NORT-29DD3C/2020 | 2020-05-07 | Europe/United Kingdom/England/ | Northumbria University / South Tees Hospitals NHS Foundation Trust / North Cumbria Integrated Care NHS Foundation Trust / North Tees and Hartlepool NHS Foundation Trust / Newcastle Hospitals NHS Foundation Trust | COVID-19 Genomics UK (COG-UK) Consortium | Darren L Smith et al |
| EPI_ISL_484388 | England/NOTT-11250B/2020 | 2020-05-30 | Europe/United Kingdom/England/ | University Hospitals Of Leicester NHS Trust and DeepSeq Nottingham | COVID-19 Genomics UK (COG-UK) Consortium | Christopher Holmes et al |
| EPI_ISL_484436 | England/SHEF-C11EA/2020 | 2020-05-13 | Europe/United Kingdom/England/ | Virology Department, Sheffield Teaching Hospitals NHS Foundation Trust/Department of Infection, Immunity and Cardiovascular Disease, The Medical School, University of Sheffield | COVID-19 Genomics UK (COG-UK) Consortium | Thushan de Silva et al |
| EPI_ISL_484439 | England/SHEF-C1E7A/2020 | 2020-06-05 | Europe/United Kingdom/England/ | Virology Department, Sheffield Teaching Hospitals NHS Foundation Trust/Department of Infection, Immunity and Cardiovascular Disease, The Medical School, University of Sheffield | COVID-19 Genomics UK (COG-UK) Consortium | Thushan de Silva et al |
| EPI_ISL_484468 | England/SHEF-C7524/2020 | 2020-05-15 | Europe/United Kingdom/England/ | Virology Department, Sheffield Teaching Hospitals NHS Foundation Trust/Department of Infection, Immunity and Cardiovascular Disease, The Medical School, University of Sheffield | COVID-19 Genomics UK (COG-UK) Consortium | Thushan de Silva et al |
| EPI_ISL_484480 | England/SHEF-C94BC/2020 | 2020-05-21 | Europe/United Kingdom/England/ | Virology Department, Sheffield Teaching Hospitals NHS Foundation Trust/Department of Infection, Immunity and Cardiovascular Disease, The Medical School, University of Sheffield | COVID-19 Genomics UK (COG-UK) Consortium | Thushan de Silva et al |

| | | | | | | |
|---|---|---|---|---|---|---|
| EPI_ISL_484517 | England/SHEF-D1E77/2020 | 2020-05-29 | Europe/United Kingdom/England/ | Virology Department, Sheffield Teaching Hospitals NHS Foundation Trust/Department of Infection, Immunity and Cardiovascular Disease, The Medical School, University of Sheffield | COVID-19 Genomics UK (COG-UK) Consortium | Thushan de Silva et al |
| EPI_ISL_484541 | Scotland/CVR2027/2020 | 2020-04-09 | Europe/United Kingdom/Scotland/ | West of Scotland Specialist Virology Centre, NHSGGC / MRC-University of Glasgow Centre for Virus Research | COVID-19 Genomics UK (COG-UK) Consortium | Ana da Silva Filipe et al |
| EPI_ISL_484547 | Scotland/CVR2045/2020 | 2020-04-09 | Europe/United Kingdom/Scotland/ | West of Scotland Specialist Virology Centre, NHSGGC / MRC-University of Glasgow Centre for Virus Research | COVID-19 Genomics UK (COG-UK) Consortium | Ana da Silva Filipe et al |
| EPI_ISL_484590 | Scotland/CVR2465/2020 | 2020-04-14 | Europe/United Kingdom/Scotland/ | West of Scotland Specialist Virology Centre, NHSGGC / MRC-University of Glasgow Centre for Virus Research | COVID-19 Genomics UK (COG-UK) Consortium | Ana da Silva Filipe et al |
| EPI_ISL_484758 | USA/MI-UM-2006S000927/2020 | 2020-03-27 | North America/USA/Michigan/ | University of Michigan Clinical Microbiology Laboratory | Lauring Lab, University of Michigan, Department of Microbiology and Immunology | Valesano et al. et al |
| EPI_ISL_484785 | USA/MI-UM-2006S000984/2020 | 2020-03-30 | North America/USA/Michigan/ | University of Michigan Clinical Microbiology Laboratory | Lauring Lab, University of Michigan, Department of Microbiology and Immunology | Valesano et al. et al |
| EPI_ISL_484791 | USA/MI-UM-2006S001015/2020 | 2020-03-29 | North America/USA/Michigan/ | University of Michigan Clinical Microbiology Laboratory | Lauring Lab, University of Michigan, Department of Microbiology and Immunology | Valesano et al. et al |
| EPI_ISL_484792 | USA/MI-UM-2006S001016/2020 | 2020-04-03 | North America/USA/Michigan/ | University of Michigan Clinical Microbiology Laboratory | Lauring Lab, University of Michigan, Department of Microbiology and Immunology | Valesano et al. et al |

| | | | | | | |
|---|---|---|---|---|---|---|
| EPI_ISL_484803 | USA/MI-UM-2006S001086/2020 | 2020-04-03 | North America/USA/Michigan/ | University of Michigan Clinical Microbiology Laboratory | Lauring Lab, University of Michigan, Department of Microbiology and Immunology | Valesano et al. et al |
| EPI_ISL_484808 | USA/WI-UW-433/2020 | 2020-06-18 | North America/USA/Wisconsin/Dane Co | University of Wisconsin-Madison AIDS Vaccine Research Laboratories | University of Wisconsin-Madison AIDS Vaccine Research Laboratories | Gage Moreno et al |
| EPI_ISL_484812 | USA/WI-UW-437/2020 | 2020-06-14 | North America/USA/Wisconsin/Dane Co | University of Wisconsin-Madison AIDS Vaccine Research Laboratories | University of Wisconsin-Madison AIDS Vaccine Research Laboratories | Gage Moreno et al |
| EPI_ISL_484816 | USA/WI-UW-441/2020 | 2020-06-18 | North America/USA/Wisconsin/Dane Co | University of Wisconsin-Madison AIDS Vaccine Research Laboratories | University of Wisconsin-Madison AIDS Vaccine Research Laboratories | Gage Moreno et al |
| EPI_ISL_484825 | USA/WI-UW-450/2020 | 2020-06-17 | North America/USA/Wisconsin/Dane Co | University of Wisconsin-Madison AIDS Vaccine Research Laboratories | University of Wisconsin-Madison AIDS Vaccine Research Laboratories | Gage Moreno et al |
| EPI_ISL_484826 | USA/WI-UW-451/2020 | 2020-06-15 | North America/USA/Wisconsin/Dane Co | University of Wisconsin-Madison AIDS Vaccine Research Laboratories | University of Wisconsin-Madison AIDS Vaccine Research Laboratories | Gage Moreno et al |
| EPI_ISL_484834 | USA/WI-UW-459/2020 | 2020-06-17 | North America/USA/Wisconsin/Dane Co | University of Wisconsin-Madison AIDS Vaccine Research Laboratories | University of Wisconsin-Madison AIDS Vaccine Research Laboratories | Gage Moreno et al |
| EPI_ISL_484835 | USA/WI-UW-460/2020 | 2020-06-17 | North America/USA/Wisconsin/Waukesl | University of Wisconsin-Madison AIDS Vaccine Research Laboratories | University of Wisconsin-Madison AIDS Vaccine Research Laboratories | Gage Moreno et al |

| | | | | | | |
|---|---|---|---|---|---|---|
| EPI_ISL_484842 | USA/WI-UW-467/2020 | 2020-06-06 | North America/USA/Wisconsin/Dane Co | University of Wisconsin-Madison AIDS Vaccine Research Laboratories | University of Wisconsin-Madison AIDS Vaccine Research Laboratories | Gage Moreno et al |
| EPI_ISL_484848 | USA/FL-UW-473/2020 | 2020-06-09 | North America/USA/Florida/Walton Cou | University of Wisconsin-Madison AIDS Vaccine Research Laboratories | University of Wisconsin-Madison AIDS Vaccine Research Laboratories | Gage Moreno et al |
| EPI_ISL_484849 | USA/WI-UW-474/2020 | 2020-06-09 | North America/USA/Wisconsin/Dane Co | University of Wisconsin-Madison AIDS Vaccine Research Laboratories | University of Wisconsin-Madison AIDS Vaccine Research Laboratories | Gage Moreno et al |
| EPI_ISL_484852 | USA/WI-UW-477/2020 | 2020-06-09 | North America/USA/Wisconsin/Dane Co | University of Wisconsin-Madison AIDS Vaccine Research Laboratories | University of Wisconsin-Madison AIDS Vaccine Research Laboratories | Gage Moreno et al |
| EPI_ISL_484865 | USA/WI-UW-490/2020 | 2020-06-04 | North America/USA/Wisconsin/Dane Co | University of Wisconsin-Madison AIDS Vaccine Research Laboratories | University of Wisconsin-Madison AIDS Vaccine Research Laboratories | Gage Moreno et al |
| EPI_ISL_484866 | USA/WI-UW-491/2020 | 2020-06-11 | North America/USA/Wisconsin/Jefferson | University of Wisconsin-Madison AIDS Vaccine Research Laboratories | University of Wisconsin-Madison AIDS Vaccine Research Laboratories | Gage Moreno et al |
| EPI_ISL_484871 | USA/WI-UW-496/2020 | 2020-06-11 | North America/USA/Wisconsin/Dane Co | University of Wisconsin-Madison AIDS Vaccine Research Laboratories | University of Wisconsin-Madison AIDS Vaccine Research Laboratories | Gage Moreno et al |
| EPI_ISL_484874 | USA/WI-UW-499/2020 | 2020-06-08 | North America/USA/Wisconsin/Dane Co | University of Wisconsin-Madison AIDS Vaccine Research Laboratories | University of Wisconsin-Madison AIDS Vaccine Research Laboratories | Gage Moreno et al |

| | | | | | | |
|---|---|---|---|---|---|---|
| EPI_ISL_484877 | USA/WI-UW-502/2020 | 2020-06-03 | North America/USA/Wisconsin/Dane Co | University of Wisconsin-Madison AIDS Vaccine Research Laboratories | University of Wisconsin-Madison AIDS Vaccine Research Laboratories | Gage Moreno et al |
| EPI_ISL_484895 | USA/WI-UW-520/2020 | 2020-06-24 | North America/USA/Wisconsin/Dane Co | University of Wisconsin-Madison AIDS Vaccine Research Laboratories | University of Wisconsin-Madison AIDS Vaccine Research Laboratories | Gage Moreno et al |
| EPI_ISL_484896 | USA/WI-UW-521/2020 | 2020-06-24 | North America/USA/Wisconsin/Dane Co | University of Wisconsin-Madison AIDS Vaccine Research Laboratories | University of Wisconsin-Madison AIDS Vaccine Research Laboratories | Gage Moreno et al |
| EPI_ISL_484900 | USA/IL-UW-525/2020 | 2020-06-24 | North America/USA/Illinois/Winnebago | University of Wisconsin-Madison AIDS Vaccine Research Laboratories | University of Wisconsin-Madison AIDS Vaccine Research Laboratories | Gage Moreno et al |
| EPI_ISL_484913 | USA/WI-UW-538/2020 | 2020-06-24 | North America/USA/Wisconsin/Dane Co | University of Wisconsin-Madison AIDS Vaccine Research Laboratories | University of Wisconsin-Madison AIDS Vaccine Research Laboratories | Gage Moreno et al |
| EPI_ISL_484915 | USA/WI-UW-540/2020 | 2020-06-22 | North America/USA/Wisconsin/Dane Co | University of Wisconsin-Madison AIDS Vaccine Research Laboratories | University of Wisconsin-Madison AIDS Vaccine Research Laboratories | Gage Moreno et al |
| EPI_ISL_484917 | USA/IL-UW-542/2020 | 2020-06-23 | North America/USA/Illinois/Winnebago | University of Wisconsin-Madison AIDS Vaccine Research Laboratories | University of Wisconsin-Madison AIDS Vaccine Research Laboratories | Gage Moreno et al |
| EPI_ISL_484934 | USA/WI-UW-559/2020 | 2020-06-25 | North America/USA/Wisconsin/Dane Co | University of Wisconsin-Madison AIDS Vaccine Research Laboratories | University of Wisconsin-Madison AIDS Vaccine Research Laboratories | Gage Moreno et al |

| | | | | | | |
|---|---|---|---|---|---|---|
| EPI_ISL_484937 | USA/WI-UW-562/2020 | 2020-06-25 | North America/USA/Wisconsin/Dane Co | University of Wisconsin-Madison AIDS Vaccine Research Laboratories | University of Wisconsin-Madison AIDS Vaccine Research Laboratories | Gage Moreno et al |
| EPI_ISL_484945 | USA/WI-UW-570/2020 | 2020-06-25 | North America/USA/Wisconsin/Dane Co | University of Wisconsin-Madison AIDS Vaccine Research Laboratories | University of Wisconsin-Madison AIDS Vaccine Research Laboratories | Gage Moreno et al |
| EPI_ISL_484949 | USA/WI-UW-574/2020 | 2020-06-23 | North America/USA/Wisconsin/Outagan | University of Wisconsin-Madison AIDS Vaccine Research Laboratories | University of Wisconsin-Madison AIDS Vaccine Research Laboratories | Gage Moreno et al |
| EPI_ISL_484959 | USA/NY-UW-584/2020 | 2020-06-23 | North America/USA/New York/Westche | University of Wisconsin-Madison AIDS Vaccine Research Laboratories | University of Wisconsin-Madison AIDS Vaccine Research Laboratories | Gage Moreno et al |
| EPI_ISL_484963 | USA/WI-UW-588/2020 | 2020-06-22 | North America/USA/Wisconsin/Dane Co | University of Wisconsin-Madison AIDS Vaccine Research Laboratories | University of Wisconsin-Madison AIDS Vaccine Research Laboratories | Gage Moreno et al |
| EPI_ISL_484964 | USA/WI-UW-589/2020 | 2020-06-19 | North America/USA/Wisconsin/Dane Co | University of Wisconsin-Madison AIDS Vaccine Research Laboratories | University of Wisconsin-Madison AIDS Vaccine Research Laboratories | Gage Moreno et al |
| EPI_ISL_484968 | USA/WI-UW-593/2020 | 2020-06-27 | North America/USA/Wisconsin/Dane Co | University of Wisconsin-Madison AIDS Vaccine Research Laboratories | University of Wisconsin-Madison AIDS Vaccine Research Laboratories | Gage Moreno et al |
| EPI_ISL_484970 | USA/WI-UW-595/2020 | 2020-06-25 | North America/USA/Wisconsin/Dane Co | University of Wisconsin-Madison AIDS Vaccine Research Laboratories | University of Wisconsin-Madison AIDS Vaccine Research Laboratories | Gage Moreno et al |

| | | | | | | |
|---|---|---|---|---|---|---|
| EPI_ISL_484973 | USA/WI-UW-598/2020 | 2020-06-27 | North America/USA/Wisconsin/Dane Co | University of Wisconsin-Madison AIDS Vaccine Research Laboratories | University of Wisconsin-Madison AIDS Vaccine Research Laboratories | Gage Moreno et al |
| EPI_ISL_484974 | USA/WI-UW-599/2020 | 2020-06-28 | North America/USA/Wisconsin/Dane Co | University of Wisconsin-Madison AIDS Vaccine Research Laboratories | University of Wisconsin-Madison AIDS Vaccine Research Laboratories | Gage Moreno et al |
| EPI_ISL_484977 | USA/WI-UW-602/2020 | 2020-06-28 | North America/USA/Wisconsin/Dane Co | University of Wisconsin-Madison AIDS Vaccine Research Laboratories | University of Wisconsin-Madison AIDS Vaccine Research Laboratories | Gage Moreno et al |
| EPI_ISL_484981 | USA/CA-UW-606/2020 | 2020-06-27 | North America/USA/California/Los Ange | University of Wisconsin-Madison AIDS Vaccine Research Laboratories | University of Wisconsin-Madison AIDS Vaccine Research Laboratories | Gage Moreno et al |
| EPI_ISL_484989 | USA/WI-UW-614/2020 | 2020-06-23 | North America/USA/Wisconsin/Dane Co | University of Wisconsin-Madison AIDS Vaccine Research Laboratories | University of Wisconsin-Madison AIDS Vaccine Research Laboratories | Gage Moreno et al |
| EPI_ISL_484992 | USA/WI-UW-617/2020 | 2020-06-23 | North America/USA/Wisconsin/Waukes | University of Wisconsin-Madison AIDS Vaccine Research Laboratories | University of Wisconsin-Madison AIDS Vaccine Research Laboratories | Gage Moreno et al |
| EPI_ISL_485001 | SouthKorea/A2/2020 | 2020-03-08 | Asia/South Korea/Seoul/ | University of Ulsan College of Medicine and Asan Medical Center | University of Ulsan College of Medicine and Asan Medical Center | Kuenyoul Park et al |
| EPI_ISL_485006 | USA/LA-GBCL-26159754/2020 | 2020-04-11 | North America/USA/Louisiana/East Bato | River Road Testing Lab | Ginkgo Bioworks Clinical Laboratory | Rebecca C. Christofferson et al |
| EPI_ISL_485013 | USA/LA-GBCL-26159763/2020 | 2020-04-11 | North America/USA/Louisiana/East Bato | River Road Testing Lab | Ginkgo Bioworks Clinical Laboratory | Rebecca C. Christofferson et al |

| | | | | | | |
|---|---|---|---|---|---|---|
| EPI_ISL_485015 | USA/LA-GBCL-26159765/2020 | 2020-04-11 | North America/USA/Louisiana/East Bato | River Road Testing Lab | Ginkgo Bioworks Clinical Laboratory | Rebecca C. Christofferson et al |
| EPI_ISL_485020 | USA/LA-GBCL-26492435/2020 | 2020-04-28 | North America/USA/Louisiana/East Bato | River Road Testing Lab | Ginkgo Bioworks Clinical Laboratory | Rebecca C. Christofferson et al |
| EPI_ISL_485042 | USA/LA-GBCL-26492470/2020 | 2020-04-27 | North America/USA/Louisiana/East Bato | River Road Testing Lab | Ginkgo Bioworks Clinical Laboratory | Rebecca C. Christofferson et al |
| EPI_ISL_485044 | USA/LA-GBCL-26492474/2020 | 2020-04-25 | North America/USA/Louisiana/East Bato | River Road Testing Lab | Ginkgo Bioworks Clinical Laboratory | Rebecca C. Christofferson et al |
| EPI_ISL_485046 | USA/LA-GBCL-26492480/2020 | 2020-04-25 | North America/USA/Louisiana/East Bato | River Road Testing Lab | Ginkgo Bioworks Clinical Laboratory | Rebecca C. Christofferson et al |
| EPI_ISL_485049 | USA/LA-GBCL-26492488/2020 | 2020-04-25 | North America/USA/Louisiana/East Bato | River Road Testing Lab | Ginkgo Bioworks Clinical Laboratory | Rebecca C. Christofferson et al |
| EPI_ISL_485051 | USA/LA-GBCL-26492495/2020 | 2020-04-24 | North America/USA/Louisiana/East Bato | River Road Testing Lab | Ginkgo Bioworks Clinical Laboratory | Rebecca C. Christofferson et al |
| EPI_ISL_485053 | USA/LA-GBCL-26492497/2020 | 2020-04-24 | North America/USA/Louisiana/East Bato | River Road Testing Lab | Ginkgo Bioworks Clinical Laboratory | Rebecca C. Christofferson et al |
| EPI_ISL_485054 | USA/LA-GBCL-26492498/2020 | 2020-04-24 | North America/USA/Louisiana/East Bato | River Road Testing Lab | Ginkgo Bioworks Clinical Laboratory | Rebecca C. Christofferson et al |
| EPI_ISL_485055 | USA/LA-GBCL-26492499/2020 | 2020-04-24 | North America/USA/Louisiana/East Bato | River Road Testing Lab | Ginkgo Bioworks Clinical Laboratory | Rebecca C. Christofferson et al |
| EPI_ISL_485056 | USA/LA-GBCL-26492500/2020 | 2020-04-24 | North America/USA/Louisiana/East Bato | River Road Testing Lab | Ginkgo Bioworks Clinical Laboratory | Rebecca C. Christofferson et al |
| EPI_ISL_485075 | USA/LA-GBCL-26513021/2020 | 2020-04-23 | North America/USA/Louisiana/East Bato | River Road Testing Lab | Ginkgo Bioworks Clinical Laboratory | Rebecca C. Christofferson et al |
| EPI_ISL_485081 | USA/LA-GBCL-26513032/2020 | 2020-04-22 | North America/USA/Louisiana/East Bato | River Road Testing Lab | Ginkgo Bioworks Clinical Laboratory | Rebecca C. Christofferson et al |

| | | | | | | |
|---|---|---|---|---|---|---|
| EPI_ISL_485098 | USA/LA-GBCL-26513066/2020 | 2020-04-21 | North America/USA/Louisiana/East Bato | River Road Testing Lab | Ginkgo Bioworks Clinical Laboratory | Rebecca C. Christofferson et al |
| EPI_ISL_485102 | USA/LA-GBCL-26544504/2020 | 2020-04-27 | North America/USA/Louisiana/East Bato | River Road Testing Lab | Ginkgo Bioworks Clinical Laboratory | Rebecca C. Christofferson et al |
| EPI_ISL_485105 | USA/LA-GBCL-26544513/2020 | 2020-04-22 | North America/USA/Louisiana/East Bato | River Road Testing Lab | Ginkgo Bioworks Clinical Laboratory | Rebecca C. Christofferson et al |
| EPI_ISL_485131 | USA/LA-GBCL-26544553/2020 | 2020-04-18 | North America/USA/Louisiana/East Bato | River Road Testing Lab | Ginkgo Bioworks Clinical Laboratory | Rebecca C. Christofferson et al |
| EPI_ISL_485137 | USA/LA-GBCL-26544563/2020 | 2020-04-18 | North America/USA/Louisiana/East Bato | River Road Testing Lab | Ginkgo Bioworks Clinical Laboratory | Rebecca C. Christofferson et al |
| EPI_ISL_485145 | USA/LA-GBCL-26544573/2020 | 2020-04-17 | North America/USA/Louisiana/East Bato | River Road Testing Lab | Ginkgo Bioworks Clinical Laboratory | Rebecca C. Christofferson et al |
| EPI_ISL_485155 | USA/LA-GBCL-26544586/2020 | 2020-04-17 | North America/USA/Louisiana/East Bato | River Road Testing Lab | Ginkgo Bioworks Clinical Laboratory | Rebecca C. Christofferson et al |
| EPI_ISL_485158 | USA/LA-GBCL-26556503/2020 | 2020-04-17 | North America/USA/Louisiana/East Bato | River Road Testing Lab | Ginkgo Bioworks Clinical Laboratory | Rebecca C. Christofferson et al |
| EPI_ISL_485160 | USA/LA-GBCL-26556508/2020 | 2020-04-16 | North America/USA/Louisiana/East Bato | River Road Testing Lab | Ginkgo Bioworks Clinical Laboratory | Rebecca C. Christofferson et al |
| EPI_ISL_485163 | USA/LA-GBCL-26556515/2020 | 2020-04-16 | North America/USA/Louisiana/East Bato | River Road Testing Lab | Ginkgo Bioworks Clinical Laboratory | Rebecca C. Christofferson et al |
| EPI_ISL_485165 | USA/LA-GBCL-26556519/2020 | 2020-04-16 | North America/USA/Louisiana/East Bato | River Road Testing Lab | Ginkgo Bioworks Clinical Laboratory | Rebecca C. Christofferson et al |
| EPI_ISL_485196 | USA/LA-GBCL-26556566/2020 | 2020-04-15 | North America/USA/Louisiana/East Bato | River Road Testing Lab | Ginkgo Bioworks Clinical Laboratory | Rebecca C. Christofferson et al |
| EPI_ISL_485199 | USA/LA-GBCL-26556570/2020 | 2020-04-15 | North America/USA/Louisiana/East Bato | River Road Testing Lab | Ginkgo Bioworks Clinical Laboratory | Rebecca C. Christofferson et al |

| | | | | | | |
|---|---|---|---|---|---|---|
| EPI_ISL_485200 | USA/LA-GBCL-26556572/2020 | 2020-04-14 | North America/USA/Louisiana/East Bat | River Road Testing Lab | Ginkgo Bioworks Clinical Laboratory | Rebecca C. Christofferson et al |
| EPI_ISL_485201 | USA/LA-GBCL-26556574/2020 | 2020-04-14 | North America/USA/Louisiana/East Bat | River Road Testing Lab | Ginkgo Bioworks Clinical Laboratory | Rebecca C. Christofferson et al |
| EPI_ISL_485207 | USA/LA-GBCL-26556585/2020 | 2020-04-14 | North America/USA/Louisiana/East Bat | River Road Testing Lab | Ginkgo Bioworks Clinical Laboratory | Rebecca C. Christofferson et al |
| EPI_ISL_485209 | USA/LA-GBCL-26556587/2020 | 2020-04-14 | North America/USA/Louisiana/East Bat | River Road Testing Lab | Ginkgo Bioworks Clinical Laboratory | Rebecca C. Christofferson et al |
| EPI_ISL_485212 | USA/LA-GBCL-26566518/2020 | 2020-04-18 | North America/USA/Louisiana/East Bat | River Road Testing Lab | Ginkgo Bioworks Clinical Laboratory | Rebecca C. Christofferson et al |
| EPI_ISL_485218 | USA/LA-GBCL-26629204/2020 | 2020-04-23 | North America/USA/Louisiana/East Bat | River Road Testing Lab | Ginkgo Bioworks Clinical Laboratory | Rebecca C. Christofferson et al |
| EPI_ISL_485224 | USA/LA-GBCL-26629220/2020 | 2020-04-23 | North America/USA/Louisiana/East Bat | River Road Testing Lab | Ginkgo Bioworks Clinical Laboratory | Rebecca C. Christofferson et al |
| EPI_ISL_485229 | USA/LA-GBCL-26629233/2020 | 2020-04-22 | North America/USA/Louisiana/East Bat | River Road Testing Lab | Ginkgo Bioworks Clinical Laboratory | Rebecca C. Christofferson et al |
| EPI_ISL_485233 | USA/LA-GBCL-26629240/2020 | 2020-04-21 | North America/USA/Louisiana/East Bat | River Road Testing Lab | Ginkgo Bioworks Clinical Laboratory | Rebecca C. Christofferson et al |
| EPI_ISL_485250 | USA/LA-GBCL-26674722/2020 | 2020-04-11 | North America/USA/Louisiana/East Bat | River Road Testing Lab | Ginkgo Bioworks Clinical Laboratory | Rebecca C. Christofferson et al |
| EPI_ISL_485254 | USA/LA-GBCL-26674728/2020 | 2020-04-10 | North America/USA/Louisiana/East Bat | River Road Testing Lab | Ginkgo Bioworks Clinical Laboratory | Rebecca C. Christofferson et al |
| EPI_ISL_485259 | USA/LA-GBCL-26674735/2020 | 2020-04-04 | North America/USA/Louisiana/East Bat | River Road Testing Lab | Ginkgo Bioworks Clinical Laboratory | Rebecca C. Christofferson et al |
| EPI_ISL_485265 | USA/LA-GBCL-26684440/2020 | 2020-04-11 | North America/USA/Louisiana/East Bat | River Road Testing Lab | Ginkgo Bioworks Clinical Laboratory | Rebecca C. Christofferson et al |

| | | | | | | |
|---|---|---|---|---|---|---|
| EPI_ISL_485279 | USA/LA-GBCL-26684458/2020 | 2020-04-04 | North America/USA/Louisiana/East Batc | River Road Testing Lab | Ginkgo Bioworks Clinical Laboratory | Rebecca C. Christofferson et al |
| EPI_ISL_485285 | USA/LA-GBCL-26684465/2020 | 2020-04-11 | North America/USA/Louisiana/East Batc | River Road Testing Lab | Ginkgo Bioworks Clinical Laboratory | Rebecca C. Christofferson et al |
| EPI_ISL_485290 | USA/LA-GBCL-26684472/2020 | 2020-04-10 | North America/USA/Louisiana/East Batc | River Road Testing Lab | Ginkgo Bioworks Clinical Laboratory | Rebecca C. Christofferson et al |
| EPI_ISL_485296 | USA/LA-GBCL-26684481/2020 | 2020-04-10 | North America/USA/Louisiana/East Batc | River Road Testing Lab | Ginkgo Bioworks Clinical Laboratory | Rebecca C. Christofferson et al |
| EPI_ISL_485304 | USA/LA-GBCL-26684496/2020 | 2020-04-10 | North America/USA/Louisiana/East Batc | River Road Testing Lab | Ginkgo Bioworks Clinical Laboratory | Rebecca C. Christofferson et al |
| EPI_ISL_485306 | USA/LA-GBCL-26684499/2020 | 2020-04-10 | North America/USA/Louisiana/East Batc | River Road Testing Lab | Ginkgo Bioworks Clinical Laboratory | Rebecca C. Christofferson et al |
| EPI_ISL_485307 | USA/LA-GBCL-26684500/2020 | 2020-04-10 | North America/USA/Louisiana/East Batc | River Road Testing Lab | Ginkgo Bioworks Clinical Laboratory | Rebecca C. Christofferson et al |
| EPI_ISL_485314 | USA/LA-GBCL-26684509/2020 | 2020-04-10 | North America/USA/Louisiana/East Batc | River Road Testing Lab | Ginkgo Bioworks Clinical Laboratory | Rebecca C. Christofferson et al |
| EPI_ISL_485324 | USA/LA-GBCL-26791700/2020 | 2020-04-10 | North America/USA/Louisiana/East Batc | River Road Testing Lab | Ginkgo Bioworks Clinical Laboratory | Rebecca C. Christofferson et al |
| EPI_ISL_485329 | USA/LA-GBCL-26791706/2020 | 2020-04-09 | North America/USA/Louisiana/East Batc | River Road Testing Lab | Ginkgo Bioworks Clinical Laboratory | Rebecca C. Christofferson et al |
| EPI_ISL_485344 | USA/LA-GBCL-26791725/2020 | 2020-04-09 | North America/USA/Louisiana/East Batc | River Road Testing Lab | Ginkgo Bioworks Clinical Laboratory | Rebecca C. Christofferson et al |
| EPI_ISL_485347 | USA/LA-GBCL-26791728/2020 | 2020-04-09 | North America/USA/Louisiana/East Batc | River Road Testing Lab | Ginkgo Bioworks Clinical Laboratory | Rebecca C. Christofferson et al |
| EPI_ISL_485356 | USA/LA-GBCL-26791739/2020 | 2020-04-06 | North America/USA/Louisiana/East Batc | River Road Testing Lab | Ginkgo Bioworks Clinical Laboratory | Rebecca C. Christofferson et al |

| | | | | | | |
|---|---|---|---|---|---|---|
| EPI_ISL_485367 | USA/LA-GBCL-26791752/2020 | 2020-04-04 | North America/USA/Louisiana/East Bato | River Road Testing Lab | Ginkgo Bioworks Clinical Laboratory | Rebecca C. Christofferson et al |
| EPI_ISL_485371 | USA/LA-GBCL-26791756/2020 | 2020-04-06 | North America/USA/Louisiana/East Bato | River Road Testing Lab | Ginkgo Bioworks Clinical Laboratory | Rebecca C. Christofferson et al |
| EPI_ISL_485376 | USA/LA-GBCL-26791764/2020 | 2020-04-06 | North America/USA/Louisiana/East Bato | River Road Testing Lab | Ginkgo Bioworks Clinical Laboratory | Rebecca C. Christofferson et al |
| EPI_ISL_485379 | USA/LA-GBCL-26791769/2020 | 2020-04-04 | North America/USA/Louisiana/East Bato | River Road Testing Lab | Ginkgo Bioworks Clinical Laboratory | Rebecca C. Christofferson et al |
| EPI_ISL_485387 | USA/LA-GBCL-26791781/2020 | 2020-04-04 | North America/USA/Louisiana/East Bato | River Road Testing Lab | Ginkgo Bioworks Clinical Laboratory | Rebecca C. Christofferson et al |
| EPI_ISL_485391 | SouthKorea/A7/2020 | 2020-04-17 | Asia/South Korea/Seoul/ | University of Ulsan College of Medicine and Asan Medical Center | University of Ulsan College of Medicine and Asan Medical Center | Kuenyoul Park et al |
| EPI_ISL_485393 | SouthKorea/S2/2020 | 2020-03-30 | Asia/South Korea/Gyeonggi Province/Ui | University of Ulsan College of Medicine and Asan Medical Center | University of Ulsan College of Medicine and Asan Medical Center | Kuenyoul Park et al |
| EPI_ISL_485398 | SouthKorea/CoV55/2020 | 2020-02-26 | Asia/South Korea/South Korea/ | Department of Internal Medicine, College of Medicine, Chosun University | Department of Internal Medicine, College of Medicine, Chosun University | Kim et al |
| EPI_ISL_485401 | Bahrain/Hu-1/2020 | 2020-06-23 | Asia/Bahrain/Capital Governorate/Man | Communicable Disease Laboratory, Public Health Directorate | Communicable Disease Laboratory, Public Health Directorate | Zaed et al |
| EPI_ISL_485604 | England/200920118/2020 | 2020-02-24 | Europe/United Kingdom/England/ | Respiratory Virus Unit, Microbiology Services Colindale, Public Health England | Respiratory Virus Unit, Microbiology Services Colindale, Public Health England | PHE Covid Sequencing Team et al |

| | | | | | | |
|---|---|---|---|---|---|---|
| EPI_ISL_485809 | Germany/BW-UT-001/2020 | 2020-05-13 | Europe/Germany/Baden-Wuerttemberg | Institut f√ºr Virologie und Epidemiologie der Viruskrankheiten, Universit√§tsklinikum T√ºbingen | NGS Competence Center T√ºbingen, Institut f√ºr Medizinische Mikrobiologie und Hygiene, Universit√§tsklinikum T√ºbingen | Angelov at al. et al |
| EPI_ISL_485818 | USA/VA-DCLS-0506/2020 | 2020-05-29 | North America/USA/Virginia/ | Virginia DCLS | Virginia DCLS | Virginia DCLS et al |
| EPI_ISL_485822 | USA/VA-DCLS-0511/2020 | 2020-05-29 | North America/USA/Virginia/ | Virginia DCLS | Virginia DCLS | Virginia DCLS et al |
| EPI_ISL_485823 | USA/VA-DCLS-0512/2020 | 2020-05-29 | North America/USA/Virginia/ | Virginia DCLS | Virginia DCLS | Virginia DCLS et al |
| EPI_ISL_485824 | USA/VA-DCLS-0513/2020 | 2020-05-29 | North America/USA/Virginia/ | Virginia DCLS | Virginia DCLS | Virginia DCLS et al |
| EPI_ISL_485829 | USA/VA-DCLS-0527/2020 | 2020-05-29 | North America/USA/Virginia/ | Virginia DCLS | Virginia DCLS | Virginia DCLS et al |
| EPI_ISL_485836 | USA/VA-DCLS-0535/2020 | 2020-03-13 | North America/USA/Virginia/ | Virginia DCLS | Virginia DCLS | Virginia DCLS et al |
| EPI_ISL_485837 | USA/VA-DCLS-0536/2020 | 2020-03-14 | North America/USA/Virginia/ | Virginia DCLS | Virginia DCLS | Virginia DCLS et al |
| EPI_ISL_485871 | USA/VA-DCLS-0620/2020 | 2020-04-01 | North America/USA/Virginia/ | Virginia DCLS | Virginia DCLS | Virginia DCLS et al |
| EPI_ISL_485873 | USA/LA-GBCL-26492436/2020 | 2020-04-28 | North America/USA/Louisiana/East Bato | River Road Testing Lab | Ginkgo Bioworks Clinical Laboratory | Rebecca C. Christofferson et al |
| EPI_ISL_485888 | USA/LA-GBCL-26544562/2020 | 2020-04-18 | North America/USA/Louisiana/East Bato | River Road Testing Lab | Ginkgo Bioworks Clinical Laboratory | Rebecca C. Christofferson et al |
| EPI_ISL_485897 | USA/LA-GBCL-26674724/2020 | 2020-04-10 | North America/USA/Louisiana/East Bato | River Road Testing Lab | Ginkgo Bioworks Clinical Laboratory | Rebecca C. Christofferson et al |
| EPI_ISL_485901 | USA/LA-GBCL-26684456/2020 | 2020-04-11 | North America/USA/Louisiana/East Bato | River Road Testing Lab | Ginkgo Bioworks Clinical Laboratory | Rebecca C. Christofferson et al |
| EPI_ISL_485903 | USA/LA-GBCL-26684476/2020 | 2020-04-10 | North America/USA/Louisiana/East Bato | River Road Testing Lab | Ginkgo Bioworks Clinical Laboratory | Rebecca C. Christofferson et al |
| EPI_ISL_485919 | USA/CT-UW-2441/2020 | 2020-03-28 | North America/USA/Connecticut/ | UW Virology Lab | UW Virology Lab | Pavitra Roychoudhury et al |
| EPI_ISL_485923 | USA/CT-UW-2475/2020 | 2020-03-30 | North America/USA/Connecticut/ | UW Virology Lab | UW Virology Lab | Pavitra Roychoudhury et al |
| EPI_ISL_485953 | USA/UN-UW-2820/2020 | 2020-04-15 | North America/USA/USA/ | UW Virology Lab | UW Virology Lab | Pavitra Roychoudhury et al |
| EPI_ISL_485955 | USA/UN-UW-2834/2020 | 2020-04-15 | North America/USA/USA/ | UW Virology Lab | UW Virology Lab | Pavitra Roychoudhury et al |
| EPI_ISL_485958 | USA/UN-UW-2849/2020 | 2020-04-15 | North America/USA/USA/ | UW Virology Lab | UW Virology Lab | Pavitra Roychoudhury et al |

| | | | | | | |
|---|---|---|---|---|---|---|
| EPI_ISL_485963 | USA/UN-UW-2859/2020 | 2020-04-15 | North America/USA/USA/ | UW Virology Lab | UW Virology Lab | Pavitra Roychoudhury et al |
| EPI_ISL_485984 | USA/WA-UW-2346/2020 | 2020-04-19 | North America/USA/Washington/ | UW Virology Lab | UW Virology Lab | Pavitra Roychoudhury et al |
| EPI_ISL_486015 | USA/WA-UW-2686/2020 | 2020-04-22 | North America/USA/Washington/ | UW Virology Lab | UW Virology Lab | Pavitra Roychoudhury et al |
| EPI_ISL_486027 | USA/WA-UW-2747/2020 | 2020-04-15 | North America/USA/Washington/ | UW Virology Lab | UW Virology Lab | Pavitra Roychoudhury et al |
| EPI_ISL_486029 | USA/WA-UW-2761/2020 | 2020-04-15 | North America/USA/Washington/ | UW Virology Lab | UW Virology Lab | Pavitra Roychoudhury et al |
| EPI_ISL_486045 | USA/WA-UW-2885/2020 | 2020-03-29 | North America/USA/Washington/ | UW Virology Lab | UW Virology Lab | Pavitra Roychoudhury et al |
| EPI_ISL_486058 | USA/WA-UW-2943/2020 | 2020-03-26 | North America/USA/Washington/ | UW Virology Lab | UW Virology Lab | Pavitra Roychoudhury et al |
| EPI_ISL_486061 | USA/WA-UW-2961/2020 | 2020-03-26 | North America/USA/Washington/ | UW Virology Lab | UW Virology Lab | Pavitra Roychoudhury et al |
| EPI_ISL_486062 | USA/WA-UW-2963/2020 | 2020-03-26 | North America/USA/Washington/ | UW Virology Lab | UW Virology Lab | Pavitra Roychoudhury et al |
| EPI_ISL_486082 | USA/WA-UW-3030/2020 | 2020-03-26 | North America/USA/Washington/ | UW Virology Lab | UW Virology Lab | Pavitra Roychoudhury et al |
| EPI_ISL_486088 | USA/WA-UW-3069/2020 | 2020-03-27 | North America/USA/Washington/ | UW Virology Lab | UW Virology Lab | Pavitra Roychoudhury et al |
| EPI_ISL_486109 | USA/WA-UW-7178/2020 | 2020-04-13 | North America/USA/Washington/ | UW Virology Lab | UW Virology Lab | Pavitra Roychoudhury et al |
| EPI_ISL_486134 | USA/CA-CZB-1817/2020 | 2020-04-22 | North America/USA/California/Orange ( | Orange County Public Health Laboratory | Chan-Zuckerberg Biohub | CZB Cliahub Consortium et al |
| EPI_ISL_486138 | USA/CA-CZB-1822/2020 | 2020-04-23 | North America/USA/California/Orange ( | Orange County Public Health Laboratory | Chan-Zuckerberg Biohub | CZB Cliahub Consortium et al |
| EPI_ISL_486152 | USA/CA-CZB-1839/2020 | 2020-04-28 | North America/USA/California/Orange ( | Orange County Public Health Laboratory | Chan-Zuckerberg Biohub | CZB Cliahub Consortium et al |
| EPI_ISL_486164 | USA/CA-CZB-1855/2020 | 2020-04-27 | North America/USA/California/Orange ( | Orange County Public Health Laboratory | Chan-Zuckerberg Biohub | CZB Cliahub Consortium et al |
| EPI_ISL_486165 | USA/CA-CZB-1857/2020 | 2020-04-27 | North America/USA/California/Orange ( | Orange County Public Health Laboratory | Chan-Zuckerberg Biohub | CZB Cliahub Consortium et al |
| EPI_ISL_486166 | USA/CA-CZB-1859/2020 | 2020-04-28 | North America/USA/California/Orange ( | Orange County Public Health Laboratory | Chan-Zuckerberg Biohub | CZB Cliahub Consortium et al |
| EPI_ISL_486171 | USA/CA-CZB-1871/2020 | 2020-04-28 | North America/USA/California/Orange ( | Orange County Public Health Laboratory | Chan-Zuckerberg Biohub | CZB Cliahub Consortium et al |
| EPI_ISL_486174 | USA/CA-CZB-1874/2020 | 2020-04-28 | North America/USA/California/Orange ( | Orange County Public Health Laboratory | Chan-Zuckerberg Biohub | CZB Cliahub Consortium et al |
| EPI_ISL_486177 | USA/CA-CZB-1878/2020 | 2020-04-28 | North America/USA/California/Orange ( | Orange County Public Health Laboratory | Chan-Zuckerberg Biohub | CZB Cliahub Consortium et al |
| EPI_ISL_486183 | USA/CA-CZB-1884/2020 | 2020-04-29 | North America/USA/California/Orange ( | Orange County Public Health Laboratory | Chan-Zuckerberg Biohub | CZB Cliahub Consortium et al |
| EPI_ISL_486193 | USA/CA-CZB-1894/2020 | 2020-04-21 | North America/USA/California/Orange ( | Orange County Public Health Laboratory | Chan-Zuckerberg Biohub | CZB Cliahub Consortium et al |
| EPI_ISL_486198 | USA/CA-CZB-1899/2020 | 2020-04-22 | North America/USA/California/Orange ( | Orange County Public Health Laboratory | Chan-Zuckerberg Biohub | CZB Cliahub Consortium et al |

| | | | | | | |
|---|---|---|---|---|---|---|
| EPI_ISL_486210 | USA/CA-CZB-1918/2020 | 2020-04-22 | North America/USA/California/Orange ( | Orange County Public Health Laboratory | Chan-Zuckerberg Biohub | CZB Cliahub Consortium et al |
| EPI_ISL_486214 | USA/CA-CZB-1922/2020 | 2020-04-23 | North America/USA/California/Orange ( | Orange County Public Health Laboratory | Chan-Zuckerberg Biohub | CZB Cliahub Consortium et al |
| EPI_ISL_486216 | USA/CA-CZB-1926/2020 | 2020-04-23 | North America/USA/California/Orange ( | Orange County Public Health Laboratory | Chan-Zuckerberg Biohub | CZB Cliahub Consortium et al |
| EPI_ISL_486219 | USA/CA-CZB-1930/2020 | 2020-04-15 | North America/USA/California/Orange ( | Orange County Public Health Laboratory | Chan-Zuckerberg Biohub | CZB Cliahub Consortium et al |
| EPI_ISL_486225 | USA/CA-CZB-1947/2020 | 2020-05-04 | North America/USA/California/Orange ( | Orange County Public Health Laboratory | Chan-Zuckerberg Biohub | CZB Cliahub Consortium et al |
| EPI_ISL_486228 | USA/CA-CZB-1955/2020 | 2020-05-06 | North America/USA/California/Orange ( | Orange County Public Health Laboratory | Chan-Zuckerberg Biohub | CZB Cliahub Consortium et al |
| EPI_ISL_486232 | USA/CA-CZB-1960/2020 | 2020-05-06 | North America/USA/California/Orange ( | Orange County Public Health Laboratory | Chan-Zuckerberg Biohub | CZB Cliahub Consortium et al |
| EPI_ISL_486234 | USA/CA-CZB-1962/2020 | 2020-05-06 | North America/USA/California/Orange ( | Orange County Public Health Laboratory | Chan-Zuckerberg Biohub | CZB Cliahub Consortium et al |
| EPI_ISL_486237 | USA/CA-CZB-1965/2020 | 2020-05-06 | North America/USA/California/Orange ( | Orange County Public Health Laboratory | Chan-Zuckerberg Biohub | CZB Cliahub Consortium et al |
| EPI_ISL_486241 | USA/CA-CZB-1969/2020 | 2020-05-06 | North America/USA/California/Orange ( | Orange County Public Health Laboratory | Chan-Zuckerberg Biohub | CZB Cliahub Consortium et al |
| EPI_ISL_486245 | USA/CA-CZB-1973/2020 | 2020-05-07 | North America/USA/California/Orange ( | Orange County Public Health Laboratory | Chan-Zuckerberg Biohub | CZB Cliahub Consortium et al |
| EPI_ISL_486250 | USA/CA-CZB-1982/2020 | 2020-05-06 | North America/USA/California/Orange ( | Orange County Public Health Laboratory | Chan-Zuckerberg Biohub | CZB Cliahub Consortium et al |
| EPI_ISL_486267 | USA/CA-CZB-2013/2020 | 2020-05-01 | North America/USA/California/Orange ( | Orange County Public Health Laboratory | Chan-Zuckerberg Biohub | CZB Cliahub Consortium et al |
| EPI_ISL_486272 | USA/CA-CZB-2020/2020 | 2020-05-03 | North America/USA/California/Orange ( | Orange County Public Health Laboratory | Chan-Zuckerberg Biohub | CZB Cliahub Consortium et al |
| EPI_ISL_486278 | USA/CA-CZB-2030/2020 | 2020-05-08 | North America/USA/California/Orange ( | Orange County Public Health Laboratory | Chan-Zuckerberg Biohub | CZB Cliahub Consortium et al |
| EPI_ISL_486281 | USA/CA-CZB-2035/2020 | 2020-05-12 | North America/USA/California/Humbold | Humboldt County Public Health Laboratory | Chan-Zuckerberg Biohub | CZB Cliahub Consortium et al |
| EPI_ISL_486285 | USA/CA-CZB-2041/2020 | 2020-05-15 | North America/USA/California/Humbold | Humboldt County Public Health Laboratory | Chan-Zuckerberg Biohub | CZB Cliahub Consortium et al |
| EPI_ISL_486294 | USA/CA-CZB-2050/2020 | 2020-04-21 | North America/USA/California/San Joac | San Joaquin County Public Health Lab | Chan-Zuckerberg Biohub | CZB Cliahub Consortium et al |
| EPI_ISL_486297 | USA/CA-CZB-2053/2020 | 2020-04-22 | North America/USA/California/San Joac | San Joaquin County Public Health Lab | Chan-Zuckerberg Biohub | CZB Cliahub Consortium et al |
| EPI_ISL_486306 | USA/CA-CZB-2063/2020 | 2020-04-22 | North America/USA/California/San Joac | San Joaquin County Public Health Lab | Chan-Zuckerberg Biohub | CZB Cliahub Consortium et al |

| | | | | | | |
|---|---|---|---|---|---|---|
| EPI_ISL_486320 | USA/CA-CZB-2077/2020 | 2020-05-04 | North America/USA/California/San Joac | San Joaquin County Public Health Lab | Chan-Zuckerberg Biohub | CZB Cliahub Consortium et al |
| EPI_ISL_486328 | USA/CA-CZB-2085/2020 | 2020-05-04 | North America/USA/California/San Joac | San Joaquin County Public Health Lab | Chan-Zuckerberg Biohub | CZB Cliahub Consortium et al |
| EPI_ISL_486334 | USA/CA-CZB-2091/2020 | 2020-05-07 | North America/USA/California/San Joac | San Joaquin County Public Health Lab | Chan-Zuckerberg Biohub | CZB Cliahub Consortium et al |
| EPI_ISL_486350 | USA/CA-CZB-2122/2020 | 2020-05-22 | North America/USA/California/Marin Co | UCSF Clinical Microbiology Laboratory | Chan-Zuckerberg Biohub | CZB Cliahub Consortium et al |
| EPI_ISL_486356 | USA/CA-CZB-2129/2020 | 2020-05-22 | North America/USA/California/Marin Co | UCSF Clinical Microbiology Laboratory | Chan-Zuckerberg Biohub | CZB Cliahub Consortium et al |
| EPI_ISL_486361 | USA/CA-CZB-2139/2020 | 2020-05-21 | North America/USA/California/Marin Co | UCSF Clinical Microbiology Laboratory | Chan-Zuckerberg Biohub | CZB Cliahub Consortium et al |
| EPI_ISL_486385 | India/KA-nimh-10102/2020 | 2020-05-01 | Asia/India/Karnataka/ | Victoria Hospital | Department of Neurovirology, National Institute of Mental Health and Neuroscience (NIMHANS) | Chitra Pattabiraman et al |
| EPI_ISL_486405 | India/KA-nimh-14709/2020 | 2020-05-10 | Asia/India/Karnataka/ | DH | Department of Neurovirology, National Institute of Mental Health and Neuroscience (NIMHANS) | Chitra Pattabiraman et al |
| EPI_ISL_486419 | Latvia/036/2020 | 2020-04-29 | Europe/Latvia/Latvia/ | CentrƒÅlƒÅ laboratorija | Latvian Biomedical Research and Study Centre | Ivars Silamiƒ∑elis et al |
| EPI_ISL_486422 | Latvia/039/2020 | 2020-03-15 | Europe/Latvia/Latvia/ | Latvijas Infektoloƒ£ijas centrs | Latvian Biomedical Research and Study Centre | Ivars Silamiƒ∑elis et al |
| EPI_ISL_486433 | Latvia/049/2020 | 2020-03-16 | Europe/Latvia/Latvia/ | Latvijas Infektoloƒ£ijas centrs | Latvian Biomedical Research and Study Centre | Ivars Silamiƒ∑elis et al |
| EPI_ISL_486438 | Latvia/054/2020 | 2020-05-28 | Europe/Latvia/Latvia/ | E. Gulbja laboratorija | Latvian Biomedical Research and Study Centre | Ivars Silamiƒ∑elis et al |

| | | | | | | |
|---|---|---|---|---|---|---|
| EPI_ISL_486519 | Switzerland/UR-ETHZ-170004/2020 | 2020-06-26 | Europe/Switzerland/Uri/ | Viollier AG | Department of Biosystems Science and Engineering, ETH Z√ºrich | Christian Beisel et al |
| EPI_ISL_486533 | Switzerland/VD-ETHZ-170021/2020 | 2020-06-24 | Europe/Switzerland/Vaud/ | Viollier AG | Department of Biosystems Science and Engineering, ETH Z√ºrich | Christian Beisel et al |
| EPI_ISL_486645 | Wuhan/ZY38-1/2020 | 2020-02-14 | Asia/China/Hubei/Wuhan | Renmin Hospital of Wuhan University Hubei General Hospital | State Key Laboratory of Agriculture Microbiology, Huazhong Agric | Zhong Zou et al |
| EPI_ISL_486646 | Italy/LOM-ASST-2/2020 | 2020-03-09 | Europe/Italy/Lombardy/ | Microbiology, Virology and Biemergency Laboratory-ASST FBF Sacco | Microbiology, Virology and Biemergency Laboratory-ASST FBF Sacco | Mancon A et al |
| EPI_ISL_486648 | Italy/LOM-ASST-11/2020 | 2020-03-02 | Europe/Italy/Lombardy/ | Microbiology, Virology and Biemergency Laboratory-ASST FBF Sacco | Microbiology, Virology and Biemergency Laboratory-ASST FBF Sacco | Micheli V et al |
| EPI_ISL_486649 | Italy/LOM-ASST-12/2020 | 2020-03-13 | Europe/Italy/Lombardy/ | Microbiology, Virology and Biemergency Laboratory-ASST FBF Sacco | Microbiology, Virology and Biemergency Laboratory-ASST FBF Sacco | Rimoldi SG et al |
| EPI_ISL_486651 | Italy/LOM-ASST-14/2020 | 2020-03-24 | Europe/Italy/Lombardy/ | Microbiology, Virology and Biemergency Laboratory-ASST FBF Sacco | Microbiology, Virology and Biemergency Laboratory-ASST FBF Sacco | Mancon A et al |
| EPI_ISL_486656 | Italy/LOM-ASST-19/2020 | 2020-02-20 | Europe/Italy/Lombardy/ | Microbiology, Virology and Biemergency Laboratory-ASST FBF Sacco | Microbiology, Virology and Biemergency Laboratory-ASST FBF Sacco | Micheli V et al |

| | | | | | | |
|---|---|---|---|---|---|---|
| EPI_ISL_486657 | Italy/LOM-ASST-20/2020 | 2020-02-26 | Europe/Italy/Lombardy/ | Microbiology, Virology and Biemergency Laboratory-ASST FBF Sacco | Microbiology, Virology and Biemergency Laboratory-ASST FBF Sacco | Rimoldi SG et al |
| EPI_ISL_486659 | Italy/LOM-ASST-3/2020 | 2020-02-22 | Europe/Italy/Lombardy/ | Microbiology, Virology and Biemergency Laboratory-ASST FBF Sacco | Microbiology, Virology and Biemergency Laboratory-ASST FBF Sacco | Micheli V et al |
| EPI_ISL_486660 | Italy/LOM-ASST-4/2020 | 2020-02-23 | Europe/Italy/Lombardy/ | Microbiology, Virology and Biemergency Laboratory-ASST FBF Sacco | Microbiology, Virology and Biemergency Laboratory-ASST FBF Sacco | Rimoldi SG et al |
| EPI_ISL_486661 | Italy/LOM-ASST-5/2020 | 2020-03-02 | Europe/Italy/Lombardy/ | Microbiology, Virology and Biemergency Laboratory-ASST FBF Sacco | Microbiology, Virology and Biemergency Laboratory-ASST FBF Sacco | Romeri F et al |
| EPI_ISL_486662 | Italy/LOM-ASST-6/2020 | 2020-03-02 | Europe/Italy/Lombardy/ | Microbiology, Virology and Biemergency Laboratory-ASST FBF Sacco | Microbiology, Virology and Biemergency Laboratory-ASST FBF Sacco | Mancon A et al |
| EPI_ISL_486663 | Italy/LOM-ASST-7/2020 | 2020-03-06 | Europe/Italy/Lombardy/ | Microbiology, Virology and Biemergency Laboratory-ASST FBF Sacco | Microbiology, Virology and Biemergency Laboratory-ASST FBF Sacco | Micheli V et al |
| EPI_ISL_486664 | Italy/LOM-ASST-8/2020 | 2020-02-26 | Europe/Italy/Lombardy/ | Microbiology, Virology and Biemergency Laboratory-ASST FBF Sacco | Microbiology, Virology and Biemergency Laboratory-ASST FBF Sacco | Rimoldi SG et al |
| EPI_ISL_486665 | Italy/LOM-AMVRC-9/2020 | 2020-03-03 | Europe/Italy/Lombardy/ | Microbiology, Virology and Biemergency Laboratory-ASST FBF Sacco | Microbiology, Virology and Biemergency Laboratory-ASST FBF Sacco | Micheli V et al |

| | | | | | | |
|---|---|---|---|---|---|---|
| EPI_ISL_486816 | Russia/CRIE159875/2020 | 2020-03-30 | Europe/Russia/Moscow Oblast/ | "Molecular diagnostic laboratory of Federal Budget Institution of Science ""Central Research Institute of Epidemiology"" of The Federal Service on Customers' Rights Protection and Human Well-being Surveillance" | Group of Genomics and Postgenomic Technologies of Central Research Institute of Epidemilology | Speranskaya AS et al |
| EPI_ISL_486817 | Russia/CRIE159932/2020 | 2020-03-30 | Europe/Russia/Moscow Oblast/ | "Molecular diagnostic laboratory of Federal Budget Institution of Science ""Central Research Institute of Epidemiology"" of The Federal Service on Customers' Rights Protection and Human Well-being Surveillance" | Group of Genomics and Postgenomic Technologies of Central Research Institute of Epidemilology | Speranskaya AS et al |
| EPI_ISL_486821 | Russia/CRIE162398/2020 | 2020-03-31 | Europe/Russia/Moscow Oblast/ | "Molecular diagnostic laboratory of Federal Budget Institution of Science ""Central Research Institute of Epidemiology"" of The Federal Service on Customers' Rights Protection and Human Well-being Surveillance" | Group of Genomics and Postgenomic Technologies of Central Research Institute of Epidemilology | Speranskaya AS et al |
| EPI_ISL_486827 | Russia/CRIE175960/2020 | 2020-04-01 | Europe/Russia/Moscow Oblast/ | "Molecular diagnostic laboratory of Federal Budget Institution of Science ""Central Research Institute of Epidemiology"" of The Federal Service on Customers' Rights Protection and Human Well-being Surveillance" | Group of Genomics and Postgenomic Technologies of Central Research Institute of Epidemilology | Speranskaya AS et al |
| EPI_ISL_486828 | Russia/CRIE176675/2020 | 2020-04-01 | Europe/Russia/Moscow Oblast/ | "Molecular diagnostic laboratory of Federal Budget Institution of Science ""Central Research Institute of Epidemiology"" of The Federal Service on Customers' Rights Protection and Human Well-being Surveillance" | Group of Genomics and Postgenomic Technologies of Central Research Institute of Epidemilology | Speranskaya AS et al |
| EPI_ISL_486842 | Ecuador/USFQ-105/2020 | 2020-07-01 | South America/Ecuador/Chimborazo/Ric | Institute of Microbiology, Universidad San Francisco de Quito | Institute of Microbiology, Universidad San Francisco de Quito | BelV©n Prado-Vivar et al |

| | | | | | | |
|---|---|---|---|---|---|---|
| EPI_ISL_486843 | Ecuador/USFQ-108/2020 | 2020-07-01 | South America/Ecuador/Chimborazo/Rio | Institute of Microbiology, Universidad San Francisco de Quito | Institute of Microbiology, Universidad San Francisco de Quito | Bel√©n Prado-Vivar et al |
| EPI_ISL_486844 | Ecuador/USFQ-109/2020 | 2020-07-01 | South America/Ecuador/Chimborazo/Rio | Institute of Microbiology, Universidad San Francisco de Quito | Institute of Microbiology, Universidad San Francisco de Quito | Bel√©n Prado-Vivar et al |
| EPI_ISL_486846 | Ecuador/USFQ-111/2020 | 2020-06-30 | South America/Ecuador/Imbabura/Ibarr | Institute of Microbiology, Universidad San Francisco de Quito | Institute of Microbiology, Universidad San Francisco de Quito | Bel√©n Prado-Vivar et al |
| EPI_ISL_486847 | Ecuador/USFQ-1112/2020 | 2020-06-30 | South America/Ecuador/Imbabura/Ibarr | Institute of Microbiology, Universidad San Francisco de Quito | Institute of Microbiology, Universidad San Francisco de Quito | Bel√©n Prado-Vivar et al |
| EPI_ISL_486848 | Ecuador/USFQ-113/2020 | 2020-06-30 | South America/Ecuador/Imbabura/Ibarr | Institute of Microbiology, Universidad San Francisco de Quito | Institute of Microbiology, Universidad San Francisco de Quito | Bel√©n Prado-Vivar et al |
| EPI_ISL_486853 | India/UN-UN2/2020 | 2020-06-17 | Asia/India/India/ | CSIR-CDRI/SGPGI | CSIR-CDRI/SGPGI | Saumya Sarkar et al |
| EPI_ISL_486854 | Romania/ROSV_12646/2020 | 2020-06-10 | Europe/Romania/Suceava/ | Emergency County Hospital Suceava | Stefan cel Mare, University Metagenomics lab | Lobiuc Andrei et al. et al |
| EPI_ISL_486855 | Romania/ROSV/2020 | 2020-06-08 | Europe/Romania/Suceava/ | Emergency county Hospital Suceava | """Stefan cel Mare"" University Metagenomics Lab" | Lobiuc Andrei et al. et al |
| EPI_ISL_486865 | Senegal/6847/2020 | 2020-04-02 | Africa/Senegal/Dakar/ | Institut Pasteur Dakar | Institut Pasteur de Dakar | Ndongo Dia et al |
| EPI_ISL_486866 | Senegal/9250/2020 | 2020-04-22 | Africa/Senegal/Dakar/ | Institut Pasteur Dakar | Institut Pasteur de Dakar | Ndongo Dia et al |
| EPI_ISL_486881 | India/KA-nimh-3975/2020 | 2020-04-23 | Asia/India/Karnataka/ | CV Raman Hospital | Department of Neurovirology, National Institute of Mental Health and Neuroscience (NIMHANS) | Chitra Pattabiraman et al |
| EPI_ISL_486887 | Bahrain/02/2020 | 2020-06-23 | Asia/Bahrain/Bahrain/ | National Influenza Center, Bahrain | National Influenza Center, Bahrain | Zaed et al |

| | | | | | | |
|---|---|---|---|---|---|---|
| EPI_ISL_486888 | Bahrain/03/2020 | 2020-06-23 | Asia/Bahrain/Bahrain/ | National Influenza Center, Bahrain | National Influenza Center, Bahrain | AlWasti et al |
| EPI_ISL_487092 | Nigeria/KW314-CV63/2020 | 2020-05-09 | Africa/Nigeria/Kwara State/ | Nigeria Centre for Disease Control (NCDC) | African Centre of Excellence for Genomics of Infectious Diseases (ACEGID), Redeemer's University, Ede, Osun State, Nigeria | Oluniyi P.E. et al |
| EPI_ISL_487096 | Nigeria/EK254-CV60/2020 | 2020-05-19 | Africa/Nigeria/Ekiti State/ | Nigeria Centre for Disease Control (NCDC) | African Centre of Excellence for Genomics of Infectious Diseases (ACEGID), Redeemer's University, Ede, Osun State, Nigeria | Oluniyi P.E. et al |
| EPI_ISL_487101 | Nigeria/OY260-CV55/2020 | 2020-05-16 | Africa/Nigeria/Oyo State/ | Nigeria Centre for Disease Control (NCDC) | African Centre of Excellence for Genomics of Infectious Diseases (ACEGID), Redeemer's University, Ede, Osun State, Nigeria | Oluniyi P.E. et al |

| | | | | | | |
|---|---|---|---|---|---|---|
| EPI_ISL_487112 | Nigeria/ON505-CV43/2020 | 2020-05-24 | Africa/Nigeria/Ondo State/ | Nigeria Centre for Disease Control (NCDC) | African Centre of Excellence for Genomics of Infectious Diseases (ACEGID), Redeemer's University, Ede, Osun State, Nigeria | Oluniyi P.E. et al |
| EPI_ISL_487197 | USA/UT-02536/2020 | 2020-04-24 | North America/USA/Utah/ | Utah Public Health Laboratory | Utah Public Health Laboratory | Heidi Butz et al |
| EPI_ISL_487200 | USA/UT-02543/2020 | 2020-04-24 | North America/USA/Utah/ | Utah Public Health Laboratory | Utah Public Health Laboratory | Heidi Butz et al |
| EPI_ISL_487211 | USA/UT-02562/2020 | 2020-04-24 | North America/USA/Utah/ | Utah Public Health Laboratory | Utah Public Health Laboratory | Heidi Butz et al |
| EPI_ISL_487217 | USA/UT-02569/2020 | 2020-04-25 | North America/USA/Utah/ | Utah Public Health Laboratory | Utah Public Health Laboratory | Heidi Butz et al |
| EPI_ISL_487247 | USA/UT-04116/2020 | 2020-05-12 | North America/USA/Utah/ | Utah Public Health Laboratory | Utah Public Health Laboratory | Heidi Butz et al |
| EPI_ISL_487248 | USA/UT-04117/2020 | 2020-05-12 | North America/USA/Utah/ | Utah Public Health Laboratory | Utah Public Health Laboratory | Heidi Butz et al |
| EPI_ISL_487249 | USA/UT-04118/2020 | 2020-05-13 | North America/USA/Utah/ | Utah Public Health Laboratory | Utah Public Health Laboratory | Heidi Butz et al |
| EPI_ISL_487251 | USA/UT-04127/2020 | 2020-05-13 | North America/USA/Utah/ | Utah Public Health Laboratory | Utah Public Health Laboratory | Heidi Butz et al |
| EPI_ISL_487253 | USA/UT-04151/2020 | 2020-05-14 | North America/USA/Utah/ | Utah Public Health Laboratory | Utah Public Health Laboratory | Heidi Butz et al |
| EPI_ISL_487259 | USA/UT-04158/2020 | 2020-05-14 | North America/USA/Utah/ | Utah Public Health Laboratory | Utah Public Health Laboratory | Heidi Butz et al |
| EPI_ISL_487262 | USA/UT-05783/2020 | 2020-06-04 | North America/USA/Utah/ | Utah Public Health Laboratory | Utah Public Health Laboratory | Heidi Butz et al |
| EPI_ISL_487270 | Bahrain/340217593/2020 | 2020-06-22 | Asia/Bahrain/Bahrain/ | unknown | Communicable Disease Laboratory, Public Health Directorate | AlWasti et al |

| | | | | | | |
|---|---|---|---|---|---|---|
| EPI_ISL_487273 | Bahrain/110003611/2020 | 2020-06-25 | Asia/Bahrain/Bahrain/ | unknown | Communicable Disease Laboratory, Public Health Directorate | Zaed et al |
| EPI_ISL_487274 | Bahrain/023232093/2020 | 2020-06-24 | Asia/Bahrain/Bahrain/ | unknown | Communicable Disease Laboratory, Public Health Directorate | AlWasti et al |
| EPI_ISL_487276 | Italy/LOM-UniMI-1/2020 | 2020-04-04 | Europe/Italy/Lombardy/Milan | Department of Food Safety, Nutrition and Veterinary public health, Istituto Superiore di Sanita' | Department of Biomedical, Surgical and Dental Sciences and Department of Biomedical Sciences for Health | Delbue et al |
| EPI_ISL_487278 | SouthAfrica/KRISP-0287/2020 | 2020-06-26 | Africa/South Africa/KwaZulu-Natal/eThe | NHLS-IALCH | KRISP, KZN Research Innovation and Sequencing Platform | Giandhari J et al |
| EPI_ISL_487279 | SouthAfrica/KRISP-0288/2020 | 2020-06-26 | Africa/South Africa/KwaZulu-Natal/eThe | NHLS-IALCH | KRISP, KZN Research Innovation and Sequencing Platform | Giandhari J et al |
| EPI_ISL_487281 | SouthAfrica/KRISP-0291/2020 | 2020-06-26 | Africa/South Africa/KwaZulu-Natal/eThe | NHLS-IALCH | KRISP, KZN Research Innovation and Sequencing Platform | Giandhari J et al |
| EPI_ISL_487282 | SouthAfrica/KRISP-0293/2020 | 2020-06-25 | Africa/South Africa/KwaZulu-Natal/eThe | NHLS-IALCH | KRISP, KZN Research Innovation and Sequencing Platform | Giandhari J et al |

| | | | | | | |
|---|---|---|---|---|---|---|
| EPI_ISL_487283 | SouthAfrica/KRISP-0294/2020 | 2020-06-26 | Africa/South Africa/KwaZulu-Natal/eThe | NHLS-IALCH | KRISP, KZN Research Innovation and Sequencing Platform | Giandhari J et al |
| EPI_ISL_487284 | SouthAfrica/KRISP-0296/2020 | 2020-06-26 | Africa/South Africa/KwaZulu-Natal/eThe | NHLS-IALCH | KRISP, KZN Research Innovation and Sequencing Platform | Giandhari J et al |
| EPI_ISL_487285 | SouthAfrica/KRISP-0297/2020 | 2020-06-26 | Africa/South Africa/KwaZulu-Natal/eThe | NHLS-IALCH | KRISP, KZN Research Innovation and Sequencing Platform | Giandhari J et al |
| EPI_ISL_487287 | SouthAfrica/KRISP-0299/2020 | 2020-06-25 | Africa/South Africa/KwaZulu-Natal/eThe | NHLS-IALCH | KRISP, KZN Research Innovation and Sequencing Platform | Giandhari J et al |
| EPI_ISL_487288 | SouthAfrica/KRISP-0300/2020 | 2020-06-25 | Africa/South Africa/KwaZulu-Natal/eThe | NHLS-IALCH | KRISP, KZN Research Innovation and Sequencing Platform | Giandhari J et al |
| EPI_ISL_487291 | SouthAfrica/KRISP-0305/2020 | 2020-06-24 | Africa/South Africa/KwaZulu-Natal/uMg | NHLS-IALCH | KRISP, KZN Research Innovation and Sequencing Platform | Giandhari J et al |
| EPI_ISL_487292 | SouthAfrica/KRISP-0308/2020 | 2020-06-24 | Africa/South Africa/KwaZulu-Natal/Zulu | NHLS-IALCH | KRISP, KZN Research Innovation and Sequencing Platform | Giandhari J et al |
| EPI_ISL_487294 | SouthAfrica/KRISP-0310/2020 | 2020-06-24 | Africa/South Africa/KwaZulu-Natal/Zulu | NHLS-IALCH | KRISP, KZN Research Innovation and Sequencing Platform | Giandhari J et al |

| | | | | | | |
|---|---|---|---|---|---|---|
| EPI_ISL_487298 | SouthAfrica/KRISP-0321/2020 | 2020-06-24 | Africa/South Africa/KwaZulu-Natal/Zulu | NHLS-IALCH | KRISP, KZN Research Innovation and Sequencing Platform | Giandhari J et al |
| EPI_ISL_487299 | SouthAfrica/KRISP-0322/2020 | 2020-06-24 | Africa/South Africa/KwaZulu-Natal/Zulu | NHLS-IALCH | KRISP, KZN Research Innovation and Sequencing Platform | Giandhari J et al |
| EPI_ISL_487302 | SouthAfrica/KRISP-0331/2020 | 2020-06-24 | Africa/South Africa/KwaZulu-Natal/Zulu | NHLS-IALCH | KRISP, KZN Research Innovation and Sequencing Platform | Giandhari J et al |
| EPI_ISL_487303 | SouthAfrica/KRISP-0332/2020 | 2020-06-24 | Africa/South Africa/KwaZulu-Natal/Zulu | NHLS-IALCH | KRISP, KZN Research Innovation and Sequencing Platform | Giandhari J et al |
| EPI_ISL_487304 | SouthAfrica/KRISP-0336/2020 | 2020-06-18 | Africa/South Africa/KwaZulu-Natal/uMg | NHLS-IALCH | KRISP, KZN Research Innovation and Sequencing Platform | Giandhari J et al |
| EPI_ISL_487305 | SouthAfrica/KRISP-0338/2020 | 2020-06-18 | Africa/South Africa/KwaZulu-Natal/Ugu | NHLS-IALCH | KRISP, KZN Research Innovation and Sequencing Platform | Giandhari J et al |
| EPI_ISL_487307 | SouthAfrica/KRISP-0340/2020 | 2020-06-17 | Africa/South Africa/KwaZulu-Natal/uThu | NHLS-IALCH | KRISP, KZN Research Innovation and Sequencing Platform | Giandhari J et al |
| EPI_ISL_487308 | SouthAfrica/KRISP-0347/2020 | 2020-06-18 | Africa/South Africa/KwaZulu-Natal/Ugu | NHLS-IALCH | KRISP, KZN Research Innovation and Sequencing Platform | Giandhari J et al |

| | | | | | | |
|---|---|---|---|---|---|---|
| EPI_ISL_487309 | SouthAfrica/KRISP-0348/2020 | 2020-06-18 | Africa/South Africa/KwaZulu-Natal/uMg | NHLS-IALCH | KRISP, KZN Research Innovation and Sequencing Platform | Giandhari J et al |
| EPI_ISL_487310 | SouthAfrica/KRISP-0349/2020 | 2020-06-17 | Africa/South Africa/KwaZulu-Natal/uThu | NHLS-IALCH | KRISP, KZN Research Innovation and Sequencing Platform | Giandhari J et al |
| EPI_ISL_487311 | SouthAfrica/KRISP-0350/2020 | 2020-06-17 | Africa/South Africa/KwaZulu-Natal/uThu | NHLS-IALCH | KRISP, KZN Research Innovation and Sequencing Platform | Giandhari J et al |
| EPI_ISL_487312 | SouthAfrica/KRISP-0352/2020 | 2020-06-17 | Africa/South Africa/KwaZulu-Natal/uThu | NHLS-IALCH | KRISP, KZN Research Innovation and Sequencing Platform | Giandhari J et al |
| EPI_ISL_487313 | SouthAfrica/KRISP-0356/2020 | 2020-06-25 | Africa/South Africa/KwaZulu-Natal/eThe | NHLS-IALCH | KRISP, KZN Research Innovation and Sequencing Platform | Giandhari J et al |
| EPI_ISL_487314 | SouthAfrica/KRISP-0360/2020 | 2020-06-19 | Africa/South Africa/KwaZulu-Natal/Ama | NHLS-IALCH | KRISP, KZN Research Innovation and Sequencing Platform | Giandhari J et al |
| EPI_ISL_487315 | SouthAfrica/KRISP-0361/2020 | 2020-06-18 | Africa/South Africa/KwaZulu-Natal/uMg | NHLS-IALCH | KRISP, KZN Research Innovation and Sequencing Platform | Giandhari J et al |
| EPI_ISL_487316 | SouthAfrica/KRISP-0365/2020 | 2020-06-25 | Africa/South Africa/KwaZulu-Natal/eThe | NHLS-IALCH | KRISP, KZN Research Innovation and Sequencing Platform | Giandhari J et al |

| | | | | | | |
|---|---|---|---|---|---|---|
| EPI_ISL_487317 | SouthAfrica/KRISP-0366/2020 | 2020-06-18 | Africa/South Africa/KwaZulu-Natal/Ama | NHLS-IALCH | KRISP, KZN Research Innovation and Sequencing Platform | Giandhari J et al |
| EPI_ISL_487318 | SouthAfrica/KRISP-0367/2020 | 2020-06-25 | Africa/South Africa/KwaZulu-Natal/eThe | NHLS-IALCH | KRISP, KZN Research Innovation and Sequencing Platform | Giandhari J et al |
| EPI_ISL_487319 | SouthAfrica/KRISP-0368/2020 | 2020-06-18 | Africa/South Africa/KwaZulu-Natal/Ama | NHLS-IALCH | KRISP, KZN Research Innovation and Sequencing Platform | Giandhari J et al |
| EPI_ISL_487320 | SouthAfrica/KRISP-0369/2020 | 2020-06-25 | Africa/South Africa/KwaZulu-Natal/eThe | NHLS-IALCH | KRISP, KZN Research Innovation and Sequencing Platform | Giandhari J et al |
| EPI_ISL_487321 | SouthAfrica/KRISP-0370/2020 | 2020-06-25 | Africa/South Africa/KwaZulu-Natal/eThe | NHLS-IALCH | KRISP, KZN Research Innovation and Sequencing Platform | Giandhari J et al |
| EPI_ISL_487323 | SouthAfrica/KRISP-0372/2020 | 2020-06-19 | Africa/South Africa/KwaZulu-Natal/eThe | NHLS-IALCH | KRISP, KZN Research Innovation and Sequencing Platform | Giandhari J et al |
| EPI_ISL_487324 | SouthAfrica/KRISP-0373/2020 | 2020-06-19 | Africa/South Africa/KwaZulu-Natal/eThe | NHLS-IALCH | KRISP, KZN Research Innovation and Sequencing Platform | Giandhari J et al |
| EPI_ISL_487325 | SouthAfrica/KRISP-0375/2020 | 2020-06-25 | Africa/South Africa/KwaZulu-Natal/eThe | NHLS-IALCH | KRISP, KZN Research Innovation and Sequencing Platform | Giandhari J et al |

| | | | | | | |
|---|---|---|---|---|---|---|
| EPI_ISL_487326 | SouthAfrica/KRISP-0376/2020 | 2020-06-22 | Africa/South Africa/KwaZulu-Natal/eThe | NHLS-IALCH | KRISP, KZN Research Innovation and Sequencing Platform | Giandhari J et al |
| EPI_ISL_487327 | SouthAfrica/KRISP-0378/2020 | 2020-06-22 | Africa/South Africa/KwaZulu-Natal/eThe | NHLS-IALCH | KRISP, KZN Research Innovation and Sequencing Platform | Giandhari J et al |
| EPI_ISL_487330 | SouthAfrica/KRISP-0387/2020 | 2020-07-01 | Africa/South Africa/KwaZulu-Natal/eThe | Molecular Diagnostics Services (MDS) | KRISP, KZN Research Innovation and Sequencing Platform | Giandhari J et al |
| EPI_ISL_487335 | SouthAfrica/KRISP-0392/2020 | 2020-07-01 | Africa/South Africa/KwaZulu-Natal/eThe | Molecular Diagnostics Services (MDS) | KRISP, KZN Research Innovation and Sequencing Platform | Giandhari J et al |
| EPI_ISL_487337 | SouthAfrica/KRISP-0394/2020 | 2020-07-01 | Africa/South Africa/KwaZulu-Natal/eThe | Molecular Diagnostics Services (MDS) | KRISP, KZN Research Innovation and Sequencing Platform | Giandhari J et al |
| EPI_ISL_487363 | Bangladesh/BCSIR-NILMRC-223/2020 | 2020-07-06 | Asia/Bangladesh/Barishal/ | National Institute of Laboratory Medicine and Referral Center | Genomic Research Lab, BCSIR | Tanjina Akhter Banu et al |
| EPI_ISL_487364 | Bangladesh/BCSIR-NILMRC-224/2020 | 2020-07-06 | Asia/Bangladesh/Barishal/ | National Institute of Laboratory Medicine and Referral Center | Genomic Research Lab, BCSIR | Tanjina Akhter Banu et al |
| EPI_ISL_487366 | Bangladesh/BCSIR-NILMRC-226/2020 | 2020-07-06 | Asia/Bangladesh/Barishal/ | National Institute of Laboratory Medicine and Referral Center | Genomic Research Lab, BCSIR | Shahina Akter et al |
| EPI_ISL_487372 | Bangladesh/BCSIR-NILMRC-230/2020 | 2020-07-06 | Asia/Bangladesh/Barishal/ | National Institute of Laboratory Medicine and Referral Center | Genomic Research Lab, BCSIR | Iffat Jahan et al |
| EPI_ISL_487380 | Bangladesh/BCSIR-NILMRC-235/2020 | 2020-07-06 | Asia/Bangladesh/Barishal/ | National Institute of Laboratory Medicine and Referral Center | Genomic Research Lab, BCSIR | Md. Murshed Hasan Sarkar et al |
| EPI_ISL_487383 | Bangladesh/BCSIR-NILMRC-243/2020 | 2020-07-06 | Asia/Bangladesh/Barishal/ | National Institute of Laboratory Medicine and Referral Center | Genomic Research Lab, BCSIR | Md. Murshed Hasan Sarkar et al |
| EPI_ISL_487392 | Bangladesh/BCSIR-NILMRC-237/2020 | 2020-07-06 | Asia/Bangladesh/Barishal/ | National Institute of Laboratory Medicine and Referral Center | Genomic Research Lab, BCSIR | Abu Sayeed Mohammad Mahmud et al |

| | | | | | | |
|---|---|---|---|---|---|---|
| EPI_ISL_487394 | Bangladesh/BCSIR-NILMRC-239/2020 | 2020-07-06 | Asia/Bangladesh/Barishal/ | National Institute of Laboratory Medicine and Referral Center | Genomic Research Lab, BCSIR | Abu Sayeed Mohammad Mahmud et al |
| EPI_ISL_487396 | Bangladesh/BCSIR-NILMRC-241/2020 | 2020-07-06 | Asia/Bangladesh/Barishal/ | National Institute of Laboratory Medicine and Referral Center | Genomic Research Lab, BCSIR | Abu Sayeed Mohammad Mahmud et al |
| EPI_ISL_487400 | Germany/NW-MPP-12/2020 | 2020-05-25 | Europe/Germany/North Rhine Westphal | Labor Kneiÿüler GmbH & Co. KG | Heinrich Pette Institute, Leibniz Institute for Experimental Virology | Thomas GŸnther et al |
| EPI_ISL_487422 | Germany/NW-MPP-32/2020 | 2020-06-17 | Europe/Germany/North Rhine Westphal | Labor Kneiÿüler GmbH & Co. KG | Heinrich Pette Institute, Leibniz Institute for Experimental Virology | Thomas GŸnther et al |
| EPI_ISL_487428 | Germany/NW-MPP-6/2020 | 2020-05-25 | Europe/Germany/North Rhine Westphal | Labor Kneiÿüler GmbH & Co. KG | Heinrich Pette Institute, Leibniz Institute for Experimental Virology | Thomas GŸnther et al |
| EPI_ISL_487433 | Belgium/ITM_C2181/2020 | 2020-05-20 | Europe/Belgium/Belgium/ | Queen Astrid Military Hospital | Institute of Tropical Medicine | Philippe Selhorst et al |
| EPI_ISL_487552 | England/BRIS-12B46C/2020 | 2020-05-25 | Europe/United Kingdom/England/ | PHE South West Regional Laboratory, National Infection Service | Wellcome Sanger Institute for the COVID-19 Genomics UK (COG-UK) consortium | Stephanie Hutchings et al |
| EPI_ISL_487609 | England/BRIS-12B569/2020 | 2020-05-22 | Europe/United Kingdom/England/ | PHE South West Regional Laboratory, National Infection Service | Wellcome Sanger Institute for the COVID-19 Genomics UK (COG-UK) Consortium | Stephanie Hutchings et al |
| EPI_ISL_487615 | England/LOND-D64BC/2020 | 2020-04-22 | Europe/United Kingdom/England/ | University College London, Great Ormond Street Hospital for Children NHS Foundation Trust, Imperial College Healthcare NHS Trust | Wellcome Sanger Institute for the COVID-19 Genomics UK (COG-UK) consortium | Sergi Castellano et al |

| | | | | | | |
|---|---|---|---|---|---|---|
| EPI_ISL_487688 | Scotland/EDB3357/2020 | 2020-04-09 | Europe/United Kingdom/Scotland/ | Virology Department, Royal Infirmary of Edinburgh, NHS Lothian / School of Biological Sciences, University of Edinburgh | Wellcome Sanger Institute for the COVID-19 Genomics UK (COG-UK) consortium | McHugh M et al |
| EPI_ISL_487691 | Scotland/EDB5883/2020 | 2020-04-29 | Europe/United Kingdom/Scotland/ | Virology Department, Royal Infirmary of Edinburgh, NHS Lothian / School of Biological Sciences, University of Edinburgh | Wellcome Sanger Institute for the COVID-19 Genomics UK (COG-UK) consortium | McHugh M et al |
| EPI_ISL_487710 | Scotland/EDB5663/2020 | 2020-03-31 | Europe/United Kingdom/Scotland/ | Virology Department, Royal Infirmary of Edinburgh, NHS Lothian / School of Biological Sciences, University of Edinburgh | Wellcome Sanger Institute for the COVID-19 Genomics UK (COG-UK) consortium | McHugh M et al |
| EPI_ISL_487803 | Scotland/EDB3521/2020 | 2020-04-13 | Europe/United Kingdom/Scotland/ | Virology Department, Royal Infirmary of Edinburgh, NHS Lothian / School of Biological Sciences, University of Edinburgh | Wellcome Sanger Institute for the COVID-19 Genomics UK (COG-UK) consortium | McHugh M et al |
| EPI_ISL_487819 | Scotland/EDB5431/2020 | 2020-04-08 | Europe/United Kingdom/Scotland/ | Virology Department, Royal Infirmary of Edinburgh, NHS Lothian / School of Biological Sciences, University of Edinburgh | Wellcome Sanger Institute for the COVID-19 Genomics UK (COG-UK) consortium | McHugh M et al |
| EPI_ISL_487867 | Scotland/EDB3452/2020 | 2020-04-10 | Europe/United Kingdom/Scotland/ | Virology Department, Royal Infirmary of Edinburgh, NHS Lothian / School of Biological Sciences, University of Edinburgh | Wellcome Sanger Institute for the COVID-19 Genomics UK (COG-UK) consortium | McHugh M et al |
| EPI_ISL_487892 | Scotland/EDB5418/2020 | 2020-04-07 | Europe/United Kingdom/Scotland/ | Virology Department, Royal Infirmary of Edinburgh, NHS Lothian / School of Biological Sciences, University of Edinburgh | Wellcome Sanger Institute for the COVID-19 Genomics UK (COG-UK) consortium | McHugh M et al |
| EPI_ISL_487908 | Scotland/EDB6414/2020 | 2020-04-12 | Europe/United Kingdom/Scotland/ | Virology Department, Royal Infirmary of Edinburgh, NHS Lothian / School of Biological Sciences, University of Edinburgh | Wellcome Sanger Institute for the COVID-19 Genomics UK (COG-UK) consortium | McHugh M et al |

| | | | | | | |
|---|---|---|---|---|---|---|
| EPI_ISL_487919 | Scotland/EDB5632/2020 | 2020-03-23 | Europe/United Kingdom/Scotland/ | Virology Department, Royal Infirmary of Edinburgh, NHS Lothian / School of Biological Sciences, University of Edinburgh | Wellcome Sanger Institute for the COVID-19 Genomics UK (COG-UK) consortium | McHugh M et al |
| EPI_ISL_487929 | Scotland/EDB3547/2020 | 2020-04-13 | Europe/United Kingdom/Scotland/ | Virology Department, Royal Infirmary of Edinburgh, NHS Lothian / School of Biological Sciences, University of Edinburgh | Wellcome Sanger Institute for the COVID-19 Genomics UK (COG-UK) consortium | McHugh M et al |
| EPI_ISL_487934 | Scotland/EDB2907/2020 | 2020-04-01 | Europe/United Kingdom/Scotland/ | Virology Department, Royal Infirmary of Edinburgh, NHS Lothian / School of Biological Sciences, University of Edinburgh | Wellcome Sanger Institute for the COVID-19 Genomics UK (COG-UK) consortium | McHugh M et al |
| EPI_ISL_487966 | Scotland/EDB4829/2020 | 2020-04-03 | Europe/United Kingdom/Scotland/ | Virology Department, Royal Infirmary of Edinburgh, NHS Lothian / School of Biological Sciences, University of Edinburgh | Wellcome Sanger Institute for the COVID-19 Genomics UK (COG-UK) consortium | McHugh M et al |
| EPI_ISL_487993 | Scotland/EDB5582/2020 | 2020-03-30 | Europe/United Kingdom/Scotland/ | Virology Department, Royal Infirmary of Edinburgh, NHS Lothian / School of Biological Sciences, University of Edinburgh | Wellcome Sanger Institute for the COVID-19 Genomics UK (COG-UK) consortium | McHugh M et al |
| EPI_ISL_488013 | England/NORT-2985EE/2020 | 2020-04-22 | Europe/United Kingdom/England/ | NU-OMICS DNA Sequencing research facility, Northumbria University | Wellcome Sanger Institute for the COVID-19 Genomics UK (COG-UK) consortium | Chris Duncan et al |
| EPI_ISL_488131 | England/NORT-29D9A1/2020 | 2020-05-07 | Europe/United Kingdom/England/ | NU-OMICS DNA Sequencing research facility, Northumbria University | Wellcome Sanger Institute for the COVID-19 Genomics UK (COG-UK) consortium | Chris Duncan et al |
| EPI_ISL_488133 | England/NORT-29DD96/2020 | 2020-05-06 | Europe/United Kingdom/England/ | NU-OMICS DNA Sequencing research facility, Northumbria University | Wellcome Sanger Institute for the COVID-19 Genomics UK (COG-UK) consortium | Chris Duncan et al |

| | | | | | | |
|---|---|---|---|---|---|---|
| EPI_ISL_488192 | England/BRIS-12F9E1/2020 | 2020-04-03 | Europe/United Kingdom/England/ | PHE South West Regional Laboratory, National Infection Service | Wellcome Sanger Institute for the COVID-19 Genomics UK (COG-UK) consortium | Stephanie Hutchings et al |
| EPI_ISL_488282 | England/BRIS-12F556/2020 | 2020-02-29 | Europe/United Kingdom/England/ | PHE South West Regional Laboratory, National Infection Service | Wellcome Sanger Institute for the COVID-19 Genomics UK (COG-UK) consortium | Stephanie Hutchings et al |
| EPI_ISL_488293 | England/BRIS-12FA0C/2020 | 2020-04-03 | Europe/United Kingdom/England/ | PHE South West Regional Laboratory, National Infection Service | Wellcome Sanger Institute for the COVID-19 Genomics UK (COG-UK) consortium | Stephanie Hutchings et al |
| EPI_ISL_488354 | England/BRIS-12E654/2020 | 2020-04-21 | Europe/United Kingdom/England/ | PHE South West Regional Laboratory, National Infection Service | Wellcome Sanger Institute for the COVID-19 Genomics UK (COG-UK) consortium | Stephanie Hutchings et al |
| EPI_ISL_488362 | England/BRIS-12F8E4/2020 | 2020-03-30 | Europe/United Kingdom/England/ | PHE South West Regional Laboratory, National Infection Service | Wellcome Sanger Institute for the COVID-19 Genomics UK (COG-UK) consortium | Stephanie Hutchings et al |
| EPI_ISL_488388 | England/BRIS-12F565/2020 | 2020-02-29 | Europe/United Kingdom/England/ | PHE South West Regional Laboratory, National Infection Service | Wellcome Sanger Institute for the COVID-19 Genomics UK (COG-UK) consortium | Stephanie Hutchings et al |
| EPI_ISL_488438 | England/BRIS-12EEC5/2020 | 2020-04-22 | Europe/United Kingdom/England/ | PHE South West Regional Laboratory, National Infection Service | Wellcome Sanger Institute for the COVID-19 Genomics UK (COG-UK) consortium | Stephanie Hutchings et al |
| EPI_ISL_488456 | England/NORT-283D0C/2020 | 2020-04-04 | Europe/United Kingdom/England/ | NU-OMICS DNA Sequencing research facility, Northumbria University | Wellcome Sanger Institute for the COVID-19 Genomics UK (COG-UK) consortium | Chris Duncan et al |

| | | | | | | |
|---|---|---|---|---|---|---|
| EPI_ISL_488464 | England/NORT-294CFC/2020 | 2020-03-27 | Europe/United Kingdom/England/ | NU-OMICS DNA Sequencing research facility, Northumbria University | Wellcome Sanger Institute for the COVID-19 Genomics UK (COG-UK) consortium | Chris Duncan et al |
| EPI_ISL_488533 | England/NORT-295648/2020 | 2020-03-31 | Europe/United Kingdom/England/ | NU-OMICS DNA Sequencing research facility, Northumbria University | Wellcome Sanger Institute for the COVID-19 Genomics UK (COG-UK) consortium | Chris Duncan et al |
| EPI_ISL_488573 | England/NORT-2826D5/2020 | 2020-03-27 | Europe/United Kingdom/England/ | NU-OMICS DNA Sequencing research facility, Northumbria University | Wellcome Sanger Institute for the COVID-19 Genomics UK (COG-UK) consortium | Chris Duncan et al |
| EPI_ISL_488593 | England/NORT-294F4E/2020 | 2020-03-29 | Europe/United Kingdom/England/ | NU-OMICS DNA Sequencing research facility, Northumbria University | Wellcome Sanger Institute for the COVID-19 Genomics UK (COG-UK) consortium | Chris Duncan et al |
| EPI_ISL_488661 | England/NORT-283DC0/2020 | 2020-04-04 | Europe/United Kingdom/England/ | NU-OMICS DNA Sequencing research facility, Northumbria University | Wellcome Sanger Institute for the COVID-19 Genomics UK (COG-UK) consortium | Chris Duncan et al |
| EPI_ISL_489002 | Scotland/EDB2745/2020 | 2020-03-27 | Europe/United Kingdom/Scotland/ | Virology Department, Royal Infirmary of Edinburgh, NHS Lothian / School of Biological Sciences, University of Edinburgh | Wellcome Sanger Institute for the COVID-19 Genomics UK (COG-UK) consortium | McHugh M et al |
| EPI_ISL_489024 | Scotland/EDB3237/2020 | 2020-04-07 | Europe/United Kingdom/Scotland/ | Virology Department, Royal Infirmary of Edinburgh, NHS Lothian / School of Biological Sciences, University of Edinburgh | Wellcome Sanger Institute for the COVID-19 Genomics UK (COG-UK) consortium | McHugh M et al |
| EPI_ISL_489048 | Scotland/EDB3175/2020 | 2020-04-06 | Europe/United Kingdom/Scotland/ | Virology Department, Royal Infirmary of Edinburgh, NHS Lothian / School of Biological Sciences, University of Edinburgh | Wellcome Sanger Institute for the COVID-19 Genomics UK (COG-UK) consortium | McHugh M et al |

| | | | | | | |
|---|---|---|---|---|---|---|
| EPI_ISL_489081 | England/NORT-291E80/2020 | 2020-05-06 | Europe/United Kingdom/England/ | NU-OMICS DNA Sequencing research facility, Northumbria University | Wellcome Sanger Institute for the COVID-19 Genomics UK (COG-UK) consortium | Chris Duncan et al |
| EPI_ISL_489083 | England/NORT-29261E/2020 | 2020-05-12 | Europe/United Kingdom/England/ | NU-OMICS DNA Sequencing research facility, Northumbria University | Wellcome Sanger Institute for the COVID-19 Genomics UK (COG-UK) consortium | Chris Duncan et al |
| EPI_ISL_489085 | England/NORT-2928AF/2020 | 2020-05-30 | Europe/United Kingdom/England/ | NU-OMICS DNA Sequencing research facility, Northumbria University | Wellcome Sanger Institute for the COVID-19 Genomics UK (COG-UK) consortium | Chris Duncan et al |
| EPI_ISL_489139 | England/NORT-2921C2/2020 | 2020-05-07 | Europe/United Kingdom/England/ | NU-OMICS DNA Sequencing research facility, Northumbria University | Wellcome Sanger Institute for the COVID-19 Genomics UK (COG-UK) consortium | Chris Duncan et al |
| EPI_ISL_489173 | NorthernIreland/NIRE-102E30/2020 | 2020-04-20 | Europe/United Kingdom/Northern Irelar | Regional Virus Laboratory, Belfast Health and Social Care Trust | Wellcome Sanger Institute for the COVID-19 Genomics UK (COG-UK) consortium | Conall McCaughey et al |
| EPI_ISL_489303 | NorthernIreland/NIRE-FD613/2020 | 2020-05-01 | Europe/United Kingdom/Northern Irelar | Regional Virus Laboratory, Belfast Health and Social Care Trust | Wellcome Sanger Institute for the COVID-19 Genomics UK (COG-UK) consortium | Conall McCaughey et al |
| EPI_ISL_489364 | NorthernIreland/NIRE-10048B/2020 | 2020-05-03 | Europe/United Kingdom/Northern Irelar | Regional Virus Laboratory, Belfast Health and Social Care Trust | Wellcome Sanger Institute for the COVID-19 Genomics UK (COG-UK) consortium | Conall McCaughey et al |
| EPI_ISL_489368 | NorthernIreland/NIRE-FCCD6/2020 | 2020-04-26 | Europe/United Kingdom/Northern Irelar | Regional Virus Laboratory, Belfast Health and Social Care Trust | Wellcome Sanger Institute for the COVID-19 Genomics UK (COG-UK) consortium | Conall McCaughey et al |

| | | | | | | |
|---|---|---|---|---|---|---|
| EPI_ISL_489376 | NorthernIreland/NIRE-FD033/2020 | 2020-04-24 | Europe/United Kingdom/Northern Irelar | Regional Virus Laboratory, Belfast Health and Social Care Trust | Wellcome Sanger Institute for the COVID-19 Genomics UK (COG-UK) consortium | Conall McCaughey et al |
| EPI_ISL_489388 | England/CAMB-1AC7A6/2020 | 2020-04-23 | Europe/United Kingdom/England/ | Department of Pathology, University of Cambridge | Wellcome Sanger Institute for the COVID-19 Genomics UK (COG-UK) consortium | Luke W Meredith et al |
| EPI_ISL_489405 | England/CAMB-1ACB31/2020 | 2020-04-22 | Europe/United Kingdom/England/ | Department of Pathology, University of Cambridge | Wellcome Sanger Institute for the COVID-19 Genomics UK (COG-UK) consortium | Luke W Meredith et al |
| EPI_ISL_489446 | England/CAMB-1AD505/2020 | 2020-04-26 | Europe/United Kingdom/England/ | Department of Pathology, University of Cambridge | Wellcome Sanger Institute for the COVID-19 Genomics UK (COG-UK) consortium | Luke W Meredith et al |
| EPI_ISL_489509 | England/CAMB-1AC6D6/2020 | 2020-04-23 | Europe/United Kingdom/England/ | Department of Pathology, University of Cambridge | Wellcome Sanger Institute for the COVID-19 Genomics UK (COG-UK) consortium | Luke W Meredith et al |
| EPI_ISL_489549 | England/CAMB-1AC928/2020 | 2020-04-23 | Europe/United Kingdom/England/ | Department of Pathology, University of Cambridge | Wellcome Sanger Institute for the COVID-19 Genomics UK (COG-UK) consortium | Luke W Meredith et al |
| EPI_ISL_489636 | Scotland/GCVR-16FD52/2020 | 2020-03-23 | Europe/United Kingdom/Scotland/ | NHSGGC West of Scotland Specialist Virology Centre / MRC-University of Glasgow Centre for Virus Research | Wellcome Sanger Institute for the COVID-19 Genomics UK (COG-UK) consortium | Ana da Silva Filipe et al |
| EPI_ISL_489712 | USA/FL-BPHL-0303/2020 | 2020-05-29 | North America/USA/Florida/ | Florida Bureau of Public Health Laboratories | Florida Bureau of Public Health Laboratories | Sarah Schmedes et al |
| EPI_ISL_489719 | USA/FL-BPHL-0310/2020 | 2020-06-01 | North America/USA/Florida/ | Florida Bureau of Public Health Laboratories | Florida Bureau of Public Health Laboratories | Sarah Schmedes et al |

| | | | | | | |
|---|---|---|---|---|---|---|
| EPI_ISL_489723 | USA/FL-BPHL-0314/2020 | 2020-06-02 | North America/USA/Florida/ | Florida Bureau of Public Health Laboratories | Florida Bureau of Public Health Laboratories | Sarah Schmedes et al |
| EPI_ISL_489735 | USA/FL-BPHL-0326/2020 | 2020-04-13 | North America/USA/Florida/ | Florida Bureau of Public Health Laboratories | Florida Bureau of Public Health Laboratories | Sarah Schmedes et al |
| EPI_ISL_489746 | USA/FL-BPHL-0337/2020 | 2020-04-19 | North America/USA/Florida/ | Florida Bureau of Public Health Laboratories | Florida Bureau of Public Health Laboratories | Sarah Schmedes et al |
| EPI_ISL_489752 | USA/FL-BPHL-0343/2020 | 2020-04-11 | North America/USA/Florida/ | Florida Bureau of Public Health Laboratories | Florida Bureau of Public Health Laboratories | Sarah Schmedes et al |
| EPI_ISL_489772 | USA/FL-BPHL-0363/2020 | 2020-04-13 | North America/USA/Florida/ | Florida Bureau of Public Health Laboratories | Florida Bureau of Public Health Laboratories | Sarah Schmedes et al |
| EPI_ISL_489773 | USA/FL-BPHL-0364/2020 | 2020-04-13 | North America/USA/Florida/ | Florida Bureau of Public Health Laboratories | Florida Bureau of Public Health Laboratories | Sarah Schmedes et al |
| EPI_ISL_489778 | USA/FL-BPHL-0369/2020 | 2020-04-21 | North America/USA/Florida/ | Florida Bureau of Public Health Laboratories | Florida Bureau of Public Health Laboratories | Sarah Schmedes et al |
| EPI_ISL_489784 | USA/FL-BPHL-0375/2020 | 2020-04-21 | North America/USA/Florida/ | Florida Bureau of Public Health Laboratories | Florida Bureau of Public Health Laboratories | Sarah Schmedes et al |
| EPI_ISL_489790 | USA/FL-BPHL-0381/2020 | 2020-04-22 | North America/USA/Florida/ | Florida Bureau of Public Health Laboratories | Florida Bureau of Public Health Laboratories | Sarah Schmedes et al |
| EPI_ISL_489792 | USA/FL-BPHL-0383/2020 | 2020-04-23 | North America/USA/Florida/ | Florida Bureau of Public Health Laboratories | Florida Bureau of Public Health Laboratories | Sarah Schmedes et al |
| EPI_ISL_489799 | USA/FL-BPHL-0390/2020 | 2020-04-29 | North America/USA/Florida/ | Florida Bureau of Public Health Laboratories | Florida Bureau of Public Health Laboratories | Sarah Schmedes et al |
| EPI_ISL_489819 | USA/FL-BPHL-0410/2020 | 2020-05-14 | North America/USA/Florida/ | Florida Bureau of Public Health Laboratories | Florida Bureau of Public Health Laboratories | Sarah Schmedes et al |

| | | | | | | |
|---|---|---|---|---|---|---|
| EPI_ISL_489839 | Peru/HUC-INS-008/2020 | 2020-03-09 | South America/Peru/Huanuco/ | Laboratorio de Referencia Nacional de Virus Respiratorio. Instituto Nacional de Salud Per√∫ | Laboratorio de Referencia Nacional de Biotecnolog√≠a y Biolog√≠a Molecular. Instituto Nacional de Salud Per√∫ | Carlos Padilla Rojas et al |
| EPI_ISL_489898 | USA/IA-GMF-08809/2020 | 2020-07-01 | North America/USA/Iowa/Fayette Coun | Gundersen Molecular Diagnostics Laboratory | Kabara Cancer Research Institute | Craig S. Richmond et al |
| EPI_ISL_489909 | USA/WI-GMF-06776/2020 | 2020-06-17 | North America/USA/Wisconsin/La Crosse | Gundersen Molecular Diagnostics Laboratory | Kabara Cancer Research Institute | Craig S. Richmond et al |
| EPI_ISL_489912 | USA/WI-GMF-06870/2020 | 2020-06-18 | North America/USA/Wisconsin/La Crosse | Gundersen Molecular Diagnostics Laboratory | Kabara Cancer Research Institute | Craig S. Richmond et al |
| EPI_ISL_489920 | USA/WI-GMF-07403/2020 | 2020-06-22 | North America/USA/Wisconsin/La Crosse | Gundersen Molecular Diagnostics Laboratory | Kabara Cancer Research Institute | Craig S. Richmond et al |
| EPI_ISL_489933 | USA/WI-GMF-08666/2020 | 2020-06-30 | North America/USA/Wisconsin/La Crosse | Gundersen Molecular Diagnostics Laboratory | Kabara Cancer Research Institute | Craig S. Richmond et al |
| EPI_ISL_489937 | USA/WI-GMF-08878/2020 | 2020-07-02 | North America/USA/Wisconsin/La Crosse | Gundersen Molecular Diagnostics Laboratory | Kabara Cancer Research Institute | Craig S. Richmond et al |
| EPI_ISL_489940 | USA/WI-GMF-09008/2020 | 2020-07-03 | North America/USA/Wisconsin/Crawford | Gundersen Molecular Diagnostics Laboratory | Kabara Cancer Research Institute | Craig S. Richmond et al |
| EPI_ISL_489942 | USA/WI-GMF-09171/2020 | 2020-07-04 | North America/USA/Wisconsin/Trempea | Gundersen Molecular Diagnostics Laboratory | Kabara Cancer Research Institute | Craig S. Richmond et al |
| EPI_ISL_489947 | USA/WI-GMF-09497/2020 | 2020-07-05 | North America/USA/Wisconsin/Trempea | Gundersen Molecular Diagnostics Laboratory | Kabara Cancer Research Institute | Craig S. Richmond et al |
| EPI_ISL_489993 | Malaysia/IMR-WC9174/2020 | 2020-03-04 | Asia/Malaysia/Selangor/ | Institute for Medical Research, Infectious Disease Research Centre, National Institutes of Health, Ministry of Health Malaysia | Institute for Medical Research, Infectious Disease Research Centre, National Institutes of Health, Ministry of Health Malaysia | Suppiah J et al |
| EPI_ISL_489996 | SaudiArabia/477/2020 | 2020-02-03 | Asia/Saudi Arabia/Saudi Arabia/ | King Fahad Medical City | King Fahad Medical City | Alosaimi et al |
| EPI_ISL_489997 | SaudiArabia/492/2020 | 2020-02-04 | Asia/Saudi Arabia/Saudi Arabia/ | King Fahad Medical City | King Fahad Medical City | Alosaimi et al |

| | | | | | | |
|---|---|---|---|---|---|---|
| EPI_ISL_489998 | SaudiArabia/509/2020 | 2020-02-09 | Asia/Saudi Arabia/Saudi Arabia/ | King Fahad Medical City | King Fahad Medical City | Alosaimi et al |
| EPI_ISL_489999 | SaudiArabia/518/2020 | 2020-02-10 | Asia/Saudi Arabia/Saudi Arabia/ | King Fahad Medical City | King Fahad Medical City | Alosaimi et al |
| EPI_ISL_490000 | SaudiArabia/523/2020 | 2020-02-10 | Asia/Saudi Arabia/Saudi Arabia/ | King Fahad Medical City | King Fahad Medical City | Alosaimi et al |
| EPI_ISL_490001 | SaudiArabia/545/2020 | 2020-02-13 | Asia/Saudi Arabia/Saudi Arabia/ | King Fahad Medical City | King Fahad Medical City | Alosaimi et al |
| EPI_ISL_490002 | SaudiArabia/553/2020 | 2020-02-13 | Asia/Saudi Arabia/Saudi Arabia/ | King Fahad Medical City | King Fahad Medical City | Alosaimi et al |
| EPI_ISL_490003 | SaudiArabia/562/2020 | 2020-02-16 | Asia/Saudi Arabia/Saudi Arabia/ | King Fahad Medical City | King Fahad Medical City | Alosaimi et al |
| EPI_ISL_490004 | SaudiArabia/578/2020 | 2020-02-17 | Asia/Saudi Arabia/Saudi Arabia/ | King Fahad Medical City | King Fahad Medical City | Alosaimi et al |
| EPI_ISL_490005 | SaudiArabia/583/2020 | 2020-02-18 | Asia/Saudi Arabia/Saudi Arabia/ | King Fahad Medical City | King Fahad Medical City | Alosaimi et al |
| EPI_ISL_490006 | SaudiArabia/596/2020 | 2020-02-18 | Asia/Saudi Arabia/Saudi Arabia/ | King Fahad Medical City | King Fahad Medical City | Alosaimi et al |
| EPI_ISL_490007 | SaudiArabia/610/2020 | 2020-02-20 | Asia/Saudi Arabia/Saudi Arabia/ | King Fahad Medical City | King Fahad Medical City | Alosaimi et al |
| EPI_ISL_490008 | SaudiArabia/627/2020 | 2020-02-23 | Asia/Saudi Arabia/Saudi Arabia/ | King Fahad Medical City | King Fahad Medical City | Alosaimi et al |
| EPI_ISL_490009 | SaudiArabia/640/2020 | 2020-02-24 | Asia/Saudi Arabia/Saudi Arabia/ | King Fahad Medical City | King Fahad Medical City | Alosaimi et al |
| EPI_ISL_490010 | SaudiArabia/656/2020 | 2020-02-26 | Asia/Saudi Arabia/Saudi Arabia/ | King Fahad Medical City | King Fahad Medical City | Alosaimi et al |
| EPI_ISL_490011 | SaudiArabia/671/2020 | 2020-02-27 | Asia/Saudi Arabia/Saudi Arabia/ | King Fahad Medical City | King Fahad Medical City | Alosaimi et al |
| EPI_ISL_490052 | Singapore/602/2020 | 2020-05-14 | Asia/Singapore/Singapore/ | National Public Health Laboratory, National Centre for Infectious Diseases | National Public Health Laboratory, National Centre for Infectious Diseases | Mak TM et al |
| EPI_ISL_490056 | Singapore/606/2020 | 2020-07-02 | Asia/Singapore/Singapore/ | National Public Health Laboratory, National Centre for Infectious Diseases | National Public Health Laboratory, National Centre for Infectious Diseases | Mak TM et al |

| | | | | | | |
|---|---|---|---|---|---|---|
| EPI_ISL_490059 | Singapore/609/2020 | 2020-05-08 | Asia/Singapore/Singapore/ | National Public Health Laboratory, National Centre for Infectious Diseases | National Public Health Laboratory, National Centre for Infectious Diseases | Mak TM et al |
| EPI_ISL_490064 | Singapore/614/2020 | 2020-05-09 | Asia/Singapore/Singapore/ | National Public Health Laboratory, National Centre for Infectious Diseases | National Public Health Laboratory, National Centre for Infectious Diseases | Mak TM et al |
| EPI_ISL_490070 | Singapore/620/2020 | 2020-05-08 | Asia/Singapore/Singapore/ | National Public Health Laboratory, National Centre for Infectious Diseases | National Public Health Laboratory, National Centre for Infectious Diseases | Mak TM et al |
| EPI_ISL_490076 | Singapore/626/2020 | 2020-05-10 | Asia/Singapore/Singapore/ | National Public Health Laboratory, National Centre for Infectious Diseases | National Public Health Laboratory, National Centre for Infectious Diseases | Mak TM et al |
| EPI_ISL_490094 | Malaysia/IMR_WC80109/2020 | 2020-05-23 | Asia/Malaysia/Malaysia/ | Institute for Medical Research, Infectious Disease Research Centre, National Institutes of Health, Ministry of Health Malaysia | Institute for Medical Research, Infectious Disease Research Centre, National Institutes of Health, Ministry of Health Malaysia | Suppiah J et al |
| EPI_ISL_490109 | Bangladesh/BCSIR-NILMRC-246/2020 | 2020-07-07 | Asia/Bangladesh/Khulna/ | National Institute of Laboratory Medicine and Referral Center | Genomic Research Lab, BCSIR | Md. Murshed Hasan Sarkar et al |
| EPI_ISL_490144 | Bangladesh/BCSIR-NILMRC-251/2020 | 2020-07-07 | Asia/Bangladesh/Khulna/ | National Institute of Laboratory Medicine and Referral Center | Genomic Research Lab, BCSIR | Barna Goswami et al |
| EPI_ISL_490165 | Bangladesh/BCSIR-NILMRC-256/2020 | 2020-07-07 | Asia/Bangladesh/Khulna/Jashore | National Institute of Laboratory Medicine and Referral Center | Genomic Research Lab, BCSIR | Md. Saddam Hossain et al |
| EPI_ISL_490168 | Bangladesh/BCSIR-NILMRC-257/2020 | 2020-07-07 | Asia/Bangladesh/Khulna/Jashore | National Institute of Laboratory Medicine and Referral Center | Genomic Research Lab, BCSIR | Abu Sayeed Mohammad Mahmud et al |

| | | | | | | |
|---|---|---|---|---|---|---|
| EPI_ISL_490203 | Spain/PV-HUB-50378855/2020 | 2020-06-10 | Europe/Spain/Basque Country/Bizkaia | Clinical Microbiology Laboratory- Basurto University Hospital | Biocruces-Bizkaia | Mikel J. Urrutikoetxea-Gutierrez et al |
| EPI_ISL_490205 | Germany/BY-MGZ-01/2020 | 2020-04-19 | Europe/Germany/Bavaria/Munich | M√ºnchen Klinik Schwabing | MGZ Medical Genetics Center | Dieter A. Wolf et al |
| EPI_ISL_490206 | Germany/BY-MGZ-02/2020 | 2020-04-21 | Europe/Germany/Bavaria/Munich | M√ºnchen Klinik Schwabing | MGZ Medical Genetics Center | Dieter A. Wolf et al |
| EPI_ISL_490210 | USA/CA-QRTP-Donor1/2020 | 2020-04-01 | North America/USA/California/ | Quest Diagnostics | Q Squared Solutions - QRTP facility | Victor J Weigman et al |
| EPI_ISL_490217 | USA/CA-QRTP-Donor13/2020 | 2020-04-01 | North America/USA/California/ | Quest Diagnostics | Q Squared Solutions - QRTP facility | Victor J Weigman et al |
| EPI_ISL_490219 | USA/CA-QRTP-Donor15/2020 | 2020-04-01 | North America/USA/California/ | Quest Diagnostics | Q Squared Solutions - QRTP facility | Victor J Weigman et al |
| EPI_ISL_490246 | England/202822190/2020 | 2020-06-16 | Europe/United Kingdom/England/ | Respiratory Virus Unit, Microbiology Services Colindale, Public Health England | Respiratory Virus Unit, Microbiology Services Colindale, Public Health England | PHE Covid Sequencing Team et al |
| EPI_ISL_490258 | SouthAfrica/R10007/2020 | 2020-05-08 | Africa/South Africa/Western Cape Provi | National Institute for Communicable Diseases of the National Health Laboratory Service | National Institute for Communicable Diseases of the National Health Laboratory Service | Allam M et al |
| EPI_ISL_490259 | SouthAfrica/R10139/2020 | 2020-05-21 | Africa/South Africa/Western Cape Provi | National Institute for Communicable Diseases of the National Health Laboratory Service | National Institute for Communicable Diseases of the National Health Laboratory Service | Allam M et al |
| EPI_ISL_490260 | SouthAfrica/R10166/2020 | 2020-05-20 | Africa/South Africa/Western Cape Provi | National Institute for Communicable Diseases of the National Health Laboratory Service | National Institute for Communicable Diseases of the National Health Laboratory Service | Allam M et al |

| | | | | | | |
|---|---|---|---|---|---|---|
| EPI_ISL_490263 | SouthAfrica/R10199/2020 | 2020-05-22 | Africa/South Africa/Western Cape Provi | National Institute for Communicable Diseases of the National Health Laboratory Service | National Institute for Communicable Diseases of the National Health Laboratory Service | Allam M et al |
| EPI_ISL_490264 | SouthAfrica/R10236/2020 | 2020-05-27 | Africa/South Africa/Western Cape Provi | National Institute for Communicable Diseases of the National Health Laboratory Service | National Institute for Communicable Diseases of the National Health Laboratory Service | Allam M et al |
| EPI_ISL_490271 | SouthAfrica/R10364/2020 | 2020-06-01 | Africa/South Africa/Gauteng/ | National Institute for Communicable Diseases of the National Health Laboratory Service | National Institute for Communicable Diseases of the National Health Laboratory Service | Allam M et al |
| EPI_ISL_490283 | SouthAfrica/R10518/2020 | 2020-06-09 | Africa/South Africa/Gauteng/ | National Institute for Communicable Diseases of the National Health Laboratory Service | National Institute for Communicable Diseases of the National Health Laboratory Service | Allam M et al |
| EPI_ISL_490287 | SouthAfrica/R10571/2020 | 2020-06-10 | Africa/South Africa/Gauteng/ | National Institute for Communicable Diseases of the National Health Laboratory Service | National Institute for Communicable Diseases of the National Health Laboratory Service | Allam M et al |
| EPI_ISL_490289 | SouthAfrica/R10608/2020 | 2020-06-08 | Africa/South Africa/Western Cape Provi | National Institute for Communicable Diseases of the National Health Laboratory Service | National Institute for Communicable Diseases of the National Health Laboratory Service | Allam M et al |

| | | | | | | |
|---|---|---|---|---|---|---|
| EPI_ISL_490295 | SouthAfrica/R10712/2020 | 2020-06-15 | Africa/South Africa/Gauteng/ | National Institute for Communicable Diseases of the National Health Laboratory Service | National Institute for Communicable Diseases of the National Health Laboratory Service | Allam M et al |
| EPI_ISL_490296 | SouthAfrica/R10713/2020 | 2020-06-15 | Africa/South Africa/Gauteng/ | National Institute for Communicable Diseases of the National Health Laboratory Service | National Institute for Communicable Diseases of the National Health Laboratory Service | Allam M et al |
| EPI_ISL_490301 | SouthAfrica/R10741/2020 | 2020-06-15 | Africa/South Africa/Free State/ | National Institute for Communicable Diseases of the National Health Laboratory Service | National Institute for Communicable Diseases of the National Health Laboratory Service | Allam M et al |
| EPI_ISL_490302 | SouthAfrica/R10752/2020 | 2020-06-12 | Africa/South Africa/Western Cape Provi | National Institute for Communicable Diseases of the National Health Laboratory Service | National Institute for Communicable Diseases of the National Health Laboratory Service | Allam M et al |
| EPI_ISL_490309 | SouthAfrica/R10836/2020 | 2020-06-18 | Africa/South Africa/Gauteng/ | National Institute for Communicable Diseases of the National Health Laboratory Service | National Institute for Communicable Diseases of the National Health Laboratory Service | Allam M et al |
| EPI_ISL_490313 | SouthAfrica/R10883/2020 | 2020-06-19 | Africa/South Africa/Gauteng/ | National Institute for Communicable Diseases of the National Health Laboratory Service | National Institute for Communicable Diseases of the National Health Laboratory Service | Allam M et al |
| EPI_ISL_490369 | England/LIVE-A1F13/2020 | 2020-05-18 | Europe/United Kingdom/England/ | Liverpool Clinical Laboratories | COVID-19 Genomics UK (COG-UK) Consortium | Sam Haldenby et al |

| | | | | | | |
|---|---|---|---|---|---|---|
| EPI_ISL_490399 | England/LIVE-A3F7B/2020 | 2020-06-29 | Europe/United Kingdom/England/ | Liverpool Clinical Laboratories | COVID-19 Genomics UK (COG-UK) Consortium | Sam Haldenby et al |
| EPI_ISL_490472 | England/NORT-29EAAD/2020 | 2020-05-17 | Europe/United Kingdom/England/ | Northumbria University / South Tees Hospitals NHS Foundation Trust / North Cumbria Integrated Care NHS Foundation Trust / North Tees and Hartlepool NHS Foundation Trust / Newcastle Hospitals NHS Foundation Trust | COVID-19 Genomics UK (COG-UK) Consortium | Darren L Smith et al |
| EPI_ISL_490473 | England/NORT-29EAE9/2020 | 2020-05-12 | Europe/United Kingdom/England/ | Northumbria University / South Tees Hospitals NHS Foundation Trust / North Cumbria Integrated Care NHS Foundation Trust / North Tees and Hartlepool NHS Foundation Trust / Newcastle Hospitals NHS Foundation Trust | COVID-19 Genomics UK (COG-UK) Consortium | Darren L Smith et al |
| EPI_ISL_490535 | England/NORW-EE3C3/2020 | 2020-04-24 | Europe/United Kingdom/England/ | Quadram Institute Bioscience | COVID-19 Genomics UK (COG-UK) Consortium | Dave J. Baker et al |
| EPI_ISL_490693 | Scotland/CVR3140/2020 | 2020-04-28 | Europe/United Kingdom/Scotland/ | West of Scotland Specialist Virology Centre, NHSGGC / MRC-University of Glasgow Centre for Virus Research | COVID-19 Genomics UK (COG-UK) Consortium | Ana da Silva Filipe et al |
| EPI_ISL_490738 | Wales/PHWC-1658E2/2020 | 2020-05-15 | Europe/United Kingdom/Wales/ | Wales Specialist Virology Centre Sequencing lab: Pathogen Genomics Unit | COVID-19 Genomics UK (COG-UK) Consortium | Catherine Moore et al |
| EPI_ISL_490993 | USA/ID-UW-3204/2020 | 2020-03-26 | North America/USA/Idaho/ | UW Virology Lab | UW Virology Lab | Pavitra Roychoudhury et al |
| EPI_ISL_491017 | USA/WA-UW-3340/2020 | 2020-03-26 | North America/USA/Washington/ | UW Virology Lab | UW Virology Lab | Pavitra Roychoudhury et al |
| EPI_ISL_491022 | USA/WA-UW-3413/2020 | 2020-03-26 | North America/USA/Washington/ | UW Virology Lab | UW Virology Lab | Pavitra Roychoudhury et al |
| EPI_ISL_491036 | Romania/ROSV_10626/2020 | 2020-05-26 | Europe/Romania/Suceava/ | Suceava County Emergency Hospital | """Stefan cel Mare"" University Metagenomics Lab" | Lobiuc Andrei et al |
| EPI_ISL_491038 | Romania/ROSV_11967/2020 | 2020-06-04 | Europe/Romania/Suceava/ | Suceava County Emergency Hospital | """Stefan cel Mare"" University Metagenomics Lab" | Lobiuc Andrei et al |
| EPI_ISL_491039 | Romania/ROSV_11990/2020 | 2020-06-05 | Europe/Romania/Suceava/ | Suceava County Emergency Hospital | """Stefan cel Mare"" University Metagenomics Lab" | Lobiuc Andrei et al. et al |

| | | | | | | |
|---|---|---|---|---|---|---|
| EPI_ISL_491040 | Romania/ROSV_12235/2020 | 2020-06-08 | Europe/Romania/Suceava/ | Suceava County Emergency Hospital | """Stefan cel Mare"" University Metagenomics Lab" | Lobiuc Andrei et al. et al |
| EPI_ISL_491042 | Romania/ROSV_12421/2020 | 2020-06-08 | Europe/Romania/Suceava/ | Suceava County Emergency Hospital | """Stefan cel Mare"" University Metagenomics Lab" | Lobiuc Andrei et al |
| EPI_ISL_491045 | Romania/ROSV_12615/2020 | 2020-06-09 | Europe/Romania/Suceava/ | Suceava County Emergency Hospital | """Stefan cel Mare"" University Metagenomics Lab" | Lobiuc Andrei et al |
| EPI_ISL_491050 | Romania/ROSV_1296/2020 | 2020-04-09 | Europe/Romania/Suceava/ | Suceava County Emergency Hospital | """Stefan cel Mare"" University Metagenomics Lab" | Lobiuc Andrei et al. et al |
| EPI_ISL_491058 | Romania/ROSV_2492/2020 | 2020-04-16 | Europe/Romania/Suceava/ | Suceava County Emergency Hospital | """Stefan cel Mare"" University Metagenomics Lab" | Lobiuc Andrei et al |
| EPI_ISL_491062 | Romania/ROSV_2874/2020 | 2020-04-17 | Europe/Romania/Suceava/ | Suceava County Emergency Hospital | """Stefan cel Mare"" University Metagenomics Lab" | Lobiuc Andrei et al |
| EPI_ISL_491069 | Romania/ROSV_3376/2020 | 2020-04-20 | Europe/Romania/Suceava/ | Suceava County Emergency Hospital | """Stefan cel Mare"" University Metagenomics Lab" | Lobiuc Andrei et al. et al |
| EPI_ISL_491073 | Romania/ROSV_4271/2020 | 2020-04-23 | Europe/Romania/Suceava/ | Suceava County Emergency Hospital | """Stefan cel Mare"" University Metagenomics Lab" | Lobiuc Andrei et al |
| EPI_ISL_491076 | Romania/ROSV_4659/2020 | 2020-04-25 | Europe/Romania/Suceava/ | Suceava County Emergency Hospital | """Stefan cel Mare"" University Metagenomics Lab" | Lobiuc Andrei et al. et al |
| EPI_ISL_491084 | Romania/ROSV_5259/2020 | 2020-04-29 | Europe/Romania/Suceava/ | Suceava County Emergency Hospital | """Stefan cel Mare"" University Metagenomics Lab" | Lobiuc Andrei et al. et al |

| | | | | | | |
|---|---|---|---|---|---|---|
| EPI_ISL_491088 | Romania/ROSV_7981/2020 | 2020-05-12 | Europe/Romania/Suceava/ | Suceava County Emergency Hospital | """Stefan cel Mare"" University Metagenomics Lab" | Lobiuc Andrei et al |
| EPI_ISL_491089 | Romania/ROSV_8235/2020 | 2020-05-14 | Europe/Romania/Suceava/ | Suceava County Emergency Hospital | """Stefan cel Mare"" University Metagenomics Lab" | Lobiuc Andrei et al. et al |
| EPI_ISL_491091 | Romania/ROSV_9826/2020 | 2020-05-22 | Europe/Romania/Suceava/ | Suceava County Emergency Hospital | """Stefan cel Mare"" University Metagenomics Lab" | Lobiuc Andrei et al. et al |
| EPI_ISL_491108 | USA/SC-DHEC-0093/2020 | 2020-04-24 | North America/USA/South Carolina/Rich | SC Department of Health and Environmental Control | SC Department of Health and Environmental Control | Flores et al |
| EPI_ISL_491133 | Oman/C-9263/2020 | 2020-04-28 | Asia/Oman/Dakhiliyah/Manah | Oman-National Influenza Center | Biotechnology & OMICs Laboratory | Samiha Al-Kharusi et al |
| EPI_ISL_491155 | Oman/11374/2020 | 2020-04-07 | Asia/Oman/South Batinah/Barka | Oman-National Influenza Center | Biotechnology & OMICs Laboratory | Abdul Latif Khan et al |
| EPI_ISL_491161 | Oman/11467/2020 | 2020-04-06 | Asia/Oman/North Sharqiyah/Ibra | Oman-National Influenza Center | Biotechnology & OMICs Laboratory | Sajjad Asaf et al |
| EPI_ISL_491180 | Portugal/IGC2991/2020 | 2020-05-19 | Europe/Portugal/Lisbon/Queluz | Instituto Gulbenkian de Civ™ncia | Instituto Gulbenkian de Civ™ncia | Jov£o Costa et al |
| EPI_ISL_491183 | Portugal/IGC3008/2020 | 2020-05-19 | Europe/Portugal/Lisbon/Queluz | Instituto Gulbenkian de Civ™ncia | Instituto Gulbenkian de Civ™ncia | Jov£o Costa et al |
| EPI_ISL_491191 | Portugal/IGC3492/2020 | 2020-05-25 | Europe/Portugal/Lisbon/Encosta de Sol | Instituto Gulbenkian de Civ™ncia | Instituto Gulbenkian de Civ™ncia | Jov£o Costa et al |
| EPI_ISL_491195 | Portugal/IGC3628/2020 | 2020-05-26 | Europe/Portugal/Lisbon/Amadora | Instituto Gulbenkian de Civ™ncia | Instituto Gulbenkian de Civ™ncia | Jov£o Costa et al |
| EPI_ISL_491197 | Portugal/IGC3632/2020 | 2020-05-26 | Europe/Portugal/Lisbon/Amadora | Instituto Gulbenkian de Civ™ncia | Instituto Gulbenkian de Civ™ncia | Jov£o Costa et al |
| EPI_ISL_491205 | Portugal/IGC3667/2020 | 2020-05-27 | Europe/Portugal/Lisbon/Agualva | Instituto Gulbenkian de Civ™ncia | Instituto Gulbenkian de Civ™ncia | Cathy Paulino et al |

| | | | | | | |
|---|---|---|---|---|---|---|
| EPI_ISL_491206 | Portugal/IGC3668/2020 | 2020-05-27 | Europe/Portugal/Lisbon/Rio de Mouro | Instituto Gulbenkian de Civ™ncia | Instituto Gulbenkian de Civ™ncia | Cathy Paulino et al |
| EPI_ISL_491221 | Portugal/IGC4011/2020 | 2020-05-31 | Europe/Portugal/Lisbon/Encosta de Sol | Instituto Gulbenkian de Civ™ncia | Instituto Gulbenkian de Civ™ncia | Cathy Paulino et al |
| EPI_ISL_491225 | Portugal/IGC4033/2020 | 2020-06-01 | Europe/Portugal/Lisbon/Massama | Instituto Gulbenkian de Civ™ncia | Instituto Gulbenkian de Civ™ncia | Cathy Paulino et al |
| EPI_ISL_491226 | Portugal/IGC4089/2020 | 2020-06-01 | Europe/Portugal/Lisbon/Cacem | Instituto Gulbenkian de Civ™ncia | Instituto Gulbenkian de Civ™ncia | Cathy Paulino et al |
| EPI_ISL_491230 | Portugal/IGC4102/2020 | 2020-06-01 | Europe/Portugal/Lisbon/Massama | Instituto Gulbenkian de Civ™ncia | Instituto Gulbenkian de Civ™ncia | Cathy Paulino et al |
| EPI_ISL_491233 | Portugal/IGC4108/2020 | 2020-06-01 | Europe/Portugal/Lisbon/Sacavem | Instituto Gulbenkian de Civ™ncia | Instituto Gulbenkian de Civ™ncia | Joao Sobral et al |
| EPI_ISL_491235 | Portugal/IGC4124/2020 | 2020-06-02 | Europe/Portugal/Lisbon/Amadora | Instituto Gulbenkian de Civ™ncia | Instituto Gulbenkian de Civ™ncia | Joao Sobral et al |
| EPI_ISL_491236 | Portugal/IGC4126/2020 | 2020-06-01 | Europe/Portugal/Lisbon/Aguas livres | Instituto Gulbenkian de Civ™ncia | Instituto Gulbenkian de Civ™ncia | Joao Sobral et al |
| EPI_ISL_491248 | Portugal/IGC4403/2020 | 2020-06-07 | Europe/Portugal/Lisbon/Massama | Instituto Gulbenkian de Civ™ncia | Instituto Gulbenkian de Civ™ncia | Joao Sobral et al |
| EPI_ISL_491264 | Portugal/IGC4570/2020 | 2020-06-12 | Europe/Portugal/Lisbon/Encosta de Sol | Instituto Gulbenkian de Civ™ncia | Instituto Gulbenkian de Civ™ncia | Joao Sobral et al |
| EPI_ISL_491271 | Portugal/R1237/2020 | 2020-06-12 | Europe/Portugal/Lisbon/Amadora | Instituto Gulbenkian de Civ™ncia | Instituto Gulbenkian de Civ™ncia | Susana Ladeiro et al |
| EPI_ISL_491280 | Portugal/R147/2020 | 2020-05-21 | Europe/Portugal/Lisbon/Massama | Instituto Gulbenkian de Civ™ncia | Instituto Gulbenkian de Civ™ncia | Susana Ladeiro et al |
| EPI_ISL_491281 | Portugal/R150/2020 | 2020-05-21 | Europe/Portugal/Lisbon/Monte Abrv£o | Instituto Gulbenkian de Civ™ncia | Instituto Gulbenkian de Civ™ncia | Susana Ladeiro et al |

| | | | | | | |
|---|---|---|---|---|---|---|
| EPI_ISL_491289 | Portugal/R473/2020 | 2020-05-28 | Europe/Portugal/Lisbon/Agualva | Instituto Gulbenkian de Civ™ncia | Instituto Gulbenkian de Civ™ncia | Susana Ladeiro et al |
| EPI_ISL_491295 | Portugal/R502/2020 | 2020-05-27 | Europe/Portugal/Lisbon/Rio de Mouro | Instituto Gulbenkian de Civ™ncia | Instituto Gulbenkian de Civ™ncia | Susana Ladeiro et al |
| EPI_ISL_491298 | Philippines/PH-RITM-0019/2020 | 2020-06-25 | Asia/Philippines/Philippines/ | Rural Health Unit - Calauan, Laguna | Research Institute for Tropical Medicine | Tujan et al |
| EPI_ISL_491306 | USA/FL-UW-741/2020 | 2020-06-30 | North America/USA/Florida/Palm Beach | University of Wisconsin-Madison AIDS Vaccine Research Laboratories | University of Wisconsin-Madison AIDS Vaccine Research Laboratories | Gage Moreno et al |
| EPI_ISL_491309 | USA/IL-UW-639/2020 | 2020-06-28 | North America/USA/Illinois/Kane County | University of Wisconsin-Madison AIDS Vaccine Research Laboratories | University of Wisconsin-Madison AIDS Vaccine Research Laboratories | Gage Moreno et al |
| EPI_ISL_491322 | USA/WI-UW-626/2020 | 2020-06-25 | North America/USA/Wisconsin/Dane Co | University of Wisconsin-Madison AIDS Vaccine Research Laboratories | University of Wisconsin-Madison AIDS Vaccine Research Laboratories | Gage Moreno et al |
| EPI_ISL_491324 | USA/WI-UW-630/2020 | 2020-06-26 | North America/USA/Wisconsin/Dane Co | University of Wisconsin-Madison AIDS Vaccine Research Laboratories | University of Wisconsin-Madison AIDS Vaccine Research Laboratories | Gage Moreno et al |
| EPI_ISL_491325 | USA/WI-UW-631/2020 | 2020-06-28 | North America/USA/Wisconsin/Dane Co | University of Wisconsin-Madison AIDS Vaccine Research Laboratories | University of Wisconsin-Madison AIDS Vaccine Research Laboratories | Gage Moreno et al |
| EPI_ISL_491335 | USA/WI-UW-644/2020 | 2020-06-26 | North America/USA/Wisconsin/Dane Co | University of Wisconsin-Madison AIDS Vaccine Research Laboratories | University of Wisconsin-Madison AIDS Vaccine Research Laboratories | Gage Moreno et al |

| | | | | | | |
|---|---|---|---|---|---|---|
| EPI_ISL_491341 | USA/WI-UW-650/2020 | 2020-06-26 | North America/USA/Wisconsin/Dane Co | University of Wisconsin-Madison AIDS Vaccine Research Laboratories | University of Wisconsin-Madison AIDS Vaccine Research Laboratories | Gage Moreno et al |
| EPI_ISL_491349 | USA/WI-UW-663/2020 | 2020-06-26 | North America/USA/Wisconsin/Dane Co | University of Wisconsin-Madison AIDS Vaccine Research Laboratories | University of Wisconsin-Madison AIDS Vaccine Research Laboratories | Gage Moreno et al |
| EPI_ISL_491356 | USA/WI-UW-675/2020 | 2020-06-26 | North America/USA/Wisconsin/Dane Co | University of Wisconsin-Madison AIDS Vaccine Research Laboratories | University of Wisconsin-Madison AIDS Vaccine Research Laboratories | Gage Moreno et al |
| EPI_ISL_491362 | USA/WI-UW-681/2020 | 2020-06-28 | North America/USA/Wisconsin/Dane Co | University of Wisconsin-Madison AIDS Vaccine Research Laboratories | University of Wisconsin-Madison AIDS Vaccine Research Laboratories | Gage Moreno et al |
| EPI_ISL_491381 | USA/WI-UW-703/2020 | 2020-06-30 | North America/USA/Wisconsin/Outagan | University of Wisconsin-Madison AIDS Vaccine Research Laboratories | University of Wisconsin-Madison AIDS Vaccine Research Laboratories | Gage Moreno et al |
| EPI_ISL_491382 | USA/WI-UW-704/2020 | 2020-07-02 | North America/USA/Wisconsin/Dane Co | University of Wisconsin-Madison AIDS Vaccine Research Laboratories | University of Wisconsin-Madison AIDS Vaccine Research Laboratories | Gage Moreno et al |
| EPI_ISL_491383 | USA/WI-UW-705/2020 | 2020-06-30 | North America/USA/Wisconsin/Dane Co | University of Wisconsin-Madison AIDS Vaccine Research Laboratories | University of Wisconsin-Madison AIDS Vaccine Research Laboratories | Gage Moreno et al |
| EPI_ISL_491385 | USA/WI-UW-708/2020 | 2020-07-02 | North America/USA/Wisconsin/Dane Co | University of Wisconsin-Madison AIDS Vaccine Research Laboratories | University of Wisconsin-Madison AIDS Vaccine Research Laboratories | Gage Moreno et al |

| | | | | | | |
|---|---|---|---|---|---|---|
| EPI_ISL_491394 | USA/WI-UW-719/2020 | 2020-06-29 | North America/USA/Wisconsin/Dane Co | University of Wisconsin-Madison AIDS Vaccine Research Laboratories | University of Wisconsin-Madison AIDS Vaccine Research Laboratories | Gage Moreno et al |
| EPI_ISL_491396 | USA/WI-UW-721/2020 | 2020-07-01 | North America/USA/Wisconsin/Dane Co | University of Wisconsin-Madison AIDS Vaccine Research Laboratories | University of Wisconsin-Madison AIDS Vaccine Research Laboratories | Gage Moreno et al |
| EPI_ISL_491405 | USA/WI-UW-730/2020 | 2020-06-30 | North America/USA/Wisconsin/Dane Co | University of Wisconsin-Madison AIDS Vaccine Research Laboratories | University of Wisconsin-Madison AIDS Vaccine Research Laboratories | Gage Moreno et al |
| EPI_ISL_491406 | USA/WI-UW-731/2020 | 2020-07-02 | North America/USA/Wisconsin/Dane Co | University of Wisconsin-Madison AIDS Vaccine Research Laboratories | University of Wisconsin-Madison AIDS Vaccine Research Laboratories | Gage Moreno et al |
| EPI_ISL_491410 | USA/WI-UW-735/2020 | 2020-07-02 | North America/USA/Wisconsin/Dane Co | University of Wisconsin-Madison AIDS Vaccine Research Laboratories | University of Wisconsin-Madison AIDS Vaccine Research Laboratories | Gage Moreno et al |
| EPI_ISL_491412 | USA/WI-UW-737/2020 | 2020-06-29 | North America/USA/Wisconsin/Dane Co | University of Wisconsin-Madison AIDS Vaccine Research Laboratories | University of Wisconsin-Madison AIDS Vaccine Research Laboratories | Gage Moreno et al |
| EPI_ISL_491414 | USA/WI-UW-740/2020 | 2020-06-30 | North America/USA/Wisconsin/Dane Co | University of Wisconsin-Madison AIDS Vaccine Research Laboratories | University of Wisconsin-Madison AIDS Vaccine Research Laboratories | Gage Moreno et al |
| EPI_ISL_491420 | USA/WI-UW-747/2020 | 2020-07-02 | North America/USA/Wisconsin/Dane Co | University of Wisconsin-Madison AIDS Vaccine Research Laboratories | University of Wisconsin-Madison AIDS Vaccine Research Laboratories | Gage Moreno et al |

| | | | | | | |
|---|---|---|---|---|---|---|
| EPI_ISL_491433 | Peru/LIM-INS-029/2020 | 2020-04-23 | South America/Peru/Lima/ | Laboratorio de Referencia Nacional de Virus Respiratorio. Instituto Nacional de Salud Perv∫ | Laboratorio de Referencia Nacional de Biotecnologv≠a y Biologv≠a Molecular. Instituto Nacional de Salud Perv∫ | Carlos Padilla Rojas et al |
| EPI_ISL_491434 | Peru/LIM-INS-030/2020 | 2020-04-23 | South America/Peru/Lima/ | Laboratorio de Referencia Nacional de Virus Respiratorio. Instituto Nacional de Salud Perv∫ | Laboratorio de Referencia Nacional de Biotecnologv≠a y Biologv≠a Molecular. Instituto Nacional de Salud Perv∫ | Carlos Padilla Rojas et al |
| EPI_ISL_491435 | Peru/LIM-INS-031/2020 | 2020-04-23 | South America/Peru/Lima/ | Laboratorio de Referencia Nacional de Virus Respiratorio. Instituto Nacional de Salud Perv∫ | Laboratorio de Referencia Nacional de Biotecnologv≠a y Biologv≠a Molecular. Instituto Nacional de Salud Perv∫ | Carlos Padilla Rojas et al |
| EPI_ISL_491458 | Peru/LIM-INS-033/2020 | 2020-04-13 | South America/Peru/Lima/ | Laboratorio de Referencia Nacional de Virus Respiratorio. Instituto Nacional de Salud Perv∫ | Laboratorio de Referencia Nacional de Biotecnologv≠a y Biologv≠a Molecular. Instituto Nacional de Salud Perv∫ | Carlos Padilla Rojas et al |
| EPI_ISL_491461 | Peru/LAM-INS-038/2020 | 2020-04-01 | South America/Peru/Lambayeque/ | Laboratorio de Referencia Nacional de Virus Respiratorio. Instituto Nacional de Salud Perv∫ | Laboratorio de Referencia Nacional de Biotecnologv≠a y Biologv≠a Molecular. Instituto Nacional de Salud Perv∫ | Carlos Padilla Rojas et al |

| | | | | | | |
|---|---|---|---|---|---|---|
| EPI_ISL_491471 | Philippines/PH-RITM-0016/2020 | 2020-01-29 | Asia/Philippines/National Capital Regior | San Lazaro Hospital | Research Institute for Tropical Medicine | Ma. Angelica Tujan et al |
| EPI_ISL_491473 | Philippines/PH-RITM-0018/2020 | 2020-03-06 | Asia/Philippines/National Capital Regior | The Medical City | Research Institute for Tropical Medicine | Ma. Angelica Tujan et al |
| EPI_ISL_491474 | Philippines/PH-RITM-0020/2020 | 2020-06-22 | Asia/Philippines/National Capital Regior | Research Institute for Tropical Medicine | Research Institute for Tropical Medicine | Ma. Angelica Tujan et al |
| EPI_ISL_491574 | Scotland/EDB6724/2020 | 2020-03-31 | Europe/United Kingdom/Scotland/ | Virology Department, Royal Infirmary of Edinburgh, NHS Lothian / School of Biological Sciences, University of Edinburgh | Wellcome Sanger Institute for the COVID-19 Genomics UK (COG-UK) consortium | McHugh M et al |
| EPI_ISL_491674 | Scotland/EDB2481/2020 | 2020-04-23 | Europe/United Kingdom/Scotland/ | Virology Department, Royal Infirmary of Edinburgh, NHS Lothian / School of Biological Sciences, University of Edinburgh | Wellcome Sanger Institute for the COVID-19 Genomics UK (COG-UK) consortium | McHugh M et al |
| EPI_ISL_491909 | USA/MD_NIDDL_3202/2020 | 2020-04-16 | North America/USA/Maryland/ | Naval Infectious Diseases Diagnostic Laboratory | Naval Medical Research Center Biological Defense Research Directorate | Logan Voegtly et al |
| EPI_ISL_491917 | USA/VA_NIDDL_2783/2020 | 2020-03-30 | North America/USA/Virginia/ | Naval Infectious Diseases Diagnostic Laboratory | Naval Medical Research Center Biological Defense Research Directorate | Logan Voegtly et al |
| EPI_ISL_491921 | USA/VA_NIDDL_2896/2020 | 2020-04-02 | North America/USA/Virginia/ | Naval Infectious Diseases Diagnostic Laboratory | Naval Medical Research Center Biological Defense Research Directorate | Logan Voegtly et al |
| EPI_ISL_491931 | USA/VA_NIDDL_3129/2020 | 2020-04-11 | North America/USA/Virginia/ | Naval Infectious Diseases Diagnostic Laboratory | Naval Medical Research Center Biological Defense Research Directorate | Logan Voegtly et al |

| | | | | | | |
|---|---|---|---|---|---|---|
| EPI_ISL_491932 | Ecuador/USFQ-120/2020 | 2020-07-03 | South America/Ecuador/Carchi/Awa Res | Institute of Microbiology, Universidad San Francisco de Quito | Institute of Microbiology, Universidad San Francisco de Quito | Bel√©n Prado-Vivar et al |
| EPI_ISL_491933 | Ecuador/USFQ-128/2020 | 2020-04-07 | South America/Ecuador/Guayas/Guayaq | Centro de Investigaciones, Universidad de Especialidades Esp√≠ritu Santo | Institute of Microbiology, Universidad San Francisco de Quito | Derly Andrade et al |
| EPI_ISL_491934 | Ecuador/USFQ-129/2020 | 2020-04-07 | South America/Ecuador/Guayas/Guayaq | Centro de Investigaciones, Universidad de Especialidades Esp√≠ritu Santo | Institute of Microbiology, Universidad San Francisco de Quito | Derly Andrade et al |
| EPI_ISL_491935 | Ecuador/USFQ-131/2020 | 2020-04-13 | South America/Ecuador/Guayas/Genera | Centro de Investigaciones, Universidad de Especialidades Esp√≠ritu Santo | Institute of Microbiology, Universidad San Francisco de Quito | Derly Andrade et al |
| EPI_ISL_491936 | Ecuador/USFQ-139/2020 | 2020-06-30 | South America/Ecuador/Manabi/Portov | Institute of Microbiology, Universidad San Francisco de Quito | Institute of Microbiology, Universidad San Francisco de Quito | Bel√©n Prado-Vivar et al |
| EPI_ISL_491937 | Ecuador/USFQ-147/2020 | 2020-06-30 | South America/Ecuador/Manabi/Calceta | Institute of Microbiology, Universidad San Francisco de Quito | Institute of Microbiology, Universidad San Francisco de Quito | Bel√©n Prado-Vivar et al |
| EPI_ISL_491938 | Ecuador/USFQ-150/2020 | 2020-06-30 | South America/Ecuador/Manabi/Manta | Institute of Microbiology, Universidad San Francisco de Quito | Institute of Microbiology, Universidad San Francisco de Quito | Bel√©n Prado-Vivar et al |
| EPI_ISL_491939 | Ecuador/USFQ-126/2020 | 2020-07-03 | South America/Ecuador/Orellana/ | Institute of Microbiology, Universidad San Francisco de Quito | Institute of Microbiology, Universidad San Francisco de Quito | Bel√©n Prado-Vivar et al |
| EPI_ISL_491941 | Ecuador/USFQ-133/2020 | 2020-05-02 | South America/Ecuador/Galapagos/Puer | Centro de Investigaciones, Universidad de Especialidades Esp√≠ritu Santo | Institute of Microbiology, Universidad San Francisco de Quito | Derly Andrade et al |
| EPI_ISL_491944 | Ecuador/23890/2020 | 2020-04-25 | South America/Ecuador/Daule/ | Instituto Nacional de Investigaci√≥n en Salud P√∫blica - INSPI | INSPI - Charit√© | Alfredo Bruno Caicedo et al |
| EPI_ISL_491945 | Ecuador/23055/2020 | 2020-04-22 | South America/Ecuador/Manabi/Puerto | Instituto Nacional de Investigaci√≥n en Salud P√∫blica - INSPI | INSPI - Charit√© | Alfredo Bruno Caicedo et al |

| | | | | | | |
|---|---|---|---|---|---|---|
| EPI_ISL_491946 | Ecuador/52872/2020 | 2020-05-28 | South America/Ecuador/Los Rios/Ventar | Instituto Nacional de Investigaci√≥n en Salud P√∫blica - INSPI | INSPI - Charit√© | Alfredo Bruno Caicedo et al |
| EPI_ISL_491947 | Ecuador/36192/2020 | 2020-05-08 | South America/Ecuador/Manabi/Jipijapa | Instituto Nacional de Investigaci√≥n en Salud P√∫blica - INSPI | INSPI - Charit√© | Alfredo Bruno Caicedo et al |
| EPI_ISL_491948 | Ecuador/30755/2020 | 2020-04-29 | South America/Ecuador/Manabi/Pajan | Instituto Nacional de Investigaci√≥n en Salud P√∫blica - INSPI | INSPI - Charit√© | Alfredo Bruno Caicedo et al |
| EPI_ISL_491949 | Ecuador/51908/2020 | 2020-05-26 | South America/Ecuador/Manabi/Sucre | Instituto Nacional de Investigaci√≥n en Salud P√∫blica - INSPI | INSPI - Charit√© | Alfredo Bruno Caicedo et al |
| EPI_ISL_491950 | Ecuador/36230/2020 | 2020-05-08 | South America/Ecuador/Los Rios/Babaho | Instituto Nacional de Investigaci√≥n en Salud P√∫blica - INSPI | INSPI - Charit√© | Alfredo Bruno Caicedo et al |
| EPI_ISL_491951 | Ecuador/52438/2020 | 2020-06-01 | South America/Ecuador/Guayas/Guayaq | Instituto Nacional de Investigaci√≥n en Salud P√∫blica - INSPI | INSPI - Charit√© | Alfredo Bruno Caicedo et al |
| EPI_ISL_491952 | Ecuador/52255/2020 | 2020-05-29 | South America/Ecuador/Guayas/San Jac | Instituto Nacional de Investigaci√≥n en Salud P√∫blica - INSPI | INSPI - Charit√© | Alfredo Bruno Caicedo et al |
| EPI_ISL_491953 | Ecuador/36226/2020 | 2020-05-08 | South America/Ecuador/Los Rios/Babaho | Instituto Nacional de Investigaci√≥n en Salud P√∫blica - INSPI | INSPI - Charit√© | Alfredo Bruno Caicedo et al |
| EPI_ISL_491954 | Ecuador/52389/2020 | 2020-05-28 | South America/Ecuador/Santa Elena/ | Instituto Nacional de Investigaci√≥n en Salud P√∫blica - INSPI | INSPI - Charit√© | Alfredo Bruno Caicedo et al |
| EPI_ISL_491968 | Oman/205035037/2020 | 2020-05-10 | Asia/Oman/Muscat/ | Oman-NIC | Department of Microbiology and Immunology-SQUH | Fahad Zadjali et al |
| EPI_ISL_491976 | Oman/205036179/2020 | 2020-05-16 | Asia/Oman/Muscat/ | Oman-NIC | Department of Microbiology and Immunology-SQUH | Fahad Zadjali et al |
| EPI_ISL_491980 | Oman/205036471/2020 | 2020-05-17 | Asia/Oman/Muscat/ | Oman-NIC | Department of Microbiology and Immunology-SQUH | Fahad Zadjali et al |
| EPI_ISL_491984 | Oman/205036714/2020 | 2020-05-17 | Asia/Oman/Muscat/ | Oman-NIC | Department of Microbiology and Immunology-SQUH | Fahad Zadjali et al |
| EPI_ISL_491985 | Oman/205037001/2020 | 2020-05-19 | Asia/Oman/Muscat/ | Oman-NIC | Department of Microbiology and Immunology-SQUH | Fahad Zadjali et al |

| | | | | | | |
|---|---|---|---|---|---|---|
| EPI_ISL_491991 | Oman/205038035/2020 | 2020-05-21 | Asia/Oman/Muscat/ | Oman-NIC | Department of Microbiology and Immunology-SQUH | Fahad Zadjali et al |
| EPI_ISL_491992 | Oman/205038041/2020 | 2020-05-21 | Asia/Oman/Muscat/ | Oman-NIC | Department of Microbiology and Immunology-SQUH | Fahad Zadjali et al |
| EPI_ISL_491995 | Oman/205038321/2020 | 2020-05-22 | Asia/Oman/Muscat/ | Oman-NIC | Department of Microbiology and Immunology-SQUH | Fahad Zadjali et al |
| EPI_ISL_491996 | Oman/205038337/2020 | 2020-05-22 | Asia/Oman/Muscat/ | Oman-NIC | Department of Microbiology and Immunology-SQUH | Fahad Zadjali et al |
| EPI_ISL_491999 | Oman/205038885/2020 | 2020-05-26 | Asia/Oman/Muscat/ | Oman-NIC | Department of Microbiology and Immunology-SQUH | Fahad Zadjali et al |
| EPI_ISL_492003 | Oman/205039149/2020 | 2020-05-27 | Asia/Oman/Muscat/ | Oman-NIC | Department of Microbiology and Immunology-SQUH | Fahad Zadjali et al |
| EPI_ISL_492004 | Oman/205039432/2020 | 2020-05-28 | Asia/Oman/Muscat/ | Oman-NIC | Department of Microbiology and Immunology-SQUH | Fahad Zadjali et al |
| EPI_ISL_492009 | Oman/205039804/2020 | 2020-05-30 | Asia/Oman/Muscat/ | Oman-NIC | Department of Microbiology and Immunology-SQUH | Fahad Zadjali et al |
| EPI_ISL_492012 | Oman/205040194/2020 | 2020-05-31 | Asia/Oman/Muscat/ | Oman-NIC | Department of Microbiology and Immunology-SQUH | Fahad Zadjali et al |
| EPI_ISL_492014 | Oman/205041214/2020 | 2020-06-03 | Asia/Oman/Muscat/ | Oman-NIC | Department of Microbiology and Immunology-SQUH | Fahad Zadjali et al |

| | | | | | | |
|---|---|---|---|---|---|---|
| EPI_ISL_492015 | Oman/205041335/2020 | 2020-06-04 | Asia/Oman/Muscat/ | Oman-NIC | Department of Microbiology and Immunology-SQUH | Fahad Zadjali et al |
| EPI_ISL_492016 | Oman/205041374/2020 | 2020-06-04 | Asia/Oman/Muscat/ | Oman-NIC | Department of Microbiology and Immunology-SQUH | Fahad Zadjali et al |
| EPI_ISL_492017 | Oman/205041592/2020 | 2020-06-05 | Asia/Oman/Muscat/ | Oman-NIC | Department of Microbiology and Immunology-SQUH | Fahad Zadjali et al |
| EPI_ISL_492018 | Oman/205042569/2020 | 2020-06-08 | Asia/Oman/Muscat/ | Oman-NIC | Department of Microbiology and Immunology-SQUH | Fahad Zadjali et al |
| EPI_ISL_492019 | Oman/205042592/2020 | 2020-06-08 | Asia/Oman/Muscat/ | Oman-NIC | Department of Microbiology and Immunology-SQUH | Fahad Zadjali et al |
| EPI_ISL_492020 | Oman/205042822/2020 | 2020-06-09 | Asia/Oman/Muscat/ | Oman-NIC | Department of Microbiology and Immunology-SQUH | Fahad Zadjali et al |
| EPI_ISL_492021 | Oman/205042904/2020 | 2020-06-09 | Asia/Oman/Muscat/ | Oman-NIC | Department of Microbiology and Immunology-SQUH | Fahad Zadjali et al |
| EPI_ISL_492022 | Oman/205042917/2020 | 2020-06-09 | Asia/Oman/Muscat/ | Oman-NIC | Department of Microbiology and Immunology-SQUH | Fahad Zadjali et al |
| EPI_ISL_492023 | Oman/205042975/2020 | 2020-06-09 | Asia/Oman/Muscat/ | Oman-NIC | Department of Microbiology and Immunology-SQUH | Fahad Zadjali et al |
| EPI_ISL_492024 | Oman/205043002/2020 | 2020-06-09 | Asia/Oman/Muscat/ | Oman-NIC | Department of Microbiology and Immunology-SQUH | Fahad Zadjali et al |

| | | | | | | |
|---|---|---|---|---|---|---|
| EPI_ISL_492025 | Oman/205043071/2020 | 2020-06-10 | Asia/Oman/Muscat/ | Oman-NIC | Department of Microbiology and Immunology-SQUH | Fahad Zadjali et al |
| EPI_ISL_492026 | Oman/205043490/2020 | 2020-06-11 | Asia/Oman/Muscat/ | Oman-NIC | Department of Microbiology and Immunology-SQUH | Fahad Zadjali et al |
| EPI_ISL_492029 | Bangladesh/CHRF-0018/2020 | 2020-05-08 | Asia/Bangladesh/Bangladesh/ | Child Health Research Foundation | Child Health Research Foundation | Senjuti Saha et al |
| EPI_ISL_492031 | Bangladesh/CHRF-0021/2020 | 2020-05-14 | Asia/Bangladesh/Bangladesh/ | Child Health Research Foundation | Child Health Research Foundation | Senjuti Saha et al |
| EPI_ISL_492032 | Brazil/RJ-UFRJ-1019/2020 | 2020-04-15 | South America/Brazil/Rio de Janeiro/Rio | Instituto de Biologia do Ex√©rcito | Laborat√≥rio Metabolismo Macromolecular FirminoTorres de Castro, Instituto de Biof√≠sica Carlos Chagas Filho, Universidade Federal do Rio de Janeiro | Bianca Catarina Azevedo Cabral et al |
| EPI_ISL_492033 | Brazil/RJ-UFRJ-1640/2020 | 2020-04-29 | South America/Brazil/Rio de Janeiro/Rio | Instituto de Biologia do Ex√©rcito | Laborat√≥rio Metabolismo Macromolecular FirminoTorres de Castro, Instituto de Biof√≠sica Carlos Chagas Filho, Universidade Federal do Rio de Janeiro | Bianca Catarina Azevedo Cabral et al |

| | | | | | | |
|---|---|---|---|---|---|---|
| EPI_ISL_492035 | Brazil/RJ-UFRJ-2517/2020 | 2020-05-29 | South America/Brazil/Rio de Janeiro/Rio | Instituto de Biologia do Ex√©rcito | Laborat√≥rio Metabolismo Macromolecular FirminoTorres de Castro, Instituto de Biof√≠sica Carlos Chagas Filho, Universidade Federal do Rio de Janeiro | Bianca Catarina Azevedo Cabral et al |
| EPI_ISL_492036 | Brazil/RJ-UFRJ-9331/2020 | 2020-06-01 | South America/Brazil/Rio de Janeiro/Res | Instituto de Biologia do Ex√©rcito | Laborat√≥rio Metabolismo Macromolecular FirminoTorres de Castro, Instituto de Biof√≠sica Carlos Chagas Filho, Universidade Federal do Rio de Janeiro | Bianca Catarina Azevedo Cabral et al |
| EPI_ISL_492043 | Brazil/RJ-UFRJ-57437/2020 | 2020-05-25 | South America/Brazil/Rio de Janeiro/Rio | Instituto de Biologia do Ex√©rcito | Laborat√≥rio Metabolismo Macromolecular FirminoTorres de Castro, Instituto de Biof√≠sica Carlos Chagas Filho, Universidade Federal do Rio de Janeiro | Bianca Catarina Azevedo Cabral et al |

| | | | | | | |
|---|---|---|---|---|---|---|
| EPI_ISL_492044 | Brazil/RJ-UFRJ-57721/2020 | 2020-05-27 | South America/Brazil/Rio de Janeiro/Rio | Instituto de Biologia do Ex√©rcito | Laborat√≥rio Metabolismo Macromolecular FirminoTorres de Castro, Instituto de Biof√≠sica Carlos Chagas Filho, Universidade Federal do Rio de Janeiro | Bianca Catarina Azevedo Cabral et al |
| EPI_ISL_492045 | Brazil/RJ-UFRJ-57722/2020 | 2020-05-27 | South America/Brazil/Rio de Janeiro/Rio | Instituto de Biologia do Ex√©rcito | Laborat√≥rio Metabolismo Macromolecular FirminoTorres de Castro, Instituto de Biof√≠sica Carlos Chagas Filho, Universidade Federal do Rio de Janeiro | Bianca Catarina Azevedo Cabral et al |
| EPI_ISL_492046 | Brazil/RJ-UFRJ-58090/2020 | 2020-05-29 | South America/Brazil/S√£o Paulo/S√£o | Instituto de Biologia do Ex√©rcito | Laborat√≥rio Metabolismo Macromolecular FirminoTorres de Castro, Instituto de Biof√≠sica Carlos Chagas Filho, Universidade Federal do Rio de Janeiro | Bianca Catarina Azevedo Cabral et al |

| | | | | | | |
|---|---|---|---|---|---|---|
| EPI_ISL_492047 | Brazil/RJ-UFRJ-58110/2020 | 2020-05-29 | South America/Brazil/S√£o Paulo/S√£o | Instituto de Biologia do Ex√©rcito | Laborat√≥rio Metabolismo Macromolecular FirminoTorres de Castro, Instituto de Biof√≠sica Carlos Chagas Filho, Universidade Federal do Rio de Janeiro | Bianca Catarina Azevedo Cabral et al |
| EPI_ISL_492048 | Brazil/RJ-UFRJ-58271/2020 | 2020-06-01 | South America/Brazil/Rio de Janeiro/Rio | Instituto de Biologia do Ex√©rcito | Laborat√≥rio Metabolismo Macromolecular FirminoTorres de Castro, Instituto de Biof√≠sica Carlos Chagas Filho, Universidade Federal do Rio de Janeiro | Bianca Catarina Azevedo Cabral et al |
| EPI_ISL_492052 | USA/AK-PHL048/2020 | 2020-03-26 | North America/USA/Alaska/ | Alaska State Virology Laboratory | Alaska State Virology Laboratory | Chen J et al with Pathogenomics group Dagdag R et al |
| EPI_ISL_492065 | Oman/810962/2020 | 2020-06-20 | Asia/Oman/Muscat/Mutrah | Oman-National Influenza Center | Department of Microbiology and Immunology-SQUH Department of Microbiology and Immunology, Sultan Qaboos University Hopsital, P.O 35, Postal code 123 | Samira Al-Maruqi et al |

| | | | | | | |
|---|---|---|---|---|---|---|
| EPI_ISL_492067 | Poland/PL_P42/2020 | 2020-03-28 | Europe/Poland/Mazowieckie/Brodno-Wa | 1. ViroGenetics - BSL3 Laboratory of Virology, Ma≈Çopolska Centre of Biotechnology, Jagiellonian University; 2. II Department of Internal Medicine, Faculty of Medicine, Jagiellonian University Medical College; 3. Narodowy Instytut Zdrowia Publicznego ‚Äì Pa≈Ñstwowy Zak≈Çad Higieny (NIZP-PZH) | 1. ViroGenetics - BSL3 Laboratory of Virology, Ma≈Çopolska Centre of Biotechnology, Jagiellonian University; 2. II Department of Internal Medicine, Faculty of Medicine, Jagiellonian University Medical College; 3. Narodowy Instytut Zdrowia Publicznego ‚Äì Pa≈Ñstwowy Zak≈Çad Higieny (NIZP-PZH). | Katarzyna Pancer et al |

| | | | | | | |
|---|---|---|---|---|---|---|
| EPI_ISL_492068 | Poland/PL_P43/2020 | 2020-04-01 | Europe/Poland/Mazowieckie/Radom | 1. ViroGenetics - BSL3 Laboratory of Virology, Ma≈Çopolska Centre of Biotechnology, Jagiellonian University; 2. II Department of Internal Medicine, Faculty of Medicine, Jagiellonian University Medical College; 3. Narodowy Instytut Zdrowia Publicznego ‚Äì Pa≈Ñstwowy Zak≈Çad Higieny (NIZP-PZH) | 1. ViroGenetics - BSL3 Laboratory of Virology, Ma≈Çopolska Centre of Biotechnology, Jagiellonian University; 2. II Department of Internal Medicine, Faculty of Medicine, Jagiellonian University Medical College; 3. Narodowy Instytut Zdrowia Publicznego ‚Äì Pa≈Ñstwowy Zak≈Çad Higieny (NIZP-PZH). | Katarzyna Pancer et al |

| | | | | | | |
|---|---|---|---|---|---|---|
| EPI_ISL_492069 | Poland/PL_P44/2020 | 2020-04-02 | Europe/Poland/Mazowieckie/Radom | 1. ViroGenetics - BSL3 Laboratory of Virology, Ma≈Çopolska Centre of Biotechnology, Jagiellonian University; 2. II Department of Internal Medicine, Faculty of Medicine, Jagiellonian University Medical College; 3. Narodowy Instytut Zdrowia Publicznego ,Äì Pa≈Ñstwowy Zak≈Çad Higieny (NIZP-PZH) | 1. ViroGenetics - BSL3 Laboratory of Virology, Ma≈Çopolska Centre of Biotechnology, Jagiellonian University; 2. II Department of Internal Medicine, Faculty of Medicine, Jagiellonian University Medical College; 3. Narodowy Instytut Zdrowia Publicznego ,Äì Pa≈Ñstwowy Zak≈Çad Higieny (NIZP-PZH). | Katarzyna Pancer et al |
| EPI_ISL_492074 | USA/SC-USC-PH00769/2020 | 2020-06-19 | North America/USA/South Carolina/Rich | Functional Genomics Core University of South Carolina / Prisma Health,ÄìMidlands | Functional Genomics Core, University of South Carolina, | Hao Ji et al |
| EPI_ISL_492081 | NorthMacedonia/ChVir7911/2020 | 2020-06-27 | Europe/North Macedonia/North Maced | Institute for Public Health of the Republic of North Macedonia | Charite Universitatsmedizin Berlin, Institute of Virology | Victor M Corman et al |
| EPI_ISL_492187 | England/BRIS-132848/2020 | 2020-04-03 | Europe/United Kingdom/England/ | PHE South West Regional Laboratory, National Infection Service | Wellcome Sanger Institute for the COVID-19 Genomics UK (COG-UK) consortium | Stephanie Hutchings et al |
| EPI_ISL_492195 | England/BRIS-1325BA/2020 | 2020-06-10 | Europe/United Kingdom/England/ | PHE South West Regional Laboratory, National Infection Service | Wellcome Sanger Institute for the COVID-19 Genomics UK (COG-UK) consortium | Stephanie Hutchings et al |

| | | | | | | |
|---|---|---|---|---|---|---|
| EPI_ISL_492222 | England/BRIS-1853AAB/2020 | 2020-04-17 | Europe/United Kingdom/England/ | PHE South West Regional Laboratory, National Infection Service | Wellcome Sanger Institute for the COVID-19 Genomics UK (COG-UK) consortium | Stephanie Hutchings et al |
| EPI_ISL_492483 | England/NORT-29ABEA/2020 | 2020-05-22 | Europe/United Kingdom/England/ | NU-OMICS DNA Sequencing research facility, Northumbria University | Wellcome Sanger Institute for the COVID-19 Genomics UK (COG-UK) consortium | Chris Duncan et al |
| EPI_ISL_492518 | England/BRIS-12FD12/2020 | 2020-04-09 | Europe/United Kingdom/England/ | PHE South West Regional Laboratory, National Infection Service | Wellcome Sanger Institute for the COVID-19 Genomics UK (COG-UK) consortium | Stephanie Hutchings et al |
| EPI_ISL_492586 | England/BRIS-1304CE/2020 | 2020-04-11 | Europe/United Kingdom/England/ | PHE South West Regional Laboratory, National Infection Service | Wellcome Sanger Institute for the COVID-19 Genomics UK (COG-UK) consortium | Stephanie Hutchings et al |
| EPI_ISL_492682 | England/BRIS-130CF3/2020 | 2020-03-24 | Europe/United Kingdom/England/ | PHE South West Regional Laboratory, National Infection Service | Wellcome Sanger Institute for the COVID-19 Genomics UK (COG-UK) consortium | Stephanie Hutchings et al |
| EPI_ISL_492690 | England/BRIS-1302B5/2020 | 2020-04-11 | Europe/United Kingdom/England/ | PHE South West Regional Laboratory, National Infection Service | Wellcome Sanger Institute for the COVID-19 Genomics UK (COG-UK) consortium | Stephanie Hutchings et al |
| EPI_ISL_492724 | England/BRIS-12FDD6/2020 | 2020-04-10 | Europe/United Kingdom/England/ | PHE South West Regional Laboratory, National Infection Service | Wellcome Sanger Institute for the COVID-19 Genomics UK (COG-UK) consortium | Stephanie Hutchings et al |
| EPI_ISL_492730 | England/BRIS-130AEA/2020 | 2020-03-13 | Europe/United Kingdom/England/ | PHE South West Regional Laboratory, National Infection Service | Wellcome Sanger Institute for the COVID-19 Genomics UK (COG-UK) consortium | Stephanie Hutchings et al |

| | | | | | | |
|---|---|---|---|---|---|---|
| EPI_ISL_492809 | England/BRIS-123D4B/2020 | 2020-04-05 | Europe/United Kingdom/England/ | PHE South West Regional Laboratory, National Infection Service | Wellcome Sanger Institute for the COVID-19 Genomics UK (COG-UK) consortium | Stephanie Hutchings et al |
| EPI_ISL_492899 | England/WSFT-25C110D/2020 | 2020-05-06 | Europe/United Kingdom/England/ | Department of Medical Microbiology, Western Sussex Hospitals NHS Foundation Trust, St Richard's Hospital | Wellcome Sanger Institute for the COVID-19 Genomics UK (COG-UK) consortium | Manasa Mutingwende et al |
| EPI_ISL_492929 | England/NORT-28C301/2020 | 2020-05-13 | Europe/United Kingdom/England/ | NU-OMICS DNA Sequencing research facility, Northumbria University | Wellcome Sanger Institute for the COVID-19 Genomics UK (COG-UK) Consortium | Chris Duncan et al |
| EPI_ISL_492973 | England/NORT-28AEA8/2020 | 2020-04-16 | Europe/United Kingdom/England/ | NU-OMICS DNA Sequencing research facility, Northumbria University | Wellcome Sanger Institute for the COVID-19 Genomics UK (COG-UK) consortium | Chris Duncan et al |
| EPI_ISL_492982 | Italy/VEN-UniVR-3/2020 | 2020-04-07 | Europe/Italy/Veneto/Verona IT | IRCCS Sacro Cuore Don Calabria Hospital, Department of Infectious, Tropical Diseases & Microbiology | University of Verona, Department of Biotechnology | Antonio Mori et al |
| EPI_ISL_492983 | Italy/VEN-UniVR-4/2020 | 2020-04-18 | Europe/Italy/Veneto/Verona IT | IRCCS Sacro Cuore Don Calabria Hospital, Department of Infectious, Tropical Diseases & Microbiology | University of Verona, Department of Biotechnology | Antonio Mori et al |
| EPI_ISL_492984 | Italy/VEN-UniVR-5/2020 | 2020-03-11 | Europe/Italy/Veneto/Verona IT | IRCCS Sacro Cuore Don Calabria Hospital, Department of Infectious, Tropical Diseases & Microbiology | University of Verona, Department of Biotechnology | Antonio Mori et al |
| EPI_ISL_492985 | Italy/VEN-UniVR-6/2020 | 2020-03-08 | Europe/Italy/Veneto/Verona IT | IRCCS Sacro Cuore Don Calabria Hospital, Department of Infectious, Tropical Diseases & Microbiology | University of Verona, Department of Biotechnology | Antonio Mori et al |
| EPI_ISL_492987 | Italy/VEN-UniVR-8/2020 | 2020-03-05 | Europe/Italy/Veneto/Verona IT | IRCCS Sacro Cuore Don Calabria Hospital, Department of Infectious, Tropical Diseases & Microbiology | University of Verona, Department of Biotechnology | Antonio Mori et al |

| | | | | | | |
|---|---|---|---|---|---|---|
| EPI_ISL_493008 | USA/UT-03268/2020 | 2020-05-08 | North America/USA/Utah/ | Utah Public Health Laboratory | Utah Public Health Laboratory | Heidi Butz et al |
| EPI_ISL_493010 | USA/UT-03272/2020 | 2020-05-08 | North America/USA/Utah/ | Utah Public Health Laboratory | Utah Public Health Laboratory | Heidi Butz et al |
| EPI_ISL_493015 | USA/UT-03288/2020 | 2020-05-09 | North America/USA/Utah/ | Utah Public Health Laboratory | Utah Public Health Laboratory | Heidi Butz et al |
| EPI_ISL_493020 | USA/UT-03305/2020 | 2020-05-09 | North America/USA/Utah/ | Utah Public Health Laboratory | Utah Public Health Laboratory | Heidi Butz et al |
| EPI_ISL_493027 | USA/UT-04016/2020 | 2020-05-11 | North America/USA/Utah/ | Utah Public Health Laboratory | Utah Public Health Laboratory | Heidi Butz et al |
| EPI_ISL_493031 | USA/UT-04132/2020 | 2020-05-13 | North America/USA/Utah/ | Utah Public Health Laboratory | Utah Public Health Laboratory | Heidi Butz et al |
| EPI_ISL_493035 | USA/UT-04136/2020 | 2020-05-13 | North America/USA/Utah/ | Utah Public Health Laboratory | Utah Public Health Laboratory | Heidi Butz et al |
| EPI_ISL_493040 | USA/UT-04141/2020 | 2020-05-13 | North America/USA/Utah/ | Utah Public Health Laboratory | Utah Public Health Laboratory | Heidi Butz et al |
| EPI_ISL_493041 | USA/UT-04143/2020 | 2020-05-13 | North America/USA/Utah/ | Utah Public Health Laboratory | Utah Public Health Laboratory | Heidi Butz et al |
| EPI_ISL_493050 | USA/UT-04172/2020 | 2020-05-14 | North America/USA/Utah/ | Utah Public Health Laboratory | Utah Public Health Laboratory | Heidi Butz et al |
| EPI_ISL_493051 | USA/UT-04176/2020 | 2020-05-14 | North America/USA/Utah/ | Utah Public Health Laboratory | Utah Public Health Laboratory | Heidi Butz et al |
| EPI_ISL_493052 | USA/UT-04186/2020 | 2020-05-14 | North America/USA/Utah/ | Utah Public Health Laboratory | Utah Public Health Laboratory | Heidi Butz et al |
| EPI_ISL_493055 | USA/UT-04195/2020 | 2020-05-14 | North America/USA/Utah/ | Utah Public Health Laboratory | Utah Public Health Laboratory | Heidi Butz et al |
| EPI_ISL_493057 | USA/UT-04199/2020 | 2020-05-14 | North America/USA/Utah/ | Utah Public Health Laboratory | Utah Public Health Laboratory | Heidi Butz et al |
| EPI_ISL_493058 | USA/UT-04204/2020 | 2020-05-14 | North America/USA/Utah/ | Utah Public Health Laboratory | Utah Public Health Laboratory | Heidi Butz et al |
| EPI_ISL_493071 | USA/MO-WUSTL036/2020 | 2020-06-19 | North America/USA/Missouri/St. Louis [illegible] | Washington University in St. Louis | Washington University in St. Louis | David Wang et al |
| EPI_ISL_493072 | USA/MO-WUSTL037/2020 | 2020-06-19 | North America/USA/Missouri/St. Louis [illegible] | Washington University in St. Louis | Washington University in St. Louis | David Wang et al |
| EPI_ISL_493075 | USA/MO-WUSTL046/2020 | 2020-06-19 | North America/USA/Missouri/St. Louis [illegible] | Washington University in St. Louis | Washington University in St. Louis | David Wang et al |

| | | | | | | |
|---|---|---|---|---|---|---|
| EPI_ISL_493080 | USA/MO-WUSTL061/2020 | 2020-06-19 | North America/USA/Missouri/St. Louis N | Washington University in St. Louis | Washington University in St. Louis | David Wang et al |
| EPI_ISL_493085 | USA/MO-WUSTL068/2020 | 2020-06-19 | North America/USA/Missouri/St. Louis N | Washington University in St. Louis | Washington University in St. Louis | David Wang et al |
| EPI_ISL_493100 | USA/UT-02512/2020 | 2020-04-23 | North America/USA/Utah/ | Utah Public Health Laboratory | Utah Public Health Laboratory | Heidi Butz et al |
| EPI_ISL_493103 | USA/UT-02515/2020 | 2020-04-23 | North America/USA/Utah/ | Utah Public Health Laboratory | Utah Public Health Laboratory | Heidi Butz et al |
| EPI_ISL_493104 | USA/UT-02516/2020 | 2020-04-23 | North America/USA/Utah/ | Utah Public Health Laboratory | Utah Public Health Laboratory | Heidi Butz et al |
| EPI_ISL_493113 | USA/UT-02528/2020 | 2020-04-23 | North America/USA/Utah/ | Utah Public Health Laboratory | Utah Public Health Laboratory | Heidi Butz et al |
| EPI_ISL_493137 | Thailand/MUMT-3/2020 | 2020-02-25 | Asia/Thailand/Bangkok/ | Center for Research and Innovation, Faculty of Medical Technology, Mahidol University | Center for Research and Innovation, Faculty of Medical Technology, Mahidol University | Kantima Sangsiriwut et al |
| EPI_ISL_493149 | Wuhan/0125-A137/2020 | 2020-01-25 | Asia/China/Hubei/Wuhan | National Virus Resource Center, Chinese Academy of Sciences, Wuhan 430071, China | Computational Virology Group, Center for Bacteria and Viruses Resources and Bioinformation, Wuhan Institute of Virology, Chinese Academy of SciencesÔºåWuhan 430071, China | Jianjun Chen et al |

| | | | | | | |
|---|---|---|---|---|---|---|
| EPI_ISL_493150 | Wuhan/0125-A142/2020 | 2020-01-25 | Asia/China/Hubei/Wuhan | National Virus Resource Center, Chinese Academy of Sciences, Wuhan 430071, China | Computational Virology Group, Center for Bacteria and Viruses Resources and Bioinformation, Wuhan Institute of Virology, Chinese Academy of SciencesÔºåWuhan 430071, China | Jianjun Chen et al |
| EPI_ISL_493151 | Wuhan/0125-A145/2020 | 2020-01-25 | Asia/China/Hubei/Wuhan | National Virus Resource Center, Chinese Academy of Sciences, Wuhan 430071, China | Computational Virology Group, Center for Bacteria and Viruses Resources and Bioinformation, Wuhan Institute of Virology, Chinese Academy of SciencesÔºåWuhan 430071, China | Jianjun Chen et al |
| EPI_ISL_493152 | Wuhan/0125-A148/2020 | 2020-01-25 | Asia/China/Hubei/Wuhan | National Virus Resource Center, Chinese Academy of Sciences, Wuhan 430071, China | Computational Virology Group, Center for Bacteria and Viruses Resources and Bioinformation, Wuhan Institute of Virology, Chinese Academy of SciencesÔºåWuhan 430071, China | Jianjun Chen et al |

| | | | | | | |
|---|---|---|---|---|---|---|
| EPI_ISL_493153 | Wuhan/0125-A152/2020 | 2020-01-25 | Asia/China/Hubei/Wuhan | National Virus Resource Center, Chinese Academy of Sciences, Wuhan 430071, China | Computational Virology Group, Center for Bacteria and Viruses Resources and Bioinformation, Wuhan Institute of Virology, Chinese Academy of SciencesÔºåWuhan 430071, China | Jianjun Chen et al |
| EPI_ISL_493154 | Wuhan/0125-A153/2020 | 2020-01-25 | Asia/China/Hubei/Wuhan | National Virus Resource Center, Chinese Academy of Sciences, Wuhan 430071, China | Computational Virology Group, Center for Bacteria and Viruses Resources and Bioinformation, Wuhan Institute of Virology, Chinese Academy of SciencesÔºåWuhan 430071, China | Jianjun Chen et al |
| EPI_ISL_493155 | Wuhan/0125-A160/2020 | 2020-01-25 | Asia/China/Hubei/Wuhan | National Virus Resource Center, Chinese Academy of Sciences, Wuhan 430071, China | Computational Virology Group, Center for Bacteria and Viruses Resources and Bioinformation, Wuhan Institute of Virology, Chinese Academy of SciencesÔºåWuhan 430071, China | Jianjun Chen et al |

| | | | | | | |
|---|---|---|---|---|---|---|
| EPI_ISL_493156 | Wuhan/0125-A163/2020 | 2020-01-25 | Asia/China/Hubei/Wuhan | National Virus Resource Center, Chinese Academy of Sciences, Wuhan 430071, China | Computational Virology Group, Center for Bacteria and Viruses Resources and Bioinformation, Wuhan Institute of Virology, Chinese Academy of SciencesÔºåWuhan 430071, China | Jianjun Chen et al |
| EPI_ISL_493157 | Wuhan/0125-A164/2020 | 2020-01-25 | Asia/China/Hubei/Wuhan | National Virus Resource Center, Chinese Academy of Sciences, Wuhan 430071, China | Computational Virology Group, Center for Bacteria and Viruses Resources and Bioinformation, Wuhan Institute of Virology, Chinese Academy of SciencesÔºåWuhan 430071, China | Jianjun Chen et al |
| EPI_ISL_493158 | Wuhan/0125-A167/2020 | 2020-01-25 | Asia/China/Hubei/Wuhan | National Virus Resource Center, Chinese Academy of Sciences, Wuhan 430071, China | Computational Virology Group, Center for Bacteria and Viruses Resources and Bioinformation, Wuhan Institute of Virology, Chinese Academy of SciencesÔºåWuhan 430071, China | Jianjun Chen et al |

| | | | | | | |
|---|---|---|---|---|---|---|
| EPI_ISL_493159 | Wuhan/0125-A168/2020 | 2020-01-25 | Asia/China/Hubei/Wuhan | National Virus Resource Center, Chinese Academy of Sciences, Wuhan 430071, China | Computational Virology Group, Center for Bacteria and Viruses Resources and Bioinformation, Wuhan Institute of Virology, Chinese Academy of SciencesÔºåWuhan 430071, China | Jianjun Chen et al |
| EPI_ISL_493160 | Wuhan/0125-A169/2020 | 2020-01-25 | Asia/China/Hubei/Wuhan | National Virus Resource Center, Chinese Academy of Sciences, Wuhan 430071, China | Computational Virology Group, Center for Bacteria and Viruses Resources and Bioinformation, Wuhan Institute of Virology, Chinese Academy of SciencesÔºåWuhan 430071, China | Jianjun Chen et al |
| EPI_ISL_493161 | Wuhan/0125-A170/2020 | 2020-01-25 | Asia/China/Hubei/Wuhan | National Virus Resource Center, Chinese Academy of Sciences, Wuhan 430071, China | Computational Virology Group, Center for Bacteria and Viruses Resources and Bioinformation, Wuhan Institute of Virology, Chinese Academy of SciencesÔºåWuhan 430071, China | Jianjun Chen et al |

| | | | | | | |
|---|---|---|---|---|---|---|
| EPI_ISL_493162 | Wuhan/0125-A171/2020 | 2020-01-25 | Asia/China/Hubei/Wuhan | National Virus Resource Center, Chinese Academy of Sciences, Wuhan 430071, China | Computational Virology Group, Center for Bacteria and Viruses Resources and Bioinformation, Wuhan Institute of Virology, Chinese Academy of SciencesÔºåWuhan 430071, China | Jianjun Chen et al |
| EPI_ISL_493163 | Wuhan/0125-A174/2020 | 2020-01-25 | Asia/China/Hubei/Wuhan | National Virus Resource Center, Chinese Academy of Sciences, Wuhan 430071, China | Computational Virology Group, Center for Bacteria and Viruses Resources and Bioinformation, Wuhan Institute of Virology, Chinese Academy of SciencesÔºåWuhan 430071, China | Jianjun Chen et al |
| EPI_ISL_493164 | Wuhan/0125-A182/2020 | 2020-01-25 | Asia/China/Hubei/Wuhan | National Virus Resource Center, Chinese Academy of Sciences, Wuhan 430071, China | Computational Virology Group, Center for Bacteria and Viruses Resources and Bioinformation, Wuhan Institute of Virology, Chinese Academy of SciencesÔºåWuhan 430071, China | Jianjun Chen et al |

| | | | | | | |
|---|---|---|---|---|---|---|
| EPI_ISL_493165 | Wuhan/0125-A191/2020 | 2020-01-25 | Asia/China/Hubei/Wuhan | National Virus Resource Center, Chinese Academy of Sciences, Wuhan 430071, China | Computational Virology Group, Center for Bacteria and Viruses Resources and Bioinformation, Wuhan Institute of Virology, Chinese Academy of SciencesÔºåWuhan 430071, China | Jianjun Chen et al |
| EPI_ISL_493166 | Wuhan/0125-A192/2020 | 2020-01-25 | Asia/China/Hubei/Wuhan | National Virus Resource Center, Chinese Academy of Sciences, Wuhan 430071, China | Computational Virology Group, Center for Bacteria and Viruses Resources and Bioinformation, Wuhan Institute of Virology, Chinese Academy of SciencesÔºåWuhan 430071, China | Jianjun Chen et al |
| EPI_ISL_493167 | Wuhan/0126-C5/2020 | 2020-01-26 | Asia/China/Hubei/Wuhan | National Virus Resource Center, Chinese Academy of Sciences, Wuhan 430071, China | Computational Virology Group, Center for Bacteria and Viruses Resources and Bioinformation, Wuhan Institute of Virology, Chinese Academy of SciencesÔºåWuhan 430071, China | Jianjun Chen et al |

| | | | | | | |
|---|---|---|---|---|---|---|
| EPI_ISL_493168 | Wuhan/0126-C7/2020 | 2020-01-26 | Asia/China/Hubei/Wuhan | National Virus Resource Center, Chinese Academy of Sciences, Wuhan 430071, China | Computational Virology Group, Center for Bacteria and Viruses Resources and Bioinformation, Wuhan Institute of Virology, Chinese Academy of SciencesÔºåWuhan 430071, China | Jianjun Chen et al |
| EPI_ISL_493169 | Wuhan/0126-C10/2020 | 2020-01-26 | Asia/China/Hubei/Wuhan | National Virus Resource Center, Chinese Academy of Sciences, Wuhan 430071, China | Computational Virology Group, Center for Bacteria and Viruses Resources and Bioinformation, Wuhan Institute of Virology, Chinese Academy of SciencesÔºåWuhan 430071, China | Jianjun Chen et al |
| EPI_ISL_493170 | Wuhan/0126-C12/2020 | 2020-01-26 | Asia/China/Hubei/Wuhan | National Virus Resource Center, Chinese Academy of Sciences, Wuhan 430071, China | Computational Virology Group, Center for Bacteria and Viruses Resources and Bioinformation, Wuhan Institute of Virology, Chinese Academy of SciencesÔºåWuhan 430071, China | Jianjun Chen et al |

| | | | | | | |
|---|---|---|---|---|---|---|
| EPI_ISL_493171 | Wuhan/0126-C13/2020 | 2020-01-26 | Asia/China/Hubei/Wuhan | National Virus Resource Center, Chinese Academy of Sciences, Wuhan 430071, China | Computational Virology Group, Center for Bacteria and Viruses Resources and Bioinformation, Wuhan Institute of Virology, Chinese Academy of SciencesÔºåWuhan 430071, China | Jianjun Chen et al |
| EPI_ISL_493172 | Wuhan/0126-C14/2020 | 2020-01-26 | Asia/China/Hubei/Wuhan | National Virus Resource Center, Chinese Academy of Sciences, Wuhan 430071, China | Computational Virology Group, Center for Bacteria and Viruses Resources and Bioinformation, Wuhan Institute of Virology, Chinese Academy of SciencesÔºåWuhan 430071, China | Jianjun Chen et al |
| EPI_ISL_493173 | Wuhan/0126-C26/2020 | 2020-01-26 | Asia/China/Hubei/Wuhan | National Virus Resource Center, Chinese Academy of Sciences, Wuhan 430071, China | Computational Virology Group, Center for Bacteria and Viruses Resources and Bioinformation, Wuhan Institute of Virology, Chinese Academy of SciencesÔºåWuhan 430071, China | Jianjun Chen et al |

| | | | | | | |
|---|---|---|---|---|---|---|
| EPI_ISL_493174 | Wuhan/0126-C31/2020 | 2020-01-26 | Asia/China/Hubei/Wuhan | National Virus Resource Center, Chinese Academy of Sciences, Wuhan 430071, China | Computational Virology Group, Center for Bacteria and Viruses Resources and Bioinformation, Wuhan Institute of Virology, Chinese Academy of SciencesÔºåWuhan 430071, China | Jianjun Chen et al |
| EPI_ISL_493175 | Wuhan/0126-C43/2020 | 2020-01-26 | Asia/China/Hubei/Wuhan | National Virus Resource Center, Chinese Academy of Sciences, Wuhan 430071, China | Computational Virology Group, Center for Bacteria and Viruses Resources and Bioinformation, Wuhan Institute of Virology, Chinese Academy of SciencesÔºåWuhan 430071, China | Jianjun Chen et al |
| EPI_ISL_493176 | Wuhan/0126-C45/2020 | 2020-01-26 | Asia/China/Hubei/Wuhan | National Virus Resource Center, Chinese Academy of Sciences, Wuhan 430071, China | Computational Virology Group, Center for Bacteria and Viruses Resources and Bioinformation, Wuhan Institute of Virology, Chinese Academy of SciencesÔºåWuhan 430071, China | Jianjun Chen et al |

| | | | | | | |
|---|---|---|---|---|---|---|
| EPI_ISL_493177 | Wuhan/0126-C47/2020 | 2020-01-26 | Asia/China/Hubei/Wuhan | National Virus Resource Center, Chinese Academy of Sciences, Wuhan 430071, China | Computational Virology Group, Center for Bacteria and Viruses Resources and Bioinformation, Wuhan Institute of Virology, Chinese Academy of SciencesÔºåWuhan 430071, China | Jianjun Chen et al |
| EPI_ISL_493178 | Wuhan/0126-C77/2020 | 2020-01-26 | Asia/China/Hubei/Wuhan | National Virus Resource Center, Chinese Academy of Sciences, Wuhan 430071, China | Computational Virology Group, Center for Bacteria and Viruses Resources and Bioinformation, Wuhan Institute of Virology, Chinese Academy of SciencesÔºåWuhan 430071, China | Jianjun Chen et al |
| EPI_ISL_493179 | Wuhan/0126-C93/2020 | 2020-01-26 | Asia/China/Hubei/Wuhan | National Virus Resource Center, Chinese Academy of Sciences, Wuhan 430071, China | Computational Virology Group, Center for Bacteria and Viruses Resources and Bioinformation, Wuhan Institute of Virology, Chinese Academy of SciencesÔºåWuhan 430071, China | Jianjun Chen et al |

| | | | | | | |
|---|---|---|---|---|---|---|
| EPI_ISL_493180 | Wuhan/0126-C94/2020 | 2020-01-26 | Asia/China/Hubei/Wuhan | National Virus Resource Center, Chinese Academy of Sciences, Wuhan 430071, China | Computational Virology Group, Center for Bacteria and Viruses Resources and Bioinformation, Wuhan Institute of Virology, Chinese Academy of SciencesÔºåWuhan 430071, China | Jianjun Chen et al |
| EPI_ISL_493181 | Wuhan/0126-C98/2020 | 2020-01-26 | Asia/China/Hubei/Wuhan | National Virus Resource Center, Chinese Academy of Sciences, Wuhan 430071, China | Computational Virology Group, Center for Bacteria and Viruses Resources and Bioinformation, Wuhan Institute of Virology, Chinese Academy of SciencesÔºåWuhan 430071, China | Jianjun Chen et al |
| EPI_ISL_493182 | Wuhan/0126-C100/2020 | 2020-01-26 | Asia/China/Hubei/Wuhan | National Virus Resource Center, Chinese Academy of Sciences, Wuhan 430071, China | Computational Virology Group, Center for Bacteria and Viruses Resources and Bioinformation, Wuhan Institute of Virology, Chinese Academy of SciencesÔºåWuhan 430071, China | Jianjun Chen et al |

| | | | | | | |
|---|---|---|---|---|---|---|
| EPI_ISL_493183 | Wuhan/0126-C101/2020 | 2020-01-26 | Asia/China/Hubei/Wuhan | National Virus Resource Center, Chinese Academy of Sciences, Wuhan 430071, China | Computational Virology Group, Center for Bacteria and Viruses Resources and Bioinformation, Wuhan Institute of Virology, Chinese Academy of SciencesÔºåWuhan 430071, China | Jianjun Chen et al |
| EPI_ISL_493184 | Wuhan/0126-C102/2020 | 2020-01-26 | Asia/China/Hubei/Wuhan | National Virus Resource Center, Chinese Academy of Sciences, Wuhan 430071, China | Computational Virology Group, Center for Bacteria and Viruses Resources and Bioinformation, Wuhan Institute of Virology, Chinese Academy of SciencesÔºåWuhan 430071, China | Jianjun Chen et al |
| EPI_ISL_493185 | Wuhan/0126-C103/2020 | 2020-01-26 | Asia/China/Hubei/Wuhan | National Virus Resource Center, Chinese Academy of Sciences, Wuhan 430071, China | Computational Virology Group, Center for Bacteria and Viruses Resources and Bioinformation, Wuhan Institute of Virology, Chinese Academy of SciencesÔºåWuhan 430071, China | Jianjun Chen et al |

| | | | | | | |
|---|---|---|---|---|---|---|
| EPI_ISL_493186 | Wuhan/0126-C106/2020 | 2020-01-26 | Asia/China/Hubei/Wuhan | National Virus Resource Center, Chinese Academy of Sciences, Wuhan 430071, China | Computational Virology Group, Center for Bacteria and Viruses Resources and Bioinformation, Wuhan Institute of Virology, Chinese Academy of SciencesÔºåWuhan 430071, China | Jianjun Chen et al |
| EPI_ISL_493187 | Wuhan/0126-C108/2020 | 2020-01-26 | Asia/China/Hubei/Wuhan | National Virus Resource Center, Chinese Academy of Sciences, Wuhan 430071, China | Computational Virology Group, Center for Bacteria and Viruses Resources and Bioinformation, Wuhan Institute of Virology, Chinese Academy of SciencesÔºåWuhan 430071, China | Jianjun Chen et al |
| EPI_ISL_493188 | Wuhan/0126-C109/2020 | 2020-01-26 | Asia/China/Hubei/Wuhan | National Virus Resource Center, Chinese Academy of Sciences, Wuhan 430071, China | Computational Virology Group, Center for Bacteria and Viruses Resources and Bioinformation, Wuhan Institute of Virology, Chinese Academy of SciencesÔºåWuhan 430071, China | Jianjun Chen et al |

| | | | | | | |
|---|---|---|---|---|---|---|
| EPI_ISL_493189 | Wuhan/0126-C111/2020 | 2020-01-26 | Asia/China/Hubei/Wuhan | National Virus Resource Center, Chinese Academy of Sciences, Wuhan 430071, China | Computational Virology Group, Center for Bacteria and Viruses Resources and Bioinformation, Wuhan Institute of Virology, Chinese Academy of SciencesÔºåWuhan 430071, China | Jianjun Chen et al |
| EPI_ISL_493190 | Wuhan/0126-C113/2020 | 2020-01-26 | Asia/China/Hubei/Wuhan | National Virus Resource Center, Chinese Academy of Sciences, Wuhan 430071, China | Computational Virology Group, Center for Bacteria and Viruses Resources and Bioinformation, Wuhan Institute of Virology, Chinese Academy of SciencesÔºåWuhan 430071, China | Jianjun Chen et al |
| EPI_ISL_493206 | Taiwan/NCKUH-013/2020 | 2020-03-28 | Asia/Taiwan/Tainan/ | Virology Lab,Department of Pathology, National Cheng Kung University Hospital | Virology Lab,Department of Pathology, National Cheng Kung University Hospital | Huey-Pin Tsai et al |
| EPI_ISL_493330 | Italy/LAZ-INMI18/2020 | 2020-05-10 | Europe/Italy/Lazio/ | INMI Lazzaro Spallanzani IRCCS | INMI Lazzaro Spallanzani IRCCS | Cesare E.M. Gruber et al |
| EPI_ISL_493332 | Italy/CAM-INMI-32798-45/2020 | 2020-03-28 | Europe/Italy/Campania/Benevento | Istituto Zooprofilattico Sperimentale del Mezzogiorno | INMI Lazzaro Spallanzani IRCCS | Cesare E.M. Gruber et al |
| EPI_ISL_493334 | Mexico/GUA-InDRE-19/2020 | 2020-03-27 | North America/Mexico/Guanajuato/ | Instituto de Diagnostico y Referencia Epidemiologicos (INDRE) | Instituto de Diagnostico y Referencia Epidemiologicos (INDRE) | Gisela Barrera-Badillo et al |

| | | | | | | |
|---|---|---|---|---|---|---|
| EPI_ISL_493338 | Mexico/CHP-InDRE-23/2020 | 2020-05-04 | North America/Mexico/Chiapas/ | Instituto de Diagnostico y Referencia Epidemiologicos (INDRE) | Instituto de Diagnostico y Referencia Epidemiologicos (INDRE) | Gisela Barrera-Badillo et al |
| EPI_ISL_493339 | Mexico/GRO-InDRE-24/2020 | 2020-05-04 | North America/Mexico/Guerrero/ | Instituto de Diagnostico y Referencia Epidemiologicos (INDRE) | Instituto de Diagnostico y Referencia Epidemiologicos (INDRE) | Gisela Barrera-Badillo et al |
| EPI_ISL_493341 | Mexico/MOR-InDRE-26/2020 | 2020-05-27 | North America/Mexico/Morelos/ | Instituto de Diagnostico y Referencia Epidemiologicos (INDRE) | Instituto de Diagnostico y Referencia Epidemiologicos (INDRE) | Gisela Barrera-Badillo et al |
| EPI_ISL_493342 | Mexico/NAY-InDRE_27/2020 | 2020-05-26 | North America/Mexico/Nayarit/ | Instituto de Diagnostico y Referencia Epidemiologicos (INDRE) | Instituto de Diagnostico y Referencia Epidemiologicos (INDRE) | Ernesto Ramirez-Gonzalez et al |
| EPI_ISL_493343 | Mexico/NAY-InDRE_28/2020 | 2020-05-29 | North America/Mexico/Nayarit/ | Instituto de Diagnostico y Referencia Epidemiologicos (INDRE) | Instituto de Diagnostico y Referencia Epidemiologicos (INDRE) | Ernesto Ramirez-Gonzalez et al |
| EPI_ISL_493344 | Mexico/SON-InDRE_29/2020 | 2020-05-15 | North America/Mexico/Sonora/ | Instituto de Diagnostico y Referencia Epidemiologicos (INDRE) | Instituto de Diagnostico y Referencia Epidemiologicos (INDRE) | Ernesto Ramirez-Gonzalez et al |
| EPI_ISL_493345 | Mexico/BCS-InDRE_30/2020 | 2020-05-29 | North America/Mexico/Baja California S | Instituto de Diagnostico y Referencia Epidemiologicos (INDRE) | Instituto de Diagnostico y Referencia Epidemiologicos (INDRE) | Ernesto Ramirez-Gonzalez et al |
| EPI_ISL_493346 | Mexico/BCS-InDRE_31/2020 | 2020-05-26 | North America/Mexico/Baja California S | Instituto de Diagnostico y Referencia Epidemiologicos (INDRE) | Instituto de Diagnostico y Referencia Epidemiologicos (INDRE) | Ernesto Ramirez-Gonzalez et al |

| | | | | | | |
|---|---|---|---|---|---|---|
| EPI_ISL_493348 | Mexico/YUC-InDRE_33/2020 | 2020-05-27 | North America/Mexico/Yucatan/ | Instituto de Diagnostico y Referencia Epidemiologicos (INDRE) | Instituto de Diagnostico y Referencia Epidemiologicos (INDRE) | Ernesto Ramirez-Gonzalez et al |
| EPI_ISL_493349 | Mexico/SON-InDRE_34/2020 | 2020-05-28 | North America/Mexico/Sonora/ | Instituto de Diagnostico y Referencia Epidemiologicos (INDRE) | Instituto de Diagnostico y Referencia Epidemiologicos (INDRE) | Ernesto Ramirez-Gonzalez et al |
| EPI_ISL_493356 | Norway/2789/2020 | 2020-06-26 | Europe/Norway/Norway/ | Furst Medical Laboratory | Norwegian Institute of Public Health, Department of Virology | Kathrine Stene-Johansen et al |
| EPI_ISL_493390 | Singapore/630/2020 | 2020-02-06 | Asia/Singapore/Singapore/ | National Public Health Laboratory, National Centre for Infectious Diseases | National Public Health Laboratory, National Centre for Infectious Diseases | Mak TM et al |
| EPI_ISL_493399 | Singapore/639/2020 | 2020-05-06 | Asia/Singapore/Singapore/ | National Public Health Laboratory, National Centre for Infectious Diseases | National Public Health Laboratory, National Centre for Infectious Diseases | Mak TM et al |
| EPI_ISL_493400 | Singapore/640/2020 | 2020-05-05 | Asia/Singapore/Singapore/ | National Public Health Laboratory, National Centre for Infectious Diseases | National Public Health Laboratory, National Centre for Infectious Diseases | Mak TM et al |
| EPI_ISL_493404 | Singapore/644/2020 | 2020-05-14 | Asia/Singapore/Singapore/ | National Public Health Laboratory, National Centre for Infectious Diseases | National Public Health Laboratory, National Centre for Infectious Diseases | Mak TM et al |

| | | | | | | |
|---|---|---|---|---|---|---|
| EPI_ISL_493406 | Singapore/646/2020 | 2020-05-18 | Asia/Singapore/Singapore/ | National Public Health Laboratory, National Centre for Infectious Diseases | National Public Health Laboratory, National Centre for Infectious Diseases | Mak TM et al |
| EPI_ISL_493411 | Singapore/651/2020 | 2020-05-08 | Asia/Singapore/Singapore/ | National Public Health Laboratory, National Centre for Infectious Diseases | National Public Health Laboratory, National Centre for Infectious Diseases | Mak TM et al |
| EPI_ISL_493414 | Singapore/654/2020 | 2020-02-18 | Asia/Singapore/Singapore/ | National Public Health Laboratory, National Centre for Infectious Diseases | National Public Health Laboratory, National Centre for Infectious Diseases | Mak TM et al |
| EPI_ISL_493418 | Singapore/658/2020 | 2020-04-14 | Asia/Singapore/Singapore/ | National Public Health Laboratory, National Centre for Infectious Diseases | National Public Health Laboratory, National Centre for Infectious Diseases | Mak TM et al |
| EPI_ISL_493421 | Singapore/661/2020 | 2020-02-07 | Asia/Singapore/Singapore/ | National Public Health Laboratory, National Centre for Infectious Diseases | National Public Health Laboratory, National Centre for Infectious Diseases | Mak TM et al |
| EPI_ISL_493520 | England/NORT-29E5BB/2020 | 2020-05-02 | Europe/United Kingdom/England/ | Northumbria University / South Tees Hospitals NHS Foundation Trust / North Cumbria Integrated Care NHS Foundation Trust / North Tees and Hartlepool NHS Foundation Trust / Newcastle Hospitals NHS Foundation Trust | COVID-19 Genomics UK (COG-UK) Consortium | Darren L Smith et al |

| | | | | | | |
|---|---|---|---|---|---|---|
| EPI_ISL_493528 | England/NORT-29E81C/2020 | 2020-05-04 | Europe/United Kingdom/England/ | Northumbria University / South Tees Hospitals NHS Foundation Trust / North Cumbria Integrated Care NHS Foundation Trust / North Tees and Hartlepool NHS Foundation Trust / Newcastle Hospitals NHS Foundation Trust | COVID-19 Genomics UK (COG-UK) Consortium | Darren L Smith et al |
| EPI_ISL_493605 | England/NOTT-112C8E/2020 | 2020-04-17 | Europe/United Kingdom/England/ | Lincolnshire Hospitals and DeepSeq Nottingham | COVID-19 Genomics UK (COG-UK) Consortium | Nichola Duckworth et al |
| EPI_ISL_493625 | England/PORT-2CFB0A/2020 | 2020-05-20 | Europe/United Kingdom/England/ | Centre for Enzyme Innovation, University of Portsmouth / Translational Research Laboratory, Portsmouth Hospitals NHS Trust | COVID-19 Genomics UK (COG-UK) Consortium | Angela Beckett et al |
| EPI_ISL_493684 | England/SHEF-C6051/2020 | 2020-07-12 | Europe/United Kingdom/England/ | Virology Department, Sheffield Teaching Hospitals NHS Foundation Trust/Department of Infection, Immunity and Cardiovascular Disease, The Medical School, University of Sheffield | COVID-19 Genomics UK (COG-UK) Consortium | Thushan de Silva et al |
| EPI_ISL_493701 | England/SHEF-CB39F/2020 | 2020-05-15 | Europe/United Kingdom/England/ | Virology Department, Sheffield Teaching Hospitals NHS Foundation Trust/Department of Infection, Immunity and Cardiovascular Disease, The Medical School, University of Sheffield | COVID-19 Genomics UK (COG-UK) Consortium | Thushan de Silva et al |
| EPI_ISL_493787 | Scotland/CVR2311/2020 | 2020-04-13 | Europe/United Kingdom/Scotland/ | West of Scotland Specialist Virology Centre, NHSGGC / MRC-University of Glasgow Centre for Virus Research | COVID-19 Genomics UK (COG-UK) Consortium | Ana da Silva Filipe et al |
| EPI_ISL_493851 | Scotland/CVR3289/2020 | 2020-05-01 | Europe/United Kingdom/Scotland/ | West of Scotland Specialist Virology Centre, NHSGGC / MRC-University of Glasgow Centre for Virus Research | COVID-19 Genomics UK (COG-UK) Consortium | Ana da Silva Filipe et al |
| EPI_ISL_493856 | Scotland/CVR3300/2020 | 2020-04-30 | Europe/United Kingdom/Scotland/ | West of Scotland Specialist Virology Centre, NHSGGC / MRC-University of Glasgow Centre for Virus Research | COVID-19 Genomics UK (COG-UK) Consortium | Ana da Silva Filipe et al |
| EPI_ISL_493887 | Scotland/CVR3338/2020 | 2020-05-01 | Europe/United Kingdom/Scotland/ | West of Scotland Specialist Virology Centre, NHSGGC / MRC-University of Glasgow Centre for Virus Research | COVID-19 Genomics UK (COG-UK) Consortium | Ana da Silva Filipe et al |
| EPI_ISL_493897 | Scotland/CVR3887/2020 | 2020-05-19 | Europe/United Kingdom/Scotland/ | West of Scotland Specialist Virology Centre, NHSGGC / MRC-University of Glasgow Centre for Virus Research | COVID-19 Genomics UK (COG-UK) Consortium | Ana da Silva Filipe et al |

| | | | | | | |
|---|---|---|---|---|---|---|
| EPI_ISL_493937 | Scotland/EDB5019/2020 | 2020-05-15 | Europe/United Kingdom/Scotland/ | Virology Department, Royal Infirmary of Edinburgh, NHS Lothian / School of Biological Sciences, University of Edinburgh / Institute of Genetics and Molecular Medicine, University of Edinburgh | COVID-19 Genomics UK (COG-UK) Consortium | McHugh M et al |
| EPI_ISL_494251 | Wales/PHWC-168515/2020 | 2020-04-17 | Europe/United Kingdom/Wales/ | Wales Specialist Virology Centre Sequencing lab: Pathogen Genomics Unit | COVID-19 Genomics UK (COG-UK) Consortium | Catherine Moore et al |
| EPI_ISL_494356 | Wales/PHWC-168D1D/2020 | 2020-04-18 | Europe/United Kingdom/Wales/ | Wales Specialist Virology Centre Sequencing lab: Pathogen Genomics Unit | COVID-19 Genomics UK (COG-UK) Consortium | Catherine Moore et al |
| EPI_ISL_494375 | USA/CA-ALSR-1123/2020 | 2020-04-28 | North America/USA/California/San Dieg | San Diego County Public Health Laboratory | Andersen lab at Scripps Research | SEARCH Alliance San Diego with Tracy Basler et al |
| EPI_ISL_494381 | USA/CA-ALSR-1130/2020 | 2020-04-04 | North America/USA/California/ | San Diego County Public Health Laboratory | Andersen lab at Scripps Research | SEARCH Alliance San Diego with Tracy Basler et al |
| EPI_ISL_494385 | USA/CA-ALSR-1140/2020 | 2020-04-02 | North America/USA/California/ | San Diego County Public Health Laboratory | Andersen lab at Scripps Research | SEARCH Alliance San Diego with Tracy Basler et al |
| EPI_ISL_494391 | USA/CA-ALSR-1146/2020 | 2020-04-08 | North America/USA/California/ | San Diego County Public Health Laboratory | Andersen lab at Scripps Research | SEARCH Alliance San Diego with Tracy Basler et al |
| EPI_ISL_494397 | USA/CA-ALSR-1154/2020 | 2020-04-27 | North America/USA/California/San Dieg | San Diego County Public Health Laboratory | Andersen lab at Scripps Research | SEARCH Alliance San Diego with Tracy Basler et al |
| EPI_ISL_494407 | USA/CA-ALSR-1165/2020 | 2020-04-05 | North America/USA/California/San Dieg | San Diego County Public Health Laboratory | Andersen lab at Scripps Research | SEARCH Alliance San Diego with Tracy Basler et al |
| EPI_ISL_494418 | USA/CA-ALSR-1182/2020 | 2020-04-02 | North America/USA/California/ | San Diego County Public Health Laboratory | Andersen lab at Scripps Research | SEARCH Alliance San Diego with Tracy Basler et al |
| EPI_ISL_494419 | USA/CA-ALSR-1183/2020 | 2020-04-02 | North America/USA/California/ | San Diego County Public Health Laboratory | Andersen lab at Scripps Research | SEARCH Alliance San Diego with Tracy Basler et al |
| EPI_ISL_494428 | USA/CA-ALSR-1192/2020 | 2020-04-28 | North America/USA/California/San Dieg | San Diego County Public Health Laboratory | Andersen lab at Scripps Research | SEARCH Alliance San Diego with Tracy Basler et al |
| EPI_ISL_494442 | USA/CA-ALSR-1214/2020 | 2020-04-24 | North America/USA/California/San Dieg | San Diego County Public Health Laboratory | Andersen lab at Scripps Research | SEARCH Alliance San Diego with Tracy Basler et al |
| EPI_ISL_494443 | USA/CA-ALSR-1217/2020 | 2020-04-24 | North America/USA/California/San Dieg | San Diego County Public Health Laboratory | Andersen lab at Scripps Research | SEARCH Alliance San Diego with Tracy Basler et al |
| EPI_ISL_494452 | USA/CA-ALSR-1231/2020 | 2020-05-06 | North America/USA/California/San Dieg | San Diego County Public Health Laboratory | Andersen lab at Scripps Research | SEARCH Alliance San Diego with Tracy Basler et al |
| EPI_ISL_494453 | USA/CA-ALSR-1239/2020 | 2020-05-05 | North America/USA/California/San Dieg | San Diego County Public Health Laboratory | Andersen lab at Scripps Research | SEARCH Alliance San Diego with Tracy Basler et al |
| EPI_ISL_494454 | USA/CA-ALSR-1240/2020 | 2020-05-04 | North America/USA/California/San Dieg | San Diego County Public Health Laboratory | Andersen lab at Scripps Research | SEARCH Alliance San Diego with Tracy Basler et al |

| | | | | | | |
|---|---|---|---|---|---|---|
| EPI_ISL_494462 | USA/CA-ALSR-1261/2020 | 2020-05-05 | North America/USA/California/San Dieg | San Diego County Public Health Laboratory | Andersen lab at Scripps Research | SEARCH Alliance San Diego with Tracy Basler et al |
| EPI_ISL_494470 | USA/CA-ALSR-1272/2020 | 2020-05-08 | North America/USA/California/San Dieg | San Diego County Public Health Laboratory | Andersen lab at Scripps Research | SEARCH Alliance San Diego with Tracy Basler et al |
| EPI_ISL_494494 | USA/CA-ALSR-1324/2020 | 2020-04-30 | North America/USA/California/San Dieg | San Diego County Public Health Laboratory | Andersen lab at Scripps Research | SEARCH Alliance San Diego with Tracy Basler et al |
| EPI_ISL_494502 | USA/CA-ALSR-1340/2020 | 2020-04-30 | North America/USA/California/San Dieg | San Diego County Public Health Laboratory | Andersen lab at Scripps Research | SEARCH Alliance San Diego with Tracy Basler et al |
| EPI_ISL_494518 | USA/IN-QDX-182/2020 | 2020-04-28 | North America/USA/Indiana/ | Quest Diagnostics | Quest Diagnostics | Anderson et al |
| EPI_ISL_494521 | USA/LA-QDX-167/2020 | 2020-04-10 | North America/USA/Louisiana/ | Quest Diagnostics | Quest Diagnostics | Anderson et al |
| EPI_ISL_494531 | USA/NM-QDX-208/2020 | 2020-04-30 | North America/USA/New Mexico/ | Quest Diagnostics | Quest Diagnostics | Anderson et al |
| EPI_ISL_494534 | USA/NM-QDX-211/2020 | 2020-04-30 | North America/USA/New Mexico/ | Quest Diagnostics | Quest Diagnostics | Anderson et al |
| EPI_ISL_494537 | USA/NM-QDX-215/2020 | 2020-05-01 | North America/USA/New Mexico/ | Quest Diagnostics | Quest Diagnostics | Anderson et al |
| EPI_ISL_494539 | USA/TX-QDX-168/2020 | 2020-04-10 | North America/USA/Texas/ | Quest Diagnostics | Quest Diagnostics | Anderson et al |
| EPI_ISL_494545 | USA/TX-QDX-190/2020 | 2020-04-29 | North America/USA/Texas/ | Quest Diagnostics | Quest Diagnostics | Anderson et al |
| EPI_ISL_494554 | USA/SC-USC-PRG505/2020 | 2020-05-30 | North America/USA/South Carolina/Rich | Functional Genomics Core University of South Carolina / Prisma Health,ÄìMidlands | Functional Genomics Core, University of South Carolina | Hao Ji et al |
| EPI_ISL_494588 | USA/CA-ALSR-1413/2020 | 2020-05-04 | North America/USA/California/San Dieg | San Diego County Public Health Laboratory | Andersen lab at Scripps Research | SEARCH Alliance San Diego with Tracy Basler et al |
| EPI_ISL_494590 | USA/CA-ALSR-1416/2020 | 2020-04-21 | North America/USA/California/San Dieg | San Diego County Public Health Laboratory | Andersen lab at Scripps Research | SEARCH Alliance San Diego with Tracy Basler et al |
| EPI_ISL_494594 | USA/CA-ALSR-1425/2020 | 2020-04-20 | North America/USA/California/San Dieg | San Diego County Public Health Laboratory | Andersen lab at Scripps Research | SEARCH Alliance San Diego with Tracy Basler et al |
| EPI_ISL_494597 | USA/CA-ALSR-1430/2020 | 2020-03-28 | North America/USA/California/San Dieg | Scripps Medical Laboratory | Andersen lab at Scripps Research | SEARCH Alliance San Diego with Michael Quigley et al |
| EPI_ISL_494603 | USA/CA-ALSR-1437/2020 | 2020-03-27 | North America/USA/California/San Dieg | Scripps Medical Laboratory | Andersen lab at Scripps Research | SEARCH Alliance San Diego with Michael Quigley et al |
| EPI_ISL_494609 | USA/CA-ALSR-1448/2020 | 2020-03-26 | North America/USA/California/San Dieg | Scripps Medical Laboratory | Andersen lab at Scripps Research | SEARCH Alliance San Diego with Michael Quigley et al |
| EPI_ISL_494616 | USA/CA-ALSR-1460/2020 | 2020-03-20 | North America/USA/California/San Dieg | Scripps Medical Laboratory | Andersen lab at Scripps Research | SEARCH Alliance San Diego with Michael Quigley et al |
| EPI_ISL_494617 | USA/CA-ALSR-1461/2020 | 2020-03-23 | North America/USA/California/San Dieg | Scripps Medical Laboratory | Andersen lab at Scripps Research | SEARCH Alliance San Diego with Michael Quigley et al |
| EPI_ISL_494622 | USA/CA-ALSR-1497/2020 | 2020-04-01 | North America/USA/California/ | San Diego County Public Health Laboratory | Andersen lab at Scripps Research | SEARCH Alliance San Diego with Tracy Basler et al |

| | | | | | | |
|---|---|---|---|---|---|---|
| EPI_ISL_494628 | USA/CA-ALSR-1505/2020 | 2020-04-17 | North America/USA/California/ | San Diego County Public Health Laboratory | Andersen lab at Scripps Research | SEARCH Alliance San Diego with Tracy Basler et al |
| EPI_ISL_494629 | USA/CA-ALSR-1506/2020 | 2020-04-02 | North America/USA/California/ | San Diego County Public Health Laboratory | Andersen lab at Scripps Research | SEARCH Alliance San Diego with Tracy Basler et al |
| EPI_ISL_494630 | USA/CA-ALSR-1508/2020 | 2020-04-02 | North America/USA/California/ | San Diego County Public Health Laboratory | Andersen lab at Scripps Research | SEARCH Alliance San Diego with Tracy Basler et al |
| EPI_ISL_494632 | USA/CA-ALSR-1510/2020 | 2020-04-02 | North America/USA/California/ | San Diego County Public Health Laboratory | Andersen lab at Scripps Research | SEARCH Alliance San Diego with Tracy Basler et al |
| EPI_ISL_494634 | USA/CA-ALSR-1512/2020 | 2020-04-21 | North America/USA/California/San Dieg | San Diego County Public Health Laboratory | Andersen lab at Scripps Research | SEARCH Alliance San Diego with Tracy Basler et al |
| EPI_ISL_494641 | USA/CA-ALSR-1522/2020 | 2020-05-03 | North America/USA/California/San Dieg | Scripps Medical Laboratory | Andersen lab at Scripps Research | SEARCH Alliance San Diego with Michael Quigley et al |
| EPI_ISL_494642 | USA/CA-ALSR-1525/2020 | 2020-05-12 | North America/USA/California/San Dieg | Scripps Medical Laboratory | Andersen lab at Scripps Research | SEARCH Alliance San Diego with Michael Quigley et al |
| EPI_ISL_494651 | USA/CA-ALSR-1538/2020 | 2020-05-01 | North America/USA/California/San Dieg | Scripps Medical Laboratory | Andersen lab at Scripps Research | SEARCH Alliance San Diego with Michael Quigley et al |
| EPI_ISL_494659 | USA/CA-ALSR-1556/2020 | 2020-05-13 | North America/USA/California/San Dieg | Scripps Medical Laboratory | Andersen lab at Scripps Research | SEARCH Alliance San Diego with Michael Quigley et al |
| EPI_ISL_494660 | USA/CA-ALSR-1558/2020 | 2020-05-12 | North America/USA/California/San Dieg | Scripps Medical Laboratory | Andersen lab at Scripps Research | SEARCH Alliance San Diego with Michael Quigley et al |
| EPI_ISL_494661 | USA/CA-ALSR-1560/2020 | 2020-05-11 | North America/USA/California/San Dieg | Scripps Medical Laboratory | Andersen lab at Scripps Research | SEARCH Alliance San Diego with Michael Quigley et al |
| EPI_ISL_494663 | USA/CA-ALSR-1567/2020 | 2020-05-03 | North America/USA/California/San Dieg | Scripps Medical Laboratory | Andersen lab at Scripps Research | SEARCH Alliance San Diego with Michael Quigley et al |
| EPI_ISL_494665 | USA/CA-ALSR-1572/2020 | 2020-04-03 | North America/USA/California/San Dieg | Scripps Medical Laboratory | Andersen lab at Scripps Research | SEARCH Alliance San Diego with Michael Quigley et al |
| EPI_ISL_494673 | USA/CA-ALSR-1585/2020 | 2020-04-23 | North America/USA/California/San Dieg | Scripps Medical Laboratory | Andersen lab at Scripps Research | SEARCH Alliance San Diego with Michael Quigley et al |
| EPI_ISL_494675 | USA/CA-ALSR-1587/2020 | 2020-04-27 | North America/USA/California/San Dieg | Scripps Medical Laboratory | Andersen lab at Scripps Research | SEARCH Alliance San Diego with Michael Quigley et al |
| EPI_ISL_494685 | USA/CA-ALSR-1610/2020 | 2020-05-20 | North America/USA/California/San Dieg | Scripps Medical Laboratory | Andersen lab at Scripps Research | SEARCH Alliance San Diego with Michael Quigley et al |
| EPI_ISL_494704 | USA/CA-ALSR-1634/2020 | 2020-04-23 | North America/USA/California/San Dieg | Scripps Medical Laboratory | Andersen lab at Scripps Research | SEARCH Alliance San Diego with Michael Quigley et al |
| EPI_ISL_494708 | USA/CA-ALSR-1641/2020 | 2020-05-19 | North America/USA/California/San Dieg | Scripps Medical Laboratory | Andersen lab at Scripps Research | SEARCH Alliance San Diego with Michael Quigley et al |
| EPI_ISL_494710 | USA/CA-ALSR-1645/2020 | 2020-04-22 | North America/USA/California/San Dieg | Scripps Medical Laboratory | Andersen lab at Scripps Research | SEARCH Alliance San Diego with Michael Quigley et al |
| EPI_ISL_494714 | USA/CA-ALSR-1649/2020 | 2020-03-23 | North America/USA/California/San Dieg | San Diego County Public Health Laboratory | Andersen lab at Scripps Research | SEARCH Alliance San Diego with Tracy Basler et al |

| | | | | | | |
|---|---|---|---|---|---|---|
| EPI_ISL_494716 | USA/CA-ALSR-1651/2020 | 2020-03-25 | North America/USA/California/San Dieg | San Diego County Public Health Laboratory | Andersen lab at Scripps Research | SEARCH Alliance San Diego with Tracy Basler et al |
| EPI_ISL_494722 | USA/CA-ALSR-1661/2020 | 2020-04-02 | North America/USA/California/San Dieg | San Diego County Public Health Laboratory | Andersen lab at Scripps Research | SEARCH Alliance San Diego with Tracy Basler et al |
| EPI_ISL_494724 | USA/CA-ALSR-1663/2020 | 2020-04-02 | North America/USA/California/San Dieg | San Diego County Public Health Laboratory | Andersen lab at Scripps Research | SEARCH Alliance San Diego with Tracy Basler et al |
| EPI_ISL_494736 | USA/CA-ALSR-1676/2020 | 2020-04-01 | North America/USA/California/San Dieg | San Diego County Public Health Laboratory | Andersen lab at Scripps Research | SEARCH Alliance San Diego with Tracy Basler et al |
| EPI_ISL_494970 | Canada/LTRI-18/2020 | 2020-03-20 | North America/Canada/Ontario/ | Dr. Tony Mazzulli Microbiologist-in-Chief | Dr. Jeff Wrana, Senior Investigator | Jeff Wrana et al |
| EPI_ISL_495012 | Canada/LTRI-23/2020 | 2020-03-22 | North America/Canada/Ontario/ | Dr. Tony Mazzulli Microbiologist-in-Chief | Dr. Jeff Wrana, Senior Investigator | Jeff Wrana et al |
| EPI_ISL_495013 | Canada/LTRI-25/2020 | 2020-03-26 | North America/Canada/Ontario/ | Dr. Tony Mazzulli Microbiologist-in-Chief | Dr. Jeff Wrana, Senior Investigator | Jeff Wrana et al |
| EPI_ISL_495015 | India/GJ-GBRC278b/2020 | 2020-06-18 | Asia/India/Gujarat/Daskroi | B.J. Medical College and Civil hospital | Gujarat Biotechnology Research Centre | Zarna Patel et al |
| EPI_ISL_495016 | India/GJ-GBRC279/2020 | 2020-06-18 | Asia/India/Gujarat/Dholka | B.J. Medical College and Civil hospital | Gujarat Biotechnology Research Centre | Monika Gandhi et al |
| EPI_ISL_495018 | India/GJ-GBRC281/2020 | 2020-06-18 | Asia/India/Gujarat/Ahmedabad | B.J. Medical College and Civil hospital | Gujarat Biotechnology Research Centre | Maharshi Pandya et al |
| EPI_ISL_495020 | India/GJ-GBRC283/2020 | 2020-06-28 | Asia/India/Gujarat/Botad | Government Medical College, Bhavnagar | Gujarat Biotechnology Research Centre | Kairavi Desai et al |
| EPI_ISL_495022 | India/GJ-GBRC284b/2020 | 2020-06-30 | Asia/India/Gujarat/Bhavnagar | Government Medical College, Bhavnagar | Gujarat Biotechnology Research Centre | Shirish Patel et al |
| EPI_ISL_495023 | India/GJ-GBRC285/2020 | 2020-07-02 | Asia/India/Gujarat/Bhavnagar | Government Medical College, Bhavnagar | Gujarat Biotechnology Research Centre | Nitin Savaliya et al |
| EPI_ISL_495024 | India/GJ-GBRC286/2020 | 2020-07-03 | Asia/India/Gujarat/Ghogha | Government Medical College, Bhavnagar | Gujarat Biotechnology Research Centre | Raghawendra Kumar et al |
| EPI_ISL_495025 | India/GJ-GBRC287/2020 | 2020-07-04 | Asia/India/Gujarat/Mahuva | Government Medical College, Bhavnagar | Gujarat Biotechnology Research Centre | Dinesh Kumar et al |

| | | | | | | |
|---|---|---|---|---|---|---|
| EPI_ISL_495026 | India/GJ-GBRC289/2020 | 2020-06-05 | Asia/India/Gujarat/Vadodara | GMERS Medical College & Hospital, Gotri, Vadodara | Gujarat Biotechnology Research Centre | Komal Patel et al |
| EPI_ISL_495027 | India/GJ-GBRC290a/2020 | 2020-06-15 | Asia/India/Gujarat/Vadodara | GMERS Medical College & Hospital, Gotri, Vadodara | Gujarat Biotechnology Research Centre | Labdhi Pandya et al |
| EPI_ISL_495028 | India/GJ-GBRC290b/2020 | 2020-06-15 | Asia/India/Gujarat/Vadodara | GMERS Medical College & Hospital, Gotri, Vadodara | Gujarat Biotechnology Research Centre | Afzal Ansari et al |
| EPI_ISL_495029 | India/GJ-GBRC291/2020 | 2020-06-15 | Asia/India/Gujarat/Vadodara | GMERS Medical College & Hospital, Gotri, Vadodara | Gujarat Biotechnology Research Centre | Nikha Trivedi et al |
| EPI_ISL_495030 | India/GJ-GBRC292a/2020 | 2020-06-15 | Asia/India/Gujarat/Vadodara | GMERS Medical College & Hospital, Gotri, Vadodara | Gujarat Biotechnology Research Centre | Meenakshi Shah et al |
| EPI_ISL_495031 | India/GJ-GBRC292b/2020 | 2020-06-15 | Asia/India/Gujarat/Vadodara | GMERS Medical College & Hospital, Gotri, Vadodara | Gujarat Biotechnology Research Centre | Neena Doshi et al |
| EPI_ISL_495032 | India/GJ-GBRC293a/2020 | 2020-06-15 | Asia/India/Gujarat/Vadodara | GMERS Medical College & Hospital, Gotri, Vadodara | Gujarat Biotechnology Research Centre | Varsha Godbole et al |
| EPI_ISL_495034 | India/GJ-GBRC294/2020 | 2020-06-15 | Asia/India/Gujarat/Vadodara | GMERS Medical College & Hospital, Gotri, Vadodara | Gujarat Biotechnology Research Centre | Janvi Raval et al |
| EPI_ISL_495035 | India/GJ-GBRC295/2020 | 2020-06-15 | Asia/India/Gujarat/Vadodara | GMERS Medical College & Hospital, Gotri, Vadodara | Gujarat Biotechnology Research Centre | Zarna Patel et al |
| EPI_ISL_495036 | India/GJ-GBRC296a/2020 | 2020-06-15 | Asia/India/Gujarat/Vadodara | GMERS Medical College & Hospital, Gotri, Vadodara | Gujarat Biotechnology Research Centre | Monika Gandhi et al |
| EPI_ISL_495038 | India/GJ-GBRC297a/2020 | 2020-06-15 | Asia/India/Gujarat/Vadodara | GMERS Medical College & Hospital, Gotri, Vadodara | Gujarat Biotechnology Research Centre | Maharshi Pandya et al |
| EPI_ISL_495040 | India/GJ-GBRC298a/2020 | 2020-06-15 | Asia/India/Gujarat/Vadodara | GMERS Medical College & Hospital, Gotri, Vadodara | Gujarat Biotechnology Research Centre | Nitin Savaliya et al |
| EPI_ISL_495041 | India/GJ-GBRC298b/2020 | 2020-06-15 | Asia/India/Gujarat/Vadodara | GMERS Medical College & Hospital, Gotri, Vadodara | Gujarat Biotechnology Research Centre | Raghawendra Kumar et al |

| | | | | | | |
|---|---|---|---|---|---|---|
| EPI_ISL_495043 | India/GJ-GBRC299b/2020 | 2020-06-15 | Asia/India/Gujarat/Vadodara | GMERS Medical College & Hospital, Gotri, Vadodara | Gujarat Biotechnology Research Centre | Zuber Saiyed et al |
| EPI_ISL_495047 | India/GJ-GBRC301b/2020 | 2020-06-15 | Asia/India/Gujarat/Vadodara | GMERS Medical College & Hospital, Gotri, Vadodara | Gujarat Biotechnology Research Centre | Nikha Trivedi et al |
| EPI_ISL_495049 | India/GJ-GBRC302b/2020 | 2020-06-15 | Asia/India/Gujarat/Vadodara | GMERS Medical College & Hospital, Gotri, Vadodara | Gujarat Biotechnology Research Centre | Neena Doshi et al |
| EPI_ISL_495052 | India/GJ-GBRC304a/2020 | 2020-06-15 | Asia/India/Gujarat/Vadodara | GMERS Medical College & Hospital, Gotri, Vadodara | Gujarat Biotechnology Research Centre | Janvi Raval et al |
| EPI_ISL_495054 | India/GJ-GBRC305a/2020 | 2020-06-15 | Asia/India/Gujarat/Vadodara | GMERS Medical College & Hospital, Gotri, Vadodara | Gujarat Biotechnology Research Centre | Monika Gandhi et al |
| EPI_ISL_495055 | India/GJ-GBRC305b/2020 | 2020-06-15 | Asia/India/Gujarat/Vadodara | GMERS Medical College & Hospital, Gotri, Vadodara | Gujarat Biotechnology Research Centre | Pinal Trivedi et al |
| EPI_ISL_495056 | India/GJ-GBRC306a/2020 | 2020-06-15 | Asia/India/Gujarat/Vadodara | GMERS Medical College & Hospital, Gotri, Vadodara | Gujarat Biotechnology Research Centre | Maharshi Pandya et al |
| EPI_ISL_495057 | India/GJ-GBRC306b/2020 | 2020-06-15 | Asia/India/Gujarat/Vadodara | GMERS Medical College & Hospital, Gotri, Vadodara | Gujarat Biotechnology Research Centre | Nidhi Patel et al |
| EPI_ISL_495058 | India/GJ-GBRC307a/2020 | 2020-06-15 | Asia/India/Gujarat/Vadodara | GMERS Medical College & Hospital, Gotri, Vadodara | Gujarat Biotechnology Research Centre | Nitin Savaliya et al |
| EPI_ISL_495059 | India/GJ-GBRC307b/2020 | 2020-06-15 | Asia/India/Gujarat/Vadodara | GMERS Medical College & Hospital, Gotri, Vadodara | Gujarat Biotechnology Research Centre | Raghawendra Kumar et al |
| EPI_ISL_495060 | India/GJ-GBRC308/2020 | 2020-06-15 | Asia/India/Gujarat/Vadodara | GMERS Medical College & Hospital, Gotri, Vadodara | Gujarat Biotechnology Research Centre | Dinesh Kumar et al |
| EPI_ISL_495062 | India/GJ-GBRC309b/2020 | 2020-06-15 | Asia/India/Gujarat/Vadodara | GMERS Medical College & Hospital, Gotri, Vadodara | Gujarat Biotechnology Research Centre | Komal Patel et al |
| EPI_ISL_495063 | India/GJ-GBRC310a/2020 | 2020-06-19 | Asia/India/Gujarat/Prantij | GMERS Medical College & Hospital, Himmatnagar | Gujarat Biotechnology Research Centre | Labdhi Pandya et al |

| | | | | | | |
|---|---|---|---|---|---|---|
| EPI_ISL_495066 | India/GJ-GBRC311b/2020 | 2020-06-20 | Asia/India/Gujarat/Mandvi | GAIMS & G K General Hospital | Gujarat Biotechnology Research Centre | Babulal Babhoria et al |
| EPI_ISL_495067 | India/GJ-GBRC312a/2020 | 2020-06-08 | Asia/India/Gujarat/Kheda | Dr. N. D. Desai Medical College & Hospital | Gujarat Biotechnology Research Centre | J G Buch et al |
| EPI_ISL_495069 | India/GJ-GBRC313/2020 | 2020-06-08 | Asia/India/Gujarat/Nadiad | Dr. N. D. Desai Medical College & Hospital | Gujarat Biotechnology Research Centre | Supreet Prabhu et al |
| EPI_ISL_495072 | India/GJ-GBRC315b/2020 | 2020-06-17 | Asia/India/Gujarat/Kamrej | Department of MicroBiology, Government Medical College, Surat | Gujarat Biotechnology Research Centre | Zarna Patel et al |
| EPI_ISL_495073 | India/GJ-GBRC316a/2020 | 2020-06-17 | Asia/India/Gujarat/Surat | Department of MicroBiology, Government Medical College, Surat | Gujarat Biotechnology Research Centre | Monika Gandhi et al |
| EPI_ISL_495075 | India/GJ-GBRC318a/2020 | 2020-06-17 | Asia/India/Gujarat/Surat | Department of MicroBiology, Government Medical College, Surat | Gujarat Biotechnology Research Centre | Nidhi Patel et al |
| EPI_ISL_495076 | India/GJ-GBRC318b/2020 | 2020-06-17 | Asia/India/Gujarat/Surat | Department of MicroBiology, Government Medical College, Surat | Gujarat Biotechnology Research Centre | Nitin Savaliya et al |
| EPI_ISL_495077 | India/GJ-GBRC319/2020 | 2020-06-17 | Asia/India/Gujarat/Surat | Department of MicroBiology, Government Medical College, Surat | Gujarat Biotechnology Research Centre | Raghawendra Kumar et al |
| EPI_ISL_495078 | India/GJ-GBRC320/2020 | 2020-06-17 | Asia/India/Gujarat/Surat | Department of MicroBiology, Government Medical College, Surat | Gujarat Biotechnology Research Centre | Dinesh Kumar et al |
| EPI_ISL_495079 | India/GJ-GBRC323a/2020 | 2020-06-12 | Asia/India/Gujarat/Surat | Department of MicroBiology, Government Medical College, Surat | Gujarat Biotechnology Research Centre | Afzal Ansari et al |
| EPI_ISL_495080 | India/GJ-GBRC323b/2020 | 2020-06-12 | Asia/India/Gujarat/Surat | Department of MicroBiology, Government Medical College, Surat | Gujarat Biotechnology Research Centre | Nikha Trivedi et al |
| EPI_ISL_495125 | USA/CA-IGI-0103/2020 | 2020-05-20 | North America/USA/California/Alameda | Innovative Genomics Institute, UC Berkeley | Innovative Genomics Institute, UC Berkeley | Stacia Wyman et al |
| EPI_ISL_495128 | USA/CA-IGI-0106/2020 | 2020-06-10 | North America/USA/California/Alameda | Innovative Genomics Institute, UC Berkeley | Innovative Genomics Institute, UC Berkeley | Stacia Wyman et al |

| | | | | | | |
|---|---|---|---|---|---|---|
| EPI_ISL_495129 | USA/CA-IGI-0107/2020 | 2020-05-22 | North America/USA/California/Alameda | Innovative Genomics Institute, UC Berkeley | Innovative Genomics Institute, UC Berkeley | Stacia Wyman et al |
| EPI_ISL_495131 | USA/CA-IGI-0109/2020 | 2020-05-22 | North America/USA/California/Alameda | Innovative Genomics Institute, UC Berkeley | Innovative Genomics Institute, UC Berkeley | Stacia Wyman et al |
| EPI_ISL_495133 | USA/CA-IGI-0111/2020 | 2020-06-19 | North America/USA/California/Alameda | Innovative Genomics Institute, UC Berkeley | Innovative Genomics Institute, UC Berkeley | Stacia Wyman et al |
| EPI_ISL_495144 | USA/CA-IGI-0122/2020 | 2020-06-08 | North America/USA/California/Alameda | Innovative Genomics Institute, UC Berkeley | Innovative Genomics Institute, UC Berkeley | Stacia Wyman et al |
| EPI_ISL_495147 | USA/CA-IGI-0125/2020 | 2020-06-08 | North America/USA/California/Alameda | Innovative Genomics Institute, UC Berkeley | Innovative Genomics Institute, UC Berkeley | Stacia Wyman et al |
| EPI_ISL_495148 | USA/CA-IGI-0126/2020 | 2020-05-22 | North America/USA/California/Alameda | Innovative Genomics Institute, UC Berkeley | Innovative Genomics Institute, UC Berkeley | Stacia Wyman et al |
| EPI_ISL_495157 | USA/CA-IGI-0136/2020 | 2020-06-03 | North America/USA/California/Alameda | Innovative Genomics Institute, UC Berkeley | Innovative Genomics Institute, UC Berkeley | Stacia Wyman et al |
| EPI_ISL_495162 | India/TG-CCMB-L1023/2020 | 2020-05-11 | Asia/India/Telangana/Hyderabad | CSIR-Centre for Cellular and Molecular Biology | CSIR-Centre for Cellular and Molecular Biology | Onkar Kulkarni et al |
| EPI_ISL_495167 | India/TG-CCMB-L1885/2020 | 2020-06-03 | Asia/India/Telangana/Hyderabad | CSIR-Centre for Cellular and Molecular Biology | CSIR-Centre for Cellular and Molecular Biology | Onkar Kulkarni et al |
| EPI_ISL_495168 | India/TG-CCMB-L1892/2020 | 2020-06-03 | Asia/India/Telangana/Hyderabad | CSIR-Centre for Cellular and Molecular Biology | CSIR-Centre for Cellular and Molecular Biology | Sofia Banu et al |
| EPI_ISL_495169 | India/TG-CCMB-L1893/2020 | 2020-06-03 | Asia/India/Telangana/Hyderabad | CSIR-Centre for Cellular and Molecular Biology | CSIR-Centre for Cellular and Molecular Biology | Onkar Kulkarni et al |

| | | | | | | |
|---|---|---|---|---|---|---|
| EPI_ISL_495170 | India/TG-CCMB-L1930/2020 | 2020-06-04 | Asia/India/Telangana/Hyderabad | CSIR-Centre for Cellular and Molecular Biology | CSIR-Centre for Cellular and Molecular Biology | Onkar Kulkarni et al |
| EPI_ISL_495172 | India/TG-CCMB-L1934/2020 | 2020-06-04 | Asia/India/Telangana/Hyderabad | CSIR-Centre for Cellular and Molecular Biology | CSIR-Centre for Cellular and Molecular Biology | Onkar Kulkarni et al |
| EPI_ISL_495173 | India/TG-CCMB-L1935/2020 | 2020-06-04 | Asia/India/Telangana/Hyderabad | CSIR-Centre for Cellular and Molecular Biology | CSIR-Centre for Cellular and Molecular Biology | Onkar Kulkarni et al |
| EPI_ISL_495174 | India/TG-CCMB-L1959/2020 | 2020-06-04 | Asia/India/Telangana/Hyderabad | CSIR-Centre for Cellular and Molecular Biology | CSIR-Centre for Cellular and Molecular Biology | M Soujanya Reddy et al |
| EPI_ISL_495175 | India/TG-CCMB-L1963/2020 | 2020-06-04 | Asia/India/Telangana/Hyderabad | CSIR-Centre for Cellular and Molecular Biology | CSIR-Centre for Cellular and Molecular Biology | Onkar Kulkarni et al |
| EPI_ISL_495177 | India/TG-CCMB-L1967/2020 | 2020-06-04 | Asia/India/Telangana/Hyderabad | CSIR-Centre for Cellular and Molecular Biology | CSIR-Centre for Cellular and Molecular Biology | Sakshi Shambhavi et al |
| EPI_ISL_495178 | India/TG-CCMB-L1973/2020 | 2020-06-04 | Asia/India/Telangana/Hyderabad | CSIR-Centre for Cellular and Molecular Biology | CSIR-Centre for Cellular and Molecular Biology | Payel Mukherjee et al |
| EPI_ISL_495179 | India/TG-CCMB-L1975/2020 | 2020-06-04 | Asia/India/Telangana/Hyderabad | CSIR-Centre for Cellular and Molecular Biology | CSIR-Centre for Cellular and Molecular Biology | Pratheusa Maccha et al |
| EPI_ISL_495182 | India/TG-CCMB-M116/2020 | 2020-06-06 | Asia/India/Telangana/Hyderabad | CSIR-Centre for Cellular and Molecular Biology | CSIR-Centre for Cellular and Molecular Biology | Sakshi Shambhavi et al |
| EPI_ISL_495183 | India/TG-CCMB-M118/2020 | 2020-06-06 | Asia/India/Telangana/Hyderabad | CSIR-Centre for Cellular and Molecular Biology | CSIR-Centre for Cellular and Molecular Biology | Namami Gaur et al |
| EPI_ISL_495184 | India/TG-CCMB-M135/2020 | 2020-06-06 | Asia/India/Telangana/Hyderabad | CSIR-Centre for Cellular and Molecular Biology | CSIR-Centre for Cellular and Molecular Biology | Namami Gaur et al |
| EPI_ISL_495185 | India/TG-CCMB-M14/2020 | 2020-06-04 | Asia/India/Telangana/Hyderabad | CSIR-Centre for Cellular and Molecular Biology | CSIR-Centre for Cellular and Molecular Biology | Shagufta Khan et al |
| EPI_ISL_495187 | India/TG-CCMB-M152/2020 | 2020-06-06 | Asia/India/Telangana/Hyderabad | CSIR-Centre for Cellular and Molecular Biology | CSIR-Centre for Cellular and Molecular Biology | Shagufta Khan et al |

| | | | | | | |
|---|---|---|---|---|---|---|
| EPI_ISL_495190 | India/TG-CCMB-M175/2020 | 2020-06-06 | Asia/India/Telangana/Hyderabad | CSIR-Centre for Cellular and Molecular Biology | CSIR-Centre for Cellular and Molecular Biology | Sakshi Shambhavi et al |
| EPI_ISL_495191 | India/TG-CCMB-M181/2020 | 2020-06-07 | Asia/India/Telangana/Hyderabad | CSIR-Centre for Cellular and Molecular Biology | CSIR-Centre for Cellular and Molecular Biology | Nikhil Hajirnis et al |
| EPI_ISL_495192 | India/TG-CCMB-M2/2020 | 2020-06-04 | Asia/India/Telangana/Hyderabad | CSIR-Centre for Cellular and Molecular Biology | CSIR-Centre for Cellular and Molecular Biology | Sofia Banu et al |
| EPI_ISL_495193 | India/TG-CCMB-M200/2020 | 2020-06-07 | Asia/India/Telangana/Hyderabad | CSIR-Centre for Cellular and Molecular Biology | CSIR-Centre for Cellular and Molecular Biology | Namami Gaur et al |
| EPI_ISL_495196 | India/TG-CCMB-M25/2020 | 2020-06-04 | Asia/India/Telangana/Hyderabad | CSIR-Centre for Cellular and Molecular Biology | CSIR-Centre for Cellular and Molecular Biology | Payel Mukherjee et al |
| EPI_ISL_495197 | India/TG-CCMB-M288/2020 | 2020-06-08 | Asia/India/Telangana/Hyderabad | CSIR-Centre for Cellular and Molecular Biology | CSIR-Centre for Cellular and Molecular Biology | Sofia Banu et al |
| EPI_ISL_495198 | India/TG-CCMB-M343/2020 | 2020-06-08 | Asia/India/Telangana/Hyderabad | CSIR-Centre for Cellular and Molecular Biology | CSIR-Centre for Cellular and Molecular Biology | Tulasi Nagabandi et al |
| EPI_ISL_495200 | India/TG-CCMB-M359/2020 | 2020-06-09 | Asia/India/Telangana/Hyderabad | CSIR-Centre for Cellular and Molecular Biology | CSIR-Centre for Cellular and Molecular Biology | Lamuk Zaveri et al |
| EPI_ISL_495203 | India/TG-CCMB-M437/2020 | 2020-06-09 | Asia/India/Telangana/Hyderabad | CSIR-Centre for Cellular and Molecular Biology | CSIR-Centre for Cellular and Molecular Biology | Tulasi Nagabandi et al |
| EPI_ISL_495204 | India/TG-CCMB-M441/2020 | 2020-06-09 | Asia/India/Telangana/Hyderabad | CSIR-Centre for Cellular and Molecular Biology | CSIR-Centre for Cellular and Molecular Biology | Pratheusa Maccha et al |
| EPI_ISL_495205 | India/TG-CCMB-M446/2020 | 2020-06-09 | Asia/India/Telangana/Hyderabad | CSIR-Centre for Cellular and Molecular Biology | CSIR-Centre for Cellular and Molecular Biology | Shagufta Khan et al |
| EPI_ISL_495207 | India/TG-CCMB-M456/2020 | 2020-06-09 | Asia/India/Telangana/Hyderabad | CSIR-Centre for Cellular and Molecular Biology | CSIR-Centre for Cellular and Molecular Biology | Namami Gaur et al |
| EPI_ISL_495208 | India/TG-CCMB-M458/2020 | 2020-06-09 | Asia/India/Telangana/Hyderabad | CSIR-Centre for Cellular and Molecular Biology | CSIR-Centre for Cellular and Molecular Biology | Shagufta Khan et al |

| | | | | | | |
|---|---|---|---|---|---|---|
| EPI_ISL_495210 | India/TG-CCMB-M466/2020 | 2020-06-09 | Asia/India/Telangana/Hyderabad | CSIR-Centre for Cellular and Molecular Biology | CSIR-Centre for Cellular and Molecular Biology | Sofia Banu et al |
| EPI_ISL_495211 | India/TG-CCMB-M471/2020 | 2020-06-09 | Asia/India/Telangana/Hyderabad | CSIR-Centre for Cellular and Molecular Biology | CSIR-Centre for Cellular and Molecular Biology | Shagufta Khan et al |
| EPI_ISL_495213 | India/TG-CCMB-M499/2020 | 2020-06-10 | Asia/India/Telangana/Hyderabad | CSIR-Centre for Cellular and Molecular Biology | CSIR-Centre for Cellular and Molecular Biology | Sakshi Shambhavi et al |
| EPI_ISL_495214 | India/TG-CCMB-M5/2020 | 2020-06-04 | Asia/India/Telangana/Hyderabad | CSIR-Centre for Cellular and Molecular Biology | CSIR-Centre for Cellular and Molecular Biology | Tulasi Nagabandi et al |
| EPI_ISL_495215 | India/TG-CCMB-M58/2020 | 2020-06-05 | Asia/India/Telangana/Hyderabad | CSIR-Centre for Cellular and Molecular Biology | CSIR-Centre for Cellular and Molecular Biology | Lamuk Zaveri et al |
| EPI_ISL_495217 | India/TG-CCMB-M588/2020 | 2020-06-10 | Asia/India/Telangana/Hyderabad | CSIR-Centre for Cellular and Molecular Biology | CSIR-Centre for Cellular and Molecular Biology | Nikhil Hajirnis et al |
| EPI_ISL_495218 | India/TG-CCMB-M64/2020 | 2020-06-05 | Asia/India/Telangana/Hyderabad | CSIR-Centre for Cellular and Molecular Biology | CSIR-Centre for Cellular and Molecular Biology | Sakshi Shambhavi et al |
| EPI_ISL_495219 | India/TG-CCMB-M651/2020 | 2020-06-11 | Asia/India/Telangana/Hyderabad | CSIR-Centre for Cellular and Molecular Biology | CSIR-Centre for Cellular and Molecular Biology | Sofia Banu et al |
| EPI_ISL_495220 | India/TG-CCMB-M652/2020 | 2020-06-11 | Asia/India/Telangana/Hyderabad | CSIR-Centre for Cellular and Molecular Biology | CSIR-Centre for Cellular and Molecular Biology | Shagufta Khan et al |
| EPI_ISL_495229 | India/TG-CCMB-M711/2020 | 2020-06-11 | Asia/India/Telangana/Hyderabad | CSIR-Centre for Cellular and Molecular Biology | CSIR-Centre for Cellular and Molecular Biology | Namami Gaur et al |
| EPI_ISL_495234 | India/TG-CCMB-M739/2020 | 2020-06-11 | Asia/India/Telangana/Hyderabad | CSIR-Centre for Cellular and Molecular Biology | CSIR-Centre for Cellular and Molecular Biology | Lamuk Zaveri et al |
| EPI_ISL_495235 | India/TG-CCMB-M740/2020 | 2020-06-11 | Asia/India/Telangana/Hyderabad | CSIR-Centre for Cellular and Molecular Biology | CSIR-Centre for Cellular and Molecular Biology | Tulasi Nagabandi et al |
| EPI_ISL_495236 | India/TG-CCMB-M750/2020 | 2020-06-12 | Asia/India/Telangana/Hyderabad | CSIR-Centre for Cellular and Molecular Biology | CSIR-Centre for Cellular and Molecular Biology | Nikhil Hajirnis et al |

| | | | | | | |
|---|---|---|---|---|---|---|
| EPI_ISL_495237 | India/TG-CCMB-M762/2020 | 2020-06-12 | Asia/India/Telangana/Hyderabad | CSIR-Centre for Cellular and Molecular Biology | CSIR-Centre for Cellular and Molecular Biology | Payel Mukherjee et al |
| EPI_ISL_495239 | India/TG-CCMB-M779/2020 | 2020-06-12 | Asia/India/Telangana/Hyderabad | CSIR-Centre for Cellular and Molecular Biology | CSIR-Centre for Cellular and Molecular Biology | Nikhil Hajirnis et al |
| EPI_ISL_495240 | India/TG-CCMB-M780/2020 | 2020-06-12 | Asia/India/Telangana/Hyderabad | CSIR-Centre for Cellular and Molecular Biology | CSIR-Centre for Cellular and Molecular Biology | M Soujanya Reddy et al |
| EPI_ISL_495241 | India/TG-CCMB-M785/2020 | 2020-06-12 | Asia/India/Telangana/Hyderabad | CSIR-Centre for Cellular and Molecular Biology | CSIR-Centre for Cellular and Molecular Biology | Shagufta Khan et al |
| EPI_ISL_495242 | India/TG-CCMB-M79/2020 | 2020-06-05 | Asia/India/Telangana/Hyderabad | CSIR-Centre for Cellular and Molecular Biology | CSIR-Centre for Cellular and Molecular Biology | Tulasi Nagabandi et al |
| EPI_ISL_495243 | India/TG-CCMB-M80/2020 | 2020-06-05 | Asia/India/Telangana/Hyderabad | CSIR-Centre for Cellular and Molecular Biology | CSIR-Centre for Cellular and Molecular Biology | Lamuk Zaveri et al |
| EPI_ISL_495244 | India/TG-CCMB-M804/2020 | 2020-06-12 | Asia/India/Telangana/Hyderabad | CSIR-Centre for Cellular and Molecular Biology | CSIR-Centre for Cellular and Molecular Biology | Payel Mukherjee et al |
| EPI_ISL_495245 | India/TG-CCMB-M806/2020 | 2020-06-12 | Asia/India/Telangana/Hyderabad | CSIR-Centre for Cellular and Molecular Biology | CSIR-Centre for Cellular and Molecular Biology | Payel Mukherjee et al |
| EPI_ISL_495248 | India/TG-CCMB-M817/2020 | 2020-06-12 | Asia/India/Telangana/Hyderabad | CSIR-Centre for Cellular and Molecular Biology | CSIR-Centre for Cellular and Molecular Biology | Sakshi Shambhavi et al |
| EPI_ISL_495249 | India/TG-CCMB-M818/2020 | 2020-06-12 | Asia/India/Telangana/Hyderabad | CSIR-Centre for Cellular and Molecular Biology | CSIR-Centre for Cellular and Molecular Biology | Payel Mukherjee et al |
| EPI_ISL_495251 | India/TG-CCMB-M830/2020 | 2020-06-12 | Asia/India/Telangana/Hyderabad | CSIR-Centre for Cellular and Molecular Biology | CSIR-Centre for Cellular and Molecular Biology | Pratheusa Maccha et al |
| EPI_ISL_495253 | India/TG-CCMB-M840/2020 | 2020-06-12 | Asia/India/Telangana/Hyderabad | CSIR-Centre for Cellular and Molecular Biology | CSIR-Centre for Cellular and Molecular Biology | M Soujanya Reddy et al |
| EPI_ISL_495256 | India/TG-CCMB-M844/2020 | 2020-06-12 | Asia/India/Telangana/Hyderabad | CSIR-Centre for Cellular and Molecular Biology | CSIR-Centre for Cellular and Molecular Biology | Sakshi Shambhavi et al |

| | | | | | | |
|---|---|---|---|---|---|---|
| EPI_ISL_495257 | India/TG-CCMB-M845/2020 | 2020-06-12 | Asia/India/Telangana/Hyderabad | CSIR-Centre for Cellular and Molecular Biology | CSIR-Centre for Cellular and Molecular Biology | M Soujanya Reddy et al |
| EPI_ISL_495262 | India/TG-CCMB-M900/2020 | 2020-06-13 | Asia/India/Telangana/Hyderabad | CSIR-Centre for Cellular and Molecular Biology | CSIR-Centre for Cellular and Molecular Biology | Sofia Banu et al |
| EPI_ISL_495263 | India/TG-CCMB-M925/2020 | 2020-06-13 | Asia/India/Telangana/Hyderabad | CSIR-Centre for Cellular and Molecular Biology | CSIR-Centre for Cellular and Molecular Biology | Tulasi Nagabandi et al |
| EPI_ISL_495265 | India/TG-CCMB-M931/2020 | 2020-06-13 | Asia/India/Telangana/Hyderabad | CSIR-Centre for Cellular and Molecular Biology | CSIR-Centre for Cellular and Molecular Biology | Lamuk Zaveri et al |
| EPI_ISL_495266 | India/TG-CCMB-M950/2020 | 2020-06-13 | Asia/India/Telangana/Hyderabad | CSIR-Centre for Cellular and Molecular Biology | CSIR-Centre for Cellular and Molecular Biology | Namami Gaur et al |
| EPI_ISL_495268 | India/TG-CCMB-M971/2020 | 2020-06-13 | Asia/India/Telangana/Hyderabad | CSIR-Centre for Cellular and Molecular Biology | CSIR-Centre for Cellular and Molecular Biology | Tulasi Nagabandi et al |
| EPI_ISL_495272 | India/TG-CCMB-N4/2020 | 2020-06-13 | Asia/India/Telangana/Hyderabad | CSIR-Centre for Cellular and Molecular Biology | CSIR-Centre for Cellular and Molecular Biology | M Soujanya Reddy et al |
| EPI_ISL_495273 | India/TG-CCMB-N7/2020 | 2020-06-13 | Asia/India/Telangana/Hyderabad | CSIR-Centre for Cellular and Molecular Biology | CSIR-Centre for Cellular and Molecular Biology | Pratheusa Maccha et al |
| EPI_ISL_495274 | India/TG-CCMB-OM1/2020 | 2020-05-13 | Asia/India/Telangana/Hyderabad | Osmania Medical College | CSIR-Centre for Cellular and Molecular Biology | Shashikala Reddy et al |
| EPI_ISL_495275 | India/TG-CCMB-OM1-ONT/2020 | 2020-05-13 | Asia/India/Telangana/Hyderabad | Osmania Medical College | CSIR-Centre for Cellular and Molecular Biology | Shashikala Reddy et al |
| EPI_ISL_495276 | India/TG-CCMB-OM10/2020 | 2020-05-15 | Asia/India/Telangana/Hyderabad | Osmania Medical College | CSIR-Centre for Cellular and Molecular Biology | Shashikala Reddy et al |
| EPI_ISL_495277 | India/TG-CCMB-OM11/2020 | 2020-05-15 | Asia/India/Telangana/Hyderabad | Osmania Medical College | CSIR-Centre for Cellular and Molecular Biology | Shashikala Reddy et al |
| EPI_ISL_495284 | India/TG-CCMB-OM18/2020 | 2020-05-17 | Asia/India/Telangana/Hyderabad | Osmania Medical College | CSIR-Centre for Cellular and Molecular Biology | Shashikala Reddy et al |

| | | | | | | |
|---|---|---|---|---|---|---|
| EPI_ISL_495285 | India/TG-CCMB-OM19/2020 | 2020-05-17 | Asia/India/Telangana/Hyderabad | Osmania Medical College | CSIR-Centre for Cellular and Molecular Biology | Shashikala Reddy et al |
| EPI_ISL_495295 | India/TG-CCMB-OM7/2020 | 2020-05-14 | Asia/India/Telangana/Hyderabad | Osmania Medical College | CSIR-Centre for Cellular and Molecular Biology | Shashikala Reddy et al |
| EPI_ISL_495304 | USA/VA-DCLS-0631/2020 | 2020-04-09 | North America/USA/Virginia/ | Virginia DCLS | Virginia DCLS | Virginia DCLS et al |
| EPI_ISL_495308 | USA/VA-DCLS-0636/2020 | 2020-04-11 | North America/USA/Virginia/ | Virginia DCLS | Virginia DCLS | Virginia DCLS et al |
| EPI_ISL_495325 | USA/VA-DCLS-0653/2020 | 2020-04-11 | North America/USA/Virginia/ | Virginia DCLS | Virginia DCLS | Virginia DCLS et al |
| EPI_ISL_495328 | USA/VA-DCLS-0657/2020 | 2020-04-11 | North America/USA/Virginia/ | Virginia DCLS | Virginia DCLS | Virginia DCLS et al |
| EPI_ISL_495351 | USA/FL-BPHL-0445/2020 | 2020-03-23 | North America/USA/Florida/ | Florida Bureau of Public Health Laboratories | Florida Bureau of Public Health Laboratories | Sarah Schmedes et al |
| EPI_ISL_495353 | USA/FL-BPHL-0447/2020 | 2020-03-23 | North America/USA/Florida/ | Florida Bureau of Public Health Laboratories | Florida Bureau of Public Health Laboratories | Sarah Schmedes et al |
| EPI_ISL_495369 | USA/FL-BPHL-0463/2020 | 2020-03-24 | North America/USA/Florida/ | Florida Bureau of Public Health Laboratories | Florida Bureau of Public Health Laboratories | Sarah Schmedes et al |
| EPI_ISL_495387 | USA/FL-BPHL-0481/2020 | 2020-05-21 | North America/USA/Florida/ | Florida Bureau of Public Health Laboratories | Florida Bureau of Public Health Laboratories | Sarah Schmedes et al |
| EPI_ISL_495401 | USA/FL-BPHL-0495/2020 | 2020-03-25 | North America/USA/Florida/ | Florida Bureau of Public Health Laboratories | Florida Bureau of Public Health Laboratories | Sarah Schmedes et al |
| EPI_ISL_495404 | USA/FL-BPHL-0498/2020 | 2020-03-25 | North America/USA/Florida/ | Florida Bureau of Public Health Laboratories | Florida Bureau of Public Health Laboratories | Sarah Schmedes et al |
| EPI_ISL_495411 | Turkey/KU-001/2020 | 2020-05-19 | Europe/Turkey/Kars Province/ | Kafkas University, Faculty of Medicine, Department of Medical Microbiology | Kafkas University, Faculty of Medicine, Department of Medical Microbiology | Murat Karamese et al |

| | | | | | | |
|---|---|---|---|---|---|---|
| EPI_ISL_495415 | Turkey/KU-005/2020 | 2020-05-28 | Europe/Turkey/Kars Province/ | Kafkas University, Faculty of Medicine, Department of Medical Microbiology | Kafkas University, Faculty of Medicine, Department of Medical Microbiology | Murat Karamese et al |
| EPI_ISL_495416 | Turkey/KU-006/2020 | 2020-05-22 | Europe/Turkey/Kars Province/ | Kafkas University, Faculty of Medicine, Department of Medical Microbiology | Kafkas University, Faculty of Medicine, Department of Medical Microbiology | Murat Karamese et al |
| EPI_ISL_495418 | Turkey/KU-008/2020 | 2020-05-16 | Europe/Turkey/Kars Province/ | Kafkas University, Faculty of Medicine, Department of Medical Microbiology | Kafkas University, Faculty of Medicine, Department of Medical Microbiology | Murat Karamese et al |
| EPI_ISL_495419 | Turkey/KU-009/2020 | 2020-04-29 | Europe/Turkey/Kars Province/ | Kafkas University, Faculty of Medicine, Department of Medical Microbiology | Kafkas University, Faculty of Medicine, Department of Medical Microbiology | Murat Karamese et al |
| EPI_ISL_495425 | Turkey/KU-015/2020 | 2020-05-27 | Europe/Turkey/Kars Province/ | Kafkas University, Faculty of Medicine, Department of Medical Microbiology | Kafkas University, Faculty of Medicine, Department of Medical Microbiology | Murat Karamese et al |
| EPI_ISL_495426 | Turkey/KU-016/2020 | 2020-05-12 | Europe/Turkey/Kars Province/ | Kafkas University, Faculty of Medicine, Department of Medical Microbiology | Kafkas University, Faculty of Medicine, Department of Medical Microbiology | Murat Karamese et al |

| | | | | | | |
|---|---|---|---|---|---|---|
| EPI_ISL_495427 | Turkey/KU-017/2020 | 2020-05-18 | Europe/Turkey/Kars Province/ | Kafkas University, Faculty of Medicine, Department of Medical Microbiology | Kafkas University, Faculty of Medicine, Department of Medical Microbiology | Murat Karamese et al |
| EPI_ISL_495428 | Turkey/KU-018/2020 | 2020-04-29 | Europe/Turkey/Kars Province/ | Kafkas University, Faculty of Medicine, Department of Medical Microbiology | Kafkas University, Faculty of Medicine, Department of Medical Microbiology | Murat Karamese et al |
| EPI_ISL_495429 | Turkey/KU-019/2020 | 2020-06-05 | Europe/Turkey/Kars Province/ | Kafkas University, Faculty of Medicine, Department of Medical Microbiology | Kafkas University, Faculty of Medicine, Department of Medical Microbiology | Murat Karamese et al |
| EPI_ISL_495432 | Turkey/KU-022/2020 | 2020-04-27 | Europe/Turkey/Kars Province/ | Kafkas University, Faculty of Medicine, Department of Medical Microbiology | Kafkas University, Faculty of Medicine, Department of Medical Microbiology | Murat Karamese et al |
| EPI_ISL_495433 | Turkey/KU-023/2020 | 2020-06-06 | Europe/Turkey/Kars Province/ | Kafkas University, Faculty of Medicine, Department of Medical Microbiology | Kafkas University, Faculty of Medicine, Department of Medical Microbiology | Murat Karamese et al |
| EPI_ISL_495435 | Turkey/KU-025/2020 | 2020-05-05 | Europe/Turkey/Kars Province/ | Kafkas University, Faculty of Medicine, Department of Medical Microbiology | Kafkas University, Faculty of Medicine, Department of Medical Microbiology | Murat Karamese et al |

| | | | | | | |
|---|---|---|---|---|---|---|
| EPI_ISL_495436 | Turkey/KU-026/2020 | 2020-06-03 | Europe/Turkey/Kars Province/ | Kafkas University, Faculty of Medicine, Department of Medical Microbiology | Kafkas University, Faculty of Medicine, Department of Medical Microbiology | Murat Karamese et al |
| EPI_ISL_495438 | Turkey/KU-028/2020 | 2020-05-28 | Europe/Turkey/Kars Province/ | Kafkas University, Faculty of Medicine, Department of Medical Microbiology | Kafkas University, Faculty of Medicine, Department of Medical Microbiology | Murat Karamese et al |
| EPI_ISL_495439 | Turkey/KU-029/2020 | 2020-05-08 | Europe/Turkey/Kars Province/ | Kafkas University, Faculty of Medicine, Department of Medical Microbiology | Kafkas University, Faculty of Medicine, Department of Medical Microbiology | Murat Karamese et al |
| EPI_ISL_495441 | Turkey/KU-031/2020 | 2020-04-28 | Europe/Turkey/Kars Province/ | Kafkas University, Faculty of Medicine, Department of Medical Microbiology | Kafkas University, Faculty of Medicine, Department of Medical Microbiology | Murat Karamese et al |
| EPI_ISL_495443 | Turkey/KU-033/2020 | 2020-04-29 | Europe/Turkey/Kars Province/ | Kafkas University, Faculty of Medicine, Department of Medical Microbiology | Kafkas University, Faculty of Medicine, Department of Medical Microbiology | Murat Karamese et al |
| EPI_ISL_495444 | Turkey/KU-034/2020 | 2020-05-20 | Europe/Turkey/Kars Province/ | Kafkas University, Faculty of Medicine, Department of Medical Microbiology | Kafkas University, Faculty of Medicine, Department of Medical Microbiology | Murat Karamese et al |

| | | | | | | |
|---|---|---|---|---|---|---|
| EPI_ISL_495445 | Turkey/KU-035/2020 | 2020-06-17 | Europe/Turkey/Kars Province/ | Kafkas University, Faculty of Medicine, Department of Medical Microbiology | Kafkas University, Faculty of Medicine, Department of Medical Microbiology | Murat Karamese et al |
| EPI_ISL_495448 | Turkey/KU-038/2020 | 2020-04-25 | Europe/Turkey/Kars Province/ | Kafkas University, Faculty of Medicine, Department of Medical Microbiology | Kafkas University, Faculty of Medicine, Department of Medical Microbiology | Murat Karamese et al |
| EPI_ISL_495449 | Turkey/KU-039/2020 | 2020-05-26 | Europe/Turkey/Kars Province/ | Kafkas University, Faculty of Medicine, Department of Medical Microbiology | Kafkas University, Faculty of Medicine, Department of Medical Microbiology | Murat Karamese et al |
| EPI_ISL_495450 | Turkey/KU-040/2020 | 2020-05-24 | Europe/Turkey/Kars Province/ | Kafkas University, Faculty of Medicine, Department of Medical Microbiology | Kafkas University, Faculty of Medicine, Department of Medical Microbiology | Murat Karamese et al |
| EPI_ISL_495453 | Turkey/KU-043/2020 | 2020-05-23 | Europe/Turkey/Kars Province/ | Kafkas University, Faculty of Medicine, Department of Medical Microbiology | Kafkas University, Faculty of Medicine, Department of Medical Microbiology | Murat Karamese et al |
| EPI_ISL_495455 | Turkey/KU-045/2020 | 2020-05-10 | Europe/Turkey/Kars Province/ | Kafkas University, Faculty of Medicine, Department of Medical Microbiology | Kafkas University, Faculty of Medicine, Department of Medical Microbiology | Murat Karamese et al |

| | | | | | | |
|---|---|---|---|---|---|---|
| EPI_ISL_495456 | Turkey/KU-046/2020 | 2020-05-26 | Europe/Turkey/Kars Province/ | Kafkas University, Faculty of Medicine, Department of Medical Microbiology | Kafkas University, Faculty of Medicine, Department of Medical Microbiology | Murat Karamese et al |
| EPI_ISL_495457 | Turkey/KU-047/2020 | 2020-05-07 | Europe/Turkey/Kars Province/ | Kafkas University, Faculty of Medicine, Department of Medical Microbiology | Kafkas University, Faculty of Medicine, Department of Medical Microbiology | Murat Karamese et al |
| EPI_ISL_495459 | Lishui/LS003/2020 | 2020-01-24 | Asia/China/Zhejiang/Lishui | Centers for Disease Control and Prevention of Lishui | Department of InspectionÔºåCenters for Disease Control and Prevention of Lishui | Wang Xiaoguang et al |
| EPI_ISL_495462 | USA/WI-UW-757/2020 | 2020-07-04 | North America/USA/Wisconsin/Dane Co | University of Wisconsin-Madison AIDS Vaccine Research Laboratories | University of Wisconsin-Madison AIDS Vaccine Research Laboratories | Gage Moreno et al |
| EPI_ISL_495475 | USA/WI-UW-770/2020 | 2020-07-07 | North America/USA/Wisconsin/Dane Co | University of Wisconsin-Madison AIDS Vaccine Research Laboratories | University of Wisconsin-Madison AIDS Vaccine Research Laboratories | Gage Moreno et al |
| EPI_ISL_495479 | USA/WI-UW-775/2020 | 2020-07-03 | North America/USA/Wisconsin/Dane Co | University of Wisconsin-Madison AIDS Vaccine Research Laboratories | University of Wisconsin-Madison AIDS Vaccine Research Laboratories | Gage Moreno et al |
| EPI_ISL_495480 | USA/WI-UW-776/2020 | 2020-07-04 | North America/USA/Wisconsin/Dane Co | University of Wisconsin-Madison AIDS Vaccine Research Laboratories | University of Wisconsin-Madison AIDS Vaccine Research Laboratories | Gage Moreno et al |

| | | | | | | |
|---|---|---|---|---|---|---|
| EPI_ISL_495483 | USA/WI-UW-779/2020 | 2020-07-03 | North America/USA/Wisconsin/Dane Co | University of Wisconsin-Madison AIDS Vaccine Research Laboratories | University of Wisconsin-Madison AIDS Vaccine Research Laboratories | Gage Moreno et al |
| EPI_ISL_495487 | USA/WI-UW-783/2020 | 2020-07-03 | North America/USA/Wisconsin/Dane Co | University of Wisconsin-Madison AIDS Vaccine Research Laboratories | University of Wisconsin-Madison AIDS Vaccine Research Laboratories | Gage Moreno et al |
| EPI_ISL_495488 | USA/WI-UW-784/2020 | 2020-07-06 | North America/USA/Wisconsin/Dane Co | University of Wisconsin-Madison AIDS Vaccine Research Laboratories | University of Wisconsin-Madison AIDS Vaccine Research Laboratories | Gage Moreno et al |
| EPI_ISL_495491 | USA/WI-UW-787/2020 | 2020-07-06 | North America/USA/Wisconsin/Rock Cou | University of Wisconsin-Madison AIDS Vaccine Research Laboratories | University of Wisconsin-Madison AIDS Vaccine Research Laboratories | Gage Moreno et al |
| EPI_ISL_495493 | USA/WI-UW-789/2020 | 2020-07-04 | North America/USA/Wisconsin/Milwauk | University of Wisconsin-Madison AIDS Vaccine Research Laboratories | University of Wisconsin-Madison AIDS Vaccine Research Laboratories | Gage Moreno et al |
| EPI_ISL_495496 | USA/WI-UW-792/2020 | 2020-07-05 | North America/USA/Wisconsin/Dane Co | University of Wisconsin-Madison AIDS Vaccine Research Laboratories | University of Wisconsin-Madison AIDS Vaccine Research Laboratories | Gage Moreno et al |
| EPI_ISL_495503 | USA/IL-UW-799/2020 | 2020-07-07 | North America/USA/Illinois/Winnebago | University of Wisconsin-Madison AIDS Vaccine Research Laboratories | University of Wisconsin-Madison AIDS Vaccine Research Laboratories | Gage Moreno et al |
| EPI_ISL_495508 | USA/IL-UW-804/2020 | 2020-07-03 | North America/USA/Illinois/Winnebago | University of Wisconsin-Madison AIDS Vaccine Research Laboratories | University of Wisconsin-Madison AIDS Vaccine Research Laboratories | Gage Moreno et al |

| | | | | | | |
|---|---|---|---|---|---|---|
| EPI_ISL_495519 | SouthAfrica/KRISP-0343/2020 | 2020-06-18 | Africa/South Africa/KwaZulu-Natal/Harr | NHLS-IALCH | KRISP, KZN Research Innovation and Sequencing Platform | Giandhari J et al |
| EPI_ISL_495527 | SouthAfrica/KRISP-0402/2020 | 2020-06-23 | Africa/South Africa/KwaZulu-Natal/Harr | NHLS-IALCH | KRISP, KZN Research Innovation and Sequencing Platform | Giandhari J et al |
| EPI_ISL_495528 | SouthAfrica/KRISP-0403/2020 | 2020-06-23 | Africa/South Africa/KwaZulu-Natal/Ama | NHLS-IALCH | KRISP, KZN Research Innovation and Sequencing Platform | Giandhari J et al |
| EPI_ISL_495529 | SouthAfrica/KRISP-0404/2020 | 2020-06-24 | Africa/South Africa/KwaZulu-Natal/Ama | NHLS-IALCH | KRISP, KZN Research Innovation and Sequencing Platform | Giandhari J et al |
| EPI_ISL_495530 | SouthAfrica/KRISP-0405/2020 | 2020-06-23 | Africa/South Africa/KwaZulu-Natal/Harr | NHLS-IALCH | KRISP, KZN Research Innovation and Sequencing Platform | Giandhari J et al |
| EPI_ISL_495531 | SouthAfrica/KRISP-0406/2020 | 2020-06-22 | Africa/South Africa/KwaZulu-Natal/uMz | NHLS-IALCH | KRISP, KZN Research Innovation and Sequencing Platform | Giandhari J et al |
| EPI_ISL_495532 | SouthAfrica/KRISP-0407/2020 | 2020-06-24 | Africa/South Africa/KwaZulu-Natal/Ama | NHLS-IALCH | KRISP, KZN Research Innovation and Sequencing Platform | Giandhari J et al |
| EPI_ISL_495534 | SouthAfrica/KRISP-0409/2020 | 2020-06-23 | Africa/South Africa/KwaZulu-Natal/Ama | NHLS-IALCH | KRISP, KZN Research Innovation and Sequencing Platform | Giandhari J et al |

| | | | | | | |
|---|---|---|---|---|---|---|
| EPI_ISL_495536 | SouthAfrica/KRISP-0411/2020 | 2020-07-02 | Africa/South Africa/KwaZulu-Natal/eThe | Medical Disagnostics Services (MDS) | KRISP, KZN Research Innovation and Sequencing Platform | Giandhari J et al |
| EPI_ISL_495537 | SouthAfrica/KRISP-0412/2020 | 2020-07-02 | Africa/South Africa/KwaZulu-Natal/eThe | Medical Disagnostics Services (MDS) | KRISP, KZN Research Innovation and Sequencing Platform | Giandhari J et al |
| EPI_ISL_495543 | SouthAfrica/KRISP-0420/2020 | 2020-07-01 | Africa/South Africa/KwaZulu-Natal/iLem | NHLS-IALCH | KRISP, KZN Research Innovation and Sequencing Platform | Giandhari J et al |
| EPI_ISL_495550 | SouthAfrica/KRISP-0460/2020 | 2020-07-02 | Africa/South Africa/KwaZulu-Natal/eThe | NHLS-IALCH | KRISP, KZN Research Innovation and Sequencing Platform | Giandhari J et al |
| EPI_ISL_495553 | SouthAfrica/KRISP-0464/2020 | 2020-06-30 | Africa/South Africa/KwaZulu-Natal/King | NHLS-IALCH | KRISP, KZN Research Innovation and Sequencing Platform | Giandhari J et al |
| EPI_ISL_495556 | SouthAfrica/KRISP-0473/2020 | 2020-07-06 | Africa/South Africa/KwaZulu-Natal/Ugu | NHLS-IALCH | KRISP, KZN Research Innovation and Sequencing Platform | Giandhari J et al |
| EPI_ISL_495563 | USA/MI-UM-2006S000956/2020 | 2020-04-13 | North America/USA/Michigan/ | University of Michigan Clinical Microbiology Laboratory | Lauring Lab, University of Michigan, Department of Microbiology and Immunology | Valesano et al. et al |

| | | | | | | |
|---|---|---|---|---|---|---|
| EPI_ISL_495587 | USA/MI-UM-2006S001116/2020 | 2020-04-20 | North America/USA/Michigan/ | University of Michigan Clinical Microbiology Laboratory | Lauring Lab, University of Michigan, Department of Microbiology and Immunology | Valesano et al. et al |
| EPI_ISL_495592 | USA/MI-UM-2006S001135/2020 | 2020-05-05 | North America/USA/Michigan/ | University of Michigan Clinical Microbiology Laboratory | Lauring Lab, University of Michigan, Department of Microbiology and Immunology | Valesano et al. et al |
| EPI_ISL_495597 | USA/MN-MDH-1298/2020 | 2020-03-30 | North America/USA/Minnesota/ | Mayo Clinic & Mayo Clinic Laboratories | Minnesota Department of Health, Public Health Laboratory | Matt Plumb et al |
| EPI_ISL_495600 | USA/MN-MDH-1301/2020 | 2020-04-15 | North America/USA/Minnesota/ | Mayo Clinic & Mayo Clinic Laboratories | Minnesota Department of Health, Public Health Laboratory | Matt Plumb et al |
| EPI_ISL_495610 | USA/MN-MDH-1311/2020 | 2020-03-15 | North America/USA/Minnesota/ | Minnesota Department of Health, Public Health Laboratory | Minnesota Department of Health, Public Health Laboratory | Matt Plumb et al |
| EPI_ISL_495623 | USA/MN-MDH-1324/2020 | 2020-03-25 | North America/USA/Minnesota/ | Minnesota Department of Health, Public Health Laboratory | Minnesota Department of Health, Public Health Laboratory | Matt Plumb et al |
| EPI_ISL_495627 | USA/MN-MDH-1328/2020 | 2020-03-25 | North America/USA/Minnesota/ | Minnesota Department of Health, Public Health Laboratory | Minnesota Department of Health, Public Health Laboratory | Matt Plumb et al |
| EPI_ISL_495642 | USA/WA-S1467/2020 | 2020-05-08 | North America/USA/Washington/Yakim | Washington State Department of Health | Seattle Flu Study | Deborah A. Nickerson et al |
| EPI_ISL_495644 | USA/WA-S1469/2020 | 2020-05-07 | North America/USA/Washington/Yakim | Washington State Department of Health | Seattle Flu Study | Deborah A. Nickerson et al |
| EPI_ISL_495646 | USA/WA-S1471/2020 | 2020-05-07 | North America/USA/Washington/Yakim | Washington State Department of Health | Seattle Flu Study | Deborah A. Nickerson et al |
| EPI_ISL_495648 | USA/WA-S1473/2020 | 2020-05-12 | North America/USA/Washington/What | Washington State Department of Health | Seattle Flu Study | Deborah A. Nickerson et al |
| EPI_ISL_495652 | USA/WA-S1477/2020 | 2020-05-11 | North America/USA/Washington/Yakim | Washington State Department of Health | Seattle Flu Study | Deborah A. Nickerson et al |
| EPI_ISL_495653 | USA/WA-S1478/2020 | 2020-05-10 | North America/USA/Washington/Yakim | Washington State Department of Health | Seattle Flu Study | Deborah A. Nickerson et al |

| | | | | | | |
|---|---|---|---|---|---|---|
| EPI_ISL_495654 | USA/WA-S1479/2020 | 2020-05-01 | North America/USA/Washington/King C | Seattle Flu Study | Seattle Flu Study | Deborah A. Nickerson et al |
| EPI_ISL_495659 | USA/WA-S1484/2020 | 2020-05-01 | North America/USA/Washington/King C | Seattle Flu Study | Seattle Flu Study | Deborah A. Nickerson et al |
| EPI_ISL_495667 | USA/WA-S1217/2020 | 2020-05-08 | North America/USA/Washington/Yakim | Washington State Department of Health | Seattle Flu Study | Deborah A. Nickerson et al |
| EPI_ISL_495668 | USA/WA-S1218/2020 | 2020-05-08 | North America/USA/Washington/Yakim | Washington State Department of Health | Seattle Flu Study | Deborah A. Nickerson et al |
| EPI_ISL_495672 | USA/WA-S1222/2020 | 2020-05-08 | North America/USA/Washington/Yakim | Washington State Department of Health | Seattle Flu Study | Deborah A. Nickerson et al |
| EPI_ISL_495676 | USA/WA-S1226/2020 | 2020-05-09 | North America/USA/Washington/Yakim | Washington State Department of Health | Seattle Flu Study | Deborah A. Nickerson et al |
| EPI_ISL_495680 | USA/WA-S1230/2020 | 2020-05-09 | North America/USA/Washington/Yakim | Washington State Department of Health | Seattle Flu Study | Deborah A. Nickerson et al |
| EPI_ISL_495686 | USA/WA-S1236/2020 | 2020-05-06 | North America/USA/Washington/King C | Washington State Department of Health | Seattle Flu Study | Deborah A. Nickerson et al |
| EPI_ISL_495687 | USA/WA-S1237/2020 | 2020-05-11 | North America/USA/Washington/Yakim | Washington State Department of Health | Seattle Flu Study | Deborah A. Nickerson et al |
| EPI_ISL_495689 | USA/WA-S1239/2020 | 2020-05-11 | North America/USA/Washington/Yakim | Washington State Department of Health | Seattle Flu Study | Deborah A. Nickerson et al |
| EPI_ISL_495690 | USA/WA-S1240/2020 | 2020-05-11 | North America/USA/Washington/Yakim | Washington State Department of Health | Seattle Flu Study | Deborah A. Nickerson et al |
| EPI_ISL_495691 | USA/WA-S1241/2020 | 2020-05-11 | North America/USA/Washington/Yakim | Washington State Department of Health | Seattle Flu Study | Deborah A. Nickerson et al |
| EPI_ISL_495692 | USA/WA-S1242/2020 | 2020-05-11 | North America/USA/Washington/Yakim | Washington State Department of Health | Seattle Flu Study | Deborah A. Nickerson et al |
| EPI_ISL_495695 | USA/WA-S1245/2020 | 2020-05-11 | North America/USA/Washington/Yakim | Washington State Department of Health | Seattle Flu Study | Deborah A. Nickerson et al |
| EPI_ISL_495698 | USA/WA-S1248/2020 | 2020-05-11 | North America/USA/Washington/Yakim | Washington State Department of Health | Seattle Flu Study | Deborah A. Nickerson et al |
| EPI_ISL_495699 | USA/WA-S1249/2020 | 2020-05-11 | North America/USA/Washington/Yakim | Washington State Department of Health | Seattle Flu Study | Deborah A. Nickerson et al |
| EPI_ISL_495703 | USA/WA-S1253/2020 | 2020-05-13 | North America/USA/Washington/Yakim | Washington State Department of Health | Seattle Flu Study | Deborah A. Nickerson et al |
| EPI_ISL_495705 | USA/WA-S1255/2020 | 2020-05-13 | North America/USA/Washington/Skagit | Washington State Department of Health | Seattle Flu Study | Deborah A. Nickerson et al |
| EPI_ISL_495706 | USA/WA-S1256/2020 | 2020-05-14 | North America/USA/Washington/Yakim | Washington State Department of Health | Seattle Flu Study | Deborah A. Nickerson et al |
| EPI_ISL_495707 | USA/WA-S1257/2020 | 2020-05-13 | North America/USA/Washington/What | Washington State Department of Health | Seattle Flu Study | Deborah A. Nickerson et al |
| EPI_ISL_495709 | USA/WA-S1259/2020 | 2020-05-13 | North America/USA/Washington/What | Washington State Department of Health | Seattle Flu Study | Deborah A. Nickerson et al |
| EPI_ISL_495711 | USA/WA-S1261/2020 | 2020-05-13 | North America/USA/Washington/What | Washington State Department of Health | Seattle Flu Study | Deborah A. Nickerson et al |
| EPI_ISL_495717 | USA/WA-S1267/2020 | 2020-05-07 | North America/USA/Washington/Yakim | Washington State Department of Health | Seattle Flu Study | Deborah A. Nickerson et al |
| EPI_ISL_495718 | USA/WA-S1268/2020 | 2020-05-11 | North America/USA/Washington/What | Washington State Department of Health | Seattle Flu Study | Deborah A. Nickerson et al |
| EPI_ISL_495720 | USA/WA-S1270/2020 | 2020-05-11 | North America/USA/Washington/Pierce | Washington State Department of Health | Seattle Flu Study | Deborah A. Nickerson et al |
| EPI_ISL_495735 | USA/WA-S1285/2020 | 2020-05-12 | North America/USA/Washington/Yakim | Washington State Department of Health | Seattle Flu Study | Deborah A. Nickerson et al |
| EPI_ISL_495736 | USA/WA-S1286/2020 | 2020-05-12 | North America/USA/Washington/Yakim | Washington State Department of Health | Seattle Flu Study | Deborah A. Nickerson et al |
| EPI_ISL_495740 | USA/WA-S1290/2020 | 2020-05-14 | North America/USA/Washington/Pierce | Washington State Department of Health | Seattle Flu Study | Deborah A. Nickerson et al |
| EPI_ISL_495743 | USA/WA-S1293/2020 | 2020-05-13 | North America/USA/Washington/Frank | Washington State Department of Health | Seattle Flu Study | Deborah A. Nickerson et al |
| EPI_ISL_495746 | USA/WA-S1296/2020 | 2020-05-15 | North America/USA/Washington/Yakim | Washington State Department of Health | Seattle Flu Study | Deborah A. Nickerson et al |
| EPI_ISL_495769 | USA/WA-S1319/2020 | 2020-05-15 | North America/USA/Washington/Snoho | Washington State Department of Health | Seattle Flu Study | Deborah A. Nickerson et al |

| | | | | | | |
|---|---|---|---|---|---|---|
| EPI_ISL_495770 | USA/WA-S1320/2020 | 2020-05-15 | North America/USA/Washington/Snoho | Washington State Department of Health | Seattle Flu Study | Deborah A. Nickerson et al |
| EPI_ISL_495773 | USA/WA-S1323/2020 | 2020-05-14 | North America/USA/Washington/Yakim | Washington State Department of Health | Seattle Flu Study | Deborah A. Nickerson et al |
| EPI_ISL_495776 | USA/WA-S1326/2020 | 2020-05-15 | North America/USA/Washington/Yakim | Washington State Department of Health | Seattle Flu Study | Deborah A. Nickerson et al |
| EPI_ISL_495777 | USA/WA-S1327/2020 | 2020-05-15 | North America/USA/Washington/Yakim | Washington State Department of Health | Seattle Flu Study | Deborah A. Nickerson et al |
| EPI_ISL_495778 | USA/WA-S1328/2020 | 2020-05-15 | North America/USA/Washington/Yakim | Washington State Department of Health | Seattle Flu Study | Deborah A. Nickerson et al |
| EPI_ISL_495779 | USA/WA-S1329/2020 | 2020-05-15 | North America/USA/Washington/Yakim | Washington State Department of Health | Seattle Flu Study | Deborah A. Nickerson et al |
| EPI_ISL_495780 | USA/WA-S1330/2020 | 2020-05-15 | North America/USA/Washington/Yakim | Washington State Department of Health | Seattle Flu Study | Deborah A. Nickerson et al |
| EPI_ISL_495786 | USA/WA-S1336/2020 | 2020-05-13 | North America/USA/Washington/Yakim | Washington State Department of Health | Seattle Flu Study | Deborah A. Nickerson et al |
| EPI_ISL_495790 | USA/WA-S1340/2020 | 2020-05-13 | North America/USA/Washington/Yakim | Washington State Department of Health | Seattle Flu Study | Deborah A. Nickerson et al |
| EPI_ISL_495809 | USA/WA-S1359/2020 | 2020-05-18 | North America/USA/Washington/Yakim | Washington State Department of Health | Seattle Flu Study | Deborah A. Nickerson et al |
| EPI_ISL_495810 | USA/WA-S1360/2020 | 2020-05-18 | North America/USA/Washington/Yakim | Washington State Department of Health | Seattle Flu Study | Deborah A. Nickerson et al |
| EPI_ISL_495812 | USA/WA-S1362/2020 | 2020-05-18 | North America/USA/Washington/Yakim | Washington State Department of Health | Seattle Flu Study | Deborah A. Nickerson et al |
| EPI_ISL_495816 | USA/WA-S1366/2020 | 2020-05-18 | North America/USA/Washington/Yakim | Washington State Department of Health | Seattle Flu Study | Deborah A. Nickerson et al |
| EPI_ISL_495818 | USA/WA-S1368/2020 | 2020-05-16 | North America/USA/Washington/Yakim | Washington State Department of Health | Seattle Flu Study | Deborah A. Nickerson et al |
| EPI_ISL_495824 | USA/WA-S1374/2020 | 2020-05-18 | North America/USA/Washington/Yakim | Washington State Department of Health | Seattle Flu Study | Deborah A. Nickerson et al |
| EPI_ISL_495825 | USA/WA-S1375/2020 | 2020-05-18 | North America/USA/Washington/Spoka | Washington State Department of Health | Seattle Flu Study | Deborah A. Nickerson et al |
| EPI_ISL_495827 | USA/WA-S1377/2020 | 2020-05-19 | North America/USA/Washington/Snoho | Washington State Department of Health | Seattle Flu Study | Deborah A. Nickerson et al |
| EPI_ISL_495835 | USA/WA-S1385/2020 | 2020-05-19 | North America/USA/Washington/Snoho | Washington State Department of Health | Seattle Flu Study | Deborah A. Nickerson et al |
| EPI_ISL_495841 | USA/WA-S1391/2020 | 2020-05-19 | North America/USA/Washington/Snoho | Washington State Department of Health | Seattle Flu Study | Deborah A. Nickerson et al |
| EPI_ISL_495843 | USA/WA-S1393/2020 | 2020-05-19 | North America/USA/Washington/Snoho | Washington State Department of Health | Seattle Flu Study | Deborah A. Nickerson et al |
| EPI_ISL_495845 | USA/WA-S1395/2020 | 2020-05-19 | North America/USA/Washington/Snoho | Washington State Department of Health | Seattle Flu Study | Deborah A. Nickerson et al |
| EPI_ISL_495851 | USA/WA-S1401/2020 | 2020-05-19 | North America/USA/Washington/Yakim | Washington State Department of Health | Seattle Flu Study | Deborah A. Nickerson et al |
| EPI_ISL_495852 | USA/WA-S1402/2020 | 2020-05-19 | North America/USA/Washington/What | Washington State Department of Health | Seattle Flu Study | Deborah A. Nickerson et al |
| EPI_ISL_495868 | USA/WA-S1418/2020 | 2020-05-19 | North America/USA/Washington/Yakim | Washington State Department of Health | Seattle Flu Study | Deborah A. Nickerson et al |
| EPI_ISL_495877 | USA/WA-S1427/2020 | 2020-05-20 | North America/USA/Washington/Yakim | Washington State Department of Health | Seattle Flu Study | Deborah A. Nickerson et al |
| EPI_ISL_495890 | USA/WA-S1440/2020 | 2020-05-21 | North America/USA/Washington/Yakim | Washington State Department of Health | Seattle Flu Study | Deborah A. Nickerson et al |
| EPI_ISL_495895 | USA/WA-S1445/2020 | 2020-05-21 | North America/USA/Washington/Yakim | Washington State Department of Health | Seattle Flu Study | Deborah A. Nickerson et al |
| EPI_ISL_495898 | USA/WA-S1448/2020 | 2020-05-21 | North America/USA/Washington/Spoka | Washington State Department of Health | Seattle Flu Study | Deborah A. Nickerson et al |
| EPI_ISL_495899 | USA/WA-S1449/2020 | 2020-05-21 | North America/USA/Washington/Spoka | Washington State Department of Health | Seattle Flu Study | Deborah A. Nickerson et al |
| EPI_ISL_495907 | USA/WA-S1457/2020 | 2020-05-21 | North America/USA/Washington/Yakim | Washington State Department of Health | Seattle Flu Study | Deborah A. Nickerson et al |
| EPI_ISL_495911 | USA/WA-S1461/2020 | 2020-05-22 | North America/USA/Washington/Chelar | Washington State Department of Health | Seattle Flu Study | Deborah A. Nickerson et al |

| | | | | | | |
|---|---|---|---|---|---|---|
| EPI_ISL_495913 | USA/WA-S1463/2020 | 2020-05-21 | North America/USA/Washington/Yakim | Washington State Department of Health | Seattle Flu Study | Deborah A. Nickerson et al |
| EPI_ISL_495920 | USA/WA-S1494/2020 | 2020-05-22 | North America/USA/Washington/Yakim | Washington State Department of Health | Seattle Flu Study | Deborah A. Nickerson et al |
| EPI_ISL_495923 | USA/WA-S1497/2020 | 2020-05-21 | North America/USA/Washington/Yakim | Washington State Department of Health | Seattle Flu Study | Deborah A. Nickerson et al |
| EPI_ISL_495929 | USA/WA-S1503/2020 | 2020-05-21 | North America/USA/Washington/Yakim | Washington State Department of Health | Seattle Flu Study | Deborah A. Nickerson et al |
| EPI_ISL_495930 | USA/WA-S1504/2020 | 2020-05-21 | North America/USA/Washington/Yakim | Washington State Department of Health | Seattle Flu Study | Deborah A. Nickerson et al |
| EPI_ISL_495932 | USA/WA-S1506/2020 | 2020-05-22 | North America/USA/Washington/Yakim | Washington State Department of Health | Seattle Flu Study | Deborah A. Nickerson et al |
| EPI_ISL_495933 | USA/WA-S1507/2020 | 2020-05-22 | North America/USA/Washington/Yakim | Washington State Department of Health | Seattle Flu Study | Deborah A. Nickerson et al |
| EPI_ISL_495935 | USA/WA-S1509/2020 | 2020-05-22 | North America/USA/Washington/Yakim | Washington State Department of Health | Seattle Flu Study | Deborah A. Nickerson et al |
| EPI_ISL_495940 | USA/WA-S1514/2020 | 2020-05-22 | North America/USA/Washington/Yakim | Washington State Department of Health | Seattle Flu Study | Deborah A. Nickerson et al |
| EPI_ISL_495944 | USA/WA-S1518/2020 | 2020-05-22 | North America/USA/Washington/Yakim | Washington State Department of Health | Seattle Flu Study | Deborah A. Nickerson et al |
| EPI_ISL_495945 | USA/WA-S1519/2020 | 2020-05-22 | North America/USA/Washington/Yakim | Washington State Department of Health | Seattle Flu Study | Deborah A. Nickerson et al |
| EPI_ISL_495952 | USA/WA-S1526/2020 | 2020-05-23 | North America/USA/Washington/Yakim | Washington State Department of Health | Seattle Flu Study | Deborah A. Nickerson et al |
| EPI_ISL_495954 | USA/WA-S1528/2020 | 2020-05-24 | North America/USA/Washington/Yakim | Washington State Department of Health | Seattle Flu Study | Deborah A. Nickerson et al |
| EPI_ISL_495956 | USA/WA-S1530/2020 | 2020-05-24 | North America/USA/Washington/Yakim | Washington State Department of Health | Seattle Flu Study | Deborah A. Nickerson et al |
| EPI_ISL_495962 | USA/WA-S1536/2020 | 2020-05-25 | North America/USA/Washington/Yakim | Washington State Department of Health | Seattle Flu Study | Deborah A. Nickerson et al |
| EPI_ISL_495965 | USA/WA-S1539/2020 | 2020-05-26 | North America/USA/Washington/Yakim | Washington State Department of Health | Seattle Flu Study | Deborah A. Nickerson et al |
| EPI_ISL_495969 | USA/WA-S1543/2020 | 2020-05-26 | North America/USA/Washington/Yakim | Washington State Department of Health | Seattle Flu Study | Deborah A. Nickerson et al |
| EPI_ISL_495974 | USA/WA-S1548/2020 | 2020-05-26 | North America/USA/Washington/Yakim | Washington State Department of Health | Seattle Flu Study | Deborah A. Nickerson et al |
| EPI_ISL_495975 | USA/WA-S1549/2020 | 2020-05-26 | North America/USA/Washington/Yakim | Washington State Department of Health | Seattle Flu Study | Deborah A. Nickerson et al |
| EPI_ISL_495984 | USA/WA-S1558/2020 | 2020-05-26 | North America/USA/Washington/Yakim | Washington State Department of Health | Seattle Flu Study | Deborah A. Nickerson et al |
| EPI_ISL_495999 | USA/WA-S1573/2020 | 2020-05-26 | North America/USA/Washington/Yakim | Washington State Department of Health | Seattle Flu Study | Deborah A. Nickerson et al |
| EPI_ISL_496010 | USA/WA-S1584/2020 | 2020-05-26 | North America/USA/Washington/Yakim | Washington State Department of Health | Seattle Flu Study | Deborah A. Nickerson et al |
| EPI_ISL_496012 | USA/WA-S1586/2020 | 2020-05-26 | North America/USA/Washington/Yakim | Washington State Department of Health | Seattle Flu Study | Deborah A. Nickerson et al |
| EPI_ISL_496016 | USA/WA-S1590/2020 | 2020-05-26 | North America/USA/Washington/Yakim | Washington State Department of Health | Seattle Flu Study | Deborah A. Nickerson et al |
| EPI_ISL_496022 | USA/WA-S1596/2020 | 2020-05-26 | North America/USA/Washington/Yakim | Washington State Department of Health | Seattle Flu Study | Deborah A. Nickerson et al |
| EPI_ISL_496024 | USA/WA-S1598/2020 | 2020-05-26 | North America/USA/Washington/Yakim | Washington State Department of Health | Seattle Flu Study | Deborah A. Nickerson et al |
| EPI_ISL_496029 | USA/WA-S1603/2020 | 2020-05-26 | North America/USA/Washington/Yakim | Washington State Department of Health | Seattle Flu Study | Deborah A. Nickerson et al |
| EPI_ISL_496031 | USA/WA-S1605/2020 | 2020-05-26 | North America/USA/Washington/Yakim | Washington State Department of Health | Seattle Flu Study | Deborah A. Nickerson et al |
| EPI_ISL_496033 | USA/WA-S1607/2020 | 2020-05-26 | North America/USA/Washington/Yakim | Washington State Department of Health | Seattle Flu Study | Deborah A. Nickerson et al |
| EPI_ISL_496035 | USA/WA-S1609/2020 | 2020-05-26 | North America/USA/Washington/Yakim | Washington State Department of Health | Seattle Flu Study | Deborah A. Nickerson et al |
| EPI_ISL_496037 | USA/WA-S1611/2020 | 2020-05-26 | North America/USA/Washington/Yakim | Washington State Department of Health | Seattle Flu Study | Deborah A. Nickerson et al |

| | | | | | | |
|---|---|---|---|---|---|---|
| EPI_ISL_496038 | USA/WA-S1612/2020 | 2020-05-26 | North America/USA/Washington/Yakim | Washington State Department of Health | Seattle Flu Study | Deborah A. Nickerson et al |
| EPI_ISL_496044 | USA/WA-S1618/2020 | 2020-05-26 | North America/USA/Washington/Yakim | Washington State Department of Health | Seattle Flu Study | Deborah A. Nickerson et al |
| EPI_ISL_496049 | USA/WA-S1623/2020 | 2020-05-27 | North America/USA/Washington/Yakim | Washington State Department of Health | Seattle Flu Study | Deborah A. Nickerson et al |
| EPI_ISL_496052 | USA/WA-S1626/2020 | 2020-05-27 | North America/USA/Washington/Yakim | Washington State Department of Health | Seattle Flu Study | Deborah A. Nickerson et al |
| EPI_ISL_496060 | USA/WA-S1634/2020 | 2020-05-27 | North America/USA/Washington/Spoka | Washington State Department of Health | Seattle Flu Study | Deborah A. Nickerson et al |
| EPI_ISL_496064 | USA/WA-S1638/2020 | 2020-05-27 | North America/USA/Washington/Spoka | Washington State Department of Health | Seattle Flu Study | Deborah A. Nickerson et al |
| EPI_ISL_496073 | USA/WA-S1647/2020 | 2020-05-27 | North America/USA/Washington/Spoka | Washington State Department of Health | Seattle Flu Study | Deborah A. Nickerson et al |
| EPI_ISL_496075 | USA/WA-S1649/2020 | 2020-05-29 | North America/USA/Washington/Frankl | Washington State Department of Health | Seattle Flu Study | Deborah A. Nickerson et al |
| EPI_ISL_496079 | USA/WA-S1653/2020 | 2020-05-29 | North America/USA/Washington/Yakim | Washington State Department of Health | Seattle Flu Study | Deborah A. Nickerson et al |
| EPI_ISL_496080 | USA/WA-S1654/2020 | 2020-05-29 | North America/USA/Washington/Yakim | Washington State Department of Health | Seattle Flu Study | Deborah A. Nickerson et al |
| EPI_ISL_496081 | USA/WA-S1655/2020 | 2020-05-29 | North America/USA/Washington/Yakim | Washington State Department of Health | Seattle Flu Study | Deborah A. Nickerson et al |
| EPI_ISL_496093 | USA/WA-S1667/2020 | 2020-05-29 | North America/USA/Washington/Yakim | Washington State Department of Health | Seattle Flu Study | Deborah A. Nickerson et al |
| EPI_ISL_496100 | USA/WA-S1674/2020 | 2020-05-29 | North America/USA/Washington/Yakim | Washington State Department of Health | Seattle Flu Study | Deborah A. Nickerson et al |
| EPI_ISL_496102 | USA/WA-S1676/2020 | 2020-05-29 | North America/USA/Washington/Yakim | Washington State Department of Health | Seattle Flu Study | Deborah A. Nickerson et al |
| EPI_ISL_496108 | USA/WA-S1682/2020 | 2020-05-29 | North America/USA/Washington/Yakim | Washington State Department of Health | Seattle Flu Study | Deborah A. Nickerson et al |
| EPI_ISL_496112 | USA/WA-S1686/2020 | 2020-05-30 | North America/USA/Washington/Yakim | Washington State Department of Health | Seattle Flu Study | Deborah A. Nickerson et al |
| EPI_ISL_496121 | USA/WA-S1695/2020 | 2020-05-31 | North America/USA/Washington/Yakim | Washington State Department of Health | Seattle Flu Study | Deborah A. Nickerson et al |
| EPI_ISL_496132 | USA/WA-S1706/2020 | 2020-06-01 | North America/USA/Washington/Yakim | Washington State Department of Health | Seattle Flu Study | Deborah A. Nickerson et al |
| EPI_ISL_496133 | USA/WA-S1707/2020 | 2020-06-01 | North America/USA/Washington/Yakim | Washington State Department of Health | Seattle Flu Study | Deborah A. Nickerson et al |
| EPI_ISL_496134 | USA/WA-S1708/2020 | 2020-05-31 | North America/USA/Washington/Yakim | Washington State Department of Health | Seattle Flu Study | Deborah A. Nickerson et al |
| EPI_ISL_496144 | USA/WA-S1718/2020 | 2020-06-01 | North America/USA/Washington/Yakim | Washington State Department of Health | Seattle Flu Study | Deborah A. Nickerson et al |
| EPI_ISL_496149 | USA/WA-S1723/2020 | 2020-06-01 | North America/USA/Washington/Spoka | Washington State Department of Health | Seattle Flu Study | Deborah A. Nickerson et al |
| EPI_ISL_496162 | USA/WA-S1736/2020 | 2020-06-01 | North America/USA/Washington/Yakim | Washington State Department of Health | Seattle Flu Study | Deborah A. Nickerson et al |
| EPI_ISL_496166 | USA/WA-S1740/2020 | 2020-06-02 | North America/USA/Washington/Yakim | Washington State Department of Health | Seattle Flu Study | Deborah A. Nickerson et al |
| EPI_ISL_496177 | USA/WA-S1751/2020 | 2020-06-01 | North America/USA/Washington/Yakim | Washington State Department of Health | Seattle Flu Study | Deborah A. Nickerson et al |
| EPI_ISL_496180 | USA/WA-S1754/2020 | 2020-06-01 | North America/USA/Washington/Yakim | Washington State Department of Health | Seattle Flu Study | Deborah A. Nickerson et al |
| EPI_ISL_496186 | USA/WA-S1760/2020 | 2020-06-01 | North America/USA/Washington/Yakim | Washington State Department of Health | Seattle Flu Study | Deborah A. Nickerson et al |
| EPI_ISL_496188 | USA/WA-S1762/2020 | 2020-06-01 | North America/USA/Washington/Yakim | Washington State Department of Health | Seattle Flu Study | Deborah A. Nickerson et al |
| EPI_ISL_496190 | USA/WA-S1764/2020 | 2020-06-01 | North America/USA/Washington/Yakim | Washington State Department of Health | Seattle Flu Study | Deborah A. Nickerson et al |
| EPI_ISL_496198 | USA/WA-S1772/2020 | 2020-06-01 | North America/USA/Washington/Yakim | Washington State Department of Health | Seattle Flu Study | Deborah A. Nickerson et al |
| EPI_ISL_496211 | USA/WA-S1785/2020 | 2020-06-02 | North America/USA/Washington/Spoka | Washington State Department of Health | Seattle Flu Study | Deborah A. Nickerson et al |

| | | | | | | |
|---|---|---|---|---|---|---|
| EPI_ISL_496219 | USA/WA-S1793/2020 | 2020-06-02 | North America/USA/Washington/Yakim | Washington State Department of Health | Seattle Flu Study | Deborah A. Nickerson et al |
| EPI_ISL_496238 | USA/WA-S1812/2020 | 2020-05-27 | North America/USA/Washington/Thurst | Washington State Department of Health | Seattle Flu Study | Deborah A. Nickerson et al |
| EPI_ISL_496242 | USA/WA-S1816/2020 | 2020-06-03 | North America/USA/Washington/Yakim | Washington State Department of Health | Seattle Flu Study | Deborah A. Nickerson et al |
| EPI_ISL_496254 | USA/WA-S1828/2020 | 2020-05-26 | North America/USA/Washington/Spoka | Washington State Department of Health | Seattle Flu Study | Deborah A. Nickerson et al |
| EPI_ISL_496259 | USA/WA-S1833/2020 | 2020-05-27 | North America/USA/Washington/Whatc | Washington State Department of Health | Seattle Flu Study | Deborah A. Nickerson et al |
| EPI_ISL_496266 | USA/WA-S1840/2020 | 2020-05-27 | North America/USA/Washington/Yakim | Washington State Department of Health | Seattle Flu Study | Deborah A. Nickerson et al |
| EPI_ISL_496269 | USA/WA-S1843/2020 | 2020-05-27 | North America/USA/Washington/Yakim | Washington State Department of Health | Seattle Flu Study | Deborah A. Nickerson et al |
| EPI_ISL_496272 | USA/WA-S1846/2020 | 2020-05-27 | North America/USA/Washington/Yakim | Washington State Department of Health | Seattle Flu Study | Deborah A. Nickerson et al |
| EPI_ISL_496274 | USA/WA-S1848/2020 | 2020-05-27 | North America/USA/Washington/Yakim | Washington State Department of Health | Seattle Flu Study | Deborah A. Nickerson et al |
| EPI_ISL_496275 | USA/WA-S1849/2020 | 2020-05-27 | North America/USA/Washington/Yakim | Washington State Department of Health | Seattle Flu Study | Deborah A. Nickerson et al |
| EPI_ISL_496278 | USA/WA-S1852/2020 | 2020-05-27 | North America/USA/Washington/Yakim | Washington State Department of Health | Seattle Flu Study | Deborah A. Nickerson et al |
| EPI_ISL_496280 | USA/WA-S1854/2020 | 2020-05-27 | North America/USA/Washington/Yakim | Washington State Department of Health | Seattle Flu Study | Deborah A. Nickerson et al |
| EPI_ISL_496282 | USA/WA-S1856/2020 | 2020-05-27 | North America/USA/Washington/Yakim | Washington State Department of Health | Seattle Flu Study | Deborah A. Nickerson et al |
| EPI_ISL_496283 | USA/WA-S1857/2020 | 2020-05-27 | North America/USA/Washington/Yakim | Washington State Department of Health | Seattle Flu Study | Deborah A. Nickerson et al |
| EPI_ISL_496285 | USA/WA-S1859/2020 | 2020-05-28 | North America/USA/Washington/Yakim | Washington State Department of Health | Seattle Flu Study | Deborah A. Nickerson et al |
| EPI_ISL_496287 | USA/WA-S1861/2020 | 2020-05-27 | North America/USA/Washington/Thurst | Washington State Department of Health | Seattle Flu Study | Deborah A. Nickerson et al |
| EPI_ISL_496288 | USA/WA-S1862/2020 | 2020-05-27 | North America/USA/Washington/Thurst | Washington State Department of Health | Seattle Flu Study | Deborah A. Nickerson et al |
| EPI_ISL_496292 | USA/WA-S1866/2020 | 2020-05-27 | North America/USA/Washington/Thurst | Washington State Department of Health | Seattle Flu Study | Deborah A. Nickerson et al |
| EPI_ISL_496306 | USA/WA-S1880/2020 | 2020-05-28 | North America/USA/Washington/Yakim | Washington State Department of Health | Seattle Flu Study | Deborah A. Nickerson et al |
| EPI_ISL_496317 | USA/WA-S1891/2020 | 2020-05-28 | North America/USA/Washington/Yakim | Washington State Department of Health | Seattle Flu Study | Deborah A. Nickerson et al |
| EPI_ISL_496324 | USA/WA-S1898/2020 | 2020-05-28 | North America/USA/Washington/Yakim | Washington State Department of Health | Seattle Flu Study | Deborah A. Nickerson et al |
| EPI_ISL_496328 | USA/WA-S1902/2020 | 2020-05-28 | North America/USA/Washington/Yakim | Washington State Department of Health | Seattle Flu Study | Deborah A. Nickerson et al |
| EPI_ISL_496329 | USA/WA-S1903/2020 | 2020-05-29 | North America/USA/Washington/Yakim | Washington State Department of Health | Seattle Flu Study | Deborah A. Nickerson et al |
| EPI_ISL_496334 | USA/WA-S1908/2020 | 2020-05-29 | North America/USA/Washington/Yakim | Washington State Department of Health | Seattle Flu Study | Deborah A. Nickerson et al |
| EPI_ISL_496340 | Mexico/BCN-ALSR-1470/2020 | 2020-04-28 | North America/Mexico/Baja California/T | Infectolab | Andersen lab at Scripps Research | SEARCH Alliance San Diego with Samuel Navarro Alvarez et al |
| EPI_ISL_496344 | Mexico/BCN-ALSR-1475/2020 | 2020-04-28 | North America/Mexico/Baja California/T | Infectolab | Andersen lab at Scripps Research | SEARCH Alliance San Diego with Samuel Navarro Alvarez et al |
| EPI_ISL_496345 | Mexico/BCN-ALSR-1476/2020 | 2020-04-28 | North America/Mexico/Baja California/T | Infectolab | Andersen lab at Scripps Research | SEARCH Alliance San Diego with Samuel Navarro Alvarez et al |

| | | | | | | |
|---|---|---|---|---|---|---|
| EPI_ISL_496347 | Mexico/BCN-ALSR-1478/2020 | 2020-04-28 | North America/Mexico/Baja California/T | Infectolab | Andersen lab at Scripps Research | SEARCH Alliance San Diego with Samuel Navarro Alvarez et al |
| EPI_ISL_496349 | Mexico/BCN-ALSR-1485/2020 | 2020-04-28 | North America/Mexico/Baja California/T | Infectolab | Andersen lab at Scripps Research | SEARCH Alliance San Diego with Samuel Navarro Alvarez et al |
| EPI_ISL_496356 | Mexico/BCN-ALSR-1695/2020 | 2020-04-25 | North America/Mexico/Baja California/T | Infectolab | Andersen lab at Scripps Research | SEARCH Alliance San Diego with Samuel Navarro Alvarez et al |
| EPI_ISL_496357 | Mexico/BCN-ALSR-1696/2020 | 2020-04-24 | North America/Mexico/Baja California/T | Infectolab | Andersen lab at Scripps Research | SEARCH Alliance San Diego with Samuel Navarro Alvarez et al |
| EPI_ISL_496365 | Mexico/BCN-ALSR-1704/2020 | 2020-04-22 | North America/Mexico/Baja California/T | Infectolab | Andersen lab at Scripps Research | SEARCH Alliance San Diego with Samuel Navarro Alvarez et al |
| EPI_ISL_496367 | Mexico/BCN-ALSR-1706/2020 | 2020-04-22 | North America/Mexico/Baja California/T | Infectolab | Andersen lab at Scripps Research | SEARCH Alliance San Diego with Samuel Navarro Alvarez et al |
| EPI_ISL_496368 | Mexico/BCN-ALSR-1707/2020 | 2020-04-21 | North America/Mexico/Baja California/T | Infectolab | Andersen lab at Scripps Research | SEARCH Alliance San Diego with Samuel Navarro Alvarez et al |
| EPI_ISL_496482 | Italy/VEN-IZSV-9/2020 | 2020-03-17 | Europe/Italy/Veneto/Verona IT | Dept. Infectious, Tropical Diseases & Microbiology, IRCCS Sacro Cuore Don Calabria Hospital | 1) Dept. Infectious, Tropical Diseases & Microbiology, IRCCS Sacro Cuore Don Calabria Hospital; 2) Centro Piattaforme Tecnologiche, University of Verona; 3) Dept. Neurosciences, Biomedicine and Movement Sciences, University of Verona. | 1) Antonio Mori et al |
| EPI_ISL_496526 | India/MH-NCCS-NR2257/2020 | 2020-05-21 | Asia/India/Maharashtra/ | National Centre For Cell Science | National Centre For Cell Science | Dhiraj Paul et al |

| | | | | | | |
|---|---|---|---|---|---|---|
| EPI_ISL_496529 | India/MH-AFMC-5458/2020 | 2020-05-23 | Asia/India/Maharashtra/ | Armed Forces Medical College | National Centre For Cell Science | Dhiraj Paul et al |
| EPI_ISL_496533 | India/MH-AFMC-5868/2020 | 2020-05-26 | Asia/India/Maharashtra/ | Armed Forces Medical College | National Centre For Cell Science | Dhiraj Paul et al |
| EPI_ISL_496539 | India/MH-AFMC-5318/2020 | 2020-05-21 | Asia/India/Maharashtra/ | Armed Forces Medical College | National Centre For Cell Science | Dhiraj Paul et al |
| EPI_ISL_496540 | India/MH-AFMC-5670/2020 | 2020-05-25 | Asia/India/Maharashtra/ | Armed Forces Medical College | National Centre For Cell Science | Dhiraj Paul et al |
| EPI_ISL_496542 | India/MH-AFMC-5694/2020 | 2020-05-25 | Asia/India/Maharashtra/ | Armed Forces Medical College | National Centre For Cell Science | Dhiraj Paul et al |
| EPI_ISL_496545 | India/MH-AFMC-5807/2020 | 2020-05-25 | Asia/India/Maharashtra/ | Armed Forces Medical College | National Centre For Cell Science | Dhiraj Paul et al |
| EPI_ISL_496562 | India/MH-NCCS-NR1553/2020 | 2020-05-17 | Asia/India/Maharashtra/ | National Centre For Cell Science | National Centre For Cell Science | Dhiraj Paul et al |
| EPI_ISL_496563 | India/MH-NCCS-NR1565/2020 | 2020-05-17 | Asia/India/Maharashtra/ | National Centre For Cell Science | National Centre For Cell Science | Dhiraj Paul et al |
| EPI_ISL_496565 | India/MH-NCCS-NR1715/2020 | 2020-05-18 | Asia/India/Maharashtra/ | National Centre For Cell Science | National Centre For Cell Science | Dhiraj Paul et al |
| EPI_ISL_496571 | India/MH-NCCS-NR2122/2020 | 2020-05-21 | Asia/India/Maharashtra/ | National Centre For Cell Science | National Centre For Cell Science | Dhiraj Paul et al |
| EPI_ISL_496572 | India/MH-NCCS-NR2245/2020 | 2020-05-21 | Asia/India/Maharashtra/ | National Centre For Cell Science | National Centre For Cell Science | Dhiraj Paul et al |
| EPI_ISL_496575 | India/MH-NCCS-NR2254/2020 | 2020-05-21 | Asia/India/Maharashtra/ | National Centre For Cell Science | National Centre For Cell Science | Dhiraj Paul et al |
| EPI_ISL_496576 | India/MH-NCCS-NR2260/2020 | 2020-05-21 | Asia/India/Maharashtra/ | National Centre For Cell Science | National Centre For Cell Science | Dhiraj Paul et al |
| EPI_ISL_496585 | India/MH-NCCS-NR2527/2020 | 2020-05-24 | Asia/India/Maharashtra/ | National Centre For Cell Science | National Centre For Cell Science | Dhiraj Paul et al |
| EPI_ISL_496605 | Panama/328709/2020 | 2020-02-28 | North America/Panama/Panama Center | Gorgas Memorial Laboratory of Health Studies | Gorgas Memorial Laboratory of Health Studies | Danilo Franco et al |
| EPI_ISL_496606 | Panama/328710/2020 | 2020-02-20 | North America/Panama/Panama Center | Gorgas Memorial Laboratory of Health Studies | Gorgas Memorial Laboratory of Health Studies | Danilo Franco et al |
| EPI_ISL_496607 | Panama/328719/2020 | 2020-02-22 | North America/Panama/Panama Center | Gorgas Memorial Laboratory of Health Studies | Gorgas Memorial Laboratory of Health Studies | Danilo Franco et al |
| EPI_ISL_496609 | Panama/328723/2020 | 2020-02-17 | North America/Panama/Panama Center | Gorgas Memorial Laboratory of Health Studies | Gorgas Memorial Laboratory of Health Studies | Danilo Franco et al |

| | | | | | | |
|---|---|---|---|---|---|---|
| EPI_ISL_496615 | Panama/328927/2020 | 2020-02-15 | North America/Panama/Panama Center | Gorgas Memorial Laboratory of Health Studies | Gorgas Memorial Laboratory of Health Studies | Danilo Franco et al |
| EPI_ISL_496617 | Panama/328941/2020 | 2020-02-27 | North America/Panama/Panama Center | Gorgas Memorial Laboratory of Health Studies | Gorgas Memorial Laboratory of Health Studies | Danilo Franco et al |
| EPI_ISL_496624 | Panama/329108/2020 | 2020-03-10 | North America/Panama/Panama Center | Gorgas Memorial Laboratory of Health Studies | Gorgas Memorial Laboratory of Health Studies | Danilo Franco et al |
| EPI_ISL_496640 | Panama/329667/2020 | 2020-03-14 | North America/Panama/Panama Center | Gorgas Memorial Laboratory of Health Studies | Gorgas Memorial Laboratory of Health Studies | Danilo Franco et al |
| EPI_ISL_496689 | Panama/331596/2020 | 2020-03-17 | North America/Panama/Panama Center | Gorgas Memorial Laboratory of Health Studies | Gorgas Memorial Laboratory of Health Studies | Danilo Franco et al |
| EPI_ISL_496699 | Panama/331954/2020 | 2020-03-20 | North America/Panama/Panama Center | Gorgas Memorial Laboratory of Health Studies | Gorgas Memorial Laboratory of Health Studies | Danilo Franco et al |
| EPI_ISL_496736 | Panama/333271/2020 | 2020-02-23 | North America/Panama/Panama Center | Gorgas Memorial Laboratory of Health Studies | Gorgas Memorial Laboratory of Health Studies | Danilo Franco et al |
| EPI_ISL_496741 | Panama/333427/2020 | 2020-03-26 | North America/Panama/Panama Oeste/ | Gorgas Memorial Laboratory of Health Studies | Gorgas Memorial Laboratory of Health Studies | Danilo Franco et al |
| EPI_ISL_496748 | Panama/333568/2020 | 2020-03-16 | North America/Panama/Cocle/ | Gorgas Memorial Laboratory of Health Studies | Gorgas Memorial Laboratory of Health Studies | Danilo Franco et al |
| EPI_ISL_496798 | Panama/334913/2020 | 2020-03-31 | North America/Panama/Panama Center | Gorgas Memorial Laboratory of Health Studies | Gorgas Memorial Laboratory of Health Studies | Danilo Franco et al |
| EPI_ISL_496813 | Panama/335473/2020 | 2020-04-02 | North America/Panama/Panama Center | Gorgas Memorial Laboratory of Health Studies | Gorgas Memorial Laboratory of Health Studies | Danilo Franco et al |
| EPI_ISL_496836 | Panama/336047/2020 | 2020-04-03 | North America/Panama/Colon/ | Gorgas Memorial Laboratory of Health Studies | Gorgas Memorial Laboratory of Health Studies | Danilo Franco et al |
| EPI_ISL_496843 | Panama/336344/2020 | 2020-04-04 | North America/Panama/Panama Center | Gorgas Memorial Laboratory of Health Studies | Gorgas Memorial Laboratory of Health Studies | Danilo Franco et al |

| | | | | | | |
|---|---|---|---|---|---|---|
| EPI_ISL_496854 | Panama/336897/2020 | 2020-06-04 | North America/Panama/Panama Center | Gorgas Memorial Laboratory of Health Studies | Gorgas Memorial Laboratory of Health Studies | Danilo Franco et al |
| EPI_ISL_496858 | Panama/337306/2020 | 2020-04-07 | North America/Panama/Los Santos/ | Gorgas Memorial Laboratory of Health Studies | Gorgas Memorial Laboratory of Health Studies | Danilo Franco et al |
| EPI_ISL_496860 | Panama/337442/2020 | 2020-07-04 | North America/Panama/Chiriqui/ | Gorgas Memorial Laboratory of Health Studies | Gorgas Memorial Laboratory of Health Studies | Danilo Franco et al |
| EPI_ISL_496861 | Panama/337467/2020 | 2020-07-04 | North America/Panama/Panama Oeste/ | Gorgas Memorial Laboratory of Health Studies | Gorgas Memorial Laboratory of Health Studies | Danilo Franco et al |
| EPI_ISL_496863 | Panama/337660/2020 | 2020-07-04 | North America/Panama/Panama Oeste/ | Gorgas Memorial Laboratory of Health Studies | Gorgas Memorial Laboratory of Health Studies | Danilo Franco et al |
| EPI_ISL_496878 | Panama/338677/2020 | 2020-04-09 | North America/Panama/Colon/ | Gorgas Memorial Laboratory of Health Studies | Gorgas Memorial Laboratory of Health Studies | Danilo Franco et al |
| EPI_ISL_496879 | Panama/338681/2020 | 2020-04-09 | North America/Panama/Cocle/ | Gorgas Memorial Laboratory of Health Studies | Gorgas Memorial Laboratory of Health Studies | Danilo Franco et al |
| EPI_ISL_496880 | Panama/338687/2020 | 2020-04-09 | North America/Panama/Colon/ | Gorgas Memorial Laboratory of Health Studies | Gorgas Memorial Laboratory of Health Studies | Danilo Franco et al |
| EPI_ISL_496897 | Panama/339631/2020 | 2020-04-12 | North America/Panama/Cocle/ | Gorgas Memorial Laboratory of Health Studies | Gorgas Memorial Laboratory of Health Studies | Danilo Franco et al |
| EPI_ISL_496908 | Panama/341001/2020 | 2020-04-14 | North America/Panama/Colon/ | Gorgas Memorial Laboratory of Health Studies | Gorgas Memorial Laboratory of Health Studies | Danilo Franco et al |
| EPI_ISL_496928 | USA/MN-MDH-1342/2020 | 2020-03-26 | North America/USA/Minnesota/ | Minnesota Department of Health, Public Health Laboratory | Minnesota Department of Health, Public Health Laboratory | Matt Plumb et al |
| EPI_ISL_496931 | USA/WA-S2299/2020 | 2020-06-12 | North America/USA/Washington/Yakim | Washington State Department of Health | Seattle Flu Study | Deborah A. Nickerson et al |
| EPI_ISL_496944 | USA/WA-S2312/2020 | 2020-06-11 | North America/USA/Washington/Snoho | Washington State Department of Health | Seattle Flu Study | Deborah A. Nickerson et al |
| EPI_ISL_496952 | USA/WA-S2320/2020 | 2020-06-11 | North America/USA/Washington/Yakim | Washington State Department of Health | Seattle Flu Study | Deborah A. Nickerson et al |
| EPI_ISL_496959 | USA/WA-S2327/2020 | 2020-06-10 | North America/USA/Washington/Yakim | Washington State Department of Health | Seattle Flu Study | Deborah A. Nickerson et al |
| EPI_ISL_496964 | USA/WA-S2332/2020 | 2020-06-11 | North America/USA/Washington/Yakim | Washington State Department of Health | Seattle Flu Study | Deborah A. Nickerson et al |

| | | | | | | |
|---|---|---|---|---|---|---|
| EPI_ISL_496970 | USA/WA-S2338/2020 | 2020-06-11 | North America/USA/Washington/Yakim | Washington State Department of Health | Seattle Flu Study | Deborah A. Nickerson et al |
| EPI_ISL_496986 | USA/WA-S2354/2020 | 2020-06-12 | North America/USA/Washington/Yakim | Washington State Department of Health | Seattle Flu Study | Deborah A. Nickerson et al |
| EPI_ISL_496990 | USA/WA-S2358/2020 | 2020-06-11 | North America/USA/Washington/Yakim | Washington State Department of Health | Seattle Flu Study | Deborah A. Nickerson et al |
| EPI_ISL_497003 | USA/WA-S2371/2020 | 2020-06-10 | North America/USA/Washington/Yakim | Washington State Department of Health | Seattle Flu Study | Deborah A. Nickerson et al |
| EPI_ISL_497008 | USA/WA-S2376/2020 | 2020-06-11 | North America/USA/Washington/Yakim | Washington State Department of Health | Seattle Flu Study | Deborah A. Nickerson et al |
| EPI_ISL_497011 | USA/WA-S2379/2020 | 2020-06-12 | North America/USA/Washington/Yakim | Washington State Department of Health | Seattle Flu Study | Deborah A. Nickerson et al |
| EPI_ISL_497016 | USA/WA-S2384/2020 | 2020-06-13 | North America/USA/Washington/Yakim | Washington State Department of Health | Seattle Flu Study | Deborah A. Nickerson et al |
| EPI_ISL_497027 | USA/WA-S2395/2020 | 2020-06-14 | North America/USA/Washington/Yakim | Washington State Department of Health | Seattle Flu Study | Deborah A. Nickerson et al |
| EPI_ISL_497028 | USA/WA-S2396/2020 | 2020-06-14 | North America/USA/Washington/Yakim | Washington State Department of Health | Seattle Flu Study | Deborah A. Nickerson et al |
| EPI_ISL_497045 | USA/WA-S2413/2020 | 2020-06-15 | North America/USA/Washington/Yakim | Washington State Department of Health | Seattle Flu Study | Deborah A. Nickerson et al |
| EPI_ISL_497054 | USA/WA-S2422/2020 | 2020-06-15 | North America/USA/Washington/Yakim | Washington State Department of Health | Seattle Flu Study | Deborah A. Nickerson et al |
| EPI_ISL_497074 | USA/WA-S2442/2020 | 2020-06-16 | North America/USA/Washington/Yakim | Washington State Department of Health | Seattle Flu Study | Deborah A. Nickerson et al |
| EPI_ISL_497090 | USA/WA-S2458/2020 | 2020-06-16 | North America/USA/Washington/Yakim | Washington State Department of Health | Seattle Flu Study | Deborah A. Nickerson et al |
| EPI_ISL_497095 | USA/WA-S2463/2020 | 2020-06-14 | North America/USA/Washington/Yakim | Washington State Department of Health | Seattle Flu Study | Deborah A. Nickerson et al |
| EPI_ISL_497101 | USA/WA-S2469/2020 | 2020-06-15 | North America/USA/Washington/Yakim | Washington State Department of Health | Seattle Flu Study | Deborah A. Nickerson et al |
| EPI_ISL_497103 | USA/WA-S2471/2020 | 2020-06-16 | North America/USA/Washington/Yakim | Washington State Department of Health | Seattle Flu Study | Deborah A. Nickerson et al |
| EPI_ISL_497105 | USA/WA-S2473/2020 | 2020-06-16 | North America/USA/Washington/Yakim | Washington State Department of Health | Seattle Flu Study | Deborah A. Nickerson et al |
| EPI_ISL_497106 | USA/WA-S2474/2020 | 2020-06-16 | North America/USA/Washington/Yakim | Washington State Department of Health | Seattle Flu Study | Deborah A. Nickerson et al |
| EPI_ISL_497108 | USA/WA-S2476/2020 | 2020-06-15 | North America/USA/Washington/Yakim | Washington State Department of Health | Seattle Flu Study | Deborah A. Nickerson et al |
| EPI_ISL_497119 | USA/WA-S2487/2020 | 2020-06-16 | North America/USA/Washington/Yakim | Washington State Department of Health | Seattle Flu Study | Deborah A. Nickerson et al |
| EPI_ISL_497122 | USA/WA-S2490/2020 | 2020-06-16 | North America/USA/Washington/Yakim | Washington State Department of Health | Seattle Flu Study | Deborah A. Nickerson et al |
| EPI_ISL_497124 | USA/WA-S2492/2020 | 2020-06-16 | North America/USA/Washington/Yakim | Washington State Department of Health | Seattle Flu Study | Deborah A. Nickerson et al |
| EPI_ISL_497125 | USA/WA-S2493/2020 | 2020-06-16 | North America/USA/Washington/Yakim | Washington State Department of Health | Seattle Flu Study | Deborah A. Nickerson et al |
| EPI_ISL_497141 | USA/WA-S2509/2020 | 2020-06-18 | North America/USA/Washington/Pierce | Washington State Department of Health | Seattle Flu Study | Deborah A. Nickerson et al |
| EPI_ISL_497142 | USA/WA-S2510/2020 | 2020-06-18 | North America/USA/Washington/Pierce | Washington State Department of Health | Seattle Flu Study | Deborah A. Nickerson et al |
| EPI_ISL_497148 | USA/WA-S2516/2020 | 2020-06-15 | North America/USA/Washington/Yakim | Washington State Department of Health | Seattle Flu Study | Deborah A. Nickerson et al |
| EPI_ISL_497151 | USA/WA-S2519/2020 | 2020-06-15 | North America/USA/Washington/Yakim | Washington State Department of Health | Seattle Flu Study | Deborah A. Nickerson et al |
| EPI_ISL_497152 | USA/WA-S2520/2020 | 2020-06-15 | North America/USA/Washington/Yakim | Washington State Department of Health | Seattle Flu Study | Deborah A. Nickerson et al |
| EPI_ISL_497155 | USA/WA-S2523/2020 | 2020-06-15 | North America/USA/Washington/Yakim | Washington State Department of Health | Seattle Flu Study | Deborah A. Nickerson et al |
| EPI_ISL_497158 | USA/WA-S2526/2020 | 2020-06-16 | North America/USA/Washington/King C | Washington State Department of Health | Seattle Flu Study | Deborah A. Nickerson et al |
| EPI_ISL_497163 | USA/WA-S2531/2020 | 2020-06-15 | North America/USA/Washington/Yakim | Washington State Department of Health | Seattle Flu Study | Deborah A. Nickerson et al |

| | | | | | | |
|---|---|---|---|---|---|---|
| EPI_ISL_497173 | USA/WA-S2541/2020 | 2020-06-16 | North America/USA/Washington/Yakim | Washington State Department of Health | Seattle Flu Study | Deborah A. Nickerson et al |
| EPI_ISL_497196 | USA/WA-S2564/2020 | 2020-06-17 | North America/USA/Washington/Yakim | Washington State Department of Health | Seattle Flu Study | Deborah A. Nickerson et al |
| EPI_ISL_497209 | USA/WA-S2577/2020 | 2020-06-19 | North America/USA/Washington/Yakim | Washington State Department of Health | Seattle Flu Study | Deborah A. Nickerson et al |
| EPI_ISL_497214 | USA/WA-S2582/2020 | 2020-06-18 | North America/USA/Washington/Yakim | Washington State Department of Health | Seattle Flu Study | Deborah A. Nickerson et al |
| EPI_ISL_497217 | USA/WA-S2585/2020 | 2020-06-19 | North America/USA/Washington/Yakim | Washington State Department of Health | Seattle Flu Study | Deborah A. Nickerson et al |
| EPI_ISL_497224 | USA/WA-S2592/2020 | 2020-06-18 | North America/USA/Washington/Yakim | Washington State Department of Health | Seattle Flu Study | Deborah A. Nickerson et al |
| EPI_ISL_497235 | USA/WA-S2603/2020 | 2020-06-18 | North America/USA/Washington/Yakim | Washington State Department of Health | Seattle Flu Study | Deborah A. Nickerson et al |
| EPI_ISL_497253 | USA/WA-S2621/2020 | 2020-06-19 | North America/USA/Washington/Yakim | Washington State Department of Health | Seattle Flu Study | Deborah A. Nickerson et al |
| EPI_ISL_497255 | USA/WA-S2623/2020 | 2020-06-19 | North America/USA/Washington/Yakim | Washington State Department of Health | Seattle Flu Study | Deborah A. Nickerson et al |
| EPI_ISL_497271 | USA/WA-S2639/2020 | 2020-06-20 | North America/USA/Washington/Snoho | Washington State Department of Health | Seattle Flu Study | Deborah A. Nickerson et al |
| EPI_ISL_497290 | USA/WA-S2658/2020 | 2020-06-21 | North America/USA/Washington/Pierce | Washington State Department of Health | Seattle Flu Study | Deborah A. Nickerson et al |
| EPI_ISL_497295 | USA/WA-S2663/2020 | 2020-06-20 | North America/USA/Washington/Yakim | Washington State Department of Health | Seattle Flu Study | Deborah A. Nickerson et al |
| EPI_ISL_497300 | USA/WA-S2668/2020 | 2020-06-22 | North America/USA/Washington/Yakim | Washington State Department of Health | Seattle Flu Study | Deborah A. Nickerson et al |
| EPI_ISL_497304 | USA/WA-S2672/2020 | 2020-06-22 | North America/USA/Washington/Yakim | Washington State Department of Health | Seattle Flu Study | Deborah A. Nickerson et al |
| EPI_ISL_497316 | USA/WA-S2684/2020 | 2020-06-22 | North America/USA/Washington/Yakim | Washington State Department of Health | Seattle Flu Study | Deborah A. Nickerson et al |
| EPI_ISL_497319 | USA/WA-S2687/2020 | 2020-06-22 | North America/USA/Washington/Yakim | Washington State Department of Health | Seattle Flu Study | Deborah A. Nickerson et al |
| EPI_ISL_497322 | USA/WA-S2690/2020 | 2020-06-22 | North America/USA/Washington/Yakim | Washington State Department of Health | Seattle Flu Study | Deborah A. Nickerson et al |
| EPI_ISL_497325 | USA/WA-S2693/2020 | 2020-06-22 | North America/USA/Washington/Yakim | Washington State Department of Health | Seattle Flu Study | Deborah A. Nickerson et al |
| EPI_ISL_497326 | USA/WA-S2694/2020 | 2020-06-22 | North America/USA/Washington/Yakim | Washington State Department of Health | Seattle Flu Study | Deborah A. Nickerson et al |
| EPI_ISL_497331 | USA/WA-S2699/2020 | 2020-06-22 | North America/USA/Washington/Yakim | Washington State Department of Health | Seattle Flu Study | Deborah A. Nickerson et al |
| EPI_ISL_497341 | USA/CA-S1920/2020 | 2020-06-03 | North America/USA/California/Los Ange | Washington State Department of Health | Seattle Flu Study | Deborah A. Nickerson et al |
| EPI_ISL_497342 | USA/WA-S1913/2020 | 2020-06-02 | North America/USA/Washington/Yakim | Washington State Department of Health | Seattle Flu Study | Deborah A. Nickerson et al |
| EPI_ISL_497349 | USA/WA-S1921/2020 | 2020-06-03 | North America/USA/Washington/Frankl | Washington State Department of Health | Seattle Flu Study | Deborah A. Nickerson et al |
| EPI_ISL_497353 | USA/WA-S1925/2020 | 2020-06-04 | North America/USA/Washington/Yakim | Washington State Department of Health | Seattle Flu Study | Deborah A. Nickerson et al |
| EPI_ISL_497365 | USA/WA-S1937/2020 | 2020-06-02 | North America/USA/Washington/Yakim | Washington State Department of Health | Seattle Flu Study | Deborah A. Nickerson et al |
| EPI_ISL_497370 | USA/WA-S1942/2020 | 2020-06-02 | North America/USA/Washington/Yakim | Washington State Department of Health | Seattle Flu Study | Deborah A. Nickerson et al |
| EPI_ISL_497374 | USA/WA-S1946/2020 | 2020-06-02 | North America/USA/Washington/Yakim | Washington State Department of Health | Seattle Flu Study | Deborah A. Nickerson et al |
| EPI_ISL_497375 | USA/WA-S1947/2020 | 2020-06-02 | North America/USA/Washington/Yakim | Washington State Department of Health | Seattle Flu Study | Deborah A. Nickerson et al |
| EPI_ISL_497379 | USA/WA-S1951/2020 | 2020-06-02 | North America/USA/Washington/Yakim | Washington State Department of Health | Seattle Flu Study | Deborah A. Nickerson et al |
| EPI_ISL_497392 | USA/WA-S1964/2020 | 2020-06-03 | North America/USA/Washington/Yakim | Washington State Department of Health | Seattle Flu Study | Deborah A. Nickerson et al |
| EPI_ISL_497402 | USA/WA-S1974/2020 | 2020-06-03 | North America/USA/Washington/Yakim | Washington State Department of Health | Seattle Flu Study | Deborah A. Nickerson et al |

| | | | | | | |
|---|---|---|---|---|---|---|
| EPI_ISL_497410 | USA/WA-S1982/2020 | 2020-06-04 | North America/USA/Washington/Yakim | Washington State Department of Health | Seattle Flu Study | Deborah A. Nickerson et al |
| EPI_ISL_497412 | USA/WA-S1984/2020 | 2020-06-04 | North America/USA/Washington/Yakim | Washington State Department of Health | Seattle Flu Study | Deborah A. Nickerson et al |
| EPI_ISL_497413 | USA/WA-S1985/2020 | 2020-06-04 | North America/USA/Washington/Yakim | Washington State Department of Health | Seattle Flu Study | Deborah A. Nickerson et al |
| EPI_ISL_497446 | USA/WA-S2018/2020 | 2020-06-05 | North America/USA/Washington/Yakim | Washington State Department of Health | Seattle Flu Study | Deborah A. Nickerson et al |
| EPI_ISL_497450 | USA/WA-S2022/2020 | 2020-06-05 | North America/USA/Washington/Yakim | Washington State Department of Health | Seattle Flu Study | Deborah A. Nickerson et al |
| EPI_ISL_497452 | USA/WA-S2024/2020 | 2020-06-05 | North America/USA/Washington/Bentoı | Washington State Department of Health | Seattle Flu Study | Deborah A. Nickerson et al |
| EPI_ISL_497456 | USA/WA-S2028/2020 | 2020-06-05 | North America/USA/Washington/Yakim | Washington State Department of Health | Seattle Flu Study | Deborah A. Nickerson et al |
| EPI_ISL_497457 | USA/WA-S2029/2020 | 2020-06-05 | North America/USA/Washington/Yakim | Washington State Department of Health | Seattle Flu Study | Deborah A. Nickerson et al |
| EPI_ISL_497459 | USA/WA-S2031/2020 | 2020-06-06 | North America/USA/Washington/Yakim | Washington State Department of Health | Seattle Flu Study | Deborah A. Nickerson et al |
| EPI_ISL_497463 | USA/WA-S2035/2020 | 2020-06-06 | North America/USA/Washington/Yakim | Washington State Department of Health | Seattle Flu Study | Deborah A. Nickerson et al |
| EPI_ISL_497466 | USA/WA-S2038/2020 | 2020-06-05 | North America/USA/Washington/Yakim | Washington State Department of Health | Seattle Flu Study | Deborah A. Nickerson et al |
| EPI_ISL_497467 | USA/WA-S2039/2020 | 2020-06-05 | North America/USA/Washington/Yakim | Washington State Department of Health | Seattle Flu Study | Deborah A. Nickerson et al |
| EPI_ISL_497468 | USA/WA-S2040/2020 | 2020-06-05 | North America/USA/Washington/Yakim | Washington State Department of Health | Seattle Flu Study | Deborah A. Nickerson et al |
| EPI_ISL_497477 | USA/WA-S2049/2020 | 2020-06-05 | North America/USA/Washington/Yakim | Washington State Department of Health | Seattle Flu Study | Deborah A. Nickerson et al |
| EPI_ISL_497480 | USA/WA-S2052/2020 | 2020-06-05 | North America/USA/Washington/Yakim | Washington State Department of Health | Seattle Flu Study | Deborah A. Nickerson et al |
| EPI_ISL_497489 | USA/WA-S2061/2020 | 2020-06-05 | North America/USA/Washington/Yakim | Washington State Department of Health | Seattle Flu Study | Deborah A. Nickerson et al |
| EPI_ISL_497495 | USA/WA-S2067/2020 | 2020-06-06 | North America/USA/Washington/Yakim | Washington State Department of Health | Seattle Flu Study | Deborah A. Nickerson et al |
| EPI_ISL_497508 | USA/WA-S2080/2020 | 2020-06-06 | North America/USA/Washington/Yakim | Washington State Department of Health | Seattle Flu Study | Deborah A. Nickerson et al |
| EPI_ISL_497514 | USA/WA-S2086/2020 | 2020-06-05 | North America/USA/Washington/Yakim | Washington State Department of Health | Seattle Flu Study | Deborah A. Nickerson et al |
| EPI_ISL_497516 | USA/WA-S2088/2020 | 2020-06-05 | North America/USA/Washington/Yakim | Washington State Department of Health | Seattle Flu Study | Deborah A. Nickerson et al |
| EPI_ISL_497532 | USA/WA-S2104/2020 | 2020-06-06 | North America/USA/Washington/Yakim | Washington State Department of Health | Seattle Flu Study | Deborah A. Nickerson et al |
| EPI_ISL_497533 | USA/WA-S2105/2020 | 2020-06-07 | North America/USA/Washington/Yakim | Washington State Department of Health | Seattle Flu Study | Deborah A. Nickerson et al |
| EPI_ISL_497537 | USA/WA-S2109/2020 | 2020-06-08 | North America/USA/Washington/Yakim | Washington State Department of Health | Seattle Flu Study | Deborah A. Nickerson et al |
| EPI_ISL_497539 | USA/WA-S2111/2020 | 2020-06-08 | North America/USA/Washington/Yakim | Washington State Department of Health | Seattle Flu Study | Deborah A. Nickerson et al |
| EPI_ISL_497555 | USA/WA-S2127/2020 | 2020-06-07 | North America/USA/Washington/Yakim | Washington State Department of Health | Seattle Flu Study | Deborah A. Nickerson et al |
| EPI_ISL_497558 | USA/WA-S2130/2020 | 2020-06-08 | North America/USA/Washington/Thurst | Washington State Department of Health | Seattle Flu Study | Deborah A. Nickerson et al |
| EPI_ISL_497602 | USA/WA-S2174/2020 | 2020-06-08 | North America/USA/Washington/Yakim | Washington State Department of Health | Seattle Flu Study | Deborah A. Nickerson et al |
| EPI_ISL_497608 | USA/WA-S2180/2020 | 2020-06-08 | North America/USA/Washington/Yakim | Washington State Department of Health | Seattle Flu Study | Deborah A. Nickerson et al |
| EPI_ISL_497614 | USA/WA-S2186/2020 | 2020-06-08 | North America/USA/Washington/Yakim | Washington State Department of Health | Seattle Flu Study | Deborah A. Nickerson et al |
| EPI_ISL_497619 | USA/WA-S2191/2020 | 2020-06-08 | North America/USA/Washington/Yakim | Washington State Department of Health | Seattle Flu Study | Deborah A. Nickerson et al |
| EPI_ISL_497627 | USA/WA-S2199/2020 | 2020-06-08 | North America/USA/Washington/Yakim | Washington State Department of Health | Seattle Flu Study | Deborah A. Nickerson et al |

| | | | | | | |
|---|---|---|---|---|---|---|
| EPI_ISL_497642 | USA/WA-S2214/2020 | 2020-06-08 | North America/USA/Washington/Yakim | Washington State Department of Health | Seattle Flu Study | Deborah A. Nickerson et al |
| EPI_ISL_497653 | USA/WA-S2225/2020 | 2020-06-08 | North America/USA/Washington/Yakim | Washington State Department of Health | Seattle Flu Study | Deborah A. Nickerson et al |
| EPI_ISL_497658 | USA/WA-S2230/2020 | 2020-06-10 | North America/USA/Washington/King C | Washington State Department of Health | Seattle Flu Study | Deborah A. Nickerson et al |
| EPI_ISL_497661 | USA/WA-S2233/2020 | 2020-06-09 | North America/USA/Washington/Yakim | Washington State Department of Health | Seattle Flu Study | Deborah A. Nickerson et al |
| EPI_ISL_497664 | USA/WA-S2236/2020 | 2020-06-09 | North America/USA/Washington/Yakim | Washington State Department of Health | Seattle Flu Study | Deborah A. Nickerson et al |
| EPI_ISL_497672 | USA/WA-S2244/2020 | 2020-06-09 | North America/USA/Washington/Yakim | Washington State Department of Health | Seattle Flu Study | Deborah A. Nickerson et al |
| EPI_ISL_497685 | USA/WA-S2257/2020 | 2020-06-09 | North America/USA/Washington/Yakim | Washington State Department of Health | Seattle Flu Study | Deborah A. Nickerson et al |
| EPI_ISL_497688 | USA/WA-S2260/2020 | 2020-06-09 | North America/USA/Washington/Yakim | Washington State Department of Health | Seattle Flu Study | Deborah A. Nickerson et al |
| EPI_ISL_497691 | USA/WA-S2263/2020 | 2020-06-09 | North America/USA/Washington/Yakim | Washington State Department of Health | Seattle Flu Study | Deborah A. Nickerson et al |
| EPI_ISL_497695 | USA/WA-S2267/2020 | 2020-06-09 | North America/USA/Washington/Yakim | Washington State Department of Health | Seattle Flu Study | Deborah A. Nickerson et al |
| EPI_ISL_497698 | USA/WA-S2270/2020 | 2020-06-09 | North America/USA/Washington/Yakim | Washington State Department of Health | Seattle Flu Study | Deborah A. Nickerson et al |
| EPI_ISL_497702 | USA/WA-S2274/2020 | 2020-06-09 | North America/USA/Washington/Frankl | Washington State Department of Health | Seattle Flu Study | Deborah A. Nickerson et al |
| EPI_ISL_497717 | USA/WA-S2289/2020 | 2020-06-09 | North America/USA/Washington/Pierce | Washington State Department of Health | Seattle Flu Study | Deborah A. Nickerson et al |
| EPI_ISL_497723 | USA/WA-S2295/2020 | 2020-06-10 | North America/USA/Washington/Yakim | Washington State Department of Health | Seattle Flu Study | Deborah A. Nickerson et al |
| EPI_ISL_497726 | USA/WA-S2298/2020 | 2020-06-10 | North America/USA/Washington/Yakim | Washington State Department of Health | Seattle Flu Study | Deborah A. Nickerson et al |
| EPI_ISL_497771 | HongKong/HKU-200723-004/2020 | 2020-01-29 | Asia/Hong Kong/Hong Kong/ | Department of Microbiology, The University of Hong Kong | Department of Microbiology, The University of Hong Kong | Kelvin K.W. To et al |
| EPI_ISL_497783 | HongKong/HKU-200723-016/2020 | 2020-02-03 | Asia/Hong Kong/Hong Kong/ | Department of Microbiology, The University of Hong Kong | Department of Microbiology, The University of Hong Kong | Kelvin K.W. To et al |
| EPI_ISL_497784 | HongKong/HKU-200723-017/2020 | 2020-02-24 | Asia/Hong Kong/Hong Kong/ | Department of Microbiology, The University of Hong Kong | Department of Microbiology, The University of Hong Kong | Kelvin K.W. To et al |
| EPI_ISL_497792 | HongKong/HKU-200723-025/2020 | 2020-02-25 | Asia/Hong Kong/Hong Kong/ | Department of Microbiology, The University of Hong Kong | Department of Microbiology, The University of Hong Kong | Kelvin K.W. To et al |
| EPI_ISL_497797 | HongKong/HKU-200723-030/2020 | 2020-01-29 | Asia/Hong Kong/Hong Kong/ | Department of Microbiology, The University of Hong Kong | Department of Microbiology, The University of Hong Kong | Kelvin K.W. To et al |

| | | | | | | |
|---|---|---|---|---|---|---|
| EPI_ISL_497812 | HongKong/HKU-200723-045/2020 | 2020-02-15 | Asia/Hong Kong/Hong Kong/ | Department of Microbiology, The University of Hong Kong | Department of Microbiology, The University of Hong Kong | Kelvin K.W. To et al |
| EPI_ISL_497827 | HongKong/HKU-200723-060/2020 | 2020-01-29 | Asia/Hong Kong/Hong Kong/ | Department of Microbiology, The University of Hong Kong | Department of Microbiology, The University of Hong Kong | Kelvin K.W. To et al |
| EPI_ISL_497848 | HongKong/HKU-200723-081/2020 | 2020-02-15 | Asia/Hong Kong/Hong Kong/ | Department of Microbiology, The University of Hong Kong | Department of Microbiology, The University of Hong Kong | Kelvin K.W. To et al |
| EPI_ISL_497860 | HongKong/HKU-200723-093/2020 | 2020-01-25 | Asia/Hong Kong/Hong Kong/ | Department of Microbiology, The University of Hong Kong | Department of Microbiology, The University of Hong Kong | Kelvin K.W. To et al |
| EPI_ISL_497879 | India/MH-NCCS-NR099/2020 | 2020-04-28 | Asia/India/Maharashtra/ | National Centre For Cell Science | National Centre For Cell Science | Dhiraj Paul et al |
| EPI_ISL_497880 | India/MH-AFMC-5897/2020 | 2020-05-26 | Asia/India/Maharashtra/ | Armed Forces Medical College | National Centre For Cell Science | Dhiraj Paul et al |
| EPI_ISL_497883 | India/MH-AFMC-5643/2020 | 2020-05-24 | Asia/India/Maharashtra/ | Armed Forces Medical College | National Centre For Cell Science | Dhiraj Paul et al |
| EPI_ISL_497884 | India/MH-AFMC-5604/2020 | 2020-05-24 | Asia/India/Maharashtra/ | Armed Forces Medical College | National Centre For Cell Science | Dhiraj Paul et al |
| EPI_ISL_497889 | India/MH-BJMC-1701/2020 | 2020-04-19 | Asia/India/Maharashtra/ | B.J. Govt. Medical College | National Centre For Cell Science | Dhiraj Paul et al |
| EPI_ISL_497951 | SouthKorea/KCDC2030/2020 | 2020-02-20 | Asia/South Korea/South Korea/ | Division of Viral Diseases, Center for Laboratory Control of Infectious Diseases, Korea Centers for Diseases Control and Prevention | Division of Viral Diseases, Center for Laboratory Control of Infectious Diseases, Korea Centers for Diseases Control and Prevention | Jeong-Min Kim et al |

| | | | | | | |
|---|---|---|---|---|---|---|
| EPI_ISL_497953 | SouthKorea/KCDC2032/2020 | 2020-02-20 | Asia/South Korea/South Korea/ | Division of Viral Diseases, Center for Laboratory Control of Infectious Diseases, Korea Centers for Diseases Control and Prevention | Division of Viral Diseases, Center for Laboratory Control of Infectious Diseases, Korea Centers for Diseases Control and Prevention | Jeong-Min Kim et al |
| EPI_ISL_497954 | SouthKorea/KCDC2033/2020 | 2020-02-20 | Asia/South Korea/South Korea/ | Division of Viral Diseases, Center for Laboratory Control of Infectious Diseases, Korea Centers for Diseases Control and Prevention | Division of Viral Diseases, Center for Laboratory Control of Infectious Diseases, Korea Centers for Diseases Control and Prevention | Jeong-Min Kim et al |
| EPI_ISL_497956 | SouthKorea/KCDC2035/2020 | 2020-02-20 | Asia/South Korea/South Korea/ | Division of Viral Diseases, Center for Laboratory Control of Infectious Diseases, Korea Centers for Diseases Control and Prevention | Division of Viral Diseases, Center for Laboratory Control of Infectious Diseases, Korea Centers for Diseases Control and Prevention | Jeong-Min Kim et al |
| EPI_ISL_497957 | SouthKorea/KCDC2036/2020 | 2020-02-20 | Asia/South Korea/South Korea/ | Division of Viral Diseases, Center for Laboratory Control of Infectious Diseases, Korea Centers for Diseases Control and Prevention | Division of Viral Diseases, Center for Laboratory Control of Infectious Diseases, Korea Centers for Diseases Control and Prevention | Jeong-Min Kim et al |

| | | | | | | |
|---|---|---|---|---|---|---|
| EPI_ISL_497958 | SouthKorea/KCDC2037/2020 | 2020-02-20 | Asia/South Korea/South Korea/ | Division of Viral Diseases, Center for Laboratory Control of Infectious Diseases, Korea Centers for Diseases Control and Prevention | Division of Viral Diseases, Center for Laboratory Control of Infectious Diseases, Korea Centers for Diseases Control and Prevention | Jeong-Min Kim et al |
| EPI_ISL_497959 | SouthKorea/KCDC2038/2020 | 2020-02-20 | Asia/South Korea/South Korea/ | Division of Viral Diseases, Center for Laboratory Control of Infectious Diseases, Korea Centers for Diseases Control and Prevention | Division of Viral Diseases, Center for Laboratory Control of Infectious Diseases, Korea Centers for Diseases Control and Prevention | Jeong-Min Kim et al |
| EPI_ISL_497960 | SouthKorea/KCDC2039/2020 | 2020-02-19 | Asia/South Korea/South Korea/ | Division of Viral Diseases, Center for Laboratory Control of Infectious Diseases, Korea Centers for Diseases Control and Prevention | Division of Viral Diseases, Center for Laboratory Control of Infectious Diseases, Korea Centers for Diseases Control and Prevention | Jeong-Min Kim et al |
| EPI_ISL_497961 | SouthKorea/KCDC2060/2020 | 2020-02-17 | Asia/South Korea/South Korea/ | Division of Viral Diseases, Center for Laboratory Control of Infectious Diseases, Korea Centers for Diseases Control and Prevention | Division of Viral Diseases, Center for Laboratory Control of Infectious Diseases, Korea Centers for Diseases Control and Prevention | Jeong-Min Kim et al |

| | | | | | | |
|---|---|---|---|---|---|---|
| EPI_ISL_497962 | SouthKorea/KCDC2061/2020 | 2020-01-30 | Asia/South Korea/South Korea/ | Division of Viral Diseases, Center for Laboratory Control of Infectious Diseases, Korea Centers for Diseases Control and Prevention | Division of Viral Diseases, Center for Laboratory Control of Infectious Diseases, Korea Centers for Diseases Control and Prevention | Jeong-Min Kim et al |
| EPI_ISL_497963 | SouthKorea/KCDC2062/2020 | 2020-01-31 | Asia/South Korea/South Korea/ | Division of Viral Diseases, Center for Laboratory Control of Infectious Diseases, Korea Centers for Diseases Control and Prevention | Division of Viral Diseases, Center for Laboratory Control of Infectious Diseases, Korea Centers for Diseases Control and Prevention | Jeong-Min Kim et al |
| EPI_ISL_497964 | SouthKorea/KCDC2063/2020 | 2020-02-06 | Asia/South Korea/South Korea/ | Division of Viral Diseases, Center for Laboratory Control of Infectious Diseases, Korea Centers for Diseases Control and Prevention | Division of Viral Diseases, Center for Laboratory Control of Infectious Diseases, Korea Centers for Diseases Control and Prevention | Jeong-Min Kim et al |
| EPI_ISL_497965 | SouthKorea/KCDC2064/2020 | 2020-02-06 | Asia/South Korea/South Korea/ | Division of Viral Diseases, Center for Laboratory Control of Infectious Diseases, Korea Centers for Diseases Control and Prevention | Division of Viral Diseases, Center for Laboratory Control of Infectious Diseases, Korea Centers for Diseases Control and Prevention | Jeong-Min Kim et al |

| | | | | | | |
|---|---|---|---|---|---|---|
| EPI_ISL_497966 | SouthKorea/KCDC2065/2020 | 2020-02-09 | Asia/South Korea/South Korea/ | Division of Viral Diseases, Center for Laboratory Control of Infectious Diseases, Korea Centers for Diseases Control and Prevention | Division of Viral Diseases, Center for Laboratory Control of Infectious Diseases, Korea Centers for Diseases Control and Prevention | Jeong-Min Kim et al |
| EPI_ISL_497967 | SouthKorea/KCDC2066/2020 | 2020-02-20 | Asia/South Korea/South Korea/ | Division of Viral Diseases, Center for Laboratory Control of Infectious Diseases, Korea Centers for Diseases Control and Prevention | Division of Viral Diseases, Center for Laboratory Control of Infectious Diseases, Korea Centers for Diseases Control and Prevention | Jeong-Min Kim et al |
| EPI_ISL_497968 | SouthKorea/KCDC2067/2020 | 2020-02-19 | Asia/South Korea/South Korea/ | Division of Viral Diseases, Center for Laboratory Control of Infectious Diseases, Korea Centers for Diseases Control and Prevention | Division of Viral Diseases, Center for Laboratory Control of Infectious Diseases, Korea Centers for Diseases Control and Prevention | Jeong-Min Kim et al |
| EPI_ISL_497969 | SouthKorea/KCDC2068/2020 | 2020-02-19 | Asia/South Korea/South Korea/ | Division of Viral Diseases, Center for Laboratory Control of Infectious Diseases, Korea Centers for Diseases Control and Prevention | Division of Viral Diseases, Center for Laboratory Control of Infectious Diseases, Korea Centers for Diseases Control and Prevention | Jeong-Min Kim et al |

| | | | | | | |
|---|---|---|---|---|---|---|
| EPI_ISL_497970 | SouthKorea/KCDC2069/2020 | 2020-02-21 | Asia/South Korea/South Korea/ | Division of Viral Diseases, Center for Laboratory Control of Infectious Diseases, Korea Centers for Diseases Control and Prevention | Division of Viral Diseases, Center for Laboratory Control of Infectious Diseases, Korea Centers for Diseases Control and Prevention | Jeong-Min Kim et al |
| EPI_ISL_497971 | SouthKorea/KCDC2070/2020 | 2020-02-20 | Asia/South Korea/South Korea/ | Division of Viral Diseases, Center for Laboratory Control of Infectious Diseases, Korea Centers for Diseases Control and Prevention | Division of Viral Diseases, Center for Laboratory Control of Infectious Diseases, Korea Centers for Diseases Control and Prevention | Jeong-Min Kim et al |
| EPI_ISL_497972 | SouthKorea/KCDC2071/2020 | 2020-02-20 | Asia/South Korea/South Korea/ | Division of Viral Diseases, Center for Laboratory Control of Infectious Diseases, Korea Centers for Diseases Control and Prevention | Division of Viral Diseases, Center for Laboratory Control of Infectious Diseases, Korea Centers for Diseases Control and Prevention | Jeong-Min Kim et al |
| EPI_ISL_497973 | SouthKorea/KCDC2072/2020 | 2020-02-20 | Asia/South Korea/South Korea/ | Division of Viral Diseases, Center for Laboratory Control of Infectious Diseases, Korea Centers for Diseases Control and Prevention | Division of Viral Diseases, Center for Laboratory Control of Infectious Diseases, Korea Centers for Diseases Control and Prevention | Jeong-Min Kim et al |

| | | | | | | |
|---|---|---|---|---|---|---|
| EPI_ISL_497974 | SouthKorea/KCDC2073/2020 | 2020-02-20 | Asia/South Korea/South Korea/ | Division of Viral Diseases, Center for Laboratory Control of Infectious Diseases, Korea Centers for Diseases Control and Prevention | Division of Viral Diseases, Center for Laboratory Control of Infectious Diseases, Korea Centers for Diseases Control and Prevention | Jeong-Min Kim et al |
| EPI_ISL_497975 | SouthKorea/KCDC2074/2020 | 2020-02-21 | Asia/South Korea/South Korea/ | Division of Viral Diseases, Center for Laboratory Control of Infectious Diseases, Korea Centers for Diseases Control and Prevention | Division of Viral Diseases, Center for Laboratory Control of Infectious Diseases, Korea Centers for Diseases Control and Prevention | Jeong-Min Kim et al |
| EPI_ISL_497976 | SouthKorea/KCDC2075/2020 | 2020-02-21 | Asia/South Korea/South Korea/ | Division of Viral Diseases, Center for Laboratory Control of Infectious Diseases, Korea Centers for Diseases Control and Prevention | Division of Viral Diseases, Center for Laboratory Control of Infectious Diseases, Korea Centers for Diseases Control and Prevention | Jeong-Min Kim et al |
| EPI_ISL_497977 | SouthKorea/KCDC2076/2020 | 2020-02-21 | Asia/South Korea/South Korea/ | Division of Viral Diseases, Center for Laboratory Control of Infectious Diseases, Korea Centers for Diseases Control and Prevention | Division of Viral Diseases, Center for Laboratory Control of Infectious Diseases, Korea Centers for Diseases Control and Prevention | Jeong-Min Kim et al |

| | | | | | | |
|---|---|---|---|---|---|---|
| EPI_ISL_497978 | SouthKorea/KCDC2077/2020 | 2020-02-21 | Asia/South Korea/South Korea/ | Division of Viral Diseases, Center for Laboratory Control of Infectious Diseases, Korea Centers for Diseases Control and Prevention | Division of Viral Diseases, Center for Laboratory Control of Infectious Diseases, Korea Centers for Diseases Control and Prevention | Jeong-Min Kim et al |
| EPI_ISL_497979 | SouthKorea/KCDC2078/2020 | 2020-02-21 | Asia/South Korea/South Korea/ | Division of Viral Diseases, Center for Laboratory Control of Infectious Diseases, Korea Centers for Diseases Control and Prevention | Division of Viral Diseases, Center for Laboratory Control of Infectious Diseases, Korea Centers for Diseases Control and Prevention | Jeong-Min Kim et al |
| EPI_ISL_497980 | SouthKorea/KCDC2079/2020 | 2020-02-21 | Asia/South Korea/South Korea/ | Division of Viral Diseases, Center for Laboratory Control of Infectious Diseases, Korea Centers for Diseases Control and Prevention | Division of Viral Diseases, Center for Laboratory Control of Infectious Diseases, Korea Centers for Diseases Control and Prevention | Jeong-Min Kim et al |
| EPI_ISL_497981 | SouthKorea/KCDC2080/2020 | 2020-02-21 | Asia/South Korea/South Korea/ | Division of Viral Diseases, Center for Laboratory Control of Infectious Diseases, Korea Centers for Diseases Control and Prevention | Division of Viral Diseases, Center for Laboratory Control of Infectious Diseases, Korea Centers for Diseases Control and Prevention | Jeong-Min Kim et al |

| | | | | | | |
|---|---|---|---|---|---|---|
| EPI_ISL_497982 | SouthKorea/KCDC2081/2020 | 2020-02-21 | Asia/South Korea/South Korea/ | Division of Viral Diseases, Center for Laboratory Control of Infectious Diseases, Korea Centers for Diseases Control and Prevention | Division of Viral Diseases, Center for Laboratory Control of Infectious Diseases, Korea Centers for Diseases Control and Prevention | Jeong-Min Kim et al |
| EPI_ISL_497983 | SouthKorea/KCDC2082/2020 | 2020-02-22 | Asia/South Korea/South Korea/ | Division of Viral Diseases, Center for Laboratory Control of Infectious Diseases, Korea Centers for Diseases Control and Prevention | Division of Viral Diseases, Center for Laboratory Control of Infectious Diseases, Korea Centers for Diseases Control and Prevention | Jeong-Min Kim et al |
| EPI_ISL_497984 | SouthKorea/KCDC2083/2020 | 2020-02-21 | Asia/South Korea/South Korea/ | Division of Viral Diseases, Center for Laboratory Control of Infectious Diseases, Korea Centers for Diseases Control and Prevention | Division of Viral Diseases, Center for Laboratory Control of Infectious Diseases, Korea Centers for Diseases Control and Prevention | Jeong-Min Kim et al |
| EPI_ISL_497985 | SouthKorea/KCDC2084/2020 | 2020-02-22 | Asia/South Korea/South Korea/ | Division of Viral Diseases, Center for Laboratory Control of Infectious Diseases, Korea Centers for Diseases Control and Prevention | Division of Viral Diseases, Center for Laboratory Control of Infectious Diseases, Korea Centers for Diseases Control and Prevention | Jeong-Min Kim et al |

| | | | | | | |
|---|---|---|---|---|---|---|
| EPI_ISL_497986 | SouthKorea/KCDC2085/2020 | 2020-02-24 | Asia/South Korea/South Korea/ | Division of Viral Diseases, Center for Laboratory Control of Infectious Diseases, Korea Centers for Diseases Control and Prevention | Division of Viral Diseases, Center for Laboratory Control of Infectious Diseases, Korea Centers for Diseases Control and Prevention | Jeong-Min Kim et al |
| EPI_ISL_497987 | SouthKorea/KCDC2086/2020 | 2020-02-22 | Asia/South Korea/South Korea/ | Division of Viral Diseases, Center for Laboratory Control of Infectious Diseases, Korea Centers for Diseases Control and Prevention | Division of Viral Diseases, Center for Laboratory Control of Infectious Diseases, Korea Centers for Diseases Control and Prevention | Jeong-Min Kim et al |
| EPI_ISL_497988 | SouthKorea/KCDC2087/2020 | 2020-02-23 | Asia/South Korea/South Korea/ | Division of Viral Diseases, Center for Laboratory Control of Infectious Diseases, Korea Centers for Diseases Control and Prevention | Division of Viral Diseases, Center for Laboratory Control of Infectious Diseases, Korea Centers for Diseases Control and Prevention | Jeong-Min Kim et al |
| EPI_ISL_497989 | SouthKorea/KCDC2088/2020 | 2020-02-22 | Asia/South Korea/South Korea/ | Division of Viral Diseases, Center for Laboratory Control of Infectious Diseases, Korea Centers for Diseases Control and Prevention | Division of Viral Diseases, Center for Laboratory Control of Infectious Diseases, Korea Centers for Diseases Control and Prevention | Jeong-Min Kim et al |

| | | | | | | |
|---|---|---|---|---|---|---|
| EPI_ISL_497990 | SouthKorea/KCDC2089/2020 | 2020-02-24 | Asia/South Korea/South Korea/ | Division of Viral Diseases, Center for Laboratory Control of Infectious Diseases, Korea Centers for Diseases Control and Prevention | Division of Viral Diseases, Center for Laboratory Control of Infectious Diseases, Korea Centers for Diseases Control and Prevention | Jeong-Min Kim et al |
| EPI_ISL_497991 | SouthKorea/KCDC2090/2020 | 2020-02-21 | Asia/South Korea/South Korea/ | Division of Viral Diseases, Center for Laboratory Control of Infectious Diseases, Korea Centers for Diseases Control and Prevention | Division of Viral Diseases, Center for Laboratory Control of Infectious Diseases, Korea Centers for Diseases Control and Prevention | Jeong-Min Kim et al |
| EPI_ISL_497992 | SouthKorea/KCDC2091/2020 | 2020-02-21 | Asia/South Korea/South Korea/ | Division of Viral Diseases, Center for Laboratory Control of Infectious Diseases, Korea Centers for Diseases Control and Prevention | Division of Viral Diseases, Center for Laboratory Control of Infectious Diseases, Korea Centers for Diseases Control and Prevention | Jeong-Min Kim et al |
| EPI_ISL_497993 | SouthKorea/KCDC2092/2020 | 2020-02-21 | Asia/South Korea/South Korea/ | Division of Viral Diseases, Center for Laboratory Control of Infectious Diseases, Korea Centers for Diseases Control and Prevention | Division of Viral Diseases, Center for Laboratory Control of Infectious Diseases, Korea Centers for Diseases Control and Prevention | Jeong-Min Kim et al |

| | | | | | | |
|---|---|---|---|---|---|---|
| EPI_ISL_497994 | SouthKorea/KCDC2093/2020 | 2020-02-23 | Asia/South Korea/South Korea/ | Division of Viral Diseases, Center for Laboratory Control of Infectious Diseases, Korea Centers for Diseases Control and Prevention | Division of Viral Diseases, Center for Laboratory Control of Infectious Diseases, Korea Centers for Diseases Control and Prevention | Jeong-Min Kim et al |
| EPI_ISL_497995 | SouthKorea/KCDC2094/2020 | 2020-02-26 | Asia/South Korea/South Korea/ | Division of Viral Diseases, Center for Laboratory Control of Infectious Diseases, Korea Centers for Diseases Control and Prevention | Division of Viral Diseases, Center for Laboratory Control of Infectious Diseases, Korea Centers for Diseases Control and Prevention | Jeong-Min Kim et al |
| EPI_ISL_497996 | SouthKorea/KCDC2095/2020 | 2020-02-27 | Asia/South Korea/South Korea/ | Division of Viral Diseases, Center for Laboratory Control of Infectious Diseases, Korea Centers for Diseases Control and Prevention | Division of Viral Diseases, Center for Laboratory Control of Infectious Diseases, Korea Centers for Diseases Control and Prevention | Jeong-Min Kim et al |
| EPI_ISL_497998 | SouthKorea/KCDC2097/2020 | 2020-02-27 | Asia/South Korea/South Korea/ | Division of Viral Diseases, Center for Laboratory Control of Infectious Diseases, Korea Centers for Diseases Control and Prevention | Division of Viral Diseases, Center for Laboratory Control of Infectious Diseases, Korea Centers for Diseases Control and Prevention | Jeong-Min Kim et al |

| | | | | | | |
|---|---|---|---|---|---|---|
| EPI_ISL_498002 | SouthKorea/KCDC2101/2020 | 2020-03-05 | Asia/South Korea/South Korea/ | Division of Viral Diseases, Center for Laboratory Control of Infectious Diseases, Korea Centers for Diseases Control and Prevention | Division of Viral Diseases, Center for Laboratory Control of Infectious Diseases, Korea Centers for Diseases Control and Prevention | Jeong-Min Kim et al |
| EPI_ISL_498008 | SouthKorea/KCDC2107/2020 | 2020-03-10 | Asia/South Korea/South Korea/ | Division of Viral Diseases, Center for Laboratory Control of Infectious Diseases, Korea Centers for Diseases Control and Prevention | Division of Viral Diseases, Center for Laboratory Control of Infectious Diseases, Korea Centers for Diseases Control and Prevention | Jeong-Min Kim et al |
| EPI_ISL_498011 | SouthKorea/KCDC2110/2020 | 2020-03-12 | Asia/South Korea/South Korea/ | Division of Viral Diseases, Center for Laboratory Control of Infectious Diseases, Korea Centers for Diseases Control and Prevention | Division of Viral Diseases, Center for Laboratory Control of Infectious Diseases, Korea Centers for Diseases Control and Prevention | Jeong-Min Kim et al |
| EPI_ISL_498014 | SouthKorea/KCDC2113/2020 | 2020-03-12 | Asia/South Korea/South Korea/ | Division of Viral Diseases, Center for Laboratory Control of Infectious Diseases, Korea Centers for Diseases Control and Prevention | Division of Viral Diseases, Center for Laboratory Control of Infectious Diseases, Korea Centers for Diseases Control and Prevention | Jeong-Min Kim et al |

| | | | | | | |
|---|---|---|---|---|---|---|
| EPI_ISL_498018 | SouthKorea/KCDC2117/2020 | 2020-03-15 | Asia/South Korea/South Korea/ | Division of Viral Diseases, Center for Laboratory Control of Infectious Diseases, Korea Centers for Diseases Control and Prevention | Division of Viral Diseases, Center for Laboratory Control of Infectious Diseases, Korea Centers for Diseases Control and Prevention | Jeong-Min Kim et al |
| EPI_ISL_498022 | SouthKorea/KCDC2121/2020 | 2020-03-15 | Asia/South Korea/South Korea/ | Division of Viral Diseases, Center for Laboratory Control of Infectious Diseases, Korea Centers for Diseases Control and Prevention | Division of Viral Diseases, Center for Laboratory Control of Infectious Diseases, Korea Centers for Diseases Control and Prevention | Jeong-Min Kim et al |
| EPI_ISL_498027 | SouthKorea/KCDC2126/2020 | 2020-03-15 | Asia/South Korea/South Korea/ | Division of Viral Diseases, Center for Laboratory Control of Infectious Diseases, Korea Centers for Diseases Control and Prevention | Division of Viral Diseases, Center for Laboratory Control of Infectious Diseases, Korea Centers for Diseases Control and Prevention | Jeong-Min Kim et al |
| EPI_ISL_498029 | SouthKorea/KCDC2128/2020 | 2020-03-15 | Asia/South Korea/South Korea/ | Division of Viral Diseases, Center for Laboratory Control of Infectious Diseases, Korea Centers for Diseases Control and Prevention | Division of Viral Diseases, Center for Laboratory Control of Infectious Diseases, Korea Centers for Diseases Control and Prevention | Jeong-Min Kim et al |

| | | | | | | |
|---|---|---|---|---|---|---|
| EPI_ISL_498031 | SouthKorea/KCDC2130/2020 | 2020-03-16 | Asia/South Korea/South Korea/ | Division of Viral Diseases, Center for Laboratory Control of Infectious Diseases, Korea Centers for Diseases Control and Prevention | Division of Viral Diseases, Center for Laboratory Control of Infectious Diseases, Korea Centers for Diseases Control and Prevention | Jeong-Min Kim et al |
| EPI_ISL_498033 | SouthKorea/KCDC2132/2020 | 2020-03-17 | Asia/South Korea/South Korea/ | Division of Viral Diseases, Center for Laboratory Control of Infectious Diseases, Korea Centers for Diseases Control and Prevention | Division of Viral Diseases, Center for Laboratory Control of Infectious Diseases, Korea Centers for Diseases Control and Prevention | Jeong-Min Kim et al |
| EPI_ISL_498035 | SouthKorea/KCDC2134/2020 | 2020-03-20 | Asia/South Korea/South Korea/ | Division of Viral Diseases, Center for Laboratory Control of Infectious Diseases, Korea Centers for Diseases Control and Prevention | Division of Viral Diseases, Center for Laboratory Control of Infectious Diseases, Korea Centers for Diseases Control and Prevention | Jeong-Min Kim et al |
| EPI_ISL_498039 | SouthKorea/KCDC2138/2020 | 2020-03-20 | Asia/South Korea/South Korea/ | Division of Viral Diseases, Center for Laboratory Control of Infectious Diseases, Korea Centers for Diseases Control and Prevention | Division of Viral Diseases, Center for Laboratory Control of Infectious Diseases, Korea Centers for Diseases Control and Prevention | Jeong-Min Kim et al |

| | | | | | | |
|---|---|---|---|---|---|---|
| EPI_ISL_498042 | SouthKorea/KCDC2141/2020 | 2020-03-23 | Asia/South Korea/South Korea/ | Division of Viral Diseases, Center for Laboratory Control of Infectious Diseases, Korea Centers for Diseases Control and Prevention | Division of Viral Diseases, Center for Laboratory Control of Infectious Diseases, Korea Centers for Diseases Control and Prevention | Jeong-Min Kim et al |
| EPI_ISL_498046 | SouthKorea/KCDC2145/2020 | 2020-03-22 | Asia/South Korea/South Korea/ | Division of Viral Diseases, Center for Laboratory Control of Infectious Diseases, Korea Centers for Diseases Control and Prevention | Division of Viral Diseases, Center for Laboratory Control of Infectious Diseases, Korea Centers for Diseases Control and Prevention | Jeong-Min Kim et al |
| EPI_ISL_498049 | SouthKorea/KCDC2148/2020 | 2020-03-22 | Asia/South Korea/South Korea/ | Division of Viral Diseases, Center for Laboratory Control of Infectious Diseases, Korea Centers for Diseases Control and Prevention | Division of Viral Diseases, Center for Laboratory Control of Infectious Diseases, Korea Centers for Diseases Control and Prevention | Jeong-Min Kim et al |
| EPI_ISL_498051 | SouthKorea/KCDC2150/2020 | 2020-03-22 | Asia/South Korea/South Korea/ | Division of Viral Diseases, Center for Laboratory Control of Infectious Diseases, Korea Centers for Diseases Control and Prevention | Division of Viral Diseases, Center for Laboratory Control of Infectious Diseases, Korea Centers for Diseases Control and Prevention | Jeong-Min Kim et al |
| EPI_ISL_498055 | SouthAfrica/KRISP-0436/2020 | 2020-07-01 | Africa/South Africa/KwaZulu-Natal/Ugu | NHLS-IALCH | KRISP, KZN Research Innovation and Sequencing Platform | Giandhari J et al |

| | | | | | | |
|---|---|---|---|---|---|---|
| EPI_ISL_498056 | SouthAfrica/KRISP-0440/2020 | 2020-07-01 | Africa/South Africa/KwaZulu-Natal/iLem | NHLS-IALCH | KRISP, KZN Research Innovation and Sequencing Platform | Giandhari J et al |
| EPI_ISL_498057 | SouthAfrica/KRISP-0441/2020 | 2020-06-30 | Africa/South Africa/KwaZulu-Natal/iLem | NHLS-IALCH | KRISP, KZN Research Innovation and Sequencing Platform | Giandhari J et al |
| EPI_ISL_498058 | SouthAfrica/KRISP-0445/2020 | 2020-06-30 | Africa/South Africa/KwaZulu-Natal/iLem | NHLS-IALCH | KRISP, KZN Research Innovation and Sequencing Platform | Giandhari J et al |
| EPI_ISL_498059 | SouthAfrica/KRISP-0446/2020 | 2020-06-30 | Africa/South Africa/KwaZulu-Natal/iLem | NHLS-IALCH | KRISP, KZN Research Innovation and Sequencing Platform | Giandhari J et al |
| EPI_ISL_498062 | SouthAfrica/KRISP-0491/2020 | 2020-07-07 | Africa/South Africa/KwaZulu-Natal/uMk | NHLS-IALCH | KRISP, KZN Research Innovation and Sequencing Platform | Giandhari J et al |
| EPI_ISL_498065 | SouthAfrica/KRISP-0494/2020 | 2020-07-09 | Africa/South Africa/KwaZulu-Natal/uMk | NHLS-IALCH | KRISP, KZN Research Innovation and Sequencing Platform | Giandhari J et al |
| EPI_ISL_498069 | SouthAfrica/KRISP-0501/2020 | 2020-07-11 | Africa/South Africa/KwaZulu-Natal/uMk | NHLS-IALCH | KRISP, KZN Research Innovation and Sequencing Platform | Giandhari J et al |
| EPI_ISL_498081 | SouthAfrica/KRISP-0539/2020 | 2020-07-07 | Africa/South Africa/KwaZulu-Natal/uMz | NHLS-IALCH | KRISP, KZN Research Innovation and Sequencing Platform | Giandhari J et al |

| | | | | | | |
|---|---|---|---|---|---|---|
| EPI_ISL_498084 | SouthAfrica/KRISP-0543/2020 | 2020-07-12 | Africa/South Africa/KwaZulu-Natal/iLem | NHLS-IALCH | KRISP, KZN Research Innovation and Sequencing Platform | Giandhari J et al |
| EPI_ISL_498085 | SouthAfrica/KRISP-0544/2020 | 2020-07-13 | Africa/South Africa/KwaZulu-Natal/uMz | NHLS-IALCH | KRISP, KZN Research Innovation and Sequencing Platform | Giandhari J et al |
| EPI_ISL_498089 | SouthAfrica/KRISP-0550/2020 | 2020-07-15 | Africa/South Africa/KwaZulu-Natal/eThe | NHLS-IALCH | KRISP, KZN Research Innovation and Sequencing Platform | Giandhari J et al |
| EPI_ISL_498094 | SouthAfrica/KRISP-0555/2020 | 2020-07-15 | Africa/South Africa/KwaZulu-Natal/eThe | NHLS-IALCH | KRISP, KZN Research Innovation and Sequencing Platform | Giandhari J et al |
| EPI_ISL_498096 | SouthAfrica/KRISP-0557/2020 | 2020-07-14 | Africa/South Africa/KwaZulu-Natal/uThu | NHLS-IALCH | KRISP, KZN Research Innovation and Sequencing Platform | Giandhari J et al |
| EPI_ISL_498099 | SouthAfrica/KRISP-0560/2020 | 2020-07-15 | Africa/South Africa/KwaZulu-Natal/uMz | NHLS-IALCH | KRISP, KZN Research Innovation and Sequencing Platform | Giandhari J et al |
| EPI_ISL_498101 | SouthAfrica/KRISP-0562/2020 | 2020-07-15 | Africa/South Africa/KwaZulu-Natal/eThe | NHLS-IALCH | KRISP, KZN Research Innovation and Sequencing Platform | Giandhari J et al |
| EPI_ISL_498107 | SouthAfrica/KRISP-0569/2020 | 2020-07-15 | Africa/South Africa/KwaZulu-Natal/eThe | NHLS-IALCH | KRISP, KZN Research Innovation and Sequencing Platform | Giandhari J et al |

| | | | | | | |
|---|---|---|---|---|---|---|
| EPI_ISL_498109 | SouthAfrica/KRISP-0571/2020 | 2020-07-14 | Africa/South Africa/KwaZulu-Natal/uMz | NHLS-IALCH | KRISP, KZN Research Innovation and Sequencing Platform | Giandhari J et al |
| EPI_ISL_498111 | SouthAfrica/KRISP-0573/2020 | 2020-07-14 | Africa/South Africa/KwaZulu-Natal/uMz | NHLS-IALCH | KRISP, KZN Research Innovation and Sequencing Platform | Giandhari J et al |
| EPI_ISL_498114 | SouthAfrica/KRISP-0576/2020 | 2020-07-07 | Africa/South Africa/KwaZulu-Natal/iLem | NHLS-IALCH | KRISP, KZN Research Innovation and Sequencing Platform | Giandhari J et al |
| EPI_ISL_498115 | SouthAfrica/KRISP-0577/2020 | 2020-07-09 | Africa/South Africa/KwaZulu-Natal/iLem | NHLS-IALCH | KRISP, KZN Research Innovation and Sequencing Platform | Giandhari J et al |
| EPI_ISL_498117 | SouthAfrica/KRISP-0579/2020 | 2020-07-08 | Africa/South Africa/KwaZulu-Natal/iLem | NHLS-IALCH | KRISP, KZN Research Innovation and Sequencing Platform | Giandhari J et al |
| EPI_ISL_498122 | SouthAfrica/KRISP-0584/2020 | 2020-07-07 | Africa/South Africa/KwaZulu-Natal/iLem | NHLS-IALCH | KRISP, KZN Research Innovation and Sequencing Platform | Giandhari J et al |
| EPI_ISL_498124 | SouthAfrica/KRISP-0586/2020 | 2020-07-08 | Africa/South Africa/KwaZulu-Natal/iLem | NHLS-IALCH | KRISP, KZN Research Innovation and Sequencing Platform | Giandhari J et al |
| EPI_ISL_498141 | Belgium/ULG-10235/2020 | 2020-06-23 | Europe/Belgium/Li√®ge/ | Department of Clinical Microbiology | GIGA Medical Genomics | Keith Durkin et al |
| EPI_ISL_498163 | Colombia/COR-INS-95319/2020 | 2020-04-01 | South America/Colombia/Cordoba CO/Ci | Instituto Nacional de Salud, Bogot√°, Colombia | Instituto Nacional de Salud, Bogot√°, Colombia | Katherine Laiton-Donato et al |

| | | | | | | |
|---|---|---|---|---|---|---|
| EPI_ISL_498228 | Bangladesh/BCSIR-NILMRC-167/2020 | 2020-06-18 | Asia/Bangladesh/Rajshahi/ | National Institute of Laboratory Medicine and Referral Center | Genomic Research Lab, BCSIR | Shahina Akter et al |
| EPI_ISL_498238 | Senegal/45436/2020 | 2020-05-27 | Africa/Senegal/Dakar/ | Institut Pasteur de Dakar | Institut Pasteur de Dakar | Ndongo Dia et al |
| EPI_ISL_498240 | Senegal/52357/2020 | 2020-06-03 | Africa/Senegal/Dakar/Mbao | Institut Pasteur de Dakar | Institut Pasteur de Dakar | Ndongo Dia et al |
| EPI_ISL_498247 | Senegal/62431/2020 | 2020-06-10 | Africa/Senegal/Dakar/ | Institut Pasteur de Dakar | Institut Pasteur de Dakar | Ndongo Dia et al |
| EPI_ISL_498252 | Senegal/83493/2020 | 2020-07-09 | Africa/Senegal/Dakar/ | Institut Pasteur de Dakar | Institut Pasteur de Dakar | Ndongo Dia et al |
| EPI_ISL_498253 | Bangladesh/BCSIR-NILMRC-258/2020 | 2020-07-07 | Asia/Bangladesh/Khulna/Jashore | National Institute of Laboratory Medicine and Referral Center | Genomic Research Lab, BCSIR | Tanjina Akhter Banu et al |
| EPI_ISL_498254 | Bangladesh/BCSIR-NILMRC-260/2020 | 2020-07-07 | Asia/Bangladesh/Khulna/Jashore | National Institute of Laboratory Medicine and Referral Center | Genomic Research Lab, BCSIR | Tanjina Akhter Banu et al |
| EPI_ISL_498417 | Bangladesh/BCSIR-NILMRC-276/2020 | 2020-07-19 | Asia/Bangladesh/Chattogram/Brahmanb | National Institute of Laboratory Medicine and Referral Center | Genomic Research Lab, BCSIR | Tasnim Nafisa et al |
| EPI_ISL_498467 | Bangladesh/BCSIR-NILMRC-285/2020 | 2020-07-21 | Asia/Bangladesh/Chattogram/Chandpur | National Institute of Laboratory Medicine and Referral Center | Genomic Research Lab, BCSIR | Mahmuda Yeasmin et al |
| EPI_ISL_498549 | Bangladesh/BCSIR-NILMRC-290/2020 | 2020-07-14 | Asia/Bangladesh/Rangpur/Nilphamari | National Institute of Laboratory Medicine and Referral Center | Genomic Research Lab, BCSIR | Asish Kumar Ghosh et al |
| EPI_ISL_498552 | Lebanon/LB-LR9/2020 | 2020-03-18 | Asia/Lebanon/Beirut/ | Lebanese American University | Lebanese American University | Abi Habib et al |
| EPI_ISL_498562 | Italy/FVG-ICGEB-S217/2020 | 2020-07-07 | Europe/Italy/Friuli Venezia Giulia/ | Laboratory of Molecular Virology International Center for Genetic Engineering and Biotechnology (ICGEB) | ARGO Open Lab Platform for Genome Sequencing | Licastro D et al |
| EPI_ISL_498571 | Singapore/673/2020 | 2020-07-02 | Asia/Singapore/Singapore/ | National Public Health Laboratory, National Centre for Infectious Diseases | National Public Health Laboratory, National Centre for Infectious Diseases | Mak TM et al |
| EPI_ISL_498576 | Singapore/678/2020 | 2020-06-23 | Asia/Singapore/Singapore/ | National Public Health Laboratory, National Centre for Infectious Diseases | National Public Health Laboratory, National Centre for Infectious Diseases | Mak TM et al |

| | | | | | | |
|---|---|---|---|---|---|---|
| EPI_ISL_498604 | Singapore/706/2020 | 2020-06-13 | Asia/Singapore/Singapore/ | National Public Health Laboratory, National Centre for Infectious Diseases | National Public Health Laboratory, National Centre for Infectious Diseases | Mak TM et al |
| EPI_ISL_498630 | USA/UT-01856/2020 | 2020-04-13 | North America/USA/Utah/ | Utah Public Health Laboratory | Utah Public Health Laboratory | Heidi Butz et al |
| EPI_ISL_498645 | USA/UT-03338/2020 | 2020-04-22 | North America/USA/Utah/ | Utah Public Health Laboratory | Utah Public Health Laboratory | Heidi Butz et al |
| EPI_ISL_498647 | USA/UT-03345/2020 | 2020-04-22 | North America/USA/Utah/ | Utah Public Health Laboratory | Utah Public Health Laboratory | Heidi Butz et al |
| EPI_ISL_498653 | USA/UT-03368/2020 | 2020-04-23 | North America/USA/Utah/ | Utah Public Health Laboratory | Utah Public Health Laboratory | Heidi Butz et al |
| EPI_ISL_498668 | USA/UT-02898/2020 | 2020-04-30 | North America/USA/Utah/ | Utah Public Health Laboratory | Utah Public Health Laboratory | Heidi Butz et al |
| EPI_ISL_498684 | USA/UT-04206/2020 | 2020-05-14 | North America/USA/Utah/ | Utah Public Health Laboratory | Utah Public Health Laboratory | Heidi Butz et al |
| EPI_ISL_498686 | USA/UT-04208/2020 | 2020-05-14 | North America/USA/Utah/ | Utah Public Health Laboratory | Utah Public Health Laboratory | Heidi Butz et al |
| EPI_ISL_498700 | USA/CA-QDX-265/2020 | 2020-05-01 | North America/USA/California/ | Quest Diagnostics | Quest Diagnostics | Rosenthal et al |
| EPI_ISL_498704 | USA/CA-QDX-269/2020 | 2020-05-01 | North America/USA/California/ | Quest Diagnostics | Quest Diagnostics | Rosenthal et al |
| EPI_ISL_498712 | USA/FL-QDX-231/2020 | 2020-04-18 | North America/USA/Florida/ | Quest Diagnostics | Quest Diagnostics | Rosenthal et al |
| EPI_ISL_498713 | USA/GA-QDX-232/2020 | 2020-04-24 | North America/USA/Georgia/ | Quest Diagnostics | Quest Diagnostics | Rosenthal et al |
| EPI_ISL_498738 | USA/TX-QDX-218/2020 | 2020-04-10 | North America/USA/Texas/ | Quest Diagnostics | Quest Diagnostics | Rosenthal et al |
| EPI_ISL_498785 | Bangladesh/BCSIR-NILMRC-298/2020 | 2020-07-18 | Asia/Bangladesh/Sylhet/ | National Institute of Laboratory Medicine and Referral Center | Genomic Research Lab, BCSIR | Md. Saddam Hossain et al |
| EPI_ISL_498786 | Bangladesh/BCSIR-NILMRC-306/2020 | 2020-07-18 | Asia/Bangladesh/Sylhet/Moulvibazar | National Institute of Laboratory Medicine and Referral Center | Genomic Research Lab, BCSIR | Md. Saddam Hossain et al |
| EPI_ISL_498788 | Bangladesh/BCSIR-NILMRC-315/2020 | 2020-05-31 | Asia/Bangladesh/Mymensingh/ | National Institute of Laboratory Medicine and Referral Center | Genomic Research Lab, BCSIR | Md. Saddam Hossain et al |
| EPI_ISL_498789 | Bangladesh/BCSIR-NILMRC-318/2020 | 2020-05-31 | Asia/Bangladesh/Mymensingh/ | National Institute of Laboratory Medicine and Referral Center | Genomic Research Lab, BCSIR | Md. Saddam Hossain et al |
| EPI_ISL_498790 | Bangladesh/BCSIR-NILMRC-322/2020 | 2020-05-31 | Asia/Bangladesh/Mymensingh/ | National Institute of Laboratory Medicine and Referral Center | Genomic Research Lab, BCSIR | Md. Saddam Hossain et al |
| EPI_ISL_498791 | Bangladesh/BCSIR-NILMRC-323/2020 | 2020-06-03 | Asia/Bangladesh/Mymensingh/ | National Institute of Laboratory Medicine and Referral Center | Genomic Research Lab, BCSIR | Md. Saddam Hossain et al |
| EPI_ISL_498792 | Bangladesh/BCSIR-NILMRC-326/2020 | 2020-06-03 | Asia/Bangladesh/Mymensingh/ | National Institute of Laboratory Medicine and Referral Center | Genomic Research Lab, BCSIR | Md. Saddam Hossain et al |

| | | | | | | |
|---|---|---|---|---|---|---|
| EPI_ISL_498793 | Bangladesh/BCSIR-NILMRC-327/2020 | 2020-06-03 | Asia/Bangladesh/Mymensingh/ | National Institute of Laboratory Medicine and Referral Center | Genomic Research Lab, BCSIR | Md. Murshed Hasan Sarkar et al |
| EPI_ISL_498795 | Bangladesh/BCSIR-NILMRC-329/2020 | 2020-06-03 | Asia/Bangladesh/Mymensingh/ | National Institute of Laboratory Medicine and Referral Center | Genomic Research Lab, BCSIR | Md. Murshed Hasan Sarkar et al |
| EPI_ISL_498796 | Bangladesh/BCSIR-NILMRC-333/2020 | 2020-06-04 | Asia/Bangladesh/Mymensingh/ | National Institute of Laboratory Medicine and Referral Center | Genomic Research Lab, BCSIR | Md. Murshed Hasan Sarkar et al |
| EPI_ISL_498801 | Bangladesh/BCSIR-NILMRC-340/2020 | 2020-07-15 | Asia/Bangladesh/Rajshahi/ | National Institute of Laboratory Medicine and Referral Center | Genomic Research Lab, BCSIR | Md. Murshed Hasan Sarkar et al |
| EPI_ISL_498802 | Bangladesh/BCSIR-NILMRC-341/2020 | 2020-07-15 | Asia/Bangladesh/Rajshahi/ | National Institute of Laboratory Medicine and Referral Center | Genomic Research Lab, BCSIR | Md. Murshed Hasan Sarkar et al |
| EPI_ISL_498804 | Bangladesh/BCSIR-NILMRC-345/2020 | 2020-07-12 | Asia/Bangladesh/Rajshahi/ | National Institute of Laboratory Medicine and Referral Center | Genomic Research Lab, BCSIR | Md. Murshed Hasan Sarkar et al |
| EPI_ISL_498805 | Bangladesh/BCSIR-NILMRC-346/2020 | 2020-07-15 | Asia/Bangladesh/Rajshahi/ | National Institute of Laboratory Medicine and Referral Center | Genomic Research Lab, BCSIR | Md. Murshed Hasan Sarkar et al |
| EPI_ISL_498806 | Bangladesh/BCSIR-NILMRC-349/2020 | 2020-07-15 | Asia/Bangladesh/Khulna/ | National Institute of Laboratory Medicine and Referral Center | Genomic Research Lab, BCSIR | Md. Murshed Hasan Sarkar et al |
| EPI_ISL_498814 | Bangladesh/BCSIR-NILMRC-281/2020 | 2020-07-21 | Asia/Bangladesh/Chattogram/Chandpur | National Institute of Laboratory Medicine and Referral Center | Genomic Research Lab, BCSIR | Abu Sayeed Mohammad Mahmud et al |
| EPI_ISL_498815 | Bangladesh/BCSIR-NILMRC-283/2020 | 2020-07-21 | Asia/Bangladesh/Chattogram/Chandpur | National Institute of Laboratory Medicine and Referral Center | Genomic Research Lab, BCSIR | Abu Sayeed Mohammad Mahmud et al |
| EPI_ISL_498816 | Bangladesh/BCSIR-NILMRC-288/2020 | 2020-07-14 | Asia/Bangladesh/Rangpur/Nilphamari | National Institute of Laboratory Medicine and Referral Center | Genomic Research Lab, BCSIR | Abu Sayeed Mohammad Mahmud et al |
| EPI_ISL_498818 | Bangladesh/BCSIR-NILMRC-305/2020 | 2020-07-18 | Asia/Bangladesh/Sylhet/Moulvibazar | National Institute of Laboratory Medicine and Referral Center | Genomic Research Lab, BCSIR | Abu Sayeed Mohammad Mahmud et al |
| EPI_ISL_498830 | Bangladesh/BCSIR-NILMRC-311/2020 | 2020-07-19 | Asia/Bangladesh/Sylhet/Moulvibazar | National Institute of Laboratory Medicine and Referral Center | Genomic Research Lab, BCSIR | Abu Sayeed Mohammad Mahmud et al |
| EPI_ISL_498892 | Bangladesh/BCSIR-NILMRC-312/2020 | 2020-07-20 | Asia/Bangladesh/Sylhet/Moulvibazar | National Institute of Laboratory Medicine and Referral Center | Genomic Research Lab, BCSIR | Abu Sayeed Mohammad Mahmud et al |
| EPI_ISL_499340 | England/SHEF-C80C8/2020 | 2020-04-04 | Europe/United Kingdom/England/ | Virology Department, Sheffield Teaching Hospitals NHS Foundation Trust/Department of Infection, Immunity and Cardiovascular Disease, The Medical School, University of Sheffield | COVID-19 Genomics UK (COG-UK) Consortium | Thushan de Silva et al |
| EPI_ISL_499358 | Wales/PHWC-168E74/2020 | 2020-04-01 | Europe/United Kingdom/Wales/ | Wales Specialist Virology Centre Sequencing lab: Pathogen Genomics Unit | COVID-19 Genomics UK (COG-UK) Consortium | Catherine Moore et al |
| EPI_ISL_499411 | Wales/PHWC-169471/2020 | 2020-05-31 | Europe/United Kingdom/Wales/ | Wales Specialist Virology Centre Sequencing lab: Pathogen Genomics Unit | COVID-19 Genomics UK (COG-UK) Consortium | Catherine Moore et al |

| | | | | | | |
|---|---|---|---|---|---|---|
| EPI_ISL_499506 | England/NORT-29C66F/2020 | 2020-04-23 | Europe/United Kingdom/England/ | Northumbria University / South Tees Hospitals NHS Foundation Trust / North Cumbria Integrated Care NHS Foundation Trust / North Tees and Hartlepool NHS Foundation Trust / Newcastle Hospitals NHS Foundation Trust | COVID-19 Genomics UK (COG-UK) Consortium | Darren L Smith et al |
| EPI_ISL_499535 | England/LIVE-A5F6A/2020 | 2020-03-21 | Europe/United Kingdom/England/ | Liverpool Clinical Laboratories | COVID-19 Genomics UK (COG-UK) Consortium | Sam Haldenby et al |
| EPI_ISL_499589 | England/LIVE-A6154/2020 | 2020-03-21 | Europe/United Kingdom/England/ | Liverpool Clinical Laboratories | COVID-19 Genomics UK (COG-UK) Consortium | Sam Haldenby et al |
| EPI_ISL_499650 | England/LIVE-A598A/2020 | 2020-03-17 | Europe/United Kingdom/England/ | Liverpool Clinical Laboratories | COVID-19 Genomics UK (COG-UK) Consortium | Sam Haldenby et al |
| EPI_ISL_499652 | England/LIVE-A59E4/2020 | 2020-03-16 | Europe/United Kingdom/England/ | Liverpool Clinical Laboratories | COVID-19 Genomics UK (COG-UK) Consortium | Sam Haldenby et al |
| EPI_ISL_499678 | England/LIVE-A6048/2020 | 2020-03-21 | Europe/United Kingdom/England/ | Liverpool Clinical Laboratories | COVID-19 Genomics UK (COG-UK) Consortium | Sam Haldenby et al |
| EPI_ISL_499807 | England/NORT-2A0413/2020 | 2020-04-25 | Europe/United Kingdom/England/ | Northumbria University / South Tees Hospitals NHS Foundation Trust / North Cumbria Integrated Care NHS Foundation Trust / North Tees and Hartlepool NHS Foundation Trust / Newcastle Hospitals NHS Foundation Trust | COVID-19 Genomics UK (COG-UK) Consortium | Darren L Smith et al |
| EPI_ISL_499843 | England/LIVE-A4F89/2020 | 2020-02-28 | Europe/United Kingdom/England/ | Liverpool Clinical Laboratories | COVID-19 Genomics UK (COG-UK) Consortium | Sam Haldenby et al |
| EPI_ISL_499900 | England/LIVE-A51DD/2020 | 2020-03-01 | Europe/United Kingdom/England/ | Liverpool Clinical Laboratories | COVID-19 Genomics UK (COG-UK) Consortium | Sam Haldenby et al |
| EPI_ISL_499916 | England/LIVE-A4E9B/2020 | 2020-03-05 | Europe/United Kingdom/England/ | Liverpool Clinical Laboratories | COVID-19 Genomics UK (COG-UK) Consortium | Sam Haldenby et al |
| EPI_ISL_499921 | England/LIVE-A2ED9/2020 | 2020-05-20 | Europe/United Kingdom/England/ | Liverpool Clinical Laboratories | COVID-19 Genomics UK (COG-UK) Consortium | Sam Haldenby et al |

| | | | | | | |
|---|---|---|---|---|---|---|
| EPI_ISL_500084 | England/LIVE-A4D16/2020 | 2020-03-12 | Europe/United Kingdom/England/ | Liverpool Clinical Laboratories | COVID-19 Genomics UK (COG-UK) Consortium | Sam Haldenby et al |
| EPI_ISL_500110 | England/LIVE-A5191/2020 | 2020-03-14 | Europe/United Kingdom/England/ | Liverpool Clinical Laboratories | COVID-19 Genomics UK (COG-UK) Consortium | Sam Haldenby et al |
| EPI_ISL_500117 | England/LIVE-A546B/2020 | 2020-03-19 | Europe/United Kingdom/England/ | Liverpool Clinical Laboratories | COVID-19 Genomics UK (COG-UK) Consortium | Sam Haldenby et al |
| EPI_ISL_500126 | England/LIVE-A5568/2020 | 2020-03-21 | Europe/United Kingdom/England/ | Liverpool Clinical Laboratories | COVID-19 Genomics UK (COG-UK) Consortium | Sam Haldenby et al |
| EPI_ISL_500133 | England/LIVE-A561A/2020 | 2020-03-19 | Europe/United Kingdom/England/ | Liverpool Clinical Laboratories | COVID-19 Genomics UK (COG-UK) Consortium | Sam Haldenby et al |
| EPI_ISL_500163 | Spain/AN-IBV-003946/2020 | 2020-04-07 | Europe/Spain/Andalusia/Atarfe | Hospital Universitario Virgen de las Nieves de Granada-SAS | SeqCOVID-SPAIN consortium/IBV(CSIC) | Mercedes P√©rez Ruiz et al |
| EPI_ISL_500166 | Spain/CM-IBV-003919/2020 | 2020-04-29 | Europe/Spain/Castilla la Mancha/Bogar | Complejo Hospitalario Universitario de Albacete | SeqCOVID-SPAIN consortium/IBV(CSIC) | Encarnacion Simarro C√≥rdoba et al |
| EPI_ISL_500167 | Spain/GA-IBV-004070/2020 | 2020-03-19 | Europe/Spain/Galicia/Boiro | Hospital Cl√≠nico Universitario de Santiago de Compostela | SeqCOVID-SPAIN consortium/IBV(CSIC) | Jos√© Javier Costa Alcalde et al |
| EPI_ISL_500173 | Spain/AN-IBV-004006/2020 | 2020-04-21 | Europe/Spain/Andalusia/Durcal | Hospital Universitario Virgen de las Nieves de Granada-SAS | SeqCOVID-SPAIN consortium/IBV(CSIC) | Mercedes P√©rez Ruiz et al |
| EPI_ISL_500185 | Spain/AN-IBV-003970/2020 | 2020-04-12 | Europe/Spain/Andalusia/Granada ES | Hospital Universitario Virgen de las Nieves de Granada-SAS | SeqCOVID-SPAIN consortium/IBV(CSIC) | Mercedes P√©rez Ruiz et al |
| EPI_ISL_500187 | Spain/AN-IBV-003984/2020 | 2020-04-16 | Europe/Spain/Andalusia/Granada ES | Hospital Universitario Virgen de las Nieves de Granada-SAS | SeqCOVID-SPAIN consortium/IBV(CSIC) | Mercedes P√©rez Ruiz et al |
| EPI_ISL_500188 | Spain/AN-IBV-003985/2020 | 2020-04-16 | Europe/Spain/Andalusia/Granada ES | Hospital Universitario Virgen de las Nieves de Granada-SAS | SeqCOVID-SPAIN consortium/IBV(CSIC) | Mercedes P√©rez Ruiz et al |
| EPI_ISL_500190 | Spain/AN-IBV-003991/2020 | 2020-04-17 | Europe/Spain/Andalusia/Granada ES | Hospital Universitario Virgen de las Nieves de Granada-SAS | SeqCOVID-SPAIN consortium/IBV(CSIC) | Mercedes P√©rez Ruiz et al |

| | | | | | | |
|---|---|---|---|---|---|---|
| EPI_ISL_500194 | Spain/AN-IBV-003998/2020 | 2020-04-20 | Europe/Spain/Andalusia/Granada ES | Hospital Universitario Virgen de las Nieves de Granada-SAS | SeqCOVID-SPAIN consortium/IBV(CSIC) | Mercedes P√©rez Ruiz et al |
| EPI_ISL_500198 | Spain/AN-IBV-004002/2020 | 2020-04-20 | Europe/Spain/Andalusia/Granada ES | Hospital Universitario Virgen de las Nieves de Granada-SAS | SeqCOVID-SPAIN consortium/IBV(CSIC) | Mercedes P√©rez Ruiz et al |
| EPI_ISL_500201 | Spain/AN-IBV-004023/2020 | 2020-04-27 | Europe/Spain/Andalusia/Granada ES | Hospital Universitario Virgen de las Nieves de Granada-SAS | SeqCOVID-SPAIN consortium/IBV(CSIC) | Mercedes P√©rez Ruiz et al |
| EPI_ISL_500202 | Spain/AN-IBV-003948/2020 | 2020-04-07 | Europe/Spain/Andalusia/Guadix | Hospital Universitario Virgen de las Nieves de Granada-SAS | SeqCOVID-SPAIN consortium/IBV(CSIC) | Mercedes P√©rez Ruiz et al |
| EPI_ISL_500207 | Spain/CM-IBV-003928/2020 | 2020-04-28 | Europe/Spain/Castilla la Mancha/Hellin | Complejo Hospitalario Universitario de Albacete | SeqCOVID-SPAIN consortium/IBV(CSIC) | Encarnacion Simarro C√≥rdoba et al |
| EPI_ISL_500211 | Spain/AN-IBV-003956/2020 | 2020-04-09 | Europe/Spain/Andalusia/Las Gabias | Hospital Universitario Virgen de las Nieves de Granada-SAS | SeqCOVID-SPAIN consortium/IBV(CSIC) | Mercedes P√©rez Ruiz et al |
| EPI_ISL_500214 | Spain/AN-IBV-003969/2020 | 2020-04-11 | Europe/Spain/Andalusia/Maracena | Hospital Universitario Virgen de las Nieves de Granada-SAS | SeqCOVID-SPAIN consortium/IBV(CSIC) | Mercedes P√©rez Ruiz et al |
| EPI_ISL_500216 | Spain/AN-IBV-003973/2020 | 2020-04-12 | Europe/Spain/Andalusia/Maracena | Hospital Universitario Virgen de las Nieves de Granada-SAS | SeqCOVID-SPAIN consortium/IBV(CSIC) | Mercedes P√©rez Ruiz et al |
| EPI_ISL_500223 | Spain/AN-IBV-004004/2020 | 2020-04-21 | Europe/Spain/Andalusia/Puebla de Don | Hospital Universitario Virgen de las Nieves de Granada-SAS | SeqCOVID-SPAIN consortium/IBV(CSIC) | Mercedes P√©rez Ruiz et al |
| EPI_ISL_500293 | Spain/PV-IBV-004229/2020 | 2020-04-05 | Europe/Spain/Basque Country/San Seba | Servicio de Microbiolog√≠a. Hospital Universitario Donostia. OSI Donostialdea. √Årea de Enfermedades Infecciosas, Grupo de Infecci√≥n Respiratoria y Resistencia Antimicrobiana. Instituto de Investigaci√≥n Sanitaria Biodonostia | SeqCOVID-SPAIN consortium/IBV(CSIC) | Gustavo Cilla et al |
| EPI_ISL_500294 | Spain/PV-IBV-004230/2020 | 2020-04-05 | Europe/Spain/Basque Country/San Seba | Servicio de Microbiolog√≠a. Hospital Universitario Donostia. OSI Donostialdea. √Årea de Enfermedades Infecciosas, Grupo de Infecci√≥n Respiratoria y Resistencia Antimicrobiana. Instituto de Investigaci√≥n Sanitaria Biodonostia | SeqCOVID-SPAIN consortium/IBV(CSIC) | Gustavo Cilla et al |

| | | | | | | |
|---|---|---|---|---|---|---|
| EPI_ISL_500310 | Spain/PV-IBV-004246/2020 | 2020-04-06 | Europe/Spain/Basque Country/San Seba | Servicio de Microbiolog√≠a. Hospital Universitario Donostia. OSI Donostialdea. √Årea de Enfermedades Infecciosas, Grupo de Infecci√≥n Respiratoria y Resistencia Antimicrobiana. Instituto de Investigaci√≥n Sanitaria Biodonostia | SeqCOVID-SPAIN consortium/IBV(CSIC) | Gustavo Cilla et al |
| EPI_ISL_500311 | Spain/PV-IBV-004247/2020 | 2020-04-06 | Europe/Spain/Basque Country/San Seba | Servicio de Microbiolog√≠a. Hospital Universitario Donostia. OSI Donostialdea. √Årea de Enfermedades Infecciosas, Grupo de Infecci√≥n Respiratoria y Resistencia Antimicrobiana. Instituto de Investigaci√≥n Sanitaria Biodonostia | SeqCOVID-SPAIN consortium/IBV(CSIC) | Gustavo Cilla et al |
| EPI_ISL_500312 | Spain/PV-IBV-004248/2020 | 2020-04-06 | Europe/Spain/Basque Country/San Seba | Servicio de Microbiolog√≠a. Hospital Universitario Donostia. OSI Donostialdea. √Årea de Enfermedades Infecciosas, Grupo de Infecci√≥n Respiratoria y Resistencia Antimicrobiana. Instituto de Investigaci√≥n Sanitaria Biodonostia | SeqCOVID-SPAIN consortium/IBV(CSIC) | Gustavo Cilla et al |
| EPI_ISL_500319 | Spain/PV-IBV-004255/2020 | 2020-04-06 | Europe/Spain/Basque Country/San Seba | Servicio de Microbiolog√≠a. Hospital Universitario Donostia. OSI Donostialdea. √Årea de Enfermedades Infecciosas, Grupo de Infecci√≥n Respiratoria y Resistencia Antimicrobiana. Instituto de Investigaci√≥n Sanitaria Biodonostia | SeqCOVID-SPAIN consortium/IBV(CSIC) | Gustavo Cilla et al |
| EPI_ISL_500333 | Spain/PV-IBV-004269/2020 | 2020-04-07 | Europe/Spain/Basque Country/San Seba | Servicio de Microbiolog√≠a. Hospital Universitario Donostia. OSI Donostialdea. √Årea de Enfermedades Infecciosas, Grupo de Infecci√≥n Respiratoria y Resistencia Antimicrobiana. Instituto de Investigaci√≥n Sanitaria Biodonostia | SeqCOVID-SPAIN consortium/IBV(CSIC) | Gustavo Cilla et al |
| EPI_ISL_500341 | Spain/AR-IBV-005046/2020 | 2020-04-21 | Europe/Spain/Aragon/Zaragoza | Servicio de Microbiolog√≠a, Hospital Miguel Servet, Zaragoza | SeqCOVID-SPAIN consortium/IBV(CSIC) | Antonio Rezusta L√≥pez et al |
| EPI_ISL_500346 | Spain/AR-IBV-005058/2020 | 2020-04-21 | Europe/Spain/Aragon/Zaragoza | Servicio de Microbiolog√≠a, Hospital Miguel Servet, Zaragoza | SeqCOVID-SPAIN consortium/IBV(CSIC) | Antonio Rezusta L√≥pez et al |
| EPI_ISL_500347 | Spain/AR-IBV-005061/2020 | 2020-04-21 | Europe/Spain/Aragon/Zaragoza | Servicio de Microbiolog√≠a, Hospital Miguel Servet, Zaragoza | SeqCOVID-SPAIN consortium/IBV(CSIC) | Antonio Rezusta L√≥pez et al |

| | | | | | | |
|---|---|---|---|---|---|---|
| EPI_ISL_500353 | Spain/AR-IBV-005078/2020 | 2020-04-22 | Europe/Spain/Aragon/Zaragoza | Servicio de Microbiolog√≠a, Hospital Miguel Servet, Zaragoza | SeqCOVID-SPAIN consortium/IBV(CSIC) | Antonio Rezusta L√≥pez et al |
| EPI_ISL_500354 | Spain/AR-IBV-005092/2020 | 2020-04-23 | Europe/Spain/Aragon/Zaragoza | Servicio de Microbiolog√≠a, Hospital Miguel Servet, Zaragoza | SeqCOVID-SPAIN consortium/IBV(CSIC) | Antonio Rezusta L√≥pez et al |
| EPI_ISL_500356 | Spain/AR-IBV-005100/2020 | 2020-04-25 | Europe/Spain/Aragon/Zaragoza | Servicio de Microbiolog√≠a, Hospital Miguel Servet, Zaragoza | SeqCOVID-SPAIN consortium/IBV(CSIC) | Antonio Rezusta L√≥pez et al |
| EPI_ISL_500367 | Spain/AR-IBV-005129/2020 | 2020-05-07 | Europe/Spain/Aragon/Zaragoza | Servicio de Microbiolog√≠a, Hospital Miguel Servet, Zaragoza | SeqCOVID-SPAIN consortium/IBV(CSIC) | Antonio Rezusta L√≥pez et al |
| EPI_ISL_500391 | Spain/RI-IBV-005957/2020 | 2020-03-05 | Europe/Spain/La Rioja/Haro | Centro de Investigaci√≥n Biom√©dica de La Rioja - Hospital San Pedro Logro√±o | SeqCOVID-SPAIN consortium/IBV(CSIC) | Mar√≠a de Toro et al |
| EPI_ISL_500393 | Spain/RI-IBV-005959/2020 | 2020-03-05 | Europe/Spain/La Rioja/Haro | Centro de Investigaci√≥n Biom√©dica de La Rioja - Hospital San Pedro Logro√±o | SeqCOVID-SPAIN consortium/IBV(CSIC) | Mar√≠a de Toro et al |
| EPI_ISL_500395 | Spain/RI-IBV-005961/2020 | 2020-03-07 | Europe/Spain/La Rioja/Haro | Centro de Investigaci√≥n Biom√©dica de La Rioja - Hospital San Pedro Logro√±o | SeqCOVID-SPAIN consortium/IBV(CSIC) | Mar√≠a de Toro et al |
| EPI_ISL_500413 | Spain/RI-IBV-005979/2020 | 2020-03-11 | Europe/Spain/La Rioja/Santo Domingo d | Centro de Investigaci√≥n Biom√©dica de La Rioja - Hospital San Pedro Logro√±o | SeqCOVID-SPAIN consortium/IBV(CSIC) | Mar√≠a de Toro et al |
| EPI_ISL_500420 | Spain/RI-IBV-005986/2020 | 2020-03-22 | Europe/Spain/La Rioja/Logrono | Centro de Investigaci√≥n Biom√©dica de La Rioja - Hospital San Pedro Logro√±o | SeqCOVID-SPAIN consortium/IBV(CSIC) | Mar√≠a de Toro et al |
| EPI_ISL_500427 | Spain/RI-IBV-005995/2020 | 2020-03-22 | Europe/Spain/La Rioja/Albelda de Iregua | Centro de Investigaci√≥n Biom√©dica de La Rioja - Hospital San Pedro Logro√±o | SeqCOVID-SPAIN consortium/IBV(CSIC) | Mar√≠a de Toro et al |
| EPI_ISL_500436 | Spain/RI-IBV-006005/2020 | 2020-03-21 | Europe/Spain/La Rioja/Casalarreina | Centro de Investigaci√≥n Biom√©dica de La Rioja - Hospital San Pedro Logro√±o | SeqCOVID-SPAIN consortium/IBV(CSIC) | Mar√≠a de Toro et al |
| EPI_ISL_500447 | Spain/RI-IBV-006017/2020 | 2020-03-30 | Europe/Spain/La Rioja/Santo Domingo d | Centro de Investigaci√≥n Biom√©dica de La Rioja - Hospital San Pedro Logro√±o | SeqCOVID-SPAIN consortium/IBV(CSIC) | Mar√≠a de Toro et al |
| EPI_ISL_500457 | Spain/RI-IBV-006027/2020 | 2020-03-31 | Europe/Spain/La Rioja/Logrono | Centro de Investigaci√≥n Biom√©dica de La Rioja - Hospital San Pedro Logro√±o | SeqCOVID-SPAIN consortium/IBV(CSIC) | Mar√≠a de Toro et al |

| | | | | | | |
|---|---|---|---|---|---|---|
| EPI_ISL_500460 | Brazil/PE-IAM08/2020 | 2020-04-07 | South America/Brazil/Pernambuco/Recif | LACEN/PE | WallauLab, Aggeu Magalhaes Institute | Marcelo Henrique Santos Paiva et al |
| EPI_ISL_500464 | Brazil/PE-IAM139/2020 | 2020-04-09 | South America/Brazil/Pernambuco/Recif | LACEN/PE | WallauLab, Aggeu Magalhaes Institute | Marcelo Henrique Santos Paiva et al |
| EPI_ISL_500466 | Brazil/PE-IAM167/2020 | 2020-04-09 | South America/Brazil/Pernambuco/Recif | LACEN/PE | WallauLab, Aggeu Magalhaes Institute | Marcelo Henrique Santos Paiva et al |
| EPI_ISL_500471 | Brazil/PE-IAM215/2020 | 2020-04-06 | South America/Brazil/Pernambuco/Olinc | LACEN/PE | WallauLab, Aggeu Magalhaes Institute | Marcelo Henrique Santos Paiva et al |
| EPI_ISL_500478 | Brazil/PE-IAM305/2020 | 2020-04-13 | South America/Brazil/Pernambuco/Cama | LACEN/PE | WallauLab, Aggeu Magalhaes Institute | Marcelo Henrique Santos Paiva et al |
| EPI_ISL_500485 | Brazil/PE-IAM87/2020 | 2020-04-08 | South America/Brazil/Pernambuco/Sao I | LACEN/PE | WallauLab, Aggeu Magalhaes Institute | Marcelo Henrique Santos Paiva et al |
| EPI_ISL_500486 | Brazil/PE-IAM89/2020 | 2020-04-07 | South America/Brazil/Pernambuco/Cama | LACEN/PE | WallauLab, Aggeu Magalhaes Institute | Marcelo Henrique Santos Paiva et al |
| EPI_ISL_500503 | USA/NY-UW-MAYO21/2020 | 2020-03-18 | North America/USA/New York/ | Mayo Clinic Laboratories | University of Washington Virology Lab | Pavitra Roychoudhury et al |
| EPI_ISL_500512 | USA/WA-UW-MAYO57/2020 | 2020-03-27 | North America/USA/Washington/ | Mayo Clinic Laboratories | University of Washington Virology Lab | Pavitra Roychoudhury et al |
| EPI_ISL_500544 | Singapore/723/2020 | 2020-06-24 | Asia/Singapore/Singapore/ | Singapore General Hospital | Department of Microbiology | Nurdyana Abdul Rahman et al |
| EPI_ISL_500550 | Singapore/729/2020 | 2020-06-22 | Asia/Singapore/Singapore/ | Singapore General Hospital | Department of Microbiology | Nurdyana Abdul Rahman et al |
| EPI_ISL_500554 | Singapore/744/2020 | 2020-06-30 | Asia/Singapore/Singapore/ | Singapore General Hospital | Department of Microbiology | Nurdyana Abdul Rahman et al |
| EPI_ISL_500556 | Singapore/742/2020 | 2020-07-04 | Asia/Singapore/Singapore/ | Singapore General Hospital | Department of Microbiology | Nurdyana Abdul Rahman et al |
| EPI_ISL_500563 | Singapore/751/2020 | 2020-06-13 | Asia/Singapore/Singapore/ | Singapore General Hospital | Department of Microbiology | Nurdyana Abdul Rahman et al |
| EPI_ISL_500569 | Singapore/755/2020 | 2020-06-06 | Asia/Singapore/Singapore/ | Singapore General Hospital | Department of Microbiology | Nurdyana Abdul Rahman et al |

| | | | | | | |
|---|---|---|---|---|---|---|
| EPI_ISL_500613 | Australia/NSW2042/2020 | 2020-03-20 | Oceania/Australia/New South Wales/ | Area of Virology, Serology and Virology Division (SAViD), New South Wales Health Pathology Randwick | Area of Virology, Serology and Virology Division (SAViD), New South Wales Health Pathology Randwick | Rawlinson et al |
| EPI_ISL_500624 | Australia/NSW2131/2020 | 2020-04-05 | Oceania/Australia/New South Wales/ | Area of Virology, Serology and Virology Division (SAViD), New South Wales Health Pathology Randwick | Area of Virology, Serology and Virology Division (SAViD), New South Wales Health Pathology Randwick | Rawlinson et al |
| EPI_ISL_500656 | Australia/NSW2195/2020 | 2020-03-21 | Oceania/Australia/New South Wales/ | Area of Virology, Serology and Virology Division (SAViD), New South Wales Health Pathology Randwick | Area of Virology, Serology and Virology Division (SAViD), New South Wales Health Pathology Randwick | Rawlinson et al |
| EPI_ISL_500716 | Turkey/Pen07-2/2020 | 2020-03-17 | Europe/Turkey/Sakarya/ | BSL3 Lab Pendik Veterinary Control Institute | Department of Medicinal Genetics, Bursa Uludaƒü University, Faculty of medicine By Sehime G√ºls√ºn Temel, Adem Alemdar, Kadir Ye≈üilbaƒü | Mustafa HASOKSUZ et al |
| EPI_ISL_500777 | Norway/2854/2020 | 2020-06-29 | Europe/Norway/Norway/ | Foerde Hospital, Department of Microbiology | Norwegian Institute of Public Health, Department of Virology | Kathrine Stene-Johansen et al |
| EPI_ISL_500834 | USA/VA-DCLS-0671/2020 | 2020-04-17 | North America/USA/Virginia/ | Virginia DCLS | Virginia DCLS | Virginia DCLS et al |
| EPI_ISL_500837 | USA/VA-DCLS-0674/2020 | 2020-04-17 | North America/USA/Virginia/ | Virginia DCLS | Virginia DCLS | Virginia DCLS et al |
| EPI_ISL_500838 | USA/VA-DCLS-0675/2020 | 2020-04-18 | North America/USA/Virginia/ | Virginia DCLS | Virginia DCLS | Virginia DCLS et al |

| | | | | | | |
|---|---|---|---|---|---|---|
| EPI_ISL_500840 | USA/VA-DCLS-0677/2020 | 2020-04-18 | North America/USA/Virginia/ | Virginia DCLS | Virginia DCLS | Virginia DCLS et al |
| EPI_ISL_500851 | USA/VA-DCLS-0690/2020 | 2020-04-18 | North America/USA/Virginia/ | Virginia DCLS | Virginia DCLS | Virginia DCLS et al |
| EPI_ISL_500852 | USA/VA-DCLS-0691/2020 | 2020-04-18 | North America/USA/Virginia/ | Virginia DCLS | Virginia DCLS | Virginia DCLS et al |
| EPI_ISL_500853 | USA/VA-DCLS-0692/2020 | 2020-04-18 | North America/USA/Virginia/ | Virginia DCLS | Virginia DCLS | Virginia DCLS et al |
| EPI_ISL_500858 | USA/VA-DCLS-0702/2020 | 2020-04-18 | North America/USA/Virginia/ | Virginia DCLS | Virginia DCLS | Virginia DCLS et al |
| EPI_ISL_500863 | USA/VA-DCLS-0711/2020 | 2020-04-25 | North America/USA/Virginia/ | Virginia DCLS | Virginia DCLS | Virginia DCLS et al |
| EPI_ISL_500872 | Brazil/PE-IAM29/2020 | 2020-04-08 | South America/Brazil/Pernambuco/Olinc | LACEN/PE | WallauLab, Aggeu Magalhaes Institute | Marcelo Henrique Santos Paiva et al |
| EPI_ISL_500873 | Brazil/PE-IAM291/2020 | 2020-04-13 | South America/Brazil/Pernambuco/Recif | LACEN/PE | WallauLab, Aggeu Magalhaes Institute | Marcelo Henrique Santos Paiva et al |
| EPI_ISL_500947 | India/GJ-GBRC317/2020 | 2020-06-17 | Asia/India/Gujarat/Surat | Department of MicroBiology, Government Medical College, Surat | Gujarat Biotechnology Research Centre | Maharshi Pandya et al |
| EPI_ISL_500948 | India/GJ-GBRC321/2020 | 2020-06-17 | Asia/India/Gujarat/Surat | Department of MicroBiology, Government Medical College, Surat | Gujarat Biotechnology Research Centre | Zuber Saiyed et al |
| EPI_ISL_500950 | India/GJ-GBRC322b/2020 | 2020-06-17 | Asia/India/Gujarat/Surat | Department of MicroBiology, Government Medical College, Surat | Gujarat Biotechnology Research Centre | Labdhi Pandya et al |
| EPI_ISL_500963 | NorthernIreland/NIRE-106F0C/2020 | 2020-05-22 | Europe/United Kingdom/Northern Irelar | Regional Virus Laboratory, Belfast Health and Social Care Trust | Wellcome Sanger Institute for the COVID-19 Genomics UK (COG-UK) consortium | Conall McCaughey et al |
| EPI_ISL_501023 | NorthernIreland/NIRE-106F57/2020 | 2020-05-30 | Europe/United Kingdom/Northern Irelar | Regional Virus Laboratory, Belfast Health and Social Care Trust | Wellcome Sanger Institute for the COVID-19 Genomics UK (COG-UK) consortium | Conall McCaughey et al |
| EPI_ISL_501066 | NorthernIreland/NIRE-10678C/2020 | 2020-05-14 | Europe/United Kingdom/Northern Irelar | Regional Virus Laboratory, Belfast Health and Social Care Trust | Wellcome Sanger Institute for the COVID-19 Genomics UK (COG-UK) consortium | Conall McCaughey et al |
| EPI_ISL_501085 | USA/UN-UW-13444/2020 | 2020-06-30 | North America/USA/USA/ | University of Washington Virology Lab | University of Washington Virology Lab | Pavitra Roychoudhury et al |

| | | | | | | |
|---|---|---|---|---|---|---|
| EPI_ISL_501087 | USA/WA-UW-10212/2020 | 2020-05-31 | North America/USA/Washington/ | University of Washington Virology Lab | University of Washington Virology Lab | Pavitra Roychoudhury et al |
| EPI_ISL_501091 | USA/WA-UW-10613/2020 | 2020-06-12 | North America/USA/Washington/ | University of Washington Virology Lab | University of Washington Virology Lab | Pavitra Roychoudhury et al |
| EPI_ISL_501095 | USA/WA-UW-11279/2020 | 2020-06-21 | North America/USA/Washington/ | University of Washington Virology Lab | University of Washington Virology Lab | Pavitra Roychoudhury et al |
| EPI_ISL_501097 | USA/WA-UW-11300/2020 | 2020-06-20 | North America/USA/Washington/ | University of Washington Virology Lab | University of Washington Virology Lab | Pavitra Roychoudhury et al |
| EPI_ISL_501107 | USA/WA-UW-12149/2020 | 2020-06-25 | North America/USA/Washington/ | University of Washington Virology Lab | University of Washington Virology Lab | Pavitra Roychoudhury et al |
| EPI_ISL_501112 | USA/WA-UW-12596/2020 | 2020-06-27 | North America/USA/Washington/ | University of Washington Virology Lab | University of Washington Virology Lab | Pavitra Roychoudhury et al |
| EPI_ISL_501113 | USA/WA-UW-12612/2020 | 2020-06-28 | North America/USA/Washington/ | University of Washington Virology Lab | University of Washington Virology Lab | Pavitra Roychoudhury et al |
| EPI_ISL_501114 | USA/WA-UW-12689/2020 | 2020-06-29 | North America/USA/Washington/ | University of Washington Virology Lab | University of Washington Virology Lab | Pavitra Roychoudhury et al |
| EPI_ISL_501120 | USA/WA-UW-13226/2020 | 2020-07-01 | North America/USA/Washington/ | University of Washington Virology Lab | University of Washington Virology Lab | Pavitra Roychoudhury et al |
| EPI_ISL_501123 | USA/WA-UW-13233/2020 | 2020-06-29 | North America/USA/Washington/ | University of Washington Virology Lab | University of Washington Virology Lab | Pavitra Roychoudhury et al |
| EPI_ISL_501128 | USA/WA-UW-13274/2020 | 2020-06-30 | North America/USA/Washington/ | University of Washington Virology Lab | University of Washington Virology Lab | Pavitra Roychoudhury et al |
| EPI_ISL_501136 | USA/WA-UW-13310/2020 | 2020-07-03 | North America/USA/Washington/ | University of Washington Virology Lab | University of Washington Virology Lab | Pavitra Roychoudhury et al |
| EPI_ISL_501137 | USA/WA-UW-13317/2020 | 2020-07-02 | North America/USA/Washington/ | University of Washington Virology Lab | University of Washington Virology Lab | Pavitra Roychoudhury et al |

| | | | | | | |
|---|---|---|---|---|---|---|
| EPI_ISL_501141 | USA/WA-UW-13356/2020 | 2020-07-01 | North America/USA/Washington/ | University of Washington Virology Lab | University of Washington Virology Lab | Pavitra Roychoudhury et al |
| EPI_ISL_501144 | USA/WA-UW-13366/2020 | 2020-07-01 | North America/USA/Washington/ | University of Washington Virology Lab | University of Washington Virology Lab | Pavitra Roychoudhury et al |
| EPI_ISL_501147 | USA/WA-UW-13395/2020 | 2020-07-02 | North America/USA/Washington/ | University of Washington Virology Lab | University of Washington Virology Lab | Pavitra Roychoudhury et al |
| EPI_ISL_501151 | USA/WA-UW-7745/2020 | 2020-04-19 | North America/USA/Washington/ | University of Washington Virology Lab | University of Washington Virology Lab | Pavitra Roychoudhury et al |
| EPI_ISL_501152 | USA/WA-UW-7764/2020 | 2020-04-20 | North America/USA/Washington/ | University of Washington Virology Lab | University of Washington Virology Lab | Pavitra Roychoudhury et al |
| EPI_ISL_501161 | USA/WA-UW-9551/2020 | 2020-05-19 | North America/USA/Washington/ | University of Washington Virology Lab | University of Washington Virology Lab | Pavitra Roychoudhury et al |
| EPI_ISL_501167 | USA/TX-GCID-192000051B/2020 | 2020-03-23 | North America/USA/Texas/Harris Count | Baylor College of Medicine | Baylor College of Medicine: HGSC | Vasanthi Avadhanula et al |
| EPI_ISL_501172 | USA/TX-GCID-192000296/2020 | 2020-04-13 | North America/USA/Texas/Harris Count | Baylor College of Medicine | Baylor College of Medicine: HGSC | Vasanthi Avadhanula et al |
| EPI_ISL_501173 | USA/TX-GCID-192000331/2020 | 2020-04-16 | North America/USA/Texas/Harris Count | Baylor College of Medicine | Baylor College of Medicine: HGSC | Vasanthi Avadhanula et al |
| EPI_ISL_501178 | Malaysia/0478/2020 | 2020-03-23 | Asia/Malaysia/Kuala Lumpur/ | Department of Medical Microbiology, University Malaya Medical Centre | Department of Medical Microbiology, Faculty of Medicine, University of Malaya | Yoong Min CHONG et al |
| EPI_ISL_501182 | Malaysia/1121/2020 | 2020-03-24 | Asia/Malaysia/Kuala Lumpur/ | Department of Medical Microbiology, University Malaya Medical Centre | Department of Medical Microbiology, Faculty of Medicine, University of Malaya | Yoong Min CHONG et al |

| | | | | | | |
|---|---|---|---|---|---|---|
| EPI_ISL_501187 | Malaysia/1798/2020 | 2020-03-25 | Asia/Malaysia/Kuala Lumpur/ | Department of Medical Microbiology, University Malaya Medical Centre | Department of Medical Microbiology, Faculty of Medicine, University of Malaya | Yoong Min CHONG et al |
| EPI_ISL_501190 | Malaysia/2079/2020 | 2020-03-26 | Asia/Malaysia/Kuala Lumpur/ | Department of Medical Microbiology, University Malaya Medical Centre | Department of Medical Microbiology, Faculty of Medicine, University of Malaya | Yoong Min CHONG et al |
| EPI_ISL_501194 | Malaysia/2363/2020 | 2020-03-27 | Asia/Malaysia/Kuala Lumpur/ | Department of Medical Microbiology, University Malaya Medical Centre | Department of Medical Microbiology, Faculty of Medicine, University of Malaya | Yoong Min CHONG et al |
| EPI_ISL_501195 | Malaysia/2735/2020 | 2020-03-28 | Asia/Malaysia/Kuala Lumpur/ | Department of Medical Microbiology, University Malaya Medical Centre | Department of Medical Microbiology, Faculty of Medicine, University of Malaya | Yoong Min CHONG et al |
| EPI_ISL_501199 | Malaysia/3012/2020 | 2020-03-29 | Asia/Malaysia/Kuala Lumpur/ | Department of Medical Microbiology, University Malaya Medical Centre | Department of Medical Microbiology, Faculty of Medicine, University of Malaya | Yoong Min CHONG et al |

| | | | | | | |
|---|---|---|---|---|---|---|
| EPI_ISL_501202 | Malaysia/3145/2020 | 2020-03-30 | Asia/Malaysia/Kuala Lumpur/ | Department of Medical Microbiology, University Malaya Medical Centre | Department of Medical Microbiology, Faculty of Medicine, University of Malaya | Yoong Min CHONG et al |
| EPI_ISL_501204 | Malaysia/3479/2020 | 2020-03-30 | Asia/Malaysia/Kuala Lumpur/ | Department of Medical Microbiology, University Malaya Medical Centre | Department of Medical Microbiology, Faculty of Medicine, University of Malaya | Yoong Min CHONG et al |
| EPI_ISL_501205 | Malaysia/3499/2020 | 2020-03-30 | Asia/Malaysia/Kuala Lumpur/ | Department of Medical Microbiology, University Malaya Medical Centre | Department of Medical Microbiology, Faculty of Medicine, University of Malaya | Yoong Min CHONG et al |
| EPI_ISL_501208 | Malaysia/3703/2020 | 2020-03-31 | Asia/Malaysia/Kuala Lumpur/ | Department of Medical Microbiology, University Malaya Medical Centre | Department of Medical Microbiology, Faculty of Medicine, University of Malaya | Yoong Min CHONG et al |
| EPI_ISL_501220 | Malaysia/6359/2020 | 2020-02-25 | Asia/Malaysia/Kuala Lumpur/ | Department of Medical Microbiology, University Malaya Medical Centre | Department of Medical Microbiology, Faculty of Medicine, University of Malaya | Yoong Min CHONG et al |

| | | | | | | |
|---|---|---|---|---|---|---|
| EPI_ISL_501221 | Malaysia/7618/2020 | 2020-03-17 | Asia/Malaysia/Kuala Lumpur/ | Department of Medical Microbiology, University Malaya Medical Centre | Department of Medical Microbiology, Faculty of Medicine, University of Malaya | Yoong Min CHONG et al |
| EPI_ISL_501224 | Malaysia/8816/2020 | 2020-03-18 | Asia/Malaysia/Kuala Lumpur/ | Department of Medical Microbiology, University Malaya Medical Centre | Department of Medical Microbiology, Faculty of Medicine, University of Malaya | Yoong Min CHONG et al |
| EPI_ISL_501230 | Greece/16034_HPI/2020 | 2020-04-24 | Europe/Greece/Central Greece/Livadeia | Hellenic Pasteur Institute, Public Health Laboratories | Hellenic Pasteur Institute, National Influenza Reference laboratory of Southern Greece & Unit of Bioinformatics and Applied Genomics | Vasiliki Pogka et al |
| EPI_ISL_501233 | Greece/19553_HPI/2020 | 2020-05-03 | Europe/Greece/Athens/ | Hellenic Pasteur Institute, National Influenza Reference laboratory of Southern Greece & Unit of Bioinformatics and Applied Genomics | Hellenic Pasteur Institute, National Influenza Reference laboratory of Southern Greece & Unit of Bioinformatics and Applied Genomics | Vasiliki Pogka et al |
| EPI_ISL_501236 | Greece/26072_HPI/2020 | 2020-05-13 | Europe/Greece/Athens/ | Hellenic Pasteur Institute, National Influenza Reference laboratory of Southern Greece & Unit of Bioinformatics and Applied Genomics | Hellenic Pasteur Institute, National Influenza Reference laboratory of Southern Greece & Unit of Bioinformatics and Applied Genomics | Vasiliki Pogka et al |

| | | | | | | |
|---|---|---|---|---|---|---|
| EPI_ISL_501253 | Ireland/LH-NVRL-20IRL22140/2020 | 2020-05-23 | Europe/Ireland/Louth/ | National Virus Reference Laboratory | National Virus Reference Laboratory | Michael Carr et al |
| EPI_ISL_501259 | Ireland/LH-NVRL-20IRL22956/2020 | 2020-05-27 | Europe/Ireland/Louth/ | National Virus Reference Laboratory | National Virus Reference Laboratory | Michael Carr et al |
| EPI_ISL_502779 | Brazil/PE-AMU0036/2020 | 2020-06-24 | South America/Brazil/Pernambuco/Caru | LACEN/PE | LABBE, Federal University of Pernambuco | WILSON JOSE DA SILVA JUNIOR et al |
| EPI_ISL_502875 | Brazil/PE-COV0260/2020 | 2020-06-24 | South America/Brazil/Pernambuco/Recif | LACEN/PE | LABBE, Federal University of Pernambuco | WILSON JOSE DA SILVA JUNIOR et al |
| EPI_ISL_504176 | Bangladesh/BCSIR-NILMRC-284/2020 | 2020-07-21 | Asia/Bangladesh/Chattogram/Chandpur | National Institute of Laboratory Medicine and Referral Center | Genomic Research Lab, BCSIR | Abu Sayeed Mohammad Mahmud et al |
| EPI_ISL_504182 | Bangladesh/BCSIR-NILMRC-330/2020 | 2020-06-03 | Asia/Bangladesh/Mymensingh/ | National Institute of Laboratory Medicine and Referral Center | Genomic Research Lab, BCSIR | Abu Sayeed Mohammad Mahmud et al |
| EPI_ISL_504190 | SouthAfrica/R12550-20/2020 | 2020-06-16 | Africa/South Africa/North-West/ | National Institute for Communicable Diseases of the National Health Laboratory Service | National Institute for Communicable Diseases of the National Health Laboratory Service | Allam M et al |
| EPI_ISL_504191 | SouthAfrica/R12566-20/2020 | 2020-06-16 | Africa/South Africa/North-West/ | National Institute for Communicable Diseases of the National Health Laboratory Service | National Institute for Communicable Diseases of the National Health Laboratory Service | Allam M et al |
| EPI_ISL_504194 | SouthAfrica/R12578-20/2020 | 2020-06-16 | Africa/South Africa/North-West/ | National Institute for Communicable Diseases of the National Health Laboratory Service | National Institute for Communicable Diseases of the National Health Laboratory Service | Allam M et al |
| EPI_ISL_504196 | SouthAfrica/R12583-20/2020 | 2020-06-16 | Africa/South Africa/North-West/ | National Institute for Communicable Diseases of the National Health Laboratory Service | National Institute for Communicable Diseases of the National Health Laboratory Service | Allam M et al |

| | | | | | | |
|---|---|---|---|---|---|---|
| EPI_ISL_504203 | SouthAfrica/R12641-20/2020 | 2020-06-16 | Africa/South Africa/North-West/ | National Institute for Communicable Diseases of the National Health Laboratory Service | National Institute for Communicable Diseases of the National Health Laboratory Service | Allam M et al |
| EPI_ISL_504205 | SouthAfrica/R12643-20/2020 | 2020-06-16 | Africa/South Africa/North-West/ | National Institute for Communicable Diseases of the National Health Laboratory Service | National Institute for Communicable Diseases of the National Health Laboratory Service | Allam M et al |
| EPI_ISL_504207 | SouthAfrica/R12647-20/2020 | 2020-06-16 | Africa/South Africa/North-West/ | National Institute for Communicable Diseases of the National Health Laboratory Service | National Institute for Communicable Diseases of the National Health Laboratory Service | Allam M et al |
| EPI_ISL_504213 | SouthAfrica/R12659-20/2020 | 2020-06-16 | Africa/South Africa/North-West/ | National Institute for Communicable Diseases of the National Health Laboratory Service | National Institute for Communicable Diseases of the National Health Laboratory Service | Allam M et al |
| EPI_ISL_504214 | SouthAfrica/R12660-20/2020 | 2020-06-16 | Africa/South Africa/North-West/ | National Institute for Communicable Diseases of the National Health Laboratory Service | National Institute for Communicable Diseases of the National Health Laboratory Service | Allam M et al |
| EPI_ISL_504220 | SouthAfrica/R12678-20/2020 | 2020-06-16 | Africa/South Africa/North-West/ | National Institute for Communicable Diseases of the National Health Laboratory Service | National Institute for Communicable Diseases of the National Health Laboratory Service | Allam M et al |

| | | | | | | |
|---|---|---|---|---|---|---|
| EPI_ISL_504223 | SouthAfrica/R12682-20/2020 | 2020-06-16 | Africa/South Africa/North-West/ | National Institute for Communicable Diseases of the National Health Laboratory Service | National Institute for Communicable Diseases of the National Health Laboratory Service | Allam M et al |
| EPI_ISL_504224 | SouthAfrica/R12683-20/2020 | 2020-06-16 | Africa/South Africa/North-West/ | National Institute for Communicable Diseases of the National Health Laboratory Service | National Institute for Communicable Diseases of the National Health Laboratory Service | Allam M et al |
| EPI_ISL_504227 | SouthAfrica/R12686-20/2020 | 2020-06-16 | Africa/South Africa/North-West/ | National Institute for Communicable Diseases of the National Health Laboratory Service | National Institute for Communicable Diseases of the National Health Laboratory Service | Allam M et al |
| EPI_ISL_504229 | SouthAfrica/R12695-20/2020 | 2020-06-16 | Africa/South Africa/North-West/ | National Institute for Communicable Diseases of the National Health Laboratory Service | National Institute for Communicable Diseases of the National Health Laboratory Service | Allam M et al |
| EPI_ISL_504231 | SouthAfrica/R12701-20/2020 | 2020-06-16 | Africa/South Africa/North-West/ | National Institute for Communicable Diseases of the National Health Laboratory Service | National Institute for Communicable Diseases of the National Health Laboratory Service | Allam M et al |
| EPI_ISL_504238 | SouthAfrica/R12711-20/2020 | 2020-06-16 | Africa/South Africa/North-West/ | National Institute for Communicable Diseases of the National Health Laboratory Service | National Institute for Communicable Diseases of the National Health Laboratory Service | Allam M et al |

| | | | | | | |
|---|---|---|---|---|---|---|
| EPI_ISL_504243 | SouthAfrica/R12723-20/2020 | 2020-06-16 | Africa/South Africa/North-West/ | National Institute for Communicable Diseases of the National Health Laboratory Service | National Institute for Communicable Diseases of the National Health Laboratory Service | Allam M et al |
| EPI_ISL_504244 | SouthAfrica/R12726-20/2020 | 2020-06-16 | Africa/South Africa/North-West/ | National Institute for Communicable Diseases of the National Health Laboratory Service | National Institute for Communicable Diseases of the National Health Laboratory Service | Allam M et al |
| EPI_ISL_506947 | SouthKorea/KCDC2153/2020 | 2020-02-24 | Asia/South Korea/South Korea/ | Division of Viral Diseases, Center for Laboratory Control of Infectious Diseases, Korea Centers for Diseases Control and Prevention | Division of Viral Diseases, Center for Laboratory Control of Infectious Diseases, Korea Centers for Diseases Control and Prevention | Jeong-Min Kim et al |
| EPI_ISL_506948 | SouthKorea/KCDC2154/2020 | 2020-02-27 | Asia/South Korea/South Korea/ | Division of Viral Diseases, Center for Laboratory Control of Infectious Diseases, Korea Centers for Diseases Control and Prevention | Division of Viral Diseases, Center for Laboratory Control of Infectious Diseases, Korea Centers for Diseases Control and Prevention | Jeong-Min Kim et al |
| EPI_ISL_506949 | SouthKorea/KCDC2155/2020 | 2020-02-27 | Asia/South Korea/South Korea/ | Division of Viral Diseases, Center for Laboratory Control of Infectious Diseases, Korea Centers for Diseases Control and Prevention | Division of Viral Diseases, Center for Laboratory Control of Infectious Diseases, Korea Centers for Diseases Control and Prevention | Jeong-Min Kim et al |

| | | | | | | |
|---|---|---|---|---|---|---|
| EPI_ISL_506950 | SouthKorea/KCDC2156/2020 | 2020-02-28 | Asia/South Korea/South Korea/ | Division of Viral Diseases, Center for Laboratory Control of Infectious Diseases, Korea Centers for Diseases Control and Prevention | Division of Viral Diseases, Center for Laboratory Control of Infectious Diseases, Korea Centers for Diseases Control and Prevention | Jeong-Min Kim et al |
| EPI_ISL_506951 | SouthKorea/KCDC2157/2020 | 2020-02-29 | Asia/South Korea/South Korea/ | Division of Viral Diseases, Center for Laboratory Control of Infectious Diseases, Korea Centers for Diseases Control and Prevention | Division of Viral Diseases, Center for Laboratory Control of Infectious Diseases, Korea Centers for Diseases Control and Prevention | Jeong-Min Kim et al |
| EPI_ISL_506952 | SouthKorea/KCDC2158/2020 | 2020-02-28 | Asia/South Korea/South Korea/ | Division of Viral Diseases, Center for Laboratory Control of Infectious Diseases, Korea Centers for Diseases Control and Prevention | Division of Viral Diseases, Center for Laboratory Control of Infectious Diseases, Korea Centers for Diseases Control and Prevention | Jeong-Min Kim et al |
| EPI_ISL_506953 | SouthKorea/KCDC2159/2020 | 2020-02-28 | Asia/South Korea/South Korea/ | Division of Viral Diseases, Center for Laboratory Control of Infectious Diseases, Korea Centers for Diseases Control and Prevention | Division of Viral Diseases, Center for Laboratory Control of Infectious Diseases, Korea Centers for Diseases Control and Prevention | Jeong-Min Kim et al |

| | | | | | | |
|---|---|---|---|---|---|---|
| EPI_ISL_506955 | SouthKorea/KCDC2161/2020 | 2020-03-06 | Asia/South Korea/South Korea/ | Division of Viral Diseases, Center for Laboratory Control of Infectious Diseases, Korea Centers for Diseases Control and Prevention | Division of Viral Diseases, Center for Laboratory Control of Infectious Diseases, Korea Centers for Diseases Control and Prevention | Jeong-Min Kim et al |
| EPI_ISL_506957 | SouthKorea/KCDC2163/2020 | 2020-03-09 | Asia/South Korea/South Korea/ | Division of Viral Diseases, Center for Laboratory Control of Infectious Diseases, Korea Centers for Diseases Control and Prevention | Division of Viral Diseases, Center for Laboratory Control of Infectious Diseases, Korea Centers for Diseases Control and Prevention | Jeong-Min Kim et al |
| EPI_ISL_506959 | SouthKorea/KCDC2165/2020 | 2020-03-09 | Asia/South Korea/South Korea/ | Division of Viral Diseases, Center for Laboratory Control of Infectious Diseases, Korea Centers for Diseases Control and Prevention | Division of Viral Diseases, Center for Laboratory Control of Infectious Diseases, Korea Centers for Diseases Control and Prevention | Jeong-Min Kim et al |
| EPI_ISL_506968 | SouthKorea/KCDC2174/2020 | 2020-03-23 | Asia/South Korea/South Korea/ | Division of Viral Diseases, Center for Laboratory Control of Infectious Diseases, Korea Centers for Diseases Control and Prevention | Division of Viral Diseases, Center for Laboratory Control of Infectious Diseases, Korea Centers for Diseases Control and Prevention | Jeong-Min Kim et al |

| | | | | | | |
|---|---|---|---|---|---|---|
| EPI_ISL_506971 | SouthKorea/KCDC2177/2020 | 2020-03-24 | Asia/South Korea/South Korea/ | Division of Viral Diseases, Center for Laboratory Control of Infectious Diseases, Korea Centers for Diseases Control and Prevention | Division of Viral Diseases, Center for Laboratory Control of Infectious Diseases, Korea Centers for Diseases Control and Prevention | Jeong-Min Kim et al |
| EPI_ISL_506974 | SouthKorea/KCDC2180/2020 | 2020-03-25 | Asia/South Korea/South Korea/ | Division of Viral Diseases, Center for Laboratory Control of Infectious Diseases, Korea Centers for Diseases Control and Prevention | Division of Viral Diseases, Center for Laboratory Control of Infectious Diseases, Korea Centers for Diseases Control and Prevention | Jeong-Min Kim et al |
| EPI_ISL_506975 | SouthKorea/KCDC2181/2020 | 2020-03-25 | Asia/South Korea/South Korea/ | Division of Viral Diseases, Center for Laboratory Control of Infectious Diseases, Korea Centers for Diseases Control and Prevention | Division of Viral Diseases, Center for Laboratory Control of Infectious Diseases, Korea Centers for Diseases Control and Prevention | Jeong-Min Kim et al |
| EPI_ISL_506981 | SouthKorea/KCDC2187/2020 | 2020-03-27 | Asia/South Korea/South Korea/ | Division of Viral Diseases, Center for Laboratory Control of Infectious Diseases, Korea Centers for Diseases Control and Prevention | Division of Viral Diseases, Center for Laboratory Control of Infectious Diseases, Korea Centers for Diseases Control and Prevention | Jeong-Min Kim et al |

| | | | | | | |
|---|---|---|---|---|---|---|
| EPI_ISL_506991 | SouthKorea/KCDC2197/2020 | 2020-04-02 | Asia/South Korea/South Korea/ | Division of Viral Diseases, Center for Laboratory Control of Infectious Diseases, Korea Centers for Diseases Control and Prevention | Division of Viral Diseases, Center for Laboratory Control of Infectious Diseases, Korea Centers for Diseases Control and Prevention | Jeong-Min Kim et al |
| EPI_ISL_506992 | SouthKorea/KCDC2198/2020 | 2020-04-04 | Asia/South Korea/South Korea/ | Division of Viral Diseases, Center for Laboratory Control of Infectious Diseases, Korea Centers for Diseases Control and Prevention | Division of Viral Diseases, Center for Laboratory Control of Infectious Diseases, Korea Centers for Diseases Control and Prevention | Jeong-Min Kim et al |
| EPI_ISL_507006 | Singapore/761/2020 | 2020-04-27 | Asia/Singapore/Singapore/ | Department of Laboratory Medicine, Tan Tock Seng Hospital | Department of Laboratory Medicine, Tan Tock Seng Hospital | Chen YYC et al |
| EPI_ISL_507011 | SaudiArabia/KAIMRC02/2020 | 2020-03-10 | Asia/Saudi Arabia/Saudi Arabia/ | unknown | Infectious Diseases Research, King Abdullah International Medical Research Center (KAIMRC) | Alghoribi et al |
| EPI_ISL_507051 | England/LOND-124D099/2020 | 2020-03-14 | Europe/United Kingdom/England/ | University College London Hospital | COVID-19 Genomics UK (COG-UK) Consortium | Judith Heaney et al |
| EPI_ISL_507061 | England/LOND-124D187/2020 | 2020-03-23 | Europe/United Kingdom/England/ | University College London Hospital | COVID-19 Genomics UK (COG-UK) Consortium | Judith Heaney et al |
| EPI_ISL_507153 | England/PORT-2D2515/2020 | 2020-03-26 | Europe/United Kingdom/England/ | Centre for Enzyme Innovation, University of Portsmouth / Translational Research Laboratory, Portsmouth Hospitals NHS Trust | COVID-19 Genomics UK (COG-UK) Consortium | Angela Beckett et al |

| | | | | | | |
|---|---|---|---|---|---|---|
| EPI_ISL_507190 | Scotland/EDB6699/2020 | 2020-05-07 | Europe/United Kingdom/Scotland/ | Virology Department, Royal Infirmary of Edinburgh, NHS Lothian / School of Biological Sciences, University of Edinburgh / Institute of Genetics and Molecular Medicine, University of Edinburgh | COVID-19 Genomics UK (COG-UK) Consortium | McHugh M et al |
| EPI_ISL_507196 | Scotland/EDB6710/2020 | 2020-05-05 | Europe/United Kingdom/Scotland/ | Virology Department, Royal Infirmary of Edinburgh, NHS Lothian / School of Biological Sciences, University of Edinburgh / Institute of Genetics and Molecular Medicine, University of Edinburgh | COVID-19 Genomics UK (COG-UK) Consortium | McHugh M et al |
| EPI_ISL_507207 | Russia/Novosibirsk-RII27313S/2020 | 2020-04-29 | Europe/Russia/Novosibirsk/ | Department of Experimental Modeling and Pathogenesis of Infectious Diseases | WHO National Influenza Centre Russian Federation | Andrey Komissarov et al |
| EPI_ISL_507208 | Russia/Novosibirsk-RII27314S/2020 | 2020-04-29 | Europe/Russia/Novosibirsk/ | Department of Experimental Modeling and Pathogenesis of Infectious Diseases | WHO National Influenza Centre Russian Federation | Andrey Komissarov et al |
| EPI_ISL_507210 | Russia/Novosibirsk-RII27316S/2020 | 2020-05-04 | Europe/Russia/Novosibirsk/ | Department of Experimental Modeling and Pathogenesis of Infectious Diseases | WHO National Influenza Centre Russian Federation | Andrey Komissarov et al |
| EPI_ISL_507211 | Russia/Novosibirsk-RII27317S/2020 | 2020-04-30 | Europe/Russia/Novosibirsk/ | Department of Experimental Modeling and Pathogenesis of Infectious Diseases | WHO National Influenza Centre Russian Federation | Andrey Komissarov et al |
| EPI_ISL_507213 | Russia/Novosibirsk-RII27320S/2020 | 2020-05-07 | Europe/Russia/Novosibirsk/ | Department of Experimental Modeling and Pathogenesis of Infectious Diseases | WHO National Influenza Centre Russian Federation | Andrey Komissarov et al |
| EPI_ISL_507214 | Russia/Novosibirsk-RII27321S/2020 | 2020-05-07 | Europe/Russia/Novosibirsk/ | Department of Experimental Modeling and Pathogenesis of Infectious Diseases | WHO National Influenza Centre Russian Federation | Andrey Komissarov et al |
| EPI_ISL_507215 | Russia/Novosibirsk-RII27322S/2020 | 2020-05-07 | Europe/Russia/Novosibirsk/ | Department of Experimental Modeling and Pathogenesis of Infectious Diseases | WHO National Influenza Centre Russian Federation | Andrey Komissarov et al |

| | | | | | | |
|---|---|---|---|---|---|---|
| EPI_ISL_507226 | Russia/StPetersburg-RII10183S/2020 | 2020-04-26 | Europe/Russia/Saint-Petersburg/ | WHO National Influenza Centre Russian Federation | WHO National Influenza Centre Russian Federation | Andrey Komissarov et al |
| EPI_ISL_507227 | Russia/StPetersburg-RII10219S/2020 | 2020-04-24 | Europe/Russia/Saint-Petersburg/ | WHO National Influenza Centre Russian Federation | WHO National Influenza Centre Russian Federation | Andrey Komissarov et al |
| EPI_ISL_507231 | Russia/StPetersburg-RII11126S/2020 | 2020-05-02 | Europe/Russia/Saint-Petersburg/ | WHO National Influenza Centre Russian Federation | WHO National Influenza Centre Russian Federation | Andrey Komissarov et al |
| EPI_ISL_507232 | Russia/StPetersburg-RII11142S/2020 | 2020-05-02 | Europe/Russia/Saint-Petersburg/ | WHO National Influenza Centre Russian Federation | WHO National Influenza Centre Russian Federation | Andrey Komissarov et al |
| EPI_ISL_507233 | Russia/StPetersburg-RII11144S/2020 | 2020-05-03 | Europe/Russia/Saint-Petersburg/ | WHO National Influenza Centre Russian Federation | WHO National Influenza Centre Russian Federation | Andrey Komissarov et al |
| EPI_ISL_507234 | Russia/StPetersburg-RII11149S/2020 | 2020-04-30 | Europe/Russia/Saint-Petersburg/ | WHO National Influenza Centre Russian Federation | WHO National Influenza Centre Russian Federation | Andrey Komissarov et al |
| EPI_ISL_507235 | Russia/StPetersburg-RII11149V/2020 | 2020-04-30 | Europe/Russia/Saint-Petersburg/ | WHO National Influenza Centre Russian Federation | WHO National Influenza Centre Russian Federation | Andrey Komissarov et al |
| EPI_ISL_507237 | Russia/StPetersburg-RII11153S/2020 | 2020-04-30 | Europe/Russia/Saint-Petersburg/ | WHO National Influenza Centre Russian Federation | WHO National Influenza Centre Russian Federation | Andrey Komissarov et al |
| EPI_ISL_507239 | Russia/StPetersburg-RII11164S/2020 | 2020-05-03 | Europe/Russia/Saint-Petersburg/ | WHO National Influenza Centre Russian Federation | WHO National Influenza Centre Russian Federation | Andrey Komissarov et al |
| EPI_ISL_507241 | Russia/StPetersburg-RII11172S/2020 | 2020-05-02 | Europe/Russia/Saint-Petersburg/ | WHO National Influenza Centre Russian Federation | WHO National Influenza Centre Russian Federation | Andrey Komissarov et al |

| | | | | | | |
|---|---|---|---|---|---|---|
| EPI_ISL_507242 | Russia/StPetersburg-RII11183S/2020 | 2020-05-02 | Europe/Russia/Saint-Petersburg/ | WHO National Influenza Centre Russian Federation | WHO National Influenza Centre Russian Federation | Andrey Komissarov et al |
| EPI_ISL_507243 | Russia/StPetersburg-RII11184S/2020 | 2020-05-02 | Europe/Russia/Saint-Petersburg/ | WHO National Influenza Centre Russian Federation | WHO National Influenza Centre Russian Federation | Andrey Komissarov et al |
| EPI_ISL_507244 | Russia/StPetersburg-RII11188S/2020 | 2020-05-02 | Europe/Russia/Saint-Petersburg/ | WHO National Influenza Centre Russian Federation | WHO National Influenza Centre Russian Federation | Andrey Komissarov et al |
| EPI_ISL_507245 | Russia/StPetersburg-RII11247S/2020 | 2020-04-30 | Europe/Russia/Saint-Petersburg/ | WHO National Influenza Centre Russian Federation | WHO National Influenza Centre Russian Federation | Andrey Komissarov et al |
| EPI_ISL_507247 | Russia/StPetersburg-RII11251S/2020 | 2020-04-30 | Europe/Russia/Saint-Petersburg/ | WHO National Influenza Centre Russian Federation | WHO National Influenza Centre Russian Federation | Andrey Komissarov et al |
| EPI_ISL_507248 | Russia/StPetersburg-RII11256S/2020 | 2020-04-30 | Europe/Russia/Saint-Petersburg/ | WHO National Influenza Centre Russian Federation | WHO National Influenza Centre Russian Federation | Andrey Komissarov et al |
| EPI_ISL_507249 | Russia/StPetersburg-RII11271S/2020 | 2020-05-02 | Europe/Russia/Saint-Petersburg/ | WHO National Influenza Centre Russian Federation | WHO National Influenza Centre Russian Federation | Andrey Komissarov et al |
| EPI_ISL_507250 | Russia/StPetersburg-RII11273S/2020 | 2020-05-02 | Europe/Russia/Saint-Petersburg/ | WHO National Influenza Centre Russian Federation | WHO National Influenza Centre Russian Federation | Andrey Komissarov et al |
| EPI_ISL_507251 | Russia/StPetersburg-RII11275S/2020 | 2020-05-02 | Europe/Russia/Saint-Petersburg/ | WHO National Influenza Centre Russian Federation | WHO National Influenza Centre Russian Federation | Andrey Komissarov et al |
| EPI_ISL_507252 | Russia/StPetersburg-RII11292S/2020 | 2020-05-01 | Europe/Russia/Saint-Petersburg/ | WHO National Influenza Centre Russian Federation | WHO National Influenza Centre Russian Federation | Andrey Komissarov et al |

| | | | | | | |
|---|---|---|---|---|---|---|
| EPI_ISL_507253 | Russia/StPetersburg-RII11299S/2020 | 2020-05-02 | Europe/Russia/Saint-Petersburg/ | WHO National Influenza Centre Russian Federation | WHO National Influenza Centre Russian Federation | Andrey Komissarov et al |
| EPI_ISL_507254 | Russia/StPetersburg-RII11306S/2020 | 2020-05-01 | Europe/Russia/Saint-Petersburg/ | WHO National Influenza Centre Russian Federation | WHO National Influenza Centre Russian Federation | Andrey Komissarov et al |
| EPI_ISL_507255 | Russia/StPetersburg-RII11311S/2020 | 2020-05-03 | Europe/Russia/Saint-Petersburg/ | WHO National Influenza Centre Russian Federation | WHO National Influenza Centre Russian Federation | Andrey Komissarov et al |
| EPI_ISL_507256 | Russia/StPetersburg-RII13532S/2020 | 2020-05-10 | Europe/Russia/Saint-Petersburg/ | WHO National Influenza Centre Russian Federation | WHO National Influenza Centre Russian Federation | Andrey Komissarov et al |
| EPI_ISL_507257 | Russia/StPetersburg-RII14570V/2020 | 2020-05-14 | Europe/Russia/Saint-Petersburg/ | WHO National Influenza Centre Russian Federation | WHO National Influenza Centre Russian Federation | Andrey Komissarov et al |
| EPI_ISL_507259 | Russia/StPetersburg-RII15065S/2020 | 2020-05-14 | Europe/Russia/Saint-Petersburg/ | WHO National Influenza Centre Russian Federation | WHO National Influenza Centre Russian Federation | Andrey Komissarov et al |
| EPI_ISL_507260 | Russia/StPetersburg-RII15163S/2020 | 2020-05-13 | Europe/Russia/Saint-Petersburg/ | WHO National Influenza Centre Russian Federation | WHO National Influenza Centre Russian Federation | Andrey Komissarov et al |
| EPI_ISL_507261 | Russia/StPetersburg-RII15221S/2020 | 2020-05-12 | Europe/Russia/Saint-Petersburg/ | WHO National Influenza Centre Russian Federation | WHO National Influenza Centre Russian Federation | Andrey Komissarov et al |
| EPI_ISL_507262 | Russia/StPetersburg-RII15228S/2020 | 2020-05-12 | Europe/Russia/Saint-Petersburg/ | WHO National Influenza Centre Russian Federation | WHO National Influenza Centre Russian Federation | Andrey Komissarov et al |
| EPI_ISL_507263 | Russia/StPetersburg-RII15229S/2020 | 2020-05-12 | Europe/Russia/Saint-Petersburg/ | WHO National Influenza Centre Russian Federation | WHO National Influenza Centre Russian Federation | Andrey Komissarov et al |

| | | | | | | |
|---|---|---|---|---|---|---|
| EPI_ISL_507264 | Russia/StPetersburg-RII15231S/2020 | 2020-05-12 | Europe/Russia/Saint-Petersburg/ | WHO National Influenza Centre Russian Federation | WHO National Influenza Centre Russian Federation | Andrey Komissarov et al |
| EPI_ISL_507265 | Russia/StPetersburg-RII15516S/2020 | 2020-05-15 | Europe/Russia/Saint-Petersburg/ | WHO National Influenza Centre Russian Federation | WHO National Influenza Centre Russian Federation | Andrey Komissarov et al |
| EPI_ISL_507266 | Russia/StPetersburg-RII15517S/2020 | 2020-05-15 | Europe/Russia/Saint-Petersburg/ | WHO National Influenza Centre Russian Federation | WHO National Influenza Centre Russian Federation | Andrey Komissarov et al |
| EPI_ISL_507267 | Russia/StPetersburg-RII15518S/2020 | 2020-05-15 | Europe/Russia/Saint-Petersburg/ | WHO National Influenza Centre Russian Federation | WHO National Influenza Centre Russian Federation | Andrey Komissarov et al |
| EPI_ISL_507268 | Russia/StPetersburg-RII15520S/2020 | 2020-05-15 | Europe/Russia/Saint-Petersburg/ | WHO National Influenza Centre Russian Federation | WHO National Influenza Centre Russian Federation | Andrey Komissarov et al |
| EPI_ISL_507269 | Russia/StPetersburg-RII15671S/2020 | 2020-05-15 | Europe/Russia/Saint-Petersburg/ | WHO National Influenza Centre Russian Federation | WHO National Influenza Centre Russian Federation | Andrey Komissarov et al |
| EPI_ISL_507270 | Russia/StPetersburg-RII15673S/2020 | 2020-05-15 | Europe/Russia/Saint-Petersburg/ | WHO National Influenza Centre Russian Federation | WHO National Influenza Centre Russian Federation | Andrey Komissarov et al |
| EPI_ISL_507271 | Russia/StPetersburg-RII15674S/2020 | 2020-05-15 | Europe/Russia/Saint-Petersburg/ | WHO National Influenza Centre Russian Federation | WHO National Influenza Centre Russian Federation | Andrey Komissarov et al |
| EPI_ISL_507272 | Russia/StPetersburg-RII15675S/2020 | 2020-05-15 | Europe/Russia/Saint-Petersburg/ | WHO National Influenza Centre Russian Federation | WHO National Influenza Centre Russian Federation | Andrey Komissarov et al |
| EPI_ISL_507273 | Russia/StPetersburg-RII15817S/2020 | 2020-05-18 | Europe/Russia/Saint-Petersburg/ | WHO National Influenza Centre Russian Federation | WHO National Influenza Centre Russian Federation | Andrey Komissarov et al |

| | | | | | | |
|---|---|---|---|---|---|---|
| EPI_ISL_507274 | Russia/StPetersburg-RII15932V/2020 | 2020-05-16 | Europe/Russia/Saint-Petersburg/ | WHO National Influenza Centre Russian Federation | WHO National Influenza Centre Russian Federation | Andrey Komissarov et al |
| EPI_ISL_507275 | Russia/StPetersburg-RII15933S/2020 | 2020-05-16 | Europe/Russia/Saint-Petersburg/ | WHO National Influenza Centre Russian Federation | WHO National Influenza Centre Russian Federation | Andrey Komissarov et al |
| EPI_ISL_507276 | Russia/StPetersburg-RII15950V/2020 | 2020-05-18 | Europe/Russia/Saint-Petersburg/ | WHO National Influenza Centre Russian Federation | WHO National Influenza Centre Russian Federation | Andrey Komissarov et al |
| EPI_ISL_507277 | Russia/StPetersburg-RII16106S/2020 | 2020-05-19 | Europe/Russia/Saint-Petersburg/ | WHO National Influenza Centre Russian Federation | WHO National Influenza Centre Russian Federation | Andrey Komissarov et al |
| EPI_ISL_507278 | Russia/StPetersburg-RII17536S/2020 | 2020-05-26 | Europe/Russia/Saint-Petersburg/ | WHO National Influenza Centre Russian Federation | WHO National Influenza Centre Russian Federation | Andrey Komissarov et al |
| EPI_ISL_507279 | Russia/StPetersburg-RII17574S/2020 | 2020-05-26 | Europe/Russia/Saint-Petersburg/ | WHO National Influenza Centre Russian Federation | WHO National Influenza Centre Russian Federation | Andrey Komissarov et al |
| EPI_ISL_507280 | Russia/StPetersburg-RII17940S/2020 | 2020-05-27 | Europe/Russia/Saint-Petersburg/ | WHO National Influenza Centre Russian Federation | WHO National Influenza Centre Russian Federation | Andrey Komissarov et al |
| EPI_ISL_507281 | Russia/StPetersburg-RII17941S/2020 | 2020-05-27 | Europe/Russia/Saint-Petersburg/ | WHO National Influenza Centre Russian Federation | WHO National Influenza Centre Russian Federation | Andrey Komissarov et al |
| EPI_ISL_507282 | Russia/StPetersburg-RII17948S/2020 | 2020-05-27 | Europe/Russia/Saint-Petersburg/ | WHO National Influenza Centre Russian Federation | WHO National Influenza Centre Russian Federation | Andrey Komissarov et al |
| EPI_ISL_507283 | Russia/StPetersburg-RII17949S/2020 | 2020-05-27 | Europe/Russia/Saint-Petersburg/ | WHO National Influenza Centre Russian Federation | WHO National Influenza Centre Russian Federation | Andrey Komissarov et al |

| | | | | | | |
|---|---|---|---|---|---|---|
| EPI_ISL_507284 | Russia/StPetersburg-RII18088S/2020 | 2020-05-27 | Europe/Russia/Saint-Petersburg/ | WHO National Influenza Centre Russian Federation | WHO National Influenza Centre Russian Federation | Andrey Komissarov et al |
| EPI_ISL_507285 | Russia/StPetersburg-RII18089S/2020 | 2020-05-27 | Europe/Russia/Saint-Petersburg/ | WHO National Influenza Centre Russian Federation | WHO National Influenza Centre Russian Federation | Andrey Komissarov et al |
| EPI_ISL_507286 | Russia/StPetersburg-RII18090S/2020 | 2020-05-27 | Europe/Russia/Saint-Petersburg/ | WHO National Influenza Centre Russian Federation | WHO National Influenza Centre Russian Federation | Andrey Komissarov et al |
| EPI_ISL_507288 | Russia/StPetersburg-RII21769S/2020 | 2020-06-06 | Europe/Russia/Saint-Petersburg/ | WHO National Influenza Centre Russian Federation | WHO National Influenza Centre Russian Federation | Andrey Komissarov et al |
| EPI_ISL_507289 | Russia/StPetersburg-RII21797S/2020 | 2020-06-07 | Europe/Russia/Saint-Petersburg/ | WHO National Influenza Centre Russian Federation | WHO National Influenza Centre Russian Federation | Andrey Komissarov et al |
| EPI_ISL_507290 | Russia/StPetersburg-RII21802S/2020 | 2020-06-06 | Europe/Russia/Saint-Petersburg/ | WHO National Influenza Centre Russian Federation | WHO National Influenza Centre Russian Federation | Andrey Komissarov et al |
| EPI_ISL_507293 | Russia/StPetersburg-RII8317V/2020 | 2020-04-21 | Europe/Russia/Saint-Petersburg/ | WHO National Influenza Centre Russian Federation | WHO National Influenza Centre Russian Federation | Andrey Komissarov et al |
| EPI_ISL_507295 | Russia/StPetersburg-RII9304V/2020 | 2020-04-24 | Europe/Russia/Saint-Petersburg/ | WHO National Influenza Centre Russian Federation | WHO National Influenza Centre Russian Federation | Andrey Komissarov et al |
| EPI_ISL_507420 | USA/MI-MDHHS-SC21163/2020 | 2020-06-24 | North America/USA/Michigan/ | Michigan Department of Health and Human Services, Bureau of Laboratories | Michigan Department of Health and Human Services, Bureau of Laboratories | Blankenship HM et al |

| | | | | | | |
|---|---|---|---|---|---|---|
| EPI_ISL_507425 | USA/MI-MDHHS-SC21168/2020 | 2020-04-04 | North America/USA/Michigan/ | Michigan Department of Health and Human Services, Bureau of Laboratories | Michigan Department of Health and Human Services, Bureau of Laboratories | Blankenship HM et al |
| EPI_ISL_507431 | USA/MI-MDHHS-SC21174/2020 | 2020-04-01 | North America/USA/Michigan/ | Michigan Department of Health and Human Services, Bureau of Laboratories | Michigan Department of Health and Human Services, Bureau of Laboratories | Blankenship HM et al |
| EPI_ISL_507433 | USA/MI-MDHHS-SC21176/2020 | 2020-04-02 | North America/USA/Michigan/ | Michigan Department of Health and Human Services, Bureau of Laboratories | Michigan Department of Health and Human Services, Bureau of Laboratories | Blankenship HM et al |
| EPI_ISL_507438 | USA/MI-MDHHS-SC21181/2020 | 2020-06-30 | North America/USA/Michigan/ | Michigan Department of Health and Human Services, Bureau of Laboratories | Michigan Department of Health and Human Services, Bureau of Laboratories | Blankenship HM et al |
| EPI_ISL_507440 | USA/MI-MDHHS-SC21183/2020 | 2020-04-03 | North America/USA/Michigan/ | Michigan Department of Health and Human Services, Bureau of Laboratories | Michigan Department of Health and Human Services, Bureau of Laboratories | Blankenship HM et al |
| EPI_ISL_507443 | USA/MI-MDHHS-SC21186/2020 | 2020-04-04 | North America/USA/Michigan/ | Michigan Department of Health and Human Services, Bureau of Laboratories | Michigan Department of Health and Human Services, Bureau of Laboratories | Blankenship HM et al |

| | | | | | | |
|---|---|---|---|---|---|---|
| EPI_ISL_507445 | USA/MI-MDHHS-SC21188/2020 | 2020-07-06 | North America/USA/Michigan/ | Michigan Department of Health and Human Services, Bureau of Laboratories | Michigan Department of Health and Human Services, Bureau of Laboratories | Blankenship HM et al |
| EPI_ISL_507446 | USA/MI-MDHHS-SC21189/2020 | 2020-04-02 | North America/USA/Michigan/ | Michigan Department of Health and Human Services, Bureau of Laboratories | Michigan Department of Health and Human Services, Bureau of Laboratories | Blankenship HM et al |
| EPI_ISL_507456 | USA/MI-MDHHS-SC21199/2020 | 2020-03-30 | North America/USA/Michigan/ | Michigan Department of Health and Human Services, Bureau of Laboratories | Michigan Department of Health and Human Services, Bureau of Laboratories | Blankenship HM et al |
| EPI_ISL_507460 | USA/MI-MDHHS-SC21203/2020 | 2020-03-31 | North America/USA/Michigan/ | Michigan Department of Health and Human Services, Bureau of Laboratories | Michigan Department of Health and Human Services, Bureau of Laboratories | Blankenship HM et al |
| EPI_ISL_507485 | USA/MI-MDHHS-SC21229/2020 | 2020-04-07 | North America/USA/Michigan/ | Michigan Department of Health and Human Services, Bureau of Laboratories | Michigan Department of Health and Human Services, Bureau of Laboratories | Blankenship HM et al |
| EPI_ISL_507486 | USA/MI-MDHHS-SC21230/2020 | 2020-04-08 | North America/USA/Michigan/ | Michigan Department of Health and Human Services, Bureau of Laboratories | Michigan Department of Health and Human Services, Bureau of Laboratories | Blankenship HM et al |

| | | | | | | |
|---|---|---|---|---|---|---|
| EPI_ISL_507497 | USA/MI-MDHHS-SC21242/2020 | 2020-04-04 | North America/USA/Michigan/ | Michigan Department of Health and Human Services, Bureau of Laboratories | Michigan Department of Health and Human Services, Bureau of Laboratories | Blankenship HM et al |
| EPI_ISL_507501 | USA/MI-MDHHS-SC21246/2020 | 2020-04-03 | North America/USA/Michigan/ | Michigan Department of Health and Human Services, Bureau of Laboratories | Michigan Department of Health and Human Services, Bureau of Laboratories | Blankenship HM et al |
| EPI_ISL_507507 | USA/MI-MDHHS-SC21252/2020 | 2020-04-04 | North America/USA/Michigan/ | Michigan Department of Health and Human Services, Bureau of Laboratories | Michigan Department of Health and Human Services, Bureau of Laboratories | Blankenship HM et al |
| EPI_ISL_507515 | USA/MI-MDHHS-SC21260/2020 | 2020-04-03 | North America/USA/Michigan/ | Michigan Department of Health and Human Services, Bureau of Laboratories | Michigan Department of Health and Human Services, Bureau of Laboratories | Blankenship HM et al |
| EPI_ISL_507552 | USA/MI-MDHHS-SC21297/2020 | 2020-04-08 | North America/USA/Michigan/ | Michigan Department of Health and Human Services, Bureau of Laboratories | Michigan Department of Health and Human Services, Bureau of Laboratories | Blankenship HM et al |
| EPI_ISL_507571 | USA/MI-MDHHS-SC21316/2020 | 2020-04-03 | North America/USA/Michigan/ | Michigan Department of Health and Human Services, Bureau of Laboratories | Michigan Department of Health and Human Services, Bureau of Laboratories | Blankenship HM et al |

| | | | | | | |
|---|---|---|---|---|---|---|
| EPI_ISL_507576 | USA/MI-MDHHS-SC21321/2020 | 2020-04-02 | North America/USA/Michigan/ | Michigan Department of Health and Human Services, Bureau of Laboratories | Michigan Department of Health and Human Services, Bureau of Laboratories | Blankenship HM et al |
| EPI_ISL_507605 | USA/MI-MDHHS-SC21351/2020 | 2020-04-02 | North America/USA/Michigan/ | Michigan Department of Health and Human Services, Bureau of Laboratories | Michigan Department of Health and Human Services, Bureau of Laboratories | Blankenship HM et al |
| EPI_ISL_507610 | USA/MI-MDHHS-SC21356/2020 | 2020-04-02 | North America/USA/Michigan/ | Michigan Department of Health and Human Services, Bureau of Laboratories | Michigan Department of Health and Human Services, Bureau of Laboratories | Blankenship HM et al |
| EPI_ISL_507650 | USA/MI-MDHHS-SC21396/2020 | 2020-03-21 | North America/USA/Michigan/ | Michigan Department of Health and Human Services, Bureau of Laboratories | Michigan Department of Health and Human Services, Bureau of Laboratories | Blankenship HM et al |
| EPI_ISL_507687 | USA/MI-MDHHS-SC21433/2020 | 2020-03-24 | North America/USA/Michigan/ | Michigan Department of Health and Human Services, Bureau of Laboratories | Michigan Department of Health and Human Services, Bureau of Laboratories | Blankenship HM et al |
| EPI_ISL_507697 | USA/MI-MDHHS-SC21443/2020 | 2020-03-25 | North America/USA/Michigan/ | Michigan Department of Health and Human Services, Bureau of Laboratories | Michigan Department of Health and Human Services, Bureau of Laboratories | Blankenship HM et al |

| | | | | | | |
|---|---|---|---|---|---|---|
| EPI_ISL_507737 | USA/MI-MDHHS-SC20948/2020 | 2020-03-23 | North America/USA/Michigan/ | Michigan Department of Health and Human Services, Bureau of Laboratories | Michigan Department of Health and Human Services, Bureau of Laboratories | Blankenship HM et al |
| EPI_ISL_507738 | USA/MI-MDHHS-SC20949/2020 | 2020-05-29 | North America/USA/Michigan/ | Michigan Department of Health and Human Services, Bureau of Laboratories | Michigan Department of Health and Human Services, Bureau of Laboratories | Blankenship HM et al |
| EPI_ISL_507745 | USA/MI-MDHHS-SC20956/2020 | 2020-03-21 | North America/USA/Michigan/ | Michigan Department of Health and Human Services, Bureau of Laboratories | Michigan Department of Health and Human Services, Bureau of Laboratories | Blankenship HM et al |
| EPI_ISL_507765 | USA/MI-MDHHS-SC20977/2020 | 2020-05-14 | North America/USA/Michigan/ | Michigan Department of Health and Human Services, Bureau of Laboratories | Michigan Department of Health and Human Services, Bureau of Laboratories | Blankenship HM et al |
| EPI_ISL_507802 | USA/MI-MDHHS-SC21026/2020 | 2020-03-27 | North America/USA/Michigan/ | Michigan Department of Health and Human Services, Bureau of Laboratories | Michigan Department of Health and Human Services, Bureau of Laboratories | Blankenship HM et al |
| EPI_ISL_507820 | USA/MI-MDHHS-SC21044/2020 | 2020-03-24 | North America/USA/Michigan/ | Michigan Department of Health and Human Services, Bureau of Laboratories | Michigan Department of Health and Human Services, Bureau of Laboratories | Blankenship HM et al |

| | | | | | | |
|---|---|---|---|---|---|---|
| EPI_ISL_507826 | USA/MI-MDHHS-SC21050/2020 | 2020-04-01 | North America/USA/Michigan/ | Michigan Department of Health and Human Services, Bureau of Laboratories | Michigan Department of Health and Human Services, Bureau of Laboratories | Blankenship HM et al |
| EPI_ISL_507828 | USA/MI-MDHHS-SC21052/2020 | 2020-03-28 | North America/USA/Michigan/ | Michigan Department of Health and Human Services, Bureau of Laboratories | Michigan Department of Health and Human Services, Bureau of Laboratories | Blankenship HM et al |
| EPI_ISL_507829 | USA/MI-MDHHS-SC21053/2020 | 2020-03-26 | North America/USA/Michigan/ | Michigan Department of Health and Human Services, Bureau of Laboratories | Michigan Department of Health and Human Services, Bureau of Laboratories | Blankenship HM et al |
| EPI_ISL_507841 | USA/MI-MDHHS-SC21065/2020 | 2020-04-01 | North America/USA/Michigan/ | Michigan Department of Health and Human Services, Bureau of Laboratories | Michigan Department of Health and Human Services, Bureau of Laboratories | Blankenship HM et al |
| EPI_ISL_507851 | USA/MI-MDHHS-SC21075/2020 | 2020-03-26 | North America/USA/Michigan/ | Michigan Department of Health and Human Services, Bureau of Laboratories | Michigan Department of Health and Human Services, Bureau of Laboratories | Blankenship HM et al |
| EPI_ISL_507863 | USA/MI-MDHHS-SC21087/2020 | 2020-04-01 | North America/USA/Michigan/ | Michigan Department of Health and Human Services, Bureau of Laboratories | Michigan Department of Health and Human Services, Bureau of Laboratories | Blankenship HM et al |

| | | | | | | |
|---|---|---|---|---|---|---|
| EPI_ISL_507865 | USA/MI-MDHHS-SC21089/2020 | 2020-03-28 | North America/USA/Michigan/ | Michigan Department of Health and Human Services, Bureau of Laboratories | Michigan Department of Health and Human Services, Bureau of Laboratories | Blankenship HM et al |
| EPI_ISL_507866 | USA/MI-MDHHS-SC21090/2020 | 2020-03-25 | North America/USA/Michigan/ | Michigan Department of Health and Human Services, Bureau of Laboratories | Michigan Department of Health and Human Services, Bureau of Laboratories | Blankenship HM et al |
| EPI_ISL_507870 | USA/MI-MDHHS-SC21094/2020 | 2020-03-26 | North America/USA/Michigan/ | Michigan Department of Health and Human Services, Bureau of Laboratories | Michigan Department of Health and Human Services, Bureau of Laboratories | Blankenship HM et al |
| EPI_ISL_507871 | USA/MI-MDHHS-SC21095/2020 | 2020-04-02 | North America/USA/Michigan/ | Michigan Department of Health and Human Services, Bureau of Laboratories | Michigan Department of Health and Human Services, Bureau of Laboratories | Blankenship HM et al |
| EPI_ISL_507874 | USA/MI-MDHHS-SC21098/2020 | 2020-03-27 | North America/USA/Michigan/ | Michigan Department of Health and Human Services, Bureau of Laboratories | Michigan Department of Health and Human Services, Bureau of Laboratories | Blankenship HM et al |
| EPI_ISL_507876 | USA/MI-MDHHS-SC21100/2020 | 2020-03-25 | North America/USA/Michigan/ | Michigan Department of Health and Human Services, Bureau of Laboratories | Michigan Department of Health and Human Services, Bureau of Laboratories | Blankenship HM et al |

| | | | | | | |
|---|---|---|---|---|---|---|
| EPI_ISL_507891 | USA/MI-MDHHS-SC21115/2020 | 2020-03-29 | North America/USA/Michigan/ | Michigan Department of Health and Human Services, Bureau of Laboratories | Michigan Department of Health and Human Services, Bureau of Laboratories | Blankenship HM et al |
| EPI_ISL_507912 | USA/MI-MDHHS-SC21136/2020 | 2020-03-30 | North America/USA/Michigan/ | Michigan Department of Health and Human Services, Bureau of Laboratories | Michigan Department of Health and Human Services, Bureau of Laboratories | Blankenship HM et al |
| EPI_ISL_507915 | USA/MI-MDHHS-SC21139/2020 | 2020-03-30 | North America/USA/Michigan/ | Michigan Department of Health and Human Services, Bureau of Laboratories | Michigan Department of Health and Human Services, Bureau of Laboratories | Blankenship HM et al |
| EPI_ISL_507918 | USA/MI-MDHHS-SC21142/2020 | 2020-03-25 | North America/USA/Michigan/ | Michigan Department of Health and Human Services, Bureau of Laboratories | Michigan Department of Health and Human Services, Bureau of Laboratories | Blankenship HM et al |
| EPI_ISL_507922 | USA/MI-MDHHS-SC21146/2020 | 2020-03-31 | North America/USA/Michigan/ | Michigan Department of Health and Human Services, Bureau of Laboratories | Michigan Department of Health and Human Services, Bureau of Laboratories | Blankenship HM et al |
| EPI_ISL_507935 | USA/MN-MDH-1346/2020 | 2020-04-13 | North America/USA/Minnesota/ | Minnesota Department of Health, Public Health Laboratory | Minnesota Department of Health, Public Health Laboratory | Matt Plumb et al |
| EPI_ISL_507942 | USA/MN-MDH-1353/2020 | 2020-04-21 | North America/USA/Minnesota/ | Minnesota Department of Health, Public Health Laboratory | Minnesota Department of Health, Public Health Laboratory | Matt Plumb et al |

| | | | | | | |
|---|---|---|---|---|---|---|
| EPI_ISL_507945 | USA/MN-MDH-1356/2020 | 2020-04-21 | North America/USA/Minnesota/ | Minnesota Department of Health, Public Health Laboratory | Minnesota Department of Health, Public Health Laboratory | Matt Plumb et al |
| EPI_ISL_507950 | USA/MN-MDH-1361/2020 | 2020-04-22 | North America/USA/Minnesota/ | Minnesota Department of Health, Public Health Laboratory | Minnesota Department of Health, Public Health Laboratory | Matt Plumb et al |
| EPI_ISL_507951 | USA/MN-MDH-1362/2020 | 2020-04-22 | North America/USA/Minnesota/ | Minnesota Department of Health, Public Health Laboratory | Minnesota Department of Health, Public Health Laboratory | Matt Plumb et al |
| EPI_ISL_507955 | USA/MN-MDH-1367/2020 | 2020-04-23 | North America/USA/Minnesota/ | Minnesota Department of Health, Public Health Laboratory | Minnesota Department of Health, Public Health Laboratory | Matt Plumb et al |
| EPI_ISL_507957 | USA/MN-MDH-1369/2020 | 2020-06-30 | North America/USA/Minnesota/ | Mayo Clinic & Mayo Clinic Laboratories | Minnesota Department of Health, Public Health Laboratory | Matt Plumb et al |
| EPI_ISL_507962 | USA/MN-MDH-1374/2020 | 2020-05-22 | North America/USA/Minnesota/ | Children's Hospitals and Clinics of Minnesota | Minnesota Department of Health, Public Health Laboratory | Matt Plumb et al |
| EPI_ISL_507964 | USA/MN-MDH-1376/2020 | 2020-07-09 | North America/USA/Minnesota/ | Avera Mckennan Laboratory | Minnesota Department of Health, Public Health Laboratory | Matt Plumb et al |
| EPI_ISL_507980 | USA/MN-MDH-1393/2020 | 2020-03-27 | North America/USA/Minnesota/ | Minnesota Department of Health, Public Health Laboratory | Minnesota Department of Health, Public Health Laboratory | Matt Plumb et al |
| EPI_ISL_507991 | USA/NM-UNM-00336/2020 | 2020-05-08 | North America/USA/New Mexico/ | New Mexico Department of Health Scientific Laboratory Division | Center for Global Health, University of New Mexico Health Sciences Center | Daryl Domman et al |

| | | | | | | |
|---|---|---|---|---|---|---|
| EPI_ISL_507992 | USA/NM-UNM-00310/2020 | 2020-05-13 | North America/USA/New Mexico/ | New Mexico Department of Health Scientific Laboratory Division | Center for Global Health, University of New Mexico Health Sciences Center | Daryl Domman et al |
| EPI_ISL_508060 | USA/NM-UNM-00398/2020 | 2020-04-17 | North America/USA/New Mexico/ | New Mexico Department of Health Scientific Laboratory Division | Center for Global Health, University of New Mexico Health Sciences Center | Daryl Domman et al |
| EPI_ISL_508069 | USA/NM-UNM-00302/2020 | 2020-05-13 | North America/USA/New Mexico/ | New Mexico Department of Health Scientific Laboratory Division | Center for Global Health, University of New Mexico Health Sciences Center | Daryl Domman et al |
| EPI_ISL_508071 | USA/NM-UNM-00277/2020 | 2020-05-13 | North America/USA/New Mexico/ | New Mexico Department of Health Scientific Laboratory Division | Center for Global Health, University of New Mexico Health Sciences Center | Daryl Domman et al |
| EPI_ISL_508075 | USA/NM-UNM-00386/2020 | 2020-04-12 | North America/USA/New Mexico/ | New Mexico Department of Health Scientific Laboratory Division | Center for Global Health, University of New Mexico Health Sciences Center | Daryl Domman et al |
| EPI_ISL_508080 | USA/NM-UNM-00396/2020 | 2020-05-04 | North America/USA/New Mexico/ | New Mexico Department of Health Scientific Laboratory Division | Center for Global Health, University of New Mexico Health Sciences Center | Daryl Domman et al |
| EPI_ISL_508091 | USA/NM-UNM-00403/2020 | 2020-05-01 | North America/USA/New Mexico/ | New Mexico Department of Health Scientific Laboratory Division | Center for Global Health, University of New Mexico Health Sciences Center | Daryl Domman et al |
| EPI_ISL_508093 | USA/NM-UNM-00341/2020 | 2020-05-07 | North America/USA/New Mexico/ | New Mexico Department of Health Scientific Laboratory Division | Center for Global Health, University of New Mexico Health Sciences Center | Daryl Domman et al |

| | | | | | | |
|---|---|---|---|---|---|---|
| EPI_ISL_508108 | USA/NM-UNM-00268/2020 | 2020-05-13 | North America/USA/New Mexico/ | New Mexico Department of Health Scientific Laboratory Division | Center for Global Health, University of New Mexico Health Sciences Center | Daryl Domman et al |
| EPI_ISL_508109 | USA/NM-UNM-00269/2020 | 2020-05-13 | North America/USA/New Mexico/ | New Mexico Department of Health Scientific Laboratory Division | Center for Global Health, University of New Mexico Health Sciences Center | Daryl Domman et al |
| EPI_ISL_508158 | India/UT-AR11/2020 | 2020-05-29 | Asia/India/Uttarakhand/Dehradun | All india institute of Medical Sciences Rishikesh | National Institute of Biomedical Genomics | Arindam Maitra et al |
| EPI_ISL_508160 | India/UT-AR13/2020 | 2020-06-06 | Asia/India/Uttarakhand/Dehradun | All india institute of Medical Sciences Rishikesh | National Institute of Biomedical Genomics | Arindam Maitra et al |
| EPI_ISL_508161 | India/UT-AR14/2020 | 2020-06-06 | Asia/India/Uttarakhand/Dehradun | All india institute of Medical Sciences Rishikesh | National Institute of Biomedical Genomics | Arindam Maitra et al |
| EPI_ISL_508162 | India/UT-AR15/2020 | 2020-05-24 | Asia/India/Uttarakhand/Haridwar | All india institute of Medical Sciences Rishikesh | National Institute of Biomedical Genomics | Arindam Maitra et al |
| EPI_ISL_508164 | India/UT-AR17/2020 | 2020-05-27 | Asia/India/Uttarakhand/Pauri Garhwal | All india institute of Medical Sciences Rishikesh | National Institute of Biomedical Genomics | Arindam Maitra et al |
| EPI_ISL_508170 | India/UT-AR22/2020 | 2020-05-22 | Asia/India/Uttarakhand/Haridwar | All india institute of Medical Sciences Rishikesh | National Institute of Biomedical Genomics | Arindam Maitra et al |
| EPI_ISL_508171 | India/UT-AR23/2020 | 2020-05-21 | Asia/India/Uttarakhand/Haridwar | All india institute of Medical Sciences Rishikesh | National Institute of Biomedical Genomics | Arindam Maitra et al |
| EPI_ISL_508174 | India/UT-AR27/2020 | 2020-06-01 | Asia/India/Uttarakhand/Tehri Garhwal | All india institute of Medical Sciences Rishikesh | National Institute of Biomedical Genomics | Arindam Maitra et al |
| EPI_ISL_508175 | India/UT-AR28/2020 | 2020-06-29 | Asia/India/Uttarakhand/Haridwar | All india institute of Medical Sciences Rishikesh | National Institute of Biomedical Genomics | Arindam Maitra et al |
| EPI_ISL_508178 | India/UT-AR30/2020 | 2020-05-29 | Asia/India/Uttarakhand/Haridwar | All india institute of Medical Sciences Rishikesh | National Institute of Biomedical Genomics | Arindam Maitra et al |

| | | | | | | |
|---|---|---|---|---|---|---|
| EPI_ISL_508180 | India/UT-AR32/2020 | 2020-06-04 | Asia/India/Uttarakhand/Uttarkashi | All india institute of Medical Sciences Rishikesh | National Institute of Biomedical Genomics | Arindam Maitra et al |
| EPI_ISL_508182 | India/UT-AR34/2020 | 2020-06-08 | Asia/India/Uttarakhand/Haridwar | All india institute of Medical Sciences Rishikesh | National Institute of Biomedical Genomics | Arindam Maitra et al |
| EPI_ISL_508184 | India/UT-AR4/2020 | 2020-06-01 | Asia/India/Uttarakhand/Dehradun | All india institute of Medical Sciences Rishikesh | National Institute of Biomedical Genomics | Arindam Maitra et al |
| EPI_ISL_508185 | India/UT-AR41/2020 | 2020-05-28 | Asia/India/Uttarakhand/Tehri Garhwal | All india institute of Medical Sciences Rishikesh | National Institute of Biomedical Genomics | Arindam Maitra et al |
| EPI_ISL_508192 | India/UT-AR53/2020 | 2020-06-07 | Asia/India/Uttarakhand/Haridwar | All india institute of Medical Sciences Rishikesh | National Institute of Biomedical Genomics | Arindam Maitra et al |
| EPI_ISL_508195 | India/UT-AR59/2020 | 2020-06-03 | Asia/India/Uttarakhand/Tehri Garhwal | All india institute of Medical Sciences Rishikesh | National Institute of Biomedical Genomics | Arindam Maitra et al |
| EPI_ISL_508199 | India/UT-AR62/2020 | 2020-06-08 | Asia/India/Uttarakhand/Tehri Garhwal | All india institute of Medical Sciences Rishikesh | National Institute of Biomedical Genomics | Arindam Maitra et al |
| EPI_ISL_508202 | India/UP-AR66/2020 | 2020-06-09 | Asia/India/Uttar Pradesh/Muzaffarnaga | All india institute of Medical Sciences Rishikesh | National Institute of Biomedical Genomics | Arindam Maitra et al |
| EPI_ISL_508204 | India/UT-AR7/2020 | 2020-05-30 | Asia/India/Uttarakhand/Dehradun | All india institute of Medical Sciences Rishikesh | National Institute of Biomedical Genomics | Arindam Maitra et al |
| EPI_ISL_508206 | India/UT-AR9/2020 | 2020-05-22 | Asia/India/Uttarakhand/Haridwar | All india institute of Medical Sciences Rishikesh | National Institute of Biomedical Genomics | Arindam Maitra et al |
| EPI_ISL_508209 | India/MH-GA13/2020 | 2020-06-17 | Asia/India/Maharashtra/Aurangabad | Government Medical College | National Institute of Biomedical Genomics | Arindam Maitra et al |
| EPI_ISL_508212 | India/MH-GA16/2020 | 2020-06-17 | Asia/India/Maharashtra/Aurangabad | Government Medical College | National Institute of Biomedical Genomics | Arindam Maitra et al |
| EPI_ISL_508214 | India/MH-GA19/2020 | 2020-06-17 | Asia/India/Maharashtra/Aurangabad | Government Medical College | National Institute of Biomedical Genomics | Arindam Maitra et al |

| | | | | | | |
|---|---|---|---|---|---|---|
| EPI_ISL_508217 | India/MH-GA21/2020 | 2020-06-17 | Asia/India/Maharashtra/Aurangabad | Government Medical College | National Institute of Biomedical Genomics | Arindam Maitra et al |
| EPI_ISL_508221 | India/MH-GA26/2020 | 2020-06-16 | Asia/India/Maharashtra/Jalna | Government Medical College | National Institute of Biomedical Genomics | Arindam Maitra et al |
| EPI_ISL_508222 | India/MH-GA27/2020 | 2020-06-16 | Asia/India/Maharashtra/Jalna | Government Medical College | National Institute of Biomedical Genomics | Arindam Maitra et al |
| EPI_ISL_508223 | India/MH-GA28/2020 | 2020-06-17 | Asia/India/Maharashtra/Aurangabad | Government Medical College | National Institute of Biomedical Genomics | Arindam Maitra et al |
| EPI_ISL_508226 | India/MH-GA32/2020 | 2020-06-20 | Asia/India/Maharashtra/Aurangabad | Government Medical College | National Institute of Biomedical Genomics | Arindam Maitra et al |
| EPI_ISL_508228 | India/MH-GA34/2020 | 2020-06-20 | Asia/India/Maharashtra/Aurangabad | Government Medical College | National Institute of Biomedical Genomics | Arindam Maitra et al |
| EPI_ISL_508230 | India/MH-GA36/2020 | 2020-06-20 | Asia/India/Maharashtra/Aurangabad | Government Medical College | National Institute of Biomedical Genomics | Arindam Maitra et al |
| EPI_ISL_508242 | India/MH-GA48/2020 | 2020-06-20 | Asia/India/Maharashtra/Aurangabad | Government Medical College | National Institute of Biomedical Genomics | Arindam Maitra et al |
| EPI_ISL_508247 | India/MH-GA52/2020 | 2020-06-20 | Asia/India/Maharashtra/Aurangabad | Government Medical College | National Institute of Biomedical Genomics | Arindam Maitra et al |
| EPI_ISL_508250 | India/MH-GA55/2020 | 2020-06-20 | Asia/India/Maharashtra/Aurangabad | Government Medical College | National Institute of Biomedical Genomics | Arindam Maitra et al |
| EPI_ISL_508251 | India/MH-GA56/2020 | 2020-06-20 | Asia/India/Maharashtra/Aurangabad | Government Medical College | National Institute of Biomedical Genomics | Arindam Maitra et al |
| EPI_ISL_508252 | India/MH-GA57/2020 | 2020-06-20 | Asia/India/Maharashtra/Aurangabad | Government Medical College | National Institute of Biomedical Genomics | Arindam Maitra et al |
| EPI_ISL_508262 | India/MH-GA68/2020 | 2020-06-18 | Asia/India/Maharashtra/Aurangabad | Government Medical College | National Institute of Biomedical Genomics | Arindam Maitra et al |

| | | | | | | |
|---|---|---|---|---|---|---|
| EPI_ISL_508265 | India/MH-GA70/2020 | 2020-06-18 | Asia/India/Maharashtra/Aurangabad | Government Medical College | National Institute of Biomedical Genomics | Arindam Maitra et al |
| EPI_ISL_508266 | India/MH-GA71/2020 | 2020-06-18 | Asia/India/Maharashtra/Aurangabad | Government Medical College | National Institute of Biomedical Genomics | Arindam Maitra et al |
| EPI_ISL_508268 | India/MH-GA74/2020 | 2020-06-18 | Asia/India/Maharashtra/Aurangabad | Government Medical College | National Institute of Biomedical Genomics | Arindam Maitra et al |
| EPI_ISL_508271 | India/MH-GA77/2020 | 2020-06-18 | Asia/India/Maharashtra/Aurangabad | Government Medical College | National Institute of Biomedical Genomics | Arindam Maitra et al |
| EPI_ISL_508272 | India/MH-GA78/2020 | 2020-06-18 | Asia/India/Maharashtra/Aurangabad | Government Medical College | National Institute of Biomedical Genomics | Arindam Maitra et al |
| EPI_ISL_508273 | India/MH-GA79/2020 | 2020-06-18 | Asia/India/Maharashtra/Aurangabad | Government Medical College | National Institute of Biomedical Genomics | Arindam Maitra et al |
| EPI_ISL_508274 | India/MH-GA8/2020 | 2020-06-17 | Asia/India/Maharashtra/Aurangabad | Government Medical College | National Institute of Biomedical Genomics | Arindam Maitra et al |
| EPI_ISL_508275 | India/MH-GA80/2020 | 2020-06-18 | Asia/India/Maharashtra/Aurangabad | Government Medical College | National Institute of Biomedical Genomics | Arindam Maitra et al |
| EPI_ISL_508277 | India/MH-GA82/2020 | 2020-06-18 | Asia/India/Maharashtra/Aurangabad | Government Medical College | National Institute of Biomedical Genomics | Arindam Maitra et al |
| EPI_ISL_508278 | India/MH-GA83/2020 | 2020-06-18 | Asia/India/Maharashtra/Aurangabad | Government Medical College | National Institute of Biomedical Genomics | Arindam Maitra et al |
| EPI_ISL_508279 | India/MH-GA84/2020 | 2020-06-18 | Asia/India/Maharashtra/Aurangabad | Government Medical College | National Institute of Biomedical Genomics | Arindam Maitra et al |
| EPI_ISL_508280 | India/MH-GA85/2020 | 2020-06-18 | Asia/India/Maharashtra/Aurangabad | Government Medical College | National Institute of Biomedical Genomics | Arindam Maitra et al |
| EPI_ISL_508282 | India/MH-GA87/2020 | 2020-06-15 | Asia/India/Maharashtra/Jalna | Government Medical College | National Institute of Biomedical Genomics | Arindam Maitra et al |

| | | | | | | |
|---|---|---|---|---|---|---|
| EPI_ISL_508283 | India/MH-GA88/2020 | 2020-06-15 | Asia/India/Maharashtra/Jalna | Government Medical College | National Institute of Biomedical Genomics | Arindam Maitra et al |
| EPI_ISL_508285 | India/MH-GA93/2020 | 2020-06-17 | Asia/India/Maharashtra/Aurangabad | Government Medical College | National Institute of Biomedical Genomics | Arindam Maitra et al |
| EPI_ISL_508286 | India/MH-GA98/2020 | 2020-06-17 | Asia/India/Maharashtra/Aurangabad | Government Medical College | National Institute of Biomedical Genomics | Arindam Maitra et al |
| EPI_ISL_508287 | India/KA-IB1/2020 | 2020-06-19 | Asia/India/Karnataka/Kolar | Indian Institute of Science | National Institute of Biomedical Genomics | Arindam Maitra et al |
| EPI_ISL_508288 | India/KA-IB10/2020 | 2020-06-20 | Asia/India/Karnataka/Kolar | Indian Institute of Science | National Institute of Biomedical Genomics | Arindam Maitra et al |
| EPI_ISL_508290 | India/KA-IB12/2020 | 2020-06-21 | Asia/India/Karnataka/Bengaluru Urban | Indian Institute of Science | National Institute of Biomedical Genomics | Arindam Maitra et al |
| EPI_ISL_508291 | India/KA-IB13/2020 | 2020-06-15 | Asia/India/Karnataka/Bengaluru Urban | Indian Institute of Science | National Institute of Biomedical Genomics | Arindam Maitra et al |
| EPI_ISL_508305 | India/KA-IB27/2020 | 2020-05-26 | Asia/India/Karnataka/Kolar | Indian Institute of Science | National Institute of Biomedical Genomics | Arindam Maitra et al |
| EPI_ISL_508306 | India/KA-IB28/2020 | 2020-06-20 | Asia/India/Karnataka/Bengaluru Urban | Indian Institute of Science | National Institute of Biomedical Genomics | Arindam Maitra et al |
| EPI_ISL_508308 | India/KA-IB3/2020 | 2020-06-17 | Asia/India/Karnataka/Bengaluru Urban | Indian Institute of Science | National Institute of Biomedical Genomics | Arindam Maitra et al |
| EPI_ISL_508310 | India/KA-IB31/2020 | 2020-06-03 | Asia/India/Karnataka/Bengaluru Urban | Indian Institute of Science | National Institute of Biomedical Genomics | Arindam Maitra et al |
| EPI_ISL_508311 | India/KA-IB32/2020 | 2020-06-18 | Asia/India/Karnataka/Bengaluru Urban | Indian Institute of Science | National Institute of Biomedical Genomics | Arindam Maitra et al |
| EPI_ISL_508312 | India/KA-IB33/2020 | 2020-06-20 | Asia/India/Karnataka/Bengaluru Urban | Indian Institute of Science | National Institute of Biomedical Genomics | Arindam Maitra et al |

| | | | | | | |
|---|---|---|---|---|---|---|
| EPI_ISL_508313 | India/KA-IB34/2020 | 2020-06-15 | Asia/India/Karnataka/Bengaluru Urban | Indian Institute of Science | National Institute of Biomedical Genomics | Arindam Maitra et al |
| EPI_ISL_508314 | India/KA-IB35/2020 | 2020-05-12 | Asia/India/Karnataka/Bengaluru Urban | Indian Institute of Science | National Institute of Biomedical Genomics | Arindam Maitra et al |
| EPI_ISL_508315 | India/KA-IB36/2020 | 2020-06-10 | Asia/India/Karnataka/Bengaluru Urban | Indian Institute of Science | National Institute of Biomedical Genomics | Arindam Maitra et al |
| EPI_ISL_508317 | India/KA-IB39/2020 | 2020-06-17 | Asia/India/Karnataka/Bengaluru Urban | Indian Institute of Science | National Institute of Biomedical Genomics | Arindam Maitra et al |
| EPI_ISL_508318 | India/KA-IB4/2020 | 2020-06-17 | Asia/India/Karnataka/Bengaluru Urban | Indian Institute of Science | National Institute of Biomedical Genomics | Arindam Maitra et al |
| EPI_ISL_508320 | India/KA-IB41/2020 | 2020-06-04 | Asia/India/Karnataka/Kolar | Indian Institute of Science | National Institute of Biomedical Genomics | Arindam Maitra et al |
| EPI_ISL_508321 | India/KA-IB42/2020 | 2020-06-15 | Asia/India/Karnataka/Bengaluru Urban | Indian Institute of Science | National Institute of Biomedical Genomics | Arindam Maitra et al |
| EPI_ISL_508322 | India/KA-IB43/2020 | 2020-06-17 | Asia/India/Karnataka/Bengaluru Urban | Indian Institute of Science | National Institute of Biomedical Genomics | Arindam Maitra et al |
| EPI_ISL_508323 | India/KA-IB44/2020 | 2020-06-15 | Asia/India/Karnataka/Bengaluru Urban | Indian Institute of Science | National Institute of Biomedical Genomics | Arindam Maitra et al |
| EPI_ISL_508329 | India/KA-IB51/2020 | 2020-06-19 | Asia/India/Karnataka/Bengaluru Urban | Indian Institute of Science | National Institute of Biomedical Genomics | Arindam Maitra et al |
| EPI_ISL_508330 | India/KA-IB53/2020 | 2020-06-03 | Asia/India/Karnataka/Bengaluru Urban | Indian Institute of Science | National Institute of Biomedical Genomics | Arindam Maitra et al |
| EPI_ISL_508331 | India/KA-IB54/2020 | 2020-06-20 | Asia/India/Karnataka/Bengaluru Urban | Indian Institute of Science | National Institute of Biomedical Genomics | Arindam Maitra et al |
| EPI_ISL_508332 | India/KA-IB58/2020 | 2020-05-27 | Asia/India/Karnataka/Chikkaballapura | Indian Institute of Science | National Institute of Biomedical Genomics | Arindam Maitra et al |

| | | | | | | |
|---|---|---|---|---|---|---|
| EPI_ISL_508335 | India/KA-IB7/2020 | 2020-06-17 | Asia/India/Karnataka/Bengaluru Urban | Indian Institute of Science | National Institute of Biomedical Genomics | Arindam Maitra et al |
| EPI_ISL_508336 | India/KA-IB8/2020 | 2020-06-20 | Asia/India/Karnataka/Bengaluru Urban | Indian Institute of Science | National Institute of Biomedical Genomics | Arindam Maitra et al |
| EPI_ISL_508337 | India/KA-IB9/2020 | 2020-06-19 | Asia/India/Karnataka/Kolar | Indian Institute of Science | National Institute of Biomedical Genomics | Arindam Maitra et al |
| EPI_ISL_508343 | India/WB-IK14/2020 | 2020-04-21 | Asia/India/West Bengal/Kolkata | Institute of Post Graduate Medical Education & Research | National Institute of Biomedical Genomics | Arindam Maitra et al |
| EPI_ISL_508348 | India/WB-IK19/2020 | 2020-04-24 | Asia/India/West Bengal/North 24 Parga | Institute of Post Graduate Medical Education & Research | National Institute of Biomedical Genomics | Arindam Maitra et al |
| EPI_ISL_508364 | India/WB-IK35/2020 | 2020-05-01 | Asia/India/West Bengal/North 24 Parga | Institute of Post Graduate Medical Education & Research | National Institute of Biomedical Genomics | Arindam Maitra et al |
| EPI_ISL_508369 | India/WB-IK41/2020 | 2020-05-04 | Asia/India/West Bengal/Kolkata | Institute of Post Graduate Medical Education & Research | National Institute of Biomedical Genomics | Arindam Maitra et al |
| EPI_ISL_508377 | India/WB-IK50/2020 | 2020-05-05 | Asia/India/West Bengal/North 24 Parga | Institute of Post Graduate Medical Education & Research | National Institute of Biomedical Genomics | Arindam Maitra et al |
| EPI_ISL_508378 | India/WB-IK51/2020 | 2020-05-05 | Asia/India/West Bengal/Kolkata | Institute of Post Graduate Medical Education & Research | National Institute of Biomedical Genomics | Arindam Maitra et al |
| EPI_ISL_508382 | India/WB-IK56/2020 | 2020-05-07 | Asia/India/West Bengal/Kolkata | Institute of Post Graduate Medical Education & Research | National Institute of Biomedical Genomics | Arindam Maitra et al |
| EPI_ISL_508389 | India/WB-IK65/2020 | 2020-05-12 | Asia/India/West Bengal/Howrah | Institute of Post Graduate Medical Education & Research | National Institute of Biomedical Genomics | Arindam Maitra et al |
| EPI_ISL_508391 | India/WB-IK67/2020 | 2020-05-12 | Asia/India/West Bengal/Kolkata | Institute of Post Graduate Medical Education & Research | National Institute of Biomedical Genomics | Arindam Maitra et al |
| EPI_ISL_508393 | India/WB-IK69/2020 | 2020-05-14 | Asia/India/West Bengal/Kolkata | Institute of Post Graduate Medical Education & Research | National Institute of Biomedical Genomics | Arindam Maitra et al |

| | | | | | | |
|---|---|---|---|---|---|---|
| EPI_ISL_508397 | India/WB-IK73/2020 | 2020-05-14 | Asia/India/West Bengal/Kolkata | Institute of Post Graduate Medical Education & Research | National Institute of Biomedical Genomics | Arindam Maitra et al |
| EPI_ISL_508398 | India/WB-IK75/2020 | 2020-05-19 | Asia/India/West Bengal/East Medinipur | Institute of Post Graduate Medical Education & Research | National Institute of Biomedical Genomics | Arindam Maitra et al |
| EPI_ISL_508401 | India/WB-IK80/2020 | 2020-05-17 | Asia/India/West Bengal/Kolkata | Institute of Post Graduate Medical Education & Research | National Institute of Biomedical Genomics | Arindam Maitra et al |
| EPI_ISL_508404 | India/WB-IK85/2020 | 2020-05-17 | Asia/India/West Bengal/Kolkata | Institute of Post Graduate Medical Education & Research | National Institute of Biomedical Genomics | Arindam Maitra et al |
| EPI_ISL_508406 | India/WB-IK88/2020 | 2020-05-19 | Asia/India/West Bengal/Kolkata | Institute of Post Graduate Medical Education & Research | National Institute of Biomedical Genomics | Arindam Maitra et al |
| EPI_ISL_508407 | India/WB-IK9/2020 | 2020-04-13 | Asia/India/West Bengal/Kolkata | Institute of Post Graduate Medical Education & Research | National Institute of Biomedical Genomics | Arindam Maitra et al |
| EPI_ISL_508409 | India/WB-IK91/2020 | 2020-05-20 | Asia/India/West Bengal/Kolkata | Institute of Post Graduate Medical Education & Research | National Institute of Biomedical Genomics | Arindam Maitra et al |
| EPI_ISL_508415 | India/DL-MN1/2020 | 2020-06-16 | Asia/India/Delhi/ | Maulana Azad Medical College | National Institute of Biomedical Genomics | Arindam Maitra et al |
| EPI_ISL_508418 | India/DL-MN26/2020 | 2020-06-20 | Asia/India/Delhi/ | Maulana Azad Medical College | National Institute of Biomedical Genomics | Arindam Maitra et al |
| EPI_ISL_508420 | India/DL-MN4/2020 | 2020-06-18 | Asia/India/Delhi/ | Maulana Azad Medical College | National Institute of Biomedical Genomics | Arindam Maitra et al |
| EPI_ISL_508424 | India/MH-MW12/2020 | 2020-05-01 | Asia/India/Maharashtra/Yavatmal | Mahatma Gandhi Institute of Medical Sciences | National Institute of Biomedical Genomics | Arindam Maitra et al |
| EPI_ISL_508437 | India/MH-MW34/2020 | 2020-06-04 | Asia/India/Maharashtra/Wardha | Mahatma Gandhi Institute of Medical Sciences | National Institute of Biomedical Genomics | Arindam Maitra et al |
| EPI_ISL_508438 | India/MH-MW38/2020 | 2020-06-08 | Asia/India/Maharashtra/Wardha | Mahatma Gandhi Institute of Medical Sciences | National Institute of Biomedical Genomics | Arindam Maitra et al |

| | | | | | | |
|---|---|---|---|---|---|---|
| EPI_ISL_508442 | India/WB-S101/2020 | 2020-06-09 | Asia/India/West Bengal/Kolkata | ICMR-National Institute of Cholera and Enteric Diseases | National Institute of Biomedical Genomics | Arindam Maitra et al |
| EPI_ISL_508443 | India/WB-S102/2020 | 2020-06-07 | Asia/India/West Bengal/North 24 Parga | ICMR-National Institute of Cholera and Enteric Diseases | National Institute of Biomedical Genomics | Arindam Maitra et al |
| EPI_ISL_508448 | India/WB-S40/2020 | 2020-05-01 | Asia/India/West Bengal/Kolkata | ICMR-National Institute of Cholera and Enteric Diseases | National Institute of Biomedical Genomics | Arindam Maitra et al |
| EPI_ISL_508461 | India/WB-S71/2020 | 2020-05-25 | Asia/India/West Bengal/North 24 Parga | ICMR-National Institute of Cholera and Enteric Diseases | National Institute of Biomedical Genomics | Arindam Maitra et al |
| EPI_ISL_508462 | India/WB-S72/2020 | 2020-05-20 | Asia/India/West Bengal/Nadia | ICMR-National Institute of Cholera and Enteric Diseases | National Institute of Biomedical Genomics | Arindam Maitra et al |
| EPI_ISL_508463 | India/WB-S73/2020 | 2020-05-26 | Asia/India/West Bengal/Birbhum | ICMR-National Institute of Cholera and Enteric Diseases | National Institute of Biomedical Genomics | Arindam Maitra et al |
| EPI_ISL_508465 | India/WB-S75/2020 | 2020-05-20 | Asia/India/West Bengal/Nadia | ICMR-National Institute of Cholera and Enteric Diseases | National Institute of Biomedical Genomics | Arindam Maitra et al |
| EPI_ISL_508469 | India/WB-S79/2020 | 2020-05-20 | Asia/India/West Bengal/Nadia | ICMR-National Institute of Cholera and Enteric Diseases | National Institute of Biomedical Genomics | Arindam Maitra et al |
| EPI_ISL_508472 | India/WB-S82/2020 | 2020-05-29 | Asia/India/West Bengal/Howrah | ICMR-National Institute of Cholera and Enteric Diseases | National Institute of Biomedical Genomics | Arindam Maitra et al |
| EPI_ISL_508480 | India/WB-S91/2020 | 2020-06-01 | Asia/India/West Bengal/Kolkata | ICMR-National Institute of Cholera and Enteric Diseases | National Institute of Biomedical Genomics | Arindam Maitra et al |
| EPI_ISL_508483 | India/WB-S94/2020 | 2020-06-03 | Asia/India/West Bengal/North 24 Parga | ICMR-National Institute of Cholera and Enteric Diseases | National Institute of Biomedical Genomics | Arindam Maitra et al |
| EPI_ISL_508484 | India/WB-S95/2020 | 2020-06-03 | Asia/India/West Bengal/North 24 Parga | ICMR-National Institute of Cholera and Enteric Diseases | National Institute of Biomedical Genomics | Arindam Maitra et al |
| EPI_ISL_508486 | India/WB-S97/2020 | 2020-06-11 | Asia/India/West Bengal/North 24 Parga | ICMR-National Institute of Cholera and Enteric Diseases | National Institute of Biomedical Genomics | Arindam Maitra et al |

| | | | | | | |
|---|---|---|---|---|---|---|
| EPI_ISL_508489 | India/HR-TF1/2020 | 2020-06-12 | Asia/India/Haryana/Faridabad | Translational Health Science and Technology Institute | National Institute of Biomedical Genomics | Arindam Maitra et al |
| EPI_ISL_508490 | India/HR-TF10/2020 | 2020-06-17 | Asia/India/Haryana/Faridabad | Translational Health Science and Technology Institute | National Institute of Biomedical Genomics | Arindam Maitra et al |
| EPI_ISL_508491 | India/HR-TF12/2020 | 2020-06-17 | Asia/India/Haryana/Faridabad | Translational Health Science and Technology Institute | National Institute of Biomedical Genomics | Arindam Maitra et al |
| EPI_ISL_508492 | India/HR-TF13/2020 | 2020-06-17 | Asia/India/Haryana/Faridabad | Translational Health Science and Technology Institute | National Institute of Biomedical Genomics | Arindam Maitra et al |
| EPI_ISL_508494 | India/HR-TF15/2020 | 2020-06-17 | Asia/India/Haryana/Faridabad | Translational Health Science and Technology Institute | National Institute of Biomedical Genomics | Arindam Maitra et al |
| EPI_ISL_508495 | India/DL-TF17/2020 | 2020-06-17 | Asia/India/Delhi/Tughlakabad | Translational Health Science and Technology Institute | National Institute of Biomedical Genomics | Arindam Maitra et al |
| EPI_ISL_508496 | India/HR-TF2/2020 | 2020-06-14 | Asia/India/Haryana/Palwal | Translational Health Science and Technology Institute | National Institute of Biomedical Genomics | Arindam Maitra et al |
| EPI_ISL_508498 | India/HR-TF24/2020 | 2020-06-19 | Asia/India/Haryana/Palwal | Translational Health Science and Technology Institute | National Institute of Biomedical Genomics | Arindam Maitra et al |
| EPI_ISL_508499 | India/HR-TF25/2020 | 2020-06-19 | Asia/India/Haryana/Faridabad | Translational Health Science and Technology Institute | National Institute of Biomedical Genomics | Arindam Maitra et al |
| EPI_ISL_508500 | India/HR-TF26/2020 | 2020-06-19 | Asia/India/Haryana/Palwal | Translational Health Science and Technology Institute | National Institute of Biomedical Genomics | Arindam Maitra et al |
| EPI_ISL_508502 | India/HR-TF3/2020 | 2020-06-14 | Asia/India/Haryana/Palwal | Translational Health Science and Technology Institute | National Institute of Biomedical Genomics | Arindam Maitra et al |
| EPI_ISL_508615 | Spain/CT-IBV-005618/2020 | 2020-03-18 | Europe/Spain/Catalunya/Badalona | Departamento de Microbiologv≠a, CDB, Hospital Clv≠nic, Barcelona | SeqCOVID-SPAIN consortium/IBV(CSIC) | Andrea Vergara et al |
| EPI_ISL_508617 | Spain/CT-IBV-005554/2020 | 2020-03-18 | Europe/Spain/Catalunya/Barcelona | Departamento de Microbiologv≠a, CDB, Hospital Clv≠nic, Barcelona | SeqCOVID-SPAIN consortium/IBV(CSIC) | Andrea Vergara et al |

| | | | | | | |
|---|---|---|---|---|---|---|
| EPI_ISL_508623 | Spain/CT-IBV-005566/2020 | 2020-03-18 | Europe/Spain/Catalunya/Barcelona | Departamento de Microbiologv≠a, CDB, Hospital Clv≠nic, Barcelona | SeqCOVID-SPAIN consortium/IBV(CSIC) | Andrea Vergara et al |
| EPI_ISL_508627 | Spain/CT-IBV-005574/2020 | 2020-03-29 | Europe/Spain/Catalunya/Barcelona | Departamento de Microbiologv≠a, CDB, Hospital Clv≠nic, Barcelona | SeqCOVID-SPAIN consortium/IBV(CSIC) | Andrea Vergara et al |
| EPI_ISL_508632 | Spain/CT-IBV-005581/2020 | 2020-03-26 | Europe/Spain/Catalunya/Barcelona | Departamento de Microbiologv≠a, CDB, Hospital Clv≠nic, Barcelona | SeqCOVID-SPAIN consortium/IBV(CSIC) | Andrea Vergara et al |
| EPI_ISL_508639 | Spain/CT-IBV-005590/2020 | 2020-03-17 | Europe/Spain/Catalunya/Barcelona | Departamento de Microbiologv≠a, CDB, Hospital Clv≠nic, Barcelona | SeqCOVID-SPAIN consortium/IBV(CSIC) | Andrea Vergara et al |
| EPI_ISL_508644 | Spain/CT-IBV-005596/2020 | 2020-03-18 | Europe/Spain/Catalunya/Barcelona | Departamento de Microbiologv≠a, CDB, Hospital Clv≠nic, Barcelona | SeqCOVID-SPAIN consortium/IBV(CSIC) | Andrea Vergara et al |
| EPI_ISL_508645 | Spain/CT-IBV-005597/2020 | 2020-03-18 | Europe/Spain/Catalunya/Barcelona | Departamento de Microbiologv≠a, CDB, Hospital Clv≠nic, Barcelona | SeqCOVID-SPAIN consortium/IBV(CSIC) | Andrea Vergara et al |
| EPI_ISL_508647 | Spain/CT-IBV-005601/2020 | 2020-03-26 | Europe/Spain/Catalunya/Barcelona | Departamento de Microbiologv≠a, CDB, Hospital Clv≠nic, Barcelona | SeqCOVID-SPAIN consortium/IBV(CSIC) | Andrea Vergara et al |
| EPI_ISL_508650 | Spain/CT-IBV-005604/2020 | 2020-03-26 | Europe/Spain/Catalunya/Barcelona | Departamento de Microbiologv≠a, CDB, Hospital Clv≠nic, Barcelona | SeqCOVID-SPAIN consortium/IBV(CSIC) | Andrea Vergara et al |
| EPI_ISL_508661 | Spain/CT-IBV-005626/2020 | 2020-03-29 | Europe/Spain/Catalunya/Barcelona | Departamento de Microbiologv≠a, CDB, Hospital Clv≠nic, Barcelona | SeqCOVID-SPAIN consortium/IBV(CSIC) | Andrea Vergara et al |
| EPI_ISL_508663 | Spain/CT-IBV-005630/2020 | 2020-04-10 | Europe/Spain/Catalunya/Barcelona | Departamento de Microbiologv≠a, CDB, Hospital Clv≠nic, Barcelona | SeqCOVID-SPAIN consortium/IBV(CSIC) | Andrea Vergara et al |
| EPI_ISL_508665 | Spain/CT-IBV-005632/2020 | 2020-04-23 | Europe/Spain/Catalunya/Barcelona | Departamento de Microbiologv≠a, CDB, Hospital Clv≠nic, Barcelona | SeqCOVID-SPAIN consortium/IBV(CSIC) | Andrea Vergara et al |
| EPI_ISL_508667 | Spain/CT-IBV-005634/2020 | 2020-04-24 | Europe/Spain/Catalunya/Barcelona | Departamento de Microbiologv≠a, CDB, Hospital Clv≠nic, Barcelona | SeqCOVID-SPAIN consortium/IBV(CSIC) | Andrea Vergara et al |
| EPI_ISL_508675 | Spain/CT-IBV-005552/2020 | 2020-03-17 | Europe/Spain/Catalunya/Barcelona | Departamento de Microbiologv≠a, CDB, Hospital Clv≠nic, Barcelona | SeqCOVID-SPAIN consortium/IBV(CSIC) | Andrea Vergara et al |

| | | | | | | |
|---|---|---|---|---|---|---|
| EPI_ISL_508677 | Spain/IB-IBV-005617/2020 | 2020-04-27 | Europe/Spain/Balear Islands/Calvia | Departamento de Microbiolog√≠a, CDB, Hospital Cl√≠nic, Barcelona | SeqCOVID-SPAIN consortium/IBV(CSIC) | Andrea Vergara et al |
| EPI_ISL_508686 | Spain/CT-IBV-005637/2020 | 2020-04-22 | Europe/Spain/Catalunya/Vic | Departamento de Microbiolog√≠a, CDB, Hospital Cl√≠nic, Barcelona | SeqCOVID-SPAIN consortium/IBV(CSIC) | Andrea Vergara et al |
| EPI_ISL_508687 | Germany/BW-UT-006/2020 | 2020-05-20 | Europe/Germany/Baden-Wuerttemberg | Institut f√ºr Virologie und Epidemiologie der Viruskrankheiten, Universit√§tsklinikum T√ºbingen | NGS Competence Center T√ºbingen, Institut f√ºr Medizinische Mikrobiologie und Hygiene, Universit√§tsklinikum T√ºbingen | Angel Angelov et al |
| EPI_ISL_508692 | Germany/BW-UT-013/2020 | 2020-05-20 | Europe/Germany/Baden-Wuerttemberg | Institut f√ºr Virologie und Epidemiologie der Viruskrankheiten, Universit√§tsklinikum T√ºbingen | NGS Competence Center T√ºbingen, Institut f√ºr Medizinische Mikrobiologie und Hygiene, Universit√§tsklinikum T√ºbingen | Angel Angelov et al |
| EPI_ISL_508694 | Germany/BW-UT-015/2020 | 2020-05-20 | Europe/Germany/Baden-Wuerttemberg | Institut f√ºr Virologie und Epidemiologie der Viruskrankheiten, Universit√§tsklinikum T√ºbingen | NGS Competence Center T√ºbingen, Institut f√ºr Medizinische Mikrobiologie und Hygiene, Universit√§tsklinikum T√ºbingen | Angel Angelov et al |
| EPI_ISL_508703 | Germany/BW-UT-028/2020 | 2020-05-20 | Europe/Germany/Baden-Wuerttemberg | Institut f√ºr Virologie und Epidemiologie der Viruskrankheiten, Universit√§tsklinikum T√ºbingen | NGS Competence Center T√ºbingen, Institut f√ºr Medizinische Mikrobiologie und Hygiene, Universit√§tsklinikum T√ºbingen | Angel Angelov et al |
| EPI_ISL_508716 | USA/FL-BPHL-0513/2020 | 2020-06-12 | North America/USA/Florida/ | Florida Bureau of Public Health Laboratories | Florida Bureau of Public Health Laboratories | Sarah Schmedes et al |

| | | | | | | |
|---|---|---|---|---|---|---|
| EPI_ISL_508720 | USA/FL-BPHL-0517/2020 | 2020-06-16 | North America/USA/Florida/ | Florida Bureau of Public Health Laboratories | Florida Bureau of Public Health Laboratories | Sarah Schmedes et al |
| EPI_ISL_508721 | USA/FL-BPHL-0518/2020 | 2020-06-15 | North America/USA/Florida/ | Florida Bureau of Public Health Laboratories | Florida Bureau of Public Health Laboratories | Sarah Schmedes et al |
| EPI_ISL_508729 | USA/FL-BPHL-0526/2020 | 2020-06-19 | North America/USA/Florida/ | Florida Bureau of Public Health Laboratories | Florida Bureau of Public Health Laboratories | Sarah Schmedes et al |
| EPI_ISL_508737 | USA/FL-BPHL-0534/2020 | 2020-06-19 | North America/USA/Florida/ | Florida Bureau of Public Health Laboratories | Florida Bureau of Public Health Laboratories | Sarah Schmedes et al |
| EPI_ISL_508773 | USA/FL-BPHL-0569/2020 | 2020-03-15 | North America/USA/Florida/ | Florida Bureau of Public Health Laboratories | Florida Bureau of Public Health Laboratories | Sarah Schmedes et al |
| EPI_ISL_508779 | USA/FL-BPHL-0575/2020 | 2020-03-21 | North America/USA/Florida/ | Florida Bureau of Public Health Laboratories | Florida Bureau of Public Health Laboratories | Sarah Schmedes et al |
| EPI_ISL_508783 | USA/FL-BPHL-0579/2020 | 2020-03-20 | North America/USA/Florida/ | Florida Bureau of Public Health Laboratories | Florida Bureau of Public Health Laboratories | Sarah Schmedes et al |
| EPI_ISL_508797 | USA/FL-BPHL-0593/2020 | 2020-03-24 | North America/USA/Florida/ | Florida Bureau of Public Health Laboratories | Florida Bureau of Public Health Laboratories | Sarah Schmedes et al |
| EPI_ISL_508811 | USA/FL-BPHL-0607/2020 | 2020-04-03 | North America/USA/Florida/ | Florida Bureau of Public Health Laboratories | Florida Bureau of Public Health Laboratories | Sarah Schmedes et al |
| EPI_ISL_508814 | USA/FL-BPHL-0610/2020 | 2020-04-03 | North America/USA/Florida/ | Florida Bureau of Public Health Laboratories | Florida Bureau of Public Health Laboratories | Sarah Schmedes et al |
| EPI_ISL_508817 | USA/FL-BPHL-0613/2020 | 2020-04-03 | North America/USA/Florida/ | Florida Bureau of Public Health Laboratories | Florida Bureau of Public Health Laboratories | Sarah Schmedes et al |
| EPI_ISL_508822 | USA/FL-BPHL-0618/2020 | 2020-04-03 | North America/USA/Florida/ | Florida Bureau of Public Health Laboratories | Florida Bureau of Public Health Laboratories | Sarah Schmedes et al |
| EPI_ISL_508843 | USA/FL-BPHL-0639/2020 | 2020-04-06 | North America/USA/Florida/ | Florida Bureau of Public Health Laboratories | Florida Bureau of Public Health Laboratories | Sarah Schmedes et al |

| | | | | | | |
|---|---|---|---|---|---|---|
| EPI_ISL_508857 | USA/FL-BPHL-0653/2020 | 2020-04-03 | North America/USA/Florida/ | Florida Bureau of Public Health Laboratories | Florida Bureau of Public Health Laboratories | Sarah Schmedes et al |
| EPI_ISL_508872 | France/ARA-07639/2020 | 2020-03-11 | Europe/France/Auvergne-Rh√¥ne-Alpes/ | Institut des Agents Infectieux (IAI), Hospices Civils de Lyon | CNR Virus des Infections Respiratoires - France SUD | Antonin Bal et al |
| EPI_ISL_508874 | France/ARA-08589/2020 | 2020-03-13 | Europe/France/Auvergne-Rh√¥ne-Alpes/ | Institut des Agents Infectieux (IAI), Hospices Civils de Lyon | CNR Virus des Infections Respiratoires - France SUD | Antonin Bal et al |
| EPI_ISL_508875 | France/ARA-0873/2020 | 2020-03-14 | Europe/France/Auvergne-Rh√¥ne-Alpes/ | Centre Hospitalier de Macon | CNR Virus des Infections Respiratoires - France SUD | Antonin Bal et al |
| EPI_ISL_508876 | France/ARA-08938/2020 | 2020-03-14 | Europe/France/Auvergne-Rh√¥ne-Alpes/ | Centre Hospitalier de Macon | CNR Virus des Infections Respiratoires - France SUD | Antonin Bal et al |
| EPI_ISL_508883 | France/ARA-10825/2020 | 2020-03-18 | Europe/France/Auvergne-Rh√¥ne-Alpes/ | Institut des Agents Infectieux (IAI), Hospices Civils de Lyon | CNR Virus des Infections Respiratoires - France SUD | Antonin Bal et al |
| EPI_ISL_508889 | France/ARA-14570/2020 | 2020-03-26 | Europe/France/Auvergne-Rh√¥ne-Alpes/ | Institut des Agents Infectieux (IAI), Hospices Civils de Lyon | CNR Virus des Infections Respiratoires - France SUD | Antonin Bal et al |
| EPI_ISL_508899 | France/ARA-14997/2020 | 2020-03-27 | Europe/France/Auvergne-Rh√¥ne-Alpes/ | Institut des Agents Infectieux (IAI), Hospices Civils de Lyon | CNR Virus des Infections Respiratoires - France SUD | Antonin Bal et al |
| EPI_ISL_508903 | France/ARA-15296/2020 | 2020-03-27 | Europe/France/Auvergne-Rh√¥ne-Alpes/ | Institut des Agents Infectieux (IAI), Hospices Civils de Lyon | CNR Virus des Infections Respiratoires - France SUD | Antonin Bal et al |
| EPI_ISL_508908 | France/ARA-16322/2020 | 2020-03-30 | Europe/France/Auvergne-Rh√¥ne-Alpes/ | Institut des Agents Infectieux (IAI), Hospices Civils de Lyon | CNR Virus des Infections Respiratoires - France SUD | Antonin Bal et al |

| | | | | | | |
|---|---|---|---|---|---|---|
| EPI_ISL_508910 | France/NAQ-NA01745/2020 | 2020-03-11 | Europe/France/Nouvelle-Aquitaine/Limo | Institut des Agents Infectieux (IAI), Hospices Civils de Lyon | CNR Virus des Infections Respiratoires - France SUD | Antonin Bal et al |
| EPI_ISL_508915 | France/ARA-20976/2020 | 2020-04-06 | Europe/France/Auvergne-Rhv¥ne-Alpes/ | CNR Virus des Infections Respiratoires - France SUD | CNR Virus des Infections Respiratoires - France SUD | Antonin Bal et al |
| EPI_ISL_508917 | France/ARA-2221/2020 | 2020-04-08 | Europe/France/Auvergne-Rhv¥ne-Alpes/ | CNR Virus des Infections Respiratoires - France SUD | CNR Virus des Infections Respiratoires - France SUD | Antonin Bal et al |
| EPI_ISL_508928 | France/ARA-23813/2020 | 2020-04-11 | Europe/France/Auvergne-Rhv¥ne-Alpes/ | CNR Virus des Infections Respiratoires - France SUD | CNR Virus des Infections Respiratoires - France SUD | Antonin Bal et al |
| EPI_ISL_508930 | France/ARA-23943/2020 | 2020-04-12 | Europe/France/Auvergne-Rhv¥ne-Alpes/ | CNR Virus des Infections Respiratoires - France SUD | CNR Virus des Infections Respiratoires - France SUD | Antonin Bal et al |
| EPI_ISL_508931 | France/ARA-24023/2020 | 2020-04-11 | Europe/France/Auvergne-Rhv¥ne-Alpes/ | L'Hv¥pital Nord-Ouest Tarare-Grandris | CNR Virus des Infections Respiratoires - France SUD | Antonin Bal et al |
| EPI_ISL_508933 | France/ARA-05288/2020 | 2020-02-28 | Europe/France/Auvergne-Rhv¥ne-Alpes/ | CNR Virus des Infections Respiratoires - France SUD | CNR Virus des Infections Respiratoires - France SUD | Antonin Bal et al |
| EPI_ISL_508941 | France/ARA-0731/2020 | 2020-03-10 | Europe/France/Auvergne-Rhv¥ne-Alpes/ | Centre Hospitalier de Macon | CNR Virus des Infections Respiratoires - France SUD | Antonin Bal et al |
| EPI_ISL_508952 | France/ARA-12638/2020 | 2020-03-23 | Europe/France/Auvergne-Rhv¥ne-Alpes/ | Centre Hospitalier de Macon | CNR Virus des Infections Respiratoires - France SUD | Antonin Bal et al |
| EPI_ISL_508955 | France/ARA-13367/2020 | 2020-03-24 | Europe/France/Auvergne-Rhv¥ne-Alpes/ | CNR Virus des Infections Respiratoires - France SUD | CNR Virus des Infections Respiratoires - France SUD | Antonin Bal et al |

| | | | | | | |
|---|---|---|---|---|---|---|
| EPI_ISL_508956 | France/ARA-13374/2020 | 2020-03-24 | Europe/France/Auvergne-Rh√¥ne-Alpes/ | CNR Virus des Infections Respiratoires - France SUD | CNR Virus des Infections Respiratoires - France SUD | Antonin Bal et al |
| EPI_ISL_508961 | France/ARA-1783/2020 | 2020-04-01 | Europe/France/Auvergne-Rh√¥ne-Alpes/ | CNR Virus des Infections Respiratoires - France SUD | CNR Virus des Infections Respiratoires - France SUD | Antonin Bal et al |
| EPI_ISL_508963 | France/ARA-18212/2020 | 2020-04-01 | Europe/France/Auvergne-Rh√¥ne-Alpes/ | CNR Virus des Infections Respiratoires - France SUD | CNR Virus des Infections Respiratoires - France SUD | Antonin Bal et al |
| EPI_ISL_508968 | France/ARA-18437/2020 | 2020-04-02 | Europe/France/Auvergne-Rh√¥ne-Alpes/ | Centre Hospitalier de Bourg en Bresse | CNR Virus des Infections Respiratoires - France SUD | Antonin Bal et al |
| EPI_ISL_508969 | France/ARA-18440/2020 | 2020-04-02 | Europe/France/Auvergne-Rh√¥ne-Alpes/ | CNR Virus des Infections Respiratoires - France SUD | CNR Virus des Infections Respiratoires - France SUD | Antonin Bal et al |
| EPI_ISL_508970 | France/ARA-18487/2020 | 2020-04-02 | Europe/France/Auvergne-Rh√¥ne-Alpes/ | CNR Virus des Infections Respiratoires - France SUD | CNR Virus des Infections Respiratoires - France SUD | Antonin Bal et al |
| EPI_ISL_508971 | France/ARA-18625/2020 | 2020-04-02 | Europe/France/Auvergne-Rh√¥ne-Alpes/ | CNR Virus des Infections Respiratoires - France SUD | CNR Virus des Infections Respiratoires - France SUD | Antonin Bal et al |
| EPI_ISL_508976 | France/ARA-19119/2020 | 2020-04-01 | Europe/France/Auvergne-Rh√¥ne-Alpes/ | CNR Virus des Infections Respiratoires - France SUD | CNR Virus des Infections Respiratoires - France SUD | Antonin Bal et al |
| EPI_ISL_508990 | France/ARA-20869/2020 | 2020-04-06 | Europe/France/Auvergne-Rh√¥ne-Alpes/ | CNR Virus des Infections Respiratoires - France SUD | CNR Virus des Infections Respiratoires - France SUD | Antonin Bal et al |
| EPI_ISL_508992 | France/ARA-21163/2020 | 2020-04-07 | Europe/France/Auvergne-Rh√¥ne-Alpes/ | CNR Virus des Infections Respiratoires - France SUD | CNR Virus des Infections Respiratoires - France SUD | Antonin Bal et al |

| | | | | | | |
|---|---|---|---|---|---|---|
| EPI_ISL_508994 | France/ARA-21399/2020 | 2020-04-07 | Europe/France/Auvergne-Rh√¥ne-Alpes/ | CNR Virus des Infections Respiratoires - France SUD | CNR Virus des Infections Respiratoires - France SUD | Antonin Bal et al |
| EPI_ISL_509008 | France/ARA-12740/2020 | 2020-03-23 | Europe/France/Auvergne-Rh√¥ne-Alpes/ | Institut des Agents Infectieux (IAI), Hospices Civils de Lyon | CNR Virus des Infections Respiratoires - France SUD | Antonin Bal et al |
| EPI_ISL_509012 | France/ARA-17181/2020 | 2020-03-30 | Europe/France/Auvergne-Rh√¥ne-Alpes/ | Centre Hospitalier Alpes Leman | CNR Virus des Infections Respiratoires - France SUD | Antonin Bal et al |
| EPI_ISL_509015 | France/ARA-18066/2020 | 2020-04-01 | Europe/France/Auvergne-Rh√¥ne-Alpes/ | Centre Hospitalier de Villefranche | CNR Virus des Infections Respiratoires - France SUD | Antonin Bal et al |
| EPI_ISL_509070 | USA/OR-OHSU-0193/2020 | 2020-03-30 | North America/USA/Oregon/ | OHSU Lab Services Molecular Microbiology Lab | Oregon SARS-CoV-2 Genome Sequencing Center | Brendan L. O'Connell et al |
| EPI_ISL_509071 | USA/OR-OHSU-0200/2020 | 2020-03-30 | North America/USA/Oregon/Multnomal | OHSU Lab Services Molecular Microbiology Lab | Oregon SARS-CoV-2 Genome Sequencing Center | Brendan L. O'Connell et al |
| EPI_ISL_509100 | USA/OR-OHSU-0309/2020 | 2020-05-09 | North America/USA/Oregon/Washingto | OHSU Lab Services Molecular Microbiology Lab | Oregon SARS-CoV-2 Genome Sequencing Center | Brendan L. O'Connell et al |
| EPI_ISL_509101 | USA/OR-OHSU-0310/2020 | 2020-05-09 | North America/USA/Oregon/Clackamas | OHSU Lab Services Molecular Microbiology Lab | Oregon SARS-CoV-2 Genome Sequencing Center | Brendan L. O'Connell et al |
| EPI_ISL_509103 | USA/WA-OHSU-0327/2020 | 2020-05-14 | North America/USA/Washington/Klickit | OHSU Lab Services Molecular Microbiology Lab | Oregon SARS-CoV-2 Genome Sequencing Center | Brendan L. O'Connell et al |
| EPI_ISL_509114 | USA/OR-OHSU-0349/2020 | 2020-05-18 | North America/USA/Oregon/Multnomal | OHSU Lab Services Molecular Microbiology Lab | Oregon SARS-CoV-2 Genome Sequencing Center | Brendan L. O'Connell et al |

| | | | | | | |
|---|---|---|---|---|---|---|
| EPI_ISL_509115 | USA/OR-OHSU-0352/2020 | 2020-05-18 | North America/USA/Oregon/Clackamas | OHSU Lab Services Molecular Microbiology Lab | Oregon SARS-CoV-2 Genome Sequencing Center | Brendan L. O'Connell et al |
| EPI_ISL_509116 | USA/OR-OHSU-0353/2020 | 2020-05-18 | North America/USA/Oregon/Washingto | OHSU Lab Services Molecular Microbiology Lab | Oregon SARS-CoV-2 Genome Sequencing Center | Brendan L. O'Connell et al |
| EPI_ISL_509118 | USA/WA-OHSU-0366/2020 | 2020-05-29 | North America/USA/Washington/Klickit | OHSU Lab Services Molecular Microbiology Lab | Oregon SARS-CoV-2 Genome Sequencing Center | Brendan L. O'Connell et al |
| EPI_ISL_509121 | USA/OR-OHSU-0380/2020 | 2020-05-08 | North America/USA/Oregon/Washingto | OHSU Lab Services Molecular Microbiology Lab | Oregon SARS-CoV-2 Genome Sequencing Center | Brendan L. O'Connell et al |
| EPI_ISL_509124 | USA/WA-OHSU-0386/2020 | 2020-05-08 | North America/USA/Washington/Klickit | OHSU Lab Services Molecular Microbiology Lab | Oregon SARS-CoV-2 Genome Sequencing Center | Brendan L. O'Connell et al |
| EPI_ISL_509131 | USA/OR-OHSU-0415/2020 | 2020-05-16 | North America/USA/Oregon/Multnomal | OHSU Lab Services Molecular Microbiology Lab | Oregon SARS-CoV-2 Genome Sequencing Center | Brendan L. O'Connell et al |
| EPI_ISL_509134 | USA/OR-OHSU-0422/2020 | 2020-05-25 | North America/USA/Oregon/Washingto | OHSU Lab Services Molecular Microbiology Lab | Oregon SARS-CoV-2 Genome Sequencing Center | Brendan L. O'Connell et al |
| EPI_ISL_509136 | USA/OR-OHSU-0433/2020 | 2020-05-26 | North America/USA/Oregon/Washingto | OHSU Lab Services Molecular Microbiology Lab | Oregon SARS-CoV-2 Genome Sequencing Center | Brendan L. O'Connell et al |
| EPI_ISL_509153 | USA/OR-OHSU-1167/2020 | 2020-07-10 | North America/USA/Oregon/Columbia C | OHSU Lab Services Molecular Microbiology Lab | Oregon SARS-CoV-2 Genome Sequencing Center | Brendan L. O'Connell et al |
| EPI_ISL_509154 | USA/OR-OHSU-1169/2020 | 2020-07-11 | North America/USA/Oregon/Clackamas | OHSU Lab Services Molecular Microbiology Lab | Oregon SARS-CoV-2 Genome Sequencing Center | Brendan L. O'Connell et al |

| | | | | | | |
|---|---|---|---|---|---|---|
| EPI_ISL_509156 | USA/OR-OHSU-1175/2020 | 2020-07-11 | North America/USA/Oregon/Washingto | OHSU Lab Services Molecular Microbiology Lab | Oregon SARS-CoV-2 Genome Sequencing Center | Brendan L. O'Connell et al |
| EPI_ISL_509159 | USA/OR-OHSU-1185/2020 | 2020-07-11 | North America/USA/Oregon/Washingto | OHSU Lab Services Molecular Microbiology Lab | Oregon SARS-CoV-2 Genome Sequencing Center | Brendan L. O'Connell et al |
| EPI_ISL_509170 | USA/OR-OHSU-1243/2020 | 2020-07-13 | North America/USA/Oregon/Multnomal | OHSU Lab Services Molecular Microbiology Lab | Oregon SARS-CoV-2 Genome Sequencing Center | Brendan L. O'Connell et al |
| EPI_ISL_509178 | USA/OR-OHSU-1285/2020 | 2020-07-14 | North America/USA/Oregon/Marion Co | OHSU Lab Services Molecular Microbiology Lab | Oregon SARS-CoV-2 Genome Sequencing Center | Brendan L. O'Connell et al |
| EPI_ISL_509184 | USA/OR-OHSU-1331/2020 | 2020-07-15 | North America/USA/Oregon/Multnomal | OHSU Lab Services Molecular Microbiology Lab | Oregon SARS-CoV-2 Genome Sequencing Center | Brendan L. O'Connell et al |
| EPI_ISL_509187 | USA/OR-OHSU-1349/2020 | 2020-07-15 | North America/USA/Oregon/Multnomal | OHSU Lab Services Molecular Microbiology Lab | Oregon SARS-CoV-2 Genome Sequencing Center | Brendan L. O'Connell et al |
| EPI_ISL_509193 | USA/OR-OHSU-1401/2020 | 2020-07-16 | North America/USA/Oregon/Washingto | OHSU Lab Services Molecular Microbiology Lab | Oregon SARS-CoV-2 Genome Sequencing Center | Brendan L. O'Connell et al |
| EPI_ISL_509200 | USA/OR-OHSU-1433/2020 | 2020-07-16 | North America/USA/Oregon/Washingto | OHSU Lab Services Molecular Microbiology Lab | Oregon SARS-CoV-2 Genome Sequencing Center | Brendan L. O'Connell et al |
| EPI_ISL_509203 | USA/OR-OHSU-1447/2020 | 2020-07-17 | North America/USA/Oregon/Washingto | OHSU Lab Services Molecular Microbiology Lab | Oregon SARS-CoV-2 Genome Sequencing Center | Brendan L. O'Connell et al |
| EPI_ISL_509208 | USA/OR-OHSU-1463/2020 | 2020-07-17 | North America/USA/Oregon/Multnomal | OHSU Lab Services Molecular Microbiology Lab | Oregon SARS-CoV-2 Genome Sequencing Center | Brendan L. O'Connell et al |

| | | | | | | |
|---|---|---|---|---|---|---|
| EPI_ISL_509209 | USA/OR-OHSU-1465/2020 | 2020-07-17 | North America/USA/Oregon/Washingto | OHSU Lab Services Molecular Microbiology Lab | Oregon SARS-CoV-2 Genome Sequencing Center | Brendan L. O'Connell et al |
| EPI_ISL_509210 | USA/OR-OHSU-1469/2020 | 2020-07-17 | North America/USA/Oregon/Multnomal | OHSU Lab Services Molecular Microbiology Lab | Oregon SARS-CoV-2 Genome Sequencing Center | Brendan L. O'Connell et al |
| EPI_ISL_509211 | USA/OR-OHSU-1475/2020 | 2020-07-17 | North America/USA/Oregon/Columbia C | OHSU Lab Services Molecular Microbiology Lab | Oregon SARS-CoV-2 Genome Sequencing Center | Brendan L. O'Connell et al |
| EPI_ISL_509223 | SouthAfrica/KRISP-K000566/2020 | 2020-07-13 | Africa/South Africa/KwaZulu-Natal/uThu | NHLS-IALCH | KRISP, KZN Research Innovation and Sequencing Platform | Giandhari J et al |
| EPI_ISL_509227 | SouthAfrica/KRISP-K000571/2020 | 2020-07-13 | Africa/South Africa/KwaZulu-Natal/uThu | NHLS-IALCH | KRISP, KZN Research Innovation and Sequencing Platform | Giandhari J et al |
| EPI_ISL_509228 | SouthAfrica/KRISP-K000572/2020 | 2020-07-13 | Africa/South Africa/KwaZulu-Natal/uThu | NHLS-IALCH | KRISP, KZN Research Innovation and Sequencing Platform | Giandhari J et al |
| EPI_ISL_509229 | SouthAfrica/KRISP-K000573/2020 | 2020-07-10 | Africa/South Africa/KwaZulu-Natal/uMz | NHLS-IALCH | KRISP, KZN Research Innovation and Sequencing Platform | Giandhari J et al |
| EPI_ISL_509231 | SouthAfrica/KRISP-K000575/2020 | 2020-07-13 | Africa/South Africa/KwaZulu-Natal/iLem | NHLS-IALCH | KRISP, KZN Research Innovation and Sequencing Platform | Giandhari J et al |

| | | | | | | |
|---|---|---|---|---|---|---|
| EPI_ISL_509233 | SouthAfrica/KRISP-K000577/2020 | 2020-07-11 | Africa/South Africa/KwaZulu-Natal/uThu | NHLS-IALCH | KRISP, KZN Research Innovation and Sequencing Platform | Giandhari J et al |
| EPI_ISL_509242 | SouthAfrica/KRISP-K000586/2020 | 2020-07-13 | Africa/South Africa/KwaZulu-Natal/uMg | NHLS-IALCH | KRISP, KZN Research Innovation and Sequencing Platform | Giandhari J et al |
| EPI_ISL_509243 | SouthAfrica/KRISP-K000588/2020 | 2020-07-13 | Africa/South Africa/KwaZulu-Natal/uThu | NHLS-IALCH | KRISP, KZN Research Innovation and Sequencing Platform | Giandhari J et al |
| EPI_ISL_509244 | SouthAfrica/KRISP-K000589/2020 | 2020-07-13 | Africa/South Africa/KwaZulu-Natal/Ugu | NHLS-IALCH | KRISP, KZN Research Innovation and Sequencing Platform | Giandhari J et al |
| EPI_ISL_509248 | SouthAfrica/KRISP-K000595/2020 | 2020-07-02 | Africa/South Africa/KwaZulu-Natal/uThu | NHLS-IALCH | KRISP, KZN Research Innovation and Sequencing Platform | Giandhari J et al |
| EPI_ISL_509249 | SouthAfrica/KRISP-K000596/2020 | 2020-07-02 | Africa/South Africa/KwaZulu-Natal/uThu | NHLS-IALCH | KRISP, KZN Research Innovation and Sequencing Platform | Giandhari J et al |
| EPI_ISL_509252 | SouthAfrica/KRISP-K000599/2020 | 2020-07-13 | Africa/South Africa/KwaZulu-Natal/iLem | NHLS-IALCH | KRISP, KZN Research Innovation and Sequencing Platform | Giandhari J et al |
| EPI_ISL_509254 | SouthAfrica/KRISP-K000601/2020 | 2020-07-13 | Africa/South Africa/KwaZulu-Natal/Ugu | NHLS-IALCH | KRISP, KZN Research Innovation and Sequencing Platform | Giandhari J et al |

| | | | | | | |
|---|---|---|---|---|---|---|
| EPI_ISL_509255 | SouthAfrica/KRISP-K000602/2020 | 2020-07-01 | Africa/South Africa/KwaZulu-Natal/uThu | NHLS-IALCH | KRISP, KZN Research Innovation and Sequencing Platform | Giandhari J et al |
| EPI_ISL_509259 | SouthAfrica/KRISP-K000606/2020 | 2020-07-02 | Africa/South Africa/KwaZulu-Natal/uThu | NHLS-IALCH | KRISP, KZN Research Innovation and Sequencing Platform | Giandhari J et al |
| EPI_ISL_509266 | SouthAfrica/KRISP-K000614/2020 | 2020-07-02 | Africa/South Africa/KwaZulu-Natal/uThu | NHLS-IALCH | KRISP, KZN Research Innovation and Sequencing Platform | Giandhari J et al |
| EPI_ISL_509267 | SouthAfrica/KRISP-K000615/2020 | 2020-07-02 | Africa/South Africa/KwaZulu-Natal/uThu | NHLS-IALCH | KRISP, KZN Research Innovation and Sequencing Platform | Giandhari J et al |
| EPI_ISL_509274 | SouthAfrica/KRISP-K000622/2020 | 2020-07-03 | Africa/South Africa/KwaZulu-Natal/uThu | NHLS-IALCH | KRISP, KZN Research Innovation and Sequencing Platform | Giandhari J et al |
| EPI_ISL_509275 | SouthAfrica/KRISP-K000623/2020 | 2020-07-04 | Africa/South Africa/KwaZulu-Natal/uThu | NHLS-IALCH | KRISP, KZN Research Innovation and Sequencing Platform | Giandhari J et al |
| EPI_ISL_509276 | SouthAfrica/KRISP-K000624/2020 | 2020-07-02 | Africa/South Africa/KwaZulu-Natal/uThu | NHLS-IALCH | KRISP, KZN Research Innovation and Sequencing Platform | Giandhari J et al |
| EPI_ISL_509277 | SouthAfrica/KRISP-K000625/2020 | 2020-07-11 | Africa/South Africa/KwaZulu-Natal/uThu | NHLS-IALCH | KRISP, KZN Research Innovation and Sequencing Platform | Giandhari J et al |

| | | | | | | |
|---|---|---|---|---|---|---|
| EPI_ISL_509282 | SouthAfrica/KRISP-K000631/2020 | 2020-07-04 | Africa/South Africa/KwaZulu-Natal/uThu | NHLS-IALCH | KRISP, KZN Research Innovation and Sequencing Platform | Giandhari J et al |
| EPI_ISL_509292 | SouthAfrica/KRISP-K000643/2020 | 2020-07-10 | Africa/South Africa/KwaZulu-Natal/uThu | NHLS-IALCH | KRISP, KZN Research Innovation and Sequencing Platform | Giandhari J et al |
| EPI_ISL_509296 | SouthAfrica/KRISP-K000652/2020 | 2020-07-13 | Africa/South Africa/KwaZulu-Natal/uMg | NHLS-IALCH | KRISP, KZN Research Innovation and Sequencing Platform | Giandhari J et al |
| EPI_ISL_509299 | SouthAfrica/KRISP-K000658/2020 | 2020-07-13 | Africa/South Africa/KwaZulu-Natal/uThu | NHLS-IALCH | KRISP, KZN Research Innovation and Sequencing Platform | Giandhari J et al |
| EPI_ISL_509301 | SouthAfrica/KRISP-K001035/2020 | 2020-07-17 | Africa/South Africa/KwaZulu-Natal/eThe | NHLS-IALCH | KRISP, KZN Research Innovation and Sequencing Platform | Giandhari J et al |
| EPI_ISL_509304 | SouthAfrica/KRISP-K001041/2020 | 2020-07-16 | Africa/South Africa/KwaZulu-Natal/eThe | NHLS-IALCH | KRISP, KZN Research Innovation and Sequencing Platform | Giandhari J et al |
| EPI_ISL_509305 | SouthAfrica/KRISP-K001043/2020 | 2020-07-02 | Africa/South Africa/KwaZulu-Natal/uMk | NHLS-IALCH | KRISP, KZN Research Innovation and Sequencing Platform | Giandhari J et al |
| EPI_ISL_509308 | SouthAfrica/KRISP-K001049/2020 | 2020-07-16 | Africa/South Africa/KwaZulu-Natal/eThe | NHLS-IALCH | KRISP, KZN Research Innovation and Sequencing Platform | Giandhari J et al |

| | | | | | | |
|---|---|---|---|---|---|---|
| EPI_ISL_509310 | SouthAfrica/KRISP-K001052/2020 | 2020-07-14 | Africa/South Africa/KwaZulu-Natal/Ama | NHLS-IALCH | KRISP, KZN Research Innovation and Sequencing Platform | Giandhari J et al |
| EPI_ISL_509313 | SouthAfrica/KRISP-K001055/2020 | 2020-07-13 | Africa/South Africa/KwaZulu-Natal/eThe | NHLS-IALCH | KRISP, KZN Research Innovation and Sequencing Platform | Giandhari J et al |
| EPI_ISL_509316 | SouthAfrica/KRISP-K001059/2020 | 2020-07-01 | Africa/South Africa/KwaZulu-Natal/uMz | NHLS-IALCH | KRISP, KZN Research Innovation and Sequencing Platform | Giandhari J et al |
| EPI_ISL_509321 | SouthAfrica/KRISP-K001065/2020 | 2020-07-03 | Africa/South Africa/KwaZulu-Natal/Zulu | NHLS-IALCH | KRISP, KZN Research Innovation and Sequencing Platform | Giandhari J et al |
| EPI_ISL_509324 | SouthAfrica/KRISP-K001068/2020 | 2020-07-14 | Africa/South Africa/KwaZulu-Natal/Ama | NHLS-IALCH | KRISP, KZN Research Innovation and Sequencing Platform | Giandhari J et al |
| EPI_ISL_509326 | SouthAfrica/KRISP-K001072/2020 | 2020-07-14 | Africa/South Africa/KwaZulu-Natal/Ama | NHLS-IALCH | KRISP, KZN Research Innovation and Sequencing Platform | Giandhari J et al |
| EPI_ISL_509331 | SouthAfrica/KRISP-K001077/2020 | 2020-07-03 | Africa/South Africa/KwaZulu-Natal/uMz | NHLS-IALCH | KRISP, KZN Research Innovation and Sequencing Platform | Giandhari J et al |
| EPI_ISL_509332 | SouthAfrica/KRISP-K001078/2020 | 2020-07-03 | Africa/South Africa/KwaZulu-Natal/Ugu | NHLS-IALCH | KRISP, KZN Research Innovation and Sequencing Platform | Giandhari J et al |

| | | | | | | |
|---|---|---|---|---|---|---|
| EPI_ISL_509333 | SouthAfrica/KRISP-K001079/2020 | 2020-07-03 | Africa/South Africa/KwaZulu-Natal/Ama | NHLS-IALCH | KRISP, KZN Research Innovation and Sequencing Platform | Giandhari J et al |
| EPI_ISL_509334 | SouthAfrica/KRISP-K001080/2020 | 2020-07-01 | Africa/South Africa/KwaZulu-Natal/uThu | NHLS-IALCH | KRISP, KZN Research Innovation and Sequencing Platform | Giandhari J et al |
| EPI_ISL_509336 | SouthAfrica/KRISP-K001083/2020 | 2020-07-03 | Africa/South Africa/KwaZulu-Natal/Ama | NHLS-IALCH | KRISP, KZN Research Innovation and Sequencing Platform | Giandhari J et al |
| EPI_ISL_509337 | SouthAfrica/KRISP-K001084/2020 | 2020-07-03 | Africa/South Africa/KwaZulu-Natal/Ama | NHLS-IALCH | KRISP, KZN Research Innovation and Sequencing Platform | Giandhari J et al |
| EPI_ISL_509340 | SouthAfrica/KRISP-K001088/2020 | 2020-07-13 | Africa/South Africa/KwaZulu-Natal/uThu | NHLS-IALCH | KRISP, KZN Research Innovation and Sequencing Platform | Giandhari J et al |
| EPI_ISL_509345 | SouthAfrica/KRISP-K001096/2020 | 2020-07-13 | Africa/South Africa/KwaZulu-Natal/uThu | NHLS-IALCH | KRISP, KZN Research Innovation and Sequencing Platform | Giandhari J et al |
| EPI_ISL_509347 | SouthAfrica/KRISP-K001098/2020 | 2020-07-10 | Africa/South Africa/KwaZulu-Natal/uThu | NHLS-IALCH | KRISP, KZN Research Innovation and Sequencing Platform | Giandhari J et al |
| EPI_ISL_509348 | SouthAfrica/KRISP-K001099/2020 | 2020-07-13 | Africa/South Africa/KwaZulu-Natal/Zulu | NHLS-IALCH | KRISP, KZN Research Innovation and Sequencing Platform | Giandhari J et al |

| | | | | | | |
|---|---|---|---|---|---|---|
| EPI_ISL_509349 | SouthAfrica/KRISP-K001100/2020 | 2020-07-10 | Africa/South Africa/KwaZulu-Natal/uThu | NHLS-IALCH | KRISP, KZN Research Innovation and Sequencing Platform | Giandhari J et al |
| EPI_ISL_509352 | SouthAfrica/KRISP-K001103/2020 | 2020-07-13 | Africa/South Africa/KwaZulu-Natal/uThu | NHLS-IALCH | KRISP, KZN Research Innovation and Sequencing Platform | Giandhari J et al |
| EPI_ISL_509354 | SouthAfrica/KRISP-K001105/2020 | 2020-07-10 | Africa/South Africa/KwaZulu-Natal/uThu | NHLS-IALCH | KRISP, KZN Research Innovation and Sequencing Platform | Giandhari J et al |
| EPI_ISL_509358 | SouthAfrica/KRISP-K001112/2020 | 2020-07-17 | Africa/South Africa/KwaZulu-Natal/iLem | NHLS-IALCH | KRISP, KZN Research Innovation and Sequencing Platform | Giandhari J et al |
| EPI_ISL_509359 | SouthAfrica/KRISP-K001113/2020 | 2020-07-14 | Africa/South Africa/KwaZulu-Natal/iLem | NHLS-IALCH | KRISP, KZN Research Innovation and Sequencing Platform | Giandhari J et al |
| EPI_ISL_509360 | SouthAfrica/KRISP-K001114/2020 | 2020-07-17 | Africa/South Africa/KwaZulu-Natal/iLem | NHLS-IALCH | KRISP, KZN Research Innovation and Sequencing Platform | Giandhari J et al |
| EPI_ISL_509366 | SouthAfrica/KRISP-K001123/2020 | 2020-07-15 | Africa/South Africa/KwaZulu-Natal/iLem | NHLS-IALCH | KRISP, KZN Research Innovation and Sequencing Platform | Giandhari J et al |
| EPI_ISL_509367 | SouthAfrica/KRISP-K001124/2020 | 2020-07-16 | Africa/South Africa/KwaZulu-Natal/iLem | NHLS-IALCH | KRISP, KZN Research Innovation and Sequencing Platform | Giandhari J et al |

| | | | | | | |
|---|---|---|---|---|---|---|
| EPI_ISL_509369 | SouthAfrica/KRISP-K001126/2020 | 2020-07-16 | Africa/South Africa/KwaZulu-Natal/iLem | NHLS-IALCH | KRISP, KZN Research Innovation and Sequencing Platform | Giandhari J et al |
| EPI_ISL_509374 | Singapore/764/2020 | 2020-06-17 | Asia/Singapore/Singapore/ | Singapore General Hospital | Department of Microbiology | Nurdyana Abdul Rahman et al |
| EPI_ISL_509393 | Singapore/783/2020 | 2020-06-13 | Asia/Singapore/Singapore/ | Singapore General Hospital | Department of Microbiology | Nurdyana Abdul Rahman et al |
| EPI_ISL_509412 | Turkey/ACUTG-1/2020 | 2020-04-13 | Europe/Turkey/Istanbul/ | Acibadem Labcell Cellular Therapy Laboratory | Acibadem Mehmet Ali Aydinlar University School of Medicine, Medical Genetics Department | Ozden Hatirnaz Ng et al |
| EPI_ISL_509413 | Turkey/ACUTG-2/2020 | 2020-04-06 | Europe/Turkey/Istanbul/ | Acibadem Labcell Cellular Therapy Laboratory | Acibadem Mehmet Ali Aydinlar University School of Medicine, Medical Genetics Department | Ozden Hatirnaz Ng et al |
| EPI_ISL_509416 | Turkey/ACUTG-5/2020 | 2020-04-22 | Europe/Turkey/Istanbul/ | Acibadem Labcell Cellular Therapy Laboratory | Acibadem Mehmet Ali Aydinlar University School of Medicine, Medical Genetics Department | Ozden Hatirnaz Ng et al |
| EPI_ISL_509427 | USA/AL-SV106V1/2020 | 2020-05-05 | North America/USA/Alabama/ | Microbiology and Immunology, University of South Alabama | Microbiology and Immunology, University of South Alabama | Wood et al |
| EPI_ISL_509430 | Brazil/RJ-DCV05/2020 | 2020-07-20 | South America/Brazil/Rio de Janeiro/ | Centro de Desenvolvimento Tecnologico em Saude, Fundacao Oswaldo Cruz | Centro de Desenvolvimento Tecnologico em Saude, Fundacao Oswaldo Cruz | Souza et al |

| | | | | | | |
|---|---|---|---|---|---|---|
| EPI_ISL_509431 | Brazil/RJ-DCV1/2020 | 2020-07-20 | South America/Brazil/Rio de Janeiro/ | Centro de Desenvolvimento Tecnologico em Saude, Fundacao Oswaldo Cruz | Centro de Desenvolvimento Tecnologico em Saude, Fundacao Oswaldo Cruz | Souza et al |
| EPI_ISL_509432 | Brazil/RJ-DCV3/2020 | 2020-07-20 | South America/Brazil/Rio de Janeiro/ | Centro de Desenvolvimento Tecnologico em Saude, Fundacao Oswaldo Cruz | Centro de Desenvolvimento Tecnologico em Saude, Fundacao Oswaldo Cruz | Souza et al |
| EPI_ISL_509433 | Brazil/RJ-DCV5/2020 | 2020-07-20 | South America/Brazil/Rio de Janeiro/ | Centro de Desenvolvimento Tecnologico em Saude, Fundacao Oswaldo Cruz | Centro de Desenvolvimento Tecnologico em Saude, Fundacao Oswaldo Cruz | Souza et al |
| EPI_ISL_509435 | Brazil/RJ-DCVTV507Q/2020 | 2020-07-20 | South America/Brazil/Rio de Janeiro/ | Centro de Desenvolvimento Tecnologico em Saude, Fundacao Oswaldo Cruz | Centro de Desenvolvimento Tecnologico em Saude, Fundacao Oswaldo Cruz | Souza et al |
| EPI_ISL_509446 | USA/MD-MDH-0042/2020 | 2020-04-03 | North America/USA/Maryland/ | Maryland Department of Health | Maryland Department of Health | Keller et al |
| EPI_ISL_509454 | USA/MD-MDH-0050/2020 | 2020-05-26 | North America/USA/Maryland/ | Maryland Department of Health | Maryland Department of Health | Keller et al |
| EPI_ISL_509455 | USA/MD-MDH-0051/2020 | 2020-05-26 | North America/USA/Maryland/ | Maryland Department of Health | Maryland Department of Health | Keller et al |
| EPI_ISL_509464 | USA/MD-MDH-0060/2020 | 2020-05-25 | North America/USA/Maryland/ | Maryland Department of Health | Maryland Department of Health | Keller et al |
| EPI_ISL_509471 | USA/MD-MDH-0067/2020 | 2020-05-25 | North America/USA/Maryland/ | Maryland Department of Health | Maryland Department of Health | Keller et al |
| EPI_ISL_509488 | USA/MD-MDH-0084/2020 | 2020-05-25 | North America/USA/Maryland/ | Maryland Department of Health | Maryland Department of Health | Keller et al |

| | | | | | | |
|---|---|---|---|---|---|---|
| EPI_ISL_509505 | Australia/NSW2153/2020 | 2020-01-25 | Oceania/Australia/New South Wales/ | Area of Virology, Serology and Virology Division (SAViD), New South Wales Health Pathology Randwick | Area of Virology, Serology and Virology Division (SAViD), New South Wales Health Pathology Randwick | Rawlinson et al |
| EPI_ISL_509527 | USA/UT-02882/2020 | 2020-04-30 | North America/USA/Utah/ | Utah Public Health Laboratory | Utah Public Health Laboratory | Heidi Butz et al |
| EPI_ISL_509542 | USA/UT-02919/2020 | 2020-04-30 | North America/USA/Utah/ | Utah Public Health Laboratory | Utah Public Health Laboratory | Heidi Butz et al |
| EPI_ISL_509550 | USA/UT-02946/2020 | 2020-04-30 | North America/USA/Utah/ | Utah Public Health Laboratory | Utah Public Health Laboratory | Heidi Butz et al |
| EPI_ISL_509551 | USA/UT-02954/2020 | 2020-05-01 | North America/USA/Utah/ | Utah Public Health Laboratory | Utah Public Health Laboratory | Heidi Butz et al |
| EPI_ISL_509554 | USA/UT-02962/2020 | 2020-05-01 | North America/USA/Utah/ | Utah Public Health Laboratory | Utah Public Health Laboratory | Heidi Butz et al |
| EPI_ISL_509557 | USA/UT-03007/2020 | 2020-04-30 | North America/USA/Utah/ | Utah Public Health Laboratory | Utah Public Health Laboratory | Heidi Butz et al |
| EPI_ISL_509561 | USA/UT-03019/2020 | 2020-05-01 | North America/USA/Utah/ | Utah Public Health Laboratory | Utah Public Health Laboratory | Heidi Butz et al |
| EPI_ISL_509573 | USA/UT-01823/2020 | 2020-04-10 | North America/USA/Utah/ | Utah Public Health Laboratory | Utah Public Health Laboratory | Heidi Butz et al |
| EPI_ISL_509579 | USA/UT-03736/2020 | 2020-04-05 | North America/USA/Utah/ | Utah Public Health Laboratory | Utah Public Health Laboratory | Heidi Butz et al |
| EPI_ISL_509584 | USA/UT-03745/2020 | 2020-04-06 | North America/USA/Utah/ | Utah Public Health Laboratory | Utah Public Health Laboratory | Heidi Butz et al |
| EPI_ISL_509603 | Spain/AN-IBV-001976/2020 | 2020-03-15 | Europe/Spain/Andalusia/Malaga | Servicio de Microbiolog√≠a. HRU de M√°laga. Servicio Andaluz de Salud | SeqCOVID-SPAIN consortium/IBV(CSIC) | Inmaculada de Toro Peinado. M¬™Concepci√≥n Mediavilla Gradolph. Begov√±a Palop Borr√°s et al |
| EPI_ISL_509604 | Spain/AN-IBV-001991/2020 | 2020-03-15 | Europe/Spain/Andalusia/Malaga | Servicio de Microbiolog√≠a. HRU de M√°laga. Servicio Andaluz de Salud | SeqCOVID-SPAIN consortium/IBV(CSIC) | Inmaculada de Toro Peinado. M¬™Concepci√≥n Mediavilla Gradolph. Begov√±a Palop Borr√°s et al |
| EPI_ISL_509606 | Spain/AN-IBV-001974/2020 | 2020-03-15 | Europe/Spain/Andalusia/Malaga | Servicio de Microbiolog√≠a. HRU de M√°laga. Servicio Andaluz de Salud | SeqCOVID-SPAIN consortium/IBV(CSIC) | Inmaculada de Toro Peinado. M¬™Concepci√≥n Mediavilla Gradolph. Begov√±a Palop Borr√°s et al |

| | | | | | | |
|---|---|---|---|---|---|---|
| EPI_ISL_509614 | Spain/PV-IBV-005178/2020 | 2020-06-29 | Europe/Spain/Basque Country/San Seba | Servicio de Microbiolog√≠a. Hospital Universitario Donostia. OSI Donostialdea. √Årea de Enfermedades Infecciosas, Grupo de Infecci√≥n Respiratoria y Resistencia Antimicrobiana. Instituto de Investigaci√≥n Sanitaria Biodonostia | SeqCOVID-SPAIN consortium/IBV(CSIC) | Gustavo Cilla et al |
| EPI_ISL_509616 | Spain/PV-IBV-001537/2020 | 2020-03-16 | Europe/Spain/Basque Country/Vitoria-G | Hospital Universitario Araba. Vitoria-Gasteiz | SeqCOVID-SPAIN consortium/IBV(CSIC) | Silvia Hern√°ez Crespo et al |
| EPI_ISL_509641 | Spain/PV-IBV-005153/2020 | 2020-06-21 | Europe/Spain/Basque Country/San Seba | Servicio de Microbiolog√≠a. Hospital Universitario Donostia. OSI Donostialdea. √Årea de Enfermedades Infecciosas, Grupo de Infecci√≥n Respiratoria y Resistencia Antimicrobiana. Instituto de Investigaci√≥n Sanitaria Biodonostia | SeqCOVID-SPAIN consortium/IBV(CSIC) | Gustavo Cilla et al |
| EPI_ISL_509642 | Spain/PV-IBV-005154/2020 | 2020-06-22 | Europe/Spain/Basque Country/San Seba | Servicio de Microbiolog√≠a. Hospital Universitario Donostia. OSI Donostialdea. √Årea de Enfermedades Infecciosas, Grupo de Infecci√≥n Respiratoria y Resistencia Antimicrobiana. Instituto de Investigaci√≥n Sanitaria Biodonostia | SeqCOVID-SPAIN consortium/IBV(CSIC) | Gustavo Cilla et al |
| EPI_ISL_509664 | USA/AR-CDC-4201/2020 | 2020-07-10 | North America/USA/Arkansas/ | AR Dept. of Health-Public Health Lab | Pathogen Discovery, Respiratory Viruses Branch, Division of Viral Diseases, Centers for Disease Control and Prevention | Yan Li et al |
| EPI_ISL_509667 | USA/AR-CDC-4174/2020 | 2020-07-07 | North America/USA/Arkansas/ | AR Dept. of Health-Public Health Lab | Pathogen Discovery, Respiratory Viruses Branch, Division of Viral Diseases, Centers for Disease Control and Prevention | Yan Li et al |

| | | | | | | |
|---|---|---|---|---|---|---|
| EPI_ISL_509672 | USA/AR-CDC-4187/2020 | 2020-07-08 | North America/USA/Arkansas/ | AR Dept. of Health-Public Health Lab | Pathogen Discovery, Respiratory Viruses Branch, Division of Viral Diseases, Centers for Disease Control and Prevention | Yan Li et al |
| EPI_ISL_509677 | USA/AR-CDC-4196/2020 | 2020-07-09 | North America/USA/Arkansas/ | AR Dept. of Health-Public Health Lab | Pathogen Discovery, Respiratory Viruses Branch, Division of Viral Diseases, Centers for Disease Control and Prevention | Yan Li et al |
| EPI_ISL_509678 | USA/AR-CDC-4195/2020 | 2020-07-09 | North America/USA/Arkansas/ | AR Dept. of Health-Public Health Lab | Pathogen Discovery, Respiratory Viruses Branch, Division of Viral Diseases, Centers for Disease Control and Prevention | Yan Li et al |
| EPI_ISL_509679 | USA/AR-CDC-4171/2020 | 2020-07-07 | North America/USA/Arkansas/ | AR Dept. of Health-Public Health Lab | Pathogen Discovery, Respiratory Viruses Branch, Division of Viral Diseases, Centers for Disease Control and Prevention | Yan Li et al |

| | | | | | | |
|---|---|---|---|---|---|---|
| EPI_ISL_509684 | USA/AR-CDC-4194/2020 | 2020-07-09 | North America/USA/Arkansas/ | AR Dept. of Health-Public Health Lab | Pathogen Discovery, Respiratory Viruses Branch, Division of Viral Diseases, Centers for Disease Control and Prevention | Jing Zhang et al |
| EPI_ISL_509691 | USA/UT-CDC-9750/2020 | 2020-04-25 | North America/USA/Utah/ | Utah Public Health Laboratory | Pathogen Discovery, Respiratory Viruses Branch, Division of Viral Diseases, Centers for Disease Control and Prevention | Ying Tao et al |
| EPI_ISL_509694 | USA/UT-CDC-9757/2020 | 2020-04-29 | North America/USA/Utah/ | Utah Public Health Laboratory | Pathogen Discovery, Respiratory Viruses Branch, Division of Viral Diseases, Centers for Disease Control and Prevention | Ying Tao et al |
| EPI_ISL_509695 | Guatemala/CDC-1227/2020 | 2020-03-13 | North America/Guatemala/Guatemala/ | Guatemala Ministry of Public Health | Pathogen Discovery, Respiratory Viruses Branch, Division of Viral Diseases, Centers for Disease Control and Prevention | Ying Tao et al |

| | | | | | | |
|---|---|---|---|---|---|---|
| EPI_ISL_509704 | USA/WI-CDC-0944/2020 | 2020-04-16 | North America/USA/Wisconsin/ | Wisconsin Department of Health Services | Pathogen Discovery, Respiratory Viruses Branch, Division of Viral Diseases, Centers for Disease Control and Prevention | Ying Tao et al |
| EPI_ISL_509709 | USA/UT-CDC-9782/2020 | 2020-05-06 | North America/USA/Utah/ | Utah Public Health Laboratory | Pathogen Discovery, Respiratory Viruses Branch, Division of Viral Diseases, Centers for Disease Control and Prevention | Jing Zhang et al |
| EPI_ISL_509722 | USA/FL-BPHL-0664/2020 | 2020-03-14 | North America/USA/Florida/ | Florida Bureau of Public Health Laboratories | Florida Bureau of Public Health Laboratories | Sarah Schmedes et al |
| EPI_ISL_509723 | USA/FL-BPHL-0665/2020 | 2020-03-15 | North America/USA/Florida/ | Florida Bureau of Public Health Laboratories | Florida Bureau of Public Health Laboratories | Sarah Schmedes et al |
| EPI_ISL_509731 | USA/FL-BPHL-0673/2020 | 2020-03-24 | North America/USA/Florida/ | Florida Bureau of Public Health Laboratories | Florida Bureau of Public Health Laboratories | Sarah Schmedes et al |
| EPI_ISL_509732 | USA/FL-BPHL-0674/2020 | 2020-03-24 | North America/USA/Florida/ | Florida Bureau of Public Health Laboratories | Florida Bureau of Public Health Laboratories | Sarah Schmedes et al |
| EPI_ISL_509747 | USA/FL-BPHL-0689/2020 | 2020-03-26 | North America/USA/Florida/ | Florida Bureau of Public Health Laboratories | Florida Bureau of Public Health Laboratories | Sarah Schmedes et al |
| EPI_ISL_509749 | USA/FL-BPHL-0691/2020 | 2020-03-26 | North America/USA/Florida/ | Florida Bureau of Public Health Laboratories | Florida Bureau of Public Health Laboratories | Sarah Schmedes et al |
| EPI_ISL_509763 | USA/FL-BPHL-0705/2020 | 2020-03-27 | North America/USA/Florida/ | Florida Bureau of Public Health Laboratories | Florida Bureau of Public Health Laboratories | Sarah Schmedes et al |

| | | | | | | |
|---|---|---|---|---|---|---|
| EPI_ISL_509779 | USA/FL-BPHL-0721/2020 | 2020-07-21 | North America/USA/Florida/ | Florida Bureau of Public Health Laboratories | Florida Bureau of Public Health Laboratories | Sarah Schmedes et al |
| EPI_ISL_509783 | USA/FL-BPHL-0725/2020 | 2020-07-14 | North America/USA/Florida/ | Florida Bureau of Public Health Laboratories | Florida Bureau of Public Health Laboratories | Sarah Schmedes et al |
| EPI_ISL_509796 | USA/FL-BPHL-0738/2020 | 2020-07-15 | North America/USA/Florida/ | Florida Bureau of Public Health Laboratories | Florida Bureau of Public Health Laboratories | Sarah Schmedes et al |
| EPI_ISL_509805 | USA/IL-UW-987/2020 | 2020-07-16 | North America/USA/Illinois/Winnebago | University of Wisconsin-Madison AIDS Vaccine Research Laboratories | University of Wisconsin-Madison AIDS Vaccine Research Laboratories | Gage Moreno et al |
| EPI_ISL_509807 | USA/OH-UW-921/2020 | 2020-07-12 | North America/USA/Ohio/Cuyahoga Cou | University of Wisconsin-Madison AIDS Vaccine Research Laboratories | University of Wisconsin-Madison AIDS Vaccine Research Laboratories | Gage Moreno et al |
| EPI_ISL_509808 | USA/SD-UW-859/2020 | 2020-07-09 | North America/USA/South Dakota/Min | University of Wisconsin-Madison AIDS Vaccine Research Laboratories | University of Wisconsin-Madison AIDS Vaccine Research Laboratories | Gage Moreno et al |
| EPI_ISL_509811 | USA/WI-UW-1002/2020 | 2020-07-16 | North America/USA/Wisconsin/Dane Co | University of Wisconsin-Madison AIDS Vaccine Research Laboratories | University of Wisconsin-Madison AIDS Vaccine Research Laboratories | Gage Moreno et al |
| EPI_ISL_509816 | USA/WI-UW-807/2020 | 2020-05-19 | North America/USA/Wisconsin/Ozaukee | University of Wisconsin-Madison AIDS Vaccine Research Laboratories | University of Wisconsin-Madison AIDS Vaccine Research Laboratories | Gage Moreno et al |
| EPI_ISL_509823 | USA/WI-UW-816/2020 | 2020-07-09 | North America/USA/Wisconsin/Milwauk | University of Wisconsin-Madison AIDS Vaccine Research Laboratories | University of Wisconsin-Madison AIDS Vaccine Research Laboratories | Gage Moreno et al |

| | | | | | | |
|---|---|---|---|---|---|---|
| EPI_ISL_509824 | USA/WI-UW-817/2020 | 2020-05-26 | North America/USA/Wisconsin/Milwauk | University of Wisconsin-Madison AIDS Vaccine Research Laboratories | University of Wisconsin-Madison AIDS Vaccine Research Laboratories | Gage Moreno et al |
| EPI_ISL_509840 | USA/WI-UW-833/2020 | 2020-05-20 | North America/USA/Wisconsin/Washing | University of Wisconsin-Madison AIDS Vaccine Research Laboratories | University of Wisconsin-Madison AIDS Vaccine Research Laboratories | Gage Moreno et al |
| EPI_ISL_509846 | USA/WI-UW-839/2020 | 2020-05-05 | North America/USA/Wisconsin/Milwauk | University of Wisconsin-Madison AIDS Vaccine Research Laboratories | University of Wisconsin-Madison AIDS Vaccine Research Laboratories | Gage Moreno et al |
| EPI_ISL_509849 | USA/WI-UW-842/2020 | 2020-05-07 | North America/USA/Wisconsin/Milwauk | University of Wisconsin-Madison AIDS Vaccine Research Laboratories | University of Wisconsin-Madison AIDS Vaccine Research Laboratories | Gage Moreno et al |
| EPI_ISL_509862 | USA/WI-UW-856/2020 | 2020-07-10 | North America/USA/Wisconsin/Dane Co | University of Wisconsin-Madison AIDS Vaccine Research Laboratories | University of Wisconsin-Madison AIDS Vaccine Research Laboratories | Gage Moreno et al |
| EPI_ISL_509865 | USA/WI-UW-862/2020 | 2020-07-15 | North America/USA/Wisconsin/Dane Co | University of Wisconsin-Madison AIDS Vaccine Research Laboratories | University of Wisconsin-Madison AIDS Vaccine Research Laboratories | Gage Moreno et al |
| EPI_ISL_509868 | USA/WI-UW-865/2020 | 2020-07-06 | North America/USA/Wisconsin/Dane Co | University of Wisconsin-Madison AIDS Vaccine Research Laboratories | University of Wisconsin-Madison AIDS Vaccine Research Laboratories | Gage Moreno et al |
| EPI_ISL_509870 | USA/WI-UW-867/2020 | 2020-07-09 | North America/USA/Wisconsin/Dane Co | University of Wisconsin-Madison AIDS Vaccine Research Laboratories | University of Wisconsin-Madison AIDS Vaccine Research Laboratories | Gage Moreno et al |

| | | | | | | |
|---|---|---|---|---|---|---|
| EPI_ISL_509872 | USA/WI-UW-870/2020 | 2020-07-08 | North America/USA/Wisconsin/Dane Co | University of Wisconsin-Madison AIDS Vaccine Research Laboratories | University of Wisconsin-Madison AIDS Vaccine Research Laboratories | Gage Moreno et al |
| EPI_ISL_509874 | USA/WI-UW-872/2020 | 2020-07-06 | North America/USA/Wisconsin/Dane Co | University of Wisconsin-Madison AIDS Vaccine Research Laboratories | University of Wisconsin-Madison AIDS Vaccine Research Laboratories | Gage Moreno et al |
| EPI_ISL_509881 | USA/WI-UW-879/2020 | 2020-07-14 | North America/USA/Wisconsin/Dane Co | University of Wisconsin-Madison AIDS Vaccine Research Laboratories | University of Wisconsin-Madison AIDS Vaccine Research Laboratories | Gage Moreno et al |
| EPI_ISL_509882 | USA/WI-UW-880/2020 | 2020-07-13 | North America/USA/Wisconsin/Marquet | University of Wisconsin-Madison AIDS Vaccine Research Laboratories | University of Wisconsin-Madison AIDS Vaccine Research Laboratories | Gage Moreno et al |
| EPI_ISL_509890 | USA/WI-UW-888/2020 | 2020-07-14 | North America/USA/Wisconsin/Dane Co | University of Wisconsin-Madison AIDS Vaccine Research Laboratories | University of Wisconsin-Madison AIDS Vaccine Research Laboratories | Gage Moreno et al |
| EPI_ISL_509894 | USA/WI-UW-892/2020 | 2020-07-10 | North America/USA/Wisconsin/Dane Co | University of Wisconsin-Madison AIDS Vaccine Research Laboratories | University of Wisconsin-Madison AIDS Vaccine Research Laboratories | Gage Moreno et al |
| EPI_ISL_509900 | USA/WI-UW-898/2020 | 2020-07-15 | North America/USA/Wisconsin/Dane Co | University of Wisconsin-Madison AIDS Vaccine Research Laboratories | University of Wisconsin-Madison AIDS Vaccine Research Laboratories | Gage Moreno et al |
| EPI_ISL_509904 | USA/WI-UW-903/2020 | 2020-07-11 | North America/USA/Wisconsin/Dane Co | University of Wisconsin-Madison AIDS Vaccine Research Laboratories | University of Wisconsin-Madison AIDS Vaccine Research Laboratories | Gage Moreno et al |

| | | | | | | |
|---|---|---|---|---|---|---|
| EPI_ISL_509907 | USA/WI-UW-906/2020 | 2020-07-15 | North America/USA/Wisconsin/Dane Co | University of Wisconsin-Madison AIDS Vaccine Research Laboratories | University of Wisconsin-Madison AIDS Vaccine Research Laboratories | Gage Moreno et al |
| EPI_ISL_509910 | USA/WI-UW-909/2020 | 2020-07-10 | North America/USA/Wisconsin/Dane Co | University of Wisconsin-Madison AIDS Vaccine Research Laboratories | University of Wisconsin-Madison AIDS Vaccine Research Laboratories | Gage Moreno et al |
| EPI_ISL_509924 | USA/WI-UW-924/2020 | 2020-07-10 | North America/USA/Wisconsin/Dane Co | University of Wisconsin-Madison AIDS Vaccine Research Laboratories | University of Wisconsin-Madison AIDS Vaccine Research Laboratories | Gage Moreno et al |
| EPI_ISL_509927 | USA/WI-UW-927/2020 | 2020-07-13 | North America/USA/Wisconsin/Dane Co | University of Wisconsin-Madison AIDS Vaccine Research Laboratories | University of Wisconsin-Madison AIDS Vaccine Research Laboratories | Gage Moreno et al |
| EPI_ISL_509929 | USA/WI-UW-929/2020 | 2020-07-10 | North America/USA/Wisconsin/Dane Co | University of Wisconsin-Madison AIDS Vaccine Research Laboratories | University of Wisconsin-Madison AIDS Vaccine Research Laboratories | Gage Moreno et al |
| EPI_ISL_509942 | USA/WI-UW-942/2020 | 2020-05-10 | North America/USA/Wisconsin/ | University of Wisconsin-Madison AIDS Vaccine Research Laboratories | University of Wisconsin-Madison AIDS Vaccine Research Laboratories | Gage Moreno et al |
| EPI_ISL_509949 | USA/WI-UW-950/2020 | 2020-05-15 | North America/USA/Wisconsin/Milwauk | University of Wisconsin-Madison AIDS Vaccine Research Laboratories | University of Wisconsin-Madison AIDS Vaccine Research Laboratories | Gage Moreno et al |
| EPI_ISL_509962 | USA/WI-UW-964/2020 | 2020-06-04 | North America/USA/Wisconsin/Ozaukee | University of Wisconsin-Madison AIDS Vaccine Research Laboratories | University of Wisconsin-Madison AIDS Vaccine Research Laboratories | Gage Moreno et al |

| | | | | | | |
|---|---|---|---|---|---|---|
| EPI_ISL_509966 | USA/WI-UW-968/2020 | 2020-07-03 | North America/USA/Wisconsin/Columbia | University of Wisconsin-Madison AIDS Vaccine Research Laboratories | University of Wisconsin-Madison AIDS Vaccine Research Laboratories | Gage Moreno et al |
| EPI_ISL_509990 | USA/WI-UW-995/2020 | 2020-07-16 | North America/USA/Wisconsin/Dane Co | University of Wisconsin-Madison AIDS Vaccine Research Laboratories | University of Wisconsin-Madison AIDS Vaccine Research Laboratories | Gage Moreno et al |
| EPI_ISL_510053 | Spain/AN-IBV-004098/2020 | 2020-04-17 | Europe/Spain/Andalusia/Malaga | Servicio de Microbiolog√≠a. HRU de M√°laga. Servicio Andaluz de Salud | SeqCOVID-SPAIN consortium/IBV(CSIC) | Inmaculada de Toro Peinado. M¬™Concepci√≥n Mediavilla Gradolph. Bego√±a Palop Borr√°s et al |
| EPI_ISL_510062 | Spain/AN-IBV-004130/2020 | 2020-04-30 | Europe/Spain/Andalusia/Nerja | Servicio de Microbiolog√≠a. HRU de M√°laga. Servicio Andaluz de Salud | SeqCOVID-SPAIN consortium/IBV(CSIC) | Inmaculada de Toro Peinado. M¬™Concepci√≥n Mediavilla Gradolph. Bego√±a Palop Borr√°s et al |
| EPI_ISL_510071 | Spain/CT-IBV-004161/2020 | 2020-04-01 | Europe/Spain/Catalunya/Barcelona | Instituto de Investigaciones Biom√©dicas de Barcelona (CSIC), Hospital Clinic i Provincial de Barcelona, Instituto de Biomedicina de Valencia (CSIC), Hospital de Sant Pau | SeqCOVID-SPAIN consortium/IBV(CSIC) | Anna M. Planas et al |
| EPI_ISL_510073 | Spain/CT-IBV-004163/2020 | 2020-04-21 | Europe/Spain/Catalunya/Barcelona | Instituto de Investigaciones Biom√©dicas de Barcelona (CSIC), Hospital Clinic i Provincial de Barcelona, Instituto de Biomedicina de Valencia (CSIC), Hospital de Sant Pau | SeqCOVID-SPAIN consortium/IBV(CSIC) | Anna M. Planas et al |
| EPI_ISL_510075 | Spain/CT-IBV-004166/2020 | 2020-04-21 | Europe/Spain/Catalunya/Barcelona | Instituto de Investigaciones Biom√©dicas de Barcelona (CSIC), Hospital Clinic i Provincial de Barcelona, Instituto de Biomedicina de Valencia (CSIC), Hospital de Sant Pau | SeqCOVID-SPAIN consortium/IBV(CSIC) | Anna M. Planas et al |
| EPI_ISL_510105 | Spain/MD-IBV-006356/2020 | 2020-03-23 | Europe/Spain/Madrid/ | Hospital General Universitario Gregorio Mara√±√≥n | SeqCOVID-SPAIN consortium/IBV(CSIC) | Laura P√©rez-Lago et al |
| EPI_ISL_510107 | Spain/MD-IBV-006368/2020 | 2020-06-01 | Europe/Spain/Madrid/Fuenlabrada | Hospital General Universitario Gregorio Mara√±√≥n | SeqCOVID-SPAIN consortium/IBV(CSIC) | Laura P√©rez-Lago et al |

| | | | | | | |
|---|---|---|---|---|---|---|
| EPI_ISL_510110 | Spain/MD-IBV-006379/2020 | 2020-06-05 | Europe/Spain/Madrid/ | Hospital General Universitario Gregorio Marav√±√≥n | SeqCOVID-SPAIN consortium/IBV(CSIC) | Laura P√©rez-Lago et al |
| EPI_ISL_510112 | Spain/MD-IBV-006382/2020 | 2020-04-08 | Europe/Spain/Madrid/ | Hospital General Universitario Gregorio Marav√±√≥n | SeqCOVID-SPAIN consortium/IBV(CSIC) | Laura P√©rez-Lago et al |
| EPI_ISL_510113 | Spain/MD-IBV-006384/2020 | 2020-03-26 | Europe/Spain/Madrid/ | Hospital General Universitario Gregorio Marav√±√≥n | SeqCOVID-SPAIN consortium/IBV(CSIC) | Laura P√©rez-Lago et al |
| EPI_ISL_510116 | Spain/MD-IBV-006393/2020 | 2020-04-29 | Europe/Spain/Madrid/ | Hospital General Universitario Gregorio Marav√±√≥n | SeqCOVID-SPAIN consortium/IBV(CSIC) | Laura P√©rez-Lago et al |
| EPI_ISL_510122 | Spain/MD-IBV-006404/2020 | 2020-03-19 | Europe/Spain/Madrid/ | Hospital General Universitario Gregorio Marav√±√≥n | SeqCOVID-SPAIN consortium/IBV(CSIC) | Laura P√©rez-Lago et al |
| EPI_ISL_510124 | Spain/MD-IBV-006408/2020 | 2020-03-23 | Europe/Spain/Madrid/ | Hospital General Universitario Gregorio Marav√±√≥n | SeqCOVID-SPAIN consortium/IBV(CSIC) | Laura P√©rez-Lago et al |
| EPI_ISL_510128 | Spain/MD-IBV-006413/2020 | 2020-04-03 | Europe/Spain/Madrid/ | Hospital General Universitario Gregorio Marav√±√≥n | SeqCOVID-SPAIN consortium/IBV(CSIC) | Laura P√©rez-Lago et al |
| EPI_ISL_510130 | Spain/MD-IBV-006415/2020 | 2020-05-11 | Europe/Spain/Madrid/ | Hospital General Universitario Gregorio Marav√±√≥n | SeqCOVID-SPAIN consortium/IBV(CSIC) | Laura P√©rez-Lago et al |
| EPI_ISL_510144 | Spain/MD-IBV-006431/2020 | 2020-04-02 | Europe/Spain/Madrid/ | Hospital General Universitario Gregorio Marav√±√≥n | SeqCOVID-SPAIN consortium/IBV(CSIC) | Laura P√©rez-Lago et al |
| EPI_ISL_510146 | Spain/MD-IBV-006434/2020 | 2020-03-09 | Europe/Spain/Madrid/ | Hospital General Universitario Gregorio Marav√±√≥n | SeqCOVID-SPAIN consortium/IBV(CSIC) | Laura P√©rez-Lago et al |
| EPI_ISL_510179 | Spain/MD-IBV-005294/2020 | 2020-03-15 | Europe/Spain/Madrid/ | Hospital General Universitario Gregorio Marav√±√≥n | SeqCOVID-SPAIN consortium/IBV(CSIC) | Laura P√©rez-Lago et al |
| EPI_ISL_510184 | Spain/MD-IBV-005300/2020 | 2020-03-15 | Europe/Spain/Madrid/ | Hospital General Universitario Gregorio Marav√±√≥n | SeqCOVID-SPAIN consortium/IBV(CSIC) | Laura P√©rez-Lago et al |
| EPI_ISL_510189 | Spain/MD-IBV-005308/2020 | 2020-03-16 | Europe/Spain/Madrid/ | Hospital General Universitario Gregorio Marav√±√≥n | SeqCOVID-SPAIN consortium/IBV(CSIC) | Laura P√©rez-Lago et al |

| | | | | | | |
|---|---|---|---|---|---|---|
| EPI_ISL_510191 | Spain/MD-IBV-005311/2020 | 2020-03-17 | Europe/Spain/Madrid/ | Hospital General Universitario Gregorio Marav√±√≥n | SeqCOVID-SPAIN consortium/IBV(CSIC) | Laura P√©rez-Lago et al |
| EPI_ISL_510193 | Spain/MD-IBV-005313/2020 | 2020-03-17 | Europe/Spain/Madrid/ | Hospital General Universitario Gregorio Marav√±√≥n | SeqCOVID-SPAIN consortium/IBV(CSIC) | Laura P√©rez-Lago et al |
| EPI_ISL_510197 | Spain/MD-IBV-005318/2020 | 2020-03-17 | Europe/Spain/Madrid/ | Hospital General Universitario Gregorio Marav√±√≥n | SeqCOVID-SPAIN consortium/IBV(CSIC) | Laura P√©rez-Lago et al |
| EPI_ISL_510199 | Spain/MD-IBV-005320/2020 | 2020-03-17 | Europe/Spain/Madrid/ | Hospital General Universitario Gregorio Marav√±√≥n | SeqCOVID-SPAIN consortium/IBV(CSIC) | Laura P√©rez-Lago et al |
| EPI_ISL_510201 | Spain/MD-IBV-005322/2020 | 2020-03-17 | Europe/Spain/Madrid/ | Hospital General Universitario Gregorio Marav√±√≥n | SeqCOVID-SPAIN consortium/IBV(CSIC) | Laura P√©rez-Lago et al |
| EPI_ISL_510205 | Spain/MD-IBV-005326/2020 | 2020-03-17 | Europe/Spain/Madrid/ | Hospital General Universitario Gregorio Marav√±√≥n | SeqCOVID-SPAIN consortium/IBV(CSIC) | Laura P√©rez-Lago et al |
| EPI_ISL_510208 | Spain/MD-IBV-005330/2020 | 2020-03-17 | Europe/Spain/Madrid/ | Hospital General Universitario Gregorio Marav√±√≥n | SeqCOVID-SPAIN consortium/IBV(CSIC) | Laura P√©rez-Lago et al |
| EPI_ISL_510226 | Spain/MD-IBV-005349/2020 | 2020-03-19 | Europe/Spain/Madrid/ | Hospital General Universitario Gregorio Marav√±√≥n | SeqCOVID-SPAIN consortium/IBV(CSIC) | Laura P√©rez-Lago et al |
| EPI_ISL_510239 | Spain/MD-IBV-005364/2020 | 2020-03-31 | Europe/Spain/Madrid/ | Hospital General Universitario Gregorio Marav√±√≥n | SeqCOVID-SPAIN consortium/IBV(CSIC) | Laura P√©rez-Lago et al |
| EPI_ISL_510241 | Spain/MD-IBV-005366/2020 | 2020-04-01 | Europe/Spain/Madrid/ | Hospital General Universitario Gregorio Marav√±√≥n | SeqCOVID-SPAIN consortium/IBV(CSIC) | Laura P√©rez-Lago et al |
| EPI_ISL_510248 | Spain/CT-IBV-003151/2020 | 2020-03-16 | Europe/Spain/Catalunya/Tiana | Hospital de la Santa Creu i Sant Pau. Servicio de Microbiolog√≠a | SeqCOVID-SPAIN consortium/IBV(CSIC) | Ferran Navarro et al |
| EPI_ISL_510249 | Spain/CT-IBV-003152/2020 | 2020-03-16 | Europe/Spain/Catalunya/Barcelona | Hospital de la Santa Creu i Sant Pau. Servicio de Microbiolog√≠a | SeqCOVID-SPAIN consortium/IBV(CSIC) | Ferran Navarro et al |
| EPI_ISL_510254 | Spain/CT-IBV-003157/2020 | 2020-03-16 | Europe/Spain/Catalunya/Barcelona | Hospital de la Santa Creu i Sant Pau. Servicio de Microbiolog√≠a | SeqCOVID-SPAIN consortium/IBV(CSIC) | Ferran Navarro et al |

| | | | | | | |
|---|---|---|---|---|---|---|
| EPI_ISL_510258 | Spain/CT-IBV-003162/2020 | 2020-03-16 | Europe/Spain/Catalunya/Barcelona | Hospital de la Santa Creu i Sant Pau. Servicio de Microbiolog√≠a | SeqCOVID-SPAIN consortium/IBV(CSIC) | Ferran Navarro et al |
| EPI_ISL_510263 | Spain/CT-IBV-003167/2020 | 2020-03-16 | Europe/Spain/Catalunya/Barcelona | Hospital de la Santa Creu i Sant Pau. Servicio de Microbiolog√≠a | SeqCOVID-SPAIN consortium/IBV(CSIC) | Ferran Navarro et al |
| EPI_ISL_510270 | Spain/GA-IBV-004028/2020 | 2020-03-13 | Europe/Spain/Galicia/Santiago de Comp | Hospital Cl√≠nico Universitario de Santiago de Compostela | SeqCOVID-SPAIN consortium/IBV(CSIC) | Jos√© Javier Costa Alcalde et al |
| EPI_ISL_510276 | Spain/GA-IBV-004037/2020 | 2020-03-14 | Europe/Spain/Galicia/Bertamirans | Hospital Cl√≠nico Universitario de Santiago de Compostela | SeqCOVID-SPAIN consortium/IBV(CSIC) | Jos√© Javier Costa Alcalde et al |
| EPI_ISL_510280 | Spain/GA-IBV-004041/2020 | 2020-03-16 | Europe/Spain/Galicia/Boiro | Hospital Cl√≠nico Universitario de Santiago de Compostela | SeqCOVID-SPAIN consortium/IBV(CSIC) | Jos√© Javier Costa Alcalde et al |
| EPI_ISL_510281 | Spain/GA-IBV-004042/2020 | 2020-03-16 | Europe/Spain/Galicia/Boiro | Hospital Cl√≠nico Universitario de Santiago de Compostela | SeqCOVID-SPAIN consortium/IBV(CSIC) | Jos√© Javier Costa Alcalde et al |
| EPI_ISL_510289 | Spain/GA-IBV-004050/2020 | 2020-03-18 | Europe/Spain/Galicia/Santiago de Comp | Hospital Cl√≠nico Universitario de Santiago de Compostela | SeqCOVID-SPAIN consortium/IBV(CSIC) | Jos√© Javier Costa Alcalde et al |
| EPI_ISL_510294 | Spain/GA-IBV-004055/2020 | 2020-03-18 | Europe/Spain/Galicia/La Estrada | Hospital Cl√≠nico Universitario de Santiago de Compostela | SeqCOVID-SPAIN consortium/IBV(CSIC) | Jos√© Javier Costa Alcalde et al |
| EPI_ISL_510295 | Spain/GA-IBV-004057/2020 | 2020-03-18 | Europe/Spain/Galicia/Ribeira | Hospital Cl√≠nico Universitario de Santiago de Compostela | SeqCOVID-SPAIN consortium/IBV(CSIC) | Jos√© Javier Costa Alcalde et al |
| EPI_ISL_510297 | Spain/GA-IBV-004059/2020 | 2020-03-18 | Europe/Spain/Galicia/Santiago de Comp | Hospital Cl√≠nico Universitario de Santiago de Compostela | SeqCOVID-SPAIN consortium/IBV(CSIC) | Jos√© Javier Costa Alcalde et al |
| EPI_ISL_510304 | Spain/GA-IBV-004066/2020 | 2020-03-18 | Europe/Spain/Galicia/Outes | Hospital Cl√≠nico Universitario de Santiago de Compostela | SeqCOVID-SPAIN consortium/IBV(CSIC) | Jos√© Javier Costa Alcalde et al |
| EPI_ISL_510308 | Spain/EX-IBV-004993/2020 | 2020-05-16 | Europe/Spain/Extremadura/Caceres | Hospital San Pedro de Alc√°ntara (C√°ceres) | SeqCOVID-SPAIN consortium/IBV(CSIC) | Cristina Mu√±oz Cuevas et al |
| EPI_ISL_510322 | Spain/EX-IBV-005013/2020 | 2020-03-12 | Europe/Spain/Extremadura/Arroyo de la | Hospital San Pedro de Alc√°ntara (C√°ceres) | SeqCOVID-SPAIN consortium/IBV(CSIC) | Cristina Mu√±oz Cuevas et al |

| | | | | | | |
|---|---|---|---|---|---|---|
| EPI_ISL_510324 | Spain/EX-IBV-005015/2020 | 2020-03-13 | Europe/Spain/Extremadura/Arroyo de la | Hospital San Pedro de Alc√°ntara (C√°ceres) | SeqCOVID-SPAIN consortium/IBV(CSIC) | Cristina Mu√±oz Cuevas et al |
| EPI_ISL_510336 | Spain/AR-IBV-005027/2020 | 2020-04-19 | Europe/Spain/Aragon/Zaragoza | Servicio de Microbiolog√≠a, Hospital Miguel Servet, Zaragoza | SeqCOVID-SPAIN consortium/IBV(CSIC) | Antonio Rezusta L√≥pez et al |
| EPI_ISL_510338 | Spain/AR-IBV-005034/2020 | 2020-04-20 | Europe/Spain/Aragon/Zaragoza | Servicio de Microbiolog√≠a, Hospital Miguel Servet, Zaragoza | SeqCOVID-SPAIN consortium/IBV(CSIC) | Antonio Rezusta L√≥pez et al |
| EPI_ISL_510340 | Spain/AR-IBV-005037/2020 | 2020-04-20 | Europe/Spain/Aragon/Zaragoza | Servicio de Microbiolog√≠a, Hospital Miguel Servet, Zaragoza | SeqCOVID-SPAIN consortium/IBV(CSIC) | Antonio Rezusta L√≥pez et al |
| EPI_ISL_510344 | Spain/AR-IBV-005041/2020 | 2020-04-20 | Europe/Spain/Aragon/Zaragoza | Servicio de Microbiolog√≠a, Hospital Miguel Servet, Zaragoza | SeqCOVID-SPAIN consortium/IBV(CSIC) | Antonio Rezusta L√≥pez et al |
| EPI_ISL_510346 | Spain/AR-IBV-005043/2020 | 2020-04-20 | Europe/Spain/Aragon/Zaragoza | Servicio de Microbiolog√≠a, Hospital Miguel Servet, Zaragoza | SeqCOVID-SPAIN consortium/IBV(CSIC) | Antonio Rezusta L√≥pez et al |
| EPI_ISL_510350 | Spain/AR-IBV-005050/2020 | 2020-04-21 | Europe/Spain/Aragon/Zaragoza | Servicio de Microbiolog√≠a, Hospital Miguel Servet, Zaragoza | SeqCOVID-SPAIN consortium/IBV(CSIC) | Antonio Rezusta L√≥pez et al |
| EPI_ISL_510369 | Spain/AR-IBV-005086/2020 | 2020-04-23 | Europe/Spain/Aragon/Zaragoza | Servicio de Microbiolog√≠a, Hospital Miguel Servet, Zaragoza | SeqCOVID-SPAIN consortium/IBV(CSIC) | Antonio Rezusta L√≥pez et al |
| EPI_ISL_510374 | Spain/AR-IBV-005091/2020 | 2020-04-23 | Europe/Spain/Aragon/Zaragoza | Servicio de Microbiolog√≠a, Hospital Miguel Servet, Zaragoza | SeqCOVID-SPAIN consortium/IBV(CSIC) | Antonio Rezusta L√≥pez et al |
| EPI_ISL_510381 | Spain/AR-IBV-005105/2020 | 2020-04-26 | Europe/Spain/Aragon/Zaragoza | Servicio de Microbiolog√≠a, Hospital Miguel Servet, Zaragoza | SeqCOVID-SPAIN consortium/IBV(CSIC) | Antonio Rezusta L√≥pez et al |
| EPI_ISL_510386 | Spain/AR-IBV-005121/2020 | 2020-04-27 | Europe/Spain/Aragon/Zaragoza | Servicio de Microbiolog√≠a, Hospital Miguel Servet, Zaragoza | SeqCOVID-SPAIN consortium/IBV(CSIC) | Antonio Rezusta L√≥pez et al |
| EPI_ISL_510388 | Spain/AR-IBV-005124/2020 | 2020-04-28 | Europe/Spain/Aragon/Zaragoza | Servicio de Microbiolog√≠a, Hospital Miguel Servet, Zaragoza | SeqCOVID-SPAIN consortium/IBV(CSIC) | Antonio Rezusta L√≥pez et al |

| | | | | | | |
|---|---|---|---|---|---|---|
| EPI_ISL_510398 | Spain/PV-IBV-004281/2020 | 2020-04-08 | Europe/Spain/Basque Country/San Seba | Servicio de Microbiolog√≠a. Hospital Universitario Donostia. OSI Donostialdea. √Årea de Enfermedades Infecciosas, Grupo de Infecci√≥n Respiratoria y Resistencia Antimicrobiana. Instituto de Investigaci√≥n Sanitaria Biodonostia | SeqCOVID-SPAIN consortium/IBV(CSIC) | Gustavo Cilla et al |
| EPI_ISL_510408 | Spain/PV-IBV-004292/2020 | 2020-04-09 | Europe/Spain/Basque Country/San Seba | Servicio de Microbiolog√≠a. Hospital Universitario Donostia. OSI Donostialdea. √Årea de Enfermedades Infecciosas, Grupo de Infecci√≥n Respiratoria y Resistencia Antimicrobiana. Instituto de Investigaci√≥n Sanitaria Biodonostia | SeqCOVID-SPAIN consortium/IBV(CSIC) | Gustavo Cilla et al |
| EPI_ISL_510426 | Spain/AN-IBV-006086/2020 | 2020-04-27 | Europe/Spain/Andalusia/Santa Fe ES | Hospital Universitario Virgen de las Nieves de Granada-SAS | SeqCOVID-SPAIN consortium/IBV(CSIC) | Mercedes P√©rez Ruiz et al |
| EPI_ISL_510435 | Spain/AN-IBV-006103/2020 | 2020-06-29 | Europe/Spain/Andalusia/Granada ES | Hospital Universitario Virgen de las Nieves de Granada-SAS | SeqCOVID-SPAIN consortium/IBV(CSIC) | Mercedes P√©rez Ruiz et al |
| EPI_ISL_510437 | Spain/AN-IBV-006105/2020 | 2020-05-26 | Europe/Spain/Andalusia/Guadix | Hospital Universitario Virgen de las Nieves de Granada-SAS | SeqCOVID-SPAIN consortium/IBV(CSIC) | Mercedes P√©rez Ruiz et al |
| EPI_ISL_510439 | Spain/AN-IBV-006108/2020 | 2020-07-02 | Europe/Spain/Andalusia/Granada ES | Hospital Universitario Virgen de las Nieves de Granada-SAS | SeqCOVID-SPAIN consortium/IBV(CSIC) | Mercedes P√©rez Ruiz et al |
| EPI_ISL_510445 | Spain/AN-IBV-006116/2020 | 2020-07-14 | Europe/Spain/Andalusia/Granada ES | Hospital Universitario Virgen de las Nieves de Granada-SAS | SeqCOVID-SPAIN consortium/IBV(CSIC) | Mercedes P√©rez Ruiz et al |
| EPI_ISL_510454 | Spain/AN-IBV-006126/2020 | 2020-07-10 | Europe/Spain/Andalusia/Granada ES | Hospital Universitario Virgen de las Nieves de Granada-SAS | SeqCOVID-SPAIN consortium/IBV(CSIC) | Mercedes P√©rez Ruiz et al |
| EPI_ISL_510455 | Spain/AN-IBV-006128/2020 | 2020-07-06 | Europe/Spain/Andalusia/Santa Fe ES | Hospital Universitario Virgen de las Nieves de Granada-SAS | SeqCOVID-SPAIN consortium/IBV(CSIC) | Mercedes P√©rez Ruiz et al |
| EPI_ISL_510456 | Spain/AN-IBV-006130/2020 | 2020-07-10 | Europe/Spain/Andalusia/Granada ES | Hospital Universitario Virgen de las Nieves de Granada-SAS | SeqCOVID-SPAIN consortium/IBV(CSIC) | Mercedes P√©rez Ruiz et al |
| EPI_ISL_510460 | Spain/AN-IBV-006136/2020 | 2020-07-16 | Europe/Spain/Andalusia/Granada ES | Hospital Universitario Virgen de las Nieves de Granada-SAS | SeqCOVID-SPAIN consortium/IBV(CSIC) | Mercedes P√©rez Ruiz et al |

| | | | | | | |
|---|---|---|---|---|---|---|
| EPI_ISL_510465 | Spain/CT-IBV-004176/2020 | 2020-04-03 | Europe/Spain/Catalunya/Barcelona | Instituto de Investigaciones Biom√©dicas de Barcelona (CSIC), Hospital Clinic i Provincial de Barcelona, Instituto de Biomedicina de Valencia (CSIC), Hospital de Sant Pau | SeqCOVID-SPAIN consortium/IBV(CSIC) | Anna M. Planas et al |
| EPI_ISL_510466 | Spain/CT-IBV-004177/2020 | 2020-04-10 | Europe/Spain/Catalunya/Barcelona | Instituto de Investigaciones Biom√©dicas de Barcelona (CSIC), Hospital Clinic i Provincial de Barcelona, Instituto de Biomedicina de Valencia (CSIC), Hospital de Sant Pau | SeqCOVID-SPAIN consortium/IBV(CSIC) | Anna M. Planas et al |
| EPI_ISL_510469 | Spain/PV-IBV-004182/2020 | 2020-04-02 | Europe/Spain/Basque Country/San Seba | Servicio de Microbiolog√≠a. Hospital Universitario Donostia. OSI Donostialdea. √Årea de Enfermedades Infecciosas, Grupo de Infecci√≥n Respiratoria y Resistencia Antimicrobiana. Instituto de Investigaci√≥n Sanitaria Biodonostia | SeqCOVID-SPAIN consortium/IBV(CSIC) | Gustavo Cilla et al |
| EPI_ISL_510470 | Spain/PV-IBV-004183/2020 | 2020-04-02 | Europe/Spain/Basque Country/San Seba | Servicio de Microbiolog√≠a. Hospital Universitario Donostia. OSI Donostialdea. √Årea de Enfermedades Infecciosas, Grupo de Infecci√≥n Respiratoria y Resistencia Antimicrobiana. Instituto de Investigaci√≥n Sanitaria Biodonostia | SeqCOVID-SPAIN consortium/IBV(CSIC) | Gustavo Cilla et al |
| EPI_ISL_510482 | Spain/PV-IBV-004197/2020 | 2020-04-03 | Europe/Spain/Basque Country/San Seba | Servicio de Microbiolog√≠a. Hospital Universitario Donostia. OSI Donostialdea. √Årea de Enfermedades Infecciosas, Grupo de Infecci√≥n Respiratoria y Resistencia Antimicrobiana. Instituto de Investigaci√≥n Sanitaria Biodonostia | SeqCOVID-SPAIN consortium/IBV(CSIC) | Gustavo Cilla et al |
| EPI_ISL_510483 | Spain/PV-IBV-004198/2020 | 2020-04-03 | Europe/Spain/Basque Country/San Seba | Servicio de Microbiolog√≠a. Hospital Universitario Donostia. OSI Donostialdea. √Årea de Enfermedades Infecciosas, Grupo de Infecci√≥n Respiratoria y Resistencia Antimicrobiana. Instituto de Investigaci√≥n Sanitaria Biodonostia | SeqCOVID-SPAIN consortium/IBV(CSIC) | Gustavo Cilla et al |
| EPI_ISL_510492 | Spain/PV-IBV-004207/2020 | 2020-04-03 | Europe/Spain/Basque Country/San Seba | Servicio de Microbiolog√≠a. Hospital Universitario Donostia. OSI Donostialdea. √Årea de Enfermedades Infecciosas, Grupo de Infecci√≥n Respiratoria y Resistencia Antimicrobiana. Instituto de Investigaci√≥n Sanitaria Biodonostia | SeqCOVID-SPAIN consortium/IBV(CSIC) | Gustavo Cilla et al |

| | | | | | | |
|---|---|---|---|---|---|---|
| EPI_ISL_510493 | Spain/PV-IBV-004208/2020 | 2020-04-03 | Europe/Spain/Basque Country/San Seba | Servicio de Microbiolog√≠a. Hospital Universitario Donostia. OSI Donostialdea. √Årea de Enfermedades Infecciosas, Grupo de Infecci√≥n Respiratoria y Resistencia Antimicrobiana. Instituto de Investigaci√≥n Sanitaria Biodonostia | SeqCOVID-SPAIN consortium/IBV(CSIC) | Gustavo Cilla et al |
| EPI_ISL_510514 | Spain/CT-IBV-005483/2020 | 2020-05-12 | Europe/Spain/Catalunya/Mollet del Val | Servicio de Microbiolog√≠a, Laboratori Cl√≠nic Metropolitana Nord. Hospital Universitari Germans Trias i Pujol. Institut d,ÄôInvestigaci√≥ en Ci√®ncies de la Salut Germans Trias i Pujol (IGTP) | SeqCOVID-SPAIN consortium/IBV(CSIC) | Elisa Mart√≥ et al |
| EPI_ISL_510516 | Spain/CT-IBV-005488/2020 | 2020-05-21 | Europe/Spain/Catalunya/Badalona | Servicio de Microbiolog√≠a, Laboratori Cl√≠nic Metropolitana Nord. Hospital Universitari Germans Trias i Pujol. Institut d,ÄôInvestigaci√≥ en Ci√®ncies de la Salut Germans Trias i Pujol (IGTP) | SeqCOVID-SPAIN consortium/IBV(CSIC) | Elisa Mart√≥ et al |
| EPI_ISL_510520 | Spain/CT-IBV-005502/2020 | 2020-05-26 | Europe/Spain/Catalunya/Mataro | Servicio de Microbiolog√≠a, Laboratori Cl√≠nic Metropolitana Nord. Hospital Universitari Germans Trias i Pujol. Institut d,ÄôInvestigaci√≥ en Ci√®ncies de la Salut Germans Trias i Pujol (IGTP) | SeqCOVID-SPAIN consortium/IBV(CSIC) | Elisa Mart√≥ et al |
| EPI_ISL_510522 | Spain/CT-IBV-005479/2020 | 2020-05-12 | Europe/Spain/Catalunya/Badalona | Servicio de Microbiolog√≠a, Laboratori Cl√≠nic Metropolitana Nord. Hospital Universitari Germans Trias i Pujol. Institut d,ÄôInvestigaci√≥ en Ci√®ncies de la Salut Germans Trias i Pujol (IGTP) | SeqCOVID-SPAIN consortium/IBV(CSIC) | Elisa Mart√≥ et al |
| EPI_ISL_510526 | Egypt/Army-MCL001/2020 | 2020-06-01 | Africa/Egypt/Faiyum/ | Biological prevention, army | Biological prevention, army | Seadawy et al |
| EPI_ISL_510528 | Bahrain/140008296/2020 | 2020-06-23 | Asia/Bahrain/Capital Governorate/Man | Communicable Disease Laboratory, Public Health Directorate | Communicable Disease Laboratory, Public Health Directorate | Al Wasti et al |
| EPI_ISL_510531 | Bahrain/920268866/2020 | 2020-06-23 | Asia/Bahrain/Capital Governorate/Man | Communicable Disease Laboratory, Public Health Directorate | Communicable Disease Laboratory, Public Health Directorate | Shehab et al |
| EPI_ISL_510532 | Egypt/HCoV2-Egy-002/2020 | 2020-06-02 | Africa/Egypt/Faiyum/ | Biological prevention, army | Biological prevention, army | Seadawy et al |

| | | | | | | |
|---|---|---|---|---|---|---|
| EPI_ISL_510535 | Brazil/PR-LRV-01/2020 | 2020-05-08 | South America/Brazil/Paranv°/Curitiba | Molecular Virology, Instituto Carlos Chagas / Fiocruz Paranv° | Universidade Federal do Parana (UFPR) | Suzukawa et al |
| EPI_ISL_510536 | Brazil/RJ-DCVN5/2020 | 2020-07-20 | South America/Brazil/Rio de Janeiro/ | Centro de Desenvolvimento Tecnologico em Saude, Fundacao Oswaldo Cruz | Centro de Desenvolvimento Tecnologico em Saude, Fundacao Oswaldo Cruz | Souza et al |
| EPI_ISL_510541 | Brazil/RJ-DCVN4/2020 | 2020-07-20 | South America/Brazil/Rio de Janeiro/ | Centro de Desenvolvimento Tecnologico em Saude, Fundacao Oswaldo Cruz | Centro de Desenvolvimento Tecnologico em Saude, Fundacao Oswaldo Cruz | Souza et al |
| EPI_ISL_510547 | SouthKorea/KCDC2203/2020 | 2020-02-26 | Asia/South Korea/South Korea/ | Division of Viral Diseases, Center for Laboratory Control of Infectious Diseases, Korea Centers for Diseases Control and Prevention | Division of Viral Diseases, Center for Laboratory Control of Infectious Diseases, Korea Centers for Diseases Control and Prevention | Jeong-Min Kim et al |
| EPI_ISL_510614 | SouthKorea/KCDC2277/2020 | 2020-05-14 | Asia/South Korea/South Korea/ | Division of Viral Diseases, Center for Laboratory Control of Infectious Diseases, Korea Centers for Diseases Control and Prevention | Division of Viral Diseases, Center for Laboratory Control of Infectious Diseases, Korea Centers for Diseases Control and Prevention | Jeong-Min Kim et al |
| EPI_ISL_510811 | Sweden/20-08904/2020 | 2020-05-23 | Europe/Sweden/Stockholm/ | NA | The Public Health Agency of Sweden | Oskar Karlsson Lindsjo et al |
| EPI_ISL_510812 | Sweden/20-08905/2020 | 2020-05-24 | Europe/Sweden/Stockholm/ | NA | The Public Health Agency of Sweden | Oskar Karlsson Lindsjo et al |
| EPI_ISL_510816 | Sweden/20-08910/2020 | 2020-05-28 | Europe/Sweden/Stockholm/ | NA | The Public Health Agency of Sweden | Oskar Karlsson Lindsjo et al |
| EPI_ISL_510821 | Sweden/20-51275/2020 | 2020-05-04 | Europe/Sweden/Varmland/ | Klinisk mikrobiologi centralsjukhuset Karlstad | The Public Health Agency of Sweden | Oskar Karlsson Lindsjo et al |
| EPI_ISL_510834 | Sweden/20-51697/2020 | 2020-05-23 | Europe/Sweden/Stockholm/ | Karolinska Universitetslaboratoriet | The Public Health Agency of Sweden | Oskar Karlsson Lindsjo et al |

| | | | | | | |
|---|---|---|---|---|---|---|
| EPI_ISL_510836 | Sweden/20-51710/2020 | 2020-05-22 | Europe/Sweden/Kalmar/ | Kalmar klinisk mikrobiologi | The Public Health Agency of Sweden | Oskar Karlsson Lindsjo et al |
| EPI_ISL_510841 | Sweden/20-51740/2020 | 2020-05-25 | Europe/Sweden/Vastra Gotaland/ | Klinisk mikrobiologi NAL Trollhattan | The Public Health Agency of Sweden | Oskar Karlsson Lindsjo et al |
| EPI_ISL_510845 | Sweden/20-51765/2020 | 2020-05-27 | Europe/Sweden/Vastra Gotaland/ | Skovde/Unilabs | The Public Health Agency of Sweden | Oskar Karlsson Lindsjo et al |
| EPI_ISL_510849 | Sweden/20-51772/2020 | 2020-05-27 | Europe/Sweden/Vastra Gotaland/ | Skovde/Unilabs | The Public Health Agency of Sweden | Oskar Karlsson Lindsjo et al |
| EPI_ISL_510858 | Sweden/20-51854/2020 | 2020-06-01 | Europe/Sweden/Vasternorrland/ | Klinisk mikrobiologi Vasternorrland | The Public Health Agency of Sweden | Oskar Karlsson Lindsjo et al |
| EPI_ISL_510859 | Sweden/20-51894/2020 | 2020-05-25 | Europe/Sweden/Kalmar/ | Kalmar klinisk mikrobiologi | The Public Health Agency of Sweden | Oskar Karlsson Lindsjo et al |
| EPI_ISL_510861 | Sweden/20-51947/2020 | 2020-06-05 | Europe/Sweden/Stockholm/ | Karolinska Universitetslaboratoriet | The Public Health Agency of Sweden | Oskar Karlsson Lindsjo et al |
| EPI_ISL_510862 | Sweden/20-51948/2020 | 2020-06-05 | Europe/Sweden/Stockholm/ | Karolinska Universitetslaboratoriet | The Public Health Agency of Sweden | Oskar Karlsson Lindsjo et al |
| EPI_ISL_510864 | Sweden/20-51997/2020 | 2020-06-05 | Europe/Sweden/Kalmar/ | Kalmar klinisk mikrobiologi | The Public Health Agency of Sweden | Oskar Karlsson Lindsjo et al |
| EPI_ISL_510865 | Sweden/20-52000/2020 | 2020-06-05 | Europe/Sweden/Kalmar/ | Kalmar klinisk mikrobiologi | The Public Health Agency of Sweden | Oskar Karlsson Lindsjo et al |
| EPI_ISL_510872 | Sweden/20-52073/2020 | 2020-06-15 | Europe/Sweden/Ostergotland/ | Klinsisk mikrobiologi Linkoping | The Public Health Agency of Sweden | Oskar Karlsson Lindsjo et al |
| EPI_ISL_510873 | Sweden/20-52074/2020 | 2020-06-15 | Europe/Sweden/Ostergotland/ | Klinsisk mikrobiologi Linkoping | The Public Health Agency of Sweden | Oskar Karlsson Lindsjo et al |
| EPI_ISL_510874 | Sweden/20-52077/2020 | 2020-06-15 | Europe/Sweden/Ostergotland/ | Klinsisk mikrobiologi Linkoping | The Public Health Agency of Sweden | Oskar Karlsson Lindsjo et al |
| EPI_ISL_510875 | Sweden/20-52078/2020 | 2020-06-15 | Europe/Sweden/Ostergotland/ | Klinsisk mikrobiologi Linkoping | The Public Health Agency of Sweden | Oskar Karlsson Lindsjo et al |
| EPI_ISL_510876 | Sweden/20-52079/2020 | 2020-06-15 | Europe/Sweden/Ostergotland/ | Klinsisk mikrobiologi Linkoping | The Public Health Agency of Sweden | Oskar Karlsson Lindsjo et al |
| EPI_ISL_510877 | Sweden/20-52080/2020 | 2020-06-15 | Europe/Sweden/Ostergotland/ | Klinsisk mikrobiologi Linkoping | The Public Health Agency of Sweden | Oskar Karlsson Lindsjo et al |
| EPI_ISL_510884 | USA/NM-NMDOH-2020242035/2020 | 2020-07-22 | North America/USA/New Mexico/ | New Mexico Department of Health Scientific Laboratory | New Mexico Department of Health Scientific Laboratory | Ellie Johnson et al |
| EPI_ISL_510888 | USA/NM-NMDOH-2020242041/2020 | 2020-07-22 | North America/USA/New Mexico/ | New Mexico Department of Health Scientific Laboratory | New Mexico Department of Health Scientific Laboratory | Ellie Johnson et al |

| | | | | | | |
|---|---|---|---|---|---|---|
| EPI_ISL_510890 | Portugal/PT1522/2020 | 2020-06-22 | Europe/Portugal/Portugal/ | Instituto Nacional de Saude (INSA) | Instituto Nacional de Saude (INSA) | Borges et al et al |
| EPI_ISL_510922 | Portugal/PT0663/2020 | 2020-04-04 | Europe/Portugal/Portugal/ | Instituto Nacional de Saude (INSA) | Instituto Nacional de Saude (INSA) | Borges et al et al |
| EPI_ISL_510942 | Portugal/PT0683/2020 | 2020-03-27 | Europe/Portugal/Portugal/ | Instituto Nacional de Saude (INSA) | Instituto Nacional de Saude (INSA) | Borges et al et al |
| EPI_ISL_510953 | Portugal/PT0694/2020 | 2020-03-14 | Europe/Portugal/Portugal/ | Instituto Nacional de Saude (INSA) | Instituto Nacional de Saude (INSA) | Borges et al et al |
| EPI_ISL_510960 | Portugal/PT0701/2020 | 2020-03-17 | Europe/Portugal/Portugal/ | Instituto Nacional de Saude (INSA) | Instituto Nacional de Saude (INSA) | Borges et al et al |
| EPI_ISL_510965 | Portugal/PT0706/2020 | 2020-03-21 | Europe/Portugal/Portugal/ | Instituto Nacional de Saude (INSA) | Instituto Nacional de Saude (INSA) | Borges et al et al |
| EPI_ISL_510984 | Portugal/PT0725/2020 | 2020-03-27 | Europe/Portugal/Portugal/ | Instituto Nacional de Saude (INSA) | Instituto Nacional de Saude (INSA) | Borges et al et al |
| EPI_ISL_511014 | Portugal/PT0755/2020 | 2020-03-23 | Europe/Portugal/Portugal/ | Instituto Nacional de Saude (INSA) | Instituto Nacional de Saude (INSA) | Borges et al et al |
| EPI_ISL_511023 | Portugal/PT0764/2020 | 2020-03-25 | Europe/Portugal/Portugal/ | Instituto Nacional de Saude (INSA) | Instituto Nacional de Saude (INSA) | Borges et al et al |
| EPI_ISL_511029 | Portugal/PT0770a/2020 | 2020-04-08 | Europe/Portugal/Portugal/ | Instituto Nacional de Saude (INSA) | Instituto Nacional de Saude (INSA) | Borges et al et al |
| EPI_ISL_511030 | Portugal/PT0771/2020 | 2020-05-05 | Europe/Portugal/Portugal/ | Instituto Nacional de Saude (INSA) | Instituto Nacional de Saude (INSA) | Borges et al et al |
| EPI_ISL_511060 | Portugal/PT0801/2020 | 2020-03-19 | Europe/Portugal/Portugal/ | Instituto Nacional de Saude (INSA) | Instituto Nacional de Saude (INSA) | Borges et al et al |
| EPI_ISL_511070 | Portugal/PT0811/2020 | 2020-03-20 | Europe/Portugal/Portugal/ | Instituto Nacional de Saude (INSA) | Instituto Nacional de Saude (INSA) | Borges et al et al |
| EPI_ISL_511077 | Portugal/PT0818/2020 | 2020-03-20 | Europe/Portugal/Portugal/ | Instituto Nacional de Saude (INSA) | Instituto Nacional de Saude (INSA) | Borges et al et al |
| EPI_ISL_511105 | Portugal/PT0846/2020 | 2020-05-05 | Europe/Portugal/Portugal/ | Instituto Nacional de Saude (INSA) | Instituto Nacional de Saude (INSA) | Borges et al et al |
| EPI_ISL_511145 | Portugal/PT0887/2020 | 2020-03-19 | Europe/Portugal/Portugal/ | Instituto Nacional de Saude (INSA) | Instituto Nacional de Saude (INSA) | Borges et al et al |
| EPI_ISL_511184 | Portugal/PT0927/2020 | 2020-04-26 | Europe/Portugal/Portugal/ | Instituto Nacional de Saude (INSA) and Instituto Gulbenkian de Ciencia (IGC) | Instituto Nacional de Saude (INSA) and Instituto Gulbenkian de Ciencia (IGC) | Borges et al et al |

| | | | | | | |
|---|---|---|---|---|---|---|
| EPI_ISL_511190 | Portugal/PT1229/2020 | 2020-03-29 | Europe/Portugal/Portugal/ | Instituto Nacional de Saude (INSA) and Instituto Gulbenkian de Ciencia (IGC) | Instituto Nacional de Saude (INSA) and Instituto Gulbenkian de Ciencia (IGC) | Borges et al et al |
| EPI_ISL_511194 | Portugal/PT1233/2020 | 2020-03-31 | Europe/Portugal/Portugal/ | Instituto Nacional de Saude (INSA) and Instituto Gulbenkian de Ciencia (IGC) | Instituto Nacional de Saude (INSA) and Instituto Gulbenkian de Ciencia (IGC) | Borges et al et al |
| EPI_ISL_511232 | Portugal/PT1271/2020 | 2020-03-20 | Europe/Portugal/Portugal/ | Instituto Nacional de Saude (INSA) and Instituto Gulbenkian de Ciencia (IGC) | Instituto Nacional de Saude (INSA) and Instituto Gulbenkian de Ciencia (IGC) | Borges et al et al |
| EPI_ISL_511321 | Portugal/PT1362/2020 | 2020-03-24 | Europe/Portugal/Portugal/ | Instituto Nacional de Saude (INSA) | Instituto Nacional de Saude (INSA) | Borges et al et al |
| EPI_ISL_511373 | Portugal/PT1414/2020 | 2020-03-26 | Europe/Portugal/Portugal/ | Instituto Nacional de Saude (INSA) | Instituto Nacional de Saude (INSA) | Borges et al et al |
| EPI_ISL_511383 | Portugal/PT1424/2020 | 2020-03-27 | Europe/Portugal/Portugal/ | Instituto Nacional de Saude (INSA) | Instituto Nacional de Saude (INSA) | Borges et al et al |
| EPI_ISL_511391 | Portugal/PT1432/2020 | 2020-03-26 | Europe/Portugal/Portugal/ | Instituto Nacional de Saude (INSA) | Instituto Nacional de Saude (INSA) | Borges et al et al |
| EPI_ISL_511408 | Portugal/PT1449/2020 | 2020-03-26 | Europe/Portugal/Portugal/ | Instituto Nacional de Saude (INSA) | Instituto Nacional de Saude (INSA) | Borges et al et al |
| EPI_ISL_511447 | Portugal/PT1488/2020 | 2020-03-17 | Europe/Portugal/Portugal/ | Instituto Nacional de Saude (INSA) | Instituto Nacional de Saude (INSA) | Borges et al et al |
| EPI_ISL_511483 | Portugal/PT0659a/2020 | 2020-03-23 | Europe/Portugal/Portugal/ | Instituto Nacional de Saude (INSA) | Instituto Nacional de Saude (INSA) | Borges et al et al |
| EPI_ISL_511512 | Portugal/PT0963/2020 | 2020-03-26 | Europe/Portugal/Portugal/ | Instituto Nacional de Saude (INSA) | Instituto Nacional de Saude (INSA) | Borges et al et al |
| EPI_ISL_511530 | Portugal/PT0981/2020 | 2020-03-22 | Europe/Portugal/Portugal/ | Instituto Nacional de Saude (INSA) | Instituto Nacional de Saude (INSA) | Borges et al et al |
| EPI_ISL_511539 | Portugal/PT0991/2020 | 2020-03-23 | Europe/Portugal/Portugal/ | Instituto Nacional de Saude (INSA) | Instituto Nacional de Saude (INSA) | Borges et al et al |
| EPI_ISL_511591 | Portugal/PT1044/2020 | 2020-03-27 | Europe/Portugal/Portugal/ | Instituto Nacional de Saude (INSA) | Instituto Nacional de Saude (INSA) | Borges et al et al |
| EPI_ISL_511637 | Portugal/PT1090/2020 | 2020-03-28 | Europe/Portugal/Portugal/ | Instituto Nacional de Saude (INSA) | Instituto Nacional de Saude (INSA) | Borges et al et al |

| | | | | | | |
|---|---|---|---|---|---|---|
| EPI_ISL_511638 | Portugal/PT1091/2020 | 2020-03-28 | Europe/Portugal/Portugal/ | Instituto Nacional de Saude (INSA) | Instituto Nacional de Saude (INSA) | Borges et al et al |
| EPI_ISL_511680 | Portugal/PT1133/2020 | 2020-03-27 | Europe/Portugal/Portugal/ | Instituto Nacional de Saude (INSA) | Instituto Nacional de Saude (INSA) | Borges et al et al |
| EPI_ISL_511683 | Portugal/PT1136/2020 | 2020-05-06 | Europe/Portugal/Portugal/ | Instituto Nacional de Saude (INSA) | Instituto Nacional de Saude (INSA) | Borges et al et al |
| EPI_ISL_511695 | Portugal/PT1148/2020 | 2020-04-08 | Europe/Portugal/Portugal/ | Instituto Nacional de Saude (INSA) | Instituto Nacional de Saude (INSA) | Borges et al et al |
| EPI_ISL_511707 | Portugal/PT1160/2020 | 2020-04-01 | Europe/Portugal/Portugal/ | Instituto Nacional de Saude (INSA) | Instituto Nacional de Saude (INSA) | Borges et al et al |
| EPI_ISL_511713 | Portugal/PT1167/2020 | 2020-04-05 | Europe/Portugal/Portugal/ | Instituto Nacional de Saude (INSA) | Instituto Nacional de Saude (INSA) | Borges et al et al |
| EPI_ISL_511715 | Portugal/PT1169/2020 | 2020-04-05 | Europe/Portugal/Portugal/ | Instituto Nacional de Saude (INSA) | Instituto Nacional de Saude (INSA) | Borges et al et al |
| EPI_ISL_511746 | Portugal/PT1203/2020 | 2020-03-31 | Europe/Portugal/Portugal/ | Instituto Nacional de Saude (INSA) | Instituto Nacional de Saude (INSA) | Borges et al et al |
| EPI_ISL_511810 | USA/CA-IGI-0055/2020 | 2020-05-21 | North America/USA/California/Alameda | Innovative Genomics Institute, UC Berkeley | Innovative Genomics Institute, UC Berkeley | Stacia Wyman et al |
| EPI_ISL_511812 | USA/CA-IGI-0057/2020 | 2020-05-20 | North America/USA/California/Alameda | Innovative Genomics Institute, UC Berkeley | Innovative Genomics Institute, UC Berkeley | Stacia Wyman et al |
| EPI_ISL_511818 | USA/CA-IGI-0063/2020 | 2020-06-12 | North America/USA/California/Alameda | Innovative Genomics Institute, UC Berkeley | Innovative Genomics Institute, UC Berkeley | Stacia Wyman et al |
| EPI_ISL_511846 | USA/CA-IGI-0092/2020 | 2020-06-18 | North America/USA/California/Alameda | Innovative Genomics Institute, UC Berkeley | Innovative Genomics Institute, UC Berkeley | Stacia Wyman et al |
| EPI_ISL_511850 | USA/CA-IGI-0096/2020 | 2020-06-03 | North America/USA/California/Alameda | Innovative Genomics Institute, UC Berkeley | Innovative Genomics Institute, UC Berkeley | Stacia Wyman et al |
| EPI_ISL_511852 | USA/WA-UW-10030/2020 | 2020-05-29 | North America/USA/Washington/ | UW Virology Lab | UW Virology Lab | Pavitra Roychoudhury et al |
| EPI_ISL_511858 | USA/WA-UW-10115/2020 | 2020-05-29 | North America/USA/Washington/ | UW Virology Lab | UW Virology Lab | Pavitra Roychoudhury et al |

| | | | | | | |
|---|---|---|---|---|---|---|
| EPI_ISL_511871 | USA/MD-HP00567/2020 | 2020-04-06 | North America/USA/Maryland/ | Johns Hopkins Hospital Department of Pathology | Johns Hopkins Hospital Department of Pathology | Peter M. Thielen et al |
| EPI_ISL_511876 | USA/MD-HP02026/2020 | 2020-04-19 | North America/USA/Maryland/ | Johns Hopkins Hospital Department of Pathology | Johns Hopkins Hospital Department of Pathology | Peter M. Thielen et al |
| EPI_ISL_511878 | Indonesia/JB-TFRIC19-R42550/2020 | 2020-07-11 | Asia/Indonesia/West Java/Bandung | Santosa Hospital Bandung Kopo | Laboratorium Kesehatan Provinsi Jawa Barat; Molecular Genetics Laboratory-Faculty of Medicine-Universitas Padjadjaran; School of Life Sciences and Technology & School of Pharmacy-Institut Teknologi Bandung | Ema Rahmawati et al |

| | | | | | | |
|---|---|---|---|---|---|---|
| EPI_ISL_511879 | Indonesia/JB-TFRIC19-L4173/2020 | 2020-07-08 | Asia/Indonesia/West Java/Bandung | Laboratorium Kesehatan Provinsi Jawa Barat | Molecular Genetics Laboratory-Faculty of Medicine-Universitas Padjadjaran; School of Life Sciences and Technology & School of Pharmacy-Institut Teknologi Bandung; Laboratorium Kesehatan Provinsi Jawa Barat | Marselina Irasonia Tan et al |
| EPI_ISL_511901 | India/WB-IK8/2020 | 2020-04-05 | Asia/India/West Bengal/Kolkata | Institute of Post Graduate Medical Education & Research | National Institute of Biomedical Genomics - DBT's PAN-INDIA 1000 SARS--CoV-2 RNA Genome Sequencing Consortium | Arindam Maitra et al |
| EPI_ISL_511905 | India/WB-IK81/2020 | 2020-05-17 | Asia/India/West Bengal/North 24 Parga | Institute of Post Graduate Medical Education & Research | National Institute of Biomedical Genomics - DBT's PAN-INDIA 1000 SARS--CoV-2 RNA Genome Sequencing Consortium | Arindam Maitra et al |

| | | | | | | |
|---|---|---|---|---|---|---|
| EPI_ISL_511909 | India/UT-AR35/2020 | 2020-05-28 | Asia/India/Uttarakhand/Tehri Garhwal | All india institute of Medical Sciences Rishikesh | National Institute of Biomedical Genomics - DBT's PAN-INDIA 1000 SARS--CoV-2 RNA Genome Sequencing Consortium | Arindam Maitra et al |
| EPI_ISL_511910 | India/UT-AR37/2020 | 2020-05-28 | Asia/India/Uttarakhand/Tehri Garhwal | All india institute of Medical Sciences Rishikesh | National Institute of Biomedical Genomics - DBT's PAN-INDIA 1000 SARS--CoV-2 RNA Genome Sequencing Consortium | Arindam Maitra et al |
| EPI_ISL_511911 | India/UT-AR38/2020 | 2020-05-28 | Asia/India/Uttarakhand/Tehri Garhwal | All india institute of Medical Sciences Rishikesh | National Institute of Biomedical Genomics - DBT's PAN-INDIA 1000 SARS--CoV-2 RNA Genome Sequencing Consortium | Arindam Maitra et al |
| EPI_ISL_511913 | India/UT-AR47/2020 | 2020-06-01 | Asia/India/Uttarakhand/Pauri Garhwal | All india institute of Medical Sciences Rishikesh | National Institute of Biomedical Genomics - DBT's PAN-INDIA 1000 SARS--CoV-2 RNA Genome Sequencing Consortium | Arindam Maitra et al |
| EPI_ISL_511914 | India/UT-AR48/2020 | 2020-06-01 | Asia/India/Uttarakhand/Tehri Garhwal | All india institute of Medical Sciences Rishikesh | National Institute of Biomedical Genomics - DBT's PAN-INDIA 1000 SARS--CoV-2 RNA Genome Sequencing Consortium | Arindam Maitra et al |

| | | | | | | |
|---|---|---|---|---|---|---|
| EPI_ISL_511921 | India/UT-AR57/2020 | 2020-06-02 | Asia/India/Uttarakhand/Tehri Garhwal | All india institute of Medical Sciences Rishikesh | National Institute of Biomedical Genomics - DBT's PAN-INDIA 1000 SARS--CoV-2 RNA Genome Sequencing Consortium | Arindam Maitra et al |
| EPI_ISL_511922 | India/UT-AR64/2020 | 2020-06-09 | Asia/India/Uttarakhand/Dehradun | All india institute of Medical Sciences Rishikesh | National Institute of Biomedical Genomics - DBT's PAN-INDIA 1000 SARS--CoV-2 RNA Genome Sequencing Consortium | Arindam Maitra et al |
| EPI_ISL_511926 | India/MH-MW35/2020 | 2020-06-06 | Asia/India/Maharashtra/Wardha | Mahatma Gandhi Institute of Medical Sciences | National Institute of Biomedical Genomics - DBT's PAN-INDIA 1000 SARS--CoV-2 RNA Genome Sequencing Consortium | Arindam Maitra et al |
| EPI_ISL_511928 | India/HR-MW39/2020 | 2020-06-10 | Asia/India/Haryana/Gurugram | Mahatma Gandhi Institute of Medical Sciences | National Institute of Biomedical Genomics - DBT's PAN-INDIA 1000 SARS--CoV-2 RNA Genome Sequencing Consortium | Arindam Maitra et al |
| EPI_ISL_511930 | India/MH-GA18/2020 | 2020-06-17 | Asia/India/Maharashtra/Aurangabad | Government Medical College | National Institute of Biomedical Genomics - DBT's PAN-INDIA 1000 SARS--CoV-2 RNA Genome Sequencing Consortium | Arindam Maitra et al |

| | | | | | | |
|---|---|---|---|---|---|---|
| EPI_ISL_511931 | India/MH-GA30/2020 | 2020-06-17 | Asia/India/Maharashtra/Aurangabad | Government Medical College | National Institute of Biomedical Genomics - DBT's PAN-INDIA 1000 SARS--CoV-2 RNA Genome Sequencing Consortium | Arindam Maitra et al |
| EPI_ISL_511934 | India/MH-GA72/2020 | 2020-06-18 | Asia/India/Maharashtra/Aurangabad | Government Medical College | National Institute of Biomedical Genomics - DBT's PAN-INDIA 1000 SARS--CoV-2 RNA Genome Sequencing Consortium | Arindam Maitra et al |
| EPI_ISL_511935 | India/MH-GA90/2020 | 2020-06-16 | Asia/India/Maharashtra/Aurangabad | Government Medical College | National Institute of Biomedical Genomics - DBT's PAN-INDIA 1000 SARS--CoV-2 RNA Genome Sequencing Consortium | Arindam Maitra et al |
| EPI_ISL_511937 | India/MH-GA92/2020 | 2020-06-17 | Asia/India/Maharashtra/Aurangabad | Government Medical College | National Institute of Biomedical Genomics - DBT's PAN-INDIA 1000 SARS--CoV-2 RNA Genome Sequencing Consortium | Arindam Maitra et al |
| EPI_ISL_511938 | India/MH-GA94/2020 | 2020-06-18 | Asia/India/Maharashtra/Aurangabad | Government Medical College | National Institute of Biomedical Genomics - DBT's PAN-INDIA 1000 SARS--CoV-2 RNA Genome Sequencing Consortium | Arindam Maitra et al |

| | | | | | | |
|---|---|---|---|---|---|---|
| EPI_ISL_511939 | India/MH-GA95/2020 | 2020-06-18 | Asia/India/Maharashtra/Aurangabad | Government Medical College | National Institute of Biomedical Genomics - DBT's PAN-INDIA 1000 SARS--CoV-2 RNA Genome Sequencing Consortium | Arindam Maitra et al |
| EPI_ISL_511941 | India/MH-GA100/2020 | 2020-06-17 | Asia/India/Maharashtra/Aurangabad | Government Medical College | National Institute of Biomedical Genomics - DBT's PAN-INDIA 1000 SARS--CoV-2 RNA Genome Sequencing Consortium | Arindam Maitra et al |
| EPI_ISL_511949 | India/KA-IB59/2020 | 2020-05-19 | Asia/India/Karnataka/Haveri | Indian Institute of Science | National Institute of Biomedical Genomics - DBT's PAN-INDIA 1000 SARS--CoV-2 RNA Genome Sequencing Consortium | Arindam Maitra et al |
| EPI_ISL_511976 | England/BRIS-13259C/2020 | 2020-06-10 | Europe/United Kingdom/England/ | PHE South West Regional Laboratory, National Infection Service | Wellcome Sanger Institute for the COVID-19 Genomics UK (COG-UK) consortium | Stephanie Hutchings et al |
| EPI_ISL_511997 | Switzerland/ZH-ETHZ-220010/2020 | 2020-07-24 | Europe/Switzerland/Zvºrich/ | Viollier AG | Department of Biosystems Science and Engineering, ETH Zvºrich | Christian Beisel et al |
| EPI_ISL_512014 | Switzerland/AG-ETHZ-220032/2020 | 2020-07-29 | Europe/Switzerland/Aargau/ | Viollier AG | Department of Biosystems Science and Engineering, ETH Zvºrich | Christian Beisel et al |

| | | | | | | |
|---|---|---|---|---|---|---|
| EPI_ISL_512060 | India/GJ-GBRC326/2020 | 2020-06-25 | Asia/India/Gujarat/Ahmedabad | B.J. Medical College and Civil hospital, Ahmedabad | Gujarat Biotechnology Research Centre | Maharshi Pandya et al |
| EPI_ISL_512063 | India/GJ-GBRC328/2020 | 2020-06-25 | Asia/India/Gujarat/Ahmedabad | B.J. Medical College and Civil hospital, Ahmedabad | Gujarat Biotechnology Research Centre | Raghawendra Kumar et al |
| EPI_ISL_512066 | India/GJ-GBRC331/2020 | 2020-07-27 | Asia/India/Gujarat/Ahmedabad | Sardar Vallabhbhai Patel Institute of Medical Sciences & Research | Gujarat Biotechnology Research Centre | Komal Patel et al |
| EPI_ISL_512067 | India/GJ-GBRC332a/2020 | 2020-07-27 | Asia/India/Gujarat/Ahmedabad | Sardar Vallabhbhai Patel Institute of Medical Sciences & Research | Gujarat Biotechnology Research Centre | Labdhi Pandya et al |
| EPI_ISL_512068 | India/GJ-GBRC332b/2020 | 2020-07-27 | Asia/India/Gujarat/Ahmedabad | Sardar Vallabhbhai Patel Institute of Medical Sciences & Research | Gujarat Biotechnology Research Centre | Afzal Ansari et al |
| EPI_ISL_512071 | India/GJ-GBRC334b/2020 | 2020-06-12 | Asia/India/Gujarat/Adajan | Department of MicroBiology, Government Medical College, Surat | Gujarat Biotechnology Research Centre | Summaiya Mullan et al |
| EPI_ISL_512072 | India/GJ-GBRC335a/2020 | 2020-07-06 | Asia/India/Gujarat/Mehsana | Saikrishna Hospital, Mehsana | Gujarat Biotechnology Research Centre | Harshadbhai Parmar et al |
| EPI_ISL_512073 | India/GJ-GBRC335b/2020 | 2020-07-06 | Asia/India/Gujarat/Mehsana | Saikrishna Hospital, Mehsana | Gujarat Biotechnology Research Centre | Apurvasinh Puvar et al |
| EPI_ISL_512074 | India/GJ-GBRC336/2020 | 2020-07-06 | Asia/India/Gujarat/Mehsana | Saikrishna Hospital, Mehsana | Gujarat Biotechnology Research Centre | Janvi Raval et al |
| EPI_ISL_512075 | India/GJ-GBRC338a/2020 | 2020-07-09 | Asia/India/Gujarat/Modasa | Dr. RSS Hospital, Modasa | Gujarat Biotechnology Research Centre | Monika Gandhi et al |
| EPI_ISL_512076 | India/GJ-GBRC338b/2020 | 2020-07-09 | Asia/India/Gujarat/Modasa | Dr. RSS Hospital, Modasa | Gujarat Biotechnology Research Centre | Pinal Trivedi et al |
| EPI_ISL_512077 | India/GJ-GBRC-339/2020 | 2020-07-09 | Asia/India/Gujarat/Modasa | Dr. RSS Hospital, Modasa | Gujarat Biotechnology Research Centre | Maharshi Pandya et al |
| EPI_ISL_512136 | USA/AK-PHL427/2020 | 2020-06-05 | North America/USA/Alaska/ | Alaska State Virology Laboratory | Alaska State Virology Laboratory | Chen J et al with Pathogenomics group Dagdag R et al |

| | | | | | | |
|---|---|---|---|---|---|---|
| EPI_ISL_512148 | USA/AK-PHL575/2020 | 2020-06-22 | North America/USA/Alaska/ | Alaska State Virology Laboratory | Alaska State Virology Laboratory | Chen J et al with Pathogenomics group Dagdag R et al |
| EPI_ISL_512152 | USA/AK-PHL587/2020 | 2020-06-22 | North America/USA/Alaska/ | Alaska State Virology Laboratory | Alaska State Virology Laboratory | Chen J et al with Pathogenomics group Dagdag R et al |
| EPI_ISL_512153 | USA/AK-PHL588/2020 | 2020-06-23 | North America/USA/Alaska/ | Alaska State Virology Laboratory | Alaska State Virology Laboratory | Chen J et al with Pathogenomics group Dagdag R et al |
| EPI_ISL_512158 | USA/CA-ALSR-1735/2020 | 2020-05-18 | North America/USA/California/San Dieg | San Diego County Public Health Laboratory | Andersen lab at Scripps Research | SEARCH Alliance San Diego with Tracy Basler et al |
| EPI_ISL_512161 | USA/CA-ALSR-1740/2020 | 2020-05-19 | North America/USA/California/San Dieg | San Diego County Public Health Laboratory | Andersen lab at Scripps Research | SEARCH Alliance San Diego with Tracy Basler et al |
| EPI_ISL_512166 | USA/CA-ALSR-1753/2020 | 2020-05-18 | North America/USA/California/San Dieg | San Diego County Public Health Laboratory | Andersen lab at Scripps Research | SEARCH Alliance San Diego with Tracy Basler et al |
| EPI_ISL_512178 | USA/CA-ALSR-1779/2020 | 2020-05-13 | North America/USA/California/San Dieg | San Diego County Public Health Laboratory | Andersen lab at Scripps Research | SEARCH Alliance San Diego with Tracy Basler et al |
| EPI_ISL_512182 | USA/CA-ALSR-1784/2020 | 2020-05-12 | North America/USA/California/San Dieg | San Diego County Public Health Laboratory | Andersen lab at Scripps Research | SEARCH Alliance San Diego with Tracy Basler et al |
| EPI_ISL_512187 | USA/CA-ALSR-1791/2020 | 2020-05-13 | North America/USA/California/San Dieg | San Diego County Public Health Laboratory | Andersen lab at Scripps Research | SEARCH Alliance San Diego with Tracy Basler et al |
| EPI_ISL_512191 | USA/CA-ALSR-1800/2020 | 2020-05-11 | North America/USA/California/San Dieg | San Diego County Public Health Laboratory | Andersen lab at Scripps Research | SEARCH Alliance San Diego with Tracy Basler et al |
| EPI_ISL_512196 | USA/CA-ALSR-1818/2020 | 2020-05-20 | North America/USA/California/San Dieg | San Diego County Public Health Laboratory | Andersen lab at Scripps Research | SEARCH Alliance San Diego with Tracy Basler et al |
| EPI_ISL_512211 | USA/CA-ALSR-1862/2020 | 2020-05-19 | North America/USA/California/San Dieg | San Diego County Public Health Laboratory | Andersen lab at Scripps Research | SEARCH Alliance San Diego with Tracy Basler et al |
| EPI_ISL_512213 | USA/CA-ALSR-1869/2020 | 2020-05-19 | North America/USA/California/San Dieg | San Diego County Public Health Laboratory | Andersen lab at Scripps Research | SEARCH Alliance San Diego with Tracy Basler et al |
| EPI_ISL_512214 | USA/CA-ALSR-1876/2020 | 2020-05-20 | North America/USA/California/San Dieg | San Diego County Public Health Laboratory | Andersen lab at Scripps Research | SEARCH Alliance San Diego with Tracy Basler et al |
| EPI_ISL_512225 | USA/CA-ALSR-1903/2020 | 2020-04-25 | North America/USA/California/San Dieg | San Diego County Public Health Laboratory | Andersen lab at Scripps Research | SEARCH Alliance San Diego with Tracy Basler et al |
| EPI_ISL_512236 | USA/CA-ALSR-1925/2020 | 2020-07-08 | North America/USA/California/San Dieg | San Diego County Public Health Laboratory | Andersen lab at Scripps Research | SEARCH Alliance San Diego with Tracy Basler et al |
| EPI_ISL_512238 | USA/CA-ALSR-1927/2020 | 2020-07-08 | North America/USA/California/San Dieg | San Diego County Public Health Laboratory | Andersen lab at Scripps Research | SEARCH Alliance San Diego with Tracy Basler et al |
| EPI_ISL_512245 | USA/CA-ALSR-1939/2020 | 2020-07-08 | North America/USA/California/San Dieg | San Diego County Public Health Laboratory | Andersen lab at Scripps Research | SEARCH Alliance San Diego with Tracy Basler et al |

| | | | | | | |
|---|---|---|---|---|---|---|
| EPI_ISL_512273 | USA/CA-ALSR-1975/2020 | 2020-04-22 | North America/USA/California/San Dieg | San Diego County Public Health Laboratory | Andersen lab at Scripps Research | SEARCH Alliance San Diego with Tracy Basler et al |
| EPI_ISL_512296 | Poland/1132916/2020 | 2020-05-08 | Europe/Poland/Pomorskie/ | Hematology Laboratory, Section of Molecular Diagnostics, University Clinical Centre, Medical University of Gdansk | Department of Virology, Faculty of Medicine, University of Helsinki, Helsinki, Finland | Maciej Grzybek et al |
| EPI_ISL_512297 | Poland/1134056/2020 | 2020-05-09 | Europe/Poland/Pomorskie/ | Hematology Laboratory, Section of Molecular Diagnostics, University Clinical Centre, Medical University of Gdansk | Department of Virology, Faculty of Medicine, University of Helsinki, Helsinki, Finland | Maciej Grzybek et al |
| EPI_ISL_512298 | Poland/1140683/2020 | 2020-05-19 | Europe/Poland/Pomorskie/ | Hematology Laboratory, Section of Molecular Diagnostics, University Clinical Centre, Medical University of Gdansk | Department of Virology, Faculty of Medicine, University of Helsinki, Helsinki, Finland | Maciej Grzybek et al |
| EPI_ISL_512299 | Poland/1154942/2020 | 2020-06-07 | Europe/Poland/Pomorskie/ | Hematology Laboratory, Section of Molecular Diagnostics, University Clinical Centre, Medical University of Gdansk | Department of Virology, Faculty of Medicine, University of Helsinki, Helsinki, Finland | Maciej Grzybek et al |
| EPI_ISL_512301 | Poland/1156085/2020 | 2020-06-08 | Europe/Poland/Pomorskie/ | Hematology Laboratory, Section of Molecular Diagnostics, University Clinical Centre, Medical University of Gdansk | Department of Virology, Faculty of Medicine, University of Helsinki, Helsinki, Finland | Maciej Grzybek et al |
| EPI_ISL_512302 | Poland/1157220/2020 | 2020-06-09 | Europe/Poland/Pomorskie/ | Hematology Laboratory, Section of Molecular Diagnostics, University Clinical Centre, Medical University of Gdansk | Department of Virology, Faculty of Medicine, University of Helsinki, Helsinki, Finland | Maciej Grzybek et al |

| | | | | | | |
|---|---|---|---|---|---|---|
| EPI_ISL_512396 | England/PORT-2D14DD/2020 | 2020-03-31 | Europe/United Kingdom/England/ | Centre for Enzyme Innovation, University of Portsmouth / Translational Research Laboratory, Portsmouth Hospitals NHS Trust | COVID-19 Genomics UK (COG-UK) Consortium | Angela Beckett et al |
| EPI_ISL_512403 | England/PORT-2D15BC/2020 | 2020-04-20 | Europe/United Kingdom/England/ | Centre for Enzyme Innovation, University of Portsmouth / Translational Research Laboratory, Portsmouth Hospitals NHS Trust | COVID-19 Genomics UK (COG-UK) Consortium | Angela Beckett et al |
| EPI_ISL_512435 | England/PORT-2F5E02/2020 | 2020-05-04 | Europe/United Kingdom/England/ | Centre for Enzyme Innovation, University of Portsmouth / Translational Research Laboratory, Portsmouth Hospitals NHS Trust | COVID-19 Genomics UK (COG-UK) Consortium | Angela Beckett et al |
| EPI_ISL_512506 | Wales/PHWC-16974B/2020 | 2020-07-20 | Europe/United Kingdom/Wales/ | Wales Specialist Virology Centre Sequencing lab: Pathogen Genomics Unit | COVID-19 Genomics UK (COG-UK) Consortium | Catherine Moore et al |
| EPI_ISL_512559 | USA/FL-BPHL-0753/2020 | 2020-06-22 | North America/USA/Florida/ | Florida Bureau of Public Health Laboratories | Florida Bureau of Public Health Laboratories | Sarah Schmedes et al |
| EPI_ISL_512579 | USA/FL-BPHL-0775/2020 | 2020-06-29 | North America/USA/Florida/ | Florida Bureau of Public Health Laboratories | Florida Bureau of Public Health Laboratories | Sarah Schmedes et al |
| EPI_ISL_512597 | Ukraine/203100317/2020 | 2020-06-23 | Europe/Ukraine/Ukraine/ | National Laboratory for Influenza/Virology reference laboratory, Public Health Center of the Ministry of Health of Ukraine | Respiratory Virus Unit, Microbiology Services Colindale, Public Health England | PHE Covid Sequencing Team et al |
| EPI_ISL_512598 | Ukraine/203100318/2020 | 2020-06-23 | Europe/Ukraine/Ukraine/ | National Laboratory for Influenza/Virology reference laboratory, Public Health Center of the Ministry of Health of Ukraine | Respiratory Virus Unit, Microbiology Services Colindale, Public Health England | PHE Covid Sequencing Team et al |
| EPI_ISL_512599 | Ukraine/203100319/2020 | 2020-06-23 | Europe/Ukraine/Ukraine/ | National Laboratory for Influenza/Virology reference laboratory, Public Health Center of the Ministry of Health of Ukraine | Respiratory Virus Unit, Microbiology Services Colindale, Public Health England | PHE Covid Sequencing Team et al |

| | | | | | | |
|---|---|---|---|---|---|---|
| EPI_ISL_512600 | Ukraine/203100320/2020 | 2020-06-23 | Europe/Ukraine/Ukraine/ | National Laboratory for Influenza/Virology reference laboratory, Public Health Center of the Ministry of Health of Ukraine | Respiratory Virus Unit, Microbiology Services Colindale, Public Health England | PHE Covid Sequencing Team et al |
| EPI_ISL_512601 | Ukraine/203100321/2020 | 2020-06-23 | Europe/Ukraine/Ukraine/ | National Laboratory for Influenza/Virology reference laboratory, Public Health Center of the Ministry of Health of Ukraine | Respiratory Virus Unit, Microbiology Services Colindale, Public Health England | PHE Covid Sequencing Team et al |
| EPI_ISL_512602 | Ukraine/203100322/2020 | 2020-05-26 | Europe/Ukraine/Ukraine/ | National Laboratory for Influenza/Virology reference laboratory, Public Health Center of the Ministry of Health of Ukraine | Respiratory Virus Unit, Microbiology Services Colindale, Public Health England | PHE Covid Sequencing Team et al |
| EPI_ISL_512603 | Ukraine/203100323/2020 | 2020-05-27 | Europe/Ukraine/Ukraine/ | National Laboratory for Influenza/Virology reference laboratory, Public Health Center of the Ministry of Health of Ukraine | Respiratory Virus Unit, Microbiology Services Colindale, Public Health England | PHE Covid Sequencing Team et al |
| EPI_ISL_512604 | Ukraine/203100324/2020 | 2020-05-27 | Europe/Ukraine/Ukraine/ | National Laboratory for Influenza/Virology reference laboratory, Public Health Center of the Ministry of Health of Ukraine | Respiratory Virus Unit, Microbiology Services Colindale, Public Health England | PHE Covid Sequencing Team et al |
| EPI_ISL_512605 | Ukraine/203100325/2020 | 2020-05-27 | Europe/Ukraine/Ukraine/ | National Laboratory for Influenza/Virology reference laboratory, Public Health Center of the Ministry of Health of Ukraine | Respiratory Virus Unit, Microbiology Services Colindale, Public Health England | PHE Covid Sequencing Team et al |

| | | | | | | |
|---|---|---|---|---|---|---|
| EPI_ISL_512607 | Ukraine/203100328/2020 | 2020-05-27 | Europe/Ukraine/Ukraine/ | National Laboratory for Influenza/Virology reference laboratory, Public Health Center of the Ministry of Health of Ukraine | Respiratory Virus Unit, Microbiology Services Colindale, Public Health England | PHE Covid Sequencing Team et al |
| EPI_ISL_512608 | Ukraine/203100329/2020 | 2020-05-27 | Europe/Ukraine/Ukraine/ | National Laboratory for Influenza/Virology reference laboratory, Public Health Center of the Ministry of Health of Ukraine | Respiratory Virus Unit, Microbiology Services Colindale, Public Health England | PHE Covid Sequencing Team et al |
| EPI_ISL_512609 | Ukraine/203100330/2020 | 2020-05-27 | Europe/Ukraine/Ukraine/ | National Laboratory for Influenza/Virology reference laboratory, Public Health Center of the Ministry of Health of Ukraine | Respiratory Virus Unit, Microbiology Services Colindale, Public Health England | PHE Covid Sequencing Team et al |
| EPI_ISL_512610 | Ukraine/203100331/2020 | 2020-05-27 | Europe/Ukraine/Ukraine/ | National Laboratory for Influenza/Virology reference laboratory, Public Health Center of the Ministry of Health of Ukraine | Respiratory Virus Unit, Microbiology Services Colindale, Public Health England | PHE Covid Sequencing Team et al |
| EPI_ISL_512611 | Ukraine/203100332/2020 | 2020-05-27 | Europe/Ukraine/Ukraine/ | National Laboratory for Influenza/Virology reference laboratory, Public Health Center of the Ministry of Health of Ukraine | Respiratory Virus Unit, Microbiology Services Colindale, Public Health England | PHE Covid Sequencing Team et al |
| EPI_ISL_512612 | Ukraine/203100333/2020 | 2020-06-24 | Europe/Ukraine/Ukraine/ | National Laboratory for Influenza/Virology reference laboratory, Public Health Center of the Ministry of Health of Ukraine | Respiratory Virus Unit, Microbiology Services Colindale, Public Health England | PHE Covid Sequencing Team et al |

| | | | | | | |
|---|---|---|---|---|---|---|
| EPI_ISL_512613 | Ukraine/203100334/2020 | 2020-06-26 | Europe/Ukraine/Ukraine/ | National Laboratory for Influenza/Virology reference laboratory, Public Health Center of the Ministry of Health of Ukraine | Respiratory Virus Unit, Microbiology Services Colindale, Public Health England | PHE Covid Sequencing Team et al |
| EPI_ISL_512614 | Ukraine/203100335/2020 | 2020-06-26 | Europe/Ukraine/Ukraine/ | National Laboratory for Influenza/Virology reference laboratory, Public Health Center of the Ministry of Health of Ukraine | Respiratory Virus Unit, Microbiology Services Colindale, Public Health England | PHE Covid Sequencing Team et al |
| EPI_ISL_512615 | Ukraine/203100336/2020 | 2020-06-23 | Europe/Ukraine/Ukraine/ | National Laboratory for Influenza/Virology reference laboratory, Public Health Center of the Ministry of Health of Ukraine | Respiratory Virus Unit, Microbiology Services Colindale, Public Health England | PHE Covid Sequencing Team et al |
| EPI_ISL_512616 | Ukraine/203100337/2020 | 2020-07-10 | Europe/Ukraine/Ukraine/ | National Laboratory for Influenza/Virology reference laboratory, Public Health Center of the Ministry of Health of Ukraine | Respiratory Virus Unit, Microbiology Services Colindale, Public Health England | PHE Covid Sequencing Team et al |
| EPI_ISL_512617 | Ukraine/203100338/2020 | 2020-07-10 | Europe/Ukraine/Ukraine/ | National Laboratory for Influenza/Virology reference laboratory, Public Health Center of the Ministry of Health of Ukraine | Respiratory Virus Unit, Microbiology Services Colindale, Public Health England | PHE Covid Sequencing Team et al |
| EPI_ISL_512618 | Ukraine/203100339/2020 | 2020-07-11 | Europe/Ukraine/Ukraine/ | National Laboratory for Influenza/Virology reference laboratory, Public Health Center of the Ministry of Health of Ukraine | Respiratory Virus Unit, Microbiology Services Colindale, Public Health England | PHE Covid Sequencing Team et al |

| | | | | | | |
|---|---|---|---|---|---|---|
| EPI_ISL_512621 | Ukraine/203100342/2020 | 2020-05-27 | Europe/Ukraine/Ukraine/ | National Laboratory for Influenza/Virology reference laboratory, Public Health Center of the Ministry of Health of Ukraine | Respiratory Virus Unit, Microbiology Services Colindale, Public Health England | PHE Covid Sequencing Team et al |
| EPI_ISL_512622 | Ukraine/203100343/2020 | 2020-05-27 | Europe/Ukraine/Ukraine/ | National Laboratory for Influenza/Virology reference laboratory, Public Health Center of the Ministry of Health of Ukraine | Respiratory Virus Unit, Microbiology Services Colindale, Public Health England | PHE Covid Sequencing Team et al |
| EPI_ISL_512624 | Ukraine/203100345/2020 | 2020-05-27 | Europe/Ukraine/Ukraine/ | National Laboratory for Influenza/Virology reference laboratory, Public Health Center of the Ministry of Health of Ukraine | Respiratory Virus Unit, Microbiology Services Colindale, Public Health England | PHE Covid Sequencing Team et al |
| EPI_ISL_512626 | Ukraine/203100347/2020 | 2020-05-16 | Europe/Ukraine/Ukraine/ | National Laboratory for Influenza/Virology reference laboratory, Public Health Center of the Ministry of Health of Ukraine | Respiratory Virus Unit, Microbiology Services Colindale, Public Health England | PHE Covid Sequencing Team et al |
| EPI_ISL_512627 | Ukraine/203100348/2020 | 2020-05-16 | Europe/Ukraine/Ukraine/ | National Laboratory for Influenza/Virology reference laboratory, Public Health Center of the Ministry of Health of Ukraine | Respiratory Virus Unit, Microbiology Services Colindale, Public Health England | PHE Covid Sequencing Team et al |
| EPI_ISL_512628 | Ukraine/203100349/2020 | 2020-06-23 | Europe/Ukraine/Ukraine/ | National Laboratory for Influenza/Virology reference laboratory, Public Health Center of the Ministry of Health of Ukraine | Respiratory Virus Unit, Microbiology Services Colindale, Public Health England | PHE Covid Sequencing Team et al |

| | | | | | | |
|---|---|---|---|---|---|---|
| EPI_ISL_512629 | Ukraine/203100350/2020 | 2020-06-24 | Europe/Ukraine/Ukraine/ | National Laboratory for Influenza/Virology reference laboratory, Public Health Center of the Ministry of Health of Ukraine | Respiratory Virus Unit, Microbiology Services Colindale, Public Health England | PHE Covid Sequencing Team et al |
| EPI_ISL_512630 | Ukraine/203100351/2020 | 2020-06-24 | Europe/Ukraine/Ukraine/ | National Laboratory for Influenza/Virology reference laboratory, Public Health Center of the Ministry of Health of Ukraine | Respiratory Virus Unit, Microbiology Services Colindale, Public Health England | PHE Covid Sequencing Team et al |
| EPI_ISL_512631 | Ukraine/203100352/2020 | 2020-06-24 | Europe/Ukraine/Ukraine/ | National Laboratory for Influenza/Virology reference laboratory, Public Health Center of the Ministry of Health of Ukraine | Respiratory Virus Unit, Microbiology Services Colindale, Public Health England | PHE Covid Sequencing Team et al |
| EPI_ISL_512632 | Ukraine/203100353/2020 | 2020-06-24 | Europe/Ukraine/Ukraine/ | National Laboratory for Influenza/Virology reference laboratory, Public Health Center of the Ministry of Health of Ukraine | Respiratory Virus Unit, Microbiology Services Colindale, Public Health England | PHE Covid Sequencing Team et al |
| EPI_ISL_512635 | Ukraine/203100356/2020 | 2020-05-28 | Europe/Ukraine/Ukraine/ | National Laboratory for Influenza/Virology reference laboratory, Public Health Center of the Ministry of Health of Ukraine | Respiratory Virus Unit, Microbiology Services Colindale, Public Health England | PHE Covid Sequencing Team et al |
| EPI_ISL_512636 | Ukraine/203100357/2020 | 2020-05-08 | Europe/Ukraine/Ukraine/ | National Laboratory for Influenza/Virology reference laboratory, Public Health Center of the Ministry of Health of Ukraine | Respiratory Virus Unit, Microbiology Services Colindale, Public Health England | PHE Covid Sequencing Team et al |

| | | | | | | |
|---|---|---|---|---|---|---|
| EPI_ISL_512639 | Ukraine/203100360/2020 | 2020-04-24 | Europe/Ukraine/Ukraine/ | National Laboratory for Influenza/Virology reference laboratory, Public Health Center of the Ministry of Health of Ukraine | Respiratory Virus Unit, Microbiology Services Colindale, Public Health England | PHE Covid Sequencing Team et al |
| EPI_ISL_512640 | Ukraine/203100361/2020 | 2020-04-24 | Europe/Ukraine/Ukraine/ | National Laboratory for Influenza/Virology reference laboratory, Public Health Center of the Ministry of Health of Ukraine | Respiratory Virus Unit, Microbiology Services Colindale, Public Health England | PHE Covid Sequencing Team et al |
| EPI_ISL_512641 | Ukraine/203100363/2020 | 2020-05-16 | Europe/Ukraine/Ukraine/ | National Laboratory for Influenza/Virology reference laboratory, Public Health Center of the Ministry of Health of Ukraine | Respiratory Virus Unit, Microbiology Services Colindale, Public Health England | PHE Covid Sequencing Team et al |
| EPI_ISL_512642 | Ukraine/203100364/2020 | 2020-05-15 | Europe/Ukraine/Ukraine/ | National Laboratory for Influenza/Virology reference laboratory, Public Health Center of the Ministry of Health of Ukraine | Respiratory Virus Unit, Microbiology Services Colindale, Public Health England | PHE Covid Sequencing Team et al |
| EPI_ISL_512643 | Ukraine/203100365/2020 | 2020-05-15 | Europe/Ukraine/Ukraine/ | National Laboratory for Influenza/Virology reference laboratory, Public Health Center of the Ministry of Health of Ukraine | Respiratory Virus Unit, Microbiology Services Colindale, Public Health England | PHE Covid Sequencing Team et al |
| EPI_ISL_512644 | Ukraine/203100366/2020 | 2020-05-16 | Europe/Ukraine/Ukraine/ | National Laboratory for Influenza/Virology reference laboratory, Public Health Center of the Ministry of Health of Ukraine | Respiratory Virus Unit, Microbiology Services Colindale, Public Health England | PHE Covid Sequencing Team et al |
| EPI_ISL_512670 | CostaRica/INC-0064/2020 | 2020-06-19 | North America/Costa Rica/Alajuela/Gre | Centro Nacional De Rehabilitacion Humberto Araya Rojas (Cenare) | Inciensa, Instituto Costarricense de Investigaci√≥n y Ense√±anza en Nutrici√≥n y Salud | Francisco Duarte et al |

| | | | | | | |
|---|---|---|---|---|---|---|
| EPI_ISL_512677 | USA/FL-BPHL-0772/2020 | 2020-06-29 | North America/USA/Florida/ | Florida Bureau of Public Health Laboratories | Florida Bureau of Public Health Laboratories | Sarah Schmedes et al |
| EPI_ISL_512737 | Australia/WA84/2020 | 2020-05-11 | Oceania/Australia/Western Australia/ | PathWest Laboratory Medicine WA | PathWest Laboratory Medicine WA Microbial Surveillance Unit | PathWest Laboratory Medicine WA Microbial Surveillance Unit et al |
| EPI_ISL_512774 | Bangladesh/BCSIR-NILMRC-254/2020 | 2020-07-07 | Asia/Bangladesh/Khulna/Jashore | Genomic Research Lab, BCSIR | Bangladesh Council of Scientific and Industrial Research | Abu Sayeed Mohammad Mahmud et al |
| EPI_ISL_512786 | USA/FL-USAFSAM-S010/2020 | 2020-03-24 | North America/USA/Florida/ | Public Health, United States Air Force School of Aerospace Medicine | Public Health, United States Air Force School of Aerospace Medicine | Fries et al |
| EPI_ISL_512789 | USA/WY-USAFSAM-S015/2020 | 2020-03-26 | North America/USA/Wyoming/ | Public Health, United States Air Force School of Aerospace Medicine | Public Health, United States Air Force School of Aerospace Medicine | Fries et al |
| EPI_ISL_512796 | USA/MS-USAFSAM-S022/2020 | 2020-03-24 | North America/USA/Mississippi/ | Public Health, United States Air Force School of Aerospace Medicine | Public Health, United States Air Force School of Aerospace Medicine | Fries et al |
| EPI_ISL_512810 | Ukraine/203100362/2020 | 2020-05-15 | Europe/Ukraine/Ukraine/ | National Laboratory for Influenza/Virology reference laboratory, Public Health Center of the Ministry of Health of Ukraine | Respiratory Virus Unit, Microbiology Services Colindale, Public Health England | PHE Covid Sequencing Team et al |
| EPI_ISL_512823 | Singapore/787/2020 | 2020-04-06 | Asia/Singapore/Singapore/ | National Public Health Laboratory, National Centre for Infectious Diseases | National Public Health Laboratory, National Centre for Infectious Diseases | Mak TM et al |

| | | | | | | |
|---|---|---|---|---|---|---|
| EPI_ISL_512825 | Singapore/789/2020 | 2020-04-06 | Asia/Singapore/Singapore/ | National Public Health Laboratory, National Centre for Infectious Diseases | National Public Health Laboratory, National Centre for Infectious Diseases | Mak TM et al |
| EPI_ISL_512829 | Singapore/793/2020 | 2020-06-04 | Asia/Singapore/Singapore/ | National Public Health Laboratory, National Centre for Infectious Diseases | National Public Health Laboratory, National Centre for Infectious Diseases | Mak TM et al |
| EPI_ISL_512836 | Singapore/800/2020 | 2020-07-17 | Asia/Singapore/Singapore/ | National Public Health Laboratory, National Centre for Infectious Diseases | National Public Health Laboratory, National Centre for Infectious Diseases | Mak TM et al |
| EPI_ISL_512847 | Thailand/Bangkok-CONI-0197/2020 | 2020-03-24 | Asia/Thailand/Bangkok/ | Ramathibodi Hospital | COVID-19 Network Investigations (CONI) Alliance | Elizabeth Batty et al |
| EPI_ISL_512874 | SaudiArabia/KAUST-JEDDAH1000/20 | 2020-06-23 | Asia/Saudi Arabia/Jeddah/ | Pathogen Genomics Lab King Abdullah University of Science and Technology(KAUST) | Pathogen Genomics Lab King Abdullah University of Science and Technology(KAUST ) | Raeece Naeem et al |
| EPI_ISL_512875 | SaudiArabia/KAUST-JEDDAH1001/20 | 2020-06-23 | Asia/Saudi Arabia/Jeddah/ | Pathogen Genomics Lab King Abdullah University of Science and Technology(KAUST) | Pathogen Genomics Lab King Abdullah University of Science and Technology(KAUST ) | Raeece Naeem et al |

| | | | | | | |
|---|---|---|---|---|---|---|
| EPI_ISL_512878 | SaudiArabia/KAUST-JEDDAH1007/20 | 2020-06-17 | Asia/Saudi Arabia/Jeddah/ | Pathogen Genomics Lab King Abdullah University of Science and Technology(KAUST) | Pathogen Genomics Lab King Abdullah University of Science and Technology(KAUST) | Raeece Naeem et al |
| EPI_ISL_512882 | SaudiArabia/KAUST-JEDDAH1011/20 | 2020-06-23 | Asia/Saudi Arabia/Jeddah/ | Pathogen Genomics Lab King Abdullah University of Science and Technology(KAUST) | Pathogen Genomics Lab King Abdullah University of Science and Technology(KAUST) | Raeece Naeem et al |
| EPI_ISL_512883 | SaudiArabia/KAUST-JEDDAH1012/20 | 2020-06-23 | Asia/Saudi Arabia/Jeddah/ | Pathogen Genomics Lab King Abdullah University of Science and Technology(KAUST) | Pathogen Genomics Lab King Abdullah University of Science and Technology(KAUST) | Raeece Naeem et al |
| EPI_ISL_512885 | SaudiArabia/KAUST-JEDDAH1014/20 | 2020-06-17 | Asia/Saudi Arabia/Jeddah/ | Pathogen Genomics Lab King Abdullah University of Science and Technology(KAUST) | Pathogen Genomics Lab King Abdullah University of Science and Technology(KAUST) | Raeece Naeem et al |
| EPI_ISL_512892 | SaudiArabia/KAUST-JEDDAH1022/20 | 2020-06-23 | Asia/Saudi Arabia/Jeddah/ | Pathogen Genomics Lab King Abdullah University of Science and Technology(KAUST) | Pathogen Genomics Lab King Abdullah University of Science and Technology(KAUST) | Raeece Naeem et al |

| | | | | | | |
|---|---|---|---|---|---|---|
| EPI_ISL_512894 | SaudiArabia/KAUST-JEDDAH1024/20 | 2020-06-23 | Asia/Saudi Arabia/Jeddah/ | Pathogen Genomics Lab King Abdullah University of Science and Technology(KAUST) | Pathogen Genomics Lab King Abdullah University of Science and Technology(KAUST) | Raeece Naeem et al |
| EPI_ISL_512896 | SaudiArabia/KAUST-JEDDAH1026/20 | 2020-06-17 | Asia/Saudi Arabia/Jeddah/ | Pathogen Genomics Lab King Abdullah University of Science and Technology(KAUST) | Pathogen Genomics Lab King Abdullah University of Science and Technology(KAUST) | Raeece Naeem et al |
| EPI_ISL_512898 | SaudiArabia/KAUST-JEDDAH1028/20 | 2020-06-23 | Asia/Saudi Arabia/Jeddah/ | Pathogen Genomics Lab King Abdullah University of Science and Technology(KAUST) | Pathogen Genomics Lab King Abdullah University of Science and Technology(KAUST) | Raeece Naeem et al |
| EPI_ISL_512899 | SaudiArabia/KAUST-JEDDAH1030/20 | 2020-06-23 | Asia/Saudi Arabia/Jeddah/ | Pathogen Genomics Lab King Abdullah University of Science and Technology(KAUST) | Pathogen Genomics Lab King Abdullah University of Science and Technology(KAUST) | Raeece Naeem et al |
| EPI_ISL_512900 | SaudiArabia/KAUST-JEDDAH1031/20 | 2020-06-23 | Asia/Saudi Arabia/Jeddah/ | Pathogen Genomics Lab King Abdullah University of Science and Technology(KAUST) | Pathogen Genomics Lab King Abdullah University of Science and Technology(KAUST) | Raeece Naeem et al |

| | | | | | | |
|---|---|---|---|---|---|---|
| EPI_ISL_512902 | SaudiArabia/KAUST-JEDDAH1033/20 | 2020-06-23 | Asia/Saudi Arabia/Jeddah/ | Pathogen Genomics Lab King Abdullah University of Science and Technology(KAUST) | Pathogen Genomics Lab King Abdullah University of Science and Technology(KAUST) | Raeece Naeem et al |
| EPI_ISL_512903 | SaudiArabia/KAUST-JEDDAH1034/20 | 2020-06-17 | Asia/Saudi Arabia/Jeddah/ | Pathogen Genomics Lab King Abdullah University of Science and Technology(KAUST) | Pathogen Genomics Lab King Abdullah University of Science and Technology(KAUST) | Raeece Naeem et al |
| EPI_ISL_512907 | SaudiArabia/KAUST-JEDDAH1058/20 | 2020-06-23 | Asia/Saudi Arabia/Jeddah/ | Pathogen Genomics Lab King Abdullah University of Science and Technology(KAUST) | Pathogen Genomics Lab King Abdullah University of Science and Technology(KAUST) | Fathia Ben Rached et al |
| EPI_ISL_512912 | SaudiArabia/KAUST-JEDDAH323/202 | 2020-04-15 | Asia/Saudi Arabia/Jeddah/ | Pathogen Genomics Lab King Abdullah University of Science and Technology(KAUST) | Pathogen Genomics Lab King Abdullah University of Science and Technology(KAUST) | Sharif Hala et al |
| EPI_ISL_512915 | SaudiArabia/KAUST-JEDDAH330/202 | 2020-04-12 | Asia/Saudi Arabia/Jeddah/ | Pathogen Genomics Lab King Abdullah University of Science and Technology(KAUST) | Pathogen Genomics Lab King Abdullah University of Science and Technology(KAUST) | Sharif Hala et al |

| | | | | | | |
|---|---|---|---|---|---|---|
| EPI_ISL_512920 | SaudiArabia/KAUST-JEDDAH462/202 | 2020-08-04 | Asia/Saudi Arabia/Jeddah/ | Pathogen Genomics Lab King Abdullah University of Science and Technology(KAUST) | Pathogen Genomics Lab King Abdullah University of Science and Technology(KAUST) | Fadwa Alofi et al |
| EPI_ISL_512921 | SaudiArabia/KAUST-JEDDAH466/202 | 2020-08-04 | Asia/Saudi Arabia/Jeddah/ | Pathogen Genomics Lab King Abdullah University of Science and Technology(KAUST) | Pathogen Genomics Lab King Abdullah University of Science and Technology(KAUST) | Fadwa Alofi et al |
| EPI_ISL_512922 | SaudiArabia/KAUST-JEDDAH467/202 | 2020-08-04 | Asia/Saudi Arabia/Jeddah/ | Pathogen Genomics Lab King Abdullah University of Science and Technology(KAUST) | Pathogen Genomics Lab King Abdullah University of Science and Technology(KAUST) | Fadwa Alofi et al |
| EPI_ISL_512923 | SaudiArabia/KAUST-JEDDAH468/202 | 2020-08-04 | Asia/Saudi Arabia/Jeddah/ | Pathogen Genomics Lab King Abdullah University of Science and Technology(KAUST) | Pathogen Genomics Lab King Abdullah University of Science and Technology(KAUST) | Fadwa Alofi et al |
| EPI_ISL_512924 | SaudiArabia/KAUST-JEDDAH469/202 | 2020-08-04 | Asia/Saudi Arabia/Jeddah/ | Pathogen Genomics Lab King Abdullah University of Science and Technology(KAUST) | Pathogen Genomics Lab King Abdullah University of Science and Technology(KAUST) | Fadwa Alofi et al |

| | | | | | | |
|---|---|---|---|---|---|---|
| EPI_ISL_512925 | SaudiArabia/KAUST-JEDDAH470/202 | 2020-08-04 | Asia/Saudi Arabia/Jeddah/ | Pathogen Genomics Lab King Abdullah University of Science and Technology(KAUST) | Pathogen Genomics Lab King Abdullah University of Science and Technology(KAUST ) | Fadwa Alofi et al |
| EPI_ISL_512926 | SaudiArabia/KAUST-JEDDAH471/202 | 2020-08-04 | Asia/Saudi Arabia/Jeddah/ | Pathogen Genomics Lab King Abdullah University of Science and Technology(KAUST) | Pathogen Genomics Lab King Abdullah University of Science and Technology(KAUST ) | Fadwa Alofi et al |
| EPI_ISL_512927 | SaudiArabia/KAUST-JEDDAH473/202 | 2020-08-04 | Asia/Saudi Arabia/Jeddah/ | Pathogen Genomics Lab King Abdullah University of Science and Technology(KAUST) | Pathogen Genomics Lab King Abdullah University of Science and Technology(KAUST ) | Fadwa Alofi et al |
| EPI_ISL_512932 | SaudiArabia/KAUST-JEDDAH487/202 | 2020-04-06 | Asia/Saudi Arabia/Jeddah/ | Pathogen Genomics Lab King Abdullah University of Science and Technology(KAUST) | Pathogen Genomics Lab King Abdullah University of Science and Technology(KAUST ) | Fadwa Alofi et al |
| EPI_ISL_512939 | SaudiArabia/KAUST-JEDDAH497/202 | 2020-04-09 | Asia/Saudi Arabia/Jeddah/ | Pathogen Genomics Lab King Abdullah University of Science and Technology(KAUST) | Pathogen Genomics Lab King Abdullah University of Science and Technology(KAUST ) | Fadwa Alofi et al |

| | | | | | | |
|---|---|---|---|---|---|---|
| EPI_ISL_512959 | SaudiArabia/KAUST-JEDDAH558/202 | 2020-04-21 | Asia/Saudi Arabia/Jeddah/ | Pathogen Genomics Lab King Abdullah University of Science and Technology(KAUST) | Pathogen Genomics Lab King Abdullah University of Science and Technology(KAUST) | Sara Mfarrej et al |
| EPI_ISL_512960 | SaudiArabia/KAUST-JEDDAH559/202 | 2020-04-21 | Asia/Saudi Arabia/Jeddah/ | Pathogen Genomics Lab King Abdullah University of Science and Technology(KAUST) | Pathogen Genomics Lab King Abdullah University of Science and Technology(KAUST) | Sara Mfarrej et al |
| EPI_ISL_512969 | SaudiArabia/KAUST-JEDDAH576/202 | 2020-04-30 | Asia/Saudi Arabia/Jeddah/ | Pathogen Genomics Lab King Abdullah University of Science and Technology(KAUST) | Pathogen Genomics Lab King Abdullah University of Science and Technology(KAUST) | Sara Mfarrej et al |
| EPI_ISL_512985 | SaudiArabia/KAUST-JEDDAH611/202 | 2020-04-30 | Asia/Saudi Arabia/Jeddah/ | Pathogen Genomics Lab King Abdullah University of Science and Technology(KAUST) | Pathogen Genomics Lab King Abdullah University of Science and Technology(KAUST) | Sara Mfarrej et al |
| EPI_ISL_512986 | SaudiArabia/KAUST-JEDDAH613/202 | 2020-04-30 | Asia/Saudi Arabia/Jeddah/ | Pathogen Genomics Lab King Abdullah University of Science and Technology(KAUST) | Pathogen Genomics Lab King Abdullah University of Science and Technology(KAUST) | Sara Mfarrej et al |

| | | | | | | |
|---|---|---|---|---|---|---|
| EPI_ISL_512990 | SaudiArabia/KAUST-JEDDAH721/202 | 2020-05-04 | Asia/Saudi Arabia/Jeddah/ | Pathogen Genomics Lab King Abdullah University of Science and Technology(KAUST) | Pathogen Genomics Lab King Abdullah University of Science and Technology(KAUST ) | Amit Kumar Subudhi et al |
| EPI_ISL_512991 | SaudiArabia/KAUST-JEDDAH722/202 | 2020-05-04 | Asia/Saudi Arabia/Jeddah/ | Pathogen Genomics Lab King Abdullah University of Science and Technology(KAUST) | Pathogen Genomics Lab King Abdullah University of Science and Technology(KAUST ) | Amit Kumar Subudhi et al |
| EPI_ISL_512992 | SaudiArabia/KAUST-JEDDAH723/202 | 2020-05-04 | Asia/Saudi Arabia/Jeddah/ | Pathogen Genomics Lab King Abdullah University of Science and Technology(KAUST) | Pathogen Genomics Lab King Abdullah University of Science and Technology(KAUST ) | Amit Kumar Subudhi et al |
| EPI_ISL_513002 | SaudiArabia/KAUST-JEDDAH826/202 | 2020-05-24 | Asia/Saudi Arabia/Jeddah/ | Pathogen Genomics Lab King Abdullah University of Science and Technology(KAUST) | Pathogen Genomics Lab King Abdullah University of Science and Technology(KAUST ) | Afrah Alsomali et al |
| EPI_ISL_513005 | SaudiArabia/KAUST-JEDDAH831/202 | 2020-05-24 | Asia/Saudi Arabia/Jeddah/ | Pathogen Genomics Lab King Abdullah University of Science and Technology(KAUST) | Pathogen Genomics Lab King Abdullah University of Science and Technology(KAUST ) | Afrah Alsomali et al |

| | | | | | | |
|---|---|---|---|---|---|---|
| EPI_ISL_513006 | SaudiArabia/KAUST-JEDDAH833/202 | 2020-05-24 | Asia/Saudi Arabia/Jeddah/ | Pathogen Genomics Lab King Abdullah University of Science and Technology(KAUST) | Pathogen Genomics Lab King Abdullah University of Science and Technology(KAUST) | Afrah Alsomali et al |
| EPI_ISL_513007 | SaudiArabia/KAUST-JEDDAH834/202 | 2020-05-24 | Asia/Saudi Arabia/Jeddah/ | Pathogen Genomics Lab King Abdullah University of Science and Technology(KAUST) | Pathogen Genomics Lab King Abdullah University of Science and Technology(KAUST) | Afrah Alsomali et al |
| EPI_ISL_513008 | SaudiArabia/KAUST-JEDDAH835/202 | 2020-05-24 | Asia/Saudi Arabia/Jeddah/ | Pathogen Genomics Lab King Abdullah University of Science and Technology(KAUST) | Pathogen Genomics Lab King Abdullah University of Science and Technology(KAUST) | Afrah Alsomali et al |
| EPI_ISL_513009 | SaudiArabia/KAUST-JEDDAH836/202 | 2020-05-24 | Asia/Saudi Arabia/Jeddah/ | Pathogen Genomics Lab King Abdullah University of Science and Technology(KAUST) | Pathogen Genomics Lab King Abdullah University of Science and Technology(KAUST) | Afrah Alsomali et al |
| EPI_ISL_513010 | SaudiArabia/KAUST-JEDDAH837/202 | 2020-05-24 | Asia/Saudi Arabia/Jeddah/ | Pathogen Genomics Lab King Abdullah University of Science and Technology(KAUST) | Pathogen Genomics Lab King Abdullah University of Science and Technology(KAUST) | Afrah Alsomali et al |

| | | | | | | |
|---|---|---|---|---|---|---|
| EPI_ISL_513011 | SaudiArabia/KAUST-JEDDAH838/202 | 2020-05-24 | Asia/Saudi Arabia/Jeddah/ | Pathogen Genomics Lab King Abdullah University of Science and Technology(KAUST) | Pathogen Genomics Lab King Abdullah University of Science and Technology(KAUST) | Afrah Alsomali et al |
| EPI_ISL_513012 | SaudiArabia/KAUST-JEDDAH840/202 | 2020-05-24 | Asia/Saudi Arabia/Jeddah/ | Pathogen Genomics Lab King Abdullah University of Science and Technology(KAUST) | Pathogen Genomics Lab King Abdullah University of Science and Technology(KAUST) | Afrah Alsomali et al |
| EPI_ISL_513013 | SaudiArabia/KAUST-JEDDAH841/202 | 2020-05-24 | Asia/Saudi Arabia/Jeddah/ | Pathogen Genomics Lab King Abdullah University of Science and Technology(KAUST) | Pathogen Genomics Lab King Abdullah University of Science and Technology(KAUST) | Afrah Alsomali et al |
| EPI_ISL_513014 | SaudiArabia/KAUST-JEDDAH842/202 | 2020-05-24 | Asia/Saudi Arabia/Jeddah/ | Pathogen Genomics Lab King Abdullah University of Science and Technology(KAUST) | Pathogen Genomics Lab King Abdullah University of Science and Technology(KAUST) | Afrah Alsomali et al |
| EPI_ISL_513015 | SaudiArabia/KAUST-JEDDAH843/202 | 2020-05-24 | Asia/Saudi Arabia/Jeddah/ | Pathogen Genomics Lab King Abdullah University of Science and Technology(KAUST) | Pathogen Genomics Lab King Abdullah University of Science and Technology(KAUST) | Afrah Alsomali et al |

| | | | | | | |
|---|---|---|---|---|---|---|
| EPI_ISL_513016 | SaudiArabia/KAUST-JEDDAH844/202 | 2020-05-24 | Asia/Saudi Arabia/Jeddah/ | Pathogen Genomics Lab King Abdullah University of Science and Technology(KAUST) | Pathogen Genomics Lab King Abdullah University of Science and Technology(KAUST) | Afrah Alsomali et al |
| EPI_ISL_513017 | SaudiArabia/KAUST-JEDDAH845/202 | 2020-05-24 | Asia/Saudi Arabia/Jeddah/ | Pathogen Genomics Lab King Abdullah University of Science and Technology(KAUST) | Pathogen Genomics Lab King Abdullah University of Science and Technology(KAUST) | Afrah Alsomali et al |
| EPI_ISL_513018 | SaudiArabia/KAUST-JEDDAH846/202 | 2020-05-24 | Asia/Saudi Arabia/Jeddah/ | Pathogen Genomics Lab King Abdullah University of Science and Technology(KAUST) | Pathogen Genomics Lab King Abdullah University of Science and Technology(KAUST) | Afrah Alsomali et al |
| EPI_ISL_513019 | SaudiArabia/KAUST-JEDDAH847/202 | 2020-05-24 | Asia/Saudi Arabia/Jeddah/ | Pathogen Genomics Lab King Abdullah University of Science and Technology(KAUST) | Pathogen Genomics Lab King Abdullah University of Science and Technology(KAUST) | Afrah Alsomali et al |
| EPI_ISL_513020 | SaudiArabia/KAUST-JEDDAH866/202 | 2020-05-24 | Asia/Saudi Arabia/Jeddah/ | Pathogen Genomics Lab King Abdullah University of Science and Technology(KAUST) | Pathogen Genomics Lab King Abdullah University of Science and Technology(KAUST) | Afrah Alsomali et al |

| | | | | | | |
|---|---|---|---|---|---|---|
| EPI_ISL_513021 | SaudiArabia/KAUST-JEDDAH867/202 | 2020-05-24 | Asia/Saudi Arabia/Jeddah/ | Pathogen Genomics Lab King Abdullah University of Science and Technology(KAUST) | Pathogen Genomics Lab King Abdullah University of Science and Technology(KAUST ) | Afrah Alsomali et al |
| EPI_ISL_513022 | SaudiArabia/KAUST-JEDDAH868/202 | 2020-05-24 | Asia/Saudi Arabia/Jeddah/ | Pathogen Genomics Lab King Abdullah University of Science and Technology(KAUST) | Pathogen Genomics Lab King Abdullah University of Science and Technology(KAUST ) | Afrah Alsomali et al |
| EPI_ISL_513023 | SaudiArabia/KAUST-JEDDAH869/202 | 2020-05-24 | Asia/Saudi Arabia/Jeddah/ | Pathogen Genomics Lab King Abdullah University of Science and Technology(KAUST) | Pathogen Genomics Lab King Abdullah University of Science and Technology(KAUST ) | Afrah Alsomali et al |
| EPI_ISL_513026 | SaudiArabia/KAUST-JEDDAH872/202 | 2020-05-24 | Asia/Saudi Arabia/Jeddah/ | Pathogen Genomics Lab King Abdullah University of Science and Technology(KAUST) | Pathogen Genomics Lab King Abdullah University of Science and Technology(KAUST ) | Afrah Alsomali et al |
| EPI_ISL_513027 | SaudiArabia/KAUST-JEDDAH873/202 | 2020-05-24 | Asia/Saudi Arabia/Jeddah/ | Pathogen Genomics Lab King Abdullah University of Science and Technology(KAUST) | Pathogen Genomics Lab King Abdullah University of Science and Technology(KAUST ) | Rahul P Salunke et al |

| | | | | | | |
|---|---|---|---|---|---|---|
| EPI_ISL_513028 | SaudiArabia/KAUST-JEDDAH875/202 | 2020-05-24 | Asia/Saudi Arabia/Jeddah/ | Pathogen Genomics Lab King Abdullah University of Science and Technology(KAUST) | Pathogen Genomics Lab King Abdullah University of Science and Technology(KAUST ) | Rahul P Salunke et al |
| EPI_ISL_513030 | SaudiArabia/KAUST-JEDDAH878/202 | 2020-05-24 | Asia/Saudi Arabia/Jeddah/ | Pathogen Genomics Lab King Abdullah University of Science and Technology(KAUST) | Pathogen Genomics Lab King Abdullah University of Science and Technology(KAUST ) | Rahul P Salunke et al |
| EPI_ISL_513032 | SaudiArabia/KAUST-JEDDAH882/202 | 2020-05-24 | Asia/Saudi Arabia/Jeddah/ | Pathogen Genomics Lab King Abdullah University of Science and Technology(KAUST) | Pathogen Genomics Lab King Abdullah University of Science and Technology(KAUST ) | Rahul P Salunke et al |
| EPI_ISL_513033 | SaudiArabia/KAUST-JEDDAH883/202 | 2020-05-24 | Asia/Saudi Arabia/Jeddah/ | Pathogen Genomics Lab King Abdullah University of Science and Technology(KAUST) | Pathogen Genomics Lab King Abdullah University of Science and Technology(KAUST ) | Rahul P Salunke et al |
| EPI_ISL_513034 | SaudiArabia/KAUST-JEDDAH884/202 | 2020-05-24 | Asia/Saudi Arabia/Jeddah/ | Pathogen Genomics Lab King Abdullah University of Science and Technology(KAUST) | Pathogen Genomics Lab King Abdullah University of Science and Technology(KAUST ) | Rahul P Salunke et al |

| | | | | | | |
|---|---|---|---|---|---|---|
| EPI_ISL_513036 | SaudiArabia/KAUST-JEDDAH887/202 | 2020-05-24 | Asia/Saudi Arabia/Jeddah/ | Pathogen Genomics Lab King Abdullah University of Science and Technology(KAUST) | Pathogen Genomics Lab King Abdullah University of Science and Technology(KAUST ) | Rahul P Salunke et al |
| EPI_ISL_513037 | SaudiArabia/KAUST-JEDDAH888/202 | 2020-05-24 | Asia/Saudi Arabia/Jeddah/ | Pathogen Genomics Lab King Abdullah University of Science and Technology(KAUST) | Pathogen Genomics Lab King Abdullah University of Science and Technology(KAUST ) | Rahul P Salunke et al |
| EPI_ISL_513038 | SaudiArabia/KAUST-JEDDAH889/202 | 2020-05-24 | Asia/Saudi Arabia/Jeddah/ | Pathogen Genomics Lab King Abdullah University of Science and Technology(KAUST) | Pathogen Genomics Lab King Abdullah University of Science and Technology(KAUST ) | Rahul P Salunke et al |
| EPI_ISL_513039 | SaudiArabia/KAUST-JEDDAH890/202 | 2020-05-24 | Asia/Saudi Arabia/Jeddah/ | Pathogen Genomics Lab King Abdullah University of Science and Technology(KAUST) | Pathogen Genomics Lab King Abdullah University of Science and Technology(KAUST ) | Rahul P Salunke et al |
| EPI_ISL_513040 | SaudiArabia/KAUST-JEDDAH891/202 | 2020-05-24 | Asia/Saudi Arabia/Jeddah/ | Pathogen Genomics Lab King Abdullah University of Science and Technology(KAUST) | Pathogen Genomics Lab King Abdullah University of Science and Technology(KAUST ) | Rahul P Salunke et al |

| | | | | | | |
|---|---|---|---|---|---|---|
| EPI_ISL_513064 | SaudiArabia/KAUST-JEDDAH990/202 | 2020-06-23 | Asia/Saudi Arabia/Jeddah/ | Pathogen Genomics Lab King Abdullah University of Science and Technology(KAUST) | Pathogen Genomics Lab King Abdullah University of Science and Technology(KAUST) | Raeece Naeem et al |
| EPI_ISL_513065 | SaudiArabia/KAUST-JEDDAH991/202 | 2020-06-23 | Asia/Saudi Arabia/Jeddah/ | Pathogen Genomics Lab King Abdullah University of Science and Technology(KAUST) | Pathogen Genomics Lab King Abdullah University of Science and Technology(KAUST) | Raeece Naeem et al |
| EPI_ISL_513066 | SaudiArabia/KAUST-JEDDAH992/202 | 2020-06-17 | Asia/Saudi Arabia/Jeddah/ | Pathogen Genomics Lab King Abdullah University of Science and Technology(KAUST) | Pathogen Genomics Lab King Abdullah University of Science and Technology(KAUST) | Raeece Naeem et al |
| EPI_ISL_513067 | SaudiArabia/KAUST-JEDDAH993/202 | 2020-06-17 | Asia/Saudi Arabia/Jeddah/ | Pathogen Genomics Lab King Abdullah University of Science and Technology(KAUST) | Pathogen Genomics Lab King Abdullah University of Science and Technology(KAUST) | Raeece Naeem et al |
| EPI_ISL_513068 | SaudiArabia/KAUST-JEDDAH995/202 | 2020-06-17 | Asia/Saudi Arabia/Jeddah/ | Pathogen Genomics Lab King Abdullah University of Science and Technology(KAUST) | Pathogen Genomics Lab King Abdullah University of Science and Technology(KAUST) | Raeece Naeem et al |

| | | | | | | |
|---|---|---|---|---|---|---|
| EPI_ISL_513070 | SaudiArabia/KAUST-JEDDAH997/202 | 2020-06-17 | Asia/Saudi Arabia/Jeddah/ | Pathogen Genomics Lab King Abdullah University of Science and Technology(KAUST) | Pathogen Genomics Lab King Abdullah University of Science and Technology(KAUST) | Raeece Naeem et al |
| EPI_ISL_513073 | SaudiArabia/KAUST-MADINAH1036/ | 2020-06-11 | Asia/Saudi Arabia/Madinah/ | Pathogen Genomics Lab King Abdullah University of Science and Technology(KAUST) | Pathogen Genomics Lab King Abdullah University of Science and Technology(KAUST) | Raeece Naeem et al |
| EPI_ISL_513074 | SaudiArabia/KAUST-MADINAH1038/ | 2020-06-13 | Asia/Saudi Arabia/Madinah/ | Pathogen Genomics Lab King Abdullah University of Science and Technology(KAUST) | Pathogen Genomics Lab King Abdullah University of Science and Technology(KAUST) | Raeece Naeem et al |
| EPI_ISL_513078 | SaudiArabia/KAUST-MADINAH1047/ | 2020-06-13 | Asia/Saudi Arabia/Madinah/ | Pathogen Genomics Lab King Abdullah University of Science and Technology(KAUST) | Pathogen Genomics Lab King Abdullah University of Science and Technology(KAUST) | Fathia Ben Rached et al |
| EPI_ISL_513080 | SaudiArabia/KAUST-MADINAH1049/ | 2020-06-13 | Asia/Saudi Arabia/Madinah/ | Pathogen Genomics Lab King Abdullah University of Science and Technology(KAUST) | Pathogen Genomics Lab King Abdullah University of Science and Technology(KAUST) | Fathia Ben Rached et al |

| | | | | | | |
|---|---|---|---|---|---|---|
| EPI_ISL_513082 | SaudiArabia/KAUST-MADINAH1052/ | 2020-06-13 | Asia/Saudi Arabia/Madinah/ | Pathogen Genomics Lab King Abdullah University of Science and Technology(KAUST) | Pathogen Genomics Lab King Abdullah University of Science and Technology(KAUST ) | Fathia Ben Rached et al |
| EPI_ISL_513083 | SaudiArabia/KAUST-MADINAH1053/ | 2020-06-13 | Asia/Saudi Arabia/Madinah/ | Pathogen Genomics Lab King Abdullah University of Science and Technology(KAUST) | Pathogen Genomics Lab King Abdullah University of Science and Technology(KAUST ) | Fathia Ben Rached et al |
| EPI_ISL_513084 | SaudiArabia/KAUST-MADINAH1059/ | 2020-06-13 | Asia/Saudi Arabia/Madinah/ | Pathogen Genomics Lab King Abdullah University of Science and Technology(KAUST) | Pathogen Genomics Lab King Abdullah University of Science and Technology(KAUST ) | Fathia Ben Rached et al |
| EPI_ISL_513085 | SaudiArabia/KAUST-MADINAH1060/ | 2020-06-13 | Asia/Saudi Arabia/Madinah/ | Pathogen Genomics Lab King Abdullah University of Science and Technology(KAUST) | Pathogen Genomics Lab King Abdullah University of Science and Technology(KAUST ) | Fathia Ben Rached et al |
| EPI_ISL_513086 | SaudiArabia/KAUST-MADINAH1061/ | 2020-06-13 | Asia/Saudi Arabia/Madinah/ | Pathogen Genomics Lab King Abdullah University of Science and Technology(KAUST) | Pathogen Genomics Lab King Abdullah University of Science and Technology(KAUST ) | Fathia Ben Rached et al |

| | | | | | | |
|---|---|---|---|---|---|---|
| EPI_ISL_513087 | SaudiArabia/KAUST-MADINAH1066/ | 2020-06-13 | Asia/Saudi Arabia/Madinah/ | Pathogen Genomics Lab King Abdullah University of Science and Technology(KAUST) | Pathogen Genomics Lab King Abdullah University of Science and Technology(KAUST) | Fathia Ben Rached et al |
| EPI_ISL_513088 | SaudiArabia/KAUST-MADINAH1070/ | 2020-06-13 | Asia/Saudi Arabia/Madinah/ | Pathogen Genomics Lab King Abdullah University of Science and Technology(KAUST) | Pathogen Genomics Lab King Abdullah University of Science and Technology(KAUST) | Fathia Ben Rached et al |
| EPI_ISL_513090 | SaudiArabia/KAUST-MADINAH1073/ | 2020-06-11 | Asia/Saudi Arabia/Madinah/ | Pathogen Genomics Lab King Abdullah University of Science and Technology(KAUST) | Pathogen Genomics Lab King Abdullah University of Science and Technology(KAUST) | Fathia Ben Rached et al |
| EPI_ISL_513092 | SaudiArabia/KAUST-MADINAH1075/ | 2020-06-11 | Asia/Saudi Arabia/Madinah/ | Pathogen Genomics Lab King Abdullah University of Science and Technology(KAUST) | Pathogen Genomics Lab King Abdullah University of Science and Technology(KAUST) | Fathia Ben Rached et al |
| EPI_ISL_513093 | SaudiArabia/KAUST-MADINAH1076/ | 2020-06-11 | Asia/Saudi Arabia/Madinah/ | Pathogen Genomics Lab King Abdullah University of Science and Technology(KAUST) | Pathogen Genomics Lab King Abdullah University of Science and Technology(KAUST) | Fathia Ben Rached et al |

| | | | | | | |
|---|---|---|---|---|---|---|
| EPI_ISL_513095 | SaudiArabia/KAUST-MADINAH1078/ | 2020-05-27 | Asia/Saudi Arabia/Madinah/ | Pathogen Genomics Lab King Abdullah University of Science and Technology(KAUST) | Pathogen Genomics Lab King Abdullah University of Science and Technology(KAUST) | Fathia Ben Rached et al |
| EPI_ISL_513097 | SaudiArabia/KAUST-MADINAH1081/ | 2020-06-11 | Asia/Saudi Arabia/Madinah/ | Pathogen Genomics Lab King Abdullah University of Science and Technology(KAUST) | Pathogen Genomics Lab King Abdullah University of Science and Technology(KAUST) | Fathia Ben Rached et al |
| EPI_ISL_513098 | SaudiArabia/KAUST-MADINAH1085/ | 2020-06-11 | Asia/Saudi Arabia/Madinah/ | Pathogen Genomics Lab King Abdullah University of Science and Technology(KAUST) | Pathogen Genomics Lab King Abdullah University of Science and Technology(KAUST) | Fathia Ben Rached et al |
| EPI_ISL_513099 | SaudiArabia/KAUST-MADINAH1086/ | 2020-06-13 | Asia/Saudi Arabia/Madinah/ | Pathogen Genomics Lab King Abdullah University of Science and Technology(KAUST) | Pathogen Genomics Lab King Abdullah University of Science and Technology(KAUST) | Fathia Ben Rached et al |
| EPI_ISL_513100 | SaudiArabia/KAUST-MADINAH1087/ | 2020-06-11 | Asia/Saudi Arabia/Madinah/ | Pathogen Genomics Lab King Abdullah University of Science and Technology(KAUST) | Pathogen Genomics Lab King Abdullah University of Science and Technology(KAUST) | Fathia Ben Rached et al |

| | | | | | | |
|---|---|---|---|---|---|---|
| EPI_ISL_513101 | SaudiArabia/KAUST-MADINAH1090/ | 2020-06-11 | Asia/Saudi Arabia/Madinah/ | Pathogen Genomics Lab King Abdullah University of Science and Technology(KAUST) | Pathogen Genomics Lab King Abdullah University of Science and Technology(KAUST) | Fathia Ben Rached et al |
| EPI_ISL_513102 | SaudiArabia/KAUST-MADINAH1093/ | 2020-06-11 | Asia/Saudi Arabia/Madinah/ | Pathogen Genomics Lab King Abdullah University of Science and Technology(KAUST) | Pathogen Genomics Lab King Abdullah University of Science and Technology(KAUST) | Fathia Ben Rached et al |
| EPI_ISL_513103 | SaudiArabia/KAUST-MADINAH1094/ | 2020-06-13 | Asia/Saudi Arabia/Madinah/ | Pathogen Genomics Lab King Abdullah University of Science and Technology(KAUST) | Pathogen Genomics Lab King Abdullah University of Science and Technology(KAUST) | Fathia Ben Rached et al |
| EPI_ISL_513107 | SaudiArabia/KAUST-MADINAH1100/ | 2020-06-13 | Asia/Saudi Arabia/Madinah/ | Pathogen Genomics Lab King Abdullah University of Science and Technology(KAUST) | Pathogen Genomics Lab King Abdullah University of Science and Technology(KAUST) | Fathia Ben Rached et al |
| EPI_ISL_513108 | SaudiArabia/KAUST-MADINAH1102/ | 2020-06-13 | Asia/Saudi Arabia/Madinah/ | Pathogen Genomics Lab King Abdullah University of Science and Technology(KAUST) | Pathogen Genomics Lab King Abdullah University of Science and Technology(KAUST) | Fathia Ben Rached et al |

| | | | | | | |
|---|---|---|---|---|---|---|
| EPI_ISL_513109 | SaudiArabia/KAUST-MADINAH1103/ | 2020-06-13 | Asia/Saudi Arabia/Madinah/ | Pathogen Genomics Lab King Abdullah University of Science and Technology(KAUST) | Pathogen Genomics Lab King Abdullah University of Science and Technology(KAUST ) | Fathia Ben Rached et al |
| EPI_ISL_513112 | SaudiArabia/KAUST-MADINAH1108/ | 2020-06-11 | Asia/Saudi Arabia/Madinah/ | Pathogen Genomics Lab King Abdullah University of Science and Technology(KAUST) | Pathogen Genomics Lab King Abdullah University of Science and Technology(KAUST ) | Fathia Ben Rached et al |
| EPI_ISL_513113 | SaudiArabia/KAUST-MADINAH1109/ | 2020-06-11 | Asia/Saudi Arabia/Madinah/ | Pathogen Genomics Lab King Abdullah University of Science and Technology(KAUST) | Pathogen Genomics Lab King Abdullah University of Science and Technology(KAUST ) | Fathia Ben Rached et al |
| EPI_ISL_513116 | SaudiArabia/KAUST-MADINAH1112/ | 2020-06-11 | Asia/Saudi Arabia/Madinah/ | Pathogen Genomics Lab King Abdullah University of Science and Technology(KAUST) | Pathogen Genomics Lab King Abdullah University of Science and Technology(KAUST ) | Fathia Ben Rached et al |
| EPI_ISL_513117 | SaudiArabia/KAUST-MADINAH1113/ | 2020-06-11 | Asia/Saudi Arabia/Madinah/ | Pathogen Genomics Lab King Abdullah University of Science and Technology(KAUST) | Pathogen Genomics Lab King Abdullah University of Science and Technology(KAUST ) | Fathia Ben Rached et al |

| | | | | | | |
|---|---|---|---|---|---|---|
| EPI_ISL_513118 | SaudiArabia/KAUST-MADINAH1114/ | 2020-06-11 | Asia/Saudi Arabia/Madinah/ | Pathogen Genomics Lab King Abdullah University of Science and Technology(KAUST) | Pathogen Genomics Lab King Abdullah University of Science and Technology(KAUST) | Fathia Ben Rached et al |
| EPI_ISL_513134 | SaudiArabia/KAUST-MADINAH354/2 | 2020-04-23 | Asia/Saudi Arabia/Madinah/ | Pathogen Genomics Lab King Abdullah University of Science and Technology(KAUST) | Pathogen Genomics Lab King Abdullah University of Science and Technology(KAUST) | Sharif Hala et al |
| EPI_ISL_513141 | SaudiArabia/KAUST-MADINAH364/2 | 2020-04-23 | Asia/Saudi Arabia/Madinah/ | Pathogen Genomics Lab King Abdullah University of Science and Technology(KAUST) | Pathogen Genomics Lab King Abdullah University of Science and Technology(KAUST) | Sharif Hala et al |
| EPI_ISL_513144 | SaudiArabia/KAUST-MADINAH380/2 | 2020-04-24 | Asia/Saudi Arabia/Madinah/ | Pathogen Genomics Lab King Abdullah University of Science and Technology(KAUST) | Pathogen Genomics Lab King Abdullah University of Science and Technology(KAUST) | Sharif Hala et al |
| EPI_ISL_513157 | SaudiArabia/KAUST-MADINAH437/2 | 2020-04-23 | Asia/Saudi Arabia/Madinah/ | Pathogen Genomics Lab King Abdullah University of Science and Technology(KAUST) | Pathogen Genomics Lab King Abdullah University of Science and Technology(KAUST) | Fadwa Alofi et al |

| | | | | | | |
|---|---|---|---|---|---|---|
| EPI_ISL_513161 | SaudiArabia/KAUST-MADINAH452/2 | 2020-04-21 | Asia/Saudi Arabia/Madinah/ | Pathogen Genomics Lab King Abdullah University of Science and Technology(KAUST) | Pathogen Genomics Lab King Abdullah University of Science and Technology(KAUST) | Fadwa Alofi et al |
| EPI_ISL_513170 | SaudiArabia/KAUST-MADINAH605/2 | 2020-05-06 | Asia/Saudi Arabia/Madinah/ | Pathogen Genomics Lab King Abdullah University of Science and Technology(KAUST) | Pathogen Genomics Lab King Abdullah University of Science and Technology(KAUST) | Sara Mfarrej et al |
| EPI_ISL_513171 | SaudiArabia/KAUST-MADINAH606/2 | 2020-05-06 | Asia/Saudi Arabia/Madinah/ | Pathogen Genomics Lab King Abdullah University of Science and Technology(KAUST) | Pathogen Genomics Lab King Abdullah University of Science and Technology(KAUST) | Sara Mfarrej et al |
| EPI_ISL_513172 | SaudiArabia/KAUST-MADINAH609/2 | 2020-05-06 | Asia/Saudi Arabia/Madinah/ | Pathogen Genomics Lab King Abdullah University of Science and Technology(KAUST) | Pathogen Genomics Lab King Abdullah University of Science and Technology(KAUST) | Sara Mfarrej et al |
| EPI_ISL_513174 | SaudiArabia/KAUST-MADINAH647/2 | 2020-05-08 | Asia/Saudi Arabia/Madinah/ | Pathogen Genomics Lab King Abdullah University of Science and Technology(KAUST) | Pathogen Genomics Lab King Abdullah University of Science and Technology(KAUST) | Sara Mfarrej et al |

| | | | | | | |
|---|---|---|---|---|---|---|
| EPI_ISL_513176 | SaudiArabia/KAUST-MADINAH684/2 | 2020-05-01 | Asia/Saudi Arabia/Madinah/ | Pathogen Genomics Lab King Abdullah University of Science and Technology(KAUST) | Pathogen Genomics Lab King Abdullah University of Science and Technology(KAUST ) | Amit Kumar Subudhi et al |
| EPI_ISL_513177 | SaudiArabia/KAUST-MADINAH687/2 | 2020-05-08 | Asia/Saudi Arabia/Madinah/ | Pathogen Genomics Lab King Abdullah University of Science and Technology(KAUST) | Pathogen Genomics Lab King Abdullah University of Science and Technology(KAUST ) | Amit Kumar Subudhi et al |
| EPI_ISL_513178 | SaudiArabia/KAUST-MADINAH688/2 | 2020-05-08 | Asia/Saudi Arabia/Madinah/ | Pathogen Genomics Lab King Abdullah University of Science and Technology(KAUST) | Pathogen Genomics Lab King Abdullah University of Science and Technology(KAUST ) | Amit Kumar Subudhi et al |
| EPI_ISL_513179 | SaudiArabia/KAUST-MADINAH691/2 | 2020-05-06 | Asia/Saudi Arabia/Madinah/ | Pathogen Genomics Lab King Abdullah University of Science and Technology(KAUST) | Pathogen Genomics Lab King Abdullah University of Science and Technology(KAUST ) | Amit Kumar Subudhi et al |
| EPI_ISL_513180 | SaudiArabia/KAUST-MADINAH694/2 | 2020-05-08 | Asia/Saudi Arabia/Madinah/ | Pathogen Genomics Lab King Abdullah University of Science and Technology(KAUST) | Pathogen Genomics Lab King Abdullah University of Science and Technology(KAUST ) | Amit Kumar Subudhi et al |

| | | | | | | |
|---|---|---|---|---|---|---|
| EPI_ISL_513182 | SaudiArabia/KAUST-MADINAH704/2 | 2020-04-01 | Asia/Saudi Arabia/Madinah/ | Pathogen Genomics Lab King Abdullah University of Science and Technology(KAUST) | Pathogen Genomics Lab King Abdullah University of Science and Technology(KAUST) | Amit Kumar Subudhi et al |
| EPI_ISL_513188 | SaudiArabia/KAUST-MADINAH716/2 | 2020-04-01 | Asia/Saudi Arabia/Madinah/ | Pathogen Genomics Lab King Abdullah University of Science and Technology(KAUST) | Pathogen Genomics Lab King Abdullah University of Science and Technology(KAUST) | Amit Kumar Subudhi et al |
| EPI_ISL_513190 | SaudiArabia/KAUST-MADINAH850/2 | 2020-05-26 | Asia/Saudi Arabia/Madinah/ | Pathogen Genomics Lab King Abdullah University of Science and Technology(KAUST) | Pathogen Genomics Lab King Abdullah University of Science and Technology(KAUST) | Afrah Alsomali et al |
| EPI_ISL_513193 | SaudiArabia/KAUST-MADINAH857/2 | 2020-05-26 | Asia/Saudi Arabia/Madinah/ | Pathogen Genomics Lab King Abdullah University of Science and Technology(KAUST) | Pathogen Genomics Lab King Abdullah University of Science and Technology(KAUST) | Afrah Alsomali et al |
| EPI_ISL_513194 | SaudiArabia/KAUST-MADINAH858/2 | 2020-05-26 | Asia/Saudi Arabia/Madinah/ | Pathogen Genomics Lab King Abdullah University of Science and Technology(KAUST) | Pathogen Genomics Lab King Abdullah University of Science and Technology(KAUST) | Afrah Alsomali et al |

| | | | | | | |
|---|---|---|---|---|---|---|
| EPI_ISL_513196 | SaudiArabia/KAUST-MADINAH860/2 | 2020-05-26 | Asia/Saudi Arabia/Madinah/ | Pathogen Genomics Lab King Abdullah University of Science and Technology(KAUST) | Pathogen Genomics Lab King Abdullah University of Science and Technology(KAUST) | Afrah Alsomali et al |
| EPI_ISL_513197 | SaudiArabia/KAUST-MADINAH862/2 | 2020-05-26 | Asia/Saudi Arabia/Madinah/ | Pathogen Genomics Lab King Abdullah University of Science and Technology(KAUST) | Pathogen Genomics Lab King Abdullah University of Science and Technology(KAUST) | Afrah Alsomali et al |
| EPI_ISL_513198 | SaudiArabia/KAUST-MADINAH892/2 | 2020-05-12 | Asia/Saudi Arabia/Madinah/ | Pathogen Genomics Lab King Abdullah University of Science and Technology(KAUST) | Pathogen Genomics Lab King Abdullah University of Science and Technology(KAUST) | Rahul P Salunke et al |
| EPI_ISL_513199 | SaudiArabia/KAUST-MADINAH896/2 | 2020-05-26 | Asia/Saudi Arabia/Madinah/ | Pathogen Genomics Lab King Abdullah University of Science and Technology(KAUST) | Pathogen Genomics Lab King Abdullah University of Science and Technology(KAUST) | Rahul P Salunke et al |
| EPI_ISL_513200 | SaudiArabia/KAUST-MADINAH898/2 | 2020-05-26 | Asia/Saudi Arabia/Madinah/ | Pathogen Genomics Lab King Abdullah University of Science and Technology(KAUST) | Pathogen Genomics Lab King Abdullah University of Science and Technology(KAUST) | Rahul P Salunke et al |

| | | | | | | |
|---|---|---|---|---|---|---|
| EPI_ISL_513201 | SaudiArabia/KAUST-MADINAH937/2 | 2020-05-26 | Asia/Saudi Arabia/Madinah/ | Pathogen Genomics Lab King Abdullah University of Science and Technology(KAUST) | Pathogen Genomics Lab King Abdullah University of Science and Technology(KAUST ) | Rahul P Salunke et al |
| EPI_ISL_513202 | SaudiArabia/KAUST-MADINAH938/2 | 2020-05-26 | Asia/Saudi Arabia/Madinah/ | Pathogen Genomics Lab King Abdullah University of Science and Technology(KAUST) | Pathogen Genomics Lab King Abdullah University of Science and Technology(KAUST ) | Rahul P Salunke et al |
| EPI_ISL_513203 | SaudiArabia/KAUST-MADINAH941/2 | 2020-05-26 | Asia/Saudi Arabia/Madinah/ | Pathogen Genomics Lab King Abdullah University of Science and Technology(KAUST) | Pathogen Genomics Lab King Abdullah University of Science and Technology(KAUST ) | Rahul P Salunke et al |
| EPI_ISL_513204 | SaudiArabia/KAUST-MADINAH947/2 | 2020-05-26 | Asia/Saudi Arabia/Madinah/ | Pathogen Genomics Lab King Abdullah University of Science and Technology(KAUST) | Pathogen Genomics Lab King Abdullah University of Science and Technology(KAUST ) | Rahul P Salunke et al |
| EPI_ISL_513205 | SaudiArabia/KAUST-MADINAH955/2 | 2020-05-12 | Asia/Saudi Arabia/Madinah/ | Pathogen Genomics Lab King Abdullah University of Science and Technology(KAUST) | Pathogen Genomics Lab King Abdullah University of Science and Technology(KAUST ) | Rahul P Salunke et al |

| | | | | | | |
|---|---|---|---|---|---|---|
| EPI_ISL_513206 | SaudiArabia/KAUST-MADINAH956/2 | 2020-05-12 | Asia/Saudi Arabia/Madinah/ | Pathogen Genomics Lab King Abdullah University of Science and Technology(KAUST) | Pathogen Genomics Lab King Abdullah University of Science and Technology(KAUST) | Rahul P Salunke et al |
| EPI_ISL_513207 | SaudiArabia/KAUST-MADINAH957/2 | 2020-05-26 | Asia/Saudi Arabia/Madinah/ | Pathogen Genomics Lab King Abdullah University of Science and Technology(KAUST) | Pathogen Genomics Lab King Abdullah University of Science and Technology(KAUST) | Rahul P Salunke et al |
| EPI_ISL_513208 | SaudiArabia/KAUST-MADINAH958/2 | 2020-05-12 | Asia/Saudi Arabia/Madinah/ | Pathogen Genomics Lab King Abdullah University of Science and Technology(KAUST) | Pathogen Genomics Lab King Abdullah University of Science and Technology(KAUST) | Raeece Naeem et al |
| EPI_ISL_513210 | SaudiArabia/KAUST-MADINAH960/2 | 2020-05-26 | Asia/Saudi Arabia/Madinah/ | Pathogen Genomics Lab King Abdullah University of Science and Technology(KAUST) | Pathogen Genomics Lab King Abdullah University of Science and Technology(KAUST) | Raeece Naeem et al |
| EPI_ISL_513211 | SaudiArabia/KAUST-MADINAH961/2 | 2020-05-12 | Asia/Saudi Arabia/Madinah/ | Pathogen Genomics Lab King Abdullah University of Science and Technology(KAUST) | Pathogen Genomics Lab King Abdullah University of Science and Technology(KAUST) | Raeece Naeem et al |

| | | | | | | |
|---|---|---|---|---|---|---|
| EPI_ISL_513222 | SaudiArabia/KAUST-MAKKAH301/20 | 2020-04-21 | Asia/Saudi Arabia/Makkah/ | Pathogen Genomics Lab King Abdullah University of Science and Technology(KAUST) | Pathogen Genomics Lab King Abdullah University of Science and Technology(KAUST) | Sharif Hala et al |
| EPI_ISL_513233 | SaudiArabia/KAUST-MAKKAH741/20 | 2020-04-01 | Asia/Saudi Arabia/Makkah/ | Pathogen Genomics Lab King Abdullah University of Science and Technology(KAUST) | Pathogen Genomics Lab King Abdullah University of Science and Technology(KAUST) | Amit Kumar Subudhi et al |
| EPI_ISL_513234 | SaudiArabia/KAUST-MAKKAH760/20 | 2020-04-18 | Asia/Saudi Arabia/Makkah/ | Pathogen Genomics Lab King Abdullah University of Science and Technology(KAUST) | Pathogen Genomics Lab King Abdullah University of Science and Technology(KAUST) | Amit Kumar Subudhi et al |
| EPI_ISL_513246 | SaudiArabia/KAUST-MAKKAH777/20 | 2020-04-18 | Asia/Saudi Arabia/Makkah/ | Pathogen Genomics Lab King Abdullah University of Science and Technology(KAUST) | Pathogen Genomics Lab King Abdullah University of Science and Technology(KAUST) | Amit Kumar Subudhi et al |
| EPI_ISL_513248 | SaudiArabia/KAUST-MAKKAH779/20 | 2020-04-24 | Asia/Saudi Arabia/Makkah/ | Pathogen Genomics Lab King Abdullah University of Science and Technology(KAUST) | Pathogen Genomics Lab King Abdullah University of Science and Technology(KAUST) | Amit Kumar Subudhi et al |
| EPI_ISL_513265 | USA/FL-9F97/2020 | 2020-04-24 | North America/USA/Florida/Miami-Dad | University of Miami Immunology and Histocompatibility Laboratory | University of Miami Immunology and Histocompatibility Laboratory | Emilio Margolles-Clark et al |

| | | | | | | |
|---|---|---|---|---|---|---|
| EPI_ISL_513271 | USA/FL-AD68/2020 | 2020-06-26 | North America/USA/Florida/Miami-Dad | University of Miami Immunology and Histocompatibility Laboratory | University of Miami Immunology and Histocompatibility Laboratory | Emilio Margolles-Clark et al |
| EPI_ISL_513276 | USA/FL-ADNQ/2020 | 2020-06-27 | North America/USA/Florida/Miami-Dad | University of Miami Immunology and Histocompatibility Laboratory | University of Miami Immunology and Histocompatibility Laboratory | Emilio Margolles-Clark et al |
| EPI_ISL_513291 | USA/FL-AJ5U/2020 | 2020-07-05 | North America/USA/Florida/Miami-Dad | University of Miami Immunology and Histocompatibility Laboratory | University of Miami Immunology and Histocompatibility Laboratory | Emilio Margolles-Clark et al |
| EPI_ISL_513292 | USA/PR-AGN5/2020 | 2020-06-29 | North America/USA/Puerto Rico/Ponce | University of Miami Immunology and Histocompatibility Laboratory | University of Miami Immunology and Histocompatibility Laboratory | Emilio Margolles-Clark et al |
| EPI_ISL_513302 | Germany/BY-UHR-5ZK/2020 | 2020-03-22 | Europe/Germany/Bavaria/ | Department of Infection Prevention and Infectious Diseases, University Hospital Regensburg | University Hospital Regensburg | Fritsch et al |
| EPI_ISL_513404 | Australia/NSW762/2020 | 2020-07-21 | Oceania/Australia/New South Wales/Sy | Laverty Pathology | NSW Health Pathology - Institute of Clinical Pathology and Medical Research; Westmead Hospital; University of Sydney | CIDM-PH et al. et al |
| EPI_ISL_513426 | USA/ME-HETL-J0013/2020 | 2020-03-14 | North America/USA/Maine/ | Maine HETL | Tewhey Lab, The Jackson Laboratory | Matluk et al |
| EPI_ISL_513445 | USA/ME-HETL-J0033/2020 | 2020-03-17 | North America/USA/Maine/ | Maine HETL | Tewhey Lab, The Jackson Laboratory | Matluk et al |

| | | | | | | |
|---|---|---|---|---|---|---|
| EPI_ISL_513465 | USA/ME-HETL-J0054/2020 | 2020-03-14 | North America/USA/Maine/ | Maine HETL | Tewhey Lab, The Jackson Laboratory | Matluk et al |
| EPI_ISL_513473 | USA/ME-HETL-J0062/2020 | 2020-03-23 | North America/USA/Maine/ | Maine HETL | Tewhey Lab, The Jackson Laboratory | Matluk et al |
| EPI_ISL_513474 | USA/ME-HETL-J0063/2020 | 2020-03-17 | North America/USA/Maine/ | Maine HETL | Tewhey Lab, The Jackson Laboratory | Matluk et al |
| EPI_ISL_513477 | USA/ME-HETL-J0066/2020 | 2020-03-19 | North America/USA/Maine/ | Maine HETL | Tewhey Lab, The Jackson Laboratory | Matluk et al |
| EPI_ISL_513484 | USA/ME-HETL-J0073/2020 | 2020-03-19 | North America/USA/Maine/ | Maine HETL | Tewhey Lab, The Jackson Laboratory | Matluk et al |
| EPI_ISL_513508 | USA/ME-HETL-J0099/2020 | 2020-03-23 | North America/USA/Maine/ | Maine HETL | Tewhey Lab, The Jackson Laboratory | Matluk et al |
| EPI_ISL_513514 | Brazil/RJ-INCA-C07/2020 | 2020-04-08 | South America/Brazil/Rio de Janeiro/ | Programa de Oncovirologia, Instituto Nacional de CV¢ncer | Programa de Oncovirologia, Instituto Nacional de CV¢ncer | Juliana D. Siqueira et al |
| EPI_ISL_513525 | Brazil/RJ-INCA-C194/2020 | 2020-04-30 | South America/Brazil/Rio de Janeiro/ | Programa de Oncovirologia, Instituto Nacional de CV¢ncer | Programa de Oncovirologia, Instituto Nacional de CV¢ncer | Juliana D. Siqueira et al |
| EPI_ISL_513556 | Brazil/RJ-INCA-C63/2020 | 2020-04-22 | South America/Brazil/Rio de Janeiro/ | Programa de Oncovirologia, Instituto Nacional de CV¢ncer | Programa de Oncovirologia, Instituto Nacional de CV¢ncer | Juliana D. Siqueira et al |
| EPI_ISL_513559 | Brazil/RJ-INCA-C67/2020 | 2020-04-22 | South America/Brazil/Rio de Janeiro/ | Programa de Oncovirologia, Instituto Nacional de CV¢ncer | Programa de Oncovirologia, Instituto Nacional de CV¢ncer | Juliana D. Siqueira et al |
| EPI_ISL_513573 | Brazil/RJ-INCA-I29/2020 | 2020-05-04 | South America/Brazil/Rio de Janeiro/ | Programa de Oncovirologia, Instituto Nacional de CV¢ncer | Programa de Oncovirologia, Instituto Nacional de CV¢ncer | Juliana D. Siqueira et al |

| | | | | | | |
|---|---|---|---|---|---|---|
| EPI_ISL_513574 | Brazil/RJ-INCA-I30/2020 | 2020-05-04 | South America/Brazil/Rio de Janeiro/ | Programa de Oncovirologia, Instituto Nacional de CV¢ncer | Programa de Oncovirologia, Instituto Nacional de CV¢ncer | Juliana D. Siqueira et al |
| EPI_ISL_513575 | Brazil/RJ-INCA-I34/2020 | 2020-05-04 | South America/Brazil/Rio de Janeiro/ | Programa de Oncovirologia, Instituto Nacional de CV¢ncer | Programa de Oncovirologia, Instituto Nacional de CV¢ncer | Juliana D. Siqueira et al |
| EPI_ISL_513576 | Brazil/RJ-INCA-I35/2020 | 2020-05-04 | South America/Brazil/Rio de Janeiro/ | Programa de Oncovirologia, Instituto Nacional de CV¢ncer | Programa de Oncovirologia, Instituto Nacional de CV¢ncer | Juliana D. Siqueira et al |
| EPI_ISL_513577 | Brazil/RJ-INCA-I37/2020 | 2020-05-04 | South America/Brazil/Rio de Janeiro/ | Programa de Oncovirologia, Instituto Nacional de CV¢ncer | Programa de Oncovirologia, Instituto Nacional de CV¢ncer | Juliana D. Siqueira et al |
| EPI_ISL_513578 | Brazil/RJ-INCA-I39/2020 | 2020-05-04 | South America/Brazil/Rio de Janeiro/ | Programa de Oncovirologia, Instituto Nacional de CV¢ncer | Programa de Oncovirologia, Instituto Nacional de CV¢ncer | Juliana D. Siqueira et al |
| EPI_ISL_513579 | Brazil/RJ-INCA-I43/2020 | 2020-05-04 | South America/Brazil/Rio de Janeiro/ | Programa de Oncovirologia, Instituto Nacional de CV¢ncer | Programa de Oncovirologia, Instituto Nacional de CV¢ncer | Juliana D. Siqueira et al |
| EPI_ISL_513580 | Brazil/RJ-INCA-I44/2020 | 2020-05-04 | South America/Brazil/Rio de Janeiro/ | Programa de Oncovirologia, Instituto Nacional de CV¢ncer | Programa de Oncovirologia, Instituto Nacional de CV¢ncer | Juliana D. Siqueira et al |
| EPI_ISL_513581 | Brazil/RJ-INCA-I51/2020 | 2020-05-05 | South America/Brazil/Rio de Janeiro/ | Programa de Oncovirologia, Instituto Nacional de CV¢ncer | Programa de Oncovirologia, Instituto Nacional de CV¢ncer | Juliana D. Siqueira et al |
| EPI_ISL_513582 | Brazil/RJ-INCA-I54/2020 | 2020-05-05 | South America/Brazil/Rio de Janeiro/ | Programa de Oncovirologia, Instituto Nacional de CV¢ncer | Programa de Oncovirologia, Instituto Nacional de CV¢ncer | Juliana D. Siqueira et al |
| EPI_ISL_513583 | Brazil/RJ-INCA-I57/2020 | 2020-05-05 | South America/Brazil/Rio de Janeiro/ | Programa de Oncovirologia, Instituto Nacional de CV¢ncer | Programa de Oncovirologia, Instituto Nacional de CV¢ncer | Juliana D. Siqueira et al |

| | | | | | | |
|---|---|---|---|---|---|---|
| EPI_ISL_513637 | USA/WA-UW-4758/2020 | 2020-03-28 | North America/USA/Washington/ | University of Washington Virology Lab | University of Washington Virology Lab | Pavitra Roychoudhury et al |
| EPI_ISL_513641 | USA/UT-02708/2020 | 2020-04-28 | North America/USA/Utah/ | Utah Public Health Laboratory | Utah Public Health Laboratory | Heidi Butz et al |
| EPI_ISL_513644 | USA/UT-02757/2020 | 2020-04-29 | North America/USA/Utah/ | Utah Public Health Laboratory | Utah Public Health Laboratory | Heidi Butz et al |
| EPI_ISL_513652 | USA/UT-02842/2020 | 2020-04-29 | North America/USA/Utah/ | Utah Public Health Laboratory | Utah Public Health Laboratory | Heidi Butz et al |
| EPI_ISL_513657 | USA/UT-02848/2020 | 2020-04-29 | North America/USA/Utah/ | Utah Public Health Laboratory | Utah Public Health Laboratory | Heidi Butz et al |
| EPI_ISL_513666 | USA/UT-02860/2020 | 2020-04-29 | North America/USA/Utah/ | Utah Public Health Laboratory | Utah Public Health Laboratory | Heidi Butz et al |
| EPI_ISL_513676 | USA/UT-02875/2020 | 2020-04-29 | North America/USA/Utah/ | Utah Public Health Laboratory | Utah Public Health Laboratory | Heidi Butz et al |
| EPI_ISL_513680 | USA/UT-02888/2020 | 2020-04-30 | North America/USA/Utah/ | Utah Public Health Laboratory | Utah Public Health Laboratory | Heidi Butz et al |
| EPI_ISL_513685 | USA/UT-02894/2020 | 2020-04-30 | North America/USA/Utah/ | Utah Public Health Laboratory | Utah Public Health Laboratory | Heidi Butz et al |
| EPI_ISL_513692 | USA/UT-02917/2020 | 2020-04-30 | North America/USA/Utah/ | Utah Public Health Laboratory | Utah Public Health Laboratory | Heidi Butz et al |
| EPI_ISL_513706 | USA/UT-03016/2020 | 2020-05-01 | North America/USA/Utah/ | Utah Public Health Laboratory | Utah Public Health Laboratory | Heidi Butz et al |
| EPI_ISL_513716 | USA/CA-CZB-2166/2020 | 2020-05-22 | North America/USA/California/Orange ( | Orange County Public Health Lab | Chan-Zuckerberg Biohub | CZB Cliahub Consortium et al |
| EPI_ISL_513730 | USA/CA-CZB-2180/2020 | 2020-05-26 | North America/USA/California/Orange ( | Orange County Public Health Lab | Chan-Zuckerberg Biohub | CZB Cliahub Consortium et al |
| EPI_ISL_513732 | USA/CA-CZB-2182/2020 | 2020-05-26 | North America/USA/California/Orange ( | Orange County Public Health Lab | Chan-Zuckerberg Biohub | CZB Cliahub Consortium et al |
| EPI_ISL_513734 | USA/CA-CZB-2184/2020 | 2020-05-26 | North America/USA/California/Orange ( | Orange County Public Health Lab | Chan-Zuckerberg Biohub | CZB Cliahub Consortium et al |
| EPI_ISL_513736 | USA/CA-CZB-2187/2020 | 2020-05-26 | North America/USA/California/Orange ( | Orange County Public Health Lab | Chan-Zuckerberg Biohub | CZB Cliahub Consortium et al |
| EPI_ISL_513737 | USA/CA-CZB-2188/2020 | 2020-05-26 | North America/USA/California/Orange ( | Orange County Public Health Lab | Chan-Zuckerberg Biohub | CZB Cliahub Consortium et al |
| EPI_ISL_513742 | USA/CA-CZB-2193/2020 | 2020-05-27 | North America/USA/California/Orange ( | Orange County Public Health Lab | Chan-Zuckerberg Biohub | CZB Cliahub Consortium et al |
| EPI_ISL_513746 | USA/CA-CZB-2197/2020 | 2020-05-27 | North America/USA/California/Orange ( | Orange County Public Health Lab | Chan-Zuckerberg Biohub | CZB Cliahub Consortium et al |

| | | | | | | |
|---|---|---|---|---|---|---|
| EPI_ISL_513747 | USA/CA-CZB-2198/2020 | 2020-05-27 | North America/USA/California/Orange ( | Orange County Public Health Lab | Chan-Zuckerberg Biohub | CZB Cliahub Consortium et al |
| EPI_ISL_513750 | USA/CA-CZB-2201/2020 | 2020-05-27 | North America/USA/California/Orange ( | Orange County Public Health Lab | Chan-Zuckerberg Biohub | CZB Cliahub Consortium et al |
| EPI_ISL_513771 | USA/CA-CZB-2223/2020 | 2020-05-31 | North America/USA/California/Orange ( | Orange County Public Health Lab | Chan-Zuckerberg Biohub | CZB Cliahub Consortium et al |
| EPI_ISL_513775 | USA/CA-CZB-2280/2020 | 2020-06-19 | North America/USA/California/Santa Cl | County of Santa Clara Public Health Department | Chan-Zuckerberg Biohub | CZB Cliahub Consortium et al |
| EPI_ISL_513784 | USA/CA-CZB-2293/2020 | 2020-06-20 | North America/USA/California/Santa Cl | County of Santa Clara Public Health Department | Chan-Zuckerberg Biohub | CZB Cliahub Consortium et al |
| EPI_ISL_513794 | USA/CA-CZB-2345/2020 | 2020-05-13 | North America/USA/California/Orange ( | Orange County Public Health Laboratory | Chan-Zuckerberg Biohub | CZB Cliahub Consortium et al |
| EPI_ISL_513799 | USA/CA-CZB-2351/2020 | 2020-05-13 | North America/USA/California/Orange ( | Orange County Public Health Laboratory | Chan-Zuckerberg Biohub | CZB Cliahub Consortium et al |
| EPI_ISL_513803 | USA/CA-CZB-2356/2020 | 2020-05-13 | North America/USA/California/Orange ( | Orange County Public Health Laboratory | Chan-Zuckerberg Biohub | CZB Cliahub Consortium et al |
| EPI_ISL_513807 | USA/CA-CZB-2366/2020 | 2020-05-15 | North America/USA/California/Orange ( | Orange County Public Health Laboratory | Chan-Zuckerberg Biohub | CZB Cliahub Consortium et al |
| EPI_ISL_513810 | USA/CA-CZB-2369/2020 | 2020-05-15 | North America/USA/California/Orange ( | Orange County Public Health Laboratory | Chan-Zuckerberg Biohub | CZB Cliahub Consortium et al |
| EPI_ISL_513817 | USA/CA-CZB-2377/2020 | 2020-05-15 | North America/USA/California/Orange ( | Orange County Public Health Laboratory | Chan-Zuckerberg Biohub | CZB Cliahub Consortium et al |
| EPI_ISL_513820 | USA/CA-CZB-2387/2020 | 2020-05-18 | North America/USA/California/Orange ( | Orange County Public Health Laboratory | Chan-Zuckerberg Biohub | CZB Cliahub Consortium et al |
| EPI_ISL_513825 | USA/CA-CZB-2396/2020 | 2020-05-19 | North America/USA/California/Orange ( | Orange County Public Health Laboratory | Chan-Zuckerberg Biohub | CZB Cliahub Consortium et al |
| EPI_ISL_513829 | USA/CA-CZB-2401/2020 | 2020-05-20 | North America/USA/California/Orange ( | Orange County Public Health Laboratory | Chan-Zuckerberg Biohub | CZB Cliahub Consortium et al |
| EPI_ISL_513833 | USA/CA-CZB-2409/2020 | 2020-05-21 | North America/USA/California/Orange ( | Orange County Public Health Laboratory | Chan-Zuckerberg Biohub | CZB Cliahub Consortium et al |
| EPI_ISL_513840 | USA/CA-CZB-2419/2020 | 2020-05-15 | North America/USA/California/Humbold | Humboldt County Public Health Laboratory | Chan-Zuckerberg Biohub | CZB Cliahub Consortium et al |
| EPI_ISL_513845 | USA/CA-CZB-2424/2020 | 2020-05-16 | North America/USA/California/Humbold | Humboldt County Public Health Laboratory | Chan-Zuckerberg Biohub | CZB Cliahub Consortium et al |
| EPI_ISL_513847 | USA/CA-CZB-2426/2020 | 2020-05-18 | North America/USA/California/Humbold | Humboldt County Public Health Laboratory | Chan-Zuckerberg Biohub | CZB Cliahub Consortium et al |
| EPI_ISL_513852 | USA/CA-CZB-2436/2020 | 2020-06-02 | North America/USA/California/Humbold | Humboldt County Public Health Laboratory | Chan-Zuckerberg Biohub | CZB Cliahub Consortium et al |
| EPI_ISL_513854 | USA/CA-CZB-2445/2020 | 2020-05-27 | North America/USA/California/Humbold | Humboldt County Public Health Laboratory | Chan-Zuckerberg Biohub | CZB Cliahub Consortium et al |

| | | | | | | |
|---|---|---|---|---|---|---|
| EPI_ISL_513862 | USA/CA-CZB-2454/2020 | 2020-05-22 | North America/USA/California/San Fran | San Francisco Public Health Laboratory | Chan-Zuckerberg Biohub | CZB Cliahub Consortium et al |
| EPI_ISL_513871 | USA/CA-CZB-2466/2020 | 2020-05-19 | North America/USA/California/San Fran | San Francisco Public Health Laboratory | Chan-Zuckerberg Biohub | CZB Cliahub Consortium et al |
| EPI_ISL_513888 | USA/CA-CZB-2503/2020 | 2020-05-26 | North America/USA/California/Sonoma | UCSF Clinical Microbiology Laboratory | Chan-Zuckerberg Biohub | CZB Cliahub Consortium et al |
| EPI_ISL_513895 | USA/CA-CZB-2520/2020 | 2020-05-19 | North America/USA/California/Sonoma | UCSF Clinical Microbiology Laboratory | Chan-Zuckerberg Biohub | CZB Cliahub Consortium et al |
| EPI_ISL_513898 | USA/CA-CZB-2523/2020 | 2020-05-01 | North America/USA/California/Sonoma | UCSF Clinical Microbiology Laboratory | Chan-Zuckerberg Biohub | CZB Cliahub Consortium et al |
| EPI_ISL_513900 | USA/CA-CZB-2525/2020 | 2020-04-29 | North America/USA/California/Sonoma | UCSF Clinical Microbiology Laboratory | Chan-Zuckerberg Biohub | CZB Cliahub Consortium et al |
| EPI_ISL_513925 | Pakistan/KPK-KUST-SJTU/2020 | 2020-05-15 | Asia/Pakistan/Pakistan/ | Microbiology & Bioinformatics and Biostatistics, Kohat University of Science and Technology (Pakistan) & Shanghai Jiao Tong University (China) | Microbiology & Bioinformatics and Biostatistics, Kohat University of Science and Technology (Pakistan) & Shanghai Jiao Tong University (China) | Khan et al |
| EPI_ISL_514063 | USA/UN-CDC-7413/2020 | 2020-04-20 | North America/USA/USA/ | Navy and Marine Corps Public Health Center | Pathogen Discovery, Respiratory Viruses Branch, Division of Viral Diseases, Centers for Disease Control and Prevention | Yan Li et al |
| EPI_ISL_514071 | USA/UN-CDC-7462/2020 | 2020-04-21 | North America/USA/USA/ | Navy and Marine Corps Public Health Center | Pathogen Discovery, Respiratory Viruses Branch, Division of Viral Diseases, Centers for Disease Control and Prevention | Krista Queen et al |

| | | | | | | |
|---|---|---|---|---|---|---|
| EPI_ISL_514072 | USA/UN-CDC-7467/2020 | 2020-04-21 | North America/USA/USA/ | Navy and Marine Corps Public Health Center | Pathogen Discovery, Respiratory Viruses Branch, Division of Viral Diseases, Centers for Disease Control and Prevention | Krista Queen et al |
| EPI_ISL_514077 | USA/UN-CDC-7493/2020 | 2020-04-22 | North America/USA/USA/ | Navy and Marine Corps Public Health Center | Pathogen Discovery, Respiratory Viruses Branch, Division of Viral Diseases, Centers for Disease Control and Prevention | Yan Li et al |
| EPI_ISL_514081 | USA/UN-CDC-7529/2020 | 2020-04-22 | North America/USA/USA/ | Navy and Marine Corps Public Health Center | Pathogen Discovery, Respiratory Viruses Branch, Division of Viral Diseases, Centers for Disease Control and Prevention | Krista Queen et al |
| EPI_ISL_514083 | USA/UN-CDC-7618/2020 | 2020-04-24 | North America/USA/USA/ | Navy and Marine Corps Public Health Center | Pathogen Discovery, Respiratory Viruses Branch, Division of Viral Diseases, Centers for Disease Control and Prevention | Krista Queen et al |
| EPI_ISL_514108 | DRC/4720/2020 | 2020-05-01 | Africa/Democratic Republic of the Congo | Viral Respiratory Lab, National Institute for Biomedical Research (INRB) | Pathogen Sequencing Lab, National Institute for Biomedical Research (INRB) | Placide Mbala-Kingebeni et al |

| | | | | | | |
|---|---|---|---|---|---|---|
| EPI_ISL_514123 | DRC/7248/2020 | 2020-05-13 | Africa/Democratic Republic of the Congo | Viral Respiratory Lab, National Institute for Biomedical Research (INRB) | Pathogen Sequencing Lab, National Institute for Biomedical Research (INRB) | Placide Mbala-Kingebeni et al |
| EPI_ISL_514129 | Bangladesh/BCSIR-NILMRC-356/2020 | 2020-07-24 | Asia/Bangladesh/Chattogram/Brahmanb | National Institute of Laboratory Medicine and Referral Center | Genomic Research Lab, BCSIR | Md. Murshed Hasan Sarkar et al |
| EPI_ISL_514131 | Brazil/RO-01/2020 | 2020-05-11 | South America/Brazil/Rondonia/Jaru | Rondv¥nia Central Public Health Laboratory (LACEN/RO), vinculated to State Health Secretariat of Rondv¥nia (SESAU/RO) | Molecular Virology Laboratory of Oswaldo Cruz Foundation of Rondv¥nia | Luan Felipo Botelho-Souza et al |
| EPI_ISL_514132 | Brazil/RO-02/2020 | 2020-05-11 | South America/Brazil/Rondonia/Porto V | Rondv¥nia Central Public Health Laboratory (LACEN/RO), vinculated to State Health Secretariat of Rondv¥nia (SESAU/RO) | Molecular Virology Laboratory of Oswaldo Cruz Foundation of Rondv¥nia | Luan Felipo Botelho-Souza et al |
| EPI_ISL_514133 | Brazil/RO-03/2020 | 2020-05-11 | South America/Brazil/Rondonia/Porto V | Rondv¥nia Central Public Health Laboratory (LACEN/RO), vinculated to State Health Secretariat of Rondv¥nia (SESAU/RO) | Molecular Virology Laboratory of Oswaldo Cruz Foundation of Rondv¥nia | Luan Felipo Botelho-Souza et al |
| EPI_ISL_514134 | Brazil/RO-04/2020 | 2020-05-11 | South America/Brazil/Rondonia/Porto V | Rondv¥nia Central Public Health Laboratory (LACEN/RO), vinculated to State Health Secretariat of Rondv¥nia (SESAU/RO) | Molecular Virology Laboratory of Oswaldo Cruz Foundation of Rondv¥nia | Luan Felipo Botelho-Souza et al |
| EPI_ISL_514135 | Brazil/RO-05/2020 | 2020-05-05 | South America/Brazil/Rondonia/Porto V | Rondv¥nia Central Public Health Laboratory (LACEN/RO), vinculated to State Health Secretariat of Rondv¥nia (SESAU/RO) | Molecular Virology Laboratory of Oswaldo Cruz Foundation of Rondv¥nia | Luan Felipo Botelho-Souza et al |

| | | | | | | |
|---|---|---|---|---|---|---|
| EPI_ISL_514136 | Brazil/RO-06/2020 | 2020-05-05 | South America/Brazil/Rondonia/Porto V | Rondv¥nia Central Public Health Laboratory (LACEN/RO), vinculated to State Health Secretariat of Rondv¥nia (SESAU/RO) | Molecular Virology Laboratory of Oswaldo Cruz Foundation of Rondv¥nia | Luan Felipo Botelho-Souza et al |
| EPI_ISL_514137 | Brazil/RO-07/2020 | 2020-05-05 | South America/Brazil/Rondonia/Porto V | Rondv¥nia Central Public Health Laboratory (LACEN/RO), vinculated to State Health Secretariat of Rondv¥nia (SESAU/RO) | Molecular Virology Laboratory of Oswaldo Cruz Foundation of Rondv¥nia | Luan Felipo Botelho-Souza et al |
| EPI_ISL_514138 | Brazil/RO-08/2020 | 2020-06-05 | South America/Brazil/Rondonia/Porto V | Rondv¥nia Central Public Health Laboratory (LACEN/RO), vinculated to State Health Secretariat of Rondv¥nia (SESAU/RO) | Molecular Virology Laboratory of Oswaldo Cruz Foundation of Rondv¥nia | Luan Felipo Botelho-Souza et al |
| EPI_ISL_514146 | USA/FL-BPHL-0800/2020 | 2020-07-01 | North America/USA/Florida/ | Florida Bureau of Public Health Laboratories | Florida Bureau of Public Health Laboratories | Sarah Schmedes et al |
| EPI_ISL_514151 | USA/FL-BPHL-0805/2020 | 2020-05-12 | North America/USA/Florida/ | Florida Bureau of Public Health Laboratories | Florida Bureau of Public Health Laboratories | Sarah Schmedes et al |
| EPI_ISL_514152 | USA/FL-BPHL-0806/2020 | 2020-05-18 | North America/USA/Florida/ | Florida Bureau of Public Health Laboratories | Florida Bureau of Public Health Laboratories | Sarah Schmedes et al |
| EPI_ISL_514170 | USA/FL-BPHL-0824/2020 | 2020-06-12 | North America/USA/Florida/ | Florida Bureau of Public Health Laboratories | Florida Bureau of Public Health Laboratories | Sarah Schmedes et al |
| EPI_ISL_514189 | USA/FL-BPHL-0843/2020 | 2020-06-22 | North America/USA/Florida/ | Florida Bureau of Public Health Laboratories | Florida Bureau of Public Health Laboratories | Sarah Schmedes et al |
| EPI_ISL_514200 | USA/FL-BPHL-0854/2020 | 2020-06-29 | North America/USA/Florida/ | Florida Bureau of Public Health Laboratories | Florida Bureau of Public Health Laboratories | Sarah Schmedes et al |
| EPI_ISL_514201 | USA/FL-BPHL-0855/2020 | 2020-06-27 | North America/USA/Florida/ | Florida Bureau of Public Health Laboratories | Florida Bureau of Public Health Laboratories | Sarah Schmedes et al |

| | | | | | | |
|---|---|---|---|---|---|---|
| EPI_ISL_514211 | USA/FL-BPHL-0865/2020 | 2020-07-02 | North America/USA/Florida/ | Florida Bureau of Public Health Laboratories | Florida Bureau of Public Health Laboratories | Sarah Schmedes et al |
| EPI_ISL_514215 | USA/FL-BPHL-0869/2020 | 2020-07-06 | North America/USA/Florida/ | Florida Bureau of Public Health Laboratories | Florida Bureau of Public Health Laboratories | Sarah Schmedes et al |
| EPI_ISL_514216 | USA/FL-BPHL-0870/2020 | 2020-07-07 | North America/USA/Florida/ | Florida Bureau of Public Health Laboratories | Florida Bureau of Public Health Laboratories | Sarah Schmedes et al |
| EPI_ISL_514226 | Peru/LIM-INS-041/2020 | 2020-04-11 | South America/Peru/Lima/ | Laboratorio de Referencia Nacional de Virus Respiratorio. Instituto Nacional de Salud Per√∫ | Laboratorio de Referencia Nacional de Biotecnolog√≠a y Biolog√≠a Molecular. Instituto Nacional de Salud Per√∫ | Carlos Padilla Rojas et al |
| EPI_ISL_514227 | Peru/LIM-INS-042/2020 | 2020-04-12 | South America/Peru/Lima/ | Laboratorio de Referencia Nacional de Virus Respiratorio. Instituto Nacional de Salud. Peru | Laboratorio de Referencia Nacional de Biotecnologia y Biologia Molecular.Instituto Nacional de Salud.Peru | Carlos Padilla Rojas et al |
| EPI_ISL_514228 | Bangladesh/BCSIR-NILMRC-350/2020 | 2020-07-15 | Asia/Bangladesh/Khulna/ | National Institute of Laboratory Medicine and Referral Center | Genomic Research Lab, BCSIR | Barna Goswami et al |
| EPI_ISL_514233 | Bangladesh/BCSIR-NILMRC-359/2020 | 2020-07-24 | Asia/Bangladesh/Chattogram/Brahmanb | National Institute of Laboratory Medicine and Referral Center | Genomic Research Lab, BCSIR | Md. Murshed Hasan Sarkar et al |
| EPI_ISL_514239 | Bangladesh/BCSIR-NILMRC-367/2020 | 2020-07-21 | Asia/Bangladesh/Chattogram/Brahmanb | National Institute of Laboratory Medicine and Referral Center | Genomic Research Lab, BCSIR | Md. Saddam Hossain et al |
| EPI_ISL_514240 | Bangladesh/BCSIR-NILMRC-368/2020 | 2020-07-21 | Asia/Bangladesh/Chattogram/Brahmanb | National Institute of Laboratory Medicine and Referral Center | Genomic Research Lab, BCSIR | Md. Saddam Hossain et al |
| EPI_ISL_514241 | Bangladesh/BCSIR-NILMRC-370/2020 | 2020-07-21 | Asia/Bangladesh/Chattogram/Brahmanb | National Institute of Laboratory Medicine and Referral Center | Genomic Research Lab, BCSIR | Md. Saddam Hossain et al |
| EPI_ISL_514244 | Bangladesh/BCSIR-NILMRC-387/2020 | 2020-07-18 | Asia/Bangladesh/Chattogram/Chandpur | National Institute of Laboratory Medicine and Referral Center | Genomic Research Lab, BCSIR | Tanjina Akhter Banu et al |
| EPI_ISL_514245 | Bangladesh/BCSIR-NILMRC-369/2020 | 2020-07-21 | Asia/Bangladesh/Chattogram/Brahmanb | National Institute of Laboratory Medicine and Referral Center | Genomic Research Lab, BCSIR | Iffat Jahan et al |

| | | | | | | |
|---|---|---|---|---|---|---|
| EPI_ISL_514247 | Bangladesh/BCSIR-NILMRC-382/2020 | 2020-07-21 | Asia/Bangladesh/Chattogram/Chandpur | National Institute of Laboratory Medicine and Referral Center | Genomic Research Lab, BCSIR | Iffat Jahan et al |
| EPI_ISL_514249 | Bangladesh/BCSIR-NILMRC-372/2020 | 2020-07-31 | Asia/Bangladesh/Dhaka/Narayanganj | National Institute of Laboratory Medicine and Referral Center | Genomic Research Lab, BCSIR | Abu Sayeed Mohammad Mahmud et al |
| EPI_ISL_514253 | Bangladesh/BCSIR-DU-16/2020 | 2020-07-12 | Asia/Bangladesh/Dhaka/Mohammadpur | Advanced Biotechnology Laboratory | Genomic Research Lab, BCSIR | Abu Sayeed Mohammad Mahmud et al |
| EPI_ISL_514269 | Israel/CVL-n25863/2020 | 2020-07-20 | Asia/Israel/Jerusalem District/ | Israel Central Virology laboratory | Israel Central Virology laboratory | Neta Zuckerman et al |
| EPI_ISL_514271 | Israel/CVL-n25870/2020 | 2020-07-20 | Asia/Israel/Jerusalem District/ | Israel Central Virology laboratory | Israel Central Virology laboratory | Neta Zuckerman et al |
| EPI_ISL_514272 | Israel/CVL-n25872/2020 | 2020-07-20 | Asia/Israel/Jerusalem District/ | Israel Central Virology laboratory | Israel Central Virology laboratory | Neta Zuckerman et al |
| EPI_ISL_514274 | Israel/CVL-n25886/2020 | 2020-07-20 | Asia/Israel/Jerusalem District/ | Israel Central Virology laboratory | Israel Central Virology laboratory | Neta Zuckerman et al |
| EPI_ISL_514275 | Israel/CVL-n25904/2020 | 2020-07-19 | Asia/Israel/Jerusalem District/ | Israeli Central Virology laboratory | Israel Central Virology laboratory | Neta Zuckerman et al |
| EPI_ISL_514276 | Israel/CVL-n25907/2020 | 2020-07-20 | Asia/Israel/Jerusalem District/ | Israel Central Virology laboratory | Israel Central Virology laboratory | Neta Zuckerman et al |
| EPI_ISL_514278 | Israel/CVL-n25922/2020 | 2020-07-20 | Asia/Israel/Jerusalem District/ | Israel Central Virology laboratory | Israel Central Virology laboratory | Neta Zuckerman et al |
| EPI_ISL_514279 | Israel/CVL-n25924/2020 | 2020-07-20 | Asia/Israel/Jerusalem District/ | Israel Central Virology laboratory | Israel Central Virology laboratory | Neta Zuckerman et al |
| EPI_ISL_514280 | Israel/CVL-n25929/2020 | 2020-07-20 | Asia/Israel/Jerusalem District/ | Israel Central Virology laboratory | Israel Central Virology laboratory | Neta Zuckerman et al |
| EPI_ISL_514282 | Israel/CVL-n25955/2020 | 2020-07-20 | Asia/Israel/Jerusalem District/ | Israel Central Virology laboratory | Israel Central Virology laboratory | Neta Zuckerman et al |
| EPI_ISL_514283 | Israel/CVL-n26099/2020 | 2020-07-20 | Asia/Israel/Jerusalem District/ | Israel Central Virology laboratory | Israel Central Virology laboratory | Neta Zuckerman et al |

| | | | | | | |
|---|---|---|---|---|---|---|
| EPI_ISL_514284 | Israel/CVL-n26111/2020 | 2020-07-20 | Asia/Israel/Jerusalem District/ | Israel Central Virology laboratory | Israel Central Virology laboratory | Neta Zuckerman et al |
| EPI_ISL_514287 | Israel/CVL-n26122/2020 | 2020-07-20 | Asia/Israel/Jerusalem District/ | Israel Central Virology laboratory | Israel Central Virology laboratory | Neta Zuckerman et al |
| EPI_ISL_514288 | Israel/CVL-n26131/2020 | 2020-07-20 | Asia/Israel/Jerusalem District/ | Israel Central Virology laboratory | Israel Central Virology laboratory | Neta Zuckerman et al |
| EPI_ISL_514289 | Israel/CVL-n26134/2020 | 2020-07-20 | Asia/Israel/Jerusalem District/ | Israel Central Virology laboratory | Israel Central Virology laboratory | Neta Zuckerman et al |
| EPI_ISL_514290 | Israel/CVL-n26140/2020 | 2020-07-20 | Asia/Israel/Jerusalem District/ | Israel Central Virology laboratory | Israel Central Virology laboratory | Neta Zuckerman et al |
| EPI_ISL_514291 | Israel/CVL-n26150/2020 | 2020-07-20 | Asia/Israel/Jerusalem District/ | Israel Central Virology laboratory | Israel Central Virology laboratory | Neta Zuckerman et al |
| EPI_ISL_514292 | Israel/CVL-n26156/2020 | 2020-07-20 | Asia/Israel/Jerusalem District/ | Israel Central Virology laboratory | Israel Central Virology laboratory | Neta Zuckerman et al |
| EPI_ISL_514293 | Israel/CVL-n26166/2020 | 2020-07-20 | Asia/Israel/Jerusalem District/ | Israel Central Virology laboratory | Israel Central Virology laboratory | Neta Zuckerman et al |
| EPI_ISL_514294 | Israel/CVL-n26167/2020 | 2020-07-20 | Asia/Israel/Jerusalem District/ | Israel Central Virology laboratory | Israel Central Virology laboratory | Neta Zuckerman et al |
| EPI_ISL_514296 | Israel/CVL-n26170/2020 | 2020-07-20 | Asia/Israel/Jerusalem District/ | Israel Central Virology laboratory | Israel Central Virology laboratory | Neta Zuckerman et al |
| EPI_ISL_514297 | Israel/CVL-n26389/2020 | 2020-07-21 | Asia/Israel/Jerusalem District/ | Israel Central Virology laboratory | Israel Central Virology laboratory | Neta Zuckerman et al |
| EPI_ISL_514298 | Israel/CVL-n26400/2020 | 2020-07-21 | Asia/Israel/Jerusalem District/ | Israel Central Virology laboratory | Israel Central Virology laboratory | Neta Zuckerman et al |
| EPI_ISL_514300 | Israel/CVL-n26425/2020 | 2020-07-21 | Asia/Israel/Jerusalem District/ | Israel Central Virology laboratory | Israel Central Virology laboratory | Neta Zuckerman et al |

| | | | | | | |
|---|---|---|---|---|---|---|
| EPI_ISL_514301 | Israel/CVL-n26427/2020 | 2020-07-21 | Asia/Israel/Jerusalem District/ | Israel Central Virology laboratory | Israel Central Virology laboratory | Neta Zuckerman et al |
| EPI_ISL_514302 | Israel/CVL-n26455/2020 | 2020-07-21 | Asia/Israel/Jerusalem District/ | Israel Central Virology laboratory | Israel Central Virology laboratory | Neta Zuckerman et al |
| EPI_ISL_514303 | Israel/CVL-n26456/2020 | 2020-07-21 | Asia/Israel/Jerusalem District/ | Israel Central Virology laboratory | Israel Central Virology laboratory | Neta Zuckerman et al |
| EPI_ISL_514305 | Israel/CVL-n26496/2020 | 2020-07-23 | Asia/Israel/Central District/ | Israel Central Virology laboratory | Israel Central Virology laboratory | Neta Zuckerman et al |
| EPI_ISL_514309 | Israel/CVL-n14269/2020 | 2020-03-17 | Asia/Israel/Central District/ | Israel Central Virology laboratory | Israel Central Virology laboratory | Neta Zuckerman et al |
| EPI_ISL_514339 | Peru/LOR-INS-079/2020 | 2020-04-22 | South America/Peru/Loreto/ | Laboratorio de Referencia Nacional de Virus Respiratorio. Instituto Nacional de Salud Perv∫ | Laboratorio de Referencia Nacional de Biotecnologv≠a y Biologv≠a Molecular. Instituto Nacional de Salud Perv∫ | Carlos Padilla Rojas et al |
| EPI_ISL_514340 | Peru/LOR-INS-080/2020 | 2020-04-22 | South America/Peru/Loreto/ | Laboratorio de Referencia Nacional de Virus Respiratorio. Instituto Nacional de Salud Perv∫ | Laboratorio de Referencia Nacional de Biotecnologv≠a y Biologv≠a Molecular. Instituto Nacional de Salud Perv∫ | Carlos Padilla Rojas et al |
| EPI_ISL_514341 | Peru/LOR-INS-081/2020 | 2020-04-22 | South America/Peru/Loreto/ | Laboratorio de Referencia Nacional de Virus Respiratorio. Instituto Nacional de Salud Perv∫ | Laboratorio de Referencia Nacional de Biotecnologv≠a y Biologv≠a Molecular. Instituto Nacional de Salud Perv∫ | Carlos Padilla Rojas et al |

| | | | | | | |
|---|---|---|---|---|---|---|
| EPI_ISL_514357 | SouthAfrica/LR00125/2020 | 2020-07-20 | Africa/South Africa/Gauteng/ | National Institute for Communicable Diseases of the National Health Laboratory Service | National Institute for Communicable Diseases of the National Health Laboratory Service | Allam M et al |
| EPI_ISL_514359 | SouthAfrica/LR00127/2020 | 2020-07-20 | Africa/South Africa/Gauteng/ | National Institute for Communicable Diseases of the National Health Laboratory Service | National Institute for Communicable Diseases of the National Health Laboratory Service | Allam M et al |
| EPI_ISL_514370 | SouthAfrica/LR00139/2020 | 2020-07-20 | Africa/South Africa/Gauteng/ | National Institute for Communicable Diseases of the National Health Laboratory Service | National Institute for Communicable Diseases of the National Health Laboratory Service | Allam M et al |
| EPI_ISL_514377 | SouthAfrica/LR00064/2020 | 2020-07-13 | Africa/South Africa/Gauteng/ | National Institute for Communicable Diseases of the National Health Laboratory Service | National Institute for Communicable Diseases of the National Health Laboratory Service | Allam M et al |
| EPI_ISL_514379 | SouthAfrica/LR00067/2020 | 2020-07-13 | Africa/South Africa/Mpumalanga/ | National Institute for Communicable Diseases of the National Health Laboratory Service | National Institute for Communicable Diseases of the National Health Laboratory Service | Allam M et al |
| EPI_ISL_514381 | SouthAfrica/LR00069/2020 | 2020-07-13 | Africa/South Africa/Gauteng/ | National Institute for Communicable Diseases of the National Health Laboratory Service | National Institute for Communicable Diseases of the National Health Laboratory Service | Allam M et al |

| | | | | | | |
|---|---|---|---|---|---|---|
| EPI_ISL_514382 | SouthAfrica/LR00070/2020 | 2020-07-13 | Africa/South Africa/Gauteng/ | National Institute for Communicable Diseases of the National Health Laboratory Service | National Institute for Communicable Diseases of the National Health Laboratory Service | Allam M et al |
| EPI_ISL_514383 | SouthAfrica/LR00071/2020 | 2020-07-13 | Africa/South Africa/Gauteng/ | National Institute for Communicable Diseases of the National Health Laboratory Service | National Institute for Communicable Diseases of the National Health Laboratory Service | Allam M et al |
| EPI_ISL_514387 | SouthAfrica/LR00076/2020 | 2020-07-13 | Africa/South Africa/Gauteng/ | National Institute for Communicable Diseases of the National Health Laboratory Service | National Institute for Communicable Diseases of the National Health Laboratory Service | Allam M et al |
| EPI_ISL_514388 | SouthAfrica/LR00077/2020 | 2020-07-13 | Africa/South Africa/Gauteng/ | National Institute for Communicable Diseases of the National Health Laboratory Service | National Institute for Communicable Diseases of the National Health Laboratory Service | Allam M et al |
| EPI_ISL_514389 | SouthAfrica/LR00078/2020 | 2020-07-13 | Africa/South Africa/Gauteng/ | National Institute for Communicable Diseases of the National Health Laboratory Service | National Institute for Communicable Diseases of the National Health Laboratory Service | Allam M et al |
| EPI_ISL_514392 | SouthAfrica/LR00081/2020 | 2020-07-13 | Africa/South Africa/Gauteng/ | National Institute for Communicable Diseases of the National Health Laboratory Service | National Institute for Communicable Diseases of the National Health Laboratory Service | Allam M et al |

| | | | | | | |
|---|---|---|---|---|---|---|
| EPI_ISL_514393 | SouthAfrica/LR00083/2020 | 2020-07-13 | Africa/South Africa/Gauteng/ | National Institute for Communicable Diseases of the National Health Laboratory Service | National Institute for Communicable Diseases of the National Health Laboratory Service | Allam M et al |
| EPI_ISL_514401 | SouthAfrica/LR00102/2020 | 2020-07-14 | Africa/South Africa/Gauteng/ | National Institute for Communicable Diseases of the National Health Laboratory Service | National Institute for Communicable Diseases of the National Health Laboratory Service | Allam M et al |
| EPI_ISL_514403 | SouthAfrica/LR00106/2020 | 2020-07-15 | Africa/South Africa/Gauteng/ | National Institute for Communicable Diseases of the National Health Laboratory Service | National Institute for Communicable Diseases of the National Health Laboratory Service | Allam M et al |
| EPI_ISL_514404 | SouthAfrica/LR00107/2020 | 2020-07-14 | Africa/South Africa/Mpumalanga/ | National Institute for Communicable Diseases of the National Health Laboratory Service | National Institute for Communicable Diseases of the National Health Laboratory Service | Allam M et al |
| EPI_ISL_514416 | SouthAfrica/LR00148/2020 | 2020-07-20 | Africa/South Africa/Gauteng/ | National Institute for Communicable Diseases of the National Health Laboratory Service | National Institute for Communicable Diseases of the National Health Laboratory Service | Allam M et al |
| EPI_ISL_514431 | Serbia/UE16-22-04/2020 | 2020-04-21 | Europe/Serbia/Serbia/ | Laboratory Diagnostic, Veterinary Specialized Institute Kraljevo | Laboratory Diagnostic, Veterinary Specialized Institute Kraljevo | Vidanovic et al |
| EPI_ISL_514432 | Italy/CAM-CEINGE-5/2020 | 2020-04-15 | Europe/Italy/Campania/Benevento | Prof. Massimo Zollo CEINGE TASK-FORCE COVID19 - Regione Campania | Prof. Massimo Zollo CEINGE TASK-FORCE COVID19 - Regione Campania | Veronica Ferrucci et al |

| | | | | | | |
|---|---|---|---|---|---|---|
| EPI_ISL_514435 | India/GJ-GBRC337a/2020 | 2020-07-09 | Asia/India/Gujarat/Modasa | Dr. RSS Hospital, Modasa | Gujarat Biotechnology Research Centre | Harsh Chaudhari et al |
| EPI_ISL_514437 | India/GJ-GBRC340/2020 | 2020-07-09 | Asia/India/Gujarat/Modasa | Dr. RSS Hospital, Modasa | Gujarat Biotechnology Research Centre | Nidhi Patel et al |
| EPI_ISL_514439 | USA/ME-HETL-J0086/2020 | 2020-03-20 | North America/USA/Maine/ | Tewhey Lab, The Jackson Laboratory | Tewhey Lab, The Jackson Laboratory | Matluk et al |
| EPI_ISL_514440 | Bangladesh/NGRI-NSTU-03/2020 | 2020-07-22 | Asia/Bangladesh/Chattogram/Noakhali | NSTU COVID-19 Diagnostic Center | NSU Genome Research Institute (NGRI), North South University | Dr. Muhammad Maqsud Hossain et al |
| EPI_ISL_514441 | Bangladesh/NGRI-NSTU-05/2020 | 2020-07-23 | Asia/Bangladesh/Chattogram/Noakhali | NSTU COVID-19 Diagnostic Center | NSU Genome Research Institute (NGRI), North South University | Dr. Muhammad Maqsud Hossain et al |
| EPI_ISL_514584 | India/GJ-GBRC343a/2020 | 2020-06-18 | Asia/India/Gujarat/Ahmedabad | B.J. Medical College and Civil hospital, Ahmedabad | Gujarat Biotechnology Research Centre | Zuber Saiyed et al |
| EPI_ISL_514587 | India/GJ-GBRC345/2020 | 2020-06-18 | Asia/India/Gujarat/Ahmedabad | B.J. Medical College and Civil hospital, Ahmedabad | Gujarat Biotechnology Research Centre | Afzal Ansari et al |
| EPI_ISL_514589 | India/GJ-GBRC347a/2020 | 2020-06-18 | Asia/India/Gujarat/Daskroi | B.J. Medical College and Civil hospital, Ahmedabad | Gujarat Biotechnology Research Centre | Pranay Shah et al |
| EPI_ISL_514591 | India/GJ-GBRC348a/2020 | 2020-06-18 | Asia/India/Gujarat/Ahmedabad | B.J. Medical College and Civil hospital, Ahmedabad | Gujarat Biotechnology Research Centre | Sanjay Kapadia et al |
| EPI_ISL_514592 | India/GJ-GBRC348b/2020 | 2020-06-18 | Asia/India/Gujarat/Ahmedabad | B.J. Medical College and Civil hospital, Ahmedabad | Gujarat Biotechnology Research Centre | Apurvasinh Puvar et al |
| EPI_ISL_514593 | India/GJ-GBRC349/2020 | 2020-06-18 | Asia/India/Gujarat/Ahmedabad | B.J. Medical College and Civil hospital, Ahmedabad | Gujarat Biotechnology Research Centre | Janvi Raval et al |
| EPI_ISL_514594 | India/GJ-GBRC350a/2020 | 2020-06-18 | Asia/India/Gujarat/Ahmedabad | B.J. Medical College and Civil hospital, Ahmedabad | Gujarat Biotechnology Research Centre | Zarna Patel et al |
| EPI_ISL_514595 | India/GJ-GBRC350b/2020 | 2020-06-18 | Asia/India/Gujarat/Ahmedabad | B.J. Medical College and Civil hospital, Ahmedabad | Gujarat Biotechnology Research Centre | Monika Gandhi et al |

| | | | | | | |
|---|---|---|---|---|---|---|
| EPI_ISL_514596 | India/GJ-GBRC351a/2020 | 2020-06-18 | Asia/India/Gujarat/Ahmedabad | B.J. Medical College and Civil hospital, Ahmedabad | Gujarat Biotechnology Research Centre | Pinal Trivedi et al |
| EPI_ISL_514599 | India/GJ-GBRC352b/2020 | 2020-06-18 | Asia/India/Gujarat/Ahmedabad | B.J. Medical College and Civil hospital, Ahmedabad | Gujarat Biotechnology Research Centre | Nitin Savaliya et al |
| EPI_ISL_514602 | India/GJ-GBRC354b/2020 | 2020-06-25 | Asia/India/Gujarat/Ahmedabad | B.J. Medical College and Civil hospital, Ahmedabad | Gujarat Biotechnology Research Centre | Zuber Saiyed et al |
| EPI_ISL_514603 | India/GJ-GBRC355a/2020 | 2020-06-25 | Asia/India/Gujarat/Ahmedabad | B.J. Medical College and Civil hospital, Ahmedabad | Gujarat Biotechnology Research Centre | Komal Patel et al |
| EPI_ISL_514605 | India/GJ-GBRC356/2020 | 2020-06-25 | Asia/India/Gujarat/Ahmedabad | B.J. Medical College and Civil hospital, Ahmedabad | Gujarat Biotechnology Research Centre | Afzal Ansari et al |
| EPI_ISL_514607 | India/GJ-GBRC357b/2020 | 2020-06-25 | Asia/India/Gujarat/Ahmedabad | B.J. Medical College and Civil hospital, Ahmedabad | Gujarat Biotechnology Research Centre | Pranay Shah et al |
| EPI_ISL_514610 | India/GJ-GBRC360/2020 | 2020-06-15 | Asia/India/Gujarat/Vadodara | GMERS Medical College & Hospital, Gotri, Vadodara | Gujarat Biotechnology Research Centre | Meenakshi Shah et al |
| EPI_ISL_514612 | India/GJ-GBRC361b/2020 | 2020-06-05 | Asia/India/Gujarat/Kalol | GMERS Medical College and Hospital, Gandhinagar | Gujarat Biotechnology Research Centre | Gaurishankar Shrimali et al |
| EPI_ISL_514613 | Bangladesh/NGRI-NSTU-18/2020 | 2020-07-21 | Asia/Bangladesh/Chattogram/Noakhali | NSTU COVID-19 Diagnostic Center | NSU Genome Research Institute (NGRI), North South University | Dr. Muhammad Maqsud Hossain et al |
| EPI_ISL_514615 | Bangladesh/NGRI-NSTU-21/2020 | 2020-07-20 | Asia/Bangladesh/Chattogram/Lakshmip | NSTU COVID-19 Diagnostic Center | NSU Genome Research Institute (NGRI), North South University | Dr. Muhammad Maqsud Hossain et al |
| EPI_ISL_514626 | USA/MN-MDH-1407/2020 | 2020-06-26 | North America/USA/Minnesota/ | Mayo Clinic & Mayo Clinic Laboratories | Minnesota Department of Health, Public Health Laboratory | Matt Plumb et al |
| EPI_ISL_514660 | USA/MN-MDH-1441/2020 | 2020-03-25 | North America/USA/Minnesota/ | Minnesota Department of Health, Public Health Laboratory | Minnesota Department of Health, Public Health Laboratory | Matt Plumb et al |

| | | | | | | |
|---|---|---|---|---|---|---|
| EPI_ISL_514750 | Russia/MOW-PMU-0417/2020 | 2020-04-17 | Europe/Russia/Moscow Oblast/ | Pirogov Russian National Research Medical University | Pirogov Russian National Research Medical University, Research and Development | Blagodatskikh et al |
| EPI_ISL_514751 | Italy/CAM-CEINGE-S66/2020 | 2020-04-15 | Europe/Italy/Campania/Benevento | CoronaNet Lab- TaskForce Regione Campania, CEINGE Biotecnologie Avanzate, Via G. Salvatore | CoronaNet Lab- TaskForce Regione Campania, CEINGE Biotecnologie Avanzate, Via G. Salvatore | Zollo et al |
| EPI_ISL_514752 | Beijing/Beijing-01/2020 | 2020-01-03 | Asia/China/Beijing/ | Infectious Disease Control Center, Center for Disease Control and Prevention of PLA | Infectious Disease Control Center, Center for Disease Control and Prevention of PLA | Li et al |
| EPI_ISL_514860 | SouthKorea/KCDC2457/2020 | 2020-06-08 | Asia/South Korea/South Korea/ | Division of Viral Diseases, Center for Laboratory Control of Infectious Diseases, Korea Centers for Diseases Control and Prevention | Division of Viral Diseases, Center for Laboratory Control of Infectious Diseases, Korea Centers for Diseases Control and Prevention | Jeong-Min Kim et al |
| EPI_ISL_514976 | SouthKorea/KCDC2573/2020 | 2020-06-26 | Asia/South Korea/South Korea/ | Division of Viral Diseases, Center for Laboratory Control of Infectious Diseases, Korea Centers for Diseases Control and Prevention | Division of Viral Diseases, Center for Laboratory Control of Infectious Diseases, Korea Centers for Diseases Control and Prevention | Jeong-Min Kim et al |
| EPI_ISL_515066 | Belgium/ULG-10259/2020 | 2020-07-11 | Europe/Belgium/Liv®ge/ | Department of Clinical Microbiology | GIGA Medical Genomics | Keith Durkin et al |
| EPI_ISL_515087 | Ghana/34928_S21/2020 | 2020-05-23 | Africa/Ghana/Ghana/ | Department of Biochemistry, Cell and Molecular Biology | WACCBIP, University of Ghana | Ngoi et al |

| | | | | | | |
|---|---|---|---|---|---|---|
| EPI_ISL_515088 | Ghana/34980_S22/2020 | 2020-05-23 | Africa/Ghana/Ghana/ | Department of Biochemistry, Cell and Molecular Biology | WACCBIP, University of Ghana | Ngoi et al |
| EPI_ISL_515089 | Ghana/34983_S40/2020 | 2020-05-23 | Africa/Ghana/Ghana/ | Department of Biochemistry, Cell and Molecular Biology | WACCBIP, University of Ghana | Ngoi et al |
| EPI_ISL_515093 | Ghana/35280_S41/2020 | 2020-05-24 | Africa/Ghana/Ghana/ | Department of Biochemistry, Cell and Molecular Biology | WACCBIP, University of Ghana | Ngoi et al |
| EPI_ISL_515094 | Ghana/35394_S49/2020 | 2020-05-24 | Africa/Ghana/Ghana/ | Department of Biochemistry, Cell and Molecular Biology | WACCBIP, University of Ghana | Ngoi et al |
| EPI_ISL_515096 | Ghana/35563_S37/2020 | 2020-05-24 | Africa/Ghana/Ghana/ | Department of Biochemistry, Cell and Molecular Biology | WACCBIP, University of Ghana | Ngoi et al |
| EPI_ISL_515101 | Ghana/83637_S29/2020 | 2020-05-22 | Africa/Ghana/Ghana/ | Department of Biochemistry, Cell and Molecular Biology | WACCBIP, University of Ghana | Ngoi et al |
| EPI_ISL_515103 | Ghana/84162_S45/2020 | 2020-04-25 | Africa/Ghana/Ghana/ | Department of Biochemistry, Cell and Molecular Biology | WACCBIP, University of Ghana | Ngoi et al |
| EPI_ISL_515106 | Ghana/84218_S47/2020 | 2020-05-25 | Africa/Ghana/Ghana/ | Department of Biochemistry, Cell and Molecular Biology | WACCBIP, University of Ghana | Ngoi et al |
| EPI_ISL_515110 | Ghana/84430_S51/2020 | 2020-05-25 | Africa/Ghana/Ghana/ | Department of Biochemistry, Cell and Molecular Biology | WACCBIP, University of Ghana | Ngoi et al |
| EPI_ISL_515111 | Ghana/84438_S50/2020 | 2020-05-24 | Africa/Ghana/Ghana/ | Department of Biochemistry, Cell and Molecular Biology | WACCBIP, University of Ghana | Ngoi et al |
| EPI_ISL_515136 | SouthAfrica/LR00005/2020 | 2020-07-03 | Africa/South Africa/Gauteng/ | National Institute for Communicable Diseases of the National Health Laboratory Service | National Institute for Communicable Diseases of the National Health Laboratory Service | Allam M et al |

| | | | | | | |
|---|---|---|---|---|---|---|
| EPI_ISL_515138 | SouthAfrica/LR00008/2020 | 2020-06-30 | Africa/South Africa/Eastern Cape/ | National Institute for Communicable Diseases of the National Health Laboratory Service | National Institute for Communicable Diseases of the National Health Laboratory Service | Allam M et al |
| EPI_ISL_515151 | SouthAfrica/LR00022/2020 | 2020-07-02 | Africa/South Africa/Gauteng/ | National Institute for Communicable Diseases of the National Health Laboratory Service | National Institute for Communicable Diseases of the National Health Laboratory Service | Allam M et al |
| EPI_ISL_515155 | SouthAfrica/LR00026/2020 | 2020-07-04 | Africa/South Africa/Gauteng/ | National Institute for Communicable Diseases of the National Health Laboratory Service | National Institute for Communicable Diseases of the National Health Laboratory Service | Allam M et al |
| EPI_ISL_515159 | SouthAfrica/LR00030/2020 | 2020-07-03 | Africa/South Africa/North-West/ | National Institute for Communicable Diseases of the National Health Laboratory Service | National Institute for Communicable Diseases of the National Health Laboratory Service | Allam M et al |
| EPI_ISL_515161 | SouthAfrica/LR00033/2020 | 2020-07-01 | Africa/South Africa/Gauteng/ | National Institute for Communicable Diseases of the National Health Laboratory Service | National Institute for Communicable Diseases of the National Health Laboratory Service | Allam M et al |
| EPI_ISL_515162 | SouthAfrica/LR00034/2020 | 2020-07-04 | Africa/South Africa/North-West/ | National Institute for Communicable Diseases of the National Health Laboratory Service | National Institute for Communicable Diseases of the National Health Laboratory Service | Allam M et al |

| | | | | | | |
|---|---|---|---|---|---|---|
| EPI_ISL_515164 | SouthAfrica/LR00037/2020 | 2020-07-05 | Africa/South Africa/Gauteng/ | National Institute for Communicable Diseases of the National Health Laboratory Service | National Institute for Communicable Diseases of the National Health Laboratory Service | Allam M et al |
| EPI_ISL_515166 | SouthAfrica/LR00039/2020 | 2020-07-05 | Africa/South Africa/Gauteng/ | National Institute for Communicable Diseases of the National Health Laboratory Service | National Institute for Communicable Diseases of the National Health Laboratory Service | Allam M et al |
| EPI_ISL_515169 | SouthAfrica/LR00042/2020 | 2020-07-06 | Africa/South Africa/Gauteng/ | National Institute for Communicable Diseases of the National Health Laboratory Service | National Institute for Communicable Diseases of the National Health Laboratory Service | Allam M et al |
| EPI_ISL_515171 | SouthAfrica/LR00044/2020 | 2020-07-05 | Africa/South Africa/Gauteng/ | National Institute for Communicable Diseases of the National Health Laboratory Service | National Institute for Communicable Diseases of the National Health Laboratory Service | Allam M et al |
| EPI_ISL_515179 | SouthAfrica/LR00059/2020 | 2020-07-07 | Africa/South Africa/North-West/ | National Institute for Communicable Diseases of the National Health Laboratory Service | National Institute for Communicable Diseases of the National Health Laboratory Service | Allam M et al |
| EPI_ISL_515180 | SouthAfrica/LR00060/2020 | 2020-07-10 | Africa/South Africa/Gauteng/ | National Institute for Communicable Diseases of the National Health Laboratory Service | National Institute for Communicable Diseases of the National Health Laboratory Service | Allam M et al |
| EPI_ISL_515197 | Canada/QC_AA1/2020 | 2020-04-05 | North America/Canada/Quebec/ | Laboratoire de microbiologie, Hopital de Verdun | Smith Laboratory, Centre de Recherche CHU Sainte-Justine | Martin Smith et al |

| | | | | | | |
|---|---|---|---|---|---|---|
| EPI_ISL_515198 | Canada/QC_AE2/2020 | 2020-04-05 | North America/Canada/Quebec/ | Laboratoire de microbiologie, Hopital de Verdun | Smith Laboratory, Centre de Recherche CHU Sainte-Justine | Martin Smith et al |
| EPI_ISL_515199 | Canada/QC_AE3/2020 | 2020-04-05 | North America/Canada/Quebec/ | Laboratoire de microbiologie, Hopital de Verdun | Smith Laboratory, Centre de Recherche CHU Sainte-Justine | Martin Smith et al |
| EPI_ISL_515200 | Canada/QC_AE7/2020 | 2020-04-05 | North America/Canada/Quebec/ | Laboratoire de microbiologie, Hopital de Verdun | Smith Laboratory, Centre de Recherche CHU Sainte-Justine | Martin Smith et al |
| EPI_ISL_515201 | Canada/QC_AF3/2020 | 2020-04-06 | North America/Canada/Quebec/ | Laboratoire de microbiologie, Hopital de Verdun | Smith Laboratory, Centre de Recherche CHU Sainte-Justine | Martin Smith et al |
| EPI_ISL_515202 | Canada/QC_AF7/2020 | 2020-04-04 | North America/Canada/Quebec/ | Laboratoire de microbiologie, Hopital de Verdun | Smith Laboratory, Centre de Recherche CHU Sainte-Justine | Martin Smith et al |
| EPI_ISL_515203 | Canada/QC_AG2/2020 | 2020-04-11 | North America/Canada/Quebec/ | Laboratoire de microbiologie, Hopital de Verdun | Smith Laboratory, Centre de Recherche CHU Sainte-Justine | Martin Smith et al |
| EPI_ISL_515204 | Canada/QC_AH3/2020 | 2020-04-19 | North America/Canada/Quebec/ | Laboratoire de microbiologie, Hopital de Verdun | Smith Laboratory, Centre de Recherche CHU Sainte-Justine | Martin Smith et al |
| EPI_ISL_515205 | Canada/QC_AH8/2020 | 2020-04-19 | North America/Canada/Quebec/ | Laboratoire de microbiologie, Hopital de Verdun | Smith Laboratory, Centre de Recherche CHU Sainte-Justine | Martin Smith et al |
| EPI_ISL_515206 | Canada/QC_AI2/2020 | 2020-04-10 | North America/Canada/Quebec/ | Laboratoire de microbiologie, Hopital de Verdun | Smith Laboratory, Centre de Recherche CHU Sainte-Justine | Martin Smith et al |
| EPI_ISL_515207 | Canada/QC_AJ1/2020 | 2020-04-19 | North America/Canada/Quebec/ | Laboratoire de microbiologie, Hopital de Verdun | Smith Laboratory, Centre de Recherche CHU Sainte-Justine | Martin Smith et al |

| | | | | | | |
|---|---|---|---|---|---|---|
| EPI_ISL_515208 | Canada/QC_AJ5/2020 | 2020-04-11 | North America/Canada/Quebec/ | Laboratoire de microbiologie, Hopital de Verdun | Smith Laboratory, Centre de Recherche CHU Sainte-Justine | Martin Smith et al |
| EPI_ISL_515209 | Canada/QC_AK6/2020 | 2020-04-17 | North America/Canada/Quebec/ | Laboratoire de microbiologie, Hopital de Verdun | Smith Laboratory, Centre de Recherche CHU Sainte-Justine | Martin Smith et al |
| EPI_ISL_515210 | Canada/QC_AK9/2020 | 2020-04-17 | North America/Canada/Quebec/ | Laboratoire de microbiologie, Hopital de Verdun | Smith Laboratory, Centre de Recherche CHU Sainte-Justine | Martin Smith et al |
| EPI_ISL_515211 | Canada/QC_AL3/2020 | 2020-04-07 | North America/Canada/Quebec/ | Laboratoire de microbiologie, Hopital de Verdun | Smith Laboratory, Centre de Recherche CHU Sainte-Justine | Martin Smith et al |
| EPI_ISL_515212 | Canada/QC_AL4/2020 | 2020-04-18 | North America/Canada/Quebec/ | Laboratoire de microbiologie, Hopital de Verdun | Smith Laboratory, Centre de Recherche CHU Sainte-Justine | Martin Smith et al |
| EPI_ISL_515213 | Canada/QC_AM2/2020 | 2020-04-13 | North America/Canada/Quebec/ | Laboratoire de microbiologie, Hopital de Verdun | Smith Laboratory, Centre de Recherche CHU Sainte-Justine | Martin Smith et al |
| EPI_ISL_515214 | Canada/QC_AM4/2020 | 2020-04-09 | North America/Canada/Quebec/ | Laboratoire de microbiologie, Hopital de Verdun | Smith Laboratory, Centre de Recherche CHU Sainte-Justine | Martin Smith et al |
| EPI_ISL_515215 | Canada/QC_AM7/2020 | 2020-04-08 | North America/Canada/Quebec/ | Laboratoire de microbiologie, Hopital de Verdun | Smith Laboratory, Centre de Recherche CHU Sainte-Justine | Martin Smith et al |
| EPI_ISL_515216 | Canada/QC_AM8/2020 | 2020-04-09 | North America/Canada/Quebec/ | Laboratoire de microbiologie, Hopital de Verdun | Smith Laboratory, Centre de Recherche CHU Sainte-Justine | Martin Smith et al |
| EPI_ISL_515217 | Canada/QC_AN3/2020 | 2020-04-08 | North America/Canada/Quebec/ | Laboratoire de microbiologie, Hopital de Verdun | Smith Laboratory, Centre de Recherche CHU Sainte-Justine | Martin Smith et al |

| | | | | | | |
|---|---|---|---|---|---|---|
| EPI_ISL_515218 | Canada/QC_AN4/2020 | 2020-04-08 | North America/Canada/Quebec/ | Laboratoire de microbiologie, Hopital de Verdun | Smith Laboratory, Centre de Recherche CHU Sainte-Justine | Martin Smith et al |
| EPI_ISL_515219 | Canada/QC_AN5/2020 | 2020-04-19 | North America/Canada/Quebec/ | Laboratoire de microbiologie, Hopital de Verdun | Smith Laboratory, Centre de Recherche CHU Sainte-Justine | Martin Smith et al |
| EPI_ISL_515220 | Canada/QC_AN6/2020 | 2020-04-08 | North America/Canada/Quebec/ | Laboratoire de microbiologie, Hopital de Verdun | Smith Laboratory, Centre de Recherche CHU Sainte-Justine | Martin Smith et al |
| EPI_ISL_515221 | Canada/QC_AN8/2020 | 2020-04-16 | North America/Canada/Quebec/ | Laboratoire de microbiologie, Hopital de Verdun | Smith Laboratory, Centre de Recherche CHU Sainte-Justine | Martin Smith et al |
| EPI_ISL_515222 | Canada/QC_AN9/2020 | 2020-04-07 | North America/Canada/Quebec/ | Laboratoire de microbiologie, Hopital de Verdun | Smith Laboratory, Centre de Recherche CHU Sainte-Justine | Martin Smith et al |
| EPI_ISL_515223 | Canada/QC_AO2/2020 | 2020-04-17 | North America/Canada/Quebec/ | Laboratoire de microbiologie, Hopital de Verdun | Smith Laboratory, Centre de Recherche CHU Sainte-Justine | Martin Smith et al |
| EPI_ISL_515224 | Canada/QC_AP4/2020 | 2020-04-18 | North America/Canada/Quebec/ | Laboratoire de microbiologie, Hopital de Verdun | Smith Laboratory, Centre de Recherche CHU Sainte-Justine | Martin Smith et al |
| EPI_ISL_515225 | Canada/QC_AQ8/2020 | 2020-04-06 | North America/Canada/Quebec/ | Laboratoire de microbiologie, Hopital de Verdun | Smith Laboratory, Centre de Recherche CHU Sainte-Justine | Martin Smith et al |
| EPI_ISL_515226 | Canada/QC_AR7/2020 | 2020-04-04 | North America/Canada/Quebec/ | Laboratoire de microbiologie, Hopital de Verdun | Smith Laboratory, Centre de Recherche CHU Sainte-Justine | Martin Smith et al |
| EPI_ISL_515228 | Canada/QC_AT4/2020 | 2020-04-04 | North America/Canada/Quebec/ | Laboratoire de microbiologie, Hopital de Verdun | Smith Laboratory, Centre de Recherche CHU Sainte-Justine | Martin Smith et al |

| | | | | | | |
|---|---|---|---|---|---|---|
| EPI_ISL_515229 | Canada/QC_AU3/2020 | 2020-04-05 | North America/Canada/Quebec/ | Laboratoire de microbiologie, Hopital de Verdun | Smith Laboratory, Centre de Recherche CHU Sainte-Justine | Martin Smith et al |
| EPI_ISL_515230 | Canada/QC_AU6/2020 | 2020-04-05 | North America/Canada/Quebec/ | Laboratoire de microbiologie, Hopital de Verdun | Smith Laboratory, Centre de Recherche CHU Sainte-Justine | Martin Smith et al |
| EPI_ISL_515231 | Canada/QC_AV5/2020 | 2020-04-02 | North America/Canada/Quebec/ | Laboratoire de microbiologie, Hopital de Verdun | Smith Laboratory, Centre de Recherche CHU Sainte-Justine | Martin Smith et al |
| EPI_ISL_515232 | Canada/QC_AW4/2020 | 2020-04-14 | North America/Canada/Quebec/ | Laboratoire de microbiologie, Hopital de Verdun | Smith Laboratory, Centre de Recherche CHU Sainte-Justine | Martin Smith et al |
| EPI_ISL_515233 | Canada/QC_AW7/2020 | 2020-04-13 | North America/Canada/Quebec/ | Laboratoire de microbiologie, Hopital de Verdun | Smith Laboratory, Centre de Recherche CHU Sainte-Justine | Martin Smith et al |
| EPI_ISL_515234 | Canada/QC_AX3/2020 | 2020-04-09 | North America/Canada/Quebec/ | Laboratoire de microbiologie, Hopital de Verdun | Smith Laboratory, Centre de Recherche CHU Sainte-Justine | Martin Smith et al |
| EPI_ISL_515235 | Canada/QC_AY6/2020 | 2020-04-07 | North America/Canada/Quebec/ | Laboratoire de microbiologie, Hopital de Verdun | Smith Laboratory, Centre de Recherche CHU Sainte-Justine | Martin Smith et al |
| EPI_ISL_515236 | Canada/QC_AZ5/2020 | 2020-04-11 | North America/Canada/Quebec/ | Laboratoire de microbiologie, Hopital de Verdun | Smith Laboratory, Centre de Recherche CHU Sainte-Justine | Martin Smith et al |
| EPI_ISL_515237 | Canada/QC_AZ6/2020 | 2020-04-14 | North America/Canada/Quebec/ | Laboratoire de microbiologie, Hopital de Verdun | Smith Laboratory, Centre de Recherche CHU Sainte-Justine | Martin Smith et al |
| EPI_ISL_515238 | Canada/QC_BA1/2020 | 2020-04-14 | North America/Canada/Quebec/ | Laboratoire de microbiologie, Hopital de Verdun | Smith Laboratory, Centre de Recherche CHU Sainte-Justine | Martin Smith et al |

| | | | | | | |
|---|---|---|---|---|---|---|
| EPI_ISL_515239 | Canada/QC_BA2/2020 | 2020-04-14 | North America/Canada/Quebec/ | Laboratoire de microbiologie, Hopital de Verdun | Smith Laboratory, Centre de Recherche CHU Sainte-Justine | Martin Smith et al |
| EPI_ISL_515240 | Canada/QC_BA3/2020 | 2020-04-13 | North America/Canada/Quebec/ | Laboratoire de microbiologie, Hopital de Verdun | Smith Laboratory, Centre de Recherche CHU Sainte-Justine | Martin Smith et al |
| EPI_ISL_515241 | Canada/QC_BB3/2020 | 2020-04-14 | North America/Canada/Quebec/ | Laboratoire de microbiologie, Hopital de Verdun | Smith Laboratory, Centre de Recherche CHU Sainte-Justine | Martin Smith et al |
| EPI_ISL_515242 | Canada/QC_BB4/2020 | 2020-04-13 | North America/Canada/Quebec/ | Laboratoire de microbiologie, Hopital de Verdun | Smith Laboratory, Centre de Recherche CHU Sainte-Justine | Martin Smith et al |
| EPI_ISL_515243 | Canada/QC_BB6/2020 | 2020-04-11 | North America/Canada/Quebec/ | Laboratoire de microbiologie, Hopital de Verdun | Smith Laboratory, Centre de Recherche CHU Sainte-Justine | Martin Smith et al |
| EPI_ISL_515244 | Canada/QC_BB9/2020 | 2020-04-13 | North America/Canada/Quebec/ | Laboratoire de microbiologie, Hopital de Verdun | Smith Laboratory, Centre de Recherche CHU Sainte-Justine | Martin Smith et al |
| EPI_ISL_515245 | Canada/QC_BC6/2020 | 2020-04-11 | North America/Canada/Quebec/ | Laboratoire de microbiologie, Hopital de Verdun | Smith Laboratory, Centre de Recherche CHU Sainte-Justine | Martin Smith et al |
| EPI_ISL_515246 | Canada/QC_BC8/2020 | 2020-04-11 | North America/Canada/Quebec/ | Laboratoire de microbiologie, Hopital de Verdun | Smith Laboratory, Centre de Recherche CHU Sainte-Justine | Martin Smith et al |
| EPI_ISL_515250 | USA/TX-DSHS-0001/2020 | 2020-05-07 | North America/USA/Texas/Parker Count | Texas Department of State Health Services | Texas Department of State Health Services | Rashmi Tuladhar et al |
| EPI_ISL_515253 | USA/TX-DSHS-0004/2020 | 2020-05-07 | North America/USA/Texas/Ochiltree Co | Texas Department of State Health Services | Texas Department of State Health Services | Rashmi Tuladhar et al |

| | | | | | | |
|---|---|---|---|---|---|---|
| EPI_ISL_515257 | USA/TX-DSHS-0011/2020 | 2020-05-08 | North America/USA/Texas/Ochiltree Co | Texas Department of State Health Services | Texas Department of State Health Services | Rashmi Tuladhar et al |
| EPI_ISL_515259 | USA/TX-DSHS-0013/2020 | 2020-05-08 | North America/USA/Texas/Ochiltree Co | Texas Department of State Health Services | Texas Department of State Health Services | Rashmi Tuladhar et al |
| EPI_ISL_515260 | USA/TX-DSHS-0015/2020 | 2020-05-08 | North America/USA/Texas/Ochiltree Co | Texas Department of State Health Services | Texas Department of State Health Services | Rashmi Tuladhar et al |
| EPI_ISL_515281 | USA/WA-UW-4615/2020 | 2020-03-26 | North America/USA/Washington/ | University of Washington Virology Lab | University of Washington Virology Lab | Pavitra Roychoudhury et al |
| EPI_ISL_515284 | USA/WA-UW-4620/2020 | 2020-03-29 | North America/USA/Washington/ | University of Washington Virology Lab | University of Washington Virology Lab | Pavitra Roychoudhury et al |
| EPI_ISL_515301 | USA/NV-NSPHL-A0014/2020 | 2020-03-15 | North America/USA/Nevada/ | Nevada State Public Health Laboratory | Nevada State Public Health Laboratory | Richard Tillett et al |
| EPI_ISL_515319 | USA/NV-NSPHL-A0033/2020 | 2020-03-21 | North America/USA/Nevada/ | Nevada State Public Health Laboratory | Nevada State Public Health Laboratory | Richard Tillett et al |
| EPI_ISL_515324 | USA/NV-NSPHL-A0041/2020 | 2020-03-23 | North America/USA/Nevada/ | Nevada State Public Health Laboratory | Nevada State Public Health Laboratory | Richard Tillett et al |
| EPI_ISL_515344 | USA/NV-NSPHL-A0062/2020 | 2020-03-30 | North America/USA/Nevada/ | Nevada State Public Health Laboratory | Nevada State Public Health Laboratory | Richard Tillett et al |
| EPI_ISL_515363 | USA/NV-NSPHL-A0086/2020 | 2020-04-13 | North America/USA/Nevada/ | Nevada State Public Health Laboratory | Nevada State Public Health Laboratory | Richard Tillett et al |
| EPI_ISL_515365 | USA/NV-NSPHL-A0088/2020 | 2020-04-13 | North America/USA/Nevada/ | Nevada State Public Health Laboratory | Nevada State Public Health Laboratory | Richard Tillett et al |
| EPI_ISL_515367 | USA/NV-NSPHL-A0090/2020 | 2020-04-13 | North America/USA/Nevada/ | Nevada State Public Health Laboratory | Nevada State Public Health Laboratory | Richard Tillett et al |
| EPI_ISL_515373 | USA/NV-NSPHL-A0097/2020 | 2020-04-14 | North America/USA/Nevada/ | Nevada State Public Health Laboratory | Nevada State Public Health Laboratory | Richard Tillett et al |

| | | | | | | |
|---|---|---|---|---|---|---|
| EPI_ISL_515374 | USA/NV-NSPHL-A0098/2020 | 2020-04-15 | North America/USA/Nevada/ | Nevada State Public Health Laboratory | Nevada State Public Health Laboratory | Richard Tillett et al |
| EPI_ISL_515382 | USA/NV-NSPHL-A0111/2020 | 2020-04-18 | North America/USA/Nevada/ | Nevada State Public Health Laboratory | Nevada State Public Health Laboratory | Richard Tillett et al |
| EPI_ISL_515384 | USA/NV-NSPHL-A0113/2020 | 2020-04-18 | North America/USA/Nevada/ | Nevada State Public Health Laboratory | Nevada State Public Health Laboratory | Richard Tillett et al |
| EPI_ISL_515390 | USA/NV-NSPHL-A0119/2020 | 2020-04-20 | North America/USA/Nevada/ | Nevada State Public Health Laboratory | Nevada State Public Health Laboratory | Richard Tillett et al |
| EPI_ISL_515394 | USA/NV-NSPHL-A0123/2020 | 2020-04-26 | North America/USA/Nevada/ | Nevada State Public Health Laboratory | Nevada State Public Health Laboratory | Richard Tillett et al |
| EPI_ISL_515399 | USA/NV-NSPHL-A0131/2020 | 2020-04-29 | North America/USA/Nevada/ | Nevada State Public Health Laboratory | Nevada State Public Health Laboratory | Richard Tillett et al |
| EPI_ISL_515400 | USA/NV-NSPHL-A0132/2020 | 2020-04-29 | North America/USA/Nevada/ | Nevada State Public Health Laboratory | Nevada State Public Health Laboratory | Richard Tillett et al |
| EPI_ISL_515405 | USA/NV-NSPHL-A0138/2020 | 2020-04-30 | North America/USA/Nevada/ | Nevada State Public Health Laboratory | Nevada State Public Health Laboratory | Richard Tillett et al |
| EPI_ISL_515417 | USA/NV-NSPHL-A0153/2020 | 2020-05-14 | North America/USA/Nevada/ | Nevada State Public Health Laboratory | Nevada State Public Health Laboratory | Richard Tillett et al |
| EPI_ISL_515419 | USA/NV-NSPHL-A0155/2020 | 2020-05-14 | North America/USA/Nevada/ | Nevada State Public Health Laboratory | Nevada State Public Health Laboratory | Richard Tillett et al |
| EPI_ISL_515422 | USA/NV-NSPHL-A0160/2020 | 2020-05-15 | North America/USA/Nevada/ | Nevada State Public Health Laboratory | Nevada State Public Health Laboratory | Richard Tillett et al |
| EPI_ISL_515427 | USA/NV-NSPHL-A0165/2020 | 2020-05-16 | North America/USA/Nevada/ | Nevada State Public Health Laboratory | Nevada State Public Health Laboratory | Richard Tillett et al |
| EPI_ISL_515440 | USA/NV-NSPHL-A0183/2020 | 2020-05-19 | North America/USA/Nevada/ | Nevada State Public Health Laboratory | Nevada State Public Health Laboratory | Richard Tillett et al |

| | | | | | | |
|---|---|---|---|---|---|---|
| EPI_ISL_515441 | USA/NV-NSPHL-A0185/2020 | 2020-05-20 | North America/USA/Nevada/ | Nevada State Public Health Laboratory | Nevada State Public Health Laboratory | Richard Tillett et al |
| EPI_ISL_515442 | USA/NV-NSPHL-A0186/2020 | 2020-05-20 | North America/USA/Nevada/ | Nevada State Public Health Laboratory | Nevada State Public Health Laboratory | Richard Tillett et al |
| EPI_ISL_515448 | USA/NV-NSPHL-A0194/2020 | 2020-05-26 | North America/USA/Nevada/ | Nevada State Public Health Laboratory | Nevada State Public Health Laboratory | Richard Tillett et al |
| EPI_ISL_515450 | USA/NV-NSPHL-A0196/2020 | 2020-05-26 | North America/USA/Nevada/ | Nevada State Public Health Laboratory | Nevada State Public Health Laboratory | Richard Tillett et al |
| EPI_ISL_515452 | USA/NV-NSPHL-A0198/2020 | 2020-05-27 | North America/USA/Nevada/ | Nevada State Public Health Laboratory | Nevada State Public Health Laboratory | Richard Tillett et al |
| EPI_ISL_515461 | USA/NV-NSPHL-A0208/2020 | 2020-05-07 | North America/USA/Nevada/ | Nevada State Public Health Laboratory | Nevada State Public Health Laboratory | Richard Tillett et al |
| EPI_ISL_515466 | Canada/AB-Z0002/2020 | 2020-07-15 | North America/Canada/Alberta/ | Discovery DNA | Discovery DNA | Dustin Hittel et al |
| EPI_ISL_515525 | Brazil/SP-600/2020 | 2020-06-30 | South America/Brazil/S√£o Paulo/S√£o | National Influenza Center - Instituto Adolfo Lutz | Instituto Adolfo Lutz, Interdiciplinary Procedures Center, Strategic Laboratory | Claudio Tavares Sacchi et al |
| EPI_ISL_515559 | Brazil/SP-155/2020 | 2020-04-18 | South America/Brazil/S√£o Paulo/S√£o | Hospital Sao Paulo de Ensino da Unifesp | Instituto Adolfo Lutz, Interdiciplinary Procedures Center, Strategic Laboratory | Claudio Tavares Sacchi et al |
| EPI_ISL_515573 | SouthAfrica/KRISP-K002171/2020 | 2020-07-08 | Africa/South Africa/KwaZulu-Natal/eThe | NHLS-IALCH | KRISP, KZN Research Innovation and Sequencing Platform | Giandhari J et al |

| | | | | | | |
|---|---|---|---|---|---|---|
| EPI_ISL_515576 | SouthAfrica/KRISP-K002174/2020 | 2020-07-09 | Africa/South Africa/KwaZulu-Natal/eThe | NHLS-IALCH | KRISP, KZN Research Innovation and Sequencing Platform | Giandhari J et al |
| EPI_ISL_515577 | SouthAfrica/KRISP-K002175/2020 | 2020-07-09 | Africa/South Africa/KwaZulu-Natal/eThe | NHLS-IALCH | KRISP, KZN Research Innovation and Sequencing Platform | Giandhari J et al |
| EPI_ISL_515578 | SouthAfrica/KRISP-K002176/2020 | 2020-06-29 | Africa/South Africa/KwaZulu-Natal/eThe | NHLS-IALCH | KRISP, KZN Research Innovation and Sequencing Platform | Giandhari J et al |
| EPI_ISL_515582 | SouthAfrica/KRISP-K002183/2020 | 2020-07-09 | Africa/South Africa/KwaZulu-Natal/eThe | NHLS-IALCH | KRISP, KZN Research Innovation and Sequencing Platform | Giandhari J et al |
| EPI_ISL_515584 | SouthAfrica/KRISP-K002185/2020 | 2020-06-30 | Africa/South Africa/KwaZulu-Natal/eThe | NHLS-IALCH | KRISP, KZN Research Innovation and Sequencing Platform | Giandhari J et al |
| EPI_ISL_515585 | SouthAfrica/KRISP-K002186/2020 | 2020-06-30 | Africa/South Africa/KwaZulu-Natal/eThe | NHLS-IALCH | KRISP, KZN Research Innovation and Sequencing Platform | Giandhari J et al |
| EPI_ISL_515586 | SouthAfrica/KRISP-K002187/2020 | 2020-07-08 | Africa/South Africa/KwaZulu-Natal/eThe | NHLS-IALCH | KRISP, KZN Research Innovation and Sequencing Platform | Giandhari J et al |
| EPI_ISL_515588 | SouthAfrica/KRISP-K002189/2020 | 2020-07-08 | Africa/South Africa/KwaZulu-Natal/eThe | NHLS-IALCH | KRISP, KZN Research Innovation and Sequencing Platform | Giandhari J et al |

| | | | | | | |
|---|---|---|---|---|---|---|
| EPI_ISL_515589 | SouthAfrica/KRISP-K002190/2020 | 2020-07-14 | Africa/South Africa/KwaZulu-Natal/eThe | NHLS-IALCH | KRISP, KZN Research Innovation and Sequencing Platform | Giandhari J et al |
| EPI_ISL_515590 | SouthAfrica/KRISP-K002191/2020 | 2020-07-06 | Africa/South Africa/KwaZulu-Natal/eThe | NHLS-IALCH | KRISP, KZN Research Innovation and Sequencing Platform | Giandhari J et al |
| EPI_ISL_515593 | SouthAfrica/KRISP-K002195/2020 | 2020-07-10 | Africa/South Africa/KwaZulu-Natal/eThe | NHLS-IALCH | KRISP, KZN Research Innovation and Sequencing Platform | Giandhari J et al |
| EPI_ISL_515594 | SouthAfrica/KRISP-K002196/2020 | 2020-07-05 | Africa/South Africa/KwaZulu-Natal/eThe | NHLS-IALCH | KRISP, KZN Research Innovation and Sequencing Platform | Giandhari J et al |
| EPI_ISL_515597 | SouthAfrica/KRISP-K002199/2020 | 2020-07-14 | Africa/South Africa/KwaZulu-Natal/eThe | NHLS-IALCH | KRISP, KZN Research Innovation and Sequencing Platform | Giandhari J et al |
| EPI_ISL_515599 | SouthAfrica/KRISP-K002201/2020 | 2020-07-01 | Africa/South Africa/KwaZulu-Natal/eThe | NHLS-IALCH | KRISP, KZN Research Innovation and Sequencing Platform | Giandhari J et al |
| EPI_ISL_515600 | SouthAfrica/KRISP-K002202/2020 | 2020-07-16 | Africa/South Africa/KwaZulu-Natal/eThe | NHLS-IALCH | KRISP, KZN Research Innovation and Sequencing Platform | Giandhari J et al |
| EPI_ISL_515601 | SouthAfrica/KRISP-K002203/2020 | 2020-07-09 | Africa/South Africa/KwaZulu-Natal/eThe | NHLS-IALCH | KRISP, KZN Research Innovation and Sequencing Platform | Giandhari J et al |

| | | | | | | |
|---|---|---|---|---|---|---|
| EPI_ISL_515603 | SouthAfrica/KRISP-K002205/2020 | 2020-07-16 | Africa/South Africa/KwaZulu-Natal/eThe | NHLS-IALCH | KRISP, KZN Research Innovation and Sequencing Platform | Giandhari J et al |
| EPI_ISL_515604 | SouthAfrica/KRISP-K002206/2020 | 2020-07-17 | Africa/South Africa/KwaZulu-Natal/eThe | NHLS-IALCH | KRISP, KZN Research Innovation and Sequencing Platform | Giandhari J et al |
| EPI_ISL_515605 | SouthAfrica/KRISP-K002208/2020 | 2020-07-24 | Africa/South Africa/KwaZulu-Natal/eThe | NHLS-IALCH | KRISP, KZN Research Innovation and Sequencing Platform | Giandhari J et al |
| EPI_ISL_515610 | SouthAfrica/KRISP-K002213/2020 | 2020-07-30 | Africa/South Africa/KwaZulu-Natal/uMg | NHLS-IALCH | KRISP, KZN Research Innovation and Sequencing Platform | Giandhari J et al |
| EPI_ISL_515618 | SouthAfrica/KRISP-K002221/2020 | 2020-07-20 | Africa/South Africa/KwaZulu-Natal/King | NHLS-IALCH | KRISP, KZN Research Innovation and Sequencing Platform | Giandhari J et al |
| EPI_ISL_515619 | SouthAfrica/KRISP-K002222/2020 | 2020-07-21 | Africa/South Africa/KwaZulu-Natal/Ama | NHLS-IALCH | KRISP, KZN Research Innovation and Sequencing Platform | Giandhari J et al |
| EPI_ISL_515620 | SouthAfrica/KRISP-K002223/2020 | 2020-07-21 | Africa/South Africa/KwaZulu-Natal/Zulu | NHLS-IALCH | KRISP, KZN Research Innovation and Sequencing Platform | Giandhari J et al |
| EPI_ISL_515626 | SouthAfrica/KRISP-K002230/2020 | 2020-07-21 | Africa/South Africa/KwaZulu-Natal/Zulu | NHLS-IALCH | KRISP, KZN Research Innovation and Sequencing Platform | Giandhari J et al |

| | | | | | | |
|---|---|---|---|---|---|---|
| EPI_ISL_515627 | SouthAfrica/KRISP-K002231/2020 | 2020-07-18 | Africa/South Africa/KwaZulu-Natal/Zulu | NHLS-IALCH | KRISP, KZN Research Innovation and Sequencing Platform | Giandhari J et al |
| EPI_ISL_515630 | SouthAfrica/KRISP-K002238/2020 | 2020-07-20 | Africa/South Africa/KwaZulu-Natal/uMz | NHLS-IALCH | KRISP, KZN Research Innovation and Sequencing Platform | Giandhari J et al |
| EPI_ISL_515631 | SouthAfrica/KRISP-K002239/2020 | 2020-07-20 | Africa/South Africa/KwaZulu-Natal/uThu | NHLS-IALCH | KRISP, KZN Research Innovation and Sequencing Platform | Giandhari J et al |
| EPI_ISL_515633 | SouthAfrica/KRISP-K002242/2020 | 2020-07-21 | Africa/South Africa/KwaZulu-Natal/Ama | NHLS-IALCH | KRISP, KZN Research Innovation and Sequencing Platform | Giandhari J et al |
| EPI_ISL_515635 | SouthAfrica/KRISP-K002244/2020 | 2020-07-20 | Africa/South Africa/KwaZulu-Natal/King | NHLS-IALCH | KRISP, KZN Research Innovation and Sequencing Platform | Giandhari J et al |
| EPI_ISL_515641 | SouthAfrica/KRISP-K002256/2020 | 2020-07-21 | Africa/South Africa/KwaZulu-Natal/Ugu | NHLS-IALCH | KRISP, KZN Research Innovation and Sequencing Platform | Giandhari J et al |
| EPI_ISL_515645 | SouthAfrica/KRISP-K002260/2020 | 2020-07-16 | Africa/South Africa/KwaZulu-Natal/uMg | NHLS-IALCH | KRISP, KZN Research Innovation and Sequencing Platform | Giandhari J et al |
| EPI_ISL_515648 | SouthAfrica/KRISP-K002265/2020 | 2020-07-20 | Africa/South Africa/KwaZulu-Natal/uMg | NHLS-IALCH | KRISP, KZN Research Innovation and Sequencing Platform | Giandhari J et al |

| | | | | | | |
|---|---|---|---|---|---|---|
| EPI_ISL_515657 | SouthAfrica/KRISP-K002277/2020 | 2020-07-18 | Africa/South Africa/KwaZulu-Natal/uMz | NHLS-IALCH | KRISP, KZN Research Innovation and Sequencing Platform | Giandhari J et al |
| EPI_ISL_515662 | SouthAfrica/KRISP-K002282/2020 | 2020-07-21 | Africa/South Africa/KwaZulu-Natal/Ama | NHLS-IALCH | KRISP, KZN Research Innovation and Sequencing Platform | Giandhari J et al |
| EPI_ISL_515666 | SouthAfrica/KRISP-K002288/2020 | 2020-07-21 | Africa/South Africa/KwaZulu-Natal/uMg | NHLS-IALCH | KRISP, KZN Research Innovation and Sequencing Platform | Giandhari J et al |
| EPI_ISL_515672 | SouthAfrica/KRISP-K002295/2020 | 2020-07-23 | Africa/South Africa/KwaZulu-Natal/Ama | NHLS-IALCH | KRISP, KZN Research Innovation and Sequencing Platform | Giandhari J et al |
| EPI_ISL_515674 | SouthAfrica/KRISP-K002297/2020 | 2020-07-20 | Africa/South Africa/KwaZulu-Natal/uMz | NHLS-IALCH | KRISP, KZN Research Innovation and Sequencing Platform | Giandhari J et al |
| EPI_ISL_515676 | SouthAfrica/KRISP-K002299/2020 | 2020-07-22 | Africa/South Africa/KwaZulu-Natal/King | NHLS-IALCH | KRISP, KZN Research Innovation and Sequencing Platform | Giandhari J et al |
| EPI_ISL_515681 | SouthAfrica/KRISP-K002304/2020 | 2020-07-23 | Africa/South Africa/KwaZulu-Natal/Ama | NHLS-IALCH | KRISP, KZN Research Innovation and Sequencing Platform | Giandhari J et al |
| EPI_ISL_515683 | SouthAfrica/KRISP-K002307/2020 | 2020-07-23 | Africa/South Africa/KwaZulu-Natal/uThu | NHLS-IALCH | KRISP, KZN Research Innovation and Sequencing Platform | Giandhari J et al |

| | | | | | | |
|---|---|---|---|---|---|---|
| EPI_ISL_515686 | SouthAfrica/KRISP-K002310/2020 | 2020-07-23 | Africa/South Africa/KwaZulu-Natal/Ama | NHLS-IALCH | KRISP, KZN Research Innovation and Sequencing Platform | Giandhari J et al |
| EPI_ISL_515687 | SouthAfrica/KRISP-K002311/2020 | 2020-07-22 | Africa/South Africa/KwaZulu-Natal/iLem | NHLS-IALCH | KRISP, KZN Research Innovation and Sequencing Platform | Giandhari J et al |
| EPI_ISL_515688 | SouthAfrica/KRISP-K002312/2020 | 2020-07-22 | Africa/South Africa/KwaZulu-Natal/King | NHLS-IALCH | KRISP, KZN Research Innovation and Sequencing Platform | Giandhari J et al |
| EPI_ISL_515696 | SouthAfrica/KRISP-K002320/2020 | 2020-07-22 | Africa/South Africa/KwaZulu-Natal/uMg | NHLS-IALCH | KRISP, KZN Research Innovation and Sequencing Platform | Giandhari J et al |
| EPI_ISL_515701 | SouthAfrica/KRISP-K002325/2020 | 2020-07-22 | Africa/South Africa/KwaZulu-Natal/uMg | NHLS-IALCH | KRISP, KZN Research Innovation and Sequencing Platform | Giandhari J et al |
| EPI_ISL_515704 | SouthAfrica/KRISP-K002328/2020 | 2020-07-22 | Africa/South Africa/KwaZulu-Natal/King | NHLS-IALCH | KRISP, KZN Research Innovation and Sequencing Platform | Giandhari J et al |
| EPI_ISL_515706 | SouthAfrica/KRISP-K002331/2020 | 2020-07-22 | Africa/South Africa/KwaZulu-Natal/uMg | NHLS-IALCH | KRISP, KZN Research Innovation and Sequencing Platform | Giandhari J et al |
| EPI_ISL_515713 | SouthAfrica/KRISP-K002340/2020 | 2020-07-22 | Africa/South Africa/KwaZulu-Natal/uMg | NHLS-IALCH | KRISP, KZN Research Innovation and Sequencing Platform | Giandhari J et al |

| | | | | | | |
|---|---|---|---|---|---|---|
| EPI_ISL_515716 | SouthAfrica/KRISP-K002343/2020 | 2020-07-23 | Africa/South Africa/KwaZulu-Natal/Zulu | NHLS-IALCH | KRISP, KZN Research Innovation and Sequencing Platform | Giandhari J et al |
| EPI_ISL_515718 | SouthAfrica/KRISP-K002346/2020 | 2020-07-23 | Africa/South Africa/KwaZulu-Natal/uMz | NHLS-IALCH | KRISP, KZN Research Innovation and Sequencing Platform | Giandhari J et al |
| EPI_ISL_515719 | SouthAfrica/KRISP-K002347/2020 | 2020-07-23 | Africa/South Africa/KwaZulu-Natal/uMz | NHLS-IALCH | KRISP, KZN Research Innovation and Sequencing Platform | Giandhari J et al |
| EPI_ISL_515723 | SouthAfrica/KRISP-K002351/2020 | 2020-07-21 | Africa/South Africa/KwaZulu-Natal/Zulu | NHLS-IALCH | KRISP, KZN Research Innovation and Sequencing Platform | Giandhari J et al |
| EPI_ISL_515727 | SouthAfrica/KRISP-K002356/2020 | 2020-07-22 | Africa/South Africa/KwaZulu-Natal/uMg | NHLS-IALCH | KRISP, KZN Research Innovation and Sequencing Platform | Giandhari J et al |
| EPI_ISL_515729 | SouthAfrica/KRISP-K002358/2020 | 2020-07-21 | Africa/South Africa/KwaZulu-Natal/uMz | NHLS-IALCH | KRISP, KZN Research Innovation and Sequencing Platform | Giandhari J et al |
| EPI_ISL_515730 | SouthAfrica/KRISP-K002359/2020 | 2020-07-22 | Africa/South Africa/KwaZulu-Natal/uThu | NHLS-IALCH | KRISP, KZN Research Innovation and Sequencing Platform | Giandhari J et al |
| EPI_ISL_515731 | SouthAfrica/KRISP-K002363/2020 | 2020-07-21 | Africa/South Africa/KwaZulu-Natal/uThu | NHLS-IALCH | KRISP, KZN Research Innovation and Sequencing Platform | Giandhari J et al |

| | | | | | | |
|---|---|---|---|---|---|---|
| EPI_ISL_515733 | SouthAfrica/KRISP-K002365/2020 | 2020-07-22 | Africa/South Africa/KwaZulu-Natal/uMk | NHLS-IALCH | KRISP, KZN Research Innovation and Sequencing Platform | Giandhari J et al |
| EPI_ISL_515735 | SouthAfrica/KRISP-K002367/2020 | 2020-07-22 | Africa/South Africa/KwaZulu-Natal/uThu | NHLS-IALCH | KRISP, KZN Research Innovation and Sequencing Platform | Giandhari J et al |
| EPI_ISL_515736 | SouthAfrica/KRISP-K002368/2020 | 2020-07-22 | Africa/South Africa/KwaZulu-Natal/uMk | NHLS-IALCH | KRISP, KZN Research Innovation and Sequencing Platform | Giandhari J et al |
| EPI_ISL_515737 | SouthAfrica/KRISP-K002369/2020 | 2020-07-22 | Africa/South Africa/KwaZulu-Natal/iLem | NHLS-IALCH | KRISP, KZN Research Innovation and Sequencing Platform | Giandhari J et al |
| EPI_ISL_515738 | SouthAfrica/KRISP-K002370/2020 | 2020-07-22 | Africa/South Africa/KwaZulu-Natal/uMg | NHLS-IALCH | KRISP, KZN Research Innovation and Sequencing Platform | Giandhari J et al |
| EPI_ISL_515739 | SouthAfrica/KRISP-K002371/2020 | 2020-07-22 | Africa/South Africa/KwaZulu-Natal/iLem | NHLS-IALCH | KRISP, KZN Research Innovation and Sequencing Platform | Giandhari J et al |
| EPI_ISL_515743 | SouthAfrica/KRISP-K002377/2020 | 2020-07-22 | Africa/South Africa/KwaZulu-Natal/King | NHLS-IALCH | KRISP, KZN Research Innovation and Sequencing Platform | Giandhari J et al |
| EPI_ISL_515744 | SouthAfrica/KRISP-K002378/2020 | 2020-07-20 | Africa/South Africa/KwaZulu-Natal/uMk | NHLS-IALCH | KRISP, KZN Research Innovation and Sequencing Platform | Giandhari J et al |

| | | | | | | |
|---|---|---|---|---|---|---|
| EPI_ISL_515746 | SouthAfrica/KRISP-K002380/2020 | 2020-07-22 | Africa/South Africa/KwaZulu-Natal/uMk | NHLS-IALCH | KRISP, KZN Research Innovation and Sequencing Platform | Giandhari J et al |
| EPI_ISL_515754 | SouthAfrica/KRISP-K002388/2020 | 2020-07-22 | Africa/South Africa/KwaZulu-Natal/uMk | NHLS-IALCH | KRISP, KZN Research Innovation and Sequencing Platform | Giandhari J et al |
| EPI_ISL_515757 | SouthAfrica/KRISP-K002392/2020 | 2020-07-27 | Africa/South Africa/KwaZulu-Natal/Ugu | NHLS-IALCH | KRISP, KZN Research Innovation and Sequencing Platform | Giandhari J et al |
| EPI_ISL_515758 | SouthAfrica/KRISP-K002393/2020 | 2020-07-28 | Africa/South Africa/KwaZulu-Natal/King | NHLS-IALCH | KRISP, KZN Research Innovation and Sequencing Platform | Giandhari J et al |
| EPI_ISL_515759 | SouthAfrica/KRISP-K002394/2020 | 2020-07-28 | Africa/South Africa/KwaZulu-Natal/King | NHLS-IALCH | KRISP, KZN Research Innovation and Sequencing Platform | Giandhari J et al |
| EPI_ISL_515760 | SouthAfrica/KRISP-K002395/2020 | 2020-07-28 | Africa/South Africa/KwaZulu-Natal/Ugu | NHLS-IALCH | KRISP, KZN Research Innovation and Sequencing Platform | Giandhari J et al |
| EPI_ISL_515762 | SouthAfrica/KRISP-K002398/2020 | 2020-07-27 | Africa/South Africa/KwaZulu-Natal/Ugu | NHLS-IALCH | KRISP, KZN Research Innovation and Sequencing Platform | Giandhari J et al |
| EPI_ISL_515763 | SouthAfrica/KRISP-K002399/2020 | 2020-07-30 | Africa/South Africa/KwaZulu-Natal/iLem | NHLS-IALCH | KRISP, KZN Research Innovation and Sequencing Platform | Giandhari J et al |

| | | | | | | |
|---|---|---|---|---|---|---|
| EPI_ISL_515768 | SouthAfrica/KRISP-K002404/2020 | 2020-07-27 | Africa/South Africa/KwaZulu-Natal/King | NHLS-IALCH | KRISP, KZN Research Innovation and Sequencing Platform | Giandhari J et al |
| EPI_ISL_515769 | SouthAfrica/KRISP-K002405/2020 | 2020-07-27 | Africa/South Africa/KwaZulu-Natal/Ugu | NHLS-IALCH | KRISP, KZN Research Innovation and Sequencing Platform | Giandhari J et al |
| EPI_ISL_515770 | SouthAfrica/KRISP-K002406/2020 | 2020-07-29 | Africa/South Africa/KwaZulu-Natal/uMk | NHLS-IALCH | KRISP, KZN Research Innovation and Sequencing Platform | Giandhari J et al |
| EPI_ISL_515775 | SouthAfrica/KRISP-K002413/2020 | 2020-07-24 | Africa/South Africa/KwaZulu-Natal/Ugu | NHLS-IALCH | KRISP, KZN Research Innovation and Sequencing Platform | Giandhari J et al |
| EPI_ISL_515779 | SouthAfrica/KRISP-K002417/2020 | 2020-07-30 | Africa/South Africa/KwaZulu-Natal/Ama | NHLS-IALCH | KRISP, KZN Research Innovation and Sequencing Platform | Giandhari J et al |
| EPI_ISL_515780 | SouthAfrica/KRISP-K002418/2020 | 2020-07-30 | Africa/South Africa/KwaZulu-Natal/Ama | NHLS-IALCH | KRISP, KZN Research Innovation and Sequencing Platform | Giandhari J et al |
| EPI_ISL_515785 | SouthAfrica/KRISP-K002424/2020 | 2020-07-30 | Africa/South Africa/KwaZulu-Natal/uMg | NHLS-IALCH | KRISP, KZN Research Innovation and Sequencing Platform | Giandhari J et al |
| EPI_ISL_515786 | SouthAfrica/KRISP-K002425/2020 | 2020-07-29 | Africa/South Africa/KwaZulu-Natal/Zulu | NHLS-IALCH | KRISP, KZN Research Innovation and Sequencing Platform | Giandhari J et al |

| | | | | | | |
|---|---|---|---|---|---|---|
| EPI_ISL_515787 | SouthAfrica/KRISP-K002426/2020 | 2020-07-29 | Africa/South Africa/KwaZulu-Natal/eThe | NHLS-IALCH | KRISP, KZN Research Innovation and Sequencing Platform | Giandhari J et al |
| EPI_ISL_515789 | SouthAfrica/KRISP-K002428/2020 | 2020-07-30 | Africa/South Africa/KwaZulu-Natal/uMg | NHLS-IALCH | KRISP, KZN Research Innovation and Sequencing Platform | Giandhari J et al |
| EPI_ISL_515790 | SouthAfrica/KRISP-K002429/2020 | 2020-07-30 | Africa/South Africa/KwaZulu-Natal/Ama | NHLS-IALCH | KRISP, KZN Research Innovation and Sequencing Platform | Giandhari J et al |
| EPI_ISL_515791 | SouthAfrica/KRISP-K002430/2020 | 2020-07-30 | Africa/South Africa/KwaZulu-Natal/uMg | NHLS-IALCH | KRISP, KZN Research Innovation and Sequencing Platform | Giandhari J et al |
| EPI_ISL_515798 | SouthAfrica/KRISP-K002442/2020 | 2020-07-30 | Africa/South Africa/KwaZulu-Natal/Ama | NHLS-IALCH | KRISP, KZN Research Innovation and Sequencing Platform | Giandhari J et al |
| EPI_ISL_515803 | SouthAfrica/KRISP-K001318/2020 | 2020-07-18 | Africa/South Africa/KwaZulu-Natal/eThe | Medical Disagnostics Services (MDS) | KRISP, KZN Research Innovation and Sequencing Platform | Giandhari J et al |
| EPI_ISL_515810 | SouthAfrica/KRISP-K001325/2020 | 2020-07-18 | Africa/South Africa/KwaZulu-Natal/eThe | Medical Disagnostics Services (MDS) | KRISP, KZN Research Innovation and Sequencing Platform | Giandhari J et al |
| EPI_ISL_515811 | SouthAfrica/KRISP-K001326/2020 | 2020-07-18 | Africa/South Africa/KwaZulu-Natal/eThe | Medical Disagnostics Services (MDS) | KRISP, KZN Research Innovation and Sequencing Platform | Giandhari J et al |

| | | | | | | |
|---|---|---|---|---|---|---|
| EPI_ISL_515813 | SouthAfrica/KRISP-K001328/2020 | 2020-07-18 | Africa/South Africa/KwaZulu-Natal/eThe | Medical Disagnostics Services (MDS) | KRISP, KZN Research Innovation and Sequencing Platform | Giandhari J et al |
| EPI_ISL_515814 | SouthAfrica/KRISP-K001329/2020 | 2020-07-18 | Africa/South Africa/KwaZulu-Natal/eThe | Medical Disagnostics Services (MDS) | KRISP, KZN Research Innovation and Sequencing Platform | Giandhari J et al |
| EPI_ISL_515816 | SouthAfrica/KRISP-K001331/2020 | 2020-07-19 | Africa/South Africa/KwaZulu-Natal/eThe | Medical Disagnostics Services (MDS) | KRISP, KZN Research Innovation and Sequencing Platform | Giandhari J et al |
| EPI_ISL_515819 | SouthAfrica/KRISP-K001334/2020 | 2020-07-19 | Africa/South Africa/KwaZulu-Natal/eThe | Medical Disagnostics Services (MDS) | KRISP, KZN Research Innovation and Sequencing Platform | Giandhari J et al |
| EPI_ISL_515821 | SouthAfrica/KRISP-K001336/2020 | 2020-07-19 | Africa/South Africa/KwaZulu-Natal/eThe | Medical Disagnostics Services (MDS) | KRISP, KZN Research Innovation and Sequencing Platform | Giandhari J et al |
| EPI_ISL_515824 | SouthAfrica/KRISP-K001339/2020 | 2020-07-10 | Africa/South Africa/KwaZulu-Natal/eThe | Medical Disagnostics Services (MDS) | KRISP, KZN Research Innovation and Sequencing Platform | Giandhari J et al |
| EPI_ISL_515825 | SouthAfrica/KRISP-K001340/2020 | 2020-07-10 | Africa/South Africa/KwaZulu-Natal/eThe | Medical Disagnostics Services (MDS) | KRISP, KZN Research Innovation and Sequencing Platform | Giandhari J et al |
| EPI_ISL_515828 | SouthAfrica/KRISP-K001343/2020 | 2020-07-10 | Africa/South Africa/KwaZulu-Natal/eThe | Medical Disagnostics Services (MDS) | KRISP, KZN Research Innovation and Sequencing Platform | Giandhari J et al |

| | | | | | | |
|---|---|---|---|---|---|---|
| EPI_ISL_515831 | SouthAfrica/KRISP-K001346/2020 | 2020-07-13 | Africa/South Africa/KwaZulu-Natal/eThe | Medical Disagnostics Services (MDS) | KRISP, KZN Research Innovation and Sequencing Platform | Giandhari J et al |
| EPI_ISL_515832 | SouthAfrica/KRISP-K001347/2020 | 2020-07-01 | Africa/South Africa/KwaZulu-Natal/Ugu | Medical Disagnostics Services (MDS) | KRISP, KZN Research Innovation and Sequencing Platform | Giandhari J et al |
| EPI_ISL_515835 | SouthAfrica/KRISP-K001350/2020 | 2020-06-30 | Africa/South Africa/KwaZulu-Natal/uMg | Medical Disagnostics Services (MDS) | KRISP, KZN Research Innovation and Sequencing Platform | Giandhari J et al |
| EPI_ISL_515837 | SouthAfrica/KRISP-K001352/2020 | 2020-06-24 | Africa/South Africa/KwaZulu-Natal/uMg | Medical Disagnostics Services (MDS) | KRISP, KZN Research Innovation and Sequencing Platform | Giandhari J et al |
| EPI_ISL_515840 | SouthAfrica/KRISP-K001355/2020 | 2020-07-17 | Africa/South Africa/KwaZulu-Natal/iLem | Medical Disagnostics Services (MDS) | KRISP, KZN Research Innovation and Sequencing Platform | Giandhari J et al |
| EPI_ISL_515850 | SouthAfrica/KRISP-K001365/2020 | 2020-07-16 | Africa/South Africa/KwaZulu-Natal/iLem | Medical Disagnostics Services (MDS) | KRISP, KZN Research Innovation and Sequencing Platform | Giandhari J et al |
| EPI_ISL_515852 | SouthAfrica/KRISP-K001367/2020 | 2020-07-16 | Africa/South Africa/KwaZulu-Natal/iLem | Medical Disagnostics Services (MDS) | KRISP, KZN Research Innovation and Sequencing Platform | Giandhari J et al |
| EPI_ISL_515856 | SouthAfrica/KRISP-K001371/2020 | 2020-07-16 | Africa/South Africa/KwaZulu-Natal/iLem | Medical Disagnostics Services (MDS) | KRISP, KZN Research Innovation and Sequencing Platform | Giandhari J et al |

| | | | | | | |
|---|---|---|---|---|---|---|
| EPI_ISL_515857 | SouthAfrica/KRISP-K001372/2020 | 2020-07-15 | Africa/South Africa/KwaZulu-Natal/iLem | Medical Disagnostics Services (MDS) | KRISP, KZN Research Innovation and Sequencing Platform | Giandhari J et al |
| EPI_ISL_515858 | SouthAfrica/KRISP-K001373/2020 | 2020-07-17 | Africa/South Africa/KwaZulu-Natal/iLem | Medical Disagnostics Services (MDS) | KRISP, KZN Research Innovation and Sequencing Platform | Giandhari J et al |
| EPI_ISL_515860 | SouthAfrica/KRISP-K001375/2020 | 2020-07-16 | Africa/South Africa/KwaZulu-Natal/iLem | Medical Disagnostics Services (MDS) | KRISP, KZN Research Innovation and Sequencing Platform | Giandhari J et al |
| EPI_ISL_515861 | SouthAfrica/KRISP-K001376/2020 | 2020-07-16 | Africa/South Africa/KwaZulu-Natal/iLem | Medical Disagnostics Services (MDS) | KRISP, KZN Research Innovation and Sequencing Platform | Giandhari J et al |
| EPI_ISL_515868 | SouthAfrica/KRISP-K001384/2020 | 2020-07-17 | Africa/South Africa/KwaZulu-Natal/Zulu | Medical Disagnostics Services (MDS) | KRISP, KZN Research Innovation and Sequencing Platform | Giandhari J et al |
| EPI_ISL_515869 | SouthAfrica/KRISP-K001385/2020 | 2020-07-09 | Africa/South Africa/KwaZulu-Natal/uThu | Medical Disagnostics Services (MDS) | KRISP, KZN Research Innovation and Sequencing Platform | Giandhari J et al |
| EPI_ISL_515875 | SouthAfrica/KRISP-K001391/2020 | 2020-07-18 | Africa/South Africa/KwaZulu-Natal/uMk | Medical Disagnostics Services (MDS) | KRISP, KZN Research Innovation and Sequencing Platform | Giandhari J et al |
| EPI_ISL_515877 | SouthAfrica/KRISP-K001393/2020 | 2020-07-16 | Africa/South Africa/KwaZulu-Natal/uMk | Medical Disagnostics Services (MDS) | KRISP, KZN Research Innovation and Sequencing Platform | Giandhari J et al |

| | | | | | | |
|---|---|---|---|---|---|---|
| EPI_ISL_515878 | SouthAfrica/KRISP-K001394/2020 | 2020-07-18 | Africa/South Africa/KwaZulu-Natal/uMk | Medical Disagnostics Services (MDS) | KRISP, KZN Research Innovation and Sequencing Platform | Giandhari J et al |
| EPI_ISL_515884 | SouthAfrica/KRISP-K001401/2020 | 2020-07-17 | Africa/South Africa/KwaZulu-Natal/uMk | Medical Disagnostics Services (MDS) | KRISP, KZN Research Innovation and Sequencing Platform | Giandhari J et al |
| EPI_ISL_515895 | USA/CA-CDPH002/2020 | 2020-03-18 | North America/USA/California/ | California Department of Public Health | California Department of Public Health | CDPH IDLB COVIDNet et al |
| EPI_ISL_515897 | USA/CA-CDPH004/2020 | 2020-03-23 | North America/USA/California/Marin Co | California Department of Public Health | California Department of Public Health | CDPH IDLB COVIDNet et al |
| EPI_ISL_515910 | USA/CA-CDPH017/2020 | 2020-02-27 | North America/USA/California/Solano C | California Department of Public Health | California Department of Public Health | CDPH IDLB COVIDNet et al |
| EPI_ISL_515911 | USA/CA-CDPH018/2020 | 2020-02-27 | North America/USA/California/Solano C | California Department of Public Health | California Department of Public Health | CDPH IDLB COVIDNet et al |
| EPI_ISL_515913 | USA/CA-CDPH020/2020 | 2020-03-07 | North America/USA/California/Sacrame | California Department of Public Health | California Department of Public Health | CDPH IDLB COVIDNet et al |
| EPI_ISL_515917 | USA/CA-CDPH025/2020 | 2020-05-19 | North America/USA/California/Los Ange | California Department of Public Health | California Department of Public Health | CDPH IDLB COVIDNet et al |
| EPI_ISL_515919 | USA/CA-CDPH027/2020 | 2020-05-22 | North America/USA/California/Madera | California Department of Public Health | California Department of Public Health | CDPH IDLB COVIDNet et al |
| EPI_ISL_515927 | USA/CA-CDPH036/2020 | 2020-06-25 | North America/USA/California/Lake Cou | California Department of Public Health | California Department of Public Health | CDPH IDLB COVIDNet et al |
| EPI_ISL_515945 | India/KA-nimh-11068/2020 | 2020-05-04 | Asia/India/Karnataka/ | DH | Department of Neurovirology, National Institute of Mental Health and Neuroscience (NIMHANS) | Chitra Pattabiraman et al |

| | | | | | | |
|---|---|---|---|---|---|---|
| EPI_ISL_515946 | India/KA-nimh-11069/2020 | 2020-05-04 | Asia/India/Karnataka/ | DH | Department of Neurovirology, National Institute of Mental Health and Neuroscience (NIMHANS) | Chitra Pattabiraman et al |
| EPI_ISL_515948 | India/KA-nimh-11074/2020 | 2020-05-04 | Asia/India/Karnataka/ | DH | Department of Neurovirology, National Institute of Mental Health and Neuroscience (NIMHANS) | Chitra Pattabiraman et al |
| EPI_ISL_515955 | India/KA-nimh-13894/2020 | 2020-05-09 | Asia/India/Karnataka/ | DH | Department of Neurovirology, National Institute of Mental Health and Neuroscience (NIMHANS) | Chitra Pattabiraman et al |
| EPI_ISL_515959 | India/KA-nimh-14835/2020 | 2020-05-08 | Asia/India/Karnataka/ | DH | Department of Neurovirology, National Institute of Mental Health and Neuroscience (NIMHANS) | Chitra Pattabiraman et al |
| EPI_ISL_515960 | India/KA-nimh-15365/2020 | 2020-05-11 | Asia/India/Karnataka/ | JGH | Department of Neurovirology, National Institute of Mental Health and Neuroscience (NIMHANS) | Chitra Pattabiraman et al |
| EPI_ISL_515963 | India/KA-nimh-17585/2020 | 2020-05-12 | Asia/India/Karnataka/ | VICTORIA HOSPITAL | Department of Neurovirology, National Institute of Mental Health and Neuroscience (NIMHANS) | Chitra Pattabiraman et al |

| | | | | | | |
|---|---|---|---|---|---|---|
| EPI_ISL_515966 | India/KA-nimh-19184/2020 | 2020-05-14 | Asia/India/Karnataka/ | VICTORIA HOSPITAL | Department of Neurovirology, National Institute of Mental Health and Neuroscience (NIMHANS) | Chitra Pattabiraman et al |
| EPI_ISL_516076 | India/KA-nimh-1114/2020 | 2020-04-15 | Asia/India/Karnataka/ | VICTORIA HOSPITAL | Department of Neurovirology, National Institute of Mental Health and Neuroscience (NIMHANS) | Chitra Pattabiraman et al |
| EPI_ISL_516079 | Italy/MAR-UnivPM-32898/2020 | 2020-03-08 | Europe/Italy/Marche/Ancona | Biomedical Sciences and Public Health, Polytechnic University of Marche | Biomedical Sciences and Public Health, Polytechnic University of Marche | Bagnarelli et al |
| EPI_ISL_516080 | Italy/MAR-UnivPM-34104/2020 | 2020-03-09 | Europe/Italy/Marche/Ancona | Biomedical Sciences and Public Health, Polytechnic University of Marche | Biomedical Sciences and Public Health, Polytechnic University of Marche | Bagnarelli et al |
| EPI_ISL_516082 | Italy/MAR-UnivPM-43950/2020 | 2020-03-18 | Europe/Italy/Marche/Ancona | Biomedical Sciences and Public Health, Polytechnic University of Marche | Biomedical Sciences and Public Health, Polytechnic University of Marche | Bagnarelli et al |
| EPI_ISL_516083 | Italy/MAR-UnivPM-71205/2020 | 2020-02-26 | Europe/Italy/Marche/Ancona | Biomedical Sciences and Public Health, Polytechnic University of Marche | Biomedical Sciences and Public Health, Polytechnic University of Marche | Bagnarelli et al |

| | | | | | | |
|---|---|---|---|---|---|---|
| EPI_ISL_516084 | Italy/MAR-UnivPM-73696/2020 | 2020-02-28 | Europe/Italy/Marche/Ancona | Biomedical Sciences and Public Health, Polytechnic University of Marche | Biomedical Sciences and Public Health, Polytechnic University of Marche | Bagnarelli et al |
| EPI_ISL_516085 | Italy/MAR-UnivPM-73702/2020 | 2020-02-28 | Europe/Italy/Marche/Ancona | Biomedical Sciences and Public Health, Polytechnic University of Marche | Biomedical Sciences and Public Health, Polytechnic University of Marche | Bagnarelli et al |
| EPI_ISL_516087 | Italy/MAR-UnivPM-78955/2020 | 2020-03-04 | Europe/Italy/Marche/Ancona | Biomedical Sciences and Public Health, Polytechnic University of Marche | Biomedical Sciences and Public Health, Polytechnic University of Marche | Bagnarelli et al |
| EPI_ISL_516088 | Italy/MAR-UnivPM-78955-2/2020 | 2020-03-04 | Europe/Italy/Marche/Ancona | Biomedical Sciences and Public Health, Polytechnic University of Marche | Biomedical Sciences and Public Health, Polytechnic University of Marche | Bagnarelli et al |
| EPI_ISL_516195 | USA/MI-MDHHS-SC21468/2020 | 2020-03-30 | North America/USA/Michigan/ | Michigan Department of Health and Human Services, Bureau of Laboratories | Michigan Department of Health and Human Services, Bureau of Laboratories | Blankenship HM et al |
| EPI_ISL_516203 | USA/MI-MDHHS-SC21476/2020 | 2020-07-16 | North America/USA/Michigan/ | Michigan Department of Health and Human Services, Bureau of Laboratories | Michigan Department of Health and Human Services, Bureau of Laboratories | Blankenship HM et al |

| | | | | | | |
|---|---|---|---|---|---|---|
| EPI_ISL_516210 | USA/MI-MDHHS-SC21483/2020 | 2020-07-20 | North America/USA/Michigan/ | Michigan Department of Health and Human Services, Bureau of Laboratories | Michigan Department of Health and Human Services, Bureau of Laboratories | Blankenship HM et al |
| EPI_ISL_516211 | USA/MI-MDHHS-SC21484/2020 | 2020-07-27 | North America/USA/Michigan/ | Michigan Department of Health and Human Services, Bureau of Laboratories | Michigan Department of Health and Human Services, Bureau of Laboratories | Blankenship HM et al |
| EPI_ISL_516212 | USA/MI-MDHHS-SC21485/2020 | 2020-07-26 | North America/USA/Michigan/ | Michigan Department of Health and Human Services, Bureau of Laboratories | Michigan Department of Health and Human Services, Bureau of Laboratories | Blankenship HM et al |
| EPI_ISL_516213 | USA/MI-MDHHS-SC21486/2020 | 2020-07-24 | North America/USA/Michigan/ | Michigan Department of Health and Human Services, Bureau of Laboratories | Michigan Department of Health and Human Services, Bureau of Laboratories | Blankenship HM et al |
| EPI_ISL_516216 | USA/MI-MDHHS-SC21489/2020 | 2020-07-24 | North America/USA/Michigan/ | Michigan Department of Health and Human Services, Bureau of Laboratories | Michigan Department of Health and Human Services, Bureau of Laboratories | Blankenship HM et al |
| EPI_ISL_516217 | USA/MI-MDHHS-SC21490/2020 | 2020-07-28 | North America/USA/Michigan/ | Michigan Department of Health and Human Services, Bureau of Laboratories | Michigan Department of Health and Human Services, Bureau of Laboratories | Blankenship HM et al |

| | | | | | | |
|---|---|---|---|---|---|---|
| EPI_ISL_516218 | USA/MI-MDHHS-SC21491/2020 | 2020-07-28 | North America/USA/Michigan/ | Michigan Department of Health and Human Services, Bureau of Laboratories | Michigan Department of Health and Human Services, Bureau of Laboratories | Blankenship HM et al |
| EPI_ISL_516221 | USA/MI-MDHHS-SC21494/2020 | 2020-07-30 | North America/USA/Michigan/ | Michigan Department of Health and Human Services, Bureau of Laboratories | Michigan Department of Health and Human Services, Bureau of Laboratories | Blankenship HM et al |
| EPI_ISL_516223 | USA/MI-MDHHS-SC21496/2020 | 2020-07-30 | North America/USA/Michigan/ | Michigan Department of Health and Human Services, Bureau of Laboratories | Michigan Department of Health and Human Services, Bureau of Laboratories | Blankenship HM et al |
| EPI_ISL_516243 | USA/MI-MDHHS-SC21516/2020 | 2020-04-03 | North America/USA/Michigan/ | Michigan Department of Health and Human Services, Bureau of Laboratories | Michigan Department of Health and Human Services, Bureau of Laboratories | Blankenship HM et al |
| EPI_ISL_516245 | USA/MI-MDHHS-SC21518/2020 | 2020-04-02 | North America/USA/Michigan/ | Michigan Department of Health and Human Services, Bureau of Laboratories | Michigan Department of Health and Human Services, Bureau of Laboratories | Blankenship HM et al |
| EPI_ISL_516256 | USA/MI-MDHHS-SC21529/2020 | 2020-04-03 | North America/USA/Michigan/ | Michigan Department of Health and Human Services, Bureau of Laboratories | Michigan Department of Health and Human Services, Bureau of Laboratories | Blankenship HM et al |

| | | | | | | |
|---|---|---|---|---|---|---|
| EPI_ISL_516261 | USA/MI-MDHHS-SC21534/2020 | 2020-04-05 | North America/USA/Michigan/ | Michigan Department of Health and Human Services, Bureau of Laboratories | Michigan Department of Health and Human Services, Bureau of Laboratories | Blankenship HM et al |
| EPI_ISL_516271 | USA/MI-MDHHS-SC21544/2020 | 2020-03-19 | North America/USA/Michigan/ | Michigan Department of Health and Human Services, Bureau of Laboratories | Michigan Department of Health and Human Services, Bureau of Laboratories | Blankenship HM et al |
| EPI_ISL_516281 | USA/MI-MDHHS-SC21554/2020 | 2020-03-18 | North America/USA/Michigan/ | Michigan Department of Health and Human Services, Bureau of Laboratories | Michigan Department of Health and Human Services, Bureau of Laboratories | Blankenship HM et al |
| EPI_ISL_516311 | USA/MI-MDHHS-SC21584/2020 | 2020-03-23 | North America/USA/Michigan/ | Michigan Department of Health and Human Services, Bureau of Laboratories | Michigan Department of Health and Human Services, Bureau of Laboratories | Blankenship HM et al |
| EPI_ISL_516363 | USA/MI-MDHHS-SC21636/2020 | 2020-03-29 | North America/USA/Michigan/ | Michigan Department of Health and Human Services, Bureau of Laboratories | Michigan Department of Health and Human Services, Bureau of Laboratories | Blankenship HM et al |
| EPI_ISL_516381 | USA/MI-MDHHS-SC21654/2020 | 2020-07-13 | North America/USA/Michigan/ | Michigan Department of Health and Human Services, Bureau of Laboratories | Michigan Department of Health and Human Services, Bureau of Laboratories | Blankenship HM et al |

| | | | | | | |
|---|---|---|---|---|---|---|
| EPI_ISL_516390 | USA/MI-MDHHS-SC21663/2020 | 2020-07-12 | North America/USA/Michigan/ | Michigan Department of Health and Human Services, Bureau of Laboratories | Michigan Department of Health and Human Services, Bureau of Laboratories | Blankenship HM et al |
| EPI_ISL_516392 | USA/MI-MDHHS-SC21665/2020 | 2020-07-14 | North America/USA/Michigan/ | Michigan Department of Health and Human Services, Bureau of Laboratories | Michigan Department of Health and Human Services, Bureau of Laboratories | Blankenship HM et al |
| EPI_ISL_516398 | USA/MI-MDHHS-SC21671/2020 | 2020-07-07 | North America/USA/Michigan/ | Michigan Department of Health and Human Services, Bureau of Laboratories | Michigan Department of Health and Human Services, Bureau of Laboratories | Blankenship HM et al |
| EPI_ISL_516401 | USA/MI-MDHHS-SC21674/2020 | 2020-07-06 | North America/USA/Michigan/ | Michigan Department of Health and Human Services, Bureau of Laboratories | Michigan Department of Health and Human Services, Bureau of Laboratories | Blankenship HM et al |
| EPI_ISL_516403 | USA/MI-MDHHS-SC21676/2020 | 2020-07-13 | North America/USA/Michigan/ | Michigan Department of Health and Human Services, Bureau of Laboratories | Michigan Department of Health and Human Services, Bureau of Laboratories | Blankenship HM et al |
| EPI_ISL_516414 | NorthMacedonia/2531/2020 | 2020-06-06 | Europe/North Macedonia/Skopje/ | Center for public health - Skopje | "Research Center for Genetic Engineering and Biotechnology ""Georgi D. Efremov,Äù , Macedonian Academy of Sciences and Arts" | RCGEB - MASA et al |

| | | | | | | |
|---|---|---|---|---|---|---|
| EPI_ISL_516421 | NorthMacedonia/3122/2020 | 2020-06-11 | Europe/North Macedonia/Skopje/ | Center for public health - Skopje | "Research Center for Genetic Engineering and Biotechnology ""Georgi D. Efremov,Äù , Macedonian Academy of Sciences and Arts" | RCGEB - MASA et al |
| EPI_ISL_516429 | NorthMacedonia/7229/2020 | 2020-07-14 | Europe/North Macedonia/Eastern Regio | Clinical Hospital - Shtip | "Research Center for Genetic Engineering and Biotechnology ""Georgi D. Efremov,Äù , Macedonian Academy of Sciences and Arts" | RCGEB - MASA et al |
| EPI_ISL_516431 | NorthMacedonia/7808/2020 | 2020-07-20 | Europe/North Macedonia/Eastern Regio | Clinical Hospital - Shtip | "Research Center for Genetic Engineering and Biotechnology ""Georgi D. Efremov,Äù , Macedonian Academy of Sciences and Arts" | RCGEB - MASA et al |
| EPI_ISL_516441 | USA/WI-UW-1015/2020 | 2020-07-22 | North America/USA/Wisconsin/Dane Co | University of Wisconsin-Madison AIDS Vaccine Research Laboratories | University of Wisconsin-Madison AIDS Vaccine Research Laboratories | Gage Moreno et al |
| EPI_ISL_516443 | USA/WI-UW-1017/2020 | 2020-07-17 | North America/USA/Wisconsin/Dane Co | University of Wisconsin-Madison AIDS Vaccine Research Laboratories | University of Wisconsin-Madison AIDS Vaccine Research Laboratories | Gage Moreno et al |

| | | | | | | |
|---|---|---|---|---|---|---|
| EPI_ISL_516446 | USA/WI-UW-1020/2020 | 2020-08-04 | North America/USA/Wisconsin/Dane Co | University of Wisconsin-Madison AIDS Vaccine Research Laboratories | University of Wisconsin-Madison AIDS Vaccine Research Laboratories | Gage Moreno et al |
| EPI_ISL_516448 | USA/WI-UW-1022/2020 | 2020-07-31 | North America/USA/Wisconsin/Dane Co | University of Wisconsin-Madison AIDS Vaccine Research Laboratories | University of Wisconsin-Madison AIDS Vaccine Research Laboratories | Gage Moreno et al |
| EPI_ISL_516449 | USA/WI-UW-1023/2020 | 2020-08-02 | North America/USA/Wisconsin/Adams C | University of Wisconsin-Madison AIDS Vaccine Research Laboratories | University of Wisconsin-Madison AIDS Vaccine Research Laboratories | Gage Moreno et al |
| EPI_ISL_516453 | USA/WI-UW-1027/2020 | 2020-08-05 | North America/USA/Wisconsin/Dane Co | University of Wisconsin-Madison AIDS Vaccine Research Laboratories | University of Wisconsin-Madison AIDS Vaccine Research Laboratories | Gage Moreno et al |
| EPI_ISL_516454 | USA/WI-UW-1028/2020 | 2020-08-04 | North America/USA/Wisconsin/Dane Co | University of Wisconsin-Madison AIDS Vaccine Research Laboratories | University of Wisconsin-Madison AIDS Vaccine Research Laboratories | Gage Moreno et al |
| EPI_ISL_516469 | USA/WI-UW-1043/2020 | 2020-08-01 | North America/USA/Wisconsin/Dane Co | University of Wisconsin-Madison AIDS Vaccine Research Laboratories | University of Wisconsin-Madison AIDS Vaccine Research Laboratories | Gage Moreno et al |
| EPI_ISL_516475 | USA/WI-UW-1049/2020 | 2020-08-01 | North America/USA/Wisconsin/Dane Co | University of Wisconsin-Madison AIDS Vaccine Research Laboratories | University of Wisconsin-Madison AIDS Vaccine Research Laboratories | Gage Moreno et al |
| EPI_ISL_516478 | USA/WI-UW-1052/2020 | 2020-08-03 | North America/USA/Wisconsin/Jefferson | University of Wisconsin-Madison AIDS Vaccine Research Laboratories | University of Wisconsin-Madison AIDS Vaccine Research Laboratories | Gage Moreno et al |

| | | | | | | |
|---|---|---|---|---|---|---|
| EPI_ISL_516479 | USA/WI-UW-1053/2020 | 2020-07-30 | North America/USA/Wisconsin/Dane Co | University of Wisconsin-Madison AIDS Vaccine Research Laboratories | University of Wisconsin-Madison AIDS Vaccine Research Laboratories | Gage Moreno et al |
| EPI_ISL_516482 | USA/WI-UW-1056/2020 | 2020-07-30 | North America/USA/Wisconsin/Dane Co | University of Wisconsin-Madison AIDS Vaccine Research Laboratories | University of Wisconsin-Madison AIDS Vaccine Research Laboratories | Gage Moreno et al |
| EPI_ISL_516488 | USA/WI-UW-1062/2020 | 2020-07-31 | North America/USA/Wisconsin/Dane Co | University of Wisconsin-Madison AIDS Vaccine Research Laboratories | University of Wisconsin-Madison AIDS Vaccine Research Laboratories | Gage Moreno et al |
| EPI_ISL_516497 | USA/WI-UW-1071/2020 | 2020-07-29 | North America/USA/Wisconsin/Dane Co | University of Wisconsin-Madison AIDS Vaccine Research Laboratories | University of Wisconsin-Madison AIDS Vaccine Research Laboratories | Gage Moreno et al |
| EPI_ISL_516500 | USA/WI-UW-1074/2020 | 2020-07-24 | North America/USA/Wisconsin/Dane Co | University of Wisconsin-Madison AIDS Vaccine Research Laboratories | University of Wisconsin-Madison AIDS Vaccine Research Laboratories | Gage Moreno et al |
| EPI_ISL_516503 | USA/WI-UW-1077/2020 | 2020-07-26 | North America/USA/Wisconsin/Dane Co | University of Wisconsin-Madison AIDS Vaccine Research Laboratories | University of Wisconsin-Madison AIDS Vaccine Research Laboratories | Gage Moreno et al |
| EPI_ISL_516511 | USA/IL-UW-1085/2020 | 2020-07-25 | North America/USA/Illinois/Winnebago | University of Wisconsin-Madison AIDS Vaccine Research Laboratories | University of Wisconsin-Madison AIDS Vaccine Research Laboratories | Gage Moreno et al |
| EPI_ISL_516512 | USA/WI-UW-1086/2020 | 2020-07-25 | North America/USA/Wisconsin/Dane Co | University of Wisconsin-Madison AIDS Vaccine Research Laboratories | University of Wisconsin-Madison AIDS Vaccine Research Laboratories | Gage Moreno et al |

| | | | | | | |
|---|---|---|---|---|---|---|
| EPI_ISL_516517 | USA/WI-UW-1092/2020 | 2020-07-27 | North America/USA/Wisconsin/Jefferson | University of Wisconsin-Madison AIDS Vaccine Research Laboratories | University of Wisconsin-Madison AIDS Vaccine Research Laboratories | Gage Moreno et al |
| EPI_ISL_516524 | USA/WI-UW-1099/2020 | 2020-08-02 | North America/USA/Wisconsin/Dane Co | University of Wisconsin-Madison AIDS Vaccine Research Laboratories | University of Wisconsin-Madison AIDS Vaccine Research Laboratories | Gage Moreno et al |
| EPI_ISL_516526 | USA/WI-UW-1101/2020 | 2020-07-24 | North America/USA/Wisconsin/Dane Co | University of Wisconsin-Madison AIDS Vaccine Research Laboratories | University of Wisconsin-Madison AIDS Vaccine Research Laboratories | Gage Moreno et al |
| EPI_ISL_516527 | USA/WI-UW-1102/2020 | 2020-07-26 | North America/USA/Wisconsin/Dane Co | University of Wisconsin-Madison AIDS Vaccine Research Laboratories | University of Wisconsin-Madison AIDS Vaccine Research Laboratories | Gage Moreno et al |
| EPI_ISL_516544 | USA/WI-UW-1119/2020 | 2020-07-27 | North America/USA/Wisconsin/Dane Co | University of Wisconsin-Madison AIDS Vaccine Research Laboratories | University of Wisconsin-Madison AIDS Vaccine Research Laboratories | Gage Moreno et al |
| EPI_ISL_516546 | USA/WI-UW-1121/2020 | 2020-07-23 | North America/USA/Wisconsin/Dane Co | University of Wisconsin-Madison AIDS Vaccine Research Laboratories | University of Wisconsin-Madison AIDS Vaccine Research Laboratories | Gage Moreno et al |
| EPI_ISL_516564 | Switzerland/BL-ETHZ-230021/2020 | 2020-08-03 | Europe/Switzerland/Basel-Land/ | Viollier AG | Department of Biosystems Science and Engineering, ETH Zvºrich | Christian Beisel et al |
| EPI_ISL_516608 | Mexico/BC-InDRE-44/2020 | 2020-06-26 | North America/Mexico/Baja California/ | Instituto de Diagnostico y Referencia Epidemiologicos (INDRE) | Instituto de Diagnostico y Referencia Epidemiologicos (INDRE) | Ernesto Ramirez-Gonzalez et al |

| | | | | | | |
|---|---|---|---|---|---|---|
| EPI_ISL_516609 | Mexico/BCN-InDRE-45/2020 | 2020-06-27 | North America/Mexico/Baja California/ | Instituto de Diagnostico y Referencia Epidemiologicos (INDRE) | Instituto de Diagnostico y Referencia Epidemiologicos (INDRE) | Ernesto Ramirez-Gonzalez et al |
| EPI_ISL_516610 | Mexico/BCN-InDRE-50/2020 | 2020-06-30 | North America/Mexico/Baja California/ | Instituto de Diagnostico y Referencia Epidemiologicos (INDRE) | Instituto de Diagnostico y Referencia Epidemiologicos (INDRE) | Ernesto Ramirez-Gonzalez et al |
| EPI_ISL_516611 | Mexico/BCN-InDRE-51/2020 | 2020-07-05 | North America/Mexico/Baja California/ | Instituto de Diagnostico y Referencia Epidemiologicos (INDRE) | Instituto de Diagnostico y Referencia Epidemiologicos (INDRE) | Gisela Barrera-Badillo et al |
| EPI_ISL_516612 | Mexico/BCN-InDRE-52/2020 | 2020-07-06 | North America/Mexico/Baja California/ | Instituto de Diagnostico y Referencia Epidemiologicos (INDRE) | Instituto de Diagnostico y Referencia Epidemiologicos (INDRE) | Ernesto Ramirez-Gonzalez et al |
| EPI_ISL_516613 | Mexico/CHH-InDRE-43/2020 | 2020-06-26 | North America/Mexico/Chihuahua/ | Instituto de Diagnostico y Referencia Epidemiologicos (INDRE) | Instituto de Diagnostico y Referencia Epidemiologicos (INDRE) | Ernesto Ramirez-Gonzalez et al |
| EPI_ISL_516614 | Mexico/CHH-InDRE-46/2020 | 2020-06-28 | North America/Mexico/Chihuahua/ | Instituto de Diagnostico y Referencia Epidemiologicos (INDRE) | Instituto de Diagnostico y Referencia Epidemiologicos (INDRE) | Ernesto Ramirez-Gonzalez et al |
| EPI_ISL_516615 | Mexico/CHH-InDRE-47/2020 | 2020-06-27 | North America/Mexico/Chihuahua/ | Instituto de Diagnostico y Referencia Epidemiologicos (INDRE) | Instituto de Diagnostico y Referencia Epidemiologicos (INDRE) | Ernesto Ramirez-Gonzalez et al |
| EPI_ISL_516616 | Mexico/CHH-InDRE-48/2020 | 2020-06-30 | North America/Mexico/Chihuahua/ | Instituto de Diagnostico y Referencia Epidemiologicos (INDRE) | Instituto de Diagnostico y Referencia Epidemiologicos (INDRE) | Ernesto Ramirez-Gonzalez et al |

| | | | | | | |
|---|---|---|---|---|---|---|
| EPI_ISL_516617 | Mexico/CHH-InDRE-49/2020 | 2020-07-01 | North America/Mexico/Chihuahua/ | Instituto de Diagnostico y Referencia Epidemiologicos (INDRE) | Instituto de Diagnostico y Referencia Epidemiologicos (INDRE) | Ernesto Ramirez-Gonzalez et al |
| EPI_ISL_516619 | Mexico/COA-InDRE-37/2020 | 2020-06-13 | North America/Mexico/Coahuila/ | Instituto de Diagnostico y Referencia Epidemiologicos (INDRE) | Instituto de Diagnostico y Referencia Epidemiologicos (INDRE) | Gisela Barrera-Badillo et al |
| EPI_ISL_516620 | Mexico/COA-InDRE_40/2020 | 2020-06-19 | North America/Mexico/Coahuila/ | Instituto de Diagnostico y Referencia Epidemiologicos (INDRE) | Instituto de Diagnostico y Referencia Epidemiologicos (INDRE) | Gisela Barrera-Badillo et al |
| EPI_ISL_516621 | Mexico/COA-InDRE_41/2020 | 2020-06-26 | North America/Mexico/Coahuila/ | Instituto de Diagnostico y Referencia Epidemiologicos (INDRE) | Instituto de Diagnostico y Referencia Epidemiologicos (INDRE) | Gisela Barrera-Badillo et al |
| EPI_ISL_516622 | Mexico/SON-InDRE_35/2020 | 2020-05-15 | North America/Mexico/Sonora/ | Instituto de Diagnostico y Referencia Epidemiologicos (INDRE) | Instituto de Diagnostico y Referencia Epidemiologicos (INDRE) | Gisela Barrera-Badillo et al |
| EPI_ISL_516623 | Mexico/SON-InDRE_42/2020 | 2020-06-06 | North America/Mexico/Sonora/ | Instituto de Diagnostico y Referencia Epidemiologicos (INDRE) | Instituto de Diagnostico y Referencia Epidemiologicos (INDRE) | Gisela Barrera-Badillo et al |
| EPI_ISL_516624 | Mexico/TAM-InDRE_38/2020 | 2020-06-19 | North America/Mexico/Tamaulipas/ | Instituto de Diagnostico y Referencia Epidemiologicos (INDRE) | Instituto de Diagnostico y Referencia Epidemiologicos (INDRE) | Gisela Barrera-Badillo et al |
| EPI_ISL_516625 | Mexico/TAM-InDRE_39/2020 | 2020-06-15 | North America/Mexico/Tamaulipas/ | Instituto de Diagnostico y Referencia Epidemiologicos (INDRE) | Instituto de Diagnostico y Referencia Epidemiologicos (INDRE) | Gisela Barrera-Badillo et al |

| | | | | | | |
|---|---|---|---|---|---|---|
| EPI_ISL_516636 | Germany/BE-ChVir-D671-8/2020 | 2020-03-07 | Europe/Germany/Berlin/ | Charit√© Universit√§tsmedizin Berlin, Institut f√ºr Virologie/Labor Berlin | Charit√© Universit√§tsmedizin Berlin, Institut f√ºr Virologie/Labor Berlin | Victor M Corman et al |
| EPI_ISL_516645 | Germany/BE-ChVir-D715-D799-17/20 | 2020-03-06 | Europe/Germany/Berlin/ | Charit√© Universit√§tsmedizin Berlin, Institut f√ºr Virologie/Labor Berlin | Charit√© Universit√§tsmedizin Berlin, Institut f√ºr Virologie/Labor Berlin | Victor M Corman et al |
| EPI_ISL_516646 | Peru/LIM-INS-046/2020 | 2020-04-12 | South America/Peru/Lima/ | Laboratorio de Referencia Nacional de Virus Respiratorio. Centro Nacional de Salud Publica. Instituto Nacional de Salud Peru. | Laboratorio de Referencia Nacional de Biotecnolog√≠a y Biolog√≠a Molecular. Centro Nacional de Salud Publica. Instituto Nacional de Salud Peru. | Carlos Padilla Rojas et al |
| EPI_ISL_516647 | Peru/JUN-INS-047/2020 | 2020-04-11 | South America/Peru/Junin/ | Laboratorio de Referencia Nacional de Virus Respiratorio. Centro Nacional de Salud Publica. Instituto Nacional de Salud Peru. | Laboratorio de Referencia Nacional de Biotecnolog√≠a y Biolog√≠a Molecular. Centro Nacional de Salud Publica. Instituto Nacional de Salud Peru. | Carlos Padilla Rojas et al |
| EPI_ISL_516648 | Ecuador/USFQ-161/2020 | 2020-07-14 | South America/Ecuador/Zamora-Chinchi | Institute of Microbiology, Universidad San Francisco de Quito | Institute of Microbiology, Universidad San Francisco de Quito | Juan Jos√© Guadalupe et al |
| EPI_ISL_516650 | Ecuador/USFQ-165/2020 | 2020-07-22 | South America/Ecuador/Pichincha/Quito | Institute of Microbiology, Universidad San Francisco de Quito | Institute of Microbiology, Universidad San Francisco de Quito | Prado-Vivar et al |

| | | | | | | |
|---|---|---|---|---|---|---|
| EPI_ISL_516651 | Ecuador/USFQ-167/2020 | 2020-07-25 | South America/Ecuador/Los Rios/Queved | Institute of Microbiology, Universidad San Francisco de Quito | Institute of Microbiology, Universidad San Francisco de Quito | Prado-Vivar et al |
| EPI_ISL_516652 | Ecuador/USFQ-171/2020 | 2020-07-29 | South America/Ecuador/Orellana/Coca | Institute of Microbiology, Universidad San Francisco de Quito | Institute of Microbiology, Universidad San Francisco de Quito | Prado-Vivar et al |
| EPI_ISL_516653 | Peru/ANC-INS-049/2020 | 2020-04-10 | South America/Peru/Ancash/ | Laboratorio de Referencia Nacional de Virus Respiratorio. Centro Nacional de Salud Publica. Instituto Nacional de Salud Peru. | Laboratorio de Referencia Nacional de Biotecnolog√≠a y Biolog√≠a Molecular. Centro Nacional de Salud Publica. Instituto Nacional de Salud Peru. | Carlos Padilla Rojas et al |
| EPI_ISL_516673 | USA/VA-DCLS-0883/2020 | 2020-04-30 | North America/USA/Virginia/ | Virginia DCLS | Virginia DCLS | Virginia DCLS et al |
| EPI_ISL_516679 | USA/VA-DCLS-0893/2020 | 2020-05-12 | North America/USA/Virginia/ | Virginia DCLS | Virginia DCLS | Virginia DCLS et al |
| EPI_ISL_516686 | Peru/ANC-INS-052/2020 | 2020-04-11 | South America/Peru/Ancash/ | Laboratorio de Referencia Nacional de Virus Respiratorio. Centro Nacional de Salud Publica. Instituto Nacional de Salud Peru. | Laboratorio de Referencia Nacional de Biotecnolog√≠a y Biolog√≠a Molecular. Centro Nacional de Salud Publica. Instituto Nacional de Salud Peru. | Carlos Padilla Rojas et al |
| EPI_ISL_516691 | USA/VA-DCLS-0362/2020 | 2020-04-27 | North America/USA/Virginia/ | Virginia DCLS | Virginia DCLS | Virginia DCLS et al |
| EPI_ISL_516701 | USA/VA-DCLS-0828/2020 | 2020-05-06 | North America/USA/Virginia/ | Virginia DCLS | Virginia DCLS | Virginia DCLS et al |
| EPI_ISL_516702 | USA/VA-DCLS-0829/2020 | 2020-05-06 | North America/USA/Virginia/ | Virginia DCLS | Virginia DCLS | Virginia DCLS et al |
| EPI_ISL_516707 | USA/VA-DCLS-0840/2020 | 2020-05-05 | North America/USA/Virginia/ | Virginia DCLS | Virginia DCLS | Virginia DCLS et al |
| EPI_ISL_516710 | USA/VA-DCLS-0845/2020 | 2020-05-06 | North America/USA/Virginia/ | Virginia DCLS | Virginia DCLS | Virginia DCLS et al |
| EPI_ISL_516711 | USA/VA-DCLS-0846/2020 | 2020-05-06 | North America/USA/Virginia/ | Virginia DCLS | Virginia DCLS | Virginia DCLS et al |
| EPI_ISL_516717 | USA/VA-DCLS-0854/2020 | 2020-04-30 | North America/USA/Virginia/ | Virginia DCLS | Virginia DCLS | Virginia DCLS et al |

| | | | | | | |
|---|---|---|---|---|---|---|
| EPI_ISL_516722 | Peru/LIM-INS-054/2020 | 2020-04-11 | South America/Peru/Lima/ | Laboratorio de Referencia Nacional de Virus Respiratorio. Centro Nacional de Salud Publica. Instituto Nacional de Salud Peru. | Laboratorio de Referencia Nacional de Biotecnolog√≠a y Biolog√≠a Molecular. Centro Nacional de Salud Publica. Instituto Nacional de Salud Peru. | Carlos Padilla Rojas et al |
| EPI_ISL_516723 | USA/CA-CZB-3648/2020 | 2020-07-07 | North America/USA/California/Madera | UCSF Clinical Microbiology Laboratory | Chan-Zuckerberg Biohub | CZB Cliahub Consortium et al |
| EPI_ISL_516731 | USA/CA-CZB-3656/2020 | 2020-07-18 | North America/USA/California/Madera | UCSF Clinical Microbiology Laboratory | Chan-Zuckerberg Biohub | CZB Cliahub Consortium et al |
| EPI_ISL_516732 | USA/CA-CZB-3657/2020 | 2020-07-18 | North America/USA/California/Madera | UCSF Clinical Microbiology Laboratory | Chan-Zuckerberg Biohub | CZB Cliahub Consortium et al |
| EPI_ISL_516745 | USA/CA-CZB-3670/2020 | 2020-08-11 | North America/USA/California/Madera | UCSF Clinical Microbiology Laboratory | Chan-Zuckerberg Biohub | CZB Cliahub Consortium et al |
| EPI_ISL_516750 | USA/SC-20-0946-T10/2020 | 2020-06-20 | North America/USA/South Carolina/ | van Bakel Laboratory, Genetics and Genomics Sciences, Icahn School of Medicine at Mount Sinai | van Bakel Laboratory, Genetics and Genomics Sciences, Icahn School of Medicine at Mount Sinai | Andrew G. Letizia et al |
| EPI_ISL_516752 | USA/SC-20-1167-T07/2020 | 2020-06-24 | North America/USA/South Carolina/ | van Bakel Laboratory, Genetics and Genomics Sciences, Icahn School of Medicine at Mount Sinai | van Bakel Laboratory, Genetics and Genomics Sciences, Icahn School of Medicine at Mount Sinai | Andrew G. Letizia et al |

| | | | | | | |
|---|---|---|---|---|---|---|
| EPI_ISL_516753 | USA/SC-20-1273-T07/2020 | 2020-06-30 | North America/USA/South Carolina/ | van Bakel Laboratory, Genetics and Genomics Sciences, Icahn School of Medicine at Mount Sinai | van Bakel Laboratory, Genetics and Genomics Sciences, Icahn School of Medicine at Mount Sinai | Andrew G. Letizia et al |
| EPI_ISL_516764 | USA/SC-20-1418-T07/2020 | 2020-07-01 | North America/USA/South Carolina/ | van Bakel Laboratory, Genetics and Genomics Sciences, Icahn School of Medicine at Mount Sinai | van Bakel Laboratory, Genetics and Genomics Sciences, Icahn School of Medicine at Mount Sinai | Andrew G. Letizia et al |
| EPI_ISL_516773 | USA/SC-20-1601-T03/2020 | 2020-07-03 | North America/USA/South Carolina/ | van Bakel Laboratory, Genetics and Genomics Sciences, Icahn School of Medicine at Mount Sinai | van Bakel Laboratory, Genetics and Genomics Sciences, Icahn School of Medicine at Mount Sinai | Andrew G. Letizia et al |
| EPI_ISL_516778 | USA/SC-20-1820-T07/2020 | 2020-07-21 | North America/USA/South Carolina/ | van Bakel Laboratory, Genetics and Genomics Sciences, Icahn School of Medicine at Mount Sinai | van Bakel Laboratory, Genetics and Genomics Sciences, Icahn School of Medicine at Mount Sinai | Andrew G. Letizia et al |
| EPI_ISL_516779 | USA/SC-20-1936-T00/2020 | 2020-07-15 | North America/USA/South Carolina/ | van Bakel Laboratory, Genetics and Genomics Sciences, Icahn School of Medicine at Mount Sinai | van Bakel Laboratory, Genetics and Genomics Sciences, Icahn School of Medicine at Mount Sinai | Andrew G. Letizia et al |

| | | | | | | |
|---|---|---|---|---|---|---|
| EPI_ISL_516781 | USA/SC-20-1963-T00/2020 | 2020-07-15 | North America/USA/South Carolina/ | van Bakel Laboratory, Genetics and Genomics Sciences, Icahn School of Medicine at Mount Sinai | van Bakel Laboratory, Genetics and Genomics Sciences, Icahn School of Medicine at Mount Sinai | Andrew G. Letizia et al |
| EPI_ISL_516800 | Indonesia/YO-UGM-202449/2020 | 2020-06-22 | Asia/Indonesia/Yogyakarta/ | Rumah Sakit Akademik Universitas Gadjah Mada | Genetics Working Group (Pokja Genetik) Faculty of Medicine, Public Health and Nursing Universitas Gadjah Mada (FK-KMK UGM); Disease Investigation Center Wates Ministry of Agriculture Indonesia; Department of Microbiology FK-KMK UGM; Laboratorium Diagnostik Yayasan Tahija World Mosquito Program (WMP) Yogyakarta Center for Tropical Medicine FK-KMK UGM; Integrated Research center FK-KMK UGM; Department of Computer Science and Electronics FMIPA UGM | Gunadi et al |

| | | | | | | |
|---|---|---|---|---|---|---|
| EPI_ISL_516806 | Indonesia/YO-UGM-200927/2020 | 2020-05-16 | Asia/Indonesia/Yogyakarta/ | Rumah Sakit PKU Gamping | Genetics Working Group (Pokja Genetik) Faculty of Medicine, Public Health and Nursing Universitas Gadjah Mada (FK-KMK UGM); Disease Investigation Center Wates Ministry of Agriculture Indonesia; Department of Microbiology FK-KMK UGM; Laboratorium Diagnostik Yayasan Tahija World Mosquito Program (WMP) Yogyakarta Center for Tropical Medicine FK-KMK UGM; Integrated Research center FK-KMK UGM; Department of Computer Science and Electronics FMIPA UGM | Gunadi et al |
| EPI_ISL_516807 | Singapore/807/2020 | 2020-04-10 | Asia/Singapore/Singapore/ | National Public Health Laboratory, National Centre for Infectious Diseases | National Public Health Laboratory, National Centre for Infectious Diseases | Mak TM et al |

| | | | | | | |
|---|---|---|---|---|---|---|
| EPI_ISL_516812 | Singapore/812/2020 | 2020-08-11 | Asia/Singapore/Singapore/ | National Public Health Laboratory, National Centre for Infectious Diseases | National Public Health Laboratory, National Centre for Infectious Diseases | Mak TM et al |
| EPI_ISL_516826 | Singapore/826/2020 | 2020-08-11 | Asia/Singapore/Singapore/ | National Public Health Laboratory, National Centre for Infectious Diseases | National Public Health Laboratory, National Centre for Infectious Diseases | Mak TM et al |

| | | | | | | |
|---|---|---|---|---|---|---|
| EPI_ISL_516829 | Indonesia/YO-UGM-781481/2020 | 2020-08-10 | Asia/Indonesia/Yogyakarta/ | RSUD Nyi Ageng Serang | Genetics Working Group (Pokja Genetik) Faculty of Medicine, Public Health and Nursing Universitas Gadjah Mada (FK-KMK UGM), Disease Investigation Center Wates Ministry of Agriculture Indonesia, Department of Microbiology FK-KMK UGM, Laboratorium Diagnostik Yayasan Tahija World Mosquito Program (WMP) Yogyakarta Center for Tropical Medicine FK-KMK UGM, Integrated Research Center FK-KMK UGM, Department of Computer Science and Electronics FMIPA UGM | Gunadi et al |
| EPI_ISL_516873 | England/LOND-125F814/2020 | 2020-04-06 | Europe/United Kingdom/England/ | North West London Pathology, Imperial College Healthcare NHS Trust | Wellcome Sanger Institute for the COVID-19 Genomics UK (COG-UK) consortium | Ling Li et al |
| EPI_ISL_516884 | Israel/CVL-n17051/2020 | 2020-06-01 | Asia/Israel/Tel Aviv District/ | Israel Central Virology laboratory | Israel Central Virology laboratory | Neta Zuckerman et al |

| | | | | | | |
|---|---|---|---|---|---|---|
| EPI_ISL_516891 | Israel/CVL-n17953/2020 | 2020-06-03 | Asia/Israel/Tel Aviv District/ | Israel Central Virology laboratory | Israel Central Virology laboratory | Neta Zuckerman et al |
| EPI_ISL_516892 | Israel/CVL-n18311/2020 | 2020-06-04 | Asia/Israel/Tel Aviv District/ | Israeli Central Virology laboratory | Israel Central Virology laboratory | Neta Zuckerman et al |
| EPI_ISL_516893 | Israel/CVL-n18346/2020 | 2020-06-04 | Asia/Israel/Tel Aviv District/ | Israel Central Virology laboratory | Israel Central Virology laboratory | Neta Zuckerman et al |
| EPI_ISL_516896 | Israel/CVL-n18467/2020 | 2020-06-04 | Asia/Israel/Tel Aviv District/ | Israel Central Virology laboratory | Israel Central Virology laboratory | Neta Zuckerman et al |
| EPI_ISL_516897 | Israel/CVL-n19018/2020 | 2020-06-05 | Asia/Israel/Tel Aviv District/ | Israel Central Virology laboratory | Israel Central Virology laboratory | Neta Zuckerman et al |
| EPI_ISL_516898 | Israel/CVL-n19042/2020 | 2020-06-05 | Asia/Israel/Tel Aviv District/ | Israel Central Virology laboratory | Israel Central Virology laboratory | Neta Zuckerman et al |
| EPI_ISL_516903 | Israel/CVL1133/2020 | 2020-06-29 | Asia/Israel/North District/ | Israel Central Virology laboratory | Israel Central Virology laboratory | Neta Zuckerman et al |
| EPI_ISL_516910 | Israel/CVL1610/2020 | 2020-07-03 | Asia/Israel/North District/ | Israel Central Virology laboratory | Israel Central Virology laboratory | Neta Zuckerman et al |
| EPI_ISL_516912 | Israel/CVL34959/2020 | 2020-07-05 | Asia/Israel/Haifa District/ | Israel Central Virology laboratory | Israel Central Virology laboratory | Neta Zuckerman et al |
| EPI_ISL_516915 | Israel/CVL4577/2020 | 2020-06-15 | Asia/Israel/Haifa District/ | Israel Central Virology laboratory | Israel Central Virology laboratory | Neta Zuckerman et al |
| EPI_ISL_516919 | Israel/CVL5786/2020 | 2020-06-25 | Asia/Israel/North District/ | Israel Central Virology laboratory | Israel Central Virology laboratory | Neta Zuckerman et al |
| EPI_ISL_516926 | Montenegro/ChVir-1794332002/202( | 2020-07-17 | Europe/Montenegro/Montenegro/ | Department for Molecular Diagnostics, Centre for Medical Microbiology, Institute of Public Health of Montenegro | Charit√© Universit√§tsmedizin Berlin, Institut f√ºr Virologie | Victor M Corman et al |

| | | | | | | |
|---|---|---|---|---|---|---|
| EPI_ISL_516934 | Moldova/ICGEB_MD1/2020 | 2020-06-18 | Europe/Moldova/Chisinau/ | Nicolae Testemitanu State University of Medicine and Pharmacy | International Centre for Genetic Engineering and Biotechnology (ICGEB) and ARGO Open Lab Platform for Genome Sequencing | Ulinici M et al |
| EPI_ISL_516935 | Moldova/ICGEB_MD4/2020 | 2020-06-18 | Europe/Moldova/Balti/ | Nicolae Testemitanu State University of Medicine and Pharmacy | International Centre for Genetic Engineering and Biotechnology (ICGEB) and ARGO Open Lab Platform for Genome Sequencing | Ulinici M et al |
| EPI_ISL_516936 | Moldova/ICGEB_MD7/2020 | 2020-06-17 | Europe/Moldova/Straseni/ | Nicolae Testemitanu State University of Medicine and Pharmacy | International Centre for Genetic Engineering and Biotechnology (ICGEB) and ARGO Open Lab Platform for Genome Sequencing | Ulinici M et al |
| EPI_ISL_516938 | Moldova/ICGEB_MD6/2020 | 2020-06-17 | Europe/Moldova/Rezina/Rabnita | Nicolae Testemitanu State University of Medicine and Pharmacy | International Centre for Genetic Engineering and Biotechnology (ICGEB) and ARGO Open Lab Platform for Genome Sequencing | Ulinici M et al |
| EPI_ISL_516942 | India/UP-NBRI-N13-TCR323/2020 | 2020-04-02 | Asia/India/Uttar Pradesh/Lucknow | King Georges Medical University | CSIR-National Botanical Research Institute | Priti Prasad et al |
| EPI_ISL_516946 | India/UP-NBRI-N17-TCP057/2020 | 2020-03-11 | Asia/India/Uttar Pradesh/Lucknow | King Georges Medical University | CSIR-National Botanical Research Institute | Priti Prasad et al |
| EPI_ISL_516947 | India/UP-NBRI-N18-TDU012/2020 | 2020-05-14 | Asia/India/Uttar Pradesh/Farrukhabad | King Georges Medical University | CSIR-National Botanical Research Institute | Priti Prasad et al |

| | | | | | | |
|---|---|---|---|---|---|---|
| EPI_ISL_516948 | India/UP-NBRI-N19-TCP645/2020 | 2020-03-18 | Asia/India/Uttar Pradesh/Lucknow | King Georges Medical University | CSIR-National Botanical Research Institute | Priti Prasad et al |
| EPI_ISL_516953 | India/UP-NBRI-N24-TDU424/2020 | 2020-05-15 | Asia/India/Uttar Pradesh/Lucknow | King Georges Medical University | CSIR-National Botanical Research Institute | Priti Prasad et al |
| EPI_ISL_516954 | India/UP-NBRI-N25-TDU890/2020 | 2020-05-15 | Asia/India/Uttar Pradesh/Hardoi | King Georges Medical University | CSIR-National Botanical Research Institute | Priti Prasad et al |
| EPI_ISL_516956 | India/UP-NBRI-N27-TDV034/2020 | 2020-05-15 | Asia/India/Uttar Pradesh/Kannauj | King Georges Medical University | CSIR-National Botanical Research Institute | Priti Prasad et al |
| EPI_ISL_516964 | India/UP-NBRI-N35-TDV640/2020 | 2020-05-15 | Asia/India/Uttar Pradesh/Shahjahanpur | King Georges Medical University | CSIR-National Botanical Research Institute | Priti Prasad et al |
| EPI_ISL_516967 | India/UP-NBRI-N38-TDT660/2020 | 2020-05-14 | Asia/India/Uttar Pradesh/Lucknow | King Georges Medical University | CSIR-National Botanical Research Institute | Priti Prasad et al |
| EPI_ISL_516968 | India/UP-NBRI-N39-TDV163/2020 | 2020-05-15 | Asia/India/Uttar Pradesh/Kanpur | King Georges Medical University | CSIR-National Botanical Research Institute | Priti Prasad et al |
| EPI_ISL_516971 | India/UP-NBRI-N42-TCX099/2020 | 2020-04-16 | Asia/India/Uttar Pradesh/Lucknow | King Georges Medical University | CSIR-National Botanical Research Institute | Priti Prasad et al |
| EPI_ISL_516973 | India/UP-NBRI-N45-TCW721/2020 | 2020-04-15 | Asia/India/Uttar Pradesh/Agra | King Georges Medical University | CSIR-National Botanical Research Institute | Priti Prasad et al |
| EPI_ISL_516979 | India/UP-NBRI-N51-TEH465/2020 | 2020-05-25 | Asia/India/Uttar Pradesh/Hardoi | King Georges Medical University | CSIR-National Botanical Research Institute | Priti Prasad et al |
| EPI_ISL_516986 | India/UP-NBRI-N58-TEG811/2020 | 2020-05-24 | Asia/India/Uttar Pradesh/Moradabad | King Georges Medical University | CSIR-National Botanical Research Institute | Priti Prasad et al |

| | | | | | | |
|---|---|---|---|---|---|---|
| EPI_ISL_516987 | Peru/ANC-INS-066/2020 | 2020-03-16 | South America/Peru/Ancash/ | Laboratorio de Referencia Nacional de Virus Respiratorio. Centro Nacional de Salud Publica. Instituto Nacional de Salud Peru. | Laboratorio de Referencia Nacional de Biotecnolog√≠a y Biolog√≠a Molecular. Centro Nacional de Salud Publica. Instituto Nacional de Salud Peru. | Carlos Padilla Rojas et al |
| EPI_ISL_517099 | England/LIVE-A7FB3/2020 | 2020-03-26 | Europe/United Kingdom/England/ | Liverpool Clinical Laboratories | COVID-19 Genomics UK (COG-UK) Consortium | Sam Haldenby et al |
| EPI_ISL_517255 | England/LIVE-A9081/2020 | 2020-03-25 | Europe/United Kingdom/England/ | Liverpool Clinical Laboratories | COVID-19 Genomics UK (COG-UK) Consortium | Sam Haldenby et al |
| EPI_ISL_517303 | England/LIVE-A99C2/2020 | 2020-03-24 | Europe/United Kingdom/England/ | Liverpool Clinical Laboratories | COVID-19 Genomics UK (COG-UK) Consortium | Sam Haldenby et al |
| EPI_ISL_517328 | England/LIVE-A9B53/2020 | 2020-03-24 | Europe/United Kingdom/England/ | Liverpool Clinical Laboratories | COVID-19 Genomics UK (COG-UK) Consortium | Sam Haldenby et al |
| EPI_ISL_517336 | England/LIVE-A9BF9/2020 | 2020-03-24 | Europe/United Kingdom/England/ | Liverpool Clinical Laboratories | COVID-19 Genomics UK (COG-UK) Consortium | Sam Haldenby et al |
| EPI_ISL_517344 | England/LIVE-A9C7E/2020 | 2020-03-24 | Europe/United Kingdom/England/ | Liverpool Clinical Laboratories | COVID-19 Genomics UK (COG-UK) Consortium | Sam Haldenby et al |
| EPI_ISL_517389 | England/LIVE-AA017/2020 | 2020-03-23 | Europe/United Kingdom/England/ | Liverpool Clinical Laboratories | COVID-19 Genomics UK (COG-UK) Consortium | Sam Haldenby et al |
| EPI_ISL_517447 | England/LIVE-AA3D2/2020 | 2020-03-23 | Europe/United Kingdom/England/ | Liverpool Clinical Laboratories | COVID-19 Genomics UK (COG-UK) Consortium | Sam Haldenby et al |
| EPI_ISL_517479 | England/LIVE-AA642/2020 | 2020-03-24 | Europe/United Kingdom/England/ | Liverpool Clinical Laboratories | COVID-19 Genomics UK (COG-UK) Consortium | Sam Haldenby et al |
| EPI_ISL_517490 | England/LIVE-AA6F7/2020 | 2020-03-24 | Europe/United Kingdom/England/ | Liverpool Clinical Laboratories | COVID-19 Genomics UK (COG-UK) Consortium | Sam Haldenby et al |

| | | | | | | |
|---|---|---|---|---|---|---|
| EPI_ISL_517493 | England/LIVE-AA721/2020 | 2020-03-24 | Europe/United Kingdom/England/ | Liverpool Clinical Laboratories | COVID-19 Genomics UK (COG-UK) Consortium | Sam Haldenby et al |
| EPI_ISL_517531 | Peru/ICA-INS-055/2020 | 2020-04-10 | South America/Peru/Ica/ | Laboratorio de Referencia Nacional de Virus Respiratorio. Centro Nacional de Salud Publica. Instituto Nacional de Salud Peru. | Laboratorio de Referencia Nacional de Biotecnolog√≠a y Biolog√≠a Molecular. Centro Nacional de Salud Publica. Instituto Nacional de Salud Peru. | Carlos Padilla Rojas et al |
| EPI_ISL_517537 | England/PORT-2D373C/2020 | 2020-04-15 | Europe/United Kingdom/England/ | Centre for Enzyme Innovation, University of Portsmouth / Translational Research Laboratory, Portsmouth Hospitals NHS Trust | COVID-19 Genomics UK (COG-UK) Consortium | Angela Beckett et al |
| EPI_ISL_517661 | Suriname/SR-69/2020 | 2020-08-07 | South America/Suriname/Suriname/ | Academic Hospital Paramaribo | Erasmus Medical Center | Bas Oude Munnink et al |
| EPI_ISL_517662 | Suriname/SR-70/2020 | 2020-08-07 | South America/Suriname/Suriname/ | Academic Hospital Paramaribo | Erasmus Medical Center | Bas Oude Munnink et al |
| EPI_ISL_517686 | Peru/LAL-INS-056/2020 | 2020-04-12 | South America/Peru/La Libertad/ | Laboratorio de Referencia Nacional de Virus Respiratorio. Centro Nacional de Salud Publica. Instituto Nacional de Salud Peru. | Laboratorio de Referencia Nacional de Biotecnolog√≠a y Biolog√≠a Molecular. Centro Nacional de Salud Publica. Instituto Nacional de Salud Peru. | Carlos Padilla Rojas et al |
| EPI_ISL_517713 | Peru/CAJ-INS-057/2020 | 2020-04-01 | South America/Peru/Cajamarca/ | Laboratorio de Referencia Nacional de Virus Respiratorio. Centro Nacional de Salud Publica. Instituto Nacional de Salud Peru. | Laboratorio de Referencia Nacional de Biotecnolog√≠a y Biolog√≠a Molecular. Centro Nacional de Salud Publica. Instituto Nacional de Salud Peru. | Carlos Padilla Rojas et al |

| | | | | | | |
|---|---|---|---|---|---|---|
| EPI_ISL_517770 | Peru/CAJ-INS-061/2020 | 2020-03-13 | South America/Peru/Cajamarca/ | Laboratorio de Referencia Nacional de Virus Respiratorio. Centro Nacional de Salud Publica. Instituto Nacional de Salud Peru | Laboratorio de Referencia Nacional de Biotecnolog√≠a y Biolog√≠a Molecular. Centro Nacional de Salud Publica. Instituto Nacional de Salud Peru. | Carlos Padilla Rojas et al |
| EPI_ISL_517778 | USA/FL-BPHL-0888/2020 | 2020-05-06 | North America/USA/Florida/ | Florida Bureau of Public Health Laboratories | Florida Bureau of Public Health Laboratories | Sarah Schmedes et al |
| EPI_ISL_517780 | USA/FL-BPHL-0890/2020 | 2020-04-14 | North America/USA/Florida/ | Florida Bureau of Public Health Laboratories | Florida Bureau of Public Health Laboratories | Sarah Schmedes et al |
| EPI_ISL_517781 | USA/FL-BPHL-0891/2020 | 2020-04-14 | North America/USA/Florida/ | Florida Bureau of Public Health Laboratories | Florida Bureau of Public Health Laboratories | Sarah Schmedes et al |
| EPI_ISL_517796 | USA/FL-BPHL-0906/2020 | 2020-06-29 | North America/USA/Florida/ | Florida Bureau of Public Health Laboratories | Florida Bureau of Public Health Laboratories | Sarah Schmedes et al |
| EPI_ISL_517804 | USA/FL-BPHL-0914/2020 | 2020-07-02 | North America/USA/Florida/ | Florida Bureau of Public Health Laboratories | Florida Bureau of Public Health Laboratories | Sarah Schmedes et al |
| EPI_ISL_517807 | USA/FL-BPHL-0917/2020 | 2020-07-02 | North America/USA/Florida/ | Florida Bureau of Public Health Laboratories | Florida Bureau of Public Health Laboratories | Sarah Schmedes et al |
| EPI_ISL_517809 | USA/FL-BPHL-0919/2020 | 2020-07-02 | North America/USA/Florida/ | Florida Bureau of Public Health Laboratories | Florida Bureau of Public Health Laboratories | Sarah Schmedes et al |
| EPI_ISL_517818 | USA/FL-BPHL-0928/2020 | 2020-07-11 | North America/USA/Florida/ | Florida Bureau of Public Health Laboratories | Florida Bureau of Public Health Laboratories | Sarah Schmedes et al |
| EPI_ISL_517819 | USA/FL-BPHL-0929/2020 | 2020-07-10 | North America/USA/Florida/ | Florida Bureau of Public Health Laboratories | Florida Bureau of Public Health Laboratories | Sarah Schmedes et al |
| EPI_ISL_517821 | USA/FL-BPHL-0931/2020 | 2020-07-13 | North America/USA/Florida/ | Florida Bureau of Public Health Laboratories | Florida Bureau of Public Health Laboratories | Sarah Schmedes et al |

| | | | | | | |
|---|---|---|---|---|---|---|
| EPI_ISL_517832 | USA/FL-BPHL-0942/2020 | 2020-07-17 | North America/USA/Florida/ | Florida Bureau of Public Health Laboratories | Florida Bureau of Public Health Laboratories | Sarah Schmedes et al |
| EPI_ISL_517833 | USA/FL-BPHL-0943/2020 | 2020-07-17 | North America/USA/Florida/ | Florida Bureau of Public Health Laboratories | Florida Bureau of Public Health Laboratories | Sarah Schmedes et al |
| EPI_ISL_517843 | USA/FL-BPHL-0953/2020 | 2020-07-07 | North America/USA/Florida/ | Florida Bureau of Public Health Laboratories | Florida Bureau of Public Health Laboratories | Sarah Schmedes et al |
| EPI_ISL_517851 | USA/FL-BPHL-0961/2020 | 2020-07-08 | North America/USA/Florida/ | Florida Bureau of Public Health Laboratories | Florida Bureau of Public Health Laboratories | Sarah Schmedes et al |
| EPI_ISL_517853 | USA/FL-BPHL-0963/2020 | 2020-07-08 | North America/USA/Florida/ | Florida Bureau of Public Health Laboratories | Florida Bureau of Public Health Laboratories | Sarah Schmedes et al |
| EPI_ISL_517859 | USA/FL-BPHL-0969/2020 | 2020-07-09 | North America/USA/Florida/ | Florida Bureau of Public Health Laboratories | Florida Bureau of Public Health Laboratories | Sarah Schmedes et al |
| EPI_ISL_517863 | USA/FL-BPHL-0973/2020 | 2020-07-29 | North America/USA/Florida/ | Florida Bureau of Public Health Laboratories | Florida Bureau of Public Health Laboratories | Sarah Schmedes et al |
| EPI_ISL_517866 | USA/FL-BPHL-0976/2020 | 2020-07-29 | North America/USA/Florida/ | Florida Bureau of Public Health Laboratories | Florida Bureau of Public Health Laboratories | Sarah Schmedes et al |
| EPI_ISL_517880 | USA/FL-BPHL-0990/2020 | 2020-07-02 | North America/USA/Florida/ | Florida Bureau of Public Health Laboratories | Florida Bureau of Public Health Laboratories | Sarah Schmedes et al |
| EPI_ISL_517883 | USA/FL-BPHL-0993/2020 | 2020-07-02 | North America/USA/Florida/ | Florida Bureau of Public Health Laboratories | Florida Bureau of Public Health Laboratories | Sarah Schmedes et al |
| EPI_ISL_517885 | USA/FL-BPHL-0995/2020 | 2020-07-10 | North America/USA/Florida/ | Florida Bureau of Public Health Laboratories | Florida Bureau of Public Health Laboratories | Sarah Schmedes et al |
| EPI_ISL_517888 | USA/FL-BPHL-0998/2020 | 2020-07-10 | North America/USA/Florida/ | Florida Bureau of Public Health Laboratories | Florida Bureau of Public Health Laboratories | Sarah Schmedes et al |
| EPI_ISL_517890 | USA/FL-BPHL-1000/2020 | 2020-07-10 | North America/USA/Florida/ | Florida Bureau of Public Health Laboratories | Florida Bureau of Public Health Laboratories | Sarah Schmedes et al |

| | | | | | | |
|---|---|---|---|---|---|---|
| EPI_ISL_517893 | USA/FL-BPHL-1003/2020 | 2020-07-13 | North America/USA/Florida/ | Florida Bureau of Public Health Laboratories | Florida Bureau of Public Health Laboratories | Sarah Schmedes et al |
| EPI_ISL_517894 | USA/FL-BPHL-1004/2020 | 2020-07-13 | North America/USA/Florida/ | Florida Bureau of Public Health Laboratories | Florida Bureau of Public Health Laboratories | Sarah Schmedes et al |
| EPI_ISL_517900 | USA/FL-BPHL-1010/2020 | 2020-07-15 | North America/USA/Florida/ | Florida Bureau of Public Health Laboratories | Florida Bureau of Public Health Laboratories | Sarah Schmedes et al |
| EPI_ISL_517903 | USA/FL-BPHL-1013/2020 | 2020-07-16 | North America/USA/Florida/ | Florida Bureau of Public Health Laboratories | Florida Bureau of Public Health Laboratories | Sarah Schmedes et al |
| EPI_ISL_517905 | USA/FL-BPHL-1015/2020 | 2020-07-17 | North America/USA/Florida/ | Florida Bureau of Public Health Laboratories | Florida Bureau of Public Health Laboratories | Sarah Schmedes et al |
| EPI_ISL_517906 | USA/FL-BPHL-1016/2020 | 2020-07-17 | North America/USA/Florida/ | Florida Bureau of Public Health Laboratories | Florida Bureau of Public Health Laboratories | Sarah Schmedes et al |
| EPI_ISL_517912 | USA/FL-BPHL-1022/2020 | 2020-07-20 | North America/USA/Florida/ | Florida Bureau of Public Health Laboratories | Florida Bureau of Public Health Laboratories | Sarah Schmedes et al |
| EPI_ISL_517915 | USA/FL-BPHL-1025/2020 | 2020-07-21 | North America/USA/Florida/ | Florida Bureau of Public Health Laboratories | Florida Bureau of Public Health Laboratories | Sarah Schmedes et al |
| EPI_ISL_517916 | USA/FL-BPHL-1026/2020 | 2020-07-21 | North America/USA/Florida/ | Florida Bureau of Public Health Laboratories | Florida Bureau of Public Health Laboratories | Sarah Schmedes et al |
| EPI_ISL_517920 | USA/FL-BPHL-1030/2020 | 2020-07-22 | North America/USA/Florida/ | Florida Bureau of Public Health Laboratories | Florida Bureau of Public Health Laboratories | Sarah Schmedes et al |
| EPI_ISL_517923 | USA/FL-BPHL-1033/2020 | 2020-07-23 | North America/USA/Florida/ | Florida Bureau of Public Health Laboratories | Florida Bureau of Public Health Laboratories | Sarah Schmedes et al |
| EPI_ISL_517926 | USA/FL-BPHL-1036/2020 | 2020-07-22 | North America/USA/Florida/ | Florida Bureau of Public Health Laboratories | Florida Bureau of Public Health Laboratories | Sarah Schmedes et al |
| EPI_ISL_517929 | USA/FL-BPHL-1039/2020 | 2020-08-01 | North America/USA/Florida/ | Florida Bureau of Public Health Laboratories | Florida Bureau of Public Health Laboratories | Sarah Schmedes et al |

| | | | | | | |
|---|---|---|---|---|---|---|
| EPI_ISL_517930 | USA/FL-BPHL-1040/2020 | 2020-08-01 | North America/USA/Florida/ | Florida Bureau of Public Health Laboratories | Florida Bureau of Public Health Laboratories | Sarah Schmedes et al |
| EPI_ISL_517942 | USA/FL-BPHL-1052/2020 | 2020-05-08 | North America/USA/Florida/ | Florida Bureau of Public Health Laboratories | Florida Bureau of Public Health Laboratories | Sarah Schmedes et al |
| EPI_ISL_517945 | USA/FL-BPHL-1055/2020 | 2020-06-03 | North America/USA/Florida/ | Florida Bureau of Public Health Laboratories | Florida Bureau of Public Health Laboratories | Sarah Schmedes et al |
| EPI_ISL_517947 | USA/FL-BPHL-1057/2020 | 2020-06-04 | North America/USA/Florida/ | Florida Bureau of Public Health Laboratories | Florida Bureau of Public Health Laboratories | Sarah Schmedes et al |
| EPI_ISL_517949 | USA/FL-BPHL-1059/2020 | 2020-07-17 | North America/USA/Florida/ | Florida Bureau of Public Health Laboratories | Florida Bureau of Public Health Laboratories | Sarah Schmedes et al |
| EPI_ISL_517952 | USA/FL-BPHL-1062/2020 | 2020-07-17 | North America/USA/Florida/ | Florida Bureau of Public Health Laboratories | Florida Bureau of Public Health Laboratories | Sarah Schmedes et al |
| EPI_ISL_517959 | Peru/LIM-INS-048/2020 | 2020-04-12 | South America/Peru/Lima/ | Laboratorio de Referencia Nacional de Virus Respiratorio. Centro Nacional de Salud Publica. Instituto Nacional de Salud Peru | Laboratorio de Referencia Nacional de Biotecnolog√≠a y Biolog√≠a Molecular. Centro Nacional de Salud Publica. Instituto Nacional de Salud Peru. | Carlos Padilla Rojas et al |
| EPI_ISL_517964 | USA/TX-DSHS-0031/2020 | 2020-05-08 | North America/USA/Texas/Bexar County | Texas Department of State Health Services | Texas Department of State Health Services | Rashmi Tuladhar et al |
| EPI_ISL_517965 | USA/TX-DSHS-0032/2020 | 2020-05-07 | North America/USA/Texas/Panola Count | Texas Department of State Health Services | Texas Department of State Health Services | Rashmi Tuladhar et al |
| EPI_ISL_517966 | USA/TX-DSHS-0033/2020 | 2020-05-08 | North America/USA/Texas/Deaf Smith C | Texas Department of State Health Services | Texas Department of State Health Services | Rashmi Tuladhar et al |
| EPI_ISL_517974 | USA/TX-DSHS-0042/2020 | 2020-05-09 | North America/USA/Texas/Titus County | Texas Department of State Health Services | Texas Department of State Health Services | Rashmi Tuladhar et al |

| | | | | | | |
|---|---|---|---|---|---|---|
| EPI_ISL_517986 | USA/TX-DSHS-0055/2020 | 2020-05-09 | North America/USA/Texas/Titus County | Texas Department of State Health Services | Texas Department of State Health Services | Rashmi Tuladhar et al |
| EPI_ISL_517997 | Singapore/834/2020 | 2020-06-15 | Asia/Singapore/Singapore/ | Singapore General Hospital | Department of Microbiology | Nurdyana Abdul Rahman et al |
| EPI_ISL_518015 | Singapore/835_2/2020 | 2020-06-17 | Asia/Singapore/Singapore/ | Singapore General Hospital | Department of Microbiology | Nurdyana Abdul Rahman et al |
| EPI_ISL_518035 | SouthAfrica/KRISP-K002445/2020 | 2020-07-07 | Africa/South Africa/KwaZulu-Natal/eThe | NHLS-IALCH | KRISP, KZN Research Innovation and Sequencing Platform | Giandhari J et al |
| EPI_ISL_518036 | SouthAfrica/KRISP-K002446/2020 | 2020-07-07 | Africa/South Africa/KwaZulu-Natal/eThe | NHLS-IALCH | KRISP, KZN Research Innovation and Sequencing Platform | Giandhari J et al |
| EPI_ISL_518047 | SouthAfrica/KRISP-K002464/2020 | 2020-07-14 | Africa/South Africa/KwaZulu-Natal/eThe | NHLS-IALCH | KRISP, KZN Research Innovation and Sequencing Platform | Giandhari J et al |
| EPI_ISL_518048 | SouthAfrica/KRISP-K002465/2020 | 2020-06-25 | Africa/South Africa/KwaZulu-Natal/eThe | NHLS-IALCH | KRISP, KZN Research Innovation and Sequencing Platform | Giandhari J et al |
| EPI_ISL_518052 | SouthAfrica/KRISP-K002474/2020 | 2020-06-29 | Africa/South Africa/KwaZulu-Natal/eThe | NHLS-IALCH | KRISP, KZN Research Innovation and Sequencing Platform | Giandhari J et al |
| EPI_ISL_518254 | Australia/VIC6263/2020 | 2020-07-24 | Oceania/Australia/Victoria/ | Microbiological Diagnostic Unit - Public Health Laboratory (MDU-PHL) | MDU-PHL | Seemann T. et al |
| EPI_ISL_518802 | Suriname/SR-53/2020 | 2020-07-16 | South America/Suriname/Suriname/ | Academic Hospital Paramaribo | Erasmus Medical Center | Bas Oude Munnink et al |

| | | | | | | |
|---|---|---|---|---|---|---|
| EPI_ISL_518819 | Indonesia/TNG-LIPI001/2020 | 2020-08-13 | Asia/Indonesia/Banten/Tangerang | Qadr Hospital, Tangerang, Banten | Biosafety Level-3 Laboratory, Indonesian Institute of Sciences (LIPI) | Anik Budhi Dharmayanthi et al |
| EPI_ISL_518820 | Indonesia/JK-LIPI002/2020 | 2020-08-13 | Asia/Indonesia/Jakarta/ | Klinik Apotek Dein, Jakarta, Indonesia | Biosafety Level-3 Laboratory, Indonesian Institute of Sciences (LIPI) | Anik Budhi Dharmayanthi et al |
| EPI_ISL_518824 | Oman/RESP-20-B-15095/2020 | 2020-05-20 | Asia/Oman/Muscat/ | Oman-National Influenza Center | Biotechnology & OMICs Laboratory, Natural & Medical Sciences Research Center, University of Nizwa | Abdul Latif Khan et al |
| EPI_ISL_518825 | Oman/RESP-20-B-15133/2020 | 2020-05-21 | Asia/Oman/Muscat/ | Oman-National Influenza Center | Biotechnology & OMICs Laboratory, Natural & Medical Sciences Research Center, University of Nizwa | Samira Al-Mahruqi et al |
| EPI_ISL_518831 | Oman/RESP-20-B-15411/2020 | 2020-05-21 | Asia/Oman/Muscat/ | Oman-National Influenza Center | Biotechnology & OMICs Laboratory, Natural & Medical Sciences Research Center, University of Nizwa | Sajjad Asaf et al |
| EPI_ISL_518832 | Oman/RESP-20-B-15719/2020 | 2020-05-22 | Asia/Oman/Muscat/ | Oman-National Influenza Center | Biotechnology & OMICs Laboratory, Natural & Medical Sciences Research Center, University of Nizwa | Sajjad Asaf et al |

| | | | | | | |
|---|---|---|---|---|---|---|
| EPI_ISL_518840 | Oman/RESP-20-B-16254/2020 | 2020-05-22 | Asia/Oman/Muscat/ | Oman-National Influenza Center | Biotechnology & OMICs Laboratory, Natural & Medical Sciences Research Center, University of Nizwa | Samiha Al-Kharusi et al |
| EPI_ISL_518868 | USA/MN-MDH-1474/2020 | 2020-03-18 | North America/USA/Minnesota/ | Mayo Clinic & Mayo Clinic Laboratories | Minnesota Department of Health, Public Health Laboratory | Matt Plumb et al |
| EPI_ISL_518883 | USA/MN-MDH-1490/2020 | 2020-03-22 | North America/USA/Minnesota/ | Mayo Clinic & Mayo Clinic Laboratories | Minnesota Department of Health, Public Health Laboratory | Matt Plumb et al |
| EPI_ISL_518889 | USA/MN-MDH-1496/2020 | 2020-03-21 | North America/USA/Minnesota/ | Mayo Clinic & Mayo Clinic Laboratories | Minnesota Department of Health, Public Health Laboratory | Matt Plumb et al |
| EPI_ISL_518893 | USA/MN-MDH-1500/2020 | 2020-03-24 | North America/USA/Minnesota/ | Mayo Clinic & Mayo Clinic Laboratories | Minnesota Department of Health, Public Health Laboratory | Matt Plumb et al |
| EPI_ISL_520066 | Australia/VIC5303/2020 | 2020-07-18 | Oceania/Australia/Victoria/ | Microbiological Diagnostic Unit - Public Health Laboratory (MDU-PHL) | MDU-PHL | Seemann T. et al |
| EPI_ISL_520664 | UnitedArabEmirates/L109275764/20 | 2020-04-18 | Asia/United Arab Emirates/United Arab | Mohammed Bin Rashid University of Medicine and Health Sciences | Al Jalila Genomics Center | Ahmad Abou Tayoun et al |
| EPI_ISL_520666 | UnitedArabEmirates/L121759318/20 | 2020-04-21 | Asia/United Arab Emirates/United Arab | Mohammed Bin Rashid University of Medicine and Health Sciences | Al Jalila Genomics Center | Ahmad Abou Tayoun et al |
| EPI_ISL_520667 | UnitedArabEmirates/L123065607/20 | 2020-04-21 | Asia/United Arab Emirates/United Arab | Mohammed Bin Rashid University of Medicine and Health Sciences | Al Jalila Genomics Center | Ahmad Abou Tayoun et al |
| EPI_ISL_520670 | UnitedArabEmirates/L2004034664/2 | 2020-04-17 | Asia/United Arab Emirates/United Arab | Mohammed Bin Rashid University of Medicine and Health Sciences | Al Jalila Genomics Center | Ahmad Abou Tayoun et al |
| EPI_ISL_520680 | UnitedArabEmirates/L3280050877/2 | 2020-04-08 | Asia/United Arab Emirates/United Arab | Mohammed Bin Rashid University of Medicine and Health Sciences | Al Jalila Genomics Center | Ahmad Abou Tayoun et al |
| EPI_ISL_520690 | UnitedArabEmirates/L3290224549/2 | 2020-05-18 | Asia/United Arab Emirates/United Arab | Mohammed Bin Rashid University of Medicine and Health Sciences | Al Jalila Genomics Center | Ahmad Abou Tayoun et al |
| EPI_ISL_520698 | UnitedArabEmirates/L3290250781/2 | 2020-05-20 | Asia/United Arab Emirates/United Arab | Mohammed Bin Rashid University of Medicine and Health Sciences | Al Jalila Genomics Center | Ahmad Abou Tayoun et al |

| | | | | | | |
|---|---|---|---|---|---|---|
| EPI_ISL_520706 | UnitedArabEmirates/L3290314014/2 | 2020-05-25 | Asia/United Arab Emirates/United Arab | Mohammed Bin Rashid University of Medicine and Health Sciences | Al Jalila Genomics Center | Ahmad Abou Tayoun et al |
| EPI_ISL_520708 | UnitedArabEmirates/L3290361549/2 | 2020-05-30 | Asia/United Arab Emirates/United Arab | Mohammed Bin Rashid University of Medicine and Health Sciences | Al Jalila Genomics Center | Ahmad Abou Tayoun et al |
| EPI_ISL_520711 | UnitedArabEmirates/L3300054450/2 | 2020-06-05 | Asia/United Arab Emirates/United Arab | Mohammed Bin Rashid University of Medicine and Health Sciences | Al Jalila Genomics Center | Ahmad Abou Tayoun et al |
| EPI_ISL_520716 | UnitedArabEmirates/L46302981/202 | 2020-04-28 | Asia/United Arab Emirates/United Arab | Mohammed Bin Rashid University of Medicine and Health Sciences | Al Jalila Genomics Center | Ahmad Abou Tayoun et al |
| EPI_ISL_520719 | UnitedArabEmirates/L920626247/20 | 2020-03-26 | Asia/United Arab Emirates/United Arab | Mohammed Bin Rashid University of Medicine and Health Sciences | Al Jalila Genomics Center | Ahmad Abou Tayoun et al |
| EPI_ISL_520735 | UnitedArabEmirates/L920667218/20 | 2020-04-15 | Asia/United Arab Emirates/United Arab | Mohammed Bin Rashid University of Medicine and Health Sciences | Al Jalila Genomics Center | Ahmad Abou Tayoun et al |
| EPI_ISL_520740 | UnitedArabEmirates/L920702211/20 | 2020-04-27 | Asia/United Arab Emirates/United Arab | Mohammed Bin Rashid University of Medicine and Health Sciences | Al Jalila Genomics Center | Ahmad Abou Tayoun et al |
| EPI_ISL_520849 | Australia/VIC3142/2020 | 2020-07-08 | Oceania/Australia/Victoria/ | Microbiological Diagnostic Unit - Public Health Laboratory (MDU-PHL) | MDU-PHL | Seemann T. et al |
| EPI_ISL_521956 | Australia/VIC2216/2020 | 2020-06-14 | Oceania/Australia/Victoria/ | Victorian Infectious Diseases Reference Laboratory (VIDRL) | VIDRL and MDU-PHL | Caly L. et al |
| EPI_ISL_522266 | USA/UT-02699/2020 | 2020-04-27 | North America/USA/Utah/ | Utah Public Health Laboratory | Utah Public Health Laboratory | Erin Young et al |
| EPI_ISL_522272 | USA/UT-02713/2020 | 2020-04-26 | North America/USA/Utah/ | Utah Public Health Laboratory | Utah Public Health Laboratory | Erin Young et al |
| EPI_ISL_522280 | USA/UT-02964/2020 | 2020-05-01 | North America/USA/Utah/ | Utah Public Health Laboratory | Utah Public Health Laboratory | Erin Young et al |
| EPI_ISL_522288 | USA/UT-02979/2020 | 2020-05-01 | North America/USA/Utah/ | Utah Public Health Laboratory | Utah Public Health Laboratory | Erin Young et al |
| EPI_ISL_522295 | USA/UT-03012/2020 | 2020-05-01 | North America/USA/Utah/ | Utah Public Health Laboratory | Utah Public Health Laboratory | Erin Young et al |
| EPI_ISL_522296 | USA/UT-03013/2020 | 2020-05-01 | North America/USA/Utah/ | Utah Public Health Laboratory | Utah Public Health Laboratory | Erin Young et al |
| EPI_ISL_522297 | USA/UT-03025/2020 | 2020-05-01 | North America/USA/Utah/ | Utah Public Health Laboratory | Utah Public Health Laboratory | Erin Young et al |
| EPI_ISL_522302 | USA/UT-03030/2020 | 2020-05-01 | North America/USA/Utah/ | Utah Public Health Laboratory | Utah Public Health Laboratory | Erin Young et al |
| EPI_ISL_522306 | USA/UT-03034/2020 | 2020-05-01 | North America/USA/Utah/ | Utah Public Health Laboratory | Utah Public Health Laboratory | Erin Young et al |
| EPI_ISL_522309 | USA/UT-03037/2020 | 2020-05-01 | North America/USA/Utah/ | Utah Public Health Laboratory | Utah Public Health Laboratory | Erin Young et al |
| EPI_ISL_522312 | USA/UT-03041/2020 | 2020-05-01 | North America/USA/Utah/ | Utah Public Health Laboratory | Utah Public Health Laboratory | Erin Young et al |

| | | | | | | |
|---|---|---|---|---|---|---|
| EPI_ISL_522323 | USA/UT-04226/2020 | 2020-05-15 | North America/USA/Utah/ | Utah Public Health Laboratory | Utah Public Health Laboratory | Erin Young et al |
| EPI_ISL_522324 | USA/UT-04229/2020 | 2020-05-15 | North America/USA/Utah/ | Utah Public Health Laboratory | Utah Public Health Laboratory | Erin Young et al |
| EPI_ISL_522325 | USA/UT-04257/2020 | 2020-05-15 | North America/USA/Utah/ | Utah Public Health Laboratory | Utah Public Health Laboratory | Erin Young et al |
| EPI_ISL_522338 | USA/UT-04292/2020 | 2020-05-15 | North America/USA/Utah/ | Utah Public Health Laboratory | Utah Public Health Laboratory | Erin Young et al |
| EPI_ISL_522339 | USA/UT-04294/2020 | 2020-05-15 | North America/USA/Utah/ | Utah Public Health Laboratory | Utah Public Health Laboratory | Erin Young et al |
| EPI_ISL_522342 | USA/UT-04490/2020 | 2020-05-19 | North America/USA/Utah/ | Utah Public Health Laboratory | Utah Public Health Laboratory | Erin Young et al |
| EPI_ISL_522343 | USA/UT-04499/2020 | 2020-05-21 | North America/USA/Utah/ | Utah Public Health Laboratory | Utah Public Health Laboratory | Erin Young et al |
| EPI_ISL_522348 | USA/UT-04521/2020 | 2020-05-22 | North America/USA/Utah/ | Utah Public Health Laboratory | Utah Public Health Laboratory | Erin Young et al |
| EPI_ISL_522352 | USA/TX-DSHS-0066/2020 | 2020-05-09 | North America/USA/Texas/Williamson C | Texas Department of State Health Services | Texas Department of State Health Services | Rashmi Tuladhar et al |
| EPI_ISL_522354 | USA/TX-DSHS-0069/2020 | 2020-05-09 | North America/USA/Texas/Nueces Coun | Texas Department of State Health Services | Texas Department of State Health Services | Rashmi Tuladhar et al |
| EPI_ISL_522362 | USA/TX-DSHS-0077/2020 | 2020-05-09 | North America/USA/Texas/Nueces Coun | Texas Department of State Health Services | Texas Department of State Health Services | Rashmi Tuladhar et al |
| EPI_ISL_522379 | USA/TX-DSHS-0096/2020 | 2020-05-24 | North America/USA/Texas/Jim Wells Co | Texas Department of State Health Services | Texas Department of State Health Services | Rashmi Tuladhar et al |
| EPI_ISL_522387 | USA/TX-DSHS-0104/2020 | 2020-05-28 | North America/USA/Texas/El Paso Coun | Texas Department of State Health Services | Texas Department of State Health Services | Rashmi Tuladhar et al |
| EPI_ISL_522389 | USA/TX-DSHS-0106/2020 | 2020-05-28 | North America/USA/Texas/El Paso Coun | Texas Department of State Health Services | Texas Department of State Health Services | Rashmi Tuladhar et al |
| EPI_ISL_522396 | USA/AK-PHL346/2020 | 2020-05-27 | North America/USA/Alaska/ | Alaska State Virology Laboratory | Alaska State Virology Laboratory | Jack Chen et al |

| | | | | | | |
|---|---|---|---|---|---|---|
| EPI_ISL_522408 | Germany/BY-UHR-3O/2020 | 2020-03-19 | Europe/Germany/Bavaria/ | Department of Infection Prevention and Infectious Diseases, University Hospital Regensburg | Department of Infection Prevention and Infectious Diseases, University Hospital Regensburg | Fritsch et al |
| EPI_ISL_522572 | Australia/TAS107/2020 | 2020-03-24 | Oceania/Australia/Tasmania/ | Royal Hobart Hospital Microbiology Department | MDU-PHL | Cooley L. et al |
| EPI_ISL_522605 | Australia/TAS143/2020 | 2020-04-16 | Oceania/Australia/Tasmania/ | Royal Hobart Hospital Microbiology Department | MDU-PHL | Cooley L. et al |
| EPI_ISL_522699 | Australia/TAS270/2020 | 2020-04-13 | Oceania/Australia/Tasmania/ | Royal Hobart Hospital Microbiology Department | MDU-PHL | Cooley L. et al |
| EPI_ISL_522783 | USA/VA-DCLS-0910/2020 | 2020-05-13 | North America/USA/Virginia/ | Virginia DCLS | Virginia DCLS | Virginia DCLS et al |
| EPI_ISL_522784 | USA/VA-DCLS-0912/2020 | 2020-05-13 | North America/USA/Virginia/ | Virginia DCLS | Virginia DCLS | Virginia DCLS et al |
| EPI_ISL_522787 | USA/VA-DCLS-0916/2020 | 2020-05-12 | North America/USA/Virginia/ | Virginia DCLS | Virginia DCLS | Virginia DCLS et al |
| EPI_ISL_522789 | USA/VA-DCLS-0918/2020 | 2020-05-12 | North America/USA/Virginia/ | Virginia DCLS | Virginia DCLS | Virginia DCLS et al |
| EPI_ISL_522791 | USA/VA-DCLS-0920/2020 | 2020-05-13 | North America/USA/Virginia/ | Virginia DCLS | Virginia DCLS | Virginia DCLS et al |
| EPI_ISL_522797 | USA/VA-DCLS-0930/2020 | 2020-05-21 | North America/USA/Virginia/ | Virginia DCLS | Virginia DCLS | Virginia DCLS et al |
| EPI_ISL_522799 | USA/VA-DCLS-0934/2020 | 2020-05-21 | North America/USA/Virginia/ | Virginia DCLS | Virginia DCLS | Virginia DCLS et al |
| EPI_ISL_522801 | USA/VA-DCLS-0936/2020 | 2020-05-21 | North America/USA/Virginia/ | Virginia DCLS | Virginia DCLS | Virginia DCLS et al |
| EPI_ISL_522802 | USA/VA-DCLS-0938/2020 | 2020-05-21 | North America/USA/Virginia/ | Virginia DCLS | Virginia DCLS | Virginia DCLS et al |
| EPI_ISL_522808 | USA/VA-DCLS-0797/2020 | 2020-07-07 | North America/USA/Virginia/ | Virginia DCLS | Virginia DCLS | Virginia DCLS et al |
| EPI_ISL_522823 | USA/VA-DCLS-1156/2020 | 2020-07-23 | North America/USA/Virginia/ | Virginia DCLS | Virginia DCLS | Virginia DCLS et al |
| EPI_ISL_522826 | USA/MD-MDH-0090/2020 | 2020-05-26 | North America/USA/Maryland/ | Maryland Department of Health | Maryland Department of Health | Maryland Department of Health Laboratories Administration et al |
| EPI_ISL_522827 | USA/MD-MDH-0091/2020 | 2020-05-26 | North America/USA/Maryland/ | Maryland Department of Health | Maryland Department of Health | Maryland Department of Health Laboratories Administration et al |
| EPI_ISL_522832 | USA/MD-MDH-0096/2020 | 2020-05-26 | North America/USA/Maryland/ | Maryland Department of Health | Maryland Department of Health | Maryland Department of Health Laboratories Administration et al |
| EPI_ISL_522833 | USA/MD-MDH-0098/2020 | 2020-05-23 | North America/USA/Maryland/ | Maryland Department of Health | Maryland Department of Health | Maryland Department of Health Laboratories Administration et al |

| | | | | | | |
|---|---|---|---|---|---|---|
| EPI_ISL_522846 | USA/MD-MDH-0112/2020 | 2020-05-26 | North America/USA/Maryland/ | Maryland Department of Health | Maryland Department of Health | Maryland Department of Health Laboratories Administration et al |
| EPI_ISL_522853 | USA/MD-MDH-0120/2020 | 2020-05-27 | North America/USA/Maryland/ | Maryland Department of Health | Maryland Department of Health | Maryland Department of Health Laboratories Administration et al |
| EPI_ISL_522858 | Italy/VEN-IZSVe-51-5/2020 | 2020-04-07 | Europe/Italy/Veneto/Verona IT | ULSS9 Distretto di San Bonifacio | Istituto Zooprofilattico Sperimentale delle Venezie | Adelaide Milani et al |
| EPI_ISL_522860 | Italy/VEN-IZSVe-54-17/2020 | 2020-04-08 | Europe/Italy/Veneto/Verona IT | ULSS9 Distretto di Bussolengo | Istituto Zooprofilattico Sperimentale delle Venezie | Adelaide Milani et al |
| EPI_ISL_522861 | Italy/VEN-IZSVe-55-111/2020 | 2020-04-08 | Europe/Italy/Veneto/Verona IT | ULSS9 Distretto di Bussolengo | Istituto Zooprofilattico Sperimentale delle Venezie | Adelaide Milani et al |
| EPI_ISL_522862 | Italy/VEN-IZSVe-55-16/2020 | 2020-04-08 | Europe/Italy/Veneto/Verona IT | ULSS9 Distretto di Bussolengo | Istituto Zooprofilattico Sperimentale delle Venezie | Adelaide Milani et al |
| EPI_ISL_522865 | Italy/VEN-IZSVe-60-40/2020 | 2020-04-08 | Europe/Italy/Veneto/Verona IT | ULSS9 Distretto di Bussolengo | Istituto Zooprofilattico Sperimentale delle Venezie | Adelaide Milani et al |
| EPI_ISL_522866 | Italy/VEN-IZSVe-63-7/2020 | 2020-04-09 | Europe/Italy/Veneto/Verona IT | ULSS9 Distretto di Bussolengo | Istituto Zooprofilattico Sperimentale delle Venezie | Adelaide Milani et al |
| EPI_ISL_522868 | Italy/VEN-IZSVe-94-28/2020 | 2020-04-17 | Europe/Italy/Veneto/Verona IT | ULSS9 Distretto di Bussolengo | Istituto Zooprofilattico Sperimentale delle Venezie | Adelaide Milani et al |
| EPI_ISL_522872 | Mexico/CMX-INMEGEN-01/2020 | 2020-07-31 | North America/Mexico/Mexico City/ | Instituto Nacional de Medicina Gen√≥mica | Instituto Nacional de Medicina Gen√≥mica | Hidalgo-Miranda A et al |
| EPI_ISL_522874 | Mexico/CMX-INMEGEN-03/2020 | 2020-07-31 | North America/Mexico/Mexico City/ | Instituto Nacional de Medicina Genomica | Instituto Nacional de Medicina Genomica | Hidalgo-Miranda A et al |

| | | | | | | |
|---|---|---|---|---|---|---|
| EPI_ISL_522875 | Mexico/CMX-INMEGEN-04/2020 | 2020-07-31 | North America/Mexico/Mexico City/ | Instituto Nacional de Medicina Genomica | Instituto Nacional de Medicina Genomica | Hidalgo-Miranda A et al |
| EPI_ISL_522876 | Mexico/CMX-INMEGEN-05/2020 | 2020-07-31 | North America/Mexico/Mexico City/ | Instituto Nacional de Medicina Genomica | Instituto Nacional de Medicina Genomica | Hidalgo-Miranda A et al |
| EPI_ISL_522877 | Mexico/CMX-INMEGEN-06/2020 | 2020-08-01 | North America/Mexico/Mexico City/ | Instituto Nacional de Medicina Genomica | Instituto Nacional de Medicina Genomica | Hidalgo-Miranda A et al |
| EPI_ISL_522878 | Mexico/CMX-INMEGEN-07/2020 | 2020-08-01 | North America/Mexico/Mexico City/ | Instituto Nacional de Medicina Genomica | Instituto Nacional de Medicina Genomica | Hidalgo-Miranda A et al |
| EPI_ISL_522879 | Mexico/CMX-INMEGEN-08/2020 | 2020-07-31 | North America/Mexico/Mexico City/ | Instituto Nacional de Medicina Genv≥mica | Instituto Nacional de Medicina Genv≥mica | Hidalgo-Miranda A et al |
| EPI_ISL_522882 | USA/MN-UW-1130/2020 | 2020-08-13 | North America/USA/Minnesota/Housto | University of Wisconsin-Madison AIDS Vaccine Research Laboratories | University of Wisconsin-Madison AIDS Vaccine Research Laboratories | Gage Moreno et al |
| EPI_ISL_522883 | USA/WI-UW-1125/2020 | 2020-08-05 | North America/USA/Wisconsin/Dodge C | University of Wisconsin-Madison AIDS Vaccine Research Laboratories | University of Wisconsin-Madison AIDS Vaccine Research Laboratories | Gage Moreno et al |
| EPI_ISL_522887 | USA/WI-UW-1129/2020 | 2020-08-13 | North America/USA/Wisconsin/Dane Co | University of Wisconsin-Madison AIDS Vaccine Research Laboratories | University of Wisconsin-Madison AIDS Vaccine Research Laboratories | Gage Moreno et al |
| EPI_ISL_522891 | USA/WI-UW-1134/2020 | 2020-08-11 | North America/USA/Wisconsin/Dane Co | University of Wisconsin-Madison AIDS Vaccine Research Laboratories | University of Wisconsin-Madison AIDS Vaccine Research Laboratories | Gage Moreno et al |
| EPI_ISL_522896 | USA/WI-UW-1139/2020 | 2020-08-13 | North America/USA/Wisconsin/Dane Co | University of Wisconsin-Madison AIDS Vaccine Research Laboratories | University of Wisconsin-Madison AIDS Vaccine Research Laboratories | Gage Moreno et al |

| | | | | | | |
|---|---|---|---|---|---|---|
| EPI_ISL_522898 | USA/WI-UW-1141/2020 | 2020-08-13 | North America/USA/Wisconsin/Green Co | University of Wisconsin-Madison AIDS Vaccine Research Laboratories | University of Wisconsin-Madison AIDS Vaccine Research Laboratories | Gage Moreno et al |
| EPI_ISL_522899 | USA/WI-UW-1142/2020 | 2020-08-13 | North America/USA/Wisconsin/Dane Co | University of Wisconsin-Madison AIDS Vaccine Research Laboratories | University of Wisconsin-Madison AIDS Vaccine Research Laboratories | Gage Moreno et al |
| EPI_ISL_522902 | USA/WI-UW-1145/2020 | 2020-08-13 | North America/USA/Wisconsin/Dane Co | University of Wisconsin-Madison AIDS Vaccine Research Laboratories | University of Wisconsin-Madison AIDS Vaccine Research Laboratories | Gage Moreno et al |
| EPI_ISL_522906 | USA/WI-UW-1150/2020 | 2020-08-07 | North America/USA/Wisconsin/Dane Co | University of Wisconsin-Madison AIDS Vaccine Research Laboratories | University of Wisconsin-Madison AIDS Vaccine Research Laboratories | Gage Moreno et al |
| EPI_ISL_522909 | USA/WI-UW-1153/2020 | 2020-08-13 | North America/USA/Wisconsin/Green Co | University of Wisconsin-Madison AIDS Vaccine Research Laboratories | University of Wisconsin-Madison AIDS Vaccine Research Laboratories | Gage Moreno et al |
| EPI_ISL_522910 | USA/WI-UW-1154/2020 | 2020-08-11 | North America/USA/Wisconsin/Dane Co | University of Wisconsin-Madison AIDS Vaccine Research Laboratories | University of Wisconsin-Madison AIDS Vaccine Research Laboratories | Gage Moreno et al |
| EPI_ISL_522912 | USA/WI-UW-1156/2020 | 2020-08-11 | North America/USA/Wisconsin/Dane Co | University of Wisconsin-Madison AIDS Vaccine Research Laboratories | University of Wisconsin-Madison AIDS Vaccine Research Laboratories | Gage Moreno et al |
| EPI_ISL_522913 | USA/WI-UW-1157/2020 | 2020-08-11 | North America/USA/Wisconsin/Dane Co | University of Wisconsin-Madison AIDS Vaccine Research Laboratories | University of Wisconsin-Madison AIDS Vaccine Research Laboratories | Gage Moreno et al |

| | | | | | | |
|---|---|---|---|---|---|---|
| EPI_ISL_522915 | USA/WI-UW-1159/2020 | 2020-08-06 | North America/USA/Wisconsin/Dane Co | University of Wisconsin-Madison AIDS Vaccine Research Laboratories | University of Wisconsin-Madison AIDS Vaccine Research Laboratories | Gage Moreno et al |
| EPI_ISL_522917 | USA/WI-UW-1161/2020 | 2020-08-05 | North America/USA/Wisconsin/Dodge C | University of Wisconsin-Madison AIDS Vaccine Research Laboratories | University of Wisconsin-Madison AIDS Vaccine Research Laboratories | Gage Moreno et al |
| EPI_ISL_522922 | USA/WI-UW-1166/2020 | 2020-08-11 | North America/USA/Wisconsin/Dane Co | University of Wisconsin-Madison AIDS Vaccine Research Laboratories | University of Wisconsin-Madison AIDS Vaccine Research Laboratories | Gage Moreno et al |
| EPI_ISL_522923 | USA/WI-UW-1167/2020 | 2020-08-07 | North America/USA/Wisconsin/Lafayett | University of Wisconsin-Madison AIDS Vaccine Research Laboratories | University of Wisconsin-Madison AIDS Vaccine Research Laboratories | Gage Moreno et al |
| EPI_ISL_522927 | USA/WI-UW-1171/2020 | 2020-08-12 | North America/USA/Wisconsin/Dane Co | University of Wisconsin-Madison AIDS Vaccine Research Laboratories | University of Wisconsin-Madison AIDS Vaccine Research Laboratories | Gage Moreno et al |
| EPI_ISL_522929 | USA/WI-UW-1174/2020 | 2020-08-07 | North America/USA/Wisconsin/Dane Co | University of Wisconsin-Madison AIDS Vaccine Research Laboratories | University of Wisconsin-Madison AIDS Vaccine Research Laboratories | Gage Moreno et al |
| EPI_ISL_522931 | USA/WI-UW-1176/2020 | 2020-08-01 | North America/USA/Wisconsin/Dane Co | University of Wisconsin-Madison AIDS Vaccine Research Laboratories | University of Wisconsin-Madison AIDS Vaccine Research Laboratories | Gage Moreno et al |
| EPI_ISL_522940 | Mexico/CMX-INMEGEN-10/2020 | 2020-08-01 | North America/Mexico/Mexico City/ | Instituto Nacional de Medicina Gen√≥mica | Instituto Nacional de Medicina Gen√≥mica | Hidalgo-Miranda A et al |
| EPI_ISL_522941 | Mexico/CMX-INMEGEN-11/2020 | 2020-07-31 | North America/Mexico/Mexico City/ | Instituto Nacional de Medicina Gen√≥mica | Instituto Nacional de Medicina Gen√≥mica | Hidalgo-Miranda A et al |

| | | | | | | |
|---|---|---|---|---|---|---|
| EPI_ISL_522942 | Mexico/CMX-INMEGEN-12/2020 | 2020-08-06 | North America/Mexico/Mexico City/ | Instituto Nacional de Medicina Gen√≥mica | Instituto Nacional de Medicina Gen√≥mica | Hidalgo-Miranda A et al |
| EPI_ISL_522944 | USA/TX-DSHS-0115/2020 | 2020-06-11 | North America/USA/Texas/El Paso Coun | Texas Department of State Health Services | Texas Department of State Health Services | Rashmi Tuladhar et al |
| EPI_ISL_522945 | USA/TX-DSHS-0116/2020 | 2020-06-11 | North America/USA/Texas/Travis Count | Texas Department of State Health Services | Texas Department of State Health Services | Rashmi Tuladhar et al |
| EPI_ISL_522946 | USA/TX-DSHS-0117/2020 | 2020-06-11 | North America/USA/Texas/El Paso Coun | Texas Department of State Health Services | Texas Department of State Health Services | Rashmi Tuladhar et al |
| EPI_ISL_522956 | USA/TX-DSHS-0129/2020 | 2020-06-11 | North America/USA/Texas/Galveston Co | Texas Department of State Health Services | Texas Department of State Health Services | Rashmi Tuladhar et al |
| EPI_ISL_522958 | USA/TX-DSHS-0131/2020 | 2020-06-11 | North America/USA/Texas/Brewster Co | Texas Department of State Health Services | Texas Department of State Health Services | Rashmi Tuladhar et al |
| EPI_ISL_522962 | USA/TX-DSHS-0135/2020 | 2020-06-06 | North America/USA/Texas/Brewster Co | Texas Department of State Health Services | Texas Department of State Health Services | Rashmi Tuladhar et al |
| EPI_ISL_522971 | USA/TX-DSHS-0145/2020 | 2020-05-28 | North America/USA/Texas/El Paso Coun | Texas Department of State Health Services | Texas Department of State Health Services | Rashmi Tuladhar et al |
| EPI_ISL_522978 | Mexico/CMX-INMEGEN-13/2020 | 2020-08-06 | North America/Mexico/Mexico City/ | Instituto Nacional de Medicina Gen√≥mica | Instituto Nacional de Medicina Gen√≥mica | Hidalgo-Miranda A et al |
| EPI_ISL_522980 | Mexico/CMX-INMEGEN-15/2020 | 2020-08-06 | North America/Mexico/Mexico City/ | Instituto Nacional de Medicina Gen√≥mica | Instituto Nacional de Medicina Gen√≥mica | Hidalgo-Miranda A et al |
| EPI_ISL_522981 | Mexico/CMX-INMEGEN-16/2020 | 2020-08-06 | North America/Mexico/Mexico City/ | Instituto Nacional de Medicina Gen√≥mica | Instituto Nacional de Medicina Gen√≥mica | Hidalgo-Miranda A et al |
| EPI_ISL_522982 | Mexico/CMX-INMEGEN-17/2020 | 2020-08-06 | North America/Mexico/Mexico City/ | Instituto Nacional de Medicina Gen√≥mica | Instituto Nacional de Medicina Gen√≥mica | Hidalgo-Miranda A et al |
| EPI_ISL_522983 | Mexico/CMX-INMEGEN-18/2020 | 2020-08-06 | North America/Mexico/Mexico City/ | Instituto Nacional de Medicina Gen√≥mica | Instituto Nacional de Medicina Gen√≥mica | Hidalgo-Miranda A et al |

| | | | | | | |
|---|---|---|---|---|---|---|
| EPI_ISL_522984 | Mexico/CMX-INMEGEN-19/2020 | 2020-08-06 | North America/Mexico/Mexico City/ | Instituto Nacional de Medicina Gen√≥mica | Instituto Nacional de Medicina Gen√≥mica | Hidalgo-Miranda A et al |
| EPI_ISL_522985 | Mexico/CMX-INMEGEN-20/2020 | 2020-08-05 | North America/Mexico/Mexico City/ | Instituto Nacional de Medicina Gen√≥mica | Instituto Nacional de Medicina Gen√≥mica | Hidalgo-Miranda A et al |
| EPI_ISL_522986 | Mexico/CMX-INMEGEN-21/2020 | 2020-08-06 | North America/Mexico/Mexico City/ | Instituto Nacional de Medicina Gen√≥mica | Instituto Nacional de Medicina Gen√≥mica | Hidalgo-Miranda A et al |
| EPI_ISL_523126 | Netherlands/DR-EMC-8/2020 | 2020-05-30 | Europe/Netherlands/Drenthe/ | Dutch COVID-19 response team | Erasmus Medical Center | Bas Oude Munnink et al |
| EPI_ISL_523127 | Netherlands/DR-EMC-9/2020 | 2020-06-03 | Europe/Netherlands/Drenthe/ | Dutch COVID-19 response team | Erasmus Medical Center | Bas Oude Munnink et al |
| EPI_ISL_523136 | Netherlands/FL-EMC-27/2020 | 2020-04-19 | Europe/Netherlands/Flevoland/ | Dutch COVID-19 response team | Erasmus Medical Center | Bas Oude Munnink et al |
| EPI_ISL_523152 | Netherlands/GE-EMC-169/2020 | 2020-05-22 | Europe/Netherlands/Gelderland/ | Dutch COVID-19 response team | Erasmus Medical Center | Bas Oude Munnink et al |
| EPI_ISL_523157 | Netherlands/GE-EMC-175/2020 | 2020-04-16 | Europe/Netherlands/Gelderland/ | Dutch COVID-19 response team | Erasmus Medical Center | Bas Oude Munnink et al |
| EPI_ISL_523165 | Netherlands/GE-EMC-183/2020 | 2020-05-26 | Europe/Netherlands/Gelderland/ | Dutch COVID-19 response team | Erasmus Medical Center | Bas Oude Munnink et al |
| EPI_ISL_523178 | Netherlands/GE-EMC-196/2020 | 2020-05-28 | Europe/Netherlands/Gelderland/ | Dutch COVID-19 response team | Erasmus Medical Center | Bas Oude Munnink et al |
| EPI_ISL_523235 | Netherlands/GE-EMC-256/2020 | 2020-04-18 | Europe/Netherlands/Gelderland/ | Dutch COVID-19 response team | Erasmus Medical Center | Bas Oude Munnink et al |
| EPI_ISL_523238 | Netherlands/GE-EMC-259/2020 | 2020-04-24 | Europe/Netherlands/Gelderland/ | Dutch COVID-19 response team | Erasmus Medical Center | Bas Oude Munnink et al |
| EPI_ISL_523260 | Netherlands/GE-EMC-281/2020 | 2020-05-27 | Europe/Netherlands/Gelderland/ | Dutch COVID-19 response team | Erasmus Medical Center | Bas Oude Munnink et al |
| EPI_ISL_523262 | Netherlands/GE-EMC-283/2020 | 2020-05-25 | Europe/Netherlands/Gelderland/ | Dutch COVID-19 response team | Erasmus Medical Center | Bas Oude Munnink et al |
| EPI_ISL_523275 | Netherlands/GE-EMC-296/2020 | 2020-08-07 | Europe/Netherlands/Gelderland/ | Dutch COVID-19 response team | Erasmus Medical Center | Bas Oude Munnink et al |
| EPI_ISL_523280 | Netherlands/GE-EMC-301/2020 | 2020-08-04 | Europe/Netherlands/Gelderland/ | Dutch COVID-19 response team | Erasmus Medical Center | Bas Oude Munnink et al |
| EPI_ISL_523285 | Netherlands/LI-EMC-80/2020 | 2020-06-03 | Europe/Netherlands/Limburg/ | Dutch COVID-19 response team | Erasmus Medical Center | Bas Oude Munnink et al |
| EPI_ISL_523306 | Netherlands/NB-EMC-183/2020 | 2020-05-08 | Europe/Netherlands/North Brabant/ | Dutch COVID-19 response team | Erasmus Medical Center | Bas Oude Munnink et al |

| | | | | | | |
|---|---|---|---|---|---|---|
| EPI_ISL_523346 | Netherlands/NB-EMC-223/2020 | 2020-07-08 | Europe/Netherlands/North Brabant/ | Dutch COVID-19 response team | Erasmus Medical Center | Bas Oude Munnink et al |
| EPI_ISL_523434 | Netherlands/OV-EMC-50/2020 | 2020-05-04 | Europe/Netherlands/Overijssel/ | Dutch COVID-19 response team | Erasmus Medical Center | Bas Oude Munnink et al |
| EPI_ISL_523442 | Netherlands/OV-EMC-58/2020 | 2020-05-14 | Europe/Netherlands/Overijssel/ | Dutch COVID-19 response team | Erasmus Medical Center | Bas Oude Munnink et al |
| EPI_ISL_523451 | Netherlands/OV-EMC-67/2020 | 2020-05-29 | Europe/Netherlands/Overijssel/ | Dutch COVID-19 response team | Erasmus Medical Center | Bas Oude Munnink et al |
| EPI_ISL_523490 | Netherlands/un-EMC-735/2020 | 2020-03-14 | Europe/Netherlands/Netherlands/ | Dutch COVID-19 response team | Erasmus Medical Center | Bas Oude Munnink et al |
| EPI_ISL_523500 | Mexico/CMX-INMEGEN-22/2020 | 2020-08-06 | North America/Mexico/Mexico City/ | Instituto Nacional de Medicina Gen√≥mica | Instituto Nacional de Medicina Gen√≥mica | Hidalgo-Miranda A et al |
| EPI_ISL_523508 | Netherlands/un-EMC-752/2020 | 2020-06-27 | Europe/Netherlands/Netherlands/ | Dutch COVID-19 response team | Erasmus Medical Center | Bas Oude Munnink et al |
| EPI_ISL_523540 | Netherlands/UT-EMC-63/2020 | 2020-08-02 | Europe/Netherlands/Utrecht/ | Dutch COVID-19 response team | Erasmus Medical Center | Bas Oude Munnink et al |
| EPI_ISL_523570 | Netherlands/ZH-EMC-228/2020 | 2020-05-17 | Europe/Netherlands/South Holland/ | Dutch COVID-19 response team | Erasmus Medical Center | Bas Oude Munnink et al |
| EPI_ISL_523607 | Netherlands/ZH-EMC-265/2020 | 2020-04-16 | Europe/Netherlands/South Holland/ | Dutch COVID-19 response team | Erasmus Medical Center | Bas Oude Munnink et al |
| EPI_ISL_523694 | Netherlands/ZH-EMC-354/2020 | 2020-05-22 | Europe/Netherlands/South Holland/ | Dutch COVID-19 response team | Erasmus Medical Center | Bas Oude Munnink et al |
| EPI_ISL_523697 | Netherlands/ZH-EMC-357/2020 | 2020-08-01 | Europe/Netherlands/South Holland/ | Dutch COVID-19 response team | Erasmus Medical Center | Bas Oude Munnink et al |
| EPI_ISL_523700 | Netherlands/ZH-EMC-360/2020 | 2020-08-02 | Europe/Netherlands/South Holland/ | Dutch COVID-19 response team | Erasmus Medical Center | Bas Oude Munnink et al |
| EPI_ISL_523739 | Netherlands/ZH-EMC-399/2020 | 2020-07-23 | Europe/Netherlands/South Holland/ | Dutch COVID-19 response team | Erasmus Medical Center | Bas Oude Munnink et al |
| EPI_ISL_523751 | Netherlands/ZH-EMC-411/2020 | 2020-08-10 | Europe/Netherlands/South Holland/ | Dutch COVID-19 response team | Erasmus Medical Center | Bas Oude Munnink et al |
| EPI_ISL_523756 | Netherlands/ZH-EMC-416/2020 | 2020-08-11 | Europe/Netherlands/South Holland/ | Dutch COVID-19 response team | Erasmus Medical Center | Bas Oude Munnink et al |
| EPI_ISL_523760 | Netherlands/ZH-EMC-421/2020 | 2020-08-11 | Europe/Netherlands/South Holland/ | Dutch COVID-19 response team | Erasmus Medical Center | Bas Oude Munnink et al |
| EPI_ISL_523776 | Netherlands/ZH-EMC-438/2020 | 2020-08-12 | Europe/Netherlands/South Holland/ | Dutch COVID-19 response team | Erasmus Medical Center | Bas Oude Munnink et al |
| EPI_ISL_523784 | Netherlands/ZH-EMC-446/2020 | 2020-08-12 | Europe/Netherlands/South Holland/ | Dutch COVID-19 response team | Erasmus Medical Center | Bas Oude Munnink et al |

| | | | | | | |
|---|---|---|---|---|---|---|
| EPI_ISL_523786 | Netherlands/ZH-EMC-448/2020 | 2020-08-12 | Europe/Netherlands/South Holland/ | Dutch COVID-19 response team | Erasmus Medical Center | Bas Oude Munnink et al |
| EPI_ISL_523811 | DominicanRepublic/ICGEB_UNIBE051/ | 2020-03-01 | North America/Dominican Republic/Dom | Universidad Iberoamericana, Instituto de Medicina Tropical & Salud Global | International Centre for Genetic Engineering and Biotechnology (ICGEB) and ARGO Open Lab Platform | Robert Paulino-Ramirez et al |
| EPI_ISL_523812 | DominicanRepublic/ICGEB_UNIBE258/ | 2020-03-04 | North America/Dominican Republic/Dom | Universidad Iberoamericana, Instituto de Medicina Tropical & Salud Global | International Centre for Genetic Engineering and Biotechnology (ICGEB) and ARGO Open Lab Platform | Robert Paulino-Ramirez et al |
| EPI_ISL_523885 | Switzerland/GE-ETHZ-240043/2020 | 2020-08-11 | Europe/Switzerland/Geneva/ | Viollier AG | Department of Biosystems Science and Engineering, ETH Zvºrich | Christian Beisel et al |
| EPI_ISL_523927 | Germany/NW-HHU-56/2020 | 2020-03-07 | Europe/Germany/North Rhine Westphal | Center of Medical Microbiology, Virology, and Hospital Hygiene, University of Duesseldorf | Center of Medical Microbiology, Virology, and Hospital Hygiene, University of Duesseldorf | Maximilian Damagnez et al |
| EPI_ISL_523928 | Germany/NW-HHU-57/2020 | 2020-04-20 | Europe/Germany/North Rhine Westphal | Center of Medical Microbiology, Virology, and Hospital Hygiene, University of Duesseldorf | Center of Medical Microbiology, Virology, and Hospital Hygiene, University of Duesseldorf | Maximilian Damagnez et al |
| EPI_ISL_523934 | Germany/NW-HHU-63/2020 | 2020-03-11 | Europe/Germany/North Rhine Westphal | Center of Medical Microbiology, Virology, and Hospital Hygiene, University of Duesseldorf | Center of Medical Microbiology, Virology, and Hospital Hygiene, University of Duesseldorf | Maximilian Damagnez et al |

| | | | | | | |
|---|---|---|---|---|---|---|
| EPI_ISL_523936 | Germany/NW-HHU-65/2020 | 2020-04-01 | Europe/Germany/North Rhine Westphal | Center of Medical Microbiology, Virology, and Hospital Hygiene, University of Duesseldorf | Center of Medical Microbiology, Virology, and Hospital Hygiene, University of Duesseldorf | Maximilian Damagnez et al |
| EPI_ISL_523944 | Germany/NW-HHU-73/2020 | 2020-03-30 | Europe/Germany/North Rhine Westphal | Center of Medical Microbiology, Virology, and Hospital Hygiene, University of Duesseldorf | Center of Medical Microbiology, Virology, and Hospital Hygiene, University of Duesseldorf | Maximilian Damagnez et al |
| EPI_ISL_523945 | Germany/NW-HHU-74/2020 | 2020-04-01 | Europe/Germany/North Rhine Westphal | Center of Medical Microbiology, Virology, and Hospital Hygiene, University of Duesseldorf | Center of Medical Microbiology, Virology, and Hospital Hygiene, University of Duesseldorf | Maximilian Damagnez et al |
| EPI_ISL_523946 | Germany/NW-HHU-75/2020 | 2020-03-31 | Europe/Germany/North Rhine Westphal | Center of Medical Microbiology, Virology, and Hospital Hygiene, University of Duesseldorf | Center of Medical Microbiology, Virology, and Hospital Hygiene, University of Duesseldorf | Maximilian Damagnez et al |
| EPI_ISL_523948 | Germany/NW-HHU-77/2020 | 2020-03-12 | Europe/Germany/North Rhine Westphal | Center of Medical Microbiology, Virology, and Hospital Hygiene, University of Duesseldorf | Center of Medical Microbiology, Virology, and Hospital Hygiene, University of Duesseldorf | Maximilian Damagnez et al |
| EPI_ISL_523958 | Brazil/SP-162/2020 | 2020-04-15 | South America/Brazil/S√£o Paulo/S√£o | Pronto Socorro Municipal de Perus | Instituto Adolfo Lutz, Interdiciplinary Procedures Center, Strategic Laboratory | Claudio Tavares Sacchi et al |

| | | | | | | |
|---|---|---|---|---|---|---|
| EPI_ISL_523959 | Brazil/SP-163/2020 | 2020-08-15 | South America/Brazil/S√£o Paulo/S√£o | Pronto Socorro Municipal de Perus | Instituto Adolfo Lutz, Interdiciplinary Procedures Center, Strategic Laboratory | Claudio Tavares Sacchi et al |
| EPI_ISL_523960 | Moldova/ChVir7768/2020 | 2020-03-13 | Europe/Italy/Soroca/ | National Agency for Public Health, Republic of Moldova | Charite Universitatsmedizin Berlin, Institute of Virology | Victor M Corman et al |
| EPI_ISL_523962 | Moldova/ChVir7770/2020 | 2020-03-14 | Europe/Moldova/Chisinau/ | National Agency for Public Health, Republic of Moldova | Charite Universitatsmedizin Berlin, Institute of Virology | Victor M Corman et al |
| EPI_ISL_523964 | Moldova/ChVir7771/2020 | 2020-03-14 | Europe/Moldova/Cahul/ | National Agency for Public Health, Republic of Moldova | Charite Universitatsmedizin Berlin, Institute of Virology | Victor M Corman et al |
| EPI_ISL_523966 | Moldova/ChVir7777/2020 | 2020-03-12 | Europe/Italy/Chisinau/ | National Agency for Public Health, Republic of Moldova | Charite Universitatsmedizin Berlin, Institute of Virology | Victor M Corman et al |
| EPI_ISL_523968 | Moldova/ChVir7776/2020 | 2020-03-11 | Europe/Italy/Chisinau/ | National Agency for Public Health, Republic of Moldova | Charite Universitatsmedizin Berlin, Institute of Virology | Victor M Corman et al |
| EPI_ISL_523972 | Brazil/SP-176/2020 | 2020-04-11 | South America/Brazil/S√£o Paulo/S√£o | Hospital do Servidor P√∫blico Estadual Francisco Morato de Oliveira | Instituto Adolfo Lutz, Interdiciplinary Procedures Center, Strategic Laboratory | Claudio Tavares Sacchi et al |
| EPI_ISL_523978 | Brazil/SP-195/2020 | 2020-04-10 | South America/Brazil/S√£o Paulo/S√£o | Hospital do Servidor P√∫blico Estadual Francisco Morato de Oliveira | Instituto Adolfo Lutz, Interdiciplinary Procedures Center, Strategic Laboratory | Claudio Tavares Sacchi et al |

| | | | | | | |
|---|---|---|---|---|---|---|
| EPI_ISL_523985 | Brazil/SP-202/2020 | 2020-04-18 | South America/Brazil/S√£o Paulo/S√£o | Hospital Municipal Dr. Benedicto Montenegro | Instituto Adolfo Lutz, Interdiciplinary Procedures Center, Strategic Laboratory | Claudio Tavares Sacchi et al |
| EPI_ISL_523986 | Brazil/SP-203/2020 | 2020-04-23 | South America/Brazil/S√£o Paulo/S√£o | Ama Dr Jose Soares Hungria | Instituto Adolfo Lutz, Interdiciplinary Procedures Center, Strategic Laboratory | Claudio Tavares Sacchi et al |
| EPI_ISL_523990 | Brazil/SP-209/2020 | 2020-04-19 | South America/Brazil/S√£o Paulo/S√£o | AMA Jardim Peri | Instituto Adolfo Lutz, Interdiciplinary Procedures Center, Strategic Laboratory | Claudio Tavares Sacchi et al |
| EPI_ISL_523991 | Brazil/SP-211/2020 | 2020-04-22 | South America/Brazil/S√£o Paulo/S√£o | Hospital Municipal Carmen Prudente | Instituto Adolfo Lutz, Interdiciplinary Procedures Center, Strategic Laboratory | Claudio Tavares Sacchi et al |
| EPI_ISL_523995 | Russia/SPE-RII-10152V/2020 | 2020-04-26 | Europe/Russia/Saint-Petersburg/ | WHO National Influenza Centre Russian Federation | WHO National Influenza Centre Russian Federation | Andrey Komissarov et al |
| EPI_ISL_523997 | Russia/SPE-RII-10325V/2020 | 2020-04-29 | Europe/Russia/Saint-Petersburg/ | WHO National Influenza Centre Russian Federation | WHO National Influenza Centre Russian Federation | Andrey Komissarov et al |
| EPI_ISL_523998 | Russia/SPE-RII-11036V/2020 | 2020-05-04 | Europe/Russia/Saint-Petersburg/ | WHO National Influenza Centre Russian Federation | WHO National Influenza Centre Russian Federation | Andrey Komissarov et al |
| EPI_ISL_523999 | Russia/SPE-RII-11688V/2020 | 2020-05-06 | Europe/Russia/Saint-Petersburg/ | WHO National Influenza Centre Russian Federation | WHO National Influenza Centre Russian Federation | Andrey Komissarov et al |

| | | | | | | |
|---|---|---|---|---|---|---|
| EPI_ISL_524000 | Russia/SPE-RII-12083V/2020 | 2020-05-08 | Europe/Russia/Saint-Petersburg/ | WHO National Influenza Centre Russian Federation | WHO National Influenza Centre Russian Federation | Andrey Komissarov et al |
| EPI_ISL_524001 | Russia/SPE-RII-12101V/2020 | 2020-05-08 | Europe/Russia/Saint-Petersburg/ | WHO National Influenza Centre Russian Federation | WHO National Influenza Centre Russian Federation | Andrey Komissarov et al |
| EPI_ISL_524002 | Russia/SPE-RII-12105V/2020 | 2020-05-08 | Europe/Russia/Saint-Petersburg/ | WHO National Influenza Centre Russian Federation | WHO National Influenza Centre Russian Federation | Andrey Komissarov et al |
| EPI_ISL_524003 | Russia/SPE-RII-13179V/2020 | 2020-05-12 | Europe/Russia/Saint-Petersburg/ | WHO National Influenza Centre Russian Federation | WHO National Influenza Centre Russian Federation | Andrey Komissarov et al |
| EPI_ISL_524004 | Russia/SPE-RII-13247V/2020 | 2020-05-12 | Europe/Russia/Saint-Petersburg/ | WHO National Influenza Centre Russian Federation | WHO National Influenza Centre Russian Federation | Andrey Komissarov et al |
| EPI_ISL_524005 | Russia/SPE-RII-13482V/2020 | 2020-05-11 | Europe/Russia/Saint-Petersburg/ | WHO National Influenza Centre Russian Federation | WHO National Influenza Centre Russian Federation | Andrey Komissarov et al |
| EPI_ISL_524006 | Russia/SPE-RII-13784V/2020 | 2020-05-12 | Europe/Russia/Saint-Petersburg/ | WHO National Influenza Centre Russian Federation | WHO National Influenza Centre Russian Federation | Andrey Komissarov et al |
| EPI_ISL_524007 | Russia/SPE-RII-14716V/2020 | 2020-05-12 | Europe/Russia/Saint-Petersburg/ | WHO National Influenza Centre Russian Federation | WHO National Influenza Centre Russian Federation | Andrey Komissarov et al |
| EPI_ISL_524008 | Russia/SPE-RII-15054S/2020 | 2020-05-13 | Europe/Russia/Saint-Petersburg/ | WHO National Influenza Centre Russian Federation | WHO National Influenza Centre Russian Federation | Andrey Komissarov et al |
| EPI_ISL_524009 | Russia/SPE-RII-15320V/2020 | 2020-05-14 | Europe/Russia/Saint-Petersburg/ | WHO National Influenza Centre Russian Federation | WHO National Influenza Centre Russian Federation | Andrey Komissarov et al |

| | | | | | | |
|---|---|---|---|---|---|---|
| EPI_ISL_524010 | Russia/SPE-RII-15911V/2020 | 2020-05-18 | Europe/Russia/Saint-Petersburg/ | WHO National Influenza Centre Russian Federation | WHO National Influenza Centre Russian Federation | Andrey Komissarov et al |
| EPI_ISL_524011 | Russia/SPE-RII-16011V/2020 | 2020-05-19 | Europe/Russia/Saint-Petersburg/ | WHO National Influenza Centre Russian Federation | WHO National Influenza Centre Russian Federation | Andrey Komissarov et al |
| EPI_ISL_524012 | Russia/SPE-RII-16850V/2020 | 2020-05-22 | Europe/Russia/Saint-Petersburg/ | WHO National Influenza Centre Russian Federation | WHO National Influenza Centre Russian Federation | Andrey Komissarov et al |
| EPI_ISL_524013 | Russia/SPE-RII-17217V/2020 | 2020-05-24 | Europe/Russia/Saint-Petersburg/ | WHO National Influenza Centre Russian Federation | WHO National Influenza Centre Russian Federation | Andrey Komissarov et al |
| EPI_ISL_524014 | Russia/SPE-RII-17291V/2020 | 2020-05-25 | Europe/Russia/Saint-Petersburg/ | WHO National Influenza Centre Russian Federation | WHO National Influenza Centre Russian Federation | Andrey Komissarov et al |
| EPI_ISL_524015 | Russia/SPE-RII-17684V/2020 | 2020-05-27 | Europe/Russia/Saint-Petersburg/ | WHO National Influenza Centre Russian Federation | WHO National Influenza Centre Russian Federation | Andrey Komissarov et al |
| EPI_ISL_524016 | Russia/SPE-RII-17942V/2020 | 2020-05-27 | Europe/Russia/Saint-Petersburg/ | WHO National Influenza Centre Russian Federation | WHO National Influenza Centre Russian Federation | Andrey Komissarov et al |
| EPI_ISL_524017 | Russia/SPE-RII-18220V/2020 | 2020-05-28 | Europe/Russia/Saint-Petersburg/ | WHO National Influenza Centre Russian Federation | WHO National Influenza Centre Russian Federation | Andrey Komissarov et al |
| EPI_ISL_524018 | Russia/SPE-RII-18357V/2020 | 2020-05-28 | Europe/Russia/Saint-Petersburg/ | WHO National Influenza Centre Russian Federation | WHO National Influenza Centre Russian Federation | Andrey Komissarov et al |
| EPI_ISL_524019 | Russia/SPE-RII-18358V/2020 | 2020-05-28 | Europe/Russia/Saint-Petersburg/ | WHO National Influenza Centre Russian Federation | WHO National Influenza Centre Russian Federation | Andrey Komissarov et al |

| | | | | | | |
|---|---|---|---|---|---|---|
| EPI_ISL_524020 | Russia/SPE-RII-18574V/2020 | 2020-05-29 | Europe/Russia/Saint-Petersburg/ | WHO National Influenza Centre Russian Federation | WHO National Influenza Centre Russian Federation | Andrey Komissarov et al |
| EPI_ISL_524021 | Russia/SPE-RII-18663V/2020 | 2020-05-29 | Europe/Russia/Saint-Petersburg/ | WHO National Influenza Centre Russian Federation | WHO National Influenza Centre Russian Federation | Andrey Komissarov et al |
| EPI_ISL_524022 | Russia/SPE-RII-18966V/2020 | 2020-06-01 | Europe/Russia/Saint-Petersburg/ | WHO National Influenza Centre Russian Federation | WHO National Influenza Centre Russian Federation | Andrey Komissarov et al |
| EPI_ISL_524023 | Russia/SPE-RII-19109V/2020 | 2020-06-01 | Europe/Russia/Saint-Petersburg/ | WHO National Influenza Centre Russian Federation | WHO National Influenza Centre Russian Federation | Andrey Komissarov et al |
| EPI_ISL_524025 | Russia/SPE-RII-19426V/2020 | 2020-06-03 | Europe/Russia/Saint-Petersburg/ | WHO National Influenza Centre Russian Federation | WHO National Influenza Centre Russian Federation | Andrey Komissarov et al |
| EPI_ISL_524026 | Russia/SPE-RII-19578V/2020 | 2020-06-03 | Europe/Russia/Saint-Petersburg/ | WHO National Influenza Centre Russian Federation | WHO National Influenza Centre Russian Federation | Andrey Komissarov et al |
| EPI_ISL_524027 | Russia/SPE-RII-19602V/2020 | 2020-06-03 | Europe/Russia/Saint-Petersburg/ | WHO National Influenza Centre Russian Federation | WHO National Influenza Centre Russian Federation | Andrey Komissarov et al |
| EPI_ISL_524028 | Russia/SPE-RII-19652V/2020 | 2020-06-03 | Europe/Russia/Saint-Petersburg/ | WHO National Influenza Centre Russian Federation | WHO National Influenza Centre Russian Federation | Andrey Komissarov et al |
| EPI_ISL_524029 | Russia/SPE-RII-20404V/2020 | 2020-06-04 | Europe/Russia/Saint-Petersburg/ | WHO National Influenza Centre Russian Federation | WHO National Influenza Centre Russian Federation | Andrey Komissarov et al |
| EPI_ISL_524030 | Russia/SPE-RII-20906V/2020 | 2020-06-04 | Europe/Russia/Saint-Petersburg/ | WHO National Influenza Centre Russian Federation | WHO National Influenza Centre Russian Federation | Andrey Komissarov et al |

| | | | | | | |
|---|---|---|---|---|---|---|
| EPI_ISL_524031 | Russia/SPE-RII-21056V/2020 | 2020-06-04 | Europe/Russia/Saint-Petersburg/ | WHO National Influenza Centre Russian Federation | WHO National Influenza Centre Russian Federation | Andrey Komissarov et al |
| EPI_ISL_524032 | Russia/SPE-RII-21119V/2020 | 2020-06-04 | Europe/Russia/Saint-Petersburg/ | WHO National Influenza Centre Russian Federation | WHO National Influenza Centre Russian Federation | Andrey Komissarov et al |
| EPI_ISL_524033 | Russia/SPE-RII-21357V/2020 | 2020-06-05 | Europe/Russia/Saint-Petersburg/ | WHO National Influenza Centre Russian Federation | WHO National Influenza Centre Russian Federation | Andrey Komissarov et al |
| EPI_ISL_524034 | Russia/SPE-RII-21358V/2020 | 2020-06-05 | Europe/Russia/Saint-Petersburg/ | WHO National Influenza Centre Russian Federation | WHO National Influenza Centre Russian Federation | Andrey Komissarov et al |
| EPI_ISL_524035 | Russia/SPE-RII-22043V/2020 | 2020-06-08 | Europe/Russia/Saint-Petersburg/ | WHO National Influenza Centre Russian Federation | WHO National Influenza Centre Russian Federation | Andrey Komissarov et al |
| EPI_ISL_524036 | Russia/SPE-RII-22068V/2020 | 2020-06-08 | Europe/Russia/Saint-Petersburg/ | WHO National Influenza Centre Russian Federation | WHO National Influenza Centre Russian Federation | Andrey Komissarov et al |
| EPI_ISL_524037 | Russia/SPE-RII-22069V/2020 | 2020-06-08 | Europe/Russia/Saint-Petersburg/ | WHO National Influenza Centre Russian Federation | WHO National Influenza Centre Russian Federation | Andrey Komissarov et al |
| EPI_ISL_524038 | Russia/SPE-RII-22086V/2020 | 2020-06-08 | Europe/Russia/Saint-Petersburg/ | WHO National Influenza Centre Russian Federation | WHO National Influenza Centre Russian Federation | Andrey Komissarov et al |
| EPI_ISL_524039 | Russia/SPE-RII-22207V/2020 | 2020-06-08 | Europe/Russia/Saint-Petersburg/ | WHO National Influenza Centre Russian Federation | WHO National Influenza Centre Russian Federation | Andrey Komissarov et al |
| EPI_ISL_524040 | Russia/SPE-RII-22266V/2020 | 2020-06-08 | Europe/Russia/Saint-Petersburg/ | WHO National Influenza Centre Russian Federation | WHO National Influenza Centre Russian Federation | Andrey Komissarov et al |

| | | | | | | |
|---|---|---|---|---|---|---|
| EPI_ISL_524041 | Russia/SPE-RII-22269V/2020 | 2020-06-08 | Europe/Russia/Saint-Petersburg/ | WHO National Influenza Centre Russian Federation | WHO National Influenza Centre Russian Federation | Andrey Komissarov et al |
| EPI_ISL_524042 | Russia/SPE-RII-22276V/2020 | 2020-06-08 | Europe/Russia/Saint-Petersburg/ | WHO National Influenza Centre Russian Federation | WHO National Influenza Centre Russian Federation | Andrey Komissarov et al |
| EPI_ISL_524043 | Russia/SPE-RII-22311V/2020 | 2020-06-08 | Europe/Russia/Saint-Petersburg/ | WHO National Influenza Centre Russian Federation | WHO National Influenza Centre Russian Federation | Andrey Komissarov et al |
| EPI_ISL_524044 | Russia/SPE-RII-22388V/2020 | 2020-06-09 | Europe/Russia/Saint-Petersburg/ | WHO National Influenza Centre Russian Federation | WHO National Influenza Centre Russian Federation | Andrey Komissarov et al |
| EPI_ISL_524045 | Russia/SPE-RII-22478V/2020 | 2020-06-09 | Europe/Russia/Saint-Petersburg/ | WHO National Influenza Centre Russian Federation | WHO National Influenza Centre Russian Federation | Andrey Komissarov et al |
| EPI_ISL_524046 | Russia/SPE-RII-23386V/2020 | 2020-06-11 | Europe/Russia/Saint-Petersburg/ | WHO National Influenza Centre Russian Federation | WHO National Influenza Centre Russian Federation | Andrey Komissarov et al |
| EPI_ISL_524047 | Russia/SPE-RII-24810V/2020 | 2020-06-16 | Europe/Russia/Saint-Petersburg/ | WHO National Influenza Centre Russian Federation | WHO National Influenza Centre Russian Federation | Andrey Komissarov et al |
| EPI_ISL_524048 | Russia/SPE-RII-6425V/2020 | 2020-04-15 | Europe/Russia/Saint-Petersburg/ | WHO National Influenza Centre Russian Federation | WHO National Influenza Centre Russian Federation | Andrey Komissarov et al |
| EPI_ISL_524049 | Russia/SPE-RII-7388V/2020 | 2020-04-20 | Europe/Russia/Saint-Petersburg/ | WHO National Influenza Centre Russian Federation | WHO National Influenza Centre Russian Federation | Andrey Komissarov et al |
| EPI_ISL_524061 | USA/TX-DSHS-0180/2020 | 2020-03-16 | North America/USA/Texas/Llano County | Texas Department of State Health Services | Texas Department of State Health Services | Rashmi Tuladhar et al |

| | | | | | | |
|---|---|---|---|---|---|---|
| EPI_ISL_524067 | USA/TX-DSHS-0187/2020 | 2020-04-10 | North America/USA/Texas/Travis Count | Texas Department of State Health Services | Texas Department of State Health Services | Rashmi Tuladhar et al |
| EPI_ISL_524077 | USA/UT-08275/2020 | 2020-05-22 | North America/USA/Utah/ | Utah Public Health Laboratory | Utah Public Health Laboratory | Erin L. Young et al |
| EPI_ISL_524084 | USA/UT-08285/2020 | 2020-05-22 | North America/USA/Utah/ | Utah Public Health Laboratory | Utah Public Health Laboratory | Erin L. Young et al |
| EPI_ISL_524086 | USA/UT-08287/2020 | 2020-05-21 | North America/USA/Utah/ | Utah Public Health Laboratory | Utah Public Health Laboratory | Erin L. Young et al |
| EPI_ISL_524087 | USA/UT-08288/2020 | 2020-05-26 | North America/USA/Utah/ | Utah Public Health Laboratory | Utah Public Health Laboratory | Erin L. Young et al |
| EPI_ISL_524088 | USA/UT-08289/2020 | 2020-05-24 | North America/USA/Utah/ | Utah Public Health Laboratory | Utah Public Health Laboratory | Erin L. Young et al |
| EPI_ISL_524098 | USA/UT-08301/2020 | 2020-05-19 | North America/USA/Utah/ | Utah Public Health Laboratory | Utah Public Health Laboratory | Erin L. Young et al |
| EPI_ISL_524105 | USA/UT-08309/2020 | 2020-05-21 | North America/USA/Utah/ | Utah Public Health Laboratory | Utah Public Health Laboratory | Erin L. Young et al |
| EPI_ISL_524121 | USA/UT-08330/2020 | 2020-05-14 | North America/USA/Utah/ | Utah Public Health Laboratory | Utah Public Health Laboratory | Erin L. Young et al |
| EPI_ISL_524131 | USA/UT-08344/2020 | 2020-05-26 | North America/USA/Utah/ | Utah Public Health Laboratory | Utah Public Health Laboratory | Erin L. Young et al |
| EPI_ISL_524138 | USA/UT-08353/2020 | 2020-05-12 | North America/USA/Utah/ | Utah Public Health Laboratory | Utah Public Health Laboratory | Erin L. Young et al |
| EPI_ISL_524147 | USA/UT-08364/2020 | 2020-05-20 | North America/USA/Utah/ | Utah Public Health Laboratory | Utah Public Health Laboratory | Erin L. Young et al |
| EPI_ISL_524153 | USA/UT-08370/2020 | 2020-05-15 | North America/USA/Utah/ | Utah Public Health Laboratory | Utah Public Health Laboratory | Erin L. Young et al |
| EPI_ISL_524157 | USA/UT-08374/2020 | 2020-05-20 | North America/USA/Utah/ | Utah Public Health Laboratory | Utah Public Health Laboratory | Erin L. Young et al |
| EPI_ISL_524158 | USA/UT-08375/2020 | 2020-05-19 | North America/USA/Utah/ | Utah Public Health Laboratory | Utah Public Health Laboratory | Erin L. Young et al |
| EPI_ISL_524166 | USA/UT-08383/2020 | 2020-05-15 | North America/USA/Utah/ | Utah Public Health Laboratory | Utah Public Health Laboratory | Erin L. Young et al |
| EPI_ISL_524171 | USA/UT-08389/2020 | 2020-05-26 | North America/USA/Utah/ | Utah Public Health Laboratory | Utah Public Health Laboratory | Erin L. Young et al |
| EPI_ISL_524174 | USA/UT-08393/2020 | 2020-05-15 | North America/USA/Utah/ | Utah Public Health Laboratory | Utah Public Health Laboratory | Erin L. Young et al |
| EPI_ISL_524175 | USA/UT-08395/2020 | 2020-05-20 | North America/USA/Utah/ | Utah Public Health Laboratory | Utah Public Health Laboratory | Erin L. Young et al |

| | | | | | | |
|---|---|---|---|---|---|---|
| EPI_ISL_524176 | USA/UT-08396/2020 | 2020-05-20 | North America/USA/Utah/ | Utah Public Health Laboratory | Utah Public Health Laboratory | Erin L. Young et al |
| EPI_ISL_524180 | USA/UT-08400/2020 | 2020-05-26 | North America/USA/Utah/ | Utah Public Health Laboratory | Utah Public Health Laboratory | Erin L. Young et al |
| EPI_ISL_524183 | USA/UT-08404/2020 | 2020-05-13 | North America/USA/Utah/ | Utah Public Health Laboratory | Utah Public Health Laboratory | Erin L. Young et al |
| EPI_ISL_524186 | USA/UT-08408/2020 | 2020-05-20 | North America/USA/Utah/ | Utah Public Health Laboratory | Utah Public Health Laboratory | Erin L. Young et al |
| EPI_ISL_524189 | USA/UT-08411/2020 | 2020-05-26 | North America/USA/Utah/ | Utah Public Health Laboratory | Utah Public Health Laboratory | Erin L. Young et al |
| EPI_ISL_524191 | USA/UT-08413/2020 | 2020-05-13 | North America/USA/Utah/ | Utah Public Health Laboratory | Utah Public Health Laboratory | Erin L. Young et al |
| EPI_ISL_524192 | USA/UT-08414/2020 | 2020-05-13 | North America/USA/Utah/ | Utah Public Health Laboratory | Utah Public Health Laboratory | Erin L. Young et al |
| EPI_ISL_524194 | USA/UT-08416/2020 | 2020-05-13 | North America/USA/Utah/ | Utah Public Health Laboratory | Utah Public Health Laboratory | Erin L. Young et al |
| EPI_ISL_524197 | USA/UT-08419/2020 | 2020-05-20 | North America/USA/Utah/ | Utah Public Health Laboratory | Utah Public Health Laboratory | Erin L. Young et al |
| EPI_ISL_524199 | USA/UT-08421/2020 | 2020-05-26 | North America/USA/Utah/ | Utah Public Health Laboratory | Utah Public Health Laboratory | Erin L. Young et al |
| EPI_ISL_524202 | USA/UT-08426/2020 | 2020-05-13 | North America/USA/Utah/ | Utah Public Health Laboratory | Utah Public Health Laboratory | Erin L. Young et al |
| EPI_ISL_524205 | USA/UT-08429/2020 | 2020-05-19 | North America/USA/Utah/ | Utah Public Health Laboratory | Utah Public Health Laboratory | Erin L. Young et al |
| EPI_ISL_524207 | USA/UT-08431/2020 | 2020-05-20 | North America/USA/Utah/ | Utah Public Health Laboratory | Utah Public Health Laboratory | Erin L. Young et al |
| EPI_ISL_524210 | USA/UT-08434/2020 | 2020-05-13 | North America/USA/Utah/ | Utah Public Health Laboratory | Utah Public Health Laboratory | Erin L. Young et al |
| EPI_ISL_524215 | USA/UT-08440/2020 | 2020-05-20 | North America/USA/Utah/ | Utah Public Health Laboratory | Utah Public Health Laboratory | Erin L. Young et al |
| EPI_ISL_524218 | USA/UT-08444/2020 | 2020-05-26 | North America/USA/Utah/ | Utah Public Health Laboratory | Utah Public Health Laboratory | Erin L. Young et al |
| EPI_ISL_524220 | USA/UT-08446/2020 | 2020-05-18 | North America/USA/Utah/ | Utah Public Health Laboratory | Utah Public Health Laboratory | Erin L. Young et al |
| EPI_ISL_524225 | USA/UT-08453/2020 | 2020-05-19 | North America/USA/Utah/ | Utah Public Health Laboratory | Utah Public Health Laboratory | Erin L. Young et al |
| EPI_ISL_524232 | USA/UT-08460/2020 | 2020-05-26 | North America/USA/Utah/ | Utah Public Health Laboratory | Utah Public Health Laboratory | Erin L. Young et al |
| EPI_ISL_524235 | USA/UT-08463/2020 | 2020-05-19 | North America/USA/Utah/ | Utah Public Health Laboratory | Utah Public Health Laboratory | Erin L. Young et al |

| | | | | | | |
|---|---|---|---|---|---|---|
| EPI_ISL_524237 | USA/UT-08466/2020 | 2020-05-19 | North America/USA/Utah/ | Utah Public Health Laboratory | Utah Public Health Laboratory | Erin L. Young et al |
| EPI_ISL_524241 | USA/UT-08470/2020 | 2020-05-22 | North America/USA/Utah/ | Utah Public Health Laboratory | Utah Public Health Laboratory | Erin L. Young et al |
| EPI_ISL_524242 | USA/UT-08471/2020 | 2020-05-26 | North America/USA/Utah/ | Utah Public Health Laboratory | Utah Public Health Laboratory | Erin L. Young et al |
| EPI_ISL_524244 | USA/UT-08474/2020 | 2020-05-20 | North America/USA/Utah/ | Utah Public Health Laboratory | Utah Public Health Laboratory | Erin L. Young et al |
| EPI_ISL_524246 | USA/UT-08476/2020 | 2020-05-20 | North America/USA/Utah/ | Utah Public Health Laboratory | Utah Public Health Laboratory | Erin L. Young et al |
| EPI_ISL_524249 | USA/UT-08480/2020 | 2020-05-21 | North America/USA/Utah/ | Utah Public Health Laboratory | Utah Public Health Laboratory | Erin L. Young et al |
| EPI_ISL_524250 | USA/UT-08481/2020 | 2020-05-21 | North America/USA/Utah/ | Utah Public Health Laboratory | Utah Public Health Laboratory | Erin L. Young et al |
| EPI_ISL_524253 | USA/UT-08484/2020 | 2020-05-19 | North America/USA/Utah/ | Utah Public Health Laboratory | Utah Public Health Laboratory | Erin L. Young et al |
| EPI_ISL_524254 | USA/UT-08485/2020 | 2020-05-20 | North America/USA/Utah/ | Utah Public Health Laboratory | Utah Public Health Laboratory | Erin L. Young et al |
| EPI_ISL_524256 | USA/UT-08487/2020 | 2020-05-20 | North America/USA/Utah/ | Utah Public Health Laboratory | Utah Public Health Laboratory | Erin L. Young et al |
| EPI_ISL_524264 | USA/UT-08497/2020 | 2020-05-18 | North America/USA/Utah/ | Utah Public Health Laboratory | Utah Public Health Laboratory | Erin L. Young et al |
| EPI_ISL_524267 | USA/UT-08500/2020 | 2020-05-18 | North America/USA/Utah/ | Utah Public Health Laboratory | Utah Public Health Laboratory | Erin L. Young et al |
| EPI_ISL_524272 | USA/UT-08507/2020 | 2020-05-18 | North America/USA/Utah/ | Utah Public Health Laboratory | Utah Public Health Laboratory | Erin L. Young et al |
| EPI_ISL_524274 | USA/UT-08510/2020 | 2020-05-18 | North America/USA/Utah/ | Utah Public Health Laboratory | Utah Public Health Laboratory | Erin L. Young et al |
| EPI_ISL_524281 | USA/UT-08518/2020 | 2020-05-18 | North America/USA/Utah/ | Utah Public Health Laboratory | Utah Public Health Laboratory | Erin L. Young et al |
| EPI_ISL_524286 | USA/UT-08523/2020 | 2020-05-18 | North America/USA/Utah/ | Utah Public Health Laboratory | Utah Public Health Laboratory | Erin L. Young et al |
| EPI_ISL_524291 | USA/UT-08528/2020 | 2020-05-17 | North America/USA/Utah/ | Utah Public Health Laboratory | Utah Public Health Laboratory | Erin L. Young et al |
| EPI_ISL_524297 | USA/UT-08536/2020 | 2020-05-18 | North America/USA/Utah/ | Utah Public Health Laboratory | Utah Public Health Laboratory | Erin L. Young et al |
| EPI_ISL_524299 | USA/UT-08538/2020 | 2020-05-18 | North America/USA/Utah/ | Utah Public Health Laboratory | Utah Public Health Laboratory | Erin L. Young et al |
| EPI_ISL_524300 | USA/UT-08539/2020 | 2020-05-18 | North America/USA/Utah/ | Utah Public Health Laboratory | Utah Public Health Laboratory | Erin L. Young et al |

| | | | | | | |
|---|---|---|---|---|---|---|
| EPI_ISL_524303 | USA/UT-08542/2020 | 2020-05-07 | North America/USA/Utah/ | Utah Public Health Laboratory | Utah Public Health Laboratory | Erin L. Young et al |
| EPI_ISL_524313 | USA/UT-08554/2020 | 2020-05-26 | North America/USA/Utah/ | Utah Public Health Laboratory | Utah Public Health Laboratory | Erin L. Young et al |
| EPI_ISL_524321 | USA/UT-08564/2020 | 2020-05-07 | North America/USA/Utah/ | Utah Public Health Laboratory | Utah Public Health Laboratory | Erin L. Young et al |
| EPI_ISL_524323 | USA/UT-08566/2020 | 2020-05-18 | North America/USA/Utah/ | Utah Public Health Laboratory | Utah Public Health Laboratory | Erin L. Young et al |
| EPI_ISL_524330 | USA/UT-08575/2020 | 2020-05-07 | North America/USA/Utah/ | Utah Public Health Laboratory | Utah Public Health Laboratory | Erin L. Young et al |
| EPI_ISL_524331 | USA/UT-08576/2020 | 2020-05-26 | North America/USA/Utah/ | Utah Public Health Laboratory | Utah Public Health Laboratory | Erin L. Young et al |
| EPI_ISL_524334 | USA/UT-08581/2020 | 2020-05-18 | North America/USA/Utah/ | Utah Public Health Laboratory | Utah Public Health Laboratory | Erin L. Young et al |
| EPI_ISL_524335 | USA/UT-08582/2020 | 2020-05-26 | North America/USA/Utah/ | Utah Public Health Laboratory | Utah Public Health Laboratory | Erin L. Young et al |
| EPI_ISL_524338 | USA/UT-08588/2020 | 2020-05-21 | North America/USA/Utah/ | Utah Public Health Laboratory | Utah Public Health Laboratory | Erin L. Young et al |
| EPI_ISL_524346 | USA/UT-08599/2020 | 2020-05-19 | North America/USA/Utah/ | Utah Public Health Laboratory | Utah Public Health Laboratory | Erin L. Young et al |
| EPI_ISL_524348 | USA/UT-08601/2020 | 2020-05-18 | North America/USA/Utah/ | Utah Public Health Laboratory | Utah Public Health Laboratory | Erin L. Young et al |
| EPI_ISL_524352 | USA/UT-08608/2020 | 2020-05-07 | North America/USA/Utah/ | Utah Public Health Laboratory | Utah Public Health Laboratory | Erin L. Young et al |
| EPI_ISL_524353 | USA/UT-08609/2020 | 2020-05-26 | North America/USA/Utah/ | Utah Public Health Laboratory | Utah Public Health Laboratory | Erin L. Young et al |
| EPI_ISL_524357 | USA/UT-08616/2020 | 2020-05-26 | North America/USA/Utah/ | Utah Public Health Laboratory | Utah Public Health Laboratory | Erin L. Young et al |
| EPI_ISL_524360 | USA/UT-08620/2020 | 2020-05-26 | North America/USA/Utah/ | Utah Public Health Laboratory | Utah Public Health Laboratory | Erin L. Young et al |
| EPI_ISL_524362 | USA/UT-08622/2020 | 2020-05-21 | North America/USA/Utah/ | Utah Public Health Laboratory | Utah Public Health Laboratory | Erin L. Young et al |
| EPI_ISL_524365 | USA/UT-08627/2020 | 2020-05-21 | North America/USA/Utah/ | Utah Public Health Laboratory | Utah Public Health Laboratory | Erin L. Young et al |
| EPI_ISL_524372 | USA/UT-08634/2020 | 2020-05-19 | North America/USA/Utah/ | Utah Public Health Laboratory | Utah Public Health Laboratory | Erin L. Young et al |
| EPI_ISL_524387 | USA/UT-08661/2020 | 2020-05-24 | North America/USA/Utah/ | Utah Public Health Laboratory | Utah Public Health Laboratory | Erin L. Young et al |
| EPI_ISL_524392 | USA/UT-08666/2020 | 2020-05-07 | North America/USA/Utah/ | Utah Public Health Laboratory | Utah Public Health Laboratory | Erin L. Young et al |

| | | | | | | |
|---|---|---|---|---|---|---|
| EPI_ISL_524395 | USA/UT-08670/2020 | 2020-05-15 | North America/USA/Utah/ | Utah Public Health Laboratory | Utah Public Health Laboratory | Erin L. Young et al |
| EPI_ISL_524397 | USA/UT-08672/2020 | 2020-05-08 | North America/USA/Utah/ | Utah Public Health Laboratory | Utah Public Health Laboratory | Erin L. Young et al |
| EPI_ISL_524411 | USA/UT-08692/2020 | 2020-05-08 | North America/USA/Utah/ | Utah Public Health Laboratory | Utah Public Health Laboratory | Erin L. Young et al |
| EPI_ISL_524413 | USA/UT-08695/2020 | 2020-05-26 | North America/USA/Utah/ | Utah Public Health Laboratory | Utah Public Health Laboratory | Erin L. Young et al |
| EPI_ISL_524414 | USA/UT-08696/2020 | 2020-05-26 | North America/USA/Utah/ | Utah Public Health Laboratory | Utah Public Health Laboratory | Erin L. Young et al |
| EPI_ISL_524427 | Egypt/CUNCI-7I026/2020 | 2020-07-19 | Africa/Egypt/Egypt/ | Egyptian National Cancer Institute (ENCI) | Egyptian National Cancer Institute (ENCI) | Zekri et al |
| EPI_ISL_524433 | USA/FL-UF-4/2020 | 2020-03-27 | North America/USA/Florida/ | Environmental and Global Health, University of Florida - Gainesville | University of Florida | Elbadry et al |
| EPI_ISL_524434 | USA/FL-UF-5/2020 | 2020-03-27 | North America/USA/Florida/ | Environmental and Global Health, University of Florida - Gainesville | University of Florida | Elbadry et al |
| EPI_ISL_524436 | USA/CA-SR-0851/2020 | 2020-05-14 | North America/USA/California/ | Department of Immunology, The Scripps Research Institute | Department of Immunology, The Scripps Research Institute | Smither et al |
| EPI_ISL_524446 | Singapore/739/2020 | 2020-07-06 | Asia/Singapore/Singapore/ | Singapore General Hospital | Department of Microbiology | Nurdyana Abdul Rahman et al |
| EPI_ISL_524468 | Brazil/SP-240/2020 | 2020-05-01 | South America/Brazil/S√£o Paulo/S√£o [illegible] | Hospital Municipal Vereador Jose Storopolli | Instituto Adolfo Lutz, Interdiciplinary Procedures Center, Strategic Laboratory | Claudio Tavares Sacchi et al |
| EPI_ISL_524469 | Brazil/SP-254/2020 | 2020-05-05 | South America/Brazil/S√£o Paulo/S√£o [illegible] | Santa Casa de Misericordia de Sao Paulo | Instituto Adolfo Lutz, Interdiciplinary Procedures Center, Strategic Laboratory | Claudio Tavares Sacchi et al |

| | | | | | | |
|---|---|---|---|---|---|---|
| EPI_ISL_524470 | Brazil/SP-259/2020 | 2020-05-06 | South America/Brazil/S√£o Paulo/S√£o | Hospital do Servidor P√∫blico Estadual Francisco Morato de Oliveira | Instituto Adolfo Lutz, Interdiciplinary Procedures Center, Strategic Laboratory | Claudio Tavares Sacchi et al |
| EPI_ISL_524481 | Switzerland/AG-UZH-1000478772/20 | 2020-03-04 | Europe/Switzerland/Aargau/ | Department of Infectious Diseases, Cantonal Hospital Baden | Institute of Medical Virology, University of Zurich | Stefan Schmutz et al |
| EPI_ISL_524493 | England/CAMB-7E3F5/2020 | 2020-04-07 | Europe/United Kingdom/England/ | Department of Pathology, University of Cambridge | Wellcome Sanger Institute for the COVID-19 Genomics UK (COG-UK) consortium | Luke W Meredith et al |
| EPI_ISL_524536 | Scotland/GCVR-171E1E/2020 | 2020-03-30 | Europe/United Kingdom/Scotland/ | NHSGGC West of Scotland Specialist Virology Centre / MRC-University of Glasgow Centre for Virus Research | Wellcome Sanger Institute for the COVID-19 Genomics UK (COG-UK) consortium | Ana da Silva Filipe et al |
| EPI_ISL_524546 | England/CAMB-80220/2020 | 2020-04-08 | Europe/United Kingdom/England/ | Department of Pathology, University of Cambridge | Wellcome Sanger Institute for the COVID-19 Genomics UK (COG-UK) consortium | Luke W Meredith et al |
| EPI_ISL_524685 | England/LOND-1260F84/2020 | 2020-04-02 | Europe/United Kingdom/England/ | North West London Pathology, Imperial College Healthcare NHS Trust | Wellcome Sanger Institute for the COVID-19 Genomics UK (COG-UK) consortium | Ling Li et al |
| EPI_ISL_524713 | India/GJ-GBRC-362a/2020 | 2020-06-18 | Asia/India/Gujarat/Ahmedabad | B.J. Medical College and Civil hospital, Ahmedabad | Gujarat Biotechnology Research Centre | Apurvasinh Puvar et al |
| EPI_ISL_524714 | India/GJ-GBRC-362b/2020 | 2020-06-18 | Asia/India/Gujarat/Ahmedabad | B.J. Medical College and Civil hospital, Ahmedabad | Gujarat Biotechnology Research Centre | Janvi Raval et al |
| EPI_ISL_524716 | India/GJ-GBRC-364a/2020 | 2020-06-18 | Asia/India/Gujarat/Daskroi | B.J. Medical College and Civil hospital, Ahmedabad | Gujarat Biotechnology Research Centre | Monika Gandhi et al |

| | | | | | | |
|---|---|---|---|---|---|---|
| EPI_ISL_524724 | India/GJ-GBRC-368b/2020 | 2020-06-18 | Asia/India/Gujarat/Dahegam | B.J. Medical College and Civil hospital, Ahmedabad | Gujarat Biotechnology Research Centre | Komal Patel et al |
| EPI_ISL_524726 | India/GJ-GBRC-370/2020 | 2020-06-18 | Asia/India/Gujarat/Ahmedabad | B.J. Medical College and Civil hospital, Ahmedabad | Gujarat Biotechnology Research Centre | Afzal Ansari et al |
| EPI_ISL_524728 | India/GJ-GBRC-372/2020 | 2020-06-18 | Asia/India/Gujarat/Ahmedabad | B.J. Medical College and Civil hospital, Ahmedabad | Gujarat Biotechnology Research Centre | Pranay Shah et al |
| EPI_ISL_524730 | India/GJ-GBRC-374a/2020 | 2020-06-15 | Asia/India/Gujarat/Vadodara | GMERS Medical College & Hospital, Gotri, Vadodara | Gujarat Biotechnology Research Centre | Meenakshi Shah et al |
| EPI_ISL_524731 | India/GJ-GBRC-374b/2020 | 2020-06-15 | Asia/India/Gujarat/Vadodara | GMERS Medical College & Hospital, Gotri, Vadodara | Gujarat Biotechnology Research Centre | Neena Doshi et al |
| EPI_ISL_524732 | India/GJ-GBRC-375/2020 | 2020-07-09 | Asia/India/Gujarat/Modasa | Dr. RSS Hospital, Modasa | Gujarat Biotechnology Research Centre | Harsh Chaudhari et al |
| EPI_ISL_524733 | India/GJ-GBRC-376a/2020 | 2020-07-06 | Asia/India/Gujarat/Patan | GMERS Medical College and Hospital, Dharpur, Patan | Gujarat Biotechnology Research Centre | A N Parmar et al |
| EPI_ISL_524734 | India/GJ-GBRC-376b/2020 | 2020-07-06 | Asia/India/Gujarat/Patan | GMERS Medical College and Hospital, Dharpur, Patan | Gujarat Biotechnology Research Centre | Apurvasinh Puvar et al |
| EPI_ISL_524735 | India/GJ-GBRC-377a/2020 | 2020-07-11 | Asia/India/Gujarat/Khambhaliya | Government General Hospital, Jam Khambhaliya, Devbhoomi Dwarka | Gujarat Biotechnology Research Centre | Harish Matani et al |
| EPI_ISL_524736 | India/GJ-GBRC-377b/2020 | 2020-07-11 | Asia/India/Gujarat/Khambhaliya | Government General Hospital, Jam Khambhaliya, Devbhoomi Dwarka | Gujarat Biotechnology Research Centre | Janvi Raval et al |
| EPI_ISL_524737 | India/GJ-GBRC-378/2020 | 2020-06-18 | Asia/India/Gujarat/Gandhinagar | GMERS Medical College and Hospital, Gandhinagar | Gujarat Biotechnology Research Centre | Zarna Patel et al |
| EPI_ISL_524739 | India/GJ-GBRC-380a/2020 | 2020-06-18 | Asia/India/Gujarat/Gandhinagar | GMERS Medical College and Hospital, Gandhinagar | Gujarat Biotechnology Research Centre | Pinal Trivedi et al |
| EPI_ISL_524741 | India/GJ-GBRC-381/2020 | 2020-06-18 | Asia/India/Gujarat/Gandhinagar | GMERS Medical College and Hospital, Gandhinagar | Gujarat Biotechnology Research Centre | Nidhi Patel et al |

| | | | | | | |
|---|---|---|---|---|---|---|
| EPI_ISL_524742 | India/GJ-GBRC-383a/2020 | 2020-06-18 | Asia/India/Gujarat/Gandhinagar | GMERS Medical College and Hospital, Gandhinagar | Gujarat Biotechnology Research Centre | Dinesh Kumar et al |
| EPI_ISL_524745 | India/GJ-GBRC-384b/2020 | 2020-06-22 | Asia/India/Gujarat/Gandhinagar | GMERS Medical College and Hospital, Gandhinagar | Gujarat Biotechnology Research Centre | Labdhi Pandya et al |
| EPI_ISL_524746 | India/GJ-GBRC-385/2020 | 2020-07-11 | Asia/India/Gujarat/Jamnagar | Sir M P Shah Government Medical College, Jamnagar | Gujarat Biotechnology Research Centre | Binita Aring et al |
| EPI_ISL_524747 | India/GJ-GBRC-386a/2020 | 2020-07-11 | Asia/India/Gujarat/Jamnagar | Sir M P Shah Government Medical College, Jamnagar | Gujarat Biotechnology Research Centre | Afzal Ansari et al |
| EPI_ISL_524748 | India/GJ-GBRC-386b/2020 | 2020-07-11 | Asia/India/Gujarat/Jamnagar | Sir M P Shah Government Medical College, Jamnagar | Gujarat Biotechnology Research Centre | Nikha Trivedi et al |
| EPI_ISL_524749 | India/GJ-GBRC-387a/2020 | 2020-07-11 | Asia/India/Gujarat/Jamnagar | Sir M P Shah Government Medical College, Jamnagar | Gujarat Biotechnology Research Centre | Apurvasinh Puvar et al |
| EPI_ISL_524750 | India/GJ-GBRC-387b/2020 | 2020-07-11 | Asia/India/Gujarat/Jamnagar | Sir M P Shah Government Medical College, Jamnagar | Gujarat Biotechnology Research Centre | Janvi Raval et al |
| EPI_ISL_524752 | India/GJ-GBRC-389a/2020 | 2020-07-06 | Asia/India/Gujarat/Patan | GMERS Medical College and Hospital, Dharpur, Patan | Gujarat Biotechnology Research Centre | Monika Gandhi et al |
| EPI_ISL_524753 | India/GJ-GBRC-389b/2020 | 2020-07-06 | Asia/India/Gujarat/Patan | GMERS Medical College and Hospital, Dharpur, Patan | Gujarat Biotechnology Research Centre | Pinal Trivedi et al |
| EPI_ISL_524755 | India/GJ-GBRC-390b/2020 | 2020-07-06 | Asia/India/Gujarat/Patan | GMERS Medical College and Hospital, Dharpur, Patan | Gujarat Biotechnology Research Centre | Nidhi Patel et al |
| EPI_ISL_524756 | India/GJ-GBRC-391a/2020 | 2020-07-06 | Asia/India/Gujarat/Patan | GMERS Medical College and Hospital, Dharpur, Patan | Gujarat Biotechnology Research Centre | Nitin Savaliya et al |
| EPI_ISL_524757 | India/GJ-GBRC-391b/2020 | 2020-07-06 | Asia/India/Gujarat/Patan | GMERS Medical College and Hospital, Dharpur, Patan | Gujarat Biotechnology Research Centre | Raghawendra Kumar et al |
| EPI_ISL_524758 | India/GJ-GBRC-392a/2020 | 2020-07-06 | Asia/India/Gujarat/Patan | GMERS Medical College and Hospital, Dharpur, Patan | Gujarat Biotechnology Research Centre | Dinesh Kumar et al |

| | | | | | | |
|---|---|---|---|---|---|---|
| EPI_ISL_524759 | India/GJ-GBRC-392b/2020 | 2020-07-06 | Asia/India/Gujarat/Patan | GMERS Medical College and Hospital, Dharpur, Patan | Gujarat Biotechnology Research Centre | Zuber Saiyed et al |
| EPI_ISL_524761 | India/GJ-GBRC-393b/2020 | 2020-07-06 | Asia/India/Gujarat/Radhanpur | GMERS Medical College and Hospital, Dharpur, Patan | Gujarat Biotechnology Research Centre | Labdhi Pandya et al |
| EPI_ISL_524763 | India/GJ-GBRC-394b/2020 | 2020-07-06 | Asia/India/Gujarat/Patan | GMERS Medical College and Hospital, Dharpur, Patan | Gujarat Biotechnology Research Centre | Nikha Trivedi et al |
| EPI_ISL_524764 | India/GJ-GBRC-395a/2020 | 2020-07-06 | Asia/India/Gujarat/Patan | GMERS Medical College and Hospital, Dharpur, Patan | Gujarat Biotechnology Research Centre | A N Parmar et al |
| EPI_ISL_524765 | India/GJ-GBRC-395b/2020 | 2020-07-06 | Asia/India/Gujarat/Patan | GMERS Medical College and Hospital, Dharpur, Patan | Gujarat Biotechnology Research Centre | Apurvasinh Puvar et al |
| EPI_ISL_524766 | India/GJ-GBRC-396/2020 | 2020-07-06 | Asia/India/Gujarat/Siddhpur | GMERS Medical College and Hospital, Dharpur, Patan | Gujarat Biotechnology Research Centre | Janvi Raval et al |
| EPI_ISL_524774 | USA/UT-08717/2020 | 2020-05-21 | North America/USA/Utah/ | Utah Public Health Laboratory | Utah Public Health Laboratory | Erin L. Young et al |
| EPI_ISL_524775 | USA/UT-08718/2020 | 2020-05-21 | North America/USA/Utah/ | Utah Public Health Laboratory | Utah Public Health Laboratory | Erin L. Young et al |
| EPI_ISL_524779 | USA/UT-08722/2020 | 2020-05-26 | North America/USA/Utah/ | Utah Public Health Laboratory | Utah Public Health Laboratory | Erin L. Young et al |
| EPI_ISL_524785 | Brazil/PA-IEC165313/2020 | 2020-05-05 | South America/Brazil/Para/ | ÔªøEvandro Chagas Institute | ÔªøEvandro Chagas Institute | Santos et al |
| EPI_ISL_524786 | Brazil/PA-IEC165302/2020 | 2020-05-01 | South America/Brazil/Para/ | ÔªøEvandro Chagas Institute | ÔªøEvandro Chagas Institute | Santos et al |
| EPI_ISL_524789 | Brazil/MA-IEC166716/2020 | 2020-05-05 | South America/Brazil/Maranh√£o/ | ÔªøEvandro Chagas Institute | ÔªøEvandro Chagas Institute | Santos et al |
| EPI_ISL_524797 | Brazil/MA-IEC162157/2020 | 2020-03-24 | South America/Brazil/Maranh√£o/ | ÔªøEvandro Chagas Institute | ÔªøEvandro Chagas Institute | Santos et al |
| EPI_ISL_524799 | Brazil/MA-IEC166867/2020 | 2020-05-08 | South America/Brazil/Maranh√£o/ | ÔªøEvandro Chagas Institute | ÔªøEvandro Chagas Institute | Santos et al |
| EPI_ISL_524800 | Brazil/PA-IEC166687/2020 | 2020-05-13 | South America/Brazil/Para/ | ÔªøEvandro Chagas Institute | ÔªøEvandro Chagas Institute | Santos et al |
| EPI_ISL_524805 | USA/UT-UPHL-2008772/2020 | 2020-05-30 | North America/USA/Utah/ | Utah Public Health Laboratory | Utah Public Health Laboratory | Erin L. Young et al |
| EPI_ISL_524807 | USA/UT-UPHL-2008775/2020 | 2020-05-20 | North America/USA/Utah/ | Utah Public Health Laboratory | Utah Public Health Laboratory | Erin L. Young et al |

| | | | | | | |
|---|---|---|---|---|---|---|
| EPI_ISL_524817 | USA/UT-UPHL-2008790/2020 | 2020-05-23 | North America/USA/Utah/ | Utah Public Health Laboratory | Utah Public Health Laboratory | Erin L. Young et al |
| EPI_ISL_524823 | USA/UT-UPHL-2008796/2020 | 2020-05-22 | North America/USA/Utah/ | Utah Public Health Laboratory | Utah Public Health Laboratory | Erin L. Young et al |
| EPI_ISL_524828 | USA/UT-UPHL-2008802/2020 | 2020-05-26 | North America/USA/Utah/ | Utah Public Health Laboratory | Utah Public Health Laboratory | Erin L. Young et al |
| EPI_ISL_524829 | USA/UT-UPHL-2008803/2020 | 2020-05-29 | North America/USA/Utah/ | Utah Public Health Laboratory | Utah Public Health Laboratory | Erin L. Young et al |
| EPI_ISL_524830 | USA/UT-UPHL-2008804/2020 | 2020-05-31 | North America/USA/Utah/ | Utah Public Health Laboratory | Utah Public Health Laboratory | Erin L. Young et al |
| EPI_ISL_524835 | USA/UT-UPHL-2008809/2020 | 2020-05-23 | North America/USA/Utah/ | Utah Public Health Laboratory | Utah Public Health Laboratory | Erin L. Young et al |
| EPI_ISL_524836 | USA/UT-UPHL-2008810/2020 | 2020-05-29 | North America/USA/Utah/ | Utah Public Health Laboratory | Utah Public Health Laboratory | Erin L. Young et al |
| EPI_ISL_524847 | USA/UT-UPHL-2008827/2020 | 2020-05-29 | North America/USA/Utah/ | Utah Public Health Laboratory | Utah Public Health Laboratory | Erin L. Young et al |
| EPI_ISL_524848 | USA/UT-UPHL-2008828/2020 | 2020-05-29 | North America/USA/Utah/ | Utah Public Health Laboratory | Utah Public Health Laboratory | Erin L. Young et al |
| EPI_ISL_524855 | USA/UT-UPHL-2008836/2020 | 2020-06-01 | North America/USA/Utah/ | Utah Public Health Laboratory | Utah Public Health Laboratory | Erin L. Young et al |
| EPI_ISL_524857 | USA/UT-UPHL-2008839/2020 | 2020-05-29 | North America/USA/Utah/ | Utah Public Health Laboratory | Utah Public Health Laboratory | Erin L. Young et al |
| EPI_ISL_524862 | USA/UT-UPHL-2008845/2020 | 2020-06-01 | North America/USA/Utah/ | Utah Public Health Laboratory | Utah Public Health Laboratory | Erin L. Young et al |
| EPI_ISL_524868 | USA/UT-UPHL-2008852/2020 | 2020-05-22 | North America/USA/Utah/ | Utah Public Health Laboratory | Utah Public Health Laboratory | Erin L. Young et al |
| EPI_ISL_524884 | USA/MD-MDH-0157/2020 | 2020-07-13 | North America/USA/Maryland/ | MD PHL | MD PHL | Maryland Department of Health Laboratories Administration et al |
| EPI_ISL_524885 | USA/MD-MDH-0158/2020 | 2020-07-14 | North America/USA/Maryland/ | MD PHL | MD PHL | Maryland Department of Health Laboratories Administration et al |
| EPI_ISL_524887 | USA/MD-MDH-0160/2020 | 2020-07-15 | North America/USA/Maryland/ | MD PHL | MD PHL | Maryland Department of Health Laboratories Administration et al |
| EPI_ISL_524891 | USA/MD-MDH-0164/2020 | 2020-07-15 | North America/USA/Maryland/ | MD PHL | MD PHL | Maryland Department of Health Laboratories Administration et al |

| | | | | | | |
|---|---|---|---|---|---|---|
| EPI_ISL_524902 | USA/MD-MDH-0139/2020 | 2020-07-09 | North America/USA/Maryland/ | MD PHL | MD PHL | Maryland Department of Health Laboratories Administration et al |
| EPI_ISL_524903 | USA/MD-MDH-0144/2020 | 2020-07-09 | North America/USA/Maryland/ | MD PHL | MD PHL | Maryland Department of Health Laboratories Administration et al |
| EPI_ISL_524907 | USA/UT-UPHL-2009298/2020 | 2020-05-16 | North America/USA/Utah/ | Utah Public Health Laboratory | Utah Public Health Laboratory | Erin L. Young et al |
| EPI_ISL_524915 | USA/UT-UPHL-2009309/2020 | 2020-05-16 | North America/USA/Utah/ | Utah Public Health Laboratory | Utah Public Health Laboratory | Erin L. Young et al |
| EPI_ISL_524918 | USA/UT-UPHL-2009313/2020 | 2020-05-17 | North America/USA/Utah/ | Utah Public Health Laboratory | Utah Public Health Laboratory | Erin L. Young et al |
| EPI_ISL_524920 | USA/UT-UPHL-2009315/2020 | 2020-05-17 | North America/USA/Utah/ | Utah Public Health Laboratory | Utah Public Health Laboratory | Erin L. Young et al |
| EPI_ISL_524925 | USA/UT-UPHL-2009324/2020 | 2020-05-16 | North America/USA/Utah/ | Utah Public Health Laboratory | Utah Public Health Laboratory | Erin L. Young et al |
| EPI_ISL_524926 | USA/UT-UPHL-2009325/2020 | 2020-05-14 | North America/USA/Utah/ | Utah Public Health Laboratory | Utah Public Health Laboratory | Erin L. Young et al |
| EPI_ISL_524928 | USA/UT-UPHL-2009327/2020 | 2020-06-22 | North America/USA/Utah/ | Utah Public Health Laboratory | Utah Public Health Laboratory | Erin L. Young et al |
| EPI_ISL_524931 | USA/UT-UPHL-2009330/2020 | 2020-05-17 | North America/USA/Utah/ | Utah Public Health Laboratory | Utah Public Health Laboratory | Erin L. Young et al |
| EPI_ISL_524942 | USA/UT-UPHL-2009344/2020 | 2020-05-16 | North America/USA/Utah/ | Utah Public Health Laboratory | Utah Public Health Laboratory | Erin L. Young et al |
| EPI_ISL_524944 | USA/UT-UPHL-2009346/2020 | 2020-05-17 | North America/USA/Utah/ | Utah Public Health Laboratory | Utah Public Health Laboratory | Erin L. Young et al |
| EPI_ISL_524954 | USA/UT-UPHL-2009359/2020 | 2020-05-17 | North America/USA/Utah/ | Utah Public Health Laboratory | Utah Public Health Laboratory | Erin L. Young et al |
| EPI_ISL_524963 | USA/UT-UPHL-2009370/2020 | 2020-05-17 | North America/USA/Utah/ | Utah Public Health Laboratory | Utah Public Health Laboratory | Erin L. Young et al |
| EPI_ISL_524964 | USA/UT-UPHL-2009371/2020 | 2020-06-22 | North America/USA/Utah/ | Utah Public Health Laboratory | Utah Public Health Laboratory | Erin L. Young et al |
| EPI_ISL_524989 | USA/UT-UPHL-2008875/2020 | 2020-05-28 | North America/USA/Utah/ | Utah Public Health Laboratory | Utah Public Health Laboratory | Erin L. Young et al |
| EPI_ISL_525002 | USA/UT-UPHL-2008888/2020 | 2020-05-26 | North America/USA/Utah/ | Utah Public Health Laboratory | Utah Public Health Laboratory | Erin L. Young et al |
| EPI_ISL_525013 | USA/UT-UPHL-2008900/2020 | 2020-05-22 | North America/USA/Utah/ | Utah Public Health Laboratory | Utah Public Health Laboratory | Erin L. Young et al |
| EPI_ISL_525015 | USA/UT-UPHL-2008904/2020 | 2020-05-26 | North America/USA/Utah/ | Utah Public Health Laboratory | Utah Public Health Laboratory | Erin L. Young et al |

| | | | | | | |
|---|---|---|---|---|---|---|
| EPI_ISL_525016 | USA/UT-UPHL-2008906/2020 | 2020-05-28 | North America/USA/Utah/ | Utah Public Health Laboratory | Utah Public Health Laboratory | Erin L. Young et al |
| EPI_ISL_525019 | USA/UT-UPHL-2008909/2020 | 2020-05-28 | North America/USA/Utah/ | Utah Public Health Laboratory | Utah Public Health Laboratory | Erin L. Young et al |
| EPI_ISL_525020 | USA/UT-UPHL-2008914/2020 | 2020-05-26 | North America/USA/Utah/ | Utah Public Health Laboratory | Utah Public Health Laboratory | Erin L. Young et al |
| EPI_ISL_525021 | USA/UT-UPHL-2008915/2020 | 2020-05-18 | North America/USA/Utah/ | Utah Public Health Laboratory | Utah Public Health Laboratory | Erin L. Young et al |
| EPI_ISL_525024 | USA/UT-UPHL-2008918/2020 | 2020-05-28 | North America/USA/Utah/ | Utah Public Health Laboratory | Utah Public Health Laboratory | Erin L. Young et al |
| EPI_ISL_525033 | USA/UT-UPHL-2008928/2020 | 2020-05-28 | North America/USA/Utah/ | Utah Public Health Laboratory | Utah Public Health Laboratory | Erin L. Young et al |
| EPI_ISL_525056 | USA/UT-UPHL-2009042/2020 | 2020-05-15 | North America/USA/Utah/ | Utah Public Health Laboratory | Utah Public Health Laboratory | Erin L. Young et al |
| EPI_ISL_525060 | USA/UT-UPHL-2009046/2020 | 2020-05-12 | North America/USA/Utah/ | Utah Public Health Laboratory | Utah Public Health Laboratory | Erin L. Young et al |
| EPI_ISL_525061 | USA/UT-UPHL-2009047/2020 | 2020-05-13 | North America/USA/Utah/ | Utah Public Health Laboratory | Utah Public Health Laboratory | Erin L. Young et al |
| EPI_ISL_525062 | USA/UT-UPHL-2009048/2020 | 2020-05-11 | North America/USA/Utah/ | Utah Public Health Laboratory | Utah Public Health Laboratory | Erin L. Young et al |
| EPI_ISL_525073 | USA/UT-UPHL-2009063/2020 | 2020-05-12 | North America/USA/Utah/ | Utah Public Health Laboratory | Utah Public Health Laboratory | Erin L. Young et al |
| EPI_ISL_525075 | USA/UT-UPHL-2009067/2020 | 2020-05-11 | North America/USA/Utah/ | Utah Public Health Laboratory | Utah Public Health Laboratory | Erin L. Young et al |
| EPI_ISL_525078 | USA/UT-UPHL-2009070/2020 | 2020-05-11 | North America/USA/Utah/ | Utah Public Health Laboratory | Utah Public Health Laboratory | Erin L. Young et al |
| EPI_ISL_525086 | USA/UT-UPHL-2009079/2020 | 2020-05-13 | North America/USA/Utah/ | Utah Public Health Laboratory | Utah Public Health Laboratory | Erin L. Young et al |
| EPI_ISL_525087 | USA/UT-UPHL-2009080/2020 | 2020-05-14 | North America/USA/Utah/ | Utah Public Health Laboratory | Utah Public Health Laboratory | Erin L. Young et al |
| EPI_ISL_525089 | USA/UT-UPHL-2009082/2020 | 2020-05-13 | North America/USA/Utah/ | Utah Public Health Laboratory | Utah Public Health Laboratory | Erin L. Young et al |
| EPI_ISL_525092 | USA/UT-UPHL-2009085/2020 | 2020-05-15 | North America/USA/Utah/ | Utah Public Health Laboratory | Utah Public Health Laboratory | Erin L. Young et al |
| EPI_ISL_525093 | USA/UT-UPHL-2009086/2020 | 2020-05-12 | North America/USA/Utah/ | Utah Public Health Laboratory | Utah Public Health Laboratory | Erin L. Young et al |
| EPI_ISL_525108 | USA/UT-UPHL-2009108/2020 | 2020-05-13 | North America/USA/Utah/ | Utah Public Health Laboratory | Utah Public Health Laboratory | Erin L. Young et al |
| EPI_ISL_525110 | USA/UT-UPHL-2009110/2020 | 2020-05-11 | North America/USA/Utah/ | Utah Public Health Laboratory | Utah Public Health Laboratory | Erin L. Young et al |

| | | | | | | |
|---|---|---|---|---|---|---|
| EPI_ISL_525112 | USA/UT-UPHL-2009113/2020 | 2020-05-11 | North America/USA/Utah/ | Utah Public Health Laboratory | Utah Public Health Laboratory | Erin L. Young et al |
| EPI_ISL_525125 | USA/UT-UPHL-2009130/2020 | 2020-05-30 | North America/USA/Utah/ | Utah Public Health Laboratory | Utah Public Health Laboratory | Erin L. Young et al |
| EPI_ISL_525126 | USA/UT-UPHL-2009131/2020 | 2020-05-31 | North America/USA/Utah/ | Utah Public Health Laboratory | Utah Public Health Laboratory | Erin L. Young et al |
| EPI_ISL_525133 | USA/UT-UPHL-2009139/2020 | 2020-05-27 | North America/USA/Utah/ | Utah Public Health Laboratory | Utah Public Health Laboratory | Erin L. Young et al |
| EPI_ISL_525134 | USA/UT-UPHL-2009140/2020 | 2020-05-30 | North America/USA/Utah/ | Utah Public Health Laboratory | Utah Public Health Laboratory | Erin L. Young et al |
| EPI_ISL_525140 | USA/UT-UPHL-2009150/2020 | 2020-05-28 | North America/USA/Utah/ | Utah Public Health Laboratory | Utah Public Health Laboratory | Erin L. Young et al |
| EPI_ISL_525143 | USA/UT-UPHL-2009153/2020 | 2020-05-30 | North America/USA/Utah/ | Utah Public Health Laboratory | Utah Public Health Laboratory | Erin L. Young et al |
| EPI_ISL_525144 | USA/UT-UPHL-2009154/2020 | 2020-05-30 | North America/USA/Utah/ | Utah Public Health Laboratory | Utah Public Health Laboratory | Erin L. Young et al |
| EPI_ISL_525146 | USA/UT-UPHL-2009156/2020 | 2020-05-22 | North America/USA/Utah/ | Utah Public Health Laboratory | Utah Public Health Laboratory | Erin L. Young et al |
| EPI_ISL_525151 | USA/UT-UPHL-2009163/2020 | 2020-05-28 | North America/USA/Utah/ | Utah Public Health Laboratory | Utah Public Health Laboratory | Erin L. Young et al |
| EPI_ISL_525157 | USA/UT-UPHL-2009171/2020 | 2020-05-28 | North America/USA/Utah/ | Utah Public Health Laboratory | Utah Public Health Laboratory | Erin L. Young et al |
| EPI_ISL_525158 | USA/UT-UPHL-2009172/2020 | 2020-05-30 | North America/USA/Utah/ | Utah Public Health Laboratory | Utah Public Health Laboratory | Erin L. Young et al |
| EPI_ISL_525159 | USA/UT-UPHL-2009173/2020 | 2020-05-30 | North America/USA/Utah/ | Utah Public Health Laboratory | Utah Public Health Laboratory | Erin L. Young et al |
| EPI_ISL_525162 | USA/UT-UPHL-2009176/2020 | 2020-05-27 | North America/USA/Utah/ | Utah Public Health Laboratory | Utah Public Health Laboratory | Erin L. Young et al |
| EPI_ISL_525169 | USA/UT-UPHL-2009186/2020 | 2020-05-30 | North America/USA/Utah/ | Utah Public Health Laboratory | Utah Public Health Laboratory | Erin L. Young et al |
| EPI_ISL_525170 | USA/UT-UPHL-2009187/2020 | 2020-05-22 | North America/USA/Utah/ | Utah Public Health Laboratory | Utah Public Health Laboratory | Erin L. Young et al |
| EPI_ISL_525172 | USA/UT-UPHL-2009189/2020 | 2020-05-26 | North America/USA/Utah/ | Utah Public Health Laboratory | Utah Public Health Laboratory | Erin L. Young et al |
| EPI_ISL_525181 | USA/UT-UPHL-2009198/2020 | 2020-05-22 | North America/USA/Utah/ | Utah Public Health Laboratory | Utah Public Health Laboratory | Erin L. Young et al |
| EPI_ISL_525183 | USA/UT-UPHL-2009201/2020 | 2020-05-11 | North America/USA/Utah/ | Utah Public Health Laboratory | Utah Public Health Laboratory | Erin L. Young et al |
| EPI_ISL_525184 | USA/UT-UPHL-2009202/2020 | 2020-05-22 | North America/USA/Utah/ | Utah Public Health Laboratory | Utah Public Health Laboratory | Erin L. Young et al |

| | | | | | | |
|---|---|---|---|---|---|---|
| EPI_ISL_525191 | USA/VA-DCLS-1161/2020 | 2020-07-07 | North America/USA/Virginia/ | Virginia DCLS | Virginia DCLS | Virginia DCLS et al |
| EPI_ISL_525192 | USA/VA-DCLS-1164/2020 | 2020-07-07 | North America/USA/Virginia/ | Virginia DCLS | Virginia DCLS | Virginia DCLS et al |
| EPI_ISL_525193 | USA/VA-DCLS-1165/2020 | 2020-07-18 | North America/USA/Virginia/ | Virginia DCLS | Virginia DCLS | Virginia DCLS et al |
| EPI_ISL_525195 | USA/VA-DCLS-1168/2020 | 2020-07-18 | North America/USA/Virginia/ | Virginia DCLS | Virginia DCLS | Virginia DCLS et al |
| EPI_ISL_525207 | USA/UT-UPHL-2009207/2020 | 2020-05-11 | North America/USA/Utah/ | Utah Public Health Laboratory | Utah Public Health Laboratory | Erin L. Young et al |
| EPI_ISL_525211 | USA/UT-UPHL-2009211/2020 | 2020-05-15 | North America/USA/Utah/ | Utah Public Health Laboratory | Utah Public Health Laboratory | Erin L. Young et al |
| EPI_ISL_525212 | USA/UT-UPHL-2009212/2020 | 2020-05-12 | North America/USA/Utah/ | Utah Public Health Laboratory | Utah Public Health Laboratory | Erin L. Young et al |
| EPI_ISL_525215 | USA/UT-UPHL-2009215/2020 | 2020-05-14 | North America/USA/Utah/ | Utah Public Health Laboratory | Utah Public Health Laboratory | Erin L. Young et al |
| EPI_ISL_525218 | USA/UT-UPHL-2009218/2020 | 2020-05-11 | North America/USA/Utah/ | Utah Public Health Laboratory | Utah Public Health Laboratory | Erin L. Young et al |
| EPI_ISL_525223 | USA/UT-UPHL-2009223/2020 | 2020-05-15 | North America/USA/Utah/ | Utah Public Health Laboratory | Utah Public Health Laboratory | Erin L. Young et al |
| EPI_ISL_525226 | USA/UT-UPHL-2009230/2020 | 2020-05-12 | North America/USA/Utah/ | Utah Public Health Laboratory | Utah Public Health Laboratory | Erin L. Young et al |
| EPI_ISL_525229 | USA/UT-UPHL-2009236/2020 | 2020-05-14 | North America/USA/Utah/ | Utah Public Health Laboratory | Utah Public Health Laboratory | Erin L. Young et al |
| EPI_ISL_525238 | USA/UT-UPHL-2009251/2020 | 2020-05-11 | North America/USA/Utah/ | Utah Public Health Laboratory | Utah Public Health Laboratory | Erin L. Young et al |
| EPI_ISL_525242 | USA/UT-UPHL-2009256/2020 | 2020-05-18 | North America/USA/Utah/ | Utah Public Health Laboratory | Utah Public Health Laboratory | Erin L. Young et al |
| EPI_ISL_525243 | USA/UT-UPHL-2009257/2020 | 2020-05-15 | North America/USA/Utah/ | Utah Public Health Laboratory | Utah Public Health Laboratory | Erin L. Young et al |
| EPI_ISL_525249 | USA/UT-UPHL-2009266/2020 | 2020-05-16 | North America/USA/Utah/ | Utah Public Health Laboratory | Utah Public Health Laboratory | Erin L. Young et al |
| EPI_ISL_525250 | USA/UT-UPHL-2009267/2020 | 2020-05-13 | North America/USA/Utah/ | Utah Public Health Laboratory | Utah Public Health Laboratory | Erin L. Young et al |
| EPI_ISL_525257 | USA/UT-UPHL-2009276/2020 | 2020-05-13 | North America/USA/Utah/ | Utah Public Health Laboratory | Utah Public Health Laboratory | Erin L. Young et al |
| EPI_ISL_525258 | USA/UT-UPHL-2009277/2020 | 2020-05-13 | North America/USA/Utah/ | Utah Public Health Laboratory | Utah Public Health Laboratory | Erin L. Young et al |
| EPI_ISL_525259 | USA/UT-UPHL-2009278/2020 | 2020-05-15 | North America/USA/Utah/ | Utah Public Health Laboratory | Utah Public Health Laboratory | Erin L. Young et al |
| EPI_ISL_525261 | USA/UT-UPHL-2009280/2020 | 2020-05-15 | North America/USA/Utah/ | Utah Public Health Laboratory | Utah Public Health Laboratory | Erin L. Young et al |

| | | | | | | |
|---|---|---|---|---|---|---|
| EPI_ISL_525267 | USA/UT-UPHL-2009289/2020 | 2020-05-15 | North America/USA/Utah/ | Utah Public Health Laboratory | Utah Public Health Laboratory | Erin L. Young et al |
| EPI_ISL_525268 | USA/UT-UPHL-2009290/2020 | 2020-05-15 | North America/USA/Utah/ | Utah Public Health Laboratory | Utah Public Health Laboratory | Erin L. Young et al |
| EPI_ISL_525283 | USA/UT-UPHL-2008956/2020 | 2020-05-11 | North America/USA/Utah/ | Utah Public Health Laboratory | Utah Public Health Laboratory | Erin L. Young et al |
| EPI_ISL_525284 | USA/UT-UPHL-2008957/2020 | 2020-05-11 | North America/USA/Utah/ | Utah Public Health Laboratory | Utah Public Health Laboratory | Erin L. Young et al |
| EPI_ISL_525288 | USA/UT-UPHL-2008962/2020 | 2020-05-15 | North America/USA/Utah/ | Utah Public Health Laboratory | Utah Public Health Laboratory | Erin L. Young et al |
| EPI_ISL_525291 | USA/UT-UPHL-2008965/2020 | 2020-05-11 | North America/USA/Utah/ | Utah Public Health Laboratory | Utah Public Health Laboratory | Erin L. Young et al |
| EPI_ISL_525302 | USA/UT-UPHL-2008985/2020 | 2020-05-11 | North America/USA/Utah/ | Utah Public Health Laboratory | Utah Public Health Laboratory | Erin L. Young et al |
| EPI_ISL_525305 | USA/UT-UPHL-2008991/2020 | 2020-05-11 | North America/USA/Utah/ | Utah Public Health Laboratory | Utah Public Health Laboratory | Erin L. Young et al |
| EPI_ISL_525310 | USA/UT-UPHL-2008999/2020 | 2020-05-11 | North America/USA/Utah/ | Utah Public Health Laboratory | Utah Public Health Laboratory | Erin L. Young et al |
| EPI_ISL_525312 | USA/UT-UPHL-2009001/2020 | 2020-05-11 | North America/USA/Utah/ | Utah Public Health Laboratory | Utah Public Health Laboratory | Erin L. Young et al |
| EPI_ISL_525313 | USA/UT-UPHL-2009003/2020 | 2020-05-27 | North America/USA/Utah/ | Utah Public Health Laboratory | Utah Public Health Laboratory | Erin L. Young et al |
| EPI_ISL_525315 | USA/UT-UPHL-2009005/2020 | 2020-05-12 | North America/USA/Utah/ | Utah Public Health Laboratory | Utah Public Health Laboratory | Erin L. Young et al |
| EPI_ISL_525317 | USA/UT-UPHL-2009008/2020 | 2020-05-15 | North America/USA/Utah/ | Utah Public Health Laboratory | Utah Public Health Laboratory | Erin L. Young et al |
| EPI_ISL_525341 | USA/UT-UPHL-04474/2020 | 2020-05-20 | North America/USA/Utah/ | Utah Public Health Laboratory | Utah Public Health Laboratory | Erin Young et al |
| EPI_ISL_525347 | USA/UT-UPHL-08760/2020 | 2020-08-13 | North America/USA/Utah/ | Utah Public Health Laboratory | Utah Public Health Laboratory | Erin Young et al |
| EPI_ISL_525348 | USA/UT-UPHL-08762/2020 | 2020-08-14 | North America/USA/Utah/ | Utah Public Health Laboratory | Utah Public Health Laboratory | Erin Young et al |
| EPI_ISL_525350 | USA/UT-UPHL-08764/2020 | 2020-08-13 | North America/USA/Utah/ | Utah Public Health Laboratory | Utah Public Health Laboratory | Erin Young et al |
| EPI_ISL_525419 | India/GJ-GBRC-382a/2020 | 2020-06-18 | Asia/India/Gujarat/Ahmedabad | GMERS Medical College and Hospital, Gandhinagar | Gujarat Biotechnology Research Centre | Nitin Savaliya et al |
| EPI_ISL_525421 | India/GJ-GBRC-397a/2020 | 2020-06-25 | Asia/India/Gujarat/Ahmedabad | B.J. Medical College and Civil hospital, Ahmedabad | Gujarat Biotechnology Research Centre | Zarna Patel et al |

| | | | | | | |
|---|---|---|---|---|---|---|
| EPI_ISL_525428 | Oman/RESP-20-B-15703/2020 | 2020-05-22 | Asia/Oman/Muscat/ | Oman-National Influenza Center | Biotechnology & OMICs Laboratory | Sajjad Asaf et al |
| EPI_ISL_525429 | Oman/RESP-20-B-6588/2020 | 2020-06-05 | Asia/Oman/Muscat/ | Oman-National Influenza Center | Biotechnology & OMICs Laboratory | Samira Al-Mahruqi et al |
| EPI_ISL_525430 | Ecuador/USFQ-163/2020 | 2020-05-20 | South America/Ecuador/Pichincha/Quito | Institute of Microbiology, Universidad San Francisco de Quito | Institute of Microbiology, Universidad San Francisco de Quito | Juan Jos√© Guadalupe et al |
| EPI_ISL_525431 | Ecuador/USFQ-177/2020 | 2020-08-12 | South America/Ecuador/Zamora-Chinchi | Institute of Microbiology, Universidad San Francisco de Quito | Institute of Microbiology, Universidad San Francisco de Quito | Juan Jos√© Guadalupe et al |
| EPI_ISL_525433 | Ecuador/USFQ-182/2020 | 2020-08-13 | South America/Ecuador/Los Rios/Queve | Institute of Microbiology, Universidad San Francisco de Quito | Institute of Microbiology, Universidad San Francisco de Quito | Juan Jos√© Guadalupe et al |
| EPI_ISL_525434 | Ecuador/USFQ-183/2020 | 2020-08-13 | South America/Ecuador/Los Rios/Queve | Institute of Microbiology, Universidad San Francisco de Quito | Institute of Microbiology, Universidad San Francisco de Quito | Juan Jos√© Guadalupe et al |
| EPI_ISL_525435 | Ecuador/USFQ-184/2020 | 2020-08-14 | South America/Ecuador/Orellana/Orella | Institute of Microbiology, Universidad San Francisco de Quito | Institute of Microbiology, Universidad San Francisco de Quito | Juan Jos√© Guadalupe et al |
| EPI_ISL_525437 | Ecuador/USFQ-188/2020 | 2020-08-12 | South America/Ecuador/Imbabura/Otav | Institute of Microbiology, Universidad San Francisco de Quito | Institute of Microbiology, Universidad San Francisco de Quito | Juan Jos√© Guadalupe et al |
| EPI_ISL_525467 | DominicanRepublic/ICGEB_UNIBE022/ | 2020-05-05 | North America/Dominican Republic/Dom | Universidad Iberoamericana | International Centre for Genetic Engineering and Biotechnology (ICGEB) and ARGO Open Lab Platform | Robert Paulino-Ramirez et al |
| EPI_ISL_525468 | DominicanRepublic/ICGEB_UNIBE032/ | 2020-05-06 | North America/Dominican Republic/Dom | Universidad Iberoamericana | International Centre for Genetic Engineering and Biotechnology (ICGEB) and ARGO Open Lab Platform | Robert Paulino-Ramirez et al |

| | | | | | | |
|---|---|---|---|---|---|---|
| EPI_ISL_525469 | DominicanRepublic/ICGEB_UNIBE032 | 2020-05-06 | North America/Dominican Republic/Dom | Universidad Iberoamericana | International Centre for Genetic Engineering and Biotechnology (ICGEB) and ARGO Open Lab Platform | Robert Paulino-Ramirez et al |
| EPI_ISL_525474 | SriLanka/CDR142/2020 | 2020-04-19 | Asia/Sri Lanka/Colombo/ | Centre for Dengue Research | Centre for Dengue Research, USJ, SL | Chandima Jeewandara et al |
| EPI_ISL_525486 | SriLanka/CDR-KK57/2020 | 2020-07-21 | Asia/Sri Lanka/Sri Lanka/ | Centre for Dengue Research | Centre for Dengue Research | Chandima Jeewandara et al |

| | | | | | | |
|---|---|---|---|---|---|---|
| EPI_ISL_525492 | Indonesia/JT-UGM-202538/2020 | 2020-06-26 | Asia/Indonesia/Central Java/ | RSUP dr. SOERADJI TIRTONEGORO | Genetics Working Group (Pokja Genetik) Faculty of Medicine, Public Health and Nursing Universitas Gadjah Mada (FK-KMK UGM), Disease Investigation Center Wates Ministry of Agriculture Indonesia, Department of Microbiology FK-KMK UGM, Laboratorium Diagnostik Yayasan Tahija World Mosquito Program (WMP) Yogyakarta Center for Tropical Medicine FK-KMK UGM, Integrated Research Center FK-KMK UGM, Department of Computer Science and Electronics FMIPA UGM | Gunadi et al |
| EPI_ISL_525541 | France/ARA-11167/2020 | 2020-03-18 | Europe/France/Auvergne-Rhv¥ne-Alpes/ | CNR Virus des Infections Respiratoires - France SUD | CNR Virus des Infections Respiratoires - France SUD | Antonin Bal et al |

| | | | | | | |
|---|---|---|---|---|---|---|
| EPI_ISL_525553 | Italy/APU-UniMI-102/2020 | 2020-03-16 | Europe/Italy/Apulia/Bari | Istituto Zooprofilattico Sperimentale Puglia e Basilicata; Dipartimento di Bioscienze, Biotecnologie e Biofarmaceutica dell,ÄôUniversit√† degli Studi di Bari ,ÄúA.Moro,Äù; Istituto di Biomembrane. Bioenergetica e Biotecnologie Molecolari del Consiglio Nazionale delle Ricerche di Bari | Beaconlab (Bioinformatics, Evolution and Comparative Genomics lab), Dept of Biosciences, University on Milan | Parisi A. et al |
| EPI_ISL_525556 | Italy/APU-UniMI-22/2020 | 2020-03-16 | Europe/Italy/Apulia/Brindisi | Istituto Zooprofilattico Sperimentale Puglia e Basilicata; Dipartimento di Bioscienze, Biotecnologie e Biofarmaceutica dell,ÄôUniversit√† degli Studi di Bari ,ÄúA.Moro,Äù; Istituto di Biomembrane. Bioenergetica e Biotecnologie Molecolari del Consiglio Nazionale delle Ricerche di Bari | Beaconlab (Bioinformatics, Evolution and Comparative Genomics lab), Dept of Biosciences, University on Milan | Parisi A. et al |
| EPI_ISL_525557 | Italy/APU-UniMI-368/2020 | 2020-03-17 | Europe/Italy/Apulia/Brindisi | Istituto Zooprofilattico Sperimentale Puglia e Basilicata; Dipartimento di Bioscienze, Biotecnologie e Biofarmaceutica dell,ÄôUniversit√† degli Studi di Bari ,ÄúA.Moro,Äù; Istituto di Biomembrane. Bioenergetica e Biotecnologie Molecolari del Consiglio Nazionale delle Ricerche di Bari | Beaconlab (Bioinformatics, Evolution and Comparative Genomics lab), Dept of Biosciences, University on Milan | Parisi A. et al |
| EPI_ISL_525559 | Italy/APU-UniMI-4431/2020 | 2020-04-09 | Europe/Italy/Apulia/Brindisi | Istituto Zooprofilattico Sperimentale Puglia e Basilicata; Dipartimento di Bioscienze, Biotecnologie e Biofarmaceutica dell,ÄôUniversit√† degli Studi di Bari ,ÄúA.Moro,Äù; Istituto di Biomembrane. Bioenergetica e Biotecnologie Molecolari del Consiglio Nazionale delle Ricerche di Bari | Beaconlab (Bioinformatics, Evolution and Comparative Genomics lab), Dept of Biosciences, University on Milan | Parisi A. et al |

| | | | | | | |
|---|---|---|---|---|---|---|
| EPI_ISL_525561 | Italy/APU-UniMI-5410/2020 | 2020-04-11 | Europe/Italy/Apulia/Brindisi | Istituto Zooprofilattico Sperimentale Puglia e Basilicata; Dipartimento di Bioscienze, Biotecnologie e Biofarmaceutica dell‚ÄôUniversit√† degli Studi di Bari ‚ÄúA.Moro‚Äù; Istituto di Biomembrane. Bioenergetica e Biotecnologie Molecolari del Consiglio Nazionale delle Ricerche di Bari | Beaconlab (Bioinformatics, Evolution and Comparative Genomics lab), Dept of Biosciences, University on Milan | Parisi A. et al |
| EPI_ISL_525563 | Italy/APU-UniMI-5425/2020 | 2020-04-11 | Europe/Italy/Apulia/Brindisi | Istituto Zooprofilattico Sperimentale Puglia e Basilicata; Dipartimento di Bioscienze, Biotecnologie e Biofarmaceutica dell‚ÄôUniversit√† degli Studi di Bari ‚ÄúA.Moro‚Äù; Istituto di Biomembrane. Bioenergetica e Biotecnologie Molecolari del Consiglio Nazionale delle Ricerche di Bari | Beaconlab (Bioinformatics, Evolution and Comparative Genomics lab), Dept of Biosciences, University on Milan | Parisi A. et al |
| EPI_ISL_525564 | Italy/APU-UniMI-5428/2020 | 2020-04-11 | Europe/Italy/Apulia/Brindisi | Istituto Zooprofilattico Sperimentale Puglia e Basilicata; Dipartimento di Bioscienze, Biotecnologie e Biofarmaceutica dell‚ÄôUniversit√† degli Studi di Bari ‚ÄúA.Moro‚Äù; Istituto di Biomembrane. Bioenergetica e Biotecnologie Molecolari del Consiglio Nazionale delle Ricerche di Bari | Beaconlab (Bioinformatics, Evolution and Comparative Genomics lab), Dept of Biosciences, University on Milan | Parisi A. et al |
| EPI_ISL_525569 | Italy/APU-UniMI-5439/2020 | 2020-04-11 | Europe/Italy/Apulia/Brindisi | Istituto Zooprofilattico Sperimentale Puglia e Basilicata; Dipartimento di Bioscienze, Biotecnologie e Biofarmaceutica dell‚ÄôUniversit√† degli Studi di Bari ‚ÄúA.Moro‚Äù; Istituto di Biomembrane. Bioenergetica e Biotecnologie Molecolari del Consiglio Nazionale delle Ricerche di Bari | Beaconlab (Bioinformatics, Evolution and Comparative Genomics lab), Dept of Biosciences, University on Milan | Parisi A. et al |

| | | | | | | |
|---|---|---|---|---|---|---|
| EPI_ISL_525570 | Italy/APU-UniMI-6/2020 | 2020-03-16 | Europe/Italy/Apulia/Brindisi | Istituto Zooprofilattico Sperimentale Puglia e Basilicata; Dipartimento di Bioscienze, Biotecnologie e Biofarmaceutica dell,ÄôUniversit√† degli Studi di Bari ,ÄúA.Moro,Äù; Istituto di Biomembrane. Bioenergetica e Biotecnologie Molecolari del Consiglio Nazionale delle Ricerche di Bari | Beaconlab (Bioinformatics, Evolution and Comparative Genomics lab), Dept of Biosciences, University on Milan | Parisi A. et al |
| EPI_ISL_525571 | Italy/APU-UniMI-62PT/2020 | 2020-03-28 | Europe/Italy/Apulia/Lecce | Istituto Zooprofilattico Sperimentale Puglia e Basilicata; Dipartimento di Bioscienze, Biotecnologie e Biofarmaceutica dell,ÄôUniversit√† degli Studi di Bari ,ÄúA.Moro,Äù; Istituto di Biomembrane. Bioenergetica e Biotecnologie Molecolari del Consiglio Nazionale delle Ricerche di Bari | Beaconlab (Bioinformatics, Evolution and Comparative Genomics lab), Dept of Biosciences, University on Milan | Parisi A. et al |
| EPI_ISL_525574 | Italy/BAS-UniMI-4160/2020 | 2020-04-08 | Europe/Italy/Basilicata/Potenza | Istituto Zooprofilattico Sperimentale Puglia e Basilicata; Dipartimento di Bioscienze, Biotecnologie e Biofarmaceutica dell,ÄôUniversit√† degli Studi di Bari ,ÄúA.Moro,Äù; Istituto di Biomembrane. Bioenergetica e Biotecnologie Molecolari del Consiglio Nazionale delle Ricerche di Bari | Beaconlab (Bioinformatics, Evolution and Comparative Genomics lab), Dept of Biosciences, University on Milan | Parisi A. et al |
| EPI_ISL_525588 | USA/NY-Wadsworth-11690-01/2020 | 2020-03-08 | North America/USA/New York/ | Wadsworth Center, New York State Department of Health | Wadsworth Center, New York State Department of Health | Kirsten St. George et al |
| EPI_ISL_525595 | USA/NY-Wadsworth-12806-01/2020 | 2020-03-10 | North America/USA/New York/ | Wadsworth Center, New York State Department of Health | Wadsworth Center, New York State Department of Health | Kirsten St. George et al |
| EPI_ISL_525633 | USA/NY-Wadsworth-30673-01/2020 | 2020-03-25 | North America/USA/New York/ | Wadsworth Center, New York State Department of Health | Wadsworth Center, New York State Department of Health | Kirsten St. George et al |

| | | | | | | |
|---|---|---|---|---|---|---|
| EPI_ISL_525648 | USA/NY-Wadsworth-36877-01/2020 | 2020-04-02 | North America/USA/New York/ | Wadsworth Center, New York State Department of Health | Wadsworth Center, New York State Department of Health | Kirsten St. George et al |
| EPI_ISL_525649 | USA/NY-Wadsworth-36878-01/2020 | 2020-04-02 | North America/USA/New York/ | Wadsworth Center, New York State Department of Health | Wadsworth Center, New York State Department of Health | Kirsten St. George et al |
| EPI_ISL_525654 | USA/NY-Wadsworth-39179-01/2020 | 2020-04-06 | North America/USA/New York/ | Wadsworth Center, New York State Department of Health | Wadsworth Center, New York State Department of Health | Kirsten St. George et al |
| EPI_ISL_525655 | USA/NY-Wadsworth-39537-01/2020 | 2020-04-06 | North America/USA/New York/ | Wadsworth Center, New York State Department of Health | Wadsworth Center, New York State Department of Health | Kirsten St. George et al |
| EPI_ISL_525664 | USA/NY-Wadsworth-43319-01/2020 | 2020-04-09 | North America/USA/New York/ | Wadsworth Center, New York State Department of Health | Wadsworth Center, New York State Department of Health | Kirsten St. George et al |
| EPI_ISL_525669 | USA/NY-Wadsworth-44600-01/2020 | 2020-04-10 | North America/USA/New York/ | Wadsworth Center, New York State Department of Health | Wadsworth Center, New York State Department of Health | Kirsten St. George et al |
| EPI_ISL_525702 | USA/WA-S2712/2020 | 2020-06-25 | North America/USA/Washington/King C | Seattle Flu Study | Seattle Flu Study | Deborah A. Nickerson et al |
| EPI_ISL_525711 | USA/WA-S2721/2020 | 2020-07-02 | North America/USA/Washington/King C | Seattle Flu Study | Seattle Flu Study | Deborah A. Nickerson et al |
| EPI_ISL_525712 | USA/WA-S2722/2020 | 2020-07-03 | North America/USA/Washington/King C | Seattle Flu Study | Seattle Flu Study | Deborah A. Nickerson et al |
| EPI_ISL_525716 | USA/WA-S2726/2020 | 2020-07-07 | North America/USA/Washington/King C | Seattle Flu Study | Seattle Flu Study | Deborah A. Nickerson et al |
| EPI_ISL_525718 | USA/WA-S2728/2020 | 2020-07-08 | North America/USA/Washington/King C | Seattle Flu Study | Seattle Flu Study | Deborah A. Nickerson et al |
| EPI_ISL_525721 | USA/WA-S2731/2020 | 2020-07-11 | North America/USA/Washington/King C | Seattle Flu Study | Seattle Flu Study | Deborah A. Nickerson et al |
| EPI_ISL_525727 | USA/WA-S2737/2020 | 2020-07-16 | North America/USA/Washington/King C | Seattle Flu Study | Seattle Flu Study | Deborah A. Nickerson et al |
| EPI_ISL_525731 | USA/WA-S2741/2020 | 2020-07-16 | North America/USA/Washington/King C | Seattle Flu Study | Seattle Flu Study | Deborah A. Nickerson et al |
| EPI_ISL_525739 | USA/WA-S2749/2020 | 2020-07-21 | North America/USA/Washington/King C | Seattle Flu Study | Seattle Flu Study | Deborah A. Nickerson et al |
| EPI_ISL_525740 | USA/WA-S2750/2020 | 2020-07-21 | North America/USA/Washington/King C | Seattle Flu Study | Seattle Flu Study | Deborah A. Nickerson et al |
| EPI_ISL_525741 | USA/WA-S2751/2020 | 2020-07-27 | North America/USA/Washington/King C | Seattle Flu Study | Seattle Flu Study | Deborah A. Nickerson et al |
| EPI_ISL_525750 | USA/WA-S2760/2020 | 2020-07-29 | North America/USA/Washington/King C | Seattle Flu Study | Seattle Flu Study | Deborah A. Nickerson et al |

| | | | | | | |
|---|---|---|---|---|---|---|
| EPI_ISL_525752 | USA/WA-S2762/2020 | 2020-07-29 | North America/USA/Washington/King C | Seattle Flu Study | Seattle Flu Study | Deborah A. Nickerson et al |
| EPI_ISL_525753 | USA/WA-S2763/2020 | 2020-07-30 | North America/USA/Washington/King C | Seattle Flu Study | Seattle Flu Study | Deborah A. Nickerson et al |
| EPI_ISL_525759 | USA/AK-PHL549/2020 | 2020-06-20 | North America/USA/Alaska/ | Alaska State Virology Laboratory | Alaska State Virology Laboratory | Jack Chen et al |
| EPI_ISL_525764 | USA/TX-DSHS-0201/2020 | 2020-03-16 | North America/USA/Texas/Williamson C | Texas Department of State Health Services | Texas Department of State Health Services | Jenny Zhang et al |
| EPI_ISL_525768 | USA/TX-DSHS-0206/2020 | 2020-03-20 | North America/USA/Texas/Webb County | Texas Department of State Health Services | Texas Department of State Health Services | Jenny Zhang et al |
| EPI_ISL_525782 | USA/TX-DSHS-0221/2020 | 2020-04-07 | North America/USA/Texas/Hidalgo Coun | Texas Department of State Health Services | Texas Department of State Health Services | Jenny Zhang et al |
| EPI_ISL_525787 | USA/TX-DSHS-0235/2020 | 2020-04-25 | North America/USA/Texas/Washington | Texas Department of State Health Services | Texas Department of State Health Services | Jenny Zhang et al |
| EPI_ISL_525806 | USA/ID-OHSU-1977/2020 | 2020-07-30 | North America/USA/Idaho/Madison Cou | OHSU Lab Services Molecular Microbiology Lab | Oregon SARS-CoV-2 Genome Sequencing Center | Brendan L. O'Connell et al |
| EPI_ISL_525807 | USA/OR-OHSU-0350/2020 | 2020-05-16 | North America/USA/Oregon/ | OHSU Lab Services Molecular Microbiology Lab | Oregon SARS-CoV-2 Genome Sequencing Center | Brendan L. O'Connell et al |
| EPI_ISL_525812 | USA/OR-OHSU-0454/2020 | 2020-06-09 | North America/USA/Oregon/Multnomal | OHSU Lab Services Molecular Microbiology Lab | Ginkgo Bioworks Clinical Laboratory | Brendan L. O'Connell et al |
| EPI_ISL_525817 | USA/OR-OHSU-0468/2020 | 2020-06-10 | North America/USA/Oregon/Multnomal | OHSU Lab Services Molecular Microbiology Lab | Ginkgo Bioworks Clinical Laboratory | Brendan L. O'Connell et al |
| EPI_ISL_525822 | USA/OR-OHSU-0478/2020 | 2020-06-11 | North America/USA/Oregon/Washingto | OHSU Lab Services Molecular Microbiology Lab | Ginkgo Bioworks Clinical Laboratory | Brendan L. O'Connell et al |
| EPI_ISL_525828 | USA/OR-OHSU-0510/2020 | 2020-06-05 | North America/USA/Oregon/Clackamas | OHSU Lab Services Molecular Microbiology Lab | Ginkgo Bioworks Clinical Laboratory | Brendan L. O'Connell et al |
| EPI_ISL_525830 | USA/OR-OHSU-0516/2020 | 2020-06-05 | North America/USA/Oregon/Yamhill Co | OHSU Lab Services Molecular Microbiology Lab | Ginkgo Bioworks Clinical Laboratory | Brendan L. O'Connell et al |

| | | | | | | |
|---|---|---|---|---|---|---|
| EPI_ISL_525832 | USA/OR-OHSU-0528/2020 | 2020-06-15 | North America/USA/Oregon/Washingto | OHSU Lab Services Molecular Microbiology Lab | Ginkgo Bioworks Clinical Laboratory | Brendan L. O'Connell et al |
| EPI_ISL_525839 | USA/OR-OHSU-0544/2020 | 2020-06-16 | North America/USA/Oregon/Multnomal | OHSU Lab Services Molecular Microbiology Lab | Ginkgo Bioworks Clinical Laboratory | Brendan L. O'Connell et al |
| EPI_ISL_525853 | USA/OR-OHSU-0622/2020 | 2020-06-13 | North America/USA/Oregon/Multnomal | OHSU Lab Services Molecular Microbiology Lab | Ginkgo Bioworks Clinical Laboratory | Brendan L. O'Connell et al |
| EPI_ISL_525855 | USA/OR-OHSU-0626/2020 | 2020-06-13 | North America/USA/Oregon/Washingto | OHSU Lab Services Molecular Microbiology Lab | Ginkgo Bioworks Clinical Laboratory | Brendan L. O'Connell et al |
| EPI_ISL_525869 | USA/OR-OHSU-0648/2020 | 2020-06-16 | North America/USA/Oregon/Multnomal | OHSU Lab Services Molecular Microbiology Lab | Ginkgo Bioworks Clinical Laboratory | Brendan L. O'Connell et al |
| EPI_ISL_525873 | USA/OR-OHSU-0660/2020 | 2020-06-15 | North America/USA/Oregon/Clackamas | OHSU Lab Services Molecular Microbiology Lab | Ginkgo Bioworks Clinical Laboratory | Brendan L. O'Connell et al |
| EPI_ISL_525883 | USA/OR-OHSU-0675/2020 | 2020-06-17 | North America/USA/Oregon/Multnomal | OHSU Lab Services Molecular Microbiology Lab | Oregon SARS-CoV-2 Genome Sequencing Center | Brendan L. O'Connell et al |
| EPI_ISL_525894 | USA/OR-OHSU-0767/2020 | 2020-06-24 | North America/USA/Oregon/Multnomal | OHSU Lab Services Molecular Microbiology Lab | Oregon SARS-CoV-2 Genome Sequencing Center | Brendan L. O'Connell et al |
| EPI_ISL_525902 | USA/OR-OHSU-0801/2020 | 2020-06-22 | North America/USA/Oregon/Multnomal | OHSU Lab Services Molecular Microbiology Lab | Oregon SARS-CoV-2 Genome Sequencing Center | Brendan L. O'Connell et al |
| EPI_ISL_525916 | USA/OR-OHSU-0869/2020 | 2020-06-29 | North America/USA/Oregon/Washingto | OHSU Lab Services Molecular Microbiology Lab | Oregon SARS-CoV-2 Genome Sequencing Center | Brendan L. O'Connell et al |
| EPI_ISL_525917 | USA/OR-OHSU-0871/2020 | 2020-06-29 | North America/USA/Oregon/Multnomal | OHSU Lab Services Molecular Microbiology Lab | Oregon SARS-CoV-2 Genome Sequencing Center | Brendan L. O'Connell et al |

| | | | | | | |
|---|---|---|---|---|---|---|
| EPI_ISL_525923 | USA/OR-OHSU-0901/2020 | 2020-06-30 | North America/USA/Oregon/Washingto | OHSU Lab Services Molecular Microbiology Lab | Oregon SARS-CoV-2 Genome Sequencing Center | Brendan L. O'Connell et al |
| EPI_ISL_525930 | USA/OR-OHSU-0929/2020 | 2020-06-30 | North America/USA/Oregon/Multnomal | OHSU Lab Services Molecular Microbiology Lab | Oregon SARS-CoV-2 Genome Sequencing Center | Brendan L. O'Connell et al |
| EPI_ISL_525935 | USA/OR-OHSU-0953/2020 | 2020-07-03 | North America/USA/Oregon/Marion Co | OHSU Lab Services Molecular Microbiology Lab | Oregon SARS-CoV-2 Genome Sequencing Center | Brendan L. O'Connell et al |
| EPI_ISL_525944 | USA/OR-OHSU-0993/2020 | 2020-07-07 | North America/USA/Oregon/ | OHSU Lab Services Molecular Microbiology Lab | Oregon SARS-CoV-2 Genome Sequencing Center | Brendan L. O'Connell et al |
| EPI_ISL_525945 | USA/OR-OHSU-1003/2020 | 2020-07-07 | North America/USA/Oregon/Multnomal | OHSU Lab Services Molecular Microbiology Lab | Oregon SARS-CoV-2 Genome Sequencing Center | Brendan L. O'Connell et al |
| EPI_ISL_525952 | USA/OR-OHSU-1027/2020 | 2020-07-07 | North America/USA/Oregon/Wasco Cou | OHSU Lab Services Molecular Microbiology Lab | Oregon SARS-CoV-2 Genome Sequencing Center | Brendan L. O'Connell et al |
| EPI_ISL_525953 | USA/OR-OHSU-1029/2020 | 2020-07-07 | North America/USA/Oregon/Wasco Cou | OHSU Lab Services Molecular Microbiology Lab | Oregon SARS-CoV-2 Genome Sequencing Center | Brendan L. O'Connell et al |
| EPI_ISL_525955 | USA/OR-OHSU-1035/2020 | 2020-07-07 | North America/USA/Oregon/Washingto | OHSU Lab Services Molecular Microbiology Lab | Oregon SARS-CoV-2 Genome Sequencing Center | Brendan L. O'Connell et al |
| EPI_ISL_525957 | USA/OR-OHSU-1043/2020 | 2020-07-08 | North America/USA/Oregon/Multnomal | OHSU Lab Services Molecular Microbiology Lab | Oregon SARS-CoV-2 Genome Sequencing Center | Brendan L. O'Connell et al |
| EPI_ISL_525958 | USA/OR-OHSU-1047/2020 | 2020-07-08 | North America/USA/Oregon/Multnomal | OHSU Lab Services Molecular Microbiology Lab | Oregon SARS-CoV-2 Genome Sequencing Center | Brendan L. O'Connell et al |

| | | | | | | |
|---|---|---|---|---|---|---|
| EPI_ISL_525961 | USA/OR-OHSU-1065/2020 | 2020-07-09 | North America/USA/Oregon/Multnomal | OHSU Lab Services Molecular Microbiology Lab | Oregon SARS-CoV-2 Genome Sequencing Center | Brendan L. O'Connell et al |
| EPI_ISL_525963 | USA/OR-OHSU-1071/2020 | 2020-07-09 | North America/USA/Oregon/Washingto | OHSU Lab Services Molecular Microbiology Lab | Oregon SARS-CoV-2 Genome Sequencing Center | Brendan L. O'Connell et al |
| EPI_ISL_525965 | USA/OR-OHSU-1079/2020 | 2020-07-09 | North America/USA/Oregon/Washingto | OHSU Lab Services Molecular Microbiology Lab | Oregon SARS-CoV-2 Genome Sequencing Center | Brendan L. O'Connell et al |
| EPI_ISL_525968 | USA/OR-OHSU-1099/2020 | 2020-06-25 | North America/USA/Oregon/Washingto | OHSU Lab Services Molecular Microbiology Lab | Oregon SARS-CoV-2 Genome Sequencing Center | Brendan L. O'Connell et al |
| EPI_ISL_525991 | USA/OR-OHSU-1643/2020 | 2020-07-22 | North America/USA/Oregon/Multnomal | OHSU Lab Services Molecular Microbiology Lab | Oregon SARS-CoV-2 Genome Sequencing Center | Brendan L. O'Connell et al |
| EPI_ISL_525995 | USA/OR-OHSU-1655/2020 | 2020-07-22 | North America/USA/Oregon/Washingto | OHSU Lab Services Molecular Microbiology Lab | Oregon SARS-CoV-2 Genome Sequencing Center | Brendan L. O'Connell et al |
| EPI_ISL_525998 | USA/OR-OHSU-1673/2020 | 2020-07-22 | North America/USA/Oregon/Multnomal | OHSU Lab Services Molecular Microbiology Lab | Oregon SARS-CoV-2 Genome Sequencing Center | Brendan L. O'Connell et al |
| EPI_ISL_526004 | USA/OR-OHSU-1689/2020 | 2020-07-23 | North America/USA/Oregon/Clackamas | OHSU Lab Services Molecular Microbiology Lab | Oregon SARS-CoV-2 Genome Sequencing Center | Brendan L. O'Connell et al |
| EPI_ISL_526007 | USA/OR-OHSU-1701/2020 | 2020-07-23 | North America/USA/Oregon/Multnomal | OHSU Lab Services Molecular Microbiology Lab | Oregon SARS-CoV-2 Genome Sequencing Center | Brendan L. O'Connell et al |
| EPI_ISL_526008 | USA/OR-OHSU-1703/2020 | 2020-07-23 | North America/USA/Oregon/Marion Co | OHSU Lab Services Molecular Microbiology Lab | Oregon SARS-CoV-2 Genome Sequencing Center | Brendan L. O'Connell et al |

| | | | | | | |
|---|---|---|---|---|---|---|
| EPI_ISL_526010 | USA/OR-OHSU-1707/2020 | 2020-07-23 | North America/USA/Oregon/Washingto | OHSU Lab Services Molecular Microbiology Lab | Oregon SARS-CoV-2 Genome Sequencing Center | Brendan L. O'Connell et al |
| EPI_ISL_526021 | USA/OR-OHSU-1757/2020 | 2020-07-24 | North America/USA/Oregon/Multnomal | OHSU Lab Services Molecular Microbiology Lab | Oregon SARS-CoV-2 Genome Sequencing Center | Brendan L. O'Connell et al |
| EPI_ISL_526029 | USA/OR-OHSU-1791/2020 | 2020-07-25 | North America/USA/Oregon/Washingto | OHSU Lab Services Molecular Microbiology Lab | Oregon SARS-CoV-2 Genome Sequencing Center | Brendan L. O'Connell et al |
| EPI_ISL_526032 | USA/OR-OHSU-1807/2020 | 2020-07-27 | North America/USA/Oregon/Multnomal | OHSU Lab Services Molecular Microbiology Lab | Oregon SARS-CoV-2 Genome Sequencing Center | Brendan L. O'Connell et al |
| EPI_ISL_526041 | USA/OR-OHSU-1833/2020 | 2020-07-27 | North America/USA/Oregon/Washingto | OHSU Lab Services Molecular Microbiology Lab | Oregon SARS-CoV-2 Genome Sequencing Center | Brendan L. O'Connell et al |
| EPI_ISL_526044 | USA/OR-OHSU-1839/2020 | 2020-07-27 | North America/USA/Oregon/Multnomal | OHSU Lab Services Molecular Microbiology Lab | Oregon SARS-CoV-2 Genome Sequencing Center | Brendan L. O'Connell et al |
| EPI_ISL_526046 | USA/OR-OHSU-1847/2020 | 2020-07-27 | North America/USA/Oregon/Clackamas | OHSU Lab Services Molecular Microbiology Lab | Oregon SARS-CoV-2 Genome Sequencing Center | Brendan L. O'Connell et al |
| EPI_ISL_526050 | USA/OR-OHSU-1859/2020 | 2020-07-27 | North America/USA/Oregon/Washingto | OHSU Lab Services Molecular Microbiology Lab | Oregon SARS-CoV-2 Genome Sequencing Center | Brendan L. O'Connell et al |
| EPI_ISL_526054 | USA/OR-OHSU-1873/2020 | 2020-07-28 | North America/USA/Oregon/Clackamas | OHSU Lab Services Molecular Microbiology Lab | Oregon SARS-CoV-2 Genome Sequencing Center | Brendan L. O'Connell et al |
| EPI_ISL_526056 | USA/OR-OHSU-1877/2020 | 2020-07-28 | North America/USA/Oregon/Multnomal | OHSU Lab Services Molecular Microbiology Lab | Oregon SARS-CoV-2 Genome Sequencing Center | Brendan L. O'Connell et al |

| | | | | | | |
|---|---|---|---|---|---|---|
| EPI_ISL_526058 | USA/OR-OHSU-1881/2020 | 2020-07-28 | North America/USA/Oregon/Washingto | OHSU Lab Services Molecular Microbiology Lab | Oregon SARS-CoV-2 Genome Sequencing Center | Brendan L. O'Connell et al |
| EPI_ISL_526060 | USA/OR-OHSU-1889/2020 | 2020-07-28 | North America/USA/Oregon/Washingto | OHSU Lab Services Molecular Microbiology Lab | Oregon SARS-CoV-2 Genome Sequencing Center | Brendan L. O'Connell et al |
| EPI_ISL_526066 | USA/OR-OHSU-1907/2020 | 2020-07-28 | North America/USA/Oregon/Multnomal | OHSU Lab Services Molecular Microbiology Lab | Oregon SARS-CoV-2 Genome Sequencing Center | Brendan L. O'Connell et al |
| EPI_ISL_526081 | USA/OR-OHSU-1979/2020 | 2020-07-30 | North America/USA/Oregon/Multnomal | OHSU Lab Services Molecular Microbiology Lab | Oregon SARS-CoV-2 Genome Sequencing Center | Brendan L. O'Connell et al |
| EPI_ISL_526084 | USA/OR-OHSU-1989/2020 | 2020-07-31 | North America/USA/Oregon/Clackamas | OHSU Lab Services Molecular Microbiology Lab | Oregon SARS-CoV-2 Genome Sequencing Center | Brendan L. O'Connell et al |
| EPI_ISL_526092 | USA/WA-OHSU-0506/2020 | 2020-06-05 | North America/USA/Washington/Clark ( | OHSU Lab Services Molecular Microbiology Lab | Ginkgo Bioworks Clinical Laboratory | Brendan L. O'Connell et al |
| EPI_ISL_526095 | USA/WA-OHSU-0515/2020 | 2020-06-05 | North America/USA/Washington/Clark ( | OHSU Lab Services Molecular Microbiology Lab | Oregon SARS-CoV-2 Genome Sequencing Center | Brendan L. O'Connell et al |
| EPI_ISL_526102 | USA/WA-OHSU-0987/2020 | 2020-07-06 | North America/USA/Washington/Clark ( | OHSU Lab Services Molecular Microbiology Lab | Oregon SARS-CoV-2 Genome Sequencing Center | Brendan L. O'Connell et al |
| EPI_ISL_526103 | USA/WA-OHSU-1571/2020 | 2020-07-20 | North America/USA/Washington/Clark ( | OHSU Lab Services Molecular Microbiology Lab | Oregon SARS-CoV-2 Genome Sequencing Center | Brendan L. O'Connell et al |
| EPI_ISL_526109 | USA/WA-OHSU-1831/2020 | 2020-07-27 | North America/USA/Washington/Clark ( | OHSU Lab Services Molecular Microbiology Lab | Oregon SARS-CoV-2 Genome Sequencing Center | Brendan L. O'Connell et al |

| | | | | | | |
|---|---|---|---|---|---|---|
| EPI_ISL_526223 | Hungary/MH-8268/2020 | 2020-05-25 | Europe/Hungary/Pest County/ | Hungarian Defence Forces Military Medical Centre | National Laboratory of Virology, Szentv°gothai Research Centre | Endre Gv°bor Tv≥th et al |
| EPI_ISL_526224 | Hungary/MH-9653/2020 | 2020-06-04 | Europe/Hungary/Pest County/ | Hungarian Defence Forces Military Medical Centre | National Laboratory of Virology, Szentv°gothai Research Centre | Endre Gv°bor Tv≥th et al |
| EPI_ISL_526225 | Hungary/MH-13211/2020 | 2020-07-06 | Europe/Hungary/Pest County/ | Hungarian Defence Forces Military Medical Centre | National Laboratory of Virology, Szentv°gothai Research Centre | Endre Gv°bor Tv≥th et al |
| EPI_ISL_526253 | Canada/ON-UHTC_0095/2020 | 2020-05-10 | North America/Canada/Ontario/ | Unity Health Toronto | Ontario Institute for Cancer Research | Ramzi Fattouh et al |
| EPI_ISL_526254 | Canada/ON-UHTC_0096/2020 | 2020-05-10 | North America/Canada/Ontario/ | Unity Health Toronto | Ontario Institute for Cancer Research | Ramzi Fattouh et al |
| EPI_ISL_526255 | Canada/ON-UHTC_0097/2020 | 2020-05-10 | North America/Canada/Ontario/ | Unity Health Toronto | Ontario Institute for Cancer Research | Ramzi Fattouh et al |
| EPI_ISL_526256 | Canada/ON-UHTC_0099/2020 | 2020-05-10 | North America/Canada/Ontario/ | Unity Health Toronto | Ontario Institute for Cancer Research | Ramzi Fattouh et al |
| EPI_ISL_526257 | Canada/ON-UHTC_0101/2020 | 2020-05-10 | North America/Canada/Ontario/ | Unity Health Toronto | Ontario Institute for Cancer Research | Ramzi Fattouh et al |
| EPI_ISL_526258 | Canada/ON-UHTC_0102/2020 | 2020-05-11 | North America/Canada/Ontario/ | Unity Health Toronto | Ontario Institute for Cancer Research | Ramzi Fattouh et al |
| EPI_ISL_526260 | Canada/ON-UHTC_0105/2020 | 2020-05-15 | North America/Canada/Ontario/ | Unity Health Toronto | Ontario Institute for Cancer Research | Ramzi Fattouh et al |
| EPI_ISL_526261 | Canada/ON-UHTC_0111/2020 | 2020-05-20 | North America/Canada/Ontario/ | Unity Health Toronto | Ontario Institute for Cancer Research | Ramzi Fattouh et al |

| | | | | | | |
|---|---|---|---|---|---|---|
| EPI_ISL_526262 | Canada/ON-UHTC_0112/2020 | 2020-05-20 | North America/Canada/Ontario/ | Unity Health Toronto | Ontario Institute for Cancer Research | Ramzi Fattouh et al |
| EPI_ISL_526263 | Canada/ON-UHTC_0115/2020 | 2020-05-20 | North America/Canada/Ontario/ | Unity Health Toronto | Ontario Institute for Cancer Research | Ramzi Fattouh et al |
| EPI_ISL_526264 | Canada/ON-UHTC_0116/2020 | 2020-05-20 | North America/Canada/Ontario/ | Unity Health Toronto | Ontario Institute for Cancer Research | Ramzi Fattouh et al |
| EPI_ISL_526265 | Canada/ON-UHTC_0117/2020 | 2020-05-20 | North America/Canada/Ontario/ | Unity Health Toronto | Ontario Institute for Cancer Research | Ramzi Fattouh et al |
| EPI_ISL_526266 | Canada/ON-UHTC_0120/2020 | 2020-05-18 | North America/Canada/Ontario/ | Unity Health Toronto | Ontario Institute for Cancer Research | Ramzi Fattouh et al |
| EPI_ISL_526267 | Canada/ON-UHTC_0122/2020 | 2020-05-20 | North America/Canada/Ontario/ | Unity Health Toronto | Ontario Institute for Cancer Research | Ramzi Fattouh et al |
| EPI_ISL_526268 | Canada/ON-UHTC_0123/2020 | 2020-05-20 | North America/Canada/Ontario/ | Unity Health Toronto | Ontario Institute for Cancer Research | Ramzi Fattouh et al |
| EPI_ISL_526269 | Canada/ON-UHTC_0124/2020 | 2020-05-20 | North America/Canada/Ontario/ | Unity Health Toronto | Ontario Institute for Cancer Research | Ramzi Fattouh et al |
| EPI_ISL_526270 | Canada/ON-UHTC_0125/2020 | 2020-05-20 | North America/Canada/Ontario/ | Unity Health Toronto | Ontario Institute for Cancer Research | Ramzi Fattouh et al |
| EPI_ISL_526271 | Canada/ON-UHTC_0129/2020 | 2020-05-21 | North America/Canada/Ontario/ | Unity Health Toronto | Ontario Institute for Cancer Research | Ramzi Fattouh et al |
| EPI_ISL_526272 | Canada/ON-UHTC_0132/2020 | 2020-05-22 | North America/Canada/Ontario/ | Unity Health Toronto | Ontario Institute for Cancer Research | Ramzi Fattouh et al |
| EPI_ISL_526273 | Canada/ON-UHTC_0133/2020 | 2020-05-23 | North America/Canada/Ontario/ | Unity Health Toronto | Ontario Institute for Cancer Research | Ramzi Fattouh et al |
| EPI_ISL_526274 | Canada/ON-UHTC_0137/2020 | 2020-05-25 | North America/Canada/Ontario/ | Unity Health Toronto | Ontario Institute for Cancer Research | Ramzi Fattouh et al |

| | | | | | | |
|---|---|---|---|---|---|---|
| EPI_ISL_526275 | Canada/ON-UHTC_0139/2020 | 2020-05-31 | North America/Canada/Ontario/ | Unity Health Toronto | Ontario Institute for Cancer Research | Ramzi Fattouh et al |
| EPI_ISL_526276 | Canada/ON-UHTC_0141/2020 | 2020-05-30 | North America/Canada/Ontario/ | Unity Health Toronto | Ontario Institute for Cancer Research | Ramzi Fattouh et al |
| EPI_ISL_526277 | Canada/ON-UHTC_0143/2020 | 2020-05-30 | North America/Canada/Ontario/ | Unity Health Toronto | Ontario Institute for Cancer Research | Ramzi Fattouh et al |
| EPI_ISL_526278 | Canada/ON-UHTC_0144/2020 | 2020-05-31 | North America/Canada/Ontario/ | Unity Health Toronto | Ontario Institute for Cancer Research | Ramzi Fattouh et al |
| EPI_ISL_526279 | Canada/ON-UHTC_0145/2020 | 2020-05-31 | North America/Canada/Ontario/ | Unity Health Toronto | Ontario Institute for Cancer Research | Ramzi Fattouh et al |
| EPI_ISL_526280 | Canada/ON-UHTC_0146/2020 | 2020-05-31 | North America/Canada/Ontario/ | Unity Health Toronto | Ontario Institute for Cancer Research | Ramzi Fattouh et al |
| EPI_ISL_526281 | Canada/ON-UHTC_0147/2020 | 2020-06-09 | North America/Canada/Ontario/ | Unity Health Toronto | Ontario Institute for Cancer Research | Ramzi Fattouh et al |
| EPI_ISL_526282 | Canada/ON-UHTC_0149/2020 | 2020-06-09 | North America/Canada/Ontario/ | Unity Health Toronto | Ontario Institute for Cancer Research | Ramzi Fattouh et al |
| EPI_ISL_526283 | Canada/ON-UHTC_0150/2020 | 2020-06-09 | North America/Canada/Ontario/ | Unity Health Toronto | Ontario Institute for Cancer Research | Ramzi Fattouh et al |
| EPI_ISL_526284 | Canada/ON-UHTC_0151/2020 | 2020-06-09 | North America/Canada/Ontario/ | Unity Health Toronto | Ontario Institute for Cancer Research | Ramzi Fattouh et al |
| EPI_ISL_526285 | Canada/ON-UHTC_0159/2020 | 2020-06-20 | North America/Canada/Ontario/ | Unity Health Toronto | Ontario Institute for Cancer Research | Ramzi Fattouh et al |
| EPI_ISL_526286 | Canada/ON-UHTC_0160/2020 | 2020-06-20 | North America/Canada/Ontario/ | Unity Health Toronto | Ontario Institute for Cancer Research | Ramzi Fattouh et al |
| EPI_ISL_526476 | Scotland/EDB4060/2020 | 2020-04-15 | Europe/United Kingdom/Scotland/ | Virology Department, Royal Infirmary of Edinburgh, NHS Lothian / School of Biological Sciences, University of Edinburgh / Institute of Genetics and Molecular Medicine, University of Edinburgh | COVID-19 Genomics UK (COG-UK) Consortium | McHugh M et al |

| | | | | | | |
|---|---|---|---|---|---|---|
| EPI_ISL_526510 | Scotland/EDB6904/2020 | 2020-05-01 | Europe/United Kingdom/Scotland/ | Virology Department, Royal Infirmary of Edinburgh, NHS Lothian / School of Biological Sciences, University of Edinburgh / Institute of Genetics and Molecular Medicine, University of Edinburgh | COVID-19 Genomics UK (COG-UK) Consortium | McHugh M et al |
| EPI_ISL_526554 | USA/FL-BPHL-1068/2020 | 2020-04-21 | North America/USA/Florida/ | Florida Bureau of Public Health Laboratories | Florida Bureau of Public Health Laboratories | Sarah Schmedes et al |
| EPI_ISL_526555 | USA/FL-BPHL-1069/2020 | 2020-04-22 | North America/USA/Florida/ | Florida Bureau of Public Health Laboratories | Florida Bureau of Public Health Laboratories | Sarah Schmedes et al |
| EPI_ISL_526563 | USA/FL-BPHL-1077/2020 | 2020-04-26 | North America/USA/Florida/ | Florida Bureau of Public Health Laboratories | Florida Bureau of Public Health Laboratories | Sarah Schmedes et al |
| EPI_ISL_526565 | USA/FL-BPHL-1079/2020 | 2020-05-15 | North America/USA/Florida/ | Florida Bureau of Public Health Laboratories | Florida Bureau of Public Health Laboratories | Sarah Schmedes et al |
| EPI_ISL_526567 | USA/FL-BPHL-1081/2020 | 2020-05-18 | North America/USA/Florida/ | Florida Bureau of Public Health Laboratories | Florida Bureau of Public Health Laboratories | Sarah Schmedes et al |
| EPI_ISL_526569 | USA/FL-BPHL-1083/2020 | 2020-05-21 | North America/USA/Florida/ | Florida Bureau of Public Health Laboratories | Florida Bureau of Public Health Laboratories | Sarah Schmedes et al |
| EPI_ISL_526573 | USA/FL-BPHL-1087/2020 | 2020-05-25 | North America/USA/Florida/ | Florida Bureau of Public Health Laboratories | Florida Bureau of Public Health Laboratories | Sarah Schmedes et al |
| EPI_ISL_526589 | USA/FL-BPHL-1103/2020 | 2020-08-03 | North America/USA/Florida/ | Florida Bureau of Public Health Laboratories | Florida Bureau of Public Health Laboratories | Sarah Schmedes et al |
| EPI_ISL_526591 | USA/FL-BPHL-1105/2020 | 2020-08-03 | North America/USA/Florida/ | Florida Bureau of Public Health Laboratories | Florida Bureau of Public Health Laboratories | Sarah Schmedes et al |
| EPI_ISL_526595 | USA/FL-BPHL-1109/2020 | 2020-08-05 | North America/USA/Florida/ | Florida Bureau of Public Health Laboratories | Florida Bureau of Public Health Laboratories | Sarah Schmedes et al |
| EPI_ISL_526599 | USA/FL-BPHL-1113/2020 | 2020-08-05 | North America/USA/Florida/ | Florida Bureau of Public Health Laboratories | Florida Bureau of Public Health Laboratories | Sarah Schmedes et al |
| EPI_ISL_526610 | USA/FL-BPHL-1124/2020 | 2020-08-11 | North America/USA/Florida/ | Florida Bureau of Public Health Laboratories | Florida Bureau of Public Health Laboratories | Sarah Schmedes et al |

| | | | | | | |
|---|---|---|---|---|---|---|
| EPI_ISL_526615 | USA/FL-BPHL-1129/2020 | 2020-08-14 | North America/USA/Florida/ | Florida Bureau of Public Health Laboratories | Florida Bureau of Public Health Laboratories | Sarah Schmedes et al |
| EPI_ISL_526616 | USA/FL-BPHL-1130/2020 | 2020-08-14 | North America/USA/Florida/ | Florida Bureau of Public Health Laboratories | Florida Bureau of Public Health Laboratories | Sarah Schmedes et al |
| EPI_ISL_526619 | USA/FL-BPHL-1133/2020 | 2020-08-14 | North America/USA/Florida/ | Florida Bureau of Public Health Laboratories | Florida Bureau of Public Health Laboratories | Sarah Schmedes et al |
| EPI_ISL_526621 | USA/FL-BPHL-1135/2020 | 2020-08-15 | North America/USA/Florida/ | Florida Bureau of Public Health Laboratories | Florida Bureau of Public Health Laboratories | Sarah Schmedes et al |
| EPI_ISL_526622 | USA/FL-BPHL-1136/2020 | 2020-08-16 | North America/USA/Florida/ | Florida Bureau of Public Health Laboratories | Florida Bureau of Public Health Laboratories | Sarah Schmedes et al |
| EPI_ISL_526623 | USA/FL-BPHL-1137/2020 | 2020-08-17 | North America/USA/Florida/ | Florida Bureau of Public Health Laboratories | Florida Bureau of Public Health Laboratories | Sarah Schmedes et al |
| EPI_ISL_526627 | USA/FL-BPHL-1141/2020 | 2020-08-17 | North America/USA/Florida/ | Florida Bureau of Public Health Laboratories | Florida Bureau of Public Health Laboratories | Sarah Schmedes et al |
| EPI_ISL_526628 | USA/FL-BPHL-1142/2020 | 2020-08-17 | North America/USA/Florida/ | Florida Bureau of Public Health Laboratories | Florida Bureau of Public Health Laboratories | Sarah Schmedes et al |
| EPI_ISL_526629 | USA/FL-BPHL-1143/2020 | 2020-08-17 | North America/USA/Florida/ | Florida Bureau of Public Health Laboratories | Florida Bureau of Public Health Laboratories | Sarah Schmedes et al |
| EPI_ISL_526630 | USA/FL-BPHL-1144/2020 | 2020-08-17 | North America/USA/Florida/ | Florida Bureau of Public Health Laboratories | Florida Bureau of Public Health Laboratories | Sarah Schmedes et al |
| EPI_ISL_526637 | USA/FL-BPHL-1151/2020 | 2020-08-18 | North America/USA/Florida/ | Florida Bureau of Public Health Laboratories | Florida Bureau of Public Health Laboratories | Sarah Schmedes et al |
| EPI_ISL_526640 | USA/FL-BPHL-1154/2020 | 2020-08-18 | North America/USA/Florida/ | Florida Bureau of Public Health Laboratories | Florida Bureau of Public Health Laboratories | Sarah Schmedes et al |
| EPI_ISL_526641 | USA/FL-BPHL-1155/2020 | 2020-08-18 | North America/USA/Florida/ | Florida Bureau of Public Health Laboratories | Florida Bureau of Public Health Laboratories | Sarah Schmedes et al |

| | | | | | | |
|---|---|---|---|---|---|---|
| EPI_ISL_526643 | USA/FL-BPHL-1157/2020 | 2020-08-18 | North America/USA/Florida/ | Florida Bureau of Public Health Laboratories | Florida Bureau of Public Health Laboratories | Sarah Schmedes et al |
| EPI_ISL_526649 | USA/FL-BPHL-1163/2020 | 2020-08-19 | North America/USA/Florida/ | Florida Bureau of Public Health Laboratories | Florida Bureau of Public Health Laboratories | Sarah Schmedes et al |
| EPI_ISL_526651 | USA/FL-BPHL-1165/2020 | 2020-08-19 | North America/USA/Florida/ | Florida Bureau of Public Health Laboratories | Florida Bureau of Public Health Laboratories | Sarah Schmedes et al |
| EPI_ISL_526655 | USA/FL-BPHL-1169/2020 | 2020-08-19 | North America/USA/Florida/ | Florida Bureau of Public Health Laboratories | Florida Bureau of Public Health Laboratories | Sarah Schmedes et al |
| EPI_ISL_526658 | USA/FL-BPHL-1172/2020 | 2020-08-19 | North America/USA/Florida/ | Florida Bureau of Public Health Laboratories | Florida Bureau of Public Health Laboratories | Sarah Schmedes et al |
| EPI_ISL_526659 | USA/FL-BPHL-1173/2020 | 2020-08-18 | North America/USA/Florida/ | Florida Bureau of Public Health Laboratories | Florida Bureau of Public Health Laboratories | Sarah Schmedes et al |
| EPI_ISL_526660 | USA/FL-BPHL-1174/2020 | 2020-08-20 | North America/USA/Florida/ | Florida Bureau of Public Health Laboratories | Florida Bureau of Public Health Laboratories | Sarah Schmedes et al |
| EPI_ISL_526661 | USA/FL-BPHL-1175/2020 | 2020-08-19 | North America/USA/Florida/ | Florida Bureau of Public Health Laboratories | Florida Bureau of Public Health Laboratories | Sarah Schmedes et al |
| EPI_ISL_526662 | USA/FL-BPHL-1176/2020 | 2020-08-20 | North America/USA/Florida/ | Florida Bureau of Public Health Laboratories | Florida Bureau of Public Health Laboratories | Sarah Schmedes et al |
| EPI_ISL_526663 | USA/FL-BPHL-1177/2020 | 2020-08-19 | North America/USA/Florida/ | Florida Bureau of Public Health Laboratories | Florida Bureau of Public Health Laboratories | Sarah Schmedes et al |
| EPI_ISL_526665 | USA/FL-BPHL-1179/2020 | 2020-08-20 | North America/USA/Florida/ | Florida Bureau of Public Health Laboratories | Florida Bureau of Public Health Laboratories | Sarah Schmedes et al |
| EPI_ISL_526667 | USA/FL-BPHL-1181/2020 | 2020-08-20 | North America/USA/Florida/ | Florida Bureau of Public Health Laboratories | Florida Bureau of Public Health Laboratories | Sarah Schmedes et al |
| EPI_ISL_526674 | USA/FL-BPHL-1188/2020 | 2020-08-20 | North America/USA/Florida/ | Florida Bureau of Public Health Laboratories | Florida Bureau of Public Health Laboratories | Sarah Schmedes et al |

| | | | | | | |
|---|---|---|---|---|---|---|
| EPI_ISL_526676 | USA/FL-BPHL-1190/2020 | 2020-08-20 | North America/USA/Florida/ | Florida Bureau of Public Health Laboratories | Florida Bureau of Public Health Laboratories | Sarah Schmedes et al |
| EPI_ISL_526677 | USA/FL-BPHL-1191/2020 | 2020-08-20 | North America/USA/Florida/ | Florida Bureau of Public Health Laboratories | Florida Bureau of Public Health Laboratories | Sarah Schmedes et al |
| EPI_ISL_526678 | USA/FL-BPHL-1192/2020 | 2020-08-21 | North America/USA/Florida/ | Florida Bureau of Public Health Laboratories | Florida Bureau of Public Health Laboratories | Sarah Schmedes et al |
| EPI_ISL_526682 | USA/FL-BPHL-1196/2020 | 2020-08-21 | North America/USA/Florida/ | Florida Bureau of Public Health Laboratories | Florida Bureau of Public Health Laboratories | Sarah Schmedes et al |
| EPI_ISL_526687 | USA/NY-MH001827/2020 | 2020-04-16 | North America/USA/New York/ | Faith Laboratory, Immunology Institute, Icahn School of Medicine at Mount Sinai | van Bakel Laboratory, Genetics and Genomics Sciences, Icahn School of Medicine at Mount Sinai | Graham J. Britton et al |
| EPI_ISL_526776 | USA/VA-DCLS-0468/2020 | 2020-05-06 | North America/USA/Virginia/ | Virginia DCLS | Virginia DCLS | Virginia DCLS et al |
| EPI_ISL_526785 | USA/VA-DCLS-0477/2020 | 2020-05-07 | North America/USA/Virginia/ | Virginia DCLS | Virginia DCLS | Virginia DCLS et al |
| EPI_ISL_526786 | USA/VA-DCLS-0478/2020 | 2020-05-07 | North America/USA/Virginia/ | Virginia DCLS | Virginia DCLS | Virginia DCLS et al |
| EPI_ISL_526788 | USA/VA-DCLS-0480/2020 | 2020-05-08 | North America/USA/Virginia/ | Virginia DCLS | Virginia DCLS | Virginia DCLS et al |
| EPI_ISL_526794 | USA/VA-DCLS-0487/2020 | 2020-05-11 | North America/USA/Virginia/ | Virginia DCLS | Virginia DCLS | Virginia DCLS et al |
| EPI_ISL_526796 | USA/VA-DCLS-0489/2020 | 2020-05-12 | North America/USA/Virginia/ | Virginia DCLS | Virginia DCLS | Virginia DCLS et al |
| EPI_ISL_526798 | USA/VA-DCLS-0577/2020 | 2020-03-17 | North America/USA/Virginia/ | Virginia DCLS | Virginia DCLS | Virginia DCLS et al |
| EPI_ISL_526831 | USA/VA-DCLS-0616/2020 | 2020-04-01 | North America/USA/Virginia/ | Virginia DCLS | Virginia DCLS | Virginia DCLS et al |
| EPI_ISL_526835 | USA/VA-DCLS-0622/2020 | 2020-04-01 | North America/USA/Virginia/ | Virginia DCLS | Virginia DCLS | Virginia DCLS et al |
| EPI_ISL_526842 | USA/VA-DCLS-0666/2020 | 2020-04-18 | North America/USA/Virginia/ | Virginia DCLS | Virginia DCLS | Virginia DCLS et al |
| EPI_ISL_526845 | USA/VA-DCLS-0722/2020 | 2020-07-06 | North America/USA/Virginia/ | Virginia DCLS | Virginia DCLS | Virginia DCLS et al |
| EPI_ISL_526846 | USA/VA-DCLS-0723/2020 | 2020-07-06 | North America/USA/Virginia/ | Virginia DCLS | Virginia DCLS | Virginia DCLS et al |
| EPI_ISL_526849 | USA/VA-DCLS-0726/2020 | 2020-07-09 | North America/USA/Virginia/ | Virginia DCLS | Virginia DCLS | Virginia DCLS et al |
| EPI_ISL_526852 | USA/VA-DCLS-0729/2020 | 2020-07-09 | North America/USA/Virginia/ | Virginia DCLS | Virginia DCLS | Virginia DCLS et al |
| EPI_ISL_526853 | USA/VA-DCLS-0731/2020 | 2020-07-08 | North America/USA/Virginia/ | Virginia DCLS | Virginia DCLS | Virginia DCLS et al |
| EPI_ISL_526855 | USA/VA-DCLS-0733/2020 | 2020-07-16 | North America/USA/Virginia/ | Virginia DCLS | Virginia DCLS | Virginia DCLS et al |

| | | | | | | |
|---|---|---|---|---|---|---|
| EPI_ISL_526856 | USA/VA-DCLS-0734/2020 | 2020-07-16 | North America/USA/Virginia/ | Virginia DCLS | Virginia DCLS | Virginia DCLS et al |
| EPI_ISL_526859 | USA/VA-DCLS-0738/2020 | 2020-07-03 | North America/USA/Virginia/ | Virginia DCLS | Virginia DCLS | Virginia DCLS et al |
| EPI_ISL_526861 | USA/VA-DCLS-0740/2020 | 2020-07-03 | North America/USA/Virginia/ | Virginia DCLS | Virginia DCLS | Virginia DCLS et al |
| EPI_ISL_526863 | USA/VA-DCLS-0743/2020 | 2020-06-12 | North America/USA/Virginia/ | Virginia DCLS | Virginia DCLS | Virginia DCLS et al |
| EPI_ISL_526867 | USA/VA-DCLS-0750/2020 | 2020-06-15 | North America/USA/Virginia/ | Virginia DCLS | Virginia DCLS | Virginia DCLS et al |
| EPI_ISL_526874 | USA/VA-DCLS-0758/2020 | 2020-07-02 | North America/USA/Virginia/ | Virginia DCLS | Virginia DCLS | Virginia DCLS et al |
| EPI_ISL_526875 | USA/VA-DCLS-0759/2020 | 2020-07-02 | North America/USA/Virginia/ | Virginia DCLS | Virginia DCLS | Virginia DCLS et al |
| EPI_ISL_526889 | USA/VA-DCLS-0775/2020 | 2020-07-01 | North America/USA/Virginia/ | Virginia DCLS | Virginia DCLS | Virginia DCLS et al |
| EPI_ISL_526896 | USA/VA-DCLS-0791/2020 | 2020-07-07 | North America/USA/Virginia/ | Virginia DCLS | Virginia DCLS | Virginia DCLS et al |
| EPI_ISL_526898 | USA/VA-DCLS-0794/2020 | 2020-07-07 | North America/USA/Virginia/ | Virginia DCLS | Virginia DCLS | Virginia DCLS et al |
| EPI_ISL_526900 | USA/VA-DCLS-0998/2020 | 2020-05-30 | North America/USA/Virginia/ | Virginia DCLS | Virginia DCLS | Virginia DCLS et al |
| EPI_ISL_526905 | USA/VA-DCLS-1014/2020 | 2020-06-03 | North America/USA/Virginia/ | Virginia DCLS | Virginia DCLS | Virginia DCLS et al |
| EPI_ISL_526919 | USA/VA-DCLS-1034/2020 | 2020-06-03 | North America/USA/Virginia/ | Virginia DCLS | Virginia DCLS | Virginia DCLS et al |
| EPI_ISL_526922 | USA/VA-DCLS-1038/2020 | 2020-06-03 | North America/USA/Virginia/ | Virginia DCLS | Virginia DCLS | Virginia DCLS et al |
| EPI_ISL_526931 | USA/VA-DCLS-1048/2020 | 2020-06-10 | North America/USA/Virginia/ | Virginia DCLS | Virginia DCLS | Virginia DCLS et al |
| EPI_ISL_526933 | Colombia/AMA-INS-105161/2020 | 2020-05-03 | South America/Colombia/Departamento | Instituto Nacional de Salud, Bogot√°, Colombia | Instituto Nacional de Salud, Bogot√°, Colombia | Katherine Laiton-Donato et al |
| EPI_ISL_526950 | Colombia/AMA-INS-105142/2020 | 2020-05-03 | South America/Colombia/Departamento | Instituto Nacional de Salud, Bogot√°, Colombia | Instituto Nacional de Salud, Bogot√°, Colombia | Katherine Laiton-Donato et al |
| EPI_ISL_526958 | Colombia/INS-CUN-AG2/2020 | 2020-07-17 | South America/Colombia/Cundinamarca | Instituto Nacional de Salud, Bogot√°, Colombia | Instituto Nacional de Salud, Bogot√°, Colombia | Katherine Laiton-Donato et al |
| EPI_ISL_526963 | Colombia/TOL-INS-0345-T20/2020 | 2020-04-14 | South America/Colombia/Tolima/Tolema | Instituto Nacional de Salud, Bogot√°, Colombia | Instituto Nacional de Salud, Bogot√°, Colombia | Katherine Laiton-Donato et al |
| EPI_ISL_526967 | Colombia/DC-INS-289/2020 | 2020-07-07 | South America/Colombia/Bogota/ | Instituto Nacional de Salud, Bogot√°, Colombia | Instituto Nacional de Salud, Bogot√°, Colombia | Katherine Laiton-Donato et al |
| EPI_ISL_526969 | Colombia/DC-INS-027/2020 | 2020-08-18 | South America/Colombia/Bogota/ | Instituto Nacional de Salud, Bogot√°, Colombia | Instituto Nacional de Salud, Bogot√°, Colombia | Katherine Laiton-Donato et al |
| EPI_ISL_526971 | Colombia/DC-INS-197/2020 | 2020-08-18 | South America/Colombia/Bogota/ | Instituto Nacional de Salud, Bogot√°, Colombia | Instituto Nacional de Salud, Bogot√°, Colombia | Katherine Laiton-Donato et al |

| | | | | | | |
|---|---|---|---|---|---|---|
| EPI_ISL_526974 | Colombia/DC-INS-05-07/2020 | 2020-06-17 | South America/Colombia/Bogota/ | Instituto Nacional de Salud, Bogot√°, Colombia | Instituto Nacional de Salud, Bogot√°, Colombia | Katherine Laiton-Donato et al |
| EPI_ISL_526976 | Egypt/EGY-021/2020 | 2020-08-14 | Africa/Egypt/Egypt/ | Biological prevention, army | Biological prevention, army | Seadawy et al |
| EPI_ISL_527382 | USA/PR-B2DR/2020 | 2020-08-04 | North America/USA/Puerto Rico/Ponce | University of Miami Immunology and Histocompatibility Laboratory | University of Miami Immunology and Histocompatibility Laboratory | Emilio Margolles-Clark et al |
| EPI_ISL_527406 | USA/CO-CSU-C0881-2/2020 | 2020-04-15 | North America/USA/Colorado/ | Colorado State University - Ebel Lab | Colorado State University - Ebel Lab | Greg Ebel et al. et al |
| EPI_ISL_527408 | USA/CO-CSU-C2673-1/2020 | 2020-04-02 | North America/USA/Colorado/ | Colorado State University - Ebel Lab | Colorado State University - Ebel Lab | Greg Ebel et al. et al |
| EPI_ISL_527424 | USA/CO-CSU-D0789-1/2020 | 2020-04-22 | North America/USA/Colorado/ | Colorado State University - Ebel Lab | Colorado State University - Ebel Lab | Greg Ebel et al. et al |
| EPI_ISL_527427 | USA/CO-CSU-D1050-2/2020 | 2020-05-06 | North America/USA/Colorado/ | Colorado State University - Ebel Lab | Colorado State University - Ebel Lab | Greg Ebel et al. et al |
| EPI_ISL_527431 | USA/CO-CSU-D1060-1/2020 | 2020-04-02 | North America/USA/Colorado/ | Colorado State University - Ebel Lab | Colorado State University - Ebel Lab | Greg Ebel et al. et al |
| EPI_ISL_527436 | USA/CO-CSU-D1085-1/2020 | 2020-04-02 | North America/USA/Colorado/ | Colorado State University - Ebel Lab | Colorado State University - Ebel Lab | Greg Ebel et al. et al |
| EPI_ISL_527449 | USA/CO-CSU-D1773/2020 | 2020-04-02 | North America/USA/Colorado/ | Colorado State University - Ebel Lab | Colorado State University - Ebel Lab | Greg Ebel et al. et al |
| EPI_ISL_527450 | USA/CO-CSU-D1862/2020 | 2020-04-02 | North America/USA/Colorado/ | Colorado State University - Ebel Lab | Colorado State University - Ebel Lab | Greg Ebel et al. et al |
| EPI_ISL_527452 | USA/CO-CSU-D1882-2/2020 | 2020-04-08 | North America/USA/Colorado/ | Colorado State University - Ebel Lab | Colorado State University - Ebel Lab | Greg Ebel et al. et al |
| EPI_ISL_527462 | USA/CO-CSU-D2282-3/2020 | 2020-04-15 | North America/USA/Colorado/ | Colorado State University - Ebel Lab | Colorado State University - Ebel Lab | Greg Ebel et al. et al |

| | | | | | | |
|---|---|---|---|---|---|---|
| EPI_ISL_527467 | USA/CO-CSU-D2575-2/2020 | 2020-05-06 | North America/USA/Colorado/ | Colorado State University - Ebel Lab | Colorado State University - Ebel Lab | Greg Ebel et al. et al |
| EPI_ISL_527476 | USA/CO-CSU-D3084-2/2020 | 2020-04-15 | North America/USA/Colorado/ | Colorado State University - Ebel Lab | Colorado State University - Ebel Lab | Greg Ebel et al. et al |
| EPI_ISL_527479 | USA/CO-CSU-E0462-1/2020 | 2020-05-06 | North America/USA/Colorado/ | Colorado State University - Ebel Lab | Colorado State University - Ebel Lab | Greg Ebel et al. et al |
| EPI_ISL_527482 | USA/CO-CSU-E0491/2020 | 2020-04-22 | North America/USA/Colorado/ | Colorado State University - Ebel Lab | Colorado State University - Ebel Lab | Greg Ebel et al. et al |
| EPI_ISL_527487 | USA/CO-CSU-E2267/2020 | 2020-04-29 | North America/USA/Colorado/ | Colorado State University - Ebel Lab | Colorado State University - Ebel Lab | Greg Ebel et al. et al |
| EPI_ISL_527516 | DRC/11450/2020 | 2020-05-25 | Africa/Democratic Republic of the Congo | Viral Respiratory Lab, National Institute for Biomedical Research (INRB) | Pathogen Sequencing Lab, National Institute for Biomedical Research (INRB) | Placide Mbala-Kingebeni et al |
| EPI_ISL_527533 | DRC/11653/2020 | 2020-05-25 | Africa/Democratic Republic of the Congo | Viral Respiratory Lab, National Institute for Biomedical Research (INRB) | Pathogen Sequencing Lab, National Institute for Biomedical Research (INRB) | Placide Mbala-Kingebeni et al |
| EPI_ISL_527555 | DRC/11041/2020 | 2020-05-24 | Africa/Democratic Republic of the Congo | Viral Respiratory Lab, National Institute for Biomedical Research (INRB) | Pathogen Sequencing Lab, National Institute for Biomedical Research (INRB) | Placide Mbala-Kingebeni et al |
| EPI_ISL_527574 | USA/MN-MDH-1509/2020 | 2020-05-21 | North America/USA/Minnesota/ | Minnesota Department of Health, Public Health Laboratory | Minnesota Department of Health, Public Health Laboratory | Matt Plumb et al |
| EPI_ISL_527576 | USA/MN-MDH-1511/2020 | 2020-07-06 | North America/USA/Minnesota/ | Minnesota Department of Health, Public Health Laboratory | Minnesota Department of Health, Public Health Laboratory | Matt Plumb et al |

| | | | | | | |
|---|---|---|---|---|---|---|
| EPI_ISL_527579 | USA/MN-MDH-1514/2020 | 2020-07-06 | North America/USA/Minnesota/ | Minnesota Department of Health, Public Health Laboratory | Minnesota Department of Health, Public Health Laboratory | Matt Plumb et al |
| EPI_ISL_527582 | USA/MN-MDH-1517/2020 | 2020-07-06 | North America/USA/Minnesota/ | Minnesota Department of Health, Public Health Laboratory | Minnesota Department of Health, Public Health Laboratory | Matt Plumb et al |
| EPI_ISL_527586 | USA/MN-MDH-1521/2020 | 2020-07-14 | North America/USA/Minnesota/ | Minnesota Department of Health, Public Health Laboratory | Minnesota Department of Health, Public Health Laboratory | Matt Plumb et al |
| EPI_ISL_527587 | USA/MN-MDH-1522/2020 | 2020-07-14 | North America/USA/Minnesota/ | Minnesota Department of Health, Public Health Laboratory | Minnesota Department of Health, Public Health Laboratory | Matt Plumb et al |
| EPI_ISL_527592 | USA/MN-MDH-1527/2020 | 2020-07-14 | North America/USA/Minnesota/ | Minnesota Department of Health, Public Health Laboratory | Minnesota Department of Health, Public Health Laboratory | Matt Plumb et al |
| EPI_ISL_527595 | USA/MN-MDH-1530/2020 | 2020-07-17 | North America/USA/Minnesota/ | Minnesota Department of Health, Public Health Laboratory | Minnesota Department of Health, Public Health Laboratory | Matt Plumb et al |
| EPI_ISL_527605 | USA/MN-MDH-1540/2020 | 2020-08-03 | North America/USA/Minnesota/ | Minnesota Department of Health, Public Health Laboratory | Minnesota Department of Health, Public Health Laboratory | Matt Plumb et al |
| EPI_ISL_527620 | USA/MN-MDH-1555/2020 | 2020-08-03 | North America/USA/Minnesota/ | Minnesota Department of Health, Public Health Laboratory | Minnesota Department of Health, Public Health Laboratory | Matt Plumb et al |

| | | | | | | |
|---|---|---|---|---|---|---|
| EPI_ISL_527640 | USA/AR-CDC-4226/2020 | 2020-07-13 | North America/USA/Arkansas/ | AR Dept. of Health-Public Health Lab | Pathogen Discovery, Respiratory Viruses Branch, Division of Viral Diseases, Centers for Disease Control and Prevention | Ying Tao et al |
| EPI_ISL_527641 | USA/AR-CDC-4227/2020 | 2020-07-13 | North America/USA/Arkansas/ | AR Dept. of Health-Public Health Lab | Pathogen Discovery, Respiratory Viruses Branch, Division of Viral Diseases, Centers for Disease Control and Prevention | Ying Tao et al |
| EPI_ISL_527648 | USA/AR-CDC-4299/2020 | 2020-07-17 | North America/USA/Arkansas/ | AR Dept. of Health-Public Health Lab | Pathogen Discovery, Respiratory Viruses Branch, Division of Viral Diseases, Centers for Disease Control and Prevention | Ying Tao et al |
| EPI_ISL_527655 | USA/AR-CDC-4380/2020 | 2020-07-03 | North America/USA/Arkansas/ | AR Dept. of Health-Public Health Lab | Pathogen Discovery, Respiratory Viruses Branch, Division of Viral Diseases, Centers for Disease Control and Prevention | Ying Tao et al |

| | | | | | | |
|---|---|---|---|---|---|---|
| EPI_ISL_527656 | USA/AR-CDC-4388/2020 | 2020-07-23 | North America/USA/Arkansas/ | AR Dept. of Health-Public Health Lab | Pathogen Discovery, Respiratory Viruses Branch, Division of Viral Diseases, Centers for Disease Control and Prevention | Ying Tao et al |
| EPI_ISL_527657 | USA/AR-CDC-6567/2020 | 2020-07-10 | North America/USA/Arkansas/ | AR Dept. of Health-Public Health Lab | Pathogen Discovery, Respiratory Viruses Branch, Division of Viral Diseases, Centers for Disease Control and Prevention | Ying Tao et al |
| EPI_ISL_527660 | USA/GA-CDC-6420/2020 | 2020-07-29 | North America/USA/Georgia/ | GA Department of Public Health Laboratory | Pathogen Discovery, Respiratory Viruses Branch, Division of Viral Diseases, Centers for Disease Control and Prevention | Yan Li et al |
| EPI_ISL_527661 | USA/GA-CDC-6424/2020 | 2020-07-29 | North America/USA/Georgia/ | GA Department of Public Health Laboratory | Pathogen Discovery, Respiratory Viruses Branch, Division of Viral Diseases, Centers for Disease Control and Prevention | Yan Li et al |

| | | | | | | |
|---|---|---|---|---|---|---|
| EPI_ISL_527662 | USA/GA-CDC-6427/2020 | 2020-07-29 | North America/USA/Georgia/ | GA Department of Public Health Laboratory | Pathogen Discovery, Respiratory Viruses Branch, Division of Viral Diseases, Centers for Disease Control and Prevention | Yan Li et al |
| EPI_ISL_527666 | USA/IN-CDC-6387/2020 | 2020-07-21 | North America/USA/Indiana/ | IN State Department of Health Laboratory Services | Pathogen Discovery, Respiratory Viruses Branch, Division of Viral Diseases, Centers for Disease Control and Prevention | Krista Queen et al |
| EPI_ISL_527690 | USA/MN-CDC-6274/2020 | 2020-05-01 | North America/USA/Minnesota/ | MN PHL Division, Minnesota Department of Health | Pathogen Discovery, Respiratory Viruses Branch, Division of Viral Diseases, Centers for Disease Control and Prevention | Yan Li et al |
| EPI_ISL_527698 | USA/MN-CDC-6305/2020 | 2020-05-08 | North America/USA/Minnesota/ | MN PHL Division, Minnesota Department of Health | Pathogen Discovery, Respiratory Viruses Branch, Division of Viral Diseases, Centers for Disease Control and Prevention | Yan Li et al |

| | | | | | | |
|---|---|---|---|---|---|---|
| EPI_ISL_527700 | USA/MN-CDC-6315/2020 | 2020-05-14 | North America/USA/Minnesota/ | MN PHL Division, Minnesota Department of Health | Pathogen Discovery, Respiratory Viruses Branch, Division of Viral Diseases, Centers for Disease Control and Prevention | Yan Li et al |
| EPI_ISL_527707 | USA/MN-CDC-6354/2020 | 2020-05-19 | North America/USA/Minnesota/ | MN PHL Division, Minnesota Department of Health | Pathogen Discovery, Respiratory Viruses Branch, Division of Viral Diseases, Centers for Disease Control and Prevention | Yan Li et al |
| EPI_ISL_527725 | USA/MN-CDC-6542/2020 | 2020-04-14 | North America/USA/Minnesota/ | MN PHL Division, Minnesota Department of Health | Pathogen Discovery, Respiratory Viruses Branch, Division of Viral Diseases, Centers for Disease Control and Prevention | Yan Li et al |
| EPI_ISL_527736 | USA/MN-CDC-6597/2020 | 2020-05-23 | North America/USA/Minnesota/ | MN PHL Division, Minnesota Department of Health | Pathogen Discovery, Respiratory Viruses Branch, Division of Viral Diseases, Centers for Disease Control and Prevention | Yan Li et al |
| EPI_ISL_527774 | USA/CT-Yale-358/2020 | 2020-08-17 | North America/USA/Connecticut/ | Connecticut State Department of Public Health | Grubaugh Lab - Yale School of Public Health | Joseph Fauver et al |

| | | | | | | |
|---|---|---|---|---|---|---|
| EPI_ISL_527776 | USA/CT-Yale-360/2020 | 2020-08-17 | North America/USA/Connecticut/ | Connecticut State Department of Public Health | Grubaugh Lab - Yale School of Public Health | Joseph Fauver et al |
| EPI_ISL_527787 | Peru/ARE-INS-075/2020 | 2020-04-16 | South America/Peru/Arequipa/ | Laboratorio de Referencia Nacional de Virus Respiratorio. Centro Nacional de Salud Publica. Instituto Nacional de Salud Peru. | Laboratorio de Referencia Nacional de Biotecnolog√≠a y Biolog√≠a Molecular. Centro Nacional de Salud Publica. Instituto Nacional de Salud Peru. | Carlos Padilla Rojas et al |
| EPI_ISL_527789 | Peru/ARE-INS-076/2020 | 2020-04-13 | South America/Peru/Arequipa/ | Laboratorio de Referencia Nacional de Virus Respiratorio. Centro Nacional de Salud Publica. Instituto Nacional de Salud Peru. | Laboratorio de Referencia Nacional de Biotecnolog√≠a y Biolog√≠a Molecular. Centro Nacional de Salud Publica. Instituto Nacional de Salud Peru. | Carlos Padilla Rojas et al |
| EPI_ISL_527799 | USA/WI-UW-1188/2020 | 2020-08-06 | North America/USA/Wisconsin/Dane Co | University of Wisconsin-Madison AIDS Vaccine Research Laboratories | University of Wisconsin-Madison AIDS Vaccine Research Laboratories | Gage Moreno et al |
| EPI_ISL_527809 | Ecuador/USFQ-193/2020 | 2020-08-04 | South America/Ecuador/Bolivar EC/Gua | Institute of Microbiology, Universidad San Francisco de Quito | Institute of Microbiology, Universidad San Francisco de Quito | Bel√©n Prado-Vivar et al |
| EPI_ISL_527810 | Ecuador/USFQ-194/2020 | 2020-08-04 | South America/Ecuador/Bolivar EC/Gua | Institute of Microbiology, Universidad San Francisco de Quito | Institute of Microbiology, Universidad San Francisco de Quito | Bel√©n Prado-Vivar et al |
| EPI_ISL_527812 | Ecuador/USFQ-196/2020 | 2020-08-20 | South America/Ecuador/Esmeraldas/Esı | Institute of Microbiology, Universidad San Francisco de Quito | Institute of Microbiology, Universidad San Francisco de Quito | Bel√©n Prado-Vivar et al |

| | | | | | | |
|---|---|---|---|---|---|---|
| EPI_ISL_527815 | Ecuador/USFQ-201/2020 | 2020-08-20 | South America/Ecuador/Esmeraldas/Esm | Institute of Microbiology, Universidad San Francisco de Quito | Institute of Microbiology, Universidad San Francisco de Quito | Bel√©n Prado-Vivar et al |
| EPI_ISL_527817 | Ecuador/USFQ-204/2020 | 2020-08-20 | South America/Ecuador/Esmeraldas/Esm | Institute of Microbiology, Universidad San Francisco de Quito | Institute of Microbiology, Universidad San Francisco de Quito | Bel√©n Prado-Vivar et al |
| EPI_ISL_527818 | Ecuador/USFQ-206/2020 | 2020-08-14 | South America/Ecuador/Guayas/Guayaq | Centro de Investigaciones, Universidad de Especialidades Esp√≠ritu Santo | Institute of Microbiology, Universidad San Francisco de Quito | Derly Andrade et al |
| EPI_ISL_527819 | Ecuador/USFQ-207/2020 | 2020-04-07 | South America/Ecuador/Guayas/Guayaq | Centro de Investigaciones, Universidad de Especialidades Esp√≠ritu Santo | Institute of Microbiology, Universidad San Francisco de Quito | Derly Andrade et al |
| EPI_ISL_527830 | USA/TX-DSHS-0257/2020 | 2020-04-26 | North America/USA/Texas/Cameron Cou | Texas Department of State Health Services | Texas Department of State Health Services | Bonnie Oh et al |
| EPI_ISL_527831 | USA/TX-DSHS-0263/2020 | 2020-06-06 | North America/USA/Texas/El Paso Coun | Texas Department of State Health Services | Texas Department of State Health Services | Bonnie Oh et al |
| EPI_ISL_527836 | USA/TX-DSHS-0269/2020 | 2020-06-13 | North America/USA/Texas/Harris Count | Texas Department of State Health Services | Texas Department of State Health Services | Bonnie Oh et al |
| EPI_ISL_527840 | USA/TX-DSHS-0273/2020 | 2020-06-13 | North America/USA/Texas/Harris Count | Texas Department of State Health Services | Texas Department of State Health Services | Bonnie Oh et al |
| EPI_ISL_527841 | USA/TX-DSHS-0274/2020 | 2020-06-13 | North America/USA/Texas/Harris Count | Texas Department of State Health Services | Texas Department of State Health Services | Bonnie Oh et al |
| EPI_ISL_527848 | USA/TX-DSHS-0281/2020 | 2020-06-13 | North America/USA/Texas/Harris Count | Texas Department of State Health Services | Texas Department of State Health Services | Bonnie Oh et al |
| EPI_ISL_527849 | USA/TX-DSHS-0282/2020 | 2020-06-13 | North America/USA/Texas/Harris Count | Texas Department of State Health Services | Texas Department of State Health Services | Bonnie Oh et al |
| EPI_ISL_527853 | USA/TX-DSHS-0286/2020 | 2020-06-13 | North America/USA/Texas/Harris Count | Texas Department of State Health Services | Texas Department of State Health Services | Bonnie Oh et al |

| | | | | | | |
|---|---|---|---|---|---|---|
| EPI_ISL_527855 | USA/TX-DSHS-0288/2020 | 2020-06-13 | North America/USA/Texas/El Paso Coun | Texas Department of State Health Services | Texas Department of State Health Services | Bonnie Oh et al |
| EPI_ISL_527869 | Brazil/SP-300/2020 | 2020-04-22 | South America/Brazil/S√£o Paulo/S√£o | Hospital Municipal Carmen Prudente | Instituto Adolfo Lutz, Interdiciplinary Procedures Center, Strategic Laboratory | Claudio Tavares Sacchi et al |
| EPI_ISL_527872 | Nigeria/AB03-CV135/2020 | 2020-05-12 | Africa/Nigeria/Ebonyi/ | Nigeria Centre for Disease Control (NCDC) | African Centre of Excellence for Genomics of Infectious Diseases (ACEGID), Redeemer's University, Ede, Osun State, Nigeria | Oluniyi P.E. et al et al |
| EPI_ISL_527882 | Nigeria/ED25-CV163/2020 | 2020-05-09 | Africa/Nigeria/Edo/ | Nigeria Centre for Disease Control (NCDC) | African Centre of Excellence for Genomics of Infectious Diseases (ACEGID), Redeemer's University, Ede, Osun State, Nigeria | Oluniyi P.E. et al et al |
| EPI_ISL_527883 | Nigeria/ED32-CV164/2020 | 2020-05-18 | Africa/Nigeria/Akwa-Ibom/ | Nigeria Centre for Disease Control (NCDC) | African Centre of Excellence for Genomics of Infectious Diseases (ACEGID), Redeemer's University, Ede, Osun State, Nigeria | Oluniyi P.E. et al et al |

| | | | | | | |
|---|---|---|---|---|---|---|
| EPI_ISL_527887 | Nigeria/EK291-CV115/2020 | 2020-06-02 | Africa/Nigeria/Ekiti State/ | Nigeria Centre for Disease Control (NCDC) | African Centre of Excellence for Genomics of Infectious Diseases (ACEGID), Redeemer's University, Ede, Osun State, Nigeria | Oluniyi P.E. et al et al |
| EPI_ISL_527889 | Nigeria/ON705-CV153/2020 | 2020-06-09 | Africa/Nigeria/Ondo State/ | Nigeria Centre for Disease Control (NCDC) | African Centre of Excellence for Genomics of Infectious Diseases (ACEGID), Redeemer's University, Ede, Osun State, Nigeria | Oluniyi P.E. et al et al |
| EPI_ISL_527898 | Nigeria/OY592-CV167/2020 | 2020-06-10 | Africa/Nigeria/Oyo State/ | Nigeria Centre for Disease Control (NCDC) | African Centre of Excellence for Genomics of Infectious Diseases (ACEGID), Redeemer's University, Ede, Osun State, Nigeria | Oluniyi P.E. et al et al |
| EPI_ISL_527899 | Nigeria/OY595-CV168/2020 | 2020-06-09 | Africa/Nigeria/Oyo State/ | Nigeria Centre for Disease Control (NCDC) | African Centre of Excellence for Genomics of Infectious Diseases (ACEGID), Redeemer's University, Ede, Osun State, Nigeria | Oluniyi P.E. et al et al |

| | | | | | | |
|---|---|---|---|---|---|---|
| EPI_ISL_527900 | Nigeria/OY598-CV169/2020 | 2020-06-09 | Africa/Nigeria/Oyo State/ | Nigeria Centre for Disease Control (NCDC) | African Centre of Excellence for Genomics of Infectious Diseases (ACEGID), Redeemer's University, Ede, Osun State, Nigeria | Oluniyi P.E. et al et al |
| EPI_ISL_527902 | Nigeria/OY607-CV171/2020 | 2020-06-12 | Africa/Nigeria/Oyo State/ | Nigeria Centre for Disease Control (NCDC) | African Centre of Excellence for Genomics of Infectious Diseases (ACEGID), Redeemer's University, Ede, Osun State, Nigeria | Oluniyi P.E. et al et al |
| EPI_ISL_527905 | Nigeria/OY616-CV174/2020 | 2020-06-12 | Africa/Nigeria/Oyo State/ | Nigeria Centre for Disease Control (NCDC) | African Centre of Excellence for Genomics of Infectious Diseases (ACEGID), Redeemer's University, Ede, Osun State, Nigeria | Oluniyi P.E. et al et al |
| EPI_ISL_527909 | Nigeria/OY625-CV182/2020 | 2020-06-13 | Africa/Nigeria/Oyo State/ | Nigeria Centre for Disease Control (NCDC) | African Centre of Excellence for Genomics of Infectious Diseases (ACEGID), Redeemer's University, Ede, Osun State, Nigeria | Oluniyi P.E. et al et al |

| | | | | | | |
|---|---|---|---|---|---|---|
| EPI_ISL_527916 | Switzerland/BS-UHB-42167340/2020 | 2020-02-26 | Europe/Italy/Basel-Stadt/ | University Hospital Basel, Clinical Virology | University Hospital Basel, Clinical Bacteriology | Madlen Stange et al |
| EPI_ISL_527917 | Switzerland/BL-UHB-42167842/2020 | 2020-02-26 | Europe/Italy/Basel-Land/ | University Hospital Basel, Clinical Virology | University Hospital Basel, Clinical Bacteriology | Madlen Stange et al |
| EPI_ISL_527918 | Switzerland/BL-UHB-42169471/2020 | 2020-02-28 | Europe/Switzerland/Basel-Land/ | University Hospital Basel, Clinical Virology | University Hospital Basel, Clinical Bacteriology | Madlen Stange et al |
| EPI_ISL_527919 | Switzerland/BL-UHB-42170345/2020 | 2020-02-28 | Europe/Switzerland/Basel-Land/ | University Hospital Basel, Clinical Virology | University Hospital Basel, Clinical Bacteriology | Madlen Stange et al |
| EPI_ISL_527926 | Switzerland/BL-UHB-42176198/2020 | 2020-03-04 | Europe/Switzerland/Basel-Land/ | University Hospital Basel, Clinical Virology | University Hospital Basel, Clinical Bacteriology | Madlen Stange et al |
| EPI_ISL_527938 | Switzerland/BS-UHB-42178000/2020 | 2020-03-05 | Europe/Switzerland/Basel-Stadt/ | University Hospital Basel, Clinical Virology | University Hospital Basel, Clinical Bacteriology | Madlen Stange et al |
| EPI_ISL_527950 | Switzerland/BL-UHB-42180271/2020 | 2020-03-06 | Europe/Switzerland/Basel-Land/ | University Hospital Basel, Clinical Virology | University Hospital Basel, Clinical Bacteriology | Madlen Stange et al |
| EPI_ISL_527991 | Switzerland/BS-UHB-42185836/2020 | 2020-03-11 | Europe/Switzerland/Basel-Stadt/ | University Hospital Basel, Clinical Virology | University Hospital Basel, Clinical Bacteriology | Madlen Stange et al |
| EPI_ISL_527996 | Switzerland/BS-UHB-42186169/2020 | 2020-03-11 | Europe/Switzerland/Basel-Stadt/ | University Hospital Basel, Clinical Virology | University Hospital Basel, Clinical Bacteriology | Madlen Stange et al |
| EPI_ISL_528037 | Switzerland/BS-UHB-42188679/2020 | 2020-03-12 | Europe/Italy/Basel-Stadt/ | University Hospital Basel, Clinical Virology | University Hospital Basel, Clinical Bacteriology | Madlen Stange et al |

| | | | | | | |
|---|---|---|---|---|---|---|
| EPI_ISL_528075 | Switzerland/BS-UHB-42190675/2020 | 2020-03-14 | Europe/Switzerland/Basel-Stadt/ | University Hospital Basel, Clinical Virology | University Hospital Basel, Clinical Bacteriology | Madlen Stange et al |
| EPI_ISL_528078 | Switzerland/BS-UHB-42190918/2020 | 2020-03-14 | Europe/Switzerland/Basel-Stadt/ | University Hospital Basel, Clinical Virology | University Hospital Basel, Clinical Bacteriology | Madlen Stange et al |
| EPI_ISL_528104 | Switzerland/BL-UHB-42191670/2020 | 2020-03-15 | Europe/Switzerland/Basel-Land/ | University Hospital Basel, Clinical Virology | University Hospital Basel, Clinical Bacteriology | Madlen Stange et al |
| EPI_ISL_528125 | Switzerland/BL-UHB-42192108/2020 | 2020-03-16 | Europe/Switzerland/Basel-Land/ | University Hospital Basel, Clinical Virology | University Hospital Basel, Clinical Bacteriology | Madlen Stange et al |
| EPI_ISL_528134 | Switzerland/BS-UHB-42192894/2020 | 2020-03-16 | Europe/Switzerland/Basel-Stadt/ | University Hospital Basel, Clinical Virology | University Hospital Basel, Clinical Bacteriology | Madlen Stange et al |
| EPI_ISL_528140 | Switzerland/VD-UHB-42192980/2020 | 2020-03-16 | Europe/Switzerland/Vaud/ | University Hospital Basel, Clinical Virology | University Hospital Basel, Clinical Bacteriology | Madlen Stange et al |
| EPI_ISL_528164 | Switzerland/BS-UHB-42194551/2020 | 2020-03-17 | Europe/Switzerland/Basel-Stadt/ | University Hospital Basel, Clinical Virology | University Hospital Basel, Clinical Bacteriology | Madlen Stange et al |
| EPI_ISL_528198 | Switzerland/BS-UHB-42196271/2020 | 2020-03-17 | Europe/Switzerland/Basel-Stadt/ | University Hospital Basel, Clinical Virology | University Hospital Basel, Clinical Bacteriology | Madlen Stange et al |
| EPI_ISL_528200 | Switzerland/BS-UHB-42196976/2020 | 2020-03-18 | Europe/Switzerland/Basel-Stadt/ | University Hospital Basel, Clinical Virology | University Hospital Basel, Clinical Bacteriology | Madlen Stange et al |
| EPI_ISL_528221 | Switzerland/BS-UHB-42198574/2020 | 2020-03-19 | Europe/Switzerland/Basel-Stadt/ | University Hospital Basel, Clinical Virology | University Hospital Basel, Clinical Bacteriology | Madlen Stange et al |

| | | | | | | |
|---|---|---|---|---|---|---|
| EPI_ISL_528224 | Switzerland/BL-UHB-42198619/2020 | 2020-03-19 | Europe/Switzerland/Basel-Land/ | University Hospital Basel, Clinical Virology | University Hospital Basel, Clinical Bacteriology | Madlen Stange et al |
| EPI_ISL_528234 | Switzerland/BS-UHB-42198938/2020 | 2020-03-19 | Europe/Italy/Basel-Stadt/ | University Hospital Basel, Clinical Virology | University Hospital Basel, Clinical Bacteriology | Madlen Stange et al |
| EPI_ISL_528251 | Switzerland/BS-UHB-42199797/2020 | 2020-03-20 | Europe/Switzerland/Basel-Stadt/ | University Hospital Basel, Clinical Virology | University Hospital Basel, Clinical Bacteriology | Madlen Stange et al |
| EPI_ISL_528270 | Switzerland/BS-UHB-42200118/2020 | 2020-03-20 | Europe/Switzerland/Basel-Stadt/ | University Hospital Basel, Clinical Virology | University Hospital Basel, Clinical Bacteriology | Madlen Stange et al |
| EPI_ISL_528310 | Switzerland/BS-UHB-42201769/2020 | 2020-03-21 | Europe/Switzerland/Basel-Stadt/ | University Hospital Basel, Clinical Virology | University Hospital Basel, Clinical Bacteriology | Madlen Stange et al |
| EPI_ISL_528321 | Switzerland/BL-UHB-42202153/2020 | 2020-03-22 | Europe/Switzerland/Basel-Land/ | University Hospital Basel, Clinical Virology | University Hospital Basel, Clinical Bacteriology | Madlen Stange et al |
| EPI_ISL_528322 | Switzerland/BL-UHB-42202165/2020 | 2020-03-22 | Europe/Switzerland/Basel-Land/ | University Hospital Basel, Clinical Virology | University Hospital Basel, Clinical Bacteriology | Madlen Stange et al |
| EPI_ISL_528349 | Switzerland/BS-UHB-42203345/2020 | 2020-03-23 | Europe/Switzerland/Basel-Stadt/ | University Hospital Basel, Clinical Virology | University Hospital Basel, Clinical Bacteriology | Madlen Stange et al |
| EPI_ISL_528354 | Switzerland/BS-UHB-42203381/2020 | 2020-03-23 | Europe/Switzerland/Basel-Stadt/ | University Hospital Basel, Clinical Virology | University Hospital Basel, Clinical Bacteriology | Madlen Stange et al |
| EPI_ISL_528365 | Switzerland/BS-UHB-42203696/2020 | 2020-03-23 | Europe/Switzerland/Basel-Stadt/ | University Hospital Basel, Clinical Virology | University Hospital Basel, Clinical Bacteriology | Madlen Stange et al |

| | | | | | | |
|---|---|---|---|---|---|---|
| EPI_ISL_528382 | India/HR-THSTI2010A/2020 | 2020-04-27 | Asia/India/Haryana/ | Translational Health Science and Technology Institute -ESIC medical college and hospital, Faridabad | THSTI Bioassay laboratory | Saurabh Kumar et al |
| EPI_ISL_528413 | England/20189182904/2020 | 2020-05-02 | Europe/United Kingdom/England/ | Respiratory Virus Unit, Microbiology Services Colindale, Public Health England | Respiratory Virus Unit, Microbiology Services Colindale, Public Health England | PHE Covid Sequencing Team et al |
| EPI_ISL_528421 | India/MH-IGIB-B1/2020 | 2020-05-15 | Asia/India/Maharashtra/Mumbai | TNMC & BYL NAIR CH. HOSPITAL | Institute of Genomics and Integrative Biology - Council of Scientific and Industrial Research | Rajesh Pandey et al |
| EPI_ISL_528426 | India/MH-IGIB-D2/2020 | 2020-07-07 | Asia/India/Maharashtra/Mumbai | P. D. Hinduja Hospital and Medical Research Centre | Institute of Genomics and Integrative Biology - Council of Scientific and Industrial Research | Rajesh Pandey et al |
| EPI_ISL_528485 | USA/AK-PHL1002/2020 | 2020-07-08 | North America/USA/Alaska/ | Alaska State Virology Laboratory | Alaska State Virology Laboratory | Chen J et al with Pathogenomics group Dagdag R et al |
| EPI_ISL_528493 | USA/AK-PHL1252/2020 | 2020-07-14 | North America/USA/Alaska/ | Alaska State Virology Laboratory | Alaska State Virology Laboratory | Chen J et al with Pathogenomics group Dagdag R et al |
| EPI_ISL_528495 | USA/AK-PHL1420/2020 | 2020-07-17 | North America/USA/Alaska/ | Alaska State Virology Laboratory | Alaska State Virology Laboratory | Chen J et al with Pathogenomics group Dagdag R et al |
| EPI_ISL_528499 | USA/AK-PHL1718/2020 | 2020-07-21 | North America/USA/Alaska/ | Alaska State Virology Laboratory | Alaska State Virology Laboratory | Chen J et al with Pathogenomics group Dagdag R et al |
| EPI_ISL_528502 | USA/AK-PHL1744/2020 | 2020-07-21 | North America/USA/Alaska/ | Alaska State Virology Laboratory | Alaska State Virology Laboratory | Chen J et al with Pathogenomics group Dagdag R et al |
| EPI_ISL_528507 | USA/AK-PHL1859/2020 | 2020-07-22 | North America/USA/Alaska/ | Alaska State Virology Laboratory | Alaska State Virology Laboratory | Chen J et al with Pathogenomics group Dagdag R et al |

| | | | | | | |
|---|---|---|---|---|---|---|
| EPI_ISL_528514 | USA/AK-PHL1896/2020 | 2020-07-22 | North America/USA/Alaska/ | Alaska State Virology Laboratory | Alaska State Virology Laboratory | Chen J et al with Pathogenomics group Dagdag R et al |
| EPI_ISL_528525 | USA/AK-PHL793/2020 | 2020-07-02 | North America/USA/Alaska/ | Alaska State Virology Laboratory | Alaska State Virology Laboratory | Chen J et al with Pathogenomics group Dagdag R et al |
| EPI_ISL_528526 | USA/AK-PHL803/2020 | 2020-07-03 | North America/USA/Alaska/ | Alaska State Virology Laboratory | Alaska State Virology Laboratory | Chen J et al with Pathogenomics group Dagdag R et al |
| EPI_ISL_528528 | USA/AK-PHL818/2020 | 2020-07-04 | North America/USA/Alaska/ | Alaska State Virology Laboratory | Alaska State Virology Laboratory | Chen J et al with Pathogenomics group Dagdag R et al |
| EPI_ISL_528531 | USA/AK-PHL868/2020 | 2020-07-06 | North America/USA/Alaska/ | Alaska State Virology Laboratory | Alaska State Virology Laboratory | Chen J et al with Pathogenomics group Dagdag R et al |
| EPI_ISL_528536 | USA/AK-PHL936/2020 | 2020-07-07 | North America/USA/Alaska/ | Alaska State Virology Laboratory | Alaska State Virology Laboratory | Chen J et al with Pathogenomics group Dagdag R et al |
| EPI_ISL_528537 | USA/AK-PHL973/2020 | 2020-07-08 | North America/USA/Alaska/ | Alaska State Virology Laboratory | Alaska State Virology Laboratory | Chen J et al with Pathogenomics group Dagdag R et al |
| EPI_ISL_528539 | Brazil/RJ-LVM161-2/2020 | 2020-03-19 | South America/Brazil/Rio de Janeiro/ | LVM/UFRJ | LNCC | Gustavo M. Romv°rio M. de Souza et al |
| EPI_ISL_528548 | India/TG-CDFD-C1132/2020 | 2020-05-30 | Asia/India/Telangana/ | National Genomics Core-Center for DNA Fingerprinting and Diagnostics | National Genomics Core- Center for DNA Fingerprinting and Diagnostics (NGC-CDFD)- DBT's PAN-INDIA-1000 Genome consortium | G Shashikanth et al |
| EPI_ISL_528552 | India/TG-CDFD-C1216/2020 | 2020-06-03 | Asia/India/Telangana/ | National Genomics Core-Center for DNA Fingerprinting and Diagnostics | National Genomics Core- Center for DNA Fingerprinting and Diagnostics (NGC-CDFD)- DBT's PAN-INDIA-1000 Genome consortium | Heena Shah et al |

| | | | | | | |
|---|---|---|---|---|---|---|
| EPI_ISL_528553 | India/TG-CDFD-C1254/2020 | 2020-06-04 | Asia/India/Telangana/ | National Genomics Core-Center for DNA Fingerprinting and Diagnostics | National Genomics Core- Center for DNA Fingerprinting and Diagnostics (NGC-CDFD)- DBT's PAN-INDIA-1000 Genome consortium | Heena Shah et al |
| EPI_ISL_528559 | India/TG-CDFD-C1359/2020 | 2020-06-06 | Asia/India/Telangana/ | National Genomics Core-Center for DNA Fingerprinting and Diagnostics | National Genomics Core- Center for DNA Fingerprinting and Diagnostics (NGC-CDFD)- DBT's PAN-INDIA-1000 Genome consortium | Heena Shah et al |
| EPI_ISL_528560 | India/TG-CDFD-C1391/2020 | 2020-06-08 | Asia/India/Telangana/ | National Genomics Core-Center for DNA Fingerprinting and Diagnostics | National Genomics Core- Center for DNA Fingerprinting and Diagnostics (NGC-CDFD)- DBT's PAN-INDIA-1000 Genome consortium | Heena Shah et al |
| EPI_ISL_528561 | India/TG-CDFD-C1399/2020 | 2020-06-08 | Asia/India/Telangana/ | National Genomics Core-Center for DNA Fingerprinting and Diagnostics | National Genomics Core- Center for DNA Fingerprinting and Diagnostics (NGC-CDFD)- DBT's PAN-INDIA-1000 Genome consortium | Heena Shah et al |

| | | | | | | |
|---|---|---|---|---|---|---|
| EPI_ISL_528562 | India/TG-CDFD-C1408/2020 | 2020-06-08 | Asia/India/Telangana/ | National Genomics Core-Center for DNA Fingerprinting and Diagnostics | National Genomics Core- Center for DNA Fingerprinting and Diagnostics (NGC-CDFD)- DBT's PAN-INDIA-1000 Genome consortium | Heena Shah et al |
| EPI_ISL_528563 | India/TG-CDFD-C1469/2020 | 2020-06-10 | Asia/India/Telangana/ | National Genomics Core-Center for DNA Fingerprinting and Diagnostics | National Genomics Core- Center for DNA Fingerprinting and Diagnostics (NGC-CDFD)- DBT's PAN-INDIA-1000 Genome consortium | Heena Shah et al |
| EPI_ISL_528569 | India/TG-CDFD-C1516/2020 | 2020-06-10 | Asia/India/Telangana/ | National Genomics Core-Center for DNA Fingerprinting and Diagnostics | National Genomics Core- Center for DNA Fingerprinting and Diagnostics (NGC-CDFD)- DBT's PAN-INDIA-1000 Genome consortium | G Shashikanth et al |
| EPI_ISL_528571 | India/TG-CDFD-C1829/2020 | 2020-06-16 | Asia/India/Telangana/ | National Genomics Core-Center for DNA Fingerprinting and Diagnostics | National Genomics Core- Center for DNA Fingerprinting and Diagnostics (NGC-CDFD)- DBT's PAN-INDIA-1000 Genome consortium | G Shashikanth et al |

| | | | | | | |
|---|---|---|---|---|---|---|
| EPI_ISL_528573 | India/TG-CDFD-C1847/2020 | 2020-06-16 | Asia/India/Telangana/ | National Genomics Core-Center for DNA Fingerprinting and Diagnostics | National Genomics Core- Center for DNA Fingerprinting and Diagnostics (NGC-CDFD)- DBT's PAN-INDIA-1000 Genome consortium | Vinay Donipadi et al |
| EPI_ISL_528577 | India/TG-CDFD-C2350/2020 | 2020-06-17 | Asia/India/Telangana/ | National Genomics Core-Center for DNA Fingerprinting and Diagnostics | National Genomics Core- Center for DNA Fingerprinting and Diagnostics (NGC-CDFD)- DBT's PAN-INDIA-1000 Genome consortium | Vinay Donipadi et al |
| EPI_ISL_528578 | India/TG-CDFD-C2520/2020 | 2020-06-18 | Asia/India/Telangana/ | National Genomics Core-Center for DNA Fingerprinting and Diagnostics | National Genomics Core- Center for DNA Fingerprinting and Diagnostics (NGC-CDFD)- DBT's PAN-INDIA-1000 Genome consortium | Vinay Donipadi et al |
| EPI_ISL_528579 | India/TG-CDFD-C2524/2020 | 2020-06-18 | Asia/India/Telangana/ | National Genomics Core-Center for DNA Fingerprinting and Diagnostics | National Genomics Core- Center for DNA Fingerprinting and Diagnostics (NGC-CDFD)- DBT's PAN-INDIA-1000 Genome consortium | Vinay Donipadi et al |

| | | | | | | |
|---|---|---|---|---|---|---|
| EPI_ISL_528581 | India/TG-CDFD-C2529/2020 | 2020-06-18 | Asia/India/Telangana/ | National Genomics Core-Center for DNA Fingerprinting and Diagnostics | National Genomics Core- Center for DNA Fingerprinting and Diagnostics (NGC-CDFD)- DBT's PAN-INDIA-1000 Genome consortium | G Shashikanth et al |
| EPI_ISL_528584 | India/TG-CDFD-C2901/2020 | 2020-06-19 | Asia/India/Telangana/ | National Genomics Core-Center for DNA Fingerprinting and Diagnostics | National Genomics Core- Center for DNA Fingerprinting and Diagnostics (NGC-CDFD)- DBT's PAN-INDIA-1000 Genome consortium | G Shashikanth et al |
| EPI_ISL_528586 | India/TG-CDFD-C2910/2020 | 2020-06-19 | Asia/India/Telangana/ | National Genomics Core-Center for DNA Fingerprinting and Diagnostics | National Genomics Core- Center for DNA Fingerprinting and Diagnostics (NGC-CDFD)- DBT's PAN-INDIA-1000 Genome consortium | G Shashikanth et al |
| EPI_ISL_528587 | India/TG-CDFD-C2916/2020 | 2020-06-19 | Asia/India/Telangana/ | National Genomics Core-Center for DNA Fingerprinting and Diagnostics | National Genomics Core- Center for DNA Fingerprinting and Diagnostics (NGC-CDFD)- DBT's PAN-INDIA-1000 Genome consortium | G Shashikanth et al |

| | | | | | | |
|---|---|---|---|---|---|---|
| EPI_ISL_528588 | India/TG-CDFD-C2926/2020 | 2020-06-19 | Asia/India/Telangana/ | National Genomics Core-Center for DNA Fingerprinting and Diagnostics | National Genomics Core- Center for DNA Fingerprinting and Diagnostics (NGC-CDFD)- DBT's PAN-INDIA-1000 Genome consortium | Heena Shah et al |
| EPI_ISL_528589 | India/TG-CDFD-C2999/2020 | 2020-06-19 | Asia/India/Telangana/ | National Genomics Core-Center for DNA Fingerprinting and Diagnostics | National Genomics Core- Center for DNA Fingerprinting and Diagnostics (NGC-CDFD)- DBT's PAN-INDIA-1000 Genome consortium | Heena Shah et al |
| EPI_ISL_528591 | India/TG-CDFD-C3145/2020 | 2020-06-20 | Asia/India/Telangana/ | National Genomics Core-Center for DNA Fingerprinting and Diagnostics | National Genomics Core- Center for DNA Fingerprinting and Diagnostics (NGC-CDFD)- DBT's PAN-INDIA-1000 Genome consortium | Heena Shah et al |
| EPI_ISL_528596 | India/TG-CDFD-C3298/2020 | 2020-06-20 | Asia/India/Telangana/ | National Genomics Core-Center for DNA Fingerprinting and Diagnostics | National Genomics Core- Center for DNA Fingerprinting and Diagnostics (NGC-CDFD)- DBT's PAN-INDIA-1000 Genome consortium | Bala Pratyusha et al |

| | | | | | | |
|---|---|---|---|---|---|---|
| EPI_ISL_528601 | India/TG-CDFD-C584/2020 | 2020-05-10 | Asia/India/Telangana/ | National Genomics Core-Center for DNA Fingerprinting and Diagnostics | National Genomics Core- Center for DNA Fingerprinting and Diagnostics (NGC-CDFD)- DBT's PAN-INDIA-1000 Genome consortium | Bala Pratyusha et al |
| EPI_ISL_528602 | India/TG-CDFD-C673/2020 | 2020-05-12 | Asia/India/Telangana/ | National Genomics Core-Center for DNA Fingerprinting and Diagnostics | National Genomics Core- Center for DNA Fingerprinting and Diagnostics (NGC-CDFD)- DBT's PAN-INDIA-1000 Genome consortium | Bala Pratyusha et al |
| EPI_ISL_528616 | India/TG-CDFD-N4853/2020 | 2020-05-27 | Asia/India/Telangana/ | National Genomics Core-Center for DNA Fingerprinting and Diagnostics | National Genomics Core- Center for DNA Fingerprinting and Diagnostics (NGC-CDFD)- DBT's PAN-INDIA-1000 Genome consortium | Divya Vashisht et al |
| EPI_ISL_528618 | India/TG-CDFD-N4895/2020 | 2020-05-28 | Asia/India/Telangana/ | National Genomics Core-Center for DNA Fingerprinting and Diagnostics | National Genomics Core- Center for DNA Fingerprinting and Diagnostics (NGC-CDFD)- DBT's PAN-INDIA-1000 Genome consortium | Heena Shah et al |

| | | | | | | |
|---|---|---|---|---|---|---|
| EPI_ISL_528620 | India/TG-CDFD-N5196/2020 | 2020-05-30 | Asia/India/Telangana/ | National Genomics Core-Center for DNA Fingerprinting and Diagnostics | National Genomics Core- Center for DNA Fingerprinting and Diagnostics (NGC-CDFD)- DBT's PAN-INDIA-1000 Genome consortium | Heena Shah et al |
| EPI_ISL_528622 | India/TG-CDFD-N5246/2020 | 2020-05-30 | Asia/India/Telangana/ | National Genomics Core-Center for DNA Fingerprinting and Diagnostics | National Genomics Core- Center for DNA Fingerprinting and Diagnostics (NGC-CDFD)- DBT's PAN-INDIA-1000 Genome consortium | Heena Shah et al |
| EPI_ISL_528628 | India/TG-CDFD-N5414/2020 | 2020-06-01 | Asia/India/Telangana/ | National Genomics Core-Center for DNA Fingerprinting and Diagnostics | National Genomics Core- Center for DNA Fingerprinting and Diagnostics (NGC-CDFD)- DBT's PAN-INDIA-1000 Genome consortium | G Shashikanth et al |
| EPI_ISL_528629 | India/TG-CDFD-N5426/2020 | 2020-06-01 | Asia/India/Telangana/ | National Genomics Core-Center for DNA Fingerprinting and Diagnostics | National Genomics Core- Center for DNA Fingerprinting and Diagnostics (NGC-CDFD)- DBT's PAN-INDIA-1000 Genome consortium | G Shashikanth et al |

| | | | | | | |
|---|---|---|---|---|---|---|
| EPI_ISL_528630 | India/TG-CDFD-N5427/2020 | 2020-06-01 | Asia/India/Telangana/ | National Genomics Core-Center for DNA Fingerprinting and Diagnostics | National Genomics Core- Center for DNA Fingerprinting and Diagnostics (NGC-CDFD)- DBT's PAN-INDIA-1000 Genome consortium | G Shashikanth et al |
| EPI_ISL_528631 | India/TG-CDFD-N5451/2020 | 2020-06-01 | Asia/India/Telangana/ | National Genomics Core-Center for DNA Fingerprinting and Diagnostics | National Genomics Core- Center for DNA Fingerprinting and Diagnostics (NGC-CDFD)- DBT's PAN-INDIA-1000 Genome consortium | Heena Shah et al |
| EPI_ISL_528637 | Brazil/RJ-MD63-1a/2020 | 2020-03-16 | South America/Brazil/Rio de Janeiro/ | LVM/UFRJ | Bioinformatics Laboratory / LNCC | Gustavo D. P. Silva et al |
| EPI_ISL_528650 | USA/VA-DCLS-1228/2020 | 2020-07-29 | North America/USA/Virginia/ | Virginia DCLS | Virginia DCLS | Virginia DCLS et al |
| EPI_ISL_528651 | USA/VA-DCLS-1232/2020 | 2020-08-04 | North America/USA/Virginia/ | Virginia DCLS | Virginia DCLS | Virginia DCLS et al |
| EPI_ISL_528659 | USA/VA-DCLS-1240/2020 | 2020-07-30 | North America/USA/Virginia/ | Virginia DCLS | Virginia DCLS | Virginia DCLS et al |
| EPI_ISL_528666 | USA/VA-DCLS-1060/2020 | 2020-06-10 | North America/USA/Virginia/ | Virginia Division of Consolidated Laboratory Services (DCLS) | Virginia Division of Consolidated Laboratory Services (DCLS) | Virginia DCLS et al |
| EPI_ISL_528668 | USA/VA-DCLS-1064/2020 | 2020-06-10 | North America/USA/Virginia/ | Virginia Division of Consolidated Laboratory Services (DCLS) | Virginia Division of Consolidated Laboratory Services (DCLS) | Virginia DCLS et al |
| EPI_ISL_528671 | USA/VA-DCLS-1072/2020 | 2020-06-10 | North America/USA/Virginia/ | Virginia Division of Consolidated Laboratory Services (DCLS) | Virginia Division of Consolidated Laboratory Services (DCLS) | Virginia DCLS et al |
| EPI_ISL_528683 | India/DL-IGIB-77797/2020 | 2020-08-24 | Asia/India/Delhi/ | NCDC Institute of Genomics and Integrative Biology | NCDC Institute of Genomics and Integrative Biology | Vivekanand A et al |

| | | | | | | |
|---|---|---|---|---|---|---|
| EPI_ISL_528684 | India/DL-IGIB-C1/2020 | 2020-06-12 | Asia/India/Delhi/ | NCDC Institute of Genomics and Integrative Biology | NCDC Institute of Genomics and Integrative Biology | Vivekanand A et al |
| EPI_ISL_528694 | UnitedArabEmirates/306/2020 | 2020-06-02 | Asia/United Arab Emirates/Dubai/ | Alsafar - Khalifa University Abu Dhabi | Alsafar - Khalifa University Abu Dhabi | Andreas Henschel et al |
| EPI_ISL_528697 | UnitedArabEmirates/31B/2020 | 2020-04-29 | Asia/United Arab Emirates/Abu Dhabi/ | Alsafar - Khalifa University Abu Dhabi | Alsafar - Khalifa University Abu Dhabi | Andreas Henschel et al |
| EPI_ISL_528703 | UnitedArabEmirates/529/2020 | 2020-06-26 | Asia/United Arab Emirates/Sharja/ | Alsafar - Khalifa University Abu Dhabi | Alsafar - Khalifa University Abu Dhabi | Andreas Henschel et al |
| EPI_ISL_528709 | UnitedArabEmirates/H1/2020 | 2020-04-30 | Asia/United Arab Emirates/Abu Dhabi/ | Alsafar - Khalifa University Abu Dhabi | Alsafar - Khalifa University Abu Dhabi | Andreas Henschel et al |
| EPI_ISL_528720 | UnitedArabEmirates/H8/2020 | 2020-04-24 | Asia/United Arab Emirates/Abu Dhabi/ | Alsafar - Khalifa University Abu Dhabi | Alsafar - Khalifa University Abu Dhabi | Andreas Henschel et al |
| EPI_ISL_528721 | UnitedArabEmirates/H9/2020 | 2020-04-24 | Asia/United Arab Emirates/Abu Dhabi/ | Alsafar - Khalifa University Abu Dhabi | Alsafar - Khalifa University Abu Dhabi | Andreas Henschel et al |
| EPI_ISL_528745 | Indonesia/JB-TFRIC19-R24754/2020 | 2020-06-22 | Asia/Indonesia/West Java/ | Laboratorium Kesehatan Provinsi Jawa Barat | School of Life Sciences and Technology & School of Pharmacy-Institut Teknologi Bandung; Molecular Genetics Laboratory-Faculty of Medicine-Universitas Padjadjaran; Laboratorium Kesehatan Provinsi Jawa Barat | Marselina Irasonia Tan et al |

| | | | | | | |
|---|---|---|---|---|---|---|
| EPI_ISL_528746 | Indonesia/JB-TFRIC19-R47609/2020 | 2020-07-21 | Asia/Indonesia/West Java/ | Immanuel Hospital | Molecular Genetics Laboratory-Faculty of Medicine-Universitas Padjadjaran; School of Life Sciences and Technology & School of Pharmacy-Institut Teknologi Bandung; Laboratorium Kesehatan Provinsi Jawa Barat | Yunia Sribudiani et al |
| EPI_ISL_528747 | Indonesia/JB-TFRIC19-R30535/2020 | 2020-06-28 | Asia/Indonesia/West Java/ | Santo Borromeus Hospital | School of Pharmacy & School of Life Sciences and Technology - Institut Teknologi Bandung; Molecular Genetics Laboratory-Faculty of Medicine-Universitas Padjadjaran; Laboratorium Kesehatan Provinsi Jawa Barat | Catur Riani et al |

| | | | | | | |
|---|---|---|---|---|---|---|
| EPI_ISL_528748 | Indonesia/JB-TFRIC19-R48562/2020 | 2020-07-23 | Asia/Indonesia/West Java/ | Dinkes Provinsi Jawa Barat | School of Life Sciences and Technology & School of Pharmacy-Institut Teknologi Bandung; Molecular Genetics Laboratory-Faculty of Medicine-Universitas Padjadjaran; Laboratorium Kesehatan Provinsi Jawa Barat | Azzania Fibriani et al |
| EPI_ISL_528749 | Indonesia/JB-TFRIC19-R4975/2020 | 2020-05-07 | Asia/Indonesia/West Java/ | Santosa Hospital Bandung Central | School of Life Sciences and Technology & School of Pharmacy-Institut Teknologi Bandung; Molecular Genetics Laboratory-Faculty of Medicine-Universitas Padjadjaran; Laboratorium Kesehatan Provinsi Jawa Barat | Husna Nugrahapraja et al |

| | | | | | | |
|---|---|---|---|---|---|---|
| EPI_ISL_528751 | Indonesia/JB-TFRIC19-R49344/2020 | 2020-07-23 | Asia/Indonesia/West Java/ | Santo Borromeus Hospital | Molecular Genetics Laboratory-Faculty of Medicine-Universitas Padjadjaran; School of Life Sciences and Technology & School of Pharmacy-Institut Teknologi Bandung; Laboratorium Kesehatan Provinsi Jawa Barat | Yunia Sribudiani et al |
| EPI_ISL_528752 | Indonesia/JB-TFRIC19-R48875/2020 | 2020-07-23 | Asia/Indonesia/West Java/ | Dr. H. A. Rotinsulu Lung Hospital | School of Pharmacy & School of Life Sciences and Technology - Institut Teknologi Bandung; Molecular Genetics Laboratory-Faculty of Medicine-Universitas Padjadjaran; Laboratorium Kesehatan Provinsi Jawa Barat | Catur Riani et al |

| | | | | | | |
|---|---|---|---|---|---|---|
| EPI_ISL_528753 | Indonesia/JB-TFRIC19-R53817/2020 | 2020-08-04 | Asia/Indonesia/West Java/ | Dinkes Kota Bogor | School of Life Sciences and Technology & School of Pharmacy-Institut Teknologi Bandung; Molecular Genetics Laboratory-Faculty of Medicine-Universitas Padjadjaran; Laboratorium Kesehatan Provinsi Jawa Barat | Azzania Fibriani et al |
| EPI_ISL_528759 | Indonesia/JB-TFRIC19-R49544/2020 | 2020-07-25 | Asia/Indonesia/West Java/ | Santo Borromeus Hospital | School of Life Sciences and Technology & School of Pharmacy-Institut Teknologi Bandung; Molecular Genetics Laboratory-Faculty of Medicine-Universitas Padjadjaran; Laboratorium Kesehatan Provinsi Jawa Barat | Husna Nugrahapraja et al |
| EPI_ISL_528795 | England/BHRT-24B38DD/2020 | 2020-05-20 | Europe/United Kingdom/England/ | Microbiology Department, Barking Havering and Redbridge University Hospitals NHS trust | Wellcome Sanger Institute for the COVID-19 Genomics UK (COG-UK) consortium | Amy Ash et al |

| | | | | | | |
|---|---|---|---|---|---|---|
| EPI_ISL_528813 | India/TG-CCMB-GC20/2020 | 2020-07-19 | Asia/India/Telangana/Hyderabad | Department of Medicine, Gandhi hospital, Hyderabad | CSIR-Centre for Cellular and Molecular Biology | Vinayasekhar Aedula et al |
| EPI_ISL_528814 | India/TG-CCMB-GC22/2020 | 2020-05-22 | Asia/India/Telangana/Hyderabad | Department of Medicine, Gandhi hospital, Hyderabad | CSIR-Centre for Cellular and Molecular Biology | Rajarao Mesipogu et al |
| EPI_ISL_528823 | India/TG-CCMB-W842/2020 | 2020-07-14 | Asia/India/Telangana/Karimnagar | CSIR-Centre for Cellular and Molecular Biology | CSIR-Centre for Cellular and Molecular Biology | Shagufta Khan et al |
| EPI_ISL_528825 | India/TG-CCMB-W849/2020 | 2020-07-14 | Asia/India/Telangana/Karimnagar | CSIR-Centre for Cellular and Molecular Biology | CSIR-Centre for Cellular and Molecular Biology | Shagufta Khan et al |
| EPI_ISL_528827 | India/TG-CCMB-W855/2020 | 2020-07-14 | Asia/India/Telangana/Karimnagar | CSIR-Centre for Cellular and Molecular Biology | CSIR-Centre for Cellular and Molecular Biology | Namami Gaur et al |
| EPI_ISL_528829 | India/TG-CCMB-W866/2020 | 2020-07-14 | Asia/India/Telangana/Karimnagar | CSIR-Centre for Cellular and Molecular Biology | CSIR-Centre for Cellular and Molecular Biology | Sakshi Shambhavi et al |
| EPI_ISL_528830 | India/TG-CCMB-W887/2020 | 2020-07-14 | Asia/India/Telangana/Karimnagar | CSIR-Centre for Cellular and Molecular Biology | CSIR-Centre for Cellular and Molecular Biology | Tulasi Nagabandi et al |
| EPI_ISL_528831 | India/TG-CCMB-W897/2020 | 2020-07-14 | Asia/India/Telangana/Karimnagar | CSIR-Centre for Cellular and Molecular Biology | CSIR-Centre for Cellular and Molecular Biology | Lamuk Zaveri et al |
| EPI_ISL_528832 | India/TG-CCMB-W899/2020 | 2020-07-14 | Asia/India/Telangana/Karimnagar | CSIR-Centre for Cellular and Molecular Biology | CSIR-Centre for Cellular and Molecular Biology | Sofia Banu et al |
| EPI_ISL_528833 | India/TG-CCMB-W900/2020 | 2020-07-14 | Asia/India/Telangana/Karimnagar | CSIR-Centre for Cellular and Molecular Biology | CSIR-Centre for Cellular and Molecular Biology | Lamuk Zaveri et al |
| EPI_ISL_528834 | India/TG-CCMB-W903/2020 | 2020-07-14 | Asia/India/Telangana/Karimnagar | CSIR-Centre for Cellular and Molecular Biology | CSIR-Centre for Cellular and Molecular Biology | Pratheusa Maccha et al |
| EPI_ISL_528835 | India/TG-CCMB-W908/2020 | 2020-07-14 | Asia/India/Telangana/Karimnagar | CSIR-Centre for Cellular and Molecular Biology | CSIR-Centre for Cellular and Molecular Biology | M Soujanya Reddy et al |
| EPI_ISL_528836 | India/TG-CCMB-W910/2020 | 2020-07-14 | Asia/India/Telangana/Karimnagar | CSIR-Centre for Cellular and Molecular Biology | CSIR-Centre for Cellular and Molecular Biology | Tulasi Nagabandi et al |

| | | | | | | |
|---|---|---|---|---|---|---|
| EPI_ISL_528837 | India/TG-CCMB-W919/2020 | 2020-07-14 | Asia/India/Telangana/Karimnagar | CSIR-Centre for Cellular and Molecular Biology | CSIR-Centre for Cellular and Molecular Biology | Sakshi Shambhavi et al |
| EPI_ISL_528838 | India/TG-CCMB-W990/2020 | 2020-07-14 | Asia/India/Telangana/Nalgonda | CSIR-Centre for Cellular and Molecular Biology | CSIR-Centre for Cellular and Molecular Biology | Payel Mukherjee et al |
| EPI_ISL_528840 | India/TG-CCMB-W998/2020 | 2020-07-14 | Asia/India/Telangana/Nalgonda | CSIR-Centre for Cellular and Molecular Biology | CSIR-Centre for Cellular and Molecular Biology | Tulasi Nagabandi et al |
| EPI_ISL_528841 | India/TG-CCMB-X10/2020 | 2020-07-14 | Asia/India/Telangana/Nalgonda | CSIR-Centre for Cellular and Molecular Biology | CSIR-Centre for Cellular and Molecular Biology | Payel Mukherjee et al |
| EPI_ISL_528842 | India/TG-CCMB-X100/2020 | 2020-07-15 | Asia/India/Telangana/Karimnagar | CSIR-Centre for Cellular and Molecular Biology | CSIR-Centre for Cellular and Molecular Biology | Payel Mukherjee et al |
| EPI_ISL_528843 | India/TG-CCMB-X102/2020 | 2020-07-15 | Asia/India/Telangana/Karimnagar | CSIR-Centre for Cellular and Molecular Biology | CSIR-Centre for Cellular and Molecular Biology | Namami Gaur et al |
| EPI_ISL_528844 | India/TG-CCMB-X103/2020 | 2020-07-15 | Asia/India/Telangana/Karimnagar | CSIR-Centre for Cellular and Molecular Biology | CSIR-Centre for Cellular and Molecular Biology | Namami Gaur et al |
| EPI_ISL_528845 | India/TG-CCMB-X110/2020 | 2020-07-15 | Asia/India/Telangana/Karimnagar | CSIR-Centre for Cellular and Molecular Biology | CSIR-Centre for Cellular and Molecular Biology | Payel Mukherjee et al |
| EPI_ISL_528846 | India/TG-CCMB-X111/2020 | 2020-07-15 | Asia/India/Telangana/Karimnagar | CSIR-Centre for Cellular and Molecular Biology | CSIR-Centre for Cellular and Molecular Biology | Namami Gaur et al |
| EPI_ISL_528847 | India/TG-CCMB-X112/2020 | 2020-07-15 | Asia/India/Telangana/Karimnagar | CSIR-Centre for Cellular and Molecular Biology | CSIR-Centre for Cellular and Molecular Biology | Shagufta Khan et al |
| EPI_ISL_528850 | India/TG-CCMB-X116/2020 | 2020-07-15 | Asia/India/Telangana/Karimnagar | CSIR-Centre for Cellular and Molecular Biology | CSIR-Centre for Cellular and Molecular Biology | Lamuk Zaveri et al |
| EPI_ISL_528851 | India/TG-CCMB-X14/2020 | 2020-07-14 | Asia/India/Telangana/Nalgonda | CSIR-Centre for Cellular and Molecular Biology | CSIR-Centre for Cellular and Molecular Biology | Payel Mukherjee et al |
| EPI_ISL_528853 | India/TG-CCMB-X16/2020 | 2020-07-14 | Asia/India/Telangana/Nalgonda | CSIR-Centre for Cellular and Molecular Biology | CSIR-Centre for Cellular and Molecular Biology | Tulasi Nagabandi et al |

| | | | | | | |
|---|---|---|---|---|---|---|
| EPI_ISL_528854 | India/TG-CCMB-X42/2020 | 2020-07-14 | Asia/India/Telangana/Nalgonda | CSIR-Centre for Cellular and Molecular Biology | CSIR-Centre for Cellular and Molecular Biology | Payel Mukherjee et al |
| EPI_ISL_528855 | India/TG-CCMB-X43/2020 | 2020-07-14 | Asia/India/Telangana/Nalgonda | CSIR-Centre for Cellular and Molecular Biology | CSIR-Centre for Cellular and Molecular Biology | Nikhil Hajirnis et al |
| EPI_ISL_528857 | India/TG-CCMB-X46/2020 | 2020-07-14 | Asia/India/Telangana/Nalgonda | CSIR-Centre for Cellular and Molecular Biology | CSIR-Centre for Cellular and Molecular Biology | Nikhil Hajirnis et al |
| EPI_ISL_528858 | India/TG-CCMB-X49/2020 | 2020-07-14 | Asia/India/Telangana/Nalgonda | CSIR-Centre for Cellular and Molecular Biology | CSIR-Centre for Cellular and Molecular Biology | Pratheusa Maccha et al |
| EPI_ISL_528859 | India/TG-CCMB-X50/2020 | 2020-07-14 | Asia/India/Telangana/Nalgonda | CSIR-Centre for Cellular and Molecular Biology | CSIR-Centre for Cellular and Molecular Biology | Pratheusa Maccha et al |
| EPI_ISL_528860 | India/TG-CCMB-X52/2020 | 2020-07-14 | Asia/India/Telangana/Nalgonda | CSIR-Centre for Cellular and Molecular Biology | CSIR-Centre for Cellular and Molecular Biology | Tulasi Nagabandi et al |
| EPI_ISL_528861 | India/TG-CCMB-X59/2020 | 2020-07-14 | Asia/India/Telangana/Nalgonda | CSIR-Centre for Cellular and Molecular Biology | CSIR-Centre for Cellular and Molecular Biology | Sakshi Shambhavi et al |
| EPI_ISL_528862 | India/TG-CCMB-X65/2020 | 2020-07-14 | Asia/India/Telangana/Nalgonda | CSIR-Centre for Cellular and Molecular Biology | CSIR-Centre for Cellular and Molecular Biology | Lamuk Zaveri et al |
| EPI_ISL_528863 | India/TG-CCMB-X67/2020 | 2020-07-14 | Asia/India/Telangana/Nalgonda | CSIR-Centre for Cellular and Molecular Biology | CSIR-Centre for Cellular and Molecular Biology | Payel Mukherjee et al |
| EPI_ISL_528864 | India/TG-CCMB-X76/2020 | 2020-07-14 | Asia/India/Telangana/Nalgonda | CSIR-Centre for Cellular and Molecular Biology | CSIR-Centre for Cellular and Molecular Biology | Sofia Banu et al |
| EPI_ISL_528865 | India/TG-CCMB-X86/2020 | 2020-07-14 | Asia/India/Telangana/Nalgonda | CSIR-Centre for Cellular and Molecular Biology | CSIR-Centre for Cellular and Molecular Biology | Shagufta Khan et al |
| EPI_ISL_528866 | India/TG-CCMB-X87/2020 | 2020-07-14 | Asia/India/Telangana/Nalgonda | CSIR-Centre for Cellular and Molecular Biology | CSIR-Centre for Cellular and Molecular Biology | Sakshi Shambhavi et al |
| EPI_ISL_528867 | India/TG-CCMB-X88/2020 | 2020-07-14 | Asia/India/Telangana/Nalgonda | CSIR-Centre for Cellular and Molecular Biology | CSIR-Centre for Cellular and Molecular Biology | M Soujanya Reddy et al |

| | | | | | | |
|---|---|---|---|---|---|---|
| EPI_ISL_528868 | India/TG-CCMB-X89/2020 | 2020-07-14 | Asia/India/Telangana/Nalgonda | CSIR-Centre for Cellular and Molecular Biology | CSIR-Centre for Cellular and Molecular Biology | Shagufta Khan et al |
| EPI_ISL_528869 | India/TG-CCMB-X94/2020 | 2020-07-14 | Asia/India/Telangana/Nalgonda | CSIR-Centre for Cellular and Molecular Biology | CSIR-Centre for Cellular and Molecular Biology | Shagufta Khan et al |
| EPI_ISL_528880 | India/TG-CDFD-C2603/2020 | 2020-06-18 | Asia/India/Telangana/ | National Genomics Core-Center for DNA Fingerprinting and Diagnostics | National Genomics Core- Center for DNA Fingerprinting and Diagnostics (NGC-CDFD)- DBT's PAN-INDIA-1000 Genome consortium | G Shashikanth et al |
| EPI_ISL_528919 | Italy/ABR-IZSGC-TE4884/2020 | 2020-03-17 | Europe/Italy/Abruzzo/ | Ospedale Civile S. Liberatore-Atri | "Istituto Zooprofilattico Sperimentale dell'Abruzzo e Molise ""G.Caporale""" | Lorusso A et al |
| EPI_ISL_528920 | Italy/ABR-IZSGC-TE4891/2020 | 2020-03-17 | Europe/Italy/Abruzzo/ | "Presidio Ospedaliero ""Santo Spirito""-Pescara" | "Istituto Zooprofilattico Sperimentale dell'Abruzzo e Molise ""G.Caporale""" | Lorusso A et al |
| EPI_ISL_528922 | Italy/ABR-IZSGC-TE5040/2020 | 2020-03-17 | Europe/Italy/Abruzzo/ | "Ospedale ""Giuseppe Mazzini""-Teramo" | "Istituto Zooprofilattico Sperimentale dell'Abruzzo e Molise ""G.Caporale""" | Lorusso A et al |
| EPI_ISL_528923 | Italy/ABR-IZSGC-TE5148/2020 | 2020-03-18 | Europe/Italy/Abruzzo/ | Ospedale Civile S. Liberatore-Atri | "Istituto Zooprofilattico Sperimentale dell'Abruzzo e Molise ""G.Caporale""" | Lorusso A et al |

| | | | | | | |
|---|---|---|---|---|---|---|
| EPI_ISL_528924 | Italy/ABR-IZSGC-TE5173/2020 | 2020-03-18 | Europe/Italy/Abruzzo/ | "Ospedale ""Giuseppe Mazzini""-Teramo" | "Istitituto Zooprofilattico Sperimentale dell'Abruzzo e Molise ""G.Caporale""" | Lorusso A et al |
| EPI_ISL_528925 | Italy/ABR-IZSGC-TE5268/2020 | 2020-03-18 | Europe/Italy/Abruzzo/ | Ospedale Regionale San Salvatore-L'Aquila | "Istitituto Zooprofilattico Sperimentale dell'Abruzzo e Molise ""G.Caporale""" | Lorusso A et al |
| EPI_ISL_528928 | Italy/ABR-IZSGC-TE6218/2020 | 2020-03-23 | Europe/Italy/Abruzzo/ | "Ospedale ""Giuseppe Mazzini""-Teramo" | "Istitituto Zooprofilattico Sperimentale dell'Abruzzo e Molise ""G.Caporale""" | Lorusso A et al |
| EPI_ISL_528929 | Italy/ABR-IZSGC-TE7097/2020 | 2020-03-26 | Europe/Italy/Abruzzo/ | Ospedale Civile S. Liberatore-Atri | "Istitituto Zooprofilattico Sperimentale dell'Abruzzo e Molise ""G.Caporale""" | Lorusso A et al |
| EPI_ISL_528938 | Italy/LOM-IZSGC-TE31751/2020 | 2020-05-02 | Europe/Italy/Lombardy/ | Agenzia di Tutela della Salute di Bergamo | "Istitituto Zooprofilattico Sperimentale dell'Abruzzo e Molise ""G.Caporale""" | Lorusso A et al |
| EPI_ISL_528941 | Italy/LOM-IZSGC-TE35432/2020 | 2020-05-08 | Europe/Italy/Lombardy/ | Agenzia di Tutela della Salute di Bergamo | "Istitituto Zooprofilattico Sperimentale dell'Abruzzo e Molise ""G.Caporale""" | Lorusso A et al |

| | | | | | | |
|---|---|---|---|---|---|---|
| EPI_ISL_528942 | Italy/LOM-IZSGC-TE35441/2020 | 2020-05-08 | Europe/Italy/Lombardy/ | Agenzia di Tutela della Salute di Bergamo | "Istitituto Zooprofilattico Sperimentale dell'Abruzzo e Molise ""G.Caporale""" | Lorusso A et al |
| EPI_ISL_528943 | Italy/LOM-IZSGC-TE35526/2020 | 2020-05-08 | Europe/Italy/Lombardy/ | Agenzia di Tutela della Salute di Bergamo | "Istitituto Zooprofilattico Sperimentale dell'Abruzzo e Molise ""G.Caporale""" | Lorusso A et al |
| EPI_ISL_528944 | Italy/LOM-IZSGC-TE35532/2020 | 2020-05-08 | Europe/Italy/Lombardy/ | Agenzia di Tutela della Salute di Bergamo | "Istitituto Zooprofilattico Sperimentale dell'Abruzzo e Molise ""G.Caporale""" | Lorusso A et al |
| EPI_ISL_528945 | Italy/LOM-IZSGC-TE35566/2020 | 2020-05-08 | Europe/Italy/Lombardy/ | Agenzia di Tutela della Salute di Bergamo | "Istitituto Zooprofilattico Sperimentale dell'Abruzzo e Molise ""G.Caporale""" | Lorusso A et al |
| EPI_ISL_528946 | Italy/LOM-IZSGC-TE35578/2020 | 2020-05-08 | Europe/Italy/Lombardy/ | Agenzia di Tutela della Salute di Bergamo | "Istitituto Zooprofilattico Sperimentale dell'Abruzzo e Molise ""G.Caporale""" | Lorusso A et al |
| EPI_ISL_528947 | Italy/LOM-IZSGC-TE38811/2020 | 2020-05-10 | Europe/Italy/Lombardy/ | Agenzia di Tutela della Salute di Bergamo | "Istitituto Zooprofilattico Sperimentale dell'Abruzzo e Molise ""G.Caporale""" | Lorusso A et al |

| | | | | | | |
|---|---|---|---|---|---|---|
| EPI_ISL_528991 | Italy/ABR-IZSGC-TE10007/2020 | 2020-04-02 | Europe/Italy/Abruzzo/ | Ospedale SS Annunziata-Sulmona | "Istitituto Zooprofilattico Sperimentale dell'Abruzzo e Molise ""G.Caporale""" | Lorusso A et al |
| EPI_ISL_528992 | Italy/ABR-IZSGC-TE32707/2020 | 2020-04-29 | Europe/Italy/Abruzzo/ | Ospedale SS Annunziata-Sulmona | "Istitituto Zooprofilattico Sperimentale dell'Abruzzo e Molise ""G.Caporale""" | Lorusso A et al |
| EPI_ISL_528993 | Italy/ABR-IZSGC-TE34742/2020 | 2020-05-04 | Europe/Italy/Abruzzo/ | Ospedale Civile S. Liberatore-Atri | "Istitituto Zooprofilattico Sperimentale dell'Abruzzo e Molise ""G.Caporale""" | Lorusso A et al |
| EPI_ISL_528996 | Italy/ABR-IZSGC-TE46428/2020 | 2020-05-14 | Europe/Italy/Abruzzo/ | Servizio di igiene epidemiologia e sanit√† pubblica (SIESP)-Chieti | "Istitituto Zooprofilattico Sperimentale dell'Abruzzo e Molise ""G.Caporale""" | Lorusso A et al |
| EPI_ISL_528999 | Italy/ABR-IZSGC-TE46435/2020 | 2020-05-14 | Europe/Italy/Abruzzo/ | Servizio di igiene epidemiologia e sanit√† pubblica (SIESP)-Chieti | "Istitituto Zooprofilattico Sperimentale dell'Abruzzo e Molise ""G.Caporale""" | Lorusso A et al |
| EPI_ISL_529001 | Italy/ABR-IZSGC-TE46439/2020 | 2020-05-14 | Europe/Italy/Abruzzo/ | Servizio di igiene epidemiologia e sanit√† pubblica (SIESP)-Chieti | "Istitituto Zooprofilattico Sperimentale dell'Abruzzo e Molise ""G.Caporale""" | Lorusso A et al |

| | | | | | | |
|---|---|---|---|---|---|---|
| EPI_ISL_529004 | Italy/ABR-IZSGC-TE49480/2020 | 2020-05-16 | Europe/Italy/Abruzzo/ | Servizio di igiene epidemiologia e sanitv† pubblica (SIESP)-Chieti | "Istitituto Zooprofilattico Sperimentale dell'Abruzzo e Molise ""G.Caporale""" | Lorusso A et al |
| EPI_ISL_529005 | Italy/ABR-IZSGC-TE49482/2020 | 2020-05-16 | Europe/Italy/Abruzzo/ | Servizio di igiene epidemiologia e sanitv† pubblica (SIESP)-Chieti | "Istitituto Zooprofilattico Sperimentale dell'Abruzzo e Molise ""G.Caporale""" | Lorusso A et al |
| EPI_ISL_529007 | Italy/ABR-IZSGC-TE79445/2020 | 2020-06-13 | Europe/Italy/Abruzzo/ | Ospedale Civile S. Liberatore-Atri | "Istitituto Zooprofilattico Sperimentale dell'Abruzzo e Molise ""G.Caporale""" | Lorusso A et al |
| EPI_ISL_529008 | Italy/ABR-IZSGC-TE82840/2020 | 2020-06-17 | Europe/Italy/Abruzzo/ | Servizio di igiene e sanitv† pubblica (SIESP)-Teramo | "Istitituto Zooprofilattico Sperimentale dell'Abruzzo e Molise ""G.Caporale""" | Lorusso A et al |
| EPI_ISL_529009 | Italy/ABR-IZSGC-TE9047/2020 | 2020-04-01 | Europe/Italy/Abruzzo/ | Ospedale Civile S. Liberatore-Atri | "Istitituto Zooprofilattico Sperimentale dell'Abruzzo e Molise ""G.Caporale""" | Lorusso A et al |
| EPI_ISL_529012 | Italy/ABR-IZSGC-TE93935/2020 | 2020-07-09 | Europe/Italy/Abruzzo/ | Servizio Igiene Epidemiologia e Sanitv† Pubblica (SIESP)-L'Aquila | "Istitituto Zooprofilattico Sperimentale dell'Abruzzo e Molise ""G.Caporale""" | Lorusso A et al |

| | | | | | | |
|---|---|---|---|---|---|---|
| EPI_ISL_529016 | Italy/ABR-IZSGC-TE10151/2020 | 2020-04-02 | Europe/Italy/Abruzzo/ | Ospedale SS Annunziata-Sulmona | "Istituto Zooprofilattico Sperimentale dell'Abruzzo e Molise ""G.Caporale""" | Lorusso A et al |
| EPI_ISL_529019 | Italy/ABR-IZSGC-TE27016/2020 | 2020-04-27 | Europe/Italy/Abruzzo/ | RSA/RP Villa San Giovanni - Gruppo Edos | "Istituto Zooprofilattico Sperimentale dell'Abruzzo e Molise ""G.Caporale""" | Lorusso A et al |
| EPI_ISL_529022 | Italy/ABR-IZSGC-TE36500/2020 | 2020-05-06 | Europe/Italy/Abruzzo/ | "Ospedale ""Ss. Annunziata""" | "Istituto Zooprofilattico Sperimentale dell'Abruzzo e Molise ""G.Caporale""" | Lorusso A et al |
| EPI_ISL_529026 | Italy/ABR-IZSGC-TE8387/2020 | 2020-03-30 | Europe/Italy/Abruzzo/ | "Ospedale ""Giuseppe Mazzini""-Teramo" | "Istituto Zooprofilattico Sperimentale dell'Abruzzo e Molise ""G.Caporale""" | Lorusso A et al |
| EPI_ISL_529030 | Canada/ON-QGLO-049/2020 | 2020-03-30 | North America/Canada/Ontario/ | Kingston Health Sciences Center | Queen's Genomics Lab at Ongwanada (Q-GLO) | Sjaarda CP et al |

| | | | | | | |
|---|---|---|---|---|---|---|
| EPI_ISL_529031 | Egypt/C-CU001a-S1/2020 | 2020-05-19 | Africa/Egypt/Cairo/ | Central Molecular Microbiology Laboratory, Clinical and Chemical Pathology Department, Faculty of Medicine, CAIRO UNIVERSITY | Next Generation Sequencing Reference Laboratory, Faculty of Medicine, Cairo University and The Center for Genome and Microbiome Research, Faculty of Pharmacy, CAIRO UNIVERSITY | May Sherif Soliman et al |
| EPI_ISL_529032 | Egypt/C-CU002b-S3/2020 | 2020-06-19 | Africa/Egypt/Cairo/ | Central Molecular Microbiology Laboratory and Next Generation Sequencing Reference Laboratory, Clinical and Chemical Pathology Department, Faculty of Medicine, CAIRO UNIVERSITY | Next Generation Sequencing Reference Laboratory, Faculty of Medicine, CAIRO UNIVERSITY and The Center for Genome and Microbiome Research, Faculty of Pharmacy, CAIRO UNIVERSITY | May Sherif Soliman et al |
| EPI_ISL_529034 | USA/NY-Wadsworth-11661-01/2020 | 2020-03-08 | North America/USA/New York/ | Wadsworth Center, New York State Department of Health | Wadsworth Center, New York State Department of Health | Kirsten St. George et al |
| EPI_ISL_529035 | USA/NY-WC-46316-01/2020 | 2020-04-13 | North America/USA/New York/ | Wadsworth Center, New York State Department of Health | Wadsworth Center, New York State Department of Health | Kirsten St. George et al |

| | | | | | | |
|---|---|---|---|---|---|---|
| EPI_ISL_529065 | Peru/LIM-UPCH-0003/2020 | 2020-06-17 | South America/Peru/Lima/Ca√±ete | Laboratorio de Referencia Nacional de Virus Respiratorios, Instituto Nacional de Salud Peru | Laboratorio de Gen√≥mica Microbiana, Universidad Peruana Cayetano Heredia | Pablo Tsukayama et al |
| EPI_ISL_529066 | Peru/LIM-UPCH-0004/2020 | 2020-06-17 | South America/Peru/Lima/Lima | Laboratorio de Referencia Nacional de Virus Respiratorios, Instituto Nacional de Salud Peru | Laboratorio de Gen√≥mica Microbiana, Universidad Peruana Cayetano Heredia | Pablo Tsukayama et al |
| EPI_ISL_529067 | Peru/LAM-UPCH-0006/2020 | 2020-06-08 | South America/Peru/Lambayeque/Chicla | Laboratorio de Referencia Nacional de Virus Respiratorios, Instituto Nacional de Salud Peru | Laboratorio de Gen√≥mica Microbiana, Universidad Peruana Cayetano Heredia | Pablo Tsukayama et al |
| EPI_ISL_529069 | Peru/LIM-UPCH-0012/2020 | 2020-06-17 | South America/Peru/Lima/Lima | Laboratorio de Referencia Nacional de Virus Respiratorios, Instituto Nacional de Salud Peru | Laboratorio de Gen√≥mica Microbiana, Universidad Peruana Cayetano Heredia | Pablo Tsukayama et al |
| EPI_ISL_529070 | Peru/CAL-UPCH-0013/2020 | 2020-06-17 | South America/Peru/Callao/ | Laboratorio de Referencia Nacional de Virus Respiratorios, Instituto Nacional de Salud Peru | Laboratorio de Gen√≥mica Microbiana, Universidad Peruana Cayetano Heredia | Pablo Tsukayama et al |
| EPI_ISL_529071 | Peru/LIM-UPCH-0014/2020 | 2020-06-16 | South America/Peru/Lima/Lima | Laboratorio de Referencia Nacional de Virus Respiratorios, Instituto Nacional de Salud Peru | Laboratorio de Gen√≥mica Microbiana, Universidad Peruana Cayetano Heredia | Pablo Tsukayama et al |

| | | | | | | |
|---|---|---|---|---|---|---|
| EPI_ISL_529073 | Peru/CAL-UPCH-0036/2020 | 2020-06-18 | South America/Peru/Callao/ | Laboratorio de Referencia Nacional de Virus Respiratorios, Instituto Nacional de Salud Peru | Laboratorio de Genv≥mica Microbiana, Universidad Peruana Cayetano Heredia | Pablo Tsukayama et al |
| EPI_ISL_529074 | Peru/CAL-UPCH-0037/2020 | 2020-06-17 | South America/Peru/Callao/ | Laboratorio de Referencia Nacional de Virus Respiratorios, Instituto Nacional de Salud Peru | Laboratorio de Genv≥mica Microbiana, Universidad Peruana Cayetano Heredia | Pablo Tsukayama et al |
| EPI_ISL_529075 | Peru/LIM-UPCH-0038/2020 | 2020-06-16 | South America/Peru/Lima/Lima | Laboratorio de Referencia Nacional de Virus Respiratorios, Instituto Nacional de Salud Peru | Laboratorio de Genv≥mica Microbiana, Universidad Peruana Cayetano Heredia | Pablo Tsukayama et al |
| EPI_ISL_529076 | Peru/LIM-UPCH-0039/2020 | 2020-06-18 | South America/Peru/Lima/Lima | Laboratorio de Referencia Nacional de Virus Respiratorios, Instituto Nacional de Salud Peru | Laboratorio de Genv≥mica Microbiana, Universidad Peruana Cayetano Heredia | Pablo Tsukayama et al |
| EPI_ISL_529077 | Peru/LIM-UPCH-0041/2020 | 2020-06-17 | South America/Peru/Lima/Lima | Laboratorio de Referencia Nacional de Virus Respiratorios, Instituto Nacional de Salud Peru | Laboratorio de Genv≥mica Microbiana, Universidad Peruana Cayetano Heredia | Pablo Tsukayama et al |
| EPI_ISL_529079 | Peru/CAL-UPCH-0044/2020 | 2020-06-16 | South America/Peru/Callao/ | Laboratorio de Referencia Nacional de Virus Respiratorios, Instituto Nacional de Salud Peru | Laboratorio de Genv≥mica Microbiana, Universidad Peruana Cayetano Heredia | Pablo Tsukayama et al |
| EPI_ISL_529081 | USA/SC-DHEC-0533/2020 | 2020-06-04 | North America/USA/South Carolina/ | Microbiology Division, SC DHEC | Microbiology Division, SC DHEC | Flores et al |
| EPI_ISL_529095 | USA/SC-DHEC-0586/2020 | 2020-06-19 | North America/USA/South Carolina/ | Microbiology Division, SC DHEC | Microbiology Division, SC DHEC | Flores et al |

| | | | | | | |
|---|---|---|---|---|---|---|
| EPI_ISL_529096 | USA/SC-DHEC-0587/2020 | 2020-06-19 | North America/USA/South Carolina/ | Microbiology Division, SC DHEC | Microbiology Division, SC DHEC | Flores et al |
| EPI_ISL_529135 | Japan/QH-329-037/2020 | 2020-03-29 | Asia/Japan/Japan/ | Department of Virology III, National Institute of Infectious Diseases | Pathogen Genomics Center, National Institute of Infectious Diseases | Tsuyoshi Sekizuka et al |
| EPI_ISL_529138 | Indonesia/EJ-ITD-8402NT/2020 | 2020-05-11 | Asia/Indonesia/East Java/Surabaya | RSAL Dr. Ramelan Surabaya | Institute of Tropical Disease, Universitas Airlangga | Jezzy R Dewantari et al |
| EPI_ISL_529139 | Brazil/RJ-DCVN2/2020 | 2020-07-20 | South America/Brazil/Rio de Janeiro/ | Centro de Desenvolvimento Tecnologico em Saude, Fundacao Oswaldo Cruz | Centro de Desenvolvimento Tecnologico em Saude, Fundacao Oswaldo Cruz | Souza et al |
| EPI_ISL_529140 | Brazil/RJ-DCVN3/2020 | 2020-07-20 | South America/Brazil/Rio de Janeiro/ | Centro de Desenvolvimento Tecnologico em Saude, Fundacao Oswaldo Cruz | Centro de Desenvolvimento Tecnologico em Saude, Fundacao Oswaldo Cruz | Souza et al |
| EPI_ISL_529141 | Egypt/CUNCI-HGC7I01/2020 | 2020-07-19 | Africa/Egypt/Cairo/ | Egyptian National Cancer Institute (ENCI) | Egyptian National Cancer Institute (ENCI) | Zekri et al |
| EPI_ISL_529142 | Egypt/CUNCI-HGC7I02/2020 | 2020-07-19 | Africa/Egypt/Cairo/ | Egyptian National Cancer Institute (ENCI) | Egyptian National Cancer Institute (ENCI) | Zekri et al |
| EPI_ISL_529149 | Guangzhou/IQTC03/2020 | 2020-01-29 | Asia/China/Guangdong/Guangzhou | Technology Centre, Guangzhou Customs | Technology Centre, Guangzhou Customs | Huang et al |
| EPI_ISL_529150 | Guangzhou/IQTC04/2020 | 2020-01-27 | Asia/China/Guangdong/Guangzhou | Technology Centre, Guangzhou Customs | Technology Centre, Guangzhou Customs | Huang et al |
| EPI_ISL_529153 | USA/CA-ALSR-0231/2020 | 2020-04-25 | North America/USA/California/ | Department of Immunology, The Scripps Research Institute | Andersen lab at Scripps Research | Quigley et al |

| | | | | | | |
|---|---|---|---|---|---|---|
| EPI_ISL_529155 | USA/CT-UW420/2020 | 2020-03-28 | North America/USA/Connecticut/ | University of Washington, Laboratory Medicine | University of Washington, Laboratory Medicine | Roychoudhury et al |
| EPI_ISL_529169 | USA/MI-MDHHS-SC2-0008/2020 | 2020-03-12 | North America/USA/Michigan/ | Michigan Department of Health and Human Services, Bureau of Laboratories | Michigan Department of Health and Human Services, Bureau of Laboratories | Blankenship et al |
| EPI_ISL_529199 | USA/SC-DHEC-0238/2020 | 2020-06-03 | North America/USA/South Carolina/ | South Carolina Department of Health and Environmental Control | South Carolina Department of Health and Environmental Control | Haley V. Flores et al |
| EPI_ISL_529213 | Wuhan/IME-WH01/2019 | 2019-12-30 | Asia/China/Hubei/Wuhan | Beijing Institute of Microbiology and Epidemiology | Beijing Institute of Microbiology and Epidemiology | Fan et al |
| EPI_ISL_529214 | Wuhan/IME-WH02/2019 | 2019-12-30 | Asia/China/Hubei/Wuhan | Beijing Institute of Microbiology and Epidemiology | Beijing Institute of Microbiology and Epidemiology | Fan et al |
| EPI_ISL_529215 | Wuhan/IME-WH03/2019 | 2019-12-30 | Asia/China/Hubei/Wuhan | Beijing Institute of Microbiology and Epidemiology | Beijing Institute of Microbiology and Epidemiology | Fan et al |
| EPI_ISL_529216 | Wuhan/IME-WH04/2019 | 2019-12-30 | Asia/China/Hubei/Wuhan | Beijing Institute of Microbiology and Epidemiology | Beijing Institute of Microbiology and Epidemiology | Fan et al |
| EPI_ISL_529217 | Wuhan/IME-WH05/2019 | 2019-12-30 | Asia/China/Hubei/Wuhan | Beijing Institute of Microbiology and Epidemiology | Beijing Institute of Microbiology and Epidemiology | Fan et al |
| EPI_ISL_529243 | England/NORW-EE730/2020 | 2020-08-24 | Europe/United Kingdom/England/ | Quadram Institute Bioscience | COVID-19 Genomics UK (COG-UK) Consortium | Dave J. Baker et al |
| EPI_ISL_529252 | England/BIRM-637E8/2020 | 2020-08-19 | Europe/United Kingdom/England/ | University of Birmingham | COVID-19 Genomics UK (COG-UK) Consortium | Institute of Microbiology et al |
| EPI_ISL_529267 | England/ALDP-985E3D/2020 | 2020-08-07 | Europe/United Kingdom/England/ | Quadram Institute Bioscience | COVID-19 Genomics UK (COG-UK) Consortium | Dave J. Baker et al |

| | | | | | | |
|---|---|---|---|---|---|---|
| EPI_ISL_529427 | England/BIRM-61C72/2020 | 2020-03-25 | Europe/United Kingdom/England/ | University of Birmingham | COVID-19 Genomics UK (COG-UK) Consortium | Institute of Microbiology et al |
| EPI_ISL_529534 | England/ALDP-97FB3F/2020 | 2020-08-05 | Europe/United Kingdom/England/ | Queens Medical Centre, Clinical Microbiology Department / DeepSeq Nottingham | COVID-19 Genomics UK (COG-UK) Consortium | Gemma Clark et al |
| EPI_ISL_529647 | England/BIRM-6377F/2020 | 2020-08-16 | Europe/United Kingdom/England/ | University of Birmingham | COVID-19 Genomics UK (COG-UK) Consortium | Institute of Microbiology et al |
| EPI_ISL_529718 | Indonesia/JB-TFRIC19-R48704/2020 | 2020-07-22 | Asia/Indonesia/West Java/ | Santo Borromeus Hospital | Laboratorium Kesehatan Provinsi Jawa Barat; School of Life Sciences and Technology & School of Pharmacy-Institut Teknologi Bandung; Molecular Genetics Laboratory-Faculty of Medicine-Universitas Padjadjaran | Ema Rahmawati et al |
| EPI_ISL_529720 | SouthAfrica/KRISP-K002717/2020 | 2020-08-14 | Africa/South Africa/KwaZulu-Natal/Ama | NHLS-IALCH | KRISP, KZN Research Innovation and Sequencing Platform | Giandhari J et al |
| EPI_ISL_529721 | SouthAfrica/KRISP-K002720/2020 | 2020-08-16 | Africa/South Africa/KwaZulu-Natal/King | NHLS-IALCH | KRISP, KZN Research Innovation and Sequencing Platform | Giandhari J et al |
| EPI_ISL_529722 | SouthAfrica/KRISP-K002721/2020 | 2020-08-14 | Africa/South Africa/KwaZulu-Natal/Ama | NHLS-IALCH | KRISP, KZN Research Innovation and Sequencing Platform | Giandhari J et al |

| | | | | | | |
|---|---|---|---|---|---|---|
| EPI_ISL_529725 | SouthAfrica/KRISP-K002726/2020 | 2020-08-14 | Africa/South Africa/KwaZulu-Natal/Ama | NHLS-IALCH | KRISP, KZN Research Innovation and Sequencing Platform | Giandhari J et al |
| EPI_ISL_529726 | SouthAfrica/KRISP-K002727/2020 | 2020-08-14 | Africa/South Africa/KwaZulu-Natal/Ama | NHLS-IALCH | KRISP, KZN Research Innovation and Sequencing Platform | Giandhari J et al |
| EPI_ISL_529727 | SouthAfrica/KRISP-K002728/2020 | 2020-08-14 | Africa/South Africa/KwaZulu-Natal/Ama | NHLS-IALCH | KRISP, KZN Research Innovation and Sequencing Platform | Giandhari J et al |
| EPI_ISL_529728 | SouthAfrica/KRISP-K002729/2020 | 2020-08-14 | Africa/South Africa/KwaZulu-Natal/Ama | NHLS-IALCH | KRISP, KZN Research Innovation and Sequencing Platform | Giandhari J et al |
| EPI_ISL_529733 | SouthAfrica/KRISP-K002735/2020 | 2020-08-16 | Africa/South Africa/KwaZulu-Natal/Harr | NHLS-IALCH | KRISP, KZN Research Innovation and Sequencing Platform | Giandhari J et al |
| EPI_ISL_529734 | SouthAfrica/KRISP-K002736/2020 | 2020-08-15 | Africa/South Africa/KwaZulu-Natal/Ugu | NHLS-IALCH | KRISP, KZN Research Innovation and Sequencing Platform | Giandhari J et al |
| EPI_ISL_529735 | SouthAfrica/KRISP-K002737/2020 | 2020-08-14 | Africa/South Africa/KwaZulu-Natal/King | NHLS-IALCH | KRISP, KZN Research Innovation and Sequencing Platform | Giandhari J et al |
| EPI_ISL_529736 | SouthAfrica/KRISP-K002738/2020 | 2020-08-14 | Africa/South Africa/KwaZulu-Natal/Ugu | NHLS-IALCH | KRISP, KZN Research Innovation and Sequencing Platform | Giandhari J et al |

| | | | | | | |
|---|---|---|---|---|---|---|
| EPI_ISL_529737 | SouthAfrica/KRISP-K002739/2020 | 2020-08-13 | Africa/South Africa/KwaZulu-Natal/Ugu | NHLS-IALCH | KRISP, KZN Research Innovation and Sequencing Platform | Giandhari J et al |
| EPI_ISL_529738 | SouthAfrica/KRISP-K002740/2020 | 2020-08-14 | Africa/South Africa/KwaZulu-Natal/King | NHLS-IALCH | KRISP, KZN Research Innovation and Sequencing Platform | Giandhari J et al |
| EPI_ISL_529740 | SouthAfrica/KRISP-K002744/2020 | 2020-08-15 | Africa/South Africa/KwaZulu-Natal/uThu | NHLS-IALCH | KRISP, KZN Research Innovation and Sequencing Platform | Giandhari J et al |
| EPI_ISL_529741 | SouthAfrica/KRISP-K002745/2020 | 2020-08-15 | Africa/South Africa/KwaZulu-Natal/uThu | NHLS-IALCH | KRISP, KZN Research Innovation and Sequencing Platform | Giandhari J et al |
| EPI_ISL_529742 | SouthAfrica/KRISP-K002746/2020 | 2020-08-13 | Africa/South Africa/KwaZulu-Natal/uMg | NHLS-IALCH | KRISP, KZN Research Innovation and Sequencing Platform | Giandhari J et al |
| EPI_ISL_529745 | SouthAfrica/KRISP-K002749/2020 | 2020-08-16 | Africa/South Africa/KwaZulu-Natal/Ugu | NHLS-IALCH | KRISP, KZN Research Innovation and Sequencing Platform | Giandhari J et al |
| EPI_ISL_529754 | SouthAfrica/KRISP-K002758/2020 | 2020-08-12 | Africa/South Africa/KwaZulu-Natal/uMk | NHLS-IALCH | KRISP, KZN Research Innovation and Sequencing Platform | Giandhari J et al |
| EPI_ISL_529755 | SouthAfrica/KRISP-K002759/2020 | 2020-08-15 | Africa/South Africa/KwaZulu-Natal/Zulu | NHLS-IALCH | KRISP, KZN Research Innovation and Sequencing Platform | Giandhari J et al |

| | | | | | | |
|---|---|---|---|---|---|---|
| EPI_ISL_529759 | SouthAfrica/KRISP-K002763/2020 | 2020-08-16 | Africa/South Africa/KwaZulu-Natal/eThe | NHLS-IALCH | KRISP, KZN Research Innovation and Sequencing Platform | Giandhari J et al |
| EPI_ISL_529760 | SouthAfrica/KRISP-K002764/2020 | 2020-08-12 | Africa/South Africa/KwaZulu-Natal/uMk | NHLS-IALCH | KRISP, KZN Research Innovation and Sequencing Platform | Giandhari J et al |
| EPI_ISL_529762 | SouthAfrica/KRISP-K002766/2020 | 2020-08-12 | Africa/South Africa/KwaZulu-Natal/eThe | NHLS-IALCH | KRISP, KZN Research Innovation and Sequencing Platform | Giandhari J et al |
| EPI_ISL_529763 | SouthAfrica/KRISP-K002768/2020 | 2020-08-21 | Africa/South Africa/KwaZulu-Natal/iLem | NHLS-IALCH | KRISP, KZN Research Innovation and Sequencing Platform | Giandhari J et al |
| EPI_ISL_529766 | SouthAfrica/KRISP-K002771/2020 | 2020-08-21 | Africa/South Africa/KwaZulu-Natal/iLem | NHLS-IALCH | KRISP, KZN Research Innovation and Sequencing Platform | Giandhari J et al |
| EPI_ISL_529768 | SouthAfrica/KRISP-K002773/2020 | 2020-08-21 | Africa/South Africa/KwaZulu-Natal/iLem | NHLS-IALCH | KRISP, KZN Research Innovation and Sequencing Platform | Giandhari J et al |
| EPI_ISL_529769 | SouthAfrica/KRISP-K002774/2020 | 2020-08-21 | Africa/South Africa/KwaZulu-Natal/uThu | NHLS-IALCH | KRISP, KZN Research Innovation and Sequencing Platform | Giandhari J et al |
| EPI_ISL_529770 | SouthAfrica/KRISP-K002775/2020 | 2020-08-21 | Africa/South Africa/KwaZulu-Natal/uThu | NHLS-IALCH | KRISP, KZN Research Innovation and Sequencing Platform | Giandhari J et al |

| | | | | | | |
|---|---|---|---|---|---|---|
| EPI_ISL_529772 | SouthAfrica/KRISP-K002777/2020 | 2020-08-21 | Africa/South Africa/KwaZulu-Natal/uThu | NHLS-IALCH | KRISP, KZN Research Innovation and Sequencing Platform | Giandhari J et al |
| EPI_ISL_529773 | SouthAfrica/KRISP-K002779/2020 | 2020-08-21 | Africa/South Africa/KwaZulu-Natal/uThu | NHLS-IALCH | KRISP, KZN Research Innovation and Sequencing Platform | Giandhari J et al |
| EPI_ISL_529774 | SouthAfrica/KRISP-K002780/2020 | 2020-08-20 | Africa/South Africa/KwaZulu-Natal/iLem | NHLS-IALCH | KRISP, KZN Research Innovation and Sequencing Platform | Giandhari J et al |
| EPI_ISL_529776 | SouthAfrica/KRISP-K002782/2020 | 2020-08-20 | Africa/South Africa/KwaZulu-Natal/iLem | NHLS-IALCH | KRISP, KZN Research Innovation and Sequencing Platform | Giandhari J et al |
| EPI_ISL_529779 | SouthAfrica/KRISP-K002785/2020 | 2020-08-24 | Africa/South Africa/KwaZulu-Natal/uThu | NHLS-IALCH | KRISP, KZN Research Innovation and Sequencing Platform | Giandhari J et al |
| EPI_ISL_529780 | SouthAfrica/KRISP-K002786/2020 | 2020-08-23 | Africa/South Africa/KwaZulu-Natal/uThu | NHLS-IALCH | KRISP, KZN Research Innovation and Sequencing Platform | Giandhari J et al |
| EPI_ISL_529782 | SouthAfrica/KRISP-K002788/2020 | 2020-08-24 | Africa/South Africa/KwaZulu-Natal/uThu | NHLS-IALCH | KRISP, KZN Research Innovation and Sequencing Platform | Giandhari J et al |
| EPI_ISL_529783 | SouthAfrica/KRISP-K002789/2020 | 2020-08-21 | Africa/South Africa/KwaZulu-Natal/uThu | NHLS-IALCH | KRISP, KZN Research Innovation and Sequencing Platform | Giandhari J et al |

| | | | | | | |
|---|---|---|---|---|---|---|
| EPI_ISL_529784 | SouthAfrica/KRISP-K002790/2020 | 2020-08-23 | Africa/South Africa/KwaZulu-Natal/uThu | NHLS-IALCH | KRISP, KZN Research Innovation and Sequencing Platform | Giandhari J et al |
| EPI_ISL_529786 | SouthAfrica/KRISP-K002792/2020 | 2020-08-23 | Africa/South Africa/KwaZulu-Natal/uThu | NHLS-IALCH | KRISP, KZN Research Innovation and Sequencing Platform | Giandhari J et al |
| EPI_ISL_529787 | SouthAfrica/KRISP-K002793/2020 | 2020-08-24 | Africa/South Africa/KwaZulu-Natal/iLem | NHLS-IALCH | KRISP, KZN Research Innovation and Sequencing Platform | Giandhari J et al |
| EPI_ISL_529789 | SouthAfrica/KRISP-K002795/2020 | 2020-08-23 | Africa/South Africa/KwaZulu-Natal/iLem | NHLS-IALCH | KRISP, KZN Research Innovation and Sequencing Platform | Giandhari J et al |
| EPI_ISL_529791 | SouthAfrica/KRISP-K002797/2020 | 2020-08-24 | Africa/South Africa/KwaZulu-Natal/iLem | NHLS-IALCH | KRISP, KZN Research Innovation and Sequencing Platform | Giandhari J et al |
| EPI_ISL_529792 | SouthAfrica/KRISP-K002798/2020 | 2020-08-24 | Africa/South Africa/KwaZulu-Natal/iLem | NHLS-IALCH | KRISP, KZN Research Innovation and Sequencing Platform | Giandhari J et al |
| EPI_ISL_529793 | SouthAfrica/KRISP-K002799/2020 | 2020-08-24 | Africa/South Africa/KwaZulu-Natal/iLem | NHLS-IALCH | KRISP, KZN Research Innovation and Sequencing Platform | Giandhari J et al |
| EPI_ISL_529794 | SouthAfrica/KRISP-K002800/2020 | 2020-08-24 | Africa/South Africa/KwaZulu-Natal/iLem | NHLS-IALCH | KRISP, KZN Research Innovation and Sequencing Platform | Giandhari J et al |

| | | | | | | |
|---|---|---|---|---|---|---|
| EPI_ISL_529795 | SouthAfrica/KRISP-K002801/2020 | 2020-08-24 | Africa/South Africa/KwaZulu-Natal/iLem | NHLS-IALCH | KRISP, KZN Research Innovation and Sequencing Platform | Giandhari J et al |
| EPI_ISL_529796 | SouthAfrica/KRISP-K002802/2020 | 2020-08-20 | Africa/South Africa/KwaZulu-Natal/Zulu | NHLS-IALCH | KRISP, KZN Research Innovation and Sequencing Platform | Giandhari J et al |
| EPI_ISL_529799 | SouthAfrica/KRISP-K002805/2020 | 2020-08-24 | Africa/South Africa/KwaZulu-Natal/uThu | NHLS-IALCH | KRISP, KZN Research Innovation and Sequencing Platform | Giandhari J et al |
| EPI_ISL_529800 | SouthAfrica/KRISP-K002806/2020 | 2020-08-25 | Africa/South Africa/KwaZulu-Natal/uThu | NHLS-IALCH | KRISP, KZN Research Innovation and Sequencing Platform | Giandhari J et al |
| EPI_ISL_529801 | SouthAfrica/KRISP-K002807/2020 | 2020-08-25 | Africa/South Africa/KwaZulu-Natal/Ama | NHLS-IALCH | KRISP, KZN Research Innovation and Sequencing Platform | Giandhari J et al |
| EPI_ISL_529802 | SouthAfrica/KRISP-K002808/2020 | 2020-08-25 | Africa/South Africa/KwaZulu-Natal/uThu | NHLS-IALCH | KRISP, KZN Research Innovation and Sequencing Platform | Giandhari J et al |
| EPI_ISL_529820 | USA/MI-MDHHS-SC21701/2020 | 2020-03-25 | North America/USA/Michigan/ | Michigan Department of Health and Human Services, Bureau of Laboratories | Michigan Department of Health and Human Services, Bureau of Laboratories | Blankenship HM et al |

| | | | | | | |
|---|---|---|---|---|---|---|
| EPI_ISL_529837 | USA/MI-MDHHS-SC21718/2020 | 2020-07-13 | North America/USA/Michigan/ | Michigan Department of Health and Human Services, Bureau of Laboratories | Michigan Department of Health and Human Services, Bureau of Laboratories | Blankenship HM et al |
| EPI_ISL_529838 | USA/MI-MDHHS-SC21719/2020 | 2020-07-14 | North America/USA/Michigan/ | Michigan Department of Health and Human Services, Bureau of Laboratories | Michigan Department of Health and Human Services, Bureau of Laboratories | Blankenship HM et al |
| EPI_ISL_529839 | USA/MI-MDHHS-SC21720/2020 | 2020-07-14 | North America/USA/Michigan/ | Michigan Department of Health and Human Services, Bureau of Laboratories | Michigan Department of Health and Human Services, Bureau of Laboratories | Blankenship HM et al |
| EPI_ISL_529845 | USA/MI-MDHHS-SC21726/2020 | 2020-08-04 | North America/USA/Michigan/ | Michigan Department of Health and Human Services, Bureau of Laboratories | Michigan Department of Health and Human Services, Bureau of Laboratories | Blankenship HM et al |
| EPI_ISL_529846 | USA/MI-MDHHS-SC21727/2020 | 2020-08-07 | North America/USA/Michigan/ | Michigan Department of Health and Human Services, Bureau of Laboratories | Michigan Department of Health and Human Services, Bureau of Laboratories | Blankenship HM et al |
| EPI_ISL_529852 | USA/MI-MDHHS-SC21733/2020 | 2020-07-08 | North America/USA/Michigan/ | Michigan Department of Health and Human Services, Bureau of Laboratories | Michigan Department of Health and Human Services, Bureau of Laboratories | Blankenship HM et al |

| | | | | | | |
|---|---|---|---|---|---|---|
| EPI_ISL_529855 | USA/MI-MDHHS-SC21736/2020 | 2020-08-10 | North America/USA/Michigan/ | Michigan Department of Health and Human Services, Bureau of Laboratories | Michigan Department of Health and Human Services, Bureau of Laboratories | Blankenship HM et al |
| EPI_ISL_529856 | USA/MI-MDHHS-SC21737/2020 | 2020-08-11 | North America/USA/Michigan/ | Michigan Department of Health and Human Services, Bureau of Laboratories | Michigan Department of Health and Human Services, Bureau of Laboratories | Blankenship HM et al |
| EPI_ISL_529860 | USA/MI-MDHHS-SC21742/2020 | 2020-07-29 | North America/USA/Michigan/ | Michigan Department of Health and Human Services, Bureau of Laboratories | Michigan Department of Health and Human Services, Bureau of Laboratories | Blankenship HM et al |
| EPI_ISL_529865 | USA/MI-MDHHS-SC21747/2020 | 2020-07-31 | North America/USA/Michigan/ | Michigan Department of Health and Human Services, Bureau of Laboratories | Michigan Department of Health and Human Services, Bureau of Laboratories | Blankenship HM et al |
| EPI_ISL_529868 | USA/MI-MDHHS-SC21750/2020 | 2020-07-31 | North America/USA/Michigan/ | Michigan Department of Health and Human Services, Bureau of Laboratories | Michigan Department of Health and Human Services, Bureau of Laboratories | Blankenship HM et al |
| EPI_ISL_529871 | USA/MI-MDHHS-SC21753/2020 | 2020-07-22 | North America/USA/Michigan/ | Michigan Department of Health and Human Services, Bureau of Laboratories | Michigan Department of Health and Human Services, Bureau of Laboratories | Blankenship HM et al |

| | | | | | | |
|---|---|---|---|---|---|---|
| EPI_ISL_529876 | USA/MI-MDHHS-SC21758/2020 | 2020-08-05 | North America/USA/Michigan/ | Michigan Department of Health and Human Services, Bureau of Laboratories | Michigan Department of Health and Human Services, Bureau of Laboratories | Blankenship HM et al |
| EPI_ISL_529886 | USA/MI-MDHHS-SC21768/2020 | 2020-08-13 | North America/USA/Michigan/ | Michigan Department of Health and Human Services, Bureau of Laboratories | Michigan Department of Health and Human Services, Bureau of Laboratories | Blankenship HM et al |
| EPI_ISL_529888 | USA/MI-MDHHS-SC21770/2020 | 2020-08-13 | North America/USA/Michigan/ | Michigan Department of Health and Human Services, Bureau of Laboratories | Michigan Department of Health and Human Services, Bureau of Laboratories | Blankenship HM et al |
| EPI_ISL_529894 | USA/MI-MDHHS-SC21776/2020 | 2020-08-13 | North America/USA/Michigan/ | Michigan Department of Health and Human Services, Bureau of Laboratories | Michigan Department of Health and Human Services, Bureau of Laboratories | Blankenship HM et al |
| EPI_ISL_529903 | USA/MI-MDHHS-SC21785/2020 | 2020-08-14 | North America/USA/Michigan/ | Michigan Department of Health and Human Services, Bureau of Laboratories | Michigan Department of Health and Human Services, Bureau of Laboratories | Blankenship HM et al |
| EPI_ISL_529906 | USA/MI-MDHHS-SC21788/2020 | 2020-08-11 | North America/USA/Michigan/ | Michigan Department of Health and Human Services, Bureau of Laboratories | Michigan Department of Health and Human Services, Bureau of Laboratories | Blankenship HM et al |
| EPI_ISL_529915 | USA/VA-DCLS-1113/2020 | 2020-06-17 | North America/USA/Virginia/ | Virginia Division of Consolidated Laboratory Services | Virginia Division of Consolidated Laboratory Services | Virginia DCLS et al |

| | | | | | | |
|---|---|---|---|---|---|---|
| EPI_ISL_529938 | USA/VA-DCLS-1245/2020 | 2020-07-30 | North America/USA/Virginia/ | Virginia DCLS | Virginia DCLS | Virginia DCLS et al |
| EPI_ISL_529944 | USA/VA-DCLS-1253/2020 | 2020-08-06 | North America/USA/Virginia/ | Virginia DCLS | Virginia DCLS | Virginia DCLS et al |
| EPI_ISL_529945 | USA/VA-DCLS-1254/2020 | 2020-08-06 | North America/USA/Virginia/ | Virginia DCLS | Virginia DCLS | Virginia DCLS et al |
| EPI_ISL_529948 | USA/VA-DCLS-1258/2020 | 2020-08-06 | North America/USA/Virginia/ | Virginia DCLS | Virginia DCLS | Virginia DCLS et al |
| EPI_ISL_529951 | USA/VA-DCLS-1261/2020 | 2020-08-06 | North America/USA/Virginia/ | Virginia DCLS | Virginia DCLS | Virginia DCLS et al |
| EPI_ISL_529952 | USA/VA-DCLS-1262/2020 | 2020-08-06 | North America/USA/Virginia/ | Virginia DCLS | Virginia DCLS | Virginia DCLS et al |
| EPI_ISL_529953 | USA/VA-DCLS-1263/2020 | 2020-08-06 | North America/USA/Virginia/ | Virginia DCLS | Virginia DCLS | Virginia DCLS et al |
| EPI_ISL_529958 | USA/VA-DCLS-1271/2020 | 2020-08-12 | North America/USA/Virginia/ | Virginia DCLS | Virginia DCLS | Virginia DCLS et al |
| EPI_ISL_529963 | Indonesia/JI-ITD-17398NT/2020 | 2020-07-18 | Asia/Indonesia/East Java/Surabaya | Universitas Airlangga Hospital | Institute of Tropical Disease, Universitas Airlangga | Rima R Prasetya et al |
| EPI_ISL_529965 | Indonesia/JI-ITD-4859V/2020 | 2020-04-22 | Asia/Indonesia/East Java/Surabaya | RSUD Dr. Soetomo | Institute of Tropical Disease, Universitas Airlangga | Krisnoadi Rahardjo et al |
| EPI_ISL_529966 | Indonesia/JI-ITD-7061V/2020 | 2020-05-05 | Asia/Indonesia/East Java/Sidoarjo | RSUD Sidoarjo | Institute of Tropical Disease, Universitas Airlangga | Kazufumi Shimizu et al |
| EPI_ISL_529967 | Indonesia/JI-ITD-12202V/2020 | 2020-06-09 | Asia/Indonesia/East Java/Sidoarjo | RSUD Sidoarjo | Institute of Tropical Disease, Universitas Airlangga | Aldise M Nastri et al |
| EPI_ISL_529991 | Spain/MD-H12-59/2020 | 2020-04-03 | Europe/Spain/Madrid/ | Hospital Universitario 12 de Octubre | Hospital Universitario 12 de Octubre | Sara Gonzv°lez et al |
| EPI_ISL_530000 | Spain/MD-H12-69/2020 | 2020-04-11 | Europe/Spain/Madrid/ | Hospital Universitario 12 de Octubre | Hospital Universitario 12 de Octubre | Sara Gonzv°lez et al |
| EPI_ISL_530001 | Spain/MD-H12-70/2020 | 2020-04-11 | Europe/Spain/Madrid/ | Hospital Universitario 12 de Octubre | Hospital Universitario 12 de Octubre | Sara Gonzv°lez et al |
| EPI_ISL_530003 | Spain/MD-H12-72/2020 | 2020-04-13 | Europe/Spain/Madrid/ | Hospital Universitario 12 de Octubre | Hospital Universitario 12 de Octubre | Sara Gonzv°lez et al |

| | | | | | | |
|---|---|---|---|---|---|---|
| EPI_ISL_530025 | Spain/MD-HLP-100-1/2020 | 2020-04-26 | Europe/Spain/Madrid/ | Hospital Universitario La Paz | Hospital Universitario La Paz | Marv≠a Rodrv≠guez et al |
| EPI_ISL_530039 | Spain/MD-HLP-110/2020 | 2020-05-21 | Europe/Spain/Madrid/ | Hospital Universitario La Paz | Hospital Universitario La Paz | Marv≠a Rodrv≠guez et al |
| EPI_ISL_530040 | Spain/MD-HLP-111/2020 | 2020-05-27 | Europe/Spain/Madrid/ | Hospital Universitario La Paz | Hospital Universitario La Paz | Marv≠a Rodrv≠guez et al |
| EPI_ISL_530042 | Spain/MD-HLP-113/2020 | 2020-02-27 | Europe/Spain/Madrid/ | Hospital Universitario La Paz | Hospital Universitario La Paz | Marv≠a Rodrv≠guez et al |
| EPI_ISL_530043 | Spain/MD-HLP-114/2020 | 2020-05-18 | Europe/Spain/Madrid/ | Hospital Universitario La Paz | Hospital Universitario La Paz | Marv≠a Rodrv≠guez et al |
| EPI_ISL_530050 | Spain/MD-HLP-46/2020 | 2020-04-12 | Europe/Spain/Madrid/ | Hospital Universitario La Paz | Hospital Universitario La Paz | Elias Dahdouh et al |
| EPI_ISL_530056 | Spain/MD-HLP-53/2020 | 2020-04-20 | Europe/Spain/Madrid/ | Hospital Universitario La Paz | Hospital Universitario La Paz | Elias Dahdouh et al |
| EPI_ISL_530063 | Spain/MD-HLP-60/2020 | 2020-04-20 | Europe/Spain/Madrid/ | Hospital Universitario La Paz | Hospital Universitario La Paz | Elias Dahdouh et al |
| EPI_ISL_530064 | Spain/MD-HLP-61/2020 | 2020-04-20 | Europe/Spain/Madrid/ | Hospital Universitario La Paz | Hospital Universitario La Paz | Elias Dahdouh et al |
| EPI_ISL_530065 | Spain/MD-HLP-62/2020 | 2020-04-21 | Europe/Spain/Madrid/ | Hospital Universitario La Paz | Hospital Universitario La Paz | Elias Dahdouh et al |
| EPI_ISL_530071 | Spain/MD-HLP-72/2020 | 2020-05-13 | Europe/Spain/Madrid/ | Hospital Universitario La Paz | Hospital Universitario La Paz | Marv≠a Rodrv≠guez et al |
| EPI_ISL_530072 | Spain/MD-HLP-73/2020 | 2020-05-22 | Europe/Spain/Madrid/ | Hospital Universitario La Paz | Hospital Universitario La Paz | Marv≠a Rodrv≠guez et al |
| EPI_ISL_530077 | Spain/MD-HLP-80/2020 | 2020-05-13 | Europe/Spain/Madrid/ | Hospital Universitario La Paz | Hospital Universitario La Paz | Marv≠a Rodrv≠guez et al |

| | | | | | | |
|---|---|---|---|---|---|---|
| EPI_ISL_530081 | Spain/MD-HLP-84/2020 | 2020-05-18 | Europe/Spain/Madrid/ | Hospital Universitario La Paz | Hospital Universitario La Paz | Mar√≠a Rodr√≠guez et al |
| EPI_ISL_530083 | Spain/MD-HLP-86/2020 | 2020-04-30 | Europe/Spain/Madrid/ | Hospital Universitario La Paz | Hospital Universitario La Paz | Mar√≠a Rodr√≠guez et al |
| EPI_ISL_530087 | Spain/MD-HLP-90/2020 | 2020-05-02 | Europe/Spain/Madrid/ | Hospital Universitario La Paz | Hospital Universitario La Paz | Mar√≠a Rodr√≠guez et al |
| EPI_ISL_530088 | Spain/MD-HLP-92/2020 | 2020-05-03 | Europe/Spain/Madrid/ | Hospital Universitario La Paz | Hospital Universitario La Paz | Mar√≠a Rodr√≠guez et al |
| EPI_ISL_530091 | Spain/MD-HLP-95/2020 | 2020-05-21 | Europe/Spain/Madrid/ | Hospital Universitario La Paz | Hospital Universitario La Paz | Mar√≠a Rodr√≠guez et al |
| EPI_ISL_530098 | Spain/MD-H12-R22/2020 | 2020-03-13 | Europe/Spain/Madrid/ | Hospital Universitario Ram√≥n y Cajal | Hospital Universitario 12 de Octubre | Ra√∫l Recio et al |
| EPI_ISL_530099 | Spain/MD-H12-R25/2020 | 2020-03-04 | Europe/Spain/Madrid/ | Hospital Universitario Ram√≥n y Cajal | Hospital Universitario 12 de Octubre | Ra√∫l Recio et al |
| EPI_ISL_530111 | Spain/MD-HLP-R65/2020 | 2020-04-16 | Europe/Spain/Madrid/ | Hospital Universitario Ram√≥n y Cajal | Hospital Universitario La Paz | Ra√∫l Recio et al |
| EPI_ISL_530114 | Spain/MD-HLP-R81/2020 | 2020-05-07 | Europe/Spain/Madrid/ | Hospital Universitario Ram√≥n y Cajal | Hospital Universitario La Paz | Mar√≠a Rodr√≠guez et al |
| EPI_ISL_530119 | Spain/MD-HLP-R90/2020 | 2020-05-21 | Europe/Spain/Madrid/ | Hospital Universitario Ram√≥n y Cajal | Hospital Universitario La Paz | Mar√≠a Rodr√≠guez et al |
| EPI_ISL_530124 | USA/WA-S2767/2020 | 2020-05-11 | North America/USA/Washington/ | Seattle Flu Study | Seattle Flu Study | Deborah A. Nickerson et al |
| EPI_ISL_530129 | USA/WA-S2772/2020 | 2020-08-03 | North America/USA/Washington/King C | Seattle Flu Study | Seattle Flu Study | Deborah A. Nickerson et al |
| EPI_ISL_530130 | USA/WA-S2773/2020 | 2020-08-04 | North America/USA/Washington/King C | Seattle Flu Study | Seattle Flu Study | Deborah A. Nickerson et al |
| EPI_ISL_530131 | USA/WA-S2774/2020 | 2020-08-05 | North America/USA/Washington/King C | Seattle Flu Study | Seattle Flu Study | Deborah A. Nickerson et al |
| EPI_ISL_530132 | USA/WA-S2775/2020 | 2020-08-06 | North America/USA/Washington/ | Seattle Flu Study | Seattle Flu Study | Deborah A. Nickerson et al |
| EPI_ISL_530135 | USA/WA-S2778/2020 | 2020-08-09 | North America/USA/Washington/King C | Seattle Flu Study | Seattle Flu Study | Deborah A. Nickerson et al |
| EPI_ISL_530139 | USA/WA-S2782/2020 | 2020-08-10 | North America/USA/Washington/King C | Seattle Flu Study | Seattle Flu Study | Deborah A. Nickerson et al |
| EPI_ISL_530142 | USA/WA-S2785/2020 | 2020-08-11 | North America/USA/Washington/King C | Seattle Flu Study | Seattle Flu Study | Deborah A. Nickerson et al |

| | | | | | | |
|---|---|---|---|---|---|---|
| EPI_ISL_530145 | USA/WA-S2788/2020 | 2020-08-12 | North America/USA/Washington/King C | Seattle Flu Study | Seattle Flu Study | Deborah A. Nickerson et al |
| EPI_ISL_530146 | USA/WA-S2789/2020 | 2020-08-13 | North America/USA/Washington/King C | Seattle Flu Study | Seattle Flu Study | Deborah A. Nickerson et al |
| EPI_ISL_530148 | USA/WA-S2791/2020 | 2020-08-14 | North America/USA/Washington/King C | Seattle Flu Study | Seattle Flu Study | Deborah A. Nickerson et al |
| EPI_ISL_530149 | USA/WA-S2792/2020 | 2020-08-14 | North America/USA/Washington/King C | Seattle Flu Study | Seattle Flu Study | Deborah A. Nickerson et al |
| EPI_ISL_530151 | USA/WA-S2794/2020 | 2020-08-14 | North America/USA/Washington/King C | Seattle Flu Study | Seattle Flu Study | Deborah A. Nickerson et al |
| EPI_ISL_530152 | USA/WA-S2795/2020 | 2020-08-16 | North America/USA/Washington/King C | Seattle Flu Study | Seattle Flu Study | Deborah A. Nickerson et al |
| EPI_ISL_530153 | USA/WA-S2796/2020 | 2020-08-19 | North America/USA/Washington/King C | Seattle Flu Study | Seattle Flu Study | Deborah A. Nickerson et al |
| EPI_ISL_530155 | USA/WA-S2798/2020 | 2020-08-22 | North America/USA/Washington/King C | Seattle Flu Study | Seattle Flu Study | Deborah A. Nickerson et al |
| EPI_ISL_530164 | USA/MN-MDH-1568/2020 | 2020-08-22 | North America/USA/Minnesota/ | M Health Fairview | Minnesota Department of Health, Public Health Laboratory | Matt Plumb et al |
| EPI_ISL_530165 | USA/MN-MDH-1569/2020 | 2020-08-21 | North America/USA/Minnesota/ | M Health Fairview | Minnesota Department of Health, Public Health Laboratory | Matt Plumb et al |
| EPI_ISL_530172 | USA/MN-MDH-1576/2020 | 2020-08-03 | North America/USA/Minnesota/ | Minnesota Department of Health, Public Health Laboratory | Minnesota Department of Health, Public Health Laboratory | Matt Plumb et al |
| EPI_ISL_530173 | USA/MN-MDH-1577/2020 | 2020-07-02 | North America/USA/Minnesota/ | Minnesota Department of Health, Public Health Laboratory | Minnesota Department of Health, Public Health Laboratory | Matt Plumb et al |
| EPI_ISL_530174 | USA/MN-MDH-1578/2020 | 2020-06-30 | North America/USA/Minnesota/ | Minnesota Department of Health, Public Health Laboratory | Minnesota Department of Health, Public Health Laboratory | Matt Plumb et al |
| EPI_ISL_530187 | USA/MN-MDH-1591/2020 | 2020-05-19 | North America/USA/Minnesota/ | Minnesota Department of Health, Public Health Laboratory | Minnesota Department of Health, Public Health Laboratory | Matt Plumb et al |
| EPI_ISL_530188 | USA/MN-MDH-1592/2020 | 2020-05-18 | North America/USA/Minnesota/ | Minnesota Department of Health, Public Health Laboratory | Minnesota Department of Health, Public Health Laboratory | Matt Plumb et al |

| | | | | | | |
|---|---|---|---|---|---|---|
| EPI_ISL_530191 | USA/MN-MDH-1595/2020 | 2020-05-13 | North America/USA/Minnesota/ | Minnesota Department of Health, Public Health Laboratory | Minnesota Department of Health, Public Health Laboratory | Matt Plumb et al |
| EPI_ISL_530197 | USA/MN-MDH-1601/2020 | 2020-05-05 | North America/USA/Minnesota/ | Minnesota Department of Health, Public Health Laboratory | Minnesota Department of Health, Public Health Laboratory | Matt Plumb et al |
| EPI_ISL_530211 | USA/MN-MDH-1615/2020 | 2020-04-25 | North America/USA/Minnesota/ | Minnesota Department of Health, Public Health Laboratory | Minnesota Department of Health, Public Health Laboratory | Matt Plumb et al |
| EPI_ISL_530267 | Australia/QLD987/2020 | 2020-03-26 | Oceania/Australia/Queensland/ | Queensland Health Forensic and Scientific Services, Public Health Virology | Public Health Virology Laboratory, Forensic and Scientific Services, Queensland Health | Son Nguyen et al et al |
| EPI_ISL_530269 | Australia/QLD1009/2020 | 2020-03-22 | Oceania/Australia/Queensland/ | Queensland Health Forensic and Scientific Services, Public Health Virology | Public Health Virology Laboratory, Forensic and Scientific Services, Queensland Health | Son Nguyen et al et al |
| EPI_ISL_530365 | England/QEUH-99045B/2020 | 2020-08-26 | Europe/United Kingdom/England/ | Lighthouse Lab in Glasgow | Wellcome Sanger Institute for the COVID-19 Genomics UK (COG-UK) consortium | Harper VanSteenhouse et al |
| EPI_ISL_530440 | Scotland/QEUH-96DFA1/2020 | 2020-08-20 | Europe/United Kingdom/Scotland/ | NHSGGC West of Scotland Specialist Virology Centre / MRC-University of Glasgow Centre for Virus Research | Wellcome Sanger Institute for the COVID-19 Genomics UK (COG-UK) consortium | Ana da Silva Filipe et al |
| EPI_ISL_530821 | NorthernIreland/QEUH-96C0FA/2020 | 2020-08-15 | Europe/United Kingdom/Northern Irelar | Lighthouse Lab in Glasgow | Wellcome Sanger Institute for the COVID-19 Genomics UK (COG-UK) consortium | Harper VanSteenhouse et al |

| | | | | | | |
|---|---|---|---|---|---|---|
| EPI_ISL_530917 | England/QEUH-969CBD/2020 | 2020-08-12 | Europe/United Kingdom/England/ | Lighthouse Lab in Glasgow | Wellcome Sanger Institute for the COVID-19 Genomics UK (COG-UK) consortium | Harper VanSteenhouse et al |
| EPI_ISL_531136 | England/QEUH-9652CE/2020 | 2020-08-12 | Europe/United Kingdom/England/ | Lighthouse Lab in Glasgow | Wellcome Sanger Institute for the COVID-19 Genomics UK (COG-UK) consortium | Harper VanSteenhouse et al |
| EPI_ISL_531186 | England/QEUH-9694A6/2020 | 2020-08-12 | Europe/United Kingdom/England/ | Lighthouse Lab in Glasgow | Wellcome Sanger Institute for the COVID-19 Genomics UK (COG-UK) consortium | Harper VanSteenhouse et al |
| EPI_ISL_531271 | England/QEUH-96B996/2020 | 2020-08-14 | Europe/United Kingdom/England/ | Lighthouse Lab in Glasgow | Wellcome Sanger Institute for the COVID-19 Genomics UK (COG-UK) consortium | Harper VanSteenhouse et al |
| EPI_ISL_531466 | England/QEUH-96A821/2020 | 2020-08-13 | Europe/United Kingdom/England/ | Lighthouse Lab in Glasgow | Wellcome Sanger Institute for the COVID-19 Genomics UK (COG-UK) consortium | Harper VanSteenhouse et al |
| EPI_ISL_531519 | England/QEUH-963011/2020 | 2020-08-11 | Europe/United Kingdom/England/ | Lighthouse Lab in Glasgow | Wellcome Sanger Institute for the COVID-19 Genomics UK (COG-UK) consortium | Harper VanSteenhouse et al |
| EPI_ISL_531582 | England/QEUH-962F60/2020 | 2020-08-10 | Europe/United Kingdom/England/ | Lighthouse Lab in Glasgow | Wellcome Sanger Institute for the COVID-19 Genomics UK (COG-UK) consortium | Harper VanSteenhouse et al |
| EPI_ISL_531632 | England/QEUH-963916/2020 | 2020-08-10 | Europe/United Kingdom/England/ | Lighthouse Lab in Glasgow | Wellcome Sanger Institute for the COVID-19 Genomics UK (COG-UK) consortium | Harper VanSteenhouse et al |

| | | | | | | |
|---|---|---|---|---|---|---|
| EPI_ISL_531692 | England/QEUH-9625E6/2020 | 2020-08-10 | Europe/United Kingdom/England/ | Lighthouse Lab in Glasgow | Wellcome Sanger Institute for the COVID-19 Genomics UK (COG-UK) consortium | Harper VanSteenhouse et al |
| EPI_ISL_532528 | England/QEUH-95EF2B/2020 | 2020-08-07 | Europe/United Kingdom/England/ | Lighthouse Lab in Glasgow | Wellcome Sanger Institute for the COVID-19 Genomics UK (COG-UK) consortium | Harper VanSteenhouse et al |
| EPI_ISL_532656 | NorthernIreland/QEUH-88F159/2020 | 2020-07-27 | Europe/United Kingdom/Northern Irelar | Lighthouse Lab in Glasgow | Wellcome Sanger Institute for the COVID-19 Genomics UK (COG-UK) consortium | Harper VanSteenhouse et al |
| EPI_ISL_532849 | England/QEUH-7BBA0F/2020 | 2020-07-23 | Europe/United Kingdom/England/ | Lighthouse Lab in Glasgow | Wellcome Sanger Institute for the COVID-19 Genomics UK (COG-UK) consortium | Harper VanSteenhouse et al |
| EPI_ISL_533180 | England/QEUH-78589B/2020 | 2020-07-15 | Europe/United Kingdom/England/ | Lighthouse Lab in Glasgow | Wellcome Sanger Institute for the COVID-19 Genomics UK (COG-UK) consortium | Harper VanSteenhouse et al |
| EPI_ISL_533218 | England/QEUH-78FEEA/2020 | 2020-07-15 | Europe/United Kingdom/England/ | Lighthouse Lab in Glasgow | Wellcome Sanger Institute for the COVID-19 Genomics UK (COG-UK) consortium | Harper VanSteenhouse et al |
| EPI_ISL_533357 | Wales/QEUH-5458A8/2020 | 2020-05-14 | Europe/United Kingdom/Wales/ | Lighthouse Lab in Glasgow | Wellcome Sanger Institute for the COVID-19 Genomics UK (COG-UK) consortium | Harper VanSteenhouse et al |
| EPI_ISL_533423 | England/QEUH-549385/2020 | 2020-05-13 | Europe/United Kingdom/England/ | Lighthouse Lab in Glasgow | Wellcome Sanger Institute for the COVID-19 Genomics UK (COG-UK) consortium | Harper VanSteenhouse et al |

| | | | | | | |
|---|---|---|---|---|---|---|
| EPI_ISL_534235 | Sweden/20-52288/2020 | 2020-07-01 | Europe/Sweden/Stockholm/ | Karolinska universitetslaboratoriet SOLNA | The Public Health Agency of Sweden | Anna-Malin Linde et al |
| EPI_ISL_534239 | Sweden/20-52384/2020 | 2020-07-02 | Europe/Sweden/Vastra Gotaland/ | Norra Alvsborgs lanssjukhus | The Public Health Agency of Sweden | Anna-Malin Linde et al |
| EPI_ISL_534247 | Sweden/20-52462/2020 | 2020-07-28 | Europe/Sweden/Ostergotland/ | Universitetssjukhuset i Linkoping | The Public Health Agency of Sweden | Anna-Malin Linde et al |
| EPI_ISL_534252 | Sweden/20-52565/2020 | 2020-08-11 | Europe/Sweden/Vasternorrland/ | Laboratoriemedicin Vasternorrland | The Public Health Agency of Sweden | Anna-Malin Linde et al |
| EPI_ISL_534253 | Sweden/20-52566/2020 | 2020-08-11 | Europe/Sweden/Vasternorrland/ | Laboratoriemedicin Vasternorrland | The Public Health Agency of Sweden | Anna-Malin Linde et al |
| EPI_ISL_534259 | Sweden/20-52647/2020 | 2020-08-06 | Europe/Sweden/Gotland/ | Karolinska universitetslaboratoriet | The Public Health Agency of Sweden | Anna-Malin Linde et al |
| EPI_ISL_534267 | USA/TX-DSHS-0299/2020 | 2020-06-13 | North America/USA/Texas/Harris Count | Texas Department of State Health Services | Texas Department of State Health Services | Rashmi Tuladhar et al |
| EPI_ISL_534270 | USA/TX-DSHS-0302/2020 | 2020-06-13 | North America/USA/Texas/Harris Count | Texas Department of State Health Services | Texas Department of State Health Services | Rashmi Tuladhar et al |
| EPI_ISL_534285 | USA/TX-DSHS-0318/2020 | 2020-06-13 | North America/USA/Texas/Harris Count | Texas Department of State Health Services | Texas Department of State Health Services | Rashmi Tuladhar et al |
| EPI_ISL_534292 | USA/TX-DSHS-0325/2020 | 2020-06-13 | North America/USA/Texas/Harris Count | Texas Department of State Health Services | Texas Department of State Health Services | Rashmi Tuladhar et al |
| EPI_ISL_534312 | Brazil/SP-315/2020 | 2020-05-01 | South America/Brazil/S√£o Paulo/Campi | Distrito Sanitario Sul | Instituto Adolfo Lutz, Interdiciplinary Procedures Center, Strategic Laboratory | Claudio Tavares Sacchi et al |
| EPI_ISL_534314 | Brazil/SP-317/2020 | 2020-05-04 | South America/Brazil/S√£o Paulo/S√£o [illegible] | Hospital Universitario da USP de SP | Instituto Adolfo Lutz, Interdiciplinary Procedures Center, Strategic Laboratory | Claudio Tavares Sacchi et al |

| | | | | | | |
|---|---|---|---|---|---|---|
| EPI_ISL_534315 | Brazil/SP-318/2020 | 2020-05-02 | South America/Brazil/S√£o Paulo/Guaru | Servi√ßo de Verifica√ß√£o de √ìbitos SVO Guaruulhos | Instituto Adolfo Lutz, Interdiciplinary Procedures Center, Strategic Laboratory | Claudio Tavares Sacchi et al |
| EPI_ISL_534316 | Brazil/SP-321/2020 | 2020-05-02 | South America/Brazil/S√£o Paulo/S√£o | OS Mun Santana Lauro Ribas Braga | Instituto Adolfo Lutz, Interdiciplinary Procedures Center, Strategic Laboratory | Claudio Tavares Sacchi et al |
| EPI_ISL_534317 | Brazil/SP-324/2020 | 2020-05-05 | South America/Brazil/S√£o Paulo/Jandir | Hospital Geral de Itapevi | Instituto Adolfo Lutz, Interdiciplinary Procedures Center, Strategic Laboratory | Claudio Tavares Sacchi et al |
| EPI_ISL_534318 | Brazil/SP-325/2020 | 2020-05-03 | South America/Brazil/S√£o Paulo/Osasc | Hospital Municipal Antonio Giglio | Instituto Adolfo Lutz, Interdiciplinary Procedures Center, Strategic Laboratory | Claudio Tavares Sacchi et al |
| EPI_ISL_534319 | Brazil/SP-255/2020 | 2020-05-06 | South America/Brazil/S√£o Paulo/S√£o | Hospital do Serv Pub ESTAFCO Morato de Oliveira | Instituto Adolfo Lutz, Interdiciplinary Procedures Center, Strategic Laboratory | Claudio Tavares Sacchi et al |
| EPI_ISL_534320 | Brazil/SP-256/2020 | 2020-05-05 | South America/Brazil/S√£o Paulo/S√£o | Hospital do Serv Pub ESTAFCO Morato de Oliveira | Instituto Adolfo Lutz, Interdiciplinary Procedures Center, Strategic Laboratory | Claudio Tavares Sacchi et al |

| | | | | | | |
|---|---|---|---|---|---|---|
| EPI_ISL_534321 | Brazil/SP-329/2020 | 2020-05-07 | South America/Brazil/S√£o Paulo/Barue | PS e Maternidade Nair Fonseca Leitao Arantes | Instituto Adolfo Lutz, Interdiciplinary Procedures Center, Strategic Laboratory | Claudio Tavares Sacchi et al |
| EPI_ISL_534325 | Brazil/SP-306/2020 | 2020-04-25 | South America/Brazil/S√£o Paulo/Guaru | Unidade de Vigilancia em Saude de Guarulhos | Instituto Adolfo Lutz, Interdiciplinary Procedures Center, Strategic Laboratory | Claudio Tavares Sacchi et al |
| EPI_ISL_534326 | Brazil/SP-340/2020 | 2020-05-01 | South America/Brazil/S√£o Paulo/Cotia | Notre Dame Intermedica Saude AS | Instituto Adolfo Lutz, Interdiciplinary Procedures Center, Strategic Laboratory | Claudio Tavares Sacchi et al |
| EPI_ISL_534329 | Spain/MD-H12-88/2020 | 2020-04-18 | Europe/Spain/Madrid/ | Hospital Universitario 12 de Octubre | Hospital Universitario 12 de Octubre | Esther Viedma et al |
| EPI_ISL_534332 | Spain/MD-HLP-105-3/2020 | 2020-05-02 | Europe/Spain/Madrid/ | Hospital Universitario La Paz | Hospital Universitario La Paz | Mar√≠a Rodr√≠guez et al |
| EPI_ISL_534334 | Spain/MD-HRC-88/2020 | 2020-05-20 | Europe/Spain/Madrid/ | Hospital Universitario Ram√≥n y Cajal | Hospital Universitario La Paz | Mar√≠a Rodr√≠guez et al |
| EPI_ISL_534337 | Russia/CRIE4924360/2020 | 2020-07-15 | Europe/Russia/Moscow Oblast/ | "Molecular diagnostic laboratory of Federal Budget Institution of Science ""Central Research Institute of Epidemiology"" of The Federal Service on Customers' Rights Protection and Human Well-being Surveillance" | Group of Genomics and Postgenomic Technologies of Central Research Institute of Epidemilology | Speranskaya AS et al |
| EPI_ISL_534338 | Russia/CRIE4955821/2020 | 2020-07-16 | Europe/Russia/Moscow Oblast/ | "Molecular diagnostic laboratory of Federal Budget Institution of Science ""Central Research Institute of Epidemiology"" of The Federal Service on Customers' Rights Protection and Human Well-being Surveillance" | Group of Genomics and Postgenomic Technologies of Central Research Institute of Epidemilology | Speranskaya AS et al |

| | | | | | | |
|---|---|---|---|---|---|---|
| EPI_ISL_534339 | Russia/CRIE4991898/2020 | 2020-07-16 | Europe/Russia/Moscow Oblast/ | "Molecular diagnostic laboratory of Federal Budget Institution of Science ""Central Research Institute of Epidemiology"" of The Federal Service on Customers' Rights Protection and Human Well-being Surveillance" | Group of Genomics and Postgenomic Technologies of Central Research Institute of Epidemilology | Speranskaya AS et al |
| EPI_ISL_534340 | Russia/CRIE5027243/2020 | 2020-07-16 | Europe/Russia/Moscow Oblast/ | "Molecular diagnostic laboratory of Federal Budget Institution of Science ""Central Research Institute of Epidemiology"" of The Federal Service on Customers' Rights Protection and Human Well-being Surveillance" | Group of Genomics and Postgenomic Technologies of Central Research Institute of Epidemilology | Speranskaya AS et al |
| EPI_ISL_534341 | Russia/CRIE5006606/2020 | 2020-07-16 | Europe/Russia/Moscow Oblast/ | "Molecular diagnostic laboratory of Federal Budget Institution of Science ""Central Research Institute of Epidemiology"" of The Federal Service on Customers' Rights Protection and Human Well-being Surveillance" | Group of Genomics and Postgenomic Technologies of Central Research Institute of Epidemilology | Speranskaya AS et al |
| EPI_ISL_534342 | Russia/CRIE4980432/2020 | 2020-07-16 | Europe/Russia/Moscow Oblast/ | "Molecular diagnostic laboratory of Federal Budget Institution of Science ""Central Research Institute of Epidemiology"" of The Federal Service on Customers' Rights Protection and Human Well-being Surveillance" | Group of Genomics and Postgenomic Technologies of Central Research Institute of Epidemilology | Speranskaya AS et al |
| EPI_ISL_534343 | Russia/CRIE5047362/2020 | 2020-07-16 | Europe/Russia/Moscow Oblast/ | "Molecular diagnostic laboratory of Federal Budget Institution of Science ""Central Research Institute of Epidemiology"" of The Federal Service on Customers' Rights Protection and Human Well-being Surveillance" | Group of Genomics and Postgenomic Technologies of Central Research Institute of Epidemilology | Speranskaya AS et al |

| | | | | | | |
|---|---|---|---|---|---|---|
| EPI_ISL_534344 | Russia/CRIE5047007/2020 | 2020-07-16 | Europe/Russia/Moscow Oblast/ | "Molecular diagnostic laboratory of Federal Budget Institution of Science ""Central Research Institute of Epidemiology"" of The Federal Service on Customers' Rights Protection and Human Well-being Surveillance" | Group of Genomics and Postgenomic Technologies of Central Research Institute of Epidemilology | Speranskaya AS et al |
| EPI_ISL_534345 | Russia/CRIE5049919/2020 | 2020-07-17 | Europe/Russia/Moscow Oblast/ | "Molecular diagnostic laboratory of Federal Budget Institution of Science ""Central Research Institute of Epidemiology"" of The Federal Service on Customers' Rights Protection and Human Well-being Surveillance" | Group of Genomics and Postgenomic Technologies of Central Research Institute of Epidemilology | Speranskaya AS et al |
| EPI_ISL_534465 | Scotland/QEUH-96EBE8/2020 | 2020-08-19 | Europe/United Kingdom/Scotland/ | NHSGGC West of Scotland Specialist Virology Centre / MRC-University of Glasgow Centre for Virus Research | Wellcome Sanger Institute for the COVID-19 Genomics UK (COG-UK) consortium | Ana da Silva Filipe et al |
| EPI_ISL_534634 | Scotland/QEUH-96D396/2020 | 2020-08-20 | Europe/United Kingdom/Scotland/ | NHSGGC West of Scotland Specialist Virology Centre / MRC-University of Glasgow Centre for Virus Research | Wellcome Sanger Institute for the COVID-19 Genomics UK (COG-UK) Consortium | Ana da Silva Filipe et al |
| EPI_ISL_534697 | USA/PR-B5XN/2020 | 2020-08-11 | North America/USA/Puerto Rico/Ponce | University of Miami Immunology and Histocompatibility Laboratory | University of Miami Immunology and Histocompatibility Laboratory | Emilio Margolles-Clark et al |
| EPI_ISL_534700 | USA/MD-MDH-0140/2020 | 2020-07-09 | North America/USA/Maryland/ | MD PHL | MD PHL | Maryland Department of Health Laboratories Administration et al |
| EPI_ISL_534701 | USA/MD-MDH-0141/2020 | 2020-07-09 | North America/USA/Maryland/ | MD PHL | MD PHL | Maryland Department of Health Laboratories Administration et al |
| EPI_ISL_534702 | USA/MD-MDH-0142/2020 | 2020-07-09 | North America/USA/Maryland/ | MD PHL | MD PHL | Maryland Department of Health Laboratories Administration et al |
| EPI_ISL_534705 | USA/MD-MDH-0166/2020 | 2020-06-02 | North America/USA/Maryland/ | MD PHL | MD PHL | Maryland Department of Health Laboratories Administration et al |

| | | | | | | |
|---|---|---|---|---|---|---|
| EPI_ISL_534710 | USA/MD-MDH-0173/2020 | 2020-06-01 | North America/USA/Maryland/ | MD PHL | MD PHL | Maryland Department of Health Laboratories Administration et al |
| EPI_ISL_534711 | USA/MD-MDH-0174/2020 | 2020-06-03 | North America/USA/Maryland/ | MD PHL | MD PHL | Maryland Department of Health Laboratories Administration et al |
| EPI_ISL_534781 | England/OXON-B294E/2020 | 2020-04-13 | Europe/United Kingdom/England/ | Oxford Viromics, NDM, University of Oxford; Oxford University Hospitals; Basingstoke and North Hampshire Hospital | COVID-19 Genomics UK (COG-UK) Consortium | Tanya Golubchik et al |
| EPI_ISL_534876 | England/OXON-B3664/2020 | 2020-04-07 | Europe/United Kingdom/England/ | Oxford Viromics, NDM, University of Oxford; Oxford University Hospitals; Basingstoke and North Hampshire Hospital | COVID-19 Genomics UK (COG-UK) Consortium | Tanya Golubchik et al |
| EPI_ISL_534925 | England/OXON-B4038/2020 | 2020-03-23 | Europe/United Kingdom/England/ | Oxford Viromics, NDM, University of Oxford; Oxford University Hospitals; Basingstoke and North Hampshire Hospital | COVID-19 Genomics UK (COG-UK) Consortium | Tanya Golubchik et al |
| EPI_ISL_534993 | England/OXON-B4AC1/2020 | 2020-04-23 | Europe/United Kingdom/England/ | Oxford Viromics, NDM, University of Oxford; Oxford University Hospitals; Basingstoke and North Hampshire Hospital | COVID-19 Genomics UK (COG-UK) Consortium | Tanya Golubchik et al |
| EPI_ISL_534999 | England/OXON-B4BFB/2020 | 2020-03-31 | Europe/United Kingdom/England/ | Oxford Viromics, NDM, University of Oxford; Oxford University Hospitals; Basingstoke and North Hampshire Hospital | COVID-19 Genomics UK (COG-UK) Consortium | Tanya Golubchik et al |
| EPI_ISL_535227 | Wales/PHWC-16A2B2/2020 | 2020-09-06 | Europe/United Kingdom/Wales/ | Wales Specialist Virology Centre Sequencing lab: Pathogen Genomics Unit | COVID-19 Genomics UK (COG-UK) Consortium | Catherine Moore et al |
| EPI_ISL_535255 | Wales/PHWC-16A48F/2020 | 2020-09-06 | Europe/United Kingdom/Wales/ | Wales Specialist Virology Centre Sequencing lab: Pathogen Genomics Unit | COVID-19 Genomics UK (COG-UK) Consortium | Catherine Moore et al |
| EPI_ISL_535263 | USA/NM-NMDOH-2020099248/2020 | 2020-06-01 | North America/USA/New Mexico/ | New Mexico Department of Health Scientific Laboratory | New Mexico Department of Health Scientific Laboratory | Ellie Johnson et al |
| EPI_ISL_535281 | USA/NM-NMDOH-2020134989/2020 | 2020-06-18 | North America/USA/New Mexico/ | New Mexico Department of Health Scientific Laboratory | New Mexico Department of Health Scientific Laboratory | Ellie Johnson et al |
| EPI_ISL_535282 | USA/NM-NMDOH-2020134990/2020 | 2020-06-18 | North America/USA/New Mexico/ | New Mexico Department of Health Scientific Laboratory | New Mexico Department of Health Scientific Laboratory | Ellie Johnson et al |

| | | | | | | |
|---|---|---|---|---|---|---|
| EPI_ISL_535300 | USA/NM-NMDOH-2020230022/2020 | 2020-07-20 | North America/USA/New Mexico/ | New Mexico Department of Health Scientific Laboratory | New Mexico Department of Health Scientific Laboratory | Ellie Johnson et al |
| EPI_ISL_535322 | USA/LA-CDC-1281/2020 | 2020-06-10 | North America/USA/Louisiana/ | LA Office of Public Health Laboratories | Pathogen Discovery, Respiratory Viruses Branch, Division of Viral Diseases, Centers for Disease Control and Prevention | Ying Tao et al |
| EPI_ISL_535330 | USA/LA-CDC-1289/2020 | 2020-06-14 | North America/USA/Louisiana/ | LA Office of Public Health Laboratories | Pathogen Discovery, Respiratory Viruses Branch, Division of Viral Diseases, Centers for Disease Control and Prevention | Ying Tao et al |
| EPI_ISL_535331 | USA/LA-CDC-1290/2020 | 2020-06-11 | North America/USA/Louisiana/ | LA Office of Public Health Laboratories | Pathogen Discovery, Respiratory Viruses Branch, Division of Viral Diseases, Centers for Disease Control and Prevention | Ying Tao et al |
| EPI_ISL_535335 | USA/LA-CDC-1294/2020 | 2020-06-11 | North America/USA/Louisiana/ | LA Office of Public Health Laboratories | Pathogen Discovery, Respiratory Viruses Branch, Division of Viral Diseases, Centers for Disease Control and Prevention | Ying Tao et al |

| | | | | | | |
|---|---|---|---|---|---|---|
| EPI_ISL_535341 | USA/LA-CDC-1905/2020 | 2020-05-07 | North America/USA/Louisiana/ | LA Office of Public Health Laboratories | Pathogen Discovery, Respiratory Viruses Branch, Division of Viral Diseases, Centers for Disease Control and Prevention | Jing Zhang et al |
| EPI_ISL_535342 | USA/LA-CDC-1915/2020 | 2020-05-07 | North America/USA/Louisiana/ | LA Office of Public Health Laboratories | Pathogen Discovery, Respiratory Viruses Branch, Division of Viral Diseases, Centers for Disease Control and Prevention | Jing Zhang et al |
| EPI_ISL_535351 | USA/LA-CDC-2119/2020 | 2020-05-29 | North America/USA/Louisiana/ | LA Office of Public Health Laboratories | Pathogen Discovery, Respiratory Viruses Branch, Division of Viral Diseases, Centers for Disease Control and Prevention | Jing Zhang et al |
| EPI_ISL_535359 | USA/RI-CDC-6616/2020 | 2020-07-30 | North America/USA/Rhode Island/ | RI State Health Laboratories | Pathogen Discovery, Respiratory Viruses Branch, Division of Viral Diseases, Centers for Disease Control and Prevention | Jing Zhang et al |
| EPI_ISL_535363 | USA/OK-ADDL-04/2020 | 2020-05-27 | North America/USA/Oklahoma/ | Oklahoma Animal Disease Diagnostic Laboratory | Oklahoma Animal Disease Diagnostic Laboratory | Sai Narayanan et al |

| | | | | | | |
|---|---|---|---|---|---|---|
| EPI_ISL_535391 | SouthAfrica/KRISP-K002483/2020 | 2020-08-07 | Africa/South Africa/KwaZulu-Natal/Ama | NHLS-IALCH | KRISP, KZN Research Innovation and Sequencing Platform | Giandhari J et al |
| EPI_ISL_535392 | SouthAfrica/KRISP-K002484/2020 | 2020-08-07 | Africa/South Africa/KwaZulu-Natal/Ama | NHLS-IALCH | KRISP, KZN Research Innovation and Sequencing Platform | Giandhari J et al |
| EPI_ISL_535394 | SouthAfrica/KRISP-K002490/2020 | 2020-08-07 | Africa/South Africa/KwaZulu-Natal/Ama | NHLS-IALCH | KRISP, KZN Research Innovation and Sequencing Platform | Giandhari J et al |
| EPI_ISL_535397 | SouthAfrica/KRISP-K002498/2020 | 2020-08-05 | Africa/South Africa/KwaZulu-Natal/iLem | NHLS-IALCH | KRISP, KZN Research Innovation and Sequencing Platform | Giandhari J et al |
| EPI_ISL_535398 | SouthAfrica/KRISP-K002501/2020 | 2020-08-07 | Africa/South Africa/KwaZulu-Natal/iLem | NHLS-IALCH | KRISP, KZN Research Innovation and Sequencing Platform | Giandhari J et al |
| EPI_ISL_535399 | SouthAfrica/KRISP-K002502/2020 | 2020-08-05 | Africa/South Africa/KwaZulu-Natal/iLem | NHLS-IALCH | KRISP, KZN Research Innovation and Sequencing Platform | Giandhari J et al |
| EPI_ISL_535400 | SouthAfrica/KRISP-K002504/2020 | 2020-08-06 | Africa/South Africa/KwaZulu-Natal/iLem | NHLS-IALCH | KRISP, KZN Research Innovation and Sequencing Platform | Giandhari J et al |
| EPI_ISL_535401 | SouthAfrica/KRISP-K002506/2020 | 2020-08-07 | Africa/South Africa/KwaZulu-Natal/uMg | NHLS-IALCH | KRISP, KZN Research Innovation and Sequencing Platform | Giandhari J et al |

| | | | | | | |
|---|---|---|---|---|---|---|
| EPI_ISL_535403 | SouthAfrica/KRISP-K002520/2020 | 2020-08-07 | Africa/South Africa/KwaZulu-Natal/uMz | NHLS-IALCH | KRISP, KZN Research Innovation and Sequencing Platform | Giandhari J et al |
| EPI_ISL_535405 | SouthAfrica/KRISP-K002532/2020 | 2020-07-30 | Africa/South Africa/KwaZulu-Natal/uMz | NHLS-IALCH | KRISP, KZN Research Innovation and Sequencing Platform | Giandhari J et al |
| EPI_ISL_535406 | SouthAfrica/KRISP-K002533/2020 | 2020-07-30 | Africa/South Africa/KwaZulu-Natal/Ama | NHLS-IALCH | KRISP, KZN Research Innovation and Sequencing Platform | Giandhari J et al |
| EPI_ISL_535407 | SouthAfrica/KRISP-K002534/2020 | 2020-07-30 | Africa/South Africa/KwaZulu-Natal/uMg | NHLS-IALCH | KRISP, KZN Research Innovation and Sequencing Platform | Giandhari J et al |
| EPI_ISL_535412 | SouthAfrica/KRISP-K002542/2020 | 2020-07-30 | Africa/South Africa/KwaZulu-Natal/uMg | NHLS-IALCH | KRISP, KZN Research Innovation and Sequencing Platform | Giandhari J et al |
| EPI_ISL_535413 | SouthAfrica/KRISP-K002543/2020 | 2020-07-30 | Africa/South Africa/KwaZulu-Natal/Zulu | NHLS-IALCH | KRISP, KZN Research Innovation and Sequencing Platform | Giandhari J et al |
| EPI_ISL_535414 | SouthAfrica/KRISP-K002544/2020 | 2020-07-30 | Africa/South Africa/KwaZulu-Natal/uThu | NHLS-IALCH | KRISP, KZN Research Innovation and Sequencing Platform | Giandhari J et al |
| EPI_ISL_535415 | SouthAfrica/KRISP-K002545/2020 | 2020-07-30 | Africa/South Africa/KwaZulu-Natal/Ama | NHLS-IALCH | KRISP, KZN Research Innovation and Sequencing Platform | Giandhari J et al |

| | | | | | | |
|---|---|---|---|---|---|---|
| EPI_ISL_535425 | SouthAfrica/KRISP-K002556/2020 | 2020-07-29 | Africa/South Africa/KwaZulu-Natal/uMg | NHLS-IALCH | KRISP, KZN Research Innovation and Sequencing Platform | Giandhari J et al |
| EPI_ISL_535427 | SouthAfrica/KRISP-K002559/2020 | 2020-07-29 | Africa/South Africa/KwaZulu-Natal/uMg | NHLS-IALCH | KRISP, KZN Research Innovation and Sequencing Platform | Giandhari J et al |
| EPI_ISL_535431 | SouthAfrica/KRISP-K002563/2020 | 2020-07-30 | Africa/South Africa/KwaZulu-Natal/King | NHLS-IALCH | KRISP, KZN Research Innovation and Sequencing Platform | Giandhari J et al |
| EPI_ISL_535435 | SouthAfrica/KRISP-K002569/2020 | 2020-07-29 | Africa/South Africa/KwaZulu-Natal/King | NHLS-IALCH | KRISP, KZN Research Innovation and Sequencing Platform | Giandhari J et al |
| EPI_ISL_535436 | SouthAfrica/KRISP-K002570/2020 | 2020-07-29 | Africa/South Africa/KwaZulu-Natal/King | NHLS-IALCH | KRISP, KZN Research Innovation and Sequencing Platform | Giandhari J et al |
| EPI_ISL_535442 | SouthAfrica/KRISP-K002577/2020 | 2020-07-31 | Africa/South Africa/KwaZulu-Natal/iLem | NHLS-IALCH | KRISP, KZN Research Innovation and Sequencing Platform | Giandhari J et al |
| EPI_ISL_535447 | SouthAfrica/KRISP-K002582/2020 | 2020-07-31 | Africa/South Africa/KwaZulu-Natal/iLem | NHLS-IALCH | KRISP, KZN Research Innovation and Sequencing Platform | Giandhari J et al |
| EPI_ISL_535449 | SouthAfrica/KRISP-K002584/2020 | 2020-08-01 | Africa/South Africa/KwaZulu-Natal/uThu | NHLS-IALCH | KRISP, KZN Research Innovation and Sequencing Platform | Giandhari J et al |

| | | | | | | |
|---|---|---|---|---|---|---|
| EPI_ISL_535456 | SouthAfrica/KRISP-K002591/2020 | 2020-07-28 | Africa/South Africa/KwaZulu-Natal/Ugu | NHLS-IALCH | KRISP, KZN Research Innovation and Sequencing Platform | Giandhari J et al |
| EPI_ISL_535459 | SouthAfrica/KRISP-K002594/2020 | 2020-07-28 | Africa/South Africa/KwaZulu-Natal/Ugu | NHLS-IALCH | KRISP, KZN Research Innovation and Sequencing Platform | Giandhari J et al |
| EPI_ISL_535463 | SouthAfrica/KRISP-K002598/2020 | 2020-07-30 | Africa/South Africa/KwaZulu-Natal/Ugu | NHLS-IALCH | KRISP, KZN Research Innovation and Sequencing Platform | Giandhari J et al |
| EPI_ISL_535467 | SouthAfrica/KRISP-K002603/2020 | 2020-07-30 | Africa/South Africa/KwaZulu-Natal/iLem | NHLS-IALCH | KRISP, KZN Research Innovation and Sequencing Platform | Giandhari J et al |
| EPI_ISL_535469 | SouthAfrica/KRISP-K002605/2020 | 2020-07-30 | Africa/South Africa/KwaZulu-Natal/iLem | NHLS-IALCH | KRISP, KZN Research Innovation and Sequencing Platform | Giandhari J et al |
| EPI_ISL_535472 | SouthAfrica/KRISP-K002608/2020 | 2020-08-01 | Africa/South Africa/KwaZulu-Natal/Ugu | NHLS-IALCH | KRISP, KZN Research Innovation and Sequencing Platform | Giandhari J et al |
| EPI_ISL_535473 | SouthAfrica/KRISP-K002609/2020 | 2020-07-30 | Africa/South Africa/KwaZulu-Natal/Ugu | NHLS-IALCH | KRISP, KZN Research Innovation and Sequencing Platform | Giandhari J et al |
| EPI_ISL_535475 | SouthAfrica/KRISP-K002611/2020 | 2020-07-29 | Africa/South Africa/KwaZulu-Natal/Ugu | NHLS-IALCH | KRISP, KZN Research Innovation and Sequencing Platform | Giandhari J et al |

| | | | | | | |
|---|---|---|---|---|---|---|
| EPI_ISL_535480 | SouthAfrica/KRISP-K002616/2020 | 2020-08-01 | Africa/South Africa/KwaZulu-Natal/iLem | NHLS-IALCH | KRISP, KZN Research Innovation and Sequencing Platform | Giandhari J et al |
| EPI_ISL_535481 | SouthAfrica/KRISP-K002617/2020 | 2020-07-29 | Africa/South Africa/KwaZulu-Natal/Ugu | NHLS-IALCH | KRISP, KZN Research Innovation and Sequencing Platform | Giandhari J et al |
| EPI_ISL_535503 | SouthAfrica/KRISP-K002633/2020 | 2020-08-03 | Africa/South Africa/KwaZulu-Natal/iLem | NHLS-IALCH | KRISP, KZN Research Innovation and Sequencing Platform | Giandhari J et al |
| EPI_ISL_535504 | SouthAfrica/KRISP-K002634/2020 | 2020-08-03 | Africa/South Africa/KwaZulu-Natal/iLem | NHLS-IALCH | KRISP, KZN Research Innovation and Sequencing Platform | Giandhari J et al |
| EPI_ISL_535505 | SouthAfrica/KRISP-K002635/2020 | 2020-08-04 | Africa/South Africa/KwaZulu-Natal/iLem | NHLS-IALCH | KRISP, KZN Research Innovation and Sequencing Platform | Giandhari J et al |
| EPI_ISL_535508 | SouthAfrica/KRISP-K002638/2020 | 2020-08-04 | Africa/South Africa/KwaZulu-Natal/iLem | NHLS-IALCH | KRISP, KZN Research Innovation and Sequencing Platform | Giandhari J et al |
| EPI_ISL_535509 | SouthAfrica/KRISP-K002639/2020 | 2020-08-03 | Africa/South Africa/KwaZulu-Natal/iLem | NHLS-IALCH | KRISP, KZN Research Innovation and Sequencing Platform | Giandhari J et al |
| EPI_ISL_535510 | SouthAfrica/KRISP-K002640/2020 | 2020-08-04 | Africa/South Africa/KwaZulu-Natal/iLem | NHLS-IALCH | KRISP, KZN Research Innovation and Sequencing Platform | Giandhari J et al |

| | | | | | | |
|---|---|---|---|---|---|---|
| EPI_ISL_535511 | SouthAfrica/KRISP-K002641/2020 | 2020-08-02 | Africa/South Africa/KwaZulu-Natal/iLem | NHLS-IALCH | KRISP, KZN Research Innovation and Sequencing Platform | Giandhari J et al |
| EPI_ISL_535512 | SouthAfrica/KRISP-K002642/2020 | 2020-08-02 | Africa/South Africa/KwaZulu-Natal/iLem | NHLS-IALCH | KRISP, KZN Research Innovation and Sequencing Platform | Giandhari J et al |
| EPI_ISL_535514 | SouthAfrica/KRISP-K002644/2020 | 2020-08-02 | Africa/South Africa/KwaZulu-Natal/iLem | NHLS-IALCH | KRISP, KZN Research Innovation and Sequencing Platform | Giandhari J et al |
| EPI_ISL_535515 | SouthAfrica/KRISP-K002645/2020 | 2020-08-01 | Africa/South Africa/KwaZulu-Natal/iLem | NHLS-IALCH | KRISP, KZN Research Innovation and Sequencing Platform | Giandhari J et al |
| EPI_ISL_535516 | SouthAfrica/KRISP-K002646/2020 | 2020-08-03 | Africa/South Africa/KwaZulu-Natal/iLem | NHLS-IALCH | KRISP, KZN Research Innovation and Sequencing Platform | Giandhari J et al |
| EPI_ISL_535518 | SouthAfrica/KRISP-K002648/2020 | 2020-08-04 | Africa/South Africa/KwaZulu-Natal/iLem | NHLS-IALCH | KRISP, KZN Research Innovation and Sequencing Platform | Giandhari J et al |
| EPI_ISL_535519 | SouthAfrica/KRISP-K002649/2020 | 2020-08-02 | Africa/South Africa/KwaZulu-Natal/iLem | NHLS-IALCH | KRISP, KZN Research Innovation and Sequencing Platform | Giandhari J et al |
| EPI_ISL_535525 | SouthAfrica/KRISP-K002657/2020 | 2020-08-03 | Africa/South Africa/KwaZulu-Natal/King | NHLS-IALCH | KRISP, KZN Research Innovation and Sequencing Platform | Giandhari J et al |

| | | | | | | |
|---|---|---|---|---|---|---|
| EPI_ISL_535526 | SouthAfrica/KRISP-K002658/2020 | 2020-08-03 | Africa/South Africa/KwaZulu-Natal/iLem | NHLS-IALCH | KRISP, KZN Research Innovation and Sequencing Platform | Giandhari J et al |
| EPI_ISL_535528 | SouthAfrica/KRISP-K002660/2020 | 2020-08-03 | Africa/South Africa/KwaZulu-Natal/iLem | NHLS-IALCH | KRISP, KZN Research Innovation and Sequencing Platform | Giandhari J et al |
| EPI_ISL_535529 | SouthAfrica/KRISP-K002661/2020 | 2020-08-03 | Africa/South Africa/KwaZulu-Natal/iLem | NHLS-IALCH | KRISP, KZN Research Innovation and Sequencing Platform | Giandhari J et al |
| EPI_ISL_535531 | SouthAfrica/KRISP-K002665/2020 | 2020-08-03 | Africa/South Africa/KwaZulu-Natal/King | NHLS-IALCH | KRISP, KZN Research Innovation and Sequencing Platform | Giandhari J et al |
| EPI_ISL_535533 | SouthAfrica/KRISP-K002667/2020 | 2020-08-04 | Africa/South Africa/KwaZulu-Natal/iLem | NHLS-IALCH | KRISP, KZN Research Innovation and Sequencing Platform | Giandhari J et al |
| EPI_ISL_535540 | SouthAfrica/KRISP-K002675/2020 | 2020-08-03 | Africa/South Africa/KwaZulu-Natal/iLem | NHLS-IALCH | KRISP, KZN Research Innovation and Sequencing Platform | Giandhari J et al |
| EPI_ISL_535541 | SouthAfrica/KRISP-K002676/2020 | 2020-08-02 | Africa/South Africa/KwaZulu-Natal/King | NHLS-IALCH | KRISP, KZN Research Innovation and Sequencing Platform | Giandhari J et al |
| EPI_ISL_535543 | SouthAfrica/KRISP-K002679/2020 | 2020-08-02 | Africa/South Africa/KwaZulu-Natal/King | NHLS-IALCH | KRISP, KZN Research Innovation and Sequencing Platform | Giandhari J et al |

| | | | | | | |
|---|---|---|---|---|---|---|
| EPI_ISL_535544 | SouthAfrica/KRISP-K002680/2020 | 2020-08-03 | Africa/South Africa/KwaZulu-Natal/King | NHLS-IALCH | KRISP, KZN Research Innovation and Sequencing Platform | Giandhari J et al |
| EPI_ISL_535546 | SouthAfrica/KRISP-K002682/2020 | 2020-08-01 | Africa/South Africa/KwaZulu-Natal/King | NHLS-IALCH | KRISP, KZN Research Innovation and Sequencing Platform | Giandhari J et al |
| EPI_ISL_535548 | SouthAfrica/KRISP-K002685/2020 | 2020-08-02 | Africa/South Africa/KwaZulu-Natal/King | NHLS-IALCH | KRISP, KZN Research Innovation and Sequencing Platform | Giandhari J et al |
| EPI_ISL_535549 | SouthAfrica/KRISP-K002686/2020 | 2020-08-04 | Africa/South Africa/KwaZulu-Natal/iLem | NHLS-IALCH | KRISP, KZN Research Innovation and Sequencing Platform | Giandhari J et al |
| EPI_ISL_535552 | SouthAfrica/KRISP-K002690/2020 | 2020-07-27 | Africa/South Africa/KwaZulu-Natal/eThe | NHLS-IALCH | KRISP, KZN Research Innovation and Sequencing Platform | Giandhari J et al |
| EPI_ISL_535555 | SouthAfrica/KRISP-K002693/2020 | 2020-07-27 | Africa/South Africa/KwaZulu-Natal/eThe | NHLS-IALCH | KRISP, KZN Research Innovation and Sequencing Platform | Giandhari J et al |
| EPI_ISL_535556 | SouthAfrica/KRISP-K002694/2020 | 2020-07-27 | Africa/South Africa/KwaZulu-Natal/eThe | NHLS-IALCH | KRISP, KZN Research Innovation and Sequencing Platform | Giandhari J et al |
| EPI_ISL_535559 | SouthAfrica/KRISP-K002698/2020 | 2020-07-30 | Africa/South Africa/KwaZulu-Natal/eThe | NHLS-IALCH | KRISP, KZN Research Innovation and Sequencing Platform | Giandhari J et al |

| | | | | | | |
|---|---|---|---|---|---|---|
| EPI_ISL_535561 | SouthAfrica/KRISP-K002700/2020 | 2020-07-30 | Africa/South Africa/KwaZulu-Natal/eThe | NHLS-IALCH | KRISP, KZN Research Innovation and Sequencing Platform | Giandhari J et al |
| EPI_ISL_535566 | SouthAfrica/KRISP-K002707/2020 | 2020-07-30 | Africa/South Africa/KwaZulu-Natal/eThe | NHLS-IALCH | KRISP, KZN Research Innovation and Sequencing Platform | Giandhari J et al |
| EPI_ISL_535567 | SouthAfrica/KRISP-K002708/2020 | 2020-07-30 | Africa/South Africa/KwaZulu-Natal/eThe | NHLS-IALCH | KRISP, KZN Research Innovation and Sequencing Platform | Giandhari J et al |
| EPI_ISL_535568 | SouthAfrica/KRISP-K002709/2020 | 2020-07-30 | Africa/South Africa/KwaZulu-Natal/eThe | NHLS-IALCH | KRISP, KZN Research Innovation and Sequencing Platform | Giandhari J et al |
| EPI_ISL_535575 | Spain/CT-IrsiCaixa-2020090134/2020 | 2020-08-31 | Europe/Spain/Catalunya/Gerona | Hospital Universitari Germans Trias i Pujol(HUGTiP)/Fundaci√≥ Lluita contra la SIDA (FLSida) | IrsiCaixa AIDS Research Lab | Marc Noguera-Julian et al |
| EPI_ISL_535603 | Switzerland/SZ-ETHZ-250084/2020 | 2020-08-19 | Europe/Switzerland/Schwyz/ | Viollier AG | Department of Biosystems Science and Engineering, ETH Z√ºrich | Christian Beisel et al |
| EPI_ISL_535615 | Switzerland/ZH-ETHZ-250099/2020 | 2020-08-20 | Europe/Switzerland/Z√ºrich/ | Viollier AG | Department of Biosystems Science and Engineering, ETH Z√ºrich | Christian Beisel et al |

| | | | | | | |
|---|---|---|---|---|---|---|
| EPI_ISL_535651 | USA/AR-CDC-0838/2020 | 2020-07-07 | North America/USA/Arkansas/ | AR Dept. of Health-Public Health Lab | Pathogen Discovery, Respiratory Viruses Branch, Division of Viral Diseases, Centers for Disease Control and Prevention | Yan Li et al |
| EPI_ISL_535654 | USA/AR-CDC-0848/2020 | 2020-07-08 | North America/USA/Arkansas/ | AR Dept. of Health-Public Health Lab | Pathogen Discovery, Respiratory Viruses Branch, Division of Viral Diseases, Centers for Disease Control and Prevention | Brian Lynch et al |
| EPI_ISL_535656 | USA/AR-CDC-0863/2020 | 2020-07-10 | North America/USA/Arkansas/ | AR Dept. of Health-Public Health Lab | Pathogen Discovery, Respiratory Viruses Branch, Division of Viral Diseases, Centers for Disease Control and Prevention | Yan Li et al |
| EPI_ISL_535671 | USA/CA-CDC-4694/2020 | 2020-06-02 | North America/USA/California/ | CDPH, Microbial Diseases Laboratory | Pathogen Discovery, Respiratory Viruses Branch, Division of Viral Diseases, Centers for Disease Control and Prevention | Yan Li et al |

| | | | | | | |
|---|---|---|---|---|---|---|
| EPI_ISL_535677 | USA/CA-CDC-4700/2020 | 2020-06-04 | North America/USA/California/ | CDPH, Microbial Diseases Laboratory | Pathogen Discovery, Respiratory Viruses Branch, Division of Viral Diseases, Centers for Disease Control and Prevention | Yan Li et al |
| EPI_ISL_535683 | USA/CA-CDC-4706/2020 | 2020-06-04 | North America/USA/California/ | CDPH, Microbial Diseases Laboratory | Pathogen Discovery, Respiratory Viruses Branch, Division of Viral Diseases, Centers for Disease Control and Prevention | Yan Li et al |
| EPI_ISL_535686 | USA/CA-CDC-4710/2020 | 2020-06-04 | North America/USA/California/ | CDPH, Microbial Diseases Laboratory | Pathogen Discovery, Respiratory Viruses Branch, Division of Viral Diseases, Centers for Disease Control and Prevention | Yan Li et al |
| EPI_ISL_535688 | USA/CA-CDC-4712/2020 | 2020-06-04 | North America/USA/California/ | CDPH, Microbial Diseases Laboratory | Pathogen Discovery, Respiratory Viruses Branch, Division of Viral Diseases, Centers for Disease Control and Prevention | Yan Li et al |

| | | | | | | |
|---|---|---|---|---|---|---|
| EPI_ISL_535696 | USA/CA-CDC-4724/2020 | 2020-06-16 | North America/USA/California/ | CDPH, Microbial Diseases Laboratory | Pathogen Discovery, Respiratory Viruses Branch, Division of Viral Diseases, Centers for Disease Control and Prevention | Yan Li et al |
| EPI_ISL_535698 | USA/CA-CDC-4727/2020 | 2020-06-15 | North America/USA/California/ | CDPH, Microbial Diseases Laboratory | Pathogen Discovery, Respiratory Viruses Branch, Division of Viral Diseases, Centers for Disease Control and Prevention | Yan Li et al |
| EPI_ISL_535699 | USA/CA-CDC-4728/2020 | 2020-06-16 | North America/USA/California/ | CDPH, Microbial Diseases Laboratory | Pathogen Discovery, Respiratory Viruses Branch, Division of Viral Diseases, Centers for Disease Control and Prevention | Yan Li et al |
| EPI_ISL_535710 | USA/CA-CDC-4748/2020 | 2020-06-17 | North America/USA/California/ | CDPH, Microbial Diseases Laboratory | Pathogen Discovery, Respiratory Viruses Branch, Division of Viral Diseases, Centers for Disease Control and Prevention | Yan Li et al |

| | | | | | | |
|---|---|---|---|---|---|---|
| EPI_ISL_535711 | USA/CA-CDC-4749/2020 | 2020-06-17 | North America/USA/California/ | CDPH, Microbial Diseases Laboratory | Pathogen Discovery, Respiratory Viruses Branch, Division of Viral Diseases, Centers for Disease Control and Prevention | Yan Li et al |
| EPI_ISL_535712 | USA/CA-CDC-4750/2020 | 2020-06-17 | North America/USA/California/ | CDPH, Microbial Diseases Laboratory | Pathogen Discovery, Respiratory Viruses Branch, Division of Viral Diseases, Centers for Disease Control and Prevention | Yan Li et al |
| EPI_ISL_535715 | USA/CA-CDC-4753/2020 | 2020-06-17 | North America/USA/California/ | CDPH, Microbial Diseases Laboratory | Pathogen Discovery, Respiratory Viruses Branch, Division of Viral Diseases, Centers for Disease Control and Prevention | Yan Li et al |
| EPI_ISL_535716 | Canada/QC-LSPQ-L00210314/2020 | 2020-03-01 | North America/Canada/Quebec/ | H√¥pital de Verdun | Laboratoire de sant√© publique du Qu√©bec | Sandrine Moreira et al |
| EPI_ISL_535717 | Canada/QC-LSPQ-L00212401/2020 | 2020-03-01 | North America/Canada/Quebec/ | H√¥pital Honor√©-Mercier | Laboratoire de sant√© publique du Qu√©bec | Sandrine Moreira et al |
| EPI_ISL_535718 | Canada/QC-LSPQ-L00213013/2020 | 2020-03-01 | North America/Canada/Quebec/ | H√¥pital Pierre-Boucher | Laboratoire de sant√© publique du Qu√©bec | Sandrine Moreira et al |
| EPI_ISL_535719 | Canada/QC-LSPQ-L00213312/2020 | 2020-03-01 | North America/Canada/Quebec/ | H√¥pital g√©n√©ral Juif | Laboratoire de sant√© publique du Qu√©bec | Sandrine Moreira et al |

| | | | | | | |
|---|---|---|---|---|---|---|
| EPI_ISL_535720 | Canada/QC-LSPQ-L00213733/2020 | 2020-03-10 | North America/Canada/Quebec/ | H√¥pital g√©n√©ral Juif | Laboratoire de sant√© publique du Qu√©bec | Sandrine Moreira et al |
| EPI_ISL_535721 | Canada/QC-LSPQ-L00213861/2020 | 2020-03-10 | North America/Canada/Quebec/ | H√¥pital Notre-Dame | Laboratoire de sant√© publique du Qu√©bec | Sandrine Moreira et al |
| EPI_ISL_535722 | Canada/QC-LSPQ-L00213981/2020 | 2020-03-10 | North America/Canada/Quebec/ | Centre de SSS de Trois-Rivi√®res | Laboratoire de sant√© publique du Qu√©bec | Sandrine Moreira et al |
| EPI_ISL_535723 | Canada/QC-LSPQ-L00214023/2020 | 2020-03-10 | North America/Canada/Quebec/ | H√¥pital de Verdun | Laboratoire de sant√© publique du Qu√©bec | Sandrine Moreira et al |
| EPI_ISL_535724 | Canada/QC-LSPQ-L00214050/2020 | 2020-03-10 | North America/Canada/Quebec/ | Centre de SSS La Pommeraie | Laboratoire de sant√© publique du Qu√©bec | Sandrine Moreira et al |
| EPI_ISL_535725 | Canada/QC-LSPQ-L00214081/2020 | 2020-03-10 | North America/Canada/Quebec/ | H√¥tel-Dieu de L√©vis | Laboratoire de sant√© publique du Qu√©bec | Sandrine Moreira et al |
| EPI_ISL_535726 | Canada/QC-LSPQ-L00214237/2020 | 2020-03-11 | North America/Canada/Quebec/ | CHUM - Microbiologie - H√¥pital Saint-Luc | Laboratoire de sant√© publique du Qu√©bec | Sandrine Moreira et al |
| EPI_ISL_535727 | Canada/QC-LSPQ-L00214488/2020 | 2020-03-11 | North America/Canada/Quebec/ | Centre de SSS de la Haute-Yamaska | Laboratoire de sant√© publique du Qu√©bec | Sandrine Moreira et al |
| EPI_ISL_535728 | Canada/QC-LSPQ-L00214517/2020 | 2020-03-11 | North America/Canada/Quebec/ | H√¥pital de Verdun | Laboratoire de sant√© publique du Qu√©bec | Sandrine Moreira et al |
| EPI_ISL_535729 | Canada/QC-LSPQ-L00214546/2020 | 2020-03-11 | North America/Canada/Quebec/ | CHUM - Microbiologie - H√¥pital Saint-Luc | Laboratoire de sant√© publique du Qu√©bec | Sandrine Moreira et al |
| EPI_ISL_535730 | Canada/QC-LSPQ-L00214634/2020 | 2020-03-12 | North America/Canada/Quebec/ | CHUM - Microbiologie - H√¥pital Saint-Luc | Laboratoire de sant√© publique du Qu√©bec | Sandrine Moreira et al |
| EPI_ISL_535731 | Canada/QC-LSPQ-L00215022/2020 | 2020-03-12 | North America/Canada/Quebec/ | H√¥pital Pierre-Le Gardeur | Laboratoire de sant√© publique du Qu√©bec | Sandrine Moreira et al |
| EPI_ISL_535732 | Canada/QC-LSPQ-L00215586/2020 | 2020-03-12 | North America/Canada/Quebec/ | CHU Sainte-Justine | Laboratoire de sant√© publique du Qu√©bec | Sandrine Moreira et al |

| | | | | | | |
|---|---|---|---|---|---|---|
| EPI_ISL_535733 | Canada/QC-LSPQ-L00215627/2020 | 2020-03-12 | North America/Canada/Quebec/ | H√¥pital Pierre-Boucher | Laboratoire de sant√© publique du Qu√©bec | Sandrine Moreira et al |
| EPI_ISL_535734 | Canada/QC-LSPQ-L00215628/2020 | 2020-03-12 | North America/Canada/Quebec/ | H√¥pital Pierre-Boucher | Laboratoire de sant√© publique du Qu√©bec | Sandrine Moreira et al |
| EPI_ISL_535735 | Canada/QC-LSPQ-L00215711/2020 | 2020-03-12 | North America/Canada/Quebec/ | CHUM - Microbiologie - H√¥pital Saint-Luc | Laboratoire de sant√© publique du Qu√©bec | Sandrine Moreira et al |
| EPI_ISL_535736 | Canada/QC-LSPQ-L00215713/2020 | 2020-03-12 | North America/Canada/Quebec/ | CHUL-LABO MULTI / MICRO | Laboratoire de sant√© publique du Qu√©bec | Sandrine Moreira et al |
| EPI_ISL_535737 | Canada/QC-LSPQ-L00215798/2020 | 2020-03-12 | North America/Canada/Quebec/ | CHUL-LABO MULTI / MICRO | Laboratoire de sant√© publique du Qu√©bec | Sandrine Moreira et al |
| EPI_ISL_535738 | Canada/QC-LSPQ-L00215812/2020 | 2020-03-12 | North America/Canada/Quebec/ | H√¥pital Charles-LeMoyne | Laboratoire de sant√© publique du Qu√©bec | Sandrine Moreira et al |
| EPI_ISL_535739 | Canada/QC-LSPQ-L00215817/2020 | 2020-03-12 | North America/Canada/Quebec/ | H√¥pital de l'Enfant-J√©sus | Laboratoire de sant√© publique du Qu√©bec | Sandrine Moreira et al |
| EPI_ISL_535740 | Canada/QC-LSPQ-L00215858/2020 | 2020-03-12 | North America/Canada/Quebec/ | CHUM - Microbiologie - H√¥pital Saint-Luc | Laboratoire de sant√© publique du Qu√©bec | Sandrine Moreira et al |
| EPI_ISL_535741 | Canada/QC-LSPQ-L00215879/2020 | 2020-03-12 | North America/Canada/Quebec/ | CHUL-LABO MULTI / MICRO | Laboratoire de sant√© publique du Qu√©bec | Sandrine Moreira et al |
| EPI_ISL_535742 | Canada/QC-LSPQ-L00215977/2020 | 2020-03-12 | North America/Canada/Quebec/ | Centre de SSS La Pommeraie | Laboratoire de sant√© publique du Qu√©bec | Sandrine Moreira et al |
| EPI_ISL_535743 | Canada/QC-LSPQ-L00215981/2020 | 2020-03-12 | North America/Canada/Quebec/ | Centre de SSS La Pommeraie | Laboratoire de sant√© publique du Qu√©bec | Sandrine Moreira et al |
| EPI_ISL_535744 | Canada/QC-LSPQ-L00216190/2020 | 2020-03-12 | North America/Canada/Quebec/ | H√¥pital de Lasalle | Laboratoire de sant√© publique du Qu√©bec | Sandrine Moreira et al |
| EPI_ISL_535745 | Canada/QC-LSPQ-L00216197/2020 | 2020-03-12 | North America/Canada/Quebec/ | H√¥pital Laurentien | Laboratoire de sant√© publique du Qu√©bec | Sandrine Moreira et al |

| | | | | | | |
|---|---|---|---|---|---|---|
| EPI_ISL_535746 | Canada/QC-LSPQ-L00216421/2020 | 2020-03-13 | North America/Canada/Quebec/ | Centre hospitalier de St-Mary | Laboratoire de santv© publique du Quv©bec | Sandrine Moreira et al |
| EPI_ISL_535747 | Canada/QC-LSPQ-L00216427/2020 | 2020-03-12 | North America/Canada/Quebec/ | Centre de SSS de Trois-Riviv®res | Laboratoire de santv© publique du Quv©bec | Sandrine Moreira et al |
| EPI_ISL_535748 | Canada/QC-LSPQ-L00216432/2020 | 2020-03-12 | North America/Canada/Quebec/ | Centre de SSS de Trois-Riviv®res | Laboratoire de santv© publique du Quv©bec | Sandrine Moreira et al |
| EPI_ISL_535749 | Canada/QC-LSPQ-L00216478/2020 | 2020-03-12 | North America/Canada/Quebec/ | CUSM-Site Glen-LAB Microbiologie | Laboratoire de santv© publique du Quv©bec | Sandrine Moreira et al |
| EPI_ISL_535750 | Canada/QC-LSPQ-L00216503/2020 | 2020-03-12 | North America/Canada/Quebec/ | Hv¥tel-Dieu de Lv©vis | Laboratoire de santv© publique du Quv©bec | Sandrine Moreira et al |
| EPI_ISL_535751 | Canada/QC-LSPQ-L00216532/2020 | 2020-03-12 | North America/Canada/Quebec/ | Hv¥tel-Dieu de Lv©vis | Laboratoire de santv© publique du Quv©bec | Sandrine Moreira et al |
| EPI_ISL_535752 | Canada/QC-LSPQ-L00216731/2020 | 2020-03-13 | North America/Canada/Quebec/ | Hv¥pital Pierre-Le Gardeur | Laboratoire de santv© publique du Quv©bec | Sandrine Moreira et al |
| EPI_ISL_535753 | Canada/QC-LSPQ-L00216909/2020 | 2020-03-13 | North America/Canada/Quebec/ | CHUM - Microbiologie - Hv¥pital Saint-Luc | Laboratoire de santv© publique du Quv©bec | Sandrine Moreira et al |
| EPI_ISL_535754 | Canada/QC-LSPQ-L00216920/2020 | 2020-03-13 | North America/Canada/Quebec/ | CHUM - Microbiologie - Hv¥pital Saint-Luc | Laboratoire de santv© publique du Quv©bec | Sandrine Moreira et al |
| EPI_ISL_535755 | Canada/QC-LSPQ-L00216993/2020 | 2020-03-13 | North America/Canada/Quebec/ | Hv¥pital Maisonneuve-Rosemont | Laboratoire de santv© publique du Quv©bec | Sandrine Moreira et al |
| EPI_ISL_535756 | Canada/QC-LSPQ-L00217024/2020 | 2020-03-13 | North America/Canada/Quebec/ | Hv¥pital gv©nv©ral Juif | Laboratoire de santv© publique du Quv©bec | Sandrine Moreira et al |
| EPI_ISL_535757 | Canada/QC-LSPQ-L00217032/2020 | 2020-03-13 | North America/Canada/Quebec/ | Hv¥pital gv©nv©ral Juif | Laboratoire de santv© publique du Quv©bec | Sandrine Moreira et al |
| EPI_ISL_535758 | Canada/QC-LSPQ-L00217092/2020 | 2020-03-13 | North America/Canada/Quebec/ | Hv¥tel-Dieu de Lv©vis | Laboratoire de santv© publique du Quv©bec | Sandrine Moreira et al |

| | | | | | | |
|---|---|---|---|---|---|---|
| EPI_ISL_535759 | Canada/QC-LSPQ-L00217098/2020 | 2020-03-13 | North America/Canada/Quebec/ | H√¥tel-Dieu de L√©vis | Laboratoire de sant√© publique du Qu√©bec | Sandrine Moreira et al |
| EPI_ISL_535760 | Canada/QC-LSPQ-L00217408/2020 | 2020-03-13 | North America/Canada/Quebec/ | CHUM - Microbiologie - H√¥pital Saint-Luc | Laboratoire de sant√© publique du Qu√©bec | Sandrine Moreira et al |
| EPI_ISL_535761 | Canada/QC-LSPQ-L00217411/2020 | 2020-03-13 | North America/Canada/Quebec/ | CUSM-Site Glen-LAB Microbiologie | Laboratoire de sant√© publique du Qu√©bec | Sandrine Moreira et al |
| EPI_ISL_535762 | Canada/QC-LSPQ-L00217527/2020 | 2020-03-13 | North America/Canada/Quebec/ | H√¥pital Pierre-Le Gardeur | Laboratoire de sant√© publique du Qu√©bec | Sandrine Moreira et al |
| EPI_ISL_535763 | Canada/QC-LSPQ-L00217534/2020 | 2020-03-13 | North America/Canada/Quebec/ | H√¥pital Pierre-Boucher | Laboratoire de sant√© publique du Qu√©bec | Sandrine Moreira et al |
| EPI_ISL_535764 | Canada/QC-LSPQ-L00217559/2020 | 2020-03-12 | North America/Canada/Quebec/ | CUSM-Site Glen-LAB Microbiologie | Laboratoire de sant√© publique du Qu√©bec | Sandrine Moreira et al |
| EPI_ISL_535765 | Canada/QC-LSPQ-L00217574/2020 | 2020-03-13 | North America/Canada/Quebec/ | CUSM-Site Glen-LAB Microbiologie | Laboratoire de sant√© publique du Qu√©bec | Sandrine Moreira et al |
| EPI_ISL_535766 | Canada/QC-LSPQ-L00217586/2020 | 2020-03-13 | North America/Canada/Quebec/ | H√¥pital Pierre-Le Gardeur | Laboratoire de sant√© publique du Qu√©bec | Sandrine Moreira et al |
| EPI_ISL_535767 | Canada/QC-LSPQ-L00217831/2020 | 2020-03-13 | North America/Canada/Quebec/ | Centre hospitalier de St-Mary | Laboratoire de sant√© publique du Qu√©bec | Sandrine Moreira et al |
| EPI_ISL_535768 | Canada/QC-LSPQ-L00217833/2020 | 2020-03-13 | North America/Canada/Quebec/ | Centre hospitalier de St-Mary | Laboratoire de sant√© publique du Qu√©bec | Sandrine Moreira et al |
| EPI_ISL_535769 | Canada/QC-LSPQ-L00218011/2020 | 2020-03-14 | North America/Canada/Quebec/ | H√¥pital g√©n√©ral Juif | Laboratoire de sant√© publique du Qu√©bec | Sandrine Moreira et al |
| EPI_ISL_535770 | Canada/QC-LSPQ-L00218056/2020 | 2020-03-14 | North America/Canada/Quebec/ | H√¥pital Pierre-Le Gardeur | Laboratoire de sant√© publique du Qu√©bec | Sandrine Moreira et al |
| EPI_ISL_535771 | Canada/QC-LSPQ-L00218090/2020 | 2020-03-14 | North America/Canada/Quebec/ | H√¥pital Pierre-Le Gardeur | Laboratoire de sant√© publique du Qu√©bec | Sandrine Moreira et al |

| | | | | | | |
|---|---|---|---|---|---|---|
| EPI_ISL_535772 | Canada/QC-LSPQ-L00218268/2020 | 2020-03-14 | North America/Canada/Quebec/ | CHUL-LABO MULTI / MICRO | Laboratoire de santv© publique du Quv©bec | Sandrine Moreira et al |
| EPI_ISL_535773 | Canada/QC-LSPQ-L00218269/2020 | 2020-03-14 | North America/Canada/Quebec/ | CHUL-LABO MULTI / MICRO | Laboratoire de santv© publique du Quv©bec | Sandrine Moreira et al |
| EPI_ISL_535774 | Canada/QC-LSPQ-L00218308/2020 | 2020-03-14 | North America/Canada/Quebec/ | CHUL-LABO MULTI / MICRO | Laboratoire de santv© publique du Quv©bec | Sandrine Moreira et al |
| EPI_ISL_535775 | Canada/QC-LSPQ-L00218310/2020 | 2020-03-14 | North America/Canada/Quebec/ | CHUL-LABO MULTI / MICRO | Laboratoire de santv© publique du Quv©bec | Sandrine Moreira et al |
| EPI_ISL_535776 | Canada/QC-LSPQ-L00218334/2020 | 2020-03-14 | North America/Canada/Quebec/ | Centre de SSS de la Haute-Yamaska | Laboratoire de santv© publique du Quv©bec | Sandrine Moreira et al |
| EPI_ISL_535777 | Canada/QC-LSPQ-L00218337/2020 | 2020-03-14 | North America/Canada/Quebec/ | Centre de SSS de la Haute-Yamaska | Laboratoire de santv© publique du Quv©bec | Sandrine Moreira et al |
| EPI_ISL_535778 | Canada/QC-LSPQ-L00218350/2020 | 2020-03-14 | North America/Canada/Quebec/ | Centre de SSS de la Haute-Yamaska | Laboratoire de santv© publique du Quv©bec | Sandrine Moreira et al |
| EPI_ISL_535779 | Canada/QC-LSPQ-L00218353/2020 | 2020-03-14 | North America/Canada/Quebec/ | Centre de SSS de la Haute-Yamaska | Laboratoire de santv© publique du Quv©bec | Sandrine Moreira et al |
| EPI_ISL_535780 | Canada/QC-LSPQ-L00218371/2020 | 2020-03-14 | North America/Canada/Quebec/ | Centre de SSS de la Haute-Yamaska | Laboratoire de santv© publique du Quv©bec | Sandrine Moreira et al |
| EPI_ISL_535781 | Canada/QC-LSPQ-L00218472/2020 | 2020-03-14 | North America/Canada/Quebec/ | CHUL-LABO MULTI / MICRO | Laboratoire de santv© publique du Quv©bec | Sandrine Moreira et al |
| EPI_ISL_535782 | Canada/QC-LSPQ-L00218475/2020 | 2020-03-14 | North America/Canada/Quebec/ | CHUL-LABO MULTI / MICRO | Laboratoire de santv© publique du Quv©bec | Sandrine Moreira et al |
| EPI_ISL_535783 | Canada/QC-LSPQ-L00218477/2020 | 2020-03-14 | North America/Canada/Quebec/ | CHUM - Microbiologie - Hv¥pital Saint-Luc | Laboratoire de santv© publique du Quv©bec | Sandrine Moreira et al |
| EPI_ISL_535784 | Canada/QC-LSPQ-L00218534/2020 | 2020-03-14 | North America/Canada/Quebec/ | CHUL-LABO MULTI / MICRO | Laboratoire de santv© publique du Quv©bec | Sandrine Moreira et al |

| | | | | | | |
|---|---|---|---|---|---|---|
| EPI_ISL_535785 | Canada/QC-LSPQ-L00218559/2020 | 2020-03-14 | North America/Canada/Quebec/ | CHUL-LABO MULTI / MICRO | Laboratoire de santv© publique du Quv©bec | Sandrine Moreira et al |
| EPI_ISL_535786 | Canada/QC-LSPQ-L00219020/2020 | 2020-03-14 | North America/Canada/Quebec/ | Hv¥pital Pierre-Boucher | Laboratoire de santv© publique du Quv©bec | Sandrine Moreira et al |
| EPI_ISL_535787 | Canada/QC-LSPQ-L00219039/2020 | 2020-03-14 | North America/Canada/Quebec/ | Centre Hospitalier Rv©gional de Lanaudiv®re | Laboratoire de santv© publique du Quv©bec | Sandrine Moreira et al |
| EPI_ISL_535788 | Canada/QC-LSPQ-L00219049/2020 | 2020-03-14 | North America/Canada/Quebec/ | Centre Hospitalier Rv©gional de Lanaudiv®re | Laboratoire de santv© publique du Quv©bec | Sandrine Moreira et al |
| EPI_ISL_535789 | Canada/QC-LSPQ-L00219066/2020 | 2020-03-14 | North America/Canada/Quebec/ | CHUM - Microbiologie - Hv¥pital Saint-Luc | Laboratoire de santv© publique du Quv©bec | Sandrine Moreira et al |
| EPI_ISL_535790 | Canada/QC-LSPQ-L00219089/2020 | 2020-03-14 | North America/Canada/Quebec/ | Centre Hospitalier Rv©gional de Lanaudiv®re | Laboratoire de santv© publique du Quv©bec | Sandrine Moreira et al |
| EPI_ISL_535791 | Canada/QC-LSPQ-L00219335/2020 | 2020-03-14 | North America/Canada/Quebec/ | Hv¥pital rv©gional de Saint-Jv©rv¥me | Laboratoire de santv© publique du Quv©bec | Sandrine Moreira et al |
| EPI_ISL_535792 | Canada/QC-LSPQ-L00219340/2020 | 2020-03-14 | North America/Canada/Quebec/ | Hv¥pital Fleurimont | Laboratoire de santv© publique du Quv©bec | Sandrine Moreira et al |
| EPI_ISL_535793 | Canada/QC-LSPQ-L00219533/2020 | 2020-03-15 | North America/Canada/Quebec/ | Hv¥pital de Saint-Eustache | Laboratoire de santv© publique du Quv©bec | Sandrine Moreira et al |
| EPI_ISL_535794 | Canada/QC-LSPQ-L00219618/2020 | 2020-03-14 | North America/Canada/Quebec/ | Hv¥pital de Hull | Laboratoire de santv© publique du Quv©bec | Sandrine Moreira et al |
| EPI_ISL_535795 | Canada/QC-LSPQ-L00219619/2020 | 2020-03-15 | North America/Canada/Quebec/ | Hv¥pital Notre-Dame | Laboratoire de santv© publique du Quv©bec | Sandrine Moreira et al |
| EPI_ISL_535796 | Canada/QC-LSPQ-L00219966/2020 | 2020-03-15 | North America/Canada/Quebec/ | Hv¥pital Charles-LeMoyne | Laboratoire de santv© publique du Quv©bec | Sandrine Moreira et al |
| EPI_ISL_535797 | Canada/QC-LSPQ-L00220289/2020 | 2020-03-15 | North America/Canada/Quebec/ | Hv¥pital Honorv©-Mercier | Laboratoire de santv© publique du Quv©bec | Sandrine Moreira et al |

| | | | | | | |
|---|---|---|---|---|---|---|
| EPI_ISL_535798 | Canada/QC-LSPQ-L00220369/2020 | 2020-03-15 | North America/Canada/Quebec/ | H√¥pital r√©gional de Saint-J√©r√¥me | Laboratoire de sant√© publique du Qu√©bec | Sandrine Moreira et al |
| EPI_ISL_535799 | Canada/QC-LSPQ-L00220546/2020 | 2020-03-15 | North America/Canada/Quebec/ | CSSS Haut-Richelieu/Rouville (H√¥pital) | Laboratoire de sant√© publique du Qu√©bec | Sandrine Moreira et al |
| EPI_ISL_535800 | Canada/QC-LSPQ-L00220634/2020 | 2020-03-15 | North America/Canada/Quebec/ | CSSS Haut-Richelieu/Rouville (H√¥pital) | Laboratoire de sant√© publique du Qu√©bec | Sandrine Moreira et al |
| EPI_ISL_535801 | Canada/QC-LSPQ-L00220733/2020 | 2020-03-15 | North America/Canada/Quebec/ | CHU Sainte-Justine | Laboratoire de sant√© publique du Qu√©bec | Sandrine Moreira et al |
| EPI_ISL_535802 | Canada/QC-LSPQ-L00220736/2020 | 2020-03-15 | North America/Canada/Quebec/ | CHUM - Microbiologie - H√¥pital Saint-Luc | Laboratoire de sant√© publique du Qu√©bec | Sandrine Moreira et al |
| EPI_ISL_535803 | Canada/QC-LSPQ-L00220827/2020 | 2020-03-14 | North America/Canada/Quebec/ | Centre de SSS de la Haute-Yamaska | Laboratoire de sant√© publique du Qu√©bec | Sandrine Moreira et al |
| EPI_ISL_535804 | Canada/QC-LSPQ-L00220832/2020 | 2020-03-14 | North America/Canada/Quebec/ | Centre de SSS de la Haute-Yamaska | Laboratoire de sant√© publique du Qu√©bec | Sandrine Moreira et al |
| EPI_ISL_535805 | Canada/QC-LSPQ-L00220897/2020 | 2020-03-14 | North America/Canada/Quebec/ | Centre de SSS de la Haute-Yamaska | Laboratoire de sant√© publique du Qu√©bec | Sandrine Moreira et al |
| EPI_ISL_535806 | Canada/QC-LSPQ-L00220933/2020 | 2020-03-15 | North America/Canada/Quebec/ | CHUM - Microbiologie - H√¥pital Saint-Luc | Laboratoire de sant√© publique du Qu√©bec | Sandrine Moreira et al |
| EPI_ISL_535807 | Canada/QC-LSPQ-L00221340/2020 | 2020-03-15 | North America/Canada/Quebec/ | Centre hospitalier r√©gional du Grand Portage | Laboratoire de sant√© publique du Qu√©bec | Sandrine Moreira et al |
| EPI_ISL_535808 | Canada/QC-LSPQ-L00221391/2020 | 2020-03-13 | North America/Canada/Quebec/ | Centre de sant√© Chibougamau | Laboratoire de sant√© publique du Qu√©bec | Sandrine Moreira et al |
| EPI_ISL_535809 | Canada/QC-LSPQ-L00221605/2020 | 2020-03-16 | North America/Canada/Quebec/ | H√¥pital de Hull | Laboratoire de sant√© publique du Qu√©bec | Sandrine Moreira et al |
| EPI_ISL_535810 | Canada/QC-LSPQ-L00221627/2020 | 2020-03-15 | North America/Canada/Quebec/ | H√¥pital Sainte-Croix | Laboratoire de sant√© publique du Qu√©bec | Sandrine Moreira et al |

| | | | | | | |
|---|---|---|---|---|---|---|
| EPI_ISL_535811 | Canada/QC-LSPQ-L00221852/2020 | 2020-03-15 | North America/Canada/Quebec/ | CUSM-Site Glen-LAB Microbiologie | Laboratoire de santv© publique du Quv©bec | Sandrine Moreira et al |
| EPI_ISL_535812 | Canada/QC-LSPQ-L00221945/2020 | 2020-03-16 | North America/Canada/Quebec/ | Hv¥pital du Sacrv©-Coeur de Montrv©al | Laboratoire de santv© publique du Quv©bec | Sandrine Moreira et al |
| EPI_ISL_535813 | Canada/QC-LSPQ-L00222020/2020 | 2020-03-16 | North America/Canada/Quebec/ | CSSS Haut-Richelieu/Rouville (Hv¥pital) | Laboratoire de santv© publique du Quv©bec | Sandrine Moreira et al |
| EPI_ISL_535814 | Canada/QC-LSPQ-L00222022/2020 | 2020-03-15 | North America/Canada/Quebec/ | CHUM - Microbiologie - Hv¥pital Saint-Luc | Laboratoire de santv© publique du Quv©bec | Sandrine Moreira et al |
| EPI_ISL_535815 | Canada/QC-LSPQ-L00222035/2020 | 2020-03-16 | North America/Canada/Quebec/ | CSSS Haut-Richelieu/Rouville (Hv¥pital) | Laboratoire de santv© publique du Quv©bec | Sandrine Moreira et al |
| EPI_ISL_535816 | Canada/QC-LSPQ-L00222448/2020 | 2020-03-14 | North America/Canada/Quebec/ | Centre de SSS de la Haute-Yamaska | Laboratoire de santv© publique du Quv©bec | Sandrine Moreira et al |
| EPI_ISL_535817 | Canada/QC-LSPQ-L00222575/2020 | 2020-03-15 | North America/Canada/Quebec/ | Hv¥pital Maisonneuve-Rosemont | Laboratoire de santv© publique du Quv©bec | Sandrine Moreira et al |
| EPI_ISL_535818 | Canada/QC-LSPQ-L00222731/2020 | 2020-03-16 | North America/Canada/Quebec/ | Hv¥pital Pierre-Boucher | Laboratoire de santv© publique du Quv©bec | Sandrine Moreira et al |
| EPI_ISL_535819 | Canada/QC-LSPQ-L00222823/2020 | 2020-03-16 | North America/Canada/Quebec/ | Hv¥pital Charles-LeMoyne | Laboratoire de santv© publique du Quv©bec | Sandrine Moreira et al |
| EPI_ISL_535820 | Canada/QC-LSPQ-L00224744/2020 | 2020-03-17 | North America/Canada/Quebec/ | Hv¥tel-Dieu de Lv©vis | Laboratoire de santv© publique du Quv©bec | Sandrine Moreira et al |
| EPI_ISL_535821 | Canada/QC-LSPQ-L00224745/2020 | 2020-03-17 | North America/Canada/Quebec/ | Hv¥tel-Dieu de Lv©vis | Laboratoire de santv© publique du Quv©bec | Sandrine Moreira et al |
| EPI_ISL_535822 | Canada/QC-LSPQ-L00224793/2020 | 2020-03-17 | North America/Canada/Quebec/ | CHUL-LABO MULTI / MICRO | Laboratoire de santv© publique du Quv©bec | Sandrine Moreira et al |
| EPI_ISL_535823 | Canada/QC-LSPQ-L00224795/2020 | 2020-03-11 | North America/Canada/Quebec/ | CHUL-LABO MULTI / MICRO | Laboratoire de santv© publique du Quv©bec | Sandrine Moreira et al |

| | | | | | | |
|---|---|---|---|---|---|---|
| EPI_ISL_535824 | Canada/QC-LSPQ-L00224812/2020 | 2020-03-17 | North America/Canada/Quebec/ | CHUL-LABO MULTI / MICRO | Laboratoire de santv© publique du Quv©bec | Sandrine Moreira et al |
| EPI_ISL_535825 | Canada/QC-LSPQ-L00224827/2020 | 2020-03-17 | North America/Canada/Quebec/ | CHUL-LABO MULTI / MICRO | Laboratoire de santv© publique du Quv©bec | Sandrine Moreira et al |
| EPI_ISL_535826 | Canada/QC-LSPQ-L00224868/2020 | 2020-03-17 | North America/Canada/Quebec/ | CHUM - Microbiologie - Hv¥pital Saint-Luc | Laboratoire de santv© publique du Quv©bec | Sandrine Moreira et al |
| EPI_ISL_535827 | Canada/QC-LSPQ-L00224967/2020 | 2020-03-17 | North America/Canada/Quebec/ | CHUM - Microbiologie - Hv¥pital Saint-Luc | Laboratoire de santv© publique du Quv©bec | Sandrine Moreira et al |
| EPI_ISL_535828 | Canada/QC-LSPQ-L00225045/2020 | 2020-03-17 | North America/Canada/Quebec/ | CHUL-LABO MULTI / MICRO | Laboratoire de santv© publique du Quv©bec | Sandrine Moreira et al |
| EPI_ISL_535829 | Canada/QC-LSPQ-L00225185/2020 | 2020-03-16 | North America/Canada/Quebec/ | Hv¥pital de Chicoutimi | Laboratoire de santv© publique du Quv©bec | Sandrine Moreira et al |
| EPI_ISL_535830 | Canada/QC-LSPQ-L00225484/2020 | 2020-03-17 | North America/Canada/Quebec/ | CHUM - Microbiologie - Hv¥pital Saint-Luc | Laboratoire de santv© publique du Quv©bec | Sandrine Moreira et al |
| EPI_ISL_535831 | Canada/QC-LSPQ-L00225500/2020 | 2020-03-18 | North America/Canada/Quebec/ | Hv¥pital Maisonneuve-Rosemont | Laboratoire de santv© publique du Quv©bec | Sandrine Moreira et al |
| EPI_ISL_535832 | Canada/QC-LSPQ-L00225515/2020 | 2020-03-17 | North America/Canada/Quebec/ | CHUM - Microbiologie - Hv¥pital Saint-Luc | Laboratoire de santv© publique du Quv©bec | Sandrine Moreira et al |
| EPI_ISL_535833 | Canada/QC-LSPQ-L00225655/2020 | 2020-03-17 | North America/Canada/Quebec/ | Hv¥tel-Dieu de Lv©vis | Laboratoire de santv© publique du Quv©bec | Sandrine Moreira et al |
| EPI_ISL_535834 | Canada/QC-LSPQ-L00225662/2020 | 2020-03-17 | North America/Canada/Quebec/ | Hv¥tel-Dieu de Lv©vis | Laboratoire de santv© publique du Quv©bec | Sandrine Moreira et al |
| EPI_ISL_535835 | Canada/QC-LSPQ-L00225667/2020 | 2020-03-17 | North America/Canada/Quebec/ | Hv¥tel-Dieu de Lv©vis | Laboratoire de santv© publique du Quv©bec | Sandrine Moreira et al |
| EPI_ISL_535836 | Canada/QC-LSPQ-L00225685/2020 | 2020-03-17 | North America/Canada/Quebec/ | Hv¥tel-Dieu de Lv©vis | Laboratoire de santv© publique du Quv©bec | Sandrine Moreira et al |

| | | | | | | |
|---|---|---|---|---|---|---|
| EPI_ISL_535837 | Canada/QC-LSPQ-L00225800/2020 | 2020-03-16 | North America/Canada/Quebec/ | H√¥pital r√©gional de Saint-J√©r√¥me | Laboratoire de sant√© publique du Qu√©bec | Sandrine Moreira et al |
| EPI_ISL_535838 | Canada/QC-LSPQ-L00225808/2020 | 2020-03-16 | North America/Canada/Quebec/ | H√¥pital r√©gional de Saint-J√©r√¥me | Laboratoire de sant√© publique du Qu√©bec | Sandrine Moreira et al |
| EPI_ISL_535839 | Canada/QC-LSPQ-L00225894/2020 | 2020-03-17 | North America/Canada/Quebec/ | H√¥pital Cit√© de la Sant√© | Laboratoire de sant√© publique du Qu√©bec | Sandrine Moreira et al |
| EPI_ISL_535840 | Canada/QC-LSPQ-L00225956/2020 | 2020-03-18 | North America/Canada/Quebec/ | H√¥pital Pierre-Boucher | Laboratoire de sant√© publique du Qu√©bec | Sandrine Moreira et al |
| EPI_ISL_535841 | Canada/QC-LSPQ-L00226107/2020 | 2020-03-17 | North America/Canada/Quebec/ | CSSS Haut-Richelieu/Rouville (H√¥pital) | Laboratoire de sant√© publique du Qu√©bec | Sandrine Moreira et al |
| EPI_ISL_535842 | Canada/QC-LSPQ-L00226142/2020 | 2020-03-17 | North America/Canada/Quebec/ | Centre Hospitalier R√©gional de Lanaudi√®re | Laboratoire de sant√© publique du Qu√©bec | Sandrine Moreira et al |
| EPI_ISL_535843 | Canada/QC-LSPQ-L00226159/2020 | 2020-03-17 | North America/Canada/Quebec/ | Centre Hospitalier R√©gional de Lanaudi√®re | Laboratoire de sant√© publique du Qu√©bec | Sandrine Moreira et al |
| EPI_ISL_535844 | Canada/QC-LSPQ-L00226167/2020 | 2020-03-17 | North America/Canada/Quebec/ | Centre Hospitalier R√©gional de Lanaudi√®re | Laboratoire de sant√© publique du Qu√©bec | Sandrine Moreira et al |
| EPI_ISL_535846 | Canada/QC-LSPQ-L00226390/2020 | 2020-03-17 | North America/Canada/Quebec/ | CSSS Haut-Richelieu/Rouville (H√¥pital) | Laboratoire de sant√© publique du Qu√©bec | Sandrine Moreira et al |
| EPI_ISL_535847 | Canada/QC-LSPQ-L00226474/2020 | 2020-03-18 | North America/Canada/Quebec/ | H√¥pital du Suro√Æt | Laboratoire de sant√© publique du Qu√©bec | Sandrine Moreira et al |
| EPI_ISL_535848 | Canada/QC-LSPQ-L00226547/2020 | 2020-03-18 | North America/Canada/Quebec/ | H√¥pital Charles-LeMoyne | Laboratoire de sant√© publique du Qu√©bec | Sandrine Moreira et al |
| EPI_ISL_535849 | Canada/QC-LSPQ-L00226589/2020 | 2020-03-18 | North America/Canada/Quebec/ | Centre de SSS de Trois-Rivi√®res | Laboratoire de sant√© publique du Qu√©bec | Sandrine Moreira et al |
| EPI_ISL_535850 | Canada/QC-LSPQ-L00226624/2020 | 2020-03-18 | North America/Canada/Quebec/ | H√¥pital Pierre-Le Gardeur | Laboratoire de sant√© publique du Qu√©bec | Sandrine Moreira et al |

| | | | | | | |
|---|---|---|---|---|---|---|
| EPI_ISL_535851 | Canada/QC-LSPQ-L00226858/2020 | 2020-03-18 | North America/Canada/Quebec/ | H√¥pital Charles-LeMoyne | Laboratoire de sant√© publique du Qu√©bec | Sandrine Moreira et al |
| EPI_ISL_535852 | Canada/QC-LSPQ-L00226861/2020 | 2020-03-18 | North America/Canada/Quebec/ | H√¥pital g√©n√©ral du Lakeshore | Laboratoire de sant√© publique du Qu√©bec | Sandrine Moreira et al |
| EPI_ISL_535853 | Canada/QC-LSPQ-L00226912/2020 | 2020-03-18 | North America/Canada/Quebec/ | H√¥pital Charles-LeMoyne | Laboratoire de sant√© publique du Qu√©bec | Sandrine Moreira et al |
| EPI_ISL_535854 | Canada/QC-LSPQ-L00226938/2020 | 2020-03-17 | North America/Canada/Quebec/ | Centre de SSS de la Haute-Yamaska | Laboratoire de sant√© publique du Qu√©bec | Sandrine Moreira et al |
| EPI_ISL_535855 | Canada/QC-LSPQ-L00226950/2020 | 2020-03-17 | North America/Canada/Quebec/ | Centre de SSS de la Haute-Yamaska | Laboratoire de sant√© publique du Qu√©bec | Sandrine Moreira et al |
| EPI_ISL_535856 | Canada/QC-LSPQ-L00227048/2020 | 2020-03-17 | North America/Canada/Quebec/ | H√¥pital de Verdun | Laboratoire de sant√© publique du Qu√©bec | Sandrine Moreira et al |
| EPI_ISL_535857 | Canada/QC-LSPQ-L00227094/2020 | 2020-03-18 | North America/Canada/Quebec/ | H√¥pital Pierre-Boucher | Laboratoire de sant√© publique du Qu√©bec | Sandrine Moreira et al |
| EPI_ISL_535858 | Canada/QC-LSPQ-L00227147/2020 | 2020-03-18 | North America/Canada/Quebec/ | H√¥pital de Gatineau | Laboratoire de sant√© publique du Qu√©bec | Sandrine Moreira et al |
| EPI_ISL_535859 | Canada/QC-LSPQ-L00227316/2020 | 2020-03-18 | North America/Canada/Quebec/ | H√¥pital Sainte-Croix | Laboratoire de sant√© publique du Qu√©bec | Sandrine Moreira et al |
| EPI_ISL_535860 | Canada/QC-LSPQ-L00227376/2020 | 2020-03-18 | North America/Canada/Quebec/ | H√¥pital Sainte-Croix | Laboratoire de sant√© publique du Qu√©bec | Sandrine Moreira et al |
| EPI_ISL_535861 | Canada/QC-LSPQ-L00227457/2020 | 2020-03-18 | North America/Canada/Quebec/ | H√¥pital Pierre-Le Gardeur | Laboratoire de sant√© publique du Qu√©bec | Sandrine Moreira et al |
| EPI_ISL_535862 | Canada/QC-LSPQ-L00227476/2020 | 2020-03-18 | North America/Canada/Quebec/ | H√¥pital Pierre-Le Gardeur | Laboratoire de sant√© publique du Qu√©bec | Sandrine Moreira et al |
| EPI_ISL_535863 | Canada/QC-LSPQ-L00227481/2020 | 2020-03-19 | North America/Canada/Quebec/ | H√¥pital Pierre-Le Gardeur | Laboratoire de sant√© publique du Qu√©bec | Sandrine Moreira et al |

| | | | | | | |
|---|---|---|---|---|---|---|
| EPI_ISL_535864 | Canada/QC-LSPQ-L00227528/2020 | 2020-03-19 | North America/Canada/Quebec/ | CSSS Haut-Richelieu/Rouville (H√¥pital) | Laboratoire de sant√© publique du Qu√©bec | Sandrine Moreira et al |
| EPI_ISL_535865 | Canada/QC-LSPQ-L00227658/2020 | 2020-03-19 | North America/Canada/Quebec/ | H√¥pital de Saint-Eustache | Laboratoire de sant√© publique du Qu√©bec | Sandrine Moreira et al |
| EPI_ISL_535866 | Canada/QC-LSPQ-L00227663/2020 | 2020-03-19 | North America/Canada/Quebec/ | H√¥pital Charles-LeMoyne | Laboratoire de sant√© publique du Qu√©bec | Sandrine Moreira et al |
| EPI_ISL_535867 | Canada/QC-LSPQ-L00227703/2020 | 2020-03-19 | North America/Canada/Quebec/ | H√¥pital de Saint-Eustache | Laboratoire de sant√© publique du Qu√©bec | Sandrine Moreira et al |
| EPI_ISL_535868 | Canada/QC-LSPQ-L00227830/2020 | 2020-03-19 | North America/Canada/Quebec/ | H√¥pital du Surо√Æt | Laboratoire de sant√© publique du Qu√©bec | Sandrine Moreira et al |
| EPI_ISL_535869 | Canada/QC-LSPQ-L00227996/2020 | 2020-03-19 | North America/Canada/Quebec/ | H√¥pital Charles-LeMoyne | Laboratoire de sant√© publique du Qu√©bec | Sandrine Moreira et al |
| EPI_ISL_535870 | Canada/QC-LSPQ-L00228138/2020 | 2020-03-18 | North America/Canada/Quebec/ | Centre Hospitalier R√©gional de Lanaudi√®re | Laboratoire de sant√© publique du Qu√©bec | Sandrine Moreira et al |
| EPI_ISL_535871 | Canada/QC-LSPQ-L00228149/2020 | 2020-03-19 | North America/Canada/Quebec/ | Centre Hospitalier R√©gional de Lanaudi√®re | Laboratoire de sant√© publique du Qu√©bec | Sandrine Moreira et al |
| EPI_ISL_535872 | Canada/QC-LSPQ-L00228225/2020 | 2020-03-18 | North America/Canada/Quebec/ | Centre hospitalier r√©gional du Grand Portage | Laboratoire de sant√© publique du Qu√©bec | Sandrine Moreira et al |
| EPI_ISL_535873 | Canada/QC-LSPQ-L00228231/2020 | 2020-03-19 | North America/Canada/Quebec/ | Centre hospitalier r√©gional du Grand Portage | Laboratoire de sant√© publique du Qu√©bec | Sandrine Moreira et al |
| EPI_ISL_535874 | Canada/QC-LSPQ-L00228636/2020 | 2020-03-20 | North America/Canada/Quebec/ | H√¥pital Charles-LeMoyne | Laboratoire de sant√© publique du Qu√©bec | Sandrine Moreira et al |
| EPI_ISL_535875 | Canada/QC-LSPQ-L00228687/2020 | 2020-03-20 | North America/Canada/Quebec/ | H√¥pital g√©n√©ral du Lakeshore | Laboratoire de sant√© publique du Qu√©bec | Sandrine Moreira et al |
| EPI_ISL_535877 | Canada/QC-LSPQ-L00228790/2020 | 2020-03-20 | North America/Canada/Quebec/ | CSSS Haut-Richelieu/Rouville (H√¥pital) | Laboratoire de sant√© publique du Qu√©bec | Sandrine Moreira et al |

| | | | | | | |
|---|---|---|---|---|---|---|
| EPI_ISL_535878 | Canada/QC-LSPQ-L00228839/2020 | 2020-03-20 | North America/Canada/Quebec/ | H√¥pital de Saint-Eustache | Laboratoire de sant√© publique du Qu√©bec | Sandrine Moreira et al |
| EPI_ISL_535879 | Canada/QC-LSPQ-L00228862/2020 | 2020-03-20 | North America/Canada/Quebec/ | H√¥pital Pierre-Boucher | Laboratoire de sant√© publique du Qu√©bec | Sandrine Moreira et al |
| EPI_ISL_535880 | Canada/QC-LSPQ-L00228918/2020 | 2020-03-20 | North America/Canada/Quebec/ | H√¥pital du Surov√Æt | Laboratoire de sant√© publique du Qu√©bec | Sandrine Moreira et al |
| EPI_ISL_535881 | Canada/QC-LSPQ-L00228950/2020 | 2020-03-20 | North America/Canada/Quebec/ | H√¥pital Pierre-Le Gardeur | Laboratoire de sant√© publique du Qu√©bec | Sandrine Moreira et al |
| EPI_ISL_535882 | Canada/QC-LSPQ-L00228954/2020 | 2020-03-20 | North America/Canada/Quebec/ | H√¥pital Pierre-Le Gardeur | Laboratoire de sant√© publique du Qu√©bec | Sandrine Moreira et al |
| EPI_ISL_535883 | Canada/QC-LSPQ-L00228964/2020 | 2020-03-20 | North America/Canada/Quebec/ | H√¥pital Pierre-Le Gardeur | Laboratoire de sant√© publique du Qu√©bec | Sandrine Moreira et al |
| EPI_ISL_535884 | Canada/QC-LSPQ-L00229065/2020 | 2020-03-20 | North America/Canada/Quebec/ | CSSS Haut-Richelieu/Rouville (H√¥pital) | Laboratoire de sant√© publique du Qu√©bec | Sandrine Moreira et al |
| EPI_ISL_535885 | Canada/QC-LSPQ-L00229136/2020 | 2020-03-19 | North America/Canada/Quebec/ | H√¥pital de Maria | Laboratoire de sant√© publique du Qu√©bec | Sandrine Moreira et al |
| EPI_ISL_535886 | Canada/QC-LSPQ-L00229172/2020 | 2020-03-20 | North America/Canada/Quebec/ | H√¥pital de Hull | Laboratoire de sant√© publique du Qu√©bec | Sandrine Moreira et al |
| EPI_ISL_535887 | Canada/QC-LSPQ-L00229212/2020 | 2020-03-20 | North America/Canada/Quebec/ | H√¥pital de Hull | Laboratoire de sant√© publique du Qu√©bec | Sandrine Moreira et al |
| EPI_ISL_535888 | Canada/QC-LSPQ-L00229288/2020 | 2020-03-20 | North America/Canada/Quebec/ | H√¥pital Charles-LeMoyne | Laboratoire de sant√© publique du Qu√©bec | Sandrine Moreira et al |
| EPI_ISL_535889 | Canada/QC-LSPQ-L00229319/2020 | 2020-03-20 | North America/Canada/Quebec/ | H√¥pital de Saint-Eustache | Laboratoire de sant√© publique du Qu√©bec | Sandrine Moreira et al |
| EPI_ISL_535890 | Canada/QC-LSPQ-L00229326/2020 | 2020-03-20 | North America/Canada/Quebec/ | H√¥pital Charles-LeMoyne | Laboratoire de sant√© publique du Qu√©bec | Sandrine Moreira et al |

| | | | | | | |
|---|---|---|---|---|---|---|
| EPI_ISL_535891 | Canada/QC-LSPQ-L00229363/2020 | 2020-03-20 | North America/Canada/Quebec/ | H√¥pital Charles-LeMoyne | Laboratoire de sant√© publique du Qu√©bec | Sandrine Moreira et al |
| EPI_ISL_535892 | Canada/QC-LSPQ-L00229384/2020 | 2020-03-20 | North America/Canada/Quebec/ | H√¥pital Pierre-Le Gardeur | Laboratoire de sant√© publique du Qu√©bec | Sandrine Moreira et al |
| EPI_ISL_535893 | Canada/QC-LSPQ-L00229410/2020 | 2020-03-20 | North America/Canada/Quebec/ | H√¥pital Pierre-Le Gardeur | Laboratoire de sant√© publique du Qu√©bec | Sandrine Moreira et al |
| EPI_ISL_535894 | Canada/QC-LSPQ-L00229443/2020 | 2020-03-20 | North America/Canada/Quebec/ | Centre Hospitalier R√©gional de Lanaudi√®re | Laboratoire de sant√© publique du Qu√©bec | Sandrine Moreira et al |
| EPI_ISL_535895 | Canada/QC-LSPQ-L00229450/2020 | 2020-03-20 | North America/Canada/Quebec/ | Centre Hospitalier R√©gional de Lanaudi√®re | Laboratoire de sant√© publique du Qu√©bec | Sandrine Moreira et al |
| EPI_ISL_535896 | Canada/QC-LSPQ-L00229554/2020 | 2020-03-20 | North America/Canada/Quebec/ | Centre Hospitalier R√©gional de Lanaudi√®re | Laboratoire de sant√© publique du Qu√©bec | Sandrine Moreira et al |
| EPI_ISL_535897 | Canada/QC-LSPQ-L00229615/2020 | 2020-03-20 | North America/Canada/Quebec/ | H√¥pital Sainte-Croix | Laboratoire de sant√© publique du Qu√©bec | Sandrine Moreira et al |
| EPI_ISL_535898 | Canada/QC-LSPQ-L00229648/2020 | 2020-03-20 | North America/Canada/Quebec/ | H√¥pital Sainte-Croix | Laboratoire de sant√© publique du Qu√©bec | Sandrine Moreira et al |
| EPI_ISL_535899 | Canada/QC-LSPQ-L00229671/2020 | 2020-03-20 | North America/Canada/Quebec/ | H√¥pital Sainte-Croix | Laboratoire de sant√© publique du Qu√©bec | Sandrine Moreira et al |
| EPI_ISL_535900 | Canada/QC-LSPQ-L00229674/2020 | 2020-03-20 | North America/Canada/Quebec/ | H√¥pital Sainte-Croix | Laboratoire de sant√© publique du Qu√©bec | Sandrine Moreira et al |
| EPI_ISL_535901 | Canada/QC-LSPQ-L00229727/2020 | 2020-03-20 | North America/Canada/Quebec/ | H√¥pital Sainte-Croix | Laboratoire de sant√© publique du Qu√©bec | Sandrine Moreira et al |
| EPI_ISL_535902 | Canada/QC-LSPQ-L00229741/2020 | 2020-03-21 | North America/Canada/Quebec/ | CSSS Haut-Richelieu/Rouville (H√¥pital) | Laboratoire de sant√© publique du Qu√©bec | Sandrine Moreira et al |
| EPI_ISL_535903 | Canada/QC-LSPQ-L00229794/2020 | 2020-03-20 | North America/Canada/Quebec/ | H√¥tel-Dieu de Sorel | Laboratoire de sant√© publique du Qu√©bec | Sandrine Moreira et al |

| | | | | | | |
|---|---|---|---|---|---|---|
| EPI_ISL_535904 | Canada/QC-LSPQ-L00229796/2020 | 2020-03-20 | North America/Canada/Quebec/ | H√¥tel-Dieu de Sorel | Laboratoire de sant√© publique du Qu√©bec | Sandrine Moreira et al |
| EPI_ISL_535905 | Canada/QC-LSPQ-L00229929/2020 | 2020-03-19 | North America/Canada/Quebec/ | H√¥pital de Chicoutimi | Laboratoire de sant√© publique du Qu√©bec | Sandrine Moreira et al |
| EPI_ISL_535906 | Canada/QC-LSPQ-L00229943/2020 | 2020-03-19 | North America/Canada/Quebec/ | H√¥pital de Chicoutimi | Laboratoire de sant√© publique du Qu√©bec | Sandrine Moreira et al |
| EPI_ISL_535907 | Canada/QC-LSPQ-L00229952/2020 | 2020-03-21 | North America/Canada/Quebec/ | Centre Hospitalier R√©gional de Lanaudi√®re | Laboratoire de sant√© publique du Qu√©bec | Sandrine Moreira et al |
| EPI_ISL_535908 | Canada/QC-LSPQ-L00230018/2020 | 2020-03-21 | North America/Canada/Quebec/ | H√¥pital Pierre-Boucher | Laboratoire de sant√© publique du Qu√©bec | Sandrine Moreira et al |
| EPI_ISL_535909 | Canada/QC-LSPQ-L00230020/2020 | 2020-03-21 | North America/Canada/Quebec/ | H√¥pital Pierre-Boucher | Laboratoire de sant√© publique du Qu√©bec | Sandrine Moreira et al |
| EPI_ISL_535910 | Canada/QC-LSPQ-L00230110/2020 | 2020-03-21 | North America/Canada/Quebec/ | CSSS Haut-Richelieu/Rouville (H√¥pital) | Laboratoire de sant√© publique du Qu√©bec | Sandrine Moreira et al |
| EPI_ISL_535911 | Canada/QC-LSPQ-L00230159/2020 | 2020-03-21 | North America/Canada/Quebec/ | H√¥pital de Hull | Laboratoire de sant√© publique du Qu√©bec | Sandrine Moreira et al |
| EPI_ISL_535912 | Canada/QC-LSPQ-L00230189/2020 | 2020-03-21 | North America/Canada/Quebec/ | H√¥pital Honor√©-Mercier | Laboratoire de sant√© publique du Qu√©bec | Sandrine Moreira et al |
| EPI_ISL_535913 | Canada/QC-LSPQ-L00230299/2020 | 2020-03-21 | North America/Canada/Quebec/ | H√¥pital du Suro√Æt | Laboratoire de sant√© publique du Qu√©bec | Sandrine Moreira et al |
| EPI_ISL_535914 | Canada/QC-LSPQ-L00230315/2020 | 2020-03-21 | North America/Canada/Quebec/ | H√¥pital du Centre-de-la-Mauricie | Laboratoire de sant√© publique du Qu√©bec | Sandrine Moreira et al |
| EPI_ISL_535915 | Canada/QC-LSPQ-L00230319/2020 | 2020-03-21 | North America/Canada/Quebec/ | H√¥pital Honor√©-Mercier | Laboratoire de sant√© publique du Qu√©bec | Sandrine Moreira et al |
| EPI_ISL_535916 | Canada/QC-LSPQ-L00230351/2020 | 2020-03-21 | North America/Canada/Quebec/ | Centre de SSS de Trois-Rivi√®res | Laboratoire de sant√© publique du Qu√©bec | Sandrine Moreira et al |

| | | | | | | |
|---|---|---|---|---|---|---|
| EPI_ISL_535917 | Canada/QC-LSPQ-L00230444/2020 | 2020-03-21 | North America/Canada/Quebec/ | H√¥pital du Surov√Æt | Laboratoire de sant√© publique du Qu√©bec | Sandrine Moreira et al |
| EPI_ISL_535918 | Canada/QC-LSPQ-L00230448/2020 | 2020-03-21 | North America/Canada/Quebec/ | H√¥pital du Surov√Æt | Laboratoire de sant√© publique du Qu√©bec | Sandrine Moreira et al |
| EPI_ISL_535919 | Canada/QC-LSPQ-L00230472/2020 | 2020-03-21 | North America/Canada/Quebec/ | H√¥pital du Surov√Æt | Laboratoire de sant√© publique du Qu√©bec | Sandrine Moreira et al |
| EPI_ISL_535920 | Canada/QC-LSPQ-L00230473/2020 | 2020-03-21 | North America/Canada/Quebec/ | H√¥pital Charles-LeMoyne | Laboratoire de sant√© publique du Qu√©bec | Sandrine Moreira et al |
| EPI_ISL_535921 | Canada/QC-LSPQ-L00230482/2020 | 2020-03-22 | North America/Canada/Quebec/ | H√¥pital Charles-LeMoyne | Laboratoire de sant√© publique du Qu√©bec | Sandrine Moreira et al |
| EPI_ISL_535922 | Canada/QC-LSPQ-L00230506/2020 | 2020-03-21 | North America/Canada/Quebec/ | H√¥pital Charles-LeMoyne | Laboratoire de sant√© publique du Qu√©bec | Sandrine Moreira et al |
| EPI_ISL_535923 | Canada/QC-LSPQ-L00230536/2020 | 2020-03-21 | North America/Canada/Quebec/ | H√¥pital Charles-LeMoyne | Laboratoire de sant√© publique du Qu√©bec | Sandrine Moreira et al |
| EPI_ISL_535924 | Canada/QC-LSPQ-L00230575/2020 | 2020-03-21 | North America/Canada/Quebec/ | H√¥pital de Saint-Eustache | Laboratoire de sant√© publique du Qu√©bec | Sandrine Moreira et al |
| EPI_ISL_535925 | Canada/QC-LSPQ-L00230637/2020 | 2020-03-21 | North America/Canada/Quebec/ | CSSS Haut-Richelieu/Rouville (H√¥pital) | Laboratoire de sant√© publique du Qu√©bec | Sandrine Moreira et al |
| EPI_ISL_535926 | Canada/QC-LSPQ-L00230647/2020 | 2020-03-21 | North America/Canada/Quebec/ | H√¥pital de Saint-Eustache | Laboratoire de sant√© publique du Qu√©bec | Sandrine Moreira et al |
| EPI_ISL_535927 | Canada/QC-LSPQ-L00230666/2020 | 2020-03-21 | North America/Canada/Quebec/ | H√¥pital Charles-LeMoyne | Laboratoire de sant√© publique du Qu√©bec | Sandrine Moreira et al |
| EPI_ISL_535928 | Canada/QC-LSPQ-L00230676/2020 | 2020-03-21 | North America/Canada/Quebec/ | CSSS Haut-Richelieu/Rouville (H√¥pital) | Laboratoire de sant√© publique du Qu√©bec | Sandrine Moreira et al |
| EPI_ISL_535929 | Canada/QC-LSPQ-L00230720/2020 | 2020-03-21 | North America/Canada/Quebec/ | H√¥pital de Saint-Eustache | Laboratoire de sant√© publique du Qu√©bec | Sandrine Moreira et al |

| | | | | | | |
|---|---|---|---|---|---|---|
| EPI_ISL_535930 | Canada/QC-LSPQ-L00230755/2020 | 2020-03-21 | North America/Canada/Quebec/ | H√¥pital de Saint-Eustache | Laboratoire de sant√© publique du Qu√©bec | Sandrine Moreira et al |
| EPI_ISL_535931 | Canada/QC-LSPQ-L00230787/2020 | 2020-03-21 | North America/Canada/Quebec/ | Centre Hospitalier R√©gional de Lanaudi√®re | Laboratoire de sant√© publique du Qu√©bec | Sandrine Moreira et al |
| EPI_ISL_535932 | Canada/QC-LSPQ-L00230796/2020 | 2020-03-21 | North America/Canada/Quebec/ | Centre Hospitalier R√©gional de Lanaudi√®re | Laboratoire de sant√© publique du Qu√©bec | Sandrine Moreira et al |
| EPI_ISL_535933 | Canada/QC-LSPQ-L00230798/2020 | 2020-03-21 | North America/Canada/Quebec/ | Centre Hospitalier R√©gional de Lanaudi√®re | Laboratoire de sant√© publique du Qu√©bec | Sandrine Moreira et al |
| EPI_ISL_535934 | Canada/QC-LSPQ-L00230813/2020 | 2020-03-21 | North America/Canada/Quebec/ | Centre Hospitalier R√©gional de Lanaudi√®re | Laboratoire de sant√© publique du Qu√©bec | Sandrine Moreira et al |
| EPI_ISL_535935 | Canada/QC-LSPQ-L00230821/2020 | 2020-03-21 | North America/Canada/Quebec/ | Centre Hospitalier R√©gional de Lanaudi√®re | Laboratoire de sant√© publique du Qu√©bec | Sandrine Moreira et al |
| EPI_ISL_535936 | Canada/QC-LSPQ-L00230881/2020 | 2020-03-21 | North America/Canada/Quebec/ | H√¥pital Pierre-Le Gardeur | Laboratoire de sant√© publique du Qu√©bec | Sandrine Moreira et al |
| EPI_ISL_535937 | Canada/QC-LSPQ-L00230882/2020 | 2020-03-21 | North America/Canada/Quebec/ | H√¥pital du Centre-de-la-Mauricie | Laboratoire de sant√© publique du Qu√©bec | Sandrine Moreira et al |
| EPI_ISL_535938 | Canada/QC-LSPQ-L00230895/2020 | 2020-03-21 | North America/Canada/Quebec/ | H√¥pital Pierre-Le Gardeur | Laboratoire de sant√© publique du Qu√©bec | Sandrine Moreira et al |
| EPI_ISL_535939 | Canada/QC-LSPQ-L00230946/2020 | 2020-03-21 | North America/Canada/Quebec/ | Centre de SSS D'Arthabaska-et-de-l'√©rable - H√¥tel-Dieu | Laboratoire de sant√© publique du Qu√©bec | Sandrine Moreira et al |
| EPI_ISL_535940 | Canada/QC-LSPQ-L00230948/2020 | 2020-03-21 | North America/Canada/Quebec/ | Centre de SSS D'Arthabaska-et-de-l'√©rable - H√¥tel-Dieu | Laboratoire de sant√© publique du Qu√©bec | Sandrine Moreira et al |
| EPI_ISL_535941 | Canada/QC-LSPQ-L00230998/2020 | 2020-03-21 | North America/Canada/Quebec/ | H√¥pital de Saint-Eustache | Laboratoire de sant√© publique du Qu√©bec | Sandrine Moreira et al |
| EPI_ISL_535942 | Canada/QC-LSPQ-L00231066/2020 | 2020-03-21 | North America/Canada/Quebec/ | H√¥pital de Saint-Eustache | Laboratoire de sant√© publique du Qu√©bec | Sandrine Moreira et al |

| | | | | | | |
|---|---|---|---|---|---|---|
| EPI_ISL_535943 | Canada/QC-LSPQ-L00231068/2020 | 2020-03-21 | North America/Canada/Quebec/ | H√¥pital de Saint-Eustache | Laboratoire de sant√© publique du Qu√©bec | Sandrine Moreira et al |
| EPI_ISL_535944 | Canada/QC-LSPQ-L00231139/2020 | 2020-03-22 | North America/Canada/Quebec/ | H√¥pital Charles-LeMoyne | Laboratoire de sant√© publique du Qu√©bec | Sandrine Moreira et al |
| EPI_ISL_535945 | Canada/QC-LSPQ-L00231156/2020 | 2020-03-21 | North America/Canada/Quebec/ | H√¥pital de Mont-Laurier | Laboratoire de sant√© publique du Qu√©bec | Sandrine Moreira et al |
| EPI_ISL_535946 | Canada/QC-LSPQ-L00231158/2020 | 2020-03-22 | North America/Canada/Quebec/ | CSSS Haut-Richelieu/Rouville (H√¥pital) | Laboratoire de sant√© publique du Qu√©bec | Sandrine Moreira et al |
| EPI_ISL_535947 | Canada/QC-LSPQ-L00231230/2020 | 2020-03-22 | North America/Canada/Quebec/ | H√¥pital de Saint-Eustache | Laboratoire de sant√© publique du Qu√©bec | Sandrine Moreira et al |
| EPI_ISL_535948 | Canada/QC-LSPQ-L00231249/2020 | 2020-03-22 | North America/Canada/Quebec/ | H√¥pital Pierre-Boucher | Laboratoire de sant√© publique du Qu√©bec | Sandrine Moreira et al |
| EPI_ISL_535949 | Canada/QC-LSPQ-L00231271/2020 | 2020-03-22 | North America/Canada/Quebec/ | H√¥pital Charles-LeMoyne | Laboratoire de sant√© publique du Qu√©bec | Sandrine Moreira et al |
| EPI_ISL_535950 | Canada/QC-LSPQ-L00231277/2020 | 2020-03-22 | North America/Canada/Quebec/ | H√¥pital Charles-LeMoyne | Laboratoire de sant√© publique du Qu√©bec | Sandrine Moreira et al |
| EPI_ISL_535951 | Canada/QC-LSPQ-L00231289/2020 | 2020-03-21 | North America/Canada/Quebec/ | H√¥pital de Saint-Eustache | Laboratoire de sant√© publique du Qu√©bec | Sandrine Moreira et al |
| EPI_ISL_535952 | Canada/QC-LSPQ-L00231296/2020 | 2020-03-22 | North America/Canada/Quebec/ | H√¥pital Charles-LeMoyne | Laboratoire de sant√© publique du Qu√©bec | Sandrine Moreira et al |
| EPI_ISL_535953 | Canada/QC-LSPQ-L00231304/2020 | 2020-03-20 | North America/Canada/Quebec/ | H√¥pital Charles-LeMoyne | Laboratoire de sant√© publique du Qu√©bec | Sandrine Moreira et al |
| EPI_ISL_535954 | Canada/QC-LSPQ-L00231415/2020 | 2020-03-22 | North America/Canada/Quebec/ | H√¥pital Pierre-Le Gardeur | Laboratoire de sant√© publique du Qu√©bec | Sandrine Moreira et al |
| EPI_ISL_535955 | Canada/QC-LSPQ-L00231416/2020 | 2020-03-22 | North America/Canada/Quebec/ | Centre Hospitalier R√©gional de Lanaudi√®re | Laboratoire de sant√© publique du Qu√©bec | Sandrine Moreira et al |

| | | | | | | |
|---|---|---|---|---|---|---|
| EPI_ISL_535956 | Canada/QC-LSPQ-L00231456/2020 | 2020-03-22 | North America/Canada/Quebec/ | H√¥pital Pierre-Le Gardeur | Laboratoire de sant√© publique du Qu√©bec | Sandrine Moreira et al |
| EPI_ISL_535957 | Canada/QC-LSPQ-L00231569/2020 | 2020-03-22 | North America/Canada/Quebec/ | H√¥pital Charles-LeMoyne | Laboratoire de sant√© publique du Qu√©bec | Sandrine Moreira et al |
| EPI_ISL_535958 | Canada/QC-LSPQ-L00231633/2020 | 2020-03-21 | North America/Canada/Quebec/ | Centre hospitalier r√©gional du Grand Portage | Laboratoire de sant√© publique du Qu√©bec | Sandrine Moreira et al |
| EPI_ISL_535959 | Canada/QC-LSPQ-L00231634/2020 | 2020-03-21 | North America/Canada/Quebec/ | Centre hospitalier r√©gional du Grand Portage | Laboratoire de sant√© publique du Qu√©bec | Sandrine Moreira et al |
| EPI_ISL_535960 | Canada/QC-LSPQ-L00231755/2020 | 2020-03-22 | North America/Canada/Quebec/ | Centre hospitalier Anna-Laberge | Laboratoire de sant√© publique du Qu√©bec | Sandrine Moreira et al |
| EPI_ISL_535961 | Canada/QC-LSPQ-L00231766/2020 | 2020-03-22 | North America/Canada/Quebec/ | H√¥pital Charles-LeMoyne | Laboratoire de sant√© publique du Qu√©bec | Sandrine Moreira et al |
| EPI_ISL_535962 | Canada/QC-LSPQ-L00231779/2020 | 2020-03-22 | North America/Canada/Quebec/ | H√¥pital Charles-LeMoyne | Laboratoire de sant√© publique du Qu√©bec | Sandrine Moreira et al |
| EPI_ISL_535963 | Canada/QC-LSPQ-L00231805/2020 | 2020-03-20 | North America/Canada/Quebec/ | H√¥pital d'Alma | Laboratoire de sant√© publique du Qu√©bec | Sandrine Moreira et al |
| EPI_ISL_535964 | Canada/QC-LSPQ-L00231814/2020 | 2020-03-20 | North America/Canada/Quebec/ | H√¥pital de Chicoutimi | Laboratoire de sant√© publique du Qu√©bec | Sandrine Moreira et al |
| EPI_ISL_535965 | Canada/QC-LSPQ-L00231860/2020 | 2020-03-20 | North America/Canada/Quebec/ | H√¥pital de Chicoutimi | Laboratoire de sant√© publique du Qu√©bec | Sandrine Moreira et al |
| EPI_ISL_535966 | Canada/QC-LSPQ-L00231946/2020 | 2020-03-22 | North America/Canada/Quebec/ | H√¥pital Charles-LeMoyne | Laboratoire de sant√© publique du Qu√©bec | Sandrine Moreira et al |
| EPI_ISL_535967 | Canada/QC-LSPQ-L00231966/2020 | 2020-03-22 | North America/Canada/Quebec/ | CSSS Haut-Richelieu/Rouville (H√¥pital) | Laboratoire de sant√© publique du Qu√©bec | Sandrine Moreira et al |
| EPI_ISL_535968 | Canada/QC-LSPQ-L00232003/2020 | 2020-03-22 | North America/Canada/Quebec/ | H√¥pital du Centre-de-la-Mauricie | Laboratoire de sant√© publique du Qu√©bec | Sandrine Moreira et al |

| | | | | | | |
|---|---|---|---|---|---|---|
| EPI_ISL_535969 | Canada/QC-LSPQ-L00232004/2020 | 2020-03-21 | North America/Canada/Quebec/ | H√¥pital Sainte-Croix | Laboratoire de sant√© publique du Qu√©bec | Sandrine Moreira et al |
| EPI_ISL_535970 | Canada/QC-LSPQ-L00232042/2020 | 2020-03-19 | North America/Canada/Quebec/ | CSSS de la Minganie | Laboratoire de sant√© publique du Qu√©bec | Sandrine Moreira et al |
| EPI_ISL_535971 | Canada/QC-LSPQ-L00232064/2020 | 2020-03-22 | North America/Canada/Quebec/ | H√¥pital du Centre-de-la-Mauricie | Laboratoire de sant√© publique du Qu√©bec | Sandrine Moreira et al |
| EPI_ISL_535972 | Canada/QC-LSPQ-L00232070/2020 | 2020-03-22 | North America/Canada/Quebec/ | H√¥pital Charles-LeMoyne | Laboratoire de sant√© publique du Qu√©bec | Sandrine Moreira et al |
| EPI_ISL_535973 | Canada/QC-LSPQ-L00232073/2020 | 2020-03-22 | North America/Canada/Quebec/ | H√¥pital Charles-LeMoyne | Laboratoire de sant√© publique du Qu√©bec | Sandrine Moreira et al |
| EPI_ISL_535974 | Canada/QC-LSPQ-L00232080/2020 | 2020-03-22 | North America/Canada/Quebec/ | H√¥pital du Centre-de-la-Mauricie | Laboratoire de sant√© publique du Qu√©bec | Sandrine Moreira et al |
| EPI_ISL_535975 | Canada/QC-LSPQ-L00232097/2020 | 2020-03-22 | North America/Canada/Quebec/ | H√¥pital du Centre-de-la-Mauricie | Laboratoire de sant√© publique du Qu√©bec | Sandrine Moreira et al |
| EPI_ISL_535976 | Canada/QC-LSPQ-L00232098/2020 | 2020-03-22 | North America/Canada/Quebec/ | Centre de SSS de Trois-Rivi√®res | Laboratoire de sant√© publique du Qu√©bec | Sandrine Moreira et al |
| EPI_ISL_535977 | Canada/QC-LSPQ-L00232101/2020 | 2020-03-22 | North America/Canada/Quebec/ | Centre de SSS de Trois-Rivi√®res | Laboratoire de sant√© publique du Qu√©bec | Sandrine Moreira et al |
| EPI_ISL_535978 | Canada/QC-LSPQ-L00232107/2020 | 2020-03-22 | North America/Canada/Quebec/ | H√¥pital du Centre-de-la-Mauricie | Laboratoire de sant√© publique du Qu√©bec | Sandrine Moreira et al |
| EPI_ISL_535979 | Canada/QC-LSPQ-L00232113/2020 | 2020-03-22 | North America/Canada/Quebec/ | Centre de SSS de Trois-Rivi√®res | Laboratoire de sant√© publique du Qu√©bec | Sandrine Moreira et al |
| EPI_ISL_535980 | Canada/QC-LSPQ-L00232148/2020 | 2020-03-22 | North America/Canada/Quebec/ | Centre hospitalier Anna-Laberge | Laboratoire de sant√© publique du Qu√©bec | Sandrine Moreira et al |
| EPI_ISL_535981 | Canada/QC-LSPQ-L00232187/2020 | 2020-03-22 | North America/Canada/Quebec/ | Centre hospitalier Anna-Laberge | Laboratoire de sant√© publique du Qu√©bec | Sandrine Moreira et al |

| | | | | | | |
|---|---|---|---|---|---|---|
| EPI_ISL_535982 | Canada/QC-LSPQ-L00232239/2020 | 2020-03-22 | North America/Canada/Quebec/ | H√¥pital de Saint-Eustache | Laboratoire de sant√© publique du Qu√©bec | Sandrine Moreira et al |
| EPI_ISL_535983 | Canada/QC-LSPQ-L00232271/2020 | 2020-03-22 | North America/Canada/Quebec/ | H√¥pital Charles-LeMoyne | Laboratoire de sant√© publique du Qu√©bec | Sandrine Moreira et al |
| EPI_ISL_535984 | Canada/QC-LSPQ-L00232310/2020 | 2020-03-22 | North America/Canada/Quebec/ | CSSS Haut-Richelieu/Rouville (H√¥pital) | Laboratoire de sant√© publique du Qu√©bec | Sandrine Moreira et al |
| EPI_ISL_535985 | Canada/QC-LSPQ-L00232405/2020 | 2020-03-22 | North America/Canada/Quebec/ | H√¥pital de Rouyn-Noranda | Laboratoire de sant√© publique du Qu√©bec | Sandrine Moreira et al |
| EPI_ISL_535986 | Canada/QC-LSPQ-L00232439/2020 | 2020-03-22 | North America/Canada/Quebec/ | H√¥pital de Rouyn-Noranda | Laboratoire de sant√© publique du Qu√©bec | Sandrine Moreira et al |
| EPI_ISL_535987 | Canada/QC-LSPQ-L00232447/2020 | 2020-03-22 | North America/Canada/Quebec/ | H√¥pital de Rouyn-Noranda | Laboratoire de sant√© publique du Qu√©bec | Sandrine Moreira et al |
| EPI_ISL_535988 | Canada/QC-LSPQ-L00232511/2020 | 2020-03-22 | North America/Canada/Quebec/ | H√¥pital du Surov√Æt | Laboratoire de sant√© publique du Qu√©bec | Sandrine Moreira et al |
| EPI_ISL_535989 | Canada/QC-LSPQ-L00232601/2020 | 2020-03-21 | North America/Canada/Quebec/ | H√¥pital de Hull | Laboratoire de sant√© publique du Qu√©bec | Sandrine Moreira et al |
| EPI_ISL_535990 | Canada/QC-LSPQ-L00232603/2020 | 2020-03-22 | North America/Canada/Quebec/ | H√¥tel-Dieu de Sorel | Laboratoire de sant√© publique du Qu√©bec | Sandrine Moreira et al |
| EPI_ISL_535991 | Canada/QC-LSPQ-L00232609/2020 | 2020-03-21 | North America/Canada/Quebec/ | H√¥pital et Centre d'H√©bergement de Sept-Iles | Laboratoire de sant√© publique du Qu√©bec | Sandrine Moreira et al |
| EPI_ISL_535992 | Canada/QC-LSPQ-L00232614/2020 | 2020-03-22 | North America/Canada/Quebec/ | H√¥tel-Dieu de Sorel | Laboratoire de sant√© publique du Qu√©bec | Sandrine Moreira et al |
| EPI_ISL_535993 | Canada/QC-LSPQ-L00232688/2020 | 2020-03-22 | North America/Canada/Quebec/ | H√¥pital de Hull | Laboratoire de sant√© publique du Qu√©bec | Sandrine Moreira et al |
| EPI_ISL_535994 | Canada/QC-LSPQ-L00232723/2020 | 2020-03-22 | North America/Canada/Quebec/ | H√¥pital Laurentien | Laboratoire de sant√© publique du Qu√©bec | Sandrine Moreira et al |

| | | | | | | |
|---|---|---|---|---|---|---|
| EPI_ISL_535995 | Canada/QC-LSPQ-L00232726/2020 | 2020-03-22 | North America/Canada/Quebec/ | H√¥pital de Hull | Laboratoire de sant√© publique du Qu√©bec | Sandrine Moreira et al |
| EPI_ISL_535996 | Canada/QC-LSPQ-L00232810/2020 | 2020-03-23 | North America/Canada/Quebec/ | H√¥pital du Centre-de-la-Mauricie | Laboratoire de sant√© publique du Qu√©bec | Sandrine Moreira et al |
| EPI_ISL_535997 | Canada/QC-LSPQ-L00232813/2020 | 2020-03-22 | North America/Canada/Quebec/ | H√¥pital du Centre-de-la-Mauricie | Laboratoire de sant√© publique du Qu√©bec | Sandrine Moreira et al |
| EPI_ISL_535998 | Canada/QC-LSPQ-L00232822/2020 | 2020-03-22 | North America/Canada/Quebec/ | H√¥pital du Centre-de-la-Mauricie | Laboratoire de sant√© publique du Qu√©bec | Sandrine Moreira et al |
| EPI_ISL_535999 | Canada/QC-LSPQ-L00232866/2020 | 2020-03-22 | North America/Canada/Quebec/ | H√¥pital du Centre-de-la-Mauricie | Laboratoire de sant√© publique du Qu√©bec | Sandrine Moreira et al |
| EPI_ISL_536000 | Canada/QC-LSPQ-L00232917/2020 | 2020-03-23 | North America/Canada/Quebec/ | Centre hospitalier Anna-Laberge | Laboratoire de sant√© publique du Qu√©bec | Sandrine Moreira et al |
| EPI_ISL_536001 | Canada/QC-LSPQ-L00232925/2020 | 2020-03-23 | North America/Canada/Quebec/ | H√¥pital du Centre-de-la-Mauricie | Laboratoire de sant√© publique du Qu√©bec | Sandrine Moreira et al |
| EPI_ISL_536002 | Canada/QC-LSPQ-L00232941/2020 | 2020-03-22 | North America/Canada/Quebec/ | Centre Hospitalier R√©gional de Lanaudi√®re | Laboratoire de sant√© publique du Qu√©bec | Sandrine Moreira et al |
| EPI_ISL_536003 | Canada/QC-LSPQ-L00232949/2020 | 2020-03-22 | North America/Canada/Quebec/ | H√¥pital Sainte-Croix | Laboratoire de sant√© publique du Qu√©bec | Sandrine Moreira et al |
| EPI_ISL_536004 | Canada/QC-LSPQ-L00232952/2020 | 2020-03-23 | North America/Canada/Quebec/ | Centre hospitalier Anna-Laberge | Laboratoire de sant√© publique du Qu√©bec | Sandrine Moreira et al |
| EPI_ISL_536005 | Canada/QC-LSPQ-L00232955/2020 | 2020-03-22 | North America/Canada/Quebec/ | Centre Hospitalier R√©gional de Lanaudi√®re | Laboratoire de sant√© publique du Qu√©bec | Sandrine Moreira et al |
| EPI_ISL_536006 | Canada/QC-LSPQ-L00233003/2020 | 2020-03-22 | North America/Canada/Quebec/ | Centre Hospitalier R√©gional de Lanaudi√®re | Laboratoire de sant√© publique du Qu√©bec | Sandrine Moreira et al |
| EPI_ISL_536007 | Canada/QC-LSPQ-L00233038/2020 | 2020-03-22 | North America/Canada/Quebec/ | Centre de SSS de Trois-Rivi√®res | Laboratoire de sant√© publique du Qu√©bec | Sandrine Moreira et al |

| | | | | | | |
|---|---|---|---|---|---|---|
| EPI_ISL_536008 | Canada/QC-LSPQ-L00233063/2020 | 2020-03-22 | North America/Canada/Quebec/ | H√¥pital de Saint-Eustache | Laboratoire de sant√© publique du Qu√©bec | Sandrine Moreira et al |
| EPI_ISL_536009 | Canada/QC-LSPQ-L00233115/2020 | 2020-03-22 | North America/Canada/Quebec/ | Centre Hospitalier R√©gional de Lanaudi√®re | Laboratoire de sant√© publique du Qu√©bec | Sandrine Moreira et al |
| EPI_ISL_536010 | Canada/QC-LSPQ-L00233169/2020 | 2020-03-22 | North America/Canada/Quebec/ | H√¥pital Sainte-Croix | Laboratoire de sant√© publique du Qu√©bec | Sandrine Moreira et al |
| EPI_ISL_536011 | Canada/QC-LSPQ-L00233171/2020 | 2020-03-22 | North America/Canada/Quebec/ | H√¥pital Sainte-Croix | Laboratoire de sant√© publique du Qu√©bec | Sandrine Moreira et al |
| EPI_ISL_536012 | Canada/QC-LSPQ-L00233222/2020 | 2020-03-22 | North America/Canada/Quebec/ | Centre Hospitalier R√©gional de Lanaudi√®re | Laboratoire de sant√© publique du Qu√©bec | Sandrine Moreira et al |
| EPI_ISL_536013 | Canada/QC-LSPQ-L00233253/2020 | 2020-03-23 | North America/Canada/Quebec/ | H√¥pital Honor√©-Mercier | Laboratoire de sant√© publique du Qu√©bec | Sandrine Moreira et al |
| EPI_ISL_536014 | Canada/QC-LSPQ-L00233281/2020 | 2020-03-22 | North America/Canada/Quebec/ | Centre Hospitalier R√©gional de Lanaudi√®re | Laboratoire de sant√© publique du Qu√©bec | Sandrine Moreira et al |
| EPI_ISL_536015 | Canada/QC-LSPQ-L00233316/2020 | 2020-03-22 | North America/Canada/Quebec/ | Centre Hospitalier R√©gional de Lanaudi√®re | Laboratoire de sant√© publique du Qu√©bec | Sandrine Moreira et al |
| EPI_ISL_536016 | Canada/QC-LSPQ-L00233323/2020 | 2020-03-22 | North America/Canada/Quebec/ | Centre Hospitalier R√©gional de Lanaudi√®re | Laboratoire de sant√© publique du Qu√©bec | Sandrine Moreira et al |
| EPI_ISL_536017 | Canada/QC-LSPQ-L00233333/2020 | 2020-03-23 | North America/Canada/Quebec/ | H√¥pital Charles-LeMoyne | Laboratoire de sant√© publique du Qu√©bec | Sandrine Moreira et al |
| EPI_ISL_536018 | Canada/QC-LSPQ-L00233340/2020 | 2020-03-23 | North America/Canada/Quebec/ | H√¥pital Charles-LeMoyne | Laboratoire de sant√© publique du Qu√©bec | Sandrine Moreira et al |
| EPI_ISL_536019 | Canada/QC-LSPQ-L00233346/2020 | 2020-03-21 | North America/Canada/Quebec/ | H√¥pital de Chicoutimi | Laboratoire de sant√© publique du Qu√©bec | Sandrine Moreira et al |
| EPI_ISL_536020 | Canada/QC-LSPQ-L00233357/2020 | 2020-03-23 | North America/Canada/Quebec/ | H√¥pital Charles-LeMoyne | Laboratoire de sant√© publique du Qu√©bec | Sandrine Moreira et al |

| | | | | | | |
|---|---|---|---|---|---|---|
| EPI_ISL_536021 | Canada/QC-LSPQ-L00233380/2020 | 2020-03-23 | North America/Canada/Quebec/ | H√¥pital Pierre-Boucher | Laboratoire de sant√© publique du Qu√©bec | Sandrine Moreira et al |
| EPI_ISL_536022 | Canada/QC-LSPQ-L00233398/2020 | 2020-03-23 | North America/Canada/Quebec/ | H√¥pital r√©gional de Saint-J√©r√¥me | Laboratoire de sant√© publique du Qu√©bec | Sandrine Moreira et al |
| EPI_ISL_536023 | Canada/QC-LSPQ-L00233422/2020 | 2020-03-23 | North America/Canada/Quebec/ | Centre hospitalier Anna-Laberge | Laboratoire de sant√© publique du Qu√©bec | Sandrine Moreira et al |
| EPI_ISL_536024 | Canada/QC-LSPQ-L00233493/2020 | 2020-03-23 | North America/Canada/Quebec/ | H√¥pital Charles-LeMoyne | Laboratoire de sant√© publique du Qu√©bec | Sandrine Moreira et al |
| EPI_ISL_536025 | Canada/QC-LSPQ-L00233496/2020 | 2020-03-23 | North America/Canada/Quebec/ | H√¥pital Charles-LeMoyne | Laboratoire de sant√© publique du Qu√©bec | Sandrine Moreira et al |
| EPI_ISL_536026 | Canada/QC-LSPQ-L00233527/2020 | 2020-03-23 | North America/Canada/Quebec/ | H√¥pital du Suro√Æt | Laboratoire de sant√© publique du Qu√©bec | Sandrine Moreira et al |
| EPI_ISL_536027 | Canada/QC-LSPQ-L00233561/2020 | 2020-03-23 | North America/Canada/Quebec/ | CSSS Haut-Richelieu/Rouville (H√¥pital) | Laboratoire de sant√© publique du Qu√©bec | Sandrine Moreira et al |
| EPI_ISL_536028 | Canada/QC-LSPQ-L00233567/2020 | 2020-03-23 | North America/Canada/Quebec/ | CSSS Haut-Richelieu/Rouville (H√¥pital) | Laboratoire de sant√© publique du Qu√©bec | Sandrine Moreira et al |
| EPI_ISL_536029 | Canada/QC-LSPQ-L00233585/2020 | 2020-03-23 | North America/Canada/Quebec/ | H√¥pital Honor√©-Mercier | Laboratoire de sant√© publique du Qu√©bec | Sandrine Moreira et al |
| EPI_ISL_536030 | Canada/QC-LSPQ-L00233633/2020 | 2020-03-23 | North America/Canada/Quebec/ | H√¥pital du Centre-de-la-Mauricie | Laboratoire de sant√© publique du Qu√©bec | Sandrine Moreira et al |
| EPI_ISL_536031 | Canada/QC-LSPQ-L00233661/2020 | 2020-03-23 | North America/Canada/Quebec/ | Centre de SSS de Trois-Rivi√®res | Laboratoire de sant√© publique du Qu√©bec | Sandrine Moreira et al |
| EPI_ISL_536032 | Canada/QC-LSPQ-L00233681/2020 | 2020-03-23 | North America/Canada/Quebec/ | H√¥pital du Centre-de-la-Mauricie | Laboratoire de sant√© publique du Qu√©bec | Sandrine Moreira et al |
| EPI_ISL_536033 | Canada/QC-LSPQ-L00233683/2020 | 2020-03-23 | North America/Canada/Quebec/ | H√¥pital du Centre-de-la-Mauricie | Laboratoire de sant√© publique du Qu√©bec | Sandrine Moreira et al |

| | | | | | | |
|---|---|---|---|---|---|---|
| EPI_ISL_536034 | Canada/QC-LSPQ-L00233695/2020 | 2020-03-23 | North America/Canada/Quebec/ | Centre de SSS de Trois-Riviv®res | Laboratoire de santv© publique du Quv©bec | Sandrine Moreira et al |
| EPI_ISL_536035 | Canada/QC-LSPQ-L00233702/2020 | 2020-03-23 | North America/Canada/Quebec/ | Centre de SSS de Trois-Riviv®res | Laboratoire de santv© publique du Quv©bec | Sandrine Moreira et al |
| EPI_ISL_536036 | Canada/QC-LSPQ-L00233714/2020 | 2020-03-23 | North America/Canada/Quebec/ | Centre de SSS de Trois-Riviv®res | Laboratoire de santv© publique du Quv©bec | Sandrine Moreira et al |
| EPI_ISL_536037 | Canada/QC-LSPQ-L00233721/2020 | 2020-03-23 | North America/Canada/Quebec/ | Hv¥pital du Centre-de-la-Mauricie | Laboratoire de santv© publique du Quv©bec | Sandrine Moreira et al |
| EPI_ISL_536038 | Canada/QC-LSPQ-L00233734/2020 | 2020-03-23 | North America/Canada/Quebec/ | Centre de SSS de Trois-Riviv®res | Laboratoire de santv© publique du Quv©bec | Sandrine Moreira et al |
| EPI_ISL_536039 | Canada/QC-LSPQ-L00233793/2020 | 2020-03-23 | North America/Canada/Quebec/ | Hv¥pital Pierre-Boucher | Laboratoire de santv© publique du Quv©bec | Sandrine Moreira et al |
| EPI_ISL_536040 | Canada/QC-LSPQ-L00233812/2020 | 2020-03-23 | North America/Canada/Quebec/ | CSSS Haut-Richelieu/Rouville (Hv¥pital) | Laboratoire de santv© publique du Quv©bec | Sandrine Moreira et al |
| EPI_ISL_536041 | Canada/QC-LSPQ-L00233835/2020 | 2020-03-23 | North America/Canada/Quebec/ | Hv¥pital Pierre-Boucher | Laboratoire de santv© publique du Quv©bec | Sandrine Moreira et al |
| EPI_ISL_536042 | Canada/QC-LSPQ-L00233849/2020 | 2020-03-23 | North America/Canada/Quebec/ | CSSS Haut-Richelieu/Rouville (Hv¥pital) | Laboratoire de santv© publique du Quv©bec | Sandrine Moreira et al |
| EPI_ISL_536043 | Canada/QC-LSPQ-L00233853/2020 | 2020-03-23 | North America/Canada/Quebec/ | Centre hospitalier Anna-Laberge | Laboratoire de santv© publique du Quv©bec | Sandrine Moreira et al |
| EPI_ISL_536044 | Canada/QC-LSPQ-L00233867/2020 | 2020-03-23 | North America/Canada/Quebec/ | CSSS Haut-Richelieu/Rouville (Hv¥pital) | Laboratoire de santv© publique du Quv©bec | Sandrine Moreira et al |
| EPI_ISL_536045 | Canada/QC-LSPQ-L00233870/2020 | 2020-03-23 | North America/Canada/Quebec/ | Centre hospitalier Anna-Laberge | Laboratoire de santv© publique du Quv©bec | Sandrine Moreira et al |
| EPI_ISL_536046 | Canada/QC-LSPQ-L00233970/2020 | 2020-03-22 | North America/Canada/Quebec/ | Hv¥pital d'Alma | Laboratoire de santv© publique du Quv©bec | Sandrine Moreira et al |

| | | | | | | |
|---|---|---|---|---|---|---|
| EPI_ISL_536047 | Canada/QC-LSPQ-L00234248/2020 | 2020-03-23 | North America/Canada/Quebec/ | Centre Hospitalier R√©gional de Lanaudi√®re | Laboratoire de sant√© publique du Qu√©bec | Sandrine Moreira et al |
| EPI_ISL_536048 | Canada/QC-LSPQ-L00234260/2020 | 2020-03-23 | North America/Canada/Quebec/ | Centre Hospitalier R√©gional de Lanaudi√®re | Laboratoire de sant√© publique du Qu√©bec | Sandrine Moreira et al |
| EPI_ISL_536049 | Canada/QC-LSPQ-L00234287/2020 | 2020-03-23 | North America/Canada/Quebec/ | H√¥pital de Saint-Eustache | Laboratoire de sant√© publique du Qu√©bec | Sandrine Moreira et al |
| EPI_ISL_536050 | Canada/QC-LSPQ-L00234302/2020 | 2020-03-23 | North America/Canada/Quebec/ | Centre Hospitalier R√©gional de Lanaudi√®re | Laboratoire de sant√© publique du Qu√©bec | Sandrine Moreira et al |
| EPI_ISL_536051 | Canada/QC-LSPQ-L00234333/2020 | 2020-03-23 | North America/Canada/Quebec/ | H√¥pital de Hull | Laboratoire de sant√© publique du Qu√©bec | Sandrine Moreira et al |
| EPI_ISL_536052 | Canada/QC-LSPQ-L00234347/2020 | 2020-03-23 | North America/Canada/Quebec/ | Centre Hospitalier R√©gional de Lanaudi√®re | Laboratoire de sant√© publique du Qu√©bec | Sandrine Moreira et al |
| EPI_ISL_536053 | Canada/QC-LSPQ-L00234397/2020 | 2020-03-23 | North America/Canada/Quebec/ | Centre Hospitalier R√©gional de Lanaudi√®re | Laboratoire de sant√© publique du Qu√©bec | Sandrine Moreira et al |
| EPI_ISL_536054 | Canada/QC-LSPQ-L00234442/2020 | 2020-03-23 | North America/Canada/Quebec/ | Centre Hospitalier R√©gional de Lanaudi√®re | Laboratoire de sant√© publique du Qu√©bec | Sandrine Moreira et al |
| EPI_ISL_536055 | Canada/QC-LSPQ-L00234460/2020 | 2020-03-23 | North America/Canada/Quebec/ | H√¥pital Honor√©-Mercier | Laboratoire de sant√© publique du Qu√©bec | Sandrine Moreira et al |
| EPI_ISL_536056 | Canada/QC-LSPQ-L00234533/2020 | 2020-03-23 | North America/Canada/Quebec/ | H√¥pital Sainte-Croix | Laboratoire de sant√© publique du Qu√©bec | Sandrine Moreira et al |
| EPI_ISL_536057 | Canada/QC-LSPQ-L00234537/2020 | 2020-03-23 | North America/Canada/Quebec/ | H√¥pital Sainte-Croix | Laboratoire de sant√© publique du Qu√©bec | Sandrine Moreira et al |
| EPI_ISL_536058 | Canada/QC-LSPQ-L00234540/2020 | 2020-03-23 | North America/Canada/Quebec/ | H√¥pital de Saint-Eustache | Laboratoire de sant√© publique du Qu√©bec | Sandrine Moreira et al |
| EPI_ISL_536059 | Canada/QC-LSPQ-L00234574/2020 | 2020-03-23 | North America/Canada/Quebec/ | Centre Hospitalier R√©gional de Lanaudi√®re | Laboratoire de sant√© publique du Qu√©bec | Sandrine Moreira et al |

| | | | | | | |
|---|---|---|---|---|---|---|
| EPI_ISL_536060 | Canada/QC-LSPQ-L00234577/2020 | 2020-03-23 | North America/Canada/Quebec/ | Centre Hospitalier R√©gional de Lanaudi√®re | Laboratoire de sant√© publique du Qu√©bec | Sandrine Moreira et al |
| EPI_ISL_536061 | Canada/QC-LSPQ-L00234602/2020 | 2020-03-23 | North America/Canada/Quebec/ | H√¥pital Pierre-Le Gardeur | Laboratoire de sant√© publique du Qu√©bec | Sandrine Moreira et al |
| EPI_ISL_536062 | Canada/QC-LSPQ-L00234611/2020 | 2020-03-23 | North America/Canada/Quebec/ | H√¥pital Pierre-Le Gardeur | Laboratoire de sant√© publique du Qu√©bec | Sandrine Moreira et al |
| EPI_ISL_536063 | Canada/QC-LSPQ-L00234621/2020 | 2020-03-23 | North America/Canada/Quebec/ | Centre Hospitalier R√©gional de Lanaudi√®re | Laboratoire de sant√© publique du Qu√©bec | Sandrine Moreira et al |
| EPI_ISL_536064 | Canada/QC-LSPQ-L00234627/2020 | 2020-03-23 | North America/Canada/Quebec/ | H√¥pital Pierre-Le Gardeur | Laboratoire de sant√© publique du Qu√©bec | Sandrine Moreira et al |
| EPI_ISL_536065 | Canada/QC-LSPQ-L00234639/2020 | 2020-03-23 | North America/Canada/Quebec/ | H√¥pital Pierre-Le Gardeur | Laboratoire de sant√© publique du Qu√©bec | Sandrine Moreira et al |
| EPI_ISL_536066 | Canada/QC-LSPQ-L00234652/2020 | 2020-03-23 | North America/Canada/Quebec/ | H√¥pital Pierre-Le Gardeur | Laboratoire de sant√© publique du Qu√©bec | Sandrine Moreira et al |
| EPI_ISL_536067 | Canada/QC-LSPQ-L00234655/2020 | 2020-03-23 | North America/Canada/Quebec/ | H√¥pital Pierre-Le Gardeur | Laboratoire de sant√© publique du Qu√©bec | Sandrine Moreira et al |
| EPI_ISL_536068 | Canada/QC-LSPQ-L00234676/2020 | 2020-03-23 | North America/Canada/Quebec/ | H√¥pital Pierre-Le Gardeur | Laboratoire de sant√© publique du Qu√©bec | Sandrine Moreira et al |
| EPI_ISL_536069 | Canada/QC-LSPQ-L00234776/2020 | 2020-03-23 | North America/Canada/Quebec/ | H√¥pital de Saint-Eustache | Laboratoire de sant√© publique du Qu√©bec | Sandrine Moreira et al |
| EPI_ISL_536070 | Canada/QC-LSPQ-L00234790/2020 | 2020-03-23 | North America/Canada/Quebec/ | H√¥pital Pierre-Le Gardeur | Laboratoire de sant√© publique du Qu√©bec | Sandrine Moreira et al |
| EPI_ISL_536071 | Canada/QC-LSPQ-L00234851/2020 | 2020-03-23 | North America/Canada/Quebec/ | H√¥pital de Saint-Eustache | Laboratoire de sant√© publique du Qu√©bec | Sandrine Moreira et al |
| EPI_ISL_536072 | Canada/QC-LSPQ-L00234853/2020 | 2020-03-24 | North America/Canada/Quebec/ | H√¥pital de Saint-Eustache | Laboratoire de sant√© publique du Qu√©bec | Sandrine Moreira et al |

| | | | | | | |
|---|---|---|---|---|---|---|
| EPI_ISL_536073 | Canada/QC-LSPQ-L00234865/2020 | 2020-03-22 | North America/Canada/Quebec/ | Centre Hospitalier R√©gional de Lanaudi√®re | Laboratoire de sant√© publique du Qu√©bec | Sandrine Moreira et al |
| EPI_ISL_536074 | Canada/QC-LSPQ-L00234871/2020 | 2020-03-23 | North America/Canada/Quebec/ | Centre Hospitalier R√©gional de Lanaudi√®re | Laboratoire de sant√© publique du Qu√©bec | Sandrine Moreira et al |
| EPI_ISL_536075 | Canada/QC-LSPQ-L00234872/2020 | 2020-03-24 | North America/Canada/Quebec/ | H√¥pital Charles-LeMoyne | Laboratoire de sant√© publique du Qu√©bec | Sandrine Moreira et al |
| EPI_ISL_536076 | Canada/QC-LSPQ-L00234877/2020 | 2020-03-23 | North America/Canada/Quebec/ | Centre Hospitalier R√©gional de Lanaudi√®re | Laboratoire de sant√© publique du Qu√©bec | Sandrine Moreira et al |
| EPI_ISL_536077 | Canada/QC-LSPQ-L00234878/2020 | 2020-03-24 | North America/Canada/Quebec/ | H√¥pital Charles-LeMoyne | Laboratoire de sant√© publique du Qu√©bec | Sandrine Moreira et al |
| EPI_ISL_536078 | Canada/QC-LSPQ-L00234883/2020 | 2020-03-24 | North America/Canada/Quebec/ | H√¥pital Charles-LeMoyne | Laboratoire de sant√© publique du Qu√©bec | Sandrine Moreira et al |
| EPI_ISL_536079 | Canada/QC-LSPQ-L00234887/2020 | 2020-03-23 | North America/Canada/Quebec/ | Centre Hospitalier R√©gional de Lanaudi√®re | Laboratoire de sant√© publique du Qu√©bec | Sandrine Moreira et al |
| EPI_ISL_536080 | Canada/QC-LSPQ-L00234905/2020 | 2020-03-24 | North America/Canada/Quebec/ | Centre hospitalier Anna-Laberge | Laboratoire de sant√© publique du Qu√©bec | Sandrine Moreira et al |
| EPI_ISL_536081 | Canada/QC-LSPQ-L00234922/2020 | 2020-03-24 | North America/Canada/Quebec/ | H√¥pital Charles-LeMoyne | Laboratoire de sant√© publique du Qu√©bec | Sandrine Moreira et al |
| EPI_ISL_536082 | Canada/QC-LSPQ-L00234990/2020 | 2020-03-24 | North America/Canada/Quebec/ | H√¥pital Charles-LeMoyne | Laboratoire de sant√© publique du Qu√©bec | Sandrine Moreira et al |
| EPI_ISL_536083 | Canada/QC-LSPQ-L00235020/2020 | 2020-03-24 | North America/Canada/Quebec/ | H√¥pital Pierre-Le Gardeur | Laboratoire de sant√© publique du Qu√©bec | Sandrine Moreira et al |
| EPI_ISL_536084 | Canada/QC-LSPQ-L00235027/2020 | 2020-03-24 | North America/Canada/Quebec/ | H√¥pital Pierre-Le Gardeur | Laboratoire de sant√© publique du Qu√©bec | Sandrine Moreira et al |
| EPI_ISL_536085 | Canada/QC-LSPQ-L00235085/2020 | 2020-03-24 | North America/Canada/Quebec/ | H√¥pital Pierre-Le Gardeur | Laboratoire de sant√© publique du Qu√©bec | Sandrine Moreira et al |

| | | | | | | |
|---|---|---|---|---|---|---|
| EPI_ISL_536086 | Canada/QC-LSPQ-L00235107/2020 | 2020-03-23 | North America/Canada/Quebec/ | H√¥pital Pierre-Le Gardeur | Laboratoire de sant√© publique du Qu√©bec | Sandrine Moreira et al |
| EPI_ISL_536087 | Canada/QC-LSPQ-L00235132/2020 | 2020-03-24 | North America/Canada/Quebec/ | H√¥pital Charles-LeMoyne | Laboratoire de sant√© publique du Qu√©bec | Sandrine Moreira et al |
| EPI_ISL_536088 | Canada/QC-LSPQ-L00235136/2020 | 2020-03-24 | North America/Canada/Quebec/ | H√¥pital Charles-LeMoyne | Laboratoire de sant√© publique du Qu√©bec | Sandrine Moreira et al |
| EPI_ISL_536089 | Canada/QC-LSPQ-L00235149/2020 | 2020-03-24 | North America/Canada/Quebec/ | H√¥pital Charles-LeMoyne | Laboratoire de sant√© publique du Qu√©bec | Sandrine Moreira et al |
| EPI_ISL_536090 | Canada/QC-LSPQ-L00235218/2020 | 2020-03-24 | North America/Canada/Quebec/ | H√¥pital Charles-LeMoyne | Laboratoire de sant√© publique du Qu√©bec | Sandrine Moreira et al |
| EPI_ISL_536091 | Canada/QC-LSPQ-L00235225/2020 | 2020-03-24 | North America/Canada/Quebec/ | H√¥pital Charles-LeMoyne | Laboratoire de sant√© publique du Qu√©bec | Sandrine Moreira et al |
| EPI_ISL_536092 | Canada/QC-LSPQ-L00235261/2020 | 2020-03-24 | North America/Canada/Quebec/ | H√¥pital Pierre-Boucher | Laboratoire de sant√© publique du Qu√©bec | Sandrine Moreira et al |
| EPI_ISL_536093 | Canada/QC-LSPQ-L00235289/2020 | 2020-03-24 | North America/Canada/Quebec/ | H√¥pital Charles-LeMoyne | Laboratoire de sant√© publique du Qu√©bec | Sandrine Moreira et al |
| EPI_ISL_536094 | Canada/QC-LSPQ-L00235295/2020 | 2020-03-24 | North America/Canada/Quebec/ | H√¥pital Charles-LeMoyne | Laboratoire de sant√© publique du Qu√©bec | Sandrine Moreira et al |
| EPI_ISL_536095 | Canada/QC-LSPQ-L00235378/2020 | 2020-03-24 | North America/Canada/Quebec/ | H√¥pital du Surou√Æt | Laboratoire de sant√© publique du Qu√©bec | Sandrine Moreira et al |
| EPI_ISL_536096 | Canada/QC-LSPQ-L00235380/2020 | 2020-03-24 | North America/Canada/Quebec/ | H√¥pital Pierre-Boucher | Laboratoire de sant√© publique du Qu√©bec | Sandrine Moreira et al |
| EPI_ISL_536097 | Canada/QC-LSPQ-L00235424/2020 | 2020-03-24 | North America/Canada/Quebec/ | H√¥pital du Surou√Æt | Laboratoire de sant√© publique du Qu√©bec | Sandrine Moreira et al |
| EPI_ISL_536098 | Canada/QC-LSPQ-L00235430/2020 | 2020-03-24 | North America/Canada/Quebec/ | H√¥pital du Surou√Æt | Laboratoire de sant√© publique du Qu√©bec | Sandrine Moreira et al |

| | | | | | | |
|---|---|---|---|---|---|---|
| EPI_ISL_536099 | Canada/QC-LSPQ-L00235552/2020 | 2020-03-24 | North America/Canada/Quebec/ | H√¥pital Pierre-Boucher | Laboratoire de sant√© publique du Qu√©bec | Sandrine Moreira et al |
| EPI_ISL_536100 | Canada/QC-LSPQ-L00235558/2020 | 2020-03-24 | North America/Canada/Quebec/ | H√¥pital Charles-LeMoyne | Laboratoire de sant√© publique du Qu√©bec | Sandrine Moreira et al |
| EPI_ISL_536101 | Canada/QC-LSPQ-L00235561/2020 | 2020-03-24 | North America/Canada/Quebec/ | H√¥pital, CLSC et Centre d'H√©bergement de Roberval | Laboratoire de sant√© publique du Qu√©bec | Sandrine Moreira et al |
| EPI_ISL_536102 | Canada/QC-LSPQ-L00235607/2020 | 2020-03-24 | North America/Canada/Quebec/ | H√¥pital du Surov√Æt | Laboratoire de sant√© publique du Qu√©bec | Sandrine Moreira et al |
| EPI_ISL_536103 | Canada/QC-LSPQ-L00235638/2020 | 2020-03-24 | North America/Canada/Quebec/ | H√¥pital Pierre-Boucher | Laboratoire de sant√© publique du Qu√©bec | Sandrine Moreira et al |
| EPI_ISL_536104 | Canada/QC-LSPQ-L00235639/2020 | 2020-03-24 | North America/Canada/Quebec/ | Centre hospitalier Anna-Laberge | Laboratoire de sant√© publique du Qu√©bec | Sandrine Moreira et al |
| EPI_ISL_536105 | Canada/QC-LSPQ-L00235671/2020 | 2020-03-24 | North America/Canada/Quebec/ | H√¥pital du Surov√Æt | Laboratoire de sant√© publique du Qu√©bec | Sandrine Moreira et al |
| EPI_ISL_536106 | Canada/QC-LSPQ-L00235753/2020 | 2020-03-24 | North America/Canada/Quebec/ | H√¥pital r√©gional de Rimouski | Laboratoire de sant√© publique du Qu√©bec | Sandrine Moreira et al |
| EPI_ISL_536107 | Canada/QC-LSPQ-L00235772/2020 | 2020-03-24 | North America/Canada/Quebec/ | H√¥pital de Chicoutimi | Laboratoire de sant√© publique du Qu√©bec | Sandrine Moreira et al |
| EPI_ISL_536108 | Canada/QC-LSPQ-L00235800/2020 | 2020-03-23 | North America/Canada/Quebec/ | H√¥pital Le Royer | Laboratoire de sant√© publique du Qu√©bec | Sandrine Moreira et al |
| EPI_ISL_536109 | Canada/QC-LSPQ-L00235802/2020 | 2020-03-24 | North America/Canada/Quebec/ | CSSS de Port-Cartier | Laboratoire de sant√© publique du Qu√©bec | Sandrine Moreira et al |
| EPI_ISL_536110 | Canada/QC-LSPQ-L00235815/2020 | 2020-03-24 | North America/Canada/Quebec/ | CSSS Haut-Richelieu/Rouville (H√¥pital) | Laboratoire de sant√© publique du Qu√©bec | Sandrine Moreira et al |
| EPI_ISL_536111 | Canada/QC-LSPQ-L00235849/2020 | 2020-03-23 | North America/Canada/Quebec/ | H√¥pital de Maria | Laboratoire de sant√© publique du Qu√©bec | Sandrine Moreira et al |

| | | | | | | |
|---|---|---|---|---|---|---|
| EPI_ISL_536112 | Canada/QC-LSPQ-L00235855/2020 | 2020-03-23 | North America/Canada/Quebec/ | H√¥pital de Maria | Laboratoire de sant√© publique du Qu√©bec | Sandrine Moreira et al |
| EPI_ISL_536113 | Canada/QC-LSPQ-L00235866/2020 | 2020-03-22 | North America/Canada/Quebec/ | H√¥pital et Centre d'H√©bergement de Sept-Iles | Laboratoire de sant√© publique du Qu√©bec | Sandrine Moreira et al |
| EPI_ISL_536114 | Canada/QC-LSPQ-L00235907/2020 | 2020-03-22 | North America/Canada/Quebec/ | H√¥pital et Centre d'H√©bergement de Sept-Iles | Laboratoire de sant√© publique du Qu√©bec | Sandrine Moreira et al |
| EPI_ISL_536115 | Canada/QC-LSPQ-L00235965/2020 | 2020-03-23 | North America/Canada/Quebec/ | Centre hospitalier r√©gional du Grand Portage | Laboratoire de sant√© publique du Qu√©bec | Sandrine Moreira et al |
| EPI_ISL_536116 | Canada/QC-LSPQ-L00236074/2020 | 2020-03-24 | North America/Canada/Quebec/ | H√¥tel-Dieu de Sorel | Laboratoire de sant√© publique du Qu√©bec | Sandrine Moreira et al |
| EPI_ISL_536117 | Canada/QC-LSPQ-L00236091/2020 | 2020-03-24 | North America/Canada/Quebec/ | H√¥pital de Hull | Laboratoire de sant√© publique du Qu√©bec | Sandrine Moreira et al |
| EPI_ISL_536118 | Canada/QC-LSPQ-L00236132/2020 | 2020-03-24 | North America/Canada/Quebec/ | H√¥pital de Hull | Laboratoire de sant√© publique du Qu√©bec | Sandrine Moreira et al |
| EPI_ISL_536119 | Canada/QC-LSPQ-L00236207/2020 | 2020-03-24 | North America/Canada/Quebec/ | Centre Hospitalier R√©gional de Lanaudi√®re | Laboratoire de sant√© publique du Qu√©bec | Sandrine Moreira et al |
| EPI_ISL_536120 | Canada/QC-LSPQ-L00236212/2020 | 2020-03-24 | North America/Canada/Quebec/ | Centre Hospitalier R√©gional de Lanaudi√®re | Laboratoire de sant√© publique du Qu√©bec | Sandrine Moreira et al |
| EPI_ISL_536121 | Canada/QC-LSPQ-L00236215/2020 | 2020-03-24 | North America/Canada/Quebec/ | H√¥tel-Dieu de Sorel | Laboratoire de sant√© publique du Qu√©bec | Sandrine Moreira et al |
| EPI_ISL_536122 | Canada/QC-LSPQ-L00236258/2020 | 2020-03-24 | North America/Canada/Quebec/ | H√¥pital de Hull | Laboratoire de sant√© publique du Qu√©bec | Sandrine Moreira et al |
| EPI_ISL_536123 | Canada/QC-LSPQ-L00236332/2020 | 2020-03-24 | North America/Canada/Quebec/ | Centre Hospitalier R√©gional de Lanaudi√®re | Laboratoire de sant√© publique du Qu√©bec | Sandrine Moreira et al |
| EPI_ISL_536124 | Canada/QC-LSPQ-L00236367/2020 | 2020-03-24 | North America/Canada/Quebec/ | H√¥pital de Hull | Laboratoire de sant√© publique du Qu√©bec | Sandrine Moreira et al |

| | | | | | | |
|---|---|---|---|---|---|---|
| EPI_ISL_536125 | Canada/QC-LSPQ-L00236385/2020 | 2020-03-24 | North America/Canada/Quebec/ | H√¥pital Pierre-Le Gardeur | Laboratoire de sant√© publique du Qu√©bec | Sandrine Moreira et al |
| EPI_ISL_536126 | Canada/QC-LSPQ-L00236394/2020 | 2020-03-24 | North America/Canada/Quebec/ | Centre Hospitalier R√©gional de Lanaudi√®re | Laboratoire de sant√© publique du Qu√©bec | Sandrine Moreira et al |
| EPI_ISL_536127 | Canada/QC-LSPQ-L00236396/2020 | 2020-03-24 | North America/Canada/Quebec/ | H√¥pital de Hull | Laboratoire de sant√© publique du Qu√©bec | Sandrine Moreira et al |
| EPI_ISL_536128 | Canada/QC-LSPQ-L00236403/2020 | 2020-03-24 | North America/Canada/Quebec/ | H√¥pital Pierre-Le Gardeur | Laboratoire de sant√© publique du Qu√©bec | Sandrine Moreira et al |
| EPI_ISL_536129 | Canada/QC-LSPQ-L00236426/2020 | 2020-03-24 | North America/Canada/Quebec/ | Centre Hospitalier R√©gional de Lanaudi√®re | Laboratoire de sant√© publique du Qu√©bec | Sandrine Moreira et al |
| EPI_ISL_536130 | Canada/QC-LSPQ-L00236430/2020 | 2020-03-24 | North America/Canada/Quebec/ | Centre Hospitalier R√©gional de Lanaudi√®re | Laboratoire de sant√© publique du Qu√©bec | Sandrine Moreira et al |
| EPI_ISL_536131 | Canada/QC-LSPQ-L00236439/2020 | 2020-03-24 | North America/Canada/Quebec/ | H√¥pital Pierre-Le Gardeur | Laboratoire de sant√© publique du Qu√©bec | Sandrine Moreira et al |
| EPI_ISL_536132 | Canada/QC-LSPQ-L00236526/2020 | 2020-03-24 | North America/Canada/Quebec/ | H√¥pital de Hull | Laboratoire de sant√© publique du Qu√©bec | Sandrine Moreira et al |
| EPI_ISL_536133 | Canada/QC-LSPQ-L00236528/2020 | 2020-03-24 | North America/Canada/Quebec/ | Centre Hospitalier R√©gional de Lanaudi√®re | Laboratoire de sant√© publique du Qu√©bec | Sandrine Moreira et al |
| EPI_ISL_536134 | Canada/QC-LSPQ-L00236536/2020 | 2020-03-24 | North America/Canada/Quebec/ | Centre Hospitalier R√©gional de Lanaudi√®re | Laboratoire de sant√© publique du Qu√©bec | Sandrine Moreira et al |
| EPI_ISL_536135 | Canada/QC-LSPQ-L00236544/2020 | 2020-03-24 | North America/Canada/Quebec/ | H√¥pital de Hull | Laboratoire de sant√© publique du Qu√©bec | Sandrine Moreira et al |
| EPI_ISL_536136 | Canada/QC-LSPQ-L00236551/2020 | 2020-03-24 | North America/Canada/Quebec/ | H√¥pital de Hull | Laboratoire de sant√© publique du Qu√©bec | Sandrine Moreira et al |
| EPI_ISL_536137 | Canada/QC-LSPQ-L00236558/2020 | 2020-03-24 | North America/Canada/Quebec/ | H√¥pital Pierre-Le Gardeur | Laboratoire de sant√© publique du Qu√©bec | Sandrine Moreira et al |

| | | | | | | |
|---|---|---|---|---|---|---|
| EPI_ISL_536138 | Canada/QC-LSPQ-L00236573/2020 | 2020-03-24 | North America/Canada/Quebec/ | Centre Hospitalier R√©gional de Lanaudi√®re | Laboratoire de sant√© publique du Qu√©bec | Sandrine Moreira et al |
| EPI_ISL_536139 | Canada/QC-LSPQ-L00236582/2020 | 2020-03-24 | North America/Canada/Quebec/ | H√¥pital Pierre-Le Gardeur | Laboratoire de sant√© publique du Qu√©bec | Sandrine Moreira et al |
| EPI_ISL_536140 | Canada/QC-LSPQ-L00236587/2020 | 2020-03-24 | North America/Canada/Quebec/ | H√¥pital Pierre-Le Gardeur | Laboratoire de sant√© publique du Qu√©bec | Sandrine Moreira et al |
| EPI_ISL_536141 | Canada/QC-LSPQ-L00236595/2020 | 2020-03-25 | North America/Canada/Quebec/ | Centre hospitalier Anna-Laberge | Laboratoire de sant√© publique du Qu√©bec | Sandrine Moreira et al |
| EPI_ISL_536142 | Canada/QC-LSPQ-L00236598/2020 | 2020-03-25 | North America/Canada/Quebec/ | Centre hospitalier Anna-Laberge | Laboratoire de sant√© publique du Qu√©bec | Sandrine Moreira et al |
| EPI_ISL_536143 | Canada/QC-LSPQ-L00236601/2020 | 2020-03-24 | North America/Canada/Quebec/ | H√¥pital Pierre-Le Gardeur | Laboratoire de sant√© publique du Qu√©bec | Sandrine Moreira et al |
| EPI_ISL_536144 | Canada/QC-LSPQ-L00236620/2020 | 2020-03-24 | North America/Canada/Quebec/ | H√¥pital Pierre-Le Gardeur | Laboratoire de sant√© publique du Qu√©bec | Sandrine Moreira et al |
| EPI_ISL_536145 | Canada/QC-LSPQ-L00236653/2020 | 2020-03-25 | North America/Canada/Quebec/ | Centre hospitalier Anna-Laberge | Laboratoire de sant√© publique du Qu√©bec | Sandrine Moreira et al |
| EPI_ISL_536146 | Canada/QC-LSPQ-L00236657/2020 | 2020-03-25 | North America/Canada/Quebec/ | H√¥pital Charles-LeMoyne | Laboratoire de sant√© publique du Qu√©bec | Sandrine Moreira et al |
| EPI_ISL_536147 | Canada/QC-LSPQ-L00236779/2020 | 2020-03-25 | North America/Canada/Quebec/ | H√¥pital Charles-LeMoyne | Laboratoire de sant√© publique du Qu√©bec | Sandrine Moreira et al |
| EPI_ISL_536148 | Canada/QC-LSPQ-L00236781/2020 | 2020-03-25 | North America/Canada/Quebec/ | H√¥pital Charles-LeMoyne | Laboratoire de sant√© publique du Qu√©bec | Sandrine Moreira et al |
| EPI_ISL_536149 | Canada/QC-LSPQ-L00236787/2020 | 2020-03-25 | North America/Canada/Quebec/ | H√¥pital Charles-LeMoyne | Laboratoire de sant√© publique du Qu√©bec | Sandrine Moreira et al |
| EPI_ISL_536150 | Canada/QC-LSPQ-L00236805/2020 | 2020-03-25 | North America/Canada/Quebec/ | CSSS Haut-Richelieu/Rouville (H√¥pital) | Laboratoire de sant√© publique du Qu√©bec | Sandrine Moreira et al |

| | | | | | | |
|---|---|---|---|---|---|---|
| EPI_ISL_536151 | Canada/QC-LSPQ-L00236905/2020 | 2020-03-25 | North America/Canada/Quebec/ | Centre Hospitalier R√©gional de Lanaudi√®re | Laboratoire de sant√© publique du Qu√©bec | Sandrine Moreira et al |
| EPI_ISL_536152 | Canada/QC-LSPQ-L00236922/2020 | 2020-03-25 | North America/Canada/Quebec/ | H√¥pital Pierre-Le Gardeur | Laboratoire de sant√© publique du Qu√©bec | Sandrine Moreira et al |
| EPI_ISL_536153 | Canada/QC-LSPQ-L00236940/2020 | 2020-03-25 | North America/Canada/Quebec/ | Centre Hospitalier R√©gional de Lanaudi√®re | Laboratoire de sant√© publique du Qu√©bec | Sandrine Moreira et al |
| EPI_ISL_536154 | Canada/QC-LSPQ-L00236943/2020 | 2020-03-25 | North America/Canada/Quebec/ | Centre Hospitalier R√©gional de Lanaudi√®re | Laboratoire de sant√© publique du Qu√©bec | Sandrine Moreira et al |
| EPI_ISL_536155 | Canada/QC-LSPQ-L00236965/2020 | 2020-03-25 | North America/Canada/Quebec/ | Centre hospitalier Anna-Laberge | Laboratoire de sant√© publique du Qu√©bec | Sandrine Moreira et al |
| EPI_ISL_536156 | Canada/QC-LSPQ-L00236973/2020 | 2020-03-25 | North America/Canada/Quebec/ | Centre hospitalier Anna-Laberge | Laboratoire de sant√© publique du Qu√©bec | Sandrine Moreira et al |
| EPI_ISL_536157 | Canada/QC-LSPQ-L00237011/2020 | 2020-03-25 | North America/Canada/Quebec/ | H√¥pital Pierre-Le Gardeur | Laboratoire de sant√© publique du Qu√©bec | Sandrine Moreira et al |
| EPI_ISL_536158 | Canada/QC-LSPQ-L00237078/2020 | 2020-03-25 | North America/Canada/Quebec/ | CSSS Haut-Richelieu/Rouville (H√¥pital) | Laboratoire de sant√© publique du Qu√©bec | Sandrine Moreira et al |
| EPI_ISL_536159 | Canada/QC-LSPQ-L00237080/2020 | 2020-03-25 | North America/Canada/Quebec/ | Centre hospitalier Anna-Laberge | Laboratoire de sant√© publique du Qu√©bec | Sandrine Moreira et al |
| EPI_ISL_536160 | Canada/QC-LSPQ-L00237091/2020 | 2020-03-25 | North America/Canada/Quebec/ | CSSS Haut-Richelieu/Rouville (H√¥pital) | Laboratoire de sant√© publique du Qu√©bec | Sandrine Moreira et al |
| EPI_ISL_536161 | Canada/QC-LSPQ-L00237107/2020 | 2020-03-25 | North America/Canada/Quebec/ | CSSS Haut-Richelieu/Rouville (H√¥pital) | Laboratoire de sant√© publique du Qu√©bec | Sandrine Moreira et al |
| EPI_ISL_536162 | Canada/QC-LSPQ-L00237108/2020 | 2020-03-25 | North America/Canada/Quebec/ | Centre hospitalier Anna-Laberge | Laboratoire de sant√© publique du Qu√©bec | Sandrine Moreira et al |
| EPI_ISL_536163 | Canada/QC-LSPQ-L00237109/2020 | 2020-03-25 | North America/Canada/Quebec/ | H√¥pital du Surov√Æt | Laboratoire de sant√© publique du Qu√©bec | Sandrine Moreira et al |

| | | | | | | |
|---|---|---|---|---|---|---|
| EPI_ISL_536164 | Canada/QC-LSPQ-L00237113/2020 | 2020-03-25 | North America/Canada/Quebec/ | CSSS Haut-Richelieu/Rouville (H√¥pital) | Laboratoire de sant√© publique du Qu√©bec | Sandrine Moreira et al |
| EPI_ISL_536165 | Canada/QC-LSPQ-L00237128/2020 | 2020-03-25 | North America/Canada/Quebec/ | H√¥pital du Surov√Æt | Laboratoire de sant√© publique du Qu√©bec | Sandrine Moreira et al |
| EPI_ISL_536166 | Canada/QC-LSPQ-L00237135/2020 | 2020-03-25 | North America/Canada/Quebec/ | CSSS Haut-Richelieu/Rouville (H√¥pital) | Laboratoire de sant√© publique du Qu√©bec | Sandrine Moreira et al |
| EPI_ISL_536167 | Canada/QC-LSPQ-L00237180/2020 | 2020-03-25 | North America/Canada/Quebec/ | H√¥pital du Surov√Æt | Laboratoire de sant√© publique du Qu√©bec | Sandrine Moreira et al |
| EPI_ISL_536168 | Canada/QC-LSPQ-L00237217/2020 | 2020-03-25 | North America/Canada/Quebec/ | H√¥pital de Hull | Laboratoire de sant√© publique du Qu√©bec | Sandrine Moreira et al |
| EPI_ISL_536169 | Canada/QC-LSPQ-L00237242/2020 | 2020-03-25 | North America/Canada/Quebec/ | H√¥pital de Hull | Laboratoire de sant√© publique du Qu√©bec | Sandrine Moreira et al |
| EPI_ISL_536170 | Canada/QC-LSPQ-L00237267/2020 | 2020-03-25 | North America/Canada/Quebec/ | H√¥pital du Surov√Æt | Laboratoire de sant√© publique du Qu√©bec | Sandrine Moreira et al |
| EPI_ISL_536171 | Canada/QC-LSPQ-L00237287/2020 | 2020-03-25 | North America/Canada/Quebec/ | H√¥pital de Papineau | Laboratoire de sant√© publique du Qu√©bec | Sandrine Moreira et al |
| EPI_ISL_536172 | Canada/QC-LSPQ-L00237309/2020 | 2020-03-25 | North America/Canada/Quebec/ | CSSS Haut-Richelieu/Rouville (H√¥pital) | Laboratoire de sant√© publique du Qu√©bec | Sandrine Moreira et al |
| EPI_ISL_536173 | Canada/QC-LSPQ-L00237312/2020 | 2020-03-25 | North America/Canada/Quebec/ | H√¥pital Pierre-Boucher | Laboratoire de sant√© publique du Qu√©bec | Sandrine Moreira et al |
| EPI_ISL_536174 | Canada/QC-LSPQ-L00237340/2020 | 2020-03-25 | North America/Canada/Quebec/ | H√¥pital Pierre-Boucher | Laboratoire de sant√© publique du Qu√©bec | Sandrine Moreira et al |
| EPI_ISL_536175 | Canada/QC-LSPQ-L00237352/2020 | 2020-03-25 | North America/Canada/Quebec/ | H√¥pital Pierre-Boucher | Laboratoire de sant√© publique du Qu√©bec | Sandrine Moreira et al |
| EPI_ISL_536176 | Canada/QC-LSPQ-L00237365/2020 | 2020-03-25 | North America/Canada/Quebec/ | H√¥pital Pierre-Boucher | Laboratoire de sant√© publique du Qu√©bec | Sandrine Moreira et al |

| | | | | | | |
|---|---|---|---|---|---|---|
| EPI_ISL_536177 | Canada/QC-LSPQ-L00237371/2020 | 2020-03-25 | North America/Canada/Quebec/ | H√¥pital Honor√©-Mercier | Laboratoire de sant√© publique du Qu√©bec | Sandrine Moreira et al |
| EPI_ISL_536178 | Canada/QC-LSPQ-L00237372/2020 | 2020-03-25 | North America/Canada/Quebec/ | H√¥pital Pierre-Boucher | Laboratoire de sant√© publique du Qu√©bec | Sandrine Moreira et al |
| EPI_ISL_536179 | Canada/QC-LSPQ-L00237374/2020 | 2020-03-25 | North America/Canada/Quebec/ | H√¥pital Pierre-Boucher | Laboratoire de sant√© publique du Qu√©bec | Sandrine Moreira et al |
| EPI_ISL_536180 | Canada/QC-LSPQ-L00237419/2020 | 2020-03-25 | North America/Canada/Quebec/ | H√¥pital Honor√©-Mercier | Laboratoire de sant√© publique du Qu√©bec | Sandrine Moreira et al |
| EPI_ISL_536181 | Canada/QC-LSPQ-L00237465/2020 | 2020-03-25 | North America/Canada/Quebec/ | CSSS Haut-Richelieu/Rouville (H√¥pital) | Laboratoire de sant√© publique du Qu√©bec | Sandrine Moreira et al |
| EPI_ISL_536182 | Canada/QC-LSPQ-L00237490/2020 | 2020-03-25 | North America/Canada/Quebec/ | Centre hospitalier Anna-Laberge | Laboratoire de sant√© publique du Qu√©bec | Sandrine Moreira et al |
| EPI_ISL_536183 | Canada/QC-LSPQ-L00237503/2020 | 2020-03-25 | North America/Canada/Quebec/ | Centre hospitalier Anna-Laberge | Laboratoire de sant√© publique du Qu√©bec | Sandrine Moreira et al |
| EPI_ISL_536184 | Canada/QC-LSPQ-L00237507/2020 | 2020-03-25 | North America/Canada/Quebec/ | Centre hospitalier Anna-Laberge | Laboratoire de sant√© publique du Qu√©bec | Sandrine Moreira et al |
| EPI_ISL_536185 | Canada/QC-LSPQ-L00237533/2020 | 2020-03-25 | North America/Canada/Quebec/ | H√¥pital Charles-LeMoyne | Laboratoire de sant√© publique du Qu√©bec | Sandrine Moreira et al |
| EPI_ISL_536186 | Canada/QC-LSPQ-L00237577/2020 | 2020-03-25 | North America/Canada/Quebec/ | H√¥pital Charles-LeMoyne | Laboratoire de sant√© publique du Qu√©bec | Sandrine Moreira et al |
| EPI_ISL_536187 | Canada/QC-LSPQ-L00237589/2020 | 2020-03-25 | North America/Canada/Quebec/ | H√¥pital Charles-LeMoyne | Laboratoire de sant√© publique du Qu√©bec | Sandrine Moreira et al |
| EPI_ISL_536188 | Canada/QC-LSPQ-L00237598/2020 | 2020-03-25 | North America/Canada/Quebec/ | H√¥pital Pierre-Boucher | Laboratoire de sant√© publique du Qu√©bec | Sandrine Moreira et al |
| EPI_ISL_536189 | Canada/QC-LSPQ-L00237611/2020 | 2020-03-25 | North America/Canada/Quebec/ | H√¥pital Pierre-Boucher | Laboratoire de sant√© publique du Qu√©bec | Sandrine Moreira et al |

| | | | | | | |
|---|---|---|---|---|---|---|
| EPI_ISL_536190 | Canada/QC-LSPQ-L00237624/2020 | 2020-03-25 | North America/Canada/Quebec/ | H√¥pital du Surov√Æt | Laboratoire de sant√© publique du Qu√©bec | Sandrine Moreira et al |
| EPI_ISL_536191 | Canada/QC-LSPQ-L00237636/2020 | 2020-03-25 | North America/Canada/Quebec/ | H√¥pital Charles-LeMoyne | Laboratoire de sant√© publique du Qu√©bec | Sandrine Moreira et al |
| EPI_ISL_536192 | Canada/QC-LSPQ-L00237727/2020 | 2020-03-25 | North America/Canada/Quebec/ | H√¥pital du Surov√Æt | Laboratoire de sant√© publique du Qu√©bec | Sandrine Moreira et al |
| EPI_ISL_536193 | Canada/QC-LSPQ-L00237734/2020 | 2020-03-25 | North America/Canada/Quebec/ | H√¥pital du Surov√Æt | Laboratoire de sant√© publique du Qu√©bec | Sandrine Moreira et al |
| EPI_ISL_536194 | Canada/QC-LSPQ-L00237747/2020 | 2020-03-25 | North America/Canada/Quebec/ | H√¥pital Charles-LeMoyne | Laboratoire de sant√© publique du Qu√©bec | Sandrine Moreira et al |
| EPI_ISL_536195 | Canada/QC-LSPQ-L00237841/2020 | 2020-03-25 | North America/Canada/Quebec/ | H√¥pital Pierre-Boucher | Laboratoire de sant√© publique du Qu√©bec | Sandrine Moreira et al |
| EPI_ISL_536196 | Canada/QC-LSPQ-L00237878/2020 | 2020-03-25 | North America/Canada/Quebec/ | H√¥pital Pierre-Boucher | Laboratoire de sant√© publique du Qu√©bec | Sandrine Moreira et al |
| EPI_ISL_536197 | Canada/QC-LSPQ-L00237883/2020 | 2020-03-25 | North America/Canada/Quebec/ | CSSS Haut-Richelieu/Rouville (H√¥pital) | Laboratoire de sant√© publique du Qu√©bec | Sandrine Moreira et al |
| EPI_ISL_536198 | Canada/QC-LSPQ-L00237886/2020 | 2020-03-25 | North America/Canada/Quebec/ | H√¥pital du Surov√Æt | Laboratoire de sant√© publique du Qu√©bec | Sandrine Moreira et al |
| EPI_ISL_536199 | Canada/QC-LSPQ-L00237888/2020 | 2020-03-25 | North America/Canada/Quebec/ | Centre hospitalier r√©gional du Grand Portage | Laboratoire de sant√© publique du Qu√©bec | Sandrine Moreira et al |
| EPI_ISL_536200 | Canada/QC-LSPQ-L00237930/2020 | 2020-03-25 | North America/Canada/Quebec/ | H√¥pital du Surov√Æt | Laboratoire de sant√© publique du Qu√©bec | Sandrine Moreira et al |
| EPI_ISL_536201 | Canada/QC-LSPQ-L00237942/2020 | 2020-03-25 | North America/Canada/Quebec/ | H√¥pital du Surov√Æt | Laboratoire de sant√© publique du Qu√©bec | Sandrine Moreira et al |
| EPI_ISL_536202 | Canada/QC-LSPQ-L00237972/2020 | 2020-03-25 | North America/Canada/Quebec/ | H√¥pital du Surov√Æt | Laboratoire de sant√© publique du Qu√©bec | Sandrine Moreira et al |

| | | | | | | |
|---|---|---|---|---|---|---|
| EPI_ISL_536203 | Canada/QC-LSPQ-L00238204/2020 | 2020-03-26 | North America/Canada/Quebec/ | H√¥pital Charles-LeMoyne | Laboratoire de sant√© publique du Qu√©bec | Sandrine Moreira et al |
| EPI_ISL_536204 | Canada/QC-LSPQ-L00238216/2020 | 2020-03-25 | North America/Canada/Quebec/ | Centre hospitalier r√©gional du Grand Portage | Laboratoire de sant√© publique du Qu√©bec | Sandrine Moreira et al |
| EPI_ISL_536205 | Canada/QC-LSPQ-L00238226/2020 | 2020-03-24 | North America/Canada/Quebec/ | H√¥pital r√©gional de Rimouski | Laboratoire de sant√© publique du Qu√©bec | Sandrine Moreira et al |
| EPI_ISL_536206 | Canada/QC-LSPQ-L00238246/2020 | 2020-03-24 | North America/Canada/Quebec/ | CSSS de la Minganie | Laboratoire de sant√© publique du Qu√©bec | Sandrine Moreira et al |
| EPI_ISL_536207 | Canada/QC-LSPQ-L00238342/2020 | 2020-03-25 | North America/Canada/Quebec/ | H√¥pital de Hull | Laboratoire de sant√© publique du Qu√©bec | Sandrine Moreira et al |
| EPI_ISL_536208 | Canada/QC-LSPQ-L00238445/2020 | 2020-03-25 | North America/Canada/Quebec/ | H√¥pital de Hull | Laboratoire de sant√© publique du Qu√©bec | Sandrine Moreira et al |
| EPI_ISL_536209 | Canada/QC-LSPQ-L00238518/2020 | 2020-03-25 | North America/Canada/Quebec/ | H√¥pital de Hull | Laboratoire de sant√© publique du Qu√©bec | Sandrine Moreira et al |
| EPI_ISL_536210 | Canada/QC-LSPQ-L00238539/2020 | 2020-03-25 | North America/Canada/Quebec/ | H√¥pital de Hull | Laboratoire de sant√© publique du Qu√©bec | Sandrine Moreira et al |
| EPI_ISL_536211 | Canada/QC-LSPQ-L00238550/2020 | 2020-03-26 | North America/Canada/Quebec/ | Centre hospitalier Anna-Laberge | Laboratoire de sant√© publique du Qu√©bec | Sandrine Moreira et al |
| EPI_ISL_536212 | Canada/QC-LSPQ-L00238674/2020 | 2020-03-26 | North America/Canada/Quebec/ | Centre hospitalier Anna-Laberge | Laboratoire de sant√© publique du Qu√©bec | Sandrine Moreira et al |
| EPI_ISL_536213 | Canada/QC-LSPQ-L00238675/2020 | 2020-03-25 | North America/Canada/Quebec/ | CSSS Haut-Richelieu/Rouville (H√¥pital) | Laboratoire de sant√© publique du Qu√©bec | Sandrine Moreira et al |
| EPI_ISL_536214 | Canada/QC-LSPQ-L00238697/2020 | 2020-03-26 | North America/Canada/Quebec/ | H√¥pital Charles-LeMoyne | Laboratoire de sant√© publique du Qu√©bec | Sandrine Moreira et al |
| EPI_ISL_536215 | Canada/QC-LSPQ-L00238722/2020 | 2020-03-26 | North America/Canada/Quebec/ | CSSS Haut-Richelieu/Rouville (H√¥pital) | Laboratoire de sant√© publique du Qu√©bec | Sandrine Moreira et al |

| | | | | | | |
|---|---|---|---|---|---|---|
| EPI_ISL_536216 | Canada/QC-LSPQ-L00238725/2020 | 2020-03-26 | North America/Canada/Quebec/ | CSSS Haut-Richelieu/Rouville (H√¥pital) | Laboratoire de sant√© publique du Qu√©bec | Sandrine Moreira et al |
| EPI_ISL_536217 | Canada/QC-LSPQ-L00238742/2020 | 2020-03-26 | North America/Canada/Quebec/ | CSSS Haut-Richelieu/Rouville (H√¥pital) | Laboratoire de sant√© publique du Qu√©bec | Sandrine Moreira et al |
| EPI_ISL_536218 | Canada/QC-LSPQ-L00238793/2020 | 2020-03-26 | North America/Canada/Quebec/ | Centre hospitalier Anna-Laberge | Laboratoire de sant√© publique du Qu√©bec | Sandrine Moreira et al |
| EPI_ISL_536219 | Canada/QC-LSPQ-L00238797/2020 | 2020-03-26 | North America/Canada/Quebec/ | Centre hospitalier Anna-Laberge | Laboratoire de sant√© publique du Qu√©bec | Sandrine Moreira et al |
| EPI_ISL_536220 | Canada/QC-LSPQ-L00238803/2020 | 2020-03-26 | North America/Canada/Quebec/ | Centre hospitalier Anna-Laberge | Laboratoire de sant√© publique du Qu√©bec | Sandrine Moreira et al |
| EPI_ISL_536221 | Canada/QC-LSPQ-L00238804/2020 | 2020-03-26 | North America/Canada/Quebec/ | Centre hospitalier Anna-Laberge | Laboratoire de sant√© publique du Qu√©bec | Sandrine Moreira et al |
| EPI_ISL_536222 | Canada/QC-LSPQ-L00238812/2020 | 2020-03-24 | North America/Canada/Quebec/ | Conseil Cri de la SSS de la Baie-James | Laboratoire de sant√© publique du Qu√©bec | Sandrine Moreira et al |
| EPI_ISL_536223 | Canada/QC-LSPQ-L00238819/2020 | 2020-03-26 | North America/Canada/Quebec/ | H√¥pital Charles-LeMoyne | Laboratoire de sant√© publique du Qu√©bec | Sandrine Moreira et al |
| EPI_ISL_536224 | Canada/QC-LSPQ-L00238862/2020 | 2020-03-26 | North America/Canada/Quebec/ | Centre hospitalier Anna-Laberge | Laboratoire de sant√© publique du Qu√©bec | Sandrine Moreira et al |
| EPI_ISL_536225 | Canada/QC-LSPQ-L00238888/2020 | 2020-03-26 | North America/Canada/Quebec/ | H√¥pital Charles-LeMoyne | Laboratoire de sant√© publique du Qu√©bec | Sandrine Moreira et al |
| EPI_ISL_536226 | Canada/QC-LSPQ-L00238906/2020 | 2020-03-26 | North America/Canada/Quebec/ | H√¥pital du Suro√Æt | Laboratoire de sant√© publique du Qu√©bec | Sandrine Moreira et al |
| EPI_ISL_536227 | Canada/QC-LSPQ-L00238920/2020 | 2020-03-26 | North America/Canada/Quebec/ | H√¥pital du Suro√Æt | Laboratoire de sant√© publique du Qu√©bec | Sandrine Moreira et al |
| EPI_ISL_536228 | Canada/QC-LSPQ-L00238922/2020 | 2020-03-26 | North America/Canada/Quebec/ | H√¥pital du Suro√Æt | Laboratoire de sant√© publique du Qu√©bec | Sandrine Moreira et al |

| | | | | | | |
|---|---|---|---|---|---|---|
| EPI_ISL_536229 | Canada/QC-LSPQ-L00238978/2020 | 2020-03-26 | North America/Canada/Quebec/ | H√¥pital Pierre-Boucher | Laboratoire de sant√© publique du Qu√©bec | Sandrine Moreira et al |
| EPI_ISL_536230 | Canada/QC-LSPQ-L00239009/2020 | 2020-03-22 | North America/Canada/Quebec/ | H√¥pital de Hull | Laboratoire de sant√© publique du Qu√©bec | Sandrine Moreira et al |
| EPI_ISL_536231 | Canada/QC-LSPQ-L00239061/2020 | 2020-03-26 | North America/Canada/Quebec/ | H√¥pital Pierre-Boucher | Laboratoire de sant√© publique du Qu√©bec | Sandrine Moreira et al |
| EPI_ISL_536232 | Canada/QC-LSPQ-L00239077/2020 | 2020-03-26 | North America/Canada/Quebec/ | Centre hospitalier Anna-Laberge | Laboratoire de sant√© publique du Qu√©bec | Sandrine Moreira et al |
| EPI_ISL_536233 | Canada/QC-LSPQ-L00239086/2020 | 2020-03-26 | North America/Canada/Quebec/ | H√¥pital Charles-LeMoyne | Laboratoire de sant√© publique du Qu√©bec | Sandrine Moreira et al |
| EPI_ISL_536234 | Canada/QC-LSPQ-L00239134/2020 | 2020-03-26 | North America/Canada/Quebec/ | H√¥pital Pierre-Boucher | Laboratoire de sant√© publique du Qu√©bec | Sandrine Moreira et al |
| EPI_ISL_536235 | Canada/QC-LSPQ-L00239182/2020 | 2020-03-26 | North America/Canada/Quebec/ | CSSS Haut-Richelieu/Rouville (H√¥pital) | Laboratoire de sant√© publique du Qu√©bec | Sandrine Moreira et al |
| EPI_ISL_536236 | Canada/QC-LSPQ-L00239192/2020 | 2020-03-26 | North America/Canada/Quebec/ | H√¥pital Pierre-Boucher | Laboratoire de sant√© publique du Qu√©bec | Sandrine Moreira et al |
| EPI_ISL_536237 | Canada/QC-LSPQ-L00239194/2020 | 2020-03-26 | North America/Canada/Quebec/ | CSSS Haut-Richelieu/Rouville (H√¥pital) | Laboratoire de sant√© publique du Qu√©bec | Sandrine Moreira et al |
| EPI_ISL_536238 | Canada/QC-LSPQ-L00239209/2020 | 2020-03-21 | North America/Canada/Quebec/ | H√¥pital de Hull | Laboratoire de sant√© publique du Qu√©bec | Sandrine Moreira et al |
| EPI_ISL_536239 | Canada/QC-LSPQ-L00239235/2020 | 2020-03-21 | North America/Canada/Quebec/ | H√¥pital de Hull | Laboratoire de sant√© publique du Qu√©bec | Sandrine Moreira et al |
| EPI_ISL_536240 | Canada/QC-LSPQ-L00239249/2020 | 2020-03-26 | North America/Canada/Quebec/ | Centre hospitalier Anna-Laberge | Laboratoire de sant√© publique du Qu√©bec | Sandrine Moreira et al |
| EPI_ISL_536241 | Canada/QC-LSPQ-L00239255/2020 | 2020-03-26 | North America/Canada/Quebec/ | Centre hospitalier Anna-Laberge | Laboratoire de sant√© publique du Qu√©bec | Sandrine Moreira et al |

| | | | | | | |
|---|---|---|---|---|---|---|
| EPI_ISL_536242 | Canada/QC-LSPQ-L00239272/2020 | 2020-03-26 | North America/Canada/Quebec/ | H√¥pital de Hull | Laboratoire de sant√© publique du Qu√©bec | Sandrine Moreira et al |
| EPI_ISL_536243 | Canada/QC-LSPQ-L00239296/2020 | 2020-03-26 | North America/Canada/Quebec/ | CSSS Haut-Richelieu/Rouville (H√¥pital) | Laboratoire de sant√© publique du Qu√©bec | Sandrine Moreira et al |
| EPI_ISL_536244 | Canada/QC-LSPQ-L00239298/2020 | 2020-03-26 | North America/Canada/Quebec/ | H√¥pital de Hull | Laboratoire de sant√© publique du Qu√©bec | Sandrine Moreira et al |
| EPI_ISL_536245 | Canada/QC-LSPQ-L00239332/2020 | 2020-03-26 | North America/Canada/Quebec/ | H√¥pital Pierre-Boucher | Laboratoire de sant√© publique du Qu√©bec | Sandrine Moreira et al |
| EPI_ISL_536246 | Canada/QC-LSPQ-L00239391/2020 | 2020-03-24 | North America/Canada/Quebec/ | H√¥pital du Centre-de-la-Mauricie | Laboratoire de sant√© publique du Qu√©bec | Sandrine Moreira et al |
| EPI_ISL_536247 | Canada/QC-LSPQ-L00239396/2020 | 2020-03-26 | North America/Canada/Quebec/ | H√¥pital Charles-LeMoyne | Laboratoire de sant√© publique du Qu√©bec | Sandrine Moreira et al |
| EPI_ISL_536248 | Canada/QC-LSPQ-L00239397/2020 | 2020-03-24 | North America/Canada/Quebec/ | H√¥pital du Centre-de-la-Mauricie | Laboratoire de sant√© publique du Qu√©bec | Sandrine Moreira et al |
| EPI_ISL_536249 | Canada/QC-LSPQ-L00239402/2020 | 2020-03-24 | North America/Canada/Quebec/ | H√¥pital du Centre-de-la-Mauricie | Laboratoire de sant√© publique du Qu√©bec | Sandrine Moreira et al |
| EPI_ISL_536250 | Canada/QC-LSPQ-L00239407/2020 | 2020-03-24 | North America/Canada/Quebec/ | H√¥pital du Centre-de-la-Mauricie | Laboratoire de sant√© publique du Qu√©bec | Sandrine Moreira et al |
| EPI_ISL_536251 | Canada/QC-LSPQ-L00239412/2020 | 2020-03-24 | North America/Canada/Quebec/ | H√¥pital du Centre-de-la-Mauricie | Laboratoire de sant√© publique du Qu√©bec | Sandrine Moreira et al |
| EPI_ISL_536252 | Canada/QC-LSPQ-L00239413/2020 | 2020-03-24 | North America/Canada/Quebec/ | H√¥pital Sainte-Croix | Laboratoire de sant√© publique du Qu√©bec | Sandrine Moreira et al |
| EPI_ISL_536253 | Canada/QC-LSPQ-L00239440/2020 | 2020-03-26 | North America/Canada/Quebec/ | H√¥pital Pierre-Boucher | Laboratoire de sant√© publique du Qu√©bec | Sandrine Moreira et al |
| EPI_ISL_536254 | Canada/QC-LSPQ-L00239501/2020 | 2020-03-26 | North America/Canada/Quebec/ | H√¥pital du Surো√Æt | Laboratoire de sant√© publique du Qu√©bec | Sandrine Moreira et al |

| | | | | | | |
|---|---|---|---|---|---|---|
| EPI_ISL_536255 | Canada/QC-LSPQ-L00239612/2020 | 2020-03-26 | North America/Canada/Quebec/ | Centre hospitalier Anna-Laberge | Laboratoire de santv© publique du Quv©bec | Sandrine Moreira et al |
| EPI_ISL_536256 | Canada/QC-LSPQ-L00239847/2020 | 2020-03-26 | North America/Canada/Quebec/ | CSSS Haut-Richelieu/Rouville (Hv¥pital) | Laboratoire de santv© publique du Quv©bec | Sandrine Moreira et al |
| EPI_ISL_536257 | Canada/QC-LSPQ-L00239865/2020 | 2020-03-26 | North America/Canada/Quebec/ | Hv¥pital du SurovÆt | Laboratoire de santv© publique du Quv©bec | Sandrine Moreira et al |
| EPI_ISL_536258 | Canada/QC-LSPQ-L00239920/2020 | 2020-03-26 | North America/Canada/Quebec/ | CSSS Haut-Richelieu/Rouville (Hv¥pital) | Laboratoire de santv© publique du Quv©bec | Sandrine Moreira et al |
| EPI_ISL_536259 | Canada/QC-LSPQ-L00239981/2020 | 2020-03-26 | North America/Canada/Quebec/ | Hv¥pital du SurovÆt | Laboratoire de santv© publique du Quv©bec | Sandrine Moreira et al |
| EPI_ISL_536260 | Canada/QC-LSPQ-L00239991/2020 | 2020-03-25 | North America/Canada/Quebec/ | CSSS de la Minganie | Laboratoire de santv© publique du Quv©bec | Sandrine Moreira et al |
| EPI_ISL_536261 | Canada/QC-LSPQ-L00240029/2020 | 2020-03-26 | North America/Canada/Quebec/ | Hv¥pital du SurovÆt | Laboratoire de santv© publique du Quv©bec | Sandrine Moreira et al |
| EPI_ISL_536262 | Canada/QC-LSPQ-L00240070/2020 | 2020-03-25 | North America/Canada/Quebec/ | Hv¥pital de Maria | Laboratoire de santv© publique du Quv©bec | Sandrine Moreira et al |
| EPI_ISL_536263 | Canada/QC-LSPQ-L00240113/2020 | 2020-03-25 | North America/Canada/Quebec/ | Point de Service de Tv©miscaming-et-Kipawa | Laboratoire de santv© publique du Quv©bec | Sandrine Moreira et al |
| EPI_ISL_536264 | Canada/QC-LSPQ-L00240115/2020 | 2020-03-25 | North America/Canada/Quebec/ | Point de Service de Tv©miscaming-et-Kipawa | Laboratoire de santv© publique du Quv©bec | Sandrine Moreira et al |
| EPI_ISL_536265 | Canada/QC-LSPQ-L00240158/2020 | 2020-03-26 | North America/Canada/Quebec/ | Hv¥pital de Chandler | Laboratoire de santv© publique du Quv©bec | Sandrine Moreira et al |
| EPI_ISL_536266 | Canada/QC-LSPQ-L00240197/2020 | 2020-03-26 | North America/Canada/Quebec/ | Hv¥pital de Gatineau | Laboratoire de santv© publique du Quv©bec | Sandrine Moreira et al |
| EPI_ISL_536267 | Canada/QC-LSPQ-L00240217/2020 | 2020-03-26 | North America/Canada/Quebec/ | Hv¥pital de Hull | Laboratoire de santv© publique du Quv©bec | Sandrine Moreira et al |

| | | | | | | |
|---|---|---|---|---|---|---|
| EPI_ISL_536268 | Canada/QC-LSPQ-L00240292/2020 | 2020-03-27 | North America/Canada/Quebec/ | H√¥pital Charles-LeMoyne | Laboratoire de sant√© publique du Qu√©bec | Sandrine Moreira et al |
| EPI_ISL_536269 | Canada/QC-LSPQ-L00240404/2020 | 2020-03-27 | North America/Canada/Quebec/ | H√¥pital Charles-LeMoyne | Laboratoire de sant√© publique du Qu√©bec | Sandrine Moreira et al |
| EPI_ISL_536270 | Canada/QC-LSPQ-L00240414/2020 | 2020-03-27 | North America/Canada/Quebec/ | H√¥pital Charles-LeMoyne | Laboratoire de sant√© publique du Qu√©bec | Sandrine Moreira et al |
| EPI_ISL_536271 | Canada/QC-LSPQ-L00240444/2020 | 2020-03-27 | North America/Canada/Quebec/ | H√¥pital Honor√©-Mercier | Laboratoire de sant√© publique du Qu√©bec | Sandrine Moreira et al |
| EPI_ISL_536272 | Canada/QC-LSPQ-L00240479/2020 | 2020-03-27 | North America/Canada/Quebec/ | CSSS Haut-Richelieu/Rouville (H√¥pital) | Laboratoire de sant√© publique du Qu√©bec | Sandrine Moreira et al |
| EPI_ISL_536273 | Canada/QC-LSPQ-L00240536/2020 | 2020-03-27 | North America/Canada/Quebec/ | H√¥pital du Suro√Æt | Laboratoire de sant√© publique du Qu√©bec | Sandrine Moreira et al |
| EPI_ISL_536274 | Canada/QC-LSPQ-L00240552/2020 | 2020-03-27 | North America/Canada/Quebec/ | H√¥pital du Suro√Æt | Laboratoire de sant√© publique du Qu√©bec | Sandrine Moreira et al |
| EPI_ISL_536275 | Canada/QC-LSPQ-L00240811/2020 | 2020-03-27 | North America/Canada/Quebec/ | H√¥pital de Hull | Laboratoire de sant√© publique du Qu√©bec | Sandrine Moreira et al |
| EPI_ISL_536276 | Canada/QC-LSPQ-L00241242/2020 | 2020-03-27 | North America/Canada/Quebec/ | H√¥pital Pierre-Boucher | Laboratoire de sant√© publique du Qu√©bec | Sandrine Moreira et al |
| EPI_ISL_536277 | Canada/QC-LSPQ-L00241376/2020 | 2020-03-27 | North America/Canada/Quebec/ | H√¥pital Pierre-Boucher | Laboratoire de sant√© publique du Qu√©bec | Sandrine Moreira et al |
| EPI_ISL_536278 | Canada/QC-LSPQ-L00241406/2020 | 2020-03-27 | North America/Canada/Quebec/ | CSSS Haut-Richelieu/Rouville (H√¥pital) | Laboratoire de sant√© publique du Qu√©bec | Sandrine Moreira et al |
| EPI_ISL_536279 | Canada/QC-LSPQ-L00241433/2020 | 2020-03-27 | North America/Canada/Quebec/ | H√¥pital Charles-LeMoyne | Laboratoire de sant√© publique du Qu√©bec | Sandrine Moreira et al |
| EPI_ISL_536280 | Canada/QC-LSPQ-L00241435/2020 | 2020-03-27 | North America/Canada/Quebec/ | H√¥pital Charles-LeMoyne | Laboratoire de sant√© publique du Qu√©bec | Sandrine Moreira et al |

| | | | | | | |
|---|---|---|---|---|---|---|
| EPI_ISL_536281 | Canada/QC-LSPQ-L00241439/2020 | 2020-03-27 | North America/Canada/Quebec/ | H√¥pital Charles-LeMoyne | Laboratoire de sant√© publique du Qu√©bec | Sandrine Moreira et al |
| EPI_ISL_536282 | Canada/QC-LSPQ-L00241444/2020 | 2020-03-27 | North America/Canada/Quebec/ | H√¥pital Charles-LeMoyne | Laboratoire de sant√© publique du Qu√©bec | Sandrine Moreira et al |
| EPI_ISL_536283 | Canada/QC-LSPQ-L00241514/2020 | 2020-03-28 | North America/Canada/Quebec/ | Centre hospitalier Anna-Laberge | Laboratoire de sant√© publique du Qu√©bec | Sandrine Moreira et al |
| EPI_ISL_536284 | Canada/QC-LSPQ-L00241602/2020 | 2020-03-28 | North America/Canada/Quebec/ | H√¥pital Charles-LeMoyne | Laboratoire de sant√© publique du Qu√©bec | Sandrine Moreira et al |
| EPI_ISL_536285 | Canada/QC-LSPQ-L00241610/2020 | 2020-03-28 | North America/Canada/Quebec/ | H√¥pital Charles-LeMoyne | Laboratoire de sant√© publique du Qu√©bec | Sandrine Moreira et al |
| EPI_ISL_536286 | Canada/QC-LSPQ-L00241648/2020 | 2020-03-28 | North America/Canada/Quebec/ | CSSS Haut-Richelieu/Rouville (H√¥pital) | Laboratoire de sant√© publique du Qu√©bec | Sandrine Moreira et al |
| EPI_ISL_536287 | Canada/QC-LSPQ-L00241659/2020 | 2020-03-28 | North America/Canada/Quebec/ | H√¥pital du Surov√Æt | Laboratoire de sant√© publique du Qu√©bec | Sandrine Moreira et al |
| EPI_ISL_536288 | Canada/QC-LSPQ-L00241670/2020 | 2020-03-26 | North America/Canada/Quebec/ | Conseil Cri de la SSS de la Baie-James | Laboratoire de sant√© publique du Qu√©bec | Sandrine Moreira et al |
| EPI_ISL_536289 | Canada/QC-LSPQ-L00241753/2020 | 2020-03-27 | North America/Canada/Quebec/ | H√¥pital de Maria | Laboratoire de sant√© publique du Qu√©bec | Sandrine Moreira et al |
| EPI_ISL_536290 | Canada/QC-LSPQ-L00241782/2020 | 2020-03-28 | North America/Canada/Quebec/ | H√¥pital Charles-LeMoyne | Laboratoire de sant√© publique du Qu√©bec | Sandrine Moreira et al |
| EPI_ISL_536291 | Canada/QC-LSPQ-L00241862/2020 | 2020-03-28 | North America/Canada/Quebec/ | H√¥pital de Hull | Laboratoire de sant√© publique du Qu√©bec | Sandrine Moreira et al |
| EPI_ISL_536292 | Canada/QC-LSPQ-L00241900/2020 | 2020-03-28 | North America/Canada/Quebec/ | H√¥pital Charles-LeMoyne | Laboratoire de sant√© publique du Qu√©bec | Sandrine Moreira et al |
| EPI_ISL_536293 | Canada/QC-LSPQ-L00241901/2020 | 2020-03-28 | North America/Canada/Quebec/ | H√¥pital Charles-LeMoyne | Laboratoire de sant√© publique du Qu√©bec | Sandrine Moreira et al |

| | | | | | | |
|---|---|---|---|---|---|---|
| EPI_ISL_536294 | Canada/QC-LSPQ-L00241907/2020 | 2020-03-28 | North America/Canada/Quebec/ | H√¥pital Charles-LeMoyne | Laboratoire de sant√© publique du Qu√©bec | Sandrine Moreira et al |
| EPI_ISL_536295 | Canada/QC-LSPQ-L00241909/2020 | 2020-03-28 | North America/Canada/Quebec/ | H√¥pital Pierre-Boucher | Laboratoire de sant√© publique du Qu√©bec | Sandrine Moreira et al |
| EPI_ISL_536296 | Canada/QC-LSPQ-L00241924/2020 | 2020-03-28 | North America/Canada/Quebec/ | H√¥pital Charles-LeMoyne | Laboratoire de sant√© publique du Qu√©bec | Sandrine Moreira et al |
| EPI_ISL_536297 | Canada/QC-LSPQ-L00241945/2020 | 2020-03-28 | North America/Canada/Quebec/ | H√¥pital Charles-LeMoyne | Laboratoire de sant√© publique du Qu√©bec | Sandrine Moreira et al |
| EPI_ISL_536298 | Canada/QC-LSPQ-L00241948/2020 | 2020-03-28 | North America/Canada/Quebec/ | H√¥pital Charles-LeMoyne | Laboratoire de sant√© publique du Qu√©bec | Sandrine Moreira et al |
| EPI_ISL_536299 | Canada/QC-LSPQ-L00241959/2020 | 2020-03-28 | North America/Canada/Quebec/ | CSSS Haut-Richelieu/Rouville (H√¥pital) | Laboratoire de sant√© publique du Qu√©bec | Sandrine Moreira et al |
| EPI_ISL_536300 | Canada/QC-LSPQ-L00241965/2020 | 2020-03-28 | North America/Canada/Quebec/ | H√¥pital Pierre-Boucher | Laboratoire de sant√© publique du Qu√©bec | Sandrine Moreira et al |
| EPI_ISL_536301 | Canada/QC-LSPQ-L00241993/2020 | 2020-03-28 | North America/Canada/Quebec/ | CSSS Haut-Richelieu/Rouville (H√¥pital) | Laboratoire de sant√© publique du Qu√©bec | Sandrine Moreira et al |
| EPI_ISL_536302 | Canada/QC-LSPQ-L00242011/2020 | 2020-03-28 | North America/Canada/Quebec/ | CSSS Haut-Richelieu/Rouville (H√¥pital) | Laboratoire de sant√© publique du Qu√©bec | Sandrine Moreira et al |
| EPI_ISL_536303 | Canada/QC-LSPQ-L00242018/2020 | 2020-03-28 | North America/Canada/Quebec/ | H√¥pital Pierre-Boucher | Laboratoire de sant√© publique du Qu√©bec | Sandrine Moreira et al |
| EPI_ISL_536304 | Canada/QC-LSPQ-L00242019/2020 | 2020-03-28 | North America/Canada/Quebec/ | CSSS Haut-Richelieu/Rouville (H√¥pital) | Laboratoire de sant√© publique du Qu√©bec | Sandrine Moreira et al |
| EPI_ISL_536305 | Canada/QC-LSPQ-L00242033/2020 | 2020-03-28 | North America/Canada/Quebec/ | H√¥pital Pierre-Boucher | Laboratoire de sant√© publique du Qu√©bec | Sandrine Moreira et al |
| EPI_ISL_536306 | Canada/QC-LSPQ-L00242035/2020 | 2020-03-28 | North America/Canada/Quebec/ | H√¥pital Pierre-Boucher | Laboratoire de sant√© publique du Qu√©bec | Sandrine Moreira et al |

| | | | | | | |
|---|---|---|---|---|---|---|
| EPI_ISL_536307 | Canada/QC-LSPQ-L00242049/2020 | 2020-03-28 | North America/Canada/Quebec/ | H√¥pital Pierre-Boucher | Laboratoire de sant√© publique du Qu√©bec | Sandrine Moreira et al |
| EPI_ISL_536308 | Canada/QC-LSPQ-L00242051/2020 | 2020-03-28 | North America/Canada/Quebec/ | H√¥pital Pierre-Boucher | Laboratoire de sant√© publique du Qu√©bec | Sandrine Moreira et al |
| EPI_ISL_536309 | Canada/QC-LSPQ-L00242105/2020 | 2020-03-28 | North America/Canada/Quebec/ | H√¥pital Pierre-Boucher | Laboratoire de sant√© publique du Qu√©bec | Sandrine Moreira et al |
| EPI_ISL_536310 | Canada/QC-LSPQ-L00242152/2020 | 2020-03-28 | North America/Canada/Quebec/ | CLSC et Centre d'H√©bergement la Petite-Nation | Laboratoire de sant√© publique du Qu√©bec | Sandrine Moreira et al |
| EPI_ISL_536311 | Canada/QC-LSPQ-L00242214/2020 | 2020-03-29 | North America/Canada/Quebec/ | H√¥pital de Gatineau | Laboratoire de sant√© publique du Qu√©bec | Sandrine Moreira et al |
| EPI_ISL_536312 | Canada/QC-LSPQ-L00242228/2020 | 2020-03-29 | North America/Canada/Quebec/ | H√¥pital de Hull | Laboratoire de sant√© publique du Qu√©bec | Sandrine Moreira et al |
| EPI_ISL_536313 | Canada/QC-LSPQ-L00242235/2020 | 2020-03-29 | North America/Canada/Quebec/ | H√¥pital Charles-LeMoyne | Laboratoire de sant√© publique du Qu√©bec | Sandrine Moreira et al |
| EPI_ISL_536314 | Canada/QC-LSPQ-L00242238/2020 | 2020-03-29 | North America/Canada/Quebec/ | H√¥pital Charles-LeMoyne | Laboratoire de sant√© publique du Qu√©bec | Sandrine Moreira et al |
| EPI_ISL_536315 | Canada/QC-LSPQ-L00242292/2020 | 2020-03-29 | North America/Canada/Quebec/ | Centre hospitalier Anna-Laberge | Laboratoire de sant√© publique du Qu√©bec | Sandrine Moreira et al |
| EPI_ISL_536316 | Canada/QC-LSPQ-L00242308/2020 | 2020-03-29 | North America/Canada/Quebec/ | H√¥pital Charles-LeMoyne | Laboratoire de sant√© publique du Qu√©bec | Sandrine Moreira et al |
| EPI_ISL_536317 | Canada/QC-LSPQ-L00242317/2020 | 2020-03-29 | North America/Canada/Quebec/ | H√¥pital Charles-LeMoyne | Laboratoire de sant√© publique du Qu√©bec | Sandrine Moreira et al |
| EPI_ISL_536318 | Canada/QC-LSPQ-L00242370/2020 | 2020-03-29 | North America/Canada/Quebec/ | H√¥pital du Surov√Æt | Laboratoire de sant√© publique du Qu√©bec | Sandrine Moreira et al |
| EPI_ISL_536319 | Canada/QC-LSPQ-L00242384/2020 | 2020-03-29 | North America/Canada/Quebec/ | H√¥pital du Surov√Æt | Laboratoire de sant√© publique du Qu√©bec | Sandrine Moreira et al |

| | | | | | | |
|---|---|---|---|---|---|---|
| EPI_ISL_536320 | Canada/QC-LSPQ-L00242401/2020 | 2020-03-29 | North America/Canada/Quebec/ | H√¥pital du Surov√Æt | Laboratoire de sant√© publique du Qu√©bec | Sandrine Moreira et al |
| EPI_ISL_536321 | Canada/QC-LSPQ-L00242447/2020 | 2020-03-29 | North America/Canada/Quebec/ | CSSS Haut-Richelieu/Rouville (H√¥pital) | Laboratoire de sant√© publique du Qu√©bec | Sandrine Moreira et al |
| EPI_ISL_536322 | Canada/QC-LSPQ-L00242487/2020 | 2020-03-29 | North America/Canada/Quebec/ | H√¥pital Pierre-Boucher | Laboratoire de sant√© publique du Qu√©bec | Sandrine Moreira et al |
| EPI_ISL_536323 | Canada/QC-LSPQ-L00242505/2020 | 2020-03-29 | North America/Canada/Quebec/ | H√¥pital Pierre-Boucher | Laboratoire de sant√© publique du Qu√©bec | Sandrine Moreira et al |
| EPI_ISL_536324 | Canada/QC-LSPQ-L00242509/2020 | 2020-03-29 | North America/Canada/Quebec/ | H√¥pital Pierre-Boucher | Laboratoire de sant√© publique du Qu√©bec | Sandrine Moreira et al |
| EPI_ISL_536325 | Canada/QC-LSPQ-L00242547/2020 | 2020-03-29 | North America/Canada/Quebec/ | Centre hospitalier Anna-Laberge | Laboratoire de sant√© publique du Qu√©bec | Sandrine Moreira et al |
| EPI_ISL_536326 | Canada/QC-LSPQ-L00242558/2020 | 2020-03-29 | North America/Canada/Quebec/ | H√¥pital Charles-LeMoyne | Laboratoire de sant√© publique du Qu√©bec | Sandrine Moreira et al |
| EPI_ISL_536327 | Canada/QC-LSPQ-L00242573/2020 | 2020-03-29 | North America/Canada/Quebec/ | H√¥pital Charles-LeMoyne | Laboratoire de sant√© publique du Qu√©bec | Sandrine Moreira et al |
| EPI_ISL_536328 | Canada/QC-LSPQ-L00242576/2020 | 2020-03-29 | North America/Canada/Quebec/ | H√¥pital Charles-LeMoyne | Laboratoire de sant√© publique du Qu√©bec | Sandrine Moreira et al |
| EPI_ISL_536329 | Canada/QC-LSPQ-L00242582/2020 | 2020-03-29 | North America/Canada/Quebec/ | Centre hospitalier Anna-Laberge | Laboratoire de sant√© publique du Qu√©bec | Sandrine Moreira et al |
| EPI_ISL_536330 | Canada/QC-LSPQ-L00242623/2020 | 2020-03-29 | North America/Canada/Quebec/ | H√¥pital Honor√©-Mercier | Laboratoire de sant√© publique du Qu√©bec | Sandrine Moreira et al |
| EPI_ISL_536331 | Canada/QC-LSPQ-L00242662/2020 | 2020-03-29 | North America/Canada/Quebec/ | H√¥pital Charles-LeMoyne | Laboratoire de sant√© publique du Qu√©bec | Sandrine Moreira et al |
| EPI_ISL_536332 | Canada/QC-LSPQ-L00242684/2020 | 2020-03-29 | North America/Canada/Quebec/ | H√¥pital Pierre-Boucher | Laboratoire de sant√© publique du Qu√©bec | Sandrine Moreira et al |

| | | | | | | |
|---|---|---|---|---|---|---|
| EPI_ISL_536333 | Canada/QC-LSPQ-L00242687/2020 | 2020-03-29 | North America/Canada/Quebec/ | H√¥pital Pierre-Boucher | Laboratoire de sant√© publique du Qu√©bec | Sandrine Moreira et al |
| EPI_ISL_536334 | Canada/QC-LSPQ-L00242697/2020 | 2020-03-29 | North America/Canada/Quebec/ | H√¥pital Pierre-Boucher | Laboratoire de sant√© publique du Qu√©bec | Sandrine Moreira et al |
| EPI_ISL_536335 | Canada/QC-LSPQ-L00242714/2020 | 2020-03-29 | North America/Canada/Quebec/ | H√¥pital Pierre-Boucher | Laboratoire de sant√© publique du Qu√©bec | Sandrine Moreira et al |
| EPI_ISL_536336 | Canada/QC-LSPQ-L00242858/2020 | 2020-03-30 | North America/Canada/Quebec/ | H√¥pital Charles-LeMoyne | Laboratoire de sant√© publique du Qu√©bec | Sandrine Moreira et al |
| EPI_ISL_536337 | Canada/QC-LSPQ-L00242867/2020 | 2020-03-27 | North America/Canada/Quebec/ | CUSM-Site Glen-LAB Microbiologie | Laboratoire de sant√© publique du Qu√©bec | Sandrine Moreira et al |
| EPI_ISL_536338 | Canada/QC-LSPQ-L00242878/2020 | 2020-03-30 | North America/Canada/Quebec/ | H√¥pital Charles-LeMoyne | Laboratoire de sant√© publique du Qu√©bec | Sandrine Moreira et al |
| EPI_ISL_536339 | Canada/QC-LSPQ-L00242891/2020 | 2020-03-30 | North America/Canada/Quebec/ | H√¥pital Charles-LeMoyne | Laboratoire de sant√© publique du Qu√©bec | Sandrine Moreira et al |
| EPI_ISL_536340 | Canada/QC-LSPQ-L00242900/2020 | 2020-03-30 | North America/Canada/Quebec/ | H√¥pital Charles-LeMoyne | Laboratoire de sant√© publique du Qu√©bec | Sandrine Moreira et al |
| EPI_ISL_536341 | Canada/QC-LSPQ-L00242917/2020 | 2020-03-30 | North America/Canada/Quebec/ | H√¥pital Charles-LeMoyne | Laboratoire de sant√© publique du Qu√©bec | Sandrine Moreira et al |
| EPI_ISL_536342 | Canada/QC-LSPQ-L00242918/2020 | 2020-03-30 | North America/Canada/Quebec/ | H√¥pital Charles-LeMoyne | Laboratoire de sant√© publique du Qu√©bec | Sandrine Moreira et al |
| EPI_ISL_536343 | Canada/QC-LSPQ-L00242991/2020 | 2020-03-30 | North America/Canada/Quebec/ | H√¥pital Charles-LeMoyne | Laboratoire de sant√© publique du Qu√©bec | Sandrine Moreira et al |
| EPI_ISL_536344 | Canada/QC-LSPQ-L00243024/2020 | 2020-03-30 | North America/Canada/Quebec/ | CSSS Haut-Richelieu/Rouville (H√¥pital) | Laboratoire de sant√© publique du Qu√©bec | Sandrine Moreira et al |
| EPI_ISL_536345 | Canada/QC-LSPQ-L00243036/2020 | 2020-03-30 | North America/Canada/Quebec/ | CSSS Haut-Richelieu/Rouville (H√¥pital) | Laboratoire de sant√© publique du Qu√©bec | Sandrine Moreira et al |

| | | | | | | |
|---|---|---|---|---|---|---|
| EPI_ISL_536346 | Canada/QC-LSPQ-L00243049/2020 | 2020-03-29 | North America/Canada/Quebec/ | H√¥pital Pierre-Boucher | Laboratoire de sant√© publique du Qu√©bec | Sandrine Moreira et al |
| EPI_ISL_536347 | Canada/QC-LSPQ-L00243054/2020 | 2020-03-30 | North America/Canada/Quebec/ | H√¥pital Pierre-Boucher | Laboratoire de sant√© publique du Qu√©bec | Sandrine Moreira et al |
| EPI_ISL_536348 | Canada/QC-LSPQ-L00243079/2020 | 2020-03-30 | North America/Canada/Quebec/ | Centre hospitalier Anna-Laberge | Laboratoire de sant√© publique du Qu√©bec | Sandrine Moreira et al |
| EPI_ISL_536349 | Canada/QC-LSPQ-L00243118/2020 | 2020-03-30 | North America/Canada/Quebec/ | H√¥pital du Surov√Æt | Laboratoire de sant√© publique du Qu√©bec | Sandrine Moreira et al |
| EPI_ISL_536350 | Canada/QC-LSPQ-L00243222/2020 | 2020-03-30 | North America/Canada/Quebec/ | H√¥pital Charles-LeMoyne | Laboratoire de sant√© publique du Qu√©bec | Sandrine Moreira et al |
| EPI_ISL_536351 | Canada/QC-LSPQ-L00243245/2020 | 2020-03-30 | North America/Canada/Quebec/ | H√¥pital Pierre-Boucher | Laboratoire de sant√© publique du Qu√©bec | Sandrine Moreira et al |
| EPI_ISL_536352 | Canada/QC-LSPQ-L00243257/2020 | 2020-03-30 | North America/Canada/Quebec/ | H√¥pital Pierre-Boucher | Laboratoire de sant√© publique du Qu√©bec | Sandrine Moreira et al |
| EPI_ISL_536353 | Canada/QC-LSPQ-L00243284/2020 | 2020-03-29 | North America/Canada/Quebec/ | Centre de sant√© Inuulitsivik | Laboratoire de sant√© publique du Qu√©bec | Sandrine Moreira et al |
| EPI_ISL_536354 | Canada/QC-LSPQ-L00243313/2020 | 2020-03-30 | North America/Canada/Quebec/ | H√¥pital du Surov√Æt | Laboratoire de sant√© publique du Qu√©bec | Sandrine Moreira et al |
| EPI_ISL_536355 | Canada/QC-LSPQ-L00243317/2020 | 2020-03-30 | North America/Canada/Quebec/ | H√¥pital du Surov√Æt | Laboratoire de sant√© publique du Qu√©bec | Sandrine Moreira et al |
| EPI_ISL_536356 | Canada/QC-LSPQ-L00243323/2020 | 2020-03-30 | North America/Canada/Quebec/ | H√¥pital du Surov√Æt | Laboratoire de sant√© publique du Qu√©bec | Sandrine Moreira et al |
| EPI_ISL_536357 | Canada/QC-LSPQ-L00243325/2020 | 2020-03-30 | North America/Canada/Quebec/ | H√¥pital du Surov√Æt | Laboratoire de sant√© publique du Qu√©bec | Sandrine Moreira et al |
| EPI_ISL_536358 | Canada/QC-LSPQ-L00243328/2020 | 2020-03-30 | North America/Canada/Quebec/ | H√¥pital du Surov√Æt | Laboratoire de sant√© publique du Qu√©bec | Sandrine Moreira et al |

| | | | | | | |
|---|---|---|---|---|---|---|
| EPI_ISL_536359 | Canada/QC-LSPQ-L00243345/2020 | 2020-03-30 | North America/Canada/Quebec/ | H√¥pital du Surov√Æt | Laboratoire de sant√© publique du Qu√©bec | Sandrine Moreira et al |
| EPI_ISL_536360 | Canada/QC-LSPQ-L00243347/2020 | 2020-03-30 | North America/Canada/Quebec/ | H√¥pital Pierre-Boucher | Laboratoire de sant√© publique du Qu√©bec | Sandrine Moreira et al |
| EPI_ISL_536361 | Canada/QC-LSPQ-L00243417/2020 | 2020-03-30 | North America/Canada/Quebec/ | CSSS Haut-Richelieu/Rouville (H√¥pital) | Laboratoire de sant√© publique du Qu√©bec | Sandrine Moreira et al |
| EPI_ISL_536362 | Canada/QC-LSPQ-L00243460/2020 | 2020-03-30 | North America/Canada/Quebec/ | H√¥pital du Surov√Æt | Laboratoire de sant√© publique du Qu√©bec | Sandrine Moreira et al |
| EPI_ISL_536363 | Canada/QC-LSPQ-L00243560/2020 | 2020-03-30 | North America/Canada/Quebec/ | H√¥pital de Hull | Laboratoire de sant√© publique du Qu√©bec | Sandrine Moreira et al |
| EPI_ISL_536364 | Canada/QC-LSPQ-L00243590/2020 | 2020-03-30 | North America/Canada/Quebec/ | H√¥pital de Hull | Laboratoire de sant√© publique du Qu√©bec | Sandrine Moreira et al |
| EPI_ISL_536365 | Canada/QC-LSPQ-L00243603/2020 | 2020-03-29 | North America/Canada/Quebec/ | CUSM-Site Glen-LAB Microbiologie | Laboratoire de sant√© publique du Qu√©bec | Sandrine Moreira et al |
| EPI_ISL_536366 | Canada/QC-LSPQ-L00243607/2020 | 2020-03-29 | North America/Canada/Quebec/ | CUSM-Site Glen-LAB Microbiologie | Laboratoire de sant√© publique du Qu√©bec | Sandrine Moreira et al |
| EPI_ISL_536367 | Canada/QC-LSPQ-L00243608/2020 | 2020-03-30 | North America/Canada/Quebec/ | H√¥pital de Hull | Laboratoire de sant√© publique du Qu√©bec | Sandrine Moreira et al |
| EPI_ISL_536368 | Canada/QC-LSPQ-L00243646/2020 | 2020-03-31 | North America/Canada/Quebec/ | Centre hospitalier Anna-Laberge | Laboratoire de sant√© publique du Qu√©bec | Sandrine Moreira et al |
| EPI_ISL_536369 | Canada/QC-LSPQ-L00243673/2020 | 2020-03-31 | North America/Canada/Quebec/ | Centre hospitalier Anna-Laberge | Laboratoire de sant√© publique du Qu√©bec | Sandrine Moreira et al |
| EPI_ISL_536370 | Canada/QC-LSPQ-L00243675/2020 | 2020-03-31 | North America/Canada/Quebec/ | Centre hospitalier Anna-Laberge | Laboratoire de sant√© publique du Qu√©bec | Sandrine Moreira et al |
| EPI_ISL_536371 | Canada/QC-LSPQ-L00243706/2020 | 2020-03-31 | North America/Canada/Quebec/ | H√¥pital Charles-LeMoyne | Laboratoire de sant√© publique du Qu√©bec | Sandrine Moreira et al |

| | | | | | | |
|---|---|---|---|---|---|---|
| EPI_ISL_536372 | Canada/QC-LSPQ-L00243708/2020 | 2020-03-31 | North America/Canada/Quebec/ | H√¥pital Charles-LeMoyne | Laboratoire de sant√© publique du Qu√©bec | Sandrine Moreira et al |
| EPI_ISL_536373 | Canada/QC-LSPQ-L00243710/2020 | 2020-03-31 | North America/Canada/Quebec/ | H√¥pital Charles-LeMoyne | Laboratoire de sant√© publique du Qu√©bec | Sandrine Moreira et al |
| EPI_ISL_536374 | Canada/QC-LSPQ-L00243735/2020 | 2020-03-31 | North America/Canada/Quebec/ | H√¥pital Pierre-Boucher | Laboratoire de sant√© publique du Qu√©bec | Sandrine Moreira et al |
| EPI_ISL_536375 | Canada/QC-LSPQ-L00243746/2020 | 2020-03-30 | North America/Canada/Quebec/ | H√¥pital Pierre-Boucher | Laboratoire de sant√© publique du Qu√©bec | Sandrine Moreira et al |
| EPI_ISL_536376 | Canada/QC-LSPQ-L00243751/2020 | 2020-03-31 | North America/Canada/Quebec/ | Centre hospitalier Anna-Laberge | Laboratoire de sant√© publique du Qu√©bec | Sandrine Moreira et al |
| EPI_ISL_536377 | Canada/QC-LSPQ-L00243755/2020 | 2020-03-31 | North America/Canada/Quebec/ | Centre hospitalier Anna-Laberge | Laboratoire de sant√© publique du Qu√©bec | Sandrine Moreira et al |
| EPI_ISL_536378 | Canada/QC-LSPQ-L00243761/2020 | 2020-03-31 | North America/Canada/Quebec/ | Centre hospitalier Anna-Laberge | Laboratoire de sant√© publique du Qu√©bec | Sandrine Moreira et al |
| EPI_ISL_536379 | Canada/QC-LSPQ-L00243797/2020 | 2020-03-31 | North America/Canada/Quebec/ | H√¥pital Honor√©-Mercier | Laboratoire de sant√© publique du Qu√©bec | Sandrine Moreira et al |
| EPI_ISL_536380 | Canada/QC-LSPQ-L00243862/2020 | 2020-03-31 | North America/Canada/Quebec/ | H√¥pital Pierre-Boucher | Laboratoire de sant√© publique du Qu√©bec | Sandrine Moreira et al |
| EPI_ISL_536381 | Canada/QC-LSPQ-L00243864/2020 | 2020-03-31 | North America/Canada/Quebec/ | Centre Hospitalier R√©gional de Lanaudi√®re | Laboratoire de sant√© publique du Qu√©bec | Sandrine Moreira et al |
| EPI_ISL_536382 | Canada/QC-LSPQ-L00243874/2020 | 2020-03-31 | North America/Canada/Quebec/ | CSSS Haut-Richelieu/Rouville (H√¥pital) | Laboratoire de sant√© publique du Qu√©bec | Sandrine Moreira et al |
| EPI_ISL_536383 | Canada/QC-LSPQ-L00243875/2020 | 2020-03-31 | North America/Canada/Quebec/ | CSSS Haut-Richelieu/Rouville (H√¥pital) | Laboratoire de sant√© publique du Qu√©bec | Sandrine Moreira et al |
| EPI_ISL_536384 | Canada/QC-LSPQ-L00243973/2020 | 2020-03-31 | North America/Canada/Quebec/ | H√¥pital de Gatineau | Laboratoire de sant√© publique du Qu√©bec | Sandrine Moreira et al |

| | | | | | | |
|---|---|---|---|---|---|---|
| EPI_ISL_536385 | Canada/QC-LSPQ-L00243976/2020 | 2020-03-31 | North America/Canada/Quebec/ | H√¥pital de Hull | Laboratoire de sant√© publique du Qu√©bec | Sandrine Moreira et al |
| EPI_ISL_536386 | Canada/QC-LSPQ-L00243994/2020 | 2020-03-31 | North America/Canada/Quebec/ | H√¥pital de Hull | Laboratoire de sant√© publique du Qu√©bec | Sandrine Moreira et al |
| EPI_ISL_536387 | Canada/QC-LSPQ-L00244008/2020 | 2020-03-31 | North America/Canada/Quebec/ | H√¥pital de Hull | Laboratoire de sant√© publique du Qu√©bec | Sandrine Moreira et al |
| EPI_ISL_536388 | Canada/QC-LSPQ-L00244068/2020 | 2020-03-31 | North America/Canada/Quebec/ | H√¥pital Pierre-Boucher | Laboratoire de sant√© publique du Qu√©bec | Sandrine Moreira et al |
| EPI_ISL_536389 | Canada/QC-LSPQ-L00244094/2020 | 2020-03-31 | North America/Canada/Quebec/ | H√¥pital Pierre-Boucher | Laboratoire de sant√© publique du Qu√©bec | Sandrine Moreira et al |
| EPI_ISL_536390 | Canada/QC-LSPQ-L00244160/2020 | 2020-03-31 | North America/Canada/Quebec/ | H√¥pital du Surov√Æt | Laboratoire de sant√© publique du Qu√©bec | Sandrine Moreira et al |
| EPI_ISL_536391 | Canada/QC-LSPQ-L00244197/2020 | 2020-03-31 | North America/Canada/Quebec/ | H√¥pital Pierre-Boucher | Laboratoire de sant√© publique du Qu√©bec | Sandrine Moreira et al |
| EPI_ISL_536392 | Canada/QC-LSPQ-L00244326/2020 | 2020-03-31 | North America/Canada/Quebec/ | H√¥pital de Hull | Laboratoire de sant√© publique du Qu√©bec | Sandrine Moreira et al |
| EPI_ISL_536393 | Canada/QC-LSPQ-L00244338/2020 | 2020-03-31 | North America/Canada/Quebec/ | H√¥pital de Hull | Laboratoire de sant√© publique du Qu√©bec | Sandrine Moreira et al |
| EPI_ISL_536394 | Canada/QC-LSPQ-L00244347/2020 | 2020-03-31 | North America/Canada/Quebec/ | H√¥pital de Hull | Laboratoire de sant√© publique du Qu√©bec | Sandrine Moreira et al |
| EPI_ISL_536395 | Canada/QC-LSPQ-L00244354/2020 | 2020-03-31 | North America/Canada/Quebec/ | H√¥pital Pierre-Boucher | Laboratoire de sant√© publique du Qu√©bec | Sandrine Moreira et al |
| EPI_ISL_536396 | Canada/QC-LSPQ-L00244358/2020 | 2020-03-31 | North America/Canada/Quebec/ | H√¥pital de Hull | Laboratoire de sant√© publique du Qu√©bec | Sandrine Moreira et al |
| EPI_ISL_536397 | Canada/QC-LSPQ-L00244359/2020 | 2020-03-31 | North America/Canada/Quebec/ | H√¥pital Pierre-Boucher | Laboratoire de sant√© publique du Qu√©bec | Sandrine Moreira et al |

| | | | | | | |
|---|---|---|---|---|---|---|
| EPI_ISL_536413 | Singapore/872/2020 | 2020-05-26 | Asia/Singapore/Singapore/ | National Public Health Laboratory, National Centre for Infectious Diseases | National Public Health Laboratory, National Centre for Infectious Diseases | Mak TM et al |
| EPI_ISL_536427 | Singapore/886/2020 | 2020-08-31 | Asia/Singapore/Singapore/ | National Public Health Laboratory, National Centre for Infectious Diseases | National Public Health Laboratory, National Centre for Infectious Diseases | Mak TM et al |
| EPI_ISL_536435 | Singapore/894/2020 | 2020-02-02 | Asia/Singapore/Singapore/ | National Public Health Laboratory, National Centre for Infectious Diseases | National Public Health Laboratory, National Centre for Infectious Diseases | Mak TM et al |
| EPI_ISL_536439 | Singapore/898/2020 | 2020-02-03 | Asia/Singapore/Singapore/ | National Public Health Laboratory, National Centre for Infectious Diseases | National Public Health Laboratory, National Centre for Infectious Diseases | Mak TM et al |
| EPI_ISL_536441 | Singapore/900/2020 | 2020-02-05 | Asia/Singapore/Singapore/ | National Public Health Laboratory, National Centre for Infectious Diseases | National Public Health Laboratory, National Centre for Infectious Diseases | Mak TM et al |
| EPI_ISL_536442 | Singapore/901/2020 | 2020-09-04 | Asia/Singapore/Singapore/ | National Public Health Laboratory, National Centre for Infectious Diseases | National Public Health Laboratory, National Centre for Infectious Diseases | Mak TM et al |

| | | | | | | |
|---|---|---|---|---|---|---|
| EPI_ISL_536447 | Singapore/906/2020 | 2020-01-25 | Asia/Singapore/Singapore/ | National Public Health Laboratory, National Centre for Infectious Diseases | National Public Health Laboratory, National Centre for Infectious Diseases | Mak TM et al |
| EPI_ISL_536452 | Singapore/911/2020 | 2020-02-03 | Asia/Singapore/Singapore/ | National Public Health Laboratory, National Centre for Infectious Diseases | National Public Health Laboratory, National Centre for Infectious Diseases | Mak TM et al |
| EPI_ISL_536477 | Peru/LIM-INS-077/2020 | 2020-07-03 | South America/Peru/Lima/ | Instituto Nacional de Salud | Laboratorio de Infecciones Respiratorias Agudas | Eduardo Juscamayta Lopez et al |
| EPI_ISL_536478 | Peru/LIM-INS-078/2020 | 2020-07-02 | South America/Peru/Lima/ | Instituto Nacional de Salud | Laboratorio de Infecciones Respiratorias Agudas | Eduardo Juscamayta Lopez et al |
| EPI_ISL_536479 | Peru/CAL-INS-079/2020 | 2020-07-01 | South America/Peru/Callao/ | Instituto Nacional de Salud | Laboratorio de Infecciones Respiratorias Agudas | Eduardo Juscamayta Lopez et al |
| EPI_ISL_536480 | Peru/CAL-INS-081/2020 | 2020-07-01 | South America/Peru/Callao/ | Instituto Nacional de Salud | Laboratorio de Infecciones Respiratorias Agudas | Eduardo Juscamayta Lopez et al |
| EPI_ISL_536481 | Peru/CAL-INS-082/2020 | 2020-07-01 | South America/Peru/Callao/ | Instituto Nacional de Salud | Laboratorio de Infecciones Respiratorias Agudas | Eduardo Juscamayta Lopez et al |
| EPI_ISL_536482 | Peru/ANC-INS-083/2020 | 2020-06-30 | South America/Peru/Ancash/ | Instituto Nacional de Salud | Laboratorio de Infecciones Respiratorias Agudas | Eduardo Juscamayta Lopez et al |
| EPI_ISL_536483 | Peru/ANC-INS-084/2020 | 2020-07-02 | South America/Peru/Ancash/ | Instituto Nacional de Salud | Laboratorio de Infecciones Respiratorias Agudas | Eduardo Juscamayta Lopez et al |

| | | | | | | |
|---|---|---|---|---|---|---|
| EPI_ISL_536484 | Peru/ANC-INS-085/2020 | 2020-07-02 | South America/Peru/Ancash/ | Instituto Nacional de Salud | Laboratorio de Infecciones Respiratorias Agudas | Eduardo Juscamayta Lopez et al |
| EPI_ISL_536485 | Peru/LIM-INS-086/2020 | 2020-07-04 | South America/Peru/Lima/ | Instituto Nacional de Salud | Laboratorio de Infecciones Respiratorias Agudas | Eduardo Juscamayta Lopez et al |
| EPI_ISL_536486 | Peru/LIM-INS-087/2020 | 2020-07-04 | South America/Peru/Lima/ | Instituto Nacional de Salud | Laboratorio de Infecciones Respiratorias Agudas | Eduardo Juscamayta Lopez et al |
| EPI_ISL_536487 | Peru/LIM-INS-088/2020 | 2020-07-04 | South America/Peru/Lima/ | Instituto Nacional de Salud | Laboratorio de Infecciones Respiratorias Agudas | Eduardo Juscamayta Lopez et al |
| EPI_ISL_536488 | Peru/LIM-INS-089/2020 | 2020-07-04 | South America/Peru/Lima/ | Instituto Nacional de Salud | Laboratorio de Infecciones Respiratorias Agudas | Eduardo Juscamayta Lopez et al |
| EPI_ISL_536489 | Peru/LIM-INS-090/2020 | 2020-07-04 | South America/Peru/Lima/ | Instituto Nacional de Salud | Laboratorio de Infecciones Respiratorias Agudas | Eduardo Juscamayta Lopez et al |
| EPI_ISL_536490 | Peru/LIM-INS-091/2020 | 2020-07-03 | South America/Peru/Lima/ | Instituto Nacional de Salud | Laboratorio de Infecciones Respiratorias Agudas | Eduardo Juscamayta Lopez et al |
| EPI_ISL_536491 | Peru/ANC-INS-092/2020 | 2020-07-03 | South America/Peru/Ancash/ | Instituto Nacional de Salud | Laboratorio de Infecciones Respiratorias Agudas | Eduardo Juscamayta Lopez et al |
| EPI_ISL_536492 | Peru/LIM-INS-093/2020 | 2020-07-04 | South America/Peru/Lima/ | Instituto Nacional de Salud | Laboratorio de Infecciones Respiratorias Agudas | Eduardo Juscamayta Lopez et al |
| EPI_ISL_536493 | Peru/LIM-INS-094/2020 | 2020-07-03 | South America/Peru/Lima/ | Instituto Nacional de Salud | Laboratorio de Infecciones Respiratorias Agudas | Eduardo Juscamayta Lopez et al |

| | | | | | | |
|---|---|---|---|---|---|---|
| EPI_ISL_536494 | Peru/LIM-INS-095/2020 | 2020-07-03 | South America/Peru/Lima/ | Instituto Nacional de Salud | Laboratorio de Infecciones Respiratorias Agudas | Eduardo Juscamayta Lopez et al |
| EPI_ISL_536495 | Peru/LIM-INS-096/2020 | 2020-07-03 | South America/Peru/Lima/ | Instituto Nacional de Salud | Laboratorio de Infecciones Respiratorias Agudas | Eduardo Juscamayta Lopez et al |
| EPI_ISL_536496 | Peru/LIM-INS-097/2020 | 2020-07-01 | South America/Peru/Lima/ | Instituto Nacional de Salud | Laboratorio de Infecciones Respiratorias Agudas | Eduardo Juscamayta Lopez et al |
| EPI_ISL_536497 | Peru/LIM-INS-098/2020 | 2020-07-04 | South America/Peru/Lima/ | Instituto Nacional de Salud | Laboratorio de Infecciones Respiratorias Agudas | Eduardo Juscamayta Lopez et al |
| EPI_ISL_536498 | Peru/LIM-INS-099/2020 | 2020-07-04 | South America/Peru/Lima/ | Instituto Nacional de Salud | Laboratorio de Infecciones Respiratorias Agudas | Eduardo Juscamayta Lopez et al |
| EPI_ISL_536499 | Peru/LIM-INS-100/2020 | 2020-07-04 | South America/Peru/Lima/ | Instituto Nacional de Salud | Laboratorio de Infecciones Respiratorias Agudas | Eduardo Juscamayta Lopez et al |
| EPI_ISL_536500 | Peru/LIM-INS-101/2020 | 2020-07-04 | South America/Peru/Lima/ | Instituto Nacional de Salud | Laboratorio de Infecciones Respiratorias Agudas | Eduardo Juscamayta Lopez et al |
| EPI_ISL_536501 | Peru/LIM-INS-102/2020 | 2020-05-02 | South America/Peru/Lima/ | Instituto Nacional de Salud | Laboratorio de Infecciones Respiratorias Agudas | Eduardo Juscamayta Lopez et al |
| EPI_ISL_536502 | Peru/CAL-INS-103/2020 | 2020-05-02 | South America/Peru/Callao/ | Instituto Nacional de Salud | Laboratorio de Infecciones Respiratorias Agudas | Eduardo Juscamayta Lopez et al |
| EPI_ISL_536503 | Peru/CAL-INS-104/2020 | 2020-05-02 | South America/Peru/Callao/ | Instituto Nacional de Salud | Laboratorio de Infecciones Respiratorias Agudas | Eduardo Juscamayta Lopez et al |

| | | | | | | |
|---|---|---|---|---|---|---|
| EPI_ISL_536504 | Peru/CAL-INS-105/2020 | 2020-05-02 | South America/Peru/Callao/ | Instituto Nacional de Salud | Laboratorio de Infecciones Respiratorias Agudas | Eduardo Juscamayta Lopez et al |
| EPI_ISL_536505 | Peru/ICA-INS-107/2020 | 2020-05-04 | South America/Peru/Ica/ | Instituto Nacional de Salud | Laboratorio de Infecciones Respiratorias Agudas | Eduardo Juscamayta Lopez et al |
| EPI_ISL_536506 | Peru/ICA-INS-108/2020 | 2020-05-04 | South America/Peru/Ica/ | Instituto Nacional de Salud | Laboratorio de Infecciones Respiratorias Agudas | Eduardo Juscamayta Lopez et al |
| EPI_ISL_536507 | Peru/ICA-INS-109/2020 | 2020-05-04 | South America/Peru/Ica/ | Instituto Nacional de Salud | Laboratorio de Infecciones Respiratorias Agudas | Eduardo Juscamayta Lopez et al |
| EPI_ISL_536508 | Peru/LIM-INS-110/2020 | 2020-05-04 | South America/Peru/Lima/ | Instituto Nacional de Salud | Laboratorio de Infecciones Respiratorias Agudas | Eduardo Juscamayta Lopez et al |
| EPI_ISL_536509 | Peru/LIM-INS-111/2020 | 2020-05-04 | South America/Peru/Lima/ | Instituto Nacional de Salud | Laboratorio de Infecciones Respiratorias Agudas | Eduardo Juscamayta Lopez et al |
| EPI_ISL_536510 | Peru/LIM-INS-112/2020 | 2020-05-04 | South America/Peru/Lima/ | Instituto Nacional de Salud | Laboratorio de Infecciones Respiratorias Agudas | Eduardo Juscamayta Lopez et al |
| EPI_ISL_536511 | Peru/LIM-INS-113/2020 | 2020-05-04 | South America/Peru/Lima/ | Instituto Nacional de Salud | Laboratorio de Infecciones Respiratorias Agudas | Eduardo Juscamayta Lopez et al |
| EPI_ISL_536512 | Peru/LIM-INS-114/2020 | 2020-05-04 | South America/Peru/Lima/ | Instituto Nacional de Salud | Laboratorio de Infecciones Respiratorias Agudas | Eduardo Juscamayta Lopez et al |
| EPI_ISL_536513 | Peru/LIM-INS-115/2020 | 2020-05-04 | South America/Peru/Lima/ | Instituto Nacional de Salud | Laboratorio de Infecciones Respiratorias Agudas | Eduardo Juscamayta Lopez et al |

| | | | | | | |
|---|---|---|---|---|---|---|
| EPI_ISL_536514 | Peru/LIM-INS-116/2020 | 2020-05-05 | South America/Peru/Lima/ | Instituto Nacional de Salud | Laboratorio de Infecciones Respiratorias Agudas | Eduardo Juscamayta Lopez et al |
| EPI_ISL_536515 | Peru/LIM-INS-117/2020 | 2020-05-04 | South America/Peru/Lima/ | Instituto Nacional de Salud | Laboratorio de Infecciones Respiratorias Agudas | Eduardo Juscamayta Lopez et al |
| EPI_ISL_536516 | Peru/LIM-INS-118/2020 | 2020-05-04 | South America/Peru/Lima/ | Instituto Nacional de Salud | Laboratorio de Infecciones Respiratorias Agudas | Eduardo Juscamayta Lopez et al |
| EPI_ISL_536517 | Peru/LIM-INS-119/2020 | 2020-05-05 | South America/Peru/Lima/ | Instituto Nacional de Salud | Laboratorio de Infecciones Respiratorias Agudas | Eduardo Juscamayta Lopez et al |
| EPI_ISL_536518 | Peru/LIM-INS-120/2020 | 2020-05-05 | South America/Peru/Lima/ | Instituto Nacional de Salud | Laboratorio de Infecciones Respiratorias Agudas | Eduardo Juscamayta Lopez et al |
| EPI_ISL_536519 | Peru/LIM-INS-121/2020 | 2020-05-04 | South America/Peru/Lima/ | Instituto Nacional de Salud | Laboratorio de Infecciones Respiratorias Agudas | Eduardo Juscamayta Lopez et al |
| EPI_ISL_536520 | Peru/LIM-INS-122/2020 | 2020-05-05 | South America/Peru/Lima/ | Instituto Nacional de Salud | Laboratorio de Infecciones Respiratorias Agudas | Eduardo Juscamayta Lopez et al |
| EPI_ISL_536521 | Peru/LIM-INS-123/2020 | 2020-05-06 | South America/Peru/Lima/ | Instituto Nacional de Salud | Laboratorio de Infecciones Respiratorias Agudas | Eduardo Juscamayta Lopez et al |
| EPI_ISL_536522 | Peru/LIM-INS-124/2020 | 2020-06-24 | South America/Peru/Lima/ | Instituto Nacional de Salud | Laboratorio de Infecciones Respiratorias Agudas | Eduardo Juscamayta Lopez et al |
| EPI_ISL_536523 | Peru/LAM-INS-125/2020 | 2020-04-01 | South America/Peru/Lambayeque/ | Instituto Nacional de Salud | Laboratorio de Infecciones Respiratorias Agudas | Eduardo Juscamayta Lopez et al |

| | | | | | | |
|---|---|---|---|---|---|---|
| EPI_ISL_536524 | Peru/LAM-INS-126/2020 | 2020-04-01 | South America/Peru/Lambayeque/ | Instituto Nacional de Salud | Laboratorio de Infecciones Respiratorias Agudas | Eduardo Juscamayta Lopez et al |
| EPI_ISL_536525 | Peru/JUN-INS-127/2020 | 2020-04-01 | South America/Peru/Junin/ | Instituto Nacional de Salud | Laboratorio de Infecciones Respiratorias Agudas | Eduardo Juscamayta Lopez et al |
| EPI_ISL_536526 | Peru/JUN-INS-128/2020 | 2020-04-01 | South America/Peru/Junin/ | Instituto Nacional de Salud | Laboratorio de Infecciones Respiratorias Agudas | Eduardo Juscamayta Lopez et al |
| EPI_ISL_536562 | Peru/LAM-INS-169/2020 | 2020-04-01 | South America/Peru/Lambayeque/ | Instituto Nacional de Salud | Laboratorio de Infecciones Respiratorias Agudas | Eduardo Juscamayta Lopez et al |
| EPI_ISL_536563 | Peru/LAM-INS-170/2020 | 2020-04-01 | South America/Peru/Lambayeque/ | Instituto Nacional de Salud | Laboratorio de Infecciones Respiratorias Agudas | Eduardo Juscamayta Lopez et al |
| EPI_ISL_536564 | Peru/LAM-INS-171/2020 | 2020-04-01 | South America/Peru/Lambayeque/ | Instituto Nacional de Salud | Laboratorio de Infecciones Respiratorias Agudas | Eduardo Juscamayta Lopez et al |
| EPI_ISL_536574 | USA/WI-UW-1201/2020 | 2020-07-28 | North America/USA/Wisconsin/Dane Co | University of Wisconsin-Madison AIDS Vaccine Research Laboratories | University of Wisconsin-Madison AIDS Vaccine Research Laboratories | Gage Moreno et al |
| EPI_ISL_536577 | USA/WI-UW-1204/2020 | 2020-08-06 | North America/USA/Wisconsin/Dane Co | University of Wisconsin-Madison AIDS Vaccine Research Laboratories | University of Wisconsin-Madison AIDS Vaccine Research Laboratories | Gage Moreno et al |
| EPI_ISL_536585 | USA/WI-UW-1212/2020 | 2020-08-19 | North America/USA/Wisconsin/Dodge C | University of Wisconsin-Madison AIDS Vaccine Research Laboratories | University of Wisconsin-Madison AIDS Vaccine Research Laboratories | Gage Moreno et al |

| | | | | | | |
|---|---|---|---|---|---|---|
| EPI_ISL_536587 | USA/WI-UW-1214/2020 | 2020-08-18 | North America/USA/Wisconsin/Dane Co | University of Wisconsin-Madison AIDS Vaccine Research Laboratories | University of Wisconsin-Madison AIDS Vaccine Research Laboratories | Gage Moreno et al |
| EPI_ISL_536590 | USA/WI-UW-1217/2020 | 2020-08-20 | North America/USA/Wisconsin/Green Co | University of Wisconsin-Madison AIDS Vaccine Research Laboratories | University of Wisconsin-Madison AIDS Vaccine Research Laboratories | Gage Moreno et al |
| EPI_ISL_536591 | USA/WI-UW-1218/2020 | 2020-08-20 | North America/USA/Wisconsin/Dane Co | University of Wisconsin-Madison AIDS Vaccine Research Laboratories | University of Wisconsin-Madison AIDS Vaccine Research Laboratories | Gage Moreno et al |
| EPI_ISL_536592 | USA/WI-UW-1219/2020 | 2020-08-19 | North America/USA/Wisconsin/Dane Co | University of Wisconsin-Madison AIDS Vaccine Research Laboratories | University of Wisconsin-Madison AIDS Vaccine Research Laboratories | Gage Moreno et al |
| EPI_ISL_536593 | USA/WI-UW-1220/2020 | 2020-08-17 | North America/USA/Wisconsin/Green Co | University of Wisconsin-Madison AIDS Vaccine Research Laboratories | University of Wisconsin-Madison AIDS Vaccine Research Laboratories | Gage Moreno et al |
| EPI_ISL_536598 | USA/WI-UW-1225/2020 | 2020-08-14 | North America/USA/Wisconsin/Dane Co | University of Wisconsin-Madison AIDS Vaccine Research Laboratories | University of Wisconsin-Madison AIDS Vaccine Research Laboratories | Gage Moreno et al |
| EPI_ISL_536601 | USA/WI-UW-1228/2020 | 2020-08-15 | North America/USA/Wisconsin/Dane Co | University of Wisconsin-Madison AIDS Vaccine Research Laboratories | University of Wisconsin-Madison AIDS Vaccine Research Laboratories | Gage Moreno et al |
| EPI_ISL_536605 | USA/WI-UW-1232/2020 | 2020-08-17 | North America/USA/Wisconsin/Dane Co | University of Wisconsin-Madison AIDS Vaccine Research Laboratories | University of Wisconsin-Madison AIDS Vaccine Research Laboratories | Gage Moreno et al |

| | | | | | | |
|---|---|---|---|---|---|---|
| EPI_ISL_536607 | USA/WI-UW-1234/2020 | 2020-08-14 | North America/USA/Wisconsin/Dane Co | University of Wisconsin-Madison AIDS Vaccine Research Laboratories | University of Wisconsin-Madison AIDS Vaccine Research Laboratories | Gage Moreno et al |
| EPI_ISL_536614 | USA/WI-UW-1241/2020 | 2020-08-17 | North America/USA/Wisconsin/Dane Co | University of Wisconsin-Madison AIDS Vaccine Research Laboratories | University of Wisconsin-Madison AIDS Vaccine Research Laboratories | Gage Moreno et al |
| EPI_ISL_536617 | USA/WI-UW-1244/2020 | 2020-08-17 | North America/USA/Wisconsin/Dane Co | University of Wisconsin-Madison AIDS Vaccine Research Laboratories | University of Wisconsin-Madison AIDS Vaccine Research Laboratories | Gage Moreno et al |
| EPI_ISL_536619 | USA/WI-UW-1246/2020 | 2020-08-17 | North America/USA/Wisconsin/Iowa Cou | University of Wisconsin-Madison AIDS Vaccine Research Laboratories | University of Wisconsin-Madison AIDS Vaccine Research Laboratories | Gage Moreno et al |
| EPI_ISL_536622 | USA/WI-UW-1249/2020 | 2020-08-17 | North America/USA/Wisconsin/Dane Co | University of Wisconsin-Madison AIDS Vaccine Research Laboratories | University of Wisconsin-Madison AIDS Vaccine Research Laboratories | Gage Moreno et al |
| EPI_ISL_536626 | USA/WI-UW-1253/2020 | 2020-08-14 | North America/USA/Wisconsin/Dane Co | University of Wisconsin-Madison AIDS Vaccine Research Laboratories | University of Wisconsin-Madison AIDS Vaccine Research Laboratories | Gage Moreno et al |
| EPI_ISL_536628 | USA/WI-UW-1255/2020 | 2020-08-20 | North America/USA/Wisconsin/Green Co | University of Wisconsin-Madison AIDS Vaccine Research Laboratories | University of Wisconsin-Madison AIDS Vaccine Research Laboratories | Gage Moreno et al |
| EPI_ISL_536632 | USA/WI-UW-1259/2020 | 2020-08-22 | North America/USA/Wisconsin/Rock Cou | University of Wisconsin-Madison AIDS Vaccine Research Laboratories | University of Wisconsin-Madison AIDS Vaccine Research Laboratories | Gage Moreno et al |

| | | | | | | |
|---|---|---|---|---|---|---|
| EPI_ISL_536633 | USA/WI-UW-1260/2020 | 2020-08-21 | North America/USA/Wisconsin/Dane Co | University of Wisconsin-Madison AIDS Vaccine Research Laboratories | University of Wisconsin-Madison AIDS Vaccine Research Laboratories | Gage Moreno et al |
| EPI_ISL_536634 | USA/WI-UW-1261/2020 | 2020-08-25 | North America/USA/Wisconsin/Dane Co | University of Wisconsin-Madison AIDS Vaccine Research Laboratories | University of Wisconsin-Madison AIDS Vaccine Research Laboratories | Gage Moreno et al |
| EPI_ISL_536643 | USA/WI-UW-1271/2020 | 2020-08-21 | North America/USA/Wisconsin/Dane Co | University of Wisconsin-Madison AIDS Vaccine Research Laboratories | University of Wisconsin-Madison AIDS Vaccine Research Laboratories | Gage Moreno et al |
| EPI_ISL_536644 | USA/WI-UW-1272/2020 | 2020-08-21 | North America/USA/Wisconsin/Columbia | University of Wisconsin-Madison AIDS Vaccine Research Laboratories | University of Wisconsin-Madison AIDS Vaccine Research Laboratories | Gage Moreno et al |
| EPI_ISL_536653 | USA/WI-UW-1281/2020 | 2020-08-22 | North America/USA/Wisconsin/Dane Co | University of Wisconsin-Madison AIDS Vaccine Research Laboratories | University of Wisconsin-Madison AIDS Vaccine Research Laboratories | Gage Moreno et al |
| EPI_ISL_536659 | USA/WI-UW-1287/2020 | 2020-09-02 | North America/USA/Wisconsin/Dane Co | University of Wisconsin-Madison AIDS Vaccine Research Laboratories | University of Wisconsin-Madison AIDS Vaccine Research Laboratories | Gage Moreno et al |
| EPI_ISL_536663 | USA/WI-UW-1291/2020 | 2020-08-31 | North America/USA/Wisconsin/Jefferson | University of Wisconsin-Madison AIDS Vaccine Research Laboratories | University of Wisconsin-Madison AIDS Vaccine Research Laboratories | Gage Moreno et al |
| EPI_ISL_536666 | USA/WI-UW-1294/2020 | 2020-09-02 | North America/USA/Wisconsin/Dane Co | University of Wisconsin-Madison AIDS Vaccine Research Laboratories | University of Wisconsin-Madison AIDS Vaccine Research Laboratories | Gage Moreno et al |

| | | | | | | |
|---|---|---|---|---|---|---|
| EPI_ISL_536667 | USA/WI-UW-1295/2020 | 2020-09-01 | North America/USA/Wisconsin/Dodge C | University of Wisconsin-Madison AIDS Vaccine Research Laboratories | University of Wisconsin-Madison AIDS Vaccine Research Laboratories | Gage Moreno et al |
| EPI_ISL_536675 | USA/WI-UW-1303/2020 | 2020-08-29 | North America/USA/Wisconsin/Dane Co | University of Wisconsin-Madison AIDS Vaccine Research Laboratories | University of Wisconsin-Madison AIDS Vaccine Research Laboratories | Gage Moreno et al |
| EPI_ISL_536681 | USA/WI-UW-1309/2020 | 2020-08-31 | North America/USA/Wisconsin/Dane Co | University of Wisconsin-Madison AIDS Vaccine Research Laboratories | University of Wisconsin-Madison AIDS Vaccine Research Laboratories | Gage Moreno et al |
| EPI_ISL_536685 | USA/WI-UW-1313/2020 | 2020-08-26 | North America/USA/Wisconsin/Dane Co | University of Wisconsin-Madison AIDS Vaccine Research Laboratories | University of Wisconsin-Madison AIDS Vaccine Research Laboratories | Gage Moreno et al |
| EPI_ISL_536687 | USA/WI-UW-1315/2020 | 2020-08-31 | North America/USA/Wisconsin/Dane Co | University of Wisconsin-Madison AIDS Vaccine Research Laboratories | University of Wisconsin-Madison AIDS Vaccine Research Laboratories | Gage Moreno et al |
| EPI_ISL_536695 | USA/WI-UW-1324/2020 | 2020-08-27 | North America/USA/Wisconsin/Iowa Co | University of Wisconsin-Madison AIDS Vaccine Research Laboratories | University of Wisconsin-Madison AIDS Vaccine Research Laboratories | Gage Moreno et al |
| EPI_ISL_536698 | USA/WI-UW-1327/2020 | 2020-08-27 | North America/USA/Wisconsin/Dane Co | University of Wisconsin-Madison AIDS Vaccine Research Laboratories | University of Wisconsin-Madison AIDS Vaccine Research Laboratories | Gage Moreno et al |
| EPI_ISL_536703 | USA/WI-UW-1332/2020 | 2020-08-31 | North America/USA/Wisconsin/Dane Co | University of Wisconsin-Madison AIDS Vaccine Research Laboratories | University of Wisconsin-Madison AIDS Vaccine Research Laboratories | Gage Moreno et al |

| | | | | | | |
|---|---|---|---|---|---|---|
| EPI_ISL_536706 | USA/WI-UW-1335/2020 | 2020-08-30 | North America/USA/Wisconsin/Dane Co | University of Wisconsin-Madison AIDS Vaccine Research Laboratories | University of Wisconsin-Madison AIDS Vaccine Research Laboratories | Gage Moreno et al |
| EPI_ISL_536708 | USA/WI-UW-1337/2020 | 2020-09-04 | North America/USA/Wisconsin/Monroe | University of Wisconsin-Madison AIDS Vaccine Research Laboratories | University of Wisconsin-Madison AIDS Vaccine Research Laboratories | Gage Moreno et al |
| EPI_ISL_536709 | USA/WI-UW-1338/2020 | 2020-09-08 | North America/USA/Wisconsin/Dane Co | University of Wisconsin-Madison AIDS Vaccine Research Laboratories | University of Wisconsin-Madison AIDS Vaccine Research Laboratories | Gage Moreno et al |
| EPI_ISL_536710 | USA/WI-UW-1339/2020 | 2020-09-07 | North America/USA/Wisconsin/Dane Co | University of Wisconsin-Madison AIDS Vaccine Research Laboratories | University of Wisconsin-Madison AIDS Vaccine Research Laboratories | Gage Moreno et al |
| EPI_ISL_536712 | USA/WI-UW-1341/2020 | 2020-09-08 | North America/USA/Wisconsin/Dodge C | University of Wisconsin-Madison AIDS Vaccine Research Laboratories | University of Wisconsin-Madison AIDS Vaccine Research Laboratories | Gage Moreno et al |
| EPI_ISL_536719 | USA/WI-UW-1348/2020 | 2020-09-08 | North America/USA/Wisconsin/Dane Co | University of Wisconsin-Madison AIDS Vaccine Research Laboratories | University of Wisconsin-Madison AIDS Vaccine Research Laboratories | Gage Moreno et al |
| EPI_ISL_536723 | USA/WI-UW-1352/2020 | 2020-09-09 | North America/USA/Wisconsin/Rock Cou | University of Wisconsin-Madison AIDS Vaccine Research Laboratories | University of Wisconsin-Madison AIDS Vaccine Research Laboratories | Gage Moreno et al |
| EPI_ISL_536725 | USA/WI-UW-1354/2020 | 2020-09-09 | North America/USA/Wisconsin/Dane Co | University of Wisconsin-Madison AIDS Vaccine Research Laboratories | University of Wisconsin-Madison AIDS Vaccine Research Laboratories | Gage Moreno et al |

| | | | | | | |
|---|---|---|---|---|---|---|
| EPI_ISL_536729 | USA/WI-UW-1358/2020 | 2020-09-08 | North America/USA/Wisconsin/Dane Co | University of Wisconsin-Madison AIDS Vaccine Research Laboratories | University of Wisconsin-Madison AIDS Vaccine Research Laboratories | Gage Moreno et al |
| EPI_ISL_536730 | USA/WI-UW-1359/2020 | 2020-09-08 | North America/USA/Wisconsin/Dane Co | University of Wisconsin-Madison AIDS Vaccine Research Laboratories | University of Wisconsin-Madison AIDS Vaccine Research Laboratories | Gage Moreno et al |
| EPI_ISL_536733 | USA/WI-UW-1362/2020 | 2020-09-07 | North America/USA/Wisconsin/Dane Co | University of Wisconsin-Madison AIDS Vaccine Research Laboratories | University of Wisconsin-Madison AIDS Vaccine Research Laboratories | Gage Moreno et al |
| EPI_ISL_536736 | USA/NJ-UW-1365/2020 | 2020-09-06 | North America/USA/New Jersey/Morris | University of Wisconsin-Madison AIDS Vaccine Research Laboratories | University of Wisconsin-Madison AIDS Vaccine Research Laboratories | Gage Moreno et al |
| EPI_ISL_536739 | USA/WI-UW-1368/2020 | 2020-09-08 | North America/USA/Wisconsin/Dane Co | University of Wisconsin-Madison AIDS Vaccine Research Laboratories | University of Wisconsin-Madison AIDS Vaccine Research Laboratories | Gage Moreno et al |
| EPI_ISL_536740 | USA/WI-UW-1369/2020 | 2020-09-08 | North America/USA/Wisconsin/Dane Co | University of Wisconsin-Madison AIDS Vaccine Research Laboratories | University of Wisconsin-Madison AIDS Vaccine Research Laboratories | Gage Moreno et al |
| EPI_ISL_536746 | USA/WI-UW-1375/2020 | 2020-09-05 | North America/USA/Wisconsin/Dane Co | University of Wisconsin-Madison AIDS Vaccine Research Laboratories | University of Wisconsin-Madison AIDS Vaccine Research Laboratories | Gage Moreno et al |
| EPI_ISL_536747 | USA/WI-UW-1376/2020 | 2020-09-06 | North America/USA/Wisconsin/Lincoln C | University of Wisconsin-Madison AIDS Vaccine Research Laboratories | University of Wisconsin-Madison AIDS Vaccine Research Laboratories | Gage Moreno et al |

| | | | | | | |
|---|---|---|---|---|---|---|
| EPI_ISL_536750 | USA/WI-UW-1379/2020 | 2020-09-04 | North America/USA/Wisconsin/Dane Co | University of Wisconsin-Madison AIDS Vaccine Research Laboratories | University of Wisconsin-Madison AIDS Vaccine Research Laboratories | Gage Moreno et al |
| EPI_ISL_536756 | USA/WI-UW-1385/2020 | 2020-09-06 | North America/USA/Wisconsin/Dane Co | University of Wisconsin-Madison AIDS Vaccine Research Laboratories | University of Wisconsin-Madison AIDS Vaccine Research Laboratories | Gage Moreno et al |
| EPI_ISL_536763 | USA/MO-UW-1392/2020 | 2020-09-07 | North America/USA/Missouri/Saint Loui | University of Wisconsin-Madison AIDS Vaccine Research Laboratories | University of Wisconsin-Madison AIDS Vaccine Research Laboratories | Gage Moreno et al |
| EPI_ISL_536768 | USA/WI-UW-1397/2020 | 2020-09-04 | North America/USA/Wisconsin/Jeffersor | University of Wisconsin-Madison AIDS Vaccine Research Laboratories | University of Wisconsin-Madison AIDS Vaccine Research Laboratories | Gage Moreno et al |
| EPI_ISL_536769 | USA/WI-UW-1398/2020 | 2020-09-03 | North America/USA/Wisconsin/Jeffersor | University of Wisconsin-Madison AIDS Vaccine Research Laboratories | University of Wisconsin-Madison AIDS Vaccine Research Laboratories | Gage Moreno et al |
| EPI_ISL_536772 | USA/WI-UW-1401/2020 | 2020-09-06 | North America/USA/Wisconsin/Dane Co | University of Wisconsin-Madison AIDS Vaccine Research Laboratories | University of Wisconsin-Madison AIDS Vaccine Research Laboratories | Gage Moreno et al |
| EPI_ISL_536774 | USA/WI-UW-1404/2020 | 2020-09-07 | North America/USA/Wisconsin/Dane Co | University of Wisconsin-Madison AIDS Vaccine Research Laboratories | University of Wisconsin-Madison AIDS Vaccine Research Laboratories | Gage Moreno et al |
| EPI_ISL_536776 | USA/WI-UW-1406/2020 | 2020-09-03 | North America/USA/Wisconsin/Sheboyg | University of Wisconsin-Madison AIDS Vaccine Research Laboratories | University of Wisconsin-Madison AIDS Vaccine Research Laboratories | Gage Moreno et al |

| | | | | | | |
|---|---|---|---|---|---|---|
| EPI_ISL_536779 | USA/WI-UW-1409/2020 | 2020-09-03 | North America/USA/Wisconsin/Dane Co | University of Wisconsin-Madison AIDS Vaccine Research Laboratories | University of Wisconsin-Madison AIDS Vaccine Research Laboratories | Gage Moreno et al |
| EPI_ISL_536782 | USA/WI-UW-1412/2020 | 2020-09-04 | North America/USA/Wisconsin/Dane Co | University of Wisconsin-Madison AIDS Vaccine Research Laboratories | University of Wisconsin-Madison AIDS Vaccine Research Laboratories | Gage Moreno et al |
| EPI_ISL_536783 | USA/WI-UW-1413/2020 | 2020-09-03 | North America/USA/Wisconsin/Dane Co | University of Wisconsin-Madison AIDS Vaccine Research Laboratories | University of Wisconsin-Madison AIDS Vaccine Research Laboratories | Gage Moreno et al |
| EPI_ISL_536784 | USA/WI-UW-1414/2020 | 2020-09-03 | North America/USA/Wisconsin/Dane Co | University of Wisconsin-Madison AIDS Vaccine Research Laboratories | University of Wisconsin-Madison AIDS Vaccine Research Laboratories | Gage Moreno et al |
| EPI_ISL_536786 | USA/WI-UW-1416/2020 | 2020-09-02 | North America/USA/Wisconsin/Dane Co | University of Wisconsin-Madison AIDS Vaccine Research Laboratories | University of Wisconsin-Madison AIDS Vaccine Research Laboratories | Gage Moreno et al |
| EPI_ISL_536933 | NorthernIreland/QEUH-9650C4/2020 | 2020-08-13 | Europe/United Kingdom/Northern Irelar | Lighthouse Lab in Glasgow | Wellcome Sanger Institute for the COVID-19 Genomics UK (COG-UK) consortium | Harper VanSteenhouse et al |
| EPI_ISL_537055 | England/QEUH-9615E7/2020 | 2020-08-03 | Europe/United Kingdom/England/ | Lighthouse Lab in Glasgow | Wellcome Sanger Institute for the COVID-19 Genomics UK (COG-UK) consortium | Harper VanSteenhouse et al |
| EPI_ISL_537121 | England/QEUH-9618A2/2020 | 2020-08-05 | Europe/United Kingdom/England/ | Lighthouse Lab in Glasgow | Wellcome Sanger Institute for the COVID-19 Genomics UK (COG-UK) consortium | Harper VanSteenhouse et al |

| | | | | | | |
|---|---|---|---|---|---|---|
| EPI_ISL_537269 | England/SHEF-10B2D30/2020 | 2020-05-15 | Europe/United Kingdom/England/ | Virology Department, Sheffield Teaching Hospitals NHS Foundation Trust / Department of Infection, Immunity and Cardiovascular Disease, The Medical School, University of Sheffield | Wellcome Sanger Institute for the COVID-19 Genomics UK (COG-UK) consortium | Thushan de Silva et al |
| EPI_ISL_537270 | England/SHEF-10B3018/2020 | 2020-05-26 | Europe/United Kingdom/England/ | Virology Department, Sheffield Teaching Hospitals NHS Foundation Trust / Department of Infection, Immunity and Cardiovascular Disease, The Medical School, University of Sheffield | Wellcome Sanger Institute for the COVID-19 Genomics UK (COG-UK) consortium | Thushan de Silva et al |
| EPI_ISL_537288 | Spain/CL-IBV-97006140/2020 | 2020-03-31 | Europe/Spain/Castilla y Leon/La Baneza | Universidad de Lev≥n | SeqCOVID-SPAIN consortium/IBV(CSIC) | Ana Carvajal et al |
| EPI_ISL_537289 | Spain/CL-IBV-97006141/2020 | 2020-03-31 | Europe/Spain/Castilla y Leon/Leon | Universidad de Lev≥n | SeqCOVID-SPAIN consortium/IBV(CSIC) | Ana Carvajal et al |
| EPI_ISL_537299 | Spain/CL-IBV-97006152/2020 | 2020-04-05 | Europe/Spain/Castilla y Leon/Leon | Universidad de Lev≥n | SeqCOVID-SPAIN consortium/IBV(CSIC) | Ana Carvajal et al |
| EPI_ISL_537302 | Spain/CL-IBV-97006155/2020 | 2020-04-05 | Europe/Spain/Castilla y Leon/Leon | Universidad de Lev≥n | SeqCOVID-SPAIN consortium/IBV(CSIC) | Ana Carvajal et al |
| EPI_ISL_537307 | Spain/CL-IBV-97006160/2020 | 2020-04-05 | Europe/Spain/Castilla y Leon/Leon | Universidad de Lev≥n | SeqCOVID-SPAIN consortium/IBV(CSIC) | Ana Carvajal et al |
| EPI_ISL_537328 | Spain/CL-IBV-97006181/2020 | 2020-04-13 | Europe/Spain/Castilla y Leon/Riego de la | Universidad de Lev≥n | SeqCOVID-SPAIN consortium/IBV(CSIC) | Ana Carvajal et al |
| EPI_ISL_537330 | Spain/CL-IBV-97006184/2020 | 2020-04-13 | Europe/Spain/Castilla y Leon/Castrillo d | Universidad de Lev≥n | SeqCOVID-SPAIN consortium/IBV(CSIC) | Ana Carvajal et al |
| EPI_ISL_537345 | Spain/CL-IBV-97006199/2020 | 2020-04-13 | Europe/Spain/Castilla y Leon/La Virgen | Universidad de Lev≥n | SeqCOVID-SPAIN consortium/IBV(CSIC) | Ana Carvajal et al |
| EPI_ISL_537349 | Spain/CL-IBV-97006203/2020 | 2020-04-13 | Europe/Spain/Castilla y Leon/Villarejo d | Universidad de Lev≥n | SeqCOVID-SPAIN consortium/IBV(CSIC) | Ana Carvajal et al |
| EPI_ISL_537356 | Spain/CL-IBV-97006210/2020 | 2020-04-13 | Europe/Spain/Castilla y Leon/San Cristo | Universidad de Lev≥n | SeqCOVID-SPAIN consortium/IBV(CSIC) | Ana Carvajal et al |

| | | | | | | |
|---|---|---|---|---|---|---|
| EPI_ISL_537366 | Spain/CL-IBV-97006220/2020 | 2020-04-13 | Europe/Spain/Castilla y Leon/Villarejo d | Universidad de Lev≥n | SeqCOVID-SPAIN consortium/IBV(CSIC) | Ana Carvajal et al |
| EPI_ISL_537373 | Spain/CL-IBV-97006227/2020 | 2020-04-13 | Europe/Spain/Castilla y Leon/Villarejo d | Universidad de Lev≥n | SeqCOVID-SPAIN consortium/IBV(CSIC) | Ana Carvajal et al |
| EPI_ISL_537374 | Spain/CL-IBV-97006228/2020 | 2020-04-13 | Europe/Spain/Castilla y Leon/Val de Sar | Universidad de Lev≥n | SeqCOVID-SPAIN consortium/IBV(CSIC) | Ana Carvajal et al |
| EPI_ISL_537376 | Spain/CL-IBV-97006230/2020 | 2020-04-14 | Europe/Spain/Castilla y Leon/Leon | Universidad de Lev≥n | SeqCOVID-SPAIN consortium/IBV(CSIC) | Ana Carvajal et al |
| EPI_ISL_537386 | Spain/RI-IBV-99005843/2020 | 2020-03-11 | Europe/Spain/La Rioja/Logrono | Centro de Investigaci√≥n Biom√©dica de La Rioja - Hospital San Pedro Logro√±o | SeqCOVID-SPAIN consortium/IBV(CSIC) | Mar√≠a de Toro et al |
| EPI_ISL_537388 | Spain/RI-IBV-99005845/2020 | 2020-03-11 | Europe/Spain/La Rioja/Alberite | Centro de Investigaci√≥n Biom√©dica de La Rioja - Hospital San Pedro Logro√±o | SeqCOVID-SPAIN consortium/IBV(CSIC) | Mar√≠a de Toro et al |
| EPI_ISL_537391 | Spain/RI-IBV-99005848/2020 | 2020-03-27 | Europe/Spain/La Rioja/Navarrete | Centro de Investigaci√≥n Biom√©dica de La Rioja - Hospital San Pedro Logro√±o | SeqCOVID-SPAIN consortium/IBV(CSIC) | Mar√≠a de Toro et al |
| EPI_ISL_537394 | Spain/RI-IBV-99005851/2020 | 2020-04-01 | Europe/Spain/La Rioja/Haro | Centro de Investigaci√≥n Biom√©dica de La Rioja - Hospital San Pedro Logro√±o | SeqCOVID-SPAIN consortium/IBV(CSIC) | Mar√≠a de Toro et al |
| EPI_ISL_537398 | Spain/RI-IBV-99005855/2020 | 2020-03-24 | Europe/Spain/La Rioja/Calahorra | Centro de Investigaci√≥n Biom√©dica de La Rioja - Hospital San Pedro Logro√±o | SeqCOVID-SPAIN consortium/IBV(CSIC) | Mar√≠a de Toro et al |
| EPI_ISL_537408 | Spain/RI-IBV-99005865/2020 | 2020-03-05 | Europe/Spain/La Rioja/Haro | Centro de Investigaci√≥n Biom√©dica de La Rioja - Hospital San Pedro Logro√±o | SeqCOVID-SPAIN consortium/IBV(CSIC) | Mar√≠a de Toro et al |
| EPI_ISL_537412 | Spain/RI-IBV-99005869/2020 | 2020-03-07 | Europe/Spain/La Rioja/Logrono | Centro de Investigaci√≥n Biom√©dica de La Rioja - Hospital San Pedro Logro√±o | SeqCOVID-SPAIN consortium/IBV(CSIC) | Mar√≠a de Toro et al |
| EPI_ISL_537413 | Spain/RI-IBV-99005870/2020 | 2020-03-07 | Europe/Spain/La Rioja/Haro | Centro de Investigaci√≥n Biom√©dica de La Rioja - Hospital San Pedro Logro√±o | SeqCOVID-SPAIN consortium/IBV(CSIC) | Mar√≠a de Toro et al |
| EPI_ISL_537430 | Spain/RI-IBV-99005888/2020 | 2020-03-11 | Europe/Spain/La Rioja/Santo Domingo d | Centro de Investigaci√≥n Biom√©dica de La Rioja - Hospital San Pedro Logro√±o | SeqCOVID-SPAIN consortium/IBV(CSIC) | Mar√≠a de Toro et al |

| | | | | | | |
|---|---|---|---|---|---|---|
| EPI_ISL_537435 | Spain/RI-IBV-99005893/2020 | 2020-03-22 | Europe/Spain/La Rioja/Logrono | Centro de Investigaci√≥n Biom√©dica de La Rioja - Hospital San Pedro Logro√±o | SeqCOVID-SPAIN consortium/IBV(CSIC) | Mar√≠a de Toro et al |
| EPI_ISL_537436 | Spain/RI-IBV-99005894/2020 | 2020-03-22 | Europe/Spain/La Rioja/Ezcaray | Centro de Investigaci√≥n Biom√©dica de La Rioja - Hospital San Pedro Logro√±o | SeqCOVID-SPAIN consortium/IBV(CSIC) | Mar√≠a de Toro et al |
| EPI_ISL_537441 | Spain/RI-IBV-99005903/2020 | 2020-04-03 | Europe/Spain/La Rioja/Arnedo | Centro de Investigaci√≥n Biom√©dica de La Rioja - Hospital San Pedro Logro√±o | SeqCOVID-SPAIN consortium/IBV(CSIC) | Mar√≠a de Toro et al |
| EPI_ISL_537448 | Spain/RI-IBV-99005910/2020 | 2020-03-25 | Europe/Spain/La Rioja/Lardero | Centro de Investigaci√≥n Biom√©dica de La Rioja - Hospital San Pedro Logro√±o | SeqCOVID-SPAIN consortium/IBV(CSIC) | Mar√≠a de Toro et al |
| EPI_ISL_537454 | Spain/RI-IBV-99005916/2020 | 2020-03-28 | Europe/Spain/La Rioja/Logrono | Centro de Investigaci√≥n Biom√©dica de La Rioja - Hospital San Pedro Logro√±o | SeqCOVID-SPAIN consortium/IBV(CSIC) | Mar√≠a de Toro et al |
| EPI_ISL_537455 | Spain/RI-IBV-99005918/2020 | 2020-03-28 | Europe/Spain/La Rioja/Torrecilla en Cam | Centro de Investigaci√≥n Biom√©dica de La Rioja - Hospital San Pedro Logro√±o | SeqCOVID-SPAIN consortium/IBV(CSIC) | Mar√≠a de Toro et al |
| EPI_ISL_537461 | Spain/RI-IBV-99005924/2020 | 2020-03-29 | Europe/Spain/La Rioja/Santo Domingo d | Centro de Investigaci√≥n Biom√©dica de La Rioja - Hospital San Pedro Logro√±o | SeqCOVID-SPAIN consortium/IBV(CSIC) | Mar√≠a de Toro et al |
| EPI_ISL_537462 | Spain/RI-IBV-99005926/2020 | 2020-03-12 | Europe/Spain/La Rioja/Alberite | Centro de Investigaci√≥n Biom√©dica de La Rioja - Hospital San Pedro Logro√±o | SeqCOVID-SPAIN consortium/IBV(CSIC) | Mar√≠a de Toro et al |
| EPI_ISL_537471 | USA/CA-UCLA-115/2020 | 2020-04-07 | North America/USA/California/Los Ange | UCLA Pathology Clinical Microbiology Lab | Kruglyak Lab | Guo et al. et al |
| EPI_ISL_537473 | USA/CA-UCLA-119/2020 | 2020-04-07 | North America/USA/California/Los Ange | UCLA Pathology Clinical Microbiology Lab | Kruglyak Lab | Guo et al. et al |
| EPI_ISL_537489 | USA/CA-UCLA-146/2020 | 2020-04-12 | North America/USA/California/Los Ange | UCLA Pathology Clinical Microbiology Lab | Kruglyak Lab | Guo et al. et al |
| EPI_ISL_537491 | USA/CA-UCLA-15/2020 | 2020-04-25 | North America/USA/California/Los Ange | UCLA Pathology Clinical Microbiology Lab | Kruglyak Lab | Guo et al. et al |
| EPI_ISL_537499 | USA/CA-UCLA-16/2020 | 2020-04-26 | North America/USA/California/Los Ange | UCLA Pathology Clinical Microbiology Lab | Kruglyak Lab | Guo et al. et al |
| EPI_ISL_537506 | USA/CA-UCLA-18/2020 | 2020-05-01 | North America/USA/California/Los Ange | UCLA Pathology Clinical Microbiology Lab | Kruglyak Lab | Guo et al. et al |
| EPI_ISL_537508 | USA/CA-UCLA-19/2020 | 2020-05-08 | North America/USA/California/Los Ange | UCLA Pathology Clinical Microbiology Lab | Kruglyak Lab | Guo et al. et al |
| EPI_ISL_537519 | USA/CA-UCLA-212/2020 | 2020-05-04 | North America/USA/California/Los Ange | UCLA Pathology Clinical Microbiology Lab | Kruglyak Lab | Guo et al. et al |
| EPI_ISL_537525 | USA/CA-UCLA-219/2020 | 2020-06-03 | North America/USA/California/Los Ange | UCLA Pathology Clinical Microbiology Lab | Kruglyak Lab | Guo et al. et al |
| EPI_ISL_537527 | USA/CA-UCLA-220/2020 | 2020-06-09 | North America/USA/California/Los Ange | UCLA Pathology Clinical Microbiology Lab | Kruglyak Lab | Guo et al. et al |
| EPI_ISL_537532 | USA/CA-UCLA-227/2020 | 2020-05-15 | North America/USA/California/Los Ange | UCLA Pathology Clinical Microbiology Lab | Kruglyak Lab | Guo et al. et al |
| EPI_ISL_537536 | USA/CA-UCLA-230/2020 | 2020-05-01 | North America/USA/California/Los Ange | UCLA Pathology Clinical Microbiology Lab | Kruglyak Lab | Guo et al. et al |
| EPI_ISL_537543 | USA/CA-UCLA-237/2020 | 2020-06-18 | North America/USA/California/Los Ange | UCLA Pathology Clinical Microbiology Lab | Kruglyak Lab | Guo et al. et al |

| | | | | | | |
|---|---|---|---|---|---|---|
| EPI_ISL_537545 | USA/CA-UCLA-24/2020 | 2020-05-02 | North America/USA/California/Los Ange | UCLA Pathology Clinical Microbiology Lab | Kruglyak Lab | Guo et al. et al |
| EPI_ISL_537546 | USA/CA-UCLA-240/2020 | 2020-06-20 | North America/USA/California/Los Ange | UCLA Pathology Clinical Microbiology Lab | Kruglyak Lab | Guo et al. et al |
| EPI_ISL_537549 | USA/CA-UCLA-244/2020 | 2020-04-22 | North America/USA/California/Los Ange | UCLA Pathology Clinical Microbiology Lab | Kruglyak Lab | Guo et al. et al |
| EPI_ISL_537558 | USA/CA-UCLA-260/2020 | 2020-02-28 | North America/USA/California/Los Ange | UCLA Pathology Clinical Microbiology Lab | Kruglyak Lab | Guo et al. et al |
| EPI_ISL_537562 | USA/CA-UCLA-3/2020 | 2020-04-13 | North America/USA/California/Los Ange | UCLA Pathology Clinical Microbiology Lab | Kruglyak Lab | Guo et al. et al |
| EPI_ISL_537573 | USA/CA-UCLA-44/2020 | 2020-03-31 | North America/USA/California/Los Ange | UCLA Pathology Clinical Microbiology Lab | Kruglyak Lab | Guo et al. et al |
| EPI_ISL_537580 | USA/CA-UCLA-51/2020 | 2020-03-14 | North America/USA/California/Los Ange | UCLA Pathology Clinical Microbiology Lab | Kruglyak Lab | Guo et al. et al |
| EPI_ISL_537584 | USA/CA-UCLA-59/2020 | 2020-03-12 | North America/USA/California/Los Ange | UCLA Pathology Clinical Microbiology Lab | Kruglyak Lab | Guo et al. et al |
| EPI_ISL_537596 | USA/CA-UCLA-71/2020 | 2020-03-17 | North America/USA/California/Los Ange | UCLA Pathology Clinical Microbiology Lab | Kruglyak Lab | Guo et al. et al |
| EPI_ISL_537609 | Spain/CL-IBV-97006234/2020 | 2020-04-14 | Europe/Spain/Castilla y Leon/Leon | Universidad de Lev≥n | SeqCOVID-SPAIN consortium/IBV(CSIC) | Ana Carvajal et al |
| EPI_ISL_537620 | Spain/CL-IBV-97006248/2020 | 2020-04-20 | Europe/Spain/Castilla y Leon/Villaobisp | Universidad de Lev≥n | SeqCOVID-SPAIN consortium/IBV(CSIC) | Ana Carvajal et al |
| EPI_ISL_537622 | Spain/CL-IBV-97006250/2020 | 2020-04-20 | Europe/Spain/Castilla y Leon/Villaquilar | Universidad de Lev≥n | SeqCOVID-SPAIN consortium/IBV(CSIC) | Ana Carvajal et al |
| EPI_ISL_537627 | Spain/CL-IBV-97006255/2020 | 2020-04-20 | Europe/Spain/Castilla y Leon/Villaquilar | Universidad de Lev≥n | SeqCOVID-SPAIN consortium/IBV(CSIC) | Ana Carvajal et al |
| EPI_ISL_537634 | Spain/CL-IBV-97006262/2020 | 2020-04-20 | Europe/Spain/Castilla y Leon/Villaobisp | Universidad de Lev≥n | SeqCOVID-SPAIN consortium/IBV(CSIC) | Ana Carvajal et al |
| EPI_ISL_537637 | Spain/CL-IBV-97006265/2020 | 2020-04-21 | Europe/Spain/Castilla y Leon/Astorga | Universidad de Lev≥n | SeqCOVID-SPAIN consortium/IBV(CSIC) | Ana Carvajal et al |
| EPI_ISL_537638 | Spain/CL-IBV-97006266/2020 | 2020-04-21 | Europe/Spain/Castilla y Leon/Astorga | Universidad de Lev≥n | SeqCOVID-SPAIN consortium/IBV(CSIC) | Ana Carvajal et al |
| EPI_ISL_537643 | Spain/CL-IBV-97006272/2020 | 2020-04-21 | Europe/Spain/Castilla y Leon/Astorga | Universidad de Lev≥n | SeqCOVID-SPAIN consortium/IBV(CSIC) | Ana Carvajal et al |
| EPI_ISL_537644 | Spain/CL-IBV-97006273/2020 | 2020-04-21 | Europe/Spain/Castilla y Leon/La Carrera | Universidad de Lev≥n | SeqCOVID-SPAIN consortium/IBV(CSIC) | Ana Carvajal et al |

| | | | | | | |
|---|---|---|---|---|---|---|
| EPI_ISL_537646 | Spain/CL-IBV-97006275/2020 | 2020-04-21 | Europe/Spain/Castilla y Leon/Castro de | Universidad de Lev≥n | SeqCOVID-SPAIN consortium/IBV(CSIC) | Ana Carvajal et al |
| EPI_ISL_537647 | Spain/CL-IBV-97006276/2020 | 2020-04-21 | Europe/Spain/Castilla y Leon/La Baneza | Universidad de Lev≥n | SeqCOVID-SPAIN consortium/IBV(CSIC) | Ana Carvajal et al |
| EPI_ISL_537651 | Spain/CL-IBV-97006280/2020 | 2020-04-21 | Europe/Spain/Castilla y Leon/Astorga | Universidad de Lev≥n | SeqCOVID-SPAIN consortium/IBV(CSIC) | Ana Carvajal et al |
| EPI_ISL_537652 | Spain/CL-IBV-97006282/2020 | 2020-04-21 | Europe/Spain/Castilla y Leon/Astorga | Universidad de Lev≥n | SeqCOVID-SPAIN consortium/IBV(CSIC) | Ana Carvajal et al |
| EPI_ISL_537654 | Spain/CL-IBV-97006284/2020 | 2020-04-21 | Europe/Spain/Castilla y Leon/Astorga | Universidad de Lev≥n | SeqCOVID-SPAIN consortium/IBV(CSIC) | Ana Carvajal et al |
| EPI_ISL_537660 | Spain/CL-IBV-97006290/2020 | 2020-04-21 | Europe/Spain/Castilla y Leon/Astorga | Universidad de Lev≥n | SeqCOVID-SPAIN consortium/IBV(CSIC) | Ana Carvajal et al |
| EPI_ISL_537665 | Spain/CL-IBV-97006295/2020 | 2020-04-21 | Europe/Spain/Castilla y Leon/La Baneza | Universidad de Lev≥n | SeqCOVID-SPAIN consortium/IBV(CSIC) | Ana Carvajal et al |
| EPI_ISL_537668 | Spain/CL-IBV-97006299/2020 | 2020-04-21 | Europe/Spain/Castilla y Leon/Bercianos | Universidad de Lev≥n | SeqCOVID-SPAIN consortium/IBV(CSIC) | Ana Carvajal et al |
| EPI_ISL_537669 | Spain/CL-IBV-97006300/2020 | 2020-04-21 | Europe/Spain/Castilla y Leon/Posada de | Universidad de Lev≥n | SeqCOVID-SPAIN consortium/IBV(CSIC) | Ana Carvajal et al |
| EPI_ISL_537682 | Spain/CL-IBV-97006326/2020 | 2020-05-08 | Europe/Spain/Castilla y Leon/La Baneza | Universidad de Lev≥n | SeqCOVID-SPAIN consortium/IBV(CSIC) | Ana Carvajal et al |
| EPI_ISL_537689 | Spain/CL-IBV-97006342/2020 | 2020-05-26 | Europe/Spain/Castilla y Leon/Astorga | Universidad de Lev≥n | SeqCOVID-SPAIN consortium/IBV(CSIC) | Ana Carvajal et al |
| EPI_ISL_537695 | Spain/CN-IBV-97006479/2020 | 2020-03-24 | Europe/Spain/Canary Islands/Las Palmas | Hospital Universitario de Gran Canaria Dr. Negr√≠n | SeqCOVID-SPAIN consortium/IBV(CSIC) | M. Carmen P√©rez Gonz√°lez et al |
| EPI_ISL_537708 | Spain/CN-IBV-97006498/2020 | 2020-03-27 | Europe/Spain/Canary Islands/Las Palmas | Hospital Universitario de Gran Canaria Dr. Negr√≠n | SeqCOVID-SPAIN consortium/IBV(CSIC) | M. Carmen P√©rez Gonz√°lez et al |

| | | | | | | |
|---|---|---|---|---|---|---|
| EPI_ISL_537709 | Spain/CN-IBV-97006499/2020 | 2020-03-27 | Europe/Spain/Canary Islands/Las Palmas | Hospital Universitario de Gran Canaria Dr. Negrv≠n | SeqCOVID-SPAIN consortium/IBV(CSIC) | M. Carmen Pv©rez Gonzv°lez et al |
| EPI_ISL_537713 | Spain/CN-IBV-97006505/2020 | 2020-03-29 | Europe/Spain/Canary Islands/Las Palmas | Hospital Universitario de Gran Canaria Dr. Negrv≠n | SeqCOVID-SPAIN consortium/IBV(CSIC) | M. Carmen Pv©rez Gonzv°lez et al |
| EPI_ISL_537727 | Spain/CN-IBV-97006524/2020 | 2020-04-12 | Europe/Spain/Canary Islands/Las Palmas | Hospital Universitario de Gran Canaria Dr. Negrv≠n | SeqCOVID-SPAIN consortium/IBV(CSIC) | M. Carmen Pv©rez Gonzv°lez et al |
| EPI_ISL_537728 | Spain/CN-IBV-97006525/2020 | 2020-06-18 | Europe/Spain/Canary Islands/Las Palmas | Hospital Universitario de Gran Canaria Dr. Negrv≠n | SeqCOVID-SPAIN consortium/IBV(CSIC) | M. Carmen Pv©rez Gonzv°lez et al |
| EPI_ISL_537752 | Spain/AR-IBV-98005704/2020 | 2020-06-25 | Europe/Spain/Aragon/Caspe | Servicio de Microbiologv≠a, Hospital Miguel Servet, Zaragoza | SeqCOVID-SPAIN consortium/IBV(CSIC) | Antonio Rezusta Lv≥pez et al |
| EPI_ISL_537777 | Spain/AR-IBV-98005765/2020 | 2020-04-30 | Europe/Spain/Aragon/Zaragoza | Servicio de Microbiologv≠a, Hospital Miguel Servet, Zaragoza | SeqCOVID-SPAIN consortium/IBV(CSIC) | Antonio Rezusta Lv≥pez et al |
| EPI_ISL_537787 | Spain/AR-IBV-98005794/2020 | 2020-05-24 | Europe/Spain/Aragon/Zaragoza | Servicio de Microbiologv≠a, Hospital Miguel Servet, Zaragoza | SeqCOVID-SPAIN consortium/IBV(CSIC) | Antonio Rezusta Lv≥pez et al |
| EPI_ISL_537794 | Spain/CN-IBV-98005809/2020 | 2020-03-20 | Europe/Spain/Canary Islands/Las Palmas | Hospital Universitario de Gran Canaria Dr. Negrv≠n | SeqCOVID-SPAIN consortium/IBV(CSIC) | M. Carmen Pv©rez Gonzv°lez et al |
| EPI_ISL_537798 | Spain/CN-IBV-98005820/2020 | 2020-03-21 | Europe/Spain/Canary Islands/Las Palmas | Hospital Universitario de Gran Canaria Dr. Negrv≠n | SeqCOVID-SPAIN consortium/IBV(CSIC) | M. Carmen Pv©rez Gonzv°lez et al |
| EPI_ISL_537807 | Spain/CN-IBV-98005835/2020 | 2020-03-24 | Europe/Spain/Canary Islands/Las Palmas | Hospital Universitario de Gran Canaria Dr. Negrv≠n | SeqCOVID-SPAIN consortium/IBV(CSIC) | M. Carmen Pv©rez Gonzv°lez et al |
| EPI_ISL_537810 | Spain/CB-IBV-98002761/2020 | 2020-03-22 | Europe/Spain/Cantabria/Zurita | Hospital Universitario Marquv©s de Valdecilla (Santander), Servicio de Microbiologv≠a | SeqCOVID-SPAIN consortium/IBV(CSIC) | Mv≥nica Gozalo Margv∫ello et al |
| EPI_ISL_537820 | Spain/RI-IBV-98002297/2020 | 2020-03-07 | Europe/Spain/La Rioja/Haro | Centro de Investigaciv≥n Biomv©dica de La Rioja - Hospital San Pedro Logrov±o | SeqCOVID-SPAIN consortium/IBV(CSIC) | Marv≠a de Toro et al |
| EPI_ISL_537834 | Spain/RI-IBV-98002312/2020 | 2020-03-11 | Europe/Spain/La Rioja/Logrono | Centro de Investigaciv≥n Biomv©dica de La Rioja - Hospital San Pedro Logrov±o | SeqCOVID-SPAIN consortium/IBV(CSIC) | Marv≠a de Toro et al |

| | | | | | | |
|---|---|---|---|---|---|---|
| EPI_ISL_537835 | Spain/RI-IBV-98002314/2020 | 2020-03-11 | Europe/Spain/La Rioja/Santo Domingo d | Centro de Investigaci√≥n Biom√©dica de La Rioja - Hospital San Pedro Logro√±o | SeqCOVID-SPAIN consortium/IBV(CSIC) | Mar√≠a de Toro et al |
| EPI_ISL_537839 | Spain/RI-IBV-98002320/2020 | 2020-03-20 | Europe/Spain/La Rioja/Ventas del Bano | Centro de Investigaci√≥n Biom√©dica de La Rioja - Hospital San Pedro Logro√±o | SeqCOVID-SPAIN consortium/IBV(CSIC) | Mar√≠a de Toro et al |
| EPI_ISL_537840 | Spain/RI-IBV-98002321/2020 | 2020-03-20 | Europe/Spain/La Rioja/Autol | Centro de Investigaci√≥n Biom√©dica de La Rioja - Hospital San Pedro Logro√±o | SeqCOVID-SPAIN consortium/IBV(CSIC) | Mar√≠a de Toro et al |
| EPI_ISL_537846 | Spain/RI-IBV-98002330/2020 | 2020-03-24 | Europe/Spain/La Rioja/Calahorra | Centro de Investigaci√≥n Biom√©dica de La Rioja - Hospital San Pedro Logro√±o | SeqCOVID-SPAIN consortium/IBV(CSIC) | Mar√≠a de Toro et al |
| EPI_ISL_537854 | Spain/RI-IBV-98002341/2020 | 2020-03-28 | Europe/Spain/La Rioja/Logrono | Centro de Investigaci√≥n Biom√©dica de La Rioja - Hospital San Pedro Logro√±o | SeqCOVID-SPAIN consortium/IBV(CSIC) | Mar√≠a de Toro et al |
| EPI_ISL_537866 | Spain/RI-IBV-98002370/2020 | 2020-03-20 | Europe/Spain/La Rioja/Haro | Centro de Investigaci√≥n Biom√©dica de La Rioja - Hospital San Pedro Logro√±o | SeqCOVID-SPAIN consortium/IBV(CSIC) | Mar√≠a de Toro et al |
| EPI_ISL_537874 | Spain/PV-IBV-98001430/2020 | 2020-03-17 | Europe/Spain/Basque Country/Vitoria-G | Hospital Universitario Araba. Vitoria-Gasteiz | SeqCOVID-SPAIN consortium/IBV(CSIC) | Silvia Hern√°ez Crespo et al |
| EPI_ISL_537950 | Spain/VC-IBV-98003505/2020 | 2020-03-23 | Europe/Spain/Comunitat Valenciana/Ca | Servicio de Microbiolog√≠a. Hospital General Universitario de Castell√≥n | SeqCOVID-SPAIN consortium/IBV(CSIC) | Rosario Moreno et al |
| EPI_ISL_537952 | Spain/VC-IBV-98003509/2020 | 2020-03-26 | Europe/Spain/Comunitat Valenciana/Ca | Servicio de Microbiolog√≠a. Hospital General Universitario de Castell√≥n | SeqCOVID-SPAIN consortium/IBV(CSIC) | Rosario Moreno et al |
| EPI_ISL_537963 | Spain/VC-IBV-98003531/2020 | 2020-03-28 | Europe/Spain/Comunitat Valenciana/Ca | Servicio de Microbiolog√≠a. Hospital General Universitario de Castell√≥n | SeqCOVID-SPAIN consortium/IBV(CSIC) | Rosario Moreno et al |
| EPI_ISL_537965 | Spain/VC-IBV-98003538/2020 | 2020-03-28 | Europe/Spain/Comunitat Valenciana/Ca | Servicio de Microbiolog√≠a. Hospital General Universitario de Castell√≥n | SeqCOVID-SPAIN consortium/IBV(CSIC) | Rosario Moreno et al |
| EPI_ISL_537976 | Spain/VC-IBV-98003554/2020 | 2020-03-30 | Europe/Spain/Comunitat Valenciana/Ca | Servicio de Microbiolog√≠a. Hospital General Universitario de Castell√≥n | SeqCOVID-SPAIN consortium/IBV(CSIC) | Rosario Moreno et al |
| EPI_ISL_537977 | Spain/VC-IBV-98003555/2020 | 2020-03-30 | Europe/Spain/Comunitat Valenciana/Ca | Servicio de Microbiolog√≠a. Hospital General Universitario de Castell√≥n | SeqCOVID-SPAIN consortium/IBV(CSIC) | Rosario Moreno et al |

| | | | | | | |
|---|---|---|---|---|---|---|
| EPI_ISL_537978 | Spain/VC-IBV-98003556/2020 | 2020-03-30 | Europe/Spain/Comunitat Valenciana/Ca | Servicio de Microbiolog√≠a. Hospital General Universitario de Castell√≥n | SeqCOVID-SPAIN consortium/IBV(CSIC) | Rosario Moreno et al |
| EPI_ISL_537982 | Spain/VC-IBV-98003565/2020 | 2020-03-14 | Europe/Spain/Comunitat Valenciana/Ca | Servicio de Microbiolog√≠a. Hospital General Universitario de Castell√≥n | SeqCOVID-SPAIN consortium/IBV(CSIC) | Rosario Moreno et al |
| EPI_ISL_537993 | Spain/VC-IBV-98003580/2020 | 2020-03-30 | Europe/Spain/Comunitat Valenciana/Ca | Servicio de Microbiolog√≠a. Hospital General Universitario de Castell√≥n | SeqCOVID-SPAIN consortium/IBV(CSIC) | Rosario Moreno et al |
| EPI_ISL_537994 | Spain/VC-IBV-98003581/2020 | 2020-03-31 | Europe/Spain/Comunitat Valenciana/Ca | Servicio de Microbiolog√≠a. Hospital General Universitario de Castell√≥n | SeqCOVID-SPAIN consortium/IBV(CSIC) | Rosario Moreno et al |
| EPI_ISL_538002 | Spain/VC-IBV-98002665/2020 | 2020-04-24 | Europe/Spain/Comunitat Valenciana/Va | Servicio de Microbiologia. Hospital Arnau de Vilanova | SeqCOVID-SPAIN consortium/IBV(CSIC) | Victoria Dominguez et al |
| EPI_ISL_538003 | Spain/VC-IBV-98003062/2020 | 2020-03-13 | Europe/Spain/Comunitat Valenciana/Va | Hospital Universitari i Polit√®cnic La Fe de Val√®ncia | SeqCOVID-SPAIN consortium/IBV(CSIC) | Mar√≠a Dolores G√≥mez Ruiz et al |
| EPI_ISL_538005 | Spain/VC-IBV-98003065/2020 | 2020-03-13 | Europe/Spain/Comunitat Valenciana/Va | Hospital Universitari i Polit√®cnic La Fe de Val√®ncia | SeqCOVID-SPAIN consortium/IBV(CSIC) | Mar√≠a Dolores G√≥mez Ruiz et al |
| EPI_ISL_538006 | Spain/VC-IBV-98003066/2020 | 2020-03-13 | Europe/Spain/Comunitat Valenciana/Va | Hospital Universitari i Polit√®cnic La Fe de Val√®ncia | SeqCOVID-SPAIN consortium/IBV(CSIC) | Mar√≠a Dolores G√≥mez Ruiz et al |
| EPI_ISL_538017 | Spain/VC-IBV-98003081/2020 | 2020-03-14 | Europe/Spain/Comunitat Valenciana/Va | Hospital Universitari i Polit√®cnic La Fe de Val√®ncia | SeqCOVID-SPAIN consortium/IBV(CSIC) | Mar√≠a Dolores G√≥mez Ruiz et al |
| EPI_ISL_538026 | Spain/VC-IBV-98003090/2020 | 2020-03-14 | Europe/Spain/Comunitat Valenciana/Va | Hospital Universitari i Polit√®cnic La Fe de Val√®ncia | SeqCOVID-SPAIN consortium/IBV(CSIC) | Mar√≠a Dolores G√≥mez Ruiz et al |
| EPI_ISL_538028 | Spain/VC-IBV-98003092/2020 | 2020-03-14 | Europe/Spain/Comunitat Valenciana/Va | Hospital Universitari i Polit√®cnic La Fe de Val√®ncia | SeqCOVID-SPAIN consortium/IBV(CSIC) | Mar√≠a Dolores G√≥mez Ruiz et al |
| EPI_ISL_538030 | Spain/VC-IBV-98003094/2020 | 2020-03-14 | Europe/Spain/Comunitat Valenciana/Va | Hospital Universitari i Polit√®cnic La Fe de Val√®ncia | SeqCOVID-SPAIN consortium/IBV(CSIC) | Mar√≠a Dolores G√≥mez Ruiz et al |
| EPI_ISL_538031 | Spain/VC-IBV-98003095/2020 | 2020-03-14 | Europe/Spain/Comunitat Valenciana/Va | Hospital Universitari i Polit√®cnic La Fe de Val√®ncia | SeqCOVID-SPAIN consortium/IBV(CSIC) | Mar√≠a Dolores G√≥mez Ruiz et al |

| | | | | | | |
|---|---|---|---|---|---|---|
| EPI_ISL_538033 | Spain/VC-IBV-98003097/2020 | 2020-03-15 | Europe/Spain/Comunitat Valenciana/Va | Hospital Universitari i Politv®cnic La Fe de Valv®ncia | SeqCOVID-SPAIN consortium/IBV(CSIC) | Marv≠a Dolores Gv≥mez Ruiz et al |
| EPI_ISL_538039 | Spain/VC-IBV-98003103/2020 | 2020-03-15 | Europe/Spain/Comunitat Valenciana/Va | Hospital Universitari i Politv®cnic La Fe de Valv®ncia | SeqCOVID-SPAIN consortium/IBV(CSIC) | Marv≠a Dolores Gv≥mez Ruiz et al |
| EPI_ISL_538040 | Spain/VC-IBV-98003104/2020 | 2020-03-15 | Europe/Spain/Comunitat Valenciana/Va | Hospital Universitari i Politv®cnic La Fe de Valv®ncia | SeqCOVID-SPAIN consortium/IBV(CSIC) | Marv≠a Dolores Gv≥mez Ruiz et al |
| EPI_ISL_538050 | Spain/VC-IBV-98003114/2020 | 2020-03-16 | Europe/Spain/Comunitat Valenciana/Va | Hospital Universitari i Politv®cnic La Fe de Valv®ncia | SeqCOVID-SPAIN consortium/IBV(CSIC) | Marv≠a Dolores Gv≥mez Ruiz et al |
| EPI_ISL_538052 | Spain/VC-IBV-98003116/2020 | 2020-03-16 | Europe/Spain/Comunitat Valenciana/Va | Hospital Universitari i Politv®cnic La Fe de Valv®ncia | SeqCOVID-SPAIN consortium/IBV(CSIC) | Marv≠a Dolores Gv≥mez Ruiz et al |
| EPI_ISL_538053 | Spain/VC-IBV-98003117/2020 | 2020-03-16 | Europe/Spain/Comunitat Valenciana/Va | Hospital Universitari i Politv®cnic La Fe de Valv®ncia | SeqCOVID-SPAIN consortium/IBV(CSIC) | Marv≠a Dolores Gv≥mez Ruiz et al |
| EPI_ISL_538079 | Spain/PV-IBV-98001040/2020 | 2020-04-01 | Europe/Spain/Basque Country/Donostia | Servicio de Microbiologv≠a. Hospital Universitario Donostia. OSI Donostialdea. vÅrea de Enfermedades Infecciosas, Grupo de Infecciv≥n Respiratoria y Resistencia Antimicrobiana. Instituto de Investigaciv≥n Sanitaria Biodonostia | SeqCOVID-SPAIN consortium/IBV(CSIC) | Gustavo Cilla et al |
| EPI_ISL_538095 | Spain/NC-IBV-98005405/2020 | 2020-03-25 | Europe/Spain/Navarra/Huarte | Clv≠nica Universidad de Navarra. Servicio de Enfermedades Infecciosas y Microbiologv≠a clv≠nica | SeqCOVID-SPAIN consortium/IBV(CSIC) | Mirian Fernv°ndez-Alonso et al |
| EPI_ISL_538097 | Spain/NC-IBV-98005411/2020 | 2020-03-25 | Europe/Spain/Navarra/Pamplona | Clv≠nica Universidad de Navarra. Servicio de Enfermedades Infecciosas y Microbiologv≠a clv≠nica | SeqCOVID-SPAIN consortium/IBV(CSIC) | Mirian Fernv°ndez-Alonso et al |
| EPI_ISL_538106 | Spain/NC-IBV-98005427/2020 | 2020-03-26 | Europe/Spain/Navarra/Villava | Clv≠nica Universidad de Navarra. Servicio de Enfermedades Infecciosas y Microbiologv≠a clv≠nica | SeqCOVID-SPAIN consortium/IBV(CSIC) | Mirian Fernv°ndez-Alonso et al |
| EPI_ISL_538111 | Spain/NC-IBV-98005449/2020 | 2020-03-27 | Europe/Spain/Navarra/Pamplona | Clv≠nica Universidad de Navarra. Servicio de Enfermedades Infecciosas y Microbiologv≠a clv≠nica | SeqCOVID-SPAIN consortium/IBV(CSIC) | Mirian Fernv°ndez-Alonso et al |
| EPI_ISL_538112 | Spain/NC-IBV-98005451/2020 | 2020-03-27 | Europe/Spain/Navarra/Artajona | Clv≠nica Universidad de Navarra. Servicio de Enfermedades Infecciosas y Microbiologv≠a clv≠nica | SeqCOVID-SPAIN consortium/IBV(CSIC) | Mirian Fernv°ndez-Alonso et al |

| | | | | | | |
|---|---|---|---|---|---|---|
| EPI_ISL_538120 | Spain/AN-IBV-98001638/2020 | 2020-04-07 | Europe/Spain/Andalusia/Sevilla | Servicio de Microbiolog√≠a y Parasitolog√≠a cl√≠nica. UCEIMP. Hospital Universitario Virgen del Roc√≠o/IBIS/CSIC/US | SeqCOVID-SPAIN consortium/IBV(CSIC) | Guillermo Mart√≠n Guti√©rrez et al |
| EPI_ISL_538133 | Spain/AN-IBV-98001705/2020 | 2020-04-13 | Europe/Spain/Andalusia/Sevilla | Servicio de Microbiolog√≠a y Parasitolog√≠a cl√≠nica. UCEIMP. Hospital Universitario Virgen del Roc√≠o/IBIS/CSIC/US | SeqCOVID-SPAIN consortium/IBV(CSIC) | Guillermo Mart√≠n Guti√©rrez et al |
| EPI_ISL_538138 | Spain/AN-IBV-98001723/2020 | 2020-04-05 | Europe/Spain/Andalusia/Sevilla | Servicio de Microbiolog√≠a y Parasitolog√≠a cl√≠nica. UCEIMP. Hospital Universitario Virgen del Roc√≠o/IBIS/CSIC/US | SeqCOVID-SPAIN consortium/IBV(CSIC) | Guillermo Mart√≠n Guti√©rrez et al |
| EPI_ISL_538153 | Spain/AN-IBV-98001780/2020 | 2020-04-04 | Europe/Spain/Andalusia/Sevilla | Servicio de Microbiolog√≠a y Parasitolog√≠a cl√≠nica. UCEIMP. Hospital Universitario Virgen del Roc√≠o/IBIS/CSIC/US | SeqCOVID-SPAIN consortium/IBV(CSIC) | Guillermo Mart√≠n Guti√©rrez et al |
| EPI_ISL_538154 | Spain/AN-IBV-98001782/2020 | 2020-04-04 | Europe/Spain/Andalusia/Sevilla | Servicio de Microbiolog√≠a y Parasitolog√≠a cl√≠nica. UCEIMP. Hospital Universitario Virgen del Roc√≠o/IBIS/CSIC/US | SeqCOVID-SPAIN consortium/IBV(CSIC) | Guillermo Mart√≠n Guti√©rrez et al |
| EPI_ISL_538162 | Spain/AN-IBV-98001806/2020 | 2020-04-03 | Europe/Spain/Andalusia/Sevilla | Servicio de Microbiolog√≠a y Parasitolog√≠a cl√≠nica. UCEIMP. Hospital Universitario Virgen del Roc√≠o/IBIS/CSIC/US | SeqCOVID-SPAIN consortium/IBV(CSIC) | Guillermo Mart√≠n Guti√©rrez et al |
| EPI_ISL_538187 | Spain/AN-IBV-98001135/2020 | 2020-03-27 | Europe/Spain/Andalusia/Sevilla | Servicio de Microbiolog√≠a y Parasitolog√≠a cl√≠nica. UCEIMP. Hospital Universitario Virgen del Roc√≠o/IBIS/CSIC/US | SeqCOVID-SPAIN consortium/IBV(CSIC) | Guillermo Mart√≠n Guti√©rrez et al |
| EPI_ISL_538188 | Spain/AN-IBV-98001138/2020 | 2020-03-27 | Europe/Spain/Andalusia/Sevilla | Servicio de Microbiolog√≠a y Parasitolog√≠a cl√≠nica. UCEIMP. Hospital Universitario Virgen del Roc√≠o/IBIS/CSIC/US | SeqCOVID-SPAIN consortium/IBV(CSIC) | Guillermo Mart√≠n Guti√©rrez et al |
| EPI_ISL_538197 | Spain/AN-IBV-98001185/2020 | 2020-03-26 | Europe/Spain/Andalusia/Sevilla | Servicio de Microbiolog√≠a y Parasitolog√≠a cl√≠nica. UCEIMP. Hospital Universitario Virgen del Roc√≠o/IBIS/CSIC/US | SeqCOVID-SPAIN consortium/IBV(CSIC) | Guillermo Mart√≠n Guti√©rrez et al |
| EPI_ISL_538198 | Spain/AN-IBV-98001186/2020 | 2020-03-26 | Europe/Spain/Andalusia/Sevilla | Servicio de Microbiolog√≠a y Parasitolog√≠a cl√≠nica. UCEIMP. Hospital Universitario Virgen del Roc√≠o/IBIS/CSIC/US | SeqCOVID-SPAIN consortium/IBV(CSIC) | Guillermo Mart√≠n Guti√©rrez et al |

| | | | | | | |
|---|---|---|---|---|---|---|
| EPI_ISL_538206 | Spain/AN-IBV-98001203/2020 | 2020-03-25 | Europe/Spain/Andalusia/Sevilla | Servicio de Microbiolog√≠a y Parasitolog√≠a cl√≠nica. UCEIMP. Hospital Universitario Virgen del Roc√≠o/IBIS/CSIC/US | SeqCOVID-SPAIN consortium/IBV(CSIC) | Guillermo Mart√≠n Guti√©rrez et al |
| EPI_ISL_538211 | Spain/AN-IBV-98001266/2020 | 2020-03-25 | Europe/Spain/Andalusia/Sevilla | Servicio de Microbiolog√≠a y Parasitolog√≠a cl√≠nica. UCEIMP. Hospital Universitario Virgen del Roc√≠o/IBIS/CSIC/US | SeqCOVID-SPAIN consortium/IBV(CSIC) | Guillermo Mart√≠n Guti√©rrez et al |
| EPI_ISL_538213 | Spain/AN-IBV-98001268/2020 | 2020-03-25 | Europe/Spain/Andalusia/Sevilla | Servicio de Microbiolog√≠a y Parasitolog√≠a cl√≠nica. UCEIMP. Hospital Universitario Virgen del Roc√≠o/IBIS/CSIC/US | SeqCOVID-SPAIN consortium/IBV(CSIC) | Guillermo Mart√≠n Guti√©rrez et al |
| EPI_ISL_538223 | Spain/AN-IBV-98001286/2020 | 2020-03-24 | Europe/Spain/Andalusia/Sevilla | Servicio de Microbiolog√≠a y Parasitolog√≠a cl√≠nica. UCEIMP. Hospital Universitario Virgen del Roc√≠o/IBIS/CSIC/US | SeqCOVID-SPAIN consortium/IBV(CSIC) | Guillermo Mart√≠n Guti√©rrez et al |
| EPI_ISL_538224 | Spain/AN-IBV-98001287/2020 | 2020-03-24 | Europe/Spain/Andalusia/Sevilla | Servicio de Microbiolog√≠a y Parasitolog√≠a cl√≠nica. UCEIMP. Hospital Universitario Virgen del Roc√≠o/IBIS/CSIC/US | SeqCOVID-SPAIN consortium/IBV(CSIC) | Guillermo Mart√≠n Guti√©rrez et al |
| EPI_ISL_538225 | Spain/AN-IBV-98001288/2020 | 2020-03-24 | Europe/Spain/Andalusia/Sevilla | Servicio de Microbiolog√≠a y Parasitolog√≠a cl√≠nica. UCEIMP. Hospital Universitario Virgen del Roc√≠o/IBIS/CSIC/US | SeqCOVID-SPAIN consortium/IBV(CSIC) | Guillermo Mart√≠n Guti√©rrez et al |
| EPI_ISL_538229 | Spain/AN-IBV-98001333/2020 | 2020-03-23 | Europe/Spain/Andalusia/Sevilla | Servicio de Microbiolog√≠a y Parasitolog√≠a cl√≠nica. UCEIMP. Hospital Universitario Virgen del Roc√≠o/IBIS/CSIC/US | SeqCOVID-SPAIN consortium/IBV(CSIC) | Guillermo Mart√≠n Guti√©rrez et al |
| EPI_ISL_538233 | USA/NM-UNM-00407/2020 | 2020-08-15 | North America/USA/New Mexico/ | TriCore Reference Laboratories | Center for Global Health, University of New Mexico Health Sciences Center | Daryl Domman et al |
| EPI_ISL_538247 | USA/NM-UNM-00439/2020 | 2020-05-04 | North America/USA/New Mexico/ | TriCore Reference Laboratories | Center for Global Health, University of New Mexico Health Sciences Center | Daryl Domman et al |

| | | | | | | |
|---|---|---|---|---|---|---|
| EPI_ISL_538254 | USA/NM-UNM-00469/2020 | 2020-05-08 | North America/USA/New Mexico/ | TriCore Reference Laboratories | Center for Global Health, University of New Mexico Health Sciences Center | Daryl Domman et al |
| EPI_ISL_538263 | USA/NM-UNM-00497/2020 | 2020-07-24 | North America/USA/New Mexico/ | TriCore Reference Laboratories | Center for Global Health, University of New Mexico Health Sciences Center | Daryl Domman et al |
| EPI_ISL_538277 | USA/TX-DSHS-0339/2020 | 2020-06-13 | North America/USA/Texas/Harris Count | Texas Department of State Health Services | Texas Department of State Health Services | Bonnie Oh et al |
| EPI_ISL_538282 | USA/TX-DSHS-0344/2020 | 2020-06-13 | North America/USA/Texas/Harris Count | Texas Department of State Health Services | Texas Department of State Health Services | Bonnie Oh et al |
| EPI_ISL_538286 | USA/TX-DSHS-0348/2020 | 2020-06-13 | North America/USA/Texas/Harris Count | Texas Department of State Health Services | Texas Department of State Health Services | Bonnie Oh et al |
| EPI_ISL_538298 | USA/TX-DSHS-0361/2020 | 2020-06-13 | North America/USA/Texas/Harris Count | Texas Department of State Health Services | Texas Department of State Health Services | Bonnie Oh et al |
| EPI_ISL_538309 | USA/TX-DSHS-0373/2020 | 2020-06-14 | North America/USA/Texas/El Paso Coun | Texas Department of State Health Services | Texas Department of State Health Services | Bonnie Oh et al |
| EPI_ISL_538313 | USA/TX-DSHS-0377/2020 | 2020-06-14 | North America/USA/Texas/El Paso Coun | Texas Department of State Health Services | Texas Department of State Health Services | Bonnie Oh et al |
| EPI_ISL_538315 | USA/TX-DSHS-0379/2020 | 2020-06-14 | North America/USA/Texas/El Paso Coun | Texas Department of State Health Services | Texas Department of State Health Services | Bonnie Oh et al |
| EPI_ISL_538321 | Canada/ON-OICR-SLB2787/2020 | 2020-03-25 | North America/Canada/Ontario/ | Kingston Health Sciences Centre / Queen's University | Ontario Institute for Cancer Research | Prameet M. Sheth et al |
| EPI_ISL_538322 | Canada/ON-OICR-SLB2788/2020 | 2020-03-25 | North America/Canada/Ontario/ | Kingston Health Sciences Centre / Queen's University | Ontario Institute for Cancer Research | Prameet M. Sheth et al |
| EPI_ISL_538323 | Canada/ON-OICR-SLB2789/2020 | 2020-03-25 | North America/Canada/Ontario/ | Kingston Health Sciences Centre / Queen's University | Ontario Institute for Cancer Research | Prameet M. Sheth et al |

| | | | | | | |
|---|---|---|---|---|---|---|
| EPI_ISL_538324 | Canada/ON-OICR-SLB2790/2020 | 2020-03-25 | North America/Canada/Ontario/ | Kingston Health Sciences Centre / Queen's University | Ontario Institute for Cancer Research | Prameet M. Sheth et al |
| EPI_ISL_538325 | Canada/ON-OICR-SLB2791/2020 | 2020-03-25 | North America/Canada/Ontario/ | Kingston Health Sciences Centre / Queen's University | Ontario Institute for Cancer Research | Prameet M. Sheth et al |
| EPI_ISL_538326 | Canada/ON-OICR-SLB2792/2020 | 2020-03-26 | North America/Canada/Ontario/ | Kingston Health Sciences Centre / Queen's University | Ontario Institute for Cancer Research | Prameet M. Sheth et al |
| EPI_ISL_538327 | Canada/ON-OICR-SLB2793/2020 | 2020-03-27 | North America/Canada/Ontario/ | Kingston Health Sciences Centre / Queen's University | Ontario Institute for Cancer Research | Prameet M. Sheth et al |
| EPI_ISL_538328 | Canada/ON-OICR-SLB2796/2020 | 2020-03-28 | North America/Canada/Ontario/ | Kingston Health Sciences Centre / Queen's University | Ontario Institute for Cancer Research | Prameet M. Sheth et al |
| EPI_ISL_538329 | Canada/ON-OICR-SLB2797/2020 | 2020-03-28 | North America/Canada/Ontario/ | Kingston Health Sciences Centre / Queen's University | Ontario Institute for Cancer Research | Prameet M. Sheth et al |
| EPI_ISL_538330 | Canada/ON-OICR-SLB2798/2020 | 2020-03-28 | North America/Canada/Ontario/ | Kingston Health Sciences Centre / Queen's University | Ontario Institute for Cancer Research | Prameet M. Sheth et al |
| EPI_ISL_538331 | Canada/ON-OICR-SLB2799/2020 | 2020-03-28 | North America/Canada/Ontario/ | Kingston Health Sciences Centre / Queen's University | Ontario Institute for Cancer Research | Prameet M. Sheth et al |
| EPI_ISL_538332 | Canada/ON-OICR-SLB2800/2020 | 2020-03-28 | North America/Canada/Ontario/ | Kingston Health Sciences Centre / Queen's University | Ontario Institute for Cancer Research | Prameet M. Sheth et al |
| EPI_ISL_538333 | Canada/ON-OICR-SLB2802/2020 | 2020-03-30 | North America/Canada/Ontario/ | Kingston Health Sciences Centre / Queen's University | Ontario Institute for Cancer Research | Prameet M. Sheth et al |
| EPI_ISL_538334 | Canada/ON-OICR-SLB2803/2020 | 2020-03-30 | North America/Canada/Ontario/ | Kingston Health Sciences Centre / Queen's University | Ontario Institute for Cancer Research | Prameet M. Sheth et al |
| EPI_ISL_538335 | Canada/ON-OICR-SLB2804/2020 | 2020-03-30 | North America/Canada/Ontario/ | Kingston Health Sciences Centre / Queen's University | Ontario Institute for Cancer Research | Prameet M. Sheth et al |
| EPI_ISL_538336 | Canada/ON-OICR-SLB2806/2020 | 2020-03-31 | North America/Canada/Ontario/ | Kingston Health Sciences Centre / Queen's University | Ontario Institute for Cancer Research | Prameet M. Sheth et al |

| | | | | | | |
|---|---|---|---|---|---|---|
| EPI_ISL_538337 | Canada/ON-OICR-SLB2807/2020 | 2020-03-31 | North America/Canada/Ontario/ | Kingston Health Sciences Centre / Queen's University | Ontario Institute for Cancer Research | Prameet M. Sheth et al |
| EPI_ISL_538338 | Canada/ON-OICR-SLB2809/2020 | 2020-03-31 | North America/Canada/Ontario/ | Kingston Health Sciences Centre / Queen's University | Ontario Institute for Cancer Research | Prameet M. Sheth et al |
| EPI_ISL_538339 | Canada/ON-OICR-SLB2811/2020 | 2020-03-31 | North America/Canada/Ontario/ | Kingston Health Sciences Centre / Queen's University | Ontario Institute for Cancer Research | Prameet M. Sheth et al |
| EPI_ISL_538340 | Canada/ON-OICR-SLB2812/2020 | 2020-04-01 | North America/Canada/Ontario/ | Kingston Health Sciences Centre / Queen's University | Ontario Institute for Cancer Research | Prameet M. Sheth et al |
| EPI_ISL_538341 | Canada/ON-OICR-SLB2813/2020 | 2020-04-01 | North America/Canada/Ontario/ | Kingston Health Sciences Centre / Queen's University | Ontario Institute for Cancer Research | Prameet M. Sheth et al |
| EPI_ISL_538342 | Canada/ON-OICR-SLB2814/2020 | 2020-04-01 | North America/Canada/Ontario/ | Kingston Health Sciences Centre / Queen's University | Ontario Institute for Cancer Research | Prameet M. Sheth et al |
| EPI_ISL_538343 | Canada/ON-OICR-SLB2815/2020 | 2020-04-01 | North America/Canada/Ontario/ | Kingston Health Sciences Centre / Queen's University | Ontario Institute for Cancer Research | Prameet M. Sheth et al |
| EPI_ISL_538344 | Canada/ON-OICR-SLB2816/2020 | 2020-04-02 | North America/Canada/Ontario/ | Kingston Health Sciences Centre / Queen's University | Ontario Institute for Cancer Research | Prameet M. Sheth et al |
| EPI_ISL_538345 | Canada/ON-OICR-SLB2817/2020 | 2020-04-02 | North America/Canada/Ontario/ | Kingston Health Sciences Centre / Queen's University | Ontario Institute for Cancer Research | Prameet M. Sheth et al |
| EPI_ISL_538346 | Canada/ON-OICR-SLB2818/2020 | 2020-04-02 | North America/Canada/Ontario/ | Kingston Health Sciences Centre / Queen's University | Ontario Institute for Cancer Research | Prameet M. Sheth et al |
| EPI_ISL_538347 | Canada/ON-OICR-SLB2819/2020 | 2020-04-02 | North America/Canada/Ontario/ | Kingston Health Sciences Centre / Queen's University | Ontario Institute for Cancer Research | Prameet M. Sheth et al |
| EPI_ISL_538348 | Canada/ON-OICR-SLB2820/2020 | 2020-04-02 | North America/Canada/Ontario/ | Kingston Health Sciences Centre / Queen's University | Ontario Institute for Cancer Research | Prameet M. Sheth et al |
| EPI_ISL_538349 | Canada/ON-OICR-SLB2821/2020 | 2020-04-02 | North America/Canada/Ontario/ | Kingston Health Sciences Centre / Queen's University | Ontario Institute for Cancer Research | Prameet M. Sheth et al |

| | | | | | | |
|---|---|---|---|---|---|---|
| EPI_ISL_538350 | Canada/ON-OICR-SLB2822/2020 | 2020-04-02 | North America/Canada/Ontario/ | Kingston Health Sciences Centre / Queen's University | Ontario Institute for Cancer Research | Prameet M. Sheth et al |
| EPI_ISL_538351 | Canada/ON-OICR-SLB2823/2020 | 2020-04-02 | North America/Canada/Ontario/ | Kingston Health Sciences Centre / Queen's University | Ontario Institute for Cancer Research | Prameet M. Sheth et al |
| EPI_ISL_538352 | Canada/ON-OICR-SLB2825/2020 | 2020-04-02 | North America/Canada/Ontario/ | Kingston Health Sciences Centre / Queen's University | Ontario Institute for Cancer Research | Prameet M. Sheth et al |
| EPI_ISL_538353 | Canada/ON-OICR-SLB2827/2020 | 2020-04-02 | North America/Canada/Ontario/ | Kingston Health Sciences Centre / Queen's University | Ontario Institute for Cancer Research | Prameet M. Sheth et al |
| EPI_ISL_538354 | Canada/ON-OICR-SLB2828/2020 | 2020-04-02 | North America/Canada/Ontario/ | Kingston Health Sciences Centre / Queen's University | Ontario Institute for Cancer Research | Prameet M. Sheth et al |
| EPI_ISL_538355 | Canada/ON-OICR-SLB2829/2020 | 2020-04-03 | North America/Canada/Ontario/ | Kingston Health Sciences Centre / Queen's University | Ontario Institute for Cancer Research | Prameet M. Sheth et al |
| EPI_ISL_538356 | Canada/ON-OICR-SLB2830/2020 | 2020-04-03 | North America/Canada/Ontario/ | Kingston Health Sciences Centre / Queen's University | Ontario Institute for Cancer Research | Prameet M. Sheth et al |
| EPI_ISL_538357 | Canada/ON-OICR-SLB2831/2020 | 2020-04-03 | North America/Canada/Ontario/ | Kingston Health Sciences Centre / Queen's University | Ontario Institute for Cancer Research | Prameet M. Sheth et al |
| EPI_ISL_538358 | Canada/ON-OICR-SLB2832/2020 | 2020-04-03 | North America/Canada/Ontario/ | Kingston Health Sciences Centre / Queen's University | Ontario Institute for Cancer Research | Prameet M. Sheth et al |
| EPI_ISL_538359 | Canada/ON-OICR-SLB2835/2020 | 2020-04-03 | North America/Canada/Ontario/ | Kingston Health Sciences Centre / Queen's University | Ontario Institute for Cancer Research | Prameet M. Sheth et al |
| EPI_ISL_538360 | Canada/ON-OICR-SLB2836/2020 | 2020-04-03 | North America/Canada/Ontario/ | Kingston Health Sciences Centre / Queen's University | Ontario Institute for Cancer Research | Prameet M. Sheth et al |
| EPI_ISL_538361 | Canada/ON-OICR-SLB2838/2020 | 2020-04-03 | North America/Canada/Ontario/ | Kingston Health Sciences Centre / Queen's University | Ontario Institute for Cancer Research | Prameet M. Sheth et al |
| EPI_ISL_538362 | Canada/ON-OICR-SLB2841/2020 | 2020-04-03 | North America/Canada/Ontario/ | Kingston Health Sciences Centre / Queen's University | Ontario Institute for Cancer Research | Prameet M. Sheth et al |

| | | | | | | |
|---|---|---|---|---|---|---|
| EPI_ISL_538363 | Canada/ON-OICR-SLB2842/2020 | 2020-04-04 | North America/Canada/Ontario/ | Kingston Health Sciences Centre / Queen's University | Ontario Institute for Cancer Research | Prameet M. Sheth et al |
| EPI_ISL_538364 | Canada/ON-OICR-SLB2843/2020 | 2020-04-04 | North America/Canada/Ontario/ | Kingston Health Sciences Centre / Queen's University | Ontario Institute for Cancer Research | Prameet M. Sheth et al |
| EPI_ISL_538365 | Canada/ON-OICR-SLB2844/2020 | 2020-04-03 | North America/Canada/Ontario/ | Kingston Health Sciences Centre / Queen's University | Ontario Institute for Cancer Research | Prameet M. Sheth et al |
| EPI_ISL_538366 | Canada/ON-OICR-SLB2846/2020 | 2020-04-04 | North America/Canada/Ontario/ | Kingston Health Sciences Centre / Queen's University | Ontario Institute for Cancer Research | Prameet M. Sheth et al |
| EPI_ISL_538367 | Canada/ON-OICR-SLB2847/2020 | 2020-04-04 | North America/Canada/Ontario/ | Kingston Health Sciences Centre / Queen's University | Ontario Institute for Cancer Research | Prameet M. Sheth et al |
| EPI_ISL_538368 | Canada/ON-OICR-SLB2848/2020 | 2020-04-04 | North America/Canada/Ontario/ | Kingston Health Sciences Centre / Queen's University | Ontario Institute for Cancer Research | Prameet M. Sheth et al |
| EPI_ISL_538369 | Canada/ON-OICR-SLB2849/2020 | 2020-04-05 | North America/Canada/Ontario/ | Kingston Health Sciences Centre / Queen's University | Ontario Institute for Cancer Research | Prameet M. Sheth et al |
| EPI_ISL_538370 | Canada/ON-OICR-SLB2851/2020 | 2020-04-05 | North America/Canada/Ontario/ | Kingston Health Sciences Centre / Queen's University | Ontario Institute for Cancer Research | Prameet M. Sheth et al |
| EPI_ISL_538371 | Canada/ON-OICR-SLB2852/2020 | 2020-04-07 | North America/Canada/Ontario/ | Kingston Health Sciences Centre / Queen's University | Ontario Institute for Cancer Research | Prameet M. Sheth et al |
| EPI_ISL_538372 | Canada/ON-OICR-SLB2853/2020 | 2020-04-07 | North America/Canada/Ontario/ | Kingston Health Sciences Centre / Queen's University | Ontario Institute for Cancer Research | Prameet M. Sheth et al |
| EPI_ISL_538373 | Canada/ON-OICR-SLB2854/2020 | 2020-04-07 | North America/Canada/Ontario/ | Kingston Health Sciences Centre / Queen's University | Ontario Institute for Cancer Research | Prameet M. Sheth et al |
| EPI_ISL_538374 | Canada/ON-OICR-SLB2855/2020 | 2020-04-07 | North America/Canada/Ontario/ | Kingston Health Sciences Centre / Queen's University | Ontario Institute for Cancer Research | Prameet M. Sheth et al |
| EPI_ISL_538375 | Canada/ON-OICR-SLB2856/2020 | 2020-04-07 | North America/Canada/Ontario/ | Kingston Health Sciences Centre / Queen's University | Ontario Institute for Cancer Research | Prameet M. Sheth et al |

| | | | | | | |
|---|---|---|---|---|---|---|
| EPI_ISL_538376 | Canada/ON-OICR-SLB2857/2020 | 2020-04-07 | North America/Canada/Ontario/ | Kingston Health Sciences Centre / Queen's University | Ontario Institute for Cancer Research | Prameet M. Sheth et al |
| EPI_ISL_538377 | Canada/ON-OICR-SLB2858/2020 | 2020-04-07 | North America/Canada/Ontario/ | Kingston Health Sciences Centre / Queen's University | Ontario Institute for Cancer Research | Prameet M. Sheth et al |
| EPI_ISL_538378 | Canada/ON-OICR-SLB2859/2020 | 2020-04-07 | North America/Canada/Ontario/ | Kingston Health Sciences Centre / Queen's University | Ontario Institute for Cancer Research | Prameet M. Sheth et al |
| EPI_ISL_538379 | Canada/ON-OICR-SLB2860/2020 | 2020-04-07 | North America/Canada/Ontario/ | Kingston Health Sciences Centre / Queen's University | Ontario Institute for Cancer Research | Prameet M. Sheth et al |
| EPI_ISL_538380 | Canada/ON-OICR-SLB2863/2020 | 2020-04-08 | North America/Canada/Ontario/ | Kingston Health Sciences Centre / Queen's University | Ontario Institute for Cancer Research | Prameet M. Sheth et al |
| EPI_ISL_538381 | Canada/ON-OICR-SLB2864/2020 | 2020-04-09 | North America/Canada/Ontario/ | Kingston Health Sciences Centre / Queen's University | Ontario Institute for Cancer Research | Prameet M. Sheth et al |
| EPI_ISL_538382 | Canada/ON-OICR-SLB2865/2020 | 2020-04-10 | North America/Canada/Ontario/ | Kingston Health Sciences Centre / Queen's University | Ontario Institute for Cancer Research | Prameet M. Sheth et al |
| EPI_ISL_538383 | Canada/ON-OICR-SLB2866/2020 | 2020-04-10 | North America/Canada/Ontario/ | Kingston Health Sciences Centre / Queen's University | Ontario Institute for Cancer Research | Prameet M. Sheth et al |
| EPI_ISL_538384 | Canada/ON-OICR-SLB2867/2020 | 2020-04-10 | North America/Canada/Ontario/ | Kingston Health Sciences Centre / Queen's University | Ontario Institute for Cancer Research | Prameet M. Sheth et al |
| EPI_ISL_538385 | Canada/ON-OICR-SLB2868/2020 | 2020-04-10 | North America/Canada/Ontario/ | Kingston Health Sciences Centre / Queen's University | Ontario Institute for Cancer Research | Prameet M. Sheth et al |
| EPI_ISL_538386 | Canada/ON-OICR-SLB2869/2020 | 2020-04-11 | North America/Canada/Ontario/ | Kingston Health Sciences Centre / Queen's University | Ontario Institute for Cancer Research | Prameet M. Sheth et al |
| EPI_ISL_538396 | USA/SC-DHEC-0777/2020 | 2020-06-26 | North America/USA/South Carolina/ | Microbiology Division, South Carolina Department of Health and Environmental Control | Microbiology Division, South Carolina Department of Health and Environmental Control | Flores et al |

| | | | | | | |
|---|---|---|---|---|---|---|
| EPI_ISL_538421 | USA/SC-DHEC-0930/2020 | 2020-06-26 | North America/USA/South Carolina/ | Microbiology Division, South Carolina Department of Health and Environmental Control | Microbiology Division, South Carolina Department of Health and Environmental Control | Flores et al |
| EPI_ISL_538425 | USA/SC-DHEC-0601/2020 | 2020-06-19 | North America/USA/South Carolina/ | Microbiology Division, South Carolina Department of Health and Environmental Control | Microbiology Division, South Carolina Department of Health and Environmental Control | Flores et al |
| EPI_ISL_538435 | Singapore/36nan/2020 | 2020-02-16 | Asia/Singapore/Singapore/ | Department of Laboratory Medicine, Tan Tock Seng Hospital | Department of Laboratory Medicine, Tan Tock Seng Hospital | Chen YYC et al |
| EPI_ISL_538436 | Singapore/913/2020 | 2020-02-20 | Asia/Singapore/Singapore/ | Department of Laboratory Medicine, Tan Tock Seng Hospital | Department of Laboratory Medicine, Tan Tock Seng Hospital | Chen YYC et al |
| EPI_ISL_538451 | Singapore/920/2020 | 2020-03-22 | Asia/Singapore/Singapore/ | Department of Laboratory Medicine, Tan Tock Seng Hospital | Department of Laboratory Medicine, Tan Tock Seng Hospital | Chen YYC et al |
| EPI_ISL_538471 | Singapore/564nan/2020 | 2020-04-02 | Asia/Singapore/Singapore/ | Department of Laboratory Medicine, Tan Tock Seng Hospital | Department of Laboratory Medicine, Tan Tock Seng Hospital | Chen YYC et al |
| EPI_ISL_538474 | Singapore/940/2020 | 2020-04-03 | Asia/Singapore/Singapore/ | Department of Laboratory Medicine, Tan Tock Seng Hospital | Department of Laboratory Medicine, Tan Tock Seng Hospital | Chen YYC et al |

| | | | | | | |
|---|---|---|---|---|---|---|
| EPI_ISL_538477 | Singapore/943/2020 | 2020-04-04 | Asia/Singapore/Singapore/ | Department of Laboratory Medicine, Tan Tock Seng Hospital | Department of Laboratory Medicine, Tan Tock Seng Hospital | Chen YYC et al |
| EPI_ISL_538481 | Singapore/947/2020 | 2020-04-06 | Asia/Singapore/Singapore/ | Department of Laboratory Medicine, Tan Tock Seng Hospital | Department of Laboratory Medicine, Tan Tock Seng Hospital | Chen YYC et al |
| EPI_ISL_538493 | Singapore/959/2020 | 2020-04-09 | Asia/Singapore/Singapore/ | Department of Laboratory Medicine, Tan Tock Seng Hospital | Department of Laboratory Medicine, Tan Tock Seng Hospital | Chen YYC et al |
| EPI_ISL_538498 | Indonesia/JI-NIHRD-PME0999/2020 | 2020-12-07 | Asia/Indonesia/East Java/ | RSUD Soediono Madiun East Java | National Institute of Health Research and Development | Pawestri et al |
| EPI_ISL_538499 | Indonesia/JI-NIHRD-PME2054/2020 | 2020-06-29 | Asia/Indonesia/East Java/ | RS Lavallete Malang East Java | National Institute of Health Research and Development | Pawestri et al |
| EPI_ISL_538500 | Indonesia/SU-NIHRD-PME4745/2020 | 2020-08-05 | Asia/Indonesia/North Sumatra/ | RS Prima Husada Cipta Medan North Sumatra | National Institute of Health Research and Development | Pawestri et al |
| EPI_ISL_538501 | Indonesia/SU-NIHRD-PME4760/2020 | 2020-08-01 | Asia/Indonesia/North Sumatra/ | RS Prima Husada Cipta Medan North Sumatra | National Institute of Health Research and Development | Pawestri et al |
| EPI_ISL_538502 | Indonesia/KT-NIHRD-PME4206/2020 | 2020-07-24 | Asia/Indonesia/Center Kalimantan/ | RSUD Sultan Imanudin Pangkalan Bun Center Kalimantan | National Institute of Health Research and Development | Pawestri et al |
| EPI_ISL_538504 | Indonesia/JK-NIHRD-MI41613/2020 | 2020-07-27 | Asia/Indonesia/Jakarta/ | National Institute of Health Research and Development | National Institute of Health Research and Development | Pawestri et al |

| | | | | | | |
|---|---|---|---|---|---|---|
| EPI_ISL_538505 | Indonesia/JK-NIHRD-MI52885/2020 | 2020-08-20 | Asia/Indonesia/Jakarta/ | National Institute of Health Research and Development | National Institute of Health Research and Development | Pawestri et al |
| EPI_ISL_538506 | Indonesia/PA-NIHRD-C0710717/2020 | 2020-06-13 | Asia/Indonesia/Papua/ | Balai Penelitian dan Pengembangan Biomedis Papua | National Institute of Health Research and Development | Pawestri et al |
| EPI_ISL_538507 | Indonesia/PA-NIHRD-C0710684/2020 | 2020-06-13 | Asia/Indonesia/Papua/ | Balai Penelitian dan Pengembangan Biomedis Papua | National Institute of Health Research and Development | Pawestri et al |
| EPI_ISL_538508 | Indonesia/JK-NIHRD-MI2851/2020 | 2020-05-17 | Asia/Indonesia/Jakarta/ | National Institute of Health Research and Development | National Institute of Health Research and Development | Pawestri et al |
| EPI_ISL_538510 | Indonesia/KS-NIHRD-PME4900/2020 | 2020-08-06 | Asia/Indonesia/South Kalimantan/ | RSUD Ulin Banjarmasin South Kalimantan | National Institute of Health Research and Development | Pawestri et al |
| EPI_ISL_538512 | Indonesia/PA-NIHRD-PME4306/2020 | 2020-07-30 | Asia/Indonesia/Papua/ | Balai Penelitian dan Pengembangan Biomedis Papua | National Institute of Health Research and Development | Pawestri et al |
| EPI_ISL_538513 | Indonesia/JB-NIHRD-PME4992/2020 | 2020-12-07 | Asia/Indonesia/West Java/ | Provincial Health Laboratory Bekasi West Java | National Institute of Health Research and Development | Pawestri et al |
| EPI_ISL_538515 | USA/NC-SLPH-0002/2020 | 2020-06-12 | North America/USA/North Carolina/ | Infectious Diseases, North Carolina State Laboratory of Public Health COVID-19 Response Team | Infectious Diseases, North Carolina State Laboratory of Public Health COVID-19 Response Team | Chase et al |

| | | | | | | |
|---|---|---|---|---|---|---|
| EPI_ISL_538518 | USA/NC-SLPH-0008/2020 | 2020-07-31 | North America/USA/North Carolina/ | Infectious Diseases, North Carolina State Laboratory of Public Health COVID-19 Response Team | Infectious Diseases, North Carolina State Laboratory of Public Health COVID-19 Response Team | Chase et al |
| EPI_ISL_538519 | USA/NC-SLPH-0009/2020 | 2020-07-31 | North America/USA/North Carolina/ | Infectious Diseases, North Carolina State Laboratory of Public Health COVID-19 Response Team | Infectious Diseases, North Carolina State Laboratory of Public Health COVID-19 Response Team | Chase et al |
| EPI_ISL_538552 | Spain/CT-IrsiCaixa-2020090134-2/202 | 2020-08-31 | Europe/Spain/Catalunya/Palamos | Hospital Universitari Germans Trias i Pujol(HUGTiP)/Fundaci√≥ Lluita contra la SIDA (FLSida) | IrsiCaixa AIDS Research Lab | Marc Noguera-Julian et al |
| EPI_ISL_538570 | Spain/AN-IBV-98001651/2020 | 2020-04-07 | Europe/Spain/Andalusia/Sevilla | Servicio de Microbiolog√≠a y Parasitolog√≠a cl√≠nica. UCEIMP. Hospital Universitario Virgen del Roc√≠o/IBIS/CSIC/US | SeqCOVID-SPAIN consortium/IBV(CSIC) | Guillermo Mart√≠n Guti√©rrez et al |
| EPI_ISL_538573 | Spain/AN-IBV-98001656/2020 | 2020-04-07 | Europe/Spain/Andalusia/Sevilla | Servicio de Microbiolog√≠a y Parasitolog√≠a cl√≠nica. UCEIMP. Hospital Universitario Virgen del Roc√≠o/IBIS/CSIC/US | SeqCOVID-SPAIN consortium/IBV(CSIC) | Guillermo Mart√≠n Guti√©rrez et al |
| EPI_ISL_538578 | Spain/AN-IBV-98001670/2020 | 2020-04-06 | Europe/Spain/Andalusia/Sevilla | Servicio de Microbiolog√≠a y Parasitolog√≠a cl√≠nica. UCEIMP. Hospital Universitario Virgen del Roc√≠o/IBIS/CSIC/US | SeqCOVID-SPAIN consortium/IBV(CSIC) | Guillermo Mart√≠n Guti√©rrez et al |
| EPI_ISL_538585 | Spain/AN-IBV-98001734/2020 | 2020-04-09 | Europe/Spain/Andalusia/Sevilla | Servicio de Microbiolog√≠a y Parasitolog√≠a cl√≠nica. UCEIMP. Hospital Universitario Virgen del Roc√≠o/IBIS/CSIC/US | SeqCOVID-SPAIN consortium/IBV(CSIC) | Guillermo Mart√≠n Guti√©rrez et al |
| EPI_ISL_538587 | Spain/AN-IBV-98001737/2020 | 2020-04-07 | Europe/Spain/Andalusia/Sevilla | Servicio de Microbiolog√≠a y Parasitolog√≠a cl√≠nica. UCEIMP. Hospital Universitario Virgen del Roc√≠o/IBIS/CSIC/US | SeqCOVID-SPAIN consortium/IBV(CSIC) | Guillermo Mart√≠n Guti√©rrez et al |

| | | | | | | |
|---|---|---|---|---|---|---|
| EPI_ISL_538588 | Spain/AN-IBV-98001740/2020 | 2020-04-06 | Europe/Spain/Andalusia/Sevilla | Servicio de Microbiolog√≠a y Parasitolog√≠a cl√≠nica. UCEIMP. Hospital Universitario Virgen del Roc√≠o/IBIS/CSIC/US | SeqCOVID-SPAIN consortium/IBV(CSIC) | Guillermo Mart√≠n Guti√©rrez et al |
| EPI_ISL_538608 | Spain/NC-IBV-98005469/2020 | 2020-03-28 | Europe/Spain/Navarra/Gartzain | Cl√≠nica Universidad de Navarra. Servicio de Enfermedades Infecciosas y Microbiolog√≠a cl√≠nica | SeqCOVID-SPAIN consortium/IBV(CSIC) | Mirian Fern√°ndez-Alonso et al |
| EPI_ISL_538610 | Spain/NC-IBV-98005472/2020 | 2020-03-28 | Europe/Spain/Navarra/Mendillorri | Cl√≠nica Universidad de Navarra. Servicio de Enfermedades Infecciosas y Microbiolog√≠a cl√≠nica | SeqCOVID-SPAIN consortium/IBV(CSIC) | Mirian Fern√°ndez-Alonso et al |
| EPI_ISL_538611 | Spain/NC-IBV-98005473/2020 | 2020-03-28 | Europe/Spain/Navarra/Pamplona | Cl√≠nica Universidad de Navarra. Servicio de Enfermedades Infecciosas y Microbiolog√≠a cl√≠nica | SeqCOVID-SPAIN consortium/IBV(CSIC) | Mirian Fern√°ndez-Alonso et al |
| EPI_ISL_538617 | Spain/VC-IBV-98003600/2020 | 2020-03-15 | Europe/Spain/Comunitat Valenciana/Va | Servicio de Microbiolog√≠a. Hospital General Universitario de Castell√≥n | SeqCOVID-SPAIN consortium/IBV(CSIC) | Rosario Moreno et al |
| EPI_ISL_538618 | Spain/VC-IBV-98003601/2020 | 2020-03-15 | Europe/Spain/Comunitat Valenciana/Va | Servicio de Microbiolog√≠a. Hospital General Universitario de Castell√≥n | SeqCOVID-SPAIN consortium/IBV(CSIC) | Rosario Moreno et al |
| EPI_ISL_538656 | Spain/VC-IBV-98006448/2020 | 2020-06-26 | Europe/Spain/Comunitat Valenciana/Ca | Servicio de Microbiolog√≠a. Hospital General Universitario de Castell√≥n | SeqCOVID-SPAIN consortium/IBV(CSIC) | Rosario Moreno et al |
| EPI_ISL_538659 | Spain/VC-IBV-98006451/2020 | 2020-06-27 | Europe/Spain/Comunitat Valenciana/Ca | Servicio de Microbiolog√≠a. Hospital General Universitario de Castell√≥n | SeqCOVID-SPAIN consortium/IBV(CSIC) | Rosario Moreno et al |
| EPI_ISL_538667 | Spain/VC-IBV-98006466/2020 | 2020-07-03 | Europe/Spain/Comunitat Valenciana/Ca | Servicio de Microbiolog√≠a. Hospital General Universitario de Castell√≥n | SeqCOVID-SPAIN consortium/IBV(CSIC) | Rosario Moreno et al |
| EPI_ISL_538671 | Spain/VC-IBV-98006471/2020 | 2020-07-06 | Europe/Spain/Comunitat Valenciana/Ca | Servicio de Microbiolog√≠a. Hospital General Universitario de Castell√≥n | SeqCOVID-SPAIN consortium/IBV(CSIC) | Rosario Moreno et al |
| EPI_ISL_538674 | Spain/AN-IBV-98002873/2020 | 2020-03-16 | Europe/Spain/Andalusia/Granada ES | Hospital Universitario Virgen de las Nieves de Granada-SAS | SeqCOVID-SPAIN consortium/IBV(CSIC) | Mercedes P√©rez Ruiz et al |
| EPI_ISL_538675 | Spain/AN-IBV-98002883/2020 | 2020-03-16 | Europe/Spain/Andalusia/Granada ES | Hospital Universitario Virgen de las Nieves de Granada-SAS | SeqCOVID-SPAIN consortium/IBV(CSIC) | Mercedes P√©rez Ruiz et al |
| EPI_ISL_538681 | Spain/AN-IBV-98002907/2020 | 2020-03-18 | Europe/Spain/Andalusia/Ogijares | Hospital Universitario Virgen de las Nieves de Granada-SAS | SeqCOVID-SPAIN consortium/IBV(CSIC) | Mercedes P√©rez Ruiz et al |

| | | | | | | |
|---|---|---|---|---|---|---|
| EPI_ISL_538686 | Spain/AN-IBV-98002954/2020 | 2020-03-16 | Europe/Spain/Andalusia/Iznalloz | Hospital Universitario Virgen de las Nieves de Granada-SAS | SeqCOVID-SPAIN consortium/IBV(CSIC) | Mercedes P√©rez Ruiz et al |
| EPI_ISL_538700 | Spain/CB-IBV-98002774/2020 | 2020-03-24 | Europe/Spain/Cantabria/Polanco | Hospital Universitario Marqu√©s de Valdecilla - IDIVAL (Santander, Cantabria) | SeqCOVID-SPAIN consortium/IBV(CSIC) | Mar√≠a Eliecer Cano Garc√≠a et al |
| EPI_ISL_538707 | Spain/CB-IBV-98002782/2020 | 2020-03-24 | Europe/Spain/Cantabria/Santander | Hospital Universitario Marqu√©s de Valdecilla - IDIVAL (Santander, Cantabria) | SeqCOVID-SPAIN consortium/IBV(CSIC) | Mar√≠a Eliecer Cano Garc√≠a et al |
| EPI_ISL_538708 | Spain/CB-IBV-98002784/2020 | 2020-03-24 | Europe/Spain/Cantabria/Santander | Hospital Universitario Marqu√©s de Valdecilla - IDIVAL (Santander, Cantabria) | SeqCOVID-SPAIN consortium/IBV(CSIC) | Mar√≠a Eliecer Cano Garc√≠a et al |
| EPI_ISL_538716 | Spain/CB-IBV-98002792/2020 | 2020-03-25 | Europe/Spain/Cantabria/Oruna | Hospital Universitario Marqu√©s de Valdecilla - IDIVAL (Santander, Cantabria) | SeqCOVID-SPAIN consortium/IBV(CSIC) | Mar√≠a Eliecer Cano Garc√≠a et al |
| EPI_ISL_538726 | Spain/CB-IBV-98002805/2020 | 2020-03-24 | Europe/Spain/Cantabria/Valderredible | Hospital Universitario Marqu√©s de Valdecilla - IDIVAL (Santander, Cantabria) | SeqCOVID-SPAIN consortium/IBV(CSIC) | Mar√≠a Eliecer Cano Garc√≠a et al |
| EPI_ISL_538731 | Spain/CB-IBV-98002812/2020 | 2020-03-24 | Europe/Spain/Cantabria/Puente San Mi | Hospital Universitario Marqu√©s de Valdecilla - IDIVAL (Santander, Cantabria) | SeqCOVID-SPAIN consortium/IBV(CSIC) | Mar√≠a Eliecer Cano Garc√≠a et al |
| EPI_ISL_538826 | England/LEED-2AA552/2020 | 2020-04-09 | Europe/United Kingdom/England/ | Leeds Teaching Hospitals NHS Trust and Public Health England, National Infection Service (Leeds laboratory) | Wellcome Sanger Institute for the COVID-19 Genomics UK (COG-UK) consortium | Louissa Macfarlane-Smith et al |
| EPI_ISL_538842 | England/LEED-2AA14F/2020 | 2020-03-12 | Europe/United Kingdom/England/ | Leeds Teaching Hospitals NHS Trust and Public Health England, National Infection Service (Leeds laboratory) | Wellcome Sanger Institute for the COVID-19 Genomics UK (COG-UK) consortium | Louissa Macfarlane-Smith et al |
| EPI_ISL_538850 | England/LEED-2A94B0/2020 | 2020-04-11 | Europe/United Kingdom/England/ | Leeds Teaching Hospitals NHS Trust and Public Health England, National Infection Service (Leeds laboratory) | Wellcome Sanger Institute for the COVID-19 Genomics UK (COG-UK) consortium | Louissa Macfarlane-Smith et al |
| EPI_ISL_538885 | England/LEED-2A9A54/2020 | 2020-04-11 | Europe/United Kingdom/England/ | Leeds Teaching Hospitals NHS Trust and Public Health England, National Infection Service (Leeds laboratory) | Wellcome Sanger Institute for the COVID-19 Genomics UK (COG-UK) consortium | Louissa Macfarlane-Smith et al |

| | | | | | | |
|---|---|---|---|---|---|---|
| EPI_ISL_538893 | England/LEED-2AA5CB/2020 | 2020-04-03 | Europe/United Kingdom/England/ | Leeds Teaching Hospitals NHS Trust and Public Health England, National Infection Service (Leeds laboratory) | Wellcome Sanger Institute for the COVID-19 Genomics UK (COG-UK) consortium | Louissa Macfarlane-Smith et al |
| EPI_ISL_538937 | England/LEED-2A9B60/2020 | 2020-04-12 | Europe/United Kingdom/England/ | Leeds Teaching Hospitals NHS Trust and Public Health England, National Infection Service (Leeds laboratory) | Wellcome Sanger Institute for the COVID-19 Genomics UK (COG-UK) consortium | Louissa Macfarlane-Smith et al |
| EPI_ISL_538955 | England/LEED-2A9CE5/2020 | 2020-03-28 | Europe/United Kingdom/England/ | Leeds Teaching Hospitals NHS Trust and Public Health England, National Infection Service (Leeds laboratory) | Wellcome Sanger Institute for the COVID-19 Genomics UK (COG-UK) consortium | Louissa Macfarlane-Smith et al |
| EPI_ISL_539088 | England/LEED-2A8B43/2020 | 2020-04-01 | Europe/United Kingdom/England/ | Leeds Teaching Hospitals NHS Trust and Public Health England, National Infection Service (Leeds laboratory) | Wellcome Sanger Institute for the COVID-19 Genomics UK (COG-UK) consortium | Louissa Macfarlane-Smith et al |
| EPI_ISL_539148 | England/LEED-2A8D6B/2020 | 2020-03-23 | Europe/United Kingdom/England/ | Leeds Teaching Hospitals NHS Trust and Public Health England, National Infection Service (Leeds laboratory) | Wellcome Sanger Institute for the COVID-19 Genomics UK (COG-UK) consortium | Louissa Macfarlane-Smith et al |
| EPI_ISL_539221 | Spain/VC-IBV-98000385/2020 | 2020-03-24 | Europe/Spain/Comunitat Valenciana/Al | Servicio de Microbiologia. Hospital Clinico Universitario de Valencia | SeqCOVID-SPAIN consortium/IBV(CSIC) | Maria Dolores Ocete et al |
| EPI_ISL_539225 | Spain/VC-IBV-98000391/2020 | 2020-03-23 | Europe/Spain/Comunitat Valenciana/Ga | Servicio de Microbiologia. Hospital Clinico Universitario de Valencia | SeqCOVID-SPAIN consortium/IBV(CSIC) | Maria Dolores Ocete et al |
| EPI_ISL_539226 | Spain/VC-IBV-98000395/2020 | 2020-03-13 | Europe/Spain/Comunitat Valenciana/Ga | Servicio de Microbiologia. Hospital Clinico Universitario de Valencia | SeqCOVID-SPAIN consortium/IBV(CSIC) | Maria Dolores Ocete et al |
| EPI_ISL_539240 | Spain/VC-IBV-98000343/2020 | 2020-03-22 | Europe/Spain/Comunitat Valenciana/Va | Servicio de Microbiolog√≠a. Consorcio Hospital General Universitario de Valencia | SeqCOVID-SPAIN consortium/IBV(CSIC) | Paula Ruiz-Hueso et al |
| EPI_ISL_539250 | Spain/VC-IBV-98003434/2020 | 2020-04-02 | Europe/Spain/Comunitat Valenciana/Be | Hospital Universitario de La Ribera (Alzira, Val√®ncia) | SeqCOVID-SPAIN consortium/IBV(CSIC) | Olalla Mart√≠nez Macias et al |

| | | | | | | |
|---|---|---|---|---|---|---|
| EPI_ISL_539253 | Spain/VC-IBV-98003444/2020 | 2020-04-02 | Europe/Spain/Comunitat Valenciana/Va | Hospital Universitario de La Ribera (Alzira, Valv®ncia) | SeqCOVID-SPAIN consortium/IBV(CSIC) | Olalla Martv≠nez Macias et al |
| EPI_ISL_539257 | Spain/VC-IBV-98003450/2020 | 2020-04-03 | Europe/Spain/Comunitat Valenciana/Ta | Hospital Universitario de La Ribera (Alzira, Valv®ncia) | SeqCOVID-SPAIN consortium/IBV(CSIC) | Olalla Martv≠nez Macias et al |
| EPI_ISL_539266 | Spain/VC-IBV-98003462/2020 | 2020-04-03 | Europe/Spain/Comunitat Valenciana/Al | Hospital Universitario de La Ribera (Alzira, Valv®ncia) | SeqCOVID-SPAIN consortium/IBV(CSIC) | Olalla Martv≠nez Macias et al |
| EPI_ISL_539272 | Spain/VC-IBV-98003473/2020 | 2020-04-07 | Europe/Spain/Comunitat Valenciana/Su | Hospital Universitario de La Ribera (Alzira, Valv®ncia) | SeqCOVID-SPAIN consortium/IBV(CSIC) | Olalla Martv≠nez Macias et al |
| EPI_ISL_539273 | Spain/VC-IBV-98003474/2020 | 2020-04-07 | Europe/Spain/Comunitat Valenciana/Fa | Hospital Universitario de La Ribera (Alzira, Valv®ncia) | SeqCOVID-SPAIN consortium/IBV(CSIC) | Olalla Martv≠nez Macias et al |
| EPI_ISL_539289 | Spain/VC-IBV-98000083/2020 | 2020-03-10 | Europe/Spain/Comunitat Valenciana/To | Servicio de Microbiologv≠a. Consorcio Hospital General Universitario de Valencia | SeqCOVID-SPAIN consortium/IBV(CSIC) | Paula Ruiz-Hueso et al |
| EPI_ISL_539299 | Russia/MOW-CRIE-160158/2020 | 2020-03-30 | Europe/Russia/Moscow Oblast/ | "Molecular diagnostic laboratory of Federal Budget Institution of Science ""Central Research Institute of Epidemiology"" of The Federal Service on Customers' Rights Protection and Human Well-being Surveillance" | Group of Genomics and Postgenomic Technologies of Central Research Institute of Epidemilology | Speranskaya AS et al |
| EPI_ISL_539333 | Hunan/HN-CIDC-P2/2020 | 2020-01-17 | Asia/China/Hunan/Huaihua | Institute of Disease Control and Prevention, People's Liberation Army | Institute of Disease Control and Prevention, People's Liberation Army | Qiu et al |
| EPI_ISL_539335 | Hunan/HN-CIDC-P6/2020 | 2020-01-28 | Asia/China/Hunan/Huaihua | Institute of Disease Control and Prevention, People's Liberation Army | Institute of Disease Control and Prevention, People's Liberation Army | Qiu et al |
| EPI_ISL_539336 | Hunan/HN-CIDC-P7/2020 | 2020-01-27 | Asia/China/Hunan/Shaoyang | Institute of Disease Control and Prevention, People's Liberation Army | Institute of Disease Control and Prevention, People's Liberation Army | Qiu et al |

| | | | | | | |
|---|---|---|---|---|---|---|
| EPI_ISL_539337 | Hunan/HN-CIDC-P8/2020 | 2020-01-26 | Asia/China/Hunan/Shaoyang | Institute of Disease Control and Prevention, People's Liberation Army | Institute of Disease Control and Prevention, People's Liberation Army | Qiu et al |
| EPI_ISL_539338 | Hunan/HN-CIDC-P10/2020 | 2020-01-29 | Asia/China/Hunan/Shaoyang | Institute of Disease Control and Prevention, People's Liberation Army | Institute of Disease Control and Prevention, People's Liberation Army | Qiu et al |
| EPI_ISL_539339 | Hunan/HN-CIDC-P11/2020 | 2020-01-29 | Asia/China/Hunan/Shaoyang | Institute of Disease Control and Prevention, People's Liberation Army | Institute of Disease Control and Prevention, People's Liberation Army | Qiu et al |
| EPI_ISL_539477 | Switzerland/SO-ETHZ-260188/2020 | 2020-08-27 | Europe/Switzerland/Solothurn/ | Viollier AG | Department of Biosystems Science and Engineering, ETH Zvºrich | Christian Beisel et al |
| EPI_ISL_539489 | India/PB-IMT-P131/2020 | 2020-05-05 | Asia/India/Punjab IN/Rupnagar | Civil Hospital, Rupnagar | CSIR-Institute of Microbial Technology | Kanika Bansal et al |
| EPI_ISL_539491 | India/UT-IMT-AO15/2020 | 2020-06-18 | Asia/India/Uttarakhand/Dehradun | IDSP unit, Dehradun | CSIR-Institute of Microbial Technology | Kanika Bansal et al |
| EPI_ISL_539494 | CzechRepublic/NRL-5635/2020 | 2020-03-31 | Europe/Czech Republic/Usti nad Labem/I | The National Institute of Public Health | State Veterinary Institute Prague | Nagy et al |
| EPI_ISL_539496 | Andorra/202552/2020 | 2020-03-13 | Europe/Andorra/Andorra/ | Hospital Nostra Senyora de Meritxell | Instituto de Salud Carlos III | Iglesias-Caballero et al |
| EPI_ISL_539497 | Spain/AN-ISCIII-201273/2020 | 2020-02-28 | Europe/Spain/Andalusia/ | Hospital Virgen de las Nieves | Instituto de Salud Carlos III | Iglesias-Caballero et al |
| EPI_ISL_539499 | Spain/AN-ISCIII-202657/2020 | 2020-03-12 | Europe/Spain/Andalusia/ | Hospital Costa del Sol | Instituto de Salud Carlos III | Iglesias-Caballero et al |
| EPI_ISL_539514 | Spain/AR-ISCIII-201780/2020 | 2020-03-07 | Europe/Spain/Aragon/ | Hospital Universitario Miguel Servet | Instituto de Salud Carlos III | Iglesias-Caballero et al |
| EPI_ISL_539517 | Spain/AR-ISCIII-201848/2020 | 2020-03-08 | Europe/Spain/Aragon/ | Hospital Universitario Miguel Servet | Instituto de Salud Carlos III | Iglesias-Caballero et al |
| EPI_ISL_539522 | Spain/CE-ISCIII-204881/2020 | 2020-03-18 | Europe/Spain/Ceuta/ | Hospital Universitario de Ceuta | Instituto de Salud Carlos III | Iglesias-Caballero et al |

| | | | | | | |
|---|---|---|---|---|---|---|
| EPI_ISL_539527 | Spain/CM-ISCIII-202018234/2020 | 2020-03-22 | Europe/Spain/Castilla la Mancha/ | Consejeria de Sanidad y Asuntos Sociales | Instituto de Salud Carlos III | Iglesias-Caballero et al |
| EPI_ISL_539531 | Spain/CN-ISCIII-201048/2020 | 2020-02-24 | Europe/Spain/Canary Islands/ | C.H.U Nuestra Se√±ora de Candelaria | Instituto de Salud Carlos III | Iglesias-Caballero et al |
| EPI_ISL_539534 | Spain/CT-ISCIII-2012563/2020 | 2020-05-13 | Europe/Spain/Catalunya/ | Hospital Cl√≠nic | Instituto de Salud Carlos III | Iglesias-Caballero et al |
| EPI_ISL_539538 | Spain/CT-ISCIII-2012571/2020 | 2020-05-20 | Europe/Spain/Catalunya/ | Hospital Cl√≠nic | Instituto de Salud Carlos III | Iglesias-Caballero et al |
| EPI_ISL_539540 | Spain/CT-ISCIII-2012573/2020 | 2020-05-21 | Europe/Spain/Catalunya/ | Hospital Cl√≠nic | Instituto de Salud Carlos III | Iglesias-Caballero et al |
| EPI_ISL_539557 | Spain/EX-ISCIII-201239/2020 | 2020-02-28 | Europe/Spain/Extremadura/ | Gerencia del √°rea de salud de Badajoz, Llerena y Zafra | Instituto de Salud Carlos III | Iglesias-Caballero et al |
| EPI_ISL_539558 | Spain/EX-ISCIII-201248/2020 | 2020-02-28 | Europe/Spain/Extremadura/ | Hospital San Pedro de Alc√°ntara | Instituto de Salud Carlos III | Iglesias-Caballero et al |
| EPI_ISL_539560 | Spain/EX-ISCIII-201655/2020 | 2020-03-07 | Europe/Spain/Extremadura/ | Hospital Campo Ara√±uelo | Instituto de Salud Carlos III | Iglesias-Caballero et al |
| EPI_ISL_539564 | Spain/GA-ISCIII-201859/2020 | 2020-03-10 | Europe/Spain/Galicia/ | Complejo Hospitalario Universitario La Coruna | Instituto de Salud Carlos III | Iglesias-Caballero et al |
| EPI_ISL_539571 | Spain/NA-ISCIII-201951/2020 | 2020-03-09 | Europe/Spain/Navarra/ | Complejo Hospitalario de Navarra | Instituto de Salud Carlos III | Iglesias-Caballero et al |
| EPI_ISL_539572 | Spain/NA-ISCIII-201952/2020 | 2020-03-09 | Europe/Spain/Navarra/ | Complejo Hospitalario de Navarra | Instituto de Salud Carlos III | Iglesias-Caballero et al |
| EPI_ISL_539581 | Germany/NW-HHU-104/2020 | 2020-08-28 | Europe/Germany/North Rhine Westphal | ZOTZ KLIMAS MVZ D√ºsseldorf-Centrum GbR √úBAG f√ºr Labormedizin, Genetik, Zytologie, Pathologie | Center of Medical Microbiology, Virology, and Hospital Hygiene, University of Duesseldorf | Maximilian Damagnez et al |
| EPI_ISL_539585 | Germany/NW-HHU-108/2020 | 2020-08-28 | Europe/Germany/North Rhine Westphal | ZOTZ KLIMAS MVZ D√ºsseldorf-Centrum GbR √úBAG f√ºr Labormedizin, Genetik, Zytologie, Pathologie | Center of Medical Microbiology, Virology, and Hospital Hygiene, University of Duesseldorf | Maximilian Damagnez et al |
| EPI_ISL_539588 | Germany/NW-HHU-111/2020 | 2020-08-27 | Europe/Germany/North Rhine Westphal | ZOTZ KLIMAS MVZ D√ºsseldorf-Centrum GbR √úBAG f√ºr Labormedizin, Genetik, Zytologie, Pathologie | Center of Medical Microbiology, Virology, and Hospital Hygiene, University of Duesseldorf | Maximilian Damagnez et al |

| | | | | | | |
|---|---|---|---|---|---|---|
| EPI_ISL_539589 | Germany/NW-HHU-112/2020 | 2020-08-26 | Europe/Germany/North Rhine Westphal | ZOTZ KLIMAS MVZ Dv∫sseldorf-Centrum GbR √úBAG fv∫r Labormedizin, Genetik, Zytologie, Pathologie | Center of Medical Microbiology, Virology, and Hospital Hygiene, University of Duesseldorf | Maximilian Damagnez et al |
| EPI_ISL_539590 | Germany/NW-HHU-113/2020 | 2020-08-26 | Europe/Germany/North Rhine Westphal | ZOTZ KLIMAS MVZ Dv∫sseldorf-Centrum GbR √úBAG fv∫r Labormedizin, Genetik, Zytologie, Pathologie | Center of Medical Microbiology, Virology, and Hospital Hygiene, University of Duesseldorf | Maximilian Damagnez et al |
| EPI_ISL_539594 | Germany/NW-HHU-117/2020 | 2020-08-24 | Europe/Germany/North Rhine Westphal | ZOTZ KLIMAS MVZ Dv∫sseldorf-Centrum GbR √úBAG fv∫r Labormedizin, Genetik, Zytologie, Pathologie | Center of Medical Microbiology, Virology, and Hospital Hygiene, University of Duesseldorf | Maximilian Damagnez et al |
| EPI_ISL_539603 | Germany/NW-HHU-87/2020 | 2020-08-17 | Europe/Germany/North Rhine Westphal | ZOTZ KLIMAS MVZ Dv∫sseldorf-Centrum GbR √úBAG fv∫r Labormedizin, Genetik, Zytologie, Pathologie | Center of Medical Microbiology, Virology, and Hospital Hygiene, University of Duesseldorf | Maximilian Damagnez et al |
| EPI_ISL_539612 | Germany/NW-HHU-96/2020 | 2020-08-19 | Europe/Germany/North Rhine Westphal | ZOTZ KLIMAS MVZ Dv∫sseldorf-Centrum GbR √úBAG fv∫r Labormedizin, Genetik, Zytologie, Pathologie | Center of Medical Microbiology, Virology, and Hospital Hygiene, University of Duesseldorf | Maximilian Damagnez et al |
| EPI_ISL_539616 | India/TG-CCMB-AA147/2020 | 2020-08-10 | Asia/India/Telangana/ | CSIR-Centre for Cellular and Molecular Biology | CSIR-Centre for Cellular and Molecular Biology | Lamuk Zaveri et al |
| EPI_ISL_539617 | India/TG-CCMB-AB465/2020 | 2020-08-14 | Asia/India/Telangana/ | CSIR-Centre for Cellular and Molecular Biology | CSIR-Centre for Cellular and Molecular Biology | Lamuk Zaveri et al |
| EPI_ISL_539618 | India/TG-CCMB-AB583/2020 | 2020-08-16 | Asia/India/Telangana/ | CSIR-Centre for Cellular and Molecular Biology | CSIR-Centre for Cellular and Molecular Biology | Lamuk Zaveri et al |

| | | | | | | |
|---|---|---|---|---|---|---|
| EPI_ISL_539619 | India/TG-CCMB-AC539/2020 | 2020-08-19 | Asia/India/Telangana/ | CSIR-Centre for Cellular and Molecular Biology | CSIR-Centre for Cellular and Molecular Biology | M Soujanya Reddy et al |
| EPI_ISL_539620 | India/TG-CCMB-R249/2020 | 2020-08-09 | Asia/India/Telangana/ | CSIR-Centre for Cellular and Molecular Biology | CSIR-Centre for Cellular and Molecular Biology | M Soujanya Reddy et al |
| EPI_ISL_539621 | India/TG-CCMB-R220/2020 | 2020-08-09 | Asia/India/Telangana/ | CSIR-Centre for Cellular and Molecular Biology | CSIR-Centre for Cellular and Molecular Biology | M Soujanya Reddy et al |
| EPI_ISL_539622 | India/TG-CCMB-R159/2020 | 2020-08-09 | Asia/India/Telangana/ | CSIR-Centre for Cellular and Molecular Biology | CSIR-Centre for Cellular and Molecular Biology | Namami Gaur et al |
| EPI_ISL_539623 | India/TG-CCMB-R178/2020 | 2020-08-09 | Asia/India/Telangana/ | CSIR-Centre for Cellular and Molecular Biology | CSIR-Centre for Cellular and Molecular Biology | Namami Gaur et al |
| EPI_ISL_539624 | India/TG-CCMB-R208/2020 | 2020-08-09 | Asia/India/Telangana/ | CSIR-Centre for Cellular and Molecular Biology | CSIR-Centre for Cellular and Molecular Biology | Namami Gaur et al |
| EPI_ISL_539625 | India/TG-CCMB-R91/2020 | 2020-08-09 | Asia/India/Telangana/ | CSIR-Centre for Cellular and Molecular Biology | CSIR-Centre for Cellular and Molecular Biology | Nikhil Hajirnis et al |
| EPI_ISL_539626 | India/TG-CCMB-R320/2020 | 2020-08-09 | Asia/India/Telangana/ | CSIR-Centre for Cellular and Molecular Biology | CSIR-Centre for Cellular and Molecular Biology | Nikhil Hajirnis et al |
| EPI_ISL_539627 | India/TG-CCMB-AB443/2020 | 2020-08-14 | Asia/India/Telangana/ | CSIR-Centre for Cellular and Molecular Biology | CSIR-Centre for Cellular and Molecular Biology | Nikhil Hajirnis et al |
| EPI_ISL_539628 | India/TG-CCMB-AA527/2020 | 2020-08-12 | Asia/India/Telangana/ | CSIR-Centre for Cellular and Molecular Biology | CSIR-Centre for Cellular and Molecular Biology | Payel Mukherjee et al |
| EPI_ISL_539629 | India/TG-CCMB-AB520/2020 | 2020-08-14 | Asia/India/Telangana/ | CSIR-Centre for Cellular and Molecular Biology | CSIR-Centre for Cellular and Molecular Biology | Payel Mukherjee et al |
| EPI_ISL_539630 | India/TG-CCMB-R394/2020 | 2020-08-09 | Asia/India/Telangana/ | CSIR-Centre for Cellular and Molecular Biology | CSIR-Centre for Cellular and Molecular Biology | Payel Mukherjee et al |
| EPI_ISL_539631 | India/TG-CCMB-AB364/2020 | 2020-08-13 | Asia/India/Telangana/ | CSIR-Centre for Cellular and Molecular Biology | CSIR-Centre for Cellular and Molecular Biology | Pratheusa Maccha et al |

| | | | | | | |
|---|---|---|---|---|---|---|
| EPI_ISL_539632 | India/TG-CCMB-AB352/2020 | 2020-08-13 | Asia/India/Telangana/ | CSIR-Centre for Cellular and Molecular Biology | CSIR-Centre for Cellular and Molecular Biology | Pratheusa Maccha et al |
| EPI_ISL_539633 | India/TG-CCMB-AA560/2020 | 2020-08-12 | Asia/India/Telangana/ | CSIR-Centre for Cellular and Molecular Biology | CSIR-Centre for Cellular and Molecular Biology | Pratheusa Maccha et al |
| EPI_ISL_539634 | India/TG-CCMB-AA562/2020 | 2020-08-12 | Asia/India/Telangana/ | CSIR-Centre for Cellular and Molecular Biology | CSIR-Centre for Cellular and Molecular Biology | Sakshi Shambhavi et al |
| EPI_ISL_539635 | India/TG-CCMB-AB395/2020 | 2020-08-13 | Asia/India/Telangana/ | CSIR-Centre for Cellular and Molecular Biology | CSIR-Centre for Cellular and Molecular Biology | Sakshi Shambhavi et al |
| EPI_ISL_539636 | India/TG-CCMB-AA972/2020 | 2020-08-13 | Asia/India/Telangana/ | CSIR-Centre for Cellular and Molecular Biology | CSIR-Centre for Cellular and Molecular Biology | Sakshi Shambhavi et al |
| EPI_ISL_539637 | India/TG-CCMB-AB358/2020 | 2020-08-13 | Asia/India/Telangana/ | CSIR-Centre for Cellular and Molecular Biology | CSIR-Centre for Cellular and Molecular Biology | Shagufta Khan et al |
| EPI_ISL_539638 | India/TG-CCMB-AA622/2020 | 2020-08-12 | Asia/India/Telangana/ | CSIR-Centre for Cellular and Molecular Biology | CSIR-Centre for Cellular and Molecular Biology | Shagufta Khan et al |
| EPI_ISL_539639 | India/TG-CCMB-AB300/2020 | 2020-08-15 | Asia/India/Telangana/ | CSIR-Centre for Cellular and Molecular Biology | CSIR-Centre for Cellular and Molecular Biology | Shagufta Khan et al |
| EPI_ISL_539640 | India/TG-CCMB-AB223/2020 | 2020-08-14 | Asia/India/Telangana/ | CSIR-Centre for Cellular and Molecular Biology | CSIR-Centre for Cellular and Molecular Biology | Sofia Banu et al |
| EPI_ISL_539641 | India/TG-CCMB-AA624/2020 | 2020-08-12 | Asia/India/Telangana/ | CSIR-Centre for Cellular and Molecular Biology | CSIR-Centre for Cellular and Molecular Biology | Sofia Banu et al |
| EPI_ISL_539642 | India/TG-CCMB-AC437/2020 | 2020-08-19 | Asia/India/Telangana/ | CSIR-Centre for Cellular and Molecular Biology | CSIR-Centre for Cellular and Molecular Biology | Sofia Banu et al |
| EPI_ISL_539644 | India/TG-CCMB-R217/2020 | 2020-08-09 | Asia/India/Telangana/ | CSIR-Centre for Cellular and Molecular Biology | CSIR-Centre for Cellular and Molecular Biology | Tulasi Nagabandi et al |
| EPI_ISL_539645 | India/TG-CCMB-AA227/2020 | 2020-08-10 | Asia/India/Telangana/ | CSIR-Centre for Cellular and Molecular Biology | CSIR-Centre for Cellular and Molecular Biology | Tulasi Nagabandi et al |

| | | | | | | |
|---|---|---|---|---|---|---|
| EPI_ISL_539646 | India/TG-CCMB-AA158/2020 | 2020-08-10 | Asia/India/Telangana/ | CSIR-Centre for Cellular and Molecular Biology | CSIR-Centre for Cellular and Molecular Biology | Lamuk Zaveri et al |
| EPI_ISL_539647 | India/TG-CCMB-AA509/2020 | 2020-08-12 | Asia/India/Telangana/ | CSIR-Centre for Cellular and Molecular Biology | CSIR-Centre for Cellular and Molecular Biology | Lamuk Zaveri et al |
| EPI_ISL_539648 | India/TG-CCMB-R372/2020 | 2020-08-09 | Asia/India/Telangana/ | CSIR-Centre for Cellular and Molecular Biology | CSIR-Centre for Cellular and Molecular Biology | Lamuk Zaveri et al |
| EPI_ISL_539649 | India/TG-CCMB-AA586/2020 | 2020-08-12 | Asia/India/Telangana/ | CSIR-Centre for Cellular and Molecular Biology | CSIR-Centre for Cellular and Molecular Biology | M Soujanya Reddy et al |
| EPI_ISL_539650 | India/TG-CCMB-AA589/2020 | 2020-08-12 | Asia/India/Telangana/ | CSIR-Centre for Cellular and Molecular Biology | CSIR-Centre for Cellular and Molecular Biology | M Soujanya Reddy et al |
| EPI_ISL_539651 | India/TG-CCMB-AA226/2020 | 2020-08-10 | Asia/India/Telangana/ | CSIR-Centre for Cellular and Molecular Biology | CSIR-Centre for Cellular and Molecular Biology | M Soujanya Reddy et al |
| EPI_ISL_539652 | India/TG-CCMB-AA661/2020 | 2020-08-12 | Asia/India/Telangana/ | CSIR-Centre for Cellular and Molecular Biology | CSIR-Centre for Cellular and Molecular Biology | Namami Gaur et al |
| EPI_ISL_539653 | India/TG-CCMB-AA785/2020 | 2020-08-13 | Asia/India/Telangana/ | CSIR-Centre for Cellular and Molecular Biology | CSIR-Centre for Cellular and Molecular Biology | Namami Gaur et al |
| EPI_ISL_539654 | India/TG-CCMB-AA407/2020 | 2020-08-11 | Asia/India/Telangana/ | CSIR-Centre for Cellular and Molecular Biology | CSIR-Centre for Cellular and Molecular Biology | Namami Gaur et al |
| EPI_ISL_539655 | India/TG-CCMB-AA669/2020 | 2020-08-12 | Asia/India/Telangana/ | CSIR-Centre for Cellular and Molecular Biology | CSIR-Centre for Cellular and Molecular Biology | Nikhil Hajirnis et al |
| EPI_ISL_539656 | India/TG-CCMB-AB314/2020 | 2020-08-15 | Asia/India/Telangana/ | CSIR-Centre for Cellular and Molecular Biology | CSIR-Centre for Cellular and Molecular Biology | Nikhil Hajirnis et al |
| EPI_ISL_539657 | India/TG-CCMB-AB143/2020 | 2020-08-14 | Asia/India/Telangana/ | CSIR-Centre for Cellular and Molecular Biology | CSIR-Centre for Cellular and Molecular Biology | Nikhil Hajirnis et al |
| EPI_ISL_539658 | India/TG-CCMB-AC284/2020 | 2020-08-19 | Asia/India/Telangana/ | CSIR-Centre for Cellular and Molecular Biology | CSIR-Centre for Cellular and Molecular Biology | Payel Mukherjee et al |

| | | | | | | |
|---|---|---|---|---|---|---|
| EPI_ISL_539659 | India/TG-CCMB-AC434/2020 | 2020-08-19 | Asia/India/Telangana/ | CSIR-Centre for Cellular and Molecular Biology | CSIR-Centre for Cellular and Molecular Biology | Payel Mukherjee et al |
| EPI_ISL_539660 | India/TG-CCMB-AC438/2020 | 2020-08-19 | Asia/India/Telangana/ | CSIR-Centre for Cellular and Molecular Biology | CSIR-Centre for Cellular and Molecular Biology | Payel Mukherjee et al |
| EPI_ISL_539661 | India/TG-CCMB-AC488/2020 | 2020-08-19 | Asia/India/Telangana/ | CSIR-Centre for Cellular and Molecular Biology | CSIR-Centre for Cellular and Molecular Biology | Pratheusa Maccha et al |
| EPI_ISL_539662 | India/TG-CCMB-AB336/2020 | 2020-08-15 | Asia/India/Telangana/ | CSIR-Centre for Cellular and Molecular Biology | CSIR-Centre for Cellular and Molecular Biology | Pratheusa Maccha et al |
| EPI_ISL_539663 | India/TG-CCMB-AA332/2020 | 2020-08-11 | Asia/India/Telangana/ | CSIR-Centre for Cellular and Molecular Biology | CSIR-Centre for Cellular and Molecular Biology | Pratheusa Maccha et al |
| EPI_ISL_539664 | India/TG-CCMB-AB203/2020 | 2020-08-14 | Asia/India/Telangana/ | CSIR-Centre for Cellular and Molecular Biology | CSIR-Centre for Cellular and Molecular Biology | Sakshi Shambhavi et al |
| EPI_ISL_539665 | India/TG-CCMB-AB578/2020 | 2020-08-16 | Asia/India/Telangana/ | CSIR-Centre for Cellular and Molecular Biology | CSIR-Centre for Cellular and Molecular Biology | Sakshi Shambhavi et al |
| EPI_ISL_539666 | India/TG-CCMB-AB397/2020 | 2020-08-13 | Asia/India/Telangana/ | CSIR-Centre for Cellular and Molecular Biology | CSIR-Centre for Cellular and Molecular Biology | Sakshi Shambhavi et al |
| EPI_ISL_539667 | India/TG-CCMB-AA01/2020 | 2020-08-10 | Asia/India/Telangana/ | CSIR-Centre for Cellular and Molecular Biology | CSIR-Centre for Cellular and Molecular Biology | Shagufta Khan et al |
| EPI_ISL_539668 | India/TG-CCMB-AA33/2020 | 2020-08-10 | Asia/India/Telangana/ | CSIR-Centre for Cellular and Molecular Biology | CSIR-Centre for Cellular and Molecular Biology | Shagufta Khan et al |
| EPI_ISL_539669 | India/TG-CCMB-AA819/2020 | 2020-08-13 | Asia/India/Telangana/ | CSIR-Centre for Cellular and Molecular Biology | CSIR-Centre for Cellular and Molecular Biology | Shagufta Khan et al |
| EPI_ISL_539670 | India/TG-CCMB-AA524/2020 | 2020-08-12 | Asia/India/Telangana/ | CSIR-Centre for Cellular and Molecular Biology | CSIR-Centre for Cellular and Molecular Biology | Sofia Banu et al |
| EPI_ISL_539671 | India/TG-CCMB-AA863/2020 | 2020-08-13 | Asia/India/Telangana/ | CSIR-Centre for Cellular and Molecular Biology | CSIR-Centre for Cellular and Molecular Biology | Sofia Banu et al |

| | | | | | | |
|---|---|---|---|---|---|---|
| EPI_ISL_539672 | India/TG-CCMB-AA320/2020 | 2020-08-11 | Asia/India/Telangana/ | CSIR-Centre for Cellular and Molecular Biology | CSIR-Centre for Cellular and Molecular Biology | Sofia Banu et al |
| EPI_ISL_539673 | India/TG-CCMB-AC377/2020 | 2020-08-19 | Asia/India/Telangana/ | CSIR-Centre for Cellular and Molecular Biology | CSIR-Centre for Cellular and Molecular Biology | Tulasi Nagabandi et al |
| EPI_ISL_539674 | India/TG-CCMB-AA900/2020 | 2020-08-13 | Asia/India/Telangana/ | CSIR-Centre for Cellular and Molecular Biology | CSIR-Centre for Cellular and Molecular Biology | Tulasi Nagabandi et al |
| EPI_ISL_539675 | India/TG-CCMB-AB559/2020 | 2020-08-16 | Asia/India/Telangana/ | CSIR-Centre for Cellular and Molecular Biology | CSIR-Centre for Cellular and Molecular Biology | Tulasi Nagabandi et al |
| EPI_ISL_539676 | India/TG-CCMB-AA04/2020 | 2020-08-10 | Asia/India/Telangana/ | CSIR-Centre for Cellular and Molecular Biology | CSIR-Centre for Cellular and Molecular Biology | Lamuk Zaveri et al |
| EPI_ISL_539677 | India/TG-CCMB-AA102/2020 | 2020-08-10 | Asia/India/Telangana/ | CSIR-Centre for Cellular and Molecular Biology | CSIR-Centre for Cellular and Molecular Biology | Lamuk Zaveri et al |
| EPI_ISL_539678 | India/TG-CCMB-AA773/2020 | 2020-08-13 | Asia/India/Telangana/ | CSIR-Centre for Cellular and Molecular Biology | CSIR-Centre for Cellular and Molecular Biology | Lamuk Zaveri et al |
| EPI_ISL_539679 | India/TG-CCMB-R187/2020 | 2020-08-09 | Asia/India/Telangana/ | CSIR-Centre for Cellular and Molecular Biology | CSIR-Centre for Cellular and Molecular Biology | M Soujanya Reddy et al |
| EPI_ISL_539680 | India/TG-CCMB-AA19/2020 | 2020-08-10 | Asia/India/Telangana/ | CSIR-Centre for Cellular and Molecular Biology | CSIR-Centre for Cellular and Molecular Biology | M Soujanya Reddy et al |
| EPI_ISL_539681 | India/TG-CCMB-AB259/2020 | 2020-08-14 | Asia/India/Telangana/ | CSIR-Centre for Cellular and Molecular Biology | CSIR-Centre for Cellular and Molecular Biology | M Soujanya Reddy et al |
| EPI_ISL_539682 | India/TG-CCMB-AB268/2020 | 2020-08-14 | Asia/India/Telangana/ | CSIR-Centre for Cellular and Molecular Biology | CSIR-Centre for Cellular and Molecular Biology | Namami Gaur et al |
| EPI_ISL_539683 | India/TG-CCMB-AA105/2020 | 2020-08-10 | Asia/India/Telangana/ | CSIR-Centre for Cellular and Molecular Biology | CSIR-Centre for Cellular and Molecular Biology | Namami Gaur et al |
| EPI_ISL_539684 | India/TG-CCMB-AA28/2020 | 2020-08-10 | Asia/India/Telangana/ | CSIR-Centre for Cellular and Molecular Biology | CSIR-Centre for Cellular and Molecular Biology | Namami Gaur et al |

| | | | | | | |
|---|---|---|---|---|---|---|
| EPI_ISL_539685 | India/TG-CCMB-AA378/2020 | 2020-08-11 | Asia/India/Telangana/ | CSIR-Centre for Cellular and Molecular Biology | CSIR-Centre for Cellular and Molecular Biology | Nikhil Hajirnis et al |
| EPI_ISL_539686 | India/TG-CCMB-AA608/2020 | 2020-08-12 | Asia/India/Telangana/ | CSIR-Centre for Cellular and Molecular Biology | CSIR-Centre for Cellular and Molecular Biology | Nikhil Hajirnis et al |
| EPI_ISL_539687 | India/TG-CCMB-AA370/2020 | 2020-08-11 | Asia/India/Telangana/ | CSIR-Centre for Cellular and Molecular Biology | CSIR-Centre for Cellular and Molecular Biology | Nikhil Hajirnis et al |
| EPI_ISL_539688 | India/TG-CCMB-AA371/2020 | 2020-08-11 | Asia/India/Telangana/ | CSIR-Centre for Cellular and Molecular Biology | CSIR-Centre for Cellular and Molecular Biology | Payel Mukherjee et al |
| EPI_ISL_539689 | India/TG-CCMB-AA375/2020 | 2020-08-11 | Asia/India/Telangana/ | CSIR-Centre for Cellular and Molecular Biology | CSIR-Centre for Cellular and Molecular Biology | Payel Mukherjee et al |
| EPI_ISL_539690 | India/TG-CCMB-AA403/2020 | 2020-08-11 | Asia/India/Telangana/ | CSIR-Centre for Cellular and Molecular Biology | CSIR-Centre for Cellular and Molecular Biology | Payel Mukherjee et al |
| EPI_ISL_539691 | India/TG-CCMB-AB927/2020 | 2020-08-17 | Asia/India/Telangana/ | CSIR-Centre for Cellular and Molecular Biology | CSIR-Centre for Cellular and Molecular Biology | Pratheusa Maccha et al |
| EPI_ISL_539692 | India/TG-CCMB-AA373/2020 | 2020-08-11 | Asia/India/Telangana/ | CSIR-Centre for Cellular and Molecular Biology | CSIR-Centre for Cellular and Molecular Biology | Pratheusa Maccha et al |
| EPI_ISL_539693 | India/TG-CCMB-AA770/2020 | 2020-08-13 | Asia/India/Telangana/ | CSIR-Centre for Cellular and Molecular Biology | CSIR-Centre for Cellular and Molecular Biology | Pratheusa Maccha et al |
| EPI_ISL_539694 | India/TG-CCMB-AA456/2020 | 2020-08-11 | Asia/India/Telangana/ | CSIR-Centre for Cellular and Molecular Biology | CSIR-Centre for Cellular and Molecular Biology | Sakshi Shambhavi et al |
| EPI_ISL_539695 | India/TG-CCMB-AA867/2020 | 2020-08-13 | Asia/India/Telangana/ | CSIR-Centre for Cellular and Molecular Biology | CSIR-Centre for Cellular and Molecular Biology | Sakshi Shambhavi et al |
| EPI_ISL_539696 | India/TG-CCMB-AA868/2020 | 2020-08-13 | Asia/India/Telangana/ | CSIR-Centre for Cellular and Molecular Biology | CSIR-Centre for Cellular and Molecular Biology | Sakshi Shambhavi et al |
| EPI_ISL_539697 | India/TG-CCMB-AA869/2020 | 2020-08-13 | Asia/India/Telangana/ | CSIR-Centre for Cellular and Molecular Biology | CSIR-Centre for Cellular and Molecular Biology | Shagufta Khan et al |

| | | | | | | |
|---|---|---|---|---|---|---|
| EPI_ISL_539698 | India/TG-CCMB-AA870/2020 | 2020-08-13 | Asia/India/Telangana/ | CSIR-Centre for Cellular and Molecular Biology | CSIR-Centre for Cellular and Molecular Biology | Shagufta Khan et al |
| EPI_ISL_539699 | India/TG-CCMB-AA789/2020 | 2020-08-13 | Asia/India/Telangana/ | CSIR-Centre for Cellular and Molecular Biology | CSIR-Centre for Cellular and Molecular Biology | Shagufta Khan et al |
| EPI_ISL_539700 | India/TG-CCMB-AC568/2020 | 2020-08-19 | Asia/India/Telangana/ | CSIR-Centre for Cellular and Molecular Biology | CSIR-Centre for Cellular and Molecular Biology | Sofia Banu et al |
| EPI_ISL_539701 | India/TG-CCMB-AB91/2020 | 2020-08-14 | Asia/India/Telangana/ | CSIR-Centre for Cellular and Molecular Biology | CSIR-Centre for Cellular and Molecular Biology | Sofia Banu et al |
| EPI_ISL_539702 | India/TG-CCMB-AC214/2020 | 2020-08-18 | Asia/India/Telangana/ | CSIR-Centre for Cellular and Molecular Biology | CSIR-Centre for Cellular and Molecular Biology | Sofia Banu et al |
| EPI_ISL_539703 | India/TG-CCMB-AC528/2020 | 2020-08-19 | Asia/India/Telangana/ | CSIR-Centre for Cellular and Molecular Biology | CSIR-Centre for Cellular and Molecular Biology | Tulasi Nagabandi et al |
| EPI_ISL_539704 | India/TG-CCMB-AC329/2020 | 2020-08-19 | Asia/India/Telangana/ | CSIR-Centre for Cellular and Molecular Biology | CSIR-Centre for Cellular and Molecular Biology | Tulasi Nagabandi et al |
| EPI_ISL_539705 | India/TG-CCMB-AB261/2020 | 2020-08-14 | Asia/India/Telangana/ | CSIR-Centre for Cellular and Molecular Biology | CSIR-Centre for Cellular and Molecular Biology | Tulasi Nagabandi et al |
| EPI_ISL_539706 | India/TG-CCMB-AB16/2020 | 2020-08-14 | Asia/India/Telangana/ | CSIR-Centre for Cellular and Molecular Biology | CSIR-Centre for Cellular and Molecular Biology | Lamuk Zaveri et al |
| EPI_ISL_539707 | India/TG-CCMB-AB401/2020 | 2020-08-13 | Asia/India/Telangana/ | CSIR-Centre for Cellular and Molecular Biology | CSIR-Centre for Cellular and Molecular Biology | Lamuk Zaveri et al |
| EPI_ISL_539708 | India/TG-CCMB-AB966/2020 | 2020-08-17 | Asia/India/Telangana/ | CSIR-Centre for Cellular and Molecular Biology | CSIR-Centre for Cellular and Molecular Biology | Lamuk Zaveri et al |
| EPI_ISL_539709 | India/TG-CCMB-AB182/2020 | 2020-08-14 | Asia/India/Telangana/ | CSIR-Centre for Cellular and Molecular Biology | CSIR-Centre for Cellular and Molecular Biology | M Soujanya Reddy et al |
| EPI_ISL_539710 | India/TG-CCMB-AB192/2020 | 2020-08-14 | Asia/India/Telangana/ | CSIR-Centre for Cellular and Molecular Biology | CSIR-Centre for Cellular and Molecular Biology | M Soujanya Reddy et al |

| | | | | | | |
|---|---|---|---|---|---|---|
| EPI_ISL_539711 | India/TG-CCMB-R356/2020 | 2020-08-09 | Asia/India/Telangana/ | CSIR-Centre for Cellular and Molecular Biology | CSIR-Centre for Cellular and Molecular Biology | M Soujanya Reddy et al |
| EPI_ISL_539712 | India/TG-CCMB-AA222/2020 | 2020-08-10 | Asia/India/Telangana/ | CSIR-Centre for Cellular and Molecular Biology | CSIR-Centre for Cellular and Molecular Biology | Namami Gaur et al |
| EPI_ISL_539713 | India/TG-CCMB-AB226/2020 | 2020-08-14 | Asia/India/Telangana/ | CSIR-Centre for Cellular and Molecular Biology | CSIR-Centre for Cellular and Molecular Biology | Namami Gaur et al |
| EPI_ISL_539714 | India/TG-CCMB-AB215/2020 | 2020-08-14 | Asia/India/Telangana/ | CSIR-Centre for Cellular and Molecular Biology | CSIR-Centre for Cellular and Molecular Biology | Namami Gaur et al |
| EPI_ISL_539715 | India/TG-CCMB-AB248/2020 | 2020-08-14 | Asia/India/Telangana/ | CSIR-Centre for Cellular and Molecular Biology | CSIR-Centre for Cellular and Molecular Biology | Nikhil Hajirnis et al |
| EPI_ISL_539716 | India/TG-CCMB-AA311/2020 | 2020-08-11 | Asia/India/Telangana/ | CSIR-Centre for Cellular and Molecular Biology | CSIR-Centre for Cellular and Molecular Biology | Nikhil Hajirnis et al |
| EPI_ISL_539717 | India/TG-CCMB-AC317/2020 | 2020-08-19 | Asia/India/Telangana/ | CSIR-Centre for Cellular and Molecular Biology | CSIR-Centre for Cellular and Molecular Biology | Nikhil Hajirnis et al |
| EPI_ISL_539718 | India/TG-CCMB-AB344/2020 | 2020-08-13 | Asia/India/Telangana/ | CSIR-Centre for Cellular and Molecular Biology | CSIR-Centre for Cellular and Molecular Biology | Payel Mukherjee et al |
| EPI_ISL_539719 | India/TG-CCMB-AA591/2020 | 2020-08-12 | Asia/India/Telangana/ | CSIR-Centre for Cellular and Molecular Biology | CSIR-Centre for Cellular and Molecular Biology | Payel Mukherjee et al |
| EPI_ISL_539720 | India/TG-CCMB-R296/2020 | 2020-08-09 | Asia/India/Telangana/ | CSIR-Centre for Cellular and Molecular Biology | CSIR-Centre for Cellular and Molecular Biology | Payel Mukherjee et al |
| EPI_ISL_539721 | India/TG-CCMB-AB415/2020 | 2020-08-14 | Asia/India/Telangana/ | CSIR-Centre for Cellular and Molecular Biology | CSIR-Centre for Cellular and Molecular Biology | Pratheusa Maccha et al |
| EPI_ISL_539722 | India/TG-CCMB-AA871/2020 | 2020-08-13 | Asia/India/Telangana/ | CSIR-Centre for Cellular and Molecular Biology | CSIR-Centre for Cellular and Molecular Biology | Pratheusa Maccha et al |
| EPI_ISL_539723 | India/TG-CCMB-AC314/2020 | 2020-08-19 | Asia/India/Telangana/ | CSIR-Centre for Cellular and Molecular Biology | CSIR-Centre for Cellular and Molecular Biology | Pratheusa Maccha et al |

| | | | | | | |
|---|---|---|---|---|---|---|
| EPI_ISL_539724 | India/TG-CCMB-R338/2020 | 2020-08-09 | Asia/India/Telangana/ | CSIR-Centre for Cellular and Molecular Biology | CSIR-Centre for Cellular and Molecular Biology | Sakshi Shambhavi et al |
| EPI_ISL_539725 | India/TG-CCMB-AA344/2020 | 2020-08-11 | Asia/India/Telangana/ | CSIR-Centre for Cellular and Molecular Biology | CSIR-Centre for Cellular and Molecular Biology | Sakshi Shambhavi et al |
| EPI_ISL_539726 | India/TG-CCMB-AB610/2020 | 2020-08-16 | Asia/India/Telangana/ | CSIR-Centre for Cellular and Molecular Biology | CSIR-Centre for Cellular and Molecular Biology | Sakshi Shambhavi et al |
| EPI_ISL_539727 | India/TG-CCMB-AC475/2020 | 2020-08-19 | Asia/India/Telangana/ | CSIR-Centre for Cellular and Molecular Biology | CSIR-Centre for Cellular and Molecular Biology | Shagufta Khan et al |
| EPI_ISL_539728 | India/TG-CCMB-AB603/2020 | 2020-08-16 | Asia/India/Telangana/ | CSIR-Centre for Cellular and Molecular Biology | CSIR-Centre for Cellular and Molecular Biology | Shagufta Khan et al |
| EPI_ISL_539729 | India/TG-CCMB-AB05/2020 | 2020-08-13 | Asia/India/Telangana/ | CSIR-Centre for Cellular and Molecular Biology | CSIR-Centre for Cellular and Molecular Biology | Shagufta Khan et al |
| EPI_ISL_539730 | India/TG-CCMB-AB509/2020 | 2020-08-14 | Asia/India/Telangana/ | CSIR-Centre for Cellular and Molecular Biology | CSIR-Centre for Cellular and Molecular Biology | Sofia Banu et al |
| EPI_ISL_539731 | India/TG-CCMB-AA902/2020 | 2020-08-13 | Asia/India/Telangana/ | CSIR-Centre for Cellular and Molecular Biology | CSIR-Centre for Cellular and Molecular Biology | Sofia Banu et al |
| EPI_ISL_539732 | India/TG-CCMB-AA903/2020 | 2020-08-13 | Asia/India/Telangana/ | CSIR-Centre for Cellular and Molecular Biology | CSIR-Centre for Cellular and Molecular Biology | Sofia Banu et al |
| EPI_ISL_539733 | India/TG-CCMB-AA616/2020 | 2020-08-12 | Asia/India/Telangana/ | CSIR-Centre for Cellular and Molecular Biology | CSIR-Centre for Cellular and Molecular Biology | Tulasi Nagabandi et al |
| EPI_ISL_539734 | India/TG-CCMB-AA713/2020 | 2020-08-12 | Asia/India/Telangana/ | CSIR-Centre for Cellular and Molecular Biology | CSIR-Centre for Cellular and Molecular Biology | Tulasi Nagabandi et al |
| EPI_ISL_539735 | India/TG-CCMB-AA818/2020 | 2020-08-13 | Asia/India/Telangana/ | CSIR-Centre for Cellular and Molecular Biology | CSIR-Centre for Cellular and Molecular Biology | Tulasi Nagabandi et al |
| EPI_ISL_539736 | India/TG-CCMB-AA620/2020 | 2020-08-12 | Asia/India/Telangana/ | CSIR-Centre for Cellular and Molecular Biology | CSIR-Centre for Cellular and Molecular Biology | Lamuk Zaveri et al |

| | | | | | | |
|---|---|---|---|---|---|---|
| EPI_ISL_539737 | India/TG-CCMB-AC338/2020 | 2020-08-19 | Asia/India/Telangana/ | CSIR-Centre for Cellular and Molecular Biology | CSIR-Centre for Cellular and Molecular Biology | Lamuk Zaveri et al |
| EPI_ISL_539739 | India/TG-CCMB-AA41/2020 | 2020-08-10 | Asia/India/Telangana/ | CSIR-Centre for Cellular and Molecular Biology | CSIR-Centre for Cellular and Molecular Biology | M Soujanya Reddy et al |
| EPI_ISL_539740 | India/TG-CCMB-AB227/2020 | 2020-08-14 | Asia/India/Telangana/ | CSIR-Centre for Cellular and Molecular Biology | CSIR-Centre for Cellular and Molecular Biology | M Soujanya Reddy et al |
| EPI_ISL_539741 | India/TG-CCMB-AC391/2020 | 2020-08-19 | Asia/India/Telangana/ | CSIR-Centre for Cellular and Molecular Biology | CSIR-Centre for Cellular and Molecular Biology | M Soujanya Reddy et al |
| EPI_ISL_539742 | India/TG-CCMB-AB955/2020 | 2020-08-17 | Asia/India/Telangana/ | CSIR-Centre for Cellular and Molecular Biology | CSIR-Centre for Cellular and Molecular Biology | Namami Gaur et al |
| EPI_ISL_539743 | India/TG-CCMB-AC222/2020 | 2020-08-18 | Asia/India/Telangana/ | CSIR-Centre for Cellular and Molecular Biology | CSIR-Centre for Cellular and Molecular Biology | Namami Gaur et al |
| EPI_ISL_539744 | India/TG-CCMB-AC511/2020 | 2020-08-19 | Asia/India/Telangana/ | CSIR-Centre for Cellular and Molecular Biology | CSIR-Centre for Cellular and Molecular Biology | Namami Gaur et al |
| EPI_ISL_539745 | India/TG-CCMB-R286/2020 | 2020-08-09 | Asia/India/Telangana/ | CSIR-Centre for Cellular and Molecular Biology | CSIR-Centre for Cellular and Molecular Biology | Nikhil Hajirnis et al |
| EPI_ISL_539746 | India/TG-CCMB-AB198/2020 | 2020-08-14 | Asia/India/Telangana/ | CSIR-Centre for Cellular and Molecular Biology | CSIR-Centre for Cellular and Molecular Biology | Nikhil Hajirnis et al |
| EPI_ISL_539747 | India/TG-CCMB-AA812/2020 | 2020-08-13 | Asia/India/Telangana/ | CSIR-Centre for Cellular and Molecular Biology | CSIR-Centre for Cellular and Molecular Biology | Nikhil Hajirnis et al |
| EPI_ISL_539749 | India/TG-CCMB-AA340/2020 | 2020-08-11 | Asia/India/Telangana/ | CSIR-Centre for Cellular and Molecular Biology | CSIR-Centre for Cellular and Molecular Biology | Payel Mukherjee et al |
| EPI_ISL_539750 | India/TG-CCMB-AA590/2020 | 2020-08-12 | Asia/India/Telangana/ | CSIR-Centre for Cellular and Molecular Biology | CSIR-Centre for Cellular and Molecular Biology | Payel Mukherjee et al |
| EPI_ISL_539751 | India/TG-CCMB-AA241/2020 | 2020-08-11 | Asia/India/Telangana/ | CSIR-Centre for Cellular and Molecular Biology | CSIR-Centre for Cellular and Molecular Biology | Pratheusa Maccha et al |

| | | | | | | |
|---|---|---|---|---|---|---|
| EPI_ISL_539752 | India/TG-CCMB-AA272/2020 | 2020-08-11 | Asia/India/Telangana/ | CSIR-Centre for Cellular and Molecular Biology | CSIR-Centre for Cellular and Molecular Biology | Pratheusa Maccha et al |
| EPI_ISL_539753 | India/TG-CCMB-AA29/2020 | 2020-08-10 | Asia/India/Telangana/ | CSIR-Centre for Cellular and Molecular Biology | CSIR-Centre for Cellular and Molecular Biology | Pratheusa Maccha et al |
| EPI_ISL_539754 | India/TG-CCMB-AA315/2020 | 2020-08-11 | Asia/India/Telangana/ | CSIR-Centre for Cellular and Molecular Biology | CSIR-Centre for Cellular and Molecular Biology | Sakshi Shambhavi et al |
| EPI_ISL_539755 | India/TG-CCMB-AA32/2020 | 2020-08-10 | Asia/India/Telangana/ | CSIR-Centre for Cellular and Molecular Biology | CSIR-Centre for Cellular and Molecular Biology | Sakshi Shambhavi et al |
| EPI_ISL_539756 | India/TG-CCMB-AA335/2020 | 2020-08-11 | Asia/India/Telangana/ | CSIR-Centre for Cellular and Molecular Biology | CSIR-Centre for Cellular and Molecular Biology | Sakshi Shambhavi et al |
| EPI_ISL_539757 | India/TG-CCMB-AA372/2020 | 2020-08-11 | Asia/India/Telangana/ | CSIR-Centre for Cellular and Molecular Biology | CSIR-Centre for Cellular and Molecular Biology | Shagufta Khan et al |
| EPI_ISL_539758 | India/TG-CCMB-AA42/2020 | 2020-08-10 | Asia/India/Telangana/ | CSIR-Centre for Cellular and Molecular Biology | CSIR-Centre for Cellular and Molecular Biology | Shagufta Khan et al |
| EPI_ISL_539759 | India/TG-CCMB-AA617/2020 | 2020-08-12 | Asia/India/Telangana/ | CSIR-Centre for Cellular and Molecular Biology | CSIR-Centre for Cellular and Molecular Biology | Shagufta Khan et al |
| EPI_ISL_539760 | India/TG-CCMB-AA636/2020 | 2020-08-12 | Asia/India/Telangana/ | CSIR-Centre for Cellular and Molecular Biology | CSIR-Centre for Cellular and Molecular Biology | Sofia Banu et al |
| EPI_ISL_539761 | India/TG-CCMB-AA68/2020 | 2020-08-10 | Asia/India/Telangana/ | CSIR-Centre for Cellular and Molecular Biology | CSIR-Centre for Cellular and Molecular Biology | Sofia Banu et al |
| EPI_ISL_539762 | India/TG-CCMB-AA833/2020 | 2020-08-13 | Asia/India/Telangana/ | CSIR-Centre for Cellular and Molecular Biology | CSIR-Centre for Cellular and Molecular Biology | Sofia Banu et al |
| EPI_ISL_539763 | India/TG-CCMB-AA979/2020 | 2020-08-13 | Asia/India/Telangana/ | CSIR-Centre for Cellular and Molecular Biology | CSIR-Centre for Cellular and Molecular Biology | Tulasi Nagabandi et al |
| EPI_ISL_539764 | India/TG-CCMB-AB229/2020 | 2020-08-14 | Asia/India/Telangana/ | CSIR-Centre for Cellular and Molecular Biology | CSIR-Centre for Cellular and Molecular Biology | Tulasi Nagabandi et al |

| | | | | | | |
|---|---|---|---|---|---|---|
| EPI_ISL_539765 | India/TG-CCMB-AB247/2020 | 2020-08-14 | Asia/India/Telangana/ | CSIR-Centre for Cellular and Molecular Biology | CSIR-Centre for Cellular and Molecular Biology | Tulasi Nagabandi et al |
| EPI_ISL_539766 | India/TG-CCMB-AB249/2020 | 2020-08-14 | Asia/India/Telangana/ | CSIR-Centre for Cellular and Molecular Biology | CSIR-Centre for Cellular and Molecular Biology | Tulasi Nagabandi et al |
| EPI_ISL_539767 | India/TG-CCMB-AB297/2020 | 2020-08-15 | Asia/India/Telangana/ | CSIR-Centre for Cellular and Molecular Biology | CSIR-Centre for Cellular and Molecular Biology | Tulasi Nagabandi et al |
| EPI_ISL_539768 | India/TG-CCMB-AB430/2020 | 2020-08-14 | Asia/India/Telangana/ | CSIR-Centre for Cellular and Molecular Biology | CSIR-Centre for Cellular and Molecular Biology | Tulasi Nagabandi et al |
| EPI_ISL_539769 | India/TG-CCMB-AB490/2020 | 2020-08-14 | Asia/India/Telangana/ | CSIR-Centre for Cellular and Molecular Biology | CSIR-Centre for Cellular and Molecular Biology | Lamuk Zaveri et al |
| EPI_ISL_539770 | India/TG-CCMB-AB519/2020 | 2020-08-14 | Asia/India/Telangana/ | CSIR-Centre for Cellular and Molecular Biology | CSIR-Centre for Cellular and Molecular Biology | Lamuk Zaveri et al |
| EPI_ISL_539771 | India/TG-CCMB-AB523/2020 | 2020-08-14 | Asia/India/Telangana/ | CSIR-Centre for Cellular and Molecular Biology | CSIR-Centre for Cellular and Molecular Biology | Pratheusa Maccha et al |
| EPI_ISL_539772 | India/TG-CCMB-AC529/2020 | 2020-08-19 | Asia/India/Telangana/ | CSIR-Centre for Cellular and Molecular Biology | CSIR-Centre for Cellular and Molecular Biology | Pratheusa Maccha et al |
| EPI_ISL_539773 | India/TG-CCMB-AC576/2020 | 2020-08-19 | Asia/India/Telangana/ | CSIR-Centre for Cellular and Molecular Biology | CSIR-Centre for Cellular and Molecular Biology | Payel Mukherjee et al |
| EPI_ISL_539774 | India/TG-CCMB-R124/2020 | 2020-08-09 | Asia/India/Telangana/ | CSIR-Centre for Cellular and Molecular Biology | CSIR-Centre for Cellular and Molecular Biology | Payel Mukherjee et al |
| EPI_ISL_539775 | India/TG-CCMB-R325/2020 | 2020-08-09 | Asia/India/Telangana/ | CSIR-Centre for Cellular and Molecular Biology | CSIR-Centre for Cellular and Molecular Biology | Payel Mukherjee et al |
| EPI_ISL_539780 | CzechRepublic/NRL-6047/2020 | 2020-04-21 | Europe/Czech Republic/Prague/ | National Institute of Public Health (Czech Republic) | State Veterinary Institute Prague | Nagy et al |
| EPI_ISL_539783 | Ecuador/USFQ-208/2020 | 2020-06-15 | South America/Ecuador/Napo/Tena | Universidad Regional Amazonica IKIAM | Institute of Microbiology, Universidad San Francisco de Quito | Fabian Aguilar et al |

| | | | | | | |
|---|---|---|---|---|---|---|
| EPI_ISL_539784 | Ecuador/USFQ-209/2020 | 2020-08-19 | South America/Ecuador/Napo/Tena | Universidad Regional Amazonica IKIAM | Institute of Microbiology, Universidad San Francisco de Quito | Fabian Aguilar et al |
| EPI_ISL_539785 | Ecuador/USFQ-212/2020 | 2020-08-04 | South America/Ecuador/Azuay/Cuenca E | Institute of Microbiology, Universidad San Francisco de Quito | Institute of Microbiology, Universidad San Francisco de Quito | Andrea Macias et al |
| EPI_ISL_539788 | Ecuador/USFQ-216/2020 | 2020-08-05 | South America/Ecuador/Azuay/Cuenca E | Institute of Microbiology, Universidad San Francisco de Quito | Institute of Microbiology, Universidad San Francisco de Quito | Andrea Macias et al |
| EPI_ISL_539789 | Ecuador/USFQ-219/2020 | 2020-08-15 | South America/Ecuador/Loja/Macara | Institute of Microbiology, Universidad San Francisco de Quito | Institute of Microbiology, Universidad San Francisco de Quito | Belv©n Prado-Vivar et al |
| EPI_ISL_539790 | Ecuador/USFQ-220/2020 | 2020-08-15 | South America/Ecuador/Loja/Macara | Institute of Microbiology, Universidad San Francisco de Quito | Institute of Microbiology, Universidad San Francisco de Quito | Belv©n Prado-Vivar et al |
| EPI_ISL_539791 | Ecuador/USFQ-222/2020 | 2020-08-15 | South America/Ecuador/Loja/Macara | Institute of Microbiology, Universidad San Francisco de Quito | Institute of Microbiology, Universidad San Francisco de Quito | Belv©n Prado-Vivar et al |
| EPI_ISL_539792 | Ecuador/USFQ-224/2020 | 2020-08-15 | South America/Ecuador/Loja/Catamayo | Institute of Microbiology, Universidad San Francisco de Quito | Institute of Microbiology, Universidad San Francisco de Quito | Belv©n Prado-Vivar et al |
| EPI_ISL_539793 | Ecuador/USFQ-226/2020 | 2020-08-15 | South America/Ecuador/Loja/Catamayo | Institute of Microbiology, Universidad San Francisco de Quito | Institute of Microbiology, Universidad San Francisco de Quito | Belv©n Prado-Vivar et al |
| EPI_ISL_539832 | USA/MN-MDH-1638/2020 | 2020-05-21 | North America/USA/Minnesota/ | Minnesota Department of Health, Public Health Laboratory | Minnesota Department of Health, Public Health Laboratory | Matt Plumb et al |
| EPI_ISL_539836 | USA/MN-MDH-1642/2020 | 2020-05-21 | North America/USA/Minnesota/ | Minnesota Department of Health, Public Health Laboratory | Minnesota Department of Health, Public Health Laboratory | Matt Plumb et al |

| | | | | | | |
|---|---|---|---|---|---|---|
| EPI_ISL_539843 | USA/MN-MDH-1649/2020 | 2020-08-18 | North America/USA/Minnesota/ | Mayo Clinic & Mayo Clinic Laboratories | Minnesota Department of Health, Public Health Laboratory | Matt Plumb et al |
| EPI_ISL_539845 | USA/MN-MDH-1651/2020 | 2020-08-17 | North America/USA/Minnesota/ | Mayo Clinic & Mayo Clinic Laboratories | Minnesota Department of Health, Public Health Laboratory | Matt Plumb et al |
| EPI_ISL_539847 | USA/MN-MDH-1653/2020 | 2020-08-12 | North America/USA/Minnesota/ | Mayo Clinic & Mayo Clinic Laboratories | Minnesota Department of Health, Public Health Laboratory | Matt Plumb et al |
| EPI_ISL_539849 | USA/MN-MDH-1655/2020 | 2020-08-19 | North America/USA/Minnesota/ | Mayo Clinic & Mayo Clinic Laboratories | Minnesota Department of Health, Public Health Laboratory | Matt Plumb et al |
| EPI_ISL_539883 | Sweden/20-52156/2020 | 2020-06-24 | Europe/Sweden/Vastra Gotaland/ | Omtanken Grimmered | The Public Health Agency of Sweden | Anna-Malin Linde et al |
| EPI_ISL_539884 | Sweden/20-52440/2020 | 2020-07-22 | Europe/Sweden/Vastra Gotaland/ | Omtanken Grimmered | The Public Health Agency of Sweden | Anna-Malin Linde et al |
| EPI_ISL_540052 | England/QEUH-990400/2020 | 2020-08-26 | Europe/United Kingdom/England/ | Lighthouse Lab in Glasgow | Wellcome Sanger Institute for the COVID-19 Genomics UK (COG-UK) consortium | Harper VanSteenhouse et al |
| EPI_ISL_540149 | England/QEUH-9A4427/2020 | 2020-09-01 | Europe/United Kingdom/England/ | Lighthouse Lab in Glasgow | Wellcome Sanger Institute for the COVID-19 Genomics UK (COG-UK) consortium | Harper VanSteenhouse et al |
| EPI_ISL_540172 | England/QEUH-9A468B/2020 | 2020-09-01 | Europe/United Kingdom/England/ | Lighthouse Lab in Glasgow | Wellcome Sanger Institute for the COVID-19 Genomics UK (COG-UK) consortium | Harper VanSteenhouse et al |
| EPI_ISL_540179 | England/QEUH-9A46A9/2020 | 2020-09-01 | Europe/United Kingdom/England/ | Lighthouse Lab in Glasgow | Wellcome Sanger Institute for the COVID-19 Genomics UK (COG-UK) consortium | Harper VanSteenhouse et al |

| | | | | | | |
|---|---|---|---|---|---|---|
| EPI_ISL_540186 | England/QEUH-9A4348/2020 | 2020-09-01 | Europe/United Kingdom/England/ | Lighthouse Lab in Glasgow | Wellcome Sanger Institute for the COVID-19 Genomics UK (COG-UK) consortium | Harper VanSteenhouse et al |
| EPI_ISL_540422 | USA/WY-WYPHL-20011656/2020 | 2020-05-01 | North America/USA/Wyoming/ | Wyoming Public Health Laboratory | Wyoming Public Health Laboratory | Noah Hull et al |
| EPI_ISL_540431 | USA/IN-CDC-0821/2020 | 2020-04-05 | North America/USA/Indiana/ | IN State Department of Health Laboratory Services | Pathogen Discovery, Respiratory Viruses Branch, Division of Viral Diseases, Centers for Disease Control and Prevention | Yan Li et al |
| EPI_ISL_540449 | Belgium/ULG-10282/2020 | 2020-07-25 | Europe/Belgium/Limburg BE/ | University of Li√®ge COVID-19 testing center | GIGA Medical Genomics | Keith Durkin et al |
| EPI_ISL_540475 | Belgium/ULG-10308/2020 | 2020-07-24 | Europe/Belgium/Li√®ge/ | Department of Clinical Microbiology | GIGA Medical Genomics | Keith Durkin et al |
| EPI_ISL_540522 | Belgium/ULG-10355/2020 | 2020-08-04 | Europe/Belgium/Li√®ge/ | Department of Clinical Microbiology | GIGA Medical Genomics | Keith Durkin et al |
| EPI_ISL_540526 | Belgium/ULG-10359/2020 | 2020-08-06 | Europe/Belgium/Li√®ge/ | Department of Clinical Microbiology | GIGA Medical Genomics | Keith Durkin et al |
| EPI_ISL_540551 | Belgium/ULG-10384/2020 | 2020-08-07 | Europe/Belgium/Li√®ge/ | Department of Clinical Microbiology | GIGA Medical Genomics | Keith Durkin et al |
| EPI_ISL_540553 | Belgium/ULG-10386/2020 | 2020-08-07 | Europe/Belgium/Li√®ge/ | Department of Clinical Microbiology | GIGA Medical Genomics | Keith Durkin et al |
| EPI_ISL_540621 | England/LIVE-AAE3B/2020 | 2020-07-28 | Europe/United Kingdom/England/ | Liverpool Clinical Laboratories | COVID-19 Genomics UK (COG-UK) Consortium | Sam Haldenby et al |
| EPI_ISL_540921 | USA/WY-WYPHL-20015627/2020 | 2020-05-18 | North America/USA/Wyoming/ | Wyoming Public Health Laboratory | Wyoming Public Health Laboratory | Noah Hull et al |
| EPI_ISL_540923 | Peru/ANC-UPCH-0010/2020 | 2020-06-17 | South America/Peru/Ancash/Huarmey | Laboratorio de Referencia Nacional de Virus Respiratorios, Instituto Nacional de Salud Peru | Laboratorio de Gen√≥mica Microbiana, Universidad Peruana Cayetano Heredia | Pablo Tsukayama et al |

| | | | | | | |
|---|---|---|---|---|---|---|
| EPI_ISL_540926 | Peru/LIM-UPCH-0025/2020 | 2020-06-17 | South America/Peru/Lima/Lima | Laboratorio de Referencia Nacional de Virus Respiratorios, Instituto Nacional de Salud Peru | Laboratorio de Genv≥mica Microbiana, Universidad Peruana Cayetano Heredia | Pablo Tsukayama et al |
| EPI_ISL_540927 | Peru/CAL-UPCH-0026/2020 | 2020-06-16 | South America/Peru/Callao/Callao | Laboratorio de Referencia Nacional de Virus Respiratorios, Instituto Nacional de Salud Peru | Laboratorio de Genv≥mica Microbiana, Universidad Peruana Cayetano Heredia | Pablo Tsukayama et al |
| EPI_ISL_540928 | Peru/CAL-UPCH-0027/2020 | 2020-06-17 | South America/Peru/Callao/Callao | Laboratorio de Referencia Nacional de Virus Respiratorios, Instituto Nacional de Salud Peru | Laboratorio de Genv≥mica Microbiana, Universidad Peruana Cayetano Heredia | Pablo Tsukayama et al |
| EPI_ISL_540929 | Peru/LIM-UPCH-0029/2020 | 2020-06-15 | South America/Peru/Lima/Lima | Laboratorio de Referencia Nacional de Virus Respiratorios, Instituto Nacional de Salud Peru | Laboratorio de Genv≥mica Microbiana, Universidad Peruana Cayetano Heredia | Pablo Tsukayama et al |
| EPI_ISL_540930 | Peru/LIM-UPCH-0030/2020 | 2020-06-17 | South America/Peru/Lima/Lima | Laboratorio de Referencia Nacional de Virus Respiratorios, Instituto Nacional de Salud Peru | Laboratorio de Genv≥mica Microbiana, Universidad Peruana Cayetano Heredia | Pablo Tsukayama et al |
| EPI_ISL_540931 | Peru/LIM-UPCH-0031/2020 | 2020-06-18 | South America/Peru/Lima/Lima | Laboratorio de Referencia Nacional de Virus Respiratorios, Instituto Nacional de Salud Peru | Laboratorio de Genv≥mica Microbiana, Universidad Peruana Cayetano Heredia | Pablo Tsukayama et al |

| | | | | | | |
|---|---|---|---|---|---|---|
| EPI_ISL_540933 | Peru/LIM-UPCH-0033/2020 | 2020-06-17 | South America/Peru/Lima/Lima | Laboratorio de Referencia Nacional de Virus Respiratorios, Instituto Nacional de Salud Peru | Laboratorio de Genv≥mica Microbiana, Universidad Peruana Cayetano Heredia | Pablo Tsukayama et al |
| EPI_ISL_540934 | Peru/LIM-UPCH-0034/2020 | 2020-06-17 | South America/Peru/Lima/Lima | Laboratorio de Referencia Nacional de Virus Respiratorios, Instituto Nacional de Salud Peru | Laboratorio de Genv≥mica Microbiana, Universidad Peruana Cayetano Heredia | Pablo Tsukayama et al |
| EPI_ISL_540935 | Peru/CAL-UPCH-0035/2020 | 2020-06-17 | South America/Peru/Callao/Callao | Laboratorio de Referencia Nacional de Virus Respiratorios, Instituto Nacional de Salud Peru | Laboratorio de Genv≥mica Microbiana, Universidad Peruana Cayetano Heredia | Pablo Tsukayama et al |
| EPI_ISL_540936 | Peru/LIM-UPCH-0045/2020 | 2020-06-17 | South America/Peru/Lima/Cav±ete | Laboratorio de Referencia Nacional de Virus Respiratorios, Instituto Nacional de Salud Peru | Laboratorio de Genv≥mica Microbiana, Universidad Peruana Cayetano Heredia | Pablo Tsukayama et al |
| EPI_ISL_540937 | Peru/CAL-UPCH-0046/2020 | 2020-06-16 | South America/Peru/Callao/Callao | Laboratorio de Referencia Nacional de Virus Respiratorios, Instituto Nacional de Salud Peru | Laboratorio de Genv≥mica Microbiana, Universidad Peruana Cayetano Heredia | Pablo Tsukayama et al |
| EPI_ISL_540938 | Peru/CAL-UPCH-0047/2020 | 2020-06-18 | South America/Peru/Callao/Callao | Laboratorio de Referencia Nacional de Virus Respiratorios, Instituto Nacional de Salud Peru | Laboratorio de Genv≥mica Microbiana, Universidad Peruana Cayetano Heredia | Pablo Tsukayama et al |

| | | | | | | |
|---|---|---|---|---|---|---|
| EPI_ISL_540939 | Peru/LIM-UPCH-0050/2020 | 2020-06-17 | South America/Peru/Lima/Lima | Laboratorio de Referencia Nacional de Virus Respiratorios, Instituto Nacional de Salud Peru | Laboratorio de Genv≥mica Microbiana, Universidad Peruana Cayetano Heredia | Pablo Tsukayama et al |
| EPI_ISL_540940 | Peru/ANC-UPCH-0051/2020 | 2020-06-16 | South America/Peru/Ancash/Santa | Laboratorio de Referencia Nacional de Virus Respiratorios, Instituto Nacional de Salud Peru | Laboratorio de Genv≥mica Microbiana, Universidad Peruana Cayetano Heredia | Pablo Tsukayama et al |
| EPI_ISL_540941 | Peru/CAL-UPCH-0053/2020 | 2020-06-16 | South America/Peru/Callao/Callao | Laboratorio de Referencia Nacional de Virus Respiratorios, Instituto Nacional de Salud Peru | Laboratorio de Genv≥mica Microbiana, Universidad Peruana Cayetano Heredia | Pablo Tsukayama et al |
| EPI_ISL_540942 | Peru/LIM-UPCH-0056/2020 | 2020-06-18 | South America/Peru/Lima/Lima | Laboratorio de Referencia Nacional de Virus Respiratorios, Instituto Nacional de Salud Peru | Laboratorio de Genv≥mica Microbiana, Universidad Peruana Cayetano Heredia | Pablo Tsukayama et al |
| EPI_ISL_540944 | Peru/LIM-UPCH-0059/2020 | 2020-06-17 | South America/Peru/Lima/Lima | Laboratorio de Referencia Nacional de Virus Respiratorios, Instituto Nacional de Salud Peru | Laboratorio de Genv≥mica Microbiana, Universidad Peruana Cayetano Heredia | Pablo Tsukayama et al |
| EPI_ISL_540945 | Peru/LIM-UPCH-0061/2020 | 2020-06-17 | South America/Peru/Lima/Lima | Laboratorio de Referencia Nacional de Virus Respiratorios, Instituto Nacional de Salud Peru | Laboratorio de Genv≥mica Microbiana, Universidad Peruana Cayetano Heredia | Pablo Tsukayama et al |

| | | | | | | |
|---|---|---|---|---|---|---|
| EPI_ISL_540946 | Peru/CAL-UPCH-0062/2020 | 2020-06-17 | South America/Peru/Callao/Callao | Laboratorio de Referencia Nacional de Virus Respiratorios, Instituto Nacional de Salud Peru | Laboratorio de Gen√≥mica Microbiana, Universidad Peruana Cayetano Heredia | Pablo Tsukayama et al |
| EPI_ISL_540947 | Peru/LIM-UPCH-0063/2020 | 2020-06-17 | South America/Peru/Lima/Ca√±ete | Laboratorio de Referencia Nacional de Virus Respiratorios, Instituto Nacional de Salud Peru | Laboratorio de Gen√≥mica Microbiana, Universidad Peruana Cayetano Heredia | Pablo Tsukayama et al |
| EPI_ISL_540948 | Peru/LIM-UPCH-0065/2020 | 2020-06-18 | South America/Peru/Lima/Lima | Laboratorio de Referencia Nacional de Virus Respiratorios, Instituto Nacional de Salud Peru | Laboratorio de Gen√≥mica Microbiana, Universidad Peruana Cayetano Heredia | Pablo Tsukayama et al |
| EPI_ISL_540949 | Peru/LIM-UPCH-0066/2020 | 2020-06-17 | South America/Peru/Lima/Lima | Laboratorio de Referencia Nacional de Virus Respiratorios, Instituto Nacional de Salud Peru | Laboratorio de Gen√≥mica Microbiana, Universidad Peruana Cayetano Heredia | Pablo Tsukayama et al |
| EPI_ISL_540950 | Peru/UCA-UPCH-0067/2020 | 2020-06-11 | South America/Peru/Ucayali/Coronel Po | Laboratorio de Referencia Nacional de Virus Respiratorios, Instituto Nacional de Salud Peru | Laboratorio de Gen√≥mica Microbiana, Universidad Peruana Cayetano Heredia | Pablo Tsukayama et al |
| EPI_ISL_540951 | Peru/LIM-UPCH-0068/2020 | 2020-06-18 | South America/Peru/Lima/Lima | Laboratorio de Referencia Nacional de Virus Respiratorios, Instituto Nacional de Salud Peru | Laboratorio de Gen√≥mica Microbiana, Universidad Peruana Cayetano Heredia | Pablo Tsukayama et al |

| | | | | | | |
|---|---|---|---|---|---|---|
| EPI_ISL_540952 | Peru/LIM-UPCH-0070/2020 | 2020-06-17 | South America/Peru/Lima/Lima | Laboratorio de Referencia Nacional de Virus Respiratorios, Instituto Nacional de Salud Peru | Laboratorio de Genv≥mica Microbiana, Universidad Peruana Cayetano Heredia | Pablo Tsukayama et al |
| EPI_ISL_540953 | Peru/CAL-UPCH-0071/2020 | 2020-06-17 | South America/Peru/Callao/Callao | Laboratorio de Referencia Nacional de Virus Respiratorios, Instituto Nacional de Salud Peru | Laboratorio de Genv≥mica Microbiana, Universidad Peruana Cayetano Heredia | Pablo Tsukayama et al |
| EPI_ISL_540955 | Peru/LIM-UPCH-0074/2020 | 2020-06-18 | South America/Peru/Lima/Lima | Laboratorio de Referencia Nacional de Virus Respiratorios, Instituto Nacional de Salud Peru | Laboratorio de Genv≥mica Microbiana, Universidad Peruana Cayetano Heredia | Pablo Tsukayama et al |
| EPI_ISL_540956 | Peru/LIM-UPCH-0078/2020 | 2020-06-17 | South America/Peru/Lima/Lima | Laboratorio de Referencia Nacional de Virus Respiratorios, Instituto Nacional de Salud Peru | Laboratorio de Genv≥mica Microbiana, Universidad Peruana Cayetano Heredia | Pablo Tsukayama et al |
| EPI_ISL_540957 | Peru/LIM-UPCH-0079/2020 | 2020-06-17 | South America/Peru/Lima/Lima | Laboratorio de Referencia Nacional de Virus Respiratorios, Instituto Nacional de Salud Peru | Laboratorio de Genv≥mica Microbiana, Universidad Peruana Cayetano Heredia | Pablo Tsukayama et al |
| EPI_ISL_540958 | Peru/CAL-UPCH-0080/2020 | 2020-06-17 | South America/Peru/Callao/Callao | Laboratorio de Referencia Nacional de Virus Respiratorios, Instituto Nacional de Salud Peru | Laboratorio de Genv≥mica Microbiana, Universidad Peruana Cayetano Heredia | Pablo Tsukayama et al |

| | | | | | | |
|---|---|---|---|---|---|---|
| EPI_ISL_540960 | Peru/LIM-UPCH-0082/2020 | 2020-06-17 | South America/Peru/Lima/Lima | Laboratorio de Referencia Nacional de Virus Respiratorios, Instituto Nacional de Salud Peru | Laboratorio de Gen√≥mica Microbiana, Universidad Peruana Cayetano Heredia | Pablo Tsukayama et al |
| EPI_ISL_540961 | Peru/LIM-UPCH-0083/2020 | 2020-06-18 | South America/Peru/Lima/Lima | Laboratorio de Referencia Nacional de Virus Respiratorios, Instituto Nacional de Salud Peru | Laboratorio de Gen√≥mica Microbiana, Universidad Peruana Cayetano Heredia | Pablo Tsukayama et al |
| EPI_ISL_540962 | Peru/LIM-UPCH-0084/2020 | 2020-06-17 | South America/Peru/Lima/Lima | Laboratorio de Referencia Nacional de Virus Respiratorios, Instituto Nacional de Salud Peru | Laboratorio de Gen√≥mica Microbiana, Universidad Peruana Cayetano Heredia | Pablo Tsukayama et al |
| EPI_ISL_540963 | Peru/LIM-UPCH-0085/2020 | 2020-06-17 | South America/Peru/Lima/Huarochiri | Laboratorio de Referencia Nacional de Virus Respiratorios, Instituto Nacional de Salud Peru | Laboratorio de Gen√≥mica Microbiana, Universidad Peruana Cayetano Heredia | Pablo Tsukayama et al |
| EPI_ISL_540964 | Peru/LIM-UPCH-0086/2020 | 2020-06-18 | South America/Peru/Lima/Lima | Laboratorio de Referencia Nacional de Virus Respiratorios, Instituto Nacional de Salud Peru | Laboratorio de Gen√≥mica Microbiana, Universidad Peruana Cayetano Heredia | Pablo Tsukayama et al |
| EPI_ISL_540965 | Peru/LIM-UPCH-0087/2020 | 2020-06-17 | South America/Peru/Lima/Lima | Laboratorio de Referencia Nacional de Virus Respiratorios, Instituto Nacional de Salud Peru | Laboratorio de Gen√≥mica Microbiana, Universidad Peruana Cayetano Heredia | Pablo Tsukayama et al |

| | | | | | | |
|---|---|---|---|---|---|---|
| EPI_ISL_540966 | Peru/LIM-UPCH-0088/2020 | 2020-06-17 | South America/Peru/Lima/Lima | Laboratorio de Referencia Nacional de Virus Respiratorios, Instituto Nacional de Salud Peru | Laboratorio de Genv≥mica Microbiana, Universidad Peruana Cayetano Heredia | Pablo Tsukayama et al |
| EPI_ISL_540968 | Peru/LIM-UPCH-0090/2020 | 2020-06-17 | South America/Peru/Lima/Lima | Laboratorio de Referencia Nacional de Virus Respiratorios, Instituto Nacional de Salud Peru | Laboratorio de Genv≥mica Microbiana, Universidad Peruana Cayetano Heredia | Pablo Tsukayama et al |
| EPI_ISL_540969 | Peru/LIM-UPCH-0091/2020 | 2020-06-17 | South America/Peru/Lima/Lima | Laboratorio de Referencia Nacional de Virus Respiratorios, Instituto Nacional de Salud Peru | Laboratorio de Genv≥mica Microbiana, Universidad Peruana Cayetano Heredia | Pablo Tsukayama et al |
| EPI_ISL_540970 | Peru/CAL-UPCH-0092/2020 | 2020-06-17 | South America/Peru/Callao/Callao | Laboratorio de Referencia Nacional de Virus Respiratorios, Instituto Nacional de Salud Peru | Laboratorio de Genv≥mica Microbiana, Universidad Peruana Cayetano Heredia | Pablo Tsukayama et al |
| EPI_ISL_540971 | Peru/LIM-UPCH-0093/2020 | 2020-06-17 | South America/Peru/Lima/Lima | Laboratorio de Referencia Nacional de Virus Respiratorios, Instituto Nacional de Salud Peru | Laboratorio de Genv≥mica Microbiana, Universidad Peruana Cayetano Heredia | Pablo Tsukayama et al |
| EPI_ISL_540972 | Peru/LIM-UPCH-0094/2020 | 2020-06-17 | South America/Peru/Lima/Lima | Laboratorio de Referencia Nacional de Virus Respiratorios, Instituto Nacional de Salud Peru | Laboratorio de Genv≥mica Microbiana, Universidad Peruana Cayetano Heredia | Pablo Tsukayama et al |

| | | | | | | |
|---|---|---|---|---|---|---|
| EPI_ISL_540973 | Peru/LIM-UPCH-0095/2020 | 2020-06-18 | South America/Peru/Lima/Lima | Laboratorio de Referencia Nacional de Virus Respiratorios, Instituto Nacional de Salud Peru | Laboratorio de Gen√≥mica Microbiana, Universidad Peruana Cayetano Heredia | Pablo Tsukayama et al |
| EPI_ISL_540974 | Peru/LIM-UPCH-0096/2020 | 2020-06-17 | South America/Peru/Lima/Lima | Laboratorio de Referencia Nacional de Virus Respiratorios, Instituto Nacional de Salud Peru | Laboratorio de Gen√≥mica Microbiana, Universidad Peruana Cayetano Heredia | Pablo Tsukayama et al |
| EPI_ISL_540975 | Peru/CAL-UPCH-0097/2020 | 2020-06-16 | South America/Peru/Callao/Callao | Laboratorio de Referencia Nacional de Virus Respiratorios, Instituto Nacional de Salud Peru | Laboratorio de Gen√≥mica Microbiana, Universidad Peruana Cayetano Heredia | Pablo Tsukayama et al |
| EPI_ISL_540976 | Peru/CAL-UPCH-0098/2020 | 2020-06-17 | South America/Peru/Callao/Callao | Laboratorio de Referencia Nacional de Virus Respiratorios, Instituto Nacional de Salud Peru | Laboratorio de Gen√≥mica Microbiana, Universidad Peruana Cayetano Heredia | Pablo Tsukayama et al |
| EPI_ISL_540977 | Peru/LIM-UPCH-0099/2020 | 2020-06-17 | South America/Peru/Lima/Lima | Laboratorio de Referencia Nacional de Virus Respiratorios, Instituto Nacional de Salud Peru | Laboratorio de Gen√≥mica Microbiana, Universidad Peruana Cayetano Heredia | Pablo Tsukayama et al |
| EPI_ISL_540978 | Peru/LIM-UPCH-0100/2020 | 2020-06-16 | South America/Peru/Lima/Lima | Laboratorio de Referencia Nacional de Virus Respiratorios, Instituto Nacional de Salud Peru | Laboratorio de Gen√≥mica Microbiana, Universidad Peruana Cayetano Heredia | Pablo Tsukayama et al |

| | | | | | | |
|---|---|---|---|---|---|---|
| EPI_ISL_540979 | Peru/LIM-UPCH-0101/2020 | 2020-06-17 | South America/Peru/Lima/Lima | Laboratorio de Referencia Nacional de Virus Respiratorios, Instituto Nacional de Salud Peru | Laboratorio de Genv≥mica Microbiana, Universidad Peruana Cayetano Heredia | Pablo Tsukayama et al |
| EPI_ISL_540980 | Peru/LIM-UPCH-0102/2020 | 2020-06-17 | South America/Peru/Lima/Lima | Laboratorio de Referencia Nacional de Virus Respiratorios, Instituto Nacional de Salud Peru | Laboratorio de Genv≥mica Microbiana, Universidad Peruana Cayetano Heredia | Pablo Tsukayama et al |
| EPI_ISL_540981 | Peru/LIM-UPCH-0103/2020 | 2020-06-17 | South America/Peru/Lima/Lima | Laboratorio de Referencia Nacional de Virus Respiratorios, Instituto Nacional de Salud Peru | Laboratorio de Genv≥mica Microbiana, Universidad Peruana Cayetano Heredia | Pablo Tsukayama et al |
| EPI_ISL_540982 | Peru/LIM-UPCH-0104/2020 | 2020-06-17 | South America/Peru/Lima/Lima | Laboratorio de Referencia Nacional de Virus Respiratorios, Instituto Nacional de Salud Peru | Laboratorio de Genv≥mica Microbiana, Universidad Peruana Cayetano Heredia | Pablo Tsukayama et al |
| EPI_ISL_540983 | Peru/ANC-UPCH-0105/2020 | 2020-06-16 | South America/Peru/Ancash/Santa | Laboratorio de Referencia Nacional de Virus Respiratorios, Instituto Nacional de Salud Peru | Laboratorio de Genv≥mica Microbiana, Universidad Peruana Cayetano Heredia | Pablo Tsukayama et al |
| EPI_ISL_540984 | Peru/ANC-UPCH-0106/2020 | 2020-06-16 | South America/Peru/Ancash/Santa | Laboratorio de Referencia Nacional de Virus Respiratorios, Instituto Nacional de Salud Peru | Laboratorio de Genv≥mica Microbiana, Universidad Peruana Cayetano Heredia | Pablo Tsukayama et al |

| | | | | | | |
|---|---|---|---|---|---|---|
| EPI_ISL_540985 | Peru/ANC-UPCH-0107/2020 | 2020-06-17 | South America/Peru/Ancash/Santa | Laboratorio de Referencia Nacional de Virus Respiratorios, Instituto Nacional de Salud Peru | Laboratorio de Gen√≥mica Microbiana, Universidad Peruana Cayetano Heredia | Pablo Tsukayama et al |
| EPI_ISL_540989 | Peru/LIM-UPCH-0112/2020 | 2020-06-18 | South America/Peru/Lima/Lima | Laboratorio de Referencia Nacional de Virus Respiratorios, Instituto Nacional de Salud Peru | Laboratorio de Gen√≥mica Microbiana, Universidad Peruana Cayetano Heredia | Pablo Tsukayama et al |
| EPI_ISL_540990 | Peru/LIM-UPCH-0113/2020 | 2020-06-18 | South America/Peru/Lima/Lima | Laboratorio de Referencia Nacional de Virus Respiratorios, Instituto Nacional de Salud Peru | Laboratorio de Gen√≥mica Microbiana, Universidad Peruana Cayetano Heredia | Pablo Tsukayama et al |
| EPI_ISL_540991 | Peru/LIM-UPCH-0114/2020 | 2020-06-15 | South America/Peru/Lima/Lima | Laboratorio de Referencia Nacional de Virus Respiratorios, Instituto Nacional de Salud Peru | Laboratorio de Gen√≥mica Microbiana, Universidad Peruana Cayetano Heredia | Pablo Tsukayama et al |
| EPI_ISL_540992 | Peru/LIM-UPCH-0115/2020 | 2020-06-18 | South America/Peru/Lima/Lima | Laboratorio de Referencia Nacional de Virus Respiratorios, Instituto Nacional de Salud Peru | Laboratorio de Gen√≥mica Microbiana, Universidad Peruana Cayetano Heredia | Pablo Tsukayama et al |
| EPI_ISL_540993 | SouthKorea/IHC597/2020 | 2020-02-20 | Asia/South Korea/Gwangju/ | Health and Environmental Research Institute of Gwangju Metropolitan city | Health and Environmental Research Institute of Gwangju Metropolitan city | Min Ji Kim et al |
| EPI_ISL_540994 | SouthKorea/IHC610/2020 | 2020-02-20 | Asia/South Korea/Gwangju/ | Health and Environmental Research Institute of Gwangju Metropolitan city | Health and Environmental Research Institute of Gwangju Metropolitan city | Min Ji Kim et al |

| | | | | | | |
|---|---|---|---|---|---|---|
| EPI_ISL_540995 | SouthKorea/IHC614/2020 | 2020-02-21 | Asia/South Korea/Gwangju/ | Health and Environmental Research Institute of Gwangju Metropolitan city | Health and Environmental Research Institute of Gwangju Metropolitan city | Min Ji Kim et al |
| EPI_ISL_540996 | SouthKorea/IHC685/2020 | 2020-02-22 | Asia/South Korea/Gwangju/ | Health and Environmental Research Institute of Gwangju Metropolitan city | Health and Environmental Research Institute of Gwangju Metropolitan city | Min Ji Kim et al |
| EPI_ISL_540997 | SouthKorea/IHC719/2020 | 2020-02-23 | Asia/South Korea/Gwangju/ | Health and Environmental Research Institute of Gwangju Metropolitan city | Health and Environmental Research Institute of Gwangju Metropolitan city | Min Ji Kim et al |
| EPI_ISL_540998 | SouthKorea/IHC1215/2020 | 2020-03-03 | Asia/South Korea/Gwangju/ | Health and Environmental Research Institute of Gwangju Metropolitan city | Health and Environmental Research Institute of Gwangju Metropolitan city | Min Ji Kim et al |
| EPI_ISL_541001 | SouthKorea/IHC3486/2020 | 2020-03-26 | Asia/South Korea/Gwangju/ | Health and Environmental Research Institute of Gwangju Metropolitan city | Health and Environmental Research Institute of Gwangju Metropolitan city | Min Ji Kim et al |
| EPI_ISL_541010 | Spain/AR-IBV-98005657/2020 | 2020-06-23 | Europe/Spain/Aragon/Zaragoza | Servicio de Microbiolog√≠a, Hospital Miguel Servet, Zaragoza | SeqCOVID-SPAIN consortium/Institute of Biomedicine of Valencia, IBV-CSIC | Antonio Rezusta L√≥pez et al |
| EPI_ISL_541021 | Spain/AR-IBV-98005745/2020 | 2020-04-28 | Europe/Spain/Aragon/Zaragoza | Servicio de Microbiolog√≠a, Hospital Miguel Servet, Zaragoza | SeqCOVID-SPAIN consortium/Institute of Biomedicine of Valencia, IBV-CSIC | Antonio Rezusta L√≥pez et al |
| EPI_ISL_541023 | Spain/AR-IBV-98005747/2020 | 2020-04-28 | Europe/Spain/Aragon/Zaragoza | Servicio de Microbiolog√≠a, Hospital Miguel Servet, Zaragoza | SeqCOVID-SPAIN consortium/Institute of Biomedicine of Valencia, IBV-CSIC | Antonio Rezusta L√≥pez et al |

| | | | | | | |
|---|---|---|---|---|---|---|
| EPI_ISL_541025 | Spain/AR-IBV-98005750/2020 | 2020-04-28 | Europe/Spain/Aragon/Zaragoza | Servicio de Microbiolog√≠a, Hospital Miguel Servet, Zaragoza | SeqCOVID-SPAIN consortium/Institute of Biomedicine of Valencia, IBV-CSIC | Antonio Rezusta L√≥pez et al |
| EPI_ISL_541029 | Spain/AR-IBV-98005757/2020 | 2020-04-28 | Europe/Spain/Aragon/Zaragoza | Servicio de Microbiolog√≠a, Hospital Miguel Servet, Zaragoza | SeqCOVID-SPAIN consortium/Institute of Biomedicine of Valencia, IBV-CSIC | Antonio Rezusta L√≥pez et al |
| EPI_ISL_541036 | Spain/AR-IBV-98005784/2020 | 2020-05-07 | Europe/Spain/Aragon/Zaragoza | Servicio de Microbiolog√≠a, Hospital Miguel Servet, Zaragoza | SeqCOVID-SPAIN consortium/Institute of Biomedicine of Valencia, IBV-CSIC | Antonio Rezusta L√≥pez et al |
| EPI_ISL_541041 | Spain/GA-IBV-98006036/2020 | 2020-05-09 | Europe/Spain/Galicia/Rois | Hospital Cl√≠nico Universitario de Santiago de Compostela | SeqCOVID-SPAIN consortium/Institute of Biomedicine of Valencia, IBV-CSIC | Jos√© Javier Costa Alcalde et al |
| EPI_ISL_541048 | Spain/GA-IBV-98006045/2020 | 2020-04-04 | Europe/Spain/Galicia/Boiro | Hospital Cl√≠nico Universitario de Santiago de Compostela | SeqCOVID-SPAIN consortium/Institute of Biomedicine of Valencia, IBV-CSIC | Jos√© Javier Costa Alcalde et al |
| EPI_ISL_541053 | Spain/GA-IBV-98006051/2020 | 2020-04-05 | Europe/Spain/Galicia/Santiago de Comp | Hospital Cl√≠nico Universitario de Santiago de Compostela | SeqCOVID-SPAIN consortium/Institute of Biomedicine of Valencia, IBV-CSIC | Jos√© Javier Costa Alcalde et al |
| EPI_ISL_541055 | Spain/GA-IBV-98006064/2020 | 2020-04-17 | Europe/Spain/Galicia/Santiago de Comp | Hospital Cl√≠nico Universitario de Santiago de Compostela | SeqCOVID-SPAIN consortium/Institute of Biomedicine of Valencia, IBV-CSIC | Jos√© Javier Costa Alcalde et al |
| EPI_ISL_541057 | Spain/GA-IBV-98006066/2020 | 2020-04-23 | Europe/Spain/Galicia/Santiago de Comp | Hospital Cl√≠nico Universitario de Santiago de Compostela | SeqCOVID-SPAIN consortium/Institute of Biomedicine of Valencia, IBV-CSIC | Jos√© Javier Costa Alcalde et al |

| | | | | | | |
|---|---|---|---|---|---|---|
| EPI_ISL_541064 | Spain/GA-IBV-98006077/2020 | 2020-05-02 | Europe/Spain/Galicia/Santiago de Comp | Hospital Cl√≠nico Universitario de Santiago de Compostela | SeqCOVID-SPAIN consortium/Institute of Biomedicine of Valencia, IBV-CSIC | Jos√© Javier Costa Alcalde et al |
| EPI_ISL_541065 | Spain/GA-IBV-98006078/2020 | 2020-04-30 | Europe/Spain/Galicia/Puebla del Carami | Hospital Cl√≠nico Universitario de Santiago de Compostela | SeqCOVID-SPAIN consortium/Institute of Biomedicine of Valencia, IBV-CSIC | Jos√© Javier Costa Alcalde et al |
| EPI_ISL_541073 | Spain/CT-IBV-98006559/2020 | 2020-03-16 | Europe/Spain/Catalunya/Cambrils | Hospital de la Santa Creu i Sant Pau. Servicio de Microbiolog√≠a | SeqCOVID-SPAIN consortium/Institute of Biomedicine of Valencia, IBV-CSIC | Ferran Navarro et al |
| EPI_ISL_541077 | Spain/CT-IBV-98006565/2020 | 2020-07-03 | Europe/Spain/Catalunya/Barcelona | Hospital de la Santa Creu i Sant Pau. Servicio de Microbiolog√≠a | SeqCOVID-SPAIN consortium/Institute of Biomedicine of Valencia, IBV-CSIC | Ferran Navarro et al |
| EPI_ISL_541087 | Spain/PV-IBV-98006688/2020 | 2020-07-08 | Europe/Spain/Basque Country/Ordizia | Servicio de Microbiolog√≠a. Hospital Universitario Donostia. OSI Donostialdea. √Årea de Enfermedades Infecciosas, Grupo de Infecci√≥n Respiratoria y Resistencia Antimicrobiana. Instituto de Investigaci√≥n Sanitaria Biodonostia | SeqCOVID-SPAIN consortium/Institute of Biomedicine of Valencia, IBV-CSIC | Gustavo Cilla et al |
| EPI_ISL_541101 | Spain/PV-IBV-98006703/2020 | 2020-07-11 | Europe/Spain/Basque Country/Irun | Servicio de Microbiolog√≠a. Hospital Universitario Donostia. OSI Donostialdea. √Årea de Enfermedades Infecciosas, Grupo de Infecci√≥n Respiratoria y Resistencia Antimicrobiana. Instituto de Investigaci√≥n Sanitaria Biodonostia | SeqCOVID-SPAIN consortium/Institute of Biomedicine of Valencia, IBV-CSIC | Gustavo Cilla et al |
| EPI_ISL_541102 | Spain/PV-IBV-98006705/2020 | 2020-07-12 | Europe/Spain/Basque Country/Zarauz | Servicio de Microbiolog√≠a. Hospital Universitario Donostia. OSI Donostialdea. √Årea de Enfermedades Infecciosas, Grupo de Infecci√≥n Respiratoria y Resistencia Antimicrobiana. Instituto de Investigaci√≥n Sanitaria Biodonostia | SeqCOVID-SPAIN consortium/Institute of Biomedicine of Valencia, IBV-CSIC | Gustavo Cilla et al |

| | | | | | | |
|---|---|---|---|---|---|---|
| EPI_ISL_541103 | Spain/PV-IBV-98006707/2020 | 2020-07-13 | Europe/Spain/Basque Country/Zumarrag | Servicio de Microbiolog√≠a. Hospital Universitario Donostia. OSI Donostialdea. √Årea de Enfermedades Infecciosas, Grupo de Infecci√≥n Respiratoria y Resistencia Antimicrobiana. Instituto de Investigaci√≥n Sanitaria Biodonostia | SeqCOVID-SPAIN consortium/Institute of Biomedicine of Valencia, IBV-CSIC | Gustavo Cilla et al |
| EPI_ISL_541111 | Spain/PV-IBV-98006720/2020 | 2020-07-15 | Europe/Spain/Basque Country/Eibar | Servicio de Microbiolog√≠a. Hospital Universitario Donostia. OSI Donostialdea. √Årea de Enfermedades Infecciosas, Grupo de Infecci√≥n Respiratoria y Resistencia Antimicrobiana. Instituto de Investigaci√≥n Sanitaria Biodonostia | SeqCOVID-SPAIN consortium/Institute of Biomedicine of Valencia, IBV-CSIC | Gustavo Cilla et al |
| EPI_ISL_541116 | Spain/PV-IBV-98006728/2020 | 2020-07-16 | Europe/Spain/Basque Country/Donostia | Servicio de Microbiolog√≠a. Hospital Universitario Donostia. OSI Donostialdea. √Årea de Enfermedades Infecciosas, Grupo de Infecci√≥n Respiratoria y Resistencia Antimicrobiana. Instituto de Investigaci√≥n Sanitaria Biodonostia | SeqCOVID-SPAIN consortium/Institute of Biomedicine of Valencia, IBV-CSIC | Gustavo Cilla et al |
| EPI_ISL_541117 | Spain/PV-IBV-98006730/2020 | 2020-07-16 | Europe/Spain/Basque Country/Eibar | Servicio de Microbiolog√≠a. Hospital Universitario Donostia. OSI Donostialdea. √Årea de Enfermedades Infecciosas, Grupo de Infecci√≥n Respiratoria y Resistencia Antimicrobiana. Instituto de Investigaci√≥n Sanitaria Biodonostia | SeqCOVID-SPAIN consortium/Institute of Biomedicine of Valencia, IBV-CSIC | Gustavo Cilla et al |
| EPI_ISL_541123 | Spain/PV-IBV-98006737/2020 | 2020-07-17 | Europe/Spain/Basque Country/Tolosa | Servicio de Microbiolog√≠a. Hospital Universitario Donostia. OSI Donostialdea. √Årea de Enfermedades Infecciosas, Grupo de Infecci√≥n Respiratoria y Resistencia Antimicrobiana. Instituto de Investigaci√≥n Sanitaria Biodonostia | SeqCOVID-SPAIN consortium/Institute of Biomedicine of Valencia, IBV-CSIC | Gustavo Cilla et al |
| EPI_ISL_541128 | Spain/PV-IBV-98006763/2020 | 2020-07-18 | Europe/Spain/Basque Country/Irun | Servicio de Microbiolog√≠a. Hospital Universitario Donostia. OSI Donostialdea. √Årea de Enfermedades Infecciosas, Grupo de Infecci√≥n Respiratoria y Resistencia Antimicrobiana. Instituto de Investigaci√≥n Sanitaria Biodonostia | SeqCOVID-SPAIN consortium/Institute of Biomedicine of Valencia, IBV-CSIC | Gustavo Cilla et al |

| | | | | | | |
|---|---|---|---|---|---|---|
| EPI_ISL_541130 | Spain/PV-IBV-98006920/2020 | 2020-08-01 | Europe/Spain/Basque Country/Donostia | Servicio de Microbiolog√≠a. Hospital Universitario Donostia. OSI Donostialdea. √Årea de Enfermedades Infecciosas, Grupo de Infecci√≥n Respiratoria y Resistencia Antimicrobiana. Instituto de Investigaci√≥n Sanitaria Biodonostia | SeqCOVID-SPAIN consortium/Institute of Biomedicine of Valencia, IBV-CSIC | Gustavo Cilla et al |
| EPI_ISL_541131 | Spain/PV-IBV-98006926/2020 | 2020-08-08 | Europe/Spain/Basque Country/Donostia | Servicio de Microbiolog√≠a. Hospital Universitario Donostia. OSI Donostialdea. √Årea de Enfermedades Infecciosas, Grupo de Infecci√≥n Respiratoria y Resistencia Antimicrobiana. Instituto de Investigaci√≥n Sanitaria Biodonostia | SeqCOVID-SPAIN consortium/Institute of Biomedicine of Valencia, IBV-CSIC | Gustavo Cilla et al |
| EPI_ISL_541133 | Spain/PV-IBV-98006932/2020 | 2020-08-05 | Europe/Spain/Basque Country/Donostia | Servicio de Microbiolog√≠a. Hospital Universitario Donostia. OSI Donostialdea. √Årea de Enfermedades Infecciosas, Grupo de Infecci√≥n Respiratoria y Resistencia Antimicrobiana. Instituto de Investigaci√≥n Sanitaria Biodonostia | SeqCOVID-SPAIN consortium/Institute of Biomedicine of Valencia, IBV-CSIC | Gustavo Cilla et al |
| EPI_ISL_541134 | Spain/PV-IBV-98006933/2020 | 2020-08-05 | Europe/Spain/Basque Country/Donostia | Servicio de Microbiolog√≠a. Hospital Universitario Donostia. OSI Donostialdea. √Årea de Enfermedades Infecciosas, Grupo de Infecci√≥n Respiratoria y Resistencia Antimicrobiana. Instituto de Investigaci√≥n Sanitaria Biodonostia | SeqCOVID-SPAIN consortium/Institute of Biomedicine of Valencia, IBV-CSIC | Gustavo Cilla et al |
| EPI_ISL_541135 | Spain/PV-IBV-98006934/2020 | 2020-08-08 | Europe/Spain/Basque Country/Donostia | Servicio de Microbiolog√≠a. Hospital Universitario Donostia. OSI Donostialdea. √Årea de Enfermedades Infecciosas, Grupo de Infecci√≥n Respiratoria y Resistencia Antimicrobiana. Instituto de Investigaci√≥n Sanitaria Biodonostia | SeqCOVID-SPAIN consortium/Institute of Biomedicine of Valencia, IBV-CSIC | Gustavo Cilla et al |
| EPI_ISL_541137 | Spain/PV-IBV-98006936/2020 | 2020-08-08 | Europe/Spain/Basque Country/Donostia | Servicio de Microbiolog√≠a. Hospital Universitario Donostia. OSI Donostialdea. √Årea de Enfermedades Infecciosas, Grupo de Infecci√≥n Respiratoria y Resistencia Antimicrobiana. Instituto de Investigaci√≥n Sanitaria Biodonostia | SeqCOVID-SPAIN consortium/Institute of Biomedicine of Valencia, IBV-CSIC | Gustavo Cilla et al |

| | | | | | | |
|---|---|---|---|---|---|---|
| EPI_ISL_541155 | USA/FL-BPHL-1249/2020 | 2020-04-20 | North America/USA/Florida/ | Florida Bureau of Public Health Laboratories, Florida Department of Health | Florida Bureau of Public Health Laboratories, Florida Department of Health | Schmedes et al |
| EPI_ISL_541169 | USA/FL-BPHL-1262/2020 | 2020-06-04 | North America/USA/Florida/ | Florida Bureau of Public Health Laboratories, Florida Department of Health | Florida Bureau of Public Health Laboratories, Florida Department of Health | Schmedes et al |
| EPI_ISL_541171 | USA/FL-BPHL-1264/2020 | 2020-06-04 | North America/USA/Florida/ | Florida Bureau of Public Health Laboratories, Florida Department of Health | Florida Bureau of Public Health Laboratories, Florida Department of Health | Schmedes et al |
| EPI_ISL_541178 | USA/FL-BPHL-1205/2020 | 2020-07-23 | North America/USA/Florida/ | Florida Bureau of Public Health Laboratories, Florida Department of Health | Florida Bureau of Public Health Laboratories, Florida Department of Health | Schmedes et al |
| EPI_ISL_541180 | USA/FL-BPHL-1207/2020 | 2020-07-24 | North America/USA/Florida/ | Florida Bureau of Public Health Laboratories, Florida Department of Health | Florida Bureau of Public Health Laboratories, Florida Department of Health | Schmedes et al |
| EPI_ISL_541187 | USA/FL-BPHL-1214/2020 | 2020-07-27 | North America/USA/Florida/ | Florida Bureau of Public Health Laboratories, Florida Department of Health | Florida Bureau of Public Health Laboratories, Florida Department of Health | Schmedes et al |

| | | | | | | |
|---|---|---|---|---|---|---|
| EPI_ISL_541188 | USA/FL-BPHL-1215/2020 | 2020-07-27 | North America/USA/Florida/ | Florida Bureau of Public Health Laboratories, Florida Department of Health | Florida Bureau of Public Health Laboratories, Florida Department of Health | Schmedes et al |
| EPI_ISL_541189 | USA/FL-BPHL-1216/2020 | 2020-07-27 | North America/USA/Florida/ | Florida Bureau of Public Health Laboratories, Florida Department of Health | Florida Bureau of Public Health Laboratories, Florida Department of Health | Schmedes et al |
| EPI_ISL_541192 | USA/FL-BPHL-1219/2020 | 2020-07-27 | North America/USA/Florida/ | Florida Bureau of Public Health Laboratories, Florida Department of Health | Florida Bureau of Public Health Laboratories, Florida Department of Health | Schmedes et al |
| EPI_ISL_541193 | USA/FL-BPHL-1220/2020 | 2020-07-27 | North America/USA/Florida/ | Florida Bureau of Public Health Laboratories, Florida Department of Health | Florida Bureau of Public Health Laboratories, Florida Department of Health | Schmedes et al |
| EPI_ISL_541196 | USA/FL-BPHL-1223/2020 | 2020-07-27 | North America/USA/Florida/ | Florida Bureau of Public Health Laboratories, Florida Department of Health | Florida Bureau of Public Health Laboratories, Florida Department of Health | Schmedes et al |
| EPI_ISL_541198 | USA/FL-BPHL-1225/2020 | 2020-07-28 | North America/USA/Florida/ | Florida Bureau of Public Health Laboratories, Florida Department of Health | Florida Bureau of Public Health Laboratories, Florida Department of Health | Schmedes et al |

| | | | | | | |
|---|---|---|---|---|---|---|
| EPI_ISL_541201 | USA/FL-BPHL-1269/2020 | 2020-08-04 | North America/USA/Florida/ | Florida Bureau of Public Health Laboratories, Florida Department of Health | Florida Bureau of Public Health Laboratories, Florida Department of Health | Schmedes et al |
| EPI_ISL_541204 | USA/FL-BPHL-1272/2020 | 2020-08-03 | North America/USA/Florida/ | Florida Bureau of Public Health Laboratories, Florida Department of Health | Florida Bureau of Public Health Laboratories, Florida Department of Health | Schmedes et al |
| EPI_ISL_541205 | USA/FL-BPHL-1273/2020 | 2020-08-03 | North America/USA/Florida/ | Florida Bureau of Public Health Laboratories, Florida Department of Health | Florida Bureau of Public Health Laboratories, Florida Department of Health | Schmedes et al |
| EPI_ISL_541209 | USA/FL-BPHL-1277/2020 | 2020-08-07 | North America/USA/Florida/ | Florida Bureau of Public Health Laboratories, Florida Department of Health | Florida Bureau of Public Health Laboratories, Florida Department of Health | Schmedes et al |
| EPI_ISL_541214 | USA/FL-BPHL-1282/2020 | 2020-08-25 | North America/USA/Florida/ | Florida Bureau of Public Health Laboratories, Florida Department of Health | Florida Bureau of Public Health Laboratories, Florida Department of Health | Schmedes et al |
| EPI_ISL_541219 | USA/FL-BPHL-1287/2020 | 2020-08-28 | North America/USA/Florida/ | Florida Bureau of Public Health Laboratories, Florida Department of Health | Florida Bureau of Public Health Laboratories, Florida Department of Health | Schmedes et al |

| | | | | | | |
|---|---|---|---|---|---|---|
| EPI_ISL_541224 | USA/FL-BPHL-1292/2020 | 2020-08-28 | North America/USA/Florida/ | Florida Bureau of Public Health Laboratories, Florida Department of Health | Florida Bureau of Public Health Laboratories, Florida Department of Health | Schmedes et al |
| EPI_ISL_541225 | USA/FL-BPHL-1293/2020 | 2020-08-28 | North America/USA/Florida/ | Florida Bureau of Public Health Laboratories, Florida Department of Health | Florida Bureau of Public Health Laboratories, Florida Department of Health | Schmedes et al |
| EPI_ISL_541228 | USA/FL-BPHL-1296/2020 | 2020-08-31 | North America/USA/Florida/ | Florida Bureau of Public Health Laboratories, Florida Department of Health | Florida Bureau of Public Health Laboratories, Florida Department of Health | Schmedes et al |
| EPI_ISL_541229 | USA/FL-BPHL-1297/2020 | 2020-08-31 | North America/USA/Florida/ | Florida Bureau of Public Health Laboratories, Florida Department of Health | Florida Bureau of Public Health Laboratories, Florida Department of Health | Schmedes et al |
| EPI_ISL_541259 | USA/FL-BPHL-1321/2020 | 2020-07-14 | North America/USA/Florida/ | Florida Bureau of Public Health Laboratories, Florida Department of Health | Florida Bureau of Public Health Laboratories, Florida Department of Health | Schmedes et al |
| EPI_ISL_541263 | USA/FL-BPHL-1325/2020 | 2020-07-14 | North America/USA/Florida/ | Florida Bureau of Public Health Laboratories, Florida Department of Health | Florida Bureau of Public Health Laboratories, Florida Department of Health | Schmedes et al |

| | | | | | | |
|---|---|---|---|---|---|---|
| EPI_ISL_541267 | USA/FL-BPHL-1329/2020 | 2020-07-14 | North America/USA/Florida/ | Florida Bureau of Public Health Laboratories, Florida Department of Health | Florida Bureau of Public Health Laboratories, Florida Department of Health | Schmedes et al |
| EPI_ISL_541268 | USA/FL-BPHL-1330/2020 | 2020-07-14 | North America/USA/Florida/ | Florida Bureau of Public Health Laboratories, Florida Department of Health | Florida Bureau of Public Health Laboratories, Florida Department of Health | Schmedes et al |
| EPI_ISL_541270 | USA/FL-BPHL-1332/2020 | 2020-07-14 | North America/USA/Florida/ | Florida Bureau of Public Health Laboratories, Florida Department of Health | Florida Bureau of Public Health Laboratories, Florida Department of Health | Schmedes et al |
| EPI_ISL_541275 | USA/FL-BPHL-1337/2020 | 2020-07-14 | North America/USA/Florida/ | Florida Bureau of Public Health Laboratories, Florida Department of Health | Florida Bureau of Public Health Laboratories, Florida Department of Health | Schmedes et al |
| EPI_ISL_541280 | USA/FL-BPHL-1342/2020 | 2020-07-15 | North America/USA/Florida/ | Florida Bureau of Public Health Laboratories, Florida Department of Health | Florida Bureau of Public Health Laboratories, Florida Department of Health | Schmedes et al |
| EPI_ISL_541287 | USA/FL-BPHL-1349/2020 | 2020-07-15 | North America/USA/Florida/ | Florida Bureau of Public Health Laboratories, Florida Department of Health | Florida Bureau of Public Health Laboratories, Florida Department of Health | Schmedes et al |

| | | | | | | |
|---|---|---|---|---|---|---|
| EPI_ISL_541300 | USA/FL-BPHL-1363/2020 | 2020-07-17 | North America/USA/Florida/ | Florida Bureau of Public Health Laboratories, Florida Department of Health | Florida Bureau of Public Health Laboratories, Florida Department of Health | Schmedes et al |
| EPI_ISL_541301 | USA/FL-BPHL-1364/2020 | 2020-07-17 | North America/USA/Florida/ | Florida Bureau of Public Health Laboratories, Florida Department of Health | Florida Bureau of Public Health Laboratories, Florida Department of Health | Schmedes et al |
| EPI_ISL_541304 | USA/FL-BPHL-1367/2020 | 2020-07-20 | North America/USA/Florida/ | Florida Bureau of Public Health Laboratories, Florida Department of Health | Florida Bureau of Public Health Laboratories, Florida Department of Health | Schmedes et al |
| EPI_ISL_541305 | USA/FL-BPHL-1368/2020 | 2020-07-20 | North America/USA/Florida/ | Florida Bureau of Public Health Laboratories, Florida Department of Health | Florida Bureau of Public Health Laboratories, Florida Department of Health | Schmedes et al |
| EPI_ISL_541314 | USA/FL-BPHL-1377/2020 | 2020-07-21 | North America/USA/Florida/ | Florida Bureau of Public Health Laboratories, Florida Department of Health | Florida Bureau of Public Health Laboratories, Florida Department of Health | Schmedes et al |
| EPI_ISL_541317 | USA/FL-BPHL-1380/2020 | 2020-07-21 | North America/USA/Florida/ | Florida Bureau of Public Health Laboratories, Florida Department of Health | Florida Bureau of Public Health Laboratories, Florida Department of Health | Schmedes et al |

| | | | | | | |
|---|---|---|---|---|---|---|
| EPI_ISL_541318 | USA/FL-BPHL-1381/2020 | 2020-07-21 | North America/USA/Florida/ | Florida Bureau of Public Health Laboratories, Florida Department of Health | Florida Bureau of Public Health Laboratories, Florida Department of Health | Schmedes et al |
| EPI_ISL_541320 | USA/FL-BPHL-1383/2020 | 2020-07-21 | North America/USA/Florida/ | Florida Bureau of Public Health Laboratories, Florida Department of Health | Florida Bureau of Public Health Laboratories, Florida Department of Health | Schmedes et al |
| EPI_ISL_541324 | USA/FL-BPHL-1387/2020 | 2020-07-22 | North America/USA/Florida/ | Florida Bureau of Public Health Laboratories, Florida Department of Health | Florida Bureau of Public Health Laboratories, Florida Department of Health | Schmedes et al |
| EPI_ISL_541330 | USA/FL-BPHL-1393/2020 | 2020-07-22 | North America/USA/Florida/ | Florida Bureau of Public Health Laboratories, Florida Department of Health | Florida Bureau of Public Health Laboratories, Florida Department of Health | Schmedes et al |
| EPI_ISL_541334 | CzechRepublic/NRL-6835/2020 | 2020-05-23 | Europe/Czech Republic/Moravian-Silesia | The National Institute of Public Health | State Veterinary Institute Prague | Nagy et al |
| EPI_ISL_541336 | CzechRepublic/NRL-7913/2020 | 2020-07-27 | Europe/Czech Republic/South Moravian | The National Institute of Public Health | State Veterinary Institute Prague | Nagy et al |
| EPI_ISL_541356 | Brazil/RJ-2231/2020 | 2020-04-17 | South America/Brazil/Rio de Janeiro/Rio | Laboratory of Respiratory Viruses and Measles, Oswaldo Cruz Institute, FIOCRUZ | Laboratory of Respiratory Viruses and Measles, Oswaldo Cruz Institute, FIOCRUZ | Paola Resende et al |
| EPI_ISL_541371 | Brazil/SC-771/2020 | 2020-03-13 | South America/Italy/Santa Catarina/Bal | LACEN/SC | Laboratory of Respiratory Viruses and Measles, Oswaldo Cruz Institute, FIOCRUZ | Paola Resende et al |

| | | | | | | |
|---|---|---|---|---|---|---|
| EPI_ISL_541389 | Brazil/SE-6583/2020 | 2020-04-20 | South America/Brazil/Sergipe/Aracaju | LACEN/SE | Laboratory of Respiratory Viruses and Measles, Oswaldo Cruz Institute, FIOCRUZ | Paola Resende et al |
| EPI_ISL_541409 | Switzerland/AG-ETHZ-270111/2020 | 2020-09-03 | Europe/Switzerland/Aargau/ | Viollier AG | Department of Biosystems Science and Engineering, ETH Zvºrich | Christian Beisel et al |
| EPI_ISL_541472 | Switzerland/VD-ETHZ-270056/2020 | 2020-09-02 | Europe/Switzerland/Vaud/ | Viollier AG | Department of Biosystems Science and Engineering, ETH Zvºrich | Christian Beisel et al |
| EPI_ISL_541479 | Switzerland/VD-ETHZ-270077/2020 | 2020-09-03 | Europe/Switzerland/Vaud/ | Viollier AG | Department of Biosystems Science and Engineering, ETH Zvºrich | Christian Beisel et al |
| EPI_ISL_541491 | Switzerland/VD-ETHZ-270123/2020 | 2020-09-03 | Europe/Switzerland/Vaud/ | Viollier AG | Department of Biosystems Science and Engineering, ETH Zvºrich | Christian Beisel et al |
| EPI_ISL_541541 | USA/IL-UW-1481/2020 | 2020-09-15 | North America/USA/Illinois/Winnebago | University of Wisconsin-Madison AIDS Vaccine Research Laboratories | University of Wisconsin-Madison AIDS Vaccine Research Laboratories | Gage Moreno et al |
| EPI_ISL_541543 | USA/IL-UW-1488/2020 | 2020-09-14 | North America/USA/Illinois/Winnebago | University of Wisconsin-Madison AIDS Vaccine Research Laboratories | University of Wisconsin-Madison AIDS Vaccine Research Laboratories | Gage Moreno et al |
| EPI_ISL_541544 | USA/IL-UW-1490/2020 | 2020-09-15 | North America/USA/Illinois/Winnebago | University of Wisconsin-Madison AIDS Vaccine Research Laboratories | University of Wisconsin-Madison AIDS Vaccine Research Laboratories | Gage Moreno et al |

| | | | | | | |
|---|---|---|---|---|---|---|
| EPI_ISL_541545 | USA/IL-UW-1494/2020 | 2020-09-16 | North America/USA/Illinois/Winnebago | University of Wisconsin-Madison AIDS Vaccine Research Laboratories | University of Wisconsin-Madison AIDS Vaccine Research Laboratories | Gage Moreno et al |
| EPI_ISL_541546 | USA/IL-UW-1503/2020 | 2020-09-16 | North America/USA/Illinois/Winnebago | University of Wisconsin-Madison AIDS Vaccine Research Laboratories | University of Wisconsin-Madison AIDS Vaccine Research Laboratories | Gage Moreno et al |
| EPI_ISL_541548 | USA/IL-UW-1521/2020 | 2020-09-14 | North America/USA/Illinois/Winnebago | University of Wisconsin-Madison AIDS Vaccine Research Laboratories | University of Wisconsin-Madison AIDS Vaccine Research Laboratories | Gage Moreno et al |
| EPI_ISL_541551 | USA/WI-UW-1418/2020 | 2020-09-11 | North America/USA/Wisconsin/Dane Co | University of Wisconsin-Madison Campus AIDS Vaccine Research Laboratories | University of Wisconsin-Madison AIDS Vaccine Research Laboratories | Gage Moreno et al |
| EPI_ISL_541552 | USA/WI-UW-1419/2020 | 2020-09-11 | North America/USA/Wisconsin/Dane Co | University of Wisconsin-Madison Campus AIDS Vaccine Research Laboratories | University of Wisconsin-Madison AIDS Vaccine Research Laboratories | Gage Moreno et al |
| EPI_ISL_541553 | USA/WI-UW-1420/2020 | 2020-09-11 | North America/USA/Wisconsin/Dane Co | University of Wisconsin-Madison Campus AIDS Vaccine Research Laboratories | University of Wisconsin-Madison AIDS Vaccine Research Laboratories | Gage Moreno et al |
| EPI_ISL_541557 | USA/WI-UW-1424/2020 | 2020-09-11 | North America/USA/Wisconsin/Dane Co | University of Wisconsin-Madison Campus AIDS Vaccine Research Laboratories | University of Wisconsin-Madison AIDS Vaccine Research Laboratories | Gage Moreno et al |
| EPI_ISL_541559 | USA/WI-UW-1426/2020 | 2020-09-11 | North America/USA/Wisconsin/Dane Co | University of Wisconsin-Madison Campus AIDS Vaccine Research Laboratories | University of Wisconsin-Madison AIDS Vaccine Research Laboratories | Gage Moreno et al |

| | | | | | | |
|---|---|---|---|---|---|---|
| EPI_ISL_541561 | USA/WI-UW-1428/2020 | 2020-09-11 | North America/USA/Wisconsin/Dane Co | University of Wisconsin-Madison Campus AIDS Vaccine Research Laboratories | University of Wisconsin-Madison AIDS Vaccine Research Laboratories | Gage Moreno et al |
| EPI_ISL_541565 | USA/WI-UW-1432/2020 | 2020-09-11 | North America/USA/Wisconsin/Dane Co | University of Wisconsin-Madison Campus AIDS Vaccine Research Laboratories | University of Wisconsin-Madison AIDS Vaccine Research Laboratories | Gage Moreno et al |
| EPI_ISL_541566 | USA/WI-UW-1433/2020 | 2020-09-11 | North America/USA/Wisconsin/Dane Co | University of Wisconsin-Madison AIDS Vaccine Research Laboratories | University of Wisconsin-Madison AIDS Vaccine Research Laboratories | Gage Moreno et al |
| EPI_ISL_541568 | USA/WI-UW-1435/2020 | 2020-09-09 | North America/USA/Wisconsin/Dane Co | University of Wisconsin-Madison Campus AIDS Vaccine Research Laboratories | University of Wisconsin-Madison AIDS Vaccine Research Laboratories | Gage Moreno et al |
| EPI_ISL_541570 | USA/WI-UW-1437/2020 | 2020-09-12 | North America/USA/Wisconsin/Dane Co | University of Wisconsin-Madison AIDS Vaccine Research Laboratories | University of Wisconsin-Madison AIDS Vaccine Research Laboratories | Gage Moreno et al |
| EPI_ISL_541571 | USA/WI-UW-1438/2020 | 2020-09-11 | North America/USA/Wisconsin/Dane Co | University of Wisconsin-Madison AIDS Vaccine Research Laboratories | University of Wisconsin-Madison AIDS Vaccine Research Laboratories | Gage Moreno et al |
| EPI_ISL_541580 | USA/WI-UW-1448/2020 | 2020-09-13 | North America/USA/Wisconsin/Dane Co | University of Wisconsin-Madison AIDS Vaccine Research Laboratories | University of Wisconsin-Madison AIDS Vaccine Research Laboratories | Gage Moreno et al |
| EPI_ISL_541591 | USA/WI-UW-1459/2020 | 2020-09-11 | North America/USA/Wisconsin/Dane Co | University of Wisconsin-Madison AIDS Vaccine Research Laboratories | University of Wisconsin-Madison AIDS Vaccine Research Laboratories | Gage Moreno et al |

| | | | | | | |
|---|---|---|---|---|---|---|
| EPI_ISL_541600 | USA/WI-UW-1468/2020 | 2020-09-12 | North America/USA/Wisconsin/Dane Co | University of Wisconsin-Madison AIDS Vaccine Research Laboratories | University of Wisconsin-Madison AIDS Vaccine Research Laboratories | Gage Moreno et al |
| EPI_ISL_541603 | USA/WI-UW-1471/2020 | 2020-09-10 | North America/USA/Wisconsin/Dane Co | University of Wisconsin-Madison AIDS Vaccine Research Laboratories | University of Wisconsin-Madison AIDS Vaccine Research Laboratories | Gage Moreno et al |
| EPI_ISL_541615 | USA/WI-UW-1485/2020 | 2020-09-15 | North America/USA/Wisconsin/Dane Co | University of Wisconsin-Madison AIDS Vaccine Research Laboratories | University of Wisconsin-Madison AIDS Vaccine Research Laboratories | Gage Moreno et al |
| EPI_ISL_541616 | USA/WI-UW-1486/2020 | 2020-09-16 | North America/USA/Wisconsin/Dane Co | University of Wisconsin-Madison AIDS Vaccine Research Laboratories | University of Wisconsin-Madison AIDS Vaccine Research Laboratories | Gage Moreno et al |
| EPI_ISL_541619 | USA/WI-UW-1491/2020 | 2020-09-16 | North America/USA/Wisconsin/Dane Co | University of Wisconsin-Madison AIDS Vaccine Research Laboratories | University of Wisconsin-Madison AIDS Vaccine Research Laboratories | Gage Moreno et al |
| EPI_ISL_541623 | USA/WI-UW-1496/2020 | 2020-09-14 | North America/USA/Wisconsin/Dane Co | University of Wisconsin-Madison AIDS Vaccine Research Laboratories | University of Wisconsin-Madison AIDS Vaccine Research Laboratories | Gage Moreno et al |
| EPI_ISL_541625 | USA/WI-UW-1498/2020 | 2020-09-15 | North America/USA/Wisconsin/Dane Co | University of Wisconsin-Madison AIDS Vaccine Research Laboratories | University of Wisconsin-Madison AIDS Vaccine Research Laboratories | Gage Moreno et al |
| EPI_ISL_541635 | USA/WI-UW-1509/2020 | 2020-09-14 | North America/USA/Wisconsin/Dodge C | University of Wisconsin-Madison AIDS Vaccine Research Laboratories | University of Wisconsin-Madison AIDS Vaccine Research Laboratories | Gage Moreno et al |

| | | | | | | |
|---|---|---|---|---|---|---|
| EPI_ISL_541636 | USA/WI-UW-1510/2020 | 2020-09-16 | North America/USA/Wisconsin/Dane Co | University of Wisconsin-Madison AIDS Vaccine Research Laboratories | University of Wisconsin-Madison AIDS Vaccine Research Laboratories | Gage Moreno et al |
| EPI_ISL_541642 | USA/WI-UW-1516/2020 | 2020-09-14 | North America/USA/Wisconsin/Dane Co | University of Wisconsin-Madison AIDS Vaccine Research Laboratories | University of Wisconsin-Madison AIDS Vaccine Research Laboratories | Gage Moreno et al |
| EPI_ISL_541648 | USA/WI-UW-1526/2020 | 2020-09-14 | North America/USA/Wisconsin/Dane Co | University of Wisconsin-Madison AIDS Vaccine Research Laboratories | University of Wisconsin-Madison AIDS Vaccine Research Laboratories | Gage Moreno et al |
| EPI_ISL_541653 | Serbia/NP-0332007/2020 | 2020-07-19 | Europe/Serbia/Ra≈°ka District/Novi Paz | Laboratory Diagnostic, Veterinary Specialized Institute Kraljevo | Laboratory Diagnostic, Veterinary Specialized Institute Kraljevo | Vidanovic et al |
| EPI_ISL_541655 | Serbia/KM-09962407/2020 | 2020-07-23 | Europe/Kosovo/Kosovska Mitrovica/ | Laboratory Diagnostic, Veterinary Specialized Institute Kraljevo | Laboratory Diagnostic, Veterinary Specialized Institute Kraljevo | Vidanovic et al |
| EPI_ISL_541658 | Serbia/KM-050707/2020 | 2020-07-06 | Europe/Kosovo/Kosovska Mitrovica/ | Laboratory Diagnostic, Veterinary Specialized Institute Kraljevo | Laboratory Diagnostic, Veterinary Specialized Institute Kraljevo | Vidanovic et al |
| EPI_ISL_541665 | USA/SC-DHEC-1081/2020 | 2020-07-06 | North America/USA/South Carolina/Hor | Microbiology Division, South Carolina Department of Health and Environmental Control | Microbiology Division, South Carolina Department of Health and Environmental Control | Flores et al |

| | | | | | | |
|---|---|---|---|---|---|---|
| EPI_ISL_541666 | USA/SC-DHEC-1083/2020 | 2020-07-07 | North America/USA/South Carolina/Hor | Microbiology Division, South Carolina Department of Health and Environmental Control | Microbiology Division, South Carolina Department of Health and Environmental Control | Flores et al |
| EPI_ISL_541668 | USA/SC-DHEC-1092/2020 | 2020-07-10 | North America/USA/South Carolina/Geo | Microbiology Division, South Carolina Department of Health and Environmental Control | Microbiology Division, South Carolina Department of Health and Environmental Control | Flores et al |
| EPI_ISL_541669 | USA/SC-DHEC-1093/2020 | 2020-07-10 | North America/USA/South Carolina/Hor | Microbiology Division, South Carolina Department of Health and Environmental Control | Microbiology Division, South Carolina Department of Health and Environmental Control | Flores et al |
| EPI_ISL_541670 | USA/SC-DHEC-1095/2020 | 2020-07-10 | North America/USA/South Carolina/Geo | Microbiology Division, South Carolina Department of Health and Environmental Control | Microbiology Division, South Carolina Department of Health and Environmental Control | Flores et al |
| EPI_ISL_541672 | USA/SC-DHEC-0932/2020 | 2020-06-26 | North America/USA/South Carolina/Hor | Microbiology Division, South Carolina Department of Health and Environmental Control | Microbiology Division, South Carolina Department of Health and Environmental Control | Flores et al |

| | | | | | | |
|---|---|---|---|---|---|---|
| EPI_ISL_541677 | USA/SC-DHEC-0942/2020 | 2020-06-26 | North America/USA/South Carolina/Hor | Microbiology Division, South Carolina Department of Health and Environmental Control | Microbiology Division, South Carolina Department of Health and Environmental Control | Flores et al |
| EPI_ISL_541681 | India/MH-NIV-101459/2020 | 2020-07-06 | Asia/India/Maharashtra/ | National Institute of Virology, NIV Influenza | National Institute of Virology, NIV Influenza | Potdar V et al |
| EPI_ISL_541682 | India/MH-NIV-10418/2020 | 2020-04-16 | Asia/India/Maharashtra/ | National Institute of Virology, NIV Influenza | National Institute of Virology, NIV Influenza | Potdar V et al |
| EPI_ISL_541684 | India/MH-NIV-112247/2020 | 2020-07-13 | Asia/India/Maharashtra/ | National Institute of Virology, NIV Influenza | National Institute of Virology, NIV Influenza | Potdar V et al |
| EPI_ISL_541685 | India/MH-NIV-113454/2020 | 2020-07-14 | Asia/India/Maharashtra/ | National Institute of Virology, NIV Influenza | National Institute of Virology, NIV Influenza | Potdar V et al |
| EPI_ISL_541687 | India/MH-NIV-116690/2020 | 2020-07-15 | Asia/India/Maharashtra/ | National Institute of Virology, NIV Influenza | National Institute of Virology, NIV Influenza | Potdar V et al |
| EPI_ISL_541688 | India/MH-NIV-119994/2020 | 2020-07-17 | Asia/India/Maharashtra/ | National Institute of Virology, NIV Influenza | National Institute of Virology, NIV Influenza | Potdar V et al |
| EPI_ISL_541689 | India/MH-NIV-121164/2020 | 2020-07-17 | Asia/India/Maharashtra/ | National Institute of Virology, NIV Influenza | National Institute of Virology, NIV Influenza | Potdar V et al |
| EPI_ISL_541690 | India/MH-NIV-122530/2020 | 2020-07-18 | Asia/India/Maharashtra/ | National Institute of Virology, NIV Influenza | National Institute of Virology, NIV Influenza | Potdar V et al |
| EPI_ISL_541691 | India/MH-NIV-123415/2020 | 2020-07-18 | Asia/India/Maharashtra/ | National Institute of Virology, NIV Influenza | National Institute of Virology, NIV Influenza | Potdar V et al |
| EPI_ISL_541692 | India/MH-NIV-124593/2020 | 2020-07-20 | Asia/India/Maharashtra/ | National Institute of Virology, NIV Influenza | National Institute of Virology, NIV Influenza | Potdar V et al |
| EPI_ISL_541694 | India/MH-NIV-126959/2020 | 2020-07-21 | Asia/India/Maharashtra/ | National Institute of Virology, NIV Influenza | National Institute of Virology, NIV Influenza | Potdar V et al |

| | | | | | | |
|---|---|---|---|---|---|---|
| EPI_ISL_541695 | India/MH-NIV-136271/2020 | 2020-07-26 | Asia/India/Maharashtra/ | National Institute of Virology, NIV Influenza | National Institute of Virology, NIV Influenza | Potdar V et al |
| EPI_ISL_541696 | India/MH-NIV-137244/2020 | 2020-07-27 | Asia/India/Maharashtra/ | National Institute of Virology, NIV Influenza | National Institute of Virology, NIV Influenza | Potdar V et al |
| EPI_ISL_541697 | India/MH-NIV-13768/2020 | 2020-04-23 | Asia/India/Maharashtra/ | National Institute of Virology, NIV Influenza | National Institute of Virology, NIV Influenza | Potdar V et al |
| EPI_ISL_541699 | India/MH-NIV-138100/2020 | 2020-07-27 | Asia/India/Maharashtra/ | National Institute of Virology, NIV Influenza | National Institute of Virology, NIV Influenza | Potdar V et al |
| EPI_ISL_541701 | India/MH-NIV-150015/2020 | 2020-08-04 | Asia/India/Maharashtra/ | National Institute of Virology, NIV Influenza | National Institute of Virology, NIV Influenza | Potdar V et al |
| EPI_ISL_541702 | India/MH-NIV-150596/2020 | 2020-08-05 | Asia/India/Maharashtra/ | National Institute of Virology, NIV Influenza | National Institute of Virology, NIV Influenza | Potdar V et al |
| EPI_ISL_541703 | India/MH-NIV-152698/2020 | 2020-08-06 | Asia/India/Maharashtra/ | National Institute of Virology, NIV Influenza | National Institute of Virology, NIV Influenza | Potdar V et al |
| EPI_ISL_541704 | India/MH-NIV-153805/2020 | 2020-08-07 | Asia/India/Maharashtra/ | National Institute of Virology, NIV Influenza | National Institute of Virology, NIV Influenza | Potdar V et al |
| EPI_ISL_541705 | India/MH-NIV-155669/2020 | 2020-08-08 | Asia/India/Maharashtra/ | National Institute of Virology, NIV Influenza | National Institute of Virology, NIV Influenza | Potdar V et al |
| EPI_ISL_541706 | India/MH-NIV-156143/2020 | 2020-08-08 | Asia/India/Maharashtra/ | National Institute of Virology, NIV Influenza | National Institute of Virology, NIV Influenza | Potdar V et al |
| EPI_ISL_541707 | India/MH-NIV-157478/2020 | 2020-08-10 | Asia/India/Maharashtra/ | National Institute of Virology, NIV Influenza | National Institute of Virology, NIV Influenza | Potdar V et al |
| EPI_ISL_541708 | India/MH-NIV-157939/2020 | 2020-08-10 | Asia/India/Maharashtra/ | National Institute of Virology, NIV Influenza | National Institute of Virology, NIV Influenza | Potdar V et al |
| EPI_ISL_541710 | India/MH-NIV-159610/2020 | 2020-08-11 | Asia/India/Maharashtra/ | National Institute of Virology, NIV Influenza | National Institute of Virology, NIV Influenza | Potdar V et al |

| | | | | | | |
|---|---|---|---|---|---|---|
| EPI_ISL_541711 | India/MH-NIV-164425/2020 | 2020-08-14 | Asia/India/Maharashtra/ | National Institute of Virology, NIV Influenza | National Institute of Virology, NIV Influenza | Potdar V et al |
| EPI_ISL_541712 | India/MH-NIV-171235/2020 | 2020-08-19 | Asia/India/Maharashtra/ | National Institute of Virology, NIV Influenza | National Institute of Virology, NIV Influenza | Potdar V et al |
| EPI_ISL_541713 | India/MH-NIV-172607/2020 | 2020-08-20 | Asia/India/Maharashtra/ | National Institute of Virology, NIV Influenza | National Institute of Virology, NIV Influenza | Potdar V et al |
| EPI_ISL_541714 | India/MH-NIV-173797/2020 | 2020-08-21 | Asia/India/Maharashtra/ | National Institute of Virology, NIV Influenza | National Institute of Virology, NIV Influenza | Potdar V et al |
| EPI_ISL_541715 | India/MH-NIV-174910/2020 | 2020-08-23 | Asia/India/Maharashtra/ | National Institute of Virology, NIV Influenza | National Institute of Virology, NIV Influenza | Potdar V et al |
| EPI_ISL_541716 | India/MH-NIV-176041/2020 | 2020-08-24 | Asia/India/Maharashtra/ | National Institute of Virology, NIV Influenza | National Institute of Virology, NIV Influenza | Potdar V et al |
| EPI_ISL_541717 | India/MH-NIV-176352/2020 | 2020-08-24 | Asia/India/Maharashtra/ | National Institute of Virology, NIV Influenza | National Institute of Virology, NIV Influenza | Potdar V et al |
| EPI_ISL_541718 | India/MH-NIV-183948/2020 | 2020-08-28 | Asia/India/Maharashtra/ | National Institute of Virology, NIV Influenza | National Institute of Virology, NIV Influenza | Potdar V et al |
| EPI_ISL_541719 | India/MH-NIV-185628/2020 | 2020-08-29 | Asia/India/Maharashtra/ | National Institute of Virology, NIV Influenza | National Institute of Virology, NIV Influenza | Potdar V et al |
| EPI_ISL_541720 | India/MH-NIV-186959/2020 | 2020-08-31 | Asia/India/Maharashtra/ | National Institute of Virology, NIV Influenza | National Institute of Virology, NIV Influenza | Potdar V et al |
| EPI_ISL_541721 | India/MH-NIV-187756/2020 | 2020-08-31 | Asia/India/Maharashtra/ | National Institute of Virology, NIV Influenza | National Institute of Virology, NIV Influenza | Potdar V et al |
| EPI_ISL_541723 | India/MH-NIV-191205/2020 | 2020-09-02 | Asia/India/Maharashtra/ | National Institute of Virology, NIV Influenza | National Institute of Virology, NIV Influenza | Potdar V et al |
| EPI_ISL_541724 | India/MH-NIV-192922/2020 | 2020-09-03 | Asia/India/Maharashtra/ | National Institute of Virology, NIV Influenza | National Institute of Virology, NIV Influenza | Potdar V et al |

| | | | | | | |
|---|---|---|---|---|---|---|
| EPI_ISL_541726 | India/MH-NIV-57622/2020 | 2020-06-10 | Asia/India/Maharashtra/ | National Institute of Virology, NIV Influenza | National Institute of Virology, NIV Influenza | Potdar V et al |
| EPI_ISL_541728 | India/MH-NIV-58210/2020 | 2020-06-10 | Asia/India/Maharashtra/ | National Institute of Virology, NIV Influenza | National Institute of Virology, NIV Influenza | Potdar V et al |
| EPI_ISL_541729 | India/MH-NIV-59437/2020 | 2020-06-11 | Asia/India/Maharashtra/ | National Institute of Virology, NIV Influenza | National Institute of Virology, NIV Influenza | Potdar V et al |
| EPI_ISL_541731 | India/MH-NIV-60033/2020 | 2020-06-12 | Asia/India/Maharashtra/ | National Institute of Virology, NIV Influenza | National Institute of Virology, NIV Influenza | Potdar V et al |
| EPI_ISL_541733 | India/MH-NIV-69925/2020 | 2020-06-19 | Asia/India/Maharashtra/ | National Institute of Virology, NIV Influenza | National Institute of Virology, NIV Influenza | Potdar V et al |
| EPI_ISL_541734 | India/MH-NIV-71548/2020 | 2020-06-20 | Asia/India/Maharashtra/ | National Institute of Virology, NIV Influenza | National Institute of Virology, NIV Influenza | Potdar V et al |
| EPI_ISL_541736 | India/MH-NIV-72638/2020 | 2020-06-20 | Asia/India/Maharashtra/ | National Institute of Virology, NIV Influenza | National Institute of Virology, NIV Influenza | Potdar V et al |
| EPI_ISL_541737 | India/MH-NIV-73012/2020 | 2020-06-22 | Asia/India/Maharashtra/ | National Institute of Virology, NIV Influenza | National Institute of Virology, NIV Influenza | Potdar V et al |
| EPI_ISL_541740 | India/MH-NIV-74916/2020 | 2020-07-03 | Asia/India/Maharashtra/ | National Institute of Virology, NIV Influenza | National Institute of Virology, NIV Influenza | Potdar V et al |
| EPI_ISL_541743 | India/MH-NIV-77702/2020 | 2020-06-24 | Asia/India/Maharashtra/ | National Institute of Virology, NIV Influenza | National Institute of Virology, NIV Influenza | Potdar V et al |
| EPI_ISL_541744 | India/MH-NIV-78532/2020 | 2020-06-24 | Asia/India/Maharashtra/ | National Institute of Virology, NIV Influenza | National Institute of Virology, NIV Influenza | Potdar V et al |
| EPI_ISL_541746 | India/MH-NIV-80896/2020 | 2020-06-25 | Asia/India/Maharashtra/ | National Institute of Virology, NIV Influenza | National Institute of Virology, NIV Influenza | Potdar V et al |
| EPI_ISL_541749 | India/MH-NIV-97052/2020 | 2020-07-03 | Asia/India/Maharashtra/ | National Institute of Virology, NIV Influenza | National Institute of Virology, NIV Influenza | Potdar V et al |

| | | | | | | |
|---|---|---|---|---|---|---|
| EPI_ISL_541750 | India/MH-NIV-99083/2020 | 2020-07-04 | Asia/India/Maharashtra/ | National Institute of Virology, NIV Influenza | National Institute of Virology, NIV Influenza | Potdar V et al |
| EPI_ISL_541751 | India/MH-NIV-99913/2020 | 2020-07-06 | Asia/India/Maharashtra/ | National Institute of Virology, NIV Influenza | National Institute of Virology, NIV Influenza | Potdar V et al |
| EPI_ISL_541879 | Spain/MD-IBV-99007091/2020 | 2020-03-23 | Europe/Spain/Madrid/Estremera | Hospital General Universitario Gregorio Marav√±√≥n | SeqCOVID-SPAIN consortium/IBV(CSIC) | Laura P√©rez-Lago et al |
| EPI_ISL_541881 | Spain/MD-IBV-99007096/2020 | 2020-04-15 | Europe/Spain/Madrid/Estremera | Hospital General Universitario Gregorio Marav√±√≥n | SeqCOVID-SPAIN consortium/IBV(CSIC) | Laura P√©rez-Lago et al |
| EPI_ISL_541884 | Spain/MD-IBV-99007102/2020 | 2020-06-12 | Europe/Spain/Madrid/Madrid | Hospital General Universitario Gregorio Marav√±√≥n | SeqCOVID-SPAIN consortium/IBV(CSIC) | Laura P√©rez-Lago et al |
| EPI_ISL_541896 | Spain/MD-IBV-99007122/2020 | 2020-07-29 | Europe/Spain/Madrid/Madrid | Hospital General Universitario Gregorio Marav√±√≥n | SeqCOVID-SPAIN consortium/IBV(CSIC) | Laura P√©rez-Lago et al |
| EPI_ISL_541897 | Spain/MD-IBV-99007123/2020 | 2020-07-30 | Europe/Spain/Madrid/Madrid | Hospital General Universitario Gregorio Marav√±√≥n | SeqCOVID-SPAIN consortium/IBV(CSIC) | Laura P√©rez-Lago et al |
| EPI_ISL_541902 | Spain/MD-IBV-99007128/2020 | 2020-07-31 | Europe/Spain/Madrid/Madrid | Hospital General Universitario Gregorio Marav√±√≥n | SeqCOVID-SPAIN consortium/IBV(CSIC) | Laura P√©rez-Lago et al |
| EPI_ISL_541907 | Spain/MD-IBV-99007134/2020 | 2020-08-03 | Europe/Spain/Madrid/Madrid | Hospital General Universitario Gregorio Marav√±√≥n | SeqCOVID-SPAIN consortium/IBV(CSIC) | Laura P√©rez-Lago et al |
| EPI_ISL_541908 | Spain/MD-IBV-99007135/2020 | 2020-08-03 | Europe/Spain/Madrid/Madrid | Hospital General Universitario Gregorio Marav√±√≥n | SeqCOVID-SPAIN consortium/IBV(CSIC) | Laura P√©rez-Lago et al |
| EPI_ISL_541909 | Spain/MD-IBV-99007136/2020 | 2020-08-04 | Europe/Spain/Madrid/Madrid | Hospital General Universitario Gregorio Marav√±√≥n | SeqCOVID-SPAIN consortium/IBV(CSIC) | Laura P√©rez-Lago et al |
| EPI_ISL_541910 | Spain/MD-IBV-99007137/2020 | 2020-08-04 | Europe/Spain/Madrid/Madrid | Hospital General Universitario Gregorio Marav√±√≥n | SeqCOVID-SPAIN consortium/IBV(CSIC) | Laura P√©rez-Lago et al |
| EPI_ISL_541914 | Spain/MD-IBV-99007141/2020 | 2020-08-04 | Europe/Spain/Madrid/Madrid | Hospital General Universitario Gregorio Marav√±√≥n | SeqCOVID-SPAIN consortium/IBV(CSIC) | Laura P√©rez-Lago et al |

| | | | | | | |
|---|---|---|---|---|---|---|
| EPI_ISL_541917 | Spain/MD-IBV-99007145/2020 | 2020-08-06 | Europe/Spain/Madrid/Madrid | Hospital General Universitario Gregorio Marav√±√≥n | SeqCOVID-SPAIN consortium/IBV(CSIC) | Laura P√©rez-Lago et al |
| EPI_ISL_541918 | Spain/MD-IBV-99007146/2020 | 2020-08-07 | Europe/Spain/Madrid/Madrid | Hospital General Universitario Gregorio Marav√±√≥n | SeqCOVID-SPAIN consortium/IBV(CSIC) | Laura P√©rez-Lago et al |
| EPI_ISL_541919 | Spain/MD-IBV-99007147/2020 | 2020-08-07 | Europe/Spain/Madrid/Madrid | Hospital General Universitario Gregorio Marav√±√≥n | SeqCOVID-SPAIN consortium/IBV(CSIC) | Laura P√©rez-Lago et al |
| EPI_ISL_541920 | Spain/MD-IBV-99007148/2020 | 2020-08-11 | Europe/Spain/Madrid/Madrid | Hospital General Universitario Gregorio Marav√±√≥n | SeqCOVID-SPAIN consortium/IBV(CSIC) | Laura P√©rez-Lago et al |
| EPI_ISL_541921 | Spain/MD-IBV-99007149/2020 | 2020-08-12 | Europe/Spain/Madrid/Madrid | Hospital General Universitario Gregorio Marav√±√≥n | SeqCOVID-SPAIN consortium/IBV(CSIC) | Laura P√©rez-Lago et al |
| EPI_ISL_541923 | Spain/MD-IBV-99007151/2020 | 2020-08-13 | Europe/Spain/Madrid/Madrid | Hospital General Universitario Gregorio Marav√±√≥n | SeqCOVID-SPAIN consortium/IBV(CSIC) | Laura P√©rez-Lago et al |
| EPI_ISL_541924 | Spain/MD-IBV-99007152/2020 | 2020-08-13 | Europe/Spain/Madrid/Madrid | Hospital General Universitario Gregorio Marav√±√≥n | SeqCOVID-SPAIN consortium/IBV(CSIC) | Laura P√©rez-Lago et al |
| EPI_ISL_541926 | Spain/MD-IBV-99007154/2020 | 2020-08-14 | Europe/Spain/Madrid/Madrid | Hospital General Universitario Gregorio Marav√±√≥n | SeqCOVID-SPAIN consortium/IBV(CSIC) | Laura P√©rez-Lago et al |
| EPI_ISL_541927 | Spain/MD-IBV-99007157/2020 | 2020-08-18 | Europe/Spain/Madrid/Madrid | Hospital General Universitario Gregorio Marav√±√≥n | SeqCOVID-SPAIN consortium/IBV(CSIC) | Laura P√©rez-Lago et al |
| EPI_ISL_541928 | Spain/MD-IBV-99007158/2020 | 2020-08-17 | Europe/Spain/Madrid/Madrid | Hospital General Universitario Gregorio Marav√±√≥n | SeqCOVID-SPAIN consortium/IBV(CSIC) | Laura P√©rez-Lago et al |
| EPI_ISL_541929 | Spain/MD-IBV-99007159/2020 | 2020-08-18 | Europe/Spain/Madrid/Madrid | Hospital General Universitario Gregorio Marav√±√≥n | SeqCOVID-SPAIN consortium/IBV(CSIC) | Laura P√©rez-Lago et al |
| EPI_ISL_541931 | Spain/MD-IBV-99007161/2020 | 2020-08-19 | Europe/Spain/Madrid/Madrid | Hospital General Universitario Gregorio Marav√±√≥n | SeqCOVID-SPAIN consortium/IBV(CSIC) | Laura P√©rez-Lago et al |
| EPI_ISL_541932 | Spain/MD-IBV-99007162/2020 | 2020-08-19 | Europe/Spain/Madrid/Madrid | Hospital General Universitario Gregorio Marav√±√≥n | SeqCOVID-SPAIN consortium/IBV(CSIC) | Laura P√©rez-Lago et al |

| | | | | | | |
|---|---|---|---|---|---|---|
| EPI_ISL_541933 | Spain/MD-IBV-99007163/2020 | 2020-08-19 | Europe/Spain/Madrid/Madrid | Hospital General Universitario Gregorio Marav±V≥n | SeqCOVID-SPAIN consortium/IBV(CSIC) | Laura PV©rez-Lago et al |
| EPI_ISL_541935 | Spain/MD-IBV-99007165/2020 | 2020-08-19 | Europe/Spain/Madrid/Madrid | Hospital General Universitario Gregorio Marav±V≥n | SeqCOVID-SPAIN consortium/IBV(CSIC) | Laura PV©rez-Lago et al |
| EPI_ISL_541936 | Spain/MD-IBV-99007166/2020 | 2020-08-23 | Europe/Spain/Madrid/Madrid | Hospital General Universitario Gregorio Marav±V≥n | SeqCOVID-SPAIN consortium/IBV(CSIC) | Laura PV©rez-Lago et al |
| EPI_ISL_541937 | Spain/MD-IBV-99007167/2020 | 2020-08-24 | Europe/Spain/Madrid/Madrid | Hospital General Universitario Gregorio Marav±V≥n | SeqCOVID-SPAIN consortium/IBV(CSIC) | Laura PV©rez-Lago et al |
| EPI_ISL_541938 | Spain/MD-IBV-99007168/2020 | 2020-08-24 | Europe/Spain/Madrid/Madrid | Hospital General Universitario Gregorio Marav±V≥n | SeqCOVID-SPAIN consortium/IBV(CSIC) | Laura PV©rez-Lago et al |
| EPI_ISL_541939 | Spain/MD-IBV-99007169/2020 | 2020-08-24 | Europe/Spain/Madrid/Madrid | Hospital General Universitario Gregorio Marav±V≥n | SeqCOVID-SPAIN consortium/IBV(CSIC) | Laura PV©rez-Lago et al |
| EPI_ISL_541949 | Spain/IB-IBV-99007181/2020 | 2020-03-14 | Europe/Spain/Balear Islands/Mahon | Servicio de MicrobiologV≠a, Hospital Universitario Son Espases | SeqCOVID-SPAIN consortium/IBV(CSIC) | Carla LV≥pez-CausapV© et al |
| EPI_ISL_541950 | Spain/IB-IBV-99007182/2020 | 2020-03-02 | Europe/Spain/Balear Islands/Pont d'Inca | Servicio de MicrobiologV≠a, Hospital Universitario Son Espases | SeqCOVID-SPAIN consortium/IBV(CSIC) | Carla LV≥pez-CausapV© et al |
| EPI_ISL_541951 | Spain/IB-IBV-99007192/2020 | 2020-03-08 | Europe/Spain/Balear Islands/Menorca | Servicio de MicrobiologV≠a, Hospital Universitario Son Espases | SeqCOVID-SPAIN consortium/IBV(CSIC) | Carla LV≥pez-CausapV© et al |
| EPI_ISL_541953 | Spain/IB-IBV-99007194/2020 | 2020-04-01 | Europe/Spain/Balear Islands/Palma | Servicio de MicrobiologV≠a, Hospital Universitario Son Espases | SeqCOVID-SPAIN consortium/IBV(CSIC) | Carla LV≥pez-CausapV© et al |
| EPI_ISL_541955 | Spain/IB-IBV-99007205/2020 | 2020-03-22 | Europe/Spain/Balear Islands/Formenter | Servicio de MicrobiologV≠a, Hospital Universitario Son Espases | SeqCOVID-SPAIN consortium/IBV(CSIC) | Carla LV≥pez-CausapV© et al |
| EPI_ISL_541968 | Spain/IB-IBV-99007264/2020 | 2020-02-28 | Europe/Spain/Balear Islands/Palma | Servicio de MicrobiologV≠a, Hospital Universitario Son Espases | SeqCOVID-SPAIN consortium/IBV(CSIC) | Carla LV≥pez-CausapV© et al |
| EPI_ISL_541969 | Spain/IB-IBV-99007265/2020 | 2020-03-24 | Europe/Spain/Balear Islands/Manacor | Servicio de MicrobiologV≠a, Hospital Universitario Son Espases | SeqCOVID-SPAIN consortium/IBV(CSIC) | Carla LV≥pez-CausapV© et al |

| | | | | | | |
|---|---|---|---|---|---|---|
| EPI_ISL_541970 | Norway/Bergen-01/2020 | 2020-03-18 | Europe/Norway/Vestland/Bergen | Influenza Centre, University of Bergen | Norwegian Institute of Public Health, Department of Virology | Fan Zhou et al |
| EPI_ISL_541980 | USA/TX-DSHS-0445/2020 | 2020-06-05 | North America/USA/Texas/Harris Count | Texas Department of State Health Services | Texas Department of State Health Services | Rashmi Tuladhar et al |
| EPI_ISL_541983 | USA/TX-DSHS-0448/2020 | 2020-06-05 | North America/USA/Texas/Harris Count | Texas Department of State Health Services | Texas Department of State Health Services | Rashmi Tuladhar et al |
| EPI_ISL_541996 | USA/TX-DSHS-0465/2020 | 2020-06-21 | North America/USA/Texas/Hidalgo Cour | Texas Department of State Health Services | Texas Department of State Health Services | Rashmi Tuladhar et al |
| EPI_ISL_541998 | USA/TX-DSHS-0467/2020 | 2020-06-21 | North America/USA/Texas/Hidalgo Cour | Texas Department of State Health Services | Texas Department of State Health Services | Rashmi Tuladhar et al |
| EPI_ISL_542000 | USA/TX-DSHS-0469/2020 | 2020-06-21 | North America/USA/Texas/Hidalgo Cour | Texas Department of State Health Services | Texas Department of State Health Services | Rashmi Tuladhar et al |
| EPI_ISL_542007 | USA/TX-DSHS-0476/2020 | 2020-06-21 | North America/USA/Texas/Hidalgo Cour | Texas Department of State Health Services | Texas Department of State Health Services | Rashmi Tuladhar et al |
| EPI_ISL_542009 | USA/TX-DSHS-0478/2020 | 2020-06-21 | North America/USA/Texas/Hidalgo Cour | Texas Department of State Health Services | Texas Department of State Health Services | Rashmi Tuladhar et al |
| EPI_ISL_542010 | USA/TX-DSHS-0479/2020 | 2020-06-21 | North America/USA/Texas/Hidalgo Cour | Texas Department of State Health Services | Texas Department of State Health Services | Rashmi Tuladhar et al |
| EPI_ISL_542021 | USA/NM-NMDOH-2020132668/2020 | 2020-06-16 | North America/USA/New Mexico/ | New Mexico Department of Health Scientific Laboratory | New Mexico Department of Health Scientific Laboratory | Ellie Johnson et al |
| EPI_ISL_542022 | USA/NMDOH-2020136739/2020 | 2020-06-18 | North America/USA/Texas/ | New Mexico Department of Health Scientific Laboratory | New Mexico Department of Health Scientific Laboratory | Ellie Johnson et al |
| EPI_ISL_542030 | USA/NMDOH-2020185052/2020 | 2020-07-07 | North America/USA/New Mexico/ | New Mexico Department of Health Scientific Laboratory | New Mexico Department of Health Scientific Laboratory | Ellie Johnson et al |

| | | | | | | |
|---|---|---|---|---|---|---|
| EPI_ISL_542032 | USA/NMDOH-2020189541/2020 | 2020-07-07 | North America/USA/New Mexico/ | New Mexico Department of Health Scientific Laboratory | New Mexico Department of Health Scientific Laboratory | Ellie Johnson et al |
| EPI_ISL_542033 | USA/NMDOH-2020189581/2020 | 2020-07-07 | North America/USA/Texas/ | New Mexico Department of Health Scientific Laboratory | New Mexico Department of Health Scientific Laboratory | Ellie Johnson et al |
| EPI_ISL_542038 | USA/NMDOH-2020239972/2020 | 2020-07-22 | North America/USA/Texas/ | New Mexico Department of Health Scientific Laboratory | New Mexico Department of Health Scientific Laboratory | Ellie Johnson et al |
| EPI_ISL_542040 | USA/NMDOH-2020245669/2020 | 2020-07-23 | North America/USA/New Mexico/ | New Mexico Department of Health Scientific Laboratory | New Mexico Department of Health Scientific Laboratory | Ellie Johnson et al |
| EPI_ISL_542042 | USA/NMDOH-2020305741/2020 | 2020-08-21 | North America/USA/New Mexico/ | New Mexico Department of Health Scientific Laboratory | New Mexico Department of Health Scientific Laboratory | Ellie Johnson et al |
| EPI_ISL_542061 | USA/NMDOH-2020305829/2020 | 2020-08-21 | North America/USA/New Mexico/ | New Mexico Department of Health Scientific Laboratory | New Mexico Department of Health Scientific Laboratory | Ellie Johnson et al |
| EPI_ISL_542062 | USA/NMDOH-2020320036/2020 | 2020-08-31 | North America/USA/New Mexico/ | New Mexico Department of Health Scientific Laboratory | New Mexico Department of Health Scientific Laboratory | Ellie Johnson et al |
| EPI_ISL_542063 | USA/NMDOH-2020320930/2020 | 2020-09-01 | North America/USA/New Mexico/ | New Mexico Department of Health Scientific Laboratory | New Mexico Department of Health Scientific Laboratory | Ellie Johnson et al |
| EPI_ISL_542066 | USA/NMDOH-2020320943/2020 | 2020-09-01 | North America/USA/Texas/ | New Mexico Department of Health Scientific Laboratory | New Mexico Department of Health Scientific Laboratory | Ellie Johnson et al |
| EPI_ISL_542070 | USA/NMDOH-2020321648/2020 | 2020-09-01 | North America/USA/New Mexico/ | New Mexico Department of Health Scientific Laboratory | New Mexico Department of Health Scientific Laboratory | Ellie Johnson et al |

| | | | | | | |
|---|---|---|---|---|---|---|
| EPI_ISL_542077 | USA/NMDOH-2020322105/2020 | 2020-09-01 | North America/USA/New Mexico/ | New Mexico Department of Health Scientific Laboratory | New Mexico Department of Health Scientific Laboratory | Ellie Johnson et al |
| EPI_ISL_542082 | USA/NMDOH-2020322173/2020 | 2020-09-01 | North America/USA/New Mexico/ | New Mexico Department of Health Scientific Laboratory | New Mexico Department of Health Scientific Laboratory | Ellie Johnson et al |
| EPI_ISL_542085 | USA/NMDOH-2020322366/2020 | 2020-09-01 | North America/USA/New Mexico/ | New Mexico Department of Health Scientific Laboratory | New Mexico Department of Health Scientific Laboratory | Ellie Johnson et al |
| EPI_ISL_542086 | USA/NMDOH-2020323484/2020 | 2020-09-01 | North America/USA/New Mexico/ | New Mexico Department of Health Scientific Laboratory | New Mexico Department of Health Scientific Laboratory | Ellie Johnson et al |
| EPI_ISL_542088 | USA/NMDOH-2020323670/2020 | 2020-09-01 | North America/USA/New Mexico/ | New Mexico Department of Health Scientific Laboratory | New Mexico Department of Health Scientific Laboratory | Ellie Johnson et al |
| EPI_ISL_542089 | USA/NMDOH-2020323679/2020 | 2020-09-01 | North America/USA/New Mexico/ | New Mexico Department of Health Scientific Laboratory | New Mexico Department of Health Scientific Laboratory | Ellie Johnson et al |
| EPI_ISL_542090 | USA/NMDOH-2020323686/2020 | 2020-09-01 | North America/USA/New Mexico/ | New Mexico Department of Health Scientific Laboratory | New Mexico Department of Health Scientific Laboratory | Ellie Johnson et al |
| EPI_ISL_542095 | USA/NMDOH-2020324359/2020 | 2020-09-02 | North America/USA/New Mexico/ | New Mexico Department of Health Scientific Laboratory | New Mexico Department of Health Scientific Laboratory | Ellie Johnson et al |
| EPI_ISL_542098 | Italy/LOM-UniMI-L100/2020 | 2020-03-08 | Europe/Italy/Lombardy/ | ASST GOM Niguarda | Dep. Of Oncology and Hemato-Oncology University of Milan | Claudia Alteri et al |

| | | | | | | |
|---|---|---|---|---|---|---|
| EPI_ISL_542099 | Italy/LOM-UniMI-L101/2020 | 2020-03-08 | Europe/Italy/Lombardy/ | ASST GOM Niguarda | Dep. Of Oncology and Hemato-Oncology University of Milan | Claudia Alteri et al |
| EPI_ISL_542105 | Italy/LOM-UniMI-L107/2020 | 2020-03-05 | Europe/Italy/Lombardy/ | ASST GOM Niguarda | Dep. Of Oncology and Hemato-Oncology University of Milan | Claudia Alteri et al |
| EPI_ISL_542106 | Italy/LOM-UniMI-L108/2020 | 2020-03-02 | Europe/Italy/Lombardy/ | ASST GOM Niguarda | Dep. Of Oncology and Hemato-Oncology University of Milan | Claudia Alteri et al |
| EPI_ISL_542107 | Italy/LOM-UniMI-L109/2020 | 2020-03-10 | Europe/Italy/Lombardy/ | ASST GOM Niguarda | Dep. Of Oncology and Hemato-Oncology University of Milan | Claudia Alteri et al |
| EPI_ISL_542111 | Italy/LOM-UniMI-L115/2020 | 2020-03-07 | Europe/Italy/Lombardy/ | ASST GOM Niguarda | Dep. Of Oncology and Hemato-Oncology University of Milan | Claudia Alteri et al |
| EPI_ISL_542118 | Italy/LOM-UniMI-L122/2020 | 2020-03-09 | Europe/Italy/Lombardy/ | ASST GOM Niguarda | Dep. Of Oncology and Hemato-Oncology University of Milan | Claudia Alteri et al |
| EPI_ISL_542119 | Italy/LOM-UniMI-L123/2020 | 2020-03-10 | Europe/Italy/Lombardy/ | ASST GOM Niguarda | Dep. Of Oncology and Hemato-Oncology University of Milan | Claudia Alteri et al |
| EPI_ISL_542120 | Italy/LOM-UniMI-L124/2020 | 2020-03-10 | Europe/Italy/Lombardy/ | ASST GOM Niguarda | Dep. Of Oncology and Hemato-Oncology University of Milan | Claudia Alteri et al |

| | | | | | | |
|---|---|---|---|---|---|---|
| EPI_ISL_542121 | Italy/LOM-UniMI-L125/2020 | 2020-03-13 | Europe/Italy/Lombardy/ | ASST GOM Niguarda | Dep. Of Oncology and Hemato-Oncology University of Milan | Claudia Alteri et al |
| EPI_ISL_542122 | Italy/LOM-UniMI-L126/2020 | 2020-03-18 | Europe/Italy/Lombardy/ | ASST GOM Niguarda | Dep. Of Oncology and Hemato-Oncology University of Milan | Claudia Alteri et al |
| EPI_ISL_542125 | Italy/LOM-UniMI-L129/2020 | 2020-02-28 | Europe/Italy/Lombardy/ | ASST GOM Niguarda | Dep. Of Oncology and Hemato-Oncology University of Milan | Claudia Alteri et al |
| EPI_ISL_542126 | Italy/LOM-UniMI-L130/2020 | 2020-03-10 | Europe/Italy/Lombardy/ | ASST GOM Niguarda | Dep. Of Oncology and Hemato-Oncology University of Milan | Claudia Alteri et al |
| EPI_ISL_542128 | Italy/LOM-UniMI-L132/2020 | 2020-03-21 | Europe/Italy/Lombardy/ | ASST GOM Niguarda | Dep. Of Oncology and Hemato-Oncology University of Milan | Claudia Alteri et al |
| EPI_ISL_542129 | Italy/LOM-UniMI-L133/2020 | 2020-03-18 | Europe/Italy/Lombardy/ | ASST GOM Niguarda | Dep. Of Oncology and Hemato-Oncology University of Milan | Claudia Alteri et al |
| EPI_ISL_542130 | Italy/LOM-UniMI-L134/2020 | 2020-03-17 | Europe/Italy/Lombardy/ | ASST GOM Niguarda | Dep. Of Oncology and Hemato-Oncology University of Milan | Claudia Alteri et al |
| EPI_ISL_542132 | Italy/LOM-UniMI-L136/2020 | 2020-03-22 | Europe/Italy/Lombardy/ | ASST GOM Niguarda | Dep. Of Oncology and Hemato-Oncology University of Milan | Claudia Alteri et al |

| | | | | | | |
|---|---|---|---|---|---|---|
| EPI_ISL_542137 | Italy/LOM-UniMI-L141/2020 | 2020-03-19 | Europe/Italy/Lombardy/ | ASST GOM Niguarda | Dep. Of Oncology and Hemato-Oncology University of Milan | Claudia Alteri et al |
| EPI_ISL_542138 | Italy/LOM-UniMI-L142/2020 | 2020-03-19 | Europe/Italy/Lombardy/ | ASST GOM Niguarda | Dep. Of Oncology and Hemato-Oncology University of Milan | Claudia Alteri et al |
| EPI_ISL_542139 | Italy/LOM-UniMI-L144/2020 | 2020-03-19 | Europe/Italy/Lombardy/ | ASST GOM Niguarda | Dep. Of Oncology and Hemato-Oncology University of Milan | Claudia Alteri et al |
| EPI_ISL_542140 | Italy/LOM-UniMI-L145/2020 | 2020-03-19 | Europe/Italy/Lombardy/ | ASST GOM Niguarda | Dep. Of Oncology and Hemato-Oncology University of Milan | Claudia Alteri et al |
| EPI_ISL_542141 | Italy/LOM-UniMI-L146/2020 | 2020-03-19 | Europe/Italy/Lombardy/ | ASST GOM Niguarda | Dep. Of Oncology and Hemato-Oncology University of Milan | Claudia Alteri et al |
| EPI_ISL_542144 | Italy/LOM-UniMI-L149/2020 | 2020-03-19 | Europe/Italy/Lombardy/ | ASST GOM Niguarda | Dep. Of Oncology and Hemato-Oncology University of Milan | Claudia Alteri et al |
| EPI_ISL_542147 | Italy/LOM-UniMI-L152/2020 | 2020-03-16 | Europe/Italy/Lombardy/ | ASST GOM Niguarda | Dep. Of Oncology and Hemato-Oncology University of Milan | Claudia Alteri et al |
| EPI_ISL_542148 | Italy/LOM-UniMI-L153/2020 | 2020-03-15 | Europe/Italy/Lombardy/ | ASST GOM Niguarda | Dep. Of Oncology and Hemato-Oncology University of Milan | Claudia Alteri et al |

| | | | | | | |
|---|---|---|---|---|---|---|
| EPI_ISL_542149 | Italy/LOM-UniMI-L154/2020 | 2020-03-21 | Europe/Italy/Lombardy/ | ASST GOM Niguarda | Dep. Of Oncology and Hemato-Oncology University of Milan | Claudia Alteri et al |
| EPI_ISL_542151 | Italy/LOM-UniMI-L156/2020 | 2020-03-19 | Europe/Italy/Lombardy/ | ASST GOM Niguarda | Dep. Of Oncology and Hemato-Oncology University of Milan | Claudia Alteri et al |
| EPI_ISL_542153 | Italy/LOM-UniMI-L158/2020 | 2020-03-19 | Europe/Italy/Lombardy/ | ASST GOM Niguarda | Dep. Of Oncology and Hemato-Oncology University of Milan | Claudia Alteri et al |
| EPI_ISL_542156 | Italy/LOM-UniMI-L162/2020 | 2020-03-08 | Europe/Italy/Lombardy/ | ASST GOM Niguarda | Dep. Of Oncology and Hemato-Oncology University of Milan | Claudia Alteri et al |
| EPI_ISL_542157 | Italy/LOM-UniMI-L163/2020 | 2020-03-03 | Europe/Italy/Lombardy/ | ASST GOM Niguarda | Dep. Of Oncology and Hemato-Oncology University of Milan | Claudia Alteri et al |
| EPI_ISL_542158 | Italy/LOM-UniMI-L164/2020 | 2020-03-21 | Europe/Italy/Lombardy/ | ASST GOM Niguarda | Dep. Of Oncology and Hemato-Oncology University of Milan | Claudia Alteri et al |
| EPI_ISL_542161 | Italy/LOM-UniMI-L167/2020 | 2020-03-15 | Europe/Italy/Lombardy/ | ASST GOM Niguarda | Dep. Of Oncology and Hemato-Oncology University of Milan | Claudia Alteri et al |
| EPI_ISL_542163 | Italy/LOM-UniMI-L169/2020 | 2020-03-14 | Europe/Italy/Lombardy/ | ASST GOM Niguarda | Dep. Of Oncology and Hemato-Oncology University of Milan | Claudia Alteri et al |

| | | | | | | |
|---|---|---|---|---|---|---|
| EPI_ISL_542166 | Italy/LOM-UniMI-L172/2020 | 2020-03-19 | Europe/Italy/Lombardy/ | ASST GOM Niguarda | Dep. Of Oncology and Hemato-Oncology University of Milan | Claudia Alteri et al |
| EPI_ISL_542167 | Italy/LOM-UniMI-L173/2020 | 2020-03-19 | Europe/Italy/Lombardy/ | ASST GOM Niguarda | Dep. Of Oncology and Hemato-Oncology University of Milan | Claudia Alteri et al |
| EPI_ISL_542168 | Italy/LOM-UniMI-L174/2020 | 2020-03-21 | Europe/Italy/Lombardy/ | ASST GOM Niguarda | Dep. Of Oncology and Hemato-Oncology University of Milan | Claudia Alteri et al |
| EPI_ISL_542169 | Italy/LOM-UniMI-L175/2020 | 2020-03-16 | Europe/Italy/Lombardy/ | ASST GOM Niguarda | Dep. Of Oncology and Hemato-Oncology University of Milan | Claudia Alteri et al |
| EPI_ISL_542172 | Italy/LOM-UniMI-L178/2020 | 2020-03-21 | Europe/Italy/Lombardy/ | ASST GOM Niguarda | Dep. Of Oncology and Hemato-Oncology University of Milan | Claudia Alteri et al |
| EPI_ISL_542174 | Italy/LOM-UniMI-L183/2020 | 2020-03-21 | Europe/Italy/Lombardy/ | ASST GOM Niguarda | Dep. Of Oncology and Hemato-Oncology University of Milan | Claudia Alteri et al |
| EPI_ISL_542177 | Italy/LOM-UniMI-L186/2020 | 2020-03-20 | Europe/Italy/Lombardy/ | ASST GOM Niguarda | Dep. Of Oncology and Hemato-Oncology University of Milan | Claudia Alteri et al |
| EPI_ISL_542180 | Italy/LOM-UniMI-L190/2020 | 2020-03-10 | Europe/Italy/Lombardy/ | ASST GOM Niguarda | Dep. Of Oncology and Hemato-Oncology University of Milan | Claudia Alteri et al |

| | | | | | | |
|---|---|---|---|---|---|---|
| EPI_ISL_542181 | Italy/LOM-UniMI-L191/2020 | 2020-03-21 | Europe/Italy/Lombardy/ | ASST GOM Niguarda | Dep. Of Oncology and Hemato-Oncology University of Milan | Claudia Alteri et al |
| EPI_ISL_542182 | Italy/LOM-UniMI-L192/2020 | 2020-03-21 | Europe/Italy/Lombardy/ | ASST GOM Niguarda | Dep. Of Oncology and Hemato-Oncology University of Milan | Claudia Alteri et al |
| EPI_ISL_542184 | Italy/LOM-UniMI-L194/2020 | 2020-03-22 | Europe/Italy/Lombardy/ | ASST GOM Niguarda | Dep. Of Oncology and Hemato-Oncology University of Milan | Claudia Alteri et al |
| EPI_ISL_542188 | Italy/LOM-UniMI-L198/2020 | 2020-03-21 | Europe/Italy/Lombardy/ | ASST GOM Niguarda | Dep. Of Oncology and Hemato-Oncology University of Milan | Claudia Alteri et al |
| EPI_ISL_542192 | Italy/LOM-UniMI-L204/2020 | 2020-03-08 | Europe/Italy/Lombardy/ | ASST GOM Niguarda | Dep. Of Oncology and Hemato-Oncology University of Milan | Claudia Alteri et al |
| EPI_ISL_542197 | Italy/LOM-UniMI-L210/2020 | 2020-03-12 | Europe/Italy/Lombardy/ | ASST GOM Niguarda | Dep. Of Oncology and Hemato-Oncology University of Milan | Claudia Alteri et al |
| EPI_ISL_542198 | Italy/LOM-UniMI-L211/2020 | 2020-03-12 | Europe/Italy/Lombardy/ | ASST GOM Niguarda | Dep. Of Oncology and Hemato-Oncology University of Milan | Claudia Alteri et al |
| EPI_ISL_542199 | Italy/LOM-UniMI-L212/2020 | 2020-03-14 | Europe/Italy/Lombardy/ | ASST GOM Niguarda | Dep. Of Oncology and Hemato-Oncology University of Milan | Claudia Alteri et al |

| | | | | | | |
|---|---|---|---|---|---|---|
| EPI_ISL_542202 | Italy/LOM-UniMI-L215/2020 | 2020-03-12 | Europe/Italy/Lombardy/ | ASST GOM Niguarda | Dep. Of Oncology and Hemato-Oncology University of Milan | Claudia Alteri et al |
| EPI_ISL_542203 | Italy/LOM-UniMI-L216/2020 | 2020-03-13 | Europe/Italy/Lombardy/ | ASST GOM Niguarda | Dep. Of Oncology and Hemato-Oncology University of Milan | Claudia Alteri et al |
| EPI_ISL_542204 | Italy/LOM-UniMI-L217/2020 | 2020-03-15 | Europe/Italy/Lombardy/ | ASST GOM Niguarda | Dep. Of Oncology and Hemato-Oncology University of Milan | Claudia Alteri et al |
| EPI_ISL_542205 | Italy/LOM-UniMI-L218/2020 | 2020-03-10 | Europe/Italy/Lombardy/ | ASST GOM Niguarda | Dep. Of Oncology and Hemato-Oncology University of Milan | Claudia Alteri et al |
| EPI_ISL_542206 | Italy/LOM-UniMI-L219/2020 | 2020-03-15 | Europe/Italy/Lombardy/ | ASST GOM Niguarda | Dep. Of Oncology and Hemato-Oncology University of Milan | Claudia Alteri et al |
| EPI_ISL_542212 | Italy/LOM-UniMI-L225/2020 | 2020-03-14 | Europe/Italy/Lombardy/ | ASST GOM Niguarda | Dep. Of Oncology and Hemato-Oncology University of Milan | Claudia Alteri et al |
| EPI_ISL_542215 | Italy/LOM-UniMI-L228/2020 | 2020-03-11 | Europe/Italy/Lombardy/ | ASST GOM Niguarda | Dep. Of Oncology and Hemato-Oncology University of Milan | Claudia Alteri et al |
| EPI_ISL_542216 | Italy/LOM-UniMI-L229/2020 | 2020-03-11 | Europe/Italy/Lombardy/ | ASST GOM Niguarda | Dep. Of Oncology and Hemato-Oncology University of Milan | Claudia Alteri et al |

| | | | | | | |
|---|---|---|---|---|---|---|
| EPI_ISL_542217 | Italy/LOM-UniMI-L230/2020 | 2020-02-23 | Europe/Italy/Lombardy/ | ASST GOM Niguarda | Dep. Of Oncology and Hemato-Oncology University of Milan | Claudia Alteri et al |
| EPI_ISL_542218 | Italy/LOM-UniMI-L239/2020 | 2020-03-04 | Europe/Italy/Lombardy/ | ASST GOM Niguarda | Dep. Of Oncology and Hemato-Oncology University of Milan | Claudia Alteri et al |
| EPI_ISL_542221 | Italy/LOM-UniMI-L249/2020 | 2020-02-22 | Europe/Italy/Lombardy/ | ASST GOM Niguarda | Dep. Of Oncology and Hemato-Oncology University of Milan | Claudia Alteri et al |
| EPI_ISL_542222 | Italy/LOM-UniMI-L250/2020 | 2020-02-22 | Europe/Italy/Lombardy/ | ASST GOM Niguarda | Dep. Of Oncology and Hemato-Oncology University of Milan | Claudia Alteri et al |
| EPI_ISL_542223 | Italy/LOM-UniMI-L251/2020 | 2020-02-22 | Europe/Italy/Lombardy/ | ASST GOM Niguarda | Dep. Of Oncology and Hemato-Oncology University of Milan | Claudia Alteri et al |
| EPI_ISL_542224 | Italy/LOM-UniMI-L252/2020 | 2020-02-22 | Europe/Italy/Lombardy/ | ASST GOM Niguarda | Dep. Of Oncology and Hemato-Oncology University of Milan | Claudia Alteri et al |
| EPI_ISL_542225 | Italy/LOM-UniMI-L253/2020 | 2020-02-23 | Europe/Italy/Lombardy/ | ASST GOM Niguarda | Dep. Of Oncology and Hemato-Oncology University of Milan | Claudia Alteri et al |
| EPI_ISL_542226 | Italy/LOM-UniMI-L254/2020 | 2020-02-23 | Europe/Italy/Lombardy/ | ASST GOM Niguarda | Dep. Of Oncology and Hemato-Oncology University of Milan | Claudia Alteri et al |

| | | | | | | |
|---|---|---|---|---|---|---|
| EPI_ISL_542227 | Italy/LOM-UniMI-L256/2020 | 2020-02-23 | Europe/Italy/Lombardy/ | ASST GOM Niguarda | Dep. Of Oncology and Hemato-Oncology University of Milan | Claudia Alteri et al |
| EPI_ISL_542228 | Italy/LOM-UniMI-L257/2020 | 2020-02-23 | Europe/Italy/Lombardy/ | ASST GOM Niguarda | Dep. Of Oncology and Hemato-Oncology University of Milan | Claudia Alteri et al |
| EPI_ISL_542229 | Italy/LOM-UniMI-L259/2020 | 2020-02-24 | Europe/Italy/Lombardy/ | ASST GOM Niguarda | Dep. Of Oncology and Hemato-Oncology University of Milan | Claudia Alteri et al |
| EPI_ISL_542230 | Italy/LOM-UniMI-L260/2020 | 2020-02-24 | Europe/Italy/Lombardy/ | ASST GOM Niguarda | Dep. Of Oncology and Hemato-Oncology University of Milan | Claudia Alteri et al |
| EPI_ISL_542231 | Italy/LOM-UniMI-L261/2020 | 2020-02-24 | Europe/Italy/Lombardy/ | ASST GOM Niguarda | Dep. Of Oncology and Hemato-Oncology University of Milan | Claudia Alteri et al |
| EPI_ISL_542232 | Italy/LOM-UniMI-L265/2020 | 2020-02-24 | Europe/Italy/Lombardy/ | ASST GOM Niguarda | Dep. Of Oncology and Hemato-Oncology University of Milan | Claudia Alteri et al |
| EPI_ISL_542233 | Italy/LOM-UniMI-L267/2020 | 2020-02-24 | Europe/Italy/Lombardy/ | ASST GOM Niguarda | Dep. Of Oncology and Hemato-Oncology University of Milan | Claudia Alteri et al |
| EPI_ISL_542234 | Italy/LOM-UniMI-L268/2020 | 2020-02-25 | Europe/Italy/Lombardy/ | ASST GOM Niguarda | Dep. Of Oncology and Hemato-Oncology University of Milan | Claudia Alteri et al |

| | | | | | | |
|---|---|---|---|---|---|---|
| EPI_ISL_542235 | Italy/LOM-UniMI-L269/2020 | 2020-02-25 | Europe/Italy/Lombardy/ | ASST GOM Niguarda | Dep. Of Oncology and Hemato-Oncology University of Milan | Claudia Alteri et al |
| EPI_ISL_542236 | Italy/LOM-UniMI-L270/2020 | 2020-02-25 | Europe/Italy/Lombardy/ | ASST GOM Niguarda | Dep. Of Oncology and Hemato-Oncology University of Milan | Claudia Alteri et al |
| EPI_ISL_542237 | Italy/LOM-UniMI-L271/2020 | 2020-02-25 | Europe/Italy/Lombardy/ | ASST GOM Niguarda | Dep. Of Oncology and Hemato-Oncology University of Milan | Claudia Alteri et al |
| EPI_ISL_542238 | Italy/LOM-UniMI-L272/2020 | 2020-02-25 | Europe/Italy/Lombardy/ | ASST GOM Niguarda | Dep. Of Oncology and Hemato-Oncology University of Milan | Claudia Alteri et al |
| EPI_ISL_542239 | Italy/LOM-UniMI-L273/2020 | 2020-02-25 | Europe/Italy/Lombardy/ | ASST GOM Niguarda | Dep. Of Oncology and Hemato-Oncology University of Milan | Claudia Alteri et al |
| EPI_ISL_542240 | Italy/LOM-UniMI-L274/2020 | 2020-02-25 | Europe/Italy/Lombardy/ | ASST GOM Niguarda | Dep. Of Oncology and Hemato-Oncology University of Milan | Claudia Alteri et al |
| EPI_ISL_542241 | Italy/LOM-UniMI-L276/2020 | 2020-02-26 | Europe/Italy/Lombardy/ | ASST GOM Niguarda | Dep. Of Oncology and Hemato-Oncology University of Milan | Claudia Alteri et al |
| EPI_ISL_542242 | Italy/LOM-UniMI-L278/2020 | 2020-02-27 | Europe/Italy/Lombardy/ | ASST GOM Niguarda | Dep. Of Oncology and Hemato-Oncology University of Milan | Claudia Alteri et al |

| | | | | | | |
|---|---|---|---|---|---|---|
| EPI_ISL_542243 | Italy/LOM-UniMI-L279/2020 | 2020-02-28 | Europe/Italy/Lombardy/ | ASST GOM Niguarda | Dep. Of Oncology and Hemato-Oncology University of Milan | Claudia Alteri et al |
| EPI_ISL_542244 | Italy/LOM-UniMI-L280/2020 | 2020-02-28 | Europe/Italy/Lombardy/ | ASST GOM Niguarda | Dep. Of Oncology and Hemato-Oncology University of Milan | Claudia Alteri et al |
| EPI_ISL_542245 | Italy/LOM-UniMI-L281/2020 | 2020-02-28 | Europe/Italy/Lombardy/ | ASST GOM Niguarda | Dep. Of Oncology and Hemato-Oncology University of Milan | Claudia Alteri et al |
| EPI_ISL_542246 | Italy/LOM-UniMI-L282/2020 | 2020-02-28 | Europe/Italy/Lombardy/ | ASST GOM Niguarda | Dep. Of Oncology and Hemato-Oncology University of Milan | Claudia Alteri et al |
| EPI_ISL_542247 | Italy/LOM-UniMI-L283/2020 | 2020-02-28 | Europe/Italy/Lombardy/ | ASST GOM Niguarda | Dep. Of Oncology and Hemato-Oncology University of Milan | Claudia Alteri et al |
| EPI_ISL_542248 | Italy/LOM-UniMI-L284/2020 | 2020-02-28 | Europe/Italy/Lombardy/ | ASST GOM Niguarda | Dep. Of Oncology and Hemato-Oncology University of Milan | Claudia Alteri et al |
| EPI_ISL_542249 | Italy/LOM-UniMI-L285/2020 | 2020-02-28 | Europe/Italy/Lombardy/ | ASST GOM Niguarda | Dep. Of Oncology and Hemato-Oncology University of Milan | Claudia Alteri et al |
| EPI_ISL_542250 | Italy/LOM-UniMI-L286/2020 | 2020-02-28 | Europe/Italy/Lombardy/ | ASST GOM Niguarda | Dep. Of Oncology and Hemato-Oncology University of Milan | Claudia Alteri et al |

| | | | | | | |
|---|---|---|---|---|---|---|
| EPI_ISL_542251 | Italy/LOM-UniMI-L287/2020 | 2020-02-28 | Europe/Italy/Lombardy/ | ASST GOM Niguarda | Dep. Of Oncology and Hemato-Oncology University of Milan | Claudia Alteri et al |
| EPI_ISL_542252 | Italy/LOM-UniMI-L288/2020 | 2020-02-28 | Europe/Italy/Lombardy/ | ASST GOM Niguarda | Dep. Of Oncology and Hemato-Oncology University of Milan | Claudia Alteri et al |
| EPI_ISL_542253 | Italy/LOM-UniMI-L289/2020 | 2020-02-29 | Europe/Italy/Lombardy/ | ASST GOM Niguarda | Dep. Of Oncology and Hemato-Oncology University of Milan | Claudia Alteri et al |
| EPI_ISL_542254 | Italy/LOM-UniMI-L290/2020 | 2020-02-29 | Europe/Italy/Lombardy/ | ASST GOM Niguarda | Dep. Of Oncology and Hemato-Oncology University of Milan | Claudia Alteri et al |
| EPI_ISL_542255 | Italy/LOM-UniMI-L292/2020 | 2020-02-29 | Europe/Italy/Lombardy/ | ASST GOM Niguarda | Dep. Of Oncology and Hemato-Oncology University of Milan | Claudia Alteri et al |
| EPI_ISL_542256 | Italy/LOM-UniMI-L293/2020 | 2020-02-29 | Europe/Italy/Lombardy/ | ASST GOM Niguarda | Dep. Of Oncology and Hemato-Oncology University of Milan | Claudia Alteri et al |
| EPI_ISL_542257 | Italy/LOM-UniMI-L294/2020 | 2020-02-29 | Europe/Italy/Lombardy/ | ASST GOM Niguarda | Dep. Of Oncology and Hemato-Oncology University of Milan | Claudia Alteri et al |
| EPI_ISL_542258 | Italy/LOM-UniMI-L295/2020 | 2020-02-29 | Europe/Italy/Lombardy/ | ASST GOM Niguarda | Dep. Of Oncology and Hemato-Oncology University of Milan | Claudia Alteri et al |

| | | | | | | |
|---|---|---|---|---|---|---|
| EPI_ISL_542259 | Italy/LOM-UniMI-L296/2020 | 2020-02-29 | Europe/Italy/Lombardy/ | ASST GOM Niguarda | Dep. Of Oncology and Hemato-Oncology University of Milan | Claudia Alteri et al |
| EPI_ISL_542260 | Italy/LOM-UniMI-L299/2020 | 2020-02-29 | Europe/Italy/Lombardy/ | ASST GOM Niguarda | Dep. Of Oncology and Hemato-Oncology University of Milan | Claudia Alteri et al |
| EPI_ISL_542261 | Italy/LOM-UniMI-L302/2020 | 2020-02-29 | Europe/Italy/Lombardy/ | ASST GOM Niguarda | Dep. Of Oncology and Hemato-Oncology University of Milan | Claudia Alteri et al |
| EPI_ISL_542262 | Italy/LOM-UniMI-L309/2020 | 2020-02-29 | Europe/Italy/Lombardy/ | ASST GOM Niguarda | Dep. Of Oncology and Hemato-Oncology University of Milan | Claudia Alteri et al |
| EPI_ISL_542263 | Italy/LOM-UniMI-L310/2020 | 2020-02-29 | Europe/Italy/Lombardy/ | ASST GOM Niguarda | Dep. Of Oncology and Hemato-Oncology University of Milan | Claudia Alteri et al |
| EPI_ISL_542267 | Italy/LOM-UniMI-L316/2020 | 2020-03-01 | Europe/Italy/Lombardy/ | ASST GOM Niguarda | Dep. Of Oncology and Hemato-Oncology University of Milan | Claudia Alteri et al |
| EPI_ISL_542269 | Italy/LOM-UniMI-L318/2020 | 2020-03-01 | Europe/Italy/Lombardy/ | ASST GOM Niguarda | Dep. Of Oncology and Hemato-Oncology University of Milan | Claudia Alteri et al |
| EPI_ISL_542271 | Italy/LOM-UniMI-L321/2020 | 2020-03-01 | Europe/Italy/Lombardy/ | ASST GOM Niguarda | Dep. Of Oncology and Hemato-Oncology University of Milan | Claudia Alteri et al |

| | | | | | | |
|---|---|---|---|---|---|---|
| EPI_ISL_542274 | Italy/LOM-UniMI-L327/2020 | 2020-03-02 | Europe/Italy/Lombardy/ | ASST GOM Niguarda | Dep. Of Oncology and Hemato-Oncology University of Milan | Claudia Alteri et al |
| EPI_ISL_542275 | Italy/LOM-UniMI-L328/2020 | 2020-03-02 | Europe/Italy/Lombardy/ | ASST GOM Niguarda | Dep. Of Oncology and Hemato-Oncology University of Milan | Claudia Alteri et al |
| EPI_ISL_542276 | Italy/LOM-UniMI-L329/2020 | 2020-03-02 | Europe/Italy/Lombardy/ | ASST GOM Niguarda | Dep. Of Oncology and Hemato-Oncology University of Milan | Claudia Alteri et al |
| EPI_ISL_542277 | Italy/LOM-UniMI-L330/2020 | 2020-03-02 | Europe/Italy/Lombardy/ | ASST GOM Niguarda | Dep. Of Oncology and Hemato-Oncology University of Milan | Claudia Alteri et al |
| EPI_ISL_542278 | Italy/LOM-UniMI-L331/2020 | 2020-03-03 | Europe/Italy/Lombardy/ | San Matteo Hospital Pavia | Dep. Of Oncology and Hemato-Oncology University of Milan | Claudia Alteri et al |
| EPI_ISL_542282 | Italy/LOM-UniMI-L343/2020 | 2020-03-04 | Europe/Italy/Lombardy/ | San Matteo Hospital Pavia | Dep. Of Oncology and Hemato-Oncology University of Milan | Claudia Alteri et al |
| EPI_ISL_542283 | Italy/LOM-UniMI-L345/2020 | 2020-03-04 | Europe/Italy/Lombardy/ | San Matteo Hospital Pavia | Dep. Of Oncology and Hemato-Oncology University of Milan | Claudia Alteri et al |
| EPI_ISL_542287 | Italy/LOM-UniMI-L350/2020 | 2020-03-05 | Europe/Italy/Lombardy/ | San Matteo Hospital Pavia | Dep. Of Oncology and Hemato-Oncology University of Milan | Claudia Alteri et al |

| | | | | | | |
|---|---|---|---|---|---|---|
| EPI_ISL_542293 | Italy/LOM-UniMI-L361/2020 | 2020-03-05 | Europe/Italy/Lombardy/ | San Matteo Hospital Pavia | Dep. Of Oncology and Hemato-Oncology University of Milan | Claudia Alteri et al |
| EPI_ISL_542295 | Italy/LOM-UniMI-L363/2020 | 2020-03-06 | Europe/Italy/Lombardy/ | San Matteo Hospital Pavia | Dep. Of Oncology and Hemato-Oncology University of Milan | Claudia Alteri et al |
| EPI_ISL_542296 | Italy/LOM-UniMI-L364/2020 | 2020-03-06 | Europe/Italy/Lombardy/ | San Matteo Hospital Pavia | Dep. Of Oncology and Hemato-Oncology University of Milan | Claudia Alteri et al |
| EPI_ISL_542298 | Italy/LOM-UniMI-L367/2020 | 2020-03-07 | Europe/Italy/Lombardy/ | San Matteo Hospital Pavia | Dep. Of Oncology and Hemato-Oncology University of Milan | Claudia Alteri et al |
| EPI_ISL_542300 | Italy/LOM-UniMI-L369/2020 | 2020-03-07 | Europe/Italy/Lombardy/ | San Matteo Hospital Pavia | Dep. Of Oncology and Hemato-Oncology University of Milan | Claudia Alteri et al |
| EPI_ISL_542301 | Italy/LOM-UniMI-L370/2020 | 2020-03-07 | Europe/Italy/Lombardy/ | San Matteo Hospital Pavia | Dep. Of Oncology and Hemato-Oncology University of Milan | Claudia Alteri et al |
| EPI_ISL_542302 | Italy/LOM-UniMI-L371/2020 | 2020-03-07 | Europe/Italy/Lombardy/ | San Matteo Hospital Pavia | Dep. Of Oncology and Hemato-Oncology University of Milan | Claudia Alteri et al |
| EPI_ISL_542304 | Italy/LOM-UniMI-L373/2020 | 2020-03-08 | Europe/Italy/Lombardy/ | San Matteo Hospital Pavia | Dep. Of Oncology and Hemato-Oncology University of Milan | Claudia Alteri et al |

| | | | | | | |
|---|---|---|---|---|---|---|
| EPI_ISL_542307 | Italy/LOM-UniMI-L384/2020 | 2020-03-09 | Europe/Italy/Lombardy/ | San Matteo Hospital Pavia | Dep. Of Oncology and Hemato-Oncology University of Milan | Claudia Alteri et al |
| EPI_ISL_542308 | Italy/LOM-UniMI-L385/2020 | 2020-03-09 | Europe/Italy/Lombardy/ | San Matteo Hospital Pavia | Dep. Of Oncology and Hemato-Oncology University of Milan | Claudia Alteri et al |
| EPI_ISL_542311 | Italy/LOM-UniMI-L388/2020 | 2020-03-09 | Europe/Italy/Lombardy/ | San Matteo Hospital Pavia | Dep. Of Oncology and Hemato-Oncology University of Milan | Claudia Alteri et al |
| EPI_ISL_542312 | Italy/LOM-UniMI-L389/2020 | 2020-03-09 | Europe/Italy/Lombardy/ | San Matteo Hospital Pavia | Dep. Of Oncology and Hemato-Oncology University of Milan | Claudia Alteri et al |
| EPI_ISL_542314 | Italy/LOM-UniMI-L391/2020 | 2020-03-09 | Europe/Italy/Lombardy/ | San Matteo Hospital Pavia | Dep. Of Oncology and Hemato-Oncology University of Milan | Claudia Alteri et al |
| EPI_ISL_542315 | Italy/LOM-UniMI-L393/2020 | 2020-03-09 | Europe/Italy/Lombardy/ | San Matteo Hospital Pavia | Dep. Of Oncology and Hemato-Oncology University of Milan | Claudia Alteri et al |
| EPI_ISL_542316 | Italy/LOM-UniMI-L394/2020 | 2020-03-09 | Europe/Italy/Lombardy/ | San Matteo Hospital Pavia | Dep. Of Oncology and Hemato-Oncology University of Milan | Claudia Alteri et al |
| EPI_ISL_542320 | Italy/LOM-UniMI-L398/2020 | 2020-03-13 | Europe/Italy/Lombardy/ | San Matteo Hospital Pavia | Dep. Of Oncology and Hemato-Oncology University of Milan | Claudia Alteri et al |

| | | | | | | |
|---|---|---|---|---|---|---|
| EPI_ISL_542324 | Italy/LOM-UniMI-L402/2020 | 2020-03-13 | Europe/Italy/Lombardy/ | San Matteo Hospital Pavia | Dep. Of Oncology and Hemato-Oncology University of Milan | Claudia Alteri et al |
| EPI_ISL_542325 | Italy/LOM-UniMI-L403/2020 | 2020-03-15 | Europe/Italy/Lombardy/ | San Matteo Hospital Pavia | Dep. Of Oncology and Hemato-Oncology University of Milan | Claudia Alteri et al |
| EPI_ISL_542326 | Italy/LOM-UniMI-L404/2020 | 2020-03-14 | Europe/Italy/Lombardy/ | San Matteo Hospital Pavia | Dep. Of Oncology and Hemato-Oncology University of Milan | Claudia Alteri et al |
| EPI_ISL_542328 | Italy/LOM-UniMI-L407/2020 | 2020-03-13 | Europe/Italy/Lombardy/ | San Matteo Hospital Pavia | Dep. Of Oncology and Hemato-Oncology University of Milan | Claudia Alteri et al |
| EPI_ISL_542330 | Italy/LOM-UniMI-L410/2020 | 2020-03-03 | Europe/Italy/Lombardy/ | San Matteo Hospital Pavia | Dep. Of Oncology and Hemato-Oncology University of Milan | Claudia Alteri et al |
| EPI_ISL_542331 | Italy/LOM-UniMI-L411/2020 | 2020-03-01 | Europe/Italy/Lombardy/ | San Matteo Hospital Pavia | Dep. Of Oncology and Hemato-Oncology University of Milan | Claudia Alteri et al |
| EPI_ISL_542332 | Italy/LOM-UniMI-L412/2020 | 2020-03-14 | Europe/Italy/Lombardy/ | San Matteo Hospital Pavia | Dep. Of Oncology and Hemato-Oncology University of Milan | Claudia Alteri et al |
| EPI_ISL_542333 | Italy/LOM-UniMI-L413/2020 | 2020-03-16 | Europe/Italy/Lombardy/ | San Matteo Hospital Pavia | Dep. Of Oncology and Hemato-Oncology University of Milan | Claudia Alteri et al |

| | | | | | | |
|---|---|---|---|---|---|---|
| EPI_ISL_542338 | Italy/LOM-UniMI-L419/2020 | 2020-03-12 | Europe/Italy/Lombardy/ | San Matteo Hospital Pavia | Dep. Of Oncology and Hemato-Oncology University of Milan | Claudia Alteri et al |
| EPI_ISL_542339 | Italy/LOM-UniMI-L420/2020 | 2020-03-18 | Europe/Italy/Lombardy/ | San Matteo Hospital Pavia | Dep. Of Oncology and Hemato-Oncology University of Milan | Claudia Alteri et al |
| EPI_ISL_542340 | Italy/LOM-UniMI-L421/2020 | 2020-03-17 | Europe/Italy/Lombardy/ | San Matteo Hospital Pavia | Dep. Of Oncology and Hemato-Oncology University of Milan | Claudia Alteri et al |
| EPI_ISL_542341 | Italy/LOM-UniMI-L422/2020 | 2020-03-18 | Europe/Italy/Lombardy/ | San Matteo Hospital Pavia | Dep. Of Oncology and Hemato-Oncology University of Milan | Claudia Alteri et al |
| EPI_ISL_542342 | Italy/LOM-UniMI-L423/2020 | 2020-03-19 | Europe/Italy/Lombardy/ | San Matteo Hospital Pavia | Dep. Of Oncology and Hemato-Oncology University of Milan | Claudia Alteri et al |
| EPI_ISL_542346 | Italy/LOM-UniMI-L427/2020 | 2020-03-15 | Europe/Italy/Lombardy/ | San Matteo Hospital Pavia | Dep. Of Oncology and Hemato-Oncology University of Milan | Claudia Alteri et al |
| EPI_ISL_542348 | Italy/LOM-UniMI-L429/2020 | 2020-03-17 | Europe/Italy/Lombardy/ | San Matteo Hospital Pavia | Dep. Of Oncology and Hemato-Oncology University of Milan | Claudia Alteri et al |
| EPI_ISL_542349 | Italy/LOM-UniMI-L430/2020 | 2020-03-19 | Europe/Italy/Lombardy/ | San Matteo Hospital Pavia | Dep. Of Oncology and Hemato-Oncology University of Milan | Claudia Alteri et al |

| | | | | | | |
|---|---|---|---|---|---|---|
| EPI_ISL_542350 | Italy/LOM-UniMI-L431/2020 | 2020-03-20 | Europe/Italy/Lombardy/ | San Matteo Hospital Pavia | Dep. Of Oncology and Hemato-Oncology University of Milan | Claudia Alteri et al |
| EPI_ISL_542353 | Italy/LOM-UniMI-L434/2020 | 2020-03-20 | Europe/Italy/Lombardy/ | San Matteo Hospital Pavia | Dep. Of Oncology and Hemato-Oncology University of Milan | Claudia Alteri et al |
| EPI_ISL_542355 | Italy/LOM-UniMI-L436/2020 | 2020-03-19 | Europe/Italy/Lombardy/ | San Matteo Hospital Pavia | Dep. Of Oncology and Hemato-Oncology University of Milan | Claudia Alteri et al |
| EPI_ISL_542357 | Italy/LOM-UniMI-L438/2020 | 2020-03-21 | Europe/Italy/Lombardy/ | San Matteo Hospital Pavia | Dep. Of Oncology and Hemato-Oncology University of Milan | Claudia Alteri et al |
| EPI_ISL_542358 | Italy/LOM-UniMI-L439/2020 | 2020-03-28 | Europe/Italy/Lombardy/ | San Matteo Hospital Pavia | Dep. Of Oncology and Hemato-Oncology University of Milan | Claudia Alteri et al |
| EPI_ISL_542359 | Italy/LOM-UniMI-L440/2020 | 2020-03-22 | Europe/Italy/Lombardy/ | San Matteo Hospital Pavia | Dep. Of Oncology and Hemato-Oncology University of Milan | Claudia Alteri et al |
| EPI_ISL_542361 | Italy/LOM-UniMI-L442/2020 | 2020-03-22 | Europe/Italy/Lombardy/ | San Matteo Hospital Pavia | Dep. Of Oncology and Hemato-Oncology University of Milan | Claudia Alteri et al |
| EPI_ISL_542363 | Italy/LOM-UniMI-L444/2020 | 2020-03-20 | Europe/Italy/Lombardy/ | San Matteo Hospital Pavia | Dep. Of Oncology and Hemato-Oncology University of Milan | Claudia Alteri et al |

| | | | | | | |
|---|---|---|---|---|---|---|
| EPI_ISL_542364 | Italy/LOM-UniMI-L445/2020 | 2020-03-14 | Europe/Italy/Lombardy/ | San Matteo Hospital Pavia | Dep. Of Oncology and Hemato-Oncology University of Milan | Claudia Alteri et al |
| EPI_ISL_542371 | Italy/LOM-UniMI-L453/2020 | 2020-03-21 | Europe/Italy/Lombardy/ | San Matteo Hospital Pavia | Dep. Of Oncology and Hemato-Oncology University of Milan | Claudia Alteri et al |
| EPI_ISL_542373 | Italy/LOM-UniMI-L455/2020 | 2020-03-25 | Europe/Italy/Lombardy/ | San Matteo Hospital Pavia | Dep. Of Oncology and Hemato-Oncology University of Milan | Claudia Alteri et al |
| EPI_ISL_542374 | Italy/LOM-UniMI-L457/2020 | 2020-03-25 | Europe/Italy/Lombardy/ | San Matteo Hospital Pavia | Dep. Of Oncology and Hemato-Oncology University of Milan | Claudia Alteri et al |
| EPI_ISL_542375 | Italy/LOM-UniMI-L458/2020 | 2020-03-25 | Europe/Italy/Lombardy/ | San Matteo Hospital Pavia | Dep. Of Oncology and Hemato-Oncology University of Milan | Claudia Alteri et al |
| EPI_ISL_542376 | Italy/LOM-UniMI-L459/2020 | 2020-03-13 | Europe/Italy/Lombardy/ | San Matteo Hospital Pavia | Dep. Of Oncology and Hemato-Oncology University of Milan | Claudia Alteri et al |
| EPI_ISL_542378 | Italy/LOM-UniMI-L462/2020 | 2020-03-16 | Europe/Italy/Lombardy/ | San Matteo Hospital Pavia | Dep. Of Oncology and Hemato-Oncology University of Milan | Claudia Alteri et al |
| EPI_ISL_542379 | Italy/LOM-UniMI-L463/2020 | 2020-03-23 | Europe/Italy/Lombardy/ | San Matteo Hospital Pavia | Dep. Of Oncology and Hemato-Oncology University of Milan | Claudia Alteri et al |

| | | | | | | |
|---|---|---|---|---|---|---|
| EPI_ISL_542380 | Italy/LOM-UniMI-L464/2020 | 2020-03-27 | Europe/Italy/Lombardy/ | San Matteo Hospital Pavia | Dep. Of Oncology and Hemato-Oncology University of Milan | Claudia Alteri et al |
| EPI_ISL_542382 | Italy/LOM-UniMI-L469/2020 | 2020-03-28 | Europe/Italy/Lombardy/ | San Matteo Hospital Pavia | Dep. Of Oncology and Hemato-Oncology University of Milan | Claudia Alteri et al |
| EPI_ISL_542389 | Italy/LOM-UniMI-L476/2020 | 2020-04-01 | Europe/Italy/Lombardy/ | San Matteo Hospital Pavia | Dep. Of Oncology and Hemato-Oncology University of Milan | Claudia Alteri et al |
| EPI_ISL_542392 | Italy/LOM-UniMI-L480/2020 | 2020-03-31 | Europe/Italy/Lombardy/ | San Matteo Hospital Pavia | Dep. Of Oncology and Hemato-Oncology University of Milan | Claudia Alteri et al |
| EPI_ISL_542396 | Italy/LOM-UniMI-L491/2020 | 2020-03-20 | Europe/Italy/Lombardy/ | San Matteo Hospital Pavia | Dep. Of Oncology and Hemato-Oncology University of Milan | Claudia Alteri et al |
| EPI_ISL_542397 | Italy/LOM-UniMI-L492/2020 | 2020-03-20 | Europe/Italy/Lombardy/ | San Matteo Hospital Pavia | Dep. Of Oncology and Hemato-Oncology University of Milan | Claudia Alteri et al |
| EPI_ISL_542399 | Italy/LOM-UniMI-L494/2020 | 2020-03-21 | Europe/Italy/Lombardy/ | San Matteo Hospital Pavia | Dep. Of Oncology and Hemato-Oncology University of Milan | Claudia Alteri et al |
| EPI_ISL_542400 | Italy/LOM-UniMI-L53/2020 | 2020-03-02 | Europe/Italy/Lombardy/ | ASST GOM Niguarda | Dep. Of Oncology and Hemato-Oncology University of Milan | Claudia Alteri et al |

| | | | | | | |
|---|---|---|---|---|---|---|
| EPI_ISL_542401 | Italy/LOM-UniMI-L54/2020 | 2020-02-26 | Europe/Italy/Lombardy/ | ASST GOM Niguarda | Dep. Of Oncology and Hemato-Oncology University of Milan | Claudia Alteri et al |
| EPI_ISL_542402 | Italy/LOM-UniMI-L55/2020 | 2020-03-02 | Europe/Italy/Lombardy/ | ASST GOM Niguarda | Dep. Of Oncology and Hemato-Oncology University of Milan | Claudia Alteri et al |
| EPI_ISL_542405 | Italy/LOM-UniMI-L58/2020 | 2020-03-04 | Europe/Italy/Lombardy/ | ASST GOM Niguarda | Dep. Of Oncology and Hemato-Oncology University of Milan | Claudia Alteri et al |
| EPI_ISL_542406 | Italy/LOM-UniMI-L60/2020 | 2020-02-27 | Europe/Italy/Lombardy/ | ASST GOM Niguarda | Dep. Of Oncology and Hemato-Oncology University of Milan | Claudia Alteri et al |
| EPI_ISL_542407 | Italy/LOM-UniMI-L61/2020 | 2020-03-02 | Europe/Italy/Lombardy/ | ASST GOM Niguarda | Dep. Of Oncology and Hemato-Oncology University of Milan | Claudia Alteri et al |
| EPI_ISL_542408 | Italy/LOM-UniMI-L62/2020 | 2020-03-04 | Europe/Italy/Lombardy/ | ASST GOM Niguarda | Dep. Of Oncology and Hemato-Oncology University of Milan | Claudia Alteri et al |
| EPI_ISL_542409 | Italy/LOM-UniMI-L63/2020 | 2020-03-03 | Europe/Italy/Lombardy/ | ASST GOM Niguarda | Dep. Of Oncology and Hemato-Oncology University of Milan | Claudia Alteri et al |
| EPI_ISL_542410 | Italy/LOM-UniMI-L64/2020 | 2020-02-25 | Europe/Italy/Lombardy/ | ASST GOM Niguarda | Dep. Of Oncology and Hemato-Oncology University of Milan | Claudia Alteri et al |

| | | | | | | |
|---|---|---|---|---|---|---|
| EPI_ISL_542411 | Italy/LOM-UniMI-L65/2020 | 2020-03-02 | Europe/Italy/Lombardy/ | ASST GOM Niguarda | Dep. Of Oncology and Hemato-Oncology University of Milan | Claudia Alteri et al |
| EPI_ISL_542412 | Italy/LOM-UniMI-L66/2020 | 2020-02-29 | Europe/Italy/Lombardy/ | ASST GOM Niguarda | Dep. Of Oncology and Hemato-Oncology University of Milan | Claudia Alteri et al |
| EPI_ISL_542413 | Italy/LOM-UniMI-L67/2020 | 2020-03-05 | Europe/Italy/Lombardy/ | ASST GOM Niguarda | Dep. Of Oncology and Hemato-Oncology University of Milan | Claudia Alteri et al |
| EPI_ISL_542414 | Italy/LOM-UniMI-L68/2020 | 2020-03-04 | Europe/Italy/Lombardy/ | ASST GOM Niguarda | Dep. Of Oncology and Hemato-Oncology University of Milan | Claudia Alteri et al |
| EPI_ISL_542415 | Italy/LOM-UniMI-L69/2020 | 2020-03-01 | Europe/Italy/Lombardy/ | ASST GOM Niguarda | Dep. Of Oncology and Hemato-Oncology University of Milan | Claudia Alteri et al |
| EPI_ISL_542416 | Italy/LOM-UniMI-L70/2020 | 2020-03-06 | Europe/Italy/Lombardy/ | ASST GOM Niguarda | Dep. Of Oncology and Hemato-Oncology University of Milan | Claudia Alteri et al |
| EPI_ISL_542421 | Italy/LOM-UniMI-L76/2020 | 2020-03-05 | Europe/Italy/Lombardy/ | ASST GOM Niguarda | Dep. Of Oncology and Hemato-Oncology University of Milan | Claudia Alteri et al |
| EPI_ISL_542423 | Italy/LOM-UniMI-L78/2020 | 2020-03-06 | Europe/Italy/Lombardy/ | ASST GOM Niguarda | Dep. Of Oncology and Hemato-Oncology University of Milan | Claudia Alteri et al |

| | | | | | | |
|---|---|---|---|---|---|---|
| EPI_ISL_542425 | Italy/LOM-UniMI-L80/2020 | 2020-03-10 | Europe/Italy/Lombardy/ | ASST GOM Niguarda | Dep. Of Oncology and Hemato-Oncology University of Milan | Claudia Alteri et al |
| EPI_ISL_542426 | Italy/LOM-UniMI-L81/2020 | 2020-03-08 | Europe/Italy/Lombardy/ | ASST GOM Niguarda | Dep. Of Oncology and Hemato-Oncology University of Milan | Claudia Alteri et al |
| EPI_ISL_542429 | Italy/LOM-UniMI-L85/2020 | 2020-03-02 | Europe/Italy/Lombardy/ | ASST GOM Niguarda | Dep. Of Oncology and Hemato-Oncology University of Milan | Claudia Alteri et al |
| EPI_ISL_542432 | Italy/LOM-UniMI-L88/2020 | 2020-03-08 | Europe/Italy/Lombardy/ | ASST GOM Niguarda | Dep. Of Oncology and Hemato-Oncology University of Milan | Claudia Alteri et al |
| EPI_ISL_542435 | Italy/LOM-UniMI-L91/2020 | 2020-03-11 | Europe/Italy/Lombardy/ | ASST GOM Niguarda | Dep. Of Oncology and Hemato-Oncology University of Milan | Claudia Alteri et al |
| EPI_ISL_542438 | Italy/LOM-UniMI-L94/2020 | 2020-03-10 | Europe/Italy/Lombardy/ | ASST GOM Niguarda | Dep. Of Oncology and Hemato-Oncology University of Milan | Claudia Alteri et al |
| EPI_ISL_542440 | Italy/LOM-UniMI-L96/2020 | 2020-03-06 | Europe/Italy/Lombardy/ | ASST GOM Niguarda | Dep. Of Oncology and Hemato-Oncology University of Milan | Claudia Alteri et al |
| EPI_ISL_542442 | Italy/LOM-UniMI-L98/2020 | 2020-02-26 | Europe/Italy/Lombardy/ | ASST GOM Niguarda | Dep. Of Oncology and Hemato-Oncology University of Milan | Claudia Alteri et al |

| | | | | | | |
|---|---|---|---|---|---|---|
| EPI_ISL_542444 | USA/TX-DSHS-0388/2020 | 2020-05-31 | North America/USA/Texas/El Paso Coun | Texas Department of State Health Services | Texas Department of State Health Services | Rashmi Tuladhar et al |
| EPI_ISL_542454 | USA/TX-DSHS-0394/2020 | 2020-05-31 | North America/USA/Texas/El Paso Coun | Texas Department of State Health Services | Texas Department of State Health Services | Rashmi Tuladhar et al |
| EPI_ISL_542458 | USA/TX-DSHS-0398/2020 | 2020-05-31 | North America/USA/Texas/El Paso Coun | Texas Department of State Health Services | Texas Department of State Health Services | Rashmi Tuladhar et al |
| EPI_ISL_542473 | USA/TX-DSHS-0414/2020 | 2020-06-16 | North America/USA/Texas/Harris Count | Texas Department of State Health Services | Texas Department of State Health Services | Rashmi Tuladhar et al |
| EPI_ISL_542485 | USA/TX-DSHS-0432/2020 | 2020-06-26 | North America/USA/Texas/Mclennan Co | Texas Department of State Health Services | Texas Department of State Health Services | Rashmi Tuladhar et al |
| EPI_ISL_542490 | USA/TX-HMH-1006/2020 | 2020-04-03 | North America/USA/Texas/Greater Hou | Houston Methodist Hospital | Houston Methodist Hospital | S. Wesley Long et al |
| EPI_ISL_542500 | USA/TX-HMH-1026/2020 | 2020-04-05 | North America/USA/Texas/Greater Hou | Houston Methodist Hospital | Houston Methodist Hospital | S. Wesley Long et al |
| EPI_ISL_542503 | USA/TX-HMH-1036/2020 | 2020-04-06 | North America/USA/Texas/Greater Hou | Houston Methodist Hospital | Houston Methodist Hospital | S. Wesley Long et al |
| EPI_ISL_542507 | USA/TX-HMH-1046/2020 | 2020-04-05 | North America/USA/Texas/Greater Hou | Houston Methodist Hospital | Houston Methodist Hospital | S. Wesley Long et al |
| EPI_ISL_542510 | USA/TX-HMH-1056/2020 | 2020-04-05 | North America/USA/Texas/Greater Hou | Houston Methodist Hospital | Houston Methodist Hospital | S. Wesley Long et al |
| EPI_ISL_542512 | USA/TX-HMH-1058/2020 | 2020-04-09 | North America/USA/Texas/Greater Hou | Houston Methodist Hospital | Houston Methodist Hospital | S. Wesley Long et al |
| EPI_ISL_542516 | USA/TX-HMH-1069/2020 | 2020-04-13 | North America/USA/Texas/Greater Hou | Houston Methodist Hospital | Houston Methodist Hospital | S. Wesley Long et al |
| EPI_ISL_542525 | USA/TX-HMH-1087/2020 | 2020-04-18 | North America/USA/Texas/Greater Hou | Houston Methodist Hospital | Houston Methodist Hospital | S. Wesley Long et al |

| | | | | | | |
|---|---|---|---|---|---|---|
| EPI_ISL_542529 | USA/TX-HMH-1096/2020 | 2020-04-20 | North America/USA/Texas/Greater Hou | Houston Methodist Hospital | Houston Methodist Hospital | S. Wesley Long et al |
| EPI_ISL_542542 | USA/TX-HMH-1120/2020 | 2020-04-22 | North America/USA/Texas/Greater Hou | Houston Methodist Hospital | Houston Methodist Hospital | S. Wesley Long et al |
| EPI_ISL_542547 | USA/TX-HMH-1139/2020 | 2020-04-26 | North America/USA/Texas/Greater Hou | Houston Methodist Hospital | Houston Methodist Hospital | S. Wesley Long et al |
| EPI_ISL_542550 | USA/TX-HMH-1143/2020 | 2020-04-18 | North America/USA/Texas/Greater Hou | Houston Methodist Hospital | Houston Methodist Hospital | S. Wesley Long et al |
| EPI_ISL_542552 | USA/TX-HMH-1147/2020 | 2020-04-27 | North America/USA/Texas/Greater Hou | Houston Methodist Hospital | Houston Methodist Hospital | S. Wesley Long et al |
| EPI_ISL_542555 | USA/TX-HMH-1154/2020 | 2020-04-19 | North America/USA/Texas/Greater Hou | Houston Methodist Hospital | Houston Methodist Hospital | S. Wesley Long et al |
| EPI_ISL_542556 | USA/TX-HMH-1155/2020 | 2020-04-27 | North America/USA/Texas/Greater Hou | Houston Methodist Hospital | Houston Methodist Hospital | S. Wesley Long et al |
| EPI_ISL_542557 | USA/TX-HMH-1156/2020 | 2020-04-27 | North America/USA/Texas/Greater Hou | Houston Methodist Hospital | Houston Methodist Hospital | S. Wesley Long et al |
| EPI_ISL_542561 | USA/TX-HMH-1162/2020 | 2020-04-18 | North America/USA/Texas/Greater Hou | Houston Methodist Hospital | Houston Methodist Hospital | S. Wesley Long et al |
| EPI_ISL_542562 | USA/TX-HMH-1164/2020 | 2020-04-26 | North America/USA/Texas/Greater Hou | Houston Methodist Hospital | Houston Methodist Hospital | S. Wesley Long et al |
| EPI_ISL_542573 | USA/TX-HMH-1185/2020 | 2020-04-01 | North America/USA/Texas/Greater Hou | Houston Methodist Hospital | Houston Methodist Hospital | S. Wesley Long et al |
| EPI_ISL_542579 | USA/TX-HMH-1194/2020 | 2020-04-19 | North America/USA/Texas/Greater Hou | Houston Methodist Hospital | Houston Methodist Hospital | S. Wesley Long et al |
| EPI_ISL_542583 | USA/TX-HMH-1200/2020 | 2020-04-24 | North America/USA/Texas/Greater Hou | Houston Methodist Hospital | Houston Methodist Hospital | S. Wesley Long et al |

| | | | | | | |
|---|---|---|---|---|---|---|
| EPI_ISL_542598 | USA/TX-HMH-1233/2020 | 2020-04-23 | North America/USA/Texas/Greater Hou | Houston Methodist Hospital | Houston Methodist Hospital | S. Wesley Long et al |
| EPI_ISL_542636 | USA/TX-HMH-1299/2020 | 2020-04-25 | North America/USA/Texas/Greater Hou | Houston Methodist Hospital | Houston Methodist Hospital | S. Wesley Long et al |
| EPI_ISL_542644 | USA/TX-HMH-1310/2020 | 2020-04-13 | North America/USA/Texas/Greater Hou | Houston Methodist Hospital | Houston Methodist Hospital | S. Wesley Long et al |
| EPI_ISL_542645 | USA/TX-HMH-1311/2020 | 2020-04-16 | North America/USA/Texas/Greater Hou | Houston Methodist Hospital | Houston Methodist Hospital | S. Wesley Long et al |
| EPI_ISL_542646 | USA/TX-HMH-1313/2020 | 2020-04-21 | North America/USA/Texas/Greater Hou | Houston Methodist Hospital | Houston Methodist Hospital | S. Wesley Long et al |
| EPI_ISL_542650 | USA/TX-HMH-1318/2020 | 2020-04-12 | North America/USA/Texas/Greater Hou | Houston Methodist Hospital | Houston Methodist Hospital | S. Wesley Long et al |
| EPI_ISL_542653 | USA/TX-HMH-1325/2020 | 2020-04-14 | North America/USA/Texas/Greater Hou | Houston Methodist Hospital | Houston Methodist Hospital | S. Wesley Long et al |
| EPI_ISL_542659 | USA/TX-HMH-1341/2020 | 2020-04-14 | North America/USA/Texas/Greater Hou | Houston Methodist Hospital | Houston Methodist Hospital | S. Wesley Long et al |
| EPI_ISL_542660 | USA/TX-HMH-1342/2020 | 2020-04-12 | North America/USA/Texas/Greater Hou | Houston Methodist Hospital | Houston Methodist Hospital | S. Wesley Long et al |
| EPI_ISL_542662 | USA/TX-HMH-1347/2020 | 2020-04-12 | North America/USA/Texas/Greater Hou | Houston Methodist Hospital | Houston Methodist Hospital | S. Wesley Long et al |
| EPI_ISL_542664 | USA/TX-HMH-1350/2020 | 2020-04-12 | North America/USA/Texas/Greater Hou | Houston Methodist Hospital | Houston Methodist Hospital | S. Wesley Long et al |
| EPI_ISL_542670 | USA/TX-HMH-1361/2020 | 2020-05-01 | North America/USA/Texas/Greater Hou | Houston Methodist Hospital | Houston Methodist Hospital | S. Wesley Long et al |
| EPI_ISL_542677 | USA/TX-HMH-1370/2020 | 2020-05-02 | North America/USA/Texas/Greater Hou | Houston Methodist Hospital | Houston Methodist Hospital | S. Wesley Long et al |

| | | | | | | |
|---|---|---|---|---|---|---|
| EPI_ISL_542684 | USA/TX-HMH-1380/2020 | 2020-05-01 | North America/USA/Texas/Greater Hou | Houston Methodist Hospital | Houston Methodist Hospital | S. Wesley Long et al |
| EPI_ISL_542692 | USA/TX-HMH-1391/2020 | 2020-04-01 | North America/USA/Texas/Greater Hou | Houston Methodist Hospital | Houston Methodist Hospital | S. Wesley Long et al |
| EPI_ISL_542701 | USA/TX-HMH-1403/2020 | 2020-04-17 | North America/USA/Texas/Greater Hou | Houston Methodist Hospital | Houston Methodist Hospital | S. Wesley Long et al |
| EPI_ISL_542711 | USA/TX-HMH-1418/2020 | 2020-04-21 | North America/USA/Texas/Greater Hou | Houston Methodist Hospital | Houston Methodist Hospital | S. Wesley Long et al |
| EPI_ISL_542717 | USA/TX-HMH-1426/2020 | 2020-04-29 | North America/USA/Texas/Greater Hou | Houston Methodist Hospital | Houston Methodist Hospital | S. Wesley Long et al |
| EPI_ISL_542718 | USA/TX-HMH-1427/2020 | 2020-05-01 | North America/USA/Texas/Greater Hou | Houston Methodist Hospital | Houston Methodist Hospital | S. Wesley Long et al |
| EPI_ISL_542721 | USA/TX-HMH-1433/2020 | 2020-04-21 | North America/USA/Texas/Greater Hou | Houston Methodist Hospital | Houston Methodist Hospital | S. Wesley Long et al |
| EPI_ISL_542722 | USA/TX-HMH-1435/2020 | 2020-05-02 | North America/USA/Texas/Greater Hou | Houston Methodist Hospital | Houston Methodist Hospital | S. Wesley Long et al |
| EPI_ISL_542723 | USA/TX-HMH-1436/2020 | 2020-04-30 | North America/USA/Texas/Greater Hou | Houston Methodist Hospital | Houston Methodist Hospital | S. Wesley Long et al |
| EPI_ISL_542725 | USA/TX-HMH-1441/2020 | 2020-05-02 | North America/USA/Texas/Greater Hou | Houston Methodist Hospital | Houston Methodist Hospital | S. Wesley Long et al |
| EPI_ISL_542727 | USA/TX-HMH-1444/2020 | 2020-04-13 | North America/USA/Texas/Greater Hou | Houston Methodist Hospital | Houston Methodist Hospital | S. Wesley Long et al |
| EPI_ISL_542730 | USA/TX-HMH-1452/2020 | 2020-05-03 | North America/USA/Texas/Greater Hou | Houston Methodist Hospital | Houston Methodist Hospital | S. Wesley Long et al |
| EPI_ISL_542734 | USA/TX-HMH-1459/2020 | 2020-04-27 | North America/USA/Texas/Greater Hou | Houston Methodist Hospital | Houston Methodist Hospital | S. Wesley Long et al |

| | | | | | | |
|---|---|---|---|---|---|---|
| EPI_ISL_542735 | USA/TX-HMH-1461/2020 | 2020-05-01 | North America/USA/Texas/Greater Hou | Houston Methodist Hospital | Houston Methodist Hospital | S. Wesley Long et al |
| EPI_ISL_542749 | USA/TX-HMH-1487/2020 | 2020-05-02 | North America/USA/Texas/Greater Hou | Houston Methodist Hospital | Houston Methodist Hospital | S. Wesley Long et al |
| EPI_ISL_542751 | USA/TX-HMH-1490/2020 | 2020-04-30 | North America/USA/Texas/Greater Hou | Houston Methodist Hospital | Houston Methodist Hospital | S. Wesley Long et al |
| EPI_ISL_542756 | USA/TX-HMH-1499/2020 | 2020-05-02 | North America/USA/Texas/Greater Hou | Houston Methodist Hospital | Houston Methodist Hospital | S. Wesley Long et al |
| EPI_ISL_542759 | USA/TX-HMH-1503/2020 | 2020-04-30 | North America/USA/Texas/Greater Hou | Houston Methodist Hospital | Houston Methodist Hospital | S. Wesley Long et al |
| EPI_ISL_542766 | USA/TX-HMH-1516/2020 | 2020-04-28 | North America/USA/Texas/Greater Hou | Houston Methodist Hospital | Houston Methodist Hospital | S. Wesley Long et al |
| EPI_ISL_542777 | USA/TX-HMH-1530/2020 | 2020-04-03 | North America/USA/Texas/Greater Hou | Houston Methodist Hospital | Houston Methodist Hospital | S. Wesley Long et al |
| EPI_ISL_542782 | USA/TX-HMH-1542/2020 | 2020-04-14 | North America/USA/Texas/Greater Hou | Houston Methodist Hospital | Houston Methodist Hospital | S. Wesley Long et al |
| EPI_ISL_542785 | USA/TX-HMH-1545/2020 | 2020-04-13 | North America/USA/Texas/Greater Hou | Houston Methodist Hospital | Houston Methodist Hospital | S. Wesley Long et al |
| EPI_ISL_542786 | USA/TX-HMH-1546/2020 | 2020-04-13 | North America/USA/Texas/Greater Hou | Houston Methodist Hospital | Houston Methodist Hospital | S. Wesley Long et al |
| EPI_ISL_542796 | USA/TX-HMH-1727/2020 | 2020-04-02 | North America/USA/Texas/Greater Hou | Houston Methodist Hospital | Houston Methodist Hospital | S. Wesley Long et al |
| EPI_ISL_542797 | USA/TX-HMH-1728/2020 | 2020-04-20 | North America/USA/Texas/Greater Hou | Houston Methodist Hospital | Houston Methodist Hospital | S. Wesley Long et al |
| EPI_ISL_542804 | USA/TX-HMH-1737/2020 | 2020-05-07 | North America/USA/Texas/Greater Hou | Houston Methodist Hospital | Houston Methodist Hospital | S. Wesley Long et al |

| | | | | | | |
|---|---|---|---|---|---|---|
| EPI_ISL_542811 | USA/TX-HMH-1746/2020 | 2020-05-12 | North America/USA/Texas/Greater Hou | Houston Methodist Hospital | Houston Methodist Hospital | S. Wesley Long et al |
| EPI_ISL_542812 | USA/TX-HMH-1747/2020 | 2020-05-12 | North America/USA/Texas/Greater Hou | Houston Methodist Hospital | Houston Methodist Hospital | S. Wesley Long et al |
| EPI_ISL_542816 | USA/TX-HMH-1756/2020 | 2020-05-12 | North America/USA/Texas/Greater Hou | Houston Methodist Hospital | Houston Methodist Hospital | S. Wesley Long et al |
| EPI_ISL_542818 | USA/TX-HMH-1758/2020 | 2020-05-16 | North America/USA/Texas/Greater Hou | Houston Methodist Hospital | Houston Methodist Hospital | S. Wesley Long et al |
| EPI_ISL_542822 | USA/TX-HMH-1763/2020 | 2020-05-14 | North America/USA/Texas/Greater Hou | Houston Methodist Hospital | Houston Methodist Hospital | S. Wesley Long et al |
| EPI_ISL_542828 | USA/TX-HMH-1770/2020 | 2020-05-07 | North America/USA/Texas/Greater Hou | Houston Methodist Hospital | Houston Methodist Hospital | S. Wesley Long et al |
| EPI_ISL_542830 | USA/TX-HMH-1774/2020 | 2020-05-14 | North America/USA/Texas/Greater Hou | Houston Methodist Hospital | Houston Methodist Hospital | S. Wesley Long et al |
| EPI_ISL_542833 | USA/TX-HMH-1778/2020 | 2020-05-17 | North America/USA/Texas/Greater Hou | Houston Methodist Hospital | Houston Methodist Hospital | S. Wesley Long et al |
| EPI_ISL_542834 | USA/TX-HMH-1780/2020 | 2020-05-07 | North America/USA/Texas/Greater Hou | Houston Methodist Hospital | Houston Methodist Hospital | S. Wesley Long et al |
| EPI_ISL_542838 | USA/TX-HMH-1786/2020 | 2020-05-17 | North America/USA/Texas/Greater Hou | Houston Methodist Hospital | Houston Methodist Hospital | S. Wesley Long et al |
| EPI_ISL_542839 | USA/TX-HMH-1788/2020 | 2020-05-12 | North America/USA/Texas/Greater Hou | Houston Methodist Hospital | Houston Methodist Hospital | S. Wesley Long et al |
| EPI_ISL_542845 | USA/TX-HMH-1796/2020 | 2020-05-16 | North America/USA/Texas/Greater Hou | Houston Methodist Hospital | Houston Methodist Hospital | S. Wesley Long et al |
| EPI_ISL_542850 | USA/TX-HMH-1802/2020 | 2020-05-16 | North America/USA/Texas/Greater Hou | Houston Methodist Hospital | Houston Methodist Hospital | S. Wesley Long et al |

| | | | | | | |
|---|---|---|---|---|---|---|
| EPI_ISL_542851 | USA/TX-HMH-1803/2020 | 2020-05-16 | North America/USA/Texas/Greater Hou | Houston Methodist Hospital | Houston Methodist Hospital | S. Wesley Long et al |
| EPI_ISL_542860 | USA/TX-HMH-1817/2020 | 2020-05-13 | North America/USA/Texas/Greater Hou | Houston Methodist Hospital | Houston Methodist Hospital | S. Wesley Long et al |
| EPI_ISL_542867 | USA/TX-HMH-1826/2020 | 2020-05-16 | North America/USA/Texas/Greater Hou | Houston Methodist Hospital | Houston Methodist Hospital | S. Wesley Long et al |
| EPI_ISL_542874 | USA/TX-HMH-1837/2020 | 2020-05-12 | North America/USA/Texas/Greater Hou | Houston Methodist Hospital | Houston Methodist Hospital | S. Wesley Long et al |
| EPI_ISL_542877 | USA/TX-HMH-1846/2020 | 2020-04-29 | North America/USA/Texas/Greater Hou | Houston Methodist Hospital | Houston Methodist Hospital | S. Wesley Long et al |
| EPI_ISL_542878 | USA/TX-HMH-1847/2020 | 2020-04-18 | North America/USA/Texas/Greater Hou | Houston Methodist Hospital | Houston Methodist Hospital | S. Wesley Long et al |
| EPI_ISL_542881 | USA/TX-HMH-1856/2020 | 2020-05-10 | North America/USA/Texas/Greater Hou | Houston Methodist Hospital | Houston Methodist Hospital | S. Wesley Long et al |
| EPI_ISL_542885 | USA/TX-HMH-1865/2020 | 2020-05-07 | North America/USA/Texas/Greater Hou | Houston Methodist Hospital | Houston Methodist Hospital | S. Wesley Long et al |
| EPI_ISL_542891 | USA/TX-HMH-1880/2020 | 2020-05-04 | North America/USA/Texas/Greater Hou | Houston Methodist Hospital | Houston Methodist Hospital | S. Wesley Long et al |
| EPI_ISL_542893 | USA/TX-HMH-1895/2020 | 2020-04-29 | North America/USA/Texas/Greater Hou | Houston Methodist Hospital | Houston Methodist Hospital | S. Wesley Long et al |
| EPI_ISL_542898 | USA/TX-HMH-1910/2020 | 2020-05-16 | North America/USA/Texas/Greater Hou | Houston Methodist Hospital | Houston Methodist Hospital | S. Wesley Long et al |
| EPI_ISL_542901 | USA/TX-HMH-1917/2020 | 2020-04-17 | North America/USA/Texas/Greater Hou | Houston Methodist Hospital | Houston Methodist Hospital | S. Wesley Long et al |
| EPI_ISL_542911 | USA/TX-HMH-1939/2020 | 2020-04-28 | North America/USA/Texas/Greater Hou | Houston Methodist Hospital | Houston Methodist Hospital | S. Wesley Long et al |

| | | | | | | |
|---|---|---|---|---|---|---|
| EPI_ISL_542912 | USA/TX-HMH-1942/2020 | 2020-05-20 | North America/USA/Texas/Greater Hou | Houston Methodist Hospital | Houston Methodist Hospital | S. Wesley Long et al |
| EPI_ISL_542922 | USA/TX-HMH-1961/2020 | 2020-05-17 | North America/USA/Texas/Greater Hou | Houston Methodist Hospital | Houston Methodist Hospital | S. Wesley Long et al |
| EPI_ISL_542928 | USA/TX-HMH-1972/2020 | 2020-05-19 | North America/USA/Texas/Greater Hou | Houston Methodist Hospital | Houston Methodist Hospital | S. Wesley Long et al |
| EPI_ISL_542929 | USA/TX-HMH-1973/2020 | 2020-05-18 | North America/USA/Texas/Greater Hou | Houston Methodist Hospital | Houston Methodist Hospital | S. Wesley Long et al |
| EPI_ISL_542930 | USA/TX-HMH-1974/2020 | 2020-05-18 | North America/USA/Texas/Greater Hou | Houston Methodist Hospital | Houston Methodist Hospital | S. Wesley Long et al |
| EPI_ISL_542949 | USA/NM-UNM-00526/2020 | 2020-04-28 | North America/USA/New Mexico/ | TriCore Reference Laboratories | Center for Global Health, University of New Mexico Health Sciences Center | Daryl Domman et al |
| EPI_ISL_542951 | USA/NM-UNM-00529/2020 | 2020-05-09 | North America/USA/New Mexico/ | TriCore Reference Laboratories | Center for Global Health, University of New Mexico Health Sciences Center | Daryl Domman et al |
| EPI_ISL_542959 | USA/NM-UNM-00537/2020 | 2020-05-11 | North America/USA/New Mexico/ | TriCore Reference Laboratories | Center for Global Health, University of New Mexico Health Sciences Center | Daryl Domman et al |
| EPI_ISL_542960 | USA/NM-UNM-00538/2020 | 2020-05-11 | North America/USA/New Mexico/ | TriCore Reference Laboratories | Center for Global Health, University of New Mexico Health Sciences Center | Daryl Domman et al |
| EPI_ISL_542967 | USA/NM-UNM-00546/2020 | 2020-05-12 | North America/USA/New Mexico/ | TriCore Reference Laboratories | Center for Global Health, University of New Mexico Health Sciences Center | Daryl Domman et al |

| | | | | | | |
|---|---|---|---|---|---|---|
| EPI_ISL_542968 | USA/NM-UNM-00547/2020 | 2020-04-27 | North America/USA/Tennessee/ | TriCore Reference Laboratories | Center for Global Health, University of New Mexico Health Sciences Center | Daryl Domman et al |
| EPI_ISL_542969 | USA/NM-UNM-00548/2020 | 2020-05-10 | North America/USA/New Mexico/ | TriCore Reference Laboratories | Center for Global Health, University of New Mexico Health Sciences Center | Daryl Domman et al |
| EPI_ISL_542970 | USA/NM-UNM-00549/2020 | 2020-04-30 | North America/USA/New Mexico/ | TriCore Reference Laboratories | Center for Global Health, University of New Mexico Health Sciences Center | Daryl Domman et al |
| EPI_ISL_542977 | USA/NM-UNM-00561/2020 | 2020-05-07 | North America/USA/New Mexico/ | TriCore Reference Laboratories | Center for Global Health, University of New Mexico Health Sciences Center | Daryl Domman et al |
| EPI_ISL_542978 | USA/NM-UNM-00562/2020 | 2020-05-07 | North America/USA/New Mexico/ | TriCore Reference Laboratories | Center for Global Health, University of New Mexico Health Sciences Center | Daryl Domman et al |
| EPI_ISL_542980 | USA/NM-UNM-00565/2020 | 2020-05-09 | North America/USA/New Mexico/ | TriCore Reference Laboratories | Center for Global Health, University of New Mexico Health Sciences Center | Daryl Domman et al |
| EPI_ISL_542982 | USA/NM-UNM-00567/2020 | 2020-05-08 | North America/USA/New Mexico/ | TriCore Reference Laboratories | Center for Global Health, University of New Mexico Health Sciences Center | Daryl Domman et al |
| EPI_ISL_542983 | USA/NM-UNM-00569/2020 | 2020-08-08 | North America/USA/New Mexico/ | TriCore Reference Laboratories | Center for Global Health, University of New Mexico Health Sciences Center | Daryl Domman et al |

| | | | | | | |
|---|---|---|---|---|---|---|
| EPI_ISL_542991 | USA/NM-UNM-00577/2020 | 2020-08-07 | North America/USA/New Mexico/ | TriCore Reference Laboratories | Center for Global Health, University of New Mexico Health Sciences Center | Daryl Domman et al |
| EPI_ISL_542993 | USA/NM-UNM-00579/2020 | 2020-07-08 | North America/USA/New Mexico/ | TriCore Reference Laboratories | Center for Global Health, University of New Mexico Health Sciences Center | Daryl Domman et al |
| EPI_ISL_542994 | USA/NM-UNM-00581/2020 | 2020-07-09 | North America/USA/Texas/ | TriCore Reference Laboratories | Center for Global Health, University of New Mexico Health Sciences Center | Daryl Domman et al |
| EPI_ISL_542995 | USA/NM-UNM-00582/2020 | 2020-07-09 | North America/USA/New Mexico/ | TriCore Reference Laboratories | Center for Global Health, University of New Mexico Health Sciences Center | Daryl Domman et al |
| EPI_ISL_543013 | USA/TX-HMH-3551/2020 | 2020-06-15 | North America/USA/Texas/Greater Hou | Houston Methodist Hospital | Houston Methodist Hospital | S. Wesley Long et al |
| EPI_ISL_543014 | USA/TX-HMH-3552/2020 | 2020-06-14 | North America/USA/Texas/Greater Hou | Houston Methodist Hospital | Houston Methodist Hospital | S. Wesley Long et al |
| EPI_ISL_543015 | USA/TX-HMH-3553/2020 | 2020-06-14 | North America/USA/Texas/Greater Hou | Houston Methodist Hospital | Houston Methodist Hospital | S. Wesley Long et al |
| EPI_ISL_543021 | USA/TX-HMH-3559/2020 | 2020-06-10 | North America/USA/Texas/Greater Hou | Houston Methodist Hospital | Houston Methodist Hospital | S. Wesley Long et al |
| EPI_ISL_543026 | USA/TX-HMH-3564/2020 | 2020-06-15 | North America/USA/Texas/Greater Hou | Houston Methodist Hospital | Houston Methodist Hospital | S. Wesley Long et al |
| EPI_ISL_543042 | USA/TX-HMH-3585/2020 | 2020-06-15 | North America/USA/Texas/Greater Hou | Houston Methodist Hospital | Houston Methodist Hospital | S. Wesley Long et al |
| EPI_ISL_543057 | USA/TX-HMH-3601/2020 | 2020-06-15 | North America/USA/Texas/Greater Hou | Houston Methodist Hospital | Houston Methodist Hospital | S. Wesley Long et al |

| | | | | | | |
|---|---|---|---|---|---|---|
| EPI_ISL_543065 | USA/TX-HMH-3610/2020 | 2020-06-15 | North America/USA/Texas/Greater Hou | Houston Methodist Hospital | Houston Methodist Hospital | S. Wesley Long et al |
| EPI_ISL_543074 | USA/TX-HMH-3621/2020 | 2020-06-15 | North America/USA/Texas/Greater Hou | Houston Methodist Hospital | Houston Methodist Hospital | S. Wesley Long et al |
| EPI_ISL_543077 | USA/TX-HMH-3624/2020 | 2020-06-15 | North America/USA/Texas/Greater Hou | Houston Methodist Hospital | Houston Methodist Hospital | S. Wesley Long et al |
| EPI_ISL_543085 | USA/TX-HMH-3635/2020 | 2020-06-15 | North America/USA/Texas/Greater Hou | Houston Methodist Hospital | Houston Methodist Hospital | S. Wesley Long et al |
| EPI_ISL_543090 | USA/TX-HMH-3644/2020 | 2020-06-15 | North America/USA/Texas/Greater Hou | Houston Methodist Hospital | Houston Methodist Hospital | S. Wesley Long et al |
| EPI_ISL_543092 | USA/TX-HMH-3646/2020 | 2020-06-15 | North America/USA/Texas/Greater Hou | Houston Methodist Hospital | Houston Methodist Hospital | S. Wesley Long et al |
| EPI_ISL_543093 | USA/TX-HMH-3647/2020 | 2020-06-15 | North America/USA/Texas/Greater Hou | Houston Methodist Hospital | Houston Methodist Hospital | S. Wesley Long et al |
| EPI_ISL_543120 | USA/TX-HMH-3702/2020 | 2020-06-15 | North America/USA/Texas/Greater Hou | Houston Methodist Hospital | Houston Methodist Hospital | S. Wesley Long et al |
| EPI_ISL_543121 | USA/TX-HMH-3703/2020 | 2020-06-15 | North America/USA/Texas/Greater Hou | Houston Methodist Hospital | Houston Methodist Hospital | S. Wesley Long et al |
| EPI_ISL_543131 | USA/TX-HMH-3714/2020 | 2020-06-16 | North America/USA/Texas/Greater Hou | Houston Methodist Hospital | Houston Methodist Hospital | S. Wesley Long et al |
| EPI_ISL_543135 | USA/TX-HMH-3718/2020 | 2020-06-16 | North America/USA/Texas/Greater Hou | Houston Methodist Hospital | Houston Methodist Hospital | S. Wesley Long et al |
| EPI_ISL_543137 | USA/TX-HMH-3721/2020 | 2020-06-16 | North America/USA/Texas/Greater Hou | Houston Methodist Hospital | Houston Methodist Hospital | S. Wesley Long et al |
| EPI_ISL_543139 | USA/TX-HMH-3724/2020 | 2020-06-16 | North America/USA/Texas/Greater Hou | Houston Methodist Hospital | Houston Methodist Hospital | S. Wesley Long et al |

| | | | | | | |
|---|---|---|---|---|---|---|
| EPI_ISL_543140 | USA/TX-HMH-3728/2020 | 2020-06-16 | North America/USA/Texas/Greater Hou | Houston Methodist Hospital | Houston Methodist Hospital | S. Wesley Long et al |
| EPI_ISL_543141 | USA/TX-HMH-3729/2020 | 2020-06-16 | North America/USA/Texas/Greater Hou | Houston Methodist Hospital | Houston Methodist Hospital | S. Wesley Long et al |
| EPI_ISL_543149 | USA/TX-HMH-3737/2020 | 2020-06-15 | North America/USA/Texas/Greater Hou | Houston Methodist Hospital | Houston Methodist Hospital | S. Wesley Long et al |
| EPI_ISL_543152 | USA/TX-HMH-3740/2020 | 2020-06-16 | North America/USA/Texas/Greater Hou | Houston Methodist Hospital | Houston Methodist Hospital | S. Wesley Long et al |
| EPI_ISL_543157 | USA/TX-HMH-3745/2020 | 2020-06-16 | North America/USA/Texas/Greater Hou | Houston Methodist Hospital | Houston Methodist Hospital | S. Wesley Long et al |
| EPI_ISL_543164 | USA/TX-HMH-3753/2020 | 2020-06-16 | North America/USA/Texas/Greater Hou | Houston Methodist Hospital | Houston Methodist Hospital | S. Wesley Long et al |
| EPI_ISL_543171 | USA/TX-HMH-3760/2020 | 2020-06-16 | North America/USA/Texas/Greater Hou | Houston Methodist Hospital | Houston Methodist Hospital | S. Wesley Long et al |
| EPI_ISL_543176 | USA/TX-HMH-3765/2020 | 2020-06-16 | North America/USA/Texas/Greater Hou | Houston Methodist Hospital | Houston Methodist Hospital | S. Wesley Long et al |
| EPI_ISL_543184 | USA/TX-HMH-3778/2020 | 2020-06-16 | North America/USA/Texas/Greater Hou | Houston Methodist Hospital | Houston Methodist Hospital | S. Wesley Long et al |
| EPI_ISL_543196 | USA/TX-HMH-3791/2020 | 2020-06-12 | North America/USA/Texas/Greater Hou | Houston Methodist Hospital | Houston Methodist Hospital | S. Wesley Long et al |
| EPI_ISL_543204 | USA/TX-HMH-3800/2020 | 2020-06-11 | North America/USA/Texas/Greater Hou | Houston Methodist Hospital | Houston Methodist Hospital | S. Wesley Long et al |
| EPI_ISL_543217 | USA/TX-HMH-3813/2020 | 2020-06-12 | North America/USA/Texas/Greater Hou | Houston Methodist Hospital | Houston Methodist Hospital | S. Wesley Long et al |
| EPI_ISL_543230 | USA/TX-HMH-3826/2020 | 2020-06-12 | North America/USA/Texas/Greater Hou | Houston Methodist Hospital | Houston Methodist Hospital | S. Wesley Long et al |

| | | | | | | |
|---|---|---|---|---|---|---|
| EPI_ISL_543232 | USA/TX-HMH-3829/2020 | 2020-06-12 | North America/USA/Texas/Greater Hou | Houston Methodist Hospital | Houston Methodist Hospital | S. Wesley Long et al |
| EPI_ISL_543237 | USA/TX-HMH-3834/2020 | 2020-06-11 | North America/USA/Texas/Greater Hou | Houston Methodist Hospital | Houston Methodist Hospital | S. Wesley Long et al |
| EPI_ISL_543238 | USA/TX-HMH-3835/2020 | 2020-06-16 | North America/USA/Texas/Greater Hou | Houston Methodist Hospital | Houston Methodist Hospital | S. Wesley Long et al |
| EPI_ISL_543243 | USA/TX-HMH-3840/2020 | 2020-06-12 | North America/USA/Texas/Greater Hou | Houston Methodist Hospital | Houston Methodist Hospital | S. Wesley Long et al |
| EPI_ISL_543246 | USA/TX-HMH-3843/2020 | 2020-06-11 | North America/USA/Texas/Greater Hou | Houston Methodist Hospital | Houston Methodist Hospital | S. Wesley Long et al |
| EPI_ISL_543255 | USA/TX-HMH-3854/2020 | 2020-06-16 | North America/USA/Texas/Greater Hou | Houston Methodist Hospital | Houston Methodist Hospital | S. Wesley Long et al |
| EPI_ISL_543263 | USA/TX-HMH-3864/2020 | 2020-06-17 | North America/USA/Texas/Greater Hou | Houston Methodist Hospital | Houston Methodist Hospital | S. Wesley Long et al |
| EPI_ISL_543265 | USA/TX-HMH-3867/2020 | 2020-06-16 | North America/USA/Texas/Greater Hou | Houston Methodist Hospital | Houston Methodist Hospital | S. Wesley Long et al |
| EPI_ISL_543273 | USA/TX-HMH-3904/2020 | 2020-06-16 | North America/USA/Texas/Greater Hou | Houston Methodist Hospital | Houston Methodist Hospital | S. Wesley Long et al |
| EPI_ISL_543274 | USA/TX-HMH-3908/2020 | 2020-06-16 | North America/USA/Texas/Greater Hou | Houston Methodist Hospital | Houston Methodist Hospital | S. Wesley Long et al |
| EPI_ISL_543284 | USA/TX-HMH-3925/2020 | 2020-06-16 | North America/USA/Texas/Greater Hou | Houston Methodist Hospital | Houston Methodist Hospital | S. Wesley Long et al |
| EPI_ISL_543295 | USA/TX-HMH-4013/2020 | 2020-06-16 | North America/USA/Texas/Greater Hou | Houston Methodist Hospital | Houston Methodist Hospital | S. Wesley Long et al |
| EPI_ISL_543303 | USA/TX-HMH-4021/2020 | 2020-06-16 | North America/USA/Texas/Greater Hou | Houston Methodist Hospital | Houston Methodist Hospital | S. Wesley Long et al |

| | | | | | | |
|---|---|---|---|---|---|---|
| EPI_ISL_543306 | USA/TX-HMH-4024/2020 | 2020-06-16 | North America/USA/Texas/Greater Hou | Houston Methodist Hospital | Houston Methodist Hospital | S. Wesley Long et al |
| EPI_ISL_543307 | USA/TX-HMH-4027/2020 | 2020-06-16 | North America/USA/Texas/Greater Hou | Houston Methodist Hospital | Houston Methodist Hospital | S. Wesley Long et al |
| EPI_ISL_543311 | USA/TX-HMH-4031/2020 | 2020-06-12 | North America/USA/Texas/Greater Hou | Houston Methodist Hospital | Houston Methodist Hospital | S. Wesley Long et al |
| EPI_ISL_543315 | USA/TX-HMH-4035/2020 | 2020-06-16 | North America/USA/Texas/Greater Hou | Houston Methodist Hospital | Houston Methodist Hospital | S. Wesley Long et al |
| EPI_ISL_543317 | USA/TX-HMH-4039/2020 | 2020-06-16 | North America/USA/Texas/Greater Hou | Houston Methodist Hospital | Houston Methodist Hospital | S. Wesley Long et al |
| EPI_ISL_543318 | USA/TX-HMH-4040/2020 | 2020-06-17 | North America/USA/Texas/Greater Hou | Houston Methodist Hospital | Houston Methodist Hospital | S. Wesley Long et al |
| EPI_ISL_543319 | USA/TX-HMH-4041/2020 | 2020-06-17 | North America/USA/Texas/Greater Hou | Houston Methodist Hospital | Houston Methodist Hospital | S. Wesley Long et al |
| EPI_ISL_543332 | USA/TX-HMH-4056/2020 | 2020-06-17 | North America/USA/Texas/Greater Hou | Houston Methodist Hospital | Houston Methodist Hospital | S. Wesley Long et al |
| EPI_ISL_543335 | USA/TX-HMH-4061/2020 | 2020-06-17 | North America/USA/Texas/Greater Hou | Houston Methodist Hospital | Houston Methodist Hospital | S. Wesley Long et al |
| EPI_ISL_543338 | USA/TX-HMH-4065/2020 | 2020-06-18 | North America/USA/Texas/Greater Hou | Houston Methodist Hospital | Houston Methodist Hospital | S. Wesley Long et al |
| EPI_ISL_543357 | USA/TX-HMH-4087/2020 | 2020-06-18 | North America/USA/Texas/Greater Hou | Houston Methodist Hospital | Houston Methodist Hospital | S. Wesley Long et al |
| EPI_ISL_543359 | USA/TX-HMH-4089/2020 | 2020-06-17 | North America/USA/Texas/Greater Hou | Houston Methodist Hospital | Houston Methodist Hospital | S. Wesley Long et al |
| EPI_ISL_543360 | USA/TX-HMH-4090/2020 | 2020-06-17 | North America/USA/Texas/Greater Hou | Houston Methodist Hospital | Houston Methodist Hospital | S. Wesley Long et al |

| | | | | | | |
|---|---|---|---|---|---|---|
| EPI_ISL_543367 | USA/TX-HMH-4097/2020 | 2020-06-17 | North America/USA/Texas/Greater Hou | Houston Methodist Hospital | Houston Methodist Hospital | S. Wesley Long et al |
| EPI_ISL_543393 | USA/TX-HMH-4127/2020 | 2020-06-18 | North America/USA/Texas/Greater Hou | Houston Methodist Hospital | Houston Methodist Hospital | S. Wesley Long et al |
| EPI_ISL_543400 | USA/TX-HMH-4136/2020 | 2020-06-18 | North America/USA/Texas/Greater Hou | Houston Methodist Hospital | Houston Methodist Hospital | S. Wesley Long et al |
| EPI_ISL_543411 | USA/TX-HMH-4174/2020 | 2020-06-18 | North America/USA/Texas/Greater Hou | Houston Methodist Hospital | Houston Methodist Hospital | S. Wesley Long et al |
| EPI_ISL_543416 | USA/TX-HMH-4180/2020 | 2020-06-18 | North America/USA/Texas/Greater Hou | Houston Methodist Hospital | Houston Methodist Hospital | S. Wesley Long et al |
| EPI_ISL_543426 | USA/TX-HMH-4192/2020 | 2020-06-19 | North America/USA/Texas/Greater Hou | Houston Methodist Hospital | Houston Methodist Hospital | S. Wesley Long et al |
| EPI_ISL_543428 | USA/TX-HMH-4196/2020 | 2020-06-18 | North America/USA/Texas/Greater Hou | Houston Methodist Hospital | Houston Methodist Hospital | S. Wesley Long et al |
| EPI_ISL_543437 | USA/TX-HMH-4206/2020 | 2020-06-19 | North America/USA/Texas/Greater Hou | Houston Methodist Hospital | Houston Methodist Hospital | S. Wesley Long et al |
| EPI_ISL_543442 | USA/TX-HMH-4212/2020 | 2020-06-19 | North America/USA/Texas/Greater Hou | Houston Methodist Hospital | Houston Methodist Hospital | S. Wesley Long et al |
| EPI_ISL_543449 | USA/TX-HMH-4220/2020 | 2020-06-19 | North America/USA/Texas/Greater Hou | Houston Methodist Hospital | Houston Methodist Hospital | S. Wesley Long et al |
| EPI_ISL_543455 | USA/TX-HMH-4242/2020 | 2020-06-18 | North America/USA/Texas/Greater Hou | Houston Methodist Hospital | Houston Methodist Hospital | S. Wesley Long et al |
| EPI_ISL_543480 | USA/TX-HMH-4268/2020 | 2020-06-18 | North America/USA/Texas/Greater Hou | Houston Methodist Hospital | Houston Methodist Hospital | S. Wesley Long et al |
| EPI_ISL_543481 | USA/TX-HMH-4269/2020 | 2020-06-18 | North America/USA/Texas/Greater Hou | Houston Methodist Hospital | Houston Methodist Hospital | S. Wesley Long et al |

| | | | | | | |
|---|---|---|---|---|---|---|
| EPI_ISL_543486 | USA/TX-HMH-4274/2020 | 2020-06-18 | North America/USA/Texas/Greater Hou | Houston Methodist Hospital | Houston Methodist Hospital | S. Wesley Long et al |
| EPI_ISL_543488 | USA/TX-HMH-4276/2020 | 2020-06-18 | North America/USA/Texas/Greater Hou | Houston Methodist Hospital | Houston Methodist Hospital | S. Wesley Long et al |
| EPI_ISL_543498 | USA/TX-HMH-4288/2020 | 2020-06-18 | North America/USA/Texas/Greater Hou | Houston Methodist Hospital | Houston Methodist Hospital | S. Wesley Long et al |
| EPI_ISL_543503 | USA/TX-HMH-4293/2020 | 2020-06-18 | North America/USA/Texas/Greater Hou | Houston Methodist Hospital | Houston Methodist Hospital | S. Wesley Long et al |
| EPI_ISL_543508 | USA/TX-HMH-4298/2020 | 2020-06-19 | North America/USA/Texas/Greater Hou | Houston Methodist Hospital | Houston Methodist Hospital | S. Wesley Long et al |
| EPI_ISL_543520 | USA/TX-HMH-4311/2020 | 2020-06-18 | North America/USA/Texas/Greater Hou | Houston Methodist Hospital | Houston Methodist Hospital | S. Wesley Long et al |
| EPI_ISL_543522 | USA/TX-HMH-4313/2020 | 2020-06-18 | North America/USA/Texas/Greater Hou | Houston Methodist Hospital | Houston Methodist Hospital | S. Wesley Long et al |
| EPI_ISL_543528 | USA/TX-HMH-4319/2020 | 2020-06-18 | North America/USA/Texas/Greater Hou | Houston Methodist Hospital | Houston Methodist Hospital | S. Wesley Long et al |
| EPI_ISL_543551 | USA/TX-HMH-4350/2020 | 2020-06-17 | North America/USA/Texas/Greater Hou | Houston Methodist Hospital | Houston Methodist Hospital | S. Wesley Long et al |
| EPI_ISL_543555 | USA/TX-HMH-4356/2020 | 2020-06-19 | North America/USA/Texas/Greater Hou | Houston Methodist Hospital | Houston Methodist Hospital | S. Wesley Long et al |
| EPI_ISL_543558 | USA/TX-HMH-4359/2020 | 2020-06-19 | North America/USA/Texas/Greater Hou | Houston Methodist Hospital | Houston Methodist Hospital | S. Wesley Long et al |
| EPI_ISL_543562 | USA/TX-HMH-4363/2020 | 2020-06-18 | North America/USA/Texas/Greater Hou | Houston Methodist Hospital | Houston Methodist Hospital | S. Wesley Long et al |
| EPI_ISL_543570 | USA/TX-HMH-4373/2020 | 2020-06-17 | North America/USA/Texas/Greater Hou | Houston Methodist Hospital | Houston Methodist Hospital | S. Wesley Long et al |

| | | | | | | |
|---|---|---|---|---|---|---|
| EPI_ISL_543585 | USA/TX-HMH-4395/2020 | 2020-06-17 | North America/USA/Texas/Greater Hou | Houston Methodist Hospital | Houston Methodist Hospital | S. Wesley Long et al |
| EPI_ISL_543591 | USA/TX-HMH-4402/2020 | 2020-06-17 | North America/USA/Texas/Greater Hou | Houston Methodist Hospital | Houston Methodist Hospital | S. Wesley Long et al |
| EPI_ISL_543595 | USA/TX-HMH-4406/2020 | 2020-06-17 | North America/USA/Texas/Greater Hou | Houston Methodist Hospital | Houston Methodist Hospital | S. Wesley Long et al |
| EPI_ISL_543597 | USA/TX-HMH-4408/2020 | 2020-06-17 | North America/USA/Texas/Greater Hou | Houston Methodist Hospital | Houston Methodist Hospital | S. Wesley Long et al |
| EPI_ISL_543605 | USA/TX-HMH-4419/2020 | 2020-06-17 | North America/USA/Texas/Greater Hou | Houston Methodist Hospital | Houston Methodist Hospital | S. Wesley Long et al |
| EPI_ISL_543636 | USA/TX-HMH-4459/2020 | 2020-06-17 | North America/USA/Texas/Greater Hou | Houston Methodist Hospital | Houston Methodist Hospital | S. Wesley Long et al |
| EPI_ISL_543648 | USA/TX-HMH-4472/2020 | 2020-06-17 | North America/USA/Texas/Greater Hou | Houston Methodist Hospital | Houston Methodist Hospital | S. Wesley Long et al |
| EPI_ISL_543660 | USA/TX-HMH-4487/2020 | 2020-06-19 | North America/USA/Texas/Greater Hou | Houston Methodist Hospital | Houston Methodist Hospital | S. Wesley Long et al |
| EPI_ISL_543664 | USA/TX-HMH-4492/2020 | 2020-06-19 | North America/USA/Texas/Greater Hou | Houston Methodist Hospital | Houston Methodist Hospital | S. Wesley Long et al |
| EPI_ISL_543684 | USA/TX-HMH-4515/2020 | 2020-06-19 | North America/USA/Texas/Greater Hou | Houston Methodist Hospital | Houston Methodist Hospital | S. Wesley Long et al |
| EPI_ISL_543708 | USA/TX-HMH-4545/2020 | 2020-06-21 | North America/USA/Texas/Greater Hou | Houston Methodist Hospital | Houston Methodist Hospital | S. Wesley Long et al |
| EPI_ISL_543710 | USA/TX-HMH-4547/2020 | 2020-06-21 | North America/USA/Texas/Greater Hou | Houston Methodist Hospital | Houston Methodist Hospital | S. Wesley Long et al |
| EPI_ISL_543711 | USA/TX-HMH-4551/2020 | 2020-06-20 | North America/USA/Texas/Greater Hou | Houston Methodist Hospital | Houston Methodist Hospital | S. Wesley Long et al |

| | | | | | | |
|---|---|---|---|---|---|---|
| EPI_ISL_543717 | USA/TX-HMH-4559/2020 | 2020-06-21 | North America/USA/Texas/Greater Hou | Houston Methodist Hospital | Houston Methodist Hospital | S. Wesley Long et al |
| EPI_ISL_543721 | USA/TX-HMH-4567/2020 | 2020-06-22 | North America/USA/Texas/Greater Hou | Houston Methodist Hospital | Houston Methodist Hospital | S. Wesley Long et al |
| EPI_ISL_543729 | USA/TX-HMH-4579/2020 | 2020-06-22 | North America/USA/Texas/Greater Hou | Houston Methodist Hospital | Houston Methodist Hospital | S. Wesley Long et al |
| EPI_ISL_543741 | USA/TX-HMH-4594/2020 | 2020-06-21 | North America/USA/Texas/Greater Hou | Houston Methodist Hospital | Houston Methodist Hospital | S. Wesley Long et al |
| EPI_ISL_543747 | USA/TX-HMH-4601/2020 | 2020-06-20 | North America/USA/Texas/Greater Hou | Houston Methodist Hospital | Houston Methodist Hospital | S. Wesley Long et al |
| EPI_ISL_543763 | USA/TX-HMH-4624/2020 | 2020-06-22 | North America/USA/Texas/Greater Hou | Houston Methodist Hospital | Houston Methodist Hospital | S. Wesley Long et al |
| EPI_ISL_543770 | USA/TX-HMH-4632/2020 | 2020-06-22 | North America/USA/Texas/Greater Hou | Houston Methodist Hospital | Houston Methodist Hospital | S. Wesley Long et al |
| EPI_ISL_543773 | USA/TX-HMH-4636/2020 | 2020-06-22 | North America/USA/Texas/Greater Hou | Houston Methodist Hospital | Houston Methodist Hospital | S. Wesley Long et al |
| EPI_ISL_543780 | USA/TX-HMH-4650/2020 | 2020-06-22 | North America/USA/Texas/Greater Hou | Houston Methodist Hospital | Houston Methodist Hospital | S. Wesley Long et al |
| EPI_ISL_543786 | USA/TX-HMH-466/2020 | 2020-04-04 | North America/USA/Texas/Greater Hou | Houston Methodist Hospital | Houston Methodist Hospital | S. Wesley Long et al |
| EPI_ISL_543788 | USA/TX-HMH-4664/2020 | 2020-06-22 | North America/USA/Texas/Greater Hou | Houston Methodist Hospital | Houston Methodist Hospital | S. Wesley Long et al |
| EPI_ISL_543793 | USA/TX-HMH-2817/2020 | 2020-06-08 | North America/USA/Texas/Greater Hou | Houston Methodist Hospital | Houston Methodist Hospital | S. Wesley Long et al |
| EPI_ISL_543794 | USA/TX-HMH-2818/2020 | 2020-06-08 | North America/USA/Texas/Greater Hou | Houston Methodist Hospital | Houston Methodist Hospital | S. Wesley Long et al |

| | | | | | | |
|---|---|---|---|---|---|---|
| EPI_ISL_543795 | USA/TX-HMH-2819/2020 | 2020-06-08 | North America/USA/Texas/Greater Hou | Houston Methodist Hospital | Houston Methodist Hospital | S. Wesley Long et al |
| EPI_ISL_543809 | USA/TX-HMH-2838/2020 | 2020-06-07 | North America/USA/Texas/Greater Hou | Houston Methodist Hospital | Houston Methodist Hospital | S. Wesley Long et al |
| EPI_ISL_543815 | USA/TX-HMH-2844/2020 | 2020-06-08 | North America/USA/Texas/Greater Hou | Houston Methodist Hospital | Houston Methodist Hospital | S. Wesley Long et al |
| EPI_ISL_543820 | USA/TX-HMH-2849/2020 | 2020-06-08 | North America/USA/Texas/Greater Hou | Houston Methodist Hospital | Houston Methodist Hospital | S. Wesley Long et al |
| EPI_ISL_543827 | USA/TX-HMH-2857/2020 | 2020-06-08 | North America/USA/Texas/Greater Hou | Houston Methodist Hospital | Houston Methodist Hospital | S. Wesley Long et al |
| EPI_ISL_543832 | USA/TX-HMH-2862/2020 | 2020-06-08 | North America/USA/Texas/Greater Hou | Houston Methodist Hospital | Houston Methodist Hospital | S. Wesley Long et al |
| EPI_ISL_543834 | USA/TX-HMH-2864/2020 | 2020-06-08 | North America/USA/Texas/Greater Hou | Houston Methodist Hospital | Houston Methodist Hospital | S. Wesley Long et al |
| EPI_ISL_543835 | USA/TX-HMH-2865/2020 | 2020-06-08 | North America/USA/Texas/Greater Hou | Houston Methodist Hospital | Houston Methodist Hospital | S. Wesley Long et al |
| EPI_ISL_543840 | USA/TX-HMH-2870/2020 | 2020-06-08 | North America/USA/Texas/Greater Hou | Houston Methodist Hospital | Houston Methodist Hospital | S. Wesley Long et al |
| EPI_ISL_543843 | USA/TX-HMH-2874/2020 | 2020-06-08 | North America/USA/Texas/Greater Hou | Houston Methodist Hospital | Houston Methodist Hospital | S. Wesley Long et al |
| EPI_ISL_543860 | USA/TX-HMH-2892/2020 | 2020-06-08 | North America/USA/Texas/Greater Hou | Houston Methodist Hospital | Houston Methodist Hospital | S. Wesley Long et al |
| EPI_ISL_543869 | USA/TX-HMH-2927/2020 | 2020-06-09 | North America/USA/Texas/Greater Hou | Houston Methodist Hospital | Houston Methodist Hospital | S. Wesley Long et al |
| EPI_ISL_543883 | USA/TX-HMH-2947/2020 | 2020-06-09 | North America/USA/Texas/Greater Hou | Houston Methodist Hospital | Houston Methodist Hospital | S. Wesley Long et al |

| | | | | | | |
|---|---|---|---|---|---|---|
| EPI_ISL_543884 | USA/TX-HMH-2948/2020 | 2020-06-09 | North America/USA/Texas/Greater Hou | Houston Methodist Hospital | Houston Methodist Hospital | S. Wesley Long et al |
| EPI_ISL_543890 | USA/TX-HMH-2956/2020 | 2020-06-09 | North America/USA/Texas/Greater Hou | Houston Methodist Hospital | Houston Methodist Hospital | S. Wesley Long et al |
| EPI_ISL_543892 | USA/TX-HMH-2958/2020 | 2020-06-08 | North America/USA/Texas/Greater Hou | Houston Methodist Hospital | Houston Methodist Hospital | S. Wesley Long et al |
| EPI_ISL_543893 | USA/TX-HMH-2959/2020 | 2020-06-08 | North America/USA/Texas/Greater Hou | Houston Methodist Hospital | Houston Methodist Hospital | S. Wesley Long et al |
| EPI_ISL_543911 | USA/TX-HMH-2983/2020 | 2020-05-31 | North America/USA/Texas/Greater Hou | Houston Methodist Hospital | Houston Methodist Hospital | S. Wesley Long et al |
| EPI_ISL_543912 | USA/TX-HMH-2984/2020 | 2020-05-30 | North America/USA/Texas/Greater Hou | Houston Methodist Hospital | Houston Methodist Hospital | S. Wesley Long et al |
| EPI_ISL_543914 | USA/TX-HMH-2987/2020 | 2020-05-31 | North America/USA/Texas/Greater Hou | Houston Methodist Hospital | Houston Methodist Hospital | S. Wesley Long et al |
| EPI_ISL_543918 | USA/TX-HMH-2994/2020 | 2020-05-31 | North America/USA/Texas/Greater Hou | Houston Methodist Hospital | Houston Methodist Hospital | S. Wesley Long et al |
| EPI_ISL_543926 | USA/TX-HMH-3004/2020 | 2020-05-30 | North America/USA/Texas/Greater Hou | Houston Methodist Hospital | Houston Methodist Hospital | S. Wesley Long et al |
| EPI_ISL_543927 | USA/TX-HMH-3006/2020 | 2020-05-29 | North America/USA/Texas/Greater Hou | Houston Methodist Hospital | Houston Methodist Hospital | S. Wesley Long et al |
| EPI_ISL_543933 | USA/TX-HMH-3013/2020 | 2020-05-30 | North America/USA/Texas/Greater Hou | Houston Methodist Hospital | Houston Methodist Hospital | S. Wesley Long et al |
| EPI_ISL_543934 | USA/TX-HMH-3014/2020 | 2020-05-31 | North America/USA/Texas/Greater Hou | Houston Methodist Hospital | Houston Methodist Hospital | S. Wesley Long et al |
| EPI_ISL_543936 | USA/TX-HMH-3016/2020 | 2020-05-30 | North America/USA/Texas/Greater Hou | Houston Methodist Hospital | Houston Methodist Hospital | S. Wesley Long et al |

| | | | | | | |
|---|---|---|---|---|---|---|
| EPI_ISL_543937 | USA/TX-HMH-3017/2020 | 2020-05-30 | North America/USA/Texas/Greater Hou | Houston Methodist Hospital | Houston Methodist Hospital | S. Wesley Long et al |
| EPI_ISL_543940 | USA/TX-HMH-3022/2020 | 2020-05-30 | North America/USA/Texas/Greater Hou | Houston Methodist Hospital | Houston Methodist Hospital | S. Wesley Long et al |
| EPI_ISL_543944 | USA/TX-HMH-3026/2020 | 2020-05-30 | North America/USA/Texas/Greater Hou | Houston Methodist Hospital | Houston Methodist Hospital | S. Wesley Long et al |
| EPI_ISL_543951 | USA/TX-HMH-3039/2020 | 2020-05-22 | North America/USA/Texas/Greater Hou | Houston Methodist Hospital | Houston Methodist Hospital | S. Wesley Long et al |
| EPI_ISL_543957 | USA/TX-HMH-3051/2020 | 2020-04-16 | North America/USA/Texas/Greater Hou | Houston Methodist Hospital | Houston Methodist Hospital | S. Wesley Long et al |
| EPI_ISL_543960 | USA/TX-HMH-3054/2020 | 2020-04-16 | North America/USA/Texas/Greater Hou | Houston Methodist Hospital | Houston Methodist Hospital | S. Wesley Long et al |
| EPI_ISL_543966 | USA/TX-HMH-3060/2020 | 2020-05-31 | North America/USA/Texas/Greater Hou | Houston Methodist Hospital | Houston Methodist Hospital | S. Wesley Long et al |
| EPI_ISL_543968 | USA/TX-HMH-3063/2020 | 2020-06-01 | North America/USA/Texas/Greater Hou | Houston Methodist Hospital | Houston Methodist Hospital | S. Wesley Long et al |
| EPI_ISL_543974 | USA/TX-HMH-3070/2020 | 2020-05-31 | North America/USA/Texas/Greater Hou | Houston Methodist Hospital | Houston Methodist Hospital | S. Wesley Long et al |
| EPI_ISL_543999 | USA/TX-HMH-3111/2020 | 2020-06-10 | North America/USA/Texas/Greater Hou | Houston Methodist Hospital | Houston Methodist Hospital | S. Wesley Long et al |
| EPI_ISL_544011 | USA/TX-HMH-3126/2020 | 2020-06-11 | North America/USA/Texas/Greater Hou | Houston Methodist Hospital | Houston Methodist Hospital | S. Wesley Long et al |
| EPI_ISL_544027 | USA/TX-HMH-3148/2020 | 2020-06-10 | North America/USA/Texas/Greater Hou | Houston Methodist Hospital | Houston Methodist Hospital | S. Wesley Long et al |
| EPI_ISL_544033 | USA/TX-HMH-3157/2020 | 2020-06-10 | North America/USA/Texas/Greater Hou | Houston Methodist Hospital | Houston Methodist Hospital | S. Wesley Long et al |

| | | | | | | |
|---|---|---|---|---|---|---|
| EPI_ISL_544034 | USA/TX-HMH-3158/2020 | 2020-06-10 | North America/USA/Texas/Greater Hou | Houston Methodist Hospital | Houston Methodist Hospital | S. Wesley Long et al |
| EPI_ISL_544036 | USA/TX-HMH-3160/2020 | 2020-06-10 | North America/USA/Texas/Greater Hou | Houston Methodist Hospital | Houston Methodist Hospital | S. Wesley Long et al |
| EPI_ISL_544037 | USA/TX-HMH-3161/2020 | 2020-06-10 | North America/USA/Texas/Greater Hou | Houston Methodist Hospital | Houston Methodist Hospital | S. Wesley Long et al |
| EPI_ISL_544039 | USA/TX-HMH-3163/2020 | 2020-06-10 | North America/USA/Texas/Greater Hou | Houston Methodist Hospital | Houston Methodist Hospital | S. Wesley Long et al |
| EPI_ISL_544061 | USA/TX-HMH-3187/2020 | 2020-06-13 | North America/USA/Texas/Greater Hou | Houston Methodist Hospital | Houston Methodist Hospital | S. Wesley Long et al |
| EPI_ISL_544072 | USA/TX-HMH-3200/2020 | 2020-06-13 | North America/USA/Texas/Greater Hou | Houston Methodist Hospital | Houston Methodist Hospital | S. Wesley Long et al |
| EPI_ISL_544074 | USA/TX-HMH-3203/2020 | 2020-06-12 | North America/USA/Texas/Greater Hou | Houston Methodist Hospital | Houston Methodist Hospital | S. Wesley Long et al |
| EPI_ISL_544078 | USA/TX-HMH-3209/2020 | 2020-06-12 | North America/USA/Texas/Greater Hou | Houston Methodist Hospital | Houston Methodist Hospital | S. Wesley Long et al |
| EPI_ISL_544081 | USA/TX-HMH-3213/2020 | 2020-06-13 | North America/USA/Texas/Greater Hou | Houston Methodist Hospital | Houston Methodist Hospital | S. Wesley Long et al |
| EPI_ISL_544118 | USA/TX-HMH-3306/2020 | 2020-06-11 | North America/USA/Texas/Greater Hou | Houston Methodist Hospital | Houston Methodist Hospital | S. Wesley Long et al |
| EPI_ISL_544130 | USA/TX-HMH-3321/2020 | 2020-06-10 | North America/USA/Texas/Greater Hou | Houston Methodist Hospital | Houston Methodist Hospital | S. Wesley Long et al |
| EPI_ISL_544145 | USA/TX-HMH-3360/2020 | 2020-06-12 | North America/USA/Texas/Greater Hou | Houston Methodist Hospital | Houston Methodist Hospital | S. Wesley Long et al |
| EPI_ISL_544153 | USA/TX-HMH-3369/2020 | 2020-06-13 | North America/USA/Texas/Greater Hou | Houston Methodist Hospital | Houston Methodist Hospital | S. Wesley Long et al |

| | | | | | | |
|---|---|---|---|---|---|---|
| EPI_ISL_544156 | USA/TX-HMH-3372/2020 | 2020-06-13 | North America/USA/Texas/Greater Hou | Houston Methodist Hospital | Houston Methodist Hospital | S. Wesley Long et al |
| EPI_ISL_544163 | USA/TX-HMH-3403/2020 | 2020-06-14 | North America/USA/Texas/Greater Hou | Houston Methodist Hospital | Houston Methodist Hospital | S. Wesley Long et al |
| EPI_ISL_544175 | USA/TX-HMH-3416/2020 | 2020-06-14 | North America/USA/Texas/Greater Hou | Houston Methodist Hospital | Houston Methodist Hospital | S. Wesley Long et al |
| EPI_ISL_544176 | USA/TX-HMH-3417/2020 | 2020-06-14 | North America/USA/Texas/Greater Hou | Houston Methodist Hospital | Houston Methodist Hospital | S. Wesley Long et al |
| EPI_ISL_544180 | USA/TX-HMH-3422/2020 | 2020-06-14 | North America/USA/Texas/Greater Hou | Houston Methodist Hospital | Houston Methodist Hospital | S. Wesley Long et al |
| EPI_ISL_544184 | USA/TX-HMH-3427/2020 | 2020-06-13 | North America/USA/Texas/Greater Hou | Houston Methodist Hospital | Houston Methodist Hospital | S. Wesley Long et al |
| EPI_ISL_544195 | USA/TX-HMH-3440/2020 | 2020-06-12 | North America/USA/Texas/Greater Hou | Houston Methodist Hospital | Houston Methodist Hospital | S. Wesley Long et al |
| EPI_ISL_544198 | USA/TX-HMH-3443/2020 | 2020-06-11 | North America/USA/Texas/Greater Hou | Houston Methodist Hospital | Houston Methodist Hospital | S. Wesley Long et al |
| EPI_ISL_544211 | USA/TX-HMH-3511/2020 | 2020-06-15 | North America/USA/Texas/Greater Hou | Houston Methodist Hospital | Houston Methodist Hospital | S. Wesley Long et al |
| EPI_ISL_544212 | USA/TX-HMH-3512/2020 | 2020-06-15 | North America/USA/Texas/Greater Hou | Houston Methodist Hospital | Houston Methodist Hospital | S. Wesley Long et al |
| EPI_ISL_544223 | USA/TX-HMH-3525/2020 | 2020-06-10 | North America/USA/Texas/Greater Hou | Houston Methodist Hospital | Houston Methodist Hospital | S. Wesley Long et al |
| EPI_ISL_544227 | USA/TX-HMH-3529/2020 | 2020-06-15 | North America/USA/Texas/Greater Hou | Houston Methodist Hospital | Houston Methodist Hospital | S. Wesley Long et al |
| EPI_ISL_544235 | USA/TX-HMH-3541/2020 | 2020-06-10 | North America/USA/Texas/Greater Hou | Houston Methodist Hospital | Houston Methodist Hospital | S. Wesley Long et al |

| | | | | | | |
|---|---|---|---|---|---|---|
| EPI_ISL_544236 | USA/TX-HMH-3542/2020 | 2020-06-15 | North America/USA/Texas/Greater Hou | Houston Methodist Hospital | Houston Methodist Hospital | S. Wesley Long et al |
| EPI_ISL_544239 | USA/TX-HMH-4666/2020 | 2020-06-22 | North America/USA/Texas/Greater Hou | Houston Methodist Hospital | Houston Methodist Hospital | S. Wesley Long et al |
| EPI_ISL_544240 | USA/TX-HMH-4667/2020 | 2020-06-22 | North America/USA/Texas/Greater Hou | Houston Methodist Hospital | Houston Methodist Hospital | S. Wesley Long et al |
| EPI_ISL_544245 | USA/TX-HMH-4671/2020 | 2020-06-22 | North America/USA/Texas/Greater Hou | Houston Methodist Hospital | Houston Methodist Hospital | S. Wesley Long et al |
| EPI_ISL_544247 | USA/TX-HMH-4673/2020 | 2020-06-19 | North America/USA/Texas/Greater Hou | Houston Methodist Hospital | Houston Methodist Hospital | S. Wesley Long et al |
| EPI_ISL_544248 | USA/TX-HMH-4674/2020 | 2020-06-19 | North America/USA/Texas/Greater Hou | Houston Methodist Hospital | Houston Methodist Hospital | S. Wesley Long et al |
| EPI_ISL_544249 | USA/TX-HMH-4678/2020 | 2020-06-21 | North America/USA/Texas/Greater Hou | Houston Methodist Hospital | Houston Methodist Hospital | S. Wesley Long et al |
| EPI_ISL_544250 | USA/TX-HMH-4679/2020 | 2020-06-21 | North America/USA/Texas/Greater Hou | Houston Methodist Hospital | Houston Methodist Hospital | S. Wesley Long et al |
| EPI_ISL_544253 | USA/TX-HMH-4681/2020 | 2020-06-20 | North America/USA/Texas/Greater Hou | Houston Methodist Hospital | Houston Methodist Hospital | S. Wesley Long et al |
| EPI_ISL_544255 | USA/TX-HMH-4684/2020 | 2020-06-20 | North America/USA/Texas/Greater Hou | Houston Methodist Hospital | Houston Methodist Hospital | S. Wesley Long et al |
| EPI_ISL_544256 | USA/TX-HMH-4685/2020 | 2020-06-20 | North America/USA/Texas/Greater Hou | Houston Methodist Hospital | Houston Methodist Hospital | S. Wesley Long et al |
| EPI_ISL_544259 | USA/TX-HMH-469/2020 | 2020-04-04 | North America/USA/Texas/Greater Hou | Houston Methodist Hospital | Houston Methodist Hospital | S. Wesley Long et al |
| EPI_ISL_544269 | USA/TX-HMH-4701/2020 | 2020-06-24 | North America/USA/Texas/Greater Hou | Houston Methodist Hospital | Houston Methodist Hospital | S. Wesley Long et al |

| | | | | | | |
|---|---|---|---|---|---|---|
| EPI_ISL_544286 | USA/TX-HMH-4720/2020 | 2020-06-21 | North America/USA/Texas/Greater Hou | Houston Methodist Hospital | Houston Methodist Hospital | S. Wesley Long et al |
| EPI_ISL_544291 | USA/TX-HMH-4730/2020 | 2020-06-20 | North America/USA/Texas/Greater Hou | Houston Methodist Hospital | Houston Methodist Hospital | S. Wesley Long et al |
| EPI_ISL_544298 | USA/TX-HMH-4741/2020 | 2020-06-20 | North America/USA/Texas/Greater Hou | Houston Methodist Hospital | Houston Methodist Hospital | S. Wesley Long et al |
| EPI_ISL_544300 | USA/TX-HMH-4743/2020 | 2020-06-22 | North America/USA/Texas/Greater Hou | Houston Methodist Hospital | Houston Methodist Hospital | S. Wesley Long et al |
| EPI_ISL_544302 | USA/TX-HMH-4748/2020 | 2020-06-22 | North America/USA/Texas/Greater Hou | Houston Methodist Hospital | Houston Methodist Hospital | S. Wesley Long et al |
| EPI_ISL_544307 | USA/TX-HMH-4753/2020 | 2020-06-22 | North America/USA/Texas/Greater Hou | Houston Methodist Hospital | Houston Methodist Hospital | S. Wesley Long et al |
| EPI_ISL_544322 | USA/TX-HMH-4775/2020 | 2020-06-22 | North America/USA/Texas/Greater Hou | Houston Methodist Hospital | Houston Methodist Hospital | S. Wesley Long et al |
| EPI_ISL_544323 | USA/TX-HMH-4777/2020 | 2020-06-22 | North America/USA/Texas/Greater Hou | Houston Methodist Hospital | Houston Methodist Hospital | S. Wesley Long et al |
| EPI_ISL_544331 | USA/TX-HMH-4789/2020 | 2020-06-22 | North America/USA/Texas/Greater Hou | Houston Methodist Hospital | Houston Methodist Hospital | S. Wesley Long et al |
| EPI_ISL_544335 | USA/TX-HMH-4792/2020 | 2020-06-22 | North America/USA/Texas/Greater Hou | Houston Methodist Hospital | Houston Methodist Hospital | S. Wesley Long et al |
| EPI_ISL_544357 | USA/TX-HMH-4825/2020 | 2020-06-22 | North America/USA/Texas/Greater Hou | Houston Methodist Hospital | Houston Methodist Hospital | S. Wesley Long et al |
| EPI_ISL_544365 | USA/TX-HMH-4836/2020 | 2020-06-22 | North America/USA/Texas/Greater Hou | Houston Methodist Hospital | Houston Methodist Hospital | S. Wesley Long et al |
| EPI_ISL_544385 | USA/TX-HMH-4863/2020 | 2020-06-22 | North America/USA/Texas/Greater Hou | Houston Methodist Hospital | Houston Methodist Hospital | S. Wesley Long et al |

| | | | | | | |
|---|---|---|---|---|---|---|
| EPI_ISL_544397 | USA/TX-HMH-4879/2020 | 2020-06-22 | North America/USA/Texas/Greater Hou | Houston Methodist Hospital | Houston Methodist Hospital | S. Wesley Long et al |
| EPI_ISL_544399 | USA/TX-HMH-4880/2020 | 2020-06-22 | North America/USA/Texas/Greater Hou | Houston Methodist Hospital | Houston Methodist Hospital | S. Wesley Long et al |
| EPI_ISL_544409 | USA/TX-HMH-492/2020 | 2020-04-05 | North America/USA/Texas/Greater Hou | Houston Methodist Hospital | Houston Methodist Hospital | S. Wesley Long et al |
| EPI_ISL_544413 | USA/TX-HMH-4937/2020 | 2020-06-21 | North America/USA/Texas/Greater Hou | Houston Methodist Hospital | Houston Methodist Hospital | S. Wesley Long et al |
| EPI_ISL_544416 | USA/TX-HMH-4940/2020 | 2020-06-22 | North America/USA/Texas/Greater Hou | Houston Methodist Hospital | Houston Methodist Hospital | S. Wesley Long et al |
| EPI_ISL_544417 | USA/TX-HMH-4941/2020 | 2020-06-21 | North America/USA/Texas/Greater Hou | Houston Methodist Hospital | Houston Methodist Hospital | S. Wesley Long et al |
| EPI_ISL_544453 | USA/TX-HMH-5006/2020 | 2020-06-18 | North America/USA/Texas/Greater Hou | Houston Methodist Hospital | Houston Methodist Hospital | S. Wesley Long et al |
| EPI_ISL_544482 | USA/TX-HMH-5045/2020 | 2020-06-24 | North America/USA/Texas/Greater Hou | Houston Methodist Hospital | Houston Methodist Hospital | S. Wesley Long et al |
| EPI_ISL_544488 | USA/TX-HMH-5052/2020 | 2020-06-24 | North America/USA/Texas/Greater Hou | Houston Methodist Hospital | Houston Methodist Hospital | S. Wesley Long et al |
| EPI_ISL_544501 | USA/TX-HMH-5069/2020 | 2020-06-23 | North America/USA/Texas/Greater Hou | Houston Methodist Hospital | Houston Methodist Hospital | S. Wesley Long et al |
| EPI_ISL_544511 | USA/TX-HMH-5082/2020 | 2020-06-24 | North America/USA/Texas/Greater Hou | Houston Methodist Hospital | Houston Methodist Hospital | S. Wesley Long et al |
| EPI_ISL_544514 | USA/TX-HMH-5086/2020 | 2020-06-21 | North America/USA/Texas/Greater Hou | Houston Methodist Hospital | Houston Methodist Hospital | S. Wesley Long et al |
| EPI_ISL_544516 | USA/TX-HMH-5088/2020 | 2020-06-24 | North America/USA/Texas/Greater Hou | Houston Methodist Hospital | Houston Methodist Hospital | S. Wesley Long et al |

| | | | | | | |
|---|---|---|---|---|---|---|
| EPI_ISL_544518 | USA/TX-HMH-5091/2020 | 2020-06-24 | North America/USA/Texas/Greater Hou | Houston Methodist Hospital | Houston Methodist Hospital | S. Wesley Long et al |
| EPI_ISL_544529 | USA/TX-HMH-5106/2020 | 2020-06-22 | North America/USA/Texas/Greater Hou | Houston Methodist Hospital | Houston Methodist Hospital | S. Wesley Long et al |
| EPI_ISL_544531 | USA/TX-HMH-5108/2020 | 2020-06-29 | North America/USA/Texas/Greater Hou | Houston Methodist Hospital | Houston Methodist Hospital | S. Wesley Long et al |
| EPI_ISL_544532 | USA/TX-HMH-511/2020 | 2020-04-02 | North America/USA/Texas/Greater Hou | Houston Methodist Hospital | Houston Methodist Hospital | S. Wesley Long et al |
| EPI_ISL_544536 | USA/TX-HMH-5118/2020 | 2020-06-29 | North America/USA/Texas/Greater Hou | Houston Methodist Hospital | Houston Methodist Hospital | S. Wesley Long et al |
| EPI_ISL_544547 | USA/TX-HMH-5131/2020 | 2020-06-21 | North America/USA/Texas/Greater Hou | Houston Methodist Hospital | Houston Methodist Hospital | S. Wesley Long et al |
| EPI_ISL_544556 | USA/TX-HMH-5145/2020 | 2020-06-22 | North America/USA/Texas/Greater Hou | Houston Methodist Hospital | Houston Methodist Hospital | S. Wesley Long et al |
| EPI_ISL_544566 | USA/TX-HMH-5157/2020 | 2020-06-21 | North America/USA/Texas/Greater Hou | Houston Methodist Hospital | Houston Methodist Hospital | S. Wesley Long et al |
| EPI_ISL_544578 | USA/TX-HMH-5179/2020 | 2020-06-21 | North America/USA/Texas/Greater Hou | Houston Methodist Hospital | Houston Methodist Hospital | S. Wesley Long et al |
| EPI_ISL_544587 | USA/TX-HMH-5192/2020 | 2020-06-20 | North America/USA/Texas/Greater Hou | Houston Methodist Hospital | Houston Methodist Hospital | S. Wesley Long et al |
| EPI_ISL_544590 | USA/TX-HMH-5201/2020 | 2020-06-26 | North America/USA/Texas/Greater Hou | Houston Methodist Hospital | Houston Methodist Hospital | S. Wesley Long et al |
| EPI_ISL_544592 | USA/TX-HMH-5206/2020 | 2020-06-21 | North America/USA/Texas/Greater Hou | Houston Methodist Hospital | Houston Methodist Hospital | S. Wesley Long et al |
| EPI_ISL_544596 | USA/TX-HMH-5210/2020 | 2020-06-22 | North America/USA/Texas/Greater Hou | Houston Methodist Hospital | Houston Methodist Hospital | S. Wesley Long et al |

| | | | | | | |
|---|---|---|---|---|---|---|
| EPI_ISL_544601 | USA/TX-HMH-5217/2020 | 2020-06-22 | North America/USA/Texas/Greater Hou | Houston Methodist Hospital | Houston Methodist Hospital | S. Wesley Long et al |
| EPI_ISL_544605 | USA/TX-HMH-5220/2020 | 2020-06-26 | North America/USA/Texas/Greater Hou | Houston Methodist Hospital | Houston Methodist Hospital | S. Wesley Long et al |
| EPI_ISL_544617 | USA/TX-HMH-524/2020 | 2020-04-04 | North America/USA/Texas/Greater Hou | Houston Methodist Hospital | Houston Methodist Hospital | S. Wesley Long et al |
| EPI_ISL_544622 | USA/TX-HMH-5245/2020 | 2020-06-22 | North America/USA/Texas/Greater Hou | Houston Methodist Hospital | Houston Methodist Hospital | S. Wesley Long et al |
| EPI_ISL_544626 | USA/TX-HMH-5249/2020 | 2020-06-25 | North America/USA/Texas/Greater Hou | Houston Methodist Hospital | Houston Methodist Hospital | S. Wesley Long et al |
| EPI_ISL_544629 | USA/TX-HMH-5254/2020 | 2020-06-21 | North America/USA/Texas/Greater Hou | Houston Methodist Hospital | Houston Methodist Hospital | S. Wesley Long et al |
| EPI_ISL_544632 | USA/TX-HMH-526/2020 | 2020-04-05 | North America/USA/Texas/Greater Hou | Houston Methodist Hospital | Houston Methodist Hospital | S. Wesley Long et al |
| EPI_ISL_544633 | USA/TX-HMH-5260/2020 | 2020-06-26 | North America/USA/Texas/Greater Hou | Houston Methodist Hospital | Houston Methodist Hospital | S. Wesley Long et al |
| EPI_ISL_544635 | USA/TX-HMH-5263/2020 | 2020-06-22 | North America/USA/Texas/Greater Hou | Houston Methodist Hospital | Houston Methodist Hospital | S. Wesley Long et al |
| EPI_ISL_544638 | USA/TX-HMH-5267/2020 | 2020-06-21 | North America/USA/Texas/Greater Hou | Houston Methodist Hospital | Houston Methodist Hospital | S. Wesley Long et al |
| EPI_ISL_544640 | USA/TX-HMH-5269/2020 | 2020-06-21 | North America/USA/Texas/Greater Hou | Houston Methodist Hospital | Houston Methodist Hospital | S. Wesley Long et al |
| EPI_ISL_544647 | USA/TX-HMH-5278/2020 | 2020-06-23 | North America/USA/Texas/Greater Hou | Houston Methodist Hospital | Houston Methodist Hospital | S. Wesley Long et al |
| EPI_ISL_544648 | USA/TX-HMH-5279/2020 | 2020-06-21 | North America/USA/Texas/Greater Hou | Houston Methodist Hospital | Houston Methodist Hospital | S. Wesley Long et al |

| | | | | | | |
|---|---|---|---|---|---|---|
| EPI_ISL_544666 | USA/TX-HMH-5305/2020 | 2020-06-18 | North America/USA/Texas/Greater Hou | Houston Methodist Hospital | Houston Methodist Hospital | S. Wesley Long et al |
| EPI_ISL_544669 | USA/TX-HMH-5308/2020 | 2020-06-18 | North America/USA/Texas/Greater Hou | Houston Methodist Hospital | Houston Methodist Hospital | S. Wesley Long et al |
| EPI_ISL_544679 | USA/TX-HMH-5322/2020 | 2020-06-25 | North America/USA/Texas/Greater Hou | Houston Methodist Hospital | Houston Methodist Hospital | S. Wesley Long et al |
| EPI_ISL_544692 | USA/TX-HMH-5336/2020 | 2020-06-25 | North America/USA/Texas/Greater Hou | Houston Methodist Hospital | Houston Methodist Hospital | S. Wesley Long et al |
| EPI_ISL_544694 | USA/TX-HMH-5338/2020 | 2020-06-24 | North America/USA/Texas/Greater Hou | Houston Methodist Hospital | Houston Methodist Hospital | S. Wesley Long et al |
| EPI_ISL_544697 | USA/TX-HMH-5341/2020 | 2020-06-24 | North America/USA/Texas/Greater Hou | Houston Methodist Hospital | Houston Methodist Hospital | S. Wesley Long et al |
| EPI_ISL_544710 | USA/TX-HMH-5355/2020 | 2020-06-25 | North America/USA/Texas/Greater Hou | Houston Methodist Hospital | Houston Methodist Hospital | S. Wesley Long et al |
| EPI_ISL_544716 | USA/TX-HMH-5362/2020 | 2020-06-26 | North America/USA/Texas/Greater Hou | Houston Methodist Hospital | Houston Methodist Hospital | S. Wesley Long et al |
| EPI_ISL_544721 | USA/TX-HMH-5367/2020 | 2020-06-29 | North America/USA/Texas/Greater Hou | Houston Methodist Hospital | Houston Methodist Hospital | S. Wesley Long et al |
| EPI_ISL_544731 | USA/TX-HMH-5379/2020 | 2020-06-26 | North America/USA/Texas/Greater Hou | Houston Methodist Hospital | Houston Methodist Hospital | S. Wesley Long et al |
| EPI_ISL_544742 | USA/TX-HMH-5396/2020 | 2020-06-25 | North America/USA/Texas/Greater Hou | Houston Methodist Hospital | Houston Methodist Hospital | S. Wesley Long et al |
| EPI_ISL_544752 | USA/TX-HMH-5407/2020 | 2020-06-29 | North America/USA/Texas/Greater Hou | Houston Methodist Hospital | Houston Methodist Hospital | S. Wesley Long et al |
| EPI_ISL_544759 | USA/TX-HMH-5415/2020 | 2020-06-29 | North America/USA/Texas/Greater Hou | Houston Methodist Hospital | Houston Methodist Hospital | S. Wesley Long et al |

| | | | | | | |
|---|---|---|---|---|---|---|
| EPI_ISL_544780 | USA/TX-HMH-5443/2020 | 2020-06-24 | North America/USA/Texas/Greater Hou | Houston Methodist Hospital | Houston Methodist Hospital | S. Wesley Long et al |
| EPI_ISL_544785 | USA/TX-HMH-5450/2020 | 2020-06-26 | North America/USA/Texas/Greater Hou | Houston Methodist Hospital | Houston Methodist Hospital | S. Wesley Long et al |
| EPI_ISL_544791 | USA/TX-HMH-5458/2020 | 2020-06-26 | North America/USA/Texas/Greater Hou | Houston Methodist Hospital | Houston Methodist Hospital | S. Wesley Long et al |
| EPI_ISL_544819 | USA/TX-HMH-5496/2020 | 2020-06-24 | North America/USA/Texas/Greater Hou | Houston Methodist Hospital | Houston Methodist Hospital | S. Wesley Long et al |
| EPI_ISL_544828 | USA/TX-HMH-5504/2020 | 2020-06-26 | North America/USA/Texas/Greater Hou | Houston Methodist Hospital | Houston Methodist Hospital | S. Wesley Long et al |
| EPI_ISL_544829 | USA/TX-HMH-5506/2020 | 2020-06-29 | North America/USA/Texas/Greater Hou | Houston Methodist Hospital | Houston Methodist Hospital | S. Wesley Long et al |
| EPI_ISL_544837 | USA/TX-HMH-5514/2020 | 2020-06-25 | North America/USA/Texas/Greater Hou | Houston Methodist Hospital | Houston Methodist Hospital | S. Wesley Long et al |
| EPI_ISL_544843 | USA/TX-HMH-5523/2020 | 2020-06-29 | North America/USA/Texas/Greater Hou | Houston Methodist Hospital | Houston Methodist Hospital | S. Wesley Long et al |
| EPI_ISL_544853 | USA/TX-HMH-5546/2020 | 2020-06-25 | North America/USA/Texas/Greater Hou | Houston Methodist Hospital | Houston Methodist Hospital | S. Wesley Long et al |
| EPI_ISL_544863 | USA/TX-HMH-5562/2020 | 2020-06-26 | North America/USA/Texas/Greater Hou | Houston Methodist Hospital | Houston Methodist Hospital | S. Wesley Long et al |
| EPI_ISL_544873 | USA/TX-HMH-5575/2020 | 2020-06-22 | North America/USA/Texas/Greater Hou | Houston Methodist Hospital | Houston Methodist Hospital | S. Wesley Long et al |
| EPI_ISL_544876 | USA/TX-HMH-5579/2020 | 2020-06-25 | North America/USA/Texas/Greater Hou | Houston Methodist Hospital | Houston Methodist Hospital | S. Wesley Long et al |
| EPI_ISL_544878 | USA/TX-HMH-5581/2020 | 2020-06-26 | North America/USA/Texas/Greater Hou | Houston Methodist Hospital | Houston Methodist Hospital | S. Wesley Long et al |

| | | | | | | |
|---|---|---|---|---|---|---|
| EPI_ISL_544886 | USA/TX-HMH-5592/2020 | 2020-06-23 | North America/USA/Texas/Greater Hou | Houston Methodist Hospital | Houston Methodist Hospital | S. Wesley Long et al |
| EPI_ISL_544895 | USA/TX-HMH-5604/2020 | 2020-06-23 | North America/USA/Texas/Greater Hou | Houston Methodist Hospital | Houston Methodist Hospital | S. Wesley Long et al |
| EPI_ISL_544913 | USA/TX-HMH-5627/2020 | 2020-06-24 | North America/USA/Texas/Greater Hou | Houston Methodist Hospital | Houston Methodist Hospital | S. Wesley Long et al |
| EPI_ISL_544917 | USA/TX-HMH-5630/2020 | 2020-06-19 | North America/USA/Texas/Greater Hou | Houston Methodist Hospital | Houston Methodist Hospital | S. Wesley Long et al |
| EPI_ISL_544931 | USA/TX-HMH-5646/2020 | 2020-06-20 | North America/USA/Texas/Greater Hou | Houston Methodist Hospital | Houston Methodist Hospital | S. Wesley Long et al |
| EPI_ISL_544947 | USA/TX-HMH-5665/2020 | 2020-06-23 | North America/USA/Texas/Greater Hou | Houston Methodist Hospital | Houston Methodist Hospital | S. Wesley Long et al |
| EPI_ISL_544950 | USA/TX-HMH-567/2020 | 2020-04-06 | North America/USA/Texas/Greater Hou | Houston Methodist Hospital | Houston Methodist Hospital | S. Wesley Long et al |
| EPI_ISL_544953 | USA/TX-HMH-5673/2020 | 2020-06-24 | North America/USA/Texas/Greater Hou | Houston Methodist Hospital | Houston Methodist Hospital | S. Wesley Long et al |
| EPI_ISL_545054 | USA/TX-HMH-1989/2020 | 2020-05-19 | North America/USA/Texas/Greater Hou | Houston Methodist Hospital | Houston Methodist Hospital | S. Wesley Long et al |
| EPI_ISL_545057 | USA/TX-HMH-1993/2020 | 2020-05-17 | North America/USA/Texas/Greater Hou | Houston Methodist Hospital | Houston Methodist Hospital | S. Wesley Long et al |
| EPI_ISL_545064 | USA/TX-HMH-2005/2020 | 2020-05-18 | North America/USA/Texas/Greater Hou | Houston Methodist Hospital | Houston Methodist Hospital | S. Wesley Long et al |
| EPI_ISL_545071 | USA/TX-HMH-2021/2020 | 2020-05-12 | North America/USA/Texas/Greater Hou | Houston Methodist Hospital | Houston Methodist Hospital | S. Wesley Long et al |
| EPI_ISL_545077 | USA/TX-HMH-2035/2020 | 2020-05-09 | North America/USA/Texas/Greater Hou | Houston Methodist Hospital | Houston Methodist Hospital | S. Wesley Long et al |

| | | | | | | |
|---|---|---|---|---|---|---|
| EPI_ISL_545079 | USA/TX-HMH-2040/2020 | 2020-05-07 | North America/USA/Texas/Greater Hou | Houston Methodist Hospital | Houston Methodist Hospital | S. Wesley Long et al |
| EPI_ISL_545088 | USA/TX-HMH-2051/2020 | 2020-05-08 | North America/USA/Texas/Greater Hou | Houston Methodist Hospital | Houston Methodist Hospital | S. Wesley Long et al |
| EPI_ISL_545091 | USA/TX-HMH-2055/2020 | 2020-05-12 | North America/USA/Texas/Greater Hou | Houston Methodist Hospital | Houston Methodist Hospital | S. Wesley Long et al |
| EPI_ISL_545092 | USA/TX-HMH-2056/2020 | 2020-04-30 | North America/USA/Texas/Greater Hou | Houston Methodist Hospital | Houston Methodist Hospital | S. Wesley Long et al |
| EPI_ISL_545095 | USA/TX-HMH-2061/2020 | 2020-05-12 | North America/USA/Texas/Greater Hou | Houston Methodist Hospital | Houston Methodist Hospital | S. Wesley Long et al |
| EPI_ISL_545105 | USA/TX-HMH-2076/2020 | 2020-05-11 | North America/USA/Texas/Greater Hou | Houston Methodist Hospital | Houston Methodist Hospital | S. Wesley Long et al |
| EPI_ISL_545107 | USA/TX-HMH-2078/2020 | 2020-05-09 | North America/USA/Texas/Greater Hou | Houston Methodist Hospital | Houston Methodist Hospital | S. Wesley Long et al |
| EPI_ISL_545111 | USA/TX-HMH-2082/2020 | 2020-05-11 | North America/USA/Texas/Greater Hou | Houston Methodist Hospital | Houston Methodist Hospital | S. Wesley Long et al |
| EPI_ISL_545118 | USA/TX-HMH-2095/2020 | 2020-05-23 | North America/USA/Texas/Greater Hou | Houston Methodist Hospital | Houston Methodist Hospital | S. Wesley Long et al |
| EPI_ISL_545119 | USA/TX-HMH-2098/2020 | 2020-05-23 | North America/USA/Texas/Greater Hou | Houston Methodist Hospital | Houston Methodist Hospital | S. Wesley Long et al |
| EPI_ISL_545142 | USA/TX-HMH-2142/2020 | 2020-05-19 | North America/USA/Texas/Greater Hou | Houston Methodist Hospital | Houston Methodist Hospital | S. Wesley Long et al |
| EPI_ISL_545146 | USA/TX-HMH-2150/2020 | 2020-05-22 | North America/USA/Texas/Greater Hou | Houston Methodist Hospital | Houston Methodist Hospital | S. Wesley Long et al |
| EPI_ISL_545158 | USA/TX-HMH-2169/2020 | 2020-05-22 | North America/USA/Texas/Greater Hou | Houston Methodist Hospital | Houston Methodist Hospital | S. Wesley Long et al |

| | | | | | | |
|---|---|---|---|---|---|---|
| EPI_ISL_545165 | USA/TX-HMH-2178/2020 | 2020-05-21 | North America/USA/Texas/Greater Hou | Houston Methodist Hospital | Houston Methodist Hospital | S. Wesley Long et al |
| EPI_ISL_545171 | USA/TX-HMH-2189/2020 | 2020-05-23 | North America/USA/Texas/Greater Hou | Houston Methodist Hospital | Houston Methodist Hospital | S. Wesley Long et al |
| EPI_ISL_545178 | USA/TX-HMH-2201/2020 | 2020-05-23 | North America/USA/Texas/Greater Hou | Houston Methodist Hospital | Houston Methodist Hospital | S. Wesley Long et al |
| EPI_ISL_545194 | USA/TX-HMH-2234/2020 | 2020-05-15 | North America/USA/Texas/Greater Hou | Houston Methodist Hospital | Houston Methodist Hospital | S. Wesley Long et al |
| EPI_ISL_545208 | USA/TX-HMH-2255/2020 | 2020-05-23 | North America/USA/Texas/Greater Hou | Houston Methodist Hospital | Houston Methodist Hospital | S. Wesley Long et al |
| EPI_ISL_545211 | USA/TX-HMH-2261/2020 | 2020-05-24 | North America/USA/Texas/Greater Hou | Houston Methodist Hospital | Houston Methodist Hospital | S. Wesley Long et al |
| EPI_ISL_545218 | USA/TX-HMH-2276/2020 | 2020-05-11 | North America/USA/Texas/Greater Hou | Houston Methodist Hospital | Houston Methodist Hospital | S. Wesley Long et al |
| EPI_ISL_545219 | USA/TX-HMH-2277/2020 | 2020-05-22 | North America/USA/Texas/Greater Hou | Houston Methodist Hospital | Houston Methodist Hospital | S. Wesley Long et al |
| EPI_ISL_545220 | USA/TX-HMH-2278/2020 | 2020-05-22 | North America/USA/Texas/Greater Hou | Houston Methodist Hospital | Houston Methodist Hospital | S. Wesley Long et al |
| EPI_ISL_545230 | USA/TX-HMH-2295/2020 | 2020-05-10 | North America/USA/Texas/Greater Hou | Houston Methodist Hospital | Houston Methodist Hospital | S. Wesley Long et al |
| EPI_ISL_545232 | USA/TX-HMH-2297/2020 | 2020-05-06 | North America/USA/Texas/Greater Hou | Houston Methodist Hospital | Houston Methodist Hospital | S. Wesley Long et al |
| EPI_ISL_545239 | USA/TX-HMH-2310/2020 | 2020-04-21 | North America/USA/Texas/Greater Hou | Houston Methodist Hospital | Houston Methodist Hospital | S. Wesley Long et al |
| EPI_ISL_545243 | USA/TX-HMH-2323/2020 | 2020-05-29 | North America/USA/Texas/Greater Hou | Houston Methodist Hospital | Houston Methodist Hospital | S. Wesley Long et al |

| | | | | | | |
|---|---|---|---|---|---|---|
| EPI_ISL_545246 | USA/TX-HMH-2327/2020 | 2020-05-31 | North America/USA/Texas/Greater Hou | Houston Methodist Hospital | Houston Methodist Hospital | S. Wesley Long et al |
| EPI_ISL_545247 | USA/TX-HMH-2328/2020 | 2020-05-31 | North America/USA/Texas/Greater Hou | Houston Methodist Hospital | Houston Methodist Hospital | S. Wesley Long et al |
| EPI_ISL_545260 | USA/TX-HMH-2352/2020 | 2020-05-27 | North America/USA/Texas/Greater Hou | Houston Methodist Hospital | Houston Methodist Hospital | S. Wesley Long et al |
| EPI_ISL_545261 | USA/TX-HMH-2356/2020 | 2020-05-27 | North America/USA/Texas/Greater Hou | Houston Methodist Hospital | Houston Methodist Hospital | S. Wesley Long et al |
| EPI_ISL_545262 | USA/TX-HMH-2357/2020 | 2020-05-27 | North America/USA/Texas/Greater Hou | Houston Methodist Hospital | Houston Methodist Hospital | S. Wesley Long et al |
| EPI_ISL_545264 | USA/TX-HMH-2361/2020 | 2020-05-31 | North America/USA/Texas/Greater Hou | Houston Methodist Hospital | Houston Methodist Hospital | S. Wesley Long et al |
| EPI_ISL_545269 | USA/TX-HMH-2371/2020 | 2020-05-29 | North America/USA/Texas/Greater Hou | Houston Methodist Hospital | Houston Methodist Hospital | S. Wesley Long et al |
| EPI_ISL_545272 | USA/TX-HMH-2377/2020 | 2020-05-28 | North America/USA/Texas/Greater Hou | Houston Methodist Hospital | Houston Methodist Hospital | S. Wesley Long et al |
| EPI_ISL_545280 | USA/TX-HMH-2388/2020 | 2020-04-24 | North America/USA/Texas/Greater Hou | Houston Methodist Hospital | Houston Methodist Hospital | S. Wesley Long et al |
| EPI_ISL_545282 | USA/TX-HMH-2390/2020 | 2020-05-29 | North America/USA/Texas/Greater Hou | Houston Methodist Hospital | Houston Methodist Hospital | S. Wesley Long et al |
| EPI_ISL_545285 | USA/TX-HMH-2397/2020 | 2020-05-25 | North America/USA/Texas/Greater Hou | Houston Methodist Hospital | Houston Methodist Hospital | S. Wesley Long et al |
| EPI_ISL_545291 | USA/TX-HMH-2404/2020 | 2020-05-25 | North America/USA/Texas/Greater Hou | Houston Methodist Hospital | Houston Methodist Hospital | S. Wesley Long et al |
| EPI_ISL_545292 | USA/TX-HMH-2407/2020 | 2020-05-25 | North America/USA/Texas/Greater Hou | Houston Methodist Hospital | Houston Methodist Hospital | S. Wesley Long et al |

| | | | | | | |
|---|---|---|---|---|---|---|
| EPI_ISL_545297 | USA/TX-HMH-2415/2020 | 2020-05-11 | North America/USA/Texas/Greater Hou | Houston Methodist Hospital | Houston Methodist Hospital | S. Wesley Long et al |
| EPI_ISL_545300 | USA/TX-HMH-2419/2020 | 2020-05-27 | North America/USA/Texas/Greater Hou | Houston Methodist Hospital | Houston Methodist Hospital | S. Wesley Long et al |
| EPI_ISL_545301 | USA/TX-HMH-2420/2020 | 2020-05-27 | North America/USA/Texas/Greater Hou | Houston Methodist Hospital | Houston Methodist Hospital | S. Wesley Long et al |
| EPI_ISL_545314 | USA/TX-HMH-2447/2020 | 2020-05-30 | North America/USA/Texas/Greater Hou | Houston Methodist Hospital | Houston Methodist Hospital | S. Wesley Long et al |
| EPI_ISL_545319 | USA/TX-HMH-2456/2020 | 2020-05-27 | North America/USA/Texas/Greater Hou | Houston Methodist Hospital | Houston Methodist Hospital | S. Wesley Long et al |
| EPI_ISL_545325 | USA/TX-HMH-2465/2020 | 2020-05-29 | North America/USA/Texas/Greater Hou | Houston Methodist Hospital | Houston Methodist Hospital | S. Wesley Long et al |
| EPI_ISL_545327 | USA/TX-HMH-2470/2020 | 2020-05-28 | North America/USA/Texas/Greater Hou | Houston Methodist Hospital | Houston Methodist Hospital | S. Wesley Long et al |
| EPI_ISL_545332 | USA/TX-HMH-2475/2020 | 2020-05-28 | North America/USA/Texas/Greater Hou | Houston Methodist Hospital | Houston Methodist Hospital | S. Wesley Long et al |
| EPI_ISL_545333 | USA/TX-HMH-2478/2020 | 2020-05-27 | North America/USA/Texas/Greater Hou | Houston Methodist Hospital | Houston Methodist Hospital | S. Wesley Long et al |
| EPI_ISL_545334 | USA/TX-HMH-2482/2020 | 2020-05-28 | North America/USA/Texas/Greater Hou | Houston Methodist Hospital | Houston Methodist Hospital | S. Wesley Long et al |
| EPI_ISL_545335 | USA/TX-HMH-2483/2020 | 2020-05-28 | North America/USA/Texas/Greater Hou | Houston Methodist Hospital | Houston Methodist Hospital | S. Wesley Long et al |
| EPI_ISL_545343 | USA/TX-HMH-2505/2020 | 2020-05-15 | North America/USA/Texas/Greater Hou | Houston Methodist Hospital | Houston Methodist Hospital | S. Wesley Long et al |
| EPI_ISL_545345 | USA/TX-HMH-2507/2020 | 2020-05-19 | North America/USA/Texas/Greater Hou | Houston Methodist Hospital | Houston Methodist Hospital | S. Wesley Long et al |

| | | | | | | |
|---|---|---|---|---|---|---|
| EPI_ISL_545351 | USA/TX-HMH-2515/2020 | 2020-06-04 | North America/USA/Texas/Greater Hou | Houston Methodist Hospital | Houston Methodist Hospital | S. Wesley Long et al |
| EPI_ISL_545358 | USA/TX-HMH-2522/2020 | 2020-06-02 | North America/USA/Texas/Greater Hou | Houston Methodist Hospital | Houston Methodist Hospital | S. Wesley Long et al |
| EPI_ISL_545371 | USA/TX-HMH-2540/2020 | 2020-06-01 | North America/USA/Texas/Greater Hou | Houston Methodist Hospital | Houston Methodist Hospital | S. Wesley Long et al |
| EPI_ISL_545376 | USA/TX-HMH-2546/2020 | 2020-06-01 | North America/USA/Texas/Greater Hou | Houston Methodist Hospital | Houston Methodist Hospital | S. Wesley Long et al |
| EPI_ISL_545378 | USA/TX-HMH-2549/2020 | 2020-06-01 | North America/USA/Texas/Greater Hou | Houston Methodist Hospital | Houston Methodist Hospital | S. Wesley Long et al |
| EPI_ISL_545379 | USA/TX-HMH-2551/2020 | 2020-06-01 | North America/USA/Texas/Greater Hou | Houston Methodist Hospital | Houston Methodist Hospital | S. Wesley Long et al |
| EPI_ISL_545380 | USA/TX-HMH-2552/2020 | 2020-06-01 | North America/USA/Texas/Greater Hou | Houston Methodist Hospital | Houston Methodist Hospital | S. Wesley Long et al |
| EPI_ISL_545402 | USA/TX-HMH-2579/2020 | 2020-06-06 | North America/USA/Texas/Greater Hou | Houston Methodist Hospital | Houston Methodist Hospital | S. Wesley Long et al |
| EPI_ISL_545406 | USA/TX-HMH-2583/2020 | 2020-06-04 | North America/USA/Texas/Greater Hou | Houston Methodist Hospital | Houston Methodist Hospital | S. Wesley Long et al |
| EPI_ISL_545414 | USA/TX-HMH-2591/2020 | 2020-06-02 | North America/USA/Texas/Greater Hou | Houston Methodist Hospital | Houston Methodist Hospital | S. Wesley Long et al |
| EPI_ISL_545416 | USA/TX-HMH-2595/2020 | 2020-06-07 | North America/USA/Texas/Greater Hou | Houston Methodist Hospital | Houston Methodist Hospital | S. Wesley Long et al |
| EPI_ISL_545417 | USA/TX-HMH-2597/2020 | 2020-06-04 | North America/USA/Texas/Greater Hou | Houston Methodist Hospital | Houston Methodist Hospital | S. Wesley Long et al |
| EPI_ISL_545428 | USA/TX-HMH-2614/2020 | 2020-06-04 | North America/USA/Texas/Greater Hou | Houston Methodist Hospital | Houston Methodist Hospital | S. Wesley Long et al |

| | | | | | | |
|---|---|---|---|---|---|---|
| EPI_ISL_545433 | USA/TX-HMH-2623/2020 | 2020-06-05 | North America/USA/Texas/Greater Hou | Houston Methodist Hospital | Houston Methodist Hospital | S. Wesley Long et al |
| EPI_ISL_545435 | USA/TX-HMH-2626/2020 | 2020-06-05 | North America/USA/Texas/Greater Hou | Houston Methodist Hospital | Houston Methodist Hospital | S. Wesley Long et al |
| EPI_ISL_545438 | USA/TX-HMH-2631/2020 | 2020-06-04 | North America/USA/Texas/Greater Hou | Houston Methodist Hospital | Houston Methodist Hospital | S. Wesley Long et al |
| EPI_ISL_545443 | USA/TX-HMH-2636/2020 | 2020-06-05 | North America/USA/Texas/Greater Hou | Houston Methodist Hospital | Houston Methodist Hospital | S. Wesley Long et al |
| EPI_ISL_545456 | USA/TX-HMH-2658/2020 | 2020-06-05 | North America/USA/Texas/Greater Hou | Houston Methodist Hospital | Houston Methodist Hospital | S. Wesley Long et al |
| EPI_ISL_545464 | USA/TX-HMH-2668/2020 | 2020-06-06 | North America/USA/Texas/Greater Hou | Houston Methodist Hospital | Houston Methodist Hospital | S. Wesley Long et al |
| EPI_ISL_545468 | USA/TX-HMH-2674/2020 | 2020-06-03 | North America/USA/Texas/Greater Hou | Houston Methodist Hospital | Houston Methodist Hospital | S. Wesley Long et al |
| EPI_ISL_545474 | USA/TX-HMH-2685/2020 | 2020-06-05 | North America/USA/Texas/Greater Hou | Houston Methodist Hospital | Houston Methodist Hospital | S. Wesley Long et al |
| EPI_ISL_545477 | USA/TX-HMH-2688/2020 | 2020-06-05 | North America/USA/Texas/Greater Hou | Houston Methodist Hospital | Houston Methodist Hospital | S. Wesley Long et al |
| EPI_ISL_545480 | USA/TX-HMH-2692/2020 | 2020-06-05 | North America/USA/Texas/Greater Hou | Houston Methodist Hospital | Houston Methodist Hospital | S. Wesley Long et al |
| EPI_ISL_545484 | USA/TX-HMH-2697/2020 | 2020-06-07 | North America/USA/Texas/Greater Hou | Houston Methodist Hospital | Houston Methodist Hospital | S. Wesley Long et al |
| EPI_ISL_545502 | USA/TX-HMH-2719/2020 | 2020-06-06 | North America/USA/Texas/Greater Hou | Houston Methodist Hospital | Houston Methodist Hospital | S. Wesley Long et al |
| EPI_ISL_545505 | USA/TX-HMH-2722/2020 | 2020-06-05 | North America/USA/Texas/Greater Hou | Houston Methodist Hospital | Houston Methodist Hospital | S. Wesley Long et al |

| | | | | | | |
|---|---|---|---|---|---|---|
| EPI_ISL_545509 | USA/TX-HMH-2727/2020 | 2020-06-07 | North America/USA/Texas/Greater Hou | Houston Methodist Hospital | Houston Methodist Hospital | S. Wesley Long et al |
| EPI_ISL_545531 | USA/TX-HMH-2756/2020 | 2020-06-04 | North America/USA/Texas/Greater Hou | Houston Methodist Hospital | Houston Methodist Hospital | S. Wesley Long et al |
| EPI_ISL_545539 | USA/TX-HMH-2765/2020 | 2020-06-05 | North America/USA/Texas/Greater Hou | Houston Methodist Hospital | Houston Methodist Hospital | S. Wesley Long et al |
| EPI_ISL_545542 | USA/TX-HMH-2769/2020 | 2020-06-04 | North America/USA/Texas/Greater Hou | Houston Methodist Hospital | Houston Methodist Hospital | S. Wesley Long et al |
| EPI_ISL_545543 | USA/TX-HMH-2771/2020 | 2020-06-03 | North America/USA/Texas/Greater Hou | Houston Methodist Hospital | Houston Methodist Hospital | S. Wesley Long et al |
| EPI_ISL_545544 | USA/TX-HMH-2772/2020 | 2020-06-08 | North America/USA/Texas/Greater Hou | Houston Methodist Hospital | Houston Methodist Hospital | S. Wesley Long et al |
| EPI_ISL_545545 | USA/TX-HMH-2775/2020 | 2020-06-03 | North America/USA/Texas/Greater Hou | Houston Methodist Hospital | Houston Methodist Hospital | S. Wesley Long et al |
| EPI_ISL_545550 | USA/TX-HMH-2780/2020 | 2020-06-03 | North America/USA/Texas/Greater Hou | Houston Methodist Hospital | Houston Methodist Hospital | S. Wesley Long et al |
| EPI_ISL_545551 | USA/TX-HMH-2782/2020 | 2020-06-08 | North America/USA/Texas/Greater Hou | Houston Methodist Hospital | Houston Methodist Hospital | S. Wesley Long et al |
| EPI_ISL_545570 | USA/TX-HMH-2808/2020 | 2020-06-08 | North America/USA/Texas/Greater Hou | Houston Methodist Hospital | Houston Methodist Hospital | S. Wesley Long et al |
| EPI_ISL_545589 | USA/TX-HMH-7697/2020 | 2020-06-26 | North America/USA/Texas/Greater Hou | Houston Methodist Hospital | Houston Methodist Hospital | S. Wesley Long et al |
| EPI_ISL_545607 | USA/TX-HMH-7733/2020 | 2020-07-02 | North America/USA/Texas/Greater Hou | Houston Methodist Hospital | Houston Methodist Hospital | S. Wesley Long et al |
| EPI_ISL_545612 | USA/TX-HMH-7742/2020 | 2020-06-30 | North America/USA/Texas/Greater Hou | Houston Methodist Hospital | Houston Methodist Hospital | S. Wesley Long et al |

| | | | | | | |
|---|---|---|---|---|---|---|
| EPI_ISL_545614 | USA/TX-HMH-7747/2020 | 2020-06-26 | North America/USA/Texas/Greater Hou | Houston Methodist Hospital | Houston Methodist Hospital | S. Wesley Long et al |
| EPI_ISL_545619 | USA/TX-HMH-7757/2020 | 2020-06-26 | North America/USA/Texas/Greater Hou | Houston Methodist Hospital | Houston Methodist Hospital | S. Wesley Long et al |
| EPI_ISL_545622 | USA/TX-HMH-7763/2020 | 2020-06-30 | North America/USA/Texas/Greater Hou | Houston Methodist Hospital | Houston Methodist Hospital | S. Wesley Long et al |
| EPI_ISL_545629 | USA/TX-HMH-7773/2020 | 2020-06-30 | North America/USA/Texas/Greater Hou | Houston Methodist Hospital | Houston Methodist Hospital | S. Wesley Long et al |
| EPI_ISL_545633 | USA/TX-HMH-7780/2020 | 2020-06-26 | North America/USA/Texas/Greater Hou | Houston Methodist Hospital | Houston Methodist Hospital | S. Wesley Long et al |
| EPI_ISL_545634 | USA/TX-HMH-7782/2020 | 2020-06-30 | North America/USA/Texas/Greater Hou | Houston Methodist Hospital | Houston Methodist Hospital | S. Wesley Long et al |
| EPI_ISL_545648 | USA/TX-HMH-782/2020 | 2020-04-09 | North America/USA/Texas/Greater Hou | Houston Methodist Hospital | Houston Methodist Hospital | S. Wesley Long et al |
| EPI_ISL_545663 | USA/TX-HMH-7845/2020 | 2020-06-30 | North America/USA/Texas/Greater Hou | Houston Methodist Hospital | Houston Methodist Hospital | S. Wesley Long et al |
| EPI_ISL_545670 | USA/TX-HMH-7864/2020 | 2020-06-30 | North America/USA/Texas/Greater Hou | Houston Methodist Hospital | Houston Methodist Hospital | S. Wesley Long et al |
| EPI_ISL_545673 | USA/TX-HMH-7870/2020 | 2020-07-01 | North America/USA/Texas/Greater Hou | Houston Methodist Hospital | Houston Methodist Hospital | S. Wesley Long et al |
| EPI_ISL_545675 | USA/TX-HMH-7873/2020 | 2020-06-30 | North America/USA/Texas/Greater Hou | Houston Methodist Hospital | Houston Methodist Hospital | S. Wesley Long et al |
| EPI_ISL_545682 | USA/TX-HMH-7886/2020 | 2020-07-01 | North America/USA/Texas/Greater Hou | Houston Methodist Hospital | Houston Methodist Hospital | S. Wesley Long et al |
| EPI_ISL_545690 | USA/TX-HMH-7899/2020 | 2020-07-07 | North America/USA/Texas/Greater Hou | Houston Methodist Hospital | Houston Methodist Hospital | S. Wesley Long et al |

| | | | | | | |
|---|---|---|---|---|---|---|
| EPI_ISL_545700 | USA/TX-HMH-7910/2020 | 2020-06-30 | North America/USA/Texas/Greater Hou | Houston Methodist Hospital | Houston Methodist Hospital | S. Wesley Long et al |
| EPI_ISL_545709 | USA/TX-HMH-7923/2020 | 2020-06-30 | North America/USA/Texas/Greater Hou | Houston Methodist Hospital | Houston Methodist Hospital | S. Wesley Long et al |
| EPI_ISL_545713 | USA/TX-HMH-7929/2020 | 2020-06-29 | North America/USA/Texas/Greater Hou | Houston Methodist Hospital | Houston Methodist Hospital | S. Wesley Long et al |
| EPI_ISL_545717 | USA/TX-HMH-7932/2020 | 2020-06-30 | North America/USA/Texas/Greater Hou | Houston Methodist Hospital | Houston Methodist Hospital | S. Wesley Long et al |
| EPI_ISL_545719 | USA/TX-HMH-7935/2020 | 2020-07-06 | North America/USA/Texas/Greater Hou | Houston Methodist Hospital | Houston Methodist Hospital | S. Wesley Long et al |
| EPI_ISL_545737 | USA/TX-HMH-7955/2020 | 2020-06-29 | North America/USA/Texas/Greater Hou | Houston Methodist Hospital | Houston Methodist Hospital | S. Wesley Long et al |
| EPI_ISL_545738 | USA/TX-HMH-7956/2020 | 2020-06-26 | North America/USA/Texas/Greater Hou | Houston Methodist Hospital | Houston Methodist Hospital | S. Wesley Long et al |
| EPI_ISL_545739 | USA/TX-HMH-7957/2020 | 2020-06-29 | North America/USA/Texas/Greater Hou | Houston Methodist Hospital | Houston Methodist Hospital | S. Wesley Long et al |
| EPI_ISL_545740 | USA/TX-HMH-7958/2020 | 2020-07-01 | North America/USA/Texas/Greater Hou | Houston Methodist Hospital | Houston Methodist Hospital | S. Wesley Long et al |
| EPI_ISL_545741 | USA/TX-HMH-7959/2020 | 2020-07-07 | North America/USA/Texas/Greater Hou | Houston Methodist Hospital | Houston Methodist Hospital | S. Wesley Long et al |
| EPI_ISL_545748 | USA/TX-HMH-7966/2020 | 2020-06-29 | North America/USA/Texas/Greater Hou | Houston Methodist Hospital | Houston Methodist Hospital | S. Wesley Long et al |
| EPI_ISL_545753 | USA/TX-HMH-7971/2020 | 2020-06-30 | North America/USA/Texas/Greater Hou | Houston Methodist Hospital | Houston Methodist Hospital | S. Wesley Long et al |
| EPI_ISL_545754 | USA/TX-HMH-7972/2020 | 2020-07-06 | North America/USA/Texas/Greater Hou | Houston Methodist Hospital | Houston Methodist Hospital | S. Wesley Long et al |

| | | | | | | |
|---|---|---|---|---|---|---|
| EPI_ISL_545763 | USA/TX-HMH-7982/2020 | 2020-07-01 | North America/USA/Texas/Greater Hou | Houston Methodist Hospital | Houston Methodist Hospital | S. Wesley Long et al |
| EPI_ISL_545765 | USA/TX-HMH-7984/2020 | 2020-07-07 | North America/USA/Texas/Greater Hou | Houston Methodist Hospital | Houston Methodist Hospital | S. Wesley Long et al |
| EPI_ISL_545771 | USA/TX-HMH-7990/2020 | 2020-07-01 | North America/USA/Texas/Greater Hou | Houston Methodist Hospital | Houston Methodist Hospital | S. Wesley Long et al |
| EPI_ISL_545774 | USA/TX-HMH-7993/2020 | 2020-07-06 | North America/USA/Texas/Greater Hou | Houston Methodist Hospital | Houston Methodist Hospital | S. Wesley Long et al |
| EPI_ISL_545777 | USA/TX-HMH-7997/2020 | 2020-07-01 | North America/USA/Texas/Greater Hou | Houston Methodist Hospital | Houston Methodist Hospital | S. Wesley Long et al |
| EPI_ISL_545780 | USA/TX-HMH-800/2020 | 2020-04-08 | North America/USA/Texas/Greater Hou | Houston Methodist Hospital | Houston Methodist Hospital | S. Wesley Long et al |
| EPI_ISL_545783 | USA/TX-HMH-8005/2020 | 2020-07-01 | North America/USA/Texas/Greater Hou | Houston Methodist Hospital | Houston Methodist Hospital | S. Wesley Long et al |
| EPI_ISL_545786 | USA/TX-HMH-8008/2020 | 2020-07-07 | North America/USA/Texas/Greater Hou | Houston Methodist Hospital | Houston Methodist Hospital | S. Wesley Long et al |
| EPI_ISL_545794 | USA/TX-HMH-8017/2020 | 2020-07-06 | North America/USA/Texas/Greater Hou | Houston Methodist Hospital | Houston Methodist Hospital | S. Wesley Long et al |
| EPI_ISL_545799 | USA/TX-HMH-8027/2020 | 2020-07-01 | North America/USA/Texas/Greater Hou | Houston Methodist Hospital | Houston Methodist Hospital | S. Wesley Long et al |
| EPI_ISL_545818 | USA/TX-HMH-8050/2020 | 2020-07-01 | North America/USA/Texas/Greater Hou | Houston Methodist Hospital | Houston Methodist Hospital | S. Wesley Long et al |
| EPI_ISL_545822 | USA/TX-HMH-8055/2020 | 2020-07-01 | North America/USA/Texas/Greater Hou | Houston Methodist Hospital | Houston Methodist Hospital | S. Wesley Long et al |
| EPI_ISL_545833 | USA/TX-HMH-8068/2020 | 2020-07-07 | North America/USA/Texas/Greater Hou | Houston Methodist Hospital | Houston Methodist Hospital | S. Wesley Long et al |

| | | | | | | |
|---|---|---|---|---|---|---|
| EPI_ISL_545834 | USA/TX-HMH-8069/2020 | 2020-07-01 | North America/USA/Texas/Greater Hou | Houston Methodist Hospital | Houston Methodist Hospital | S. Wesley Long et al |
| EPI_ISL_545835 | USA/TX-HMH-807/2020 | 2020-04-10 | North America/USA/Texas/Greater Hou | Houston Methodist Hospital | Houston Methodist Hospital | S. Wesley Long et al |
| EPI_ISL_545839 | USA/TX-HMH-8073/2020 | 2020-07-01 | North America/USA/Texas/Greater Hou | Houston Methodist Hospital | Houston Methodist Hospital | S. Wesley Long et al |
| EPI_ISL_545848 | USA/TX-HMH-814/2020 | 2020-04-10 | North America/USA/Texas/Greater Hou | Houston Methodist Hospital | Houston Methodist Hospital | S. Wesley Long et al |
| EPI_ISL_545851 | USA/TX-HMH-817/2020 | 2020-04-08 | North America/USA/Texas/Greater Hou | Houston Methodist Hospital | Houston Methodist Hospital | S. Wesley Long et al |
| EPI_ISL_545861 | USA/TX-HMH-838/2020 | 2020-04-10 | North America/USA/Texas/Greater Hou | Houston Methodist Hospital | Houston Methodist Hospital | S. Wesley Long et al |
| EPI_ISL_545869 | USA/TX-HMH-849/2020 | 2020-04-03 | North America/USA/Texas/Greater Hou | Houston Methodist Hospital | Houston Methodist Hospital | S. Wesley Long et al |
| EPI_ISL_545873 | USA/TX-HMH-853/2020 | 2020-04-03 | North America/USA/Texas/Greater Hou | Houston Methodist Hospital | Houston Methodist Hospital | S. Wesley Long et al |
| EPI_ISL_545887 | USA/TX-HMH-870/2020 | 2020-04-08 | North America/USA/Texas/Greater Hou | Houston Methodist Hospital | Houston Methodist Hospital | S. Wesley Long et al |
| EPI_ISL_545900 | USA/TX-HMH-887/2020 | 2020-04-15 | North America/USA/Texas/Greater Hou | Houston Methodist Hospital | Houston Methodist Hospital | S. Wesley Long et al |
| EPI_ISL_545902 | USA/TX-HMH-889/2020 | 2020-04-16 | North America/USA/Texas/Greater Hou | Houston Methodist Hospital | Houston Methodist Hospital | S. Wesley Long et al |
| EPI_ISL_545910 | USA/TX-HMH-908/2020 | 2020-04-14 | North America/USA/Texas/Greater Hou | Houston Methodist Hospital | Houston Methodist Hospital | S. Wesley Long et al |
| EPI_ISL_545917 | USA/TX-HMH-916/2020 | 2020-04-17 | North America/USA/Texas/Greater Hou | Houston Methodist Hospital | Houston Methodist Hospital | S. Wesley Long et al |

| | | | | | | |
|---|---|---|---|---|---|---|
| EPI_ISL_545938 | USA/TX-HMH-969/2020 | 2020-04-11 | North America/USA/Texas/Greater Hou | Houston Methodist Hospital | Houston Methodist Hospital | S. Wesley Long et al |
| EPI_ISL_545939 | USA/TX-HMH-970/2020 | 2020-04-10 | North America/USA/Texas/Greater Hou | Houston Methodist Hospital | Houston Methodist Hospital | S. Wesley Long et al |
| EPI_ISL_545944 | USA/TX-HMH-979/2020 | 2020-04-09 | North America/USA/Texas/Greater Hou | Houston Methodist Hospital | Houston Methodist Hospital | S. Wesley Long et al |
| EPI_ISL_545947 | USA/TX-HMH-985/2020 | 2020-04-10 | North America/USA/Texas/Greater Hou | Houston Methodist Hospital | Houston Methodist Hospital | S. Wesley Long et al |
| EPI_ISL_545955 | Peru/LIM-INS-001/2020 | 2020-04-01 | South America/Peru/Lima/ | Laboratorio de Infecciones Respiratorias Agudas. Centro Nacional de Salud Publica,Instituto Nacional de Salud | Laboratorio de Infecciones Respiratorias Agudas. Centro Nacional de Salud Publica,Instituto Nacional de Salud | Juscamayta et al |
| EPI_ISL_545956 | CzechRepublic/NRL_7942/2020 | 2020-07-28 | Europe/Czech Republic/Olomouc Region/ | The National Institute of Public Health | State Veterinary Institute Prague | Nagy et al |
| EPI_ISL_545962 | USA/TX-HMH-6389/2020 | 2020-07-05 | North America/USA/Texas/Greater Hou | Houston Methodist Hospital | Houston Methodist Hospital | S. Wesley Long et al |
| EPI_ISL_545966 | USA/TX-HMH-6393/2020 | 2020-07-05 | North America/USA/Texas/Greater Hou | Houston Methodist Hospital | Houston Methodist Hospital | S. Wesley Long et al |
| EPI_ISL_545972 | USA/TX-HMH-6400/2020 | 2020-07-06 | North America/USA/Texas/Greater Hou | Houston Methodist Hospital | Houston Methodist Hospital | S. Wesley Long et al |
| EPI_ISL_545978 | USA/TX-HMH-6407/2020 | 2020-07-05 | North America/USA/Texas/Greater Hou | Houston Methodist Hospital | Houston Methodist Hospital | S. Wesley Long et al |
| EPI_ISL_545979 | USA/TX-HMH-6408/2020 | 2020-07-06 | North America/USA/Texas/Greater Hou | Houston Methodist Hospital | Houston Methodist Hospital | S. Wesley Long et al |
| EPI_ISL_545982 | USA/TX-HMH-6412/2020 | 2020-07-05 | North America/USA/Texas/Greater Hou | Houston Methodist Hospital | Houston Methodist Hospital | S. Wesley Long et al |

| | | | | | | |
|---|---|---|---|---|---|---|
| EPI_ISL_545989 | USA/TX-HMH-6426/2020 | 2020-07-05 | North America/USA/Texas/Greater Hou | Houston Methodist Hospital | Houston Methodist Hospital | S. Wesley Long et al |
| EPI_ISL_545994 | USA/TX-HMH-6435/2020 | 2020-07-04 | North America/USA/Texas/Greater Hou | Houston Methodist Hospital | Houston Methodist Hospital | S. Wesley Long et al |
| EPI_ISL_545997 | USA/TX-HMH-6440/2020 | 2020-06-23 | North America/USA/Texas/Greater Hou | Houston Methodist Hospital | Houston Methodist Hospital | S. Wesley Long et al |
| EPI_ISL_545999 | USA/TX-HMH-6442/2020 | 2020-07-05 | North America/USA/Texas/Greater Hou | Houston Methodist Hospital | Houston Methodist Hospital | S. Wesley Long et al |
| EPI_ISL_546002 | USA/TX-HMH-6447/2020 | 2020-06-27 | North America/USA/Texas/Greater Hou | Houston Methodist Hospital | Houston Methodist Hospital | S. Wesley Long et al |
| EPI_ISL_546006 | USA/TX-HMH-6453/2020 | 2020-06-27 | North America/USA/Texas/Greater Hou | Houston Methodist Hospital | Houston Methodist Hospital | S. Wesley Long et al |
| EPI_ISL_546019 | USA/TX-HMH-647/2020 | 2020-04-10 | North America/USA/Texas/Greater Hou | Houston Methodist Hospital | Houston Methodist Hospital | S. Wesley Long et al |
| EPI_ISL_546043 | USA/TX-HMH-6504/2020 | 2020-06-27 | North America/USA/Texas/Greater Hou | Houston Methodist Hospital | Houston Methodist Hospital | S. Wesley Long et al |
| EPI_ISL_546045 | USA/TX-HMH-6506/2020 | 2020-06-27 | North America/USA/Texas/Greater Hou | Houston Methodist Hospital | Houston Methodist Hospital | S. Wesley Long et al |
| EPI_ISL_546046 | USA/TX-HMH-6507/2020 | 2020-06-27 | North America/USA/Texas/Greater Hou | Houston Methodist Hospital | Houston Methodist Hospital | S. Wesley Long et al |
| EPI_ISL_546066 | USA/TX-HMH-6540/2020 | 2020-06-26 | North America/USA/Texas/Greater Hou | Houston Methodist Hospital | Houston Methodist Hospital | S. Wesley Long et al |
| EPI_ISL_546070 | USA/TX-HMH-6544/2020 | 2020-06-18 | North America/USA/Texas/Greater Hou | Houston Methodist Hospital | Houston Methodist Hospital | S. Wesley Long et al |
| EPI_ISL_546071 | USA/TX-HMH-6545/2020 | 2020-06-24 | North America/USA/Texas/Greater Hou | Houston Methodist Hospital | Houston Methodist Hospital | S. Wesley Long et al |

| | | | | | | |
|---|---|---|---|---|---|---|
| EPI_ISL_546074 | USA/TX-HMH-6548/2020 | 2020-06-24 | North America/USA/Texas/Greater Hou | Houston Methodist Hospital | Houston Methodist Hospital | S. Wesley Long et al |
| EPI_ISL_546081 | USA/TX-HMH-6556/2020 | 2020-06-23 | North America/USA/Texas/Greater Hou | Houston Methodist Hospital | Houston Methodist Hospital | S. Wesley Long et al |
| EPI_ISL_546098 | USA/TX-HMH-6577/2020 | 2020-06-24 | North America/USA/Texas/Greater Hou | Houston Methodist Hospital | Houston Methodist Hospital | S. Wesley Long et al |
| EPI_ISL_546107 | USA/TX-HMH-6589/2020 | 2020-06-24 | North America/USA/Texas/Greater Hou | Houston Methodist Hospital | Houston Methodist Hospital | S. Wesley Long et al |
| EPI_ISL_546110 | USA/TX-HMH-6592/2020 | 2020-06-24 | North America/USA/Texas/Greater Hou | Houston Methodist Hospital | Houston Methodist Hospital | S. Wesley Long et al |
| EPI_ISL_546111 | USA/TX-HMH-6593/2020 | 2020-06-24 | North America/USA/Texas/Greater Hou | Houston Methodist Hospital | Houston Methodist Hospital | S. Wesley Long et al |
| EPI_ISL_546112 | USA/TX-HMH-6594/2020 | 2020-06-24 | North America/USA/Texas/Greater Hou | Houston Methodist Hospital | Houston Methodist Hospital | S. Wesley Long et al |
| EPI_ISL_546122 | USA/TX-HMH-6619/2020 | 2020-06-24 | North America/USA/Texas/Greater Hou | Houston Methodist Hospital | Houston Methodist Hospital | S. Wesley Long et al |
| EPI_ISL_546127 | USA/TX-HMH-6625/2020 | 2020-06-24 | North America/USA/Texas/Greater Hou | Houston Methodist Hospital | Houston Methodist Hospital | S. Wesley Long et al |
| EPI_ISL_546135 | USA/TX-HMH-6637/2020 | 2020-06-24 | North America/USA/Texas/Greater Hou | Houston Methodist Hospital | Houston Methodist Hospital | S. Wesley Long et al |
| EPI_ISL_546136 | USA/TX-HMH-6638/2020 | 2020-06-24 | North America/USA/Texas/Greater Hou | Houston Methodist Hospital | Houston Methodist Hospital | S. Wesley Long et al |
| EPI_ISL_546143 | USA/TX-HMH-6647/2020 | 2020-06-24 | North America/USA/Texas/Greater Hou | Houston Methodist Hospital | Houston Methodist Hospital | S. Wesley Long et al |
| EPI_ISL_546154 | USA/TX-HMH-6661/2020 | 2020-06-24 | North America/USA/Texas/Greater Hou | Houston Methodist Hospital | Houston Methodist Hospital | S. Wesley Long et al |

| | | | | | | |
|---|---|---|---|---|---|---|
| EPI_ISL_546158 | USA/TX-HMH-6667/2020 | 2020-06-24 | North America/USA/Texas/Greater Hou | Houston Methodist Hospital | Houston Methodist Hospital | S. Wesley Long et al |
| EPI_ISL_546168 | USA/TX-HMH-6680/2020 | 2020-06-24 | North America/USA/Texas/Greater Hou | Houston Methodist Hospital | Houston Methodist Hospital | S. Wesley Long et al |
| EPI_ISL_546173 | USA/TX-HMH-6686/2020 | 2020-06-24 | North America/USA/Texas/Greater Hou | Houston Methodist Hospital | Houston Methodist Hospital | S. Wesley Long et al |
| EPI_ISL_546178 | USA/TX-HMH-6699/2020 | 2020-06-24 | North America/USA/Texas/Greater Hou | Houston Methodist Hospital | Houston Methodist Hospital | S. Wesley Long et al |
| EPI_ISL_546182 | USA/TX-HMH-6703/2020 | 2020-06-24 | North America/USA/Texas/Greater Hou | Houston Methodist Hospital | Houston Methodist Hospital | S. Wesley Long et al |
| EPI_ISL_546198 | USA/TX-HMH-673/2020 | 2020-04-11 | North America/USA/Texas/Greater Hou | Houston Methodist Hospital | Houston Methodist Hospital | S. Wesley Long et al |
| EPI_ISL_546199 | USA/TX-HMH-6730/2020 | 2020-06-25 | North America/USA/Texas/Greater Hou | Houston Methodist Hospital | Houston Methodist Hospital | S. Wesley Long et al |
| EPI_ISL_546201 | USA/TX-HMH-6733/2020 | 2020-06-25 | North America/USA/Texas/Greater Hou | Houston Methodist Hospital | Houston Methodist Hospital | S. Wesley Long et al |
| EPI_ISL_546202 | USA/TX-HMH-6735/2020 | 2020-06-24 | North America/USA/Texas/Greater Hou | Houston Methodist Hospital | Houston Methodist Hospital | S. Wesley Long et al |
| EPI_ISL_546203 | USA/TX-HMH-6736/2020 | 2020-06-25 | North America/USA/Texas/Greater Hou | Houston Methodist Hospital | Houston Methodist Hospital | S. Wesley Long et al |
| EPI_ISL_546209 | USA/TX-HMH-6742/2020 | 2020-06-25 | North America/USA/Texas/Greater Hou | Houston Methodist Hospital | Houston Methodist Hospital | S. Wesley Long et al |
| EPI_ISL_546211 | USA/TX-HMH-6746/2020 | 2020-06-24 | North America/USA/Texas/Greater Hou | Houston Methodist Hospital | Houston Methodist Hospital | S. Wesley Long et al |
| EPI_ISL_546225 | USA/TX-HMH-6762/2020 | 2020-06-24 | North America/USA/Texas/Greater Hou | Houston Methodist Hospital | Houston Methodist Hospital | S. Wesley Long et al |

| | | | | | | |
|---|---|---|---|---|---|---|
| EPI_ISL_546227 | USA/TX-HMH-6764/2020 | 2020-06-25 | North America/USA/Texas/Greater Hou | Houston Methodist Hospital | Houston Methodist Hospital | S. Wesley Long et al |
| EPI_ISL_546234 | USA/TX-HMH-6771/2020 | 2020-06-25 | North America/USA/Texas/Greater Hou | Houston Methodist Hospital | Houston Methodist Hospital | S. Wesley Long et al |
| EPI_ISL_546245 | USA/TX-HMH-6791/2020 | 2020-06-25 | North America/USA/Texas/Greater Hou | Houston Methodist Hospital | Houston Methodist Hospital | S. Wesley Long et al |
| EPI_ISL_546247 | USA/TX-HMH-6793/2020 | 2020-06-24 | North America/USA/Texas/Greater Hou | Houston Methodist Hospital | Houston Methodist Hospital | S. Wesley Long et al |
| EPI_ISL_546260 | USA/TX-HMH-6810/2020 | 2020-06-24 | North America/USA/Texas/Greater Hou | Houston Methodist Hospital | Houston Methodist Hospital | S. Wesley Long et al |
| EPI_ISL_546263 | USA/TX-HMH-6816/2020 | 2020-06-25 | North America/USA/Texas/Greater Hou | Houston Methodist Hospital | Houston Methodist Hospital | S. Wesley Long et al |
| EPI_ISL_546264 | USA/TX-HMH-6819/2020 | 2020-06-25 | North America/USA/Texas/Greater Hou | Houston Methodist Hospital | Houston Methodist Hospital | S. Wesley Long et al |
| EPI_ISL_546265 | USA/TX-HMH-6820/2020 | 2020-06-25 | North America/USA/Texas/Greater Hou | Houston Methodist Hospital | Houston Methodist Hospital | S. Wesley Long et al |
| EPI_ISL_546271 | USA/TX-HMH-6828/2020 | 2020-06-24 | North America/USA/Texas/Greater Hou | Houston Methodist Hospital | Houston Methodist Hospital | S. Wesley Long et al |
| EPI_ISL_546297 | USA/TX-HMH-6861/2020 | 2020-06-24 | North America/USA/Texas/Greater Hou | Houston Methodist Hospital | Houston Methodist Hospital | S. Wesley Long et al |
| EPI_ISL_546303 | USA/TX-HMH-6868/2020 | 2020-06-25 | North America/USA/Texas/Greater Hou | Houston Methodist Hospital | Houston Methodist Hospital | S. Wesley Long et al |
| EPI_ISL_546311 | USA/TX-HMH-6876/2020 | 2020-06-25 | North America/USA/Texas/Greater Hou | Houston Methodist Hospital | Houston Methodist Hospital | S. Wesley Long et al |
| EPI_ISL_546314 | USA/TX-HMH-6879/2020 | 2020-06-25 | North America/USA/Texas/Greater Hou | Houston Methodist Hospital | Houston Methodist Hospital | S. Wesley Long et al |

| | | | | | | |
|---|---|---|---|---|---|---|
| EPI_ISL_546338 | USA/TX-HMH-6903/2020 | 2020-06-25 | North America/USA/Texas/Greater Hou | Houston Methodist Hospital | Houston Methodist Hospital | S. Wesley Long et al |
| EPI_ISL_546339 | USA/TX-HMH-6905/2020 | 2020-06-25 | North America/USA/Texas/Greater Hou | Houston Methodist Hospital | Houston Methodist Hospital | S. Wesley Long et al |
| EPI_ISL_546344 | USA/TX-HMH-6911/2020 | 2020-06-25 | North America/USA/Texas/Greater Hou | Houston Methodist Hospital | Houston Methodist Hospital | S. Wesley Long et al |
| EPI_ISL_546354 | USA/TX-HMH-6922/2020 | 2020-06-25 | North America/USA/Texas/Greater Hou | Houston Methodist Hospital | Houston Methodist Hospital | S. Wesley Long et al |
| EPI_ISL_546367 | USA/TX-HMH-6939/2020 | 2020-06-27 | North America/USA/Texas/Greater Hou | Houston Methodist Hospital | Houston Methodist Hospital | S. Wesley Long et al |
| EPI_ISL_546372 | USA/TX-HMH-6943/2020 | 2020-06-28 | North America/USA/Texas/Greater Hou | Houston Methodist Hospital | Houston Methodist Hospital | S. Wesley Long et al |
| EPI_ISL_546380 | USA/TX-HMH-6952/2020 | 2020-06-28 | North America/USA/Texas/Greater Hou | Houston Methodist Hospital | Houston Methodist Hospital | S. Wesley Long et al |
| EPI_ISL_546387 | USA/TX-HMH-6960/2020 | 2020-06-28 | North America/USA/Texas/Greater Hou | Houston Methodist Hospital | Houston Methodist Hospital | S. Wesley Long et al |
| EPI_ISL_546394 | USA/TX-HMH-6967/2020 | 2020-06-29 | North America/USA/Texas/Greater Hou | Houston Methodist Hospital | Houston Methodist Hospital | S. Wesley Long et al |
| EPI_ISL_546422 | USA/TX-HMH-7001/2020 | 2020-06-28 | North America/USA/Texas/Greater Hou | Houston Methodist Hospital | Houston Methodist Hospital | S. Wesley Long et al |
| EPI_ISL_546424 | USA/TX-HMH-7003/2020 | 2020-06-28 | North America/USA/Texas/Greater Hou | Houston Methodist Hospital | Houston Methodist Hospital | S. Wesley Long et al |
| EPI_ISL_546429 | USA/TX-HMH-7010/2020 | 2020-06-28 | North America/USA/Texas/Greater Hou | Houston Methodist Hospital | Houston Methodist Hospital | S. Wesley Long et al |
| EPI_ISL_546445 | USA/TX-HMH-7026/2020 | 2020-06-28 | North America/USA/Texas/Greater Hou | Houston Methodist Hospital | Houston Methodist Hospital | S. Wesley Long et al |

| | | | | | | |
|---|---|---|---|---|---|---|
| EPI_ISL_546458 | USA/TX-HMH-7042/2020 | 2020-06-28 | North America/USA/Texas/Greater Hou | Houston Methodist Hospital | Houston Methodist Hospital | S. Wesley Long et al |
| EPI_ISL_546462 | USA/TX-HMH-705/2020 | 2020-04-18 | North America/USA/Texas/Greater Hou | Houston Methodist Hospital | Houston Methodist Hospital | S. Wesley Long et al |
| EPI_ISL_546463 | USA/TX-HMH-7050/2020 | 2020-06-28 | North America/USA/Texas/Greater Hou | Houston Methodist Hospital | Houston Methodist Hospital | S. Wesley Long et al |
| EPI_ISL_546477 | USA/TX-HMH-7068/2020 | 2020-06-28 | North America/USA/Texas/Greater Hou | Houston Methodist Hospital | Houston Methodist Hospital | S. Wesley Long et al |
| EPI_ISL_546493 | USA/TX-HMH-7087/2020 | 2020-06-29 | North America/USA/Texas/Greater Hou | Houston Methodist Hospital | Houston Methodist Hospital | S. Wesley Long et al |
| EPI_ISL_546494 | USA/TX-HMH-7089/2020 | 2020-06-28 | North America/USA/Texas/Greater Hou | Houston Methodist Hospital | Houston Methodist Hospital | S. Wesley Long et al |
| EPI_ISL_546496 | USA/TX-HMH-7091/2020 | 2020-06-28 | North America/USA/Texas/Greater Hou | Houston Methodist Hospital | Houston Methodist Hospital | S. Wesley Long et al |
| EPI_ISL_546497 | USA/TX-HMH-7092/2020 | 2020-06-28 | North America/USA/Texas/Greater Hou | Houston Methodist Hospital | Houston Methodist Hospital | S. Wesley Long et al |
| EPI_ISL_546501 | USA/TX-HMH-7098/2020 | 2020-06-28 | North America/USA/Texas/Greater Hou | Houston Methodist Hospital | Houston Methodist Hospital | S. Wesley Long et al |
| EPI_ISL_546505 | USA/TX-HMH-7103/2020 | 2020-06-28 | North America/USA/Texas/Greater Hou | Houston Methodist Hospital | Houston Methodist Hospital | S. Wesley Long et al |
| EPI_ISL_546518 | USA/TX-HMH-7117/2020 | 2020-06-28 | North America/USA/Texas/Greater Hou | Houston Methodist Hospital | Houston Methodist Hospital | S. Wesley Long et al |
| EPI_ISL_546527 | USA/TX-HMH-7126/2020 | 2020-06-29 | North America/USA/Texas/Greater Hou | Houston Methodist Hospital | Houston Methodist Hospital | S. Wesley Long et al |
| EPI_ISL_546533 | USA/TX-HMH-7133/2020 | 2020-06-29 | North America/USA/Texas/Greater Hou | Houston Methodist Hospital | Houston Methodist Hospital | S. Wesley Long et al |

| | | | | | | |
|---|---|---|---|---|---|---|
| EPI_ISL_546534 | USA/TX-HMH-7134/2020 | 2020-06-29 | North America/USA/Texas/Greater Hou | Houston Methodist Hospital | Houston Methodist Hospital | S. Wesley Long et al |
| EPI_ISL_546538 | USA/TX-HMH-7139/2020 | 2020-06-29 | North America/USA/Texas/Greater Hou | Houston Methodist Hospital | Houston Methodist Hospital | S. Wesley Long et al |
| EPI_ISL_546541 | USA/TX-HMH-7143/2020 | 2020-06-29 | North America/USA/Texas/Greater Hou | Houston Methodist Hospital | Houston Methodist Hospital | S. Wesley Long et al |
| EPI_ISL_546546 | USA/TX-HMH-7148/2020 | 2020-06-29 | North America/USA/Texas/Greater Hou | Houston Methodist Hospital | Houston Methodist Hospital | S. Wesley Long et al |
| EPI_ISL_546558 | USA/TX-HMH-7162/2020 | 2020-06-29 | North America/USA/Texas/Greater Hou | Houston Methodist Hospital | Houston Methodist Hospital | S. Wesley Long et al |
| EPI_ISL_546583 | USA/TX-HMH-7187/2020 | 2020-06-30 | North America/USA/Texas/Greater Hou | Houston Methodist Hospital | Houston Methodist Hospital | S. Wesley Long et al |
| EPI_ISL_546594 | USA/TX-HMH-7203/2020 | 2020-06-29 | North America/USA/Texas/Greater Hou | Houston Methodist Hospital | Houston Methodist Hospital | S. Wesley Long et al |
| EPI_ISL_546602 | USA/TX-HMH-7213/2020 | 2020-06-29 | North America/USA/Texas/Greater Hou | Houston Methodist Hospital | Houston Methodist Hospital | S. Wesley Long et al |
| EPI_ISL_546603 | USA/TX-HMH-7214/2020 | 2020-06-29 | North America/USA/Texas/Greater Hou | Houston Methodist Hospital | Houston Methodist Hospital | S. Wesley Long et al |
| EPI_ISL_546604 | USA/TX-HMH-7215/2020 | 2020-06-29 | North America/USA/Texas/Greater Hou | Houston Methodist Hospital | Houston Methodist Hospital | S. Wesley Long et al |
| EPI_ISL_546607 | USA/TX-HMH-7218/2020 | 2020-06-29 | North America/USA/Texas/Greater Hou | Houston Methodist Hospital | Houston Methodist Hospital | S. Wesley Long et al |
| EPI_ISL_546625 | USA/TX-HMH-7248/2020 | 2020-06-28 | North America/USA/Texas/Greater Hou | Houston Methodist Hospital | Houston Methodist Hospital | S. Wesley Long et al |
| EPI_ISL_546626 | USA/TX-HMH-7250/2020 | 2020-06-29 | North America/USA/Texas/Greater Hou | Houston Methodist Hospital | Houston Methodist Hospital | S. Wesley Long et al |

| | | | | | | |
|---|---|---|---|---|---|---|
| EPI_ISL_546627 | USA/TX-HMH-7252/2020 | 2020-06-29 | North America/USA/Texas/Greater Hou | Houston Methodist Hospital | Houston Methodist Hospital | S. Wesley Long et al |
| EPI_ISL_546628 | USA/TX-HMH-7254/2020 | 2020-06-29 | North America/USA/Texas/Greater Hou | Houston Methodist Hospital | Houston Methodist Hospital | S. Wesley Long et al |
| EPI_ISL_546631 | USA/TX-HMH-7259/2020 | 2020-06-29 | North America/USA/Texas/Greater Hou | Houston Methodist Hospital | Houston Methodist Hospital | S. Wesley Long et al |
| EPI_ISL_546634 | USA/TX-HMH-7263/2020 | 2020-06-29 | North America/USA/Texas/Greater Hou | Houston Methodist Hospital | Houston Methodist Hospital | S. Wesley Long et al |
| EPI_ISL_546637 | USA/TX-HMH-7267/2020 | 2020-06-29 | North America/USA/Texas/Greater Hou | Houston Methodist Hospital | Houston Methodist Hospital | S. Wesley Long et al |
| EPI_ISL_546638 | USA/TX-HMH-7269/2020 | 2020-06-29 | North America/USA/Texas/Greater Hou | Houston Methodist Hospital | Houston Methodist Hospital | S. Wesley Long et al |
| EPI_ISL_546643 | USA/TX-HMH-7274/2020 | 2020-06-29 | North America/USA/Texas/Greater Hou | Houston Methodist Hospital | Houston Methodist Hospital | S. Wesley Long et al |
| EPI_ISL_546645 | USA/TX-HMH-7277/2020 | 2020-06-29 | North America/USA/Texas/Greater Hou | Houston Methodist Hospital | Houston Methodist Hospital | S. Wesley Long et al |
| EPI_ISL_546649 | USA/TX-HMH-7290/2020 | 2020-06-29 | North America/USA/Texas/Greater Hou | Houston Methodist Hospital | Houston Methodist Hospital | S. Wesley Long et al |
| EPI_ISL_546657 | USA/TX-HMH-7300/2020 | 2020-06-29 | North America/USA/Texas/Greater Hou | Houston Methodist Hospital | Houston Methodist Hospital | S. Wesley Long et al |
| EPI_ISL_546658 | USA/TX-HMH-7301/2020 | 2020-06-29 | North America/USA/Texas/Greater Hou | Houston Methodist Hospital | Houston Methodist Hospital | S. Wesley Long et al |
| EPI_ISL_546660 | USA/TX-HMH-7303/2020 | 2020-06-29 | North America/USA/Texas/Greater Hou | Houston Methodist Hospital | Houston Methodist Hospital | S. Wesley Long et al |
| EPI_ISL_546668 | USA/TX-HMH-7311/2020 | 2020-06-29 | North America/USA/Texas/Greater Hou | Houston Methodist Hospital | Houston Methodist Hospital | S. Wesley Long et al |

| | | | | | | |
|---|---|---|---|---|---|---|
| EPI_ISL_546676 | USA/TX-HMH-7327/2020 | 2020-06-29 | North America/USA/Texas/Greater Hou | Houston Methodist Hospital | Houston Methodist Hospital | S. Wesley Long et al |
| EPI_ISL_546680 | USA/TX-HMH-7331/2020 | 2020-06-29 | North America/USA/Texas/Greater Hou | Houston Methodist Hospital | Houston Methodist Hospital | S. Wesley Long et al |
| EPI_ISL_546687 | USA/TX-HMH-7340/2020 | 2020-06-29 | North America/USA/Texas/Greater Hou | Houston Methodist Hospital | Houston Methodist Hospital | S. Wesley Long et al |
| EPI_ISL_546689 | USA/TX-HMH-7342/2020 | 2020-06-29 | North America/USA/Texas/Greater Hou | Houston Methodist Hospital | Houston Methodist Hospital | S. Wesley Long et al |
| EPI_ISL_546694 | USA/TX-HMH-7349/2020 | 2020-06-29 | North America/USA/Texas/Greater Hou | Houston Methodist Hospital | Houston Methodist Hospital | S. Wesley Long et al |
| EPI_ISL_546699 | USA/TX-HMH-7354/2020 | 2020-06-29 | North America/USA/Texas/Greater Hou | Houston Methodist Hospital | Houston Methodist Hospital | S. Wesley Long et al |
| EPI_ISL_546700 | USA/TX-HMH-7355/2020 | 2020-06-29 | North America/USA/Texas/Greater Hou | Houston Methodist Hospital | Houston Methodist Hospital | S. Wesley Long et al |
| EPI_ISL_546716 | USA/TX-HMH-7376/2020 | 2020-06-29 | North America/USA/Texas/Greater Hou | Houston Methodist Hospital | Houston Methodist Hospital | S. Wesley Long et al |
| EPI_ISL_546717 | USA/TX-HMH-7378/2020 | 2020-06-29 | North America/USA/Texas/Greater Hou | Houston Methodist Hospital | Houston Methodist Hospital | S. Wesley Long et al |
| EPI_ISL_546719 | USA/TX-HMH-738/2020 | 2020-04-04 | North America/USA/Texas/Greater Hou | Houston Methodist Hospital | Houston Methodist Hospital | S. Wesley Long et al |
| EPI_ISL_546733 | USA/TX-HMH-7395/2020 | 2020-06-29 | North America/USA/Texas/Greater Hou | Houston Methodist Hospital | Houston Methodist Hospital | S. Wesley Long et al |
| EPI_ISL_546742 | USA/TX-HMH-7403/2020 | 2020-06-29 | North America/USA/Texas/Greater Hou | Houston Methodist Hospital | Houston Methodist Hospital | S. Wesley Long et al |
| EPI_ISL_546745 | USA/TX-HMH-7408/2020 | 2020-06-30 | North America/USA/Texas/Greater Hou | Houston Methodist Hospital | Houston Methodist Hospital | S. Wesley Long et al |

| | | | | | | |
|---|---|---|---|---|---|---|
| EPI_ISL_546751 | USA/TX-HMH-7414/2020 | 2020-06-30 | North America/USA/Texas/Greater Hou | Houston Methodist Hospital | Houston Methodist Hospital | S. Wesley Long et al |
| EPI_ISL_546756 | USA/TX-HMH-7423/2020 | 2020-06-30 | North America/USA/Texas/Greater Hou | Houston Methodist Hospital | Houston Methodist Hospital | S. Wesley Long et al |
| EPI_ISL_546761 | USA/TX-HMH-7429/2020 | 2020-06-30 | North America/USA/Texas/Greater Hou | Houston Methodist Hospital | Houston Methodist Hospital | S. Wesley Long et al |
| EPI_ISL_546764 | USA/TX-HMH-7432/2020 | 2020-06-30 | North America/USA/Texas/Greater Hou | Houston Methodist Hospital | Houston Methodist Hospital | S. Wesley Long et al |
| EPI_ISL_546765 | USA/TX-HMH-7433/2020 | 2020-06-30 | North America/USA/Texas/Greater Hou | Houston Methodist Hospital | Houston Methodist Hospital | S. Wesley Long et al |
| EPI_ISL_546767 | USA/TX-HMH-7436/2020 | 2020-06-30 | North America/USA/Texas/Greater Hou | Houston Methodist Hospital | Houston Methodist Hospital | S. Wesley Long et al |
| EPI_ISL_546770 | USA/TX-HMH-7440/2020 | 2020-06-26 | North America/USA/Texas/Greater Hou | Houston Methodist Hospital | Houston Methodist Hospital | S. Wesley Long et al |
| EPI_ISL_546784 | USA/TX-HMH-7466/2020 | 2020-06-26 | North America/USA/Texas/Greater Hou | Houston Methodist Hospital | Houston Methodist Hospital | S. Wesley Long et al |
| EPI_ISL_546789 | USA/TX-HMH-7473/2020 | 2020-06-26 | North America/USA/Texas/Greater Hou | Houston Methodist Hospital | Houston Methodist Hospital | S. Wesley Long et al |
| EPI_ISL_546795 | USA/TX-HMH-7487/2020 | 2020-06-26 | North America/USA/Texas/Greater Hou | Houston Methodist Hospital | Houston Methodist Hospital | S. Wesley Long et al |
| EPI_ISL_546796 | USA/TX-HMH-7488/2020 | 2020-06-26 | North America/USA/Texas/Greater Hou | Houston Methodist Hospital | Houston Methodist Hospital | S. Wesley Long et al |
| EPI_ISL_546802 | USA/TX-HMH-7496/2020 | 2020-06-26 | North America/USA/Texas/Greater Hou | Houston Methodist Hospital | Houston Methodist Hospital | S. Wesley Long et al |
| EPI_ISL_546809 | USA/TX-HMH-7509/2020 | 2020-06-25 | North America/USA/Texas/Greater Hou | Houston Methodist Hospital | Houston Methodist Hospital | S. Wesley Long et al |

| | | | | | | |
|---|---|---|---|---|---|---|
| EPI_ISL_546821 | USA/TX-HMH-7526/2020 | 2020-06-25 | North America/USA/Texas/Greater Hou | Houston Methodist Hospital | Houston Methodist Hospital | S. Wesley Long et al |
| EPI_ISL_546826 | USA/TX-HMH-7531/2020 | 2020-06-25 | North America/USA/Texas/Greater Hou | Houston Methodist Hospital | Houston Methodist Hospital | S. Wesley Long et al |
| EPI_ISL_546833 | USA/TX-HMH-7539/2020 | 2020-06-25 | North America/USA/Texas/Greater Hou | Houston Methodist Hospital | Houston Methodist Hospital | S. Wesley Long et al |
| EPI_ISL_546836 | USA/TX-HMH-7541/2020 | 2020-06-25 | North America/USA/Texas/Greater Hou | Houston Methodist Hospital | Houston Methodist Hospital | S. Wesley Long et al |
| EPI_ISL_546842 | USA/TX-HMH-7547/2020 | 2020-06-25 | North America/USA/Texas/Greater Hou | Houston Methodist Hospital | Houston Methodist Hospital | S. Wesley Long et al |
| EPI_ISL_546845 | USA/TX-HMH-7552/2020 | 2020-06-25 | North America/USA/Texas/Greater Hou | Houston Methodist Hospital | Houston Methodist Hospital | S. Wesley Long et al |
| EPI_ISL_546846 | USA/TX-HMH-7553/2020 | 2020-06-25 | North America/USA/Texas/Greater Hou | Houston Methodist Hospital | Houston Methodist Hospital | S. Wesley Long et al |
| EPI_ISL_546848 | USA/TX-HMH-7556/2020 | 2020-06-25 | North America/USA/Texas/Greater Hou | Houston Methodist Hospital | Houston Methodist Hospital | S. Wesley Long et al |
| EPI_ISL_546863 | USA/TX-HMH-7573/2020 | 2020-06-25 | North America/USA/Texas/Greater Hou | Houston Methodist Hospital | Houston Methodist Hospital | S. Wesley Long et al |
| EPI_ISL_546866 | USA/TX-HMH-7578/2020 | 2020-06-25 | North America/USA/Texas/Greater Hou | Houston Methodist Hospital | Houston Methodist Hospital | S. Wesley Long et al |
| EPI_ISL_546869 | USA/TX-HMH-7580/2020 | 2020-06-26 | North America/USA/Texas/Greater Hou | Houston Methodist Hospital | Houston Methodist Hospital | S. Wesley Long et al |
| EPI_ISL_546876 | USA/TX-HMH-7591/2020 | 2020-06-25 | North America/USA/Texas/Greater Hou | Houston Methodist Hospital | Houston Methodist Hospital | S. Wesley Long et al |
| EPI_ISL_546886 | USA/TX-HMH-7604/2020 | 2020-06-26 | North America/USA/Texas/Greater Hou | Houston Methodist Hospital | Houston Methodist Hospital | S. Wesley Long et al |

| | | | | | | |
|---|---|---|---|---|---|---|
| EPI_ISL_546887 | USA/TX-HMH-7605/2020 | 2020-06-26 | North America/USA/Texas/Greater Hou | Houston Methodist Hospital | Houston Methodist Hospital | S. Wesley Long et al |
| EPI_ISL_546896 | USA/TX-HMH-7626/2020 | 2020-06-26 | North America/USA/Texas/Greater Hou | Houston Methodist Hospital | Houston Methodist Hospital | S. Wesley Long et al |
| EPI_ISL_546904 | USA/TX-HMH-7635/2020 | 2020-06-26 | North America/USA/Texas/Greater Hou | Houston Methodist Hospital | Houston Methodist Hospital | S. Wesley Long et al |
| EPI_ISL_546906 | USA/TX-HMH-7639/2020 | 2020-06-26 | North America/USA/Texas/Greater Hou | Houston Methodist Hospital | Houston Methodist Hospital | S. Wesley Long et al |
| EPI_ISL_546912 | USA/TX-HMH-7649/2020 | 2020-06-26 | North America/USA/Texas/Greater Hou | Houston Methodist Hospital | Houston Methodist Hospital | S. Wesley Long et al |
| EPI_ISL_546928 | USA/TX-HMH-7670/2020 | 2020-06-30 | North America/USA/Texas/Greater Hou | Houston Methodist Hospital | Houston Methodist Hospital | S. Wesley Long et al |
| EPI_ISL_546936 | CzechRepublic/NRL-8118-2/2020 | 2020-08-07 | Europe/Czech Republic/Usti nad Labem/ | The National Institute of Public Health | State Veterinary Institute Prague | Nagy et al |
| EPI_ISL_546939 | USA/TX-HMH-5678/2020 | 2020-06-23 | North America/USA/Texas/Greater Hou | Houston Methodist Hospital | Houston Methodist Hospital | S. Wesley Long et al |
| EPI_ISL_546941 | USA/TX-HMH-5680/2020 | 2020-06-23 | North America/USA/Texas/Greater Hou | Houston Methodist Hospital | Houston Methodist Hospital | S. Wesley Long et al |
| EPI_ISL_546944 | USA/TX-HMH-5684/2020 | 2020-06-20 | North America/USA/Texas/Greater Hou | Houston Methodist Hospital | Houston Methodist Hospital | S. Wesley Long et al |
| EPI_ISL_546945 | USA/TX-HMH-5685/2020 | 2020-06-26 | North America/USA/Texas/Greater Hou | Houston Methodist Hospital | Houston Methodist Hospital | S. Wesley Long et al |
| EPI_ISL_546950 | USA/TX-HMH-5697/2020 | 2020-06-30 | North America/USA/Texas/Greater Hou | Houston Methodist Hospital | Houston Methodist Hospital | S. Wesley Long et al |
| EPI_ISL_546952 | USA/TX-HMH-570/2020 | 2020-04-06 | North America/USA/Texas/Greater Hou | Houston Methodist Hospital | Houston Methodist Hospital | S. Wesley Long et al |
| EPI_ISL_546971 | USA/TX-HMH-5728/2020 | 2020-06-24 | North America/USA/Texas/Greater Hou | Houston Methodist Hospital | Houston Methodist Hospital | S. Wesley Long et al |

| | | | | | | |
|---|---|---|---|---|---|---|
| EPI_ISL_546974 | USA/TX-HMH-5732/2020 | 2020-06-23 | North America/USA/Texas/Greater Hou | Houston Methodist Hospital | Houston Methodist Hospital | S. Wesley Long et al |
| EPI_ISL_546979 | USA/TX-HMH-5738/2020 | 2020-06-24 | North America/USA/Texas/Greater Hou | Houston Methodist Hospital | Houston Methodist Hospital | S. Wesley Long et al |
| EPI_ISL_546982 | USA/TX-HMH-5742/2020 | 2020-06-29 | North America/USA/Texas/Greater Hou | Houston Methodist Hospital | Houston Methodist Hospital | S. Wesley Long et al |
| EPI_ISL_546991 | USA/TX-HMH-5755/2020 | 2020-06-18 | North America/USA/Texas/Greater Hou | Houston Methodist Hospital | Houston Methodist Hospital | S. Wesley Long et al |
| EPI_ISL_547000 | USA/TX-HMH-5768/2020 | 2020-06-26 | North America/USA/Texas/Greater Hou | Houston Methodist Hospital | Houston Methodist Hospital | S. Wesley Long et al |
| EPI_ISL_547005 | USA/TX-HMH-5773/2020 | 2020-06-24 | North America/USA/Texas/Greater Hou | Houston Methodist Hospital | Houston Methodist Hospital | S. Wesley Long et al |
| EPI_ISL_547014 | USA/TX-HMH-5787/2020 | 2020-06-28 | North America/USA/Texas/Greater Hou | Houston Methodist Hospital | Houston Methodist Hospital | S. Wesley Long et al |
| EPI_ISL_547015 | USA/TX-HMH-5788/2020 | 2020-07-01 | North America/USA/Texas/Greater Hou | Houston Methodist Hospital | Houston Methodist Hospital | S. Wesley Long et al |
| EPI_ISL_547018 | USA/TX-HMH-5791/2020 | 2020-06-30 | North America/USA/Texas/Greater Hou | Houston Methodist Hospital | Houston Methodist Hospital | S. Wesley Long et al |
| EPI_ISL_547021 | USA/TX-HMH-5795/2020 | 2020-06-30 | North America/USA/Texas/Greater Hou | Houston Methodist Hospital | Houston Methodist Hospital | S. Wesley Long et al |
| EPI_ISL_547024 | USA/TX-HMH-5801/2020 | 2020-06-30 | North America/USA/Texas/Greater Hou | Houston Methodist Hospital | Houston Methodist Hospital | S. Wesley Long et al |
| EPI_ISL_547034 | USA/TX-HMH-5812/2020 | 2020-06-29 | North America/USA/Texas/Greater Hou | Houston Methodist Hospital | Houston Methodist Hospital | S. Wesley Long et al |
| EPI_ISL_547038 | USA/TX-HMH-5816/2020 | 2020-06-23 | North America/USA/Texas/Greater Hou | Houston Methodist Hospital | Houston Methodist Hospital | S. Wesley Long et al |

| | | | | | | |
|---|---|---|---|---|---|---|
| EPI_ISL_547039 | USA/TX-HMH-5817/2020 | 2020-06-28 | North America/USA/Texas/Greater Hou | Houston Methodist Hospital | Houston Methodist Hospital | S. Wesley Long et al |
| EPI_ISL_547045 | USA/TX-HMH-5829/2020 | 2020-06-30 | North America/USA/Texas/Greater Hou | Houston Methodist Hospital | Houston Methodist Hospital | S. Wesley Long et al |
| EPI_ISL_547053 | USA/TX-HMH-5839/2020 | 2020-06-30 | North America/USA/Texas/Greater Hou | Houston Methodist Hospital | Houston Methodist Hospital | S. Wesley Long et al |
| EPI_ISL_547069 | USA/TX-HMH-586/2020 | 2020-04-02 | North America/USA/Texas/Greater Hou | Houston Methodist Hospital | Houston Methodist Hospital | S. Wesley Long et al |
| EPI_ISL_547083 | USA/TX-HMH-5880/2020 | 2020-06-30 | North America/USA/Texas/Greater Hou | Houston Methodist Hospital | Houston Methodist Hospital | S. Wesley Long et al |
| EPI_ISL_547088 | USA/TX-HMH-5890/2020 | 2020-06-30 | North America/USA/Texas/Greater Hou | Houston Methodist Hospital | Houston Methodist Hospital | S. Wesley Long et al |
| EPI_ISL_547091 | USA/TX-HMH-5895/2020 | 2020-06-30 | North America/USA/Texas/Greater Hou | Houston Methodist Hospital | Houston Methodist Hospital | S. Wesley Long et al |
| EPI_ISL_547092 | USA/TX-HMH-5896/2020 | 2020-06-30 | North America/USA/Texas/Greater Hou | Houston Methodist Hospital | Houston Methodist Hospital | S. Wesley Long et al |
| EPI_ISL_547095 | USA/TX-HMH-5900/2020 | 2020-06-30 | North America/USA/Texas/Greater Hou | Houston Methodist Hospital | Houston Methodist Hospital | S. Wesley Long et al |
| EPI_ISL_547097 | USA/TX-HMH-5902/2020 | 2020-06-30 | North America/USA/Texas/Greater Hou | Houston Methodist Hospital | Houston Methodist Hospital | S. Wesley Long et al |
| EPI_ISL_547112 | USA/TX-HMH-5945/2020 | 2020-06-30 | North America/USA/Texas/Greater Hou | Houston Methodist Hospital | Houston Methodist Hospital | S. Wesley Long et al |
| EPI_ISL_547143 | USA/TX-HMH-5983/2020 | 2020-06-30 | North America/USA/Texas/Greater Hou | Houston Methodist Hospital | Houston Methodist Hospital | S. Wesley Long et al |
| EPI_ISL_547146 | USA/TX-HMH-5986/2020 | 2020-06-30 | North America/USA/Texas/Greater Hou | Houston Methodist Hospital | Houston Methodist Hospital | S. Wesley Long et al |

| | | | | | | |
|---|---|---|---|---|---|---|
| EPI_ISL_547150 | USA/TX-HMH-5991/2020 | 2020-06-30 | North America/USA/Texas/Greater Hou | Houston Methodist Hospital | Houston Methodist Hospital | S. Wesley Long et al |
| EPI_ISL_547157 | USA/TX-HMH-6000/2020 | 2020-06-30 | North America/USA/Texas/Greater Hou | Houston Methodist Hospital | Houston Methodist Hospital | S. Wesley Long et al |
| EPI_ISL_547159 | USA/TX-HMH-6003/2020 | 2020-06-23 | North America/USA/Texas/Greater Hou | Houston Methodist Hospital | Houston Methodist Hospital | S. Wesley Long et al |
| EPI_ISL_547161 | USA/TX-HMH-6005/2020 | 2020-06-18 | North America/USA/Texas/Greater Hou | Houston Methodist Hospital | Houston Methodist Hospital | S. Wesley Long et al |
| EPI_ISL_547163 | USA/TX-HMH-6008/2020 | 2020-06-30 | North America/USA/Texas/Greater Hou | Houston Methodist Hospital | Houston Methodist Hospital | S. Wesley Long et al |
| EPI_ISL_547165 | USA/TX-HMH-601/2020 | 2020-04-08 | North America/USA/Texas/Greater Hou | Houston Methodist Hospital | Houston Methodist Hospital | S. Wesley Long et al |
| EPI_ISL_547170 | USA/TX-HMH-6016/2020 | 2020-06-28 | North America/USA/Texas/Greater Hou | Houston Methodist Hospital | Houston Methodist Hospital | S. Wesley Long et al |
| EPI_ISL_547174 | USA/TX-HMH-6022/2020 | 2020-06-30 | North America/USA/Texas/Greater Hou | Houston Methodist Hospital | Houston Methodist Hospital | S. Wesley Long et al |
| EPI_ISL_547175 | USA/TX-HMH-6025/2020 | 2020-06-30 | North America/USA/Texas/Greater Hou | Houston Methodist Hospital | Houston Methodist Hospital | S. Wesley Long et al |
| EPI_ISL_547181 | USA/TX-HMH-6035/2020 | 2020-06-27 | North America/USA/Texas/Greater Hou | Houston Methodist Hospital | Houston Methodist Hospital | S. Wesley Long et al |
| EPI_ISL_547184 | USA/TX-HMH-6038/2020 | 2020-06-28 | North America/USA/Texas/Greater Hou | Houston Methodist Hospital | Houston Methodist Hospital | S. Wesley Long et al |
| EPI_ISL_547185 | USA/TX-HMH-6040/2020 | 2020-07-01 | North America/USA/Texas/Greater Hou | Houston Methodist Hospital | Houston Methodist Hospital | S. Wesley Long et al |
| EPI_ISL_547209 | USA/TX-HMH-6067/2020 | 2020-07-02 | North America/USA/Texas/Greater Hou | Houston Methodist Hospital | Houston Methodist Hospital | S. Wesley Long et al |

| | | | | | | |
|---|---|---|---|---|---|---|
| EPI_ISL_547214 | USA/TX-HMH-6075/2020 | 2020-07-02 | North America/USA/Texas/Greater Hou | Houston Methodist Hospital | Houston Methodist Hospital | S. Wesley Long et al |
| EPI_ISL_547215 | USA/TX-HMH-6078/2020 | 2020-06-23 | North America/USA/Texas/Greater Hou | Houston Methodist Hospital | Houston Methodist Hospital | S. Wesley Long et al |
| EPI_ISL_547220 | USA/TX-HMH-6082/2020 | 2020-06-22 | North America/USA/Texas/Greater Hou | Houston Methodist Hospital | Houston Methodist Hospital | S. Wesley Long et al |
| EPI_ISL_547238 | USA/TX-HMH-6102/2020 | 2020-06-22 | North America/USA/Texas/Greater Hou | Houston Methodist Hospital | Houston Methodist Hospital | S. Wesley Long et al |
| EPI_ISL_547239 | USA/TX-HMH-6103/2020 | 2020-06-30 | North America/USA/Texas/Greater Hou | Houston Methodist Hospital | Houston Methodist Hospital | S. Wesley Long et al |
| EPI_ISL_547246 | USA/TX-HMH-6110/2020 | 2020-07-04 | North America/USA/Texas/Greater Hou | Houston Methodist Hospital | Houston Methodist Hospital | S. Wesley Long et al |
| EPI_ISL_547260 | USA/TX-HMH-6127/2020 | 2020-07-07 | North America/USA/Texas/Greater Hou | Houston Methodist Hospital | Houston Methodist Hospital | S. Wesley Long et al |
| EPI_ISL_547266 | USA/TX-HMH-6139/2020 | 2020-07-07 | North America/USA/Texas/Greater Hou | Houston Methodist Hospital | Houston Methodist Hospital | S. Wesley Long et al |
| EPI_ISL_547275 | USA/TX-HMH-6152/2020 | 2020-06-22 | North America/USA/Texas/Greater Hou | Houston Methodist Hospital | Houston Methodist Hospital | S. Wesley Long et al |
| EPI_ISL_547279 | USA/TX-HMH-6158/2020 | 2020-06-24 | North America/USA/Texas/Greater Hou | Houston Methodist Hospital | Houston Methodist Hospital | S. Wesley Long et al |
| EPI_ISL_547293 | USA/TX-HMH-6176/2020 | 2020-06-27 | North America/USA/Texas/Greater Hou | Houston Methodist Hospital | Houston Methodist Hospital | S. Wesley Long et al |
| EPI_ISL_547295 | USA/TX-HMH-6181/2020 | 2020-07-03 | North America/USA/Texas/Greater Hou | Houston Methodist Hospital | Houston Methodist Hospital | S. Wesley Long et al |
| EPI_ISL_547296 | USA/TX-HMH-6183/2020 | 2020-07-04 | North America/USA/Texas/Greater Hou | Houston Methodist Hospital | Houston Methodist Hospital | S. Wesley Long et al |

| | | | | | | |
|---|---|---|---|---|---|---|
| EPI_ISL_547304 | USA/TX-HMH-6195/2020 | 2020-07-04 | North America/USA/Texas/Greater Hou | Houston Methodist Hospital | Houston Methodist Hospital | S. Wesley Long et al |
| EPI_ISL_547308 | USA/TX-HMH-6201/2020 | 2020-07-04 | North America/USA/Texas/Greater Hou | Houston Methodist Hospital | Houston Methodist Hospital | S. Wesley Long et al |
| EPI_ISL_547310 | USA/TX-HMH-6205/2020 | 2020-07-04 | North America/USA/Texas/Greater Hou | Houston Methodist Hospital | Houston Methodist Hospital | S. Wesley Long et al |
| EPI_ISL_547316 | USA/TX-HMH-6213/2020 | 2020-07-07 | North America/USA/Texas/Greater Hou | Houston Methodist Hospital | Houston Methodist Hospital | S. Wesley Long et al |
| EPI_ISL_547324 | USA/TX-HMH-6223/2020 | 2020-06-22 | North America/USA/Texas/Greater Hou | Houston Methodist Hospital | Houston Methodist Hospital | S. Wesley Long et al |
| EPI_ISL_547336 | USA/TX-HMH-6243/2020 | 2020-07-02 | North America/USA/Texas/Greater Hou | Houston Methodist Hospital | Houston Methodist Hospital | S. Wesley Long et al |
| EPI_ISL_547340 | USA/TX-HMH-625/2020 | 2020-04-11 | North America/USA/Texas/Greater Hou | Houston Methodist Hospital | Houston Methodist Hospital | S. Wesley Long et al |
| EPI_ISL_547343 | USA/TX-HMH-6255/2020 | 2020-06-22 | North America/USA/Texas/Greater Hou | Houston Methodist Hospital | Houston Methodist Hospital | S. Wesley Long et al |
| EPI_ISL_547349 | USA/TX-HMH-6264/2020 | 2020-06-23 | North America/USA/Texas/Greater Hou | Houston Methodist Hospital | Houston Methodist Hospital | S. Wesley Long et al |
| EPI_ISL_547350 | USA/TX-HMH-6265/2020 | 2020-06-22 | North America/USA/Texas/Greater Hou | Houston Methodist Hospital | Houston Methodist Hospital | S. Wesley Long et al |
| EPI_ISL_547352 | USA/TX-HMH-6267/2020 | 2020-07-03 | North America/USA/Texas/Greater Hou | Houston Methodist Hospital | Houston Methodist Hospital | S. Wesley Long et al |
| EPI_ISL_547356 | USA/TX-HMH-6271/2020 | 2020-06-24 | North America/USA/Texas/Greater Hou | Houston Methodist Hospital | Houston Methodist Hospital | S. Wesley Long et al |
| EPI_ISL_547361 | USA/TX-HMH-6281/2020 | 2020-06-27 | North America/USA/Texas/Greater Hou | Houston Methodist Hospital | Houston Methodist Hospital | S. Wesley Long et al |

| | | | | | | |
|---|---|---|---|---|---|---|
| EPI_ISL_547362 | USA/TX-HMH-6283/2020 | 2020-06-30 | North America/USA/Texas/Greater Hou | Houston Methodist Hospital | Houston Methodist Hospital | S. Wesley Long et al |
| EPI_ISL_547363 | USA/TX-HMH-6284/2020 | 2020-06-28 | North America/USA/Texas/Greater Hou | Houston Methodist Hospital | Houston Methodist Hospital | S. Wesley Long et al |
| EPI_ISL_547365 | USA/TX-HMH-6286/2020 | 2020-06-21 | North America/USA/Texas/Greater Hou | Houston Methodist Hospital | Houston Methodist Hospital | S. Wesley Long et al |
| EPI_ISL_547369 | USA/TX-HMH-6294/2020 | 2020-07-04 | North America/USA/Texas/Greater Hou | Houston Methodist Hospital | Houston Methodist Hospital | S. Wesley Long et al |
| EPI_ISL_547370 | USA/TX-HMH-6296/2020 | 2020-06-28 | North America/USA/Texas/Greater Hou | Houston Methodist Hospital | Houston Methodist Hospital | S. Wesley Long et al |
| EPI_ISL_547374 | USA/TX-HMH-6300/2020 | 2020-07-04 | North America/USA/Texas/Greater Hou | Houston Methodist Hospital | Houston Methodist Hospital | S. Wesley Long et al |
| EPI_ISL_547378 | USA/TX-HMH-6304/2020 | 2020-07-03 | North America/USA/Texas/Greater Hou | Houston Methodist Hospital | Houston Methodist Hospital | S. Wesley Long et al |
| EPI_ISL_547382 | USA/TX-HMH-6311/2020 | 2020-07-04 | North America/USA/Texas/Greater Hou | Houston Methodist Hospital | Houston Methodist Hospital | S. Wesley Long et al |
| EPI_ISL_547385 | USA/TX-HMH-6316/2020 | 2020-07-02 | North America/USA/Texas/Greater Hou | Houston Methodist Hospital | Houston Methodist Hospital | S. Wesley Long et al |
| EPI_ISL_547386 | USA/TX-HMH-6317/2020 | 2020-07-04 | North America/USA/Texas/Greater Hou | Houston Methodist Hospital | Houston Methodist Hospital | S. Wesley Long et al |
| EPI_ISL_547396 | USA/TX-HMH-6332/2020 | 2020-07-04 | North America/USA/Texas/Greater Hou | Houston Methodist Hospital | Houston Methodist Hospital | S. Wesley Long et al |
| EPI_ISL_547397 | USA/TX-HMH-6333/2020 | 2020-06-21 | North America/USA/Texas/Greater Hou | Houston Methodist Hospital | Houston Methodist Hospital | S. Wesley Long et al |
| EPI_ISL_547403 | USA/TX-HMH-6341/2020 | 2020-07-03 | North America/USA/Texas/Greater Hou | Houston Methodist Hospital | Houston Methodist Hospital | S. Wesley Long et al |

| | | | | | | |
|---|---|---|---|---|---|---|
| EPI_ISL_547414 | USA/TX-HMH-6357/2020 | 2020-07-05 | North America/USA/Texas/Greater Hou | Houston Methodist Hospital | Houston Methodist Hospital | S. Wesley Long et al |
| EPI_ISL_547418 | USA/TX-HMH-6361/2020 | 2020-07-05 | North America/USA/Texas/Greater Hou | Houston Methodist Hospital | Houston Methodist Hospital | S. Wesley Long et al |
| EPI_ISL_547495 | Netherlands/NH-RIVM-10062/2020 | 2020-04-03 | Europe/Netherlands/North Holland/ | Dutch COVID-19 response team | National Institute for Public Health and the Environment (RIVM) | Adam Meijer et al |
| EPI_ISL_547499 | Netherlands/NH-RIVM-10073/2020 | 2020-02-27 | Europe/Netherlands/North Holland/ | Dutch COVID-19 response team | National Institute for Public Health and the Environment (RIVM) | Adam Meijer et al |
| EPI_ISL_547513 | Netherlands/OV-RIVM-10152/2020 | 2020-07-21 | Europe/Netherlands/Overijssel/ | Dutch COVID-19 response team | National Institute for Public Health and the Environment (RIVM) | Adam Meijer et al |
| EPI_ISL_547559 | Netherlands/ZH-RIVM-10076/2020 | 2020-03-05 | Europe/Netherlands/South Holland/ | Dutch COVID-19 response team | National Institute for Public Health and the Environment (RIVM) | Adam Meijer et al |
| EPI_ISL_547571 | Brazil/SP-341/2020 | 2020-06-01 | South America/Brazil/S√£o Paulo/Maua | Hospital Municipal Ant√¥nio Giglio | Instituto Adolfo Lutz, Interdiciplinary Procedures Center, Strategic Laboratory | Claudio Tavares Sacchi et al |
| EPI_ISL_547573 | Brazil/SP-342/2020 | 2020-06-01 | South America/Brazil/S√£o Paulo/Cajam | Vigil√¢ncia em Sa√∫de de Cajamar | Instituto Adolfo Lutz, Interdiciplinary Procedures Center, Strategic Laboratory | Claudio Tavares Sacchi et al |

| | | | | | | |
|---|---|---|---|---|---|---|
| EPI_ISL_547574 | Brazil/SP-354/2020 | 2020-06-19 | South America/Brazil/S√£o Paulo/Taboa | Hospital Universitario da USP | Instituto Adolfo Lutz, Interdiciplinary Procedures Center, Strategic Laboratory | Claudio Tavares Sacchi et al |
| EPI_ISL_547575 | Brazil/SP-356/2020 | 2020-06-16 | South America/Brazil/S√£o Paulo/Jundia | SVO Jundia√≠ | Instituto Adolfo Lutz, Interdiciplinary Procedures Center, Strategic Laboratory | Claudio Tavares Sacchi et al |
| EPI_ISL_547576 | Brazil/SP-358/2020 | 2020-06-23 | South America/Brazil/S√£o Paulo/Birigu | Secretaria Municipal de Sa√∫de | Instituto Adolfo Lutz, Interdiciplinary Procedures Center, Strategic Laboratory | Claudio Tavares Sacchi et al |
| EPI_ISL_547577 | Brazil/SP-405/2020 | 2020-07-06 | South America/Brazil/S√£o Paulo/Presid | Hospital e Maternidade Nossa Senhora das Gra√ßas | Instituto Adolfo Lutz, Interdiciplinary Procedures Center, Strategic Laboratory | Claudio Tavares Sacchi et al |
| EPI_ISL_547578 | Brazil/SP-406/2020 | 2020-07-05 | South America/Brazil/S√£o Paulo/Presid | Hospital Doutor Domingos Leonardo Cer√°volo | Instituto Adolfo Lutz, Interdiciplinary Procedures Center, Strategic Laboratory | Claudio Tavares Sacchi et al |
| EPI_ISL_547579 | Brazil/SP-417/2020 | 2020-07-07 | South America/Brazil/S√£o Paulo/Ara√ß | Santa Casa de Miseric√≥rdia de Ara√ßatuba | Instituto Adolfo Lutz, Interdiciplinary Procedures Center, Strategic Laboratory | Claudio Tavares Sacchi et al |

| | | | | | | |
|---|---|---|---|---|---|---|
| EPI_ISL_547580 | Brazil/SP-418/2020 | 2020-07-09 | South America/Brazil/S√£o Paulo/Euclid | Santa Casa da Miseric√≥rdia de Presidente Prudente | Instituto Adolfo Lutz, Interdiciplinary Procedures Center, Strategic Laboratory | Claudio Tavares Sacchi et al |
| EPI_ISL_547581 | USA/WI-UW-1265/2020 | 2020-08-22 | North America/USA/Wisconsin/Dane Co | University of Wisconsin-Madison AIDS Vaccine Research Laboratories | University of Wisconsin-Madison AIDS Vaccine Research Laboratories | Gage Moreno et al |
| EPI_ISL_547584 | India/UT-IMT-BY60/2020 | 2020-08-24 | Asia/India/Uttarakhand/ | CMS, Roorkee | CSIR-Institute of Microbial Technology | Kanika Bansal et al |
| EPI_ISL_547585 | India/HR-IMT-BZ16/2020 | 2020-08-25 | Asia/India/Haryana/Panchkula | Civil Hospital, Panchkula | CSIR-Institute of Microbial Technology | Kanika Bansal et al |
| EPI_ISL_547586 | India/HR-IMT-BZ102/2020 | 2020-08-25 | Asia/India/Haryana/Panchkula | Civil Hospital, Panchkula | CSIR-Institute of Microbial Technology | Kanika Bansal et al |
| EPI_ISL_547588 | India/HR-IMT-BZ140/2020 | 2020-08-25 | Asia/India/Haryana/Panchkula | Civil Hospital, Panchkula | CSIR-Institute of Microbial Technology | Kanika Bansal et al |
| EPI_ISL_547589 | India/HR-IMT-CB29/2020 | 2020-08-27 | Asia/India/Haryana/Panchkula | Civil Hospital, Panchkula | CSIR-Institute of Microbial Technology | Kanika Bansal et al |
| EPI_ISL_547590 | India/HR-IMT-CB44/2020 | 2020-08-27 | Asia/India/Haryana/Panchkula | Civil Hospital, Panchkula | CSIR-Institute of Microbial Technology | Kanika Bansal et al |
| EPI_ISL_547591 | India/HR-IMT-CB45/2020 | 2020-08-27 | Asia/India/Haryana/Panchkula | Civil Hospital, Panchkula | CSIR-Institute of Microbial Technology | Kanika Bansal et al |
| EPI_ISL_547592 | India/HR-IMT-CB130/2020 | 2020-08-27 | Asia/India/Haryana/Panchkula | Civil Hospital, Panchkula | CSIR-Institute of Microbial Technology | Kanika Bansal et al |
| EPI_ISL_547593 | India/HR-IMT-CB193/2020 | 2020-08-27 | Asia/India/Haryana/Panchkula | Civil Hospital, Panchkula | CSIR-Institute of Microbial Technology | Kanika Bansal et al |
| EPI_ISL_547606 | USA/WI-GMF-09934/2020 | 2020-07-08 | North America/USA/Wisconsin/Monroe | Gundersen Molecular Diagnostics Laboratory | Kabara Cancer Research Institute | Craig S. Richmond et al |

| | | | | | | |
|---|---|---|---|---|---|---|
| EPI_ISL_547610 | USA/WI-GMF-10571/2020 | 2020-07-13 | North America/USA/Wisconsin/Trempea | Gundersen Clinical Microbiology Laboratory | Kabara Cancer Research Institute | Craig S. Richmond et al |
| EPI_ISL_547617 | USA/WI-GMF-10885/2020 | 2020-07-15 | North America/USA/Wisconsin/La Cross | Gundersen Molecular Diagnostics Laboratory | Kabara Cancer Research Institute | Craig S. Richmond et al |
| EPI_ISL_547618 | USA/WI-GMF-10889/2020 | 2020-07-15 | North America/USA/Wisconsin/Monroe | Gundersen Molecular Diagnostics Laboratory | Kabara Cancer Research Institute | Craig S. Richmond et al |
| EPI_ISL_547619 | USA/WI-GMF-10900/2020 | 2020-07-15 | North America/USA/Wisconsin/Vernon | Gundersen Molecular Diagnostics Laboratory | Kabara Cancer Research Institute | Craig S. Richmond et al |
| EPI_ISL_547633 | USA/WI-GMF-11818/2020 | 2020-07-21 | North America/USA/Wisconsin/La Cross | Gundersen Molecular Diagnostics Laboratory | Kabara Cancer Research Institute | Craig S. Richmond et al |
| EPI_ISL_547638 | USA/WI-GMF-12045/2020 | 2020-07-23 | North America/USA/Wisconsin/La Cross | Gundersen Molecular Diagnostics Laboratory | Kabara Cancer Research Institute | Craig S. Richmond et al |
| EPI_ISL_547647 | USA/WI-GMF-12930/2020 | 2020-07-28 | North America/USA/Wisconsin/Juneau | Gundersen Molecular Diagnostics Laboratory | Kabara Cancer Research Institute | Craig S. Richmond et al |
| EPI_ISL_547648 | USA/WI-GMF-12945/2020 | 2020-07-28 | North America/USA/Wisconsin/Vernon | Gundersen Molecular Diagnostics Laboratory | Kabara Cancer Research Institute | Craig S. Richmond et al |
| EPI_ISL_547649 | USA/WI-GMF-13006/2020 | 2020-07-29 | North America/USA/Wisconsin/Vernon | Gundersen Molecular Diagnostics Laboratory | Kabara Cancer Research Institute | Craig S. Richmond et al |
| EPI_ISL_547652 | USA/WI-GMF-13033/2020 | 2020-07-29 | North America/USA/Wisconsin/Trempea | Gundersen Molecular Diagnostics Laboratory | Kabara Cancer Research Institute | Craig S. Richmond et al |
| EPI_ISL_547656 | USA/WI-GMF-13352/2020 | 2020-08-01 | North America/USA/Wisconsin/La Cross | Gundersen Molecular Diagnostics Laboratory | Kabara Cancer Research Institute | Craig S. Richmond et al |
| EPI_ISL_547661 | USA/MN-GMF-13847/2020 | 2020-08-04 | North America/USA/Minnesota/Winona | Gundersen Molecular Diagnostics Laboratory | Kabara Cancer Research Institute | Craig S. Richmond et al |
| EPI_ISL_547664 | USA/WI-GMF-13999/2020 | 2020-08-05 | North America/USA/Wisconsin/Monroe | Gundersen Molecular Diagnostics Laboratory | Kabara Cancer Research Institute | Craig S. Richmond et al |
| EPI_ISL_547666 | USA/WI-GMF-M00015/2020 | 2020-07-11 | North America/USA/Wisconsin/Monroe | Gundersen Clinical Microbiology Laboratory | Kabara Cancer Research Institute | Craig S. Richmond et al |
| EPI_ISL_547680 | USA/WI-GMF-14557/2020 | 2020-08-09 | North America/USA/Wisconsin/Vernon | Gundersen Molecular Diagnostics Laboratory | Kabara Cancer Research Institute | Craig S. Richmond et al |
| EPI_ISL_547685 | USA/WI-GMF-14621/2020 | 2020-08-10 | North America/USA/Wisconsin/La Cross | Gundersen Molecular Diagnostics Laboratory | Kabara Cancer Research Institute | Craig S. Richmond et al |
| EPI_ISL_547689 | USA/WI-GMF-14849/2020 | 2020-08-11 | North America/USA/Wisconsin/Jackson | Gundersen Molecular Diagnostics Laboratory | Kabara Cancer Research Institute | Craig S. Richmond et al |
| EPI_ISL_547699 | USA/WI-GMF-15113/2020 | 2020-08-13 | North America/USA/Wisconsin/La Cross | Gundersen Molecular Diagnostics Laboratory | Kabara Cancer Research Institute | Craig S. Richmond et al |
| EPI_ISL_547701 | USA/WI-GMF-15214/2020 | 2020-08-14 | North America/USA/Wisconsin/La Cross | Gundersen Molecular Diagnostics Laboratory | Kabara Cancer Research Institute | Craig S. Richmond et al |
| EPI_ISL_547703 | USA/MN-GMF-15330/2020 | 2020-08-14 | North America/USA/Minnesota/Housto | Gundersen Molecular Diagnostics Laboratory | Kabara Cancer Research Institute | Craig S. Richmond et al |

| | | | | | | |
|---|---|---|---|---|---|---|
| EPI_ISL_547705 | USA/MN-GMF-M00029/2020 | 2020-08-12 | North America/USA/Minnesota/Winona | Gundersen Clinical Microbiology Laboratory | Kabara Cancer Research Institute | Craig S. Richmond et al |
| EPI_ISL_547706 | USA/WI-GMF-15609/2020 | 2020-08-15 | North America/USA/Wisconsin/Trempea | Gundersen Molecular Diagnostics Laboratory | Kabara Cancer Research Institute | Craig S. Richmond et al |
| EPI_ISL_547710 | USA/MN-GMF-15765/2020 | 2020-08-17 | North America/USA/Minnesota/Housto | Gundersen Molecular Diagnostics Laboratory | Kabara Cancer Research Institute | Craig S. Richmond et al |
| EPI_ISL_547714 | USA/WI-GMF-15972/2020 | 2020-08-18 | North America/USA/Wisconsin/La Cross | Gundersen Molecular Diagnostics Laboratory | Kabara Cancer Research Institute | Craig S. Richmond et al |
| EPI_ISL_547720 | USA/WI-GMF-16214/2020 | 2020-08-20 | North America/USA/Wisconsin/La Cross | Gundersen Molecular Diagnostics Laboratory | Kabara Cancer Research Institute | Craig S. Richmond et al |
| EPI_ISL_547721 | USA/WI-GMF-16244/2020 | 2020-08-20 | North America/USA/Wisconsin/Monroe | Gundersen Molecular Diagnostics Laboratory | Kabara Cancer Research Institute | Craig S. Richmond et al |
| EPI_ISL_547723 | USA/WI-GMF-16264/2020 | 2020-08-20 | North America/USA/Wisconsin/La Cross | Gundersen Molecular Diagnostics Laboratory | Kabara Cancer Research Institute | Craig S. Richmond et al |
| EPI_ISL_547725 | USA/WI-GMF-16473/2020 | 2020-08-21 | North America/USA/Wisconsin/Shawan | Gundersen Molecular Diagnostics Laboratory | Kabara Cancer Research Institute | Craig S. Richmond et al |
| EPI_ISL_547727 | USA/WI-GMF-16632/2020 | 2020-08-22 | North America/USA/Wisconsin/La Cross | Gundersen Molecular Diagnostics Laboratory | Kabara Cancer Research Institute | Craig S. Richmond et al |
| EPI_ISL_547733 | USA/MN-GMF-16946/2020 | 2020-08-24 | North America/USA/Minnesota/Housto | Gundersen Molecular Diagnostics Laboratory | Kabara Cancer Research Institute | Craig S. Richmond et al |
| EPI_ISL_547736 | USA/WI-GMF-17327/2020 | 2020-08-26 | North America/USA/Wisconsin/Trempea | Gundersen Molecular Diagnostics Laboratory | Kabara Cancer Research Institute | Craig S. Richmond et al |
| EPI_ISL_547738 | USA/WI-GMF-17378/2020 | 2020-08-27 | North America/USA/Wisconsin/Vernon | Gundersen Molecular Diagnostics Laboratory | Kabara Cancer Research Institute | Craig S. Richmond et al |
| EPI_ISL_547739 | USA/WI-GMF-17385/2020 | 2020-08-28 | North America/USA/Wisconsin/Crawfor | Gundersen Molecular Diagnostics Laboratory | Kabara Cancer Research Institute | Craig S. Richmond et al |
| EPI_ISL_547743 | USA/WI-GMF-17507/2020 | 2020-08-28 | North America/USA/Wisconsin/La Cross | Gundersen Molecular Diagnostics Laboratory | Kabara Cancer Research Institute | Craig S. Richmond et al |
| EPI_ISL_547754 | USA/IA-GMF-18219/2020 | 2020-08-31 | North America/USA/Iowa/Fayette Coun | Gundersen Molecular Diagnostics Laboratory | Kabara Cancer Research Institute | Craig S. Richmond et al |
| EPI_ISL_547755 | USA/WI-GMF-18264/2020 | 2020-09-01 | North America/USA/Wisconsin/La Cross | Gundersen Molecular Diagnostics Laboratory | Kabara Cancer Research Institute | Craig S. Richmond et al |
| EPI_ISL_547767 | USA/WI-GMF-18563/2020 | 2020-09-03 | North America/USA/Wisconsin/Monroe | Gundersen Molecular Diagnostics Laboratory | Kabara Cancer Research Institute | Craig S. Richmond et al |
| EPI_ISL_547768 | USA/WI-GMF-18574/2020 | 2020-09-03 | North America/USA/Wisconsin/Vernon | Gundersen Molecular Diagnostics Laboratory | Kabara Cancer Research Institute | Craig S. Richmond et al |
| EPI_ISL_547773 | USA/WI-GMF-18660/2020 | 2020-09-02 | North America/USA/Wisconsin/Adams | Gundersen Molecular Diagnostics Laboratory | Kabara Cancer Research Institute | Craig S. Richmond et al |
| EPI_ISL_547778 | USA/WI-GMF-18724/2020 | 2020-09-04 | North America/USA/Wisconsin/La Cross | Gundersen Molecular Diagnostics Laboratory | Kabara Cancer Research Institute | Craig S. Richmond et al |

| | | | | | | |
|---|---|---|---|---|---|---|
| EPI_ISL_547781 | USA/WI-GMF-18735/2020 | 2020-09-04 | North America/USA/Wisconsin/Juneau C | Gundersen Molecular Diagnostics Laboratory | Kabara Cancer Research Institute | Craig S. Richmond et al |
| EPI_ISL_547784 | USA/WI-GMF-18789/2020 | 2020-09-04 | North America/USA/Wisconsin/La Crosse | Gundersen Molecular Diagnostics Laboratory | Kabara Cancer Research Institute | Craig S. Richmond et al |
| EPI_ISL_547791 | USA/MN-GMF-19171/2020 | 2020-09-08 | North America/USA/Minnesota/Winona | Gundersen Molecular Diagnostics Laboratory | Kabara Cancer Research Institute | Craig S. Richmond et al |
| EPI_ISL_547793 | USA/WI-GMF-19195/2020 | 2020-09-08 | North America/USA/Wisconsin/La Crosse | Gundersen Molecular Diagnostics Laboratory | Kabara Cancer Research Institute | Craig S. Richmond et al |
| EPI_ISL_547795 | USA/WI-GMF-19225/2020 | 2020-09-08 | North America/USA/Wisconsin/La Crosse | Gundersen Molecular Diagnostics Laboratory | Kabara Cancer Research Institute | Craig S. Richmond et al |
| EPI_ISL_547798 | USA/WI-GMF-19240/2020 | 2020-09-08 | North America/USA/Wisconsin/La Crosse | Gundersen Molecular Diagnostics Laboratory | Kabara Cancer Research Institute | Craig S. Richmond et al |
| EPI_ISL_547800 | USA/MN-GMF-19243/2020 | 2020-09-08 | North America/USA/Minnesota/Winona | Gundersen Molecular Diagnostics Laboratory | Kabara Cancer Research Institute | Craig S. Richmond et al |
| EPI_ISL_547828 | USA/WI-GMF-19599/2020 | 2020-09-10 | North America/USA/Wisconsin/La Crosse | Gundersen Molecular Diagnostics Laboratory | Kabara Cancer Research Institute | Craig S. Richmond et al |
| EPI_ISL_547829 | USA/WI-GMF-19600/2020 | 2020-09-10 | North America/USA/Wisconsin/La Crosse | Gundersen Molecular Diagnostics Laboratory | Kabara Cancer Research Institute | Craig S. Richmond et al |
| EPI_ISL_547831 | USA/WI-GMF-19604/2020 | 2020-09-10 | North America/USA/Wisconsin/Grant Co | Gundersen Molecular Diagnostics Laboratory | Kabara Cancer Research Institute | Craig S. Richmond et al |
| EPI_ISL_547832 | USA/WI-GMF-19612/2020 | 2020-09-10 | North America/USA/Wisconsin/La Crosse | Gundersen Molecular Diagnostics Laboratory | Kabara Cancer Research Institute | Craig S. Richmond et al |
| EPI_ISL_547839 | USA/WI-GMF-19714/2020 | 2020-09-11 | North America/USA/Wisconsin/La Crosse | Gundersen Molecular Diagnostics Laboratory | Kabara Cancer Research Institute | Craig S. Richmond et al |
| EPI_ISL_547842 | USA/WI-GMF-19732/2020 | 2020-09-11 | North America/USA/Wisconsin/La Crosse | Gundersen Molecular Diagnostics Laboratory | Kabara Cancer Research Institute | Craig S. Richmond et al |
| EPI_ISL_547843 | USA/WI-GMF-19743/2020 | 2020-09-11 | North America/USA/Wisconsin/La Crosse | Gundersen Molecular Diagnostics Laboratory | Kabara Cancer Research Institute | Craig S. Richmond et al |
| EPI_ISL_547848 | USA/WI-GMF-19968/2020 | 2020-09-12 | North America/USA/Wisconsin/La Crosse | Gundersen Molecular Diagnostics Laboratory | Kabara Cancer Research Institute | Craig S. Richmond et al |
| EPI_ISL_547851 | USA/WI-GMF-20122/2020 | 2020-09-14 | North America/USA/Wisconsin/Vernon C | Gundersen Molecular Diagnostics Laboratory | Kabara Cancer Research Institute | Craig S. Richmond et al |
| EPI_ISL_547853 | USA/WI-GMF-20149/2020 | 2020-09-14 | North America/USA/Wisconsin/Trempea | Gundersen Molecular Diagnostics Laboratory | Kabara Cancer Research Institute | Craig S. Richmond et al |
| EPI_ISL_547854 | USA/WI-GMF-20170/2020 | 2020-09-14 | North America/USA/Wisconsin/La Crosse | Gundersen Molecular Diagnostics Laboratory | Kabara Cancer Research Institute | Craig S. Richmond et al |
| EPI_ISL_547857 | USA/IA-GMF-20217/2020 | 2020-09-14 | North America/USA/Iowa/Allamakee Co | Gundersen Molecular Diagnostics Laboratory | Kabara Cancer Research Institute | Craig S. Richmond et al |
| EPI_ISL_547858 | USA/WI-GMF-20228/2020 | 2020-09-14 | North America/USA/Wisconsin/Vernon C | Gundersen Molecular Diagnostics Laboratory | Kabara Cancer Research Institute | Craig S. Richmond et al |

| | | | | | | |
|---|---|---|---|---|---|---|
| EPI_ISL_547860 | USA/WI-GMF-20273/2020 | 2020-09-14 | North America/USA/Wisconsin/La Cross | Gundersen Molecular Diagnostics Laboratory | Kabara Cancer Research Institute | Craig S. Richmond et al |
| EPI_ISL_547862 | USA/WI-GMF-20295/2020 | 2020-09-15 | North America/USA/Wisconsin/La Cross | Gundersen Molecular Diagnostics Laboratory | Kabara Cancer Research Institute | Craig S. Richmond et al |
| EPI_ISL_547863 | USA/WI-GMF-20333/2020 | 2020-09-14 | North America/USA/Wisconsin/La Cross | Gundersen Molecular Diagnostics Laboratory | Kabara Cancer Research Institute | Craig S. Richmond et al |
| EPI_ISL_547867 | USA/WI-GMF-20407/2020 | 2020-09-15 | North America/USA/Wisconsin/Trempea | Gundersen Molecular Diagnostics Laboratory | Kabara Cancer Research Institute | Craig S. Richmond et al |
| EPI_ISL_547968 | CzechRepublic/NRL_8118-5/2020 | 2020-08-07 | Europe/Czech Republic/Usti nad Labem/ | The National Institute of Public Health | State Veterinary Institute Prague | Nagy et al |
| EPI_ISL_548156 | Switzerland/GE-SNRCI-2616/2020 | 2020-03-14 | Europe/Switzerland/Geneva/ | Laboratoire de Virologie, HUG | Swiss National Reference Centre for Influenza | LAUBSCHER F. et al |
| EPI_ISL_548173 | Switzerland/GE-SNRCI-5671/2020 | 2020-03-14 | Europe/Switzerland/Geneva/ | Laboratoire de Virologie, HUG | Swiss National Reference Centre for Influenza | LAUBSCHER F. et al |
| EPI_ISL_548186 | Switzerland/GE-SNRCI-7201/2020 | 2020-03-14 | Europe/Switzerland/Geneva/ | Laboratoire de Virologie, HUG | Swiss National Reference Centre for Influenza | LAUBSCHER F. et al |
| EPI_ISL_548203 | Switzerland/GE-SNRCI-7821/2020 | 2020-03-14 | Europe/Switzerland/Geneva/ | Laboratoire de Virologie, HUG | Swiss National Reference Centre for Influenza | LAUBSCHER F. et al |
| EPI_ISL_548204 | Switzerland/GE-SNRCI-7836/2020 | 2020-03-14 | Europe/Switzerland/Geneva/ | Laboratoire de Virologie, HUG | Swiss National Reference Centre for Influenza | LAUBSCHER F. et al |
| EPI_ISL_548239 | Switzerland/GE-SNRCI-9268/2020 | 2020-03-14 | Europe/Switzerland/Geneva/ | Laboratoire de Virologie, HUG | Swiss National Reference Centre for Influenza | LAUBSCHER F. et al |
| EPI_ISL_548246 | Sweden/20-08186/2020 | 2020-03-13 | Europe/Sweden/Vastra Gotaland/ | Skovde/Unilabs | The Public Health Agency of Sweden | Anna-Malin Linde et al |
| EPI_ISL_548252 | Sweden/20-52034/2020 | 2020-06-14 | Europe/Sweden/Varmland/ | Klinisk mikrobiologi centralsjukhuset Karlstad | The Public Health Agency of Sweden | Anna-Malin Linde et al |
| EPI_ISL_548253 | Sweden/20-52051/2020 | 2020-06-05 | Europe/Sweden/Vastra Gotaland/ | Klinisk mikrobiologi NAL Trollhattan | The Public Health Agency of Sweden | Anna-Malin Linde et al |
| EPI_ISL_548267 | USA/CA-CZB-3269/2020 | 2020-07-15 | North America/USA/California/Santa Cl | County of Santa Clara Public Health Department | Chan-Zuckerberg Biohub | CZB Cliahub Consortium et al |
| EPI_ISL_548272 | USA/CA-CZB-3333/2020 | 2020-07-10 | North America/USA/California/Orange ( | Orange County Public Health Laboratory | Chan-Zuckerberg Biohub | CZB Cliahub Consortium et al |
| EPI_ISL_548273 | USA/CA-CZB-3351/2020 | 2020-07-13 | North America/USA/California/Orange ( | Orange County Public Health Laboratory | Chan-Zuckerberg Biohub | CZB Cliahub Consortium et al |

| | | | | | | |
|---|---|---|---|---|---|---|
| EPI_ISL_548279 | USA/CA-CZB-3279/2020 | 2020-07-16 | North America/USA/California/Santa Cla | County of Santa Clara Public Health Department | Chan-Zuckerberg Biohub | CZB Cliahub Consortium et al |
| EPI_ISL_548280 | USA/CA-CZB-3318/2020 | 2020-07-17 | North America/USA/California/San Luis | County of San Luis Obispo Public Health Laboratory | Chan-Zuckerberg Biohub | CZB Cliahub Consortium et al |
| EPI_ISL_548282 | USA/CA-CZB-3280/2020 | 2020-07-16 | North America/USA/California/Santa Cla | County of Santa Clara Public Health Department | Chan-Zuckerberg Biohub | CZB Cliahub Consortium et al |
| EPI_ISL_548284 | USA/CA-CZB-3296/2020 | 2020-07-21 | North America/USA/California/San Luis | County of San Luis Obispo Public Health Laboratory | Chan-Zuckerberg Biohub | CZB Cliahub Consortium et al |
| EPI_ISL_548291 | USA/CA-CZB-3357/2020 | 2020-07-15 | North America/USA/California/Orange C | Orange County Public Health Laboratory | Chan-Zuckerberg Biohub | CZB Cliahub Consortium et al |
| EPI_ISL_548292 | USA/CA-CZB-3302/2020 | 2020-07-17 | North America/USA/California/San Luis | County of San Luis Obispo Public Health Laboratory | Chan-Zuckerberg Biohub | CZB Cliahub Consortium et al |
| EPI_ISL_548294 | USA/CA-CZB-3325/2020 | 2020-07-21 | North America/USA/California/San Luis | County of San Luis Obispo Public Health Laboratory | Chan-Zuckerberg Biohub | CZB Cliahub Consortium et al |
| EPI_ISL_548299 | USA/CA-CZB-3286/2020 | 2020-07-17 | North America/USA/California/Santa Cla | County of Santa Clara Public Health Department | Chan-Zuckerberg Biohub | CZB Cliahub Consortium et al |
| EPI_ISL_548301 | USA/CA-CZB-3361/2020 | 2020-07-14 | North America/USA/California/Orange C | Orange County Public Health Laboratory | Chan-Zuckerberg Biohub | CZB Cliahub Consortium et al |
| EPI_ISL_548302 | USA/CA-CZB-3292/2020 | 2020-07-19 | North America/USA/California/Santa Cla | County of Santa Clara Public Health Department | Chan-Zuckerberg Biohub | CZB Cliahub Consortium et al |
| EPI_ISL_548309 | USA/CA-CZB-3353/2020 | 2020-07-13 | North America/USA/California/Orange C | Orange County Public Health Laboratory | Chan-Zuckerberg Biohub | CZB Cliahub Consortium et al |
| EPI_ISL_548312 | USA/CA-CZB-3356/2020 | 2020-07-14 | North America/USA/California/Orange C | Orange County Public Health Laboratory | Chan-Zuckerberg Biohub | CZB Cliahub Consortium et al |
| EPI_ISL_548325 | USA/CA-CZB-3321/2020 | 2020-07-17 | North America/USA/California/San Luis | County of San Luis Obispo Public Health Laboratory | Chan-Zuckerberg Biohub | CZB Cliahub Consortium et al |
| EPI_ISL_548330 | USA/CA-CZB-3310/2020 | 2020-07-17 | North America/USA/California/San Luis | County of San Luis Obispo Public Health Laboratory | Chan-Zuckerberg Biohub | CZB Cliahub Consortium et al |
| EPI_ISL_548333 | USA/CA-CZB-3283/2020 | 2020-07-17 | North America/USA/California/Santa Cla | County of Santa Clara Public Health Department | Chan-Zuckerberg Biohub | CZB Cliahub Consortium et al |
| EPI_ISL_548339 | USA/CA-CZB-3616/2020 | 2020-07-28 | North America/USA/California/Santa Cla | County of Santa Clara Public Health Department | Chan-Zuckerberg Biohub | CZB Cliahub Consortium et al |
| EPI_ISL_548344 | USA/CA-CZB-3617/2020 | 2020-07-28 | North America/USA/California/Santa Cla | County of Santa Clara Public Health Department | Chan-Zuckerberg Biohub | CZB Cliahub Consortium et al |
| EPI_ISL_548347 | USA/CA-CZB-3634/2020 | 2020-08-01 | North America/USA/California/San Luis | County of San Luis Obispo Public Health Laboratory | Chan-Zuckerberg Biohub | CZB Cliahub Consortium et al |
| EPI_ISL_548363 | USA/CA-CZB-3673/2020 | 2020-07-13 | North America/USA/California/Tuolumn | Tuolumne County Public Health | Chan-Zuckerberg Biohub | CZB Cliahub Consortium et al |
| EPI_ISL_548365 | USA/CA-CZB-3627/2020 | 2020-07-27 | North America/USA/California/San Luis | County of San Luis Obispo Public Health Laboratory | Chan-Zuckerberg Biohub | CZB Cliahub Consortium et al |

| | | | | | | |
|---|---|---|---|---|---|---|
| EPI_ISL_548370 | USA/CA-CZB-3626/2020 | 2020-07-27 | North America/USA/California/San Luis | County of San Luis Obispo Public Health Laboratory | Chan-Zuckerberg Biohub | CZB Cliahub Consortium et al |
| EPI_ISL_548373 | USA/CA-CZB-3739/2020 | 2020-07-22 | North America/USA/California/Orange ( | Orange County Public Health Laboratory | Chan-Zuckerberg Biohub | CZB Cliahub Consortium et al |
| EPI_ISL_548375 | USA/CA-CZB-3757/2020 | 2020-07-27 | North America/USA/California/Orange ( | Orange County Public Health Laboratory | Chan-Zuckerberg Biohub | CZB Cliahub Consortium et al |
| EPI_ISL_548378 | USA/CA-CZB-3851/2020 | 2020-07-24 | North America/USA/California/Orange ( | Orange County Public Health Laboratory | Chan-Zuckerberg Biohub | CZB Cliahub Consortium et al |
| EPI_ISL_548383 | USA/CA-CZB-3754/2020 | 2020-07-27 | North America/USA/California/Orange ( | Orange County Public Health Laboratory | Chan-Zuckerberg Biohub | CZB Cliahub Consortium et al |
| EPI_ISL_548386 | USA/CA-CZB-3725/2020 | 2020-07-21 | North America/USA/California/Orange ( | Orange County Public Health Laboratory | Chan-Zuckerberg Biohub | CZB Cliahub Consortium et al |
| EPI_ISL_548388 | USA/CA-CZB-3831/2020 | 2020-07-21 | North America/USA/California/Santa Cl | County of Santa Clara Public Health Department | Chan-Zuckerberg Biohub | CZB Cliahub Consortium et al |
| EPI_ISL_548393 | USA/CA-CZB-3743/2020 | 2020-07-24 | North America/USA/California/Orange ( | Orange County Public Health Laboratory | Chan-Zuckerberg Biohub | CZB Cliahub Consortium et al |
| EPI_ISL_548404 | USA/CA-CZB-3738/2020 | 2020-07-21 | North America/USA/California/Orange ( | Orange County Public Health Laboratory | Chan-Zuckerberg Biohub | CZB Cliahub Consortium et al |
| EPI_ISL_548419 | USA/CA-CZB-3707/2020 | 2020-07-22 | North America/USA/California/Santa Cl | County of Santa Clara Public Health Department | Chan-Zuckerberg Biohub | CZB Cliahub Consortium et al |
| EPI_ISL_548424 | USA/CA-CZB-3853/2020 | 2020-07-26 | North America/USA/California/Orange ( | Orange County Public Health Laboratory | Chan-Zuckerberg Biohub | CZB Cliahub Consortium et al |
| EPI_ISL_548427 | USA/CA-CZB-3767/2020 | 2020-06-18 | North America/USA/California/Ventura | Ventura County Public Health Lab | Chan-Zuckerberg Biohub | CZB Cliahub Consortium et al |
| EPI_ISL_548436 | USA/CA-CZB-3722/2020 | 2020-07-24 | North America/USA/California/San Luis | County of San Luis Obispo Public Health Laboratory | Chan-Zuckerberg Biohub | CZB Cliahub Consortium et al |
| EPI_ISL_548438 | USA/CA-CZB-3735/2020 | 2020-07-22 | North America/USA/California/Orange ( | Orange County Public Health Laboratory | Chan-Zuckerberg Biohub | CZB Cliahub Consortium et al |
| EPI_ISL_548439 | USA/CA-CZB-3724/2020 | 2020-07-20 | North America/USA/California/Orange ( | Orange County Public Health Laboratory | Chan-Zuckerberg Biohub | CZB Cliahub Consortium et al |
| EPI_ISL_548444 | USA/CA-CZB-3726/2020 | 2020-07-21 | North America/USA/California/Orange ( | Orange County Public Health Laboratory | Chan-Zuckerberg Biohub | CZB Cliahub Consortium et al |
| EPI_ISL_548445 | USA/CA-CZB-3848/2020 | 2020-07-23 | North America/USA/California/Orange ( | Orange County Public Health Laboratory | Chan-Zuckerberg Biohub | CZB Cliahub Consortium et al |
| EPI_ISL_548450 | USA/CA-CZB-3784/2020 | 2020-06-04 | North America/USA/California/Ventura | Ventura County Public Health Lab | Chan-Zuckerberg Biohub | CZB Cliahub Consortium et al |
| EPI_ISL_548451 | USA/CA-CZB-3830/2020 | 2020-07-21 | North America/USA/California/Santa Cl | County of Santa Clara Public Health Department | Chan-Zuckerberg Biohub | CZB Cliahub Consortium et al |
| EPI_ISL_548456 | USA/CA-CZB-3740/2020 | 2020-07-23 | North America/USA/California/Orange ( | Orange County Public Health Laboratory | Chan-Zuckerberg Biohub | CZB Cliahub Consortium et al |

| | | | | | | |
|---|---|---|---|---|---|---|
| EPI_ISL_548459 | USA/CA-CZB-3775/2020 | 2020-05-30 | North America/USA/California/Ventura | Ventura County Public Health Lab | Chan-Zuckerberg Biohub | CZB Cliahub Consortium et al |
| EPI_ISL_548464 | USA/CA-CZB-3733/2020 | 2020-07-21 | North America/USA/California/Orange ( | Orange County Public Health Laboratory | Chan-Zuckerberg Biohub | CZB Cliahub Consortium et al |
| EPI_ISL_548465 | USA/CA-CZB-3750/2020 | 2020-07-27 | North America/USA/California/Orange ( | Orange County Public Health Laboratory | Chan-Zuckerberg Biohub | CZB Cliahub Consortium et al |
| EPI_ISL_548467 | USA/CA-CZB-3755/2020 | 2020-07-27 | North America/USA/California/Orange ( | Orange County Public Health Laboratory | Chan-Zuckerberg Biohub | CZB Cliahub Consortium et al |
| EPI_ISL_548470 | USA/CA-CZB-3719/2020 | 2020-08-01 | North America/USA/California/Santa Cl | County of Santa Clara Public Health Department | Chan-Zuckerberg Biohub | CZB Cliahub Consortium et al |
| EPI_ISL_548488 | USA/CA-CZB-3741/2020 | 2020-07-24 | North America/USA/California/Orange ( | Orange County Public Health Laboratory | Chan-Zuckerberg Biohub | CZB Cliahub Consortium et al |
| EPI_ISL_548497 | USA/CA-CZB-3866/2020 | 2020-07-20 | North America/USA/California/Santa Cl | County of Santa Clara Public Health Department | Chan-Zuckerberg Biohub | CZB Cliahub Consortium et al |
| EPI_ISL_548500 | USA/CA-CZB-3884/2020 | 2020-07-21 | North America/USA/California/Santa Cl | County of Santa Clara Public Health Department | Chan-Zuckerberg Biohub | CZB Cliahub Consortium et al |
| EPI_ISL_548502 | USA/CA-CZB-3904/2020 | 2020-07-27 | North America/USA/California/San Luis | County of San Luis Obispo Public Health Laboratory | Chan-Zuckerberg Biohub | CZB Cliahub Consortium et al |
| EPI_ISL_548504 | USA/CA-CZB-3908/2020 | 2020-07-27 | North America/USA/California/San Luis | County of San Luis Obispo Public Health Laboratory | Chan-Zuckerberg Biohub | CZB Cliahub Consortium et al |
| EPI_ISL_548512 | USA/CA-CZB-3886/2020 | 2020-07-21 | North America/USA/California/Santa Cl | County of Santa Clara Public Health Department | Chan-Zuckerberg Biohub | CZB Cliahub Consortium et al |
| EPI_ISL_548515 | USA/CA-CZB-3935/2020 | 2020-07-30 | North America/USA/California/Contra C | Contra Costa Public Health Lab | Chan-Zuckerberg Biohub | CZB Cliahub Consortium et al |
| EPI_ISL_548516 | USA/CA-CZB-3929/2020 | 2020-07-29 | North America/USA/California/Contra C | Contra Costa Public Health Lab | Chan-Zuckerberg Biohub | CZB Cliahub Consortium et al |
| EPI_ISL_548523 | USA/CA-CZB-3888/2020 | 2020-07-21 | North America/USA/California/Santa Cl | County of Santa Clara Public Health Department | Chan-Zuckerberg Biohub | CZB Cliahub Consortium et al |
| EPI_ISL_548532 | USA/CA-CZB-3906/2020 | 2020-07-27 | North America/USA/California/San Luis | County of San Luis Obispo Public Health Laboratory | Chan-Zuckerberg Biohub | CZB Cliahub Consortium et al |
| EPI_ISL_548535 | USA/CA-CZB-3923/2020 | 2020-07-25 | North America/USA/California/Contra C | Contra Costa Public Health Lab | Chan-Zuckerberg Biohub | CZB Cliahub Consortium et al |
| EPI_ISL_548539 | USA/CA-CZB-3911/2020 | 2020-07-23 | North America/USA/California/Contra C | Contra Costa Public Health Lab | Chan-Zuckerberg Biohub | CZB Cliahub Consortium et al |
| EPI_ISL_548540 | USA/CA-CZB-3912/2020 | 2020-07-23 | North America/USA/California/Contra C | Contra Costa Public Health Lab | Chan-Zuckerberg Biohub | CZB Cliahub Consortium et al |
| EPI_ISL_548546 | USA/CA-CZB-3881/2020 | 2020-07-17 | North America/USA/California/Santa Cl | County of Santa Clara Public Health Department | Chan-Zuckerberg Biohub | CZB Cliahub Consortium et al |
| EPI_ISL_548547 | USA/CA-CZB-3893/2020 | 2020-07-21 | North America/USA/California/Santa Cl | County of Santa Clara Public Health Department | Chan-Zuckerberg Biohub | CZB Cliahub Consortium et al |

| | | | | | | |
|---|---|---|---|---|---|---|
| EPI_ISL_548549 | USA/CA-CZB-3927/2020 | 2020-07-26 | North America/USA/California/Contra C | Contra Costa Public Health Lab | Chan-Zuckerberg Biohub | CZB Cliahub Consortium et al |
| EPI_ISL_548552 | USA/CA-CZB-3932/2020 | 2020-07-29 | North America/USA/California/Contra C | Contra Costa Public Health Lab | Chan-Zuckerberg Biohub | CZB Cliahub Consortium et al |
| EPI_ISL_548560 | USA/CA-CZB-3903/2020 | 2020-07-24 | North America/USA/California/San Luis | County of San Luis Obispo Public Health Laboratory | Chan-Zuckerberg Biohub | CZB Cliahub Consortium et al |
| EPI_ISL_548562 | USA/CA-CZB-3879/2020 | 2020-07-17 | North America/USA/California/Santa Cl | County of Santa Clara Public Health Department | Chan-Zuckerberg Biohub | CZB Cliahub Consortium et al |
| EPI_ISL_548567 | USA/CA-CZB-3910/2020 | 2020-07-22 | North America/USA/California/Contra C | Contra Costa Public Health Lab | Chan-Zuckerberg Biohub | CZB Cliahub Consortium et al |
| EPI_ISL_548569 | USA/CA-CZB-3943/2020 | 2020-07-31 | North America/USA/California/Contra C | Contra Costa Public Health Lab | Chan-Zuckerberg Biohub | CZB Cliahub Consortium et al |
| EPI_ISL_548570 | USA/CA-CZB-3871/2020 | 2020-07-17 | North America/USA/California/Santa Cl | County of Santa Clara Public Health Department | Chan-Zuckerberg Biohub | CZB Cliahub Consortium et al |
| EPI_ISL_548576 | USA/CA-CZB-3870/2020 | 2020-07-17 | North America/USA/California/Santa Cl | County of Santa Clara Public Health Department | Chan-Zuckerberg Biohub | CZB Cliahub Consortium et al |
| EPI_ISL_548582 | USA/CA-CZB-4029/2020 | 2020-08-05 | North America/USA/California/Santa Cl | County of Santa Clara Public Health Department | Chan-Zuckerberg Biohub | CZB Cliahub Consortium et al |
| EPI_ISL_548583 | USA/CA-CZB-4044/2020 | 2020-08-12 | North America/USA/California/Santa Cl | County of Santa Clara Public Health Department | Chan-Zuckerberg Biohub | CZB Cliahub Consortium et al |
| EPI_ISL_548585 | USA/CA-CZB-4076/2020 | 2020-08-05 | North America/USA/California/Orange C | Orange County Public Health Laboratory | Chan-Zuckerberg Biohub | CZB Cliahub Consortium et al |
| EPI_ISL_548598 | USA/CA-CZB-3995/2020 | 2020-07-29 | North America/USA/California/Santa Cl | County of Santa Clara Public Health Department | Chan-Zuckerberg Biohub | CZB Cliahub Consortium et al |
| EPI_ISL_548605 | USA/CA-CZB-3997/2020 | 2020-07-30 | North America/USA/California/Santa Cl | County of Santa Clara Public Health Department | Chan-Zuckerberg Biohub | CZB Cliahub Consortium et al |
| EPI_ISL_548606 | USA/CA-CZB-4033/2020 | 2020-08-10 | North America/USA/California/Santa Cl | County of Santa Clara Public Health Department | Chan-Zuckerberg Biohub | CZB Cliahub Consortium et al |
| EPI_ISL_548609 | USA/CA-CZB-4087/2020 | 2020-08-10 | North America/USA/California/Orange C | Orange County Public Health Laboratory | Chan-Zuckerberg Biohub | CZB Cliahub Consortium et al |
| EPI_ISL_548610 | USA/CA-CZB-4041/2020 | 2020-08-12 | North America/USA/California/Santa Cl | County of Santa Clara Public Health Department | Chan-Zuckerberg Biohub | CZB Cliahub Consortium et al |
| EPI_ISL_548612 | USA/CA-CZB-4009/2020 | 2020-08-01 | North America/USA/California/Santa Cl | County of Santa Clara Public Health Department | Chan-Zuckerberg Biohub | CZB Cliahub Consortium et al |
| EPI_ISL_548613 | USA/CA-CZB-4083/2020 | 2020-08-11 | North America/USA/California/Orange C | Orange County Public Health Laboratory | Chan-Zuckerberg Biohub | CZB Cliahub Consortium et al |
| EPI_ISL_548620 | USA/CA-CZB-4089/2020 | 2020-08-12 | North America/USA/California/Orange C | Orange County Public Health Laboratory | Chan-Zuckerberg Biohub | CZB Cliahub Consortium et al |
| EPI_ISL_548622 | USA/CA-CZB-4042/2020 | 2020-08-12 | North America/USA/California/Santa Cl | County of Santa Clara Public Health Department | Chan-Zuckerberg Biohub | CZB Cliahub Consortium et al |

| | | | | | | |
|---|---|---|---|---|---|---|
| EPI_ISL_548623 | USA/CA-CZB-4086/2020 | 2020-08-10 | North America/USA/California/Orange ( | Orange County Public Health Laboratory | Chan-Zuckerberg Biohub | CZB Cliahub Consortium et al |
| EPI_ISL_548632 | USA/CA-CZB-3996/2020 | 2020-07-29 | North America/USA/California/Santa Cl | County of Santa Clara Public Health Department | Chan-Zuckerberg Biohub | CZB Cliahub Consortium et al |
| EPI_ISL_548633 | USA/CA-CZB-4035/2020 | 2020-08-10 | North America/USA/California/Santa Cl | County of Santa Clara Public Health Department | Chan-Zuckerberg Biohub | CZB Cliahub Consortium et al |
| EPI_ISL_548634 | USA/CA-CZB-4091/2020 | 2020-08-13 | North America/USA/California/Orange ( | Orange County Public Health Laboratory | Chan-Zuckerberg Biohub | CZB Cliahub Consortium et al |
| EPI_ISL_548641 | USA/CA-CZB-4098/2020 | 2020-08-14 | North America/USA/California/Orange ( | Orange County Public Health Laboratory | Chan-Zuckerberg Biohub | CZB Cliahub Consortium et al |
| EPI_ISL_548644 | USA/CA-CZB-4054/2020 | 2020-07-16 | North America/USA/California/San Luis | County of San Luis Obispo Public Health Laboratory | Chan-Zuckerberg Biohub | CZB Cliahub Consortium et al |
| EPI_ISL_548648 | USA/CA-CZB-4065/2020 | 2020-08-04 | North America/USA/California/Orange ( | Orange County Public Health Laboratory | Chan-Zuckerberg Biohub | CZB Cliahub Consortium et al |
| EPI_ISL_548651 | USA/CA-CZB-4081/2020 | 2020-08-10 | North America/USA/California/Orange ( | Orange County Public Health Laboratory | Chan-Zuckerberg Biohub | CZB Cliahub Consortium et al |
| EPI_ISL_548653 | USA/CA-CZB-4064/2020 | 2020-08-04 | North America/USA/California/Orange ( | Orange County Public Health Laboratory | Chan-Zuckerberg Biohub | CZB Cliahub Consortium et al |
| EPI_ISL_548655 | USA/CA-CZB-4004/2020 | 2020-08-03 | North America/USA/California/Santa Cl | County of Santa Clara Public Health Department | Chan-Zuckerberg Biohub | CZB Cliahub Consortium et al |
| EPI_ISL_548662 | USA/CA-CZB-4062/2020 | 2020-08-04 | North America/USA/California/Orange ( | Orange County Public Health Laboratory | Chan-Zuckerberg Biohub | CZB Cliahub Consortium et al |
| EPI_ISL_548666 | USA/CA-CZB-4099/2020 | 2020-08-15 | North America/USA/California/Orange ( | Orange County Public Health Laboratory | Chan-Zuckerberg Biohub | CZB Cliahub Consortium et al |
| EPI_ISL_548671 | USA/CA-CZB-4056/2020 | 2020-07-16 | North America/USA/California/San Luis | County of San Luis Obispo Public Health Laboratory | Chan-Zuckerberg Biohub | CZB Cliahub Consortium et al |
| EPI_ISL_548675 | USA/CA-CZB-4074/2020 | 2020-08-05 | North America/USA/California/Orange ( | Orange County Public Health Laboratory | Chan-Zuckerberg Biohub | CZB Cliahub Consortium et al |
| EPI_ISL_548762 | Canada/ON-PHL-20-00169/2020 | 20-Mar | North America/Canada/Ontario/ | Public Health Ontario Laboratory | Public Health Ontario Laboratory | Vanessa G Allen et al |
| EPI_ISL_548763 | Canada/ON-PHL-20-00170/2020 | 20-Mar | North America/Canada/Ontario/ | Public Health Ontario Laboratory | Public Health Ontario Laboratory | Vanessa G Allen et al |
| EPI_ISL_548764 | Canada/ON-PHL-20-00171/2020 | 20-Mar | North America/Canada/Ontario/ | Public Health Ontario Laboratory | Public Health Ontario Laboratory | Vanessa G Allen et al |
| EPI_ISL_548765 | Canada/ON-PHL-20-00172/2020 | 20-Mar | North America/Canada/Ontario/ | Public Health Ontario Laboratory | Public Health Ontario Laboratory | Vanessa G Allen et al |

| | | | | | | |
|---|---|---|---|---|---|---|
| EPI_ISL_548766 | Canada/ON-PHL-20-00173/2020 | 20-Mar | North America/Canada/Ontario/ | Public Health Ontario Laboratory | Public Health Ontario Laboratory | Vanessa G Allen et al |
| EPI_ISL_548767 | Canada/ON-PHL-20-00174/2020 | 20-Mar | North America/Canada/Ontario/ | Public Health Ontario Laboratory | Public Health Ontario Laboratory | Vanessa G Allen et al |
| EPI_ISL_548768 | Canada/ON-PHL-20-00233/2020 | 20-Mar | North America/Canada/Ontario/ | Public Health Ontario Laboratory | Public Health Ontario Laboratory | Vanessa G Allen et al |
| EPI_ISL_548769 | Canada/ON-PHL-20-00234/2020 | 20-Mar | North America/Canada/Ontario/ | Public Health Ontario Laboratory | Public Health Ontario Laboratory | Vanessa G Allen et al |
| EPI_ISL_548770 | Canada/ON-PHL-20-00235/2020 | 20-Mar | North America/Canada/Ontario/ | Public Health Ontario Laboratory | Public Health Ontario Laboratory | Vanessa G Allen et al |
| EPI_ISL_548771 | Canada/ON-PHL-20-00236/2020 | 20-Mar | North America/Canada/Ontario/ | Public Health Ontario Laboratory | Public Health Ontario Laboratory | Vanessa G Allen et al |
| EPI_ISL_548772 | Canada/ON-PHL-20-00237/2020 | 20-Mar | North America/Canada/Ontario/ | Public Health Ontario Laboratory | Public Health Ontario Laboratory | Vanessa G Allen et al |
| EPI_ISL_548773 | Canada/ON-PHL-20-00238/2020 | 20-Mar | North America/Canada/Ontario/ | Public Health Ontario Laboratory | Public Health Ontario Laboratory | Vanessa G Allen et al |
| EPI_ISL_548774 | Canada/ON-PHL-20-00320/2020 | 20-Mar | North America/Canada/Ontario/ | Public Health Ontario Laboratory | Public Health Ontario Laboratory | Vanessa G Allen et al |
| EPI_ISL_548775 | Canada/ON-PHL-20-00384/2020 | 20-Mar | North America/Canada/Ontario/ | Public Health Ontario Laboratory | Public Health Ontario Laboratory | Vanessa G Allen et al |
| EPI_ISL_548776 | Canada/ON-PHL-20-00385/2020 | 20-Mar | North America/Canada/Ontario/ | Public Health Ontario Laboratory | Public Health Ontario Laboratory | Vanessa G Allen et al |
| EPI_ISL_548777 | Canada/ON-PHL-20-00386/2020 | 20-Mar | North America/Canada/Ontario/ | Public Health Ontario Laboratory | Public Health Ontario Laboratory | Vanessa G Allen et al |
| EPI_ISL_548778 | Canada/ON-PHL-20-00397/2020 | 20-Mar | North America/Canada/Ontario/ | Public Health Ontario Laboratory | Public Health Ontario Laboratory | Vanessa G Allen et al |

| | | | | | | |
|---|---|---|---|---|---|---|
| EPI_ISL_548779 | Canada/ON-PHL-20-00820/2020 | 20-Jun | North America/Canada/Ontario/ | Public Health Ontario Laboratory | Public Health Ontario Laboratory | Vanessa G Allen et al |
| EPI_ISL_548780 | Canada/ON-PHL-20-00987/2020 | 20-Mar | North America/Canada/Ontario/ | Public Health Ontario Laboratory | Public Health Ontario Laboratory | Vanessa G Allen et al |
| EPI_ISL_548781 | Canada/ON-PHL-20-01158/2020 | 20-Apr | North America/Canada/Ontario/ | Public Health Ontario Laboratory | Public Health Ontario Laboratory | Vanessa G Allen et al |
| EPI_ISL_548782 | Canada/ON-PHL-20-01160/2020 | 20-Jun | North America/Canada/Ontario/ | Public Health Ontario Laboratory | Public Health Ontario Laboratory | Vanessa G Allen et al |
| EPI_ISL_548783 | Canada/ON-PHL-20-01161/2020 | 20-Jun | North America/Canada/Ontario/ | Public Health Ontario Laboratory | Public Health Ontario Laboratory | Vanessa G Allen et al |
| EPI_ISL_548784 | Canada/ON-PHL-20-01162/2020 | 20-Jun | North America/Canada/Ontario/ | Public Health Ontario Laboratory | Public Health Ontario Laboratory | Vanessa G Allen et al |
| EPI_ISL_548785 | Canada/ON-PHL-20-01163/2020 | 20-Jun | North America/Canada/Ontario/ | Public Health Ontario Laboratory | Public Health Ontario Laboratory | Vanessa G Allen et al |
| EPI_ISL_548786 | Canada/ON-PHL-20-01164/2020 | 20-Jun | North America/Canada/Ontario/ | Public Health Ontario Laboratory | Public Health Ontario Laboratory | Vanessa G Allen et al |
| EPI_ISL_548787 | Canada/ON-PHL-20-01165/2020 | 20-Jun | North America/Canada/Ontario/ | Public Health Ontario Laboratory | Public Health Ontario Laboratory | Vanessa G Allen et al |
| EPI_ISL_548788 | Canada/ON-PHL-20-01166/2020 | 20-Jun | North America/Canada/Ontario/ | Public Health Ontario Laboratory | Public Health Ontario Laboratory | Vanessa G Allen et al |
| EPI_ISL_548789 | Canada/ON-PHL-20-01167/2020 | 20-Jun | North America/Canada/Ontario/ | Public Health Ontario Laboratory | Public Health Ontario Laboratory | Vanessa G Allen et al |
| EPI_ISL_548790 | Canada/ON-PHL-20-01168/2020 | 20-Jun | North America/Canada/Ontario/ | Public Health Ontario Laboratory | Public Health Ontario Laboratory | Vanessa G Allen et al |
| EPI_ISL_548791 | Canada/ON-PHL-20-01169/2020 | 20-Jun | North America/Canada/Ontario/ | Public Health Ontario Laboratory | Public Health Ontario Laboratory | Vanessa G Allen et al |

| | | | | | | |
|---|---|---|---|---|---|---|
| EPI_ISL_548792 | Canada/ON-PHL-20-01170/2020 | 20-Jun | North America/Canada/Ontario/ | Public Health Ontario Laboratory | Public Health Ontario Laboratory | Vanessa G Allen et al |
| EPI_ISL_548793 | Canada/ON-PHL-20-01171/2020 | 20-Jun | North America/Canada/Ontario/ | Public Health Ontario Laboratory | Public Health Ontario Laboratory | Vanessa G Allen et al |
| EPI_ISL_548794 | Canada/ON-PHL-20-01172/2020 | 20-Jun | North America/Canada/Ontario/ | Public Health Ontario Laboratory | Public Health Ontario Laboratory | Vanessa G Allen et al |
| EPI_ISL_548795 | Canada/ON-PHL-20-01173/2020 | 20-Jun | North America/Canada/Ontario/ | Public Health Ontario Laboratory | Public Health Ontario Laboratory | Vanessa G Allen et al |
| EPI_ISL_548796 | Canada/ON-PHL-20-01174/2020 | 20-Jun | North America/Canada/Ontario/ | Public Health Ontario Laboratory | Public Health Ontario Laboratory | Vanessa G Allen et al |
| EPI_ISL_548797 | Canada/ON-PHL-20-01175/2020 | 20-Jun | North America/Canada/Ontario/ | Public Health Ontario Laboratory | Public Health Ontario Laboratory | Vanessa G Allen et al |
| EPI_ISL_548798 | Canada/ON-PHL-20-01176/2020 | 20-Jun | North America/Canada/Ontario/ | Public Health Ontario Laboratory | Public Health Ontario Laboratory | Vanessa G Allen et al |
| EPI_ISL_548799 | Canada/ON-PHL-20-01177/2020 | 20-Jun | North America/Canada/Ontario/ | Public Health Ontario Laboratory | Public Health Ontario Laboratory | Vanessa G Allen et al |
| EPI_ISL_548800 | Canada/ON-PHL-20-01178/2020 | 20-Jun | North America/Canada/Ontario/ | Public Health Ontario Laboratory | Public Health Ontario Laboratory | Vanessa G Allen et al |
| EPI_ISL_548801 | Canada/ON-PHL-20-01179/2020 | 20-Jun | North America/Canada/Ontario/ | Public Health Ontario Laboratory | Public Health Ontario Laboratory | Vanessa G Allen et al |
| EPI_ISL_548802 | Canada/ON-PHL-20-01180/2020 | 20-Jun | North America/Canada/Ontario/ | Public Health Ontario Laboratory | Public Health Ontario Laboratory | Vanessa G Allen et al |
| EPI_ISL_548803 | Canada/ON-PHL-20-01181/2020 | 20-Jun | North America/Canada/Ontario/ | Public Health Ontario Laboratory | Public Health Ontario Laboratory | Vanessa G Allen et al |
| EPI_ISL_548804 | Canada/ON-PHL-20-01182/2020 | 20-Jun | North America/Canada/Ontario/ | Public Health Ontario Laboratory | Public Health Ontario Laboratory | Vanessa G Allen et al |

| | | | | | | |
|---|---|---|---|---|---|---|
| EPI_ISL_548805 | Canada/ON-PHL-20-01183/2020 | 20-Jun | North America/Canada/Ontario/ | Public Health Ontario Laboratory | Public Health Ontario Laboratory | Vanessa G Allen et al |
| EPI_ISL_548806 | Canada/ON-PHL-20-01184/2020 | 20-Jun | North America/Canada/Ontario/ | Public Health Ontario Laboratory | Public Health Ontario Laboratory | Vanessa G Allen et al |
| EPI_ISL_548807 | Canada/ON-PHL-20-01185/2020 | 20-Jun | North America/Canada/Ontario/ | Public Health Ontario Laboratory | Public Health Ontario Laboratory | Vanessa G Allen et al |
| EPI_ISL_548808 | Canada/ON-PHL-20-01186/2020 | 20-Jun | North America/Canada/Ontario/ | Public Health Ontario Laboratory | Public Health Ontario Laboratory | Vanessa G Allen et al |
| EPI_ISL_548809 | Canada/ON-PHL-20-01187/2020 | 20-Jun | North America/Canada/Ontario/ | Public Health Ontario Laboratory | Public Health Ontario Laboratory | Vanessa G Allen et al |
| EPI_ISL_548810 | Canada/ON-PHL-20-01188/2020 | 20-Jun | North America/Canada/Ontario/ | Public Health Ontario Laboratory | Public Health Ontario Laboratory | Vanessa G Allen et al |
| EPI_ISL_548811 | Canada/ON-PHL-20-01189/2020 | 20-Jun | North America/Canada/Ontario/ | Public Health Ontario Laboratory | Public Health Ontario Laboratory | Vanessa G Allen et al |
| EPI_ISL_548812 | Canada/ON-PHL-20-01190/2020 | 20-Jun | North America/Canada/Ontario/ | Public Health Ontario Laboratory | Public Health Ontario Laboratory | Vanessa G Allen et al |
| EPI_ISL_548813 | Canada/ON-PHL-20-01191/2020 | 20-Jun | North America/Canada/Ontario/ | Public Health Ontario Laboratory | Public Health Ontario Laboratory | Vanessa G Allen et al |
| EPI_ISL_548814 | Canada/ON-PHL-20-01192/2020 | 20-Jun | North America/Canada/Ontario/ | Public Health Ontario Laboratory | Public Health Ontario Laboratory | Vanessa G Allen et al |
| EPI_ISL_548815 | Canada/ON-PHL-20-01193/2020 | 20-Jun | North America/Canada/Ontario/ | Public Health Ontario Laboratory | Public Health Ontario Laboratory | Vanessa G Allen et al |
| EPI_ISL_548816 | Canada/ON-PHL-20-01194/2020 | 20-Jun | North America/Canada/Ontario/ | Public Health Ontario Laboratory | Public Health Ontario Laboratory | Vanessa G Allen et al |
| EPI_ISL_548817 | Canada/ON-PHL-20-01195/2020 | 20-Jun | North America/Canada/Ontario/ | Public Health Ontario Laboratory | Public Health Ontario Laboratory | Vanessa G Allen et al |

| | | | | | | |
|---|---|---|---|---|---|---|
| EPI_ISL_548818 | Canada/ON-PHL-20-01196/2020 | 20-Jun | North America/Canada/Ontario/ | Public Health Ontario Laboratory | Public Health Ontario Laboratory | Vanessa G Allen et al |
| EPI_ISL_548819 | Canada/ON-PHL-20-01197/2020 | 20-Jun | North America/Canada/Ontario/ | Public Health Ontario Laboratory | Public Health Ontario Laboratory | Vanessa G Allen et al |
| EPI_ISL_548820 | Canada/ON-PHL-20-01198/2020 | 20-Jun | North America/Canada/Ontario/ | Public Health Ontario Laboratory | Public Health Ontario Laboratory | Vanessa G Allen et al |
| EPI_ISL_548821 | Canada/ON-PHL-20-01199/2020 | 20-Jun | North America/Canada/Ontario/ | Public Health Ontario Laboratory | Public Health Ontario Laboratory | Vanessa G Allen et al |
| EPI_ISL_548822 | Canada/ON-PHL-20-01200/2020 | 20-Jun | North America/Canada/Ontario/ | Public Health Ontario Laboratory | Public Health Ontario Laboratory | Vanessa G Allen et al |
| EPI_ISL_548823 | Canada/ON-PHL-20-01201/2020 | 20-Jun | North America/Canada/Ontario/ | Public Health Ontario Laboratory | Public Health Ontario Laboratory | Vanessa G Allen et al |
| EPI_ISL_548824 | Canada/ON-PHL-20-01202/2020 | 20-Jun | North America/Canada/Ontario/ | Public Health Ontario Laboratory | Public Health Ontario Laboratory | Vanessa G Allen et al |
| EPI_ISL_548825 | Canada/ON-PHL-20-01203/2020 | 20-Jun | North America/Canada/Ontario/ | Public Health Ontario Laboratory | Public Health Ontario Laboratory | Vanessa G Allen et al |
| EPI_ISL_548826 | Canada/ON-PHL-20-01204/2020 | 20-Jun | North America/Canada/Ontario/ | Public Health Ontario Laboratory | Public Health Ontario Laboratory | Vanessa G Allen et al |
| EPI_ISL_548827 | Canada/ON-PHL-20-01205/2020 | 20-Jun | North America/Canada/Ontario/ | Public Health Ontario Laboratory | Public Health Ontario Laboratory | Vanessa G Allen et al |
| EPI_ISL_548828 | Canada/ON-PHL-20-01206/2020 | 20-Jun | North America/Canada/Ontario/ | Public Health Ontario Laboratory | Public Health Ontario Laboratory | Vanessa G Allen et al |
| EPI_ISL_548829 | Canada/ON-PHL-20-01207/2020 | 20-Jun | North America/Canada/Ontario/ | Public Health Ontario Laboratory | Public Health Ontario Laboratory | Vanessa G Allen et al |
| EPI_ISL_548830 | Canada/ON-PHL-20-01208/2020 | 20-Jun | North America/Canada/Ontario/ | Public Health Ontario Laboratory | Public Health Ontario Laboratory | Vanessa G Allen et al |

| | | | | | | |
|---|---|---|---|---|---|---|
| EPI_ISL_548831 | Canada/ON-PHL-20-01209/2020 | 20-Jun | North America/Canada/Ontario/ | Public Health Ontario Laboratory | Public Health Ontario Laboratory | Vanessa G Allen et al |
| EPI_ISL_548832 | Canada/ON-PHL-20-01210/2020 | 20-Jun | North America/Canada/Ontario/ | Public Health Ontario Laboratory | Public Health Ontario Laboratory | Vanessa G Allen et al |
| EPI_ISL_548833 | Canada/ON-PHL-20-01211/2020 | 20-Jun | North America/Canada/Ontario/ | Public Health Ontario Laboratory | Public Health Ontario Laboratory | Vanessa G Allen et al |
| EPI_ISL_548834 | Canada/ON-PHL-20-01212/2020 | 20-Jun | North America/Canada/Ontario/ | Public Health Ontario Laboratory | Public Health Ontario Laboratory | Vanessa G Allen et al |
| EPI_ISL_548835 | Canada/ON-PHL-20-01213/2020 | 20-Jun | North America/Canada/Ontario/ | Public Health Ontario Laboratory | Public Health Ontario Laboratory | Vanessa G Allen et al |
| EPI_ISL_548836 | Canada/ON-PHL-20-01214/2020 | 20-Jun | North America/Canada/Ontario/ | Public Health Ontario Laboratory | Public Health Ontario Laboratory | Vanessa G Allen et al |
| EPI_ISL_548837 | Canada/ON-PHL-20-01215/2020 | 20-Jun | North America/Canada/Ontario/ | Public Health Ontario Laboratory | Public Health Ontario Laboratory | Vanessa G Allen et al |
| EPI_ISL_548838 | Canada/ON-PHL-20-01216/2020 | 20-Jun | North America/Canada/Ontario/ | Public Health Ontario Laboratory | Public Health Ontario Laboratory | Vanessa G Allen et al |
| EPI_ISL_548839 | Canada/ON-PHL-20-01217/2020 | 20-Jun | North America/Canada/Ontario/ | Public Health Ontario Laboratory | Public Health Ontario Laboratory | Vanessa G Allen et al |
| EPI_ISL_548840 | Canada/ON-PHL-20-01218/2020 | 20-Jun | North America/Canada/Ontario/ | Public Health Ontario Laboratory | Public Health Ontario Laboratory | Vanessa G Allen et al |
| EPI_ISL_548841 | Canada/ON-PHL-20-01219/2020 | 20-Jun | North America/Canada/Ontario/ | Public Health Ontario Laboratory | Public Health Ontario Laboratory | Vanessa G Allen et al |
| EPI_ISL_548842 | Canada/ON-PHL-20-01220/2020 | 20-Jun | North America/Canada/Ontario/ | Public Health Ontario Laboratory | Public Health Ontario Laboratory | Vanessa G Allen et al |
| EPI_ISL_548843 | Canada/ON-PHL-20-01221/2020 | 20-Jun | North America/Canada/Ontario/ | Public Health Ontario Laboratory | Public Health Ontario Laboratory | Vanessa G Allen et al |

| | | | | | | |
|---|---|---|---|---|---|---|
| EPI_ISL_548844 | Canada/ON-PHL-20-01222/2020 | 20-Jun | North America/Canada/Ontario/ | Public Health Ontario Laboratory | Public Health Ontario Laboratory | Vanessa G Allen et al |
| EPI_ISL_548845 | Canada/ON-PHL-20-01223/2020 | 20-Jun | North America/Canada/Ontario/ | Public Health Ontario Laboratory | Public Health Ontario Laboratory | Vanessa G Allen et al |
| EPI_ISL_548846 | Canada/ON-PHL-20-01224/2020 | 20-Jun | North America/Canada/Ontario/ | Public Health Ontario Laboratory | Public Health Ontario Laboratory | Vanessa G Allen et al |
| EPI_ISL_548847 | Canada/ON-PHL-20-01225/2020 | 20-Jun | North America/Canada/Ontario/ | Public Health Ontario Laboratory | Public Health Ontario Laboratory | Vanessa G Allen et al |
| EPI_ISL_548848 | Canada/ON-PHL-20-01226/2020 | 20-Jun | North America/Canada/Ontario/ | Public Health Ontario Laboratory | Public Health Ontario Laboratory | Vanessa G Allen et al |
| EPI_ISL_548849 | Canada/ON-PHL-20-01227/2020 | 20-Jun | North America/Canada/Ontario/ | Public Health Ontario Laboratory | Public Health Ontario Laboratory | Vanessa G Allen et al |
| EPI_ISL_548850 | Canada/ON-PHL-20-01228/2020 | 20-Jun | North America/Canada/Ontario/ | Public Health Ontario Laboratory | Public Health Ontario Laboratory | Vanessa G Allen et al |
| EPI_ISL_548851 | Canada/ON-PHL-20-01229/2020 | 20-Jun | North America/Canada/Ontario/ | Public Health Ontario Laboratory | Public Health Ontario Laboratory | Vanessa G Allen et al |
| EPI_ISL_548852 | Canada/ON-PHL-20-01230/2020 | 20-Jun | North America/Canada/Ontario/ | Public Health Ontario Laboratory | Public Health Ontario Laboratory | Vanessa G Allen et al |
| EPI_ISL_548853 | Canada/ON-PHL-20-01231/2020 | 20-Jun | North America/Canada/Ontario/ | Public Health Ontario Laboratory | Public Health Ontario Laboratory | Vanessa G Allen et al |
| EPI_ISL_548854 | Canada/ON-PHL-20-01232/2020 | 20-Jun | North America/Canada/Ontario/ | Public Health Ontario Laboratory | Public Health Ontario Laboratory | Vanessa G Allen et al |
| EPI_ISL_548855 | Canada/ON-PHL-20-01233/2020 | 20-Jun | North America/Canada/Ontario/ | Public Health Ontario Laboratory | Public Health Ontario Laboratory | Vanessa G Allen et al |
| EPI_ISL_548856 | Canada/ON-PHL-20-01234/2020 | 20-Jun | North America/Canada/Ontario/ | Public Health Ontario Laboratory | Public Health Ontario Laboratory | Vanessa G Allen et al |

| | | | | | | |
|---|---|---|---|---|---|---|
| EPI_ISL_548857 | Canada/ON-PHL-20-01235/2020 | 20-Jun | North America/Canada/Ontario/ | Public Health Ontario Laboratory | Public Health Ontario Laboratory | Vanessa G Allen et al |
| EPI_ISL_548858 | Canada/ON-PHL-20-01236/2020 | 20-Jun | North America/Canada/Ontario/ | Public Health Ontario Laboratory | Public Health Ontario Laboratory | Vanessa G Allen et al |
| EPI_ISL_548859 | Canada/ON-PHL-20-01237/2020 | 20-Jun | North America/Canada/Ontario/ | Public Health Ontario Laboratory | Public Health Ontario Laboratory | Vanessa G Allen et al |
| EPI_ISL_548860 | Canada/ON-PHL-20-01238/2020 | 20-Jun | North America/Canada/Ontario/ | Public Health Ontario Laboratory | Public Health Ontario Laboratory | Vanessa G Allen et al |
| EPI_ISL_548861 | Canada/ON-PHL-20-01239/2020 | 20-Jun | North America/Canada/Ontario/ | Public Health Ontario Laboratory | Public Health Ontario Laboratory | Vanessa G Allen et al |
| EPI_ISL_548862 | Canada/ON-PHL-20-01240/2020 | 20-Jun | North America/Canada/Ontario/ | Public Health Ontario Laboratory | Public Health Ontario Laboratory | Vanessa G Allen et al |
| EPI_ISL_548863 | Canada/ON-PHL-20-01241/2020 | 20-Jun | North America/Canada/Ontario/ | Public Health Ontario Laboratory | Public Health Ontario Laboratory | Vanessa G Allen et al |
| EPI_ISL_548864 | Canada/ON-PHL-20-01242/2020 | 20-Jun | North America/Canada/Ontario/ | Public Health Ontario Laboratory | Public Health Ontario Laboratory | Vanessa G Allen et al |
| EPI_ISL_548865 | Canada/ON-PHL-20-01243/2020 | 20-Jun | North America/Canada/Ontario/ | Public Health Ontario Laboratory | Public Health Ontario Laboratory | Vanessa G Allen et al |
| EPI_ISL_548866 | Canada/ON-PHL-20-01244/2020 | 20-Jun | North America/Canada/Ontario/ | Public Health Ontario Laboratory | Public Health Ontario Laboratory | Vanessa G Allen et al |
| EPI_ISL_548867 | Canada/ON-PHL-20-01245/2020 | 20-Jun | North America/Canada/Ontario/ | Public Health Ontario Laboratory | Public Health Ontario Laboratory | Vanessa G Allen et al |
| EPI_ISL_548868 | Canada/ON-PHL-20-01246/2020 | 20-Jun | North America/Canada/Ontario/ | Public Health Ontario Laboratory | Public Health Ontario Laboratory | Vanessa G Allen et al |
| EPI_ISL_548869 | Canada/ON-PHL-20-01247/2020 | 20-Jun | North America/Canada/Ontario/ | Public Health Ontario Laboratory | Public Health Ontario Laboratory | Vanessa G Allen et al |

| | | | | | | |
|---|---|---|---|---|---|---|
| EPI_ISL_548870 | Canada/ON-PHL-20-01248/2020 | 20-Jun | North America/Canada/Ontario/ | Public Health Ontario Laboratory | Public Health Ontario Laboratory | Vanessa G Allen et al |
| EPI_ISL_548871 | Canada/ON-PHL-20-01249/2020 | 20-Jun | North America/Canada/Ontario/ | Public Health Ontario Laboratory | Public Health Ontario Laboratory | Vanessa G Allen et al |
| EPI_ISL_548872 | Canada/ON-PHL-20-01250/2020 | 20-Jun | North America/Canada/Ontario/ | Public Health Ontario Laboratory | Public Health Ontario Laboratory | Vanessa G Allen et al |
| EPI_ISL_548873 | Canada/ON-PHL-20-01255/2020 | 20-Jun | North America/Canada/Ontario/ | Public Health Ontario Laboratory | Public Health Ontario Laboratory | Vanessa G Allen et al |
| EPI_ISL_548874 | Canada/ON-PHL-20-01256/2020 | 20-Jun | North America/Canada/Ontario/ | Public Health Ontario Laboratory | Public Health Ontario Laboratory | Vanessa G Allen et al |
| EPI_ISL_548875 | Canada/ON-PHL-20-01260/2020 | 20-Jun | North America/Canada/Ontario/ | Public Health Ontario Laboratory | Public Health Ontario Laboratory | Vanessa G Allen et al |
| EPI_ISL_548876 | Canada/ON-PHL-20-01261/2020 | 20-Jun | North America/Canada/Ontario/ | Public Health Ontario Laboratory | Public Health Ontario Laboratory | Vanessa G Allen et al |
| EPI_ISL_548878 | Canada/ON-PHL-20-01264/2020 | 20-Jun | North America/Canada/Ontario/ | Public Health Ontario Laboratory | Public Health Ontario Laboratory | Vanessa G Allen et al |
| EPI_ISL_548879 | Canada/ON-PHL-20-01265/2020 | 20-Jun | North America/Canada/Ontario/ | Public Health Ontario Laboratory | Public Health Ontario Laboratory | Vanessa G Allen et al |
| EPI_ISL_548880 | Canada/ON-PHL-20-01272/2020 | 20-Jun | North America/Canada/Ontario/ | Public Health Ontario Laboratory | Public Health Ontario Laboratory | Vanessa G Allen et al |
| EPI_ISL_548881 | Canada/ON-PHL-20-01273/2020 | 20-Jun | North America/Canada/Ontario/ | Public Health Ontario Laboratory | Public Health Ontario Laboratory | Vanessa G Allen et al |
| EPI_ISL_548882 | Canada/ON-PHL-20-01274/2020 | 20-Jun | North America/Canada/Ontario/ | Public Health Ontario Laboratory | Public Health Ontario Laboratory | Vanessa G Allen et al |
| EPI_ISL_548883 | Canada/ON-PHL-20-01276/2020 | 20-Jun | North America/Canada/Ontario/ | Public Health Ontario Laboratory | Public Health Ontario Laboratory | Vanessa G Allen et al |

| | | | | | | |
|---|---|---|---|---|---|---|
| EPI_ISL_548884 | Canada/ON-PHL-20-01277/2020 | 20-Jun | North America/Canada/Ontario/ | Public Health Ontario Laboratory | Public Health Ontario Laboratory | Vanessa G Allen et al |
| EPI_ISL_548885 | Canada/ON-PHL-20-01278/2020 | 20-Jun | North America/Canada/Ontario/ | Public Health Ontario Laboratory | Public Health Ontario Laboratory | Vanessa G Allen et al |
| EPI_ISL_548886 | Canada/ON-PHL-20-01280/2020 | 20-Jun | North America/Canada/Ontario/ | Public Health Ontario Laboratory | Public Health Ontario Laboratory | Vanessa G Allen et al |
| EPI_ISL_548887 | Canada/ON-PHL-20-01282/2020 | 20-Jun | North America/Canada/Ontario/ | Public Health Ontario Laboratory | Public Health Ontario Laboratory | Vanessa G Allen et al |
| EPI_ISL_548888 | Canada/ON-PHL-20-01283/2020 | 20-Jun | North America/Canada/Ontario/ | Public Health Ontario Laboratory | Public Health Ontario Laboratory | Vanessa G Allen et al |
| EPI_ISL_548889 | Canada/ON-PHL-20-01284/2020 | 20-Jun | North America/Canada/Ontario/ | Public Health Ontario Laboratory | Public Health Ontario Laboratory | Vanessa G Allen et al |
| EPI_ISL_548890 | Canada/ON-PHL-20-01285/2020 | 20-Jun | North America/Canada/Ontario/ | Public Health Ontario Laboratory | Public Health Ontario Laboratory | Vanessa G Allen et al |
| EPI_ISL_548891 | Canada/ON-PHL-20-01286/2020 | 20-Jun | North America/Canada/Ontario/ | Public Health Ontario Laboratory | Public Health Ontario Laboratory | Vanessa G Allen et al |
| EPI_ISL_548892 | Canada/ON-PHL-20-01287/2020 | 20-Jun | North America/Canada/Ontario/ | Public Health Ontario Laboratory | Public Health Ontario Laboratory | Vanessa G Allen et al |
| EPI_ISL_548893 | Canada/ON-PHL-20-01288/2020 | 20-Jun | North America/Canada/Ontario/ | Public Health Ontario Laboratory | Public Health Ontario Laboratory | Vanessa G Allen et al |
| EPI_ISL_548894 | Canada/ON-PHL-20-01290/2020 | 20-Jun | North America/Canada/Ontario/ | Public Health Ontario Laboratory | Public Health Ontario Laboratory | Vanessa G Allen et al |
| EPI_ISL_548895 | Canada/ON-PHL-20-01291/2020 | 20-Jun | North America/Canada/Ontario/ | Public Health Ontario Laboratory | Public Health Ontario Laboratory | Vanessa G Allen et al |
| EPI_ISL_548896 | Canada/ON-PHL-20-01292/2020 | 20-Jun | North America/Canada/Ontario/ | Public Health Ontario Laboratory | Public Health Ontario Laboratory | Vanessa G Allen et al |

| | | | | | | |
|---|---|---|---|---|---|---|
| EPI_ISL_548897 | Canada/ON-PHL-20-01293/2020 | 20-Jun | North America/Canada/Ontario/ | Public Health Ontario Laboratory | Public Health Ontario Laboratory | Vanessa G Allen et al |
| EPI_ISL_548898 | Canada/ON-PHL-20-01294/2020 | 20-Jun | North America/Canada/Ontario/ | Public Health Ontario Laboratory | Public Health Ontario Laboratory | Vanessa G Allen et al |
| EPI_ISL_548899 | Canada/ON-PHL-20-01295/2020 | 20-Jun | North America/Canada/Ontario/ | Public Health Ontario Laboratory | Public Health Ontario Laboratory | Vanessa G Allen et al |
| EPI_ISL_548900 | Canada/ON-PHL-20-01296/2020 | 20-Jun | North America/Canada/Ontario/ | Public Health Ontario Laboratory | Public Health Ontario Laboratory | Vanessa G Allen et al |
| EPI_ISL_548901 | Canada/ON-PHL-20-01297/2020 | 20-Jun | North America/Canada/Ontario/ | Public Health Ontario Laboratory | Public Health Ontario Laboratory | Vanessa G Allen et al |
| EPI_ISL_548902 | Canada/ON-PHL-20-01298/2020 | 20-Jun | North America/Canada/Ontario/ | Public Health Ontario Laboratory | Public Health Ontario Laboratory | Vanessa G Allen et al |
| EPI_ISL_548903 | Canada/ON-PHL-20-01299/2020 | 20-Jun | North America/Canada/Ontario/ | Public Health Ontario Laboratory | Public Health Ontario Laboratory | Vanessa G Allen et al |
| EPI_ISL_548904 | Canada/ON-PHL-20-01300/2020 | 20-Jun | North America/Canada/Ontario/ | Public Health Ontario Laboratory | Public Health Ontario Laboratory | Vanessa G Allen et al |
| EPI_ISL_548905 | Canada/ON-PHL-20-01302/2020 | 20-Jun | North America/Canada/Ontario/ | Public Health Ontario Laboratory | Public Health Ontario Laboratory | Vanessa G Allen et al |
| EPI_ISL_548906 | Canada/ON-PHL-20-01303/2020 | 20-Jun | North America/Canada/Ontario/ | Public Health Ontario Laboratory | Public Health Ontario Laboratory | Vanessa G Allen et al |
| EPI_ISL_548908 | Canada/ON-PHL-20-01305/2020 | 20-Jun | North America/Canada/Ontario/ | Public Health Ontario Laboratory | Public Health Ontario Laboratory | Vanessa G Allen et al |
| EPI_ISL_548909 | Canada/ON-PHL-20-01306/2020 | 20-Jun | North America/Canada/Ontario/ | Public Health Ontario Laboratory | Public Health Ontario Laboratory | Vanessa G Allen et al |
| EPI_ISL_548910 | Canada/ON-PHL-20-01307/2020 | 20-Jun | North America/Canada/Ontario/ | Public Health Ontario Laboratory | Public Health Ontario Laboratory | Vanessa G Allen et al |

| | | | | | | |
|---|---|---|---|---|---|---|
| EPI_ISL_548911 | Canada/ON-PHL-20-01308/2020 | 20-Jun | North America/Canada/Ontario/ | Public Health Ontario Laboratory | Public Health Ontario Laboratory | Vanessa G Allen et al |
| EPI_ISL_548913 | Canada/ON-PHL-20-01310/2020 | 20-Jun | North America/Canada/Ontario/ | Public Health Ontario Laboratory | Public Health Ontario Laboratory | Vanessa G Allen et al |
| EPI_ISL_548914 | Canada/ON-PHL-20-01311/2020 | 20-Jun | North America/Canada/Ontario/ | Public Health Ontario Laboratory | Public Health Ontario Laboratory | Vanessa G Allen et al |
| EPI_ISL_548915 | Canada/ON-PHL-20-01312/2020 | 20-Jun | North America/Canada/Ontario/ | Public Health Ontario Laboratory | Public Health Ontario Laboratory | Vanessa G Allen et al |
| EPI_ISL_548916 | Canada/ON-PHL-20-01313/2020 | 20-Jun | North America/Canada/Ontario/ | Public Health Ontario Laboratory | Public Health Ontario Laboratory | Vanessa G Allen et al |
| EPI_ISL_548917 | Canada/ON-PHL-20-01314/2020 | 20-Jun | North America/Canada/Ontario/ | Public Health Ontario Laboratory | Public Health Ontario Laboratory | Vanessa G Allen et al |
| EPI_ISL_548918 | Canada/ON-PHL-20-01315/2020 | 20-Jun | North America/Canada/Ontario/ | Public Health Ontario Laboratory | Public Health Ontario Laboratory | Vanessa G Allen et al |
| EPI_ISL_548919 | Canada/ON-PHL-20-01316/2020 | 20-Jun | North America/Canada/Ontario/ | Public Health Ontario Laboratory | Public Health Ontario Laboratory | Vanessa G Allen et al |
| EPI_ISL_548920 | Canada/ON-PHL-20-01317/2020 | 20-Jun | North America/Canada/Ontario/ | Public Health Ontario Laboratory | Public Health Ontario Laboratory | Vanessa G Allen et al |
| EPI_ISL_548921 | Canada/ON-PHL-20-01318/2020 | 20-Jun | North America/Canada/Ontario/ | Public Health Ontario Laboratory | Public Health Ontario Laboratory | Vanessa G Allen et al |
| EPI_ISL_548922 | Canada/ON-PHL-20-01319/2020 | 20-Jun | North America/Canada/Ontario/ | Public Health Ontario Laboratory | Public Health Ontario Laboratory | Vanessa G Allen et al |
| EPI_ISL_548923 | Canada/ON-PHL-20-01320/2020 | 20-Jun | North America/Canada/Ontario/ | Public Health Ontario Laboratory | Public Health Ontario Laboratory | Vanessa G Allen et al |
| EPI_ISL_548924 | Canada/ON-PHL-20-01321/2020 | 20-Jun | North America/Canada/Ontario/ | Public Health Ontario Laboratory | Public Health Ontario Laboratory | Vanessa G Allen et al |

| | | | | | | |
|---|---|---|---|---|---|---|
| EPI_ISL_548925 | Canada/ON-PHL-20-01322/2020 | 20-Jun | North America/Canada/Ontario/ | Public Health Ontario Laboratory | Public Health Ontario Laboratory | Vanessa G Allen et al |
| EPI_ISL_548926 | Canada/ON-PHL-20-01323/2020 | 20-Jun | North America/Canada/Ontario/ | Public Health Ontario Laboratory | Public Health Ontario Laboratory | Vanessa G Allen et al |
| EPI_ISL_548927 | Canada/ON-PHL-20-01324/2020 | 20-Jun | North America/Canada/Ontario/ | Public Health Ontario Laboratory | Public Health Ontario Laboratory | Vanessa G Allen et al |
| EPI_ISL_548928 | Canada/ON-PHL-20-01325/2020 | 20-Jun | North America/Canada/Ontario/ | Public Health Ontario Laboratory | Public Health Ontario Laboratory | Vanessa G Allen et al |
| EPI_ISL_548929 | Canada/ON-PHL-20-01326/2020 | 20-Jun | North America/Canada/Ontario/ | Public Health Ontario Laboratory | Public Health Ontario Laboratory | Vanessa G Allen et al |
| EPI_ISL_548930 | Canada/ON-PHL-20-01327/2020 | 20-Jun | North America/Canada/Ontario/ | Public Health Ontario Laboratory | Public Health Ontario Laboratory | Vanessa G Allen et al |
| EPI_ISL_548931 | Canada/ON-PHL-20-01328/2020 | 20-Apr | North America/Canada/Ontario/ | Public Health Ontario Laboratory | Public Health Ontario Laboratory | Vanessa G Allen et al |
| EPI_ISL_548932 | Canada/ON-PHL-20-90069/2020 | 20-Mar | North America/Canada/Ontario/ | Public Health Ontario Laboratory | Public Health Ontario Laboratory | Vanessa G Allen et al |
| EPI_ISL_548933 | Canada/ON-PHL-20-90070/2020 | 20-Mar | North America/Canada/Ontario/ | Public Health Ontario Laboratory | Public Health Ontario Laboratory | Vanessa G Allen et al |
| EPI_ISL_548934 | Canada/ON-PHL-20-90084/2020 | 20-Mar | North America/Canada/Ontario/ | Public Health Ontario Laboratory | Public Health Ontario Laboratory | Vanessa G Allen et al |
| EPI_ISL_548935 | Canada/ON-PHL-20-90085/2020 | 20-Mar | North America/Canada/Ontario/ | Public Health Ontario Laboratory | Public Health Ontario Laboratory | Vanessa G Allen et al |
| EPI_ISL_548936 | Canada/ON-PHL-20-90086/2020 | 20-Mar | North America/Canada/Ontario/ | Public Health Ontario Laboratory | Public Health Ontario Laboratory | Vanessa G Allen et al |
| EPI_ISL_548937 | Canada/ON-PHL-20-90087/2020 | 20-Mar | North America/Canada/Ontario/ | Public Health Ontario Laboratory | Public Health Ontario Laboratory | Vanessa G Allen et al |

| | | | | | | |
|---|---|---|---|---|---|---|
| EPI_ISL_548938 | Canada/ON-PHL-20-90089/2020 | 20-Mar | North America/Canada/Ontario/ | Public Health Ontario Laboratory | Public Health Ontario Laboratory | Vanessa G Allen et al |
| EPI_ISL_548939 | Canada/ON-PHL-20-90090/2020 | 20-Mar | North America/Canada/Ontario/ | Public Health Ontario Laboratory | Public Health Ontario Laboratory | Vanessa G Allen et al |
| EPI_ISL_548940 | Canada/ON-PHL-20-90107/2020 | 20-Mar | North America/Canada/Ontario/ | Public Health Ontario Laboratory | Public Health Ontario Laboratory | Vanessa G Allen et al |
| EPI_ISL_548941 | Canada/ON-PHL-20-90108/2020 | 20-Mar | North America/Canada/Ontario/ | Public Health Ontario Laboratory | Public Health Ontario Laboratory | Vanessa G Allen et al |
| EPI_ISL_548942 | Pakistan/UN-UVAS-Lahore-I/2020 | 2020-05-11 | Asia/Pakistan/Pakistan/ | Institute of Microbiology, University of Veterinary and Animal sciences | Institute of Microbiology, University of Veterinary and Animal sciences | Yaqub et al |
| EPI_ISL_548943 | Pakistan/UN-UVAS-Lahore-II/2020 | 2020-05-11 | Asia/Pakistan/Pakistan/ | Institute of Microbiology, University of Veterinary and Animal sciences | Institute of Microbiology, University of Veterinary and Animal sciences | Yaqub et al |
| EPI_ISL_548944 | Pakistan/UN-UVAS-Lahore-III/2020 | 2020-05-11 | Asia/Pakistan/Pakistan/ | Institute of Microbiology, University of Veterinary and Animal sciences | Institute of Microbiology, University of Veterinary and Animal sciences | Yaqub et al |
| EPI_ISL_548945 | Pakistan/UN-UVAS-Lahore-IV/2020 | 2020-05-11 | Asia/Pakistan/Pakistan/ | Institute of Microbiology, University of Veterinary and Animal sciences | Institute of Microbiology, University of Veterinary and Animal sciences | Yaqub et al |
| EPI_ISL_548946 | Pakistan/UN-UVAS-Sialkot/2020 | 2020-05-11 | Asia/Pakistan/Pakistan/ | Institute of Microbiology, University of Veterinary and Animal sciences | Institute of Microbiology, University of Veterinary and Animal sciences | Yaqub et al |

| | | | | | | |
|---|---|---|---|---|---|---|
| EPI_ISL_548956 | Germany/BAV-MVP0264/2020 | 2020-05-12 | Europe/Germany/Bavaria/Munich | Max von Pettenkofer Institute, Virology, National Reference Center for Retroviruses, LMU M√ºnchen | Laboratory for Functional Genome Analysis, Dept. Genomics, Gene Center of the LMU Munich | Max Muenchhoff et al |
| EPI_ISL_548957 | Germany/BAV-MVP0292/2020 | 2020-05-25 | Europe/Germany/Bavaria/Munich | Max von Pettenkofer Institute, Virology, National Reference Center for Retroviruses, LMU M√ºnchen | Laboratory for Functional Genome Analysis, Dept. Genomics, Gene Center of the LMU Munich | Max Muenchhoff et al |
| EPI_ISL_548958 | Germany/BAV-MVP0295/2020 | 2020-05-26 | Europe/Germany/Bavaria/Munich | Max von Pettenkofer Institute, Virology, National Reference Center for Retroviruses, LMU M√ºnchen | Laboratory for Functional Genome Analysis, Dept. Genomics, Gene Center of the LMU Munich | Max Muenchhoff et al |
| EPI_ISL_548960 | Germany/BAV-V2010517/2020 | 2020-03-11 | Europe/Germany/Bavaria/Munich | Max von Pettenkofer Institute, Virology, National Reference Center for Retroviruses, LMU M√ºnchen | Laboratory for Functional Genome Analysis, Dept. Genomics, Gene Center of the LMU Munich | Max Muenchhoff et al |
| EPI_ISL_548961 | Germany/BAV-V2011674/2020 | 2020-03-16 | Europe/Germany/Bavaria/Munich | Max von Pettenkofer Institute, Virology, National Reference Center for Retroviruses, LMU M√ºnchen | Laboratory for Functional Genome Analysis, Dept. Genomics, Gene Center of the LMU Munich | Max Muenchhoff et al |
| EPI_ISL_548968 | UnitedArabEmirates/DU-A-S3-3_S9/2 | 2020-04-23 | Asia/United Arab Emirates/Dubai/ | Expo2020 Emergency Center | Agiomix | Walaa Allam et al |
| EPI_ISL_548974 | Singapore/965/2020 | 2020-09-13 | Asia/Singapore/Singapore/ | National Public Health Laboratory, National Centre for Infectious Diseases | National Public Health Laboratory, National Centre for Infectious Diseases | Mak TM et al |
| EPI_ISL_549184 | USA/FL-BPHL-1402/2020 | 2020-07-30 | North America/USA/Florida/ | Florida Bureau of Public Health Laboratories | Florida Bureau of Public Health Laboratories | Sarah Schmedes et al |

| | | | | | | |
|---|---|---|---|---|---|---|
| EPI_ISL_549186 | USA/FL-BPHL-1404/2020 | 2020-05-12 | North America/USA/Florida/ | Florida Bureau of Public Health Laboratories | Florida Bureau of Public Health Laboratories | Sarah Schmedes et al |
| EPI_ISL_549200 | USA/FL-BPHL-1418/2020 | 2020-08-13 | North America/USA/Florida/ | Florida Bureau of Public Health Laboratories | Florida Bureau of Public Health Laboratories | Sarah Schmedes et al |
| EPI_ISL_549201 | USA/FL-BPHL-1419/2020 | 2020-09-01 | North America/USA/Florida/ | Florida Bureau of Public Health Laboratories | Florida Bureau of Public Health Laboratories | Sarah Schmedes et al |
| EPI_ISL_549205 | USA/FL-BPHL-1423/2020 | 2020-08-31 | North America/USA/Florida/ | Florida Bureau of Public Health Laboratories | Florida Bureau of Public Health Laboratories | Sarah Schmedes et al |
| EPI_ISL_549211 | USA/FL-BPHL-1429/2020 | 2020-09-01 | North America/USA/Florida/ | Florida Bureau of Public Health Laboratories | Florida Bureau of Public Health Laboratories | Sarah Schmedes et al |
| EPI_ISL_549215 | USA/FL-BPHL-1433/2020 | 2020-09-03 | North America/USA/Florida/ | Florida Bureau of Public Health Laboratories | Florida Bureau of Public Health Laboratories | Sarah Schmedes et al |
| EPI_ISL_549218 | USA/FL-BPHL-1436/2020 | 2020-09-03 | North America/USA/Florida/ | Florida Bureau of Public Health Laboratories | Florida Bureau of Public Health Laboratories | Sarah Schmedes et al |
| EPI_ISL_549226 | USA/FL-BPHL-1444/2020 | 2020-09-08 | North America/USA/Florida/ | Florida Bureau of Public Health Laboratories | Florida Bureau of Public Health Laboratories | Sarah Schmedes et al |
| EPI_ISL_549227 | USA/FL-BPHL-1445/2020 | 2020-09-08 | North America/USA/Florida/ | Florida Bureau of Public Health Laboratories | Florida Bureau of Public Health Laboratories | Sarah Schmedes et al |
| EPI_ISL_549244 | USA/FL-BPHL-1462/2020 | 2020-04-20 | North America/USA/Florida/ | Florida Bureau of Public Health Laboratories | Florida Bureau of Public Health Laboratories | Sarah Schmedes et al |
| EPI_ISL_549268 | USA/FL-BPHL-1486/2020 | 2020-06-29 | North America/USA/Florida/ | Florida Bureau of Public Health Laboratories | Florida Bureau of Public Health Laboratories | Sarah Schmedes et al |
| EPI_ISL_549491 | England/NOTT-11329A/2020 | 2020-09-14 | Europe/United Kingdom/England/ | Queens Medical Centre, Clinical Microbiology Department / DeepSeq Nottingham | COVID-19 Genomics UK (COG-UK) Consortium | Gemma Clark et al |
| EPI_ISL_549502 | England/NOTT-1133C4/2020 | 2020-09-16 | Europe/United Kingdom/England/ | Queens Medical Centre, Clinical Microbiology Department / DeepSeq Nottingham | COVID-19 Genomics UK (COG-UK) Consortium | Gemma Clark et al |

| | | | | | | |
|---|---|---|---|---|---|---|
| EPI_ISL_549539 | England/QEUH-9979CE/2020 | 2020-08-28 | Europe/United Kingdom/England/ | Oxford Viromics, NDM, University of Oxford; Oxford University Hospitals; Basingstoke and North Hampshire Hospital | COVID-19 Genomics UK (COG-UK) Consortium | Tanya Golubchik et al |
| EPI_ISL_549599 | England/QEUH-9B83AB/2020 | 2020-09-07 | Europe/United Kingdom/England/ | Lighthouse Lab in Glasgow | Wellcome Sanger Institute for the COVID-19 Genomics UK (COG-UK) consortium | Harper VanSteenhouse et al |
| EPI_ISL_549609 | England/QEUH-9B8DF8/2020 | 2020-09-07 | Europe/United Kingdom/England/ | Lighthouse Lab in Glasgow | Wellcome Sanger Institute for the COVID-19 Genomics UK (COG-UK) consortium | Harper VanSteenhouse et al |
| EPI_ISL_549677 | England/QEUH-9B935F/2020 | 2020-09-07 | Europe/United Kingdom/England/ | Lighthouse Lab in Glasgow | Wellcome Sanger Institute for the COVID-19 Genomics UK (COG-UK) consortium | Harper VanSteenhouse et al |
| EPI_ISL_549724 | England/QEUH-9B87AF/2020 | 2020-09-10 | Europe/United Kingdom/England/ | Lighthouse Lab in Glasgow | Wellcome Sanger Institute for the COVID-19 Genomics UK (COG-UK) consortium | Harper VanSteenhouse et al |
| EPI_ISL_549768 | England/QEUH-9B9D51/2020 | 2020-09-09 | Europe/United Kingdom/England/ | Lighthouse Lab in Glasgow | Wellcome Sanger Institute for the COVID-19 Genomics UK (COG-UK) consortium | Harper VanSteenhouse et al |
| EPI_ISL_549809 | England/MILK-99CE06/2020 | 2020-08-30 | Europe/United Kingdom/England/ | Lighthouse Lab in Milton Keynes | Wellcome Sanger Institute for the COVID-19 Genomics UK (COG-UK) consortium | The Lighthouse Lab in Milton Keynes et al |
| EPI_ISL_549889 | England/MILK-99D46D/2020 | 2020-08-30 | Europe/United Kingdom/England/ | Lighthouse Lab in Milton Keynes | Wellcome Sanger Institute for the COVID-19 Genomics UK (COG-UK) consortium | The Lighthouse Lab in Milton Keynes et al |

| | | | | | | |
|---|---|---|---|---|---|---|
| EPI_ISL_550355 | England/MILK-99A4D9/2020 | 2020-08-29 | Europe/United Kingdom/England/ | Lighthouse Lab in Milton Keynes | Wellcome Sanger Institute for the COVID-19 Genomics UK (COG-UK) consortium | The Lighthouse Lab in Milton Keynes et al |
| EPI_ISL_550393 | England/MILK-99A1A5/2020 | 2020-08-30 | Europe/United Kingdom/England/ | Lighthouse Lab in Milton Keynes | Wellcome Sanger Institute for the COVID-19 Genomics UK (COG-UK) consortium | The Lighthouse Lab in Milton Keynes et al |
| EPI_ISL_550435 | England/MILK-977566/2020 | 2020-07-24 | Europe/United Kingdom/England/ | Lighthouse Lab in Milton Keynes | Wellcome Sanger Institute for the COVID-19 Genomics UK (COG-UK) consortium | The Lighthouse Lab in Milton Keynes et al |
| EPI_ISL_550538 | England/MILK-99C413/2020 | 2020-08-23 | Europe/United Kingdom/England/ | Lighthouse Lab in Milton Keynes | Wellcome Sanger Institute for the COVID-19 Genomics UK (COG-UK) consortium | The Lighthouse Lab in Milton Keynes et al |
| EPI_ISL_550610 | England/MILK-99E45D/2020 | 2020-08-30 | Europe/United Kingdom/England/ | Lighthouse Lab in Milton Keynes | Wellcome Sanger Institute for the COVID-19 Genomics UK (COG-UK) consortium | The Lighthouse Lab in Milton Keynes et al |
| EPI_ISL_550748 | England/MILK-99E0C2/2020 | 2020-08-31 | Europe/United Kingdom/England/ | Lighthouse Lab in Milton Keynes | Wellcome Sanger Institute for the COVID-19 Genomics UK (COG-UK) consortium | The Lighthouse Lab in Milton Keynes et al |
| EPI_ISL_551234 | England/MILK-9A8F40/2020 | 2020-09-01 | Europe/United Kingdom/England/ | Lighthouse Lab in Milton Keynes | Wellcome Sanger Institute for the COVID-19 Genomics UK (COG-UK) consortium | The Lighthouse Lab in Milton Keynes et al |
| EPI_ISL_551305 | England/ALDP-951DD3/2020 | 2020-06-10 | Europe/United Kingdom/England/ | Lighthouse Lab in Alderley Park | Wellcome Sanger Institute for the COVID-19 Genomics UK (COG-UK) consortium | The Lighthouse Lab in Alderley Park et al |

| | | | | | | |
|---|---|---|---|---|---|---|
| EPI_ISL_551409 | England/ALDP-90459B/2020 | 2020-06-24 | Europe/United Kingdom/England/ | Lighthouse Lab in Alderley Park | Wellcome Sanger Institute for the COVID-19 Genomics UK (COG-UK) consortium | The Lighthouse Lab in Alderley Park et al |
| EPI_ISL_551543 | England/ALDP-90E573/2020 | 2020-06-23 | Europe/United Kingdom/England/ | Lighthouse Lab in Alderley Park | Wellcome Sanger Institute for the COVID-19 Genomics UK (COG-UK) consortium | The Lighthouse Lab in Alderley Park et al |
| EPI_ISL_551635 | England/MILK-99581E/2020 | 2020-08-23 | Europe/United Kingdom/England/ | Lighthouse Lab in Milton Keynes | Wellcome Sanger Institute for the COVID-19 Genomics UK (COG-UK) consortium | The Lighthouse Lab in Milton Keynes et al |
| EPI_ISL_551878 | England/CAMC-99128D/2020 | 2020-08-26 | Europe/United Kingdom/England/ | Lighthouse Lab in Cambridge | Wellcome Sanger Institute for the COVID-19 Genomics UK (COG-UK) consortium | Rob Howes et al |
| EPI_ISL_551938 | Wales/MILK-993142/2020 | 2020-08-27 | Europe/United Kingdom/Wales/ | Lighthouse Lab in Milton Keynes | Wellcome Sanger Institute for the COVID-19 Genomics UK (COG-UK) consortium | The Lighthouse Lab in Alderley Park et al |
| EPI_ISL_552121 | England/MILK-99319D/2020 | 2020-08-27 | Europe/United Kingdom/England/ | Lighthouse Lab in Milton Keynes | Wellcome Sanger Institute for the COVID-19 Genomics UK (COG-UK) consortium | The Lighthouse Lab in Milton Keynes et al |
| EPI_ISL_552254 | England/MILK-995395/2020 | 2020-08-24 | Europe/United Kingdom/England/ | Lighthouse Lab in Milton Keynes | Wellcome Sanger Institute for the COVID-19 Genomics UK (COG-UK) consortium | The Lighthouse Lab in Milton Keynes et al |
| EPI_ISL_552828 | England/ALDP-925567/2020 | 2020-06-23 | Europe/United Kingdom/England/ | Lighthouse Lab in Alderley Park | Wellcome Sanger Institute for the COVID-19 Genomics UK (COG-UK) consortium | The Lighthouse Lab in Alderley Park et al |

| | | | | | | |
|---|---|---|---|---|---|---|
| EPI_ISL_552896 | England/ALDP-937016/2020 | 2020-06-21 | Europe/United Kingdom/England/ | Lighthouse Lab in Alderley Park | Wellcome Sanger Institute for the COVID-19 Genomics UK (COG-UK) consortium | The Lighthouse Lab in Alderley Park et al |
| EPI_ISL_552976 | England/ALDP-913CFE/2020 | 2020-06-23 | Europe/United Kingdom/England/ | Lighthouse Lab in Alderley Park | Wellcome Sanger Institute for the COVID-19 Genomics UK (COG-UK) consortium | The Lighthouse Lab in Alderley Park et al |
| EPI_ISL_553281 | England/ALDP-9221DF/2020 | 2020-06-23 | Europe/United Kingdom/England/ | Lighthouse Lab in Alderley Park | Wellcome Sanger Institute for the COVID-19 Genomics UK (COG-UK) consortium | The Lighthouse Lab in Alderley Park et al |
| EPI_ISL_553336 | England/ALDP-90C6BE/2020 | 2020-06-22 | Europe/United Kingdom/England/ | Lighthouse Lab in Alderley Park | Wellcome Sanger Institute for the COVID-19 Genomics UK (COG-UK) consortium | The Lighthouse Lab in Alderley Park et al |
| EPI_ISL_553344 | England/ALDP-923095/2020 | 2020-06-23 | Europe/United Kingdom/England/ | Lighthouse Lab in Alderley Park | Wellcome Sanger Institute for the COVID-19 Genomics UK (COG-UK) consortium | The Lighthouse Lab in Alderley Park et al |
| EPI_ISL_553409 | England/ALDP-955540/2020 | 2020-06-12 | Europe/United Kingdom/England/ | Lighthouse Lab in Alderley Park | Wellcome Sanger Institute for the COVID-19 Genomics UK (COG-UK) consortium | The Lighthouse Lab in Alderley Park et al |
| EPI_ISL_553416 | England/ALDP-954FBC/2020 | 2020-06-12 | Europe/United Kingdom/England/ | Lighthouse Lab in Alderley Park | Wellcome Sanger Institute for the COVID-19 Genomics UK (COG-UK) consortium | The Lighthouse Lab in Alderley Park et al |
| EPI_ISL_553525 | England/ALDP-955328/2020 | 2020-06-12 | Europe/United Kingdom/England/ | Lighthouse Lab in Alderley Park | Wellcome Sanger Institute for the COVID-19 Genomics UK (COG-UK) consortium | The Lighthouse Lab in Alderley Park et al |

| | | | | | | |
|---|---|---|---|---|---|---|
| EPI_ISL_553549 | England/ALDP-95A4B7/2020 | 2020-06-12 | Europe/United Kingdom/England/ | Lighthouse Lab in Alderley Park | Wellcome Sanger Institute for the COVID-19 Genomics UK (COG-UK) consortium | The Lighthouse Lab in Alderley Park et al |
| EPI_ISL_553640 | England/ALDP-952FEB/2020 | 2020-06-11 | Europe/United Kingdom/England/ | Lighthouse Lab in Alderley Park | Wellcome Sanger Institute for the COVID-19 Genomics UK (COG-UK) consortium | The Lighthouse Lab in Alderley Park et al |
| EPI_ISL_553989 | England/MILK-95747E/2020 | 2020-06-12 | Europe/United Kingdom/England/ | Lighthouse Lab in Milton Keynes | Wellcome Sanger Institute for the COVID-19 Genomics UK (COG-UK) consortium | The Lighthouse Lab in Milton Keynes et al |
| EPI_ISL_554060 | England/ALDP-952516/2020 | 2020-06-09 | Europe/United Kingdom/England/ | Lighthouse Lab in Alderley Park | Wellcome Sanger Institute for the COVID-19 Genomics UK (COG-UK) consortium | The Lighthouse Lab in Alderley Park et al |
| EPI_ISL_554282 | England/ALDP-95109E/2020 | 2020-06-13 | Europe/United Kingdom/England/ | Lighthouse Lab in Alderley Park | Wellcome Sanger Institute for the COVID-19 Genomics UK (COG-UK) consortium | The Lighthouse Lab in Alderley Park et al |
| EPI_ISL_554387 | Wales/ALDP-95572C/2020 | 2020-06-12 | Europe/United Kingdom/Wales/ | Lighthouse Lab in Alderley Park | Wellcome Sanger Institute for the COVID-19 Genomics UK (COG-UK) consortium | The Lighthouse Lab in Alderley Park et al |
| EPI_ISL_554438 | England/ALDP-953FF9/2020 | 2020-06-18 | Europe/United Kingdom/England/ | Lighthouse Lab in Alderley Park | Wellcome Sanger Institute for the COVID-19 Genomics UK (COG-UK) consortium | The Lighthouse Lab in Alderley Park et al |
| EPI_ISL_554563 | England/ALDP-950888/2020 | 2020-06-30 | Europe/United Kingdom/England/ | Lighthouse Lab in Alderley Park | Wellcome Sanger Institute for the COVID-19 Genomics UK (COG-UK) consortium | The Lighthouse Lab in Alderley Park et al |

| | | | | | | |
|---|---|---|---|---|---|---|
| EPI_ISL_554585 | England/ALDP-955980/2020 | 2020-06-12 | Europe/United Kingdom/England/ | Lighthouse Lab in Alderley Park | Wellcome Sanger Institute for the COVID-19 Genomics UK (COG-UK) consortium | The Lighthouse Lab in Alderley Park et al |
| EPI_ISL_554641 | England/MILK-95853E/2020 | 2020-06-09 | Europe/United Kingdom/England/ | Lighthouse Lab in Milton Keynes | Wellcome Sanger Institute for the COVID-19 Genomics UK (COG-UK) consortium | The Lighthouse Lab in Milton Keynes et al |
| EPI_ISL_554649 | England/MILK-9580C4/2020 | 2020-06-15 | Europe/United Kingdom/England/ | Lighthouse Lab in Milton Keynes | Wellcome Sanger Institute for the COVID-19 Genomics UK (COG-UK) consortium | The Lighthouse Lab in Milton Keynes et al |
| EPI_ISL_554909 | England/ALDP-95CF78/2020 | 2020-06-18 | Europe/United Kingdom/England/ | Lighthouse Lab in Alderley Park | Wellcome Sanger Institute for the COVID-19 Genomics UK (COG-UK) consortium | The Lighthouse Lab in Alderley Park et al |
| EPI_ISL_554950 | England/ALDP-94D7DC/2020 | 2020-06-14 | Europe/United Kingdom/England/ | Lighthouse Lab in Alderley Park | Wellcome Sanger Institute for the COVID-19 Genomics UK (COG-UK) consortium | The Lighthouse Lab in Alderley Park et al |
| EPI_ISL_555242 | England/ALDP-94FD32/2020 | 2020-06-15 | Europe/United Kingdom/England/ | Lighthouse Lab in Alderley Park | Wellcome Sanger Institute for the COVID-19 Genomics UK (COG-UK) consortium | The Lighthouse Lab in Alderley Park et al |
| EPI_ISL_555526 | England/ALDP-95B137/2020 | 2020-06-10 | Europe/United Kingdom/England/ | Lighthouse Lab in Alderley Park | Wellcome Sanger Institute for the COVID-19 Genomics UK (COG-UK) consortium | The Lighthouse Lab in Alderley Park et al |
| EPI_ISL_555556 | England/ALDP-944A09/2020 | 2020-06-07 | Europe/United Kingdom/England/ | Lighthouse Lab in Alderley Park | Wellcome Sanger Institute for the COVID-19 Genomics UK (COG-UK) consortium | The Lighthouse Lab in Alderley Park et al |

| | | | | | | |
|---|---|---|---|---|---|---|
| EPI_ISL_555606 | England/ALDP-94D94F/2020 | 2020-06-18 | Europe/United Kingdom/England/ | Lighthouse Lab in Alderley Park | Wellcome Sanger Institute for the COVID-19 Genomics UK (COG-UK) consortium | The Lighthouse Lab in Alderley Park et al |
| EPI_ISL_555665 | England/ALDP-65EAC3/2020 | 2020-07-03 | Europe/United Kingdom/England/ | Lighthouse Lab in Alderley Park | Wellcome Sanger Institute for the COVID-19 Genomics UK (COG-UK) consortium | The Lighthouse Lab in Alderley Park et al |
| EPI_ISL_555783 | England/ALDP-94B880/2020 | 2020-06-11 | Europe/United Kingdom/England/ | Lighthouse Lab in Alderley Park | Wellcome Sanger Institute for the COVID-19 Genomics UK (COG-UK) consortium | The Lighthouse Lab in Alderley Park et al |
| EPI_ISL_555792 | England/ALDP-94BB96/2020 | 2020-06-13 | Europe/United Kingdom/England/ | Lighthouse Lab in Alderley Park | Wellcome Sanger Institute for the COVID-19 Genomics UK (COG-UK) consortium | The Lighthouse Lab in Alderley Park et al |
| EPI_ISL_556353 | England/ALDP-94EDF7/2020 | 2020-06-17 | Europe/United Kingdom/England/ | Lighthouse Lab in Alderley Park | Wellcome Sanger Institute for the COVID-19 Genomics UK (COG-UK) consortium | The Lighthouse Lab in Alderley Park et al |
| EPI_ISL_556569 | England/CAMC-9473DE/2020 | 2020-07-21 | Europe/United Kingdom/England/ | Lighthouse Lab in Cambridge | Wellcome Sanger Institute for the COVID-19 Genomics UK (COG-UK) consortium | Rob Howes et al |
| EPI_ISL_556598 | England/ALDP-8C12B2/2020 | 2020-06-25 | Europe/United Kingdom/England/ | Lighthouse Lab in Alderley Park | Wellcome Sanger Institute for the COVID-19 Genomics UK (COG-UK) consortium | The Lighthouse Lab in Alderley Park et al |
| EPI_ISL_556706 | England/CAMC-94AA9A/2020 | 2020-07-18 | Europe/United Kingdom/England/ | Lighthouse Lab in Cambridge | Wellcome Sanger Institute for the COVID-19 Genomics UK (COG-UK) consortium | Rob Howes et al |

| | | | | | | |
|---|---|---|---|---|---|---|
| EPI_ISL_556710 | England/CAMC-94B237/2020 | 2020-07-16 | Europe/United Kingdom/England/ | Lighthouse Lab in Cambridge | Wellcome Sanger Institute for the COVID-19 Genomics UK (COG-UK) consortium | Rob Howes et al |
| EPI_ISL_556835 | England/CAMC-941A66/2020 | 2020-07-23 | Europe/United Kingdom/England/ | Lighthouse Lab in Cambridge | Wellcome Sanger Institute for the COVID-19 Genomics UK (COG-UK) consortium | Rob Howes et al |
| EPI_ISL_557143 | England/ALDP-8E25EF/2020 | 2020-06-25 | Europe/United Kingdom/England/ | Lighthouse Lab in Alderley Park | Wellcome Sanger Institute for the COVID-19 Genomics UK (COG-UK) consortium | The Lighthouse Lab in Alderley Park et al |
| EPI_ISL_557230 | England/MILK-8249F4/2020 | 2020-07-15 | Europe/United Kingdom/England/ | Lighthouse Lab in Milton Keynes | Wellcome Sanger Institute for the COVID-19 Genomics UK (COG-UK) consortium | The Lighthouse Lab in Milton Keynes et al |
| EPI_ISL_557340 | England/MILK-879E1F/2020 | 2020-07-18 | Europe/United Kingdom/England/ | Lighthouse Lab in Milton Keynes | Wellcome Sanger Institute for the COVID-19 Genomics UK (COG-UK) consortium | The Lighthouse Lab in Milton Keynes et al |
| EPI_ISL_557515 | England/MILK-85C54A/2020 | 2020-07-12 | Europe/United Kingdom/England/ | Lighthouse Lab in Milton Keynes | Wellcome Sanger Institute for the COVID-19 Genomics UK (COG-UK) consortium | The Lighthouse Lab in Milton Keynes et al |
| EPI_ISL_557591 | England/ALDP-70414E/2020 | 2020-06-30 | Europe/United Kingdom/England/ | Lighthouse Lab in Alderley Park | Wellcome Sanger Institute for the COVID-19 Genomics UK (COG-UK) consortium | The Lighthouse Lab in Alderley Park et al |
| EPI_ISL_557592 | England/ALDP-6E9C6D/2020 | 2020-06-30 | Europe/United Kingdom/England/ | Lighthouse Lab in Alderley Park | Wellcome Sanger Institute for the COVID-19 Genomics UK (COG-UK) consortium | The Lighthouse Lab in Alderley Park et al |

| | | | | | | |
|---|---|---|---|---|---|---|
| EPI_ISL_557742 | England/MILK-6285D0/2020 | 2020-07-05 | Europe/United Kingdom/England/ | Lighthouse Lab in Milton Keynes | Wellcome Sanger Institute for the COVID-19 Genomics UK (COG-UK) consortium | The Lighthouse Lab in Milton Keynes et al |
| EPI_ISL_557927 | England/MILK-6160F2/2020 | 2020-07-06 | Europe/United Kingdom/England/ | Lighthouse Lab in Milton Keynes | Wellcome Sanger Institute for the COVID-19 Genomics UK (COG-UK) consortium | The Lighthouse Lab in Milton Keynes et al |
| EPI_ISL_557932 | England/ALDP-6B5D76/2020 | 2020-07-01 | Europe/United Kingdom/England/ | Lighthouse Lab in Alderley Park | Wellcome Sanger Institute for the COVID-19 Genomics UK (COG-UK) consortium | The Lighthouse Lab in Alderley Park et al |
| EPI_ISL_557934 | England/ALDP-6B663E/2020 | 2020-06-29 | Europe/United Kingdom/England/ | Lighthouse Lab in Alderley Park | Wellcome Sanger Institute for the COVID-19 Genomics UK (COG-UK) consortium | The Lighthouse Lab in Alderley Park et al |
| EPI_ISL_558065 | England/ALDP-64C4D9/2020 | 2020-07-04 | Europe/United Kingdom/England/ | Lighthouse Lab in Alderley Park | Wellcome Sanger Institute for the COVID-19 Genomics UK (COG-UK) consortium | The Lighthouse Lab in Alderley Park et al |
| EPI_ISL_558243 | England/MILK-559ABA/2020 | 2020-06-10 | Europe/United Kingdom/England/ | Lighthouse Lab in Milton Keynes | Wellcome Sanger Institute for the COVID-19 Genomics UK (COG-UK) consortium | The Lighthouse Lab in Milton Keynes et al |
| EPI_ISL_558244 | England/ALDP-59BB9B/2020 | 2020-06-13 | Europe/United Kingdom/England/ | Lighthouse Lab in Alderley Park | Wellcome Sanger Institute for the COVID-19 Genomics UK (COG-UK) consortium | The Lighthouse Lab in Alderley Park et al |
| EPI_ISL_558274 | England/ALDP-59A419/2020 | 2020-06-15 | Europe/United Kingdom/England/ | Lighthouse Lab in Alderley Park | Wellcome Sanger Institute for the COVID-19 Genomics UK (COG-UK) consortium | The Lighthouse Lab in Alderley Park et al |

| | | | | | | |
|---|---|---|---|---|---|---|
| EPI_ISL_558337 | England/MILK-565D2B/2020 | 2020-06-10 | Europe/United Kingdom/England/ | Lighthouse Lab in Milton Keynes | Wellcome Sanger Institute for the COVID-19 Genomics UK (COG-UK) consortium | The Lighthouse Lab in Milton Keynes et al |
| EPI_ISL_558776 | England/ALDP-5A0629/2020 | 2020-06-14 | Europe/United Kingdom/England/ | Lighthouse Lab in Alderley Park | Wellcome Sanger Institute for the COVID-19 Genomics UK (COG-UK) consortium | The Lighthouse Lab in Alderley Park et al |
| EPI_ISL_558913 | England/ALDP-529E6C/2020 | 2020-05-31 | Europe/United Kingdom/England/ | Lighthouse Lab in Alderley Park | Wellcome Sanger Institute for the COVID-19 Genomics UK (COG-UK) consortium | The Lighthouse Lab in Alderley Park et al |
| EPI_ISL_559020 | England/ALDP-5249BB/2020 | 2020-06-02 | Europe/United Kingdom/England/ | Lighthouse Lab in Alderley Park | Wellcome Sanger Institute for the COVID-19 Genomics UK (COG-UK) consortium | The Lighthouse Lab in Alderley Park et al |
| EPI_ISL_559032 | England/ALDP-51F142/2020 | 2020-06-02 | Europe/United Kingdom/England/ | Lighthouse Lab in Alderley Park | Wellcome Sanger Institute for the COVID-19 Genomics UK (COG-UK) consortium | The Lighthouse Lab in Alderley Park et al |
| EPI_ISL_559169 | England/ALDP-50085E/2020 | 2020-06-27 | Europe/United Kingdom/England/ | Lighthouse Lab in Alderley Park | Wellcome Sanger Institute for the COVID-19 Genomics UK (COG-UK) consortium | The Lighthouse Lab in Alderley Park et al |
| EPI_ISL_559183 | England/ALDP-4F4092/2020 | 2020-06-24 | Europe/United Kingdom/England/ | Lighthouse Lab in Alderley Park | Wellcome Sanger Institute for the COVID-19 Genomics UK (COG-UK) consortium | The Lighthouse Lab in Alderley Park et al |
| EPI_ISL_559381 | England/ALDP-4997C7/2020 | 2020-05-27 | Europe/United Kingdom/England/ | Lighthouse Lab in Alderley Park | Wellcome Sanger Institute for the COVID-19 Genomics UK (COG-UK) consortium | The Lighthouse Lab in Alderley Park et al |

| | | | | | | |
|---|---|---|---|---|---|---|
| EPI_ISL_559403 | England/ALDP-4A22AC/2020 | 2020-05-21 | Europe/United Kingdom/England/ | Lighthouse Lab in Alderley Park | Wellcome Sanger Institute for the COVID-19 Genomics UK (COG-UK) consortium | The Lighthouse Lab in Alderley Park et al |
| EPI_ISL_559433 | England/ALDP-499B9E/2020 | 2020-05-26 | Europe/United Kingdom/England/ | Lighthouse Lab in Alderley Park | Wellcome Sanger Institute for the COVID-19 Genomics UK (COG-UK) consortium | The Lighthouse Lab in Alderley Park et al |
| EPI_ISL_559444 | England/ALDP-49F169/2020 | 2020-05-24 | Europe/United Kingdom/England/ | Lighthouse Lab in Alderley Park | Wellcome Sanger Institute for the COVID-19 Genomics UK (COG-UK) consortium | The Lighthouse Lab in Alderley Park et al |
| EPI_ISL_559509 | England/ALDP-4A5B53/2020 | 2020-06-09 | Europe/United Kingdom/England/ | Lighthouse Lab in Alderley Park | Wellcome Sanger Institute for the COVID-19 Genomics UK (COG-UK) consortium | The Lighthouse Lab in Alderley Park et al |
| EPI_ISL_559569 | England/ALDP-4AD863/2020 | 2020-06-09 | Europe/United Kingdom/England/ | Lighthouse Lab in Alderley Park | Wellcome Sanger Institute for the COVID-19 Genomics UK (COG-UK) consortium | The Lighthouse Lab in Alderley Park et al |
| EPI_ISL_559704 | England/MILK-45F6DF/2020 | 2020-05-13 | Europe/United Kingdom/England/ | Lighthouse Lab in Milton Keynes | Wellcome Sanger Institute for the COVID-19 Genomics UK (COG-UK) consortium | The Lighthouse Lab in Milton Keynes et al |
| EPI_ISL_560056 | Scotland/QEUH-9B3A50/2020 | 2020-09-08 | Europe/United Kingdom/Scotland/ | Virology Department, Sheffield Teaching Hospitals NHS Foundation Trust/Department of Infection, Immunity and Cardiovascular Disease, The Medical School, University of Sheffield | COVID-19 Genomics UK (COG-UK) Consortium | Thushan de Silva et al |
| EPI_ISL_560266 | Wales/PHWC-16B275/2020 | 2020-09-13 | Europe/United Kingdom/Wales/ | Wales Specialist Virology Centre Sequencing lab: Pathogen Genomics Unit | COVID-19 Genomics UK (COG-UK) Consortium | Catherine Moore et al |

| | | | | | | |
|---|---|---|---|---|---|---|
| EPI_ISL_560308 | Russia/SVE-RII-MH182S/2020 | 2020-06-26 | Europe/Russia/Sverdlovsk/Yekaterinbur | UMMC-Health | WHO National Influenza Centre Russian Federation | Andrey Komissarov et al |
| EPI_ISL_560309 | Russia/SVE-RII-MH184S/2020 | 2020-07-08 | Europe/Russia/Sverdlovsk/Verkhnyaya | UMMC-Health | WHO National Influenza Centre Russian Federation | Andrey Komissarov et al |
| EPI_ISL_560310 | Russia/SVE-RII-MH187S/2020 | 2020-07-08 | Europe/Russia/Sverdlovsk/Yekaterinbur | UMMC-Health | WHO National Influenza Centre Russian Federation | Andrey Komissarov et al |
| EPI_ISL_560311 | Russia/SVE-RII-MH197S/2020 | 2020-07-16 | Europe/Russia/Sverdlovsk/Yekaterinbur | UMMC-Health | WHO National Influenza Centre Russian Federation | Andrey Komissarov et al |
| EPI_ISL_560312 | Russia/SVE-RII-MH198S/2020 | 2020-07-17 | Europe/Russia/Sverdlovsk/Yekaterinbur | UMMC-Health | WHO National Influenza Centre Russian Federation | Andrey Komissarov et al |
| EPI_ISL_560313 | Russia/SVE-RII-MH199S/2020 | 2020-07-16 | Europe/Russia/Sverdlovsk/Yekaterinbur | UMMC-Health | WHO National Influenza Centre Russian Federation | Andrey Komissarov et al |
| EPI_ISL_560314 | Russia/SVE-RII-MH200S/2020 | 2020-07-17 | Europe/Russia/Sverdlovsk/Yekaterinbur | UMMC-Health | WHO National Influenza Centre Russian Federation | Andrey Komissarov et al |
| EPI_ISL_560315 | Russia/SVE-RII-MH202S/2020 | 2020-07-17 | Europe/Russia/Sverdlovsk/Yekaterinbur | UMMC-Health | WHO National Influenza Centre Russian Federation | Andrey Komissarov et al |
| EPI_ISL_560316 | Russia/SVE-RII-MH210S/2020 | 2020-07-22 | Europe/Russia/Sverdlovsk/Yekaterinbur | UMMC-Health | WHO National Influenza Centre Russian Federation | Andrey Komissarov et al |
| EPI_ISL_560317 | Russia/SVE-RII-MH231S/2020 | 2020-08-08 | Europe/Russia/Sverdlovsk/Yekaterinbur | UMMC-Health | WHO National Influenza Centre Russian Federation | Andrey Komissarov et al |

| | | | | | | |
|---|---|---|---|---|---|---|
| EPI_ISL_560320 | India/UT-IMT-AT52/2020 | 2020-07-03 | Asia/India/Uttarakhand/Roorkee | CMS, Roorkee | CSIR-Institute of Microbial Technology | Kanika Bansal et al |
| EPI_ISL_560322 | India/HR-IMT-BZ115/2020 | 2020-08-25 | Asia/India/Haryana/Panchkula | Civil Hospital, Panchkula | CSIR-Institute of Microbial Technology | Kanika Bansal et al |
| EPI_ISL_560323 | India/HR-IMT-BZ119/2020 | 2020-08-25 | Asia/India/Haryana/Panchkula | Civil Hospital, Panchkula | CSIR-Institute of Microbial Technology | Kanika Bansal et al |
| EPI_ISL_560324 | India/HR-IMT-CB117/2020 | 2020-08-27 | Asia/India/Haryana/Panchkula | Civil Hospital, Panchkula | CSIR-Institute of Microbial Technology | Kanika Bansal et al |
| EPI_ISL_560329 | USA/NM-UNM-00661/2020 | 2020-06-29 | North America/USA/New Mexico/ | TriCore Reference Laboratories | Center for Global Health, University of New Mexico Health Sciences Center | Daryl Domman et al |
| EPI_ISL_560334 | USA/NM-UNM-00646/2020 | 2020-05-09 | North America/USA/New Mexico/ | TriCore Reference Laboratories | Center for Global Health, University of New Mexico Health Sciences Center | Daryl Domman et al |
| EPI_ISL_560354 | USA/NM-UNM-00599/2020 | 2020-06-03 | North America/USA/New Mexico/ | TriCore Reference Laboratories | Center for Global Health, University of New Mexico Health Sciences Center | Daryl Domman et al |
| EPI_ISL_560356 | USA/NM-UNM-00688/2020 | 2020-05-07 | North America/USA/New Mexico/ | TriCore Reference Laboratories | Center for Global Health, University of New Mexico Health Sciences Center | Daryl Domman et al |
| EPI_ISL_560407 | Italy/CAM-IZSM-52479-129/2020 | 2020-05-13 | Europe/Italy/Campania/Benevento | Istituto Zooprofilattico Sperimentale del Mezzogiorno | INMI Lazzaro Spallanzani IRCCS | Barbara Bartolini et al |
| EPI_ISL_560413 | Canada/QC-LSPQ-L00232672/2020 | 2020-03-22 | North America/Canada/Quebec/ | Centre de SSS D'Arthabaska-et-de-l'√©rable - H√¥tel-Dieu | Laboratoire de sant√© publique du Qu√©bec | Sandrine Moreira et al |
| EPI_ISL_560414 | Canada/QC-LSPQ-L00232710/2020 | 2020-03-22 | North America/Canada/Quebec/ | Centre de SSS D'Arthabaska-et-de-l'√©rable - H√¥tel-Dieu | Laboratoire de sant√© publique du Qu√©bec | Sandrine Moreira et al |

| | | | | | | |
|---|---|---|---|---|---|---|
| EPI_ISL_560415 | Canada/QC-LSPQ-L00234169/2020 | 2020-03-23 | North America/Canada/Quebec/ | Centre Hospitalier R√©gional de Lanaudi√®re | Laboratoire de sant√© publique du Qu√©bec | Sandrine Moreira et al |
| EPI_ISL_560430 | Switzerland/BE-ETHZ-280094/2020 | 2020-09-07 | Europe/Switzerland/Bern/ | Viollier AG | Department of Biosystems Science and Engineering, ETH Z√ºrich | Christian Beisel et al |
| EPI_ISL_560434 | Switzerland/BE-ETHZ-280116/2020 | 2020-09-08 | Europe/Switzerland/Bern/ | Viollier AG | Department of Biosystems Science and Engineering, ETH Z√ºrich | Christian Beisel et al |
| EPI_ISL_560449 | Switzerland/BE-ETHZ-280236/2020 | 2020-09-10 | Europe/Switzerland/Bern/ | Viollier AG | Department of Biosystems Science and Engineering, ETH Z√ºrich | Christian Beisel et al |
| EPI_ISL_560472 | Switzerland/BS-ETHZ-280162/2020 | 2020-09-09 | Europe/Switzerland/Basel-Stadt/ | Viollier AG | Department of Biosystems Science and Engineering, ETH Z√ºrich | Christian Beisel et al |
| EPI_ISL_560486 | Switzerland/VD-ETHZ-280075/2020 | 2020-09-07 | Europe/Switzerland/Vaud/ | Viollier AG | Department of Biosystems Science and Engineering, ETH Z√ºrich | Christian Beisel et al |
| EPI_ISL_560525 | Switzerland/VD-ETHZ-280207/2020 | 2020-09-09 | Europe/Switzerland/Vaud/ | Viollier AG | Department of Biosystems Science and Engineering, ETH Z√ºrich | Christian Beisel et al |
| EPI_ISL_560543 | Switzerland/ZH-ETHZ-280088/2020 | 2020-09-07 | Europe/Switzerland/Z√ºrich/ | Viollier AG | Department of Biosystems Science and Engineering, ETH Z√ºrich | Christian Beisel et al |
| EPI_ISL_560558 | USA/AK-PHL448/2020 | 2020-06-08 | North America/USA/Alaska/Anchorage | Alaska State Virology Laboratory | Alaska State Virology Laboratory | Jack Chen et al |

| | | | | | | |
|---|---|---|---|---|---|---|
| EPI_ISL_560561 | USA/AK-PHL473/2020 | 2020-06-11 | North America/USA/Alaska/Anchorage | Alaska State Virology Laboratory | Alaska State Virology Laboratory | Jack Chen et al |
| EPI_ISL_560566 | USA/AK-PHL534/2020 | 2020-06-18 | North America/USA/Alaska/Anchorage | Alaska State Virology Laboratory | Alaska State Virology Laboratory | Jack Chen et al |
| EPI_ISL_560568 | France/CVL-8260/2020 | 2020-04-07 | Europe/France/Pays de la Loire/Angers | h√¥pital | National Reference Center for Viruses of Respiratory Infections, Institut Pasteur, Paris | Sylvie Behillil et al |
| EPI_ISL_560573 | France/CVL-8271/2020 | 2020-03-27 | Europe/France/Pays de la Loire/Angers | h√¥pital | National Reference Center for Viruses of Respiratory Infections, Institut Pasteur, Paris | Sylvie Behillil et al |
| EPI_ISL_560580 | France/CVL-8297/2020 | 2020-04-21 | Europe/France/Pays de la Loire/Angers | h√¥pital | National Reference Center for Viruses of Respiratory Infections, Institut Pasteur, Paris | Sylvie Behillil et al |
| EPI_ISL_560583 | France/GES-8509/2020 | 2020-03-31 | Europe/France/Grand Est/Munster | Hopital | National Reference Center for Viruses of Respiratory Infections, Institut Pasteur, Paris | Sylvie Behillil et al |
| EPI_ISL_560586 | France/GES-8524/2020 | 2020-04-23 | Europe/France/Grand Est/Guebwiller | Hopital | National Reference Center for Viruses of Respiratory Infections, Institut Pasteur, Paris | Sylvie Behillil et al |

| | | | | | | |
|---|---|---|---|---|---|---|
| EPI_ISL_560593 | France/HDF-8236/2020 | 2020-03-24 | Europe/France/Hauts de France/Lille FR | hopital | National Reference Center for Viruses of Respiratory Infections, Institut Pasteur, Paris | Sylvie Behillil et al |
| EPI_ISL_560607 | France/BRE-8939/2020 | 2020-08-01 | Europe/France/Bretagne/Brest | Hospital | National Reference Center for Viruses of Respiratory Infections, Institut Pasteur, Paris | Sylvie Behillil et al |
| EPI_ISL_560608 | France/BRE-8940/2020 | 2020-08-01 | Europe/France/Bretagne/Vannes | Hospital | National Reference Center for Viruses of Respiratory Infections, Institut Pasteur, Paris | Sylvie Behillil et al |
| EPI_ISL_560616 | France/BRE-8949/2020 | 2020-08-07 | Europe/France/Bretagne/Lannilis | Hospital | National Reference Center for Viruses of Respiratory Infections, Institut Pasteur, Paris | Sylvie Behillil et al |
| EPI_ISL_560618 | France/BRE-8951/2020 | 2020-08-07 | Europe/France/Bretagne/Rennes | Hospital | National Reference Center for Viruses of Respiratory Infections, Institut Pasteur, Paris | Sylvie Behillil et al |
| EPI_ISL_560624 | France/GES-8782/2020 | 2020-03-21 | Europe/France/Grand Est/Metz | Hospital | National Reference Center for Viruses of Respiratory Infections, Institut Pasteur, Paris | Sylvie Behillil et al |

| | | | | | | |
|---|---|---|---|---|---|---|
| EPI_ISL_560627 | France/GES-8789/2020 | 2020-03-28 | Europe/France/Grand Est/Ottonville | Hospital | National Reference Center for Viruses of Respiratory Infections, Institut Pasteur, Paris | Sylvie Behillil et al |
| EPI_ISL_560629 | France/GES-8799/2020 | 2020-04-08 | Europe/France/Grand Est/Fameck | Hospital | National Reference Center for Viruses of Respiratory Infections, Institut Pasteur, Paris | Sylvie Behillil et al |
| EPI_ISL_560630 | France/GES-8802/2020 | 2020-04-14 | Europe/France/Grand Est/Metz | Hospital | National Reference Center for Viruses of Respiratory Infections, Institut Pasteur, Paris | Sylvie Behillil et al |
| EPI_ISL_560634 | France/GES-8809/2020 | 2020-04-27 | Europe/France/Grand Est/Metz | Hospital | National Reference Center for Viruses of Respiratory Infections, Institut Pasteur, Paris | Sylvie Behillil et al |
| EPI_ISL_560636 | France/IDF-8589/2020 | 2020-06-29 | Europe/France/Ile de France/Argenteuil | Hospital | National Reference Center for Viruses of Respiratory Infections, Institut Pasteur, Paris | Sylvie Behillil et al |
| EPI_ISL_560637 | France/IDF-8679/2020 | 2020-06-25 | Europe/France/Ile de France/Sarcelles | Labo Analyses Med | National Reference Center for Viruses of Respiratory Infections, Institut Pasteur, Paris | Sylvie Behillil et al |

| | | | | | | |
|---|---|---|---|---|---|---|
| EPI_ISL_560638 | France/IDF-8683/2020 | 2020-06-25 | Europe/France/Ile de France/Sarcelles | Labo Analyses Med | National Reference Center for Viruses of Respiratory Infections, Institut Pasteur, Paris | Sylvie Behillil et al |
| EPI_ISL_560642 | France/IDF-8707/2020 | 2020-06-29 | Europe/France/Ile de France/Sarcelles | Labo Analyses Med | National Reference Center for Viruses of Respiratory Infections, Institut Pasteur, Paris | Sylvie Behillil et al |
| EPI_ISL_560644 | France/IDF-8763/2020 | 2020-05-09 | Europe/France/Ile de France/Le Chesnay | Hospital | National Reference Center for Viruses of Respiratory Infections, Institut Pasteur, Paris | Sylvie Behillil et al |
| EPI_ISL_560651 | England/GSTT-5152/2020 | 2020-03-18 | Europe/United Kingdom/England/London | Centre for Clinical Infection and Diagnostics Research and Genomics Innovation Unit, Guy's and St. Thomas' NHS Trust | Centre for Clinical Infection and Diagnostics Research and Genomics Innovation Unit, Guy's and St. Thomas' NHS Trust | Chloe Fisher et al |
| EPI_ISL_560734 | England/GSTT-6773/2020 | 2020-03-30 | Europe/United Kingdom/England/London | Centre for Clinical Infection and Diagnostics Research and Genomics Innovation Unit, Guy's and St. Thomas' NHS Trust | Centre for Clinical Infection and Diagnostics Research and Genomics Innovation Unit, Guy's and St. Thomas' NHS Trust | Chloe Fisher et al |
| EPI_ISL_560744 | USA/MN-MDH-1657/2020 | 2020-04-01 | North America/USA/Minnesota/ | Minnesota Department of Health, Public Health Laboratory | Minnesota Department of Health, Public Health Laboratory | Matt Plumb et al |

| | | | | | | |
|---|---|---|---|---|---|---|
| EPI_ISL_560753 | USA/MN-MDH-1666/2020 | 2020-04-04 | North America/USA/Minnesota/ | Minnesota Department of Health, Public Health Laboratory | Minnesota Department of Health, Public Health Laboratory | Matt Plumb et al |
| EPI_ISL_560754 | USA/MN-MDH-1667/2020 | 2020-04-04 | North America/USA/Minnesota/ | Minnesota Department of Health, Public Health Laboratory | Minnesota Department of Health, Public Health Laboratory | Matt Plumb et al |
| EPI_ISL_560758 | USA/MN-MDH-1671/2020 | 2020-04-05 | North America/USA/Minnesota/ | Minnesota Department of Health, Public Health Laboratory | Minnesota Department of Health, Public Health Laboratory | Matt Plumb et al |
| EPI_ISL_560768 | USA/MN-MDH-1681/2020 | 2020-04-07 | North America/USA/Minnesota/ | Minnesota Department of Health, Public Health Laboratory | Minnesota Department of Health, Public Health Laboratory | Matt Plumb et al |
| EPI_ISL_560784 | USA/MN-MDH-1697/2020 | 2020-04-08 | North America/USA/Minnesota/ | Minnesota Department of Health, Public Health Laboratory | Minnesota Department of Health, Public Health Laboratory | Matt Plumb et al |
| EPI_ISL_560794 | USA/MN-MDH-1707/2020 | 2020-08-24 | North America/USA/Minnesota/ | Mayo Clinic & Mayo Clinic Laboratories | Minnesota Department of Health, Public Health Laboratory | Matt Plumb et al |
| EPI_ISL_560800 | USA/MN-MDH-1713/2020 | 2020-08-23 | North America/USA/Minnesota/ | M Health Fairview | Minnesota Department of Health, Public Health Laboratory | Matt Plumb et al |
| EPI_ISL_560803 | USA/MN-MDH-1716/2020 | 2020-08-25 | North America/USA/Minnesota/ | M Health Fairview | Minnesota Department of Health, Public Health Laboratory | Matt Plumb et al |
| EPI_ISL_560804 | USA/MN-MDH-1717/2020 | 2020-08-26 | North America/USA/Minnesota/ | M Health Fairview | Minnesota Department of Health, Public Health Laboratory | Matt Plumb et al |
| EPI_ISL_560812 | USA/MD-MDH-0199/2020 | 2020-06-08 | North America/USA/Maryland/ | Maryland Public Health Laboratory | Maryland Public Health Laboratory | Maryland Department of Health Laboratories Administration et al |

| | | | | | | |
|---|---|---|---|---|---|---|
| EPI_ISL_560814 | USA/MD-MDH-0201/2020 | 2020-06-08 | North America/USA/Maryland/ | Maryland Public Health Laboratory | Maryland Public Health Laboratory | Maryland Department of Health Laboratories Administration et al |
| EPI_ISL_560819 | USA/MD-MDH-0206/2020 | 2020-09-16 | North America/USA/Maryland/ | Maryland Public Health Laboratory | Maryland Public Health Laboratory | Maryland Department of Health Laboratories Administration et al |
| EPI_ISL_560820 | USA/MD-MDH-0207/2020 | 2020-09-15 | North America/USA/Maryland/ | Maryland Public Health Laboratory | Maryland Public Health Laboratory | Maryland Department of Health Laboratories Administration et al |
| EPI_ISL_560824 | USA/MD-MDH-0211/2020 | 2020-09-17 | North America/USA/Maryland/ | Maryland Public Health Laboratory | Maryland Public Health Laboratory | Maryland Department of Health Laboratories Administration et al |
| EPI_ISL_560825 | USA/MD-MDH-0212/2020 | 2020-09-17 | North America/USA/Maryland/ | Maryland Public Health Laboratory | Maryland Public Health Laboratory | Maryland Department of Health Laboratories Administration et al |
| EPI_ISL_560829 | USA/MD-MDH-0216/2020 | 2020-09-17 | North America/USA/Maryland/ | Maryland Public Health Laboratory | Maryland Public Health Laboratory | Maryland Department of Health Laboratories Administration et al |
| EPI_ISL_560837 | USA/UT-UPHL-04427/2020 | 2020-05-18 | North America/USA/Utah/ | Utah Public Health Laboratory | Utah Public Health Laboratory | Erin Young et al |
| EPI_ISL_560838 | USA/UT-UPHL-04426/2020 | 2020-05-19 | North America/USA/Utah/ | Utah Public Health Laboratory | Utah Public Health Laboratory | Erin Young et al |
| EPI_ISL_560844 | USA/UT-UPHL-03938/2020 | 2020-04-03 | North America/USA/Utah/ | Utah Public Health Laboratory | Utah Public Health Laboratory | Erin Young et al |
| EPI_ISL_560850 | USA/UT-UPHL-01648/2020 | 2020-04-07 | North America/USA/Utah/ | Utah Public Health Laboratory | Utah Public Health Laboratory | Erin Young et al |
| EPI_ISL_560861 | USA/UT-UPHL-03843/2020 | 2020-04-01 | North America/USA/Utah/ | Utah Public Health Laboratory | Utah Public Health Laboratory | Erin Young et al |
| EPI_ISL_560869 | USA/UT-UPHL-05273/2020 | 2020-05-30 | North America/USA/Utah/ | Utah Public Health Laboratory | Utah Public Health Laboratory | Erin Young et al |
| EPI_ISL_560871 | USA/UT-UPHL-05313/2020 | 2020-05-31 | North America/USA/Utah/ | Utah Public Health Laboratory | Utah Public Health Laboratory | Erin Young et al |
| EPI_ISL_560875 | USA/UT-UPHL-05411/2020 | 2020-05-30 | North America/USA/Utah/ | Utah Public Health Laboratory | Utah Public Health Laboratory | Erin Young et al |
| EPI_ISL_560903 | USA/UT-UPHL-03057/2020 | 2020-05-01 | North America/USA/Utah/ | Utah Public Health Laboratory | Utah Public Health Laboratory | Erin Young et al |
| EPI_ISL_560905 | USA/UT-UPHL-03056/2020 | 2020-04-30 | North America/USA/Utah/ | Utah Public Health Laboratory | Utah Public Health Laboratory | Erin Young et al |
| EPI_ISL_560907 | USA/UT-UPHL-03068/2020 | 2020-05-04 | North America/USA/Utah/ | Utah Public Health Laboratory | Utah Public Health Laboratory | Erin Young et al |

| | | | | | | |
|---|---|---|---|---|---|---|
| EPI_ISL_560917 | USA/UT-UPHL-03080/2020 | 2020-05-04 | North America/USA/Utah/ | Utah Public Health Laboratory | Utah Public Health Laboratory | Erin Young et al |
| EPI_ISL_560919 | USA/UT-UPHL-03083/2020 | 2020-05-04 | North America/USA/Utah/ | Utah Public Health Laboratory | Utah Public Health Laboratory | Erin Young et al |
| EPI_ISL_560921 | USA/UT-UPHL-03791/2020 | 2020-04-03 | North America/USA/Utah/ | Utah Public Health Laboratory | Utah Public Health Laboratory | Erin Young et al |
| EPI_ISL_560927 | USA/TX-DSHS-0483/2020 | 2020-05-14 | North America/USA/Texas/Harris Count | Texas Department of State Health Services | Texas Department of State Health Services | Rashmi Tuladhar et al |
| EPI_ISL_560928 | USA/TX-DSHS-0484/2020 | 2020-05-16 | North America/USA/Texas/Comanche Co | Texas Department of State Health Services | Texas Department of State Health Services | Rashmi Tuladhar et al |
| EPI_ISL_560929 | USA/TX-DSHS-0485/2020 | 2020-06-19 | North America/USA/Texas/Wharton Cou | Texas Department of State Health Services | Texas Department of State Health Services | Rashmi Tuladhar et al |
| EPI_ISL_560931 | USA/TX-DSHS-0487/2020 | 2020-06-21 | North America/USA/Texas/Hidalgo Cour | Texas Department of State Health Services | Texas Department of State Health Services | Rashmi Tuladhar et al |
| EPI_ISL_560936 | USA/TX-DSHS-0492/2020 | 2020-06-21 | North America/USA/Texas/Hidalgo Cour | Texas Department of State Health Services | Texas Department of State Health Services | Rashmi Tuladhar et al |
| EPI_ISL_560940 | USA/TX-DSHS-0496/2020 | 2020-06-21 | North America/USA/Texas/Hidalgo Cour | Texas Department of State Health Services | Texas Department of State Health Services | Rashmi Tuladhar et al |
| EPI_ISL_560941 | USA/TX-DSHS-0497/2020 | 2020-06-21 | North America/USA/Texas/Hidalgo Cour | Texas Department of State Health Services | Texas Department of State Health Services | Rashmi Tuladhar et al |
| EPI_ISL_560949 | USA/TX-DSHS-0505/2020 | 2020-06-21 | North America/USA/Texas/Hidalgo Cour | Texas Department of State Health Services | Texas Department of State Health Services | Rashmi Tuladhar et al |
| EPI_ISL_560951 | USA/TX-DSHS-0508/2020 | 2020-06-21 | North America/USA/Texas/Hidalgo Cour | Texas Department of State Health Services | Texas Department of State Health Services | Rashmi Tuladhar et al |
| EPI_ISL_560952 | USA/TX-DSHS-0509/2020 | 2020-06-21 | North America/USA/Texas/Hidalgo Cour | Texas Department of State Health Services | Texas Department of State Health Services | Rashmi Tuladhar et al |
| EPI_ISL_560972 | Sweden/20-50303/2020 | 2020-03-10 | Europe/Sweden/Vastmanland/ | Orebro klinisk mikrobiologi | The Public Health Agency of Sweden | Anna-Malin Linde et al |
| EPI_ISL_560974 | Sweden/20-51952/2020 | 2020-06-05 | Europe/Sweden/Stockholm/ | Karolinska Universitetslaboratoriet | The Public Health Agency of Sweden | Anna-Malin Linde et al |

| | | | | | | |
|---|---|---|---|---|---|---|
| EPI_ISL_560975 | Sweden/20-52037/2020 | 2020-06-13 | Europe/Sweden/Varmland/ | Klinisk mikrobiologi centralsjukhuset Karlstad | The Public Health Agency of Sweden | Anna-Malin Linde et al |
| EPI_ISL_561118 | Gambia/GC192579/2020 | 2020-07-21 | Africa/Gambia/Kombo/ | MRCG at LSHTM Genomics lab | MRCG at LSHTM Genomics lab | Abdul Karim sesay et al |
| EPI_ISL_561288 | Gambia/NPHL-2712/2020 | 2020-08-03 | Africa/Gambia/Kombo/ | MRCG at LSHTM Genomics lab | MRCG at LSHTM Genomics lab | Abdul Karim sesay et al |
| EPI_ISL_561292 | Gambia/NPHL-2759/2020 | 2020-08-07 | Africa/Gambia/Kombo/ | MRCG at LSHTM Genomics lab | MRCG at LSHTM Genomics lab | Abdul Karim sesay et al |
| EPI_ISL_561339 | India/HR-IMT-BZ124/2020 | 2020-08-25 | Asia/India/Haryana/Panchkula | Civil Hospital, Panchkula | CSIR-Institute of Microbial Technology | Kanika Bansal et al |
| EPI_ISL_561340 | India/HR-IMT-BZ144/2020 | 2020-08-25 | Asia/India/Haryana/Panchkula | Civil Hospital, Panchkula | CSIR-Institute of Microbial Technology | Kanika Bansal et al |
| EPI_ISL_561341 | India/HR-IMT-BZ164/2020 | 2020-08-25 | Asia/India/Haryana/Panchkula | Civil Hospital, Panchkula | CSIR-Institute of Microbial Technology | Kanika Bansal et al |
| EPI_ISL_561342 | India/HR-IMT-BZ98/2020 | 2020-08-25 | Asia/India/Haryana/Panchkula | Civil Hospital, Panchkula | CSIR-Institute of Microbial Technology | Kanika Bansal et al |
| EPI_ISL_561343 | India/HR-IMT-CB126/2020 | 2020-08-27 | Asia/India/Haryana/Panchkula | Civil Hospital, Panchkula | CSIR-Institute of Microbial Technology | Kanika Bansal et al |
| EPI_ISL_561371 | Spain/CN-IBV-97006522/2020 | 2020-04-06 | Europe/Spain/Canary Islands/Las Palmas | Hospital Universitario de Gran Canaria Dr. Negr√≠n | SeqCOVID-SPAIN consortium/IBV(CSIC) | M. Carmen P√©rez Gonz√°lez et al |
| EPI_ISL_562063 | Australia/VIC12043/2020 | 2020-07-26 | Oceania/Australia/Victoria/ | Microbiological Diagnostic Unit - Public Health Laboratory (MDU-PHL) | MDU-PHL | Seemann et al |
| EPI_ISL_565086 | Australia/VIC8549/2020 | 2020-08-18 | Oceania/Australia/Victoria/ | Microbiological Diagnostic Unit - Public Health Laboratory (MDU-PHL) | MDU-PHL | Seemann et al |
| EPI_ISL_565151 | Australia/VIC8711/2020 | 2020-07-26 | Oceania/Australia/Victoria/ | Victorian Infectious Diseases Reference Laboratory (VIDRL) | VIDRL and MDU-PHL | Caly et al |
| EPI_ISL_565518 | Australia/VIC9354/2020 | 2020-07-26 | Oceania/Australia/Victoria/ | Microbiological Diagnostic Unit - Public Health Laboratory (MDU-PHL) | MDU-PHL | Seemann et al |
| EPI_ISL_565843 | USA/MI-MDHHS-SC21800/2020 | 2020-07-06 | North America/USA/Michigan/ | Michigan Department of Health and Human Services, Bureau of Laboratories | Michigan Department of Health and Human Services, Bureau of Laboratories | Blankenship HM et al |

| | | | | | | |
|---|---|---|---|---|---|---|
| EPI_ISL_565847 | USA/MI-MDHHS-SC21804/2020 | 2020-07-09 | North America/USA/Michigan/ | Michigan Department of Health and Human Services, Bureau of Laboratories | Michigan Department of Health and Human Services, Bureau of Laboratories | Blankenship HM et al |
| EPI_ISL_565849 | USA/MI-MDHHS-SC21806/2020 | 2020-07-07 | North America/USA/Michigan/ | Michigan Department of Health and Human Services, Bureau of Laboratories | Michigan Department of Health and Human Services, Bureau of Laboratories | Blankenship HM et al |
| EPI_ISL_565853 | USA/MI-MDHHS-SC21810/2020 | 2020-07-15 | North America/USA/Michigan/ | Michigan Department of Health and Human Services, Bureau of Laboratories | Michigan Department of Health and Human Services, Bureau of Laboratories | Blankenship HM et al |
| EPI_ISL_565854 | USA/MI-MDHHS-SC21811/2020 | 2020-07-21 | North America/USA/Michigan/ | Michigan Department of Health and Human Services, Bureau of Laboratories | Michigan Department of Health and Human Services, Bureau of Laboratories | Blankenship HM et al |
| EPI_ISL_565856 | USA/MI-MDHHS-SC21813/2020 | 2020-06-29 | North America/USA/Michigan/ | Michigan Department of Health and Human Services, Bureau of Laboratories | Michigan Department of Health and Human Services, Bureau of Laboratories | Blankenship HM et al |
| EPI_ISL_565861 | USA/MI-MDHHS-SC21818/2020 | 2020-03-13 | North America/USA/Michigan/ | Michigan Department of Health and Human Services, Bureau of Laboratories | Michigan Department of Health and Human Services, Bureau of Laboratories | Blankenship HM et al |

| | | | | | | |
|---|---|---|---|---|---|---|
| EPI_ISL_565881 | USA/MI-MDHHS-SC21838/2020 | 2020-04-16 | North America/USA/Michigan/ | Michigan Department of Health and Human Services, Bureau of Laboratories | Michigan Department of Health and Human Services, Bureau of Laboratories | Blankenship HM et al |
| EPI_ISL_565893 | USA/MI-MDHHS-SC21850/2020 | 2020-04-27 | North America/USA/Michigan/ | Michigan Department of Health and Human Services, Bureau of Laboratories | Michigan Department of Health and Human Services, Bureau of Laboratories | Blankenship HM et al |
| EPI_ISL_565895 | USA/MI-MDHHS-SC21852/2020 | 2020-04-27 | North America/USA/Michigan/ | Michigan Department of Health and Human Services, Bureau of Laboratories | Michigan Department of Health and Human Services, Bureau of Laboratories | Blankenship HM et al |
| EPI_ISL_565904 | USA/MI-MDHHS-SC21861/2020 | 2020-05-11 | North America/USA/Michigan/ | Michigan Department of Health and Human Services, Bureau of Laboratories | Michigan Department of Health and Human Services, Bureau of Laboratories | Blankenship HM et al |
| EPI_ISL_565907 | USA/MI-MDHHS-SC21864/2020 | 2020-04-11 | North America/USA/Michigan/ | Michigan Department of Health and Human Services, Bureau of Laboratories | Michigan Department of Health and Human Services, Bureau of Laboratories | Blankenship HM et al |
| EPI_ISL_565911 | Spain/AN-IBV-98001966/2020 | 2020-03-14 | Europe/Spain/Andalusia/Malaga | Servicio de Microbiolog√≠a. HRU de M√°laga. Servicio Andaluz de Salud | SeqCOVID-SPAIN consortium/IBV(CSIC) | Inmaculada de Toro Peinado et al |
| EPI_ISL_565923 | Spain/PV-IBV-98005146/2020 | 2020-06-19 | Europe/Spain/Basque Country/San Seba | Servicio de Microbiolog√≠a. Hospital Universitario Donostia. OSI Donostialdea. √Årea de Enfermedades Infecciosas, Grupo de Infecci√≥n Respiratoria y Resistencia Antimicrobiana. Instituto de Investigaci√≥n Sanitaria Biodonostia | SeqCOVID-SPAIN consortium/IBV(CSIC) | Gustavo Cilla et al |

| | | | | | | |
|---|---|---|---|---|---|---|
| EPI_ISL_565937 | USA/MI-MDHHS-SC21876/2020 | 2020-04-06 | North America/USA/Michigan/ | Michigan Department of Health and Human Services, Bureau of Laboratories | Michigan Department of Health and Human Services, Bureau of Laboratories | Blankenship HM et al |
| EPI_ISL_565947 | USA/MI-MDHHS-SC21886/2020 | 2020-04-14 | North America/USA/Michigan/ | Michigan Department of Health and Human Services, Bureau of Laboratories | Michigan Department of Health and Human Services, Bureau of Laboratories | Blankenship HM et al |
| EPI_ISL_565975 | USA/MI-MDHHS-SC21914/2020 | 2020-04-06 | North America/USA/Michigan/ | Michigan Department of Health and Human Services, Bureau of Laboratories | Michigan Department of Health and Human Services, Bureau of Laboratories | Blankenship HM et al |
| EPI_ISL_566006 | USA/MI-MDHHS-SC21955/2020 | 2020-08-12 | North America/USA/Michigan/ | Michigan Department of Health and Human Services, Bureau of Laboratories | Michigan Department of Health and Human Services, Bureau of Laboratories | Blankenship HM et al |
| EPI_ISL_566017 | USA/MI-MDHHS-SC21966/2020 | 2020-08-20 | North America/USA/Michigan/ | Michigan Department of Health and Human Services, Bureau of Laboratories | Michigan Department of Health and Human Services, Bureau of Laboratories | Blankenship HM et al |
| EPI_ISL_566018 | USA/MI-MDHHS-SC21967/2020 | 2020-08-18 | North America/USA/Michigan/ | Michigan Department of Health and Human Services, Bureau of Laboratories | Michigan Department of Health and Human Services, Bureau of Laboratories | Blankenship HM et al |

| | | | | | | |
|---|---|---|---|---|---|---|
| EPI_ISL_566020 | USA/MI-MDHHS-SC21969/2020 | 2020-08-23 | North America/USA/Michigan/ | Michigan Department of Health and Human Services, Bureau of Laboratories | Michigan Department of Health and Human Services, Bureau of Laboratories | Blankenship HM et al |
| EPI_ISL_566022 | USA/MI-MDHHS-SC21971/2020 | 2020-08-17 | North America/USA/Michigan/ | Michigan Department of Health and Human Services, Bureau of Laboratories | Michigan Department of Health and Human Services, Bureau of Laboratories | Blankenship HM et al |
| EPI_ISL_566024 | USA/MI-MDHHS-SC21973/2020 | 2020-03-15 | North America/USA/Michigan/ | Michigan Department of Health and Human Services, Bureau of Laboratories | Michigan Department of Health and Human Services, Bureau of Laboratories | Blankenship HM et al |
| EPI_ISL_566025 | USA/MI-MDHHS-SC21974/2020 | 2020-06-26 | North America/USA/Michigan/ | Michigan Department of Health and Human Services, Bureau of Laboratories | Michigan Department of Health and Human Services, Bureau of Laboratories | Blankenship HM et al |
| EPI_ISL_566035 | USA/MI-MDHHS-SC21985/2020 | 2020-07-31 | North America/USA/Michigan/ | Michigan Department of Health and Human Services, Bureau of Laboratories | Michigan Department of Health and Human Services, Bureau of Laboratories | Blankenship HM et al |
| EPI_ISL_566077 | Russia/MOW-RII-MH249S/2020 | 2020-06-16 | Europe/Russia/Moscow Oblast/ | Pathogenic Microorganisms Variability Laboratory | WHO National Influenza Centre Russian Federation | Andrey Komissarov et al |
| EPI_ISL_566078 | Russia/MOW-RII-MH250S/2020 | 2020-06-22 | Europe/Russia/Moscow Oblast/ | Pathogenic Microorganisms Variability Laboratory | WHO National Influenza Centre Russian Federation | Andrey Komissarov et al |

| | | | | | | |
|---|---|---|---|---|---|---|
| EPI_ISL_566079 | Russia/MOW-RII-MH251S/2020 | 2020-06-19 | Europe/Russia/Moscow Oblast/ | Pathogenic Microorganisms Variability Laboratory | WHO National Influenza Centre Russian Federation | Andrey Komissarov et al |
| EPI_ISL_566080 | Russia/MOW-RII-MH252S/2020 | 2020-06-25 | Europe/Russia/Moscow Oblast/ | Pathogenic Microorganisms Variability Laboratory | WHO National Influenza Centre Russian Federation | Andrey Komissarov et al |
| EPI_ISL_566081 | Russia/MOW-RII-MH254S/2020 | 2020-06-08 | Europe/Russia/Moscow Oblast/ | Pathogenic Microorganisms Variability Laboratory | WHO National Influenza Centre Russian Federation | Andrey Komissarov et al |
| EPI_ISL_566082 | Russia/MOW-RII-MH255S/2020 | 2020-06-18 | Europe/Russia/Moscow Oblast/ | Pathogenic Microorganisms Variability Laboratory | WHO National Influenza Centre Russian Federation | Andrey Komissarov et al |
| EPI_ISL_566083 | Russia/MOW-RII-MH256S/2020 | 2020-06-18 | Europe/Russia/Moscow Oblast/ | Pathogenic Microorganisms Variability Laboratory | WHO National Influenza Centre Russian Federation | Andrey Komissarov et al |
| EPI_ISL_566091 | USA/GA-CDC-5103/2020 | 2020-07-01 | North America/USA/Georgia/ | GA Department of Public Health Laboratory | Pathogen Discovery, Respiratory Viruses Branch, Division of Viral Diseases, Centers for Disease Control and Prevention | Jing Zhang et al |
| EPI_ISL_566095 | USA/GA-CDC-5110/2020 | 2020-07-03 | North America/USA/Georgia/ | GA Department of Public Health Laboratory | Pathogen Discovery, Respiratory Viruses Branch, Division of Viral Diseases, Centers for Disease Control and Prevention | Jing Zhang et al |

| | | | | | | |
|---|---|---|---|---|---|---|
| EPI_ISL_566103 | USA/GA-CDC-5123/2020 | 2020-07-01 | North America/USA/Georgia/ | GA Department of Public Health Laboratory | Pathogen Discovery, Respiratory Viruses Branch, Division of Viral Diseases, Centers for Disease Control and Prevention | Jing Zhang et al |
| EPI_ISL_566204 | England/ALDP-9CD042/2020 | 2020-09-16 | Europe/United Kingdom/England/ | Lighthouse Lab in Alderley Park | Wellcome Sanger Institute for the COVID-19 Genomics UK (COG-UK) consortium | Jacquelyn Wynn et al |
| EPI_ISL_566278 | England/ALDP-9CDE84/2020 | 2020-09-16 | Europe/United Kingdom/England/ | Lighthouse Lab in Alderley Park | Wellcome Sanger Institute for the COVID-19 Genomics UK (COG-UK) consortium | Jacquelyn Wynn et al |
| EPI_ISL_566280 | England/ALDP-9CD75C/2020 | 2020-09-15 | Europe/United Kingdom/England/ | Lighthouse Lab in Alderley Park | Wellcome Sanger Institute for the COVID-19 Genomics UK (COG-UK) consortium | Jacquelyn Wynn et al |
| EPI_ISL_566331 | England/ALDP-9CD859/2020 | 2020-09-16 | Europe/United Kingdom/England/ | Lighthouse Lab in Alderley Park | Wellcome Sanger Institute for the COVID-19 Genomics UK (COG-UK) consortium | Jacquelyn Wynn et al |
| EPI_ISL_566427 | England/MILK-9C23F9/2020 | 2020-09-07 | Europe/United Kingdom/England/ | Lighthouse Lab in Milton Keynes | Wellcome Sanger Institute for the COVID-19 Genomics UK (COG-UK) consortium | The Lighthouse Lab in Milton Keynes et al |
| EPI_ISL_566493 | England/CAMC-9B1ADA/2020 | 2020-09-04 | Europe/United Kingdom/England/ | Lighthouse Lab in Cambridge | Wellcome Sanger Institute for the COVID-19 Genomics UK (COG-UK) consortium | Rob Howes et al |

| | | | | | | |
|---|---|---|---|---|---|---|
| EPI_ISL_566504 | England/MILK-9C23EA/2020 | 2020-09-07 | Europe/United Kingdom/England/ | Lighthouse Lab in Milton Keynes | Wellcome Sanger Institute for the COVID-19 Genomics UK (COG-UK) consortium | The Lighthouse Lab in Milton Keynes et al |
| EPI_ISL_566540 | England/MILK-9C206B/2020 | 2020-09-10 | Europe/United Kingdom/England/ | Lighthouse Lab in Milton Keynes | Wellcome Sanger Institute for the COVID-19 Genomics UK (COG-UK) consortium | The Lighthouse Lab in Milton Keynes et al |
| EPI_ISL_566634 | England/MILK-9C2809/2020 | 2020-09-10 | Europe/United Kingdom/England/ | Lighthouse Lab in Milton Keynes | Wellcome Sanger Institute for the COVID-19 Genomics UK (COG-UK) consortium | The Lighthouse Lab in Milton Keynes et al |
| EPI_ISL_566737 | England/MILK-9C3176/2020 | 2020-09-10 | Europe/United Kingdom/England/ | Lighthouse Lab in Milton Keynes | Wellcome Sanger Institute for the COVID-19 Genomics UK (COG-UK) consortium | The Lighthouse Lab in Milton Keynes et al |
| EPI_ISL_566927 | England/MILK-9C3246/2020 | 2020-09-10 | Europe/United Kingdom/England/ | Lighthouse Lab in Milton Keynes | Wellcome Sanger Institute for the COVID-19 Genomics UK (COG-UK) consortium | The Lighthouse Lab in Milton Keynes et al |
| EPI_ISL_566935 | England/MILK-9C30F1/2020 | 2020-09-07 | Europe/United Kingdom/England/ | Lighthouse Lab in Milton Keynes | Wellcome Sanger Institute for the COVID-19 Genomics UK (COG-UK) consortium | The Lighthouse Lab in Milton Keynes et al |
| EPI_ISL_567146 | England/QEUH-9C843C/2020 | 2020-09-11 | Europe/United Kingdom/England/ | Lighthouse Lab in Glasgow | Wellcome Sanger Institute for the COVID-19 Genomics UK (COG-UK) consortium | Harper VanSteenhouse et al |
| EPI_ISL_567272 | England/QEUH-9CA528/2020 | 2020-09-10 | Europe/United Kingdom/England/ | Lighthouse Lab in Glasgow | Wellcome Sanger Institute for the COVID-19 Genomics UK (COG-UK) consortium | Harper VanSteenhouse et al |

| | | | | | | |
|---|---|---|---|---|---|---|
| EPI_ISL_567310 | Wales/QEUH-9C98D5/2020 | 2020-09-10 | Europe/United Kingdom/Wales/ | Lighthouse Lab in Glasgow | Wellcome Sanger Institute for the COVID-19 Genomics UK (COG-UK) consortium | Harper VanSteenhouse et al |
| EPI_ISL_567313 | England/QEUH-9C9CCA/2020 | 2020-09-10 | Europe/United Kingdom/England/ | Lighthouse Lab in Glasgow | Wellcome Sanger Institute for the COVID-19 Genomics UK (COG-UK) consortium | Harper VanSteenhouse et al |
| EPI_ISL_567635 | England/ALDP-9C5DCB/2020 | 2020-09-10 | Europe/United Kingdom/England/ | Lighthouse Lab in Alderley Park | Wellcome Sanger Institute for the COVID-19 Genomics UK (COG-UK) consortium | Jacquelyn Wynn et al |
| EPI_ISL_567675 | England/ALDP-9C58D9/2020 | 2020-09-11 | Europe/United Kingdom/England/ | Lighthouse Lab in Alderley Park | Wellcome Sanger Institute for the COVID-19 Genomics UK (COG-UK) consortium | Jacquelyn Wynn et al |
| EPI_ISL_567792 | Wales/ALDP-9C5138/2020 | 2020-09-10 | Europe/United Kingdom/Wales/ | Lighthouse Lab in Alderley Park | Wellcome Sanger Institute for the COVID-19 Genomics UK (COG-UK) consortium | Jacquelyn Wynn et al |
| EPI_ISL_567899 | England/ALDP-9BE5FA/2020 | 2020-09-10 | Europe/United Kingdom/England/ | Lighthouse Lab in Alderley Park | Wellcome Sanger Institute for the COVID-19 Genomics UK (COG-UK) consortium | Jacquelyn Wynn et al |
| EPI_ISL_567900 | England/ALDP-9BED6B/2020 | 2020-09-10 | Europe/United Kingdom/England/ | Lighthouse Lab in Alderley Park | Wellcome Sanger Institute for the COVID-19 Genomics UK (COG-UK) consortium | Jacquelyn Wynn et al |
| EPI_ISL_568154 | England/ALDP-9BC8D5/2020 | 2020-09-09 | Europe/United Kingdom/England/ | Lighthouse Lab in Alderley Park | Wellcome Sanger Institute for the COVID-19 Genomics UK (COG-UK) consortium | Jacquelyn Wynn et al |

| | | | | | | |
|---|---|---|---|---|---|---|
| EPI_ISL_568160 | England/ALDP-9BC8B7/2020 | 2020-09-09 | Europe/United Kingdom/England/ | Lighthouse Lab in Alderley Park | Wellcome Sanger Institute for the COVID-19 Genomics UK (COG-UK) consortium | Jacquelyn Wynn et al |
| EPI_ISL_568188 | England/ALDP-9BC6F9/2020 | 2020-09-09 | Europe/United Kingdom/England/ | Lighthouse Lab in Alderley Park | Wellcome Sanger Institute for the COVID-19 Genomics UK (COG-UK) consortium | Jacquelyn Wynn et al |
| EPI_ISL_568243 | Scotland/QEUH-9B9B93/2020 | 2020-09-09 | Europe/United Kingdom/Scotland/ | Lighthouse Lab in Glasgow | Wellcome Sanger Institute for the COVID-19 Genomics UK (COG-UK) consortium | Harper VanSteenhouse et al |
| EPI_ISL_568382 | Scotland/QEUH-9A44CD/2020 | 2020-09-01 | Europe/United Kingdom/Scotland/ | Lighthouse Lab in Glasgow | Wellcome Sanger Institute for the COVID-19 Genomics UK (COG-UK) consortium | Harper VanSteenhouse et al |
| EPI_ISL_568510 | Peru/LIM-UPCH-0028/2020 | 2020-06-17 | South America/Peru/Lima/ | Laboratorio de Referencia Nacional de Virus Respiratorios, Instituto Nacional de Salud Peru | Laboratorio de Gen√≥mica Microbiana, Universidad Peruana Cayetano Heredia | Pablo Tsukayama et al |
| EPI_ISL_568511 | Peru/LIM-UPCH-0054/2020 | 2020-06-17 | South America/Peru/Lima/Ca√±ete | Laboratorio de Referencia Nacional de Virus Respiratorios, Instituto Nacional de Salud Peru | Laboratorio de Gen√≥mica Microbiana, Universidad Peruana Cayetano Heredia | Pablo Tsukayama et al |
| EPI_ISL_568512 | Peru/LIM-UPCH-0077/2020 | 2020-06-18 | South America/Peru/Lima/ | Laboratorio de Referencia Nacional de Virus Respiratorios, Instituto Nacional de Salud Peru | Laboratorio de Gen√≥mica Microbiana, Universidad Peruana Cayetano Heredia | Pablo Tsukayama et al |

| | | | | | | |
|---|---|---|---|---|---|---|
| EPI_ISL_568513 | Peru/LIM-UPCH-0116/2020 | 2020-06-18 | South America/Peru/Lima/ | Laboratorio de Referencia Nacional de Virus Respiratorios, Instituto Nacional de Salud Peru | Laboratorio de Genv≥mica Microbiana, Universidad Peruana Cayetano Heredia | Pablo Tsukayama et al |
| EPI_ISL_568514 | Peru/LIM-UPCH-0117/2020 | 2020-06-09 | South America/Peru/Lima/ | Laboratorio de Referencia Nacional de Virus Respiratorios, Instituto Nacional de Salud Peru | Laboratorio de Genv≥mica Microbiana, Universidad Peruana Cayetano Heredia | Pablo Tsukayama et al |
| EPI_ISL_568517 | Peru/LIM-UPCH-0120/2020 | 2020-06-18 | South America/Peru/Lima/ | Laboratorio de Referencia Nacional de Virus Respiratorios, Instituto Nacional de Salud Peru | Laboratorio de Genv≥mica Microbiana, Universidad Peruana Cayetano Heredia | Pablo Tsukayama et al |
| EPI_ISL_568518 | Peru/LIM-UPCH-0122/2020 | 2020-08-27 | South America/Peru/Lima/ | Laboratorio de Referencia Nacional de Virus Respiratorios, Instituto Nacional de Salud Peru | Laboratorio de Genv≥mica Microbiana, Universidad Peruana Cayetano Heredia | Pablo Tsukayama et al |
| EPI_ISL_568519 | Peru/LIM-UPCH-0123/2020 | 2020-08-27 | South America/Peru/Lima/ | Laboratorio de Referencia Nacional de Virus Respiratorios, Instituto Nacional de Salud Peru | Laboratorio de Genv≥mica Microbiana, Universidad Peruana Cayetano Heredia | Pablo Tsukayama et al |
| EPI_ISL_568520 | Peru/LIM-UPCH-0124/2020 | 2020-08-27 | South America/Peru/Lima/ | Laboratorio de Referencia Nacional de Virus Respiratorios, Instituto Nacional de Salud Peru | Laboratorio de Genv≥mica Microbiana, Universidad Peruana Cayetano Heredia | Pablo Tsukayama et al |

| | | | | | | |
|---|---|---|---|---|---|---|
| EPI_ISL_568521 | Peru/LIM-UPCH-0125/2020 | 2020-08-27 | South America/Peru/Lima/ | Laboratorio de Referencia Nacional de Virus Respiratorios, Instituto Nacional de Salud Peru | Laboratorio de Genv≥mica Microbiana, Universidad Peruana Cayetano Heredia | Pablo Tsukayama et al |
| EPI_ISL_568522 | Peru/LIM-UPCH-0126/2020 | 2020-08-27 | South America/Peru/Lima/ | Laboratorio de Referencia Nacional de Virus Respiratorios, Instituto Nacional de Salud Peru | Laboratorio de Genv≥mica Microbiana, Universidad Peruana Cayetano Heredia | Pablo Tsukayama et al |
| EPI_ISL_568523 | Peru/LIM-UPCH-0127/2020 | 2020-08-28 | South America/Peru/Lima/ | Laboratorio de Referencia Nacional de Virus Respiratorios, Instituto Nacional de Salud Peru | Laboratorio de Genv≥mica Microbiana, Universidad Peruana Cayetano Heredia | Pablo Tsukayama et al |
| EPI_ISL_568524 | Peru/LIM-UPCH-0128/2020 | 2020-08-28 | South America/Peru/Lima/ | Laboratorio de Referencia Nacional de Virus Respiratorios, Instituto Nacional de Salud Peru | Laboratorio de Genv≥mica Microbiana, Universidad Peruana Cayetano Heredia | Pablo Tsukayama et al |
| EPI_ISL_568525 | Peru/LIM-UPCH-0129/2020 | 2020-08-28 | South America/Peru/Lima/ | Laboratorio de Referencia Nacional de Virus Respiratorios, Instituto Nacional de Salud Peru | Laboratorio de Genv≥mica Microbiana, Universidad Peruana Cayetano Heredia | Pablo Tsukayama et al |
| EPI_ISL_568526 | Peru/LIM-UPCH-0130/2020 | 2020-08-28 | South America/Peru/Lima/ | Laboratorio de Referencia Nacional de Virus Respiratorios, Instituto Nacional de Salud Peru | Laboratorio de Genv≥mica Microbiana, Universidad Peruana Cayetano Heredia | Pablo Tsukayama et al |

| | | | | | | |
|---|---|---|---|---|---|---|
| EPI_ISL_568527 | Peru/LIM-UPCH-0131/2020 | 2020-08-28 | South America/Peru/Lima/ | Laboratorio de Referencia Nacional de Virus Respiratorios, Instituto Nacional de Salud Peru | Laboratorio de Genv≥mica Microbiana, Universidad Peruana Cayetano Heredia | Pablo Tsukayama et al |
| EPI_ISL_568528 | Peru/LIM-UPCH-0132/2020 | 2020-08-27 | South America/Peru/Lima/ | Laboratorio de Referencia Nacional de Virus Respiratorios, Instituto Nacional de Salud Peru | Laboratorio de Genv≥mica Microbiana, Universidad Peruana Cayetano Heredia | Pablo Tsukayama et al |
| EPI_ISL_568529 | Peru/LIM-UPCH-0133/2020 | 2020-08-27 | South America/Peru/Lima/ | Laboratorio de Referencia Nacional de Virus Respiratorios, Instituto Nacional de Salud Peru | Laboratorio de Genv≥mica Microbiana, Universidad Peruana Cayetano Heredia | Pablo Tsukayama et al |
| EPI_ISL_568530 | Peru/LIM-UPCH-0134/2020 | 2020-08-27 | South America/Peru/Lima/ | Laboratorio de Referencia Nacional de Virus Respiratorios, Instituto Nacional de Salud Peru | Laboratorio de Genv≥mica Microbiana, Universidad Peruana Cayetano Heredia | Pablo Tsukayama et al |
| EPI_ISL_568531 | Peru/LIM-UPCH-0135/2020 | 2020-08-27 | South America/Peru/Lima/ | Laboratorio de Referencia Nacional de Virus Respiratorios, Instituto Nacional de Salud Peru | Laboratorio de Genv≥mica Microbiana, Universidad Peruana Cayetano Heredia | Pablo Tsukayama et al |
| EPI_ISL_568532 | Peru/LIM-UPCH-0136/2020 | 2020-08-27 | South America/Peru/Lima/ | Laboratorio de Referencia Nacional de Virus Respiratorios, Instituto Nacional de Salud Peru | Laboratorio de Genv≥mica Microbiana, Universidad Peruana Cayetano Heredia | Pablo Tsukayama et al |

| | | | | | | |
|---|---|---|---|---|---|---|
| EPI_ISL_568533 | Peru/LIM-UPCH-0137/2020 | 2020-08-28 | South America/Peru/Lima/ | Laboratorio de Referencia Nacional de Virus Respiratorios, Instituto Nacional de Salud Peru | Laboratorio de Genv≥mica Microbiana, Universidad Peruana Cayetano Heredia | Pablo Tsukayama et al |
| EPI_ISL_568534 | Peru/LIM-UPCH-0138/2020 | 2020-08-27 | South America/Peru/Lima/ | Laboratorio de Referencia Nacional de Virus Respiratorios, Instituto Nacional de Salud Peru | Laboratorio de Genv≥mica Microbiana, Universidad Peruana Cayetano Heredia | Pablo Tsukayama et al |
| EPI_ISL_568535 | Peru/LIM-UPCH-0141/2020 | 2020-08-27 | South America/Peru/Lima/ | Laboratorio de Referencia Nacional de Virus Respiratorios, Instituto Nacional de Salud Peru | Laboratorio de Genv≥mica Microbiana, Universidad Peruana Cayetano Heredia | Pablo Tsukayama et al |
| EPI_ISL_568536 | Peru/LIM-UPCH-0142/2020 | 2020-08-28 | South America/Peru/Lima/ | Laboratorio de Referencia Nacional de Virus Respiratorios, Instituto Nacional de Salud Peru | Laboratorio de Genv≥mica Microbiana, Universidad Peruana Cayetano Heredia | Pablo Tsukayama et al |
| EPI_ISL_568537 | Peru/LIM-UPCH-0143/2020 | 2020-08-27 | South America/Peru/Lima/ | Laboratorio de Referencia Nacional de Virus Respiratorios, Instituto Nacional de Salud Peru | Laboratorio de Genv≥mica Microbiana, Universidad Peruana Cayetano Heredia | Pablo Tsukayama et al |
| EPI_ISL_568538 | Peru/LIM-UPCH-0144/2020 | 2020-08-27 | South America/Peru/Lima/ | Laboratorio de Referencia Nacional de Virus Respiratorios, Instituto Nacional de Salud Peru | Laboratorio de Genv≥mica Microbiana, Universidad Peruana Cayetano Heredia | Pablo Tsukayama et al |

| | | | | | | |
|---|---|---|---|---|---|---|
| EPI_ISL_568539 | Peru/LIM-UPCH-0145/2020 | 2020-08-27 | South America/Peru/Lima/ | Laboratorio de Referencia Nacional de Virus Respiratorios, Instituto Nacional de Salud Peru | Laboratorio de Genv≥mica Microbiana, Universidad Peruana Cayetano Heredia | Pablo Tsukayama et al |
| EPI_ISL_568540 | Peru/LIM-UPCH-0146/2020 | 2020-08-28 | South America/Peru/Lima/ | Laboratorio de Referencia Nacional de Virus Respiratorios, Instituto Nacional de Salud Peru | Laboratorio de Genv≥mica Microbiana, Universidad Peruana Cayetano Heredia | Pablo Tsukayama et al |
| EPI_ISL_568541 | Peru/LIM-UPCH-0147/2020 | 2020-08-28 | South America/Peru/Lima/ | Laboratorio de Referencia Nacional de Virus Respiratorios, Instituto Nacional de Salud Peru | Laboratorio de Genv≥mica Microbiana, Universidad Peruana Cayetano Heredia | Pablo Tsukayama et al |
| EPI_ISL_568542 | Peru/LIM-UPCH-0148/2020 | 2020-08-28 | South America/Peru/Lima/ | Laboratorio de Referencia Nacional de Virus Respiratorios, Instituto Nacional de Salud Peru | Laboratorio de Genv≥mica Microbiana, Universidad Peruana Cayetano Heredia | Pablo Tsukayama et al |
| EPI_ISL_568543 | Peru/LIM-UPCH-0149/2020 | 2020-08-27 | South America/Peru/Lima/ | Laboratorio de Referencia Nacional de Virus Respiratorios, Instituto Nacional de Salud Peru | Laboratorio de Genv≥mica Microbiana, Universidad Peruana Cayetano Heredia | Pablo Tsukayama et al |
| EPI_ISL_568544 | Peru/LIM-UPCH-0150/2020 | 2020-08-27 | South America/Peru/Lima/ | Laboratorio de Referencia Nacional de Virus Respiratorios, Instituto Nacional de Salud Peru | Laboratorio de Genv≥mica Microbiana, Universidad Peruana Cayetano Heredia | Pablo Tsukayama et al |

| | | | | | | |
|---|---|---|---|---|---|---|
| EPI_ISL_568545 | Peru/LIM-UPCH-0151/2020 | 2020-08-28 | South America/Peru/Lima/ | Laboratorio de Referencia Nacional de Virus Respiratorios, Instituto Nacional de Salud Peru | Laboratorio de Genv≥mica Microbiana, Universidad Peruana Cayetano Heredia | Pablo Tsukayama et al |
| EPI_ISL_568546 | Peru/LIM-UPCH-0152/2020 | 2020-08-28 | South America/Peru/Lima/ | Laboratorio de Referencia Nacional de Virus Respiratorios, Instituto Nacional de Salud Peru | Laboratorio de Genv≥mica Microbiana, Universidad Peruana Cayetano Heredia | Pablo Tsukayama et al |
| EPI_ISL_568547 | Peru/LIM-UPCH-0154/2020 | 2020-08-28 | South America/Peru/Lima/ | Laboratorio de Referencia Nacional de Virus Respiratorios, Instituto Nacional de Salud Peru | Laboratorio de Genv≥mica Microbiana, Universidad Peruana Cayetano Heredia | Pablo Tsukayama et al |
| EPI_ISL_568548 | Peru/LIM-UPCH-0155/2020 | 2020-08-27 | South America/Peru/Lima/ | Laboratorio de Referencia Nacional de Virus Respiratorios, Instituto Nacional de Salud Peru | Laboratorio de Genv≥mica Microbiana, Universidad Peruana Cayetano Heredia | Pablo Tsukayama et al |
| EPI_ISL_568549 | Peru/LIM-UPCH-0156/2020 | 2020-08-28 | South America/Peru/Lima/ | Laboratorio de Referencia Nacional de Virus Respiratorios, Instituto Nacional de Salud Peru | Laboratorio de Genv≥mica Microbiana, Universidad Peruana Cayetano Heredia | Pablo Tsukayama et al |
| EPI_ISL_568550 | Peru/LIM-UPCH-0157/2020 | 2020-08-28 | South America/Peru/Lima/ | Laboratorio de Referencia Nacional de Virus Respiratorios, Instituto Nacional de Salud Peru | Laboratorio de Genv≥mica Microbiana, Universidad Peruana Cayetano Heredia | Pablo Tsukayama et al |

| | | | | | | |
|---|---|---|---|---|---|---|
| EPI_ISL_568551 | Peru/LIM-UPCH-0158/2020 | 2020-08-28 | South America/Peru/Lima/ | Laboratorio de Referencia Nacional de Virus Respiratorios, Instituto Nacional de Salud Peru | Laboratorio de Genv≥mica Microbiana, Universidad Peruana Cayetano Heredia | Pablo Tsukayama et al |
| EPI_ISL_568552 | Peru/LIM-UPCH-0159/2020 | 2020-08-27 | South America/Peru/Lima/ | Laboratorio de Referencia Nacional de Virus Respiratorios, Instituto Nacional de Salud Peru | Laboratorio de Genv≥mica Microbiana, Universidad Peruana Cayetano Heredia | Pablo Tsukayama et al |
| EPI_ISL_568553 | Peru/LIM-UPCH-0160/2020 | 2020-08-28 | South America/Peru/Lima/ | Laboratorio de Referencia Nacional de Virus Respiratorios, Instituto Nacional de Salud Peru | Laboratorio de Genv≥mica Microbiana, Universidad Peruana Cayetano Heredia | Pablo Tsukayama et al |
| EPI_ISL_568554 | Peru/LIM-UPCH-0161/2020 | 2020-08-28 | South America/Peru/Lima/ | Laboratorio de Referencia Nacional de Virus Respiratorios, Instituto Nacional de Salud Peru | Laboratorio de Genv≥mica Microbiana, Universidad Peruana Cayetano Heredia | Pablo Tsukayama et al |
| EPI_ISL_568555 | Peru/LIM-UPCH-0162/2020 | 2020-08-28 | South America/Peru/Lima/ | Laboratorio de Referencia Nacional de Virus Respiratorios, Instituto Nacional de Salud Peru | Laboratorio de Genv≥mica Microbiana, Universidad Peruana Cayetano Heredia | Pablo Tsukayama et al |
| EPI_ISL_568577 | CzechRepublic/NRL-8631/2020 | 2020-09-04 | Europe/Czech Republic/Prague/ | The National Institute of Public Health | State Veterinary Institute Prague | Nagy et al |
| EPI_ISL_568579 | Italy/LOM-INMI-10734/2020 | 2020-02-24 | Europe/Italy/Lombardy/Pavia | Virus Molecular Laboratory of the Microbiology and Virology Department | INMI Lazzaro Spallanzani IRCCS | Cesare E.M. Gruber et al |
| EPI_ISL_568588 | USA/FL-BPHL-1490/2020 | 2020-08-10 | North America/USA/Florida/ | Florida Bureau of Public Health Laboratories | Florida Bureau of Public Health Laboratories | Sarah Schmedes et al |
| EPI_ISL_568598 | USA/FL-BPHL-1500/2020 | 2020-07-23 | North America/USA/Florida/ | Florida Bureau of Public Health Laboratories | Florida Bureau of Public Health Laboratories | Sarah Schmedes et al |

| | | | | | | |
|---|---|---|---|---|---|---|
| EPI_ISL_568602 | USA/FL-BPHL-1504/2020 | 2020-07-23 | North America/USA/Florida/ | Florida Bureau of Public Health Laboratories | Florida Bureau of Public Health Laboratories | Sarah Schmedes et al |
| EPI_ISL_568619 | USA/FL-BPHL-1521/2020 | 2020-07-28 | North America/USA/Florida/ | Florida Bureau of Public Health Laboratories | Florida Bureau of Public Health Laboratories | Sarah Schmedes et al |
| EPI_ISL_568621 | USA/FL-BPHL-1523/2020 | 2020-07-28 | North America/USA/Florida/ | Florida Bureau of Public Health Laboratories | Florida Bureau of Public Health Laboratories | Sarah Schmedes et al |
| EPI_ISL_568638 | USA/FL-BPHL-1540/2020 | 2020-08-28 | North America/USA/Florida/ | Florida Bureau of Public Health Laboratories | Florida Bureau of Public Health Laboratories | Sarah Schmedes et al |
| EPI_ISL_568650 | USA/FL-BPHL-1552/2020 | 2020-09-09 | North America/USA/Florida/ | Florida Bureau of Public Health Laboratories | Florida Bureau of Public Health Laboratories | Sarah Schmedes et al |
| EPI_ISL_568651 | USA/FL-BPHL-1553/2020 | 2020-09-09 | North America/USA/Florida/ | Florida Bureau of Public Health Laboratories | Florida Bureau of Public Health Laboratories | Sarah Schmedes et al |
| EPI_ISL_568652 | USA/FL-BPHL-1554/2020 | 2020-09-09 | North America/USA/Florida/ | Florida Bureau of Public Health Laboratories | Florida Bureau of Public Health Laboratories | Sarah Schmedes et al |
| EPI_ISL_568654 | USA/FL-BPHL-1556/2020 | 2020-09-09 | North America/USA/Florida/ | Florida Bureau of Public Health Laboratories | Florida Bureau of Public Health Laboratories | Sarah Schmedes et al |
| EPI_ISL_568657 | USA/FL-BPHL-1559/2020 | 2020-09-09 | North America/USA/Florida/ | Florida Bureau of Public Health Laboratories | Florida Bureau of Public Health Laboratories | Sarah Schmedes et al |
| EPI_ISL_568659 | USA/FL-BPHL-1561/2020 | 2020-09-10 | North America/USA/Florida/ | Florida Bureau of Public Health Laboratories | Florida Bureau of Public Health Laboratories | Sarah Schmedes et al |
| EPI_ISL_568674 | USA/FL-BPHL-1576/2020 | 2020-09-16 | North America/USA/Florida/ | Florida Bureau of Public Health Laboratories | Florida Bureau of Public Health Laboratories | Sarah Schmedes et al |
| EPI_ISL_568677 | USA/FL-BPHL-1579/2020 | 2020-09-16 | North America/USA/Florida/ | Florida Bureau of Public Health Laboratories | Florida Bureau of Public Health Laboratories | Sarah Schmedes et al |
| EPI_ISL_568683 | USA/FL-BPHL-1585/2020 | 2020-03-21 | North America/USA/Florida/ | Florida Bureau of Public Health Laboratories | Florida Bureau of Public Health Laboratories | Sarah Schmedes et al |

| | | | | | | |
|---|---|---|---|---|---|---|
| EPI_ISL_568686 | Indonesia/JT-EIJK-10/2020 | 2020-03-26 | Asia/Indonesia/Central Java/Purwokert | RSUD Prof. DR. Margono Soekarjo | Eijkman Institute for Molecular Biology, Ministry of Research and Technology/National Agency for Research and Innovation | Frilasita A Yudhaputri et al |
| EPI_ISL_568687 | Indonesia/JK-EIJK-23/2020 | 2020-04-11 | Asia/Indonesia/Jakarta/ | RSUP Fatmawati | Eijkman Institute for Molecular Biology, Ministry of Research and Technology/National Agency for Research and Innovation | Frilasita A Yudhaputri et al |
| EPI_ISL_568688 | Indonesia/JK-EIJK-32/2020 | 2020-04-22 | Asia/Indonesia/Jakarta/ | RS Pelni | Eijkman Institute for Molecular Biology, Ministry of Research and Technology/National Agency for Research and Innovation | Frilasita A Yudhaputri et al |
| EPI_ISL_568689 | Indonesia/JK-EIJK-38/2020 | 2020-05-20 | Asia/Indonesia/Jakarta/ | RS Pondok Indah Puri Indah | Eijkman Institute for Molecular Biology, Ministry of Research and Technology/National Agency for Research and Innovation | Frilasita A Yudhaputri et al |
| EPI_ISL_568690 | Indonesia/JK-EIJK-39/2020 | 2020-05-26 | Asia/Indonesia/Jakarta/ | RSUP Fatmawati | Eijkman Institute for Molecular Biology, Ministry of Research and Technology/National Agency for Research and Innovation | Frilasita A Yudhaputri et al |

| | | | | | | |
|---|---|---|---|---|---|---|
| EPI_ISL_568691 | Indonesia/JK-EIJK-40/2020 | 2020-05-27 | Asia/Indonesia/Jakarta/ | RSUP Fatmawati | Eijkman Institute for Molecular Biology, Ministry of Research and Technology/National Agency for Research and Innovation | Frilasita A Yudhaputri et al |
| EPI_ISL_568692 | Indonesia/JK-EIJK-43/2020 | 2020-06-04 | Asia/Indonesia/Jakarta/ | RS Kramat 128 | Eijkman Institute for Molecular Biology, Ministry of Research and Technology/National Agency for Research and Innovation | Frilasita A Yudhaputri et al |
| EPI_ISL_568693 | Indonesia/PP-EIJK-11/2020 | 2020-03-28 | Asia/Indonesia/Papua/Timika | RS Freeport Tembagapura | Eijkman Institute for Molecular Biology, Ministry of Research and Technology/National Agency for Research and Innovation | Frilasita A Yudhaputri et al |
| EPI_ISL_568723 | Kenya/C4195/2020 | 2020-06-05 | Africa/Kenya/Mombasa/ | KEMRI-Wellcome Trust Research Programme/KEMRI-CGMR-C Kilifi | KEMRI-Wellcome Trust Research Programme/KEMRI-CGMR-C Kilifi | Githinji et al 2020 et al |
| EPI_ISL_568725 | Kenya/C4214/2020 | 2020-06-05 | Africa/Kenya/Mombasa/ | KEMRI-Wellcome Trust Research Programme/KEMRI-CGMR-C Kilifi | KEMRI-Wellcome Trust Research Programme/KEMRI-CGMR-C Kilifi | Githinji et al 2020 et al |
| EPI_ISL_568755 | Kenya/C5288/2020 | 2020-05-10 | Africa/Kenya/Mombasa/ | KEMRI-Wellcome Trust Research Programme/KEMRI-CGMR-C Kilifi | KEMRI-Wellcome Trust Research Programme/KEMRI-CGMR-C Kilifi | Githinji et al 2020 et al |
| EPI_ISL_568873 | Malaysia/MGI-M23/2020 | 2020-04-02 | Asia/Malaysia/Selangor/ | Malaysia Genome Institute | Malaysia Genome Institute | Mohd Noor Mat Isa et al |

| | | | | | | |
|---|---|---|---|---|---|---|
| EPI_ISL_568881 | France/PAC-IHU-0601/2020 | 2020-04-10 | Europe/France/Provence-Alpes-CV¥te d' | MEPHI, Aix Marseille University | MEPHI, Aix Marseille University | Anthony LEVASSEUR et al |
| EPI_ISL_568882 | France/PAC-IHU-0602/2020 | 2020-04-10 | Europe/France/Provence-Alpes-CV¥te d' | MEPHI, Aix Marseille University | MEPHI, Aix Marseille University | Anthony LEVASSEUR et al |
| EPI_ISL_568885 | France/PAC-IHU-0609/2020 | 2020-04-11 | Europe/France/Provence-Alpes-CV¥te d' | MEPHI, Aix Marseille University | MEPHI, Aix Marseille University | Anthony LEVASSEUR et al |
| EPI_ISL_568888 | France/PAC-IHU-0612/2020 | 2020-04-11 | Europe/France/Provence-Alpes-CV¥te d' | MEPHI, Aix Marseille University | MEPHI, Aix Marseille University | Anthony LEVASSEUR et al |
| EPI_ISL_568893 | France/PAC-IHU-0620/2020 | 2020-04-12 | Europe/France/Provence-Alpes-CV¥te d' | MEPHI, Aix Marseille University | MEPHI, Aix Marseille University | Anthony LEVASSEUR et al |
| EPI_ISL_568898 | France/PAC-IHU-0627/2020 | 2020-04-13 | Europe/France/Provence-Alpes-CV¥te d' | MEPHI, Aix Marseille University | MEPHI, Aix Marseille University | Anthony LEVASSEUR et al |
| EPI_ISL_568909 | France/PAC-IHU-0641/2020 | 2020-04-02 | Europe/France/Provence-Alpes-CV¥te d' | MEPHI, Aix Marseille University | MEPHI, Aix Marseille University | Anthony LEVASSEUR et al |
| EPI_ISL_568913 | France/PAC-IHU-0649/2020 | 2020-03-06 | Europe/France/Provence-Alpes-CV¥te d' | MEPHI, Aix Marseille University | MEPHI, Aix Marseille University | Anthony LEVASSEUR et al |
| EPI_ISL_568921 | France/PAC-IHU-0668/2020 | 2020-03-17 | Europe/France/Provence-Alpes-CV¥te d' | MEPHI, Aix Marseille University | MEPHI, Aix Marseille University | Anthony LEVASSEUR et al |
| EPI_ISL_568922 | France/PAC-IHU-0670/2020 | 2020-03-16 | Europe/France/Provence-Alpes-CV¥te d' | MEPHI, Aix Marseille University | MEPHI, Aix Marseille University | Anthony LEVASSEUR et al |
| EPI_ISL_568929 | France/PAC-IHU-0677/2020 | 2020-04-15 | Europe/France/Provence-Alpes-CV¥te d' | MEPHI, Aix Marseille University | MEPHI, Aix Marseille University | Anthony LEVASSEUR et al |
| EPI_ISL_568937 | France/PAC-IHU-0691/2020 | 2020-04-14 | Europe/France/Provence-Alpes-CV¥te d' | MEPHI, Aix Marseille University | MEPHI, Aix Marseille University | Anthony LEVASSEUR et al |
| EPI_ISL_568955 | France/PAC-IHU-0743/2020 | 2020-04-24 | Europe/France/Provence-Alpes-CV¥te d' | MEPHI, Aix Marseille University | MEPHI, Aix Marseille University | Anthony LEVASSEUR et al |

| | | | | | | |
|---|---|---|---|---|---|---|
| EPI_ISL_568956 | France/PAC-IHU-0756/2020 | 2020-03-15 | Europe/France/Provence-Alpes-CV¥te d' | MEPHI, Aix Marseille University | MEPHI, Aix Marseille University | Anthony LEVASSEUR et al |
| EPI_ISL_568959 | France/PAC-IHU-0783/2020 | 2020-04-27 | Europe/France/Provence-Alpes-CV¥te d' | MEPHI, Aix Marseille University | MEPHI, Aix Marseille University | Anthony LEVASSEUR et al |
| EPI_ISL_568961 | France/PAC-IHU-0785/2020 | 2020-04-29 | Europe/France/Provence-Alpes-CV¥te d' | MEPHI, Aix Marseille University | MEPHI, Aix Marseille University | Anthony LEVASSEUR et al |
| EPI_ISL_568963 | France/PAC-IHU-0787/2020 | 2020-05-01 | Europe/France/Provence-Alpes-CV¥te d' | MEPHI, Aix Marseille University | MEPHI, Aix Marseille University | Anthony LEVASSEUR et al |
| EPI_ISL_568964 | France/PAC-IHU-0788/2020 | 2020-05-03 | Europe/France/Provence-Alpes-CV¥te d' | MEPHI, Aix Marseille University | MEPHI, Aix Marseille University | Anthony LEVASSEUR et al |
| EPI_ISL_568968 | France/PAC-IHU-0796/2020 | 2020-05-03 | Europe/France/Provence-Alpes-CV¥te d' | MEPHI, Aix Marseille University | MEPHI, Aix Marseille University | Anthony LEVASSEUR et al |
| EPI_ISL_568969 | France/PAC-IHU-0798/2020 | 2020-05-02 | Europe/France/Provence-Alpes-CV¥te d' | MEPHI, Aix Marseille University | MEPHI, Aix Marseille University | Anthony LEVASSEUR et al |
| EPI_ISL_568970 | France/PAC-IHU-0799/2020 | 2020-04-30 | Europe/France/Provence-Alpes-CV¥te d' | MEPHI, Aix Marseille University | MEPHI, Aix Marseille University | Anthony LEVASSEUR et al |
| EPI_ISL_568972 | France/PAC-IHU-0808/2020 | 2020-05-06 | Europe/France/Provence-Alpes-CV¥te d' | MEPHI, Aix Marseille University | MEPHI, Aix Marseille University | Anthony LEVASSEUR et al |
| EPI_ISL_568976 | France/PAC-IHU-0814/2020 | 2020-05-07 | Europe/France/Provence-Alpes-CV¥te d' | MEPHI, Aix Marseille University | MEPHI, Aix Marseille University | Anthony LEVASSEUR et al |
| EPI_ISL_568977 | France/PAC-IHU-0817/2020 | 2020-05-05 | Europe/France/Provence-Alpes-CV¥te d' | MEPHI, Aix Marseille University | MEPHI, Aix Marseille University | Anthony LEVASSEUR et al |
| EPI_ISL_568983 | France/PAC-IHU-0830/2020 | 2020-05-20 | Europe/France/Provence-Alpes-CV¥te d' | MEPHI, Aix Marseille University | MEPHI, Aix Marseille University | Anthony LEVASSEUR et al |
| EPI_ISL_568985 | France/PAC-IHU-0851/2020 | 2020-03-11 | Europe/France/Provence-Alpes-CV¥te d' | MEPHI, Aix Marseille University | MEPHI, Aix Marseille University | Anthony LEVASSEUR et al |

| | | | | | | |
|---|---|---|---|---|---|---|
| EPI_ISL_568986 | France/PAC-IHU-0852/2020 | 2020-03-11 | Europe/France/Provence-Alpes-CV¥te d' | MEPHI, Aix Marseille University | MEPHI, Aix Marseille University | Anthony LEVASSEUR et al |
| EPI_ISL_568988 | France/PAC-IHU-0871/2020 | 2020-03-12 | Europe/France/Provence-Alpes-CV¥te d' | MEPHI, Aix Marseille University | MEPHI, Aix Marseille University | Anthony LEVASSEUR et al |
| EPI_ISL_569012 | France/PAC-IHU-0962/2020 | 2020-07-13 | Europe/France/Provence-Alpes-CV¥te d' | MEPHI, Aix Marseille University | MEPHI, Aix Marseille University | Anthony LEVASSEUR et al |
| EPI_ISL_569019 | France/PAC-IHU-0971/2020 | 2020-07-20 | Europe/France/Provence-Alpes-CV¥te d' | MEPHI, Aix Marseille University | MEPHI, Aix Marseille University | Anthony LEVASSEUR et al |
| EPI_ISL_569027 | France/PAC-IHU-0980/2020 | 2020-07-27 | Europe/France/Provence-Alpes-CV¥te d' | MEPHI, Aix Marseille University | MEPHI, Aix Marseille University | Anthony LEVASSEUR et al |
| EPI_ISL_569031 | France/PAC-IHU-0985/2020 | 2020-08-07 | Europe/France/Provence-Alpes-CV¥te d' | MEPHI, Aix Marseille University | MEPHI, Aix Marseille University | Anthony LEVASSEUR et al |
| EPI_ISL_569042 | France/PAC-IHU-1056/2020 | 2020-08-04 | Europe/France/Provence-Alpes-CV¥te d' | MEPHI, Aix Marseille University | MEPHI, Aix Marseille University | Anthony LEVASSEUR et al |
| EPI_ISL_569046 | France/PAC-IHU-1068/2020 | 2020-08-05 | Europe/France/Provence-Alpes-CV¥te d' | MEPHI, Aix Marseille University | MEPHI, Aix Marseille University | Anthony LEVASSEUR et al |
| EPI_ISL_569051 | France/PAC-IHU-1097/2020 | 2020-08-05 | Europe/France/Provence-Alpes-CV¥te d' | MEPHI, Aix Marseille University | MEPHI, Aix Marseille University | Anthony LEVASSEUR et al |
| EPI_ISL_569053 | France/PAC-IHU-1126/2020 | 2020-08-07 | Europe/France/Provence-Alpes-CV¥te d' | MEPHI, Aix Marseille University | MEPHI, Aix Marseille University | Anthony LEVASSEUR et al |
| EPI_ISL_569055 | France/PAC-IHU-1131/2020 | 2020-08-07 | Europe/France/Provence-Alpes-CV¥te d' | MEPHI, Aix Marseille University | MEPHI, Aix Marseille University | Anthony LEVASSEUR et al |
| EPI_ISL_569061 | France/PAC-IHU-1151/2020 | 2020-08-07 | Europe/France/Provence-Alpes-CV¥te d' | MEPHI, Aix Marseille University | MEPHI, Aix Marseille University | Anthony LEVASSEUR et al |
| EPI_ISL_569063 | France/PAC-IHU-1157/2020 | 2020-08-08 | Europe/France/Provence-Alpes-CV¥te d' | MEPHI, Aix Marseille University | MEPHI, Aix Marseille University | Anthony LEVASSEUR et al |

| | | | | | | |
|---|---|---|---|---|---|---|
| EPI_ISL_569065 | France/PAC-IHU-1160/2020 | 2020-08-12 | Europe/France/Provence-Alpes-CV¥te d' | MEPHI, Aix Marseille University | MEPHI, Aix Marseille University | Anthony LEVASSEUR et al |
| EPI_ISL_569069 | France/PAC-IHU-1165/2020 | 2020-08-14 | Europe/France/Provence-Alpes-CV¥te d' | MEPHI, Aix Marseille University | MEPHI, Aix Marseille University | Anthony LEVASSEUR et al |
| EPI_ISL_569071 | France/PAC-IHU-1167/2020 | 2020-08-14 | Europe/France/Provence-Alpes-CV¥te d' | MEPHI, Aix Marseille University | MEPHI, Aix Marseille University | Anthony LEVASSEUR et al |
| EPI_ISL_569073 | France/PAC-IHU-1171/2020 | 2020-08-14 | Europe/France/Provence-Alpes-CV¥te d' | MEPHI, Aix Marseille University | MEPHI, Aix Marseille University | Anthony LEVASSEUR et al |
| EPI_ISL_569076 | France/PAC-IHU-1175/2020 | 2020-08-14 | Europe/France/Provence-Alpes-CV¥te d' | MEPHI, Aix Marseille University | MEPHI, Aix Marseille University | Anthony LEVASSEUR et al |
| EPI_ISL_569077 | France/PAC-IHU-1176/2020 | 2020-08-15 | Europe/France/Provence-Alpes-CV¥te d' | MEPHI, Aix Marseille University | MEPHI, Aix Marseille University | Anthony LEVASSEUR et al |
| EPI_ISL_569081 | France/PAC-IHU-1193/2020 | 2020-08-17 | Europe/France/Provence-Alpes-CV¥te d' | MEPHI, Aix Marseille University | MEPHI, Aix Marseille University | Anthony LEVASSEUR et al |
| EPI_ISL_569083 | France/PAC-IHU-1203/2020 | 2020-08-17 | Europe/France/Provence-Alpes-CV¥te d' | MEPHI, Aix Marseille University | MEPHI, Aix Marseille University | Anthony LEVASSEUR et al |
| EPI_ISL_569091 | France/PAC-IHU-1223/2020 | 2020-08-17 | Europe/France/Provence-Alpes-CV¥te d' | MEPHI, Aix Marseille University | MEPHI, Aix Marseille University | Anthony LEVASSEUR et al |
| EPI_ISL_569095 | France/PAC-IHU-1236/2020 | 2020-08-17 | Europe/France/Provence-Alpes-CV¥te d' | MEPHI, Aix Marseille University | MEPHI, Aix Marseille University | Anthony LEVASSEUR et al |
| EPI_ISL_569102 | France/PAC-IHU-1268/2020 | 2020-08-17 | Europe/France/Provence-Alpes-CV¥te d' | MEPHI, Aix Marseille University | MEPHI, Aix Marseille University | Anthony LEVASSEUR et al |
| EPI_ISL_569103 | France/PAC-IHU-1270/2020 | 2020-08-18 | Europe/France/Provence-Alpes-CV¥te d' | MEPHI, Aix Marseille University | MEPHI, Aix Marseille University | Anthony LEVASSEUR et al |
| EPI_ISL_569113 | France/PAC-IHU-1307/2020 | 2020-08-18 | Europe/France/Provence-Alpes-CV¥te d' | MEPHI, Aix Marseille University | MEPHI, Aix Marseille University | Anthony LEVASSEUR et al |

| | | | | | | |
|---|---|---|---|---|---|---|
| EPI_ISL_569114 | France/PAC-IHU-1315/2020 | 2020-08-18 | Europe/France/Provence-Alpes-CV¥te d' | MEPHI, Aix Marseille University | MEPHI, Aix Marseille University | Anthony LEVASSEUR et al |
| EPI_ISL_569115 | France/PAC-IHU-1316/2020 | 2020-08-18 | Europe/France/Provence-Alpes-CV¥te d' | MEPHI, Aix Marseille University | MEPHI, Aix Marseille University | Anthony LEVASSEUR et al |
| EPI_ISL_569122 | France/PAC-IHU-1335/2020 | 2020-08-19 | Europe/France/Provence-Alpes-CV¥te d' | MEPHI, Aix Marseille University | MEPHI, Aix Marseille University | Anthony LEVASSEUR et al |
| EPI_ISL_569126 | France/PAC-IHU-1341/2020 | 2020-08-19 | Europe/France/Provence-Alpes-CV¥te d' | MEPHI, Aix Marseille University | MEPHI, Aix Marseille University | Anthony LEVASSEUR et al |
| EPI_ISL_569129 | France/PAC-IHU-1345/2020 | 2020-08-19 | Europe/France/Provence-Alpes-CV¥te d' | MEPHI, Aix Marseille University | MEPHI, Aix Marseille University | Anthony LEVASSEUR et al |
| EPI_ISL_569133 | France/PAC-IHU-1349/2020 | 2020-08-19 | Europe/France/Provence-Alpes-CV¥te d' | MEPHI, Aix Marseille University | MEPHI, Aix Marseille University | Anthony LEVASSEUR et al |
| EPI_ISL_569137 | France/PAC-IHU-1355/2020 | 2020-08-20 | Europe/France/Provence-Alpes-CV¥te d' | MEPHI, Aix Marseille University | MEPHI, Aix Marseille University | Anthony LEVASSEUR et al |
| EPI_ISL_569138 | France/PAC-IHU-1357/2020 | 2020-08-20 | Europe/France/Provence-Alpes-CV¥te d' | MEPHI, Aix Marseille University | MEPHI, Aix Marseille University | Anthony LEVASSEUR et al |
| EPI_ISL_569142 | France/PAC-IHU-1363/2020 | 2020-08-20 | Europe/France/Provence-Alpes-CV¥te d' | MEPHI, Aix Marseille University | MEPHI, Aix Marseille University | Anthony LEVASSEUR et al |
| EPI_ISL_569146 | France/PAC-IHU-1377/2020 | 2020-08-08 | Europe/France/Provence-Alpes-CV¥te d' | MEPHI, Aix Marseille University | MEPHI, Aix Marseille University | Anthony LEVASSEUR et al |
| EPI_ISL_569147 | France/PAC-IHU-1382/2020 | 2020-08-08 | Europe/France/Provence-Alpes-CV¥te d' | MEPHI, Aix Marseille University | MEPHI, Aix Marseille University | Anthony LEVASSEUR et al |
| EPI_ISL_569148 | France/PAC-IHU-1383/2020 | 2020-08-08 | Europe/France/Provence-Alpes-CV¥te d' | MEPHI, Aix Marseille University | MEPHI, Aix Marseille University | Anthony LEVASSEUR et al |
| EPI_ISL_569154 | France/PAC-IHU-1407/2020 | 2020-08-10 | Europe/France/Provence-Alpes-CV¥te d' | MEPHI, Aix Marseille University | MEPHI, Aix Marseille University | Anthony LEVASSEUR et al |

| | | | | | | |
|---|---|---|---|---|---|---|
| EPI_ISL_569156 | France/PAC-IHU-1414/2020 | 2020-08-10 | Europe/France/Provence-Alpes-CV¥te d' | MEPHI, Aix Marseille University | MEPHI, Aix Marseille University | Anthony LEVASSEUR et al |
| EPI_ISL_569160 | France/PAC-IHU-1426/2020 | 2020-08-10 | Europe/France/Provence-Alpes-CV¥te d' | MEPHI, Aix Marseille University | MEPHI, Aix Marseille University | Anthony LEVASSEUR et al |
| EPI_ISL_569165 | France/PAC-IHU-1431/2020 | 2020-08-10 | Europe/France/Provence-Alpes-CV¥te d' | MEPHI, Aix Marseille University | MEPHI, Aix Marseille University | Anthony LEVASSEUR et al |
| EPI_ISL_569170 | France/PAC-IHU-1446/2020 | 2020-08-10 | Europe/France/Provence-Alpes-CV¥te d' | MEPHI, Aix Marseille University | MEPHI, Aix Marseille University | Anthony LEVASSEUR et al |
| EPI_ISL_569175 | France/PAC-IHU-1475/2020 | 2020-08-11 | Europe/France/Provence-Alpes-CV¥te d' | MEPHI, Aix Marseille University | MEPHI, Aix Marseille University | Anthony LEVASSEUR et al |
| EPI_ISL_569176 | France/PAC-IHU-1476/2020 | 2020-08-11 | Europe/France/Provence-Alpes-CV¥te d' | MEPHI, Aix Marseille University | MEPHI, Aix Marseille University | Anthony LEVASSEUR et al |
| EPI_ISL_569180 | France/PAC-IHU-1502/2020 | 2020-08-10 | Europe/France/Provence-Alpes-CV¥te d' | MEPHI, Aix Marseille University | MEPHI, Aix Marseille University | Anthony LEVASSEUR et al |
| EPI_ISL_569181 | France/PAC-IHU-1505/2020 | 2020-08-12 | Europe/France/Provence-Alpes-CV¥te d' | MEPHI, Aix Marseille University | MEPHI, Aix Marseille University | Anthony LEVASSEUR et al |
| EPI_ISL_569184 | France/PAC-IHU-1513/2020 | 2020-08-12 | Europe/France/Provence-Alpes-CV¥te d' | MEPHI, Aix Marseille University | MEPHI, Aix Marseille University | Anthony LEVASSEUR et al |
| EPI_ISL_569185 | France/PAC-IHU-1519/2020 | 2020-08-12 | Europe/France/Provence-Alpes-CV¥te d' | MEPHI, Aix Marseille University | MEPHI, Aix Marseille University | Anthony LEVASSEUR et al |
| EPI_ISL_569189 | France/PAC-IHU-1525/2020 | 2020-08-13 | Europe/France/Provence-Alpes-CV¥te d' | MEPHI, Aix Marseille University | MEPHI, Aix Marseille University | Anthony LEVASSEUR et al |
| EPI_ISL_569191 | France/PAC-IHU-1529/2020 | 2020-08-13 | Europe/France/Provence-Alpes-CV¥te d' | MEPHI, Aix Marseille University | MEPHI, Aix Marseille University | Anthony LEVASSEUR et al |
| EPI_ISL_569198 | France/PAC-IHU-1565/2020 | 2020-09-01 | Europe/France/Provence-Alpes-CV¥te d' | MEPHI, Aix Marseille University | MEPHI, Aix Marseille University | Anthony LEVASSEUR et al |

| | | | | | | |
|---|---|---|---|---|---|---|
| EPI_ISL_569199 | France/PAC-IHU-1566/2020 | 2020-09-02 | Europe/France/Provence-Alpes-CV¥te d' | MEPHI, Aix Marseille University | MEPHI, Aix Marseille University | Anthony LEVASSEUR et al |
| EPI_ISL_569200 | France/PAC-IHU-1567/2020 | 2020-09-01 | Europe/France/Provence-Alpes-CV¥te d' | MEPHI, Aix Marseille University | MEPHI, Aix Marseille University | Anthony LEVASSEUR et al |
| EPI_ISL_569202 | France/PAC-IHU-1569/2020 | 2020-09-02 | Europe/France/Provence-Alpes-CV¥te d' | MEPHI, Aix Marseille University | MEPHI, Aix Marseille University | Anthony LEVASSEUR et al |
| EPI_ISL_569206 | France/PAC-IHU-1573/2020 | 2020-09-02 | Europe/France/Provence-Alpes-CV¥te d' | MEPHI, Aix Marseille University | MEPHI, Aix Marseille University | Anthony LEVASSEUR et al |
| EPI_ISL_569207 | France/PAC-IHU-1574/2020 | 2020-09-02 | Europe/France/Provence-Alpes-CV¥te d' | MEPHI, Aix Marseille University | MEPHI, Aix Marseille University | Anthony LEVASSEUR et al |
| EPI_ISL_569208 | France/PAC-IHU-1575/2020 | 2020-09-08 | Europe/France/Provence-Alpes-CV¥te d' | MEPHI, Aix Marseille University | MEPHI, Aix Marseille University | Anthony LEVASSEUR et al |
| EPI_ISL_569209 | France/PAC-IHU-1579/2020 | 2020-09-01 | Europe/France/Provence-Alpes-CV¥te d' | MEPHI, Aix Marseille University | MEPHI, Aix Marseille University | Anthony LEVASSEUR et al |
| EPI_ISL_569210 | France/PAC-IHU-1581/2020 | 2020-09-01 | Europe/France/Provence-Alpes-CV¥te d' | MEPHI, Aix Marseille University | MEPHI, Aix Marseille University | Anthony LEVASSEUR et al |
| EPI_ISL_569211 | France/PAC-IHU-1582/2020 | 2020-09-09 | Europe/France/Provence-Alpes-CV¥te d' | MEPHI, Aix Marseille University | MEPHI, Aix Marseille University | Anthony LEVASSEUR et al |
| EPI_ISL_569212 | France/PAC-IHU-1583/2020 | 2020-09-09 | Europe/France/Provence-Alpes-CV¥te d' | MEPHI, Aix Marseille University | MEPHI, Aix Marseille University | Anthony LEVASSEUR et al |
| EPI_ISL_569213 | France/PAC-IHU-1584/2020 | 2020-09-01 | Europe/France/Provence-Alpes-CV¥te d' | MEPHI, Aix Marseille University | MEPHI, Aix Marseille University | Anthony LEVASSEUR et al |
| EPI_ISL_569214 | France/PAC-IHU-1585/2020 | 2020-09-08 | Europe/France/Provence-Alpes-CV¥te d' | MEPHI, Aix Marseille University | MEPHI, Aix Marseille University | Anthony LEVASSEUR et al |
| EPI_ISL_569215 | France/PAC-IHU-1586/2020 | 2020-09-01 | Europe/France/Provence-Alpes-CV¥te d' | MEPHI, Aix Marseille University | MEPHI, Aix Marseille University | Anthony LEVASSEUR et al |

| | | | | | | |
|---|---|---|---|---|---|---|
| EPI_ISL_569216 | France/PAC-IHU-1587/2020 | 2020-09-01 | Europe/France/Provence-Alpes-CV¥te d' | MEPHI, Aix Marseille University | MEPHI, Aix Marseille University | Anthony LEVASSEUR et al |
| EPI_ISL_569218 | France/PAC-IHU-1589/2020 | 2020-09-01 | Europe/France/Provence-Alpes-CV¥te d' | MEPHI, Aix Marseille University | MEPHI, Aix Marseille University | Anthony LEVASSEUR et al |
| EPI_ISL_569219 | France/PAC-IHU-1590/2020 | 2020-09-08 | Europe/France/Provence-Alpes-CV¥te d' | MEPHI, Aix Marseille University | MEPHI, Aix Marseille University | Anthony LEVASSEUR et al |
| EPI_ISL_569220 | France/PAC-IHU-1591/2020 | 2020-09-10 | Europe/France/Provence-Alpes-CV¥te d' | MEPHI, Aix Marseille University | MEPHI, Aix Marseille University | Anthony LEVASSEUR et al |
| EPI_ISL_569221 | France/PAC-IHU-1592/2020 | 2020-09-01 | Europe/France/Provence-Alpes-CV¥te d' | MEPHI, Aix Marseille University | MEPHI, Aix Marseille University | Anthony LEVASSEUR et al |
| EPI_ISL_569222 | France/PAC-IHU-1597/2020 | 2020-09-02 | Europe/France/Provence-Alpes-CV¥te d' | MEPHI, Aix Marseille University | MEPHI, Aix Marseille University | Anthony LEVASSEUR et al |
| EPI_ISL_569223 | France/PAC-IHU-1599/2020 | 2020-09-02 | Europe/France/Provence-Alpes-CV¥te d' | MEPHI, Aix Marseille University | MEPHI, Aix Marseille University | Anthony LEVASSEUR et al |
| EPI_ISL_569225 | France/PAC-IHU-1607/2020 | 2020-09-04 | Europe/France/Provence-Alpes-CV¥te d' | MEPHI, Aix Marseille University | MEPHI, Aix Marseille University | Anthony LEVASSEUR et al |
| EPI_ISL_569226 | France/PAC-IHU-1609/2020 | 2020-09-04 | Europe/France/Provence-Alpes-CV¥te d' | MEPHI, Aix Marseille University | MEPHI, Aix Marseille University | Anthony LEVASSEUR et al |
| EPI_ISL_569227 | France/PAC-IHU-1610/2020 | 2020-09-04 | Europe/France/Provence-Alpes-CV¥te d' | MEPHI, Aix Marseille University | MEPHI, Aix Marseille University | Anthony LEVASSEUR et al |
| EPI_ISL_569228 | France/PAC-IHU-1615/2020 | 2020-09-04 | Europe/France/Provence-Alpes-CV¥te d' | MEPHI, Aix Marseille University | MEPHI, Aix Marseille University | Anthony LEVASSEUR et al |
| EPI_ISL_569229 | France/PAC-IHU-1618/2020 | 2020-09-09 | Europe/France/Provence-Alpes-CV¥te d' | MEPHI, Aix Marseille University | MEPHI, Aix Marseille University | Anthony LEVASSEUR et al |
| EPI_ISL_569230 | France/PAC-IHU-1620/2020 | 2020-09-10 | Europe/France/Provence-Alpes-CV¥te d' | MEPHI, Aix Marseille University | MEPHI, Aix Marseille University | Anthony LEVASSEUR et al |

| | | | | | | |
|---|---|---|---|---|---|---|
| EPI_ISL_569231 | France/PAC-IHU-1622/2020 | 2020-09-10 | Europe/France/Provence-Alpes-CV¥te d' | MEPHI, Aix Marseille University | MEPHI, Aix Marseille University | Anthony LEVASSEUR et al |
| EPI_ISL_569232 | France/PAC-IHU-1623/2020 | 2020-09-10 | Europe/France/Provence-Alpes-CV¥te d' | MEPHI, Aix Marseille University | MEPHI, Aix Marseille University | Anthony LEVASSEUR et al |
| EPI_ISL_569233 | France/PAC-IHU-1624/2020 | 2020-09-10 | Europe/France/Provence-Alpes-CV¥te d' | MEPHI, Aix Marseille University | MEPHI, Aix Marseille University | Anthony LEVASSEUR et al |
| EPI_ISL_569234 | France/PAC-IHU-1625/2020 | 2020-09-10 | Europe/France/Provence-Alpes-CV¥te d' | MEPHI, Aix Marseille University | MEPHI, Aix Marseille University | Anthony LEVASSEUR et al |
| EPI_ISL_569236 | France/PAC-IHU-1627/2020 | 2020-09-10 | Europe/France/Provence-Alpes-CV¥te d' | MEPHI, Aix Marseille University | MEPHI, Aix Marseille University | Anthony LEVASSEUR et al |
| EPI_ISL_569238 | France/PAC-IHU-1629/2020 | 2020-09-11 | Europe/France/Provence-Alpes-CV¥te d' | MEPHI, Aix Marseille University | MEPHI, Aix Marseille University | Anthony LEVASSEUR et al |
| EPI_ISL_569239 | France/PAC-IHU-1630/2020 | 2020-09-11 | Europe/France/Provence-Alpes-CV¥te d' | MEPHI, Aix Marseille University | MEPHI, Aix Marseille University | Anthony LEVASSEUR et al |
| EPI_ISL_569240 | France/PAC-IHU-1631/2020 | 2020-09-11 | Europe/France/Provence-Alpes-CV¥te d' | MEPHI, Aix Marseille University | MEPHI, Aix Marseille University | Anthony LEVASSEUR et al |
| EPI_ISL_569241 | France/PAC-IHU-1632/2020 | 2020-09-12 | Europe/France/Provence-Alpes-CV¥te d' | MEPHI, Aix Marseille University | MEPHI, Aix Marseille University | Anthony LEVASSEUR et al |
| EPI_ISL_569243 | France/PAC-IHU-0012/2020 | 2020-03-06 | Europe/France/Provence-Alpes-CV¥te d' | MEPHI, Aix Marseille University | MEPHI, Aix Marseille University | Anthony LEVASSEUR et al |
| EPI_ISL_569246 | France/PAC-IHU-0017/2020 | 2020-03-07 | Europe/France/Provence-Alpes-CV¥te d' | MEPHI, Aix Marseille University | MEPHI, Aix Marseille University | Anthony LEVASSEUR et al |
| EPI_ISL_569255 | France/PAC-IHU-0029/2020 | 2020-03-17 | Europe/France/Provence-Alpes-CV¥te d' | MEPHI, Aix Marseille University | MEPHI, Aix Marseille University | Anthony LEVASSEUR et al |
| EPI_ISL_569257 | France/PAC-IHU-0031/2020 | 2020-03-18 | Europe/France/Provence-Alpes-CV¥te d' | MEPHI, Aix Marseille University | MEPHI, Aix Marseille University | Anthony LEVASSEUR et al |

| | | | | | | |
|---|---|---|---|---|---|---|
| EPI_ISL_569258 | France/PAC-IHU-0032/2020 | 2020-03-18 | Europe/France/Provence-Alpes-CV¥te d' | MEPHI, Aix Marseille University | MEPHI, Aix Marseille University | Anthony LEVASSEUR et al |
| EPI_ISL_569268 | France/PAC-IHU-0047/2020 | 2020-03-18 | Europe/France/Provence-Alpes-CV¥te d' | MEPHI, Aix Marseille University | MEPHI, Aix Marseille University | Anthony LEVASSEUR et al |
| EPI_ISL_569269 | France/PAC-IHU-0049/2020 | 2020-03-18 | Europe/France/Provence-Alpes-CV¥te d' | MEPHI, Aix Marseille University | MEPHI, Aix Marseille University | Anthony LEVASSEUR et al |
| EPI_ISL_569285 | France/PAC-IHU-0066/2020 | 2020-03-19 | Europe/France/Provence-Alpes-CV¥te d' | MEPHI, Aix Marseille University | MEPHI, Aix Marseille University | Anthony LEVASSEUR et al |
| EPI_ISL_569287 | France/PAC-IHU-0070/2020 | 2020-03-20 | Europe/France/Provence-Alpes-CV¥te d' | MEPHI, Aix Marseille University | MEPHI, Aix Marseille University | Anthony LEVASSEUR et al |
| EPI_ISL_569292 | France/PAC-IHU-0075/2020 | 2020-03-20 | Europe/France/Provence-Alpes-CV¥te d' | MEPHI, Aix Marseille University | MEPHI, Aix Marseille University | Anthony LEVASSEUR et al |
| EPI_ISL_569293 | France/PAC-IHU-0076/2020 | 2020-03-20 | Europe/France/Provence-Alpes-CV¥te d' | MEPHI, Aix Marseille University | MEPHI, Aix Marseille University | Anthony LEVASSEUR et al |
| EPI_ISL_569305 | France/PAC-IHU-0094/2020 | 2020-03-22 | Europe/France/Provence-Alpes-CV¥te d' | MEPHI, Aix Marseille University | MEPHI, Aix Marseille University | Anthony LEVASSEUR et al |
| EPI_ISL_569306 | France/PAC-IHU-0095/2020 | 2020-03-22 | Europe/France/Provence-Alpes-CV¥te d' | MEPHI, Aix Marseille University | MEPHI, Aix Marseille University | Anthony LEVASSEUR et al |
| EPI_ISL_569319 | France/PAC-IHU-0119/2020 | 2020-03-24 | Europe/France/Provence-Alpes-CV¥te d' | MEPHI, Aix Marseille University | MEPHI, Aix Marseille University | Anthony LEVASSEUR et al |
| EPI_ISL_569321 | France/PAC-IHU-0122/2020 | 2020-03-24 | Europe/France/Provence-Alpes-CV¥te d' | MEPHI, Aix Marseille University | MEPHI, Aix Marseille University | Anthony LEVASSEUR et al |
| EPI_ISL_569323 | France/PAC-IHU-0126/2020 | 2020-03-24 | Europe/France/Provence-Alpes-CV¥te d' | MEPHI, Aix Marseille University | MEPHI, Aix Marseille University | Anthony LEVASSEUR et al |
| EPI_ISL_569328 | France/PAC-IHU-0132/2020 | 2020-03-13 | Europe/France/Provence-Alpes-CV¥te d' | MEPHI, Aix Marseille University | MEPHI, Aix Marseille University | Anthony LEVASSEUR et al |

| | | | | | | |
|---|---|---|---|---|---|---|
| EPI_ISL_569335 | France/PAC-IHU-0146/2020 | 2020-03-21 | Europe/France/Provence-Alpes-CV¥te d' | MEPHI, Aix Marseille University | MEPHI, Aix Marseille University | Anthony LEVASSEUR et al |
| EPI_ISL_569336 | France/PAC-IHU-0149/2020 | 2020-03-21 | Europe/France/Provence-Alpes-CV¥te d' | MEPHI, Aix Marseille University | MEPHI, Aix Marseille University | Anthony LEVASSEUR et al |
| EPI_ISL_569337 | France/PAC-IHU-0151/2020 | 2020-03-22 | Europe/France/Provence-Alpes-CV¥te d' | MEPHI, Aix Marseille University | MEPHI, Aix Marseille University | Anthony LEVASSEUR et al |
| EPI_ISL_569342 | France/PAC-IHU-0160/2020 | 2020-03-24 | Europe/France/Provence-Alpes-CV¥te d' | MEPHI, Aix Marseille University | MEPHI, Aix Marseille University | Anthony LEVASSEUR et al |
| EPI_ISL_569343 | France/PAC-IHU-0162/2020 | 2020-03-24 | Europe/France/Provence-Alpes-CV¥te d' | MEPHI, Aix Marseille University | MEPHI, Aix Marseille University | Anthony LEVASSEUR et al |
| EPI_ISL_569344 | France/PAC-IHU-0167/2020 | 2020-03-25 | Europe/France/Provence-Alpes-CV¥te d' | MEPHI, Aix Marseille University | MEPHI, Aix Marseille University | Anthony LEVASSEUR et al |
| EPI_ISL_569357 | France/PAC-IHU-0189/2020 | 2020-03-27 | Europe/France/Provence-Alpes-CV¥te d' | MEPHI, Aix Marseille University | MEPHI, Aix Marseille University | Anthony LEVASSEUR et al |
| EPI_ISL_569358 | France/PAC-IHU-0190/2020 | 2020-03-27 | Europe/France/Provence-Alpes-CV¥te d' | MEPHI, Aix Marseille University | MEPHI, Aix Marseille University | Anthony LEVASSEUR et al |
| EPI_ISL_569359 | France/PAC-IHU-0191/2020 | 2020-03-27 | Europe/France/Provence-Alpes-CV¥te d' | MEPHI, Aix Marseille University | MEPHI, Aix Marseille University | Anthony LEVASSEUR et al |
| EPI_ISL_569363 | France/PAC-IHU-0196/2020 | 2020-03-27 | Europe/France/Provence-Alpes-CV¥te d' | MEPHI, Aix Marseille University | MEPHI, Aix Marseille University | Anthony LEVASSEUR et al |
| EPI_ISL_569366 | France/PAC-IHU-0199/2020 | 2020-03-27 | Europe/France/Provence-Alpes-CV¥te d' | MEPHI, Aix Marseille University | MEPHI, Aix Marseille University | Anthony LEVASSEUR et al |
| EPI_ISL_569367 | France/PAC-IHU-0200/2020 | 2020-03-27 | Europe/France/Provence-Alpes-CV¥te d' | MEPHI, Aix Marseille University | MEPHI, Aix Marseille University | Anthony LEVASSEUR et al |
| EPI_ISL_569379 | France/PAC-IHU-0214/2020 | 2020-03-28 | Europe/France/Provence-Alpes-CV¥te d' | MEPHI, Aix Marseille University | MEPHI, Aix Marseille University | Anthony LEVASSEUR et al |

| | | | | | | |
|---|---|---|---|---|---|---|
| EPI_ISL_569383 | France/PAC-IHU-0218/2020 | 2020-03-28 | Europe/France/Provence-Alpes-CV¥te d' | MEPHI, Aix Marseille University | MEPHI, Aix Marseille University | Anthony LEVASSEUR et al |
| EPI_ISL_569384 | France/PAC-IHU-0219/2020 | 2020-03-28 | Europe/France/Provence-Alpes-CV¥te d' | MEPHI, Aix Marseille University | MEPHI, Aix Marseille University | Anthony LEVASSEUR et al |
| EPI_ISL_569387 | France/PAC-IHU-0224/2020 | 2020-03-29 | Europe/France/Provence-Alpes-CV¥te d' | MEPHI, Aix Marseille University | MEPHI, Aix Marseille University | Anthony LEVASSEUR et al |
| EPI_ISL_569389 | France/PAC-IHU-0226/2020 | 2020-03-29 | Europe/France/Provence-Alpes-CV¥te d' | MEPHI, Aix Marseille University | MEPHI, Aix Marseille University | Anthony LEVASSEUR et al |
| EPI_ISL_569391 | France/PAC-IHU-0228/2020 | 2020-03-29 | Europe/France/Provence-Alpes-CV¥te d' | MEPHI, Aix Marseille University | MEPHI, Aix Marseille University | Anthony LEVASSEUR et al |
| EPI_ISL_569398 | France/PAC-IHU-0235/2020 | 2020-03-29 | Europe/France/Provence-Alpes-CV¥te d' | MEPHI, Aix Marseille University | MEPHI, Aix Marseille University | Anthony LEVASSEUR et al |
| EPI_ISL_569411 | France/PAC-IHU-0251/2020 | 2020-03-30 | Europe/France/Provence-Alpes-CV¥te d' | MEPHI, Aix Marseille University | MEPHI, Aix Marseille University | Anthony LEVASSEUR et al |
| EPI_ISL_569414 | France/PAC-IHU-0256/2020 | 2020-03-30 | Europe/France/Provence-Alpes-CV¥te d' | MEPHI, Aix Marseille University | MEPHI, Aix Marseille University | Anthony LEVASSEUR et al |
| EPI_ISL_569418 | France/PAC-IHU-0261/2020 | 2020-03-31 | Europe/France/Provence-Alpes-CV¥te d' | MEPHI, Aix Marseille University | MEPHI, Aix Marseille University | Anthony LEVASSEUR et al |
| EPI_ISL_569419 | France/PAC-IHU-0262/2020 | 2020-03-31 | Europe/France/Provence-Alpes-CV¥te d' | MEPHI, Aix Marseille University | MEPHI, Aix Marseille University | Anthony LEVASSEUR et al |
| EPI_ISL_569420 | France/PAC-IHU-0263/2020 | 2020-03-31 | Europe/France/Provence-Alpes-CV¥te d' | MEPHI, Aix Marseille University | MEPHI, Aix Marseille University | Anthony LEVASSEUR et al |
| EPI_ISL_569422 | France/PAC-IHU-0268/2020 | 2020-03-30 | Europe/France/Provence-Alpes-CV¥te d' | MEPHI, Aix Marseille University | MEPHI, Aix Marseille University | Anthony LEVASSEUR et al |
| EPI_ISL_569427 | France/PAC-IHU-0280/2020 | 2020-03-31 | Europe/France/Provence-Alpes-CV¥te d' | MEPHI, Aix Marseille University | MEPHI, Aix Marseille University | Anthony LEVASSEUR et al |

| | | | | | | |
|---|---|---|---|---|---|---|
| EPI_ISL_569429 | France/PAC-IHU-0282/2020 | 2020-03-31 | Europe/France/Provence-Alpes-CV¥te d' | MEPHI, Aix Marseille University | MEPHI, Aix Marseille University | Anthony LEVASSEUR et al |
| EPI_ISL_569443 | France/PAC-IHU-0300/2020 | 2020-03-31 | Europe/France/Provence-Alpes-CV¥te d' | MEPHI, Aix Marseille University | MEPHI, Aix Marseille University | Anthony LEVASSEUR et al |
| EPI_ISL_569445 | France/PAC-IHU-0304/2020 | 2020-03-31 | Europe/France/Provence-Alpes-CV¥te d' | MEPHI, Aix Marseille University | MEPHI, Aix Marseille University | Anthony LEVASSEUR et al |
| EPI_ISL_569447 | France/PAC-IHU-0308/2020 | 2020-03-31 | Europe/France/Provence-Alpes-CV¥te d' | MEPHI, Aix Marseille University | MEPHI, Aix Marseille University | Anthony LEVASSEUR et al |
| EPI_ISL_569450 | France/PAC-IHU-0318/2020 | 2020-03-23 | Europe/France/Provence-Alpes-CV¥te d' | MEPHI, Aix Marseille University | MEPHI, Aix Marseille University | Anthony LEVASSEUR et al |
| EPI_ISL_569460 | France/PAC-IHU-0333/2020 | 2020-04-01 | Europe/France/Provence-Alpes-CV¥te d' | MEPHI, Aix Marseille University | MEPHI, Aix Marseille University | Anthony LEVASSEUR et al |
| EPI_ISL_569461 | France/PAC-IHU-0334/2020 | 2020-04-01 | Europe/France/Provence-Alpes-CV¥te d' | MEPHI, Aix Marseille University | MEPHI, Aix Marseille University | Anthony LEVASSEUR et al |
| EPI_ISL_569462 | France/PAC-IHU-0336/2020 | 2020-04-01 | Europe/France/Provence-Alpes-CV¥te d' | MEPHI, Aix Marseille University | MEPHI, Aix Marseille University | Anthony LEVASSEUR et al |
| EPI_ISL_569464 | France/PAC-IHU-0338/2020 | 2020-04-01 | Europe/France/Provence-Alpes-CV¥te d' | MEPHI, Aix Marseille University | MEPHI, Aix Marseille University | Anthony LEVASSEUR et al |
| EPI_ISL_569465 | France/PAC-IHU-0340/2020 | 2020-03-19 | Europe/France/Provence-Alpes-CV¥te d' | MEPHI, Aix Marseille University | MEPHI, Aix Marseille University | Anthony LEVASSEUR et al |
| EPI_ISL_569466 | France/PAC-IHU-0342/2020 | 2020-03-20 | Europe/France/Provence-Alpes-CV¥te d' | MEPHI, Aix Marseille University | MEPHI, Aix Marseille University | Anthony LEVASSEUR et al |
| EPI_ISL_569468 | France/PAC-IHU-0345/2020 | 2020-03-21 | Europe/France/Provence-Alpes-CV¥te d' | MEPHI, Aix Marseille University | MEPHI, Aix Marseille University | Anthony LEVASSEUR et al |
| EPI_ISL_569470 | France/PAC-IHU-0349/2020 | 2020-03-21 | Europe/France/Provence-Alpes-CV¥te d' | MEPHI, Aix Marseille University | MEPHI, Aix Marseille University | Anthony LEVASSEUR et al |

| | | | | | | |
|---|---|---|---|---|---|---|
| EPI_ISL_569471 | France/PAC-IHU-0351/2020 | 2020-03-21 | Europe/France/Provence-Alpes-CV¥te d' | MEPHI, Aix Marseille University | MEPHI, Aix Marseille University | Anthony LEVASSEUR et al |
| EPI_ISL_569472 | France/PAC-IHU-0354/2020 | 2020-03-21 | Europe/France/Provence-Alpes-CV¥te d' | MEPHI, Aix Marseille University | MEPHI, Aix Marseille University | Anthony LEVASSEUR et al |
| EPI_ISL_569478 | France/PAC-IHU-0379/2020 | 2020-03-27 | Europe/France/Provence-Alpes-CV¥te d' | MEPHI, Aix Marseille University | MEPHI, Aix Marseille University | Anthony LEVASSEUR et al |
| EPI_ISL_569480 | France/PAC-IHU-0382/2020 | 2020-03-28 | Europe/France/Provence-Alpes-CV¥te d' | MEPHI, Aix Marseille University | MEPHI, Aix Marseille University | Anthony LEVASSEUR et al |
| EPI_ISL_569483 | France/PAC-IHU-0388/2020 | 2020-03-28 | Europe/France/Provence-Alpes-CV¥te d' | MEPHI, Aix Marseille University | MEPHI, Aix Marseille University | Anthony LEVASSEUR et al |
| EPI_ISL_569487 | France/PAC-IHU-0396/2020 | 2020-03-29 | Europe/France/Provence-Alpes-CV¥te d' | MEPHI, Aix Marseille University | MEPHI, Aix Marseille University | Anthony LEVASSEUR et al |
| EPI_ISL_569489 | France/PAC-IHU-0400/2020 | 2020-03-29 | Europe/France/Provence-Alpes-CV¥te d' | MEPHI, Aix Marseille University | MEPHI, Aix Marseille University | Anthony LEVASSEUR et al |
| EPI_ISL_569497 | France/PAC-IHU-0419/2020 | 2020-04-02 | Europe/France/Provence-Alpes-CV¥te d' | MEPHI, Aix Marseille University | MEPHI, Aix Marseille University | Anthony LEVASSEUR et al |
| EPI_ISL_569501 | France/PAC-IHU-0429/2020 | 2020-04-02 | Europe/France/Provence-Alpes-CV¥te d' | MEPHI, Aix Marseille University | MEPHI, Aix Marseille University | Anthony LEVASSEUR et al |
| EPI_ISL_569503 | France/PAC-IHU-0431/2020 | 2020-04-02 | Europe/France/Provence-Alpes-CV¥te d' | MEPHI, Aix Marseille University | MEPHI, Aix Marseille University | Anthony LEVASSEUR et al |
| EPI_ISL_569510 | France/PAC-IHU-0440/2020 | 2020-04-03 | Europe/France/Provence-Alpes-CV¥te d' | MEPHI, Aix Marseille University | MEPHI, Aix Marseille University | Anthony LEVASSEUR et al |
| EPI_ISL_569513 | France/PAC-IHU-0445/2020 | 2020-04-04 | Europe/France/Provence-Alpes-CV¥te d' | MEPHI, Aix Marseille University | MEPHI, Aix Marseille University | Anthony LEVASSEUR et al |
| EPI_ISL_569518 | France/PAC-IHU-0451/2020 | 2020-04-04 | Europe/France/Provence-Alpes-CV¥te d' | MEPHI, Aix Marseille University | MEPHI, Aix Marseille University | Anthony LEVASSEUR et al |

| | | | | | | |
|---|---|---|---|---|---|---|
| EPI_ISL_569527 | France/PAC-IHU-0462/2020 | 2020-04-05 | Europe/France/Provence-Alpes-CV¥te d' | MEPHI, Aix Marseille University | MEPHI, Aix Marseille University | Anthony LEVASSEUR et al |
| EPI_ISL_569528 | France/PAC-IHU-0464/2020 | 2020-04-05 | Europe/France/Provence-Alpes-CV¥te d' | MEPHI, Aix Marseille University | MEPHI, Aix Marseille University | Anthony LEVASSEUR et al |
| EPI_ISL_569535 | France/PAC-IHU-0471/2020 | 2020-04-05 | Europe/France/Provence-Alpes-CV¥te d' | MEPHI, Aix Marseille University | MEPHI, Aix Marseille University | Anthony LEVASSEUR et al |
| EPI_ISL_569537 | France/PAC-IHU-0473/2020 | 2020-04-05 | Europe/France/Provence-Alpes-CV¥te d' | MEPHI, Aix Marseille University | MEPHI, Aix Marseille University | Anthony LEVASSEUR et al |
| EPI_ISL_569541 | France/PAC-IHU-0483/2020 | 2020-04-03 | Europe/France/Provence-Alpes-CV¥te d' | MEPHI, Aix Marseille University | MEPHI, Aix Marseille University | Anthony LEVASSEUR et al |
| EPI_ISL_569552 | France/PAC-IHU-0497/2020 | 2020-04-03 | Europe/France/Provence-Alpes-CV¥te d' | MEPHI, Aix Marseille University | MEPHI, Aix Marseille University | Anthony LEVASSEUR et al |
| EPI_ISL_569553 | France/PAC-IHU-0498/2020 | 2020-04-03 | Europe/France/Provence-Alpes-CV¥te d' | MEPHI, Aix Marseille University | MEPHI, Aix Marseille University | Anthony LEVASSEUR et al |
| EPI_ISL_569555 | France/PAC-IHU-0500/2020 | 2020-04-03 | Europe/France/Provence-Alpes-CV¥te d' | MEPHI, Aix Marseille University | MEPHI, Aix Marseille University | Anthony LEVASSEUR et al |
| EPI_ISL_569561 | France/PAC-IHU-0507/2020 | 2020-04-06 | Europe/France/Provence-Alpes-CV¥te d' | MEPHI, Aix Marseille University | MEPHI, Aix Marseille University | Anthony LEVASSEUR et al |
| EPI_ISL_569563 | France/PAC-IHU-0510/2020 | 2020-04-06 | Europe/France/Provence-Alpes-CV¥te d' | MEPHI, Aix Marseille University | MEPHI, Aix Marseille University | Anthony LEVASSEUR et al |
| EPI_ISL_569568 | France/PAC-IHU-0520/2020 | 2020-04-06 | Europe/France/Provence-Alpes-CV¥te d' | MEPHI, Aix Marseille University | MEPHI, Aix Marseille University | Anthony LEVASSEUR et al |
| EPI_ISL_569571 | France/PAC-IHU-0525/2020 | 2020-04-07 | Europe/France/Provence-Alpes-CV¥te d' | MEPHI, Aix Marseille University | MEPHI, Aix Marseille University | Anthony LEVASSEUR et al |
| EPI_ISL_569572 | France/PAC-IHU-0526/2020 | 2020-04-07 | Europe/France/Provence-Alpes-CV¥te d' | MEPHI, Aix Marseille University | MEPHI, Aix Marseille University | Anthony LEVASSEUR et al |

| | | | | | | |
|---|---|---|---|---|---|---|
| EPI_ISL_569573 | France/PAC-IHU-0527/2020 | 2020-04-07 | Europe/France/Provence-Alpes-CV¥te d' | MEPHI, Aix Marseille University | MEPHI, Aix Marseille University | Anthony LEVASSEUR et al |
| EPI_ISL_569576 | France/PAC-IHU-0532/2020 | 2020-04-07 | Europe/France/Provence-Alpes-CV¥te d' | MEPHI, Aix Marseille University | MEPHI, Aix Marseille University | Anthony LEVASSEUR et al |
| EPI_ISL_569579 | France/PAC-IHU-0536/2020 | 2020-04-07 | Europe/France/Provence-Alpes-CV¥te d' | MEPHI, Aix Marseille University | MEPHI, Aix Marseille University | Anthony LEVASSEUR et al |
| EPI_ISL_569583 | France/PAC-IHU-0542/2020 | 2020-04-07 | Europe/France/Provence-Alpes-CV¥te d' | MEPHI, Aix Marseille University | MEPHI, Aix Marseille University | Anthony LEVASSEUR et al |
| EPI_ISL_569584 | France/PAC-IHU-0545/2020 | 2020-04-06 | Europe/France/Provence-Alpes-CV¥te d' | MEPHI, Aix Marseille University | MEPHI, Aix Marseille University | Anthony LEVASSEUR et al |
| EPI_ISL_569588 | France/PAC-IHU-0550/2020 | 2020-04-08 | Europe/France/Provence-Alpes-CV¥te d' | MEPHI, Aix Marseille University | MEPHI, Aix Marseille University | Anthony LEVASSEUR et al |
| EPI_ISL_569593 | France/PAC-IHU-0558/2020 | 2020-04-08 | Europe/France/Provence-Alpes-CV¥te d' | MEPHI, Aix Marseille University | MEPHI, Aix Marseille University | Anthony LEVASSEUR et al |
| EPI_ISL_569596 | France/PAC-IHU-0562/2020 | 2020-04-09 | Europe/France/Provence-Alpes-CV¥te d' | MEPHI, Aix Marseille University | MEPHI, Aix Marseille University | Anthony LEVASSEUR et al |
| EPI_ISL_569597 | France/PAC-IHU-0573/2020 | 2020-04-09 | Europe/France/Provence-Alpes-CV¥te d' | MEPHI, Aix Marseille University | MEPHI, Aix Marseille University | Anthony LEVASSEUR et al |
| EPI_ISL_569600 | France/PAC-IHU-0578/2020 | 2020-04-10 | Europe/France/Provence-Alpes-CV¥te d' | MEPHI, Aix Marseille University | MEPHI, Aix Marseille University | Anthony LEVASSEUR et al |
| EPI_ISL_569614 | USA/SD-SDPHL-0006/2020 | 2020-07-10 | North America/USA/South Dakota/ | IHS Fort Yates Hospital | South Dakota Public Health Laboratory | Matt Plumb et al |
| EPI_ISL_569616 | USA/SD-SDPHL-0008/2020 | 2020-07-12 | North America/USA/South Dakota/ | Aurora County Clinic | South Dakota Public Health Laboratory | Matt Plumb et al |
| EPI_ISL_569627 | USA/NM-NMDOH-2020321224/2020 | 2020-09-03 | North America/USA/New Mexico/ | New Mexico Department of Health Scientific Laboratory | New Mexico Department of Health Scientific Laboratory | Ellie Johnson et al |

| | | | | | | |
|---|---|---|---|---|---|---|
| EPI_ISL_569634 | USA/NM-NMDOH-2020321223/2020 | 2020-09-03 | North America/USA/New Mexico/ | New Mexico Department of Health Scientific Laboratory | New Mexico Department of Health Scientific Laboratory | Ellie Johnson et al |
| EPI_ISL_569636 | USA/NM-NMDOH-2020322592/2020 | 2020-09-03 | North America/USA/New Mexico/ | New Mexico Department of Health Scientific Laboratory | New Mexico Department of Health Scientific Laboratory | Ellie Johnson et al |
| EPI_ISL_569637 | USA/NM-NMDOH-2020324092/2020 | 2020-09-03 | North America/USA/New Mexico/ | New Mexico Department of Health Scientific Laboratory | New Mexico Department of Health Scientific Laboratory | Ellie Johnson et al |
| EPI_ISL_569639 | USA/NM-NMDOH-2020324378/2020 | 2020-09-03 | North America/USA/New Mexico/ | New Mexico Department of Health Scientific Laboratory | New Mexico Department of Health Scientific Laboratory | Ellie Johnson et al |
| EPI_ISL_569644 | USA/NM-NMDOH-2020327064/2020 | 2020-09-03 | North America/USA/New Mexico/ | New Mexico Department of Health Scientific Laboratory | New Mexico Department of Health Scientific Laboratory | Ellie Johnson et al |
| EPI_ISL_569648 | USA/NM-NMDOH-2020327510/2020 | 2020-09-03 | North America/USA/New Mexico/ | New Mexico Department of Health Scientific Laboratory | New Mexico Department of Health Scientific Laboratory | Ellie Johnson et al |
| EPI_ISL_569655 | USA/NM-NMDOH-2020328416/2020 | 2020-09-04 | North America/USA/New Mexico/ | New Mexico Department of Health Scientific Laboratory | New Mexico Department of Health Scientific Laboratory | Ellie Johnson et al |
| EPI_ISL_569656 | USA/NM-NMDOH-2020328523/2020 | 2020-09-03 | North America/USA/New Mexico/ | New Mexico Department of Health Scientific Laboratory | New Mexico Department of Health Scientific Laboratory | Ellie Johnson et al |
| EPI_ISL_569658 | USA/NM-NMDOH-2020328640/2020 | 2020-09-02 | North America/USA/New Mexico/ | New Mexico Department of Health Scientific Laboratory | New Mexico Department of Health Scientific Laboratory | Ellie Johnson et al |
| EPI_ISL_569684 | USA/CA-LAC-USC21/2020 | 2020-06-11 | North America/USA/California/Los Ange | Lee Lab | Lee Lab | Sung Yong Park et al |
| EPI_ISL_569689 | USA/TX-DSHS-0577/2020 | 2020-06-02 | North America/USA/Texas/El Paso Coun | Texas Department of State Health Services | Texas Department of State Health Services | Rashmi Tuladhar et al |

| | | | | | | |
|---|---|---|---|---|---|---|
| EPI_ISL_569693 | USA/TX-DSHS-0581/2020 | 2020-06-02 | North America/USA/Texas/El Paso Coun | Texas Department of State Health Services | Texas Department of State Health Services | Rashmi Tuladhar et al |
| EPI_ISL_569695 | USA/TX-DSHS-0583/2020 | 2020-06-03 | North America/USA/Texas/Harris Count | Texas Department of State Health Services | Texas Department of State Health Services | Rashmi Tuladhar et al |
| EPI_ISL_569702 | USA/TX-DSHS-0591/2020 | 2020-06-03 | North America/USA/Texas/El Paso Coun | Texas Department of State Health Services | Texas Department of State Health Services | Rashmi Tuladhar et al |
| EPI_ISL_569716 | USA/TX-DSHS-0606/2020 | 2020-06-04 | North America/USA/Texas/El Paso Coun | Texas Department of State Health Services | Texas Department of State Health Services | Rashmi Tuladhar et al |
| EPI_ISL_569721 | USA/TX-DSHS-0611/2020 | 2020-06-04 | North America/USA/Texas/El Paso Coun | Texas Department of State Health Services | Texas Department of State Health Services | Rashmi Tuladhar et al |
| EPI_ISL_569731 | USA/TX-DSHS-0621/2020 | 2020-06-07 | North America/USA/Texas/El Paso Coun | Texas Department of State Health Services | Texas Department of State Health Services | Rashmi Tuladhar et al |
| EPI_ISL_569732 | USA/TX-DSHS-0622/2020 | 2020-06-07 | North America/USA/Texas/El Paso Coun | Texas Department of State Health Services | Texas Department of State Health Services | Rashmi Tuladhar et al |
| EPI_ISL_569735 | Russia/OMS-ORINFI-5710S/2020 | 2020-05-08 | Europe/Russia/Omsk/Omsk | Omsk Research Institute of Natural Focal Infections | WHO National Influenza Centre Russian Federation | Artem Fadeev et al |
| EPI_ISL_569737 | Russia/OMS-ORINFI-5835S/2020 | 2020-05-08 | Europe/Russia/Omsk/Omsk | Omsk Research Institute of Natural Focal Infections | WHO National Influenza Centre Russian Federation | Artem Fadeev et al |
| EPI_ISL_569738 | Russia/OMS-ORINFI-6215S/2020 | 2020-05-10 | Europe/Russia/Omsk/Kalachinsk | Omsk Research Institute of Natural Focal Infections | WHO National Influenza Centre Russian Federation | Artem Fadeev et al |
| EPI_ISL_569739 | Russia/OMS-ORINFI-6673S/2020 | 2020-05-11 | Europe/Russia/Omsk/Omsk | Omsk Research Institute of Natural Focal Infections | WHO National Influenza Centre Russian Federation | Artem Fadeev et al |

| | | | | | | |
|---|---|---|---|---|---|---|
| EPI_ISL_569740 | Russia/OMS-ORINFI-6675S/2020 | 2020-05-12 | Europe/Russia/Omsk/Omsk | Omsk Research Institute of Natural Focal Infections | WHO National Influenza Centre Russian Federation | Artem Fadeev et al |
| EPI_ISL_569741 | Russia/OMS-ORINFI-6889S/2020 | 2020-05-13 | Europe/Russia/Omsk/Omsk | Omsk Research Institute of Natural Focal Infections | WHO National Influenza Centre Russian Federation | Artem Fadeev et al |
| EPI_ISL_569742 | Russia/OMS-ORINFI-7121S/2020 | 2020-05-14 | Europe/Russia/Omsk/Omsk | Omsk Research Institute of Natural Focal Infections | WHO National Influenza Centre Russian Federation | Artem Fadeev et al |
| EPI_ISL_569743 | Russia/OMS-ORINFI-7126S/2020 | 2020-05-14 | Europe/Russia/Omsk/Omsk | Omsk Research Institute of Natural Focal Infections | WHO National Influenza Centre Russian Federation | Artem Fadeev et al |
| EPI_ISL_569744 | Russia/OMS-ORINFI-7132S/2020 | 2020-05-14 | Europe/Russia/Omsk/Omsk | Omsk Research Institute of Natural Focal Infections | WHO National Influenza Centre Russian Federation | Artem Fadeev et al |
| EPI_ISL_569745 | Russia/OMS-ORINFI-7146S/2020 | 2020-05-14 | Europe/Russia/Omsk/Omsk | Omsk Research Institute of Natural Focal Infections | WHO National Influenza Centre Russian Federation | Artem Fadeev et al |
| EPI_ISL_569746 | Russia/OMS-ORINFI-9080S/2020 | 2020-05-19 | Europe/Russia/Omsk/Omsk | Omsk Research Institute of Natural Focal Infections | WHO National Influenza Centre Russian Federation | Artem Fadeev et al |
| EPI_ISL_569747 | Russia/OMS-ORINFI-9431S/2020 | 2020-05-21 | Europe/Russia/Omsk/Omsk | Omsk Research Institute of Natural Focal Infections | WHO National Influenza Centre Russian Federation | Artem Fadeev et al |
| EPI_ISL_569748 | Russia/OMS-ORINFI-9433S/2020 | 2020-05-21 | Europe/Russia/Omsk/Omsk | Omsk Research Institute of Natural Focal Infections | WHO National Influenza Centre Russian Federation | Artem Fadeev et al |
| EPI_ISL_569749 | Russia/OMS-ORINFI-9440S/2020 | 2020-05-21 | Europe/Russia/Omsk/Omsk | Omsk Research Institute of Natural Focal Infections | WHO National Influenza Centre Russian Federation | Artem Fadeev et al |

| | | | | | | |
|---|---|---|---|---|---|---|
| EPI_ISL_569750 | Russia/OMS-ORINFI-9064S/2020 | 2020-05-19 | Europe/Russia/Omsk/Moskalenki | Omsk Research Institute of Natural Focal Infections | WHO National Influenza Centre Russian Federation | Artem Fadeev et al |
| EPI_ISL_569751 | Russia/OMS-ORINFI-543S/2020 | 2020-05-23 | Europe/Russia/Omsk/Omsk | Omsk Research Institute of Natural Focal Infections | WHO National Influenza Centre Russian Federation | Artem Fadeev et al |
| EPI_ISL_569752 | Russia/OMS-ORINFI-1129S/2020 | 2020-05-24 | Europe/Russia/Omsk/Russkaya Polyana | Omsk Research Institute of Natural Focal Infections | WHO National Influenza Centre Russian Federation | Artem Fadeev et al |
| EPI_ISL_569753 | Russia/OMS-ORINFI-9761S/2020 | 2020-05-21 | Europe/Russia/Omsk/Omsk | Omsk Research Institute of Natural Focal Infections | WHO National Influenza Centre Russian Federation | Artem Fadeev et al |
| EPI_ISL_569754 | Russia/OMS-ORINFI-9785S/2020 | 2020-05-21 | Europe/Russia/Omsk/Omsk | Omsk Research Institute of Natural Focal Infections | WHO National Influenza Centre Russian Federation | Artem Fadeev et al |
| EPI_ISL_569755 | Russia/OMS-ORINFI-9809S/2020 | 2020-05-21 | Europe/Russia/Omsk/Omsk | Omsk Research Institute of Natural Focal Infections | WHO National Influenza Centre Russian Federation | Artem Fadeev et al |
| EPI_ISL_569756 | Russia/OMS-ORINFI-9815S/2020 | 2020-05-21 | Europe/Russia/Omsk/Omsk | Omsk Research Institute of Natural Focal Infections | WHO National Influenza Centre Russian Federation | Artem Fadeev et al |
| EPI_ISL_569757 | Russia/OMS-ORINFI-9820S/2020 | 2020-05-21 | Europe/Russia/Omsk/Omsk | Omsk Research Institute of Natural Focal Infections | WHO National Influenza Centre Russian Federation | Artem Fadeev et al |
| EPI_ISL_569758 | Russia/OMS-ORINFI-1296S/2020 | 2020-05-24 | Europe/Russia/Omsk/Moskalenki | Omsk Research Institute of Natural Focal Infections | WHO National Influenza Centre Russian Federation | Artem Fadeev et al |
| EPI_ISL_569759 | Russia/OMS-ORINFI-2022S/2020 | 2020-05-26 | Europe/Russia/Omsk/Omsk | Omsk Research Institute of Natural Focal Infections | WHO National Influenza Centre Russian Federation | Artem Fadeev et al |

| | | | | | | |
|---|---|---|---|---|---|---|
| EPI_ISL_569760 | Russia/OMS-ORINFI-2306S/2020 | 2020-05-28 | Europe/Russia/Omsk/Omsk | Omsk Research Institute of Natural Focal Infections | WHO National Influenza Centre Russian Federation | Artem Fadeev et al |
| EPI_ISL_569762 | Russia/OMS-ORINFI-2990S/2020 | 2020-05-27 | Europe/Russia/Omsk/Omsk | Omsk Research Institute of Natural Focal Infections | WHO National Influenza Centre Russian Federation | Artem Fadeev et al |
| EPI_ISL_569763 | Russia/OMS-ORINFI-3807S/2020 | 2020-05-13 | Europe/Russia/Omsk/Omsk | Omsk Research Institute of Natural Focal Infections | WHO National Influenza Centre Russian Federation | Artem Fadeev et al |
| EPI_ISL_569764 | Russia/OMS-ORINFI-3811S/2020 | 2020-06-02 | Europe/Russia/Omsk/Omsk | Omsk Research Institute of Natural Focal Infections | WHO National Influenza Centre Russian Federation | Artem Fadeev et al |
| EPI_ISL_569765 | Russia/OMS-ORINFI-3846S/2020 | 2020-06-02 | Europe/Russia/Omsk/Omsk | Omsk Research Institute of Natural Focal Infections | WHO National Influenza Centre Russian Federation | Artem Fadeev et al |
| EPI_ISL_569766 | Russia/OMS-ORINFI-4143S/2020 | 2020-05-28 | Europe/Russia/Omsk/Omsk | Omsk Research Institute of Natural Focal Infections | WHO National Influenza Centre Russian Federation | Artem Fadeev et al |
| EPI_ISL_569767 | Russia/OMS-ORINFI-4507S/2020 | 2020-06-03 | Europe/Russia/Omsk/Omsk | Omsk Research Institute of Natural Focal Infections | WHO National Influenza Centre Russian Federation | Artem Fadeev et al |
| EPI_ISL_569768 | Russia/OMS-ORINFI-5350S/2020 | 2020-06-07 | Europe/Russia/Omsk/Nizhnyaya Omka | Omsk Research Institute of Natural Focal Infections | WHO National Influenza Centre Russian Federation | Artem Fadeev et al |
| EPI_ISL_569769 | Russia/OMS-ORINFI-5363S/2020 | 2020-06-05 | Europe/Russia/Omsk/Omsk | Omsk Research Institute of Natural Focal Infections | WHO National Influenza Centre Russian Federation | Artem Fadeev et al |
| EPI_ISL_569770 | Russia/OMS-ORINFI-5372S/2020 | 2020-06-05 | Europe/Russia/Omsk/Omsk | Omsk Research Institute of Natural Focal Infections | WHO National Influenza Centre Russian Federation | Artem Fadeev et al |

| | | | | | | |
|---|---|---|---|---|---|---|
| EPI_ISL_569771 | Russia/OMS-ORINFI-5782S/2020 | 2020-06-05 | Europe/Russia/Omsk/Omsk | Omsk Research Institute of Natural Focal Infections | WHO National Influenza Centre Russian Federation | Artem Fadeev et al |
| EPI_ISL_569772 | Russia/OMS-ORINFI-5900S/2020 | 2020-06-01 | Europe/Russia/Omsk/Omsk | Omsk Research Institute of Natural Focal Infections | WHO National Influenza Centre Russian Federation | Artem Fadeev et al |
| EPI_ISL_569773 | Russia/OMS-ORINFI-5948S/2020 | 2020-06-07 | Europe/Russia/Omsk/Omsk | Omsk Research Institute of Natural Focal Infections | WHO National Influenza Centre Russian Federation | Artem Fadeev et al |
| EPI_ISL_569774 | Russia/OMS-ORINFI-6331S/2020 | 2020-06-07 | Europe/Russia/Omsk/Omsk | Omsk Research Institute of Natural Focal Infections | WHO National Influenza Centre Russian Federation | Artem Fadeev et al |
| EPI_ISL_569775 | Russia/OMS-ORINFI-9033S/2020 | 2020-06-17 | Europe/Russia/Omsk/Omsk | Omsk Research Institute of Natural Focal Infections | WHO National Influenza Centre Russian Federation | Artem Fadeev et al |
| EPI_ISL_569776 | Russia/OMS-ORINFI-1157S/2020 | 2020-06-26 | Europe/Russia/Omsk/Novovarshavka | Omsk Research Institute of Natural Focal Infections | WHO National Influenza Centre Russian Federation | Artem Fadeev et al |
| EPI_ISL_569778 | Russia/OMS-ORINFI-2911S/2020 | 2020-07-04 | Europe/Russia/Omsk/Moskalenki | Omsk Research Institute of Natural Focal Infections | WHO National Influenza Centre Russian Federation | Artem Fadeev et al |
| EPI_ISL_569779 | Russia/OMS-ORINFI-3012S/2020 | 2020-07-06 | Europe/Russia/Omsk/Azovo | Omsk Research Institute of Natural Focal Infections | WHO National Influenza Centre Russian Federation | Artem Fadeev et al |
| EPI_ISL_569781 | Russia/OMS-ORINFI-3868S/2020 | 2020-07-08 | Europe/Russia/Omsk/Moskalenki | Omsk Research Institute of Natural Focal Infections | WHO National Influenza Centre Russian Federation | Artem Fadeev et al |
| EPI_ISL_569782 | Russia/OMS-ORINFI-4361S/2020 | 2020-07-08 | Europe/Russia/Omsk/Novovarshavka | Omsk Research Institute of Natural Focal Infections | WHO National Influenza Centre Russian Federation | Artem Fadeev et al |

| | | | | | | |
|---|---|---|---|---|---|---|
| EPI_ISL_569783 | Russia/OMS-ORINFI-4761S/2020 | 2020-06-30 | Europe/Russia/Omsk/Sedelnikovo | Omsk Research Institute of Natural Focal Infections | WHO National Influenza Centre Russian Federation | Artem Fadeev et al |
| EPI_ISL_569786 | Russia/OMS-ORINFI-5207S/2020 | 2020-07-13 | Europe/Russia/Omsk/Azovo | Omsk Research Institute of Natural Focal Infections | WHO National Influenza Centre Russian Federation | Artem Fadeev et al |
| EPI_ISL_569787 | Russia/OMS-ORINFI-5762S/2020 | 2020-07-13 | Europe/Russia/Omsk/Moskalenki | Omsk Research Institute of Natural Focal Infections | WHO National Influenza Centre Russian Federation | Artem Fadeev et al |
| EPI_ISL_569788 | Russia/OMS-ORINFI-5463S/2020 | 2020-07-13 | Europe/Russia/Omsk/Novovarshavka | Omsk Research Institute of Natural Focal Infections | WHO National Influenza Centre Russian Federation | Artem Fadeev et al |
| EPI_ISL_569789 | Russia/OMS-ORINFI-7899S/2020 | 2020-07-21 | Europe/Russia/Omsk/Muromtsevo | Omsk Research Institute of Natural Focal Infections | WHO National Influenza Centre Russian Federation | Artem Fadeev et al |
| EPI_ISL_569790 | Russia/OMS-ORINFI-8583S/2020 | 2020-07-22 | Europe/Russia/Omsk/Novovarshavka | Omsk Research Institute of Natural Focal Infections | WHO National Influenza Centre Russian Federation | Artem Fadeev et al |
| EPI_ISL_569791 | Russia/OMS-ORINFI-8803S/2020 | 2020-07-23 | Europe/Russia/Omsk/Maryanovka | Omsk Research Institute of Natural Focal Infections | WHO National Influenza Centre Russian Federation | Artem Fadeev et al |
| EPI_ISL_569792 | Russia/OMS-ORINFI-8945S/2020 | 2020-07-23 | Europe/Russia/Omsk/Moskalenki | Omsk Research Institute of Natural Focal Infections | WHO National Influenza Centre Russian Federation | Artem Fadeev et al |
| EPI_ISL_569793 | Russia/OMS-ORINFI-9295S/2020 | 2020-07-25 | Europe/Russia/Omsk/Omsk | Omsk Research Institute of Natural Focal Infections | WHO National Influenza Centre Russian Federation | Artem Fadeev et al |
| EPI_ISL_569794 | Russia/OMS-ORINFI-9642S/2020 | 2020-07-27 | Europe/Russia/Omsk/Lyubinsky | Omsk Research Institute of Natural Focal Infections | WHO National Influenza Centre Russian Federation | Artem Fadeev et al |

| | | | | | | |
|---|---|---|---|---|---|---|
| EPI_ISL_569795 | Russia/OMS-ORINFI-9674S/2020 | 2020-07-27 | Europe/Russia/Omsk/Ust-Ishim | Omsk Research Institute of Natural Focal Infections | WHO National Influenza Centre Russian Federation | Artem Fadeev et al |
| EPI_ISL_569796 | Russia/OMS-ORINFI-4764S/2020 | 2020-05-02 | Europe/Russia/Omsk/Omsk | Omsk Research Institute of Natural Focal Infections | WHO National Influenza Centre Russian Federation | Artem Fadeev et al |
| EPI_ISL_569797 | Russia/OMS-ORINFI-4760S/2020 | 2020-05-02 | Europe/Russia/Omsk/Omsk | Omsk Research Institute of Natural Focal Infections | WHO National Influenza Centre Russian Federation | Artem Fadeev et al |
| EPI_ISL_569798 | Russia/OMS-ORINFI-4370S/2020 | 2020-07-08 | Europe/Russia/Omsk/Novovarshavka | Omsk Research Institute of Natural Focal Infections | WHO National Influenza Centre Russian Federation | Artem Fadeev et al |
| EPI_ISL_569800 | Russia/OMS-ORINFI-4822S/2020 | 2020-07-10 | Europe/Russia/Omsk/Azovo | Omsk Research Institute of Natural Focal Infections | WHO National Influenza Centre Russian Federation | Artem Fadeev et al |
| EPI_ISL_569801 | Russia/OMS-ORINFI-8941S/2020 | 2020-07-23 | Europe/Russia/Omsk/Moskalenki | Omsk Research Institute of Natural Focal Infections | WHO National Influenza Centre Russian Federation | Artem Fadeev et al |
| EPI_ISL_569802 | Russia/OMS-ORINFI-61S/2020 | 2020-05-14 | Europe/Russia/Omsk/Omsk | Omsk Research Institute of Natural Focal Infections | WHO National Influenza Centre Russian Federation | Artem Fadeev et al |
| EPI_ISL_569803 | Russia/OMS-ORINFI-5912S/2020 | 2020-06-07 | Europe/Russia/Omsk/Azovo | Omsk Research Institute of Natural Focal Infections | WHO National Influenza Centre Russian Federation | Artem Fadeev et al |
| EPI_ISL_569804 | Russia/OMS-ORINFI-4882S/2020 | 2020-07-10 | Europe/Russia/Omsk/Novovarshavka | Omsk Research Institute of Natural Focal Infections | WHO National Influenza Centre Russian Federation | Artem Fadeev et al |
| EPI_ISL_569805 | Russia/OMS-ORINFI-3019S/2020 | 2020-07-06 | Europe/Russia/Omsk/Azovo | Omsk Research Institute of Natural Focal Infections | WHO National Influenza Centre Russian Federation | Artem Fadeev et al |

| | | | | | | |
|---|---|---|---|---|---|---|
| EPI_ISL_569806 | Russia/OMS-ORINFI-4827S/2020 | 2020-07-10 | Europe/Russia/Omsk/Azovo | Omsk Research Institute of Natural Focal Infections | WHO National Influenza Centre Russian Federation | Artem Fadeev et al |
| EPI_ISL_569807 | Russia/OMS-ORINFI-6240S/2020 | 2020-07-15 | Europe/Russia/Omsk/Azovo | Omsk Research Institute of Natural Focal Infections | WHO National Influenza Centre Russian Federation | Artem Fadeev et al |
| EPI_ISL_569808 | Russia/OMS-ORINFI-8942S/2020 | 2020-07-23 | Europe/Russia/Omsk/Moskalenki | Omsk Research Institute of Natural Focal Infections | WHO National Influenza Centre Russian Federation | Artem Fadeev et al |
| EPI_ISL_569809 | Russia/OMS-ORINFI-70S/2020 | 2020-05-14 | Europe/Russia/Omsk/Omsk | Omsk Research Institute of Natural Focal Infections | WHO National Influenza Centre Russian Federation | Artem Fadeev et al |
| EPI_ISL_569810 | Russia/OMS-ORINFI-56S-2/2020 | 2020-06-17 | Europe/Russia/Omsk/Omsk | Omsk Research Institute of Natural Focal Infections | WHO National Influenza Centre Russian Federation | Artem Fadeev et al |
| EPI_ISL_569811 | Russia/OMS-ORINFI-4877S/2020 | 2020-05-03 | Europe/Russia/Omsk/Omsk | Omsk Research Institute of Natural Focal Infections | WHO National Influenza Centre Russian Federation | Artem Fadeev et al |
| EPI_ISL_569812 | Russia/OMS-ORINFI-66S/2020 | 2020-05-14 | Europe/Russia/Omsk/Omsk | Omsk Research Institute of Natural Focal Infections | WHO National Influenza Centre Russian Federation | Artem Fadeev et al |
| EPI_ISL_569813 | Russia/OMS-ORINFI-5286S/2020 | 2020-07-13 | Europe/Russia/Omsk/Omsk | Omsk Research Institute of Natural Focal Infections | WHO National Influenza Centre Russian Federation | Artem Fadeev et al |
| EPI_ISL_569815 | Russia/OMS-ORINFI-3873S/2020 | 2020-07-08 | Europe/Russia/Omsk/Moskalenki | Omsk Research Institute of Natural Focal Infections | WHO National Influenza Centre Russian Federation | Artem Fadeev et al |
| EPI_ISL_569816 | Russia/OMS-ORINFI-4828S/2020 | 2020-07-10 | Europe/Russia/Omsk/Azovo | Omsk Research Institute of Natural Focal Infections | WHO National Influenza Centre Russian Federation | Artem Fadeev et al |

| | | | | | | |
|---|---|---|---|---|---|---|
| EPI_ISL_569817 | Russia/OMS-ORINFI-47S/2020 | 2020-05-14 | Europe/Russia/Omsk/Omsk | Omsk Research Institute of Natural Focal Infections | WHO National Influenza Centre Russian Federation | Artem Fadeev et al |
| EPI_ISL_569818 | Russia/OMS-ORINFI-120S/2020 | 2020-05-14 | Europe/Russia/Omsk/Omsk | Omsk Research Institute of Natural Focal Infections | WHO National Influenza Centre Russian Federation | Artem Fadeev et al |
| EPI_ISL_569819 | Russia/OMS-ORINFI-1630S/2020 | 2020-06-29 | Europe/Russia/Omsk/Moskalenki | Omsk Research Institute of Natural Focal Infections | WHO National Influenza Centre Russian Federation | Artem Fadeev et al |
| EPI_ISL_569820 | Russia/OMS-ORINFI-4791S/2020 | 2020-07-10 | Europe/Russia/Omsk/Omsk | Omsk Research Institute of Natural Focal Infections | WHO National Influenza Centre Russian Federation | Artem Fadeev et al |
| EPI_ISL_569821 | Russia/OMS-ORINFI-3S/2020 | 2020-05-13 | Europe/Russia/Omsk/Omsk | Omsk Research Institute of Natural Focal Infections | WHO National Influenza Centre Russian Federation | Artem Fadeev et al |
| EPI_ISL_569822 | Russia/OMS-ORINFI-78S/2020 | 2020-05-13 | Europe/Russia/Omsk/Omsk | Omsk Research Institute of Natural Focal Infections | WHO National Influenza Centre Russian Federation | Artem Fadeev et al |
| EPI_ISL_569823 | Russia/OMS-ORINFI-5195S/2020 | 2020-07-13 | Europe/Russia/Omsk/Azovo | Omsk Research Institute of Natural Focal Infections | WHO National Influenza Centre Russian Federation | Artem Fadeev et al |
| EPI_ISL_569824 | Russia/OMS-ORINFI-6957S/2020 | 2020-07-17 | Europe/Russia/Omsk/Moskalenki | Omsk Research Institute of Natural Focal Infections | WHO National Influenza Centre Russian Federation | Artem Fadeev et al |
| EPI_ISL_569825 | Russia/OMS-ORINFI-55S/2020 | 2020-05-14 | Europe/Russia/Omsk/Omsk | Omsk Research Institute of Natural Focal Infections | WHO National Influenza Centre Russian Federation | Artem Fadeev et al |
| EPI_ISL_569826 | Russia/OMS-ORINFI-2535S/2020 | 2020-04-08 | Europe/Russia/Omsk/Omsk | Omsk Research Institute of Natural Focal Infections | WHO National Influenza Centre Russian Federation | Artem Fadeev et al |

| | | | | | | |
|---|---|---|---|---|---|---|
| EPI_ISL_569827 | Russia/OMS-ORINFI-1631S/2020 | 2020-06-29 | Europe/Russia/Omsk/Moskalenki | Omsk Research Institute of Natural Focal Infections | WHO National Influenza Centre Russian Federation | Artem Fadeev et al |
| EPI_ISL_569828 | Russia/OMS-ORINFI-2112S/2020 | 2020-07-02 | Europe/Russia/Omsk/Omsk | Omsk Research Institute of Natural Focal Infections | WHO National Influenza Centre Russian Federation | Artem Fadeev et al |
| EPI_ISL_569829 | Russia/OMS-ORINFI-4814S/2020 | 2020-07-10 | Europe/Russia/Omsk/Azovo | Omsk Research Institute of Natural Focal Infections | WHO National Influenza Centre Russian Federation | Artem Fadeev et al |
| EPI_ISL_569830 | Russia/OMS-ORINFI-5199S/2020 | 2020-07-13 | Europe/Russia/Omsk/Azovo | Omsk Research Institute of Natural Focal Infections | WHO National Influenza Centre Russian Federation | Artem Fadeev et al |
| EPI_ISL_569831 | Russia/OMS-ORINFI-8436S/2020 | 2020-07-22 | Europe/Russia/Omsk/Moskalenki | Omsk Research Institute of Natural Focal Infections | WHO National Influenza Centre Russian Federation | Artem Fadeev et al |
| EPI_ISL_569832 | Russia/OMS-ORINFI-56S/2020 | 2020-05-14 | Europe/Russia/Omsk/Omsk | Omsk Research Institute of Natural Focal Infections | WHO National Influenza Centre Russian Federation | Artem Fadeev et al |
| EPI_ISL_569833 | Russia/OMS-ORINFI-1632S/2020 | 2020-06-29 | Europe/Russia/Omsk/Moskalenki | Omsk Research Institute of Natural Focal Infections | WHO National Influenza Centre Russian Federation | Artem Fadeev et al |
| EPI_ISL_569834 | Russia/OMS-ORINFI-3961S/2020 | 2020-07-08 | Europe/Russia/Omsk/Novovarshavka | Omsk Research Institute of Natural Focal Infections | WHO National Influenza Centre Russian Federation | Artem Fadeev et al |
| EPI_ISL_569835 | Russia/OMS-ORINFI-2427S/2020 | 2020-07-03 | Europe/Russia/Omsk/Omsk | Omsk Research Institute of Natural Focal Infections | WHO National Influenza Centre Russian Federation | Artem Fadeev et al |
| EPI_ISL_569836 | Russia/OMS-ORINFI-1141S/2020 | 2020-08-18 | Europe/Russia/Omsk/Omsk | Omsk Research Institute of Natural Focal Infections | WHO National Influenza Centre Russian Federation | Artem Fadeev et al |

| | | | | | | |
|---|---|---|---|---|---|---|
| EPI_ISL_569837 | Russia/OMS-ORINFI-106S/2020 | 2020-05-13 | Europe/Russia/Omsk/Omsk | Omsk Research Institute of Natural Focal Infections | WHO National Influenza Centre Russian Federation | Artem Fadeev et al |
| EPI_ISL_569839 | Russia/OMS-ORINFI-8474S/2020 | 2020-07-22 | Europe/Russia/Omsk/Azovo | Omsk Research Institute of Natural Focal Infections | WHO National Influenza Centre Russian Federation | Artem Fadeev et al |
| EPI_ISL_569840 | Russia/OMS-ORINFI-57S/2020 | 2020-05-14 | Europe/Russia/Omsk/Omsk | Omsk Research Institute of Natural Focal Infections | WHO National Influenza Centre Russian Federation | Artem Fadeev et al |
| EPI_ISL_569842 | Russia/OMS-ORINFI-1639S/2020 | 2020-06-17 | Europe/Russia/Omsk/Moskalenki | Omsk Research Institute of Natural Focal Infections | WHO National Influenza Centre Russian Federation | Artem Fadeev et al |
| EPI_ISL_569843 | Russia/OMS-ORINFI-9661S/2020 | 2020-06-22 | Europe/Russia/Omsk/Omsk | Omsk Research Institute of Natural Focal Infections | WHO National Influenza Centre Russian Federation | Artem Fadeev et al |
| EPI_ISL_569844 | Russia/OMS-ORINFI-7593S/2020 | 2020-07-20 | Europe/Russia/Omsk/Omsk | Omsk Research Institute of Natural Focal Infections | WHO National Influenza Centre Russian Federation | Artem Fadeev et al |
| EPI_ISL_569845 | Russia/OMS-ORINFI-4818S/2020 | 2020-07-10 | Europe/Russia/Omsk/Azovo | Omsk Research Institute of Natural Focal Infections | WHO National Influenza Centre Russian Federation | Artem Fadeev et al |
| EPI_ISL_569846 | Russia/OMS-ORINFI-5208S/2020 | 2020-07-13 | Europe/Russia/Omsk/Azovo | Omsk Research Institute of Natural Focal Infections | WHO National Influenza Centre Russian Federation | Artem Fadeev et al |
| EPI_ISL_569847 | Russia/OMS-ORINFI-8499S/2020 | 2020-07-22 | Europe/Russia/Omsk/Azovo | Omsk Research Institute of Natural Focal Infections | WHO National Influenza Centre Russian Federation | Artem Fadeev et al |
| EPI_ISL_569848 | Russia/OMS-ORINFI-58S/2020 | 2020-05-14 | Europe/Russia/Omsk/Omsk | Omsk Research Institute of Natural Focal Infections | WHO National Influenza Centre Russian Federation | Artem Fadeev et al |

| | | | | | | |
|---|---|---|---|---|---|---|
| EPI_ISL_569849 | Russia/OMS-ORINFI-3401S/2020 | 2020-04-20 | Europe/Russia/Omsk/Cherlak | Omsk Research Institute of Natural Focal Infections | WHO National Influenza Centre Russian Federation | Artem Fadeev et al |
| EPI_ISL_569850 | Russia/OMS-ORINFI-3951S/2020 | 2020-07-08 | Europe/Russia/Omsk/Novovarshavka | Omsk Research Institute of Natural Focal Infections | WHO National Influenza Centre Russian Federation | Artem Fadeev et al |
| EPI_ISL_569851 | Russia/OMS-ORINFI-7598S/2020 | 2020-06-12 | Europe/Russia/Omsk/Omsk | Omsk Research Institute of Natural Focal Infections | WHO National Influenza Centre Russian Federation | Artem Fadeev et al |
| EPI_ISL_569852 | Russia/OMS-ORINFI-9975S/2020 | 2020-06-22 | Europe/Russia/Omsk/Omsk | Omsk Research Institute of Natural Focal Infections | WHO National Influenza Centre Russian Federation | Artem Fadeev et al |
| EPI_ISL_569853 | Russia/OMS-ORINFI-4368S/2020 | 2020-07-08 | Europe/Russia/Omsk/Novovarshavka | Omsk Research Institute of Natural Focal Infections | WHO National Influenza Centre Russian Federation | Artem Fadeev et al |
| EPI_ISL_569854 | Russia/OMS-ORINFI-4890S/2020 | 2020-05-03 | Europe/Russia/Omsk/Omsk | Omsk Research Institute of Natural Focal Infections | WHO National Influenza Centre Russian Federation | Artem Fadeev et al |
| EPI_ISL_569855 | Russia/OMS-ORINFI-60S/2020 | 2020-05-13 | Europe/Russia/Omsk/Omsk | Omsk Research Institute of Natural Focal Infections | WHO National Influenza Centre Russian Federation | Artem Fadeev et al |
| EPI_ISL_569856 | Russia/OMS-ORINFI-9060S/2020 | 2020-05-19 | Europe/Russia/Omsk/Moskalenki | Omsk Research Institute of Natural Focal Infections | WHO National Influenza Centre Russian Federation | Artem Fadeev et al |
| EPI_ISL_569857 | Russia/OMS-ORINFI-4895S/2020 | 2020-05-03 | Europe/Russia/Omsk/Omsk | Omsk Research Institute of Natural Focal Infections | WHO National Influenza Centre Russian Federation | Artem Fadeev et al |

| | | | | | | |
|---|---|---|---|---|---|---|
| EPI_ISL_569859 | India/WB-IICB-001/2020 | 2020-09-10 | Asia/India/West Bengal/ | CSIR-Indian Institute of Chemical Biology, MEDICA Supercpecialty Hospital Kolkata | CSIR-Indian Institute of Chemical Biology, MEDICA Supercpecialty Hospital Kolkata | Sujay Krishna Maity et al |
| EPI_ISL_569860 | India/WB-IICB-002/2020 | 2020-09-10 | Asia/India/West Bengal/ | CSIR-Indian Institute of Chemical Biology, MEDICA Supercpecialty Hospital Kolkata | CSIR-Indian Institute of Chemical Biology, MEDICA Supercpecialty Hospital Kolkata | Sujay Krishna Maity et al |
| EPI_ISL_569861 | India/WB-IICB-003/2020 | 2020-09-10 | Asia/India/West Bengal/ | CSIR-Indian Institute of Chemical Biology, MEDICA Supercpecialty Hospital Kolkata | CSIR-Indian Institute of Chemical Biology, MEDICA Supercpecialty Hospital Kolkata | Sujay Krishna Maity et al |
| EPI_ISL_569862 | India/WB-IICB-004/2020 | 2020-09-10 | Asia/India/West Bengal/ | CSIR-Indian Institute of Chemical Biology, MEDICA Supercpecialty Hospital Kolkata | CSIR-Indian Institute of Chemical Biology, MEDICA Supercpecialty Hospital Kolkata | Sujay Krishna Maity et al |
| EPI_ISL_569863 | India/WB-IICB-005/2020 | 2020-08-12 | Asia/India/West Bengal/ | CSIR-Indian Institute of Chemical Biology, MEDICA Supercpecialty Hospital Kolkata | CSIR-Indian Institute of Chemical Biology, MEDICA Supercpecialty Hospital Kolkata | Sujay Krishna Maity et al |
| EPI_ISL_569866 | Italy/PIE-SLL-MS45/2020 | 2020-02-25 | Europe/Italy/Piedmont/Turin | Amedeo di savoia | Crosetto lab, Karolinska Institutet, SciLifeLab | Michele Simonetti et al |
| EPI_ISL_569867 | Italy/PIE-SLL-MS46/2020 | 2020-02-22 | Europe/Italy/Piedmont/Turin | Amedeo di savoia | Crosetto lab, Karolinska Institutet, SciLifeLab | Michele Simonetti et al |

| | | | | | | |
|---|---|---|---|---|---|---|
| EPI_ISL_569870 | Italy/PIE-SLL-MS54/2020 | 2020-03-31 | Europe/Italy/Piedmont/Turin | Amedeo di savoia | Crosetto lab, Karolinska Institutet, SciLifeLab | Michele Simonetti et al |
| EPI_ISL_569877 | Italy/PIE-SLL-MS62/2020 | 2020-03-31 | Europe/Italy/Piedmont/Turin | Amedeo di savoia | Crosetto lab, Karolinska Institutet, SciLifeLab | Michele Simonetti et al |
| EPI_ISL_569878 | Italy/PIE-SLL-MS63/2020 | 2020-04-01 | Europe/Italy/Piedmont/Turin | Amedeo di savoia | Crosetto lab, Karolinska Institutet, SciLifeLab | Michele Simonetti et al |
| EPI_ISL_569879 | Italy/PIE-SLL-NZ138/2020 | 2020-04-01 | Europe/Italy/Piedmont/Turin | Amedeo di savoia | Crosetto lab, Karolinska Institutet, SciLifeLab | Michele Simonetti et al |
| EPI_ISL_569883 | Italy/PIE-SLL-NZ144/2020 | 2020-04-08 | Europe/Italy/Piedmont/Turin | Amedeo di savoia | Crosetto lab, Karolinska Institutet, SciLifeLab | Michele Simonetti et al |
| EPI_ISL_569884 | Italy/PIE-SLL-NZ146/2020 | 2020-04-08 | Europe/Italy/Piedmont/Turin | Amedeo di savoia | Crosetto lab, Karolinska Institutet, SciLifeLab | Michele Simonetti et al |
| EPI_ISL_569886 | Italy/PIE-SLL-NZ148/2020 | 2020-04-08 | Europe/Italy/Piedmont/Turin | Amedeo di savoia | Crosetto lab, Karolinska Institutet, SciLifeLab | Michele Simonetti et al |
| EPI_ISL_569888 | USA/CA-IGI-0203/2020 | 2020-07-02 | North America/USA/California/Contra C | Innovative Genomics Institute, UC Berkeley | Innovative Genomics Institute, UC Berkeley | Stacia Wyman et al |
| EPI_ISL_569889 | USA/CA-IGI-0205/2020 | 2020-07-09 | North America/USA/California/Contra C | Innovative Genomics Institute, UC Berkeley | Innovative Genomics Institute, UC Berkeley | Stacia Wyman et al |
| EPI_ISL_569891 | USA/CA-IGI-0210/2020 | 2020-07-02 | North America/USA/California/Alameda | Innovative Genomics Institute, UC Berkeley | Innovative Genomics Institute, UC Berkeley | Stacia Wyman et al |

| | | | | | | |
|---|---|---|---|---|---|---|
| EPI_ISL_569892 | USA/CA-IGI-0211/2020 | 2020-07-02 | North America/USA/California/Alameda | Innovative Genomics Institute, UC Berkeley | Innovative Genomics Institute, UC Berkeley | Stacia Wyman et al |
| EPI_ISL_569896 | USA/CA-IGI-0220/2020 | 2020-07-01 | North America/USA/California/Alameda | Innovative Genomics Institute, UC Berkeley | Innovative Genomics Institute, UC Berkeley | Stacia Wyman et al |
| EPI_ISL_569907 | USA/CA-IGI-0240/2020 | 2020-07-06 | North America/USA/California/Alameda | Innovative Genomics Institute, UC Berkeley | Innovative Genomics Institute, UC Berkeley | Stacia Wyman et al |
| EPI_ISL_569909 | USA/CA-IGI-0245/2020 | 2020-07-01 | North America/USA/California/Alameda | Innovative Genomics Institute, UC Berkeley | Innovative Genomics Institute, UC Berkeley | Stacia Wyman et al |
| EPI_ISL_569915 | USA/CA-IGI-0253/2020 | 2020-06-30 | North America/USA/California/Alameda | Innovative Genomics Institute, UC Berkeley | Innovative Genomics Institute, UC Berkeley | Stacia Wyman et al |
| EPI_ISL_569919 | USA/CA-IGI-0263/2020 | 2020-07-01 | North America/USA/California/Contra C | Innovative Genomics Institute, UC Berkeley | Innovative Genomics Institute, UC Berkeley | Stacia Wyman et al |
| EPI_ISL_569920 | USA/CA-IGI-0265/2020 | 2020-07-13 | North America/USA/California/Alameda | Innovative Genomics Institute, UC Berkeley | Innovative Genomics Institute, UC Berkeley | Stacia Wyman et al |
| EPI_ISL_569923 | USA/CA-IGI-0271/2020 | 2020-07-06 | North America/USA/California/Contra C | Innovative Genomics Institute, UC Berkeley | Innovative Genomics Institute, UC Berkeley | Stacia Wyman et al |
| EPI_ISL_569928 | USA/CA-IGI-0281/2020 | 2020-07-08 | North America/USA/California/Alameda | Innovative Genomics Institute, UC Berkeley | Innovative Genomics Institute, UC Berkeley | Stacia Wyman et al |
| EPI_ISL_569941 | USA/CA-IGI-0223/2020 | 2020-07-01 | North America/USA/California/Alameda | Innovative Genomics Institute, UC Berkeley | Innovative Genomics Institute, UC Berkeley | Stacia Wyman et al |

| | | | | | | |
|---|---|---|---|---|---|---|
| EPI_ISL_569947 | USA/CA-IGI-0243/2020 | 2020-07-01 | North America/USA/California/Alameda | Innovative Genomics Institute, UC Berkeley | Innovative Genomics Institute, UC Berkeley | Stacia Wyman et al |
| EPI_ISL_569949 | Canada/ON-UHTC-0174/2020 | 2020-05-23 | North America/Canada/Ontario/ | Unity Health Toronto | Ontario Institute for Cancer Research | Ramzi Fattouh et al |
| EPI_ISL_569950 | Canada/ON-UHTC-0167/2020 | 2020-05-19 | North America/Canada/Ontario/ | Unity Health Toronto | Ontario Institute for Cancer Research | Ramzi Fattouh et al |
| EPI_ISL_569951 | Canada/ON-UHTC-0169/2020 | 2020-05-18 | North America/Canada/Ontario/ | Unity Health Toronto | Ontario Institute for Cancer Research | Ramzi Fattouh et al |
| EPI_ISL_569952 | Canada/ON-UHTC-0173/2020 | 2020-05-23 | North America/Canada/Ontario/ | Unity Health Toronto | Ontario Institute for Cancer Research | Ramzi Fattouh et al |
| EPI_ISL_569953 | Canada/ON-UHTC-0166/2020 | 2020-05-18 | North America/Canada/Ontario/ | Unity Health Toronto | Ontario Institute for Cancer Research | Ramzi Fattouh et al |
| EPI_ISL_569954 | Canada/ON-UHTC-0188/2020 | 2020-05-25 | North America/Canada/Ontario/ | Unity Health Toronto | Ontario Institute for Cancer Research | Ramzi Fattouh et al |
| EPI_ISL_569955 | Canada/ON-UHTC-0186/2020 | 2020-05-26 | North America/Canada/Ontario/ | Unity Health Toronto | Ontario Institute for Cancer Research | Ramzi Fattouh et al |
| EPI_ISL_569956 | Canada/ON-UHTC-0204/2020 | 2020-06-11 | North America/Canada/Ontario/ | Unity Health Toronto | Ontario Institute for Cancer Research | Ramzi Fattouh et al |
| EPI_ISL_569957 | Canada/ON-UHTC-0168/2020 | 2020-05-18 | North America/Canada/Ontario/ | Unity Health Toronto | Ontario Institute for Cancer Research | Ramzi Fattouh et al |
| EPI_ISL_569958 | Canada/ON-UHTC-0194/2020 | 2020-05-27 | North America/Canada/Ontario/ | Unity Health Toronto | Ontario Institute for Cancer Research | Ramzi Fattouh et al |
| EPI_ISL_569959 | Canada/ON-UHTC-0171/2020 | 2020-05-18 | North America/Canada/Ontario/ | Unity Health Toronto | Ontario Institute for Cancer Research | Ramzi Fattouh et al |
| EPI_ISL_569960 | Canada/ON-UHTC-0196/2020 | 2020-05-27 | North America/Canada/Ontario/ | Unity Health Toronto | Ontario Institute for Cancer Research | Ramzi Fattouh et al |

| | | | | | | |
|---|---|---|---|---|---|---|
| EPI_ISL_569961 | Canada/ON-UHTC-0200/2020 | 2020-06-10 | North America/Canada/Ontario/ | Unity Health Toronto | Ontario Institute for Cancer Research | Ramzi Fattouh et al |
| EPI_ISL_569962 | Canada/ON-UHTC-0193/2020 | 2020-05-27 | North America/Canada/Ontario/ | Unity Health Toronto | Ontario Institute for Cancer Research | Ramzi Fattouh et al |
| EPI_ISL_569963 | Canada/ON-UHTC-0187/2020 | 2020-05-25 | North America/Canada/Ontario/ | Unity Health Toronto | Ontario Institute for Cancer Research | Ramzi Fattouh et al |
| EPI_ISL_569964 | Canada/ON-UHTC-0181/2020 | 2020-05-25 | North America/Canada/Ontario/ | Unity Health Toronto | Ontario Institute for Cancer Research | Ramzi Fattouh et al |
| EPI_ISL_569965 | Canada/ON-UHTC-0189/2020 | 2020-05-26 | North America/Canada/Ontario/ | Unity Health Toronto | Ontario Institute for Cancer Research | Ramzi Fattouh et al |
| EPI_ISL_569966 | Canada/ON-UHTC-0180/2020 | 2020-05-24 | North America/Canada/Ontario/ | Unity Health Toronto | Ontario Institute for Cancer Research | Ramzi Fattouh et al |
| EPI_ISL_569967 | Canada/ON-UHTC-0206/2020 | 2020-05-08 | North America/Canada/Ontario/ | Unity Health Toronto | Ontario Institute for Cancer Research | Ramzi Fattouh et al |
| EPI_ISL_569968 | Canada/ON-UHTC-0177/2020 | 2020-05-23 | North America/Canada/Ontario/ | Unity Health Toronto | Ontario Institute for Cancer Research | Ramzi Fattouh et al |
| EPI_ISL_569969 | Canada/ON-UHTC-0199/2020 | 2020-06-10 | North America/Canada/Ontario/ | Unity Health Toronto | Ontario Institute for Cancer Research | Ramzi Fattouh et al |
| EPI_ISL_569970 | Canada/ON-UHTC-0176/2020 | 2020-05-23 | North America/Canada/Ontario/ | Unity Health Toronto | Ontario Institute for Cancer Research | Ramzi Fattouh et al |
| EPI_ISL_569971 | Canada/ON-UHTC-0178/2020 | 2020-05-23 | North America/Canada/Ontario/ | Unity Health Toronto | Ontario Institute for Cancer Research | Ramzi Fattouh et al |
| EPI_ISL_569972 | Canada/ON-UHTC-0202/2020 | 2020-06-10 | North America/Canada/Ontario/ | Unity Health Toronto | Ontario Institute for Cancer Research | Ramzi Fattouh et al |
| EPI_ISL_569973 | Canada/ON-UHTC-0175/2020 | 2020-05-23 | North America/Canada/Ontario/ | Unity Health Toronto | Ontario Institute for Cancer Research | Ramzi Fattouh et al |

| | | | | | | |
|---|---|---|---|---|---|---|
| EPI_ISL_569974 | Canada/ON-UHTC-0205/2020 | 2020-05-04 | North America/Canada/Ontario/ | Unity Health Toronto | Ontario Institute for Cancer Research | Ramzi Fattouh et al |
| EPI_ISL_569975 | Canada/ON-UHTC-0203/2020 | 2020-06-10 | North America/Canada/Ontario/ | Unity Health Toronto | Ontario Institute for Cancer Research | Ramzi Fattouh et al |
| EPI_ISL_569976 | Canada/ON-UHTC-0179/2020 | 2020-05-24 | North America/Canada/Ontario/ | Unity Health Toronto | Ontario Institute for Cancer Research | Ramzi Fattouh et al |
| EPI_ISL_569977 | Canada/ON-UHTC-0170/2020 | 2020-05-18 | North America/Canada/Ontario/ | Unity Health Toronto | Ontario Institute for Cancer Research | Ramzi Fattouh et al |
| EPI_ISL_569978 | Canada/ON-UHTC-0197/2020 | 2020-05-29 | North America/Canada/Ontario/ | Unity Health Toronto | Ontario Institute for Cancer Research | Ramzi Fattouh et al |
| EPI_ISL_569979 | Canada/ON-UHTC-0185/2020 | 2020-05-25 | North America/Canada/Ontario/ | Unity Health Toronto | Ontario Institute for Cancer Research | Ramzi Fattouh et al |
| EPI_ISL_569980 | Canada/ON-UHTC-0293/2020 | 2020-07-30 | North America/Canada/Ontario/ | Unity Health Toronto | Ontario Institute for Cancer Research | Ramzi Fattouh et al |
| EPI_ISL_569981 | Canada/ON-UHTC-0283/2020 | 2020-07-06 | North America/Canada/Ontario/ | Unity Health Toronto | Ontario Institute for Cancer Research | Ramzi Fattouh et al |
| EPI_ISL_569982 | Canada/ON-UHTC-0285/2020 | 2020-07-16 | North America/Canada/Ontario/ | Unity Health Toronto | Ontario Institute for Cancer Research | Ramzi Fattouh et al |
| EPI_ISL_569983 | Canada/ON-UHTC-0284/2020 | 2020-07-14 | North America/Canada/Ontario/ | Unity Health Toronto | Ontario Institute for Cancer Research | Ramzi Fattouh et al |
| EPI_ISL_569984 | Canada/ON-UHTC-0287/2020 | 2020-07-22 | North America/Canada/Ontario/ | Unity Health Toronto | Ontario Institute for Cancer Research | Ramzi Fattouh et al |
| EPI_ISL_569985 | Canada/ON-UHTC-0299/2020 | 2020-05-22 | North America/Canada/Ontario/ | Unity Health Toronto | Ontario Institute for Cancer Research | Ramzi Fattouh et al |
| EPI_ISL_569986 | Canada/ON-UHTC-0296/2020 | 2020-05-20 | North America/Canada/Ontario/ | Unity Health Toronto | Ontario Institute for Cancer Research | Ramzi Fattouh et al |

| | | | | | | |
|---|---|---|---|---|---|---|
| EPI_ISL_569987 | Canada/ON-UHTC-0286/2020 | 2020-07-22 | North America/Canada/Ontario/ | Unity Health Toronto | Ontario Institute for Cancer Research | Ramzi Fattouh et al |
| EPI_ISL_569988 | Canada/ON-UHTC-0298/2020 | 2020-05-22 | North America/Canada/Ontario/ | Unity Health Toronto | Ontario Institute for Cancer Research | Ramzi Fattouh et al |
| EPI_ISL_569989 | Canada/ON-UHTC-0295/2020 | 2020-05-19 | North America/Canada/Ontario/ | Unity Health Toronto | Ontario Institute for Cancer Research | Ramzi Fattouh et al |
| EPI_ISL_569990 | Canada/ON-UHTC-0289/2020 | 2020-07-28 | North America/Canada/Ontario/ | Unity Health Toronto | Ontario Institute for Cancer Research | Ramzi Fattouh et al |
| EPI_ISL_569991 | Canada/ON-UHTC-0291/2020 | 2020-07-29 | North America/Canada/Ontario/ | Unity Health Toronto | Ontario Institute for Cancer Research | Ramzi Fattouh et al |
| EPI_ISL_569992 | Canada/ON-UHTC-0282/2020 | 2020-07-06 | North America/Canada/Ontario/ | Unity Health Toronto | Ontario Institute for Cancer Research | Ramzi Fattouh et al |
| EPI_ISL_569994 | Canada/ON-UHTC-0292/2020 | 2020-07-30 | North America/Canada/Ontario/ | Unity Health Toronto | Ontario Institute for Cancer Research | Ramzi Fattouh et al |
| EPI_ISL_569995 | Canada/ON-UHTC-0258/2020 | 2020-06-14 | North America/Canada/Ontario/ | Unity Health Toronto | Ontario Institute for Cancer Research | Ramzi Fattouh et al |
| EPI_ISL_569996 | Canada/ON-UHTC-0267/2020 | 2020-06-21 | North America/Canada/Ontario/ | Unity Health Toronto | Ontario Institute for Cancer Research | Ramzi Fattouh et al |
| EPI_ISL_569997 | Canada/ON-UHTC-0256/2020 | 2020-06-15 | North America/Canada/Ontario/ | Unity Health Toronto | Ontario Institute for Cancer Research | Ramzi Fattouh et al |
| EPI_ISL_569998 | Canada/ON-UHTC-0272/2020 | 2020-08-17 | North America/Canada/Ontario/ | Unity Health Toronto | Ontario Institute for Cancer Research | Ramzi Fattouh et al |
| EPI_ISL_569999 | Canada/ON-UHTC-0127/2020 | 2020-05-21 | North America/Canada/Ontario/ | Unity Health Toronto | Ontario Institute for Cancer Research | Ramzi Fattouh et al |
| EPI_ISL_570000 | Canada/ON-UHTC-0271/2020 | 2020-08-10 | North America/Canada/Ontario/ | Unity Health Toronto | Ontario Institute for Cancer Research | Ramzi Fattouh et al |

| | | | | | | |
|---|---|---|---|---|---|---|
| EPI_ISL_570001 | Canada/ON-UHTC-0261/2020 | 2020-06-16 | North America/Canada/Ontario/ | Unity Health Toronto | Ontario Institute for Cancer Research | Ramzi Fattouh et al |
| EPI_ISL_570002 | Canada/ON-UHTC-0270/2020 | 2020-06-19 | North America/Canada/Ontario/ | Unity Health Toronto | Ontario Institute for Cancer Research | Ramzi Fattouh et al |
| EPI_ISL_570003 | Canada/ON-UHTC-0277/2020 | 2020-06-23 | North America/Canada/Ontario/ | Unity Health Toronto | Ontario Institute for Cancer Research | Ramzi Fattouh et al |
| EPI_ISL_570004 | Canada/ON-UHTC-0257/2020 | 2020-06-14 | North America/Canada/Ontario/ | Unity Health Toronto | Ontario Institute for Cancer Research | Ramzi Fattouh et al |
| EPI_ISL_570005 | Canada/ON-UHTC-0275/2020 | 2020-06-22 | North America/Canada/Ontario/ | Unity Health Toronto | Ontario Institute for Cancer Research | Ramzi Fattouh et al |
| EPI_ISL_570006 | Canada/ON-UHTC-0262/2020 | 2020-06-16 | North America/Canada/Ontario/ | Unity Health Toronto | Ontario Institute for Cancer Research | Ramzi Fattouh et al |
| EPI_ISL_570007 | Canada/ON-UHTC-0253/2020 | 2020-06-13 | North America/Canada/Ontario/ | Unity Health Toronto | Ontario Institute for Cancer Research | Ramzi Fattouh et al |
| EPI_ISL_570008 | Canada/ON-UHTC-0278/2020 | 2020-06-29 | North America/Canada/Ontario/ | Unity Health Toronto | Ontario Institute for Cancer Research | Ramzi Fattouh et al |
| EPI_ISL_570009 | Canada/ON-UHTC-0241/2020 | 2020-06-08 | North America/Canada/Ontario/ | Unity Health Toronto | Ontario Institute for Cancer Research | Ramzi Fattouh et al |
| EPI_ISL_570010 | Canada/ON-UHTC-0209/2020 | 2020-05-04 | North America/Canada/Ontario/ | Unity Health Toronto | Ontario Institute for Cancer Research | Ramzi Fattouh et al |
| EPI_ISL_570011 | Canada/ON-UHTC-0228/2020 | 2020-06-04 | North America/Canada/Ontario/ | Unity Health Toronto | Ontario Institute for Cancer Research | Ramzi Fattouh et al |
| EPI_ISL_570012 | Canada/ON-UHTC-0239/2020 | 2020-06-08 | North America/Canada/Ontario/ | Unity Health Toronto | Ontario Institute for Cancer Research | Ramzi Fattouh et al |
| EPI_ISL_570013 | Canada/ON-UHTC-0211/2020 | 2020-05-05 | North America/Canada/Ontario/ | Unity Health Toronto | Ontario Institute for Cancer Research | Ramzi Fattouh et al |

| | | | | | | |
|---|---|---|---|---|---|---|
| EPI_ISL_570014 | Canada/ON-UHTC-0240/2020 | 2020-06-08 | North America/Canada/Ontario/ | Unity Health Toronto | Ontario Institute for Cancer Research | Ramzi Fattouh et al |
| EPI_ISL_570015 | Canada/ON-UHTC-0232/2020 | 2020-06-06 | North America/Canada/Ontario/ | Unity Health Toronto | Ontario Institute for Cancer Research | Ramzi Fattouh et al |
| EPI_ISL_570016 | Canada/ON-UHTC-0243/2020 | 2020-06-12 | North America/Canada/Ontario/ | Unity Health Toronto | Ontario Institute for Cancer Research | Ramzi Fattouh et al |
| EPI_ISL_570017 | Canada/ON-UHTC-0222/2020 | 2020-06-01 | North America/Canada/Ontario/ | Unity Health Toronto | Ontario Institute for Cancer Research | Ramzi Fattouh et al |
| EPI_ISL_570018 | Canada/ON-UHTC-0229/2020 | 2020-06-04 | North America/Canada/Ontario/ | Unity Health Toronto | Ontario Institute for Cancer Research | Ramzi Fattouh et al |
| EPI_ISL_570019 | Canada/ON-UHTC-0238/2020 | 2020-06-07 | North America/Canada/Ontario/ | Unity Health Toronto | Ontario Institute for Cancer Research | Ramzi Fattouh et al |
| EPI_ISL_570020 | Canada/ON-UHTC-0230/2020 | 2020-06-05 | North America/Canada/Ontario/ | Unity Health Toronto | Ontario Institute for Cancer Research | Ramzi Fattouh et al |
| EPI_ISL_570021 | Canada/ON-UHTC-0225/2020 | 2020-06-02 | North America/Canada/Ontario/ | Unity Health Toronto | Ontario Institute for Cancer Research | Ramzi Fattouh et al |
| EPI_ISL_570022 | Canada/ON-UHTC-0219/2020 | 2020-06-01 | North America/Canada/Ontario/ | Unity Health Toronto | Ontario Institute for Cancer Research | Ramzi Fattouh et al |
| EPI_ISL_570023 | Canada/ON-UHTC-0218/2020 | 2020-06-01 | North America/Canada/Ontario/ | Unity Health Toronto | Ontario Institute for Cancer Research | Ramzi Fattouh et al |
| EPI_ISL_570024 | Canada/ON-UHTC-0226/2020 | 2020-06-03 | North America/Canada/Ontario/ | Unity Health Toronto | Ontario Institute for Cancer Research | Ramzi Fattouh et al |
| EPI_ISL_570025 | Canada/ON-UHTC-0245/2020 | 2020-06-12 | North America/Canada/Ontario/ | Unity Health Toronto | Ontario Institute for Cancer Research | Ramzi Fattouh et al |
| EPI_ISL_570026 | Canada/ON-UHTC-0210/2020 | 2020-05-05 | North America/Canada/Ontario/ | Unity Health Toronto | Ontario Institute for Cancer Research | Ramzi Fattouh et al |
| EPI_ISL_570048 | USA/ID-UW-4417/2020 | 2020-03-28 | North America/USA/Idaho/ | UW Virology Lab | UW Virology Lab | Pavitra Roychoudhury et al |

| | | | | | | |
|---|---|---|---|---|---|---|
| EPI_ISL_570052 | USA/WA-UW-4630/2020 | 2020-03-28 | North America/USA/Washington/ | UW Virology Lab | UW Virology Lab | Pavitra Roychoudhury et al |
| EPI_ISL_570072 | USA/WA-UW-5670/2020 | 2020-04-09 | North America/USA/Washington/ | UW Virology Lab | UW Virology Lab | Pavitra Roychoudhury et al |
| EPI_ISL_570085 | USA/WA-UW-6187/2020 | 2020-04-01 | North America/USA/Washington/ | UW Virology Lab | UW Virology Lab | Pavitra Roychoudhury et al |
| EPI_ISL_570086 | USA/WA-UW-6190/2020 | 2020-04-01 | North America/USA/Washington/ | UW Virology Lab | UW Virology Lab | Pavitra Roychoudhury et al |
| EPI_ISL_570095 | USA/CT-UW-6288/2020 | 2020-04-01 | North America/USA/Connecticut/ | UW Virology Lab | UW Virology Lab | Pavitra Roychoudhury et al |
| EPI_ISL_570106 | USA/ID-UW-6367/2020 | 2020-04-01 | North America/USA/Idaho/ | UW Virology Lab | UW Virology Lab | Pavitra Roychoudhury et al |
| EPI_ISL_570109 | USA/CT-UW-6379/2020 | 2020-04-01 | North America/USA/Connecticut/ | UW Virology Lab | UW Virology Lab | Pavitra Roychoudhury et al |
| EPI_ISL_570119 | USA/ID-UW-6464/2020 | 2020-04-02 | North America/USA/Idaho/ | UW Virology Lab | UW Virology Lab | Pavitra Roychoudhury et al |
| EPI_ISL_570123 | USA/WA-UW-6496/2020 | 2020-04-03 | North America/USA/Washington/ | UW Virology Lab | UW Virology Lab | Pavitra Roychoudhury et al |
| EPI_ISL_570146 | USA/CT-UW-6790/2020 | 2020-04-08 | North America/USA/Connecticut/ | UW Virology Lab | UW Virology Lab | Pavitra Roychoudhury et al |
| EPI_ISL_570169 | USA/WA-UW-7115/2020 | 2020-04-12 | North America/USA/Washington/ | UW Virology Lab | UW Virology Lab | Pavitra Roychoudhury et al |
| EPI_ISL_570172 | USA/WA-UW-7160/2020 | 2020-04-11 | North America/USA/Washington/ | UW Virology Lab | UW Virology Lab | Pavitra Roychoudhury et al |
| EPI_ISL_570175 | USA/WA-UW-7236/2020 | 2020-04-10 | North America/USA/Washington/ | UW Virology Lab | UW Virology Lab | Pavitra Roychoudhury et al |
| EPI_ISL_570192 | USA/WA-UW-7763/2020 | 2020-04-20 | North America/USA/Washington/ | UW Virology Lab | UW Virology Lab | Pavitra Roychoudhury et al |
| EPI_ISL_570197 | USA/WA-UW-8065/2020 | 2020-04-24 | North America/USA/Washington/ | UW Virology Lab | UW Virology Lab | Pavitra Roychoudhury et al |
| EPI_ISL_570207 | USA/WA-UW-8422/2020 | 2020-05-01 | North America/USA/Washington/ | UW Virology Lab | UW Virology Lab | Pavitra Roychoudhury et al |
| EPI_ISL_570241 | USA/WA-UW-9266/2020 | 2020-05-15 | North America/USA/Washington/ | UW Virology Lab | UW Virology Lab | Pavitra Roychoudhury et al |
| EPI_ISL_570244 | USA/WA-UW-9333/2020 | 2020-05-13 | North America/USA/Washington/ | UW Virology Lab | UW Virology Lab | Pavitra Roychoudhury et al |
| EPI_ISL_570246 | USA/WA-UW-9339/2020 | 2020-05-13 | North America/USA/Washington/ | UW Virology Lab | UW Virology Lab | Pavitra Roychoudhury et al |
| EPI_ISL_570248 | USA/WA-UW-9361/2020 | 2020-05-15 | North America/USA/Washington/ | UW Virology Lab | UW Virology Lab | Pavitra Roychoudhury et al |
| EPI_ISL_570250 | USA/WA-UW-9374/2020 | 2020-05-16 | North America/USA/Washington/ | UW Virology Lab | UW Virology Lab | Pavitra Roychoudhury et al |
| EPI_ISL_570253 | USA/WA-UW-9422/2020 | 2020-05-18 | North America/USA/Washington/ | UW Virology Lab | UW Virology Lab | Pavitra Roychoudhury et al |
| EPI_ISL_570257 | USA/WA-UW-9504/2020 | 2020-05-18 | North America/USA/Washington/ | UW Virology Lab | UW Virology Lab | Pavitra Roychoudhury et al |
| EPI_ISL_570258 | USA/WA-UW-9520/2020 | 2020-05-18 | North America/USA/Washington/ | UW Virology Lab | UW Virology Lab | Pavitra Roychoudhury et al |
| EPI_ISL_570264 | USA/WA-UW-9578/2020 | 2020-05-20 | North America/USA/Washington/ | UW Virology Lab | UW Virology Lab | Pavitra Roychoudhury et al |
| EPI_ISL_570279 | USA/WA-UW-10206/2020 | 2020-05-31 | North America/USA/Washington/ | UW Virology Lab | UW Virology Lab | Pavitra Roychoudhury et al |
| EPI_ISL_570281 | USA/WA-UW-10216/2020 | 2020-05-31 | North America/USA/Washington/ | UW Virology Lab | UW Virology Lab | Pavitra Roychoudhury et al |
| EPI_ISL_570300 | USA/WA-UW-10394/2020 | 2020-06-05 | North America/USA/Washington/ | UW Virology Lab | UW Virology Lab | Pavitra Roychoudhury et al |
| EPI_ISL_570318 | USA/WA-UW-10472/2020 | 2020-06-07 | North America/USA/Washington/ | UW Virology Lab | UW Virology Lab | Pavitra Roychoudhury et al |
| EPI_ISL_570325 | USA/WA-UW-10510/2020 | 2020-05-29 | North America/USA/Washington/ | UW Virology Lab | UW Virology Lab | Pavitra Roychoudhury et al |
| EPI_ISL_570332 | USA/WA-UW-10533/2020 | 2020-06-09 | North America/USA/Washington/ | UW Virology Lab | UW Virology Lab | Pavitra Roychoudhury et al |

| | | | | | | |
|---|---|---|---|---|---|---|
| EPI_ISL_570333 | USA/WA-UW-10552/2020 | 2020-06-11 | North America/USA/Washington/ | UW Virology Lab | UW Virology Lab | Pavitra Roychoudhury et al |
| EPI_ISL_570345 | USA/WA-UW-10663/2020 | 2020-06-09 | North America/USA/Washington/ | UW Virology Lab | UW Virology Lab | Pavitra Roychoudhury et al |
| EPI_ISL_570351 | USA/WA-UW-10691/2020 | 2020-06-09 | North America/USA/Washington/ | UW Virology Lab | UW Virology Lab | Pavitra Roychoudhury et al |
| EPI_ISL_570354 | USA/WA-UW-10698/2020 | 2020-06-10 | North America/USA/Washington/ | UW Virology Lab | UW Virology Lab | Pavitra Roychoudhury et al |
| EPI_ISL_570359 | USA/WA-UW-10826/2020 | 2020-06-12 | North America/USA/Washington/ | UW Virology Lab | UW Virology Lab | Pavitra Roychoudhury et al |
| EPI_ISL_570361 | USA/WA-UW-10834/2020 | 2020-06-12 | North America/USA/Washington/ | UW Virology Lab | UW Virology Lab | Pavitra Roychoudhury et al |
| EPI_ISL_570364 | USA/WA-UW-11002/2020 | 2020-06-15 | North America/USA/Washington/ | UW Virology Lab | UW Virology Lab | Pavitra Roychoudhury et al |
| EPI_ISL_570378 | USA/WA-UW-11165/2020 | 2020-06-19 | North America/USA/Washington/ | UW Virology Lab | UW Virology Lab | Pavitra Roychoudhury et al |
| EPI_ISL_570381 | USA/WA-UW-11196/2020 | 2020-06-19 | North America/USA/Washington/ | UW Virology Lab | UW Virology Lab | Pavitra Roychoudhury et al |
| EPI_ISL_570386 | USA/WA-UW-11280/2020 | 2020-06-19 | North America/USA/Washington/ | UW Virology Lab | UW Virology Lab | Pavitra Roychoudhury et al |
| EPI_ISL_570392 | USA/WA-UW-11309/2020 | 2020-06-22 | North America/USA/Washington/ | UW Virology Lab | UW Virology Lab | Pavitra Roychoudhury et al |
| EPI_ISL_570394 | USA/WA-UW-11325/2020 | 2020-06-22 | North America/USA/Washington/ | UW Virology Lab | UW Virology Lab | Pavitra Roychoudhury et al |
| EPI_ISL_570402 | USA/WA-UW-11394/2020 | 2020-06-23 | North America/USA/Washington/ | UW Virology Lab | UW Virology Lab | Pavitra Roychoudhury et al |
| EPI_ISL_570403 | USA/WA-UW-11401/2020 | 2020-06-23 | North America/USA/Washington/ | UW Virology Lab | UW Virology Lab | Pavitra Roychoudhury et al |
| EPI_ISL_570409 | USA/WA-UW-11443/2020 | 2020-06-23 | North America/USA/Washington/ | UW Virology Lab | UW Virology Lab | Pavitra Roychoudhury et al |
| EPI_ISL_570419 | USA/WA-UW-11484/2020 | 2020-06-24 | North America/USA/Washington/ | UW Virology Lab | UW Virology Lab | Pavitra Roychoudhury et al |
| EPI_ISL_570427 | USA/WA-UW-11519/2020 | 2020-06-20 | North America/USA/Washington/ | UW Virology Lab | UW Virology Lab | Pavitra Roychoudhury et al |
| EPI_ISL_570430 | USA/WA-UW-11529/2020 | 2020-06-21 | North America/USA/Washington/ | UW Virology Lab | UW Virology Lab | Pavitra Roychoudhury et al |
| EPI_ISL_570432 | USA/WA-UW-11533/2020 | 2020-06-21 | North America/USA/Washington/ | UW Virology Lab | UW Virology Lab | Pavitra Roychoudhury et al |
| EPI_ISL_570453 | USA/WA-UW-11645/2020 | 2020-06-19 | North America/USA/Washington/ | UW Virology Lab | UW Virology Lab | Pavitra Roychoudhury et al |
| EPI_ISL_570463 | USA/WA-UW-11722/2020 | 2020-06-16 | North America/USA/Washington/ | UW Virology Lab | UW Virology Lab | Pavitra Roychoudhury et al |
| EPI_ISL_570478 | USA/WA-UW-11909/2020 | 2020-06-24 | North America/USA/Washington/ | UW Virology Lab | UW Virology Lab | Pavitra Roychoudhury et al |
| EPI_ISL_570480 | USA/WA-UW-11919/2020 | 2020-06-24 | North America/USA/Washington/ | UW Virology Lab | UW Virology Lab | Pavitra Roychoudhury et al |
| EPI_ISL_570491 | USA/WA-UW-12127/2020 | 2020-06-25 | North America/USA/Washington/ | UW Virology Lab | UW Virology Lab | Pavitra Roychoudhury et al |
| EPI_ISL_570493 | USA/WA-UW-12140/2020 | 2020-06-26 | North America/USA/Washington/ | UW Virology Lab | UW Virology Lab | Pavitra Roychoudhury et al |
| EPI_ISL_570511 | USA/WA-UW-12356/2020 | 2020-06-27 | North America/USA/Washington/ | UW Virology Lab | UW Virology Lab | Pavitra Roychoudhury et al |
| EPI_ISL_570513 | USA/WA-UW-12360/2020 | 2020-06-27 | North America/USA/Washington/ | UW Virology Lab | UW Virology Lab | Pavitra Roychoudhury et al |
| EPI_ISL_570519 | USA/WA-UW-12371/2020 | 2020-06-27 | North America/USA/Washington/ | UW Virology Lab | UW Virology Lab | Pavitra Roychoudhury et al |
| EPI_ISL_570543 | USA/WA-UW-12539/2020 | 2020-06-27 | North America/USA/Washington/ | UW Virology Lab | UW Virology Lab | Pavitra Roychoudhury et al |
| EPI_ISL_570552 | USA/WA-UW-22375/2020 | 2020-08-07 | North America/USA/Washington/ | UW Virology Lab | UW Virology Lab | Pavitra Roychoudhury et al |
| EPI_ISL_570557 | USA/WA-UW-26866/2020 | 2020-09-02 | North America/USA/Washington/ | UW Virology Lab | UW Virology Lab | Pavitra Roychoudhury et al |

| | | | | | | |
|---|---|---|---|---|---|---|
| EPI_ISL_570559 | USA/WA-UW-22090/2020 | 2020-08-04 | North America/USA/Washington/ | UW Virology Lab | UW Virology Lab | Pavitra Roychoudhury et al |
| EPI_ISL_570563 | USA/WA-UW-22060/2020 | 2020-08-03 | North America/USA/Washington/ | UW Virology Lab | UW Virology Lab | Pavitra Roychoudhury et al |
| EPI_ISL_570568 | USA/WA-UW-22172/2020 | 2020-08-06 | North America/USA/Washington/ | UW Virology Lab | UW Virology Lab | Pavitra Roychoudhury et al |
| EPI_ISL_570571 | USA/WA-UW-22256/2020 | 2020-08-03 | North America/USA/Washington/ | UW Virology Lab | UW Virology Lab | Pavitra Roychoudhury et al |
| EPI_ISL_570573 | USA/WA-UW-24887/2020 | 2020-08-10 | North America/USA/Washington/ | UW Virology Lab | UW Virology Lab | Pavitra Roychoudhury et al |
| EPI_ISL_570576 | USA/WA-UW-22855/2020 | 2020-08-05 | North America/USA/Washington/ | UW Virology Lab | UW Virology Lab | Pavitra Roychoudhury et al |
| EPI_ISL_570581 | USA/WA-UW-24809/2020 | 2020-08-12 | North America/USA/Washington/ | UW Virology Lab | UW Virology Lab | Pavitra Roychoudhury et al |
| EPI_ISL_570585 | USA/WA-UW-22056/2020 | 2020-08-03 | North America/USA/Washington/ | UW Virology Lab | UW Virology Lab | Pavitra Roychoudhury et al |
| EPI_ISL_570591 | USA/WA-UW-22415/2020 | 2020-08-07 | North America/USA/Washington/ | UW Virology Lab | UW Virology Lab | Pavitra Roychoudhury et al |
| EPI_ISL_570592 | USA/WA-UW-22521/2020 | 2020-08-04 | North America/USA/Washington/ | UW Virology Lab | UW Virology Lab | Pavitra Roychoudhury et al |
| EPI_ISL_570594 | USA/WA-UW-23335/2020 | 2020-08-08 | North America/USA/Washington/ | UW Virology Lab | UW Virology Lab | Pavitra Roychoudhury et al |
| EPI_ISL_570595 | USA/WA-UW-23555/2020 | 2020-08-11 | North America/USA/Washington/ | UW Virology Lab | UW Virology Lab | Pavitra Roychoudhury et al |
| EPI_ISL_570607 | USA/WA-UW-22451/2020 | 2020-08-04 | North America/USA/Washington/ | UW Virology Lab | UW Virology Lab | Pavitra Roychoudhury et al |
| EPI_ISL_570611 | USA/WA-UW-24891/2020 | 2020-08-10 | North America/USA/Washington/ | UW Virology Lab | UW Virology Lab | Pavitra Roychoudhury et al |
| EPI_ISL_570614 | USA/OR-UW-22416/2020 | 2020-08-06 | North America/USA/Oregon/ | UW Virology Lab | UW Virology Lab | Pavitra Roychoudhury et al |
| EPI_ISL_570615 | USA/OR-UW-22446/2020 | 2020-08-03 | North America/USA/Oregon/ | UW Virology Lab | UW Virology Lab | Pavitra Roychoudhury et al |
| EPI_ISL_570618 | USA/OR-UW-22871/2020 | 2020-08-04 | North America/USA/Oregon/ | UW Virology Lab | UW Virology Lab | Pavitra Roychoudhury et al |
| EPI_ISL_570619 | USA/OR-UW-22918/2020 | 2020-08-09 | North America/USA/Oregon/ | UW Virology Lab | UW Virology Lab | Pavitra Roychoudhury et al |
| EPI_ISL_570623 | USA/WA-UW-22080/2020 | 2020-08-03 | North America/USA/Washington/ | UW Virology Lab | UW Virology Lab | Pavitra Roychoudhury et al |
| EPI_ISL_570630 | USA/WA-UW-22148/2020 | 2020-08-05 | North America/USA/Washington/ | UW Virology Lab | UW Virology Lab | Pavitra Roychoudhury et al |
| EPI_ISL_570631 | USA/WA-UW-22149/2020 | 2020-08-06 | North America/USA/Washington/ | UW Virology Lab | UW Virology Lab | Pavitra Roychoudhury et al |
| EPI_ISL_570633 | USA/WA-UW-22157/2020 | 2020-08-05 | North America/USA/Washington/ | UW Virology Lab | UW Virology Lab | Pavitra Roychoudhury et al |
| EPI_ISL_570636 | USA/WA-UW-22177/2020 | 2020-08-06 | North America/USA/Washington/ | UW Virology Lab | UW Virology Lab | Pavitra Roychoudhury et al |
| EPI_ISL_570638 | USA/WA-UW-22186/2020 | 2020-08-06 | North America/USA/Washington/ | UW Virology Lab | UW Virology Lab | Pavitra Roychoudhury et al |
| EPI_ISL_570639 | USA/WA-UW-22201/2020 | 2020-08-03 | North America/USA/Washington/ | UW Virology Lab | UW Virology Lab | Pavitra Roychoudhury et al |
| EPI_ISL_570640 | USA/WA-UW-22212/2020 | 2020-08-03 | North America/USA/Washington/ | UW Virology Lab | UW Virology Lab | Pavitra Roychoudhury et al |
| EPI_ISL_570641 | USA/WA-UW-22217/2020 | 2020-08-01 | North America/USA/Washington/ | UW Virology Lab | UW Virology Lab | Pavitra Roychoudhury et al |
| EPI_ISL_570642 | USA/WA-UW-22228/2020 | 2020-08-02 | North America/USA/Washington/ | UW Virology Lab | UW Virology Lab | Pavitra Roychoudhury et al |
| EPI_ISL_570645 | USA/WA-UW-22253/2020 | 2020-08-03 | North America/USA/Washington/ | UW Virology Lab | UW Virology Lab | Pavitra Roychoudhury et al |
| EPI_ISL_570646 | USA/WA-UW-22258/2020 | 2020-08-03 | North America/USA/Washington/ | UW Virology Lab | UW Virology Lab | Pavitra Roychoudhury et al |
| EPI_ISL_570652 | USA/WA-UW-22293/2020 | 2020-08-06 | North America/USA/Washington/ | UW Virology Lab | UW Virology Lab | Pavitra Roychoudhury et al |

| | | | | | | |
|---|---|---|---|---|---|---|
| EPI_ISL_570663 | USA/WA-UW-22331/2020 | 2020-08-06 | North America/USA/Washington/ | UW Virology Lab | UW Virology Lab | Pavitra Roychoudhury et al |
| EPI_ISL_570664 | USA/WA-UW-22339/2020 | 2020-08-05 | North America/USA/Washington/ | UW Virology Lab | UW Virology Lab | Pavitra Roychoudhury et al |
| EPI_ISL_570666 | USA/WA-UW-22358/2020 | 2020-08-07 | North America/USA/Washington/ | UW Virology Lab | UW Virology Lab | Pavitra Roychoudhury et al |
| EPI_ISL_570669 | USA/WA-UW-22365/2020 | 2020-08-08 | North America/USA/Washington/ | UW Virology Lab | UW Virology Lab | Pavitra Roychoudhury et al |
| EPI_ISL_570671 | USA/WA-UW-22376/2020 | 2020-08-08 | North America/USA/Washington/ | UW Virology Lab | UW Virology Lab | Pavitra Roychoudhury et al |
| EPI_ISL_570679 | USA/WA-UW-22412/2020 | 2020-08-08 | North America/USA/Washington/ | UW Virology Lab | UW Virology Lab | Pavitra Roychoudhury et al |
| EPI_ISL_570680 | USA/WA-UW-22418/2020 | 2020-08-07 | North America/USA/Washington/ | UW Virology Lab | UW Virology Lab | Pavitra Roychoudhury et al |
| EPI_ISL_570682 | USA/WA-UW-22422/2020 | 2020-08-07 | North America/USA/Washington/ | UW Virology Lab | UW Virology Lab | Pavitra Roychoudhury et al |
| EPI_ISL_570683 | USA/WA-UW-22426/2020 | 2020-08-07 | North America/USA/Washington/ | UW Virology Lab | UW Virology Lab | Pavitra Roychoudhury et al |
| EPI_ISL_570685 | USA/WA-UW-22431/2020 | 2020-08-07 | North America/USA/Washington/ | UW Virology Lab | UW Virology Lab | Pavitra Roychoudhury et al |
| EPI_ISL_570687 | USA/WA-UW-22436/2020 | 2020-08-06 | North America/USA/Washington/ | UW Virology Lab | UW Virology Lab | Pavitra Roychoudhury et al |
| EPI_ISL_570693 | USA/WA-UW-22458/2020 | 2020-08-04 | North America/USA/Washington/ | UW Virology Lab | UW Virology Lab | Pavitra Roychoudhury et al |
| EPI_ISL_570695 | USA/WA-UW-22476/2020 | 2020-08-05 | North America/USA/Washington/ | UW Virology Lab | UW Virology Lab | Pavitra Roychoudhury et al |
| EPI_ISL_570697 | USA/WA-UW-22493/2020 | 2020-08-04 | North America/USA/Washington/ | UW Virology Lab | UW Virology Lab | Pavitra Roychoudhury et al |
| EPI_ISL_570703 | USA/WA-UW-22720/2020 | 2020-08-07 | North America/USA/Washington/ | UW Virology Lab | UW Virology Lab | Pavitra Roychoudhury et al |
| EPI_ISL_570709 | USA/WA-UW-22752/2020 | 2020-08-07 | North America/USA/Washington/ | UW Virology Lab | UW Virology Lab | Pavitra Roychoudhury et al |
| EPI_ISL_570713 | USA/WA-UW-22758/2020 | 2020-08-06 | North America/USA/Washington/ | UW Virology Lab | UW Virology Lab | Pavitra Roychoudhury et al |
| EPI_ISL_570716 | USA/WA-UW-22774/2020 | 2020-08-06 | North America/USA/Washington/ | UW Virology Lab | UW Virology Lab | Pavitra Roychoudhury et al |
| EPI_ISL_570719 | USA/WA-UW-22791/2020 | 2020-08-07 | North America/USA/Washington/ | UW Virology Lab | UW Virology Lab | Pavitra Roychoudhury et al |
| EPI_ISL_570727 | USA/WA-UW-22859/2020 | 2020-08-05 | North America/USA/Washington/ | UW Virology Lab | UW Virology Lab | Pavitra Roychoudhury et al |
| EPI_ISL_570729 | USA/WA-UW-22885/2020 | 2020-08-06 | North America/USA/Washington/ | UW Virology Lab | UW Virology Lab | Pavitra Roychoudhury et al |
| EPI_ISL_570747 | USA/WA-UW-23004/2020 | 2020-08-10 | North America/USA/Washington/ | UW Virology Lab | UW Virology Lab | Pavitra Roychoudhury et al |
| EPI_ISL_570748 | USA/WA-UW-23124/2020 | 2020-08-08 | North America/USA/Washington/ | UW Virology Lab | UW Virology Lab | Pavitra Roychoudhury et al |
| EPI_ISL_570749 | USA/WA-UW-23126/2020 | 2020-08-07 | North America/USA/Washington/ | UW Virology Lab | UW Virology Lab | Pavitra Roychoudhury et al |
| EPI_ISL_570751 | USA/WA-UW-23152/2020 | 2020-08-03 | North America/USA/Washington/ | UW Virology Lab | UW Virology Lab | Pavitra Roychoudhury et al |
| EPI_ISL_570752 | USA/WA-UW-23164/2020 | 2020-08-03 | North America/USA/Washington/ | UW Virology Lab | UW Virology Lab | Pavitra Roychoudhury et al |
| EPI_ISL_570757 | USA/WA-UW-23206/2020 | 2020-08-07 | North America/USA/Washington/ | UW Virology Lab | UW Virology Lab | Pavitra Roychoudhury et al |
| EPI_ISL_570758 | USA/WA-UW-23213/2020 | 2020-08-07 | North America/USA/Washington/ | UW Virology Lab | UW Virology Lab | Pavitra Roychoudhury et al |
| EPI_ISL_570759 | USA/WA-UW-23483/2020 | 2020-08-10 | North America/USA/Washington/ | UW Virology Lab | UW Virology Lab | Pavitra Roychoudhury et al |
| EPI_ISL_570762 | USA/WA-UW-24020/2020 | 2020-08-13 | North America/USA/Washington/ | UW Virology Lab | UW Virology Lab | Pavitra Roychoudhury et al |
| EPI_ISL_570773 | USA/WA-UW-24930/2020 | 2020-08-12 | North America/USA/Washington/ | UW Virology Lab | UW Virology Lab | Pavitra Roychoudhury et al |

| | | | | | | |
|---|---|---|---|---|---|---|
| EPI_ISL_570780 | USA/WA-UW-26858/2020 | 2020-09-01 | North America/USA/Washington/ | UW Virology Lab | UW Virology Lab | Pavitra Roychoudhury et al |
| EPI_ISL_570781 | USA/WA-UW-26859/2020 | 2020-09-01 | North America/USA/Washington/ | UW Virology Lab | UW Virology Lab | Pavitra Roychoudhury et al |
| EPI_ISL_570790 | USA/WA-UW-27038/2020 | 2020-09-06 | North America/USA/Washington/ | UW Virology Lab | UW Virology Lab | Pavitra Roychoudhury et al |
| EPI_ISL_570803 | USA/WA-UW-19463/2020 | 2020-07-31 | North America/USA/Washington/ | UW Virology Lab | UW Virology Lab | Pavitra Roychoudhury et al |
| EPI_ISL_570812 | USA/WA-UW-19249/2020 | 2020-08-01 | North America/USA/Washington/ | UW Virology Lab | UW Virology Lab | Pavitra Roychoudhury et al |
| EPI_ISL_570820 | USA/WA-UW-21969/2020 | 2020-08-06 | North America/USA/Washington/ | UW Virology Lab | UW Virology Lab | Pavitra Roychoudhury et al |
| EPI_ISL_570821 | USA/WA-UW-19536/2020 | 2020-08-01 | North America/USA/Washington/ | UW Virology Lab | UW Virology Lab | Pavitra Roychoudhury et al |
| EPI_ISL_570822 | USA/WA-UW-21712/2020 | 2020-08-05 | North America/USA/Washington/ | UW Virology Lab | UW Virology Lab | Pavitra Roychoudhury et al |
| EPI_ISL_570828 | USA/WA-UW-22020/2020 | 2020-08-06 | North America/USA/Washington/ | UW Virology Lab | UW Virology Lab | Pavitra Roychoudhury et al |
| EPI_ISL_570829 | USA/WA-UW-22027/2020 | 2020-08-06 | North America/USA/Washington/ | UW Virology Lab | UW Virology Lab | Pavitra Roychoudhury et al |
| EPI_ISL_570853 | USA/WA-UW-19262/2020 | 2020-08-01 | North America/USA/Washington/ | UW Virology Lab | UW Virology Lab | Pavitra Roychoudhury et al |
| EPI_ISL_570854 | USA/WA-UW-19376/2020 | 2020-08-01 | North America/USA/Washington/ | UW Virology Lab | UW Virology Lab | Pavitra Roychoudhury et al |
| EPI_ISL_570855 | USA/WA-UW-21726/2020 | 2020-08-05 | North America/USA/Washington/ | UW Virology Lab | UW Virology Lab | Pavitra Roychoudhury et al |
| EPI_ISL_570867 | USA/WA-UW-12578/2020 | 2020-06-27 | North America/USA/Washington/ | UW Virology Lab | UW Virology Lab | Pavitra Roychoudhury et al |
| EPI_ISL_570872 | USA/WA-UW-12602/2020 | 2020-06-28 | North America/USA/Washington/ | UW Virology Lab | UW Virology Lab | Pavitra Roychoudhury et al |
| EPI_ISL_570875 | USA/WA-UW-12613/2020 | 2020-06-28 | North America/USA/Washington/ | UW Virology Lab | UW Virology Lab | Pavitra Roychoudhury et al |
| EPI_ISL_570888 | USA/WA-UW-12671/2020 | 2020-06-29 | North America/USA/Washington/ | UW Virology Lab | UW Virology Lab | Pavitra Roychoudhury et al |
| EPI_ISL_570893 | USA/WA-UW-12716/2020 | 2020-06-30 | North America/USA/Washington/ | UW Virology Lab | UW Virology Lab | Pavitra Roychoudhury et al |
| EPI_ISL_570894 | USA/WA-UW-12728/2020 | 2020-06-29 | North America/USA/Washington/ | UW Virology Lab | UW Virology Lab | Pavitra Roychoudhury et al |
| EPI_ISL_570919 | USA/WA-UW-12878/2020 | 2020-06-29 | North America/USA/Washington/ | UW Virology Lab | UW Virology Lab | Pavitra Roychoudhury et al |
| EPI_ISL_570925 | USA/WA-UW-12901/2020 | 2020-06-29 | North America/USA/Washington/ | UW Virology Lab | UW Virology Lab | Pavitra Roychoudhury et al |
| EPI_ISL_570927 | USA/WA-UW-12908/2020 | 2020-06-29 | North America/USA/Washington/ | UW Virology Lab | UW Virology Lab | Pavitra Roychoudhury et al |
| EPI_ISL_570939 | USA/WA-UW-12978/2020 | 2020-06-30 | North America/USA/Washington/ | UW Virology Lab | UW Virology Lab | Pavitra Roychoudhury et al |
| EPI_ISL_570949 | USA/WA-UW-13082/2020 | 2020-06-30 | North America/USA/Washington/ | UW Virology Lab | UW Virology Lab | Pavitra Roychoudhury et al |
| EPI_ISL_570956 | USA/WA-UW-13196/2020 | 2020-07-01 | North America/USA/Washington/ | UW Virology Lab | UW Virology Lab | Pavitra Roychoudhury et al |
| EPI_ISL_570964 | USA/WA-UW-13311/2020 | 2020-07-03 | North America/USA/Washington/ | UW Virology Lab | UW Virology Lab | Pavitra Roychoudhury et al |
| EPI_ISL_570965 | USA/WA-UW-13319/2020 | 2020-06-30 | North America/USA/Washington/ | UW Virology Lab | UW Virology Lab | Pavitra Roychoudhury et al |
| EPI_ISL_570967 | USA/WA-UW-13370/2020 | 2020-07-01 | North America/USA/Washington/ | UW Virology Lab | UW Virology Lab | Pavitra Roychoudhury et al |
| EPI_ISL_570969 | USA/WA-UW-13397/2020 | 2020-07-02 | North America/USA/Washington/ | UW Virology Lab | UW Virology Lab | Pavitra Roychoudhury et al |
| EPI_ISL_570973 | USA/WA-UW-182/2020 | 2020-03-08 | North America/USA/Washington/ | UW Virology Lab | UW Virology Lab | Pavitra Roychoudhury et al |
| EPI_ISL_570978 | USA/WA-UW-19384/2020 | 2020-08-01 | North America/USA/Washington/ | UW Virology Lab | UW Virology Lab | Pavitra Roychoudhury et al |

| | | | | | | |
|---|---|---|---|---|---|---|
| EPI_ISL_570982 | USA/WA-UW-19488/2020 | 2020-07-31 | North America/USA/Washington/ | UW Virology Lab | UW Virology Lab | Pavitra Roychoudhury et al |
| EPI_ISL_570984 | USA/WA-UW-19676/2020 | 2020-08-02 | North America/USA/Washington/ | UW Virology Lab | UW Virology Lab | Pavitra Roychoudhury et al |
| EPI_ISL_570989 | USA/WA-UW-21600/2020 | 2020-08-04 | North America/USA/Washington/ | UW Virology Lab | UW Virology Lab | Pavitra Roychoudhury et al |
| EPI_ISL_570995 | USA/WA-UW-21732/2020 | 2020-08-05 | North America/USA/Washington/ | UW Virology Lab | UW Virology Lab | Pavitra Roychoudhury et al |
| EPI_ISL_570998 | USA/WA-UW-21736/2020 | 2020-08-05 | North America/USA/Washington/ | UW Virology Lab | UW Virology Lab | Pavitra Roychoudhury et al |
| EPI_ISL_570999 | USA/WA-UW-21742/2020 | 2020-08-05 | North America/USA/Washington/ | UW Virology Lab | UW Virology Lab | Pavitra Roychoudhury et al |
| EPI_ISL_571002 | USA/WA-UW-21753/2020 | 2020-08-05 | North America/USA/Washington/ | UW Virology Lab | UW Virology Lab | Pavitra Roychoudhury et al |
| EPI_ISL_571004 | USA/WA-UW-21771/2020 | 2020-08-04 | North America/USA/Washington/ | UW Virology Lab | UW Virology Lab | Pavitra Roychoudhury et al |
| EPI_ISL_571005 | USA/WA-UW-21784/2020 | 2020-08-05 | North America/USA/Washington/ | UW Virology Lab | UW Virology Lab | Pavitra Roychoudhury et al |
| EPI_ISL_571010 | USA/WA-UW-21987/2020 | 2020-08-06 | North America/USA/Washington/ | UW Virology Lab | UW Virology Lab | Pavitra Roychoudhury et al |
| EPI_ISL_571011 | USA/WA-UW-21998/2020 | 2020-08-05 | North America/USA/Washington/ | UW Virology Lab | UW Virology Lab | Pavitra Roychoudhury et al |
| EPI_ISL_571012 | USA/WA-UW-22003/2020 | 2020-08-05 | North America/USA/Washington/ | UW Virology Lab | UW Virology Lab | Pavitra Roychoudhury et al |
| EPI_ISL_571013 | USA/WA-UW-22016/2020 | 2020-08-06 | North America/USA/Washington/ | UW Virology Lab | UW Virology Lab | Pavitra Roychoudhury et al |
| EPI_ISL_571015 | USA/WA-UW-22039/2020 | 2020-08-03 | North America/USA/Washington/ | UW Virology Lab | UW Virology Lab | Pavitra Roychoudhury et al |
| EPI_ISL_571039 | USA/MI-QDX-295/2020 | 2020-03-20 | North America/USA/Michigan/ | Quest Diagnostics | Quest Diagnostics | Rosenthal et al |
| EPI_ISL_571077 | USA/MI-QDX-333/2020 | 2020-03-19 | North America/USA/Michigan/ | Quest Diagnostics | Quest Diagnostics | Rosenthal et al |
| EPI_ISL_571083 | USA/MI-QDX-339/2020 | 2020-03-23 | North America/USA/Michigan/ | Quest Diagnostics | Quest Diagnostics | Rosenthal et al |
| EPI_ISL_571084 | USA/MI-QDX-340/2020 | 2020-03-22 | North America/USA/Michigan/ | Quest Diagnostics | Quest Diagnostics | Rosenthal et al |
| EPI_ISL_571096 | USA/MI-QDX-352/2020 | 2020-03-23 | North America/USA/Michigan/ | Quest Diagnostics | Quest Diagnostics | Rosenthal et al |
| EPI_ISL_571098 | USA/MI-QDX-354/2020 | 2020-03-23 | North America/USA/Michigan/ | Quest Diagnostics | Quest Diagnostics | Rosenthal et al |
| EPI_ISL_571113 | USA/MI-QDX-370/2020 | 2020-03-23 | North America/USA/Michigan/ | Quest Diagnostics | Quest Diagnostics | Rosenthal et al |
| EPI_ISL_571114 | USA/MI-QDX-371/2020 | 2020-03-23 | North America/USA/Michigan/ | Quest Diagnostics | Quest Diagnostics | Rosenthal et al |
| EPI_ISL_571129 | USA/MI-QDX-388/2020 | 2020-03-23 | North America/USA/Michigan/ | Quest Diagnostics | Quest Diagnostics | Rosenthal et al |
| EPI_ISL_571137 | USA/MI-QDX-398/2020 | 2020-03-24 | North America/USA/Michigan/ | Quest Diagnostics | Quest Diagnostics | Rosenthal et al |
| EPI_ISL_571138 | USA/MI-QDX-399/2020 | 2020-03-24 | North America/USA/Michigan/ | Quest Diagnostics | Quest Diagnostics | Rosenthal et al |
| EPI_ISL_571146 | USA/MI-QDX-407/2020 | 2020-03-24 | North America/USA/Michigan/ | Quest Diagnostics | Quest Diagnostics | Rosenthal et al |
| EPI_ISL_571147 | USA/MI-QDX-408/2020 | 2020-03-24 | North America/USA/Michigan/ | Quest Diagnostics | Quest Diagnostics | Rosenthal et al |
| EPI_ISL_571162 | USA/CA-QDX-437/2020 | 2020-03-21 | North America/USA/California/ | Quest Diagnostics | Quest Diagnostics | Rosenthal et al |
| EPI_ISL_571181 | USA/CT-QDX-429/2020 | 2020-03-17 | North America/USA/Connecticut/ | Quest Diagnostics | Quest Diagnostics | Rosenthal et al |
| EPI_ISL_571193 | USA/MI-QDX-458/2020 | 2020-03-21 | North America/USA/Michigan/ | Quest Diagnostics | Quest Diagnostics | Rosenthal et al |
| EPI_ISL_571203 | USA/MI-QDX-468/2020 | 2020-03-23 | North America/USA/Michigan/ | Quest Diagnostics | Quest Diagnostics | Rosenthal et al |

| | | | | | | |
|---|---|---|---|---|---|---|
| EPI_ISL_571214 | USA/MI-QDX-480/2020 | 2020-03-24 | North America/USA/Michigan/ | Quest Diagnostics | Quest Diagnostics | Rosenthal et al |
| EPI_ISL_571217 | USA/MI-QDX-483/2020 | 2020-03-23 | North America/USA/Michigan/ | Quest Diagnostics | Quest Diagnostics | Rosenthal et al |
| EPI_ISL_571228 | USA/MI-QDX-494/2020 | 2020-03-23 | North America/USA/Michigan/ | Quest Diagnostics | Quest Diagnostics | Rosenthal et al |
| EPI_ISL_571243 | USA/MI-QDX-509/2020 | 2020-03-22 | North America/USA/Michigan/ | Quest Diagnostics | Quest Diagnostics | Rosenthal et al |
| EPI_ISL_571288 | USA/GA-QDX-577/2020 | 2020-03-18 | North America/USA/Georgia/ | Quest Diagnostics | Quest Diagnostics | Rosenthal et al |
| EPI_ISL_571325 | USA/MI-QDX-587/2020 | 2020-03-18 | North America/USA/Michigan/ | Quest Diagnostics | Quest Diagnostics | Rosenthal et al |
| EPI_ISL_571336 | USA/MI-QDX-601/2020 | 2020-03-19 | North America/USA/Michigan/ | Quest Diagnostics | Quest Diagnostics | Rosenthal et al |
| EPI_ISL_571345 | USA/MI-QDX-616/2020 | 2020-03-18 | North America/USA/Michigan/ | Quest Diagnostics | Quest Diagnostics | Rosenthal et al |
| EPI_ISL_571349 | USA/MI-QDX-622/2020 | 2020-03-17 | North America/USA/Michigan/ | Quest Diagnostics | Quest Diagnostics | Rosenthal et al |
| EPI_ISL_571355 | USA/MI-QDX-628/2020 | 2020-03-19 | North America/USA/Michigan/ | Quest Diagnostics | Quest Diagnostics | Rosenthal et al |
| EPI_ISL_571381 | USA/MO-QDX-524/2020 | 2020-03-18 | North America/USA/Missouri/ | Quest Diagnostics | Quest Diagnostics | Rosenthal et al |
| EPI_ISL_571386 | USA/MO-QDX-535/2020 | 2020-03-18 | North America/USA/Missouri/ | Quest Diagnostics | Quest Diagnostics | Rosenthal et al |
| EPI_ISL_571397 | USA/TX-QDX-663/2020 | 2020-03-27 | North America/USA/Texas/ | Quest Diagnostics | Quest Diagnostics | Rosenthal et al |
| EPI_ISL_571401 | USA/WA-QDX-528/2020 | 2020-03-18 | North America/USA/Washington/ | Quest Diagnostics | Quest Diagnostics | Rosenthal et al |
| EPI_ISL_571448 | USA/MI-QDX-715/2020 | 2020-03-21 | North America/USA/Michigan/ | Quest Diagnostics | Quest Diagnostics | Rosenthal et al |
| EPI_ISL_571452 | USA/MI-QDX-719/2020 | 2020-03-21 | North America/USA/Michigan/ | Quest Diagnostics | Quest Diagnostics | Rosenthal et al |
| EPI_ISL_571456 | USA/MI-QDX-723/2020 | 2020-03-20 | North America/USA/Michigan/ | Quest Diagnostics | Quest Diagnostics | Rosenthal et al |
| EPI_ISL_571458 | USA/MI-QDX-725/2020 | 2020-03-21 | North America/USA/Michigan/ | Quest Diagnostics | Quest Diagnostics | Rosenthal et al |
| EPI_ISL_571469 | USA/MI-QDX-736/2020 | 2020-03-20 | North America/USA/Michigan/ | Quest Diagnostics | Quest Diagnostics | Rosenthal et al |
| EPI_ISL_571486 | USA/CA-QDX-776/2020 | 2020-03-20 | North America/USA/California/ | Quest Diagnostics | Quest Diagnostics | Rosenthal et al |
| EPI_ISL_571488 | USA/CA-QDX-794/2020 | 2020-03-21 | North America/USA/California/ | Quest Diagnostics | Quest Diagnostics | Rosenthal et al |
| EPI_ISL_571495 | USA/CA-QDX-802/2020 | 2020-03-21 | North America/USA/California/ | Quest Diagnostics | Quest Diagnostics | Rosenthal et al |
| EPI_ISL_571499 | USA/CA-QDX-806/2020 | 2020-03-23 | North America/USA/California/ | Quest Diagnostics | Quest Diagnostics | Rosenthal et al |
| EPI_ISL_571536 | USA/MI-QDX-819/2020 | 2020-03-20 | North America/USA/Michigan/ | Quest Diagnostics | Quest Diagnostics | Rosenthal et al |
| EPI_ISL_571547 | USA/MN-QDX-753/2020 | 2020-03-19 | North America/USA/Minnesota/ | Quest Diagnostics | Quest Diagnostics | Rosenthal et al |
| EPI_ISL_571568 | USA/WI-QDX-783/2020 | 2020-03-16 | North America/USA/Wisconsin/ | Quest Diagnostics | Quest Diagnostics | Rosenthal et al |
| EPI_ISL_571572 | USA/AL-QDX-949/2020 | 2020-03-23 | North America/USA/Alabama/ | Quest Diagnostics | Quest Diagnostics | Rosenthal et al |
| EPI_ISL_571584 | USA/CA-QDX-939/2020 | 2020-03-23 | North America/USA/California/ | Quest Diagnostics | Quest Diagnostics | Rosenthal et al |
| EPI_ISL_571593 | USA/FL-QDX-909/2020 | 2020-03-21 | North America/USA/Florida/ | Quest Diagnostics | Quest Diagnostics | Rosenthal et al |
| EPI_ISL_571602 | USA/IN-QDX-830/2020 | 2020-03-16 | North America/USA/Indiana/ | Quest Diagnostics | Quest Diagnostics | Rosenthal et al |
| EPI_ISL_571608 | USA/MI-QDX-887/2020 | 2020-03-18 | North America/USA/Michigan/ | Quest Diagnostics | Quest Diagnostics | Rosenthal et al |

| | | | | | | |
|---|---|---|---|---|---|---|
| EPI_ISL_571628 | USA/MI-QDX-918/2020 | 2020-03-22 | North America/USA/Michigan/ | Quest Diagnostics | Quest Diagnostics | Rosenthal et al |
| EPI_ISL_571632 | USA/MI-QDX-927/2020 | 2020-03-22 | North America/USA/Michigan/ | Quest Diagnostics | Quest Diagnostics | Rosenthal et al |
| EPI_ISL_571640 | USA/MI-QDX-947/2020 | 2020-03-23 | North America/USA/Michigan/ | Quest Diagnostics | Quest Diagnostics | Rosenthal et al |
| EPI_ISL_571674 | USA/NY-QDX-868/2020 | 2020-03-16 | North America/USA/New York/ | Quest Diagnostics | Quest Diagnostics | Rosenthal et al |
| EPI_ISL_571679 | USA/NY-QDX-875/2020 | 2020-03-17 | North America/USA/New York/ | Quest Diagnostics | Quest Diagnostics | Rosenthal et al |
| EPI_ISL_571711 | USA/CA-QDX-1049/2020 | 2020-03-17 | North America/USA/California/ | Quest Diagnostics | Quest Diagnostics | Rosenthal et al |
| EPI_ISL_571723 | USA/MI-QDX-1047/2020 | 2020-03-17 | North America/USA/Michigan/ | Quest Diagnostics | Quest Diagnostics | Rosenthal et al |
| EPI_ISL_571735 | USA/MN-QDX-1009/2020 | 2020-03-15 | North America/USA/Minnesota/ | Quest Diagnostics | Quest Diagnostics | Rosenthal et al |
| EPI_ISL_571737 | USA/MN-QDX-1011/2020 | 2020-03-14 | North America/USA/Minnesota/ | Quest Diagnostics | Quest Diagnostics | Rosenthal et al |
| EPI_ISL_571753 | USA/MN-QDX-952/2020 | 2020-03-16 | North America/USA/Minnesota/ | Quest Diagnostics | Quest Diagnostics | Rosenthal et al |
| EPI_ISL_571765 | USA/MN-QDX-966/2020 | 2020-03-15 | North America/USA/Minnesota/ | Quest Diagnostics | Quest Diagnostics | Rosenthal et al |
| EPI_ISL_571790 | USA/MN-QDX-992/2020 | 2020-03-13 | North America/USA/Minnesota/ | Quest Diagnostics | Quest Diagnostics | Rosenthal et al |
| EPI_ISL_571795 | USA/MN-QDX-998/2020 | 2020-03-14 | North America/USA/Minnesota/ | Quest Diagnostics | Quest Diagnostics | Rosenthal et al |
| EPI_ISL_571802 | USA/WA-QDX-1060/2020 | 2020-03-17 | North America/USA/Washington/ | Quest Diagnostics | Quest Diagnostics | Rosenthal et al |
| EPI_ISL_571814 | USA/CA-QDX-1119/2020 | 2020-03-20 | North America/USA/California/ | Quest Diagnostics | Quest Diagnostics | Rosenthal et al |
| EPI_ISL_571818 | USA/CA-QDX-1171/2020 | 2020-03-28 | North America/USA/California/ | Quest Diagnostics | Quest Diagnostics | Rosenthal et al |
| EPI_ISL_571860 | USA/MI-QDX-1130/2020 | 2020-03-21 | North America/USA/Michigan/ | Quest Diagnostics | Quest Diagnostics | Rosenthal et al |
| EPI_ISL_571867 | USA/MI-QDX-1137/2020 | 2020-03-21 | North America/USA/Michigan/ | Quest Diagnostics | Quest Diagnostics | Rosenthal et al |
| EPI_ISL_571872 | USA/MI-QDX-1142/2020 | 2020-03-19 | North America/USA/Michigan/ | Quest Diagnostics | Quest Diagnostics | Rosenthal et al |
| EPI_ISL_571880 | USA/MI-QDX-1150/2020 | 2020-03-23 | North America/USA/Michigan/ | Quest Diagnostics | Quest Diagnostics | Rosenthal et al |
| EPI_ISL_571886 | USA/MI-QDX-1156/2020 | 2020-03-24 | North America/USA/Michigan/ | Quest Diagnostics | Quest Diagnostics | Rosenthal et al |
| EPI_ISL_571899 | USA/PR-QDX-1074/2020 | 2020-03-20 | North America/USA/Puerto Rico/ | Quest Diagnostics | Quest Diagnostics | Rosenthal et al |
| EPI_ISL_571908 | USA/WA-QDX-1104/2020 | 2020-03-19 | North America/USA/Washington/ | Quest Diagnostics | Quest Diagnostics | Rosenthal et al |
| EPI_ISL_571919 | USA/CA-QDX-1246/2020 | 2020-03-16 | North America/USA/California/ | Quest Diagnostics | Quest Diagnostics | Rosenthal et al |
| EPI_ISL_571925 | USA/CO-QDX-1312/2020 | 2020-03-20 | North America/USA/Colorado/ | Quest Diagnostics | Quest Diagnostics | Rosenthal et al |
| EPI_ISL_571940 | USA/LA-QDX-1194/2020 | 2020-03-16 | North America/USA/Louisiana/ | Quest Diagnostics | Quest Diagnostics | Rosenthal et al |
| EPI_ISL_571958 | USA/MI-QDX-1303/2020 | 2020-03-17 | North America/USA/Michigan/ | Quest Diagnostics | Quest Diagnostics | Rosenthal et al |
| EPI_ISL_571967 | USA/MI-QDX-1313/2020 | 2020-03-20 | North America/USA/Michigan/ | Quest Diagnostics | Quest Diagnostics | Rosenthal et al |
| EPI_ISL_571968 | USA/MS-QDX-1243/2020 | 2020-03-16 | North America/USA/Mississippi/ | Quest Diagnostics | Quest Diagnostics | Rosenthal et al |
| EPI_ISL_571974 | USA/NJ-QDX-1213/2020 | 2020-03-16 | North America/USA/New Jersey/ | Quest Diagnostics | Quest Diagnostics | Rosenthal et al |
| EPI_ISL_571975 | USA/NJ-QDX-1216/2020 | 2020-03-17 | North America/USA/New Jersey/ | Quest Diagnostics | Quest Diagnostics | Rosenthal et al |

| | | | | | | |
|---|---|---|---|---|---|---|
| EPI_ISL_571992 | USA/NY-QDX-1182/2020 | 2020-03-13 | North America/USA/New York/ | Quest Diagnostics | Quest Diagnostics | Rosenthal et al |
| EPI_ISL_571999 | USA/NY-QDX-1222/2020 | 2020-03-16 | North America/USA/New York/ | Quest Diagnostics | Quest Diagnostics | Rosenthal et al |
| EPI_ISL_572004 | USA/NY-QDX-1228/2020 | 2020-03-16 | North America/USA/New York/ | Quest Diagnostics | Quest Diagnostics | Rosenthal et al |
| EPI_ISL_572010 | USA/NY-QDX-1234/2020 | 2020-03-17 | North America/USA/New York/ | Quest Diagnostics | Quest Diagnostics | Rosenthal et al |
| EPI_ISL_572038 | USA/TX-QDX-1201/2020 | 2020-03-16 | North America/USA/Texas/ | Quest Diagnostics | Quest Diagnostics | Rosenthal et al |
| EPI_ISL_572052 | USA/WA-QDX-1273/2020 | 2020-03-16 | North America/USA/Washington/ | Quest Diagnostics | Quest Diagnostics | Rosenthal et al |
| EPI_ISL_572054 | USA/AR-QDX-1315/2020 | 2020-03-15 | North America/USA/Arkansas/ | Quest Diagnostics | Quest Diagnostics | Rosenthal et al |
| EPI_ISL_572104 | USA/MI-QDX-1439/2020 | 2020-03-16 | North America/USA/Michigan/ | Quest Diagnostics | Quest Diagnostics | Rosenthal et al |
| EPI_ISL_572112 | USA/MN-QDX-1323/2020 | 2020-03-15 | North America/USA/Minnesota/ | Quest Diagnostics | Quest Diagnostics | Rosenthal et al |
| EPI_ISL_572120 | USA/MN-QDX-1331/2020 | 2020-03-15 | North America/USA/Minnesota/ | Quest Diagnostics | Quest Diagnostics | Rosenthal et al |
| EPI_ISL_572124 | USA/MN-QDX-1335/2020 | 2020-03-14 | North America/USA/Minnesota/ | Quest Diagnostics | Quest Diagnostics | Rosenthal et al |
| EPI_ISL_572126 | USA/MN-QDX-1337/2020 | 2020-03-14 | North America/USA/Minnesota/ | Quest Diagnostics | Quest Diagnostics | Rosenthal et al |
| EPI_ISL_572127 | USA/MN-QDX-1338/2020 | 2020-03-15 | North America/USA/Minnesota/ | Quest Diagnostics | Quest Diagnostics | Rosenthal et al |
| EPI_ISL_572128 | USA/MN-QDX-1339/2020 | 2020-03-16 | North America/USA/Minnesota/ | Quest Diagnostics | Quest Diagnostics | Rosenthal et al |
| EPI_ISL_572133 | USA/MN-QDX-1344/2020 | 2020-03-15 | North America/USA/Minnesota/ | Quest Diagnostics | Quest Diagnostics | Rosenthal et al |
| EPI_ISL_572138 | USA/MN-QDX-1349/2020 | 2020-03-14 | North America/USA/Minnesota/ | Quest Diagnostics | Quest Diagnostics | Rosenthal et al |
| EPI_ISL_572140 | USA/MN-QDX-1351/2020 | 2020-03-14 | North America/USA/Minnesota/ | Quest Diagnostics | Quest Diagnostics | Rosenthal et al |
| EPI_ISL_572177 | USA/MN-QDX-1388/2020 | 2020-03-14 | North America/USA/Minnesota/ | Quest Diagnostics | Quest Diagnostics | Rosenthal et al |
| EPI_ISL_572185 | USA/UT-QDX-1391/2020 | 2020-03-14 | North America/USA/Utah/ | Quest Diagnostics | Quest Diagnostics | Rosenthal et al |
| EPI_ISL_572199 | USA/VA-DCLS-1456/2020 | 2020-08-19 | North America/USA/Virginia/ | Virginia DCLS | Virginia DCLS | Virginia DCLS et al |
| EPI_ISL_572201 | USA/VA-DCLS-1460/2020 | 2020-08-21 | North America/USA/Virginia/ | Virginia DCLS | Virginia DCLS | Virginia DCLS et al |
| EPI_ISL_572203 | USA/VA-DCLS-1462/2020 | 2020-08-21 | North America/USA/Virginia/ | Virginia DCLS | Virginia DCLS | Virginia DCLS et al |
| EPI_ISL_572208 | USA/VA-DCLS-1469/2020 | 2020-08-21 | North America/USA/Virginia/ | Virginia DCLS | Virginia DCLS | Virginia DCLS et al |
| EPI_ISL_572211 | USA/VA-DCLS-1472/2020 | 2020-08-22 | North America/USA/Virginia/ | Virginia DCLS | Virginia DCLS | Virginia DCLS et al |
| EPI_ISL_572212 | USA/VA-DCLS-1473/2020 | 2020-08-22 | North America/USA/Virginia/ | Virginia DCLS | Virginia DCLS | Virginia DCLS et al |
| EPI_ISL_572213 | USA/VA-DCLS-1475/2020 | 2020-08-22 | North America/USA/Virginia/ | Virginia DCLS | Virginia DCLS | Virginia DCLS et al |
| EPI_ISL_572217 | USA/VA-DCLS-1479/2020 | 2020-08-22 | North America/USA/Virginia/ | Virginia DCLS | Virginia DCLS | Virginia DCLS et al |
| EPI_ISL_572219 | USA/VA-DCLS-1678/2020 | 2020-09-03 | North America/USA/Virginia/ | Virginia DCLS | Virginia DCLS | Virginia DCLS et al |
| EPI_ISL_572220 | USA/VA-DCLS-1376/2020 | 2020-07-05 | North America/USA/Virginia/ | Virginia DCLS | Virginia DCLS | Virginia DCLS et al |
| EPI_ISL_572221 | USA/VA-DCLS-1362/2020 | 2020-07-07 | North America/USA/Virginia/ | Virginia DCLS | Virginia DCLS | Virginia DCLS et al |
| EPI_ISL_572222 | USA/VA-DCLS-1363/2020 | 2020-07-07 | North America/USA/Virginia/ | Virginia DCLS | Virginia DCLS | Virginia DCLS et al |

| | | | | | | |
|---|---|---|---|---|---|---|
| EPI_ISL_572238 | USA/VA-DCLS-1398/2020 | 2020-07-13 | North America/USA/Virginia/ | Virginia DCLS | Virginia DCLS | Virginia DCLS et al |
| EPI_ISL_572241 | USA/VA-DCLS-1404/2020 | 2020-07-13 | North America/USA/Virginia/ | Virginia DCLS | Virginia DCLS | Virginia DCLS et al |
| EPI_ISL_572246 | USA/VA-DCLS-1309/2020 | 2020-07-23 | North America/USA/Virginia/ | Virginia DCLS | Virginia DCLS | Virginia DCLS et al |
| EPI_ISL_572248 | USA/VA-DCLS-1308/2020 | 2020-07-22 | North America/USA/Virginia/ | Virginia DCLS | Virginia DCLS | Virginia DCLS et al |
| EPI_ISL_572250 | USA/VA-DCLS-1315/2020 | 2020-07-23 | North America/USA/Virginia/ | Virginia DCLS | Virginia DCLS | Virginia DCLS et al |
| EPI_ISL_572270 | USA/VA-DCLS-1709/2020 | 2020-04-02 | North America/USA/Virginia/ | Virginia DCLS | Virginia DCLS | Virginia DCLS et al |
| EPI_ISL_572276 | USA/VA-DCLS-1200/2020 | 2020-06-22 | North America/USA/Virginia/ | Virginia DCLS | Virginia DCLS | Virginia DCLS et al |
| EPI_ISL_572277 | USA/VA-DCLS-1203/2020 | 2020-06-23 | North America/USA/Virginia/ | Virginia DCLS | Virginia DCLS | Virginia DCLS et al |
| EPI_ISL_572279 | USA/VA-DCLS-1208/2020 | 2020-06-23 | North America/USA/Virginia/ | Virginia DCLS | Virginia DCLS | Virginia DCLS et al |
| EPI_ISL_572280 | USA/VA-DCLS-1209/2020 | 2020-06-23 | North America/USA/Virginia/ | Virginia DCLS | Virginia DCLS | Virginia DCLS et al |
| EPI_ISL_572293 | USA/VA-DCLS-1277/2020 | 2020-08-12 | North America/USA/Virginia/ | Virginia DCLS | Virginia DCLS | Virginia DCLS et al |
| EPI_ISL_572298 | USA/VA-DCLS-1284/2020 | 2020-08-14 | North America/USA/Virginia/ | Virginia DCLS | Virginia DCLS | Virginia DCLS et al |
| EPI_ISL_572307 | USA/VA-DCLS-1440/2020 | 2020-08-18 | North America/USA/Virginia/ | Virginia DCLS | Virginia DCLS | Virginia DCLS et al |
| EPI_ISL_572310 | USA/VA-DCLS-1443/2020 | 2020-08-18 | North America/USA/Virginia/ | Virginia DCLS | Virginia DCLS | Virginia DCLS et al |
| EPI_ISL_572311 | USA/VA-DCLS-1444/2020 | 2020-08-18 | North America/USA/Virginia/ | Virginia DCLS | Virginia DCLS | Virginia DCLS et al |
| EPI_ISL_572314 | USA/VA-DCLS-1447/2020 | 2020-08-19 | North America/USA/Virginia/ | Virginia DCLS | Virginia DCLS | Virginia DCLS et al |
| EPI_ISL_572320 | Italy/CAM-IZSM-112/2020 | 2020-09-12 | Europe/Italy/Campania/ | IZSM | IZSM | Maurizio Viscardi et al |
| EPI_ISL_572329 | Slovakia/UKBA-101/2020 | 2020-04-03 | Europe/Slovakia/Bratislava/ | Public Health Authority of the Slovak Republic, Bratislava | Faculty of Natural Sciences, Comenius University in Bratislava | Dominika Frifçovv° et al |
| EPI_ISL_572334 | Brazil/PE-IAM109/2020 | 2020-04-08 | South America/Brazil/Pernambuco/Olinc | LACEN/PE | WallauLab, Aggeu Magalhaes Institute | Marcelo Henrique Santos Paiva et al |
| EPI_ISL_572355 | Brazil/PE-IAM766/2020 | 2020-04-26 | South America/Brazil/Pernambuco/Pauli | LACEN/PE | WallauLab, Aggeu Magalhaes Institute | Marcelo Henrique Santos Paiva et al |
| EPI_ISL_572358 | Brazil/PE-IAM889/2020 | 2020-05-03 | South America/Brazil/Pernambuco/Pauli | LACEN/PE | WallauLab, Aggeu Magalhaes Institute | Marcelo Henrique Santos Paiva et al |
| EPI_ISL_572359 | Brazil/PE-IAM900/2020 | 2020-05-03 | South America/Brazil/Pernambuco/Recif | LACEN/PE | WallauLab, Aggeu Magalhaes Institute | Marcelo Henrique Santos Paiva et al |

| | | | | | | |
|---|---|---|---|---|---|---|
| EPI_ISL_572360 | Brazil/PE-IAM914/2020 | 2020-05-03 | South America/Brazil/Pernambuco/Carp | LACEN/PE | WallauLab, Aggeu Magalhaes Institute | Marcelo Henrique Santos Paiva et al |
| EPI_ISL_572361 | Brazil/PE-IAM939/2020 | 2020-05-03 | South America/Brazil/Pernambuco/Cabo | LACEN/PE | WallauLab, Aggeu Magalhaes Institute | Marcelo Henrique Santos Paiva et al |
| EPI_ISL_572363 | Brazil/PE-IAM958/2020 | 2020-05-03 | South America/Brazil/Pernambuco/Ipoju | LACEN/PE | WallauLab, Aggeu Magalhaes Institute | Marcelo Henrique Santos Paiva et al |
| EPI_ISL_572366 | Brazil/PE-IAM961/2020 | 2020-05-02 | South America/Brazil/Pernambuco/Carp | LACEN/PE | WallauLab, Aggeu Magalhaes Institute | Marcelo Henrique Santos Paiva et al |
| EPI_ISL_572367 | Brazil/PE-IAM965/2020 | 2020-05-02 | South America/Brazil/Pernambuco/Caru | LACEN/PE | WallauLab, Aggeu Magalhaes Institute | Marcelo Henrique Santos Paiva et al |
| EPI_ISL_572371 | Brazil/PE-IAM991/2020 | 2020-05-06 | South America/Brazil/Pernambuco/Recif | LACEN/PE | WallauLab, Aggeu Magalhaes Institute | Marcelo Henrique Santos Paiva et al |
| EPI_ISL_572372 | Brazil/PE-IAM992/2020 | 2020-05-06 | South America/Brazil/Pernambuco/Olind | LACEN/PE | WallauLab, Aggeu Magalhaes Institute | Marcelo Henrique Santos Paiva et al |
| EPI_ISL_572373 | Brazil/PE-IAM1026/2020 | 2020-05-06 | South America/Brazil/Pernambuco/Cama | LACEN/PE | WallauLab, Aggeu Magalhaes Institute | Marcelo Henrique Santos Paiva et al |
| EPI_ISL_572375 | Brazil/PE-IAM1082/2020 | 2020-05-03 | South America/Brazil/Pernambuco/Jaqu | LACEN/PE | WallauLab, Aggeu Magalhaes Institute | Marcelo Henrique Santos Paiva et al |
| EPI_ISL_572379 | Brazil/PE-IAM1126/2020 | 2020-05-07 | South America/Brazil/Pernambuco/Carp | LACEN/PE | WallauLab, Aggeu Magalhaes Institute | Marcelo Henrique Santos Paiva et al |
| EPI_ISL_572380 | Brazil/PE-IAM1127/2020 | 2020-05-07 | South America/Brazil/Pernambuco/Recif | LACEN/PE | WallauLab, Aggeu Magalhaes Institute | Marcelo Henrique Santos Paiva et al |
| EPI_ISL_572381 | Brazil/PE-IAM1148/2020 | 2020-05-06 | South America/Brazil/Pernambuco/Vitor | LACEN/PE | WallauLab, Aggeu Magalhaes Institute | Marcelo Henrique Santos Paiva et al |
| EPI_ISL_572382 | Brazil/PE-IAM1149/2020 | 2020-05-06 | South America/Brazil/Pernambuco/Cha ( | LACEN/PE | WallauLab, Aggeu Magalhaes Institute | Marcelo Henrique Santos Paiva et al |

| | | | | | | |
|---|---|---|---|---|---|---|
| EPI_ISL_572384 | Brazil/PE-IAM1254/2020 | 2020-05-09 | South America/Brazil/Pernambuco/Caru | LACEN/PE | WallauLab, Aggeu Magalhaes Institute | Marcelo Henrique Santos Paiva et al |
| EPI_ISL_572385 | Brazil/PE-IAM1264/2020 | 2020-05-10 | South America/Brazil/Pernambuco/Caru | LACEN/PE | WallauLab, Aggeu Magalhaes Institute | Marcelo Henrique Santos Paiva et al |
| EPI_ISL_572386 | Brazil/PE-IAM1309/2020 | 2020-05-10 | South America/Brazil/Pernambuco/Ipoju | LACEN/PE | WallauLab, Aggeu Magalhaes Institute | Marcelo Henrique Santos Paiva et al |
| EPI_ISL_572388 | Brazil/PE-IAM1315/2020 | 2020-05-11 | South America/Brazil/Pernambuco/Prim | LACEN/PE | WallauLab, Aggeu Magalhaes Institute | Marcelo Henrique Santos Paiva et al |
| EPI_ISL_572394 | Brazil/PE-IAM1447/2020 | 2020-05-13 | South America/Brazil/Pernambuco/Goia | LACEN/PE | WallauLab, Aggeu Magalhaes Institute | Marcelo Henrique Santos Paiva et al |
| EPI_ISL_572396 | Brazil/PE-IAM1468/2020 | 2020-05-14 | South America/Brazil/Pernambuco/Pauli | LACEN/PE | WallauLab, Aggeu Magalhaes Institute | Marcelo Henrique Santos Paiva et al |
| EPI_ISL_572398 | Russia/Moscow-PMVL-12/2020 | 2020-05-18 | Europe/Russia/Moscow Oblast/ | Laboratory of Biology and Identification of Arboviruses | Pathogenic Microorganisms Variability Laboratory | Alexey Shchetinin et al |
| EPI_ISL_572814 | England/NORW-EF5CC/2020 | 2020-09-25 | Europe/United Kingdom/England/ | Quadram Institute Bioscience | COVID-19 Genomics UK (COG-UK) Consortium | Dave J. Baker et al |
| EPI_ISL_572826 | England/ALDP-9CCBBB/2020 | 2020-09-16 | Europe/United Kingdom/England/ | Quadram Institute Bioscience | COVID-19 Genomics UK (COG-UK) Consortium | Dave J. Baker et al |
| EPI_ISL_572943 | England/NOTT-113889/2020 | 2020-09-15 | Europe/United Kingdom/England/ | Lincolnshire Hospitals and DeepSeq Nottingham | COVID-19 Genomics UK (COG-UK) Consortium | Nichola Duckworth et al |
| EPI_ISL_572991 | England/ALDP-9B61D0/2020 | 2020-08-28 | Europe/United Kingdom/England/ | Oxford Viromics, NDM, University of Oxford; Oxford University Hospitals; Basingstoke and North Hampshire Hospital | COVID-19 Genomics UK (COG-UK) Consortium | Tanya Golubchik et al |
| EPI_ISL_573058 | England/ALDP-9BCBAF/2020 | 2020-09-09 | Europe/United Kingdom/England/ | Oxford Viromics, NDM, University of Oxford; Oxford University Hospitals; Basingstoke and North Hampshire Hospital | COVID-19 Genomics UK (COG-UK) Consortium | Tanya Golubchik et al |
| EPI_ISL_573083 | England/ALDP-9BCF58/2020 | 2020-09-10 | Europe/United Kingdom/England/ | Oxford Viromics, NDM, University of Oxford; Oxford University Hospitals; Basingstoke and North Hampshire Hospital | COVID-19 Genomics UK (COG-UK) Consortium | Tanya Golubchik et al |

| | | | | | | |
|---|---|---|---|---|---|---|
| EPI_ISL_573114 | England/ALDP-9CC5AE/2020 | 2020-09-16 | Europe/United Kingdom/England/ | Quadram Institute Bioscience | COVID-19 Genomics UK (COG-UK) Consortium | Dave J. Baker et al |
| EPI_ISL_573179 | England/CAMC-9C0319/2020 | 2020-09-08 | Europe/United Kingdom/England/ | Oxford Viromics, NDM, University of Oxford; Oxford University Hospitals; Basingstoke and North Hampshire Hospital | COVID-19 Genomics UK (COG-UK) Consortium | Tanya Golubchik et al |
| EPI_ISL_573234 | England/CAMC-9C1D92/2020 | 2020-09-09 | Europe/United Kingdom/England/ | Oxford Viromics, NDM, University of Oxford; Oxford University Hospitals; Basingstoke and North Hampshire Hospital | COVID-19 Genomics UK (COG-UK) Consortium | Tanya Golubchik et al |
| EPI_ISL_573423 | England/NOTT-113722/2020 | 2020-09-21 | Europe/United Kingdom/England/ | Queens Medical Centre, Clinical Microbiology Department / DeepSeq Nottingham | COVID-19 Genomics UK (COG-UK) Consortium | Gemma Clark et al |
| EPI_ISL_573481 | England/QEUH-9AEB46/2020 | 2020-09-03 | Europe/United Kingdom/England/ | University College London, Great Ormond Street Hospital for Children NHS Foundation Trust, Imperial College Healthcare NHS Trust | COVID-19 Genomics UK (COG-UK) Consortium | Sergi Castellano et al |
| EPI_ISL_573519 | England/QEUH-9AEF86/2020 | 2020-09-03 | Europe/United Kingdom/England/ | University College London, Great Ormond Street Hospital for Children NHS Foundation Trust, Imperial College Healthcare NHS Trust | COVID-19 Genomics UK (COG-UK) Consortium | Sergi Castellano et al |
| EPI_ISL_573581 | England/QEUH-9BA136/2020 | 2020-09-07 | Europe/United Kingdom/England/ | Oxford Viromics, NDM, University of Oxford; Oxford University Hospitals; Basingstoke and North Hampshire Hospital | COVID-19 Genomics UK (COG-UK) Consortium | Tanya Golubchik et al |
| EPI_ISL_573687 | England/SHEF-C0AFF/2020 | 2020-09-27 | Europe/United Kingdom/England/ | Virology Department, Sheffield Teaching Hospitals NHS Foundation Trust/Department of Infection, Immunity and Cardiovascular Disease, The Medical School, University of Sheffield | COVID-19 Genomics UK (COG-UK) Consortium | Thushan de Silva et al |
| EPI_ISL_573901 | Wales/PHWC-16B7A3/2020 | 2020-09-15 | Europe/United Kingdom/Wales/ | Wales Specialist Virology Centre Sequencing lab: Pathogen Genomics Unit | COVID-19 Genomics UK (COG-UK) Consortium | Catherine Moore et al |
| EPI_ISL_573918 | Wales/PHWC-16B925/2020 | 2020-09-17 | Europe/United Kingdom/Wales/ | Wales Specialist Virology Centre Sequencing lab: Pathogen Genomics Unit | COVID-19 Genomics UK (COG-UK) Consortium | Catherine Moore et al |
| EPI_ISL_574261 | USA/NM-DOH-2020313047/2020 | 2020-08-26 | North America/USA/New Mexico/ | New Mexico Department of Health Scientific Laboratory | New Mexico Department of Health Scientific Laboratory | Ellie Johnson et al |

| | | | | | | |
|---|---|---|---|---|---|---|
| EPI_ISL_574267 | USA/NM-DOH-2020313314/2020 | 2020-08-26 | North America/USA/New Mexico/ | New Mexico Department of Health Scientific Laboratory | New Mexico Department of Health Scientific Laboratory | Ellie Johnson et al |
| EPI_ISL_574269 | USA/NM-DOH-2020313398/2020 | 2020-08-26 | North America/USA/New Mexico/ | New Mexico Department of Health Scientific Laboratory | New Mexico Department of Health Scientific Laboratory | Ellie Johnson et al |
| EPI_ISL_574270 | USA/NM-DOH-2020313467/2020 | 2020-08-26 | North America/USA/New Mexico/ | New Mexico Department of Health Scientific Laboratory | New Mexico Department of Health Scientific Laboratory | Ellie Johnson et al |
| EPI_ISL_574273 | USA/NM-DOH-2020313625/2020 | 2020-08-26 | North America/USA/New Mexico/ | New Mexico Department of Health Scientific Laboratory | New Mexico Department of Health Scientific Laboratory | Ellie Johnson et al |
| EPI_ISL_574274 | USA/NM-DOH-2020313776/2020 | 2020-08-26 | North America/USA/New Mexico/ | New Mexico Department of Health Scientific Laboratory | New Mexico Department of Health Scientific Laboratory | Ellie Johnson et al |
| EPI_ISL_574276 | USA/NM-DOH-2020314787/2020 | 2020-08-26 | North America/USA/New Mexico/ | New Mexico Department of Health Scientific Laboratory | New Mexico Department of Health Scientific Laboratory | Ellie Johnson et al |
| EPI_ISL_574277 | USA/NM-DOH-2020314819/2020 | 2020-08-25 | North America/USA/New Mexico/ | New Mexico Department of Health Scientific Laboratory | New Mexico Department of Health Scientific Laboratory | Ellie Johnson et al |
| EPI_ISL_574280 | USA/NM-DOH-2020314850/2020 | 2020-08-25 | North America/USA/New Mexico/ | New Mexico Department of Health Scientific Laboratory | New Mexico Department of Health Scientific Laboratory | Ellie Johnson et al |
| EPI_ISL_574281 | USA/NM-DOH-2020314875/2020 | 2020-08-26 | North America/USA/New Mexico/ | New Mexico Department of Health Scientific Laboratory | New Mexico Department of Health Scientific Laboratory | Ellie Johnson et al |
| EPI_ISL_574282 | USA/NM-DOH-2020314888/2020 | 2020-08-25 | North America/USA/New Mexico/ | New Mexico Department of Health Scientific Laboratory | New Mexico Department of Health Scientific Laboratory | Ellie Johnson et al |

| | | | | | | |
|---|---|---|---|---|---|---|
| EPI_ISL_574288 | USA/NM-DOH-2020315317/2020 | 2020-08-27 | North America/USA/New Mexico/ | New Mexico Department of Health Scientific Laboratory | New Mexico Department of Health Scientific Laboratory | Ellie Johnson et al |
| EPI_ISL_574289 | USA/NM-DOH-2020315508/2020 | 2020-08-27 | North America/USA/New Mexico/ | New Mexico Department of Health Scientific Laboratory | New Mexico Department of Health Scientific Laboratory | Ellie Johnson et al |
| EPI_ISL_574292 | Ecuador/2343/2020 | 2020-03-16 | South America/Ecuador/Guayas/Guayac | Instituto Nacional de Investigaci√≥n en Salud P√∫blica | Instituto Nacional de Investigaci√≥n en Salud P√∫blica | Leandro Patino Patino et al |
| EPI_ISL_574293 | Ecuador/2512/2020 | 2020-03-22 | South America/Ecuador/Guayas/Guayac | Instituto Nacional de Investigaci√≥n en Salud P√∫blica | Instituto Nacional de Investigaci√≥n en Salud Publica | Leandro Patino Patino et al |
| EPI_ISL_574294 | Ecuador/2479/2020 | 2020-03-23 | South America/Ecuador/Guayas/Guayac | Instituto Nacional de Investigacion en Salud P√∫blica | Instituto Nacional de Investigaci√≥n en Salud P√∫blica | Leandro Patino Patino et al |
| EPI_ISL_574295 | USA/LA-EVTL160/2020 | 2020-05-11 | North America/USA/Louisiana/Caddo Pa | LSUHS Emerging Viral Threat Laboratory | Microbial Genome Sequencing Center | Jeremy P. Kamil et al |
| EPI_ISL_574299 | USA/LA-EVTL164/2020 | 2020-05-11 | North America/USA/Louisiana/Caddo Pa | LSUHS Emerging Viral Threat Laboratory | Microbial Genome Sequencing Center | Jeremy P. Kamil et al |
| EPI_ISL_574314 | USA/LA-EVTL179/2020 | 2020-05-14 | North America/USA/Louisiana/Rapides P | LSUHS Emerging Viral Threat Laboratory | Microbial Genome Sequencing Center | Rona S. Scott et al |
| EPI_ISL_574318 | USA/LA-EVTL183/2020 | 2020-07-19 | North America/USA/Louisiana/Caddo Pa | LSUHS Emerging Viral Threat Laboratory | Microbial Genome Sequencing Center | Rona S. Scott et al |
| EPI_ISL_574319 | USA/LA-EVTL184/2020 | 2020-07-20 | North America/USA/Louisiana/Webster | LSUHS Emerging Viral Threat Laboratory | Microbial Genome Sequencing Center | Rona S. Scott et al |
| EPI_ISL_574329 | USA/LA-EVTL195/2020 | 2020-08-11 | North America/USA/Louisiana/Bienville | LSUHS Emerging Viral Threat Laboratory | Microbial Genome Sequencing Center | Jeremy P. Kamil et al |
| EPI_ISL_574331 | USA/LA-EVTL197/2020 | 2020-08-11 | North America/USA/Louisiana/Red Rive | LSUHS Emerging Viral Threat Laboratory | Microbial Genome Sequencing Center | Jeremy P. Kamil et al |
| EPI_ISL_574332 | USA/LA-EVTL198/2020 | 2020-08-11 | North America/USA/Louisiana/Webster | LSUHS Emerging Viral Threat Laboratory | Microbial Genome Sequencing Center | Jeremy P. Kamil et al |

| | | | | | | |
|---|---|---|---|---|---|---|
| EPI_ISL_574334 | USA/LA-EVTL200/2020 | 2020-08-11 | North America/USA/Louisiana/Webster | LSUHS Emerging Viral Threat Laboratory | Microbial Genome Sequencing Center | Jeremy P. Kamil et al |
| EPI_ISL_574335 | USA/LA-EVTL201/2020 | 2020-08-11 | North America/USA/Louisiana/Webster | LSUHS Emerging Viral Threat Laboratory | Microbial Genome Sequencing Center | Jeremy P. Kamil et al |
| EPI_ISL_574339 | USA/LA-EVTL205/2020 | 2020-08-11 | North America/USA/Louisiana/Webster | LSUHS Emerging Viral Threat Laboratory | Microbial Genome Sequencing Center | Jeremy P. Kamil et al |
| EPI_ISL_574340 | USA/LA-EVTL206/2020 | 2020-08-11 | North America/USA/Louisiana/Webster | LSUHS Emerging Viral Threat Laboratory | Microbial Genome Sequencing Center | Jeremy P. Kamil et al |
| EPI_ISL_574347 | USA/LA-EVTL213/2020 | 2020-08-15 | North America/USA/Louisiana/Webster | LSUHS Emerging Viral Threat Laboratory | Microbial Genome Sequencing Center | Jeremy P. Kamil et al |
| EPI_ISL_574348 | USA/LA-EVTL214/2020 | 2020-08-15 | North America/USA/Louisiana/Webster | LSUHS Emerging Viral Threat Laboratory | Microbial Genome Sequencing Center | Jeremy P. Kamil et al |
| EPI_ISL_574354 | USA/LA-EVTL220/2020 | 2020-08-22 | North America/USA/Louisiana/Madison | LSUHS Emerging Viral Threat Laboratory | Microbial Genome Sequencing Center | Jeremy P. Kamil et al |
| EPI_ISL_574357 | USA/LA-EVTL223/2020 | 2020-08-22 | North America/USA/Louisiana/Madison | LSUHS Emerging Viral Threat Laboratory | Microbial Genome Sequencing Center | Jeremy P. Kamil et al |
| EPI_ISL_574359 | USA/LA-EVTL225/2020 | 2020-08-22 | North America/USA/Louisiana/Madison | LSUHS Emerging Viral Threat Laboratory | Microbial Genome Sequencing Center | Jeremy P. Kamil et al |
| EPI_ISL_574360 | USA/LA-EVTL226/2020 | 2020-08-22 | North America/USA/Louisiana/Madison | LSUHS Emerging Viral Threat Laboratory | Microbial Genome Sequencing Center | Jeremy P. Kamil et al |
| EPI_ISL_574361 | USA/LA-EVTL227/2020 | 2020-08-22 | North America/USA/Louisiana/Madison | LSUHS Emerging Viral Threat Laboratory | Microbial Genome Sequencing Center | Jeremy P. Kamil et al |
| EPI_ISL_574365 | USA/LA-EVTL231/2020 | 2020-08-22 | North America/USA/Louisiana/Madison | LSUHS Emerging Viral Threat Laboratory | Microbial Genome Sequencing Center | Jeremy P. Kamil et al |
| EPI_ISL_574370 | USA/LA-EVTL236/2020 | 2020-08-22 | North America/USA/Louisiana/Madison | LSUHS Emerging Viral Threat Laboratory | Microbial Genome Sequencing Center | Jeremy P. Kamil et al |

| | | | | | | |
|---|---|---|---|---|---|---|
| EPI_ISL_574371 | USA/LA-EVTL237/2020 | 2020-08-22 | North America/USA/Louisiana/Madison | LSUHS Emerging Viral Threat Laboratory | Microbial Genome Sequencing Center | Jeremy P. Kamil et al |
| EPI_ISL_574372 | USA/LA-EVTL238/2020 | 2020-08-22 | North America/USA/Louisiana/Madison | LSUHS Emerging Viral Threat Laboratory | Microbial Genome Sequencing Center | Jeremy P. Kamil et al |
| EPI_ISL_574377 | USA/LA-EVTL243/2020 | 2020-08-22 | North America/USA/Louisiana/Madison | LSUHS Emerging Viral Threat Laboratory | Microbial Genome Sequencing Center | Jeremy P. Kamil et al |
| EPI_ISL_574379 | USA/LA-EVTL245/2020 | 2020-08-22 | North America/USA/Louisiana/Madison | LSUHS Emerging Viral Threat Laboratory | Microbial Genome Sequencing Center | Jeremy P. Kamil et al |
| EPI_ISL_574380 | USA/LA-EVTL246/2020 | 2020-08-22 | North America/USA/Louisiana/Madison | LSUHS Emerging Viral Threat Laboratory | Microbial Genome Sequencing Center | Jeremy P. Kamil et al |
| EPI_ISL_574381 | USA/LA-EVTL247/2020 | 2020-08-22 | North America/USA/Louisiana/Madison | LSUHS Emerging Viral Threat Laboratory | Microbial Genome Sequencing Center | Jeremy P. Kamil et al |
| EPI_ISL_574386 | USA/LA-EVTL252/2020 | 2020-08-22 | North America/USA/Louisiana/Madison | LSUHS Emerging Viral Threat Laboratory | Microbial Genome Sequencing Center | Jeremy P. Kamil et al |
| EPI_ISL_574390 | USA/LA-EVTL256/2020 | 2020-08-22 | North America/USA/Louisiana/Madison | LSUHS Emerging Viral Threat Laboratory | Microbial Genome Sequencing Center | Jeremy P. Kamil et al |
| EPI_ISL_574392 | USA/LA-EVTL258/2020 | 2020-08-22 | North America/USA/Louisiana/Madison | LSUHS Emerging Viral Threat Laboratory | Microbial Genome Sequencing Center | Jeremy P. Kamil et al |
| EPI_ISL_574394 | USA/LA-EVTL260/2020 | 2020-08-22 | North America/USA/Louisiana/Madison | LSUHS Emerging Viral Threat Laboratory | Microbial Genome Sequencing Center | Jeremy P. Kamil et al |
| EPI_ISL_574396 | USA/LA-EVTL262/2020 | 2020-08-22 | North America/USA/Louisiana/Madison | LSUHS Emerging Viral Threat Laboratory | Microbial Genome Sequencing Center | Jeremy P. Kamil et al |
| EPI_ISL_574398 | USA/LA-EVTL264/2020 | 2020-08-22 | North America/USA/Louisiana/Madison | LSUHS Emerging Viral Threat Laboratory | Microbial Genome Sequencing Center | Jeremy P. Kamil et al |
| EPI_ISL_574403 | USA/LA-EVTL269/2020 | 2020-08-22 | North America/USA/Louisiana/Madison | LSUHS Emerging Viral Threat Laboratory | Microbial Genome Sequencing Center | Jeremy P. Kamil et al |

| | | | | | | |
|---|---|---|---|---|---|---|
| EPI_ISL_574412 | USA/LA-EVTL278/2020 | 2020-08-26 | North America/USA/Louisiana/Madison | LSUHS Emerging Viral Threat Laboratory | Microbial Genome Sequencing Center | Jeremy P. Kamil et al |
| EPI_ISL_574414 | USA/LA-EVTL280/2020 | 2020-08-26 | North America/USA/Louisiana/Madison | LSUHS Emerging Viral Threat Laboratory | Microbial Genome Sequencing Center | Jeremy P. Kamil et al |
| EPI_ISL_574416 | USA/LA-EVTL282/2020 | 2020-08-26 | North America/USA/Louisiana/Madison | LSUHS Emerging Viral Threat Laboratory | Microbial Genome Sequencing Center | Jeremy P. Kamil et al |
| EPI_ISL_574421 | USA/LA-EVTL287/2020 | 2020-08-26 | North America/USA/Louisiana/Madison | LSUHS Emerging Viral Threat Laboratory | Microbial Genome Sequencing Center | Jeremy P. Kamil et al |
| EPI_ISL_574424 | USA/LA-EVTL290/2020 | 2020-08-26 | North America/USA/Louisiana/Madison | LSUHS Emerging Viral Threat Laboratory | Microbial Genome Sequencing Center | Jeremy P. Kamil et al |
| EPI_ISL_574429 | USA/LA-EVTL295/2020 | 2020-08-26 | North America/USA/Louisiana/Madison | LSUHS Emerging Viral Threat Laboratory | Microbial Genome Sequencing Center | Jeremy P. Kamil et al |
| EPI_ISL_574430 | USA/LA-EVTL296/2020 | 2020-08-26 | North America/USA/Louisiana/Madison | LSUHS Emerging Viral Threat Laboratory | Microbial Genome Sequencing Center | Jeremy P. Kamil et al |
| EPI_ISL_574433 | Indonesia/JK-NIHRD-C0027475/2020 | 2020-07-03 | Asia/Indonesia/Jakarta/ | National Institute of Health Research and Development | National Institute of Health Research and Development | Pawestri et al |
| EPI_ISL_574437 | USA/TX-DSHS-0532/2020 | 2020-05-14 | North America/USA/Texas/Harris Count | Texas Department of State Health Services | Texas Department of State Health Services | Rashmi Tuladhar et al |
| EPI_ISL_574441 | USA/TX-DSHS-0536/2020 | 2020-05-14 | North America/USA/Texas/Harris Count | Texas Department of State Health Services | Texas Department of State Health Services | Rashmi Tuladhar et al |
| EPI_ISL_574449 | USA/TX-DSHS-0545/2020 | 2020-05-14 | North America/USA/Texas/Harris Count | Texas Department of State Health Services | Texas Department of State Health Services | Rashmi Tuladhar et al |
| EPI_ISL_574452 | USA/TX-DSHS-0548/2020 | 2020-05-14 | North America/USA/Texas/Harris Count | Texas Department of State Health Services | Texas Department of State Health Services | Rashmi Tuladhar et al |
| EPI_ISL_574458 | USA/TX-DSHS-0555/2020 | 2020-05-14 | North America/USA/Texas/Harris Count | Texas Department of State Health Services | Texas Department of State Health Services | Rashmi Tuladhar et al |

| | | | | | | |
|---|---|---|---|---|---|---|
| EPI_ISL_574461 | USA/TX-DSHS-0558/2020 | 2020-05-14 | North America/USA/Texas/Harris Count | Texas Department of State Health Services | Texas Department of State Health Services | Rashmi Tuladhar et al |
| EPI_ISL_574466 | USA/TX-DSHS-0564/2020 | 2020-05-14 | North America/USA/Texas/Harris Count | Texas Department of State Health Services | Texas Department of State Health Services | Rashmi Tuladhar et al |
| EPI_ISL_574506 | Singapore/1021/2020 | 2020-03-24 | Asia/Singapore/Singapore/ | National Public Health Laboratory, National Centre for Infectious Diseases | National Public Health Laboratory, National Centre for Infectious Diseases | Tze Minn Mak et al |
| EPI_ISL_574519 | Singapore/1034/2020 | 2020-09-28 | Asia/Singapore/Singapore/ | National Public Health Laboratory, National Centre for Infectious Diseases | National Public Health Laboratory, National Centre for Infectious Diseases | Tze Minn Mak et al |
| EPI_ISL_574544 | USA/SC-DHEC-1150/2020 | 2020-07-10 | North America/USA/South Carolina/Hor | Microbiology Division, South Carolina Department of Health and Environmental Control | Microbiology Division, South Carolina Department of Health and Environmental Control | Flores et al |
| EPI_ISL_574550 | USA/SC-DHEC-0543/2020 | 2020-06-19 | North America/USA/South Carolina/Geo | Microbiology Division, South Carolina Department of Health and Environmental Control | Microbiology Division, South Carolina Department of Health and Environmental Control | Flores et al |
| EPI_ISL_574561 | USA/SC-DHEC-1138/2020 | 2020-07-10 | North America/USA/South Carolina/Geo | Microbiology Division, South Carolina Department of Health and Environmental Control | Microbiology Division, South Carolina Department of Health and Environmental Control | Flores et al |

| | | | | | | |
|---|---|---|---|---|---|---|
| EPI_ISL_574562 | USA/SC-DHEC-1139/2020 | 2020-07-10 | North America/USA/South Carolina/Geo | Microbiology Division, South Carolina Department of Health and Environmental Control | Microbiology Division, South Carolina Department of Health and Environmental Control | Flores et al |
| EPI_ISL_574567 | USA/SC-DHEC-1109/2020 | 2020-07-10 | North America/USA/South Carolina/Geo | Microbiology Division, South Carolina Department of Health and Environmental Control | Microbiology Division, South Carolina Department of Health and Environmental Control | Flores et al |
| EPI_ISL_574586 | USA/DE-DHSS-F917680/2020 | 2020-03-23 | North America/USA/Delaware/New Cas | Delaware Public Health Lab | Delaware Public Health Lab | Gregory Hovan et al |
| EPI_ISL_574592 | Brazil/SP-560/2020 | 2020-03-31 | South America/Brazil/S√£o Paulo/Presid | Hospital Domingos Leonardo Ceravolo Presidente Prudente | Instituto Adolfo Lutz, Interdiciplinary Procedures Center, Strategic Laboratory | Claudio Tavares Sacchi et al |
| EPI_ISL_574594 | Brazil/SP-425/2020 | 2020-07-10 | South America/Brazil/S√£o Paulo/Tauba | Hospital Escola da Universidade de Taubate | Instituto Adolfo Lutz, Interdiciplinary Procedures Center, Strategic Laboratory | Claudio Tavares Sacchi et al |
| EPI_ISL_574596 | Brazil/SP-388/2020 | 2020-06-19 | South America/Brazil/S√£o Paulo/Sumar | CS II Dr. Antonio Vicoso Moreira de Rezende Sumare | Instituto Adolfo Lutz, Interdiciplinary Procedures Center, Strategic Laboratory | Claudio Tavares Sacchi et al |
| EPI_ISL_574597 | Brazil/SP-389/2020 | 2020-06-18 | South America/Brazil/S√£o Paulo/Jarinu | Secretaria Municipal de Saude de Jarinu | Instituto Adolfo Lutz, Interdiciplinary Procedures Center, Strategic Laboratory | Claudio Tavares Sacchi et al |

| | | | | | | |
|---|---|---|---|---|---|---|
| EPI_ISL_574598 | Brazil/SP-391/2020 | 2020-06-20 | South America/Brazil/S√£o Paulo/Jundia | Servico de Verificacao de Obito SVO | Instituto Adolfo Lutz, Interdiciplinary Procedures Center, Strategic Laboratory | Claudio Tavares Sacchi et al |
| EPI_ISL_574604 | Indonesia/SA-EIJK13/2020 | 2020-03-24 | Asia/Indonesia/North Sulawesi/Manado | RSUP Prof. Dr. R. Kandou Manado | Eijkman Institute for Molecular Biology, Ministry of Research and Technology/National Agency for Research and Innovation | Frilasita A Yudhaputri et al |
| EPI_ISL_574611 | Indonesia/BT-EIJK36/2020 | 2020-05-06 | Asia/Indonesia/Banten/South Tangerang | RS Premier Bintaro | Eijkman Institute for Molecular Biology, Ministry of Research and Technology/National Agency for Research and Innovation | Frilasita A Yudhaputri et al |
| EPI_ISL_574612 | Indonesia/JB-EIJK41/2020 | 2020-05-31 | Asia/Indonesia/West Java/Bogor Regen | RS Hermina Mekarsari | Eijkman Institute for Molecular Biology, Ministry of Research and Technology/National Agency for Research and Innovation | Frilasita A Yudhaputri et al |
| EPI_ISL_574613 | Indonesia/JK-EIJK42/2020 | 2020-06-03 | Asia/Indonesia/Jakarta/ | RS Harapan Bunda | Eijkman Institute for Molecular Biology, Ministry of Research and Technology/National Agency for Research and Innovation | Frilasita A Yudhaputri et al |

| | | | | | | |
|---|---|---|---|---|---|---|
| EPI_ISL_574615 | Indonesia/JK-EIJK46/2020 | 2020-06-05 | Asia/Indonesia/Jakarta/ | RS Harapan Bunda | Eijkman Institute for Molecular Biology, Ministry of Research and Technology/National Agency for Research and Innovation | Frilasita A Yudhaputri et al |
| EPI_ISL_574617 | Indonesia/JK-EIJK48/2020 | 2020-06-08 | Asia/Indonesia/Jakarta/ | RS Kramat 128 | Eijkman Institute for Molecular Biology, Ministry of Research and Technology/National Agency for Research and Innovation | Frilasita A Yudhaputri et al |
| EPI_ISL_574619 | Indonesia/SA-EIJK50/2020 | 2020-06-16 | Asia/Indonesia/North Sulawesi/Manado | BTKLPP Kelas I Manado | Eijkman Institute for Molecular Biology, Ministry of Research and Technology/National Agency for Research and Innovation | Hidayat Trimarsanto et al |
| EPI_ISL_574620 | Indonesia/SA-EIJK51/2020 | 2020-06-17 | Asia/Indonesia/North Sulawesi/Manado | BTKLPP Kelas I Manado | Eijkman Institute for Molecular Biology, Ministry of Research and Technology/National Agency for Research and Innovation | Hidayat Trimarsanto et al |
| EPI_ISL_574621 | Indonesia/SA-EIJK52/2020 | 2020-06-16 | Asia/Indonesia/North Sulawesi/Manado | BTKLPP Kelas I Manado | Eijkman Institute for Molecular Biology, Ministry of Research and Technology/National Agency for Research and Innovation | Hidayat Trimarsanto et al |

| | | | | | | |
|---|---|---|---|---|---|---|
| EPI_ISL_574623 | Indonesia/SA-EIJK54/2020 | 2020-06-18 | Asia/Indonesia/North Sulawesi/Manado | BTKLPP Kelas I Manado | Eijkman Institute for Molecular Biology, Ministry of Research and Technology/National Agency for Research and Innovation | Hidayat Trimarsanto et al |
| EPI_ISL_574657 | USA/WA-S2815/2020 | 2020-09-05 | North America/USA/Washington/King C | Seattle Flu Study | Seattle Flu Study | Deborah A. Nickerson et al |
| EPI_ISL_574659 | USA/WA-S2817/2020 | 2020-09-05 | North America/USA/Washington/King C | Seattle Flu Study | Seattle Flu Study | Deborah A. Nickerson et al |
| EPI_ISL_574662 | USA/WA-S2820/2020 | 2020-09-09 | North America/USA/Washington/King C | Seattle Flu Study | Seattle Flu Study | Deborah A. Nickerson et al |
| EPI_ISL_574663 | USA/WA-S2821/2020 | 2020-09-09 | North America/USA/Washington/ | Seattle Flu Study | Seattle Flu Study | Deborah A. Nickerson et al |
| EPI_ISL_574664 | USA/WA-S2822/2020 | 2020-09-09 | North America/USA/Washington/ | Seattle Flu Study | Seattle Flu Study | Deborah A. Nickerson et al |
| EPI_ISL_574666 | USA/WA-S2824/2020 | 2020-09-09 | North America/USA/Washington/ | Seattle Flu Study | Seattle Flu Study | Deborah A. Nickerson et al |
| EPI_ISL_574668 | USA/WA-S2826/2020 | 2020-09-09 | North America/USA/Washington/King C | Seattle Flu Study | Seattle Flu Study | Deborah A. Nickerson et al |
| EPI_ISL_574669 | USA/WA-S2827/2020 | 2020-09-07 | North America/USA/Washington/King C | Seattle Flu Study | Seattle Flu Study | Deborah A. Nickerson et al |
| EPI_ISL_574673 | USA/WA-S2831/2020 | 2020-09-08 | North America/USA/Washington/King C | Seattle Flu Study | Seattle Flu Study | Deborah A. Nickerson et al |
| EPI_ISL_574687 | USA/WA-S2845/2020 | 2020-09-16 | North America/USA/Washington/King C | Seattle Flu Study | Seattle Flu Study | Deborah A. Nickerson et al |
| EPI_ISL_574689 | USA/WA-S2847/2020 | 2020-09-18 | North America/USA/Washington/King C | Seattle Flu Study | Seattle Flu Study | Deborah A. Nickerson et al |
| EPI_ISL_574690 | USA/WA-S2848/2020 | 2020-09-18 | North America/USA/Washington/King C | Seattle Flu Study | Seattle Flu Study | Deborah A. Nickerson et al |
| EPI_ISL_574876 | Switzerland/AG-ETHZ-100193/2020 | 2020-03-17 | Europe/Switzerland/Aargau/ | Viollier AG | Department of Biosystems Science and Engineering, ETH Zvºrich | Christian Beisel et al |
| EPI_ISL_574886 | Switzerland/BL-ETHZ-100236/2020 | 2020-03-18 | Europe/Switzerland/Basel-Land/ | Viollier AG | Department of Biosystems Science and Engineering, ETH Zvºrich | Christian Beisel et al |
| EPI_ISL_574917 | Switzerland/BE-ETHZ-100216/2020 | 2020-03-17 | Europe/Switzerland/Bern/ | Viollier AG | Department of Biosystems Science and Engineering, ETH Zvºrich | Christian Beisel et al |

| | | | | | | |
|---|---|---|---|---|---|---|
| EPI_ISL_574934 | Switzerland/GE-ETHZ-100189/2020 | 2020-03-16 | Europe/Switzerland/Geneva/ | Viollier AG | Department of Biosystems Science and Engineering, ETH Z√ºrich | Christian Beisel et al |
| EPI_ISL_574950 | Switzerland/LU-ETHZ-100312/2020 | 2020-03-18 | Europe/Switzerland/Lucerne/ | Viollier AG | Department of Biosystems Science and Engineering, ETH Z√ºrich | Christian Beisel et al |
| EPI_ISL_574958 | Switzerland/TI-ETHZ-100170/2020 | 2020-03-16 | Europe/Switzerland/Ticino/ | Viollier AG | Department of Biosystems Science and Engineering, ETH Z√ºrich | Christian Beisel et al |
| EPI_ISL_574988 | Switzerland/VD-ETHZ-100181/2020 | 2020-03-16 | Europe/Switzerland/Vaud/ | Viollier AG | Department of Biosystems Science and Engineering, ETH Z√ºrich | Christian Beisel et al |
| EPI_ISL_574993 | Switzerland/VD-ETHZ-100198/2020 | 2020-03-17 | Europe/Switzerland/Vaud/ | Viollier AG | Department of Biosystems Science and Engineering, ETH Z√ºrich | Christian Beisel et al |
| EPI_ISL_574995 | Switzerland/VD-ETHZ-100220/2020 | 2020-03-17 | Europe/Switzerland/Vaud/ | Viollier AG | Department of Biosystems Science and Engineering, ETH Z√ºrich | Christian Beisel et al |
| EPI_ISL_574999 | Switzerland/VD-ETHZ-100232/2020 | 2020-03-17 | Europe/Switzerland/Vaud/ | Viollier AG | Department of Biosystems Science and Engineering, ETH Z√ºrich | Christian Beisel et al |
| EPI_ISL_575028 | USA/WA-S2850/2020 | 2020-09-01 | North America/USA/Washington/King C | Seattle Flu Study | Seattle Flu Study | Deborah A. Nickerson et al |
| EPI_ISL_575029 | USA/WA-S2851/2020 | 2020-08-31 | North America/USA/Washington/King C | Seattle Flu Study | Seattle Flu Study | Deborah A. Nickerson et al |
| EPI_ISL_575031 | USA/WA-S2853/2020 | 2020-08-31 | North America/USA/Washington/King C | Seattle Flu Study | Seattle Flu Study | Deborah A. Nickerson et al |
| EPI_ISL_575045 | USA/UT-UPHL-01382/2020 | 2020-03-31 | North America/USA/Utah/ | Utah Public Health Laboratory | Utah Public Health Laboratory | Erin Young et al |

| | | | | | | |
|---|---|---|---|---|---|---|
| EPI_ISL_575049 | USA/UT-UPHL-01396/2020 | 2020-04-04 | North America/USA/Utah/ | Utah Public Health Laboratory | Utah Public Health Laboratory | Erin Young et al |
| EPI_ISL_575054 | USA/UT-UPHL-01407/2020 | 2020-04-03 | North America/USA/Utah/ | Utah Public Health Laboratory | Utah Public Health Laboratory | Erin Young et al |
| EPI_ISL_575057 | USA/UT-UPHL-01415/2020 | 2020-04-01 | North America/USA/Utah/ | Utah Public Health Laboratory | Utah Public Health Laboratory | Erin Young et al |
| EPI_ISL_575063 | USA/UT-UPHL-01637/2020 | 2020-04-07 | North America/USA/Utah/ | Utah Public Health Laboratory | Utah Public Health Laboratory | Erin Young et al |
| EPI_ISL_575078 | USA/UT-UPHL-02799/2020 | 2020-04-25 | North America/USA/Utah/ | Utah Public Health Laboratory | Utah Public Health Laboratory | Erin Young et al |
| EPI_ISL_575085 | USA/UT-UPHL-02822/2020 | 2020-04-25 | North America/USA/Utah/ | Utah Public Health Laboratory | Utah Public Health Laboratory | Erin Young et al |
| EPI_ISL_575092 | USA/UT-UPHL-02832/2020 | 2020-04-23 | North America/USA/Utah/ | Utah Public Health Laboratory | Utah Public Health Laboratory | Erin Young et al |
| EPI_ISL_575107 | USA/UT-UPHL-03757/2020 | 2020-04-01 | North America/USA/Utah/ | Utah Public Health Laboratory | Utah Public Health Laboratory | Erin Young et al |
| EPI_ISL_575113 | USA/UT-UPHL-03768/2020 | 2020-04-05 | North America/USA/Utah/ | Utah Public Health Laboratory | Utah Public Health Laboratory | Erin Young et al |
| EPI_ISL_575125 | USA/UT-UPHL-04262/2020 | 2020-05-15 | North America/USA/Utah/ | Utah Public Health Laboratory | Utah Public Health Laboratory | Erin Young et al |
| EPI_ISL_575127 | USA/UT-UPHL-04282/2020 | 2020-05-14 | North America/USA/Utah/ | Utah Public Health Laboratory | Utah Public Health Laboratory | Erin Young et al |
| EPI_ISL_575133 | USA/UT-UPHL-04301/2020 | 2020-05-14 | North America/USA/Utah/ | Utah Public Health Laboratory | Utah Public Health Laboratory | Erin Young et al |
| EPI_ISL_575134 | USA/UT-UPHL-04302/2020 | 2020-05-14 | North America/USA/Utah/ | Utah Public Health Laboratory | Utah Public Health Laboratory | Erin Young et al |
| EPI_ISL_575139 | USA/UT-UPHL-04309/2020 | 2020-05-15 | North America/USA/Utah/ | Utah Public Health Laboratory | Utah Public Health Laboratory | Erin Young et al |
| EPI_ISL_575146 | USA/UT-UPHL-04316/2020 | 2020-05-17 | North America/USA/Utah/ | Utah Public Health Laboratory | Utah Public Health Laboratory | Erin Young et al |
| EPI_ISL_575151 | USA/UT-UPHL-04322/2020 | 2020-05-18 | North America/USA/Utah/ | Utah Public Health Laboratory | Utah Public Health Laboratory | Erin Young et al |
| EPI_ISL_575155 | USA/UT-UPHL-04525/2020 | 2020-05-22 | North America/USA/Utah/ | Utah Public Health Laboratory | Utah Public Health Laboratory | Erin Young et al |
| EPI_ISL_575158 | USA/UT-UPHL-04529/2020 | 2020-05-22 | North America/USA/Utah/ | Utah Public Health Laboratory | Utah Public Health Laboratory | Erin Young et al |
| EPI_ISL_575160 | USA/UT-UPHL-04534/2020 | 2020-05-22 | North America/USA/Utah/ | Utah Public Health Laboratory | Utah Public Health Laboratory | Erin Young et al |
| EPI_ISL_575161 | USA/UT-UPHL-04535/2020 | 2020-05-22 | North America/USA/Utah/ | Utah Public Health Laboratory | Utah Public Health Laboratory | Erin Young et al |

| | | | | | | |
|---|---|---|---|---|---|---|
| EPI_ISL_575162 | USA/UT-UPHL-04536/2020 | 2020-05-22 | North America/USA/Utah/ | Utah Public Health Laboratory | Utah Public Health Laboratory | Erin Young et al |
| EPI_ISL_575166 | USA/UT-UPHL-04840/2020 | 2020-05-22 | North America/USA/Utah/ | Utah Public Health Laboratory | Utah Public Health Laboratory | Erin Young et al |
| EPI_ISL_575167 | USA/UT-UPHL-04841/2020 | 2020-05-22 | North America/USA/Utah/ | Utah Public Health Laboratory | Utah Public Health Laboratory | Erin Young et al |
| EPI_ISL_575168 | USA/UT-UPHL-04843/2020 | 2020-05-22 | North America/USA/Utah/ | Utah Public Health Laboratory | Utah Public Health Laboratory | Erin Young et al |
| EPI_ISL_575170 | USA/UT-UPHL-04845/2020 | 2020-05-23 | North America/USA/Utah/ | Utah Public Health Laboratory | Utah Public Health Laboratory | Erin Young et al |
| EPI_ISL_575181 | USA/UT-UPHL-04861/2020 | 2020-05-23 | North America/USA/Utah/ | Utah Public Health Laboratory | Utah Public Health Laboratory | Erin Young et al |
| EPI_ISL_575189 | USA/UT-UPHL-04888/2020 | 2020-05-26 | North America/USA/Utah/ | Utah Public Health Laboratory | Utah Public Health Laboratory | Erin Young et al |
| EPI_ISL_575198 | USA/UT-UPHL-04907/2020 | 2020-05-26 | North America/USA/Utah/ | Utah Public Health Laboratory | Utah Public Health Laboratory | Erin Young et al |
| EPI_ISL_575206 | USA/UT-UPHL-04921/2020 | 2020-05-26 | North America/USA/Utah/ | Utah Public Health Laboratory | Utah Public Health Laboratory | Erin Young et al |
| EPI_ISL_575224 | USA/UT-UPHL-05565/2020 | 2020-06-02 | North America/USA/Utah/ | Utah Public Health Laboratory | Utah Public Health Laboratory | Erin Young et al |
| EPI_ISL_575227 | USA/UT-UPHL-05574/2020 | 2020-06-01 | North America/USA/Utah/ | Utah Public Health Laboratory | Utah Public Health Laboratory | Erin Young et al |
| EPI_ISL_575231 | USA/UT-UPHL-05585/2020 | 2020-06-01 | North America/USA/Utah/ | Utah Public Health Laboratory | Utah Public Health Laboratory | Erin Young et al |
| EPI_ISL_575232 | USA/UT-UPHL-05586/2020 | 2020-06-01 | North America/USA/Utah/ | Utah Public Health Laboratory | Utah Public Health Laboratory | Erin Young et al |
| EPI_ISL_575235 | USA/UT-UPHL-05589/2020 | 2020-06-01 | North America/USA/Utah/ | Utah Public Health Laboratory | Utah Public Health Laboratory | Erin Young et al |
| EPI_ISL_575236 | USA/UT-UPHL-05590/2020 | 2020-06-01 | North America/USA/Utah/ | Utah Public Health Laboratory | Utah Public Health Laboratory | Erin Young et al |
| EPI_ISL_575246 | USA/UT-UPHL-03084/2020 | 2020-05-04 | North America/USA/Utah/ | Utah Public Health Laboratory | Utah Public Health Laboratory | Erin Young et al |
| EPI_ISL_575252 | USA/UT-UPHL-03102/2020 | 2020-05-04 | North America/USA/Utah/ | Utah Public Health Laboratory | Utah Public Health Laboratory | Erin Young et al |
| EPI_ISL_575256 | USA/UT-UPHL-03107/2020 | 2020-05-04 | North America/USA/Utah/ | Utah Public Health Laboratory | Utah Public Health Laboratory | Erin Young et al |
| EPI_ISL_575259 | USA/UT-UPHL-03111/2020 | 2020-05-04 | North America/USA/Utah/ | Utah Public Health Laboratory | Utah Public Health Laboratory | Erin Young et al |
| EPI_ISL_575283 | USA/UT-UPHL-03469/2020 | 2020-05-09 | North America/USA/Utah/ | Utah Public Health Laboratory | Utah Public Health Laboratory | Erin Young et al |

| | | | | | | |
|---|---|---|---|---|---|---|
| EPI_ISL_575286 | USA/UT-UPHL-03828/2020 | 2020-05-11 | North America/USA/Utah/ | Utah Public Health Laboratory | Utah Public Health Laboratory | Erin Young et al |
| EPI_ISL_575287 | USA/UT-UPHL-04008/2020 | 2020-05-07 | North America/USA/Utah/ | Utah Public Health Laboratory | Utah Public Health Laboratory | Erin Young et al |
| EPI_ISL_575292 | USA/NM-DOH-2020321221/2020 | 2020-09-03 | North America/USA/New Mexico/ | Molecular Biology, New Mexico Department of Health Scientific Laboratory | Molecular Biology, New Mexico Department of Health Scientific Laboratory | Johnson et al |
| EPI_ISL_575295 | USA/NM-DOH-2020322589/2020 | 2020-09-03 | North America/USA/New Mexico/ | Molecular Biology, New Mexico Department of Health Scientific Laboratory | Molecular Biology, New Mexico Department of Health Scientific Laboratory | Johnson et al |
| EPI_ISL_575296 | USA/NM-DOH-2020322592/2020 | 2020-09-03 | North America/USA/New Mexico/ | Molecular Biology, New Mexico Department of Health Scientific Laboratory | Molecular Biology, New Mexico Department of Health Scientific Laboratory | Johnson et al |
| EPI_ISL_575297 | USA/NM-DOH-2020324092/2020 | 2020-09-03 | North America/USA/New Mexico/ | Molecular Biology, New Mexico Department of Health Scientific Laboratory | Molecular Biology, New Mexico Department of Health Scientific Laboratory | Johnson et al |
| EPI_ISL_575301 | USA/NM-DOH-2020326795/2020 | 2020-09-03 | North America/USA/New Mexico/ | Molecular Biology, New Mexico Department of Health Scientific Laboratory | Molecular Biology, New Mexico Department of Health Scientific Laboratory | Johnson et al |
| EPI_ISL_575305 | USA/NM-DOH-2020327026/2020 | 2020-09-03 | North America/USA/New Mexico/ | Molecular Biology, New Mexico Department of Health Scientific Laboratory | Molecular Biology, New Mexico Department of Health Scientific Laboratory | Johnson et al |

| | | | | | | |
|---|---|---|---|---|---|---|
| EPI_ISL_575309 | USA/NM-DOH-2020327163/2020 | 2020-09-03 | North America/USA/New Mexico/ | Molecular Biology, New Mexico Department of Health Scientific Laboratory | Molecular Biology, New Mexico Department of Health Scientific Laboratory | Johnson et al |
| EPI_ISL_575329 | Gansu/NPRC0003/2020 | 2020-02-24 | Asia/China/Gansu/ | National Institute for Viral Disease Control and Prevention, China CDC | National Institute for Viral Disease Control and Prevention, China CDC | Rongbao Gao et al |
| EPI_ISL_575330 | Beijing/NPRC0004/2020 | 2020-04-15 | Asia/China/Beijing/ | National Institute for Viral Disease Control and Prevention, China CDC | National Institute for Viral Disease Control and Prevention, China CDC | Rongbao Gao et al |

| | | | | | | |
|---|---|---|---|---|---|---|
| EPI_ISL_575331 | Indonesia/YO-UGM-00061/2020 | 2020-06-16 | Asia/Indonesia/Yogyakarta/ | RSUD Wates | Genetics Working Group (Pokja Genetik) Faculty of Medicine, Public Health and Nursing Universitas Gadjah Mada (FK-KMK UGM); Disease Investigation Center Wates Ministry of Agriculture Indonesia; Department of Microbiology FK-KMK UGM; Laboratorium Diagnostik Yayasan Tahija World Mosquito Program (WMP) Yogyakarta Center for Tropical Medicine FK-KMK UGM; Integrated Research Center FK-KMK UGM; Department of Computer Science and Electronics FMIPA UGM | Gunadi et al |
| EPI_ISL_575332 | Israel/CVL-s2049/2020 | 2020-09-01 | Asia/Israel/Central District/ | Israel Central Virology laboratory | Israel Central Virology laboratory | Neta Zuckerman et al |
| EPI_ISL_575333 | Israel/CVL-s2057/2020 | 2020-09-01 | Asia/Israel/Central District/ | Israel Central Virology laboratory | Israel Central Virology laboratory | Neta Zuckerman et al |

| | | | | | | |
|---|---|---|---|---|---|---|
| EPI_ISL_575334 | Israel/CVL-s2064/2020 | 2020-09-01 | Asia/Israel/North District/ | Israel Central Virology laboratory | Israel Central Virology laboratory | Neta Zuckerman et al |
| EPI_ISL_575396 | Scotland/QEUH-9DC0D7/2020 | 2020-09-22 | Europe/United Kingdom/Scotland/ | Lighthouse Lab in Glasgow | Wellcome Sanger Institute for the COVID-19 Genomics UK (COG-UK) consortium | Harper VanSteenhouse et al |
| EPI_ISL_575595 | England/ALDP-9CBA28/2020 | 2020-09-16 | Europe/United Kingdom/England/ | Lighthouse Lab in Alderley Park | Wellcome Sanger Institute for the COVID-19 Genomics UK (COG-UK) consortium | Jacquelyn Wynn et al |
| EPI_ISL_575605 | England/ALDP-9CB6E8/2020 | 2020-09-16 | Europe/United Kingdom/England/ | Lighthouse Lab in Alderley Park | Wellcome Sanger Institute for the COVID-19 Genomics UK (COG-UK) consortium | Jacquelyn Wynn et al |
| EPI_ISL_575723 | England/ALDP-9CB1BA/2020 | 2020-09-16 | Europe/United Kingdom/England/ | Lighthouse Lab in Alderley Park | Wellcome Sanger Institute for the COVID-19 Genomics UK (COG-UK) consortium | Jacquelyn Wynn et al |
| EPI_ISL_575732 | England/ALDP-9CB8B5/2020 | 2020-09-16 | Europe/United Kingdom/England/ | Lighthouse Lab in Alderley Park | Wellcome Sanger Institute for the COVID-19 Genomics UK (COG-UK) consortium | Jacquelyn Wynn et al |
| EPI_ISL_575924 | England/MILK-97338E/2020 | 2020-08-18 | Europe/United Kingdom/England/ | Lighthouse Lab in Milton Keynes | Wellcome Sanger Institute for the COVID-19 Genomics UK (COG-UK) consortium | The Lighthouse Lab in Milton Keynes et al |

| | | | | | | |
|---|---|---|---|---|---|---|
| EPI_ISL_576113 | Indonesia/YO-UGM-10001/2020 | 2020-08-18 | Asia/Indonesia/Yogyakarta/ | RSUP Dr. Sardjito | Genetics Working Group (Pokja Genetik) Faculty of Medicine, Public Health and Nursing Universitas Gadjah Mada (FK-KMK UGM); Disease Investigation Center Wates Ministry of Agriculture Indonesia; Department of Microbiology FK-KMK UGM; Laboratorium Diagnostik Yayasan Tahija World Mosquito Program (WMP) Yogyakarta Center for Tropical Medicine FK-KMK UGM; Integrated Research Center FK-KMK UGM; Department of Computer Science and Electronics FMIPA UGM | Gunadi et al |

| | | | | | | |
|---|---|---|---|---|---|---|
| EPI_ISL_576114 | Indonesia/YO-UGM-10004/2020 | 2020-08-20 | Asia/Indonesia/Yogyakarta/ | RSUP Dr. Sardjito | Genetics Working Group (Pokja Genetik) Faculty of Medicine, Public Health and Nursing Universitas Gadjah Mada (FK-KMK UGM); Disease Investigation Center Wates Ministry of Agriculture Indonesia; Department of Microbiology FK-KMK UGM; Laboratorium Diagnostik Yayasan Tahija World Mosquito Program (WMP) Yogyakarta Center for Tropical Medicine FK-KMK UGM; Integrated Research Center FK-KMK UGM; Department of Computer Science and Electronics FMIPA UGM | Gunadi et al |

| | | | | | | |
|---|---|---|---|---|---|---|
| EPI_ISL_576115 | Indonesia/YO-UGM-10003/2020 | 2020-08-16 | Asia/Indonesia/Yogyakarta/ | RSUP Dr Sardjito | Genetics Working Group (Pokja Genetik) Faculty of Medicine, Public Health and Nursing Universitas Gadjah Mada (FK-KMK UGM); Disease Investigation Center Wates Ministry of Agriculture Indonesia; Department of Microbiology FK-KMK UGM; Laboratorium Diagnostik Yayasan Tahija World Mosquito Program (WMP) Yogyakarta Center for Tropical Medicine FK-KMK UGM; Integrated Research Center FK-KMK UGM; Department of Computer Science and Electronics FMIPA UGM | Gunadi et al |

| | | | | | | |
|---|---|---|---|---|---|---|
| EPI_ISL_576116 | Indonesia/YO-UGM-10002/2020 | 2020-08-13 | Asia/Indonesia/Yogyakarta/ | RSUP Dr. Sardjito | Genetics Working Group (Pokja Genetik) Faculty of Medicine, Public Health and Nursing Universitas Gadjah Mada (FK-KMK UGM); Disease Investigation Center Wates Ministry of Agriculture Indonesia; Department of Microbiology FK-KMK UGM; Laboratorium Diagnostik Yayasan Tahija World Mosquito Program (WMP) Yogyakarta Center for Tropical Medicine FK-KMK UGM; Integrated Research Center FK-KMK UGM; Department of Computer Science and Electronics FMIPA UGM | Gunadi et al |
| EPI_ISL_576119 | Malta/BAL-Sliema-3/2020 | 2020-08-14 | Europe/Malta/Malta/ | Laboratory, The Bio Arte Limited | Laboratory, The Bio Arte Limited | Biazzo et al |
| EPI_ISL_576121 | Malta/BAL-Sliema-5/2020 | 2020-08-20 | Europe/Malta/Malta/ | Laboratory, The Bio Arte Limited | Laboratory, The Bio Arte Limited | Biazzo et al |
| EPI_ISL_576124 | Malta/BAL-Sliema-8/2020 | 2020-09-07 | Europe/Malta/Malta/ | Laboratory, The Bio Arte Limited | Laboratory, The Bio Arte Limited | Biazzo et al |
| EPI_ISL_576125 | Malta/BAL-Sliema-9/2020 | 2020-09-07 | Europe/Malta/Malta/ | Laboratory, The Bio Arte Limited | Laboratory, The Bio Arte Limited | Biazzo et al |

| | | | | | | |
|---|---|---|---|---|---|---|
| EPI_ISL_576128 | Indonesia/YO-UGM-10005/2020 | 2020-08-21 | Asia/Indonesia/Yogyakarta/ | RSUP Dr. Sardjito | Genetics Working Group (Pokja Genetik) Faculty of Medicine, Public Health and Nursing Universitas Gadjah Mada (FK-KMK UGM); Disease Investigation Center Wates Ministry of Agriculture Indonesia; Department of Microbiology FK-KMK UGM; Laboratorium Diagnostik Yayasan Tahija World Mosquito Program (WMP) Yogyakarta Center for Tropical Medicine FK-KMK UGM; Integrated Research Center FK-KMK UGM; Department of Computer Science and Electronics FMIPA UGM | Gunadi et al |

| | | | | | | |
|---|---|---|---|---|---|---|
| EPI_ISL_576130 | Indonesia/YO-UGM-10006/2020 | 2020-08-18 | Asia/Indonesia/Yogyakarta/ | RSUP Dr. Sardjito | Genetics Working Group (Pokja Genetik) Faculty of Medicine, Public Health and Nursing Universitas Gadjah Mada (FK-KMK UGM); Disease Investigation Center Wates Ministry of Agriculture Indonesia; Department of Microbiology FK-KMK UGM; Laboratorium Diagnostik Yayasan Tahija World Mosquito Program (WMP) Yogyakarta Center for Tropical Medicine FK-KMK UGM; Integrated Research Center FK-KMK UGM; Department of Computer Science and Electronics FMIPA UGM | Gunadi et al |

| | | | | | | |
|---|---|---|---|---|---|---|
| EPI_ISL_576145 | Indonesia/YO-UGM-107727/2020 | 2020-07-30 | Asia/Indonesia/Yogyakarta/ | RSA Universitas Gadjah Mada | Genetics Working Group (Pokja Genetik) Faculty of Medicine, Public Health and Nursing Universitas Gadjah Mada (FK-KMK UGM); Disease Investigation Center Wates Ministry of Agriculture Indonesia; Department of Microbiology FK-KMK UGM; Laboratorium Diagnostik Yayasan Tahija World Mosquito Program (WMP) Yogyakarta Center for Tropical Medicine FK-KMK UGM; Integrated Research Center FK-KMK UGM; Department of Computer Science and Electronics FMIPA UGM | Gunadi et al |

| | | | | | | |
|---|---|---|---|---|---|---|
| EPI_ISL_576146 | Ukraine/Kharkiv-705/2020 | 2020-07-28 | Europe/Ukraine/Kharkiv/ | Department of Respiratory & Other Viral Infections of L.V. Gromashevsky Institute of Epidemiology & Infectious Diseases NAMS of Ukraine | "Department of Respiratory & Other Viral Infections of L.V. Gromashevsky Institute of Epidemiology & Infectious Diseases NAMS of Ukraine, JSC ""Farmak""" | Alla Mironenko et al |
| EPI_ISL_576147 | Ukraine/Ivano-Frankivsk-706/2020 | 2020-07-28 | Europe/Ukraine/Ivano-Frankivsk/ | Department of Respiratory & Other Viral Infections of L.V. Gromashevsky Institute of Epidemiology & Infectious Diseases NAMS of Ukraine | "Department of Respiratory & Other Viral Infections of L.V. Gromashevsky Institute of Epidemiology & Infectious Diseases NAMS of Ukraine, JSC ""Farmak""" | Alla Mironenko et al |
| EPI_ISL_576148 | Ukraine/Kyiv-785/2020 | 2020-08-03 | Europe/Ukraine/Kyiv/ | Department of Respiratory & Other Viral Infections of L.V. Gromashevsky Institute of Epidemiology & Infectious Diseases NAMS of Ukraine | "Department of Respiratory & Other Viral Infections of L.V. Gromashevsky Institute of Epidemiology & Infectious Diseases NAMS of Ukraine, JSC ""Farmak""" | Alla Mironenko et al |

| | | | | | | |
|---|---|---|---|---|---|---|
| EPI_ISL_576149 | Ukraine/Lutsk-889/2020 | 2020-08-06 | Europe/Ukraine/Volyn/Lutsk | Department of Respiratory & Other Viral Infections of L.V. Gromashevsky Institute of Epidemiology & Infectious Diseases NAMS of Ukraine | "Department of Respiratory & Other Viral Infections of L.V. Gromashevsky Institute of Epidemiology & Infectious Diseases NAMS of Ukraine, JSC ""Farmak""" | Alla Mironenko et al |
| EPI_ISL_576169 | USA/DE-DHSS-F917785/2020 | 2020-03-24 | North America/USA/Delaware/Kent Cou | Delaware Public Health Lab | Delaware Public Health Lab | Gregory Hovan et al |
| EPI_ISL_576178 | USA/DC-CDC-8704/2020 | 2020-04-21 | North America/USA/Washington DC/ | DC Public Health Lab/ Dept. of Forensic Sciences | Pathogen Discovery, Respiratory Viruses Branch, Division of Viral Diseases, Centers for Disease Control and Prevention | Ying Tao et al |
| EPI_ISL_576180 | USA/WI-CDC-6231/2020 | 2020-03-25 | North America/USA/Wisconsin/ | WI State Laboratory of Hygiene | Pathogen Discovery, Respiratory Viruses Branch, Division of Viral Diseases, Centers for Disease Control and Prevention | Ying Tao et al |
| EPI_ISL_576187 | USA/WI-CDC-6238/2020 | 2020-03-26 | North America/USA/Wisconsin/ | WI State Laboratory of Hygiene | Pathogen Discovery, Respiratory Viruses Branch, Division of Viral Diseases, Centers for Disease Control and Prevention | Ying Tao et al |

| | | | | | | |
|---|---|---|---|---|---|---|
| EPI_ISL_576190 | USA/GA-CDC-4459/2020 | 2020-09-11 | North America/USA/Georgia/ | GA Department of Public Health Laboratory | Pathogen Discovery, Respiratory Viruses Branch, Division of Viral Diseases, Centers for Disease Control and Prevention | Ying Tao et al |
| EPI_ISL_576201 | USA/MI-MDHHS-SC21934/2020 | 2020-09-26 | North America/USA/Michigan/ | Michigan Department of Health and Human Services, Bureau of Laboratories | Michigan Department of Health and Human Services, Bureau of Laboratories | Blankenship HM et al |
| EPI_ISL_576202 | USA/MI-MDHHS-SC21936/2020 | 2020-09-26 | North America/USA/Michigan/ | Michigan Department of Health and Human Services, Bureau of Laboratories | Michigan Department of Health and Human Services, Bureau of Laboratories | Blankenship HM et al |
| EPI_ISL_576204 | USA/MI-MDHHS-SC21938/2020 | 2020-09-26 | North America/USA/Michigan/ | Michigan Department of Health and Human Services, Bureau of Laboratories | Michigan Department of Health and Human Services, Bureau of Laboratories | Blankenship HM et al |
| EPI_ISL_576206 | USA/MI-MDHHS-SC21940/2020 | 2020-09-26 | North America/USA/Michigan/ | Michigan Department of Health and Human Services, Bureau of Laboratories | Michigan Department of Health and Human Services, Bureau of Laboratories | Blankenship HM et al |
| EPI_ISL_576219 | USA/MI-MDHHS-SC21996/2020 | 2020-09-26 | North America/USA/Michigan/ | Michigan Department of Health and Human Services, Bureau of Laboratories | Michigan Department of Health and Human Services, Bureau of Laboratories | Blankenship HM et al |

| | | | | | | |
|---|---|---|---|---|---|---|
| EPI_ISL_576220 | USA/MI-MDHHS-SC21997/2020 | 2020-09-26 | North America/USA/Michigan/ | Michigan Department of Health and Human Services, Bureau of Laboratories | Michigan Department of Health and Human Services, Bureau of Laboratories | Blankenship HM et al |
| EPI_ISL_576230 | USA/MN-MDH-1721/2020 | 2020-09-16 | North America/USA/Minnesota/ | Minnesota Department of Health, Public Health Laboratory | Minnesota Department of Health, Public Health Laboratory | Matt Plumb et al |
| EPI_ISL_576235 | USA/MN-MDH-1726/2020 | 2020-09-16 | North America/USA/Minnesota/ | Minnesota Department of Health, Public Health Laboratory | Minnesota Department of Health, Public Health Laboratory | Matt Plumb et al |
| EPI_ISL_576244 | USA/MN-MDH-1735/2020 | 2020-09-16 | North America/USA/Minnesota/ | Minnesota Department of Health, Public Health Laboratory | Minnesota Department of Health, Public Health Laboratory | Matt Plumb et al |
| EPI_ISL_576248 | USA/MN-MDH-1739/2020 | 2020-09-16 | North America/USA/Minnesota/ | Minnesota Department of Health, Public Health Laboratory | Minnesota Department of Health, Public Health Laboratory | Matt Plumb et al |
| EPI_ISL_576251 | USA/MN-MDH-1742/2020 | 2020-09-16 | North America/USA/Minnesota/ | Minnesota Department of Health, Public Health Laboratory | Minnesota Department of Health, Public Health Laboratory | Matt Plumb et al |
| EPI_ISL_576254 | USA/MN-MDH-1745/2020 | 2020-04-10 | North America/USA/Minnesota/ | Minnesota Department of Health, Public Health Laboratory | Minnesota Department of Health, Public Health Laboratory | Matt Plumb et al |
| EPI_ISL_576256 | USA/MN-MDH-1747/2020 | 2020-09-16 | North America/USA/Minnesota/ | Minnesota Department of Health, Public Health Laboratory | Minnesota Department of Health, Public Health Laboratory | Matt Plumb et al |
| EPI_ISL_576257 | Mexico/HID-InDRE-54/2020 | 2020-07-22 | North America/Mexico/Hidalgo/ | Instituto de Diagnostico y Referencia Epidemiologicos (INDRE) | Instituto de Diagnostico y Referencia Epidemiologicos (INDRE) | Gisela Barrera-Badillo et al |

| | | | | | | |
|---|---|---|---|---|---|---|
| EPI_ISL_576258 | Mexico/AGU-InDRE-55/2020 | 2020-06-15 | North America/Mexico/Aguascalientes/ | Instituto de Diagnostico y Referencia Epidemiologicos (INDRE) | Instituto de Diagnostico y Referencia Epidemiologicos (INDRE) | Gisela Barrera-Badillo et al |
| EPI_ISL_576259 | Mexico/TLA-InDRE-56/2020 | 2020-08-13 | North America/Mexico/Tlaxcala/ | Instituto de Diagnostico y Referencia Epidemiologicos (INDRE) | Instituto de Diagnostico y Referencia Epidemiologicos (INDRE) | Gisela Barrera-Badillo et al |
| EPI_ISL_576260 | Mexico/TLA-InDRE-57/2020 | 2020-08-13 | North America/Mexico/Tlaxcala/ | Instituto de Diagnostico y Referencia Epidemiologicos (INDRE) | Instituto de Diagnostico y Referencia Epidemiologicos (INDRE) | Gisela Barrera-Badillo et al |
| EPI_ISL_576261 | Mexico/AGU-InDRE-58/2020 | 2020-08-07 | North America/Mexico/Aguascalientes/ | Instituto de Diagnostico y Referencia Epidemiologicos (INDRE) | Instituto de Diagnostico y Referencia Epidemiologicos (INDRE) | Gisela Barrera-Badillo et al |
| EPI_ISL_576262 | Mexico/NLE-InDRE-59/2020 | 2020-08-14 | North America/Mexico/Nuevo Leon/ | Instituto de Diagnostico y Referencia Epidemiologicos (INDRE) | Instituto de Diagnostico y Referencia Epidemiologicos (INDRE) | Gisela Barrera-Badillo et al |
| EPI_ISL_576263 | Mexico/NLE-InDRE-60/2020 | 2020-08-14 | North America/Mexico/Nuevo Leon/ | Instituto de Diagnostico y Referencia Epidemiologicos (INDRE) | Instituto de Diagnostico y Referencia Epidemiologicos (INDRE) | Gisela Barrera-Badillo et al |
| EPI_ISL_576264 | Mexico/OAX-InDRE-61/2020 | 2020-08-18 | North America/Mexico/Oaxaca/ | Instituto de Diagnostico y Referencia Epidemiologicos (INDRE) | Instituto de Diagnostico y Referencia Epidemiologicos (INDRE) | Ernesto Ramirez-Gonzalez et al |
| EPI_ISL_576265 | Mexico/CAM-InDRE-62/2020 | 2020-08-21 | North America/Mexico/Campeche/ | Instituto de Diagnostico y Referencia Epidemiologicos (INDRE) | Instituto de Diagnostico y Referencia Epidemiologicos (INDRE) | Ernesto Ramirez-Gonzalez et al |

| | | | | | | |
|---|---|---|---|---|---|---|
| EPI_ISL_576266 | Mexico/CAM-InDRE-63/2020 | 2020-08-21 | North America/Mexico/Campeche/ | Instituto de Diagnostico y Referencia Epidemiologicos (INDRE) | Instituto de Diagnostico y Referencia Epidemiologicos (INDRE) | Ernesto Ramirez-Gonzalez et al |
| EPI_ISL_576267 | Mexico/CAM-InDRE-64/2020 | 2020-08-21 | North America/Mexico/Campeche/ | Instituto de Diagnostico y Referencia Epidemiologicos (INDRE) | Instituto de Diagnostico y Referencia Epidemiologicos (INDRE) | Ernesto Ramirez-Gonzalez et al |
| EPI_ISL_576268 | Mexico/HID-InDRE-65/2020 | 2020-08-25 | North America/Mexico/Hidalgo/ | Instituto de Diagnostico y Referencia Epidemiologicos (INDRE) | Instituto de Diagnostico y Referencia Epidemiologicos (INDRE) | Ernesto Ramirez-Gonzalez et al |
| EPI_ISL_576269 | Mexico/VER-InDRE-66/2020 | 2020-08-16 | North America/Mexico/Veracruz/ | Instituto de Diagnostico y Referencia Epidemiologicos (INDRE) | Instituto de Diagnostico y Referencia Epidemiologicos (INDRE) | Ernesto Ramirez-Gonzalez et al |
| EPI_ISL_576270 | Mexico/VER-InDRE-67/2020 | 2020-08-14 | North America/Mexico/Veracruz/ | Instituto de Diagnostico y Referencia Epidemiologicos (INDRE) | Instituto de Diagnostico y Referencia Epidemiologicos (INDRE) | Ernesto Ramirez-Gonzalez et al |
| EPI_ISL_576271 | Mexico/MIC-InDRE-68/2020 | 2020-08-17 | North America/Mexico/Michoacan/ | Instituto de Diagnostico y Referencia Epidemiologicos (INDRE) | Instituto de Diagnostico y Referencia Epidemiologicos (INDRE) | Ernesto Ramirez-Gonzalez et al |
| EPI_ISL_576272 | Mexico/SLP-InDRE-69/2020 | 2020-08-21 | North America/Mexico/San Luis Potosi/ | Instituto de Diagnostico y Referencia Epidemiologicos (INDRE) | Instituto de Diagnostico y Referencia Epidemiologicos (INDRE) | Gisela Barrera-Badillo et al |
| EPI_ISL_576273 | Mexico/ZAC-InDRE-70/2020 | 2020-08-21 | North America/Mexico/Zacatecas/ | Instituto de Diagnostico y Referencia Epidemiologicos (INDRE) | Instituto de Diagnostico y Referencia Epidemiologicos (INDRE) | Ernesto Ramirez-Gonzalez et al |

| | | | | | | |
|---|---|---|---|---|---|---|
| EPI_ISL_576274 | Mexico/ZAC-InDRE-71/2020 | 2020-08-21 | North America/Mexico/Zacatecas/ | Instituto de Diagnostico y Referencia Epidemiologicos (INDRE) | Instituto de Diagnostico y Referencia Epidemiologicos (INDRE) | Gisela Barrera-Badillo et al |
| EPI_ISL_576275 | Mexico/ZAC-InDRE-72/2020 | 2020-08-14 | North America/Mexico/Zacatecas/ | Instituto de Diagnostico y Referencia Epidemiologicos (INDRE) | Instituto de Diagnostico y Referencia Epidemiologicos (INDRE) | Gisela Barrera-Badillo et al |
| EPI_ISL_576276 | Mexico/MIC-InDRE-73/2020 | 2020-08-20 | North America/Mexico/Michoacan/ | Instituto de Diagnostico y Referencia Epidemiologicos (INDRE) | Instituto de Diagnostico y Referencia Epidemiologicos (INDRE) | Gisela Barrera-Badillo et al |
| EPI_ISL_576277 | Mexico/MIC-InDRE-74/2020 | 2020-08-20 | North America/Mexico/Michoacan/ | Instituto de Diagnostico y Referencia Epidemiologicos (INDRE) | Instituto de Diagnostico y Referencia Epidemiologicos (INDRE) | Ernesto Ramirez-Gonzalez et al |
| EPI_ISL_576278 | Mexico/COL-InDRE-75/2020 | 2020-08-24 | North America/Mexico/Colima/ | Instituto de Diagnostico y Referencia Epidemiologicos (INDRE) | Instituto de Diagnostico y Referencia Epidemiologicos (INDRE) | Ernesto Ramirez-Gonzalez et al |
| EPI_ISL_576279 | Mexico/COL-InDRE-76/2020 | 2020-08-24 | North America/Mexico/Colima/ | Instituto de Diagnostico y Referencia Epidemiologicos (INDRE) | Instituto de Diagnostico y Referencia Epidemiologicos (INDRE) | Ernesto Ramirez-Gonzalez et al |
| EPI_ISL_576287 | USA/TX-DSHS-0631/2020 | 2020-06-26 | North America/USA/Texas/Dallas Count | Texas Department of State Health Services | Texas Department of State Health Services | Rashmi Tuladhar et al |
| EPI_ISL_576292 | USA/TX-DSHS-0637/2020 | 2020-07-16 | North America/USA/Texas/Dallas Count | Texas Department of State Health Services | Texas Department of State Health Services | Rashmi Tuladhar et al |
| EPI_ISL_576293 | USA/TX-DSHS-0638/2020 | 2020-07-16 | North America/USA/Texas/Dallas Count | Texas Department of State Health Services | Texas Department of State Health Services | Rashmi Tuladhar et al |

| | | | | | | |
|---|---|---|---|---|---|---|
| EPI_ISL_576299 | USA/TX-DSHS-0644/2020 | 2020-07-16 | North America/USA/Texas/Dallas Count | Texas Department of State Health Services | Texas Department of State Health Services | Rashmi Tuladhar et al |
| EPI_ISL_576305 | USA/TX-DSHS-0651/2020 | 2020-07-16 | North America/USA/Texas/Dallas Count | Texas Department of State Health Services | Texas Department of State Health Services | Rashmi Tuladhar et al |
| EPI_ISL_576308 | USA/TX-DSHS-0654/2020 | 2020-07-19 | North America/USA/Texas/Parker Count | Texas Department of State Health Services | Texas Department of State Health Services | Rashmi Tuladhar et al |
| EPI_ISL_576311 | USA/TX-DSHS-0657/2020 | 2020-07-19 | North America/USA/Texas/Parker Count | Texas Department of State Health Services | Texas Department of State Health Services | Rashmi Tuladhar et al |
| EPI_ISL_576313 | USA/TX-DSHS-0659/2020 | 2020-07-19 | North America/USA/Texas/Parker Count | Texas Department of State Health Services | Texas Department of State Health Services | Rashmi Tuladhar et al |
| EPI_ISL_576317 | USA/TX-DSHS-0663/2020 | 2020-07-19 | North America/USA/Texas/Parker Count | Texas Department of State Health Services | Texas Department of State Health Services | Rashmi Tuladhar et al |
| EPI_ISL_576324 | USA/TX-DSHS-0671/2020 | 2020-07-19 | North America/USA/Texas/Parker Count | Texas Department of State Health Services | Texas Department of State Health Services | Rashmi Tuladhar et al |
| EPI_ISL_576325 | USA/TX-DSHS-0672/2020 | 2020-07-19 | North America/USA/Texas/Parker Count | Texas Department of State Health Services | Texas Department of State Health Services | Rashmi Tuladhar et al |
| EPI_ISL_576349 | USA/TX-DSHS-0706/2020 | 2020-09-08 | North America/USA/Texas/Mclennan Co | Texas Department of State Health Services | Texas Department of State Health Services | Rashmi Tuladhar et al |
| EPI_ISL_576354 | USA/TX-DSHS-0711/2020 | 2020-09-08 | North America/USA/Texas/Mclennan Co | Texas Department of State Health Services | Texas Department of State Health Services | Rashmi Tuladhar et al |
| EPI_ISL_576358 | USA/TX-DSHS-0715/2020 | 2020-09-10 | North America/USA/Texas/Mclennan Co | Texas Department of State Health Services | Texas Department of State Health Services | Rashmi Tuladhar et al |
| EPI_ISL_576361 | USA/TX-DSHS-0719/2020 | 2020-09-10 | North America/USA/Texas/Mclennan Co | Texas Department of State Health Services | Texas Department of State Health Services | Rashmi Tuladhar et al |
| EPI_ISL_576372 | Egypt/CUNCI-HGC7I023/2020 | 2020-07-19 | Africa/Egypt/Egypt/ | Cancer Biology Department, National Cancer Institute | Cancer Biology Department, National Cancer Institute | Zekri et al |

| | | | | | | |
|---|---|---|---|---|---|---|
| EPI_ISL_576383 | Indonesia/JT-UGM-47906/2020 | 2020-09-01 | Asia/Indonesia/Central Java/ | RSUD Budi Rahayu Kota Magelang | Genetics Working Group (Pokja Genetik) Faculty of Medicine, Public Health and Nursing Universitas Gadjah Mada (FK-KMK UGM); Disease Investigation Center Wates Ministry of Agriculture Indonesia; Department of Microbiology FK-KMK UGM; Laboratorium Diagnostik Yayasan Tahija World Mosquito Program (WMP) Yogyakarta Center for Tropical Medicine FK-KMK UGM; Integrated Research Center FK-KMK UGM; Department of Computer Science and Electronics FMIPA UGM; Balai Besar Teknik Kesehatan Lingkungan dan | Gunadi et al |
| EPI_ISL_576387 | Indonesia/JK-NIHRD-MI52948/2020 | 2020-08-20 | Asia/Indonesia/Jakarta/ | National Institute of Health Research and Development | National Institute of Health Research and Development | Pawestri et al |
| EPI_ISL_576414 | USA/WA-UW-23276/2020 | 2020-08-07 | North America/USA/Washington/ | UW Virology Lab | UW Virology Lab | Pavitra Roychoudhury et al |
| EPI_ISL_576418 | USA/WA-UW-23293/2020 | 2020-08-07 | North America/USA/Washington/ | UW Virology Lab | UW Virology Lab | Pavitra Roychoudhury et al |

| | | | | | | |
|---|---|---|---|---|---|---|
| EPI_ISL_576421 | USA/WA-UW-23302/2020 | 2020-08-07 | North America/USA/Washington/ | UW Virology Lab | UW Virology Lab | Pavitra Roychoudhury et al |
| EPI_ISL_576422 | USA/WA-UW-23305/2020 | 2020-08-07 | North America/USA/Washington/ | UW Virology Lab | UW Virology Lab | Pavitra Roychoudhury et al |
| EPI_ISL_576425 | USA/WA-UW-23313/2020 | 2020-08-07 | North America/USA/Washington/ | UW Virology Lab | UW Virology Lab | Pavitra Roychoudhury et al |
| EPI_ISL_576427 | USA/OR-UW-23339/2020 | 2020-08-06 | North America/USA/Oregon/ | UW Virology Lab | UW Virology Lab | Pavitra Roychoudhury et al |
| EPI_ISL_576430 | USA/WA-UW-23349/2020 | 2020-08-03 | North America/USA/Washington/ | UW Virology Lab | UW Virology Lab | Pavitra Roychoudhury et al |
| EPI_ISL_576433 | USA/WA-UW-23365/2020 | 2020-08-02 | North America/USA/Washington/ | UW Virology Lab | UW Virology Lab | Pavitra Roychoudhury et al |
| EPI_ISL_576440 | USA/WA-UW-23389/2020 | 2020-08-10 | North America/USA/Washington/ | UW Virology Lab | UW Virology Lab | Pavitra Roychoudhury et al |
| EPI_ISL_576441 | USA/WA-UW-23390/2020 | 2020-08-11 | North America/USA/Washington/ | UW Virology Lab | UW Virology Lab | Pavitra Roychoudhury et al |
| EPI_ISL_576442 | USA/WA-UW-23391/2020 | 2020-08-11 | North America/USA/Washington/ | UW Virology Lab | UW Virology Lab | Pavitra Roychoudhury et al |
| EPI_ISL_576443 | USA/WA-UW-23394/2020 | 2020-08-11 | North America/USA/Washington/ | UW Virology Lab | UW Virology Lab | Pavitra Roychoudhury et al |
| EPI_ISL_576448 | USA/WA-UW-23414/2020 | 2020-08-09 | North America/USA/Washington/ | UW Virology Lab | UW Virology Lab | Pavitra Roychoudhury et al |
| EPI_ISL_576450 | USA/WA-UW-23422/2020 | 2020-08-10 | North America/USA/Washington/ | UW Virology Lab | UW Virology Lab | Pavitra Roychoudhury et al |
| EPI_ISL_576456 | USA/WA-UW-23461/2020 | 2020-08-10 | North America/USA/Washington/ | UW Virology Lab | UW Virology Lab | Pavitra Roychoudhury et al |
| EPI_ISL_576471 | USA/WA-UW-23530/2020 | 2020-08-10 | North America/USA/Washington/ | UW Virology Lab | UW Virology Lab | Pavitra Roychoudhury et al |
| EPI_ISL_576473 | USA/WA-UW-23538/2020 | 2020-08-12 | North America/USA/Washington/ | UW Virology Lab | UW Virology Lab | Pavitra Roychoudhury et al |
| EPI_ISL_576477 | USA/WA-UW-23576/2020 | 2020-08-10 | North America/USA/Washington/ | UW Virology Lab | UW Virology Lab | Pavitra Roychoudhury et al |
| EPI_ISL_576478 | USA/WA-UW-23579/2020 | 2020-08-11 | North America/USA/Washington/ | UW Virology Lab | UW Virology Lab | Pavitra Roychoudhury et al |
| EPI_ISL_576481 | USA/WA-UW-23596/2020 | 2020-08-11 | North America/USA/Washington/ | UW Virology Lab | UW Virology Lab | Pavitra Roychoudhury et al |
| EPI_ISL_576482 | USA/WA-UW-23597/2020 | 2020-08-10 | North America/USA/Washington/ | UW Virology Lab | UW Virology Lab | Pavitra Roychoudhury et al |
| EPI_ISL_576490 | USA/UN-UW-5251/2020 | 2020-04-09 | North America/USA/USA/ | UW Virology Lab | UW Virology Lab | Pavitra Roychoudhury et al |
| EPI_ISL_576504 | USA/UN-UW-7227/2020 | 2020-04-13 | North America/USA/USA/ | UW Virology Lab | UW Virology Lab | Pavitra Roychoudhury et al |
| EPI_ISL_576510 | USA/UN-UW-7434/2020 | 2020-04-14 | North America/USA/USA/ | UW Virology Lab | UW Virology Lab | Pavitra Roychoudhury et al |
| EPI_ISL_576513 | USA/UN-UW-7473/2020 | 2020-04-14 | North America/USA/USA/ | UW Virology Lab | UW Virology Lab | Pavitra Roychoudhury et al |
| EPI_ISL_576515 | USA/UN-UW-7770/2020 | 2020-04-20 | North America/USA/USA/ | UW Virology Lab | UW Virology Lab | Pavitra Roychoudhury et al |
| EPI_ISL_576519 | USA/CA-IGI-0321/2020 | 2020-07-17 | North America/USA/California/Alameda | Innovative Genomics Institute, UC Berkeley | Innovative Genomics Institute, UC Berkeley | Stacia Wyman et al |
| EPI_ISL_576522 | USA/CA-IGI-0330/2020 | 2020-06-26 | North America/USA/California/Alameda | Innovative Genomics Institute, UC Berkeley | Innovative Genomics Institute, UC Berkeley | Stacia Wyman et al |

| | | | | | | |
|---|---|---|---|---|---|---|
| EPI_ISL_576523 | USA/CA-IGI-0331/2020 | 2020-07-16 | North America/USA/California/Alameda | Innovative Genomics Institute, UC Berkeley | Innovative Genomics Institute, UC Berkeley | Stacia Wyman et al |
| EPI_ISL_576525 | USA/CA-IGI-0335/2020 | 2020-07-15 | North America/USA/California/Alameda | Innovative Genomics Institute, UC Berkeley | Innovative Genomics Institute, UC Berkeley | Stacia Wyman et al |
| EPI_ISL_576526 | USA/CA-IGI-0336/2020 | 2020-07-09 | North America/USA/California/Contra C | Innovative Genomics Institute, UC Berkeley | Innovative Genomics Institute, UC Berkeley | Stacia Wyman et al |
| EPI_ISL_576528 | USA/CA-IGI-0339/2020 | 2020-07-09 | North America/USA/California/Alameda | Innovative Genomics Institute, UC Berkeley | Innovative Genomics Institute, UC Berkeley | Stacia Wyman et al |
| EPI_ISL_576532 | USA/CA-IGI-0344/2020 | 2020-07-15 | North America/USA/California/Alameda | Innovative Genomics Institute, UC Berkeley | Innovative Genomics Institute, UC Berkeley | Stacia Wyman et al |
| EPI_ISL_576538 | USA/CA-IGI-0352/2020 | 2020-07-15 | North America/USA/California/Contra C | Innovative Genomics Institute, UC Berkeley | Innovative Genomics Institute, UC Berkeley | Stacia Wyman et al |
| EPI_ISL_576550 | USA/CA-IGI-0368/2020 | 2020-07-16 | North America/USA/California/Contra C | Innovative Genomics Institute, UC Berkeley | Innovative Genomics Institute, UC Berkeley | Stacia Wyman et al |
| EPI_ISL_576551 | USA/CA-IGI-0369/2020 | 2020-06-26 | North America/USA/California/Alameda | Innovative Genomics Institute, UC Berkeley | Innovative Genomics Institute, UC Berkeley | Stacia Wyman et al |
| EPI_ISL_576557 | USA/WA-UW-29526/2020 | 2020-09-29 | North America/USA/Washington/ | UW Virology Lab | UW Virology Lab | Pavitra Roychoudhury et al |
| EPI_ISL_576634 | England/ALDP-9D7E2D/2020 | 2020-09-17 | Europe/United Kingdom/England/ | Oxford Viromics, NDM, University of Oxford; Oxford University Hospitals; Basingstoke and North Hampshire Hospital | COVID-19 Genomics UK (COG-UK) Consortium | Tanya Golubchik et al |
| EPI_ISL_576682 | England/ALDP-9D8EE0/2020 | 2020-09-18 | Europe/United Kingdom/England/ | Oxford Viromics, NDM, University of Oxford; Oxford University Hospitals; Basingstoke and North Hampshire Hospital | COVID-19 Genomics UK (COG-UK) Consortium | Tanya Golubchik et al |

| | | | | | | |
|---|---|---|---|---|---|---|
| EPI_ISL_576748 | England/ALDP-9D9614/2020 | 2020-09-18 | Europe/United Kingdom/England/ | Oxford Viromics, NDM, University of Oxford; Oxford University Hospitals; Basingstoke and North Hampshire Hospital | COVID-19 Genomics UK (COG-UK) Consortium | Tanya Golubchik et al |
| EPI_ISL_576825 | England/ALDP-9D9EFE/2020 | 2020-09-18 | Europe/United Kingdom/England/ | Oxford Viromics, NDM, University of Oxford; Oxford University Hospitals; Basingstoke and North Hampshire Hospital | COVID-19 Genomics UK (COG-UK) Consortium | Tanya Golubchik et al |
| EPI_ISL_576828 | England/ALDP-9D9F46/2020 | 2020-09-18 | Europe/United Kingdom/England/ | Oxford Viromics, NDM, University of Oxford; Oxford University Hospitals; Basingstoke and North Hampshire Hospital | COVID-19 Genomics UK (COG-UK) Consortium | Tanya Golubchik et al |
| EPI_ISL_576893 | England/BIRM-63E98/2020 | 2020-09-04 | Europe/United Kingdom/England/ | University of Birmingham | COVID-19 Genomics UK (COG-UK) Consortium | Institute of Microbiology et al |
| EPI_ISL_576912 | England/CAMB-1B6475/2020 | 2020-09-22 | Europe/United Kingdom/England/ | Department of Pathology, University of Cambridge | COVID-19 Genomics UK (COG-UK) Consortium | Aminu S. Jahun et al |
| EPI_ISL_576956 | England/CAMC-9E0085/2020 | 2020-09-21 | Europe/United Kingdom/England/ | Oxford Viromics, NDM, University of Oxford; Oxford University Hospitals; Basingstoke and North Hampshire Hospital | COVID-19 Genomics UK (COG-UK) Consortium | Tanya Golubchik et al |
| EPI_ISL_577072 | England/MILK-9E134F/2020 | 2020-09-21 | Europe/United Kingdom/England/ | Oxford Viromics, NDM, University of Oxford; Oxford University Hospitals; Basingstoke and North Hampshire Hospital | COVID-19 Genomics UK (COG-UK) Consortium | Tanya Golubchik et al |
| EPI_ISL_577123 | England/NOTT-113F39/2020 | 2020-10-01 | Europe/United Kingdom/England/ | Queens Medical Centre, Clinical Microbiology Department / DeepSeq Nottingham | COVID-19 Genomics UK (COG-UK) Consortium | Gemma Clark et al |
| EPI_ISL_577145 | England/NOTT-11418D/2020 | 2020-10-04 | Europe/United Kingdom/England/ | Queens Medical Centre, Clinical Microbiology Department / DeepSeq Nottingham | COVID-19 Genomics UK (COG-UK) Consortium | Gemma Clark et al |
| EPI_ISL_577368 | Scotland/QEUH-9AE715/2020 | 2020-09-05 | Europe/United Kingdom/Scotland/ | University of Exeter | COVID-19 Genomics UK (COG-UK) Consortium | Ben Temperton et al |
| EPI_ISL_577437 | Wales/PHWC-16D71A/2020 | 2020-09-21 | Europe/United Kingdom/Wales/ | Wales Specialist Virology Centre Sequencing lab: Pathogen Genomics Unit | COVID-19 Genomics UK (COG-UK) Consortium | Catherine Moore et al |
| EPI_ISL_577467 | Wales/PHWC-16D932/2020 | 2020-09-21 | Europe/United Kingdom/Wales/ | Wales Specialist Virology Centre Sequencing lab: Pathogen Genomics Unit | COVID-19 Genomics UK (COG-UK) Consortium | Catherine Moore et al |

| | | | | | | |
|---|---|---|---|---|---|---|
| EPI_ISL_577557 | USA/MI-MDHHS-SC22004/2020 | 2020-08-21 | North America/USA/Michigan/ | Michigan Department of Health and Human Services, Bureau of Laboratories | Michigan Department of Health and Human Services, Bureau of Laboratories | Blankenship HM et al |
| EPI_ISL_577568 | USA/MI-MDHHS-SC22006/2020 | 2020-08-26 | North America/USA/Michigan/ | Michigan Department of Health and Human Services, Bureau of Laboratories | Michigan Department of Health and Human Services, Bureau of Laboratories | Blankenship HM et al |
| EPI_ISL_577569 | USA/MI-MDHHS-SC22007/2020 | 2020-08-26 | North America/USA/Michigan/ | Michigan Department of Health and Human Services, Bureau of Laboratories | Michigan Department of Health and Human Services, Bureau of Laboratories | Blankenship HM et al |
| EPI_ISL_577572 | USA/MI-MDHHS-SC22010/2020 | 2020-09-01 | North America/USA/Michigan/ | Michigan Department of Health and Human Services, Bureau of Laboratories | Michigan Department of Health and Human Services, Bureau of Laboratories | Blankenship HM et al |
| EPI_ISL_577576 | USA/MI-MDHHS-SC22016/2020 | 2020-09-02 | North America/USA/Michigan/ | Michigan Department of Health and Human Services, Bureau of Laboratories | Michigan Department of Health and Human Services, Bureau of Laboratories | Blankenship HM et al |
| EPI_ISL_577584 | USA/MI-MDHHS-SC22028/2020 | 2020-09-09 | North America/USA/Michigan/ | Michigan Department of Health and Human Services, Bureau of Laboratories | Michigan Department of Health and Human Services, Bureau of Laboratories | Blankenship HM et al |

| | | | | | | |
|---|---|---|---|---|---|---|
| EPI_ISL_577590 | USA/MI-MDHHS-SC22039/2020 | 2020-09-01 | North America/USA/Michigan/ | Michigan Department of Health and Human Services, Bureau of Laboratories | Michigan Department of Health and Human Services, Bureau of Laboratories | Blankenship HM et al |
| EPI_ISL_577591 | USA/MI-MDHHS-SC22041/2020 | 2020-09-15 | North America/USA/Michigan/ | Michigan Department of Health and Human Services, Bureau of Laboratories | Michigan Department of Health and Human Services, Bureau of Laboratories | Blankenship HM et al |
| EPI_ISL_577592 | USA/MI-MDHHS-SC22042/2020 | 2020-09-15 | North America/USA/Michigan/ | Michigan Department of Health and Human Services, Bureau of Laboratories | Michigan Department of Health and Human Services, Bureau of Laboratories | Blankenship HM et al |
| EPI_ISL_577627 | CzechRepublic/NRL_8118-6/2020 | 2020-08-06 | Europe/Czech Republic/Usti nad Labem/ | The National Institute of Public Health | State Veterinary Institute Prague | Nagy et al |
| EPI_ISL_577630 | CzechRepublic/NRL_6849-7/2020 | 2020-09-15 | Europe/Czech Republic/Prague/ | The National Institute of Public Health | State Veterinary Institute Prague | Nagy et al |
| EPI_ISL_577639 | India/MH-NIV-191373/2020 | 2020-09-02 | Asia/India/Maharashtra/ | NIV Influenza | NIV Influenza | Potdar V et al |
| EPI_ISL_577640 | India/MH-NIV-226874/2020 | 2020-09-25 | Asia/India/Maharashtra/ | NIV Influenza | NIV Influenza | Potdar V et al |
| EPI_ISL_577641 | India/MH-NIV-206486/2020 | 2020-09-10 | Asia/India/Maharashtra/ | NIV Influenza | NIV Influenza | Potdar V et al |
| EPI_ISL_577642 | India/MH-NIV-206956/2020 | 2020-09-11 | Asia/India/Maharashtra/ | NIV Influenza | NIV Influenza | Potdar V et al |
| EPI_ISL_577643 | India/MH-NIV-212454/2020 | 2020-09-15 | Asia/India/Maharashtra/ | NIV Influenza | NIV Influenza | Potdar V et al |
| EPI_ISL_577644 | India/MH-NIV-213875/2020 | 2020-09-16 | Asia/India/Maharashtra/ | NIV Influenza | NIV Influenza | Potdar V et al |
| EPI_ISL_577645 | India/MH-NIV-215370/2020 | 2020-09-16 | Asia/India/Maharashtra/ | NIV Influenza | NIV Influenza | Potdar V et al |
| EPI_ISL_577646 | India/MH-NIV-214788/2020 | 2020-09-15 | Asia/India/Maharashtra/ | NIV Influenza | NIV Influenza | Potdar V et al |
| EPI_ISL_577647 | India/MH-NIV-223127/2020 | 2020-09-23 | Asia/India/Maharashtra/ | NIV Influenza | NIV Influenza | Potdar V et al |
| EPI_ISL_577648 | India/MH-NIV-210557/2020 | 2020-09-14 | Asia/India/Maharashtra/ | NIV Influenza | NIV Influenza | Potdar V et al |
| EPI_ISL_577649 | India/MH-NIV-223537/2020 | 2020-09-23 | Asia/India/Maharashtra/ | NIV Influenza | NIV Influenza | Potdar V et al |
| EPI_ISL_577650 | India/MH-NIV-221309/2020 | 2020-09-22 | Asia/India/Maharashtra/ | NIV Influenza | NIV Influenza | Potdar V et al |
| EPI_ISL_577651 | India/MH-NIV-221624/2020 | 2020-09-21 | Asia/India/Maharashtra/ | NIV Influenza | NIV Influenza | Potdar V et al |
| EPI_ISL_577652 | India/MH-NIV-222131/2020 | 2020-09-22 | Asia/India/Maharashtra/ | NIV Influenza | NIV Influenza | Potdar V et al |

| | | | | | | |
|---|---|---|---|---|---|---|
| EPI_ISL_577654 | India/MH-NIV-219744/2020 | 2020-09-20 | Asia/India/Maharashtra/ | NIV Influenza | NIV Influenza | Potdar V et al |
| EPI_ISL_577655 | India/MH-NIV-220301/2020 | 2020-09-21 | Asia/India/Maharashtra/ | NIV Influenza | NIV Influenza | Potdar V et al |
| EPI_ISL_577656 | India/MH-NIV-218275/2020 | 2020-09-19 | Asia/India/Maharashtra/ | NIV Influenza | NIV Influenza | Potdar V et al |
| EPI_ISL_577657 | India/MH-NIV-217910/2020 | 2020-09-18 | Asia/India/Maharashtra/ | NIV Influenza | NIV Influenza | Potdar V et al |
| EPI_ISL_577658 | India/MH-NIV-223126/2020 | 2020-09-23 | Asia/India/Maharashtra/ | NIV Influenza | NIV Influenza | Potdar V et al |
| EPI_ISL_577659 | India/MH-NIV-216133/2020 | 2020-09-18 | Asia/India/Maharashtra/ | NIV Influenza | NIV Influenza | Potdar V et al |
| EPI_ISL_577660 | India/MH-NIV-216500/2020 | 2020-09-18 | Asia/India/Maharashtra/ | NIV Influenza | NIV Influenza | Potdar V et al |
| EPI_ISL_577661 | India/MH-NIV-207705/2020 | 2020-09-11 | Asia/India/Maharashtra/ | NIV Influenza | NIV Influenza | Potdar V et al |
| EPI_ISL_577662 | India/MH-NIV-216641/2020 | 2020-09-18 | Asia/India/Maharashtra/ | NIV Influenza | NIV Influenza | Potdar V et al |
| EPI_ISL_577663 | India/MH-NIV-209697/2020 | 2020-09-12 | Asia/India/Maharashtra/ | NIV Influenza | NIV Influenza | Potdar V et al |
| EPI_ISL_577664 | India/MH-NIV-216244/2020 | 2020-09-17 | Asia/India/Maharashtra/ | NIV Influenza | NIV Influenza | Potdar V et al |
| EPI_ISL_577665 | India/MH-NIV-216880/2020 | 2020-09-18 | Asia/India/Maharashtra/ | NIV Influenza | NIV Influenza | Potdar V et al |
| EPI_ISL_577666 | India/MH-NIV-206415/2020 | 2020-09-10 | Asia/India/Maharashtra/ | NIV Influenza | NIV Influenza | Potdar V et al |
| EPI_ISL_577667 | India/MH-NIV-206926/2020 | 2020-09-10 | Asia/India/Maharashtra/ | NIV Influenza | NIV Influenza | Potdar V et al |
| EPI_ISL_577668 | India/MH-NIV-207312/2020 | 2020-09-11 | Asia/India/Maharashtra/ | NIV Influenza | NIV Influenza | Potdar V et al |
| EPI_ISL_577669 | India/MH-NIV-209852/2020 | 2020-09-13 | Asia/India/Maharashtra/ | NIV Influenza | NIV Influenza | Potdar V et al |
| EPI_ISL_577670 | India/MH-NIV-210934/2020 | 2020-09-14 | Asia/India/Maharashtra/ | NIV Influenza | NIV Influenza | Potdar V et al |
| EPI_ISL_577671 | India/MH-NIV-185989/2020 | 2020-08-29 | Asia/India/Maharashtra/ | NIV Influenza | NIV Influenza | Potdar V et al |
| EPI_ISL_577672 | India/MH-NIV-197096/2020 | 2020-09-05 | Asia/India/Maharashtra/ | NIV Influenza | NIV Influenza | Potdar V et al |
| EPI_ISL_577673 | India/MH-NIV-203951/2020 | 2020-09-09 | Asia/India/Maharashtra/ | NIV Influenza | NIV Influenza | Potdar V et al |
| EPI_ISL_577674 | India/MH-NIV-208140/2020 | 2020-09-11 | Asia/India/Maharashtra/ | NIV Influenza | NIV Influenza | Potdar V et al |
| EPI_ISL_577675 | India/MH-NIV-212999/2020 | 2020-09-15 | Asia/India/Maharashtra/ | NIV Influenza | NIV Influenza | Potdar V et al |
| EPI_ISL_577676 | India/MH-NIV-214342/2020 | 2020-09-16 | Asia/India/Maharashtra/ | NIV Influenza | NIV Influenza | Potdar V et al |
| EPI_ISL_577677 | India/MH-NIV-229804/2020 | 2020-09-29 | Asia/India/Maharashtra/ | NIV Influenza | NIV Influenza | Potdar V et al |
| EPI_ISL_577678 | India/MH-NIV-217242/2020 | 2020-09-17 | Asia/India/Maharashtra/ | NIV Influenza | NIV Influenza | Potdar V et al |
| EPI_ISL_577679 | India/MH-NIV-229467/2020 | 2020-09-28 | Asia/India/Maharashtra/ | NIV Influenza | NIV Influenza | Potdar V et al |
| EPI_ISL_577680 | India/MH-NIV-229468/2020 | 2020-09-28 | Asia/India/Maharashtra/ | NIV Influenza | NIV Influenza | Potdar V et al |
| EPI_ISL_577681 | India/MH-NIV-229461/2020 | 2020-09-28 | Asia/India/Maharashtra/ | NIV Influenza | NIV Influenza | Potdar V et al |
| EPI_ISL_577682 | India/MH-NIV-229428/2020 | 2020-09-29 | Asia/India/Maharashtra/ | NIV Influenza | NIV Influenza | Potdar V et al |
| EPI_ISL_577683 | India/MH-NIV-229512/2020 | 2020-09-28 | Asia/India/Maharashtra/ | NIV Influenza | NIV Influenza | Potdar V et al |
| EPI_ISL_577684 | India/MH-NIV-229539/2020 | 2020-09-28 | Asia/India/Maharashtra/ | NIV Influenza | NIV Influenza | Potdar V et al |

| | | | | | | |
|---|---|---|---|---|---|---|
| EPI_ISL_577685 | India/MH-NIV-229542/2020 | 2020-09-28 | Asia/India/Maharashtra/ | NIV Influenza | NIV Influenza | Potdar V et al |
| EPI_ISL_577686 | India/MH-NIV-229545/2020 | 2020-09-28 | Asia/India/Maharashtra/ | NIV Influenza | NIV Influenza | Potdar V et al |
| EPI_ISL_577687 | India/MH-NIV-229555/2020 | 2020-09-28 | Asia/India/Maharashtra/ | NIV Influenza | NIV Influenza | Potdar V et al |
| EPI_ISL_577688 | India/MH-NIV-229566/2020 | 2020-09-28 | Asia/India/Maharashtra/ | NIV Influenza | NIV Influenza | Potdar V et al |
| EPI_ISL_577689 | India/MH-NIV-229576/2020 | 2020-09-28 | Asia/India/Maharashtra/ | NIV Influenza | NIV Influenza | Potdar V et al |
| EPI_ISL_577690 | India/MH-NIV-229597/2020 | 2020-09-28 | Asia/India/Maharashtra/ | NIV Influenza | NIV Influenza | Potdar V et al |
| EPI_ISL_577691 | India/MH-NIV-229622/2020 | 2020-09-28 | Asia/India/Maharashtra/ | NIV Influenza | NIV Influenza | Potdar V et al |
| EPI_ISL_577692 | India/MH-NIV-229643/2020 | 2020-09-29 | Asia/India/Maharashtra/ | NIV Influenza | NIV Influenza | Potdar V et al |
| EPI_ISL_577693 | India/MH-NIV-229644/2020 | 2020-09-29 | Asia/India/Maharashtra/ | NIV Influenza | NIV Influenza | Potdar V et al |
| EPI_ISL_577694 | India/MH-NIV-229656/2020 | 2020-09-29 | Asia/India/Maharashtra/ | NIV Influenza | NIV Influenza | Potdar V et al |
| EPI_ISL_577695 | India/MH-NIV-229665/2020 | 2020-09-29 | Asia/India/Maharashtra/ | NIV Influenza | NIV Influenza | Potdar V et al |
| EPI_ISL_577696 | India/MH-NIV-229684/2020 | 2020-09-28 | Asia/India/Maharashtra/ | NIV Influenza | NIV Influenza | Potdar V et al |
| EPI_ISL_577697 | India/MH-NIV-229699/2020 | 2020-09-29 | Asia/India/Maharashtra/ | NIV Influenza | NIV Influenza | Potdar V et al |
| EPI_ISL_577698 | India/MH-NIV-229715/2020 | 2020-09-29 | Asia/India/Maharashtra/ | NIV Influenza | NIV Influenza | Potdar V et al |
| EPI_ISL_577699 | India/MH-NIV-229717/2020 | 2020-09-29 | Asia/India/Maharashtra/ | NIV Influenza | NIV Influenza | Potdar V et al |
| EPI_ISL_577700 | India/MH-NIV-229720/2020 | 2020-09-29 | Asia/India/Maharashtra/ | NIV Influenza | NIV Influenza | Potdar V et al |
| EPI_ISL_577701 | India/MH-NIV-229771/2020 | 2020-09-29 | Asia/India/Maharashtra/ | NIV Influenza | NIV Influenza | Potdar V et al |
| EPI_ISL_577702 | India/MH-NIV-229912/2020 | 2020-09-28 | Asia/India/Maharashtra/ | NIV Influenza | NIV Influenza | Potdar V et al |
| EPI_ISL_577704 | India/MH-NIV-206715/2020 | 2020-09-11 | Asia/India/Maharashtra/ | NIV Influenza | NIV Influenza | Potdar V et al |
| EPI_ISL_577705 | India/MH-NIV-211414/2020 | 2020-09-14 | Asia/India/Maharashtra/ | NIV Influenza | NIV Influenza | Potdar V et al |
| EPI_ISL_577706 | India/MH-NIV-227343/2020 | 2020-09-26 | Asia/India/Maharashtra/ | NIV Influenza | NIV Influenza | Potdar V et al |
| EPI_ISL_577707 | India/MH-NIV-QC-1459/2020 | 2020-08-14 | Asia/India/Maharashtra/ | NIV Influenza | NIV Influenza | Potdar V et al |
| EPI_ISL_577708 | India/MH-NIV-206457-1/2020 | 2020-09-10 | Asia/India/Maharashtra/ | NIV Influenza | NIV Influenza | Potdar V et al |
| EPI_ISL_577709 | India/MH-NIV-229898/2020 | 2020-09-28 | Asia/India/Maharashtra/ | NIV Influenza | NIV Influenza | Potdar V et al |
| EPI_ISL_577710 | India/MH-NIV-229297/2020 | 2020-09-28 | Asia/India/Maharashtra/ | NIV Influenza | NIV Influenza | Potdar V et al |
| EPI_ISL_577711 | India/MH-NIV-229360/2020 | 2020-09-29 | Asia/India/Maharashtra/ | NIV Influenza | NIV Influenza | Potdar V et al |
| EPI_ISL_577712 | India/MH-NIV-229453/2020 | 2020-09-29 | Asia/India/Maharashtra/ | NIV Influenza | NIV Influenza | Potdar V et al |
| EPI_ISL_577713 | India/MH-NIV-229460/2020 | 2020-09-28 | Asia/India/Maharashtra/ | NIV Influenza | NIV Influenza | Potdar V et al |
| EPI_ISL_577716 | India/MH-NIV-206457/2020 | 2020-09-10 | Asia/India/Maharashtra/ | NIV Influenza | NIV Influenza | Potdar V et al |
| EPI_ISL_577717 | India/MH-NIV-229388/2020 | 2020-09-29 | Asia/India/Maharashtra/ | NIV Influenza | NIV Influenza | Potdar V et al |
| EPI_ISL_577720 | India/MH-NIV-226709/2020 | 2020-09-25 | Asia/India/Maharashtra/ | NIV Influenza | NIV Influenza | Potdar V et al |

| | | | | | | |
|---|---|---|---|---|---|---|
| EPI_ISL_577721 | India/MH-NIV-227244/2020 | 2020-09-26 | Asia/India/Maharashtra/ | NIV Influenza | NIV Influenza | Potdar V et al |
| EPI_ISL_577722 | India/MH-NIV-227243/2020 | 2020-09-26 | Asia/India/Maharashtra/ | NIV Influenza | NIV Influenza | Potdar V et al |
| EPI_ISL_577723 | India/MH-NIV-227512/2020 | 2020-09-26 | Asia/India/Maharashtra/ | NIV Influenza | NIV Influenza | Potdar V et al |
| EPI_ISL_577725 | India/MH-NIV-QC-1379/2020 | 2020-07-21 | Asia/India/Maharashtra/ | NIV Influenza | NIV Influenza | Potdar V et al |
| EPI_ISL_577726 | India/MH-NIV-QC-1381/2020 | 2020-07-22 | Asia/India/Maharashtra/ | NIV Influenza | NIV Influenza | Potdar V et al |
| EPI_ISL_577727 | India/MH-NIV-QC-1444/2020 | 2020-09-08 | Asia/India/Maharashtra/ | NIV Influenza | NIV Influenza | Potdar V et al |
| EPI_ISL_577728 | India/MH-NIV-QC-1450/2020 | 2020-09-08 | Asia/India/Maharashtra/ | NIV Influenza | NIV Influenza | Potdar V et al |
| EPI_ISL_577729 | India/MH-NIV-QC-1405/2020 | 2020-07-25 | Asia/India/Maharashtra/ | NIV Influenza | NIV Influenza | Potdar V et al |
| EPI_ISL_577730 | India/MH-NIV-QC-1844/2020 | 2020-09-15 | Asia/India/Maharashtra/ | NIV Influenza | NIV Influenza | Potdar V et al |
| EPI_ISL_577731 | India/MH-NIV-QC-1848/2020 | 2020-09-13 | Asia/India/Maharashtra/ | NIV Influenza | NIV Influenza | Potdar V et al |
| EPI_ISL_577735 | Slovakia/UKBA-203/2020 | 2020-04-06 | Europe/Slovakia/Kosice/Zehra | Institute of Virology, Biomedical Research Center of the Slovak Academy of Sciences, Bratislava | Faculty of Natural Sciences, Comenius University, Bratislava | Kristv≠na Bor≈°ov√° et al |
| EPI_ISL_577736 | Slovakia/UKBA-204/2020 | 2020-04-06 | Europe/Slovakia/Bratislava/ | Institute of Virology, Biomedical Research Center of the Slovak Academy of Sciences, Bratislava | Faculty of Natural Sciences, Comenius University, Bratislava | Viktv≥ria Hodorov√° et al |
| EPI_ISL_577737 | Slovakia/UKBA-205/2020 | 2020-04-13 | Europe/Slovakia/Kosice/Zehra | Institute of Virology, Biomedical Research Center of the Slovak Academy of Sciences, Bratislava | Faculty of Natural Sciences, Comenius University, Bratislava | Kristv≠na Bor≈°ov√° et al |
| EPI_ISL_577738 | Slovakia/UKBA-207/2020 | 2020-04-29 | Europe/Slovakia/Pezinok/ | Institute of Virology, Biomedical Research Center of the Slovak Academy of Sciences, Bratislava | Faculty of Natural Sciences, Comenius University, Bratislava | Viktv≥ria Hodorov√° et al |
| EPI_ISL_577742 | Slovakia/UKBA-212/2020 | 2020-03-31 | Europe/Slovakia/Bratislava/ | Institute of Virology, Biomedical Research Center of the Slovak Academy of Sciences, Bratislava | Faculty of Natural Sciences, Comenius University, Bratislava | Bro≈àa Brejov√° et al |
| EPI_ISL_577829 | Netherlands/GE-EMC-302/2020 | 2020-09-19 | Europe/Netherlands/Gelderland/ | Dutch COVID-19 response team | Erasmus Medical Center | Bas Oude Munnink et al |

| | | | | | | |
|---|---|---|---|---|---|---|
| EPI_ISL_577833 | Netherlands/LI-EMC-87/2020 | 2020-09-22 | Europe/Netherlands/Limburg/ | Dutch COVID-19 response team | Erasmus Medical Center | Bas Oude Munnink et al |
| EPI_ISL_577836 | Netherlands/LI-EMC-90/2020 | 2020-09-27 | Europe/Netherlands/Limburg/ | Dutch COVID-19 response team | Erasmus Medical Center | Bas Oude Munnink et al |
| EPI_ISL_577857 | Netherlands/NH-EMC-61/2020 | 2020-08-20 | Europe/Netherlands/North Holland/ | Dutch COVID-19 response team | Erasmus Medical Center | Bas Oude Munnink et al |
| EPI_ISL_577897 | Netherlands/UT-EMC-81/2020 | 2020-08-26 | Europe/Netherlands/Utrecht/ | Dutch COVID-19 response team | Erasmus Medical Center | Bas Oude Munnink et al |
| EPI_ISL_577905 | Netherlands/UT-EMC-89/2020 | 2020-08-30 | Europe/Netherlands/Utrecht/ | Dutch COVID-19 response team | Erasmus Medical Center | Bas Oude Munnink et al |
| EPI_ISL_577909 | Netherlands/ZH-EMC-479/2020 | 2020-08-15 | Europe/Netherlands/South Holland/ | Dutch COVID-19 response team | Erasmus Medical Center | Bas Oude Munnink et al |
| EPI_ISL_577911 | Netherlands/ZH-EMC-481/2020 | 2020-08-10 | Europe/Netherlands/South Holland/ | Dutch COVID-19 response team | Erasmus Medical Center | Bas Oude Munnink et al |
| EPI_ISL_577914 | Netherlands/ZH-EMC-484/2020 | 2020-08-14 | Europe/Netherlands/South Holland/ | Dutch COVID-19 response team | Erasmus Medical Center | Bas Oude Munnink et al |
| EPI_ISL_577937 | Netherlands/ZH-EMC-508/2020 | 2020-08-26 | Europe/Netherlands/South Holland/ | Dutch COVID-19 response team | Erasmus Medical Center | Bas Oude Munnink et al |
| EPI_ISL_577950 | Netherlands/ZH-EMC-522/2020 | 2020-09-01 | Europe/Netherlands/South Holland/ | Dutch COVID-19 response team | Erasmus Medical Center | Bas Oude Munnink et al |
| EPI_ISL_577969 | Netherlands/ZH-EMC-541/2020 | 2020-09-16 | Europe/Netherlands/South Holland/ | Dutch COVID-19 response team | Erasmus Medical Center | Bas Oude Munnink et al |
| EPI_ISL_577974 | Netherlands/ZH-EMC-546/2020 | 2020-09-16 | Europe/Netherlands/South Holland/ | Dutch COVID-19 response team | Erasmus Medical Center | Bas Oude Munnink et al |
| EPI_ISL_577981 | Netherlands/ZH-EMC-553/2020 | 2020-09-08 | Europe/Netherlands/South Holland/ | Dutch COVID-19 response team | Erasmus Medical Center | Bas Oude Munnink et al |
| EPI_ISL_577983 | Netherlands/ZH-EMC-555/2020 | 2020-09-20 | Europe/Netherlands/South Holland/ | Dutch COVID-19 response team | Erasmus Medical Center | Bas Oude Munnink et al |
| EPI_ISL_577990 | Netherlands/ZH-EMC-562/2020 | 2020-09-20 | Europe/Netherlands/South Holland/ | Dutch COVID-19 response team | Erasmus Medical Center | Bas Oude Munnink et al |
| EPI_ISL_577993 | Netherlands/ZH-EMC-565/2020 | 2020-09-20 | Europe/Netherlands/South Holland/ | Dutch COVID-19 response team | Erasmus Medical Center | Bas Oude Munnink et al |
| EPI_ISL_577997 | Netherlands/ZH-EMC-569/2020 | 2020-09-20 | Europe/Netherlands/South Holland/ | Dutch COVID-19 response team | Erasmus Medical Center | Bas Oude Munnink et al |
| EPI_ISL_578005 | Netherlands/ZH-EMC-577/2020 | 2020-09-19 | Europe/Netherlands/South Holland/ | Dutch COVID-19 response team | Erasmus Medical Center | Bas Oude Munnink et al |
| EPI_ISL_578009 | Netherlands/ZH-EMC-581/2020 | 2020-09-18 | Europe/Netherlands/South Holland/ | Dutch COVID-19 response team | Erasmus Medical Center | Bas Oude Munnink et al |
| EPI_ISL_578019 | Netherlands/ZH-EMC-591/2020 | 2020-09-18 | Europe/Netherlands/South Holland/ | Dutch COVID-19 response team | Erasmus Medical Center | Bas Oude Munnink et al |

| | | | | | | |
|---|---|---|---|---|---|---|
| EPI_ISL_578022 | Netherlands/ZH-EMC-594/2020 | 2020-09-18 | Europe/Netherlands/South Holland/ | Dutch COVID-19 response team | Erasmus Medical Center | Bas Oude Munnink et al |
| EPI_ISL_578023 | Netherlands/ZH-EMC-595/2020 | 2020-09-18 | Europe/Netherlands/South Holland/ | Dutch COVID-19 response team | Erasmus Medical Center | Bas Oude Munnink et al |
| EPI_ISL_578024 | Netherlands/ZH-EMC-596/2020 | 2020-09-18 | Europe/Netherlands/South Holland/ | Dutch COVID-19 response team | Erasmus Medical Center | Bas Oude Munnink et al |
| EPI_ISL_578030 | Netherlands/ZH-EMC-602/2020 | 2020-09-18 | Europe/Netherlands/South Holland/ | Dutch COVID-19 response team | Erasmus Medical Center | Bas Oude Munnink et al |
| EPI_ISL_578031 | Netherlands/ZH-EMC-603/2020 | 2020-09-18 | Europe/Netherlands/South Holland/ | Dutch COVID-19 response team | Erasmus Medical Center | Bas Oude Munnink et al |
| EPI_ISL_578037 | Netherlands/ZH-EMC-609/2020 | 2020-09-19 | Europe/Netherlands/South Holland/ | Dutch COVID-19 response team | Erasmus Medical Center | Bas Oude Munnink et al |
| EPI_ISL_578040 | Netherlands/ZH-EMC-612/2020 | 2020-09-22 | Europe/Netherlands/South Holland/ | Dutch COVID-19 response team | Erasmus Medical Center | Bas Oude Munnink et al |
| EPI_ISL_578046 | Netherlands/ZH-EMC-618/2020 | 2020-09-22 | Europe/Netherlands/South Holland/ | Dutch COVID-19 response team | Erasmus Medical Center | Bas Oude Munnink et al |
| EPI_ISL_578049 | Netherlands/ZH-EMC-621/2020 | 2020-09-18 | Europe/Netherlands/South Holland/ | Dutch COVID-19 response team | Erasmus Medical Center | Bas Oude Munnink et al |
| EPI_ISL_578050 | Netherlands/ZH-EMC-622/2020 | 2020-09-24 | Europe/Netherlands/South Holland/ | Dutch COVID-19 response team | Erasmus Medical Center | Bas Oude Munnink et al |
| EPI_ISL_578060 | Netherlands/ZH-EMC-632/2020 | 2020-09-21 | Europe/Netherlands/South Holland/ | Dutch COVID-19 response team | Erasmus Medical Center | Bas Oude Munnink et al |
| EPI_ISL_578065 | Netherlands/ZH-EMC-637/2020 | 2020-09-21 | Europe/Netherlands/South Holland/ | Dutch COVID-19 response team | Erasmus Medical Center | Bas Oude Munnink et al |
| EPI_ISL_578079 | Netherlands/ZH-EMC-651/2020 | 2020-09-27 | Europe/Netherlands/South Holland/ | Dutch COVID-19 response team | Erasmus Medical Center | Bas Oude Munnink et al |
| EPI_ISL_578080 | India/WB-IICB-006/2020 | 2020-09-10 | Asia/India/West Bengal/ | CSIR-Indian Institute of Chemical Biology, MEDICA Supercpecialty Hospital Kolkata | CSIR-Indian Institute of Chemical Biology, MEDICA Supercpecialty Hospital Kolkata | Sujay Krishna Maity et al |
| EPI_ISL_578081 | India/WB-IICB-007/2020 | 2020-09-10 | Asia/India/West Bengal/ | CSIR-Indian Institute of Chemical Biology, MEDICA Supercpecialty Hospital Kolkata | CSIR-Indian Institute of Chemical Biology, MEDICA Supercpecialty Hospital Kolkata | Sujay Krishna Maity et al |

| | | | | | | |
|---|---|---|---|---|---|---|
| EPI_ISL_578100 | USA/MI-UM-10033768108/2020 | 2020-03-24 | North America/USA/Michigan/ | University of Michigan Clinical Microbiology Laboratory | Lauring Lab, University of Michigan, Department of Microbiology and Immunology | Valesano et al |
| EPI_ISL_578148 | USA/MI-UM-10033872707/2020 | 2020-04-07 | North America/USA/Michigan/ | University of Michigan Clinical Microbiology Laboratory | Lauring Lab, University of Michigan, Department of Microbiology and Immunology | Valesano et al |
| EPI_ISL_578151 | USA/MI-UM-10033882288/2020 | 2020-04-08 | North America/USA/Michigan/ | University of Michigan Clinical Microbiology Laboratory | Lauring Lab, University of Michigan, Department of Microbiology and Immunology | Valesano et al |
| EPI_ISL_578160 | USA/MI-UM-10033971920/2020 | 2020-04-20 | North America/USA/Michigan/ | University of Michigan Clinical Microbiology Laboratory | Lauring Lab, University of Michigan, Department of Microbiology and Immunology | Valesano et al |
| EPI_ISL_578167 | USA/MI-UM-10034012043/2020 | 2020-04-24 | North America/USA/Michigan/ | University of Michigan Clinical Microbiology Laboratory | Lauring Lab, University of Michigan, Department of Microbiology and Immunology | Valesano et al |
| EPI_ISL_578168 | India/WB-IICB-008/2020 | 2020-09-10 | Asia/India/West Bengal/ | CSIR-Indian Institute of Chemical Biology, MEDICA Supercpecialty Hospital Kolkata | CSIR-Indian Institute of Chemical Biology, MEDICA Supercpecialty Hospital Kolkata | Sujay Krishna Maity et al |

| | | | | | | |
|---|---|---|---|---|---|---|
| EPI_ISL_578172 | USA/MI-UM-10034148754/2020 | 2020-05-08 | North America/USA/Michigan/ | University of Michigan Clinical Microbiology Laboratory | Lauring Lab, University of Michigan, Department of Microbiology and Immunology | Valesano et al |
| EPI_ISL_578175 | India/WB-IICB-009/2020 | 2020-09-10 | Asia/India/West Bengal/ | CSIR-Indian Institute of Chemical Biology, MEDICA Supercpecialty Hospital Kolkata | CSIR-Indian Institute of Chemical Biology, MEDICA Supercpecialty Hospital Kolkata | Sujay Krishna Maity et al |
| EPI_ISL_578178 | India/WB-IICB-010/2020 | 2020-09-10 | Asia/India/West Bengal/ | CSIR-Indian Institute of Chemical Biology, MEDICA Supercpecialty Hospital Kolkata | CSIR-Indian Institute of Chemical Biology, MEDICA Supercpecialty Hospital Kolkata | Sujay Krishna Maity et al |
| EPI_ISL_578179 | India/WB-IICB-011/2020 | 2020-09-10 | Asia/India/West Bengal/ | CSIR-Indian Institute of Chemical Biology, MEDICA Supercpecialty Hospital Kolkata | CSIR-Indian Institute of Chemical Biology, MEDICA Supercpecialty Hospital Kolkata | Sujay Krishna Maity et al |
| EPI_ISL_578180 | India/WB-IICB-012/2020 | 2020-09-10 | Asia/India/West Bengal/ | CSIR-Indian Institute of Chemical Biology, MEDICA Supercpecialty Hospital Kolkata | CSIR-Indian Institute of Chemical Biology, MEDICA Supercpecialty Hospital Kolkata | Sujay Krishna Maity et al |
| EPI_ISL_578181 | India/WB-IICB-013/2020 | 2020-09-10 | Asia/India/West Bengal/ | CSIR-Indian Institute of Chemical Biology, MEDICA Supercpecialty Hospital Kolkata | CSIR-Indian Institute of Chemical Biology, MEDICA Supercpecialty Hospital Kolkata | Sujay Krishna Maity et al |

| | | | | | | |
|---|---|---|---|---|---|---|
| EPI_ISL_578182 | India/WB-IICB-014/2020 | 2020-09-10 | Asia/India/West Bengal/ | CSIR-Indian Institute of Chemical Biology, MEDICA Supercpecialty Hospital Kolkata | CSIR-Indian Institute of Chemical Biology, MEDICA Supercpecialty Hospital Kolkata | Sujay Krishna Maity et al |
| EPI_ISL_578183 | India/WB-IICB-015/2020 | 2020-09-10 | Asia/India/West Bengal/ | CSIR-Indian Institute of Chemical Biology, MEDICA Supercpecialty Hospital Kolkata | CSIR-Indian Institute of Chemical Biology, MEDICA Supercpecialty Hospital Kolkata | Sujay Krishna Maity et al |
| EPI_ISL_578184 | India/WB-IICB-016/2020 | 2020-09-10 | Asia/India/West Bengal/ | CSIR-Indian Institute of Chemical Biology, MEDICA Supercpecialty Hospital Kolkata | CSIR-Indian Institute of Chemical Biology, MEDICA Supercpecialty Hospital Kolkata | Sujay Krishna Maity et al |
| EPI_ISL_578186 | Spain/AN-ISCIII-2013382/2020 | 2020-05-05 | Europe/Spain/Andalusia/ | Hospital General Juan Ramv≥n Jimv©nez | Instituto de Salud Carlos III | Iglesias-Caballero et al |
| EPI_ISL_578193 | Spain/AR-ISCIII-201781/2020 | 2020-03-08 | Europe/Spain/Aragon/ | Hospital Universitario Miguel Servet | Instituto de Salud Carlos III | Iglesias-Caballero et al |
| EPI_ISL_578197 | Spain/RI-ISCIII-201566/2020 | 2020-03-04 | Europe/Spain/La Rioja/ | Hospital San Pedro | Instituto de Salud Carlos III | Iglesias-Caballero et al |
| EPI_ISL_578206 | Ireland/D-NVRL-73IRL38288/2020 | 2020-09-08 | Europe/Ireland/Dublin/ | National Virus Reference Laboratory | National Virus Reference Laboratory | Michael Carr et al |
| EPI_ISL_578261 | Ireland/D-NVRL-73IRL22431/2020 | 2020-09-16 | Europe/Ireland/Dublin/ | National Virus Reference Laboratory | National Virus Reference Laboratory | Michael Carr et al |
| EPI_ISL_578295 | Ireland/KE-NVRL-73IRL25972/2020 | 2020-09-05 | Europe/Ireland/Kildare/ | National Virus Reference Laboratory | National Virus Reference Laboratory | Michael Carr et al |
| EPI_ISL_578319 | Ireland/RN-NVRL-73IRL30636/2020 | 2020-09-16 | Europe/Ireland/Roscommon/ | National Virus Reference Laboratory | National Virus Reference Laboratory | Michael Carr et al |
| EPI_ISL_578339 | USA/WI-WSLH-200108/2020 | 2020-04-03 | North America/USA/Wisconsin/Racine C | Wisconsin State Laboratory of Hygiene Communicable Disease Division | Wisconsin State Laboratory of Hygiene Communicable Disease Division | Kelsey R. Florek et al |

| | | | | | | |
|---|---|---|---|---|---|---|
| EPI_ISL_578352 | USA/WI-WSLH-200123/2020 | 2020-04-14 | North America/USA/Wisconsin/Grant Co | Wisconsin State Laboratory of Hygiene Communicable Disease Division | Wisconsin State Laboratory of Hygiene Communicable Disease Division | Kelsey R. Florek et al |
| EPI_ISL_578374 | USA/WI-WSLH-200147/2020 | 2020-03-13 | North America/USA/Wisconsin/Fond Du | Wisconsin State Laboratory of Hygiene Communicable Disease Division | Wisconsin State Laboratory of Hygiene Communicable Disease Division | Kelsey R. Florek et al |
| EPI_ISL_578380 | USA/WI-WSLH-200153/2020 | 2020-03-29 | North America/USA/Wisconsin/Columbia | Wisconsin State Laboratory of Hygiene Communicable Disease Division | Wisconsin State Laboratory of Hygiene Communicable Disease Division | Kelsey R. Florek et al |
| EPI_ISL_578385 | USA/WI-WSLH-200158/2020 | 2020-03-31 | North America/USA/Wisconsin/Waukesh | Wisconsin State Laboratory of Hygiene Communicable Disease Division | Wisconsin State Laboratory of Hygiene Communicable Disease Division | Kelsey R. Florek et al |
| EPI_ISL_578391 | USA/WI-WSLH-200165/2020 | 2020-04-01 | North America/USA/Wisconsin/Racine C | Wisconsin State Laboratory of Hygiene Communicable Disease Division | Wisconsin State Laboratory of Hygiene Communicable Disease Division | Kelsey R. Florek et al |
| EPI_ISL_578407 | USA/WI-WSLH-200182/2020 | 2020-04-06 | North America/USA/Wisconsin/Waukesh | Wisconsin State Laboratory of Hygiene Communicable Disease Division | Wisconsin State Laboratory of Hygiene Communicable Disease Division | Kelsey R. Florek et al |
| EPI_ISL_578410 | USA/WI-WSLH-200185/2020 | 2020-04-07 | North America/USA/Wisconsin/Rock Cou | Wisconsin State Laboratory of Hygiene Communicable Disease Division | Wisconsin State Laboratory of Hygiene Communicable Disease Division | Kelsey R. Florek et al |
| EPI_ISL_578414 | USA/WI-WSLH-200189/2020 | 2020-04-08 | North America/USA/Wisconsin/Walwort | Wisconsin State Laboratory of Hygiene Communicable Disease Division | Wisconsin State Laboratory of Hygiene Communicable Disease Division | Kelsey R. Florek et al |

| | | | | | | |
|---|---|---|---|---|---|---|
| EPI_ISL_578415 | USA/WI-WSLH-200190/2020 | 2020-04-08 | North America/USA/Wisconsin/Walwort | Wisconsin State Laboratory of Hygiene Communicable Disease Division | Wisconsin State Laboratory of Hygiene Communicable Disease Division | Kelsey R. Florek et al |
| EPI_ISL_578420 | USA/WI-WSLH-200196/2020 | 2020-03-15 | North America/USA/Wisconsin/Milwauk | Wisconsin State Laboratory of Hygiene Communicable Disease Division | Wisconsin State Laboratory of Hygiene Communicable Disease Division | Kelsey R. Florek et al |
| EPI_ISL_578443 | USA/WI-WSLH-200223/2020 | 2020-04-11 | North America/USA/Wisconsin/Grant Co | Wisconsin State Laboratory of Hygiene Communicable Disease Division | Wisconsin State Laboratory of Hygiene Communicable Disease Division | Kelsey R. Florek et al |
| EPI_ISL_578447 | USA/WI-WSLH-200227/2020 | 2020-04-13 | North America/USA/Wisconsin/Walwort | Wisconsin State Laboratory of Hygiene Communicable Disease Division | Wisconsin State Laboratory of Hygiene Communicable Disease Division | Kelsey R. Florek et al |
| EPI_ISL_578448 | USA/WI-WSLH-200228/2020 | 2020-04-13 | North America/USA/Wisconsin/Walwort | Wisconsin State Laboratory of Hygiene Communicable Disease Division | Wisconsin State Laboratory of Hygiene Communicable Disease Division | Kelsey R. Florek et al |
| EPI_ISL_578459 | USA/WI-WSLH-200239/2020 | 2020-04-13 | North America/USA/Wisconsin/Racine C | Wisconsin State Laboratory of Hygiene Communicable Disease Division | Wisconsin State Laboratory of Hygiene Communicable Disease Division | Kelsey R. Florek et al |
| EPI_ISL_578461 | USA/WI-WSLH-200241/2020 | 2020-04-14 | North America/USA/Wisconsin/Walwort | Wisconsin State Laboratory of Hygiene Communicable Disease Division | Wisconsin State Laboratory of Hygiene Communicable Disease Division | Kelsey R. Florek et al |
| EPI_ISL_578469 | USA/WI-WSLH-200256/2020 | 2020-04-02 | North America/USA/Wisconsin/Brown C | Wisconsin State Laboratory of Hygiene Communicable Disease Division | Wisconsin State Laboratory of Hygiene Communicable Disease Division | Kelsey R. Florek et al |

| | | | | | | |
|---|---|---|---|---|---|---|
| EPI_ISL_578473 | USA/WI-WSLH-200263/2020 | 2020-03-20 | North America/USA/Wisconsin/Milwauk | Wisconsin State Laboratory of Hygiene Communicable Disease Division | Wisconsin State Laboratory of Hygiene Communicable Disease Division | Kelsey R. Florek et al |
| EPI_ISL_578475 | USA/WI-WSLH-200288/2020 | 2020-04-18 | North America/USA/Wisconsin/Walwort | Wisconsin State Laboratory of Hygiene Communicable Disease Division | Wisconsin State Laboratory of Hygiene Communicable Disease Division | Kelsey R. Florek et al |
| EPI_ISL_578480 | USA/WI-WSLH-200295/2020 | 2020-04-17 | North America/USA/Wisconsin/Brown C | Wisconsin State Laboratory of Hygiene Communicable Disease Division | Wisconsin State Laboratory of Hygiene Communicable Disease Division | Kelsey R. Florek et al |
| EPI_ISL_578484 | USA/WI-WSLH-200300/2020 | 2020-04-17 | North America/USA/Wisconsin/Milwauk | Wisconsin State Laboratory of Hygiene Communicable Disease Division | Wisconsin State Laboratory of Hygiene Communicable Disease Division | Kelsey R. Florek et al |
| EPI_ISL_578485 | USA/WI-WSLH-200301/2020 | 2020-04-20 | North America/USA/Wisconsin/Dane Co | Wisconsin State Laboratory of Hygiene Communicable Disease Division | Wisconsin State Laboratory of Hygiene Communicable Disease Division | Kelsey R. Florek et al |
| EPI_ISL_578486 | USA/WI-WSLH-200302/2020 | 2020-04-19 | North America/USA/Wisconsin/Dane Co | Wisconsin State Laboratory of Hygiene Communicable Disease Division | Wisconsin State Laboratory of Hygiene Communicable Disease Division | Kelsey R. Florek et al |
| EPI_ISL_578487 | USA/WI-WSLH-200303/2020 | 2020-04-18 | North America/USA/Wisconsin/Milwauk | Wisconsin State Laboratory of Hygiene Communicable Disease Division | Wisconsin State Laboratory of Hygiene Communicable Disease Division | Kelsey R. Florek et al |
| EPI_ISL_578495 | USA/WI-WSLH-200314/2020 | 2020-04-20 | North America/USA/Wisconsin/Brown C | Wisconsin State Laboratory of Hygiene Communicable Disease Division | Wisconsin State Laboratory of Hygiene Communicable Disease Division | Kelsey R. Florek et al |

| | | | | | | |
|---|---|---|---|---|---|---|
| EPI_ISL_578532 | USA/WI-WSLH-200362/2020 | 2020-04-22 | North America/USA/Wisconsin/Brown C | Wisconsin State Laboratory of Hygiene Communicable Disease Division | Wisconsin State Laboratory of Hygiene Communicable Disease Division | Kelsey R. Florek et al |
| EPI_ISL_578537 | USA/WI-WSLH-200368/2020 | 2020-04-21 | North America/USA/Wisconsin/Brown C | Wisconsin State Laboratory of Hygiene Communicable Disease Division | Wisconsin State Laboratory of Hygiene Communicable Disease Division | Kelsey R. Florek et al |
| EPI_ISL_578538 | USA/WI-WSLH-200369/2020 | 2020-04-21 | North America/USA/Wisconsin/Brown C | Wisconsin State Laboratory of Hygiene Communicable Disease Division | Wisconsin State Laboratory of Hygiene Communicable Disease Division | Kelsey R. Florek et al |
| EPI_ISL_578542 | USA/WI-WSLH-200373/2020 | 2020-04-22 | North America/USA/Wisconsin/Brown C | Wisconsin State Laboratory of Hygiene Communicable Disease Division | Wisconsin State Laboratory of Hygiene Communicable Disease Division | Kelsey R. Florek et al |
| EPI_ISL_578552 | USA/WI-WSLH-200386/2020 | 2020-04-22 | North America/USA/Wisconsin/Milwauk | Wisconsin State Laboratory of Hygiene Communicable Disease Division | Wisconsin State Laboratory of Hygiene Communicable Disease Division | Kelsey R. Florek et al |
| EPI_ISL_578559 | USA/WI-WSLH-200393/2020 | 2020-04-23 | North America/USA/Wisconsin/Brown C | Wisconsin State Laboratory of Hygiene Communicable Disease Division | Wisconsin State Laboratory of Hygiene Communicable Disease Division | Kelsey R. Florek et al |
| EPI_ISL_578562 | USA/WI-WSLH-200396/2020 | 2020-04-23 | North America/USA/Wisconsin/Milwauk | Wisconsin State Laboratory of Hygiene Communicable Disease Division | Wisconsin State Laboratory of Hygiene Communicable Disease Division | Kelsey R. Florek et al |
| EPI_ISL_578570 | USA/WI-WSLH-200405/2020 | 2020-07-27 | North America/USA/Wisconsin/Barron C | Wisconsin State Laboratory of Hygiene Communicable Disease Division | Wisconsin State Laboratory of Hygiene Communicable Disease Division | Kelsey R. Florek et al |

| | | | | | | |
|---|---|---|---|---|---|---|
| EPI_ISL_578572 | USA/WI-WSLH-200408/2020 | 2020-07-28 | North America/USA/Wisconsin/Barron C | Wisconsin State Laboratory of Hygiene Communicable Disease Division | Wisconsin State Laboratory of Hygiene Communicable Disease Division | Kelsey R. Florek et al |
| EPI_ISL_578577 | USA/WI-WSLH-200415/2020 | 2020-05-19 | North America/USA/Wisconsin/Rock Cou | Wisconsin State Laboratory of Hygiene Communicable Disease Division | Wisconsin State Laboratory of Hygiene Communicable Disease Division | Kelsey R. Florek et al |
| EPI_ISL_578578 | USA/WI-WSLH-200416/2020 | 2020-05-18 | North America/USA/Wisconsin/Racine C | Wisconsin State Laboratory of Hygiene Communicable Disease Division | Wisconsin State Laboratory of Hygiene Communicable Disease Division | Kelsey R. Florek et al |
| EPI_ISL_578581 | USA/WI-WSLH-200419/2020 | 2020-05-19 | North America/USA/Wisconsin/Polk Cou | Wisconsin State Laboratory of Hygiene Communicable Disease Division | Wisconsin State Laboratory of Hygiene Communicable Disease Division | Kelsey R. Florek et al |
| EPI_ISL_578589 | USA/WI-WSLH-200440/2020 | 2020-05-18 | North America/USA/Wisconsin/Milwauk | Wisconsin State Laboratory of Hygiene Communicable Disease Division | Wisconsin State Laboratory of Hygiene Communicable Disease Division | Kelsey R. Florek et al |
| EPI_ISL_578590 | USA/WI-WSLH-200442/2020 | 2020-05-18 | North America/USA/Wisconsin/Brown C | Wisconsin State Laboratory of Hygiene Communicable Disease Division | Wisconsin State Laboratory of Hygiene Communicable Disease Division | Kelsey R. Florek et al |
| EPI_ISL_578592 | USA/WI-WSLH-200446/2020 | 2020-05-19 | North America/USA/Wisconsin/Rock Cou | Wisconsin State Laboratory of Hygiene Communicable Disease Division | Wisconsin State Laboratory of Hygiene Communicable Disease Division | Kelsey R. Florek et al |
| EPI_ISL_578594 | USA/WI-WSLH-200449/2020 | 2020-05-19 | North America/USA/Wisconsin/Rock Cou | Wisconsin State Laboratory of Hygiene Communicable Disease Division | Wisconsin State Laboratory of Hygiene Communicable Disease Division | Kelsey R. Florek et al |

| | | | | | | |
|---|---|---|---|---|---|---|
| EPI_ISL_578596 | USA/WI-WSLH-200451/2020 | 2020-05-19 | North America/USA/Wisconsin/Rock Cou | Wisconsin State Laboratory of Hygiene Communicable Disease Division | Wisconsin State Laboratory of Hygiene Communicable Disease Division | Kelsey R. Florek et al |
| EPI_ISL_578607 | USA/WI-WSLH-200465/2020 | 2020-05-18 | North America/USA/Wisconsin/Racine C | Wisconsin State Laboratory of Hygiene Communicable Disease Division | Wisconsin State Laboratory of Hygiene Communicable Disease Division | Kelsey R. Florek et al |
| EPI_ISL_578612 | USA/WI-WSLH-200471/2020 | 2020-05-19 | North America/USA/Wisconsin/Milwauk | Wisconsin State Laboratory of Hygiene Communicable Disease Division | Wisconsin State Laboratory of Hygiene Communicable Disease Division | Kelsey R. Florek et al |
| EPI_ISL_578613 | USA/WI-WSLH-200472/2020 | 2020-05-19 | North America/USA/Wisconsin/Milwauk | Wisconsin State Laboratory of Hygiene Communicable Disease Division | Wisconsin State Laboratory of Hygiene Communicable Disease Division | Kelsey R. Florek et al |
| EPI_ISL_578614 | USA/WI-WSLH-200473/2020 | 2020-05-19 | North America/USA/Wisconsin/Milwauk | Wisconsin State Laboratory of Hygiene Communicable Disease Division | Wisconsin State Laboratory of Hygiene Communicable Disease Division | Kelsey R. Florek et al |
| EPI_ISL_578615 | USA/WI-WSLH-200474/2020 | 2020-05-19 | North America/USA/Wisconsin/Milwauk | Wisconsin State Laboratory of Hygiene Communicable Disease Division | Wisconsin State Laboratory of Hygiene Communicable Disease Division | Kelsey R. Florek et al |
| EPI_ISL_578618 | USA/WI-WSLH-200477/2020 | 2020-05-19 | North America/USA/Wisconsin/Milwauk | Wisconsin State Laboratory of Hygiene Communicable Disease Division | Wisconsin State Laboratory of Hygiene Communicable Disease Division | Kelsey R. Florek et al |
| EPI_ISL_578621 | USA/WI-WSLH-200480/2020 | 2020-05-19 | North America/USA/Wisconsin/Racine C | Wisconsin State Laboratory of Hygiene Communicable Disease Division | Wisconsin State Laboratory of Hygiene Communicable Disease Division | Kelsey R. Florek et al |

| | | | | | | |
|---|---|---|---|---|---|---|
| EPI_ISL_578625 | USA/WI-WSLH-200484/2020 | 2020-05-20 | North America/USA/Wisconsin/Walwort | Wisconsin State Laboratory of Hygiene Communicable Disease Division | Wisconsin State Laboratory of Hygiene Communicable Disease Division | Kelsey R. Florek et al |
| EPI_ISL_578627 | USA/WI-WSLH-200486/2020 | 2020-05-20 | North America/USA/Wisconsin/Milwauk | Wisconsin State Laboratory of Hygiene Communicable Disease Division | Wisconsin State Laboratory of Hygiene Communicable Disease Division | Kelsey R. Florek et al |
| EPI_ISL_578629 | USA/WI-WSLH-200488/2020 | 2020-05-20 | North America/USA/Wisconsin/Milwauk | Wisconsin State Laboratory of Hygiene Communicable Disease Division | Wisconsin State Laboratory of Hygiene Communicable Disease Division | Kelsey R. Florek et al |
| EPI_ISL_578633 | USA/WI-WSLH-200492/2020 | 2020-05-20 | North America/USA/Wisconsin/Brown C | Wisconsin State Laboratory of Hygiene Communicable Disease Division | Wisconsin State Laboratory of Hygiene Communicable Disease Division | Kelsey R. Florek et al |
| EPI_ISL_578638 | USA/WI-WSLH-200497/2020 | 2020-05-20 | North America/USA/Wisconsin/Green C | Wisconsin State Laboratory of Hygiene Communicable Disease Division | Wisconsin State Laboratory of Hygiene Communicable Disease Division | Kelsey R. Florek et al |
| EPI_ISL_578640 | USA/WI-WSLH-200499/2020 | 2020-05-21 | North America/USA/Wisconsin/Milwauk | Wisconsin State Laboratory of Hygiene Communicable Disease Division | Wisconsin State Laboratory of Hygiene Communicable Disease Division | Kelsey R. Florek et al |
| EPI_ISL_578641 | USA/WI-WSLH-200500/2020 | 2020-05-21 | North America/USA/Wisconsin/Milwauk | Wisconsin State Laboratory of Hygiene Communicable Disease Division | Wisconsin State Laboratory of Hygiene Communicable Disease Division | Kelsey R. Florek et al |
| EPI_ISL_578643 | USA/WI-WSLH-200502/2020 | 2020-05-22 | North America/USA/Wisconsin/Brown C | Wisconsin State Laboratory of Hygiene Communicable Disease Division | Wisconsin State Laboratory of Hygiene Communicable Disease Division | Kelsey R. Florek et al |

| | | | | | | |
|---|---|---|---|---|---|---|
| EPI_ISL_578644 | USA/WI-WSLH-200503/2020 | 2020-05-22 | North America/USA/Wisconsin/Milwauk | Wisconsin State Laboratory of Hygiene Communicable Disease Division | Wisconsin State Laboratory of Hygiene Communicable Disease Division | Kelsey R. Florek et al |
| EPI_ISL_578660 | USA/WI-WSLH-200520/2020 | 2020-05-27 | North America/USA/Wisconsin/Winneba | Wisconsin State Laboratory of Hygiene Communicable Disease Division | Wisconsin State Laboratory of Hygiene Communicable Disease Division | Kelsey R. Florek et al |
| EPI_ISL_578662 | USA/WI-WSLH-200522/2020 | 2020-05-26 | North America/USA/Wisconsin/Jefferson | Wisconsin State Laboratory of Hygiene Communicable Disease Division | Wisconsin State Laboratory of Hygiene Communicable Disease Division | Kelsey R. Florek et al |
| EPI_ISL_578666 | USA/WI-WSLH-200527/2020 | 2020-05-27 | North America/USA/Wisconsin/Racine C | Wisconsin State Laboratory of Hygiene Communicable Disease Division | Wisconsin State Laboratory of Hygiene Communicable Disease Division | Kelsey R. Florek et al |
| EPI_ISL_578673 | USA/WI-WSLH-200536/2020 | 2020-05-29 | North America/USA/Wisconsin/Winneba | Wisconsin State Laboratory of Hygiene Communicable Disease Division | Wisconsin State Laboratory of Hygiene Communicable Disease Division | Kelsey R. Florek et al |
| EPI_ISL_578682 | USA/WI-WSLH-200570/2020 | 2020-06-01 | North America/USA/Wisconsin/Racine C | Wisconsin State Laboratory of Hygiene Communicable Disease Division | Wisconsin State Laboratory of Hygiene Communicable Disease Division | Kelsey R. Florek et al |
| EPI_ISL_578688 | USA/WI-WSLH-200577/2020 | 2020-06-07 | North America/USA/Wisconsin/Lafayett | Wisconsin State Laboratory of Hygiene Communicable Disease Division | Wisconsin State Laboratory of Hygiene Communicable Disease Division | Kelsey R. Florek et al |
| EPI_ISL_578704 | USA/LA-EVTL297/2020 | 2020-05-09 | North America/USA/Louisiana/Caddo Pa | LSUHS Emerging Viral Threat Laboratory | Microbial Genome Sequencing Center | Maarten Van Diest et al |
| EPI_ISL_578707 | USA/LA-EVTL300/2020 | 2020-05-09 | North America/USA/Louisiana/Caddo Pa | LSUHS Emerging Viral Threat Laboratory | Microbial Genome Sequencing Center | Maarten Van Diest et al |

| | | | | | | |
|---|---|---|---|---|---|---|
| EPI_ISL_578710 | USA/LA-EVTL303/2020 | 2020-05-12 | North America/USA/Louisiana/Caddo Pa | LSUHS Emerging Viral Threat Laboratory | Microbial Genome Sequencing Center | Maarten Van Diest et al |
| EPI_ISL_578712 | USA/LA-EVTL305/2020 | 2020-05-12 | North America/USA/Louisiana/Caddo Pa | LSUHS Emerging Viral Threat Laboratory | Microbial Genome Sequencing Center | Maarten Van Diest et al |
| EPI_ISL_578714 | USA/LA-EVTL307/2020 | 2020-05-12 | North America/USA/Louisiana/Caddo Pa | LSUHS Emerging Viral Threat Laboratory | Microbial Genome Sequencing Center | Maarten Van Diest et al |
| EPI_ISL_578715 | USA/LA-EVTL308/2020 | 2020-05-12 | North America/USA/Louisiana/Caddo Pa | LSUHS Emerging Viral Threat Laboratory | Microbial Genome Sequencing Center | Maarten Van Diest et al |
| EPI_ISL_578716 | USA/LA-EVTL309/2020 | 2020-05-12 | North America/USA/Louisiana/Caddo Pa | LSUHS Emerging Viral Threat Laboratory | Microbial Genome Sequencing Center | Maarten Van Diest et al |
| EPI_ISL_578722 | USA/LA-EVTL315/2020 | 2020-05-15 | North America/USA/Louisiana/Caddo Pa | LSUHS Emerging Viral Threat Laboratory | Microbial Genome Sequencing Center | Maarten Van Diest et al |
| EPI_ISL_578724 | USA/LA-EVTL317/2020 | 2020-05-15 | North America/USA/Louisiana/Caddo Pa | LSUHS Emerging Viral Threat Laboratory | Microbial Genome Sequencing Center | Maarten Van Diest et al |
| EPI_ISL_578726 | USA/LA-EVTL319/2020 | 2020-05-15 | North America/USA/Louisiana/Caddo Pa | LSUHS Emerging Viral Threat Laboratory | Microbial Genome Sequencing Center | Maarten Van Diest et al |
| EPI_ISL_578728 | USA/LA-EVTL322/2020 | 2020-05-18 | North America/USA/Louisiana/Webster | LSUHS Emerging Viral Threat Laboratory | Microbial Genome Sequencing Center | Rona S. Scott et al |
| EPI_ISL_578732 | USA/LA-EVTL326/2020 | 2020-05-19 | North America/USA/Louisiana/Rapides P | LSUHS Emerging Viral Threat Laboratory | Microbial Genome Sequencing Center | Rona S. Scott et al |
| EPI_ISL_578736 | USA/LA-EVTL330/2020 | 2020-05-19 | North America/USA/Louisiana/Rapides P | LSUHS Emerging Viral Threat Laboratory | Microbial Genome Sequencing Center | Rona S. Scott et al |
| EPI_ISL_578738 | USA/LA-EVTL332/2020 | 2020-05-19 | North America/USA/Louisiana/Caddo Pa | LSUHS Emerging Viral Threat Laboratory | Microbial Genome Sequencing Center | Rona S. Scott et al |
| EPI_ISL_578745 | USA/LA-EVTL339/2020 | 2020-05-20 | North America/USA/Louisiana/Rapides P | LSUHS Emerging Viral Threat Laboratory | Microbial Genome Sequencing Center | Rona S. Scott et al |

| | | | | | | |
|---|---|---|---|---|---|---|
| EPI_ISL_578746 | USA/LA-EVTL340/2020 | 2020-05-20 | North America/USA/Louisiana/Caddo Pa | LSUHS Emerging Viral Threat Laboratory | Microbial Genome Sequencing Center | Rona S. Scott et al |
| EPI_ISL_578755 | USA/LA-EVTL349/2020 | 2020-05-25 | North America/USA/Louisiana/Rapides P | LSUHS Emerging Viral Threat Laboratory | Microbial Genome Sequencing Center | Jeremy P. Kamil et al |
| EPI_ISL_578757 | USA/LA-EVTL351/2020 | 2020-05-25 | North America/USA/Louisiana/Rapides P | LSUHS Emerging Viral Threat Laboratory | Microbial Genome Sequencing Center | Jeremy P. Kamil et al |
| EPI_ISL_578758 | USA/LA-EVTL352/2020 | 2020-05-25 | North America/USA/Louisiana/Rapides P | LSUHS Emerging Viral Threat Laboratory | Microbial Genome Sequencing Center | Jeremy P. Kamil et al |
| EPI_ISL_578759 | USA/LA-EVTL353/2020 | 2020-05-25 | North America/USA/Louisiana/Rapides P | LSUHS Emerging Viral Threat Laboratory | Microbial Genome Sequencing Center | Jeremy P. Kamil et al |
| EPI_ISL_578763 | USA/LA-EVTL357/2020 | 2020-05-21 | North America/USA/Louisiana/Rapides P | LSUHS Emerging Viral Threat Laboratory | Microbial Genome Sequencing Center | Jeremy P. Kamil et al |
| EPI_ISL_578769 | USA/LA-EVTL363/2020 | 2020-05-28 | North America/USA/Louisiana/Webster | LSUHS Emerging Viral Threat Laboratory | Microbial Genome Sequencing Center | Jeremy P. Kamil et al |
| EPI_ISL_578771 | USA/LA-EVTL365/2020 | 2020-05-28 | North America/USA/Louisiana/Caddo Pa | LSUHS Emerging Viral Threat Laboratory | Microbial Genome Sequencing Center | Jeremy P. Kamil et al |
| EPI_ISL_578773 | USA/LA-EVTL367/2020 | 2020-05-29 | North America/USA/Louisiana/Ouachita | LSUHS Emerging Viral Threat Laboratory | Microbial Genome Sequencing Center | Jeremy P. Kamil et al |
| EPI_ISL_578776 | USA/LA-EVTL370/2020 | 2020-06-01 | North America/USA/Louisiana/Madison | LSUHS Emerging Viral Threat Laboratory | Microbial Genome Sequencing Center | Jeremy P. Kamil et al |
| EPI_ISL_578779 | USA/LA-EVTL373/2020 | 2020-06-01 | North America/USA/Louisiana/Union Pa | LSUHS Emerging Viral Threat Laboratory | Microbial Genome Sequencing Center | Jeremy P. Kamil et al |
| EPI_ISL_578784 | USA/LA-EVTL378/2020 | 2020-06-02 | North America/USA/Louisiana/Ouachita | LSUHS Emerging Viral Threat Laboratory | Microbial Genome Sequencing Center | Jeremy P. Kamil et al |
| EPI_ISL_578786 | USA/LA-EVTL380/2020 | 2020-06-03 | North America/USA/Louisiana/Webster | LSUHS Emerging Viral Threat Laboratory | Microbial Genome Sequencing Center | Jeremy P. Kamil et al |

| | | | | | | |
|---|---|---|---|---|---|---|
| EPI_ISL_578788 | USA/LA-EVTL382/2020 | 2020-06-04 | North America/USA/Louisiana/Madison | LSUHS Emerging Viral Threat Laboratory | Microbial Genome Sequencing Center | Jeremy P. Kamil et al |
| EPI_ISL_578795 | USA/LA-EVTL389/2020 | 2020-06-04 | North America/USA/Louisiana/Madison | LSUHS Emerging Viral Threat Laboratory | Microbial Genome Sequencing Center | Jeremy P. Kamil et al |
| EPI_ISL_578800 | USA/LA-EVTL394/2020 | 2020-06-03 | North America/USA/Louisiana/Avoyelle | LSUHS Emerging Viral Threat Laboratory | Microbial Genome Sequencing Center | Jeremy P. Kamil et al |
| EPI_ISL_578802 | USA/LA-EVTL396/2020 | 2020-06-01 | North America/USA/Louisiana/Union Pa | LSUHS Emerging Viral Threat Laboratory | Microbial Genome Sequencing Center | Jeremy P. Kamil et al |
| EPI_ISL_578806 | USA/LA-EVTL400/2020 | 2020-06-01 | North America/USA/Louisiana/Union Pa | LSUHS Emerging Viral Threat Laboratory | Microbial Genome Sequencing Center | Jeremy P. Kamil et al |
| EPI_ISL_578821 | USA/LA-EVTL415/2020 | 2020-06-08 | North America/USA/Louisiana/Union Pa | LSUHS Emerging Viral Threat Laboratory | Microbial Genome Sequencing Center | Jeremy P. Kamil et al |
| EPI_ISL_578823 | USA/LA-EVTL417/2020 | 2020-06-08 | North America/USA/Louisiana/Union Pa | LSUHS Emerging Viral Threat Laboratory | Microbial Genome Sequencing Center | Jeremy P. Kamil et al |
| EPI_ISL_578827 | USA/LA-EVTL421/2020 | 2020-06-10 | North America/USA/Louisiana/Madison | LSUHS Emerging Viral Threat Laboratory | Microbial Genome Sequencing Center | Jeremy P. Kamil et al |
| EPI_ISL_578829 | USA/LA-EVTL423/2020 | 2020-06-10 | North America/USA/Louisiana/Madison | LSUHS Emerging Viral Threat Laboratory | Microbial Genome Sequencing Center | Jeremy P. Kamil et al |
| EPI_ISL_578833 | USA/LA-EVTL427/2020 | 2020-06-12 | North America/USA/Louisiana/Caddo Pa | LSUHS Emerging Viral Threat Laboratory | Microbial Genome Sequencing Center | Jeremy P. Kamil et al |
| EPI_ISL_578854 | USA/LA-EVTL450/2020 | 2020-07-30 | North America/USA/Louisiana/Caddo Pa | LSUHS Emerging Viral Threat Laboratory | Microbial Genome Sequencing Center | Jeremy P. Kamil et al |
| EPI_ISL_578859 | USA/LA-EVTL455/2020 | 2020-06-20 | North America/USA/Louisiana/Webster | LSUHS Emerging Viral Threat Laboratory | Microbial Genome Sequencing Center | Jeremy P. Kamil et al |
| EPI_ISL_578860 | USA/LA-EVTL456/2020 | 2020-06-22 | North America/USA/Louisiana/Webster | LSUHS Emerging Viral Threat Laboratory | Microbial Genome Sequencing Center | Jeremy P. Kamil et al |

| | | | | | | |
|---|---|---|---|---|---|---|
| EPI_ISL_578861 | USA/LA-EVTL457/2020 | 2020-07-30 | North America/USA/Louisiana/Caddo Pa | LSUHS Emerging Viral Threat Laboratory | Microbial Genome Sequencing Center | Jeremy P. Kamil et al |
| EPI_ISL_578877 | USA/LA-EVTL473/2020 | 2020-06-23 | North America/USA/Louisiana/Bossier P | LSUHS Emerging Viral Threat Laboratory | Microbial Genome Sequencing Center | Jeremy P. Kamil et al |
| EPI_ISL_578907 | USA/LA-EVTL503/2020 | 2020-06-24 | North America/USA/Louisiana/Bossier P | LSUHS Emerging Viral Threat Laboratory | Microbial Genome Sequencing Center | Jeremy P. Kamil et al |
| EPI_ISL_578920 | USA/LA-EVTL516/2020 | 2020-06-25 | North America/USA/Louisiana/Caddo Pa | LSUHS Emerging Viral Threat Laboratory | Microbial Genome Sequencing Center | Jeremy P. Kamil et al |
| EPI_ISL_578922 | USA/LA-EVTL518/2020 | 2020-06-25 | North America/USA/Louisiana/Caddo Pa | LSUHS Emerging Viral Threat Laboratory | Microbial Genome Sequencing Center | Jeremy P. Kamil et al |
| EPI_ISL_578923 | USA/LA-EVTL519/2020 | 2020-08-03 | North America/USA/Louisiana/Bossier P | LSUHS Emerging Viral Threat Laboratory | Microbial Genome Sequencing Center | Jeremy P. Kamil et al |
| EPI_ISL_578924 | USA/LA-EVTL520/2020 | 2020-06-25 | North America/USA/Louisiana/Caddo Pa | LSUHS Emerging Viral Threat Laboratory | Microbial Genome Sequencing Center | Jeremy P. Kamil et al |
| EPI_ISL_578929 | USA/LA-EVTL525/2020 | 2020-08-03 | North America/USA/Louisiana/Sabine Pa | LSUHS Emerging Viral Threat Laboratory | Microbial Genome Sequencing Center | Jeremy P. Kamil et al |
| EPI_ISL_578933 | USA/LA-EVTL529/2020 | 2020-08-04 | North America/USA/Louisiana/Sabine Pa | LSUHS Emerging Viral Threat Laboratory | Microbial Genome Sequencing Center | Jeremy P. Kamil et al |
| EPI_ISL_578934 | USA/LA-EVTL530/2020 | 2020-08-04 | North America/USA/Louisiana/Bossier P | LSUHS Emerging Viral Threat Laboratory | Microbial Genome Sequencing Center | Jeremy P. Kamil et al |
| EPI_ISL_578937 | USA/LA-EVTL533/2020 | 2020-08-04 | North America/USA/Louisiana/Sabine Pa | LSUHS Emerging Viral Threat Laboratory | Microbial Genome Sequencing Center | Jeremy P. Kamil et al |
| EPI_ISL_578944 | USA/LA-EVTL540/2020 | 2020-06-26 | North America/USA/Louisiana/Caddo Pa | LSUHS Emerging Viral Threat Laboratory | Microbial Genome Sequencing Center | Jeremy P. Kamil et al |
| EPI_ISL_578946 | USA/LA-EVTL542/2020 | 2020-06-26 | North America/USA/Louisiana/Caddo Pa | LSUHS Emerging Viral Threat Laboratory | Microbial Genome Sequencing Center | Jeremy P. Kamil et al |

| | | | | | | |
|---|---|---|---|---|---|---|
| EPI_ISL_578947 | USA/LA-EVTL543/2020 | 2020-06-29 | North America/USA/Louisiana/Caddo Pa | LSUHS Emerging Viral Threat Laboratory | Microbial Genome Sequencing Center | Jeremy P. Kamil et al |
| EPI_ISL_578949 | USA/LA-EVTL545/2020 | 2020-08-05 | North America/USA/Louisiana/Red Rive | LSUHS Emerging Viral Threat Laboratory | Microbial Genome Sequencing Center | Jeremy P. Kamil et al |
| EPI_ISL_578958 | USA/LA-EVTL554/2020 | 2020-08-05 | North America/USA/Louisiana/Red Rive | LSUHS Emerging Viral Threat Laboratory | Microbial Genome Sequencing Center | Jeremy P. Kamil et al |
| EPI_ISL_578960 | USA/LA-EVTL556/2020 | 2020-08-05 | North America/USA/Louisiana/Red Rive | LSUHS Emerging Viral Threat Laboratory | Microbial Genome Sequencing Center | Jeremy P. Kamil et al |
| EPI_ISL_578965 | USA/LA-EVTL561/2020 | 2020-08-05 | North America/USA/Louisiana/Red Rive | LSUHS Emerging Viral Threat Laboratory | Microbial Genome Sequencing Center | Jeremy P. Kamil et al |
| EPI_ISL_578969 | USA/LA-EVTL565/2020 | 2020-06-30 | North America/USA/Louisiana/Bossier P | LSUHS Emerging Viral Threat Laboratory | Microbial Genome Sequencing Center | Jeremy P. Kamil et al |
| EPI_ISL_578980 | USA/LA-EVTL576/2020 | 2020-06-30 | North America/USA/Louisiana/Bossier P | LSUHS Emerging Viral Threat Laboratory | Microbial Genome Sequencing Center | Jeremy P. Kamil et al |
| EPI_ISL_578989 | USA/LA-EVTL585/2020 | 2020-06-30 | North America/USA/Louisiana/Bossier P | LSUHS Emerging Viral Threat Laboratory | Microbial Genome Sequencing Center | Jeremy P. Kamil et al |
| EPI_ISL_578996 | USA/LA-EVTL592/2020 | 2020-06-30 | North America/USA/Louisiana/Bossier P | LSUHS Emerging Viral Threat Laboratory | Microbial Genome Sequencing Center | Jeremy P. Kamil et al |
| EPI_ISL_578999 | USA/LA-EVTL595/2020 | 2020-06-30 | North America/USA/Louisiana/Bossier P | LSUHS Emerging Viral Threat Laboratory | Microbial Genome Sequencing Center | Jeremy P. Kamil et al |
| EPI_ISL_579007 | USA/LA-EVTL603/2020 | 2020-06-30 | North America/USA/Louisiana/Bossier P | LSUHS Emerging Viral Threat Laboratory | Microbial Genome Sequencing Center | Jeremy P. Kamil et al |
| EPI_ISL_579010 | USA/LA-EVTL606/2020 | 2020-07-01 | North America/USA/Louisiana/Bossier P | LSUHS Emerging Viral Threat Laboratory | Microbial Genome Sequencing Center | Jeremy P. Kamil et al |
| EPI_ISL_579013 | USA/LA-EVTL609/2020 | 2020-07-01 | North America/USA/Louisiana/Caddo Pa | LSUHS Emerging Viral Threat Laboratory | Microbial Genome Sequencing Center | Jeremy P. Kamil et al |

| | | | | | | |
|---|---|---|---|---|---|---|
| EPI_ISL_579018 | USA/LA-EVTL614/2020 | 2020-07-02 | North America/USA/Louisiana/Caddo Pa | LSUHS Emerging Viral Threat Laboratory | Microbial Genome Sequencing Center | Jeremy P. Kamil et al |
| EPI_ISL_579035 | USA/LA-EVTL631/2020 | 2020-08-06 | North America/USA/Louisiana/Bossier P | LSUHS Emerging Viral Threat Laboratory | Microbial Genome Sequencing Center | Jeremy P. Kamil et al |
| EPI_ISL_579042 | USA/LA-EVTL638/2020 | 2020-08-06 | North America/USA/Louisiana/Bossier P | LSUHS Emerging Viral Threat Laboratory | Microbial Genome Sequencing Center | Jeremy P. Kamil et al |
| EPI_ISL_579043 | USA/LA-EVTL639/2020 | 2020-08-06 | North America/USA/Louisiana/Bossier P | LSUHS Emerging Viral Threat Laboratory | Microbial Genome Sequencing Center | Jeremy P. Kamil et al |
| EPI_ISL_579055 | USA/LA-EVTL651/2020 | 2020-08-06 | North America/USA/Louisiana/Caddo Pa | LSUHS Emerging Viral Threat Laboratory | Microbial Genome Sequencing Center | Jeremy P. Kamil et al |
| EPI_ISL_579220 | NewZealand/20VR2060/2020 | 2020-04-04 | Oceania/New Zealand/Canterbury/ | Canterbury Health Laboratories | Institute of Environmental Science and Research (ESR) | Xiaoyun Ren et al |
| EPI_ISL_579225 | NewZealand/20VR2080/2020 | 2020-04-08 | Oceania/New Zealand/Canterbury/ | Canterbury Health Laboratories | Institute of Environmental Science and Research (ESR) | Xiaoyun Ren et al |
| EPI_ISL_579464 | NewZealand/20VR3175/2020 | 2020-03-25 | Oceania/New Zealand/Canterbury/ | Canterbury Health Laboratories | Institute of Environmental Science and Research (ESR) | Xiaoyun Ren et al |
| EPI_ISL_579515 | Canada/NS-NML-7/2020 | 2020-03-13 | North America/Canada/Nova Scotia/ | QEII Health Sciences Centre | National Microbiology Laboratory (NML) | Anna Majer et al |
| EPI_ISL_579516 | Canada/NS-NML-12/2020 | 2020-03-13 | North America/Canada/Nova Scotia/ | QEII Health Sciences Centre | National Microbiology Laboratory (NML) | Anna Majer et al |
| EPI_ISL_579517 | Canada/NS-NML-41/2020 | 2020-03-16 | North America/Canada/Nova Scotia/ | QEII Health Sciences Centre | National Microbiology Laboratory (NML) | Anna Majer et al |
| EPI_ISL_579519 | Canada/NS-NML-142/2020 | 2020-04-13 | North America/Canada/Nova Scotia/ | QEII Health Sciences Centre | National Microbiology Laboratory (NML) | Anna Majer et al |

| | | | | | | |
|---|---|---|---|---|---|---|
| EPI_ISL_579520 | Canada/NS-NML-143/2020 | 2020-04-14 | North America/Canada/Nova Scotia/ | QEII Health Sciences Centre | National Microbiology Laboratory (NML) | Anna Majer et al |
| EPI_ISL_579521 | Canada/NS-NML-144/2020 | 2020-04-13 | North America/Canada/Nova Scotia/ | QEII Health Sciences Centre | National Microbiology Laboratory (NML) | Anna Majer et al |
| EPI_ISL_579523 | Canada/NS-NML-152/2020 | 2020-04-18 | North America/Canada/Nova Scotia/ | QEII Health Sciences Centre | National Microbiology Laboratory (NML) | Anna Majer et al |
| EPI_ISL_579524 | Canada/NS-NML-153/2020 | 2020-04-18 | North America/Canada/Nova Scotia/ | QEII Health Sciences Centre | National Microbiology Laboratory (NML) | Anna Majer et al |
| EPI_ISL_579525 | Canada/NS-NML-154/2020 | 2020-04-14 | North America/Canada/Nova Scotia/ | QEII Health Sciences Centre | National Microbiology Laboratory (NML) | Anna Majer et al |
| EPI_ISL_579526 | Canada/NS-NML-156/2020 | 2020-04-18 | North America/Canada/Nova Scotia/ | QEII Health Sciences Centre | National Microbiology Laboratory (NML) | Anna Majer et al |
| EPI_ISL_579527 | Canada/NS-NML-157/2020 | 2020-04-18 | North America/Canada/Nova Scotia/ | QEII Health Sciences Centre | National Microbiology Laboratory (NML) | Anna Majer et al |
| EPI_ISL_579528 | Canada/NS-NML-158/2020 | 2020-04-20 | North America/Canada/Nova Scotia/ | QEII Health Sciences Centre | National Microbiology Laboratory (NML) | Anna Majer et al |
| EPI_ISL_579529 | Canada/NS-NML-159/2020 | 2020-04-17 | North America/Canada/Nova Scotia/ | QEII Health Sciences Centre | National Microbiology Laboratory (NML) | Anna Majer et al |
| EPI_ISL_579530 | Canada/NS-NML-160/2020 | 2020-04-18 | North America/Canada/Nova Scotia/ | QEII Health Sciences Centre | National Microbiology Laboratory (NML) | Anna Majer et al |
| EPI_ISL_579531 | Canada/NS-NML-193/2020 | 2020-04-19 | North America/Canada/Nova Scotia/ | QEII Health Sciences Centre | National Microbiology Laboratory (NML) | Anna Majer et al |
| EPI_ISL_579532 | Canada/NS-NML-196/2020 | 2020-04-22 | North America/Canada/Nova Scotia/ | QEII Health Sciences Centre | National Microbiology Laboratory (NML) | Anna Majer et al |
| EPI_ISL_579533 | Canada/NS-NML-197/2020 | 2020-04-22 | North America/Canada/Nova Scotia/ | QEII Health Sciences Centre | National Microbiology Laboratory (NML) | Anna Majer et al |

| | | | | | | |
|---|---|---|---|---|---|---|
| EPI_ISL_579534 | Canada/NS-NML-198/2020 | 2020-04-24 | North America/Canada/Nova Scotia/ | QEII Health Sciences Centre | National Microbiology Laboratory (NML) | Anna Majer et al |
| EPI_ISL_579536 | Canada/NS-NML-201/2020 | 2020-04-28 | North America/Canada/Nova Scotia/ | QEII Health Sciences Centre | National Microbiology Laboratory (NML) | Anna Majer et al |
| EPI_ISL_579537 | Canada/NS-NML-884/2020 | 2020-03-18 | North America/Canada/Nova Scotia/ | QEII Health Sciences Centre | National Microbiology Laboratory (NML) | Anna Majer et al |
| EPI_ISL_579538 | Canada/NS-NML-886/2020 | 2020-03-19 | North America/Canada/Nova Scotia/ | QEII Health Sciences Centre | National Microbiology Laboratory (NML) | Anna Majer et al |
| EPI_ISL_579539 | Canada/NS-NML-887/2020 | 2020-03-19 | North America/Canada/Nova Scotia/ | QEII Health Sciences Centre | National Microbiology Laboratory (NML) | Anna Majer et al |
| EPI_ISL_579540 | Canada/NS-NML-888/2020 | 2020-03-19 | North America/Canada/Nova Scotia/ | QEII Health Sciences Centre | National Microbiology Laboratory (NML) | Anna Majer et al |
| EPI_ISL_579541 | Canada/NS-NML-889/2020 | 2020-03-19 | North America/Canada/Nova Scotia/ | QEII Health Sciences Centre | National Microbiology Laboratory (NML) | Anna Majer et al |
| EPI_ISL_579542 | Canada/NS-NML-891/2020 | 2020-03-20 | North America/Canada/Nova Scotia/ | QEII Health Sciences Centre | National Microbiology Laboratory (NML) | Anna Majer et al |
| EPI_ISL_579543 | Canada/NS-NML-893/2020 | 2020-03-20 | North America/Canada/Nova Scotia/ | QEII Health Sciences Centre | National Microbiology Laboratory (NML) | Anna Majer et al |
| EPI_ISL_579544 | Canada/NS-NML-895/2020 | 2020-03-20 | North America/Canada/Nova Scotia/ | QEII Health Sciences Centre | National Microbiology Laboratory (NML) | Anna Majer et al |
| EPI_ISL_579545 | Canada/NS-NML-896/2020 | 2020-03-21 | North America/Canada/Nova Scotia/ | QEII Health Sciences Centre | National Microbiology Laboratory (NML) | Anna Majer et al |
| EPI_ISL_579546 | Canada/NS-NML-897/2020 | 2020-03-21 | North America/Canada/Nova Scotia/ | QEII Health Sciences Centre | National Microbiology Laboratory (NML) | Anna Majer et al |
| EPI_ISL_579547 | Canada/NS-NML-898/2020 | 2020-03-21 | North America/Canada/Nova Scotia/ | QEII Health Sciences Centre | National Microbiology Laboratory (NML) | Anna Majer et al |

| | | | | | | |
|---|---|---|---|---|---|---|
| EPI_ISL_579548 | Canada/NS-NML-899/2020 | 2020-03-21 | North America/Canada/Nova Scotia/ | QEII Health Sciences Centre | National Microbiology Laboratory (NML) | Anna Majer et al |
| EPI_ISL_579549 | Canada/NS-NML-900/2020 | 2020-03-21 | North America/Canada/Nova Scotia/ | QEII Health Sciences Centre | National Microbiology Laboratory (NML) | Anna Majer et al |
| EPI_ISL_579550 | Canada/NS-NML-901/2020 | 2020-03-21 | North America/Canada/Nova Scotia/ | QEII Health Sciences Centre | National Microbiology Laboratory (NML) | Anna Majer et al |
| EPI_ISL_579551 | Canada/NS-NML-902/2020 | 2020-03-21 | North America/Canada/Nova Scotia/ | QEII Health Sciences Centre | National Microbiology Laboratory (NML) | Anna Majer et al |
| EPI_ISL_579552 | Canada/NS-NML-903/2020 | 2020-03-21 | North America/Canada/Nova Scotia/ | QEII Health Sciences Centre | National Microbiology Laboratory (NML) | Anna Majer et al |
| EPI_ISL_579553 | Canada/NS-NML-904/2020 | 2020-03-21 | North America/Canada/Nova Scotia/ | QEII Health Sciences Centre | National Microbiology Laboratory (NML) | Anna Majer et al |
| EPI_ISL_579554 | Canada/NS-NML-905/2020 | 2020-03-21 | North America/Canada/Nova Scotia/ | QEII Health Sciences Centre | National Microbiology Laboratory (NML) | Anna Majer et al |
| EPI_ISL_579555 | Canada/NS-NML-906/2020 | 2020-03-22 | North America/Canada/Nova Scotia/ | QEII Health Sciences Centre | National Microbiology Laboratory (NML) | Anna Majer et al |
| EPI_ISL_579556 | Canada/NS-NML-907/2020 | 2020-03-22 | North America/Canada/Nova Scotia/ | QEII Health Sciences Centre | National Microbiology Laboratory (NML) | Anna Majer et al |
| EPI_ISL_579557 | Canada/NS-NML-908/2020 | 2020-03-22 | North America/Canada/Nova Scotia/ | QEII Health Sciences Centre | National Microbiology Laboratory (NML) | Anna Majer et al |
| EPI_ISL_579558 | Canada/NS-NML-909/2020 | 2020-03-22 | North America/Canada/Nova Scotia/ | QEII Health Sciences Centre | National Microbiology Laboratory (NML) | Anna Majer et al |
| EPI_ISL_579559 | Canada/NS-NML-910/2020 | 2020-03-22 | North America/Canada/Nova Scotia/ | QEII Health Sciences Centre | National Microbiology Laboratory (NML) | Anna Majer et al |
| EPI_ISL_579560 | Canada/NS-NML-917/2020 | 2020-03-23 | North America/Canada/Nova Scotia/ | QEII Health Sciences Centre | National Microbiology Laboratory (NML) | Anna Majer et al |

| | | | | | | |
|---|---|---|---|---|---|---|
| EPI_ISL_579561 | Canada/NS-NML-918/2020 | 2020-03-23 | North America/Canada/Nova Scotia/ | QEII Health Sciences Centre | National Microbiology Laboratory (NML) | Anna Majer et al |
| EPI_ISL_579562 | Canada/NS-NML-919/2020 | 2020-03-23 | North America/Canada/Nova Scotia/ | QEII Health Sciences Centre | National Microbiology Laboratory (NML) | Anna Majer et al |
| EPI_ISL_579563 | Canada/NS-NML-920/2020 | 2020-03-23 | North America/Canada/Nova Scotia/ | QEII Health Sciences Centre | National Microbiology Laboratory (NML) | Anna Majer et al |
| EPI_ISL_579564 | Canada/NS-NML-922/2020 | 2020-03-23 | North America/Canada/Nova Scotia/ | QEII Health Sciences Centre | National Microbiology Laboratory (NML) | Anna Majer et al |
| EPI_ISL_579565 | Canada/NS-NML-924/2020 | 2020-03-23 | North America/Canada/Nova Scotia/ | QEII Health Sciences Centre | National Microbiology Laboratory (NML) | Anna Majer et al |
| EPI_ISL_579567 | Canada/NS-NML-927/2020 | 2020-03-23 | North America/Canada/Nova Scotia/ | QEII Health Sciences Centre | National Microbiology Laboratory (NML) | Anna Majer et al |
| EPI_ISL_579568 | Canada/NS-NML-928/2020 | 2020-03-24 | North America/Canada/Nova Scotia/ | QEII Health Sciences Centre | National Microbiology Laboratory (NML) | Anna Majer et al |
| EPI_ISL_579569 | Canada/NS-NML-929/2020 | 2020-03-24 | North America/Canada/Nova Scotia/ | QEII Health Sciences Centre | National Microbiology Laboratory (NML) | Anna Majer et al |
| EPI_ISL_579570 | Canada/NS-NML-930/2020 | 2020-03-25 | North America/Canada/Nova Scotia/ | QEII Health Sciences Centre | National Microbiology Laboratory (NML) | Anna Majer et al |
| EPI_ISL_579571 | Canada/NS-NML-939/2020 | 2020-03-25 | North America/Canada/Nova Scotia/ | QEII Health Sciences Centre | National Microbiology Laboratory (NML) | Anna Majer et al |
| EPI_ISL_579572 | Canada/NS-NML-940/2020 | 2020-03-25 | North America/Canada/Nova Scotia/ | QEII Health Sciences Centre | National Microbiology Laboratory (NML) | Anna Majer et al |
| EPI_ISL_579573 | Canada/NS-NML-941/2020 | 2020-03-25 | North America/Canada/Nova Scotia/ | QEII Health Sciences Centre | National Microbiology Laboratory (NML) | Anna Majer et al |
| EPI_ISL_579574 | Canada/NS-NML-943/2020 | 2020-03-26 | North America/Canada/Nova Scotia/ | QEII Health Sciences Centre | National Microbiology Laboratory (NML) | Anna Majer et al |

| | | | | | | |
|---|---|---|---|---|---|---|
| EPI_ISL_579575 | Canada/NS-NML-944/2020 | 2020-03-26 | North America/Canada/Nova Scotia/ | QEII Health Sciences Centre | National Microbiology Laboratory (NML) | Anna Majer et al |
| EPI_ISL_579576 | Canada/NS-NML-946/2020 | 2020-03-26 | North America/Canada/Nova Scotia/ | QEII Health Sciences Centre | National Microbiology Laboratory (NML) | Anna Majer et al |
| EPI_ISL_579577 | Canada/NS-NML-947/2020 | 2020-03-26 | North America/Canada/Nova Scotia/ | QEII Health Sciences Centre | National Microbiology Laboratory (NML) | Anna Majer et al |
| EPI_ISL_579578 | Canada/NS-NML-948/2020 | 2020-03-26 | North America/Canada/Nova Scotia/ | QEII Health Sciences Centre | National Microbiology Laboratory (NML) | Anna Majer et al |
| EPI_ISL_579579 | Canada/NS-NML-949/2020 | 2020-03-26 | North America/Canada/Nova Scotia/ | QEII Health Sciences Centre | National Microbiology Laboratory (NML) | Anna Majer et al |
| EPI_ISL_579580 | Canada/NS-NML-950/2020 | 2020-03-26 | North America/Canada/Nova Scotia/ | QEII Health Sciences Centre | National Microbiology Laboratory (NML) | Anna Majer et al |
| EPI_ISL_579581 | Canada/NS-NML-951/2020 | 2020-03-26 | North America/Canada/Nova Scotia/ | QEII Health Sciences Centre | National Microbiology Laboratory (NML) | Anna Majer et al |
| EPI_ISL_579582 | Canada/NS-NML-952/2020 | 2020-03-26 | North America/Canada/Nova Scotia/ | QEII Health Sciences Centre | National Microbiology Laboratory (NML) | Anna Majer et al |
| EPI_ISL_579583 | Canada/NS-NML-953/2020 | 2020-03-27 | North America/Canada/Nova Scotia/ | QEII Health Sciences Centre | National Microbiology Laboratory (NML) | Anna Majer et al |
| EPI_ISL_579584 | Canada/NS-NML-954/2020 | 2020-03-27 | North America/Canada/Nova Scotia/ | QEII Health Sciences Centre | National Microbiology Laboratory (NML) | Anna Majer et al |
| EPI_ISL_579585 | Canada/NS-NML-955/2020 | 2020-03-27 | North America/Canada/Nova Scotia/ | QEII Health Sciences Centre | National Microbiology Laboratory (NML) | Anna Majer et al |
| EPI_ISL_579586 | Canada/NS-NML-957/2020 | 2020-03-28 | North America/Canada/Nova Scotia/ | QEII Health Sciences Centre | National Microbiology Laboratory (NML) | Anna Majer et al |
| EPI_ISL_579587 | Canada/NS-NML-960/2020 | 2020-03-29 | North America/Canada/Nova Scotia/ | QEII Health Sciences Centre | National Microbiology Laboratory (NML) | Anna Majer et al |

| | | | | | | |
|---|---|---|---|---|---|---|
| EPI_ISL_579588 | Canada/NS-NML-961/2020 | 2020-03-29 | North America/Canada/Nova Scotia/ | QEII Health Sciences Centre | National Microbiology Laboratory (NML) | Anna Majer et al |
| EPI_ISL_579589 | Canada/NS-NML-963/2020 | 2020-03-29 | North America/Canada/Nova Scotia/ | QEII Health Sciences Centre | National Microbiology Laboratory (NML) | Anna Majer et al |
| EPI_ISL_579590 | Canada/NS-NML-964/2020 | 2020-03-29 | North America/Canada/Nova Scotia/ | QEII Health Sciences Centre | National Microbiology Laboratory (NML) | Anna Majer et al |
| EPI_ISL_579591 | Canada/NS-NML-965/2020 | 2020-03-30 | North America/Canada/Nova Scotia/ | QEII Health Sciences Centre | National Microbiology Laboratory (NML) | Anna Majer et al |
| EPI_ISL_579592 | Canada/NS-NML-966/2020 | 2020-03-30 | North America/Canada/Nova Scotia/ | QEII Health Sciences Centre | National Microbiology Laboratory (NML) | Anna Majer et al |
| EPI_ISL_579593 | Canada/NS-NML-968/2020 | 2020-03-30 | North America/Canada/Nova Scotia/ | QEII Health Sciences Centre | National Microbiology Laboratory (NML) | Anna Majer et al |
| EPI_ISL_579594 | Canada/NS-NML-971/2020 | 2020-03-31 | North America/Canada/Nova Scotia/ | QEII Health Sciences Centre | National Microbiology Laboratory (NML) | Anna Majer et al |
| EPI_ISL_579595 | Canada/NS-NML-972/2020 | 2020-03-31 | North America/Canada/Nova Scotia/ | QEII Health Sciences Centre | National Microbiology Laboratory (NML) | Anna Majer et al |
| EPI_ISL_579596 | Canada/NS-NML-974/2020 | 2020-04-01 | North America/Canada/Nova Scotia/ | QEII Health Sciences Centre | National Microbiology Laboratory (NML) | Anna Majer et al |
| EPI_ISL_579597 | Canada/NS-NML-1196/2020 | 2020-03-31 | North America/Canada/Nova Scotia/ | QEII Health Sciences Centre | National Microbiology Laboratory (NML) | Anna Majer et al |
| EPI_ISL_579598 | Canada/NS-NML-1197/2020 | 2020-03-31 | North America/Canada/Nova Scotia/ | QEII Health Sciences Centre | National Microbiology Laboratory (NML) | Anna Majer et al |
| EPI_ISL_579599 | Canada/NS-NML-1198/2020 | 2020-04-02 | North America/Canada/Nova Scotia/ | QEII Health Sciences Centre | National Microbiology Laboratory (NML) | Anna Majer et al |
| EPI_ISL_579600 | Canada/NS-NML-1199/2020 | 2020-04-02 | North America/Canada/Nova Scotia/ | QEII Health Sciences Centre | National Microbiology Laboratory (NML) | Anna Majer et al |

| | | | | | | |
|---|---|---|---|---|---|---|
| EPI_ISL_579601 | Canada/NS-NML-1200/2020 | 2020-04-02 | North America/Canada/Nova Scotia/ | QEII Health Sciences Centre | National Microbiology Laboratory (NML) | Anna Majer et al |
| EPI_ISL_579602 | Canada/NS-NML-1201/2020 | 2020-04-02 | North America/Canada/Nova Scotia/ | QEII Health Sciences Centre | National Microbiology Laboratory (NML) | Anna Majer et al |
| EPI_ISL_579603 | Canada/NS-NML-1202/2020 | 2020-04-02 | North America/Canada/Nova Scotia/ | QEII Health Sciences Centre | National Microbiology Laboratory (NML) | Anna Majer et al |
| EPI_ISL_579604 | Canada/NS-NML-1203/2020 | 2020-04-02 | North America/Canada/Nova Scotia/ | QEII Health Sciences Centre | National Microbiology Laboratory (NML) | Anna Majer et al |
| EPI_ISL_579605 | Canada/NS-NML-1204/2020 | 2020-04-02 | North America/Canada/Nova Scotia/ | QEII Health Sciences Centre | National Microbiology Laboratory (NML) | Anna Majer et al |
| EPI_ISL_579606 | Canada/NS-NML-1205/2020 | 2020-04-02 | North America/Canada/Nova Scotia/ | QEII Health Sciences Centre | National Microbiology Laboratory (NML) | Anna Majer et al |
| EPI_ISL_579607 | Canada/NS-NML-1207/2020 | 2020-04-02 | North America/Canada/Nova Scotia/ | QEII Health Sciences Centre | National Microbiology Laboratory (NML) | Anna Majer et al |
| EPI_ISL_579608 | Canada/NS-NML-1210/2020 | 2020-04-03 | North America/Canada/Nova Scotia/ | QEII Health Sciences Centre | National Microbiology Laboratory (NML) | Anna Majer et al |
| EPI_ISL_579609 | Canada/NS-NML-1211/2020 | 2020-04-03 | North America/Canada/Nova Scotia/ | QEII Health Sciences Centre | National Microbiology Laboratory (NML) | Anna Majer et al |
| EPI_ISL_579610 | Canada/NS-NML-1212/2020 | 2020-04-03 | North America/Canada/Nova Scotia/ | QEII Health Sciences Centre | National Microbiology Laboratory (NML) | Anna Majer et al |
| EPI_ISL_579611 | Canada/NS-NML-1213/2020 | 2020-04-03 | North America/Canada/Nova Scotia/ | QEII Health Sciences Centre | National Microbiology Laboratory (NML) | Anna Majer et al |
| EPI_ISL_579612 | Canada/NS-NML-1214/2020 | 2020-04-03 | North America/Canada/Nova Scotia/ | QEII Health Sciences Centre | National Microbiology Laboratory (NML) | Anna Majer et al |
| EPI_ISL_579613 | Canada/NS-NML-1216/2020 | 2020-04-03 | North America/Canada/Nova Scotia/ | QEII Health Sciences Centre | National Microbiology Laboratory (NML) | Anna Majer et al |

| | | | | | | |
|---|---|---|---|---|---|---|
| EPI_ISL_579614 | Canada/NS-NML-1217/2020 | 2020-04-03 | North America/Canada/Nova Scotia/ | QEII Health Sciences Centre | National Microbiology Laboratory (NML) | Anna Majer et al |
| EPI_ISL_579615 | Canada/NS-NML-1218/2020 | 2020-04-03 | North America/Canada/Nova Scotia/ | QEII Health Sciences Centre | National Microbiology Laboratory (NML) | Anna Majer et al |
| EPI_ISL_579616 | Canada/NS-NML-1219/2020 | 2020-04-03 | North America/Canada/Nova Scotia/ | QEII Health Sciences Centre | National Microbiology Laboratory (NML) | Anna Majer et al |
| EPI_ISL_579617 | Canada/NS-NML-1221/2020 | 2020-04-04 | North America/Canada/Nova Scotia/ | QEII Health Sciences Centre | National Microbiology Laboratory (NML) | Anna Majer et al |
| EPI_ISL_579618 | Canada/NS-NML-1222/2020 | 2020-04-04 | North America/Canada/Nova Scotia/ | QEII Health Sciences Centre | National Microbiology Laboratory (NML) | Anna Majer et al |
| EPI_ISL_579619 | Canada/NS-NML-1223/2020 | 2020-04-04 | North America/Canada/Nova Scotia/ | QEII Health Sciences Centre | National Microbiology Laboratory (NML) | Anna Majer et al |
| EPI_ISL_579620 | Canada/NS-NML-1224/2020 | 2020-04-04 | North America/Canada/Nova Scotia/ | QEII Health Sciences Centre | National Microbiology Laboratory (NML) | Anna Majer et al |
| EPI_ISL_579621 | Canada/NS-NML-1225/2020 | 2020-04-04 | North America/Canada/Nova Scotia/ | QEII Health Sciences Centre | National Microbiology Laboratory (NML) | Anna Majer et al |
| EPI_ISL_579622 | Canada/NS-NML-1226/2020 | 2020-04-04 | North America/Canada/Nova Scotia/ | QEII Health Sciences Centre | National Microbiology Laboratory (NML) | Anna Majer et al |
| EPI_ISL_579623 | Canada/NS-NML-1227/2020 | 2020-04-04 | North America/Canada/Nova Scotia/ | QEII Health Sciences Centre | National Microbiology Laboratory (NML) | Anna Majer et al |
| EPI_ISL_579624 | Canada/NS-NML-1228/2020 | 2020-04-04 | North America/Canada/Nova Scotia/ | QEII Health Sciences Centre | National Microbiology Laboratory (NML) | Anna Majer et al |
| EPI_ISL_579625 | Canada/NS-NML-1230/2020 | 2020-04-04 | North America/Canada/Nova Scotia/ | QEII Health Sciences Centre | National Microbiology Laboratory (NML) | Anna Majer et al |
| EPI_ISL_579626 | Canada/NS-NML-1231/2020 | 2020-04-04 | North America/Canada/Nova Scotia/ | QEII Health Sciences Centre | National Microbiology Laboratory (NML) | Anna Majer et al |

| | | | | | | |
|---|---|---|---|---|---|---|
| EPI_ISL_579627 | Canada/NS-NML-1232/2020 | 2020-04-04 | North America/Canada/Nova Scotia/ | QEII Health Sciences Centre | National Microbiology Laboratory (NML) | Anna Majer et al |
| EPI_ISL_579628 | Canada/NS-NML-1234/2020 | 2020-04-04 | North America/Canada/Nova Scotia/ | QEII Health Sciences Centre | National Microbiology Laboratory (NML) | Anna Majer et al |
| EPI_ISL_579629 | Canada/NS-NML-1235/2020 | 2020-04-04 | North America/Canada/Nova Scotia/ | QEII Health Sciences Centre | National Microbiology Laboratory (NML) | Anna Majer et al |
| EPI_ISL_579630 | Canada/NS-NML-1236/2020 | 2020-04-05 | North America/Canada/Nova Scotia/ | QEII Health Sciences Centre | National Microbiology Laboratory (NML) | Anna Majer et al |
| EPI_ISL_579631 | Canada/NS-NML-1238/2020 | 2020-04-05 | North America/Canada/Nova Scotia/ | QEII Health Sciences Centre | National Microbiology Laboratory (NML) | Anna Majer et al |
| EPI_ISL_579632 | Canada/NS-NML-1239/2020 | 2020-04-05 | North America/Canada/Nova Scotia/ | QEII Health Sciences Centre | National Microbiology Laboratory (NML) | Anna Majer et al |
| EPI_ISL_579633 | Canada/NS-NML-1241/2020 | 2020-04-05 | North America/Canada/Nova Scotia/ | QEII Health Sciences Centre | National Microbiology Laboratory (NML) | Anna Majer et al |
| EPI_ISL_579634 | Canada/NS-NML-1242/2020 | 2020-04-05 | North America/Canada/Nova Scotia/ | QEII Health Sciences Centre | National Microbiology Laboratory (NML) | Anna Majer et al |
| EPI_ISL_579635 | Canada/NS-NML-1243/2020 | 2020-04-05 | North America/Canada/Nova Scotia/ | QEII Health Sciences Centre | National Microbiology Laboratory (NML) | Anna Majer et al |
| EPI_ISL_579636 | Canada/NS-NML-1244/2020 | 2020-04-05 | North America/Canada/Nova Scotia/ | QEII Health Sciences Centre | National Microbiology Laboratory (NML) | Anna Majer et al |
| EPI_ISL_579637 | Canada/NS-NML-1245/2020 | 2020-04-05 | North America/Canada/Nova Scotia/ | QEII Health Sciences Centre | National Microbiology Laboratory (NML) | Anna Majer et al |
| EPI_ISL_579638 | Canada/NS-NML-1246/2020 | 2020-04-05 | North America/Canada/Nova Scotia/ | QEII Health Sciences Centre | National Microbiology Laboratory (NML) | Anna Majer et al |
| EPI_ISL_579639 | Canada/NS-NML-1248/2020 | 2020-04-05 | North America/Canada/Nova Scotia/ | QEII Health Sciences Centre | National Microbiology Laboratory (NML) | Anna Majer et al |

| | | | | | | |
|---|---|---|---|---|---|---|
| EPI_ISL_579640 | Canada/NS-NML-1249/2020 | 2020-04-06 | North America/Canada/Nova Scotia/ | QEII Health Sciences Centre | National Microbiology Laboratory (NML) | Anna Majer et al |
| EPI_ISL_579641 | Canada/NS-NML-1250/2020 | 2020-04-06 | North America/Canada/Nova Scotia/ | QEII Health Sciences Centre | National Microbiology Laboratory (NML) | Anna Majer et al |
| EPI_ISL_579642 | Canada/NS-NML-1251/2020 | 2020-04-06 | North America/Canada/Nova Scotia/ | QEII Health Sciences Centre | National Microbiology Laboratory (NML) | Anna Majer et al |
| EPI_ISL_579643 | Canada/NS-NML-1252/2020 | 2020-04-06 | North America/Canada/Nova Scotia/ | QEII Health Sciences Centre | National Microbiology Laboratory (NML) | Anna Majer et al |
| EPI_ISL_579644 | Canada/NS-NML-1253/2020 | 2020-04-06 | North America/Canada/Nova Scotia/ | QEII Health Sciences Centre | National Microbiology Laboratory (NML) | Anna Majer et al |
| EPI_ISL_579645 | Canada/NS-NML-1255/2020 | 2020-04-06 | North America/Canada/Nova Scotia/ | QEII Health Sciences Centre | National Microbiology Laboratory (NML) | Anna Majer et al |
| EPI_ISL_579646 | Canada/NS-NML-1256/2020 | 2020-04-06 | North America/Canada/Nova Scotia/ | QEII Health Sciences Centre | National Microbiology Laboratory (NML) | Anna Majer et al |
| EPI_ISL_579647 | Canada/NS-NML-1257/2020 | 2020-04-06 | North America/Canada/Nova Scotia/ | QEII Health Sciences Centre | National Microbiology Laboratory (NML) | Anna Majer et al |
| EPI_ISL_579648 | Canada/NS-NML-1259/2020 | 2020-04-06 | North America/Canada/Nova Scotia/ | QEII Health Sciences Centre | National Microbiology Laboratory (NML) | Anna Majer et al |
| EPI_ISL_579649 | Canada/NS-NML-1261/2020 | 2020-04-06 | North America/Canada/Nova Scotia/ | QEII Health Sciences Centre | National Microbiology Laboratory (NML) | Anna Majer et al |
| EPI_ISL_579650 | Canada/NS-NML-1262/2020 | 2020-04-06 | North America/Canada/Nova Scotia/ | QEII Health Sciences Centre | National Microbiology Laboratory (NML) | Anna Majer et al |
| EPI_ISL_579651 | Canada/NS-NML-1263/2020 | 2020-04-07 | North America/Canada/Nova Scotia/ | QEII Health Sciences Centre | National Microbiology Laboratory (NML) | Anna Majer et al |
| EPI_ISL_579652 | Canada/NS-NML-1264/2020 | 2020-04-07 | North America/Canada/Nova Scotia/ | QEII Health Sciences Centre | National Microbiology Laboratory (NML) | Anna Majer et al |

| | | | | | | |
|---|---|---|---|---|---|---|
| EPI_ISL_579653 | Canada/NS-NML-1265/2020 | 2020-04-07 | North America/Canada/Nova Scotia/ | QEII Health Sciences Centre | National Microbiology Laboratory (NML) | Anna Majer et al |
| EPI_ISL_579654 | Canada/NS-NML-1266/2020 | 2020-04-07 | North America/Canada/Nova Scotia/ | QEII Health Sciences Centre | National Microbiology Laboratory (NML) | Anna Majer et al |
| EPI_ISL_579655 | Canada/NS-NML-1267/2020 | 2020-04-16 | North America/Canada/Nova Scotia/ | QEII Health Sciences Centre | National Microbiology Laboratory (NML) | Anna Majer et al |
| EPI_ISL_579656 | Canada/NS-NML-1268/2020 | 2020-04-16 | North America/Canada/Nova Scotia/ | QEII Health Sciences Centre | National Microbiology Laboratory (NML) | Anna Majer et al |
| EPI_ISL_579657 | Canada/NS-NML-1269/2020 | 2020-04-07 | North America/Canada/Nova Scotia/ | QEII Health Sciences Centre | National Microbiology Laboratory (NML) | Anna Majer et al |
| EPI_ISL_579658 | Canada/NS-NML-1270/2020 | 2020-04-07 | North America/Canada/Nova Scotia/ | QEII Health Sciences Centre | National Microbiology Laboratory (NML) | Anna Majer et al |
| EPI_ISL_579659 | Canada/NS-NML-1271/2020 | 2020-04-07 | North America/Canada/Nova Scotia/ | QEII Health Sciences Centre | National Microbiology Laboratory (NML) | Anna Majer et al |
| EPI_ISL_579660 | Canada/NS-NML-1272/2020 | 2020-04-07 | North America/Canada/Nova Scotia/ | QEII Health Sciences Centre | National Microbiology Laboratory (NML) | Anna Majer et al |
| EPI_ISL_579661 | Canada/NS-NML-1273/2020 | 2020-04-07 | North America/Canada/Nova Scotia/ | QEII Health Sciences Centre | National Microbiology Laboratory (NML) | Anna Majer et al |
| EPI_ISL_579662 | Canada/NS-NML-1275/2020 | 2020-04-16 | North America/Canada/Nova Scotia/ | QEII Health Sciences Centre | National Microbiology Laboratory (NML) | Anna Majer et al |
| EPI_ISL_579663 | Canada/NS-NML-1276/2020 | 2020-04-16 | North America/Canada/Nova Scotia/ | QEII Health Sciences Centre | National Microbiology Laboratory (NML) | Anna Majer et al |
| EPI_ISL_579664 | Canada/NS-NML-1277/2020 | 2020-04-07 | North America/Canada/Nova Scotia/ | QEII Health Sciences Centre | National Microbiology Laboratory (NML) | Anna Majer et al |
| EPI_ISL_579665 | Canada/NS-NML-1280/2020 | 2020-04-16 | North America/Canada/Nova Scotia/ | QEII Health Sciences Centre | National Microbiology Laboratory (NML) | Anna Majer et al |

| | | | | | | |
|---|---|---|---|---|---|---|
| EPI_ISL_579666 | Canada/NS-NML-1281/2020 | 2020-04-07 | North America/Canada/Nova Scotia/ | QEII Health Sciences Centre | National Microbiology Laboratory (NML) | Anna Majer et al |
| EPI_ISL_579667 | Canada/NS-NML-1282/2020 | 2020-04-07 | North America/Canada/Nova Scotia/ | QEII Health Sciences Centre | National Microbiology Laboratory (NML) | Anna Majer et al |
| EPI_ISL_579668 | Canada/NS-NML-1284/2020 | 2020-04-07 | North America/Canada/Nova Scotia/ | QEII Health Sciences Centre | National Microbiology Laboratory (NML) | Anna Majer et al |
| EPI_ISL_579669 | Canada/NS-NML-1285/2020 | 2020-04-07 | North America/Canada/Nova Scotia/ | QEII Health Sciences Centre | National Microbiology Laboratory (NML) | Anna Majer et al |
| EPI_ISL_579670 | Canada/NS-NML-1286/2020 | 2020-04-07 | North America/Canada/Nova Scotia/ | QEII Health Sciences Centre | National Microbiology Laboratory (NML) | Anna Majer et al |
| EPI_ISL_579671 | Canada/NS-NML-1287/2020 | 2020-04-07 | North America/Canada/Nova Scotia/ | QEII Health Sciences Centre | National Microbiology Laboratory (NML) | Anna Majer et al |
| EPI_ISL_579672 | Canada/NS-NML-1288/2020 | 2020-04-08 | North America/Canada/Nova Scotia/ | QEII Health Sciences Centre | National Microbiology Laboratory (NML) | Anna Majer et al |
| EPI_ISL_579673 | Canada/NS-NML-1289/2020 | 2020-04-08 | North America/Canada/Nova Scotia/ | QEII Health Sciences Centre | National Microbiology Laboratory (NML) | Anna Majer et al |
| EPI_ISL_579674 | Canada/NS-NML-1290/2020 | 2020-04-08 | North America/Canada/Nova Scotia/ | QEII Health Sciences Centre | National Microbiology Laboratory (NML) | Anna Majer et al |
| EPI_ISL_579675 | Canada/NS-NML-1291/2020 | 2020-04-08 | North America/Canada/Nova Scotia/ | QEII Health Sciences Centre | National Microbiology Laboratory (NML) | Anna Majer et al |
| EPI_ISL_579676 | Canada/NS-NML-1292/2020 | 2020-04-08 | North America/Canada/Nova Scotia/ | QEII Health Sciences Centre | National Microbiology Laboratory (NML) | Anna Majer et al |
| EPI_ISL_579677 | Canada/NS-NML-1294/2020 | 2020-04-08 | North America/Canada/Nova Scotia/ | QEII Health Sciences Centre | National Microbiology Laboratory (NML) | Anna Majer et al |
| EPI_ISL_579678 | Canada/NS-NML-1295/2020 | 2020-04-08 | North America/Canada/Nova Scotia/ | QEII Health Sciences Centre | National Microbiology Laboratory (NML) | Anna Majer et al |

| | | | | | | |
|---|---|---|---|---|---|---|
| EPI_ISL_579679 | Canada/NS-NML-1296/2020 | 2020-04-08 | North America/Canada/Nova Scotia/ | QEII Health Sciences Centre | National Microbiology Laboratory (NML) | Anna Majer et al |
| EPI_ISL_579680 | Canada/NS-NML-1297/2020 | 2020-04-08 | North America/Canada/Nova Scotia/ | QEII Health Sciences Centre | National Microbiology Laboratory (NML) | Anna Majer et al |
| EPI_ISL_579681 | Canada/NS-NML-1299/2020 | 2020-04-09 | North America/Canada/Nova Scotia/ | QEII Health Sciences Centre | National Microbiology Laboratory (NML) | Anna Majer et al |
| EPI_ISL_579682 | Canada/NS-NML-1301/2020 | 2020-04-09 | North America/Canada/Nova Scotia/ | QEII Health Sciences Centre | National Microbiology Laboratory (NML) | Anna Majer et al |
| EPI_ISL_579683 | Canada/NS-NML-1302/2020 | 2020-04-09 | North America/Canada/Nova Scotia/ | QEII Health Sciences Centre | National Microbiology Laboratory (NML) | Anna Majer et al |
| EPI_ISL_579684 | Canada/NS-NML-1303/2020 | 2020-04-09 | North America/Canada/Nova Scotia/ | QEII Health Sciences Centre | National Microbiology Laboratory (NML) | Anna Majer et al |
| EPI_ISL_579685 | Canada/NS-NML-1304/2020 | 2020-04-09 | North America/Canada/Nova Scotia/ | QEII Health Sciences Centre | National Microbiology Laboratory (NML) | Anna Majer et al |
| EPI_ISL_579686 | Canada/NS-NML-1305/2020 | 2020-04-09 | North America/Canada/Nova Scotia/ | QEII Health Sciences Centre | National Microbiology Laboratory (NML) | Anna Majer et al |
| EPI_ISL_579687 | Canada/NS-NML-1306/2020 | 2020-04-09 | North America/Canada/Nova Scotia/ | QEII Health Sciences Centre | National Microbiology Laboratory (NML) | Anna Majer et al |
| EPI_ISL_579688 | Canada/NS-NML-1307/2020 | 2020-04-09 | North America/Canada/Nova Scotia/ | QEII Health Sciences Centre | National Microbiology Laboratory (NML) | Anna Majer et al |
| EPI_ISL_579689 | Canada/NS-NML-1308/2020 | 2020-04-09 | North America/Canada/Nova Scotia/ | QEII Health Sciences Centre | National Microbiology Laboratory (NML) | Anna Majer et al |
| EPI_ISL_579690 | Canada/NS-NML-1309/2020 | 2020-04-09 | North America/Canada/Nova Scotia/ | QEII Health Sciences Centre | National Microbiology Laboratory (NML) | Anna Majer et al |
| EPI_ISL_579691 | Canada/NS-NML-1310/2020 | 2020-04-09 | North America/Canada/Nova Scotia/ | QEII Health Sciences Centre | National Microbiology Laboratory (NML) | Anna Majer et al |

| | | | | | | |
|---|---|---|---|---|---|---|
| EPI_ISL_579692 | Canada/NS-NML-1311/2020 | 2020-04-09 | North America/Canada/Nova Scotia/ | QEII Health Sciences Centre | National Microbiology Laboratory (NML) | Anna Majer et al |
| EPI_ISL_579693 | Canada/NS-NML-1312/2020 | 2020-04-09 | North America/Canada/Nova Scotia/ | QEII Health Sciences Centre | National Microbiology Laboratory (NML) | Anna Majer et al |
| EPI_ISL_579694 | Canada/NS-NML-1313/2020 | 2020-04-09 | North America/Canada/Nova Scotia/ | QEII Health Sciences Centre | National Microbiology Laboratory (NML) | Anna Majer et al |
| EPI_ISL_579695 | Canada/NS-NML-1314/2020 | 2020-04-09 | North America/Canada/Nova Scotia/ | QEII Health Sciences Centre | National Microbiology Laboratory (NML) | Anna Majer et al |
| EPI_ISL_579696 | Canada/NS-NML-1316/2020 | 2020-04-09 | North America/Canada/Nova Scotia/ | QEII Health Sciences Centre | National Microbiology Laboratory (NML) | Anna Majer et al |
| EPI_ISL_579697 | Canada/NS-NML-1317/2020 | 2020-04-10 | North America/Canada/Nova Scotia/ | QEII Health Sciences Centre | National Microbiology Laboratory (NML) | Anna Majer et al |
| EPI_ISL_579698 | Canada/NS-NML-1318/2020 | 2020-04-10 | North America/Canada/Nova Scotia/ | QEII Health Sciences Centre | National Microbiology Laboratory (NML) | Anna Majer et al |
| EPI_ISL_579699 | Canada/NS-NML-1319/2020 | 2020-04-10 | North America/Canada/Nova Scotia/ | QEII Health Sciences Centre | National Microbiology Laboratory (NML) | Anna Majer et al |
| EPI_ISL_579700 | Canada/NS-NML-1320/2020 | 2020-04-10 | North America/Canada/Nova Scotia/ | QEII Health Sciences Centre | National Microbiology Laboratory (NML) | Anna Majer et al |
| EPI_ISL_579701 | Canada/NS-NML-1321/2020 | 2020-04-10 | North America/Canada/Nova Scotia/ | QEII Health Sciences Centre | National Microbiology Laboratory (NML) | Anna Majer et al |
| EPI_ISL_579702 | Canada/NS-NML-1322/2020 | 2020-04-10 | North America/Canada/Nova Scotia/ | QEII Health Sciences Centre | National Microbiology Laboratory (NML) | Anna Majer et al |
| EPI_ISL_579703 | Canada/NS-NML-1323/2020 | 2020-04-10 | North America/Canada/Nova Scotia/ | QEII Health Sciences Centre | National Microbiology Laboratory (NML) | Anna Majer et al |
| EPI_ISL_579704 | Canada/NS-NML-1324/2020 | 2020-04-10 | North America/Canada/Nova Scotia/ | QEII Health Sciences Centre | National Microbiology Laboratory (NML) | Anna Majer et al |

| | | | | | | |
|---|---|---|---|---|---|---|
| EPI_ISL_579705 | Canada/NS-NML-1325/2020 | 2020-04-10 | North America/Canada/Nova Scotia/ | QEII Health Sciences Centre | National Microbiology Laboratory (NML) | Anna Majer et al |
| EPI_ISL_579706 | Canada/NS-NML-1326/2020 | 2020-04-10 | North America/Canada/Nova Scotia/ | QEII Health Sciences Centre | National Microbiology Laboratory (NML) | Anna Majer et al |
| EPI_ISL_579707 | Canada/NS-NML-1327/2020 | 2020-04-10 | North America/Canada/Nova Scotia/ | QEII Health Sciences Centre | National Microbiology Laboratory (NML) | Anna Majer et al |
| EPI_ISL_579708 | Canada/NS-NML-1328/2020 | 2020-04-10 | North America/Canada/Nova Scotia/ | QEII Health Sciences Centre | National Microbiology Laboratory (NML) | Anna Majer et al |
| EPI_ISL_579709 | Canada/NS-NML-1329/2020 | 2020-04-10 | North America/Canada/Nova Scotia/ | QEII Health Sciences Centre | National Microbiology Laboratory (NML) | Anna Majer et al |
| EPI_ISL_579710 | Canada/NS-NML-1331/2020 | 2020-04-11 | North America/Canada/Nova Scotia/ | QEII Health Sciences Centre | National Microbiology Laboratory (NML) | Anna Majer et al |
| EPI_ISL_579711 | Canada/NS-NML-1332/2020 | 2020-04-11 | North America/Canada/Nova Scotia/ | QEII Health Sciences Centre | National Microbiology Laboratory (NML) | Anna Majer et al |
| EPI_ISL_579712 | Canada/NS-NML-1333/2020 | 2020-04-11 | North America/Canada/Nova Scotia/ | QEII Health Sciences Centre | National Microbiology Laboratory (NML) | Anna Majer et al |
| EPI_ISL_579713 | Canada/NS-NML-1334/2020 | 2020-04-11 | North America/Canada/Nova Scotia/ | QEII Health Sciences Centre | National Microbiology Laboratory (NML) | Anna Majer et al |
| EPI_ISL_579714 | Canada/NS-NML-1335/2020 | 2020-04-11 | North America/Canada/Nova Scotia/ | QEII Health Sciences Centre | National Microbiology Laboratory (NML) | Anna Majer et al |
| EPI_ISL_579715 | Canada/NS-NML-1336/2020 | 2020-04-11 | North America/Canada/Nova Scotia/ | QEII Health Sciences Centre | National Microbiology Laboratory (NML) | Anna Majer et al |
| EPI_ISL_579716 | Canada/NS-NML-1338/2020 | 2020-04-11 | North America/Canada/Nova Scotia/ | QEII Health Sciences Centre | National Microbiology Laboratory (NML) | Anna Majer et al |
| EPI_ISL_579717 | Canada/NS-NML-1340/2020 | 2020-04-12 | North America/Canada/Nova Scotia/ | QEII Health Sciences Centre | National Microbiology Laboratory (NML) | Anna Majer et al |

| | | | | | | |
|---|---|---|---|---|---|---|
| EPI_ISL_579718 | Canada/NS-NML-1341/2020 | 2020-04-12 | North America/Canada/Nova Scotia/ | QEII Health Sciences Centre | National Microbiology Laboratory (NML) | Anna Majer et al |
| EPI_ISL_579719 | Canada/NS-NML-1342/2020 | 2020-04-12 | North America/Canada/Nova Scotia/ | QEII Health Sciences Centre | National Microbiology Laboratory (NML) | Anna Majer et al |
| EPI_ISL_579720 | Canada/NS-NML-1343/2020 | 2020-04-12 | North America/Canada/Nova Scotia/ | QEII Health Sciences Centre | National Microbiology Laboratory (NML) | Anna Majer et al |
| EPI_ISL_579721 | Canada/NS-NML-1344/2020 | 2020-04-13 | North America/Canada/Nova Scotia/ | QEII Health Sciences Centre | National Microbiology Laboratory (NML) | Anna Majer et al |
| EPI_ISL_579722 | Canada/NS-NML-1345/2020 | 2020-04-13 | North America/Canada/Nova Scotia/ | QEII Health Sciences Centre | National Microbiology Laboratory (NML) | Anna Majer et al |
| EPI_ISL_579723 | Canada/NS-NML-1346/2020 | 2020-04-13 | North America/Canada/Nova Scotia/ | QEII Health Sciences Centre | National Microbiology Laboratory (NML) | Anna Majer et al |
| EPI_ISL_579724 | Canada/NS-NML-1347/2020 | 2020-04-13 | North America/Canada/Nova Scotia/ | QEII Health Sciences Centre | National Microbiology Laboratory (NML) | Anna Majer et al |
| EPI_ISL_579725 | Canada/NS-NML-1348/2020 | 2020-04-13 | North America/Canada/Nova Scotia/ | QEII Health Sciences Centre | National Microbiology Laboratory (NML) | Anna Majer et al |
| EPI_ISL_579726 | Canada/NS-NML-1349/2020 | 2020-04-13 | North America/Canada/Nova Scotia/ | QEII Health Sciences Centre | National Microbiology Laboratory (NML) | Anna Majer et al |
| EPI_ISL_579727 | Canada/NS-NML-1350/2020 | 2020-04-13 | North America/Canada/Nova Scotia/ | QEII Health Sciences Centre | National Microbiology Laboratory (NML) | Anna Majer et al |
| EPI_ISL_579728 | Canada/NS-NML-1351/2020 | 2020-04-13 | North America/Canada/Nova Scotia/ | QEII Health Sciences Centre | National Microbiology Laboratory (NML) | Anna Majer et al |
| EPI_ISL_579729 | Canada/NS-NML-1352/2020 | 2020-04-13 | North America/Canada/Nova Scotia/ | QEII Health Sciences Centre | National Microbiology Laboratory (NML) | Anna Majer et al |
| EPI_ISL_579730 | Canada/NS-NML-1353/2020 | 2020-04-13 | North America/Canada/Nova Scotia/ | QEII Health Sciences Centre | National Microbiology Laboratory (NML) | Anna Majer et al |

| | | | | | | |
|---|---|---|---|---|---|---|
| EPI_ISL_579731 | Canada/NS-NML-1354/2020 | 2020-04-13 | North America/Canada/Nova Scotia/ | QEII Health Sciences Centre | National Microbiology Laboratory (NML) | Anna Majer et al |
| EPI_ISL_579733 | Canada/NS-NML-1356/2020 | 2020-04-14 | North America/Canada/Nova Scotia/ | QEII Health Sciences Centre | National Microbiology Laboratory (NML) | Anna Majer et al |
| EPI_ISL_579734 | Canada/NS-NML-1357/2020 | 2020-04-14 | North America/Canada/Nova Scotia/ | QEII Health Sciences Centre | National Microbiology Laboratory (NML) | Anna Majer et al |
| EPI_ISL_579735 | Canada/NS-NML-1358/2020 | 2020-04-14 | North America/Canada/Nova Scotia/ | QEII Health Sciences Centre | National Microbiology Laboratory (NML) | Anna Majer et al |
| EPI_ISL_579736 | Canada/NS-NML-1359/2020 | 2020-04-14 | North America/Canada/Nova Scotia/ | QEII Health Sciences Centre | National Microbiology Laboratory (NML) | Anna Majer et al |
| EPI_ISL_579737 | Canada/NS-NML-1361/2020 | 2020-04-14 | North America/Canada/Nova Scotia/ | QEII Health Sciences Centre | National Microbiology Laboratory (NML) | Anna Majer et al |
| EPI_ISL_579738 | Canada/NS-NML-1362/2020 | 2020-04-14 | North America/Canada/Nova Scotia/ | QEII Health Sciences Centre | National Microbiology Laboratory (NML) | Anna Majer et al |
| EPI_ISL_579739 | Canada/NS-NML-1363/2020 | 2020-04-14 | North America/Canada/Nova Scotia/ | QEII Health Sciences Centre | National Microbiology Laboratory (NML) | Anna Majer et al |
| EPI_ISL_579740 | Canada/NS-NML-1364/2020 | 2020-04-14 | North America/Canada/Nova Scotia/ | QEII Health Sciences Centre | National Microbiology Laboratory (NML) | Anna Majer et al |
| EPI_ISL_579741 | Canada/NS-NML-1365/2020 | 2020-04-14 | North America/Canada/Nova Scotia/ | QEII Health Sciences Centre | National Microbiology Laboratory (NML) | Anna Majer et al |
| EPI_ISL_579742 | Canada/NS-NML-1366/2020 | 2020-04-14 | North America/Canada/Nova Scotia/ | QEII Health Sciences Centre | National Microbiology Laboratory (NML) | Anna Majer et al |
| EPI_ISL_579743 | Canada/NS-NML-1367/2020 | 2020-04-16 | North America/Canada/Nova Scotia/ | QEII Health Sciences Centre | National Microbiology Laboratory (NML) | Anna Majer et al |
| EPI_ISL_579744 | Canada/NS-NML-1368/2020 | 2020-04-14 | North America/Canada/Nova Scotia/ | QEII Health Sciences Centre | National Microbiology Laboratory (NML) | Anna Majer et al |

| | | | | | | |
|---|---|---|---|---|---|---|
| EPI_ISL_579745 | Canada/NS-NML-1369/2020 | 2020-04-14 | North America/Canada/Nova Scotia/ | QEII Health Sciences Centre | National Microbiology Laboratory (NML) | Anna Majer et al |
| EPI_ISL_579746 | Canada/NS-NML-1370/2020 | 2020-04-14 | North America/Canada/Nova Scotia/ | QEII Health Sciences Centre | National Microbiology Laboratory (NML) | Anna Majer et al |
| EPI_ISL_579747 | Canada/NS-NML-1371/2020 | 2020-04-15 | North America/Canada/Nova Scotia/ | QEII Health Sciences Centre | National Microbiology Laboratory (NML) | Anna Majer et al |
| EPI_ISL_579748 | Canada/NS-NML-1372/2020 | 2020-04-15 | North America/Canada/Nova Scotia/ | QEII Health Sciences Centre | National Microbiology Laboratory (NML) | Anna Majer et al |
| EPI_ISL_579749 | Canada/NS-NML-1373/2020 | 2020-04-15 | North America/Canada/Nova Scotia/ | QEII Health Sciences Centre | National Microbiology Laboratory (NML) | Anna Majer et al |
| EPI_ISL_579750 | Canada/NS-NML-1374/2020 | 2020-04-15 | North America/Canada/Nova Scotia/ | QEII Health Sciences Centre | National Microbiology Laboratory (NML) | Anna Majer et al |
| EPI_ISL_579751 | Canada/NS-NML-1376/2020 | 2020-04-15 | North America/Canada/Nova Scotia/ | QEII Health Sciences Centre | National Microbiology Laboratory (NML) | Anna Majer et al |
| EPI_ISL_579752 | Canada/NS-NML-1377/2020 | 2020-04-15 | North America/Canada/Nova Scotia/ | QEII Health Sciences Centre | National Microbiology Laboratory (NML) | Anna Majer et al |
| EPI_ISL_579753 | Canada/NS-NML-1378/2020 | 2020-04-15 | North America/Canada/Nova Scotia/ | QEII Health Sciences Centre | National Microbiology Laboratory (NML) | Anna Majer et al |
| EPI_ISL_579754 | Canada/NS-NML-1379/2020 | 2020-04-15 | North America/Canada/Nova Scotia/ | QEII Health Sciences Centre | National Microbiology Laboratory (NML) | Anna Majer et al |
| EPI_ISL_579755 | Canada/NS-NML-1380/2020 | 2020-04-15 | North America/Canada/Nova Scotia/ | QEII Health Sciences Centre | National Microbiology Laboratory (NML) | Anna Majer et al |
| EPI_ISL_579756 | Canada/NS-NML-1381/2020 | 2020-04-15 | North America/Canada/Nova Scotia/ | QEII Health Sciences Centre | National Microbiology Laboratory (NML) | Anna Majer et al |
| EPI_ISL_579757 | Canada/NS-NML-1382/2020 | 2020-04-16 | North America/Canada/Nova Scotia/ | QEII Health Sciences Centre | National Microbiology Laboratory (NML) | Anna Majer et al |

| | | | | | | |
|---|---|---|---|---|---|---|
| EPI_ISL_579758 | Canada/NS-NML-1383/2020 | 2020-04-16 | North America/Canada/Nova Scotia/ | QEII Health Sciences Centre | National Microbiology Laboratory (NML) | Anna Majer et al |
| EPI_ISL_579834 | England/QEUH-9E3EE2/2020 | 2020-09-24 | Europe/United Kingdom/England/ | Lighthouse Lab in Glasgow | Wellcome Sanger Institute for the COVID-19 Genomics UK (COG-UK) consortium | Harper VanSteenhouse et al |
| EPI_ISL_579931 | England/ALDP-9EA6E3/2020 | 2020-09-24 | Europe/United Kingdom/England/ | Lighthouse Lab in Alderley Park | Wellcome Sanger Institute for the COVID-19 Genomics UK (COG-UK) consortium | Jacquelyn Wynn et al |
| EPI_ISL_580048 | England/ALDP-9E9CB5/2020 | 2020-09-24 | Europe/United Kingdom/England/ | Lighthouse Lab in Alderley Park | Wellcome Sanger Institute for the COVID-19 Genomics UK (COG-UK) consortium | Jacquelyn Wynn et al |
| EPI_ISL_580080 | England/ALDP-9EAF06/2020 | 2020-09-24 | Europe/United Kingdom/England/ | Lighthouse Lab in Alderley Park | Wellcome Sanger Institute for the COVID-19 Genomics UK (COG-UK) consortium | Jacquelyn Wynn et al |
| EPI_ISL_580175 | England/ALDP-9EA258/2020 | 2020-09-24 | Europe/United Kingdom/England/ | Lighthouse Lab in Alderley Park | Wellcome Sanger Institute for the COVID-19 Genomics UK (COG-UK) consortium | Jacquelyn Wynn et al |
| EPI_ISL_580520 | England/ALDP-9E6CA9/2020 | 2020-09-24 | Europe/United Kingdom/England/ | Lighthouse Lab in Alderley Park | Wellcome Sanger Institute for the COVID-19 Genomics UK (COG-UK) consortium | Jacquelyn Wynn et al |
| EPI_ISL_580543 | England/ALDP-9E6E49/2020 | 2020-09-24 | Europe/United Kingdom/England/ | Lighthouse Lab in Alderley Park | Wellcome Sanger Institute for the COVID-19 Genomics UK (COG-UK) consortium | Jacquelyn Wynn et al |

| | | | | | | |
|---|---|---|---|---|---|---|
| EPI_ISL_580702 | England/MILK-9F043B/2020 | 2020-09-24 | Europe/United Kingdom/England/ | Lighthouse Lab in Milton Keynes | Wellcome Sanger Institute for the COVID-19 Genomics UK (COG-UK) consortium | The Lighthouse Lab in Milton Keynes et al |
| EPI_ISL_580829 | England/MILK-9F04FF/2020 | 2020-09-24 | Europe/United Kingdom/England/ | Lighthouse Lab in Milton Keynes | Wellcome Sanger Institute for the COVID-19 Genomics UK (COG-UK) consortium | The Lighthouse Lab in Milton Keynes et al |
| EPI_ISL_580841 | Wales/MILK-9EE940/2020 | 2020-09-23 | Europe/United Kingdom/Wales/ | Lighthouse Lab in Milton Keynes | Wellcome Sanger Institute for the COVID-19 Genomics UK (COG-UK) consortium | The Lighthouse Lab in Milton Keynes et al |
| EPI_ISL_581039 | England/QEUH-9DC65D/2020 | 2020-09-21 | Europe/United Kingdom/England/ | Lighthouse Lab in Glasgow | Wellcome Sanger Institute for the COVID-19 Genomics UK (COG-UK) consortium | Harper VanSteenhouse et al |
| EPI_ISL_581137 | England/MILK-9E3073/2020 | 2020-09-21 | Europe/United Kingdom/England/ | Lighthouse Lab in Milton Keynes | Wellcome Sanger Institute for the COVID-19 Genomics UK (COG-UK) consortium | The Lighthouse Lab in Milton Keynes et al |
| EPI_ISL_581151 | England/MILK-9E21EA/2020 | 2020-09-20 | Europe/United Kingdom/England/ | Lighthouse Lab in Milton Keynes | Wellcome Sanger Institute for the COVID-19 Genomics UK (COG-UK) consortium | The Lighthouse Lab in Milton Keynes et al |
| EPI_ISL_581162 | England/MILK-9E2C34/2020 | 2020-09-20 | Europe/United Kingdom/England/ | Lighthouse Lab in Milton Keynes | Wellcome Sanger Institute for the COVID-19 Genomics UK (COG-UK) consortium | The Lighthouse Lab in Milton Keynes et al |
| EPI_ISL_581334 | England/QEUH-9DE7EF/2020 | 2020-09-21 | Europe/United Kingdom/England/ | Lighthouse Lab in Glasgow | Wellcome Sanger Institute for the COVID-19 Genomics UK (COG-UK) consortium | Harper VanSteenhouse et al |

| | | | | | | |
|---|---|---|---|---|---|---|
| EPI_ISL_581449 | India/WB-IICB-017/2020 | 2020-08-04 | Asia/India/West Bengal/ | CSIR-Indian Institute of Chemical Biology, MEDICA Supercpecialty Hospital Kolkata | CSIR-Indian Institute of Chemical Biology, MEDICA Supercpecialty Hospital Kolkata | Sujay Krishna Maity et al |
| EPI_ISL_581450 | India/WB-IICB-018/2020 | 2020-08-12 | Asia/India/West Bengal/ | CSIR-Indian Institute of Chemical Biology, MEDICA Supercpecialty Hospital Kolkata | CSIR-Indian Institute of Chemical Biology, MEDICA Supercpecialty Hospital Kolkata | Sujay Krishna Maity et al |
| EPI_ISL_581451 | India/WB-IICB-019/2020 | 2020-08-12 | Asia/India/West Bengal/ | CSIR-Indian Institute of Chemical Biology, MEDICA Supercpecialty Hospital Kolkata | CSIR-Indian Institute of Chemical Biology, MEDICA Supercpecialty Hospital Kolkata | Sujay Krishna Maity et al |
| EPI_ISL_581457 | Germany/BW-UKT-049/2020 | 2020-04-02 | Europe/Germany/Baden-Wuerttemberg | Medizinische Klinik Innere Medizin I, Universit√§tsklinikum T√ºbingen | NGS Competence Center T√ºbingen, Institut f√ºr Medizinische Mikrobiologie und Hygiene, Universit√§tsklinikum T√ºbingen | Angel Angelov et al |
| EPI_ISL_581458 | Germany/BW-UKT-030/2020 | 2020-05-13 | Europe/Germany/Baden-Wuerttemberg | Medizinische Klinik Innere Medizin I, Universit√§tsklinikum T√ºbingen | NGS Competence Center T√ºbingen, Institut f√ºr Medizinische Mikrobiologie und Hygiene, Universit√§tsklinikum T√ºbingen | Angel Angelov et al |

| | | | | | | |
|---|---|---|---|---|---|---|
| EPI_ISL_581463 | Germany/BW-UKT-046/2020 | 2020-03-30 | Europe/Germany/Baden-Wuerttemberg | Medizinische Klinik Innere Medizin I, Universit√§tsklinikum T√ºbingen | NGS Competence Center T√ºbingen, Institut f√ºr Medizinische Mikrobiologie und Hygiene, Universit√§tsklinikum T√ºbingen | Angel Angelov et al |
| EPI_ISL_581464 | Germany/BW-UKT-047/2020 | 2020-03-30 | Europe/Germany/Baden-Wuerttemberg | Medizinische Klinik Innere Medizin I, Universit√§tsklinikum T√ºbingen | NGS Competence Center T√ºbingen, Institut f√ºr Medizinische Mikrobiologie und Hygiene, Universit√§tsklinikum T√ºbingen | Angel Angelov et al |
| EPI_ISL_581467 | Germany/BW-UKT-066/2020 | 2020-04-23 | Europe/Germany/Baden-Wuerttemberg | Medizinische Klinik Innere Medizin I, Universit√§tsklinikum T√ºbingen | NGS Competence Center T√ºbingen, Institut f√ºr Medizinische Mikrobiologie und Hygiene, Universit√§tsklinikum T√ºbingen | Angel Angelov et al |
| EPI_ISL_581473 | Germany/BW-UKT-041/2020 | 2020-03-27 | Europe/Germany/Baden-Wuerttemberg | Medizinische Klinik Innere Medizin I, Universit√§tsklinikum T√ºbingen | NGS Competence Center T√ºbingen, Institut f√ºr Medizinische Mikrobiologie und Hygiene, Universit√§tsklinikum T√ºbingen | Angel Angelov et al |
| EPI_ISL_581476 | Germany/BW-UKT-043/2020 | 2020-03-29 | Europe/Germany/Baden-Wuerttemberg | Medizinische Klinik Innere Medizin I, Universit√§tsklinikum T√ºbingen | NGS Competence Center T√ºbingen, Institut f√ºr Medizinische Mikrobiologie und Hygiene, Universit√§tsklinikum T√ºbingen | Angel Angelov et al |

| | | | | | | |
|---|---|---|---|---|---|---|
| EPI_ISL_581487 | Congo/UKT-002/2020 | 2020-07-04 | Africa/Democratic Republic of the Congo | Fondation Congolaise pour la recherche medicale (FCRM) | NGS Competence Center T√ºbingen, Institut f√ºr Medizinische Mikrobiologie und Hygiene, Universit√§tsklinikum T√ºbingen | Angel Angelov et al |
| EPI_ISL_581492 | Congo/UKT-014/2020 | 2020-07-03 | Africa/Democratic Republic of the Congo | Fondation Congolaise pour la recherche medicale (FCRM) | NGS Competence Center T√ºbingen, Institut f√ºr Medizinische Mikrobiologie und Hygiene, Universit√§tsklinikum T√ºbingen | Angel Angelov et al |
| EPI_ISL_581493 | Congo/UKT-015/2020 | 2020-06-25 | Africa/Democratic Republic of the Congo | Fondation Congolaise pour la recherche medicale (FCRM) | NGS Competence Center T√ºbingen, Institut f√ºr Medizinische Mikrobiologie und Hygiene, Universit√§tsklinikum T√ºbingen | Angel Angelov et al |
| EPI_ISL_581494 | India/WB-IICB-020/2020 | 2020-08-12 | Asia/India/West Bengal/ | CSIR-Indian Institute of Chemical Biology, MEDICA Supercpecialty Hospital Kolkata | CSIR-Indian Institute of Chemical Biology, MEDICA Supercpecialty Hospital Kolkata | Sujay Krishna Maity et al |
| EPI_ISL_581495 | India/WB-IICB-021/2020 | 2020-08-12 | Asia/India/West Bengal/ | CSIR-Indian Institute of Chemical Biology, MEDICA Supercpecialty Hospital Kolkata | CSIR-Indian Institute of Chemical Biology, MEDICA Supercpecialty Hospital Kolkata | Sujay Krishna Maity et al |

| | | | | | | |
|---|---|---|---|---|---|---|
| EPI_ISL_581496 | India/WB-IICB-022/2020 | 2020-08-12 | Asia/India/West Bengal/ | CSIR-Indian Institute of Chemical Biology, MEDICA Supercpecialty Hospital Kolkata | CSIR-Indian Institute of Chemical Biology, MEDICA Supercpecialty Hospital Kolkata | Sujay Krishna Maity et al |
| EPI_ISL_581497 | India/WB-IICB-023/2020 | 2020-08-04 | Asia/India/West Bengal/ | CSIR-Indian Institute of Chemical Biology, MEDICA Supercpecialty Hospital Kolkata | CSIR-Indian Institute of Chemical Biology, MEDICA Supercpecialty Hospital Kolkata | Sujay Krishna Maity et al |
| EPI_ISL_581499 | India/WB-IICB-024/2020 | 2020-08-12 | Asia/India/West Bengal/ | CSIR-Indian Institute of Chemical Biology, MEDICA Supercpecialty Hospital Kolkata | CSIR-Indian Institute of Chemical Biology, MEDICA Supercpecialty Hospital Kolkata | Sujay Krishna Maity et al |
| EPI_ISL_581500 | India/WB-IICB-025/2020 | 2020-08-12 | Asia/India/West Bengal/ | CSIR-Indian Institute of Chemical Biology, MEDICA Supercpecialty Hospital Kolkata | CSIR-Indian Institute of Chemical Biology, MEDICA Supercpecialty Hospital Kolkata | Sujay Krishna Maity et al |
| EPI_ISL_581501 | India/WB-IICB-026/2020 | 2020-08-12 | Asia/India/West Bengal/ | CSIR-Indian Institute of Chemical Biology, MEDICA Supercpecialty Hospital Kolkata | CSIR-Indian Institute of Chemical Biology, MEDICA Supercpecialty Hospital Kolkata | Sujay Krishna Maity et al |
| EPI_ISL_581502 | India/WB-IICB-027/2020 | 2020-08-12 | Asia/India/West Bengal/ | CSIR-Indian Institute of Chemical Biology, MEDICA Supercpecialty Hospital Kolkata | CSIR-Indian Institute of Chemical Biology, MEDICA Supercpecialty Hospital Kolkata | Sujay Krishna Maity et al |

| | | | | | | |
|---|---|---|---|---|---|---|
| EPI_ISL_581503 | India/WB-IICB-028/2020 | 2020-08-12 | Asia/India/West Bengal/ | CSIR-Indian Institute of Chemical Biology, MEDICA Supercpecialty Hospital Kolkata | CSIR-Indian Institute of Chemical Biology, MEDICA Supercpecialty Hospital Kolkata | Sujay Krishna Maity et al |
| EPI_ISL_581504 | India/WB-IICB-029/2020 | 2020-08-04 | Asia/India/West Bengal/ | CSIR-Indian Institute of Chemical Biology, MEDICA Supercpecialty Hospital Kolkata | CSIR-Indian Institute of Chemical Biology, MEDICA Supercpecialty Hospital Kolkata | Sujay Krishna Maity et al |
| EPI_ISL_581506 | India/DL-NCDC-IGIB-77797A/2020 | 2020-08-24 | Asia/India/Delhi/ | NCDC/IGIB | NCDC/IGIB | Vivekanand A et al |
| EPI_ISL_581507 | India/DL-NCDC-IGIB-D1/2020 | 2020-08-25 | Asia/India/Delhi/ | NCDC/IGIB | NCDC/IGIB | Vivekanand A et al |
| EPI_ISL_581509 | USA/VA-DCLS-1507/2020 | 2020-07-28 | North America/USA/Virginia/ | Virginia DCLS | Virginia DCLS | Virginia DCLS et al |
| EPI_ISL_581527 | USA/VA-DCLS-1531/2020 | 2020-08-07 | North America/USA/Virginia/ | Virginia DCLS | Virginia DCLS | Virginia DCLS et al |
| EPI_ISL_581528 | USA/VA-DCLS-1532/2020 | 2020-08-08 | North America/USA/Virginia/ | Virginia DCLS | Virginia DCLS | Virginia DCLS et al |
| EPI_ISL_581535 | USA/VA-DCLS-1540/2020 | 2020-08-07 | North America/USA/Virginia/ | Virginia DCLS | Virginia DCLS | Virginia DCLS et al |
| EPI_ISL_581537 | USA/VA-DCLS-1546/2020 | 2020-08-07 | North America/USA/Virginia/ | Virginia DCLS | Virginia DCLS | Virginia DCLS et al |
| EPI_ISL_581538 | USA/VA-DCLS-1547/2020 | 2020-08-07 | North America/USA/Virginia/ | Virginia DCLS | Virginia DCLS | Virginia DCLS et al |
| EPI_ISL_581544 | USA/VA-DCLS-1421/2020 | 2020-07-13 | North America/USA/Virginia/ | Virginia DCLS | Virginia DCLS | Virginia DCLS et al |
| EPI_ISL_581545 | USA/VA-DCLS-1422/2020 | 2020-07-13 | North America/USA/Virginia/ | Virginia DCLS | Virginia DCLS | Virginia DCLS et al |
| EPI_ISL_581547 | USA/VA-DCLS-1424/2020 | 2020-07-12 | North America/USA/Virginia/ | Virginia DCLS | Virginia DCLS | Virginia DCLS et al |
| EPI_ISL_581550 | USA/VA-DCLS-1427/2020 | 2020-07-12 | North America/USA/Virginia/ | Virginia DCLS | Virginia DCLS | Virginia DCLS et al |
| EPI_ISL_581566 | USA/VA-DCLS-1492/2020 | 2020-07-28 | North America/USA/Virginia/ | Virginia DCLS | Virginia DCLS | Virginia DCLS et al |
| EPI_ISL_581569 | USA/VA-DCLS-1497/2020 | 2020-07-27 | North America/USA/Virginia/ | Virginia DCLS | Virginia DCLS | Virginia DCLS et al |
| EPI_ISL_581571 | USA/VA-DCLS-1502/2020 | 2020-07-28 | North America/USA/Virginia/ | Virginia DCLS | Virginia DCLS | Virginia DCLS et al |
| EPI_ISL_581576 | Belgium/ULG-10466/2020 | 2020-08-31 | Europe/Belgium/Li√®ge/ | Department of Clinical Microbiology | GIGA Medical Genomics | Keith Durkin et al |
| EPI_ISL_581618 | Belgium/ULG-10441/2020 | 2020-08-17 | Europe/Belgium/Li√®ge/ | Department of Clinical Microbiology | GIGA Medical Genomics | Keith Durkin et al |
| EPI_ISL_581623 | Belgium/ULG-10447/2020 | 2020-08-26 | Europe/Belgium/Li√®ge/ | Department of Clinical Microbiology | GIGA Medical Genomics | Keith Durkin et al |
| EPI_ISL_581625 | Belgium/ULG-10450/2020 | 2020-08-26 | Europe/Belgium/Li√®ge/ | Department of Clinical Microbiology | GIGA Medical Genomics | Keith Durkin et al |

| | | | | | | |
|---|---|---|---|---|---|---|
| EPI_ISL_581627 | Belgium/ULG-10453/2020 | 2020-08-26 | Europe/Belgium/Li√®ge/ | Department of Clinical Microbiology | GIGA Medical Genomics | Keith Durkin et al |
| EPI_ISL_581630 | Belgium/ULG-10457/2020 | 2020-08-28 | Europe/Belgium/Li√®ge/ | Department of Clinical Microbiology | GIGA Medical Genomics | Keith Durkin et al |
| EPI_ISL_581634 | Belgium/ULG-10463/2020 | 2020-08-31 | Europe/Belgium/Li√®ge/ | Department of Clinical Microbiology | GIGA Medical Genomics | Keith Durkin et al |
| EPI_ISL_581639 | Belgium/ULG-10469/2020 | 2020-09-22 | Europe/Belgium/Li√®ge/ | Department of Clinical Microbiology | GIGA Medical Genomics | Keith Durkin et al |
| EPI_ISL_581641 | Belgium/ULG-10471/2020 | 2020-09-22 | Europe/Belgium/Li√®ge/ | Department of Clinical Microbiology | GIGA Medical Genomics | Keith Durkin et al |
| EPI_ISL_581646 | Belgium/ULG-10478/2020 | 2020-09-23 | Europe/Belgium/Li√®ge/ | Department of Clinical Microbiology | GIGA Medical Genomics | Keith Durkin et al |
| EPI_ISL_581651 | Belgium/ULG-10488/2020 | 2020-09-23 | Europe/Belgium/Li√®ge/ | Department of Clinical Microbiology | GIGA Medical Genomics | Keith Durkin et al |
| EPI_ISL_581652 | Belgium/ULG-10489/2020 | 2020-09-23 | Europe/Belgium/Li√®ge/ | Department of Clinical Microbiology | GIGA Medical Genomics | Keith Durkin et al |
| EPI_ISL_581659 | Belgium/ULG-10497/2020 | 2020-09-23 | Europe/Belgium/Li√®ge/ | Department of Clinical Microbiology | GIGA Medical Genomics | Keith Durkin et al |
| EPI_ISL_581661 | Belgium/ULG-10499/2020 | 2020-09-23 | Europe/Belgium/Li√®ge/ | Department of Clinical Microbiology | GIGA Medical Genomics | Keith Durkin et al |
| EPI_ISL_581664 | Belgium/ULG-10502/2020 | 2020-09-25 | Europe/Belgium/Li√®ge/ | Department of Clinical Microbiology | GIGA Medical Genomics | Keith Durkin et al |
| EPI_ISL_581680 | Switzerland/BS-42221953/2020 | 2020-04-06 | Europe/Switzerland/Basel-Stadt/ | University Hospital Basel, Clinical Virology | University Hospital Basel, Clinical Bacteriology | Madlen Stange et al |
| EPI_ISL_581685 | Switzerland/BL-42222531/2020 | 2020-04-06 | Europe/Switzerland/Basel-Land/ | University Hospital Basel, Clinical Virology | University Hospital Basel, Clinical Bacteriology | Madlen Stange et al |
| EPI_ISL_581705 | Switzerland/BS-42232843/2020 | 2020-04-15 | Europe/Switzerland/Basel-Stadt/ | University Hospital Basel, Clinical Virology | University Hospital Basel, Clinical Bacteriology | Madlen Stange et al |
| EPI_ISL_581714 | Switzerland/BL-42237708/2020 | 2020-04-19 | Europe/Switzerland/Basel-Land/ | University Hospital Basel, Clinical Virology | University Hospital Basel, Clinical Bacteriology | Madlen Stange et al |

| | | | | | | |
|---|---|---|---|---|---|---|
| EPI_ISL_581732 | Switzerland/BS-42204952/2020 | 2020-03-24 | Europe/Switzerland/Basel-Stadt/ | University Hospital Basel, Clinical Virology | University Hospital Basel, Clinical Bacteriology | Madlen Stange et al |
| EPI_ISL_581738 | Switzerland/BS-42205161/2020 | 2020-03-24 | Europe/Switzerland/Basel-Stadt/ | University Hospital Basel, Clinical Virology | University Hospital Basel, Clinical Bacteriology | Madlen Stange et al |
| EPI_ISL_581742 | Switzerland/BS-42206107/2020 | 2020-03-25 | Europe/Switzerland/Basel-Stadt/ | University Hospital Basel, Clinical Virology | University Hospital Basel, Clinical Bacteriology | Madlen Stange et al |
| EPI_ISL_581757 | Switzerland/BS-42207989/2020 | 2020-03-26 | Europe/Switzerland/Basel-Stadt/ | University Hospital Basel, Clinical Virology | University Hospital Basel, Clinical Bacteriology | Madlen Stange et al |
| EPI_ISL_581759 | Switzerland/BS-42208046/2020 | 2020-03-26 | Europe/Switzerland/Basel-Stadt/ | University Hospital Basel, Clinical Virology | University Hospital Basel, Clinical Bacteriology | Madlen Stange et al |
| EPI_ISL_581773 | Switzerland/BS-42208738/2020 | 2020-03-26 | Europe/Switzerland/Basel-Stadt/ | University Hospital Basel, Clinical Virology | University Hospital Basel, Clinical Bacteriology | Madlen Stange et al |
| EPI_ISL_581808 | Switzerland/BS-42210903/2020 | 2020-03-28 | Europe/Switzerland/Basel-Stadt/ | University Hospital Basel, Clinical Virology | University Hospital Basel, Clinical Bacteriology | Madlen Stange et al |
| EPI_ISL_581843 | Switzerland/NW-42219445/2020 | 2020-04-03 | Europe/Switzerland/Nidwalden/ | University Hospital Basel, Clinical Virology | University Hospital Basel, Clinical Bacteriology | Madlen Stange et al |
| EPI_ISL_581844 | Switzerland/BS-42219570/2020 | 2020-04-03 | Europe/Switzerland/Basel-Stadt/ | University Hospital Basel, Clinical Virology | University Hospital Basel, Clinical Bacteriology | Madlen Stange et al |
| EPI_ISL_581868 | Switzerland/BS-42222016/2020 | 2020-04-06 | Europe/Switzerland/Basel-Stadt/ | University Hospital Basel, Clinical Virology | University Hospital Basel, Clinical Bacteriology | Madlen Stange et al |

| | | | | | | |
|---|---|---|---|---|---|---|
| EPI_ISL_581869 | Switzerland/BL-65239904/2020 | 2020-04-03 | Europe/Switzerland/Basel-Land/ | University Hospital Basel, Clinical Virology | University Hospital Basel, Clinical Bacteriology | Madlen Stange et al |
| EPI_ISL_581950 | Switzerland/BS-42391102/2020 | 2020-08-12 | Europe/Switzerland/Basel-Stadt/ | University Hospital Basel, Clinical Virology | University Hospital Basel, Clinical Bacteriology | Madlen Stange et al |
| EPI_ISL_581960 | Switzerland/BS-65242351/2020 | 2020-04-29 | Europe/Switzerland/Basel-Stadt/ | University Hospital Basel, Clinical Virology | University Hospital Basel, Clinical Bacteriology | Madlen Stange et al |
| EPI_ISL_582028 | India/WB-IICB-030/2020 | 2020-08-12 | Asia/India/West Bengal/ | CSIR-Indian Institute of Chemical Biology, MEDICA Supercpecialty Hospital Kolkata | CSIR-Indian Institute of Chemical Biology, MEDICA Supercpecialty Hospital Kolkata | Sujay Krishna Maity et al |
| EPI_ISL_582029 | Iraq/ICGEB-2T/2020 | 2020-06-30 | Asia/Iraq/Al-Muthanna Province/ | Biology Department, College of Science, Al-Muthanna University | International Centre for Genetic Engineering and Biotechnology (ICGEB) and ARGO Open Lab Platform | Nihad Al-Rashedi et al |
| EPI_ISL_582030 | Iraq/ICGEB-5T/2020 | 2020-06-30 | Asia/Iraq/Al-Muthanna Province/ | Biology Department, College of Science, Al-Muthanna University | International Centre for Genetic Engineering and Biotechnology (ICGEB) and ARGO Open Lab Platform | Nihad Al-Rashedi et al |
| EPI_ISL_582031 | Poland/IHG-PAS-8-29/2020 | 2020-09-05 | Europe/Poland/Dolno≈õlƒÖskie/ | Institute of Human Genetics, Polish Academy of Sciences | Institute of Human Genetics, Polish Academy of Sciences | Szymon Hryhorowicz et al |
| EPI_ISL_582032 | Poland/IHG-PAS-8-30/2020 | 2020-09-05 | Europe/Poland/Wielkopolskie/ | Institute of Human Genetics, Polish Academy of Sciences | Institute of Human Genetics, Polish Academy of Sciences | Szymon Hryhorowicz et al |

| | | | | | | |
|---|---|---|---|---|---|---|
| EPI_ISL_582033 | Iran/1600/2020 | 2020-04-03 | Asia/Iran/Tehran/ | Department of Pathology, School of Medicine, Imam Khomeini Hospital, Tehran University of Medical Sciences | Genetics Research Center. University Of Social Welfare And Rehabilitation Sciences | Zohreh Fattahi et al |
| EPI_ISL_582035 | Spain/PV-IBV-98007685/2020 | 2020-09-14 | Europe/Spain/Basque Country/San Seba | Servicio de Microbiolog√≠a. Hospital Universitario Donostia. OSI Donostialdea. √Årea de Enfermedades Infecciosas, Grupo de Infecci√≥n Respiratoria y Resistencia Antimicrobiana. Instituto de Investigaci√≥n Sanitaria Biodonostia | SeqCOVID-SPAIN consortium/IBV(CSIC) | Gustavo Cilla et al |
| EPI_ISL_582038 | Spain/PV-IBV-98007711/2020 | 2020-09-17 | Europe/Spain/Basque Country/San Seba | Servicio de Microbiolog√≠a. Hospital Universitario Donostia. OSI Donostialdea. √Årea de Enfermedades Infecciosas, Grupo de Infecci√≥n Respiratoria y Resistencia Antimicrobiana. Instituto de Investigaci√≥n Sanitaria Biodonostia | SeqCOVID-SPAIN consortium/IBV(CSIC) | Gustavo Cilla et al |
| EPI_ISL_582043 | Spain/PV-IBV-98007594/2020 | 2020-09-02 | Europe/Spain/Basque Country/San Seba | Servicio de Microbiolog√≠a. Hospital Universitario Donostia. OSI Donostialdea. √Årea de Enfermedades Infecciosas, Grupo de Infecci√≥n Respiratoria y Resistencia Antimicrobiana. Instituto de Investigaci√≥n Sanitaria Biodonostia | SeqCOVID-SPAIN consortium/IBV(CSIC) | Gustavo Cilla et al |
| EPI_ISL_582045 | Spain/PV-IBV-98007666/2020 | 2020-09-11 | Europe/Spain/Basque Country/San Seba | Servicio de Microbiolog√≠a. Hospital Universitario Donostia. OSI Donostialdea. √Årea de Enfermedades Infecciosas, Grupo de Infecci√≥n Respiratoria y Resistencia Antimicrobiana. Instituto de Investigaci√≥n Sanitaria Biodonostia | SeqCOVID-SPAIN consortium/IBV(CSIC) | Gustavo Cilla et al |
| EPI_ISL_582048 | Spain/PV-IBV-98007621/2020 | 2020-09-05 | Europe/Spain/Basque Country/San Seba | Servicio de Microbiolog√≠a. Hospital Universitario Donostia. OSI Donostialdea. √Årea de Enfermedades Infecciosas, Grupo de Infecci√≥n Respiratoria y Resistencia Antimicrobiana. Instituto de Investigaci√≥n Sanitaria Biodonostia | SeqCOVID-SPAIN consortium/IBV(CSIC) | Gustavo Cilla et al |

| | | | | | | |
|---|---|---|---|---|---|---|
| EPI_ISL_582052 | Spain/PV-IBV-98007720/2020 | 2020-09-18 | Europe/Spain/Basque Country/San Seba | Servicio de Microbiolog√≠a. Hospital Universitario Donostia. OSI Donostialdea. √Årea de Enfermedades Infecciosas, Grupo de Infecci√≥n Respiratoria y Resistencia Antimicrobiana. Instituto de Investigaci√≥n Sanitaria Biodonostia | SeqCOVID-SPAIN consortium/IBV(CSIC) | Gustavo Cilla et al |
| EPI_ISL_582053 | Spain/PV-IBV-98007364/2020 | 2020-09-20 | Europe/Spain/Basque Country/San Seba | Servicio de Microbiolog√≠a. Hospital Universitario Donostia. OSI Donostialdea. √Årea de Enfermedades Infecciosas, Grupo de Infecci√≥n Respiratoria y Resistencia Antimicrobiana. Instituto de Investigaci√≥n Sanitaria Biodonostia | SeqCOVID-SPAIN consortium/IBV(CSIC) | Gustavo Cilla et al |
| EPI_ISL_582054 | Spain/PV-IBV-98007584/2020 | 2020-09-01 | Europe/Spain/Basque Country/San Seba | Servicio de Microbiolog√≠a. Hospital Universitario Donostia. OSI Donostialdea. √Årea de Enfermedades Infecciosas, Grupo de Infecci√≥n Respiratoria y Resistencia Antimicrobiana. Instituto de Investigaci√≥n Sanitaria Biodonostia | SeqCOVID-SPAIN consortium/IBV(CSIC) | Gustavo Cilla et al |
| EPI_ISL_582057 | Spain/PV-IBV-98007600/2020 | 2020-09-03 | Europe/Spain/Basque Country/San Seba | Servicio de Microbiolog√≠a. Hospital Universitario Donostia. OSI Donostialdea. √Årea de Enfermedades Infecciosas, Grupo de Infecci√≥n Respiratoria y Resistencia Antimicrobiana. Instituto de Investigaci√≥n Sanitaria Biodonostia | SeqCOVID-SPAIN consortium/IBV(CSIC) | Gustavo Cilla et al |
| EPI_ISL_582059 | Spain/PV-IBV-98007609/2020 | 2020-09-04 | Europe/Spain/Basque Country/San Seba | Servicio de Microbiolog√≠a. Hospital Universitario Donostia. OSI Donostialdea. √Årea de Enfermedades Infecciosas, Grupo de Infecci√≥n Respiratoria y Resistencia Antimicrobiana. Instituto de Investigaci√≥n Sanitaria Biodonostia | SeqCOVID-SPAIN consortium/IBV(CSIC) | Gustavo Cilla et al |
| EPI_ISL_582062 | Spain/PV-IBV-98007620/2020 | 2020-09-05 | Europe/Spain/Basque Country/San Seba | Servicio de Microbiolog√≠a. Hospital Universitario Donostia. OSI Donostialdea. √Årea de Enfermedades Infecciosas, Grupo de Infecci√≥n Respiratoria y Resistencia Antimicrobiana. Instituto de Investigaci√≥n Sanitaria Biodonostia | SeqCOVID-SPAIN consortium/IBV(CSIC) | Gustavo Cilla et al |

| | | | | | | |
|---|---|---|---|---|---|---|
| EPI_ISL_582068 | Spain/PV-IBV-98007636/2020 | 2020-09-07 | Europe/Spain/Basque Country/San Seba | Servicio de Microbiolog√≠a. Hospital Universitario Donostia. OSI Donostialdea. √Årea de Enfermedades Infecciosas, Grupo de Infecci√≥n Respiratoria y Resistencia Antimicrobiana. Instituto de Investigaci√≥n Sanitaria Biodonostia | SeqCOVID-SPAIN consortium/IBV(CSIC) | Gustavo Cilla et al |
| EPI_ISL_582069 | Spain/PV-IBV-98007644/2020 | 2020-09-09 | Europe/Spain/Basque Country/San Seba | Servicio de Microbiolog√≠a. Hospital Universitario Donostia. OSI Donostialdea. √Årea de Enfermedades Infecciosas, Grupo de Infecci√≥n Respiratoria y Resistencia Antimicrobiana. Instituto de Investigaci√≥n Sanitaria Biodonostia | SeqCOVID-SPAIN consortium/IBV(CSIC) | Gustavo Cilla et al |
| EPI_ISL_582072 | Spain/PV-IBV-98007655/2020 | 2020-09-10 | Europe/Spain/Basque Country/San Seba | Servicio de Microbiolog√≠a. Hospital Universitario Donostia. OSI Donostialdea. √Årea de Enfermedades Infecciosas, Grupo de Infecci√≥n Respiratoria y Resistencia Antimicrobiana. Instituto de Investigaci√≥n Sanitaria Biodonostia | SeqCOVID-SPAIN consortium/IBV(CSIC) | Gustavo Cilla et al |
| EPI_ISL_582074 | Spain/PV-IBV-98007668/2020 | 2020-09-12 | Europe/Spain/Basque Country/San Seba | Servicio de Microbiolog√≠a. Hospital Universitario Donostia. OSI Donostialdea. √Årea de Enfermedades Infecciosas, Grupo de Infecci√≥n Respiratoria y Resistencia Antimicrobiana. Instituto de Investigaci√≥n Sanitaria Biodonostia | SeqCOVID-SPAIN consortium/IBV(CSIC) | Gustavo Cilla et al |
| EPI_ISL_582075 | Spain/PV-IBV-98007669/2020 | 2020-09-12 | Europe/Spain/Basque Country/San Seba | Servicio de Microbiolog√≠a. Hospital Universitario Donostia. OSI Donostialdea. √Årea de Enfermedades Infecciosas, Grupo de Infecci√≥n Respiratoria y Resistencia Antimicrobiana. Instituto de Investigaci√≥n Sanitaria Biodonostia | SeqCOVID-SPAIN consortium/IBV(CSIC) | Gustavo Cilla et al |
| EPI_ISL_582076 | Spain/PV-IBV-98007670/2020 | 2020-09-12 | Europe/Spain/Basque Country/San Seba | Servicio de Microbiolog√≠a. Hospital Universitario Donostia. OSI Donostialdea. √Årea de Enfermedades Infecciosas, Grupo de Infecci√≥n Respiratoria y Resistencia Antimicrobiana. Instituto de Investigaci√≥n Sanitaria Biodonostia | SeqCOVID-SPAIN consortium/IBV(CSIC) | Gustavo Cilla et al |

| | | | | | | |
|---|---|---|---|---|---|---|
| EPI_ISL_582077 | Spain/PV-IBV-98007677/2020 | 2020-09-13 | Europe/Spain/Basque Country/San Seba | Servicio de Microbiolog√≠a. Hospital Universitario Donostia. OSI Donostialdea. √Årea de Enfermedades Infecciosas, Grupo de Infecci√≥n Respiratoria y Resistencia Antimicrobiana. Instituto de Investigaci√≥n Sanitaria Biodonostia | SeqCOVID-SPAIN consortium/IBV(CSIC) | Gustavo Cilla et al |
| EPI_ISL_582078 | Spain/PV-IBV-98007679/2020 | 2020-09-13 | Europe/Spain/Basque Country/San Seba | Servicio de Microbiolog√≠a. Hospital Universitario Donostia. OSI Donostialdea. √Årea de Enfermedades Infecciosas, Grupo de Infecci√≥n Respiratoria y Resistencia Antimicrobiana. Instituto de Investigaci√≥n Sanitaria Biodonostia | SeqCOVID-SPAIN consortium/IBV(CSIC) | Gustavo Cilla et al |
| EPI_ISL_582080 | Spain/PV-IBV-98007689/2020 | 2020-09-14 | Europe/Spain/Basque Country/San Seba | Servicio de Microbiolog√≠a. Hospital Universitario Donostia. OSI Donostialdea. √Årea de Enfermedades Infecciosas, Grupo de Infecci√≥n Respiratoria y Resistencia Antimicrobiana. Instituto de Investigaci√≥n Sanitaria Biodonostia | SeqCOVID-SPAIN consortium/IBV(CSIC) | Gustavo Cilla et al |
| EPI_ISL_582081 | Spain/PV-IBV-98007694/2020 | 2020-09-15 | Europe/Spain/Basque Country/San Seba | Servicio de Microbiolog√≠a. Hospital Universitario Donostia. OSI Donostialdea. √Årea de Enfermedades Infecciosas, Grupo de Infecci√≥n Respiratoria y Resistencia Antimicrobiana. Instituto de Investigaci√≥n Sanitaria Biodonostia | SeqCOVID-SPAIN consortium/IBV(CSIC) | Gustavo Cilla et al |
| EPI_ISL_582082 | Spain/PV-IBV-98007696/2020 | 2020-09-15 | Europe/Spain/Basque Country/San Seba | Servicio de Microbiolog√≠a. Hospital Universitario Donostia. OSI Donostialdea. √Årea de Enfermedades Infecciosas, Grupo de Infecci√≥n Respiratoria y Resistencia Antimicrobiana. Instituto de Investigaci√≥n Sanitaria Biodonostia | SeqCOVID-SPAIN consortium/IBV(CSIC) | Gustavo Cilla et al |
| EPI_ISL_582083 | Spain/PV-IBV-98007702/2020 | 2020-09-16 | Europe/Spain/Basque Country/San Seba | Servicio de Microbiolog√≠a. Hospital Universitario Donostia. OSI Donostialdea. √Årea de Enfermedades Infecciosas, Grupo de Infecci√≥n Respiratoria y Resistencia Antimicrobiana. Instituto de Investigaci√≥n Sanitaria Biodonostia | SeqCOVID-SPAIN consortium/IBV(CSIC) | Gustavo Cilla et al |

| | | | | | | |
|---|---|---|---|---|---|---|
| EPI_ISL_582084 | Spain/PV-IBV-98007704/2020 | 2020-09-16 | Europe/Spain/Basque Country/San Seba | Servicio de Microbiolog√≠a. Hospital Universitario Donostia. OSI Donostialdea. √Årea de Enfermedades Infecciosas, Grupo de Infecci√≥n Respiratoria y Resistencia Antimicrobiana. Instituto de Investigaci√≥n Sanitaria Biodonostia | SeqCOVID-SPAIN consortium/IBV(CSIC) | Gustavo Cilla et al |
| EPI_ISL_582086 | Spain/PV-IBV-98007709/2020 | 2020-09-17 | Europe/Spain/Basque Country/San Seba | Servicio de Microbiolog√≠a. Hospital Universitario Donostia. OSI Donostialdea. √Årea de Enfermedades Infecciosas, Grupo de Infecci√≥n Respiratoria y Resistencia Antimicrobiana. Instituto de Investigaci√≥n Sanitaria Biodonostia | SeqCOVID-SPAIN consortium/IBV(CSIC) | Gustavo Cilla et al |
| EPI_ISL_582087 | Spain/PV-IBV-98007710/2020 | 2020-09-17 | Europe/Spain/Basque Country/San Seba | Servicio de Microbiolog√≠a. Hospital Universitario Donostia. OSI Donostialdea. √Årea de Enfermedades Infecciosas, Grupo de Infecci√≥n Respiratoria y Resistencia Antimicrobiana. Instituto de Investigaci√≥n Sanitaria Biodonostia | SeqCOVID-SPAIN consortium/IBV(CSIC) | Gustavo Cilla et al |
| EPI_ISL_582088 | Spain/PV-IBV-98007716/2020 | 2020-09-18 | Europe/Spain/Basque Country/San Seba | Servicio de Microbiolog√≠a. Hospital Universitario Donostia. OSI Donostialdea. √Årea de Enfermedades Infecciosas, Grupo de Infecci√≥n Respiratoria y Resistencia Antimicrobiana. Instituto de Investigaci√≥n Sanitaria Biodonostia | SeqCOVID-SPAIN consortium/IBV(CSIC) | Gustavo Cilla et al |
| EPI_ISL_582089 | Spain/PV-IBV-98007717/2020 | 2020-09-18 | Europe/Spain/Basque Country/San Seba | Servicio de Microbiolog√≠a. Hospital Universitario Donostia. OSI Donostialdea. √Årea de Enfermedades Infecciosas, Grupo de Infecci√≥n Respiratoria y Resistencia Antimicrobiana. Instituto de Investigaci√≥n Sanitaria Biodonostia | SeqCOVID-SPAIN consortium/IBV(CSIC) | Gustavo Cilla et al |
| EPI_ISL_582090 | Spain/PV-IBV-98007725/2020 | 2020-09-19 | Europe/Spain/Basque Country/San Seba | Servicio de Microbiolog√≠a. Hospital Universitario Donostia. OSI Donostialdea. √Årea de Enfermedades Infecciosas, Grupo de Infecci√≥n Respiratoria y Resistencia Antimicrobiana. Instituto de Investigaci√≥n Sanitaria Biodonostia | SeqCOVID-SPAIN consortium/IBV(CSIC) | Gustavo Cilla et al |
| EPI_ISL_582094 | Spain/CB-IBV-98007077/2020 | 2020-09-03 | Europe/Spain/Cantabria/Santander | Hospital Universitario Marqu√©s de Valdecilla - IDIVAL (Santander, Cantabria) | SeqCOVID-SPAIN consortium/IBV(CSIC) | Mar√≠a Eliecer Cano Garc√≠a et al |

| | | | | | | |
|---|---|---|---|---|---|---|
| EPI_ISL_582096 | Spain/CB-IBV-98007071/2020 | 2020-09-03 | Europe/Spain/Cantabria/Santander | Hospital Universitario Marqu√©s de Valdecilla - IDIVAL (Santander, Cantabria) | SeqCOVID-SPAIN consortium/IBV(CSIC) | Mar√≠a Eliecer Cano Garc√≠a et al |
| EPI_ISL_582098 | Spain/CB-IBV-98007082/2020 | 2020-07-03 | Europe/Spain/Cantabria/Santander | Hospital Universitario Marqu√©s de Valdecilla - IDIVAL (Santander, Cantabria) | SeqCOVID-SPAIN consortium/IBV(CSIC) | Mar√≠a Eliecer Cano Garc√≠a et al |
| EPI_ISL_582100 | Spain/CB-IBV-98007068/2020 | 2020-09-03 | Europe/Spain/Cantabria/Santander | Hospital Universitario Marqu√©s de Valdecilla - IDIVAL (Santander, Cantabria) | SeqCOVID-SPAIN consortium/IBV(CSIC) | Mar√≠a Eliecer Cano Garc√≠a et al |
| EPI_ISL_582102 | Spain/CB-IBV-98007070/2020 | 2020-09-03 | Europe/Spain/Cantabria/Laredo | Hospital Universitario Marqu√©s de Valdecilla - IDIVAL (Santander, Cantabria) | SeqCOVID-SPAIN consortium/IBV(CSIC) | Mar√≠a Eliecer Cano Garc√≠a et al |
| EPI_ISL_582104 | Spain/CB-IBV-98007074/2020 | 2020-09-03 | Europe/Spain/Cantabria/Solares | Hospital Universitario Marqu√©s de Valdecilla - IDIVAL (Santander, Cantabria) | SeqCOVID-SPAIN consortium/IBV(CSIC) | Mar√≠a Eliecer Cano Garc√≠a et al |
| EPI_ISL_582105 | Spain/CB-IBV-98007075/2020 | 2020-09-03 | Europe/Spain/Cantabria/Ampuero | Hospital Universitario Marqu√©s de Valdecilla - IDIVAL (Santander, Cantabria) | SeqCOVID-SPAIN consortium/IBV(CSIC) | Mar√≠a Eliecer Cano Garc√≠a et al |
| EPI_ISL_582107 | Spain/CB-IBV-98007079/2020 | 2020-09-03 | Europe/Spain/Cantabria/Santander | Hospital Universitario Marqu√©s de Valdecilla - IDIVAL (Santander, Cantabria) | SeqCOVID-SPAIN consortium/IBV(CSIC) | Mar√≠a Eliecer Cano Garc√≠a et al |
| EPI_ISL_582111 | France/ARA-CDP81878/2020 | 2020-09-16 | Europe/France/Auvergne-Rh√¥ne-Alpes/ | CNR Virus des Infections Respiratoires - France SUD | CNR Virus des Infections Respiratoires - France SUD | Antonin Bal et al |
| EPI_ISL_582112 | France/ARA-CDP29560/2020 | 2020-09-01 | Europe/France/Auvergne-Rh√¥ne-Alpes/ | CNR Virus des Infections Respiratoires - France SUD | CNR Virus des Infections Respiratoires - France SUD | Antonin Bal et al |
| EPI_ISL_582116 | France/ARA-CDP81800/2020 | 2020-09-16 | Europe/France/Auvergne-Rh√¥ne-Alpes/ | CNR Virus des Infections Respiratoires - France SUD | CNR Virus des Infections Respiratoires - France SUD | Antonin Bal et al |
| EPI_ISL_582118 | France/ARA-CDP82238/2020 | 2020-09-09 | Europe/France/Auvergne-Rh√¥ne-Alpes/ | CNR Virus des Infections Respiratoires - France SUD | CNR Virus des Infections Respiratoires - France SUD | Antonin Bal et al |

| | | | | | | |
|---|---|---|---|---|---|---|
| EPI_ISL_582120 | France/ARA-CDP78128/2020 | 2020-09-07 | Europe/France/Auvergne-Rhv¥ne-Alpes/ | CNR Virus des Infections Respiratoires - France SUD | CNR Virus des Infections Respiratoires - France SUD | Antonin Bal et al |
| EPI_ISL_582122 | France/ARA-71991/2020 | 2020-09-13 | Europe/France/Auvergne-Rhv¥ne-Alpes/ | CNR Virus des Infections Respiratoires - France SUD | CNR Virus des Infections Respiratoires - France SUD | Antonin Bal et al |
| EPI_ISL_582123 | Italy/TUS-DBM-1/2020 | 2020-03-01 | Europe/Italy/Tuscany/ | UOC Microbiologia e Virologia, Azienda Ospedaliera Universitaria Senese, Siena, Italy | Dipartimento di Biotecnologie Mediche | Maria Grazia Cusi et al |
| EPI_ISL_582125 | UnitedArabEmirates/skmc-92016811 | 2020-01-28 | Asia/United Arab Emirates/United Arab | Sheikh Khalifa Medical City | Molecular Surveillance lab Sheikh Khalifa Medical City | Amirtharaj Francis et al |
| EPI_ISL_582126 | UnitedArabEmirates/skmc-92056911 | 2020-01-28 | Asia/United Arab Emirates/United Arab | Sheikh Khalifa Medical City | Molecular Surveillance lab Sheikh Khalifa Medical City | Amirtharaj Francis et al |
| EPI_ISL_582134 | Germany/BAV-PL-virotum-nacq/2020 | 2020-02-15 | Europe/Germany/Bavaria/Munich | Protzer Lab | Protzer Lab | Ulrike Protzer et al |
| EPI_ISL_582155 | USA/NM-UNM-00764/2020 | 2020-05-02 | North America/USA/New Mexico/ | TriCore Reference Laboratories | Center for Global Health, University of New Mexico Health Sciences Center | Daryl Domman et al |
| EPI_ISL_582157 | USA/NM-UNM-00737/2020 | 2020-05-09 | North America/USA/New Mexico/ | TriCore Reference Laboratories | Center for Global Health, University of New Mexico Health Sciences Center | Daryl Domman et al |
| EPI_ISL_582158 | USA/NM-UNM-00714/2020 | 2020-05-08 | North America/USA/New Mexico/ | TriCore Reference Laboratories | Center for Global Health, University of New Mexico Health Sciences Center | Daryl Domman et al |
| EPI_ISL_582161 | USA/NM-UNM-00725/2020 | 2020-05-04 | North America/USA/New Mexico/ | TriCore Reference Laboratories | Center for Global Health, University of New Mexico Health Sciences Center | Daryl Domman et al |

| | | | | | | |
|---|---|---|---|---|---|---|
| EPI_ISL_582162 | USA/NM-UNM-00757/2020 | 2020-05-05 | North America/USA/New Mexico/ | TriCore Reference Laboratories | Center for Global Health, University of New Mexico Health Sciences Center | Daryl Domman et al |
| EPI_ISL_582163 | USA/NM-UNM-00749/2020 | 2020-05-13 | North America/USA/New Mexico/ | TriCore Reference Laboratories | Center for Global Health, University of New Mexico Health Sciences Center | Daryl Domman et al |
| EPI_ISL_582167 | USA/NM-UNM-00750/2020 | 2020-05-12 | North America/USA/Arizona/ | TriCore Reference Laboratories | Center for Global Health, University of New Mexico Health Sciences Center | Daryl Domman et al |
| EPI_ISL_582171 | USA/NM-UNM-00742/2020 | 2020-05-13 | North America/USA/New Mexico/ | TriCore Reference Laboratories | Center for Global Health, University of New Mexico Health Sciences Center | Daryl Domman et al |
| EPI_ISL_582184 | USA/NM-UNM-00776/2020 | 2020-05-06 | North America/USA/Arizona/ | TriCore Reference Laboratories | Center for Global Health, University of New Mexico Health Sciences Center | Daryl Domman et al |
| EPI_ISL_582202 | USA/NM-UNM-00723/2020 | 2020-05-05 | North America/USA/New Mexico/ | TriCore Reference Laboratories | Center for Global Health, University of New Mexico Health Sciences Center | Daryl Domman et al |
| EPI_ISL_582223 | USA/WY-UNM-WY00105/2020 | 2020-07-28 | North America/USA/Wyoming/ | Wyoming Public Health Laboratory | Center for Global Health, University of New Mexico Health Sciences Center | Daryl Domman et al |
| EPI_ISL_582229 | USA/WY-UNM-WY00111/2020 | 2020-07-30 | North America/USA/Wyoming/ | Wyoming Public Health Laboratory | Center for Global Health, University of New Mexico Health Sciences Center | Daryl Domman et al |

| | | | | | | |
|---|---|---|---|---|---|---|
| EPI_ISL_582241 | India/MH-NIV-229543/2020 | 2020-09-28 | Asia/India/Maharashtra/ | NIV Influenza | NIV Influenza | Potdar V et al |
| EPI_ISL_582242 | Canada/MB-NML-163/2020 | 2020-03-31 | North America/Canada/Manitoba/ | Cadham Provincial Laboratory | National Microbiology Laboratory (NML) | Anna Majer et al |
| EPI_ISL_582243 | Canada/MB-NML-166/2020 | 2020-03-30 | North America/Canada/Manitoba/ | Cadham Provincial Laboratory | National Microbiology Laboratory (NML) | Anna Majer et al |
| EPI_ISL_582244 | Canada/MB-NML-167/2020 | 2020-03-31 | North America/Canada/Manitoba/ | Cadham Provincial Laboratory | National Microbiology Laboratory (NML) | Anna Majer et al |
| EPI_ISL_582245 | Canada/MB-NML-168/2020 | 2020-03-28 | North America/Canada/Manitoba/ | Cadham Provincial Laboratory | National Microbiology Laboratory (NML) | Anna Majer et al |
| EPI_ISL_582246 | Canada/MB-NML-409/2020 | 2020-03-22 | North America/Canada/Manitoba/ | Cadham Provincial Laboratory | National Microbiology Laboratory (NML) | Anna Majer et al |
| EPI_ISL_582247 | Canada/MB-NML-410/2020 | 2020-04-07 | North America/Canada/Manitoba/ | Cadham Provincial Laboratory | National Microbiology Laboratory (NML) | Anna Majer et al |
| EPI_ISL_582248 | Canada/MB-NML-411/2020 | 2020-04-07 | North America/Canada/Manitoba/ | Cadham Provincial Laboratory | National Microbiology Laboratory (NML) | Anna Majer et al |
| EPI_ISL_582249 | Canada/MB-NML-412/2020 | 2020-04-07 | North America/Canada/Manitoba/ | Cadham Provincial Laboratory | National Microbiology Laboratory (NML) | Anna Majer et al |
| EPI_ISL_582251 | Canada/MB-NML-415/2020 | 2020-04-03 | North America/Canada/Manitoba/ | Cadham Provincial Laboratory | National Microbiology Laboratory (NML) | Anna Majer et al |
| EPI_ISL_582252 | Canada/MB-NML-416/2020 | 2020-04-02 | North America/Canada/Manitoba/ | Cadham Provincial Laboratory | National Microbiology Laboratory (NML) | Anna Majer et al |
| EPI_ISL_582253 | Canada/MB-NML-418/2020 | 2020-03-30 | North America/Canada/Manitoba/ | Cadham Provincial Laboratory | National Microbiology Laboratory (NML) | Anna Majer et al |
| EPI_ISL_582254 | Canada/MB-NML-420/2020 | 2020-03-29 | North America/Canada/Manitoba/ | Cadham Provincial Laboratory | National Microbiology Laboratory (NML) | Anna Majer et al |
| EPI_ISL_582255 | Canada/MB-NML-421/2020 | 2020-03-29 | North America/Canada/Manitoba/ | Cadham Provincial Laboratory | National Microbiology Laboratory (NML) | Anna Majer et al |

| | | | | | | |
|---|---|---|---|---|---|---|
| EPI_ISL_582256 | Canada/MB-NML-422/2020 | 2020-03-31 | North America/Canada/Manitoba/ | Cadham Provincial Laboratory | National Microbiology Laboratory (NML) | Anna Majer et al |
| EPI_ISL_582257 | Canada/MB-NML-423/2020 | 2020-04-01 | North America/Canada/Manitoba/ | Cadham Provincial Laboratory | National Microbiology Laboratory (NML) | Anna Majer et al |
| EPI_ISL_582258 | Canada/MB-NML-425/2020 | 2020-04-01 | North America/Canada/Manitoba/ | Cadham Provincial Laboratory | National Microbiology Laboratory (NML) | Anna Majer et al |
| EPI_ISL_582259 | Canada/MB-NML-426/2020 | 2020-03-30 | North America/Canada/Manitoba/ | Cadham Provincial Laboratory | National Microbiology Laboratory (NML) | Anna Majer et al |
| EPI_ISL_582260 | Canada/MB-NML-559/2020 | 2020-03-23 | North America/Canada/Manitoba/ | Cadham Provincial Laboratory | National Microbiology Laboratory (NML) | Anna Majer et al |
| EPI_ISL_582261 | Canada/MB-NML-560/2020 | 2020-03-26 | North America/Canada/Manitoba/ | Cadham Provincial Laboratory | National Microbiology Laboratory (NML) | Anna Majer et al |
| EPI_ISL_582262 | Canada/MB-NML-561/2020 | 2020-04-28 | North America/Canada/Manitoba/ | Cadham Provincial Laboratory | National Microbiology Laboratory (NML) | Anna Majer et al |
| EPI_ISL_582263 | Canada/MB-NML-565/2020 | 2020-05-03 | North America/Canada/Manitoba/ | Cadham Provincial Laboratory | National Microbiology Laboratory (NML) | Anna Majer et al |
| EPI_ISL_582264 | Canada/MB-NML-568/2020 | 2020-03-26 | North America/Canada/Manitoba/ | Cadham Provincial Laboratory | National Microbiology Laboratory (NML) | Anna Majer et al |
| EPI_ISL_582265 | Canada/MB-NML-569/2020 | 2020-03-27 | North America/Canada/Manitoba/ | Cadham Provincial Laboratory | National Microbiology Laboratory (NML) | Anna Majer et al |
| EPI_ISL_582266 | Canada/MB-NML-571/2020 | 2020-03-24 | North America/Canada/Manitoba/ | Cadham Provincial Laboratory | National Microbiology Laboratory (NML) | Anna Majer et al |
| EPI_ISL_582267 | Canada/MB-NML-572/2020 | 2020-03-27 | North America/Canada/Manitoba/ | Cadham Provincial Laboratory | National Microbiology Laboratory (NML) | Anna Majer et al |
| EPI_ISL_582268 | Canada/MB-NML-573/2020 | 2020-03-29 | North America/Canada/Manitoba/ | Cadham Provincial Laboratory | National Microbiology Laboratory (NML) | Anna Majer et al |

| | | | | | | |
|---|---|---|---|---|---|---|
| EPI_ISL_582269 | Canada/MB-NML-574/2020 | 2020-03-28 | North America/Canada/Manitoba/ | Cadham Provincial Laboratory | National Microbiology Laboratory (NML) | Anna Majer et al |
| EPI_ISL_582270 | Canada/MB-NML-576/2020 | 2020-03-24 | North America/Canada/Manitoba/ | Cadham Provincial Laboratory | National Microbiology Laboratory (NML) | Anna Majer et al |
| EPI_ISL_582271 | Canada/MB-NML-580/2020 | 2020-04-07 | North America/Canada/Manitoba/ | Cadham Provincial Laboratory | National Microbiology Laboratory (NML) | Anna Majer et al |
| EPI_ISL_582272 | Canada/MB-NML-581/2020 | 2020-03-27 | North America/Canada/Manitoba/ | Cadham Provincial Laboratory | National Microbiology Laboratory (NML) | Anna Majer et al |
| EPI_ISL_582273 | Canada/MB-NML-583/2020 | 2020-03-31 | North America/Canada/Manitoba/ | Cadham Provincial Laboratory | National Microbiology Laboratory (NML) | Anna Majer et al |
| EPI_ISL_582274 | Canada/MB-NML-584/2020 | 2020-03-25 | North America/Canada/Manitoba/ | Cadham Provincial Laboratory | National Microbiology Laboratory (NML) | Anna Majer et al |
| EPI_ISL_582275 | Canada/MB-NML-585/2020 | 2020-03-19 | North America/Canada/Manitoba/ | Cadham Provincial Laboratory | National Microbiology Laboratory (NML) | Anna Majer et al |
| EPI_ISL_582276 | Canada/MB-NML-587/2020 | 2020-03-27 | North America/Canada/Manitoba/ | Cadham Provincial Laboratory | National Microbiology Laboratory (NML) | Anna Majer et al |
| EPI_ISL_582277 | Canada/MB-NML-590/2020 | 2020-03-29 | North America/Canada/Manitoba/ | Cadham Provincial Laboratory | National Microbiology Laboratory (NML) | Anna Majer et al |
| EPI_ISL_582278 | Canada/MB-NML-591/2020 | 2020-04-01 | North America/Canada/Manitoba/ | Cadham Provincial Laboratory | National Microbiology Laboratory (NML) | Anna Majer et al |
| EPI_ISL_582279 | Canada/MB-NML-592/2020 | 2020-03-28 | North America/Canada/Manitoba/ | Cadham Provincial Laboratory | National Microbiology Laboratory (NML) | Anna Majer et al |
| EPI_ISL_582280 | Canada/MB-NML-593/2020 | 2020-03-28 | North America/Canada/Manitoba/ | Cadham Provincial Laboratory | National Microbiology Laboratory (NML) | Anna Majer et al |
| EPI_ISL_582281 | Canada/MB-NML-594/2020 | 2020-03-25 | North America/Canada/Manitoba/ | Cadham Provincial Laboratory | National Microbiology Laboratory (NML) | Anna Majer et al |

| | | | | | | |
|---|---|---|---|---|---|---|
| EPI_ISL_582282 | Canada/MB-NML-597/2020 | 2020-03-27 | North America/Canada/Manitoba/ | Cadham Provincial Laboratory | National Microbiology Laboratory (NML) | Anna Majer et al |
| EPI_ISL_582283 | Canada/MB-NML-598/2020 | 2020-05-07 | North America/Canada/Manitoba/ | Cadham Provincial Laboratory | National Microbiology Laboratory (NML) | Anna Majer et al |
| EPI_ISL_582284 | Canada/MB-NML-600/2020 | 2020-03-23 | North America/Canada/Manitoba/ | Cadham Provincial Laboratory | National Microbiology Laboratory (NML) | Anna Majer et al |
| EPI_ISL_582285 | Canada/MB-NML-603/2020 | 2020-03-23 | North America/Canada/Manitoba/ | Cadham Provincial Laboratory | National Microbiology Laboratory (NML) | Anna Majer et al |
| EPI_ISL_582286 | Canada/MB-NML-612/2020 | 2020-03-28 | North America/Canada/Manitoba/ | Cadham Provincial Laboratory | National Microbiology Laboratory (NML) | Anna Majer et al |
| EPI_ISL_582288 | Canada/MB-NML-616/2020 | 2020-03-27 | North America/Canada/Manitoba/ | Cadham Provincial Laboratory | National Microbiology Laboratory (NML) | Anna Majer et al |
| EPI_ISL_582289 | Canada/MB-NML-617/2020 | 2020-03-25 | North America/Canada/Manitoba/ | Cadham Provincial Laboratory | National Microbiology Laboratory (NML) | Anna Majer et al |
| EPI_ISL_582290 | Canada/MB-NML-619/2020 | 2020-05-30 | North America/Canada/Manitoba/ | Cadham Provincial Laboratory | National Microbiology Laboratory (NML) | Anna Majer et al |
| EPI_ISL_582291 | Canada/MB-NML-621/2020 | 2020-03-27 | North America/Canada/Manitoba/ | Cadham Provincial Laboratory | National Microbiology Laboratory (NML) | Anna Majer et al |
| EPI_ISL_582292 | Canada/MB-NML-623/2020 | 2020-03-29 | North America/Canada/Manitoba/ | Cadham Provincial Laboratory | National Microbiology Laboratory (NML) | Anna Majer et al |
| EPI_ISL_582293 | Canada/MB-NML-626/2020 | 2020-03-23 | North America/Canada/Manitoba/ | Cadham Provincial Laboratory | National Microbiology Laboratory (NML) | Anna Majer et al |
| EPI_ISL_582294 | Canada/MB-NML-628/2020 | 2020-04-02 | North America/Canada/Manitoba/ | Cadham Provincial Laboratory | National Microbiology Laboratory (NML) | Anna Majer et al |
| EPI_ISL_582295 | Canada/MB-NML-629/2020 | 2020-03-27 | North America/Canada/Manitoba/ | Cadham Provincial Laboratory | National Microbiology Laboratory (NML) | Anna Majer et al |

| | | | | | | |
|---|---|---|---|---|---|---|
| EPI_ISL_582296 | Canada/MB-NML-636/2020 | 2020-04-02 | North America/Canada/Manitoba/ | Cadham Provincial Laboratory | National Microbiology Laboratory (NML) | Anna Majer et al |
| EPI_ISL_582297 | Canada/MB-NML-640/2020 | 2020-03-23 | North America/Canada/Manitoba/ | Cadham Provincial Laboratory | National Microbiology Laboratory (NML) | Anna Majer et al |
| EPI_ISL_582298 | Canada/MB-NML-641/2020 | 2020-03-24 | North America/Canada/Manitoba/ | Cadham Provincial Laboratory | National Microbiology Laboratory (NML) | Anna Majer et al |
| EPI_ISL_582299 | Canada/MB-NML-642/2020 | 2020-03-20 | North America/Canada/Manitoba/ | Cadham Provincial Laboratory | National Microbiology Laboratory (NML) | Anna Majer et al |
| EPI_ISL_582300 | Canada/MB-NML-656/2020 | 2020-04-03 | North America/Canada/Manitoba/ | Cadham Provincial Laboratory | National Microbiology Laboratory (NML) | Anna Majer et al |
| EPI_ISL_582301 | Canada/MB-NML-657/2020 | 2020-03-28 | North America/Canada/Manitoba/ | Cadham Provincial Laboratory | National Microbiology Laboratory (NML) | Anna Majer et al |
| EPI_ISL_582302 | Canada/MB-NML-658/2020 | 2020-04-03 | North America/Canada/Manitoba/ | Cadham Provincial Laboratory | National Microbiology Laboratory (NML) | Anna Majer et al |
| EPI_ISL_582303 | Canada/MB-NML-766/2020 | 2020-03-18 | North America/Canada/Manitoba/ | Cadham Provincial Laboratory | National Microbiology Laboratory (NML) | Anna Majer et al |
| EPI_ISL_582305 | Canada/MB-NML-769/2020 | 2020-03-18 | North America/Canada/Manitoba/ | Cadham Provincial Laboratory | National Microbiology Laboratory (NML) | Anna Majer et al |
| EPI_ISL_582306 | Canada/MB-NML-779/2020 | 2020-03-28 | North America/Canada/Manitoba/ | Cadham Provincial Laboratory | National Microbiology Laboratory (NML) | Anna Majer et al |
| EPI_ISL_582308 | Canada/MB-NML-782/2020 | 2020-04-01 | North America/Canada/Manitoba/ | Cadham Provincial Laboratory | National Microbiology Laboratory (NML) | Anna Majer et al |
| EPI_ISL_582311 | Canada/MB-NML-788/2020 | 2020-04-02 | North America/Canada/Manitoba/ | Cadham Provincial Laboratory | National Microbiology Laboratory (NML) | Anna Majer et al |
| EPI_ISL_582312 | Canada/MB-NML-789/2020 | 2020-04-03 | North America/Canada/Manitoba/ | Cadham Provincial Laboratory | National Microbiology Laboratory (NML) | Anna Majer et al |

| | | | | | | |
|---|---|---|---|---|---|---|
| EPI_ISL_582313 | Canada/MB-NML-794/2020 | 2020-05-30 | North America/Canada/Manitoba/ | Cadham Provincial Laboratory | National Microbiology Laboratory (NML) | Anna Majer et al |
| EPI_ISL_582314 | Canada/MB-NML-795/2020 | 2020-06-14 | North America/Canada/Manitoba/ | Cadham Provincial Laboratory | National Microbiology Laboratory (NML) | Anna Majer et al |
| EPI_ISL_582315 | Canada/MB-NML-796/2020 | 2020-06-15 | North America/Canada/Manitoba/ | Cadham Provincial Laboratory | National Microbiology Laboratory (NML) | Anna Majer et al |
| EPI_ISL_582316 | Canada/MB-NML-797/2020 | 2020-06-16 | North America/Canada/Manitoba/ | Cadham Provincial Laboratory | National Microbiology Laboratory (NML) | Anna Majer et al |
| EPI_ISL_582317 | Canada/MB-NML-798/2020 | 2020-06-16 | North America/Canada/Manitoba/ | Cadham Provincial Laboratory | National Microbiology Laboratory (NML) | Anna Majer et al |
| EPI_ISL_582318 | Canada/MB-NML-808/2020 | 2020-04-15 | North America/Canada/Manitoba/ | Cadham Provincial Laboratory | National Microbiology Laboratory (NML) | Anna Majer et al |
| EPI_ISL_582319 | Canada/MB-NML-818/2020 | 2020-07-10 | North America/Canada/Manitoba/ | Cadham Provincial Laboratory | National Microbiology Laboratory (NML) | Anna Majer et al |
| EPI_ISL_582320 | Canada/MB-NML-823/2020 | 2020-07-14 | North America/Canada/Manitoba/ | Cadham Provincial Laboratory | National Microbiology Laboratory (NML) | Anna Majer et al |
| EPI_ISL_582321 | Canada/MB-NML-824/2020 | 2020-07-13 | North America/Canada/Manitoba/ | Cadham Provincial Laboratory | National Microbiology Laboratory (NML) | Anna Majer et al |
| EPI_ISL_582322 | Canada/MB-NML-826/2020 | 2020-07-13 | North America/Canada/Manitoba/ | Cadham Provincial Laboratory | National Microbiology Laboratory (NML) | Anna Majer et al |
| EPI_ISL_582323 | Canada/MB-NML-827/2020 | 2020-07-13 | North America/Canada/Manitoba/ | Cadham Provincial Laboratory | National Microbiology Laboratory (NML) | Anna Majer et al |
| EPI_ISL_582324 | Canada/MB-NML-828/2020 | 2020-07-15 | North America/Canada/Manitoba/ | Cadham Provincial Laboratory | National Microbiology Laboratory (NML) | Anna Majer et al |
| EPI_ISL_582325 | Canada/MB-NML-830/2020 | 2020-04-21 | North America/Canada/Manitoba/ | Cadham Provincial Laboratory | National Microbiology Laboratory (NML) | Anna Majer et al |

| | | | | | | |
|---|---|---|---|---|---|---|
| EPI_ISL_582326 | Canada/MB-NML-831/2020 | 2020-06-09 | North America/Canada/Manitoba/ | Cadham Provincial Laboratory | National Microbiology Laboratory (NML) | Anna Majer et al |
| EPI_ISL_582327 | Canada/MB-NML-832/2020 | 2020-06-09 | North America/Canada/Manitoba/ | Cadham Provincial Laboratory | National Microbiology Laboratory (NML) | Anna Majer et al |
| EPI_ISL_582328 | Canada/MB-NML-837/2020 | 2020-03-28 | North America/Canada/Manitoba/ | Cadham Provincial Laboratory | National Microbiology Laboratory (NML) | Anna Majer et al |
| EPI_ISL_582329 | Canada/MB-NML-838/2020 | 2020-03-26 | North America/Canada/Manitoba/ | Cadham Provincial Laboratory | National Microbiology Laboratory (NML) | Anna Majer et al |
| EPI_ISL_582330 | Canada/MB-NML-839/2020 | 2020-03-28 | North America/Canada/Manitoba/ | Cadham Provincial Laboratory | National Microbiology Laboratory (NML) | Anna Majer et al |
| EPI_ISL_582331 | Canada/MB-NML-840/2020 | 2020-03-27 | North America/Canada/Manitoba/ | Cadham Provincial Laboratory | National Microbiology Laboratory (NML) | Anna Majer et al |
| EPI_ISL_582332 | Canada/MB-NML-841/2020 | 2020-03-26 | North America/Canada/Manitoba/ | Cadham Provincial Laboratory | National Microbiology Laboratory (NML) | Anna Majer et al |
| EPI_ISL_582333 | Canada/MB-NML-843/2020 | 2020-03-19 | North America/Canada/Manitoba/ | Cadham Provincial Laboratory | National Microbiology Laboratory (NML) | Anna Majer et al |
| EPI_ISL_582334 | Canada/MB-NML-845/2020 | 2020-03-20 | North America/Canada/Manitoba/ | Cadham Provincial Laboratory | National Microbiology Laboratory (NML) | Anna Majer et al |
| EPI_ISL_582335 | Canada/MB-NML-846/2020 | 2020-03-20 | North America/Canada/Manitoba/ | Cadham Provincial Laboratory | National Microbiology Laboratory (NML) | Anna Majer et al |
| EPI_ISL_582336 | Canada/MB-NML-847/2020 | 2020-03-20 | North America/Canada/Manitoba/ | Cadham Provincial Laboratory | National Microbiology Laboratory (NML) | Anna Majer et al |
| EPI_ISL_582337 | Canada/MB-NML-848/2020 | 2020-03-24 | North America/Canada/Manitoba/ | Cadham Provincial Laboratory | National Microbiology Laboratory (NML) | Anna Majer et al |
| EPI_ISL_582338 | Canada/MB-NML-853/2020 | 2020-03-26 | North America/Canada/Manitoba/ | Cadham Provincial Laboratory | National Microbiology Laboratory (NML) | Anna Majer et al |

| | | | | | | |
|---|---|---|---|---|---|---|
| EPI_ISL_582339 | Canada/MB-NML-854/2020 | 2020-03-26 | North America/Canada/Manitoba/ | Cadham Provincial Laboratory | National Microbiology Laboratory (NML) | Anna Majer et al |
| EPI_ISL_582340 | Canada/MB-NML-857/2020 | 2020-03-27 | North America/Canada/Manitoba/ | Cadham Provincial Laboratory | National Microbiology Laboratory (NML) | Anna Majer et al |
| EPI_ISL_582341 | Canada/MB-NML-858/2020 | 2020-03-19 | North America/Canada/Manitoba/ | Cadham Provincial Laboratory | National Microbiology Laboratory (NML) | Anna Majer et al |
| EPI_ISL_582342 | Canada/MB-NML-859/2020 | 2020-03-30 | North America/Canada/Manitoba/ | Cadham Provincial Laboratory | National Microbiology Laboratory (NML) | Anna Majer et al |
| EPI_ISL_582343 | Canada/MB-NML-861/2020 | 2020-03-28 | North America/Canada/Manitoba/ | Cadham Provincial Laboratory | National Microbiology Laboratory (NML) | Anna Majer et al |
| EPI_ISL_582345 | Canada/MB-NML-864/2020 | 2020-03-30 | North America/Canada/Manitoba/ | Cadham Provincial Laboratory | National Microbiology Laboratory (NML) | Anna Majer et al |
| EPI_ISL_582346 | Canada/MB-NML-865/2020 | 2020-03-23 | North America/Canada/Manitoba/ | Cadham Provincial Laboratory | National Microbiology Laboratory (NML) | Anna Majer et al |
| EPI_ISL_582347 | Canada/MB-NML-866/2020 | 2020-03-30 | North America/Canada/Manitoba/ | Cadham Provincial Laboratory | National Microbiology Laboratory (NML) | Anna Majer et al |
| EPI_ISL_582349 | Canada/MB-NML-872/2020 | 2020-03-21 | North America/Canada/Manitoba/ | Cadham Provincial Laboratory | National Microbiology Laboratory (NML) | Anna Majer et al |
| EPI_ISL_582350 | Canada/MB-NML-873/2020 | 2020-03-30 | North America/Canada/Manitoba/ | Cadham Provincial Laboratory | National Microbiology Laboratory (NML) | Anna Majer et al |
| EPI_ISL_582351 | Canada/MB-NML-874/2020 | 2020-03-30 | North America/Canada/Manitoba/ | Cadham Provincial Laboratory | National Microbiology Laboratory (NML) | Anna Majer et al |
| EPI_ISL_582352 | Canada/MB-NML-876/2020 | 2020-03-19 | North America/Canada/Manitoba/ | Cadham Provincial Laboratory | National Microbiology Laboratory (NML) | Anna Majer et al |
| EPI_ISL_582353 | Canada/MB-NML-877/2020 | 2020-03-30 | North America/Canada/Manitoba/ | Cadham Provincial Laboratory | National Microbiology Laboratory (NML) | Anna Majer et al |

| | | | | | | |
|---|---|---|---|---|---|---|
| EPI_ISL_582354 | Canada/MB-NML-878/2020 | 2020-03-31 | North America/Canada/Manitoba/ | Cadham Provincial Laboratory | National Microbiology Laboratory (NML) | Anna Majer et al |
| EPI_ISL_582355 | Canada/MB-NML-880/2020 | 2020-04-15 | North America/Canada/Manitoba/ | Cadham Provincial Laboratory | National Microbiology Laboratory (NML) | Anna Majer et al |
| EPI_ISL_582356 | Canada/MB-NML-979/2020 | 2020-07-19 | North America/Canada/Manitoba/ | Cadham Provincial Laboratory | National Microbiology Laboratory (NML) | Anna Majer et al |
| EPI_ISL_582357 | Canada/MB-NML-980/2020 | 2020-07-16 | North America/Canada/Manitoba/ | Cadham Provincial Laboratory | National Microbiology Laboratory (NML) | Anna Majer et al |
| EPI_ISL_582358 | Canada/MB-NML-981/2020 | 2020-07-17 | North America/Canada/Manitoba/ | Cadham Provincial Laboratory | National Microbiology Laboratory (NML) | Anna Majer et al |
| EPI_ISL_582359 | Canada/MB-NML-983/2020 | 2020-07-29 | North America/Canada/Manitoba/ | Cadham Provincial Laboratory | National Microbiology Laboratory (NML) | Anna Majer et al |
| EPI_ISL_582360 | Canada/MB-NML-984/2020 | 2020-07-30 | North America/Canada/Manitoba/ | Cadham Provincial Laboratory | National Microbiology Laboratory (NML) | Anna Majer et al |
| EPI_ISL_582361 | Canada/MB-NML-985/2020 | 2020-07-30 | North America/Canada/Manitoba/ | Cadham Provincial Laboratory | National Microbiology Laboratory (NML) | Anna Majer et al |
| EPI_ISL_582362 | Canada/MB-NML-987/2020 | 2020-04-18 | North America/Canada/Manitoba/ | Cadham Provincial Laboratory | National Microbiology Laboratory (NML) | Anna Majer et al |
| EPI_ISL_582363 | Canada/MB-NML-989/2020 | 2020-07-17 | North America/Canada/Manitoba/ | Cadham Provincial Laboratory | National Microbiology Laboratory (NML) | Anna Majer et al |
| EPI_ISL_582364 | Canada/MB-NML-990/2020 | 2020-07-16 | North America/Canada/Manitoba/ | Cadham Provincial Laboratory | National Microbiology Laboratory (NML) | Anna Majer et al |
| EPI_ISL_582365 | Canada/MB-NML-991/2020 | 2020-07-18 | North America/Canada/Manitoba/ | Cadham Provincial Laboratory | National Microbiology Laboratory (NML) | Anna Majer et al |
| EPI_ISL_582366 | Canada/MB-NML-992/2020 | 2020-07-29 | North America/Canada/Manitoba/ | Cadham Provincial Laboratory | National Microbiology Laboratory (NML) | Anna Majer et al |

| | | | | | | |
|---|---|---|---|---|---|---|
| EPI_ISL_582367 | Canada/MB-NML-997/2020 | 2020-07-19 | North America/Canada/Manitoba/ | Cadham Provincial Laboratory | National Microbiology Laboratory (NML) | Anna Majer et al |
| EPI_ISL_582368 | Canada/MB-NML-999/2020 | 2020-07-16 | North America/Canada/Manitoba/ | Cadham Provincial Laboratory | National Microbiology Laboratory (NML) | Anna Majer et al |
| EPI_ISL_582369 | Canada/MB-NML-1001/2020 | 2020-07-29 | North America/Canada/Manitoba/ | Cadham Provincial Laboratory | National Microbiology Laboratory (NML) | Anna Majer et al |
| EPI_ISL_582370 | Canada/MB-NML-1002/2020 | 2020-07-28 | North America/Canada/Manitoba/ | Cadham Provincial Laboratory | National Microbiology Laboratory (NML) | Anna Majer et al |
| EPI_ISL_582371 | Canada/MB-NML-1003/2020 | 2020-07-30 | North America/Canada/Manitoba/ | Cadham Provincial Laboratory | National Microbiology Laboratory (NML) | Anna Majer et al |
| EPI_ISL_582372 | Canada/MB-NML-1005/2020 | 2020-07-27 | North America/Canada/Manitoba/ | Cadham Provincial Laboratory | National Microbiology Laboratory (NML) | Anna Majer et al |
| EPI_ISL_582373 | Canada/MB-NML-1006/2020 | 2020-07-18 | North America/Canada/Manitoba/ | Cadham Provincial Laboratory | National Microbiology Laboratory (NML) | Anna Majer et al |
| EPI_ISL_582374 | Canada/MB-NML-1007/2020 | 2020-07-17 | North America/Canada/Manitoba/ | Cadham Provincial Laboratory | National Microbiology Laboratory (NML) | Anna Majer et al |
| EPI_ISL_582375 | Canada/MB-NML-1008/2020 | 2020-07-16 | North America/Canada/Manitoba/ | Cadham Provincial Laboratory | National Microbiology Laboratory (NML) | Anna Majer et al |
| EPI_ISL_582376 | Canada/MB-NML-1009/2020 | 2020-07-15 | North America/Canada/Manitoba/ | Cadham Provincial Laboratory | National Microbiology Laboratory (NML) | Anna Majer et al |
| EPI_ISL_582377 | Canada/MB-NML-1010/2020 | 2020-07-29 | North America/Canada/Manitoba/ | Cadham Provincial Laboratory | National Microbiology Laboratory (NML) | Anna Majer et al |
| EPI_ISL_582378 | Canada/MB-NML-1011/2020 | 2020-07-30 | North America/Canada/Manitoba/ | Cadham Provincial Laboratory | National Microbiology Laboratory (NML) | Anna Majer et al |
| EPI_ISL_582379 | Canada/MB-NML-1012/2020 | 2020-03-27 | North America/Canada/Manitoba/ | Cadham Provincial Laboratory | National Microbiology Laboratory (NML) | Anna Majer et al |

| | | | | | | |
|---|---|---|---|---|---|---|
| EPI_ISL_582380 | Canada/MB-NML-1014/2020 | 2020-07-29 | North America/Canada/Manitoba/ | Cadham Provincial Laboratory | National Microbiology Laboratory (NML) | Anna Majer et al |
| EPI_ISL_582381 | Canada/MB-NML-1015/2020 | 2020-07-18 | North America/Canada/Manitoba/ | Cadham Provincial Laboratory | National Microbiology Laboratory (NML) | Anna Majer et al |
| EPI_ISL_582382 | Canada/MB-NML-1016/2020 | 2020-07-17 | North America/Canada/Manitoba/ | Cadham Provincial Laboratory | National Microbiology Laboratory (NML) | Anna Majer et al |
| EPI_ISL_582383 | Canada/MB-NML-1017/2020 | 2020-07-16 | North America/Canada/Manitoba/ | Cadham Provincial Laboratory | National Microbiology Laboratory (NML) | Anna Majer et al |
| EPI_ISL_582384 | Canada/MB-NML-1018/2020 | 2020-07-15 | North America/Canada/Manitoba/ | Cadham Provincial Laboratory | National Microbiology Laboratory (NML) | Anna Majer et al |
| EPI_ISL_582385 | Canada/MB-NML-1019/2020 | 2020-07-29 | North America/Canada/Manitoba/ | Cadham Provincial Laboratory | National Microbiology Laboratory (NML) | Anna Majer et al |
| EPI_ISL_582386 | Canada/MB-NML-1020/2020 | 2020-07-30 | North America/Canada/Manitoba/ | Cadham Provincial Laboratory | National Microbiology Laboratory (NML) | Anna Majer et al |
| EPI_ISL_582387 | Canada/MB-NML-1021/2020 | 2020-03-28 | North America/Canada/Manitoba/ | Cadham Provincial Laboratory | National Microbiology Laboratory (NML) | Anna Majer et al |
| EPI_ISL_582388 | Canada/MB-NML-1023/2020 | 2020-04-18 | North America/Canada/Manitoba/ | Cadham Provincial Laboratory | National Microbiology Laboratory (NML) | Anna Majer et al |
| EPI_ISL_582389 | Canada/MB-NML-1024/2020 | 2020-07-18 | North America/Canada/Manitoba/ | Cadham Provincial Laboratory | National Microbiology Laboratory (NML) | Anna Majer et al |
| EPI_ISL_582390 | Canada/MB-NML-1025/2020 | 2020-07-17 | North America/Canada/Manitoba/ | Cadham Provincial Laboratory | National Microbiology Laboratory (NML) | Anna Majer et al |
| EPI_ISL_582391 | Canada/MB-NML-1026/2020 | 2020-07-17 | North America/Canada/Manitoba/ | Cadham Provincial Laboratory | National Microbiology Laboratory (NML) | Anna Majer et al |
| EPI_ISL_582392 | Canada/MB-NML-1027/2020 | 2020-07-15 | North America/Canada/Manitoba/ | Cadham Provincial Laboratory | National Microbiology Laboratory (NML) | Anna Majer et al |

| | | | | | | |
|---|---|---|---|---|---|---|
| EPI_ISL_582393 | Canada/MB-NML-1029/2020 | 2020-07-30 | North America/Canada/Manitoba/ | Cadham Provincial Laboratory | National Microbiology Laboratory (NML) | Anna Majer et al |
| EPI_ISL_582394 | Canada/MB-NML-1030/2020 | 2020-03-27 | North America/Canada/Manitoba/ | Cadham Provincial Laboratory | National Microbiology Laboratory (NML) | Anna Majer et al |
| EPI_ISL_582395 | Canada/MB-NML-1033/2020 | 2020-07-19 | North America/Canada/Manitoba/ | Cadham Provincial Laboratory | National Microbiology Laboratory (NML) | Anna Majer et al |
| EPI_ISL_582396 | Canada/MB-NML-1034/2020 | 2020-07-17 | North America/Canada/Manitoba/ | Cadham Provincial Laboratory | National Microbiology Laboratory (NML) | Anna Majer et al |
| EPI_ISL_582397 | Canada/MB-NML-1035/2020 | 2020-07-17 | North America/Canada/Manitoba/ | Cadham Provincial Laboratory | National Microbiology Laboratory (NML) | Anna Majer et al |
| EPI_ISL_582399 | Canada/MB-NML-1038/2020 | 2020-07-28 | North America/Canada/Manitoba/ | Cadham Provincial Laboratory | National Microbiology Laboratory (NML) | Anna Majer et al |
| EPI_ISL_582400 | Canada/MB-NML-1039/2020 | 2020-03-30 | North America/Canada/Manitoba/ | Cadham Provincial Laboratory | National Microbiology Laboratory (NML) | Anna Majer et al |
| EPI_ISL_582401 | Canada/MB-NML-1041/2020 | 2020-04-28 | North America/Canada/Manitoba/ | Cadham Provincial Laboratory | National Microbiology Laboratory (NML) | Anna Majer et al |
| EPI_ISL_582402 | Canada/MB-NML-1042/2020 | 2020-07-17 | North America/Canada/Manitoba/ | Cadham Provincial Laboratory | National Microbiology Laboratory (NML) | Anna Majer et al |
| EPI_ISL_582403 | Canada/MB-NML-1043/2020 | 2020-07-17 | North America/Canada/Manitoba/ | Cadham Provincial Laboratory | National Microbiology Laboratory (NML) | Anna Majer et al |
| EPI_ISL_582404 | Canada/MB-NML-1044/2020 | 2020-07-17 | North America/Canada/Manitoba/ | Cadham Provincial Laboratory | National Microbiology Laboratory (NML) | Anna Majer et al |
| EPI_ISL_582405 | Canada/MB-NML-1045/2020 | 2020-07-29 | North America/Canada/Manitoba/ | Cadham Provincial Laboratory | National Microbiology Laboratory (NML) | Anna Majer et al |
| EPI_ISL_582406 | Canada/MB-NML-1046/2020 | 2020-07-30 | North America/Canada/Manitoba/ | Cadham Provincial Laboratory | National Microbiology Laboratory (NML) | Anna Majer et al |

| | | | | | | |
|---|---|---|---|---|---|---|
| EPI_ISL_582407 | Canada/MB-NML-1048/2020 | 2020-04-18 | North America/Canada/Manitoba/ | Cadham Provincial Laboratory | National Microbiology Laboratory (NML) | Anna Majer et al |
| EPI_ISL_582408 | Canada/MB-NML-1049/2020 | 2020-04-25 | North America/Canada/Manitoba/ | Cadham Provincial Laboratory | National Microbiology Laboratory (NML) | Anna Majer et al |
| EPI_ISL_582409 | Canada/MB-NML-1050/2020 | 2020-07-17 | North America/Canada/Manitoba/ | Cadham Provincial Laboratory | National Microbiology Laboratory (NML) | Anna Majer et al |
| EPI_ISL_582410 | Canada/MB-NML-1051/2020 | 2020-07-17 | North America/Canada/Manitoba/ | Cadham Provincial Laboratory | National Microbiology Laboratory (NML) | Anna Majer et al |
| EPI_ISL_582411 | Canada/MB-NML-1052/2020 | 2020-07-17 | North America/Canada/Manitoba/ | Cadham Provincial Laboratory | National Microbiology Laboratory (NML) | Anna Majer et al |
| EPI_ISL_582412 | Canada/MB-NML-1053/2020 | 2020-07-29 | North America/Canada/Manitoba/ | Cadham Provincial Laboratory | National Microbiology Laboratory (NML) | Anna Majer et al |
| EPI_ISL_582413 | Canada/MB-NML-1054/2020 | 2020-03-22 | North America/Canada/Manitoba/ | Cadham Provincial Laboratory | National Microbiology Laboratory (NML) | Anna Majer et al |
| EPI_ISL_582414 | Canada/MB-NML-1055/2020 | 2020-07-30 | North America/Canada/Manitoba/ | Cadham Provincial Laboratory | National Microbiology Laboratory (NML) | Anna Majer et al |
| EPI_ISL_582416 | Canada/MB-NML-1060/2020 | 2020-08-05 | North America/Canada/Manitoba/ | Cadham Provincial Laboratory | National Microbiology Laboratory (NML) | Anna Majer et al |
| EPI_ISL_582417 | Canada/MB-NML-1061/2020 | 2020-08-07 | North America/Canada/Manitoba/ | Cadham Provincial Laboratory | National Microbiology Laboratory (NML) | Anna Majer et al |
| EPI_ISL_582418 | Canada/MB-NML-1063/2020 | 2020-08-05 | North America/Canada/Manitoba/ | Cadham Provincial Laboratory | National Microbiology Laboratory (NML) | Anna Majer et al |
| EPI_ISL_582419 | Canada/MB-NML-1064/2020 | 2020-08-06 | North America/Canada/Manitoba/ | Cadham Provincial Laboratory | National Microbiology Laboratory (NML) | Anna Majer et al |
| EPI_ISL_582420 | Canada/MB-NML-1065/2020 | 2020-08-05 | North America/Canada/Manitoba/ | Cadham Provincial Laboratory | National Microbiology Laboratory (NML) | Anna Majer et al |

| | | | | | | |
|---|---|---|---|---|---|---|
| EPI_ISL_582422 | Canada/MB-NML-1067/2020 | 2020-08-07 | North America/Canada/Manitoba/ | Cadham Provincial Laboratory | National Microbiology Laboratory (NML) | Anna Majer et al |
| EPI_ISL_582423 | Canada/MB-NML-1069/2020 | 2020-07-20 | North America/Canada/Manitoba/ | Cadham Provincial Laboratory | National Microbiology Laboratory (NML) | Anna Majer et al |
| EPI_ISL_582424 | Canada/MB-NML-1070/2020 | 2020-08-04 | North America/Canada/Manitoba/ | Cadham Provincial Laboratory | National Microbiology Laboratory (NML) | Anna Majer et al |
| EPI_ISL_582425 | Canada/MB-NML-1071/2020 | 2020-08-06 | North America/Canada/Manitoba/ | Cadham Provincial Laboratory | National Microbiology Laboratory (NML) | Anna Majer et al |
| EPI_ISL_582426 | Canada/MB-NML-1073/2020 | 2020-08-04 | North America/Canada/Manitoba/ | Cadham Provincial Laboratory | National Microbiology Laboratory (NML) | Anna Majer et al |
| EPI_ISL_582427 | Canada/MB-NML-1075/2020 | 2020-08-07 | North America/Canada/Manitoba/ | Cadham Provincial Laboratory | National Microbiology Laboratory (NML) | Anna Majer et al |
| EPI_ISL_582428 | Canada/MB-NML-1076/2020 | 2020-08-05 | North America/Canada/Manitoba/ | Cadham Provincial Laboratory | National Microbiology Laboratory (NML) | Anna Majer et al |
| EPI_ISL_582429 | Canada/MB-NML-1077/2020 | 2020-08-04 | North America/Canada/Manitoba/ | Cadham Provincial Laboratory | National Microbiology Laboratory (NML) | Anna Majer et al |
| EPI_ISL_582430 | Canada/MB-NML-1078/2020 | 2020-08-04 | North America/Canada/Manitoba/ | Cadham Provincial Laboratory | National Microbiology Laboratory (NML) | Anna Majer et al |
| EPI_ISL_582431 | Canada/MB-NML-1079/2020 | 2020-08-03 | North America/Canada/Manitoba/ | Cadham Provincial Laboratory | National Microbiology Laboratory (NML) | Anna Majer et al |
| EPI_ISL_582432 | Canada/MB-NML-1080/2020 | 2020-08-04 | North America/Canada/Manitoba/ | Cadham Provincial Laboratory | National Microbiology Laboratory (NML) | Anna Majer et al |
| EPI_ISL_582433 | Canada/MB-NML-1082/2020 | 2020-08-06 | North America/Canada/Manitoba/ | Cadham Provincial Laboratory | National Microbiology Laboratory (NML) | Anna Majer et al |
| EPI_ISL_582434 | Canada/MB-NML-1083/2020 | 2020-08-05 | North America/Canada/Manitoba/ | Cadham Provincial Laboratory | National Microbiology Laboratory (NML) | Anna Majer et al |

| | | | | | | |
|---|---|---|---|---|---|---|
| EPI_ISL_582435 | Canada/MB-NML-1085/2020 | 2020-08-08 | North America/Canada/Manitoba/ | Cadham Provincial Laboratory | National Microbiology Laboratory (NML) | Anna Majer et al |
| EPI_ISL_582436 | Canada/MB-NML-1086/2020 | 2020-08-06 | North America/Canada/Manitoba/ | Cadham Provincial Laboratory | National Microbiology Laboratory (NML) | Anna Majer et al |
| EPI_ISL_582437 | Canada/MB-NML-1087/2020 | 2020-08-09 | North America/Canada/Manitoba/ | Cadham Provincial Laboratory | National Microbiology Laboratory (NML) | Anna Majer et al |
| EPI_ISL_582438 | Canada/MB-NML-1089/2020 | 2020-07-23 | North America/Canada/Manitoba/ | Cadham Provincial Laboratory | National Microbiology Laboratory (NML) | Anna Majer et al |
| EPI_ISL_582439 | Canada/MB-NML-1090/2020 | 2020-08-05 | North America/Canada/Manitoba/ | Cadham Provincial Laboratory | National Microbiology Laboratory (NML) | Anna Majer et al |
| EPI_ISL_582440 | Canada/MB-NML-1091/2020 | 2020-08-04 | North America/Canada/Manitoba/ | Cadham Provincial Laboratory | National Microbiology Laboratory (NML) | Anna Majer et al |
| EPI_ISL_582441 | Canada/MB-NML-1092/2020 | 2020-08-04 | North America/Canada/Manitoba/ | Cadham Provincial Laboratory | National Microbiology Laboratory (NML) | Anna Majer et al |
| EPI_ISL_582442 | Canada/MB-NML-1093/2020 | 2020-08-04 | North America/Canada/Manitoba/ | Cadham Provincial Laboratory | National Microbiology Laboratory (NML) | Anna Majer et al |
| EPI_ISL_582443 | Canada/MB-NML-1096/2020 | 2020-08-04 | North America/Canada/Manitoba/ | Cadham Provincial Laboratory | National Microbiology Laboratory (NML) | Anna Majer et al |
| EPI_ISL_582445 | Canada/MB-NML-1100/2020 | 2020-08-05 | North America/Canada/Manitoba/ | Cadham Provincial Laboratory | National Microbiology Laboratory (NML) | Anna Majer et al |
| EPI_ISL_582446 | Canada/MB-NML-1101/2020 | 2020-08-03 | North America/Canada/Manitoba/ | Cadham Provincial Laboratory | National Microbiology Laboratory (NML) | Anna Majer et al |
| EPI_ISL_582447 | Canada/MB-NML-1102/2020 | 2020-08-06 | North America/Canada/Manitoba/ | Cadham Provincial Laboratory | National Microbiology Laboratory (NML) | Anna Majer et al |
| EPI_ISL_582448 | Canada/MB-NML-1104/2020 | 2020-08-05 | North America/Canada/Manitoba/ | Cadham Provincial Laboratory | National Microbiology Laboratory (NML) | Anna Majer et al |

| | | | | | | |
|---|---|---|---|---|---|---|
| EPI_ISL_582449 | Canada/MB-NML-1107/2020 | 2020-08-04 | North America/Canada/Manitoba/ | Cadham Provincial Laboratory | National Microbiology Laboratory (NML) | Anna Majer et al |
| EPI_ISL_582450 | Canada/MB-NML-1110/2020 | 2020-08-04 | North America/Canada/Manitoba/ | Cadham Provincial Laboratory | National Microbiology Laboratory (NML) | Anna Majer et al |
| EPI_ISL_582451 | Canada/MB-NML-1111/2020 | 2020-08-04 | North America/Canada/Manitoba/ | Cadham Provincial Laboratory | National Microbiology Laboratory (NML) | Anna Majer et al |
| EPI_ISL_582452 | Canada/MB-NML-1112/2020 | 2020-08-05 | North America/Canada/Manitoba/ | Cadham Provincial Laboratory | National Microbiology Laboratory (NML) | Anna Majer et al |
| EPI_ISL_582453 | Canada/MB-NML-1116/2020 | 2020-08-04 | North America/Canada/Manitoba/ | Cadham Provincial Laboratory | National Microbiology Laboratory (NML) | Anna Majer et al |
| EPI_ISL_582454 | Canada/MB-NML-1117/2020 | 2020-08-04 | North America/Canada/Manitoba/ | Cadham Provincial Laboratory | National Microbiology Laboratory (NML) | Anna Majer et al |
| EPI_ISL_582455 | Canada/MB-NML-1119/2020 | 2020-08-03 | North America/Canada/Manitoba/ | Cadham Provincial Laboratory | National Microbiology Laboratory (NML) | Anna Majer et al |
| EPI_ISL_582456 | Canada/MB-NML-1121/2020 | 2020-08-04 | North America/Canada/Manitoba/ | Cadham Provincial Laboratory | National Microbiology Laboratory (NML) | Anna Majer et al |
| EPI_ISL_582457 | Canada/MB-NML-1122/2020 | 2020-08-03 | North America/Canada/Manitoba/ | Cadham Provincial Laboratory | National Microbiology Laboratory (NML) | Anna Majer et al |
| EPI_ISL_582459 | Canada/MB-NML-1125/2020 | 2020-08-04 | North America/Canada/Manitoba/ | Cadham Provincial Laboratory | National Microbiology Laboratory (NML) | Anna Majer et al |
| EPI_ISL_582460 | Canada/MB-NML-1128/2020 | 2020-08-03 | North America/Canada/Manitoba/ | Cadham Provincial Laboratory | National Microbiology Laboratory (NML) | Anna Majer et al |
| EPI_ISL_582461 | Canada/MB-NML-1130/2020 | 2020-08-03 | North America/Canada/Manitoba/ | Cadham Provincial Laboratory | National Microbiology Laboratory (NML) | Anna Majer et al |
| EPI_ISL_582462 | Canada/MB-NML-1131/2020 | 2020-08-03 | North America/Canada/Manitoba/ | Cadham Provincial Laboratory | National Microbiology Laboratory (NML) | Anna Majer et al |

| | | | | | | |
|---|---|---|---|---|---|---|
| EPI_ISL_582463 | Canada/MB-NML-1132/2020 | 2020-07-20 | North America/Canada/Manitoba/ | Cadham Provincial Laboratory | National Microbiology Laboratory (NML) | Anna Majer et al |
| EPI_ISL_582464 | Canada/MB-NML-1133/2020 | 2020-07-21 | North America/Canada/Manitoba/ | Cadham Provincial Laboratory | National Microbiology Laboratory (NML) | Anna Majer et al |
| EPI_ISL_582465 | Canada/MB-NML-1134/2020 | 2020-07-20 | North America/Canada/Manitoba/ | Cadham Provincial Laboratory | National Microbiology Laboratory (NML) | Anna Majer et al |
| EPI_ISL_582466 | Canada/MB-NML-1135/2020 | 2020-07-20 | North America/Canada/Manitoba/ | Cadham Provincial Laboratory | National Microbiology Laboratory (NML) | Anna Majer et al |
| EPI_ISL_582467 | Canada/MB-NML-1136/2020 | 2020-08-03 | North America/Canada/Manitoba/ | Cadham Provincial Laboratory | National Microbiology Laboratory (NML) | Anna Majer et al |
| EPI_ISL_582468 | Canada/MB-NML-1138/2020 | 2020-07-20 | North America/Canada/Manitoba/ | Cadham Provincial Laboratory | National Microbiology Laboratory (NML) | Anna Majer et al |
| EPI_ISL_582469 | Canada/MB-NML-1139/2020 | 2020-08-06 | North America/Canada/Manitoba/ | Cadham Provincial Laboratory | National Microbiology Laboratory (NML) | Anna Majer et al |
| EPI_ISL_582470 | Canada/MB-NML-1140/2020 | 2020-08-03 | North America/Canada/Manitoba/ | Cadham Provincial Laboratory | National Microbiology Laboratory (NML) | Anna Majer et al |
| EPI_ISL_582471 | Canada/MB-NML-1141/2020 | 2020-08-03 | North America/Canada/Manitoba/ | Cadham Provincial Laboratory | National Microbiology Laboratory (NML) | Anna Majer et al |
| EPI_ISL_582472 | Canada/MB-NML-1142/2020 | 2020-08-03 | North America/Canada/Manitoba/ | Cadham Provincial Laboratory | National Microbiology Laboratory (NML) | Anna Majer et al |
| EPI_ISL_582473 | Canada/MB-NML-1144/2020 | 2020-08-04 | North America/Canada/Manitoba/ | Cadham Provincial Laboratory | National Microbiology Laboratory (NML) | Anna Majer et al |
| EPI_ISL_582474 | Canada/MB-NML-1146/2020 | 2020-07-22 | North America/Canada/Manitoba/ | Cadham Provincial Laboratory | National Microbiology Laboratory (NML) | Anna Majer et al |
| EPI_ISL_582475 | Canada/MB-NML-1147/2020 | 2020-07-21 | North America/Canada/Manitoba/ | Cadham Provincial Laboratory | National Microbiology Laboratory (NML) | Anna Majer et al |

| | | | | | | |
|---|---|---|---|---|---|---|
| EPI_ISL_582476 | Canada/MB-NML-1148/2020 | 2020-07-22 | North America/Canada/Manitoba/ | Cadham Provincial Laboratory | National Microbiology Laboratory (NML) | Anna Majer et al |
| EPI_ISL_582477 | Canada/MB-NML-1150/2020 | 2020-07-19 | North America/Canada/Manitoba/ | Cadham Provincial Laboratory | National Microbiology Laboratory (NML) | Anna Majer et al |
| EPI_ISL_582478 | Canada/MB-NML-1152/2020 | 2020-08-01 | North America/Canada/Manitoba/ | Cadham Provincial Laboratory | National Microbiology Laboratory (NML) | Anna Majer et al |
| EPI_ISL_582479 | Canada/MB-NML-1153/2020 | 2020-07-31 | North America/Canada/Manitoba/ | Cadham Provincial Laboratory | National Microbiology Laboratory (NML) | Anna Majer et al |
| EPI_ISL_582480 | Canada/MB-NML-1154/2020 | 2020-07-31 | North America/Canada/Manitoba/ | Cadham Provincial Laboratory | National Microbiology Laboratory (NML) | Anna Majer et al |
| EPI_ISL_582481 | Canada/MB-NML-1155/2020 | 2020-08-03 | North America/Canada/Manitoba/ | Cadham Provincial Laboratory | National Microbiology Laboratory (NML) | Anna Majer et al |
| EPI_ISL_582482 | Canada/MB-NML-1156/2020 | 2020-08-03 | North America/Canada/Manitoba/ | Cadham Provincial Laboratory | National Microbiology Laboratory (NML) | Anna Majer et al |
| EPI_ISL_582483 | Canada/MB-NML-1157/2020 | 2020-08-03 | North America/Canada/Manitoba/ | Cadham Provincial Laboratory | National Microbiology Laboratory (NML) | Anna Majer et al |
| EPI_ISL_582484 | Canada/MB-NML-1162/2020 | 2020-07-24 | North America/Canada/Manitoba/ | Cadham Provincial Laboratory | National Microbiology Laboratory (NML) | Anna Majer et al |
| EPI_ISL_582485 | Canada/MB-NML-1164/2020 | 2020-07-24 | North America/Canada/Manitoba/ | Cadham Provincial Laboratory | National Microbiology Laboratory (NML) | Anna Majer et al |
| EPI_ISL_582486 | Canada/MB-NML-1165/2020 | 2020-07-22 | North America/Canada/Manitoba/ | Cadham Provincial Laboratory | National Microbiology Laboratory (NML) | Anna Majer et al |
| EPI_ISL_582487 | Canada/MB-NML-1166/2020 | 2020-07-21 | North America/Canada/Manitoba/ | Cadham Provincial Laboratory | National Microbiology Laboratory (NML) | Anna Majer et al |
| EPI_ISL_582488 | Canada/MB-NML-1167/2020 | 2020-08-03 | North America/Canada/Manitoba/ | Cadham Provincial Laboratory | National Microbiology Laboratory (NML) | Anna Majer et al |

| | | | | | | |
|---|---|---|---|---|---|---|
| EPI_ISL_582489 | Canada/MB-NML-1168/2020 | 2020-08-03 | North America/Canada/Manitoba/ | Cadham Provincial Laboratory | National Microbiology Laboratory (NML) | Anna Majer et al |
| EPI_ISL_582490 | Canada/MB-NML-1169/2020 | 2020-08-03 | North America/Canada/Manitoba/ | Cadham Provincial Laboratory | National Microbiology Laboratory (NML) | Anna Majer et al |
| EPI_ISL_582491 | Canada/MB-NML-1170/2020 | 2020-08-03 | North America/Canada/Manitoba/ | Cadham Provincial Laboratory | National Microbiology Laboratory (NML) | Anna Majer et al |
| EPI_ISL_582492 | Canada/MB-NML-1171/2020 | 2020-08-03 | North America/Canada/Manitoba/ | Cadham Provincial Laboratory | National Microbiology Laboratory (NML) | Anna Majer et al |
| EPI_ISL_582493 | Canada/MB-NML-1174/2020 | 2020-07-23 | North America/Canada/Manitoba/ | Cadham Provincial Laboratory | National Microbiology Laboratory (NML) | Anna Majer et al |
| EPI_ISL_582494 | Canada/MB-NML-1176/2020 | 2020-07-03 | North America/Canada/Manitoba/ | Cadham Provincial Laboratory | National Microbiology Laboratory (NML) | Anna Majer et al |
| EPI_ISL_582495 | Canada/MB-NML-1178/2020 | 2020-08-03 | North America/Canada/Manitoba/ | Cadham Provincial Laboratory | National Microbiology Laboratory (NML) | Anna Majer et al |
| EPI_ISL_582496 | Canada/MB-NML-1179/2020 | 2020-07-24 | North America/Canada/Manitoba/ | Cadham Provincial Laboratory | National Microbiology Laboratory (NML) | Anna Majer et al |
| EPI_ISL_582497 | Canada/MB-NML-1180/2020 | 2020-07-28 | North America/Canada/Manitoba/ | Cadham Provincial Laboratory | National Microbiology Laboratory (NML) | Anna Majer et al |
| EPI_ISL_582498 | Canada/MB-NML-1182/2020 | 2020-07-28 | North America/Canada/Manitoba/ | Cadham Provincial Laboratory | National Microbiology Laboratory (NML) | Anna Majer et al |
| EPI_ISL_582499 | Canada/MB-NML-1183/2020 | 2020-07-28 | North America/Canada/Manitoba/ | Cadham Provincial Laboratory | National Microbiology Laboratory (NML) | Anna Majer et al |
| EPI_ISL_582500 | Canada/MB-NML-1186/2020 | 2020-08-10 | North America/Canada/Manitoba/ | Cadham Provincial Laboratory | National Microbiology Laboratory (NML) | Anna Majer et al |
| EPI_ISL_582501 | Canada/MB-NML-1187/2020 | 2020-08-10 | North America/Canada/Manitoba/ | Cadham Provincial Laboratory | National Microbiology Laboratory (NML) | Anna Majer et al |

| | | | | | | |
|---|---|---|---|---|---|---|
| EPI_ISL_582502 | Canada/MB-NML-1188/2020 | 2020-08-10 | North America/Canada/Manitoba/ | Cadham Provincial Laboratory | National Microbiology Laboratory (NML) | Anna Majer et al |
| EPI_ISL_582503 | Canada/MB-NML-1189/2020 | 2020-08-08 | North America/Canada/Manitoba/ | Cadham Provincial Laboratory | National Microbiology Laboratory (NML) | Anna Majer et al |
| EPI_ISL_582504 | Canada/MB-NML-1190/2020 | 2020-08-09 | North America/Canada/Manitoba/ | Cadham Provincial Laboratory | National Microbiology Laboratory (NML) | Anna Majer et al |
| EPI_ISL_582505 | Canada/MB-NML-1191/2020 | 2020-07-20 | North America/Canada/Manitoba/ | Cadham Provincial Laboratory | National Microbiology Laboratory (NML) | Anna Majer et al |
| EPI_ISL_582506 | Canada/MB-NML-1193/2020 | 2020-04-09 | North America/Canada/Manitoba/ | Cadham Provincial Laboratory | National Microbiology Laboratory (NML) | Anna Majer et al |
| EPI_ISL_582507 | Canada/MB-NML-1195/2020 | 2020-04-25 | North America/Canada/Manitoba/ | Cadham Provincial Laboratory | National Microbiology Laboratory (NML) | Anna Majer et al |
| EPI_ISL_582508 | France/ARA-CDP29289/2020 | 2020-09-11 | Europe/France/Auvergne-Rhv¥ne-Alpes/ | CNR Virus des Infections Respiratoires - France SUD | CNR Virus des Infections Respiratoires - France SUD | Antonin Bal et al |
| EPI_ISL_582509 | Ukraine/Kharkiv-817/2020 | 2020-08-04 | Europe/Ukraine/Kharkiv/ | Department of Respiratory and other Viral Infections of L.V.Gromashevsky Institute of Epidemiology & Infectious Diseases NAMS of Ukraine | "Department of Respiratory and other Viral Infections of L.V.Gromashevsky Institute of Epidemiology & Infectious Diseases NAMS of Ukraine, JSC ""Farmak""" | Alla Mironenko et al |

| | | | | | | |
|---|---|---|---|---|---|---|
| EPI_ISL_582510 | Ukraine/Kharkiv-877/2020 | 2020-08-07 | Europe/Ukraine/Kharkiv/ | Department of Respiratory and other Viral Infections of L.V.Gromashevsky Institute of Epidemiology & Infectious Diseases NAMS of Ukraine | "Department of Respiratory and other Viral Infections of L.V.Gromashevsky Institute of Epidemiology & Infectious Diseases NAMS of Ukrain–µ, JSC ""Farmak""" | Alla Mironenko et al |
| EPI_ISL_582511 | Ukraine/Kharkiv-879/2020 | 2020-08-07 | Europe/Ukraine/Kharkiv/ | Department of Respiratory and other Viral Infections of L.V.Gromashevsky Institute of Epidemiology & Infectious Diseases NAMS of Ukrain–µ | "Department of Respiratory and other Viral Infections of L.V.Gromashevsky Institute of Epidemiology & Infectious Diseases NAMS of Ukrain–µ, JSC ""Farmak""" | Alla Mironenko et al |
| EPI_ISL_582512 | Ukraine/Kharkiv-882/2020 | 2020-08-07 | Europe/Ukraine/Kharkiv/ | Department of Respiratory and other Viral Infections of L.V.Gromashevsky Institute of Epidemiology & Infectious Diseases NAMS of Ukrain–µ | "Department of Respiratory and other Viral Infections of L.V.Gromashevsky Institute of Epidemiology & Infectious Diseases NAMS of Ukrain–µ, JSC ""Farmak""" | Alla Mironenko et al |

| | | | | | | |
|---|---|---|---|---|---|---|
| EPI_ISL_582513 | Ukraine/Kharkiv-782/2020 | 2020-07-31 | Europe/Ukraine/Kharkiv/ | Department of Respiratory and other Viral Infections of L.V.Gromashevsky Institute of Epidemiology & Infectious Diseases NAMS of Ukrain–µ | "Department of Respiratory and other Viral Infections of L.V.Gromashevsky Institute of Epidemiology & Infectious Diseases NAMS of Ukrain–µ, JSC ""Farmak""" | Alla Mironenko et al |
| EPI_ISL_582522 | Portugal/PT1295/2020 | 2020-03-24 | Europe/Portugal/Portugal/ | unknown | Instituto Nacional de Saude (INSA) and Instituto Gulbenkian de Ciencia (IGC) | Borges et al et al |
| EPI_ISL_582528 | Serbia/Krusevac-07961604/2020 | 2020-04-15 | Europe/Serbia/Rasina District/Krusevac | "Veterinary Specialized Institute ""Kraljevo"", Serbia" | "Veterinary Specialized Institute ""Kraljevo"", Serbia" | Vidanovic et al |
| EPI_ISL_582608 | UnitedArabEmirates/skmc-92056837 | 2020-01-30 | Asia/United Arab Emirates/United Arab | Sheikh Khalifa Medical City | Molecular/Surveillance lab Sheikh Khalifa Medical City | Amirtharaj Francis et al |
| EPI_ISL_582609 | UnitedArabEmirates/skmc-0381517/2 | 2020-02-07 | Asia/United Arab Emirates/United Arab | Sheikh Khalifa Medical City | Molecular/Surveillance lab Sheikh Khalifa Medical City | Amirtharaj Francis et al |
| EPI_ISL_582610 | UnitedArabEmirates/skmc-0442954/2 | 2020-02-13 | Asia/United Arab Emirates/United Arab | Sheikh Khalifa Medical City | Molecular/Surveillance lab Sheikh Khalifa Medical City | Amirtharaj Francis et al |
| EPI_ISL_582611 | UnitedArabEmirates/skmc-22005221 | 2020-02-21 | Asia/United Arab Emirates/United Arab | Sheikh Khalifa Medical City | Molecular/Surveillance lab Sheikh Khalifa Medical City | Amirtharaj Francis et al |

| | | | | | | |
|---|---|---|---|---|---|---|
| EPI_ISL_582612 | UnitedArabEmirates/skmc-002/2020 | 2020-02-21 | Asia/United Arab Emirates/United Arab | Sheikh Khalifa Medical City | Molecular/Surveillance lab Sheikh Khalifa Medical City | Amirtharaj Francis et al |
| EPI_ISL_582613 | UnitedArabEmirates/skmc-004/2020 | 2020-02-21 | Asia/United Arab Emirates/United Arab | Sheikh Khalifa Medical City | Molecular/Surveillance lab Sheikh Khalifa Medical City | Amirtharaj Francis et al |
| EPI_ISL_582614 | UnitedArabEmirates/skmc-005/2020 | 2020-02-21 | Asia/United Arab Emirates/United Arab | Sheikh Khalifa Medical City | Molecular/Surveillance lab Sheikh Khalifa Medical City | Amirtharaj Francis et al |
| EPI_ISL_582615 | UnitedArabEmirates/skmc-006/2020 | 2020-02-21 | Asia/United Arab Emirates/United Arab | Sheikh Khalifa Medical City | Molecular/Surveillance lab Sheikh Khalifa Medical City | Amirtharaj Francis et al |
| EPI_ISL_582616 | UnitedArabEmirates/skmc-007/2020 | 2020-02-21 | Asia/United Arab Emirates/United Arab | Sheikh Khalifa Medical City | Molecular/Surveillance lab Sheikh Khalifa Medical City | Amirtharaj Francis et al |
| EPI_ISL_582617 | UnitedArabEmirates/skmc-008/2020 | 2020-02-21 | Asia/United Arab Emirates/United Arab | Sheikh Khalifa Medical City | Molecular/Surveillance lab Sheikh Khalifa Medical City | Amirtharaj Francis et al |
| EPI_ISL_582618 | UnitedArabEmirates/skmc-009/2020 | 2020-02-21 | Asia/United Arab Emirates/United Arab | Sheikh Khalifa Medical City | Molecular/Surveillance lab Sheikh Khalifa Medical City | Amirtharaj Francis et al |
| EPI_ISL_582619 | UnitedArabEmirates/skmc-0532493/2 | 2020-02-21 | Asia/United Arab Emirates/United Arab | Sheikh Khalifa Medical City | Molecular/Surveillance lab Sheikh Khalifa Medical City | Amirtharaj Francis et al |
| EPI_ISL_582620 | UnitedArabEmirates/skmc-0532497/2 | 2020-02-22 | Asia/United Arab Emirates/United Arab | Sheikh Khalifa Medical City | Molecular/Surveillance lab Sheikh Khalifa Medical City | Amirtharaj Francis et al |
| EPI_ISL_582621 | UnitedArabEmirates/skmc-22005431 | 2020-02-23 | Asia/United Arab Emirates/United Arab | Sheikh Khalifa Medical City | Molecular/Surveillance lab Sheikh Khalifa Medical City | Amirtharaj Francis et al |

| | | | | | | |
|---|---|---|---|---|---|---|
| EPI_ISL_582622 | UnitedArabEmirates/skmc-22005431 | 2020-02-23 | Asia/United Arab Emirates/United Arab | Sheikh Khalifa Medical City | Molecular/Surveillance lab Sheikh Khalifa Medical City | Amirtharaj Francis et al |
| EPI_ISL_582623 | UnitedArabEmirates/skmc-22005431 | 2020-02-23 | Asia/United Arab Emirates/United Arab | Sheikh Khalifa Medical City | Molecular/Surveillance lab Sheikh Khalifa Medical City | Amirtharaj Francis et al |
| EPI_ISL_582624 | UnitedArabEmirates/skmc-001/2020 | 2020-02-27 | Asia/United Arab Emirates/United Arab | Sheikh Khalifa Medical City | Molecular/Surveillance lab Sheikh Khalifa Medical City | Amirtharaj Francis et al |
| EPI_ISL_582625 | UnitedArabEmirates/skmc-003/2020 | 2020-02-27 | Asia/United Arab Emirates/United Arab | Sheikh Khalifa Medical City | Molecular/Surveillance lab Sheikh Khalifa Medical City | Amirtharaj Francis et al |
| EPI_ISL_582626 | UnitedArabEmirates/skmc-01319674 | 2020-02-28 | Asia/United Arab Emirates/United Arab | Sheikh Khalifa Medical City | Molecular/Surveillance lab Sheikh Khalifa Medical City | Amirtharaj Francis et al |
| EPI_ISL_582628 | UnitedArabEmirates/skmc-72664/20 | 2020-04-03 | Asia/United Arab Emirates/United Arab | Sheikh Khalifa Medical City | Molecular/Surveillance lab Sheikh Khalifa Medical City | Amirtharaj Francis et al |
| EPI_ISL_582636 | UnitedArabEmirates/skmc-227578/2 | 2020-04-20 | Asia/United Arab Emirates/United Arab | Sheikh Khalifa Medical City | Molecular/Surveillance lab Sheikh Khalifa Medical City | Amirtharaj Francis et al |
| EPI_ISL_582641 | UnitedArabEmirates/skmc-359399/2 | 2020-04-29 | Asia/United Arab Emirates/United Arab | Sheikh Khalifa Medical City | Molecular/Surveillance lab Sheikh Khalifa Medical City | Amirtharaj Francis et al |
| EPI_ISL_582643 | UnitedArabEmirates/skmc-3079344/ | 2020-08-26 | Asia/United Arab Emirates/United Arab | Sheikh Khalifa Medical City | Molecular/Surveillance lab Sheikh Khalifa Medical City | Amirtharaj Francis et al |
| EPI_ISL_582650 | UnitedArabEmirates/skmc-3131248/ | 2020-08-29 | Asia/United Arab Emirates/United Arab | Sheikh Khalifa Medical City | Molecular/Surveillance lab Sheikh Khalifa Medical City | Amirtharaj Francis et al |

| | | | | | | |
|---|---|---|---|---|---|---|
| EPI_ISL_582652 | UnitedArabEmirates/skmc-3131941/2 | 2020-08-29 | Asia/United Arab Emirates/United Arab | Sheikh Khalifa Medical City | Molecular/Surveillance lab Sheikh Khalifa Medical City | Amirtharaj Francis et al |
| EPI_ISL_582657 | UnitedArabEmirates/skmc-3124949/2 | 2020-08-29 | Asia/United Arab Emirates/United Arab | Sheikh Khalifa Medical City | Molecular/Surveillance lab Sheikh Khalifa Medical City | Amirtharaj Francis et al |
| EPI_ISL_582663 | UnitedArabEmirates/skmc-3759960/2 | 2020-09-09 | Asia/United Arab Emirates/United Arab | Sheikh Khalifa Medical City | Molecular/Surveillance lab Sheikh Khalifa Medical City | Amirtharaj Francis et al |
| EPI_ISL_582667 | UnitedArabEmirates/skmc-3595813/2 | 2020-09-13 | Asia/United Arab Emirates/United Arab | Sheikh Khalifa Medical City | Molecular/Surveillance lab Sheikh Khalifa Medical City | Amirtharaj Francis et al |
| EPI_ISL_582668 | UnitedArabEmirates/skmc-3594464/2 | 2020-09-13 | Asia/United Arab Emirates/United Arab | Sheikh Khalifa Medical City | Molecular/Surveillance lab Sheikh Khalifa Medical City | Amirtharaj Francis et al |
| EPI_ISL_582669 | UnitedArabEmirates/skmc-3594255/2 | 2020-09-13 | Asia/United Arab Emirates/United Arab | Sheikh Khalifa Medical City | Molecular/Surveillance lab Sheikh Khalifa Medical City | Amirtharaj Francis et al |
| EPI_ISL_582672 | UnitedArabEmirates/skmc-3607928/2 | 2020-09-14 | Asia/United Arab Emirates/United Arab | Sheikh Khalifa Medical City | Molecular/Surveillance lab Sheikh Khalifa Medical City | Amirtharaj Francis et al |
| EPI_ISL_582674 | UnitedArabEmirates/skmc-3819699/2 | 2020-09-16 | Asia/United Arab Emirates/United Arab | Sheikh Khalifa Medical City | Molecular/Surveillance lab Sheikh Khalifa Medical City | Amirtharaj Francis et al |
| EPI_ISL_582676 | UnitedArabEmirates/skmc-3780553/2 | 2020-09-17 | Asia/United Arab Emirates/United Arab | Sheikh Khalifa Medical City | Molecular/Surveillance lab Sheikh Khalifa Medical City | Amirtharaj Francis et al |
| EPI_ISL_582677 | UnitedArabEmirates/skmc-3820378/2 | 2020-09-20 | Asia/United Arab Emirates/United Arab | Sheikh Khalifa Medical City | Molecular/Surveillance lab Sheikh Khalifa Medical City | Amirtharaj Francis et al |

| | | | | | | |
|---|---|---|---|---|---|---|
| EPI_ISL_582678 | UnitedArabEmirates/skmc-3806331/: | 2020-09-21 | Asia/United Arab Emirates/United Arab | Sheikh Khalifa Medical City | Molecular/Surveillance lab Sheikh Khalifa Medical City | Amirtharaj Francis et al |
| EPI_ISL_582690 | UnitedArabEmirates/skmc-3945800/: | 2020-09-25 | Asia/United Arab Emirates/United Arab | Sheikh Khalifa Medical City | Molecular/Surveillance lab Sheikh Khalifa Medical City | Amirtharaj Francis et al |
| EPI_ISL_582696 | Spain/PV-IBV-98005180/2020 | 2020-03-23 | Europe/Spain/Basque Country/Derio | Servicio de Microbiolog√≠a, Hospital Universitario Cruces, Bilbao | SeqCOVID-SPAIN consortium/IBV(CSIC) | Maitane Aranzamendi Zaldumbide et al |
| EPI_ISL_582700 | Spain/PV-IBV-98005184/2020 | 2020-03-23 | Europe/Spain/Basque Country/Santurtzi | Servicio de Microbiolog√≠a, Hospital Universitario Cruces, Bilbao | SeqCOVID-SPAIN consortium/IBV(CSIC) | Maitane Aranzamendi Zaldumbide et al |
| EPI_ISL_582721 | Spain/PV-IBV-98005209/2020 | 2020-03-23 | Europe/Spain/Basque Country/Baracald | Servicio de Microbiolog√≠a, Hospital Universitario Cruces, Bilbao | SeqCOVID-SPAIN consortium/IBV(CSIC) | Maitane Aranzamendi Zaldumbide et al |
| EPI_ISL_582725 | Spain/PV-IBV-98005215/2020 | 2020-03-23 | Europe/Spain/Basque Country/Baracald | Servicio de Microbiolog√≠a, Hospital Universitario Cruces, Bilbao | SeqCOVID-SPAIN consortium/IBV(CSIC) | Maitane Aranzamendi Zaldumbide et al |
| EPI_ISL_582738 | Spain/PV-IBV-98005234/2020 | 2020-03-24 | Europe/Spain/Basque Country/Baracald | Servicio de Microbiolog√≠a, Hospital Universitario Cruces, Bilbao | SeqCOVID-SPAIN consortium/IBV(CSIC) | Maitane Aranzamendi Zaldumbide et al |
| EPI_ISL_582739 | Spain/PV-IBV-98005235/2020 | 2020-03-24 | Europe/Spain/Basque Country/Baracald | Servicio de Microbiolog√≠a, Hospital Universitario Cruces, Bilbao | SeqCOVID-SPAIN consortium/IBV(CSIC) | Maitane Aranzamendi Zaldumbide et al |
| EPI_ISL_582749 | Spain/PV-IBV-98005249/2020 | 2020-03-28 | Europe/Spain/Basque Country/Baracald | Servicio de Microbiolog√≠a, Hospital Universitario Cruces, Bilbao | SeqCOVID-SPAIN consortium/IBV(CSIC) | Maitane Aranzamendi Zaldumbide et al |
| EPI_ISL_582758 | Spain/PV-IBV-98005261/2020 | 2020-03-26 | Europe/Spain/Basque Country/Baracald | Servicio de Microbiolog√≠a, Hospital Universitario Cruces, Bilbao | SeqCOVID-SPAIN consortium/IBV(CSIC) | Maitane Aranzamendi Zaldumbide et al |
| EPI_ISL_582760 | Spain/PV-IBV-98005263/2020 | 2020-03-28 | Europe/Spain/Basque Country/Baracald | Servicio de Microbiolog√≠a, Hospital Universitario Cruces, Bilbao | SeqCOVID-SPAIN consortium/IBV(CSIC) | Maitane Aranzamendi Zaldumbide et al |
| EPI_ISL_582761 | Spain/PV-IBV-98005268/2020 | 2020-03-27 | Europe/Spain/Basque Country/Baracald | Servicio de Microbiolog√≠a, Hospital Universitario Cruces, Bilbao | SeqCOVID-SPAIN consortium/IBV(CSIC) | Maitane Aranzamendi Zaldumbide et al |
| EPI_ISL_582771 | Sweden/20-08985/2020 | 2020-09-16 | Europe/Sweden/Ostergotland/ | Klinisk mikrobiologi Linkoping | The Public Health Agency of Sweden | Anna-Malin Linde et al |

| | | | | | | |
|---|---|---|---|---|---|---|
| EPI_ISL_582778 | Sweden/20-21500/2020 | 2020-06-04 | Europe/Sweden/Uppsala/ | Uppsala klinisk mikrobiologi | The Public Health Agency of Sweden | Anna-Malin Linde et al |
| EPI_ISL_582785 | Sweden/20-22107/2020 | 2020-06-01 | Europe/Sweden/Orebro/ | Orebro klinisk mikrobiologi | The Public Health Agency of Sweden | Anna-Malin Linde et al |
| EPI_ISL_582789 | Sweden/20-22115/2020 | 2020-06-04 | Europe/Sweden/Orebro/ | Orebro klinisk mikrobiologi | The Public Health Agency of Sweden | Anna-Malin Linde et al |
| EPI_ISL_582813 | Spain/MD-IBV-99007325/2020 | 2020-08-17 | Europe/Spain/Madrid/Madrid | Hospital General Universitario Gregorio Marav±√≥n | SeqCOVID-SPAIN consortium/IBV(CSIC) | Dar√≠o Garc√≠a de Viedma et al |
| EPI_ISL_582814 | Spain/MD-IBV-99007326/2020 | 2020-08-17 | Europe/Spain/Madrid/Madrid | Hospital General Universitario Gregorio Marav±√≥n | SeqCOVID-SPAIN consortium/IBV(CSIC) | Dar√≠o Garc√≠a de Viedma et al |
| EPI_ISL_582818 | Spain/MD-IBV-99007331/2020 | 2020-08-20 | Europe/Spain/Madrid/Madrid | Hospital General Universitario Gregorio Marav±√≥n | SeqCOVID-SPAIN consortium/IBV(CSIC) | Dar√≠o Garc√≠a de Viedma et al |
| EPI_ISL_582819 | Spain/MD-IBV-99007332/2020 | 2020-08-19 | Europe/Spain/Madrid/Madrid | Hospital General Universitario Gregorio Marav±√≥n | SeqCOVID-SPAIN consortium/IBV(CSIC) | Dar√≠o Garc√≠a de Viedma et al |
| EPI_ISL_582821 | Spain/MD-IBV-99007335/2020 | 2020-08-22 | Europe/Spain/Madrid/Madrid | Hospital General Universitario Gregorio Marav±√≥n | SeqCOVID-SPAIN consortium/IBV(CSIC) | Dar√≠o Garc√≠a de Viedma et al |
| EPI_ISL_582822 | Spain/MD-IBV-99007336/2020 | 2020-08-24 | Europe/Spain/Madrid/Madrid | Hospital General Universitario Gregorio Marav±√≥n | SeqCOVID-SPAIN consortium/IBV(CSIC) | Dar√≠o Garc√≠a de Viedma et al |
| EPI_ISL_582823 | Spain/MD-IBV-99007337/2020 | 2020-08-25 | Europe/Spain/Madrid/Madrid | Hospital General Universitario Gregorio Marav±√≥n | SeqCOVID-SPAIN consortium/IBV(CSIC) | Dar√≠o Garc√≠a de Viedma et al |
| EPI_ISL_582826 | Spain/MD-IBV-99007342/2020 | 2020-08-25 | Europe/Spain/Madrid/Madrid | Hospital General Universitario Gregorio Marav±√≥n | SeqCOVID-SPAIN consortium/IBV(CSIC) | Dar√≠o Garc√≠a de Viedma et al |
| EPI_ISL_582827 | Spain/MD-IBV-99007343/2020 | 2020-08-27 | Europe/Spain/Madrid/Madrid | Hospital General Universitario Gregorio Marav±√≥n | SeqCOVID-SPAIN consortium/IBV(CSIC) | Dar√≠o Garc√≠a de Viedma et al |
| EPI_ISL_582828 | Spain/MD-IBV-99007344/2020 | 2020-08-27 | Europe/Spain/Madrid/Madrid | Hospital General Universitario Gregorio Marav±√≥n | SeqCOVID-SPAIN consortium/IBV(CSIC) | Dar√≠o Garc√≠a de Viedma et al |
| EPI_ISL_582830 | Spain/MD-IBV-99007346/2020 | 2020-08-28 | Europe/Spain/Madrid/Madrid | Hospital General Universitario Gregorio Marav±√≥n | SeqCOVID-SPAIN consortium/IBV(CSIC) | Dar√≠o Garc√≠a de Viedma et al |

| | | | | | | |
|---|---|---|---|---|---|---|
| EPI_ISL_582832 | Spain/MD-IBV-99007348/2020 | 2020-09-02 | Europe/Spain/Madrid/Madrid | Hospital General Universitario Gregorio Marav√±√≥n | SeqCOVID-SPAIN consortium/IBV(CSIC) | Dar√≠o Garc√≠a de Viedma et al |
| EPI_ISL_582835 | Sweden/20-52115/2020 | 2020-06-22 | Europe/Sweden/Vasternorrland/ | Klinisk mikrobiologi Vasternorrland | The Public Health Agency of Sweden | Anna-Malin Linde et al |
| EPI_ISL_582838 | Sweden/20-52185/2020 | 2020-06-28 | Europe/Sweden/Vasternorrland/ | Klinisk mikrobiologi Vasternorrland | The Public Health Agency of Sweden | Anna-Malin Linde et al |
| EPI_ISL_582843 | Spain/MD-IBV-99007733/2020 | 2020-08-11 | Europe/Spain/Madrid/Madrid | Hospital General Universitario Gregorio Marav√±√≥n | SeqCOVID-SPAIN consortium/IBV(CSIC) | Dar√≠o Garc√≠a de Viedma et al |
| EPI_ISL_582844 | Spain/MD-IBV-99007734/2020 | 2020-08-22 | Europe/Spain/Madrid/Madrid | Hospital General Universitario Gregorio Marav√±√≥n | SeqCOVID-SPAIN consortium/IBV(CSIC) | Dar√≠o Garc√≠a de Viedma et al |
| EPI_ISL_582857 | USA/CA-CZB-2750/2020 | 2020-06-27 | North America/USA/California/Santa Cl | County of Santa Clara Public Health Department | Chan-Zuckerberg Biohub | CZB Cliahub Consortium et al |
| EPI_ISL_582858 | USA/CA-CZB-2751/2020 | 2020-06-27 | North America/USA/California/Santa Cl | County of Santa Clara Public Health Department | Chan-Zuckerberg Biohub | CZB Cliahub Consortium et al |
| EPI_ISL_582867 | USA/CA-CZB-2763/2020 | 2020-06-27 | North America/USA/California/Santa Cl | County of Santa Clara Public Health Department | Chan-Zuckerberg Biohub | CZB Cliahub Consortium et al |
| EPI_ISL_582870 | USA/CA-CZB-2771/2020 | 2020-06-27 | North America/USA/California/Santa Cl | County of Santa Clara Public Health Department | Chan-Zuckerberg Biohub | CZB Cliahub Consortium et al |
| EPI_ISL_582871 | USA/CA-CZB-2794/2020 | 2020-07-01 | North America/USA/California/Santa Cl | County of Santa Clara Public Health Department | Chan-Zuckerberg Biohub | CZB Cliahub Consortium et al |
| EPI_ISL_582873 | USA/CA-CZB-2807/2020 | 2020-07-06 | North America/USA/California/Santa Cl | County of Santa Clara Public Health Department | Chan-Zuckerberg Biohub | CZB Cliahub Consortium et al |
| EPI_ISL_582879 | USA/CA-CZB-2829/2020 | 2020-07-06 | North America/USA/California/Santa Cl | County of Santa Clara Public Health Department | Chan-Zuckerberg Biohub | CZB Cliahub Consortium et al |
| EPI_ISL_582880 | USA/CA-CZB-6802/2020 | 2020-07-14 | North America/USA/California/Santa Cl | County of Santa Clara Public Health Department | Chan-Zuckerberg Biohub | CZB Cliahub Consortium et al |
| EPI_ISL_582881 | USA/CA-CZB-6804/2020 | 2020-07-14 | North America/USA/California/Santa Cl | County of Santa Clara Public Health Department | Chan-Zuckerberg Biohub | CZB Cliahub Consortium et al |
| EPI_ISL_582882 | USA/CA-CZB-6805/2020 | 2020-07-14 | North America/USA/California/Santa Cl | County of Santa Clara Public Health Department | Chan-Zuckerberg Biohub | CZB Cliahub Consortium et al |
| EPI_ISL_582883 | USA/CA-CZB-6807/2020 | 2020-07-14 | North America/USA/California/Santa Cl | County of Santa Clara Public Health Department | Chan-Zuckerberg Biohub | CZB Cliahub Consortium et al |
| EPI_ISL_582890 | USA/CA-CZB-6932/2020 | 2020-08-22 | North America/USA/California/Santa Cl | County of Santa Clara Public Health Department | Chan-Zuckerberg Biohub | CZB Cliahub Consortium et al |
| EPI_ISL_582891 | USA/CA-CZB-6933/2020 | 2020-08-25 | North America/USA/California/Santa Cl | County of Santa Clara Public Health Department | Chan-Zuckerberg Biohub | CZB Cliahub Consortium et al |

| | | | | | | |
|---|---|---|---|---|---|---|
| EPI_ISL_582894 | USA/CA-CZB-4714/2020 | 2020-08-27 | North America/USA/California/Santa Cl | County of Santa Clara Public Health Department | Chan-Zuckerberg Biohub | CZB Cliahub Consortium et al |
| EPI_ISL_582895 | USA/CA-CZB-4730/2020 | 2020-08-26 | North America/USA/California/Santa Cl | County of Santa Clara Public Health Department | Chan-Zuckerberg Biohub | CZB Cliahub Consortium et al |
| EPI_ISL_582897 | USA/CA-CZB-4812/2020 | 2020-09-01 | North America/USA/California/Santa Cl | County of Santa Clara Public Health Department | Chan-Zuckerberg Biohub | CZB Cliahub Consortium et al |
| EPI_ISL_582898 | USA/CA-CZB-4860/2020 | 2020-09-01 | North America/USA/California/Santa Cl | County of Santa Clara Public Health Department | Chan-Zuckerberg Biohub | CZB Cliahub Consortium et al |
| EPI_ISL_582901 | USA/CA-CZB-6966/2020 | 2020-09-02 | North America/USA/California/Santa Cl | County of Santa Clara Public Health Department | Chan-Zuckerberg Biohub | CZB Cliahub Consortium et al |
| EPI_ISL_582905 | USA/CA-CZB-6973/2020 | 2020-09-02 | North America/USA/California/Santa Cl | County of Santa Clara Public Health Department | Chan-Zuckerberg Biohub | CZB Cliahub Consortium et al |
| EPI_ISL_582907 | USA/CA-CZB-6975/2020 | 2020-09-02 | North America/USA/California/Santa Cl | County of Santa Clara Public Health Department | Chan-Zuckerberg Biohub | CZB Cliahub Consortium et al |
| EPI_ISL_582914 | USA/CA-CZB-6987/2020 | 2020-09-01 | North America/USA/California/Santa Cl | County of Santa Clara Public Health Department | Chan-Zuckerberg Biohub | CZB Cliahub Consortium et al |
| EPI_ISL_582927 | USA/CA-CZB-7001/2020 | 2020-09-04 | North America/USA/California/Santa Cl | County of Santa Clara Public Health Department | Chan-Zuckerberg Biohub | CZB Cliahub Consortium et al |
| EPI_ISL_582929 | USA/CA-CZB-7145/2020 | 2020-08-31 | North America/USA/California/Santa Cl | County of Santa Clara Public Health Department | Chan-Zuckerberg Biohub | CZB Cliahub Consortium et al |
| EPI_ISL_582933 | USA/CA-CZB-7149/2020 | 2020-09-09 | North America/USA/California/Santa Cl | County of Santa Clara Public Health Department | Chan-Zuckerberg Biohub | CZB Cliahub Consortium et al |
| EPI_ISL_582934 | USA/CA-CZB-7152/2020 | 2020-09-09 | North America/USA/California/Santa Cl | County of Santa Clara Public Health Department | Chan-Zuckerberg Biohub | CZB Cliahub Consortium et al |
| EPI_ISL_582937 | USA/CA-CZB-7199/2020 | 2020-09-06 | North America/USA/California/Santa Cl | County of Santa Clara Public Health Department | Chan-Zuckerberg Biohub | CZB Cliahub Consortium et al |
| EPI_ISL_582939 | USA/CA-CZB-7203/2020 | 2020-08-25 | North America/USA/California/Santa Cl | County of Santa Clara Public Health Department | Chan-Zuckerberg Biohub | CZB Cliahub Consortium et al |
| EPI_ISL_582941 | USA/CA-CZB-7205/2020 | 2020-08-04 | North America/USA/California/Santa Cl | County of Santa Clara Public Health Department | Chan-Zuckerberg Biohub | CZB Cliahub Consortium et al |
| EPI_ISL_582943 | USA/CA-CZB-10967/2020 | 2020-09-16 | North America/USA/California/Santa Cl | County of Santa Clara Public Health Department | Chan-Zuckerberg Biohub | CZB Cliahub Consortium et al |
| EPI_ISL_582944 | USA/CA-CZB-10968/2020 | 2020-09-16 | North America/USA/California/Santa Cl | County of Santa Clara Public Health Department | Chan-Zuckerberg Biohub | CZB Cliahub Consortium et al |
| EPI_ISL_582947 | USA/CA-CZB-10978/2020 | 2020-09-17 | North America/USA/California/Santa Cl | County of Santa Clara Public Health Department | Chan-Zuckerberg Biohub | CZB Cliahub Consortium et al |
| EPI_ISL_582950 | USA/CA-CZB-11008/2020 | 2020-08-20 | North America/USA/California/Santa Cl | County of Santa Clara Public Health Department | Chan-Zuckerberg Biohub | CZB Cliahub Consortium et al |
| EPI_ISL_582953 | USA/CA-CZB-11011/2020 | 2020-09-01 | North America/USA/California/Santa Cl | County of Santa Clara Public Health Department | Chan-Zuckerberg Biohub | CZB Cliahub Consortium et al |

| | | | | | | |
|---|---|---|---|---|---|---|
| EPI_ISL_582955 | USA/CA-CZB-11015/2020 | 2020-08-20 | North America/USA/California/Santa Cl | County of Santa Clara Public Health Department | Chan-Zuckerberg Biohub | CZB Cliahub Consortium et al |
| EPI_ISL_582959 | USA/CA-CZB-5834/2020 | 2020-07-16 | North America/USA/California/San Luis | San Luis Obispo Public Health Department | Chan-Zuckerberg Biohub | CZB Cliahub Consortium et al |
| EPI_ISL_582962 | USA/CA-CZB-5780/2020 | 2020-07-12 | North America/USA/California/San Luis | San Luis Obispo Public Health Department | Chan-Zuckerberg Biohub | CZB Cliahub Consortium et al |
| EPI_ISL_582963 | USA/CA-CZB-5769/2020 | 2020-07-12 | North America/USA/California/San Luis | San Luis Obispo Public Health Department | Chan-Zuckerberg Biohub | CZB Cliahub Consortium et al |
| EPI_ISL_582971 | USA/CA-CZB-5698/2020 | 2020-07-23 | North America/USA/California/San Luis | San Luis Obispo Public Health Department | Chan-Zuckerberg Biohub | CZB Cliahub Consortium et al |
| EPI_ISL_582972 | USA/CA-CZB-5682/2020 | 2020-07-23 | North America/USA/California/San Luis | San Luis Obispo Public Health Department | Chan-Zuckerberg Biohub | CZB Cliahub Consortium et al |
| EPI_ISL_582975 | USA/CA-CZB-6711/2020 | 2020-08-17 | North America/USA/California/Orange ( | Orange County Public Health Lab | Chan-Zuckerberg Biohub | CZB Cliahub Consortium et al |
| EPI_ISL_582976 | USA/CA-CZB-6715/2020 | 2020-08-18 | North America/USA/California/Orange ( | Orange County Public Health Lab | Chan-Zuckerberg Biohub | CZB Cliahub Consortium et al |
| EPI_ISL_582977 | USA/CA-CZB-6716/2020 | 2020-08-17 | North America/USA/California/Orange ( | Orange County Public Health Lab | Chan-Zuckerberg Biohub | CZB Cliahub Consortium et al |
| EPI_ISL_582980 | USA/CA-CZB-6719/2020 | 2020-08-18 | North America/USA/California/Orange ( | Orange County Public Health Lab | Chan-Zuckerberg Biohub | CZB Cliahub Consortium et al |
| EPI_ISL_582986 | USA/CA-CZB-6727/2020 | 2020-08-19 | North America/USA/California/Orange ( | Orange County Public Health Lab | Chan-Zuckerberg Biohub | CZB Cliahub Consortium et al |
| EPI_ISL_582989 | USA/CA-CZB-6731/2020 | 2020-08-19 | North America/USA/California/Orange ( | Orange County Public Health Lab | Chan-Zuckerberg Biohub | CZB Cliahub Consortium et al |
| EPI_ISL_582992 | USA/CA-CZB-6734/2020 | 2020-08-19 | North America/USA/California/Orange ( | Orange County Public Health Lab | Chan-Zuckerberg Biohub | CZB Cliahub Consortium et al |
| EPI_ISL_582995 | USA/CA-CZB-6739/2020 | 2020-08-19 | North America/USA/California/Orange ( | Orange County Public Health Lab | Chan-Zuckerberg Biohub | CZB Cliahub Consortium et al |
| EPI_ISL_582998 | USA/CA-CZB-6742/2020 | 2020-08-20 | North America/USA/California/Orange ( | Orange County Public Health Lab | Chan-Zuckerberg Biohub | CZB Cliahub Consortium et al |
| EPI_ISL_582999 | USA/CA-CZB-6745/2020 | 2020-08-21 | North America/USA/California/Orange ( | Orange County Public Health Lab | Chan-Zuckerberg Biohub | CZB Cliahub Consortium et al |
| EPI_ISL_583002 | USA/CA-CZB-216/2020 | 2020-08-26 | North America/USA/California/Orange ( | Orange County Public Health Lab | Chan-Zuckerberg Biohub | CZB Cliahub Consortium et al |
| EPI_ISL_583008 | USA/CA-CZB-7128/2020 | 2020-08-31 | North America/USA/California/Orange ( | Orange County Public Health Lab | Chan-Zuckerberg Biohub | CZB Cliahub Consortium et al |
| EPI_ISL_583015 | USA/CA-CZB-7140/2020 | 2020-09-02 | North America/USA/California/Orange ( | Orange County Public Health Lab | Chan-Zuckerberg Biohub | CZB Cliahub Consortium et al |
| EPI_ISL_583016 | USA/CA-CZB-10285/2020 | 2020-09-04 | North America/USA/California/Orange ( | Orange County Public Health Lab | Chan-Zuckerberg Biohub | CZB Cliahub Consortium et al |

| | | | | | | |
|---|---|---|---|---|---|---|
| EPI_ISL_583024 | USA/CA-CZB-10300/2020 | 2020-09-09 | North America/USA/California/Orange ( | Orange County Public Health Lab | Chan-Zuckerberg Biohub | CZB Cliahub Consortium et al |
| EPI_ISL_583029 | USA/CA-CZB-10311/2020 | 2020-09-11 | North America/USA/California/Orange ( | Orange County Public Health Lab | Chan-Zuckerberg Biohub | CZB Cliahub Consortium et al |
| EPI_ISL_583035 | USA/CA-CZB-10951/2020 | 2020-09-15 | North America/USA/California/Orange ( | Orange County Public Health Lab | Chan-Zuckerberg Biohub | CZB Cliahub Consortium et al |
| EPI_ISL_583060 | USA/CA-CZB-10802/2020 | 2020-07-01 | North America/USA/California/Humbold | Humboldt County Public Health Laboratory | Chan-Zuckerberg Biohub | CZB Cliahub Consortium et al |
| EPI_ISL_583064 | USA/CA-CZB-10811/2020 | 2020-07-06 | North America/USA/California/Humbold | Humboldt County Public Health Laboratory | Chan-Zuckerberg Biohub | CZB Cliahub Consortium et al |
| EPI_ISL_583066 | USA/CA-CZB-10813/2020 | 2020-07-06 | North America/USA/California/Humbold | Humboldt County Public Health Laboratory | Chan-Zuckerberg Biohub | CZB Cliahub Consortium et al |
| EPI_ISL_583072 | USA/CA-CZB-10822/2020 | 2020-07-13 | North America/USA/California/Humbold | Humboldt County Public Health Laboratory | Chan-Zuckerberg Biohub | CZB Cliahub Consortium et al |
| EPI_ISL_583074 | USA/CA-CZB-10824/2020 | 2020-07-13 | North America/USA/California/Humbold | Humboldt County Public Health Laboratory | Chan-Zuckerberg Biohub | CZB Cliahub Consortium et al |
| EPI_ISL_583075 | USA/CA-CZB-10825/2020 | 2020-07-13 | North America/USA/California/Humbold | Humboldt County Public Health Laboratory | Chan-Zuckerberg Biohub | CZB Cliahub Consortium et al |
| EPI_ISL_583084 | USA/CA-CZB-10837/2020 | 2020-07-20 | North America/USA/California/Humbold | Humboldt County Public Health Laboratory | Chan-Zuckerberg Biohub | CZB Cliahub Consortium et al |
| EPI_ISL_583089 | USA/CA-CZB-10844/2020 | 2020-07-23 | North America/USA/California/Humbold | Humboldt County Public Health Laboratory | Chan-Zuckerberg Biohub | CZB Cliahub Consortium et al |
| EPI_ISL_583095 | USA/CA-CZB-10850/2020 | 2020-07-27 | North America/USA/California/Humbold | Humboldt County Public Health Laboratory | Chan-Zuckerberg Biohub | CZB Cliahub Consortium et al |
| EPI_ISL_583100 | USA/CA-CZB-10856/2020 | 2020-07-28 | North America/USA/California/Humbold | Humboldt County Public Health Laboratory | Chan-Zuckerberg Biohub | CZB Cliahub Consortium et al |
| EPI_ISL_583105 | USA/CA-CZB-10863/2020 | 2020-08-02 | North America/USA/California/Humbold | Humboldt County Public Health Laboratory | Chan-Zuckerberg Biohub | CZB Cliahub Consortium et al |
| EPI_ISL_583113 | USA/CA-CZB-10873/2020 | 2020-08-06 | North America/USA/California/Humbold | Humboldt County Public Health Laboratory | Chan-Zuckerberg Biohub | CZB Cliahub Consortium et al |
| EPI_ISL_583116 | USA/CA-CZB-10878/2020 | 2020-08-03 | North America/USA/California/Humbold | Humboldt County Public Health Laboratory | Chan-Zuckerberg Biohub | CZB Cliahub Consortium et al |
| EPI_ISL_583117 | USA/CA-CZB-10879/2020 | 2020-08-06 | North America/USA/California/Humbold | Humboldt County Public Health Laboratory | Chan-Zuckerberg Biohub | CZB Cliahub Consortium et al |
| EPI_ISL_583119 | USA/CA-CZB-10881/2020 | 2020-08-07 | North America/USA/California/Humbold | Humboldt County Public Health Laboratory | Chan-Zuckerberg Biohub | CZB Cliahub Consortium et al |
| EPI_ISL_583120 | USA/CA-CZB-10885/2020 | 2020-08-11 | North America/USA/California/Humbold | Humboldt County Public Health Laboratory | Chan-Zuckerberg Biohub | CZB Cliahub Consortium et al |
| EPI_ISL_583124 | USA/CA-CZB-10892/2020 | 2020-08-11 | North America/USA/California/Humbold | Humboldt County Public Health Laboratory | Chan-Zuckerberg Biohub | CZB Cliahub Consortium et al |

| | | | | | | |
|---|---|---|---|---|---|---|
| EPI_ISL_583127 | USA/CA-CZB-10897/2020 | 2020-08-17 | North America/USA/California/Humbold | Humboldt County Public Health Laboratory | Chan-Zuckerberg Biohub | CZB Cliahub Consortium et al |
| EPI_ISL_583129 | USA/CA-CZB-10899/2020 | 2020-08-18 | North America/USA/California/Humbold | Humboldt County Public Health Laboratory | Chan-Zuckerberg Biohub | CZB Cliahub Consortium et al |
| EPI_ISL_583136 | USA/CA-CZB-10907/2020 | 2020-08-25 | North America/USA/California/Humbold | Humboldt County Public Health Laboratory | Chan-Zuckerberg Biohub | CZB Cliahub Consortium et al |
| EPI_ISL_583143 | USA/CA-CZB-10914/2020 | 2020-08-27 | North America/USA/California/Humbold | Humboldt County Public Health Laboratory | Chan-Zuckerberg Biohub | CZB Cliahub Consortium et al |
| EPI_ISL_583144 | USA/CA-CZB-10915/2020 | 2020-08-27 | North America/USA/California/Humbold | Humboldt County Public Health Laboratory | Chan-Zuckerberg Biohub | CZB Cliahub Consortium et al |
| EPI_ISL_583146 | USA/CA-CZB-10917/2020 | 2020-08-27 | North America/USA/California/Humbold | Humboldt County Public Health Laboratory | Chan-Zuckerberg Biohub | CZB Cliahub Consortium et al |
| EPI_ISL_583147 | USA/CA-CZB-10918/2020 | 2020-08-28 | North America/USA/California/Humbold | Humboldt County Public Health Laboratory | Chan-Zuckerberg Biohub | CZB Cliahub Consortium et al |
| EPI_ISL_583150 | USA/CA-CZB-10921/2020 | 2020-08-28 | North America/USA/California/Humbold | Humboldt County Public Health Laboratory | Chan-Zuckerberg Biohub | CZB Cliahub Consortium et al |
| EPI_ISL_583165 | USA/CA-CZB-10936/2020 | 2020-09-04 | North America/USA/California/Humbold | Humboldt County Public Health Laboratory | Chan-Zuckerberg Biohub | CZB Cliahub Consortium et al |
| EPI_ISL_583166 | USA/CA-CZB-10937/2020 | 2020-09-04 | North America/USA/California/Humbold | Humboldt County Public Health Laboratory | Chan-Zuckerberg Biohub | CZB Cliahub Consortium et al |
| EPI_ISL_583168 | USA/CA-CZB-10939/2020 | 2020-09-04 | North America/USA/California/Humbold | Humboldt County Public Health Laboratory | Chan-Zuckerberg Biohub | CZB Cliahub Consortium et al |
| EPI_ISL_583170 | USA/CA-CZB-10941/2020 | 2020-09-08 | North America/USA/California/Humbold | Humboldt County Public Health Laboratory | Chan-Zuckerberg Biohub | CZB Cliahub Consortium et al |
| EPI_ISL_583174 | USA/CA-CZB-176/2020 | 2020-05-08 | North America/USA/California/San Bern | San Bernardino County Public Health Lab | Chan-Zuckerberg Biohub | CZB Cliahub Consortium et al |
| EPI_ISL_583190 | USA/CA-CZB-10526/2020 | 2020-09-07 | North America/USA/California/San Fran | San Francisco Public Health Laboratory | Chan-Zuckerberg Biohub | CZB Cliahub Consortium et al |
| EPI_ISL_583192 | USA/CA-CZB-10542/2020 | 2020-09-07 | North America/USA/California/San Fran | San Francisco Public Health Laboratory | Chan-Zuckerberg Biohub | CZB Cliahub Consortium et al |
| EPI_ISL_583193 | USA/CA-CZB-10558/2020 | 2020-09-07 | North America/USA/California/San Fran | San Francisco Public Health Laboratory | Chan-Zuckerberg Biohub | CZB Cliahub Consortium et al |
| EPI_ISL_583194 | USA/CA-CZB-10566/2020 | 2020-09-07 | North America/USA/California/San Fran | San Francisco Public Health Laboratory | Chan-Zuckerberg Biohub | CZB Cliahub Consortium et al |
| EPI_ISL_583195 | USA/CA-CZB-10582/2020 | 2020-09-07 | North America/USA/California/San Fran | San Francisco Public Health Laboratory | Chan-Zuckerberg Biohub | CZB Cliahub Consortium et al |
| EPI_ISL_583198 | USA/CA-CZB-7028/2020 | 2020-09-02 | North America/USA/California/Marin Co | Marin County Public Health Department | Chan-Zuckerberg Biohub | CZB Cliahub Consortium et al |
| EPI_ISL_583199 | USA/CA-CZB-193/2020 | 2020-08-26 | North America/USA/California/Marin Co | UCSF Clinical Microbiology Laboratory | Chan-Zuckerberg Biohub | CZB Cliahub Consortium et al |

| | | | | | | |
|---|---|---|---|---|---|---|
| EPI_ISL_583202 | USA/CA-CZB-169/2020 | 2020-08-31 | North America/USA/California/Marin Co | UCSF Clinical Microbiology Laboratory | Chan-Zuckerberg Biohub | CZB Cliahub Consortium et al |
| EPI_ISL_583203 | USA/CA-CZB-168/2020 | 2020-08-31 | North America/USA/California/Marin Co | UCSF Clinical Microbiology Laboratory | Chan-Zuckerberg Biohub | CZB Cliahub Consortium et al |
| EPI_ISL_583206 | USA/CA-CZB-163/2020 | 2020-09-01 | North America/USA/California/Marin Co | UCSF Clinical Microbiology Laboratory | Chan-Zuckerberg Biohub | CZB Cliahub Consortium et al |
| EPI_ISL_583207 | USA/CA-CZB-162/2020 | 2020-09-01 | North America/USA/California/Marin Co | UCSF Clinical Microbiology Laboratory | Chan-Zuckerberg Biohub | CZB Cliahub Consortium et al |
| EPI_ISL_583210 | USA/CA-CZB-5851/2020 | 2020-07-22 | North America/USA/California/Marin Co | UCSF Clinical Microbiology Laboratory | Chan-Zuckerberg Biohub | CZB Cliahub Consortium et al |
| EPI_ISL_583211 | USA/CA-CZB-5842/2020 | 2020-07-22 | North America/USA/California/Marin Co | UCSF Clinical Microbiology Laboratory | Chan-Zuckerberg Biohub | CZB Cliahub Consortium et al |
| EPI_ISL_583215 | USA/CA-CZB-5691/2020 | 2020-07-17 | North America/USA/California/Marin Co | UCSF Clinical Microbiology Laboratory | Chan-Zuckerberg Biohub | CZB Cliahub Consortium et al |
| EPI_ISL_583222 | USA/CA-CZB-5168/2020 | 2020-08-12 | North America/USA/California/Marin Co | UCSF Clinical Microbiology Laboratory | Chan-Zuckerberg Biohub | CZB Cliahub Consortium et al |
| EPI_ISL_583225 | USA/CA-CZB-7161/2020 | 2020-08-26 | North America/USA/California/Marin Co | UCSF Clinical Microbiology Laboratory | Chan-Zuckerberg Biohub | CZB Cliahub Consortium et al |
| EPI_ISL_583227 | USA/CA-CZB-7158/2020 | 2020-08-15 | North America/USA/California/Marin Co | UCSF Clinical Microbiology Laboratory | Chan-Zuckerberg Biohub | CZB Cliahub Consortium et al |
| EPI_ISL_583229 | USA/CA-CZB-10625/2020 | 2020-09-09 | North America/USA/California/Marin Co | UCSF Clinical Microbiology Laboratory | Chan-Zuckerberg Biohub | CZB Cliahub Consortium et al |
| EPI_ISL_583230 | USA/CA-CZB-10624/2020 | 2020-09-09 | North America/USA/California/Marin Co | UCSF Clinical Microbiology Laboratory | Chan-Zuckerberg Biohub | CZB Cliahub Consortium et al |
| EPI_ISL_583231 | USA/CA-CZB-10623/2020 | 2020-09-09 | North America/USA/California/Marin Co | UCSF Clinical Microbiology Laboratory | Chan-Zuckerberg Biohub | CZB Cliahub Consortium et al |
| EPI_ISL_583259 | USA/MI-UM-10035683340/2020 | 2020-08-27 | North America/USA/Michigan/ | University of Michigan Clinical Microbiology Laboratory | Lauring Lab, University of Michigan, Department of Microbiology and Immunology | Valesano et al |
| EPI_ISL_583261 | USA/MI-UM-10035638988/2020 | 2020-08-25 | North America/USA/Michigan/ | University of Michigan Clinical Microbiology Laboratory | Lauring Lab, University of Michigan, Department of Microbiology and Immunology | Valesano et al |

| | | | | | | |
|---|---|---|---|---|---|---|
| EPI_ISL_583264 | USA/MI-UM-10035555295/2020 | 2020-08-19 | North America/USA/Michigan/ | University of Michigan Clinical Microbiology Laboratory | Lauring Lab, University of Michigan, Department of Microbiology and Immunology | Valesano et al |
| EPI_ISL_583267 | USA/MI-UM-10035665912/2020 | 2020-08-26 | North America/USA/Michigan/ | University of Michigan Clinical Microbiology Laboratory | Lauring Lab, University of Michigan, Department of Microbiology and Immunology | Valesano et al |
| EPI_ISL_583269 | USA/MI-UM-10035702585/2020 | 2020-08-28 | North America/USA/Michigan/ | University of Michigan Clinical Microbiology Laboratory | Lauring Lab, University of Michigan, Department of Microbiology and Immunology | Valesano et al |
| EPI_ISL_583271 | USA/MI-UM-10035638276/2020 | 2020-08-25 | North America/USA/Michigan/ | University of Michigan Clinical Microbiology Laboratory | Lauring Lab, University of Michigan, Department of Microbiology and Immunology | Valesano et al |
| EPI_ISL_583273 | USA/MI-UM-10035701578/2020 | 2020-08-28 | North America/USA/Michigan/ | University of Michigan Clinical Microbiology Laboratory | Lauring Lab, University of Michigan, Department of Microbiology and Immunology | Valesano et al |
| EPI_ISL_583279 | USA/MI-UM-10035692627/2020 | 2020-08-28 | North America/USA/Michigan/ | University of Michigan Clinical Microbiology Laboratory | Lauring Lab, University of Michigan, Department of Microbiology and Immunology | Valesano et al |

| | | | | | | |
|---|---|---|---|---|---|---|
| EPI_ISL_583289 | USA/MI-UM-10035739693/2020 | 2020-08-31 | North America/USA/Michigan/ | University of Michigan Clinical Microbiology Laboratory | Lauring Lab, University of Michigan, Department of Microbiology and Immunology | Valesano et al |
| EPI_ISL_583299 | USA/MI-UM-10035733673/2020 | 2020-08-31 | North America/USA/Michigan/ | University of Michigan Clinical Microbiology Laboratory | Lauring Lab, University of Michigan, Department of Microbiology and Immunology | Valesano et al |
| EPI_ISL_583301 | USA/MI-UM-10035752975/2020 | 2020-09-01 | North America/USA/Michigan/ | University of Michigan Clinical Microbiology Laboratory | Lauring Lab, University of Michigan, Department of Microbiology and Immunology | Valesano et al |
| EPI_ISL_583305 | USA/MI-UM-10035711854/2020 | 2020-08-29 | North America/USA/Michigan/ | University of Michigan Clinical Microbiology Laboratory | Lauring Lab, University of Michigan, Department of Microbiology and Immunology | Valesano et al |
| EPI_ISL_583306 | USA/MI-UM-10035750305/2020 | 2020-09-01 | North America/USA/Michigan/ | University of Michigan Clinical Microbiology Laboratory | Lauring Lab, University of Michigan, Department of Microbiology and Immunology | Valesano et al |
| EPI_ISL_583307 | USA/MI-UM-10035747436/2020 | 2020-09-01 | North America/USA/Michigan/ | University of Michigan Clinical Microbiology Laboratory | Lauring Lab, University of Michigan, Department of Microbiology and Immunology | Valesano et al |

| | | | | | | |
|---|---|---|---|---|---|---|
| EPI_ISL_583317 | USA/MI-UM-10035803894/2020 | 2020-09-04 | North America/USA/Michigan/ | University of Michigan Clinical Microbiology Laboratory | Lauring Lab, University of Michigan, Department of Microbiology and Immunology | Valesano et al |
| EPI_ISL_583319 | USA/MI-UM-10035790102/2020 | 2020-09-03 | North America/USA/Michigan/ | University of Michigan Clinical Microbiology Laboratory | Lauring Lab, University of Michigan, Department of Microbiology and Immunology | Valesano et al |
| EPI_ISL_583322 | USA/MI-UM-10035824100/2020 | 2020-09-05 | North America/USA/Michigan/ | University of Michigan Clinical Microbiology Laboratory | Lauring Lab, University of Michigan, Department of Microbiology and Immunology | Valesano et al |
| EPI_ISL_583325 | USA/MI-UM-10035768201/2020 | 2020-09-02 | North America/USA/Michigan/ | University of Michigan Clinical Microbiology Laboratory | Lauring Lab, University of Michigan, Department of Microbiology and Immunology | Valesano et al |
| EPI_ISL_583327 | USA/MI-UM-10035806702/2020 | 2020-09-04 | North America/USA/Michigan/ | University of Michigan Clinical Microbiology Laboratory | Lauring Lab, University of Michigan, Department of Microbiology and Immunology | Valesano et al |
| EPI_ISL_583331 | USA/MI-UM-10035886189/2020 | 2020-09-10 | North America/USA/Michigan/ | University of Michigan Clinical Microbiology Laboratory | Lauring Lab, University of Michigan, Department of Microbiology and Immunology | Valesano et al |

| | | | | | | |
|---|---|---|---|---|---|---|
| EPI_ISL_583332 | USA/MI-UM-10035854187/2020 | 2020-09-08 | North America/USA/Michigan/ | University of Michigan Clinical Microbiology Laboratory | Lauring Lab, University of Michigan, Department of Microbiology and Immunology | Valesano et al |
| EPI_ISL_583334 | USA/MI-UM-10035868153/2020 | 2020-09-09 | North America/USA/Michigan/ | University of Michigan Clinical Microbiology Laboratory | Lauring Lab, University of Michigan, Department of Microbiology and Immunology | Valesano et al |
| EPI_ISL_583341 | USA/MI-UM-10035843632/2020 | 2020-09-08 | North America/USA/Michigan/ | University of Michigan Clinical Microbiology Laboratory | Lauring Lab, University of Michigan, Department of Microbiology and Immunology | Valesano et al |
| EPI_ISL_583346 | USA/MI-UM-10035872711/2020 | 2020-09-09 | North America/USA/Michigan/ | University of Michigan Clinical Microbiology Laboratory | Lauring Lab, University of Michigan, Department of Microbiology and Immunology | Valesano et al |
| EPI_ISL_583348 | USA/MI-UM-10035958075/2020 | 2020-09-15 | North America/USA/Michigan/ | University of Michigan Clinical Microbiology Laboratory | Lauring Lab, University of Michigan, Department of Microbiology and Immunology | Valesano et al |
| EPI_ISL_583351 | USA/MI-UM-10035956808/2020 | 2020-09-15 | North America/USA/Michigan/ | University of Michigan Clinical Microbiology Laboratory | Lauring Lab, University of Michigan, Department of Microbiology and Immunology | Valesano et al |

| | | | | | | |
|---|---|---|---|---|---|---|
| EPI_ISL_583359 | USA/MI-UM-10035987814/2020 | 2020-09-16 | North America/USA/Michigan/ | University of Michigan Clinical Microbiology Laboratory | Lauring Lab, University of Michigan, Department of Microbiology and Immunology | Valesano et al |
| EPI_ISL_583361 | USA/MI-UM-10035908382/2020 | 2020-09-11 | North America/USA/Michigan/ | University of Michigan Clinical Microbiology Laboratory | Lauring Lab, University of Michigan, Department of Microbiology and Immunology | Valesano et al |
| EPI_ISL_583363 | USA/MI-UM-10036001658/2020 | 2020-09-17 | North America/USA/Michigan/ | University of Michigan Clinical Microbiology Laboratory | Lauring Lab, University of Michigan, Department of Microbiology and Immunology | Valesano et al |
| EPI_ISL_583365 | USA/MI-UM-10035986324/2020 | 2020-09-16 | North America/USA/Michigan/ | University of Michigan Clinical Microbiology Laboratory | Lauring Lab, University of Michigan, Department of Microbiology and Immunology | Valesano et al |
| EPI_ISL_583369 | USA/MI-UM-10035963882/2020 | 2020-09-15 | North America/USA/Michigan/ | University of Michigan Clinical Microbiology Laboratory | Lauring Lab, University of Michigan, Department of Microbiology and Immunology | Valesano et al |
| EPI_ISL_583371 | USA/MI-UM-10036001045/2020 | 2020-09-17 | North America/USA/Michigan/ | University of Michigan Clinical Microbiology Laboratory | Lauring Lab, University of Michigan, Department of Microbiology and Immunology | Valesano et al |

| | | | | | | |
|---|---|---|---|---|---|---|
| EPI_ISL_583373 | USA/MI-UM-10035994708/2020 | 2020-09-17 | North America/USA/Michigan/ | University of Michigan Clinical Microbiology Laboratory | Lauring Lab, University of Michigan, Department of Microbiology and Immunology | Valesano et al |
| EPI_ISL_583376 | USA/MI-UM-10036033491/2020 | 2020-09-19 | North America/USA/Michigan/ | University of Michigan Clinical Microbiology Laboratory | Lauring Lab, University of Michigan, Department of Microbiology and Immunology | Valesano et al |
| EPI_ISL_583380 | USA/MI-UM-10036021679/2020 | 2020-09-18 | North America/USA/Michigan/ | University of Michigan Clinical Microbiology Laboratory | Lauring Lab, University of Michigan, Department of Microbiology and Immunology | Valesano et al |
| EPI_ISL_583381 | USA/MI-UM-10036024406/2020 | 2020-09-18 | North America/USA/Michigan/ | University of Michigan Clinical Microbiology Laboratory | Lauring Lab, University of Michigan, Department of Microbiology and Immunology | Valesano et al |
| EPI_ISL_583384 | USA/MI-UM-10036005973/2020 | 2020-09-17 | North America/USA/Michigan/ | University of Michigan Clinical Microbiology Laboratory | Lauring Lab, University of Michigan, Department of Microbiology and Immunology | Valesano et al |
| EPI_ISL_583389 | USA/MI-UM-10035945885/2020 | 2020-09-14 | North America/USA/Michigan/ | University of Michigan Clinical Microbiology Laboratory | Lauring Lab, University of Michigan, Department of Microbiology and Immunology | Valesano et al |

| | | | | | | |
|---|---|---|---|---|---|---|
| EPI_ISL_583392 | USA/MI-UM-10035907994/2020 | 2020-09-11 | North America/USA/Michigan/ | University of Michigan Clinical Microbiology Laboratory | Lauring Lab, University of Michigan, Department of Microbiology and Immunology | Valesano et al |
| EPI_ISL_583401 | USA/MI-UM-10035884804/2020 | 2020-09-10 | North America/USA/Michigan/ | University of Michigan Clinical Microbiology Laboratory | Lauring Lab, University of Michigan, Department of Microbiology and Immunology | Valesano et al |
| EPI_ISL_583408 | USA/MI-UM-10036079833/2020 | 2020-09-22 | North America/USA/Michigan/ | University of Michigan Clinical Microbiology Laboratory | Lauring Lab, University of Michigan, Department of Microbiology and Immunology | Valesano et al |
| EPI_ISL_583410 | USA/MI-UM-10036092704/2020 | 2020-09-23 | North America/USA/Michigan/ | University of Michigan Clinical Microbiology Laboratory | Lauring Lab, University of Michigan, Department of Microbiology and Immunology | Valesano et al |
| EPI_ISL_583412 | USA/MI-UM-10036095408/2020 | 2020-09-23 | North America/USA/Michigan/ | University of Michigan Clinical Microbiology Laboratory | Lauring Lab, University of Michigan, Department of Microbiology and Immunology | Valesano et al |
| EPI_ISL_583415 | USA/MI-UM-10036095605/2020 | 2020-09-23 | North America/USA/Michigan/ | University of Michigan Clinical Microbiology Laboratory | Lauring Lab, University of Michigan, Department of Microbiology and Immunology | Valesano et al |

| | | | | | | |
|---|---|---|---|---|---|---|
| EPI_ISL_583422 | USA/MI-UM-10036138856/2020 | 2020-09-25 | North America/USA/Michigan/ | University of Michigan Clinical Microbiology Laboratory | Lauring Lab, University of Michigan, Department of Microbiology and Immunology | Valesano et al |
| EPI_ISL_583423 | USA/MI-UM-10036138458/2020 | 2020-09-25 | North America/USA/Michigan/ | University of Michigan Clinical Microbiology Laboratory | Lauring Lab, University of Michigan, Department of Microbiology and Immunology | Valesano et al |
| EPI_ISL_583427 | USA/NY-MSK-1A/2020 | 2020-04-14 | North America/USA/New York/ | Memorial Sloan Kettering Cancer Center | van Bakel Laboratory, Genetics and Genomics Sciences, Icahn School of Medicine at Mount Sinai | Teresa Aydillo et al |
| EPI_ISL_583439 | USA/NY-MSK-4A/2020 | 2020-04-13 | North America/USA/New York/ | Memorial Sloan Kettering Cancer Center | van Bakel Laboratory, Genetics and Genomics Sciences, Icahn School of Medicine at Mount Sinai | Teresa Aydillo et al |
| EPI_ISL_583456 | USA/NY-MSK-8A/2020 | 2020-04-20 | North America/USA/New York/ | Memorial Sloan Kettering Cancer Center | van Bakel Laboratory, Genetics and Genomics Sciences, Icahn School of Medicine at Mount Sinai | Teresa Aydillo et al |

| | | | | | | |
|---|---|---|---|---|---|---|
| EPI_ISL_583478 | USA/NY-MSK-17B/2020 | 2020-05-02 | North America/USA/New York/ | Memorial Sloan Kettering Cancer Center | van Bakel Laboratory, Genetics and Genomics Sciences, Icahn School of Medicine at Mount Sinai | Teresa Aydillo et al |
| EPI_ISL_583481 | Slovakia/UKBA-313/2020 | 2020-09-10 | Europe/Slovakia/Tvrdosovce/ | Institute of Virology, Biomedical Research Center of the Slovak Academy of Sciences, Bratislava | Faculty of Natural Sciences, Comenius University, Bratislava | Viktv≥ria Hodorovv° et al |
| EPI_ISL_583483 | Slovakia/UKBA-315/2020 | 2020-09-10 | Europe/Slovakia/Kamenicna/ | Institute of Virology, Biomedical Research Center of the Slovak Academy of Sciences, Bratislava | Faculty of Natural Sciences, Comenius University, Bratislava | Viktv≥ria Hodorovv° et al |
| EPI_ISL_583484 | Slovakia/UKBA-316/2020 | 2020-09-10 | Europe/Slovakia/Bratislava/ | Institute of Virology, Biomedical Research Center of the Slovak Academy of Sciences, Bratislava | Faculty of Natural Sciences, Comenius University, Bratislava | Kristv≠na Bor≈°ovv° et al |
| EPI_ISL_583485 | Slovakia/UKBA-317/2020 | 2020-09-16 | Europe/Slovakia/Surany/ | Institute of Virology, Biomedical Research Center of the Slovak Academy of Sciences, Bratislava | Faculty of Natural Sciences, Comenius University, Bratislava | Viktv≥ria Hodorovv° et al |
| EPI_ISL_583486 | Slovakia/UKBA-318/2020 | 2020-09-16 | Europe/Slovakia/Surany/ | Institute of Virology, Biomedical Research Center of the Slovak Academy of Sciences, Bratislava | Faculty of Natural Sciences, Comenius University, Bratislava | Kristv≠na Bor≈°ovv° et al |
| EPI_ISL_583488 | Slovakia/UKBA-321/2020 | 2020-09-16 | Europe/Slovakia/Bratislava/ | Institute of Virology, Biomedical Research Center of the Slovak Academy of Sciences, Bratislava | Faculty of Natural Sciences, Comenius University, Bratislava | Bro≈àa Brejovv° et al |

| | | | | | | |
|---|---|---|---|---|---|---|
| EPI_ISL_583489 | Slovakia/UKBA-322/2020 | 2020-09-16 | Europe/Slovakia/Levice/ | Institute of Virology, Biomedical Research Center of the Slovak Academy of Sciences, Bratislava | Faculty of Natural Sciences, Comenius University, Bratislava | Viktv≥ria Hodorovv° et al |
| EPI_ISL_583495 | Brazil/SP-345/2020 | 2020-06-13 | South America/Brazil/Sv£o Paulo/Guaru | Serviv߸o de Verificav߸v£o de v̀bitos SVO Guaruulhos | Instituto Adolfo Lutz, Interdiciplinary Procedures Center, Strategic Laboratory | Claudio Tavares Sacchi et al |
| EPI_ISL_583496 | Brazil/SP-353/2020 | 2020-06-20 | South America/Brazil/Sv£o Paulo/Jandir | UPA Jandira | Instituto Adolfo Lutz, Interdiciplinary Procedures Center, Strategic Laboratory | Claudio Tavares Sacchi et al |
| EPI_ISL_583497 | Brazil/SP-359/2020 | 2020-06-17 | South America/Brazil/Sv£o Paulo/Campi | Complexo Hospitalar Ouro Verde de Campinas | Instituto Adolfo Lutz, Interdiciplinary Procedures Center, Strategic Laboratory | Claudio Tavares Sacchi et al |
| EPI_ISL_583498 | Brazil/SP-360/2020 | 2020-06-22 | South America/Brazil/Sv£o Paulo/Ameri | Hospital Municipal Dr. Waldemar Tebaldi | Instituto Adolfo Lutz, Interdiciplinary Procedures Center, Strategic Laboratory | Claudio Tavares Sacchi et al |
| EPI_ISL_583499 | Brazil/SP-368/2020 | 2020-06-24 | South America/Brazil/Sv£o Paulo/Campi | Distrito Sanitario Sul Campinas | Instituto Adolfo Lutz, Interdiciplinary Procedures Center, Strategic Laboratory | Claudio Tavares Sacchi et al |
| EPI_ISL_583500 | Brazil/SP-370/2020 | 2020-06-29 | South America/Brazil/Sv£o Paulo/Presid | Centro de Saude I Tacito Leite de Carvalho e Silva | Instituto Adolfo Lutz, Interdiciplinary Procedures Center, Strategic Laboratory | Claudio Tavares Sacchi et al |

| | | | | | | |
|---|---|---|---|---|---|---|
| EPI_ISL_583501 | Brazil/SP-407/2020 | 2020-07-08 | South America/Brazil/S√£o Paulo/S√£o | Hospital Estadual de CampanhaCOVID 19 Barradas | Instituto Adolfo Lutz, Interdiciplinary Procedures Center, Strategic Laboratory | Claudio Tavares Sacchi et al |
| EPI_ISL_583502 | Brazil/SP-393/2020 | 2020-06-21 | South America/Brazil/S√£o Paulo/Guaru | Serv de Vig Sanitaria Epidemio e CTRL de Zoonoses Guaruja | Instituto Adolfo Lutz, Interdiciplinary Procedures Center, Strategic Laboratory | Claudio Tavares Sacchi et al |
| EPI_ISL_583503 | Brazil/SP-394/2020 | 2020-06-22 | South America/Brazil/S√£o Paulo/Caieir | CTA Centro de Testagem e Aconselhamento | Instituto Adolfo Lutz, Interdiciplinary Procedures Center, Strategic Laboratory | Claudio Tavares Sacchi et al |
| EPI_ISL_583504 | Brazil/SP-395/2020 | 2020-06-20 | South America/Brazil/S√£o Paulo/Carag | Casa de Saude Stella Maris | Instituto Adolfo Lutz, Interdiciplinary Procedures Center, Strategic Laboratory | Claudio Tavares Sacchi et al |
| EPI_ISL_583505 | Brazil/SP-397/2020 | 2020-06-26 | South America/Brazil/S√£o Paulo/Carag | Casa de Saude Stella Maris | Instituto Adolfo Lutz, Interdiciplinary Procedures Center, Strategic Laboratory | Claudio Tavares Sacchi et al |
| EPI_ISL_583507 | USA/MI-MDHHS-SC22050/2020 | 2020-09-21 | North America/USA/Michigan/ | Michigan Department of Health and Human Services, Bureau of Laboratories | Michigan Department of Health and Human Services, Bureau of Laboratories | Blankenship HM et al |

| | | | | | | |
|---|---|---|---|---|---|---|
| EPI_ISL_583511 | USA/MI-MDHHS-SC22054/2020 | 2020-09-24 | North America/USA/Michigan/ | Michigan Department of Health and Human Services, Bureau of Laboratories | Michigan Department of Health and Human Services, Bureau of Laboratories | Blankenship HM et al |
| EPI_ISL_583515 | USA/MI-MDHHS-SC22058/2020 | 2020-09-28 | North America/USA/Michigan/ | Michigan Department of Health and Human Services, Bureau of Laboratories | Michigan Department of Health and Human Services, Bureau of Laboratories | Blankenship HM et al |
| EPI_ISL_583516 | USA/MI-MDHHS-SC22059/2020 | 2020-09-28 | North America/USA/Michigan/ | Michigan Department of Health and Human Services, Bureau of Laboratories | Michigan Department of Health and Human Services, Bureau of Laboratories | Blankenship HM et al |
| EPI_ISL_583517 | USA/MI-MDHHS-SC22060/2020 | 2020-09-28 | North America/USA/Michigan/ | Michigan Department of Health and Human Services, Bureau of Laboratories | Michigan Department of Health and Human Services, Bureau of Laboratories | Blankenship HM et al |
| EPI_ISL_583526 | USA/MI-MDHHS-SC22069/2020 | 2020-09-29 | North America/USA/Michigan/ | Michigan Department of Health and Human Services, Bureau of Laboratories | Michigan Department of Health and Human Services, Bureau of Laboratories | Blankenship HM et al |
| EPI_ISL_583527 | USA/MI-MDHHS-SC22070/2020 | 2020-09-29 | North America/USA/Michigan/ | Michigan Department of Health and Human Services, Bureau of Laboratories | Michigan Department of Health and Human Services, Bureau of Laboratories | Blankenship HM et al |

| | | | | | | |
|---|---|---|---|---|---|---|
| EPI_ISL_583528 | USA/MI-MDHHS-SC22071/2020 | 2020-09-22 | North America/USA/Michigan/ | Michigan Department of Health and Human Services, Bureau of Laboratories | Michigan Department of Health and Human Services, Bureau of Laboratories | Blankenship HM et al |
| EPI_ISL_583529 | USA/MI-MDHHS-SC22072/2020 | 2020-09-29 | North America/USA/Michigan/ | Michigan Department of Health and Human Services, Bureau of Laboratories | Michigan Department of Health and Human Services, Bureau of Laboratories | Blankenship HM et al |
| EPI_ISL_583530 | USA/MI-MDHHS-SC22073/2020 | 2020-09-23 | North America/USA/Michigan/ | Michigan Department of Health and Human Services, Bureau of Laboratories | Michigan Department of Health and Human Services, Bureau of Laboratories | Blankenship HM et al |
| EPI_ISL_583534 | USA/MI-MDHHS-SC22077/2020 | 2020-10-05 | North America/USA/Michigan/ | Michigan Department of Health and Human Services, Bureau of Laboratories | Michigan Department of Health and Human Services, Bureau of Laboratories | Blankenship HM et al |
| EPI_ISL_583558 | Austria/CeMM0069/2020 | 2020-03-17 | Europe/Austria/Tyrol/Innsbruck | Universitaetsklinik fVºr Innere Medizin II Innsbruck | Bergthaler laboratory, CeMM Research Center for Molecular Medicine of the Austrian Academy of Sciences | Alexandra Popa et al |
| EPI_ISL_583567 | Austria/CeMM0253/2020 | 2020-03-21 | Europe/Austria/Vienna/ | Center for Virology, Medical University of Vienna | Bergthaler laboratory, CeMM Research Center for Molecular Medicine of the Austrian Academy of Sciences | Alexandra Popa et al |

| | | | | | | |
|---|---|---|---|---|---|---|
| EPI_ISL_583576 | Austria/CeMM0453/2020 | 2020-03-22 | Europe/Austria/Austria/ | Austrian Agency for Health and Food Safety (AGES) | Bergthaler laboratory, CeMM Research Center for Molecular Medicine of the Austrian Academy of Sciences | Alexandra Popa et al |
| EPI_ISL_583594 | Austria/CeMM0508/2020 | 2020-03-28 | Europe/Austria/Upper Austria/Linz | Institute for Medical and Chemical Laboratory Diagnostics, Kepler Universit√§tsklinikum | Bergthaler laboratory, CeMM Research Center for Molecular Medicine of the Austrian Academy of Sciences | Alexandra Popa et al |
| EPI_ISL_583595 | Austria/CeMM0509/2020 | 2020-03-23 | Europe/Austria/Austria/ | Institute for Medical and Chemical Laboratory Diagnostics, Kepler Universit√§tsklinikum | Bergthaler laboratory, CeMM Research Center for Molecular Medicine of the Austrian Academy of Sciences | Alexandra Popa et al |
| EPI_ISL_583600 | Austria/CeMM0574/2020 | 2020-03-27 | Europe/Austria/Tyrol/Innsbruck | Institut f√ºr Virologie am Department f√ºr Hygiene, Mikrobiologie und Public Health | Bergthaler laboratory, CeMM Research Center for Molecular Medicine of the Austrian Academy of Sciences | Alexandra Popa et al |
| EPI_ISL_583611 | Austria/CeMM0600/2020 | 2020-03-17 | Europe/Austria/Tyrol/Kufstein | Institut f√ºr Virologie am Department f√ºr Hygiene, Mikrobiologie und Public Health | Bergthaler laboratory, CeMM Research Center for Molecular Medicine of the Austrian Academy of Sciences | Alexandra Popa et al |

| | | | | | | |
|---|---|---|---|---|---|---|
| EPI_ISL_583613 | Austria/CeMM0607/2020 | 2020-03-27 | Europe/Austria/Tyrol/Kufstein | Institut f√ºr Virologie am Department f√ºr Hygiene, Mikrobiologie und Public Health | Bergthaler laboratory, CeMM Research Center for Molecular Medicine of the Austrian Academy of Sciences | Alexandra Popa et al |
| EPI_ISL_583614 | Austria/CeMM0608/2020 | 2020-03-27 | Europe/Austria/Tyrol/Kufstein | Institut f√ºr Virologie am Department f√ºr Hygiene, Mikrobiologie und Public Health | Bergthaler laboratory, CeMM Research Center for Molecular Medicine of the Austrian Academy of Sciences | Alexandra Popa et al |
| EPI_ISL_583622 | Austria/CeMM0648/2020 | 2020-03-24 | Europe/Austria/Tyrol/S√∂lden | Pathologie-Labor Dr. Obrist- Dr. Brunhuber | Bergthaler laboratory, CeMM Research Center for Molecular Medicine of the Austrian Academy of Sciences | Alexandra Popa et al |
| EPI_ISL_583665 | Austria/CeMM0714/2020 | 2020-05-17 | Europe/Austria/Austria/ | Austrian Agency for Health and Food Safety (AGES) | Bergthaler laboratory, CeMM Research Center for Molecular Medicine of the Austrian Academy of Sciences | Alexandra Popa et al |
| EPI_ISL_583718 | Austria/CeMM0822/2020 | 2020-04-29 | Europe/Austria/Austria/ | Center for Virology, Medical University of Vienna | Bergthaler laboratory, CeMM Research Center for Molecular Medicine of the Austrian Academy of Sciences | Alexandra Popa et al |

| | | | | | | |
|---|---|---|---|---|---|---|
| EPI_ISL_583734 | Austria/CeMM0869/2020 | 2020-04-29 | Europe/Austria/Austria/ | Dr. Gernot Walder GmbH | Bergthaler laboratory, CeMM Research Center for Molecular Medicine of the Austrian Academy of Sciences | Alexandra Popa et al |
| EPI_ISL_583760 | Austria/CeMM0907/2020 | 2020-03-31 | Europe/Austria/Austria/ | Dr. Gernot Walder GmbH | Bergthaler laboratory, CeMM Research Center for Molecular Medicine of the Austrian Academy of Sciences | Alexandra Popa et al |
| EPI_ISL_583776 | Austria/CeMM0924/2020 | 2020-03-18 | Europe/Austria/Austria/ | Dr. Gernot Walder GmbH | Bergthaler laboratory, CeMM Research Center for Molecular Medicine of the Austrian Academy of Sciences | Alexandra Popa et al |
| EPI_ISL_583832 | Austria/CeMM0988/2020 | 2020-04-09 | Europe/Austria/Austria/ | Dr. Gernot Walder GmbH | Bergthaler laboratory, CeMM Research Center for Molecular Medicine of the Austrian Academy of Sciences | Alexandra Popa et al |
| EPI_ISL_583840 | Austria/CeMM0996/2020 | 2020-04-13 | Europe/Austria/Austria/ | Dr. Gernot Walder GmbH | Bergthaler laboratory, CeMM Research Center for Molecular Medicine of the Austrian Academy of Sciences | Alexandra Popa et al |

| | | | | | | |
|---|---|---|---|---|---|---|
| EPI_ISL_583841 | Austria/CeMM0997/2020 | 2020-04-13 | Europe/Austria/Austria/ | Dr. Gernot Walder GmbH | Bergthaler laboratory, CeMM Research Center for Molecular Medicine of the Austrian Academy of Sciences | Alexandra Popa et al |
| EPI_ISL_583846 | Austria/CeMM1005/2020 | 2020-04-25 | Europe/Austria/Austria/ | Dr. Gernot Walder GmbH | Bergthaler laboratory, CeMM Research Center for Molecular Medicine of the Austrian Academy of Sciences | Alexandra Popa et al |
| EPI_ISL_583865 | Austria/CeMM1048/2020 | 2020-04-14 | Europe/Austria/Austria/ | Institute for Laboratory Diagnostics and Microbiology, Klinikum Klagenfurt am Worthersee | Bergthaler laboratory, CeMM Research Center for Molecular Medicine of the Austrian Academy of Sciences | Alexandra Popa et al |
| EPI_ISL_583881 | Austria/CeMM1068/2020 | 2020-03-16 | Europe/Austria/Austria/ | Center for Virology, Medical University of Vienna | Bergthaler laboratory, CeMM Research Center for Molecular Medicine of the Austrian Academy of Sciences | Alexandra Popa et al |
| EPI_ISL_583884 | Austria/CeMM1078/2020 | 2020-06-30 | Europe/Austria/Austria/ | Austrian Agency for Health and Food Safety (AGES) | Bergthaler laboratory, CeMM Research Center for Molecular Medicine of the Austrian Academy of Sciences | Alexandra Popa et al |

| | | | | | | |
|---|---|---|---|---|---|---|
| EPI_ISL_583888 | Austria/CeMM1092/2020 | 2020-08-03 | Europe/Austria/Austria/ | Austrian Agency for Health and Food Safety (AGES) | Bergthaler laboratory, CeMM Research Center for Molecular Medicine of the Austrian Academy of Sciences | Alexandra Popa et al |
| EPI_ISL_583892 | Austria/CeMM1096/2020 | 2020-08-03 | Europe/Austria/Austria/ | Austrian Agency for Health and Food Safety (AGES) | Bergthaler laboratory, CeMM Research Center for Molecular Medicine of the Austrian Academy of Sciences | Alexandra Popa et al |
| EPI_ISL_583893 | Singapore/1058/2020 | 2020-09-25 | Asia/Singapore/Singapore/ | Singapore General Hospital | Department of Microbiology | Nurdyana Abdul Rahman et al |
| EPI_ISL_583918 | Spain/VC-IBV-98004412/2020 | 2020-04-13 | Europe/Spain/Comunitat Valenciana/Ca | Servicio de Microbiolog√≠a. Hospital General Universitario de Castell√≥n | SeqCOVID-SPAIN consortium/IBV(CSIC) | Rosario Moreno et al |
| EPI_ISL_583935 | Spain/VC-IBV-98004459/2020 | 2020-03-24 | Europe/Spain/Comunitat Valenciana/Ca | Servicio de Microbiolog√≠a. Hospital General Universitario de Castell√≥n | SeqCOVID-SPAIN consortium/IBV(CSIC) | Rosario Moreno et al |
| EPI_ISL_583940 | Spain/VC-IBV-98004478/2020 | 2020-03-25 | Europe/Spain/Comunitat Valenciana/Ca | Servicio de Microbiolog√≠a. Hospital General Universitario de Castell√≥n | SeqCOVID-SPAIN consortium/IBV(CSIC) | Rosario Moreno et al |
| EPI_ISL_583955 | Italy/TUS-DBM-11/2020 | 2020-03-21 | Europe/Italy/Tuscany/Siena | UOC Microbiologia e Virologia, Azienda Ospedaliera Universitaria Senese, Siena, Italy | Dipartimento di Biotecnologie Mediche | Maria Grazia Cusi et al |
| EPI_ISL_583958 | Italy/TUS-DBM-21/2020 | 2020-04-27 | Europe/Italy/Tuscany/Siena | UOC Microbiologia e Virologia, Azienda Ospedaliera Universitaria Senese, Siena, Italy | Dipartimento di Biotecnologie Mediche | Maria Grazia Cusi et al |
| EPI_ISL_583960 | Italy/TUS-DBM-6/2020 | 2020-03-21 | Europe/Italy/Tuscany/Siena | UOC Microbiologia e Virologia, Azienda Ospedaliera Universitaria Senese, Siena, Italy | Dipartimento di Biotecnologie Mediche | Maria Grazia Cusi et al |
| EPI_ISL_583961 | Italy/TUS-DBM-7/2020 | 2020-03-20 | Europe/Italy/Tuscany/Siena | UOC Microbiologia e Virologia, Azienda Ospedaliera Universitaria Senese, Siena, Italy | Dipartimento di Biotecnologie Mediche | Maria Grazia Cusi et al |

| | | | | | | |
|---|---|---|---|---|---|---|
| EPI_ISL_583962 | Italy/TUS-DBM-8/2020 | 2020-03-20 | Europe/Italy/Tuscany/Siena | UOC Microbiologia e Virologia, Azienda Ospedaliera Universitaria Senese, Siena, Italy | Dipartimento di Biotecnologie Mediche | Maria Grazia Cusi et al |
| EPI_ISL_584014 | USA/DE-DHSS-F918060/2020 | 2020-03-27 | North America/USA/Delaware/Sussex C | Delaware Public Health Lab | Delaware Public Health Lab | Gregory Hovan et al |
| EPI_ISL_584048 | Italy/LOM-UniMI-ICCS1/2020 | 2020-03-21 | Europe/Italy/Lombardy/ | Laboratory of Molecular Virology, Department of Biomedical, Surgical and Dental Sciences University of Milano | Laboratory of Molecular Virology, Department of Biomedical, Surgical and Dental Sciences University of Milano | Delbue et al |
| EPI_ISL_584049 | Italy/LOM-UniMI-ICCS2/2020 | 2020-03-24 | Europe/Italy/Lombardy/ | Laboratory of Molecular Virology, Department of Biomedical, Surgical and Dental Sciences University of Milano | Laboratory of Molecular Virology, Department of Biomedical, Surgical and Dental Sciences University of Milano | Delbue et al |
| EPI_ISL_584051 | Italy/LOM-UniMI-vir1/2020 | 2020-04-05 | Europe/Italy/Lombardy/ | Laboratory of Molecular Virology, Department of Biomedical, Surgical and Dental Sciences University of Milano | Laboratory of Molecular Virology, Department of Biomedical, Surgical and Dental Sciences University of Milano | Delbue et al |
| EPI_ISL_584052 | Italy/LOM-UniMI-ICCS3/2020 | 2020-04-05 | Europe/Italy/Lombardy/ | Laboratory of Molecular Virology, Department of Biomedical, Surgical and Dental Sciences University of Milano | Laboratory of Molecular Virology, Department of Biomedical, Surgical and Dental Sciences University of Milano | Delbue et al |

| | | | | | | |
|---|---|---|---|---|---|---|
| EPI_ISL_584076 | CzechRepublic/NRL-7322-3/2020 | 2020-06-21 | Europe/Czech Republic/Liberec Region/Li | The National Institute of Public Health | State Veterinary Institute Prague | Nagy et al |
| EPI_ISL_584081 | CzechRepublic/NRL-8598/2020 | 2020-09-03 | Europe/Czech Republic/Prague/ | The National Institute of Public Health | State Veterinary Institute Prague | Nagy et al |
| EPI_ISL_584082 | CzechRepublic/NRL-8609/2020 | 2020-09-04 | Europe/Czech Republic/Prague/ | The National Institute of Public Health | State Veterinary Institute Prague | Nagy et al |
| EPI_ISL_584123 | USA/TX-DSHS-0666/2020 | 2020-05-12 | North America/USA/Texas/El Paso Coun | Texas Department of State Health Services | Texas Department of State Health Services | Rashmi Tuladhar et al |
| EPI_ISL_584129 | USA/TX-DSHS-0727/2020 | 2020-05-13 | North America/USA/Texas/El Paso Coun | Texas Department of State Health Services | Texas Department of State Health Services | Rashmi Tuladhar et al |
| EPI_ISL_584136 | USA/TX-DSHS-0734/2020 | 2020-05-14 | North America/USA/Texas/Harris Count | Texas Department of State Health Services | Texas Department of State Health Services | Rashmi Tuladhar et al |
| EPI_ISL_584147 | USA/TX-DSHS-0746/2020 | 2020-05-14 | North America/USA/Texas/Harris Count | Texas Department of State Health Services | Texas Department of State Health Services | Rashmi Tuladhar et al |
| EPI_ISL_584152 | USA/TX-DSHS-0751/2020 | 2020-05-14 | North America/USA/Texas/Harris Count | Texas Department of State Health Services | Texas Department of State Health Services | Rashmi Tuladhar et al |
| EPI_ISL_584178 | England/ALDP-9E8F17/2020 | 2020-09-24 | Europe/United Kingdom/England/ | Centre for Enzyme Innovation, University of Portsmouth / Translational Research Laboratory, Portsmouth Hospitals NHS Trust | COVID-19 Genomics UK (COG-UK) Consortium | Angela Beckett et al |
| EPI_ISL_584214 | England/ALDP-9E9435/2020 | 2020-09-24 | Europe/United Kingdom/England/ | Centre for Enzyme Innovation, University of Portsmouth / Translational Research Laboratory, Portsmouth Hospitals NHS Trust | COVID-19 Genomics UK (COG-UK) Consortium | Angela Beckett et al |
| EPI_ISL_584241 | England/ALDP-9E972D/2020 | 2020-09-24 | Europe/United Kingdom/England/ | Virology Department, Sheffield Teaching Hospitals NHS Foundation Trust/Department of Infection, Immunity and Cardiovascular Disease, The Medical School, University of Sheffield | COVID-19 Genomics UK (COG-UK) Consortium | Thushan de Silva et al |
| EPI_ISL_584244 | England/ALDP-9E975A/2020 | 2020-09-24 | Europe/United Kingdom/England/ | Virology Department, Sheffield Teaching Hospitals NHS Foundation Trust/Department of Infection, Immunity and Cardiovascular Disease, The Medical School, University of Sheffield | COVID-19 Genomics UK (COG-UK) Consortium | Thushan de Silva et al |

| | | | | | | |
|---|---|---|---|---|---|---|
| EPI_ISL_584580 | England/CWAR-24FE180/2020 | 2020-05-02 | Europe/United Kingdom/England/ | UHCW / University of Warwick | COVID-19 Genomics UK (COG-UK) Consortium | Richard Stark et al |
| EPI_ISL_584779 | England/NOTT-114EFF/2020 | 2020-10-12 | Europe/United Kingdom/England/ | Queens Medical Centre, Clinical Microbiology Department / DeepSeq Nottingham | COVID-19 Genomics UK (COG-UK) Consortium | Gemma Clark et al |
| EPI_ISL_584808 | England/NOTT-115386/2020 | 2020-10-12 | Europe/United Kingdom/England/ | Queens Medical Centre, Clinical Microbiology Department / DeepSeq Nottingham | COVID-19 Genomics UK (COG-UK) Consortium | Gemma Clark et al |
| EPI_ISL_584835 | England/QEUH-9DEF7E/2020 | 2020-09-21 | Europe/United Kingdom/England/ | University College London, Great Ormond Street Hospital for Children NHS Foundation Trust, Imperial College Healthcare NHS Trust | COVID-19 Genomics UK (COG-UK) Consortium | Sergi Castellano et al |
| EPI_ISL_585098 | NorthernIreland/NIRE-107008/2020 | 2020-03-30 | Europe/United Kingdom/Northern Irelar | Regional Virus Laboratory, Belfast Health and Social Care Trust | COVID-19 Genomics UK (COG-UK) Consortium | Conall McCaughey et al |
| EPI_ISL_585136 | NorthernIreland/NIRE-10730F/2020 | 2020-03-30 | Europe/United Kingdom/Northern Irelar | Regional Virus Laboratory, Belfast Health and Social Care Trust | COVID-19 Genomics UK (COG-UK) Consortium | Conall McCaughey et al |
| EPI_ISL_585319 | Scotland/CVR4454/2020 | 2020-09-20 | Europe/United Kingdom/Scotland/ | Lighthouse Lab in Glasgow / MRC-University of Glasgow Centre for Virus Research | COVID-19 Genomics UK (COG-UK) Consortium | Ana da Silva Filipe et al |
| EPI_ISL_585423 | Scotland/CVR4623/2020 | 2020-09-05 | Europe/United Kingdom/Scotland/ | West of Scotland Specialist Virology Centre, NHSGGC / MRC-University of Glasgow Centre for Virus Research | COVID-19 Genomics UK (COG-UK) Consortium | Ana da Silva Filipe et al |
| EPI_ISL_585432 | Scotland/EDB7879/2020 | 2020-09-22 | Europe/United Kingdom/Scotland/ | Virology Department, Royal Infirmary of Edinburgh, NHS Lothian / School of Biological Sciences, University of Edinburgh / Institute of Genetics and Molecular Medicine, University of Edinburgh | COVID-19 Genomics UK (COG-UK) Consortium | McHugh M et al |
| EPI_ISL_585437 | Scotland/EDB7884/2020 | 2020-09-23 | Europe/United Kingdom/Scotland/ | Virology Department, Royal Infirmary of Edinburgh, NHS Lothian / School of Biological Sciences, University of Edinburgh / Institute of Genetics and Molecular Medicine, University of Edinburgh | COVID-19 Genomics UK (COG-UK) Consortium | McHugh M et al |
| EPI_ISL_585494 | Scotland/EDB8243/2020 | 2020-10-02 | Europe/United Kingdom/Scotland/ | Virology Department, Royal Infirmary of Edinburgh, NHS Lothian / School of Biological Sciences, University of Edinburgh / Institute of Genetics and Molecular Medicine, University of Edinburgh | COVID-19 Genomics UK (COG-UK) Consortium | McHugh M et al |

| | | | | | | |
|---|---|---|---|---|---|---|
| EPI_ISL_585570 | Scotland/QEUH-A00CD5/2020 | 2020-10-02 | Europe/United Kingdom/Scotland/ | University of Exeter | COVID-19 Genomics UK (COG-UK) Consortium | Ben Temperton et al |
| EPI_ISL_585699 | Wales/PHWC-16E2FA/2020 | 2020-09-29 | Europe/United Kingdom/Wales/ | Wales Specialist Virology Centre Sequencing lab: Pathogen Genomics Unit | COVID-19 Genomics UK (COG-UK) Consortium | Catherine Moore et al |
| EPI_ISL_585820 | Wales/PHWC-16EAF2/2020 | 2020-10-06 | Europe/United Kingdom/Wales/ | Wales Specialist Virology Centre Sequencing lab: Pathogen Genomics Unit | COVID-19 Genomics UK (COG-UK) Consortium | Catherine Moore et al |
| EPI_ISL_585855 | Wales/PHWC-16ED26/2020 | 2020-10-05 | Europe/United Kingdom/Wales/ | Wales Specialist Virology Centre Sequencing lab: Pathogen Genomics Unit | COVID-19 Genomics UK (COG-UK) Consortium | Catherine Moore et al |
| EPI_ISL_585971 | Wales/PHWC-16F499/2020 | 2020-10-03 | Europe/United Kingdom/Wales/ | Wales Specialist Virology Centre Sequencing lab: Pathogen Genomics Unit | COVID-19 Genomics UK (COG-UK) Consortium | Catherine Moore et al |
| EPI_ISL_586015 | Wales/PHWC-16F763/2020 | 2020-10-03 | Europe/United Kingdom/Wales/ | Wales Specialist Virology Centre Sequencing lab: Pathogen Genomics Unit | COVID-19 Genomics UK (COG-UK) Consortium | Catherine Moore et al |
| EPI_ISL_586036 | Wales/PHWC-16F8E8/2020 | 2020-10-07 | Europe/United Kingdom/Wales/ | Wales Specialist Virology Centre Sequencing lab: Pathogen Genomics Unit | COVID-19 Genomics UK (COG-UK) Consortium | Catherine Moore et al |
| EPI_ISL_586045 | Wales/PHWC-47CD7D/2020 | 2020-10-09 | Europe/United Kingdom/Wales/ | Wales Specialist Virology Centre Sequencing lab: Pathogen Genomics Unit | COVID-19 Genomics UK (COG-UK) Consortium | Catherine Moore et al |
| EPI_ISL_586060 | Wales/PHWC-47CE7A/2020 | 2020-10-09 | Europe/United Kingdom/Wales/ | Wales Specialist Virology Centre Sequencing lab: Pathogen Genomics Unit | COVID-19 Genomics UK (COG-UK) Consortium | Catherine Moore et al |
| EPI_ISL_586133 | Wales/PHWC-47D37A/2020 | 2020-10-05 | Europe/United Kingdom/Wales/ | Wales Specialist Virology Centre Sequencing lab: Pathogen Genomics Unit | COVID-19 Genomics UK (COG-UK) Consortium | Catherine Moore et al |
| EPI_ISL_586220 | Wales/PHWC-47D9C3/2020 | 2020-10-08 | Europe/United Kingdom/Wales/ | Wales Specialist Virology Centre Sequencing lab: Pathogen Genomics Unit | COVID-19 Genomics UK (COG-UK) Consortium | Catherine Moore et al |
| EPI_ISL_586253 | USA/AK-PHL672/2020 | 2020-06-27 | North America/USA/Alaska/Matanuska | Alaska State Virology Laboratory | Alaska State Virology Laboratory | Jack Chen et al |
| EPI_ISL_586255 | USA/AK-PHL683/2020 | 2020-06-29 | North America/USA/Alaska/Matanuska | Alaska State Virology Laboratory | Alaska State Virology Laboratory | Jack Chen et al |

| | | | | | | |
|---|---|---|---|---|---|---|
| EPI_ISL_586256 | USA/AK-PHL689/2020 | 2020-06-29 | North America/USA/Alaska/Matanuska | Alaska State Virology Laboratory | Alaska State Virology Laboratory | Jack Chen et al |
| EPI_ISL_586261 | USA/AK-PHL734/2020 | 2020-06-30 | North America/USA/Alaska/ | Alaska State Virology Laboratory | Alaska State Virology Laboratory | Jack Chen et al |
| EPI_ISL_586264 | USA/AK-PHL767/2020 | 2020-07-02 | North America/USA/Alaska/ | Alaska State Virology Laboratory | Alaska State Virology Laboratory | Jack Chen et al |
| EPI_ISL_586265 | USA/AK-PHL796/2020 | 2020-07-02 | North America/USA/Alaska/Matanuska | Alaska State Virology Laboratory | Alaska State Virology Laboratory | Jack Chen et al |
| EPI_ISL_586267 | Canada/ON-CVD00053/2020 | 2020-03-23 | North America/Canada/Ontario/Toronto | Toronto Invasive Bacterial Diseases Network | McMaster University | Allison McGeer et al |
| EPI_ISL_586268 | Canada/ON-CVD00060/2020 | 2020-03-23 | North America/Canada/Ontario/Toronto | Toronto Invasive Bacterial Diseases Network | McMaster University | Allison McGeer et al |
| EPI_ISL_586270 | Canada/ON-CVD00173/2020 | 2020-03-26 | North America/Canada/Ontario/Toronto | Toronto Invasive Bacterial Diseases Network | McMaster University | Allison McGeer et al |
| EPI_ISL_586271 | Canada/ON-CVD00726/2020 | 2020-04-09 | North America/Canada/Ontario/Toronto | Toronto Invasive Bacterial Diseases Network | McMaster University | Allison McGeer et al |
| EPI_ISL_586272 | Canada/ON-CVD00784/2020 | 2020-04-14 | North America/Canada/Ontario/Toronto | Toronto Invasive Bacterial Diseases Network | McMaster University | Allison McGeer et al |
| EPI_ISL_586274 | Canada/ON-CVD01006/2020 | 2020-04-20 | North America/Canada/Ontario/Toronto | Toronto Invasive Bacterial Diseases Network | McMaster University | Allison McGeer et al |
| EPI_ISL_586275 | Canada/ON-CVD01196/2020 | 2020-04-24 | North America/Canada/Ontario/Toronto | Toronto Invasive Bacterial Diseases Network | McMaster University | Allison McGeer et al |
| EPI_ISL_586276 | Canada/ON-CVD01345/2020 | 2020-04-29 | North America/Canada/Ontario/Toronto | Toronto Invasive Bacterial Diseases Network | McMaster University | Allison McGeer et al |
| EPI_ISL_586278 | Canada/ON-S10/2020 | 2020-03-14 | North America/Canada/Ontario/Toronto | Toronto Invasive Bacterial Diseases Network | McMaster University | Allison McGeer et al |
| EPI_ISL_586279 | Canada/ON-S100/2020 | 2020-03-21 | North America/Canada/Ontario/Toronto | Toronto Invasive Bacterial Diseases Network | McMaster University | Allison McGeer et al |
| EPI_ISL_586280 | Canada/ON-S1012/2020 | 2020-04-16 | North America/Canada/Ontario/Toronto | Toronto Invasive Bacterial Diseases Network | McMaster University | Allison McGeer et al |
| EPI_ISL_586281 | Canada/ON-S1013/2020 | 2020-04-16 | North America/Canada/Ontario/Toronto | Toronto Invasive Bacterial Diseases Network | McMaster University | Allison McGeer et al |
| EPI_ISL_586282 | Canada/ON-S104/2020 | 2020-03-21 | North America/Canada/Ontario/Toronto | Toronto Invasive Bacterial Diseases Network | McMaster University | Allison McGeer et al |
| EPI_ISL_586283 | Canada/ON-S106/2020 | 2020-03-21 | North America/Canada/Ontario/Toronto | Toronto Invasive Bacterial Diseases Network | McMaster University | Allison McGeer et al |

| | | | | | | |
|---|---|---|---|---|---|---|
| EPI_ISL_586284 | Canada/ON-S107/2020 | 2020-03-21 | North America/Canada/Ontario/Toront | Toronto Invasive Bacterial Diseases Network | McMaster University | Allison McGeer et al |
| EPI_ISL_586285 | Canada/ON-S108/2020 | 2020-03-21 | North America/Canada/Ontario/Toront | Toronto Invasive Bacterial Diseases Network | McMaster University | Allison McGeer et al |
| EPI_ISL_586286 | Canada/ON-S110/2020 | 2020-03-22 | North America/Canada/Ontario/Toront | Toronto Invasive Bacterial Diseases Network | McMaster University | Allison McGeer et al |
| EPI_ISL_586287 | Canada/ON-S111/2020 | 2020-03-22 | North America/Canada/Ontario/Toront | Toronto Invasive Bacterial Diseases Network | McMaster University | Allison McGeer et al |
| EPI_ISL_586288 | Canada/ON-S112/2020 | 2020-03-21 | North America/Canada/Ontario/Toront | Toronto Invasive Bacterial Diseases Network | McMaster University | Allison McGeer et al |
| EPI_ISL_586289 | Canada/ON-S113/2020 | 2020-03-22 | North America/Canada/Ontario/Toront | Toronto Invasive Bacterial Diseases Network | McMaster University | Allison McGeer et al |
| EPI_ISL_586290 | Canada/ON-S116/2020 | 2020-03-22 | North America/Canada/Ontario/Toront | Toronto Invasive Bacterial Diseases Network | McMaster University | Allison McGeer et al |
| EPI_ISL_586291 | Canada/ON-S117/2020 | 2020-03-22 | North America/Canada/Ontario/Toront | Toronto Invasive Bacterial Diseases Network | McMaster University | Allison McGeer et al |
| EPI_ISL_586292 | Canada/ON-S12/2020 | 2020-03-14 | North America/Canada/Ontario/Toront | Toronto Invasive Bacterial Diseases Network | McMaster University | Allison McGeer et al |
| EPI_ISL_586293 | Canada/ON-S120/2020 | 2020-03-22 | North America/Canada/Ontario/Toront | Toronto Invasive Bacterial Diseases Network | McMaster University | Allison McGeer et al |
| EPI_ISL_586294 | Canada/ON-S121/2020 | 2020-03-23 | North America/Canada/Ontario/Toront | Toronto Invasive Bacterial Diseases Network | McMaster University | Allison McGeer et al |
| EPI_ISL_586295 | Canada/ON-S1219/2020 | 2020-03-14 | North America/Canada/Ontario/Toront | Toronto Invasive Bacterial Diseases Network | McMaster University | Allison McGeer et al |
| EPI_ISL_586296 | Canada/ON-S122/2020 | 2020-03-23 | North America/Canada/Ontario/Toront | Toronto Invasive Bacterial Diseases Network | McMaster University | Allison McGeer et al |
| EPI_ISL_586297 | Canada/ON-S1220/2020 | 2020-03-16 | North America/Canada/Ontario/Toront | Toronto Invasive Bacterial Diseases Network | McMaster University | Allison McGeer et al |
| EPI_ISL_586298 | Canada/ON-S1221/2020 | 2020-03-18 | North America/Canada/Ontario/Toront | Toronto Invasive Bacterial Diseases Network | McMaster University | Allison McGeer et al |
| EPI_ISL_586299 | Canada/ON-S1222/2020 | 2020-03-18 | North America/Canada/Ontario/Toront | Toronto Invasive Bacterial Diseases Network | McMaster University | Allison McGeer et al |
| EPI_ISL_586300 | Canada/ON-S1224/2020 | 2020-03-27 | North America/Canada/Ontario/Toront | Toronto Invasive Bacterial Diseases Network | McMaster University | Allison McGeer et al |
| EPI_ISL_586301 | Canada/ON-S1225/2020 | 2020-03-28 | North America/Canada/Ontario/Toront | Toronto Invasive Bacterial Diseases Network | McMaster University | Allison McGeer et al |
| EPI_ISL_586302 | Canada/ON-S1227/2020 | 2020-04-02 | North America/Canada/Ontario/Toront | Toronto Invasive Bacterial Diseases Network | McMaster University | Allison McGeer et al |
| EPI_ISL_586303 | Canada/ON-S1229/2020 | 2020-04-08 | North America/Canada/Ontario/Toront | Toronto Invasive Bacterial Diseases Network | McMaster University | Allison McGeer et al |

| | | | | | | |
|---|---|---|---|---|---|---|
| EPI_ISL_586304 | Canada/ON-S123/2020 | 2020-03-23 | North America/Canada/Ontario/Toronto | Toronto Invasive Bacterial Diseases Network | McMaster University | Allison McGeer et al |
| EPI_ISL_586305 | Canada/ON-S1230/2020 | 2020-03-22 | North America/Canada/Ontario/Toronto | Toronto Invasive Bacterial Diseases Network | McMaster University | Allison McGeer et al |
| EPI_ISL_586306 | Canada/ON-S1231/2020 | 2020-04-12 | North America/Canada/Ontario/Toronto | Toronto Invasive Bacterial Diseases Network | McMaster University | Allison McGeer et al |
| EPI_ISL_586307 | Canada/ON-S1232/2020 | 2020-04-10 | North America/Canada/Ontario/Toronto | Toronto Invasive Bacterial Diseases Network | McMaster University | Allison McGeer et al |
| EPI_ISL_586308 | Canada/ON-S1237/2020 | 2020-04-16 | North America/Canada/Ontario/Toronto | Toronto Invasive Bacterial Diseases Network | McMaster University | Allison McGeer et al |
| EPI_ISL_586309 | Canada/ON-S1238/2020 | 2020-04-20 | North America/Canada/Ontario/Toronto | Toronto Invasive Bacterial Diseases Network | McMaster University | Allison McGeer et al |
| EPI_ISL_586310 | Canada/ON-S1242/2020 | 2020-04-27 | North America/Canada/Ontario/Toronto | Toronto Invasive Bacterial Diseases Network | McMaster University | Allison McGeer et al |
| EPI_ISL_586311 | Canada/ON-S1243/2020 | 2020-04-16 | North America/Canada/Ontario/Toronto | Toronto Invasive Bacterial Diseases Network | McMaster University | Allison McGeer et al |
| EPI_ISL_586312 | Canada/ON-S1245/2020 | 2020-05-01 | North America/Canada/Ontario/Toronto | Toronto Invasive Bacterial Diseases Network | McMaster University | Allison McGeer et al |
| EPI_ISL_586314 | Canada/ON-S1250/2020 | 2020-05-13 | North America/Canada/Ontario/Toronto | Toronto Invasive Bacterial Diseases Network | McMaster University | Allison McGeer et al |
| EPI_ISL_586317 | Canada/ON-S1264/2020 | 2020-05-06 | North America/Canada/Ontario/Toronto | Toronto Invasive Bacterial Diseases Network | McMaster University | Allison McGeer et al |
| EPI_ISL_586318 | Canada/ON-S1265/2020 | 2020-05-05 | North America/Canada/Ontario/Toronto | Toronto Invasive Bacterial Diseases Network | McMaster University | Allison McGeer et al |
| EPI_ISL_586319 | Canada/ON-S1266/2020 | 2020-05-04 | North America/Canada/Ontario/Toronto | Toronto Invasive Bacterial Diseases Network | McMaster University | Allison McGeer et al |
| EPI_ISL_586320 | Canada/ON-S1268/2020 | 2020-05-02 | North America/Canada/Ontario/Toronto | Toronto Invasive Bacterial Diseases Network | McMaster University | Allison McGeer et al |
| EPI_ISL_586321 | Canada/ON-S127/2020 | 2020-03-23 | North America/Canada/Ontario/Toronto | Toronto Invasive Bacterial Diseases Network | McMaster University | Allison McGeer et al |
| EPI_ISL_586322 | Canada/ON-S1270/2020 | 2020-05-01 | North America/Canada/Ontario/Toronto | Toronto Invasive Bacterial Diseases Network | McMaster University | Allison McGeer et al |
| EPI_ISL_586323 | Canada/ON-S1271/2020 | 2020-04-30 | North America/Canada/Ontario/Toronto | Toronto Invasive Bacterial Diseases Network | McMaster University | Allison McGeer et al |
| EPI_ISL_586324 | Canada/ON-S1272/2020 | 2020-04-29 | North America/Canada/Ontario/Toronto | Toronto Invasive Bacterial Diseases Network | McMaster University | Allison McGeer et al |
| EPI_ISL_586325 | Canada/ON-S1274/2020 | 2020-04-29 | North America/Canada/Ontario/Toronto | Toronto Invasive Bacterial Diseases Network | McMaster University | Allison McGeer et al |
| EPI_ISL_586326 | Canada/ON-S1275/2020 | 2020-04-29 | North America/Canada/Ontario/Toronto | Toronto Invasive Bacterial Diseases Network | McMaster University | Allison McGeer et al |

| | | | | | | |
|---|---|---|---|---|---|---|
| EPI_ISL_586327 | Canada/ON-S1276/2020 | 2020-04-28 | North America/Canada/Ontario/Toronto | Toronto Invasive Bacterial Diseases Network | McMaster University | Allison McGeer et al |
| EPI_ISL_586328 | Canada/ON-S1277/2020 | 2020-04-18 | North America/Canada/Ontario/Toronto | Toronto Invasive Bacterial Diseases Network | McMaster University | Allison McGeer et al |
| EPI_ISL_586329 | Canada/ON-S1278/2020 | 2020-04-14 | North America/Canada/Ontario/Toronto | Toronto Invasive Bacterial Diseases Network | McMaster University | Allison McGeer et al |
| EPI_ISL_586330 | Canada/ON-S1280/2020 | 2020-04-10 | North America/Canada/Ontario/Toronto | Toronto Invasive Bacterial Diseases Network | McMaster University | Allison McGeer et al |
| EPI_ISL_586331 | Canada/ON-S1282/2020 | 2020-03-21 | North America/Canada/Ontario/Toronto | Toronto Invasive Bacterial Diseases Network | McMaster University | Allison McGeer et al |
| EPI_ISL_586332 | Canada/ON-S1283/2020 | 2020-04-28 | North America/Canada/Ontario/Toronto | Toronto Invasive Bacterial Diseases Network | McMaster University | Allison McGeer et al |
| EPI_ISL_586333 | Canada/ON-S1285/2020 | 2020-05-08 | North America/Canada/Ontario/Toronto | Toronto Invasive Bacterial Diseases Network | McMaster University | Allison McGeer et al |
| EPI_ISL_586334 | Canada/ON-S1286/2020 | 2020-04-15 | North America/Canada/Ontario/Toronto | Toronto Invasive Bacterial Diseases Network | McMaster University | Allison McGeer et al |
| EPI_ISL_586335 | Canada/ON-S1288/2020 | 2020-05-13 | North America/Canada/Ontario/Toronto | Toronto Invasive Bacterial Diseases Network | McMaster University | Allison McGeer et al |
| EPI_ISL_586336 | Canada/ON-S129/2020 | 2020-03-23 | North America/Canada/Ontario/Toronto | Toronto Invasive Bacterial Diseases Network | McMaster University | Allison McGeer et al |
| EPI_ISL_586337 | Canada/ON-S1290/2020 | 2020-05-17 | North America/Canada/Ontario/Toronto | Toronto Invasive Bacterial Diseases Network | McMaster University | Allison McGeer et al |
| EPI_ISL_586338 | Canada/ON-S1292/2020 | 2020-05-16 | North America/Canada/Ontario/Toronto | Toronto Invasive Bacterial Diseases Network | McMaster University | Allison McGeer et al |
| EPI_ISL_586339 | Canada/ON-S1294/2020 | 2020-05-21 | North America/Canada/Ontario/Toronto | Toronto Invasive Bacterial Diseases Network | McMaster University | Allison McGeer et al |
| EPI_ISL_586340 | Canada/ON-S1296/2020 | 2020-05-24 | North America/Canada/Ontario/Toronto | Toronto Invasive Bacterial Diseases Network | McMaster University | Allison McGeer et al |
| EPI_ISL_586341 | Canada/ON-S1297/2020 | 2020-05-22 | North America/Canada/Ontario/Toronto | Toronto Invasive Bacterial Diseases Network | McMaster University | Allison McGeer et al |
| EPI_ISL_586342 | Canada/ON-S1298/2020 | 2020-05-18 | North America/Canada/Ontario/Toronto | Toronto Invasive Bacterial Diseases Network | McMaster University | Allison McGeer et al |
| EPI_ISL_586344 | Canada/ON-S1306/2020 | 2020-04-08 | North America/Canada/Ontario/Toronto | Toronto Invasive Bacterial Diseases Network | McMaster University | Allison McGeer et al |
| EPI_ISL_586345 | Canada/ON-S1307/2020 | 2020-04-11 | North America/Canada/Ontario/Toronto | Toronto Invasive Bacterial Diseases Network | McMaster University | Allison McGeer et al |
| EPI_ISL_586346 | Canada/ON-S1309/2020 | 2020-05-26 | North America/Canada/Ontario/Toronto | Toronto Invasive Bacterial Diseases Network | McMaster University | Allison McGeer et al |
| EPI_ISL_586347 | Canada/ON-S1310/2020 | 2020-05-27 | North America/Canada/Ontario/Toronto | Toronto Invasive Bacterial Diseases Network | McMaster University | Allison McGeer et al |

| | | | | | | |
|---|---|---|---|---|---|---|
| EPI_ISL_586348 | Canada/ON-S1311/2020 | 2020-05-25 | North America/Canada/Ontario/Toronto | Toronto Invasive Bacterial Diseases Network | McMaster University | Allison McGeer et al |
| EPI_ISL_586349 | Canada/ON-S1312/2020 | 2020-04-11 | North America/Canada/Ontario/Toronto | Toronto Invasive Bacterial Diseases Network | McMaster University | Allison McGeer et al |
| EPI_ISL_586350 | Canada/ON-S1313/2020 | 2020-04-10 | North America/Canada/Ontario/Toronto | Toronto Invasive Bacterial Diseases Network | McMaster University | Allison McGeer et al |
| EPI_ISL_586351 | Canada/ON-S1316/2020 | 2020-05-27 | North America/Canada/Ontario/Toronto | Toronto Invasive Bacterial Diseases Network | McMaster University | Allison McGeer et al |
| EPI_ISL_586352 | Canada/ON-S1317/2020 | 2020-05-27 | North America/Canada/Ontario/Toronto | Toronto Invasive Bacterial Diseases Network | McMaster University | Allison McGeer et al |
| EPI_ISL_586353 | Canada/ON-S1320/2020 | 2020-06-04 | North America/Canada/Ontario/Toronto | Toronto Invasive Bacterial Diseases Network | McMaster University | Allison McGeer et al |
| EPI_ISL_586354 | Canada/ON-S135/2020 | 2020-03-23 | North America/Canada/Ontario/Toronto | Toronto Invasive Bacterial Diseases Network | McMaster University | Allison McGeer et al |
| EPI_ISL_586355 | Canada/ON-S139/2020 | 2020-03-24 | North America/Canada/Ontario/Toronto | Toronto Invasive Bacterial Diseases Network | McMaster University | Allison McGeer et al |
| EPI_ISL_586356 | Canada/ON-S14/2020 | 2020-03-16 | North America/Canada/Ontario/Toronto | Toronto Invasive Bacterial Diseases Network | McMaster University | Allison McGeer et al |
| EPI_ISL_586357 | Canada/ON-S140/2020 | 2020-03-24 | North America/Canada/Ontario/Toronto | Toronto Invasive Bacterial Diseases Network | McMaster University | Allison McGeer et al |
| EPI_ISL_586358 | Canada/ON-S144/2020 | 2020-03-24 | North America/Canada/Ontario/Toronto | Toronto Invasive Bacterial Diseases Network | McMaster University | Allison McGeer et al |
| EPI_ISL_586359 | Canada/ON-S147/2020 | 2020-03-24 | North America/Canada/Ontario/Toronto | Toronto Invasive Bacterial Diseases Network | McMaster University | Allison McGeer et al |
| EPI_ISL_586361 | Canada/ON-S15/2020 | 2020-03-16 | North America/Canada/Ontario/Toronto | Toronto Invasive Bacterial Diseases Network | McMaster University | Allison McGeer et al |
| EPI_ISL_586362 | Canada/ON-S1507/2020 | 2020-08-30 | North America/Canada/Ontario/Toronto | Toronto Invasive Bacterial Diseases Network | McMaster University | Allison McGeer et al |
| EPI_ISL_586363 | Canada/ON-S152/2020 | 2020-03-24 | North America/Canada/Ontario/Toronto | Toronto Invasive Bacterial Diseases Network | McMaster University | Allison McGeer et al |
| EPI_ISL_586364 | Canada/ON-S153/2020 | 2020-03-24 | North America/Canada/Ontario/Toronto | Toronto Invasive Bacterial Diseases Network | McMaster University | Allison McGeer et al |
| EPI_ISL_586365 | Canada/ON-S1530/2020 | 2020-08-31 | North America/Canada/Ontario/Toronto | Toronto Invasive Bacterial Diseases Network | McMaster University | Allison McGeer et al |
| EPI_ISL_586366 | Canada/ON-S154/2020 | 2020-03-24 | North America/Canada/Ontario/Toronto | Toronto Invasive Bacterial Diseases Network | McMaster University | Allison McGeer et al |
| EPI_ISL_586367 | Canada/ON-S156/2020 | 2020-03-24 | North America/Canada/Ontario/Toronto | Toronto Invasive Bacterial Diseases Network | McMaster University | Allison McGeer et al |
| EPI_ISL_586368 | Canada/ON-S159/2020 | 2020-03-24 | North America/Canada/Ontario/Toronto | Toronto Invasive Bacterial Diseases Network | McMaster University | Allison McGeer et al |

| | | | | | | |
|---|---|---|---|---|---|---|
| EPI_ISL_586369 | Canada/ON-S16/2020 | 2020-03-15 | North America/Canada/Ontario/Toronto | Toronto Invasive Bacterial Diseases Network | McMaster University | Allison McGeer et al |
| EPI_ISL_586370 | Canada/ON-S17/2020 | 2020-03-16 | North America/Canada/Ontario/Toronto | Toronto Invasive Bacterial Diseases Network | McMaster University | Allison McGeer et al |
| EPI_ISL_586371 | Canada/ON-S171/2020 | 2020-03-24 | North America/Canada/Ontario/Toronto | Toronto Invasive Bacterial Diseases Network | McMaster University | Allison McGeer et al |
| EPI_ISL_586372 | Canada/ON-S178/2020 | 2020-03-24 | North America/Canada/Ontario/Toronto | Toronto Invasive Bacterial Diseases Network | McMaster University | Allison McGeer et al |
| EPI_ISL_586373 | Canada/ON-S18/2020 | 2020-03-16 | North America/Canada/Ontario/Toronto | Toronto Invasive Bacterial Diseases Network | McMaster University | Allison McGeer et al |
| EPI_ISL_586374 | Canada/ON-S180/2020 | 2020-03-24 | North America/Canada/Ontario/Toronto | Toronto Invasive Bacterial Diseases Network | McMaster University | Allison McGeer et al |
| EPI_ISL_586375 | Canada/ON-S188/2020 | 2020-03-25 | North America/Canada/Ontario/Toronto | Toronto Invasive Bacterial Diseases Network | McMaster University | Allison McGeer et al |
| EPI_ISL_586376 | Canada/ON-S189/2020 | 2020-03-25 | North America/Canada/Ontario/Toronto | Toronto Invasive Bacterial Diseases Network | McMaster University | Allison McGeer et al |
| EPI_ISL_586377 | Canada/ON-S190/2020 | 2020-03-25 | North America/Canada/Ontario/Toronto | Toronto Invasive Bacterial Diseases Network | McMaster University | Allison McGeer et al |
| EPI_ISL_586378 | Canada/ON-S197/2020 | 2020-03-25 | North America/Canada/Ontario/Toronto | Toronto Invasive Bacterial Diseases Network | McMaster University | Allison McGeer et al |
| EPI_ISL_586379 | Canada/ON-S201/2020 | 2020-03-25 | North America/Canada/Ontario/Toronto | Toronto Invasive Bacterial Diseases Network | McMaster University | Allison McGeer et al |
| EPI_ISL_586380 | Canada/ON-S215/2020 | 2020-03-26 | North America/Canada/Ontario/Toronto | Toronto Invasive Bacterial Diseases Network | McMaster University | Allison McGeer et al |
| EPI_ISL_586381 | Canada/ON-S22/2020 | 2020-03-16 | North America/Canada/Ontario/Toronto | Toronto Invasive Bacterial Diseases Network | McMaster University | Allison McGeer et al |
| EPI_ISL_586382 | Canada/ON-S222/2020 | 2020-03-26 | North America/Canada/Ontario/Toronto | Toronto Invasive Bacterial Diseases Network | McMaster University | Allison McGeer et al |
| EPI_ISL_586383 | Canada/ON-S223/2020 | 2020-03-26 | North America/Canada/Ontario/Toronto | Toronto Invasive Bacterial Diseases Network | McMaster University | Allison McGeer et al |
| EPI_ISL_586384 | Canada/ON-S226/2020 | 2020-03-26 | North America/Canada/Ontario/Toronto | Toronto Invasive Bacterial Diseases Network | McMaster University | Allison McGeer et al |
| EPI_ISL_586385 | Canada/ON-S230/2020 | 2020-03-26 | North America/Canada/Ontario/Toronto | Toronto Invasive Bacterial Diseases Network | McMaster University | Allison McGeer et al |
| EPI_ISL_586386 | Canada/ON-S233/2020 | 2020-03-27 | North America/Canada/Ontario/Toronto | Toronto Invasive Bacterial Diseases Network | McMaster University | Allison McGeer et al |
| EPI_ISL_586387 | Canada/ON-S234/2020 | 2020-03-27 | North America/Canada/Ontario/Toronto | Toronto Invasive Bacterial Diseases Network | McMaster University | Allison McGeer et al |
| EPI_ISL_586388 | Canada/ON-S264/2020 | 2020-03-29 | North America/Canada/Ontario/Toronto | Toronto Invasive Bacterial Diseases Network | McMaster University | Allison McGeer et al |

| | | | | | | |
|---|---|---|---|---|---|---|
| EPI_ISL_586389 | Canada/ON-S266/2020 | 2020-03-27 | North America/Canada/Ontario/Toronto | Toronto Invasive Bacterial Diseases Network | McMaster University | Allison McGeer et al |
| EPI_ISL_586390 | Canada/ON-S3/2020 | 2020-03-13 | North America/Canada/Ontario/Toronto | Toronto Invasive Bacterial Diseases Network | McMaster University | Allison McGeer et al |
| EPI_ISL_586391 | Canada/ON-S307/2020 | 2020-03-29 | North America/Canada/Ontario/Toronto | Toronto Invasive Bacterial Diseases Network | McMaster University | Allison McGeer et al |
| EPI_ISL_586392 | Canada/ON-S309/2020 | 2020-03-28 | North America/Canada/Ontario/Toronto | Toronto Invasive Bacterial Diseases Network | McMaster University | Allison McGeer et al |
| EPI_ISL_586393 | Canada/ON-S31/2020 | 2020-03-16 | North America/Canada/Ontario/Toronto | Toronto Invasive Bacterial Diseases Network | McMaster University | Allison McGeer et al |
| EPI_ISL_586394 | Canada/ON-S310/2020 | 2020-03-28 | North America/Canada/Ontario/Toronto | Toronto Invasive Bacterial Diseases Network | McMaster University | Allison McGeer et al |
| EPI_ISL_586395 | Canada/ON-S318/2020 | 2020-03-29 | North America/Canada/Ontario/Toronto | Toronto Invasive Bacterial Diseases Network | McMaster University | Allison McGeer et al |
| EPI_ISL_586396 | Canada/ON-S322/2020 | 2020-03-29 | North America/Canada/Ontario/Toronto | Toronto Invasive Bacterial Diseases Network | McMaster University | Allison McGeer et al |
| EPI_ISL_586397 | Canada/ON-S325/2020 | 2020-03-29 | North America/Canada/Ontario/Toronto | Toronto Invasive Bacterial Diseases Network | McMaster University | Allison McGeer et al |
| EPI_ISL_586398 | Canada/ON-S326/2020 | 2020-03-29 | North America/Canada/Ontario/Toronto | Toronto Invasive Bacterial Diseases Network | McMaster University | Allison McGeer et al |
| EPI_ISL_586399 | Canada/ON-S327/2020 | 2020-03-29 | North America/Canada/Ontario/Toronto | Toronto Invasive Bacterial Diseases Network | McMaster University | Allison McGeer et al |
| EPI_ISL_586400 | Canada/ON-S330/2020 | 2020-03-29 | North America/Canada/Ontario/Toronto | Toronto Invasive Bacterial Diseases Network | McMaster University | Allison McGeer et al |
| EPI_ISL_586401 | Canada/ON-S332/2020 | 2020-03-29 | North America/Canada/Ontario/Toronto | Toronto Invasive Bacterial Diseases Network | McMaster University | Allison McGeer et al |
| EPI_ISL_586402 | Canada/ON-S34/2020 | 2020-03-17 | North America/Canada/Ontario/Toronto | Toronto Invasive Bacterial Diseases Network | McMaster University | Allison McGeer et al |
| EPI_ISL_586403 | Canada/ON-S348/2020 | 2020-03-29 | North America/Canada/Ontario/Toronto | Toronto Invasive Bacterial Diseases Network | McMaster University | Allison McGeer et al |
| EPI_ISL_586405 | Canada/ON-S357/2020 | 2020-03-30 | North America/Canada/Ontario/Toronto | Toronto Invasive Bacterial Diseases Network | McMaster University | Allison McGeer et al |
| EPI_ISL_586406 | Canada/ON-S36/2020 | 2020-03-17 | North America/Canada/Ontario/Toronto | Toronto Invasive Bacterial Diseases Network | McMaster University | Allison McGeer et al |
| EPI_ISL_586407 | Canada/ON-S37/2020 | 2020-03-17 | North America/Canada/Ontario/Toronto | Toronto Invasive Bacterial Diseases Network | McMaster University | Allison McGeer et al |
| EPI_ISL_586408 | Canada/ON-S396/2020 | 2020-03-31 | North America/Canada/Ontario/Toronto | Toronto Invasive Bacterial Diseases Network | McMaster University | Allison McGeer et al |
| EPI_ISL_586409 | Canada/ON-S41/2020 | 2020-03-17 | North America/Canada/Ontario/Toronto | Toronto Invasive Bacterial Diseases Network | McMaster University | Allison McGeer et al |

| | | | | | | |
|---|---|---|---|---|---|---|
| EPI_ISL_586410 | Canada/ON-S44/2020 | 2020-03-17 | North America/Canada/Ontario/Toronto | Toronto Invasive Bacterial Diseases Network | McMaster University | Allison McGeer et al |
| EPI_ISL_586411 | Canada/ON-S46/2020 | 2020-03-17 | North America/Canada/Ontario/Toronto | Toronto Invasive Bacterial Diseases Network | McMaster University | Allison McGeer et al |
| EPI_ISL_586412 | Canada/ON-S47/2020 | 2020-03-18 | North America/Canada/Ontario/Toronto | Toronto Invasive Bacterial Diseases Network | McMaster University | Allison McGeer et al |
| EPI_ISL_586413 | Canada/ON-S48/2020 | 2020-03-17 | North America/Canada/Ontario/Toronto | Toronto Invasive Bacterial Diseases Network | McMaster University | Allison McGeer et al |
| EPI_ISL_586415 | Canada/ON-S497/2020 | 2020-04-03 | North America/Canada/Ontario/Toronto | Toronto Invasive Bacterial Diseases Network | McMaster University | Allison McGeer et al |
| EPI_ISL_586416 | Canada/ON-S50/2020 | 2020-03-18 | North America/Canada/Ontario/Toronto | Toronto Invasive Bacterial Diseases Network | McMaster University | Allison McGeer et al |
| EPI_ISL_586417 | Canada/ON-S52/2020 | 2020-03-19 | North America/Canada/Ontario/Toronto | Toronto Invasive Bacterial Diseases Network | McMaster University | Allison McGeer et al |
| EPI_ISL_586418 | Canada/ON-S54/2020 | 2020-03-18 | North America/Canada/Ontario/Toronto | Toronto Invasive Bacterial Diseases Network | McMaster University | Allison McGeer et al |
| EPI_ISL_586419 | Canada/ON-S55/2020 | 2020-03-18 | North America/Canada/Ontario/Toronto | Toronto Invasive Bacterial Diseases Network | McMaster University | Allison McGeer et al |
| EPI_ISL_586420 | Canada/ON-S566/2020 | 2020-04-06 | North America/Canada/Ontario/Toronto | Toronto Invasive Bacterial Diseases Network | McMaster University | Allison McGeer et al |
| EPI_ISL_586421 | Canada/ON-S6/2020 | 2020-03-13 | North America/Canada/Ontario/Toronto | Toronto Invasive Bacterial Diseases Network | McMaster University | Allison McGeer et al |
| EPI_ISL_586422 | Canada/ON-S60/2020 | 2020-03-18 | North America/Canada/Ontario/Toronto | Toronto Invasive Bacterial Diseases Network | McMaster University | Allison McGeer et al |
| EPI_ISL_586423 | Canada/ON-S61/2020 | 2020-03-19 | North America/Canada/Ontario/Toronto | Toronto Invasive Bacterial Diseases Network | McMaster University | Allison McGeer et al |
| EPI_ISL_586424 | Canada/ON-S62/2020 | 2020-03-19 | North America/Canada/Ontario/Toronto | Toronto Invasive Bacterial Diseases Network | McMaster University | Allison McGeer et al |
| EPI_ISL_586425 | Canada/ON-S65/2020 | 2020-03-19 | North America/Canada/Ontario/Toronto | Toronto Invasive Bacterial Diseases Network | McMaster University | Allison McGeer et al |
| EPI_ISL_586426 | Canada/ON-S67/2020 | 2020-03-19 | North America/Canada/Ontario/Toronto | Toronto Invasive Bacterial Diseases Network | McMaster University | Allison McGeer et al |
| EPI_ISL_586427 | Canada/ON-S69/2020 | 2020-03-19 | North America/Canada/Ontario/Toronto | Toronto Invasive Bacterial Diseases Network | McMaster University | Allison McGeer et al |
| EPI_ISL_586428 | Canada/ON-S70/2020 | 2020-03-19 | North America/Canada/Ontario/Toronto | Toronto Invasive Bacterial Diseases Network | McMaster University | Allison McGeer et al |
| EPI_ISL_586429 | Canada/ON-S719/2020 | 20-Apr | North America/Canada/Ontario/Toronto | Toronto Invasive Bacterial Diseases Network | McMaster University | Allison McGeer et al |
| EPI_ISL_586430 | Canada/ON-S73/2020 | 2020-03-19 | North America/Canada/Ontario/Toronto | Toronto Invasive Bacterial Diseases Network | McMaster University | Allison McGeer et al |

| | | | | | | |
|---|---|---|---|---|---|---|
| EPI_ISL_586431 | Canada/ON-S738/2020 | 2020-04-09 | North America/Canada/Ontario/Toronto | Toronto Invasive Bacterial Diseases Network | McMaster University | Allison McGeer et al |
| EPI_ISL_586432 | Canada/ON-S739/2020 | 2020-04-09 | North America/Canada/Ontario/Toronto | Toronto Invasive Bacterial Diseases Network | McMaster University | Allison McGeer et al |
| EPI_ISL_586433 | Canada/ON-S74/2020 | 2020-03-19 | North America/Canada/Ontario/Toronto | Toronto Invasive Bacterial Diseases Network | McMaster University | Allison McGeer et al |
| EPI_ISL_586434 | Canada/ON-S741/2020 | 2020-04-09 | North America/Canada/Ontario/Toronto | Toronto Invasive Bacterial Diseases Network | McMaster University | Allison McGeer et al |
| EPI_ISL_586435 | Canada/ON-S749/2020 | 2020-04-10 | North America/Canada/Ontario/Toronto | Toronto Invasive Bacterial Diseases Network | McMaster University | Allison McGeer et al |
| EPI_ISL_586436 | Canada/ON-S75/2020 | 2020-03-19 | North America/Canada/Ontario/Toronto | Toronto Invasive Bacterial Diseases Network | McMaster University | Allison McGeer et al |
| EPI_ISL_586437 | Canada/ON-S750/2020 | 2020-04-10 | North America/Canada/Ontario/Toronto | Toronto Invasive Bacterial Diseases Network | McMaster University | Allison McGeer et al |
| EPI_ISL_586438 | Canada/ON-S757/2020 | 2020-04-10 | North America/Canada/Ontario/Toronto | Toronto Invasive Bacterial Diseases Network | McMaster University | Allison McGeer et al |
| EPI_ISL_586439 | Canada/ON-S763/2020 | 20-Apr | North America/Canada/Ontario/Toronto | Toronto Invasive Bacterial Diseases Network | McMaster University | Allison McGeer et al |
| EPI_ISL_586440 | Canada/ON-S77/2020 | 2020-03-19 | North America/Canada/Ontario/Toronto | Toronto Invasive Bacterial Diseases Network | McMaster University | Allison McGeer et al |
| EPI_ISL_586441 | Canada/ON-S78/2020 | 2020-03-19 | North America/Canada/Ontario/Toronto | Toronto Invasive Bacterial Diseases Network | McMaster University | Allison McGeer et al |
| EPI_ISL_586442 | Canada/ON-S781/2020 | 2020-04-10 | North America/Canada/Ontario/Toronto | Toronto Invasive Bacterial Diseases Network | McMaster University | Allison McGeer et al |
| EPI_ISL_586443 | Canada/ON-S782/2020 | 2020-04-10 | North America/Canada/Ontario/Toronto | Toronto Invasive Bacterial Diseases Network | McMaster University | Allison McGeer et al |
| EPI_ISL_586444 | Canada/ON-S784/2020 | 2020-04-10 | North America/Canada/Ontario/Toronto | Toronto Invasive Bacterial Diseases Network | McMaster University | Allison McGeer et al |
| EPI_ISL_586445 | Canada/ON-S788/2020 | 2020-04-10 | North America/Canada/Ontario/Toronto | Toronto Invasive Bacterial Diseases Network | McMaster University | Allison McGeer et al |
| EPI_ISL_586446 | Canada/ON-S791/2020 | 2020-04-10 | North America/Canada/Ontario/Toronto | Toronto Invasive Bacterial Diseases Network | McMaster University | Allison McGeer et al |
| EPI_ISL_586448 | Canada/ON-S80/2020 | 2020-03-19 | North America/Canada/Ontario/Toronto | Toronto Invasive Bacterial Diseases Network | McMaster University | Allison McGeer et al |
| EPI_ISL_586449 | Canada/ON-S81/2020 | 2020-03-19 | North America/Canada/Ontario/Toronto | Toronto Invasive Bacterial Diseases Network | McMaster University | Allison McGeer et al |
| EPI_ISL_586450 | Canada/ON-S817/2020 | 20-Apr | North America/Canada/Ontario/Toronto | Toronto Invasive Bacterial Diseases Network | McMaster University | Allison McGeer et al |
| EPI_ISL_586451 | Canada/ON-S818/2020 | 20-Apr | North America/Canada/Ontario/Toronto | Toronto Invasive Bacterial Diseases Network | McMaster University | Allison McGeer et al |

| | | | | | | |
|---|---|---|---|---|---|---|
| EPI_ISL_586452 | Canada/ON-S824/2020 | 2020-04-11 | North America/Canada/Ontario/Toronto | Toronto Invasive Bacterial Diseases Network | McMaster University | Allison McGeer et al |
| EPI_ISL_586453 | Canada/ON-S831/2020 | 2020-04-11 | North America/Canada/Ontario/Toronto | Toronto Invasive Bacterial Diseases Network | McMaster University | Allison McGeer et al |
| EPI_ISL_586454 | Canada/ON-S834/2020 | 20-Apr | North America/Canada/Ontario/Toronto | Toronto Invasive Bacterial Diseases Network | McMaster University | Allison McGeer et al |
| EPI_ISL_586455 | Canada/ON-S835/2020 | 2020-04-11 | North America/Canada/Ontario/Toronto | Toronto Invasive Bacterial Diseases Network | McMaster University | Allison McGeer et al |
| EPI_ISL_586456 | Canada/ON-S84/2020 | 2020-03-20 | North America/Canada/Ontario/Toronto | Toronto Invasive Bacterial Diseases Network | McMaster University | Allison McGeer et al |
| EPI_ISL_586457 | Canada/ON-S842/2020 | 20-Apr | North America/Canada/Ontario/Toronto | Toronto Invasive Bacterial Diseases Network | McMaster University | Allison McGeer et al |
| EPI_ISL_586458 | Canada/ON-S85/2020 | 2020-03-20 | North America/Canada/Ontario/Toronto | Toronto Invasive Bacterial Diseases Network | McMaster University | Allison McGeer et al |
| EPI_ISL_586459 | Canada/ON-S854/2020 | 2020-04-11 | North America/Canada/Ontario/Toronto | Toronto Invasive Bacterial Diseases Network | McMaster University | Allison McGeer et al |
| EPI_ISL_586460 | Canada/ON-S855/2020 | 2020-04-11 | North America/Canada/Ontario/Toronto | Toronto Invasive Bacterial Diseases Network | McMaster University | Allison McGeer et al |
| EPI_ISL_586461 | Canada/ON-S856/2020 | 2020-04-11 | North America/Canada/Ontario/Toronto | Toronto Invasive Bacterial Diseases Network | McMaster University | Allison McGeer et al |
| EPI_ISL_586462 | Canada/ON-S860/2020 | 20-Apr | North America/Canada/Ontario/Toronto | Toronto Invasive Bacterial Diseases Network | McMaster University | Allison McGeer et al |
| EPI_ISL_586463 | Canada/ON-S861/2020 | 2020-04-11 | North America/Canada/Ontario/Toronto | Toronto Invasive Bacterial Diseases Network | McMaster University | Allison McGeer et al |
| EPI_ISL_586464 | Canada/ON-S864/2020 | 20-Apr | North America/Canada/Ontario/Toronto | Toronto Invasive Bacterial Diseases Network | McMaster University | Allison McGeer et al |
| EPI_ISL_586465 | Canada/ON-S867/2020 | 20-Apr | North America/Canada/Ontario/Toronto | Toronto Invasive Bacterial Diseases Network | McMaster University | Allison McGeer et al |
| EPI_ISL_586466 | Canada/ON-S87/2020 | 2020-03-19 | North America/Canada/Ontario/Toronto | Toronto Invasive Bacterial Diseases Network | McMaster University | Allison McGeer et al |
| EPI_ISL_586467 | Canada/ON-S877/2020 | 2020-04-12 | North America/Canada/Ontario/Toronto | Toronto Invasive Bacterial Diseases Network | McMaster University | Allison McGeer et al |
| EPI_ISL_586468 | Canada/ON-S88/2020 | 2020-03-20 | North America/Canada/Ontario/Toronto | Toronto Invasive Bacterial Diseases Network | McMaster University | Allison McGeer et al |
| EPI_ISL_586469 | Canada/ON-S882/2020 | 2020-04-12 | North America/Canada/Ontario/Toronto | Toronto Invasive Bacterial Diseases Network | McMaster University | Allison McGeer et al |
| EPI_ISL_586470 | Canada/ON-S884/2020 | 2020-04-12 | North America/Canada/Ontario/Toronto | Toronto Invasive Bacterial Diseases Network | McMaster University | Allison McGeer et al |
| EPI_ISL_586471 | Canada/ON-S889/2020 | 2020-04-12 | North America/Canada/Ontario/Toronto | Toronto Invasive Bacterial Diseases Network | McMaster University | Allison McGeer et al |

| | | | | | | |
|---|---|---|---|---|---|---|
| EPI_ISL_586472 | Canada/ON-S89/2020 | 2020-03-20 | North America/Canada/Ontario/Toronto | Toronto Invasive Bacterial Diseases Network | McMaster University | Allison McGeer et al |
| EPI_ISL_586473 | Canada/ON-S898/2020 | 2020-04-12 | North America/Canada/Ontario/Toronto | Toronto Invasive Bacterial Diseases Network | McMaster University | Allison McGeer et al |
| EPI_ISL_586474 | Canada/ON-S901/2020 | 2020-04-12 | North America/Canada/Ontario/Toronto | Toronto Invasive Bacterial Diseases Network | McMaster University | Allison McGeer et al |
| EPI_ISL_586475 | Canada/ON-S902/2020 | 2020-04-12 | North America/Canada/Ontario/Toronto | Toronto Invasive Bacterial Diseases Network | McMaster University | Allison McGeer et al |
| EPI_ISL_586476 | Canada/ON-S903/2020 | 2020-04-12 | North America/Canada/Ontario/Toronto | Toronto Invasive Bacterial Diseases Network | McMaster University | Allison McGeer et al |
| EPI_ISL_586477 | Canada/ON-S904/2020 | 2020-04-12 | North America/Canada/Ontario/Toronto | Toronto Invasive Bacterial Diseases Network | McMaster University | Allison McGeer et al |
| EPI_ISL_586478 | Canada/ON-S905/2020 | 2020-04-12 | North America/Canada/Ontario/Toronto | Toronto Invasive Bacterial Diseases Network | McMaster University | Allison McGeer et al |
| EPI_ISL_586479 | Canada/ON-S906/2020 | 2020-04-12 | North America/Canada/Ontario/Toronto | Toronto Invasive Bacterial Diseases Network | McMaster University | Allison McGeer et al |
| EPI_ISL_586480 | Canada/ON-S907/2020 | 2020-04-13 | North America/Canada/Ontario/Toronto | Toronto Invasive Bacterial Diseases Network | McMaster University | Allison McGeer et al |
| EPI_ISL_586481 | Canada/ON-S91/2020 | 2020-03-20 | North America/Canada/Ontario/Toronto | Toronto Invasive Bacterial Diseases Network | McMaster University | Allison McGeer et al |
| EPI_ISL_586482 | Canada/ON-S911/2020 | 2020-04-13 | North America/Canada/Ontario/Toronto | Toronto Invasive Bacterial Diseases Network | McMaster University | Allison McGeer et al |
| EPI_ISL_586483 | Canada/ON-S915/2020 | 2020-04-13 | North America/Canada/Ontario/Toronto | Toronto Invasive Bacterial Diseases Network | McMaster University | Allison McGeer et al |
| EPI_ISL_586484 | Canada/ON-S916/2020 | 2020-04-13 | North America/Canada/Ontario/Toronto | Toronto Invasive Bacterial Diseases Network | McMaster University | Allison McGeer et al |
| EPI_ISL_586485 | Canada/ON-S917/2020 | 2020-04-13 | North America/Canada/Ontario/Toronto | Toronto Invasive Bacterial Diseases Network | McMaster University | Allison McGeer et al |
| EPI_ISL_586486 | Canada/ON-S918/2020 | 2020-04-13 | North America/Canada/Ontario/Toronto | Toronto Invasive Bacterial Diseases Network | McMaster University | Allison McGeer et al |
| EPI_ISL_586487 | Canada/ON-S919/2020 | 2020-04-13 | North America/Canada/Ontario/Toronto | Toronto Invasive Bacterial Diseases Network | McMaster University | Allison McGeer et al |
| EPI_ISL_586488 | Canada/ON-S920/2020 | 2020-04-13 | North America/Canada/Ontario/Toronto | Toronto Invasive Bacterial Diseases Network | McMaster University | Allison McGeer et al |
| EPI_ISL_586489 | Canada/ON-S93/2020 | 2020-03-20 | North America/Canada/Ontario/Toronto | Toronto Invasive Bacterial Diseases Network | McMaster University | Allison McGeer et al |
| EPI_ISL_586490 | Canada/ON-S932/2020 | 2020-04-13 | North America/Canada/Ontario/Toronto | Toronto Invasive Bacterial Diseases Network | McMaster University | Allison McGeer et al |
| EPI_ISL_586491 | Canada/ON-S933/2020 | 2020-04-13 | North America/Canada/Ontario/Toronto | Toronto Invasive Bacterial Diseases Network | McMaster University | Allison McGeer et al |

| | | | | | | |
|---|---|---|---|---|---|---|
| EPI_ISL_586492 | Canada/ON-S934/2020 | 2020-04-13 | North America/Canada/Ontario/Toronto | Toronto Invasive Bacterial Diseases Network | McMaster University | Allison McGeer et al |
| EPI_ISL_586493 | Canada/ON-S935/2020 | 2020-04-13 | North America/Canada/Ontario/Toronto | Toronto Invasive Bacterial Diseases Network | McMaster University | Allison McGeer et al |
| EPI_ISL_586494 | Canada/ON-S936/2020 | 2020-04-13 | North America/Canada/Ontario/Toronto | Toronto Invasive Bacterial Diseases Network | McMaster University | Allison McGeer et al |
| EPI_ISL_586495 | Canada/ON-S938/2020 | 2020-04-13 | North America/Canada/Ontario/Toronto | Toronto Invasive Bacterial Diseases Network | McMaster University | Allison McGeer et al |
| EPI_ISL_586496 | Canada/ON-S939/2020 | 2020-04-13 | North America/Canada/Ontario/Toronto | Toronto Invasive Bacterial Diseases Network | McMaster University | Allison McGeer et al |
| EPI_ISL_586497 | Canada/ON-S94/2020 | 2020-03-20 | North America/Canada/Ontario/Toronto | Toronto Invasive Bacterial Diseases Network | McMaster University | Allison McGeer et al |
| EPI_ISL_586498 | Canada/ON-S945/2020 | 2020-04-14 | North America/Canada/Ontario/Toronto | Toronto Invasive Bacterial Diseases Network | McMaster University | Allison McGeer et al |
| EPI_ISL_586499 | Canada/ON-S946/2020 | 2020-04-14 | North America/Canada/Ontario/Toronto | Toronto Invasive Bacterial Diseases Network | McMaster University | Allison McGeer et al |
| EPI_ISL_586500 | Canada/ON-S947/2020 | 2020-04-14 | North America/Canada/Ontario/Toronto | Toronto Invasive Bacterial Diseases Network | McMaster University | Allison McGeer et al |
| EPI_ISL_586501 | Canada/ON-S948/2020 | 2020-04-14 | North America/Canada/Ontario/Toronto | Toronto Invasive Bacterial Diseases Network | McMaster University | Allison McGeer et al |
| EPI_ISL_586502 | Canada/ON-S949/2020 | 2020-04-14 | North America/Canada/Ontario/Toronto | Toronto Invasive Bacterial Diseases Network | McMaster University | Allison McGeer et al |
| EPI_ISL_586503 | Canada/ON-S95/2020 | 2020-03-21 | North America/Canada/Ontario/Toronto | Toronto Invasive Bacterial Diseases Network | McMaster University | Allison McGeer et al |
| EPI_ISL_586504 | Canada/ON-S952/2020 | 2020-04-14 | North America/Canada/Ontario/Toronto | Toronto Invasive Bacterial Diseases Network | McMaster University | Allison McGeer et al |
| EPI_ISL_586505 | Canada/ON-S96/2020 | 2020-03-21 | North America/Canada/Ontario/Toronto | Toronto Invasive Bacterial Diseases Network | McMaster University | Allison McGeer et al |
| EPI_ISL_586506 | Canada/ON-S97/2020 | 2020-03-21 | North America/Canada/Ontario/Toronto | Toronto Invasive Bacterial Diseases Network | McMaster University | Allison McGeer et al |
| EPI_ISL_586507 | Canada/ON-S98/2020 | 2020-03-21 | North America/Canada/Ontario/Toronto | Toronto Invasive Bacterial Diseases Network | McMaster University | Allison McGeer et al |
| EPI_ISL_586508 | Canada/ON-S99/2020 | 2020-03-21 | North America/Canada/Ontario/Toronto | Toronto Invasive Bacterial Diseases Network | McMaster University | Allison McGeer et al |
| EPI_ISL_586515 | India/GJ-GBRC-399a/2020 | 2020-06-25 | Asia/India/Gujarat/Daskroi | B.J. Medical College and Civil hospital, Ahmedabad | Gujarat Biotechnology Research Centre | Nidhi Patel et al |
| EPI_ISL_586517 | India/GJ-GBRC-400a/2020 | 2020-06-25 | Asia/India/Gujarat/Ahmedabad | B.J. Medical College and Civil hospital, Ahmedabad | Gujarat Biotechnology Research Centre | Raghawendra Kumar et al |

| | | | | | | |
|---|---|---|---|---|---|---|
| EPI_ISL_586518 | India/GJ-GBRC-400b/2020 | 2020-06-25 | Asia/India/Gujarat/Ahmedabad | B.J. Medical College and Civil hospital, Ahmedabad | Gujarat Biotechnology Research Centre | Dinesh Kumar et al |
| EPI_ISL_586519 | India/GJ-GBRC-401/2020 | 2020-06-25 | Asia/India/Gujarat/Ahmedabad | B.J. Medical College and Civil hospital, Ahmedabad | Gujarat Biotechnology Research Centre | Zuber Saiyed et al |
| EPI_ISL_586521 | India/GJ-GBRC-402b/2020 | 2020-06-25 | Asia/India/Gujarat/Ahmedabad | B.J. Medical College and Civil hospital, Ahmedabad | Gujarat Biotechnology Research Centre | Labdhi Pandya et al |
| EPI_ISL_586522 | India/GJ-GBRC-403a/2020 | 2020-06-25 | Asia/India/Gujarat/Ahmedabad | B.J. Medical College and Civil hospital, Ahmedabad | Gujarat Biotechnology Research Centre | Afzal Ansari et al |
| EPI_ISL_586523 | India/GJ-GBRC-403b/2020 | 2020-06-25 | Asia/India/Gujarat/Ahmedabad | B.J. Medical College and Civil hospital, Ahmedabad | Gujarat Biotechnology Research Centre | Nikha Trivedi et al |
| EPI_ISL_586531 | India/GJ-GBRC-408a/2020 | 2020-07-18 | Asia/India/Gujarat/Bodeli | General Hospital,Chhotaudepur | Gujarat Biotechnology Research Centre | Yogesh Parmar et al |
| EPI_ISL_586533 | India/GJ-GBRC-409a/2020 | 2020-07-18 | Asia/India/Gujarat/Bodeli | General Hospital,Chhotaudepur | Gujarat Biotechnology Research Centre | Dharak Pandya et al |
| EPI_ISL_586534 | India/GJ-GBRC-409b/2020 | 2020-07-18 | Asia/India/Gujarat/Bodeli | General Hospital,Chhotaudepur | Gujarat Biotechnology Research Centre | Maharshi Pandya et al |
| EPI_ISL_586535 | India/GJ-GBRC-410/2020 | 2020-07-18 | Asia/India/Gujarat/Bodeli | General Hospital,Chhotaudepur | Gujarat Biotechnology Research Centre | Nidhi Patel et al |
| EPI_ISL_586536 | India/GJ-GBRC-411/2020 | 2020-07-18 | Asia/India/Gujarat/Limkheda | Zydus Medical Hospital,Dahod | Gujarat Biotechnology Research Centre | Kalpesh Labana et al |
| EPI_ISL_586537 | India/GJ-GBRC-412a/2020 | 2020-07-18 | Asia/India/Gujarat/Fatepura | Zydus Medical Hospital,Dahod | Gujarat Biotechnology Research Centre | Mohit Desai et al |
| EPI_ISL_586538 | India/GJ-GBRC-412b/2020 | 2020-07-18 | Asia/India/Gujarat/Fatepura | Zydus Medical Hospital,Dahod | Gujarat Biotechnology Research Centre | Nitin Savaliya et al |
| EPI_ISL_586539 | India/GJ-GBRC-413a/2020 | 2020-07-18 | Asia/India/Gujarat/Dahod | Zydus Medical Hospital,Dahod | Gujarat Biotechnology Research Centre | Raghawendra Kumar et al |

| | | | | | | |
|---|---|---|---|---|---|---|
| EPI_ISL_586541 | India/GJ-GBRC-414/2020 | 2020-07-18 | Asia/India/Gujarat/Dahod | Zydus Medical Hospital,Dahod | Gujarat Biotechnology Research Centre | Zuber Saiyed et al |
| EPI_ISL_586542 | India/GJ-GBRC-415a/2020 | 2020-07-11 | Asia/India/Gujarat/Khambhaliya | Government General Hospital, Jam Khambhaliya, Devbhoomi Dwarka | Gujarat Biotechnology Research Centre | Komal Patel et al |
| EPI_ISL_586543 | India/GJ-GBRC-415b/2020 | 2020-07-11 | Asia/India/Gujarat/Khambhaliya | Government General Hospital, Jam Khambhaliya, Devbhoomi Dwarka | Gujarat Biotechnology Research Centre | Labdhi Pandya et al |
| EPI_ISL_586546 | India/GJ-GBRC-417a/2020 | 2020-07-18 | Asia/India/Gujarat/Godhra | General Hospital,Godhra | Gujarat Biotechnology Research Centre | Neela Katara et al |
| EPI_ISL_586547 | India/GJ-GBRC-417b/2020 | 2020-07-18 | Asia/India/Gujarat/Godhra | General Hospital,Godhra | Gujarat Biotechnology Research Centre | Pinal Gandhi et al |
| EPI_ISL_586549 | India/GJ-GBRC-419a/2020 | 2020-07-14 | Asia/India/Gujarat/Sutrapada | CCC,Veraval | Gujarat Biotechnology Research Centre | Jignesh Parmar et al |
| EPI_ISL_586550 | India/GJ-GBRC-419b/2020 | 2020-07-14 | Asia/India/Gujarat/Sutrapada | CCC,Veraval | Gujarat Biotechnology Research Centre | Jitendra Bamrotia et al |
| EPI_ISL_586551 | India/GJ-GBRC-422a/2020 | 2020-07-14 | Asia/India/Gujarat/Sutrapada | Government Hospital,Veraval | Gujarat Biotechnology Research Centre | Monika Gandhi et al |
| EPI_ISL_586553 | India/GJ-GBRC-423a/2020 | 2020-07-14 | Asia/India/Gujarat/Veraval | Government Hospital,Veraval | Gujarat Biotechnology Research Centre | Maharshi Pandya et al |
| EPI_ISL_586554 | India/GJ-GBRC-423b/2020 | 2020-07-14 | Asia/India/Gujarat/Veraval | Government Hospital,Veraval | Gujarat Biotechnology Research Centre | Nidhi Patel et al |
| EPI_ISL_586555 | India/GJ-GBRC-424a/2020 | 2020-07-14 | Asia/India/Gujarat/Talala | CCC,Veraval | Gujarat Biotechnology Research Centre | Nitin Savaliya et al |
| EPI_ISL_586556 | India/GJ-GBRC-424b/2020 | 2020-07-14 | Asia/India/Gujarat/Talala | CCC,Veraval | Gujarat Biotechnology Research Centre | Raghawendra Kumar et al |
| EPI_ISL_586557 | India/GJ-GBRC-425/2020 | 2020-07-14 | Asia/India/Gujarat/Una | CCC,Veraval | Gujarat Biotechnology Research Centre | Dinesh Kumar et al |

| | | | | | | |
|---|---|---|---|---|---|---|
| EPI_ISL_586558 | India/GJ-GBRC-426a/2020 | 2020-07-14 | Asia/India/Gujarat/Sutrapada | CCC,Veraval | Gujarat Biotechnology Research Centre | Zuber Saiyed et al |
| EPI_ISL_586559 | India/GJ-GBRC-426b/2020 | 2020-07-14 | Asia/India/Gujarat/Sutrapada | CCC,Veraval | Gujarat Biotechnology Research Centre | Komal Patel et al |
| EPI_ISL_586560 | India/GJ-GBRC-427a/2020 | 2020-07-14 | Asia/India/Gujarat/Kodinar | CCC,Veraval | Gujarat Biotechnology Research Centre | Labdhi Pandya et al |
| EPI_ISL_586561 | India/GJ-GBRC-427b/2020 | 2020-07-14 | Asia/India/Gujarat/Kodinar | CCC,Veraval | Gujarat Biotechnology Research Centre | Afzal Ansari et al |
| EPI_ISL_586562 | India/GJ-GBRC-428a/2020 | 2020-07-15 | Asia/India/Gujarat/Mangrol | GMERS GMERS Medical College and Hospital, Junagadh | Gujarat Biotechnology Research Centre | Nikha Trivedi et al |
| EPI_ISL_586564 | India/GJ-GBRC-429/2020 | 2020-07-15 | Asia/India/Gujarat/Junagadh | GMERS GMERS Medical College and Hospital, Junagadh | Gujarat Biotechnology Research Centre | Ravi Dedaniya et al |
| EPI_ISL_586565 | India/GJ-GBRC-430a/2020 | 2020-07-15 | Asia/India/Gujarat/Mangrol | GMERS GMERS Medical College and Hospital, Junagadh | Gujarat Biotechnology Research Centre | Apurvasinh Puvar et al |
| EPI_ISL_586566 | India/GJ-GBRC-430b/2020 | 2020-07-15 | Asia/India/Gujarat/Mangrol | GMERS GMERS Medical College and Hospital, Junagadh | Gujarat Biotechnology Research Centre | Janvi Raval et al |
| EPI_ISL_586567 | India/GJ-GBRC-431a/2020 | 2020-07-06 | Asia/India/Gujarat/Mehsana | Saikrishna Hospital,Mehsana | Gujarat Biotechnology Research Centre | Zarna Patel et al |
| EPI_ISL_586568 | India/GJ-GBRC-431b/2020 | 2020-07-06 | Asia/India/Gujarat/Mehsana | Saikrishna Hospital,Mehsana | Gujarat Biotechnology Research Centre | Zarna Patel et al |
| EPI_ISL_586569 | India/GJ-GBRC-432/2020 | 2020-09-03 | Asia/India/Gujarat/Ahmedabad | GCRI,Ahmedabad | Gujarat Biotechnology Research Centre | Shashank Pandya et al |
| EPI_ISL_586570 | India/GJ-GBRC-433/2020 | 2020-09-01 | Asia/India/Gujarat/Ahmedabad | GCRI,Ahmedabad | Gujarat Biotechnology Research Centre | Harsha Panchal et al |
| EPI_ISL_586718 | England/ALDP-A2E47F/2020 | 2020-10-06 | Europe/United Kingdom/England/ | Lighthouse Lab in Alderley Park | Wellcome Sanger Institute for the COVID-19 Genomics UK (COG-UK) consortium | Jacquelyn Wynn et al |

| | | | | | | |
|---|---|---|---|---|---|---|
| EPI_ISL_586821 | England/ALDP-A2D2D0/2020 | 2020-10-06 | Europe/United Kingdom/England/ | Lighthouse Lab in Alderley Park | Wellcome Sanger Institute for the COVID-19 Genomics UK (COG-UK) consortium | Jacquelyn Wynn et al |
| EPI_ISL_586972 | England/ALDP-A2ED47/2020 | 2020-10-05 | Europe/United Kingdom/England/ | Lighthouse Lab in Alderley Park | Wellcome Sanger Institute for the COVID-19 Genomics UK (COG-UK) consortium | Jacquelyn Wynn et al |
| EPI_ISL_587007 | England/ALDP-A2F915/2020 | 2020-10-06 | Europe/United Kingdom/England/ | Lighthouse Lab in Alderley Park | Wellcome Sanger Institute for the COVID-19 Genomics UK (COG-UK) consortium | Jacquelyn Wynn et al |
| EPI_ISL_587105 | England/ALDP-A2FA12/2020 | 2020-10-06 | Europe/United Kingdom/England/ | Lighthouse Lab in Alderley Park | Wellcome Sanger Institute for the COVID-19 Genomics UK (COG-UK) consortium | Jacquelyn Wynn et al |
| EPI_ISL_587399 | England/QEUH-A21F7B/2020 | 2020-10-01 | Europe/United Kingdom/England/ | Lighthouse Lab in Glasgow | Wellcome Sanger Institute for the COVID-19 Genomics UK (COG-UK) consortium | Harper VanSteenhouse et al |
| EPI_ISL_587476 | England/QEUH-A1EFBD/2020 | 2020-09-29 | Europe/United Kingdom/England/ | Lighthouse Lab in Glasgow | Wellcome Sanger Institute for the COVID-19 Genomics UK (COG-UK) consortium | Harper VanSteenhouse et al |
| EPI_ISL_587621 | England/QEUH-A20237/2020 | 2020-10-01 | Europe/United Kingdom/England/ | Lighthouse Lab in Glasgow | Wellcome Sanger Institute for the COVID-19 Genomics UK (COG-UK) consortium | Harper VanSteenhouse et al |
| EPI_ISL_587664 | England/QEUH-A21E9C/2020 | 2020-10-01 | Europe/United Kingdom/England/ | Lighthouse Lab in Glasgow | Wellcome Sanger Institute for the COVID-19 Genomics UK (COG-UK) consortium | Harper VanSteenhouse et al |

| | | | | | | |
|---|---|---|---|---|---|---|
| EPI_ISL_587665 | England/QEUH-A2161C/2020 | 2020-10-01 | Europe/United Kingdom/England/ | Lighthouse Lab in Glasgow | Wellcome Sanger Institute for the COVID-19 Genomics UK (COG-UK) consortium | Harper VanSteenhouse et al |
| EPI_ISL_587666 | England/QEUH-A208F9/2020 | 2020-10-01 | Europe/United Kingdom/England/ | Lighthouse Lab in Glasgow | Wellcome Sanger Institute for the COVID-19 Genomics UK (COG-UK) consortium | Harper VanSteenhouse et al |
| EPI_ISL_587711 | England/ALDP-9FCE31/2020 | 2020-10-01 | Europe/United Kingdom/England/ | Lighthouse Lab in Alderley Park | Wellcome Sanger Institute for the COVID-19 Genomics UK (COG-UK) consortium | Jacquelyn Wynn et al |
| EPI_ISL_587898 | England/ALDP-9FCB67/2020 | 2020-10-01 | Europe/United Kingdom/England/ | Lighthouse Lab in Alderley Park | Wellcome Sanger Institute for the COVID-19 Genomics UK (COG-UK) consortium | Jacquelyn Wynn et al |
| EPI_ISL_587947 | England/ALDP-9FCEE6/2020 | 2020-10-01 | Europe/United Kingdom/England/ | Lighthouse Lab in Alderley Park | Wellcome Sanger Institute for the COVID-19 Genomics UK (COG-UK) consortium | Jacquelyn Wynn et al |
| EPI_ISL_587966 | England/ALDP-9FCE9B/2020 | 2020-10-01 | Europe/United Kingdom/England/ | Lighthouse Lab in Alderley Park | Wellcome Sanger Institute for the COVID-19 Genomics UK (COG-UK) consortium | Jacquelyn Wynn et al |
| EPI_ISL_588078 | England/QEUH-9FA4E6/2020 | 2020-09-29 | Europe/United Kingdom/England/ | Lighthouse Lab in Glasgow | Wellcome Sanger Institute for the COVID-19 Genomics UK (COG-UK) consortium | Harper VanSteenhouse et al |
| EPI_ISL_588208 | England/QEUH-9FAEE8/2020 | 2020-09-27 | Europe/United Kingdom/England/ | Lighthouse Lab in Glasgow | Wellcome Sanger Institute for the COVID-19 Genomics UK (COG-UK) consortium | Harper VanSteenhouse et al |

| | | | | | | |
|---|---|---|---|---|---|---|
| EPI_ISL_588302 | England/QEUH-9F4EEE/2020 | 2020-09-25 | Europe/United Kingdom/England/ | Lighthouse Lab in Glasgow | Wellcome Sanger Institute for the COVID-19 Genomics UK (COG-UK) consortium | Harper VanSteenhouse et al |
| EPI_ISL_588845 | England/QEUH-9F8600/2020 | 2020-09-25 | Europe/United Kingdom/England/ | Lighthouse Lab in Glasgow | Wellcome Sanger Institute for the COVID-19 Genomics UK (COG-UK) Consortium | Harper VanSteenhouse et al |
| EPI_ISL_588901 | England/QEUH-9F9371/2020 | 2020-09-25 | Europe/United Kingdom/England/ | Lighthouse Lab in Glasgow | Wellcome Sanger Institute for the COVID-19 Genomics UK (COG-UK) consortium | Harper VanSteenhouse et al |
| EPI_ISL_588957 | England/QEUH-9F8D47/2020 | 2020-09-27 | Europe/United Kingdom/England/ | Lighthouse Lab in Glasgow | Wellcome Sanger Institute for the COVID-19 Genomics UK (COG-UK) consortium | Harper VanSteenhouse et al |
| EPI_ISL_589019 | England/MILK-9F25BE/2020 | 2020-09-24 | Europe/United Kingdom/England/ | Lighthouse Lab in Milton Keynes | Wellcome Sanger Institute for the COVID-19 Genomics UK (COG-UK) consortium | The Lighthouse Lab in Milton Keynes et al |
| EPI_ISL_589030 | England/MILK-9F267F/2020 | 2020-09-24 | Europe/United Kingdom/England/ | Lighthouse Lab in Milton Keynes | Wellcome Sanger Institute for the COVID-19 Genomics UK (COG-UK) consortium | The Lighthouse Lab in Milton Keynes et al |
| EPI_ISL_589079 | England/MILK-9F2369/2020 | 2020-09-24 | Europe/United Kingdom/England/ | Lighthouse Lab in Milton Keynes | Wellcome Sanger Institute for the COVID-19 Genomics UK (COG-UK) consortium | The Lighthouse Lab in Milton Keynes et al |
| EPI_ISL_589102 | NorthernIreland/QEUH-9F9915/2020 | 2020-09-25 | Europe/United Kingdom/Northern Irelar | Lighthouse Lab in Glasgow | Wellcome Sanger Institute for the COVID-19 Genomics UK (COG-UK) consortium | Harper VanSteenhouse et al |

| | | | | | | |
|---|---|---|---|---|---|---|
| EPI_ISL_589319 | England/ALDP-9ED6D1/2020 | 2020-05-08 | Europe/United Kingdom/England/ | Lighthouse Lab in Alderley Park | Wellcome Sanger Institute for the COVID-19 Genomics UK (COG-UK) consortium | Jacquelyn Wynn et al |
| EPI_ISL_589379 | England/ALDP-9EE0B4/2020 | 2020-05-06 | Europe/United Kingdom/England/ | Lighthouse Lab in Alderley Park | Wellcome Sanger Institute for the COVID-19 Genomics UK (COG-UK) consortium | Jacquelyn Wynn et al |
| EPI_ISL_589384 | England/ALDP-9EE078/2020 | 2020-05-08 | Europe/United Kingdom/England/ | Lighthouse Lab in Alderley Park | Wellcome Sanger Institute for the COVID-19 Genomics UK (COG-UK) consortium | Jacquelyn Wynn et al |
| EPI_ISL_589403 | England/ALDP-9ED853/2020 | 2020-05-05 | Europe/United Kingdom/England/ | Lighthouse Lab in Alderley Park | Wellcome Sanger Institute for the COVID-19 Genomics UK (COG-UK) consortium | Jacquelyn Wynn et al |
| EPI_ISL_589410 | England/ALDP-9EE1C0/2020 | 2020-05-06 | Europe/United Kingdom/England/ | Lighthouse Lab in Alderley Park | Wellcome Sanger Institute for the COVID-19 Genomics UK (COG-UK) consortium | Jacquelyn Wynn et al |
| EPI_ISL_589440 | England/MILK-9ECBC4/2020 | 2020-04-26 | Europe/United Kingdom/England/ | Lighthouse Lab in Milton Keynes | Wellcome Sanger Institute for the COVID-19 Genomics UK (COG-UK) consortium | The Lighthouse Lab in Milton Keynes et al |
| EPI_ISL_589480 | England/ALDP-9EDDCD/2020 | 2020-05-06 | Europe/United Kingdom/England/ | Lighthouse Lab in Alderley Park | Wellcome Sanger Institute for the COVID-19 Genomics UK (COG-UK) consortium | Jacquelyn Wynn et al |
| EPI_ISL_589485 | England/ALDP-9ED64A/2020 | 2020-05-18 | Europe/United Kingdom/England/ | Lighthouse Lab in Alderley Park | Wellcome Sanger Institute for the COVID-19 Genomics UK (COG-UK) consortium | Jacquelyn Wynn et al |

| | | | | | | |
|---|---|---|---|---|---|---|
| EPI_ISL_589524 | England/ALDP-9EDEF7/2020 | 2020-05-03 | Europe/United Kingdom/England/ | Lighthouse Lab in Alderley Park | Wellcome Sanger Institute for the COVID-19 Genomics UK (COG-UK) consortium | Jacquelyn Wynn et al |
| EPI_ISL_589558 | England/ALDP-9EE0E1/2020 | 2020-05-06 | Europe/United Kingdom/England/ | Lighthouse Lab in Alderley Park | Wellcome Sanger Institute for the COVID-19 Genomics UK (COG-UK) consortium | Jacquelyn Wynn et al |
| EPI_ISL_589561 | England/ALDP-9ECF04/2020 | 2020-05-04 | Europe/United Kingdom/England/ | Lighthouse Lab in Alderley Park | Wellcome Sanger Institute for the COVID-19 Genomics UK (COG-UK) consortium | Jacquelyn Wynn et al |
| EPI_ISL_589678 | England/MILK-9F1898/2020 | 2020-09-25 | Europe/United Kingdom/England/ | Lighthouse Lab in Milton Keynes | Wellcome Sanger Institute for the COVID-19 Genomics UK (COG-UK) consortium | The Lighthouse Lab in Milton Keynes et al |
| EPI_ISL_589985 | England/QEUH-9D0668/2020 | 2020-09-15 | Europe/United Kingdom/England/ | Lighthouse Lab in Glasgow | Wellcome Sanger Institute for the COVID-19 Genomics UK (COG-UK) consortium | Harper VanSteenhouse et al |
| EPI_ISL_589989 | England/QEUH-9D0FC7/2020 | 2020-09-19 | Europe/United Kingdom/England/ | Lighthouse Lab in Glasgow | Wellcome Sanger Institute for the COVID-19 Genomics UK (COG-UK) consortium | Harper VanSteenhouse et al |
| EPI_ISL_590068 | England/QEUH-9CEB5F/2020 | 2020-09-17 | Europe/United Kingdom/England/ | Lighthouse Lab in Glasgow | Wellcome Sanger Institute for the COVID-19 Genomics UK (COG-UK) consortium | Harper VanSteenhouse et al |
| EPI_ISL_590356 | England/QEUH-9B4916/2020 | 2020-09-03 | Europe/United Kingdom/England/ | Lighthouse Lab in Glasgow | Wellcome Sanger Institute for the COVID-19 Genomics UK (COG-UK) consortium | Harper VanSteenhouse et al |

| | | | | | | |
|---|---|---|---|---|---|---|
| EPI_ISL_590421 | England/QEUH-9B4767/2020 | 2020-09-04 | Europe/United Kingdom/England/ | Lighthouse Lab in Glasgow | Wellcome Sanger Institute for the COVID-19 Genomics UK (COG-UK) consortium | Harper VanSteenhouse et al |
| EPI_ISL_590521 | England/QEUH-9B7315/2020 | 2020-09-05 | Europe/United Kingdom/England/ | Lighthouse Lab in Glasgow | Wellcome Sanger Institute for the COVID-19 Genomics UK (COG-UK) consortium | Harper VanSteenhouse et al |
| EPI_ISL_590546 | England/QEUH-9B763A/2020 | 2020-09-08 | Europe/United Kingdom/England/ | Lighthouse Lab in Glasgow | Wellcome Sanger Institute for the COVID-19 Genomics UK (COG-UK) consortium | Harper VanSteenhouse et al |
| EPI_ISL_590689 | India/GJ-GBRC-406a/2020 | 2020-06-25 | Asia/India/Gujarat/Ahmedabad | B.J. Medical College and Civil hospital, Ahmedabad | Gujarat Biotechnology Research Centre | Janvi Raval et al |
| EPI_ISL_590690 | India/GJ-GBRC-420/2020 | 2020-07-14 | Asia/India/Gujarat/Veraval | CCC,Veraval | Gujarat Biotechnology Research Centre | Apurvasinh Puvar et al |
| EPI_ISL_590692 | India/GJ-GBRC-421b/2020 | 2020-07-14 | Asia/India/Gujarat/Veraval | Government Hospital,Veraval | Gujarat Biotechnology Research Centre | Zarna Patel et al |
| EPI_ISL_590701 | USA/MI-UM-10036119854/2020 | 2020-09-24 | North America/USA/Michigan/ | University of Michigan Clinical Microbiology Laboratory | Lauring Lab, University of Michigan, Department of Microbiology and Immunology | Valesano et al |
| EPI_ISL_590706 | USA/MI-UM-10036117092/2020 | 2020-09-24 | North America/USA/Michigan/ | University of Michigan Clinical Microbiology Laboratory | Lauring Lab, University of Michigan, Department of Microbiology and Immunology | Valesano et al |

| | | | | | | |
|---|---|---|---|---|---|---|
| EPI_ISL_590708 | USA/MI-UM-10036114941/2020 | 2020-09-24 | North America/USA/Michigan/ | University of Michigan Clinical Microbiology Laboratory | Lauring Lab, University of Michigan, Department of Microbiology and Immunology | Valesano et al |
| EPI_ISL_590709 | USA/MI-UM-10036133006/2020 | 2020-09-25 | North America/USA/Michigan/ | University of Michigan Clinical Microbiology Laboratory | Lauring Lab, University of Michigan, Department of Microbiology and Immunology | Valesano et al |
| EPI_ISL_590711 | USA/MI-UM-10036138820/2020 | 2020-09-25 | North America/USA/Michigan/ | University of Michigan Clinical Microbiology Laboratory | Lauring Lab, University of Michigan, Department of Microbiology and Immunology | Valesano et al |
| EPI_ISL_590713 | USA/MI-UM-10036099219/2020 | 2020-09-23 | North America/USA/Michigan/ | University of Michigan Clinical Microbiology Laboratory | Lauring Lab, University of Michigan, Department of Microbiology and Immunology | Valesano et al |
| EPI_ISL_590725 | USA/MI-UM-10036147209/2020 | 2020-09-26 | North America/USA/Michigan/ | University of Michigan Clinical Microbiology Laboratory | Lauring Lab, University of Michigan, Department of Microbiology and Immunology | Valesano et al |
| EPI_ISL_590726 | USA/MI-UM-10036150967/2020 | 2020-09-26 | North America/USA/Michigan/ | University of Michigan Clinical Microbiology Laboratory | Lauring Lab, University of Michigan, Department of Microbiology and Immunology | Valesano et al |

| | | | | | | |
|---|---|---|---|---|---|---|
| EPI_ISL_590728 | USA/MI-UM-10036150256/2020 | 2020-09-26 | North America/USA/Michigan/ | University of Michigan Clinical Microbiology Laboratory | Lauring Lab, University of Michigan, Department of Microbiology and Immunology | Valesano et al |
| EPI_ISL_590732 | USA/MI-UM-10036150579/2020 | 2020-09-26 | North America/USA/Michigan/ | University of Michigan Clinical Microbiology Laboratory | Lauring Lab, University of Michigan, Department of Microbiology and Immunology | Valesano et al |
| EPI_ISL_590742 | USA/MI-UM-10036212950/2020 | 2020-09-30 | North America/USA/Michigan/ | University of Michigan Clinical Microbiology Laboratory | Lauring Lab, University of Michigan, Department of Microbiology and Immunology | Valesano et al |
| EPI_ISL_590746 | USA/MI-UM-10036182091/2020 | 2020-09-29 | North America/USA/Michigan/ | University of Michigan Clinical Microbiology Laboratory | Lauring Lab, University of Michigan, Department of Microbiology and Immunology | Valesano et al |
| EPI_ISL_590758 | USA/MN-MDH-1753/2020 | 2020-04-10 | North America/USA/Minnesota/ | Minnesota Department of Health, Public Health Laboratory | Minnesota Department of Health, Public Health Laboratory | Matt Plumb et al |
| EPI_ISL_590772 | USA/MN-MDH-1768/2020 | 2020-04-11 | North America/USA/Minnesota/ | Minnesota Department of Health, Public Health Laboratory | Minnesota Department of Health, Public Health Laboratory | Matt Plumb et al |
| EPI_ISL_590779 | USA/MN-MDH-1776/2020 | 2020-04-13 | North America/USA/Minnesota/ | Minnesota Department of Health, Public Health Laboratory | Minnesota Department of Health, Public Health Laboratory | Matt Plumb et al |
| EPI_ISL_590797 | USA/MN-MDH-1795/2020 | 2020-04-14 | North America/USA/Minnesota/ | Minnesota Department of Health, Public Health Laboratory | Minnesota Department of Health, Public Health Laboratory | Matt Plumb et al |

| | | | | | | |
|---|---|---|---|---|---|---|
| EPI_ISL_590798 | USA/MN-MDH-1796/2020 | 2020-04-14 | North America/USA/Minnesota/ | Minnesota Department of Health, Public Health Laboratory | Minnesota Department of Health, Public Health Laboratory | Matt Plumb et al |
| EPI_ISL_590809 | USA/MN-MDH-1807/2020 | 2020-04-14 | North America/USA/Minnesota/ | Minnesota Department of Health, Public Health Laboratory | Minnesota Department of Health, Public Health Laboratory | Matt Plumb et al |
| EPI_ISL_590811 | USA/MN-MDH-1809/2020 | 2020-04-14 | North America/USA/Minnesota/ | Minnesota Department of Health, Public Health Laboratory | Minnesota Department of Health, Public Health Laboratory | Matt Plumb et al |
| EPI_ISL_590828 | Switzerland/ZH-UZH-IMV6/2020 | 2020-03-29 | Europe/Switzerland/Z√ºrich/ | Institute of Medical Virology, University of Zurich | Institute of Medical Virology, University of Zurich | Marie O. Pohl et al |
| EPI_ISL_590837 | USA/MN-MDH-1777/2020 | 2020-04-13 | North America/USA/Minnesota/ | Minnesota Department of Health, Public Health Laboratory | Minnesota Department of Health, Public Health Laboratory | Matt Plumb et al |
| EPI_ISL_590860 | USA/TX-DSHS-0795/2020 | 2020-06-02 | North America/USA/Texas/El Paso Coun | Texas Department of State Health Services | Texas Department of State Health Services | Rashmi Tuladhar et al |
| EPI_ISL_590872 | USA/TX-DSHS-0811/2020 | 2020-06-03 | North America/USA/Texas/Harris Count | Texas Department of State Health Services | Texas Department of State Health Services | Rashmi Tuladhar et al |
| EPI_ISL_590915 | Norway/1986/2020 | 2020-03-10 | Europe/Norway/Agder/ | Hospital of Southern Norway - Kristiansand, Department of Medical Microbiology | Norwegian Institute of Public Health, Department of Virology | Kathrine Stene-Johansen et al |
| EPI_ISL_590916 | Norway/2086/2020 | 2020-03-09 | Europe/Norway/Oslo/ | Oslo University Hospital, Department of Medical Microbiology | Norwegian Institute of Public Health, Department of Virology | Kathrine Stene-Johansen et al |

| | | | | | | |
|---|---|---|---|---|---|---|
| EPI_ISL_590918 | Norway/2358/2020 | 2020-03-30 | Europe/Norway/Oslo/ | Oslo University Hospital, Department of Medical Microbiology | Norwegian Institute of Public Health, Department of Virology | Kathrine Stene-Johansen et al |
| EPI_ISL_590948 | Norway/3624/2020 | 2020-09-21 | Europe/Norway/Viken/ | Ostfold Hospital Trust - Kalnes, Centre for Laboratory Medicine, Section for gene technology and infection serology | Norwegian Institute of Public Health, Department of Virology | Kathrine Stene-Johansen et al |
| EPI_ISL_590972 | Norway/4025/2020 | 2020-09-21 | Europe/Norway/Rogaland/ | Dept. of Medical Microbiology, Stavanger University Hospital, Helse Stavanger HF | Norwegian Institute of Public Health, Department of Virology | Kathrine Stene-Johansen et al |
| EPI_ISL_591022 | USA/MD-MDH-0220/2020 | 2020-09-21 | North America/USA/Maryland/ | MD PHL | MD PHL | Maryland Department of Health Laboratories Administration et al |
| EPI_ISL_591032 | USA/MD-MDH-0230/2020 | 2020-09-23 | North America/USA/Maryland/ | MD PHL | MD PHL | Maryland Department of Health Laboratories Administration et al |
| EPI_ISL_591035 | USA/MD-MDH-0233/2020 | 2020-09-28 | North America/USA/Maryland/ | MD PHL | MD PHL | Maryland Department of Health Laboratories Administration et al |
| EPI_ISL_591036 | USA/MD-MDH-0234/2020 | 2020-09-28 | North America/USA/Maryland/ | MD PHL | MD PHL | Maryland Department of Health Laboratories Administration et al |
| EPI_ISL_591085 | DRC/16145/2020 | 2020-06-01 | Africa/Democratic Republic of the Congo | Viral Respiratory Lab, National Institute for Biomedical Research (INRB) | Pathogen Sequencing Lab, National Institute for Biomedical Research (INRB) | Placide Mbala-Kingebeni et al |
| EPI_ISL_591086 | DRC/16191/2020 | 2020-06-03 | Africa/Democratic Republic of the Congo | Viral Respiratory Lab, National Institute for Biomedical Research (INRB) | Pathogen Sequencing Lab, National Institute for Biomedical Research (INRB) | Placide Mbala-Kingebeni et al |

| | | | | | | |
|---|---|---|---|---|---|---|
| EPI_ISL_591099 | France/OCC-32/2020 | 2020-06-08 | Europe/France/Occitanie/ | CHU Purpan - Laboratoire de Virologie - Institut F√©d√©ratif de Biologie | CHU Purpan - Laboratoire de Virologie - Institut F√©d√©ratif de Biologie | Latour J. et al |
| EPI_ISL_591100 | Canada/ON-S1429/2020 | 2020-08-24 | North America/Canada/Ontario/Toronto | Toronto Invasive Bacterial Diseases Network | McMaster University | Allison McGeer et al |
| EPI_ISL_591101 | Canada/ON-S1430/2020 | 2020-08-24 | North America/Canada/Ontario/Toronto | Toronto Invasive Bacterial Diseases Network | McMaster University | Allison McGeer et al |
| EPI_ISL_591102 | Canada/ON-S1455/2020 | 2020-08-26 | North America/Canada/Ontario/Toronto | Toronto Invasive Bacterial Diseases Network | McMaster University | Allison McGeer et al |
| EPI_ISL_591103 | Canada/ON-S1465/2020 | 2020-08-26 | North America/Canada/Ontario/Toronto | Toronto Invasive Bacterial Diseases Network | McMaster University | Allison McGeer et al |
| EPI_ISL_591104 | Canada/ON-S1468/2020 | 2020-08-27 | North America/Canada/Ontario/Toronto | Toronto Invasive Bacterial Diseases Network | McMaster University | Allison McGeer et al |
| EPI_ISL_591105 | Canada/ON-S1469/2020 | 2020-08-26 | North America/Canada/Ontario/Toronto | Toronto Invasive Bacterial Diseases Network | McMaster University | Allison McGeer et al |
| EPI_ISL_591106 | Canada/ON-S1470/2020 | 2020-08-26 | North America/Canada/Ontario/Toronto | Toronto Invasive Bacterial Diseases Network | McMaster University | Allison McGeer et al |
| EPI_ISL_591107 | Canada/ON-S1471/2020 | 2020-08-28 | North America/Canada/Ontario/Toronto | Toronto Invasive Bacterial Diseases Network | McMaster University | Allison McGeer et al |
| EPI_ISL_591108 | Canada/ON-S1472/2020 | 2020-08-28 | North America/Canada/Ontario/Toronto | Toronto Invasive Bacterial Diseases Network | McMaster University | Allison McGeer et al |
| EPI_ISL_591109 | Canada/ON-S1473/2020 | 2020-08-28 | North America/Canada/Ontario/Toronto | Toronto Invasive Bacterial Diseases Network | McMaster University | Allison McGeer et al |
| EPI_ISL_591110 | Canada/ON-S1474/2020 | 2020-08-27 | North America/Canada/Ontario/Toronto | Toronto Invasive Bacterial Diseases Network | McMaster University | Allison McGeer et al |
| EPI_ISL_591111 | Canada/ON-S1475/2020 | 2020-08-27 | North America/Canada/Ontario/Toronto | Toronto Invasive Bacterial Diseases Network | McMaster University | Allison McGeer et al |
| EPI_ISL_591112 | Canada/ON-S1476/2020 | 2020-08-27 | North America/Canada/Ontario/Toronto | Toronto Invasive Bacterial Diseases Network | McMaster University | Allison McGeer et al |
| EPI_ISL_591113 | Canada/ON-S1477/2020 | 2020-08-27 | North America/Canada/Ontario/Toronto | Toronto Invasive Bacterial Diseases Network | McMaster University | Allison McGeer et al |
| EPI_ISL_591114 | Canada/ON-S1479/2020 | 2020-08-28 | North America/Canada/Ontario/Toronto | Toronto Invasive Bacterial Diseases Network | McMaster University | Allison McGeer et al |
| EPI_ISL_591115 | Canada/ON-S1481/2020 | 2020-08-28 | North America/Canada/Ontario/Toronto | Toronto Invasive Bacterial Diseases Network | McMaster University | Allison McGeer et al |
| EPI_ISL_591116 | Canada/ON-S1482/2020 | 2020-08-28 | North America/Canada/Ontario/Toronto | Toronto Invasive Bacterial Diseases Network | McMaster University | Allison McGeer et al |

| | | | | | | |
|---|---|---|---|---|---|---|
| EPI_ISL_591117 | Canada/ON-S1483/2020 | 2020-08-29 | North America/Canada/Ontario/Toronto | Toronto Invasive Bacterial Diseases Network | McMaster University | Allison McGeer et al |
| EPI_ISL_591118 | Canada/ON-S160/2020 | 2020-03-24 | North America/Canada/Ontario/Toronto | Toronto Invasive Bacterial Diseases Network | McMaster University | Allison McGeer et al |
| EPI_ISL_591119 | Canada/ON-S167/2020 | 2020-03-25 | North America/Canada/Ontario/Toronto | Toronto Invasive Bacterial Diseases Network | McMaster University | Allison McGeer et al |
| EPI_ISL_591120 | Canada/ON-S177/2020 | 2020-03-24 | North America/Canada/Ontario/Toronto | Toronto Invasive Bacterial Diseases Network | McMaster University | Allison McGeer et al |
| EPI_ISL_591121 | Canada/ON-S184/2020 | 2020-03-25 | North America/Canada/Ontario/Toronto | Toronto Invasive Bacterial Diseases Network | McMaster University | Allison McGeer et al |
| EPI_ISL_591122 | Canada/ON-S192/2020 | 2020-03-26 | North America/Canada/Ontario/Toronto | Toronto Invasive Bacterial Diseases Network | McMaster University | Allison McGeer et al |
| EPI_ISL_591123 | Canada/ON-S202/2020 | 2020-04-07 | North America/Canada/Ontario/Toronto | Toronto Invasive Bacterial Diseases Network | McMaster University | Allison McGeer et al |
| EPI_ISL_591124 | Canada/ON-S21/2020 | 2020-03-16 | North America/Canada/Ontario/Toronto | Toronto Invasive Bacterial Diseases Network | McMaster University | Allison McGeer et al |
| EPI_ISL_591125 | Canada/ON-S235/2020 | 2020-03-27 | North America/Canada/Ontario/Toronto | Toronto Invasive Bacterial Diseases Network | McMaster University | Allison McGeer et al |
| EPI_ISL_591126 | Canada/ON-S245/2020 | 2020-03-27 | North America/Canada/Ontario/Toronto | Toronto Invasive Bacterial Diseases Network | McMaster University | Allison McGeer et al |
| EPI_ISL_591127 | Canada/ON-S27/2020 | 2020-03-16 | North America/Canada/Ontario/Toronto | Toronto Invasive Bacterial Diseases Network | McMaster University | Allison McGeer et al |
| EPI_ISL_591128 | Canada/ON-S28/2020 | 2020-03-16 | North America/Canada/Ontario/Toronto | Toronto Invasive Bacterial Diseases Network | McMaster University | Allison McGeer et al |
| EPI_ISL_591129 | Canada/ON-S302/2020 | 2020-03-28 | North America/Canada/Ontario/Toronto | Toronto Invasive Bacterial Diseases Network | McMaster University | Allison McGeer et al |
| EPI_ISL_591130 | Canada/ON-S32/2020 | 2020-03-16 | North America/Canada/Ontario/Toronto | Toronto Invasive Bacterial Diseases Network | McMaster University | Allison McGeer et al |
| EPI_ISL_591131 | Canada/ON-S323/2020 | 2020-03-28 | North America/Canada/Ontario/Toronto | Toronto Invasive Bacterial Diseases Network | McMaster University | Allison McGeer et al |
| EPI_ISL_591132 | Canada/ON-S337/2020 | 2020-03-29 | North America/Canada/Ontario/Toronto | Toronto Invasive Bacterial Diseases Network | McMaster University | Allison McGeer et al |
| EPI_ISL_591133 | Canada/ON-S344/2020 | 2020-03-29 | North America/Canada/Ontario/Toronto | Toronto Invasive Bacterial Diseases Network | McMaster University | Allison McGeer et al |
| EPI_ISL_591134 | Canada/ON-S363/2020 | 2020-03-30 | North America/Canada/Ontario/Toronto | Toronto Invasive Bacterial Diseases Network | McMaster University | Allison McGeer et al |
| EPI_ISL_591135 | Canada/ON-S366/2020 | 2020-03-31 | North America/Canada/Ontario/Toronto | Toronto Invasive Bacterial Diseases Network | McMaster University | Allison McGeer et al |
| EPI_ISL_591136 | Canada/ON-S376/2020 | 2020-03-30 | North America/Canada/Ontario/Toronto | Toronto Invasive Bacterial Diseases Network | McMaster University | Allison McGeer et al |

| | | | | | | |
|---|---|---|---|---|---|---|
| EPI_ISL_591137 | Canada/ON-S377/2020 | 2020-03-30 | North America/Canada/Ontario/Toronto | Toronto Invasive Bacterial Diseases Network | McMaster University | Allison McGeer et al |
| EPI_ISL_591138 | Canada/ON-S38/2020 | 2020-03-17 | North America/Canada/Ontario/Toronto | Toronto Invasive Bacterial Diseases Network | McMaster University | Allison McGeer et al |
| EPI_ISL_591139 | Canada/ON-S383/2020 | 2020-03-30 | North America/Canada/Ontario/Toronto | Toronto Invasive Bacterial Diseases Network | McMaster University | Allison McGeer et al |
| EPI_ISL_591140 | Canada/ON-S390/2020 | 2020-03-30 | North America/Canada/Ontario/Toronto | Toronto Invasive Bacterial Diseases Network | McMaster University | Allison McGeer et al |
| EPI_ISL_591141 | Canada/ON-S707/2020 | 2020-04-09 | North America/Canada/Ontario/Toronto | Toronto Invasive Bacterial Diseases Network | McMaster University | Allison McGeer et al |
| EPI_ISL_591142 | Canada/ON-S708/2020 | 20-Apr | North America/Canada/Ontario/Toronto | Toronto Invasive Bacterial Diseases Network | McMaster University | Allison McGeer et al |
| EPI_ISL_591143 | Canada/ON-S71/2020 | 2020-03-19 | North America/Canada/Ontario/Toronto | Toronto Invasive Bacterial Diseases Network | McMaster University | Allison McGeer et al |
| EPI_ISL_591144 | Canada/ON-S710/2020 | 20-Apr | North America/Canada/Ontario/Toronto | Toronto Invasive Bacterial Diseases Network | McMaster University | Allison McGeer et al |
| EPI_ISL_591145 | Canada/ON-S720/2020 | 20-Apr | North America/Canada/Ontario/Toronto | Toronto Invasive Bacterial Diseases Network | McMaster University | Allison McGeer et al |
| EPI_ISL_591146 | Canada/ON-S721/2020 | 20-Apr | North America/Canada/Ontario/Toronto | Toronto Invasive Bacterial Diseases Network | McMaster University | Allison McGeer et al |
| EPI_ISL_591147 | Canada/ON-S722/2020 | 20-Apr | North America/Canada/Ontario/Toronto | Toronto Invasive Bacterial Diseases Network | McMaster University | Allison McGeer et al |
| EPI_ISL_591148 | Canada/ON-S724/2020 | 2020-04-09 | North America/Canada/Ontario/Toronto | Toronto Invasive Bacterial Diseases Network | McMaster University | Allison McGeer et al |
| EPI_ISL_591149 | Canada/ON-S725/2020 | 20-Apr | North America/Canada/Ontario/Toronto | Toronto Invasive Bacterial Diseases Network | McMaster University | Allison McGeer et al |
| EPI_ISL_591150 | Canada/ON-S726/2020 | 2020-04-09 | North America/Canada/Ontario/Toronto | Toronto Invasive Bacterial Diseases Network | McMaster University | Allison McGeer et al |
| EPI_ISL_591151 | Canada/ON-S727/2020 | 20-Apr | North America/Canada/Ontario/Toronto | Toronto Invasive Bacterial Diseases Network | McMaster University | Allison McGeer et al |
| EPI_ISL_591152 | Canada/ON-S728/2020 | 2020-04-09 | North America/Canada/Ontario/Toronto | Toronto Invasive Bacterial Diseases Network | McMaster University | Allison McGeer et al |
| EPI_ISL_591153 | Canada/ON-S729/2020 | 20-Apr | North America/Canada/Ontario/Toronto | Toronto Invasive Bacterial Diseases Network | McMaster University | Allison McGeer et al |
| EPI_ISL_591154 | Canada/ON-S730/2020 | 2020-04-09 | North America/Canada/Ontario/Toronto | Toronto Invasive Bacterial Diseases Network | McMaster University | Allison McGeer et al |
| EPI_ISL_591155 | Canada/ON-S732/2020 | 2020-04-09 | North America/Canada/Ontario/Toronto | Toronto Invasive Bacterial Diseases Network | McMaster University | Allison McGeer et al |
| EPI_ISL_591156 | Canada/ON-S733/2020 | 2020-04-09 | North America/Canada/Ontario/Toronto | Toronto Invasive Bacterial Diseases Network | McMaster University | Allison McGeer et al |

| | | | | | | |
|---|---|---|---|---|---|---|
| EPI_ISL_591157 | Canada/ON-S736/2020 | 2020-04-09 | North America/Canada/Ontario/Toronto | Toronto Invasive Bacterial Diseases Network | McMaster University | Allison McGeer et al |
| EPI_ISL_591158 | Canada/ON-S744/2020 | 2020-04-09 | North America/Canada/Ontario/Toronto | Toronto Invasive Bacterial Diseases Network | McMaster University | Allison McGeer et al |
| EPI_ISL_591159 | Canada/ON-S754/2020 | 2020-04-10 | North America/Canada/Ontario/Toronto | Toronto Invasive Bacterial Diseases Network | McMaster University | Allison McGeer et al |
| EPI_ISL_591160 | Canada/ON-S761/2020 | 2020-04-10 | North America/Canada/Ontario/Toronto | Toronto Invasive Bacterial Diseases Network | McMaster University | Allison McGeer et al |
| EPI_ISL_591161 | Canada/ON-S797/2020 | 20-Apr | North America/Canada/Ontario/Toronto | Toronto Invasive Bacterial Diseases Network | McMaster University | Allison McGeer et al |
| EPI_ISL_591162 | Canada/ON-S803/2020 | 20-Apr | North America/Canada/Ontario/Toronto | Toronto Invasive Bacterial Diseases Network | McMaster University | Allison McGeer et al |
| EPI_ISL_591163 | Canada/ON-S804/2020 | 20-Apr | North America/Canada/Ontario/Toronto | Toronto Invasive Bacterial Diseases Network | McMaster University | Allison McGeer et al |
| EPI_ISL_591164 | Canada/ON-S811/2020 | 2020-04-10 | North America/Canada/Ontario/Toronto | Toronto Invasive Bacterial Diseases Network | McMaster University | Allison McGeer et al |
| EPI_ISL_591165 | Canada/ON-S813/2020 | 2020-04-10 | North America/Canada/Ontario/Toronto | Toronto Invasive Bacterial Diseases Network | McMaster University | Allison McGeer et al |
| EPI_ISL_591166 | Canada/ON-S815/2020 | 2020-04-10 | North America/Canada/Ontario/Toronto | Toronto Invasive Bacterial Diseases Network | McMaster University | Allison McGeer et al |
| EPI_ISL_591167 | Canada/ON-S816/2020 | 20-Apr | North America/Canada/Ontario/Toronto | Toronto Invasive Bacterial Diseases Network | McMaster University | Allison McGeer et al |
| EPI_ISL_591168 | Canada/ON-S83/2020 | 2020-03-20 | North America/Canada/Ontario/Toronto | Toronto Invasive Bacterial Diseases Network | McMaster University | Allison McGeer et al |
| EPI_ISL_591169 | Canada/ON-S846/2020 | 2020-04-11 | North America/Canada/Ontario/Toronto | Toronto Invasive Bacterial Diseases Network | McMaster University | Allison McGeer et al |
| EPI_ISL_591170 | Canada/ON-S851/2020 | 2020-04-11 | North America/Canada/Ontario/Toronto | Toronto Invasive Bacterial Diseases Network | McMaster University | Allison McGeer et al |
| EPI_ISL_591171 | Canada/ON-S857/2020 | 20-Apr | North America/Canada/Ontario/Toronto | Toronto Invasive Bacterial Diseases Network | McMaster University | Allison McGeer et al |
| EPI_ISL_591172 | Canada/ON-S859/2020 | 20-Apr | North America/Canada/Ontario/Toronto | Toronto Invasive Bacterial Diseases Network | McMaster University | Allison McGeer et al |
| EPI_ISL_591173 | Canada/ON-S870/2020 | 20-Apr | North America/Canada/Ontario/Toronto | Toronto Invasive Bacterial Diseases Network | McMaster University | Allison McGeer et al |
| EPI_ISL_591174 | Canada/ON-S873/2020 | 20-Apr | North America/Canada/Ontario/Toronto | Toronto Invasive Bacterial Diseases Network | McMaster University | Allison McGeer et al |
| EPI_ISL_591175 | Canada/ON-S878/2020 | 2020-04-12 | North America/Canada/Ontario/Toronto | Toronto Invasive Bacterial Diseases Network | McMaster University | Allison McGeer et al |
| EPI_ISL_591176 | Canada/ON-S879/2020 | 2020-04-12 | North America/Canada/Ontario/Toronto | Toronto Invasive Bacterial Diseases Network | McMaster University | Allison McGeer et al |

| | | | | | | |
|---|---|---|---|---|---|---|
| EPI_ISL_591177 | Canada/ON-S895/2020 | 2020-04-12 | North America/Canada/Ontario/Toronto | Toronto Invasive Bacterial Diseases Network | McMaster University | Allison McGeer et al |
| EPI_ISL_591178 | Canada/ON-S896/2020 | 2020-04-12 | North America/Canada/Ontario/Toronto | Toronto Invasive Bacterial Diseases Network | McMaster University | Allison McGeer et al |
| EPI_ISL_591179 | Canada/ON-S921/2020 | 2020-04-13 | North America/Canada/Ontario/Toronto | Toronto Invasive Bacterial Diseases Network | McMaster University | Allison McGeer et al |
| EPI_ISL_591180 | Canada/ON-S105/2020 | 2020-03-22 | North America/Canada/Ontario/Toronto | Toronto Invasive Bacterial Diseases Network | McMaster University | Allison McGeer et al |
| EPI_ISL_591181 | Canada/ON-S957/2020 | 2020-04-14 | North America/Canada/Ontario/Toronto | Toronto Invasive Bacterial Diseases Network | McMaster University | Allison McGeer et al |
| EPI_ISL_591182 | Canada/ON-S125/2020 | 2020-03-23 | North America/Canada/Ontario/Toronto | Toronto Invasive Bacterial Diseases Network | McMaster University | Allison McGeer et al |
| EPI_ISL_591183 | Canada/ON-S126/2020 | 2020-03-24 | North America/Canada/Ontario/Toronto | Toronto Invasive Bacterial Diseases Network | McMaster University | Allison McGeer et al |
| EPI_ISL_591184 | Canada/ON-S128/2020 | 2020-03-23 | North America/Canada/Ontario/Toronto | Toronto Invasive Bacterial Diseases Network | McMaster University | Allison McGeer et al |
| EPI_ISL_591185 | Canada/ON-S1414/2020 | 2020-08-23 | North America/Canada/Ontario/Toronto | Toronto Invasive Bacterial Diseases Network | McMaster University | Allison McGeer et al |
| EPI_ISL_591186 | Canada/ON-S1415/2020 | 2020-08-23 | North America/Canada/Ontario/Toronto | Toronto Invasive Bacterial Diseases Network | McMaster University | Allison McGeer et al |
| EPI_ISL_591187 | Canada/ON-S1416/2020 | 2020-08-23 | North America/Canada/Ontario/Toronto | Toronto Invasive Bacterial Diseases Network | McMaster University | Allison McGeer et al |
| EPI_ISL_591188 | Canada/ON-S1419/2020 | 2020-08-23 | North America/Canada/Ontario/Toronto | Toronto Invasive Bacterial Diseases Network | McMaster University | Allison McGeer et al |
| EPI_ISL_591189 | Canada/ON-S1420/2020 | 2020-08-22 | North America/Canada/Ontario/Toronto | Toronto Invasive Bacterial Diseases Network | McMaster University | Allison McGeer et al |
| EPI_ISL_591190 | Canada/ON-S1421/2020 | 2020-08-22 | North America/Canada/Ontario/Toronto | Toronto Invasive Bacterial Diseases Network | McMaster University | Allison McGeer et al |
| EPI_ISL_591191 | Canada/ON-S1422/2020 | 2020-08-22 | North America/Canada/Ontario/Toronto | Toronto Invasive Bacterial Diseases Network | McMaster University | Allison McGeer et al |
| EPI_ISL_591192 | Canada/ON-S1423/2020 | 2020-08-23 | North America/Canada/Ontario/Toronto | Toronto Invasive Bacterial Diseases Network | McMaster University | Allison McGeer et al |
| EPI_ISL_591193 | Canada/ON-S1427/2020 | 2020-08-24 | North America/Canada/Ontario/Toronto | Toronto Invasive Bacterial Diseases Network | McMaster University | Allison McGeer et al |
| EPI_ISL_591194 | Canada/ON-S1431/2020 | 2020-08-24 | North America/Canada/Ontario/Toronto | Toronto Invasive Bacterial Diseases Network | McMaster University | Allison McGeer et al |
| EPI_ISL_591195 | Canada/ON-S1432/2020 | 2020-08-24 | North America/Canada/Ontario/Toronto | Toronto Invasive Bacterial Diseases Network | McMaster University | Allison McGeer et al |
| EPI_ISL_591196 | Canada/ON-S1433/2020 | 2020-08-25 | North America/Canada/Ontario/Toronto | Toronto Invasive Bacterial Diseases Network | McMaster University | Allison McGeer et al |

| | | | | | | |
|---|---|---|---|---|---|---|
| EPI_ISL_591197 | Canada/ON-S1434/2020 | 2020-08-25 | North America/Canada/Ontario/Toronto | Toronto Invasive Bacterial Diseases Network | McMaster University | Allison McGeer et al |
| EPI_ISL_591198 | Canada/ON-S1435/2020 | 2020-08-24 | North America/Canada/Ontario/Toronto | Toronto Invasive Bacterial Diseases Network | McMaster University | Allison McGeer et al |
| EPI_ISL_591199 | Canada/ON-S1436/2020 | 2020-08-22 | North America/Canada/Ontario/Toronto | Toronto Invasive Bacterial Diseases Network | McMaster University | Allison McGeer et al |
| EPI_ISL_591200 | Canada/ON-S1438/2020 | 2020-08-26 | North America/Canada/Ontario/Toronto | Toronto Invasive Bacterial Diseases Network | McMaster University | Allison McGeer et al |
| EPI_ISL_591201 | Canada/ON-S1439/2020 | 2020-08-25 | North America/Canada/Ontario/Toronto | Toronto Invasive Bacterial Diseases Network | McMaster University | Allison McGeer et al |
| EPI_ISL_591202 | Canada/ON-S1440/2020 | 2020-08-25 | North America/Canada/Ontario/Toronto | Toronto Invasive Bacterial Diseases Network | McMaster University | Allison McGeer et al |
| EPI_ISL_591203 | Canada/ON-S1443/2020 | 2020-08-26 | North America/Canada/Ontario/Toronto | Toronto Invasive Bacterial Diseases Network | McMaster University | Allison McGeer et al |
| EPI_ISL_591204 | Canada/ON-S1444/2020 | 2020-08-26 | North America/Canada/Ontario/Toronto | Toronto Invasive Bacterial Diseases Network | McMaster University | Allison McGeer et al |
| EPI_ISL_591205 | Canada/ON-S1446/2020 | 2020-08-26 | North America/Canada/Ontario/Toronto | Toronto Invasive Bacterial Diseases Network | McMaster University | Allison McGeer et al |
| EPI_ISL_591206 | Canada/ON-S1447/2020 | 2020-08-26 | North America/Canada/Ontario/Toronto | Toronto Invasive Bacterial Diseases Network | McMaster University | Allison McGeer et al |
| EPI_ISL_591207 | Canada/ON-S1456/2020 | 2020-08-26 | North America/Canada/Ontario/Toronto | Toronto Invasive Bacterial Diseases Network | McMaster University | Allison McGeer et al |
| EPI_ISL_591208 | Canada/ON-S256/2020 | 2020-03-27 | North America/Canada/Ontario/Toronto | Toronto Invasive Bacterial Diseases Network | McMaster University | Allison McGeer et al |
| EPI_ISL_591209 | Canada/ON-S706/2020 | 2020-04-09 | North America/Canada/Ontario/Toronto | Toronto Invasive Bacterial Diseases Network | McMaster University | Allison McGeer et al |
| EPI_ISL_591210 | Canada/ON-S737/2020 | 2020-04-09 | North America/Canada/Ontario/Toronto | Toronto Invasive Bacterial Diseases Network | McMaster University | Allison McGeer et al |
| EPI_ISL_591211 | Canada/ON-S743/2020 | 2020-04-10 | North America/Canada/Ontario/Toronto | Toronto Invasive Bacterial Diseases Network | McMaster University | Allison McGeer et al |
| EPI_ISL_591212 | Canada/ON-S746/2020 | 2020-04-09 | North America/Canada/Ontario/Toronto | Toronto Invasive Bacterial Diseases Network | McMaster University | Allison McGeer et al |
| EPI_ISL_591213 | Canada/ON-S747/2020 | 20-Apr | North America/Canada/Ontario/Toronto | Toronto Invasive Bacterial Diseases Network | McMaster University | Allison McGeer et al |
| EPI_ISL_591214 | Canada/ON-S751/2020 | 20-Apr | North America/Canada/Ontario/Toronto | Toronto Invasive Bacterial Diseases Network | McMaster University | Allison McGeer et al |
| EPI_ISL_591215 | Canada/ON-S752/2020 | 20-Apr | North America/Canada/Ontario/Toronto | Toronto Invasive Bacterial Diseases Network | McMaster University | Allison McGeer et al |
| EPI_ISL_591216 | Canada/ON-S753/2020 | 2020-04-10 | North America/Canada/Ontario/Toronto | Toronto Invasive Bacterial Diseases Network | McMaster University | Allison McGeer et al |

| | | | | | | |
|---|---|---|---|---|---|---|
| EPI_ISL_591217 | Canada/ON-S756/2020 | 20-Apr | North America/Canada/Ontario/Toronto | Toronto Invasive Bacterial Diseases Network | McMaster University | Allison McGeer et al |
| EPI_ISL_591218 | Canada/ON-S760/2020 | 20-Apr | North America/Canada/Ontario/Toronto | Toronto Invasive Bacterial Diseases Network | McMaster University | Allison McGeer et al |
| EPI_ISL_591219 | Canada/ON-S762/2020 | 20-Apr | North America/Canada/Ontario/Toronto | Toronto Invasive Bacterial Diseases Network | McMaster University | Allison McGeer et al |
| EPI_ISL_591220 | Canada/ON-S764/2020 | 2020-04-10 | North America/Canada/Ontario/Toronto | Toronto Invasive Bacterial Diseases Network | McMaster University | Allison McGeer et al |
| EPI_ISL_591221 | Canada/ON-S767/2020 | 20-Apr | North America/Canada/Ontario/Toronto | Toronto Invasive Bacterial Diseases Network | McMaster University | Allison McGeer et al |
| EPI_ISL_591222 | Canada/ON-S773/2020 | 2020-04-10 | North America/Canada/Ontario/Toronto | Toronto Invasive Bacterial Diseases Network | McMaster University | Allison McGeer et al |
| EPI_ISL_591223 | Canada/ON-S775/2020 | 2020-04-10 | North America/Canada/Ontario/Toronto | Toronto Invasive Bacterial Diseases Network | McMaster University | Allison McGeer et al |
| EPI_ISL_591224 | Canada/ON-S776/2020 | 2020-04-10 | North America/Canada/Ontario/Toronto | Toronto Invasive Bacterial Diseases Network | McMaster University | Allison McGeer et al |
| EPI_ISL_591225 | Canada/ON-S778/2020 | 2020-04-10 | North America/Canada/Ontario/Toronto | Toronto Invasive Bacterial Diseases Network | McMaster University | Allison McGeer et al |
| EPI_ISL_591226 | Canada/ON-S780/2020 | 20-Apr | North America/Canada/Ontario/Toronto | Toronto Invasive Bacterial Diseases Network | McMaster University | Allison McGeer et al |
| EPI_ISL_591227 | Canada/ON-S786/2020 | 20-Apr | North America/Canada/Ontario/Toronto | Toronto Invasive Bacterial Diseases Network | McMaster University | Allison McGeer et al |
| EPI_ISL_591228 | Canada/ON-S787/2020 | 20-Apr | North America/Canada/Ontario/Toronto | Toronto Invasive Bacterial Diseases Network | McMaster University | Allison McGeer et al |
| EPI_ISL_591229 | Canada/ON-S789/2020 | 20-Apr | North America/Canada/Ontario/Toronto | Toronto Invasive Bacterial Diseases Network | McMaster University | Allison McGeer et al |
| EPI_ISL_591230 | Canada/ON-S793/2020 | 20-Apr | North America/Canada/Ontario/Toronto | Toronto Invasive Bacterial Diseases Network | McMaster University | Allison McGeer et al |
| EPI_ISL_591231 | Canada/ON-S796/2020 | 20-Apr | North America/Canada/Ontario/Toronto | Toronto Invasive Bacterial Diseases Network | McMaster University | Allison McGeer et al |
| EPI_ISL_591232 | Canada/ON-S798/2020 | 20-Apr | North America/Canada/Ontario/Toronto | Toronto Invasive Bacterial Diseases Network | McMaster University | Allison McGeer et al |
| EPI_ISL_591233 | Canada/ON-S799/2020 | 20-Apr | North America/Canada/Ontario/Toronto | Toronto Invasive Bacterial Diseases Network | McMaster University | Allison McGeer et al |
| EPI_ISL_591234 | Canada/ON-S805/2020 | 20-Apr | North America/Canada/Ontario/Toronto | Toronto Invasive Bacterial Diseases Network | McMaster University | Allison McGeer et al |
| EPI_ISL_591235 | Canada/ON-S806/2020 | 20-Apr | North America/Canada/Ontario/Toronto | Toronto Invasive Bacterial Diseases Network | McMaster University | Allison McGeer et al |
| EPI_ISL_591236 | Canada/ON-S808/2020 | 20-Apr | North America/Canada/Ontario/Toronto | Toronto Invasive Bacterial Diseases Network | McMaster University | Allison McGeer et al |

| | | | | | | |
|---|---|---|---|---|---|---|
| EPI_ISL_591237 | Canada/ON-S810/2020 | 2020-04-10 | North America/Canada/Ontario/Toronto | Toronto Invasive Bacterial Diseases Network | McMaster University | Allison McGeer et al |
| EPI_ISL_591238 | Canada/ON-S812/2020 | 2020-04-10 | North America/Canada/Ontario/Toronto | Toronto Invasive Bacterial Diseases Network | McMaster University | Allison McGeer et al |
| EPI_ISL_591239 | Canada/ON-S821/2020 | 20-Apr | North America/Canada/Ontario/Toronto | Toronto Invasive Bacterial Diseases Network | McMaster University | Allison McGeer et al |
| EPI_ISL_591240 | Canada/ON-S826/2020 | 2020-04-11 | North America/Canada/Ontario/Toronto | Toronto Invasive Bacterial Diseases Network | McMaster University | Allison McGeer et al |
| EPI_ISL_591241 | Canada/ON-S836/2020 | 20-Apr | North America/Canada/Ontario/Toronto | Toronto Invasive Bacterial Diseases Network | McMaster University | Allison McGeer et al |
| EPI_ISL_591242 | Canada/ON-S839/2020 | 20-Apr | North America/Canada/Ontario/Toronto | Toronto Invasive Bacterial Diseases Network | McMaster University | Allison McGeer et al |
| EPI_ISL_591243 | Canada/ON-S840/2020 | 20-Apr | North America/Canada/Ontario/Toronto | Toronto Invasive Bacterial Diseases Network | McMaster University | Allison McGeer et al |
| EPI_ISL_591244 | Canada/ON-S843/2020 | 20-Apr | North America/Canada/Ontario/Toronto | Toronto Invasive Bacterial Diseases Network | McMaster University | Allison McGeer et al |
| EPI_ISL_591245 | Canada/ON-S848/2020 | 2020-04-11 | North America/Canada/Ontario/Toronto | Toronto Invasive Bacterial Diseases Network | McMaster University | Allison McGeer et al |
| EPI_ISL_591246 | Canada/ON-S849/2020 | 2020-04-11 | North America/Canada/Ontario/Toronto | Toronto Invasive Bacterial Diseases Network | McMaster University | Allison McGeer et al |
| EPI_ISL_591247 | Canada/ON-S850/2020 | 2020-04-11 | North America/Canada/Ontario/Toronto | Toronto Invasive Bacterial Diseases Network | McMaster University | Allison McGeer et al |
| EPI_ISL_591248 | Canada/ON-S862/2020 | 20-Apr | North America/Canada/Ontario/Toronto | Toronto Invasive Bacterial Diseases Network | McMaster University | Allison McGeer et al |
| EPI_ISL_591249 | Canada/ON-S863/2020 | 2020-04-12 | North America/Canada/Ontario/Toronto | Toronto Invasive Bacterial Diseases Network | McMaster University | Allison McGeer et al |
| EPI_ISL_591250 | Canada/ON-S868/2020 | 20-Apr | North America/Canada/Ontario/Toronto | Toronto Invasive Bacterial Diseases Network | McMaster University | Allison McGeer et al |
| EPI_ISL_591251 | Canada/ON-S869/2020 | 20-Apr | North America/Canada/Ontario/Toronto | Toronto Invasive Bacterial Diseases Network | McMaster University | Allison McGeer et al |
| EPI_ISL_591252 | Canada/ON-S871/2020 | 20-Apr | North America/Canada/Ontario/Toronto | Toronto Invasive Bacterial Diseases Network | McMaster University | Allison McGeer et al |
| EPI_ISL_591253 | Canada/ON-S881/2020 | 2020-04-12 | North America/Canada/Ontario/Toronto | Toronto Invasive Bacterial Diseases Network | McMaster University | Allison McGeer et al |
| EPI_ISL_591254 | Canada/ON-S883/2020 | 2020-04-12 | North America/Canada/Ontario/Toronto | Toronto Invasive Bacterial Diseases Network | McMaster University | Allison McGeer et al |
| EPI_ISL_591255 | Canada/ON-S888/2020 | 2020-04-12 | North America/Canada/Ontario/Toronto | Toronto Invasive Bacterial Diseases Network | McMaster University | Allison McGeer et al |
| EPI_ISL_591256 | Canada/ON-S894/2020 | 2020-04-12 | North America/Canada/Ontario/Toronto | Toronto Invasive Bacterial Diseases Network | McMaster University | Allison McGeer et al |

| | | | | | | |
|---|---|---|---|---|---|---|
| EPI_ISL_591257 | Canada/ON-S899/2020 | 2020-04-12 | North America/Canada/Ontario/Toronto | Toronto Invasive Bacterial Diseases Network | McMaster University | Allison McGeer et al |
| EPI_ISL_591258 | Canada/ON-S908/2020 | 2020-04-13 | North America/Canada/Ontario/Toronto | Toronto Invasive Bacterial Diseases Network | McMaster University | Allison McGeer et al |
| EPI_ISL_591259 | Canada/ON-S910/2020 | 2020-04-13 | North America/Canada/Ontario/Toronto | Toronto Invasive Bacterial Diseases Network | McMaster University | Allison McGeer et al |
| EPI_ISL_591260 | Canada/ON-S926/2020 | 2020-04-13 | North America/Canada/Ontario/Toronto | Toronto Invasive Bacterial Diseases Network | McMaster University | Allison McGeer et al |
| EPI_ISL_591261 | Canada/ON-S927/2020 | 2020-04-13 | North America/Canada/Ontario/Toronto | Toronto Invasive Bacterial Diseases Network | McMaster University | Allison McGeer et al |
| EPI_ISL_591262 | Canada/ON-S930/2020 | 2020-04-13 | North America/Canada/Ontario/Toronto | Toronto Invasive Bacterial Diseases Network | McMaster University | Allison McGeer et al |
| EPI_ISL_591263 | Canada/ON-S931/2020 | 2020-04-13 | North America/Canada/Ontario/Toronto | Toronto Invasive Bacterial Diseases Network | McMaster University | Allison McGeer et al |
| EPI_ISL_591264 | Canada/ON-S942/2020 | 2020-04-14 | North America/Canada/Ontario/Toronto | Toronto Invasive Bacterial Diseases Network | McMaster University | Allison McGeer et al |
| EPI_ISL_591265 | Canada/ON-S943/2020 | 2020-04-14 | North America/Canada/Ontario/Toronto | Toronto Invasive Bacterial Diseases Network | McMaster University | Allison McGeer et al |
| EPI_ISL_591266 | Canada/ON-S944/2020 | 2020-04-14 | North America/Canada/Ontario/Toronto | Toronto Invasive Bacterial Diseases Network | McMaster University | Allison McGeer et al |
| EPI_ISL_591267 | Canada/ON-S950/2020 | 2020-04-14 | North America/Canada/Ontario/Toronto | Toronto Invasive Bacterial Diseases Network | McMaster University | Allison McGeer et al |
| EPI_ISL_591268 | Canada/ON-S951/2020 | 2020-04-14 | North America/Canada/Ontario/Toronto | Toronto Invasive Bacterial Diseases Network | McMaster University | Allison McGeer et al |
| EPI_ISL_591269 | Canada/ON-S119/2020 | 2020-03-22 | North America/Canada/Ontario/Toronto | Toronto Invasive Bacterial Diseases Network | McMaster University | Allison McGeer et al |
| EPI_ISL_591284 | USA/UT-UPHL-04235_2/2020 | 2020-05-15 | North America/USA/Utah/ | Utah Public Health Laboratory | Utah Public Health Laboratory | Erin Young et al |
| EPI_ISL_591287 | USA/UT-UPHL-04262_2/2020 | 2020-05-15 | North America/USA/Utah/ | Utah Public Health Laboratory | Utah Public Health Laboratory | Erin Young et al |
| EPI_ISL_591299 | USA/UT-UPHL-04308_2/2020 | 2020-05-15 | North America/USA/Utah/ | Utah Public Health Laboratory | Utah Public Health Laboratory | Erin Young et al |
| EPI_ISL_591327 | Italy/TUS-UOS-10/2020 | 2020-03-21 | Europe/Italy/Tuscany/Siena | Dipartimento di Biotecnologie Mediche, University of Siena | Dipartimento di Biotecnologie Mediche, University of Siena | Cusi et al |

| | | | | | | |
|---|---|---|---|---|---|---|
| EPI_ISL_591328 | Italy/TUS-UOS-11/2020 | 2020-03-21 | Europe/Italy/Tuscany/Siena | Dipartimento di Biotecnologie Mediche, University of Siena | Dipartimento di Biotecnologie Mediche, University of Siena | Cusi et al |
| EPI_ISL_591332 | Italy/TUS-UOS-4/2020 | 2020-03-23 | Europe/Italy/Tuscany/Siena | Dipartimento di Biotecnologie Mediche, University of Siena | Dipartimento di Biotecnologie Mediche, University of Siena | Cusi et al |
| EPI_ISL_591333 | Italy/TUS-UOS-6/2020 | 2020-03-21 | Europe/Italy/Tuscany/Siena | Dipartimento di Biotecnologie Mediche, University of Siena | Dipartimento di Biotecnologie Mediche, University of Siena | Cusi et al |
| EPI_ISL_591334 | Italy/TUS-UOS-07/2020 | 2020-03-20 | Europe/Italy/Tuscany/Siena | Dipartimento di Biotecnologie Mediche, University of Siena | Dipartimento di Biotecnologie Mediche, University of Siena | Cusi et al |
| EPI_ISL_591354 | Japan/IC-0008/2020 | 20-Mar | Asia/Italy/Japan/ | Pathogen Genomics Center, National Institute of Infectious Diseases | Pathogen Genomics Center, National Institute of Infectious Diseases | Tsuyoshi Sekizuka et al |
| EPI_ISL_591356 | Japan/IC-0010/2020 | 20-Mar | Asia/Italy/Japan/ | Pathogen Genomics Center, National Institute of Infectious Diseases | Pathogen Genomics Center, National Institute of Infectious Diseases | Tsuyoshi Sekizuka et al |
| EPI_ISL_591394 | Japan/IC-0052/2020 | 20-Jun | Asia/Japan/Japan/ | Pathogen Genomics Center, National Institute of Infectious Diseases | Pathogen Genomics Center, National Institute of Infectious Diseases | Tsuyoshi Sekizuka et al |
| EPI_ISL_591397 | Japan/IC-0055/2020 | 20-Jun | Asia/Japan/Japan/ | Pathogen Genomics Center, National Institute of Infectious Diseases | Pathogen Genomics Center, National Institute of Infectious Diseases | Tsuyoshi Sekizuka et al |

| | | | | | | |
|---|---|---|---|---|---|---|
| EPI_ISL_591400 | Japan/IC-0058/2020 | 20-Jun | Asia/Japan/Japan/ | Pathogen Genomics Center, National Institute of Infectious Diseases | Pathogen Genomics Center, National Institute of Infectious Diseases | Tsuyoshi Sekizuka et al |
| EPI_ISL_591433 | Japan/IC-0092/2020 | 20-Jul | Asia/Iran/Japan/ | Pathogen Genomics Center, National Institute of Infectious Diseases | Pathogen Genomics Center, National Institute of Infectious Diseases | Tsuyoshi Sekizuka et al |
| EPI_ISL_591444 | Japan/IC-0103/2020 | 20-Jul | Asia/Iran/Japan/ | Pathogen Genomics Center, National Institute of Infectious Diseases | Pathogen Genomics Center, National Institute of Infectious Diseases | Tsuyoshi Sekizuka et al |
| EPI_ISL_591520 | Chile/RM-CMM-ADF564K_42/2020 | 2020-04-17 | South America/Chile/Region Metropolit | Programa de Virolog√≠a, Facultad de Medicina UC | Center for Mathematical Modeling and Center for Genome Regulation. Santiago, Chile | Gaggero A et al |
| EPI_ISL_591521 | Chile/RM-CMM-ARP6337/2020 | 2020-05-11 | South America/Chile/Region Metropolit | Programa de Virolog√≠a, Facultad de Medicina UC | Center for Mathematical Modeling and Center for Genome Regulation. Santiago, Chile | Gaggero A et al |
| EPI_ISL_591522 | Chile/RM-CMM-LRF3924/2020 | 2020-05-29 | South America/Chile/Region Metropolit | Medicina Norte U Chile - Servicio Medico Legal | Center for Mathematical Modeling and Center for Genome Regulation. Santiago, Chile | Gaggero A et al |

| | | | | | | |
|---|---|---|---|---|---|---|
| EPI_ISL_591523 | Chile/RM-CMM-NA4204/2020 | 2020-05-26 | South America/Chile/Region Metropolit | Medicina Norte U Chile - Servicio Medico Legal | Center for Mathematical Modeling and Center for Genome Regulation. Santiago, Chile | Gaggero A et al |
| EPI_ISL_591526 | Chile/RM-CMM-HRG4342/2020 | 2020-05-29 | South America/Chile/Region Metropolit | Medicina Norte U Chile - Servicio Medico Legal | Center for Mathematical Modeling and Center for Genome Regulation. Santiago, Chile | Gaggero A et al |
| EPI_ISL_591527 | Chile/RM-CMM-LAE7918/2020 | 2020-07-28 | South America/Chile/Region Metropolit | Medicina Norte U Chile - Servicio Medico Legal | Center for Mathematical Modeling and Center for Genome Regulation. Santiago, Chile | Gaggero A et al |
| EPI_ISL_591528 | Chile/RM-CMM-ADC1858/2020 | 2020-06-08 | South America/Chile/Region Metropolit | Medicina Norte U Chile - Servicio Medico Legal | Center for Mathematical Modeling and Center for Genome Regulation. Santiago, Chile | Gaggero A et al |
| EPI_ISL_591529 | Chile/RM-CMM-MCD9529/2020 | 2020-06-11 | South America/Chile/Region Metropolit | Medicina Norte U Chile - Servicio Medico Legal | Center for Mathematical Modeling and Center for Genome Regulation. Santiago, Chile | Gaggero A et al |

| | | | | | | |
|---|---|---|---|---|---|---|
| EPI_ISL_591530 | Chile/RM-CMM-EDB244K/2020 | 2020-06-16 | South America/Chile/Region Metropolit | Medicina Norte U Chile - Servicio Medico Legal | Center for Mathematical Modeling and Center for Genome Regulation. Santiago, Chile | Gaggero A et al |
| EPI_ISL_591531 | Chile/RM-CMM-IN233883987/2020 | 2020-08-24 | South America/Chile/Region Metropolit | Laboratorio de Infectologia y virologia molecular | Center for Mathematical Modeling and Center for Genome Regulation. Santiago, Chile | Valiente F et al |
| EPI_ISL_591533 | Chile/RM-CMM-A2P533884024/2020 | 2020-08-25 | South America/Chile/Region Metropolit | Laboratorio de Infectologia y virologia molecular | Center for Mathematical Modeling and Center for Genome Regulation. Santiago, Chile | Valiente F et al |
| EPI_ISL_591541 | France/OCC-33/2020 | 2020-06-08 | Europe/France/Occitanie/ | CHU Purpan - Laboratoire de Virologie - Institut F√©d√©ratif de Biologie | CHU Purpan - Laboratoire de Virologie - Institut F√©d√©ratif de Biologie | Latour J. et al |
| EPI_ISL_591546 | France/OCC-38/2020 | 2020-06-12 | Europe/France/Occitanie/ | CHU Purpan - Laboratoire de Virologie - Institut F√©d√©ratif de Biologie | CHU Purpan - Laboratoire de Virologie - Institut F√©d√©ratif de Biologie | Latour J. et al |
| EPI_ISL_592639 | Australia/VIC14526/2020 | 2020-08-09 | Oceania/Australia/Victoria/ | Microbiological Diagnostic Unit - Public Health Laboratory (MDU-PHL) | MDU-PHL | Seemann T. et al |
| EPI_ISL_593481 | Canada/ON-E1/2020 | 2020-04-11 | North America/Canada/Ontario/Ottawa | Eastern Ontario Regional Laboratory Association | McMaster University | Leanne Mortimer et al |
| EPI_ISL_593483 | Canada/ON-E3/2020 | 2020-04-23 | North America/Canada/Ontario/Ottawa | Eastern Ontario Regional Laboratory Association | McMaster University | Leanne Mortimer et al |
| EPI_ISL_593486 | Canada/ON-E6/2020 | 2020-04-24 | North America/Canada/Ontario/Ottawa | Eastern Ontario Regional Laboratory Association | McMaster University | Leanne Mortimer et al |
| EPI_ISL_593487 | Canada/ON-E7/2020 | 2020-04-25 | North America/Canada/Ontario/Ottawa | Eastern Ontario Regional Laboratory Association | McMaster University | Leanne Mortimer et al |

| | | | | | | |
|---|---|---|---|---|---|---|
| EPI_ISL_593489 | Canada/ON-E9/2020 | 2020-04-26 | North America/Canada/Ontario/Ottawa | Eastern Ontario Regional Laboratory Association | McMaster University | Leanne Mortimer et al |
| EPI_ISL_593490 | Canada/ON-E10/2020 | 2020-04-30 | North America/Canada/Ontario/Ottawa | Eastern Ontario Regional Laboratory Association | McMaster University | Leanne Mortimer et al |
| EPI_ISL_593491 | Canada/ON-E11/2020 | 2020-05-04 | North America/Canada/Ontario/Ottawa | Eastern Ontario Regional Laboratory Association | McMaster University | Leanne Mortimer et al |
| EPI_ISL_593492 | Canada/ON-E12/2020 | 2020-05-04 | North America/Canada/Ontario/Ottawa | Eastern Ontario Regional Laboratory Association | McMaster University | Leanne Mortimer et al |
| EPI_ISL_593493 | Canada/ON-E13/2020 | 2020-05-05 | North America/Canada/Ontario/Ottawa | Eastern Ontario Regional Laboratory Association | McMaster University | Leanne Mortimer et al |
| EPI_ISL_593494 | Canada/ON-E14/2020 | 2020-05-07 | North America/Canada/Ontario/Ottawa | Eastern Ontario Regional Laboratory Association | McMaster University | Leanne Mortimer et al |
| EPI_ISL_593495 | Canada/ON-E15/2020 | 2020-05-08 | North America/Canada/Ontario/Ottawa | Eastern Ontario Regional Laboratory Association | McMaster University | Leanne Mortimer et al |
| EPI_ISL_593496 | Canada/ON-E16/2020 | 2020-05-09 | North America/Canada/Ontario/Ottawa | Eastern Ontario Regional Laboratory Association | McMaster University | Leanne Mortimer et al |
| EPI_ISL_593497 | Canada/ON-E18/2020 | 2020-05-09 | North America/Canada/Ontario/Ottawa | Eastern Ontario Regional Laboratory Association | McMaster University | Leanne Mortimer et al |
| EPI_ISL_593498 | Canada/ON-E19/2020 | 2020-05-11 | North America/Canada/Ontario/Ottawa | Eastern Ontario Regional Laboratory Association | McMaster University | Leanne Mortimer et al |
| EPI_ISL_593499 | Canada/ON-E20/2020 | 2020-05-13 | North America/Canada/Ontario/Ottawa | Eastern Ontario Regional Laboratory Association | McMaster University | Leanne Mortimer et al |
| EPI_ISL_593501 | Canada/ON-E22/2020 | 2020-05-13 | North America/Canada/Ontario/Ottawa | Eastern Ontario Regional Laboratory Association | McMaster University | Leanne Mortimer et al |
| EPI_ISL_593502 | Canada/ON-E24/2020 | 2020-05-14 | North America/Canada/Ontario/Ottawa | Eastern Ontario Regional Laboratory Association | McMaster University | Leanne Mortimer et al |
| EPI_ISL_593503 | Canada/ON-E25/2020 | 2020-05-14 | North America/Canada/Ontario/Ottawa | Eastern Ontario Regional Laboratory Association | McMaster University | Leanne Mortimer et al |
| EPI_ISL_593504 | Canada/ON-E26/2020 | 2020-05-23 | North America/Canada/Ontario/Ottawa | Eastern Ontario Regional Laboratory Association | McMaster University | Leanne Mortimer et al |
| EPI_ISL_593506 | Canada/ON-E28/2020 | 2020-05-24 | North America/Canada/Ontario/Ottawa | Eastern Ontario Regional Laboratory Association | McMaster University | Leanne Mortimer et al |
| EPI_ISL_593507 | Canada/ON-E29/2020 | 2020-05-25 | North America/Canada/Ontario/Ottawa | Eastern Ontario Regional Laboratory Association | McMaster University | Leanne Mortimer et al |
| EPI_ISL_593508 | Canada/ON-E30/2020 | 2020-05-25 | North America/Canada/Ontario/Ottawa | Eastern Ontario Regional Laboratory Association | McMaster University | Leanne Mortimer et al |
| EPI_ISL_593509 | Canada/ON-E31/2020 | 2020-05-25 | North America/Canada/Ontario/Ottawa | Eastern Ontario Regional Laboratory Association | McMaster University | Leanne Mortimer et al |
| EPI_ISL_593511 | Canada/ON-E33/2020 | 2020-05-25 | North America/Canada/Ontario/Ottawa | Eastern Ontario Regional Laboratory Association | McMaster University | Leanne Mortimer et al |

| | | | | | | |
|---|---|---|---|---|---|---|
| EPI_ISL_593512 | Canada/ON-E34/2020 | 2020-05-25 | North America/Canada/Ontario/Ottawa | Eastern Ontario Regional Laboratory Association | McMaster University | Leanne Mortimer et al |
| EPI_ISL_593513 | Canada/ON-E35/2020 | 2020-05-26 | North America/Canada/Ontario/Ottawa | Eastern Ontario Regional Laboratory Association | McMaster University | Leanne Mortimer et al |
| EPI_ISL_593514 | Canada/ON-E36/2020 | 2020-05-26 | North America/Canada/Ontario/Ottawa | Eastern Ontario Regional Laboratory Association | McMaster University | Leanne Mortimer et al |
| EPI_ISL_593515 | Canada/ON-E37/2020 | 2020-05-26 | North America/Canada/Ontario/Ottawa | Eastern Ontario Regional Laboratory Association | McMaster University | Leanne Mortimer et al |
| EPI_ISL_593517 | Canada/ON-E39/2020 | 2020-05-26 | North America/Canada/Ontario/Ottawa | Eastern Ontario Regional Laboratory Association | McMaster University | Leanne Mortimer et al |
| EPI_ISL_593518 | Canada/ON-E40/2020 | 2020-05-27 | North America/Canada/Ontario/Ottawa | Eastern Ontario Regional Laboratory Association | McMaster University | Leanne Mortimer et al |
| EPI_ISL_593519 | Canada/ON-E41/2020 | 2020-05-28 | North America/Canada/Ontario/Ottawa | Eastern Ontario Regional Laboratory Association | McMaster University | Leanne Mortimer et al |
| EPI_ISL_593523 | Canada/ON-E46/2020 | 2020-05-28 | North America/Canada/Ontario/Ottawa | Eastern Ontario Regional Laboratory Association | McMaster University | Leanne Mortimer et al |
| EPI_ISL_593524 | Canada/ON-E47/2020 | 2020-05-28 | North America/Canada/Ontario/Ottawa | Eastern Ontario Regional Laboratory Association | McMaster University | Leanne Mortimer et al |
| EPI_ISL_593525 | Canada/ON-E50/2020 | 2020-05-29 | North America/Canada/Ontario/Ottawa | Eastern Ontario Regional Laboratory Association | McMaster University | Leanne Mortimer et al |
| EPI_ISL_593526 | Canada/ON-E51/2020 | 2020-05-29 | North America/Canada/Ontario/Ottawa | Eastern Ontario Regional Laboratory Association | McMaster University | Leanne Mortimer et al |
| EPI_ISL_593527 | Canada/ON-E52/2020 | 2020-05-29 | North America/Canada/Ontario/Ottawa | Eastern Ontario Regional Laboratory Association | McMaster University | Leanne Mortimer et al |
| EPI_ISL_593528 | Canada/ON-E53/2020 | 2020-05-29 | North America/Canada/Ontario/Ottawa | Eastern Ontario Regional Laboratory Association | McMaster University | Leanne Mortimer et al |
| EPI_ISL_593529 | Canada/ON-E55/2020 | 2020-05-29 | North America/Canada/Ontario/Ottawa | Eastern Ontario Regional Laboratory Association | McMaster University | Leanne Mortimer et al |
| EPI_ISL_593531 | Canada/ON-E57/2020 | 2020-05-31 | North America/Canada/Ontario/Ottawa | Eastern Ontario Regional Laboratory Association | McMaster University | Leanne Mortimer et al |
| EPI_ISL_593532 | Canada/ON-E58/2020 | 2020-06-01 | North America/Canada/Ontario/Ottawa | Eastern Ontario Regional Laboratory Association | McMaster University | Leanne Mortimer et al |
| EPI_ISL_593533 | Canada/ON-E59/2020 | 2020-06-01 | North America/Canada/Ontario/Ottawa | Eastern Ontario Regional Laboratory Association | McMaster University | Leanne Mortimer et al |
| EPI_ISL_593534 | Canada/ON-E60/2020 | 2020-06-01 | North America/Canada/Ontario/Ottawa | Eastern Ontario Regional Laboratory Association | McMaster University | Leanne Mortimer et al |
| EPI_ISL_593536 | Canada/ON-E62/2020 | 2020-06-02 | North America/Canada/Ontario/Ottawa | Eastern Ontario Regional Laboratory Association | McMaster University | Leanne Mortimer et al |
| EPI_ISL_593537 | Canada/ON-E64/2020 | 2020-06-06 | North America/Canada/Ontario/Ottawa | Eastern Ontario Regional Laboratory Association | McMaster University | Leanne Mortimer et al |

| | | | | | | |
|---|---|---|---|---|---|---|
| EPI_ISL_593538 | Canada/ON-E65/2020 | 2020-06-07 | North America/Canada/Ontario/Ottawa | Eastern Ontario Regional Laboratory Association | McMaster University | Leanne Mortimer et al |
| EPI_ISL_593539 | Canada/ON-E66/2020 | 2020-06-09 | North America/Canada/Ontario/Ottawa | Eastern Ontario Regional Laboratory Association | McMaster University | Leanne Mortimer et al |
| EPI_ISL_593540 | Canada/ON-E67/2020 | 2020-06-10 | North America/Canada/Ontario/Ottawa | Eastern Ontario Regional Laboratory Association | McMaster University | Leanne Mortimer et al |
| EPI_ISL_593541 | Canada/ON-E68/2020 | 2020-06-11 | North America/Canada/Ontario/Ottawa | Eastern Ontario Regional Laboratory Association | McMaster University | Leanne Mortimer et al |
| EPI_ISL_593543 | Canada/ON-E70/2020 | 2020-03-28 | North America/Canada/Ontario/Ottawa | Eastern Ontario Regional Laboratory Association | McMaster University | Leanne Mortimer et al |
| EPI_ISL_593544 | Canada/ON-E71/2020 | 2020-04-03 | North America/Canada/Ontario/Ottawa | Eastern Ontario Regional Laboratory Association | McMaster University | Leanne Mortimer et al |
| EPI_ISL_593545 | Canada/ON-E72/2020 | 2020-04-03 | North America/Canada/Ontario/Ottawa | Eastern Ontario Regional Laboratory Association | McMaster University | Leanne Mortimer et al |
| EPI_ISL_593546 | Canada/ON-E73/2020 | 2020-04-03 | North America/Canada/Ontario/Ottawa | Eastern Ontario Regional Laboratory Association | McMaster University | Leanne Mortimer et al |
| EPI_ISL_593547 | Canada/ON-E74/2020 | 2020-04-10 | North America/Canada/Ontario/Ottawa | Eastern Ontario Regional Laboratory Association | McMaster University | Leanne Mortimer et al |
| EPI_ISL_593548 | Canada/ON-E75/2020 | 2020-04-13 | North America/Canada/Ontario/Ottawa | Eastern Ontario Regional Laboratory Association | McMaster University | Leanne Mortimer et al |
| EPI_ISL_593549 | Canada/ON-E76/2020 | 2020-05-06 | North America/Canada/Ontario/Ottawa | Eastern Ontario Regional Laboratory Association | McMaster University | Leanne Mortimer et al |
| EPI_ISL_593550 | Canada/ON-E77/2020 | 2020-05-13 | North America/Canada/Ontario/Ottawa | Eastern Ontario Regional Laboratory Association | McMaster University | Leanne Mortimer et al |
| EPI_ISL_593551 | Canada/ON-E78/2020 | 2020-05-29 | North America/Canada/Ontario/Ottawa | Eastern Ontario Regional Laboratory Association | McMaster University | Leanne Mortimer et al |
| EPI_ISL_593578 | USA/TX-DSHS-0836/2020 | 2020-06-07 | North America/USA/Texas/El Paso Coun | Texas Department of State Health Services | Texas Department of State Health Services | Rashmi Tuladhar et al |
| EPI_ISL_593589 | USA/TX-DSHS-0857/2020 | 2020-06-10 | North America/USA/Texas/Harris Count | Texas Department of State Health Services | Texas Department of State Health Services | Rashmi Tuladhar et al |
| EPI_ISL_593591 | USA/TX-DSHS-0859/2020 | 2020-06-16 | North America/USA/Texas/Harris Count | Texas Department of State Health Services | Texas Department of State Health Services | Rashmi Tuladhar et al |
| EPI_ISL_593594 | USA/TX-DSHS-0862/2020 | 2020-06-16 | North America/USA/Texas/Harris Count | Texas Department of State Health Services | Texas Department of State Health Services | Rashmi Tuladhar et al |
| EPI_ISL_593772 | Peru/LAM-HRL-235/2020 | 2020-04-12 | South America/Peru/Lambayeque/ | HOSPITAL REGIONAL LAMBAYEQUE | GENOMA MAYOR | Franklin R. Aguilar-Gamboa et al |

| | | | | | | |
|---|---|---|---|---|---|---|
| EPI_ISL_593773 | Peru/LAM-HRL-239/2020 | 2020-04-10 | South America/Peru/Lambayeque/Chicla | HOSPITAL REGIONAL LAMBAYEQUE | GENOMA MAYOR | Franklin R. Aguilar-Gamboa et al |
| EPI_ISL_593775 | Peru/LAM-HRL-223/2020 | 2020-04-08 | South America/Peru/Lambayeque/Chicla | HOSPITAL REGIONAL LAMBAYEQUE | GENOMA MAYOR | Franklin R. Aguilar-Gamboa et al |
| EPI_ISL_593776 | Peru/LAM-HRL-224/2020 | 2020-04-10 | South America/Peru/Lambayeque/Chicla | HOSPITAL REGIONAL LAMBAYEQUE | GENOMA MAYOR | Franklin R. Aguilar-Gamboa et al |
| EPI_ISL_593858 | France/OCC-43/2020 | 2020-06-19 | Europe/France/Occitanie/ | CHU Purpan - Laboratoire de Virologie - Institut F√©d√©ratif de Biologie | CHU Purpan - Laboratoire de Virologie - Institut F√©d√©ratif de Biologie | Latour J. et al |
| EPI_ISL_593860 | France/OCC-45/2020 | 2020-06-19 | Europe/France/Occitanie/ | CHU Purpan - Laboratoire de Virologie - Institut F√©d√©ratif de Biologie | CHU Purpan - Laboratoire de Virologie - Institut F√©d√©ratif de Biologie | Latour J. et al |
| EPI_ISL_593863 | France/OCC-48/2020 | 2020-06-19 | Europe/France/Occitanie/ | CHU Purpan - Laboratoire de Virologie - Institut F√©d√©ratif de Biologie | CHU Purpan - Laboratoire de Virologie - Institut F√©d√©ratif de Biologie | Latour J. et al |
| EPI_ISL_593864 | France/OCC-49/2020 | 2020-06-19 | Europe/France/Occitanie/ | CHU Purpan - Laboratoire de Virologie - Institut F√©d√©ratif de Biologie | CHU Purpan - Laboratoire de Virologie - Institut F√©d√©ratif de Biologie | Latour J. et al |
| EPI_ISL_593871 | France/OCC-56/2020 | 2020-07-14 | Europe/France/Occitanie/ | CHU Purpan - Laboratoire de Virologie - Institut F√©d√©ratif de Biologie | CHU Purpan - Laboratoire de Virologie - Institut F√©d√©ratif de Biologie | Latour J. et al |
| EPI_ISL_593880 | France/OCC-65/2020 | 2020-08-02 | Europe/France/Occitanie/ | CHU Purpan - Laboratoire de Virologie - Institut F√©d√©ratif de Biologie | CHU Purpan - Laboratoire de Virologie - Institut F√©d√©ratif de Biologie | Latour J. et al |

| | | | | | | |
|---|---|---|---|---|---|---|
| EPI_ISL_593883 | France/OCC-69/2020 | 2020-07-22 | Europe/France/Occitanie/ | CHU Purpan - Laboratoire de Virologie - Institut F√©d√©ratif de Biologie | CHU Purpan - Laboratoire de Virologie - Institut F√©d√©ratif de Biologie | Latour J. et al |
| EPI_ISL_593888 | France/OCC-74/2020 | 2020-09-02 | Europe/France/Occitanie/ | CHU Purpan - Laboratoire de Virologie - Institut F√©d√©ratif de Biologie | CHU Purpan - Laboratoire de Virologie - Institut F√©d√©ratif de Biologie | Latour J. et al |
| EPI_ISL_593893 | France/OCC-85/2020 | 2020-09-30 | Europe/France/Occitanie/ | CHU Purpan - Laboratoire de Virologie - Institut F√©d√©ratif de Biologie | CHU Purpan - Laboratoire de Virologie - Institut F√©d√©ratif de Biologie | Latour J. et al |
| EPI_ISL_593895 | France/OCC-87/2020 | 2020-10-05 | Europe/France/Occitanie/ | CHU Purpan - Laboratoire de Virologie - Institut F√©d√©ratif de Biologie | CHU Purpan - Laboratoire de Virologie - Institut F√©d√©ratif de Biologie | Latour J. et al |
| EPI_ISL_593896 | France/OCC-88/2020 | 2020-10-04 | Europe/France/Occitanie/ | CHU Purpan - Laboratoire de Virologie - Institut F√©d√©ratif de Biologie | CHU Purpan - Laboratoire de Virologie - Institut F√©d√©ratif de Biologie | Latour J. et al |
| EPI_ISL_593897 | France/OCC-89/2020 | 2020-10-03 | Europe/France/Occitanie/ | CHU Purpan - Laboratoire de Virologie - Institut F√©d√©ratif de Biologie | CHU Purpan - Laboratoire de Virologie - Institut F√©d√©ratif de Biologie | Latour J. et al |
| EPI_ISL_593898 | France/OCC-90/2020 | 2020-10-03 | Europe/France/Occitanie/ | CHU Purpan - Laboratoire de Virologie - Institut F√©d√©ratif de Biologie | CHU Purpan - Laboratoire de Virologie - Institut F√©d√©ratif de Biologie | Latour J. et al |
| EPI_ISL_593899 | France/OCC-91/2020 | 2020-10-03 | Europe/France/Occitanie/ | CHU Purpan - Laboratoire de Virologie - Institut F√©d√©ratif de Biologie | CHU Purpan - Laboratoire de Virologie - Institut F√©d√©ratif de Biologie | Latour J. et al |

| | | | | | | |
|---|---|---|---|---|---|---|
| EPI_ISL_593900 | France/OCC-92/2020 | 2020-10-02 | Europe/France/Occitanie/ | CHU Purpan - Laboratoire de Virologie - Institut F√©d√©ratif de Biologie | CHU Purpan - Laboratoire de Virologie - Institut F√©d√©ratif de Biologie | Latour J. et al |
| EPI_ISL_593901 | France/OCC-93/2020 | 2020-09-30 | Europe/France/Occitanie/ | CHU Purpan - Laboratoire de Virologie - Institut F√©d√©ratif de Biologie | CHU Purpan - Laboratoire de Virologie - Institut F√©d√©ratif de Biologie | Latour J. et al |
| EPI_ISL_593905 | France/IDF-9137/2020 | 2020-09-14 | Europe/France/Ile de France/Puteaux | Labo Analyses Med, Sarcelles | National Reference Center for Viruses of Respiratory Infections, Institut Pasteur, Paris | Sylvie Behillil et al |
| EPI_ISL_593906 | France/IDF-9139/2020 | 2020-09-15 | Europe/France/Ile de France/Puteaux | Labo Analyses Med, Sarcelles | National Reference Center for Viruses of Respiratory Infections, Institut Pasteur, Paris | Sylvie Behillil et al |
| EPI_ISL_593907 | France/IDF-9141/2020 | 2020-09-15 | Europe/France/Ile de France/Puteaux | Labo Analyses Med, Sarcelles | National Reference Center for Viruses of Respiratory Infections, Institut Pasteur, Paris | Sylvie Behillil et al |
| EPI_ISL_593909 | France/IDF-9143/2020 | 2020-09-14 | Europe/France/Ile de France/Puteaux | Labo Analyses Med, Sarcelles | National Reference Center for Viruses of Respiratory Infections, Institut Pasteur, Paris | Sylvie Behillil et al |
| EPI_ISL_593910 | France/IDF-9144/2020 | 2020-09-14 | Europe/France/Ile de France/Puteaux | Hospital, Boulogne-Billancourt | National Reference Center for Viruses of Respiratory Infections, Institut Pasteur, Paris | Sylvie Behillil et al |

| | | | | | | |
|---|---|---|---|---|---|---|
| EPI_ISL_593913 | France/IDF-9155/2020 | 2020-09-02 | Europe/France/Ile de France/Puteaux | Hospital, Talange | National Reference Center for Viruses of Respiratory Infections, Institut Pasteur, Paris | Sylvie Behillil et al |
| EPI_ISL_593915 | France/IDF-9163/2020 | 2020-09-02 | Europe/France/Ile de France/Puteaux | Hospital, Amneville | National Reference Center for Viruses of Respiratory Infections, Institut Pasteur, Paris | Sylvie Behillil et al |
| EPI_ISL_593916 | France/IDF-9164/2020 | 2020-09-01 | Europe/France/Ile de France/Puteaux | Hospital, Sarreguemines | National Reference Center for Viruses of Respiratory Infections, Institut Pasteur, Paris | Sylvie Behillil et al |
| EPI_ISL_593917 | France/IDF-9171/2020 | 2020-08-31 | Europe/France/Ile de France/Puteaux | Hospital, Montigny les Metz | National Reference Center for Viruses of Respiratory Infections, Institut Pasteur, Paris | Sylvie Behillil et al |
| EPI_ISL_593919 | France/IDF-9175/2020 | 2020-09-01 | Europe/France/Ile de France/Puteaux | Hospital, Ottonville | National Reference Center for Viruses of Respiratory Infections, Institut Pasteur, Paris | Sylvie Behillil et al |
| EPI_ISL_593920 | France/IDF-9177/2020 | 2020-09-01 | Europe/France/Ile de France/Puteaux | Hospital, Metz | National Reference Center for Viruses of Respiratory Infections, Institut Pasteur, Paris | Sylvie Behillil et al |

| | | | | | | |
|---|---|---|---|---|---|---|
| EPI_ISL_593921 | France/IDF-9183/2020 | 2020-09-01 | Europe/France/Ile de France/Puteaux | Hospital, Fameck | National Reference Center for Viruses of Respiratory Infections, Institut Pasteur, Paris | Sylvie Behillil et al |
| EPI_ISL_593922 | France/IDF-9190/2020 | 2020-09-01 | Europe/France/Ile de France/Puteaux | Hospital, Maiziv®res les Metz | National Reference Center for Viruses of Respiratory Infections, Institut Pasteur, Paris | Sylvie Behillil et al |
| EPI_ISL_593923 | France/IDF-9191/2020 | 2020-09-01 | Europe/France/Ile de France/Puteaux | Hospital, Maiziv®res les Metz | National Reference Center for Viruses of Respiratory Infections, Institut Pasteur, Paris | Sylvie Behillil et al |
| EPI_ISL_593927 | France/IDF-9138/2020 | 2020-09-14 | Europe/France/Ile de France/Puteaux | Labo Analyses Med, Puteaux | National Reference Center for Viruses of Respiratory Infections, Institut Pasteur, Paris | Sylvie Behillil et al |
| EPI_ISL_593928 | France/IDF-9140/2020 | 2020-09-15 | Europe/France/Ile de France/Puteaux | Labo Analyses Med, Puteaux | National Reference Center for Viruses of Respiratory Infections, Institut Pasteur, Paris | Sylvie Behillil et al |
| EPI_ISL_593932 | France/IDF-9199/2020 | 2020-08-31 | Europe/France/Ile de France/Puteaux | Labo Analyses Med, Puteaux | National Reference Center for Viruses of Respiratory Infections, Institut Pasteur, Paris | Sylvie Behillil et al |

| | | | | | | |
|---|---|---|---|---|---|---|
| EPI_ISL_593935 | France/IDF-9320/2020 | 2020-09-29 | Europe/France/Ile de France/Plessis Tre | Sentinelles, Plessis-Trevise | National Reference Center for Viruses of Respiratory Infections, Institut Pasteur, Paris | Sylvie Behillil et al |
| EPI_ISL_593936 | France/CVL-9321/2020 | 2020-10-01 | Europe/France/Centre-Val de Loire/Fon | Sentinelles, Fondettes | National Reference Center for Viruses of Respiratory Infections, Institut Pasteur, Paris | Sylvie Behillil et al |
| EPI_ISL_593937 | USA/DE-DHSS-F918017/2020 | 2020-03-27 | North America/USA/Illinois/Winnebago | Delaware Public Health Lab | Delaware Public Health Lab | Gregory Hovan et al |
| EPI_ISL_593944 | USA/DE-DHSS-F918133/2020 | 2020-03-27 | North America/USA/Delaware/New Cas | Delaware Public Health Lab | Delaware Public Health Lab | Gregory Hovan et al |
| EPI_ISL_593972 | USA/DE-DHSS-F918481/2020 | 2020-04-02 | North America/USA/Delaware/Kent Cou | Delaware Public Health Lab | Delaware Public Health Lab | Gregory Hovan et al |
| EPI_ISL_593977 | USA/DE-DHSS-F952951/2020 | 2020-04-04 | North America/USA/Delaware/Sussex C | Delaware Public Health Lab | Delaware Public Health Lab | Gregory Hovan et al |
| EPI_ISL_594008 | USA/UT-UPHL-201027142/2020 | 2020-05-31 | North America/USA/Utah/ | Utah Public Health Laboratory | Utah Public Health Laboratory | Erin Young et al |
| EPI_ISL_594009 | USA/UT-UPHL-201004118/2020 | 2020-05-30 | North America/USA/Utah/ | Utah Public Health Laboratory | Utah Public Health Laboratory | Erin Young et al |
| EPI_ISL_594010 | USA/UT-UPHL-201014531/2020 | 2020-05-31 | North America/USA/Utah/ | Utah Public Health Laboratory | Utah Public Health Laboratory | Erin Young et al |
| EPI_ISL_594011 | USA/UT-UPHL-201018212/2020 | 2020-05-31 | North America/USA/Utah/ | Utah Public Health Laboratory | Utah Public Health Laboratory | Erin Young et al |
| EPI_ISL_594017 | USA/UT-UPHL-201016782/2020 | 2020-05-30 | North America/USA/Utah/ | Utah Public Health Laboratory | Utah Public Health Laboratory | Erin Young et al |
| EPI_ISL_594019 | USA/UT-UPHL-201006744/2020 | 2020-05-31 | North America/USA/Utah/ | Utah Public Health Laboratory | Utah Public Health Laboratory | Erin Young et al |
| EPI_ISL_594020 | USA/UT-UPHL-201032430/2020 | 2020-05-29 | North America/USA/Utah/ | Utah Public Health Laboratory | Utah Public Health Laboratory | Erin Young et al |
| EPI_ISL_594021 | USA/UT-UPHL-201016722/2020 | 2020-05-29 | North America/USA/Utah/ | Utah Public Health Laboratory | Utah Public Health Laboratory | Erin Young et al |
| EPI_ISL_594024 | USA/UT-UPHL-201023411/2020 | 2020-06-01 | North America/USA/Utah/ | Utah Public Health Laboratory | Utah Public Health Laboratory | Erin Young et al |
| EPI_ISL_594032 | USA/UT-UPHL-201015212/2020 | 2020-06-02 | North America/USA/Utah/ | Utah Public Health Laboratory | Utah Public Health Laboratory | Erin Young et al |

| | | | | | | |
|---|---|---|---|---|---|---|
| EPI_ISL_594033 | USA/UT-UPHL-201023501/2020 | 2020-06-01 | North America/USA/Utah/ | Utah Public Health Laboratory | Utah Public Health Laboratory | Erin Young et al |
| EPI_ISL_594044 | USA/UT-UPHL-201026758/2020 | 2020-07-06 | North America/USA/Utah/ | Utah Public Health Laboratory | Utah Public Health Laboratory | Erin Young et al |
| EPI_ISL_594050 | USA/UT-UPHL-201025257/2020 | 2020-07-06 | North America/USA/Utah/ | Utah Public Health Laboratory | Utah Public Health Laboratory | Erin Young et al |
| EPI_ISL_594054 | USA/UT-UPHL-201019360/2020 | 2020-07-06 | North America/USA/Utah/ | Utah Public Health Laboratory | Utah Public Health Laboratory | Erin Young et al |
| EPI_ISL_594065 | USA/UT-UPHL-201023062/2020 | 2020-05-29 | North America/USA/Utah/ | Utah Public Health Laboratory | Utah Public Health Laboratory | Erin Young et al |
| EPI_ISL_594066 | USA/UT-UPHL-201011705/2020 | 2020-05-29 | North America/USA/Utah/ | Utah Public Health Laboratory | Utah Public Health Laboratory | Erin Young et al |
| EPI_ISL_594069 | USA/UT-UPHL-201025899/2020 | 2020-05-29 | North America/USA/Utah/ | Utah Public Health Laboratory | Utah Public Health Laboratory | Erin Young et al |
| EPI_ISL_594070 | USA/UT-UPHL-201030995/2020 | 2020-05-29 | North America/USA/Utah/ | Utah Public Health Laboratory | Utah Public Health Laboratory | Erin Young et al |
| EPI_ISL_594082 | USA/UT-UPHL-201018660/2020 | 2020-05-29 | North America/USA/Utah/ | Utah Public Health Laboratory | Utah Public Health Laboratory | Erin Young et al |
| EPI_ISL_594093 | USA/UT-UPHL-201026271/2020 | 2020-04-09 | North America/USA/Utah/ | Utah Public Health Laboratory | Utah Public Health Laboratory | Erin Young et al |
| EPI_ISL_594120 | USA/CT-Yale-376/2020 | 2020-04-21 | North America/USA/Delaware/New Cas | Yale COVID-19 Biorepository | Grubaugh Lab - Yale School of Public Health | Joseph Fauver et al |
| EPI_ISL_594131 | USA/CT-Yale-389/2020 | 2020-05-29 | North America/USA/Delaware/New Cas | Yale COVID-19 Biorepository | Grubaugh Lab - Yale School of Public Health | Joseph Fauver et al |
| EPI_ISL_594132 | USA/CT-Yale-390/2020 | 2020-06-22 | North America/USA/Delaware/New Cas | Yale COVID-19 Biorepository | Grubaugh Lab - Yale School of Public Health | Joseph Fauver et al |
| EPI_ISL_594155 | Israel/ISR_NH_M1_0614/2020 | 2020-06-14 | Asia/Israel/Israel/ | Israel Institute for Biological Research | Israel Institute for Biological Research | Galia Zaide et al |
| EPI_ISL_594156 | Israel/ISR_NH_M2_0614/2020 | 2020-06-14 | Asia/Israel/Israel/ | Israel Institute for Biological Research | Israel Institute for Biological Research | Galia Zaide et al |
| EPI_ISL_594158 | Israel/ISR_NH_MA_0727/2020 | 2020-07-27 | Asia/Israel/Israel/ | Israel Institute for Biological Research | Israel Institute for Biological Research | Galia Zaide et al |

| | | | | | | |
|---|---|---|---|---|---|---|
| EPI_ISL_594159 | Israel/ISR_NH_GD1_0714/2020 | 2020-07-14 | Asia/Israel/Israel/ | Israel Institute for Biological Research | Israel Institute for Biological Research | Galia Zaide et al |
| EPI_ISL_594161 | Israel/ISR_NH_NM_0720/2020 | 2020-07-20 | Asia/Israel/Israel/ | Israel Institute for Biological Research | Israel Institute for Biological Research | Galia Zaide et al |
| EPI_ISL_594185 | Iran/2297/2020 | 2020-04-13 | Asia/Iran/Tehran/ | Department of Pathology, School of Medicine, Imam Khomeini Hospital, Tehran University of Medical Sciences | Genetics Research Center, University of Social Welfare and Rehabilitation Sciences | Zohreh Fattahi et al |
| EPI_ISL_594186 | Iran/1325/2020 | 2020-03-28 | Asia/Iran/Tehran/ | Department of Pathology, School of Medicine, Imam Khomeini Hospital, Tehran University of Medical Sciences | Genetics Research Center, University of Social Welfare and Rehabilitation Sciences | Zohreh Fattahi et al |
| EPI_ISL_594187 | Iran/2024/2020 | 2020-04-11 | Asia/Iran/Tehran/ | Department of Pathology, School of Medicine, Imam Khomeini Hospital, Tehran University of Medical Sciences | Genetics Research Center, University of Social Welfare and Rehabilitation Sciences | Zohreh Fattahi et al |
| EPI_ISL_594188 | Iran/7650/2020 | 2020-06-26 | Asia/Iran/Tehran/ | Department of Pathology, School of Medicine, Imam Khomeini Hospital, Tehran University of Medical Sciences | Genetics Research Center, University of Social Welfare and Rehabilitation Sciences | Zohreh Fattahi et al |
| EPI_ISL_594197 | USA/MI-MDHHS-SC22085/2020 | 2020-03-25 | North America/USA/Michigan/ | Michigan Department of Health and Human Services, Bureau of Laboratories | Michigan Department of Health and Human Services, Bureau of Laboratories | Blankenship HM et al |
| EPI_ISL_594212 | USA/MI-MDHHS-SC22100/2020 | 2020-04-29 | North America/USA/Michigan/ | Michigan Department of Health and Human Services, Bureau of Laboratories | Michigan Department of Health and Human Services, Bureau of Laboratories | Blankenship HM et al |
| EPI_ISL_594248 | USA/UT-UPHL-201024487/2020 | 2020-05-28 | North America/USA/Utah/ | Utah Public Health Laboratory | Utah Public Health Laboratory | Erin Young et al |

| | | | | | | |
|---|---|---|---|---|---|---|
| EPI_ISL_594249 | USA/UT-UPHL-201027132/2020 | 2020-05-28 | North America/USA/Utah/ | Utah Public Health Laboratory | Utah Public Health Laboratory | Erin Young et al |
| EPI_ISL_594251 | USA/UT-UPHL-201031106/2020 | 2020-05-28 | North America/USA/Utah/ | Utah Public Health Laboratory | Utah Public Health Laboratory | Erin Young et al |
| EPI_ISL_594256 | USA/UT-UPHL-201016621/2020 | 2020-06-22 | North America/USA/Utah/ | Utah Public Health Laboratory | Utah Public Health Laboratory | Erin Young et al |
| EPI_ISL_594280 | USA/FL-BPHL-1590/2020 | 2020-05-21 | North America/USA/Florida/ | Florida Bureau of Public Health Laboratories | Florida Bureau of Public Health Laboratories | Sarah Schmedes et al |
| EPI_ISL_594284 | USA/FL-BPHL-1594/2020 | 2020-05-21 | North America/USA/Florida/ | Florida Bureau of Public Health Laboratories | Florida Bureau of Public Health Laboratories | Sarah Schmedes et al |
| EPI_ISL_594286 | USA/FL-BPHL-1596/2020 | 2020-06-05 | North America/USA/Florida/ | Florida Bureau of Public Health Laboratories | Florida Bureau of Public Health Laboratories | Sarah Schmedes et al |
| EPI_ISL_594296 | USA/FL-BPHL-1606/2020 | 2020-07-05 | North America/USA/Florida/ | Florida Bureau of Public Health Laboratories | Florida Bureau of Public Health Laboratories | Sarah Schmedes et al |
| EPI_ISL_594305 | USA/FL-BPHL-1615/2020 | 2020-09-04 | North America/USA/Florida/ | Florida Bureau of Public Health Laboratories | Florida Bureau of Public Health Laboratories | Sarah Schmedes et al |
| EPI_ISL_594313 | USA/FL-BPHL-1623/2020 | 2020-06-03 | North America/USA/Florida/ | Florida Bureau of Public Health Laboratories | Florida Bureau of Public Health Laboratories | Sarah Schmedes et al |
| EPI_ISL_594322 | USA/FL-BPHL-1633/2020 | 2020-09-11 | North America/USA/Florida/ | Florida Bureau of Public Health Laboratories | Florida Bureau of Public Health Laboratories | Sarah Schmedes et al |
| EPI_ISL_594324 | USA/FL-BPHL-1635/2020 | 2020-09-11 | North America/USA/Florida/ | Florida Bureau of Public Health Laboratories | Florida Bureau of Public Health Laboratories | Sarah Schmedes et al |
| EPI_ISL_594325 | USA/FL-BPHL-1636/2020 | 2020-09-11 | North America/USA/Florida/ | Florida Bureau of Public Health Laboratories | Florida Bureau of Public Health Laboratories | Sarah Schmedes et al |
| EPI_ISL_594339 | USA/FL-BPHL-1650/2020 | 2020-07-03 | North America/USA/Florida/ | Florida Bureau of Public Health Laboratories | Florida Bureau of Public Health Laboratories | Sarah Schmedes et al |
| EPI_ISL_594347 | USA/FL-BPHL-1658/2020 | 2020-07-05 | North America/USA/Florida/ | Florida Bureau of Public Health Laboratories | Florida Bureau of Public Health Laboratories | Sarah Schmedes et al |

| | | | | | | |
|---|---|---|---|---|---|---|
| EPI_ISL_594357 | USA/FL-BPHL-1668/2020 | 2020-07-06 | North America/USA/Florida/ | Florida Bureau of Public Health Laboratories | Florida Bureau of Public Health Laboratories | Sarah Schmedes et al |
| EPI_ISL_594363 | USA/FL-BPHL-1674/2020 | 2020-07-07 | North America/USA/Florida/ | Florida Bureau of Public Health Laboratories | Florida Bureau of Public Health Laboratories | Sarah Schmedes et al |
| EPI_ISL_594371 | USA/FL-BPHL-1682/2020 | 2020-08-11 | North America/USA/Florida/ | Florida Bureau of Public Health Laboratories | Florida Bureau of Public Health Laboratories | Sarah Schmedes et al |
| EPI_ISL_594376 | USA/FL-BPHL-1687/2020 | 2020-08-13 | North America/USA/Florida/ | Florida Bureau of Public Health Laboratories | Florida Bureau of Public Health Laboratories | Sarah Schmedes et al |
| EPI_ISL_594379 | USA/FL-BPHL-1690/2020 | 2020-08-17 | North America/USA/Florida/ | Florida Bureau of Public Health Laboratories | Florida Bureau of Public Health Laboratories | Sarah Schmedes et al |
| EPI_ISL_594382 | USA/FL-BPHL-1693/2020 | 2020-08-17 | North America/USA/Florida/ | Florida Bureau of Public Health Laboratories | Florida Bureau of Public Health Laboratories | Sarah Schmedes et al |
| EPI_ISL_594386 | USA/FL-BPHL-1697/2020 | 2020-08-20 | North America/USA/Florida/ | Florida Bureau of Public Health Laboratories | Florida Bureau of Public Health Laboratories | Sarah Schmedes et al |
| EPI_ISL_594391 | USA/FL-BPHL-1702/2020 | 2020-08-22 | North America/USA/Florida/ | Florida Bureau of Public Health Laboratories | Florida Bureau of Public Health Laboratories | Sarah Schmedes et al |
| EPI_ISL_594400 | USA/UT-UPHL-201025176/2020 | 2020-04-04 | North America/USA/Utah/ | Utah Public Health Laboratory | Utah Public Health Laboratory | Erin Young et al |
| EPI_ISL_594410 | USA/UT-UPHL-201024883/2020 | 2020-04-12 | North America/USA/Utah/ | Utah Public Health Laboratory | Utah Public Health Laboratory | Erin Young et al |
| EPI_ISL_594422 | USA/UT-UPHL-201008164/2020 | 2020-04-24 | North America/USA/Utah/ | Utah Public Health Laboratory | Utah Public Health Laboratory | Erin Young et al |
| EPI_ISL_594436 | USA/UT-UPHL-201020974/2020 | 2020-06-22 | North America/USA/Utah/ | Utah Public Health Laboratory | Utah Public Health Laboratory | Erin Young et al |
| EPI_ISL_594459 | USA/WA-CDC-8384/2020 | 2020-02-26 | North America/USA/Washington/ | Washington State Public Health Laboratories | Pathogen Discovery, Respiratory Viruses Branch, Division of Viral Diseases, Centers for Disease Control and Prevention | Ying Tao et al |

| | | | | | | |
|---|---|---|---|---|---|---|
| EPI_ISL_594464 | USA/TX-CDC-3902/2020 | 2020-07-02 | North America/USA/Texas/ | Texas Department of State Health Services Public Health Laboratory | Pathogen Discovery, Respiratory Viruses Branch, Division of Viral Diseases, Centers for Disease Control and Prevention | Ying Tao et al |
| EPI_ISL_594488 | England/BIRM-6435C/2020 | 2020-09-19 | Europe/United Kingdom/England/ | University of Birmingham | COVID-19 Genomics UK (COG-UK) Consortium | Institute of Microbiology et al |
| EPI_ISL_594500 | England/BIRM-64635/2020 | 2020-09-22 | Europe/United Kingdom/England/ | University of Birmingham | COVID-19 Genomics UK (COG-UK) Consortium | Institute of Microbiology et al |
| EPI_ISL_594549 | England/BIRM-6533D/2020 | 2020-09-29 | Europe/United Kingdom/England/ | University of Birmingham | COVID-19 Genomics UK (COG-UK) Consortium | Institute of Microbiology et al |
| EPI_ISL_594591 | England/BIRM-658B6/2020 | 2020-10-02 | Europe/United Kingdom/England/ | University of Birmingham | COVID-19 Genomics UK (COG-UK) Consortium | Institute of Microbiology et al |
| EPI_ISL_594593 | England/BIRM-658D4/2020 | 2020-10-02 | Europe/United Kingdom/England/ | University of Birmingham | COVID-19 Genomics UK (COG-UK) Consortium | Institute of Microbiology et al |
| EPI_ISL_594801 | Scotland/CVR5068/2020 | 2020-10-12 | Europe/United Kingdom/Scotland/ | West of Scotland Specialist Virology Centre, NHSGGC / MRC-University of Glasgow Centre for Virus Research | COVID-19 Genomics UK (COG-UK) Consortium | Ana da Silva Filipe et al |
| EPI_ISL_594813 | Scotland/CVR5080/2020 | 2020-10-11 | Europe/United Kingdom/Scotland/ | West of Scotland Specialist Virology Centre, NHSGGC / MRC-University of Glasgow Centre for Virus Research | COVID-19 Genomics UK (COG-UK) Consortium | Ana da Silva Filipe et al |
| EPI_ISL_594901 | England/MILK-9EF10B/2020 | 2020-09-25 | Europe/United Kingdom/England/ | Oxford Viromics, NDM, University of Oxford; Oxford University Hospitals; Basingstoke and North Hampshire Hospital | COVID-19 Genomics UK (COG-UK) Consortium | Tanya Golubchik et al |
| EPI_ISL_594945 | England/MILK-9EF806/2020 | 2020-09-23 | Europe/United Kingdom/England/ | Queens Medical Centre, Clinical Microbiology Department / DeepSeq Nottingham | COVID-19 Genomics UK (COG-UK) Consortium | Gemma Clark et al |
| EPI_ISL_594952 | England/MILK-9EF89D/2020 | 2020-09-23 | Europe/United Kingdom/England/ | Queens Medical Centre, Clinical Microbiology Department / DeepSeq Nottingham | COVID-19 Genomics UK (COG-UK) Consortium | Gemma Clark et al |

| | | | | | | |
|---|---|---|---|---|---|---|
| EPI_ISL_594961 | England/MILK-9EF96D/2020 | 2020-09-23 | Europe/United Kingdom/England/ | Queens Medical Centre, Clinical Microbiology Department / DeepSeq Nottingham | COVID-19 Genomics UK (COG-UK) Consortium | Gemma Clark et al |
| EPI_ISL_595052 | England/MILK-A1D349/2020 | 2020-10-03 | Europe/United Kingdom/England/ | Queens Medical Centre, Clinical Microbiology Department / DeepSeq Nottingham | COVID-19 Genomics UK (COG-UK) Consortium | Gemma Clark et al |
| EPI_ISL_595059 | England/MILK-A1D473/2020 | 2020-10-03 | Europe/United Kingdom/England/ | Queens Medical Centre, Clinical Microbiology Department / DeepSeq Nottingham | COVID-19 Genomics UK (COG-UK) Consortium | Gemma Clark et al |
| EPI_ISL_595090 | England/MILK-A1D81D/2020 | 2020-10-03 | Europe/United Kingdom/England/ | Queens Medical Centre, Clinical Microbiology Department / DeepSeq Nottingham | COVID-19 Genomics UK (COG-UK) Consortium | Gemma Clark et al |
| EPI_ISL_595094 | England/MILK-A1D895/2020 | 2020-10-03 | Europe/United Kingdom/England/ | Queens Medical Centre, Clinical Microbiology Department / DeepSeq Nottingham | COVID-19 Genomics UK (COG-UK) Consortium | Gemma Clark et al |
| EPI_ISL_595104 | England/MILK-A23BC0/2020 | 2020-10-03 | Europe/United Kingdom/England/ | Centre for Enzyme Innovation, University of Portsmouth / Translational Research Laboratory, Portsmouth Hospitals NHS Trust | COVID-19 Genomics UK (COG-UK) Consortium | Angela Beckett et al |
| EPI_ISL_595280 | England/NOTT-115A27/2020 | 2020-10-16 | Europe/United Kingdom/England/ | Queens Medical Centre, Clinical Microbiology Department / DeepSeq Nottingham | COVID-19 Genomics UK (COG-UK) Consortium | Gemma Clark et al |
| EPI_ISL_595314 | England/NOTT-1162B5/2020 | 2020-10-19 | Europe/United Kingdom/England/ | Queens Medical Centre, Clinical Microbiology Department / DeepSeq Nottingham | COVID-19 Genomics UK (COG-UK) Consortium | Gemma Clark et al |
| EPI_ISL_595398 | Wales/PHWC-47E063/2020 | 2020-10-09 | Europe/United Kingdom/Wales/ | Wales Specialist Virology Centre Sequencing lab: Pathogen Genomics Unit | COVID-19 Genomics UK (COG-UK) Consortium | Catherine Moore et al |
| EPI_ISL_595541 | Wales/PHWC-47EA47/2020 | 2020-10-11 | Europe/United Kingdom/Wales/ | Wales Specialist Virology Centre Sequencing lab: Pathogen Genomics Unit | COVID-19 Genomics UK (COG-UK) Consortium | Catherine Moore et al |
| EPI_ISL_595549 | Wales/PHWC-47EACF/2020 | 2020-10-08 | Europe/United Kingdom/Wales/ | Wales Specialist Virology Centre Sequencing lab: Pathogen Genomics Unit | COVID-19 Genomics UK (COG-UK) Consortium | Catherine Moore et al |
| EPI_ISL_595587 | Wales/PHWC-47ED8A/2020 | 2020-10-13 | Europe/United Kingdom/Wales/ | Wales Specialist Virology Centre Sequencing lab: Pathogen Genomics Unit | COVID-19 Genomics UK (COG-UK) Consortium | Catherine Moore et al |
| EPI_ISL_595596 | Wales/PHWC-47EE1E/2020 | 2020-10-07 | Europe/United Kingdom/Wales/ | Wales Specialist Virology Centre Sequencing lab: Pathogen Genomics Unit | COVID-19 Genomics UK (COG-UK) Consortium | Catherine Moore et al |

| | | | | | | |
|---|---|---|---|---|---|---|
| EPI_ISL_595623 | Scotland/QEUH-9F6CB5/2020 | 2020-09-27 | Europe/United Kingdom/Scotland/ | Oxford Viromics, NDM, University of Oxford; Oxford University Hospitals; Basingstoke and North Hampshire Hospital | COVID-19 Genomics UK (COG-UK) Consortium | Tanya Golubchik et al |
| EPI_ISL_595766 | Scotland/QEUH-9F9DCE/2020 | 2020-09-25 | Europe/United Kingdom/Scotland/ | Oxford Viromics, NDM, University of Oxford; Oxford University Hospitals; Basingstoke and North Hampshire Hospital | COVID-19 Genomics UK (COG-UK) Consortium | Tanya Golubchik et al |
| EPI_ISL_595817 | England/QEUH-A016F4/2020 | 2020-09-30 | Europe/United Kingdom/England/ | Virology Department, Sheffield Teaching Hospitals NHS Foundation Trust/Department of Infection, Immunity and Cardiovascular Disease, The Medical School, University of Sheffield | COVID-19 Genomics UK (COG-UK) Consortium | Thushan de Silva et al |
| EPI_ISL_595827 | England/QEUH-A01BC8/2020 | 2020-10-01 | Europe/United Kingdom/England/ | Virology Department, Sheffield Teaching Hospitals NHS Foundation Trust/Department of Infection, Immunity and Cardiovascular Disease, The Medical School, University of Sheffield | COVID-19 Genomics UK (COG-UK) Consortium | Thushan de Silva et al |
| EPI_ISL_595874 | England/QEUH-A31260/2020 | 2020-10-05 | Europe/United Kingdom/England/ | Quadram Institute Bioscience | COVID-19 Genomics UK (COG-UK) Consortium | Dave J. Baker et al |
| EPI_ISL_595878 | England/QEUH-A31312/2020 | 2020-10-04 | Europe/United Kingdom/England/ | Quadram Institute Bioscience | COVID-19 Genomics UK (COG-UK) Consortium | Dave J. Baker et al |
| EPI_ISL_595889 | England/QEUH-A3145B/2020 | 2020-10-05 | Europe/United Kingdom/England/ | Quadram Institute Bioscience | COVID-19 Genomics UK (COG-UK) Consortium | Dave J. Baker et al |
| EPI_ISL_595906 | England/QEUH-A31752/2020 | 2020-10-05 | Europe/United Kingdom/England/ | Quadram Institute Bioscience | COVID-19 Genomics UK (COG-UK) Consortium | Dave J. Baker et al |
| EPI_ISL_596004 | Scotland/QEUH-A4C824/2020 | 2020-10-08 | Europe/United Kingdom/Scotland/ | Quadram Institute Bioscience | COVID-19 Genomics UK (COG-UK) Consortium | Dave J. Baker et al |
| EPI_ISL_596010 | Scotland/QEUH-A4CA3D/2020 | 2020-10-07 | Europe/United Kingdom/Scotland/ | Quadram Institute Bioscience | COVID-19 Genomics UK (COG-UK) Consortium | Dave J. Baker et al |
| EPI_ISL_596124 | England/QEUH-A6361E/2020 | 2020-10-12 | Europe/United Kingdom/England/ | Quadram Institute Bioscience | COVID-19 Genomics UK (COG-UK) Consortium | Dave J. Baker et al |
| EPI_ISL_596136 | Scotland/QEUH-A6370C/2020 | 2020-10-12 | Europe/United Kingdom/Scotland/ | Quadram Institute Bioscience | COVID-19 Genomics UK (COG-UK) Consortium | Dave J. Baker et al |

| | | | | | | |
|---|---|---|---|---|---|---|
| EPI_ISL_596204 | England/SHEF-C8031/2020 | 2020-10-05 | Europe/United Kingdom/England/ | Virology Department, Sheffield Teaching Hospitals NHS Foundation Trust/Department of Infection, Immunity and Cardiovascular Disease, The Medical School, University of Sheffield | COVID-19 Genomics UK (COG-UK) Consortium | Thushan de Silva et al |
| EPI_ISL_596212 | England/SHEF-CD11B/2020 | 2020-10-06 | Europe/United Kingdom/England/ | Virology Department, Sheffield Teaching Hospitals NHS Foundation Trust/Department of Infection, Immunity and Cardiovascular Disease, The Medical School, University of Sheffield | COVID-19 Genomics UK (COG-UK) Consortium | Thushan de Silva et al |
| EPI_ISL_596228 | Russia/SPE-RII-30769S/2020 | 2020-08-20 | Europe/Russia/Saint-Petersburg/ | WHO National Influenza Centre Russian Federation | WHO National Influenza Centre Russian Federation | Andrey Komissarov et al |
| EPI_ISL_596230 | Russia/SPE-RII-32307S/2020 | 2020-08-31 | Europe/Russia/Saint-Petersburg/ | WHO National Influenza Centre Russian Federation | WHO National Influenza Centre Russian Federation | Andrey Komissarov et al |
| EPI_ISL_596231 | Russia/SPE-RII-32995S/2020 | 2020-09-04 | Europe/Russia/Saint-Petersburg/ | WHO National Influenza Centre Russian Federation | WHO National Influenza Centre Russian Federation | Andrey Komissarov et al |
| EPI_ISL_596235 | Russia/SPE-RII-34611S/2020 | 2020-09-17 | Europe/Russia/Saint-Petersburg/ | WHO National Influenza Centre Russian Federation | WHO National Influenza Centre Russian Federation | Andrey Komissarov et al |
| EPI_ISL_596236 | Russia/SPE-RII-34613S/2020 | 2020-09-17 | Europe/Russia/Saint-Petersburg/ | WHO National Influenza Centre Russian Federation | WHO National Influenza Centre Russian Federation | Andrey Komissarov et al |
| EPI_ISL_596239 | Russia/SPE-RII-34755S/2020 | 2020-09-18 | Europe/Russia/Saint-Petersburg/ | WHO National Influenza Centre Russian Federation | WHO National Influenza Centre Russian Federation | Andrey Komissarov et al |
| EPI_ISL_596240 | Russia/SPE-RII-34948S/2020 | 2020-09-22 | Europe/Russia/Saint-Petersburg/ | WHO National Influenza Centre Russian Federation | WHO National Influenza Centre Russian Federation | Andrey Komissarov et al |

| | | | | | | |
|---|---|---|---|---|---|---|
| EPI_ISL_596242 | Russia/SPE-RII-35144S/2020 | 2020-09-24 | Europe/Russia/Saint-Petersburg/ | WHO National Influenza Centre Russian Federation | WHO National Influenza Centre Russian Federation | Andrey Komissarov et al |
| EPI_ISL_596243 | Russia/SPE-RII-35177S/2020 | 2020-09-24 | Europe/Russia/Saint-Petersburg/ | WHO National Influenza Centre Russian Federation | WHO National Influenza Centre Russian Federation | Andrey Komissarov et al |
| EPI_ISL_596244 | Russia/SPE-RII-35178S/2020 | 2020-09-24 | Europe/Russia/Saint-Petersburg/ | WHO National Influenza Centre Russian Federation | WHO National Influenza Centre Russian Federation | Andrey Komissarov et al |
| EPI_ISL_596246 | Russia/SPE-RII-35217S/2020 | 2020-09-25 | Europe/Russia/Saint-Petersburg/ | WHO National Influenza Centre Russian Federation | WHO National Influenza Centre Russian Federation | Andrey Komissarov et al |
| EPI_ISL_596250 | Russia/SPE-RII-MH1007S/2020 | 2020-09-04 | Europe/Russia/Saint-Petersburg/ | HELIX LCC | WHO National Influenza Centre Russian Federation | Andrey Komissarov et al |
| EPI_ISL_596251 | Russia/SPE-RII-MH1008S/2020 | 2020-09-04 | Europe/Russia/Saint-Petersburg/ | HELIX LCC | WHO National Influenza Centre Russian Federation | Andrey Komissarov et al |
| EPI_ISL_596252 | Russia/SPE-RII-MH1012S/2020 | 2020-09-05 | Europe/Russia/Saint-Petersburg/ | HELIX LCC | WHO National Influenza Centre Russian Federation | Andrey Komissarov et al |
| EPI_ISL_596253 | Russia/SVE-RII-MH1028S/2020 | 2020-09-05 | Europe/Russia/Sverdlovsk/Revda | HELIX LCC | WHO National Influenza Centre Russian Federation | Andrey Komissarov et al |
| EPI_ISL_596255 | Russia/SPE-RII-MH1043S/2020 | 2020-09-06 | Europe/Russia/Saint-Petersburg/ | HELIX LCC | WHO National Influenza Centre Russian Federation | Andrey Komissarov et al |
| EPI_ISL_596256 | Russia/SPE-RII-MH1049S/2020 | 2020-09-06 | Europe/Russia/Saint-Petersburg/ | HELIX LCC | WHO National Influenza Centre Russian Federation | Andrey Komissarov et al |

| | | | | | | |
|---|---|---|---|---|---|---|
| EPI_ISL_596259 | Russia/SPE-RII-MH1058S/2020 | 2020-09-06 | Europe/Russia/Saint-Petersburg/ | HELIX LCC | WHO National Influenza Centre Russian Federation | Andrey Komissarov et al |
| EPI_ISL_596260 | Russia/SPE-RII-MH1059S/2020 | 2020-09-06 | Europe/Russia/Saint-Petersburg/ | HELIX LCC | WHO National Influenza Centre Russian Federation | Andrey Komissarov et al |
| EPI_ISL_596261 | Russia/SPE-RII-MH1061S/2020 | 2020-09-06 | Europe/Russia/Saint-Petersburg/ | HELIX LCC | WHO National Influenza Centre Russian Federation | Andrey Komissarov et al |
| EPI_ISL_596262 | Russia/SPE-RII-MH1062S/2020 | 2020-09-06 | Europe/Russia/Saint-Petersburg/ | HELIX LCC | WHO National Influenza Centre Russian Federation | Andrey Komissarov et al |
| EPI_ISL_596263 | Russia/SPE-RII-MH1064S/2020 | 2020-09-06 | Europe/Russia/Saint-Petersburg/ | HELIX LCC | WHO National Influenza Centre Russian Federation | Andrey Komissarov et al |
| EPI_ISL_596265 | Russia/NVS-RII-MH1097V/2020 | 2020-07-29 | Europe/Russia/Novosibirsk/Novosibirsk | WHO National Influenza Centre Russian Federation | WHO National Influenza Centre Russian Federation | Andrey Komissarov et al |
| EPI_ISL_596266 | Russia/NVS-RII-MH1137S/2020 | 2020-07-30 | Europe/Russia/Novosibirsk/Novosibirsk | WHO National Influenza Centre Russian Federation | WHO National Influenza Centre Russian Federation | Andrey Komissarov et al |
| EPI_ISL_596267 | Russia/NVS-RII-MH1142S/2020 | 2020-08-09 | Europe/Russia/Novosibirsk/Novosibirsk | WHO National Influenza Centre Russian Federation | WHO National Influenza Centre Russian Federation | Andrey Komissarov et al |
| EPI_ISL_596268 | Russia/SPE-RII-MH1400S/2020 | 2020-09-12 | Europe/Russia/Saint-Petersburg/ | HELIX LCC | WHO National Influenza Centre Russian Federation | Andrey Komissarov et al |
| EPI_ISL_596270 | Russia/SVE-RII-MH1411S/2020 | 2020-09-03 | Europe/Russia/Sverdlovsk/Yekaterinbur | HELIX LCC | WHO National Influenza Centre Russian Federation | Andrey Komissarov et al |

| | | | | | | |
|---|---|---|---|---|---|---|
| EPI_ISL_596271 | Russia/SVE-RII-MH1412S/2020 | 2020-09-07 | Europe/Russia/Sverdlovsk/Yekaterinbur | HELIX LCC | WHO National Influenza Centre Russian Federation | Andrey Komissarov et al |
| EPI_ISL_596272 | Russia/SVE-RII-MH1430S/2020 | 2020-09-11 | Europe/Russia/Sverdlovsk/Yekaterinbur | HELIX LCC | WHO National Influenza Centre Russian Federation | Andrey Komissarov et al |
| EPI_ISL_596273 | Russia/SVE-RII-MH1433S/2020 | 2020-09-10 | Europe/Russia/Sverdlovsk/Yekaterinbur | HELIX LCC | WHO National Influenza Centre Russian Federation | Andrey Komissarov et al |
| EPI_ISL_596274 | Russia/SVE-RII-MH1436S/2020 | 2020-09-12 | Europe/Russia/Sverdlovsk/Yekaterinbur | HELIX LCC | WHO National Influenza Centre Russian Federation | Andrey Komissarov et al |
| EPI_ISL_596275 | Russia/SVE-RII-MH1438S/2020 | 2020-09-06 | Europe/Russia/Sverdlovsk/Yekaterinbur | HELIX LCC | WHO National Influenza Centre Russian Federation | Andrey Komissarov et al |
| EPI_ISL_596281 | Russia/SPE-RII-MH1489S/2020 | 2020-09-11 | Europe/Russia/Saint-Petersburg/ | HELIX LCC | WHO National Influenza Centre Russian Federation | Andrey Komissarov et al |
| EPI_ISL_596282 | Russia/SPE-RII-MH1500S/2020 | 2020-09-11 | Europe/Russia/Saint-Petersburg/ | HELIX LCC | WHO National Influenza Centre Russian Federation | Andrey Komissarov et al |
| EPI_ISL_596284 | Russia/SPE-RII-MH1509S/2020 | 2020-09-10 | Europe/Russia/Saint-Petersburg/ | HELIX LCC | WHO National Influenza Centre Russian Federation | Andrey Komissarov et al |
| EPI_ISL_596286 | Russia/SPE-RII-MH1515S/2020 | 2020-09-11 | Europe/Russia/Saint-Petersburg/ | HELIX LCC | WHO National Influenza Centre Russian Federation | Andrey Komissarov et al |
| EPI_ISL_596289 | Russia/SPE-RII-MH1520S/2020 | 2020-09-12 | Europe/Russia/Saint-Petersburg/ | HELIX LCC | WHO National Influenza Centre Russian Federation | Andrey Komissarov et al |

| | | | | | | |
|---|---|---|---|---|---|---|
| EPI_ISL_596290 | Russia/SPE-RII-MH1521S/2020 | 2020-09-12 | Europe/Russia/Saint-Petersburg/ | HELIX LCC | WHO National Influenza Centre Russian Federation | Andrey Komissarov et al |
| EPI_ISL_596293 | Russia/SPE-RII-MH1568S/2020 | 2020-09-10 | Europe/Russia/Saint-Petersburg/ | HELIX LCC | WHO National Influenza Centre Russian Federation | Andrey Komissarov et al |
| EPI_ISL_596294 | Russia/SPE-RII-MH1609S/2020 | 2020-09-13 | Europe/Russia/Saint-Petersburg/ | HELIX LCC | WHO National Influenza Centre Russian Federation | Andrey Komissarov et al |
| EPI_ISL_596295 | Russia/SVE-RII-MH1612S/2020 | 2020-09-08 | Europe/Russia/Sverdlovsk/Yekaterinbur | HELIX LCC | WHO National Influenza Centre Russian Federation | Andrey Komissarov et al |
| EPI_ISL_596296 | Russia/SVE-RII-MH1613S/2020 | 2020-09-07 | Europe/Russia/Sverdlovsk/Yekaterinbur | HELIX LCC | WHO National Influenza Centre Russian Federation | Andrey Komissarov et al |
| EPI_ISL_596297 | Russia/SPE-RII-MH1616S/2020 | 2020-09-07 | Europe/Russia/Saint-Petersburg/ | HELIX LCC | WHO National Influenza Centre Russian Federation | Andrey Komissarov et al |
| EPI_ISL_596301 | Russia/SPE-RII-MH1628S/2020 | 2020-09-07 | Europe/Russia/Saint-Petersburg/ | HELIX LCC | WHO National Influenza Centre Russian Federation | Andrey Komissarov et al |
| EPI_ISL_596302 | Russia/SVE-RII-MH1632S/2020 | 2020-09-08 | Europe/Russia/Sverdlovsk/Revda | HELIX LCC | WHO National Influenza Centre Russian Federation | Andrey Komissarov et al |
| EPI_ISL_596303 | Russia/SVE-RII-MH1633S/2020 | 2020-09-12 | Europe/Russia/Sverdlovsk/Revda | HELIX LCC | WHO National Influenza Centre Russian Federation | Andrey Komissarov et al |
| EPI_ISL_596304 | Russia/SVE-RII-MH1634S/2020 | 2020-09-11 | Europe/Russia/Sverdlovsk/Kirovgrad | HELIX LCC | WHO National Influenza Centre Russian Federation | Andrey Komissarov et al |

| | | | | | | |
|---|---|---|---|---|---|---|
| EPI_ISL_596306 | Russia/SVE-RII-MH1637S/2020 | 2020-09-03 | Europe/Russia/Sverdlovsk/Yekaterinbur | HELIX LCC | WHO National Influenza Centre Russian Federation | Andrey Komissarov et al |
| EPI_ISL_596307 | Russia/SVE-RII-MH1638S/2020 | 2020-09-08 | Europe/Russia/Sverdlovsk/Shalya | HELIX LCC | WHO National Influenza Centre Russian Federation | Andrey Komissarov et al |
| EPI_ISL_596310 | Russia/PER-RII-MH1646S/2020 | 2020-09-06 | Europe/Russia/Perm Krai/Perm | HELIX LCC | WHO National Influenza Centre Russian Federation | Andrey Komissarov et al |
| EPI_ISL_596311 | Russia/KK-RII-MH1680S/2020 | 2020-09-03 | Europe/Russia/Khakassia/Abakan | HELIX LCC | WHO National Influenza Centre Russian Federation | Andrey Komissarov et al |
| EPI_ISL_596312 | Russia/KK-RII-MH1681S/2020 | 2020-09-03 | Europe/Russia/Khakassia/Abakan | HELIX LCC | WHO National Influenza Centre Russian Federation | Andrey Komissarov et al |
| EPI_ISL_596314 | Russia/NIZ-RII-MH1683S/2020 | 2020-09-04 | Europe/Russia/Nizhny Novgorod/Nizhny | HELIX LCC | WHO National Influenza Centre Russian Federation | Andrey Komissarov et al |
| EPI_ISL_596315 | Russia/AD-RII-MH1684S/2020 | 2020-09-09 | Europe/Russia/Adygea/Maykop | HELIX LCC | WHO National Influenza Centre Russian Federation | Andrey Komissarov et al |
| EPI_ISL_596316 | Russia/KDA-RII-MH1685S/2020 | 2020-09-04 | Europe/Russia/Krasnodar/Sochi | HELIX LCC | WHO National Influenza Centre Russian Federation | Andrey Komissarov et al |
| EPI_ISL_596317 | Russia/KL-RII-MH1686S/2020 | 2020-09-03 | Europe/Russia/Kalmykia/Elista | HELIX LCC | WHO National Influenza Centre Russian Federation | Andrey Komissarov et al |
| EPI_ISL_596318 | Russia/MOW-RII-MH1970S/2020 | 2020-05-15 | Europe/Russia/Moscow Oblast/ | Pathogenic Microorganisms Variability Laboratory | WHO National Influenza Centre Russian Federation | Andrey Komissarov et al |

| | | | | | | |
|---|---|---|---|---|---|---|
| EPI_ISL_596319 | Russia/MOW-RII-MH1971S/2020 | 2020-05-18 | Europe/Russia/Moscow Oblast/ | Pathogenic Microorganisms Variability Laboratory | WHO National Influenza Centre Russian Federation | Andrey Komissarov et al |
| EPI_ISL_596321 | Russia/MOW-RII-MH1973S/2020 | 2020-05-20 | Europe/Russia/Moscow Oblast/ | Pathogenic Microorganisms Variability Laboratory | WHO National Influenza Centre Russian Federation | Andrey Komissarov et al |
| EPI_ISL_596322 | Russia/MOW-RII-MH1974S/2020 | 2020-05-21 | Europe/Russia/Moscow Oblast/ | Pathogenic Microorganisms Variability Laboratory | WHO National Influenza Centre Russian Federation | Andrey Komissarov et al |
| EPI_ISL_596323 | Russia/MOW-RII-MH1975S/2020 | 2020-05-28 | Europe/Russia/Moscow Oblast/ | Pathogenic Microorganisms Variability Laboratory | WHO National Influenza Centre Russian Federation | Andrey Komissarov et al |
| EPI_ISL_596324 | Russia/MOW-RII-MH1976S/2020 | 2020-05-28 | Europe/Russia/Moscow Oblast/ | Pathogenic Microorganisms Variability Laboratory | WHO National Influenza Centre Russian Federation | Andrey Komissarov et al |
| EPI_ISL_596325 | Russia/MOW-RII-MH1979S/2020 | 2020-05-28 | Europe/Russia/Moscow Oblast/ | Pathogenic Microorganisms Variability Laboratory | WHO National Influenza Centre Russian Federation | Andrey Komissarov et al |
| EPI_ISL_596326 | Russia/MOW-RII-MH1980S/2020 | 2020-05-29 | Europe/Russia/Moscow Oblast/ | Pathogenic Microorganisms Variability Laboratory | WHO National Influenza Centre Russian Federation | Andrey Komissarov et al |
| EPI_ISL_596327 | Russia/MOW-RII-MH1982S/2020 | 2020-05-30 | Europe/Russia/Moscow Oblast/ | Pathogenic Microorganisms Variability Laboratory | WHO National Influenza Centre Russian Federation | Andrey Komissarov et al |
| EPI_ISL_596328 | Russia/MOW-RII-MH1983S/2020 | 2020-06-01 | Europe/Russia/Moscow Oblast/ | Pathogenic Microorganisms Variability Laboratory | WHO National Influenza Centre Russian Federation | Andrey Komissarov et al |
| EPI_ISL_596329 | Russia/MOW-RII-MH1984S/2020 | 2020-06-01 | Europe/Russia/Moscow Oblast/ | Pathogenic Microorganisms Variability Laboratory | WHO National Influenza Centre Russian Federation | Andrey Komissarov et al |

| | | | | | | |
|---|---|---|---|---|---|---|
| EPI_ISL_596330 | Russia/MOW-RII-MH1985S/2020 | 2020-06-01 | Europe/Russia/Moscow Oblast/ | Pathogenic Microorganisms Variability Laboratory | WHO National Influenza Centre Russian Federation | Andrey Komissarov et al |
| EPI_ISL_596331 | Russia/MOW-RII-MH1986S/2020 | 2020-06-01 | Europe/Russia/Moscow Oblast/ | Pathogenic Microorganisms Variability Laboratory | WHO National Influenza Centre Russian Federation | Andrey Komissarov et al |
| EPI_ISL_596332 | Russia/MOW-RII-MH1988S/2020 | 2020-05-25 | Europe/Russia/Moscow Oblast/ | Pathogenic Microorganisms Variability Laboratory | WHO National Influenza Centre Russian Federation | Andrey Komissarov et al |
| EPI_ISL_596333 | Russia/MOW-RII-MH1989S/2020 | 2020-05-25 | Europe/Russia/Moscow Oblast/ | Pathogenic Microorganisms Variability Laboratory | WHO National Influenza Centre Russian Federation | Andrey Komissarov et al |
| EPI_ISL_596334 | Russia/MOW-RII-MH1990S/2020 | 2020-05-25 | Europe/Russia/Moscow Oblast/ | Pathogenic Microorganisms Variability Laboratory | WHO National Influenza Centre Russian Federation | Andrey Komissarov et al |
| EPI_ISL_596335 | Russia/MOW-RII-MH1991S/2020 | 2020-05-25 | Europe/Russia/Moscow Oblast/ | Pathogenic Microorganisms Variability Laboratory | WHO National Influenza Centre Russian Federation | Andrey Komissarov et al |
| EPI_ISL_596336 | Russia/MOW-RII-MH1992S/2020 | 2020-06-05 | Europe/Russia/Moscow Oblast/ | Pathogenic Microorganisms Variability Laboratory | WHO National Influenza Centre Russian Federation | Andrey Komissarov et al |
| EPI_ISL_596337 | Russia/MOW-RII-MH1993S/2020 | 2020-06-05 | Europe/Russia/Moscow Oblast/ | Pathogenic Microorganisms Variability Laboratory | WHO National Influenza Centre Russian Federation | Andrey Komissarov et al |
| EPI_ISL_596338 | Russia/MOW-RII-MH1994S/2020 | 2020-06-08 | Europe/Russia/Moscow Oblast/ | Pathogenic Microorganisms Variability Laboratory | WHO National Influenza Centre Russian Federation | Andrey Komissarov et al |
| EPI_ISL_596339 | Russia/MOW-RII-MH1995S/2020 | 2020-06-06 | Europe/Russia/Moscow Oblast/ | Pathogenic Microorganisms Variability Laboratory | WHO National Influenza Centre Russian Federation | Andrey Komissarov et al |

| | | | | | | |
|---|---|---|---|---|---|---|
| EPI_ISL_596340 | Russia/MOW-RII-MH1996S/2020 | 2020-06-09 | Europe/Russia/Moscow Oblast/ | Pathogenic Microorganisms Variability Laboratory | WHO National Influenza Centre Russian Federation | Andrey Komissarov et al |
| EPI_ISL_596341 | Russia/MOW-RII-MH1997S/2020 | 2020-06-10 | Europe/Russia/Moscow Oblast/ | Pathogenic Microorganisms Variability Laboratory | WHO National Influenza Centre Russian Federation | Andrey Komissarov et al |
| EPI_ISL_596342 | Russia/MOW-RII-MH1998S/2020 | 2020-06-10 | Europe/Russia/Moscow Oblast/ | Pathogenic Microorganisms Variability Laboratory | WHO National Influenza Centre Russian Federation | Andrey Komissarov et al |
| EPI_ISL_596343 | Russia/MOW-RII-MH1999S/2020 | 2020-06-11 | Europe/Russia/Moscow Oblast/ | Pathogenic Microorganisms Variability Laboratory | WHO National Influenza Centre Russian Federation | Andrey Komissarov et al |
| EPI_ISL_596344 | Russia/MOW-RII-MH2001S/2020 | 2020-06-15 | Europe/Russia/Moscow Oblast/ | Pathogenic Microorganisms Variability Laboratory | WHO National Influenza Centre Russian Federation | Andrey Komissarov et al |
| EPI_ISL_596345 | Russia/MOW-RII-MH2003S/2020 | 2020-06-19 | Europe/Russia/Moscow Oblast/ | Pathogenic Microorganisms Variability Laboratory | WHO National Influenza Centre Russian Federation | Andrey Komissarov et al |
| EPI_ISL_596346 | Russia/MOW-RII-MH2004S/2020 | 2020-05-27 | Europe/Russia/Moscow Oblast/ | Pathogenic Microorganisms Variability Laboratory | WHO National Influenza Centre Russian Federation | Andrey Komissarov et al |
| EPI_ISL_596347 | Russia/MOW-RII-MH2005S/2020 | 2020-05-25 | Europe/Russia/Moscow Oblast/ | Pathogenic Microorganisms Variability Laboratory | WHO National Influenza Centre Russian Federation | Andrey Komissarov et al |
| EPI_ISL_596349 | Russia/MOW-RII-MH2007S/2020 | 2020-05-15 | Europe/Russia/Moscow Oblast/ | Pathogenic Microorganisms Variability Laboratory | WHO National Influenza Centre Russian Federation | Andrey Komissarov et al |
| EPI_ISL_596350 | Russia/MOW-RII-MH2008S/2020 | 2020-05-18 | Europe/Russia/Moscow Oblast/ | Pathogenic Microorganisms Variability Laboratory | WHO National Influenza Centre Russian Federation | Andrey Komissarov et al |

| | | | | | | |
|---|---|---|---|---|---|---|
| EPI_ISL_596351 | Russia/MOW-RII-MH2010S/2020 | 2020-06-05 | Europe/Russia/Moscow Oblast/ | Pathogenic Microorganisms Variability Laboratory | WHO National Influenza Centre Russian Federation | Andrey Komissarov et al |
| EPI_ISL_596352 | Russia/MOW-RII-MH2014S/2020 | 2020-05-15 | Europe/Russia/Moscow Oblast/ | Pathogenic Microorganisms Variability Laboratory | WHO National Influenza Centre Russian Federation | Andrey Komissarov et al |
| EPI_ISL_596354 | Russia/PRI-RII-MH744S/2020 | 2020-08-07 | Europe/Russia/Primorsky/Vladivostok | WHO National Influenza Centre Russian Federation | WHO National Influenza Centre Russian Federation | Andrey Komissarov et al |
| EPI_ISL_596355 | Russia/PRI-RII-MH747S/2020 | 2020-08-07 | Europe/Russia/Primorsky/Vladivostok | WHO National Influenza Centre Russian Federation | WHO National Influenza Centre Russian Federation | Andrey Komissarov et al |
| EPI_ISL_596356 | Russia/PRI-RII-MH748S/2020 | 2020-08-08 | Europe/Russia/Primorsky/Vladivostok | WHO National Influenza Centre Russian Federation | WHO National Influenza Centre Russian Federation | Andrey Komissarov et al |
| EPI_ISL_596358 | Russia/SPE-RII-MH943S/2020 | 2020-09-06 | Europe/Russia/Saint-Petersburg/ | HELIX LCC | WHO National Influenza Centre Russian Federation | Andrey Komissarov et al |
| EPI_ISL_596362 | Russia/SPE-RII-MH952S/2020 | 2020-09-03 | Europe/Russia/Saint-Petersburg/ | HELIX LCC | WHO National Influenza Centre Russian Federation | Andrey Komissarov et al |
| EPI_ISL_596363 | Russia/SPE-RII-MH953S/2020 | 2020-09-03 | Europe/Russia/Saint-Petersburg/ | HELIX LCC | WHO National Influenza Centre Russian Federation | Andrey Komissarov et al |
| EPI_ISL_596365 | Russia/SPE-RII-MH958S/2020 | 2020-09-03 | Europe/Russia/Saint-Petersburg/ | HELIX LCC | WHO National Influenza Centre Russian Federation | Andrey Komissarov et al |
| EPI_ISL_596369 | Russia/SPE-RII-MH977S/2020 | 2020-09-04 | Europe/Russia/Saint-Petersburg/ | HELIX LCC | WHO National Influenza Centre Russian Federation | Andrey Komissarov et al |

| | | | | | | |
|---|---|---|---|---|---|---|
| EPI_ISL_596370 | Russia/SPE-RII-MH981S/2020 | 2020-09-04 | Europe/Russia/Saint-Petersburg/ | HELIX LCC | WHO National Influenza Centre Russian Federation | Andrey Komissarov et al |
| EPI_ISL_596373 | Russia/SPE-RII-MH984S/2020 | 2020-09-04 | Europe/Russia/Saint-Petersburg/ | HELIX LCC | WHO National Influenza Centre Russian Federation | Andrey Komissarov et al |
| EPI_ISL_596376 | Russia/SPE-RII-MH996S/2020 | 2020-09-05 | Europe/Russia/Saint-Petersburg/ | HELIX LCC | WHO National Influenza Centre Russian Federation | Andrey Komissarov et al |
| EPI_ISL_596377 | Russia/SPE-RII-MH998S/2020 | 2020-09-05 | Europe/Russia/Saint-Petersburg/ | HELIX LCC | WHO National Influenza Centre Russian Federation | Andrey Komissarov et al |
| EPI_ISL_596387 | USA/WA-S2856/2020 | 2020-09-11 | North America/USA/Washington/ | Seattle Flu Study | Seattle Flu Study | Deborah A. Nickerson et al |
| EPI_ISL_596389 | USA/WA-S2858/2020 | 2020-09-16 | North America/USA/Washington/King C | Seattle Flu Study | Seattle Flu Study | Deborah A. Nickerson et al |
| EPI_ISL_596393 | USA/WA-S2862/2020 | 2020-09-18 | North America/USA/Washington/ | Seattle Flu Study | Seattle Flu Study | Deborah A. Nickerson et al |
| EPI_ISL_596395 | USA/WA-S2864/2020 | 2020-09-19 | North America/USA/Washington/King C | Seattle Flu Study | Seattle Flu Study | Deborah A. Nickerson et al |
| EPI_ISL_596396 | USA/WA-S2865/2020 | 2020-09-19 | North America/USA/Washington/King C | Seattle Flu Study | Seattle Flu Study | Deborah A. Nickerson et al |
| EPI_ISL_596402 | USA/WA-S2871/2020 | 2020-09-22 | North America/USA/Washington/King C | Seattle Flu Study | Seattle Flu Study | Deborah A. Nickerson et al |
| EPI_ISL_596405 | USA/WA-S2874/2020 | 2020-09-22 | North America/USA/Washington/King C | Seattle Flu Study | Seattle Flu Study | Deborah A. Nickerson et al |
| EPI_ISL_596406 | USA/WA-S2875/2020 | 2020-09-22 | North America/USA/Washington/King C | Seattle Flu Study | Seattle Flu Study | Deborah A. Nickerson et al |
| EPI_ISL_596407 | USA/WA-S2876/2020 | 2020-09-24 | North America/USA/Washington/King C | Seattle Flu Study | Seattle Flu Study | Deborah A. Nickerson et al |
| EPI_ISL_596412 | USA/WA-S2881/2020 | 2020-09-28 | North America/USA/Washington/King C | Seattle Flu Study | Seattle Flu Study | Deborah A. Nickerson et al |
| EPI_ISL_596413 | USA/WA-S2882/2020 | 2020-09-28 | North America/USA/Washington/King C | Seattle Flu Study | Seattle Flu Study | Deborah A. Nickerson et al |
| EPI_ISL_596420 | USA/WA-S2889/2020 | 2020-10-01 | North America/USA/Washington/King C | Seattle Flu Study | Seattle Flu Study | Deborah A. Nickerson et al |
| EPI_ISL_596422 | USA/WA-S2891/2020 | 2020-10-01 | North America/USA/Washington/King C | Seattle Flu Study | Seattle Flu Study | Deborah A. Nickerson et al |
| EPI_ISL_596423 | USA/WA-S2892/2020 | 2020-10-01 | North America/USA/Washington/King C | Seattle Flu Study | Seattle Flu Study | Deborah A. Nickerson et al |
| EPI_ISL_596425 | USA/WA-S2894/2020 | 2020-10-01 | North America/USA/Washington/King C | Seattle Flu Study | Seattle Flu Study | Deborah A. Nickerson et al |
| EPI_ISL_596433 | USA/WA-S2902/2020 | 2020-10-03 | North America/USA/Washington/King C | Seattle Flu Study | Seattle Flu Study | Deborah A. Nickerson et al |
| EPI_ISL_596440 | USA/WA-S2909/2020 | 2020-10-05 | North America/USA/Washington/King C | Seattle Flu Study | Seattle Flu Study | Deborah A. Nickerson et al |

| | | | | | | |
|---|---|---|---|---|---|---|
| EPI_ISL_596451 | Iran/GRC-10582/2020 | 2020-06-16 | Asia/Iran/Tehran/ | Department of Pathology, School of Medicine, Imam Khomeini Hospital, Tehran University of Medical Sciences | Genetics Research Center, University of Social Welfare and Rehabilitation Sciences | Zohreh Fattahi et al |
| EPI_ISL_596452 | Iran/GRC-9673/2020 | 2020-05-18 | Asia/Iran/Tehran/ | Department of Pathology, School of Medicine, Imam Khomeini Hospital, Tehran University of Medical Sciences | Genetics Research Center, University of Social Welfare and Rehabilitation Sciences | Zohreh Fattahi et al |
| EPI_ISL_596453 | Iran/GRC-43/2020 | 2020-03-02 | Asia/Iran/Qom/ | Booali laboratory, Qom, Iran. Department of Virology, School of Public Health, Tehran University of Medical Sciences, Tehran, Iran. | Genetics Research Center, University of Social Welfare and Rehabilitation Sciences | Zohreh Fattahi et al |
| EPI_ISL_596454 | Iran/GRC-70/2020 | 2020-03-08 | Asia/Iran/Sistan and Baluchestan/Zahed | Infectious Disease and Tropical Medicine Research Center, Resistant Tuberculosis Institute, Zahedan University of Medical Sciences, Zahedan, Iran. | Genetics Research Center, University of Social Welfare and Rehabilitation Sciences | Zohreh Fattahi et al |
| EPI_ISL_596455 | Iran/GRC-9695/2020 | 2020-06-11 | Asia/Iran/Tehran/ | Department of Pathology, School of Medicine, Imam Khomeini Hospital, Tehran University of Medical Sciences | Genetics Research Center, University of Social Welfare and Rehabilitation Sciences | Zohreh Fattahi et al |
| EPI_ISL_596472 | Singapore/1082/2020 | 2020-04-09 | Asia/Singapore/Singapore/ | National Public Health Laboratory, National Centre for Infectious Diseases | National Public Health Laboratory, National Centre for Infectious Diseases | Tze Minn Mak et al |
| EPI_ISL_596577 | USA/MI-UM-10036242970/2020 | 2020-10-02 | North America/USA/Michigan/ | University of Michigan Clinical Microbiology Laboratory | Lauring Lab, University of Michigan, Department of Microbiology and Immunology | Valesano et al |

| | | | | | | |
|---|---|---|---|---|---|---|
| EPI_ISL_596585 | USA/MI-UM-10036297467/2020 | 2020-10-06 | North America/USA/Michigan/ | University of Michigan Clinical Microbiology Laboratory | Lauring Lab, University of Michigan, Department of Microbiology and Immunology | Valesano et al |
| EPI_ISL_596587 | USA/MI-UM-10036310581/2020 | 2020-10-06 | North America/USA/Michigan/ | University of Michigan Clinical Microbiology Laboratory | Lauring Lab, University of Michigan, Department of Microbiology and Immunology | Valesano et al |
| EPI_ISL_596591 | USA/MI-UM-10036277659/2020 | 2020-10-05 | North America/USA/Michigan/ | University of Michigan Clinical Microbiology Laboratory | Lauring Lab, University of Michigan, Department of Microbiology and Immunology | Valesano et al |
| EPI_ISL_596597 | USA/MI-UM-10036284170/2020 | 2020-10-05 | North America/USA/Michigan/ | University of Michigan Clinical Microbiology Laboratory | Lauring Lab, University of Michigan, Department of Microbiology and Immunology | Valesano et al |
| EPI_ISL_596598 | USA/MI-UM-10036306472/2020 | 2020-10-06 | North America/USA/Michigan/ | University of Michigan Clinical Microbiology Laboratory | Lauring Lab, University of Michigan, Department of Microbiology and Immunology | Valesano et al |
| EPI_ISL_596605 | USA/MI-UM-10036309295/2020 | 2020-10-06 | North America/USA/Michigan/ | University of Michigan Clinical Microbiology Laboratory | Lauring Lab, University of Michigan, Department of Microbiology and Immunology | Valesano et al |

| | | | | | | |
|---|---|---|---|---|---|---|
| EPI_ISL_596612 | USA/MI-UM-10036367756/2020 | 2020-10-09 | North America/USA/Michigan/ | University of Michigan Clinical Microbiology Laboratory | Lauring Lab, University of Michigan, Department of Microbiology and Immunology | Valesano et al |
| EPI_ISL_596613 | USA/MI-UM-10036371442/2020 | 2020-10-09 | North America/USA/Michigan/ | University of Michigan Clinical Microbiology Laboratory | Lauring Lab, University of Michigan, Department of Microbiology and Immunology | Valesano et al |
| EPI_ISL_596614 | USA/MI-UM-10036324614/2020 | 2020-10-07 | North America/USA/Michigan/ | University of Michigan Clinical Microbiology Laboratory | Lauring Lab, University of Michigan, Department of Microbiology and Immunology | Valesano et al |
| EPI_ISL_596617 | USA/MI-UM-10036339633/2020 | 2020-10-08 | North America/USA/Michigan/ | University of Michigan Clinical Microbiology Laboratory | Lauring Lab, University of Michigan, Department of Microbiology and Immunology | Valesano et al |
| EPI_ISL_596618 | USA/MI-UM-10036370482/2020 | 2020-10-09 | North America/USA/Michigan/ | University of Michigan Clinical Microbiology Laboratory | Lauring Lab, University of Michigan, Department of Microbiology and Immunology | Valesano et al |
| EPI_ISL_596619 | USA/MI-UM-10036361683/2020 | 2020-10-09 | North America/USA/Michigan/ | University of Michigan Clinical Microbiology Laboratory | Lauring Lab, University of Michigan, Department of Microbiology and Immunology | Valesano et al |

| | | | | | | |
|---|---|---|---|---|---|---|
| EPI_ISL_596622 | USA/MI-UM-10036322844/2020 | 2020-10-07 | North America/USA/Michigan/ | University of Michigan Clinical Microbiology Laboratory | Lauring Lab, University of Michigan, Department of Microbiology and Immunology | Valesano et al |
| EPI_ISL_596624 | USA/MI-UM-10036440713/2020 | 2020-10-14 | North America/USA/Michigan/ | University of Michigan Clinical Microbiology Laboratory | Lauring Lab, University of Michigan, Department of Microbiology and Immunology | Valesano et al |
| EPI_ISL_596650 | Ireland/D-SVUH-R6519/2020 | 2020-04-04 | Europe/Ireland/Dublin/ | St.Vincent's University Hospital | St.Vincent's University Hospital | Mary Lucey et al |
| EPI_ISL_596654 | Ireland/D-SVUH-R6370/2020 | 2020-04-03 | Europe/Ireland/Dublin/ | St.Vincent's University Hospital | St.Vincent's University Hospital | Mary Lucey et al |
| EPI_ISL_596663 | Ireland/D-SVUH-R7377/2020 | 2020-04-11 | Europe/Ireland/Dublin/ | St.Vincent's University Hospital | St.Vincent's University Hospital | Mary Lucey et al |
| EPI_ISL_596894 | Ireland/D-NVRL-73IRL38401/2020 | 2020-09-08 | Europe/Ireland/Dublin/ | National Virus Reference Laboratory | National Virus Reference Laboratory | Michael Carr et al |
| EPI_ISL_596901 | Ireland/D-NVRL-73IRL76166/2020 | 2020-09-15 | Europe/Ireland/Dublin/ | National Virus Reference Laboratory | National Virus Reference Laboratory | Michael Carr et al |
| EPI_ISL_596915 | Ireland/D-NVRL-73IRL86995/2020 | 2020-09-16 | Europe/Ireland/Dublin/ | National Virus Reference Laboratory | National Virus Reference Laboratory | Michael Carr et al |
| EPI_ISL_597081 | England/CAMC-A57D18/2020 | 2020-10-09 | Europe/United Kingdom/England/ | Lighthouse Lab in Cambridge | Wellcome Sanger Institute for the COVID-19 Genomics UK (COG-UK) consortium | Rob Howes et al |
| EPI_ISL_597159 | England/CAMC-A584E5/2020 | 2020-10-07 | Europe/United Kingdom/England/ | Lighthouse Lab in Cambridge | Wellcome Sanger Institute for the COVID-19 Genomics UK (COG-UK) consortium | Rob Howes et al |

| | | | | | | |
|---|---|---|---|---|---|---|
| EPI_ISL_597163 | England/CAMC-A58AE3/2020 | 2020-10-09 | Europe/United Kingdom/England/ | Lighthouse Lab in Cambridge | Wellcome Sanger Institute for the COVID-19 Genomics UK (COG-UK) consortium | Rob Howes et al |
| EPI_ISL_597226 | England/CAMC-A592CC/2020 | 2020-10-07 | Europe/United Kingdom/England/ | Lighthouse Lab in Cambridge | Wellcome Sanger Institute for the COVID-19 Genomics UK (COG-UK) consortium | Rob Howes et al |
| EPI_ISL_597240 | England/QEUH-A5E124/2020 | 2020-10-12 | Europe/United Kingdom/England/ | Lighthouse Lab in Glasgow | Wellcome Sanger Institute for the COVID-19 Genomics UK (COG-UK) consortium | Harper VanSteenhouse et al |
| EPI_ISL_597322 | England/QEUH-A5E3E2/2020 | 2020-10-12 | Europe/United Kingdom/England/ | Lighthouse Lab in Glasgow | Wellcome Sanger Institute for the COVID-19 Genomics UK (COG-UK) consortium | Harper VanSteenhouse et al |
| EPI_ISL_597637 | England/ALDP-A4FB19/2020 | 2020-10-07 | Europe/United Kingdom/England/ | Lighthouse Lab in Alderley Park | Wellcome Sanger Institute for the COVID-19 Genomics UK (COG-UK) consortium | Jacquelyn Wynn et al |
| EPI_ISL_597885 | England/ALDP-A521F3/2020 | 2020-10-07 | Europe/United Kingdom/England/ | Lighthouse Lab in Alderley Park | Wellcome Sanger Institute for the COVID-19 Genomics UK (COG-UK) consortium | Jacquelyn Wynn et al |
| EPI_ISL_597938 | England/ALDP-A5274C/2020 | 2020-10-07 | Europe/United Kingdom/England/ | Lighthouse Lab in Alderley Park | Wellcome Sanger Institute for the COVID-19 Genomics UK (COG-UK) consortium | Jacquelyn Wynn et al |
| EPI_ISL_597969 | England/ALDP-A52199/2020 | 2020-10-07 | Europe/United Kingdom/England/ | Lighthouse Lab in Alderley Park | Wellcome Sanger Institute for the COVID-19 Genomics UK (COG-UK) consortium | Jacquelyn Wynn et al |

| | | | | | | |
|---|---|---|---|---|---|---|
| EPI_ISL_597979 | England/ALDP-A517E3/2020 | 2020-10-07 | Europe/United Kingdom/England/ | Lighthouse Lab in Alderley Park | Wellcome Sanger Institute for the COVID-19 Genomics UK (COG-UK) consortium | Jacquelyn Wynn et al |
| EPI_ISL_597985 | England/ALDP-A52542/2020 | 2020-10-07 | Europe/United Kingdom/England/ | Lighthouse Lab in Alderley Park | Wellcome Sanger Institute for the COVID-19 Genomics UK (COG-UK) consortium | Jacquelyn Wynn et al |
| EPI_ISL_598013 | England/ALDP-A5259D/2020 | 2020-10-07 | Europe/United Kingdom/England/ | Lighthouse Lab in Alderley Park | Wellcome Sanger Institute for the COVID-19 Genomics UK (COG-UK) consortium | Jacquelyn Wynn et al |
| EPI_ISL_598034 | England/ALDP-A5290A/2020 | 2020-10-07 | Europe/United Kingdom/England/ | Lighthouse Lab in Alderley Park | Wellcome Sanger Institute for the COVID-19 Genomics UK (COG-UK) consortium | Jacquelyn Wynn et al |
| EPI_ISL_598121 | England/ALDP-A516AA/2020 | 2020-10-07 | Europe/United Kingdom/England/ | Lighthouse Lab in Alderley Park | Wellcome Sanger Institute for the COVID-19 Genomics UK (COG-UK) consortium | Jacquelyn Wynn et al |
| EPI_ISL_598151 | England/ALDP-A52393/2020 | 2020-10-07 | Europe/United Kingdom/England/ | Lighthouse Lab in Alderley Park | Wellcome Sanger Institute for the COVID-19 Genomics UK (COG-UK) consortium | Jacquelyn Wynn et al |
| EPI_ISL_598287 | England/MILK-A47C78/2020 | 2020-10-06 | Europe/United Kingdom/England/ | Lighthouse Lab in Milton Keynes | Wellcome Sanger Institute for the COVID-19 Genomics UK (COG-UK) consortium | The Lighthouse Lab in Milton Keynes et al |
| EPI_ISL_598407 | England/MILK-A47786/2020 | 2020-10-05 | Europe/United Kingdom/England/ | Lighthouse Lab in Milton Keynes | Wellcome Sanger Institute for the COVID-19 Genomics UK (COG-UK) consortium | The Lighthouse Lab in Milton Keynes et al |

| | | | | | | |
|---|---|---|---|---|---|---|
| EPI_ISL_598443 | England/MILK-A4706D/2020 | 2020-10-06 | Europe/United Kingdom/England/ | Lighthouse Lab in Milton Keynes | Wellcome Sanger Institute for the COVID-19 Genomics UK (COG-UK) consortium | The Lighthouse Lab in Milton Keynes et al |
| EPI_ISL_598471 | England/MILK-A47425/2020 | 2020-10-06 | Europe/United Kingdom/England/ | Lighthouse Lab in Milton Keynes | Wellcome Sanger Institute for the COVID-19 Genomics UK (COG-UK) consortium | The Lighthouse Lab in Milton Keynes et al |
| EPI_ISL_598577 | England/MILK-A4613E/2020 | 2020-10-06 | Europe/United Kingdom/England/ | Lighthouse Lab in Milton Keynes | Wellcome Sanger Institute for the COVID-19 Genomics UK (COG-UK) consortium | The Lighthouse Lab in Milton Keynes et al |
| EPI_ISL_598728 | England/MILK-A44F63/2020 | 2020-10-06 | Europe/United Kingdom/England/ | Lighthouse Lab in Milton Keynes | Wellcome Sanger Institute for the COVID-19 Genomics UK (COG-UK) consortium | The Lighthouse Lab in Milton Keynes et al |
| EPI_ISL_599011 | Scotland/QEUH-A22569/2020 | 2020-10-01 | Europe/United Kingdom/Scotland/ | Lighthouse Lab in Glasgow | Wellcome Sanger Institute for the COVID-19 Genomics UK (COG-UK) consortium | Harper VanSteenhouse et al |
| EPI_ISL_599060 | Scotland/QEUH-A20E51/2020 | 2020-10-01 | Europe/United Kingdom/Scotland/ | Lighthouse Lab in Glasgow | Wellcome Sanger Institute for the COVID-19 Genomics UK (COG-UK) consortium | Harper VanSteenhouse et al |
| EPI_ISL_599105 | Scotland/QEUH-A1FBF4/2020 | 2020-10-01 | Europe/United Kingdom/Scotland/ | Lighthouse Lab in Glasgow | Wellcome Sanger Institute for the COVID-19 Genomics UK (COG-UK) consortium | Harper VanSteenhouse et al |
| EPI_ISL_599229 | England/QEUH-A0B618/2020 | 2020-09-30 | Europe/United Kingdom/England/ | Lighthouse Lab in Glasgow | Wellcome Sanger Institute for the COVID-19 Genomics UK (COG-UK) consortium | Harper VanSteenhouse et al |

| | | | | | | |
|---|---|---|---|---|---|---|
| EPI_ISL_599330 | England/QEUH-A0BF68/2020 | 2020-10-03 | Europe/United Kingdom/England/ | Lighthouse Lab in Glasgow | Wellcome Sanger Institute for the COVID-19 Genomics UK (COG-UK) consortium | Harper VanSteenhouse et al |
| EPI_ISL_599489 | Scotland/QEUH-9DD9F9/2020 | 2020-09-21 | Europe/United Kingdom/Scotland/ | Lighthouse Lab in Glasgow | Wellcome Sanger Institute for the COVID-19 Genomics UK (COG-UK) consortium | Harper VanSteenhouse et al |
| EPI_ISL_599540 | Scotland/QEUH-9DD698/2020 | 2020-09-21 | Europe/United Kingdom/Scotland/ | Lighthouse Lab in Glasgow | Wellcome Sanger Institute for the COVID-19 Genomics UK (COG-UK) consortium | Harper VanSteenhouse et al |
| EPI_ISL_599592 | Wales/QEUH-9DD0A9/2020 | 2020-09-21 | Europe/United Kingdom/Wales/ | Lighthouse Lab in Glasgow | Wellcome Sanger Institute for the COVID-19 Genomics UK (COG-UK) consortium | Harper VanSteenhouse et al |
| EPI_ISL_599693 | England/QEUH-9DDA41/2020 | 2020-09-21 | Europe/United Kingdom/England/ | Lighthouse Lab in Glasgow | Wellcome Sanger Institute for the COVID-19 Genomics UK (COG-UK) consortium | Harper VanSteenhouse et al |
| EPI_ISL_599823 | England/MILK-A05CFE/2020 | 2020-09-30 | Europe/United Kingdom/England/ | Lighthouse Lab in Milton Keynes | Wellcome Sanger Institute for the COVID-19 Genomics UK (COG-UK) consortium | The Lighthouse Lab in Milton Keynes et al |
| EPI_ISL_599940 | England/MILK-A06264/2020 | 2020-10-01 | Europe/United Kingdom/England/ | Lighthouse Lab in Milton Keynes | Wellcome Sanger Institute for the COVID-19 Genomics UK (COG-UK) consortium | The Lighthouse Lab in Milton Keynes et al |
| EPI_ISL_599954 | England/MILK-A05A8B/2020 | 2020-09-29 | Europe/United Kingdom/England/ | Lighthouse Lab in Milton Keynes | Wellcome Sanger Institute for the COVID-19 Genomics UK (COG-UK) consortium | The Lighthouse Lab in Milton Keynes et al |

| | | | | | | |
|---|---|---|---|---|---|---|
| EPI_ISL_600110 | England/MILK-A05BB5/2020 | 2020-09-30 | Europe/United Kingdom/England/ | Lighthouse Lab in Milton Keynes | Wellcome Sanger Institute for the COVID-19 Genomics UK (COG-UK) consortium | The Lighthouse Lab in Milton Keynes et al |
| EPI_ISL_600277 | England/QEUH-A002C4/2020 | 2020-10-02 | Europe/United Kingdom/England/ | Lighthouse Lab in Glasgow | Wellcome Sanger Institute for the COVID-19 Genomics UK (COG-UK) consortium | Harper VanSteenhouse et al |
| EPI_ISL_600381 | England/QEUH-9FF9B5/2020 | 2020-09-30 | Europe/United Kingdom/England/ | Lighthouse Lab in Glasgow | Wellcome Sanger Institute for the COVID-19 Genomics UK (COG-UK) consortium | Harper VanSteenhouse et al |
| EPI_ISL_600471 | Bangladesh/G-127/2020 | 2020-07-01 | Asia/Bangladesh/Dhaka/ | Institute of Epidemiology Disease Control And Research | Institute for Developing Science and Health Initiatives | Lauren Cowley et al |
| EPI_ISL_600480 | Bangladesh/G-134/2020 | 2020-06-19 | Asia/Bangladesh/Dhaka/ | Institute of Epidemiology Disease Control And Research | Institute for Developing Science and Health Initiatives | Lauren Cowley et al |
| EPI_ISL_600498 | Bangladesh/G-156/2020 | 2020-06-18 | Asia/Bangladesh/Chattogram/ | Institute of Epidemiology Disease Control And Research | Institute for Developing Science and Health Initiatives | Lauren Cowley et al |
| EPI_ISL_600741 | Scotland/QEUH-9F4507/2020 | 2020-09-27 | Europe/United Kingdom/Scotland/ | Lighthouse Lab in Glasgow | Wellcome Sanger Institute for the COVID-19 Genomics UK (COG-UK) consortium | Harper VanSteenhouse et al |
| EPI_ISL_600875 | Scotland/QEUH-9F61C5/2020 | 2020-09-27 | Europe/United Kingdom/Scotland/ | Lighthouse Lab in Glasgow | Wellcome Sanger Institute for the COVID-19 Genomics UK (COG-UK) consortium | Harper VanSteenhouse et al |

| | | | | | | |
|---|---|---|---|---|---|---|
| EPI_ISL_600933 | Scotland/QEUH-9E55BE/2020 | 2020-09-24 | Europe/United Kingdom/Scotland/ | Lighthouse Lab in Glasgow | Wellcome Sanger Institute for the COVID-19 Genomics UK (COG-UK) consortium | Harper VanSteenhouse et al |
| EPI_ISL_600996 | Scotland/QEUH-9E4A74/2020 | 2020-09-23 | Europe/United Kingdom/Scotland/ | Lighthouse Lab in Glasgow | Wellcome Sanger Institute for the COVID-19 Genomics UK (COG-UK) consortium | Harper VanSteenhouse et al |
| EPI_ISL_601215 | Scotland/QEUH-9DEAD7/2020 | 2020-09-21 | Europe/United Kingdom/Scotland/ | Lighthouse Lab in Glasgow | Wellcome Sanger Institute for the COVID-19 Genomics UK (COG-UK) consortium | Harper VanSteenhouse et al |
| EPI_ISL_601258 | England/MILK-9E0586/2020 | 2020-09-21 | Europe/United Kingdom/England/ | Lighthouse Lab in Milton Keynes | Wellcome Sanger Institute for the COVID-19 Genomics UK (COG-UK) consortium | The Lighthouse Lab in Milton Keynes et al |
| EPI_ISL_601264 | Wales/MILK-9E04F2/2020 | 2020-09-20 | Europe/United Kingdom/Wales/ | Lighthouse Lab in Milton Keynes | Wellcome Sanger Institute for the COVID-19 Genomics UK (COG-UK) consortium | The Lighthouse Lab in Milton Keynes et al |
| EPI_ISL_601308 | England/QEUH-9DA8D1/2020 | 2020-09-21 | Europe/United Kingdom/England/ | Lighthouse Lab in Glasgow | Wellcome Sanger Institute for the COVID-19 Genomics UK (COG-UK) consortium | Harper VanSteenhouse et al |
| EPI_ISL_601768 | England/CAMC-9D1F7B/2020 | 2020-09-18 | Europe/United Kingdom/England/ | Lighthouse Lab in Cambridge | Wellcome Sanger Institute for the COVID-19 Genomics UK (COG-UK) consortium | Rob Howes et al |
| EPI_ISL_602165 | USA/NM-DOH-2020307857/2020 | 2020-08-24 | North America/USA/New Mexico/ | New Mexico Department of Health Scientific Laboratory | New Mexico Department of Health Scientific Laboratory | Ellie Johnson et al |

| | | | | | | |
|---|---|---|---|---|---|---|
| EPI_ISL_602166 | USA/NM-DOH-2020307874/2020 | 2020-08-24 | North America/USA/New Mexico/ | New Mexico Department of Health Scientific Laboratory | New Mexico Department of Health Scientific Laboratory | Ellie Johnson et al |
| EPI_ISL_602183 | USA/NM-DOH-2020308662/2020 | 2020-08-24 | North America/USA/New Mexico/ | New Mexico Department of Health Scientific Laboratory | New Mexico Department of Health Scientific Laboratory | Ellie Johnson et al |
| EPI_ISL_602185 | USA/NM-DOH-2020308703/2020 | 2020-08-24 | North America/USA/New Mexico/ | New Mexico Department of Health Scientific Laboratory | New Mexico Department of Health Scientific Laboratory | Ellie Johnson et al |
| EPI_ISL_602186 | USA/NM-DOH-2020308708/2020 | 2020-08-24 | North America/USA/New Mexico/ | New Mexico Department of Health Scientific Laboratory | New Mexico Department of Health Scientific Laboratory | Ellie Johnson et al |
| EPI_ISL_602196 | USA/NM-DOH-2020309719/2020 | 2020-08-24 | North America/USA/New Mexico/ | New Mexico Department of Health Scientific Laboratory | New Mexico Department of Health Scientific Laboratory | Ellie Johnson et al |
| EPI_ISL_602198 | USA/NM-DOH-2020309736/2020 | 2020-08-24 | North America/USA/New Mexico/ | New Mexico Department of Health Scientific Laboratory | New Mexico Department of Health Scientific Laboratory | Ellie Johnson et al |
| EPI_ISL_602199 | USA/NM-DOH-2020309744/2020 | 2020-08-24 | North America/USA/New Mexico/ | New Mexico Department of Health Scientific Laboratory | New Mexico Department of Health Scientific Laboratory | Ellie Johnson et al |
| EPI_ISL_602210 | USA/TX-DSHS-0912/2020 | 2020-05-20 | North America/USA/Texas/El Paso Coun | Texas Department of State Health Services | Texas Department of State Health Services | Rashmi Tuladhar et al |
| EPI_ISL_602211 | USA/TX-DSHS-0913/2020 | 2020-05-20 | North America/USA/Texas/El Paso Coun | Texas Department of State Health Services | Texas Department of State Health Services | Rashmi Tuladhar et al |
| EPI_ISL_602223 | USA/TX-DSHS-0925/2020 | 2020-06-15 | North America/USA/Texas/Harris Count | Texas Department of State Health Services | Texas Department of State Health Services | Rashmi Tuladhar et al |
| EPI_ISL_602238 | USA/TX-DSHS-0940/2020 | 2020-06-20 | North America/USA/Texas/Travis Count | Texas Department of State Health Services | Texas Department of State Health Services | Rashmi Tuladhar et al |

| | | | | | | |
|---|---|---|---|---|---|---|
| EPI_ISL_602244 | USA/TX-DSHS-0946/2020 | 2020-07-06 | North America/USA/Texas/Travis Count | Texas Department of State Health Services | Texas Department of State Health Services | Rashmi Tuladhar et al |
| EPI_ISL_602249 | USA/TX-DSHS-0952/2020 | 2020-07-16 | North America/USA/Texas/Dallas Count | Texas Department of State Health Services | Texas Department of State Health Services | Rashmi Tuladhar et al |
| EPI_ISL_602260 | USA/MD-MDH-0237/2020 | 2020-09-29 | North America/USA/Maryland/ | MD PHL | MD PHL | Maryland Department of Health Laboratories Administration et al |
| EPI_ISL_602262 | USA/MD-MDH-0239/2020 | 2020-09-29 | North America/USA/Maryland/ | MD PHL | MD PHL | Maryland Department of Health Laboratories Administration et al |
| EPI_ISL_602267 | USA/MD-MDH-0244/2020 | 2020-10-02 | North America/USA/Maryland/ | MD PHL | MD PHL | Maryland Department of Health Laboratories Administration et al |
| EPI_ISL_602270 | USA/MD-MDH-0247/2020 | 2020-10-02 | North America/USA/Maryland/ | MD PHL | MD PHL | Maryland Department of Health Laboratories Administration et al |
| EPI_ISL_602271 | USA/MD-MDH-0248/2020 | 2020-10-03 | North America/USA/Maryland/ | MD PHL | MD PHL | Maryland Department of Health Laboratories Administration et al |
| EPI_ISL_602274 | USA/MD-MDH-0251/2020 | 2020-10-05 | North America/USA/Maryland/ | MD PHL | MD PHL | Maryland Department of Health Laboratories Administration et al |
| EPI_ISL_602276 | USA/MD-MDH-0253/2020 | 2020-10-05 | North America/USA/Maryland/ | MD PHL | MD PHL | Maryland Department of Health Laboratories Administration et al |
| EPI_ISL_602284 | Germany/NW-UBI-3/2020 | 2020-03-26 | Europe/Germany/North Rhine Westphal | Evangelisches Klinikum Bethel, Institut f√ºr Laboratoriumsmedizin, Mikrobiologie und Hygiene | Bielefeld University | David Brandt et al |
| EPI_ISL_602292 | Germany/NW-UBI-11/2020 | 2020-03-27 | Europe/Germany/North Rhine Westphal | Evangelisches Klinikum Bethel, Institut f√ºr Laboratoriumsmedizin, Mikrobiologie und Hygiene | Bielefeld University | David Brandt et al |
| EPI_ISL_602294 | Germany/NW-UBI-13/2020 | 2020-04-14 | Europe/Germany/North Rhine Westphal | Evangelisches Klinikum Bethel, Institut f√ºr Laboratoriumsmedizin, Mikrobiologie und Hygiene | Bielefeld University | David Brandt et al |
| EPI_ISL_602298 | Germany/NW-UBI-17/2020 | 2020-04-17 | Europe/Germany/North Rhine Westphal | Evangelisches Klinikum Bethel, Institut f√ºr Laboratoriumsmedizin, Mikrobiologie und Hygiene | Bielefeld University | David Brandt et al |

| | | | | | | |
|---|---|---|---|---|---|---|
| EPI_ISL_602299 | Germany/NW-UBI-18/2020 | 2020-04-08 | Europe/Germany/North Rhine Westphal | Evangelisches Klinikum Bethel, Institut f√ºr Laboratoriumsmedizin, Mikrobiologie und Hygiene | Bielefeld University | David Brandt et al |
| EPI_ISL_602302 | Germany/NW-UBI-21/2020 | 2020-04-07 | Europe/Germany/North Rhine Westphal | Evangelisches Klinikum Bethel, Institut f√ºr Laboratoriumsmedizin, Mikrobiologie und Hygiene | Bielefeld University | David Brandt et al |
| EPI_ISL_602309 | USA/FL-BA0B/2020 | 2020-08-14 | North America/USA/Florida/Miami-Dad | University of Miami Immunology and Histocompatibility Laboratory | University of Miami Immunology and Histocompatibility Laboratory | Emilio Margolles-Clark et al |
| EPI_ISL_602312 | USA/FL-B2TM/2020 | 2020-08-02 | North America/USA/Florida/Miami-Dad | University of Miami Immunology and Histocompatibility Laboratory | University of Miami Immunology and Histocompatibility Laboratory | Emilio Margolles-Clark et al |
| EPI_ISL_602321 | USA/FL-BD6J/2020 | 2020-08-20 | North America/USA/Florida/Miami-Dad | University of Miami Immunology and Histocompatibility Laboratory | University of Miami Immunology and Histocompatibility Laboratory | Emilio Margolles-Clark et al |
| EPI_ISL_602322 | USA/FL-B4R9/2020 | 2020-08-05 | North America/USA/Florida/Miami-Dad | University of Miami Immunology and Histocompatibility Laboratory | University of Miami Immunology and Histocompatibility Laboratory | Emilio Margolles-Clark et al |
| EPI_ISL_602329 | Russia/SPE-RII-MH1014S/2020 | 2020-09-05 | Europe/Russia/Saint-Petersburg/ | HELIX LLC | WHO National Influenza Centre Russian Federation | Andrey Komissarov et al |
| EPI_ISL_602330 | Russia/SPE-RII-MH1057S/2020 | 2020-09-06 | Europe/Russia/Saint-Petersburg/ | HELIX LLC | WHO National Influenza Centre Russian Federation | Andrey Komissarov et al |
| EPI_ISL_602332 | Russia/SVE-RII-MH1446S/2020 | 2020-09-08 | Europe/Russia/Sverdlovsk/Yekaterinbur | HELIX LLC | WHO National Influenza Centre Russian Federation | Andrey Komissarov et al |

| | | | | | | |
|---|---|---|---|---|---|---|
| EPI_ISL_602334 | Russia/SVE-RII-MH1448S/2020 | 2020-09-08 | Europe/Russia/Sverdlovsk/Yekaterinbur | HELIX LLC | WHO National Influenza Centre Russian Federation | Andrey Komissarov et al |
| EPI_ISL_602335 | Russia/SVE-RII-MH1449S/2020 | 2020-09-08 | Europe/Russia/Sverdlovsk/Yekaterinbur | HELIX LLC | WHO National Influenza Centre Russian Federation | Andrey Komissarov et al |
| EPI_ISL_602339 | Russia/SPE-RII-MH1597S/2020 | 2020-09-12 | Europe/Russia/Saint-Petersburg/ | HELIX LLC | WHO National Influenza Centre Russian Federation | Andrey Komissarov et al |
| EPI_ISL_602341 | Russia/SPE-RII-MH1624S/2020 | 2020-09-09 | Europe/Russia/Saint-Petersburg/ | HELIX LLC | WHO National Influenza Centre Russian Federation | Andrey Komissarov et al |
| EPI_ISL_602343 | Russia/SVE-RII-MH1697S/2020 | 2020-09-20 | Europe/Russia/Sverdlovsk/Yekaterinbur | HELIX LLC | WHO National Influenza Centre Russian Federation | Andrey Komissarov et al |
| EPI_ISL_602344 | Russia/MUR-RII-MH1700S/2020 | 2020-09-17 | Europe/Russia/Murmansk/Murmansk | HELIX LLC | WHO National Influenza Centre Russian Federation | Andrey Komissarov et al |
| EPI_ISL_602345 | Russia/MUR-RII-MH1701S/2020 | 2020-09-17 | Europe/Russia/Murmansk/Murmansk | HELIX LLC | WHO National Influenza Centre Russian Federation | Andrey Komissarov et al |
| EPI_ISL_602346 | Russia/NGR-RII-MH1702S/2020 | 2020-09-18 | Europe/Russia/Novgorod/Veliky Novgor | HELIX LLC | WHO National Influenza Centre Russian Federation | Andrey Komissarov et al |
| EPI_ISL_602347 | Russia/NGR-RII-MH1705S/2020 | 2020-09-19 | Europe/Russia/Novgorod/Veliky Novgor | HELIX LLC | WHO National Influenza Centre Russian Federation | Andrey Komissarov et al |
| EPI_ISL_602350 | Russia/SVE-RII-MH1713S/2020 | 2020-09-17 | Europe/Russia/Sverdlovsk/Yekaterinbur | HELIX LLC | WHO National Influenza Centre Russian Federation | Andrey Komissarov et al |

| | | | | | | |
|---|---|---|---|---|---|---|
| EPI_ISL_602353 | Russia/SVE-RII-MH1716S/2020 | 2020-09-17 | Europe/Russia/Sverdlovsk/Yekaterinbur | HELIX LLC | WHO National Influenza Centre Russian Federation | Andrey Komissarov et al |
| EPI_ISL_602358 | Russia/SVE-RII-MH1724S/2020 | 2020-09-18 | Europe/Russia/Sverdlovsk/Yekaterinbur | HELIX LLC | WHO National Influenza Centre Russian Federation | Andrey Komissarov et al |
| EPI_ISL_602363 | Russia/SVE-RII-MH1910S/2020 | 2020-09-12 | Europe/Russia/Sverdlovsk/Yekaterinbur | HELIX LLC | WHO National Influenza Centre Russian Federation | Andrey Komissarov et al |
| EPI_ISL_602364 | Russia/KHM-RII-MH1914S/2020 | 2020-09-11 | Europe/Russia/Khanty-Mansi/Khanty-Ma | HELIX LLC | WHO National Influenza Centre Russian Federation | Andrey Komissarov et al |
| EPI_ISL_602365 | Russia/PER-RII-MH1915S/2020 | 2020-09-16 | Europe/Russia/Perm Krai/Perm | HELIX LLC | WHO National Influenza Centre Russian Federation | Andrey Komissarov et al |
| EPI_ISL_602366 | Russia/SVE-RII-MH1918S/2020 | 2020-09-17 | Europe/Russia/Sverdlovsk/Pervouralsk | HELIX LLC | WHO National Influenza Centre Russian Federation | Andrey Komissarov et al |
| EPI_ISL_602370 | Russia/SVE-RII-MH1930S/2020 | 2020-09-17 | Europe/Russia/Sverdlovsk/Nizhny Tagil | HELIX LLC | WHO National Influenza Centre Russian Federation | Andrey Komissarov et al |
| EPI_ISL_602371 | Russia/SVE-RII-MH1931S/2020 | 2020-09-17 | Europe/Russia/Sverdlovsk/Krasnouralsk | HELIX LLC | WHO National Influenza Centre Russian Federation | Andrey Komissarov et al |
| EPI_ISL_602372 | Russia/SVE-RII-MH1932S/2020 | 2020-09-16 | Europe/Russia/Sverdlovsk/Nizhny Tagil | HELIX LLC | WHO National Influenza Centre Russian Federation | Andrey Komissarov et al |
| EPI_ISL_602374 | Russia/SVE-RII-MH1941S/2020 | 2020-09-17 | Europe/Russia/Sverdlovsk/Kachkanar | HELIX LLC | WHO National Influenza Centre Russian Federation | Andrey Komissarov et al |

| | | | | | | |
|---|---|---|---|---|---|---|
| EPI_ISL_602376 | Russia/PER-RII-MH1951S/2020 | 2020-09-16 | Europe/Russia/Perm Krai/Perm | HELIX LLC | WHO National Influenza Centre Russian Federation | Andrey Komissarov et al |
| EPI_ISL_602381 | Russia/SVE-RII-MH2022S/2020 | 2020-09-18 | Europe/Russia/Sverdlovsk/Yekaterinbur | HELIX LLC | WHO National Influenza Centre Russian Federation | Andrey Komissarov et al |
| EPI_ISL_602382 | Russia/SVE-RII-MH2024S/2020 | 2020-09-18 | Europe/Russia/Sverdlovsk/Yekaterinbur | HELIX LLC | WHO National Influenza Centre Russian Federation | Andrey Komissarov et al |
| EPI_ISL_602384 | Russia/SVE-RII-MH2033S/2020 | 2020-09-20 | Europe/Russia/Sverdlovsk/Yekaterinbur | HELIX LLC | WHO National Influenza Centre Russian Federation | Andrey Komissarov et al |
| EPI_ISL_602385 | Russia/SVE-RII-MH2034S/2020 | 2020-09-21 | Europe/Russia/Sverdlovsk/Yekaterinbur | HELIX LLC | WHO National Influenza Centre Russian Federation | Andrey Komissarov et al |
| EPI_ISL_602386 | Russia/SVE-RII-MH2038S/2020 | 2020-09-21 | Europe/Russia/Sverdlovsk/Yekaterinbur | HELIX LLC | WHO National Influenza Centre Russian Federation | Andrey Komissarov et al |
| EPI_ISL_602387 | Russia/SVE-RII-MH2039S/2020 | 2020-09-21 | Europe/Russia/Sverdlovsk/Yekaterinbur | HELIX LLC | WHO National Influenza Centre Russian Federation | Andrey Komissarov et al |
| EPI_ISL_602388 | Russia/SVE-RII-MH2045S/2020 | 2020-09-21 | Europe/Russia/Sverdlovsk/Yekaterinbur | HELIX LLC | WHO National Influenza Centre Russian Federation | Andrey Komissarov et al |
| EPI_ISL_602389 | Russia/LEN-RII-MH2050S/2020 | 2020-09-18 | Europe/Russia/Leningrad Oblast/Kirishi | HELIX LLC | WHO National Influenza Centre Russian Federation | Andrey Komissarov et al |
| EPI_ISL_602390 | Russia/LEN-RII-MH2053S/2020 | 2020-09-28 | Europe/Russia/Leningrad Oblast/Kirovsk | HELIX LLC | WHO National Influenza Centre Russian Federation | Andrey Komissarov et al |

| | | | | | | |
|---|---|---|---|---|---|---|
| EPI_ISL_602391 | Russia/MOW-RII-MH2055S/2020 | 2020-09-24 | Europe/Russia/Moscow Oblast/ | HELIX LLC | WHO National Influenza Centre Russian Federation | Andrey Komissarov et al |
| EPI_ISL_602392 | Russia/MOW-RII-MH2056S/2020 | 2020-09-24 | Europe/Russia/Moscow Oblast/ | HELIX LLC | WHO National Influenza Centre Russian Federation | Andrey Komissarov et al |
| EPI_ISL_602393 | Russia/MOW-RII-MH2058S/2020 | 2020-09-24 | Europe/Russia/Moscow Oblast/ | HELIX LLC | WHO National Influenza Centre Russian Federation | Andrey Komissarov et al |
| EPI_ISL_602400 | Russia/MUR-RII-MH2065S/2020 | 2020-09-24 | Europe/Russia/Murmansk/Murmansk | HELIX LLC | WHO National Influenza Centre Russian Federation | Andrey Komissarov et al |
| EPI_ISL_602403 | Russia/SVE-RII-MH2073S/2020 | 2020-09-22 | Europe/Russia/Sverdlovsk/Nizhny Tagil | HELIX LLC | WHO National Influenza Centre Russian Federation | Andrey Komissarov et al |
| EPI_ISL_602405 | Russia/KR-RII-MH2076S/2020 | 2020-09-25 | Europe/Russia/Karelia/Petrozavodsk | HELIX LLC | WHO National Influenza Centre Russian Federation | Andrey Komissarov et al |
| EPI_ISL_602407 | Russia/MUR-RII-MH2078S/2020 | 2020-09-25 | Europe/Russia/Murmansk/Severomorsk | HELIX LLC | WHO National Influenza Centre Russian Federation | Andrey Komissarov et al |
| EPI_ISL_602408 | Russia/MUR-RII-MH2079S/2020 | 2020-09-25 | Europe/Russia/Murmansk/Severomorsk | HELIX LLC | WHO National Influenza Centre Russian Federation | Andrey Komissarov et al |
| EPI_ISL_602409 | Russia/KO-RII-MH2080S/2020 | 2020-09-21 | Europe/Russia/Komi/Ukhta | HELIX LLC | WHO National Influenza Centre Russian Federation | Andrey Komissarov et al |
| EPI_ISL_602410 | Russia/LEN-RII-MH2081S/2020 | 2020-09-20 | Europe/Russia/Leningrad Oblast/Yanino- | HELIX LLC | WHO National Influenza Centre Russian Federation | Andrey Komissarov et al |

| | | | | | | |
|---|---|---|---|---|---|---|
| EPI_ISL_602415 | Russia/CE-RII-MH2709S/2020 | 2020-09-11 | Europe/Russia/Chechen Republic/Grozny | WHO National Influenza Centre Russian Federation | WHO National Influenza Centre Russian Federation | Andrey Komissarov et al |
| EPI_ISL_602416 | Russia/CE-RII-MH2728S/2020 | 2020-09-11 | Europe/Russia/Chechen Republic/Grozny | WHO National Influenza Centre Russian Federation | WHO National Influenza Centre Russian Federation | Andrey Komissarov et al |
| EPI_ISL_602417 | Russia/AD-RII-MH2738S/2020 | 2020-09-17 | Europe/Russia/Adygea/Maykop | HELIX LLC | WHO National Influenza Centre Russian Federation | Andrey Komissarov et al |
| EPI_ISL_602418 | Russia/DA-RII-MH2739S/2020 | 2020-09-17 | Europe/Russia/Dagestan/Makhachkala | HELIX LLC | WHO National Influenza Centre Russian Federation | Andrey Komissarov et al |
| EPI_ISL_602419 | Russia/DA-RII-MH2741S/2020 | 2020-09-26 | Europe/Russia/Dagestan/Derbent | HELIX LLC | WHO National Influenza Centre Russian Federation | Andrey Komissarov et al |
| EPI_ISL_602421 | Russia/DA-RII-MH2743S/2020 | 2020-09-25 | Europe/Russia/Dagestan/Makhachkala | HELIX LLC | WHO National Influenza Centre Russian Federation | Andrey Komissarov et al |
| EPI_ISL_602422 | Russia/DA-RII-MH2744S/2020 | 2020-09-25 | Europe/Russia/Dagestan/Makhachkala | HELIX LLC | WHO National Influenza Centre Russian Federation | Andrey Komissarov et al |
| EPI_ISL_602424 | Russia/KC-RII-MH2746S/2020 | 2020-09-17 | Europe/Russia/Karachay-Cherkess/Cherk | HELIX LLC | WHO National Influenza Centre Russian Federation | Andrey Komissarov et al |
| EPI_ISL_602426 | Russia/KC-RII-MH2751S/2020 | 2020-09-21 | Europe/Russia/Karachay-Cherkess/Zelen | HELIX LLC | WHO National Influenza Centre Russian Federation | Andrey Komissarov et al |
| EPI_ISL_602427 | Russia/KC-RII-MH2752S/2020 | 2020-09-17 | Europe/Russia/Karachay-Cherkess/Cherk | HELIX LLC | WHO National Influenza Centre Russian Federation | Andrey Komissarov et al |

| | | | | | | |
|---|---|---|---|---|---|---|
| EPI_ISL_602429 | Russia/KO-RII-MH2755S/2020 | 2020-09-17 | Europe/Russia/Komi/Sosnogorsk | HELIX LLC | WHO National Influenza Centre Russian Federation | Andrey Komissarov et al |
| EPI_ISL_602430 | Russia/ME-RII-MH2757S/2020 | 2020-09-23 | Europe/Russia/Mari El/Kozmodemyansk | HELIX LLC | WHO National Influenza Centre Russian Federation | Andrey Komissarov et al |
| EPI_ISL_602431 | Russia/KK-RII-MH2758S/2020 | 2020-09-25 | Europe/Russia/Khakassia/Abakan | HELIX LLC | WHO National Influenza Centre Russian Federation | Andrey Komissarov et al |
| EPI_ISL_602432 | Russia/KK-RII-MH2759S/2020 | 2020-09-25 | Europe/Russia/Khakassia/Abakan | HELIX LLC | WHO National Influenza Centre Russian Federation | Andrey Komissarov et al |
| EPI_ISL_602436 | Russia/ALT-RII-MH2764S/2020 | 2020-09-25 | Europe/Russia/Altai Krai/Barnaul | HELIX LLC | WHO National Influenza Centre Russian Federation | Andrey Komissarov et al |
| EPI_ISL_602437 | Russia/ALT-RII-MH2765S/2020 | 2020-09-26 | Europe/Russia/Altai Krai/Barnaul | HELIX LLC | WHO National Influenza Centre Russian Federation | Andrey Komissarov et al |
| EPI_ISL_602438 | Russia/KDA-RII-MH2766S/2020 | 2020-09-22 | Europe/Russia/Krasnodar/Krasnodar | HELIX LLC | WHO National Influenza Centre Russian Federation | Andrey Komissarov et al |
| EPI_ISL_602439 | Russia/KDA-RII-MH2768S/2020 | 2020-09-21 | Europe/Russia/Krasnodar/Anapa | HELIX LLC | WHO National Influenza Centre Russian Federation | Andrey Komissarov et al |
| EPI_ISL_602440 | Russia/KDA-RII-MH2769S/2020 | 2020-09-23 | Europe/Russia/Krasnodar/Krasnodar | HELIX LLC | WHO National Influenza Centre Russian Federation | Andrey Komissarov et al |
| EPI_ISL_602442 | Russia/KDA-RII-MH2772S/2020 | 2020-09-17 | Europe/Russia/Krasnodar/Anapa | HELIX LLC | WHO National Influenza Centre Russian Federation | Andrey Komissarov et al |

| | | | | | | |
|---|---|---|---|---|---|---|
| EPI_ISL_602444 | Russia/KDA-RII-MH2774S/2020 | 2020-09-20 | Europe/Russia/Krasnodar/Krasnodar | HELIX LLC | WHO National Influenza Centre Russian Federation | Andrey Komissarov et al |
| EPI_ISL_602445 | Russia/STA-RII-MH2777S/2020 | 2020-09-21 | Europe/Russia/Stavropol/Stavropol | HELIX LLC | WHO National Influenza Centre Russian Federation | Andrey Komissarov et al |
| EPI_ISL_602447 | Russia/STA-RII-MH2782S/2020 | 2020-09-23 | Europe/Russia/Stavropol/Stavropol | HELIX LLC | WHO National Influenza Centre Russian Federation | Andrey Komissarov et al |
| EPI_ISL_602448 | Russia/STA-RII-MH2783S/2020 | 2020-09-28 | Europe/Russia/Stavropol/Yessentuki | HELIX LLC | WHO National Influenza Centre Russian Federation | Andrey Komissarov et al |
| EPI_ISL_602450 | Russia/STA-RII-MH2785S/2020 | 2020-09-20 | Europe/Russia/Stavropol/Yessentuki | HELIX LLC | WHO National Influenza Centre Russian Federation | Andrey Komissarov et al |
| EPI_ISL_602451 | Russia/STA-RII-MH2786S/2020 | 2020-09-19 | Europe/Russia/Stavropol/Stavropol | HELIX LLC | WHO National Influenza Centre Russian Federation | Andrey Komissarov et al |
| EPI_ISL_602453 | Russia/BRY-RII-MH2790S/2020 | 2020-09-17 | Europe/Russia/Bryansk Oblast/Bryansk | HELIX LLC | WHO National Influenza Centre Russian Federation | Andrey Komissarov et al |
| EPI_ISL_602454 | Russia/VOR-RII-MH2792S/2020 | 2020-09-26 | Europe/Russia/Voronezh/Voronezh | HELIX LLC | WHO National Influenza Centre Russian Federation | Andrey Komissarov et al |
| EPI_ISL_602455 | Russia/VOR-RII-MH2793S/2020 | 2020-09-28 | Europe/Russia/Voronezh/Voronezh | HELIX LLC | WHO National Influenza Centre Russian Federation | Andrey Komissarov et al |
| EPI_ISL_602456 | Russia/IVA-RII-MH2794S/2020 | 2020-09-23 | Europe/Russia/Ivanovo/Ivanovo | HELIX LLC | WHO National Influenza Centre Russian Federation | Andrey Komissarov et al |

| | | | | | | |
|---|---|---|---|---|---|---|
| EPI_ISL_602457 | Russia/KLU-RII-MH2795S/2020 | 2020-09-16 | Europe/Russia/Kaluga/Tovarkovo | HELIX LLC | WHO National Influenza Centre Russian Federation | Andrey Komissarov et al |
| EPI_ISL_602459 | Russia/MOS-RII-MH2797S/2020 | 2020-09-20 | Europe/Russia/Moscow Oblast/Lapino | HELIX LLC | WHO National Influenza Centre Russian Federation | Andrey Komissarov et al |
| EPI_ISL_602461 | Russia/MOS-RII-MH2799S/2020 | 2020-09-23 | Europe/Russia/Moscow Oblast/Bronnits | HELIX LLC | WHO National Influenza Centre Russian Federation | Andrey Komissarov et al |
| EPI_ISL_602463 | Germany/NW-HHU-189/2020 | 2020-03-29 | Europe/Germany/North Rhine Westphal | Institute for Virology, University Hospital Essen | Center of Medical Microbiology, Virology, and Hospital Hygiene, University of Duesseldorf | Olympia E. Anastasiou et al |
| EPI_ISL_602464 | Germany/NW-HHU-190/2020 | 2020-03-29 | Europe/Germany/North Rhine Westphal | Institute for Virology, University Hospital Essen | Center of Medical Microbiology, Virology, and Hospital Hygiene, University of Duesseldorf | Olympia E. Anastasiou et al |
| EPI_ISL_602466 | Germany/NW-HHU-192/2020 | 2020-03-30 | Europe/Germany/North Rhine Westphal | Institute for Virology, University Hospital Essen | Center of Medical Microbiology, Virology, and Hospital Hygiene, University of Duesseldorf | Olympia E. Anastasiou et al |
| EPI_ISL_602467 | Germany/NW-HHU-193/2020 | 2020-03-31 | Europe/Germany/North Rhine Westphal | Institute for Virology, University Hospital Essen | Center of Medical Microbiology, Virology, and Hospital Hygiene, University of Duesseldorf | Olympia E. Anastasiou et al |

| | | | | | | |
|---|---|---|---|---|---|---|
| EPI_ISL_602469 | Germany/NW-HHU-195/2020 | 2020-04-02 | Europe/Germany/North Rhine Westphal | Institute for Virology, University Hospital Essen | Center of Medical Microbiology, Virology, and Hospital Hygiene, University of Duesseldorf | Olympia E. Anastasiou et al |
| EPI_ISL_602480 | Germany/NW-HHU-206/2020 | 2020-04-09 | Europe/Germany/North Rhine Westphal | Institute for Virology, University Hospital Essen | Center of Medical Microbiology, Virology, and Hospital Hygiene, University of Duesseldorf | Olympia E. Anastasiou et al |
| EPI_ISL_602494 | Germany/NW-HHU-172/2020 | 2020-03-25 | Europe/Germany/North Rhine Westphal | Institute for Virology, University Hospital Essen | Center of Medical Microbiology, Virology, and Hospital Hygiene, University of Duesseldorf | Olympia E. Anastasiou et al |
| EPI_ISL_602495 | Germany/NW-HHU-173/2020 | 2020-03-25 | Europe/Germany/North Rhine Westphal | Institute for Virology, University Hospital Essen | Center of Medical Microbiology, Virology, and Hospital Hygiene, University of Duesseldorf | Olympia E. Anastasiou et al |
| EPI_ISL_602499 | Germany/NW-HHU-177/2020 | 2020-03-26 | Europe/Germany/North Rhine Westphal | Institute for Virology, University Hospital Essen | Center of Medical Microbiology, Virology, and Hospital Hygiene, University of Duesseldorf | Olympia E. Anastasiou et al |
| EPI_ISL_602502 | Germany/NW-HHU-180/2020 | 2020-03-26 | Europe/Germany/North Rhine Westphal | Institute for Virology, University Hospital Essen | Center of Medical Microbiology, Virology, and Hospital Hygiene, University of Duesseldorf | Olympia E. Anastasiou et al |

| | | | | | | |
|---|---|---|---|---|---|---|
| EPI_ISL_602509 | Germany/NW-HHU-187/2020 | 2020-03-28 | Europe/Germany/North Rhine Westphal | Institute for Virology, University Hospital Essen | Center of Medical Microbiology, Virology, and Hospital Hygiene, University of Duesseldorf | Olympia E. Anastasiou et al |
| EPI_ISL_602511 | Germany/NW-HHU-141/2020 | 2020-03-14 | Europe/Germany/North Rhine Westphal | Center of Medical Microbiology, Virology, and Hospital Hygiene, University of Duesseldorf | Center of Medical Microbiology, Virology, and Hospital Hygiene, University of Duesseldorf | Maximilian Damagnez et al |
| EPI_ISL_602512 | Germany/NW-HHU-142/2020 | 2020-03-26 | Europe/Germany/North Rhine Westphal | Center of Medical Microbiology, Virology, and Hospital Hygiene, University of Duesseldorf | Center of Medical Microbiology, Virology, and Hospital Hygiene, University of Duesseldorf | Maximilian Damagnez et al |
| EPI_ISL_602513 | Germany/NW-HHU-143/2020 | 2020-03-19 | Europe/Germany/North Rhine Westphal | Center of Medical Microbiology, Virology, and Hospital Hygiene, University of Duesseldorf | Center of Medical Microbiology, Virology, and Hospital Hygiene, University of Duesseldorf | Maximilian Damagnez et al |
| EPI_ISL_602530 | Germany/NW-HHU-161/2020 | 2020-03-14 | Europe/Germany/North Rhine Westphal | Institute for Virology, University Hospital Essen | Center of Medical Microbiology, Virology, and Hospital Hygiene, University of Duesseldorf | Olympia E. Anastasiou et al |
| EPI_ISL_602538 | Germany/NW-HHU-125/2020 | 2020-03-13 | Europe/Germany/North Rhine Westphal | Center of Medical Microbiology, Virology, and Hospital Hygiene, University of Duesseldorf | Center of Medical Microbiology, Virology, and Hospital Hygiene, University of Duesseldorf | Maximilian Damagnez et al |

| | | | | | | |
|---|---|---|---|---|---|---|
| EPI_ISL_602539 | Germany/NW-HHU-126/2020 | 2020-04-07 | Europe/Germany/North Rhine Westphal | Center of Medical Microbiology, Virology, and Hospital Hygiene, University of Duesseldorf | Center of Medical Microbiology, Virology, and Hospital Hygiene, University of Duesseldorf | Maximilian Damagnez et al |
| EPI_ISL_602540 | Germany/NW-HHU-127/2020 | 2020-04-15 | Europe/Germany/North Rhine Westphal | Center of Medical Microbiology, Virology, and Hospital Hygiene, University of Duesseldorf | Center of Medical Microbiology, Virology, and Hospital Hygiene, University of Duesseldorf | Maximilian Damagnez et al |
| EPI_ISL_602543 | Germany/NW-HHU-130/2020 | 2020-03-19 | Europe/Germany/North Rhine Westphal | Center of Medical Microbiology, Virology, and Hospital Hygiene, University of Duesseldorf | Center of Medical Microbiology, Virology, and Hospital Hygiene, University of Duesseldorf | Maximilian Damagnez et al |
| EPI_ISL_602546 | Germany/NW-HHU-133/2020 | 2020-04-16 | Europe/Germany/North Rhine Westphal | Center of Medical Microbiology, Virology, and Hospital Hygiene, University of Duesseldorf | Center of Medical Microbiology, Virology, and Hospital Hygiene, University of Duesseldorf | Maximilian Damagnez et al |
| EPI_ISL_602576 | SriLanka/BIN44/2020 | 2020-10-22 | Asia/Sri Lanka/Gampaha/ | Centre for Dengue Research, Department of Immunology and Molecular Medicine | Centre for Dengue Research, Department of Immunology and Molecular Medicine | Chandima Jeewandara et al |
| EPI_ISL_602626 | SouthAfrica/KRISP-K003670/2020 | 2020-07-01 | Africa/South Africa/KwaZulu-Natal/eThe | AHRI-Sigal | KRISP, KZN Research Innovation and Sequencing Platform | Gazy I et al |
| EPI_ISL_602627 | SouthAfrica/KRISP-K003671/2020 | 2020-06-26 | Africa/South Africa/KwaZulu-Natal/eThe | AHRI-Sigal | KRISP, KZN Research Innovation and Sequencing Platform | Gazy I et al |

| | | | | | | |
|---|---|---|---|---|---|---|
| EPI_ISL_602628 | SouthAfrica/KRISP-K003672/2020 | 2020-06-29 | Africa/South Africa/KwaZulu-Natal/eThe | AHRI-Sigal | KRISP, KZN Research Innovation and Sequencing Platform | Gazy I et al |
| EPI_ISL_602629 | SouthAfrica/KRISP-K003673/2020 | 2020-07-06 | Africa/South Africa/KwaZulu-Natal/eThe | AHRI-Sigal | KRISP, KZN Research Innovation and Sequencing Platform | Gazy I et al |
| EPI_ISL_602630 | SouthAfrica/KRISP-K003674/2020 | 2020-07-27 | Africa/South Africa/KwaZulu-Natal/eThe | AHRI-Sigal | KRISP, KZN Research Innovation and Sequencing Platform | Gazy I et al |
| EPI_ISL_602635 | SouthAfrica/KRISP-K002814/2020 | 2020-08-20 | Africa/South Africa/KwaZulu-Natal/uThu | NHLS-IALCH | KRISP, KZN Research Innovation and Sequencing Platform | Giandhari J et al |
| EPI_ISL_602637 | SouthAfrica/KRISP-K002816/2020 | 2020-08-21 | Africa/South Africa/KwaZulu-Natal/iLem | NHLS-IALCH | KRISP, KZN Research Innovation and Sequencing Platform | Giandhari J et al |
| EPI_ISL_602642 | SouthAfrica/KRISP-K002821/2020 | 2020-09-01 | Africa/South Africa/KwaZulu-Natal/Ugu | NHLS-IALCH | KRISP, KZN Research Innovation and Sequencing Platform | Giandhari J et al |
| EPI_ISL_602645 | SouthAfrica/KRISP-K002825/2020 | 2020-08-31 | Africa/South Africa/KwaZulu-Natal/Ugu | NHLS-IALCH | KRISP, KZN Research Innovation and Sequencing Platform | Giandhari J et al |
| EPI_ISL_602646 | SouthAfrica/KRISP-K002826/2020 | 2020-09-01 | Africa/South Africa/KwaZulu-Natal/uMk | NHLS-IALCH | KRISP, KZN Research Innovation and Sequencing Platform | Giandhari J et al |

| | | | | | | |
|---|---|---|---|---|---|---|
| EPI_ISL_602650 | SouthAfrica/KRISP-K002830/2020 | 2020-08-28 | Africa/South Africa/KwaZulu-Natal/uMz | NHLS-IALCH | KRISP, KZN Research Innovation and Sequencing Platform | Giandhari J et al |
| EPI_ISL_602651 | SouthAfrica/KRISP-K002831/2020 | 2020-09-02 | Africa/South Africa/KwaZulu-Natal/uMz | NHLS-IALCH | KRISP, KZN Research Innovation and Sequencing Platform | Giandhari J et al |
| EPI_ISL_602654 | SouthAfrica/KRISP-K002837/2020 | 2020-08-31 | Africa/South Africa/KwaZulu-Natal/uMk | NHLS-IALCH | KRISP, KZN Research Innovation and Sequencing Platform | Giandhari J et al |
| EPI_ISL_602659 | SouthAfrica/KRISP-K002843/2020 | 2020-08-30 | Africa/South Africa/KwaZulu-Natal/uMk | NHLS-IALCH | KRISP, KZN Research Innovation and Sequencing Platform | Giandhari J et al |
| EPI_ISL_602661 | SouthAfrica/KRISP-K002845/2020 | 2020-08-31 | Africa/South Africa/KwaZulu-Natal/Ugu | NHLS-IALCH | KRISP, KZN Research Innovation and Sequencing Platform | Giandhari J et al |
| EPI_ISL_602667 | SouthAfrica/KRISP-K002851/2020 | 2020-08-31 | Africa/South Africa/KwaZulu-Natal/King | NHLS-IALCH | KRISP, KZN Research Innovation and Sequencing Platform | Giandhari J et al |
| EPI_ISL_602669 | SouthAfrica/KRISP-K002853/2020 | 2020-08-31 | Africa/South Africa/KwaZulu-Natal/Harr | NHLS-IALCH | KRISP, KZN Research Innovation and Sequencing Platform | Giandhari J et al |
| EPI_ISL_602671 | SouthAfrica/KRISP-K002856/2020 | 2020-08-31 | Africa/South Africa/KwaZulu-Natal/King | NHLS-IALCH | KRISP, KZN Research Innovation and Sequencing Platform | Giandhari J et al |

| | | | | | | |
|---|---|---|---|---|---|---|
| EPI_ISL_602674 | SouthAfrica/KRISP-K002859/2020 | 2020-08-31 | Africa/South Africa/KwaZulu-Natal/King | NHLS-IALCH | KRISP, KZN Research Innovation and Sequencing Platform | Giandhari J et al |
| EPI_ISL_602676 | SouthAfrica/KRISP-K002862/2020 | 2020-08-31 | Africa/South Africa/KwaZulu-Natal/Zulu | NHLS-IALCH | KRISP, KZN Research Innovation and Sequencing Platform | Giandhari J et al |
| EPI_ISL_602699 | SouthAfrica/KRISP-K002887/2020 | 2020-09-04 | Africa/South Africa/KwaZulu-Natal/eThe | NHLS-IALCH | KRISP, KZN Research Innovation and Sequencing Platform | Giandhari J et al |
| EPI_ISL_602703 | SouthAfrica/KRISP-K002891/2020 | 2020-09-02 | Africa/South Africa/KwaZulu-Natal/eThe | NHLS-IALCH | KRISP, KZN Research Innovation and Sequencing Platform | Giandhari J et al |
| EPI_ISL_602705 | SouthAfrica/KRISP-K002893/2020 | 2020-09-02 | Africa/South Africa/KwaZulu-Natal/eThe | NHLS-IALCH | KRISP, KZN Research Innovation and Sequencing Platform | Giandhari J et al |
| EPI_ISL_602708 | SouthAfrica/KRISP-K002896/2020 | 2020-09-03 | Africa/South Africa/KwaZulu-Natal/eThe | NHLS-IALCH | KRISP, KZN Research Innovation and Sequencing Platform | Giandhari J et al |
| EPI_ISL_602711 | SouthAfrica/KRISP-K002899/2020 | 2020-09-03 | Africa/South Africa/KwaZulu-Natal/eThe | NHLS-IALCH | KRISP, KZN Research Innovation and Sequencing Platform | Giandhari J et al |
| EPI_ISL_602712 | SouthAfrica/KRISP-K002901/2020 | 2020-09-04 | Africa/South Africa/KwaZulu-Natal/eThe | NHLS-IALCH | KRISP, KZN Research Innovation and Sequencing Platform | Giandhari J et al |

| | | | | | | |
|---|---|---|---|---|---|---|
| EPI_ISL_602717 | SouthAfrica/KRISP-K003324/2020 | 2020-09-07 | Africa/South Africa/KwaZulu-Natal/uMz | NHLS-IALCH | KRISP, KZN Research Innovation and Sequencing Platform | Giandhari J et al |
| EPI_ISL_602725 | SouthAfrica/KRISP-K003332/2020 | 2020-09-07 | Africa/South Africa/KwaZulu-Natal/uMg | NHLS-IALCH | KRISP, KZN Research Innovation and Sequencing Platform | Giandhari J et al |
| EPI_ISL_602726 | SouthAfrica/KRISP-K003334/2020 | 2020-09-05 | Africa/South Africa/KwaZulu-Natal/uThu | NHLS-IALCH | KRISP, KZN Research Innovation and Sequencing Platform | Giandhari J et al |
| EPI_ISL_602735 | SouthAfrica/KRISP-K003343/2020 | 2020-09-05 | Africa/South Africa/KwaZulu-Natal/King | NHLS-IALCH | KRISP, KZN Research Innovation and Sequencing Platform | Giandhari J et al |
| EPI_ISL_602736 | SouthAfrica/KRISP-K003344/2020 | 2020-09-06 | Africa/South Africa/KwaZulu-Natal/uMk | NHLS-IALCH | KRISP, KZN Research Innovation and Sequencing Platform | Giandhari J et al |
| EPI_ISL_602738 | SouthAfrica/KRISP-K003346/2020 | 2020-09-07 | Africa/South Africa/KwaZulu-Natal/Zulu | NHLS-IALCH | KRISP, KZN Research Innovation and Sequencing Platform | Giandhari J et al |
| EPI_ISL_602745 | SouthAfrica/KRISP-K003354/2020 | 2020-09-07 | Africa/South Africa/KwaZulu-Natal/Ugu | NHLS-IALCH | KRISP, KZN Research Innovation and Sequencing Platform | Giandhari J et al |
| EPI_ISL_602751 | SouthAfrica/KRISP-K003360/2020 | 2020-09-04 | Africa/South Africa/KwaZulu-Natal/King | NHLS-IALCH | KRISP, KZN Research Innovation and Sequencing Platform | Giandhari J et al |

| | | | | | | |
|---|---|---|---|---|---|---|
| EPI_ISL_602754 | SouthAfrica/KRISP-K003363/2020 | 2020-09-04 | Africa/South Africa/KwaZulu-Natal/iLem | NHLS-IALCH | KRISP, KZN Research Innovation and Sequencing Platform | Giandhari J et al |
| EPI_ISL_602758 | SouthAfrica/KRISP-K003367/2020 | 2020-09-04 | Africa/South Africa/KwaZulu-Natal/iLem | NHLS-IALCH | KRISP, KZN Research Innovation and Sequencing Platform | Giandhari J et al |
| EPI_ISL_602759 | SouthAfrica/KRISP-K003369/2020 | 2020-09-04 | Africa/South Africa/KwaZulu-Natal/uMk | NHLS-IALCH | KRISP, KZN Research Innovation and Sequencing Platform | Giandhari J et al |
| EPI_ISL_602763 | SouthAfrica/KRISP-K003374/2020 | 2020-09-10 | Africa/South Africa/KwaZulu-Natal/uMg | NHLS-IALCH | KRISP, KZN Research Innovation and Sequencing Platform | Giandhari J et al |
| EPI_ISL_602765 | SouthAfrica/KRISP-K003376/2020 | 2020-09-10 | Africa/South Africa/KwaZulu-Natal/Harr | NHLS-IALCH | KRISP, KZN Research Innovation and Sequencing Platform | Giandhari J et al |
| EPI_ISL_602768 | SouthAfrica/KRISP-K003380/2020 | 2020-09-09 | Africa/South Africa/KwaZulu-Natal/uMk | NHLS-IALCH | KRISP, KZN Research Innovation and Sequencing Platform | Giandhari J et al |
| EPI_ISL_602771 | SouthAfrica/KRISP-K003383/2020 | 2020-09-10 | Africa/South Africa/KwaZulu-Natal/uMk | NHLS-IALCH | KRISP, KZN Research Innovation and Sequencing Platform | Giandhari J et al |
| EPI_ISL_602773 | SouthAfrica/KRISP-K003385/2020 | 2020-09-11 | Africa/South Africa/KwaZulu-Natal/eThe | NHLS-IALCH | KRISP, KZN Research Innovation and Sequencing Platform | Giandhari J et al |

| | | | | | | |
|---|---|---|---|---|---|---|
| EPI_ISL_602779 | SouthAfrica/KRISP-K003391/2020 | 2020-09-09 | Africa/South Africa/KwaZulu-Natal/iLem | NHLS-IALCH | KRISP, KZN Research Innovation and Sequencing Platform | Giandhari J et al |
| EPI_ISL_602784 | SouthAfrica/KRISP-K003396/2020 | 2020-09-11 | Africa/South Africa/KwaZulu-Natal/Harr | NHLS-IALCH | KRISP, KZN Research Innovation and Sequencing Platform | Giandhari J et al |
| EPI_ISL_602787 | SouthAfrica/KRISP-K003399/2020 | 2020-09-11 | Africa/South Africa/KwaZulu-Natal/Harr | NHLS-IALCH | KRISP, KZN Research Innovation and Sequencing Platform | Giandhari J et al |
| EPI_ISL_602797 | SouthAfrica/KRISP-K003463/2020 | 2020-09-13 | Africa/South Africa/KwaZulu-Natal/uMk | NHLS-IALCH | KRISP, KZN Research Innovation and Sequencing Platform | Giandhari J et al |
| EPI_ISL_602800 | SouthAfrica/KRISP-K003466/2020 | 2020-09-13 | Africa/South Africa/KwaZulu-Natal/uMk | NHLS-IALCH | KRISP, KZN Research Innovation and Sequencing Platform | Giandhari J et al |
| EPI_ISL_602802 | SouthAfrica/KRISP-K003469/2020 | 2020-09-11 | Africa/South Africa/KwaZulu-Natal/uMk | NHLS-IALCH | KRISP, KZN Research Innovation and Sequencing Platform | Giandhari J et al |
| EPI_ISL_602803 | SouthAfrica/KRISP-K003470/2020 | 2020-09-10 | Africa/South Africa/KwaZulu-Natal/uMk | NHLS-IALCH | KRISP, KZN Research Innovation and Sequencing Platform | Giandhari J et al |
| EPI_ISL_602806 | SouthAfrica/KRISP-K003473/2020 | 2020-09-11 | Africa/South Africa/KwaZulu-Natal/uMk | NHLS-IALCH | KRISP, KZN Research Innovation and Sequencing Platform | Giandhari J et al |

| | | | | | | |
|---|---|---|---|---|---|---|
| EPI_ISL_602807 | SouthAfrica/KRISP-K003474/2020 | 2020-09-14 | Africa/South Africa/KwaZulu-Natal/uMg | NHLS-IALCH | KRISP, KZN Research Innovation and Sequencing Platform | Giandhari J et al |
| EPI_ISL_602812 | SouthAfrica/KRISP-K003480/2020 | 2020-09-14 | Africa/South Africa/KwaZulu-Natal/eThe | NHLS-IALCH | KRISP, KZN Research Innovation and Sequencing Platform | Giandhari J et al |
| EPI_ISL_602813 | SouthAfrica/KRISP-K003481/2020 | 2020-09-13 | Africa/South Africa/KwaZulu-Natal/Ugu | NHLS-IALCH | KRISP, KZN Research Innovation and Sequencing Platform | Giandhari J et al |
| EPI_ISL_602814 | SouthAfrica/KRISP-K003482/2020 | 2020-09-13 | Africa/South Africa/KwaZulu-Natal/Ugu | NHLS-IALCH | KRISP, KZN Research Innovation and Sequencing Platform | Giandhari J et al |
| EPI_ISL_602815 | SouthAfrica/KRISP-K003483/2020 | 2020-09-13 | Africa/South Africa/KwaZulu-Natal/Ugu | NHLS-IALCH | KRISP, KZN Research Innovation and Sequencing Platform | Giandhari J et al |
| EPI_ISL_602816 | SouthAfrica/KRISP-K003484/2020 | 2020-09-14 | Africa/South Africa/KwaZulu-Natal/uMk | NHLS-IALCH | KRISP, KZN Research Innovation and Sequencing Platform | Giandhari J et al |
| EPI_ISL_602821 | SouthAfrica/KRISP-K003491/2020 | 2020-09-14 | Africa/South Africa/KwaZulu-Natal/Zulu | NHLS-IALCH | KRISP, KZN Research Innovation and Sequencing Platform | Giandhari J et al |
| EPI_ISL_602827 | SouthAfrica/KRISP-K003503/2020 | 2020-09-16 | Africa/South Africa/KwaZulu-Natal/uMk | NHLS-IALCH | KRISP, KZN Research Innovation and Sequencing Platform | Giandhari J et al |

| | | | | | | |
|---|---|---|---|---|---|---|
| EPI_ISL_602835 | SouthAfrica/KRISP-K003514/2020 | 2020-09-28 | Africa/South Africa/KwaZulu-Natal/Ugu | NHLS-IALCH | KRISP, KZN Research Innovation and Sequencing Platform | Giandhari J et al |
| EPI_ISL_602836 | SouthAfrica/KRISP-K003515/2020 | 2020-09-28 | Africa/South Africa/KwaZulu-Natal/eThe | NHLS-IALCH | KRISP, KZN Research Innovation and Sequencing Platform | Giandhari J et al |
| EPI_ISL_602838 | SouthAfrica/KRISP-K003519/2020 | 2020-09-26 | Africa/South Africa/KwaZulu-Natal/Ugu | NHLS-IALCH | KRISP, KZN Research Innovation and Sequencing Platform | Giandhari J et al |
| EPI_ISL_602848 | SouthAfrica/KRISP-K003530/2020 | 2020-09-23 | Africa/South Africa/KwaZulu-Natal/Zulu | NHLS-IALCH | KRISP, KZN Research Innovation and Sequencing Platform | Giandhari J et al |
| EPI_ISL_602856 | SouthAfrica/KRISP-K003539/2020 | 2020-09-26 | Africa/South Africa/KwaZulu-Natal/eThe | NHLS-IALCH | KRISP, KZN Research Innovation and Sequencing Platform | Giandhari J et al |
| EPI_ISL_602860 | SouthAfrica/KRISP-K003544/2020 | 2020-09-25 | Africa/South Africa/KwaZulu-Natal/eThe | NHLS-IALCH | KRISP, KZN Research Innovation and Sequencing Platform | Giandhari J et al |
| EPI_ISL_602871 | SouthAfrica/KRISP-K003689/2020 | 2020-10-06 | Africa/South Africa/KwaZulu-Natal/Zulu | NHLS-IALCH | KRISP, KZN Research Innovation and Sequencing Platform | Giandhari J et al |
| EPI_ISL_602875 | SouthAfrica/KRISP-K003694/2020 | 2020-10-05 | Africa/South Africa/KwaZulu-Natal/Ugu | NHLS-IALCH | KRISP, KZN Research Innovation and Sequencing Platform | Giandhari J et al |

| | | | | | | |
|---|---|---|---|---|---|---|
| EPI_ISL_602897 | SouthAfrica/KRISP-K003717/2020 | 2020-10-07 | Africa/South Africa/KwaZulu-Natal/iLem | NHLS-IALCH | KRISP, KZN Research Innovation and Sequencing Platform | Giandhari J et al |
| EPI_ISL_602915 | SouthAfrica/KRISP-K003738/2020 | 2020-09-29 | Africa/South Africa/KwaZulu-Natal/iLem | NHLS-IALCH | KRISP, KZN Research Innovation and Sequencing Platform | Giandhari J et al |
| EPI_ISL_602922 | SouthAfrica/KRISP-K003748/2020 | 2020-09-30 | Africa/South Africa/KwaZulu-Natal/eThe | NHLS-IALCH | KRISP, KZN Research Innovation and Sequencing Platform | Giandhari J et al |
| EPI_ISL_602933 | USA/UT-UPHL-201011725/2020 | 2020-06-19 | North America/USA/Utah/ | Utah Public Health Laboratory | Utah Public Health Laboratory | Erin Young et al |
| EPI_ISL_602935 | USA/UT-UPHL-201010022/2020 | 2020-06-19 | North America/USA/Utah/ | Utah Public Health Laboratory | Utah Public Health Laboratory | Erin Young et al |
| EPI_ISL_602950 | USA/UT-UPHL-201005271/2020 | 2020-06-23 | North America/USA/Utah/ | Utah Public Health Laboratory | Utah Public Health Laboratory | Erin Young et al |
| EPI_ISL_602951 | USA/UT-UPHL-201018562/2020 | 2020-06-23 | North America/USA/Utah/ | Utah Public Health Laboratory | Utah Public Health Laboratory | Erin Young et al |
| EPI_ISL_602957 | USA/MN-MDH-1822/2020 | 2020-08-19 | North America/USA/Minnesota/ | Minnesota Department of Health, Public Health Laboratory | Minnesota Department of Health, Public Health Laboratory | Matt Plumb et al |
| EPI_ISL_602962 | USA/MN-MDH-1827/2020 | 2020-09-18 | North America/USA/Minnesota/ | Minnesota Department of Health, Public Health Laboratory | Minnesota Department of Health, Public Health Laboratory | Matt Plumb et al |
| EPI_ISL_602964 | USA/MN-MDH-1829/2020 | 2020-09-18 | North America/USA/Minnesota/ | Minnesota Department of Health, Public Health Laboratory | Minnesota Department of Health, Public Health Laboratory | Matt Plumb et al |
| EPI_ISL_602972 | USA/MN-MDH-1837/2020 | 2020-09-21 | North America/USA/Minnesota/ | Minnesota Department of Health, Public Health Laboratory | Minnesota Department of Health, Public Health Laboratory | Matt Plumb et al |

| | | | | | | |
|---|---|---|---|---|---|---|
| EPI_ISL_602981 | USA/MN-MDH-1846/2020 | 2020-09-30 | North America/USA/Minnesota/ | Minnesota Department of Health, Public Health Laboratory | Minnesota Department of Health, Public Health Laboratory | Matt Plumb et al |
| EPI_ISL_602984 | USA/MN-MDH-1849/2020 | 2020-10-01 | North America/USA/Minnesota/ | Minnesota Department of Health, Public Health Laboratory | Minnesota Department of Health, Public Health Laboratory | Matt Plumb et al |
| EPI_ISL_602988 | USA/MN-MDH-1853/2020 | 2020-10-05 | North America/USA/Minnesota/ | Minnesota Department of Health, Public Health Laboratory | Minnesota Department of Health, Public Health Laboratory | Matt Plumb et al |
| EPI_ISL_602989 | USA/MN-MDH-1854/2020 | 2020-10-06 | North America/USA/Minnesota/ | Minnesota Department of Health, Public Health Laboratory | Minnesota Department of Health, Public Health Laboratory | Matt Plumb et al |
| EPI_ISL_603004 | USA/MN-MDH-1869/2020 | 2020-09-28 | North America/USA/Minnesota/ | Essentia Health-St. Mary's Medical Center | Minnesota Department of Health, Public Health Laboratory | Matt Plumb et al |
| EPI_ISL_603008 | USA/MN-MDH-1873/2020 | 2020-10-14 | North America/USA/Minnesota/ | Minnesota Department of Health, Public Health Laboratory | Minnesota Department of Health, Public Health Laboratory | Matt Plumb et al |
| EPI_ISL_603015 | USA/MN-MDH-1880/2020 | 2020-10-15 | North America/USA/Minnesota/ | Minnesota Department of Health, Public Health Laboratory | Minnesota Department of Health, Public Health Laboratory | Matt Plumb et al |
| EPI_ISL_603021 | Brazil/SP-427/2020 | 2020-07-15 | South America/Brazil/S√£o Paulo/Osasc | Pronto Socorro Dr. Conrado Cesarino Nuvolini | Instituto Adolfo Lutz, Interdiciplinary Procedures Center, Strategic Laboratory | Claudio Tavares Sacchi et al |
| EPI_ISL_603022 | Brazil/SP-429/2020 | 2020-07-16 | South America/Brazil/S√£o Paulo/Diade | Departamento de Vigil√¢ncia √† Sa√∫de | Instituto Adolfo Lutz, Interdiciplinary Procedures Center, Strategic Laboratory | Claudio Tavares Sacchi et al |

| | | | | | | |
|---|---|---|---|---|---|---|
| EPI_ISL_603023 | Brazil/SP-376/2020 | 2020-06-07 | South America/Brazil/S√£o Paulo/Campi | Vigil√¢ncia em Sa√∫de Visa Sul | Instituto Adolfo Lutz, Interdiciplinary Procedures Center, Strategic Laboratory | Claudio Tavares Sacchi et al |
| EPI_ISL_603024 | Brazil/SP-383/2020 | 2020-06-20 | South America/Brazil/S√£o Paulo/Arav√ß | Santa Casa de Misericordia de Arav√ßatuba | Instituto Adolfo Lutz, Interdiciplinary Procedures Center, Strategic Laboratory | Claudio Tavares Sacchi et al |
| EPI_ISL_603025 | Brazil/SP-384/2020 | 2020-06-11 | South America/Brazil/S√£o Paulo/Carag | UPA Central de Caraguatatuba | Instituto Adolfo Lutz, Interdiciplinary Procedures Center, Strategic Laboratory | Claudio Tavares Sacchi et al |
| EPI_ISL_603026 | Brazil/SP-385/2020 | 2020-06-21 | South America/Brazil/S√£o Paulo/Emilia | Santa Casa da Miseric√≥rdia de Presidente Prudente | Instituto Adolfo Lutz, Interdiciplinary Procedures Center, Strategic Laboratory | Claudio Tavares Sacchi et al |
| EPI_ISL_603027 | Brazil/SP-387/2020 | 2020-06-20 | South America/Brazil/S√£o Paulo/Arav√ß | Santa Casa de Misericordia de Arav√ßatuba | Instituto Adolfo Lutz, Interdiciplinary Procedures Center, Strategic Laboratory | Claudio Tavares Sacchi et al |
| EPI_ISL_603028 | Brazil/SP-398/2020 | 2020-06-30 | South America/Brazil/Bahia/Teixeira de | Hospital Municipal Santa Ana | Instituto Adolfo Lutz, Interdiciplinary Procedures Center, Strategic Laboratory | Claudio Tavares Sacchi et al |

| | | | | | | |
|---|---|---|---|---|---|---|
| EPI_ISL_603029 | Brazil/SP-399/2020 | 2020-06-22 | South America/Brazil/S√£o Paulo/Campi | Hospital Municipal M√°rio Gatti | Instituto Adolfo Lutz, Interdiciplinary Procedures Center, Strategic Laboratory | Claudio Tavares Sacchi et al |
| EPI_ISL_603030 | Brazil/SP-433/2020 | 2020-07-11 | South America/Brazil/S√£o Paulo/Presid | Hospital Domingos Leonardo Ceravolo Presidente Prudente | Instituto Adolfo Lutz, Interdiciplinary Procedures Center, Strategic Laboratory | Claudio Tavares Sacchi et al |
| EPI_ISL_603031 | Brazil/SP-434/2020 | 2020-07-12 | South America/Brazil/S√£o Paulo/Presid | Santa Casa de Presidente Epit√°cio | Instituto Adolfo Lutz, Interdiciplinary Procedures Center, Strategic Laboratory | Claudio Tavares Sacchi et al |
| EPI_ISL_603032 | Brazil/SP-435/2020 | 2020-07-13 | South America/Brazil/S√£o Paulo/Santo | Santa Casa da Miseric√≥rdia de Presidente Prudente | Instituto Adolfo Lutz, Interdiciplinary Procedures Center, Strategic Laboratory | Claudio Tavares Sacchi et al |
| EPI_ISL_603033 | Brazil/SP-436/2020 | 2020-07-13 | South America/Brazil/S√£o Paulo/Sao B | Vigilancia Epidemiologica de S√£o Bernardo do Campo | Instituto Adolfo Lutz, Interdiciplinary Procedures Center, Strategic Laboratory | Claudio Tavares Sacchi et al |
| EPI_ISL_603034 | Brazil/SP-437/2020 | 2020-07-11 | South America/Brazil/S√£o Paulo/Diade | Departamento de Vigil√¢ncia √† Sa√∫de | Instituto Adolfo Lutz, Interdiciplinary Procedures Center, Strategic Laboratory | Claudio Tavares Sacchi et al |

| | | | | | | |
|---|---|---|---|---|---|---|
| EPI_ISL_603035 | Brazil/SP-438/2020 | 2020-07-10 | South America/Brazil/S√£o Paulo/Birigu | Secretaria Municipal de Sa√∫de | Instituto Adolfo Lutz, Interdiciplinary Procedures Center, Strategic Laboratory | Claudio Tavares Sacchi et al |
| EPI_ISL_603036 | Brazil/SP-439/2020 | 2020-07-10 | South America/Brazil/S√£o Paulo/Santo | Hospital Santa Ana | Instituto Adolfo Lutz, Interdiciplinary Procedures Center, Strategic Laboratory | Claudio Tavares Sacchi et al |
| EPI_ISL_603037 | Brazil/SP-440/2020 | 2020-07-20 | South America/Brazil/S√£o Paulo/S√£o | Hospital Geral de Pedreira | Instituto Adolfo Lutz, Interdiciplinary Procedures Center, Strategic Laboratory | Claudio Tavares Sacchi et al |
| EPI_ISL_603038 | Brazil/SP-441/2020 | 2020-07-20 | South America/Brazil/S√£o Paulo/Ara√ß | Santa Casa de Misericordia de Ara√ßatuba | Instituto Adolfo Lutz, Interdiciplinary Procedures Center, Strategic Laboratory | Claudio Tavares Sacchi et al |
| EPI_ISL_603039 | Brazil/SP-442/2020 | 2020-07-17 | South America/Brazil/S√£o Paulo/Campi | Hospital Municipal M√°rio Gatti | Instituto Adolfo Lutz, Interdiciplinary Procedures Center, Strategic Laboratory | Claudio Tavares Sacchi et al |
| EPI_ISL_603144 | Italy/LAZ-INMI-37/2020 | 2020-06-12 | Europe/Italy/Lazio/ | INMI Lazzaro Spallanzani IRCCS | INMI Lazzaro Spallanzani IRCCS | Cesare E.M. Gruber et al |
| EPI_ISL_603154 | Italy/LAZ-INMI-47/2020 | 2020-06-12 | Europe/Italy/Lazio/ | INMI Lazzaro Spallanzani IRCCS | INMI Lazzaro Spallanzani IRCCS | Martina Rueca et al |
| EPI_ISL_603157 | Italy/LAZ-INMI-50/2020 | 2020-06-02 | Europe/Italy/Lazio/ | INMI Lazzaro Spallanzani IRCCS | INMI Lazzaro Spallanzani IRCCS | Francesco Messina et al |
| EPI_ISL_603179 | Italy/LAZ-INMI-72/2020 | 2020-05-17 | Europe/Italy/Lazio/ | INMI Lazzaro Spallanzani IRCCS | INMI Lazzaro Spallanzani IRCCS | Cesare E.M. Gruber et al |
| EPI_ISL_603180 | Italy/LAZ-INMI-73/2020 | 2020-05-17 | Europe/Italy/Lazio/ | INMI Lazzaro Spallanzani IRCCS | INMI Lazzaro Spallanzani IRCCS | Barbara Bartolini et al |

| | | | | | | |
|---|---|---|---|---|---|---|
| EPI_ISL_603181 | Italy/LAZ-INMI-74/2020 | 2020-05-23 | Europe/Italy/Lazio/ | INMI Lazzaro Spallanzani IRCCS | INMI Lazzaro Spallanzani IRCCS | Francesco Messina et al |
| EPI_ISL_603184 | Italy/LAZ-INMI-77/2020 | 2020-05-23 | Europe/Italy/Lazio/ | INMI Lazzaro Spallanzani IRCCS | INMI Lazzaro Spallanzani IRCCS | Martina Rueca et al |
| EPI_ISL_603186 | Italy/LAZ-INMI-79/2020 | 2020-05-19 | Europe/Italy/Lazio/ | INMI Lazzaro Spallanzani IRCCS | INMI Lazzaro Spallanzani IRCCS | Francesco Messina et al |
| EPI_ISL_603218 | France/OCC-82/2020 | 2020-09-30 | Europe/France/Occitanie/ | CHU Purpan - Laboratoire de Virologie - Institut F√©d√©ratif de Biologie | CHU Purpan - Laboratoire de Virologie - Institut F√©d√©ratif de Biologie | Latour J. et al |
| EPI_ISL_603219 | France/OCC-83/2020 | 2020-09-30 | Europe/France/Occitanie/ | CHU Purpan - Laboratoire de Virologie - Institut F√©d√©ratif de Biologie | CHU Purpan - Laboratoire de Virologie - Institut F√©d√©ratif de Biologie | Latour J. et al |
| EPI_ISL_603220 | France/OCC-84/2020 | 2020-09-30 | Europe/France/Occitanie/ | CHU Purpan - Laboratoire de Virologie - Institut F√©d√©ratif de Biologie | CHU Purpan - Laboratoire de Virologie - Institut F√©d√©ratif de Biologie | Latour J. et al |
| EPI_ISL_603221 | Bangladesh/BCSIR-NILMRC_392/2020 | 2020-10-14 | Asia/Bangladesh/Dhaka/ | National Institute of Laboratory Medicine and Referral Center | Genomic Research Lab, BCSIR | Abu Sayeed Mohammad Mahmud et al |
| EPI_ISL_603222 | Bangladesh/BCSIR-NILMRC_398/2020 | 2020-10-08 | Asia/Bangladesh/Dhaka/ | National Institute of Laboratory Medicine and Referral Center | Genomic Research Lab, BCSIR | Abu Sayeed Mohammad Mahmud et al |
| EPI_ISL_603223 | Bangladesh/BCSIR-NILMRC_376A/202 | 2020-07-31 | Asia/Bangladesh/Dhaka/Narayanganj | National Institute of Laboratory Medicine and Referral Center | Genomic Research Lab, BCSIR | Md. Murshed Hasan Sarkar et al |
| EPI_ISL_603225 | Bangladesh/BCSIR-NILMRC_391/2020 | 2020-07-31 | Asia/Bangladesh/Dhaka/ | National Institute of Laboratory Medicine and Referral Center | Genomic Research Lab, BCSIR | Md. Murshed Hasan Sarkar et al |
| EPI_ISL_603238 | Bangladesh/BCSIR-NILMRC_393/2020 | 2020-10-14 | Asia/Bangladesh/Dhaka/ | National Institute of Laboratory Medicine and Referral Center | Genomic Research Lab, BCSIR | Shahina Akter et al |
| EPI_ISL_603239 | Bangladesh/BCSIR-NILMRC_396/2020 | 2020-10-08 | Asia/Bangladesh/Dhaka/ | National Institute of Laboratory Medicine and Referral Center | Genomic Research Lab, BCSIR | Shahina Akter et al |
| EPI_ISL_603240 | Bangladesh/BCSIR-NILMRC_397/2020 | 2020-10-08 | Asia/Bangladesh/Dhaka/ | National Institute of Laboratory Medicine and Referral Center | Genomic Research Lab, BCSIR | Tanjina Akhter Banu et al |
| EPI_ISL_603241 | Bangladesh/BCSIR-NILMRC_411/2020 | 2020-10-18 | Asia/Bangladesh/Dhaka/Narayanganj | National Institute of Laboratory Medicine and Referral Center | Genomic Research Lab, BCSIR | Tanjina Akhter Banu et al |
| EPI_ISL_603242 | Bangladesh/BCSIR-NILMRC_420/2020 | 2020-10-10 | Asia/Bangladesh/Dhaka/ | National Institute of Laboratory Medicine and Referral Center | Genomic Research Lab, BCSIR | Barna Goswami et al |

| | | | | | | |
|---|---|---|---|---|---|---|
| EPI_ISL_603243 | Bangladesh/BCSIR-NILMRC_421/2020 | 2020-10-10 | Asia/Bangladesh/Dhaka/ | National Institute of Laboratory Medicine and Referral Center | Genomic Research Lab, BCSIR | Barna Goswami et al |
| EPI_ISL_603244 | Bangladesh/BCSIR-NILMRC_422/2020 | 2020-10-10 | Asia/Bangladesh/Dhaka/ | National Institute of Laboratory Medicine and Referral Center | Genomic Research Lab, BCSIR | Iffat Jahan et al |
| EPI_ISL_603245 | Bangladesh/BCSIR-NILMRC_423/2020 | 2020-10-10 | Asia/Bangladesh/Dhaka/ | National Institute of Laboratory Medicine and Referral Center | Genomic Research Lab, BCSIR | Iffat Jahan et al |
| EPI_ISL_603247 | Bangladesh/BCSIR-NILMRC_426/2020 | 2020-10-05 | Asia/Bangladesh/Dhaka/ | National Institute of Laboratory Medicine and Referral Center | Genomic Research Lab, BCSIR | Md. Saddam Hossain et al |
| EPI_ISL_603249 | Bangladesh/BCSIR-NILMRC_427/2020 | 2020-10-05 | Asia/Bangladesh/Dhaka/ | National Institute of Laboratory Medicine and Referral Center | Genomic Research Lab, BCSIR | Md. Ahashan Habib et al |
| EPI_ISL_603250 | Bangladesh/BCSIR-NILMRC_428/2020 | 2020-10-05 | Asia/Bangladesh/Dhaka/ | National Institute of Laboratory Medicine and Referral Center | Genomic Research Lab, BCSIR | Md. Ahashan Habib et al |
| EPI_ISL_603251 | USA/WA-UW-23621/2020 | 2020-08-10 | North America/USA/Washington/ | UW Virology Lab | UW Virology Lab | Pavitra Roychoudhury et al |
| EPI_ISL_603254 | USA/WA-UW-29740/2020 | 2020-09-28 | North America/USA/Washington/ | UW Virology Lab | UW Virology Lab | Pavitra Roychoudhury et al |
| EPI_ISL_603255 | USA/WA-UW-29702/2020 | 2020-09-28 | North America/USA/Washington/ | UW Virology Lab | UW Virology Lab | Pavitra Roychoudhury et al |
| EPI_ISL_603305 | Switzerland/VD-ETHZ-300273/2020 | 2020-10-01 | Europe/Switzerland/Vaud/ | Viollier AG | Department of Biosystems Science and Engineering, ETH Zvºrich | Christian Beisel et al |
| EPI_ISL_603310 | Switzerland/BE-ETHZ-300278/2020 | 2020-10-01 | Europe/Switzerland/Bern/ | Viollier AG | Department of Biosystems Science and Engineering, ETH Zvºrich | Christian Beisel et al |
| EPI_ISL_603316 | Switzerland/ZH-ETHZ-300284/2020 | 2020-10-01 | Europe/Switzerland/Zvºrich/ | Viollier AG | Department of Biosystems Science and Engineering, ETH Zvºrich | Christian Beisel et al |
| EPI_ISL_603321 | Switzerland/BE-ETHZ-300289/2020 | 2020-10-01 | Europe/Switzerland/Bern/ | Viollier AG | Department of Biosystems Science and Engineering, ETH Zvºrich | Christian Beisel et al |

| | | | | | | |
|---|---|---|---|---|---|---|
| EPI_ISL_603358 | Switzerland/ZH-ETHZ-300389/2020 | 2020-09-29 | Europe/Switzerland/Zvºrich/ | Viollier AG | Department of Biosystems Science and Engineering, ETH Zvºrich | Christian Beisel et al |
| EPI_ISL_603386 | Switzerland/BE-ETHZ-300431/2020 | 2020-09-30 | Europe/Switzerland/Bern/ | Viollier AG | Department of Biosystems Science and Engineering, ETH Zvºrich | Christian Beisel et al |
| EPI_ISL_603437 | Switzerland/VD-ETHZ-310363/2020 | 2020-10-07 | Europe/Switzerland/Vaud/ | Viollier AG | Department of Biosystems Science and Engineering, ETH Zvºrich | Christian Beisel et al |
| EPI_ISL_603453 | Switzerland/VD-ETHZ-310379/2020 | 2020-10-08 | Europe/Switzerland/Vaud/ | Viollier AG | Department of Biosystems Science and Engineering, ETH Zvºrich | Christian Beisel et al |
| EPI_ISL_603471 | Switzerland/LU-ETHZ-310397/2020 | 2020-10-08 | Europe/Switzerland/Lucerne/ | Viollier AG | Department of Biosystems Science and Engineering, ETH Zvºrich | Christian Beisel et al |
| EPI_ISL_603498 | Switzerland/ZH-ETHZ-310424/2020 | 2020-10-08 | Europe/Switzerland/Zvºrich/ | Viollier AG | Department of Biosystems Science and Engineering, ETH Zvºrich | Christian Beisel et al |
| EPI_ISL_603499 | Switzerland/ZH-ETHZ-310425/2020 | 2020-10-08 | Europe/Switzerland/Zvºrich/ | Viollier AG | Department of Biosystems Science and Engineering, ETH Zvºrich | Christian Beisel et al |
| EPI_ISL_603537 | Switzerland/BE-ETHZ-310465/2020 | 2020-10-07 | Europe/Switzerland/Bern/ | Viollier AG | Department of Biosystems Science and Engineering, ETH Zvºrich | Christian Beisel et al |

| | | | | | | |
|---|---|---|---|---|---|---|
| EPI_ISL_603559 | Switzerland/SZ-ETHZ-310489/2020 | 2020-10-08 | Europe/Switzerland/Schwyz/ | Viollier AG | Department of Biosystems Science and Engineering, ETH Zv¨rich | Christian Beisel et al |
| EPI_ISL_603564 | Switzerland/VD-ETHZ-310495/2020 | 2020-10-08 | Europe/Switzerland/Vaud/ | Viollier AG | Department of Biosystems Science and Engineering, ETH Zv¨rich | Christian Beisel et al |
| EPI_ISL_603569 | Switzerland/BL-ETHZ-310501/2020 | 2020-10-08 | Europe/Switzerland/Basel-Land/ | Viollier AG | Department of Biosystems Science and Engineering, ETH Zv¨rich | Christian Beisel et al |
| EPI_ISL_603597 | Switzerland/GE-ETHZ-310529/2020 | 2020-10-08 | Europe/Switzerland/Geneva/ | Viollier AG | Department of Biosystems Science and Engineering, ETH Zv¨rich | Christian Beisel et al |
| EPI_ISL_603598 | Switzerland/BE-ETHZ-310530/2020 | 2020-10-08 | Europe/Switzerland/Bern/ | Viollier AG | Department of Biosystems Science and Engineering, ETH Zv¨rich | Christian Beisel et al |
| EPI_ISL_603606 | Switzerland/AG-ETHZ-321356/2020 | 2020-10-13 | Europe/Switzerland/Aargau/ | Viollier AG | Department of Biosystems Science and Engineering, ETH Zv¨rich | Christian Beisel et al |
| EPI_ISL_603658 | Switzerland/ZH-ETHZ-321420/2020 | 2020-10-14 | Europe/Switzerland/Zv¨rich/ | Viollier AG | Department of Biosystems Science and Engineering, ETH Zv¨rich | Christian Beisel et al |
| EPI_ISL_603699 | Switzerland/BE-ETHZ-321487/2020 | 2020-10-15 | Europe/Switzerland/Bern/ | Viollier AG | Department of Biosystems Science and Engineering, ETH Zv¨rich | Christian Beisel et al |

| | | | | | | |
|---|---|---|---|---|---|---|
| EPI_ISL_603711 | Switzerland/VD-ETHZ-321500/2020 | 2020-10-15 | Europe/Switzerland/Vaud/ | Viollier AG | Department of Biosystems Science and Engineering, ETH Zv∫rich | Christian Beisel et al |
| EPI_ISL_603715 | Switzerland/VD-ETHZ-321505/2020 | 2020-10-15 | Europe/Switzerland/Vaud/ | Viollier AG | Department of Biosystems Science and Engineering, ETH Zv∫rich | Christian Beisel et al |
| EPI_ISL_603720 | Switzerland/ZH-ETHZ-321510/2020 | 2020-10-15 | Europe/Switzerland/Zv∫rich/ | Viollier AG | Department of Biosystems Science and Engineering, ETH Zv∫rich | Christian Beisel et al |
| EPI_ISL_603723 | Switzerland/AG-ETHZ-321513/2020 | 2020-10-15 | Europe/Switzerland/Aargau/ | Viollier AG | Department of Biosystems Science and Engineering, ETH Zv∫rich | Christian Beisel et al |
| EPI_ISL_603753 | Switzerland/VD-ETHZ-321544/2020 | 2020-10-15 | Europe/Switzerland/Vaud/ | Viollier AG | Department of Biosystems Science and Engineering, ETH Zv∫rich | Christian Beisel et al |
| EPI_ISL_603761 | USA/CA-QDX-1491/2020 | 2020-03-17 | North America/USA/California/ | Quest Diagnostics | Quest Diagnostics | Rosenthal et al |
| EPI_ISL_603762 | USA/CA-QDX-1492/2020 | 2020-03-17 | North America/USA/California/ | Quest Diagnostics | Quest Diagnostics | Rosenthal et al |
| EPI_ISL_603786 | USA/CA-QDX-1538/2020 | 2020-03-18 | North America/USA/California/ | Quest Diagnostics | Quest Diagnostics | Rosenthal et al |
| EPI_ISL_603803 | USA/CA-QDX-1558/2020 | 2020-03-18 | North America/USA/California/ | Quest Diagnostics | Quest Diagnostics | Rosenthal et al |
| EPI_ISL_603808 | USA/CT-QDX-1454/2020 | 2020-03-16 | North America/USA/Connecticut/ | Quest Diagnostics | Quest Diagnostics | Rosenthal et al |
| EPI_ISL_603814 | USA/ID-QDX-1531/2020 | 2020-03-17 | North America/USA/Idaho/ | Quest Diagnostics | Quest Diagnostics | Rosenthal et al |
| EPI_ISL_603825 | USA/MA-QDX-1458/2020 | 2020-03-12 | North America/USA/Massachusetts/ | Quest Diagnostics | Quest Diagnostics | Rosenthal et al |
| EPI_ISL_603827 | USA/MA-QDX-1486/2020 | 2020-03-17 | North America/USA/Massachusetts/ | Quest Diagnostics | Quest Diagnostics | Rosenthal et al |
| EPI_ISL_603828 | USA/MA-QDX-1487/2020 | 2020-03-17 | North America/USA/Massachusetts/ | Quest Diagnostics | Quest Diagnostics | Rosenthal et al |
| EPI_ISL_603846 | USA/UT-QDX-1461/2020 | 2020-03-14 | North America/USA/Utah/ | Quest Diagnostics | Quest Diagnostics | Rosenthal et al |
| EPI_ISL_603864 | USA/UT-QDX-1479/2020 | 2020-03-14 | North America/USA/Utah/ | Quest Diagnostics | Quest Diagnostics | Rosenthal et al |
| EPI_ISL_603865 | USA/UT-QDX-1480/2020 | 2020-03-14 | North America/USA/Utah/ | Quest Diagnostics | Quest Diagnostics | Rosenthal et al |

| | | | | | | |
|---|---|---|---|---|---|---|
| EPI_ISL_603876 | USA/AZ-QDX-1626/2020 | 2020-03-19 | North America/USA/Arizona/ | Quest Diagnostics | Quest Diagnostics | Rosenthal et al |
| EPI_ISL_603889 | USA/CA-QDX-1572/2020 | 2020-03-18 | North America/USA/California/ | Quest Diagnostics | Quest Diagnostics | Rosenthal et al |
| EPI_ISL_603896 | USA/CA-QDX-1586/2020 | 2020-03-19 | North America/USA/California/ | Quest Diagnostics | Quest Diagnostics | Rosenthal et al |
| EPI_ISL_603906 | USA/CA-QDX-1624/2020 | 2020-03-18 | North America/USA/California/ | Quest Diagnostics | Quest Diagnostics | Rosenthal et al |
| EPI_ISL_603925 | USA/CA-QDX-1676/2020 | 2020-03-19 | North America/USA/California/ | Quest Diagnostics | Quest Diagnostics | Rosenthal et al |
| EPI_ISL_603933 | USA/CA-QDX-1693/2020 | 2020-03-19 | North America/USA/California/ | Quest Diagnostics | Quest Diagnostics | Rosenthal et al |
| EPI_ISL_603956 | USA/MI-QDX-1600/2020 | 2020-03-17 | North America/USA/Michigan/ | Quest Diagnostics | Quest Diagnostics | Rosenthal et al |
| EPI_ISL_603957 | USA/MI-QDX-1612/2020 | 2020-03-16 | North America/USA/Michigan/ | Quest Diagnostics | Quest Diagnostics | Rosenthal et al |
| EPI_ISL_603958 | USA/MI-QDX-1613/2020 | 2020-03-18 | North America/USA/Michigan/ | Quest Diagnostics | Quest Diagnostics | Rosenthal et al |
| EPI_ISL_603970 | USA/MI-QDX-1683/2020 | 2020-03-17 | North America/USA/Michigan/ | Quest Diagnostics | Quest Diagnostics | Rosenthal et al |
| EPI_ISL_603998 | USA/MN-QDX-1652/2020 | 2020-03-16 | North America/USA/Minnesota/ | Quest Diagnostics | Quest Diagnostics | Rosenthal et al |
| EPI_ISL_604020 | USA/CA-QDX-1773/2020 | 2020-03-15 | North America/USA/California/ | Quest Diagnostics | Quest Diagnostics | Rosenthal et al |
| EPI_ISL_604031 | USA/ID-QDX-1727/2020 | 2020-03-17 | North America/USA/Idaho/ | Quest Diagnostics | Quest Diagnostics | Rosenthal et al |
| EPI_ISL_604041 | USA/MI-QDX-1723/2020 | 2020-03-18 | North America/USA/Michigan/ | Quest Diagnostics | Quest Diagnostics | Rosenthal et al |
| EPI_ISL_604042 | USA/MI-QDX-1726/2020 | 2020-03-18 | North America/USA/Michigan/ | Quest Diagnostics | Quest Diagnostics | Rosenthal et al |
| EPI_ISL_604055 | USA/MI-QDX-1767/2020 | 2020-03-17 | North America/USA/Michigan/ | Quest Diagnostics | Quest Diagnostics | Rosenthal et al |
| EPI_ISL_604057 | USA/MI-QDX-1770/2020 | 2020-03-17 | North America/USA/Michigan/ | Quest Diagnostics | Quest Diagnostics | Rosenthal et al |
| EPI_ISL_604058 | USA/MI-QDX-1772/2020 | 2020-03-17 | North America/USA/Michigan/ | Quest Diagnostics | Quest Diagnostics | Rosenthal et al |
| EPI_ISL_604072 | USA/MN-QDX-1724/2020 | 2020-03-15 | North America/USA/Minnesota/ | Quest Diagnostics | Quest Diagnostics | Rosenthal et al |
| EPI_ISL_604109 | USA/MA-QDX-1817/2020 | 2020-03-17 | North America/USA/Massachusetts/ | Quest Diagnostics | Quest Diagnostics | Rosenthal et al |
| EPI_ISL_604123 | USA/UT-QDX-1793/2020 | 2020-03-13 | North America/USA/Utah/ | Quest Diagnostics | Quest Diagnostics | Rosenthal et al |
| EPI_ISL_604149 | USA/UT-QDX-1831/2020 | 2020-03-13 | North America/USA/Utah/ | Quest Diagnostics | Quest Diagnostics | Rosenthal et al |
| EPI_ISL_604154 | USA/UT-QDX-1836/2020 | 2020-03-14 | North America/USA/Utah/ | Quest Diagnostics | Quest Diagnostics | Rosenthal et al |
| EPI_ISL_604177 | USA/UT-QDX-1864/2020 | 2020-03-15 | North America/USA/Utah/ | Quest Diagnostics | Quest Diagnostics | Rosenthal et al |
| EPI_ISL_604194 | USA/CA-QDX-1899/2020 | 2020-03-16 | North America/USA/California/ | Quest Diagnostics | Quest Diagnostics | Rosenthal et al |
| EPI_ISL_604214 | USA/CA-QDX-1961/2020 | 2020-03-15 | North America/USA/California/ | Quest Diagnostics | Quest Diagnostics | Rosenthal et al |
| EPI_ISL_604244 | USA/NY-QDX-1893/2020 | 2020-03-15 | North America/USA/New York/ | Quest Diagnostics | Quest Diagnostics | Rosenthal et al |
| EPI_ISL_604260 | USA/NY-QDX-1935/2020 | 2020-03-16 | North America/USA/New York/ | Quest Diagnostics | Quest Diagnostics | Rosenthal et al |
| EPI_ISL_604317 | USA/CA-QDX-2037/2020 | 2020-03-18 | North America/USA/California/ | Quest Diagnostics | Quest Diagnostics | Rosenthal et al |
| EPI_ISL_604338 | USA/LA-QDX-2000/2020 | 2020-03-16 | North America/USA/Louisiana/ | Quest Diagnostics | Quest Diagnostics | Rosenthal et al |
| EPI_ISL_604354 | USA/MN-QDX-2053/2020 | 2020-03-15 | North America/USA/Minnesota/ | Quest Diagnostics | Quest Diagnostics | Rosenthal et al |

| | | | | | | |
|---|---|---|---|---|---|---|
| EPI_ISL_604363 | USA/NY-QDX-2034/2020 | 2020-03-18 | North America/USA/New York/ | Quest Diagnostics | Quest Diagnostics | Rosenthal et al |
| EPI_ISL_604383 | USA/UT-QDX-2028/2020 | 2020-03-14 | North America/USA/Utah/ | Quest Diagnostics | Quest Diagnostics | Rosenthal et al |
| EPI_ISL_604388 | USA/AZ-QDX-2135/2020 | 2020-03-18 | North America/USA/Arizona/ | Quest Diagnostics | Quest Diagnostics | Rosenthal et al |
| EPI_ISL_604404 | USA/CA-QDX-2092/2020 | 2020-03-19 | North America/USA/California/ | Quest Diagnostics | Quest Diagnostics | Rosenthal et al |
| EPI_ISL_604415 | USA/CA-QDX-2113/2020 | 2020-03-18 | North America/USA/California/ | Quest Diagnostics | Quest Diagnostics | Rosenthal et al |
| EPI_ISL_604418 | USA/CA-QDX-2120/2020 | 2020-03-18 | North America/USA/California/ | Quest Diagnostics | Quest Diagnostics | Rosenthal et al |
| EPI_ISL_604423 | USA/CA-QDX-2134/2020 | 2020-03-18 | North America/USA/California/ | Quest Diagnostics | Quest Diagnostics | Rosenthal et al |
| EPI_ISL_604471 | USA/MI-QDX-2164/2020 | 2020-03-17 | North America/USA/Michigan/ | Quest Diagnostics | Quest Diagnostics | Rosenthal et al |
| EPI_ISL_604479 | USA/MI-QDX-2187/2020 | 2020-03-18 | North America/USA/Michigan/ | Quest Diagnostics | Quest Diagnostics | Rosenthal et al |
| EPI_ISL_604551 | USA/MN-QDX-2258/2020 | 2020-03-14 | North America/USA/Minnesota/ | Quest Diagnostics | Quest Diagnostics | Rosenthal et al |
| EPI_ISL_604552 | USA/MN-QDX-2265/2020 | 2020-03-13 | North America/USA/Minnesota/ | Quest Diagnostics | Quest Diagnostics | Rosenthal et al |
| EPI_ISL_604562 | USA/NJ-QDX-2280/2020 | 2020-03-14 | North America/USA/New Jersey/ | Quest Diagnostics | Quest Diagnostics | Rosenthal et al |
| EPI_ISL_604565 | USA/NY-QDX-2188/2020 | 2020-03-15 | North America/USA/New York/ | Quest Diagnostics | Quest Diagnostics | Rosenthal et al |
| EPI_ISL_604572 | USA/NY-QDX-2243/2020 | 2020-03-13 | North America/USA/New York/ | Quest Diagnostics | Quest Diagnostics | Rosenthal et al |
| EPI_ISL_604595 | USA/UT-QDX-2240/2020 | 2020-03-13 | North America/USA/Utah/ | Quest Diagnostics | Quest Diagnostics | Rosenthal et al |
| EPI_ISL_604602 | USA/UT-QDX-2262/2020 | 2020-03-13 | North America/USA/Utah/ | Quest Diagnostics | Quest Diagnostics | Rosenthal et al |
| EPI_ISL_604620 | USA/UT-QDX-2313/2020 | 2020-03-13 | North America/USA/Utah/ | Quest Diagnostics | Quest Diagnostics | Rosenthal et al |
| EPI_ISL_604635 | USA/CA-QDX-2331/2020 | 2020-03-16 | North America/USA/California/ | Quest Diagnostics | Quest Diagnostics | Rosenthal et al |
| EPI_ISL_604641 | USA/CA-QDX-2340/2020 | 2020-03-14 | North America/USA/California/ | Quest Diagnostics | Quest Diagnostics | Rosenthal et al |
| EPI_ISL_604643 | USA/CA-QDX-2342/2020 | 2020-03-16 | North America/USA/California/ | Quest Diagnostics | Quest Diagnostics | Rosenthal et al |
| EPI_ISL_604647 | USA/CA-QDX-2346/2020 | 2020-03-16 | North America/USA/California/ | Quest Diagnostics | Quest Diagnostics | Rosenthal et al |
| EPI_ISL_604652 | USA/CA-QDX-2355/2020 | 2020-03-17 | North America/USA/California/ | Quest Diagnostics | Quest Diagnostics | Rosenthal et al |
| EPI_ISL_604660 | USA/CA-QDX-2381/2020 | 2020-03-17 | North America/USA/California/ | Quest Diagnostics | Quest Diagnostics | Rosenthal et al |
| EPI_ISL_604684 | USA/CO-QDX-2314/2020 | 2020-03-16 | North America/USA/Colorado/ | Quest Diagnostics | Quest Diagnostics | Rosenthal et al |
| EPI_ISL_604725 | USA/UT-QDX-2356/2020 | 2020-03-14 | North America/USA/Utah/ | Quest Diagnostics | Quest Diagnostics | Rosenthal et al |
| EPI_ISL_604729 | USA/UT-QDX-2415/2020 | 2020-03-16 | North America/USA/Utah/ | Quest Diagnostics | Quest Diagnostics | Rosenthal et al |
| EPI_ISL_604737 | USA/WA-QDX-2376/2020 | 2020-03-16 | North America/USA/Washington/ | Quest Diagnostics | Quest Diagnostics | Rosenthal et al |
| EPI_ISL_604763 | USA/CA-QDX-2519/2020 | 2020-03-15 | North America/USA/California/ | Quest Diagnostics | Quest Diagnostics | Rosenthal et al |
| EPI_ISL_604792 | USA/NJ-QDX-2544/2020 | 2020-03-16 | North America/USA/New Jersey/ | Quest Diagnostics | Quest Diagnostics | Rosenthal et al |
| EPI_ISL_604795 | USA/NY-QDX-2431/2020 | 2020-03-14 | North America/USA/New York/ | Quest Diagnostics | Quest Diagnostics | Rosenthal et al |
| EPI_ISL_604800 | USA/NY-QDX-2446/2020 | 2020-03-14 | North America/USA/New York/ | Quest Diagnostics | Quest Diagnostics | Rosenthal et al |

| | | | | | | |
|---|---|---|---|---|---|---|
| EPI_ISL_604826 | USA/NY-QDX-2530/2020 | 2020-03-16 | North America/USA/New York/ | Quest Diagnostics | Quest Diagnostics | Rosenthal et al |
| EPI_ISL_604829 | USA/NY-QDX-2535/2020 | 2020-03-14 | North America/USA/New York/ | Quest Diagnostics | Quest Diagnostics | Rosenthal et al |
| EPI_ISL_604836 | USA/PA-QDX-2436/2020 | 2020-03-14 | North America/USA/Pennsylvania/ | Quest Diagnostics | Quest Diagnostics | Rosenthal et al |
| EPI_ISL_604837 | USA/PA-QDX-2437/2020 | 2020-03-14 | North America/USA/Pennsylvania/ | Quest Diagnostics | Quest Diagnostics | Rosenthal et al |
| EPI_ISL_604843 | USA/TX-QDX-2554/2020 | 2020-03-14 | North America/USA/Texas/ | Quest Diagnostics | Quest Diagnostics | Rosenthal et al |
| EPI_ISL_604844 | USA/UT-QDX-2434/2020 | 2020-03-13 | North America/USA/Utah/ | Quest Diagnostics | Quest Diagnostics | Rosenthal et al |
| EPI_ISL_604864 | USA/UT-QDX-2549/2020 | 2020-03-12 | North America/USA/Utah/ | Quest Diagnostics | Quest Diagnostics | Rosenthal et al |
| EPI_ISL_604871 | USA/WA-QDX-2542/2020 | 2020-03-16 | North America/USA/Washington/ | Quest Diagnostics | Quest Diagnostics | Rosenthal et al |
| EPI_ISL_604873 | USA/AZ-QDX-2673/2020 | 2020-03-16 | North America/USA/Arizona/ | Quest Diagnostics | Quest Diagnostics | Rosenthal et al |
| EPI_ISL_604877 | USA/CA-QDX-2581/2020 | 2020-03-15 | North America/USA/California/ | Quest Diagnostics | Quest Diagnostics | Rosenthal et al |
| EPI_ISL_604904 | USA/CA-QDX-2640/2020 | 2020-03-16 | North America/USA/California/ | Quest Diagnostics | Quest Diagnostics | Rosenthal et al |
| EPI_ISL_604908 | USA/CA-QDX-2646/2020 | 2020-03-16 | North America/USA/California/ | Quest Diagnostics | Quest Diagnostics | Rosenthal et al |
| EPI_ISL_604919 | USA/CT-QDX-2579/2020 | 2020-03-12 | North America/USA/Connecticut/ | Quest Diagnostics | Quest Diagnostics | Rosenthal et al |
| EPI_ISL_604925 | USA/FL-QDX-2590/2020 | 2020-03-15 | North America/USA/Florida/ | Quest Diagnostics | Quest Diagnostics | Rosenthal et al |
| EPI_ISL_604986 | USA/UT-QDX-2668/2020 | 2020-03-14 | North America/USA/Utah/ | Quest Diagnostics | Quest Diagnostics | Rosenthal et al |
| EPI_ISL_604991 | USA/UT-QDX-2681/2020 | 2020-03-13 | North America/USA/Utah/ | Quest Diagnostics | Quest Diagnostics | Rosenthal et al |
| EPI_ISL_605038 | USA/NY-QDX-2682/2020 | 2020-03-15 | North America/USA/New York/ | Quest Diagnostics | Quest Diagnostics | Rosenthal et al |
| EPI_ISL_605039 | USA/NY-QDX-2689/2020 | 2020-03-15 | North America/USA/New York/ | Quest Diagnostics | Quest Diagnostics | Rosenthal et al |
| EPI_ISL_605049 | USA/NY-QDX-2714/2020 | 2020-03-14 | North America/USA/New York/ | Quest Diagnostics | Quest Diagnostics | Rosenthal et al |
| EPI_ISL_605050 | USA/NY-QDX-2722/2020 | 2020-03-16 | North America/USA/New York/ | Quest Diagnostics | Quest Diagnostics | Rosenthal et al |
| EPI_ISL_605057 | USA/TN-QDX-2686/2020 | 2020-03-17 | North America/USA/Tennessee/ | Quest Diagnostics | Quest Diagnostics | Rosenthal et al |
| EPI_ISL_605099 | Ireland/D-NVRL-20G44441/2020 | 2020-03-29 | Europe/Ireland/Dublin/ | National Virus Reference Laboratory | Irish Coronavirus Sequencing Consortium - Helixworks | Sachin Chalapati et al |
| EPI_ISL_605126 | Ireland/CO-NVRL-20G32584/2020 | 2020-03-17 | Europe/Ireland/Cork/ | National Virus Reference Laboratory | Irish Coronavirus Sequencing Consortium - Helixworks | Sachin Chalapati et al |
| EPI_ISL_605148 | USA/UT-UPHL-2009383/2020 | 2020-05-27 | North America/USA/Utah/ | Utah Public Health Laboratory | Utah Public Health Laboratory | Erin L. Young et al |
| EPI_ISL_605151 | USA/UT-UPHL-2009388/2020 | 2020-05-29 | North America/USA/Utah/ | Utah Public Health Laboratory | Utah Public Health Laboratory | Erin L. Young et al |

| | | | | | | |
|---|---|---|---|---|---|---|
| EPI_ISL_605155 | USA/UT-UPHL-2009399/2020 | 2020-06-01 | North America/USA/Utah/ | Utah Public Health Laboratory | Utah Public Health Laboratory | Erin L. Young et al |
| EPI_ISL_605158 | USA/UT-UPHL-2009472/2020 | 2020-07-18 | North America/USA/Utah/ | Utah Public Health Laboratory | Utah Public Health Laboratory | Erin L. Young et al |
| EPI_ISL_605165 | USA/UT-UPHL-2009481/2020 | 2020-07-11 | North America/USA/Utah/ | Utah Public Health Laboratory | Utah Public Health Laboratory | Erin L. Young et al |
| EPI_ISL_605181 | USA/UT-UPHL-2009497/2020 | 2020-07-14 | North America/USA/Utah/ | Utah Public Health Laboratory | Utah Public Health Laboratory | Erin L. Young et al |
| EPI_ISL_605182 | USA/UT-UPHL-2009498/2020 | 2020-07-15 | North America/USA/Utah/ | Utah Public Health Laboratory | Utah Public Health Laboratory | Erin L. Young et al |
| EPI_ISL_605183 | USA/UT-UPHL-2009499/2020 | 2020-07-19 | North America/USA/Utah/ | Utah Public Health Laboratory | Utah Public Health Laboratory | Erin L. Young et al |
| EPI_ISL_605184 | USA/UT-UPHL-2009500/2020 | 2020-07-19 | North America/USA/Utah/ | Utah Public Health Laboratory | Utah Public Health Laboratory | Erin L. Young et al |
| EPI_ISL_605192 | USA/UT-UPHL-2009509/2020 | 2020-07-19 | North America/USA/Utah/ | Utah Public Health Laboratory | Utah Public Health Laboratory | Erin L. Young et al |
| EPI_ISL_605193 | USA/UT-UPHL-2009510/2020 | 2020-07-19 | North America/USA/Utah/ | Utah Public Health Laboratory | Utah Public Health Laboratory | Erin L. Young et al |
| EPI_ISL_605194 | USA/UT-UPHL-2009511/2020 | 2020-07-19 | North America/USA/Utah/ | Utah Public Health Laboratory | Utah Public Health Laboratory | Erin L. Young et al |
| EPI_ISL_605196 | USA/UT-UPHL-2009515/2020 | 2020-07-18 | North America/USA/Utah/ | Utah Public Health Laboratory | Utah Public Health Laboratory | Erin L. Young et al |
| EPI_ISL_605227 | USA/UT-UPHL-2009547/2020 | 2020-07-18 | North America/USA/Utah/ | Utah Public Health Laboratory | Utah Public Health Laboratory | Erin L. Young et al |
| EPI_ISL_605230 | USA/UT-UPHL-2009551/2020 | 2020-07-18 | North America/USA/Utah/ | Utah Public Health Laboratory | Utah Public Health Laboratory | Erin L. Young et al |
| EPI_ISL_605237 | USA/UT-UPHL-2009558/2020 | 2020-07-18 | North America/USA/Utah/ | Utah Public Health Laboratory | Utah Public Health Laboratory | Erin L. Young et al |
| EPI_ISL_605239 | USA/UT-UPHL-2009560/2020 | 2020-07-18 | North America/USA/Utah/ | Utah Public Health Laboratory | Utah Public Health Laboratory | Erin L. Young et al |
| EPI_ISL_605245 | USA/UT-UPHL-2009566/2020 | 2020-07-19 | North America/USA/Utah/ | Utah Public Health Laboratory | Utah Public Health Laboratory | Erin L. Young et al |
| EPI_ISL_605249 | USA/UT-UPHL-2009570/2020 | 2020-07-18 | North America/USA/Utah/ | Utah Public Health Laboratory | Utah Public Health Laboratory | Erin L. Young et al |
| EPI_ISL_605250 | USA/UT-UPHL-2009571/2020 | 2020-07-18 | North America/USA/Utah/ | Utah Public Health Laboratory | Utah Public Health Laboratory | Erin L. Young et al |
| EPI_ISL_605253 | USA/UT-UPHL-2009575/2020 | 2020-07-18 | North America/USA/Utah/ | Utah Public Health Laboratory | Utah Public Health Laboratory | Erin L. Young et al |
| EPI_ISL_605256 | USA/UT-UPHL-2009578/2020 | 2020-07-19 | North America/USA/Utah/ | Utah Public Health Laboratory | Utah Public Health Laboratory | Erin L. Young et al |

| | | | | | | |
|---|---|---|---|---|---|---|
| EPI_ISL_605260 | USA/UT-UPHL-2009582/2020 | 2020-07-18 | North America/USA/Utah/ | Utah Public Health Laboratory | Utah Public Health Laboratory | Erin L. Young et al |
| EPI_ISL_605261 | USA/UT-UPHL-2009583/2020 | 2020-07-18 | North America/USA/Utah/ | Utah Public Health Laboratory | Utah Public Health Laboratory | Erin L. Young et al |
| EPI_ISL_605274 | USA/UT-UPHL-2009596/2020 | 2020-07-19 | North America/USA/Utah/ | Utah Public Health Laboratory | Utah Public Health Laboratory | Erin L. Young et al |
| EPI_ISL_605278 | USA/UT-UPHL-2009600/2020 | 2020-07-19 | North America/USA/Utah/ | Utah Public Health Laboratory | Utah Public Health Laboratory | Erin L. Young et al |
| EPI_ISL_605286 | USA/UT-UPHL-2009608/2020 | 2020-07-19 | North America/USA/Utah/ | Utah Public Health Laboratory | Utah Public Health Laboratory | Erin L. Young et al |
| EPI_ISL_605294 | USA/UT-UPHL-2009616/2020 | 2020-07-18 | North America/USA/Utah/ | Utah Public Health Laboratory | Utah Public Health Laboratory | Erin L. Young et al |
| EPI_ISL_605300 | USA/UT-UPHL-2009623/2020 | 2020-07-19 | North America/USA/Utah/ | Utah Public Health Laboratory | Utah Public Health Laboratory | Erin L. Young et al |
| EPI_ISL_605314 | USA/UT-UPHL-2009639/2020 | 2020-07-19 | North America/USA/Utah/ | Utah Public Health Laboratory | Utah Public Health Laboratory | Erin L. Young et al |
| EPI_ISL_605317 | USA/UT-UPHL-2009642/2020 | 2020-07-18 | North America/USA/Utah/ | Utah Public Health Laboratory | Utah Public Health Laboratory | Erin L. Young et al |
| EPI_ISL_605321 | USA/UT-UPHL-2009646/2020 | 2020-07-26 | North America/USA/Utah/ | Utah Public Health Laboratory | Utah Public Health Laboratory | Erin L. Young et al |
| EPI_ISL_605327 | USA/UT-UPHL-2009652/2020 | 2020-07-26 | North America/USA/Utah/ | Utah Public Health Laboratory | Utah Public Health Laboratory | Erin L. Young et al |
| EPI_ISL_605328 | USA/UT-UPHL-2009653/2020 | 2020-07-27 | North America/USA/Utah/ | Utah Public Health Laboratory | Utah Public Health Laboratory | Erin L. Young et al |
| EPI_ISL_605331 | USA/UT-UPHL-2009656/2020 | 2020-07-26 | North America/USA/Utah/ | Utah Public Health Laboratory | Utah Public Health Laboratory | Erin L. Young et al |
| EPI_ISL_605337 | USA/UT-UPHL-2009662/2020 | 2020-07-26 | North America/USA/Utah/ | Utah Public Health Laboratory | Utah Public Health Laboratory | Erin L. Young et al |
| EPI_ISL_605354 | USA/UT-UPHL-2009679/2020 | 2020-07-26 | North America/USA/Utah/ | Utah Public Health Laboratory | Utah Public Health Laboratory | Erin L. Young et al |
| EPI_ISL_605376 | USA/UT-UPHL-2009703/2020 | 2020-07-25 | North America/USA/Utah/ | Utah Public Health Laboratory | Utah Public Health Laboratory | Erin L. Young et al |
| EPI_ISL_605380 | USA/UT-UPHL-2009707/2020 | 2020-07-27 | North America/USA/Utah/ | Utah Public Health Laboratory | Utah Public Health Laboratory | Erin L. Young et al |
| EPI_ISL_605386 | USA/UT-UPHL-2009713/2020 | 2020-07-26 | North America/USA/Utah/ | Utah Public Health Laboratory | Utah Public Health Laboratory | Erin L. Young et al |
| EPI_ISL_605389 | USA/UT-UPHL-2009717/2020 | 2020-07-26 | North America/USA/Utah/ | Utah Public Health Laboratory | Utah Public Health Laboratory | Erin L. Young et al |
| EPI_ISL_605397 | USA/UT-UPHL-2009725/2020 | 2020-07-25 | North America/USA/Utah/ | Utah Public Health Laboratory | Utah Public Health Laboratory | Erin L. Young et al |

| | | | | | | |
|---|---|---|---|---|---|---|
| EPI_ISL_605398 | USA/UT-UPHL-2009726/2020 | 2020-07-25 | North America/USA/Utah/ | Utah Public Health Laboratory | Utah Public Health Laboratory | Erin L. Young et al |
| EPI_ISL_605403 | USA/UT-UPHL-2009731/2020 | 2020-07-27 | North America/USA/Utah/ | Utah Public Health Laboratory | Utah Public Health Laboratory | Erin L. Young et al |
| EPI_ISL_605409 | USA/WI-UW-1531/2020 | 2020-09-10 | North America/USA/Wisconsin/Milwauk | University of Wisconsin-Madison AIDS Vaccine Research Laboratories | University of Wisconsin-Madison AIDS Vaccine Research Laboratories | Gage Moreno et al |
| EPI_ISL_605414 | USA/WI-UW-1536/2020 | 2020-09-14 | North America/USA/Wisconsin/Milwauk | University of Wisconsin-Madison AIDS Vaccine Research Laboratories | University of Wisconsin-Madison AIDS Vaccine Research Laboratories | Gage Moreno et al |
| EPI_ISL_605415 | USA/WI-UW-1537/2020 | 2020-09-14 | North America/USA/Wisconsin/Milwauk | University of Wisconsin-Madison AIDS Vaccine Research Laboratories | University of Wisconsin-Madison AIDS Vaccine Research Laboratories | Gage Moreno et al |
| EPI_ISL_605417 | USA/WI-UW-1539/2020 | 2020-09-14 | North America/USA/Wisconsin/Milwauk | University of Wisconsin-Madison AIDS Vaccine Research Laboratories | University of Wisconsin-Madison AIDS Vaccine Research Laboratories | Gage Moreno et al |
| EPI_ISL_605419 | USA/WI-UW-1541/2020 | 2020-09-14 | North America/USA/Wisconsin/Milwauk | University of Wisconsin-Madison AIDS Vaccine Research Laboratories | University of Wisconsin-Madison AIDS Vaccine Research Laboratories | Gage Moreno et al |
| EPI_ISL_605422 | USA/WI-UW-1544/2020 | 2020-09-15 | North America/USA/Wisconsin/Milwauk | University of Wisconsin-Madison AIDS Vaccine Research Laboratories | University of Wisconsin-Madison AIDS Vaccine Research Laboratories | Gage Moreno et al |
| EPI_ISL_605425 | USA/WI-UW-1547/2020 | 2020-09-15 | North America/USA/Wisconsin/Milwauk | University of Wisconsin-Madison AIDS Vaccine Research Laboratories | University of Wisconsin-Madison AIDS Vaccine Research Laboratories | Gage Moreno et al |

| | | | | | | |
|---|---|---|---|---|---|---|
| EPI_ISL_605428 | USA/WI-UW-1550/2020 | 2020-09-15 | North America/USA/Wisconsin/Milwauk | University of Wisconsin-Madison AIDS Vaccine Research Laboratories | University of Wisconsin-Madison AIDS Vaccine Research Laboratories | Gage Moreno et al |
| EPI_ISL_605433 | USA/WI-UW-1555/2020 | 2020-09-15 | North America/USA/Wisconsin/Milwauk | University of Wisconsin-Madison AIDS Vaccine Research Laboratories | University of Wisconsin-Madison AIDS Vaccine Research Laboratories | Gage Moreno et al |
| EPI_ISL_605434 | USA/WI-UW-1556/2020 | 2020-09-15 | North America/USA/Wisconsin/Milwauk | University of Wisconsin-Madison AIDS Vaccine Research Laboratories | University of Wisconsin-Madison AIDS Vaccine Research Laboratories | Gage Moreno et al |
| EPI_ISL_605442 | USA/WI-UW-1564/2020 | 2020-09-16 | North America/USA/Wisconsin/Milwauk | University of Wisconsin-Madison AIDS Vaccine Research Laboratories | University of Wisconsin-Madison AIDS Vaccine Research Laboratories | Gage Moreno et al |
| EPI_ISL_605443 | USA/WI-UW-1565/2020 | 2020-09-16 | North America/USA/Wisconsin/Milwauk | University of Wisconsin-Madison AIDS Vaccine Research Laboratories | University of Wisconsin-Madison AIDS Vaccine Research Laboratories | Gage Moreno et al |
| EPI_ISL_605444 | USA/WI-UW-1566/2020 | 2020-09-16 | North America/USA/Wisconsin/Milwauk | University of Wisconsin-Madison AIDS Vaccine Research Laboratories | University of Wisconsin-Madison AIDS Vaccine Research Laboratories | Gage Moreno et al |
| EPI_ISL_605445 | USA/WI-UW-1567/2020 | 2020-09-16 | North America/USA/Wisconsin/Milwauk | University of Wisconsin-Madison AIDS Vaccine Research Laboratories | University of Wisconsin-Madison AIDS Vaccine Research Laboratories | Gage Moreno et al |
| EPI_ISL_605460 | USA/WI-UW-1583/2020 | 2020-09-17 | North America/USA/Wisconsin/Milwauk | University of Wisconsin-Madison AIDS Vaccine Research Laboratories | University of Wisconsin-Madison AIDS Vaccine Research Laboratories | Gage Moreno et al |

| | | | | | | |
|---|---|---|---|---|---|---|
| EPI_ISL_605461 | USA/WI-UW-1584/2020 | 2020-09-17 | North America/USA/Wisconsin/Milwauk | University of Wisconsin-Madison AIDS Vaccine Research Laboratories | University of Wisconsin-Madison AIDS Vaccine Research Laboratories | Gage Moreno et al |
| EPI_ISL_605468 | USA/WI-UW-1591/2020 | 2020-09-18 | North America/USA/Wisconsin/Milwauk | University of Wisconsin-Madison AIDS Vaccine Research Laboratories | University of Wisconsin-Madison AIDS Vaccine Research Laboratories | Gage Moreno et al |
| EPI_ISL_605469 | USA/WI-UW-1592/2020 | 2020-09-18 | North America/USA/Wisconsin/Milwauk | University of Wisconsin-Madison AIDS Vaccine Research Laboratories | University of Wisconsin-Madison AIDS Vaccine Research Laboratories | Gage Moreno et al |
| EPI_ISL_605470 | USA/WI-UW-1593/2020 | 2020-09-18 | North America/USA/Wisconsin/Milwauk | University of Wisconsin-Madison AIDS Vaccine Research Laboratories | University of Wisconsin-Madison AIDS Vaccine Research Laboratories | Gage Moreno et al |
| EPI_ISL_605473 | USA/WI-UW-1596/2020 | 2020-09-21 | North America/USA/Wisconsin/Milwauk | University of Wisconsin-Madison AIDS Vaccine Research Laboratories | University of Wisconsin-Madison AIDS Vaccine Research Laboratories | Gage Moreno et al |
| EPI_ISL_605475 | USA/WI-UW-1598/2020 | 2020-09-21 | North America/USA/Wisconsin/Milwauk | University of Wisconsin-Madison AIDS Vaccine Research Laboratories | University of Wisconsin-Madison AIDS Vaccine Research Laboratories | Gage Moreno et al |
| EPI_ISL_605476 | USA/WI-UW-1599/2020 | 2020-09-21 | North America/USA/Wisconsin/Milwauk | University of Wisconsin-Madison AIDS Vaccine Research Laboratories | University of Wisconsin-Madison AIDS Vaccine Research Laboratories | Gage Moreno et al |
| EPI_ISL_605477 | USA/WI-UW-1600/2020 | 2020-09-23 | North America/USA/Wisconsin/Milwauk | University of Wisconsin-Madison AIDS Vaccine Research Laboratories | University of Wisconsin-Madison AIDS Vaccine Research Laboratories | Gage Moreno et al |

| | | | | | | |
|---|---|---|---|---|---|---|
| EPI_ISL_605482 | USA/WI-UW-1605/2020 | 2020-09-24 | North America/USA/Wisconsin/Milwauk | University of Wisconsin-Madison AIDS Vaccine Research Laboratories | University of Wisconsin-Madison AIDS Vaccine Research Laboratories | Gage Moreno et al |
| EPI_ISL_605485 | USA/WI-UW-1608/2020 | 2020-09-17 | North America/USA/Wisconsin/Dane Co | University of Wisconsin-Madison AIDS Vaccine Research Laboratories | University of Wisconsin-Madison AIDS Vaccine Research Laboratories | Gage Moreno et al |
| EPI_ISL_605487 | USA/WI-UW-1610/2020 | 2020-09-17 | North America/USA/Wisconsin/Dane Co | University of Wisconsin-Madison AIDS Vaccine Research Laboratories | University of Wisconsin-Madison AIDS Vaccine Research Laboratories | Gage Moreno et al |
| EPI_ISL_605490 | USA/WI-UW-1620/2020 | 2020-09-17 | North America/USA/Wisconsin/Jefferson | University of Wisconsin-Madison AIDS Vaccine Research Laboratories | University of Wisconsin-Madison AIDS Vaccine Research Laboratories | Gage Moreno et al |
| EPI_ISL_605497 | USA/WI-UW-1623/2020 | 2020-09-24 | North America/USA/Wisconsin/Columbia | University of Wisconsin-Madison AIDS Vaccine Research Laboratories | University of Wisconsin-Madison AIDS Vaccine Research Laboratories | Gage Moreno et al |
| EPI_ISL_605498 | USA/IL-UW-1624/2020 | 2020-09-25 | North America/USA/Illinois/Winnebago | University of Wisconsin-Madison AIDS Vaccine Research Laboratories | University of Wisconsin-Madison AIDS Vaccine Research Laboratories | Gage Moreno et al |
| EPI_ISL_605508 | USA/WI-UW-1634/2020 | 2020-09-09 | North America/USA/Wisconsin/Dane Co | University of Wisconsin-Madison AIDS Vaccine Research Laboratories | University of Wisconsin-Madison AIDS Vaccine Research Laboratories | Gage Moreno et al |
| EPI_ISL_605512 | USA/WI-UW-1638/2020 | 2020-09-11 | North America/USA/Wisconsin/Dane Co | University of Wisconsin-Madison AIDS Vaccine Research Laboratories | University of Wisconsin-Madison AIDS Vaccine Research Laboratories | Gage Moreno et al |

| | | | | | | |
|---|---|---|---|---|---|---|
| EPI_ISL_605519 | USA/WI-UW-1645/2020 | 2020-09-10 | North America/USA/Wisconsin/Dane Co | University of Wisconsin-Madison AIDS Vaccine Research Laboratories | University of Wisconsin-Madison AIDS Vaccine Research Laboratories | Gage Moreno et al |
| EPI_ISL_605522 | USA/WI-UW-1648/2020 | 2020-09-09 | North America/USA/Wisconsin/Dane Co | University of Wisconsin-Madison AIDS Vaccine Research Laboratories | University of Wisconsin-Madison AIDS Vaccine Research Laboratories | Gage Moreno et al |
| EPI_ISL_605523 | USA/WI-UW-1649/2020 | 2020-09-09 | North America/USA/Wisconsin/Dane Co | University of Wisconsin-Madison AIDS Vaccine Research Laboratories | University of Wisconsin-Madison AIDS Vaccine Research Laboratories | Gage Moreno et al |
| EPI_ISL_605524 | USA/WI-UW-1650/2020 | 2020-09-10 | North America/USA/Wisconsin/Dane Co | University of Wisconsin-Madison AIDS Vaccine Research Laboratories | University of Wisconsin-Madison AIDS Vaccine Research Laboratories | Gage Moreno et al |
| EPI_ISL_605525 | USA/WI-UW-1651/2020 | 2020-09-10 | North America/USA/Wisconsin/Dane Co | University of Wisconsin-Madison AIDS Vaccine Research Laboratories | University of Wisconsin-Madison AIDS Vaccine Research Laboratories | Gage Moreno et al |
| EPI_ISL_605526 | USA/WI-UW-1652/2020 | 2020-09-10 | North America/USA/Wisconsin/Dane Co | University of Wisconsin-Madison AIDS Vaccine Research Laboratories | University of Wisconsin-Madison AIDS Vaccine Research Laboratories | Gage Moreno et al |
| EPI_ISL_605527 | USA/WI-UW-1653/2020 | 2020-09-10 | North America/USA/Wisconsin/Dane Co | University of Wisconsin-Madison AIDS Vaccine Research Laboratories | University of Wisconsin-Madison AIDS Vaccine Research Laboratories | Gage Moreno et al |
| EPI_ISL_605529 | USA/WI-UW-1655/2020 | 2020-09-10 | North America/USA/Wisconsin/Dane Co | University of Wisconsin-Madison AIDS Vaccine Research Laboratories | University of Wisconsin-Madison AIDS Vaccine Research Laboratories | Gage Moreno et al |

| | | | | | | |
|---|---|---|---|---|---|---|
| EPI_ISL_605530 | USA/WI-UW-1656/2020 | 2020-09-10 | North America/USA/Wisconsin/Dane Co | University of Wisconsin-Madison AIDS Vaccine Research Laboratories | University of Wisconsin-Madison AIDS Vaccine Research Laboratories | Gage Moreno et al |
| EPI_ISL_605532 | USA/WI-UW-1658/2020 | 2020-09-10 | North America/USA/Wisconsin/Dane Co | University of Wisconsin-Madison AIDS Vaccine Research Laboratories | University of Wisconsin-Madison AIDS Vaccine Research Laboratories | Gage Moreno et al |
| EPI_ISL_605536 | USA/WI-UW-1662/2020 | 2020-09-16 | North America/USA/Wisconsin/Dane Co | University of Wisconsin-Madison AIDS Vaccine Research Laboratories | University of Wisconsin-Madison AIDS Vaccine Research Laboratories | Gage Moreno et al |
| EPI_ISL_605540 | USA/WI-UW-1666/2020 | 2020-09-16 | North America/USA/Wisconsin/Dane Co | University of Wisconsin-Madison AIDS Vaccine Research Laboratories | University of Wisconsin-Madison AIDS Vaccine Research Laboratories | Gage Moreno et al |
| EPI_ISL_605545 | USA/WI-UW-1671/2020 | 2020-09-17 | North America/USA/Wisconsin/Dane Co | University of Wisconsin-Madison AIDS Vaccine Research Laboratories | University of Wisconsin-Madison AIDS Vaccine Research Laboratories | Gage Moreno et al |
| EPI_ISL_605547 | USA/WI-UW-1673/2020 | 2020-09-11 | North America/USA/Wisconsin/Dane Co | University of Wisconsin-Madison AIDS Vaccine Research Laboratories | University of Wisconsin-Madison AIDS Vaccine Research Laboratories | Gage Moreno et al |
| EPI_ISL_605548 | USA/WI-UW-1674/2020 | 2020-09-11 | North America/USA/Wisconsin/Dane Co | University of Wisconsin-Madison AIDS Vaccine Research Laboratories | University of Wisconsin-Madison AIDS Vaccine Research Laboratories | Gage Moreno et al |
| EPI_ISL_605549 | USA/WI-UW-1675/2020 | 2020-09-11 | North America/USA/Wisconsin/Dane Co | University of Wisconsin-Madison AIDS Vaccine Research Laboratories | University of Wisconsin-Madison AIDS Vaccine Research Laboratories | Gage Moreno et al |

| | | | | | | |
|---|---|---|---|---|---|---|
| EPI_ISL_605553 | USA/WI-UW-1680/2020 | 2020-09-11 | North America/USA/Wisconsin/Dane Co | University of Wisconsin-Madison AIDS Vaccine Research Laboratories | University of Wisconsin-Madison AIDS Vaccine Research Laboratories | Gage Moreno et al |
| EPI_ISL_605556 | USA/WI-UW-1683/2020 | 2020-09-11 | North America/USA/Wisconsin/Dane Co | University of Wisconsin-Madison AIDS Vaccine Research Laboratories | University of Wisconsin-Madison AIDS Vaccine Research Laboratories | Gage Moreno et al |
| EPI_ISL_605557 | USA/WI-UW-1684/2020 | 2020-09-11 | North America/USA/Wisconsin/Dane Co | University of Wisconsin-Madison AIDS Vaccine Research Laboratories | University of Wisconsin-Madison AIDS Vaccine Research Laboratories | Gage Moreno et al |
| EPI_ISL_605561 | USA/WI-UW-1688/2020 | 2020-09-11 | North America/USA/Wisconsin/Dane Co | University of Wisconsin-Madison AIDS Vaccine Research Laboratories | University of Wisconsin-Madison AIDS Vaccine Research Laboratories | Gage Moreno et al |
| EPI_ISL_605563 | USA/WI-UW-1690/2020 | 2020-09-11 | North America/USA/Wisconsin/Dane Co | University of Wisconsin-Madison AIDS Vaccine Research Laboratories | University of Wisconsin-Madison AIDS Vaccine Research Laboratories | Gage Moreno et al |
| EPI_ISL_605564 | USA/WI-UW-1691/2020 | 2020-09-11 | North America/USA/Wisconsin/Dane Co | University of Wisconsin-Madison AIDS Vaccine Research Laboratories | University of Wisconsin-Madison AIDS Vaccine Research Laboratories | Gage Moreno et al |
| EPI_ISL_605566 | USA/WI-UW-1693/2020 | 2020-09-11 | North America/USA/Wisconsin/Dane Co | University of Wisconsin-Madison AIDS Vaccine Research Laboratories | University of Wisconsin-Madison AIDS Vaccine Research Laboratories | Gage Moreno et al |
| EPI_ISL_605571 | USA/WI-UW-1698/2020 | 2020-09-11 | North America/USA/Wisconsin/Dane Co | University of Wisconsin-Madison AIDS Vaccine Research Laboratories | University of Wisconsin-Madison AIDS Vaccine Research Laboratories | Gage Moreno et al |

| | | | | | | |
|---|---|---|---|---|---|---|
| EPI_ISL_605577 | USA/WI-UW-1704/2020 | 2020-09-11 | North America/USA/Wisconsin/Dane Co | University of Wisconsin-Madison AIDS Vaccine Research Laboratories | University of Wisconsin-Madison AIDS Vaccine Research Laboratories | Gage Moreno et al |
| EPI_ISL_605578 | USA/WI-UW-1705/2020 | 2020-09-09 | North America/USA/Wisconsin/Dane Co | University of Wisconsin-Madison AIDS Vaccine Research Laboratories | University of Wisconsin-Madison AIDS Vaccine Research Laboratories | Gage Moreno et al |
| EPI_ISL_605581 | USA/WI-UW-1708/2020 | 2020-09-11 | North America/USA/Wisconsin/Dane Co | University of Wisconsin-Madison AIDS Vaccine Research Laboratories | University of Wisconsin-Madison AIDS Vaccine Research Laboratories | Gage Moreno et al |
| EPI_ISL_605582 | USA/WI-UW-1709/2020 | 2020-09-11 | North America/USA/Wisconsin/Dane Co | University of Wisconsin-Madison AIDS Vaccine Research Laboratories | University of Wisconsin-Madison AIDS Vaccine Research Laboratories | Gage Moreno et al |
| EPI_ISL_605584 | USA/WI-UW-1711/2020 | 2020-09-11 | North America/USA/Wisconsin/Dane Co | University of Wisconsin-Madison AIDS Vaccine Research Laboratories | University of Wisconsin-Madison AIDS Vaccine Research Laboratories | Gage Moreno et al |
| EPI_ISL_605592 | USA/WI-UW-1719/2020 | 2020-09-11 | North America/USA/Wisconsin/Dane Co | University of Wisconsin-Madison AIDS Vaccine Research Laboratories | University of Wisconsin-Madison AIDS Vaccine Research Laboratories | Gage Moreno et al |
| EPI_ISL_605594 | USA/WI-UW-1721/2020 | 2020-09-09 | North America/USA/Wisconsin/Dane Co | University of Wisconsin-Madison AIDS Vaccine Research Laboratories | University of Wisconsin-Madison AIDS Vaccine Research Laboratories | Gage Moreno et al |
| EPI_ISL_605596 | USA/WI-UW-1723/2020 | 2020-09-09 | North America/USA/Wisconsin/Dane Co | University of Wisconsin-Madison AIDS Vaccine Research Laboratories | University of Wisconsin-Madison AIDS Vaccine Research Laboratories | Gage Moreno et al |

| | | | | | | |
|---|---|---|---|---|---|---|
| EPI_ISL_605600 | USA/WI-UW-1727/2020 | 2020-09-11 | North America/USA/Wisconsin/Dane Co | University of Wisconsin-Madison AIDS Vaccine Research Laboratories | University of Wisconsin-Madison AIDS Vaccine Research Laboratories | Gage Moreno et al |
| EPI_ISL_605610 | USA/WI-UW-1737/2020 | 2020-09-08 | North America/USA/Wisconsin/Dane Co | University of Wisconsin-Madison AIDS Vaccine Research Laboratories | University of Wisconsin-Madison AIDS Vaccine Research Laboratories | Gage Moreno et al |
| EPI_ISL_605611 | USA/WI-UW-1738/2020 | 2020-09-10 | North America/USA/Wisconsin/Dane Co | University of Wisconsin-Madison AIDS Vaccine Research Laboratories | University of Wisconsin-Madison AIDS Vaccine Research Laboratories | Gage Moreno et al |
| EPI_ISL_605616 | USA/WI-UW-1743/2020 | 2020-09-22 | North America/USA/Wisconsin/Dane Co | University of Wisconsin-Madison AIDS Vaccine Research Laboratories | University of Wisconsin-Madison AIDS Vaccine Research Laboratories | Gage Moreno et al |
| EPI_ISL_605617 | USA/WI-UW-1744/2020 | 2020-09-16 | North America/USA/Wisconsin/Dane Co | University of Wisconsin-Madison AIDS Vaccine Research Laboratories | University of Wisconsin-Madison AIDS Vaccine Research Laboratories | Gage Moreno et al |
| EPI_ISL_605620 | USA/WI-UW-1747/2020 | 2020-09-16 | North America/USA/Wisconsin/Dane Co | University of Wisconsin-Madison AIDS Vaccine Research Laboratories | University of Wisconsin-Madison AIDS Vaccine Research Laboratories | Gage Moreno et al |
| EPI_ISL_605625 | USA/WI-UW-1752/2020 | 2020-09-17 | North America/USA/Wisconsin/Dane Co | University of Wisconsin-Madison AIDS Vaccine Research Laboratories | University of Wisconsin-Madison AIDS Vaccine Research Laboratories | Gage Moreno et al |
| EPI_ISL_605626 | USA/WI-UW-1753/2020 | 2020-09-17 | North America/USA/Wisconsin/Dane Co | University of Wisconsin-Madison AIDS Vaccine Research Laboratories | University of Wisconsin-Madison AIDS Vaccine Research Laboratories | Gage Moreno et al |

| | | | | | | |
|---|---|---|---|---|---|---|
| EPI_ISL_605627 | USA/WI-UW-1754/2020 | 2020-09-16 | North America/USA/Wisconsin/Dane Co | University of Wisconsin-Madison AIDS Vaccine Research Laboratories | University of Wisconsin-Madison AIDS Vaccine Research Laboratories | Gage Moreno et al |
| EPI_ISL_605629 | USA/WI-UW-1756/2020 | 2020-09-16 | North America/USA/Wisconsin/Dane Co | University of Wisconsin-Madison AIDS Vaccine Research Laboratories | University of Wisconsin-Madison AIDS Vaccine Research Laboratories | Gage Moreno et al |
| EPI_ISL_605630 | USA/WI-UW-1757/2020 | 2020-09-17 | North America/USA/Wisconsin/Dane Co | University of Wisconsin-Madison AIDS Vaccine Research Laboratories | University of Wisconsin-Madison AIDS Vaccine Research Laboratories | Gage Moreno et al |
| EPI_ISL_605641 | USA/WI-UW-1769/2020 | 2020-09-16 | North America/USA/Wisconsin/Dane Co | University of Wisconsin-Madison AIDS Vaccine Research Laboratories | University of Wisconsin-Madison AIDS Vaccine Research Laboratories | Gage Moreno et al |
| EPI_ISL_605647 | USA/WI-UW-1775/2020 | 2020-09-14 | North America/USA/Wisconsin/Dane Co | University of Wisconsin-Madison AIDS Vaccine Research Laboratories | University of Wisconsin-Madison AIDS Vaccine Research Laboratories | Gage Moreno et al |
| EPI_ISL_605648 | USA/WI-UW-1776/2020 | 2020-09-14 | North America/USA/Wisconsin/Dane Co | University of Wisconsin-Madison AIDS Vaccine Research Laboratories | University of Wisconsin-Madison AIDS Vaccine Research Laboratories | Gage Moreno et al |
| EPI_ISL_605649 | USA/WI-UW-1777/2020 | 2020-09-14 | North America/USA/Wisconsin/Dane Co | University of Wisconsin-Madison AIDS Vaccine Research Laboratories | University of Wisconsin-Madison AIDS Vaccine Research Laboratories | Gage Moreno et al |
| EPI_ISL_605654 | USA/WI-UW-1782/2020 | 2020-09-16 | North America/USA/Wisconsin/Dane Co | University of Wisconsin-Madison AIDS Vaccine Research Laboratories | University of Wisconsin-Madison AIDS Vaccine Research Laboratories | Gage Moreno et al |

| | | | | | | |
|---|---|---|---|---|---|---|
| EPI_ISL_605662 | USA/WI-UW-1790/2020 | 2020-09-14 | North America/USA/Wisconsin/Dane Co | University of Wisconsin-Madison AIDS Vaccine Research Laboratories | University of Wisconsin-Madison AIDS Vaccine Research Laboratories | Gage Moreno et al |
| EPI_ISL_605663 | USA/WI-UW-1791/2020 | 2020-09-14 | North America/USA/Wisconsin/Dane Co | University of Wisconsin-Madison AIDS Vaccine Research Laboratories | University of Wisconsin-Madison AIDS Vaccine Research Laboratories | Gage Moreno et al |
| EPI_ISL_605665 | USA/WI-UW-1793/2020 | 2020-09-15 | North America/USA/Wisconsin/Dane Co | University of Wisconsin-Madison AIDS Vaccine Research Laboratories | University of Wisconsin-Madison AIDS Vaccine Research Laboratories | Gage Moreno et al |
| EPI_ISL_605669 | USA/WI-UW-1797/2020 | 2020-09-14 | North America/USA/Wisconsin/Dane Co | University of Wisconsin-Madison AIDS Vaccine Research Laboratories | University of Wisconsin-Madison AIDS Vaccine Research Laboratories | Gage Moreno et al |
| EPI_ISL_605679 | USA/WI-UW-1807/2020 | 2020-09-11 | North America/USA/Wisconsin/Dane Co | University of Wisconsin-Madison AIDS Vaccine Research Laboratories | University of Wisconsin-Madison AIDS Vaccine Research Laboratories | Gage Moreno et al |
| EPI_ISL_605685 | USA/WI-UW-1813/2020 | 2020-09-09 | North America/USA/Wisconsin/Dane Co | University of Wisconsin-Madison AIDS Vaccine Research Laboratories | University of Wisconsin-Madison AIDS Vaccine Research Laboratories | Gage Moreno et al |
| EPI_ISL_605687 | USA/WI-UW-1815/2020 | 2020-09-09 | North America/USA/Wisconsin/Dane Co | University of Wisconsin-Madison AIDS Vaccine Research Laboratories | University of Wisconsin-Madison AIDS Vaccine Research Laboratories | Gage Moreno et al |
| EPI_ISL_605689 | USA/WI-UW-1817/2020 | 2020-09-09 | North America/USA/Wisconsin/Dane Co | University of Wisconsin-Madison AIDS Vaccine Research Laboratories | University of Wisconsin-Madison AIDS Vaccine Research Laboratories | Gage Moreno et al |

| | | | | | | |
|---|---|---|---|---|---|---|
| EPI_ISL_605694 | USA/WI-UW-1822/2020 | 2020-09-09 | North America/USA/Wisconsin/Dane Co | University of Wisconsin-Madison AIDS Vaccine Research Laboratories | University of Wisconsin-Madison AIDS Vaccine Research Laboratories | Gage Moreno et al |
| EPI_ISL_605705 | USA/WI-UW-1833/2020 | 2020-09-09 | North America/USA/Wisconsin/Dane Co | University of Wisconsin-Madison AIDS Vaccine Research Laboratories | University of Wisconsin-Madison AIDS Vaccine Research Laboratories | Gage Moreno et al |
| EPI_ISL_605709 | USA/WI-UW-1837/2020 | 2020-09-11 | North America/USA/Wisconsin/Dane Co | University of Wisconsin-Madison AIDS Vaccine Research Laboratories | University of Wisconsin-Madison AIDS Vaccine Research Laboratories | Gage Moreno et al |
| EPI_ISL_605714 | USA/WI-UW-1842/2020 | 2020-09-08 | North America/USA/Wisconsin/Dane Co | University of Wisconsin-Madison AIDS Vaccine Research Laboratories | University of Wisconsin-Madison AIDS Vaccine Research Laboratories | Gage Moreno et al |
| EPI_ISL_605717 | USA/WI-UW-1845/2020 | 2020-09-10 | North America/USA/Wisconsin/Dane Co | University of Wisconsin-Madison AIDS Vaccine Research Laboratories | University of Wisconsin-Madison AIDS Vaccine Research Laboratories | Gage Moreno et al |
| EPI_ISL_605720 | USA/WI-UW-1848/2020 | 2020-09-10 | North America/USA/Wisconsin/Dane Co | University of Wisconsin-Madison AIDS Vaccine Research Laboratories | University of Wisconsin-Madison AIDS Vaccine Research Laboratories | Gage Moreno et al |
| EPI_ISL_605721 | USA/WI-UW-1849/2020 | 2020-09-09 | North America/USA/Wisconsin/Dane Co | University of Wisconsin-Madison AIDS Vaccine Research Laboratories | University of Wisconsin-Madison AIDS Vaccine Research Laboratories | Gage Moreno et al |
| EPI_ISL_605723 | USA/WI-UW-1851/2020 | 2020-09-16 | North America/USA/Wisconsin/Dane Co | University of Wisconsin-Madison AIDS Vaccine Research Laboratories | University of Wisconsin-Madison AIDS Vaccine Research Laboratories | Gage Moreno et al |

| | | | | | | |
|---|---|---|---|---|---|---|
| EPI_ISL_605724 | USA/WI-UW-1852/2020 | 2020-09-17 | North America/USA/Wisconsin/Dane Co | University of Wisconsin-Madison AIDS Vaccine Research Laboratories | University of Wisconsin-Madison AIDS Vaccine Research Laboratories | Gage Moreno et al |
| EPI_ISL_605726 | USA/WI-UW-1854/2020 | 2020-09-17 | North America/USA/Wisconsin/Dane Co | University of Wisconsin-Madison AIDS Vaccine Research Laboratories | University of Wisconsin-Madison AIDS Vaccine Research Laboratories | Gage Moreno et al |
| EPI_ISL_605727 | USA/WI-UW-1855/2020 | 2020-09-16 | North America/USA/Wisconsin/Dane Co | University of Wisconsin-Madison AIDS Vaccine Research Laboratories | University of Wisconsin-Madison AIDS Vaccine Research Laboratories | Gage Moreno et al |
| EPI_ISL_605729 | USA/WI-UW-1857/2020 | 2020-09-17 | North America/USA/Wisconsin/Dane Co | University of Wisconsin-Madison AIDS Vaccine Research Laboratories | University of Wisconsin-Madison AIDS Vaccine Research Laboratories | Gage Moreno et al |
| EPI_ISL_605732 | USA/WI-UW-1860/2020 | 2020-09-14 | North America/USA/Wisconsin/Dane Co | University of Wisconsin-Madison AIDS Vaccine Research Laboratories | University of Wisconsin-Madison AIDS Vaccine Research Laboratories | Gage Moreno et al |
| EPI_ISL_605735 | USA/WI-UW-1863/2020 | 2020-09-15 | North America/USA/Wisconsin/Dane Co | University of Wisconsin-Madison AIDS Vaccine Research Laboratories | University of Wisconsin-Madison AIDS Vaccine Research Laboratories | Gage Moreno et al |
| EPI_ISL_605740 | USA/WI-UW-1868/2020 | 2020-09-09 | North America/USA/Wisconsin/Dane Co | University of Wisconsin-Madison AIDS Vaccine Research Laboratories | University of Wisconsin-Madison AIDS Vaccine Research Laboratories | Gage Moreno et al |
| EPI_ISL_605743 | USA/WI-UW-1871/2020 | 2020-09-11 | North America/USA/Wisconsin/Dane Co | University of Wisconsin-Madison AIDS Vaccine Research Laboratories | University of Wisconsin-Madison AIDS Vaccine Research Laboratories | Gage Moreno et al |

| | | | | | | |
|---|---|---|---|---|---|---|
| EPI_ISL_605745 | USA/WI-UW-1873/2020 | 2020-09-10 | North America/USA/Wisconsin/Milwauk | University of Wisconsin-Madison AIDS Vaccine Research Laboratories | University of Wisconsin-Madison AIDS Vaccine Research Laboratories | Gage Moreno et al |
| EPI_ISL_605751 | USA/WI-UW-1880/2020 | 2020-09-11 | North America/USA/Wisconsin/Milwauk | University of Wisconsin-Madison AIDS Vaccine Research Laboratories | University of Wisconsin-Madison AIDS Vaccine Research Laboratories | Gage Moreno et al |
| EPI_ISL_605764 | USA/WI-UW-1895/2020 | 2020-09-17 | North America/USA/Wisconsin/Dane Co | University of Wisconsin-Madison AIDS Vaccine Research Laboratories | University of Wisconsin-Madison AIDS Vaccine Research Laboratories | Gage Moreno et al |
| EPI_ISL_605775 | USA/WI-UW-1906/2020 | 2020-09-17 | North America/USA/Wisconsin/Dane Co | University of Wisconsin-Madison AIDS Vaccine Research Laboratories | University of Wisconsin-Madison AIDS Vaccine Research Laboratories | Gage Moreno et al |
| EPI_ISL_605780 | Egypt/C-CEIRS-19MOH/2020 | 2020-03-13 | Africa/Egypt/Cairo/ | CEIRS Data Processing and Coordinating Center, St. Jude Center of Excellence for Influenza Research and Surveillance (CEIRS) | CEIRS Data Processing and Coordinating Center, St. Jude Center of Excellence for Influenza Research and Surveillance (CEIRS) | Roshdy et al |
| EPI_ISL_605782 | Egypt/C-CEIRS-8MOH/2020 | 2020-05-17 | Africa/Egypt/Cairo/ | CEIRS Data Processing and Coordinating Center, St. Jude Center of Excellence for Influenza Research and Surveillance (CEIRS) | CEIRS Data Processing and Coordinating Center, St. Jude Center of Excellence for Influenza Research and Surveillance (CEIRS) | Roshdy et al |
| EPI_ISL_605783 | Bangladesh/JUST-GC91.124/2020 | 2020-08-08 | Asia/Bangladesh/Khulna/Jashore | Genome Center | Genome Center | Md. Shazid Hasan et al |

| | | | | | | |
|---|---|---|---|---|---|---|
| EPI_ISL_605786 | SouthAfrica/KRISP-K003682/2020 | 2020-09-26 | Africa/South Africa/KwaZulu-Natal/iLem | NHLS-IALCH | KRISP, KZN Research Innovation and Sequencing Platform | Giandhari J et al |
| EPI_ISL_605790 | SouthAfrica/KRISP-K003744/2020 | 2020-10-01 | Africa/South Africa/KwaZulu-Natal/Zulu | NHLS-IALCH | KRISP, KZN Research Innovation and Sequencing Platform | Giandhari J et al |
| EPI_ISL_605797 | Ireland/KY-NVRL-20G35628/2020 | 2020-02-20 | Europe/Ireland/Ireland/ | National Virus Reference Laboratory | Irish Coronavirus Sequencing Consortium - Helixworks | Sachin Chalapati et al |
| EPI_ISL_605804 | India/DL-ILBS-9306T/2020 | 2020-05-01 | Asia/India/Delhi/New Delhi | Clinical Virology Laboratory, Institute of Liver and Biliary Sciences | ILBS - IGIB | Ekta Gupta et al |
| EPI_ISL_605806 | India/DL-ILBS-1929M/2020 | 2020-05-01 | Asia/India/Delhi/New Delhi | Clinical Virology Laboratory, Institute of Liver and Biliary Sciences | ILBS - IGIB | Ekta Gupta et al |
| EPI_ISL_605891 | Bangladesh/DNAS-CPH-isl-486/2020 | 2020-04-29 | Asia/Bangladesh/Bangladesh/ | NGS Lab, DNA SOLUTION LTD. | NGS Lab, DNA SOLUTION LTD. | Khan et al |
| EPI_ISL_605897 | Bangladesh/DNAS-CPH-isl-515/2020 | 2020-04-29 | Asia/Bangladesh/Bangladesh/ | NGS Lab, DNA SOLUTION LTD. | NGS Lab, DNA SOLUTION LTD. | Khan et al |
| EPI_ISL_605900 | Bangladesh/DNAS-RAB-isl-86/2020 | 2020-04-29 | Asia/Bangladesh/Bangladesh/ | NGS Lab, DNA SOLUTION LTD. | NGS Lab, DNA SOLUTION LTD. | Khan et al |
| EPI_ISL_605904 | Bangladesh/DNAS-RAB-isl-114/2020 | 2020-04-29 | Asia/Bangladesh/Bangladesh/ | NGS Lab, DNA SOLUTION LTD. | NGS Lab, DNA SOLUTION LTD. | Khan et al |
| EPI_ISL_605908 | Bangladesh/DNAS-RAB-isl-82/2020 | 2020-04-29 | Asia/Bangladesh/Bangladesh/ | NGS Lab, DNA SOLUTION LTD. | NGS Lab, DNA SOLUTION LTD. | Khan et al |
| EPI_ISL_605909 | Bangladesh/DNAS-isl-113/2020 | 2020-05-24 | Asia/Bangladesh/Bangladesh/ | NGS Lab, DNA SOLUTION LTD. | NGS Lab, DNA SOLUTION LTD. | Khan et al |
| EPI_ISL_605910 | Bangladesh/DNAS-isl-1-003/2020 | 2020-05-24 | Asia/Bangladesh/Bangladesh/ | NGS Lab, DNA SOLUTION LTD. | NGS Lab, DNA SOLUTION LTD. | Khan et al |
| EPI_ISL_605911 | Bangladesh/DNAS-isl-75/2020 | 2020-05-24 | Asia/Bangladesh/Bangladesh/ | NGS Lab, DNA SOLUTION LTD. | NGS Lab, DNA SOLUTION LTD. | Khan et al |
| EPI_ISL_605912 | Bangladesh/DNAS-isl-95/2020 | 2020-05-24 | Asia/Bangladesh/Bangladesh/ | NGS Lab, DNA SOLUTION LTD. | NGS Lab, DNA SOLUTION LTD. | Khan et al |
| EPI_ISL_605913 | Bangladesh/DNAS-isl-116/2020 | 2020-05-24 | Asia/Bangladesh/Bangladesh/ | NGS Lab, DNA SOLUTION LTD. | NGS Lab, DNA SOLUTION LTD. | Khan et al |
| EPI_ISL_605915 | Bangladesh/DNAS-isl-6840/2020 | 2020-05-18 | Asia/Bangladesh/Bangladesh/ | NGS Lab, DNA SOLUTION LTD. | NGS Lab, DNA SOLUTION LTD. | Khan et al |

| | | | | | | |
|---|---|---|---|---|---|---|
| EPI_ISL_605917 | Bangladesh/DNAS-isl-8225/2020 | 2020-05-18 | Asia/Bangladesh/Bangladesh/ | NGS Lab, DNA SOLUTION LTD. | NGS Lab, DNA SOLUTION LTD. | Khan et al |
| EPI_ISL_605918 | Bangladesh/DNAS-isl-6300/2020 | 2020-05-18 | Asia/Bangladesh/Bangladesh/ | NGS Lab, DNA SOLUTION LTD. | NGS Lab, DNA SOLUTION LTD. | Khan et al |
| EPI_ISL_605922 | Bangladesh/DNAS-isl-6301/2020 | 2020-05-18 | Asia/Bangladesh/Bangladesh/ | NGS Lab, DNA SOLUTION LTD. | NGS Lab, DNA SOLUTION LTD. | Khan et al |
| EPI_ISL_605923 | Bangladesh/DNAS-isl-4/2020 | 2020-05-18 | Asia/Bangladesh/Bangladesh/ | NGS Lab, DNA SOLUTION LTD. | NGS Lab, DNA SOLUTION LTD. | Khan et al |
| EPI_ISL_605929 | Henan/HN01/2020 | 2020-02-24 | Asia/China/Henan/ | Department of Infectious Disease Prevention and Control, Henan Provincial Center for Disease Control and Prevention | Department of Infectious Disease Prevention and Control, Henan Provincial Center for Disease Control and Prevention | Li et al |
| EPI_ISL_605930 | Henan/HN04/2020 | 2020-02-24 | Asia/China/Henan/ | Department of Infectious Disease Prevention and Control, Henan Provincial Center for Disease Control and Prevention | Department of Infectious Disease Prevention and Control, Henan Provincial Center for Disease Control and Prevention | Li et al |
| EPI_ISL_606043 | England/ALDP-A87C9A/2020 | 2020-10-17 | Europe/United Kingdom/England/ | Lighthouse Lab in Alderley Park | Wellcome Sanger Institute for the COVID-19 Genomics UK (COG-UK) consortium | Jacquelyn Wynn et al |
| EPI_ISL_606148 | England/ALDP-A8792A/2020 | 2020-10-17 | Europe/United Kingdom/England/ | Lighthouse Lab in Alderley Park | Wellcome Sanger Institute for the COVID-19 Genomics UK (COG-UK) consortium | Jacquelyn Wynn et al |
| EPI_ISL_606257 | England/ALDP-A8777B/2020 | 2020-10-16 | Europe/United Kingdom/England/ | Lighthouse Lab in Alderley Park | Wellcome Sanger Institute for the COVID-19 Genomics UK (COG-UK) consortium | Jacquelyn Wynn et al |

| | | | | | | |
|---|---|---|---|---|---|---|
| EPI_ISL_606346 | England/MILK-A7DC1F/2020 | 2020-10-13 | Europe/United Kingdom/England/ | Lighthouse Lab in Milton Keynes | Wellcome Sanger Institute for the COVID-19 Genomics UK (COG-UK) consortium | The Lighthouse Lab in Milton Keynes et al |
| EPI_ISL_606416 | England/MILK-A7DBF4/2020 | 2020-10-13 | Europe/United Kingdom/England/ | Lighthouse Lab in Milton Keynes | Wellcome Sanger Institute for the COVID-19 Genomics UK (COG-UK) consortium | The Lighthouse Lab in Milton Keynes et al |
| EPI_ISL_606487 | England/MILK-A7AEC1/2020 | 2020-10-15 | Europe/United Kingdom/England/ | Lighthouse Lab in Milton Keynes | Wellcome Sanger Institute for the COVID-19 Genomics UK (COG-UK) consortium | The Lighthouse Lab in Milton Keynes et al |
| EPI_ISL_606602 | England/MILK-A7DBE5/2020 | 2020-10-13 | Europe/United Kingdom/England/ | Lighthouse Lab in Milton Keynes | Wellcome Sanger Institute for the COVID-19 Genomics UK (COG-UK) consortium | The Lighthouse Lab in Milton Keynes et al |
| EPI_ISL_606729 | England/MILK-A7A298/2020 | 2020-10-15 | Europe/United Kingdom/England/ | Lighthouse Lab in Milton Keynes | Wellcome Sanger Institute for the COVID-19 Genomics UK (COG-UK) consortium | The Lighthouse Lab in Milton Keynes et al |
| EPI_ISL_606932 | England/ALDP-A6D5AE/2020 | 2020-10-14 | Europe/United Kingdom/England/ | Lighthouse Lab in Alderley Park | Wellcome Sanger Institute for the COVID-19 Genomics UK (COG-UK) consortium | Jacquelyn Wynn et al |
| EPI_ISL_606957 | England/ALDP-A6EC5D/2020 | 2020-10-15 | Europe/United Kingdom/England/ | Lighthouse Lab in Alderley Park | Wellcome Sanger Institute for the COVID-19 Genomics UK (COG-UK) consortium | Jacquelyn Wynn et al |
| EPI_ISL_607046 | England/ALDP-A6E376/2020 | 2020-10-15 | Europe/United Kingdom/England/ | Lighthouse Lab in Alderley Park | Wellcome Sanger Institute for the COVID-19 Genomics UK (COG-UK) consortium | Jacquelyn Wynn et al |

| | | | | | | |
|---|---|---|---|---|---|---|
| EPI_ISL_607062 | England/ALDP-A6F73D/2020 | 2020-10-15 | Europe/United Kingdom/England/ | Lighthouse Lab in Alderley Park | Wellcome Sanger Institute for the COVID-19 Genomics UK (COG-UK) consortium | Jacquelyn Wynn et al |
| EPI_ISL_607374 | England/MILK-A7BCF3/2020 | 2020-10-15 | Europe/United Kingdom/England/ | Lighthouse Lab in Milton Keynes | Wellcome Sanger Institute for the COVID-19 Genomics UK (COG-UK) consortium | The Lighthouse Lab in Milton Keynes et al |
| EPI_ISL_607390 | England/MILK-A7C560/2020 | 2020-10-15 | Europe/United Kingdom/England/ | Lighthouse Lab in Milton Keynes | Wellcome Sanger Institute for the COVID-19 Genomics UK (COG-UK) consortium | The Lighthouse Lab in Milton Keynes et al |
| EPI_ISL_607408 | England/MILK-A7B81D/2020 | 2020-10-15 | Europe/United Kingdom/England/ | Lighthouse Lab in Milton Keynes | Wellcome Sanger Institute for the COVID-19 Genomics UK (COG-UK) consortium | The Lighthouse Lab in Milton Keynes et al |
| EPI_ISL_607474 | England/MILK-A7B72F/2020 | 2020-10-15 | Europe/United Kingdom/England/ | Lighthouse Lab in Milton Keynes | Wellcome Sanger Institute for the COVID-19 Genomics UK (COG-UK) consortium | The Lighthouse Lab in Milton Keynes et al |
| EPI_ISL_607546 | England/MILK-A7B6D7/2020 | 2020-10-15 | Europe/United Kingdom/England/ | Lighthouse Lab in Milton Keynes | Wellcome Sanger Institute for the COVID-19 Genomics UK (COG-UK) consortium | The Lighthouse Lab in Milton Keynes et al |
| EPI_ISL_607559 | England/MILK-A7C937/2020 | 2020-10-15 | Europe/United Kingdom/England/ | Lighthouse Lab in Milton Keynes | Wellcome Sanger Institute for the COVID-19 Genomics UK (COG-UK) consortium | The Lighthouse Lab in Milton Keynes et al |
| EPI_ISL_607600 | England/MILK-A7B3FE/2020 | 2020-10-14 | Europe/United Kingdom/England/ | Lighthouse Lab in Milton Keynes | Wellcome Sanger Institute for the COVID-19 Genomics UK (COG-UK) consortium | The Lighthouse Lab in Milton Keynes et al |

| | | | | | | |
|---|---|---|---|---|---|---|
| EPI_ISL_607761 | England/ALDP-A6F9EC/2020 | 2020-10-14 | Europe/United Kingdom/England/ | Lighthouse Lab in Alderley Park | Wellcome Sanger Institute for the COVID-19 Genomics UK (COG-UK) consortium | Jacquelyn Wynn et al |
| EPI_ISL_607828 | England/ALDP-A6EFBE/2020 | 2020-10-15 | Europe/United Kingdom/England/ | Lighthouse Lab in Alderley Park | Wellcome Sanger Institute for the COVID-19 Genomics UK (COG-UK) consortium | Jacquelyn Wynn et al |
| EPI_ISL_607854 | England/ALDP-A6E947/2020 | 2020-10-15 | Europe/United Kingdom/England/ | Lighthouse Lab in Alderley Park | Wellcome Sanger Institute for the COVID-19 Genomics UK (COG-UK) consortium | Jacquelyn Wynn et al |
| EPI_ISL_607858 | England/ALDP-A70696/2020 | 2020-10-15 | Europe/United Kingdom/England/ | Lighthouse Lab in Alderley Park | Wellcome Sanger Institute for the COVID-19 Genomics UK (COG-UK) consortium | Jacquelyn Wynn et al |
| EPI_ISL_607875 | England/ALDP-A6DBCA/2020 | 2020-10-14 | Europe/United Kingdom/England/ | Lighthouse Lab in Alderley Park | Wellcome Sanger Institute for the COVID-19 Genomics UK (COG-UK) consortium | Jacquelyn Wynn et al |
| EPI_ISL_607901 | England/ALDP-A6FA07/2020 | 2020-10-15 | Europe/United Kingdom/England/ | Lighthouse Lab in Alderley Park | Wellcome Sanger Institute for the COVID-19 Genomics UK (COG-UK) consortium | Jacquelyn Wynn et al |
| EPI_ISL_607963 | England/ALDP-A6F17B/2020 | 2020-10-15 | Europe/United Kingdom/England/ | Lighthouse Lab in Alderley Park | Wellcome Sanger Institute for the COVID-19 Genomics UK (COG-UK) consortium | Jacquelyn Wynn et al |
| EPI_ISL_608003 | England/ALDP-A6D3C2/2020 | 2020-10-15 | Europe/United Kingdom/England/ | Lighthouse Lab in Alderley Park | Wellcome Sanger Institute for the COVID-19 Genomics UK (COG-UK) consortium | Jacquelyn Wynn et al |

| | | | | | | |
|---|---|---|---|---|---|---|
| EPI_ISL_608095 | England/ALDP-A6D77B/2020 | 2020-10-15 | Europe/United Kingdom/England/ | Lighthouse Lab in Alderley Park | Wellcome Sanger Institute for the COVID-19 Genomics UK (COG-UK) consortium | Jacquelyn Wynn et al |
| EPI_ISL_608120 | England/ALDP-A70195/2020 | 2020-10-15 | Europe/United Kingdom/England/ | Lighthouse Lab in Alderley Park | Wellcome Sanger Institute for the COVID-19 Genomics UK (COG-UK) consortium | Jacquelyn Wynn et al |
| EPI_ISL_608160 | England/ALDP-A6D3E0/2020 | 2020-10-15 | Europe/United Kingdom/England/ | Lighthouse Lab in Alderley Park | Wellcome Sanger Institute for the COVID-19 Genomics UK (COG-UK) consortium | Jacquelyn Wynn et al |
| EPI_ISL_608244 | England/ALDP-A6CB9E/2020 | 2020-10-15 | Europe/United Kingdom/England/ | Lighthouse Lab in Alderley Park | Wellcome Sanger Institute for the COVID-19 Genomics UK (COG-UK) consortium | Jacquelyn Wynn et al |
| EPI_ISL_608317 | England/ALDP-A6C3F0/2020 | 2020-10-14 | Europe/United Kingdom/England/ | Lighthouse Lab in Alderley Park | Wellcome Sanger Institute for the COVID-19 Genomics UK (COG-UK) consortium | Jacquelyn Wynn et al |
| EPI_ISL_608429 | England/CAMC-A64CDF/2020 | 2020-10-09 | Europe/United Kingdom/England/ | Lighthouse Lab in Cambridge | Wellcome Sanger Institute for the COVID-19 Genomics UK (COG-UK) consortium | Rob Howes et al |
| EPI_ISL_608434 | England/ALDP-A6885F/2020 | 2020-10-12 | Europe/United Kingdom/England/ | Lighthouse Lab in Alderley Park | Wellcome Sanger Institute for the COVID-19 Genomics UK (COG-UK) consortium | Jacquelyn Wynn et al |
| EPI_ISL_608830 | England/MILK-A6B5BF/2020 | 2020-10-09 | Europe/United Kingdom/England/ | Lighthouse Lab in Milton Keynes | Wellcome Sanger Institute for the COVID-19 Genomics UK (COG-UK) consortium | The Lighthouse Lab in Milton Keynes et al |

| | | | | | | |
|---|---|---|---|---|---|---|
| EPI_ISL_608967 | England/CAMC-A64826/2020 | 2020-10-10 | Europe/United Kingdom/England/ | Lighthouse Lab in Cambridge | Wellcome Sanger Institute for the COVID-19 Genomics UK (COG-UK) consortium | Rob Howes et al |
| EPI_ISL_608968 | England/CAMC-A64677/2020 | 2020-10-11 | Europe/United Kingdom/England/ | Lighthouse Lab in Cambridge | Wellcome Sanger Institute for the COVID-19 Genomics UK (COG-UK) consortium | Rob Howes et al |
| EPI_ISL_608989 | England/ALDP-A67D8E/2020 | 2020-10-10 | Europe/United Kingdom/England/ | Lighthouse Lab in Alderley Park | Wellcome Sanger Institute for the COVID-19 Genomics UK (COG-UK) consortium | Jacquelyn Wynn et al |
| EPI_ISL_609016 | England/ALDP-A694C3/2020 | 2020-10-12 | Europe/United Kingdom/England/ | Lighthouse Lab in Alderley Park | Wellcome Sanger Institute for the COVID-19 Genomics UK (COG-UK) consortium | Jacquelyn Wynn et al |
| EPI_ISL_609021 | England/MILK-A6BF66/2020 | 2020-10-09 | Europe/United Kingdom/England/ | Lighthouse Lab in Milton Keynes | Wellcome Sanger Institute for the COVID-19 Genomics UK (COG-UK) consortium | The Lighthouse Lab in Milton Keynes et al |
| EPI_ISL_609043 | England/ALDP-A68109/2020 | 2020-10-10 | Europe/United Kingdom/England/ | Lighthouse Lab in Alderley Park | Wellcome Sanger Institute for the COVID-19 Genomics UK (COG-UK) consortium | Jacquelyn Wynn et al |
| EPI_ISL_609299 | Wales/MILK-A44DA5/2020 | 2020-10-06 | Europe/United Kingdom/Wales/ | Lighthouse Lab in Milton Keynes | Wellcome Sanger Institute for the COVID-19 Genomics UK (COG-UK) consortium | The Lighthouse Lab in Milton Keynes et al |
| EPI_ISL_609348 | England/CAMC-A42071/2020 | 2020-10-05 | Europe/United Kingdom/England/ | Lighthouse Lab in Cambridge | Wellcome Sanger Institute for the COVID-19 Genomics UK (COG-UK) consortium | Rob Howes et al |

| | | | | | | |
|---|---|---|---|---|---|---|
| EPI_ISL_609544 | England/CAMC-A3FB3A/2020 | 2020-10-05 | Europe/United Kingdom/England/ | Lighthouse Lab in Cambridge | Wellcome Sanger Institute for the COVID-19 Genomics UK (COG-UK) consortium | Rob Howes et al |
| EPI_ISL_609558 | England/CAMC-A3F772/2020 | 2020-10-05 | Europe/United Kingdom/England/ | Lighthouse Lab in Cambridge | Wellcome Sanger Institute for the COVID-19 Genomics UK (COG-UK) consortium | Rob Howes et al |
| EPI_ISL_609604 | England/CAMC-A3F4D5/2020 | 2020-10-06 | Europe/United Kingdom/England/ | Lighthouse Lab in Cambridge | Wellcome Sanger Institute for the COVID-19 Genomics UK (COG-UK) consortium | Rob Howes et al |
| EPI_ISL_609652 | England/CAMC-A3E70A/2020 | 2020-10-05 | Europe/United Kingdom/England/ | Lighthouse Lab in Cambridge | Wellcome Sanger Institute for the COVID-19 Genomics UK (COG-UK) consortium | Rob Howes et al |
| EPI_ISL_609740 | England/CAMC-A3F727/2020 | 2020-10-05 | Europe/United Kingdom/England/ | Lighthouse Lab in Cambridge | Wellcome Sanger Institute for the COVID-19 Genomics UK (COG-UK) consortium | Rob Howes et al |
| EPI_ISL_609801 | Canada/ON-UHTC_0321/2020 | 2020-09-06 | North America/Canada/Ontario/ | Unity Health Toronto | Ontario Institute for Cancer Research | Ramzi Fattouh et al |
| EPI_ISL_609802 | Canada/ON-UHTC_0335/2020 | 2020-09-09 | North America/Canada/Ontario/ | Unity Health Toronto | Ontario Institute for Cancer Research | Ramzi Fattouh et al |
| EPI_ISL_609803 | Canada/ON-UHTC_0344/2020 | 2020-09-11 | North America/Canada/Ontario/ | Unity Health Toronto | Ontario Institute for Cancer Research | Ramzi Fattouh et al |
| EPI_ISL_609804 | Canada/ON-UHTC_0345/2020 | 2020-09-11 | North America/Canada/Ontario/ | Unity Health Toronto | Ontario Institute for Cancer Research | Ramzi Fattouh et al |
| EPI_ISL_609805 | Canada/ON-UHTC_0326/2020 | 2020-09-07 | North America/Canada/Ontario/ | Unity Health Toronto | Ontario Institute for Cancer Research | Ramzi Fattouh et al |

| | | | | | | |
|---|---|---|---|---|---|---|
| EPI_ISL_609806 | Canada/ON-UHTC_0327/2020 | 2020-09-07 | North America/Canada/Ontario/ | Unity Health Toronto | Ontario Institute for Cancer Research | Ramzi Fattouh et al |
| EPI_ISL_609807 | Canada/ON-UHTC_0330/2020 | 2020-09-08 | North America/Canada/Ontario/ | Unity Health Toronto | Ontario Institute for Cancer Research | Ramzi Fattouh et al |
| EPI_ISL_609808 | Canada/ON-UHTC_0341/2020 | 2020-09-10 | North America/Canada/Ontario/ | Unity Health Toronto | Ontario Institute for Cancer Research | Ramzi Fattouh et al |
| EPI_ISL_609809 | Canada/ON-UHTC_0328/2020 | 2020-09-07 | North America/Canada/Ontario/ | Unity Health Toronto | Ontario Institute for Cancer Research | Ramzi Fattouh et al |
| EPI_ISL_609810 | Canada/ON-UHTC_0342/2020 | 2020-09-11 | North America/Canada/Ontario/ | Unity Health Toronto | Ontario Institute for Cancer Research | Ramzi Fattouh et al |
| EPI_ISL_609811 | Canada/ON-UHTC_0356/2020 | 2020-10-03 | North America/Canada/Ontario/ | Unity Health Toronto | Ontario Institute for Cancer Research | Ramzi Fattouh et al |
| EPI_ISL_609812 | Canada/ON-UHTC_0340/2020 | 2020-09-10 | North America/Canada/Ontario/ | Unity Health Toronto | Ontario Institute for Cancer Research | Ramzi Fattouh et al |
| EPI_ISL_609813 | Canada/ON-UHTC_0331/2020 | 2020-09-08 | North America/Canada/Ontario/ | Unity Health Toronto | Ontario Institute for Cancer Research | Ramzi Fattouh et al |
| EPI_ISL_609814 | Canada/ON-UHTC_0347/2020 | 2020-09-12 | North America/Canada/Ontario/ | Unity Health Toronto | Ontario Institute for Cancer Research | Ramzi Fattouh et al |
| EPI_ISL_609815 | Canada/ON-UHTC_0339/2020 | 2020-09-10 | North America/Canada/Ontario/ | Unity Health Toronto | Ontario Institute for Cancer Research | Ramzi Fattouh et al |
| EPI_ISL_609816 | Canada/ON-UHTC_0323/2020 | 2020-09-06 | North America/Canada/Ontario/ | Unity Health Toronto | Ontario Institute for Cancer Research | Ramzi Fattouh et al |
| EPI_ISL_609817 | Canada/ON-UHTC_0336/2020 | 2020-09-09 | North America/Canada/Ontario/ | Unity Health Toronto | Ontario Institute for Cancer Research | Ramzi Fattouh et al |
| EPI_ISL_609818 | Canada/ON-UHTC_0319/2020 | 2020-09-02 | North America/Canada/Ontario/ | Unity Health Toronto | Ontario Institute for Cancer Research | Ramzi Fattouh et al |

| | | | | | | |
|---|---|---|---|---|---|---|
| EPI_ISL_609819 | Canada/ON-UHTC_0346/2020 | 2020-09-12 | North America/Canada/Ontario/ | Unity Health Toronto | Ontario Institute for Cancer Research | Ramzi Fattouh et al |
| EPI_ISL_609820 | Canada/ON-UHTC_0324/2020 | 2020-09-06 | North America/Canada/Ontario/ | Unity Health Toronto | Ontario Institute for Cancer Research | Ramzi Fattouh et al |
| EPI_ISL_609821 | Canada/ON-UHTC_0318/2020 | 2020-09-01 | North America/Canada/Ontario/ | Unity Health Toronto | Ontario Institute for Cancer Research | Ramzi Fattouh et al |
| EPI_ISL_609822 | Canada/ON-UHTC_0355/2020 | 2020-10-03 | North America/Canada/Ontario/ | Unity Health Toronto | Ontario Institute for Cancer Research | Ramzi Fattouh et al |
| EPI_ISL_609823 | Canada/ON-UHTC_0338/2020 | 2020-09-10 | North America/Canada/Ontario/ | Unity Health Toronto | Ontario Institute for Cancer Research | Ramzi Fattouh et al |
| EPI_ISL_609824 | Canada/ON-UHTC_0325/2020 | 2020-09-07 | North America/Canada/Ontario/ | Unity Health Toronto | Ontario Institute for Cancer Research | Ramzi Fattouh et al |
| EPI_ISL_609825 | Canada/ON-UHTC_0317/2020 | 2020-09-01 | North America/Canada/Ontario/ | Unity Health Toronto | Ontario Institute for Cancer Research | Ramzi Fattouh et al |
| EPI_ISL_609826 | Canada/ON-UHTC_0322/2020 | 2020-09-06 | North America/Canada/Ontario/ | Unity Health Toronto | Ontario Institute for Cancer Research | Ramzi Fattouh et al |
| EPI_ISL_609971 | USA/VA-DCLS-1681/2020 | 2020-09-03 | North America/USA/Virginia/ | Virginia DCLS | Virginia DCLS | Virginia DCLS et al |
| EPI_ISL_609972 | USA/VA-DCLS-1682/2020 | 2020-09-03 | North America/USA/Virginia/ | Virginia DCLS | Virginia DCLS | Virginia DCLS et al |
| EPI_ISL_609977 | USA/VA-DCLS-1691/2020 | 2020-09-08 | North America/USA/Virginia/ | Virginia DCLS | Virginia DCLS | Virginia DCLS et al |
| EPI_ISL_609988 | USA/VA-DCLS-1832/2020 | 2020-09-23 | North America/USA/Virginia/ | Virginia DCLS | Virginia DCLS | Virginia DCLS et al |
| EPI_ISL_609992 | Italy/LAZ-INMI-84/2020 | 2020-08-20 | Europe/Italy/Lazio/ | INMI Lazzaro Spallanzani IRCCS | INMI Lazzaro Spallanzani IRCCS | F Messina et al |
| EPI_ISL_609993 | Italy/LAZ-INMI-85/2020 | 2020-08-23 | Europe/Italy/Lazio/ | INMI Lazzaro Spallanzani IRCCS | INMI Lazzaro Spallanzani IRCCS | E Giombini et al |
| EPI_ISL_609996 | Italy/LAZ-INMI-88/2020 | 2020-10-17 | Europe/Italy/Lazio/ | INMI Lazzaro Spallanzani IRCCS | INMI Lazzaro Spallanzani IRCCS | F Messina et al |
| EPI_ISL_609997 | Italy/LAZ-INMI-89/2020 | 2020-10-16 | Europe/Italy/Lazio/ | INMI Lazzaro Spallanzani IRCCS | INMI Lazzaro Spallanzani IRCCS | M Rueca et al |
| EPI_ISL_609998 | Italy/LAZ-INMI-90/2020 | 2020-10-19 | Europe/Italy/Lazio/ | INMI Lazzaro Spallanzani IRCCS | INMI Lazzaro Spallanzani IRCCS | F Messina et al |

| | | | | | | |
|---|---|---|---|---|---|---|
| EPI_ISL_610010 | USA/TX-DSHS-0874/2020 | 2020-06-16 | North America/USA/Texas/El Paso Coun | Texas Department of State Health Services | Texas Department of State Health Services | Rashmi Tuladhar et al |
| EPI_ISL_610016 | USA/TX-DSHS-0880/2020 | 2020-06-16 | North America/USA/Texas/Harris Count | Texas Department of State Health Services | Texas Department of State Health Services | Rashmi Tuladhar et al |
| EPI_ISL_610024 | USA/TX-DSHS-0888/2020 | 2020-06-16 | North America/USA/Texas/Harris Count | Texas Department of State Health Services | Texas Department of State Health Services | Rashmi Tuladhar et al |
| EPI_ISL_610029 | USA/TX-DSHS-0893/2020 | 2020-06-16 | North America/USA/Texas/Harris Count | Texas Department of State Health Services | Texas Department of State Health Services | Rashmi Tuladhar et al |
| EPI_ISL_610031 | USA/TX-DSHS-0895/2020 | 2020-06-16 | North America/USA/Texas/Harris Count | Texas Department of State Health Services | Texas Department of State Health Services | Rashmi Tuladhar et al |
| EPI_ISL_610032 | USA/TX-DSHS-0896/2020 | 2020-06-16 | North America/USA/Texas/Harris Count | Texas Department of State Health Services | Texas Department of State Health Services | Rashmi Tuladhar et al |
| EPI_ISL_610048 | USA/MI-UM-10036399817/2020 | 2020-10-12 | North America/USA/Michigan/ | University of Michigan Clinical Microbiology Laboratory | Lauring Lab, University of Michigan, Department of Microbiology and Immunology | Valesano et al |
| EPI_ISL_610057 | USA/MI-UM-10036381452/2020 | 2020-10-10 | North America/USA/Michigan/ | University of Michigan Clinical Microbiology Laboratory | Lauring Lab, University of Michigan, Department of Microbiology and Immunology | Valesano et al |
| EPI_ISL_610064 | USA/MI-UM-10036381059/2020 | 2020-10-10 | North America/USA/Michigan/ | University of Michigan Clinical Microbiology Laboratory | Lauring Lab, University of Michigan, Department of Microbiology and Immunology | Valesano et al |

| | | | | | | |
|---|---|---|---|---|---|---|
| EPI_ISL_610067 | USA/MI-UM-10036322730/2020 | 2020-10-07 | North America/USA/Michigan/ | University of Michigan Clinical Microbiology Laboratory | Lauring Lab, University of Michigan, Department of Microbiology and Immunology | Valesano et al |
| EPI_ISL_610068 | USA/MI-UM-10036394525/2020 | 2020-10-12 | North America/USA/Michigan/ | University of Michigan Clinical Microbiology Laboratory | Lauring Lab, University of Michigan, Department of Microbiology and Immunology | Valesano et al |
| EPI_ISL_610098 | USA/MI-UM-10036422360/2020 | 2020-10-13 | North America/USA/Michigan/ | University of Michigan Clinical Microbiology Laboratory | Lauring Lab, University of Michigan, Department of Microbiology and Immunology | Valesano et al |
| EPI_ISL_610100 | USA/MI-UM-10036397118/2020 | 2020-10-12 | North America/USA/Michigan/ | University of Michigan Clinical Microbiology Laboratory | Lauring Lab, University of Michigan, Department of Microbiology and Immunology | Valesano et al |
| EPI_ISL_610103 | USA/MI-UM-10036448245/2020 | 2020-10-14 | North America/USA/Michigan/ | University of Michigan Clinical Microbiology Laboratory | Lauring Lab, University of Michigan, Department of Microbiology and Immunology | Valesano et al |
| EPI_ISL_610105 | USA/MI-UM-10036467524/2020 | 2020-10-15 | North America/USA/Michigan/ | University of Michigan Clinical Microbiology Laboratory | Lauring Lab, University of Michigan, Department of Microbiology and Immunology | Valesano et al |

| | | | | | | |
|---|---|---|---|---|---|---|
| EPI_ISL_610113 | USA/MI-UM-10036454655/2020 | 2020-10-15 | North America/USA/Michigan/ | University of Michigan Clinical Microbiology Laboratory | Lauring Lab, University of Michigan, Department of Microbiology and Immunology | Valesano et al |
| EPI_ISL_610114 | USA/MI-UM-10036444607/2020 | 2020-10-14 | North America/USA/Michigan/ | University of Michigan Clinical Microbiology Laboratory | Lauring Lab, University of Michigan, Department of Microbiology and Immunology | Valesano et al |
| EPI_ISL_610117 | USA/MI-UM-10036447294/2020 | 2020-10-14 | North America/USA/Michigan/ | University of Michigan Clinical Microbiology Laboratory | Lauring Lab, University of Michigan, Department of Microbiology and Immunology | Valesano et al |
| EPI_ISL_610122 | USA/VA-DCLS-1558/2020 | 2020-08-07 | North America/USA/Virginia/ | Virginia DCLS | Virginia DCLS | Virginia DCLS et al |
| EPI_ISL_610130 | USA/VA-DCLS-1567/2020 | 2020-08-14 | North America/USA/Virginia/ | Virginia DCLS | Virginia DCLS | Virginia DCLS et al |
| EPI_ISL_610131 | USA/VA-DCLS-1568/2020 | 2020-08-14 | North America/USA/Virginia/ | Virginia DCLS | Virginia DCLS | Virginia DCLS et al |
| EPI_ISL_610133 | USA/VA-DCLS-1572/2020 | 2020-08-14 | North America/USA/Virginia/ | Virginia DCLS | Virginia DCLS | Virginia DCLS et al |
| EPI_ISL_610141 | USA/VA-DCLS-1586/2020 | 2020-08-14 | North America/USA/Virginia/ | Virginia DCLS | Virginia DCLS | Virginia DCLS et al |
| EPI_ISL_610148 | USA/VA-DCLS-1595/2020 | 2020-08-14 | North America/USA/Virginia/ | Virginia DCLS | Virginia DCLS | Virginia DCLS et al |
| EPI_ISL_610159 | USA/MO-WUSTL_D/2020 | 2020-08-16 | North America/USA/Missouri/St. Louis [illegible] | Washington University in St. Louis | Washington University in St. Louis | David Wang et al |
| EPI_ISL_610160 | USA/MO-WUSTL_E/2020 | 2020-08-16 | North America/USA/Missouri/St. Louis [illegible] | Washington University in St. Louis | Washington University in St. Louis | David Wang et al |

| | | | | | | |
|---|---|---|---|---|---|---|
| EPI_ISL_610161 | Indonesia/JT-UGM-48660/2020 | 2020-09-01 | Asia/Indonesia/Central Java/ | RSUD Dr. Tjitrowardojo | Genetics Working Group (Pokja Genetik) Faculty of Medicine, Public Health and Nursing Universitas Gadjah Mada (FK-KMK UGM); Disease Investigation Center Wates Ministry of Agriculture Indonesia; Department of Microbiology FK-KMK UGM; Laboratorium Diagnostik Yayasan Tahija World Mosquito Program (WMP) Yogyakarta Center for Tropical Medicine FK-KMK UGM; Integrated Research Center FK-KMK UGM; Department of Computer Science and Electronics FMIPA UGM; Balai Besar Teknik Kesehatan Lingkungan dan | Gunadi et al |

| | | | | | | |
|---|---|---|---|---|---|---|
| EPI_ISL_610162 | Indonesia/JT-UGM-47964/2020 | 2020-08-31 | Asia/Indonesia/Central Java/ | RSUD Dr. Tjitrowardojo | Genetics Working Group (Pokja Genetik) Faculty of Medicine, Public Health and Nursing Universitas Gadjah Mada (FK-KMK UGM); Disease Investigation Center Wates Ministry of Agriculture Indonesia; Department of Microbiology FK-KMK UGM; Laboratorium Diagnostik Yayasan Tahija World Mosquito Program (WMP) Yogyakarta Center for Tropical Medicine FK-KMK UGM; Integrated Research Center FK-KMK UGM; Department of Computer Science and Electronics FMIPA UGM; Balai Besar Teknik Kesehatan Lingkungan dan | Gunadi et al |
| EPI_ISL_610227 | Russia/CRIE4907775/2020 | 2020-07-14 | Europe/Russia/Moscow Oblast/ | "Molecular diagnostic laboratory of Federal Budget Institution of Science ""Central Research Institute of Epidemiology"" of The Federal Service on Customers' Rights Protection and Human Well-being Surveillance" | Group of Genomics and Postgenomic Technologies of Central Research Institute of Epidemilology | Samoilov AE et al |

| | | | | | | |
|---|---|---|---|---|---|---|
| EPI_ISL_610228 | Russia/CRIE4978885/2020 | 2020-07-15 | Europe/Russia/Moscow Oblast/ | "Molecular diagnostic laboratory of Federal Budget Institution of Science ""Central Research Institute of Epidemiology"" of The Federal Service on Customers' Rights Protection and Human Well-being Surveillance" | Group of Genomics and Postgenomic Technologies of Central Research Institute of Epidemilology | Samoilov AE et al |
| EPI_ISL_610231 | Russia/CRIE5031464/2020 | 2020-07-16 | Europe/Russia/Moscow Oblast/ | "Molecular diagnostic laboratory of Federal Budget Institution of Science ""Central Research Institute of Epidemiology"" of The Federal Service on Customers' Rights Protection and Human Well-being Surveillance" | Group of Genomics and Postgenomic Technologies of Central Research Institute of Epidemilology | Samoilov AE et al |
| EPI_ISL_610233 | Russia/CRIE4990266/2020 | 2020-07-16 | Europe/Russia/Moscow Oblast/ | "Molecular diagnostic laboratory of Federal Budget Institution of Science ""Central Research Institute of Epidemiology"" of The Federal Service on Customers' Rights Protection and Human Well-being Surveillance" | Group of Genomics and Postgenomic Technologies of Central Research Institute of Epidemilology | Samoilov AE et al |
| EPI_ISL_610234 | Russia/CRIE4988935/2020 | 2020-07-16 | Europe/Russia/Moscow Oblast/ | "Molecular diagnostic laboratory of Federal Budget Institution of Science ""Central Research Institute of Epidemiology"" of The Federal Service on Customers' Rights Protection and Human Well-being Surveillance" | Group of Genomics and Postgenomic Technologies of Central Research Institute of Epidemilology | Samoilov AE et al |
| EPI_ISL_610236 | Russia/CRIE5047165/2020 | 2020-07-16 | Europe/Russia/Moscow Oblast/ | "Molecular diagnostic laboratory of Federal Budget Institution of Science ""Central Research Institute of Epidemiology"" of The Federal Service on Customers' Rights Protection and Human Well-being Surveillance" | Group of Genomics and Postgenomic Technologies of Central Research Institute of Epidemilology | Samoilov AE et al |

| | | | | | | |
|---|---|---|---|---|---|---|
| EPI_ISL_610239 | Russia/CRIE7158978/2020 | 2020-08-24 | Europe/Russia/Moscow Oblast/ | "Molecular diagnostic laboratory of Federal Budget Institution of Science ""Central Research Institute of Epidemiology"" of The Federal Service on Customers' Rights Protection and Human Well-being Surveillance" | Group of Genomics and Postgenomic Technologies of Central Research Institute of Epidemilology | Samoilov AE et al |
| EPI_ISL_610240 | Russia/CRIE7159118/2020 | 2020-08-24 | Europe/Russia/Moscow Oblast/ | "Molecular diagnostic laboratory of Federal Budget Institution of Science ""Central Research Institute of Epidemiology"" of The Federal Service on Customers' Rights Protection and Human Well-being Surveillance" | Group of Genomics and Postgenomic Technologies of Central Research Institute of Epidemilology | Samoilov AE et al |
| EPI_ISL_610241 | Russia/CRIE7171114/2020 | 2020-08-24 | Europe/Russia/Moscow Oblast/ | "Molecular diagnostic laboratory of Federal Budget Institution of Science ""Central Research Institute of Epidemiology"" of The Federal Service on Customers' Rights Protection and Human Well-being Surveillance" | Group of Genomics and Postgenomic Technologies of Central Research Institute of Epidemilology | Samoilov AE et al |
| EPI_ISL_610242 | Russia/CRIE7175362/2020 | 2020-08-24 | Europe/Russia/Moscow Oblast/ | "Molecular diagnostic laboratory of Federal Budget Institution of Science ""Central Research Institute of Epidemiology"" of The Federal Service on Customers' Rights Protection and Human Well-being Surveillance" | Group of Genomics and Postgenomic Technologies of Central Research Institute of Epidemilology | Samoilov AE et al |
| EPI_ISL_610243 | Russia/CRIE7158368/2020 | 2020-08-24 | Europe/Russia/Moscow Oblast/ | "Molecular diagnostic laboratory of Federal Budget Institution of Science ""Central Research Institute of Epidemiology"" of The Federal Service on Customers' Rights Protection and Human Well-being Surveillance" | Group of Genomics and Postgenomic Technologies of Central Research Institute of Epidemilology | Samoilov AE et al |

| | | | | | | |
|---|---|---|---|---|---|---|
| EPI_ISL_610244 | Russia/CRIE7249896/2020 | 2020-08-25 | Europe/Russia/Moscow Oblast/ | "Molecular diagnostic laboratory of Federal Budget Institution of Science ""Central Research Institute of Epidemiology"" of The Federal Service on Customers' Rights Protection and Human Well-being Surveillance" | Group of Genomics and Postgenomic Technologies of Central Research Institute of Epidemilology | Samoilov AE et al |
| EPI_ISL_610245 | England/2/2020 | 2020-01-29 | Europe/United Kingdom/England/ | Virology, National Institute for Biological Standards and Control | Virology, National Institute for Biological Standards and Control | Mee et al |
| EPI_ISL_610247 | USA/MI-UM-10036331190/2020 | 2020-10-07 | North America/USA/Michigan/ | University of Michigan Clinical Microbiology Laboratory | Lauring Lab, University of Michigan, Department of Microbiology and Immunology | Valesano et al |
| EPI_ISL_610265 | USA/NM-UNM-00815/2020 | 2020-05-13 | North America/USA/New Mexico/ | TriCore Reference Laboratories | Center for Global Health, University of New Mexico Health Sciences Center | Daryl Domman et al |
| EPI_ISL_610276 | USA/NM-UNM-00846/2020 | 2020-05-04 | North America/USA/New Mexico/ | New Mexico Department of Health Scientific Laboratory Division | Center for Global Health, University of New Mexico Health Sciences Center | Daryl Domman et al |
| EPI_ISL_610280 | USA/NM-UNM-00854/2020 | 2020-05-04 | North America/USA/New Mexico/ | New Mexico Department of Health Scientific Laboratory Division | Center for Global Health, University of New Mexico Health Sciences Center | Daryl Domman et al |
| EPI_ISL_610282 | USA/NM-UNM-00860/2020 | 2020-05-03 | North America/USA/New Mexico/ | TriCore Reference Laboratories | Center for Global Health, University of New Mexico Health Sciences Center | Daryl Domman et al |
| EPI_ISL_610284 | USA/WA-UW-16407/2020 | 2020-07-10 | North America/USA/Washington/ | UW Virology Lab | UW Virology Lab | Pavitra Roychoudhury et al |

| | | | | | | |
|---|---|---|---|---|---|---|
| EPI_ISL_610393 | England/ALDP-A85212/2020 | 2020-10-16 | Europe/United Kingdom/England/ | Lighthouse Lab in Alderley Park | Wellcome Sanger Institute for the COVID-19 Genomics UK (COG-UK) consortium | Jacquelyn Wynn et al |
| EPI_ISL_610445 | England/ALDP-A85713/2020 | 2020-10-17 | Europe/United Kingdom/England/ | Lighthouse Lab in Alderley Park | Wellcome Sanger Institute for the COVID-19 Genomics UK (COG-UK) consortium | Jacquelyn Wynn et al |
| EPI_ISL_610450 | England/ALDP-A84D30/2020 | 2020-10-17 | Europe/United Kingdom/England/ | Lighthouse Lab in Alderley Park | Wellcome Sanger Institute for the COVID-19 Genomics UK (COG-UK) consortium | Jacquelyn Wynn et al |
| EPI_ISL_610551 | England/ALDP-A84F67/2020 | 2020-10-18 | Europe/United Kingdom/England/ | Lighthouse Lab in Alderley Park | Wellcome Sanger Institute for the COVID-19 Genomics UK (COG-UK) consortium | Jacquelyn Wynn et al |
| EPI_ISL_610631 | England/CAMC-A82F4B/2020 | 2020-10-13 | Europe/United Kingdom/England/ | Lighthouse Lab in Cambridge | Wellcome Sanger Institute for the COVID-19 Genomics UK (COG-UK) consortium | Rob Howes et al |
| EPI_ISL_610658 | England/CAMC-A82E5D/2020 | 2020-10-13 | Europe/United Kingdom/England/ | Lighthouse Lab in Cambridge | Wellcome Sanger Institute for the COVID-19 Genomics UK (COG-UK) consortium | Rob Howes et al |
| EPI_ISL_610698 | England/CAMC-A821DC/2020 | 2020-10-13 | Europe/United Kingdom/England/ | Lighthouse Lab in Cambridge | Wellcome Sanger Institute for the COVID-19 Genomics UK (COG-UK) consortium | Rob Howes et al |
| EPI_ISL_610705 | England/CAMC-A80F98/2020 | 2020-10-14 | Europe/United Kingdom/England/ | Lighthouse Lab in Cambridge | Wellcome Sanger Institute for the COVID-19 Genomics UK (COG-UK) consortium | Rob Howes et al |

| | | | | | | |
|---|---|---|---|---|---|---|
| EPI_ISL_610925 | England/ALDP-A93A55/2020 | 2020-10-19 | Europe/United Kingdom/England/ | Lighthouse Lab in Alderley Park | Wellcome Sanger Institute for the COVID-19 Genomics UK (COG-UK) consortium | Jacquelyn Wynn et al |
| EPI_ISL_610979 | England/ALDP-A92397/2020 | 2020-10-19 | Europe/United Kingdom/England/ | Lighthouse Lab in Alderley Park | Wellcome Sanger Institute for the COVID-19 Genomics UK (COG-UK) consortium | Jacquelyn Wynn et al |
| EPI_ISL_611017 | England/ALDP-A93BF8/2020 | 2020-10-19 | Europe/United Kingdom/England/ | Lighthouse Lab in Alderley Park | Wellcome Sanger Institute for the COVID-19 Genomics UK (COG-UK) consortium | Jacquelyn Wynn et al |
| EPI_ISL_611091 | England/ALDP-A92537/2020 | 2020-10-19 | Europe/United Kingdom/England/ | Lighthouse Lab in Alderley Park | Wellcome Sanger Institute for the COVID-19 Genomics UK (COG-UK) consortium | Jacquelyn Wynn et al |
| EPI_ISL_611103 | England/ALDP-A92B26/2020 | 2020-10-19 | Europe/United Kingdom/England/ | Lighthouse Lab in Alderley Park | Wellcome Sanger Institute for the COVID-19 Genomics UK (COG-UK) consortium | Jacquelyn Wynn et al |
| EPI_ISL_611107 | England/ALDP-A93DE3/2020 | 2020-10-19 | Europe/United Kingdom/England/ | Lighthouse Lab in Alderley Park | Wellcome Sanger Institute for the COVID-19 Genomics UK (COG-UK) consortium | Jacquelyn Wynn et al |
| EPI_ISL_611273 | England/ALDP-A928A7/2020 | 2020-10-20 | Europe/United Kingdom/England/ | Lighthouse Lab in Alderley Park | Wellcome Sanger Institute for the COVID-19 Genomics UK (COG-UK) consortium | Jacquelyn Wynn et al |
| EPI_ISL_611335 | England/QEUH-A9E9D5/2020 | 2020-10-20 | Europe/United Kingdom/England/ | Lighthouse Lab in Glasgow | Wellcome Sanger Institute for the COVID-19 Genomics UK (COG-UK) consortium | Harper VanSteenhouse et al |

| | | | | | | |
|---|---|---|---|---|---|---|
| EPI_ISL_611446 | England/QEUH-A9E8BA/2020 | 2020-10-20 | Europe/United Kingdom/England/ | Lighthouse Lab in Glasgow | Wellcome Sanger Institute for the COVID-19 Genomics UK (COG-UK) consortium | Harper VanSteenhouse et al |
| EPI_ISL_611474 | England/QEUH-A9ED51/2020 | 2020-10-20 | Europe/United Kingdom/England/ | Lighthouse Lab in Glasgow | Wellcome Sanger Institute for the COVID-19 Genomics UK (COG-UK) consortium | Harper VanSteenhouse et al |
| EPI_ISL_611501 | USA/UT-UPHL-201026960/2020 | 2020-06-22 | North America/USA/Utah/ | Utah Public Health Laboratory | Utah Public Health Laboratory | Erin Young et al |
| EPI_ISL_611507 | USA/UT-UPHL-201020648/2020 | 2020-06-22 | North America/USA/Utah/ | Utah Public Health Laboratory | Utah Public Health Laboratory | Erin Young et al |
| EPI_ISL_611510 | USA/UT-UPHL-201012724/2020 | 2020-06-22 | North America/USA/Utah/ | Utah Public Health Laboratory | Utah Public Health Laboratory | Erin Young et al |
| EPI_ISL_611692 | England/BIRM-65BEA/2020 | 2020-10-10 | Europe/United Kingdom/England/ | University of Birmingham | COVID-19 Genomics UK (COG-UK) Consortium | Institute of Microbiology et al |
| EPI_ISL_611711 | Wales/PHWC-480CB5/2020 | 2020-10-18 | Europe/United Kingdom/Wales/ | Wales Specialist Virology Centre Sequencing lab: Pathogen Genomics Unit | COVID-19 Genomics UK (COG-UK) Consortium | Catherine Moore et al |
| EPI_ISL_611803 | England/SHEF-C88EE/2020 | 2020-10-20 | Europe/United Kingdom/England/ | Virology Department, Sheffield Teaching Hospitals NHS Foundation Trust/Department of Infection, Immunity and Cardiovascular Disease, The Medical School, University of Sheffield | COVID-19 Genomics UK (COG-UK) Consortium | Thushan de Silva et al |
| EPI_ISL_611840 | England/NOTT-11773D/2020 | 2020-10-23 | Europe/United Kingdom/England/ | Queens Medical Centre, Clinical Microbiology Department / DeepSeq Nottingham | COVID-19 Genomics UK (COG-UK) Consortium | Gemma Clark et al |
| EPI_ISL_611874 | Scotland/CVR4655/2020 | 2020-09-23 | Europe/United Kingdom/Scotland/ | West of Scotland Specialist Virology Centre, NHSGGC / MRC-University of Glasgow Centre for Virus Research | COVID-19 Genomics UK (COG-UK) Consortium | Ana da Silva Filipe et al |
| EPI_ISL_611994 | Wales/PHWC-480022/2020 | 2020-10-22 | Europe/United Kingdom/Wales/ | Wales Specialist Virology Centre Sequencing lab: Pathogen Genomics Unit | COVID-19 Genomics UK (COG-UK) Consortium | Catherine Moore et al |
| EPI_ISL_612039 | England/LIVE-1DD584/2020 | 2020-10-01 | Europe/United Kingdom/England/ | Liverpool Clinical Laboratories | COVID-19 Genomics UK (COG-UK) Consortium | Sam Haldenby et al |

| | | | | | | |
|---|---|---|---|---|---|---|
| EPI_ISL_612150 | Wales/PHWC-480E46/2020 | 2020-10-17 | Europe/United Kingdom/Wales/ | Wales Specialist Virology Centre Sequencing lab: Pathogen Genomics Unit | COVID-19 Genomics UK (COG-UK) Consortium | Catherine Moore et al |
| EPI_ISL_612325 | Scotland/EDB8332/2020 | 2020-09-24 | Europe/United Kingdom/Scotland/ | Virology Department, Royal Infirmary of Edinburgh, NHS Lothian / School of Biological Sciences, University of Edinburgh / Institute of Genetics and Molecular Medicine, University of Edinburgh | COVID-19 Genomics UK (COG-UK) Consortium | McHugh M et al |
| EPI_ISL_612440 | England/LIVE-AB76C/2020 | 2020-06-15 | Europe/United Kingdom/England/ | Liverpool Clinical Laboratories | COVID-19 Genomics UK (COG-UK) Consortium | Sam Haldenby et al |
| EPI_ISL_612446 | England/LIVE-AB9C0/2020 | 2020-05-31 | Europe/United Kingdom/England/ | Liverpool Clinical Laboratories | COVID-19 Genomics UK (COG-UK) Consortium | Sam Haldenby et al |
| EPI_ISL_612536 | England/MILK-A229E5/2020 | 2020-10-03 | Europe/United Kingdom/England/ | Northumbria University / South Tees Hospitals NHS Foundation Trust / North Cumbria Integrated Care NHS Foundation Trust / North Tees and Hartlepool NHS Foundation Trust / Newcastle Hospitals NHS Foundation Trust | COVID-19 Genomics UK (COG-UK) Consortium | Darren L Smith et al |
| EPI_ISL_612566 | England/NORT-1B6C14D/2020 | 2020-10-07 | Europe/United Kingdom/England/ | Northumbria University / South Tees Hospitals NHS Foundation Trust / North Cumbria Integrated Care NHS Foundation Trust / North Tees and Hartlepool NHS Foundation Trust / Newcastle Hospitals NHS Foundation Trust | COVID-19 Genomics UK (COG-UK) Consortium | Darren L Smith et al |
| EPI_ISL_612597 | England/NOTT-118189/2020 | 2020-10-26 | Europe/United Kingdom/England/ | Queens Medical Centre, Clinical Microbiology Department / DeepSeq Nottingham | COVID-19 Genomics UK (COG-UK) Consortium | Gemma Clark et al |
| EPI_ISL_612615 | Wales/PHWC-47EFEE/2020 | 2020-10-11 | Europe/United Kingdom/Wales/ | Wales Specialist Virology Centre Sequencing lab: Pathogen Genomics Unit | COVID-19 Genomics UK (COG-UK) Consortium | Catherine Moore et al |
| EPI_ISL_612653 | Wales/PHWC-47F28A/2020 | 2020-10-16 | Europe/United Kingdom/Wales/ | Wales Specialist Virology Centre Sequencing lab: Pathogen Genomics Unit | COVID-19 Genomics UK (COG-UK) Consortium | Catherine Moore et al |
| EPI_ISL_612673 | Wales/PHWC-47F40C/2020 | 2020-10-16 | Europe/United Kingdom/Wales/ | Wales Specialist Virology Centre Sequencing lab: Pathogen Genomics Unit | COVID-19 Genomics UK (COG-UK) Consortium | Catherine Moore et al |
| EPI_ISL_612707 | Wales/PHWC-47F660/2020 | 2020-10-04 | Europe/United Kingdom/Wales/ | Wales Specialist Virology Centre Sequencing lab: Pathogen Genomics Unit | COVID-19 Genomics UK (COG-UK) Consortium | Catherine Moore et al |

| | | | | | | |
|---|---|---|---|---|---|---|
| EPI_ISL_612715 | Wales/PHWC-47F6F7/2020 | 2020-10-12 | Europe/United Kingdom/Wales/ | Wales Specialist Virology Centre Sequencing lab: Pathogen Genomics Unit | COVID-19 Genomics UK (COG-UK) Consortium | Catherine Moore et al |
| EPI_ISL_612840 | Wales/PHWC-47FFED/2020 | 2020-10-10 | Europe/United Kingdom/Wales/ | Wales Specialist Virology Centre Sequencing lab: Pathogen Genomics Unit | COVID-19 Genomics UK (COG-UK) Consortium | Catherine Moore et al |
| EPI_ISL_613095 | Wales/PHWC-48163E/2020 | 2020-10-16 | Europe/United Kingdom/Wales/ | Wales Specialist Virology Centre Sequencing lab: Pathogen Genomics Unit | COVID-19 Genomics UK (COG-UK) Consortium | Catherine Moore et al |
| EPI_ISL_613179 | Wales/PHWC-481E54/2020 | 2020-10-16 | Europe/United Kingdom/Wales/ | Wales Specialist Virology Centre Sequencing lab: Pathogen Genomics Unit | COVID-19 Genomics UK (COG-UK) Consortium | Catherine Moore et al |
| EPI_ISL_613211 | Wales/PHWC-4822FD/2020 | 2020-10-20 | Europe/United Kingdom/Wales/ | Wales Specialist Virology Centre Sequencing lab: Pathogen Genomics Unit | COVID-19 Genomics UK (COG-UK) Consortium | Catherine Moore et al |
| EPI_ISL_613297 | England/PORT-2D4908/2020 | 2020-10-13 | Europe/United Kingdom/England/ | Centre for Enzyme Innovation, University of Portsmouth / Translational Research Laboratory, Portsmouth Hospitals NHS Trust | COVID-19 Genomics UK (COG-UK) Consortium | Angela Beckett et al |
| EPI_ISL_613361 | England/SHEF-C340D/2020 | 2020-10-15 | Europe/United Kingdom/England/ | Virology Department, Sheffield Teaching Hospitals NHS Foundation Trust/Department of Infection, Immunity and Cardiovascular Disease, The Medical School, University of Sheffield | COVID-19 Genomics UK (COG-UK) Consortium | Thushan de Silva et al |
| EPI_ISL_613410 | Spain/EX-IBV-97004996/2020 | 2020-05-28 | Europe/Spain/Extremadura/Caceres | Hospital San Pedro de Alc√°ntara (C√°ceres) | SeqCOVID-SPAIN consortium/IBV(CSIC) | Cristina Mu√±oz Cuevas et al |
| EPI_ISL_613446 | SaintBarthelemy/IPG-7358/2020 | 2020-02-28 | North America/Saint Barth√©lemy/Sair | Institut Pasteur de la Guadeloupe | Institut Pasteur de la Guadeloupe | Marion Barbet et al |
| EPI_ISL_613460 | Turkey/KOC-IST-OD5/2020 | 2020-06-21 | Europe/Turkey/Turkey/ | Microbiology, Koc University | Microbiology, Koc University | Ozer et al |
| EPI_ISL_613461 | Turkey/KOC-IST-B288/2020 | 2020-03-26 | Europe/Turkey/Turkey/ | Microbiology, Koc University | Microbiology, Koc University | Nurtop et al |
| EPI_ISL_613464 | USA/MN-MDH-1962/2020 | 2020-08-18 | North America/USA/Minnesota/ | Public Health Laboratory - Infectious Disease Lab, Minnesota Department of Health Infectious Disease Laboratory Submission Group | Minnesota Department of Health, Public Health Laboratory | Plumb et al |

| | | | | | | |
|---|---|---|---|---|---|---|
| EPI_ISL_613466 | USA/MN-MDH-1960/2020 | 2020-08-20 | North America/USA/Minnesota/ | Public Health Laboratory - Infectious Disease Lab, Minnesota Department of Health Infectious Disease Laboratory Submission Group | Minnesota Department of Health, Public Health Laboratory | Plumb et al |
| EPI_ISL_613468 | USA/MN-MDH-1958/2020 | 2020-08-23 | North America/USA/Minnesota/ | Public Health Laboratory - Infectious Disease Lab, Minnesota Department of Health Infectious Disease Laboratory Submission Group | Minnesota Department of Health, Public Health Laboratory | Plumb et al |
| EPI_ISL_613473 | USA/MN-MDH-1953/2020 | 2020-08-31 | North America/USA/Minnesota/ | Public Health Laboratory - Infectious Disease Lab, Minnesota Department of Health Infectious Disease Laboratory Submission Group | Minnesota Department of Health, Public Health Laboratory | Plumb et al |
| EPI_ISL_613478 | USA/MN-MDH-1948/2020 | 2020-09-18 | North America/USA/Minnesota/ | Public Health Laboratory - Infectious Disease Lab, Minnesota Department of Health Infectious Disease Laboratory Submission Group | Minnesota Department of Health, Public Health Laboratory | Plumb et al |
| EPI_ISL_613481 | USA/MN-MDH-1945/2020 | 2020-09-21 | North America/USA/Minnesota/ | Public Health Laboratory - Infectious Disease Lab, Minnesota Department of Health Infectious Disease Laboratory Submission Group | Minnesota Department of Health, Public Health Laboratory | Plumb et al |
| EPI_ISL_613483 | USA/MN-MDH-1943/2020 | 2020-09-21 | North America/USA/Minnesota/ | Public Health Laboratory - Infectious Disease Lab, Minnesota Department of Health Infectious Disease Laboratory Submission Group | Minnesota Department of Health, Public Health Laboratory | Plumb et al |
| EPI_ISL_613487 | USA/MN-MDH-1939/2020 | 2020-09-23 | North America/USA/Minnesota/ | Public Health Laboratory - Infectious Disease Lab, Minnesota Department of Health Infectious Disease Laboratory Submission Group | Minnesota Department of Health, Public Health Laboratory | Plumb et al |
| EPI_ISL_613490 | USA/MN-MDH-1936/2020 | 2020-09-30 | North America/USA/Minnesota/ | Public Health Laboratory - Infectious Disease Lab, Minnesota Department of Health Infectious Disease Laboratory Submission Group | Minnesota Department of Health, Public Health Laboratory | Plumb et al |
| EPI_ISL_613497 | USA/MN-MDH-1929/2020 | 2020-10-09 | North America/USA/Minnesota/ | Public Health Laboratory - Infectious Disease Lab, Minnesota Department of Health Infectious Disease Laboratory Submission Group | Minnesota Department of Health, Public Health Laboratory | Plumb et al |
| EPI_ISL_613498 | USA/MN-MDH-1928/2020 | 2020-10-10 | North America/USA/Minnesota/ | Public Health Laboratory - Infectious Disease Lab, Minnesota Department of Health Infectious Disease Laboratory Submission Group | Minnesota Department of Health, Public Health Laboratory | Plumb et al |

| | | | | | | |
|---|---|---|---|---|---|---|
| EPI_ISL_613502 | USA/MN-MDH-1924/2020 | 2020-10-12 | North America/USA/Minnesota/ | Public Health Laboratory - Infectious Disease Lab, Minnesota Department of Health Infectious Disease Laboratory Submission Group | Minnesota Department of Health, Public Health Laboratory | Plumb et al |
| EPI_ISL_613503 | USA/MN-MDH-1923/2020 | 2020-10-13 | North America/USA/Minnesota/ | Public Health Laboratory - Infectious Disease Lab, Minnesota Department of Health Infectious Disease Laboratory Submission Group | Minnesota Department of Health, Public Health Laboratory | Plumb et al |
| EPI_ISL_613504 | USA/MN-MDH-1922/2020 | 2020-10-14 | North America/USA/Minnesota/ | Public Health Laboratory - Infectious Disease Lab, Minnesota Department of Health Infectious Disease Laboratory Submission Group | Minnesota Department of Health, Public Health Laboratory | Plumb et al |
| EPI_ISL_613505 | USA/MN-MDH-1921/2020 | 2020-10-14 | North America/USA/Minnesota/ | Public Health Laboratory - Infectious Disease Lab, Minnesota Department of Health Infectious Disease Laboratory Submission Group | Minnesota Department of Health, Public Health Laboratory | Plumb et al |
| EPI_ISL_613508 | USA/MN-MDH-1918/2020 | 2020-10-16 | North America/USA/Minnesota/ | Public Health Laboratory - Infectious Disease Lab, Minnesota Department of Health Infectious Disease Laboratory Submission Group | Minnesota Department of Health, Public Health Laboratory | Plumb et al |
| EPI_ISL_613512 | USA/MN-MDH-1914/2020 | 2020-10-18 | North America/USA/Minnesota/ | Public Health Laboratory - Infectious Disease Lab, Minnesota Department of Health Infectious Disease Laboratory Submission Group | Minnesota Department of Health, Public Health Laboratory | Plumb et al |
| EPI_ISL_613518 | USA/MN-MDH-1908/2020 | 2020-10-18 | North America/USA/Minnesota/ | Public Health Laboratory - Infectious Disease Lab, Minnesota Department of Health Infectious Disease Laboratory Submission Group | Minnesota Department of Health, Public Health Laboratory | Plumb et al |
| EPI_ISL_613540 | USA/MN-MDH-1886/2020 | 2020-10-23 | North America/USA/Minnesota/ | Public Health Laboratory - Infectious Disease Lab, Minnesota Department of Health Infectious Disease Laboratory Submission Group | Minnesota Department of Health, Public Health Laboratory | Plumb et al |
| EPI_ISL_613546 | France/BRE-BR9056/2020 | 2020-08-15 | Europe/France/Bretagne/Rennes | CHRU Pontchaillou - Laboratoire de Virologie 2, rue Henri Le Guilloux | National Reference Center for Viruses of Respiratory Infections, Institut Pasteur, Paris | Marion Barbet et al |

| | | | | | | |
|---|---|---|---|---|---|---|
| EPI_ISL_613549 | France/BRE-BR9060/2020 | 2020-09-01 | Europe/France/Bretagne/Fougv®res | CHRU Pontchaillou - Laboratoire de Virologie 2, rue Henri Le Guilloux | National Reference Center for Viruses of Respiratory Infections, Institut Pasteur, Paris | Marion Barbet et al |
| EPI_ISL_613552 | France/BRE-BR9063/2020 | 2020-09-02 | Europe/France/Bretagne/Rennes | CHRU Pontchaillou - Laboratoire de Virologie 2, rue Henri Le Guilloux | National Reference Center for Viruses of Respiratory Infections, Institut Pasteur, Paris | Marion Barbet et al |
| EPI_ISL_613553 | France/BRE-BR9064/2020 | 2020-09-03 | Europe/France/Bretagne/Rennes | CHRU Pontchaillou - Laboratoire de Virologie 2, rue Henri Le Guilloux | National Reference Center for Viruses of Respiratory Infections, Institut Pasteur, Paris | Marion Barbet et al |
| EPI_ISL_613556 | France/BRE-BR9067/2020 | 2020-09-05 | Europe/France/Bretagne/Rennes | CHRU Pontchaillou - Laboratoire de Virologie 2, rue Henri Le Guilloux | National Reference Center for Viruses of Respiratory Infections, Institut Pasteur, Paris | Marion Barbet et al |
| EPI_ISL_613557 | France/BRE-BR9068/2020 | 2020-09-06 | Europe/France/Bretagne/Rennes | CHRU Pontchaillou - Laboratoire de Virologie 2, rue Henri Le Guilloux | National Reference Center for Viruses of Respiratory Infections, Institut Pasteur, Paris | Marion Barbet et al |
| EPI_ISL_613558 | France/BRE-BR9070/2020 | 2020-09-06 | Europe/France/Bretagne/Betton | CHRU Pontchaillou - Laboratoire de Virologie 2, rue Henri Le Guilloux | National Reference Center for Viruses of Respiratory Infections, Institut Pasteur, Paris | Marion Barbet et al |

| | | | | | | |
|---|---|---|---|---|---|---|
| EPI_ISL_613559 | France/BRE-BR9071/2020 | 2020-09-06 | Europe/France/Bretagne/Rennes | CHRU Pontchaillou - Laboratoire de Virologie 2, rue Henri Le Guilloux | National Reference Center for Viruses of Respiratory Infections, Institut Pasteur, Paris | Marion Barbet et al |
| EPI_ISL_613560 | Italy/SAR-ASSL-01/2020 | 2020-04-04 | Europe/Italy/Sardinia/Nuoro | "Laboratorio Biologia Molecolare Sars Cov2 - UOC Laboratorio Analisi - Servizio Medicina di Laboratorio, Ospedale ""San Francesco"" - ATS-ASSL Nuoro Via Mannironi 1, 08100 Nuoro" | "Laboratorio specialistico UOC Ematologia - Ospedale ""San Francesco"" - ATS-ASSL Nuoro Nuoro" | Piras Giovanna et al |
| EPI_ISL_613710 | Italy/SAR-ASSL-11/2020 | 2020-04-08 | Europe/Italy/Sardinia/ | "Laboratorio Biologia Molecolare Sars Cov2 - UOC Laboratorio Analisi - Servizio Medicina di Laboratorio, Ospedale ""San Francesco"" - ATS-ASSL Nuoro" | "Laboratorio specialistico UOC Ematologia - Ospedale ""San Francesco"" - ATS-ASSL Nuoro" | Piras Giovanna et al |
| EPI_ISL_613715 | USA/FL-BPHL-1891/2020 | 2020-07-14 | North America/USA/Florida/ | Florida Bureau of Public Health Laboratories | Florida Bureau of Public Health Laboratories | Sarah Schmedes et al |
| EPI_ISL_613718 | USA/FL-BPHL-1732/2020 | 2020-10-01 | North America/USA/Florida/ | Florida Bureau of Public Health Laboratories | Florida Bureau of Public Health Laboratories | Sarah Schmedes et al |
| EPI_ISL_613724 | USA/FL-BPHL-1927/2020 | 2020-07-29 | North America/USA/Florida/ | Florida Bureau of Public Health Laboratories | Florida Bureau of Public Health Laboratories | Sarah Schmedes et al |
| EPI_ISL_613726 | USA/FL-BPHL-1911/2020 | 2020-07-17 | North America/USA/Florida/ | Florida Bureau of Public Health Laboratories | Florida Bureau of Public Health Laboratories | Sarah Schmedes et al |
| EPI_ISL_613727 | USA/FL-BPHL-1909/2020 | 2020-07-17 | North America/USA/Florida/ | Florida Bureau of Public Health Laboratories | Florida Bureau of Public Health Laboratories | Sarah Schmedes et al |
| EPI_ISL_613732 | USA/FL-BPHL-1904/2020 | 2020-07-17 | North America/USA/Florida/ | Florida Bureau of Public Health Laboratories | Florida Bureau of Public Health Laboratories | Sarah Schmedes et al |
| EPI_ISL_613751 | USA/FL-BPHL-1737/2020 | 2020-10-02 | North America/USA/Florida/ | Florida Bureau of Public Health Laboratories | Florida Bureau of Public Health Laboratories | Sarah Schmedes et al |

| | | | | | | |
|---|---|---|---|---|---|---|
| EPI_ISL_613752 | USA/FL-BPHL-1709/2020 | 2020-09-30 | North America/USA/Florida/ | Florida Bureau of Public Health Laboratories | Florida Bureau of Public Health Laboratories | Sarah Schmedes et al |
| EPI_ISL_613759 | USA/FL-BPHL-1908/2020 | 2020-07-17 | North America/USA/Florida/ | Florida Bureau of Public Health Laboratories | Florida Bureau of Public Health Laboratories | Sarah Schmedes et al |
| EPI_ISL_613760 | USA/FL-BPHL-1711/2020 | 2020-09-30 | North America/USA/Florida/ | Florida Bureau of Public Health Laboratories | Florida Bureau of Public Health Laboratories | Sarah Schmedes et al |
| EPI_ISL_613762 | USA/FL-BPHL-1808/2020 | 2020-08-03 | North America/USA/Florida/ | Florida Bureau of Public Health Laboratories | Florida Bureau of Public Health Laboratories | Sarah Schmedes et al |
| EPI_ISL_613770 | USA/FL-BPHL-1896/2020 | 2020-07-15 | North America/USA/Florida/ | Florida Bureau of Public Health Laboratories | Florida Bureau of Public Health Laboratories | Sarah Schmedes et al |
| EPI_ISL_613771 | USA/FL-BPHL-1915/2020 | 2020-07-20 | North America/USA/Florida/ | Florida Bureau of Public Health Laboratories | Florida Bureau of Public Health Laboratories | Sarah Schmedes et al |
| EPI_ISL_613772 | USA/FL-BPHL-1912/2020 | 2020-07-17 | North America/USA/Florida/ | Florida Bureau of Public Health Laboratories | Florida Bureau of Public Health Laboratories | Sarah Schmedes et al |
| EPI_ISL_613787 | USA/FL-BPHL-1917/2020 | 2020-07-21 | North America/USA/Florida/ | Florida Bureau of Public Health Laboratories | Florida Bureau of Public Health Laboratories | Sarah Schmedes et al |
| EPI_ISL_613788 | USA/FL-BPHL-1936/2020 | 2020-04-08 | North America/USA/Florida/ | Florida Bureau of Public Health Laboratories | Florida Bureau of Public Health Laboratories | Sarah Schmedes et al |
| EPI_ISL_613789 | USA/FL-BPHL-1934/2020 | 2020-07-31 | North America/USA/Florida/ | Florida Bureau of Public Health Laboratories | Florida Bureau of Public Health Laboratories | Sarah Schmedes et al |
| EPI_ISL_613822 | USA/FL-BPHL-1733/2020 | 2020-10-01 | North America/USA/Florida/ | Florida Bureau of Public Health Laboratories | Florida Bureau of Public Health Laboratories | Sarah Schmedes et al |
| EPI_ISL_613829 | USA/FL-BPHL-1897/2020 | 2020-07-14 | North America/USA/Florida/ | Florida Bureau of Public Health Laboratories | Florida Bureau of Public Health Laboratories | Sarah Schmedes et al |
| EPI_ISL_613836 | USA/FL-BPHL-1918/2020 | 2020-07-21 | North America/USA/Florida/ | Florida Bureau of Public Health Laboratories | Florida Bureau of Public Health Laboratories | Sarah Schmedes et al |

| | | | | | | |
|---|---|---|---|---|---|---|
| EPI_ISL_613838 | USA/FL-BPHL-1892/2020 | 2020-07-14 | North America/USA/Florida/ | Florida Bureau of Public Health Laboratories | Florida Bureau of Public Health Laboratories | Sarah Schmedes et al |
| EPI_ISL_613840 | USA/FL-BPHL-1926/2020 | 2020-07-28 | North America/USA/Florida/ | Florida Bureau of Public Health Laboratories | Florida Bureau of Public Health Laboratories | Sarah Schmedes et al |
| EPI_ISL_613841 | USA/FL-BPHL-1932/2020 | 2020-07-31 | North America/USA/Florida/ | Florida Bureau of Public Health Laboratories | Florida Bureau of Public Health Laboratories | Sarah Schmedes et al |
| EPI_ISL_613846 | USA/FL-BPHL-1714/2020 | 2020-09-30 | North America/USA/Florida/ | Florida Bureau of Public Health Laboratories | Florida Bureau of Public Health Laboratories | Sarah Schmedes et al |
| EPI_ISL_613849 | USA/FL-BPHL-1719/2020 | 2020-09-30 | North America/USA/Florida/ | Florida Bureau of Public Health Laboratories | Florida Bureau of Public Health Laboratories | Sarah Schmedes et al |
| EPI_ISL_613854 | USA/FL-BPHL-1728/2020 | 2020-10-01 | North America/USA/Florida/ | Florida Bureau of Public Health Laboratories | Florida Bureau of Public Health Laboratories | Sarah Schmedes et al |
| EPI_ISL_613856 | USA/FL-BPHL-1734/2020 | 2020-10-02 | North America/USA/Florida/ | Florida Bureau of Public Health Laboratories | Florida Bureau of Public Health Laboratories | Sarah Schmedes et al |
| EPI_ISL_613865 | USA/FL-BPHL-1750/2020 | 2020-10-02 | North America/USA/Florida/ | Florida Bureau of Public Health Laboratories | Florida Bureau of Public Health Laboratories | Sarah Schmedes et al |
| EPI_ISL_613876 | USA/FL-BPHL-1771/2020 | 2020-07-29 | North America/USA/Florida/ | Florida Bureau of Public Health Laboratories | Florida Bureau of Public Health Laboratories | Sarah Schmedes et al |
| EPI_ISL_613880 | USA/FL-BPHL-1775/2020 | 2020-07-29 | North America/USA/Florida/ | Florida Bureau of Public Health Laboratories | Florida Bureau of Public Health Laboratories | Sarah Schmedes et al |
| EPI_ISL_613882 | USA/FL-BPHL-1779/2020 | 2020-07-30 | North America/USA/Florida/ | Florida Bureau of Public Health Laboratories | Florida Bureau of Public Health Laboratories | Sarah Schmedes et al |
| EPI_ISL_613887 | USA/FL-BPHL-1784/2020 | 2020-07-30 | North America/USA/Florida/ | Florida Bureau of Public Health Laboratories | Florida Bureau of Public Health Laboratories | Sarah Schmedes et al |
| EPI_ISL_613894 | USA/FL-BPHL-1792/2020 | 2020-07-30 | North America/USA/Florida/ | Florida Bureau of Public Health Laboratories | Florida Bureau of Public Health Laboratories | Sarah Schmedes et al |

| | | | | | | |
|---|---|---|---|---|---|---|
| EPI_ISL_613901 | USA/FL-BPHL-1804/2020 | 2020-08-03 | North America/USA/Florida/ | Florida Bureau of Public Health Laboratories | Florida Bureau of Public Health Laboratories | Sarah Schmedes et al |
| EPI_ISL_613904 | USA/FL-BPHL-1815/2020 | 2020-04-07 | North America/USA/Florida/ | Florida Bureau of Public Health Laboratories | Florida Bureau of Public Health Laboratories | Sarah Schmedes et al |
| EPI_ISL_613909 | USA/FL-BPHL-1822/2020 | 2020-04-08 | North America/USA/Florida/ | Florida Bureau of Public Health Laboratories | Florida Bureau of Public Health Laboratories | Sarah Schmedes et al |
| EPI_ISL_613912 | USA/FL-BPHL-1825/2020 | 2020-04-08 | North America/USA/Florida/ | Florida Bureau of Public Health Laboratories | Florida Bureau of Public Health Laboratories | Sarah Schmedes et al |
| EPI_ISL_613913 | USA/FL-BPHL-1827/2020 | 2020-04-08 | North America/USA/Florida/ | Florida Bureau of Public Health Laboratories | Florida Bureau of Public Health Laboratories | Sarah Schmedes et al |
| EPI_ISL_613926 | USA/FL-BPHL-1846/2020 | 2020-04-09 | North America/USA/Florida/ | Florida Bureau of Public Health Laboratories | Florida Bureau of Public Health Laboratories | Sarah Schmedes et al |
| EPI_ISL_613935 | USA/FL-BPHL-1855/2020 | 2020-04-09 | North America/USA/Florida/ | Florida Bureau of Public Health Laboratories | Florida Bureau of Public Health Laboratories | Sarah Schmedes et al |
| EPI_ISL_613936 | USA/FL-BPHL-1858/2020 | 2020-04-10 | North America/USA/Florida/ | Florida Bureau of Public Health Laboratories | Florida Bureau of Public Health Laboratories | Sarah Schmedes et al |
| EPI_ISL_613937 | USA/FL-BPHL-1859/2020 | 2020-04-10 | North America/USA/Florida/ | Florida Bureau of Public Health Laboratories | Florida Bureau of Public Health Laboratories | Sarah Schmedes et al |
| EPI_ISL_613947 | USA/FL-BPHL-1871/2020 | 2020-04-09 | North America/USA/Florida/ | Florida Bureau of Public Health Laboratories | Florida Bureau of Public Health Laboratories | Sarah Schmedes et al |
| EPI_ISL_613949 | USA/FL-BPHL-1889/2020 | 2020-07-14 | North America/USA/Florida/ | Florida Bureau of Public Health Laboratories | Florida Bureau of Public Health Laboratories | Sarah Schmedes et al |

| | | | | | | |
|---|---|---|---|---|---|---|
| EPI_ISL_613951 | Brazil/SP-RP9/2020 | 2020-04-01 | South America/Brazil/S√£o Paulo/Ribeir | Laboratory of Molecular Biology, Blood Center of Ribeir√£o Preto, Faculty of Medicine of Ribeir√£o Preto, University of S√£o Paulo | Laboratory of Molecular Biology, Blood Center of Ribeir√£o Preto, Faculty of Medicine of Ribeir√£o Preto, University of S√£o Paulo | Svetoslav N Slavov et al |
| EPI_ISL_613953 | Italy/SAR-ASSL-03/2020 | 2020-04-10 | Europe/Italy/Sardinia/ | "Laboratorio Biologia Molecolare Sars Cov2 - UOC Laboratorio Analisi - Servizio Medicina di Laboratorio, Ospedale ""San Francesco"" - ATS-ASSL Nuoro" | "Laboratorio specialistico UOC Ematologia - Ospedale ""San Francesco"" - ATS-ASSL Nuoro" | Piras Giovanna et al |
| EPI_ISL_613976 | USA/TX-DSHS-1006/2020 | 2020-07-03 | North America/USA/Texas/Hidalgo Coun | Texas Department of State Health Services | Texas Department of State Health Services | Rashmi Tuladhar et al |
| EPI_ISL_613979 | USA/TX-DSHS-1001/2020 | 2020-07-03 | North America/USA/Texas/Hidalgo Coun | Texas Department of State Health Services | Texas Department of State Health Services | Rashmi Tuladhar et al |
| EPI_ISL_613983 | USA/TX-DSHS-1003/2020 | 2020-07-03 | North America/USA/Texas/Hidalgo Coun | Texas Department of State Health Services | Texas Department of State Health Services | Rashmi Tuladhar et al |
| EPI_ISL_613986 | USA/TX-DSHS-0980/2020 | 2020-06-16 | North America/USA/Texas/Harris Count | Texas Department of State Health Services | Texas Department of State Health Services | Rashmi Tuladhar et al |
| EPI_ISL_613990 | USA/TX-DSHS-1002/2020 | 2020-07-03 | North America/USA/Texas/Hidalgo Coun | Texas Department of State Health Services | Texas Department of State Health Services | Rashmi Tuladhar et al |
| EPI_ISL_613991 | USA/TX-DSHS-0975/2020 | 2020-06-16 | North America/USA/Texas/Harris Count | Texas Department of State Health Services | Texas Department of State Health Services | Rashmi Tuladhar et al |
| EPI_ISL_613993 | USA/TX-DSHS-0963/2020 | 2020-06-15 | North America/USA/Texas/Harris Count | Texas Department of State Health Services | Texas Department of State Health Services | Rashmi Tuladhar et al |
| EPI_ISL_613996 | USA/TX-DSHS-0962/2020 | 2020-06-15 | North America/USA/Texas/Harris Count | Texas Department of State Health Services | Texas Department of State Health Services | Rashmi Tuladhar et al |

| | | | | | | |
|---|---|---|---|---|---|---|
| EPI_ISL_614007 | USA/TX-DSHS-0987/2020 | 2020-06-16 | North America/USA/Texas/El Paso Coun | Texas Department of State Health Services | Texas Department of State Health Services | Rashmi Tuladhar et al |
| EPI_ISL_614014 | USA/VA-DCLS-1604/2020 | 2020-08-14 | North America/USA/Virginia/ | Virginia DCLS | Virginia DCLS | Virginia DCLS et al |
| EPI_ISL_614016 | USA/VA-DCLS-1606/2020 | 2020-08-20 | North America/USA/Virginia/ | Virginia DCLS | Virginia DCLS | Virginia DCLS et al |
| EPI_ISL_614017 | USA/VA-DCLS-1607/2020 | 2020-08-18 | North America/USA/Virginia/ | Virginia DCLS | Virginia DCLS | Virginia DCLS et al |
| EPI_ISL_614024 | USA/VA-DCLS-1617/2020 | 2020-08-20 | North America/USA/Virginia/ | Virginia DCLS | Virginia DCLS | Virginia DCLS et al |
| EPI_ISL_614028 | USA/VA-DCLS-1621/2020 | 2020-08-19 | North America/USA/Virginia/ | Virginia DCLS | Virginia DCLS | Virginia DCLS et al |
| EPI_ISL_614041 | USA/VA-DCLS-1638/2020 | 2020-08-20 | North America/USA/Virginia/ | Virginia DCLS | Virginia DCLS | Virginia DCLS et al |
| EPI_ISL_614042 | USA/VA-DCLS-1639/2020 | 2020-08-18 | North America/USA/Virginia/ | Virginia DCLS | Virginia DCLS | Virginia DCLS et al |
| EPI_ISL_614047 | USA/VA-DCLS-1647/2020 | 2020-08-29 | North America/USA/Virginia/ | Virginia DCLS | Virginia DCLS | Virginia DCLS et al |
| EPI_ISL_614052 | USA/VA-DCLS-1729/2020 | 2020-09-02 | North America/USA/Virginia/ | Virginia DCLS | Virginia DCLS | Virginia DCLS et al |
| EPI_ISL_614053 | USA/VA-DCLS-1733/2020 | 2020-09-02 | North America/USA/Virginia/ | Virginia DCLS | Virginia DCLS | Virginia DCLS et al |
| EPI_ISL_614055 | USA/VA-DCLS-1803/2020 | 2020-09-16 | North America/USA/Virginia/ | Virginia DCLS | Virginia DCLS | Virginia DCLS et al |
| EPI_ISL_614058 | USA/VA-DCLS-1807/2020 | 2020-09-15 | North America/USA/Virginia/ | Virginia DCLS | Virginia DCLS | Virginia DCLS et al |
| EPI_ISL_614061 | USA/VA-DCLS-1810/2020 | 2020-09-16 | North America/USA/Virginia/ | Virginia DCLS | Virginia DCLS | Virginia DCLS et al |
| EPI_ISL_614064 | USA/VA-DCLS-1917/2020 | 2020-10-05 | North America/USA/Virginia/ | Virginia DCLS | Virginia DCLS | Virginia DCLS et al |
| EPI_ISL_614065 | USA/VA-DCLS-1918/2020 | 2020-09-28 | North America/USA/Virginia/ | Virginia DCLS | Virginia DCLS | Virginia DCLS et al |
| EPI_ISL_614072 | USA/VA-DCLS-1897/2020 | 2020-10-01 | North America/USA/Virginia/ | Virginia DCLS | Virginia DCLS | Virginia DCLS et al |
| EPI_ISL_614075 | USA/VA-DCLS-1849/2020 | 2020-09-01 | North America/USA/Virginia/ | Virginia DCLS | Virginia DCLS | Virginia DCLS et al |
| EPI_ISL_614079 | USA/VA-DCLS-1855/2020 | 2020-09-23 | North America/USA/Virginia/ | Virginia DCLS | Virginia DCLS | Virginia DCLS et al |
| EPI_ISL_614086 | USA/VA-DCLS-1867/2020 | 2020-09-23 | North America/USA/Virginia/ | Virginia DCLS | Virginia DCLS | Virginia DCLS et al |
| EPI_ISL_614087 | USA/VA-DCLS-1869/2020 | 2020-09-23 | North America/USA/Virginia/ | Virginia DCLS | Virginia DCLS | Virginia DCLS et al |
| EPI_ISL_614089 | USA/VA-DCLS-1900/2020 | 2020-10-05 | North America/USA/Virginia/ | Virginia DCLS | Virginia DCLS | Virginia DCLS et al |
| EPI_ISL_614092 | USA/VA-DCLS-1905/2020 | 2020-10-05 | North America/USA/Virginia/ | Virginia DCLS | Virginia DCLS | Virginia DCLS et al |
| EPI_ISL_614093 | USA/VA-DCLS-1924/2020 | 2020-10-12 | North America/USA/Virginia/ | Virginia DCLS | Virginia DCLS | Virginia DCLS et al |
| EPI_ISL_614100 | USA/VA-DCLS-0311/2020 | 2020-05-15 | North America/USA/Virginia/ | Virginia DCLS | Virginia DCLS | Virginia DCLS et al |
| EPI_ISL_614107 | USA/VA-DCLS-0326/2020 | 2020-05-15 | North America/USA/Virginia/ | Virginia DCLS | Virginia DCLS | Virginia DCLS et al |
| EPI_ISL_614111 | USA/VA-DCLS-0501/2020 | 2020-05-25 | North America/USA/Virginia/ | Virginia DCLS | Virginia DCLS | Virginia DCLS et al |
| EPI_ISL_614113 | USA/VA-DCLS-0504/2020 | 2020-05-29 | North America/USA/Virginia/ | Virginia DCLS | Virginia DCLS | Virginia DCLS et al |
| EPI_ISL_614114 | USA/VA-DCLS-0505/2020 | 2020-05-29 | North America/USA/Virginia/ | Virginia DCLS | Virginia DCLS | Virginia DCLS et al |

| | | | | | | |
|---|---|---|---|---|---|---|
| EPI_ISL_614122 | USA/VA-DCLS-0522/2020 | 2020-04-08 | North America/USA/Virginia/ | Virginia DCLS | Virginia DCLS | Virginia DCLS et al |
| EPI_ISL_614123 | USA/VA-DCLS-1713/2020 | 2020-09-02 | North America/USA/Virginia/ | Virginia DCLS | Virginia DCLS | Virginia DCLS et al |
| EPI_ISL_614125 | USA/VA-DCLS-1715/2020 | 2020-09-03 | North America/USA/Virginia/ | Virginia DCLS | Virginia DCLS | Virginia DCLS et al |
| EPI_ISL_614128 | USA/VA-DCLS-1836/2020 | 2020-09-14 | North America/USA/Virginia/ | Virginia DCLS | Virginia DCLS | Virginia DCLS et al |
| EPI_ISL_614130 | USA/VA-DCLS-1838/2020 | 2020-09-17 | North America/USA/Virginia/ | Virginia DCLS | Virginia DCLS | Virginia DCLS et al |
| EPI_ISL_614131 | USA/VA-DCLS-1839/2020 | 2020-09-10 | North America/USA/Virginia/ | Virginia DCLS | Virginia DCLS | Virginia DCLS et al |
| EPI_ISL_614139 | USA/VA-DCLS-1819/2020 | 2020-09-22 | North America/USA/Virginia/ | Virginia DCLS | Virginia DCLS | Virginia DCLS et al |
| EPI_ISL_614141 | USA/VA-DCLS-1823/2020 | 2020-09-24 | North America/USA/Virginia/ | Virginia DCLS | Virginia DCLS | Virginia DCLS et al |
| EPI_ISL_614142 | USA/VA-DCLS-1824/2020 | 2020-09-24 | North America/USA/Virginia/ | Virginia DCLS | Virginia DCLS | Virginia DCLS et al |
| EPI_ISL_614144 | USA/VA-DCLS-1829/2020 | 2020-09-24 | North America/USA/Virginia/ | Virginia DCLS | Virginia DCLS | Virginia DCLS et al |
| EPI_ISL_614150 | USA/VA-DCLS-1907/2020 | 2020-10-06 | North America/USA/Virginia/ | Virginia DCLS | Virginia DCLS | Virginia DCLS et al |
| EPI_ISL_614151 | USA/VA-DCLS-1908/2020 | 2020-10-05 | North America/USA/Virginia/ | Virginia DCLS | Virginia DCLS | Virginia DCLS et al |
| EPI_ISL_614152 | USA/VA-DCLS-1909/2020 | 2020-10-05 | North America/USA/Virginia/ | Virginia DCLS | Virginia DCLS | Virginia DCLS et al |
| EPI_ISL_614153 | USA/VA-DCLS-1910/2020 | 2020-10-05 | North America/USA/Virginia/ | Virginia DCLS | Virginia DCLS | Virginia DCLS et al |
| EPI_ISL_614158 | USA/MI-MDHHS-SC22107/2020 | 2020-10-14 | North America/USA/Michigan/ | Michigan Department of Health and Human Services, Bureau of Laboratories | Michigan Department of Health and Human Services, Bureau of Laboratories | Blankenship HM et al |
| EPI_ISL_614173 | USA/MI-MDHHS-SC22122/2020 | 2020-10-14 | North America/USA/Michigan/ | Michigan Department of Health and Human Services, Bureau of Laboratories | Michigan Department of Health and Human Services, Bureau of Laboratories | Blankenship HM et al |
| EPI_ISL_614174 | USA/MI-MDHHS-SC22123/2020 | 2020-10-02 | North America/USA/Michigan/ | Michigan Department of Health and Human Services, Bureau of Laboratories | Michigan Department of Health and Human Services, Bureau of Laboratories | Blankenship HM et al |

| | | | | | | |
|---|---|---|---|---|---|---|
| EPI_ISL_614191 | USA/MI-MDHHS-SC22140/2020 | 2020-10-06 | North America/USA/Michigan/ | Michigan Department of Health and Human Services, Bureau of Laboratories | Michigan Department of Health and Human Services, Bureau of Laboratories | Blankenship HM et al |
| EPI_ISL_614196 | USA/MI-MDHHS-SC22145/2020 | 2020-10-06 | North America/USA/Michigan/ | Michigan Department of Health and Human Services, Bureau of Laboratories | Michigan Department of Health and Human Services, Bureau of Laboratories | Blankenship HM et al |
| EPI_ISL_614203 | USA/MI-MDHHS-SC22152/2020 | 2020-10-18 | North America/USA/Michigan/ | Michigan Department of Health and Human Services, Bureau of Laboratories | Michigan Department of Health and Human Services, Bureau of Laboratories | Blankenship HM et al |
| EPI_ISL_614210 | USA/MI-MDHHS-SC22159/2020 | 2020-10-12 | North America/USA/Michigan/ | Michigan Department of Health and Human Services, Bureau of Laboratories | Michigan Department of Health and Human Services, Bureau of Laboratories | Blankenship HM et al |
| EPI_ISL_614211 | USA/MI-MDHHS-SC22160/2020 | 2020-10-13 | North America/USA/Michigan/ | Michigan Department of Health and Human Services, Bureau of Laboratories | Michigan Department of Health and Human Services, Bureau of Laboratories | Blankenship HM et al |
| EPI_ISL_614221 | USA/MI-MDHHS-SC22170/2020 | 2020-10-20 | North America/USA/Michigan/ | Michigan Department of Health and Human Services, Bureau of Laboratories | Michigan Department of Health and Human Services, Bureau of Laboratories | Blankenship HM et al |

| | | | | | | |
|---|---|---|---|---|---|---|
| EPI_ISL_614230 | USA/MI-MDHHS-SC22179/2020 | 2020-10-19 | North America/USA/Michigan/ | Michigan Department of Health and Human Services, Bureau of Laboratories | Michigan Department of Health and Human Services, Bureau of Laboratories | Blankenship HM et al |
| EPI_ISL_614234 | USA/MI-MDHHS-SC22183/2020 | 2020-10-06 | North America/USA/Michigan/ | Michigan Department of Health and Human Services, Bureau of Laboratories | Michigan Department of Health and Human Services, Bureau of Laboratories | Blankenship HM et al |
| EPI_ISL_614235 | USA/MI-MDHHS-SC22184/2020 | 2020-10-13 | North America/USA/Michigan/ | Michigan Department of Health and Human Services, Bureau of Laboratories | Michigan Department of Health and Human Services, Bureau of Laboratories | Blankenship HM et al |
| EPI_ISL_614237 | USA/MI-MDHHS-SC22186/2020 | 2020-10-20 | North America/USA/Michigan/ | Michigan Department of Health and Human Services, Bureau of Laboratories | Michigan Department of Health and Human Services, Bureau of Laboratories | Blankenship HM et al |
| EPI_ISL_614238 | USA/MI-MDHHS-SC22187/2020 | 2020-10-18 | North America/USA/Michigan/ | Michigan Department of Health and Human Services, Bureau of Laboratories | Michigan Department of Health and Human Services, Bureau of Laboratories | Blankenship HM et al |
| EPI_ISL_614242 | USA/MI-MDHHS-SC22191/2020 | 2020-10-17 | North America/USA/Michigan/ | Michigan Department of Health and Human Services, Bureau of Laboratories | Michigan Department of Health and Human Services, Bureau of Laboratories | Blankenship HM et al |

| | | | | | | |
|---|---|---|---|---|---|---|
| EPI_ISL_614244 | USA/MI-MDHHS-SC22193/2020 | 2020-10-20 | North America/USA/Michigan/ | Michigan Department of Health and Human Services, Bureau of Laboratories | Michigan Department of Health and Human Services, Bureau of Laboratories | Blankenship HM et al |
| EPI_ISL_614255 | USA/WY-UNM-00135/2020 | 2020-06-16 | North America/USA/Wyoming/ | Wyoming Public Health Laboratory | Center for Global Health, University of New Mexico Health Sciences Center | Daryl Domman et al |
| EPI_ISL_614265 | France/BRE-9325/2020 | 2020-08-09 | Europe/France/Bretagne/Chateaulin | Eurofins | National Reference Center for Viruses of Respiratory Infections, Institut Pasteur, Paris | Marion Barbet et al |
| EPI_ISL_614267 | France/BRE-9327/2020 | 2020-08-11 | Europe/France/Bretagne/Chateaulin | Eurofins | National Reference Center for Viruses of Respiratory Infections, Institut Pasteur, Paris | Marion Barbet et al |
| EPI_ISL_614268 | France/BRE-9329/2020 | 2020-08-13 | Europe/France/Bretagne/Chateaulin | Eurofins | National Reference Center for Viruses of Respiratory Infections, Institut Pasteur, Paris | Marion Barbet et al |
| EPI_ISL_614270 | France/BRE-9331/2020 | 2020-08-13 | Europe/France/Bretagne/Chateaulin | Eurofins | National Reference Center for Viruses of Respiratory Infections, Institut Pasteur, Paris | Marion Barbet et al |
| EPI_ISL_614275 | France/BRE-9343/2020 | 2020-08-13 | Europe/France/Bretagne/Chateaulin | Eurofins | National Reference Center for Viruses of Respiratory Infections, Institut Pasteur, Paris | Marion Barbet et al |

| | | | | | | |
|---|---|---|---|---|---|---|
| EPI_ISL_614277 | France/BRE-9346/2020 | 2020-08-13 | Europe/France/Bretagne/Chateaulin | Eurofins | National Reference Center for Viruses of Respiratory Infections, Institut Pasteur, Paris | Marion Barbet et al |
| EPI_ISL_614278 | France/BRE-9348/2020 | 2020-08-13 | Europe/France/Bretagne/Chateaulin | Eurofins | National Reference Center for Viruses of Respiratory Infections, Institut Pasteur, Paris | Marion Barbet et al |
| EPI_ISL_614282 | France/GES-9388/2020 | 2020-10-05 | Europe/France/Grand Est/Bischheim | General practitioner | National Reference Center for Viruses of Respiratory Infections, Institut Pasteur, Paris | Marion Barbet et al |
| EPI_ISL_614283 | France/BFC-9406/2020 | 2020-10-06 | Europe/France/Bourgogne-France-Comt | General practitioner | National Reference Center for Viruses of Respiratory Infections, Institut Pasteur, Paris | Marion Barbet et al |
| EPI_ISL_614284 | France/HDF-9440/2020 | 2020-10-12 | Europe/France/Hauts de France/Compiv | General practitioner | National Reference Center for Viruses of Respiratory Infections, Institut Pasteur, Paris | Marion Barbet et al |
| EPI_ISL_614285 | France/IDF-9449/2020 | 2020-10-12 | Europe/France/Ile de France/Verriv®res | General practitioner | National Reference Center for Viruses of Respiratory Infections, Institut Pasteur, Paris | Marion Barbet et al |

| | | | | | | |
|---|---|---|---|---|---|---|
| EPI_ISL_614288 | France/CVL-9469/2020 | 2020-10-19 | Europe/France/Centre-Val de Loire/Blois | General practitioner | National Reference Center for Viruses of Respiratory Infections, Institut Pasteur, Paris | Marion Barbet et al |
| EPI_ISL_614289 | France/CVL-9471/2020 | 2020-10-19 | Europe/France/Centre-Val de Loire/Rou: | General practitioner | National Reference Center for Viruses of Respiratory Infections, Institut Pasteur, Paris | Marion Barbet et al |
| EPI_ISL_614290 | France/CVL-9474/2020 | 2020-10-16 | Europe/France/Centre-Val de Loire/Blois | General practitioner | National Reference Center for Viruses of Respiratory Infections, Institut Pasteur, Paris | Marion Barbet et al |
| EPI_ISL_614291 | France/IDF-9475/2020 | 2020-10-19 | Europe/France/Ile de France/Moussy Le | General practitioner | National Reference Center for Viruses of Respiratory Infections, Institut Pasteur, Paris | Marion Barbet et al |
| EPI_ISL_614292 | France/GES-9488/2020 | 2020-10-22 | Europe/France/Grand Est/Thann | General practitioner | National Reference Center for Viruses of Respiratory Infections, Institut Pasteur, Paris | Marion Barbet et al |
| EPI_ISL_614396 | Italy/SAR-ASSL-07/2020 | 2020-04-23 | Europe/Italy/Sardinia/ | "Laboratorio Biologia Molecolare Sars Cov2 - UOC Laboratorio Analisi - Servizio Medicina di Laboratorio, Ospedale ""San Francesco"" - ATS-ASSL Nuoro" | "Laboratorio specialistico UOC Ematologia - Ospedale ""San Francesco"" - ATS-ASSL Nuoro" | Piras Giovanna et al |

| | | | | | | |
|---|---|---|---|---|---|---|
| EPI_ISL_614397 | Italy/SAR-ASSL-06/2020 | 2020-04-28 | Europe/Italy/Sardinia/ | "Laboratorio Biologia Molecolare Sars Cov2 - UOC Laboratorio Analisi - Servizio Medicina di Laboratorio, Ospedale ""San Francesco"" - ATS-ASSL Nuoro" | "Laboratorio specialistico UOC Ematologia - Ospedale ""San Francesco"" - ATS-ASSL Nuoro" | Piras Giovanna et al |
| EPI_ISL_614502 | Denmark/DCGC-1077/2020 | 2020-05-11 | Europe/Denmark/Hovedstaden/ | Department of Virus and Microbiological Special Diagnostics, Statens Serum Institut, Denmark | Albertsen lab, Department of Chemistry and Bioscience, Aalborg University, Denmark | Danish Covid-19 Genome Consortia et al |
| EPI_ISL_614564 | Denmark/DCGC-954/2020 | 2020-03-09 | Europe/Denmark/Sjaelland/ | Department of Virus and Microbiological Special Diagnostics, Statens Serum Institut, Denmark | Albertsen lab, Department of Chemistry and Bioscience, Aalborg University, Denmark | Danish Covid-19 Genome Consortia et al |
| EPI_ISL_614664 | Denmark/DCGC-1170/2020 | 2020-03-16 | Europe/Denmark/Hovedstaden/ | Department of Virus and Microbiological Special Diagnostics, Statens Serum Institut, Denmark | Albertsen lab, Department of Chemistry and Bioscience, Aalborg University, Denmark | Danish Covid-19 Genome Consortia et al |
| EPI_ISL_614732 | Denmark/DCGC-807/2020 | 2020-04-20 | Europe/Denmark/Nordjylland/ | Department of Virus and Microbiological Special Diagnostics, Statens Serum Institut, Denmark | Albertsen lab, Department of Chemistry and Bioscience, Aalborg University, Denmark | Danish Covid-19 Genome Consortia et al |
| EPI_ISL_614748 | Denmark/DCGC-837/2020 | 2020-04-20 | Europe/Denmark/Nordjylland/ | Department of Virus and Microbiological Special Diagnostics, Statens Serum Institut, Denmark | Albertsen lab, Department of Chemistry and Bioscience, Aalborg University, Denmark | Danish Covid-19 Genome Consortia et al |

| | | | | | | |
|---|---|---|---|---|---|---|
| EPI_ISL_614777 | Denmark/DCGC-1006/2020 | 2020-04-20 | Europe/Denmark/Syddanmark/ | Department of Virus and Microbiological Special Diagnostics, Statens Serum Institut, Denmark | Albertsen lab, Department of Chemistry and Bioscience, Aalborg University, Denmark | Danish Covid-19 Genome Consortia et al |
| EPI_ISL_614902 | Switzerland/BS-ETHZ-330428/2020 | 2020-10-18 | Europe/Switzerland/Basel-Stadt/ | Viollier AG | Department of Biosystems Science and Engineering, ETH Zv°rich | Christian Beisel et al |
| EPI_ISL_614954 | Switzerland/VD-ETHZ-330248/2020 | 2020-10-17 | Europe/Switzerland/Vaud/ | Viollier AG | Department of Biosystems Science and Engineering, ETH Zv°rich | Christian Beisel et al |
| EPI_ISL_614959 | Switzerland/VD-ETHZ-330255/2020 | 2020-10-17 | Europe/Switzerland/Vaud/ | Viollier AG | Department of Biosystems Science and Engineering, ETH Zv°rich | Christian Beisel et al |
| EPI_ISL_615076 | Sweden/20-21494/2020 | 2020-05-15 | Europe/Sweden/Uppsala/ | Uppsala klinisk mikrobiologi | The Public Health Agency of Sweden | Anna-Malin Linde et al |
| EPI_ISL_615078 | Sweden/20-21496/2020 | 2020-05-09 | Europe/Sweden/Uppsala/ | Uppsala klinisk mikrobiologi | The Public Health Agency of Sweden | Anna-Malin Linde et al |
| EPI_ISL_615085 | Sweden/20-22146/2020 | 2020-05-22 | Europe/Sweden/Skane/ | Klinisk mikrobiologi, Skanes universitetssjukhus, Lund | The Public Health Agency of Sweden | Anna-Malin Linde et al |
| EPI_ISL_615086 | Sweden/20-22736/2020 | 2020-05-06 | Europe/Sweden/Stockholm/ | Karolinska Universitetslaboratoriet | The Public Health Agency of Sweden | Anna-Malin Linde et al |
| EPI_ISL_615091 | Sweden/20-51747/2020 | 2020-05-22 | Europe/Sweden/Vastra Gotaland/ | Klinisk mikrobiologi NAL Trollhattan | The Public Health Agency of Sweden | Anna-Malin Linde et al |
| EPI_ISL_615093 | Sweden/20-52112/2020 | 2020-05-22 | Europe/Sweden/Vasterbotten/ | Umea klinisk mikrobiologi | The Public Health Agency of Sweden | Anna-Malin Linde et al |
| EPI_ISL_615094 | Sweden/20-52114/2020 | 2020-05-15 | Europe/Sweden/Vasterbotten/ | Umea klinisk mikrobiologi | The Public Health Agency of Sweden | Anna-Malin Linde et al |
| EPI_ISL_615098 | Sweden/20-52699/2020 | 2020-05-12 | Europe/Sweden/Jonkoping/ | Klinisk mikrobiologi Lanssjukhuset Ryhov, Jonkoping | The Public Health Agency of Sweden | Anna-Malin Linde et al |
| EPI_ISL_615102 | Sweden/20-52698/2020 | 2020-08-24 | Europe/Sweden/Gavleborgs Lan/ | Gavle klinisk mikrobiologi | The Public Health Agency of Sweden | Anna-Malin Linde et al |

| | | | | | | |
|---|---|---|---|---|---|---|
| EPI_ISL_615108 | Sweden/20-52779/2020 | 2020-08-25 | Europe/Sweden/Vastmanland/ | Klinisk Mikrobiologi | The Public Health Agency of Sweden | Anna-Malin Linde et al |
| EPI_ISL_615116 | Sweden/20-52830/2020 | 2020-09-07 | Europe/Sweden/Vastra Gotaland/ | Skovde/Unilabs | The Public Health Agency of Sweden | Anna-Malin Linde et al |
| EPI_ISL_615122 | USA/TX-DSHS-1081/2020 | 2020-04-25 | North America/USA/Texas/Waller Coun | Texas Department of State Health Services | Texas Department of State Health Services | Rashmi Tuladhar et al |
| EPI_ISL_615127 | USA/TX-DSHS-1087/2020 | 2020-05-14 | North America/USA/Texas/Harris Count | Texas Department of State Health Services | Texas Department of State Health Services | Rashmi Tuladhar et al |
| EPI_ISL_615128 | USA/TX-DSHS-1089/2020 | 2020-05-14 | North America/USA/Texas/Harris Count | Texas Department of State Health Services | Texas Department of State Health Services | Rashmi Tuladhar et al |
| EPI_ISL_615132 | USA/TX-DSHS-1093/2020 | 2020-05-21 | North America/USA/Texas/El Paso Coun | Texas Department of State Health Services | Texas Department of State Health Services | Rashmi Tuladhar et al |
| EPI_ISL_615133 | USA/TX-DSHS-1094/2020 | 2020-05-22 | North America/USA/Texas/El Paso Coun | Texas Department of State Health Services | Texas Department of State Health Services | Rashmi Tuladhar et al |
| EPI_ISL_615136 | USA/TX-DSHS-1097/2020 | 2020-06-12 | North America/USA/Texas/El Paso Coun | Texas Department of State Health Services | Texas Department of State Health Services | Rashmi Tuladhar et al |
| EPI_ISL_615143 | USA/TX-DSHS-1106/2020 | 2020-06-15 | North America/USA/Texas/Harris Count | Texas Department of State Health Services | Texas Department of State Health Services | Rashmi Tuladhar et al |
| EPI_ISL_615146 | USA/TX-DSHS-1109/2020 | 2020-06-15 | North America/USA/Texas/Harris Count | Texas Department of State Health Services | Texas Department of State Health Services | Rashmi Tuladhar et al |
| EPI_ISL_615154 | USA/TX-DSHS-1117/2020 | 2020-06-21 | North America/USA/Texas/Hidalgo Cour | Texas Department of State Health Services | Texas Department of State Health Services | Rashmi Tuladhar et al |
| EPI_ISL_615160 | USA/TX-DSHS-1123/2020 | 2020-06-21 | North America/USA/Texas/Hidalgo Cour | Texas Department of State Health Services | Texas Department of State Health Services | Rashmi Tuladhar et al |
| EPI_ISL_615182 | Denmark/DCGC-5251/2020 | 2020-09-14 | Europe/Denmark/Sjaelland/ | Department of Virus and Microbiological Special Diagnostics, Statens Serum Institut, Denmark | Albertsen lab, Department of Chemistry and Bioscience, Aalborg University, Denmark | Danish Covid-19 Genome Consortia et al |

| | | | | | | |
|---|---|---|---|---|---|---|
| EPI_ISL_615317 | Denmark/DCGC-5385/2020 | 2020-09-14 | Europe/Denmark/Syddanmark/ | Department of Virus and Microbiological Special Diagnostics, Statens Serum Institut, Denmark | Albertsen lab, Department of Chemistry and Bioscience, Aalborg University, Denmark | Danish Covid-19 Genome Consortia et al |
| EPI_ISL_615369 | Denmark/DCGC-5437/2020 | 2020-09-14 | Europe/Denmark/Midtjylland/ | Department of Virus and Microbiological Special Diagnostics, Statens Serum Institut, Denmark | Albertsen lab, Department of Chemistry and Bioscience, Aalborg University, Denmark | Danish Covid-19 Genome Consortia et al |
| EPI_ISL_615557 | Denmark/DCGC-4355/2020 | 2020-09-07 | Europe/Denmark/Nordjylland/ | Department of Virus and Microbiological Special Diagnostics, Statens Serum Institut, Denmark | Albertsen lab, Department of Chemistry and Bioscience, Aalborg University, Denmark | Danish Covid-19 Genome Consortia et al |
| EPI_ISL_616037 | Denmark/DCGC-3285/2020 | 2020-08-31 | Europe/Denmark/Hovedstaden/ | Department of Virus and Microbiological Special Diagnostics, Statens Serum Institut, Denmark | Albertsen lab, Department of Chemistry and Bioscience, Aalborg University, Denmark | Danish Covid-19 Genome Consortia et al |
| EPI_ISL_616339 | Denmark/DCGC-3546/2020 | 2020-08-31 | Europe/Denmark/Hovedstaden/ | Department of Virus and Microbiological Special Diagnostics, Statens Serum Institut, Denmark | Albertsen lab, Department of Chemistry and Bioscience, Aalborg University, Denmark | Danish Covid-19 Genome Consortia et al |

| | | | | | | |
|---|---|---|---|---|---|---|
| EPI_ISL_616411 | Denmark/DCGC-2895/2020 | 2020-08-17 | Europe/Denmark/Denmark/ | Department of Virus and Microbiological Special Diagnostics, Statens Serum Institut, Denmark | Albertsen lab, Department of Chemistry and Bioscience, Aalborg University, Denmark | Danish Covid-19 Genome Consortia et al |
| EPI_ISL_617057 | Denmark/DCGC-8068/2020 | 2020-10-26 | Europe/Denmark/Hovedstaden/ | Department of Virus and Microbiological Special Diagnostics, Statens Serum Institut, Denmark | Albertsen lab, Department of Chemistry and Bioscience, Aalborg University, Denmark | Danish Covid-19 Genome Consortia et al |
| EPI_ISL_617962 | Denmark/DCGC-1541/2020 | 2020-05-25 | Europe/Denmark/Hovedstaden/ | Department of Virus and Microbiological Special Diagnostics, Statens Serum Institut, Denmark | Albertsen lab, Department of Chemistry and Bioscience, Aalborg University, Denmark | Danish Covid-19 Genome Consortia et al |
| EPI_ISL_618040 | Denmark/DCGC-1263/2020 | 2020-06-08 | Europe/Denmark/Nordjylland/ | Department of Virus and Microbiological Special Diagnostics, Statens Serum Institut, Denmark | Albertsen lab, Department of Chemistry and Bioscience, Aalborg University, Denmark | Danish Covid-19 Genome Consortia et al |
| EPI_ISL_618158 | Denmark/DCGC-1690/2020 | 2020-06-29 | Europe/Denmark/Nordjylland/ | Department of Virus and Microbiological Special Diagnostics, Statens Serum Institut, Denmark | Albertsen lab, Department of Chemistry and Bioscience, Aalborg University, Denmark | Danish Covid-19 Genome Consortia et al |

| | | | | | | |
|---|---|---|---|---|---|---|
| EPI_ISL_618363 | Denmark/DCGC-2471/2020 | 2020-08-03 | Europe/Denmark/Nordjylland/ | Department of Virus and Microbiological Special Diagnostics, Statens Serum Institut, Denmark | Albertsen lab, Department of Chemistry and Bioscience, Aalborg University, Denmark | Danish Covid-19 Genome Consortia et al |
| EPI_ISL_618636 | Denmark/DCGC-1592/2020 | 2020-04-13 | Europe/Denmark/Syddanmark/ | Department of Virus and Microbiological Special Diagnostics, Statens Serum Institut, Denmark | Albertsen lab, Department of Chemistry and Bioscience, Aalborg University, Denmark | Danish Covid-19 Genome Consortia et al |
| EPI_ISL_618846 | Denmark/DCGC-7874/2020 | 2020-10-19 | Europe/Denmark/Midtjylland/ | Department of Virus and Microbiological Special Diagnostics, Statens Serum Institut, Denmark | Albertsen lab, Department of Chemistry and Bioscience, Aalborg University, Denmark | Danish Covid-19 Genome Consortia et al |
| EPI_ISL_618978 | Denmark/DCGC-1615/2020 | 2020-04-06 | Europe/Denmark/Hovedstaden/ | Department of Virus and Microbiological Special Diagnostics, Statens Serum Institut, Denmark | Albertsen lab, Department of Chemistry and Bioscience, Aalborg University, Denmark | Danish Covid-19 Genome Consortia et al |
| EPI_ISL_618980 | Denmark/DCGC-2050/2020 | 2020-04-06 | Europe/Denmark/Hovedstaden/ | Department of Virus and Microbiological Special Diagnostics, Statens Serum Institut, Denmark | Albertsen lab, Department of Chemistry and Bioscience, Aalborg University, Denmark | Danish Covid-19 Genome Consortia et al |

| | | | | | | |
|---|---|---|---|---|---|---|
| EPI_ISL_619145 | Denmark/DCGC-1640/2020 | 2020-04-06 | Europe/Denmark/Hovedstaden/ | Department of Virus and Microbiological Special Diagnostics, Statens Serum Institut, Denmark | Albertsen lab, Department of Chemistry and Bioscience, Aalborg University, Denmark | Danish Covid-19 Genome Consortia et al |
| EPI_ISL_619735 | Denmark/DCGC-6890/2020 | 2020-10-12 | Europe/Denmark/Midtjylland/ | Department of Virus and Microbiological Special Diagnostics, Statens Serum Institut, Denmark | Albertsen lab, Department of Chemistry and Bioscience, Aalborg University, Denmark | Danish Covid-19 Genome Consortia et al |
| EPI_ISL_619908 | Denmark/DCGC-5759/2020 | 2020-09-28 | Europe/Denmark/Hovedstaden/ | Department of Virus and Microbiological Special Diagnostics, Statens Serum Institut, Denmark | Albertsen lab, Department of Chemistry and Bioscience, Aalborg University, Denmark | Danish Covid-19 Genome Consortia et al |
| EPI_ISL_620223 | Denmark/DCGC-5958/2020 | 2020-10-05 | Europe/Denmark/Midtjylland/ | Department of Virus and Microbiological Special Diagnostics, Statens Serum Institut, Denmark | Albertsen lab, Department of Chemistry and Bioscience, Aalborg University, Denmark | Danish Covid-19 Genome Consortia et al |
| EPI_ISL_620776 | Denmark/DCGC-5451/2020 | 2020-09-21 | Europe/Denmark/Syddanmark/ | Department of Virus and Microbiological Special Diagnostics, Statens Serum Institut, Denmark | Albertsen lab, Department of Chemistry and Bioscience, Aalborg University, Denmark | Danish Covid-19 Genome Consortia et al |

| | | | | | | |
|---|---|---|---|---|---|---|
| EPI_ISL_620778 | Denmark/DCGC-5453/2020 | 2020-09-21 | Europe/Denmark/Syddanmark/ | Department of Virus and Microbiological Special Diagnostics, Statens Serum Institut, Denmark | Albertsen lab, Department of Chemistry and Bioscience, Aalborg University, Denmark | Danish Covid-19 Genome Consortia et al |
| EPI_ISL_620901 | Denmark/DCGC-5576/2020 | 2020-09-28 | Europe/Denmark/Hovedstaden/ | Department of Virus and Microbiological Special Diagnostics, Statens Serum Institut, Denmark | Albertsen lab, Department of Chemistry and Bioscience, Aalborg University, Denmark | Danish Covid-19 Genome Consortia et al |
| EPI_ISL_620978 | Denmark/DCGC-5791/2020 | 2020-09-28 | Europe/Denmark/Sjaelland/ | Department of Virus and Microbiological Special Diagnostics, Statens Serum Institut, Denmark | Albertsen lab, Department of Chemistry and Bioscience, Aalborg University, Denmark | Danish Covid-19 Genome Consortia et al |
| EPI_ISL_621504 | Denmark/DCGC-4966/2020 | 2020-09-21 | Europe/Denmark/Hovedstaden/ | Department of Virus and Microbiological Special Diagnostics, Statens Serum Institut, Denmark | Albertsen lab, Department of Chemistry and Bioscience, Aalborg University, Denmark | Danish Covid-19 Genome Consortia et al |
| EPI_ISL_621659 | Denmark/DCGC-5121/2020 | 2020-09-21 | Europe/Denmark/Syddanmark/ | Department of Virus and Microbiological Special Diagnostics, Statens Serum Institut, Denmark | Albertsen lab, Department of Chemistry and Bioscience, Aalborg University, Denmark | Danish Covid-19 Genome Consortia et al |

| | | | | | | |
|---|---|---|---|---|---|---|
| EPI_ISL_621777 | Denmark/DCGC-4980/2020 | 2020-09-14 | Europe/Denmark/Denmark/ | Department of Virus and Microbiological Special Diagnostics, Statens Serum Institut, Denmark | Albertsen lab, Department of Chemistry and Bioscience, Aalborg University, Denmark | Danish Covid-19 Genome Consortia et al |
| EPI_ISL_621811 | Denmark/DCGC-5032/2020 | 2020-09-21 | Europe/Denmark/Hovedstaden/ | Department of Virus and Microbiological Special Diagnostics, Statens Serum Institut, Denmark | Albertsen lab, Department of Chemistry and Bioscience, Aalborg University, Denmark | Danish Covid-19 Genome Consortia et al |
| EPI_ISL_621929 | Denmark/DCGC-4697/2020 | 2020-09-21 | Europe/Denmark/Hovedstaden/ | Department of Virus and Microbiological Special Diagnostics, Statens Serum Institut, Denmark | Albertsen lab, Department of Chemistry and Bioscience, Aalborg University, Denmark | Danish Covid-19 Genome Consortia et al |
| EPI_ISL_621963 | Denmark/DCGC-4143/2020 | 2020-09-21 | Europe/Denmark/Hovedstaden/ | Department of Virus and Microbiological Special Diagnostics, Statens Serum Institut, Denmark | Albertsen lab, Department of Chemistry and Bioscience, Aalborg University, Denmark | Danish Covid-19 Genome Consortia et al |
| EPI_ISL_622055 | Denmark/DCGC-4099/2020 | 2020-09-14 | Europe/Denmark/Midtjylland/ | Department of Virus and Microbiological Special Diagnostics, Statens Serum Institut, Denmark | Albertsen lab, Department of Chemistry and Bioscience, Aalborg University, Denmark | Danish Covid-19 Genome Consortia et al |

| | | | | | | |
|---|---|---|---|---|---|---|
| EPI_ISL_622075 | Denmark/DCGC-4119/2020 | 2020-09-14 | Europe/Denmark/Hovedstaden/ | Department of Virus and Microbiological Special Diagnostics, Statens Serum Institut, Denmark | Albertsen lab, Department of Chemistry and Bioscience, Aalborg University, Denmark | Danish Covid-19 Genome Consortia et al |
| EPI_ISL_622104 | Denmark/DCGC-4211/2020 | 2020-09-14 | Europe/Denmark/Sjaelland/ | Department of Virus and Microbiological Special Diagnostics, Statens Serum Institut, Denmark | Albertsen lab, Department of Chemistry and Bioscience, Aalborg University, Denmark | Danish Covid-19 Genome Consortia et al |
| EPI_ISL_622116 | Denmark/DCGC-4223/2020 | 2020-09-14 | Europe/Denmark/Midtjylland/ | Department of Virus and Microbiological Special Diagnostics, Statens Serum Institut, Denmark | Albertsen lab, Department of Chemistry and Bioscience, Aalborg University, Denmark | Danish Covid-19 Genome Consortia et al |
| EPI_ISL_622175 | Denmark/DCGC-4736/2020 | 2020-09-21 | Europe/Denmark/Nordjylland/ | Department of Virus and Microbiological Special Diagnostics, Statens Serum Institut, Denmark | Albertsen lab, Department of Chemistry and Bioscience, Aalborg University, Denmark | Danish Covid-19 Genome Consortia et al |
| EPI_ISL_622221 | Denmark/DCGC-1374/2020 | 2020-03-30 | Europe/Denmark/Sjaelland/ | Department of Virus and Microbiological Special Diagnostics, Statens Serum Institut, Denmark | Albertsen lab, Department of Chemistry and Bioscience, Aalborg University, Denmark | Danish Covid-19 Genome Consortia et al |

| | | | | | | |
|---|---|---|---|---|---|---|
| EPI_ISL_622313 | Denmark/DCGC-6377/2020 | 2020-09-28 | Europe/Denmark/Hovedstaden/ | Department of Virus and Microbiological Special Diagnostics, Statens Serum Institut, Denmark | Albertsen lab, Department of Chemistry and Bioscience, Aalborg University, Denmark | Danish Covid-19 Genome Consortia et al |
| EPI_ISL_622354 | Denmark/DCGC-6730/2020 | 2020-04-27 | Europe/Denmark/Hovedstaden/ | Department of Virus and Microbiological Special Diagnostics, Statens Serum Institut, Denmark | Albertsen lab, Department of Chemistry and Bioscience, Aalborg University, Denmark | Danish Covid-19 Genome Consortia et al |
| EPI_ISL_622435 | Denmark/DCGC-1436/2020 | 2020-04-27 | Europe/Denmark/Hovedstaden/ | Department of Virus and Microbiological Special Diagnostics, Statens Serum Institut, Denmark | Albertsen lab, Department of Chemistry and Bioscience, Aalborg University, Denmark | Danish Covid-19 Genome Consortia et al |
| EPI_ISL_622438 | Denmark/DCGC-1456/2020 | 2020-03-30 | Europe/Denmark/Hovedstaden/ | Department of Virus and Microbiological Special Diagnostics, Statens Serum Institut, Denmark | Albertsen lab, Department of Chemistry and Bioscience, Aalborg University, Denmark | Danish Covid-19 Genome Consortia et al |
| EPI_ISL_622464 | Denmark/DCGC-1439/2020 | 2020-04-27 | Europe/Denmark/Hovedstaden/ | Department of Virus and Microbiological Special Diagnostics, Statens Serum Institut, Denmark | Albertsen lab, Department of Chemistry and Bioscience, Aalborg University, Denmark | Danish Covid-19 Genome Consortia et al |

| | | | | | | |
|---|---|---|---|---|---|---|
| EPI_ISL_622552 | Denmark/DCGC-1895/2020 | 2020-03-30 | Europe/Denmark/Hovedstaden/ | Department of Virus and Microbiological Special Diagnostics, Statens Serum Institut, Denmark | Albertsen lab, Department of Chemistry and Bioscience, Aalborg University, Denmark | Danish Covid-19 Genome Consortia et al |
| EPI_ISL_622580 | Denmark/DCGC-1871/2020 | 2020-04-06 | Europe/Denmark/Hovedstaden/ | Department of Virus and Microbiological Special Diagnostics, Statens Serum Institut, Denmark | Albertsen lab, Department of Chemistry and Bioscience, Aalborg University, Denmark | Danish Covid-19 Genome Consortia et al |
| EPI_ISL_622653 | Denmark/DCGC-3759/2020 | 2020-04-20 | Europe/Denmark/Hovedstaden/ | Department of Virus and Microbiological Special Diagnostics, Statens Serum Institut, Denmark | Albertsen lab, Department of Chemistry and Bioscience, Aalborg University, Denmark | Danish Covid-19 Genome Consortia et al |
| EPI_ISL_622798 | NewZealand/20CV0303/2020 | 2020-03-30 | Oceania/New Zealand/Hawkes Bay/ | Canterbury Health Laboratories | Institute of Environmental Science and Research (ESR) | Xiaoyun Ren et al |
| EPI_ISL_622906 | SouthAfrica/R13512-20/2020 | 2020-07-10 | Africa/South Africa/North-West/ | National Institute for Communicable Diseases of the National Health Laboratory Service | National Institute for Communicable Diseases of the National Health Laboratory Service | Allam M et al |
| EPI_ISL_622909 | SouthAfrica/R13300-20/2020 | 2020-06-30 | Africa/South Africa/KwaZulu-Natal/ | National Institute for Communicable Diseases of the National Health Laboratory Service | National Institute for Communicable Diseases of the National Health Laboratory Service | Allam M et al |

| | | | | | | |
|---|---|---|---|---|---|---|
| EPI_ISL_622911 | SouthAfrica/R13566-20/2020 | 2020-07-14 | Africa/South Africa/North-West/ | National Institute for Communicable Diseases of the National Health Laboratory Service | National Institute for Communicable Diseases of the National Health Laboratory Service | Allam M et al |
| EPI_ISL_622913 | SouthAfrica/R13061-20/2020 | 2020-06-23 | Africa/South Africa/Western Cape Provi | National Institute for Communicable Diseases of the National Health Laboratory Service | National Institute for Communicable Diseases of the National Health Laboratory Service | Allam M et al |
| EPI_ISL_622914 | SouthAfrica/R13472-20/2020 | 2020-07-06 | Africa/South Africa/KwaZulu-Natal/ | National Institute for Communicable Diseases of the National Health Laboratory Service | National Institute for Communicable Diseases of the National Health Laboratory Service | Allam M et al |
| EPI_ISL_622916 | SouthAfrica/R13509-20/2020 | 2020-07-10 | Africa/South Africa/North-West/ | National Institute for Communicable Diseases of the National Health Laboratory Service | National Institute for Communicable Diseases of the National Health Laboratory Service | Allam M et al |
| EPI_ISL_622919 | SouthAfrica/R13436-20/2020 | 2020-07-06 | Africa/South Africa/Mpumalanga/ | National Institute for Communicable Diseases of the National Health Laboratory Service | National Institute for Communicable Diseases of the National Health Laboratory Service | Allam M et al |
| EPI_ISL_622932 | SouthAfrica/R13276-20/2020 | 2020-07-02 | Africa/South Africa/KwaZulu-Natal/ | National Institute for Communicable Diseases of the National Health Laboratory Service | National Institute for Communicable Diseases of the National Health Laboratory Service | Allam M et al |

| | | | | | | |
|---|---|---|---|---|---|---|
| EPI_ISL_622934 | SouthAfrica/R13067-20/2020 | 2020-06-23 | Africa/South Africa/Western Cape Provi | National Institute for Communicable Diseases of the National Health Laboratory Service | National Institute for Communicable Diseases of the National Health Laboratory Service | Allam M et al |
| EPI_ISL_622935 | SouthAfrica/R13147-20/2020 | 2020-06-26 | Africa/South Africa/Western Cape Provi | National Institute for Communicable Diseases of the National Health Laboratory Service | National Institute for Communicable Diseases of the National Health Laboratory Service | Allam M et al |
| EPI_ISL_622936 | SouthAfrica/R13159-20/2020 | 2020-06-30 | Africa/South Africa/Gauteng/ | National Institute for Communicable Diseases of the National Health Laboratory Service | National Institute for Communicable Diseases of the National Health Laboratory Service | Allam M et al |
| EPI_ISL_622937 | SouthAfrica/R13274-20/2020 | 2020-06-29 | Africa/South Africa/KwaZulu-Natal/ | National Institute for Communicable Diseases of the National Health Laboratory Service | National Institute for Communicable Diseases of the National Health Laboratory Service | Allam M et al |
| EPI_ISL_622938 | SouthAfrica/R13278-20/2020 | 2020-06-29 | Africa/South Africa/KwaZulu-Natal/ | National Institute for Communicable Diseases of the National Health Laboratory Service | National Institute for Communicable Diseases of the National Health Laboratory Service | Allam M et al |
| EPI_ISL_622939 | SouthAfrica/R13317-20/2020 | 2020-07-02 | Africa/South Africa/Western Cape Provi | National Institute for Communicable Diseases of the National Health Laboratory Service | National Institute for Communicable Diseases of the National Health Laboratory Service | Allam M et al |

| | | | | | | |
|---|---|---|---|---|---|---|
| EPI_ISL_622940 | SouthAfrica/R13452-20/2020 | 2020-07-07 | Africa/South Africa/KwaZulu-Natal/ | National Institute for Communicable Diseases of the National Health Laboratory Service | National Institute for Communicable Diseases of the National Health Laboratory Service | Allam M et al |
| EPI_ISL_622942 | SouthAfrica/N00095/2020 | 2020-08-15 | Africa/South Africa/Eastern Cape/ | National Health Laboratory Service | National Institute for Communicable Diseases of the National Health Laboratory Service | Allam M et al |
| EPI_ISL_622943 | SouthAfrica/L00170/2020 | 2020-07-27 | Africa/South Africa/Gauteng/ | Lancet Laboratories | National Institute for Communicable Diseases of the National Health Laboratory Service | Allam M et al |
| EPI_ISL_622945 | SouthAfrica/N00009/2020 | 2020-08-13 | Africa/South Africa/Eastern Cape/ | National Health Laboratory Service | National Institute for Communicable Diseases of the National Health Laboratory Service | Allam M et al |
| EPI_ISL_622946 | SouthAfrica/L00174/2020 | 2020-07-27 | Africa/South Africa/Gauteng/ | Lancet Laboratories | National Institute for Communicable Diseases of the National Health Laboratory Service | Allam M et al |
| EPI_ISL_622949 | SouthAfrica/L00196/2020 | 2020-07-27 | Africa/South Africa/Limpopo/ | Lancet Laboratories | National Institute for Communicable Diseases of the National Health Laboratory Service | Allam M et al |

| | | | | | | |
|---|---|---|---|---|---|---|
| EPI_ISL_622959 | SouthAfrica/L00181/2020 | 2020-07-27 | Africa/South Africa/Gauteng/ | Lancet Laboratories | National Institute for Communicable Diseases of the National Health Laboratory Service | Allam M et al |
| EPI_ISL_622963 | SouthAfrica/L00160/2020 | 2020-07-26 | Africa/South Africa/Gauteng/ | Lancet Laboratories | National Institute for Communicable Diseases of the National Health Laboratory Service | Allam M et al |
| EPI_ISL_622967 | SouthAfrica/L00194/2020 | 2020-07-28 | Africa/South Africa/Gauteng/ | Lancet Laboratories | National Institute for Communicable Diseases of the National Health Laboratory Service | Allam M et al |
| EPI_ISL_622970 | SouthAfrica/N00131/2020 | 2020-07-10 | Africa/South Africa/Mpumalanga/ | National Health Laboratory Service | National Institute for Communicable Diseases of the National Health Laboratory Service | Allam M et al |
| EPI_ISL_622979 | SouthAfrica/N00107/2020 | 2020-06-11 | Africa/South Africa/Mpumalanga/ | National Health Laboratory Service | National Institute for Communicable Diseases of the National Health Laboratory Service | Allam M et al |
| EPI_ISL_622981 | SouthAfrica/N00042/2020 | 2020-08-12 | Africa/South Africa/Eastern Cape/ | National Health Laboratory Service | National Institute for Communicable Diseases of the National Health Laboratory Service | Allam M et al |

| | | | | | | |
|---|---|---|---|---|---|---|
| EPI_ISL_622983 | SouthAfrica/L00183/2020 | 2020-07-27 | Africa/South Africa/Gauteng/ | Lancet Laboratories | National Institute for Communicable Diseases of the National Health Laboratory Service | Allam M et al |
| EPI_ISL_622990 | SouthAfrica/L00190/2020 | 2020-07-28 | Africa/South Africa/Mpumalanga/ | Lancet Laboratories | National Institute for Communicable Diseases of the National Health Laboratory Service | Allam M et al |
| EPI_ISL_622992 | SouthAfrica/N00034/2020 | 2020-08-11 | Africa/South Africa/Eastern Cape/ | National Health Laboratory Service | National Institute for Communicable Diseases of the National Health Laboratory Service | Allam M et al |
| EPI_ISL_622994 | SouthAfrica/L00165/2020 | 2020-07-27 | Africa/South Africa/Gauteng/ | Lancet Laboratories | National Institute for Communicable Diseases of the National Health Laboratory Service | Allam M et al |
| EPI_ISL_623080 | Sweden/20-22147/2020 | 2020-06-26 | Europe/Sweden/Skane/ | Klinisk mikrobiologi, Skanes universitetssjukhus, Lund | The Public Health Agency of Sweden | Anna-Malin Linde et al |
| EPI_ISL_623081 | Sweden/20-22148/2020 | 2020-06-26 | Europe/Sweden/Skane/ | Klinisk mikrobiologi, Skanes universitetssjukhus, Lund | The Public Health Agency of Sweden | Anna-Malin Linde et al |
| EPI_ISL_623084 | Sweden/20-22153/2020 | 2020-06-02 | Europe/Sweden/Skane/ | Klinisk mikrobiologi, Skanes universitetssjukhus, Lund | The Public Health Agency of Sweden | Anna-Malin Linde et al |
| EPI_ISL_623086 | Sweden/20-52028/2020 | 2020-06-10 | Europe/Sweden/Halland/ | Halmstad klinisk mikrobiologi | The Public Health Agency of Sweden | Anna-Malin Linde et al |
| EPI_ISL_623088 | Sweden/20-52054/2020 | 2020-06-04 | Europe/Sweden/Vastra Gotaland/ | Klinisk mikrobiologi NAL Trollhattan | The Public Health Agency of Sweden | Anna-Malin Linde et al |
| EPI_ISL_623091 | Sweden/20-52162/2020 | 2020-06-16 | Europe/Sweden/Kalmar/ | Kalmar klinisk mikrobiologi | The Public Health Agency of Sweden | Anna-Malin Linde et al |
| EPI_ISL_623092 | Sweden/20-52703/2020 | 2020-06-03 | Europe/Sweden/Jonkoping/ | Klinisk mikrobiologi Lanssjukhuset Ryhov, Jonkoping | The Public Health Agency of Sweden | Anna-Malin Linde et al |
| EPI_ISL_623093 | Sweden/20-52704/2020 | 2020-06-03 | Europe/Sweden/Jonkoping/ | Klinisk mikrobiologi Lanssjukhuset Ryhov, Jonkoping | The Public Health Agency of Sweden | Anna-Malin Linde et al |

| | | | | | | |
|---|---|---|---|---|---|---|
| EPI_ISL_623096 | France/BRE-9495/2020 | 2020-10-22 | Europe/France/Bretagne/Saint-Brieuc | General practitioner | National Reference Center for Viruses of Respiratory Infections, Institut Pasteur, Paris | Marion Barbet et al |
| EPI_ISL_623097 | France/GES-9483/2020 | 2020-10-20 | Europe/France/Grand Est/Tucquegnieux | General practitioner | National Reference Center for Viruses of Respiratory Infections, Institut Pasteur, Paris | Marion Barbet et al |
| EPI_ISL_623098 | France/ARA-CDP82137/2020 | 2020-09-08 | Europe/France/Auvergne-Rhv¥ne-Alpes/ | CNR Virus des Infections Respiratoires - France SUD | CNR Virus des Infections Respiratoires - France SUD | Antonin Bal et al |
| EPI_ISL_623099 | France/ARA-CDP24988/2020 | 2020-09-23 | Europe/France/Auvergne-Rhv¥ne-Alpes/ | CNR Virus des Infections Respiratoires - France SUD | CNR Virus des Infections Respiratoires - France SUD | Antonin Bal et al |
| EPI_ISL_623100 | France/ARA-CDP82132/2020 | 2020-09-08 | Europe/France/Auvergne-Rhv¥ne-Alpes/ | CNR Virus des Infections Respiratoires - France SUD | CNR Virus des Infections Respiratoires - France SUD | Antonin Bal et al |
| EPI_ISL_623102 | France/ARA-CDP82284/2020 | 2020-09-09 | Europe/France/Auvergne-Rhv¥ne-Alpes/ | CNR Virus des Infections Respiratoires - France SUD | CNR Virus des Infections Respiratoires - France SUD | Antonin Bal et al |
| EPI_ISL_623107 | Brazil/RJ-00304/2020 | 2020-05-22 | South America/Brazil/Rio de Janeiro/ | Laboratorio de Virologia Molecular / UFRJ | Bioinformatics Laboratory / LNCC | Carolina M Voloch et al |
| EPI_ISL_623110 | Brazil/RJ-00307/2020 | 2020-04-24 | South America/Brazil/Rio de Janeiro/ | Laboratorio de Virologia Molecular / UFRJ | Bioinformatics Laboratory / LNCC | Carolina M Voloch et al |
| EPI_ISL_623111 | Brazil/RJ-00308/2020 | 2020-05-21 | South America/Brazil/Rio de Janeiro/ | Laboratorio de Virologia Molecular / UFRJ | Bioinformatics Laboratory / LNCC | Carolina M Voloch et al |
| EPI_ISL_623112 | Brazil/RJ-00309/2020 | 2020-04-20 | South America/Brazil/Rio de Janeiro/ | Laboratorio de Virologia Molecular / UFRJ | Bioinformatics Laboratory / LNCC | Carolina M Voloch et al |
| EPI_ISL_623113 | Brazil/RJ-00310/2020 | 2020-05-13 | South America/Brazil/Rio de Janeiro/ | Laboratorio de Virologia Molecular / UFRJ | Bioinformatics Laboratory / LNCC | Carolina M Voloch et al |
| EPI_ISL_623116 | Brazil/RJ-00313/2020 | 2020-04-07 | South America/Brazil/Rio de Janeiro/ | Laboratorio de Virologia Molecular / UFRJ | Bioinformatics Laboratory / LNCC | Carolina M Voloch et al |

| | | | | | | |
|---|---|---|---|---|---|---|
| EPI_ISL_623119 | Brazil/RJ-00316/2020 | 2020-05-04 | South America/Brazil/Rio de Janeiro/ | Laboratorio de Virologia Molecular / UFRJ | Bioinformatics Laboratory / LNCC | Carolina M Voloch et al |
| EPI_ISL_623121 | Brazil/RJ-00318/2020 | 2020-05-12 | South America/Brazil/Rio de Janeiro/ | Laboratorio de Virologia Molecular / UFRJ | Bioinformatics Laboratory / LNCC | Carolina M Voloch et al |
| EPI_ISL_623131 | Brazil/RJ-00328/2020 | 2020-05-14 | South America/Brazil/Rio de Janeiro/ | Laboratorio de Virologia Molecular / UFRJ | Bioinformatics Laboratory / LNCC | Carolina M Voloch et al |
| EPI_ISL_623135 | Brazil/RJ-00332/2020 | 2020-05-18 | South America/Brazil/Rio de Janeiro/ | Laboratorio de Virologia Molecular / UFRJ | Bioinformatics Laboratory / LNCC | Carolina M Voloch et al |
| EPI_ISL_623137 | Brazil/RJ-00334/2020 | 2020-05-18 | South America/Brazil/Rio de Janeiro/ | Laboratorio de Virologia Molecular / UFRJ | Bioinformatics Laboratory / LNCC | Carolina M Voloch et al |
| EPI_ISL_623139 | Brazil/RJ-00336/2020 | 2020-04-20 | South America/Brazil/Rio de Janeiro/ | Laboratorio de Virologia Molecular / UFRJ | Bioinformatics Laboratory / LNCC | Carolina M Voloch et al |
| EPI_ISL_623141 | Brazil/RJ-00338/2020 | 2020-05-04 | South America/Brazil/Rio de Janeiro/ | Laboratorio de Virologia Molecular / UFRJ | Bioinformatics Laboratory / LNCC | Carolina M Voloch et al |
| EPI_ISL_623142 | Brazil/RJ-00339/2020 | 2020-04-22 | South America/Brazil/Rio de Janeiro/ | Laboratorio de Virologia Molecular / UFRJ | Bioinformatics Laboratory / LNCC | Carolina M Voloch et al |
| EPI_ISL_623144 | Brazil/RJ-00341/2020 | 2020-05-04 | South America/Brazil/Rio de Janeiro/ | Laboratorio de Virologia Molecular / UFRJ | Bioinformatics Laboratory / LNCC | Carolina M Voloch et al |
| EPI_ISL_623145 | Brazil/RJ-00342/2020 | 2020-05-18 | South America/Brazil/Rio de Janeiro/ | Laboratorio de Virologia Molecular / UFRJ | Bioinformatics Laboratory / LNCC | Carolina M Voloch et al |
| EPI_ISL_623146 | Brazil/RJ-00343/2020 | 2020-04-29 | South America/Brazil/Rio de Janeiro/ | Laboratorio de Virologia Molecular / UFRJ | Bioinformatics Laboratory / LNCC | Carolina M Voloch et al |
| EPI_ISL_623147 | Brazil/RJ-00344/2020 | 2020-05-18 | South America/Brazil/Rio de Janeiro/ | Laboratorio de Virologia Molecular / UFRJ | Bioinformatics Laboratory / LNCC | Carolina M Voloch et al |
| EPI_ISL_623149 | Brazil/RJ-00346/2020 | 2020-05-08 | South America/Brazil/Rio de Janeiro/ | Laboratorio de Virologia Molecular / UFRJ | Bioinformatics Laboratory / LNCC | Carolina M Voloch et al |
| EPI_ISL_623151 | Brazil/RJ-00348/2020 | 2020-05-05 | South America/Brazil/Rio de Janeiro/ | Laboratorio de Virologia Molecular / UFRJ | Bioinformatics Laboratory / LNCC | Carolina M Voloch et al |
| EPI_ISL_623152 | Brazil/RJ-00349/2020 | 2020-04-16 | South America/Brazil/Rio de Janeiro/ | Laboratorio de Virologia Molecular / UFRJ | Bioinformatics Laboratory / LNCC | Carolina M Voloch et al |
| EPI_ISL_623156 | Brazil/RJ-00353/2020 | 2020-04-13 | South America/Brazil/Rio de Janeiro/ | Laboratorio de Virologia Molecular / UFRJ | Bioinformatics Laboratory / LNCC | Carolina M Voloch et al |
| EPI_ISL_623158 | Brazil/RJ-00355/2020 | 2020-05-07 | South America/Brazil/Rio de Janeiro/ | Laboratorio de Virologia Molecular / UFRJ | Bioinformatics Laboratory / LNCC | Carolina M Voloch et al |
| EPI_ISL_623159 | Brazil/RJ-00356/2020 | 2020-05-27 | South America/Brazil/Rio de Janeiro/ | Laboratorio de Virologia Molecular / UFRJ | Bioinformatics Laboratory / LNCC | Carolina M Voloch et al |
| EPI_ISL_623160 | Brazil/RJ-00357/2020 | 2020-04-22 | South America/Brazil/Rio de Janeiro/ | Laboratorio de Virologia Molecular / UFRJ | Bioinformatics Laboratory / LNCC | Carolina M Voloch et al |
| EPI_ISL_623161 | Brazil/RJ-00358/2020 | 2020-05-04 | South America/Brazil/Rio de Janeiro/ | Laboratorio de Virologia Molecular / UFRJ | Bioinformatics Laboratory / LNCC | Carolina M Voloch et al |

| | | | | | | |
|---|---|---|---|---|---|---|
| EPI_ISL_623163 | Brazil/RJ-00360/2020 | 2020-05-04 | South America/Brazil/Rio de Janeiro/ | Laboratorio de Virologia Molecular / UFRJ | Bioinformatics Laboratory / LNCC | Carolina M Voloch et al |
| EPI_ISL_623164 | Brazil/RJ-00361/2020 | 2020-04-29 | South America/Brazil/Rio de Janeiro/ | Laboratorio de Virologia Molecular / UFRJ | Bioinformatics Laboratory / LNCC | Carolina M Voloch et al |
| EPI_ISL_623165 | Brazil/RJ-00362/2020 | 2020-05-05 | South America/Brazil/Rio de Janeiro/ | Laboratorio de Virologia Molecular / UFRJ | Bioinformatics Laboratory / LNCC | Carolina M Voloch et al |
| EPI_ISL_623166 | Brazil/RJ-00363/2020 | 2020-04-20 | South America/Brazil/Rio de Janeiro/ | Laboratorio de Virologia Molecular / UFRJ | Bioinformatics Laboratory / LNCC | Carolina M Voloch et al |
| EPI_ISL_623167 | Brazil/RJ-00364/2020 | 2020-05-04 | South America/Brazil/Rio de Janeiro/ | Laboratorio de Virologia Molecular / UFRJ | Bioinformatics Laboratory / LNCC | Carolina M Voloch et al |
| EPI_ISL_623168 | Brazil/RJ-00365/2020 | 2020-04-08 | South America/Brazil/Rio de Janeiro/ | Laboratorio de Virologia Molecular / UFRJ | Bioinformatics Laboratory / LNCC | Carolina M Voloch et al |
| EPI_ISL_623172 | USA/UT-UPHL-201131371/2020 | 2020-04-03 | North America/USA/Utah/ | Utah Public Health Laboratory | Utah Public Health Laboratory | Erin Young et al |
| EPI_ISL_623177 | USA/UT-UPHL-201132611/2020 | 2020-04-04 | North America/USA/Utah/ | Utah Public Health Laboratory | Utah Public Health Laboratory | Erin Young et al |
| EPI_ISL_623181 | USA/UT-UPHL-201117349/2020 | 2020-04-01 | North America/USA/Utah/ | Utah Public Health Laboratory | Utah Public Health Laboratory | Erin Young et al |
| EPI_ISL_623186 | USA/UT-UPHL-201108812/2020 | 2020-04-09 | North America/USA/Utah/ | Utah Public Health Laboratory | Utah Public Health Laboratory | Erin Young et al |
| EPI_ISL_623192 | USA/UT-UPHL-201111436/2020 | 2020-04-09 | North America/USA/Utah/ | Utah Public Health Laboratory | Utah Public Health Laboratory | Erin Young et al |
| EPI_ISL_623203 | USA/UT-UPHL-201130116/2020 | 2020-04-09 | North America/USA/Utah/ | Utah Public Health Laboratory | Utah Public Health Laboratory | Erin Young et al |
| EPI_ISL_623208 | USA/UT-UPHL-201110993/2020 | 2020-04-20 | North America/USA/Utah/ | Utah Public Health Laboratory | Utah Public Health Laboratory | Erin Young et al |
| EPI_ISL_623237 | England/MILK-AB7718/2020 | 2020-10-19 | Europe/United Kingdom/England/ | Lighthouse Lab in Milton Keynes | Wellcome Sanger Institute for the COVID-19 Genomics UK (COG-UK) consortium | The Lighthouse Lab in Milton Keynes et al |
| EPI_ISL_623321 | England/CAMC-AAFCF6/2020 | 2020-10-21 | Europe/United Kingdom/England/ | Lighthouse Lab in Cambridge | Wellcome Sanger Institute for the COVID-19 Genomics UK (COG-UK) consortium | Rob Howes et al |

| | | | | | | |
|---|---|---|---|---|---|---|
| EPI_ISL_623386 | England/CAMC-AAFE0F/2020 | 2020-10-21 | Europe/United Kingdom/England/ | Lighthouse Lab in Cambridge | Wellcome Sanger Institute for the COVID-19 Genomics UK (COG-UK) consortium | Rob Howes et al |
| EPI_ISL_623476 | England/MILK-ABB0FB/2020 | 2020-10-21 | Europe/United Kingdom/England/ | Lighthouse Lab in Milton Keynes | Wellcome Sanger Institute for the COVID-19 Genomics UK (COG-UK) consortium | The Lighthouse Lab in Milton Keynes et al |
| EPI_ISL_623513 | England/MILK-AB86FC/2020 | 2020-10-21 | Europe/United Kingdom/England/ | Lighthouse Lab in Milton Keynes | Wellcome Sanger Institute for the COVID-19 Genomics UK (COG-UK) consortium | The Lighthouse Lab in Milton Keynes et al |
| EPI_ISL_623601 | England/MILK-AB88F6/2020 | 2020-10-22 | Europe/United Kingdom/England/ | Lighthouse Lab in Milton Keynes | Wellcome Sanger Institute for the COVID-19 Genomics UK (COG-UK) consortium | The Lighthouse Lab in Milton Keynes et al |
| EPI_ISL_623680 | England/CAMC-AB1CD3/2020 | 2020-10-22 | Europe/United Kingdom/England/ | Lighthouse Lab in Cambridge | Wellcome Sanger Institute for the COVID-19 Genomics UK (COG-UK) consortium | Rob Howes et al |
| EPI_ISL_623741 | England/CAMC-AB209A/2020 | 2020-10-23 | Europe/United Kingdom/England/ | Lighthouse Lab in Cambridge | Wellcome Sanger Institute for the COVID-19 Genomics UK (COG-UK) consortium | Rob Howes et al |
| EPI_ISL_623758 | England/CAMC-AB1347/2020 | 2020-10-23 | Europe/United Kingdom/England/ | Lighthouse Lab in Cambridge | Wellcome Sanger Institute for the COVID-19 Genomics UK (COG-UK) consortium | Rob Howes et al |
| EPI_ISL_623813 | England/MILK-AB88BA/2020 | 2020-10-22 | Europe/United Kingdom/England/ | Lighthouse Lab in Milton Keynes | Wellcome Sanger Institute for the COVID-19 Genomics UK (COG-UK) consortium | The Lighthouse Lab in Milton Keynes et al |

| | | | | | | |
|---|---|---|---|---|---|---|
| EPI_ISL_623860 | England/ALDP-AA141A/2020 | 2020-10-22 | Europe/United Kingdom/England/ | Lighthouse Lab in Alderley Park | Wellcome Sanger Institute for the COVID-19 Genomics UK (COG-UK) consortium | Jacquelyn Wynn et al |
| EPI_ISL_623932 | England/ALDP-AA330C/2020 | 2020-10-22 | Europe/United Kingdom/England/ | Lighthouse Lab in Alderley Park | Wellcome Sanger Institute for the COVID-19 Genomics UK (COG-UK) consortium | Jacquelyn Wynn et al |
| EPI_ISL_623973 | England/ALDP-AA413E/2020 | 2020-10-22 | Europe/United Kingdom/England/ | Lighthouse Lab in Alderley Park | Wellcome Sanger Institute for the COVID-19 Genomics UK (COG-UK) consortium | Jacquelyn Wynn et al |
| EPI_ISL_624119 | England/ALDP-AA5197/2020 | 2020-10-22 | Europe/United Kingdom/England/ | Lighthouse Lab in Alderley Park | Wellcome Sanger Institute for the COVID-19 Genomics UK (COG-UK) consortium | Jacquelyn Wynn et al |
| EPI_ISL_624167 | England/ALDP-AA130E/2020 | 2020-10-22 | Europe/United Kingdom/England/ | Lighthouse Lab in Alderley Park | Wellcome Sanger Institute for the COVID-19 Genomics UK (COG-UK) consortium | Jacquelyn Wynn et al |
| EPI_ISL_624300 | England/ALDP-AA2060/2020 | 2020-10-22 | Europe/United Kingdom/England/ | Lighthouse Lab in Alderley Park | Wellcome Sanger Institute for the COVID-19 Genomics UK (COG-UK) consortium | Jacquelyn Wynn et al |
| EPI_ISL_624392 | England/ALDP-AA2455/2020 | 2020-10-22 | Europe/United Kingdom/England/ | Lighthouse Lab in Alderley Park | Wellcome Sanger Institute for the COVID-19 Genomics UK (COG-UK) consortium | Jacquelyn Wynn et al |
| EPI_ISL_624399 | Wales/ALDP-AA4918/2020 | 2020-10-21 | Europe/United Kingdom/Wales/ | Lighthouse Lab in Alderley Park | Wellcome Sanger Institute for the COVID-19 Genomics UK (COG-UK) consortium | Jacquelyn Wynn et al |

| | | | | | | |
|---|---|---|---|---|---|---|
| EPI_ISL_624494 | England/ALDP-AA1F64/2020 | 2020-10-22 | Europe/United Kingdom/England/ | Lighthouse Lab in Alderley Park | Wellcome Sanger Institute for the COVID-19 Genomics UK (COG-UK) consortium | Jacquelyn Wynn et al |
| EPI_ISL_624608 | England/ALDP-A95FCD/2020 | 2020-10-19 | Europe/United Kingdom/England/ | Lighthouse Lab in Alderley Park | Wellcome Sanger Institute for the COVID-19 Genomics UK (COG-UK) consortium | Jacquelyn Wynn et al |
| EPI_ISL_624809 | England/ALDP-A95D4B/2020 | 2020-10-20 | Europe/United Kingdom/England/ | Lighthouse Lab in Alderley Park | Wellcome Sanger Institute for the COVID-19 Genomics UK (COG-UK) consortium | Jacquelyn Wynn et al |
| EPI_ISL_624921 | England/ALDP-AA35BB/2020 | 2020-10-22 | Europe/United Kingdom/England/ | Lighthouse Lab in Alderley Park | Wellcome Sanger Institute for the COVID-19 Genomics UK (COG-UK) consortium | Jacquelyn Wynn et al |
| EPI_ISL_625082 | England/ALDP-AA1887/2020 | 2020-10-22 | Europe/United Kingdom/England/ | Lighthouse Lab in Alderley Park | Wellcome Sanger Institute for the COVID-19 Genomics UK (COG-UK) consortium | Jacquelyn Wynn et al |
| EPI_ISL_625096 | England/ALDP-AA2C11/2020 | 2020-10-22 | Europe/United Kingdom/England/ | Lighthouse Lab in Alderley Park | Wellcome Sanger Institute for the COVID-19 Genomics UK (COG-UK) consortium | Jacquelyn Wynn et al |
| EPI_ISL_625456 | Madagascar/VIRO-5815/2020 | 2020-05-04 | Africa/Madagascar/Toamasina/ | Virology Unit, Institut Pasteur de Madagascar | Virology Unit, Institut Pasteur de Madagascar | Christian Ranaivoson et al |
| EPI_ISL_625458 | Bangladesh/CHRF-0044/2020 | 2020-06-09 | Asia/Bangladesh/Bangladesh/ | Child Health Research Foundation | Child Health Research Foundation | Senjuti Saha et al |
| EPI_ISL_625459 | Bangladesh/CHRF-0045/2020 | 2020-05-16 | Asia/Bangladesh/Bangladesh/ | Child Health Research Foundation | Child Health Research Foundation | Senjuti Saha et al |

| | | | | | | |
|---|---|---|---|---|---|---|
| EPI_ISL_625460 | Bangladesh/CHRF-0046/2020 | 2020-05-23 | Asia/Bangladesh/Bangladesh/ | Child Health Research Foundation | Child Health Research Foundation | Senjuti Saha et al |
| EPI_ISL_625461 | Bangladesh/CHRF-0047/2020 | 2020-05-24 | Asia/Bangladesh/Bangladesh/ | Child Health Research Foundation | Child Health Research Foundation | Senjuti Saha et al |
| EPI_ISL_625464 | Bangladesh/CHRF-0050/2020 | 2020-06-02 | Asia/Bangladesh/Bangladesh/ | Child Health Research Foundation | Child Health Research Foundation | Senjuti Saha et al |
| EPI_ISL_625465 | Bangladesh/CHRF-0051/2020 | 2020-06-14 | Asia/Bangladesh/Bangladesh/ | Child Health Research Foundation | Child Health Research Foundation | Senjuti Saha et al |
| EPI_ISL_625470 | Bangladesh/CHRF-0056/2020 | 2020-07-02 | Asia/Bangladesh/Bangladesh/ | Child Health Research Foundation | Child Health Research Foundation | Senjuti Saha et al |
| EPI_ISL_625472 | Bangladesh/CHRF-0059/2020 | 2020-08-04 | Asia/Bangladesh/Bangladesh/ | Child Health Research Foundation | Child Health Research Foundation | Senjuti Saha et al |
| EPI_ISL_625473 | Bangladesh/CHRF-0060/2020 | 2020-07-31 | Asia/Bangladesh/Bangladesh/ | Child Health Research Foundation | Child Health Research Foundation | Senjuti Saha et al |
| EPI_ISL_625474 | Bangladesh/CHRF-0061/2020 | 2020-07-17 | Asia/Bangladesh/Bangladesh/ | Child Health Research Foundation | Child Health Research Foundation | Senjuti Saha et al |
| EPI_ISL_625476 | Bangladesh/CHRF-0063/2020 | 2020-07-18 | Asia/Bangladesh/Bangladesh/ | Child Health Research Foundation | Child Health Research Foundation | Senjuti Saha et al |
| EPI_ISL_625478 | USA/CA-CZB-11644/2020 | 2020-09-23 | North America/USA/California/Santa Cl | Santa Clara County Public Health Laboratory | Chan-Zuckerberg Biohub | CZB Cliahub Consortium et al |
| EPI_ISL_625482 | USA/CA-CZB-11656/2020 | 2020-09-23 | North America/USA/California/Santa Cl | Santa Clara County Public Health Laboratory | Chan-Zuckerberg Biohub | CZB Cliahub Consortium et al |
| EPI_ISL_625490 | USA/CA-CZB-11678/2020 | 2020-09-24 | North America/USA/California/Santa Cl | Santa Clara County Public Health Laboratory | Chan-Zuckerberg Biohub | CZB Cliahub Consortium et al |
| EPI_ISL_625491 | USA/CA-CZB-11679/2020 | 2020-09-24 | North America/USA/California/Santa Cl | Santa Clara County Public Health Laboratory | Chan-Zuckerberg Biohub | CZB Cliahub Consortium et al |
| EPI_ISL_625492 | USA/CA-CZB-11681/2020 | 2020-09-25 | North America/USA/California/Santa Cl | Santa Clara County Public Health Laboratory | Chan-Zuckerberg Biohub | CZB Cliahub Consortium et al |
| EPI_ISL_625494 | USA/CA-CZB-11683/2020 | 2020-09-24 | North America/USA/California/Santa Cl | Santa Clara County Public Health Laboratory | Chan-Zuckerberg Biohub | CZB Cliahub Consortium et al |
| EPI_ISL_625498 | USA/CA-CZB-11692/2020 | 2020-09-28 | North America/USA/California/Santa Cl | Santa Clara County Public Health Laboratory | Chan-Zuckerberg Biohub | CZB Cliahub Consortium et al |

| | | | | | | |
|---|---|---|---|---|---|---|
| EPI_ISL_625501 | USA/CA-CZB-11105/2020 | 2020-05-31 | North America/USA/California/Alameda | Alameda County Public Health Lab | Chan-Zuckerberg Biohub | CZB Cliahub Consortium et al |
| EPI_ISL_625506 | USA/CA-CZB-11113/2020 | 2020-05-28 | North America/USA/California/Alameda | Alameda County Public Health Lab | Chan-Zuckerberg Biohub | CZB Cliahub Consortium et al |
| EPI_ISL_625507 | USA/CA-CZB-11114/2020 | 2020-05-28 | North America/USA/California/Alameda | Alameda County Public Health Lab | Chan-Zuckerberg Biohub | CZB Cliahub Consortium et al |
| EPI_ISL_625513 | USA/CA-CZB-11121/2020 | 2020-05-29 | North America/USA/California/Alameda | Alameda County Public Health Lab | Chan-Zuckerberg Biohub | CZB Cliahub Consortium et al |
| EPI_ISL_625527 | USA/CA-CZB-11170/2020 | 2020-09-15 | North America/USA/California/Alameda | Alameda County Public Health Lab | Chan-Zuckerberg Biohub | CZB Cliahub Consortium et al |
| EPI_ISL_625528 | USA/CA-CZB-11171/2020 | 2020-09-15 | North America/USA/California/Alameda | Alameda County Public Health Lab | Chan-Zuckerberg Biohub | CZB Cliahub Consortium et al |
| EPI_ISL_625530 | USA/CA-CZB-11174/2020 | 2020-09-15 | North America/USA/California/Alameda | Alameda County Public Health Lab | Chan-Zuckerberg Biohub | CZB Cliahub Consortium et al |
| EPI_ISL_625533 | USA/CA-CZB-11178/2020 | 2020-09-15 | North America/USA/California/Alameda | Alameda County Public Health Lab | Chan-Zuckerberg Biohub | CZB Cliahub Consortium et al |
| EPI_ISL_625535 | USA/CA-CZB-11181/2020 | 2020-09-15 | North America/USA/California/Alameda | Alameda County Public Health Lab | Chan-Zuckerberg Biohub | CZB Cliahub Consortium et al |
| EPI_ISL_625543 | USA/CA-CZB-11189/2020 | 2020-09-19 | North America/USA/California/Alameda | Alameda County Public Health Lab | Chan-Zuckerberg Biohub | CZB Cliahub Consortium et al |
| EPI_ISL_625544 | USA/CA-CZB-11190/2020 | 2020-09-16 | North America/USA/California/Alameda | Alameda County Public Health Lab | Chan-Zuckerberg Biohub | CZB Cliahub Consortium et al |
| EPI_ISL_625545 | USA/CA-CZB-11191/2020 | 2020-09-16 | North America/USA/California/Alameda | Alameda County Public Health Lab | Chan-Zuckerberg Biohub | CZB Cliahub Consortium et al |
| EPI_ISL_625555 | USA/CA-CZB-11575/2020 | 2020-07-16 | North America/USA/California/Marin Co | UCSF Clinical Microbiology Laboratory | Chan-Zuckerberg Biohub | CZB Cliahub Consortium et al |
| EPI_ISL_625570 | USA/CA-CZB-11542/2020 | 2020-07-24 | North America/USA/California/Madera | NaN | Chan-Zuckerberg Biohub | CZB Cliahub Consortium et al |
| EPI_ISL_625572 | USA/CA-CZB-11539/2020 | 2020-07-24 | North America/USA/California/Madera | NaN | Chan-Zuckerberg Biohub | CZB Cliahub Consortium et al |
| EPI_ISL_625582 | USA/CA-CZB-11514/2020 | 2020-08-02 | North America/USA/California/Madera | NaN | Chan-Zuckerberg Biohub | CZB Cliahub Consortium et al |
| EPI_ISL_625585 | USA/CA-CZB-11510/2020 | 2020-08-02 | North America/USA/California/Madera | NaN | Chan-Zuckerberg Biohub | CZB Cliahub Consortium et al |
| EPI_ISL_625587 | USA/CA-CZB-11508/2020 | 2020-08-02 | North America/USA/California/Madera | NaN | Chan-Zuckerberg Biohub | CZB Cliahub Consortium et al |
| EPI_ISL_625591 | USA/CA-CZB-11502/2020 | 2020-08-02 | North America/USA/California/Madera | NaN | Chan-Zuckerberg Biohub | CZB Cliahub Consortium et al |
| EPI_ISL_625594 | USA/CA-CZB-11872/2020 | 2020-04-10 | North America/USA/California/Santa Cl | Santa Clara County Public Health Laboratory | Chan-Zuckerberg Biohub | CZB Cliahub Consortium et al |

| | | | | | | |
|---|---|---|---|---|---|---|
| EPI_ISL_625609 | USA/CA-CZB-11933/2020 | 2020-10-06 | North America/USA/California/Santa Cl | Santa Clara County Public Health Laboratory | Chan-Zuckerberg Biohub | CZB Cliahub Consortium et al |
| EPI_ISL_625612 | USA/CA-CZB-11852/2020 | 2020-10-01 | North America/USA/California/Alameda | Alameda County Public Health Lab | Chan-Zuckerberg Biohub | CZB Cliahub Consortium et al |
| EPI_ISL_625616 | USA/CA-CZB-11859/2020 | 2020-10-04 | North America/USA/California/Alameda | Alameda County Public Health Lab | Chan-Zuckerberg Biohub | CZB Cliahub Consortium et al |
| EPI_ISL_625622 | USA/CA-CZB-11870/2020 | 2020-09-08 | North America/USA/California/Alameda | Alameda County Public Health Lab | Chan-Zuckerberg Biohub | CZB Cliahub Consortium et al |
| EPI_ISL_625625 | USA/CA-CZB-11962/2020 | 2020-06-17 | North America/USA/California/San Luis | County of San Luis Obispo Public Health Laboratory | Chan-Zuckerberg Biohub | CZB Cliahub Consortium et al |
| EPI_ISL_625628 | USA/CA-CZB-11784/2020 | 2020-09-19 | North America/USA/California/Orange C | Orange County Public Health Lab | Chan-Zuckerberg Biohub | CZB Cliahub Consortium et al |
| EPI_ISL_625630 | USA/CA-CZB-11786/2020 | 2020-09-21 | North America/USA/California/Orange C | Orange County Public Health Lab | Chan-Zuckerberg Biohub | CZB Cliahub Consortium et al |
| EPI_ISL_625633 | USA/CA-CZB-11790/2020 | 2020-09-22 | North America/USA/California/Orange C | Orange County Public Health Lab | Chan-Zuckerberg Biohub | CZB Cliahub Consortium et al |
| EPI_ISL_625636 | USA/CA-CZB-11793/2020 | 2020-09-22 | North America/USA/California/Orange C | Orange County Public Health Lab | Chan-Zuckerberg Biohub | CZB Cliahub Consortium et al |
| EPI_ISL_625638 | USA/CA-CZB-11795/2020 | 2020-09-22 | North America/USA/California/Orange C | Orange County Public Health Lab | Chan-Zuckerberg Biohub | CZB Cliahub Consortium et al |
| EPI_ISL_625640 | USA/CA-CZB-11797/2020 | 2020-09-23 | North America/USA/California/Orange C | Orange County Public Health Lab | Chan-Zuckerberg Biohub | CZB Cliahub Consortium et al |
| EPI_ISL_625642 | USA/CA-CZB-11799/2020 | 2020-09-23 | North America/USA/California/Orange C | Orange County Public Health Lab | Chan-Zuckerberg Biohub | CZB Cliahub Consortium et al |
| EPI_ISL_625643 | USA/CA-CZB-11800/2020 | 2020-09-23 | North America/USA/California/Orange C | Orange County Public Health Lab | Chan-Zuckerberg Biohub | CZB Cliahub Consortium et al |
| EPI_ISL_625646 | USA/CA-CZB-11803/2020 | 2020-09-24 | North America/USA/California/Orange C | Orange County Public Health Lab | Chan-Zuckerberg Biohub | CZB Cliahub Consortium et al |
| EPI_ISL_625647 | USA/CA-CZB-11804/2020 | 2020-09-26 | North America/USA/California/Orange C | Orange County Public Health Lab | Chan-Zuckerberg Biohub | CZB Cliahub Consortium et al |
| EPI_ISL_625648 | USA/CA-CZB-11806/2020 | 2020-09-28 | North America/USA/California/Orange C | Orange County Public Health Lab | Chan-Zuckerberg Biohub | CZB Cliahub Consortium et al |
| EPI_ISL_625652 | USA/CA-CZB-11811/2020 | 2020-09-29 | North America/USA/California/Orange C | Orange County Public Health Lab | Chan-Zuckerberg Biohub | CZB Cliahub Consortium et al |
| EPI_ISL_625653 | USA/CA-CZB-11813/2020 | 2020-09-29 | North America/USA/California/Orange C | Orange County Public Health Lab | Chan-Zuckerberg Biohub | CZB Cliahub Consortium et al |
| EPI_ISL_625657 | USA/CA-CZB-11818/2020 | 2020-10-01 | North America/USA/California/Orange C | Orange County Public Health Lab | Chan-Zuckerberg Biohub | CZB Cliahub Consortium et al |
| EPI_ISL_625660 | USA/CA-CZB-11781/2020 | 2020-09-15 | North America/USA/California/San Joac | San Joaquin County Public Health Lab | Chan-Zuckerberg Biohub | CZB Cliahub Consortium et al |

| | | | | | | |
|---|---|---|---|---|---|---|
| EPI_ISL_625664 | USA/CA-CZB-11956/2020 | 2020-10-07 | North America/USA/California/Marin Co | UCSF Clinical Microbiology Laboratory | Chan-Zuckerberg Biohub | CZB Cliahub Consortium et al |
| EPI_ISL_625670 | USA/CA-CZB-11778/2020 | 2020-09-21 | North America/USA/California/San Fran | UCSF Clinical Microbiology Laboratory | Chan-Zuckerberg Biohub | CZB Cliahub Consortium et al |
| EPI_ISL_625674 | Chile/MA-UMAG-Catg-6/2020 | 2020-09-27 | South America/Chile/Magallanes/Puerto | Laboratory of Molecular Medicine, University of Magallanes | Centro Asistencial Docente y de Investigacion, Universidad de Magallanes | Jorge Gonzalez et al |
| EPI_ISL_625675 | Chile/MA-UMAG-Catg-7/2020 | 2020-09-03 | South America/Chile/Magallanes/Porve | Laboratory of Molecular Medicine, University of Magallanes | Centro Asistencial Docente y de Investigacion, Universidad de Magallanes | Jorge Gonzalez et al |
| EPI_ISL_625677 | Chile/MA-UMAG-Catg-21/2020 | 2020-09-19 | South America/Chile/Magallanes/Punta | Laboratory of Molecular Medicine, University of Magallanes | Centro Asistencial Docente y de Investigacion, Universidad de Magallanes | Jorge Gonzalez et al |
| EPI_ISL_625681 | Chile/MA-UMAG-Catg-2/2020 | 2020-08-10 | South America/Chile/Magallanes/Porve | Laboratory of Molecular Medicine, University of Magallanes | Centro Asistencial Docente y de Investigacion, Universidad de Magallanes | Jorge Gonzalez et al |
| EPI_ISL_626216 | Germany/NW-HHU-217/2020 | 2020-04-22 | Europe/Germany/North Rhine Westphal | Institute for Virology, University Hospital Essen | Center of Medical Microbiology, Virology, and Hospital Hygiene, University of Duesseldorf | Olympia E. Anastasiou et al |
| EPI_ISL_626217 | Germany/NW-HHU-218/2020 | 2020-04-23 | Europe/Germany/North Rhine Westphal | Institute for Virology, University Hospital Essen | Center of Medical Microbiology, Virology, and Hospital Hygiene, University of Duesseldorf | Olympia E. Anastasiou et al |

| | | | | | | |
|---|---|---|---|---|---|---|
| EPI_ISL_626222 | Germany/NW-HHU-223/2020 | 2020-05-05 | Europe/Germany/North Rhine Westphal | Institute for Virology, University Hospital Essen | Center of Medical Microbiology, Virology, and Hospital Hygiene, University of Duesseldorf | Olympia E. Anastasiou et al |
| EPI_ISL_626229 | Germany/NW-HHU-230/2020 | 2020-04-01 | Europe/Germany/North Rhine Westphal | Institute for Virology, University Hospital Essen | Center of Medical Microbiology, Virology, and Hospital Hygiene, University of Duesseldorf | Olympia E. Anastasiou et al |
| EPI_ISL_626230 | Germany/NW-HHU-231/2020 | 2020-04-02 | Europe/Germany/North Rhine Westphal | Institute for Virology, University Hospital Essen | Center of Medical Microbiology, Virology, and Hospital Hygiene, University of Duesseldorf | Olympia E. Anastasiou et al |
| EPI_ISL_626232 | Belgium/ULG-10624/2020 | 2020-08-17 | Europe/Belgium/Liv®ge/ | Department of Clinical Microbiology | GIGA Medical Genomics | Keith Durkin et al |
| EPI_ISL_626249 | Belgium/ULG-10604/2020 | 2020-09-04 | Europe/Belgium/Liv®ge/ | Department of Clinical Microbiology | GIGA Medical Genomics | Keith Durkin et al |
| EPI_ISL_626252 | Belgium/ULG-10619/2020 | 2020-09-05 | Europe/Belgium/Liv®ge/ | Department of Clinical Microbiology | GIGA Medical Genomics | Keith Durkin et al |
| EPI_ISL_626256 | Belgium/ULG-10507/2020 | 2020-10-23 | Europe/Belgium/Liv®ge/ | Department of Clinical Microbiology | GIGA Medical Genomics | Keith Durkin et al |
| EPI_ISL_626259 | Belgium/ULG-10508/2020 | 2020-10-23 | Europe/Belgium/Liv®ge/ | Department of Clinical Microbiology | GIGA Medical Genomics | Keith Durkin et al |
| EPI_ISL_626260 | Belgium/ULG-10516/2020 | 2020-10-23 | Europe/Belgium/Liv®ge/ | Department of Clinical Microbiology | GIGA Medical Genomics | Keith Durkin et al |
| EPI_ISL_626263 | Belgium/ULG-10520/2020 | 2020-10-23 | Europe/Belgium/Liv®ge/ | Department of Clinical Microbiology | GIGA Medical Genomics | Keith Durkin et al |
| EPI_ISL_626264 | Belgium/ULG-10582/2020 | 2020-10-23 | Europe/Belgium/Liv®ge/ | Department of Clinical Microbiology | GIGA Medical Genomics | Keith Durkin et al |
| EPI_ISL_626270 | Belgium/ULG-10593/2020 | 2020-10-23 | Europe/Belgium/Liv®ge/ | Department of Clinical Microbiology | GIGA Medical Genomics | Keith Durkin et al |
| EPI_ISL_626276 | Belgium/ULG-10517/2020 | 2020-10-23 | Europe/Belgium/Liv®ge/ | Department of Clinical Microbiology | GIGA Medical Genomics | Keith Durkin et al |
| EPI_ISL_626281 | Belgium/ULG-10571/2020 | 2020-10-23 | Europe/Belgium/Liv®ge/ | Department of Clinical Microbiology | GIGA Medical Genomics | Keith Durkin et al |

| | | | | | | |
|---|---|---|---|---|---|---|
| EPI_ISL_626285 | Belgium/ULG-10511/2020 | 2020-10-23 | Europe/Belgium/Liv®ge/ | Department of Clinical Microbiology | GIGA Medical Genomics | Keith Durkin et al |
| EPI_ISL_626289 | Belgium/ULG-10596/2020 | 2020-10-23 | Europe/Belgium/Liv®ge/ | Department of Clinical Microbiology | GIGA Medical Genomics | Keith Durkin et al |
| EPI_ISL_626292 | Belgium/ULG-10599/2020 | 2020-10-23 | Europe/Belgium/Liv®ge/ | Department of Clinical Microbiology | GIGA Medical Genomics | Keith Durkin et al |
| EPI_ISL_626297 | Belgium/ULG-10506/2020 | 2020-10-23 | Europe/Belgium/Liv®ge/ | Department of Clinical Microbiology | GIGA Medical Genomics | Keith Durkin et al |
| EPI_ISL_626300 | Belgium/ULG-10529/2020 | 2020-10-23 | Europe/Belgium/Liv®ge/ | Department of Clinical Microbiology | GIGA Medical Genomics | Keith Durkin et al |
| EPI_ISL_626301 | Belgium/ULG-10526/2020 | 2020-10-23 | Europe/Belgium/Liv®ge/ | Department of Clinical Microbiology | GIGA Medical Genomics | Keith Durkin et al |
| EPI_ISL_626302 | Belgium/ULG-10587/2020 | 2020-10-23 | Europe/Belgium/Liv®ge/ | Department of Clinical Microbiology | GIGA Medical Genomics | Keith Durkin et al |
| EPI_ISL_626305 | Belgium/ULG-10547/2020 | 2020-10-23 | Europe/Belgium/Liv®ge/ | Department of Clinical Microbiology | GIGA Medical Genomics | Keith Durkin et al |
| EPI_ISL_626307 | Belgium/ULG-10512/2020 | 2020-10-23 | Europe/Belgium/Liv®ge/ | Department of Clinical Microbiology | GIGA Medical Genomics | Keith Durkin et al |
| EPI_ISL_626309 | Belgium/ULG-10564/2020 | 2020-10-23 | Europe/Belgium/Liv®ge/ | Department of Clinical Microbiology | GIGA Medical Genomics | Keith Durkin et al |
| EPI_ISL_626310 | Belgium/ULG-10534/2020 | 2020-10-23 | Europe/Belgium/Liv®ge/ | Department of Clinical Microbiology | GIGA Medical Genomics | Keith Durkin et al |
| EPI_ISL_626311 | Belgium/ULG-10592/2020 | 2020-10-23 | Europe/Belgium/Liv®ge/ | Department of Clinical Microbiology | GIGA Medical Genomics | Keith Durkin et al |
| EPI_ISL_626317 | Belgium/ULG-10555/2020 | 2020-10-23 | Europe/Belgium/Liv®ge/ | Department of Clinical Microbiology | GIGA Medical Genomics | Keith Durkin et al |
| EPI_ISL_626318 | Belgium/ULG-10543/2020 | 2020-10-23 | Europe/Belgium/Liv®ge/ | Department of Clinical Microbiology | GIGA Medical Genomics | Keith Durkin et al |
| EPI_ISL_626326 | Belgium/ULG-10538/2020 | 2020-10-23 | Europe/Belgium/Liv®ge/ | Department of Clinical Microbiology | GIGA Medical Genomics | Keith Durkin et al |
| EPI_ISL_626329 | Belgium/ULG-10550/2020 | 2020-10-23 | Europe/Belgium/Liv®ge/ | Department of Clinical Microbiology | GIGA Medical Genomics | Keith Durkin et al |
| EPI_ISL_626332 | Belgium/ULG-10563/2020 | 2020-10-23 | Europe/Belgium/Liv®ge/ | Department of Clinical Microbiology | GIGA Medical Genomics | Keith Durkin et al |
| EPI_ISL_626337 | Belgium/ULG-10591/2020 | 2020-10-23 | Europe/Belgium/Liv®ge/ | Department of Clinical Microbiology | GIGA Medical Genomics | Keith Durkin et al |
| EPI_ISL_626360 | USA/IL-NM-0052/2020 | 2020-03-15 | North America/USA/Illinois/Chicago | Northwestern Memorial Hospital | Ozer Lab | Ramon Lorenzo-Redondo et al |
| EPI_ISL_626379 | USA/IL-NM-0329/2020 | 2020-03-26 | North America/USA/Illinois/Chicago | Northwestern Memorial Hospital | Ozer Lab | Ramon Lorenzo-Redondo et al |
| EPI_ISL_626388 | USA/IL-NM-0398/2020 | 2020-04-15 | North America/USA/Illinois/Chicago | Northwestern Memorial Hospital | Ozer Lab | Ramon Lorenzo-Redondo et al |

| | | | | | | |
|---|---|---|---|---|---|---|
| EPI_ISL_626391 | USA/IL-NM-0405/2020 | 2020-04-14 | North America/USA/Illinois/Chicago | Northwestern Memorial Hospital | Ozer Lab | Ramon Lorenzo-Redondo et al |
| EPI_ISL_626393 | USA/IL-NM-0411/2020 | 2020-04-13 | North America/USA/Illinois/Chicago | Northwestern Memorial Hospital | Ozer Lab | Ramon Lorenzo-Redondo et al |
| EPI_ISL_626394 | USA/IL-NM-0413/2020 | 2020-04-13 | North America/USA/Illinois/Chicago | Northwestern Memorial Hospital | Ozer Lab | Ramon Lorenzo-Redondo et al |
| EPI_ISL_626400 | USA/IL-NM-0420/2020 | 2020-04-13 | North America/USA/Illinois/Chicago | Northwestern Memorial Hospital | Ozer Lab | Ramon Lorenzo-Redondo et al |
| EPI_ISL_626401 | USA/IL-NM-0422/2020 | 2020-04-13 | North America/USA/Illinois/Chicago | Northwestern Memorial Hospital | Ozer Lab | Ramon Lorenzo-Redondo et al |
| EPI_ISL_626405 | USA/IL-NM-0432/2020 | 2020-04-11 | North America/USA/Illinois/Chicago | Northwestern Memorial Hospital | Ozer Lab | Ramon Lorenzo-Redondo et al |
| EPI_ISL_626407 | USA/IL-NM-0436/2020 | 2020-04-11 | North America/USA/Illinois/Chicago | Northwestern Memorial Hospital | Ozer Lab | Ramon Lorenzo-Redondo et al |
| EPI_ISL_626414 | USA/IL-NM-0466/2020 | 2020-04-21 | North America/USA/Illinois/Chicago | Northwestern Memorial Hospital | Ozer Lab | Ramon Lorenzo-Redondo et al |
| EPI_ISL_626437 | USA/IL-NM-0514/2020 | 2020-04-26 | North America/USA/Illinois/Chicago | Northwestern Memorial Hospital | Ozer Lab | Ramon Lorenzo-Redondo et al |
| EPI_ISL_626450 | USA/IL-NM-0542/2020 | 2020-04-29 | North America/USA/Illinois/Chicago | Northwestern Memorial Hospital | Ozer Lab | Ramon Lorenzo-Redondo et al |
| EPI_ISL_626458 | USA/IL-NM-0590/2020 | 2020-05-02 | North America/USA/Illinois/Chicago | Northwestern Memorial Hospital | Ozer Lab | Ramon Lorenzo-Redondo et al |
| EPI_ISL_626461 | USA/IL-NM-0642/2020 | 2020-05-06 | North America/USA/Illinois/Chicago | Northwestern Memorial Hospital | Ozer Lab | Ramon Lorenzo-Redondo et al |
| EPI_ISL_626462 | USA/IL-NM-0656/2020 | 2020-05-07 | North America/USA/Illinois/Chicago | Northwestern Memorial Hospital | Ozer Lab | Ramon Lorenzo-Redondo et al |
| EPI_ISL_626469 | USA/IL-NM-0683/2020 | 2020-05-09 | North America/USA/Illinois/Chicago | Northwestern Memorial Hospital | Ozer Lab | Ramon Lorenzo-Redondo et al |
| EPI_ISL_626473 | USA/IL-NM-0746/2020 | 2020-05-13 | North America/USA/Illinois/Chicago | Northwestern Memorial Hospital | Ozer Lab | Ramon Lorenzo-Redondo et al |
| EPI_ISL_626478 | USA/IL-NM-0816/2020 | 2020-05-18 | North America/USA/Illinois/Chicago | Northwestern Memorial Hospital | Ozer Lab | Ramon Lorenzo-Redondo et al |
| EPI_ISL_626484 | USA/IL-NM-0924/2020 | 2020-05-25 | North America/USA/Illinois/Chicago | Northwestern Memorial Hospital | Ozer Lab | Ramon Lorenzo-Redondo et al |
| EPI_ISL_626485 | USA/IL-NM-0937/2020 | 2020-05-26 | North America/USA/Illinois/Chicago | Northwestern Memorial Hospital | Ozer Lab | Ramon Lorenzo-Redondo et al |
| EPI_ISL_626487 | USA/IL-NM-0982/2020 | 2020-05-28 | North America/USA/Illinois/Chicago | Northwestern Memorial Hospital | Ozer Lab | Ramon Lorenzo-Redondo et al |
| EPI_ISL_626490 | USA/IL-NM-1012/2020 | 2020-05-31 | North America/USA/Illinois/Chicago | Northwestern Memorial Hospital | Ozer Lab | Ramon Lorenzo-Redondo et al |
| EPI_ISL_626491 | USA/IL-NM-1021/2020 | 2020-06-02 | North America/USA/Illinois/Chicago | Northwestern Memorial Hospital | Ozer Lab | Ramon Lorenzo-Redondo et al |
| EPI_ISL_626503 | USA/IL-NM-1157/2020 | 2020-06-15 | North America/USA/Illinois/Chicago | Northwestern Memorial Hospital | Ozer Lab | Ramon Lorenzo-Redondo et al |
| EPI_ISL_626528 | USA/IL-NM-1521/2020 | 2020-07-11 | North America/USA/Illinois/Chicago | Northwestern Memorial Hospital | Ozer Lab | Ramon Lorenzo-Redondo et al |
| EPI_ISL_626530 | USA/IL-NM-1530/2020 | 2020-07-13 | North America/USA/Illinois/Chicago | Northwestern Memorial Hospital | Ozer Lab | Ramon Lorenzo-Redondo et al |
| EPI_ISL_626531 | USA/IL-NM-1550/2020 | 2020-07-14 | North America/USA/Illinois/Chicago | Northwestern Memorial Hospital | Ozer Lab | Ramon Lorenzo-Redondo et al |
| EPI_ISL_626533 | USA/IL-NM-1560/2020 | 2020-07-14 | North America/USA/Illinois/Chicago | Northwestern Memorial Hospital | Ozer Lab | Ramon Lorenzo-Redondo et al |
| EPI_ISL_626534 | USA/IL-NM-1594/2020 | 2020-07-16 | North America/USA/Illinois/Chicago | Northwestern Memorial Hospital | Ozer Lab | Ramon Lorenzo-Redondo et al |
| EPI_ISL_626538 | USA/IL-NM-1650/2020 | 2020-07-21 | North America/USA/Illinois/Chicago | Northwestern Memorial Hospital | Ozer Lab | Ramon Lorenzo-Redondo et al |
| EPI_ISL_626542 | USA/IL-NM-1703/2020 | 2020-07-24 | North America/USA/Illinois/Chicago | Northwestern Memorial Hospital | Ozer Lab | Ramon Lorenzo-Redondo et al |
| EPI_ISL_626545 | USA/IL-NM-1718/2020 | 2020-07-25 | North America/USA/Illinois/Chicago | Northwestern Memorial Hospital | Ozer Lab | Ramon Lorenzo-Redondo et al |
| EPI_ISL_626548 | USA/IL-NM-1722/2020 | 2020-07-26 | North America/USA/Illinois/Chicago | Northwestern Memorial Hospital | Ozer Lab | Ramon Lorenzo-Redondo et al |

| | | | | | | |
|---|---|---|---|---|---|---|
| EPI_ISL_626549 | Chile/AT-CMM-ATAC02/2020 | 2020-07-29 | South America/Chile/Atacama/Copiapo | Laboratorio de Biologv≠a Molecular, Facultad de Medicina, Universidad de Atacama, Copiapo, Chile/ FONDAP CRG, Universidad Andrv©s Bello, Santiago, Chile | Center for Mathematical Modeling and Center for Genome Regulation. Santiago, Chile | Echeverrv≠a C et al |
| EPI_ISL_626550 | Chile/AT-CMM-ATAC03/2020 | 2020-07-20 | South America/Chile/Atacama/Copiapo | Laboratorio de Biologv≠a Molecular, Facultad de Medicina, Universidad de Atacama, Copiapo, Chile/ FONDAP CRG, Universidad Andrv©s Bello, Santiago, Chile | Center for Mathematical Modeling and Center for Genome Regulation. Santiago, Chile | Echeverrv≠a C et al |
| EPI_ISL_626552 | Chile/AT-CMM-ATAC06/2020 | 2020-07-22 | South America/Chile/Atacama/Tierra A | Laboratorio de Biologv≠a Molecular, Facultad de Medicina, Universidad de Atacama, Copiapo, Chile/ FONDAP CRG, Universidad Andrv©s Bello, Santiago, Chile | Center for Mathematical Modeling and Center for Genome Regulation. Santiago, Chile | Echeverrv≠a C et al |
| EPI_ISL_626555 | Chile/AT-CMM-ATAC09/2020 | 2020-07-25 | South America/Chile/Atacama/Paipote | Laboratorio de Biologv≠a Molecular, Facultad de Medicina, Universidad de Atacama, Copiapo, Chile/ FONDAP CRG, Universidad Andrv©s Bello, Santiago, Chile | Center for Mathematical Modeling and Center for Genome Regulation. Santiago, Chile | Echeverrv≠a C et al |
| EPI_ISL_626556 | Chile/AT-CMM-ATAC10/2020 | 2020-08-07 | South America/Chile/Atacama/Caldera | Laboratorio de Biologv≠a Molecular, Facultad de Medicina, Universidad de Atacama, Copiapo, Chile/ FONDAP CRG, Universidad Andrv©s Bello, Santiago, Chile | Center for Mathematical Modeling and Center for Genome Regulation. Santiago, Chile | Echeverrv≠a C et al |

| | | | | | | |
|---|---|---|---|---|---|---|
| EPI_ISL_626557 | Chile/AT-CMM-ATAC11/2020 | 2020-08-07 | South America/Chile/Atacama/Caldera | Laboratorio de Biologv≠a Molecular, Facultad de Medicina, Universidad de Atacama, Copiapo, Chile/ FONDAP CRG, Universidad Andrv©s Bello, Santiago, Chile | Center for Mathematical Modeling and Center for Genome Regulation. Santiago, Chile | Echeverrv≠a C et al |
| EPI_ISL_626559 | Chile/AT-CMM-ATAC13/2020 | 2020-08-07 | South America/Chile/Atacama/Caldera | Laboratorio de Biologv≠a Molecular, Facultad de Medicina, Universidad de Atacama, Copiapo, Chile/ FONDAP CRG, Universidad Andrv©s Bello, Santiago, Chile | Center for Mathematical Modeling and Center for Genome Regulation. Santiago, Chile | Echeverrv≠a C et al |
| EPI_ISL_626561 | Chile/AT-CMM-ATAC15/2020 | 2020-08-07 | South America/Chile/Atacama/Huasco | Laboratorio de Biologv≠a Molecular, Facultad de Medicina, Universidad de Atacama, Copiapo, Chile/ FONDAP CRG, Universidad Andrv©s Bello, Santiago, Chile | Center for Mathematical Modeling and Center for Genome Regulation. Santiago, Chile | Echeverrv≠a C et al |
| EPI_ISL_626562 | Chile/AT-CMM-ATAC16/2020 | 2020-08-07 | South America/Chile/Atacama/Freirina | Laboratorio de Biologv≠a Molecular, Facultad de Medicina, Universidad de Atacama, Copiapo, Chile/ FONDAP CRG, Universidad Andrv©s Bello, Santiago, Chile | Center for Mathematical Modeling and Center for Genome Regulation. Santiago, Chile | Echeverrv≠a C et al |
| EPI_ISL_626564 | Chile/AT-CMM-ATAC19/2020 | 2020-07-30 | South America/Chile/Atacama/Copiapo | Laboratorio de Biologv≠a Molecular, Facultad de Medicina, Universidad de Atacama, Copiapo, Chile/ FONDAP CRG, Universidad Andrv©s Bello, Santiago, Chile | Center for Mathematical Modeling and Center for Genome Regulation. Santiago, Chile | Echeverrv≠a C et al |

| | | | | | | |
|---|---|---|---|---|---|---|
| EPI_ISL_626565 | Chile/AT-CMM-ATAC20/2020 | 2020-07-26 | South America/Chile/Atacama/Copiapo | Laboratorio de Biologv≠a Molecular, Facultad de Medicina, Universidad de Atacama, Copiapo, Chile/ FONDAP CRG, Universidad Andrv©s Bello, Santiago, Chile | Center for Mathematical Modeling and Center for Genome Regulation. Santiago, Chile | Echeverrv≠a C et al |
| EPI_ISL_626571 | CzechRepublic/NRL_9179-1/2020 | 2020-09-19 | Europe/Czech Republic/Central Bohemia | The National Institute of Public Health | State Veterinary Institute Prague | Nagy et al |
| EPI_ISL_626572 | CzechRepublic/NRL_9179-4/2020 | 2020-09-20 | Europe/Czech Republic/Prague/ | The National Institute of Public Health | State Veterinary Institute Prague | Nagy et al |
| EPI_ISL_626573 | CzechRepublic/NRL_9179-5/2020 | 2020-09-20 | Europe/Czech Republic/Prague/ | The National Institute of Public Health | State Veterinary Institute Prague | Nagy et al |
| EPI_ISL_626574 | CzechRepublic/NRL_8779/2020 | 2020-09-10 | Europe/Czech Republic/Southern Bohem | The National Institute of Public Health | State Veterinary Institute Prague | Nagy et al |
| EPI_ISL_626575 | CzechRepublic/NRL_8798/2020 | 2020-09-09 | Europe/Czech Republic/Central Bohemia | The National Institute of Public Health | State Veterinary Institute Prague | Nagy et al |
| EPI_ISL_626577 | CzechRepublic/NRL_8884/2020 | 2020-09-14 | Europe/Czech Republic/Prague/ | The National Institute of Public Health | State Veterinary Institute Prague | Nagy et al |
| EPI_ISL_626579 | CzechRepublic/NRL_8927/2020 | 2020-09-15 | Europe/Czech Republic/Prague/ | The National Institute of Public Health | State Veterinary Institute Prague | Nagy et al |
| EPI_ISL_626581 | CzechRepublic/NRL_8972-2/2020 | 2020-09-15 | Europe/Czech Republic/Prague/ | The National Institute of Public Health | State Veterinary Institute Prague | Nagy et al |
| EPI_ISL_626582 | CzechRepublic/NRL_8972-3/2020 | 2020-09-15 | Europe/Czech Republic/Prague/ | The National Institute of Public Health | State Veterinary Institute Prague | Nagy et al |
| EPI_ISL_626583 | CzechRepublic/NRL_8972-5/2020 | 2020-09-15 | Europe/Czech Republic/Prague/ | The National Institute of Public Health | State Veterinary Institute Prague | Nagy et al |
| EPI_ISL_626584 | CzechRepublic/NRL_8997/2020 | 2020-09-16 | Europe/Czech Republic/Prague/ | The National Institute of Public Health | State Veterinary Institute Prague | Nagy et al |
| EPI_ISL_626585 | CzechRepublic/NRL_9075/2020 | 2020-09-18 | Europe/Czech Republic/Prague/ | The National Institute of Public Health | State Veterinary Institute Prague | Nagy et al |
| EPI_ISL_626588 | CzechRepublic/NRL_9429/2020 | 2020-09-29 | Europe/Czech Republic/Prague/ | The National Institute of Public Health | State Veterinary Institute Prague | Nagy et al |
| EPI_ISL_626589 | CzechRepublic/NRL_9448/2020 | 2020-09-29 | Europe/Czech Republic/Prague/ | The National Institute of Public Health | State Veterinary Institute Prague | Nagy et al |
| EPI_ISL_626592 | CzechRepublic/NRL_9630/2020 | 2020-10-06 | Europe/Czech Republic/Prague/ | The National Institute of Public Health | State Veterinary Institute Prague | Nagy et al |
| EPI_ISL_626594 | CzechRepublic/NRL_9642/2020 | 2020-10-07 | Europe/Czech Republic/Prague/ | The National Institute of Public Health | State Veterinary Institute Prague | Nagy et al |

| | | | | | | |
|---|---|---|---|---|---|---|
| EPI_ISL_626596 | CzechRepublic/NRL_9663/2020 | 2020-10-07 | Europe/Czech Republic/Prague/ | The National Institute of Public Health | State Veterinary Institute Prague | Nagy et al |
| EPI_ISL_626599 | CzechRepublic/NRL_9677/2020 | 2020-09-30 | Europe/Czech Republic/Southern Bohem | The National Institute of Public Health | State Veterinary Institute Prague | Nagy et al |
| EPI_ISL_626600 | CzechRepublic/NRL_9678/2020 | 2020-09-30 | Europe/Czech Republic/Southern Bohem | The National Institute of Public Health | State Veterinary Institute Prague | Nagy et al |
| EPI_ISL_626601 | CzechRepublic/NRL_9679/2020 | 2020-09-30 | Europe/Czech Republic/Southern Bohem | The National Institute of Public Health | State Veterinary Institute Prague | Nagy et al |
| EPI_ISL_626602 | CzechRepublic/NRL_9681/2020 | 2020-10-06 | Europe/Czech Republic/Southern Bohem | The National Institute of Public Health | State Veterinary Institute Prague | Nagy et al |
| EPI_ISL_626603 | CzechRepublic/NRL_9682/2020 | 2020-10-07 | Europe/Czech Republic/Prague/ | The National Institute of Public Health | State Veterinary Institute Prague | Nagy et al |
| EPI_ISL_626604 | CzechRepublic/NRL_9703/2020 | 2020-10-08 | Europe/Czech Republic/Central Bohemia | The National Institute of Public Health | State Veterinary Institute Prague | Nagy et al |
| EPI_ISL_626605 | CzechRepublic/NRL_9713/2020 | 2020-10-07 | Europe/Czech Republic/Plze≈à Region/D | The National Institute of Public Health | State Veterinary Institute Prague | Nagy et al |
| EPI_ISL_626607 | CzechRepublic/NRL_9818/2020 | 2020-10-12 | Europe/Czech Republic/Prague/ | The National Institute of Public Health | State Veterinary Institute Prague | Nagy et al |
| EPI_ISL_626609 | CzechRepublic/NRL_9840/2020 | 2020-10-12 | Europe/Czech Republic/Prague/ | The National Institute of Public Health | State Veterinary Institute Prague | Nagy et al |
| EPI_ISL_626610 | CzechRepublic/NRL_9875/2020 | 2020-10-12 | Europe/Czech Republic/Prague/ | The National Institute of Public Health | State Veterinary Institute Prague | Nagy et al |
| EPI_ISL_626611 | CzechRepublic/NRL_9880/2020 | 2020-10-06 | Europe/Czech Republic/Prague/ | The National Institute of Public Health | State Veterinary Institute Prague | Nagy et al |
| EPI_ISL_626612 | CzechRepublic/NRL_9881/2020 | 2020-10-11 | Europe/Czech Republic/Central Bohemia | The National Institute of Public Health | State Veterinary Institute Prague | Nagy et al |
| EPI_ISL_626613 | CzechRepublic/NRL_9882/2020 | 2020-10-11 | Europe/Czech Republic/Prague/Prague-N | The National Institute of Public Health | State Veterinary Institute Prague | Nagy et al |
| EPI_ISL_626615 | CzechRepublic/NRL_9885/2020 | 2020-10-11 | Europe/Czech Republic/Central Bohemia | The National Institute of Public Health | State Veterinary Institute Prague | Nagy et al |
| EPI_ISL_626617 | CzechRepublic/NRL_9888/2020 | 2020-10-06 | Europe/Czech Republic/Prague/ | The National Institute of Public Health | State Veterinary Institute Prague | Nagy et al |
| EPI_ISL_626618 | CzechRepublic/NRL_9889/2020 | 2020-10-06 | Europe/Czech Republic/Prague/ | The National Institute of Public Health | State Veterinary Institute Prague | Nagy et al |
| EPI_ISL_626619 | CzechRepublic/NRL_9891/2020 | 2020-10-12 | Europe/Czech Republic/Central Bohemia | The National Institute of Public Health | State Veterinary Institute Prague | Nagy et al |
| EPI_ISL_626622 | CzechRepublic/NRL_9899/2020 | 2020-10-12 | Europe/Czech Republic/Plze≈à Region/ | The National Institute of Public Health | State Veterinary Institute Prague | Nagy et al |
| EPI_ISL_626623 | CzechRepublic/NRL_9903/2020 | 2020-10-14 | Europe/Czech Republic/Prague/ | The National Institute of Public Health | State Veterinary Institute Prague | Nagy et al |

| | | | | | | |
|---|---|---|---|---|---|---|
| EPI_ISL_626624 | CzechRepublic/NRL_10178/2020 | 2020-10-20 | Europe/Czech Republic/Prague/ | The National Institute of Public Health | State Veterinary Institute Prague | Nagy et al |
| EPI_ISL_626628 | Singapore/1117/2020 | 2020-10-26 | Asia/Singapore/Singapore/ | National Public Health Laboratory, National Centre for Infectious Diseases | National Public Health Laboratory, National Centre for Infectious Diseases | Tze Minn Mak et al |
| EPI_ISL_626778 | England/PORT-2D73DA/2020 | 2020-10-22 | Europe/United Kingdom/England/ | Centre for Enzyme Innovation, University of Portsmouth / Translational Research Laboratory, Portsmouth Hospitals NHS Trust | COVID-19 Genomics UK (COG-UK) Consortium | Angela Beckett et al |
| EPI_ISL_626814 | England/NORW-F214A/2020 | 2020-10-22 | Europe/United Kingdom/England/ | Quadram Institute Bioscience | COVID-19 Genomics UK (COG-UK) Consortium | Dave J. Baker et al |
| EPI_ISL_626900 | England/MILK-A6A37A/2020 | 2020-10-08 | Europe/United Kingdom/England/ | Virology Department, Sheffield Teaching Hospitals NHS Foundation Trust/Department of Infection, Immunity and Cardiovascular Disease, The Medical School, University of Sheffield | COVID-19 Genomics UK (COG-UK) Consortium | Thushan de Silva et al |
| EPI_ISL_626906 | England/MILK-A6A27D/2020 | 2020-10-08 | Europe/United Kingdom/England/ | Virology Department, Sheffield Teaching Hospitals NHS Foundation Trust/Department of Infection, Immunity and Cardiovascular Disease, The Medical School, University of Sheffield | COVID-19 Genomics UK (COG-UK) Consortium | Thushan de Silva et al |
| EPI_ISL_627363 | Scotland/CVR4837/2020 | 2020-09-21 | Europe/United Kingdom/Scotland/ | West of Scotland Specialist Virology Centre, NHSGGC / MRC-University of Glasgow Centre for Virus Research | COVID-19 Genomics UK (COG-UK) Consortium | Ana da Silva Filipe et al |
| EPI_ISL_627422 | Scotland/CVR5263/2020 | 2020-10-15 | Europe/United Kingdom/Scotland/ | West of Scotland Specialist Virology Centre, NHSGGC / MRC-University of Glasgow Centre for Virus Research | COVID-19 Genomics UK (COG-UK) Consortium | Ana da Silva Filipe et al |
| EPI_ISL_627427 | England/EXET-138790/2020 | 2020-10-15 | Europe/United Kingdom/England/ | University of Exeter | COVID-19 Genomics UK (COG-UK) Consortium | Ben Temperton et al |
| EPI_ISL_627441 | England/EXET-138C73/2020 | 2020-09-11 | Europe/United Kingdom/England/ | University of Exeter | COVID-19 Genomics UK (COG-UK) Consortium | Ben Temperton et al |
| EPI_ISL_627536 | NorthernIreland/NIRE-108B06/2020 | 2020-10-09 | Europe/United Kingdom/Northern Irelar | Regional Virus Laboratory, Belfast Health and Social Care Trust | COVID-19 Genomics UK (COG-UK) Consortium | Conall McCaughey et al |

| | | | | | | |
|---|---|---|---|---|---|---|
| EPI_ISL_627695 | England/NOTT-118D76/2020 | 2020-10-29 | Europe/United Kingdom/England/ | Queens Medical Centre, Clinical Microbiology Department / DeepSeq Nottingham | COVID-19 Genomics UK (COG-UK) Consortium | Gemma Clark et al |
| EPI_ISL_627706 | England/NOTT-1190A9/2020 | 2020-10-30 | Europe/United Kingdom/England/ | Queens Medical Centre, Clinical Microbiology Department / DeepSeq Nottingham | COVID-19 Genomics UK (COG-UK) Consortium | Gemma Clark et al |
| EPI_ISL_627745 | Wales/PHWC-482E17/2020 | 2020-10-21 | Europe/United Kingdom/Wales/ | Wales Specialist Virology Centre Sequencing lab: Pathogen Genomics Unit | COVID-19 Genomics UK (COG-UK) Consortium | Catherine Moore et al |
| EPI_ISL_627786 | Wales/PHWC-483177/2020 | 2020-10-13 | Europe/United Kingdom/Wales/ | Wales Specialist Virology Centre Sequencing lab: Pathogen Genomics Unit | COVID-19 Genomics UK (COG-UK) Consortium | Catherine Moore et al |
| EPI_ISL_627841 | Wales/PHWC-48365A/2020 | 2020-10-22 | Europe/United Kingdom/Wales/ | Wales Specialist Virology Centre Sequencing lab: Pathogen Genomics Unit | COVID-19 Genomics UK (COG-UK) Consortium | Catherine Moore et al |
| EPI_ISL_627929 | Wales/PHWC-483DA0/2020 | 2020-10-25 | Europe/United Kingdom/Wales/ | Wales Specialist Virology Centre Sequencing lab: Pathogen Genomics Unit | COVID-19 Genomics UK (COG-UK) Consortium | Catherine Moore et al |
| EPI_ISL_627970 | Wales/PHWC-484149/2020 | 2020-10-21 | Europe/United Kingdom/Wales/ | Wales Specialist Virology Centre Sequencing lab: Pathogen Genomics Unit | COVID-19 Genomics UK (COG-UK) Consortium | Catherine Moore et al |
| EPI_ISL_628064 | Wales/PHWC-48498D/2020 | 2020-10-18 | Europe/United Kingdom/Wales/ | Wales Specialist Virology Centre Sequencing lab: Pathogen Genomics Unit | COVID-19 Genomics UK (COG-UK) Consortium | Catherine Moore et al |
| EPI_ISL_628131 | Wales/PHWC-484F9A/2020 | 2020-10-24 | Europe/United Kingdom/Wales/ | Wales Specialist Virology Centre Sequencing lab: Pathogen Genomics Unit | COVID-19 Genomics UK (COG-UK) Consortium | Catherine Moore et al |
| EPI_ISL_628171 | Wales/PHWC-485351/2020 | 2020-10-21 | Europe/United Kingdom/Wales/ | Wales Specialist Virology Centre Sequencing lab: Pathogen Genomics Unit | COVID-19 Genomics UK (COG-UK) Consortium | Catherine Moore et al |
| EPI_ISL_628214 | Wales/PHWC-485755/2020 | 2020-10-25 | Europe/United Kingdom/Wales/ | Wales Specialist Virology Centre Sequencing lab: Pathogen Genomics Unit | COVID-19 Genomics UK (COG-UK) Consortium | Catherine Moore et al |
| EPI_ISL_628406 | England/QEUH-A33705/2020 | 2020-10-04 | Europe/United Kingdom/England/ | Virology Department, Sheffield Teaching Hospitals NHS Foundation Trust/Department of Infection, Immunity and Cardiovascular Disease, The Medical School, University of Sheffield | COVID-19 Genomics UK (COG-UK) Consortium | Thushan de Silva et al |
| EPI_ISL_628499 | Scotland/QEUH-A4B9B9/2020 | 2020-10-08 | Europe/United Kingdom/Scotland/ | Oxford Viromics, NDM, University of Oxford; Oxford University Hospitals; Basingstoke and North Hampshire Hospital | COVID-19 Genomics UK (COG-UK) Consortium | Tanya Golubchik et al |

| | | | | | | |
|---|---|---|---|---|---|---|
| EPI_ISL_628666 | England/QEUH-A63BE2/2020 | 2020-10-12 | Europe/United Kingdom/England/ | Oxford Viromics, NDM, University of Oxford; Oxford University Hospitals; Basingstoke and North Hampshire Hospital | COVID-19 Genomics UK (COG-UK) Consortium | Tanya Golubchik et al |
| EPI_ISL_628675 | England/QEUH-A63EE9/2020 | 2020-10-12 | Europe/United Kingdom/England/ | Oxford Viromics, NDM, University of Oxford; Oxford University Hospitals; Basingstoke and North Hampshire Hospital | COVID-19 Genomics UK (COG-UK) Consortium | Tanya Golubchik et al |
| EPI_ISL_628718 | USA/TX-DSHS-1075/2020 | 2020-06-16 | North America/USA/Texas/El Paso Coun | Texas Department of State Health Services | Texas Department of State Health Services | Rashmi Tuladhar et al |
| EPI_ISL_628739 | USA/TX-DSHS-1069/2020 | 2020-06-15 | North America/USA/Texas/Harris Count | Texas Department of State Health Services | Texas Department of State Health Services | Rashmi Tuladhar et al |
| EPI_ISL_628751 | Ghana/UHAS-I034/2020 | 2020-06-18 | Africa/Ghana/Volta/ | UHAS COVID-19 Lab | UHAS COVID-19 Lab | Kwabena O. Duedu et al |
| EPI_ISL_628761 | Ghana/UHAS-H009/2020 | 2020-06-16 | Africa/Ghana/Volta/ | UHAS COVID-19 Lab | UHAS COVID-19 Lab | Kwabena O. Duedu et al |
| EPI_ISL_628804 | USA/MI-UM-10036468985/2020 | 2020-10-15 | North America/USA/Michigan/ | University of Michigan Clinical Microbiology Laboratory | Lauring Lab, University of Michigan, Department of Microbiology and Immunology | Valesano et al |
| EPI_ISL_628805 | USA/MI-UM-10036437656/2020 | 2020-10-14 | North America/USA/Michigan/ | University of Michigan Clinical Microbiology Laboratory | Lauring Lab, University of Michigan, Department of Microbiology and Immunology | Valesano et al |
| EPI_ISL_628812 | USA/MI-UM-10036462976/2020 | 2020-10-15 | North America/USA/Michigan/ | University of Michigan Clinical Microbiology Laboratory | Lauring Lab, University of Michigan, Department of Microbiology and Immunology | Valesano et al |
| EPI_ISL_628815 | USA/MI-UM-10036482555/2020 | 2020-10-16 | North America/USA/Michigan/ | University of Michigan Clinical Microbiology Laboratory | Lauring Lab, University of Michigan, Department of Microbiology and Immunology | Valesano et al |

| | | | | | | |
|---|---|---|---|---|---|---|
| EPI_ISL_628819 | USA/MI-UM-10036466026/2020 | 2020-10-15 | North America/USA/Michigan/ | University of Michigan Clinical Microbiology Laboratory | Lauring Lab, University of Michigan, Department of Microbiology and Immunology | Valesano et al |
| EPI_ISL_628820 | USA/MI-UM-10036483429/2020 | 2020-10-16 | North America/USA/Michigan/ | University of Michigan Clinical Microbiology Laboratory | Lauring Lab, University of Michigan, Department of Microbiology and Immunology | Valesano et al |
| EPI_ISL_628821 | USA/MI-UM-10036483559/2020 | 2020-10-16 | North America/USA/Michigan/ | University of Michigan Clinical Microbiology Laboratory | Lauring Lab, University of Michigan, Department of Microbiology and Immunology | Valesano et al |
| EPI_ISL_628824 | USA/MI-UM-10036478916/2020 | 2020-10-16 | North America/USA/Michigan/ | University of Michigan Clinical Microbiology Laboratory | Lauring Lab, University of Michigan, Department of Microbiology and Immunology | Valesano et al |
| EPI_ISL_628828 | USA/MI-UM-10036485871/2020 | 2020-10-16 | North America/USA/Michigan/ | University of Michigan Clinical Microbiology Laboratory | Lauring Lab, University of Michigan, Department of Microbiology and Immunology | Valesano et al |
| EPI_ISL_628836 | USA/MI-UM-10036455942/2020 | 2020-10-15 | North America/USA/Michigan/ | University of Michigan Clinical Microbiology Laboratory | Lauring Lab, University of Michigan, Department of Microbiology and Immunology | Valesano et al |

| | | | | | | |
|---|---|---|---|---|---|---|
| EPI_ISL_628839 | USA/MI-UM-10036488294/2020 | 2020-10-16 | North America/USA/Michigan/ | University of Michigan Clinical Microbiology Laboratory | Lauring Lab, University of Michigan, Department of Microbiology and Immunology | Valesano et al |
| EPI_ISL_628844 | USA/MI-UM-10036496672/2020 | 2020-10-17 | North America/USA/Michigan/ | University of Michigan Clinical Microbiology Laboratory | Lauring Lab, University of Michigan, Department of Microbiology and Immunology | Valesano et al |
| EPI_ISL_628855 | USA/MI-UM-10036496560/2020 | 2020-10-17 | North America/USA/Michigan/ | University of Michigan Clinical Microbiology Laboratory | Lauring Lab, University of Michigan, Department of Microbiology and Immunology | Valesano et al |
| EPI_ISL_628864 | USA/MI-UM-10036496188/2020 | 2020-10-17 | North America/USA/Michigan/ | University of Michigan Clinical Microbiology Laboratory | Lauring Lab, University of Michigan, Department of Microbiology and Immunology | Valesano et al |
| EPI_ISL_628866 | USA/MI-UM-10036496278/2020 | 2020-10-17 | North America/USA/Michigan/ | University of Michigan Clinical Microbiology Laboratory | Lauring Lab, University of Michigan, Department of Microbiology and Immunology | Valesano et al |
| EPI_ISL_628867 | USA/MI-UM-10036524225/2020 | 2020-10-19 | North America/USA/Michigan/ | University of Michigan Clinical Microbiology Laboratory | Lauring Lab, University of Michigan, Department of Microbiology and Immunology | Valesano et al |

| | | | | | | |
|---|---|---|---|---|---|---|
| EPI_ISL_628868 | USA/MI-UM-10036518489/2020 | 2020-10-19 | North America/USA/Michigan/ | University of Michigan Clinical Microbiology Laboratory | Lauring Lab, University of Michigan, Department of Microbiology and Immunology | Valesano et al |
| EPI_ISL_628872 | USA/MI-UM-10036511106/2020 | 2020-10-19 | North America/USA/Michigan/ | University of Michigan Clinical Microbiology Laboratory | Lauring Lab, University of Michigan, Department of Microbiology and Immunology | Valesano et al |
| EPI_ISL_628874 | USA/MI-UM-10036515267/2020 | 2020-10-19 | North America/USA/Michigan/ | University of Michigan Clinical Microbiology Laboratory | Lauring Lab, University of Michigan, Department of Microbiology and Immunology | Valesano et al |
| EPI_ISL_628878 | USA/MI-UM-10036524330/2020 | 2020-10-19 | North America/USA/Michigan/ | University of Michigan Clinical Microbiology Laboratory | Lauring Lab, University of Michigan, Department of Microbiology and Immunology | Valesano et al |
| EPI_ISL_628882 | USA/MI-UM-10036524165/2020 | 2020-10-19 | North America/USA/Michigan/ | University of Michigan Clinical Microbiology Laboratory | Lauring Lab, University of Michigan, Department of Microbiology and Immunology | Valesano et al |
| EPI_ISL_628938 | USA/UT-UPHL-201110857/2020 | 2020-10-05 | North America/USA/Utah/ | Utah Public Health Laboratory | Utah Public Health Laboratory | Erin Young et al |
| EPI_ISL_628944 | USA/UT-UPHL-201103251/2020 | 2020-10-08 | North America/USA/Utah/ | Utah Public Health Laboratory | Utah Public Health Laboratory | Erin Young et al |
| EPI_ISL_628946 | USA/UT-UPHL-201109190/2020 | 2020-10-01 | North America/USA/Utah/ | Utah Public Health Laboratory | Utah Public Health Laboratory | Erin Young et al |
| EPI_ISL_628950 | USA/UT-UPHL-201123731/2020 | 2020-10-09 | North America/USA/Utah/ | Utah Public Health Laboratory | Utah Public Health Laboratory | Erin Young et al |
| EPI_ISL_628952 | USA/UT-UPHL-201109604/2020 | 2020-10-12 | North America/USA/Utah/ | Utah Public Health Laboratory | Utah Public Health Laboratory | Erin Young et al |

| | | | | | | |
|---|---|---|---|---|---|---|
| EPI_ISL_628954 | USA/UT-UPHL-201124588/2020 | 2020-10-07 | North America/USA/Utah/ | Utah Public Health Laboratory | Utah Public Health Laboratory | Erin Young et al |
| EPI_ISL_628959 | USA/UT-UPHL-201108594/2020 | 2020-10-11 | North America/USA/Utah/ | Utah Public Health Laboratory | Utah Public Health Laboratory | Erin Young et al |
| EPI_ISL_628969 | USA/UT-UPHL-201118881/2020 | 2020-05-12 | North America/USA/Utah/ | Utah Public Health Laboratory | Utah Public Health Laboratory | Erin Young et al |
| EPI_ISL_628976 | USA/UT-UPHL-201126930/2020 | 2020-05-13 | North America/USA/Utah/ | Utah Public Health Laboratory | Utah Public Health Laboratory | Erin Young et al |
| EPI_ISL_628981 | USA/UT-UPHL-201101119/2020 | 2020-05-12 | North America/USA/Utah/ | Utah Public Health Laboratory | Utah Public Health Laboratory | Erin Young et al |
| EPI_ISL_628982 | USA/UT-UPHL-201104003/2020 | 2020-05-12 | North America/USA/Utah/ | Utah Public Health Laboratory | Utah Public Health Laboratory | Erin Young et al |
| EPI_ISL_629013 | Chile/RM-CMM-0184M/2020 | 2020-05-20 | South America/Chile/Region Metropolit | Centro de Biotecnolog√≠a Vegetal, Universidad Andr√©s Bello, Center for Genome Regulation | Center for Mathematical Modeling and Center for Genome Regulation. Santiago, Chile | Bastias M et al |
| EPI_ISL_629019 | Chile/RM-CMM-0071M/2020 | 2020-05-01 | South America/Chile/Region Metropolit | Centro de Biotecnolog√≠a Vegetal, Universidad Andr√©s Bello, Center for Genome Regulation | Center for Mathematical Modeling and Center for Genome Regulation. Santiago, Chile | Bastias M et al |
| EPI_ISL_629020 | Chile/RM-CMM-0007M/2020 | 2020-05-01 | South America/Chile/Region Metropolit | Centro de Biotecnolog√≠a Vegetal, Universidad Andr√©s Bello, Center for Genome Regulation | Center for Mathematical Modeling and Center for Genome Regulation. Santiago, Chile | Bastias M et al |
| EPI_ISL_629021 | Chile/RM-CMM-0089M/2020 | 2020-05-01 | South America/Chile/Region Metropolit | Centro de Biotecnolog√≠a Vegetal, Universidad Andr√©s Bello, Center for Genome Regulation | Center for Mathematical Modeling and Center for Genome Regulation. Santiago, Chile | Bastias M et al |

| | | | | | | |
|---|---|---|---|---|---|---|
| EPI_ISL_629022 | Chile/RM-CMM-0096M/2020 | 2020-05-02 | South America/Chile/Region Metropolit | Centro de Biotecnolog√≠a Vegetal, Universidad Andr√©s Bello, Center for Genome Regulation | Center for Mathematical Modeling and Center for Genome Regulation. Santiago, Chile | Bastias M et al |
| EPI_ISL_629031 | Russia/OMS-ORINFI-4761V-10K/2020 | 2020-05-03 | Europe/Russia/Omsk/Omsk | Omsk Research Institute of Natural Focal Infections | WHO National Influenza Centre Russian Federation | Artem Fadeev et al |
| EPI_ISL_629032 | Russia/OMS-ORINFI-4761V-11K/2020 | 2020-05-03 | Europe/Russia/Omsk/Omsk | Omsk Research Institute of Natural Focal Infections | WHO National Influenza Centre Russian Federation | Artem Fadeev et al |
| EPI_ISL_629033 | Russia/OMS-ORINFI-4768V-12K/2020 | 2020-05-03 | Europe/Russia/Omsk/Omsk | Omsk Research Institute of Natural Focal Infections | WHO National Influenza Centre Russian Federation | Artem Fadeev et al |
| EPI_ISL_629034 | Russia/OMS-ORINFI-4761V/2020 | 2020-05-03 | Europe/Russia/Omsk/Omsk | Omsk Research Institute of Natural Focal Infections | WHO National Influenza Centre Russian Federation | Artem Fadeev et al |
| EPI_ISL_629069 | USA/MD-MDH-0289/2020 | 2020-10-12 | North America/USA/Maryland/ | Maryland Public Health Laboratory | Maryland Public Health Laboratory | Maryland Department of Health Laboratories Administration et al |
| EPI_ISL_629081 | France/ARA-SC001/2020 | 2020-02-28 | Europe/France/Auvergne-Rh√¥ne-Alpes/ | Laboratoire du Centre Hospitalier Annecy Genevois | CNR Virus des Infections Respiratoires - France SUD | Antonin Bal et al |
| EPI_ISL_629082 | France/ARA-SC002/2020 | 2020-02-28 | Europe/France/Auvergne-Rh√¥ne-Alpes/ | Laboratoire du Centre Hospitalier Annecy Genevois | CNR Virus des Infections Respiratoires - France SUD | Antonin Bal et al |
| EPI_ISL_629083 | France/ARA-SC003/2020 | 2020-02-29 | Europe/France/Auvergne-Rh√¥ne-Alpes/ | Laboratoire du Centre Hospitalier Annecy Genevois | CNR Virus des Infections Respiratoires - France SUD | Antonin Bal et al |

| | | | | | | |
|---|---|---|---|---|---|---|
| EPI_ISL_629084 | France/ARA-SC004/2020 | 2020-02-19 | Europe/France/Auvergne-Rh√¥ne-Alpes/ | Laboratoire du Centre Hospitalier Annecy Genevois | CNR Virus des Infections Respiratoires - France SUD | Antonin Bal et al |
| EPI_ISL_629085 | France/ARA-SC005/2020 | 2020-02-29 | Europe/France/Auvergne-Rh√¥ne-Alpes/ | Laboratoire du Centre Hospitalier Annecy Genevois | CNR Virus des Infections Respiratoires - France SUD | Antonin Bal et al |
| EPI_ISL_629087 | France/ARA-SC016/2020 | 2020-03-03 | Europe/France/Auvergne-Rh√¥ne-Alpes/ | Laboratoire du Centre Hospitalier Annecy Genevois | CNR Virus des Infections Respiratoires - France SUD | Antonin Bal et al |
| EPI_ISL_629088 | France/ARA-SC020/2020 | 2020-03-18 | Europe/France/Auvergne-Rh√¥ne-Alpes/ | Laboratoire du Centre Hospitalier Annecy Genevois | CNR Virus des Infections Respiratoires - France SUD | Antonin Bal et al |
| EPI_ISL_629091 | France/ARA-SC027/2020 | 2020-03-22 | Europe/France/Auvergne-Rh√¥ne-Alpes/ | Laboratoire du Centre Hospitalier Annecy Genevois | CNR Virus des Infections Respiratoires - France SUD | Antonin Bal et al |
| EPI_ISL_629095 | France/ARA-SC040/2020 | 2020-03-30 | Europe/France/Auvergne-Rh√¥ne-Alpes/ | Laboratoire du Centre Hospitalier Annecy Genevois | CNR Virus des Infections Respiratoires - France SUD | Antonin Bal et al |
| EPI_ISL_629102 | France/ARA-SC064/2020 | 2020-04-16 | Europe/France/Auvergne-Rh√¥ne-Alpes/ | Laboratoire du Centre Hospitalier Annecy Genevois | CNR Virus des Infections Respiratoires - France SUD | Antonin Bal et al |
| EPI_ISL_629104 | France/ARA-SC068/2020 | 2020-04-17 | Europe/France/Auvergne-Rh√¥ne-Alpes/ | Laboratoire du Centre Hospitalier Annecy Genevois | CNR Virus des Infections Respiratoires - France SUD | Antonin Bal et al |
| EPI_ISL_629105 | France/ARA-SC070/2020 | 2020-04-18 | Europe/France/Auvergne-Rh√¥ne-Alpes/ | Laboratoire du Centre Hospitalier Annecy Genevois | CNR Virus des Infections Respiratoires - France SUD | Antonin Bal et al |
| EPI_ISL_629108 | France/ARA-SC078/2020 | 2020-04-30 | Europe/France/Auvergne-Rh√¥ne-Alpes/ | Laboratoire du Centre Hospitalier Annecy Genevois | CNR Virus des Infections Respiratoires - France SUD | Antonin Bal et al |

| | | | | | | |
|---|---|---|---|---|---|---|
| EPI_ISL_629131 | England/MILK-AC837A/2020 | 2020-10-21 | Europe/United Kingdom/England/ | Lighthouse Lab in Milton Keynes | Wellcome Sanger Institute for the COVID-19 Genomics UK (COG-UK) consortium | The Lighthouse Lab in Milton Keynes et al |
| EPI_ISL_629202 | England/MILK-ACE981/2020 | 2020-10-21 | Europe/United Kingdom/England/ | Lighthouse Lab in Milton Keynes | Wellcome Sanger Institute for the COVID-19 Genomics UK (COG-UK) consortium | The Lighthouse Lab in Milton Keynes et al |
| EPI_ISL_629223 | England/MILK-ACC121/2020 | 2020-10-21 | Europe/United Kingdom/England/ | Lighthouse Lab in Milton Keynes | Wellcome Sanger Institute for the COVID-19 Genomics UK (COG-UK) consortium | The Lighthouse Lab in Milton Keynes et al |
| EPI_ISL_629242 | England/MILK-AC86AE/2020 | 2020-10-21 | Europe/United Kingdom/England/ | Lighthouse Lab in Milton Keynes | Wellcome Sanger Institute for the COVID-19 Genomics UK (COG-UK) consortium | The Lighthouse Lab in Milton Keynes et al |
| EPI_ISL_629248 | England/MILK-ACC886/2020 | 2020-10-21 | Europe/United Kingdom/England/ | Lighthouse Lab in Milton Keynes | Wellcome Sanger Institute for the COVID-19 Genomics UK (COG-UK) consortium | The Lighthouse Lab in Milton Keynes et al |
| EPI_ISL_629250 | England/MILK-AC835C/2020 | 2020-10-21 | Europe/United Kingdom/England/ | Lighthouse Lab in Milton Keynes | Wellcome Sanger Institute for the COVID-19 Genomics UK (COG-UK) consortium | The Lighthouse Lab in Milton Keynes et al |
| EPI_ISL_629363 | England/MILK-ACC385/2020 | 2020-10-21 | Europe/United Kingdom/England/ | Lighthouse Lab in Milton Keynes | Wellcome Sanger Institute for the COVID-19 Genomics UK (COG-UK) consortium | The Lighthouse Lab in Milton Keynes et al |
| EPI_ISL_629567 | England/MILK-AC8EC4/2020 | 2020-10-22 | Europe/United Kingdom/England/ | Lighthouse Lab in Milton Keynes | Wellcome Sanger Institute for the COVID-19 Genomics UK (COG-UK) consortium | The Lighthouse Lab in Milton Keynes et al |

| | | | | | | |
|---|---|---|---|---|---|---|
| EPI_ISL_629631 | England/MILK-ACE66C/2020 | 2020-10-21 | Europe/United Kingdom/England/ | Lighthouse Lab in Milton Keynes | Wellcome Sanger Institute for the COVID-19 Genomics UK (COG-UK) consortium | The Lighthouse Lab in Milton Keynes et al |
| EPI_ISL_629652 | England/MILK-AC89D2/2020 | 2020-10-22 | Europe/United Kingdom/England/ | Lighthouse Lab in Milton Keynes | Wellcome Sanger Institute for the COVID-19 Genomics UK (COG-UK) consortium | The Lighthouse Lab in Milton Keynes et al |
| EPI_ISL_629743 | England/MILK-ACF7B3/2020 | 2020-10-22 | Europe/United Kingdom/England/ | Lighthouse Lab in Milton Keynes | Wellcome Sanger Institute for the COVID-19 Genomics UK (COG-UK) consortium | The Lighthouse Lab in Milton Keynes et al |
| EPI_ISL_629798 | England/QEUH-AD2E4F/2020 | 2020-10-26 | Europe/United Kingdom/England/ | Lighthouse Lab in Glasgow | Wellcome Sanger Institute for the COVID-19 Genomics UK (COG-UK) consortium | Harper VanSteenhouse et al |
| EPI_ISL_629806 | England/QEUH-AD1DBC/2020 | 2020-10-24 | Europe/United Kingdom/England/ | Lighthouse Lab in Glasgow | Wellcome Sanger Institute for the COVID-19 Genomics UK (COG-UK) consortium | Harper VanSteenhouse et al |
| EPI_ISL_629964 | England/QEUH-AD1DCB/2020 | 2020-10-24 | Europe/United Kingdom/England/ | Lighthouse Lab in Glasgow | Wellcome Sanger Institute for the COVID-19 Genomics UK (COG-UK) consortium | Harper VanSteenhouse et al |
| EPI_ISL_629985 | England/QEUH-AD17AF/2020 | 2020-10-24 | Europe/United Kingdom/England/ | Lighthouse Lab in Glasgow | Wellcome Sanger Institute for the COVID-19 Genomics UK (COG-UK) consortium | Harper VanSteenhouse et al |
| EPI_ISL_630128 | England/MILK-ACE031/2020 | 2020-10-21 | Europe/United Kingdom/England/ | Lighthouse Lab in Milton Keynes | Wellcome Sanger Institute for the COVID-19 Genomics UK (COG-UK) consortium | The Lighthouse Lab in Milton Keynes et al |

| | | | | | | |
|---|---|---|---|---|---|---|
| EPI_ISL_630246 | England/MILK-ACCD4B/2020 | 2020-10-21 | Europe/United Kingdom/England/ | Lighthouse Lab in Milton Keynes | Wellcome Sanger Institute for the COVID-19 Genomics UK (COG-UK) consortium | The Lighthouse Lab in Milton Keynes et al |
| EPI_ISL_630343 | England/MILK-ACA879/2020 | 2020-10-21 | Europe/United Kingdom/England/ | Lighthouse Lab in Milton Keynes | Wellcome Sanger Institute for the COVID-19 Genomics UK (COG-UK) consortium | The Lighthouse Lab in Milton Keynes et al |
| EPI_ISL_630346 | England/MILK-AC734E/2020 | 2020-10-22 | Europe/United Kingdom/England/ | Lighthouse Lab in Milton Keynes | Wellcome Sanger Institute for the COVID-19 Genomics UK (COG-UK) consortium | The Lighthouse Lab in Milton Keynes et al |
| EPI_ISL_630395 | England/ALDP-A66AA6/2020 | 2020-10-12 | Europe/United Kingdom/England/ | Lighthouse Lab in Alderley Park | Wellcome Sanger Institute for the COVID-19 Genomics UK (COG-UK) consortium | Jacquelyn Wynn et al |
| EPI_ISL_630413 | England/ALDP-A671FB/2020 | 2020-10-12 | Europe/United Kingdom/England/ | Lighthouse Lab in Alderley Park | Wellcome Sanger Institute for the COVID-19 Genomics UK (COG-UK) consortium | Jacquelyn Wynn et al |
| EPI_ISL_630414 | England/ALDP-A67234/2020 | 2020-10-09 | Europe/United Kingdom/England/ | Lighthouse Lab in Alderley Park | Wellcome Sanger Institute for the COVID-19 Genomics UK (COG-UK) consortium | Jacquelyn Wynn et al |
| EPI_ISL_630646 | England/ALDP-A6761A/2020 | 2020-10-12 | Europe/United Kingdom/England/ | Lighthouse Lab in Alderley Park | Wellcome Sanger Institute for the COVID-19 Genomics UK (COG-UK) consortium | Jacquelyn Wynn et al |
| EPI_ISL_630650 | England/ALDP-A66CEC/2020 | 2020-10-12 | Europe/United Kingdom/England/ | Lighthouse Lab in Alderley Park | Wellcome Sanger Institute for the COVID-19 Genomics UK (COG-UK) consortium | Jacquelyn Wynn et al |

| | | | | | | |
|---|---|---|---|---|---|---|
| EPI_ISL_630702 | England/QEUH-AAF2B8/2020 | 2020-10-19 | Europe/United Kingdom/England/ | Lighthouse Lab in Glasgow | Wellcome Sanger Institute for the COVID-19 Genomics UK (COG-UK) consortium | Harper VanSteenhouse et al |
| EPI_ISL_630721 | England/ALDP-AA320F/2020 | 2020-10-22 | Europe/United Kingdom/England/ | Lighthouse Lab in Alderley Park | Wellcome Sanger Institute for the COVID-19 Genomics UK (COG-UK) consortium | Jacquelyn Wynn et al |
| EPI_ISL_630865 | England/ALDP-AA0976/2020 | 2020-10-22 | Europe/United Kingdom/England/ | Lighthouse Lab in Alderley Park | Wellcome Sanger Institute for the COVID-19 Genomics UK (COG-UK) consortium | Jacquelyn Wynn et al |
| EPI_ISL_630907 | England/ALDP-AA2613/2020 | 2020-10-22 | Europe/United Kingdom/England/ | Lighthouse Lab in Alderley Park | Wellcome Sanger Institute for the COVID-19 Genomics UK (COG-UK) consortium | Jacquelyn Wynn et al |
| EPI_ISL_630912 | England/ALDP-AA5030/2020 | 2020-10-21 | Europe/United Kingdom/England/ | Lighthouse Lab in Alderley Park | Wellcome Sanger Institute for the COVID-19 Genomics UK (COG-UK) consortium | Jacquelyn Wynn et al |
| EPI_ISL_630967 | England/ALDP-AA4D76/2020 | 2020-10-21 | Europe/United Kingdom/England/ | Lighthouse Lab in Alderley Park | Wellcome Sanger Institute for the COVID-19 Genomics UK (COG-UK) consortium | Jacquelyn Wynn et al |
| EPI_ISL_631212 | England/ALDP-ABD2C6/2020 | 2020-10-04 | Europe/United Kingdom/England/ | Lighthouse Lab in Alderley Park | Wellcome Sanger Institute for the COVID-19 Genomics UK (COG-UK) consortium | Jacquelyn Wynn et al |
| EPI_ISL_631221 | England/CAMC-AAF977/2020 | 2020-10-23 | Europe/United Kingdom/England/ | Lighthouse Lab in Cambridge | Wellcome Sanger Institute for the COVID-19 Genomics UK (COG-UK) consortium | Rob Howes et al |

| | | | | | | |
|---|---|---|---|---|---|---|
| EPI_ISL_631297 | Germany/NW-UBI-43/2020 | 2020-06-17 | Europe/Germany/North Rhine Westphal | MVZ DIAMEDIS Diagnostische Medizin Sennestadt GmbH | Bielefeld University | David Brandt et al |
| EPI_ISL_631308 | Germany/NW-HHU-305/2020 | 2020-10-05 | Europe/Germany/North Rhine Westphal | ZOTZ KLIMAS MVZ Dvºsseldorf-Centrum GbR √úBAG fvºr Labormedizin, Genetik, Zytologie, Pathologie | Center of Medical Microbiology, Virology, and Hospital Hygiene, University of Duesseldorf | Maximilian Damagnez et al |
| EPI_ISL_631311 | Germany/NW-HHU-308/2020 | 2020-10-05 | Europe/Germany/North Rhine Westphal | ZOTZ KLIMAS MVZ Dvºsseldorf-Centrum GbR √úBAG fvºr Labormedizin, Genetik, Zytologie, Pathologie | Center of Medical Microbiology, Virology, and Hospital Hygiene, University of Duesseldorf | Maximilian Damagnez et al |
| EPI_ISL_631316 | Germany/NW-HHU-313/2020 | 2020-10-06 | Europe/Germany/North Rhine Westphal | ZOTZ KLIMAS MVZ Dvºsseldorf-Centrum GbR √úBAG fvºr Labormedizin, Genetik, Zytologie, Pathologie | Center of Medical Microbiology, Virology, and Hospital Hygiene, University of Duesseldorf | Maximilian Damagnez et al |
| EPI_ISL_631317 | Germany/NW-HHU-314/2020 | 2020-10-06 | Europe/Germany/North Rhine Westphal | ZOTZ KLIMAS MVZ Dvºsseldorf-Centrum GbR √úBAG fvºr Labormedizin, Genetik, Zytologie, Pathologie | Center of Medical Microbiology, Virology, and Hospital Hygiene, University of Duesseldorf | Maximilian Damagnez et al |
| EPI_ISL_631320 | Germany/NW-HHU-320/2020 | 2020-10-06 | Europe/Germany/North Rhine Westphal | ZOTZ KLIMAS MVZ Dvºsseldorf-Centrum GbR √úBAG fvºr Labormedizin, Genetik, Zytologie, Pathologie | Center of Medical Microbiology, Virology, and Hospital Hygiene, University of Duesseldorf | Maximilian Damagnez et al |
| EPI_ISL_631344 | Germany/NW-HHU-256/2020 | 2020-09-21 | Europe/Germany/North Rhine Westphal | ZOTZ KLIMAS MVZ Dvºsseldorf-Centrum GbR √úBAG fvºr Labormedizin, Genetik, Zytologie, Pathologie | Center of Medical Microbiology, Virology, and Hospital Hygiene, University of Duesseldorf | Maximilian Damagnez et al |

| | | | | | | |
|---|---|---|---|---|---|---|
| EPI_ISL_631364 | Germany/NW-HHU-276/2020 | 2020-09-26 | Europe/Germany/North Rhine Westphal | ZOTZ KLIMAS MVZ Dv∫sseldorf-Centrum GbR √úBAG fv∫r Labormedizin, Genetik, Zytologie, Pathologie | Center of Medical Microbiology, Virology, and Hospital Hygiene, University of Duesseldorf | Maximilian Damagnez et al |
| EPI_ISL_631367 | Germany/NW-HHU-234/2020 | 2020-09-14 | Europe/Germany/North Rhine Westphal | ZOTZ KLIMAS MVZ Dv∫sseldorf-Centrum GbR √úBAG fv∫r Labormedizin, Genetik, Zytologie, Pathologie | Center of Medical Microbiology, Virology, and Hospital Hygiene, University of Duesseldorf | Maximilian Damagnez et al |
| EPI_ISL_631370 | Germany/NW-HHU-237/2020 | 2020-09-15 | Europe/Germany/North Rhine Westphal | ZOTZ KLIMAS MVZ Dv∫sseldorf-Centrum GbR √úBAG fv∫r Labormedizin, Genetik, Zytologie, Pathologie | Center of Medical Microbiology, Virology, and Hospital Hygiene, University of Duesseldorf | Maximilian Damagnez et al |
| EPI_ISL_631374 | Germany/NW-HHU-241/2020 | 2020-09-15 | Europe/Germany/North Rhine Westphal | ZOTZ KLIMAS MVZ Dv∫sseldorf-Centrum GbR √úBAG fv∫r Labormedizin, Genetik, Zytologie, Pathologie | Center of Medical Microbiology, Virology, and Hospital Hygiene, University of Duesseldorf | Maximilian Damagnez et al |
| EPI_ISL_631397 | USA/WI-WSLH-200600/2020 | 2020-06-26 | North America/USA/Wisconsin/Waushar | Wisconsin State Laboratory of Hygiene Communicable Disease Division | Wisconsin State Laboratory of Hygiene Communicable Disease Division | Kelsey R. Florek et al |
| EPI_ISL_631401 | USA/WI-WSLH-200608/2020 | 2020-08-19 | North America/USA/Wisconsin/Sheboyg | Wisconsin State Laboratory of Hygiene Communicable Disease Division | Wisconsin State Laboratory of Hygiene Communicable Disease Division | Kelsey R. Florek et al |
| EPI_ISL_631403 | USA/WI-WSLH-200634/2020 | 2020-06-09 | North America/USA/Wisconsin/Eau Clai | Wisconsin State Laboratory of Hygiene Communicable Disease Division | Wisconsin State Laboratory of Hygiene Communicable Disease Division | Kelsey R. Florek et al |

| | | | | | | |
|---|---|---|---|---|---|---|
| EPI_ISL_631411 | USA/WI-WSLH-200642/2020 | 2020-06-15 | North America/USA/Wisconsin/Milwauk | Wisconsin State Laboratory of Hygiene Communicable Disease Division | Wisconsin State Laboratory of Hygiene Communicable Disease Division | Kelsey R. Florek et al |
| EPI_ISL_631416 | USA/WI-WSLH-200647/2020 | 2020-06-16 | North America/USA/Wisconsin/Barron C | Wisconsin State Laboratory of Hygiene Communicable Disease Division | Wisconsin State Laboratory of Hygiene Communicable Disease Division | Kelsey R. Florek et al |
| EPI_ISL_631421 | USA/WI-WSLH-200654/2020 | 2020-06-18 | North America/USA/Wisconsin/Brown C | Wisconsin State Laboratory of Hygiene Communicable Disease Division | Wisconsin State Laboratory of Hygiene Communicable Disease Division | Kelsey R. Florek et al |
| EPI_ISL_631424 | USA/WI-WSLH-200658/2020 | 2020-06-19 | North America/USA/Wisconsin/Lafayett | Wisconsin State Laboratory of Hygiene Communicable Disease Division | Wisconsin State Laboratory of Hygiene Communicable Disease Division | Kelsey R. Florek et al |
| EPI_ISL_631432 | USA/WI-WSLH-200674/2020 | 2020-06-23 | North America/USA/Wisconsin/Winneba | Wisconsin State Laboratory of Hygiene Communicable Disease Division | Wisconsin State Laboratory of Hygiene Communicable Disease Division | Kelsey R. Florek et al |
| EPI_ISL_631436 | USA/WI-WSLH-200679/2020 | 2020-08-19 | North America/USA/Wisconsin/Sheboyg | Wisconsin State Laboratory of Hygiene Communicable Disease Division | Wisconsin State Laboratory of Hygiene Communicable Disease Division | Kelsey R. Florek et al |
| EPI_ISL_631451 | USA/WI-WSLH-200694/2020 | 2020-09-14 | North America/USA/Wisconsin/Sheboyg | Wisconsin State Laboratory of Hygiene Communicable Disease Division | Wisconsin State Laboratory of Hygiene Communicable Disease Division | Kelsey R. Florek et al |
| EPI_ISL_631453 | USA/WI-WSLH-200696/2020 | 2020-09-17 | North America/USA/Wisconsin/Sheboyg | Wisconsin State Laboratory of Hygiene Communicable Disease Division | Wisconsin State Laboratory of Hygiene Communicable Disease Division | Kelsey R. Florek et al |

| | | | | | | |
|---|---|---|---|---|---|---|
| EPI_ISL_631455 | USA/WI-WSLH-200698/2020 | 2020-09-18 | North America/USA/Wisconsin/Sheboyg | Wisconsin State Laboratory of Hygiene Communicable Disease Division | Wisconsin State Laboratory of Hygiene Communicable Disease Division | Kelsey R. Florek et al |
| EPI_ISL_631460 | USA/WI-WSLH-200703/2020 | 2020-06-25 | North America/USA/Wisconsin/Rock Cou | Wisconsin State Laboratory of Hygiene Communicable Disease Division | Wisconsin State Laboratory of Hygiene Communicable Disease Division | Kelsey R. Florek et al |
| EPI_ISL_631463 | USA/WI-WSLH-200706/2020 | 2020-06-26 | North America/USA/Wisconsin/Green Co | Wisconsin State Laboratory of Hygiene Communicable Disease Division | Wisconsin State Laboratory of Hygiene Communicable Disease Division | Kelsey R. Florek et al |
| EPI_ISL_631467 | USA/WI-WSLH-200710/2020 | 2020-06-27 | North America/USA/Wisconsin/Milwauk | Wisconsin State Laboratory of Hygiene Communicable Disease Division | Wisconsin State Laboratory of Hygiene Communicable Disease Division | Kelsey R. Florek et al |
| EPI_ISL_631475 | USA/WI-WSLH-200719/2020 | 2020-06-29 | North America/USA/Wisconsin/Brown C | Wisconsin State Laboratory of Hygiene Communicable Disease Division | Wisconsin State Laboratory of Hygiene Communicable Disease Division | Kelsey R. Florek et al |
| EPI_ISL_631481 | USA/WI-WSLH-200725/2020 | 2020-07-01 | North America/USA/Wisconsin/Dane Co | Wisconsin State Laboratory of Hygiene Communicable Disease Division | Wisconsin State Laboratory of Hygiene Communicable Disease Division | Kelsey R. Florek et al |
| EPI_ISL_631483 | USA/WI-WSLH-200727/2020 | 2020-07-02 | North America/USA/Wisconsin/Dane Co | Wisconsin State Laboratory of Hygiene Communicable Disease Division | Wisconsin State Laboratory of Hygiene Communicable Disease Division | Kelsey R. Florek et al |
| EPI_ISL_631490 | USA/WI-WSLH-200734/2020 | 2020-08-06 | North America/USA/Wisconsin/Dane Co | Wisconsin State Laboratory of Hygiene Communicable Disease Division | Wisconsin State Laboratory of Hygiene Communicable Disease Division | Kelsey R. Florek et al |

| | | | | | | |
|---|---|---|---|---|---|---|
| EPI_ISL_631491 | USA/WI-WSLH-200735/2020 | 2020-08-28 | North America/USA/Wisconsin/Dane Co | Wisconsin State Laboratory of Hygiene Communicable Disease Division | Wisconsin State Laboratory of Hygiene Communicable Disease Division | Kelsey R. Florek et al |
| EPI_ISL_631495 | USA/WI-WSLH-200739/2020 | 2020-09-14 | North America/USA/Wisconsin/Dane Co | Wisconsin State Laboratory of Hygiene Communicable Disease Division | Wisconsin State Laboratory of Hygiene Communicable Disease Division | Kelsey R. Florek et al |
| EPI_ISL_631496 | USA/WI-WSLH-200741/2020 | 2020-09-17 | North America/USA/Wisconsin/Dane Co | Wisconsin State Laboratory of Hygiene Communicable Disease Division | Wisconsin State Laboratory of Hygiene Communicable Disease Division | Kelsey R. Florek et al |
| EPI_ISL_631501 | USA/WI-WSLH-200746/2020 | 2020-06-30 | North America/USA/Wisconsin/Rock Cou | Wisconsin State Laboratory of Hygiene Communicable Disease Division | Wisconsin State Laboratory of Hygiene Communicable Disease Division | Kelsey R. Florek et al |
| EPI_ISL_631506 | USA/NY-NYCPHL-000006/2020 | 2020-03-06 | North America/USA/New York/New Yor | NYC Department Of Health And Mental Hygiene | New York City Public Health Laboratory | Jade Wang et al |
| EPI_ISL_631522 | USA/NY-NYCPHL-000026/2020 | 2020-03-10 | North America/USA/New York/Nassau C | Robert Kutnick | New York City Public Health Laboratory | Jade Wang et al |
| EPI_ISL_631559 | USA/NY-NYCPHL-000066/2020 | 2020-03-11 | North America/USA/New York/New Yor | NYU Langone Health | New York City Public Health Laboratory | Jade Wang et al |
| EPI_ISL_631562 | USA/NY-NYCPHL-000069/2020 | 2020-03-14 | North America/USA/New York/New Yor | Brookdale University Hospital Medical Center | New York City Public Health Laboratory | Jade Wang et al |
| EPI_ISL_631574 | USA/NY-NYCPHL-000081/2020 | 2020-03-17 | North America/USA/New York/New Yor | Jamaica Hospital Medical Center | New York City Public Health Laboratory | Jade Wang et al |
| EPI_ISL_631579 | USA/NY-NYCPHL-000086/2020 | 2020-03-19 | North America/USA/New York/New Yor | Flushing Hospital Medical Center | New York City Public Health Laboratory | Jade Wang et al |
| EPI_ISL_631582 | USA/NY-NYCPHL-000089/2020 | 2020-03-21 | North America/USA/New York/New Yor | Jamaica Hospital Medical Center | New York City Public Health Laboratory | Jade Wang et al |

| | | | | | | |
|---|---|---|---|---|---|---|
| EPI_ISL_631598 | USA/NY-NYCPHL-000106/2020 | 2020-03-26 | North America/USA/New York/New Yor | OCME Office Of Chief Medical Examiner | New York City Public Health Laboratory | Jade Wang et al |
| EPI_ISL_631625 | USA/NY-NYCPHL-000134/2020 | 2020-04-02 | North America/USA/New York/New Yor | Richmond University Medical Center | New York City Public Health Laboratory | Jade Wang et al |
| EPI_ISL_631636 | USA/NY-NYCPHL-000145/2020 | 2020-04-03 | North America/USA/New York/New Yor | Jamaica Hospital Medical Center | New York City Public Health Laboratory | Jade Wang et al |
| EPI_ISL_631640 | USA/TX-DSHS-1177/2020 | 2020-06-16 | North America/USA/Texas/Harris Count | Texas Department of State Health Services | Texas Department of State Health Services | Rashmi Tuladhar et al |
| EPI_ISL_631646 | USA/TX-DSHS-1184/2020 | 2020-06-16 | North America/USA/Texas/Harris Count | Texas Department of State Health Services | Texas Department of State Health Services | Rashmi Tuladhar et al |
| EPI_ISL_631661 | USA/TX-DSHS-1199/2020 | 2020-03-17 | North America/USA/Texas/Bexar County | Texas Department of State Health Services | Texas Department of State Health Services | Rashmi Tuladhar et al |
| EPI_ISL_631664 | USA/TX-DSHS-1202/2020 | 2020-03-17 | North America/USA/Texas/Bexar County | Texas Department of State Health Services | Texas Department of State Health Services | Rashmi Tuladhar et al |
| EPI_ISL_631695 | USA/NY-NYCPHL-000159/2020 | 2020-04-12 | North America/USA/New York/New Yor | OCME Office Of Chief Medical Examiner | New York City Public Health Laboratory | Jade Wang et al |
| EPI_ISL_631698 | USA/NY-NYCPHL-000162/2020 | 2020-04-08 | North America/USA/New York/New Yor | Jamaica Hospital Medical Center | New York City Public Health Laboratory | Jade Wang et al |
| EPI_ISL_631699 | USA/NY-NYCPHL-000163/2020 | 2020-04-09 | North America/USA/New York/New Yor | Flushing Hospital Medical Center | New York City Public Health Laboratory | Jade Wang et al |
| EPI_ISL_631702 | USA/NY-NYCPHL-000166/2020 | 2020-04-10 | North America/USA/New York/New Yor | Jamaica Hospital Medical Center | New York City Public Health Laboratory | Jade Wang et al |
| EPI_ISL_631715 | USA/NY-NYCPHL-000180/2020 | 2020-04-15 | North America/USA/New York/New Yor | Flushing Hospital Medical Center | New York City Public Health Laboratory | Jade Wang et al |
| EPI_ISL_631719 | USA/NY-NYCPHL-000184/2020 | 2020-04-21 | North America/USA/New York/New Yor | Jamaica Hospital Medical Center | New York City Public Health Laboratory | Jade Wang et al |

| | | | | | | |
|---|---|---|---|---|---|---|
| EPI_ISL_631723 | USA/NY-NYCPHL-000188/2020 | 2020-04-13 | North America/USA/New York/New Yor | Jamaica Hospital Medical Center | New York City Public Health Laboratory | Jade Wang et al |
| EPI_ISL_631727 | USA/NY-NYCPHL-000192/2020 | 2020-04-14 | North America/USA/New York/New Yor | Flushing Hospital Medical Center | New York City Public Health Laboratory | Jade Wang et al |
| EPI_ISL_631730 | USA/NY-NYCPHL-000195/2020 | 2020-04-15 | North America/USA/New York/New Yor | Jamaica Hospital Medical Center | New York City Public Health Laboratory | Jade Wang et al |
| EPI_ISL_631744 | USA/NY-NYCPHL-000211/2020 | 2020-04-20 | North America/USA/New York/New Yor | Jamaica Hospital Medical Center | New York City Public Health Laboratory | Jade Wang et al |
| EPI_ISL_631746 | USA/NY-NYCPHL-000214/2020 | 2020-04-21 | North America/USA/New York/New Yor | Jamaica Hospital Medical Center | New York City Public Health Laboratory | Jade Wang et al |
| EPI_ISL_631751 | USA/NY-NYCPHL-000220/2020 | 2020-04-29 | North America/USA/New York/New Yor | St Barnabas Hospital | New York City Public Health Laboratory | Jade Wang et al |
| EPI_ISL_631752 | USA/NY-NYCPHL-000222/2020 | 2020-03-18 | North America/USA/New York/New Yor | NYC HH Lincoln Medical And Mental Health Center | New York City Public Health Laboratory | Jade Wang et al |
| EPI_ISL_631755 | USA/NY-NYCPHL-000226/2020 | 2020-03-23 | North America/USA/New York/New Yor | Richmond University Medical Center | New York City Public Health Laboratory | Jade Wang et al |
| EPI_ISL_631761 | USA/NY-NYCPHL-000232/2020 | 2020-03-27 | North America/USA/New York/New Yor | OCME Office Of Chief Medical Examiner | New York City Public Health Laboratory | Jade Wang et al |
| EPI_ISL_631772 | USA/NY-NYCPHL-000246/2020 | 2020-04-13 | North America/USA/New York/New Yor | Jamaica Hospital Medical Center | New York City Public Health Laboratory | Jade Wang et al |
| EPI_ISL_631810 | USA/NY-NYCPHL-000284/2020 | 2020-03-20 | North America/USA/New York/New Yor | Jamaica Hospital Medical Center | New York City Public Health Laboratory | Jade Wang et al |
| EPI_ISL_631814 | USA/NY-NYCPHL-000288/2020 | 2020-03-20 | North America/USA/New York/New Yor | Richmond University Medical Center | New York City Public Health Laboratory | Jade Wang et al |
| EPI_ISL_631819 | USA/NY-NYCPHL-000293/2020 | 2020-03-22 | North America/USA/New York/New Yor | Brookdale University Hospital Medical Center | New York City Public Health Laboratory | Jade Wang et al |

| | | | | | | |
|---|---|---|---|---|---|---|
| EPI_ISL_631822 | USA/NY-NYCPHL-000296/2020 | 2020-03-23 | North America/USA/New York/New Yor | Jamaica Hospital Medical Center | New York City Public Health Laboratory | Jade Wang et al |
| EPI_ISL_631830 | USA/NY-NYCPHL-000303/2020 | 2020-03-14 | North America/USA/New York/New Yor | Mount Sinai Hospital | New York City Public Health Laboratory | Jade Wang et al |
| EPI_ISL_631832 | USA/NY-NYCPHL-000305/2020 | 2020-03-15 | North America/USA/New York/New Yor | Metro Urgicare | New York City Public Health Laboratory | Jade Wang et al |
| EPI_ISL_631834 | USA/NY-NYCPHL-000307/2020 | 2020-03-15 | North America/USA/New York/New Yor | Mount Sinai Hospital | New York City Public Health Laboratory | Jade Wang et al |
| EPI_ISL_631842 | USA/NY-NYCPHL-000317/2020 | 2020-04-28 | North America/USA/New York/New Yor | OCME Office Of Chief Medical Examiner | New York City Public Health Laboratory | Jade Wang et al |
| EPI_ISL_631847 | USA/NY-NYCPHL-000334/2020 | 2020-04-07 | North America/USA/New York/New Yor | Jamaica Hospital Medical Center | New York City Public Health Laboratory | Jade Wang et al |
| EPI_ISL_631848 | USA/NY-NYCPHL-000335/2020 | 2020-04-08 | North America/USA/New York/New Yor | Flushing Hospital Medical Center | New York City Public Health Laboratory | Jade Wang et al |
| EPI_ISL_631851 | USA/NY-NYCPHL-000339/2020 | 2020-04-09 | North America/USA/New York/New Yor | Flushing Hospital Medical Center | New York City Public Health Laboratory | Jade Wang et al |
| EPI_ISL_631861 | USA/NY-NYCPHL-000354/2020 | 2020-03-10 | North America/USA/New York/New Yor | New York Presbyterian Queens | New York City Public Health Laboratory | Jade Wang et al |
| EPI_ISL_631864 | USA/NY-NYCPHL-000360/2020 | 2020-03-12 | North America/USA/New York/New Yor | Brookdale University Hospital Medical Center | New York City Public Health Laboratory | Jade Wang et al |
| EPI_ISL_631865 | USA/NY-NYCPHL-000361/2020 | 2020-03-12 | North America/USA/New York/New Yor | NYC HH Elmhurst Hospital Medical Center | New York City Public Health Laboratory | Jade Wang et al |
| EPI_ISL_631876 | USA/NY-NYCPHL-000390/2020 | 2020-04-05 | North America/USA/New York/New Yor | Jamaica Hospital Medical Center | New York City Public Health Laboratory | Jade Wang et al |
| EPI_ISL_631879 | USA/NY-NYCPHL-000394/2020 | 2020-04-08 | North America/USA/New York/New Yor | Jamaica Hospital Medical Center | New York City Public Health Laboratory | Jade Wang et al |

| | | | | | | |
|---|---|---|---|---|---|---|
| EPI_ISL_631882 | USA/NY-NYCPHL-000398/2020 | 2020-04-04 | North America/USA/New York/New Yor | Jamaica Hospital Medical Center | New York City Public Health Laboratory | Jade Wang et al |
| EPI_ISL_631892 | USA/NY-NYCPHL-000408/2020 | 2020-03-25 | North America/USA/New York/New Yor | Richmond University Medical Center | New York City Public Health Laboratory | Jade Wang et al |
| EPI_ISL_631903 | USA/NY-NYCPHL-000420/2020 | 2020-03-30 | North America/USA/New York/New Yor | Richmond University Medical Center | New York City Public Health Laboratory | Jade Wang et al |
| EPI_ISL_631909 | USA/NY-NYCPHL-000426/2020 | 2020-04-14 | North America/USA/New York/New Yor | Jamaica Hospital Medical Center | New York City Public Health Laboratory | Jade Wang et al |
| EPI_ISL_631921 | USA/NY-NYCPHL-000447/2020 | 2020-04-03 | North America/USA/New York/New Yor | Jamaica Hospital Medical Center | New York City Public Health Laboratory | Jade Wang et al |
| EPI_ISL_631928 | USA/NY-NYCPHL-000454/2020 | 2020-04-05 | North America/USA/New York/New Yor | Jamaica Hospital Medical Center | New York City Public Health Laboratory | Jade Wang et al |
| EPI_ISL_631933 | USA/NY-NYCPHL-000463/2020 | 2020-05-07 | North America/USA/New York/New Yor | St Barnabas Hospital | New York City Public Health Laboratory | Jade Wang et al |
| EPI_ISL_631936 | USA/NY-NYCPHL-000466/2020 | 2020-05-08 | North America/USA/New York/New Yor | OCME Office Of Chief Medical Examiner | New York City Public Health Laboratory | Jade Wang et al |
| EPI_ISL_631941 | USA/NY-NYCPHL-000472/2020 | 2020-05-01 | North America/USA/New York/New Yor | St Barnabas Hospital | New York City Public Health Laboratory | Jade Wang et al |
| EPI_ISL_631947 | USA/NY-NYCPHL-000480/2020 | 2020-05-06 | North America/USA/New York/New Yor | St Barnabas Hospital | New York City Public Health Laboratory | Jade Wang et al |
| EPI_ISL_631950 | USA/NY-NYCPHL-000485/2020 | 2020-04-10 | North America/USA/New York/New Yor | Jamaica Hospital Medical Center | New York City Public Health Laboratory | Jade Wang et al |
| EPI_ISL_631951 | USA/NY-NYCPHL-000486/2020 | 2020-04-11 | North America/USA/New York/New Yor | Jamaica Hospital Medical Center | New York City Public Health Laboratory | Jade Wang et al |
| EPI_ISL_631956 | USA/NY-NYCPHL-000494/2020 | 2020-05-14 | North America/USA/New York/New Yor | OCME Office Of Chief Medical Examiner | New York City Public Health Laboratory | Jade Wang et al |

| | | | | | | |
|---|---|---|---|---|---|---|
| EPI_ISL_631971 | USA/NY-NYCPHL-000510/2020 | 2020-03-24 | North America/USA/New York/New Yor | St Barnabas Hospital | New York City Public Health Laboratory | Jade Wang et al |
| EPI_ISL_631977 | USA/NY-NYCPHL-000516/2020 | 2020-03-20 | North America/USA/New York/New Yor | SUNY Downstate Medical Center | New York City Public Health Laboratory | Jade Wang et al |
| EPI_ISL_631983 | USA/NY-NYCPHL-000522/2020 | 2020-03-08 | North America/USA/New York/New Yor | Mount Sinai Hospital | New York City Public Health Laboratory | Jade Wang et al |
| EPI_ISL_631991 | USA/NY-NYCPHL-000530/2020 | 2020-04-02 | North America/USA/New York/New Yor | Flushing Hospital Medical Center | New York City Public Health Laboratory | Jade Wang et al |
| EPI_ISL_631998 | USA/NY-NYCPHL-000538/2020 | 2020-03-18 | North America/USA/New York/New Yor | Richmond University Medical Center | New York City Public Health Laboratory | Jade Wang et al |
| EPI_ISL_632004 | USA/NY-NYCPHL-000544/2020 | 2020-03-22 | North America/USA/New York/New Yor | Flushing Hospital Medical Center | New York City Public Health Laboratory | Jade Wang et al |
| EPI_ISL_632025 | USA/NY-NYCPHL-000566/2020 | 2020-03-29 | North America/USA/New York/New Yor | Jamaica Hospital Medical Center | New York City Public Health Laboratory | Jade Wang et al |
| EPI_ISL_632030 | USA/NY-NYCPHL-000571/2020 | 2020-03-30 | North America/USA/New York/New Yor | Richmond University Medical Center | New York City Public Health Laboratory | Jade Wang et al |
| EPI_ISL_632032 | USA/NY-NYCPHL-000573/2020 | 2020-04-01 | North America/USA/New York/New Yor | Richmond University Medical Center | New York City Public Health Laboratory | Jade Wang et al |
| EPI_ISL_632038 | USA/NY-NYCPHL-000580/2020 | 2020-03-08 | North America/USA/New York/New Yor | New York Presbyterian Lower Manhattan | New York City Public Health Laboratory | Jade Wang et al |
| EPI_ISL_632050 | USA/NY-NYCPHL-000592/2020 | 2020-03-10 | North America/USA/New York/New Hy | Long Island Jewish Medical Center | New York City Public Health Laboratory | Jade Wang et al |
| EPI_ISL_632055 | USA/NY-NYCPHL-000597/2020 | 2020-03-10 | North America/USA/New York/New Yor | New York Community Hospital | New York City Public Health Laboratory | Jade Wang et al |
| EPI_ISL_632056 | USA/NY-NYCPHL-000598/2020 | 2020-03-10 | North America/USA/New York/New Yor | New York Community Hospital | New York City Public Health Laboratory | Jade Wang et al |

| | | | | | | |
|---|---|---|---|---|---|---|
| EPI_ISL_632060 | USA/NY-NYCPHL-000602/2020 | 2020-03-10 | North America/USA/New York/New Yor | NYC HH Queens Hospital Center | New York City Public Health Laboratory | Jade Wang et al |
| EPI_ISL_632087 | USA/NY-NYCPHL-000669/2020 | 2020-03-18 | North America/USA/New York/New Yor | Flushing Hospital Medical Center | New York City Public Health Laboratory | Jade Wang et al |
| EPI_ISL_632095 | USA/NY-NYCPHL-000677/2020 | 2020-03-13 | North America/USA/New York/New Yor | Jamaica Hospital Medical Center | New York City Public Health Laboratory | Jade Wang et al |
| EPI_ISL_632104 | USA/NY-NYCPHL-000687/2020 | 2020-03-14 | North America/USA/New York/New Yor | Flushing Hospital Medical Center | New York City Public Health Laboratory | Jade Wang et al |
| EPI_ISL_632106 | USA/NY-NYCPHL-000690/2020 | 2020-03-14 | North America/USA/New York/New Yor | Jamaica Hospital Medical Center | New York City Public Health Laboratory | Jade Wang et al |
| EPI_ISL_632118 | USA/NY-NYCPHL-000737/2020 | 2020-03-22 | North America/USA/New York/New Yor | Richmond University Medical Center | New York City Public Health Laboratory | Jade Wang et al |
| EPI_ISL_632126 | USA/NY-NYCPHL-000745/2020 | 2020-03-19 | North America/USA/New York/New Yor | Flushing Hospital Medical Center | New York City Public Health Laboratory | Jade Wang et al |
| EPI_ISL_632127 | USA/NY-NYCPHL-000746/2020 | 2020-03-20 | North America/USA/New York/New Yor | Flushing Hospital Medical Center | New York City Public Health Laboratory | Jade Wang et al |
| EPI_ISL_632145 | USA/NY-NYCPHL-000765/2020 | 2020-03-19 | North America/USA/New York/New Yor | Jamaica Hospital Medical Center | New York City Public Health Laboratory | Jade Wang et al |
| EPI_ISL_632149 | USA/NY-NYCPHL-000769/2020 | 2020-03-20 | North America/USA/New York/New Yor | Jamaica Hospital Medical Center | New York City Public Health Laboratory | Jade Wang et al |
| EPI_ISL_632161 | USA/NY-NYCPHL-000781/2020 | 2020-03-20 | North America/USA/New York/New Yor | SUNY Downstate Medical Center | New York City Public Health Laboratory | Jade Wang et al |
| EPI_ISL_632162 | USA/NY-NYCPHL-000782/2020 | 2020-03-21 | North America/USA/New York/New Yor | Jamaica Hospital Medical Center | New York City Public Health Laboratory | Jade Wang et al |
| EPI_ISL_632176 | USA/NY-NYCPHL-000796/2020 | 2020-03-25 | North America/USA/New York/New Yor | OCME Office Of Chief Medical Examiner | New York City Public Health Laboratory | Jade Wang et al |

| | | | | | | |
|---|---|---|---|---|---|---|
| EPI_ISL_632185 | USA/NY-NYCPHL-000806/2020 | 2020-03-23 | North America/USA/New York/New Yor | Flushing Hospital Medical Center | New York City Public Health Laboratory | Jade Wang et al |
| EPI_ISL_632189 | USA/NY-NYCPHL-000810/2020 | 2020-03-24 | North America/USA/New York/New Yor | Jamaica Hospital Medical Center | New York City Public Health Laboratory | Jade Wang et al |
| EPI_ISL_632202 | USA/NY-NYCPHL-000825/2020 | 2020-03-26 | North America/USA/New York/New Yor | OCME Office Of Chief Medical Examiner | New York City Public Health Laboratory | Jade Wang et al |
| EPI_ISL_632203 | USA/NY-NYCPHL-000826/2020 | 2020-03-24 | North America/USA/New York/New Yor | Richmond University Medical Center | New York City Public Health Laboratory | Jade Wang et al |
| EPI_ISL_632217 | USA/NY-NYCPHL-000840/2020 | 2020-03-29 | North America/USA/New York/New Yor | Richmond University Medical Center | New York City Public Health Laboratory | Jade Wang et al |
| EPI_ISL_632228 | USA/NY-NYCPHL-000851/2020 | 2020-03-29 | North America/USA/New York/New Yor | Jamaica Hospital Medical Center | New York City Public Health Laboratory | Jade Wang et al |
| EPI_ISL_632253 | Bahrain/340861254_S20/2020 | 2020-10-14 | Asia/Bahrain/Bahrain/ | Communicable Disease Laboratory, Public Health Directorate | Communicable Disease Laboratory, Public Health Directorate | AlWasti et al |
| EPI_ISL_632261 | Bahrain/340286642_S20_L001/2020 | 2020-07-05 | Asia/Bahrain/Bahrain/ | Communicable Disease Laboratory, Public Health Directorate | Communicable Disease Laboratory, Public Health Directorate | AlWasti et al |
| EPI_ISL_632262 | Bahrain/340513774_S1_L001/2020 | 2020-08-17 | Asia/Bahrain/Bahrain/ | Communicable Disease Laboratory, Public Health Directorate | Communicable Disease Laboratory, Public Health Directorate | AlWasti et al |
| EPI_ISL_632263 | Bahrain/340564387_S8_L001/2020 | 2020-08-25 | Asia/Bahrain/Bahrain/ | Communicable Disease Laboratory, Public Health Directorate | Communicable Disease Laboratory, Public Health Directorate | AlWasti et al |

| | | | | | | |
|---|---|---|---|---|---|---|
| EPI_ISL_632265 | Bahrain/920262933_S19_L001/2020 | 2020-06-19 | Asia/Bahrain/Bahrain/ | Communicable Disease Laboratory, Public Health Directorate | Communicable Disease Laboratory, Public Health Directorate | AlWasti et al |
| EPI_ISL_632266 | Bahrain/920265546_S18_L001/2020 | 2020-06-21 | Asia/Bahrain/Bahrain/ | Communicable Disease Laboratory, Public Health Directorate | Communicable Disease Laboratory, Public Health Directorate | AlWasti et al |
| EPI_ISL_632267 | Bahrain/920319237_S17_L001/2020 | 2020-07-26 | Asia/Bahrain/Bahrain/ | Communicable Disease Laboratory, Public Health Directorate | Communicable Disease Laboratory, Public Health Directorate | AlWasti et al |
| EPI_ISL_632274 | Bahrain/340859839_S13/2020 | 2020-10-14 | Asia/Bahrain/Bahrain/ | Communicable Disease Laboratory, Public Health Directorate | Communicable Disease Laboratory, Public Health Directorate | AlWasti et al |
| EPI_ISL_632279 | Bahrain/340859926_S12/2020 | 2020-10-14 | Asia/Bahrain/Bahrain/ | Communicable Disease Laboratory, Public Health Directorate | Communicable Disease Laboratory, Public Health Directorate | AlWasti et al |
| EPI_ISL_632284 | Bahrain/023287015_S14_L001/2020 | 2020-07-25 | Asia/Bahrain/Bahrain/ | Communicable Disease Laboratory, Public Health Directorate | Communicable Disease Laboratory, Public Health Directorate | AlWasti et al |
| EPI_ISL_632285 | Bahrain/090000572_S16_L001/2020 | 2020-05-31 | Asia/Bahrain/Bahrain/ | Communicable Disease Laboratory, Public Health Directorate | Communicable Disease Laboratory, Public Health Directorate | AlWasti et al |

| | | | | | | |
|---|---|---|---|---|---|---|
| EPI_ISL_632310 | Tunisia/55006/2020 | 2020-10-31 | Africa/Tunisia/Ben Arous/ | 1-Laboratory of Microbiology, National Reference Lab, Charles Nicolle Hospital; 2-University of Tunis ElManar, Faculty of Medicine of Tunis, LR99ES09, Tunis, Tunisia | 1-Clinical and Experimental Pharmacology Lab, LR16SP02, National Center of Pharmacovigilance, University of Tunis El Manar, Tunis, Tunisia. 2-Neurodegenerative diseases and psychiatric troubles, LR18SP03, Razi Hospital, University of Tunis El Manar, Tunis, Tunisia. 3-Ministry of Health, National Observatory of New and Emerging Diseases, 1006, Tunis, Tunisia | Ilhem Boutiba-Ben Boubaker et al |
| EPI_ISL_632322 | Netherlands/GE-EMC-333/2020 | 2020-10-20 | Europe/Netherlands/Gelderland/ | Dutch COVID-19 response team | Erasmus Medical Center | Bas Oude Munnink et al |
| EPI_ISL_632329 | Netherlands/NB-EMC-303/2020 | 2020-10-03 | Europe/Netherlands/North Brabant/ | Dutch COVID-19 response team | Erasmus Medical Center | Bas Oude Munnink et al |
| EPI_ISL_632331 | Netherlands/ZH-EMC-871/2020 | 2020-10-27 | Europe/Netherlands/South Holland/ | Dutch COVID-19 response team | Erasmus Medical Center | Bas Oude Munnink et al |
| EPI_ISL_632333 | Netherlands/ZH-EMC-684/2020 | 2020-10-05 | Europe/Netherlands/South Holland/ | Dutch COVID-19 response team | Erasmus Medical Center | Bas Oude Munnink et al |
| EPI_ISL_632340 | Netherlands/NH-EMC-91/2020 | 2020-10-17 | Europe/Netherlands/North Holland/ | Dutch COVID-19 response team | Erasmus Medical Center | Bas Oude Munnink et al |
| EPI_ISL_632344 | Netherlands/ZH-EMC-838/2020 | 2020-10-05 | Europe/Netherlands/South Holland/ | Dutch COVID-19 response team | Erasmus Medical Center | Bas Oude Munnink et al |
| EPI_ISL_632345 | Netherlands/ZH-EMC-682/2020 | 2020-09-27 | Europe/Netherlands/South Holland/ | Dutch COVID-19 response team | Erasmus Medical Center | Bas Oude Munnink et al |
| EPI_ISL_632356 | Netherlands/ZH-EMC-757/2020 | 2020-09-28 | Europe/Netherlands/South Holland/ | Dutch COVID-19 response team | Erasmus Medical Center | Bas Oude Munnink et al |

| | | | | | | |
|---|---|---|---|---|---|---|
| EPI_ISL_632370 | Netherlands/ZH-EMC-766/2020 | 2020-10-08 | Europe/Netherlands/South Holland/ | Dutch COVID-19 response team | Erasmus Medical Center | Bas Oude Munnink et al |
| EPI_ISL_632372 | Netherlands/ZH-EMC-859/2020 | 2020-10-08 | Europe/Netherlands/South Holland/ | Dutch COVID-19 response team | Erasmus Medical Center | Bas Oude Munnink et al |
| EPI_ISL_632373 | Netherlands/ZH-EMC-873/2020 | 2020-10-04 | Europe/Netherlands/South Holland/ | Dutch COVID-19 response team | Erasmus Medical Center | Bas Oude Munnink et al |
| EPI_ISL_632379 | Netherlands/UT-EMC-148/2020 | 2020-10-09 | Europe/Netherlands/Utrecht/ | Dutch COVID-19 response team | Erasmus Medical Center | Bas Oude Munnink et al |
| EPI_ISL_632390 | Netherlands/un-EMC-792/2020 | 2020-09-25 | Europe/Netherlands/Netherlands/ | Dutch COVID-19 response team | Erasmus Medical Center | Bas Oude Munnink et al |
| EPI_ISL_632403 | Netherlands/ZH-EMC-830/2020 | 2020-09-27 | Europe/Netherlands/South Holland/ | Dutch COVID-19 response team | Erasmus Medical Center | Bas Oude Munnink et al |
| EPI_ISL_632411 | Netherlands/ZH-EMC-861/2020 | 2020-10-09 | Europe/Netherlands/South Holland/ | Dutch COVID-19 response team | Erasmus Medical Center | Bas Oude Munnink et al |
| EPI_ISL_632416 | Netherlands/ZH-EMC-825/2020 | 2020-10-14 | Europe/Netherlands/South Holland/ | Dutch COVID-19 response team | Erasmus Medical Center | Bas Oude Munnink et al |
| EPI_ISL_632447 | Netherlands/ZH-EMC-836/2020 | 2020-10-20 | Europe/Netherlands/South Holland/ | Dutch COVID-19 response team | Erasmus Medical Center | Bas Oude Munnink et al |
| EPI_ISL_632448 | Netherlands/ZH-EMC-866/2020 | 2020-10-14 | Europe/Netherlands/South Holland/ | Dutch COVID-19 response team | Erasmus Medical Center | Bas Oude Munnink et al |
| EPI_ISL_632480 | Netherlands/GE-EMC-319/2020 | 2020-10-05 | Europe/Netherlands/Gelderland/ | Dutch COVID-19 response team | Erasmus Medical Center | Bas Oude Munnink et al |
| EPI_ISL_632482 | Netherlands/GE-EMC-334/2020 | 2020-10-21 | Europe/Netherlands/Gelderland/ | Dutch COVID-19 response team | Erasmus Medical Center | Bas Oude Munnink et al |
| EPI_ISL_632498 | Netherlands/UT-EMC-94/2020 | 2020-09-12 | Europe/Netherlands/Utrecht/ | Dutch COVID-19 response team | Erasmus Medical Center | Bas Oude Munnink et al |
| EPI_ISL_632500 | Netherlands/UT-EMC-96/2020 | 2020-09-13 | Europe/Netherlands/Utrecht/ | Dutch COVID-19 response team | Erasmus Medical Center | Bas Oude Munnink et al |
| EPI_ISL_632505 | Netherlands/ZH-EMC-664/2020 | 2020-09-24 | Europe/Netherlands/South Holland/ | Dutch COVID-19 response team | Erasmus Medical Center | Bas Oude Munnink et al |
| EPI_ISL_632506 | Netherlands/ZH-EMC-665/2020 | 2020-09-25 | Europe/Netherlands/South Holland/ | Dutch COVID-19 response team | Erasmus Medical Center | Bas Oude Munnink et al |
| EPI_ISL_632516 | Netherlands/ZH-EMC-676/2020 | 2020-10-02 | Europe/Netherlands/South Holland/ | Dutch COVID-19 response team | Erasmus Medical Center | Bas Oude Munnink et al |
| EPI_ISL_632523 | Netherlands/NH-EMC-75/2020 | 2020-09-09 | Europe/Netherlands/North Holland/ | Dutch COVID-19 response team | Erasmus Medical Center | Bas Oude Munnink et al |
| EPI_ISL_632530 | Netherlands/ZH-EMC-681/2020 | 2020-09-26 | Europe/Netherlands/South Holland/ | Dutch COVID-19 response team | Erasmus Medical Center | Bas Oude Munnink et al |
| EPI_ISL_632531 | Netherlands/UT-EMC-106/2020 | 2020-09-28 | Europe/Netherlands/Utrecht/ | Dutch COVID-19 response team | Erasmus Medical Center | Bas Oude Munnink et al |

| | | | | | | |
|---|---|---|---|---|---|---|
| EPI_ISL_632534 | Netherlands/NH-EMC-88/2020 | 2020-09-09 | Europe/Netherlands/North Holland/ | Dutch COVID-19 response team | Erasmus Medical Center | Bas Oude Munnink et al |
| EPI_ISL_632535 | Netherlands/NB-EMC-300/2020 | 2020-10-01 | Europe/Netherlands/North Brabant/ | Dutch COVID-19 response team | Erasmus Medical Center | Bas Oude Munnink et al |
| EPI_ISL_632556 | Netherlands/ZH-EMC-708/2020 | 2020-09-28 | Europe/Netherlands/South Holland/ | Dutch COVID-19 response team | Erasmus Medical Center | Bas Oude Munnink et al |
| EPI_ISL_632558 | Netherlands/ZH-EMC-711/2020 | 2020-09-30 | Europe/Netherlands/South Holland/ | Dutch COVID-19 response team | Erasmus Medical Center | Bas Oude Munnink et al |
| EPI_ISL_632560 | Netherlands/ZH-EMC-713/2020 | 2020-10-02 | Europe/Netherlands/South Holland/ | Dutch COVID-19 response team | Erasmus Medical Center | Bas Oude Munnink et al |
| EPI_ISL_632561 | Netherlands/ZH-EMC-714/2020 | 2020-10-05 | Europe/Netherlands/South Holland/ | Dutch COVID-19 response team | Erasmus Medical Center | Bas Oude Munnink et al |
| EPI_ISL_632562 | Netherlands/ZH-EMC-715/2020 | 2020-10-05 | Europe/Netherlands/South Holland/ | Dutch COVID-19 response team | Erasmus Medical Center | Bas Oude Munnink et al |
| EPI_ISL_632567 | Netherlands/ZH-EMC-720/2020 | 2020-10-02 | Europe/Netherlands/South Holland/ | Dutch COVID-19 response team | Erasmus Medical Center | Bas Oude Munnink et al |
| EPI_ISL_632568 | Netherlands/ZH-EMC-721/2020 | 2020-09-29 | Europe/Netherlands/South Holland/ | Dutch COVID-19 response team | Erasmus Medical Center | Bas Oude Munnink et al |
| EPI_ISL_632569 | Netherlands/ZH-EMC-722/2020 | 2020-09-29 | Europe/Netherlands/South Holland/ | Dutch COVID-19 response team | Erasmus Medical Center | Bas Oude Munnink et al |
| EPI_ISL_632572 | Netherlands/ZH-EMC-726/2020 | 2020-10-01 | Europe/Netherlands/South Holland/ | Dutch COVID-19 response team | Erasmus Medical Center | Bas Oude Munnink et al |
| EPI_ISL_632576 | Netherlands/ZH-EMC-733/2020 | 2020-10-11 | Europe/Netherlands/South Holland/ | Dutch COVID-19 response team | Erasmus Medical Center | Bas Oude Munnink et al |
| EPI_ISL_632581 | Netherlands/ZH-EMC-739/2020 | 2020-09-16 | Europe/Netherlands/South Holland/ | Dutch COVID-19 response team | Erasmus Medical Center | Bas Oude Munnink et al |
| EPI_ISL_632585 | Netherlands/ZH-EMC-744/2020 | 2020-10-04 | Europe/Netherlands/South Holland/ | Dutch COVID-19 response team | Erasmus Medical Center | Bas Oude Munnink et al |
| EPI_ISL_632594 | Netherlands/ZH-EMC-753/2020 | 2020-10-03 | Europe/Netherlands/South Holland/ | Dutch COVID-19 response team | Erasmus Medical Center | Bas Oude Munnink et al |
| EPI_ISL_632595 | Netherlands/ZH-EMC-754/2020 | 2020-10-04 | Europe/Netherlands/South Holland/ | Dutch COVID-19 response team | Erasmus Medical Center | Bas Oude Munnink et al |
| EPI_ISL_632596 | Netherlands/ZH-EMC-759/2020 | 2020-09-30 | Europe/Netherlands/South Holland/ | Dutch COVID-19 response team | Erasmus Medical Center | Bas Oude Munnink et al |
| EPI_ISL_632599 | Netherlands/ZH-EMC-762/2020 | 2020-10-01 | Europe/Netherlands/South Holland/ | Dutch COVID-19 response team | Erasmus Medical Center | Bas Oude Munnink et al |
| EPI_ISL_632600 | Netherlands/ZH-EMC-763/2020 | 2020-10-02 | Europe/Netherlands/South Holland/ | Dutch COVID-19 response team | Erasmus Medical Center | Bas Oude Munnink et al |
| EPI_ISL_632601 | Netherlands/ZH-EMC-764/2020 | 2020-10-05 | Europe/Netherlands/South Holland/ | Dutch COVID-19 response team | Erasmus Medical Center | Bas Oude Munnink et al |

| | | | | | | |
|---|---|---|---|---|---|---|
| EPI_ISL_632607 | Netherlands/ZH-EMC-771/2020 | 2020-10-04 | Europe/Netherlands/South Holland/ | Dutch COVID-19 response team | Erasmus Medical Center | Bas Oude Munnink et al |
| EPI_ISL_632609 | Netherlands/NB-EMC-328/2020 | 2020-10-04 | Europe/Netherlands/North Brabant/ | Dutch COVID-19 response team | Erasmus Medical Center | Bas Oude Munnink et al |
| EPI_ISL_632612 | Netherlands/ZH-EMC-775/2020 | 2020-09-26 | Europe/Netherlands/South Holland/ | Dutch COVID-19 response team | Erasmus Medical Center | Bas Oude Munnink et al |
| EPI_ISL_632614 | Netherlands/ZH-EMC-777/2020 | 2020-09-30 | Europe/Netherlands/South Holland/ | Dutch COVID-19 response team | Erasmus Medical Center | Bas Oude Munnink et al |
| EPI_ISL_632615 | Netherlands/ZH-EMC-778/2020 | 2020-09-26 | Europe/Netherlands/South Holland/ | Dutch COVID-19 response team | Erasmus Medical Center | Bas Oude Munnink et al |
| EPI_ISL_632623 | Netherlands/UT-EMC-107/2020 | 2020-10-07 | Europe/Netherlands/Utrecht/ | Dutch COVID-19 response team | Erasmus Medical Center | Bas Oude Munnink et al |
| EPI_ISL_632627 | Netherlands/GE-EMC-308/2020 | 2020-10-08 | Europe/Netherlands/Gelderland/ | Dutch COVID-19 response team | Erasmus Medical Center | Bas Oude Munnink et al |
| EPI_ISL_632628 | Netherlands/UT-EMC-109/2020 | 2020-10-08 | Europe/Netherlands/Utrecht/ | Dutch COVID-19 response team | Erasmus Medical Center | Bas Oude Munnink et al |
| EPI_ISL_632649 | Netherlands/GE-EMC-317/2020 | 2020-10-05 | Europe/Netherlands/Gelderland/ | Dutch COVID-19 response team | Erasmus Medical Center | Bas Oude Munnink et al |
| EPI_ISL_632651 | Netherlands/GE-EMC-320/2020 | 2020-10-05 | Europe/Netherlands/Gelderland/ | Dutch COVID-19 response team | Erasmus Medical Center | Bas Oude Munnink et al |
| EPI_ISL_632655 | Netherlands/GE-EMC-323/2020 | 2020-10-07 | Europe/Netherlands/Gelderland/ | Dutch COVID-19 response team | Erasmus Medical Center | Bas Oude Munnink et al |
| EPI_ISL_632656 | Netherlands/ZH-EMC-787/2020 | 2020-10-14 | Europe/Netherlands/South Holland/ | Dutch COVID-19 response team | Erasmus Medical Center | Bas Oude Munnink et al |
| EPI_ISL_632657 | Netherlands/ZH-EMC-788/2020 | 2020-10-14 | Europe/Netherlands/South Holland/ | Dutch COVID-19 response team | Erasmus Medical Center | Bas Oude Munnink et al |
| EPI_ISL_632658 | Netherlands/NB-EMC-305/2020 | 2020-10-04 | Europe/Netherlands/North Brabant/ | Dutch COVID-19 response team | Erasmus Medical Center | Bas Oude Munnink et al |
| EPI_ISL_632660 | Netherlands/NB-EMC-307/2020 | 2020-10-07 | Europe/Netherlands/North Brabant/ | Dutch COVID-19 response team | Erasmus Medical Center | Bas Oude Munnink et al |
| EPI_ISL_632662 | Netherlands/NB-EMC-309/2020 | 2020-10-10 | Europe/Netherlands/North Brabant/ | Dutch COVID-19 response team | Erasmus Medical Center | Bas Oude Munnink et al |
| EPI_ISL_632666 | Netherlands/NB-EMC-314/2020 | 2020-10-06 | Europe/Netherlands/North Brabant/ | Dutch COVID-19 response team | Erasmus Medical Center | Bas Oude Munnink et al |
| EPI_ISL_632669 | Netherlands/GE-EMC-326/2020 | 2020-10-06 | Europe/Netherlands/Gelderland/ | Dutch COVID-19 response team | Erasmus Medical Center | Bas Oude Munnink et al |
| EPI_ISL_632680 | Netherlands/ZH-EMC-801/2020 | 2020-10-14 | Europe/Netherlands/South Holland/ | Dutch COVID-19 response team | Erasmus Medical Center | Bas Oude Munnink et al |
| EPI_ISL_632681 | Netherlands/ZH-EMC-802/2020 | 2020-10-14 | Europe/Netherlands/South Holland/ | Dutch COVID-19 response team | Erasmus Medical Center | Bas Oude Munnink et al |

| | | | | | | |
|---|---|---|---|---|---|---|
| EPI_ISL_632683 | Netherlands/ZH-EMC-804/2020 | 2020-10-14 | Europe/Netherlands/South Holland/ | Dutch COVID-19 response team | Erasmus Medical Center | Bas Oude Munnink et al |
| EPI_ISL_632685 | Netherlands/ZH-EMC-806/2020 | 2020-10-14 | Europe/Netherlands/South Holland/ | Dutch COVID-19 response team | Erasmus Medical Center | Bas Oude Munnink et al |
| EPI_ISL_632686 | Netherlands/ZH-EMC-807/2020 | 2020-10-14 | Europe/Netherlands/South Holland/ | Dutch COVID-19 response team | Erasmus Medical Center | Bas Oude Munnink et al |
| EPI_ISL_632687 | Netherlands/ZH-EMC-808/2020 | 2020-10-15 | Europe/Netherlands/South Holland/ | Dutch COVID-19 response team | Erasmus Medical Center | Bas Oude Munnink et al |
| EPI_ISL_632688 | Netherlands/ZH-EMC-809/2020 | 2020-10-15 | Europe/Netherlands/South Holland/ | Dutch COVID-19 response team | Erasmus Medical Center | Bas Oude Munnink et al |
| EPI_ISL_632694 | Netherlands/ZH-EMC-818/2020 | 2020-10-15 | Europe/Netherlands/South Holland/ | Dutch COVID-19 response team | Erasmus Medical Center | Bas Oude Munnink et al |
| EPI_ISL_632695 | Netherlands/ZH-EMC-820/2020 | 2020-10-16 | Europe/Netherlands/South Holland/ | Dutch COVID-19 response team | Erasmus Medical Center | Bas Oude Munnink et al |
| EPI_ISL_632699 | Netherlands/ZH-EMC-827/2020 | 2020-10-06 | Europe/Netherlands/South Holland/ | Dutch COVID-19 response team | Erasmus Medical Center | Bas Oude Munnink et al |
| EPI_ISL_632704 | Netherlands/ZH-EMC-834/2020 | 2020-09-21 | Europe/Netherlands/South Holland/ | Dutch COVID-19 response team | Erasmus Medical Center | Bas Oude Munnink et al |
| EPI_ISL_632706 | Netherlands/ZH-EMC-841/2020 | 2020-10-16 | Europe/Netherlands/South Holland/ | Dutch COVID-19 response team | Erasmus Medical Center | Bas Oude Munnink et al |
| EPI_ISL_632717 | Netherlands/ZH-EMC-849/2020 | 2020-09-27 | Europe/Netherlands/South Holland/ | Dutch COVID-19 response team | Erasmus Medical Center | Bas Oude Munnink et al |
| EPI_ISL_632718 | Netherlands/ZH-EMC-850/2020 | 2020-09-25 | Europe/Netherlands/South Holland/ | Dutch COVID-19 response team | Erasmus Medical Center | Bas Oude Munnink et al |
| EPI_ISL_632721 | Netherlands/UT-EMC-127/2020 | 2020-10-21 | Europe/Netherlands/Utrecht/ | Dutch COVID-19 response team | Erasmus Medical Center | Bas Oude Munnink et al |
| EPI_ISL_632723 | Netherlands/UT-EMC-129/2020 | 2020-10-22 | Europe/Netherlands/Utrecht/ | Dutch COVID-19 response team | Erasmus Medical Center | Bas Oude Munnink et al |
| EPI_ISL_632726 | Netherlands/UT-EMC-132/2020 | 2020-10-14 | Europe/Netherlands/Utrecht/ | Dutch COVID-19 response team | Erasmus Medical Center | Bas Oude Munnink et al |
| EPI_ISL_632738 | Netherlands/ZH-EMC-857/2020 | 2020-10-26 | Europe/Netherlands/South Holland/ | Dutch COVID-19 response team | Erasmus Medical Center | Bas Oude Munnink et al |
| EPI_ISL_632741 | Netherlands/OV-EMC-83/2020 | 2020-10-18 | Europe/Netherlands/Overijssel/ | Dutch COVID-19 response team | Erasmus Medical Center | Bas Oude Munnink et al |
| EPI_ISL_632747 | Netherlands/ZH-EMC-862/2020 | 2020-10-17 | Europe/Netherlands/South Holland/ | Dutch COVID-19 response team | Erasmus Medical Center | Bas Oude Munnink et al |
| EPI_ISL_632754 | Netherlands/ZH-EMC-872/2020 | 2020-09-25 | Europe/Netherlands/South Holland/ | Dutch COVID-19 response team | Erasmus Medical Center | Bas Oude Munnink et al |
| EPI_ISL_632762 | Netherlands/ZH-EMC-883/2020 | 2020-09-11 | Europe/Netherlands/South Holland/ | Dutch COVID-19 response team | Erasmus Medical Center | Bas Oude Munnink et al |

| | | | | | | |
|---|---|---|---|---|---|---|
| EPI_ISL_632764 | Netherlands/GE-EMC-348/2020 | 2020-10-27 | Europe/Netherlands/Gelderland/ | Dutch COVID-19 response team | Erasmus Medical Center | Bas Oude Munnink et al |
| EPI_ISL_632767 | Netherlands/ZH-EMC-888/2020 | 2020-10-30 | Europe/Netherlands/South Holland/ | Dutch COVID-19 response team | Erasmus Medical Center | Bas Oude Munnink et al |
| EPI_ISL_632768 | Netherlands/ZH-EMC-890/2020 | 2020-10-29 | Europe/Netherlands/South Holland/ | Dutch COVID-19 response team | Erasmus Medical Center | Bas Oude Munnink et al |
| EPI_ISL_632769 | Netherlands/ZH-EMC-891/2020 | 2020-10-30 | Europe/Netherlands/South Holland/ | Dutch COVID-19 response team | Erasmus Medical Center | Bas Oude Munnink et al |
| EPI_ISL_632781 | Netherlands/FL-EMC-39/2020 | 2020-10-15 | Europe/Netherlands/Flevoland/ | Dutch COVID-19 response team | Erasmus Medical Center | Bas Oude Munnink et al |
| EPI_ISL_632782 | Netherlands/UT-EMC-143/2020 | 2020-10-16 | Europe/Netherlands/Utrecht/ | Dutch COVID-19 response team | Erasmus Medical Center | Bas Oude Munnink et al |
| EPI_ISL_632786 | Netherlands/ZH-EMC-896/2020 | 2020-10-04 | Europe/Netherlands/South Holland/ | Dutch COVID-19 response team | Erasmus Medical Center | Bas Oude Munnink et al |
| EPI_ISL_632787 | Netherlands/ZH-EMC-897/2020 | 2020-10-16 | Europe/Netherlands/South Holland/ | Dutch COVID-19 response team | Erasmus Medical Center | Bas Oude Munnink et al |
| EPI_ISL_632792 | Netherlands/ZH-EMC-902/2020 | 2020-10-27 | Europe/Netherlands/South Holland/ | Dutch COVID-19 response team | Erasmus Medical Center | Bas Oude Munnink et al |
| EPI_ISL_632816 | Gibraltar/204462918/2020 | 2020-10-29 | Europe/United Kingdom/Gibraltar/Gibra | Microbiology, Department of Pathology, St. Bernard's Hospital, Gibraltar Health Authority | Respiratory Virus Unit, Microbiology Services Colindale, Public Health England | PHE Covid Sequencing Team et al |
| EPI_ISL_632827 | USA/NM-NMDOH-2020324222/2020 | 2020-09-02 | North America/USA/New Mexico/ | New Mexico Department of Health Scientific Laboratory | New Mexico Department of Health Scientific Laboratory | Ellie Johnson et al |
| EPI_ISL_632829 | USA/NM-NMDOH-2020324363/2020 | 2020-09-02 | North America/USA/New Mexico/ | New Mexico Department of Health Scientific Laboratory | New Mexico Department of Health Scientific Laboratory | Ellie Johnson et al |
| EPI_ISL_632830 | USA/NM-NMDOH-2020324382/2020 | 2020-09-02 | North America/USA/New Mexico/ | New Mexico Department of Health Scientific Laboratory | New Mexico Department of Health Scientific Laboratory | Ellie Johnson et al |
| EPI_ISL_632834 | USA/NM-NMDOH-2020324645/2020 | 2020-09-02 | North America/USA/New Mexico/ | New Mexico Department of Health Scientific Laboratory | New Mexico Department of Health Scientific Laboratory | Ellie Johnson et al |

| | | | | | | |
|---|---|---|---|---|---|---|
| EPI_ISL_632838 | USA/NM-NMDOH-2020325105/2020 | 2020-09-02 | North America/USA/New Mexico/ | New Mexico Department of Health Scientific Laboratory | New Mexico Department of Health Scientific Laboratory | Ellie Johnson et al |
| EPI_ISL_632850 | USA/ID-UNM-IBL030/2020 | 2020-03-15 | North America/USA/Idaho/ | Idaho Bureau of Laboratories | Center for Global Health, University of New Mexico Health Sciences Center | Daryl Domman et al |
| EPI_ISL_632851 | USA/ID-UNM-IBL312/2020 | 2020-03-28 | North America/USA/Idaho/ | Idaho Bureau of Laboratories | Center for Global Health, University of New Mexico Health Sciences Center | Daryl Domman et al |
| EPI_ISL_632855 | USA/ID-UNM-IBL158/2020 | 2020-03-22 | North America/USA/Idaho/ | Idaho Bureau of Laboratories | Center for Global Health, University of New Mexico Health Sciences Center | Daryl Domman et al |
| EPI_ISL_632857 | USA/ID-UNM-IBL021/2020 | 2020-03-12 | North America/USA/Idaho/ | Idaho Bureau of Laboratories | Center for Global Health, University of New Mexico Health Sciences Center | Daryl Domman et al |
| EPI_ISL_632859 | USA/ID-UNM-IBL022/2020 | 2020-03-12 | North America/USA/Idaho/ | Idaho Bureau of Laboratories | Center for Global Health, University of New Mexico Health Sciences Center | Daryl Domman et al |
| EPI_ISL_632866 | USA/ID-UNM-IBL359/2020 | 2020-04-01 | North America/USA/Idaho/ | Idaho Bureau of Laboratories | Center for Global Health, University of New Mexico Health Sciences Center | Daryl Domman et al |
| EPI_ISL_632880 | USA/ID-UNM-IBL067/2020 | 2020-03-20 | North America/USA/Idaho/ | Idaho Bureau of Laboratories | Center for Global Health, University of New Mexico Health Sciences Center | Daryl Domman et al |

| | | | | | | |
|---|---|---|---|---|---|---|
| EPI_ISL_632892 | USA/WI-UW-1613/2020 | 2020-09-17 | North America/USA/Wisconsin/Dane Co | University of Wisconsin-Madison AIDS Vaccine Research Laboratories | University of Wisconsin-Madison AIDS Vaccine Research Laboratories | Gage Moreno et al |
| EPI_ISL_632894 | USA/WI-UW-1678/2020 | 2020-09-11 | North America/USA/Wisconsin/Dane Co | University of Wisconsin-Madison AIDS Vaccine Research Laboratories | University of Wisconsin-Madison AIDS Vaccine Research Laboratories | Gage Moreno et al |
| EPI_ISL_632898 | USA/WI-UW-1885/2020 | 2020-09-16 | North America/USA/Wisconsin/Milwauk | University of Wisconsin-Madison AIDS Vaccine Research Laboratories | University of Wisconsin-Madison AIDS Vaccine Research Laboratories | Gage Moreno et al |
| EPI_ISL_632899 | Bahrain/920457301_S8/2020 | 2020-10-14 | Asia/Bahrain/Bahrain/ | Communicable Disease Laboratory, Public Health Directorate | Communicable Disease Laboratory, Public Health Directorate | AlAbbas et al |
| EPI_ISL_632908 | Pakistan/KP-RMI-01/2020 | 2020-04-01 | Asia/Pakistan/Pakistan/ | Genomic Sciences, Rehman Medical Institute | Genomic Sciences, Rehman Medical Institute | Ali et al |
| EPI_ISL_632909 | Canada/MB-NML-577/2020 | 2020-04-28 | North America/Canada/Manitoba/ | Cadham Provincial laboratory | Cadham Provincial laboratory | Anna Majer et al |
| EPI_ISL_632910 | Canada/MB-NML-601/2020 | 2020-03-25 | North America/Canada/Manitoba/ | Cadham Provincial laboratory | Cadham Provincial laboratory | Anna Majer et al |
| EPI_ISL_632911 | Canada/MB-NML-602/2020 | 2020-03-22 | North America/Canada/Manitoba/ | Cadham Provincial laboratory | Cadham Provincial laboratory | Anna Majer et al |
| EPI_ISL_632912 | Canada/MB-NML-606/2020 | 2020-03-28 | North America/Canada/Manitoba/ | Cadham Provincial laboratory | Cadham Provincial laboratory | Anna Majer et al |
| EPI_ISL_632913 | Canada/MB-NML-608/2020 | 2020-04-03 | North America/Canada/Manitoba/ | Cadham Provincial laboratory | Cadham Provincial laboratory | Anna Majer et al |
| EPI_ISL_632914 | Canada/MB-NML-610/2020 | 2020-03-27 | North America/Canada/Manitoba/ | Cadham Provincial laboratory | Cadham Provincial laboratory | Anna Majer et al |
| EPI_ISL_632915 | Canada/MB-NML-613/2020 | 2020-03-23 | North America/Canada/Manitoba/ | Cadham Provincial laboratory | Cadham Provincial laboratory | Anna Majer et al |
| EPI_ISL_632916 | Canada/MB-NML-614/2020 | 2020-03-23 | North America/Canada/Manitoba/ | Cadham Provincial laboratory | Cadham Provincial laboratory | Anna Majer et al |

| | | | | | | |
|---|---|---|---|---|---|---|
| EPI_ISL_632917 | Canada/MB-NML-620/2020 | 2020-04-03 | North America/Canada/Manitoba/ | Cadham Provincial laboratory | Cadham Provincial laboratory | Anna Majer et al |
| EPI_ISL_632918 | Canada/MB-NML-633/2020 | 2020-03-21 | North America/Canada/Manitoba/ | Cadham Provincial laboratory | Cadham Provincial laboratory | Anna Majer et al |
| EPI_ISL_632919 | Canada/MB-NML-634/2020 | 2020-03-20 | North America/Canada/Manitoba/ | Cadham Provincial laboratory | Cadham Provincial laboratory | Anna Majer et al |
| EPI_ISL_632920 | Canada/MB-NML-638/2020 | 2020-03-23 | North America/Canada/Manitoba/ | Cadham Provincial laboratory | Cadham Provincial laboratory | Anna Majer et al |
| EPI_ISL_632921 | Canada/MB-NML-639/2020 | 2020-03-23 | North America/Canada/Manitoba/ | Cadham Provincial laboratory | Cadham Provincial laboratory | Anna Majer et al |
| EPI_ISL_632922 | Canada/MB-NML-643/2020 | 2020-03-21 | North America/Canada/Manitoba/ | Cadham Provincial laboratory | Cadham Provincial laboratory | Anna Majer et al |
| EPI_ISL_632923 | Canada/MB-NML-644/2020 | 2020-03-25 | North America/Canada/Manitoba/ | Cadham Provincial laboratory | Cadham Provincial laboratory | Anna Majer et al |
| EPI_ISL_632924 | Canada/MB-NML-645/2020 | 2020-03-23 | North America/Canada/Manitoba/ | Cadham Provincial laboratory | Cadham Provincial laboratory | Anna Majer et al |
| EPI_ISL_632925 | Canada/MB-NML-646/2020 | 2020-03-23 | North America/Canada/Manitoba/ | Cadham Provincial laboratory | Cadham Provincial laboratory | Anna Majer et al |
| EPI_ISL_632926 | Canada/MB-NML-647/2020 | 2020-03-16 | North America/Canada/Manitoba/ | Cadham Provincial laboratory | Cadham Provincial laboratory | Anna Majer et al |
| EPI_ISL_632927 | Canada/MB-NML-648/2020 | 2020-03-16 | North America/Canada/Manitoba/ | Cadham Provincial laboratory | Cadham Provincial laboratory | Anna Majer et al |
| EPI_ISL_632928 | Canada/MB-NML-649/2020 | 2020-03-16 | North America/Canada/Manitoba/ | Cadham Provincial laboratory | Cadham Provincial laboratory | Anna Majer et al |
| EPI_ISL_632929 | Canada/MB-NML-650/2020 | 2020-03-20 | North America/Canada/Manitoba/ | Cadham Provincial laboratory | Cadham Provincial laboratory | Anna Majer et al |
| EPI_ISL_632930 | Canada/MB-NML-651/2020 | 2020-03-13 | North America/Canada/Manitoba/ | Cadham Provincial laboratory | Cadham Provincial laboratory | Anna Majer et al |
| EPI_ISL_632931 | Canada/MB-NML-652/2020 | 2020-03-12 | North America/Canada/Manitoba/ | Cadham Provincial laboratory | Cadham Provincial laboratory | Anna Majer et al |
| EPI_ISL_632932 | Canada/MB-NML-653/2020 | 2020-03-11 | North America/Canada/Manitoba/ | Cadham Provincial laboratory | Cadham Provincial laboratory | Anna Majer et al |
| EPI_ISL_632933 | Canada/MB-NML-654/2020 | 2020-03-13 | North America/Canada/Manitoba/ | Cadham Provincial laboratory | Cadham Provincial laboratory | Anna Majer et al |

| | | | | | | |
|---|---|---|---|---|---|---|
| EPI_ISL_632935 | Yunnan/03/2020 | 2020-09-13 | Asia/China/Yunnan/ | Department of Acute Infectious Diseases Control and Prevention, Yunnan Provincial Center for Disease Control and Prevention | Department of Acute Infectious Diseases Control and Prevention, Yunnan Provincial Center for Disease Control and Prevention | Meiling Zhang et al |

| | | | | | | |
|---|---|---|---|---|---|---|
| EPI_ISL_632936 | Indonesia/YO-UGM-10007/2020 | 2020-08-22 | Asia/Indonesia/Yogyakarta/ | Puskesmas Mlati 1 Sleman | Genetics Working Group (Pokja Genetik) Faculty of Medicine, Public Health and Nursing Universitas Gadjah Mada (FK-KMK UGM); Disease Investigation Center Wates Ministry of Agriculture Indonesia; Department of Microbiology FK-KMK UGM; Laboratorium Diagnostik Yayasan Tahija World Mosquito Program (WMP) Yogyakarta Center for Tropical Medicine FK-KMK UGM; Integrated Research Center FK-KMK UGM; Department of Computer Science and Electronics FMIPA UGM; RSUP Dr. Sardjito | Gunadi et al |

| | | | | | | |
|---|---|---|---|---|---|---|
| EPI_ISL_632937 | Indonesia/YO-UGM-48651/2020 | 2020-09-09 | Asia/Indonesia/Yogyakarta/ | RSUD Saptosari Gunung Kidul | Genetics Working Group (Pokja Genetik) Faculty of Medicine, Public Health and Nursing Universitas Gadjah Mada (FK-KMK UGM); Disease Investigation Center Wates Ministry of Agriculture Indonesia; Department of Microbiology FK-KMK UGM; Laboratorium Diagnostik Yayasan Tahija World Mosquito Program (WMP) Yogyakarta Center for Tropical Medicine FK-KMK UGM; Integrated Research Center FK-KMK UGM; Department of Computer Science and Electronics FMIPA UGM; Balai Besar Teknik Kesehatan Lingkungan dan | Gunadi et al |
| EPI_ISL_632938 | Germany/TH-IIMK-CaSe-1/2020 | 2020-04-19 | Europe/Germany/Thuringia/ | Jena University Hospital; Institute for Infectious Diseases and Infection Control | Institute of infectious medicine & hospital hygiene, CaSe-Group | Spott et al |

| | | | | | | |
|---|---|---|---|---|---|---|
| EPI_ISL_632943 | USA/NY-NYCPHL-000870/2020 | 2020-03-25 | North America/USA/New York/New Yor | Richmond University Medical Center | New York City Public Health Laboratory | Jade Wang et al |
| EPI_ISL_632947 | USA/NY-NYCPHL-000875/2020 | 2020-03-26 | North America/USA/New York/New Yor | Flushing Hospital Medical Center | New York City Public Health Laboratory | Jade Wang et al |
| EPI_ISL_632949 | USA/NY-NYCPHL-000877/2020 | 2020-03-26 | North America/USA/New York/New Yor | Jamaica Hospital Medical Center | New York City Public Health Laboratory | Jade Wang et al |
| EPI_ISL_632964 | USA/NY-NYCPHL-000898/2020 | 2020-07-10 | North America/USA/New York/New Yor | Department of Homeless Services | New York City Public Health Laboratory | Jade Wang et al |
| EPI_ISL_632967 | USA/NY-NYCPHL-000906/2020 | 2020-03-18 | North America/USA/New York/New Yor | Flushing Hospital Medical Center | New York City Public Health Laboratory | Jade Wang et al |
| EPI_ISL_632975 | USA/NY-NYCPHL-000914/2020 | 2020-03-20 | North America/USA/New York/New Yor | Richmond University Medical Center | New York City Public Health Laboratory | Jade Wang et al |
| EPI_ISL_632981 | USA/NY-NYCPHL-000940/2020 | 2020-09-11 | North America/USA/New York/New Yor | DOHMH Corona | New York City Public Health Laboratory | Jade Wang et al |
| EPI_ISL_632982 | USA/NY-NYCPHL-000943/2020 | 2020-09-24 | North America/USA/New York/New Yor | DOHMH Corona | New York City Public Health Laboratory | Jade Wang et al |
| EPI_ISL_632983 | USA/NY-NYCPHL-000944/2020 | 2020-09-25 | North America/USA/New York/Nassau ( | DOHMH Corona | New York City Public Health Laboratory | Jade Wang et al |
| EPI_ISL_632985 | USA/NY-NYCPHL-000947/2020 | 2020-09-29 | North America/USA/New York/New Yor | DOHMH Corona | New York City Public Health Laboratory | Jade Wang et al |
| EPI_ISL_632991 | USA/NY-NYCPHL-000954/2020 | 2020-10-01 | North America/USA/New York/New Yor | DOHMH Corona | New York City Public Health Laboratory | Jade Wang et al |
| EPI_ISL_632994 | USA/NY-NYCPHL-000959/2020 | 2020-09-14 | North America/USA/New York/New Yor | DOHMH Crown Heights | New York City Public Health Laboratory | Jade Wang et al |
| EPI_ISL_632998 | USA/NY-NYCPHL-000966/2020 | 2020-10-06 | North America/USA/New York/New Yor | DOHMH Crown Heights | New York City Public Health Laboratory | Jade Wang et al |

| | | | | | | |
|---|---|---|---|---|---|---|
| EPI_ISL_633000 | USA/NY-NYCPHL-000969/2020 | 2020-08-25 | North America/USA/New York/New Yor | DOHMH Jamaica | New York City Public Health Laboratory | Jade Wang et al |
| EPI_ISL_633002 | USA/NY-NYCPHL-000971/2020 | 2020-08-31 | North America/USA/New York/New Yor | DOHMH Jamaica | New York City Public Health Laboratory | Jade Wang et al |
| EPI_ISL_633013 | USA/NY-NYCPHL-000987/2020 | 2020-08-24 | North America/USA/New York/New Yor | DOHMH PHL | New York City Public Health Laboratory | Jade Wang et al |
| EPI_ISL_633014 | USA/NY-NYCPHL-000988/2020 | 2020-08-26 | North America/USA/New York/New Yor | DOHMH Chelsea | New York City Public Health Laboratory | Jade Wang et al |
| EPI_ISL_633015 | USA/NY-NYCPHL-000989/2020 | 2020-08-27 | North America/USA/New York/New Yor | DOHMH Morrisania | New York City Public Health Laboratory | Jade Wang et al |
| EPI_ISL_633017 | USA/NY-NYCPHL-000992/2020 | 2020-08-31 | North America/USA/New York/New Yor | DOHMH Chelsea | New York City Public Health Laboratory | Jade Wang et al |
| EPI_ISL_633032 | USA/NY-NYCPHL-001013/2020 | 2020-10-07 | North America/USA/New York/Orange ( | DOHMH Morrisania | New York City Public Health Laboratory | Jade Wang et al |
| EPI_ISL_633034 | USA/NY-NYCPHL-001015/2020 | 2020-10-08 | North America/USA/New York/New Yor | DOHMH Riverside | New York City Public Health Laboratory | Jade Wang et al |
| EPI_ISL_633035 | USA/NY-NYCPHL-001016/2020 | 2020-09-10 | North America/USA/New York/New Yor | DOHMH PHL | New York City Public Health Laboratory | Jade Wang et al |
| EPI_ISL_633037 | USA/NY-NYCPHL-001019/2020 | 2020-09-16 | North America/USA/New York/New Yor | DOHMH Central Harlem | New York City Public Health Laboratory | Jade Wang et al |
| EPI_ISL_633039 | USA/NY-NYCPHL-001021/2020 | 2020-09-18 | North America/USA/New York/New Yor | DOHMH Chelsea | New York City Public Health Laboratory | Jade Wang et al |
| EPI_ISL_633042 | USA/NY-NYCPHL-001025/2020 | 2020-09-24 | North America/USA/New York/New Yor | DOHMH Riverside | New York City Public Health Laboratory | Jade Wang et al |
| EPI_ISL_633045 | USA/NY-NYCPHL-001029/2020 | 2020-09-29 | North America/USA/New York/New Yor | DOHMH PHL | New York City Public Health Laboratory | Jade Wang et al |

| | | | | | | |
|---|---|---|---|---|---|---|
| EPI_ISL_633051 | USA/NY-NYCPHL-001038/2020 | 2020-09-30 | North America/USA/New York/New Yor | DOHMH PHL | New York City Public Health Laboratory | Jade Wang et al |
| EPI_ISL_633052 | USA/NY-NYCPHL-001039/2020 | 2020-09-30 | North America/USA/New York/New Yor | DOHMH Chelsea | New York City Public Health Laboratory | Jade Wang et al |
| EPI_ISL_633059 | USA/NY-NYCPHL-001047/2020 | 2020-10-08 | North America/USA/New York/New Yor | DOHMH Corona | New York City Public Health Laboratory | Jade Wang et al |
| EPI_ISL_633069 | USA/NY-NYCPHL-001064/2020 | 2020-10-13 | North America/USA/New York/New Yor | DOHMH Chelsea | New York City Public Health Laboratory | Jade Wang et al |
| EPI_ISL_633077 | USA/NY-NYCPHL-001076/2020 | 2020-10-15 | North America/USA/New York/New Yor | DOHMH Chelsea | New York City Public Health Laboratory | Jade Wang et al |
| EPI_ISL_633078 | USA/NY-NYCPHL-001077/2020 | 2020-10-16 | North America/USA/New York/New Yor | DOHMH Chelsea | New York City Public Health Laboratory | Jade Wang et al |
| EPI_ISL_633098 | USA/MD-MDH-0323/2020 | 2020-10-20 | North America/USA/Maryland/ | MD PHL | MD PHL | Maryland Department of Health Laboratories Administration et al |
| EPI_ISL_633099 | USA/MD-MDH-0324/2020 | 2020-10-20 | North America/USA/Maryland/ | MD PHL | MD PHL | Maryland Department of Health Laboratories Administration et al |
| EPI_ISL_633155 | England/CAMC-AEA758/2020 | 2020-10-26 | Europe/United Kingdom/England/ | Lighthouse Lab in Cambridge | Wellcome Sanger Institute for the COVID-19 Genomics UK (COG-UK) consortium | Rob Howes et al |
| EPI_ISL_633194 | England/CAMC-AEB159/2020 | 2020-10-24 | Europe/United Kingdom/England/ | Lighthouse Lab in Cambridge | Wellcome Sanger Institute for the COVID-19 Genomics UK (COG-UK) consortium | Rob Howes et al |
| EPI_ISL_633373 | England/CAMC-AEACD1/2020 | 2020-10-25 | Europe/United Kingdom/England/ | Lighthouse Lab in Cambridge | Wellcome Sanger Institute for the COVID-19 Genomics UK (COG-UK) consortium | Rob Howes et al |

| | | | | | | |
|---|---|---|---|---|---|---|
| EPI_ISL_633376 | England/QEUH-AE4C22/2020 | 2020-10-27 | Europe/United Kingdom/England/ | Lighthouse Lab in Glasgow | Wellcome Sanger Institute for the COVID-19 Genomics UK (COG-UK) consortium | Harper VanSteenhouse et al |
| EPI_ISL_633832 | England/QEUH-AD289C/2020 | 2020-10-26 | Europe/United Kingdom/England/ | Lighthouse Lab in Glasgow | Wellcome Sanger Institute for the COVID-19 Genomics UK (COG-UK) consortium | Harper VanSteenhouse et al |
| EPI_ISL_633917 | England/QEUH-AD5D30/2020 | 2020-10-26 | Europe/United Kingdom/England/ | Lighthouse Lab in Glasgow | Wellcome Sanger Institute for the COVID-19 Genomics UK (COG-UK) consortium | Harper VanSteenhouse et al |
| EPI_ISL_634286 | England/MILK-AC789A/2020 | 2020-10-21 | Europe/United Kingdom/England/ | Lighthouse Lab in Milton Keynes | Wellcome Sanger Institute for the COVID-19 Genomics UK (COG-UK) consortium | The Lighthouse Lab in Milton Keynes et al |
| EPI_ISL_634374 | England/MILK-AC75C0/2020 | 2020-10-21 | Europe/United Kingdom/England/ | Lighthouse Lab in Milton Keynes | Wellcome Sanger Institute for the COVID-19 Genomics UK (COG-UK) consortium | The Lighthouse Lab in Milton Keynes et al |
| EPI_ISL_634375 | England/MILK-AC8143/2020 | 2020-10-21 | Europe/United Kingdom/England/ | Lighthouse Lab in Milton Keynes | Wellcome Sanger Institute for the COVID-19 Genomics UK (COG-UK) consortium | The Lighthouse Lab in Milton Keynes et al |
| EPI_ISL_634499 | England/MILK-ACD199/2020 | 2020-10-22 | Europe/United Kingdom/England/ | Lighthouse Lab in Milton Keynes | Wellcome Sanger Institute for the COVID-19 Genomics UK (COG-UK) Consortium | The Lighthouse Lab in Milton Keynes et al |
| EPI_ISL_634569 | England/QEUH-AD4663/2020 | 2020-10-26 | Europe/United Kingdom/England/ | Lighthouse Lab in Glasgow | Wellcome Sanger Institute for the COVID-19 Genomics UK (COG-UK) consortium | Harper VanSteenhouse et al |

| | | | | | | |
|---|---|---|---|---|---|---|
| EPI_ISL_634636 | England/QEUH-AD104A/2020 | 2020-10-26 | Europe/United Kingdom/England/ | Lighthouse Lab in Glasgow | Wellcome Sanger Institute for the COVID-19 Genomics UK (COG-UK) consortium | Harper VanSteenhouse et al |
| EPI_ISL_634692 | England/QEUH-AD3F78/2020 | 2020-10-26 | Europe/United Kingdom/England/ | Lighthouse Lab in Glasgow | Wellcome Sanger Institute for the COVID-19 Genomics UK (COG-UK) consortium | Harper VanSteenhouse et al |
| EPI_ISL_634818 | France/ARA-SC150/2020 | 2020-03-09 | Europe/France/Auvergne-Rhv¥ne-Alpes/ | Laboratoire de virologie, CHU de Grenoble - CS 10217 - 38043 Grenoble cedex 10 | CNR Virus des Infections Respiratoires - France SUD | Antonin Bal et al |
| EPI_ISL_634823 | France/ARA-SC156/2020 | 2020-03-10 | Europe/France/Auvergne-Rhv¥ne-Alpes/ | Laboratoire de virologie, CHU de Grenoble - CS 10217 - 38043 Grenoble cedex 15 | CNR Virus des Infections Respiratoires - France SUD | Antonin Bal et al |
| EPI_ISL_634825 | France/ARA-SC160/2020 | 2020-04-08 | Europe/France/Auvergne-Rhv¥ne-Alpes/ | Laboratoire de virologie, CHU de Grenoble - CS 10217 - 38043 Grenoble cedex 17 | CNR Virus des Infections Respiratoires - France SUD | Antonin Bal et al |
| EPI_ISL_634830 | France/ARA-SC170/2020 | 2020-04-01 | Europe/France/Auvergne-Rhv¥ne-Alpes/ | Laboratoire de virologie, CHU de Grenoble - CS 10217 - 38043 Grenoble cedex 22 | CNR Virus des Infections Respiratoires - France SUD | Antonin Bal et al |
| EPI_ISL_634831 | France/ARA-SC173/2020 | 2020-04-04 | Europe/France/Auvergne-Rhv¥ne-Alpes/ | Laboratoire de virologie, CHU de Grenoble - CS 10217 - 38043 Grenoble cedex 23 | CNR Virus des Infections Respiratoires - France SUD | Antonin Bal et al |
| EPI_ISL_634834 | France/ARA-SC180/2020 | 2020-03-29 | Europe/France/Auvergne-Rhv¥ne-Alpes/ | Laboratoire de virologie, CHU de Grenoble - CS 10217 - 38043 Grenoble cedex 26 | CNR Virus des Infections Respiratoires - France SUD | Antonin Bal et al |
| EPI_ISL_634835 | France/ARA-SC183/2020 | 2020-03-30 | Europe/France/Auvergne-Rhv¥ne-Alpes/ | Laboratoire de virologie, CHU de Grenoble - CS 10217 - 38043 Grenoble cedex 27 | CNR Virus des Infections Respiratoires - France SUD | Antonin Bal et al |

| | | | | | | |
|---|---|---|---|---|---|---|
| EPI_ISL_634837 | France/ARA-SC192/2020 | 2020-04-28 | Europe/France/Auvergne-Rhv¥ne-Alpes/ | Laboratoire de virologie, CHU de Grenoble - CS 10217 - 38043 Grenoble cedex 29 | CNR Virus des Infections Respiratoires - France SUD | Antonin Bal et al |
| EPI_ISL_634846 | USA/MN-MDH-1972/2020 | 2020-10-25 | North America/USA/Minnesota/ | Minnesota Department of Health, Public Health Laboratory | Minnesota Department of Health, Public Health Laboratory | Matt Plumb et al |
| EPI_ISL_634861 | USA/MN-MDH-1987/2020 | 2020-10-27 | North America/USA/Minnesota/ | Minnesota Department of Health, Public Health Laboratory | Minnesota Department of Health, Public Health Laboratory | Matt Plumb et al |
| EPI_ISL_634872 | USA/MN-MDH-1998/2020 | 2020-10-29 | North America/USA/Minnesota/ | Minnesota Department of Health, Public Health Laboratory | Minnesota Department of Health, Public Health Laboratory | Matt Plumb et al |
| EPI_ISL_634875 | USA/MN-MDH-2001/2020 | 2020-10-29 | North America/USA/Minnesota/ | Minnesota Department of Health, Public Health Laboratory | Minnesota Department of Health, Public Health Laboratory | Matt Plumb et al |
| EPI_ISL_634883 | Belgium/UGent-167/2020 | 2020-10-19 | Europe/Belgium/Gent/ | Lab voor klinische biologie | Onderzoeksgroep Virologie | Laurens Lambrechts et al |
| EPI_ISL_634884 | Belgium/UGent-168/2020 | 2020-10-01 | Europe/Belgium/Gent/ | Lab voor klinische biologie | Onderzoeksgroep Virologie | Laurens Lambrechts et al |
| EPI_ISL_634890 | Belgium/UGent-179/2020 | 2020-10-18 | Europe/Belgium/Gent/ | Lab voor klinische biologie | Onderzoeksgroep Virologie | Nick Vereecke et al |
| EPI_ISL_634891 | Belgium/UGent-180/2020 | 2020-10-18 | Europe/Belgium/Gent/ | Lab voor klinische biologie | Onderzoeksgroep Virologie | Nick Vereecke et al |
| EPI_ISL_634900 | USA/UT-UPHL-201130637/2020 | 2020-04-22 | North America/USA/Utah/ | Utah Public Health Laboratory | Utah Public Health Laboratory | Erin L. Young et al |
| EPI_ISL_634903 | USA/UT-UPHL-201108842/2020 | 2020-05-01 | North America/USA/Utah/ | Utah Public Health Laboratory | Utah Public Health Laboratory | Erin L. Young et al |
| EPI_ISL_634904 | USA/UT-UPHL-201119439/2020 | 2020-05-01 | North America/USA/Utah/ | Utah Public Health Laboratory | Utah Public Health Laboratory | Erin L. Young et al |
| EPI_ISL_634908 | USA/UT-UPHL-201131260/2020 | 2020-05-02 | North America/USA/Utah/ | Utah Public Health Laboratory | Utah Public Health Laboratory | Erin L. Young et al |
| EPI_ISL_634909 | USA/UT-UPHL-201113837/2020 | 2020-05-01 | North America/USA/Utah/ | Utah Public Health Laboratory | Utah Public Health Laboratory | Erin L. Young et al |
| EPI_ISL_634921 | USA/UT-UPHL-201130645/2020 | 2020-04-30 | North America/USA/Utah/ | Utah Public Health Laboratory | Utah Public Health Laboratory | Erin L. Young et al |

| | | | | | | |
|---|---|---|---|---|---|---|
| EPI_ISL_634928 | USA/UT-UPHL-201131032/2020 | 2020-10-06 | North America/USA/Utah/ | Utah Public Health Laboratory | Utah Public Health Laboratory | Erin L. Young et al |
| EPI_ISL_634931 | USA/UT-UPHL-201122052/2020 | 2020-10-06 | North America/USA/Utah/ | Utah Public Health Laboratory | Utah Public Health Laboratory | Erin L. Young et al |
| EPI_ISL_634948 | USA/TX-DSHS-1144/2020 | 2020-06-17 | North America/USA/Texas/El Paso Coun | Texas Department of State Health Services | Texas Department of State Health Services | Rashmi Tuladhar et al |
| EPI_ISL_634954 | USA/TX-DSHS-1151/2020 | 2020-07-03 | North America/USA/Texas/Hidalgo Coun | Texas Department of State Health Services | Texas Department of State Health Services | Rashmi Tuladhar et al |
| EPI_ISL_634971 | USA/TX-DSHS-1171/2020 | 2020-07-03 | North America/USA/Texas/Hidalgo Coun | Texas Department of State Health Services | Texas Department of State Health Services | Rashmi Tuladhar et al |
| EPI_ISL_634972 | USA/TX-DSHS-1172/2020 | 2020-07-03 | North America/USA/Texas/Hidalgo Coun | Texas Department of State Health Services | Texas Department of State Health Services | Rashmi Tuladhar et al |
| EPI_ISL_634975 | USA/TX-DSHS-1176/2020 | 2020-07-03 | North America/USA/Texas/Hidalgo Coun | Texas Department of State Health Services | Texas Department of State Health Services | Rashmi Tuladhar et al |

| | | | | | | |
|---|---|---|---|---|---|---|
| EPI_ISL_634977 | Tunisia/55153/2020 | 2020-10-31 | Africa/Tunisia/Ben Arous/ | 1-Laboratory of Microbiology, National Reference Lab, Charles Nicolle Hospital; 2-University of Tunis ElManar, Faculty of Medicine of Tunis, LR99ES09, Tunis, Tunisia | 1-Clinical and Experimental Pharmacology Lab, LR16SP02, National Center of Pharmacovigilance, University of Tunis El Manar, Tunis, Tunisia. 2-Neurodegenerative diseases and psychiatric troubles, LR18SP03, Razi Hospital, University of Tunis El Manar, Tunis, Tunisia. 3-Ministry of Health, National Observatory of New and Emerging Diseases, 1006, Tunis, Tunisia | Ilhem Boutiba-Ben Boubaker et al |
| EPI_ISL_634978 | SouthAfrica/KRISP-K003990/2020 | 2020-09-25 | Africa/South Africa/KwaZulu-Natal/uMk | National Health Laboratory Service - Inkosi Albert Luthuli Central Hospital (NHLS-IALCH) | KRISP, KZN Research Innovation and Sequencing Platform | Giandhari J et al |
| EPI_ISL_634987 | SouthAfrica/KRISP-K004002/2020 | 2020-09-24 | Africa/South Africa/KwaZulu-Natal/Zulu | National Health Laboratory Service - Inkosi Albert Luthuli Central Hospital (NHLS-IALCH) | KRISP, KZN Research Innovation and Sequencing Platform | Giandhari J et al |
| EPI_ISL_634997 | SouthAfrica/KRISP-K004012/2020 | 2020-10-12 | Africa/South Africa/KwaZulu-Natal/iLem | National Health Laboratory Service - Inkosi Albert Luthuli Central Hospital (NHLS-IALCH) | KRISP, KZN Research Innovation and Sequencing Platform | Giandhari J et al |

| | | | | | | |
|---|---|---|---|---|---|---|
| EPI_ISL_635001 | SouthAfrica/KRISP-K004017/2020 | 2020-10-14 | Africa/South Africa/KwaZulu-Natal/uMk | National Health Laboratory Service - Inkosi Albert Luthuli Central Hospital (NHLS-IALCH) | KRISP, KZN Research Innovation and Sequencing Platform | Giandhari J et al |
| EPI_ISL_635027 | SouthAfrica/KRISP-K004048/2020 | 2020-10-19 | Africa/South Africa/KwaZulu-Natal/eThe | National Health Laboratory Service - Inkosi Albert Luthuli Central Hospital (NHLS-IALCH) | KRISP, KZN Research Innovation and Sequencing Platform | Giandhari J et al |
| EPI_ISL_635040 | SouthAfrica/KRISP-K004061/2020 | 2020-10-18 | Africa/South Africa/KwaZulu-Natal/Ugu | National Health Laboratory Service - Inkosi Albert Luthuli Central Hospital (NHLS-IALCH) | KRISP, KZN Research Innovation and Sequencing Platform | Giandhari J et al |
| EPI_ISL_635060 | Tunisia/55319/2020 | 2020-11-02 | Africa/Tunisia/Tunis/ | 1-Laboratory of Microbiology, National Reference Lab, Charles Nicolle Hospital; 2-University of Tunis ElManar, Faculty of Medicine of Tunis, LR99ES09, Tunis, Tunisia | 1-Clinical and Experimental Pharmacology Lab, LR16SP02, National Center of Pharmacovigilance, University of Tunis El Manar, Tunis, Tunisia. 2-Neurodegenerative diseases and psychiatric troubles, LR18SP03, Razi Hospital, University of Tunis El Manar, Tunis, Tunisia. 3-Ministry of Health, National Observatory of New and Emerging Diseases, 1006, Tunis, Tunisia | Ilhem Boutiba-Ben Boubaker et al |

| | | | | | | |
|---|---|---|---|---|---|---|
| EPI_ISL_635062 | Tunisia/55439/2020 | 2020-11-02 | Africa/Tunisia/Tunis/ | 1-Laboratory of Microbiology, National Reference Lab, Charles Nicolle Hospital; 2-University of Tunis ElManar, Faculty of Medicine of Tunis, LR99ES09, Tunis, Tunisia | 1-Clinical and Experimental Pharmacology Lab, LR16SP02, National Center of Pharmacovigilance, University of Tunis El Manar, Tunis, Tunisia. 2-Neurodegenerative diseases and psychiatric troubles, LR18SP03, Razi Hospital, University of Tunis El Manar, Tunis, Tunisia. 3-Ministry of Health, National Observatory of New and Emerging Diseases, 1006, Tunis, Tunisia | Ilhem Boutiba-Ben Boubaker et al |
| EPI_ISL_635220 | Slovenia/5677/2020 | 2020-03-18 | Europe/Slovenia/Slovenia/ | Institute of Microbiology and Immunology, Faculty of Medicine, University of Ljubljana | Institute of Microbiology and Immunology, Faculty of Medicine, University of Ljubljana | Samo Zakotnik et al |
| EPI_ISL_635223 | Slovenia/5716/2020 | 2020-03-11 | Europe/Slovenia/Slovenia/ | Institute of Microbiology and Immunology, Faculty of Medicine, University of Ljubljana | Institute of Microbiology and Immunology, Faculty of Medicine, University of Ljubljana | Samo Zakotnik et al |

| | | | | | | |
|---|---|---|---|---|---|---|
| EPI_ISL_635241 | Slovenia/7455/2020 | 2020-03-14 | Europe/Slovenia/Slovenia/ | Institute of Microbiology and Immunology, Faculty of Medicine, University of Ljubljana | Institute of Microbiology and Immunology, Faculty of Medicine, University of Ljubljana | Samo Zakotnik et al |
| EPI_ISL_635285 | Slovenia/V0-3610/2020 | 2020-04-02 | Europe/Slovenia/Slovenia/ | Institute of Microbiology and Immunology, Faculty of Medicine, University of Ljubljana | Institute of Microbiology and Immunology, Faculty of Medicine, University of Ljubljana | Toma≈æ Mark Zorec et al |
| EPI_ISL_635315 | USA/CA-ALSR-2043/2020 | 2020-05-09 | North America/USA/California/San Dieg | San Diego County Public Health Laboratory | Andersen lab at Scripps Research | SEARCH Alliance San Diego with Tracy Basler et al |
| EPI_ISL_635319 | USA/CA-ALSR-2052/2020 | 2020-05-08 | North America/USA/California/San Dieg | San Diego County Public Health Laboratory | Andersen lab at Scripps Research | SEARCH Alliance San Diego with Tracy Basler et al |
| EPI_ISL_635327 | USA/CA-ALSR-2062/2020 | 2020-04-22 | North America/USA/California/San Dieg | San Diego County Public Health Laboratory | Andersen lab at Scripps Research | SEARCH Alliance San Diego with Tracy Basler et al |
| EPI_ISL_635330 | USA/CA-ALSR-2065/2020 | 2020-04-22 | North America/USA/California/San Dieg | San Diego County Public Health Laboratory | Andersen lab at Scripps Research | SEARCH Alliance San Diego with Tracy Basler et al |
| EPI_ISL_635347 | USA/CA-ALSR-2099/2020 | 2020-06-01 | North America/USA/California/San Dieg | San Diego County Public Health Laboratory | Andersen lab at Scripps Research | SEARCH Alliance San Diego with Tracy Basler et al |
| EPI_ISL_635359 | USA/CA-ALSR-2129/2020 | 2020-05-21 | North America/USA/California/San Dieg | San Diego County Public Health Laboratory | Andersen lab at Scripps Research | SEARCH Alliance San Diego with Tracy Basler et al |
| EPI_ISL_635363 | USA/CA-ALSR-2158/2020 | 2020-06-01 | North America/USA/California/San Dieg | San Diego County Public Health Laboratory | Andersen lab at Scripps Research | SEARCH Alliance San Diego with Tracy Basler et al |
| EPI_ISL_635368 | USA/CA-ALSR-2167/2020 | 2020-06-01 | North America/USA/California/San Dieg | San Diego County Public Health Laboratory | Andersen lab at Scripps Research | SEARCH Alliance San Diego with Tracy Basler et al |
| EPI_ISL_635369 | USA/CA-ALSR-2168/2020 | 2020-06-02 | North America/USA/California/San Dieg | San Diego County Public Health Laboratory | Andersen lab at Scripps Research | SEARCH Alliance San Diego with Tracy Basler et al |
| EPI_ISL_635372 | USA/CA-ALSR-2171/2020 | 2020-06-04 | North America/USA/California/San Dieg | San Diego County Public Health Laboratory | Andersen lab at Scripps Research | SEARCH Alliance San Diego with Tracy Basler et al |
| EPI_ISL_635374 | USA/CA-ALSR-2174/2020 | 2020-05-12 | North America/USA/California/San Dieg | San Diego County Public Health Laboratory | Andersen lab at Scripps Research | SEARCH Alliance San Diego with Tracy Basler et al |
| EPI_ISL_635388 | USA/CA-ALSR-2213/2020 | 2020-06-25 | North America/USA/California/San Dieg | San Diego County Public Health Laboratory | Andersen lab at Scripps Research | SEARCH Alliance San Diego with Tracy Basler et al |
| EPI_ISL_635406 | USA/CA-ALSR-2279/2020 | 2020-07-07 | North America/USA/California/San Dieg | San Diego County Public Health Laboratory | Andersen lab at Scripps Research | SEARCH Alliance San Diego with Tracy Basler et al |

| | | | | | | |
|---|---|---|---|---|---|---|
| EPI_ISL_635407 | USA/CA-ALSR-2280/2020 | 2020-07-08 | North America/USA/California/San Dieg | San Diego County Public Health Laboratory | Andersen lab at Scripps Research | SEARCH Alliance San Diego with Tracy Basler et al |
| EPI_ISL_635412 | USA/CA-ALSR-2287/2020 | 2020-07-08 | North America/USA/California/San Dieg | San Diego County Public Health Laboratory | Andersen lab at Scripps Research | SEARCH Alliance San Diego with Tracy Basler et al |
| EPI_ISL_635417 | USA/CA-ALSR-2294/2020 | 2020-07-08 | North America/USA/California/San Dieg | San Diego County Public Health Laboratory | Andersen lab at Scripps Research | SEARCH Alliance San Diego with Tracy Basler et al |
| EPI_ISL_635421 | USA/CA-ALSR-2302/2020 | 2020-07-09 | North America/USA/California/San Dieg | San Diego County Public Health Laboratory | Andersen lab at Scripps Research | SEARCH Alliance San Diego with Tracy Basler et al |
| EPI_ISL_635451 | USA/CA-ALSR-2382/2020 | 2020-07-06 | North America/USA/California/San Dieg | San Diego County Public Health Laboratory | Andersen lab at Scripps Research | SEARCH Alliance San Diego with Tracy Basler et al |
| EPI_ISL_635452 | USA/CA-ALSR-2384/2020 | 2020-07-02 | North America/USA/California/San Dieg | San Diego County Public Health Laboratory | Andersen lab at Scripps Research | SEARCH Alliance San Diego with Tracy Basler et al |
| EPI_ISL_635455 | USA/CA-ALSR-2388/2020 | 2020-07-01 | North America/USA/California/San Dieg | San Diego County Public Health Laboratory | Andersen lab at Scripps Research | SEARCH Alliance San Diego with Tracy Basler et al |
| EPI_ISL_635457 | USA/CA-ALSR-2393/2020 | 2020-07-08 | North America/USA/California/San Dieg | San Diego County Public Health Laboratory | Andersen lab at Scripps Research | SEARCH Alliance San Diego with Tracy Basler et al |
| EPI_ISL_635464 | USA/CA-ALSR-2408/2020 | 2020-06-30 | North America/USA/California/San Dieg | San Diego County Public Health Laboratory | Andersen lab at Scripps Research | SEARCH Alliance San Diego with Tracy Basler et al |
| EPI_ISL_635467 | USA/CA-ALSR-2412/2020 | 2020-07-02 | North America/USA/California/San Dieg | San Diego County Public Health Laboratory | Andersen lab at Scripps Research | SEARCH Alliance San Diego with Tracy Basler et al |
| EPI_ISL_635475 | USA/CA-ALSR-2429/2020 | 2020-07-02 | North America/USA/California/San Dieg | San Diego County Public Health Laboratory | Andersen lab at Scripps Research | SEARCH Alliance San Diego with Tracy Basler et al |
| EPI_ISL_635478 | USA/CA-ALSR-2437/2020 | 2020-07-02 | North America/USA/California/San Dieg | San Diego County Public Health Laboratory | Andersen lab at Scripps Research | SEARCH Alliance San Diego with Tracy Basler et al |
| EPI_ISL_635479 | Mexico/BCN-ALSR-2444/2020 | 2020-05-13 | North America/Mexico/Baja California/T | Centro de Diagn,àö,â•stico COVID-19 UABC Tijuana | Andersen lab at Scripps Research | SEARCH Alliance San Diego with Idanya Rub√≠ Seraf√≠n Higuera et al |
| EPI_ISL_635480 | Mexico/BCN-ALSR-2446/2020 | 2020-05-14 | North America/Mexico/Baja California/T | Centro de Diagn,àö,â•stico COVID-19 UABC Tijuana | Andersen lab at Scripps Research | SEARCH Alliance San Diego with Idanya Rub√≠ Seraf√≠n Higuera et al |
| EPI_ISL_635481 | Mexico/BCN-ALSR-2447/2020 | 2020-05-14 | North America/Mexico/Baja California/T | Centro de Diagn,àö,â•stico COVID-19 UABC Tijuana | Andersen lab at Scripps Research | SEARCH Alliance San Diego with Idanya Rub√≠ Seraf√≠n Higuera et al |
| EPI_ISL_635482 | Mexico/BCN-ALSR-2448/2020 | 2020-05-14 | North America/Mexico/Baja California/T | Centro de Diagn,àö,â•stico COVID-19 UABC Tijuana | Andersen lab at Scripps Research | SEARCH Alliance San Diego with Idanya Rub√≠ Seraf√≠n Higuera et al |
| EPI_ISL_635483 | Mexico/BCN-ALSR-2451/2020 | 2020-05-15 | North America/Mexico/Baja California/T | Centro de Diagn,àö,â•stico COVID-19 UABC Tijuana | Andersen lab at Scripps Research | SEARCH Alliance San Diego with Idanya Rub√≠ Seraf√≠n Higuera et al |

| | | | | | | |
|---|---|---|---|---|---|---|
| EPI_ISL_635484 | Mexico/BCN-ALSR-2452/2020 | 2020-05-15 | North America/Mexico/Baja California/T | Centro de Diagn‚àö‚â•stico COVID-19 UABC Tijuana | Andersen lab at Scripps Research | SEARCH Alliance San Diego with Idanya Rub√≠ Seraf√≠n Higuera et al |
| EPI_ISL_635485 | Mexico/BCN-ALSR-2457/2020 | 2020-05-16 | North America/Mexico/Baja California/T | Centro de Diagn‚àö‚â•stico COVID-19 UABC Tijuana | Andersen lab at Scripps Research | SEARCH Alliance San Diego with Idanya Rub√≠ Seraf√≠n Higuera et al |
| EPI_ISL_635486 | Mexico/BCN-ALSR-2462/2020 | 2020-05-19 | North America/Mexico/Baja California/T | Centro de Diagn‚àö‚â•stico COVID-19 UABC Tijuana | Andersen lab at Scripps Research | SEARCH Alliance San Diego with Idanya Rub√≠ Seraf√≠n Higuera et al |
| EPI_ISL_635487 | Mexico/BCN-ALSR-2466/2020 | 2020-05-19 | North America/Mexico/Baja California/T | Centro de Diagn‚àö‚â•stico COVID-19 UABC Tijuana | Andersen lab at Scripps Research | SEARCH Alliance San Diego with Idanya Rub√≠ Seraf√≠n Higuera et al |
| EPI_ISL_635488 | Mexico/BCN-ALSR-2468/2020 | 2020-05-20 | North America/Mexico/Baja California/T | Centro de Diagn‚àö‚â•stico COVID-19 UABC Tijuana | Andersen lab at Scripps Research | SEARCH Alliance San Diego with Idanya Rub√≠ Seraf√≠n Higuera et al |
| EPI_ISL_635489 | Mexico/BCN-ALSR-2475/2020 | 2020-05-22 | North America/Mexico/Baja California/T | Centro de Diagn‚àö‚â•stico COVID-19 UABC Tijuana | Andersen lab at Scripps Research | SEARCH Alliance San Diego with Idanya Rub√≠ Seraf√≠n Higuera et al |
| EPI_ISL_635490 | Mexico/BCN-ALSR-2477/2020 | 2020-05-22 | North America/Mexico/Baja California/T | Centro de Diagn‚àö‚â•stico COVID-19 UABC Tijuana | Andersen lab at Scripps Research | SEARCH Alliance San Diego with Idanya Rub√≠ Seraf√≠n Higuera et al |
| EPI_ISL_635491 | Mexico/BCN-ALSR-2479/2020 | 2020-05-26 | North America/Mexico/Baja California/T | Centro de Diagn‚àö‚â•stico COVID-19 UABC Tijuana | Andersen lab at Scripps Research | SEARCH Alliance San Diego with Idanya Rub√≠ Seraf√≠n Higuera et al |
| EPI_ISL_635492 | Mexico/BCN-ALSR-2480/2020 | 2020-05-26 | North America/Mexico/Baja California/T | Centro de Diagn‚àö‚â•stico COVID-19 UABC Tijuana | Andersen lab at Scripps Research | SEARCH Alliance San Diego with Idanya Rub√≠ Seraf√≠n Higuera et al |
| EPI_ISL_635493 | Mexico/BCN-ALSR-2481/2020 | 2020-05-26 | North America/Mexico/Baja California/T | Centro de Diagn‚àö‚â•stico COVID-19 UABC Tijuana | Andersen lab at Scripps Research | SEARCH Alliance San Diego with Idanya Rub√≠ Seraf√≠n Higuera et al |
| EPI_ISL_635495 | Mexico/BCN-ALSR-2484/2020 | 2020-05-26 | North America/Mexico/Baja California/T | Centro de Diagn‚àö‚â•stico COVID-19 UABC Tijuana | Andersen lab at Scripps Research | SEARCH Alliance San Diego with Idanya Rub√≠ Seraf√≠n Higuera et al |
| EPI_ISL_635498 | Mexico/BCN-ALSR-2489/2020 | 2020-05-29 | North America/Mexico/Baja California/T | Centro de Diagn‚àö‚â•stico COVID-19 UABC Tijuana | Andersen lab at Scripps Research | SEARCH Alliance San Diego with Idanya Rub√≠ Seraf√≠n Higuera et al |
| EPI_ISL_635499 | Mexico/BCN-ALSR-2490/2020 | 2020-05-29 | North America/Mexico/Baja California/T | Centro de Diagn‚àö‚â•stico COVID-19 UABC Tijuana | Andersen lab at Scripps Research | SEARCH Alliance San Diego with Idanya Rub√≠ Seraf√≠n Higuera et al |

| | | | | | | |
|---|---|---|---|---|---|---|
| EPI_ISL_635500 | Mexico/BCN-ALSR-2491/2020 | 2020-05-29 | North America/Mexico/Baja California/T | Centro de Diagn,àö,â•stico COVID-19 UABC Tijuana | Andersen lab at Scripps Research | SEARCH Alliance San Diego with Idanya Rub√≠ Seraf√≠n Higuera et al |
| EPI_ISL_635501 | Mexico/BCN-ALSR-2492/2020 | 2020-05-29 | North America/Mexico/Baja California/T | Centro de Diagn,àö,â•stico COVID-19 UABC Tijuana | Andersen lab at Scripps Research | SEARCH Alliance San Diego with Idanya Rub√≠ Seraf√≠n Higuera et al |
| EPI_ISL_635503 | Mexico/BCN-ALSR-2494/2020 | 2020-05-29 | North America/Mexico/Baja California/T | Centro de Diagn,àö,â•stico COVID-19 UABC Tijuana | Andersen lab at Scripps Research | SEARCH Alliance San Diego with Idanya Rub√≠ Seraf√≠n Higuera et al |
| EPI_ISL_635504 | Mexico/BCN-ALSR-2495/2020 | 2020-05-30 | North America/Mexico/Baja California/T | Centro de Diagn,àö,â•stico COVID-19 UABC Tijuana | Andersen lab at Scripps Research | SEARCH Alliance San Diego with Idanya Rub√≠ Seraf√≠n Higuera et al |
| EPI_ISL_635505 | Mexico/BCN-ALSR-2496/2020 | 2020-05-30 | North America/Mexico/Baja California/T | Centro de Diagn,àö,â•stico COVID-19 UABC Tijuana | Andersen lab at Scripps Research | SEARCH Alliance San Diego with Idanya Rub√≠ Seraf√≠n Higuera et al |
| EPI_ISL_635506 | Mexico/BCN-ALSR-2497/2020 | 2020-06-01 | North America/Mexico/Baja California/T | Centro de Diagn,àö,â•stico COVID-19 UABC Tijuana | Andersen lab at Scripps Research | SEARCH Alliance San Diego with Idanya Rub√≠ Seraf√≠n Higuera et al |
| EPI_ISL_635507 | Mexico/BCN-ALSR-2499/2020 | 2020-06-01 | North America/Mexico/Baja California/T | Centro de Diagn,àö,â•stico COVID-19 UABC Tijuana | Andersen lab at Scripps Research | SEARCH Alliance San Diego with Idanya Rub√≠ Seraf√≠n Higuera et al |
| EPI_ISL_635508 | Mexico/BCN-ALSR-2500/2020 | 2020-06-03 | North America/Mexico/Baja California/T | Centro de Diagn,àö,â•stico COVID-19 UABC Tijuana | Andersen lab at Scripps Research | SEARCH Alliance San Diego with Idanya Rub√≠ Seraf√≠n Higuera et al |
| EPI_ISL_635509 | Mexico/BCN-ALSR-2502/2020 | 2020-06-03 | North America/Mexico/Baja California/T | Centro de Diagn,àö,â•stico COVID-19 UABC Tijuana | Andersen lab at Scripps Research | SEARCH Alliance San Diego with Idanya Rub√≠ Seraf√≠n Higuera et al |
| EPI_ISL_635510 | Mexico/BCN-ALSR-2504/2020 | 2020-06-05 | North America/Mexico/Baja California/T | Centro de Diagn,àö,â•stico COVID-19 UABC Tijuana | Andersen lab at Scripps Research | SEARCH Alliance San Diego with Idanya Rub√≠ Seraf√≠n Higuera et al |
| EPI_ISL_635511 | Mexico/BCN-ALSR-2506/2020 | 2020-06-06 | North America/Mexico/Baja California/T | Centro de Diagn,àö,â•stico COVID-19 UABC Tijuana | Andersen lab at Scripps Research | SEARCH Alliance San Diego with Idanya Rub√≠ Seraf√≠n Higuera et al |
| EPI_ISL_635512 | Mexico/BCN-ALSR-2507/2020 | 2020-06-08 | North America/Mexico/Baja California/T | Centro de Diagn,àö,â•stico COVID-19 UABC Tijuana | Andersen lab at Scripps Research | SEARCH Alliance San Diego with Idanya Rub√≠ Seraf√≠n Higuera et al |
| EPI_ISL_635513 | Mexico/BCN-ALSR-2508/2020 | 2020-06-08 | North America/Mexico/Baja California/T | Centro de Diagn,àö,â•stico COVID-19 UABC Tijuana | Andersen lab at Scripps Research | SEARCH Alliance San Diego with Idanya Rub√≠ Seraf√≠n Higuera et al |

| | | | | | | |
|---|---|---|---|---|---|---|
| EPI_ISL_635514 | Mexico/BCN-ALSR-2509/2020 | 2020-06-08 | North America/Mexico/Baja California/T | Centro de Diagn‚àö‚â•stico COVID-19 UABC Tijuana | Andersen lab at Scripps Research | SEARCH Alliance San Diego with Idanya Rub√≠ Seraf√≠n Higuera et al |
| EPI_ISL_635515 | Mexico/BCN-ALSR-2510/2020 | 2020-06-10 | North America/Mexico/Baja California/T | Centro de Diagn‚àö‚â•stico COVID-19 UABC Tijuana | Andersen lab at Scripps Research | SEARCH Alliance San Diego with Idanya Rub√≠ Seraf√≠n Higuera et al |
| EPI_ISL_635516 | Mexico/BCN-ALSR-2511/2020 | 2020-06-12 | North America/Mexico/Baja California/T | Centro de Diagn‚àö‚â•stico COVID-19 UABC Tijuana | Andersen lab at Scripps Research | SEARCH Alliance San Diego with Idanya Rub√≠ Seraf√≠n Higuera et al |
| EPI_ISL_635517 | Mexico/BCN-ALSR-2512/2020 | 2020-06-12 | North America/Mexico/Baja California/T | Centro de Diagn‚àö‚â•stico COVID-19 UABC Tijuana | Andersen lab at Scripps Research | SEARCH Alliance San Diego with Idanya Rub√≠ Seraf√≠n Higuera et al |
| EPI_ISL_635518 | Mexico/BCN-ALSR-2513/2020 | 2020-06-15 | North America/Mexico/Baja California/T | Centro de Diagn‚àö‚â•stico COVID-19 UABC Tijuana | Andersen lab at Scripps Research | SEARCH Alliance San Diego with Idanya Rub√≠ Seraf√≠n Higuera et al |
| EPI_ISL_635519 | Mexico/BCN-ALSR-2514/2020 | 2020-06-15 | North America/Mexico/Baja California/T | Centro de Diagn‚àö‚â•stico COVID-19 UABC Tijuana | Andersen lab at Scripps Research | SEARCH Alliance San Diego with Idanya Rub√≠ Seraf√≠n Higuera et al |
| EPI_ISL_635520 | Mexico/BCN-ALSR-2515/2020 | 2020-06-15 | North America/Mexico/Baja California/T | Centro de Diagn‚àö‚â•stico COVID-19 UABC Tijuana | Andersen lab at Scripps Research | SEARCH Alliance San Diego with Idanya Rub√≠ Seraf√≠n Higuera et al |
| EPI_ISL_635521 | Mexico/BCN-ALSR-2516/2020 | 2020-06-15 | North America/Mexico/Baja California/T | Centro de Diagn‚àö‚â•stico COVID-19 UABC Tijuana | Andersen lab at Scripps Research | SEARCH Alliance San Diego with Idanya Rub√≠ Seraf√≠n Higuera et al |
| EPI_ISL_635522 | Mexico/BCN-ALSR-2517/2020 | 2020-06-15 | North America/Mexico/Baja California/T | Centro de Diagn‚àö‚â•stico COVID-19 UABC Tijuana | Andersen lab at Scripps Research | SEARCH Alliance San Diego with Idanya Rub√≠ Seraf√≠n Higuera et al |
| EPI_ISL_635523 | Mexico/BCN-ALSR-2519/2020 | 2020-06-15 | North America/Mexico/Baja California/T | Centro de Diagn‚àö‚â•stico COVID-19 UABC Tijuana | Andersen lab at Scripps Research | SEARCH Alliance San Diego with Idanya Rub√≠ Seraf√≠n Higuera et al |
| EPI_ISL_635524 | Mexico/BCN-ALSR-2520/2020 | 2020-06-16 | North America/Mexico/Baja California/T | Centro de Diagn‚àö‚â•stico COVID-19 UABC Tijuana | Andersen lab at Scripps Research | SEARCH Alliance San Diego with Idanya Rub√≠ Seraf√≠n Higuera et al |
| EPI_ISL_635525 | Mexico/BCN-ALSR-2521/2020 | 2020-06-16 | North America/Mexico/Baja California/T | Centro de Diagn‚àö‚â•stico COVID-19 UABC Tijuana | Andersen lab at Scripps Research | SEARCH Alliance San Diego with Idanya Rub√≠ Seraf√≠n Higuera et al |
| EPI_ISL_635526 | Mexico/BCN-ALSR-2522/2020 | 2020-06-16 | North America/Mexico/Baja California/T | Centro de Diagn‚àö‚â•stico COVID-19 UABC Tijuana | Andersen lab at Scripps Research | SEARCH Alliance San Diego with Idanya Rub√≠ Seraf√≠n Higuera et al |

| | | | | | | |
|---|---|---|---|---|---|---|
| EPI_ISL_635527 | Mexico/BCN-ALSR-2523/2020 | 2020-06-16 | North America/Mexico/Baja California/T | Centro de Diagn‚àö¬∞stico COVID-19 UABC Tijuana | Andersen lab at Scripps Research | SEARCH Alliance San Diego with Idanya Rubv≠ Serafv≠n Higuera et al |
| EPI_ISL_635528 | Mexico/BCN-ALSR-2524/2020 | 2020-06-16 | North America/Mexico/Baja California/T | Centro de Diagn‚àö¬∞stico COVID-19 UABC Tijuana | Andersen lab at Scripps Research | SEARCH Alliance San Diego with Idanya Rubv≠ Serafv≠n Higuera et al |
| EPI_ISL_635529 | Mexico/BCN-ALSR-2525/2020 | 2020-06-17 | North America/Mexico/Baja California/T | Centro de Diagn‚àö¬∞stico COVID-19 UABC Tijuana | Andersen lab at Scripps Research | SEARCH Alliance San Diego with Idanya Rubv≠ Serafv≠n Higuera et al |
| EPI_ISL_635530 | Mexico/BCN-ALSR-2526/2020 | 2020-06-17 | North America/Mexico/Baja California/T | Centro de Diagn‚àö¬∞stico COVID-19 UABC Tijuana | Andersen lab at Scripps Research | SEARCH Alliance San Diego with Idanya Rubv≠ Serafv≠n Higuera et al |
| EPI_ISL_635531 | Mexico/BCN-ALSR-2527/2020 | 2020-06-17 | North America/Mexico/Baja California/T | Centro de Diagn‚àö¬∞stico COVID-19 UABC Tijuana | Andersen lab at Scripps Research | SEARCH Alliance San Diego with Idanya Rubv≠ Serafv≠n Higuera et al |
| EPI_ISL_635532 | Mexico/BCN-ALSR-2528/2020 | 2020-06-18 | North America/Mexico/Baja California/T | Centro de Diagn‚àö¬∞stico COVID-19 UABC Tijuana | Andersen lab at Scripps Research | SEARCH Alliance San Diego with Idanya Rubv≠ Serafv≠n Higuera et al |
| EPI_ISL_635533 | Mexico/BCN-ALSR-2529/2020 | 2020-06-22 | North America/Mexico/Baja California/T | Centro de Diagn‚àö¬∞stico COVID-19 UABC Tijuana | Andersen lab at Scripps Research | SEARCH Alliance San Diego with Idanya Rubv≠ Serafv≠n Higuera et al |
| EPI_ISL_635534 | Mexico/BCN-ALSR-2530/2020 | 2020-06-22 | North America/Mexico/Baja California/T | Centro de Diagn‚àö¬∞stico COVID-19 UABC Tijuana | Andersen lab at Scripps Research | SEARCH Alliance San Diego with Idanya Rubv≠ Serafv≠n Higuera et al |
| EPI_ISL_635535 | Mexico/BCN-ALSR-2531/2020 | 2020-06-22 | North America/Mexico/Baja California/T | Centro de Diagn‚àö¬∞stico COVID-19 UABC Tijuana | Andersen lab at Scripps Research | SEARCH Alliance San Diego with Idanya Rubv≠ Serafv≠n Higuera et al |
| EPI_ISL_635536 | Mexico/BCN-ALSR-2533/2020 | 2020-06-23 | North America/Mexico/Baja California/T | Centro de Diagn‚àö¬∞stico COVID-19 UABC Tijuana | Andersen lab at Scripps Research | SEARCH Alliance San Diego with Idanya Rubv≠ Serafv≠n Higuera et al |
| EPI_ISL_635537 | Mexico/BCN-ALSR-2534/2020 | 2020-06-23 | North America/Mexico/Baja California/T | Centro de Diagn‚àö¬∞stico COVID-19 UABC Tijuana | Andersen lab at Scripps Research | SEARCH Alliance San Diego with Idanya Rubv≠ Serafv≠n Higuera et al |
| EPI_ISL_635538 | Mexico/BCN-ALSR-2535/2020 | 2020-06-23 | North America/Mexico/Baja California/T | Centro de Diagn‚àö¬∞stico COVID-19 UABC Tijuana | Andersen lab at Scripps Research | SEARCH Alliance San Diego with Idanya Rubv≠ Serafv≠n Higuera et al |
| EPI_ISL_635539 | Mexico/BCN-ALSR-2536/2020 | 2020-06-23 | North America/Mexico/Baja California/T | Centro de Diagn‚àö¬∞stico COVID-19 UABC Tijuana | Andersen lab at Scripps Research | SEARCH Alliance San Diego with Idanya Rubv≠ Serafv≠n Higuera et al |

| | | | | | | |
|---|---|---|---|---|---|---|
| EPI_ISL_635540 | Mexico/BCN-ALSR-2537/2020 | 2020-06-24 | North America/Mexico/Baja California/T | Centro de Diagn,àö,â•stico COVID-19 UABC Tijuana | Andersen lab at Scripps Research | SEARCH Alliance San Diego with Idanya RubV≠ SerafV≠n Higuera et al |
| EPI_ISL_635542 | Mexico/BCN-ALSR-2539/2020 | 2020-06-24 | North America/Mexico/Baja California/T | Centro de Diagn,àö,â•stico COVID-19 UABC Tijuana | Andersen lab at Scripps Research | SEARCH Alliance San Diego with Idanya Rubv≠ SerafV≠n Higuera et al |
| EPI_ISL_635543 | Mexico/BCN-ALSR-2540/2020 | 2020-06-24 | North America/Mexico/Baja California/T | Centro de Diagn,àö,â•stico COVID-19 UABC Tijuana | Andersen lab at Scripps Research | SEARCH Alliance San Diego with Idanya RubV≠ SerafV≠n Higuera et al |
| EPI_ISL_635544 | Mexico/BCN-ALSR-2541/2020 | 2020-06-25 | North America/Mexico/Baja California/T | Centro de Diagn,àö,â•stico COVID-19 UABC Tijuana | Andersen lab at Scripps Research | SEARCH Alliance San Diego with Idanya RubV≠ SerafV≠n Higuera et al |
| EPI_ISL_635545 | Mexico/BCN-ALSR-2542/2020 | 2020-06-25 | North America/Mexico/Baja California/T | Centro de Diagn,àö,â•stico COVID-19 UABC Tijuana | Andersen lab at Scripps Research | SEARCH Alliance San Diego with Idanya RubV≠ SerafV≠n Higuera et al |
| EPI_ISL_635547 | Mexico/BCN-ALSR-2544/2020 | 2020-06-26 | North America/Mexico/Baja California/T | Centro de Diagn,àö,â•stico COVID-19 UABC Tijuana | Andersen lab at Scripps Research | SEARCH Alliance San Diego with Idanya RubV≠ SerafV≠n Higuera et al |
| EPI_ISL_635548 | Mexico/BCN-ALSR-2545/2020 | 2020-06-27 | North America/Mexico/Baja California/T | Centro de Diagn,àö,â•stico COVID-19 UABC Tijuana | Andersen lab at Scripps Research | SEARCH Alliance San Diego with Idanya RubV≠ SerafV≠n Higuera et al |
| EPI_ISL_635549 | Mexico/BCN-ALSR-2546/2020 | 2020-06-29 | North America/Mexico/Baja California/T | Centro de Diagn,àö,â•stico COVID-19 UABC Tijuana | Andersen lab at Scripps Research | SEARCH Alliance San Diego with Idanya RubV≠ SerafV≠n Higuera et al |
| EPI_ISL_635550 | Mexico/BCN-ALSR-2547/2020 | 2020-06-29 | North America/Mexico/Baja California/T | Centro de Diagn,àö,â•stico COVID-19 UABC Tijuana | Andersen lab at Scripps Research | SEARCH Alliance San Diego with Idanya RubV≠ SerafV≠n Higuera et al |
| EPI_ISL_635551 | Mexico/BCN-ALSR-2548/2020 | 2020-06-30 | North America/Mexico/Baja California/T | Centro de Diagn,àö,â•stico COVID-19 UABC Tijuana | Andersen lab at Scripps Research | SEARCH Alliance San Diego with Idanya RubV≠ SerafV≠n Higuera et al |
| EPI_ISL_635552 | Mexico/BCN-ALSR-2549/2020 | 2020-07-01 | North America/Mexico/Baja California/T | Centro de Diagn,àö,â•stico COVID-19 UABC Tijuana | Andersen lab at Scripps Research | SEARCH Alliance San Diego with Idanya RubV≠ SerafV≠n Higuera et al |
| EPI_ISL_635553 | Mexico/BCN-ALSR-2550/2020 | 2020-07-01 | North America/Mexico/Baja California/T | Centro de Diagn,àö,â•stico COVID-19 UABC Tijuana | Andersen lab at Scripps Research | SEARCH Alliance San Diego with Idanya RubV≠ SerafV≠n Higuera et al |
| EPI_ISL_635554 | Mexico/BCN-ALSR-2551/2020 | 2020-07-01 | North America/Mexico/Baja California/T | Centro de Diagn,àö,â•stico COVID-19 UABC Tijuana | Andersen lab at Scripps Research | SEARCH Alliance San Diego with Idanya RubV≠ SerafV≠n Higuera et al |

| | | | | | | |
|---|---|---|---|---|---|---|
| EPI_ISL_635556 | Mexico/BCN-ALSR-2553/2020 | 2020-07-02 | North America/Mexico/Baja California/T | Centro de Diagn‚àö‚â•stico COVID-19 UABC Tijuana | Andersen lab at Scripps Research | SEARCH Alliance San Diego with Idanya Rub√≠ Seraf√≠n Higuera et al |
| EPI_ISL_635557 | Mexico/BCN-ALSR-2554/2020 | 2020-07-03 | North America/Mexico/Baja California/T | Centro de Diagn‚àö‚â•stico COVID-19 UABC Tijuana | Andersen lab at Scripps Research | SEARCH Alliance San Diego with Idanya Rub√≠ Seraf√≠n Higuera et al |
| EPI_ISL_635558 | Mexico/BCN-ALSR-2555/2020 | 2020-07-03 | North America/Mexico/Baja California/T | Centro de Diagn‚àö‚â•stico COVID-19 UABC Tijuana | Andersen lab at Scripps Research | SEARCH Alliance San Diego with Idanya Rub√≠ Seraf√≠n Higuera et al |
| EPI_ISL_635560 | Mexico/BCN-ALSR-2557/2020 | 2020-07-04 | North America/Mexico/Baja California/T | Centro de Diagn‚àö‚â•stico COVID-19 UABC Tijuana | Andersen lab at Scripps Research | SEARCH Alliance San Diego with Idanya Rub√≠ Seraf√≠n Higuera et al |
| EPI_ISL_635561 | Mexico/BCN-ALSR-2558/2020 | 2020-07-04 | North America/Mexico/Baja California/T | Centro de Diagn‚àö‚â•stico COVID-19 UABC Tijuana | Andersen lab at Scripps Research | SEARCH Alliance San Diego with Idanya Rub√≠ Seraf√≠n Higuera et al |
| EPI_ISL_635563 | Mexico/BCN-ALSR-2560/2020 | 2020-07-06 | North America/Mexico/Baja California/T | Centro de Diagn‚àö‚â•stico COVID-19 UABC Tijuana | Andersen lab at Scripps Research | SEARCH Alliance San Diego with Idanya Rub√≠ Seraf√≠n Higuera et al |
| EPI_ISL_635564 | Mexico/BCN-ALSR-2561/2020 | 2020-07-06 | North America/Mexico/Baja California/T | Centro de Diagn‚àö‚â•stico COVID-19 UABC Tijuana | Andersen lab at Scripps Research | SEARCH Alliance San Diego with Idanya Rub√≠ Seraf√≠n Higuera et al |
| EPI_ISL_635565 | Mexico/BCN-ALSR-2562/2020 | 2020-07-06 | North America/Mexico/Baja California/T | Centro de Diagn‚àö‚â•stico COVID-19 UABC Tijuana | Andersen lab at Scripps Research | SEARCH Alliance San Diego with Idanya Rub√≠ Seraf√≠n Higuera et al |
| EPI_ISL_635566 | Mexico/BCN-ALSR-2563/2020 | 2020-07-06 | North America/Mexico/Baja California/T | Centro de Diagn‚àö‚â•stico COVID-19 UABC Tijuana | Andersen lab at Scripps Research | SEARCH Alliance San Diego with Idanya Rub√≠ Seraf√≠n Higuera et al |
| EPI_ISL_635567 | Mexico/BCN-ALSR-2565/2020 | 2020-07-06 | North America/Mexico/Baja California/T | Centro de Diagn‚àö‚â•stico COVID-19 UABC Tijuana | Andersen lab at Scripps Research | SEARCH Alliance San Diego with Idanya Rub√≠ Seraf√≠n Higuera et al |
| EPI_ISL_635568 | Mexico/BCN-ALSR-2566/2020 | 2020-07-07 | North America/Mexico/Baja California/T | Centro de Diagn‚àö‚â•stico COVID-19 UABC Tijuana | Andersen lab at Scripps Research | SEARCH Alliance San Diego with Idanya Rub√≠ Seraf√≠n Higuera et al |
| EPI_ISL_635569 | Mexico/BCN-ALSR-2568/2020 | 2020-07-07 | North America/Mexico/Baja California/T | Centro de Diagn‚àö‚â•stico COVID-19 UABC Tijuana | Andersen lab at Scripps Research | SEARCH Alliance San Diego with Idanya Rub√≠ Seraf√≠n Higuera et al |
| EPI_ISL_635572 | Mexico/BCN-ALSR-2571/2020 | 2020-07-08 | North America/Mexico/Baja California/T | Centro de Diagn‚àö‚â•stico COVID-19 UABC Tijuana | Andersen lab at Scripps Research | SEARCH Alliance San Diego with Idanya Rub√≠ Seraf√≠n Higuera et al |

| | | | | | | |
|---|---|---|---|---|---|---|
| EPI_ISL_635573 | Mexico/BCN-ALSR-2572/2020 | 2020-07-20 | North America/Mexico/Baja California/T | Centro de Diagn‚àö‚â•stico COVID-19 UABC Tijuana | Andersen lab at Scripps Research | SEARCH Alliance San Diego with Idanya Rub√≠ Seraf√≠n Higuera et al |
| EPI_ISL_635574 | Mexico/BCN-ALSR-2573/2020 | 2020-07-16 | North America/Mexico/Baja California/T | Centro de Diagn‚àö‚â•stico COVID-19 UABC Tijuana | Andersen lab at Scripps Research | SEARCH Alliance San Diego with Idanya Rub√≠ Seraf√≠n Higuera et al |
| EPI_ISL_635576 | Mexico/BCN-ALSR-2576/2020 | 2020-07-24 | North America/Mexico/Baja California/T | Centro de Diagn‚àö‚â•stico COVID-19 UABC Tijuana | Andersen lab at Scripps Research | SEARCH Alliance San Diego with Idanya Rub√≠ Seraf√≠n Higuera et al |
| EPI_ISL_635584 | USA/CA-ALSR-2607/2020 | 2020-07-11 | North America/USA/California/San Dieg | San Diego County Public Health Laboratory | Andersen lab at Scripps Research | SEARCH Alliance San Diego with Tracy Basler et al |
| EPI_ISL_635585 | USA/CA-ALSR-2610/2020 | 2020-07-12 | North America/USA/California/San Dieg | San Diego County Public Health Laboratory | Andersen lab at Scripps Research | SEARCH Alliance San Diego with Tracy Basler et al |
| EPI_ISL_635587 | USA/CA-ALSR-2614/2020 | 2020-07-11 | North America/USA/California/San Dieg | San Diego County Public Health Laboratory | Andersen lab at Scripps Research | SEARCH Alliance San Diego with Tracy Basler et al |
| EPI_ISL_635588 | USA/CA-ALSR-2615/2020 | 2020-07-11 | North America/USA/California/San Dieg | San Diego County Public Health Laboratory | Andersen lab at Scripps Research | SEARCH Alliance San Diego with Tracy Basler et al |
| EPI_ISL_635589 | USA/CA-ALSR-2616/2020 | 2020-07-11 | North America/USA/California/San Dieg | San Diego County Public Health Laboratory | Andersen lab at Scripps Research | SEARCH Alliance San Diego with Tracy Basler et al |
| EPI_ISL_635591 | USA/CA-ALSR-2620/2020 | 2020-07-10 | North America/USA/California/San Dieg | San Diego County Public Health Laboratory | Andersen lab at Scripps Research | SEARCH Alliance San Diego with Tracy Basler et al |
| EPI_ISL_635593 | USA/CA-ALSR-2622/2020 | 2020-07-10 | North America/USA/California/San Dieg | San Diego County Public Health Laboratory | Andersen lab at Scripps Research | SEARCH Alliance San Diego with Tracy Basler et al |
| EPI_ISL_635594 | USA/CA-ALSR-2624/2020 | 2020-07-10 | North America/USA/California/San Dieg | San Diego County Public Health Laboratory | Andersen lab at Scripps Research | SEARCH Alliance San Diego with Tracy Basler et al |
| EPI_ISL_635595 | USA/CA-ALSR-2627/2020 | 2020-07-11 | North America/USA/California/San Dieg | San Diego County Public Health Laboratory | Andersen lab at Scripps Research | SEARCH Alliance San Diego with Tracy Basler et al |
| EPI_ISL_635600 | USA/CA-ALSR-2634/2020 | 2020-07-09 | North America/USA/California/San Dieg | San Diego County Public Health Laboratory | Andersen lab at Scripps Research | SEARCH Alliance San Diego with Tracy Basler et al |
| EPI_ISL_635601 | USA/CA-ALSR-2635/2020 | 2020-07-11 | North America/USA/California/San Dieg | San Diego County Public Health Laboratory | Andersen lab at Scripps Research | SEARCH Alliance San Diego with Tracy Basler et al |
| EPI_ISL_635607 | USA/CA-ALSR-2642/2020 | 2020-07-12 | North America/USA/California/San Dieg | San Diego County Public Health Laboratory | Andersen lab at Scripps Research | SEARCH Alliance San Diego with Tracy Basler et al |
| EPI_ISL_635612 | USA/CA-ALSR-2654/2020 | 2020-07-11 | North America/USA/California/San Dieg | San Diego County Public Health Laboratory | Andersen lab at Scripps Research | SEARCH Alliance San Diego with Tracy Basler et al |
| EPI_ISL_635613 | USA/CA-ALSR-2657/2020 | 2020-07-09 | North America/USA/California/San Dieg | San Diego County Public Health Laboratory | Andersen lab at Scripps Research | SEARCH Alliance San Diego with Tracy Basler et al |
| EPI_ISL_635614 | USA/CA-ALSR-2659/2020 | 2020-07-12 | North America/USA/California/San Dieg | San Diego County Public Health Laboratory | Andersen lab at Scripps Research | SEARCH Alliance San Diego with Tracy Basler et al |

| | | | | | | |
|---|---|---|---|---|---|---|
| EPI_ISL_635615 | USA/CA-ALSR-2660/2020 | 2020-07-10 | North America/USA/California/San Dieg | San Diego County Public Health Laboratory | Andersen lab at Scripps Research | SEARCH Alliance San Diego with Tracy Basler et al |
| EPI_ISL_635616 | USA/CA-ALSR-2662/2020 | 2020-07-12 | North America/USA/California/San Dieg | San Diego County Public Health Laboratory | Andersen lab at Scripps Research | SEARCH Alliance San Diego with Tracy Basler et al |
| EPI_ISL_635617 | USA/CA-ALSR-2666/2020 | 2020-07-09 | North America/USA/California/San Dieg | San Diego County Public Health Laboratory | Andersen lab at Scripps Research | SEARCH Alliance San Diego with Tracy Basler et al |
| EPI_ISL_635619 | USA/CA-ALSR-2668/2020 | 2020-07-12 | North America/USA/California/San Dieg | San Diego County Public Health Laboratory | Andersen lab at Scripps Research | SEARCH Alliance San Diego with Tracy Basler et al |
| EPI_ISL_635623 | USA/CA-ALSR-2674/2020 | 2020-07-10 | North America/USA/California/San Dieg | San Diego County Public Health Laboratory | Andersen lab at Scripps Research | SEARCH Alliance San Diego with Tracy Basler et al |
| EPI_ISL_635632 | USA/CA-ALSR-2689/2020 | 2020-07-22 | North America/USA/California/San Dieg | San Diego County Public Health Laboratory | Andersen lab at Scripps Research | SEARCH Alliance San Diego with Tracy Basler et al |
| EPI_ISL_635639 | USA/CA-ALSR-2707/2020 | 2020-07-15 | North America/USA/California/San Dieg | San Diego County Public Health Laboratory | Andersen lab at Scripps Research | SEARCH Alliance San Diego with Tracy Basler et al |
| EPI_ISL_635645 | USA/CA-ALSR-2726/2020 | 2020-07-15 | North America/USA/California/San Dieg | San Diego County Public Health Laboratory | Andersen lab at Scripps Research | SEARCH Alliance San Diego with Tracy Basler et al |
| EPI_ISL_635657 | USA/CA-ALSR-2747/2020 | 2020-07-16 | North America/USA/California/San Dieg | San Diego County Public Health Laboratory | Andersen lab at Scripps Research | SEARCH Alliance San Diego with Tracy Basler et al |
| EPI_ISL_635659 | USA/CA-ALSR-2749/2020 | 2020-07-14 | North America/USA/California/San Dieg | San Diego County Public Health Laboratory | Andersen lab at Scripps Research | SEARCH Alliance San Diego with Tracy Basler et al |
| EPI_ISL_635670 | USA/CA-ALSR-2769/2020 | 2020-07-14 | North America/USA/California/San Dieg | San Diego County Public Health Laboratory | Andersen lab at Scripps Research | SEARCH Alliance San Diego with Tracy Basler et al |
| EPI_ISL_635673 | USA/CA-ALSR-2775/2020 | 2020-07-15 | North America/USA/California/San Dieg | San Diego County Public Health Laboratory | Andersen lab at Scripps Research | SEARCH Alliance San Diego with Tracy Basler et al |
| EPI_ISL_635676 | USA/CA-ALSR-2781/2020 | 2020-07-15 | North America/USA/California/San Dieg | San Diego County Public Health Laboratory | Andersen lab at Scripps Research | SEARCH Alliance San Diego with Tracy Basler et al |
| EPI_ISL_635682 | USA/CA-ALSR-2795/2020 | 2020-07-23 | North America/USA/California/San Dieg | San Diego County Public Health Laboratory | Andersen lab at Scripps Research | SEARCH Alliance San Diego with Tracy Basler et al |
| EPI_ISL_635683 | USA/CA-ALSR-2800/2020 | 2020-07-24 | North America/USA/California/San Dieg | San Diego County Public Health Laboratory | Andersen lab at Scripps Research | SEARCH Alliance San Diego with Tracy Basler et al |
| EPI_ISL_635686 | USA/CA-ALSR-2803/2020 | 2020-07-22 | North America/USA/California/San Dieg | San Diego County Public Health Laboratory | Andersen lab at Scripps Research | SEARCH Alliance San Diego with Tracy Basler et al |
| EPI_ISL_635687 | USA/CA-ALSR-2805/2020 | 2020-07-24 | North America/USA/California/San Dieg | San Diego County Public Health Laboratory | Andersen lab at Scripps Research | SEARCH Alliance San Diego with Tracy Basler et al |
| EPI_ISL_635689 | USA/CA-ALSR-2808/2020 | 2020-07-22 | North America/USA/California/San Dieg | San Diego County Public Health Laboratory | Andersen lab at Scripps Research | SEARCH Alliance San Diego with Tracy Basler et al |
| EPI_ISL_635710 | USA/CA-ALSR-2844/2020 | 2020-07-24 | North America/USA/California/San Dieg | San Diego County Public Health Laboratory | Andersen lab at Scripps Research | SEARCH Alliance San Diego with Tracy Basler et al |
| EPI_ISL_635711 | USA/CA-ALSR-2849/2020 | 2020-07-28 | North America/USA/California/San Dieg | San Diego County Public Health Laboratory | Andersen lab at Scripps Research | SEARCH Alliance San Diego with Tracy Basler et al |

| | | | | | | |
|---|---|---|---|---|---|---|
| EPI_ISL_635713 | USA/CA-ALSR-2854/2020 | 2020-07-29 | North America/USA/California/San Dieg | San Diego County Public Health Laboratory | Andersen lab at Scripps Research | SEARCH Alliance San Diego with Tracy Basler et al |
| EPI_ISL_635715 | USA/CA-ALSR-2856/2020 | 2020-07-24 | North America/USA/California/San Dieg | San Diego County Public Health Laboratory | Andersen lab at Scripps Research | SEARCH Alliance San Diego with Tracy Basler et al |
| EPI_ISL_635722 | USA/CA-ALSR-2880/2020 | 2020-07-13 | North America/USA/California/San Dieg | San Diego County Public Health Laboratory | Andersen lab at Scripps Research | SEARCH Alliance San Diego with Tracy Basler et al |
| EPI_ISL_635724 | USA/CA-ALSR-2890/2020 | 2020-07-16 | North America/USA/California/San Dieg | San Diego County Public Health Laboratory | Andersen lab at Scripps Research | SEARCH Alliance San Diego with Tracy Basler et al |
| EPI_ISL_635726 | USA/CA-ALSR-2896/2020 | 2020-07-11 | North America/USA/California/San Dieg | San Diego County Public Health Laboratory | Andersen lab at Scripps Research | SEARCH Alliance San Diego with Tracy Basler et al |
| EPI_ISL_635730 | USA/CA-ALSR-2905/2020 | 2020-07-16 | North America/USA/California/San Dieg | San Diego County Public Health Laboratory | Andersen lab at Scripps Research | SEARCH Alliance San Diego with Tracy Basler et al |
| EPI_ISL_635731 | USA/CA-ALSR-2909/2020 | 2020-08-03 | North America/USA/California/San Dieg | San Diego County Public Health Laboratory | Andersen lab at Scripps Research | SEARCH Alliance San Diego with Tracy Basler et al |
| EPI_ISL_635739 | USA/CA-ALSR-2936/2020 | 2020-07-16 | North America/USA/California/San Dieg | San Diego County Public Health Laboratory | Andersen lab at Scripps Research | SEARCH Alliance San Diego with Tracy Basler et al |
| EPI_ISL_635743 | USA/CA-ALSR-2941/2020 | 2020-07-21 | North America/USA/California/San Dieg | San Diego County Public Health Laboratory | Andersen lab at Scripps Research | SEARCH Alliance San Diego with Tracy Basler et al |
| EPI_ISL_635748 | USA/CA-ALSR-2958/2020 | 2020-07-18 | North America/USA/California/San Dieg | San Diego County Public Health Laboratory | Andersen lab at Scripps Research | SEARCH Alliance San Diego with Tracy Basler et al |
| EPI_ISL_635749 | USA/CA-ALSR-2959/2020 | 2020-09-01 | North America/USA/California/San Dieg | San Diego County Public Health Laboratory | Andersen lab at Scripps Research | SEARCH Alliance San Diego with Tracy Basler et al |
| EPI_ISL_635754 | USA/CA-ALSR-2964/2020 | 2020-09-01 | North America/USA/California/San Dieg | San Diego County Public Health Laboratory | Andersen lab at Scripps Research | SEARCH Alliance San Diego with Tracy Basler et al |
| EPI_ISL_635756 | USA/CA-ALSR-2966/2020 | 2020-09-01 | North America/USA/California/San Dieg | San Diego County Public Health Laboratory | Andersen lab at Scripps Research | SEARCH Alliance San Diego with Tracy Basler et al |
| EPI_ISL_635761 | USA/CA-ALSR-2971/2020 | 2020-09-02 | North America/USA/California/San Dieg | San Diego County Public Health Laboratory | Andersen lab at Scripps Research | SEARCH Alliance San Diego with Tracy Basler et al |
| EPI_ISL_635763 | USA/CA-ALSR-2973/2020 | 2020-09-02 | North America/USA/California/San Dieg | San Diego County Public Health Laboratory | Andersen lab at Scripps Research | SEARCH Alliance San Diego with Tracy Basler et al |
| EPI_ISL_635765 | USA/CA-ALSR-2975/2020 | 2020-09-02 | North America/USA/California/San Dieg | San Diego County Public Health Laboratory | Andersen lab at Scripps Research | SEARCH Alliance San Diego with Tracy Basler et al |
| EPI_ISL_635766 | USA/CA-ALSR-2976/2020 | 2020-09-02 | North America/USA/California/San Dieg | San Diego County Public Health Laboratory | Andersen lab at Scripps Research | SEARCH Alliance San Diego with Tracy Basler et al |
| EPI_ISL_635767 | USA/CA-ALSR-2977/2020 | 2020-09-02 | North America/USA/California/San Dieg | San Diego County Public Health Laboratory | Andersen lab at Scripps Research | SEARCH Alliance San Diego with Tracy Basler et al |
| EPI_ISL_635771 | USA/CA-ALSR-2982/2020 | 2020-09-02 | North America/USA/California/San Dieg | San Diego County Public Health Laboratory | Andersen lab at Scripps Research | SEARCH Alliance San Diego with Tracy Basler et al |
| EPI_ISL_635772 | USA/CA-ALSR-2984/2020 | 2020-08-30 | North America/USA/California/San Dieg | San Diego County Public Health Laboratory | Andersen lab at Scripps Research | SEARCH Alliance San Diego with Tracy Basler et al |

| | | | | | | |
|---|---|---|---|---|---|---|
| EPI_ISL_635774 | USA/CA-ALSR-2987/2020 | 2020-08-30 | North America/USA/California/San Dieg | San Diego County Public Health Laboratory | Andersen lab at Scripps Research | SEARCH Alliance San Diego with Tracy Basler et al |
| EPI_ISL_635783 | USA/CA-ALSR-3009/2020 | 2020-07-17 | North America/USA/California/San Dieg | San Diego County Public Health Laboratory | Andersen lab at Scripps Research | SEARCH Alliance San Diego with Tracy Basler et al |
| EPI_ISL_635786 | USA/CA-ALSR-3012/2020 | 2020-07-12 | North America/USA/California/San Dieg | San Diego County Public Health Laboratory | Andersen lab at Scripps Research | SEARCH Alliance San Diego with Tracy Basler et al |
| EPI_ISL_635796 | USA/CA-ALSR-3026/2020 | 2020-09-04 | North America/USA/California/San Dieg | San Diego County Public Health Laboratory | Andersen lab at Scripps Research | SEARCH Alliance San Diego with Tracy Basler et al |
| EPI_ISL_635801 | USA/CA-ALSR-3032/2020 | 2020-08-27 | North America/USA/California/San Dieg | San Diego County Public Health Laboratory | Andersen lab at Scripps Research | SEARCH Alliance San Diego with Tracy Basler et al |
| EPI_ISL_635806 | USA/CA-ALSR-3037/2020 | 2020-09-03 | North America/USA/California/San Dieg | San Diego County Public Health Laboratory | Andersen lab at Scripps Research | SEARCH Alliance San Diego with Tracy Basler et al |
| EPI_ISL_635807 | USA/CA-ALSR-3038/2020 | 2020-09-03 | North America/USA/California/San Dieg | San Diego County Public Health Laboratory | Andersen lab at Scripps Research | SEARCH Alliance San Diego with Tracy Basler et al |
| EPI_ISL_635808 | USA/CA-ALSR-3040/2020 | 2020-08-28 | North America/USA/California/San Dieg | San Diego County Public Health Laboratory | Andersen lab at Scripps Research | SEARCH Alliance San Diego with Tracy Basler et al |
| EPI_ISL_635809 | USA/CA-ALSR-3043/2020 | 2020-07-31 | North America/USA/California/San Dieg | San Diego County Public Health Laboratory | Andersen lab at Scripps Research | SEARCH Alliance San Diego with Tracy Basler et al |
| EPI_ISL_635817 | USA/CA-ALSR-3053/2020 | 2020-09-04 | North America/USA/California/San Dieg | San Diego County Public Health Laboratory | Andersen lab at Scripps Research | SEARCH Alliance San Diego with Tracy Basler et al |
| EPI_ISL_635818 | USA/CA-ALSR-3054/2020 | 2020-09-04 | North America/USA/California/San Dieg | San Diego County Public Health Laboratory | Andersen lab at Scripps Research | SEARCH Alliance San Diego with Tracy Basler et al |
| EPI_ISL_635825 | USA/CA-ALSR-3066/2020 | 2020-09-01 | North America/USA/California/San Dieg | San Diego County Public Health Laboratory | Andersen lab at Scripps Research | SEARCH Alliance San Diego with Tracy Basler et al |
| EPI_ISL_635826 | USA/CA-ALSR-3067/2020 | 2020-08-27 | North America/USA/California/San Dieg | San Diego County Public Health Laboratory | Andersen lab at Scripps Research | SEARCH Alliance San Diego with Tracy Basler et al |
| EPI_ISL_635830 | USA/CA-ALSR-3076/2020 | 2020-09-02 | North America/USA/California/San Dieg | San Diego County Public Health Laboratory | Andersen lab at Scripps Research | SEARCH Alliance San Diego with Tracy Basler et al |
| EPI_ISL_635832 | USA/CA-ALSR-3079/2020 | 2020-08-27 | North America/USA/California/San Dieg | San Diego County Public Health Laboratory | Andersen lab at Scripps Research | SEARCH Alliance San Diego with Tracy Basler et al |
| EPI_ISL_635834 | USA/CA-ALSR-3082/2020 | 2020-09-04 | North America/USA/California/San Dieg | San Diego County Public Health Laboratory | Andersen lab at Scripps Research | SEARCH Alliance San Diego with Tracy Basler et al |
| EPI_ISL_635838 | USA/CA-ALSR-3086/2020 | 2020-09-02 | North America/USA/California/San Dieg | San Diego County Public Health Laboratory | Andersen lab at Scripps Research | SEARCH Alliance San Diego with Tracy Basler et al |
| EPI_ISL_635840 | USA/CA-ALSR-3088/2020 | 2020-09-04 | North America/USA/California/San Dieg | San Diego County Public Health Laboratory | Andersen lab at Scripps Research | SEARCH Alliance San Diego with Tracy Basler et al |
| EPI_ISL_635851 | USA/CA-ALSR-3104/2020 | 2020-07-14 | North America/USA/California/San Dieg | San Diego County Public Health Laboratory | Andersen lab at Scripps Research | SEARCH Alliance San Diego with Tracy Basler et al |
| EPI_ISL_635858 | USA/CA-ALSR-3120/2020 | 2020-07-14 | North America/USA/California/San Dieg | San Diego County Public Health Laboratory | Andersen lab at Scripps Research | SEARCH Alliance San Diego with Tracy Basler et al |

| | | | | | | |
|---|---|---|---|---|---|---|
| EPI_ISL_635862 | USA/CA-ALSR-3129/2020 | 2020-08-24 | North America/USA/California/San Dieg | San Diego County Public Health Laboratory | Andersen lab at Scripps Research | SEARCH Alliance San Diego with Tracy Basler et al |
| EPI_ISL_635868 | USA/CA-ALSR-3137/2020 | 2020-08-21 | North America/USA/California/San Dieg | San Diego County Public Health Laboratory | Andersen lab at Scripps Research | SEARCH Alliance San Diego with Tracy Basler et al |
| EPI_ISL_635871 | USA/CA-ALSR-3141/2020 | 2020-08-19 | North America/USA/California/San Dieg | San Diego County Public Health Laboratory | Andersen lab at Scripps Research | SEARCH Alliance San Diego with Tracy Basler et al |
| EPI_ISL_635878 | USA/CA-ALSR-3148/2020 | 2020-08-20 | North America/USA/California/San Dieg | San Diego County Public Health Laboratory | Andersen lab at Scripps Research | SEARCH Alliance San Diego with Tracy Basler et al |
| EPI_ISL_635884 | USA/CA-ALSR-3156/2020 | 2020-08-21 | North America/USA/California/San Dieg | San Diego County Public Health Laboratory | Andersen lab at Scripps Research | SEARCH Alliance San Diego with Tracy Basler et al |
| EPI_ISL_635886 | USA/CA-ALSR-3158/2020 | 2020-08-19 | North America/USA/California/San Dieg | San Diego County Public Health Laboratory | Andersen lab at Scripps Research | SEARCH Alliance San Diego with Tracy Basler et al |
| EPI_ISL_635887 | USA/CA-ALSR-3159/2020 | 2020-08-19 | North America/USA/California/San Dieg | San Diego County Public Health Laboratory | Andersen lab at Scripps Research | SEARCH Alliance San Diego with Tracy Basler et al |
| EPI_ISL_635888 | USA/CA-ALSR-3160/2020 | 2020-08-19 | North America/USA/California/San Dieg | San Diego County Public Health Laboratory | Andersen lab at Scripps Research | SEARCH Alliance San Diego with Tracy Basler et al |
| EPI_ISL_635890 | USA/CA-ALSR-3162/2020 | 2020-07-13 | North America/USA/California/San Dieg | San Diego County Public Health Laboratory | Andersen lab at Scripps Research | SEARCH Alliance San Diego with Tracy Basler et al |
| EPI_ISL_635891 | USA/CA-ALSR-3163/2020 | 2020-08-26 | North America/USA/California/San Dieg | San Diego County Public Health Laboratory | Andersen lab at Scripps Research | SEARCH Alliance San Diego with Tracy Basler et al |
| EPI_ISL_635897 | USA/CA-ALSR-3170/2020 | 2020-06-03 | North America/USA/California/San Dieg | San Diego County Public Health Laboratory | Andersen lab at Scripps Research | SEARCH Alliance San Diego with Tracy Basler et al |
| EPI_ISL_635903 | USA/CA-ALSR-3181/2020 | 2020-06-01 | North America/USA/California/San Dieg | San Diego County Public Health Laboratory | Andersen lab at Scripps Research | SEARCH Alliance San Diego with Tracy Basler et al |
| EPI_ISL_635913 | USA/CA-ALSR-3212/2020 | 2020-07-31 | North America/USA/California/San Dieg | San Diego County Public Health Laboratory | Andersen lab at Scripps Research | SEARCH Alliance San Diego with Tracy Basler et al |
| EPI_ISL_635924 | USA/CA-ALSR-3228/2020 | 2020-07-29 | North America/USA/California/San Dieg | San Diego County Public Health Laboratory | Andersen lab at Scripps Research | SEARCH Alliance San Diego with Tracy Basler et al |
| EPI_ISL_635925 | USA/CA-ALSR-3229/2020 | 2020-08-19 | North America/USA/California/San Dieg | San Diego County Public Health Laboratory | Andersen lab at Scripps Research | SEARCH Alliance San Diego with Tracy Basler et al |
| EPI_ISL_635926 | USA/CA-ALSR-3232/2020 | 2020-07-29 | North America/USA/California/San Dieg | San Diego County Public Health Laboratory | Andersen lab at Scripps Research | SEARCH Alliance San Diego with Tracy Basler et al |
| EPI_ISL_635928 | USA/CA-ALSR-3237/2020 | 2020-07-30 | North America/USA/California/San Dieg | San Diego County Public Health Laboratory | Andersen lab at Scripps Research | SEARCH Alliance San Diego with Tracy Basler et al |
| EPI_ISL_635931 | USA/CA-ALSR-3243/2020 | 2020-07-14 | North America/USA/California/San Dieg | San Diego County Public Health Laboratory | Andersen lab at Scripps Research | SEARCH Alliance San Diego with Tracy Basler et al |
| EPI_ISL_635938 | USA/CA-ALSR-3263/2020 | 2020-09-07 | North America/USA/California/San Dieg | San Diego County Public Health Laboratory | Andersen lab at Scripps Research | SEARCH Alliance San Diego with Tracy Basler et al |
| EPI_ISL_635940 | USA/CA-ALSR-3265/2020 | 2020-09-06 | North America/USA/California/San Dieg | San Diego County Public Health Laboratory | Andersen lab at Scripps Research | SEARCH Alliance San Diego with Tracy Basler et al |

| | | | | | | |
|---|---|---|---|---|---|---|
| EPI_ISL_635941 | USA/CA-ALSR-3266/2020 | 2020-09-06 | North America/USA/California/San Dieg | San Diego County Public Health Laboratory | Andersen lab at Scripps Research | SEARCH Alliance San Diego with Tracy Basler et al |
| EPI_ISL_635945 | USA/CA-ALSR-3270/2020 | 2020-09-08 | North America/USA/California/San Dieg | San Diego County Public Health Laboratory | Andersen lab at Scripps Research | SEARCH Alliance San Diego with Tracy Basler et al |
| EPI_ISL_635948 | USA/CA-ALSR-3281/2020 | 2020-09-17 | North America/USA/California/San Dieg | San Diego County Public Health Laboratory | Andersen lab at Scripps Research | SEARCH Alliance San Diego with Tracy Basler et al |
| EPI_ISL_635951 | USA/CA-ALSR-3289/2020 | 2020-09-17 | North America/USA/California/San Dieg | San Diego County Public Health Laboratory | Andersen lab at Scripps Research | SEARCH Alliance San Diego with Tracy Basler et al |
| EPI_ISL_635954 | USA/CA-ALSR-3304/2020 | 2020-09-21 | North America/USA/California/San Dieg | San Diego County Public Health Laboratory | Andersen lab at Scripps Research | SEARCH Alliance San Diego with Tracy Basler et al |
| EPI_ISL_635956 | USA/CA-ALSR-3310/2020 | 2020-09-22 | North America/USA/California/San Dieg | San Diego County Public Health Laboratory | Andersen lab at Scripps Research | SEARCH Alliance San Diego with Tracy Basler et al |
| EPI_ISL_635963 | USA/CA-ALSR-3323/2020 | 2020-09-22 | North America/USA/California/San Dieg | San Diego County Public Health Laboratory | Andersen lab at Scripps Research | SEARCH Alliance San Diego with Tracy Basler et al |
| EPI_ISL_635967 | USA/CA-ALSR-3331/2020 | 2020-09-17 | North America/USA/California/San Dieg | San Diego County Public Health Laboratory | Andersen lab at Scripps Research | SEARCH Alliance San Diego with Tracy Basler et al |
| EPI_ISL_635969 | USA/CA-ALSR-3337/2020 | 2020-09-17 | North America/USA/California/San Dieg | San Diego County Public Health Laboratory | Andersen lab at Scripps Research | SEARCH Alliance San Diego with Tracy Basler et al |
| EPI_ISL_635972 | USA/CA-ALSR-3341/2020 | 2020-09-17 | North America/USA/California/San Dieg | San Diego County Public Health Laboratory | Andersen lab at Scripps Research | SEARCH Alliance San Diego with Tracy Basler et al |
| EPI_ISL_635975 | USA/CA-ALSR-3351/2020 | 2020-09-18 | North America/USA/California/San Dieg | San Diego County Public Health Laboratory | Andersen lab at Scripps Research | SEARCH Alliance San Diego with Tracy Basler et al |
| EPI_ISL_635978 | USA/CA-ALSR-3355/2020 | 2020-09-18 | North America/USA/California/San Dieg | San Diego County Public Health Laboratory | Andersen lab at Scripps Research | SEARCH Alliance San Diego with Tracy Basler et al |
| EPI_ISL_635979 | USA/CA-ALSR-3357/2020 | 2020-09-18 | North America/USA/California/San Dieg | San Diego County Public Health Laboratory | Andersen lab at Scripps Research | SEARCH Alliance San Diego with Tracy Basler et al |
| EPI_ISL_635983 | USA/CA-ALSR-3366/2020 | 2020-05-30 | North America/USA/California/San Dieg | San Diego County Public Health Laboratory | Andersen lab at Scripps Research | SEARCH Alliance San Diego with Tracy Basler et al |
| EPI_ISL_635984 | USA/CA-ALSR-3369/2020 | 2020-05-28 | North America/USA/California/San Dieg | San Diego County Public Health Laboratory | Andersen lab at Scripps Research | SEARCH Alliance San Diego with Tracy Basler et al |
| EPI_ISL_635985 | USA/CA-ALSR-3370/2020 | 2020-08-19 | North America/USA/California/San Dieg | San Diego County Public Health Laboratory | Andersen lab at Scripps Research | SEARCH Alliance San Diego with Tracy Basler et al |
| EPI_ISL_635988 | USA/CA-ALSR-3375/2020 | 2020-07-08 | North America/USA/California/San Dieg | San Diego County Public Health Laboratory | Andersen lab at Scripps Research | SEARCH Alliance San Diego with Tracy Basler et al |
| EPI_ISL_635999 | USA/CA-ALSR-3392/2020 | 2020-07-09 | North America/USA/California/San Dieg | San Diego County Public Health Laboratory | Andersen lab at Scripps Research | SEARCH Alliance San Diego with Tracy Basler et al |
| EPI_ISL_636002 | USA/CA-ALSR-3396/2020 | 2020-08-10 | North America/USA/California/San Dieg | San Diego County Public Health Laboratory | Andersen lab at Scripps Research | SEARCH Alliance San Diego with Tracy Basler et al |
| EPI_ISL_636006 | USA/CA-ALSR-3404/2020 | 2020-07-07 | North America/USA/California/San Dieg | San Diego County Public Health Laboratory | Andersen lab at Scripps Research | SEARCH Alliance San Diego with Tracy Basler et al |

| | | | | | | |
|---|---|---|---|---|---|---|
| EPI_ISL_636009 | USA/CA-ALSR-3407/2020 | 2020-07-07 | North America/USA/California/San Dieg | San Diego County Public Health Laboratory | Andersen lab at Scripps Research | SEARCH Alliance San Diego with Tracy Basler et al |
| EPI_ISL_636010 | USA/CA-ALSR-3408/2020 | 2020-07-07 | North America/USA/California/San Dieg | San Diego County Public Health Laboratory | Andersen lab at Scripps Research | SEARCH Alliance San Diego with Tracy Basler et al |
| EPI_ISL_636024 | USA/CA-ALSR-3430/2020 | 2020-05-30 | North America/USA/California/San Dieg | San Diego County Public Health Laboratory | Andersen lab at Scripps Research | SEARCH Alliance San Diego with Tracy Basler et al |
| EPI_ISL_636026 | USA/CA-ALSR-3434/2020 | 2020-05-30 | North America/USA/California/San Dieg | San Diego County Public Health Laboratory | Andersen lab at Scripps Research | SEARCH Alliance San Diego with Tracy Basler et al |
| EPI_ISL_636028 | USA/CA-ALSR-3436/2020 | 2020-05-31 | North America/USA/California/San Dieg | San Diego County Public Health Laboratory | Andersen lab at Scripps Research | SEARCH Alliance San Diego with Tracy Basler et al |
| EPI_ISL_636035 | USA/CA-ALSR-3448/2020 | 2020-08-12 | North America/USA/California/San Dieg | San Diego County Public Health Laboratory | Andersen lab at Scripps Research | SEARCH Alliance San Diego with Tracy Basler et al |
| EPI_ISL_636036 | USA/CA-ALSR-3451/2020 | 2020-09-17 | North America/USA/California/San Dieg | San Diego County Public Health Laboratory | Andersen lab at Scripps Research | SEARCH Alliance San Diego with Tracy Basler et al |
| EPI_ISL_636038 | USA/CA-ALSR-3458/2020 | 2020-09-15 | North America/USA/California/San Dieg | San Diego County Public Health Laboratory | Andersen lab at Scripps Research | SEARCH Alliance San Diego with Tracy Basler et al |
| EPI_ISL_636039 | USA/CA-ALSR-3460/2020 | 2020-09-15 | North America/USA/California/San Dieg | San Diego County Public Health Laboratory | Andersen lab at Scripps Research | SEARCH Alliance San Diego with Tracy Basler et al |
| EPI_ISL_636044 | USA/CA-ALSR-3472/2020 | 2020-08-18 | North America/USA/California/San Dieg | San Diego County Public Health Laboratory | Andersen lab at Scripps Research | SEARCH Alliance San Diego with Tracy Basler et al |
| EPI_ISL_636052 | USA/CA-ALSR-3496/2020 | 2020-09-10 | North America/USA/California/San Dieg | San Diego County Public Health Laboratory | Andersen lab at Scripps Research | SEARCH Alliance San Diego with Tracy Basler et al |
| EPI_ISL_636054 | USA/CA-ALSR-3498/2020 | 2020-09-08 | North America/USA/California/San Dieg | San Diego County Public Health Laboratory | Andersen lab at Scripps Research | SEARCH Alliance San Diego with Tracy Basler et al |
| EPI_ISL_636056 | USA/CA-ALSR-3501/2020 | 2020-09-08 | North America/USA/California/San Dieg | San Diego County Public Health Laboratory | Andersen lab at Scripps Research | SEARCH Alliance San Diego with Tracy Basler et al |
| EPI_ISL_636057 | USA/CA-ALSR-3505/2020 | 2020-09-17 | North America/USA/California/San Dieg | San Diego County Public Health Laboratory | Andersen lab at Scripps Research | SEARCH Alliance San Diego with Tracy Basler et al |
| EPI_ISL_636061 | USA/CA-ALSR-3512/2020 | 2020-09-08 | North America/USA/California/San Dieg | San Diego County Public Health Laboratory | Andersen lab at Scripps Research | SEARCH Alliance San Diego with Tracy Basler et al |
| EPI_ISL_636064 | USA/CA-ALSR-3518/2020 | 2020-07-15 | North America/USA/California/San Dieg | San Diego County Public Health Laboratory | Andersen lab at Scripps Research | SEARCH Alliance San Diego with Tracy Basler et al |
| EPI_ISL_636067 | USA/CA-ALSR-3522/2020 | 2020-09-15 | North America/USA/California/San Dieg | San Diego County Public Health Laboratory | Andersen lab at Scripps Research | SEARCH Alliance San Diego with Tracy Basler et al |
| EPI_ISL_636079 | USA/CA-ALSR-3551/2020 | 2020-06-03 | North America/USA/California/San Dieg | San Diego County Public Health Laboratory | Andersen lab at Scripps Research | SEARCH Alliance San Diego with Tracy Basler et al |
| EPI_ISL_636085 | USA/CA-ALSR-3563/2020 | 2020-06-15 | North America/USA/California/San Dieg | San Diego County Public Health Laboratory | Andersen lab at Scripps Research | SEARCH Alliance San Diego with Tracy Basler et al |
| EPI_ISL_636097 | USA/CA-ALSR-3585/2020 | 2020-06-24 | North America/USA/California/San Dieg | San Diego County Public Health Laboratory | Andersen lab at Scripps Research | SEARCH Alliance San Diego with Tracy Basler et al |

| | | | | | | |
|---|---|---|---|---|---|---|
| EPI_ISL_636113 | Mexico/BCN-ALSR-3618/2020 | 2020-06-03 | North America/Mexico/Baja California/T | Sharp HealthCare Laboratory | Andersen lab at Scripps Research | SEARCH Alliance San Diego with Aaron Harding et al |
| EPI_ISL_636114 | Mexico/BCN-ALSR-3619/2020 | 2020-06-04 | North America/Mexico/Baja California/T | Sharp HealthCare Laboratory | Andersen lab at Scripps Research | SEARCH Alliance San Diego with Aaron Harding et al |
| EPI_ISL_636115 | Mexico/BCN-ALSR-3622/2020 | 2020-05-29 | North America/Mexico/Baja California/T | Sharp HealthCare Laboratory | Andersen lab at Scripps Research | SEARCH Alliance San Diego with Aaron Harding et al |
| EPI_ISL_636116 | Mexico/BCN-ALSR-3624/2020 | 2020-06-12 | North America/Mexico/Sonora/San Luis | Sharp HealthCare Laboratory | Andersen lab at Scripps Research | SEARCH Alliance San Diego with Aaron Harding et al |
| EPI_ISL_636117 | Mexico/BCN-ALSR-3630/2020 | 2020-06-12 | North America/Mexico/Baja California/N | Sharp HealthCare Laboratory | Andersen lab at Scripps Research | SEARCH Alliance San Diego with Aaron Harding et al |
| EPI_ISL_636118 | Mexico/BCN-ALSR-3634/2020 | 2020-05-27 | North America/Mexico/Baja California/T | Sharp HealthCare Laboratory | Andersen lab at Scripps Research | SEARCH Alliance San Diego with Aaron Harding et al |
| EPI_ISL_636119 | Mexico/BCN-ALSR-3636/2020 | 2020-05-28 | North America/Mexico/Baja California/T | Sharp HealthCare Laboratory | Andersen lab at Scripps Research | SEARCH Alliance San Diego with Aaron Harding et al |
| EPI_ISL_636122 | USA/CA-ALSR-3644/2020 | 2020-09-28 | North America/USA/California/San Dieg | San Diego County Public Health Laboratory | Andersen lab at Scripps Research | SEARCH Alliance San Diego with Tracy Basler et al |
| EPI_ISL_636123 | USA/CA-ALSR-3645/2020 | 2020-09-28 | North America/USA/California/San Dieg | San Diego County Public Health Laboratory | Andersen lab at Scripps Research | SEARCH Alliance San Diego with Tracy Basler et al |
| EPI_ISL_636130 | USA/CA-ALSR-3655/2020 | 2020-09-25 | North America/USA/California/San Dieg | San Diego County Public Health Laboratory | Andersen lab at Scripps Research | SEARCH Alliance San Diego with Tracy Basler et al |
| EPI_ISL_636133 | USA/CA-ALSR-3660/2020 | 2020-05-27 | North America/USA/California/San Dieg | San Diego County Public Health Laboratory | Andersen lab at Scripps Research | SEARCH Alliance San Diego with Tracy Basler et al |
| EPI_ISL_636141 | USA/CA-ALSR-3670/2020 | 2020-09-28 | North America/USA/California/San Dieg | San Diego County Public Health Laboratory | Andersen lab at Scripps Research | SEARCH Alliance San Diego with Tracy Basler et al |
| EPI_ISL_636143 | USA/CA-ALSR-3676/2020 | 2020-05-28 | North America/USA/California/San Dieg | San Diego County Public Health Laboratory | Andersen lab at Scripps Research | SEARCH Alliance San Diego with Tracy Basler et al |
| EPI_ISL_636145 | USA/CA-ALSR-3680/2020 | 2020-09-23 | North America/USA/California/San Dieg | San Diego County Public Health Laboratory | Andersen lab at Scripps Research | SEARCH Alliance San Diego with Tracy Basler et al |
| EPI_ISL_636146 | USA/CA-ALSR-3684/2020 | 2020-09-25 | North America/USA/California/San Dieg | San Diego County Public Health Laboratory | Andersen lab at Scripps Research | SEARCH Alliance San Diego with Tracy Basler et al |
| EPI_ISL_636150 | USA/CA-ALSR-3691/2020 | 2020-09-28 | North America/USA/California/San Dieg | San Diego County Public Health Laboratory | Andersen lab at Scripps Research | SEARCH Alliance San Diego with Tracy Basler et al |
| EPI_ISL_636154 | USA/CA-ALSR-3700/2020 | 2020-09-23 | North America/USA/California/San Dieg | San Diego County Public Health Laboratory | Andersen lab at Scripps Research | SEARCH Alliance San Diego with Tracy Basler et al |
| EPI_ISL_636156 | USA/CA-ALSR-3705/2020 | 2020-09-25 | North America/USA/California/San Dieg | San Diego County Public Health Laboratory | Andersen lab at Scripps Research | SEARCH Alliance San Diego with Tracy Basler et al |
| EPI_ISL_636157 | USA/CA-ALSR-3706/2020 | 2020-09-28 | North America/USA/California/San Dieg | San Diego County Public Health Laboratory | Andersen lab at Scripps Research | SEARCH Alliance San Diego with Tracy Basler et al |
| EPI_ISL_636158 | USA/CA-ALSR-3707/2020 | 2020-09-28 | North America/USA/California/San Dieg | San Diego County Public Health Laboratory | Andersen lab at Scripps Research | SEARCH Alliance San Diego with Tracy Basler et al |

| | | | | | | |
|---|---|---|---|---|---|---|
| EPI_ISL_636159 | USA/CA-ALSR-3708/2020 | 2020-09-23 | North America/USA/California/San Dieg | San Diego County Public Health Laboratory | Andersen lab at Scripps Research | SEARCH Alliance San Diego with Tracy Basler et al |
| EPI_ISL_636168 | USA/CA-ALSR-3725/2020 | 2020-05-27 | North America/USA/California/San Dieg | San Diego County Public Health Laboratory | Andersen lab at Scripps Research | SEARCH Alliance San Diego with Tracy Basler et al |
| EPI_ISL_636172 | USA/CA-ALSR-3731/2020 | 2020-06-05 | North America/USA/California/San Dieg | San Diego County Public Health Laboratory | Andersen lab at Scripps Research | SEARCH Alliance San Diego with Tracy Basler et al |
| EPI_ISL_636176 | USA/CA-ALSR-3738/2020 | 2020-06-05 | North America/USA/California/San Dieg | San Diego County Public Health Laboratory | Andersen lab at Scripps Research | SEARCH Alliance San Diego with Tracy Basler et al |
| EPI_ISL_636177 | USA/CA-ALSR-3739/2020 | 2020-06-05 | North America/USA/California/San Dieg | San Diego County Public Health Laboratory | Andersen lab at Scripps Research | SEARCH Alliance San Diego with Tracy Basler et al |
| EPI_ISL_636180 | USA/CA-ALSR-3749/2020 | 2020-07-02 | North America/USA/California/San Dieg | San Diego County Public Health Laboratory | Andersen lab at Scripps Research | SEARCH Alliance San Diego with Tracy Basler et al |
| EPI_ISL_636183 | USA/CA-ALSR-3753/2020 | 2020-06-04 | North America/USA/California/San Dieg | San Diego County Public Health Laboratory | Andersen lab at Scripps Research | SEARCH Alliance San Diego with Tracy Basler et al |
| EPI_ISL_636191 | USA/CA-ALSR-3771/2020 | 2020-05-27 | North America/USA/California/San Dieg | San Diego County Public Health Laboratory | Andersen lab at Scripps Research | SEARCH Alliance San Diego with Tracy Basler et al |
| EPI_ISL_636197 | USA/CA-ALSR-3781/2020 | 2020-05-23 | North America/USA/California/San Dieg | San Diego County Public Health Laboratory | Andersen lab at Scripps Research | SEARCH Alliance San Diego with Tracy Basler et al |
| EPI_ISL_636202 | USA/CA-ALSR-3789/2020 | 2020-05-23 | North America/USA/California/San Dieg | San Diego County Public Health Laboratory | Andersen lab at Scripps Research | SEARCH Alliance San Diego with Tracy Basler et al |
| EPI_ISL_636213 | USA/CA-ALSR-3807/2020 | 2020-06-05 | North America/USA/California/San Dieg | San Diego County Public Health Laboratory | Andersen lab at Scripps Research | SEARCH Alliance San Diego with Tracy Basler et al |
| EPI_ISL_636214 | USA/CA-ALSR-3808/2020 | 2020-05-27 | North America/USA/California/San Dieg | San Diego County Public Health Laboratory | Andersen lab at Scripps Research | SEARCH Alliance San Diego with Tracy Basler et al |
| EPI_ISL_636215 | USA/CA-ALSR-3809/2020 | 2020-05-27 | North America/USA/California/San Dieg | San Diego County Public Health Laboratory | Andersen lab at Scripps Research | SEARCH Alliance San Diego with Tracy Basler et al |
| EPI_ISL_636229 | USA/CA-ALSR-3827/2020 | 2020-06-13 | North America/USA/California/San Dieg | San Diego County Public Health Laboratory | Andersen lab at Scripps Research | SEARCH Alliance San Diego with Tracy Basler et al |
| EPI_ISL_636231 | USA/CA-ALSR-3831/2020 | 2020-06-15 | North America/USA/California/San Dieg | San Diego County Public Health Laboratory | Andersen lab at Scripps Research | SEARCH Alliance San Diego with Tracy Basler et al |
| EPI_ISL_636235 | USA/CA-ALSR-3838/2020 | 2020-06-11 | North America/USA/California/San Dieg | San Diego County Public Health Laboratory | Andersen lab at Scripps Research | SEARCH Alliance San Diego with Tracy Basler et al |
| EPI_ISL_636240 | USA/CA-ALSR-3847/2020 | 2020-06-11 | North America/USA/California/San Dieg | San Diego County Public Health Laboratory | Andersen lab at Scripps Research | SEARCH Alliance San Diego with Tracy Basler et al |
| EPI_ISL_636246 | USA/CA-ALSR-3856/2020 | 2020-06-29 | North America/USA/California/San Dieg | San Diego County Public Health Laboratory | Andersen lab at Scripps Research | SEARCH Alliance San Diego with Tracy Basler et al |
| EPI_ISL_636247 | USA/CA-ALSR-3857/2020 | 2020-06-29 | North America/USA/California/San Dieg | San Diego County Public Health Laboratory | Andersen lab at Scripps Research | SEARCH Alliance San Diego with Tracy Basler et al |
| EPI_ISL_636249 | USA/CA-ALSR-3862/2020 | 2020-07-01 | North America/USA/California/San Dieg | San Diego County Public Health Laboratory | Andersen lab at Scripps Research | SEARCH Alliance San Diego with Tracy Basler et al |

| | | | | | | |
|---|---|---|---|---|---|---|
| EPI_ISL_636264 | USA/CA-ALSR-3895/2020 | 2020-07-01 | North America/USA/California/San Dieg | San Diego County Public Health Laboratory | Andersen lab at Scripps Research | SEARCH Alliance San Diego with Tracy Basler et al |
| EPI_ISL_636265 | USA/CA-ALSR-3898/2020 | 2020-07-01 | North America/USA/California/San Dieg | San Diego County Public Health Laboratory | Andersen lab at Scripps Research | SEARCH Alliance San Diego with Tracy Basler et al |
| EPI_ISL_636462 | Italy/VEN-IZSVe-10599/2020 | 2020-04-15 | Europe/Italy/Veneto/Padova | ULSS6 Euganea | Istituto Zooprofilattico Sperimentale delle Venezie | Adelaide Milani et al |
| EPI_ISL_636464 | Italy/VEN-IZSVe-1-1/2020 | 2020-03-21 | Europe/Italy/Veneto/Padova | ULSS6 Piove di Sacco | Istituto Zooprofilattico Sperimentale delle Venezie | Adelaide Milani et al |
| EPI_ISL_636471 | Italy/VEN-IZSVe-20-25/2020 | 2020-03-24 | Europe/Italy/Veneto/Padova | ULSS6 Piove di Sacco | Istituto Zooprofilattico Sperimentale delle Venezie | Adelaide Milani et al |
| EPI_ISL_636475 | Italy/VEN-IZSVe-6-55/2020 | 2020-03-22 | Europe/Italy/Veneto/Padova | ULSS6 Distretto Padova Terme Colli | Istituto Zooprofilattico Sperimentale delle Venezie | Adelaide Milani et al |
| EPI_ISL_636478 | France/ARA-33485/2020 | 2020-05-03 | Europe/France/Auvergne-Rhv¥ne-Alpes/ | Centre Hospitalier Pierre Oudot | CNR Virus des Infections Respiratoires - France SUD | Antonin Bal et al |
| EPI_ISL_636479 | France/ARA-33853/2020 | 2020-05-04 | Europe/France/Auvergne-Rhv¥ne-Alpes/ | CNR Virus des Infections Respiratoires - France SUD | CNR Virus des Infections Respiratoires - France SUD | Antonin Bal et al |
| EPI_ISL_636480 | France/ARA-34732/2020 | 2020-05-06 | Europe/France/Auvergne-Rhv¥ne-Alpes/ | Centre Hospitalier Pierre Oudot | CNR Virus des Infections Respiratoires - France SUD | Antonin Bal et al |
| EPI_ISL_636483 | France/ARA-36390/2020 | 2020-05-11 | Europe/France/Auvergne-Rhv¥ne-Alpes/ | Centre Hospitalier Pierre Oudot | CNR Virus des Infections Respiratoires - France SUD | Antonin Bal et al |
| EPI_ISL_636484 | France/ARA-36541/2020 | 2020-05-11 | Europe/France/Auvergne-Rhv¥ne-Alpes/ | CNR Virus des Infections Respiratoires - France SUD | CNR Virus des Infections Respiratoires - France SUD | Antonin Bal et al |

| | | | | | | |
|---|---|---|---|---|---|---|
| EPI_ISL_636486 | France/ARA-37154/2020 | 2020-05-13 | Europe/France/Auvergne-Rhv¥ne-Alpes/ | Centre Hospitalier Pierre Oudot | CNR Virus des Infections Respiratoires - France SUD | Antonin Bal et al |
| EPI_ISL_636489 | France/ARA33145/2020 | 2020-05-02 | Europe/France/Auvergne-Rhv¥ne-Alpes/ | CNR Virus des Infections Respiratoires - France SUD | CNR Virus des Infections Respiratoires - France SUD | Antonin Bal et al |
| EPI_ISL_636490 | France/ARA33273/2020 | 2020-05-02 | Europe/France/Auvergne-Rhv¥ne-Alpes/ | CNR Virus des Infections Respiratoires - France SUD | CNR Virus des Infections Respiratoires - France SUD | Antonin Bal et al |
| EPI_ISL_636498 | Netherlands/GE-RIVM-10328/2020 | 2020-03-21 | Europe/Netherlands/Gelderland/ | Dutch COVID-19 response team | National Institute for Public Health and the Environment (RIVM) | Adam Meijer et al |
| EPI_ISL_636504 | Netherlands/NB-RIVM-10300/2020 | 2020-02-27 | Europe/Netherlands/North Brabant/ | Dutch COVID-19 response team | National Institute for Public Health and the Environment (RIVM) | Adam Meijer et al |
| EPI_ISL_636533 | Netherlands/GE-RIVM-10374/2020 | 2020-09-23 | Europe/Netherlands/Gelderland/ | Dutch COVID-19 response team | National Institute for Public Health and the Environment (RIVM) | Adam Meijer et al |
| EPI_ISL_636553 | Netherlands/NB-RIVM-10318/2020 | 2020-03-11 | Europe/Netherlands/North Brabant/ | Dutch COVID-19 response team | National Institute for Public Health and the Environment (RIVM) | Adam Meijer et al |
| EPI_ISL_636559 | Netherlands/NB-RIVM-10397/2020 | 2020-10-19 | Europe/Netherlands/North Brabant/ | Dutch COVID-19 response team | National Institute for Public Health and the Environment (RIVM) | Adam Meijer et al |

| | | | | | | |
|---|---|---|---|---|---|---|
| EPI_ISL_636569 | Netherlands/NH-RIVM-10394/2020 | 2020-10-14 | Europe/Netherlands/North Holland/ | Dutch COVID-19 response team | National Institute for Public Health and the Environment (RIVM) | Adam Meijer et al |
| EPI_ISL_636580 | Netherlands/UT-RIVM-10380/2020 | 2020-10-05 | Europe/Netherlands/Utrecht/ | Dutch COVID-19 response team | National Institute for Public Health and the Environment (RIVM) | Adam Meijer et al |
| EPI_ISL_636582 | Netherlands/UT-RIVM-10384/2020 | 2020-10-08 | Europe/Netherlands/Utrecht/ | Dutch COVID-19 response team | National Institute for Public Health and the Environment (RIVM) | Adam Meijer et al |
| EPI_ISL_636583 | Netherlands/UT-RIVM-10400/2020 | 2020-10-22 | Europe/Netherlands/Utrecht/ | Dutch COVID-19 response team | National Institute for Public Health and the Environment (RIVM) | Adam Meijer et al |
| EPI_ISL_636590 | Netherlands/ZE-RIVM-10376/2020 | 2020-09-07 | Europe/Netherlands/Zeeland/ | Dutch COVID-19 response team | National Institute for Public Health and the Environment (RIVM) | Adam Meijer et al |
| EPI_ISL_636606 | Belgium/ULG-10627/2020 | 2020-10-27 | Europe/Belgium/Li√®ge/ | Department of Clinical Microbiology | GIGA Medical Genomics | Keith Durkin et al |
| EPI_ISL_636608 | Belgium/ULG-10629/2020 | 2020-10-26 | Europe/Belgium/Li√®ge/ | Department of Clinical Microbiology | GIGA Medical Genomics | Keith Durkin et al |
| EPI_ISL_636610 | Belgium/ULG-10631/2020 | 2020-10-25 | Europe/Belgium/Li√®ge/ | Department of Clinical Microbiology | GIGA Medical Genomics | Keith Durkin et al |
| EPI_ISL_636612 | Belgium/ULG-10633/2020 | 2020-10-26 | Europe/Belgium/Li√®ge/ | Department of Clinical Microbiology | GIGA Medical Genomics | Keith Durkin et al |
| EPI_ISL_636617 | Belgium/ULG-10639/2020 | 2020-10-25 | Europe/Belgium/Li√®ge/ | Department of Clinical Microbiology | GIGA Medical Genomics | Keith Durkin et al |
| EPI_ISL_636619 | Belgium/ULG-10641/2020 | 2020-10-26 | Europe/Belgium/Li√®ge/ | Department of Clinical Microbiology | GIGA Medical Genomics | Keith Durkin et al |
| EPI_ISL_636620 | Belgium/ULG-10642/2020 | 2020-10-26 | Europe/Belgium/Li√®ge/ | Department of Clinical Microbiology | GIGA Medical Genomics | Keith Durkin et al |

| | | | | | | |
|---|---|---|---|---|---|---|
| EPI_ISL_636623 | Belgium/ULG-10645/2020 | 2020-10-26 | Europe/Belgium/Liv®ge/ | Department of Clinical Microbiology | GIGA Medical Genomics | Keith Durkin et al |
| EPI_ISL_636625 | Belgium/ULG-10647/2020 | 2020-10-26 | Europe/Belgium/Liv®ge/ | Department of Clinical Microbiology | GIGA Medical Genomics | Keith Durkin et al |
| EPI_ISL_636628 | Belgium/ULG-10650/2020 | 2020-10-26 | Europe/Belgium/Liv®ge/ | Department of Clinical Microbiology | GIGA Medical Genomics | Keith Durkin et al |
| EPI_ISL_636632 | Belgium/ULG-10655/2020 | 2020-10-26 | Europe/Belgium/Liv®ge/ | Department of Clinical Microbiology | GIGA Medical Genomics | Keith Durkin et al |
| EPI_ISL_636633 | Belgium/ULG-10656/2020 | 2020-10-26 | Europe/Belgium/Liv®ge/ | Department of Clinical Microbiology | GIGA Medical Genomics | Keith Durkin et al |
| EPI_ISL_636634 | Belgium/ULG-10657/2020 | 2020-10-26 | Europe/Belgium/Liv®ge/ | Department of Clinical Microbiology | GIGA Medical Genomics | Keith Durkin et al |
| EPI_ISL_636636 | Belgium/ULG-10659/2020 | 2020-10-26 | Europe/Belgium/Liv®ge/ | Department of Clinical Microbiology | GIGA Medical Genomics | Keith Durkin et al |
| EPI_ISL_636637 | Belgium/ULG-10660/2020 | 2020-10-26 | Europe/Belgium/Liv®ge/ | Department of Clinical Microbiology | GIGA Medical Genomics | Keith Durkin et al |
| EPI_ISL_636638 | Belgium/ULG-10661/2020 | 2020-10-26 | Europe/Belgium/Liv®ge/ | Department of Clinical Microbiology | GIGA Medical Genomics | Keith Durkin et al |
| EPI_ISL_636639 | Belgium/ULG-10662/2020 | 2020-10-26 | Europe/Belgium/Liv®ge/ | Department of Clinical Microbiology | GIGA Medical Genomics | Keith Durkin et al |
| EPI_ISL_636640 | Belgium/ULG-10663/2020 | 2020-10-26 | Europe/Belgium/Liv®ge/ | Department of Clinical Microbiology | GIGA Medical Genomics | Keith Durkin et al |
| EPI_ISL_636644 | Belgium/ULG-10668/2020 | 2020-10-26 | Europe/Belgium/Liv®ge/ | Department of Clinical Microbiology | GIGA Medical Genomics | Keith Durkin et al |
| EPI_ISL_636647 | Belgium/ULG-10672/2020 | 2020-10-26 | Europe/Belgium/Liv®ge/ | Department of Clinical Microbiology | GIGA Medical Genomics | Keith Durkin et al |
| EPI_ISL_636649 | Belgium/ULG-10675/2020 | 2020-10-25 | Europe/Belgium/Liv®ge/ | Department of Clinical Microbiology | GIGA Medical Genomics | Keith Durkin et al |
| EPI_ISL_636652 | Belgium/ULG-10680/2020 | 2020-10-26 | Europe/Belgium/Liv®ge/ | Department of Clinical Microbiology | GIGA Medical Genomics | Keith Durkin et al |
| EPI_ISL_636654 | Belgium/ULG-10682/2020 | 2020-10-26 | Europe/Belgium/Liv®ge/ | Department of Clinical Microbiology | GIGA Medical Genomics | Keith Durkin et al |
| EPI_ISL_636661 | Belgium/ULG-10689/2020 | 2020-10-26 | Europe/Belgium/Liv®ge/ | Department of Clinical Microbiology | GIGA Medical Genomics | Keith Durkin et al |
| EPI_ISL_636662 | Belgium/ULG-10690/2020 | 2020-10-26 | Europe/Belgium/Liv®ge/ | Department of Clinical Microbiology | GIGA Medical Genomics | Keith Durkin et al |
| EPI_ISL_636663 | Belgium/ULG-10691/2020 | 2020-10-26 | Europe/Belgium/Liv®ge/ | Department of Clinical Microbiology | GIGA Medical Genomics | Keith Durkin et al |
| EPI_ISL_636667 | Belgium/ULG-10695/2020 | 2020-10-26 | Europe/Belgium/Liv®ge/ | Department of Clinical Microbiology | GIGA Medical Genomics | Keith Durkin et al |

| | | | | | | |
|---|---|---|---|---|---|---|
| EPI_ISL_636674 | Belgium/ULG-10702/2020 | 2020-10-26 | Europe/Belgium/Liv®ge/ | Department of Clinical Microbiology | GIGA Medical Genomics | Keith Durkin et al |
| EPI_ISL_636675 | Belgium/ULG-10703/2020 | 2020-10-26 | Europe/Belgium/Liv®ge/ | Department of Clinical Microbiology | GIGA Medical Genomics | Keith Durkin et al |
| EPI_ISL_636679 | Belgium/ULG-10707/2020 | 2020-10-26 | Europe/Belgium/Liv®ge/ | Department of Clinical Microbiology | GIGA Medical Genomics | Keith Durkin et al |
| EPI_ISL_636680 | Belgium/ULG-10709/2020 | 2020-10-26 | Europe/Belgium/Liv®ge/ | Department of Clinical Microbiology | GIGA Medical Genomics | Keith Durkin et al |
| EPI_ISL_636685 | Belgium/ULG-10714/2020 | 2020-10-26 | Europe/Belgium/Liv®ge/ | Department of Clinical Microbiology | GIGA Medical Genomics | Keith Durkin et al |
| EPI_ISL_636689 | England/C200024019/2020 | 2020-09-28 | Europe/United Kingdom/England/ | Respiratory Virus Unit, Microbiology Services Colindale, Public Health England | Respiratory Virus Unit, Microbiology Services Colindale, Public Health England | PHE Covid Sequencing Team et al |
| EPI_ISL_636737 | Brazil/RJ-06020/2020 | 2020-05-29 | South America/Brazil/Rio de Janeiro/ | Laboratv≥rio de Imunofarmacologia - Instituto Oswaldo Cruz | Laboratv≥rio de Imunofarmacologia - Instituto Oswaldo Cruz | Souza et al |
| EPI_ISL_636739 | India/UT-1504381-CSIR-IGIB/2020 | 2020-07-29 | Asia/India/Uttarakhand/ | National Centre for Disease control (NCDC) | NCDC/CSIR-IGIB | Mahesh S. Dhar1* et al |
| EPI_ISL_636740 | India/DL-57012-CSIR-IGIB/2020 | 2020-08-02 | Asia/India/Delhi/ | National Centre for Disease control (NCDC) | NCDC/CSIR-IGIB | Mahesh S. Dhar1* et al |
| EPI_ISL_636741 | India/DL-69865-CSIR-IGIB/2020 | 2020-08-14 | Asia/India/Delhi/ | National Centre for Disease control (NCDC) | NCDC/CSIR-IGIB | Mahesh S. Dhar1* et al |
| EPI_ISL_636742 | India/UT-67339-CSIR-IGIB/2020 | 2020-08-09 | Asia/India/Uttarakhand/ | National Centre for Disease control (NCDC) | NCDC/CSIR-IGIB | Mahesh S. Dhar1* et al |
| EPI_ISL_636743 | India/UT-67341-CSIR-IGIB/2020 | 2020-08-09 | Asia/India/Uttarakhand/ | National Centre for Disease control (NCDC) | NCDC/CSIR-IGIB | Mahesh S. Dhar1* et al |
| EPI_ISL_636746 | India/UT-67340-CSIR-IGIB/2020 | 2020-08-09 | Asia/India/Uttarakhand/ | National Centre for Disease control (NCDC) | NCDC/CSIR-IGIB | Mahesh S. Dhar1* et al |
| EPI_ISL_636747 | India/UT-67330-CSIR-IGIB/2020 | 2020-08-09 | Asia/India/Uttarakhand/ | National Centre for Disease control (NCDC) | NCDC/CSIR-IGIB | Mahesh S. Dhar1* et al |
| EPI_ISL_636748 | India/UT-50438-CSIR-IGIB/2020 | 2020-07-26 | Asia/India/Uttarakhand/ | National Centre for Disease control (NCDC) | NCDC/CSIR-IGIB | Mahesh S. Dhar1* et al |
| EPI_ISL_636749 | India/DL-69779-CSIR-IGIB/2020 | 2020-08-16 | Asia/India/Delhi/ | National Centre for Disease control (NCDC) | NCDC/CSIR-IGIB | Mahesh S. Dhar1* et al |
| EPI_ISL_636750 | India/UT-67177-CSIR-IGIB/2020 | 2020-08-09 | Asia/India/Uttarakhand/ | National Centre for Disease control (NCDC) | NCDC/CSIR-IGIB | Mahesh S. Dhar1* et al |
| EPI_ISL_636752 | India/DL-59946-CSIR-IGIB/2020 | 2020-08-02 | Asia/India/Delhi/ | National Centre for Disease control (NCDC) | NCDC/CSIR-IGIB | Mahesh S. Dhar1* et al |
| EPI_ISL_636753 | India/UT-67134-CSIR-IGIB/2020 | 2020-08-09 | Asia/India/Uttarakhand/ | National Centre for Disease control (NCDC) | NCDC/CSIR-IGIB | Mahesh S. Dhar1* et al |
| EPI_ISL_636754 | India/DL-69833-CSIR-IGIB/2020 | 2020-08-14 | Asia/India/Delhi/ | National Centre for Disease control (NCDC) | NCDC/CSIR-IGIB | Mahesh S. Dhar1* et al |
| EPI_ISL_636755 | India/DL-69831-CSIR-IGIB/2020 | 2020-08-14 | Asia/India/Delhi/ | National Centre for Disease control (NCDC) | NCDC/CSIR-IGIB | Mahesh S. Dhar1* et al |
| EPI_ISL_636756 | India/UT-67165-CSIR-IGIB/2020 | 2020-08-09 | Asia/India/Uttarakhand/ | National Centre for Disease control (NCDC) | NCDC/CSIR-IGIB | Mahesh S. Dhar1* et al |

| | | | | | | |
|---|---|---|---|---|---|---|
| EPI_ISL_636757 | India/DL-69826-CSIR-IGIB/2020 | 2020-08-14 | Asia/India/Delhi/ | National Centre for Disease control (NCDC) | NCDC/CSIR-IGIB | Mahesh S. Dhar1* et al |
| EPI_ISL_636758 | India/DL-60944-CSIR-IGIB/2020 | 2020-08-06 | Asia/India/Delhi/ | National Centre for Disease control (NCDC) | NCDC/CSIR-IGIB | Mahesh S. Dhar1* et al |
| EPI_ISL_636759 | India/UT-50464-CSIR-IGIB/2020 | 2020-07-26 | Asia/India/Uttarakhand/ | National Centre for Disease control (NCDC) | NCDC/CSIR-IGIB | Mahesh S. Dhar1* et al |
| EPI_ISL_636760 | India/DL-69787-CSIR-IGIB/2020 | 2020-08-14 | Asia/India/Delhi/ | National Centre for Disease control (NCDC) | NCDC/CSIR-IGIB | Mahesh S. Dhar1* et al |
| EPI_ISL_636761 | India/DL-69794-CSIR-IGIB/2020 | 2020-08-14 | Asia/India/Delhi/ | National Centre for Disease control (NCDC) | NCDC/CSIR-IGIB | Mahesh S. Dhar1* et al |
| EPI_ISL_636762 | India/DL-69813-CSIR-IGIB/2020 | 2020-08-14 | Asia/India/Delhi/ | National Centre for Disease control (NCDC) | NCDC/CSIR-IGIB | Mahesh S. Dhar1* et al |
| EPI_ISL_636765 | India/DL-60949-CSIR-IGIB/2020 | 2020-08-06 | Asia/India/Delhi/ | National Centre for Disease control (NCDC) | NCDC/CSIR-IGIB | Mahesh S. Dhar1* et al |
| EPI_ISL_636766 | India/DL-61026-CSIR-IGIB/2020 | 2020-08-06 | Asia/India/Delhi/ | National Centre for Disease control (NCDC) | NCDC/CSIR-IGIB | Mahesh S. Dhar1* et al |
| EPI_ISL_636768 | India/DL-69843-CSIR-IGIB/2020 | 2020-08-14 | Asia/India/Delhi/ | National Centre for Disease control (NCDC) | NCDC/CSIR-IGIB | Mahesh S. Dhar1* et al |
| EPI_ISL_636770 | India/DL-50480-CSIR-IGIB/2020 | 2020-07-27 | Asia/India/Delhi/ | National Centre for Disease control (NCDC) | NCDC/CSIR-IGIB | Mahesh S. Dhar1* et al |
| EPI_ISL_636774 | India/DL-60920-CSIR-IGIB/2020 | 2020-08-06 | Asia/India/Delhi/ | National Centre for Disease control (NCDC) | NCDC/CSIR-IGIB | Mahesh S. Dhar1* et al |
| EPI_ISL_636776 | India/DL-62368-CSIR-IGIB/2020 | 2020-08-08 | Asia/India/Delhi/ | National Centre for Disease control (NCDC) | NCDC/CSIR-IGIB | Mahesh S. Dhar1* et al |
| EPI_ISL_636777 | India/DL-69775-CSIR-IGIB/2020 | 2020-08-16 | Asia/India/Delhi/ | National Centre for Disease control (NCDC) | NCDC/CSIR-IGIB | Mahesh S. Dhar1* et al |
| EPI_ISL_636778 | India/DL-60909-CSIR-IGIB/2020 | 2020-08-06 | Asia/India/Delhi/ | National Centre for Disease control (NCDC) | NCDC/CSIR-IGIB | Mahesh S. Dhar1* et al |
| EPI_ISL_636779 | India/DL-60907-CSIR-IGIB/2020 | 2020-08-06 | Asia/India/Delhi/ | National Centre for Disease control (NCDC) | NCDC/CSIR-IGIB | Mahesh S. Dhar1* et al |
| EPI_ISL_636780 | India/DL-60932-CSIR-IGIB/2020 | 2020-08-06 | Asia/India/Delhi/ | National Centre for Disease control (NCDC) | NCDC/CSIR-IGIB | Mahesh S. Dhar1* et al |
| EPI_ISL_636782 | India/UT-58384-CSIR-IGIB/2020 | 2020-08-03 | Asia/India/Uttarakhand/ | National Centre for Disease control (NCDC) | NCDC/CSIR-IGIB | Mahesh S. Dhar1* et al |
| EPI_ISL_636784 | India/UT-58366-CSIR-IGIB/2020 | 2020-08-03 | Asia/India/Uttarakhand/ | National Centre for Disease control (NCDC) | NCDC/CSIR-IGIB | Mahesh S. Dhar1* et al |
| EPI_ISL_636785 | India/UT-58373-CSIR-IGIB/2020 | 2020-08-03 | Asia/India/Uttarakhand/ | National Centre for Disease control (NCDC) | NCDC/CSIR-IGIB | Mahesh S. Dhar1* et al |
| EPI_ISL_636787 | India/DL-5504375-CSIR-IGIB/2020 | 2020-08-06 | Asia/India/Delhi/ | National Centre for Disease control (NCDC) | NCDC/CSIR-IGIB | Mahesh S. Dhar1* et al |
| EPI_ISL_636788 | India/DL-60935-CSIR-IGIB/2020 | 2020-08-06 | Asia/India/Delhi/ | National Centre for Disease control (NCDC) | NCDC/CSIR-IGIB | Mahesh S. Dhar1* et al |
| EPI_ISL_636789 | India/DL-60939-CSIR-IGIB/2020 | 2020-08-06 | Asia/India/Delhi/ | National Centre for Disease control (NCDC) | NCDC/CSIR-IGIB | Mahesh S. Dhar1* et al |
| EPI_ISL_636790 | India/DL-60943-CSIR-IGIB/2020 | 2020-08-06 | Asia/India/Delhi/ | National Centre for Disease control (NCDC) | NCDC/CSIR-IGIB | Mahesh S. Dhar1* et al |
| EPI_ISL_636791 | India/UT-59241-CSIR-IGIB/2020 | 2020-08-02 | Asia/India/Uttarakhand/ | National Centre for Disease control (NCDC) | NCDC/CSIR-IGIB | Mahesh S. Dhar1* et al |
| EPI_ISL_636793 | India/UT-58396-CSIR-IGIB/2020 | 2020-08-03 | Asia/India/Uttarakhand/ | National Centre for Disease control (NCDC) | NCDC/CSIR-IGIB | Mahesh S. Dhar1* et al |
| EPI_ISL_636795 | India/DL-60072-CSIR-IGIB/2020 | 2020-08-05 | Asia/India/Delhi/ | National Centre for Disease control (NCDC) | NCDC/CSIR-IGIB | Mahesh S. Dhar1* et al |
| EPI_ISL_636796 | India/UT-58411-CSIR-IGIB/2020 | 2020-08-03 | Asia/India/Uttarakhand/ | National Centre for Disease control (NCDC) | NCDC/CSIR-IGIB | Mahesh S. Dhar1* et al |
| EPI_ISL_636797 | India/UT-65187-CSIR-IGIB/2020 | 2020-08-07 | Asia/India/Uttarakhand/ | National Centre for Disease control (NCDC) | NCDC/CSIR-IGIB | Mahesh S. Dhar1* et al |
| EPI_ISL_636798 | India/UT-58264-CSIR-IGIB/2020 | 2020-08-03 | Asia/India/Uttarakhand/ | National Centre for Disease control (NCDC) | NCDC/CSIR-IGIB | Mahesh S. Dhar1* et al |
| EPI_ISL_636800 | India/UT-58418-CSIR-IGIB/2020 | 2020-08-03 | Asia/India/Uttarakhand/ | National Centre for Disease control (NCDC) | NCDC/CSIR-IGIB | Mahesh S. Dhar1* et al |
| EPI_ISL_636801 | India/UT-58319-CSIR-IGIB/2020 | 2020-08-03 | Asia/India/Uttarakhand/ | National Centre for Disease control (NCDC) | NCDC/CSIR-IGIB | Mahesh S. Dhar1* et al |

| | | | | | | |
|---|---|---|---|---|---|---|
| EPI_ISL_636802 | India/DL-59608-CSIR-IGIB/2020 | 2020-08-05 | Asia/India/Delhi/ | National Centre for Disease control (NCDC) | NCDC/CSIR-IGIB | Mahesh S. Dhar1* et al |
| EPI_ISL_636804 | India/UT-65173-CSIR-IGIB/2020 | 2020-08-07 | Asia/India/Uttarakhand/ | National Centre for Disease control (NCDC) | NCDC/CSIR-IGIB | Mahesh S. Dhar1* et al |
| EPI_ISL_636805 | India/UT-59267-CSIR-IGIB/2020 | 2020-08-02 | Asia/India/Uttarakhand/ | National Centre for Disease control (NCDC) | NCDC/CSIR-IGIB | Mahesh S. Dhar1* et al |
| EPI_ISL_636806 | India/UT-63142-CSIR-IGIB/2020 | 2020-08-05 | Asia/India/Uttarakhand/ | National Centre for Disease control (NCDC) | NCDC/CSIR-IGIB | Mahesh S. Dhar1* et al |
| EPI_ISL_636809 | India/DL-59108-CSIR-IGIB/2020 | 2020-08-04 | Asia/India/Delhi/ | National Centre for Disease control (NCDC) | NCDC/CSIR-IGIB | Mahesh S. Dhar1* et al |
| EPI_ISL_636810 | India/UT-63138-CSIR-IGIB/2020 | 2020-08-05 | Asia/India/Uttarakhand/ | National Centre for Disease control (NCDC) | NCDC/CSIR-IGIB | Mahesh S. Dhar1* et al |
| EPI_ISL_636811 | India/UT-7570807-CSIR-IGIB/2020 | 2020-08-07 | Asia/India/Uttarakhand/ | National Centre for Disease control (NCDC) | NCDC/CSIR-IGIB | Mahesh S. Dhar1* et al |
| EPI_ISL_636812 | India/UT-63072-CSIR-IGIB/2020 | 2020-08-05 | Asia/India/Uttarakhand/ | National Centre for Disease control (NCDC) | NCDC/CSIR-IGIB | Mahesh S. Dhar1* et al |
| EPI_ISL_636813 | India/UT-59149-CSIR-IGIB/2020 | 2020-08-02 | Asia/India/Uttarakhand/ | National Centre for Disease control (NCDC) | NCDC/CSIR-IGIB | Mahesh S. Dhar1* et al |
| EPI_ISL_636815 | India/DL-59113-CSIR-IGIB/2020 | 2020-08-04 | Asia/India/Delhi/ | National Centre for Disease control (NCDC) | NCDC/CSIR-IGIB | Mahesh S. Dhar1* et al |
| EPI_ISL_636816 | India/DL-59597-CSIR-IGIB/2020 | 2020-08-05 | Asia/India/Delhi/ | National Centre for Disease control (NCDC) | NCDC/CSIR-IGIB | Mahesh S. Dhar1* et al |
| EPI_ISL_636817 | India/DL-65029-CSIR-IGIB/2020 | 2020-08-11 | Asia/India/Delhi/ | National Centre for Disease control (NCDC) | NCDC/CSIR-IGIB | Mahesh S. Dhar1* et al |
| EPI_ISL_636818 | India/DL-65025-CSIR-IGIB/2020 | 2020-08-11 | Asia/India/Delhi/ | National Centre for Disease control (NCDC) | NCDC/CSIR-IGIB | Mahesh S. Dhar1* et al |
| EPI_ISL_636819 | India/DL-65054-CSIR-IGIB/2020 | 2020-08-11 | Asia/India/Delhi/ | National Centre for Disease control (NCDC) | NCDC/CSIR-IGIB | Mahesh S. Dhar1* et al |
| EPI_ISL_636820 | India/DL-65092-CSIR-IGIB/2020 | 2020-08-11 | Asia/India/Delhi/ | National Centre for Disease control (NCDC) | NCDC/CSIR-IGIB | Mahesh S. Dhar1* et al |
| EPI_ISL_636821 | India/DL-65112-CSIR-IGIB/2020 | 2020-08-11 | Asia/India/Delhi/ | National Centre for Disease control (NCDC) | NCDC/CSIR-IGIB | Mahesh S. Dhar1* et al |
| EPI_ISL_636822 | India/UT-65121-CSIR-IGIB/2020 | 2020-08-07 | Asia/India/Uttarakhand/ | National Centre for Disease control (NCDC) | NCDC/CSIR-IGIB | Mahesh S. Dhar1* et al |
| EPI_ISL_636823 | India/UT-8570838-CSIR-IGIB/2020 | 2020-08-07 | Asia/India/Uttarakhand/ | National Centre for Disease control (NCDC) | NCDC/CSIR-IGIB | Mahesh S. Dhar1* et al |
| EPI_ISL_636824 | India/UT-65137-CSIR-IGIB/2020 | 2020-08-07 | Asia/India/Uttarakhand/ | National Centre for Disease control (NCDC) | NCDC/CSIR-IGIB | Mahesh S. Dhar1* et al |
| EPI_ISL_636825 | India/DL-57083-CSIR-IGIB/2020 | 2020-08-02 | Asia/India/Delhi/ | National Centre for Disease control (NCDC) | NCDC/CSIR-IGIB | Mahesh S. Dhar1* et al |
| EPI_ISL_636828 | India/UT-65158-CSIR-IGIB/2020 | 2020-08-07 | Asia/India/Uttarakhand/ | National Centre for Disease control (NCDC) | NCDC/CSIR-IGIB | Mahesh S. Dhar1* et al |
| EPI_ISL_636829 | India/UT-63116-CSIR-IGIB/2020 | 2020-08-05 | Asia/India/Uttarakhand/ | National Centre for Disease control (NCDC) | NCDC/CSIR-IGIB | Mahesh S. Dhar1* et al |
| EPI_ISL_636830 | India/UT-59223-CSIR-IGIB/2020 | 2020-08-02 | Asia/India/Uttarakhand/ | National Centre for Disease control (NCDC) | NCDC/CSIR-IGIB | Mahesh S. Dhar1* et al |
| EPI_ISL_636831 | India/UT-59189-CSIR-IGIB/2020 | 2020-08-02 | Asia/India/Uttarakhand/ | National Centre for Disease control (NCDC) | NCDC/CSIR-IGIB | Mahesh S. Dhar1* et al |
| EPI_ISL_636835 | Brazil/RJ-01020R2/2020 | 2020-05-29 | South America/Brazil/Rio de Janeiro/ | Laboratv≥rio de Imunofarmacologia - Instituto Oswaldo Cruz | Laboratv≥rio de Imunofarmacologia - Instituto Oswaldo Cruz | Souza et al |
| EPI_ISL_636860 | Lithuania/MR-LUHS-Eilnr285/2020 | 2020-08-11 | Europe/Lithuania/Kauno Apskritis/ | Lithuanian University of Health Sciences Hospital, Department of Laboratory Medicine | Lithuanian University of Health Sciences, Molecular cardiology lab. | Lukas Zemaitis et al |

| | | | | | | |
|---|---|---|---|---|---|---|
| EPI_ISL_636900 | Lithuania/MR-LUHS-Eilnr470/2020 | 2020-09-12 | Europe/Lithuania/Kauno Apskritis/ | Lithuanian University of Health Sciences Hospital, Department of Laboratory Medicine | Lithuanian University of Health Sciences, Molecular cardiology lab. | Lukas Zemaitis et al |
| EPI_ISL_636907 | Canada/ON-PHL-20-01754/2020 | 20-Jul | North America/Canada/Ontario/ | Public Health Ontario Laboratory | Public Health Ontario Laboratory | Vanessa G Allen et al |
| EPI_ISL_636908 | Canada/ON-PHL-20-01755/2020 | 20-Jul | North America/Canada/Ontario/ | Public Health Ontario Laboratory | Public Health Ontario Laboratory | Vanessa G Allen et al |
| EPI_ISL_636909 | Canada/ON-PHL-20-01756/2020 | 20-Jul | North America/Canada/Ontario/ | Public Health Ontario Laboratory | Public Health Ontario Laboratory | Vanessa G Allen et al |
| EPI_ISL_636910 | Canada/ON-PHL-20-01757/2020 | 20-Jul | North America/Canada/Ontario/ | Public Health Ontario Laboratory | Public Health Ontario Laboratory | Vanessa G Allen et al |
| EPI_ISL_636911 | Canada/ON-PHL-20-01758/2020 | 20-Jul | North America/Canada/Ontario/ | Public Health Ontario Laboratory | Public Health Ontario Laboratory | Vanessa G Allen et al |
| EPI_ISL_636912 | Canada/ON-PHL-20-01759/2020 | 20-Jul | North America/Canada/Ontario/ | Public Health Ontario Laboratory | Public Health Ontario Laboratory | Vanessa G Allen et al |
| EPI_ISL_636913 | Canada/ON-PHL-20-01760/2020 | 20-Jul | North America/Canada/Ontario/ | Public Health Ontario Laboratory | Public Health Ontario Laboratory | Vanessa G Allen et al |
| EPI_ISL_636914 | Canada/ON-PHL-20-01761/2020 | 20-Jul | North America/Canada/Ontario/ | Public Health Ontario Laboratory | Public Health Ontario Laboratory | Vanessa G Allen et al |
| EPI_ISL_636915 | Canada/ON-PHL-20-01762/2020 | 20-Jul | North America/Canada/Ontario/ | Public Health Ontario Laboratory | Public Health Ontario Laboratory | Vanessa G Allen et al |
| EPI_ISL_636916 | Canada/ON-PHL-20-01763/2020 | 20-Jul | North America/Canada/Ontario/ | Public Health Ontario Laboratory | Public Health Ontario Laboratory | Vanessa G Allen et al |
| EPI_ISL_636917 | Canada/ON-PHL-20-01764/2020 | 20-Jul | North America/Canada/Ontario/ | Public Health Ontario Laboratory | Public Health Ontario Laboratory | Vanessa G Allen et al |
| EPI_ISL_636918 | Canada/ON-PHL-20-01765/2020 | 20-Jul | North America/Canada/Ontario/ | Public Health Ontario Laboratory | Public Health Ontario Laboratory | Vanessa G Allen et al |

| | | | | | | |
|---|---|---|---|---|---|---|
| EPI_ISL_636919 | Canada/ON-PHL-20-01766/2020 | 20-Jul | North America/Canada/Ontario/ | Public Health Ontario Laboratory | Public Health Ontario Laboratory | Vanessa G Allen et al |
| EPI_ISL_636920 | Canada/ON-PHL-20-01767/2020 | 20-Jul | North America/Canada/Ontario/ | Public Health Ontario Laboratory | Public Health Ontario Laboratory | Vanessa G Allen et al |
| EPI_ISL_636921 | Canada/ON-PHL-20-01768/2020 | 20-Jul | North America/Canada/Ontario/ | Public Health Ontario Laboratory | Public Health Ontario Laboratory | Vanessa G Allen et al |
| EPI_ISL_636922 | Canada/ON-PHL-20-01769/2020 | 20-Jul | North America/Canada/Ontario/ | Public Health Ontario Laboratory | Public Health Ontario Laboratory | Vanessa G Allen et al |
| EPI_ISL_636923 | Canada/ON-PHL-20-01770/2020 | 20-Jul | North America/Canada/Ontario/ | Public Health Ontario Laboratory | Public Health Ontario Laboratory | Vanessa G Allen et al |
| EPI_ISL_636924 | Canada/ON-PHL-20-01771/2020 | 20-Jul | North America/Canada/Ontario/ | Public Health Ontario Laboratory | Public Health Ontario Laboratory | Vanessa G Allen et al |
| EPI_ISL_636925 | Canada/ON-PHL-20-01772/2020 | 20-Jul | North America/Canada/Ontario/ | Public Health Ontario Laboratory | Public Health Ontario Laboratory | Vanessa G Allen et al |
| EPI_ISL_636926 | Canada/ON-PHL-20-01774/2020 | 20-Jul | North America/Canada/Ontario/ | Public Health Ontario Laboratory | Public Health Ontario Laboratory | Vanessa G Allen et al |
| EPI_ISL_636927 | Canada/ON-PHL-20-01775/2020 | 20-Jul | North America/Canada/Ontario/ | Public Health Ontario Laboratory | Public Health Ontario Laboratory | Vanessa G Allen et al |
| EPI_ISL_636928 | Canada/ON-PHL-20-01776/2020 | 20-Jul | North America/Canada/Ontario/ | Public Health Ontario Laboratory | Public Health Ontario Laboratory | Vanessa G Allen et al |
| EPI_ISL_636929 | Canada/ON-PHL-20-01777/2020 | 20-Jul | North America/Canada/Ontario/ | Public Health Ontario Laboratory | Public Health Ontario Laboratory | Vanessa G Allen et al |
| EPI_ISL_636930 | Canada/ON-PHL-20-01778/2020 | 20-Jul | North America/Canada/Ontario/ | Public Health Ontario Laboratory | Public Health Ontario Laboratory | Vanessa G Allen et al |
| EPI_ISL_636931 | Canada/ON-PHL-20-01780/2020 | 20-Jul | North America/Canada/Ontario/ | Public Health Ontario Laboratory | Public Health Ontario Laboratory | Vanessa G Allen et al |

| | | | | | | |
|---|---|---|---|---|---|---|
| EPI_ISL_636932 | Canada/ON-PHL-20-01781/2020 | 20-Jul | North America/Canada/Ontario/ | Public Health Ontario Laboratory | Public Health Ontario Laboratory | Vanessa G Allen et al |
| EPI_ISL_636933 | Canada/ON-PHL-20-01782/2020 | 20-Jul | North America/Canada/Ontario/ | Public Health Ontario Laboratory | Public Health Ontario Laboratory | Vanessa G Allen et al |
| EPI_ISL_636934 | Canada/ON-PHL-20-01783/2020 | 20-Jul | North America/Canada/Ontario/ | Public Health Ontario Laboratory | Public Health Ontario Laboratory | Vanessa G Allen et al |
| EPI_ISL_636935 | Canada/ON-PHL-20-01785/2020 | 20-Jul | North America/Canada/Ontario/ | Public Health Ontario Laboratory | Public Health Ontario Laboratory | Vanessa G Allen et al |
| EPI_ISL_636936 | Canada/ON-PHL-20-01786/2020 | 20-Jul | North America/Canada/Ontario/ | Public Health Ontario Laboratory | Public Health Ontario Laboratory | Vanessa G Allen et al |
| EPI_ISL_636937 | Canada/ON-PHL-20-01787/2020 | 20-Jul | North America/Canada/Ontario/ | Public Health Ontario Laboratory | Public Health Ontario Laboratory | Vanessa G Allen et al |
| EPI_ISL_636938 | Canada/ON-PHL-20-01788/2020 | 20-Jul | North America/Canada/Ontario/ | Public Health Ontario Laboratory | Public Health Ontario Laboratory | Vanessa G Allen et al |
| EPI_ISL_636939 | Canada/ON-PHL-20-01789/2020 | 20-Jul | North America/Canada/Ontario/ | Public Health Ontario Laboratory | Public Health Ontario Laboratory | Vanessa G Allen et al |
| EPI_ISL_636940 | Canada/ON-PHL-20-01790/2020 | 20-Jul | North America/Canada/Ontario/ | Public Health Ontario Laboratory | Public Health Ontario Laboratory | Vanessa G Allen et al |
| EPI_ISL_636941 | Canada/ON-PHL-20-01791/2020 | 20-Jul | North America/Canada/Ontario/ | Public Health Ontario Laboratory | Public Health Ontario Laboratory | Vanessa G Allen et al |
| EPI_ISL_636942 | Canada/ON-PHL-20-01792/2020 | 20-Jul | North America/Canada/Ontario/ | Public Health Ontario Laboratory | Public Health Ontario Laboratory | Vanessa G Allen et al |
| EPI_ISL_636943 | Canada/ON-PHL-20-01793/2020 | 20-Jul | North America/Canada/Ontario/ | Public Health Ontario Laboratory | Public Health Ontario Laboratory | Vanessa G Allen et al |
| EPI_ISL_636944 | Canada/ON-PHL-20-01794/2020 | 20-Jul | North America/Canada/Ontario/ | Public Health Ontario Laboratory | Public Health Ontario Laboratory | Vanessa G Allen et al |

| | | | | | | |
|---|---|---|---|---|---|---|
| EPI_ISL_636945 | Canada/ON-PHL-20-01795/2020 | 20-Jul | North America/Canada/Ontario/ | Public Health Ontario Laboratory | Public Health Ontario Laboratory | Vanessa G Allen et al |
| EPI_ISL_636946 | Canada/ON-PHL-20-01796/2020 | 20-Apr | North America/Canada/Ontario/ | Public Health Ontario Laboratory | Public Health Ontario Laboratory | Vanessa G Allen et al |
| EPI_ISL_636947 | Canada/ON-PHL-20-01797/2020 | 20-Apr | North America/Canada/Ontario/ | Public Health Ontario Laboratory | Public Health Ontario Laboratory | Vanessa G Allen et al |
| EPI_ISL_636949 | Canada/ON-PHL-20-01805/2020 | 20-Jun | North America/Canada/Ontario/ | Public Health Ontario Laboratory | Public Health Ontario Laboratory | Vanessa G Allen et al |
| EPI_ISL_636950 | Canada/ON-PHL-20-01806/2020 | 20-Jun | North America/Canada/Ontario/ | Public Health Ontario Laboratory | Public Health Ontario Laboratory | Vanessa G Allen et al |
| EPI_ISL_636951 | Canada/ON-PHL-20-01807/2020 | 20-Jun | North America/Canada/Ontario/ | Public Health Ontario Laboratory | Public Health Ontario Laboratory | Vanessa G Allen et al |
| EPI_ISL_636952 | Canada/ON-PHL-20-01808/2020 | 20-Jun | North America/Canada/Ontario/ | Public Health Ontario Laboratory | Public Health Ontario Laboratory | Vanessa G Allen et al |
| EPI_ISL_636953 | Canada/ON-PHL-20-01809/2020 | 20-Jun | North America/Canada/Ontario/ | Public Health Ontario Laboratory | Public Health Ontario Laboratory | Vanessa G Allen et al |
| EPI_ISL_636954 | Canada/ON-PHL-20-01810/2020 | 20-Jun | North America/Canada/Ontario/ | Public Health Ontario Laboratory | Public Health Ontario Laboratory | Vanessa G Allen et al |
| EPI_ISL_636955 | Canada/ON-PHL-20-01811/2020 | 20-Jun | North America/Canada/Ontario/ | Public Health Ontario Laboratory | Public Health Ontario Laboratory | Vanessa G Allen et al |
| EPI_ISL_636956 | Canada/ON-PHL-20-01812/2020 | 20-Jun | North America/Canada/Ontario/ | Public Health Ontario Laboratory | Public Health Ontario Laboratory | Vanessa G Allen et al |
| EPI_ISL_636957 | Canada/ON-PHL-20-01813/2020 | 20-Jun | North America/Canada/Ontario/ | Public Health Ontario Laboratory | Public Health Ontario Laboratory | Vanessa G Allen et al |
| EPI_ISL_636958 | Canada/ON-PHL-20-01814/2020 | 20-Jul | North America/Canada/Ontario/ | Public Health Ontario Laboratory | Public Health Ontario Laboratory | Vanessa G Allen et al |

| | | | | | | |
|---|---|---|---|---|---|---|
| EPI_ISL_636959 | Canada/ON-PHL-20-01815/2020 | 20-May | North America/Canada/Ontario/ | Public Health Ontario Laboratory | Public Health Ontario Laboratory | Vanessa G Allen et al |
| EPI_ISL_636962 | SaudiArabia/KAUST-JEDDAH876/202 | 2020-05-24 | Asia/Saudi Arabia/Jeddah/ | Pathogen Genomics Lab King Abdullah University of Science and Technology(KAUST) | Pathogen Genomics Lab King Abdullah University of Science and Technology(KAUST) | Rahul P Salunke et al |
| EPI_ISL_636963 | SaudiArabia/KAUST-JEDDAH880/202 | 2020-05-24 | Asia/Saudi Arabia/Jeddah/ | Pathogen Genomics Lab King Abdullah University of Science and Technology(KAUST) | Pathogen Genomics Lab King Abdullah University of Science and Technology(KAUST) | Rahul P Salunke et al |
| EPI_ISL_636964 | SaudiArabia/KAUST-MADINAH893/2 | 2020-05-12 | Asia/Saudi Arabia/Madinah/ | Pathogen Genomics Lab King Abdullah University of Science and Technology(KAUST) | Pathogen Genomics Lab King Abdullah University of Science and Technology(KAUST) | Rahul P Salunke et al |
| EPI_ISL_636972 | SaudiArabia/KAUST-jeddah839/2020 | 2020-05-24 | Asia/Saudi Arabia/Jeddah/ | Pathogen Genomics Lab King Abdullah University of Science and Technology (KAUST) | Pathogen Genomics Lab King Abdullah University of Science and Technology (KAUST) | Afrah Alsomali et al |
| EPI_ISL_636973 | Bahrain/023341084/2020 | 2020-08-26 | Asia/Bahrain/Bahrain/ | Public Health Lab | Public Health Lab | Alwasti et al |
| EPI_ISL_637099 | Iran/Q100/2020 | 2020-03-10 | Asia/Iran/Iran/ | Indian Council of Medical Research-National Institute of Virology, Microbial Containment Complex | Indian Council of Medical Research-National Institute of Virology, Microbial Containment Complex | Pragya D. Yadav et al |

| | | | | | | |
|---|---|---|---|---|---|---|
| EPI_ISL_637102 | Iran/Q111/2020 | 2020-03-10 | Asia/Iran/Iran/ | Indian Council of Medical Research-National Institute of Virology, Microbial Containment Complex | Indian Council of Medical Research-National Institute of Virology, Microbial Containment Complex | Pragya D. Yadav et al |
| EPI_ISL_637109 | Italy/SAR-ASSL-12/2020 | 2020-04-23 | Europe/Italy/Sardinia/ | "Laboratorio Biologia Molecolare Sars Cov2 - UOC Laboratorio Analisi - Servizio Medicina di Laboratorio, Ospedale ""San Francesco"" - ATS-ASSL Nuoro" | "Laboratorio specialistico UOC Ematologia - Ospedale ""San Francesco"" - ATS-ASSL Nuoro" | Piras Giovanna et al |
| EPI_ISL_637110 | Lebanon/LAU1-53249/2020 | 2020-08-13 | Asia/Lebanon/Beirut/ | Rafik Hariri University Hospital | Microbial Pathogenomics Lab | Georgi Merhi et al |
| EPI_ISL_637112 | Lebanon/LAU3-53451/2020 | 2020-08-14 | Asia/Lebanon/Beirut/ | Rafik Hariri University Hospital | Microbial Pathogenomics Lab | Georgi Merhi et al |
| EPI_ISL_637113 | Lebanon/LAU4-53460/2020 | 2020-08-14 | Asia/Lebanon/Beirut/ | Rafik Hariri University Hospital | Microbial Pathogenomics Lab | Georgi Merhi et al |
| EPI_ISL_637314 | England/CAMC-A652CD/2020 | 2020-10-11 | Europe/United Kingdom/England/ | Department of Pathology, University of Cambridge | COVID-19 Genomics UK (COG-UK) Consortium | Aminu S. Jahun et al |
| EPI_ISL_637334 | England/QEUH-9AE108/2020 | 2020-09-04 | Europe/United Kingdom/England/ | Department of Pathology, University of Cambridge | COVID-19 Genomics UK (COG-UK) Consortium | Aminu S. Jahun et al |
| EPI_ISL_637338 | England/LIVE-DAE78A/2020 | 2020-10-23 | Europe/United Kingdom/England/ | Liverpool Clinical Laboratories | COVID-19 Genomics UK (COG-UK) Consortium | Sam Haldenby et al |
| EPI_ISL_637404 | England/CAMB-1B6FB0/2020 | 2020-10-13 | Europe/United Kingdom/England/ | Department of Pathology, University of Cambridge | COVID-19 Genomics UK (COG-UK) Consortium | Aminu S. Jahun et al |
| EPI_ISL_637492 | England/MILK-9D51F9/2020 | 2020-09-18 | Europe/United Kingdom/England/ | Centre for Enzyme Innovation, University of Portsmouth / Translational Research Laboratory, Portsmouth Hospitals NHS Trust | COVID-19 Genomics UK (COG-UK) Consortium | Angela Beckett et al |

| | | | | | | |
|---|---|---|---|---|---|---|
| EPI_ISL_637502 | Wales/MILK-9D529C/2020 | 2020-09-18 | Europe/United Kingdom/Wales/ | Centre for Enzyme Innovation, University of Portsmouth / Translational Research Laboratory, Portsmouth Hospitals NHS Trust | COVID-19 Genomics UK (COG-UK) Consortium | Angela Beckett et al |
| EPI_ISL_637566 | England/CAMB-1B776C/2020 | 2020-10-22 | Europe/United Kingdom/England/ | Department of Pathology, University of Cambridge | COVID-19 Genomics UK (COG-UK) Consortium | Aminu S. Jahun et al |
| EPI_ISL_637702 | England/CAMB-1B8349/2020 | 2020-10-19 | Europe/United Kingdom/England/ | Department of Pathology, University of Cambridge | COVID-19 Genomics UK (COG-UK) Consortium | Aminu S. Jahun et al |
| EPI_ISL_637820 | England/QEUH-A4B1FD/2020 | 2020-10-05 | Europe/United Kingdom/England/ | Department of Pathology, University of Cambridge | COVID-19 Genomics UK (COG-UK) Consortium | Aminu S. Jahun et al |
| EPI_ISL_637892 | England/ALDP-9E8524/2020 | 2020-09-24 | Europe/United Kingdom/England/ | Centre for Enzyme Innovation, University of Portsmouth / Translational Research Laboratory, Portsmouth Hospitals NHS Trust | COVID-19 Genomics UK (COG-UK) Consortium | Angela Beckett et al |
| EPI_ISL_637982 | England/CAMC-A65B3B/2020 | 2020-10-11 | Europe/United Kingdom/England/ | Department of Pathology, University of Cambridge | COVID-19 Genomics UK (COG-UK) Consortium | Aminu S. Jahun et al |
| EPI_ISL_638077 | England/MILK-A91459/2020 | 2020-10-16 | Europe/United Kingdom/England/ | Oxford Viromics, NDM, University of Oxford; Oxford University Hospitals; Basingstoke and North Hampshire Hospital | COVID-19 Genomics UK (COG-UK) Consortium | Tanya Golubchik et al |
| EPI_ISL_638136 | England/ALDP-9E8B8C/2020 | 2020-09-24 | Europe/United Kingdom/England/ | Centre for Enzyme Innovation, University of Portsmouth / Translational Research Laboratory, Portsmouth Hospitals NHS Trust | COVID-19 Genomics UK (COG-UK) Consortium | Angela Beckett et al |
| EPI_ISL_638178 | England/CAMB-1B7D79/2020 | 2020-10-14 | Europe/United Kingdom/England/ | Department of Pathology, University of Cambridge | COVID-19 Genomics UK (COG-UK) Consortium | Aminu S. Jahun et al |
| EPI_ISL_638322 | England/CAMB-1B7492/2020 | 2020-10-19 | Europe/United Kingdom/England/ | Department of Pathology, University of Cambridge | COVID-19 Genomics UK (COG-UK) Consortium | Aminu S. Jahun et al |
| EPI_ISL_638360 | England/CAMB-1B7B15/2020 | 2020-10-21 | Europe/United Kingdom/England/ | Department of Pathology, University of Cambridge | COVID-19 Genomics UK (COG-UK) Consortium | Aminu S. Jahun et al |
| EPI_ISL_638516 | England/MILK-A9126E/2020 | 2020-10-16 | Europe/United Kingdom/England/ | Oxford Viromics, NDM, University of Oxford; Oxford University Hospitals; Basingstoke and North Hampshire Hospital | COVID-19 Genomics UK (COG-UK) Consortium | Tanya Golubchik et al |

| | | | | | | |
|---|---|---|---|---|---|---|
| EPI_ISL_638555 | England/NORT-1B61139/2020 | 2020-10-14 | Europe/United Kingdom/England/ | Northumbria University / South Tees Hospitals NHS Foundation Trust / North Cumbria Integrated Care NHS Foundation Trust / North Tees and Hartlepool NHS Foundation Trust / Newcastle Hospitals NHS Foundation Trust | COVID-19 Genomics UK (COG-UK) Consortium | Darren L Smith et al |
| EPI_ISL_638630 | England/QEUH-9AE232/2020 | 2020-09-02 | Europe/United Kingdom/England/ | Department of Pathology, University of Cambridge | COVID-19 Genomics UK (COG-UK) Consortium | Aminu S. Jahun et al |
| EPI_ISL_638648 | England/QEUH-9F5023/2020 | 2020-09-27 | Europe/United Kingdom/England/ | Department of Pathology, University of Cambridge | COVID-19 Genomics UK (COG-UK) Consortium | Aminu S. Jahun et al |
| EPI_ISL_638733 | England/QEUH-A4B11B/2020 | 2020-10-03 | Europe/United Kingdom/England/ | Department of Pathology, University of Cambridge | COVID-19 Genomics UK (COG-UK) Consortium | Aminu S. Jahun et al |
| EPI_ISL_638899 | Scotland/QEUH-9ADF96/2020 | 2020-09-05 | Europe/United Kingdom/Scotland/ | Department of Pathology, University of Cambridge | COVID-19 Genomics UK (COG-UK) Consortium | Aminu S. Jahun et al |
| EPI_ISL_638914 | Scotland/QEUH-9F4EFD/2020 | 2020-09-27 | Europe/United Kingdom/Scotland/ | Department of Pathology, University of Cambridge | COVID-19 Genomics UK (COG-UK) Consortium | Aminu S. Jahun et al |
| EPI_ISL_638931 | Scotland/QEUH-A369C0/2020 | 2020-10-04 | Europe/United Kingdom/Scotland/ | Wales Specialist Virology Centre Sequencing lab: Pathogen Genomics Unit | COVID-19 Genomics UK (COG-UK) Consortium | Catherine Moore et al |
| EPI_ISL_639209 | Wales/PHWC-48754A/2020 | 2020-10-26 | Europe/United Kingdom/Wales/ | Wales Specialist Virology Centre Sequencing lab: Pathogen Genomics Unit | COVID-19 Genomics UK (COG-UK) Consortium | Catherine Moore et al |
| EPI_ISL_639264 | Wales/PHWC-487A3C/2020 | 2020-10-26 | Europe/United Kingdom/Wales/ | Wales Specialist Virology Centre Sequencing lab: Pathogen Genomics Unit | COVID-19 Genomics UK (COG-UK) Consortium | Catherine Moore et al |
| EPI_ISL_639279 | Wales/PHWC-487B66/2020 | 2020-10-27 | Europe/United Kingdom/Wales/ | Wales Specialist Virology Centre Sequencing lab: Pathogen Genomics Unit | COVID-19 Genomics UK (COG-UK) Consortium | Catherine Moore et al |
| EPI_ISL_639576 | Wales/PHWC-48945A/2020 | 2020-10-31 | Europe/United Kingdom/Wales/ | Wales Specialist Virology Centre Sequencing lab: Pathogen Genomics Unit | COVID-19 Genomics UK (COG-UK) Consortium | Catherine Moore et al |
| EPI_ISL_639682 | Latvia/185/2020 | 2020-10-01 | Europe/Latvia/Latvia/ | E. Gulbja Laboratorija | Latvian Biomedical Research and Study Centre | Ivars Silamiƒ∑elis et al |

| | | | | | | |
|---|---|---|---|---|---|---|
| EPI_ISL_639693 | Latvia/196/2020 | 2020-10-07 | Europe/Latvia/Latvia/ | E. Gulbja Laboratorija | Latvian Biomedical Research and Study Centre | Ivars Silamiƒ∑elis et al |
| EPI_ISL_639727 | Canada/ON-PHL-20-01817/2020 | 20-Apr | North America/Canada/Ontario/ | Public Health Ontario Laboratory | Public Health Ontario Laboratory | Vanessa G Allen et al |
| EPI_ISL_639728 | Canada/ON-PHL-20-01818/2020 | 20-Apr | North America/Canada/Ontario/ | Public Health Ontario Laboratory | Public Health Ontario Laboratory | Vanessa G Allen et al |
| EPI_ISL_639739 | Romania/BUCHAREST-IMT-1/2020 | 2020-07-20 | Europe/Romania/Romania/ | Centre of Nanotechnologies, INCD IMT-Bucuresti (National Institute for Research and Development in Microtechnologies - Bucharest) | Centre of Nanotechnologies, INCD IMT-Bucuresti (National Institute for Research and Development in Microtechnologies - Bucharest) | Salceanu et al |
| EPI_ISL_639800 | Australia/QLD1209/2020 | 2020-03-22 | Oceania/Australia/Australia/ | unknown | Public Health Virology Laboratory, Forensic and Scientific Services (PHV-FSS) | Son Nguyen et al. et al |
| EPI_ISL_639875 | Ireland/MH-NVRL-20IRL82653/2020 | 2020-10-05 | Europe/Ireland/Meath/ | National Virus Reference Laboratory | National Virus Reference Laboratory | Michael Carr et al |
| EPI_ISL_639881 | Ireland/LH-NVRL-20IRL82333/2020 | 2020-10-05 | Europe/Ireland/Louth/ | National Virus Reference Laboratory | National Virus Reference Laboratory | Michael Carr et al |
| EPI_ISL_639882 | Ireland/CN-NVRL-20IRL82841/2020 | 2020-10-05 | Europe/Ireland/Cavan/ | National Virus Reference Laboratory | National Virus Reference Laboratory | Michael Carr et al |
| EPI_ISL_639895 | Ireland/LH-NVRL-20IRL52389/2020 | 2020-10-01 | Europe/Ireland/Louth/ | National Virus Reference Laboratory | National Virus Reference Laboratory | Michael Carr et al |
| EPI_ISL_639905 | Ireland/CN-NVRL-20IRL51448/2020 | 2020-09-30 | Europe/Ireland/Cavan/ | National Virus Reference Laboratory | National Virus Reference Laboratory | Michael Carr et al |

| | | | | | | |
|---|---|---|---|---|---|---|
| EPI_ISL_639912 | Russia/OMS-ORINFI-1123S/2020 | 2020-06-26 | Europe/Russia/Omsk/Omsk | Omsk Research Institute of Natural Focal Infections | WHO National Influenza Centre Russian Federation | Artem Fadeev et al |
| EPI_ISL_639913 | Russia/OMS-ORINFI-1564S/2020 | 2020-06-30 | Europe/Russia/Omsk/Omsk | Omsk Research Institute of Natural Focal Infections | WHO National Influenza Centre Russian Federation | Artem Fadeev et al |
| EPI_ISL_639914 | Russia/OMS-ORINFI-1612S/2020 | 2020-07-06 | Europe/Russia/Omsk/Omsk | Omsk Research Institute of Natural Focal Infections | WHO National Influenza Centre Russian Federation | Artem Fadeev et al |
| EPI_ISL_639915 | Russia/OMS-ORINFI-1068S/2020 | 2020-06-26 | Europe/Russia/Omsk/Omsk | Omsk Research Institute of Natural Focal Infections | WHO National Influenza Centre Russian Federation | Artem Fadeev et al |
| EPI_ISL_639917 | Russia/OMS-ORINFI-2096S/2020 | 2020-07-02 | Europe/Russia/Omsk/Omsk | Omsk Research Institute of Natural Focal Infections | WHO National Influenza Centre Russian Federation | Artem Fadeev et al |
| EPI_ISL_639921 | Russia/OMS-ORINFI-2292S/2020 | 2020-06-30 | Europe/Russia/Omsk/Omsk | Omsk Research Institute of Natural Focal Infections | WHO National Influenza Centre Russian Federation | Artem Fadeev et al |
| EPI_ISL_639922 | Russia/OMS-ORINFI-2307S/2020 | 2020-06-23 | Europe/Russia/Omsk/Omsk | Omsk Research Institute of Natural Focal Infections | WHO National Influenza Centre Russian Federation | Artem Fadeev et al |
| EPI_ISL_639923 | Russia/OMS-ORINFI-3769S/2020 | 2020-05-28 | Europe/Russia/Omsk/Omsk | Omsk Research Institute of Natural Focal Infections | WHO National Influenza Centre Russian Federation | Artem Fadeev et al |
| EPI_ISL_639924 | Russia/OMS-ORINFI-3801S/2020 | 2020-06-01 | Europe/Russia/Omsk/Omsk | Omsk Research Institute of Natural Focal Infections | WHO National Influenza Centre Russian Federation | Artem Fadeev et al |
| EPI_ISL_639925 | Russia/OMS-ORINFI-3816S/2020 | 2020-06-02 | Europe/Russia/Omsk/Omsk | Omsk Research Institute of Natural Focal Infections | WHO National Influenza Centre Russian Federation | Artem Fadeev et al |

| | | | | | | |
|---|---|---|---|---|---|---|
| EPI_ISL_639926 | Russia/OMS-ORINFI-3818S/2020 | 2020-06-02 | Europe/Russia/Omsk/Omsk | Omsk Research Institute of Natural Focal Infections | WHO National Influenza Centre Russian Federation | Artem Fadeev et al |
| EPI_ISL_639927 | Russia/OMS-ORINFI-9916S/2020 | 2020-05-21 | Europe/Russia/Omsk/Omsk | Omsk Research Institute of Natural Focal Infections | WHO National Influenza Centre Russian Federation | Artem Fadeev et al |
| EPI_ISL_639928 | Russia/OMS-ORINFI-5311S/2020 | 2020-05-05 | Europe/Russia/Omsk/Omsk | Omsk Research Institute of Natural Focal Infections | WHO National Influenza Centre Russian Federation | Artem Fadeev et al |
| EPI_ISL_639929 | Russia/OMS-ORINFI-5312S/2020 | 2020-05-05 | Europe/Russia/Omsk/Omsk | Omsk Research Institute of Natural Focal Infections | WHO National Influenza Centre Russian Federation | Artem Fadeev et al |
| EPI_ISL_639930 | Russia/OMS-ORINFI-5368S/2020 | 2020-06-05 | Europe/Russia/Omsk/Omsk | Omsk Research Institute of Natural Focal Infections | WHO National Influenza Centre Russian Federation | Artem Fadeev et al |
| EPI_ISL_639931 | Russia/OMS-ORINFI-5753S/2020 | 2020-05-25 | Europe/Russia/Omsk/Omsk | Omsk Research Institute of Natural Focal Infections | WHO National Influenza Centre Russian Federation | Artem Fadeev et al |
| EPI_ISL_639932 | Russia/OMS-ORINFI-5774S/2020 | 2020-06-07 | Europe/Russia/Omsk/Omsk | Omsk Research Institute of Natural Focal Infections | WHO National Influenza Centre Russian Federation | Artem Fadeev et al |
| EPI_ISL_639933 | Russia/OMS-ORINFI-6212S/2020 | 2020-05-10 | Europe/Russia/Omsk/Omsk | Omsk Research Institute of Natural Focal Infections | WHO National Influenza Centre Russian Federation | Artem Fadeev et al |
| EPI_ISL_639934 | Russia/OMS-ORINFI-6390S/2020 | 2020-06-05 | Europe/Russia/Omsk/Omsk | Omsk Research Institute of Natural Focal Infections | WHO National Influenza Centre Russian Federation | Artem Fadeev et al |
| EPI_ISL_639935 | Russia/OMS-ORINFI-6680S/2020 | 2020-05-12 | Europe/Russia/Omsk/Omsk | Omsk Research Institute of Natural Focal Infections | WHO National Influenza Centre Russian Federation | Artem Fadeev et al |

| | | | | | | |
|---|---|---|---|---|---|---|
| EPI_ISL_639936 | Russia/OMS-ORINFI-7131S/2020 | 2020-05-14 | Europe/Russia/Omsk/Omsk | Omsk Research Institute of Natural Focal Infections | WHO National Influenza Centre Russian Federation | Artem Fadeev et al |
| EPI_ISL_639937 | Russia/OMS-ORINFI-7139S/2020 | 2020-05-14 | Europe/Russia/Omsk/Omsk | Omsk Research Institute of Natural Focal Infections | WHO National Influenza Centre Russian Federation | Artem Fadeev et al |
| EPI_ISL_639938 | Russia/OMS-ORINFI-7871S/2020 | 2020-05-25 | Europe/Russia/Omsk/Omsk | Omsk Research Institute of Natural Focal Infections | WHO National Influenza Centre Russian Federation | Artem Fadeev et al |
| EPI_ISL_639939 | Russia/OMS-ORINFI-8293S/2020 | 2020-05-17 | Europe/Russia/Omsk/Omsk | Omsk Research Institute of Natural Focal Infections | WHO National Influenza Centre Russian Federation | Artem Fadeev et al |
| EPI_ISL_639940 | Russia/OMS-ORINFI-8296S/2020 | 2020-05-17 | Europe/Russia/Omsk/Omsk | Omsk Research Institute of Natural Focal Infections | WHO National Influenza Centre Russian Federation | Artem Fadeev et al |
| EPI_ISL_639941 | Russia/OMS-ORINFI-858S/2020 | 2020-06-26 | Europe/Russia/Omsk/Omsk | Omsk Research Institute of Natural Focal Infections | WHO National Influenza Centre Russian Federation | Artem Fadeev et al |
| EPI_ISL_639942 | Russia/OMS-ORINFI-848S/2020 | 2020-06-26 | Europe/Russia/Omsk/Omsk | Omsk Research Institute of Natural Focal Infections | WHO National Influenza Centre Russian Federation | Artem Fadeev et al |
| EPI_ISL_639943 | Russia/OMS-ORINFI-864S/2020 | 2020-06-26 | Europe/Russia/Omsk/Omsk | Omsk Research Institute of Natural Focal Infections | WHO National Influenza Centre Russian Federation | Artem Fadeev et al |
| EPI_ISL_639944 | Russia/OMS-ORINFI-865S/2020 | 2020-06-26 | Europe/Russia/Omsk/Omsk | Omsk Research Institute of Natural Focal Infections | WHO National Influenza Centre Russian Federation | Artem Fadeev et al |
| EPI_ISL_639945 | Russia/OMS-ORINFI-916S/2020 | 2020-06-26 | Europe/Russia/Omsk/Omsk | Omsk Research Institute of Natural Focal Infections | WHO National Influenza Centre Russian Federation | Artem Fadeev et al |

| | | | | | | |
|---|---|---|---|---|---|---|
| EPI_ISL_639946 | Russia/OMS-ORINFI-9781S/2020 | 2020-05-21 | Europe/Russia/Omsk/Omsk | Omsk Research Institute of Natural Focal Infections | WHO National Influenza Centre Russian Federation | Artem Fadeev et al |
| EPI_ISL_639947 | Russia/OMS-ORINFI-9981S/2020 | 2020-06-17 | Europe/Russia/Omsk/Omsk | Omsk Research Institute of Natural Focal Infections | WHO National Influenza Centre Russian Federation | Artem Fadeev et al |
| EPI_ISL_639951 | Russia/MOS-RII-MH2800S/2020 | 2020-09-22 | Europe/Russia/Moscow Oblast/Lapino | HELIX LLC | WHO National Influenza Centre Russian Federation | Andrey Komissarov et al |
| EPI_ISL_639952 | Russia/MOS-RII-MH2802S/2020 | 2020-09-19 | Europe/Russia/Moscow Oblast/Korolyov | HELIX LLC | WHO National Influenza Centre Russian Federation | Andrey Komissarov et al |
| EPI_ISL_639955 | Russia/MOS-RII-MH2806S/2020 | 2020-09-27 | Europe/Russia/Moscow Oblast/Lyuberts | HELIX LLC | WHO National Influenza Centre Russian Federation | Andrey Komissarov et al |
| EPI_ISL_639956 | Russia/MOS-RII-MH2807S/2020 | 2020-09-27 | Europe/Russia/Moscow Oblast/Lapino | HELIX LLC | WHO National Influenza Centre Russian Federation | Andrey Komissarov et al |
| EPI_ISL_639958 | Russia/NVS-RII-MH2809S/2020 | 2020-09-25 | Europe/Russia/Novosibirsk/Berdsk | HELIX LLC | WHO National Influenza Centre Russian Federation | Andrey Komissarov et al |
| EPI_ISL_639959 | Russia/NVS-RII-MH2810S/2020 | 2020-09-25 | Europe/Russia/Novosibirsk/Berdsk | HELIX LLC | WHO National Influenza Centre Russian Federation | Andrey Komissarov et al |
| EPI_ISL_639961 | Russia/ORE-RII-MH2814S/2020 | 2020-09-21 | Europe/Russia/Orenburg/Orenburg | HELIX LLC | WHO National Influenza Centre Russian Federation | Andrey Komissarov et al |
| EPI_ISL_639963 | Russia/PER-RII-MH2816S/2020 | 2020-09-24 | Europe/Russia/Perm Krai/Perm | HELIX LLC | WHO National Influenza Centre Russian Federation | Andrey Komissarov et al |

| | | | | | | |
|---|---|---|---|---|---|---|
| EPI_ISL_639964 | Russia/PER-RII-MH2818S/2020 | 2020-09-22 | Europe/Russia/Perm Krai/Perm | HELIX LLC | WHO National Influenza Centre Russian Federation | Andrey Komissarov et al |
| EPI_ISL_639965 | Russia/ROS-RII-MH2820S/2020 | 2020-09-24 | Europe/Russia/Rostov/Kamensk-Shakhti | HELIX LLC | WHO National Influenza Centre Russian Federation | Andrey Komissarov et al |
| EPI_ISL_639967 | Russia/RYA-RII-MH2823S/2020 | 2020-09-24 | Europe/Russia/Ryazan/Ryazan | HELIX LLC | WHO National Influenza Centre Russian Federation | Andrey Komissarov et al |
| EPI_ISL_639968 | Russia/RYA-RII-MH2824S/2020 | 2020-09-26 | Europe/Russia/Ryazan/Ryazan | HELIX LLC | WHO National Influenza Centre Russian Federation | Andrey Komissarov et al |
| EPI_ISL_639969 | Russia/SVE-RII-MH2824S/2020 | 2020-09-18 | Europe/Russia/Sverdlovsk/Asbest | HELIX LLC | WHO National Influenza Centre Russian Federation | Andrey Komissarov et al |
| EPI_ISL_639970 | Russia/SVE-RII-MH2828S/2020 | 2020-09-24 | Europe/Russia/Sverdlovsk/Yekaterinbur | HELIX LLC | WHO National Influenza Centre Russian Federation | Andrey Komissarov et al |
| EPI_ISL_639973 | Russia/SVE-RII-MH2831S/2020 | 2020-09-24 | Europe/Russia/Sverdlovsk/Yekaterinbur | HELIX LLC | WHO National Influenza Centre Russian Federation | Andrey Komissarov et al |
| EPI_ISL_639975 | France/PAC-NRC2718/2020 | 2020-10-05 | Europe/France/Provence-Alpes-C√¥te d' | CNR Virus des Infections Respiratoires - France SUD | CNR Virus des Infections Respiratoires - France SUD | Antonin Bal et al |
| EPI_ISL_639976 | France/ARA75790/2020 | 2020-09-21 | Europe/France/Auvergne-Rh√¥ne-Alpes/ | CNR Virus des Infections Respiratoires - France SUD | CNR Virus des Infections Respiratoires - France SUD | Antonin Bal et al |
| EPI_ISL_639977 | France/PAC-NRC02798/2020 | 2020-10-14 | Europe/France/Provence-Alpes-C√¥te d' | CNR Virus des Infections Respiratoires - France SUD | CNR Virus des Infections Respiratoires - France SUD | Antonin Bal et al |

| | | | | | | |
|---|---|---|---|---|---|---|
| EPI_ISL_639978 | France/ARA74567/2020 | 2020-09-17 | Europe/France/Auvergne-Rhv¥ne-Alpes/ | Centre Hospitalier Saint Joseph Saint Luc | CNR Virus des Infections Respiratoires - France SUD | Antonin Bal et al |
| EPI_ISL_639979 | France/ARA75377/2020 | 2020-09-19 | Europe/France/Auvergne-Rhv¥ne-Alpes/ | CNR Virus des Infections Respiratoires - France SUD | CNR Virus des Infections Respiratoires - France SUD | Antonin Bal et al |
| EPI_ISL_639980 | France/ARA76733/2020 | 2020-09-22 | Europe/France/Auvergne-Rhv¥ne-Alpes/ | CNR Virus des Infections Respiratoires - France SUD | CNR Virus des Infections Respiratoires - France SUD | Antonin Bal et al |
| EPI_ISL_639981 | France/ARA-NRC02745/2020 | 2020-10-13 | Europe/France/Auvergne-Rhv¥ne-Alpes/ | CNR Virus des Infections Respiratoires - France SUD | CNR Virus des Infections Respiratoires - France SUD | Antonin Bal et al |
| EPI_ISL_639982 | France/PAC-NRC02780/2020 | 2020-10-24 | Europe/France/Provence-Alpes-Cv¥te d' | CNR Virus des Infections Respiratoires - France SUD | CNR Virus des Infections Respiratoires - France SUD | Antonin Bal et al |
| EPI_ISL_639983 | France/ARA-NRC2813/2020 | 2020-11-02 | Europe/France/Auvergne-Rhv¥ne-Alpes/ | CNR Virus des Infections Respiratoires - France SUD | CNR Virus des Infections Respiratoires - France SUD | Antonin Bal et al |
| EPI_ISL_639984 | France/ARA75924/2020 | 2020-09-20 | Europe/France/Auvergne-Rhv¥ne-Alpes/ | Centre Hospitalier de Bourg en Bresse | CNR Virus des Infections Respiratoires - France SUD | Antonin Bal et al |
| EPI_ISL_639986 | France/ARA97420/2020 | 2020-10-30 | Europe/France/Auvergne-Rhv¥ne-Alpes/ | CNR Virus des Infections Respiratoires - France SUD | CNR Virus des Infections Respiratoires - France SUD | Antonin Bal et al |
| EPI_ISL_639987 | France/ARA-NRC02726/2020 | 2020-10-05 | Europe/France/Auvergne-Rhv¥ne-Alpes/ | CNR Virus des Infections Respiratoires - France SUD | CNR Virus des Infections Respiratoires - France SUD | Antonin Bal et al |
| EPI_ISL_639991 | France/ARA-NRC02706/2020 | 2020-09-30 | Europe/France/Auvergne-Rhv¥ne-Alpes/ | CNR Virus des Infections Respiratoires - France SUD | CNR Virus des Infections Respiratoires - France SUD | Antonin Bal et al |

| | | | | | | |
|---|---|---|---|---|---|---|
| EPI_ISL_639992 | France/ARA77081/2020 | 2020-09-23 | Europe/France/Auvergne-Rh√¥ne-Alpes/ | CNR Virus des Infections Respiratoires - France SUD | CNR Virus des Infections Respiratoires - France SUD | Antonin Bal et al |
| EPI_ISL_639999 | France/ARA68158/2020 | 2020-09-05 | Europe/France/Auvergne-Rh√¥ne-Alpes/ | CNR Virus des Infections Respiratoires - France SUD | CNR Virus des Infections Respiratoires - France SUD | Antonin Bal et al |
| EPI_ISL_640000 | France/ARA93756/2020 | 2020-10-24 | Europe/France/Auvergne-Rh√¥ne-Alpes/ | CNR Virus des Infections Respiratoires - France SUD | CNR Virus des Infections Respiratoires - France SUD | Antonin Bal et al |
| EPI_ISL_640002 | France/OCC-NRC02765/2020 | 2020-10-19 | Europe/France/Occitanie/Montpellier | CNR Virus des Infections Respiratoires - France SUD | CNR Virus des Infections Respiratoires - France SUD | Antonin Bal et al |
| EPI_ISL_640003 | France/ARA93309/2020 | 2020-10-24 | Europe/France/Auvergne-Rh√¥ne-Alpes/ | CNR Virus des Infections Respiratoires - France SUD | CNR Virus des Infections Respiratoires - France SUD | Antonin Bal et al |
| EPI_ISL_640004 | France/ARA93996/2020 | 2020-10-25 | Europe/France/Auvergne-Rh√¥ne-Alpes/ | CNR Virus des Infections Respiratoires - France SUD | CNR Virus des Infections Respiratoires - France SUD | Antonin Bal et al |
| EPI_ISL_640005 | France/ARA-NRC02786/2020 | 2020-10-26 | Europe/France/Auvergne-Rh√¥ne-Alpes/ | CNR Virus des Infections Respiratoires - France SUD | CNR Virus des Infections Respiratoires - France SUD | Antonin Bal et al |
| EPI_ISL_640006 | France/OCC-NRC02755/2020 | 2020-10-15 | Europe/France/Occitanie/Saint-Gely-Du- | CNR Virus des Infections Respiratoires - France SUD | CNR Virus des Infections Respiratoires - France SUD | Antonin Bal et al |
| EPI_ISL_640007 | France/ARA97048/2020 | 2020-10-29 | Europe/France/Auvergne-Rh√¥ne-Alpes/ | CNR Virus des Infections Respiratoires - France SUD | CNR Virus des Infections Respiratoires - France SUD | Antonin Bal et al |
| EPI_ISL_640008 | France/ARA97376/2020 | 2020-10-30 | Europe/France/Auvergne-Rh√¥ne-Alpes/ | CNR Virus des Infections Respiratoires - France SUD | CNR Virus des Infections Respiratoires - France SUD | Antonin Bal et al |

| | | | | | | |
|---|---|---|---|---|---|---|
| EPI_ISL_640011 | France/ARA97193/2020 | 2020-10-29 | Europe/France/Auvergne-Rhv¥ne-Alpes/ | CNR Virus des Infections Respiratoires - France SUD | CNR Virus des Infections Respiratoires - France SUD | Antonin Bal et al |
| EPI_ISL_640012 | France/ARA92942/2020 | 2020-10-23 | Europe/France/Auvergne-Rhv¥ne-Alpes/ | CNR Virus des Infections Respiratoires - France SUD | CNR Virus des Infections Respiratoires - France SUD | Antonin Bal et al |
| EPI_ISL_640013 | France/ARA94189/2020 | 2020-10-25 | Europe/France/Auvergne-Rhv¥ne-Alpes/ | CNR Virus des Infections Respiratoires - France SUD | CNR Virus des Infections Respiratoires - France SUD | Antonin Bal et al |
| EPI_ISL_640014 | France/ARA93236/2020 | 2020-10-23 | Europe/France/Auvergne-Rhv¥ne-Alpes/ | CNR Virus des Infections Respiratoires - France SUD | CNR Virus des Infections Respiratoires - France SUD | Antonin Bal et al |
| EPI_ISL_640016 | SouthAfrica/NHLS-UCT-GS-0014/2020 | 2020-08-10 | Africa/South Africa/Western Cape Provi | George Hospital wc GRH | NHLS/UCT | Arash Iranzadeh et al |
| EPI_ISL_640018 | SouthAfrica/NHLS-UCT-GS-0016/2020 | 2020-08-11 | Africa/South Africa/Northern Cape/ | Heidelberg Clinic wc HBC | NHLS/UCT | Arash Iranzadeh et al |
| EPI_ISL_640023 | SouthAfrica/NHLS-UCT-GS-0022/2020 | 2020-08-11 | Africa/South Africa/Western Cape Provi | Oudtshoorn Hospital wc OUD | NHLS/UCT | Arash Iranzadeh et al |
| EPI_ISL_640027 | SouthAfrica/NHLS-UCT-GS-0026/2020 | 2020-08-16 | Africa/South Africa/Western Cape Provi | George Hospital wc GRH | NHLS/UCT | Arash Iranzadeh et al |
| EPI_ISL_640028 | SouthAfrica/NHLS-UCT-GS-0027/2020 | 2020-08-18 | Africa/South Africa/Western Cape Provi | Mitchells Plain Hospital wc MPH | NHLS/UCT | Arash Iranzadeh et al |
| EPI_ISL_640029 | SouthAfrica/NHLS-UCT-GS-0028/2020 | 2020-08-18 | Africa/South Africa/Western Cape Provi | D'Almeida Clinic wc DAL | NHLS/UCT | Arash Iranzadeh et al |
| EPI_ISL_640032 | SouthAfrica/NHLS-UCT-GS-0031/2020 | 2020-08-18 | Africa/South Africa/Western Cape Provi | Victoria Hospital wc VHW | NHLS/UCT | Arash Iranzadeh et al |
| EPI_ISL_640035 | SouthAfrica/NHLS-UCT-GS-0034/2020 | 2020-08-20 | Africa/South Africa/Western Cape Provi | Riversdale Clinic wc RAV | NHLS/UCT | Arash Iranzadeh et al |
| EPI_ISL_640036 | SouthAfrica/NHLS-UCT-GS-0035/2020 | 2020-08-21 | Africa/South Africa/Western Cape Provi | Bridgeton CDC wc BTC | NHLS/UCT | Arash Iranzadeh et al |
| EPI_ISL_640037 | SouthAfrica/NHLS-UCT-GS-0036/2020 | 2020-08-22 | Africa/South Africa/Western Cape Provi | Valkenberg Hospital wc VBH | NHLS/UCT | Arash Iranzadeh et al |
| EPI_ISL_640046 | SouthAfrica/NHLS-UCT-GS-1947/2020 | 2020-08-29 | Africa/South Africa/Western Cape Provi | Valkenberg Hospital wc VBH | NHLS/UCT | Arash Iranzadeh et al |
| EPI_ISL_640048 | SouthAfrica/NHLS-UCT-GS-1984/2020 | 2020-08-25 | Africa/South Africa/Western Cape Provi | D'Almeida Clinic wc DAL | NHLS/UCT | Arash Iranzadeh et al |
| EPI_ISL_640051 | SouthAfrica/NHLS-UCT-GS-0045/2020 | 2020-03-22 | Africa/South Africa/Western Cape Provi | Groote Schuur Hospital wc GSH | NHLS/UCT | Arash Iranzadeh et al |
| EPI_ISL_640052 | SouthAfrica/NHLS-UCT-GS-0047/2020 | 2020-03-22 | Africa/South Africa/Western Cape Provi | Groote Schuur Hospital wc GSH | NHLS/UCT | Arash Iranzadeh et al |
| EPI_ISL_640065 | SouthAfrica/NHLS-UCT-GS-0146/2020 | 2020-06-29 | Africa/South Africa/Western Cape Provi | Mitchells Plain Hospital wc MPH | NHLS/UCT | Arash Iranzadeh et al |
| EPI_ISL_640066 | SouthAfrica/NHLS-UCT-GS-0149/2020 | 2020-06-29 | Africa/South Africa/Western Cape Provi | Groote Schuur Hospital wc GSH | NHLS/UCT | Arash Iranzadeh et al |
| EPI_ISL_640068 | SouthAfrica/NHLS-UCT-GS-0289/2020 | 2020-07-08 | Africa/South Africa/Western Cape Provi | Clinic-in-Asla | NHLS/UCT | Arash Iranzadeh et al |
| EPI_ISL_640073 | SouthAfrica/NHLS-UCT-LA-9007/2020 | 2020-03-19 | Africa/South Africa/Western Cape Provi | unknown | NHLS/UCT | Arash Iranzadeh et al |

| | | | | | | |
|---|---|---|---|---|---|---|
| EPI_ISL_640077 | SouthAfrica/NHLS-UCT-GS-1827/2020 | 2020-08-29 | Africa/South Africa/Western Cape Provi | Oudtshoorn Hospital wc OUD | NHLS/UCT | Arash Iranzadeh et al |
| EPI_ISL_640079 | SouthAfrica/NHLS-UCT-GS-2426/2020 | 2020-06-06 | Africa/South Africa/Western Cape Provi | Stellenbosch Hospital wc STB | NHLS/UCT | Arash Iranzadeh et al |
| EPI_ISL_640081 | SouthAfrica/NHLS-UCT-GS-3159/2020 | 2020-05-14 | Africa/South Africa/Western Cape Provi | Vanguard CHC wc VGC | NHLS/UCT | Arash Iranzadeh et al |
| EPI_ISL_640085 | SouthAfrica/NHLS-UCT-GS-2093/2020 | 2020-09-25 | Africa/South Africa/Western Cape Provi | Victoria Hospital wc VHW | NHLS/UCT | Arash Iranzadeh et al |
| EPI_ISL_640089 | SouthAfrica/NHLS-UCT-GS-3517/2020 | 2020-10-09 | Africa/South Africa/Western Cape Provi | 2 Military Hospital wc MAA | NHLS/UCT | Arash Iranzadeh et al |
| EPI_ISL_640093 | SouthAfrica/NHLS-UCT-GS-3555/2020 | 2020-10-10 | Africa/South Africa/Western Cape Provi | Kensington CDC wc KSC | NHLS/UCT | Arash Iranzadeh et al |
| EPI_ISL_640095 | SouthAfrica/NHLS-UCT-GS-3559/2020 | 2020-10-09 | Africa/South Africa/Western Cape Provi | Du Noon CDC wc DNC | NHLS/UCT | Arash Iranzadeh et al |
| EPI_ISL_640108 | SouthAfrica/NHLS-UCT-GS-1772/2020 | 2020-09-02 | Africa/South Africa/Western Cape Provi | Victoria Hospital wc VHW | NHLS/UCT | Arash Iranzadeh et al |
| EPI_ISL_640112 | SouthAfrica/NHLS-UCT-GS-1779/2020 | 2020-09-04 | Africa/South Africa/Western Cape Provi | Groote Schuur Hospital wc GSH | NHLS/UCT | Arash Iranzadeh et al |
| EPI_ISL_640114 | SouthAfrica/NHLS-UCT-GS-1782/2020 | 2020-09-04 | Africa/South Africa/Western Cape Provi | Valkenberg Hospital wc VBH | NHLS/UCT | Arash Iranzadeh et al |
| EPI_ISL_640126 | SouthAfrica/NHLS-UCT-GS-2078/2020 | 2020-09-21 | Africa/South Africa/Western Cape Provi | Victoria Hospital wc VHW | NHLS/UCT | Arash Iranzadeh et al |
| EPI_ISL_640136 | SouthAfrica/NHLS-UCT-GS-3472/2020 | 2020-09-28 | Africa/South Africa/Western Cape Provi | Red Cross Children's Hospital wc RXH | NHLS/UCT | Arash Iranzadeh et al |
| EPI_ISL_640140 | SouthAfrica/NHLS-UCT-GS-3519/2020 | 2020-10-10 | Africa/South Africa/Western Cape Provi | Groote Schuur Hospital wc GSH | NHLS/UCT | Arash Iranzadeh et al |
| EPI_ISL_640147 | USA/MI-UM-10036530676/2020 | 2020-10-20 | North America/USA/Michigan/ | University of Michigan Clinical Microbiology Laboratory | Lauring Lab, University of Michigan, Department of Microbiology and Immunology | Valesano et al |
| EPI_ISL_640149 | USA/MI-UM-10036539730/2020 | 2020-10-20 | North America/USA/Michigan/ | University of Michigan Clinical Microbiology Laboratory | Lauring Lab, University of Michigan, Department of Microbiology and Immunology | Valesano et al |
| EPI_ISL_640159 | USA/MI-UM-10036541938/2020 | 2020-10-20 | North America/USA/Michigan/ | University of Michigan Clinical Microbiology Laboratory | Lauring Lab, University of Michigan, Department of Microbiology and Immunology | Valesano et al |
| EPI_ISL_640162 | USA/MI-UM-10036532868/2020 | 2020-10-20 | North America/USA/Michigan/ | University of Michigan Clinical Microbiology Laboratory | Lauring Lab, University of Michigan, Department of Microbiology and Immunology | Valesano et al |

| | | | | | | |
|---|---|---|---|---|---|---|
| EPI_ISL_640163 | USA/MI-UM-10036578565/2020 | 2020-10-22 | North America/USA/Michigan/ | University of Michigan Clinical Microbiology Laboratory | Lauring Lab, University of Michigan, Department of Microbiology and Immunology | Valesano et al |
| EPI_ISL_640164 | USA/MI-UM-10036557572/2020 | 2020-10-21 | North America/USA/Michigan/ | University of Michigan Clinical Microbiology Laboratory | Lauring Lab, University of Michigan, Department of Microbiology and Immunology | Valesano et al |
| EPI_ISL_640172 | USA/MI-UM-10036554113/2020 | 2020-10-21 | North America/USA/Michigan/ | University of Michigan Clinical Microbiology Laboratory | Lauring Lab, University of Michigan, Department of Microbiology and Immunology | Valesano et al |
| EPI_ISL_640173 | USA/MI-UM-10036562497/2020 | 2020-10-21 | North America/USA/Michigan/ | University of Michigan Clinical Microbiology Laboratory | Lauring Lab, University of Michigan, Department of Microbiology and Immunology | Valesano et al |
| EPI_ISL_640185 | USA/MI-UM-10036548240/2020 | 2020-10-21 | North America/USA/Michigan/ | University of Michigan Clinical Microbiology Laboratory | Lauring Lab, University of Michigan, Department of Microbiology and Immunology | Valesano et al |
| EPI_ISL_640187 | USA/MI-UM-10036561188/2020 | 2020-10-21 | North America/USA/Michigan/ | University of Michigan Clinical Microbiology Laboratory | Lauring Lab, University of Michigan, Department of Microbiology and Immunology | Valesano et al |

| | | | | | | |
|---|---|---|---|---|---|---|
| EPI_ISL_640198 | USA/MI-UM-10036575544/2020 | 2020-10-22 | North America/USA/Michigan/ | University of Michigan Clinical Microbiology Laboratory | Lauring Lab, University of Michigan, Department of Microbiology and Immunology | Valesano et al |
| EPI_ISL_640205 | USA/MI-UM-10036558900/2020 | 2020-10-21 | North America/USA/Michigan/ | University of Michigan Clinical Microbiology Laboratory | Lauring Lab, University of Michigan, Department of Microbiology and Immunology | Valesano et al |
| EPI_ISL_640217 | USA/MI-UM-10036598396/2020 | 2020-10-23 | North America/USA/Michigan/ | University of Michigan Clinical Microbiology Laboratory | Lauring Lab, University of Michigan, Department of Microbiology and Immunology | Valesano et al |
| EPI_ISL_640219 | Germany/BW-MVZ-LS-01/2020 | 2020-04-29 | Europe/Germany/Baden-Wuerttemberg | MVZ Laborv§rzte Singen | MVZ Laborv§rzte Singen | Jonas Schmidt et al |
| EPI_ISL_640229 | Germany/BW-MVZ-LS-11/2020 | 2020-04-15 | Europe/Germany/Baden-Wuerttemberg | MVZ Laborv§rzte Singen | MVZ Laborv§rzte Singen | Jonas Schmidt et al |
| EPI_ISL_640230 | Germany/BW-MVZ-LS-12/2020 | 2020-04-16 | Europe/Germany/Baden-Wuerttemberg | MVZ Laborv§rzte Singen | MVZ Laborv§rzte Singen | Jonas Schmidt et al |
| EPI_ISL_640232 | Germany/BW-MVZ-LS-14/2020 | 2020-04-14 | Europe/Germany/Baden-Wuerttemberg | MVZ Laborv§rzte Singen | MVZ Laborv§rzte Singen | Jonas Schmidt et al |
| EPI_ISL_640237 | Germany/BW-MVZ-LS-19/2020 | 2020-04-14 | Europe/Germany/Baden-Wuerttemberg | MVZ Laborv§rzte Singen | MVZ Laborv§rzte Singen | Jonas Schmidt et al |
| EPI_ISL_640241 | Germany/BW-MVZ-LS-23/2020 | 2020-04-16 | Europe/Germany/Baden-Wuerttemberg | MVZ Laborv§rzte Singen | MVZ Laborv§rzte Singen | Jonas Schmidt et al |
| EPI_ISL_640242 | Germany/BW-MVZ-LS-24/2020 | 2020-04-07 | Europe/Germany/Baden-Wuerttemberg | MVZ Laborv§rzte Singen | MVZ Laborv§rzte Singen | Jonas Schmidt et al |
| EPI_ISL_640246 | Germany/BW-MVZ-LS-28/2020 | 2020-04-21 | Europe/Germany/Baden-Wuerttemberg | MVZ Laborv§rzte Singen | MVZ Laborv§rzte Singen | Jonas Schmidt et al |
| EPI_ISL_640248 | Germany/BW-MVZ-LS-30/2020 | 2020-04-16 | Europe/Germany/Baden-Wuerttemberg | MVZ Laborv§rzte Singen | MVZ Laborv§rzte Singen | Jonas Schmidt et al |
| EPI_ISL_640250 | Germany/BW-MVZ-LS-32/2020 | 2020-04-15 | Europe/Germany/Baden-Wuerttemberg | MVZ Laborv§rzte Singen | MVZ Laborv§rzte Singen | Jonas Schmidt et al |
| EPI_ISL_640254 | Germany/BW-MVZ-LS-37/2020 | 2020-04-08 | Europe/Germany/Baden-Wuerttemberg | MVZ Laborv§rzte Singen | MVZ Laborv§rzte Singen | Jonas Schmidt et al |

| | | | | | | |
|---|---|---|---|---|---|---|
| EPI_ISL_640256 | Germany/BW-MVZ-LS-39/2020 | 2020-04-16 | Europe/Germany/Baden-Wuerttemberg | MVZ Labor√§rzte Singen | MVZ Labor√§rzte Singen | Jonas Schmidt et al |
| EPI_ISL_640257 | Germany/BW-MVZ-LS-40/2020 | 2020-04-21 | Europe/Germany/Baden-Wuerttemberg | MVZ Labor√§rzte Singen | MVZ Labor√§rzte Singen | Jonas Schmidt et al |
| EPI_ISL_640258 | Germany/BW-MVZ-LS-41/2020 | 2020-04-08 | Europe/Germany/Baden-Wuerttemberg | MVZ Labor√§rzte Singen | MVZ Labor√§rzte Singen | Jonas Schmidt et al |
| EPI_ISL_640259 | Germany/BW-MVZ-LS-42/2020 | 2020-04-14 | Europe/Germany/Baden-Wuerttemberg | MVZ Labor√§rzte Singen | MVZ Labor√§rzte Singen | Jonas Schmidt et al |
| EPI_ISL_640263 | Germany/BW-MVZ-LS-46/2020 | 2020-04-09 | Europe/Germany/Baden-Wuerttemberg | MVZ Labor√§rzte Singen | MVZ Labor√§rzte Singen | Jonas Schmidt et al |
| EPI_ISL_640264 | Germany/BW-MVZ-LS-47/2020 | 2020-04-13 | Europe/Germany/Baden-Wuerttemberg | MVZ Labor√§rzte Singen | MVZ Labor√§rzte Singen | Jonas Schmidt et al |
| EPI_ISL_640267 | Germany/BW-MVZ-LS-50/2020 | 2020-04-29 | Europe/Germany/Baden-Wuerttemberg | MVZ Labor√§rzte Singen | MVZ Labor√§rzte Singen | Jonas Schmidt et al |
| EPI_ISL_640270 | Germany/BW-MVZ-LS-53/2020 | 2020-05-07 | Europe/Germany/Baden-Wuerttemberg | MVZ Labor√§rzte Singen | MVZ Labor√§rzte Singen | Jonas Schmidt et al |
| EPI_ISL_640271 | Germany/BW-MVZ-LS-54/2020 | 2020-04-07 | Europe/Germany/Baden-Wuerttemberg | MVZ Labor√§rzte Singen | MVZ Labor√§rzte Singen | Jonas Schmidt et al |
| EPI_ISL_640273 | USA/MI-UM-10036576506/2020 | 2020-10-22 | North America/USA/Michigan/ | University of Michigan Clinical Microbiology Laboratory | Lauring Lab, University of Michigan, Department of Microbiology and Immunology | Valesano et al |
| EPI_ISL_641522 | France/PAC-SC236/2020 | 2020-03-28 | Europe/France/Provence-Alpes-C√¥te d' | CHU de Nice - H√¥pital Archet 3 | CNR Virus des Infections Respiratoires - France SUD | Antonin Bal et al |
| EPI_ISL_641531 | France/PAC-SC246/2020 | 2020-04-24 | Europe/France/Provence-Alpes-C√¥te d' | CHU de Nice - H√¥pital Archet 13 | CNR Virus des Infections Respiratoires - France SUD | Antonin Bal et al |
| EPI_ISL_641532 | France/PAC-SC247/2020 | 2020-04-24 | Europe/France/Provence-Alpes-C√¥te d' | CHU de Nice - H√¥pital Archet 14 | CNR Virus des Infections Respiratoires - France SUD | Antonin Bal et al |
| EPI_ISL_641535 | France/ARA-SC310/2020 | 2020-04-07 | Europe/France/Auvergne-Rh√¥ne-Alpes/ | CHU de Saint-√âtienne H√¥pital Nord | CNR Virus des Infections Respiratoires - France SUD | Antonin Bal et al |

| | | | | | | |
|---|---|---|---|---|---|---|
| EPI_ISL_641539 | France/ARA-SC322/2020 | 2020-03-23 | Europe/France/Auvergne-Rh√¥ne-Alpes/ | CHU de Saint-√âtienne H√¥pital Nord | CNR Virus des Infections Respiratoires - France SUD | Antonin Bal et al |
| EPI_ISL_641540 | France/ARA-SC324/2020 | 2020-03-24 | Europe/France/Auvergne-Rh√¥ne-Alpes/ | CHU de Saint-√âtienne H√¥pital Nord | CNR Virus des Infections Respiratoires - France SUD | Antonin Bal et al |
| EPI_ISL_641542 | France/ARA-SC340/2020 | 2020-03-12 | Europe/France/Auvergne-Rh√¥ne-Alpes/ | CHU de Saint-√âtienne H√¥pital Nord | CNR Virus des Infections Respiratoires - France SUD | Antonin Bal et al |
| EPI_ISL_641548 | France/ARA-SC404/2020 | 2020-04-20 | Europe/France/Auvergne-Rh√¥ne-Alpes/ | CHU Clermont-Ferrand | CNR Virus des Infections Respiratoires - France SUD | Antonin Bal et al |
| EPI_ISL_641551 | France/OCC-SC415/2020 | 2020-03-14 | Europe/France/Occitanie/Toulouse | CHU Toulouse | CNR Virus des Infections Respiratoires - France SUD | Antonin Bal et al |
| EPI_ISL_641554 | France/OCC-SC427/2020 | 2020-04-13 | Europe/France/Occitanie/Toulouse | CHU Toulouse | CNR Virus des Infections Respiratoires - France SUD | Antonin Bal et al |
| EPI_ISL_641555 | France/OCC-SC432/2020 | 2020-05-24 | Europe/France/Occitanie/Toulouse | CHU Toulouse | CNR Virus des Infections Respiratoires - France SUD | Antonin Bal et al |
| EPI_ISL_641556 | France/OCC-SC465/2020 | 2020-03-03 | Europe/France/Occitanie/Montpellier | CHU Montpellier | CNR Virus des Infections Respiratoires - France SUD | Antonin Bal et al |
| EPI_ISL_641558 | Belgium/ULG-10716/2020 | 2020-09-14 | Europe/Belgium/Li√®ge/ | Department of Clinical Microbiology | GIGA Medical Genomics | Keith Durkin et al |
| EPI_ISL_641562 | Belgium/ULG-10720/2020 | 2020-09-14 | Europe/Belgium/Li√®ge/ | Department of Clinical Microbiology | GIGA Medical Genomics | Keith Durkin et al |
| EPI_ISL_641563 | Belgium/ULG-10721/2020 | 2020-09-14 | Europe/Belgium/Li√®ge/ | Department of Clinical Microbiology | GIGA Medical Genomics | Keith Durkin et al |
| EPI_ISL_641568 | Belgium/ULG-10726/2020 | 2020-09-15 | Europe/Belgium/Li√®ge/ | Department of Clinical Microbiology | GIGA Medical Genomics | Keith Durkin et al |

| | | | | | | |
|---|---|---|---|---|---|---|
| EPI_ISL_641591 | Belgium/ULG-10749/2020 | 2020-09-19 | Europe/Belgium/Li√®ge/ | Department of Clinical Microbiology | GIGA Medical Genomics | Keith Durkin et al |
| EPI_ISL_641636 | England/QEUH-B14596/2020 | 2020-11-03 | Europe/United Kingdom/England/ | Lighthouse Lab in Glasgow | Wellcome Sanger Institute for the COVID-19 Genomics UK (COG-UK) Consortium | Harper VanSteenhouse et al |
| EPI_ISL_641666 | England/QEUH-B0F7B6/2020 | 2020-11-02 | Europe/United Kingdom/England/ | Lighthouse Lab in Glasgow | Wellcome Sanger Institute for the COVID-19 Genomics UK (COG-UK) Consortium | Harper VanSteenhouse et al |
| EPI_ISL_641692 | England/QEUH-B13E8D/2020 | 2020-11-02 | Europe/United Kingdom/England/ | Lighthouse Lab in Glasgow | Wellcome Sanger Institute for the COVID-19 Genomics UK (COG-UK) Consortium | Harper VanSteenhouse et al |
| EPI_ISL_641745 | England/QEUH-B1356A/2020 | 2020-11-01 | Europe/United Kingdom/England/ | Lighthouse Lab in Glasgow | Wellcome Sanger Institute for the COVID-19 Genomics UK (COG-UK) Consortium | Harper VanSteenhouse et al |
| EPI_ISL_641858 | England/QEUH-B106C4/2020 | 2020-11-02 | Europe/United Kingdom/England/ | Lighthouse Lab in Glasgow | Wellcome Sanger Institute for the COVID-19 Genomics UK (COG-UK) Consortium | Harper VanSteenhouse et al |
| EPI_ISL_641859 | England/QEUH-B0F710/2020 | 2020-11-02 | Europe/United Kingdom/England/ | Lighthouse Lab in Glasgow | Wellcome Sanger Institute for the COVID-19 Genomics UK (COG-UK) Consortium | Harper VanSteenhouse et al |
| EPI_ISL_641864 | England/QEUH-B13A4D/2020 | 2020-10-31 | Europe/United Kingdom/England/ | Lighthouse Lab in Glasgow | Wellcome Sanger Institute for the COVID-19 Genomics UK (COG-UK) Consortium | Harper VanSteenhouse et al |

| | | | | | | |
|---|---|---|---|---|---|---|
| EPI_ISL_641984 | England/QEUH-B0E9A2/2020 | 2020-11-02 | Europe/United Kingdom/England/ | Lighthouse Lab in Glasgow | Wellcome Sanger Institute for the COVID-19 Genomics UK (COG-UK) Consortium | Harper VanSteenhouse et al |
| EPI_ISL_642027 | England/QEUH-B0EB42/2020 | 2020-11-02 | Europe/United Kingdom/England/ | Lighthouse Lab in Glasgow | Wellcome Sanger Institute for the COVID-19 Genomics UK (COG-UK) Consortium | Harper VanSteenhouse et al |
| EPI_ISL_642039 | England/QEUH-B0EA81/2020 | 2020-11-02 | Europe/United Kingdom/England/ | Lighthouse Lab in Glasgow | Wellcome Sanger Institute for the COVID-19 Genomics UK (COG-UK) Consortium | Harper VanSteenhouse et al |
| EPI_ISL_642060 | England/QEUH-B0DE3B/2020 | 2020-11-02 | Europe/United Kingdom/England/ | Lighthouse Lab in Glasgow | Wellcome Sanger Institute for the COVID-19 Genomics UK (COG-UK) Consortium | Harper VanSteenhouse et al |
| EPI_ISL_642182 | England/QEUH-B0FCB7/2020 | 2020-11-02 | Europe/United Kingdom/England/ | Lighthouse Lab in Glasgow | Wellcome Sanger Institute for the COVID-19 Genomics UK (COG-UK) Consortium | Harper VanSteenhouse et al |
| EPI_ISL_642225 | England/QEUH-B1298D/2020 | 2020-11-02 | Europe/United Kingdom/England/ | Lighthouse Lab in Glasgow | Wellcome Sanger Institute for the COVID-19 Genomics UK (COG-UK) Consortium | Harper VanSteenhouse et al |
| EPI_ISL_642858 | England/MILK-B0263E/2020 | 2020-10-24 | Europe/United Kingdom/England/ | Lighthouse Lab in Milton Keynes | Wellcome Sanger Institute for the COVID-19 Genomics UK (COG-UK) Consortium | The Lighthouse Lab in Milton Keynes et al |
| EPI_ISL_642878 | England/MILK-B02425/2020 | 2020-10-23 | Europe/United Kingdom/England/ | Lighthouse Lab in Milton Keynes | Wellcome Sanger Institute for the COVID-19 Genomics UK (COG-UK) Consortium | The Lighthouse Lab in Milton Keynes et al |

| | | | | | | |
|---|---|---|---|---|---|---|
| EPI_ISL_643206 | England/QEUH-A8CA6E/2020 | 2020-10-16 | Europe/United Kingdom/England/ | Lighthouse Lab in Glasgow | Wellcome Sanger Institute for the COVID-19 Genomics UK (COG-UK) Consortium | Harper VanSteenhouse et al |
| EPI_ISL_643312 | England/QEUH-A8C0B7/2020 | 2020-10-16 | Europe/United Kingdom/England/ | Lighthouse Lab in Glasgow | Wellcome Sanger Institute for the COVID-19 Genomics UK (COG-UK) Consortium | Harper VanSteenhouse et al |
| EPI_ISL_643327 | England/CAMC-B0C449/2020 | 2020-10-30 | Europe/United Kingdom/England/ | Lighthouse Lab in Cambridge | Wellcome Sanger Institute for the COVID-19 Genomics UK (COG-UK) Consortium | Rob Howes et al |
| EPI_ISL_643403 | England/CAMC-B080FE/2020 | 2020-10-29 | Europe/United Kingdom/England/ | Lighthouse Lab in Cambridge | Wellcome Sanger Institute for the COVID-19 Genomics UK (COG-UK) Consortium | Rob Howes et al |
| EPI_ISL_643448 | England/CAMC-B0B79C/2020 | 2020-10-29 | Europe/United Kingdom/England/ | Lighthouse Lab in Cambridge | Wellcome Sanger Institute for the COVID-19 Genomics UK (COG-UK) Consortium | Rob Howes et al |
| EPI_ISL_643569 | England/CAMC-B0C634/2020 | 2020-10-30 | Europe/United Kingdom/England/ | Lighthouse Lab in Cambridge | Wellcome Sanger Institute for the COVID-19 Genomics UK (COG-UK) Consortium | Rob Howes et al |
| EPI_ISL_643575 | England/CAMC-B0BAA2/2020 | 2020-10-29 | Europe/United Kingdom/England/ | Lighthouse Lab in Cambridge | Wellcome Sanger Institute for the COVID-19 Genomics UK (COG-UK) Consortium | Rob Howes et al |
| EPI_ISL_643689 | England/CAMC-B07138/2020 | 2020-10-29 | Europe/United Kingdom/England/ | Lighthouse Lab in Cambridge | Wellcome Sanger Institute for the COVID-19 Genomics UK (COG-UK) Consortium | Rob Howes et al |

| | | | | | | |
|---|---|---|---|---|---|---|
| EPI_ISL_643735 | England/CAMC-B07C55/2020 | 2020-10-29 | Europe/United Kingdom/England/ | Lighthouse Lab in Cambridge | Wellcome Sanger Institute for the COVID-19 Genomics UK (COG-UK) Consortium | Rob Howes et al |
| EPI_ISL_643756 | England/CAMC-B0736F/2020 | 2020-10-30 | Europe/United Kingdom/England/ | Lighthouse Lab in Cambridge | Wellcome Sanger Institute for the COVID-19 Genomics UK (COG-UK) Consortium | Rob Howes et al |
| EPI_ISL_643778 | England/CAMC-B073E7/2020 | 2020-10-30 | Europe/United Kingdom/England/ | Lighthouse Lab in Cambridge | Wellcome Sanger Institute for the COVID-19 Genomics UK (COG-UK) Consortium | Rob Howes et al |
| EPI_ISL_644170 | USA/TX-DSHS-1016/2020 | 2020-04-21 | North America/USA/Texas/Hidalgo Coun | Texas Department of State Health Services | Texas Department of State Health Services | Rashmi Tuladhar et al |
| EPI_ISL_644193 | USA/TX-DSHS-1252/2020 | 2020-05-30 | North America/USA/Texas/El Paso Coun | Texas Department of State Health Services | Texas Department of State Health Services | Rashmi Tuladhar et al |
| EPI_ISL_644200 | USA/TX-DSHS-1259/2020 | 2020-06-01 | North America/USA/Texas/El Paso Coun | Texas Department of State Health Services | Texas Department of State Health Services | Rashmi Tuladhar et al |
| EPI_ISL_644201 | USA/TX-DSHS-1260/2020 | 2020-06-12 | North America/USA/Texas/El Paso Coun | Texas Department of State Health Services | Texas Department of State Health Services | Rashmi Tuladhar et al |
| EPI_ISL_644226 | Ireland/D-NVRL-AIIDV0192v1/2020 | 2020-04-14 | Europe/Ireland/Dublin/ | CEPHR / Vincent's Hospital | Irish Coronavirus Sequencing Consortium - National Virus Reference Laboratory | Michael Carr et al |
| EPI_ISL_644231 | Ireland/CE-NVRL-AIIDV1105v3/2020 | 2020-04-26 | Europe/Ireland/Clare/ | CEPHR / Vincent's Hospital | Irish Coronavirus Sequencing Consortium - National Virus Reference Laboratory | Michael Carr et al |

| | | | | | | |
|---|---|---|---|---|---|---|
| EPI_ISL_644247 | Ireland/D-NVRL-AIIDV1124v1/2020 | 2020-04-24 | Europe/Ireland/Dublin/ | CEPHR / Vincent's Hospital | Irish Coronavirus Sequencing Consortium - National Virus Reference Laboratory | Michael Carr et al |
| EPI_ISL_644269 | Ireland/CE-NVRL-AIIDV1105v2/2020 | 2020-04-26 | Europe/Ireland/Clare/ | CEPHR / Vincent's Hospital | Irish Coronavirus Sequencing Consortium - National Virus Reference Laboratory | Michael Carr et al |
| EPI_ISL_644302 | Ireland/WW-NVRL-AIIDV0351v2/2020 | 2020-05-04 | Europe/Ireland/Wicklow/ | CEPHR / Vincent's Hospital | Irish Coronavirus Sequencing Consortium - National Virus Reference Laboratory | Michael Carr et al |
| EPI_ISL_644321 | Ireland/D-NVRL-AIIDV0002v1/2020 | 2020-03-26 | Europe/Ireland/Dublin/ | CEPHR / Vincent's Hospital | Irish Coronavirus Sequencing Consortium - National Virus Reference Laboratory | Michael Carr et al |
| EPI_ISL_644323 | Ireland/D-NVRL-AIIDV0363v1/2020 | 2020-05-05 | Europe/Ireland/Dublin/ | CEPHR / Vincent's Hospital | Irish Coronavirus Sequencing Consortium - National Virus Reference Laboratory | Michael Carr et al |
| EPI_ISL_644335 | Ireland/D-NVRL-AIIDV0053v2/2020 | 2020-04-09 | Europe/Ireland/Dublin/ | CEPHR / Vincent's Hospital | Irish Coronavirus Sequencing Consortium - National Virus Reference Laboratory | Michael Carr et al |

| | | | | | | |
|---|---|---|---|---|---|---|
| EPI_ISL_644356 | USA/MI-MDHHS-SC22211/2020 | 2020-10-26 | North America/USA/Michigan/ | Michigan Department of Health and Human Services, Bureau of Laboratories | Michigan Department of Health and Human Services, Bureau of Laboratories | Blankenship HM et al |
| EPI_ISL_644357 | USA/MI-MDHHS-SC22214/2020 | 2020-10-21 | North America/USA/Michigan/ | Michigan Department of Health and Human Services, Bureau of Laboratories | Michigan Department of Health and Human Services, Bureau of Laboratories | Blankenship HM et al |
| EPI_ISL_644362 | USA/MI-MDHHS-SC22207/2020 | 2020-10-26 | North America/USA/Michigan/ | Michigan Department of Health and Human Services, Bureau of Laboratories | Michigan Department of Health and Human Services, Bureau of Laboratories | Blankenship HM et al |
| EPI_ISL_644366 | USA/MI-MDHHS-SC22198/2020 | 2020-10-27 | North America/USA/Michigan/ | Michigan Department of Health and Human Services, Bureau of Laboratories | Michigan Department of Health and Human Services, Bureau of Laboratories | Blankenship HM et al |
| EPI_ISL_644372 | USA/MI-MDHHS-SC22204/2020 | 2020-10-21 | North America/USA/Michigan/ | Michigan Department of Health and Human Services, Bureau of Laboratories | Michigan Department of Health and Human Services, Bureau of Laboratories | Blankenship HM et al |
| EPI_ISL_644375 | France/PAC-MEPHI-1928/2020 | 2020-08-27 | Europe/France/Provence-Alpes-C√¥te d' | MEPHI, Aix Marseille University | MEPHI, Aix Marseille University | Anthony LEVASSEUR et al |
| EPI_ISL_644376 | France/PAC-MEPHI-2391/2020 | 2020-10-12 | Europe/France/Provence-Alpes-C√¥te d' | MEPHI, Aix Marseille University | MEPHI, Aix Marseille University | Anthony LEVASSEUR et al |
| EPI_ISL_644377 | France/PAC-MEPHI-2324/2020 | 2020-10-08 | Europe/France/Provence-Alpes-C√¥te d' | MEPHI, Aix Marseille University | MEPHI, Aix Marseille University | Anthony LEVASSEUR et al |

| | | | | | | |
|---|---|---|---|---|---|---|
| EPI_ISL_644378 | France/PAC-MEPHI-2214/2020 | 2020-10-02 | Europe/France/Provence-Alpes-CV¥te d' | MEPHI, Aix Marseille University | MEPHI, Aix Marseille University | Anthony LEVASSEUR et al |
| EPI_ISL_644380 | France/PAC-MEPHI-1750/2020 | 2020-09-18 | Europe/France/Provence-Alpes-CV¥te d' | MEPHI, Aix Marseille University | MEPHI, Aix Marseille University | Anthony LEVASSEUR et al |
| EPI_ISL_644381 | France/PAC-MEPHI-1776/2020 | 2020-09-19 | Europe/France/Provence-Alpes-CV¥te d' | MEPHI, Aix Marseille University | MEPHI, Aix Marseille University | Anthony LEVASSEUR et al |
| EPI_ISL_644382 | France/PAC-MEPHI-1680/2020 | 2020-09-14 | Europe/France/Provence-Alpes-CV¥te d' | MEPHI, Aix Marseille University | MEPHI, Aix Marseille University | Anthony LEVASSEUR et al |
| EPI_ISL_644383 | France/PAC-MEPHI-2133/2020 | 2020-09-28 | Europe/France/Provence-Alpes-CV¥te d' | MEPHI, Aix Marseille University | MEPHI, Aix Marseille University | Anthony LEVASSEUR et al |
| EPI_ISL_644385 | France/PAC-MEPHI-2328/2020 | 2020-10-08 | Europe/France/Provence-Alpes-CV¥te d' | MEPHI, Aix Marseille University | MEPHI, Aix Marseille University | Anthony LEVASSEUR et al |
| EPI_ISL_644386 | France/PAC-MEPHI-2196/2020 | 2020-09-30 | Europe/France/Provence-Alpes-CV¥te d' | MEPHI, Aix Marseille University | MEPHI, Aix Marseille University | Anthony LEVASSEUR et al |
| EPI_ISL_644387 | France/PAC-MEPHI-1913/2020 | 2020-08-27 | Europe/France/Provence-Alpes-CV¥te d' | MEPHI, Aix Marseille University | MEPHI, Aix Marseille University | Anthony LEVASSEUR et al |
| EPI_ISL_644388 | France/PAC-MEPHI-1983/2020 | 2020-08-27 | Europe/France/Provence-Alpes-CV¥te d' | MEPHI, Aix Marseille University | MEPHI, Aix Marseille University | Anthony LEVASSEUR et al |
| EPI_ISL_644389 | France/PAC-MEPHI-2162/2020 | 2020-09-30 | Europe/France/Provence-Alpes-CV¥te d' | MEPHI, Aix Marseille University | MEPHI, Aix Marseille University | Anthony LEVASSEUR et al |
| EPI_ISL_644390 | France/PAC-MEPHI-2312/2020 | 2020-10-07 | Europe/France/Provence-Alpes-CV¥te d' | MEPHI, Aix Marseille University | MEPHI, Aix Marseille University | Anthony LEVASSEUR et al |
| EPI_ISL_644393 | France/PAC-MEPHI-1665/2020 | 2020-09-15 | Europe/France/Provence-Alpes-CV¥te d' | MEPHI, Aix Marseille University | MEPHI, Aix Marseille University | Anthony LEVASSEUR et al |
| EPI_ISL_644394 | France/PAC-MEPHI-1666/2020 | 2020-09-15 | Europe/France/Provence-Alpes-CV¥te d' | MEPHI, Aix Marseille University | MEPHI, Aix Marseille University | Anthony LEVASSEUR et al |

| | | | | | | |
|---|---|---|---|---|---|---|
| EPI_ISL_644395 | France/PAC-MEPHI-1672/2020 | 2020-09-14 | Europe/France/Provence-Alpes-CV¥te d' | MEPHI, Aix Marseille University | MEPHI, Aix Marseille University | Anthony LEVASSEUR et al |
| EPI_ISL_644398 | France/PAC-MEPHI-1770/2020 | 2020-09-19 | Europe/France/Provence-Alpes-CV¥te d' | MEPHI, Aix Marseille University | MEPHI, Aix Marseille University | Anthony LEVASSEUR et al |
| EPI_ISL_644399 | France/PAC-MEPHI-2361/2020 | 2020-10-09 | Europe/France/Provence-Alpes-CV¥te d' | MEPHI, Aix Marseille University | MEPHI, Aix Marseille University | Anthony LEVASSEUR et al |
| EPI_ISL_644400 | France/PAC-MEPHI-1682/2020 | 2020-09-14 | Europe/France/Provence-Alpes-CV¥te d' | MEPHI, Aix Marseille University | MEPHI, Aix Marseille University | Anthony LEVASSEUR et al |
| EPI_ISL_644401 | France/PAC-MEPHI-1731/2020 | 2020-09-16 | Europe/France/Provence-Alpes-CV¥te d' | MEPHI, Aix Marseille University | MEPHI, Aix Marseille University | Anthony LEVASSEUR et al |
| EPI_ISL_644409 | France/PAC-MEPHI-0752/2020 | 2020-03-17 | Europe/France/Provence-Alpes-CV¥te d' | MEPHI, Aix Marseille University | MEPHI, Aix Marseille University | Anthony LEVASSEUR et al |
| EPI_ISL_644410 | France/PAC-MEPHI-0762/2020 | 2020-03-16 | Europe/France/Provence-Alpes-CV¥te d' | MEPHI, Aix Marseille University | MEPHI, Aix Marseille University | Anthony LEVASSEUR et al |
| EPI_ISL_644411 | France/PAC-MEPHI-1613/2020 | 2020-09-04 | Europe/France/Provence-Alpes-CV¥te d' | MEPHI, Aix Marseille University | MEPHI, Aix Marseille University | Anthony LEVASSEUR et al |
| EPI_ISL_644412 | France/PAC-MEPHI-1614/2020 | 2020-09-04 | Europe/France/Provence-Alpes-CV¥te d' | MEPHI, Aix Marseille University | MEPHI, Aix Marseille University | Anthony LEVASSEUR et al |
| EPI_ISL_644413 | France/PAC-MEPHI-1667/2020 | 2020-09-14 | Europe/France/Provence-Alpes-CV¥te d' | MEPHI, Aix Marseille University | MEPHI, Aix Marseille University | Anthony LEVASSEUR et al |
| EPI_ISL_644414 | France/PAC-MEPHI-1669/2020 | 2020-09-15 | Europe/France/Provence-Alpes-CV¥te d' | MEPHI, Aix Marseille University | MEPHI, Aix Marseille University | Anthony LEVASSEUR et al |
| EPI_ISL_644416 | France/PAC-MEPHI-1671/2020 | 2020-09-14 | Europe/France/Provence-Alpes-CV¥te d' | MEPHI, Aix Marseille University | MEPHI, Aix Marseille University | Anthony LEVASSEUR et al |
| EPI_ISL_644419 | France/PAC-MEPHI-1675/2020 | 2020-09-15 | Europe/France/Provence-Alpes-CV¥te d' | MEPHI, Aix Marseille University | MEPHI, Aix Marseille University | Anthony LEVASSEUR et al |

| | | | | | | |
|---|---|---|---|---|---|---|
| EPI_ISL_644420 | France/PAC-MEPHI-1676/2020 | 2020-09-14 | Europe/France/Provence-Alpes-CV¥te d' | MEPHI, Aix Marseille University | MEPHI, Aix Marseille University | Anthony LEVASSEUR et al |
| EPI_ISL_644422 | France/PAC-MEPHI-1678/2020 | 2020-09-14 | Europe/France/Provence-Alpes-CV¥te d' | MEPHI, Aix Marseille University | MEPHI, Aix Marseille University | Anthony LEVASSEUR et al |
| EPI_ISL_644426 | France/PAC-MEPHI-1702/2020 | 2020-09-17 | Europe/France/Provence-Alpes-CV¥te d' | MEPHI, Aix Marseille University | MEPHI, Aix Marseille University | Anthony LEVASSEUR et al |
| EPI_ISL_644427 | France/PAC-MEPHI-1703/2020 | 2020-09-17 | Europe/France/Provence-Alpes-CV¥te d' | MEPHI, Aix Marseille University | MEPHI, Aix Marseille University | Anthony LEVASSEUR et al |
| EPI_ISL_644429 | France/PAC-MEPHI-1718/2020 | 2020-09-17 | Europe/France/Provence-Alpes-CV¥te d' | MEPHI, Aix Marseille University | MEPHI, Aix Marseille University | Anthony LEVASSEUR et al |
| EPI_ISL_644430 | France/PAC-MEPHI-1719/2020 | 2020-09-17 | Europe/France/Provence-Alpes-CV¥te d' | MEPHI, Aix Marseille University | MEPHI, Aix Marseille University | Anthony LEVASSEUR et al |
| EPI_ISL_644433 | France/PAC-MEPHI-1724/2020 | 2020-09-17 | Europe/France/Provence-Alpes-CV¥te d' | MEPHI, Aix Marseille University | MEPHI, Aix Marseille University | Anthony LEVASSEUR et al |
| EPI_ISL_644434 | France/PAC-MEPHI-1729/2020 | 2020-09-17 | Europe/France/Provence-Alpes-CV¥te d' | MEPHI, Aix Marseille University | MEPHI, Aix Marseille University | Anthony LEVASSEUR et al |
| EPI_ISL_644435 | France/PAC-MEPHI-1730/2020 | 2020-09-16 | Europe/France/Provence-Alpes-CV¥te d' | MEPHI, Aix Marseille University | MEPHI, Aix Marseille University | Anthony LEVASSEUR et al |
| EPI_ISL_644439 | France/PAC-MEPHI-1741/2020 | 2020-09-18 | Europe/France/Provence-Alpes-CV¥te d' | MEPHI, Aix Marseille University | MEPHI, Aix Marseille University | Anthony LEVASSEUR et al |
| EPI_ISL_644440 | France/PAC-MEPHI-1746/2020 | 2020-09-18 | Europe/France/Provence-Alpes-CV¥te d' | MEPHI, Aix Marseille University | MEPHI, Aix Marseille University | Anthony LEVASSEUR et al |
| EPI_ISL_644442 | France/PAC-MEPHI-1755/2020 | 2020-09-18 | Europe/France/Provence-Alpes-CV¥te d' | MEPHI, Aix Marseille University | MEPHI, Aix Marseille University | Anthony LEVASSEUR et al |
| EPI_ISL_644443 | France/PAC-MEPHI-1758/2020 | 2020-09-19 | Europe/France/Provence-Alpes-CV¥te d' | MEPHI, Aix Marseille University | MEPHI, Aix Marseille University | Anthony LEVASSEUR et al |

| | | | | | | |
|---|---|---|---|---|---|---|
| EPI_ISL_644444 | France/PAC-MEPHI-1762/2020 | 2020-09-18 | Europe/France/Provence-Alpes-CV¥te d' | MEPHI, Aix Marseille University | MEPHI, Aix Marseille University | Anthony LEVASSEUR et al |
| EPI_ISL_644445 | France/PAC-MEPHI-1764/2020 | 2020-09-19 | Europe/France/Provence-Alpes-CV¥te d' | MEPHI, Aix Marseille University | MEPHI, Aix Marseille University | Anthony LEVASSEUR et al |
| EPI_ISL_644447 | France/PAC-MEPHI-1839/2020 | 2020-08-25 | Europe/France/Provence-Alpes-CV¥te d' | MEPHI, Aix Marseille University | MEPHI, Aix Marseille University | Anthony LEVASSEUR et al |
| EPI_ISL_644448 | France/PAC-MEPHI-1882/2020 | 2020-09-21 | Europe/France/Provence-Alpes-CV¥te d' | MEPHI, Aix Marseille University | MEPHI, Aix Marseille University | Anthony LEVASSEUR et al |
| EPI_ISL_644449 | France/PAC-MEPHI-1883/2020 | 2020-09-21 | Europe/France/Provence-Alpes-CV¥te d' | MEPHI, Aix Marseille University | MEPHI, Aix Marseille University | Anthony LEVASSEUR et al |
| EPI_ISL_644450 | France/PAC-MEPHI-1884/2020 | 2020-09-21 | Europe/France/Provence-Alpes-CV¥te d' | MEPHI, Aix Marseille University | MEPHI, Aix Marseille University | Anthony LEVASSEUR et al |
| EPI_ISL_644457 | France/PAC-MEPHI-1908/2020 | 2020-08-27 | Europe/France/Provence-Alpes-CV¥te d' | MEPHI, Aix Marseille University | MEPHI, Aix Marseille University | Anthony LEVASSEUR et al |
| EPI_ISL_644458 | France/PAC-MEPHI-1911/2020 | 2020-08-26 | Europe/France/Provence-Alpes-CV¥te d' | MEPHI, Aix Marseille University | MEPHI, Aix Marseille University | Anthony LEVASSEUR et al |
| EPI_ISL_644459 | France/PAC-MEPHI-1912/2020 | 2020-08-27 | Europe/France/Provence-Alpes-CV¥te d' | MEPHI, Aix Marseille University | MEPHI, Aix Marseille University | Anthony LEVASSEUR et al |
| EPI_ISL_644465 | France/PAC-MEPHI-1940/2020 | 2020-08-28 | Europe/France/Provence-Alpes-CV¥te d' | MEPHI, Aix Marseille University | MEPHI, Aix Marseille University | Anthony LEVASSEUR et al |
| EPI_ISL_644470 | France/PAC-MEPHI-1999/2020 | 2020-08-28 | Europe/France/Provence-Alpes-CV¥te d' | MEPHI, Aix Marseille University | MEPHI, Aix Marseille University | Anthony LEVASSEUR et al |
| EPI_ISL_644471 | France/PAC-MEPHI-2010/2020 | 2020-08-28 | Europe/France/Provence-Alpes-CV¥te d' | MEPHI, Aix Marseille University | MEPHI, Aix Marseille University | Anthony LEVASSEUR et al |
| EPI_ISL_644475 | France/PAC-MEPHI-2018/2020 | 2020-08-28 | Europe/France/Provence-Alpes-CV¥te d' | MEPHI, Aix Marseille University | MEPHI, Aix Marseille University | Anthony LEVASSEUR et al |

| | | | | | | |
|---|---|---|---|---|---|---|
| EPI_ISL_644478 | France/PAC-MEPHI-2033/2020 | 2020-08-29 | Europe/France/Provence-Alpes-CV¥te d' | MEPHI, Aix Marseille University | MEPHI, Aix Marseille University | Anthony LEVASSEUR et al |
| EPI_ISL_644479 | France/PAC-MEPHI-2038/2020 | 2020-08-29 | Europe/France/Provence-Alpes-CV¥te d' | MEPHI, Aix Marseille University | MEPHI, Aix Marseille University | Anthony LEVASSEUR et al |
| EPI_ISL_644480 | France/PAC-MEPHI-2041/2020 | 2020-08-29 | Europe/France/Provence-Alpes-CV¥te d' | MEPHI, Aix Marseille University | MEPHI, Aix Marseille University | Anthony LEVASSEUR et al |
| EPI_ISL_644483 | France/PAC-MEPHI-2051/2020 | 2020-08-31 | Europe/France/Provence-Alpes-CV¥te d' | MEPHI, Aix Marseille University | MEPHI, Aix Marseille University | Anthony LEVASSEUR et al |
| EPI_ISL_644484 | France/PAC-MEPHI-2054/2020 | 2020-08-31 | Europe/France/Provence-Alpes-CV¥te d' | MEPHI, Aix Marseille University | MEPHI, Aix Marseille University | Anthony LEVASSEUR et al |
| EPI_ISL_644486 | France/PAC-MEPHI-2057/2020 | 2020-09-02 | Europe/France/Provence-Alpes-CV¥te d' | MEPHI, Aix Marseille University | MEPHI, Aix Marseille University | Anthony LEVASSEUR et al |
| EPI_ISL_644490 | France/PAC-MEPHI-2071/2020 | 2020-09-23 | Europe/France/Provence-Alpes-CV¥te d' | MEPHI, Aix Marseille University | MEPHI, Aix Marseille University | Anthony LEVASSEUR et al |
| EPI_ISL_644494 | France/PAC-MEPHI-2094/2020 | 2020-09-23 | Europe/France/Provence-Alpes-CV¥te d' | MEPHI, Aix Marseille University | MEPHI, Aix Marseille University | Anthony LEVASSEUR et al |
| EPI_ISL_644495 | France/PAC-MEPHI-2099/2020 | 2020-09-27 | Europe/France/Provence-Alpes-CV¥te d' | MEPHI, Aix Marseille University | MEPHI, Aix Marseille University | Anthony LEVASSEUR et al |
| EPI_ISL_644496 | France/PAC-MEPHI-2101/2020 | 2020-09-23 | Europe/France/Provence-Alpes-CV¥te d' | MEPHI, Aix Marseille University | MEPHI, Aix Marseille University | Anthony LEVASSEUR et al |
| EPI_ISL_644497 | France/PAC-MEPHI-2103/2020 | 2020-09-26 | Europe/France/Provence-Alpes-CV¥te d' | MEPHI, Aix Marseille University | MEPHI, Aix Marseille University | Anthony LEVASSEUR et al |
| EPI_ISL_644498 | France/PAC-MEPHI-2104/2020 | 2020-09-29 | Europe/France/Provence-Alpes-CV¥te d' | MEPHI, Aix Marseille University | MEPHI, Aix Marseille University | Anthony LEVASSEUR et al |
| EPI_ISL_644499 | France/PAC-MEPHI-2105/2020 | 2020-09-29 | Europe/France/Provence-Alpes-CV¥te d' | MEPHI, Aix Marseille University | MEPHI, Aix Marseille University | Anthony LEVASSEUR et al |

| | | | | | | |
|---|---|---|---|---|---|---|
| EPI_ISL_644500 | France/PAC-MEPHI-2112/2020 | 2020-09-28 | Europe/France/Provence-Alpes-CV¥te d' | MEPHI, Aix Marseille University | MEPHI, Aix Marseille University | Anthony LEVASSEUR et al |
| EPI_ISL_644503 | France/PAC-MEPHI-2118/2020 | 2020-09-25 | Europe/France/Provence-Alpes-CV¥te d' | MEPHI, Aix Marseille University | MEPHI, Aix Marseille University | Anthony LEVASSEUR et al |
| EPI_ISL_644504 | France/PAC-MEPHI-2128/2020 | 2020-09-27 | Europe/France/Provence-Alpes-CV¥te d' | MEPHI, Aix Marseille University | MEPHI, Aix Marseille University | Anthony LEVASSEUR et al |
| EPI_ISL_644508 | France/PAC-MEPHI-2139/2020 | 2020-09-24 | Europe/France/Provence-Alpes-CV¥te d' | MEPHI, Aix Marseille University | MEPHI, Aix Marseille University | Anthony LEVASSEUR et al |
| EPI_ISL_644509 | France/PAC-MEPHI-2140/2020 | 2020-09-25 | Europe/France/Provence-Alpes-CV¥te d' | MEPHI, Aix Marseille University | MEPHI, Aix Marseille University | Anthony LEVASSEUR et al |
| EPI_ISL_644510 | France/PAC-MEPHI-2144/2020 | 2020-09-28 | Europe/France/Provence-Alpes-CV¥te d' | MEPHI, Aix Marseille University | MEPHI, Aix Marseille University | Anthony LEVASSEUR et al |
| EPI_ISL_644511 | France/PAC-MEPHI-2147/2020 | 2020-09-24 | Europe/France/Provence-Alpes-CV¥te d' | MEPHI, Aix Marseille University | MEPHI, Aix Marseille University | Anthony LEVASSEUR et al |
| EPI_ISL_644512 | France/PAC-MEPHI-2148/2020 | 2020-09-25 | Europe/France/Provence-Alpes-CV¥te d' | MEPHI, Aix Marseille University | MEPHI, Aix Marseille University | Anthony LEVASSEUR et al |
| EPI_ISL_644513 | France/PAC-MEPHI-2151/2020 | 2020-09-23 | Europe/France/Provence-Alpes-CV¥te d' | MEPHI, Aix Marseille University | MEPHI, Aix Marseille University | Anthony LEVASSEUR et al |
| EPI_ISL_644514 | France/PAC-MEPHI-2153/2020 | 2020-09-30 | Europe/France/Provence-Alpes-CV¥te d' | MEPHI, Aix Marseille University | MEPHI, Aix Marseille University | Anthony LEVASSEUR et al |
| EPI_ISL_644516 | France/PAC-MEPHI-2155/2020 | 2020-09-25 | Europe/France/Provence-Alpes-CV¥te d' | MEPHI, Aix Marseille University | MEPHI, Aix Marseille University | Anthony LEVASSEUR et al |
| EPI_ISL_644517 | France/PAC-MEPHI-2157/2020 | 2020-09-28 | Europe/France/Provence-Alpes-CV¥te d' | MEPHI, Aix Marseille University | MEPHI, Aix Marseille University | Anthony LEVASSEUR et al |
| EPI_ISL_644519 | France/PAC-MEPHI-2165/2020 | 2020-09-28 | Europe/France/Provence-Alpes-CV¥te d' | MEPHI, Aix Marseille University | MEPHI, Aix Marseille University | Anthony LEVASSEUR et al |

| | | | | | | |
|---|---|---|---|---|---|---|
| EPI_ISL_644523 | France/PAC-MEPHI-2180/2020 | 2020-09-26 | Europe/France/Provence-Alpes-CV¥te d' | MEPHI, Aix Marseille University | MEPHI, Aix Marseille University | Anthony LEVASSEUR et al |
| EPI_ISL_644525 | France/PAC-MEPHI-2195/2020 | 2020-10-01 | Europe/France/Provence-Alpes-CV¥te d' | MEPHI, Aix Marseille University | MEPHI, Aix Marseille University | Anthony LEVASSEUR et al |
| EPI_ISL_644526 | France/PAC-MEPHI-2207/2020 | 2020-10-01 | Europe/France/Provence-Alpes-CV¥te d' | MEPHI, Aix Marseille University | MEPHI, Aix Marseille University | Anthony LEVASSEUR et al |
| EPI_ISL_644527 | France/PAC-MEPHI-2211/2020 | 2020-10-01 | Europe/France/Provence-Alpes-CV¥te d' | MEPHI, Aix Marseille University | MEPHI, Aix Marseille University | Anthony LEVASSEUR et al |
| EPI_ISL_644529 | France/PAC-MEPHI-2227/2020 | 2020-10-02 | Europe/France/Provence-Alpes-CV¥te d' | MEPHI, Aix Marseille University | MEPHI, Aix Marseille University | Anthony LEVASSEUR et al |
| EPI_ISL_644531 | France/PAC-MEPHI-2272/2020 | 2020-10-05 | Europe/France/Provence-Alpes-CV¥te d' | MEPHI, Aix Marseille University | MEPHI, Aix Marseille University | Anthony LEVASSEUR et al |
| EPI_ISL_644533 | France/PAC-MEPHI-2288/2020 | 2020-10-06 | Europe/France/Provence-Alpes-CV¥te d' | MEPHI, Aix Marseille University | MEPHI, Aix Marseille University | Anthony LEVASSEUR et al |
| EPI_ISL_644534 | France/PAC-MEPHI-2289/2020 | 2020-10-06 | Europe/France/Provence-Alpes-CV¥te d' | MEPHI, Aix Marseille University | MEPHI, Aix Marseille University | Anthony LEVASSEUR et al |
| EPI_ISL_644535 | France/PAC-MEPHI-2295/2020 | 2020-10-06 | Europe/France/Provence-Alpes-CV¥te d' | MEPHI, Aix Marseille University | MEPHI, Aix Marseille University | Anthony LEVASSEUR et al |
| EPI_ISL_644537 | France/PAC-MEPHI-2308/2020 | 2020-10-07 | Europe/France/Provence-Alpes-CV¥te d' | MEPHI, Aix Marseille University | MEPHI, Aix Marseille University | Anthony LEVASSEUR et al |
| EPI_ISL_644540 | France/PAC-MEPHI-2326/2020 | 2020-10-08 | Europe/France/Provence-Alpes-CV¥te d' | MEPHI, Aix Marseille University | MEPHI, Aix Marseille University | Anthony LEVASSEUR et al |
| EPI_ISL_644541 | France/PAC-MEPHI-2341/2020 | 2020-10-08 | Europe/France/Provence-Alpes-CV¥te d' | MEPHI, Aix Marseille University | MEPHI, Aix Marseille University | Anthony LEVASSEUR et al |
| EPI_ISL_644542 | France/PAC-MEPHI-2344/2020 | 2020-10-08 | Europe/France/Provence-Alpes-CV¥te d' | MEPHI, Aix Marseille University | MEPHI, Aix Marseille University | Anthony LEVASSEUR et al |

| | | | | | | |
|---|---|---|---|---|---|---|
| EPI_ISL_644544 | France/PAC-MEPHI-2369/2020 | 2020-10-09 | Europe/France/Provence-Alpes-CV¥te d' | MEPHI, Aix Marseille University | MEPHI, Aix Marseille University | Anthony LEVASSEUR et al |
| EPI_ISL_644545 | France/PAC-MEPHI-2371/2020 | 2020-10-10 | Europe/France/Provence-Alpes-CV¥te d' | MEPHI, Aix Marseille University | MEPHI, Aix Marseille University | Anthony LEVASSEUR et al |
| EPI_ISL_644546 | France/PAC-MEPHI-2374/2020 | 2020-10-10 | Europe/France/Provence-Alpes-CV¥te d' | MEPHI, Aix Marseille University | MEPHI, Aix Marseille University | Anthony LEVASSEUR et al |
| EPI_ISL_644547 | France/PAC-MEPHI-2375/2020 | 2020-10-10 | Europe/France/Provence-Alpes-CV¥te d' | MEPHI, Aix Marseille University | MEPHI, Aix Marseille University | Anthony LEVASSEUR et al |
| EPI_ISL_644548 | France/PAC-MEPHI-2376/2020 | 2020-10-10 | Europe/France/Provence-Alpes-CV¥te d' | MEPHI, Aix Marseille University | MEPHI, Aix Marseille University | Anthony LEVASSEUR et al |
| EPI_ISL_644549 | France/PAC-MEPHI-2382/2020 | 2020-10-11 | Europe/France/Provence-Alpes-CV¥te d' | MEPHI, Aix Marseille University | MEPHI, Aix Marseille University | Anthony LEVASSEUR et al |
| EPI_ISL_644550 | France/PAC-MEPHI-2385/2020 | 2020-10-11 | Europe/France/Provence-Alpes-CV¥te d' | MEPHI, Aix Marseille University | MEPHI, Aix Marseille University | Anthony LEVASSEUR et al |
| EPI_ISL_644551 | France/PAC-MEPHI-2388/2020 | 2020-10-12 | Europe/France/Provence-Alpes-CV¥te d' | MEPHI, Aix Marseille University | MEPHI, Aix Marseille University | Anthony LEVASSEUR et al |
| EPI_ISL_644552 | France/PAC-MEPHI-2389/2020 | 2020-10-12 | Europe/France/Provence-Alpes-CV¥te d' | MEPHI, Aix Marseille University | MEPHI, Aix Marseille University | Anthony LEVASSEUR et al |
| EPI_ISL_644554 | France/PAC-MEPHI-2393/2020 | 2020-10-12 | Europe/France/Provence-Alpes-CV¥te d' | MEPHI, Aix Marseille University | MEPHI, Aix Marseille University | Anthony LEVASSEUR et al |
| EPI_ISL_644555 | France/PAC-MEPHI-2394/2020 | 2020-10-12 | Europe/France/Provence-Alpes-CV¥te d' | MEPHI, Aix Marseille University | MEPHI, Aix Marseille University | Anthony LEVASSEUR et al |
| EPI_ISL_644556 | France/PAC-MEPHI-2400/2020 | 2020-10-13 | Europe/France/Provence-Alpes-CV¥te d' | MEPHI, Aix Marseille University | MEPHI, Aix Marseille University | Anthony LEVASSEUR et al |
| EPI_ISL_644558 | France/PAC-MEPHI-2402/2020 | 2020-10-13 | Europe/France/Provence-Alpes-CV¥te d' | MEPHI, Aix Marseille University | MEPHI, Aix Marseille University | Anthony LEVASSEUR et al |

| | | | | | | |
|---|---|---|---|---|---|---|
| EPI_ISL_644559 | France/PAC-MEPHI-2410/2020 | 2020-10-13 | Europe/France/Provence-Alpes-CV¥te d' | MEPHI, Aix Marseille University | MEPHI, Aix Marseille University | Anthony LEVASSEUR et al |
| EPI_ISL_644561 | France/PAC-MEPHI-2413/2020 | 2020-10-14 | Europe/France/Provence-Alpes-CV¥te d' | MEPHI, Aix Marseille University | MEPHI, Aix Marseille University | Anthony LEVASSEUR et al |
| EPI_ISL_644562 | France/PAC-MEPHI-2414/2020 | 2020-10-14 | Europe/France/Provence-Alpes-CV¥te d' | MEPHI, Aix Marseille University | MEPHI, Aix Marseille University | Anthony LEVASSEUR et al |
| EPI_ISL_644563 | France/PAC-MEPHI-2416/2020 | 2020-10-14 | Europe/France/Provence-Alpes-CV¥te d' | MEPHI, Aix Marseille University | MEPHI, Aix Marseille University | Anthony LEVASSEUR et al |
| EPI_ISL_644564 | France/PAC-MEPHI-2417/2020 | 2020-10-14 | Europe/France/Provence-Alpes-CV¥te d' | MEPHI, Aix Marseille University | MEPHI, Aix Marseille University | Anthony LEVASSEUR et al |
| EPI_ISL_644566 | Serbia/KosovskaMitrovica-08372107 | 2020-07-20 | Europe/Kosovo/Kosovska Mitrovica/ | "Veterinary Specialized Institute ""Kraljevo"", Serbia" | "Veterinary Specialized Institute ""Kraljevo"", Serbia" | Vidanovic et al |
| EPI_ISL_644572 | Serbia/Kraljevo-00521205/2020 | 2020-05-11 | Europe/Serbia/Ra≈°ka District/Kraljevo | "Veterinary Specialized Institute ""Kraljevo"", Serbia" | "Veterinary Specialized Institute ""Kraljevo"", Serbia" | Vidanovic et al |
| EPI_ISL_644577 | Serbia/Kraljevo-03102507/2020 | 2020-07-24 | Europe/Serbia/Ra≈°ka District/Kraljevo | "Veterinary Specialized Institute ""Kraljevo"", Serbia" | "Veterinary Specialized Institute ""Kraljevo"", Serbia" | Vidanovic et al |
| EPI_ISL_644580 | Serbia/NoviPazar-17611405/2020 | 2020-05-13 | Europe/Serbia/Ra≈°ka District/Novi Paz | "Veterinary Specialized Institute ""Kraljevo"", Serbia" | "Veterinary Specialized Institute ""Kraljevo"", Serbia" | Vidanovic et al |
| EPI_ISL_644581 | Serbia/NoviPazar-17641405/2020 | 2020-05-13 | Europe/Serbia/Ra≈°ka District/Novi Paz | "Veterinary Specialized Institute ""Kraljevo"", Serbia" | "Veterinary Specialized Institute ""Kraljevo"", Serbia" | Vidanovic et al |

| | | | | | | |
|---|---|---|---|---|---|---|
| EPI_ISL_644582 | Serbia/Uzice-00200704/2020 | 2020-04-06 | Europe/Serbia/Zlatibor District/Uzice | "Veterinary Specialized Institute ""Kraljevo"", Serbia" | "Veterinary Specialized Institute ""Kraljevo"", Serbia" | Vidanovic et al |
| EPI_ISL_644587 | USA/NM-NMDOH-2020327015/2020 | 2020-09-03 | North America/USA/New Mexico/ | New Mexico Department of Health Scientific Laboratory | New Mexico Department of Health Scientific Laboratory | Ellie Johnson et al |
| EPI_ISL_644593 | USA/NM-NMDOH-2020328776/2020 | 2020-09-04 | North America/USA/New Mexico/ | New Mexico Department of Health Scientific Laboratory | New Mexico Department of Health Scientific Laboratory | Ellie Johnson et al |
| EPI_ISL_644598 | USA/NM-NMDOH-2020329076/2020 | 2020-09-04 | North America/USA/New Mexico/ | New Mexico Department of Health Scientific Laboratory | New Mexico Department of Health Scientific Laboratory | Ellie Johnson et al |
| EPI_ISL_644600 | USA/NM-NMDOH-2020329989/2020 | 2020-09-04 | North America/USA/New Mexico/ | New Mexico Department of Health Scientific Laboratory | New Mexico Department of Health Scientific Laboratory | Ellie Johnson et al |
| EPI_ISL_644601 | USA/NM-NMDOH-2020329992/2020 | 2020-09-04 | North America/USA/New Mexico/ | New Mexico Department of Health Scientific Laboratory | New Mexico Department of Health Scientific Laboratory | Ellie Johnson et al |
| EPI_ISL_644602 | USA/NM-NMDOH-2020330052/2020 | 2020-09-05 | North America/USA/New Mexico/ | New Mexico Department of Health Scientific Laboratory | New Mexico Department of Health Scientific Laboratory | Ellie Johnson et al |
| EPI_ISL_644603 | USA/NM-NMDOH-2020330065/2020 | 2020-09-05 | North America/USA/New Mexico/ | New Mexico Department of Health Scientific Laboratory | New Mexico Department of Health Scientific Laboratory | Ellie Johnson et al |
| EPI_ISL_644608 | USA/NM-NMDOH-2020330343/2020 | 2020-09-05 | North America/USA/New Mexico/ | New Mexico Department of Health Scientific Laboratory | New Mexico Department of Health Scientific Laboratory | Ellie Johnson et al |
| EPI_ISL_644610 | Ireland/LK-NVRL-73IRL84268/2020 | 2020-09-15 | Europe/Ireland/Limerick/ | National Virus Reference Laboratory | National Virus Reference Laboratory | Michael Carr et al |

| | | | | | | |
|---|---|---|---|---|---|---|
| EPI_ISL_644611 | Belgium/ULG-10765/2020 | 2020-10-31 | Europe/Belgium/Liv®ge/ | Department of Clinical Microbiology | GIGA Medical Genomics | Keith Durkin et al |
| EPI_ISL_644612 | Belgium/ULG-10766/2020 | 2020-10-31 | Europe/Belgium/Liv®ge/ | Department of Clinical Microbiology | GIGA Medical Genomics | Keith Durkin et al |
| EPI_ISL_644617 | Belgium/ULG-10771/2020 | 2020-10-31 | Europe/Belgium/Liv®ge/ | Department of Clinical Microbiology | GIGA Medical Genomics | Keith Durkin et al |
| EPI_ISL_644619 | Belgium/ULG-10773/2020 | 2020-10-31 | Europe/Belgium/Liv®ge/ | Department of Clinical Microbiology | GIGA Medical Genomics | Keith Durkin et al |
| EPI_ISL_644673 | France/NAQ-SC280/2020 | 2020-03-30 | Europe/France/Nouvelle-Aquitaine/Limo | CHU de Limoges | CNR Virus des Infections Respiratoires - France SUD | Antonin Bal et al |
| EPI_ISL_644675 | France/NAQ-SC282/2020 | 2020-03-31 | Europe/France/Nouvelle-Aquitaine/Limo | CHU de Limoges | CNR Virus des Infections Respiratoires - France SUD | Antonin Bal et al |
| EPI_ISL_644677 | France/NAQ-SC284/2020 | 2020-04-06 | Europe/France/Nouvelle-Aquitaine/Limo | CHU de Limoges | CNR Virus des Infections Respiratoires - France SUD | Antonin Bal et al |
| EPI_ISL_644678 | France/NAQ-SC288/2020 | 2020-04-10 | Europe/France/Nouvelle-Aquitaine/Limo | CHU de Limoges | CNR Virus des Infections Respiratoires - France SUD | Antonin Bal et al |
| EPI_ISL_644681 | France/OCC-SC463/2020 | 2020-02-26 | Europe/France/Occitanie/Montpellier | CHU Montpellier | CNR Virus des Infections Respiratoires - France SUD | Antonin Bal et al |
| EPI_ISL_644684 | France/OCC-SC468/2020 | 2020-03-06 | Europe/France/Occitanie/Montpellier | CHU Montpellier | CNR Virus des Infections Respiratoires - France SUD | Antonin Bal et al |
| EPI_ISL_644687 | France/OCC-SC474/2020 | 2020-03-17 | Europe/France/Occitanie/Montpellier | CHU Montpellier | CNR Virus des Infections Respiratoires - France SUD | Antonin Bal et al |
| EPI_ISL_644689 | France/OCC-SC486/2020 | 2020-03-24 | Europe/France/Occitanie/Montpellier | CHU Montpellier | CNR Virus des Infections Respiratoires - France SUD | Antonin Bal et al |

| | | | | | | |
|---|---|---|---|---|---|---|
| EPI_ISL_644693 | France/OCC-SC496/2020 | 2020-03-26 | Europe/France/Occitanie/Montpellier | CHU Montpellier | CNR Virus des Infections Respiratoires - France SUD | Antonin Bal et al |
| EPI_ISL_644701 | France/OCC-SC532/2020 | 2020-03-13 | Europe/France/Occitanie/N√Æmes | CHU N√Æmes | CNR Virus des Infections Respiratoires - France SUD | Antonin Bal et al |
| EPI_ISL_644703 | France/PAC-SC435/2020 | 2020-04-23 | Europe/France/Provence-Alpes-C√¥te d' | Unit√© des Virus √âmergents | CNR Virus des Infections Respiratoires - France SUD | Antonin Bal et al |
| EPI_ISL_644704 | France/PAC-SC436/2020 | 2020-04-23 | Europe/France/Provence-Alpes-C√¥te d' | Unit√© des Virus √âmergents | CNR Virus des Infections Respiratoires - France SUD | Antonin Bal et al |
| EPI_ISL_644705 | France/PAC-SC437/2020 | 2020-04-24 | Europe/France/Provence-Alpes-C√¥te d' | Unit√© des Virus √âmergents | CNR Virus des Infections Respiratoires - France SUD | Antonin Bal et al |
| EPI_ISL_644707 | France/PAC-SC442/2020 | 2020-04-29 | Europe/France/Provence-Alpes-C√¥te d' | Unit√© des Virus √âmergents | CNR Virus des Infections Respiratoires - France SUD | Antonin Bal et al |
| EPI_ISL_644710 | France/PAC-SC446/2020 | 2020-03-18 | Europe/France/Provence-Alpes-C√¥te d' | Unit√© des Virus √âmergents | CNR Virus des Infections Respiratoires - France SUD | Antonin Bal et al |
| EPI_ISL_644711 | France/PAC-SC448/2020 | 2020-03-10 | Europe/France/Provence-Alpes-C√¥te d' | Unit√© des Virus √âmergents | CNR Virus des Infections Respiratoires - France SUD | Antonin Bal et al |
| EPI_ISL_644715 | India/TG-CCMB-OM21/2020 | 2020-09-20 | Asia/India/Telangana/Hyderabad | Osmania Medical College | CSIR-Centre for Cellular and Molecular Biology | Dr.V.Sudha Rani et al |
| EPI_ISL_644716 | India/TG-CCMB-OM22/2020 | 2020-09-20 | Asia/India/Telangana/Hyderabad | Osmania Medical College | CSIR-Centre for Cellular and Molecular Biology | Dr.V.Sudha Rani et al |

| | | | | | | |
|---|---|---|---|---|---|---|
| EPI_ISL_644717 | India/TG-CCMB-OM23/2020 | 2020-09-20 | Asia/India/Telangana/Hyderabad | Osmania Medical College | CSIR-Centre for Cellular and Molecular Biology | Dr.V.Sudha Rani et al |
| EPI_ISL_644718 | India/TG-CCMB-OM24/2020 | 2020-09-20 | Asia/India/Telangana/Hyderabad | Osmania Medical College | CSIR-Centre for Cellular and Molecular Biology | Dr.V.Sudha Rani et al |
| EPI_ISL_644719 | India/TG-CCMB-OM25/2020 | 2020-09-20 | Asia/India/Telangana/Hyderabad | Osmania Medical College | CSIR-Centre for Cellular and Molecular Biology | Dr.V.Sudha Rani et al |
| EPI_ISL_644720 | India/TG-CCMB-OM26/2020 | 2020-09-20 | Asia/India/Telangana/Hyderabad | Osmania Medical College | CSIR-Centre for Cellular and Molecular Biology | Dr.V.Sudha Rani et al |
| EPI_ISL_644721 | India/TG-CCMB-OM27/2020 | 2020-09-20 | Asia/India/Telangana/Hyderabad | Osmania Medical College | CSIR-Centre for Cellular and Molecular Biology | Dr.V.Sudha Rani et al |
| EPI_ISL_644722 | India/TG-CCMB-OM28/2020 | 2020-09-20 | Asia/India/Telangana/Hyderabad | Osmania Medical College | CSIR-Centre for Cellular and Molecular Biology | Dr.V.Sudha Rani et al |
| EPI_ISL_644723 | India/TG-CCMB-OM29/2020 | 2020-09-20 | Asia/India/Telangana/Hyderabad | Osmania Medical College | CSIR-Centre for Cellular and Molecular Biology | Dr.V.Sudha Rani et al |
| EPI_ISL_644724 | India/TG-CCMB-OM30/2020 | 2020-09-20 | Asia/India/Telangana/Hyderabad | Osmania Medical College | CSIR-Centre for Cellular and Molecular Biology | Dr.V.Sudha Rani et al |
| EPI_ISL_644729 | India/TG-CCMB-OM35/2020 | 2020-04-15 | Asia/India/Telangana/Hyderabad | Osmania Medical College | CSIR-Centre for Cellular and Molecular Biology | Dr.V.Sudha Rani et al |
| EPI_ISL_644823 | USA/WA-USAFSAM-S060/2020 | 2020-02-22 | North America/USA/Washington/ | Madigan Army Medical Center | U.S. Air Force School of Aerospace Medicine | Emily Parsons et al |
| EPI_ISL_644826 | USA/VA-DCLS-1651/2020 | 2020-08-28 | North America/USA/Virginia/ | Virginia DCLS | Virginia DCLS | Virginia DCLS et al |
| EPI_ISL_644829 | USA/VA-DCLS-1657/2020 | 2020-08-29 | North America/USA/Virginia/ | Virginia DCLS | Virginia DCLS | Virginia DCLS et al |
| EPI_ISL_644833 | USA/VA-DCLS-1662/2020 | 2020-08-29 | North America/USA/Virginia/ | Virginia DCLS | Virginia DCLS | Virginia DCLS et al |
| EPI_ISL_644835 | USA/VA-DCLS-1664/2020 | 2020-08-29 | North America/USA/Virginia/ | Virginia DCLS | Virginia DCLS | Virginia DCLS et al |
| EPI_ISL_644846 | USA/VA-DCLS-1743/2020 | 2020-09-10 | North America/USA/Virginia/ | Virginia DCLS | Virginia DCLS | Virginia DCLS et al |
| EPI_ISL_644847 | USA/VA-DCLS-1744/2020 | 2020-09-10 | North America/USA/Virginia/ | Virginia DCLS | Virginia DCLS | Virginia DCLS et al |
| EPI_ISL_644848 | USA/VA-DCLS-1745/2020 | 2020-09-10 | North America/USA/Virginia/ | Virginia DCLS | Virginia DCLS | Virginia DCLS et al |

| | | | | | | |
|---|---|---|---|---|---|---|
| EPI_ISL_644853 | USA/VA-DCLS-1753/2020 | 2020-09-10 | North America/USA/Virginia/ | Virginia DCLS | Virginia DCLS | Virginia DCLS et al |
| EPI_ISL_644854 | USA/VA-DCLS-1755/2020 | 2020-09-10 | North America/USA/Virginia/ | Virginia DCLS | Virginia DCLS | Virginia DCLS et al |
| EPI_ISL_644857 | USA/VA-DCLS-1762/2020 | 2020-09-09 | North America/USA/Virginia/ | Virginia DCLS | Virginia DCLS | Virginia DCLS et al |
| EPI_ISL_644858 | USA/VA-DCLS-1764/2020 | 2020-09-10 | North America/USA/Virginia/ | Virginia DCLS | Virginia DCLS | Virginia DCLS et al |
| EPI_ISL_644861 | USA/VA-DCLS-1767/2020 | 2020-09-09 | North America/USA/Virginia/ | Virginia DCLS | Virginia DCLS | Virginia DCLS et al |
| EPI_ISL_644862 | USA/VA-DCLS-1769/2020 | 2020-09-09 | North America/USA/Virginia/ | Virginia DCLS | Virginia DCLS | Virginia DCLS et al |
| EPI_ISL_644872 | USA/VA-DCLS-1786/2020 | 2020-09-16 | North America/USA/Virginia/ | Virginia DCLS | Virginia DCLS | Virginia DCLS et al |
| EPI_ISL_644878 | USA/VA-DCLS-1795/2020 | 2020-09-16 | North America/USA/Virginia/ | Virginia DCLS | Virginia DCLS | Virginia DCLS et al |
| EPI_ISL_644882 | USA/VA-DCLS-1956/2020 | 2020-10-07 | North America/USA/Virginia/ | Virginia DCLS | Virginia DCLS | Virginia DCLS et al |
| EPI_ISL_644912 | USA/VA-DCLS-1872/2020 | 2020-09-23 | North America/USA/Virginia/ | Virginia DCLS | Virginia DCLS | Virginia DCLS et al |
| EPI_ISL_644915 | USA/VA-DCLS-1877/2020 | 2020-09-23 | North America/USA/Virginia/ | Virginia DCLS | Virginia DCLS | Virginia DCLS et al |
| EPI_ISL_644918 | USA/VA-DCLS-1885/2020 | 2020-09-23 | North America/USA/Virginia/ | Virginia DCLS | Virginia DCLS | Virginia DCLS et al |
| EPI_ISL_644919 | USA/VA-DCLS-1886/2020 | 2020-09-23 | North America/USA/Virginia/ | Virginia DCLS | Virginia DCLS | Virginia DCLS et al |
| EPI_ISL_644930 | USA/VA-DCLS-1948/2020 | 2020-09-29 | North America/USA/Virginia/ | Virginia DCLS | Virginia DCLS | Virginia DCLS et al |
| EPI_ISL_644935 | USA/MN-MDH-2004/2020 | 2020-10-14 | North America/USA/Minnesota/ | Essentia Health-St. Mary's Medical Center | Minnesota Department of Health, Public Health Laboratory | Matt Plumb et al |
| EPI_ISL_644936 | USA/MN-MDH-2005/2020 | 2020-10-01 | North America/USA/Minnesota/ | Mayo Clinic & Mayo Clinic Laboratories | Minnesota Department of Health, Public Health Laboratory | Matt Plumb et al |
| EPI_ISL_644940 | USA/MN-MDH-2009/2020 | 2020-09-21 | North America/USA/Minnesota/ | Mayo Clinic & Mayo Clinic Laboratories | Minnesota Department of Health, Public Health Laboratory | Matt Plumb et al |
| EPI_ISL_644943 | USA/MN-MDH-2012/2020 | 2020-10-05 | North America/USA/Minnesota/ | Essentia Health-St. Mary's Medical Center | Minnesota Department of Health, Public Health Laboratory | Matt Plumb et al |
| EPI_ISL_644987 | Japan/Donner83/2020 | 2020-08-04 | Asia/Japan/Kanto/ | Department of Infectious Diseases, Keio University School of Medicine, Tokyo, Japan | Center for Medical Genetics, Keio University School of Medicine, Tokyo, Japan | Kenjiro Kosaki et al |

| | | | | | | |
|---|---|---|---|---|---|---|
| EPI_ISL_645001 | Poland/PL_MCB_2/2020 | 2020-04-22 | Europe/Poland/Poland/ | Human Genome Variation Research Group, Malopolska Centre of Biotechnology | Human Genome Variation Research Group, Malopolska Centre of Biotechnology | Kowalski et al |
| EPI_ISL_645002 | Poland/PL_MCB_3/2020 | 2020-04-22 | Europe/Poland/Poland/ | Human Genome Variation Research Group, Malopolska Centre of Biotechnology | Human Genome Variation Research Group, Malopolska Centre of Biotechnology | Kowalski et al |
| EPI_ISL_645003 | Poland/PL_MCB_4/2020 | 2020-04-22 | Europe/Poland/Poland/ | Human Genome Variation Research Group, Malopolska Centre of Biotechnology | Human Genome Variation Research Group, Malopolska Centre of Biotechnology | Kowalski et al |
| EPI_ISL_645017 | Poland/PL_MCB_18/2020 | 2020-03-28 | Europe/Poland/Poland/ | Human Genome Variation Research Group, Malopolska Centre of Biotechnology | Human Genome Variation Research Group, Malopolska Centre of Biotechnology | Kowalski et al |
| EPI_ISL_645021 | Poland/PL_MCB_22/2020 | 2020-03-29 | Europe/Poland/Poland/ | Human Genome Variation Research Group, Malopolska Centre of Biotechnology | Human Genome Variation Research Group, Malopolska Centre of Biotechnology | Kowalski et al |
| EPI_ISL_645023 | Poland/PL_MCB_24/2020 | 2020-03-28 | Europe/Poland/Poland/ | Human Genome Variation Research Group, Malopolska Centre of Biotechnology | Human Genome Variation Research Group, Malopolska Centre of Biotechnology | Kowalski et al |

| | | | | | | |
|---|---|---|---|---|---|---|
| EPI_ISL_645028 | Poland/PL_MCB_29/2020 | 2020-05-12 | Europe/Poland/Poland/ | Human Genome Variation Research Group, Malopolska Centre of Biotechnology | Human Genome Variation Research Group, Malopolska Centre of Biotechnology | Kowalski et al |
| EPI_ISL_645033 | Poland/PL_MCB_34/2020 | 2020-05-06 | Europe/Poland/Poland/ | Human Genome Variation Research Group, Malopolska Centre of Biotechnology | Human Genome Variation Research Group, Malopolska Centre of Biotechnology | Kowalski et al |
| EPI_ISL_645035 | Poland/PL_MCB_36/2020 | 2020-04-21 | Europe/Poland/Poland/ | Human Genome Variation Research Group, Malopolska Centre of Biotechnology | Human Genome Variation Research Group, Malopolska Centre of Biotechnology | Kowalski et al |
| EPI_ISL_645036 | Poland/PL_MCB_37/2020 | 2020-04-07 | Europe/Poland/Poland/ | Human Genome Variation Research Group, Malopolska Centre of Biotechnology | Human Genome Variation Research Group, Malopolska Centre of Biotechnology | Kowalski et al |
| EPI_ISL_645040 | Poland/PL_MCB_41/2020 | 2020-04-28 | Europe/Poland/Poland/ | Human Genome Variation Research Group, Malopolska Centre of Biotechnology | Human Genome Variation Research Group, Malopolska Centre of Biotechnology | Kowalski et al |
| EPI_ISL_645041 | Poland/PL_MCB_42/2020 | 2020-04-28 | Europe/Poland/Poland/ | Human Genome Variation Research Group, Malopolska Centre of Biotechnology | Human Genome Variation Research Group, Malopolska Centre of Biotechnology | Kowalski et al |

| | | | | | | |
|---|---|---|---|---|---|---|
| EPI_ISL_645045 | Poland/PL_MCB_46/2020 | 2020-04-21 | Europe/Poland/Poland/ | Human Genome Variation Research Group, Malopolska Centre of Biotechnology | Human Genome Variation Research Group, Malopolska Centre of Biotechnology | Kowalski et al |
| EPI_ISL_645055 | Poland/PL_MCB_56/2020 | 2020-06-06 | Europe/Poland/Poland/ | Human Genome Variation Research Group, Malopolska Centre of Biotechnology | Human Genome Variation Research Group, Malopolska Centre of Biotechnology | Kowalski et al |
| EPI_ISL_645056 | Poland/PL_MCB_57/2020 | 2020-06-06 | Europe/Poland/Poland/ | Human Genome Variation Research Group, Malopolska Centre of Biotechnology | Human Genome Variation Research Group, Malopolska Centre of Biotechnology | Kowalski et al |
| EPI_ISL_645057 | Poland/PL_MCB_58/2020 | 2020-06-06 | Europe/Poland/Poland/ | Human Genome Variation Research Group, Malopolska Centre of Biotechnology | Human Genome Variation Research Group, Malopolska Centre of Biotechnology | Kowalski et al |
| EPI_ISL_645058 | Poland/PL_MCB_59/2020 | 2020-06-06 | Europe/Poland/Poland/ | Human Genome Variation Research Group, Malopolska Centre of Biotechnology | Human Genome Variation Research Group, Malopolska Centre of Biotechnology | Kowalski et al |
| EPI_ISL_645060 | Poland/PL_MCB_61/2020 | 2020-06-06 | Europe/Poland/Poland/ | Human Genome Variation Research Group, Malopolska Centre of Biotechnology | Human Genome Variation Research Group, Malopolska Centre of Biotechnology | Kowalski et al |

| | | | | | | |
|---|---|---|---|---|---|---|
| EPI_ISL_645062 | Poland/PL_MCB_63/2020 | 2020-06-06 | Europe/Poland/Poland/ | Human Genome Variation Research Group, Malopolska Centre of Biotechnology | Human Genome Variation Research Group, Malopolska Centre of Biotechnology | Kowalski et al |
| EPI_ISL_645069 | Poland/PL_MCB_70/2020 | 2020-05-10 | Europe/Poland/Poland/ | Human Genome Variation Research Group, Malopolska Centre of Biotechnology | Human Genome Variation Research Group, Malopolska Centre of Biotechnology | Kowalski et al |
| EPI_ISL_645071 | Poland/PL_MCB_72/2020 | 2020-05-10 | Europe/Poland/Poland/ | Human Genome Variation Research Group, Malopolska Centre of Biotechnology | Human Genome Variation Research Group, Malopolska Centre of Biotechnology | Kowalski et al |
| EPI_ISL_645073 | Poland/PL_MCB_74/2020 | 2020-04-16 | Europe/Poland/Poland/ | Human Genome Variation Research Group, Malopolska Centre of Biotechnology | Human Genome Variation Research Group, Malopolska Centre of Biotechnology | Kowalski et al |
| EPI_ISL_645077 | Poland/PL_MCB_78/2020 | 2020-04-07 | Europe/Poland/Poland/ | Human Genome Variation Research Group, Malopolska Centre of Biotechnology | Human Genome Variation Research Group, Malopolska Centre of Biotechnology | Kowalski et al |
| EPI_ISL_645082 | Poland/PL_MCB_83/2020 | 2020-06-06 | Europe/Poland/Poland/ | Human Genome Variation Research Group, Malopolska Centre of Biotechnology | Human Genome Variation Research Group, Malopolska Centre of Biotechnology | Kowalski et al |

| | | | | | | |
|---|---|---|---|---|---|---|
| EPI_ISL_645083 | Poland/PL_MCB_84/2020 | 2020-05-10 | Europe/Poland/Poland/ | Human Genome Variation Research Group, Malopolska Centre of Biotechnology | Human Genome Variation Research Group, Malopolska Centre of Biotechnology | Kowalski et al |
| EPI_ISL_645085 | Poland/PL_MCB_86/2020 | 2020-05-10 | Europe/Poland/Poland/ | Human Genome Variation Research Group, Malopolska Centre of Biotechnology | Human Genome Variation Research Group, Malopolska Centre of Biotechnology | Kowalski et al |
| EPI_ISL_645086 | Poland/PL_MCB_87/2020 | 2020-05-10 | Europe/Poland/Poland/ | Human Genome Variation Research Group, Malopolska Centre of Biotechnology | Human Genome Variation Research Group, Malopolska Centre of Biotechnology | Kowalski et al |
| EPI_ISL_645091 | Poland/PL_MCB_92/2020 | 2020-05-09 | Europe/Poland/Poland/ | Human Genome Variation Research Group, Malopolska Centre of Biotechnology | Human Genome Variation Research Group, Malopolska Centre of Biotechnology | Kowalski et al |
| EPI_ISL_645093 | Poland/PL_MCB_94/2020 | 2020-05-10 | Europe/Poland/Poland/ | Human Genome Variation Research Group, Malopolska Centre of Biotechnology | Human Genome Variation Research Group, Malopolska Centre of Biotechnology | Kowalski et al |
| EPI_ISL_645096 | Poland/PL_MCB_97/2020 | 2020-05-10 | Europe/Poland/Poland/ | Human Genome Variation Research Group, Malopolska Centre of Biotechnology | Human Genome Variation Research Group, Malopolska Centre of Biotechnology | Kowalski et al |

| | | | | | | |
|---|---|---|---|---|---|---|
| EPI_ISL_645106 | Poland/PL_MCB_107/2020 | 2020-05-12 | Europe/Poland/Poland/ | Human Genome Variation Research Group, Malopolska Centre of Biotechnology | Human Genome Variation Research Group, Malopolska Centre of Biotechnology | Kowalski et al |
| EPI_ISL_645108 | Poland/PL_MCB_109/2020 | 2020-04-21 | Europe/Poland/Poland/ | Human Genome Variation Research Group, Malopolska Centre of Biotechnology | Human Genome Variation Research Group, Malopolska Centre of Biotechnology | Kowalski et al |
| EPI_ISL_645109 | Poland/PL_MCB_110/2020 | 2020-06-06 | Europe/Poland/Poland/ | Human Genome Variation Research Group, Malopolska Centre of Biotechnology | Human Genome Variation Research Group, Malopolska Centre of Biotechnology | Kowalski et al |
| EPI_ISL_645171 | France/NAQ-SC220/2020 | 2020-03-28 | Europe/France/Nouvelle-Aquitaine/Bord | CHU Bordeaux | CNR Virus des Infections Respiratoires - France SUD | Antonin Bal et al |
| EPI_ISL_645174 | France/NAQ-SC251/2020 | 2020-03-16 | Europe/France/Nouvelle-Aquitaine/Limo | CHU de Limoges | CNR Virus des Infections Respiratoires - France SUD | Antonin Bal et al |
| EPI_ISL_645175 | France/NAQ-SC254/2020 | 2020-03-11 | Europe/France/Nouvelle-Aquitaine/Limo | CHU de Limoges | CNR Virus des Infections Respiratoires - France SUD | Antonin Bal et al |
| EPI_ISL_645180 | France/NAQ-SC274/2020 | 2020-03-25 | Europe/France/Nouvelle-Aquitaine/Limo | CHU de Limoges | CNR Virus des Infections Respiratoires - France SUD | Antonin Bal et al |
| EPI_ISL_645185 | France/ARA-SC369/2020 | 2020-05-01 | Europe/France/Auvergne-Rh√¥ne-Alpes/ | CHU de Saint-√âtienne H√¥pital Nord | CNR Virus des Infections Respiratoires - France SUD | Antonin Bal et al |

| | | | | | | |
|---|---|---|---|---|---|---|
| EPI_ISL_645189 | France/ARA-SC379/2020 | 2020-03-13 | Europe/France/Auvergne-Rhv¥ne-Alpes/ | CHU Clermont-Ferrand | CNR Virus des Infections Respiratoires - France SUD | Antonin Bal et al |
| EPI_ISL_645192 | France/ARA-SC388/2020 | 2020-03-20 | Europe/France/Auvergne-Rhv¥ne-Alpes/ | CHU Clermont-Ferrand | CNR Virus des Infections Respiratoires - France SUD | Antonin Bal et al |
| EPI_ISL_645194 | France/ARA-SC392/2020 | 2020-03-30 | Europe/France/Auvergne-Rhv¥ne-Alpes/ | CHU Clermont-Ferrand | CNR Virus des Infections Respiratoires - France SUD | Antonin Bal et al |
| EPI_ISL_645205 | France/OCC-SC553/2020 | 2020-04-03 | Europe/France/Occitanie/NvÆmes | CHU NvÆmes | CNR Virus des Infections Respiratoires - France SUD | Antonin Bal et al |
| EPI_ISL_645206 | France/OCC-SC554/2020 | 2020-04-03 | Europe/France/Occitanie/NvÆmes | CHU NvÆmes | CNR Virus des Infections Respiratoires - France SUD | Antonin Bal et al |
| EPI_ISL_645207 | France/OCC-SC559/2020 | 2020-04-20 | Europe/France/Occitanie/NvÆmes | CHU NvÆmes | CNR Virus des Infections Respiratoires - France SUD | Antonin Bal et al |
| EPI_ISL_645209 | France/OCC-SC572/2020 | 2020-05-29 | Europe/France/Occitanie/NvÆmes | CHU NvÆmes | CNR Virus des Infections Respiratoires - France SUD | Antonin Bal et al |
| EPI_ISL_645238 | England/CAMC-B36A26/2020 | 2020-11-05 | Europe/United Kingdom/England/ | Lighthouse Lab in Cambridge | Wellcome Sanger Institute for the COVID-19 Genomics UK (COG-UK) Consortium | Rob Howes et al |
| EPI_ISL_645258 | England/CAMC-B367C5/2020 | 2020-11-05 | Europe/United Kingdom/England/ | Lighthouse Lab in Cambridge | Wellcome Sanger Institute for the COVID-19 Genomics UK (COG-UK) Consortium | Rob Howes et al |

| | | | | | | |
|---|---|---|---|---|---|---|
| EPI_ISL_645340 | England/CAMC-B361F4/2020 | 2020-11-05 | Europe/United Kingdom/England/ | Lighthouse Lab in Cambridge | Wellcome Sanger Institute for the COVID-19 Genomics UK (COG-UK) Consortium | Rob Howes et al |
| EPI_ISL_645489 | England/MILK-B3D694/2020 | 2020-11-02 | Europe/United Kingdom/England/ | Lighthouse Lab in Milton Keynes | Wellcome Sanger Institute for the COVID-19 Genomics UK (COG-UK) Consortium | The Lighthouse Lab in Milton Keynes et al |
| EPI_ISL_645531 | England/ALDP-B1AF65/2020 | 2020-11-01 | Europe/United Kingdom/England/ | Lighthouse Lab in Alderley Park | Wellcome Sanger Institute for the COVID-19 Genomics UK (COG-UK) Consortium | Jacquelyn Wynn et al |
| EPI_ISL_645588 | England/ALDP-B1C349/2020 | 2020-11-01 | Europe/United Kingdom/England/ | Lighthouse Lab in Alderley Park | Wellcome Sanger Institute for the COVID-19 Genomics UK (COG-UK) Consortium | Jacquelyn Wynn et al |
| EPI_ISL_645712 | England/ALDP-B1CEDF/2020 | 2020-11-01 | Europe/United Kingdom/England/ | Lighthouse Lab in Alderley Park | Wellcome Sanger Institute for the COVID-19 Genomics UK (COG-UK) Consortium | Jacquelyn Wynn et al |
| EPI_ISL_645842 | England/ALDP-B1F65C/2020 | 2020-11-01 | Europe/United Kingdom/England/ | Lighthouse Lab in Alderley Park | Wellcome Sanger Institute for the COVID-19 Genomics UK (COG-UK) Consortium | Jacquelyn Wynn et al |
| EPI_ISL_645894 | England/ALDP-B1AD4D/2020 | 2020-11-01 | Europe/United Kingdom/England/ | Lighthouse Lab in Alderley Park | Wellcome Sanger Institute for the COVID-19 Genomics UK (COG-UK) Consortium | Jacquelyn Wynn et al |
| EPI_ISL_645971 | England/ALDP-B1ADF2/2020 | 2020-11-01 | Europe/United Kingdom/England/ | Lighthouse Lab in Alderley Park | Wellcome Sanger Institute for the COVID-19 Genomics UK (COG-UK) Consortium | Jacquelyn Wynn et al |

| | | | | | | |
|---|---|---|---|---|---|---|
| EPI_ISL_646002 | England/ALDP-B1AC22/2020 | 2020-11-01 | Europe/United Kingdom/England/ | Lighthouse Lab in Alderley Park | Wellcome Sanger Institute for the COVID-19 Genomics UK (COG-UK) Consortium | Jacquelyn Wynn et al |
| EPI_ISL_646076 | England/CAMC-B225B3/2020 | 2020-11-01 | Europe/United Kingdom/England/ | Lighthouse Lab in Cambridge | Wellcome Sanger Institute for the COVID-19 Genomics UK (COG-UK) Consortium | Rob Howes et al |
| EPI_ISL_646096 | England/ALDP-B1B1B9/2020 | 2020-11-02 | Europe/United Kingdom/England/ | Lighthouse Lab in Alderley Park | Wellcome Sanger Institute for the COVID-19 Genomics UK (COG-UK) Consortium | Jacquelyn Wynn et al |
| EPI_ISL_646104 | England/ALDP-B1ADE3/2020 | 2020-11-01 | Europe/United Kingdom/England/ | Lighthouse Lab in Alderley Park | Wellcome Sanger Institute for the COVID-19 Genomics UK (COG-UK) Consortium | Jacquelyn Wynn et al |
| EPI_ISL_646113 | England/ALDP-B1D603/2020 | 2020-11-01 | Europe/United Kingdom/England/ | Lighthouse Lab in Alderley Park | Wellcome Sanger Institute for the COVID-19 Genomics UK (COG-UK) Consortium | Jacquelyn Wynn et al |
| EPI_ISL_646239 | England/QEUH-B31B46/2020 | 2020-11-04 | Europe/United Kingdom/England/ | Lighthouse Lab in Glasgow | Wellcome Sanger Institute for the COVID-19 Genomics UK (COG-UK) Consortium | Harper VanSteenhouse et al |
| EPI_ISL_646312 | England/QEUH-B2F7FC/2020 | 2020-11-04 | Europe/United Kingdom/England/ | Lighthouse Lab in Glasgow | Wellcome Sanger Institute for the COVID-19 Genomics UK (COG-UK) Consortium | Harper VanSteenhouse et al |
| EPI_ISL_646403 | England/QEUH-B31F77/2020 | 2020-11-04 | Europe/United Kingdom/England/ | Lighthouse Lab in Glasgow | Wellcome Sanger Institute for the COVID-19 Genomics UK (COG-UK) Consortium | Harper VanSteenhouse et al |

| | | | | | | |
|---|---|---|---|---|---|---|
| EPI_ISL_646605 | England/ALDP-B26106/2020 | 2020-11-04 | Europe/United Kingdom/England/ | Lighthouse Lab in Alderley Park | Wellcome Sanger Institute for the COVID-19 Genomics UK (COG-UK) Consortium | Jacquelyn Wynn et al |
| EPI_ISL_647133 | England/CAMC-B21F2F/2020 | 2020-11-02 | Europe/United Kingdom/England/ | Lighthouse Lab in Cambridge | Wellcome Sanger Institute for the COVID-19 Genomics UK (COG-UK) Consortium | Rob Howes et al |
| EPI_ISL_647275 | England/CAMC-B22DF7/2020 | 2020-11-01 | Europe/United Kingdom/England/ | Lighthouse Lab in Cambridge | Wellcome Sanger Institute for the COVID-19 Genomics UK (COG-UK) Consortium | Rob Howes et al |
| EPI_ISL_647292 | England/CAMC-B216DF/2020 | 2020-11-01 | Europe/United Kingdom/England/ | Lighthouse Lab in Cambridge | Wellcome Sanger Institute for the COVID-19 Genomics UK (COG-UK) Consortium | Rob Howes et al |
| EPI_ISL_647366 | England/CAMC-B22F79/2020 | 2020-11-02 | Europe/United Kingdom/England/ | Lighthouse Lab in Cambridge | Wellcome Sanger Institute for the COVID-19 Genomics UK (COG-UK) Consortium | Rob Howes et al |
| EPI_ISL_647391 | England/CAMC-B214C6/2020 | 2020-11-03 | Europe/United Kingdom/England/ | Lighthouse Lab in Cambridge | Wellcome Sanger Institute for the COVID-19 Genomics UK (COG-UK) Consortium | Rob Howes et al |
| EPI_ISL_647412 | England/CAMC-B22EA9/2020 | 2020-11-02 | Europe/United Kingdom/England/ | Lighthouse Lab in Cambridge | Wellcome Sanger Institute for the COVID-19 Genomics UK (COG-UK) Consortium | Rob Howes et al |
| EPI_ISL_647452 | England/CAMC-B22911/2020 | 2020-11-01 | Europe/United Kingdom/England/ | Lighthouse Lab in Cambridge | Wellcome Sanger Institute for the COVID-19 Genomics UK (COG-UK) Consortium | Rob Howes et al |

| | | | | | | |
|---|---|---|---|---|---|---|
| EPI_ISL_647643 | England/ALDP-B1AB34/2020 | 2020-11-01 | Europe/United Kingdom/England/ | Lighthouse Lab in Alderley Park | Wellcome Sanger Institute for the COVID-19 Genomics UK (COG-UK) Consortium | Jacquelyn Wynn et al |
| EPI_ISL_647853 | Wales/MILK-B025AA/2020 | 2020-10-24 | Europe/United Kingdom/Wales/ | Lighthouse Lab in Milton Keynes | Wellcome Sanger Institute for the COVID-19 Genomics UK (COG-UK) Consortium | The Lighthouse Lab in Milton Keynes et al |
| EPI_ISL_647889 | Wales/QEUH-AE2091/2020 | 2020-10-24 | Europe/United Kingdom/Wales/ | Lighthouse Lab in Glasgow | Wellcome Sanger Institute for the COVID-19 Genomics UK (COG-UK) Consortium | Harper VanSteenhouse et al |
| EPI_ISL_647999 | USA/IA-CDC-4053/2020 | 2020-03-09 | North America/USA/Iowa/ | IA State Hygienic Laboratory | Pathogen Discovery, Respiratory Viruses Branch, Division of Viral Diseases, Centers for Disease Control and Prevention | Ying Tao et al |
| EPI_ISL_648007 | USA/MS-CDC-6552/2020 | 2020-09-03 | North America/USA/Mississippi/ | MS Public Health Laboratory | Pathogen Discovery, Respiratory Viruses Branch, Division of Viral Diseases, Centers for Disease Control and Prevention | Yan Li et al |

| | | | | | | |
|---|---|---|---|---|---|---|
| EPI_ISL_648010 | USA/MS-CDC-6557/2020 | 2020-09-03 | North America/USA/Mississippi/ | MS Public Health Laboratory | Pathogen Discovery, Respiratory Viruses Branch, Division of Viral Diseases, Centers for Disease Control and Prevention | Yan Li et al |
| EPI_ISL_648012 | USA/MS-CDC-6560/2020 | 2020-09-03 | North America/USA/Mississippi/ | MS Public Health Laboratory | Pathogen Discovery, Respiratory Viruses Branch, Division of Viral Diseases, Centers for Disease Control and Prevention | Yan Li et al |
| EPI_ISL_648013 | USA/MS-CDC-6561/2020 | 2020-09-03 | North America/USA/Mississippi/ | MS Public Health Laboratory | Pathogen Discovery, Respiratory Viruses Branch, Division of Viral Diseases, Centers for Disease Control and Prevention | Yan Li et al |
| EPI_ISL_648014 | USA/MS-CDC-6562/2020 | 2020-09-03 | North America/USA/Mississippi/ | MS Public Health Laboratory | Pathogen Discovery, Respiratory Viruses Branch, Division of Viral Diseases, Centers for Disease Control and Prevention | Yan Li et al |

| | | | | | | |
|---|---|---|---|---|---|---|
| EPI_ISL_648016 | USA/MS-CDC-6564/2020 | 2020-09-15 | North America/USA/Mississippi/ | MS Public Health Laboratory | Pathogen Discovery, Respiratory Viruses Branch, Division of Viral Diseases, Centers for Disease Control and Prevention | Yan Li et al |
| EPI_ISL_648024 | USA/MS-CDC-6572/2020 | 2020-03-15 | North America/USA/Mississippi/ | MS Public Health Laboratory | Pathogen Discovery, Respiratory Viruses Branch, Division of Viral Diseases, Centers for Disease Control and Prevention | Yan Li et al |
| EPI_ISL_648034 | USA/MS-CDC-6585/2020 | 2020-08-22 | North America/USA/Mississippi/ | MS Public Health Laboratory | Pathogen Discovery, Respiratory Viruses Branch, Division of Viral Diseases, Centers for Disease Control and Prevention | Yan Li et al |
| EPI_ISL_648038 | USA/MS-CDC-6515/2020 | 2020-08-22 | North America/USA/Mississippi/ | MS Public Health Laboratory | Pathogen Discovery, Respiratory Viruses Branch, Division of Viral Diseases, Centers for Disease Control and Prevention | Yan Li et al |

| | | | | | | |
|---|---|---|---|---|---|---|
| EPI_ISL_648039 | USA/MS-CDC-6517/2020 | 2020-08-22 | North America/USA/Mississippi/ | MS Public Health Laboratory | Pathogen Discovery, Respiratory Viruses Branch, Division of Viral Diseases, Centers for Disease Control and Prevention | Yan Li et al |
| EPI_ISL_648040 | USA/MS-CDC-6556/2020 | 2020-09-03 | North America/USA/Mississippi/ | MS Public Health Laboratory | Pathogen Discovery, Respiratory Viruses Branch, Division of Viral Diseases, Centers for Disease Control and Prevention | Yan Li et al |
| EPI_ISL_648043 | Singapore/1135/2020 | 2020-02-07 | Asia/Singapore/Singapore/ | Department of Laboratory Medicine, Tan Tock Seng Hospital | Department of Laboratory Medicine, Tan Tock Seng Hospital | Chen YYC et al |
| EPI_ISL_648071 | Singapore/17nan/2020 | 2020-02-08 | Asia/Singapore/Singapore/ | Department of Laboratory Medicine, Tan Tock Seng Hospital | Department of Laboratory Medicine, Tan Tock Seng Hospital | Chen YYC et al |
| EPI_ISL_648090 | Singapore/560nan/2020 | 2020-04-04 | Asia/Singapore/Singapore/ | Department of Laboratory Medicine, Tan Tock Seng Hospital | Department of Laboratory Medicine, Tan Tock Seng Hospital | Chen YYC et al |
| EPI_ISL_648119 | Singapore/1191/2020 | 2020-04-14 | Asia/Singapore/Singapore/ | Department of Laboratory Medicine, Tan Tock Seng Hospital | Department of Laboratory Medicine, Tan Tock Seng Hospital | Chen YYC et al |

| | | | | | | |
|---|---|---|---|---|---|---|
| EPI_ISL_648121 | Singapore/1193/2020 | 2020-04-14 | Asia/Singapore/Singapore/ | Department of Laboratory Medicine, Tan Tock Seng Hospital | Department of Laboratory Medicine, Tan Tock Seng Hospital | Chen YYC et al |
| EPI_ISL_648124 | Ghana/UHAS-G441/2020 | 2020-06-12 | Africa/Ghana/Volta/ | UHAS COVID-19 Lab | UHAS COVID-19 Lab | Kwabena O. Duedu et al |
| EPI_ISL_648125 | Ghana/UHAS-G541/2020 | 2020-06-12 | Africa/Ghana/Volta/ | UHAS COVID-19 Lab | UHAS COVID-19 Lab | Kwabena O. Duedu et al |
| EPI_ISL_648147 | Sweden/20-53078/2020 | 2020-10-02 | Europe/Sweden/Sweden/ | The Public Health Agency of Sweden | The Public Health Agency of Sweden | Anna-Malin Linde et al |
| EPI_ISL_648150 | Sweden/20-53076/2020 | 2020-10-05 | Europe/Sweden/Sweden/ | The Public Health Agency of Sweden | The Public Health Agency of Sweden | Anna-Malin Linde et al |
| EPI_ISL_648156 | Sweden/20-23682/2020 | 2020-10-06 | Europe/Sweden/Sweden/ | The Public Health Agency of Sweden | The Public Health Agency of Sweden | Anna-Malin Linde et al |
| EPI_ISL_648159 | Sweden/20-53276/2020 | 2020-10-29 | Europe/Sweden/Sweden/ | Sundsvall | The Public Health Agency of Sweden | Anna-Malin Linde et al |
| EPI_ISL_648161 | Sweden/20-53269/2020 | 2020-10-26 | Europe/Sweden/Sweden/ | Sundsvall | The Public Health Agency of Sweden | Anna-Malin Linde et al |
| EPI_ISL_648163 | Sweden/20-52820/2020 | 2020-08-17 | Europe/Sweden/Gavleborgs Lan/ | Gavle klinisk mikrobiologi | The Public Health Agency of Sweden | Anna-Malin Linde et al |
| EPI_ISL_648168 | Sweden/20-53189/2020 | 2020-10-13 | Europe/Sweden/Sweden/ | Eskilstuna | The Public Health Agency of Sweden | Anna-Malin Linde et al |
| EPI_ISL_648175 | Sweden/20-53131/2020 | 2020-09-28 | Europe/Sweden/Sweden/ | The Public Health Agency of Sweden | The Public Health Agency of Sweden | Anna-Malin Linde et al |
| EPI_ISL_648182 | Sweden/20-08977/2020 | 2020-08-09 | Europe/Sweden/Skane/ | The Public Health Agency of Sweden | The Public Health Agency of Sweden | Anna-Malin Linde et al |
| EPI_ISL_648200 | Sweden/20-53238/2020 | 2020-10-26 | Europe/Sweden/Sweden/ | Linkoping | The Public Health Agency of Sweden | Anna-Malin Linde et al |
| EPI_ISL_648203 | Sweden/20-53239/2020 | 2020-10-26 | Europe/Sweden/Sweden/ | Linkoping | The Public Health Agency of Sweden | Anna-Malin Linde et al |
| EPI_ISL_648208 | Sweden/20-53233/2020 | 2020-10-26 | Europe/Sweden/Sweden/ | Orebro | The Public Health Agency of Sweden | Anna-Malin Linde et al |
| EPI_ISL_648209 | Argentina/argenTAG-12C0A026/2020 | 2020-07-22 | South America/Argentina/Buenos Aires/ | INBIRS-UBA | Laboratorio Mixto de Biotecnolog√≠a Acu√°tica (LMBA) | Joaqu√≠n Ezpeleta et al |
| EPI_ISL_648210 | Argentina/argenTAG-12BBC055/2020 | 2020-07-22 | South America/Argentina/Buenos Aires/ | INBIRS-UBA | Laboratorio Mixto de Biotecnolog√≠a Acu√°tica (LMBA) | Joaqu√≠n Ezpeleta et al |

| | | | | | | |
|---|---|---|---|---|---|---|
| EPI_ISL_648211 | Argentina/argenTAG-1283F02A/202( | 2020-07-22 | South America/Argentina/Buenos Aires/ | INBIRS-UBA | Laboratorio Mixto de Biotecnolog√≠a Acu√°tica (LMBA) | Joaqu√≠n Ezpeleta et al |
| EPI_ISL_648213 | Argentina/argenTAG-12568054/202( | 2020-07-22 | South America/Argentina/Buenos Aires/ | INBIRS-UBA | Laboratorio Mixto de Biotecnolog√≠a Acu√°tica (LMBA) | Joaqu√≠n Ezpeleta et al |
| EPI_ISL_648214 | Argentina/argenTAG-12E7E043/202( | 2020-07-22 | South America/Argentina/Buenos Aires/ | INBIRS-UBA | Laboratorio Mixto de Biotecnolog√≠a Acu√°tica (LMBA) | Joaqu√≠n Ezpeleta et al |
| EPI_ISL_648216 | Argentina/argenTAG-12926008/202( | 2020-07-22 | South America/Argentina/Buenos Aires/ | INBIRS-UBA | Laboratorio Mixto de Biotecnolog√≠a Acu√°tica (LMBA) | Joaqu√≠n Ezpeleta et al |
| EPI_ISL_648217 | Argentina/argenTAG-126BE00A/202( | 2020-07-20 | South America/Argentina/Santa Fe/Tot | SILAB | Laboratorio Mixto de Biotecnolog√≠a Acu√°tica (LMBA) | Joaqu√≠n Ezpeleta et al |
| EPI_ISL_648218 | Argentina/argenTAG-12CB103F/202( | 2020-07-22 | South America/Argentina/Buenos Aires/ | INBIRS-UBA | Laboratorio Mixto de Biotecnolog√≠a Acu√°tica (LMBA) | Joaqu√≠n Ezpeleta et al |
| EPI_ISL_648223 | USA/TX-DSHS-1303/2020 | 2020-03-31 | North America/USA/Texas/Bexar County | Texas Department of State Health Services | Texas Department of State Health Services | Rashmi Tuladhar et al |
| EPI_ISL_648272 | USA/MD-MDH-0327/2020 | 2020-10-13 | North America/USA/Maryland/ | MD PHL | MD PHL | Maryland Department of Health Laboratories Administration et al |
| EPI_ISL_648276 | USA/MD-MDH-0331/2020 | 2020-10-14 | North America/USA/Maryland/ | MD PHL | MD PHL | Maryland Department of Health Laboratories Administration et al |
| EPI_ISL_648289 | USA/MD-MDH-0350/2020 | 2020-10-20 | North America/USA/Maryland/ | MD PHL | MD PHL | Maryland Department of Health Laboratories Administration et al |
| EPI_ISL_648293 | USA/MD-MDH-0354/2020 | 2020-10-20 | North America/USA/Maryland/ | MD PHL | MD PHL | Maryland Department of Health Laboratories Administration et al |
| EPI_ISL_648299 | USA/MD-MDH-0364/2020 | 2020-10-22 | North America/USA/Maryland/ | MD PHL | MD PHL | Maryland Department of Health Laboratories Administration et al |
| EPI_ISL_648302 | USA/MD-MDH-0367/2020 | 2020-10-26 | North America/USA/Maryland/ | MD PHL | MD PHL | Maryland Department of Health Laboratories Administration et al |

| | | | | | | |
|---|---|---|---|---|---|---|
| EPI_ISL_648311 | EquatorialGuinea/12462/2020 | 2020-05-29 | Africa/Equatorial Guinea/Bioko Norte/B | Laboratorio de Investigaciones de Baney | University Hospital Basel, Clinical Bacteriology | Carlos Cortes et al |
| EPI_ISL_648330 | EquatorialGuinea/38087/2020 | 2020-07-16 | Africa/Equatorial Guinea/Bioko Norte/M | Laboratorio de Investigaciones de Baney | University Hospital Basel, Clinical Bacteriology | Carlos Cortes et al |
| EPI_ISL_648350 | EquatorialGuinea/8844/2020 | 2020-05-17 | Africa/Equatorial Guinea/Litoral/Bata | Laboratorio de Investigaciones de Baney | University Hospital Basel, Clinical Bacteriology | Carlos Cortes et al |
| EPI_ISL_648369 | EquatorialGuinea/23901/2020 | 2020-06-23 | Africa/Equatorial Guinea/Bioko Norte/M | Laboratorio de Investigaciones de Baney | University Hospital Basel, Clinical Bacteriology | Carlos Cortes et al |
| EPI_ISL_648376 | EquatorialGuinea/35052/2020 | 2020-07-11 | Africa/Equatorial Guinea/Litoral/Bata | Laboratorio de Investigaciones de Baney | University Hospital Basel, Clinical Bacteriology | Carlos Cortes et al |
| EPI_ISL_648383 | USA/CA-CZB-7206/2020 | 2020-08-04 | North America/USA/California/Santa Cl | Santa Clara County Public Health Laboratory | Chan-Zuckerberg Biohub | CZB Cliahub Consortium et al |
| EPI_ISL_648384 | USA/CA-CZB-12289/2020 | 2020-10-07 | North America/USA/California/Santa Cl | Santa Clara County Public Health Laboratory | Chan-Zuckerberg Biohub | CZB Cliahub Consortium et al |
| EPI_ISL_648388 | USA/CA-CZB-12295/2020 | 2020-10-07 | North America/USA/California/Santa Cl | Santa Clara County Public Health Laboratory | Chan-Zuckerberg Biohub | CZB Cliahub Consortium et al |
| EPI_ISL_648390 | USA/CA-CZB-12298/2020 | 2020-10-07 | North America/USA/California/Santa Cl | Santa Clara County Public Health Laboratory | Chan-Zuckerberg Biohub | CZB Cliahub Consortium et al |
| EPI_ISL_648427 | USA/CA-CZB-12355/2020 | 2020-10-12 | North America/USA/California/Santa Cl | Santa Clara County Public Health Laboratory | Chan-Zuckerberg Biohub | CZB Cliahub Consortium et al |
| EPI_ISL_648428 | USA/CA-CZB-12356/2020 | 2020-10-12 | North America/USA/California/Santa Cl | Santa Clara County Public Health Laboratory | Chan-Zuckerberg Biohub | CZB Cliahub Consortium et al |
| EPI_ISL_648436 | USA/CA-CZB-12366/2020 | 2020-10-12 | North America/USA/California/Santa Cl | Santa Clara County Public Health Laboratory | Chan-Zuckerberg Biohub | CZB Cliahub Consortium et al |
| EPI_ISL_648439 | USA/CA-CZB-12776/2020 | 2020-10-14 | North America/USA/California/Santa Cl | Santa Clara County Public Health Laboratory | Chan-Zuckerberg Biohub | CZB Cliahub Consortium et al |
| EPI_ISL_648442 | USA/CA-CZB-12781/2020 | 2020-10-14 | North America/USA/California/Santa Cl | Santa Clara County Public Health Laboratory | Chan-Zuckerberg Biohub | CZB Cliahub Consortium et al |
| EPI_ISL_648446 | USA/CA-CZB-12789/2020 | 2020-10-14 | North America/USA/California/Santa Cl | Santa Clara County Public Health Laboratory | Chan-Zuckerberg Biohub | CZB Cliahub Consortium et al |

| | | | | | | |
|---|---|---|---|---|---|---|
| EPI_ISL_648452 | USA/CA-CZB-12798/2020 | 2020-10-15 | North America/USA/California/Santa Cl | Santa Clara County Public Health Laboratory | Chan-Zuckerberg Biohub | CZB Cliahub Consortium et al |
| EPI_ISL_648456 | USA/CA-CZB-12805/2020 | 2020-10-16 | North America/USA/California/Santa Cl | Santa Clara County Public Health Laboratory | Chan-Zuckerberg Biohub | CZB Cliahub Consortium et al |
| EPI_ISL_648466 | USA/CA-CZB-12815/2020 | 2020-09-14 | North America/USA/California/Santa Cl | Santa Clara County Public Health Laboratory | Chan-Zuckerberg Biohub | CZB Cliahub Consortium et al |
| EPI_ISL_648469 | USA/CA-CZB-12818/2020 | 2020-09-30 | North America/USA/California/Santa Cl | Santa Clara County Public Health Laboratory | Chan-Zuckerberg Biohub | CZB Cliahub Consortium et al |
| EPI_ISL_648475 | USA/CA-CZB-12824/2020 | 2020-09-23 | North America/USA/California/Santa Cl | Santa Clara County Public Health Laboratory | Chan-Zuckerberg Biohub | CZB Cliahub Consortium et al |
| EPI_ISL_648484 | USA/CA-CZB-12838/2020 | 2020-10-20 | North America/USA/California/Santa Cl | Santa Clara County Public Health Laboratory | Chan-Zuckerberg Biohub | CZB Cliahub Consortium et al |
| EPI_ISL_648486 | USA/CA-CZB-12840/2020 | 2020-10-20 | North America/USA/California/Santa Cl | Santa Clara County Public Health Laboratory | Chan-Zuckerberg Biohub | CZB Cliahub Consortium et al |
| EPI_ISL_648495 | USA/CA-CZB-12855/2020 | 2020-10-19 | North America/USA/California/Santa Cl | Santa Clara County Public Health Laboratory | Chan-Zuckerberg Biohub | CZB Cliahub Consortium et al |
| EPI_ISL_648504 | USA/CA-CZB-12381/2020 | 2020-10-04 | North America/USA/California/Orange C | Orange County Public Health Lab | Chan-Zuckerberg Biohub | CZB Cliahub Consortium et al |
| EPI_ISL_648505 | USA/CA-CZB-12383/2020 | 2020-10-05 | North America/USA/California/Orange C | Orange County Public Health Lab | Chan-Zuckerberg Biohub | CZB Cliahub Consortium et al |
| EPI_ISL_648522 | USA/CA-CZB-12867/2020 | 2020-10-15 | North America/USA/California/Orange C | Orange County Public Health Lab | Chan-Zuckerberg Biohub | CZB Cliahub Consortium et al |
| EPI_ISL_648525 | USA/CA-CZB-12870/2020 | 2020-10-15 | North America/USA/California/Orange C | Orange County Public Health Lab | Chan-Zuckerberg Biohub | CZB Cliahub Consortium et al |
| EPI_ISL_648530 | USA/CA-CZB-12875/2020 | 2020-10-17 | North America/USA/California/Orange C | Orange County Public Health Lab | Chan-Zuckerberg Biohub | CZB Cliahub Consortium et al |
| EPI_ISL_648533 | USA/CA-CZB-12372/2020 | 2020-05-14 | North America/USA/California/Tulare C | Tulare County Public Health Lab | Chan-Zuckerberg Biohub | CZB Cliahub Consortium et al |
| EPI_ISL_648546 | USA/CA-CZB-10362/2020 | 2020-09-12 | North America/USA/California/Madera | Madera County Department of Public Health | Chan-Zuckerberg Biohub | CZB Cliahub Consortium et al |
| EPI_ISL_648547 | USA/CA-CZB-10361/2020 | 2020-09-12 | North America/USA/California/Madera | Madera County Department of Public Health | Chan-Zuckerberg Biohub | CZB Cliahub Consortium et al |
| EPI_ISL_648550 | USA/CA-CZB-10622/2020 | 2020-09-03 | North America/USA/California/Madera | Madera County Department of Public Health | Chan-Zuckerberg Biohub | CZB Cliahub Consortium et al |
| EPI_ISL_648551 | USA/CA-CZB-10621/2020 | 2020-09-03 | North America/USA/California/Madera | Madera County Department of Public Health | Chan-Zuckerberg Biohub | CZB Cliahub Consortium et al |
| EPI_ISL_648553 | USA/CA-CZB-10619/2020 | 2020-09-03 | North America/USA/California/Madera | Madera County Department of Public Health | Chan-Zuckerberg Biohub | CZB Cliahub Consortium et al |
| EPI_ISL_648554 | USA/CA-CZB-10617/2020 | 2020-09-03 | North America/USA/California/Madera | Madera County Department of Public Health | Chan-Zuckerberg Biohub | CZB Cliahub Consortium et al |

| | | | | | | |
|---|---|---|---|---|---|---|
| EPI_ISL_648559 | USA/UT-UPHL-201117542/2020 | 2020-04-09 | North America/USA/Utah/ | Utah Public Health Laboratory | Utah Public Health Laboratory | Erin Young et al |
| EPI_ISL_648569 | USA/UT-UPHL-201126219/2020 | 2020-04-07 | North America/USA/Utah/ | Utah Public Health Laboratory | Utah Public Health Laboratory | Erin Young et al |
| EPI_ISL_648571 | USA/UT-UPHL-201108622/2020 | 2020-04-06 | North America/USA/Utah/ | Utah Public Health Laboratory | Utah Public Health Laboratory | Erin Young et al |
| EPI_ISL_648577 | USA/UT-UPHL-201122426/2020 | 2020-04-06 | North America/USA/Utah/ | Utah Public Health Laboratory | Utah Public Health Laboratory | Erin Young et al |
| EPI_ISL_648578 | USA/UT-UPHL-201112041/2020 | 2020-04-20 | North America/USA/Utah/ | Utah Public Health Laboratory | Utah Public Health Laboratory | Erin Young et al |
| EPI_ISL_648581 | USA/UT-UPHL-201108802/2020 | 2020-05-13 | North America/USA/Utah/ | Utah Public Health Laboratory | Utah Public Health Laboratory | Erin Young et al |
| EPI_ISL_648588 | USA/UT-UPHL-201116393/2020 | 2020-05-13 | North America/USA/Utah/ | Utah Public Health Laboratory | Utah Public Health Laboratory | Erin Young et al |
| EPI_ISL_648590 | USA/UT-UPHL-201131465/2020 | 2020-05-13 | North America/USA/Utah/ | Utah Public Health Laboratory | Utah Public Health Laboratory | Erin Young et al |
| EPI_ISL_648591 | USA/UT-UPHL-201104262/2020 | 2020-05-13 | North America/USA/Utah/ | Utah Public Health Laboratory | Utah Public Health Laboratory | Erin Young et al |
| EPI_ISL_648594 | USA/UT-UPHL-201128144/2020 | 2020-05-12 | North America/USA/Utah/ | Utah Public Health Laboratory | Utah Public Health Laboratory | Erin Young et al |
| EPI_ISL_648602 | Mexico/NLE-UANL-2098/2020 | 2020-05-23 | North America/Mexico/Nuevo Leon/ | Laboratorio de Infectolog√≠a Servicio de Infectolog√≠a Hospital Universitario Dr. Jos√© Eleuterio Gonz√°lez - Universidad Aut√≥noma de Nuevo Le√≥n | Laboratorio de Infectolog√≠a Molecular Departamento de Bioqu√≠mica y Medicina Molecular Facultad de Medicina - Universidad Aut√≥noma de Nuevo Le√≥n | Kame A. Gal√°n-Huerta et al |

| | | | | | | |
|---|---|---|---|---|---|---|
| EPI_ISL_648603 | Mexico/NLE-UANL-2088/2020 | 2020-05-23 | North America/Mexico/Nuevo Leon/ | Laboratorio de Infectologv≠a Servicio de Infectologv≠a Hospital Universitario Dr. Josv© Eleuterio Gonzv°lez - Universidad Autv≥noma de Nuevo Lev≥n | Laboratorio de Infectologv≠a Molecular Departamento de Bioquv≠mica y Medicina Molecular Facultad de Medicina - Universidad Autv≥noma de Nuevo Lev≥n | Kame A. Galv°n-Huerta et al |
| EPI_ISL_648607 | Mexico/NLE-UANL-4479/2020 | 2020-06-17 | North America/Mexico/Nuevo Leon/ | Laboratorio de Infectologv≠a, Servicio de Infectologv≠a, Hospital Universitario Dr. Josv© Eleuterio Gonzv°lez - Universidad Autv≥noma de Nuevo Lev≥n | Laboratorio de Infectologv≠a Molecular, Departamento de Bioquv≠mica y Medicina Molecular,Facultad de Medicina - Universidad Autv≥noma de Nuevo Lev≥n | Kame A. Galv°n-Huerta et al |
| EPI_ISL_648608 | Mexico/NLE-UANL-4485/2020 | 2020-06-17 | North America/Mexico/Nuevo Leon/ | Laboratorio de Infectologv≠a, Servicio de Infectologv≠a, Hospital Universitario Dr. Josv© Eleuterio Gonzv°lez - Universidad Autv≥noma de Nuevo Lev≥n | Laboratorio de Infectologv≠a Molecular, Departamento de Bioquv≠mica y Medicina Molecular,Facultad de Medicina - Universidad Autv≥noma de Nuevo Lev≥n | Kame A. Galv°n-Huerta et al |

| | | | | | | |
|---|---|---|---|---|---|---|
| EPI_ISL_648609 | Mexico/NLE-UANL-7088/2020 | 2020-07-05 | North America/Mexico/Nuevo Leon/ | Laboratorio de Infectolog√≠a, Servicio de Infectolog√≠a, Hospital Universitario Dr. Jos√© Eleuterio Gonz√°lez - Universidad Aut√≥noma de Nuevo Le√≥n | Laboratorio de Infectolog√≠a Molecular, Departamento de Bioqu√≠mica y Medicina Molecular,Facultad de Medicina - Universidad Aut√≥noma de Nuevo Le√≥n | Kame A. Gal√°n-Huerta et al |
| EPI_ISL_648621 | Singapore/1205/2020 | 2020-04-22 | Asia/Singapore/Singapore/ | Department of Laboratory Medicine, Tan Tock Seng Hospital | Department of Laboratory Medicine, Tan Tock Seng Hospital | Chen YYC et al |
| EPI_ISL_648622 | Singapore/1206/2020 | 2020-04-22 | Asia/Singapore/Singapore/ | Department of Laboratory Medicine, Tan Tock Seng Hospital | Department of Laboratory Medicine, Tan Tock Seng Hospital | Chen YYC et al |
| EPI_ISL_648634 | Singapore/1218/2020 | 2020-04-23 | Asia/Singapore/Singapore/ | Department of Laboratory Medicine, Tan Tock Seng Hospital | Department of Laboratory Medicine, Tan Tock Seng Hospital | Chen YYC et al |
| EPI_ISL_648650 | Singapore/1233/2020 | 2020-04-25 | Asia/Singapore/Singapore/ | Department of Laboratory Medicine, Tan Tock Seng Hospital | Department of Laboratory Medicine, Tan Tock Seng Hospital | Chen YYC et al |
| EPI_ISL_648661 | Singapore/1408/2020 | 2020-05-01 | Asia/Singapore/Singapore/ | Department of Laboratory Medicine, Tan Tock Seng Hospital | Department of Laboratory Medicine, Tan Tock Seng Hospital | Chen YYC et al |
| EPI_ISL_648669 | Singapore/1251/2020 | 2020-05-02 | Asia/Singapore/Singapore/ | Department of Laboratory Medicine, Tan Tock Seng Hospital | Department of Laboratory Medicine, Tan Tock Seng Hospital | Chen YYC et al |

| | | | | | | |
|---|---|---|---|---|---|---|
| EPI_ISL_648677 | Argentina/argenTAG-1252C02E/2020 | 2020-07-22 | South America/Argentina/Buenos Aires/ | INBIRS-UBA | Laboratorio Mixto de Biotecnolog√≠a Acu√°tica (LMBA) | Joaqu√≠n Ezpeleta et al |
| EPI_ISL_648682 | Singapore/1333/2020 | 2020-06-02 | Asia/Singapore/Singapore/ | Department of Laboratory Medicine, Tan Tock Seng Hospital | Department of Laboratory Medicine, Tan Tock Seng Hospital | Chen YYC et al |
| EPI_ISL_648692 | Singapore/1343/2020 | 2020-06-09 | Asia/Singapore/Singapore/ | Department of Laboratory Medicine, Tan Tock Seng Hospital | Department of Laboratory Medicine, Tan Tock Seng Hospital | Chen YYC et al |
| EPI_ISL_648696 | Singapore/1347/2020 | 2020-06-10 | Asia/Singapore/Singapore/ | Department of Laboratory Medicine, Tan Tock Seng Hospital | Department of Laboratory Medicine, Tan Tock Seng Hospital | Chen YYC et al |
| EPI_ISL_648715 | Singapore/1364/2020 | 2020-06-25 | Asia/Singapore/Singapore/ | Department of Laboratory Medicine, Tan Tock Seng Hospital | Department of Laboratory Medicine, Tan Tock Seng Hospital | Chen YYC et al |
| EPI_ISL_648723 | Singapore/1372/2020 | 2020-07-01 | Asia/Singapore/Singapore/ | Department of Laboratory Medicine, Tan Tock Seng Hospital | Department of Laboratory Medicine, Tan Tock Seng Hospital | Chen YYC et al |
| EPI_ISL_648730 | Singapore/1377/2020 | 2020-07-06 | Asia/Singapore/Singapore/ | Department of Laboratory Medicine, Tan Tock Seng Hospital | Department of Laboratory Medicine, Tan Tock Seng Hospital | Chen YYC et al |
| EPI_ISL_648740 | Singapore/1386/2020 | 2020-07-21 | Asia/Singapore/Singapore/ | Department of Laboratory Medicine, Tan Tock Seng Hospital | Department of Laboratory Medicine, Tan Tock Seng Hospital | Chen YYC et al |

| | | | | | | |
|---|---|---|---|---|---|---|
| EPI_ISL_648742 | Singapore/1259/2020 | 2020-04-25 | Asia/Singapore/Singapore/ | Department of Laboratory Medicine, Tan Tock Seng Hospital | Department of Laboratory Medicine, Tan Tock Seng Hospital | Chen YYC et al |
| EPI_ISL_648746 | Singapore/1262/2020 | 2020-05-04 | Asia/Singapore/Singapore/ | Department of Laboratory Medicine, Tan Tock Seng Hospital | Department of Laboratory Medicine, Tan Tock Seng Hospital | Chen YYC et al |
| EPI_ISL_648749 | Singapore/1265/2020 | 2020-05-05 | Asia/Singapore/Singapore/ | Department of Laboratory Medicine, Tan Tock Seng Hospital | Department of Laboratory Medicine, Tan Tock Seng Hospital | Chen YYC et al |
| EPI_ISL_648757 | Singapore/1273/2020 | 2020-05-05 | Asia/Singapore/Singapore/ | Department of Laboratory Medicine, Tan Tock Seng Hospital | Department of Laboratory Medicine, Tan Tock Seng Hospital | Chen YYC et al |
| EPI_ISL_648759 | Singapore/885nan/2020 | 2020-05-05 | Asia/Singapore/Singapore/ | Department of Laboratory Medicine, Tan Tock Seng Hospital | Department of Laboratory Medicine, Tan Tock Seng Hospital | Chen YYC et al |
| EPI_ISL_648762 | Singapore/1277/2020 | 2020-05-06 | Asia/Singapore/Singapore/ | Department of Laboratory Medicine, Tan Tock Seng Hospital | Department of Laboratory Medicine, Tan Tock Seng Hospital | Chen YYC et al |
| EPI_ISL_648767 | Singapore/1282/2020 | 2020-05-08 | Asia/Singapore/Singapore/ | Department of Laboratory Medicine, Tan Tock Seng Hospital | Department of Laboratory Medicine, Tan Tock Seng Hospital | Chen YYC et al |
| EPI_ISL_648768 | Singapore/1283/2020 | 2020-05-09 | Asia/Singapore/Singapore/ | Department of Laboratory Medicine, Tan Tock Seng Hospital | Department of Laboratory Medicine, Tan Tock Seng Hospital | Chen YYC et al |

| | | | | | | |
|---|---|---|---|---|---|---|
| EPI_ISL_648771 | Singapore/1286/2020 | 2020-05-09 | Asia/Singapore/Singapore/ | Department of Laboratory Medicine, Tan Tock Seng Hospital | Department of Laboratory Medicine, Tan Tock Seng Hospital | Chen YYC et al |
| EPI_ISL_648772 | Singapore/1287/2020 | 2020-05-09 | Asia/Singapore/Singapore/ | Department of Laboratory Medicine, Tan Tock Seng Hospital | Department of Laboratory Medicine, Tan Tock Seng Hospital | Chen YYC et al |
| EPI_ISL_648776 | Singapore/1291/2020 | 2020-05-11 | Asia/Singapore/Singapore/ | Department of Laboratory Medicine, Tan Tock Seng Hospital | Department of Laboratory Medicine, Tan Tock Seng Hospital | Chen YYC et al |
| EPI_ISL_648781 | Singapore/1296/2020 | 2020-05-12 | Asia/Singapore/Singapore/ | Department of Laboratory Medicine, Tan Tock Seng Hospital | Department of Laboratory Medicine, Tan Tock Seng Hospital | Chen YYC et al |
| EPI_ISL_648782 | Singapore/1297/2020 | 2020-05-12 | Asia/Singapore/Singapore/ | Department of Laboratory Medicine, Tan Tock Seng Hospital | Department of Laboratory Medicine, Tan Tock Seng Hospital | Chen YYC et al |
| EPI_ISL_648790 | Singapore/1303/2020 | 2020-05-15 | Asia/Singapore/Singapore/ | Department of Laboratory Medicine, Tan Tock Seng Hospital | Department of Laboratory Medicine, Tan Tock Seng Hospital | Chen YYC et al |
| EPI_ISL_648792 | Singapore/1305/2020 | 2020-05-16 | Asia/Singapore/Singapore/ | Department of Laboratory Medicine, Tan Tock Seng Hospital | Department of Laboratory Medicine, Tan Tock Seng Hospital | Chen YYC et al |
| EPI_ISL_648793 | Singapore/1306/2020 | 2020-05-16 | Asia/Singapore/Singapore/ | Department of Laboratory Medicine, Tan Tock Seng Hospital | Department of Laboratory Medicine, Tan Tock Seng Hospital | Chen YYC et al |

| | | | | | | |
|---|---|---|---|---|---|---|
| EPI_ISL_648797 | Singapore/1310/2020 | 2020-05-17 | Asia/Singapore/Singapore/ | Department of Laboratory Medicine, Tan Tock Seng Hospital | Department of Laboratory Medicine, Tan Tock Seng Hospital | Chen YYC et al |
| EPI_ISL_648801 | Singapore/1314/2020 | 2020-05-19 | Asia/Singapore/Singapore/ | Department of Laboratory Medicine, Tan Tock Seng Hospital | Department of Laboratory Medicine, Tan Tock Seng Hospital | Chen YYC et al |
| EPI_ISL_648804 | Singapore/320nan/2020 | 2020-05-20 | Asia/Singapore/Singapore/ | Department of Laboratory Medicine, Tan Tock Seng Hospital | Department of Laboratory Medicine, Tan Tock Seng Hospital | Chen YYC et al |
| EPI_ISL_648810 | Singapore/1322/2020 | 2020-05-22 | Asia/Singapore/Singapore/ | Department of Laboratory Medicine, Tan Tock Seng Hospital | Department of Laboratory Medicine, Tan Tock Seng Hospital | Chen YYC et al |
| EPI_ISL_648814 | Singapore/1326/2020 | 2020-05-27 | Asia/Singapore/Singapore/ | Department of Laboratory Medicine, Tan Tock Seng Hospital | Department of Laboratory Medicine, Tan Tock Seng Hospital | Chen YYC et al |
| EPI_ISL_648815 | Singapore/547nan/2020 | 2020-05-27 | Asia/Singapore/Singapore/ | Department of Laboratory Medicine, Tan Tock Seng Hospital | Department of Laboratory Medicine, Tan Tock Seng Hospital | Chen YYC et al |
| EPI_ISL_648817 | Singapore/1328/2020 | 2020-05-31 | Asia/Singapore/Singapore/ | Department of Laboratory Medicine, Tan Tock Seng Hospital | Department of Laboratory Medicine, Tan Tock Seng Hospital | Chen YYC et al |
| EPI_ISL_648822 | USA/MT-RML-22/2020 | 2020-08-30 | North America/USA/Montana/Ravalli C | Virus Ecology Section, RML | Virus Ecology Section, RML | van Doremalen et al |
| EPI_ISL_648839 | USA/CA-ALSR-4131/2020 | 2020-07-02 | North America/USA/California/San Dieg | San Diego County Public Health Laboratory | Andersen lab at Scripps Research | SEARCH Alliance San Diego with Tracy Basler et al |

| | | | | | | |
|---|---|---|---|---|---|---|
| EPI_ISL_648841 | USA/CA-ALSR-4134/2020 | 2020-07-06 | North America/USA/California/San Dieg | San Diego County Public Health Laboratory | Andersen lab at Scripps Research | SEARCH Alliance San Diego with Tracy Basler et al |
| EPI_ISL_648843 | USA/CA-ALSR-4139/2020 | 2020-07-02 | North America/USA/California/San Dieg | San Diego County Public Health Laboratory | Andersen lab at Scripps Research | SEARCH Alliance San Diego with Tracy Basler et al |
| EPI_ISL_648845 | USA/CA-ALSR-4141/2020 | 2020-06-27 | North America/USA/California/San Dieg | San Diego County Public Health Laboratory | Andersen lab at Scripps Research | SEARCH Alliance San Diego with Tracy Basler et al |
| EPI_ISL_648847 | USA/CA-ALSR-4143/2020 | 2020-06-27 | North America/USA/California/San Dieg | San Diego County Public Health Laboratory | Andersen lab at Scripps Research | SEARCH Alliance San Diego with Tracy Basler et al |
| EPI_ISL_648848 | USA/CA-ALSR-4144/2020 | 2020-06-26 | North America/USA/California/San Dieg | San Diego County Public Health Laboratory | Andersen lab at Scripps Research | SEARCH Alliance San Diego with Tracy Basler et al |
| EPI_ISL_648864 | USA/CA-ALSR-4173/2020 | 2020-07-02 | North America/USA/California/San Dieg | San Diego County Public Health Laboratory | Andersen lab at Scripps Research | SEARCH Alliance San Diego with Tracy Basler et al |
| EPI_ISL_648866 | USA/CA-ALSR-4177/2020 | 2020-07-02 | North America/USA/California/San Dieg | San Diego County Public Health Laboratory | Andersen lab at Scripps Research | SEARCH Alliance San Diego with Tracy Basler et al |
| EPI_ISL_648873 | USA/CA-ALSR-4187/2020 | 2020-08-21 | North America/USA/California/San Dieg | San Diego County Public Health Laboratory | Andersen lab at Scripps Research | SEARCH Alliance San Diego with Tracy Basler et al |
| EPI_ISL_648890 | USA/CA-ALSR-4213/2020 | 2020-08-04 | North America/USA/California/San Dieg | San Diego County Public Health Laboratory | Andersen lab at Scripps Research | SEARCH Alliance San Diego with Tracy Basler et al |
| EPI_ISL_648898 | USA/CA-ALSR-4225/2020 | 2020-08-19 | North America/USA/California/San Dieg | San Diego County Public Health Laboratory | Andersen lab at Scripps Research | SEARCH Alliance San Diego with Tracy Basler et al |
| EPI_ISL_648911 | USA/CA-ALSR-4249/2020 | 2020-08-19 | North America/USA/California/San Dieg | San Diego County Public Health Laboratory | Andersen lab at Scripps Research | SEARCH Alliance San Diego with Tracy Basler et al |
| EPI_ISL_648913 | USA/CA-ALSR-4251/2020 | 2020-10-07 | North America/USA/California/San Dieg | San Diego County Public Health Laboratory | Andersen lab at Scripps Research | SEARCH Alliance San Diego with Tracy Basler et al |
| EPI_ISL_648915 | USA/CA-ALSR-4253/2020 | 2020-10-05 | North America/USA/California/San Dieg | San Diego County Public Health Laboratory | Andersen lab at Scripps Research | SEARCH Alliance San Diego with Tracy Basler et al |
| EPI_ISL_648921 | USA/CA-ALSR-4261/2020 | 2020-08-21 | North America/USA/California/San Dieg | San Diego County Public Health Laboratory | Andersen lab at Scripps Research | SEARCH Alliance San Diego with Tracy Basler et al |
| EPI_ISL_648922 | USA/CA-ALSR-4262/2020 | 2020-09-30 | North America/USA/California/San Dieg | San Diego County Public Health Laboratory | Andersen lab at Scripps Research | SEARCH Alliance San Diego with Tracy Basler et al |
| EPI_ISL_648930 | USA/CA-ALSR-4274/2020 | 2020-10-05 | North America/USA/California/San Dieg | San Diego County Public Health Laboratory | Andersen lab at Scripps Research | SEARCH Alliance San Diego with Tracy Basler et al |
| EPI_ISL_648938 | USA/CA-ALSR-4286/2020 | 2020-10-01 | North America/USA/California/San Dieg | San Diego County Public Health Laboratory | Andersen lab at Scripps Research | SEARCH Alliance San Diego with Tracy Basler et al |
| EPI_ISL_648940 | USA/CA-ALSR-4288/2020 | 2020-10-20 | North America/USA/California/San Dieg | San Diego County Public Health Laboratory | Andersen lab at Scripps Research | SEARCH Alliance San Diego with Tracy Basler et al |
| EPI_ISL_648944 | USA/CA-ALSR-4292/2020 | 2020-10-08 | North America/USA/California/Riverside | San Diego County Public Health Laboratory | Andersen lab at Scripps Research | SEARCH Alliance San Diego with Tracy Basler et al |
| EPI_ISL_648954 | USA/CA-ALSR-4304/2020 | 2020-10-06 | North America/USA/California/San Dieg | San Diego County Public Health Laboratory | Andersen lab at Scripps Research | SEARCH Alliance San Diego with Tracy Basler et al |

| | | | | | | |
|---|---|---|---|---|---|---|
| EPI_ISL_648958 | USA/CA-ALSR-4308/2020 | 2020-09-29 | North America/USA/California/San Dieg | San Diego County Public Health Laboratory | Andersen lab at Scripps Research | SEARCH Alliance San Diego with Tracy Basler et al |
| EPI_ISL_648963 | USA/CA-ALSR-4314/2020 | 2020-09-30 | North America/USA/California/San Dieg | San Diego County Public Health Laboratory | Andersen lab at Scripps Research | SEARCH Alliance San Diego with Tracy Basler et al |
| EPI_ISL_648979 | USA/CA-ALSR-4337/2020 | 2020-10-06 | North America/USA/California/San Dieg | San Diego County Public Health Laboratory | Andersen lab at Scripps Research | SEARCH Alliance San Diego with Tracy Basler et al |
| EPI_ISL_648981 | USA/CA-ALSR-4342/2020 | 2020-07-24 | North America/USA/California/San Dieg | San Diego County Public Health Laboratory | Andersen lab at Scripps Research | SEARCH Alliance San Diego with Tracy Basler et al |
| EPI_ISL_648991 | USA/CA-ALSR-4365/2020 | 2020-07-22 | North America/USA/California/San Dieg | San Diego County Public Health Laboratory | Andersen lab at Scripps Research | SEARCH Alliance San Diego with Tracy Basler et al |
| EPI_ISL_648993 | USA/CA-ALSR-4369/2020 | 2020-07-31 | North America/USA/California/San Dieg | San Diego County Public Health Laboratory | Andersen lab at Scripps Research | SEARCH Alliance San Diego with Tracy Basler et al |
| EPI_ISL_648995 | USA/CA-ALSR-4376/2020 | 2020-07-21 | North America/USA/California/San Dieg | San Diego County Public Health Laboratory | Andersen lab at Scripps Research | SEARCH Alliance San Diego with Tracy Basler et al |
| EPI_ISL_649010 | USA/CA-ALSR-4404/2020 | 2020-10-19 | North America/USA/California/San Dieg | San Diego County Public Health Laboratory | Andersen lab at Scripps Research | SEARCH Alliance San Diego with Tracy Basler et al |
| EPI_ISL_649012 | USA/CA-ALSR-4407/2020 | 2020-10-12 | North America/USA/California/San Dieg | San Diego County Public Health Laboratory | Andersen lab at Scripps Research | SEARCH Alliance San Diego with Tracy Basler et al |
| EPI_ISL_649015 | USA/CA-ALSR-4412/2020 | 2020-10-27 | North America/USA/California/San Dieg | San Diego County Public Health Laboratory | Andersen lab at Scripps Research | SEARCH Alliance San Diego with Tracy Basler et al |
| EPI_ISL_649016 | USA/CA-ALSR-4413/2020 | 2020-06-22 | North America/USA/California/San Dieg | San Diego County Public Health Laboratory | Andersen lab at Scripps Research | SEARCH Alliance San Diego with Tracy Basler et al |
| EPI_ISL_649019 | USA/CA-ALSR-4420/2020 | 2020-06-22 | North America/USA/California/San Dieg | San Diego County Public Health Laboratory | Andersen lab at Scripps Research | SEARCH Alliance San Diego with Tracy Basler et al |
| EPI_ISL_649020 | USA/CA-ALSR-4425/2020 | 2020-10-26 | North America/USA/California/San Dieg | San Diego County Public Health Laboratory | Andersen lab at Scripps Research | SEARCH Alliance San Diego with Tracy Basler et al |
| EPI_ISL_649026 | USA/CA-ALSR-4432/2020 | 2020-09-25 | North America/USA/California/San Dieg | San Diego County Public Health Laboratory | Andersen lab at Scripps Research | SEARCH Alliance San Diego with Tracy Basler et al |
| EPI_ISL_649028 | USA/CA-ALSR-4435/2020 | 2020-09-25 | North America/USA/California/San Dieg | San Diego County Public Health Laboratory | Andersen lab at Scripps Research | SEARCH Alliance San Diego with Tracy Basler et al |
| EPI_ISL_649029 | USA/CA-ALSR-4436/2020 | 2020-10-12 | North America/USA/California/San Dieg | San Diego County Public Health Laboratory | Andersen lab at Scripps Research | SEARCH Alliance San Diego with Tracy Basler et al |
| EPI_ISL_649031 | USA/CA-ALSR-4439/2020 | 2020-10-21 | North America/USA/California/San Dieg | San Diego County Public Health Laboratory | Andersen lab at Scripps Research | SEARCH Alliance San Diego with Tracy Basler et al |
| EPI_ISL_649035 | USA/CA-ALSR-4449/2020 | 2020-09-25 | North America/USA/California/San Dieg | San Diego County Public Health Laboratory | Andersen lab at Scripps Research | SEARCH Alliance San Diego with Tracy Basler et al |
| EPI_ISL_649039 | USA/CA-ALSR-4453/2020 | 2020-09-25 | North America/USA/California/San Dieg | San Diego County Public Health Laboratory | Andersen lab at Scripps Research | SEARCH Alliance San Diego with Tracy Basler et al |
| EPI_ISL_649040 | USA/CA-ALSR-4455/2020 | 2020-08-13 | North America/USA/California/San Dieg | San Diego County Public Health Laboratory | Andersen lab at Scripps Research | SEARCH Alliance San Diego with Tracy Basler et al |

| | | | | | | |
|---|---|---|---|---|---|---|
| EPI_ISL_649043 | USA/CA-ALSR-4461/2020 | 2020-07-08 | North America/USA/California/San Dieg | San Diego County Public Health Laboratory | Andersen lab at Scripps Research | SEARCH Alliance San Diego with Tracy Basler et al |
| EPI_ISL_649044 | USA/CA-ALSR-4462/2020 | 2020-10-21 | North America/USA/California/San Dieg | San Diego County Public Health Laboratory | Andersen lab at Scripps Research | SEARCH Alliance San Diego with Tracy Basler et al |
| EPI_ISL_649048 | USA/CA-ALSR-4468/2020 | 2020-09-16 | North America/USA/California/San Dieg | San Diego County Public Health Laboratory | Andersen lab at Scripps Research | SEARCH Alliance San Diego with Tracy Basler et al |
| EPI_ISL_649050 | USA/CA-ALSR-4473/2020 | 2020-10-19 | North America/USA/California/San Dieg | San Diego County Public Health Laboratory | Andersen lab at Scripps Research | SEARCH Alliance San Diego with Tracy Basler et al |
| EPI_ISL_649051 | USA/CA-ALSR-4474/2020 | 2020-09-24 | North America/USA/California/San Dieg | San Diego County Public Health Laboratory | Andersen lab at Scripps Research | SEARCH Alliance San Diego with Tracy Basler et al |
| EPI_ISL_649052 | USA/CA-ALSR-4475/2020 | 2020-07-08 | North America/USA/California/San Dieg | San Diego County Public Health Laboratory | Andersen lab at Scripps Research | SEARCH Alliance San Diego with Tracy Basler et al |
| EPI_ISL_649054 | USA/CA-ALSR-4479/2020 | 2020-09-25 | North America/USA/California/San Dieg | San Diego County Public Health Laboratory | Andersen lab at Scripps Research | SEARCH Alliance San Diego with Tracy Basler et al |
| EPI_ISL_649060 | Israel/CVL-n4295/2020 | 2020-03-12 | Asia/Israel/Tel Aviv District/Tel-Aviv | Israel Central Virology laboratory | Israel Central Virology laboratory | Neta Zuckerman et al |
| EPI_ISL_649064 | Israel/CVL-n1541/2020 | 2020-03-02 | Asia/Israel/South District/Ashdod | Israel Central Virology laboratory | Israel Central Virology laboratory | Neta Zuckerman et al |
| EPI_ISL_649088 | Israel/CVL-n2971/2020 | 2020-03-08 | Asia/Israel/Central District/Raanana | Israel Central Virology laboratory | Israel Central Virology laboratory | Neta Zuckerman et al |
| EPI_ISL_649090 | Israel/CVL-n1467/2020 | 2020-03-02 | Asia/Israel/Tel Aviv District/Ramat Gan | Israel Central Virology laboratory | Israel Central Virology laboratory | Neta Zuckerman et al |
| EPI_ISL_649094 | Israel/CVL-n896/2020 | 2020-02-27 | Asia/Israel/Central District/Rishon-Letsi | Israel Central Virology laboratory | Israel Central Virology laboratory | Neta Zuckerman et al |
| EPI_ISL_649099 | Israel/CVL-n1750/2020 | 2020-03-04 | Asia/Israel/Central District/Sdema | Israel Central Virology laboratory | Israel Central Virology laboratory | Neta Zuckerman et al |
| EPI_ISL_649101 | Israel/CVL-n2775/2020 | 2020-03-08 | Asia/Israel/Tel Aviv District/Tel-Aviv | Israel Central Virology laboratory | Israel Central Virology laboratory | Neta Zuckerman et al |
| EPI_ISL_649104 | Israel/CVL-n3707/2020 | 2020-03-10 | Asia/Israel/Israel/ | Israel Central Virology laboratory | Israel Central Virology laboratory | Neta Zuckerman et al |
| EPI_ISL_649107 | Israel/CVL-n3975/2020 | 2020-03-11 | Asia/Israel/North District/Yuvalim | Israel Central Virology laboratory | Israel Central Virology laboratory | Neta Zuckerman et al |

| | | | | | | |
|---|---|---|---|---|---|---|
| EPI_ISL_649152 | USA/CA-CZB-6808/2020 | 2020-07-14 | North America/USA/California/Santa Cl | Infectious diseases, Chan Zuckerberg Biohub | Infectious diseases, Chan Zuckerberg Biohub | Bhatt et al |
| EPI_ISL_649162 | EquatorialGuinea/21093/2020 | 2020-06-18 | Africa/Equatorial Guinea/Bioko Norte/M | Laboratorio de Investigaciones de Baney | University Hospital Basel, Clinical Bacteriology | Carlos Cortes et al |
| EPI_ISL_649164 | EquatorialGuinea/27213/2020 | 2020-06-28 | Africa/Equatorial Guinea/Bioko Norte/B | Laboratorio de Investigaciones de Baney | University Hospital Basel, Clinical Bacteriology | Carlos Cortes et al |
| EPI_ISL_649173 | France/PDL-SC600/2020 | 2020-03-11 | Europe/France/Pays de la Loire/Nantes | CHU Nantes | CNR Virus des Infections Respiratoires - France SUD | Antonin Bal et al |
| EPI_ISL_649178 | France/PDL-SC627/2020 | 2020-04-01 | Europe/France/Pays de la Loire/Nantes | CHU Nantes | CNR Virus des Infections Respiratoires - France SUD | Antonin Bal et al |
| EPI_ISL_649189 | Italy/APU-IZSPB-98APT/2020 | 2020-04-10 | Europe/Italy/Apulia/ | Istituto Zooprofilattico Sperimentale della Puglia e della Basilicata | Istituto Zooprofilattico Sperimentale della Puglia e della Basilicata | Parisi A. et al |
| EPI_ISL_649219 | England/ALDP-B50419/2020 | 2020-10-09 | Europe/United Kingdom/England/ | Lighthouse Lab in Alderley Park | Wellcome Sanger Institute for the COVID-19 Genomics UK (COG-UK) Consortium | Jacquelyn Wynn et al |
| EPI_ISL_649322 | England/ALDP-B490F1/2020 | 2020-11-07 | Europe/United Kingdom/England/ | Lighthouse Lab in Alderley Park | Wellcome Sanger Institute for the COVID-19 Genomics UK (COG-UK) Consortium | Jacquelyn Wynn et al |
| EPI_ISL_649326 | England/ALDP-B4913A/2020 | 2020-11-07 | Europe/United Kingdom/England/ | Lighthouse Lab in Alderley Park | Wellcome Sanger Institute for the COVID-19 Genomics UK (COG-UK) Consortium | Jacquelyn Wynn et al |

| | | | | | | |
|---|---|---|---|---|---|---|
| EPI_ISL_649404 | England/ALDP-B47F05/2020 | 2020-11-07 | Europe/United Kingdom/England/ | Lighthouse Lab in Alderley Park | Wellcome Sanger Institute for the COVID-19 Genomics UK (COG-UK) Consortium | Jacquelyn Wynn et al |
| EPI_ISL_649461 | England/ALDP-B4811D/2020 | 2020-11-07 | Europe/United Kingdom/England/ | Lighthouse Lab in Alderley Park | Wellcome Sanger Institute for the COVID-19 Genomics UK (COG-UK) Consortium | Jacquelyn Wynn et al |
| EPI_ISL_649679 | Scotland/QEUH-AD5477/2020 | 2020-10-26 | Europe/United Kingdom/Scotland/ | Lighthouse Lab in Glasgow | Wellcome Sanger Institute for the COVID-19 Genomics UK (COG-UK) Consortium | Harper VanSteenhouse et al |
| EPI_ISL_649734 | Scotland/QEUH-AD051F/2020 | 2020-10-26 | Europe/United Kingdom/Scotland/ | Lighthouse Lab in Glasgow | Wellcome Sanger Institute for the COVID-19 Genomics UK (COG-UK) Consortium | Harper VanSteenhouse et al |
| EPI_ISL_649938 | Italy/APU-IZSPB-190PT/2020 | 2020-08-28 | Europe/Italy/Apulia/Bari | "I.R.C.C.S. ""S. De Bellis"" - Ente Ospedaliero" | Istituto Zooprofilattico Sperimentale della Puglia e della Basilicata | Parisi A. et al |
| EPI_ISL_649940 | Italy/APU-IZSPB-193APT/2020 | 2020-08-28 | Europe/Italy/Apulia/Foggia | Istituto Zooprofilattico Sperimentale della Puglia e della Basilicata | Istituto Zooprofilattico Sperimentale della Puglia e della Basilicata | Parisi A. et al |
| EPI_ISL_649945 | France/ARA-SC307/2020 | 2020-03-24 | Europe/France/Auvergne-Rh√¥ne-Alpes/ | CHU de Saint-√âtienne H√¥pital Nord | CNR Virus des Infections Respiratoires - France SUD | Antonin Bal et al |
| EPI_ISL_649946 | France/ARA-SC308/2020 | 2020-03-24 | Europe/France/Auvergne-Rh√¥ne-Alpes/ | CHU de Saint-√âtienne H√¥pital Nord | CNR Virus des Infections Respiratoires - France SUD | Antonin Bal et al |

| | | | | | | |
|---|---|---|---|---|---|---|
| EPI_ISL_649947 | France/ARA-SC312/2020 | 2020-04-09 | Europe/France/Auvergne-Rh√¥ne-Alpes/ | CHU de Saint-√âtienne H√¥pital Nord | CNR Virus des Infections Respiratoires - France SUD | Antonin Bal et al |
| EPI_ISL_649951 | France/ARA-SC317/2020 | 2020-04-20 | Europe/France/Auvergne-Rh√¥ne-Alpes/ | CHU de Saint-√âtienne H√¥pital Nord | CNR Virus des Infections Respiratoires - France SUD | Antonin Bal et al |
| EPI_ISL_649960 | France/NAQ-SC206/2020 | 2020-03-07 | Europe/France/Nouvelle-Aquitaine/Bord | CHU Bordeaux | CNR Virus des Infections Respiratoires - France SUD | Antonin Bal et al |
| EPI_ISL_649963 | France/NAQ-SC211/2020 | 2020-03-27 | Europe/France/Nouvelle-Aquitaine/Bord | CHU Bordeaux | CNR Virus des Infections Respiratoires - France SUD | Antonin Bal et al |
| EPI_ISL_649964 | France/NAQ-SC212/2020 | 2020-03-27 | Europe/France/Nouvelle-Aquitaine/Bord | CHU Bordeaux | CNR Virus des Infections Respiratoires - France SUD | Antonin Bal et al |
| EPI_ISL_649978 | USA/MI-UM-10036596238/2020 | 2020-10-23 | North America/USA/Michigan/ | University of Michigan Clinical Microbiology Laboratory | Lauring Lab, University of Michigan, Department of Microbiology and Immunology | Valesano et al |
| EPI_ISL_649981 | USA/MI-UM-10036578395/2020 | 2020-10-22 | North America/USA/Michigan/ | University of Michigan Clinical Microbiology Laboratory | Lauring Lab, University of Michigan, Department of Microbiology and Immunology | Valesano et al |
| EPI_ISL_649983 | USA/MI-UM-10036598438/2020 | 2020-10-23 | North America/USA/Michigan/ | University of Michigan Clinical Microbiology Laboratory | Lauring Lab, University of Michigan, Department of Microbiology and Immunology | Valesano et al |

| | | | | | | |
|---|---|---|---|---|---|---|
| EPI_ISL_649985 | USA/MI-UM-10036593588/2020 | 2020-10-23 | North America/USA/Michigan/ | University of Michigan Clinical Microbiology Laboratory | Lauring Lab, University of Michigan, Department of Microbiology and Immunology | Valesano et al |
| EPI_ISL_649991 | USA/MI-UM-10036587103/2020 | 2020-10-23 | North America/USA/Michigan/ | University of Michigan Clinical Microbiology Laboratory | Lauring Lab, University of Michigan, Department of Microbiology and Immunology | Valesano et al |
| EPI_ISL_650003 | USA/MI-UM-10036614249/2020 | 2020-10-25 | North America/USA/Michigan/ | University of Michigan Clinical Microbiology Laboratory | Lauring Lab, University of Michigan, Department of Microbiology and Immunology | Valesano et al |
| EPI_ISL_650021 | USA/MI-UM-10036619785/2020 | 2020-10-26 | North America/USA/Michigan/ | University of Michigan Clinical Microbiology Laboratory | Lauring Lab, University of Michigan, Department of Microbiology and Immunology | Valesano et al |
| EPI_ISL_650035 | USA/MI-UM-10036619926/2020 | 2020-10-26 | North America/USA/Michigan/ | University of Michigan Clinical Microbiology Laboratory | Lauring Lab, University of Michigan, Department of Microbiology and Immunology | Valesano et al |
| EPI_ISL_650047 | USA/MI-UM-10036672506/2020 | 2020-10-28 | North America/USA/Michigan/ | University of Michigan Clinical Microbiology Laboratory | Lauring Lab, University of Michigan, Department of Microbiology and Immunology | Valesano et al |

| | | | | | | |
|---|---|---|---|---|---|---|
| EPI_ISL_650053 | USA/MI-UM-10036671739/2020 | 2020-10-28 | North America/USA/Michigan/ | University of Michigan Clinical Microbiology Laboratory | Lauring Lab, University of Michigan, Department of Microbiology and Immunology | Valesano et al |
| EPI_ISL_650056 | USA/MI-UM-10036672505/2020 | 2020-10-28 | North America/USA/Michigan/ | University of Michigan Clinical Microbiology Laboratory | Lauring Lab, University of Michigan, Department of Microbiology and Immunology | Valesano et al |
| EPI_ISL_650068 | USA/MI-UM-10036660941/2020 | 2020-10-28 | North America/USA/Michigan/ | University of Michigan Clinical Microbiology Laboratory | Lauring Lab, University of Michigan, Department of Microbiology and Immunology | Valesano et al |
| EPI_ISL_650069 | USA/MI-UM-10036652026/2020 | 2020-10-27 | North America/USA/Michigan/ | University of Michigan Clinical Microbiology Laboratory | Lauring Lab, University of Michigan, Department of Microbiology and Immunology | Valesano et al |
| EPI_ISL_650071 | USA/MI-UM-10036648964/2020 | 2020-10-27 | North America/USA/Michigan/ | University of Michigan Clinical Microbiology Laboratory | Lauring Lab, University of Michigan, Department of Microbiology and Immunology | Valesano et al |
| EPI_ISL_650072 | USA/MI-UM-10036645387/2020 | 2020-10-27 | North America/USA/Michigan/ | University of Michigan Clinical Microbiology Laboratory | Lauring Lab, University of Michigan, Department of Microbiology and Immunology | Valesano et al |

| | | | | | | |
|---|---|---|---|---|---|---|
| EPI_ISL_650074 | USA/MI-UM-10036653484/2020 | 2020-10-27 | North America/USA/Michigan/ | University of Michigan Clinical Microbiology Laboratory | Lauring Lab, University of Michigan, Department of Microbiology and Immunology | Valesano et al |
| EPI_ISL_650075 | USA/MI-UM-10036673508/2020 | 2020-10-28 | North America/USA/Michigan/ | University of Michigan Clinical Microbiology Laboratory | Lauring Lab, University of Michigan, Department of Microbiology and Immunology | Valesano et al |
| EPI_ISL_650080 | USA/MI-UM-10036648956/2020 | 2020-10-27 | North America/USA/Michigan/ | University of Michigan Clinical Microbiology Laboratory | Lauring Lab, University of Michigan, Department of Microbiology and Immunology | Valesano et al |
| EPI_ISL_650083 | USA/MI-UM-10036660976/2020 | 2020-10-28 | North America/USA/Michigan/ | University of Michigan Clinical Microbiology Laboratory | Lauring Lab, University of Michigan, Department of Microbiology and Immunology | Valesano et al |
| EPI_ISL_650086 | USA/MI-UM-10036690817/2020 | 2020-10-29 | North America/USA/Michigan/ | University of Michigan Clinical Microbiology Laboratory | Lauring Lab, University of Michigan, Department of Microbiology and Immunology | Valesano et al |
| EPI_ISL_650092 | USA/MI-UM-10036671254/2020 | 2020-10-28 | North America/USA/Michigan/ | University of Michigan Clinical Microbiology Laboratory | Lauring Lab, University of Michigan, Department of Microbiology and Immunology | Valesano et al |

| | | | | | | |
|---|---|---|---|---|---|---|
| EPI_ISL_650094 | USA/MI-UM-10036696400/2020 | 2020-10-29 | North America/USA/Michigan/ | University of Michigan Clinical Microbiology Laboratory | Lauring Lab, University of Michigan, Department of Microbiology and Immunology | Valesano et al |
| EPI_ISL_650095 | USA/MI-UM-10036688546/2020 | 2020-10-29 | North America/USA/Michigan/ | University of Michigan Clinical Microbiology Laboratory | Lauring Lab, University of Michigan, Department of Microbiology and Immunology | Valesano et al |
| EPI_ISL_650109 | USA/MI-UM-10036631636/2020 | 2020-10-26 | North America/USA/Michigan/ | University of Michigan Clinical Microbiology Laboratory | Lauring Lab, University of Michigan, Department of Microbiology and Immunology | Valesano et al |
| EPI_ISL_650111 | USA/MI-UM-10036671252/2020 | 2020-10-28 | North America/USA/Michigan/ | University of Michigan Clinical Microbiology Laboratory | Lauring Lab, University of Michigan, Department of Microbiology and Immunology | Valesano et al |
| EPI_ISL_650153 | England/BIRM-67E67/2020 | 2020-10-30 | Europe/United Kingdom/England/ | University of Birmingham | COVID-19 Genomics UK (COG-UK) Consortium | Institute of Microbiology et al |
| EPI_ISL_650247 | England/MILK-A8EF9A/2020 | 2020-10-16 | Europe/United Kingdom/England/ | Northumbria University / South Tees Hospitals NHS Foundation Trust / North Cumbria Integrated Care NHS Foundation Trust / North Tees and Hartlepool NHS Foundation Trust / Newcastle Hospitals NHS Foundation Trust | COVID-19 Genomics UK (COG-UK) Consortium | Darren L Smith et al |
| EPI_ISL_650248 | England/PORT-2D754D/2020 | 2020-10-27 | Europe/United Kingdom/England/ | Centre for Enzyme Innovation, University of Portsmouth / Translational Research Laboratory, Portsmouth Hospitals NHS Trust | COVID-19 Genomics UK (COG-UK) Consortium | Angela Beckett et al |

| | | | | | | |
|---|---|---|---|---|---|---|
| EPI_ISL_650355 | England/MILK-A8F00F/2020 | 2020-10-16 | Europe/United Kingdom/England/ | Northumbria University / South Tees Hospitals NHS Foundation Trust / North Cumbria Integrated Care NHS Foundation Trust / North Tees and Hartlepool NHS Foundation Trust / Newcastle Hospitals NHS Foundation Trust | COVID-19 Genomics UK (COG-UK) Consortium | Darren L Smith et al |
| EPI_ISL_650450 | England/NOTT-11B775/2020 | 2020-11-12 | Europe/United Kingdom/England/ | Queens Medical Centre, Clinical Microbiology Department / DeepSeq Nottingham | COVID-19 Genomics UK (COG-UK) Consortium | Gemma Clark et al |
| EPI_ISL_650507 | Scotland/CVR5500/2020 | 2020-10-30 | Europe/United Kingdom/Scotland/ | West of Scotland Specialist Virology Centre, NHSGGC / MRC-University of Glasgow Centre for Virus Research | COVID-19 Genomics UK (COG-UK) Consortium | Ana da Silva Filipe et al |
| EPI_ISL_650521 | Scotland/CVR5345/2020 | 2020-10-26 | Europe/United Kingdom/Scotland/ | West of Scotland Specialist Virology Centre, NHSGGC / MRC-University of Glasgow Centre for Virus Research | COVID-19 Genomics UK (COG-UK) Consortium | Ana da Silva Filipe et al |
| EPI_ISL_650603 | England/PORT-2D753E/2020 | 2020-10-27 | Europe/United Kingdom/England/ | Centre for Enzyme Innovation, University of Portsmouth / Translational Research Laboratory, Portsmouth Hospitals NHS Trust | COVID-19 Genomics UK (COG-UK) Consortium | Angela Beckett et al |
| EPI_ISL_650840 | Scotland/CVR5304/2020 | 2020-10-21 | Europe/United Kingdom/Scotland/ | West of Scotland Specialist Virology Centre, NHSGGC / MRC-University of Glasgow Centre for Virus Research | COVID-19 Genomics UK (COG-UK) Consortium | Ana da Silva Filipe et al |
| EPI_ISL_650926 | England/LIVE-DB3C65/2020 | 2020-10-21 | Europe/United Kingdom/England/ | Liverpool Clinical Laboratories | COVID-19 Genomics UK (COG-UK) Consortium | Sam Haldenby et al |
| EPI_ISL_651583 | Scotland/CVR4877/2020 | 2020-10-01 | Europe/United Kingdom/Scotland/ | West of Scotland Specialist Virology Centre, NHSGGC / MRC-University of Glasgow Centre for Virus Research | COVID-19 Genomics UK (COG-UK) Consortium | Ana da Silva Filipe et al |
| EPI_ISL_652351 | England/NORW-F6920/2020 | 2020-11-09 | Europe/United Kingdom/England/ | Quadram Institute Bioscience | COVID-19 Genomics UK (COG-UK) Consortium | Dave J. Baker et al |
| EPI_ISL_652437 | Wales/PHWC-4899E2/2020 | 2020-10-24 | Europe/United Kingdom/Wales/ | Wales Specialist Virology Centre Sequencing lab: Pathogen Genomics Unit | COVID-19 Genomics UK (COG-UK) Consortium | Catherine Moore et al |
| EPI_ISL_652513 | Wales/PHWC-489FC2/2020 | 2020-11-02 | Europe/United Kingdom/Wales/ | Wales Specialist Virology Centre Sequencing lab: Pathogen Genomics Unit | COVID-19 Genomics UK (COG-UK) Consortium | Catherine Moore et al |
| EPI_ISL_652632 | Scotland/QEUH-A9C360/2020 | 2020-10-20 | Europe/United Kingdom/Scotland/ | Queens Medical Centre, Clinical Microbiology Department / DeepSeq Nottingham | COVID-19 Genomics UK (COG-UK) Consortium | Gemma Clark et al |

| | | | | | | |
|---|---|---|---|---|---|---|
| EPI_ISL_652652 | Scotland/QEUH-A9D0D1/2020 | 2020-10-20 | Europe/United Kingdom/Scotland/ | Queens Medical Centre, Clinical Microbiology Department / DeepSeq Nottingham | COVID-19 Genomics UK (COG-UK) Consortium | Gemma Clark et al |
| EPI_ISL_652665 | England/QEUH-A9D9A9/2020 | 2020-10-20 | Europe/United Kingdom/England/ | Queens Medical Centre, Clinical Microbiology Department / DeepSeq Nottingham | COVID-19 Genomics UK (COG-UK) Consortium | Gemma Clark et al |
| EPI_ISL_652799 | Scotland/QEUH-AAB5A4/2020 | 2020-10-21 | Europe/United Kingdom/Scotland/ | Oxford Viromics, NDM, University of Oxford; Oxford University Hospitals; Basingstoke and North Hampshire Hospital | COVID-19 Genomics UK (COG-UK) Consortium | Tanya Golubchik et al |
| EPI_ISL_652812 | England/QEUH-AAB780/2020 | 2020-10-21 | Europe/United Kingdom/England/ | Oxford Viromics, NDM, University of Oxford; Oxford University Hospitals; Basingstoke and North Hampshire Hospital | COVID-19 Genomics UK (COG-UK) Consortium | Tanya Golubchik et al |
| EPI_ISL_652864 | England/QEUH-AAC655/2020 | 2020-10-21 | Europe/United Kingdom/England/ | Oxford Viromics, NDM, University of Oxford; Oxford University Hospitals; Basingstoke and North Hampshire Hospital | COVID-19 Genomics UK (COG-UK) Consortium | Tanya Golubchik et al |
| EPI_ISL_652978 | England/QEUH-AAEA2A/2020 | 2020-10-20 | Europe/United Kingdom/England/ | Oxford Viromics, NDM, University of Oxford; Oxford University Hospitals; Basingstoke and North Hampshire Hospital | COVID-19 Genomics UK (COG-UK) Consortium | Tanya Golubchik et al |
| EPI_ISL_652989 | England/QEUH-AAEBBE/2020 | 2020-10-20 | Europe/United Kingdom/England/ | Oxford Viromics, NDM, University of Oxford; Oxford University Hospitals; Basingstoke and North Hampshire Hospital | COVID-19 Genomics UK (COG-UK) Consortium | Tanya Golubchik et al |
| EPI_ISL_652999 | England/QEUH-AAECF7/2020 | 2020-10-20 | Europe/United Kingdom/England/ | Oxford Viromics, NDM, University of Oxford; Oxford University Hospitals; Basingstoke and North Hampshire Hospital | COVID-19 Genomics UK (COG-UK) Consortium | Tanya Golubchik et al |
| EPI_ISL_653024 | Scotland/QEUH-AAF449/2020 | 2020-10-22 | Europe/United Kingdom/Scotland/ | Oxford Viromics, NDM, University of Oxford; Oxford University Hospitals; Basingstoke and North Hampshire Hospital | COVID-19 Genomics UK (COG-UK) Consortium | Tanya Golubchik et al |
| EPI_ISL_653111 | USA/FL-BPHL-1958/2020 | 2020-04-13 | North America/USA/Florida/ | Florida Bureau of Public Health Laboratories | Florida Bureau of Public Health Laboratories | Sarah Schmedes et al |
| EPI_ISL_653128 | USA/FL-BPHL-1975/2020 | 2020-04-13 | North America/USA/Florida/ | Florida Bureau of Public Health Laboratories | Florida Bureau of Public Health Laboratories | Sarah Schmedes et al |
| EPI_ISL_653129 | USA/FL-BPHL-1976/2020 | 2020-04-16 | North America/USA/Florida/ | Florida Bureau of Public Health Laboratories | Florida Bureau of Public Health Laboratories | Sarah Schmedes et al |
| EPI_ISL_653141 | USA/FL-BPHL-1988/2020 | 2020-04-18 | North America/USA/Florida/ | Florida Bureau of Public Health Laboratories | Florida Bureau of Public Health Laboratories | Sarah Schmedes et al |

| | | | | | | |
|---|---|---|---|---|---|---|
| EPI_ISL_653157 | USA/FL-BPHL-2004/2020 | 2020-03-26 | North America/USA/Florida/ | Florida Bureau of Public Health Laboratories | Florida Bureau of Public Health Laboratories | Sarah Schmedes et al |
| EPI_ISL_653180 | USA/FL-BPHL-2027/2020 | 2020-03-25 | North America/USA/Florida/ | Florida Bureau of Public Health Laboratories | Florida Bureau of Public Health Laboratories | Sarah Schmedes et al |
| EPI_ISL_653194 | USA/FL-BPHL-2041/2020 | 2020-07-21 | North America/USA/Florida/ | Florida Bureau of Public Health Laboratories | Florida Bureau of Public Health Laboratories | Sarah Schmedes et al |
| EPI_ISL_653195 | USA/FL-BPHL-2042/2020 | 2020-07-14 | North America/USA/Florida/ | Florida Bureau of Public Health Laboratories | Florida Bureau of Public Health Laboratories | Sarah Schmedes et al |
| EPI_ISL_653216 | USA/FL-BPHL-2063/2020 | 2020-06-22 | North America/USA/Florida/ | Florida Bureau of Public Health Laboratories | Florida Bureau of Public Health Laboratories | Sarah Schmedes et al |
| EPI_ISL_653222 | USA/FL-BPHL-2069/2020 | 2020-07-09 | North America/USA/Florida/ | Florida Bureau of Public Health Laboratories | Florida Bureau of Public Health Laboratories | Sarah Schmedes et al |
| EPI_ISL_653231 | USA/FL-BPHL-2078/2020 | 2020-07-08 | North America/USA/Florida/ | Florida Bureau of Public Health Laboratories | Florida Bureau of Public Health Laboratories | Sarah Schmedes et al |
| EPI_ISL_653236 | USA/FL-BPHL-2083/2020 | 2020-07-29 | North America/USA/Florida/ | Florida Bureau of Public Health Laboratories | Florida Bureau of Public Health Laboratories | Sarah Schmedes et al |
| EPI_ISL_653240 | USA/FL-BPHL-2087/2020 | 2020-07-16 | North America/USA/Florida/ | Florida Bureau of Public Health Laboratories | Florida Bureau of Public Health Laboratories | Sarah Schmedes et al |
| EPI_ISL_653247 | USA/FL-BPHL-2094/2020 | 2020-07-31 | North America/USA/Florida/ | Florida Bureau of Public Health Laboratories | Florida Bureau of Public Health Laboratories | Sarah Schmedes et al |
| EPI_ISL_653252 | USA/FL-BPHL-2099/2020 | 2020-07-20 | North America/USA/Florida/ | Florida Bureau of Public Health Laboratories | Florida Bureau of Public Health Laboratories | Sarah Schmedes et al |
| EPI_ISL_653257 | USA/FL-BPHL-2104/2020 | 2020-03-05 | North America/USA/Florida/ | Florida Bureau of Public Health Laboratories | Florida Bureau of Public Health Laboratories | Sarah Schmedes et al |
| EPI_ISL_653280 | USA/FL-BPHL-2127/2020 | 2020-04-07 | North America/USA/Florida/ | Florida Bureau of Public Health Laboratories | Florida Bureau of Public Health Laboratories | Sarah Schmedes et al |

| | | | | | | |
|---|---|---|---|---|---|---|
| EPI_ISL_653313 | USA/FL-BPHL-2160/2020 | 2020-05-21 | North America/USA/Florida/ | Florida Bureau of Public Health Laboratories | Florida Bureau of Public Health Laboratories | Sarah Schmedes et al |
| EPI_ISL_653314 | USA/FL-BPHL-2161/2020 | 2020-06-09 | North America/USA/Florida/ | Florida Bureau of Public Health Laboratories | Florida Bureau of Public Health Laboratories | Sarah Schmedes et al |
| EPI_ISL_653326 | USA/FL-BPHL-2173/2020 | 2020-08-17 | North America/USA/Florida/ | Florida Bureau of Public Health Laboratories | Florida Bureau of Public Health Laboratories | Sarah Schmedes et al |
| EPI_ISL_653329 | USA/FL-BPHL-2176/2020 | 2020-08-18 | North America/USA/Florida/ | Florida Bureau of Public Health Laboratories | Florida Bureau of Public Health Laboratories | Sarah Schmedes et al |
| EPI_ISL_653333 | USA/FL-BPHL-2180/2020 | 2020-08-19 | North America/USA/Florida/ | Florida Bureau of Public Health Laboratories | Florida Bureau of Public Health Laboratories | Sarah Schmedes et al |
| EPI_ISL_653334 | USA/FL-BPHL-2181/2020 | 2020-08-20 | North America/USA/Florida/ | Florida Bureau of Public Health Laboratories | Florida Bureau of Public Health Laboratories | Sarah Schmedes et al |
| EPI_ISL_653336 | USA/FL-BPHL-2183/2020 | 2020-08-20 | North America/USA/Florida/ | Florida Bureau of Public Health Laboratories | Florida Bureau of Public Health Laboratories | Sarah Schmedes et al |
| EPI_ISL_653337 | USA/FL-BPHL-2184/2020 | 2020-08-20 | North America/USA/Florida/ | Florida Bureau of Public Health Laboratories | Florida Bureau of Public Health Laboratories | Sarah Schmedes et al |
| EPI_ISL_653345 | USA/LA-EVTL653/2020 | 2020-07-12 | North America/USA/Louisiana/Bossier P | LSUHS Emerging Viral Threat Laboratory | Microbial Genome Sequencing Center | Maarten Van Diest et al |
| EPI_ISL_653350 | USA/LA-EVTL658/2020 | 2020-07-13 | North America/USA/Louisiana/Union Pa | LSUHS Emerging Viral Threat Laboratory | Microbial Genome Sequencing Center | Maarten Van Diest et al |
| EPI_ISL_653360 | USA/LA-EVTL668/2020 | 2020-07-13 | North America/USA/Louisiana/Caddo Pa | LSUHS Emerging Viral Threat Laboratory | Microbial Genome Sequencing Center | Maarten Van Diest et al |
| EPI_ISL_653377 | USA/LA-EVTL685/2020 | 2020-07-15 | North America/USA/Louisiana/Bossier P | LSUHS Emerging Viral Threat Laboratory | Microbial Genome Sequencing Center | Rona S. Scott et al |
| EPI_ISL_653378 | USA/LA-EVTL686/2020 | 2020-07-15 | North America/USA/Louisiana/Bossier P | LSUHS Emerging Viral Threat Laboratory | Microbial Genome Sequencing Center | Rona S. Scott et al |

| | | | | | | |
|---|---|---|---|---|---|---|
| EPI_ISL_653379 | USA/LA-EVTL687/2020 | 2020-07-14 | North America/USA/Louisiana/Caddo Pa | LSUHS Emerging Viral Threat Laboratory | Microbial Genome Sequencing Center | Rona S. Scott et al |
| EPI_ISL_653381 | USA/LA-EVTL689/2020 | 2020-07-14 | North America/USA/Louisiana/Caddo Pa | LSUHS Emerging Viral Threat Laboratory | Microbial Genome Sequencing Center | Rona S. Scott et al |
| EPI_ISL_653383 | USA/LA-EVTL691/2020 | 2020-07-14 | North America/USA/Louisiana/Caddo Pa | LSUHS Emerging Viral Threat Laboratory | Microbial Genome Sequencing Center | Rona S. Scott et al |
| EPI_ISL_653389 | USA/LA-EVTL697/2020 | 2020-07-14 | North America/USA/Louisiana/Red River | LSUHS Emerging Viral Threat Laboratory | Microbial Genome Sequencing Center | Rona S. Scott et al |
| EPI_ISL_653396 | USA/LA-EVTL704/2020 | 2020-07-14 | North America/USA/Louisiana/Bossier P | LSUHS Emerging Viral Threat Laboratory | Microbial Genome Sequencing Center | Jeremy P. Kamil et al |
| EPI_ISL_653399 | USA/LA-EVTL707/2020 | 2020-07-14 | North America/USA/Louisiana/Bossier P | LSUHS Emerging Viral Threat Laboratory | Microbial Genome Sequencing Center | Jeremy P. Kamil et al |
| EPI_ISL_653402 | USA/LA-EVTL710/2020 | 2020-07-15 | North America/USA/Louisiana/Webster | LSUHS Emerging Viral Threat Laboratory | Microbial Genome Sequencing Center | Jeremy P. Kamil et al |
| EPI_ISL_653403 | USA/LA-EVTL711/2020 | 2020-07-15 | North America/USA/Louisiana/Caddo Pa | LSUHS Emerging Viral Threat Laboratory | Microbial Genome Sequencing Center | Jeremy P. Kamil et al |
| EPI_ISL_653404 | USA/LA-EVTL712/2020 | 2020-07-15 | North America/USA/Louisiana/Webster | LSUHS Emerging Viral Threat Laboratory | Microbial Genome Sequencing Center | Jeremy P. Kamil et al |
| EPI_ISL_653405 | USA/LA-EVTL713/2020 | 2020-07-14 | North America/USA/Louisiana/Webster | LSUHS Emerging Viral Threat Laboratory | Microbial Genome Sequencing Center | Jeremy P. Kamil et al |
| EPI_ISL_653406 | USA/LA-EVTL714/2020 | 2020-07-15 | North America/USA/Louisiana/Webster | LSUHS Emerging Viral Threat Laboratory | Microbial Genome Sequencing Center | Jeremy P. Kamil et al |
| EPI_ISL_653410 | USA/LA-EVTL718/2020 | 2020-07-16 | North America/USA/Louisiana/Caddo Pa | LSUHS Emerging Viral Threat Laboratory | Microbial Genome Sequencing Center | Jeremy P. Kamil et al |
| EPI_ISL_653411 | USA/LA-EVTL719/2020 | 2020-07-16 | North America/USA/Louisiana/Caddo Pa | LSUHS Emerging Viral Threat Laboratory | Microbial Genome Sequencing Center | Jeremy P. Kamil et al |

| | | | | | | |
|---|---|---|---|---|---|---|
| EPI_ISL_653413 | USA/LA-EVTL721/2020 | 2020-07-16 | North America/USA/Louisiana/Caddo Pa | LSUHS Emerging Viral Threat Laboratory | Microbial Genome Sequencing Center | Jeremy P. Kamil et al |
| EPI_ISL_653421 | USA/LA-EVTL730/2020 | 2020-07-16 | North America/USA/Louisiana/Caddo Pa | LSUHS Emerging Viral Threat Laboratory | Microbial Genome Sequencing Center | Jeremy P. Kamil et al |
| EPI_ISL_653435 | USA/LA-EVTL744/2020 | 2020-07-18 | North America/USA/Louisiana/Bossier P | LSUHS Emerging Viral Threat Laboratory | Microbial Genome Sequencing Center | Jeremy P. Kamil et al |
| EPI_ISL_653445 | USA/LA-EVTL754/2020 | 2020-07-20 | North America/USA/Louisiana/Claiborne | LSUHS Emerging Viral Threat Laboratory | Microbial Genome Sequencing Center | Jeremy P. Kamil et al |
| EPI_ISL_653449 | USA/LA-EVTL758/2020 | 2020-07-20 | North America/USA/Louisiana/Webster | LSUHS Emerging Viral Threat Laboratory | Microbial Genome Sequencing Center | Jeremy P. Kamil et al |
| EPI_ISL_653452 | USA/LA-EVTL761/2020 | 2020-07-21 | North America/USA/Louisiana/Lincoln Pa | LSUHS Emerging Viral Threat Laboratory | Microbial Genome Sequencing Center | Jeremy P. Kamil et al |
| EPI_ISL_653454 | USA/LA-EVTL763/2020 | 2020-07-21 | North America/USA/Louisiana/Lincoln Pa | LSUHS Emerging Viral Threat Laboratory | Microbial Genome Sequencing Center | Jeremy P. Kamil et al |
| EPI_ISL_653455 | USA/LA-EVTL764/2020 | 2020-07-21 | North America/USA/Louisiana/Claiborne | LSUHS Emerging Viral Threat Laboratory | Microbial Genome Sequencing Center | Jeremy P. Kamil et al |
| EPI_ISL_653457 | USA/LA-EVTL766/2020 | 2020-07-21 | North America/USA/Louisiana/Bossier P | LSUHS Emerging Viral Threat Laboratory | Microbial Genome Sequencing Center | Jeremy P. Kamil et al |
| EPI_ISL_653459 | USA/LA-EVTL768/2020 | 2020-07-21 | North America/USA/Louisiana/Caddo Pa | LSUHS Emerging Viral Threat Laboratory | Microbial Genome Sequencing Center | Jeremy P. Kamil et al |
| EPI_ISL_653462 | USA/LA-EVTL771/2020 | 2020-07-21 | North America/USA/Louisiana/Caddo Pa | LSUHS Emerging Viral Threat Laboratory | Microbial Genome Sequencing Center | Jeremy P. Kamil et al |
| EPI_ISL_653472 | USA/LA-EVTL781/2020 | 2020-07-22 | North America/USA/Louisiana/Webster | LSUHS Emerging Viral Threat Laboratory | Microbial Genome Sequencing Center | Jeremy P. Kamil et al |
| EPI_ISL_653474 | USA/LA-EVTL783/2020 | 2020-07-21 | North America/USA/Louisiana/Caddo Pa | LSUHS Emerging Viral Threat Laboratory | Microbial Genome Sequencing Center | Jeremy P. Kamil et al |

| | | | | | | |
|---|---|---|---|---|---|---|
| EPI_ISL_653475 | USA/LA-EVTL784/2020 | 2020-07-23 | North America/USA/Louisiana/Webster | LSUHS Emerging Viral Threat Laboratory | Microbial Genome Sequencing Center | Jeremy P. Kamil et al |
| EPI_ISL_653476 | USA/LA-EVTL785/2020 | 2020-07-23 | North America/USA/Louisiana/Webster | LSUHS Emerging Viral Threat Laboratory | Microbial Genome Sequencing Center | Jeremy P. Kamil et al |
| EPI_ISL_653479 | USA/LA-EVTL788/2020 | 2020-07-23 | North America/USA/Louisiana/DeSoto P | LSUHS Emerging Viral Threat Laboratory | Microbial Genome Sequencing Center | Jeremy P. Kamil et al |
| EPI_ISL_653484 | USA/LA-EVTL793/2020 | 2020-07-23 | North America/USA/Louisiana/DeSoto P | LSUHS Emerging Viral Threat Laboratory | Microbial Genome Sequencing Center | Jeremy P. Kamil et al |
| EPI_ISL_653490 | USA/LA-EVTL799/2020 | 2020-07-24 | North America/USA/Louisiana/Bossier P | LSUHS Emerging Viral Threat Laboratory | Microbial Genome Sequencing Center | Jeremy P. Kamil et al |
| EPI_ISL_653493 | USA/LA-EVTL802/2020 | 2020-07-24 | North America/USA/Louisiana/Bossier P | LSUHS Emerging Viral Threat Laboratory | Microbial Genome Sequencing Center | Jeremy P. Kamil et al |
| EPI_ISL_653495 | USA/LA-EVTL804/2020 | 2020-07-24 | North America/USA/Louisiana/Bossier P | LSUHS Emerging Viral Threat Laboratory | Microbial Genome Sequencing Center | Jeremy P. Kamil et al |
| EPI_ISL_653496 | USA/LA-EVTL805/2020 | 2020-07-24 | North America/USA/Louisiana/Bossier P | LSUHS Emerging Viral Threat Laboratory | Microbial Genome Sequencing Center | Jeremy P. Kamil et al |
| EPI_ISL_653505 | USA/LA-EVTL814/2020 | 2020-07-27 | North America/USA/Louisiana/Claiborne | LSUHS Emerging Viral Threat Laboratory | Microbial Genome Sequencing Center | Jeremy P. Kamil et al |
| EPI_ISL_653510 | USA/LA-EVTL819/2020 | 2020-07-27 | North America/USA/Louisiana/Bossier P | LSUHS Emerging Viral Threat Laboratory | Microbial Genome Sequencing Center | Jeremy P. Kamil et al |
| EPI_ISL_653521 | USA/LA-EVTL830/2020 | 2020-07-28 | North America/USA/Louisiana/Caddo Pa | LSUHS Emerging Viral Threat Laboratory | Microbial Genome Sequencing Center | Jeremy P. Kamil et al |
| EPI_ISL_653523 | USA/LA-EVTL832/2020 | 2020-07-28 | North America/USA/Louisiana/Caddo Pa | LSUHS Emerging Viral Threat Laboratory | Microbial Genome Sequencing Center | Jeremy P. Kamil et al |
| EPI_ISL_653540 | USA/LA-EVTL849/2020 | 2020-07-28 | North America/USA/Louisiana/Caddo Pa | LSUHS Emerging Viral Threat Laboratory | Microbial Genome Sequencing Center | Jeremy P. Kamil et al |

| | | | | | | |
|---|---|---|---|---|---|---|
| EPI_ISL_653553 | USA/LA-EVTL862/2020 | 2020-07-01 | North America/USA/Louisiana/Caddo Pa | LSUHS Emerging Viral Threat Laboratory | Microbial Genome Sequencing Center | Jeremy P. Kamil et al |
| EPI_ISL_653555 | USA/LA-EVTL864/2020 | 2020-07-02 | North America/USA/Louisiana/Caddo Pa | LSUHS Emerging Viral Threat Laboratory | Microbial Genome Sequencing Center | Jeremy P. Kamil et al |
| EPI_ISL_653563 | USA/LA-EVTL872/2020 | 2020-07-03 | North America/USA/Louisiana/Caddo Pa | LSUHS Emerging Viral Threat Laboratory | Microbial Genome Sequencing Center | Jeremy P. Kamil et al |
| EPI_ISL_653567 | USA/LA-EVTL876/2020 | 2020-07-06 | North America/USA/Louisiana/Bossier P | LSUHS Emerging Viral Threat Laboratory | Microbial Genome Sequencing Center | Jeremy P. Kamil et al |
| EPI_ISL_653569 | USA/LA-EVTL878/2020 | 2020-07-06 | North America/USA/Louisiana/Bossier P | LSUHS Emerging Viral Threat Laboratory | Microbial Genome Sequencing Center | Jeremy P. Kamil et al |
| EPI_ISL_653571 | USA/LA-EVTL880/2020 | 2020-07-06 | North America/USA/Louisiana/Caddo Pa | LSUHS Emerging Viral Threat Laboratory | Microbial Genome Sequencing Center | Jeremy P. Kamil et al |
| EPI_ISL_653588 | USA/LA-EVTL897/2020 | 2020-07-08 | North America/USA/Louisiana/Webster | LSUHS Emerging Viral Threat Laboratory | Microbial Genome Sequencing Center | Jeremy P. Kamil et al |
| EPI_ISL_653589 | USA/LA-EVTL898/2020 | 2020-07-08 | North America/USA/Louisiana/Caddo Pa | LSUHS Emerging Viral Threat Laboratory | Microbial Genome Sequencing Center | Jeremy P. Kamil et al |
| EPI_ISL_653593 | USA/LA-EVTL902/2020 | 2020-07-08 | North America/USA/Louisiana/Webster | LSUHS Emerging Viral Threat Laboratory | Microbial Genome Sequencing Center | Jeremy P. Kamil et al |
| EPI_ISL_653594 | USA/LA-EVTL903/2020 | 2020-08-10 | North America/USA/Louisiana/DeSoto P | LSUHS Emerging Viral Threat Laboratory | Microbial Genome Sequencing Center | Jeremy P. Kamil et al |
| EPI_ISL_653600 | USA/LA-EVTL909/2020 | 2020-08-10 | North America/USA/Louisiana/Bossier P | LSUHS Emerging Viral Threat Laboratory | Microbial Genome Sequencing Center | Jeremy P. Kamil et al |
| EPI_ISL_653602 | USA/LA-EVTL911/2020 | 2020-07-09 | North America/USA/Louisiana/Caddo Pa | LSUHS Emerging Viral Threat Laboratory | Microbial Genome Sequencing Center | Jeremy P. Kamil et al |
| EPI_ISL_653605 | USA/LA-EVTL914/2020 | 2020-07-09 | North America/USA/Louisiana/Caddo Pa | LSUHS Emerging Viral Threat Laboratory | Microbial Genome Sequencing Center | Jeremy P. Kamil et al |

| | | | | | | |
|---|---|---|---|---|---|---|
| EPI_ISL_653607 | USA/LA-EVTL916/2020 | 2020-08-10 | North America/USA/Louisiana/DeSoto P | LSUHS Emerging Viral Threat Laboratory | Microbial Genome Sequencing Center | Jeremy P. Kamil et al |
| EPI_ISL_653608 | USA/LA-EVTL917/2020 | 2020-07-08 | North America/USA/Louisiana/Lincoln Pa | LSUHS Emerging Viral Threat Laboratory | Microbial Genome Sequencing Center | Jeremy P. Kamil et al |
| EPI_ISL_653613 | USA/LA-EVTL922/2020 | 2020-08-11 | North America/USA/Louisiana/Red Rive | LSUHS Emerging Viral Threat Laboratory | Microbial Genome Sequencing Center | Jeremy P. Kamil et al |
| EPI_ISL_653616 | USA/LA-EVTL925/2020 | 2020-08-11 | North America/USA/Louisiana/Red Rive | LSUHS Emerging Viral Threat Laboratory | Microbial Genome Sequencing Center | Jeremy P. Kamil et al |
| EPI_ISL_653620 | USA/LA-EVTL929/2020 | 2020-08-11 | North America/USA/Louisiana/Red Rive | LSUHS Emerging Viral Threat Laboratory | Microbial Genome Sequencing Center | Jeremy P. Kamil et al |
| EPI_ISL_653623 | USA/LA-EVTL932/2020 | 2020-08-11 | North America/USA/Louisiana/Red Rive | LSUHS Emerging Viral Threat Laboratory | Microbial Genome Sequencing Center | Jeremy P. Kamil et al |
| EPI_ISL_653625 | USA/LA-EVTL934/2020 | 2020-08-11 | North America/USA/Louisiana/Red Rive | LSUHS Emerging Viral Threat Laboratory | Microbial Genome Sequencing Center | Jeremy P. Kamil et al |
| EPI_ISL_653631 | USA/LA-EVTL940/2020 | 2020-08-11 | North America/USA/Louisiana/Caddo Pa | LSUHS Emerging Viral Threat Laboratory | Microbial Genome Sequencing Center | Jeremy P. Kamil et al |
| EPI_ISL_653640 | USA/LA-EVTL949/2020 | 2020-08-13 | North America/USA/Louisiana/Bossier P | LSUHS Emerging Viral Threat Laboratory | Microbial Genome Sequencing Center | Jeremy P. Kamil et al |
| EPI_ISL_653641 | USA/LA-EVTL950/2020 | 2020-08-13 | North America/USA/Louisiana/Bossier P | LSUHS Emerging Viral Threat Laboratory | Microbial Genome Sequencing Center | Jeremy P. Kamil et al |
| EPI_ISL_653644 | USA/LA-EVTL953/2020 | 2020-08-06 | North America/USA/Louisiana/Bossier P | LSUHS Emerging Viral Threat Laboratory | Microbial Genome Sequencing Center | Jeremy P. Kamil et al |
| EPI_ISL_653651 | USA/LA-EVTL960/2020 | 2020-08-13 | North America/USA/Louisiana/Webster | LSUHS Emerging Viral Threat Laboratory | Microbial Genome Sequencing Center | Jeremy P. Kamil et al |
| EPI_ISL_653654 | USA/LA-EVTL963/2020 | 2020-08-14 | North America/USA/Louisiana/Red Rive | LSUHS Emerging Viral Threat Laboratory | Microbial Genome Sequencing Center | Jeremy P. Kamil et al |

| | | | | | | |
|---|---|---|---|---|---|---|
| EPI_ISL_653661 | USA/LA-EVTL970/2020 | 2020-08-17 | North America/USA/Louisiana/Webster | LSUHS Emerging Viral Threat Laboratory | Microbial Genome Sequencing Center | Jeremy P. Kamil et al |
| EPI_ISL_653662 | USA/LA-EVTL971/2020 | 2020-08-18 | North America/USA/Louisiana/Claiborne | LSUHS Emerging Viral Threat Laboratory | Microbial Genome Sequencing Center | Jeremy P. Kamil et al |
| EPI_ISL_653664 | USA/LA-EVTL973/2020 | 2020-08-18 | North America/USA/Louisiana/Bossier P | LSUHS Emerging Viral Threat Laboratory | Microbial Genome Sequencing Center | Jeremy P. Kamil et al |
| EPI_ISL_653665 | USA/LA-EVTL974/2020 | 2020-08-18 | North America/USA/Louisiana/Claiborne | LSUHS Emerging Viral Threat Laboratory | Microbial Genome Sequencing Center | Jeremy P. Kamil et al |
| EPI_ISL_653670 | USA/LA-EVTL979/2020 | 2020-08-18 | North America/USA/Louisiana/Red River | LSUHS Emerging Viral Threat Laboratory | Microbial Genome Sequencing Center | Jeremy P. Kamil et al |
| EPI_ISL_653678 | USA/LA-EVTL987/2020 | 2020-08-18 | North America/USA/Louisiana/Caddo Pa | LSUHS Emerging Viral Threat Laboratory | Microbial Genome Sequencing Center | Jeremy P. Kamil et al |
| EPI_ISL_653681 | USA/LA-EVTL990/2020 | 2020-08-18 | North America/USA/Louisiana/Caddo Pa | LSUHS Emerging Viral Threat Laboratory | Microbial Genome Sequencing Center | Jeremy P. Kamil et al |
| EPI_ISL_653685 | USA/LA-EVTL994/2020 | 2020-08-20 | North America/USA/Louisiana/Webster | LSUHS Emerging Viral Threat Laboratory | Microbial Genome Sequencing Center | Jeremy P. Kamil et al |
| EPI_ISL_653688 | USA/LA-EVTL997/2020 | 2020-08-20 | North America/USA/Louisiana/Claiborne | LSUHS Emerging Viral Threat Laboratory | Microbial Genome Sequencing Center | Jeremy P. Kamil et al |
| EPI_ISL_653691 | USA/LA-EVTL1000/2020 | 2020-08-20 | North America/USA/Louisiana/Claiborne | LSUHS Emerging Viral Threat Laboratory | Microbial Genome Sequencing Center | Jeremy P. Kamil et al |
| EPI_ISL_653704 | USA/LA-EVTL1013/2020 | 2020-08-25 | North America/USA/Louisiana/Caddo Pa | LSUHS Emerging Viral Threat Laboratory | Microbial Genome Sequencing Center | Jeremy P. Kamil et al |
| EPI_ISL_653709 | USA/LA-EVTL1018/2020 | 2020-08-25 | North America/USA/Louisiana/Claiborne | LSUHS Emerging Viral Threat Laboratory | Microbial Genome Sequencing Center | Jeremy P. Kamil et al |
| EPI_ISL_653712 | USA/LA-EVTL1021/2020 | 2020-08-25 | North America/USA/Louisiana/Claiborne | LSUHS Emerging Viral Threat Laboratory | Microbial Genome Sequencing Center | Jeremy P. Kamil et al |

| | | | | | | |
|---|---|---|---|---|---|---|
| EPI_ISL_653713 | USA/LA-EVTL1022/2020 | 2020-08-25 | North America/USA/Louisiana/Claiborne | LSUHS Emerging Viral Threat Laboratory | Microbial Genome Sequencing Center | Jeremy P. Kamil et al |
| EPI_ISL_653715 | USA/LA-EVTL1024/2020 | 2020-08-25 | North America/USA/Louisiana/Red River | LSUHS Emerging Viral Threat Laboratory | Microbial Genome Sequencing Center | Jeremy P. Kamil et al |
| EPI_ISL_653721 | USA/LA-EVTL1030/2020 | 2020-08-25 | North America/USA/Louisiana/Caddo Pa | LSUHS Emerging Viral Threat Laboratory | Microbial Genome Sequencing Center | Jeremy P. Kamil et al |
| EPI_ISL_653724 | USA/LA-EVTL1034/2020 | 2020-08-25 | North America/USA/Louisiana/Caddo Pa | LSUHS Emerging Viral Threat Laboratory | Microbial Genome Sequencing Center | Jeremy P. Kamil et al |
| EPI_ISL_653728 | USA/LA-EVTL1038/2020 | 2020-08-25 | North America/USA/Louisiana/Caddo Pa | LSUHS Emerging Viral Threat Laboratory | Microbial Genome Sequencing Center | Jeremy P. Kamil et al |
| EPI_ISL_653730 | USA/LA-EVTL1040/2020 | 2020-08-25 | North America/USA/Louisiana/Caddo Pa | LSUHS Emerging Viral Threat Laboratory | Microbial Genome Sequencing Center | Jeremy P. Kamil et al |
| EPI_ISL_653731 | USA/LA-EVTL1041/2020 | 2020-08-25 | North America/USA/Louisiana/Caddo Pa | LSUHS Emerging Viral Threat Laboratory | Microbial Genome Sequencing Center | Jeremy P. Kamil et al |
| EPI_ISL_653732 | USA/LA-EVTL1042/2020 | 2020-08-25 | North America/USA/Louisiana/Caddo Pa | LSUHS Emerging Viral Threat Laboratory | Microbial Genome Sequencing Center | Jeremy P. Kamil et al |
| EPI_ISL_653736 | USA/LA-EVTL1046/2020 | 2020-08-25 | North America/USA/Louisiana/Caddo Pa | LSUHS Emerging Viral Threat Laboratory | Microbial Genome Sequencing Center | Jeremy P. Kamil et al |
| EPI_ISL_653740 | USA/LA-EVTL1050/2020 | 2020-08-27 | North America/USA/Louisiana/Webster | LSUHS Emerging Viral Threat Laboratory | Microbial Genome Sequencing Center | Jeremy P. Kamil et al |
| EPI_ISL_653743 | USA/LA-EVTL1053/2020 | 2020-08-28 | North America/USA/Louisiana/Caddo Pa | LSUHS Emerging Viral Threat Laboratory | Microbial Genome Sequencing Center | Jeremy P. Kamil et al |
| EPI_ISL_653745 | Colombia/DC-INS-2553/2020 | 2020-08-06 | South America/Colombia/Bogota/ | Instituto Nacional de Salud, Bogot√°, Colombia | Instituto Nacional de Salud, Bogot√°, Colombia | Katherine Laiton-Donato et al |
| EPI_ISL_653746 | Colombia/DC-INS-2624/2020 | 2020-08-07 | South America/Colombia/Bogota/ | Instituto Nacional de Salud, Bogot√°, Colombia | Instituto Nacional de Salud, Bogot√°, Colombia | Katherine Laiton-Donato et al |

| | | | | | | |
|---|---|---|---|---|---|---|
| EPI_ISL_653747 | Colombia/DC-INS-2640/2020 | 2020-08-07 | South America/Colombia/Bogota/ | Instituto Nacional de Salud, Bogotv°, Colombia | Instituto Nacional de Salud, Bogotv°, Colombia | Katherine Laiton-Donato et al |
| EPI_ISL_653748 | Colombia/ANT-INS-02/2020 | 2020-06-04 | South America/Colombia/Antioquia/Me | Instituto Nacional de Salud, Bogotv°, Colombia | Instituto Nacional de Salud, Bogotv°, Colombia | Katherine Laiton-Donato et al |
| EPI_ISL_653754 | Colombia/DC-INS-3334/2020 | 2020-09-02 | South America/Colombia/Bogota/ | Instituto Nacional de Salud, Bogotv°, Colombia | Instituto Nacional de Salud, Bogotv°, Colombia | Katherine Laiton-Donato et al |
| EPI_ISL_653755 | Colombia/DC-INS-3569/2020 | 2020-06-30 | South America/Colombia/Bogota/ | Instituto Nacional de Salud, Bogotv°, Colombia | Instituto Nacional de Salud, Bogotv°, Colombia | Katherine Laiton-Donato et al |
| EPI_ISL_653756 | Colombia/ANT-INS-01/2020 | 2020-07-07 | South America/Colombia/Antioquia/Me | Instituto Nacional de Salud, Bogotv°, Colombia | Instituto Nacional de Salud, Bogotv°, Colombia | Katherine Laiton-Donato et al |
| EPI_ISL_653757 | Colombia/ATL-INS-06/2020 | 2020-06-26 | South America/Colombia/Atlantico/Barr | Instituto Nacional de Salud, Bogotv°, Colombia | Instituto Nacional de Salud, Bogotv°, Colombia | Katherine Laiton-Donato et al |
| EPI_ISL_653758 | Colombia/ANT-INS-5/2020 | 2020-08-26 | South America/Colombia/Antioquia/Me | Instituto Nacional de Salud, Bogotv°, Colombia | Instituto Nacional de Salud, Bogotv°, Colombia | Katherine Laiton-Donato et al |
| EPI_ISL_653759 | Colombia/ATL-INS-04/2020 | 2020-06-06 | South America/Colombia/Atlantico/Barr | Instituto Nacional de Salud, Bogotv°, Colombia | Instituto Nacional de Salud, Bogotv°, Colombia | Katherine Laiton-Donato et al |
| EPI_ISL_653762 | Colombia/ATL-INS-10/2020 | 2020-08-20 | South America/Colombia/Atlantico/Barr | Instituto Nacional de Salud, Bogotv°, Colombia | Instituto Nacional de Salud, Bogotv°, Colombia | Katherine Laiton-Donato et al |
| EPI_ISL_653765 | Italy/APU-IZSPB_269PT/2020 | 2020-11-02 | Europe/Italy/Apulia/Bari | "I.R.C.C.S. ""S. De Bellis"" - Ente Ospedaliero" | Istituto Zooprofilattico Sperimentale della Puglia e della Basilicata | Parisi A. et al |
| EPI_ISL_653766 | Italy/APU-IZSPB_271PT/2020 | 2020-11-02 | Europe/Italy/Apulia/Brindisi | "I.R.C.C.S. ""S. De Bellis"" - Ente Ospedaliero" | Istituto Zooprofilattico Sperimentale della Puglia e della Basilicata | Parisi A. et al |

| | | | | | | |
|---|---|---|---|---|---|---|
| EPI_ISL_653767 | Italy/APU-IZSPB_272PT/2020 | 2020-11-02 | Europe/Italy/Apulia/Bari | "I.R.C.C.S. ""S. De Bellis"" - Ente Ospedaliero" | Istituto Zooprofilattico Sperimentale della Puglia e della Basilicata | Parisi A. et al |
| EPI_ISL_653768 | Italy/APU-IZSPB_274PT/2020 | 2020-11-02 | Europe/Italy/Apulia/Brindisi | "I.R.C.C.S. ""S. De Bellis"" - Ente Ospedaliero" | Istituto Zooprofilattico Sperimentale della Puglia e della Basilicata | Parisi A. et al |
| EPI_ISL_653769 | Italy/APU-IZSPB_275PT/2020 | 2020-11-02 | Europe/Italy/Apulia/Brindisi | "I.R.C.C.S. ""S. De Bellis"" - Ente Ospedaliero" | Istituto Zooprofilattico Sperimentale della Puglia e della Basilicata | Parisi A. et al |
| EPI_ISL_653770 | Italy/APU-IZSPB_276PT/2020 | 2020-11-02 | Europe/Italy/Apulia/Brindisi | "I.R.C.C.S. ""S. De Bellis"" - Ente Ospedaliero" | Istituto Zooprofilattico Sperimentale della Puglia e della Basilicata | Parisi A. et al |
| EPI_ISL_653771 | Italy/APU-IZSPB_277PT/2020 | 2020-11-02 | Europe/Italy/Apulia/Brindisi | "I.R.C.C.S. ""S. De Bellis"" - Ente Ospedaliero" | Istituto Zooprofilattico Sperimentale della Puglia e della Basilicata | Parisi A. et al |
| EPI_ISL_653772 | Italy/APU-IZSPB_278PT/2020 | 2020-11-02 | Europe/Italy/Apulia/Brindisi | "I.R.C.C.S. ""S. De Bellis"" - Ente Ospedaliero" | Istituto Zooprofilattico Sperimentale della Puglia e della Basilicata | Parisi A. et al |
| EPI_ISL_653773 | Italy/APU-IZSPB_280PT/2020 | 2020-11-02 | Europe/Italy/Apulia/Brindisi | "I.R.C.C.S. ""S. De Bellis"" - Ente Ospedaliero" | Istituto Zooprofilattico Sperimentale della Puglia e della Basilicata | Parisi A. et al |
| EPI_ISL_653774 | Italy/APU-IZSPB_281PT/2020 | 2020-11-02 | Europe/Italy/Apulia/Brindisi | "I.R.C.C.S. ""S. De Bellis"" - Ente Ospedaliero" | Istituto Zooprofilattico Sperimentale della Puglia e della Basilicata | Parisi A. et al |

| | | | | | | |
|---|---|---|---|---|---|---|
| EPI_ISL_653775 | Italy/APU-IZSPB_282PT/2020 | 2020-11-02 | Europe/Italy/Apulia/Brindisi | "I.R.C.C.S. ""S. De Bellis"" - Ente Ospedaliero" | Istituto Zooprofilattico Sperimentale della Puglia e della Basilicata | Parisi A. et al |
| EPI_ISL_653776 | Italy/APU-IZSPB_284PT/2020 | 2020-11-02 | Europe/Italy/Apulia/Brindisi | "I.R.C.C.S. ""S. De Bellis"" - Ente Ospedaliero" | Istituto Zooprofilattico Sperimentale della Puglia e della Basilicata | Parisi A. et al |
| EPI_ISL_653777 | Italy/APU-IZSPB_285PT/2020 | 2020-11-02 | Europe/Italy/Apulia/Brindisi | "I.R.C.C.S. ""S. De Bellis"" - Ente Ospedaliero" | Istituto Zooprofilattico Sperimentale della Puglia e della Basilicata | Parisi A. et al |
| EPI_ISL_653778 | Italy/APU-IZSPB_286PT/2020 | 2020-11-02 | Europe/Italy/Apulia/Brindisi | "I.R.C.C.S. ""S. De Bellis"" - Ente Ospedaliero" | Istituto Zooprofilattico Sperimentale della Puglia e della Basilicata | Parisi A. et al |
| EPI_ISL_653781 | Italy/APU-IZSPB_290PT/2020 | 2020-11-02 | Europe/Italy/Apulia/Brindisi | "I.R.C.C.S. ""S. De Bellis"" - Ente Ospedaliero" | Istituto Zooprofilattico Sperimentale della Puglia e della Basilicata | Parisi A. et al |
| EPI_ISL_653782 | Italy/APU-IZSPB_291PT/2020 | 2020-10-30 | Europe/Italy/Apulia/Brindisi | "I.R.C.C.S. ""S. De Bellis"" - Ente Ospedaliero" | Istituto Zooprofilattico Sperimentale della Puglia e della Basilicata | Parisi A. et al |
| EPI_ISL_653783 | Italy/APU-IZSPB_292PT/2020 | 2020-11-02 | Europe/Italy/Apulia/Brindisi | "I.R.C.C.S. ""S. De Bellis"" - Ente Ospedaliero" | Istituto Zooprofilattico Sperimentale della Puglia e della Basilicata | Parisi A. et al |
| EPI_ISL_653786 | Italy/APU-IZSPB_267PT/2020 | 2020-09-30 | Europe/Italy/Apulia/Bari | Istituto Zooprofilattico Sperimentale della Puglia e della Basilicata | Istituto Zooprofilattico Sperimentale della Puglia e della Basilicata | Parisi A. et al |

| | | | | | | |
|---|---|---|---|---|---|---|
| EPI_ISL_653787 | Italy/APU-IZSPB_293PT/2020 | 2020-11-03 | Europe/Italy/Apulia/Taranto | "I.R.C.C.S. ""S. De Bellis"" - Ente Ospedaliero" | Istituto Zooprofilattico Sperimentale della Puglia e della Basilicata | Parisi A. et al |
| EPI_ISL_653788 | Italy/APU-IZSPB_294PT/2020 | 2020-11-03 | Europe/Italy/Apulia/Bari | "I.R.C.C.S. ""S. De Bellis"" - Ente Ospedaliero" | Istituto Zooprofilattico Sperimentale della Puglia e della Basilicata | Parisi A. et al |
| EPI_ISL_653791 | Italy/APU-IZSPB_297PT/2020 | 2020-11-03 | Europe/Italy/Apulia/Bari | "I.R.C.C.S. ""S. De Bellis"" - Ente Ospedaliero" | Istituto Zooprofilattico Sperimentale della Puglia e della Basilicata | Parisi A. et al |
| EPI_ISL_653793 | Italy/APU-IZSPB_299PT/2020 | 2020-11-03 | Europe/Italy/Apulia/Bari | "I.R.C.C.S. ""S. De Bellis"" - Ente Ospedaliero" | Istituto Zooprofilattico Sperimentale della Puglia e della Basilicata | Parisi A. et al |
| EPI_ISL_653794 | Italy/APU-IZSPB_300PT/2020 | 2020-11-03 | Europe/Italy/Apulia/Bari | "I.R.C.C.S. ""S. De Bellis"" - Ente Ospedaliero" | Istituto Zooprofilattico Sperimentale della Puglia e della Basilicata | Parisi A. et al |
| EPI_ISL_653795 | Italy/APU-IZSPB_301PT/2020 | 2020-11-03 | Europe/Italy/Apulia/Bari | "I.R.C.C.S. ""S. De Bellis"" - Ente Ospedaliero" | Istituto Zooprofilattico Sperimentale della Puglia e della Basilicata | Parisi A. et al |
| EPI_ISL_653797 | Italy/APU-IZSPB_303PT/2020 | 2020-11-03 | Europe/Italy/Apulia/Bari | "I.R.C.C.S. ""S. De Bellis"" - Ente Ospedaliero" | Istituto Zooprofilattico Sperimentale della Puglia e della Basilicata | Parisi A. et al |
| EPI_ISL_653798 | Italy/APU-IZSPB_304PT/2020 | 2020-11-03 | Europe/Italy/Apulia/Bari | "I.R.C.C.S. ""S. De Bellis"" - Ente Ospedaliero" | Istituto Zooprofilattico Sperimentale della Puglia e della Basilicata | Parisi A. et al |

| | | | | | | |
|---|---|---|---|---|---|---|
| EPI_ISL_653799 | Italy/APU-IZSPB_305PT/2020 | 2020-11-03 | Europe/Italy/Apulia/Bari | "I.R.C.C.S. ""S. De Bellis"" - Ente Ospedaliero" | Istituto Zooprofilattico Sperimentale della Puglia e della Basilicata | Parisi A. et al |
| EPI_ISL_653800 | Italy/APU-IZSPB_306PT/2020 | 2020-11-03 | Europe/Italy/Apulia/Bari | "I.R.C.C.S. ""S. De Bellis"" - Ente Ospedaliero" | Istituto Zooprofilattico Sperimentale della Puglia e della Basilicata | Parisi A. et al |
| EPI_ISL_653801 | Italy/APU-IZSPB_307PT/2020 | 2020-11-02 | Europe/Italy/Apulia/Brindisi | "I.R.C.C.S. ""S. De Bellis"" - Ente Ospedaliero" | Istituto Zooprofilattico Sperimentale della Puglia e della Basilicata | Parisi A. et al |
| EPI_ISL_653803 | Italy/APU-IZSPB_309PT/2020 | 2020-10-24 | Europe/Italy/Apulia/Bari | "I.R.C.C.S. ""S. De Bellis"" - Ente Ospedaliero" | Istituto Zooprofilattico Sperimentale della Puglia e della Basilicata | Parisi A. et al |
| EPI_ISL_653804 | Italy/APU-IZSPB_310PT/2020 | 2020-11-03 | Europe/Italy/Apulia/Bari | "I.R.C.C.S. ""S. De Bellis"" - Ente Ospedaliero" | Istituto Zooprofilattico Sperimentale della Puglia e della Basilicata | Parisi A. et al |
| EPI_ISL_653805 | Italy/APU-IZSPB_311PT/2020 | 2020-11-03 | Europe/Italy/Apulia/Bari | "I.R.C.C.S. ""S. De Bellis"" - Ente Ospedaliero" | Istituto Zooprofilattico Sperimentale della Puglia e della Basilicata | Parisi A. et al |
| EPI_ISL_653806 | Italy/APU-IZSPB_312PT/2020 | 2020-11-03 | Europe/Italy/Apulia/Bari | "I.R.C.C.S. ""S. De Bellis"" - Ente Ospedaliero" | Istituto Zooprofilattico Sperimentale della Puglia e della Basilicata | Parisi A. et al |
| EPI_ISL_653807 | Italy/APU-IZSPB_313PT/2020 | 2020-11-02 | Europe/Italy/Apulia/Brindisi | "I.R.C.C.S. ""S. De Bellis"" - Ente Ospedaliero" | Istituto Zooprofilattico Sperimentale della Puglia e della Basilicata | Parisi A. et al |

| | | | | | | |
|---|---|---|---|---|---|---|
| EPI_ISL_653808 | Italy/APU-IZSPB_314PT/2020 | 2020-11-02 | Europe/Italy/Apulia/Brindisi | "I.R.C.C.S. ""S. De Bellis"" - Ente Ospedaliero" | Istituto Zooprofilattico Sperimentale della Puglia e della Basilicata | Parisi A. et al |
| EPI_ISL_653809 | Italy/APU-IZSPB_315PT/2020 | 2020-11-02 | Europe/Italy/Apulia/Brindisi | "I.R.C.C.S. ""S. De Bellis"" - Ente Ospedaliero" | Istituto Zooprofilattico Sperimentale della Puglia e della Basilicata | Parisi A. et al |
| EPI_ISL_653812 | Italy/APU-IZSPB_318PT/2020 | 2020-11-02 | Europe/Italy/Apulia/Brindisi | "I.R.C.C.S. ""S. De Bellis"" - Ente Ospedaliero" | Istituto Zooprofilattico Sperimentale della Puglia e della Basilicata | Parisi A. et al |
| EPI_ISL_653817 | Italy/APU-IZSPB_324PT/2020 | 2020-04-11 | Europe/Italy/Apulia/Brindisi | "I.R.C.C.S. ""S. De Bellis"" - Ente Ospedaliero" | Istituto Zooprofilattico Sperimentale della Puglia e della Basilicata | Parisi A. et al |
| EPI_ISL_653818 | Italy/APU-IZSPB_325PT/2020 | 2020-04-11 | Europe/Italy/Apulia/Brindisi | "I.R.C.C.S. ""S. De Bellis"" - Ente Ospedaliero" | Istituto Zooprofilattico Sperimentale della Puglia e della Basilicata | Parisi A. et al |
| EPI_ISL_653819 | Italy/APU-IZSPB_326PT/2020 | 2020-11-04 | Europe/Italy/Apulia/Bari | "I.R.C.C.S. ""S. De Bellis"" - Ente Ospedaliero" | Istituto Zooprofilattico Sperimentale della Puglia e della Basilicata | Parisi A. et al |
| EPI_ISL_653820 | Italy/APU-IZSPB_327PT/2020 | 2020-11-06 | Europe/Italy/Apulia/Bari | "I.R.C.C.S. ""S. De Bellis"" - Ente Ospedaliero" | Istituto Zooprofilattico Sperimentale della Puglia e della Basilicata | Parisi A. et al |
| EPI_ISL_653821 | Italy/APU-IZSPB_328PT/2020 | 2020-10-27 | Europe/Italy/Apulia/Bari | "I.R.C.C.S. ""S. De Bellis"" - Ente Ospedaliero" | Istituto Zooprofilattico Sperimentale della Puglia e della Basilicata | Parisi A. et al |

| | | | | | | |
|---|---|---|---|---|---|---|
| EPI_ISL_653823 | Italy/APU-IZSPB_171PT/2020 | 2020-08-17 | Europe/Italy/Apulia/Foggia | "I.R.C.C.S. ""S. De Bellis"" - Ente Ospedaliero" | Istituto Zooprofilattico Sperimentale della Puglia e della Basilicata | Parisi A. et al |
| EPI_ISL_653830 | India/WB-NIBG-IK94/2020 | 2020-05-24 | Asia/India/West Bengal/Kolkata | Institute of Post Graduate Medical Education & Research | National Institute of Biomedical Genomics | Arindam Maitra et al |
| EPI_ISL_653836 | India/DL-NIBG-MN7/2020 | 2020-06-18 | Asia/India/Delhi/ | Maulana Azad Medical College | National Institute of Biomedical Genomics | Arindam Maitra et al |
| EPI_ISL_653838 | India/DL-NIBG-MN12/2020 | 2020-06-18 | Asia/India/Delhi/ | Maulana Azad Medical College | National Institute of Biomedical Genomics | Arindam Maitra et al |
| EPI_ISL_653839 | India/DL-NIBG-MN13/2020 | 2020-06-18 | Asia/India/Delhi/ | Maulana Azad Medical College | National Institute of Biomedical Genomics | Arindam Maitra et al |
| EPI_ISL_653840 | India/DL-NIBG-MN14/2020 | 2020-06-18 | Asia/India/Delhi/ | Maulana Azad Medical College | National Institute of Biomedical Genomics | Arindam Maitra et al |
| EPI_ISL_653842 | India/DL-NIBG-MN16/2020 | 2020-06-18 | Asia/India/Delhi/ | Maulana Azad Medical College | National Institute of Biomedical Genomics | Arindam Maitra et al |
| EPI_ISL_653845 | India/DL-NIBG-MN19/2020 | 2020-06-19 | Asia/India/Delhi/ | Maulana Azad Medical College | National Institute of Biomedical Genomics | Arindam Maitra et al |
| EPI_ISL_653846 | India/DL-NIBG-MN20/2020 | 2020-06-19 | Asia/India/Delhi/ | Maulana Azad Medical College | National Institute of Biomedical Genomics | Arindam Maitra et al |
| EPI_ISL_653848 | India/DL-NIBG-MN22/2020 | 2020-06-19 | Asia/India/Delhi/ | Maulana Azad Medical College | National Institute of Biomedical Genomics | Arindam Maitra et al |
| EPI_ISL_653849 | India/DL-NIBG-MN23/2020 | 2020-06-20 | Asia/India/Delhi/ | Maulana Azad Medical College | National Institute of Biomedical Genomics | Arindam Maitra et al |
| EPI_ISL_653852 | India/DL-NIBG-MN27/2020 | 2020-06-21 | Asia/India/Delhi/ | Maulana Azad Medical College | National Institute of Biomedical Genomics | Arindam Maitra et al |
| EPI_ISL_653853 | India/DL-NIBG-MN28/2020 | 2020-06-21 | Asia/India/Delhi/ | Maulana Azad Medical College | National Institute of Biomedical Genomics | Arindam Maitra et al |

| | | | | | | |
|---|---|---|---|---|---|---|
| EPI_ISL_653854 | India/DL-NIBG-MN30/2020 | 2020-06-23 | Asia/India/Delhi/ | Maulana Azad Medical College | National Institute of Biomedical Genomics | Arindam Maitra et al |
| EPI_ISL_653855 | India/DL-NIBG-MN31/2020 | 2020-06-26 | Asia/India/Delhi/ | Maulana Azad Medical College | National Institute of Biomedical Genomics | Arindam Maitra et al |
| EPI_ISL_653856 | India/DL-NIBG-MN33/2020 | 2020-06-26 | Asia/India/Delhi/ | Maulana Azad Medical College | National Institute of Biomedical Genomics | Arindam Maitra et al |
| EPI_ISL_653859 | India/HR-NIBG-TF5/2020 | 2020-06-14 | Asia/India/Haryana/Palwal | Translational Health Science and Technology Institute | National Institute of Biomedical Genomics | Arindam Maitra et al |
| EPI_ISL_653863 | India/HR-NIBG-TF29/2020 | 2020-06-22 | Asia/India/Haryana/Faridabad | Translational Health Science and Technology Institute | National Institute of Biomedical Genomics | Arindam Maitra et al |
| EPI_ISL_653864 | India/HR-NIBG-TF31/2020 | 2020-06-22 | Asia/India/Haryana/Faridabad | Translational Health Science and Technology Institute | National Institute of Biomedical Genomics | Arindam Maitra et al |
| EPI_ISL_653865 | India/HR-NIBG-TF32/2020 | 2020-06-22 | Asia/India/Haryana/Palwal | Translational Health Science and Technology Institute | National Institute of Biomedical Genomics | Arindam Maitra et al |
| EPI_ISL_653867 | India/HR-NIBG-TF35/2020 | 2020-06-26 | Asia/India/Haryana/Palwal | Translational Health Science and Technology Institute | National Institute of Biomedical Genomics | Arindam Maitra et al |
| EPI_ISL_653868 | India/HR-NIBG-TF36/2020 | 2020-06-26 | Asia/India/Haryana/Palwal | Translational Health Science and Technology Institute | National Institute of Biomedical Genomics | Arindam Maitra et al |
| EPI_ISL_653869 | India/HR-NIBG-TF40/2020 | 2020-05-01 | Asia/India/Haryana/Palwal | Translational Health Science and Technology Institute | National Institute of Biomedical Genomics | Arindam Maitra et al |
| EPI_ISL_653870 | India/HR-NIBG-TF42/2020 | 2020-05-12 | Asia/India/Haryana/Gurugram | Translational Health Science and Technology Institute | National Institute of Biomedical Genomics | Arindam Maitra et al |
| EPI_ISL_653878 | India/HR-NIBG-TF51/2020 | 2020-05-20 | Asia/India/Haryana/Gurugram | Translational Health Science and Technology Institute | National Institute of Biomedical Genomics | Arindam Maitra et al |
| EPI_ISL_653882 | India/HR-NIBG-TF58/2020 | 2020-05-21 | Asia/India/Haryana/Gurugram | Translational Health Science and Technology Institute | National Institute of Biomedical Genomics | Arindam Maitra et al |

| | | | | | | |
|---|---|---|---|---|---|---|
| EPI_ISL_653886 | India/HR-NIBG-TF66/2020 | 2020-05-19 | Asia/India/Haryana/Gurugram | Translational Health Science and Technology Institute | National Institute of Biomedical Genomics | Arindam Maitra et al |
| EPI_ISL_653887 | India/HR-NIBG-TF69/2020 | 2020-06-19 | Asia/India/Haryana/Faridabad | Translational Health Science and Technology Institute | National Institute of Biomedical Genomics | Arindam Maitra et al |
| EPI_ISL_653888 | India/HR-NIBG-TF70/2020 | 2020-06-19 | Asia/India/Haryana/Faridabad | Translational Health Science and Technology Institute | National Institute of Biomedical Genomics | Arindam Maitra et al |
| EPI_ISL_653891 | India/HR-NIBG-TF74/2020 | 2020-05-25 | Asia/India/Haryana/Gurugram | Translational Health Science and Technology Institute | National Institute of Biomedical Genomics | Arindam Maitra et al |
| EPI_ISL_653896 | India/HR-NIBG-TF80/2020 | 2020-05-25 | Asia/India/Haryana/Gurugram | Translational Health Science and Technology Institute | National Institute of Biomedical Genomics | Arindam Maitra et al |
| EPI_ISL_653901 | India/HR-NIBG-TF85/2020 | 2020-05-25 | Asia/India/Haryana/Gurugram | Translational Health Science and Technology Institute | National Institute of Biomedical Genomics | Arindam Maitra et al |
| EPI_ISL_653903 | India/HR-NIBG-TF87/2020 | 2020-05-14 | Asia/India/Haryana/Gurugram | Translational Health Science and Technology Institute | National Institute of Biomedical Genomics | Arindam Maitra et al |
| EPI_ISL_653904 | India/HR-NIBG-TF88/2020 | 2020-05-26 | Asia/India/Haryana/Palwal | Translational Health Science and Technology Institute | National Institute of Biomedical Genomics | Arindam Maitra et al |
| EPI_ISL_653905 | India/HR-NIBG-TF89/2020 | 2020-05-26 | Asia/India/Haryana/Palwal | Translational Health Science and Technology Institute | National Institute of Biomedical Genomics | Arindam Maitra et al |
| EPI_ISL_653909 | India/HR-NIBG-TF94/2020 | 2020-06-19 | Asia/India/Haryana/Faridabad | Translational Health Science and Technology Institute | National Institute of Biomedical Genomics | Arindam Maitra et al |
| EPI_ISL_653913 | India/HR-NIBG-TF98/2020 | 2020-06-19 | Asia/India/Haryana/Faridabad | Translational Health Science and Technology Institute | National Institute of Biomedical Genomics | Arindam Maitra et al |
| EPI_ISL_653914 | India/HR-NIBG-TF99/2020 | 2020-06-19 | Asia/India/Haryana/Faridabad | Translational Health Science and Technology Institute | National Institute of Biomedical Genomics | Arindam Maitra et al |
| EPI_ISL_653915 | India/HR-NIBG-TF100/2020 | 2020-06-19 | Asia/India/Haryana/Faridabad | Translational Health Science and Technology Institute | National Institute of Biomedical Genomics | Arindam Maitra et al |

| | | | | | | |
|---|---|---|---|---|---|---|
| EPI_ISL_653916 | Austria/Graz-MUG20/2020 | 2020-09-15 | Europe/Austria/Styria/ | Diagnostic- and Research Institute of Pathology, Medical University of Graz | Diagnostic- and Research Institute of Pathology, Medical University of Graz | Karl Kashofer et al |
| EPI_ISL_653922 | Russia/MOS-CRIE7162018/2020 | 2020-08-24 | Europe/Russia/Moscow Oblast/ | "Molecular diagnostic laboratory of Federal Budget Institution of Science ""Central Research Institute of Epidemiology"" of The Federal Service on Customers' Rights Protection and Human Well-being Surveillance" | Group of Genomics and Postgenomic Technologies of Central Research Institute of Epidemilology | Samoilov AE et al |
| EPI_ISL_653923 | Russia/MOS-CRIE7160676/2020 | 2020-08-24 | Europe/Russia/Moscow Oblast/ | "Molecular diagnostic laboratory of Federal Budget Institution of Science ""Central Research Institute of Epidemiology"" of The Federal Service on Customers' Rights Protection and Human Well-being Surveillance" | Group of Genomics and Postgenomic Technologies of Central Research Institute of Epidemilology | Samoilov AE et al |
| EPI_ISL_653926 | Russia/MOS-CRIE7169887/2020 | 2020-08-23 | Europe/Russia/Moscow Oblast/ | "Molecular diagnostic laboratory of Federal Budget Institution of Science ""Central Research Institute of Epidemiology"" of The Federal Service on Customers' Rights Protection and Human Well-being Surveillance" | Group of Genomics and Postgenomic Technologies of Central Research Institute of Epidemilology | Samoilov AE et al |
| EPI_ISL_653928 | Russia/MOS-CRIE7122594/2020 | 2020-08-24 | Europe/Russia/Moscow Oblast/ | "Molecular diagnostic laboratory of Federal Budget Institution of Science ""Central Research Institute of Epidemiology"" of The Federal Service on Customers' Rights Protection and Human Well-being Surveillance" | Group of Genomics and Postgenomic Technologies of Central Research Institute of Epidemilology | Samoilov AE et al |
| EPI_ISL_653929 | Russia/MOS-CRIE7150932/2020 | 2020-08-24 | Europe/Russia/Moscow Oblast/ | "Molecular diagnostic laboratory of Federal Budget Institution of Science ""Central Research Institute of Epidemiology"" of The Federal Service on Customers' Rights Protection and Human Well-being Surveillance" | Group of Genomics and Postgenomic Technologies of Central Research Institute of Epidemilology | Samoilov AE et al |

| | | | | | | |
|---|---|---|---|---|---|---|
| EPI_ISL_653930 | Russia/MOS-CRIE7165818/2020 | 2020-08-24 | Europe/Russia/Moscow Oblast/ | "Molecular diagnostic laboratory of Federal Budget Institution of Science ""Central Research Institute of Epidemiology"" of The Federal Service on Customers' Rights Protection and Human Well-being Surveillance" | Group of Genomics and Postgenomic Technologies of Central Research Institute of Epidemilology | Samoilov AE et al |
| EPI_ISL_653932 | Russia/MOS-CRIE7141819/2020 | 2020-08-24 | Europe/Russia/Moscow Oblast/ | "Molecular diagnostic laboratory of Federal Budget Institution of Science ""Central Research Institute of Epidemiology"" of The Federal Service on Customers' Rights Protection and Human Well-being Surveillance" | Group of Genomics and Postgenomic Technologies of Central Research Institute of Epidemilology | Samoilov AE et al |
| EPI_ISL_653949 | England/204421023/2020 | 2020-10-27 | Europe/United Kingdom/England/ | Respiratory Virus Unit, Microbiology Services Colindale, Public Health England | COVID-19 Genomics UK (COG-UK) Consortium | PHE Covid Sequencing Team et al |

| | | | | | | |
|---|---|---|---|---|---|---|
| EPI_ISL_654017 | Tunisia/55262/2020 | 2020-11-02 | Africa/Tunisia/Nabeul/ | Laboratory of Microbiology, National Reference Lab, Charles Nicolle Hospital; 2-University of Tunis ElManar, Faculty of Medicine of Tunis, LR99ES09, Tunis, Tunisia | 1-Clinical and Experimental Pharmacology Lab, LR16SP02, National Center of Pharmacovigilance, University of Tunis El Manar, Tunis, Tunisia. 2-Neurodegenerative diseases and psychiatric troubles, LR18SP03, Razi Hospital, University of Tunis El Manar, Tunis, Tunisia. 3-Ministry of Health, National Observatory of New and Emerging Diseases, 1006, Tunis, Tunisia | Ilhem Boutiba-Ben Boubaker et al |

| | | | | | | |
|---|---|---|---|---|---|---|
| EPI_ISL_654018 | Tunisia/58412/2020 | 2020-11-02 | Africa/Tunisia/Ben Arous/ | Laboratory of Microbiology, National Reference Lab, Charles Nicolle Hospital; 2-University of Tunis ElManar, Faculty of Medicine of Tunis, LR99ES09, Tunis, Tunisia | 1-Clinical and Experimental Pharmacology Lab, LR16SP02, National Center of Pharmacovigilance, University of Tunis El Manar, Tunis, Tunisia. 2-Neurodegenerative diseases and psychiatric troubles, LR18SP03, Razi Hospital, University of Tunis El Manar, Tunis, Tunisia. 3-Ministry of Health, National Observatory of New and Emerging Diseases, 1006, Tunis, Tunisia | Ilhem Boutiba-Ben Boubaker et al |

| | | | | | | |
|---|---|---|---|---|---|---|
| EPI_ISL_654020 | Tunisia/53873/2020 | 2020-10-26 | Africa/Tunisia/Ben Arous/ | Laboratory of Microbiology, National Reference Lab, Charles Nicolle Hospital; 2-University of Tunis ElManar, Faculty of Medicine of Tunis, LR99ES09, Tunis, Tunisia | 1-Clinical and Experimental Pharmacology Lab, LR16SP02, National Center of Pharmacovigilance, University of Tunis El Manar, Tunis, Tunisia. 2-Neurodegenerative diseases and psychiatric troubles, LR18SP03, Razi Hospital, University of Tunis El Manar, Tunis, Tunisia. 3-Ministry of Health, National Observatory of New and Emerging Diseases, 1006, Tunis, Tunisia | Ilhem Boutiba-Ben Boubaker et al |
| EPI_ISL_654021 | Spain/MD-IBV-99008488/2020 | 2020-09-10 | Europe/Spain/Madrid/Madrid | Hospital General Universitario Gregorio Marav√±√≥n | SeqCOVID-SPAIN consortium/IBV(CSIC) | Dar√≠o Garc√≠a de Viedma et al |
| EPI_ISL_654022 | Spain/MD-IBV-99008512/2020 | 2020-09-10 | Europe/Spain/Madrid/Madrid | Hospital General Universitario Gregorio Marav√±√≥n | SeqCOVID-SPAIN consortium/IBV(CSIC) | Dar√≠o Garc√≠a de Viedma et al |
| EPI_ISL_654023 | Spain/MD-IBV-99008515/2020 | 2020-09-10 | Europe/Spain/Madrid/Madrid | Hospital General Universitario Gregorio Marav√±√≥n | SeqCOVID-SPAIN consortium/IBV(CSIC) | Dar√≠o Garc√≠a de Viedma et al |
| EPI_ISL_654024 | Spain/MD-IBV-99008520/2020 | 2020-09-10 | Europe/Spain/Madrid/Madrid | Hospital General Universitario Gregorio Marav√±√≥n | SeqCOVID-SPAIN consortium/IBV(CSIC) | Dar√≠o Garc√≠a de Viedma et al |
| EPI_ISL_654025 | Spain/MD-IBV-99008513/2020 | 2020-09-10 | Europe/Spain/Madrid/Madrid | Hospital General Universitario Gregorio Marav√±√≥n | SeqCOVID-SPAIN consortium/IBV(CSIC) | Dar√≠o Garc√≠a de Viedma et al |

| | | | | | | |
|---|---|---|---|---|---|---|
| EPI_ISL_654027 | Spain/MD-IBV-99008531/2020 | 2020-09-10 | Europe/Spain/Madrid/Madrid | Hospital General Universitario Gregorio Marav±V≥n | SeqCOVID-SPAIN consortium/IBV(CSIC) | DarV≠o GarcV≠a de Viedma et al |
| EPI_ISL_654029 | Spain/MD-IBV-99008491/2020 | 2020-09-10 | Europe/Spain/Madrid/Madrid | Hospital General Universitario Gregorio Marav±V≥n | SeqCOVID-SPAIN consortium/IBV(CSIC) | DarV≠o GarcV≠a de Viedma et al |
| EPI_ISL_654033 | Spain/MD-IBV-99008487/2020 | 2020-09-10 | Europe/Spain/Madrid/Madrid | Hospital General Universitario Gregorio Marav±V≥n | SeqCOVID-SPAIN consortium/IBV(CSIC) | DarV≠o GarcV≠a de Viedma et al |
| EPI_ISL_654034 | Spain/MD-IBV-99008516/2020 | 2020-09-10 | Europe/Spain/Madrid/Madrid | Hospital General Universitario Gregorio Marav±V≥n | SeqCOVID-SPAIN consortium/IBV(CSIC) | DarV≠o GarcV≠a de Viedma et al |
| EPI_ISL_654035 | Spain/MD-IBV-99008482/2020 | 2020-08-11 | Europe/Spain/Madrid/Madrid | Hospital General Universitario Gregorio Marav±V≥n | SeqCOVID-SPAIN consortium/IBV(CSIC) | DarV≠o GarcV≠a de Viedma et al |
| EPI_ISL_654036 | Spain/MD-IBV-99008518/2020 | 2020-09-10 | Europe/Spain/Madrid/Madrid | Hospital General Universitario Gregorio Marav±V≥n | SeqCOVID-SPAIN consortium/IBV(CSIC) | DarV≠o GarcV≠a de Viedma et al |
| EPI_ISL_654037 | Spain/MD-IBV-99008486/2020 | 2020-09-10 | Europe/Spain/Madrid/Madrid | Hospital General Universitario Gregorio Marav±V≥n | SeqCOVID-SPAIN consortium/IBV(CSIC) | DarV≠o GarcV≠a de Viedma et al |
| EPI_ISL_654041 | Spain/MD-IBV-99008530/2020 | 2020-09-10 | Europe/Spain/Madrid/Madrid | Hospital General Universitario Gregorio Marav±V≥n | SeqCOVID-SPAIN consortium/IBV(CSIC) | DarV≠o GarcV≠a de Viedma et al |
| EPI_ISL_654044 | Spain/MD-IBV-99008508/2020 | 2020-09-10 | Europe/Spain/Madrid/Madrid | Hospital General Universitario Gregorio Marav±V≥n | SeqCOVID-SPAIN consortium/IBV(CSIC) | DarV≠o GarcV≠a de Viedma et al |
| EPI_ISL_654045 | Spain/MD-IBV-99008505/2020 | 2020-09-10 | Europe/Spain/Madrid/Madrid | Hospital General Universitario Gregorio Marav±V≥n | SeqCOVID-SPAIN consortium/IBV(CSIC) | DarV≠o GarcV≠a de Viedma et al |
| EPI_ISL_654046 | Spain/MD-IBV-99008523/2020 | 2020-09-10 | Europe/Spain/Madrid/Madrid | Hospital General Universitario Gregorio Marav±V≥n | SeqCOVID-SPAIN consortium/IBV(CSIC) | DarV≠o GarcV≠a de Viedma et al |
| EPI_ISL_654048 | Spain/MD-IBV-99008481/2020 | 2020-08-11 | Europe/Spain/Madrid/Madrid | Hospital General Universitario Gregorio Marav±V≥n | SeqCOVID-SPAIN consortium/IBV(CSIC) | DarV≠o GarcV≠a de Viedma et al |
| EPI_ISL_654057 | Spain/MD-IBV-99008473/2020 | 2020-08-08 | Europe/Spain/Madrid/Madrid | Hospital General Universitario Gregorio Marav±V≥n | SeqCOVID-SPAIN consortium/IBV(CSIC) | DarV≠o GarcV≠a de Viedma et al |

| | | | | | | |
|---|---|---|---|---|---|---|
| EPI_ISL_654059 | Spain/MD-IBV-99008476/2020 | 2020-08-09 | Europe/Spain/Madrid/Madrid | Hospital General Universitario Gregorio Marav√±√≥n | SeqCOVID-SPAIN consortium/IBV(CSIC) | Dar√≠o Garc√≠a de Viedma et al |
| EPI_ISL_654060 | Spain/MD-IBV-99008477/2020 | 2020-08-09 | Europe/Spain/Madrid/Madrid | Hospital General Universitario Gregorio Marav√±√≥n | SeqCOVID-SPAIN consortium/IBV(CSIC) | Dar√≠o Garc√≠a de Viedma et al |
| EPI_ISL_654061 | Spain/MD-IBV-99008479/2020 | 2020-08-10 | Europe/Spain/Madrid/Madrid | Hospital General Universitario Gregorio Marav√±√≥n | SeqCOVID-SPAIN consortium/IBV(CSIC) | Dar√≠o Garc√≠a de Viedma et al |
| EPI_ISL_654064 | Spain/MD-IBV-99008490/2020 | 2020-09-10 | Europe/Spain/Madrid/Madrid | Hospital General Universitario Gregorio Marav√±√≥n | SeqCOVID-SPAIN consortium/IBV(CSIC) | Dar√≠o Garc√≠a de Viedma et al |
| EPI_ISL_654065 | Spain/MD-IBV-99008494/2020 | 2020-09-10 | Europe/Spain/Madrid/Madrid | Hospital General Universitario Gregorio Marav√±√≥n | SeqCOVID-SPAIN consortium/IBV(CSIC) | Dar√≠o Garc√≠a de Viedma et al |
| EPI_ISL_654066 | Spain/MD-IBV-99008495/2020 | 2020-09-10 | Europe/Spain/Madrid/Madrid | Hospital General Universitario Gregorio Marav√±√≥n | SeqCOVID-SPAIN consortium/IBV(CSIC) | Dar√≠o Garc√≠a de Viedma et al |
| EPI_ISL_654067 | Spain/MD-IBV-99008497/2020 | 2020-09-10 | Europe/Spain/Madrid/Madrid | Hospital General Universitario Gregorio Marav√±√≥n | SeqCOVID-SPAIN consortium/IBV(CSIC) | Dar√≠o Garc√≠a de Viedma et al |
| EPI_ISL_654069 | Spain/MD-IBV-99008500/2020 | 2020-09-10 | Europe/Spain/Madrid/Madrid | Hospital General Universitario Gregorio Marav√±√≥n | SeqCOVID-SPAIN consortium/IBV(CSIC) | Dar√≠o Garc√≠a de Viedma et al |
| EPI_ISL_654072 | Spain/MD-IBV-99008507/2020 | 2020-09-10 | Europe/Spain/Madrid/Madrid | Hospital General Universitario Gregorio Marav√±√≥n | SeqCOVID-SPAIN consortium/IBV(CSIC) | Dar√≠o Garc√≠a de Viedma et al |
| EPI_ISL_654073 | Spain/MD-IBV-99008510/2020 | 2020-09-10 | Europe/Spain/Madrid/Madrid | Hospital General Universitario Gregorio Marav√±√≥n | SeqCOVID-SPAIN consortium/IBV(CSIC) | Dar√≠o Garc√≠a de Viedma et al |
| EPI_ISL_654075 | Spain/MD-IBV-99008517/2020 | 2020-09-10 | Europe/Spain/Madrid/Madrid | Hospital General Universitario Gregorio Marav√±√≥n | SeqCOVID-SPAIN consortium/IBV(CSIC) | Dar√≠o Garc√≠a de Viedma et al |
| EPI_ISL_654080 | Spain/MD-IBV-99008532/2020 | 2020-09-10 | Europe/Spain/Madrid/Madrid | Hospital General Universitario Gregorio Marav√±√≥n | SeqCOVID-SPAIN consortium/IBV(CSIC) | Dar√≠o Garc√≠a de Viedma et al |
| EPI_ISL_654081 | Spain/MD-IBV-99008533/2020 | 2020-09-21 | Europe/Spain/Madrid/Madrid | Hospital General Universitario Gregorio Marav√±√≥n | SeqCOVID-SPAIN consortium/IBV(CSIC) | Dar√≠o Garc√≠a de Viedma et al |

| | | | | | | |
|---|---|---|---|---|---|---|
| EPI_ISL_654086 | Spain/MD-IBV-99008540/2020 | 2020-09-05 | Europe/Spain/Madrid/Madrid | Hospital General Universitario Gregorio Marav√±√≥n | SeqCOVID-SPAIN consortium/IBV(CSIC) | Dar√≠o Garc√≠a de Viedma et al |
| EPI_ISL_654088 | Spain/MD-IBV-99008542/2020 | 2020-09-21 | Europe/Spain/Madrid/Madrid | Hospital General Universitario Gregorio Marav√±√≥n | SeqCOVID-SPAIN consortium/IBV(CSIC) | Dar√≠o Garc√≠a de Viedma et al |
| EPI_ISL_654089 | Spain/MD-IBV-99008543/2020 | 2020-09-08 | Europe/Spain/Madrid/Madrid | Hospital General Universitario Gregorio Marav√±√≥n | SeqCOVID-SPAIN consortium/IBV(CSIC) | Dar√≠o Garc√≠a de Viedma et al |
| EPI_ISL_654090 | Spain/MD-IBV-99008545/2020 | 2020-09-09 | Europe/Spain/Madrid/Madrid | Hospital General Universitario Gregorio Marav√±√≥n | SeqCOVID-SPAIN consortium/IBV(CSIC) | Dar√≠o Garc√≠a de Viedma et al |
| EPI_ISL_654091 | Spain/MD-IBV-99008546/2020 | 2020-09-10 | Europe/Spain/Madrid/Madrid | Hospital General Universitario Gregorio Marav√±√≥n | SeqCOVID-SPAIN consortium/IBV(CSIC) | Dar√≠o Garc√≠a de Viedma et al |
| EPI_ISL_654092 | Spain/MD-IBV-99008550/2020 | 2020-09-10 | Europe/Spain/Madrid/Madrid | Hospital General Universitario Gregorio Marav√±√≥n | SeqCOVID-SPAIN consortium/IBV(CSIC) | Dar√≠o Garc√≠a de Viedma et al |
| EPI_ISL_654093 | Spain/MD-IBV-99008552/2020 | 2020-09-13 | Europe/Spain/Madrid/Madrid | Hospital General Universitario Gregorio Marav√±√≥n | SeqCOVID-SPAIN consortium/IBV(CSIC) | Dar√≠o Garc√≠a de Viedma et al |
| EPI_ISL_654094 | Spain/MD-IBV-99008553/2020 | 2020-09-15 | Europe/Spain/Madrid/Madrid | Hospital General Universitario Gregorio Marav√±√≥n | SeqCOVID-SPAIN consortium/IBV(CSIC) | Dar√≠o Garc√≠a de Viedma et al |
| EPI_ISL_654095 | Spain/MD-IBV-99008554/2020 | 2020-09-17 | Europe/Spain/Madrid/Madrid | Hospital General Universitario Gregorio Marav√±√≥n | SeqCOVID-SPAIN consortium/IBV(CSIC) | Dar√≠o Garc√≠a de Viedma et al |
| EPI_ISL_654097 | Spain/MD-IBV-99008556/2020 | 2020-09-19 | Europe/Spain/Madrid/Madrid | Hospital General Universitario Gregorio Marav√±√≥n | SeqCOVID-SPAIN consortium/IBV(CSIC) | Dar√≠o Garc√≠a de Viedma et al |
| EPI_ISL_654101 | Spain/MD-IBV-99008561/2020 | 2020-09-21 | Europe/Spain/Madrid/Madrid | Hospital General Universitario Gregorio Marav√±√≥n | SeqCOVID-SPAIN consortium/IBV(CSIC) | Dar√≠o Garc√≠a de Viedma et al |
| EPI_ISL_654103 | Spain/MD-IBV-99008563/2020 | 2020-09-21 | Europe/Spain/Madrid/Madrid | Hospital General Universitario Gregorio Marav√±√≥n | SeqCOVID-SPAIN consortium/IBV(CSIC) | Dar√≠o Garc√≠a de Viedma et al |
| EPI_ISL_654104 | Spain/MD-IBV-99008564/2020 | 2020-09-21 | Europe/Spain/Madrid/Madrid | Hospital General Universitario Gregorio Marav√±√≥n | SeqCOVID-SPAIN consortium/IBV(CSIC) | Dar√≠o Garc√≠a de Viedma et al |

| | | | | | | |
|---|---|---|---|---|---|---|
| EPI_ISL_654105 | Spain/MD-IBV-99008565/2020 | 2020-09-21 | Europe/Spain/Madrid/Madrid | Hospital General Universitario Gregorio Marav√±√≥n | SeqCOVID-SPAIN consortium/IBV(CSIC) | Dar√≠o Garc√≠a de Viedma et al |
| EPI_ISL_654106 | Spain/MD-IBV-99008566/2020 | 2020-09-21 | Europe/Spain/Madrid/Madrid | Hospital General Universitario Gregorio Marav√±√≥n | SeqCOVID-SPAIN consortium/IBV(CSIC) | Dar√≠o Garc√≠a de Viedma et al |
| EPI_ISL_654107 | Spain/MD-IBV-99008567/2020 | 2020-09-21 | Europe/Spain/Madrid/Madrid | Hospital General Universitario Gregorio Marav√±√≥n | SeqCOVID-SPAIN consortium/IBV(CSIC) | Dar√≠o Garc√≠a de Viedma et al |
| EPI_ISL_654108 | Spain/MD-IBV-99008568/2020 | 2020-09-21 | Europe/Spain/Madrid/Madrid | Hospital General Universitario Gregorio Marav√±√≥n | SeqCOVID-SPAIN consortium/IBV(CSIC) | Dar√≠o Garc√≠a de Viedma et al |
| EPI_ISL_654109 | Spain/MD-IBV-99008569/2020 | 2020-09-21 | Europe/Spain/Madrid/Madrid | Hospital General Universitario Gregorio Marav√±√≥n | SeqCOVID-SPAIN consortium/IBV(CSIC) | Dar√≠o Garc√≠a de Viedma et al |
| EPI_ISL_654110 | Spain/MD-IBV-99008570/2020 | 2020-09-21 | Europe/Spain/Madrid/Madrid | Hospital General Universitario Gregorio Marav√±√≥n | SeqCOVID-SPAIN consortium/IBV(CSIC) | Dar√≠o Garc√≠a de Viedma et al |
| EPI_ISL_654111 | Spain/MD-IBV-99008571/2020 | 2020-09-21 | Europe/Spain/Madrid/Madrid | Hospital General Universitario Gregorio Marav√±√≥n | SeqCOVID-SPAIN consortium/IBV(CSIC) | Dar√≠o Garc√≠a de Viedma et al |
| EPI_ISL_654117 | Spain/MD-IBV-99008577/2020 | 2020-09-21 | Europe/Spain/Madrid/Madrid | Hospital General Universitario Gregorio Marav√±√≥n | SeqCOVID-SPAIN consortium/IBV(CSIC) | Dar√≠o Garc√≠a de Viedma et al |
| EPI_ISL_654118 | Spain/MD-IBV-99008578/2020 | 2020-09-21 | Europe/Spain/Madrid/Madrid | Hospital General Universitario Gregorio Marav√±√≥n | SeqCOVID-SPAIN consortium/IBV(CSIC) | Dar√≠o Garc√≠a de Viedma et al |
| EPI_ISL_654119 | Spain/MD-IBV-99008579/2020 | 2020-09-21 | Europe/Spain/Madrid/Madrid | Hospital General Universitario Gregorio Marav√±√≥n | SeqCOVID-SPAIN consortium/IBV(CSIC) | Dar√≠o Garc√≠a de Viedma et al |
| EPI_ISL_654120 | Spain/MD-IBV-99008580/2020 | 2020-09-21 | Europe/Spain/Madrid/Madrid | Hospital General Universitario Gregorio Marav√±√≥n | SeqCOVID-SPAIN consortium/IBV(CSIC) | Dar√≠o Garc√≠a de Viedma et al |
| EPI_ISL_654121 | Spain/MD-IBV-99008581/2020 | 2020-09-21 | Europe/Spain/Madrid/Madrid | Hospital General Universitario Gregorio Marav√±√≥n | SeqCOVID-SPAIN consortium/IBV(CSIC) | Dar√≠o Garc√≠a de Viedma et al |
| EPI_ISL_654122 | Spain/MD-IBV-99008582/2020 | 2020-09-21 | Europe/Spain/Madrid/Madrid | Hospital General Universitario Gregorio Marav√±√≥n | SeqCOVID-SPAIN consortium/IBV(CSIC) | Dar√≠o Garc√≠a de Viedma et al |

| | | | | | | |
|---|---|---|---|---|---|---|
| EPI_ISL_654123 | Spain/MD-IBV-99008583/2020 | 2020-09-21 | Europe/Spain/Madrid/Madrid | Hospital General Universitario Gregorio Marav±√≥n | SeqCOVID-SPAIN consortium/IBV(CSIC) | Dar√≠o Garc√≠a de Viedma et al |
| EPI_ISL_654124 | Spain/MD-IBV-99008584/2020 | 2020-09-21 | Europe/Spain/Madrid/Madrid | Hospital General Universitario Gregorio Marav±√≥n | SeqCOVID-SPAIN consortium/IBV(CSIC) | Dar√≠o Garc√≠a de Viedma et al |
| EPI_ISL_654125 | Spain/MD-IBV-99008585/2020 | 2020-09-21 | Europe/Spain/Madrid/Madrid | Hospital General Universitario Gregorio Marav±√≥n | SeqCOVID-SPAIN consortium/IBV(CSIC) | Dar√≠o Garc√≠a de Viedma et al |
| EPI_ISL_654126 | Spain/MD-IBV-99008586/2020 | 2020-09-21 | Europe/Spain/Madrid/Madrid | Hospital General Universitario Gregorio Marav±√≥n | SeqCOVID-SPAIN consortium/IBV(CSIC) | Dar√≠o Garc√≠a de Viedma et al |
| EPI_ISL_654128 | Spain/MD-IBV-99008588/2020 | 2020-09-21 | Europe/Spain/Madrid/Madrid | Hospital General Universitario Gregorio Marav±√≥n | SeqCOVID-SPAIN consortium/IBV(CSIC) | Dar√≠o Garc√≠a de Viedma et al |
| EPI_ISL_654135 | Spain/MD-IBV-99008595/2020 | 2020-09-21 | Europe/Spain/Madrid/Madrid | Hospital General Universitario Gregorio Marav±√≥n | SeqCOVID-SPAIN consortium/IBV(CSIC) | Dar√≠o Garc√≠a de Viedma et al |
| EPI_ISL_654137 | Spain/MD-IBV-99008597/2020 | 2020-09-21 | Europe/Spain/Madrid/Madrid | Hospital General Universitario Gregorio Marav±√≥n | SeqCOVID-SPAIN consortium/IBV(CSIC) | Dar√≠o Garc√≠a de Viedma et al |
| EPI_ISL_654138 | Spain/MD-IBV-99008599/2020 | 2020-09-21 | Europe/Spain/Madrid/Madrid | Hospital General Universitario Gregorio Marav±√≥n | SeqCOVID-SPAIN consortium/IBV(CSIC) | Dar√≠o Garc√≠a de Viedma et al |
| EPI_ISL_654139 | Spain/MD-IBV-99008600/2020 | 2020-09-21 | Europe/Spain/Madrid/Madrid | Hospital General Universitario Gregorio Marav±√≥n | SeqCOVID-SPAIN consortium/IBV(CSIC) | Dar√≠o Garc√≠a de Viedma et al |
| EPI_ISL_654141 | Spain/MD-IBV-99008790/2020 | 2020-09-21 | Europe/Spain/Madrid/Madrid | Hospital General Universitario Gregorio Marav±√≥n | SeqCOVID-SPAIN consortium/IBV(CSIC) | Dar√≠o Garc√≠a de Viedma et al |
| EPI_ISL_654143 | Spain/MD-IBV-99008792/2020 | 2020-09-21 | Europe/Spain/Madrid/Madrid | Hospital General Universitario Gregorio Marav±√≥n | SeqCOVID-SPAIN consortium/IBV(CSIC) | Dar√≠o Garc√≠a de Viedma et al |
| EPI_ISL_654147 | Spain/MD-IBV-99008796/2020 | 2020-09-21 | Europe/Spain/Madrid/Madrid | Hospital General Universitario Gregorio Marav±√≥n | SeqCOVID-SPAIN consortium/IBV(CSIC) | Dar√≠o Garc√≠a de Viedma et al |
| EPI_ISL_654148 | Spain/MD-IBV-99008797/2020 | 2020-09-21 | Europe/Spain/Madrid/Getafe | Hospital General Universitario Gregorio Marav±√≥n | SeqCOVID-SPAIN consortium/IBV(CSIC) | Dar√≠o Garc√≠a de Viedma et al |

| | | | | | | |
|---|---|---|---|---|---|---|
| EPI_ISL_654149 | Spain/MD-IBV-99008798/2020 | 2020-09-21 | Europe/Spain/Madrid/Madrid | Hospital General Universitario Gregorio Marav±V≥n | SeqCOVID-SPAIN consortium/IBV(CSIC) | DarV≠o GarcV≠a de Viedma et al |
| EPI_ISL_654150 | Spain/MD-IBV-99008799/2020 | 2020-09-21 | Europe/Spain/Madrid/Madrid | Hospital General Universitario Gregorio Marav±V≥n | SeqCOVID-SPAIN consortium/IBV(CSIC) | DarV≠o GarcV≠a de Viedma et al |
| EPI_ISL_654152 | Spain/MD-IBV-99008801/2020 | 2020-09-21 | Europe/Spain/Madrid/Madrid | Hospital General Universitario Gregorio Marav±V≥n | SeqCOVID-SPAIN consortium/IBV(CSIC) | DarV≠o GarcV≠a de Viedma et al |
| EPI_ISL_654153 | Spain/MD-IBV-99008802/2020 | 2020-09-21 | Europe/Spain/Madrid/Madrid | Hospital General Universitario Gregorio Marav±V≥n | SeqCOVID-SPAIN consortium/IBV(CSIC) | DarV≠o GarcV≠a de Viedma et al |
| EPI_ISL_654154 | Spain/MD-IBV-99008803/2020 | 2020-09-21 | Europe/Spain/Madrid/Madrid | Hospital General Universitario Gregorio Marav±V≥n | SeqCOVID-SPAIN consortium/IBV(CSIC) | DarV≠o GarcV≠a de Viedma et al |
| EPI_ISL_654155 | Spain/MD-IBV-99008804/2020 | 2020-09-21 | Europe/Spain/Madrid/Madrid | Hospital General Universitario Gregorio Marav±V≥n | SeqCOVID-SPAIN consortium/IBV(CSIC) | DarV≠o GarcV≠a de Viedma et al |
| EPI_ISL_654156 | Spain/MD-IBV-99008805/2020 | 2020-09-21 | Europe/Spain/Madrid/Madrid | Hospital General Universitario Gregorio Marav±V≥n | SeqCOVID-SPAIN consortium/IBV(CSIC) | DarV≠o GarcV≠a de Viedma et al |
| EPI_ISL_654157 | Spain/MD-IBV-99008806/2020 | 2020-09-21 | Europe/Spain/Madrid/Madrid | Hospital General Universitario Gregorio Marav±V≥n | SeqCOVID-SPAIN consortium/IBV(CSIC) | DarV≠o GarcV≠a de Viedma et al |
| EPI_ISL_654159 | Spain/MD-IBV-99008809/2020 | 2020-09-21 | Europe/Spain/Madrid/Madrid | Hospital General Universitario Gregorio Marav±V≥n | SeqCOVID-SPAIN consortium/IBV(CSIC) | DarV≠o GarcV≠a de Viedma et al |
| EPI_ISL_654163 | Spain/MD-IBV-99008634/2020 | 2020-09-29 | Europe/Spain/Madrid/Madrid | Hospital General Universitario Gregorio Marav±V≥n | SeqCOVID-SPAIN consortium/IBV(CSIC) | DarV≠o GarcV≠a de Viedma et al |
| EPI_ISL_654164 | Spain/MD-IBV-99008672/2020 | 2020-10-01 | Europe/Spain/Madrid/Madrid | Hospital General Universitario Gregorio Marav±V≥n | SeqCOVID-SPAIN consortium/IBV(CSIC) | DarV≠o GarcV≠a de Viedma et al |
| EPI_ISL_654165 | Spain/MD-IBV-99008640/2020 | 2020-09-29 | Europe/Spain/Madrid/Madrid | Hospital General Universitario Gregorio Marav±V≥n | SeqCOVID-SPAIN consortium/IBV(CSIC) | DarV≠o GarcV≠a de Viedma et al |
| EPI_ISL_654167 | Spain/MD-IBV-99008626/2020 | 2020-09-18 | Europe/Spain/Madrid/Madrid | Hospital General Universitario Gregorio Marav±V≥n | SeqCOVID-SPAIN consortium/IBV(CSIC) | DarV≠o GarcV≠a de Viedma et al |

| | | | | | | |
|---|---|---|---|---|---|---|
| EPI_ISL_654170 | Spain/MD-IBV-99008619/2020 | 2020-09-06 | Europe/Spain/Madrid/Madrid | Hospital General Universitario Gregorio Marav√±√≥n | SeqCOVID-SPAIN consortium/IBV(CSIC) | Dar√≠o Garc√≠a de Viedma et al |
| EPI_ISL_654171 | Spain/MD-IBV-99008639/2020 | 2020-09-29 | Europe/Spain/Madrid/Madrid | Hospital General Universitario Gregorio Marav√±√≥n | SeqCOVID-SPAIN consortium/IBV(CSIC) | Dar√≠o Garc√≠a de Viedma et al |
| EPI_ISL_654174 | Spain/MD-IBV-99008673/2020 | 2020-10-01 | Europe/Spain/Madrid/Madrid | Hospital General Universitario Gregorio Marav√±√≥n | SeqCOVID-SPAIN consortium/IBV(CSIC) | Dar√≠o Garc√≠a de Viedma et al |
| EPI_ISL_654175 | Spain/MD-IBV-99008641/2020 | 2020-09-29 | Europe/Spain/Madrid/Madrid | Hospital General Universitario Gregorio Marav√±√≥n | SeqCOVID-SPAIN consortium/IBV(CSIC) | Dar√≠o Garc√≠a de Viedma et al |
| EPI_ISL_654176 | Spain/MD-IBV-99008631/2020 | 2020-09-25 | Europe/Spain/Madrid/Madrid | Hospital General Universitario Gregorio Marav√±√≥n | SeqCOVID-SPAIN consortium/IBV(CSIC) | Dar√≠o Garc√≠a de Viedma et al |
| EPI_ISL_654179 | Spain/MD-IBV-99008695/2020 | 2020-10-09 | Europe/Spain/Madrid/Madrid | Hospital General Universitario Gregorio Marav√±√≥n | SeqCOVID-SPAIN consortium/IBV(CSIC) | Dar√≠o Garc√≠a de Viedma et al |
| EPI_ISL_654180 | Spain/MD-IBV-99008659/2020 | 2020-09-29 | Europe/Spain/Madrid/Madrid | Hospital General Universitario Gregorio Marav√±√≥n | SeqCOVID-SPAIN consortium/IBV(CSIC) | Dar√≠o Garc√≠a de Viedma et al |
| EPI_ISL_654183 | Spain/MD-IBV-99008646/2020 | 2020-09-29 | Europe/Spain/Madrid/Madrid | Hospital General Universitario Gregorio Marav√±√≥n | SeqCOVID-SPAIN consortium/IBV(CSIC) | Dar√≠o Garc√≠a de Viedma et al |
| EPI_ISL_654184 | Spain/MD-IBV-99008687/2020 | 2020-10-07 | Europe/Spain/Madrid/Madrid | Hospital General Universitario Gregorio Marav√±√≥n | SeqCOVID-SPAIN consortium/IBV(CSIC) | Dar√≠o Garc√≠a de Viedma et al |
| EPI_ISL_654185 | Spain/MD-IBV-99008650/2020 | 2020-09-29 | Europe/Spain/Madrid/Madrid | Hospital General Universitario Gregorio Marav√±√≥n | SeqCOVID-SPAIN consortium/IBV(CSIC) | Dar√≠o Garc√≠a de Viedma et al |
| EPI_ISL_654193 | Spain/MD-IBV-99008614/2020 | 2020-08-11 | Europe/Spain/Madrid/Madrid | Hospital General Universitario Gregorio Marav√±√≥n | SeqCOVID-SPAIN consortium/IBV(CSIC) | Dar√≠o Garc√≠a de Viedma et al |
| EPI_ISL_654194 | Spain/MD-IBV-99008615/2020 | 2020-09-28 | Europe/Spain/Madrid/Madrid | Hospital General Universitario Gregorio Marav√±√≥n | SeqCOVID-SPAIN consortium/IBV(CSIC) | Dar√≠o Garc√≠a de Viedma et al |
| EPI_ISL_654195 | Spain/MD-IBV-99008617/2020 | 2020-08-17 | Europe/Spain/Madrid/Madrid | Hospital General Universitario Gregorio Marav√±√≥n | SeqCOVID-SPAIN consortium/IBV(CSIC) | Dar√≠o Garc√≠a de Viedma et al |

| | | | | | | |
|---|---|---|---|---|---|---|
| EPI_ISL_654197 | Spain/MD-IBV-99008621/2020 | 2020-09-14 | Europe/Spain/Madrid/Madrid | Hospital General Universitario Gregorio Marav√±√≥n | SeqCOVID-SPAIN consortium/IBV(CSIC) | Dar√≠o Garc√≠a de Viedma et al |
| EPI_ISL_654198 | Spain/MD-IBV-99008622/2020 | 2020-09-13 | Europe/Spain/Madrid/Madrid | Hospital General Universitario Gregorio Marav√±√≥n | SeqCOVID-SPAIN consortium/IBV(CSIC) | Dar√≠o Garc√≠a de Viedma et al |
| EPI_ISL_654200 | Spain/MD-IBV-99008625/2020 | 2020-09-17 | Europe/Spain/Madrid/Madrid | Hospital General Universitario Gregorio Marav√±√≥n | SeqCOVID-SPAIN consortium/IBV(CSIC) | Dar√≠o Garc√≠a de Viedma et al |
| EPI_ISL_654201 | Spain/MD-IBV-99008627/2020 | 2020-10-11 | Europe/Spain/Madrid/Madrid | Hospital General Universitario Gregorio Marav√±√≥n | SeqCOVID-SPAIN consortium/IBV(CSIC) | Dar√≠o Garc√≠a de Viedma et al |
| EPI_ISL_654202 | Spain/MD-IBV-99008628/2020 | 2020-09-22 | Europe/Spain/Madrid/Madrid | Hospital General Universitario Gregorio Marav√±√≥n | SeqCOVID-SPAIN consortium/IBV(CSIC) | Dar√≠o Garc√≠a de Viedma et al |
| EPI_ISL_654203 | Spain/MD-IBV-99008629/2020 | 2020-09-23 | Europe/Spain/Madrid/Madrid | Hospital General Universitario Gregorio Marav√±√≥n | SeqCOVID-SPAIN consortium/IBV(CSIC) | Dar√≠o Garc√≠a de Viedma et al |
| EPI_ISL_654204 | Spain/MD-IBV-99008630/2020 | 2020-09-24 | Europe/Spain/Madrid/Madrid | Hospital General Universitario Gregorio Marav√±√≥n | SeqCOVID-SPAIN consortium/IBV(CSIC) | Dar√≠o Garc√≠a de Viedma et al |
| EPI_ISL_654205 | Spain/MD-IBV-99008633/2020 | 2020-09-29 | Europe/Spain/Madrid/Madrid | Hospital General Universitario Gregorio Marav√±√≥n | SeqCOVID-SPAIN consortium/IBV(CSIC) | Dar√≠o Garc√≠a de Viedma et al |
| EPI_ISL_654206 | Spain/MD-IBV-99008635/2020 | 2020-09-29 | Europe/Spain/Madrid/Madrid | Hospital General Universitario Gregorio Marav√±√≥n | SeqCOVID-SPAIN consortium/IBV(CSIC) | Dar√≠o Garc√≠a de Viedma et al |
| EPI_ISL_654208 | Spain/MD-IBV-99008637/2020 | 2020-09-29 | Europe/Spain/Madrid/Madrid | Hospital General Universitario Gregorio Marav√±√≥n | SeqCOVID-SPAIN consortium/IBV(CSIC) | Dar√≠o Garc√≠a de Viedma et al |
| EPI_ISL_654209 | Spain/MD-IBV-99008638/2020 | 2020-09-29 | Europe/Spain/Madrid/Madrid | Hospital General Universitario Gregorio Marav√±√≥n | SeqCOVID-SPAIN consortium/IBV(CSIC) | Dar√≠o Garc√≠a de Viedma et al |
| EPI_ISL_654212 | Spain/MD-IBV-99008644/2020 | 2020-09-29 | Europe/Spain/Madrid/Madrid | Hospital General Universitario Gregorio Marav√±√≥n | SeqCOVID-SPAIN consortium/IBV(CSIC) | Dar√≠o Garc√≠a de Viedma et al |
| EPI_ISL_654213 | Spain/MD-IBV-99008647/2020 | 2020-09-29 | Europe/Spain/Madrid/Madrid | Hospital General Universitario Gregorio Marav√±√≥n | SeqCOVID-SPAIN consortium/IBV(CSIC) | Dar√≠o Garc√≠a de Viedma et al |

| | | | | | | |
|---|---|---|---|---|---|---|
| EPI_ISL_654215 | Spain/MD-IBV-99008651/2020 | 2020-09-29 | Europe/Spain/Madrid/Madrid | Hospital General Universitario Gregorio Marav√±√≥n | SeqCOVID-SPAIN consortium/IBV(CSIC) | Dar√≠o Garc√≠a de Viedma et al |
| EPI_ISL_654218 | Spain/MD-IBV-99008656/2020 | 2020-09-29 | Europe/Spain/Madrid/Madrid | Hospital General Universitario Gregorio Marav√±√≥n | SeqCOVID-SPAIN consortium/IBV(CSIC) | Dar√≠o Garc√≠a de Viedma et al |
| EPI_ISL_654219 | Spain/MD-IBV-99008657/2020 | 2020-09-29 | Europe/Spain/Madrid/Madrid | Hospital General Universitario Gregorio Marav√±√≥n | SeqCOVID-SPAIN consortium/IBV(CSIC) | Dar√≠o Garc√≠a de Viedma et al |
| EPI_ISL_654220 | Spain/MD-IBV-99008658/2020 | 2020-09-29 | Europe/Spain/Madrid/Madrid | Hospital General Universitario Gregorio Marav√±√≥n | SeqCOVID-SPAIN consortium/IBV(CSIC) | Dar√≠o Garc√≠a de Viedma et al |
| EPI_ISL_654221 | Spain/MD-IBV-99008660/2020 | 2020-09-29 | Europe/Spain/Madrid/Madrid | Hospital General Universitario Gregorio Marav√±√≥n | SeqCOVID-SPAIN consortium/IBV(CSIC) | Dar√≠o Garc√≠a de Viedma et al |
| EPI_ISL_654222 | Spain/MD-IBV-99008661/2020 | 2020-09-29 | Europe/Spain/Madrid/Madrid | Hospital General Universitario Gregorio Marav√±√≥n | SeqCOVID-SPAIN consortium/IBV(CSIC) | Dar√≠o Garc√≠a de Viedma et al |
| EPI_ISL_654223 | Spain/MD-IBV-99008662/2020 | 2020-09-29 | Europe/Spain/Madrid/Madrid | Hospital General Universitario Gregorio Marav√±√≥n | SeqCOVID-SPAIN consortium/IBV(CSIC) | Dar√≠o Garc√≠a de Viedma et al |
| EPI_ISL_654224 | Spain/MD-IBV-99008663/2020 | 2020-09-30 | Europe/Spain/Madrid/Madrid | Hospital General Universitario Gregorio Marav√±√≥n | SeqCOVID-SPAIN consortium/IBV(CSIC) | Dar√≠o Garc√≠a de Viedma et al |
| EPI_ISL_654225 | Spain/MD-IBV-99008664/2020 | 2020-09-30 | Europe/Spain/Madrid/Madrid | Hospital General Universitario Gregorio Marav√±√≥n | SeqCOVID-SPAIN consortium/IBV(CSIC) | Dar√≠o Garc√≠a de Viedma et al |
| EPI_ISL_654226 | Spain/MD-IBV-99008665/2020 | 2020-09-30 | Europe/Spain/Madrid/Madrid | Hospital General Universitario Gregorio Marav√±√≥n | SeqCOVID-SPAIN consortium/IBV(CSIC) | Dar√≠o Garc√≠a de Viedma et al |
| EPI_ISL_654230 | Spain/MD-IBV-99008671/2020 | 2020-10-01 | Europe/Spain/Madrid/Madrid | Hospital General Universitario Gregorio Marav√±√≥n | SeqCOVID-SPAIN consortium/IBV(CSIC) | Dar√≠o Garc√≠a de Viedma et al |
| EPI_ISL_654232 | Spain/MD-IBV-99008677/2020 | 2020-10-05 | Europe/Spain/Madrid/Getafe | Hospital General Universitario Gregorio Marav√±√≥n | SeqCOVID-SPAIN consortium/IBV(CSIC) | Dar√≠o Garc√≠a de Viedma et al |
| EPI_ISL_654237 | Spain/MD-IBV-99008682/2020 | 2020-10-06 | Europe/Spain/Madrid/Madrid | Hospital General Universitario Gregorio Marav√±√≥n | SeqCOVID-SPAIN consortium/IBV(CSIC) | Dar√≠o Garc√≠a de Viedma et al |

| | | | | | | |
|---|---|---|---|---|---|---|
| EPI_ISL_654238 | Spain/MD-IBV-99008683/2020 | 2020-10-06 | Europe/Spain/Madrid/Madrid | Hospital General Universitario Gregorio Marav√±√≥n | SeqCOVID-SPAIN consortium/IBV(CSIC) | Dar√≠o Garc√≠a de Viedma et al |
| EPI_ISL_654241 | Spain/MD-IBV-99008686/2020 | 2020-10-07 | Europe/Spain/Madrid/Madrid | Hospital General Universitario Gregorio Marav√±√≥n | SeqCOVID-SPAIN consortium/IBV(CSIC) | Dar√≠o Garc√≠a de Viedma et al |
| EPI_ISL_654242 | Spain/MD-IBV-99008688/2020 | 2020-10-07 | Europe/Spain/Madrid/Madrid | Hospital General Universitario Gregorio Marav√±√≥n | SeqCOVID-SPAIN consortium/IBV(CSIC) | Dar√≠o Garc√≠a de Viedma et al |
| EPI_ISL_654243 | Spain/MD-IBV-99008689/2020 | 2020-10-07 | Europe/Spain/Madrid/Madrid | Hospital General Universitario Gregorio Marav√±√≥n | SeqCOVID-SPAIN consortium/IBV(CSIC) | Dar√≠o Garc√≠a de Viedma et al |
| EPI_ISL_654244 | Spain/MD-IBV-99008690/2020 | 2020-10-07 | Europe/Spain/Madrid/Madrid | Hospital General Universitario Gregorio Marav√±√≥n | SeqCOVID-SPAIN consortium/IBV(CSIC) | Dar√≠o Garc√≠a de Viedma et al |
| EPI_ISL_654245 | Spain/MD-IBV-99008691/2020 | 2020-10-08 | Europe/Spain/Madrid/Madrid | Hospital General Universitario Gregorio Marav√±√≥n | SeqCOVID-SPAIN consortium/IBV(CSIC) | Dar√≠o Garc√≠a de Viedma et al |
| EPI_ISL_654247 | Spain/MD-IBV-99008694/2020 | 2020-10-08 | Europe/Spain/Madrid/Madrid | Hospital General Universitario Gregorio Marav√±√≥n | SeqCOVID-SPAIN consortium/IBV(CSIC) | Dar√≠o Garc√≠a de Viedma et al |
| EPI_ISL_654263 | Spain/MD-IBV-99008737/2020 | 2020-10-16 | Europe/Spain/Madrid/Madrid | Hospital General Universitario Gregorio Marav√±√≥n | SeqCOVID-SPAIN consortium/IBV(CSIC) | Dar√≠o Garc√≠a de Viedma et al |
| EPI_ISL_654266 | Spain/MD-IBV-99008740/2020 | 2020-10-16 | Europe/Spain/Madrid/Madrid | Hospital General Universitario Gregorio Marav√±√≥n | SeqCOVID-SPAIN consortium/IBV(CSIC) | Dar√≠o Garc√≠a de Viedma et al |
| EPI_ISL_654267 | Spain/MD-IBV-99008764/2020 | 2020-10-20 | Europe/Spain/Madrid/Madrid | Hospital General Universitario Gregorio Marav√±√≥n | SeqCOVID-SPAIN consortium/IBV(CSIC) | Dar√≠o Garc√≠a de Viedma et al |
| EPI_ISL_654268 | Spain/MD-IBV-99008733/2020 | 2020-10-16 | Europe/Spain/Madrid/Madrid | Hospital General Universitario Gregorio Marav√±√≥n | SeqCOVID-SPAIN consortium/IBV(CSIC) | Dar√≠o Garc√≠a de Viedma et al |
| EPI_ISL_654270 | Spain/MD-IBV-99008759/2020 | 2020-10-20 | Europe/Spain/Madrid/Madrid | Hospital General Universitario Gregorio Marav√±√≥n | SeqCOVID-SPAIN consortium/IBV(CSIC) | Dar√≠o Garc√≠a de Viedma et al |
| EPI_ISL_654275 | Spain/MD-IBV-99008756/2020 | 2020-10-20 | Europe/Spain/Madrid/Madrid | Hospital General Universitario Gregorio Marav√±√≥n | SeqCOVID-SPAIN consortium/IBV(CSIC) | Dar√≠o Garc√≠a de Viedma et al |

| | | | | | | |
|---|---|---|---|---|---|---|
| EPI_ISL_654276 | Spain/MD-IBV-99008720/2020 | 2020-10-16 | Europe/Spain/Madrid/Madrid | Hospital General Universitario Gregorio Marav±V≥n | SeqCOVID-SPAIN consortium/IBV(CSIC) | Dar√≠o Garc√≠a de Viedma et al |
| EPI_ISL_654279 | Spain/MD-IBV-99008699/2020 | 2020-08-13 | Europe/Spain/Madrid/Madrid | Hospital General Universitario Gregorio Marav±V≥n | SeqCOVID-SPAIN consortium/IBV(CSIC) | Dar√≠o Garc√≠a de Viedma et al |
| EPI_ISL_654282 | Spain/MD-IBV-99008702/2020 | 2020-09-08 | Europe/Spain/Madrid/Madrid | Hospital General Universitario Gregorio Marav±V≥n | SeqCOVID-SPAIN consortium/IBV(CSIC) | Dar√≠o Garc√≠a de Viedma et al |
| EPI_ISL_654285 | Spain/MD-IBV-99008707/2020 | 2020-10-07 | Europe/Spain/Madrid/Madrid | Hospital General Universitario Gregorio Marav±V≥n | SeqCOVID-SPAIN consortium/IBV(CSIC) | Dar√≠o Garc√≠a de Viedma et al |
| EPI_ISL_654290 | Spain/MD-IBV-99008712/2020 | 2020-10-09 | Europe/Spain/Madrid/Madrid | Hospital General Universitario Gregorio Marav±V≥n | SeqCOVID-SPAIN consortium/IBV(CSIC) | Dar√≠o Garc√≠a de Viedma et al |
| EPI_ISL_654294 | Spain/MD-IBV-99008717/2020 | 2020-10-12 | Europe/Spain/Madrid/Madrid | Hospital General Universitario Gregorio Marav±V≥n | SeqCOVID-SPAIN consortium/IBV(CSIC) | Dar√≠o Garc√≠a de Viedma et al |
| EPI_ISL_654298 | Spain/MD-IBV-99008723/2020 | 2020-10-16 | Europe/Spain/Madrid/Madrid | Hospital General Universitario Gregorio Marav±V≥n | SeqCOVID-SPAIN consortium/IBV(CSIC) | Dar√≠o Garc√≠a de Viedma et al |
| EPI_ISL_654300 | Spain/MD-IBV-99008726/2020 | 2020-10-16 | Europe/Spain/Madrid/Madrid | Hospital General Universitario Gregorio Marav±V≥n | SeqCOVID-SPAIN consortium/IBV(CSIC) | Dar√≠o Garc√≠a de Viedma et al |
| EPI_ISL_654301 | Spain/MD-IBV-99008728/2020 | 2020-10-16 | Europe/Spain/Madrid/Madrid | Hospital General Universitario Gregorio Marav±V≥n | SeqCOVID-SPAIN consortium/IBV(CSIC) | Dar√≠o Garc√≠a de Viedma et al |
| EPI_ISL_654303 | Spain/MD-IBV-99008734/2020 | 2020-10-16 | Europe/Spain/Madrid/Madrid | Hospital General Universitario Gregorio Marav±V≥n | SeqCOVID-SPAIN consortium/IBV(CSIC) | Dar√≠o Garc√≠a de Viedma et al |
| EPI_ISL_654304 | Spain/MD-IBV-99008736/2020 | 2020-10-16 | Europe/Spain/Madrid/Madrid | Hospital General Universitario Gregorio Marav±V≥n | SeqCOVID-SPAIN consortium/IBV(CSIC) | Dar√≠o Garc√≠a de Viedma et al |
| EPI_ISL_654305 | Spain/MD-IBV-99008738/2020 | 2020-10-16 | Europe/Spain/Madrid/Madrid | Hospital General Universitario Gregorio Marav±V≥n | SeqCOVID-SPAIN consortium/IBV(CSIC) | Dar√≠o Garc√≠a de Viedma et al |
| EPI_ISL_654308 | Spain/MD-IBV-99008744/2020 | 2020-10-16 | Europe/Spain/Madrid/Madrid | Hospital General Universitario Gregorio Marav±V≥n | SeqCOVID-SPAIN consortium/IBV(CSIC) | Dar√≠o Garc√≠a de Viedma et al |

| | | | | | | |
|---|---|---|---|---|---|---|
| EPI_ISL_654309 | Spain/MD-IBV-99008747/2020 | 2020-10-16 | Europe/Spain/Madrid/Madrid | Hospital General Universitario Gregorio Marav√±√≥n | SeqCOVID-SPAIN consortium/IBV(CSIC) | Dar√≠o Garc√≠a de Viedma et al |
| EPI_ISL_654312 | Spain/MD-IBV-99008754/2020 | 2020-10-20 | Europe/Spain/Madrid/Madrid | Hospital General Universitario Gregorio Marav√±√≥n | SeqCOVID-SPAIN consortium/IBV(CSIC) | Dar√≠o Garc√≠a de Viedma et al |
| EPI_ISL_654313 | Spain/MD-IBV-99008755/2020 | 2020-10-20 | Europe/Spain/Madrid/Madrid | Hospital General Universitario Gregorio Marav√±√≥n | SeqCOVID-SPAIN consortium/IBV(CSIC) | Dar√≠o Garc√≠a de Viedma et al |
| EPI_ISL_654314 | Spain/MD-IBV-99008757/2020 | 2020-10-20 | Europe/Spain/Madrid/Madrid | Hospital General Universitario Gregorio Marav√±√≥n | SeqCOVID-SPAIN consortium/IBV(CSIC) | Dar√≠o Garc√≠a de Viedma et al |
| EPI_ISL_654319 | Spain/MD-IBV-99008776/2020 | 2020-10-20 | Europe/Spain/Madrid/Madrid | Hospital General Universitario Gregorio Marav√±√≥n | SeqCOVID-SPAIN consortium/IBV(CSIC) | Dar√≠o Garc√≠a de Viedma et al |
| EPI_ISL_654324 | Spain/MD-IBV-99008781/2020 | 2020-10-20 | Europe/Spain/Madrid/Madrid | Hospital General Universitario Gregorio Marav√±√≥n | SeqCOVID-SPAIN consortium/IBV(CSIC) | Dar√≠o Garc√≠a de Viedma et al |
| EPI_ISL_654327 | Spain/MD-IBV-99008787/2020 | 2020-10-20 | Europe/Spain/Madrid/Madrid | Hospital General Universitario Gregorio Marav√±√≥n | SeqCOVID-SPAIN consortium/IBV(CSIC) | Dar√≠o Garc√≠a de Viedma et al |
| EPI_ISL_654328 | Spain/MD-IBV-99009646/2020 | 2020-10-27 | Europe/Spain/Madrid/Mostoles | Hospital General Universitario Gregorio Marav√±√≥n | SeqCOVID-SPAIN consortium/IBV(CSIC) | Dar√≠o Garc√≠a de Viedma et al |
| EPI_ISL_654330 | Spain/MD-IBV-99009616/2020 | 2020-08-07 | Europe/Spain/Madrid/Madrid | Hospital General Universitario Gregorio Marav√±√≥n | SeqCOVID-SPAIN consortium/IBV(CSIC) | Dar√≠o Garc√≠a de Viedma et al |
| EPI_ISL_654331 | Spain/MD-IBV-99009659/2020 | 2020-10-23 | Europe/Spain/Madrid/Madrid | Hospital General Universitario Gregorio Marav√±√≥n | SeqCOVID-SPAIN consortium/IBV(CSIC) | Dar√≠o Garc√≠a de Viedma et al |
| EPI_ISL_654335 | Spain/MD-IBV-99009621/2020 | 2020-08-06 | Europe/Spain/Madrid/Madrid | Hospital General Universitario Gregorio Marav√±√≥n | SeqCOVID-SPAIN consortium/IBV(CSIC) | Dar√≠o Garc√≠a de Viedma et al |
| EPI_ISL_654337 | Spain/MD-IBV-99009625/2020 | 2020-06-15 | Europe/Spain/Madrid/Madrid | Hospital General Universitario Gregorio Marav√±√≥n | SeqCOVID-SPAIN consortium/IBV(CSIC) | Dar√≠o Garc√≠a de Viedma et al |
| EPI_ISL_654339 | Spain/MD-IBV-99009630/2020 | 2020-08-08 | Europe/Spain/Madrid/Madrid | Hospital General Universitario Gregorio Marav√±√≥n | SeqCOVID-SPAIN consortium/IBV(CSIC) | Dar√≠o Garc√≠a de Viedma et al |

| | | | | | | |
|---|---|---|---|---|---|---|
| EPI_ISL_654341 | Spain/MD-IBV-99009663/2020 | 2020-10-23 | Europe/Spain/Madrid/Madrid | Hospital General Universitario Gregorio Marav√±√≥n | SeqCOVID-SPAIN consortium/IBV(CSIC) | Dar√≠o Garc√≠a de Viedma et al |
| EPI_ISL_654342 | Spain/MD-IBV-99009590/2020 | 2020-10-20 | Europe/Spain/Madrid/Madrid | Hospital General Universitario Gregorio Marav√±√≥n | SeqCOVID-SPAIN consortium/IBV(CSIC) | Dar√≠o Garc√≠a de Viedma et al |
| EPI_ISL_654344 | Spain/MD-IBV-99009592/2020 | 2020-10-27 | Europe/Spain/Madrid/Madrid | Hospital General Universitario Gregorio Marav√±√≥n | SeqCOVID-SPAIN consortium/IBV(CSIC) | Dar√≠o Garc√≠a de Viedma et al |
| EPI_ISL_654345 | Spain/MD-IBV-99009593/2020 | 2020-09-24 | Europe/Spain/Madrid/Madrid | Hospital General Universitario Gregorio Marav√±√≥n | SeqCOVID-SPAIN consortium/IBV(CSIC) | Dar√≠o Garc√≠a de Viedma et al |
| EPI_ISL_654346 | Spain/MD-IBV-99009594/2020 | 2020-08-26 | Europe/Spain/Madrid/Madrid | Hospital General Universitario Gregorio Marav√±√≥n | SeqCOVID-SPAIN consortium/IBV(CSIC) | Dar√≠o Garc√≠a de Viedma et al |
| EPI_ISL_654347 | Spain/MD-IBV-99009595/2020 | 2020-10-27 | Europe/Spain/Madrid/Madrid | Hospital General Universitario Gregorio Marav√±√≥n | SeqCOVID-SPAIN consortium/IBV(CSIC) | Dar√≠o Garc√≠a de Viedma et al |
| EPI_ISL_654348 | Spain/MD-IBV-99009596/2020 | 2020-09-16 | Europe/Spain/Madrid/Madrid | Hospital General Universitario Gregorio Marav√±√≥n | SeqCOVID-SPAIN consortium/IBV(CSIC) | Dar√≠o Garc√≠a de Viedma et al |
| EPI_ISL_654349 | Spain/MD-IBV-99009597/2020 | 2020-10-10 | Europe/Spain/Madrid/Madrid | Hospital General Universitario Gregorio Marav√±√≥n | SeqCOVID-SPAIN consortium/IBV(CSIC) | Dar√≠o Garc√≠a de Viedma et al |
| EPI_ISL_654350 | Spain/MD-IBV-99009601/2020 | 2020-10-28 | Europe/Spain/Madrid/Madrid | Hospital General Universitario Gregorio Marav√±√≥n | SeqCOVID-SPAIN consortium/IBV(CSIC) | Dar√≠o Garc√≠a de Viedma et al |
| EPI_ISL_654352 | Spain/MD-IBV-99009603/2020 | 2020-10-27 | Europe/Spain/Madrid/Madrid | Hospital General Universitario Gregorio Marav√±√≥n | SeqCOVID-SPAIN consortium/IBV(CSIC) | Dar√≠o Garc√≠a de Viedma et al |
| EPI_ISL_654353 | Spain/MD-IBV-99009604/2020 | 2020-10-14 | Europe/Spain/Madrid/Madrid | Hospital General Universitario Gregorio Marav√±√≥n | SeqCOVID-SPAIN consortium/IBV(CSIC) | Dar√≠o Garc√≠a de Viedma et al |
| EPI_ISL_654354 | Spain/MD-IBV-99009605/2020 | 2020-10-22 | Europe/Spain/Madrid/Madrid | Hospital General Universitario Gregorio Marav√±√≥n | SeqCOVID-SPAIN consortium/IBV(CSIC) | Dar√≠o Garc√≠a de Viedma et al |
| EPI_ISL_654355 | Spain/MD-IBV-99009606/2020 | 2020-10-26 | Europe/Spain/Madrid/Madrid | Hospital General Universitario Gregorio Marav√±√≥n | SeqCOVID-SPAIN consortium/IBV(CSIC) | Dar√≠o Garc√≠a de Viedma et al |

| | | | | | | |
|---|---|---|---|---|---|---|
| EPI_ISL_654358 | Spain/MD-IBV-99009609/2020 | 2020-10-16 | Europe/Spain/Madrid/Madrid | Hospital General Universitario Gregorio Marav√±√≥n | SeqCOVID-SPAIN consortium/IBV(CSIC) | Dar√≠o Garc√≠a de Viedma et al |
| EPI_ISL_654360 | Spain/MD-IBV-99009611/2020 | 2020-10-22 | Europe/Spain/Madrid/Madrid | Hospital General Universitario Gregorio Marav√±√≥n | SeqCOVID-SPAIN consortium/IBV(CSIC) | Dar√≠o Garc√≠a de Viedma et al |
| EPI_ISL_654372 | Spain/MD-IBV-99009636/2020 | 2020-10-27 | Europe/Spain/Madrid/Madrid | Hospital General Universitario Gregorio Marav√±√≥n | SeqCOVID-SPAIN consortium/IBV(CSIC) | Dar√≠o Garc√≠a de Viedma et al |
| EPI_ISL_654373 | Spain/MD-IBV-99009637/2020 | 2020-10-27 | Europe/Spain/Madrid/Madrid | Hospital General Universitario Gregorio Marav√±√≥n | SeqCOVID-SPAIN consortium/IBV(CSIC) | Dar√≠o Garc√≠a de Viedma et al |
| EPI_ISL_654374 | Spain/MD-IBV-99009638/2020 | 2020-10-27 | Europe/Spain/Madrid/Madrid | Hospital General Universitario Gregorio Marav√±√≥n | SeqCOVID-SPAIN consortium/IBV(CSIC) | Dar√≠o Garc√≠a de Viedma et al |
| EPI_ISL_654376 | Spain/MD-IBV-99009640/2020 | 2020-10-27 | Europe/Spain/Madrid/Madrid | Hospital General Universitario Gregorio Marav√±√≥n | SeqCOVID-SPAIN consortium/IBV(CSIC) | Dar√≠o Garc√≠a de Viedma et al |
| EPI_ISL_654379 | Spain/MD-IBV-99009644/2020 | 2020-10-27 | Europe/Spain/Madrid/Galapagar | Hospital General Universitario Gregorio Marav√±√≥n | SeqCOVID-SPAIN consortium/IBV(CSIC) | Dar√≠o Garc√≠a de Viedma et al |
| EPI_ISL_654383 | Spain/MD-IBV-99009649/2020 | 2020-10-27 | Europe/Spain/Madrid/Madrid | Hospital General Universitario Gregorio Marav√±√≥n | SeqCOVID-SPAIN consortium/IBV(CSIC) | Dar√≠o Garc√≠a de Viedma et al |
| EPI_ISL_654388 | Spain/MD-IBV-99009660/2020 | 2020-10-23 | Europe/Spain/Madrid/Madrid | Hospital General Universitario Gregorio Marav√±√≥n | SeqCOVID-SPAIN consortium/IBV(CSIC) | Dar√≠o Garc√≠a de Viedma et al |
| EPI_ISL_654389 | Spain/MD-IBV-99009661/2020 | 2020-10-23 | Europe/Spain/Madrid/Madrid | Hospital General Universitario Gregorio Marav√±√≥n | SeqCOVID-SPAIN consortium/IBV(CSIC) | Dar√≠o Garc√≠a de Viedma et al |
| EPI_ISL_654391 | Spain/MD-IBV-99009664/2020 | 2020-10-24 | Europe/Spain/Madrid/Madrid | Hospital General Universitario Gregorio Marav√±√≥n | SeqCOVID-SPAIN consortium/IBV(CSIC) | Dar√≠o Garc√≠a de Viedma et al |
| EPI_ISL_654395 | Spain/MD-IBV-99009670/2020 | 2020-10-22 | Europe/Spain/Madrid/Madrid | Hospital General Universitario Gregorio Marav√±√≥n | SeqCOVID-SPAIN consortium/IBV(CSIC) | Dar√≠o Garc√≠a de Viedma et al |
| EPI_ISL_654397 | Spain/MD-IBV-99009672/2020 | 2020-10-22 | Europe/Spain/Madrid/Madrid | Hospital General Universitario Gregorio Marav√±√≥n | SeqCOVID-SPAIN consortium/IBV(CSIC) | Dar√≠o Garc√≠a de Viedma et al |

| | | | | | | |
|---|---|---|---|---|---|---|
| EPI_ISL_654398 | Spain/MD-IBV-99009673/2020 | 2020-10-22 | Europe/Spain/Madrid/Madrid | Hospital General Universitario Gregorio Marav±√≥n | SeqCOVID-SPAIN consortium/IBV(CSIC) | Dar√≠o Garc√≠a de Viedma et al |
| EPI_ISL_654399 | Spain/MD-IBV-99009674/2020 | 2020-10-22 | Europe/Spain/Madrid/Madrid | Hospital General Universitario Gregorio Marav±√≥n | SeqCOVID-SPAIN consortium/IBV(CSIC) | Dar√≠o Garc√≠a de Viedma et al |
| EPI_ISL_654400 | Spain/MD-IBV-99009675/2020 | 2020-10-31 | Europe/Spain/Madrid/Madrid | Hospital General Universitario Gregorio Marav±√≥n | SeqCOVID-SPAIN consortium/IBV(CSIC) | Dar√≠o Garc√≠a de Viedma et al |
| EPI_ISL_654406 | Spain/AR-IBV-98008084/2020 | 2020-10-01 | Europe/Spain/Aragon/Zaragoza | Servicio de Microbiolog√≠a, Hospital Miguel Servet, Zaragoza | SeqCOVID-SPAIN consortium/IBV(CSIC) | Antonio Rezusta L√≥pez et al |
| EPI_ISL_654410 | Spain/AR-IBV-98008155/2020 | 2020-10-14 | Europe/Spain/Aragon/Zaragoza | Servicio de Microbiolog√≠a, Hospital Miguel Servet, Zaragoza | SeqCOVID-SPAIN consortium/IBV(CSIC) | Antonio Rezusta L√≥pez et al |
| EPI_ISL_654411 | Spain/AR-IBV-98008129/2020 | 2020-10-13 | Europe/Spain/Aragon/Zaragoza | Servicio de Microbiolog√≠a, Hospital Miguel Servet, Zaragoza | SeqCOVID-SPAIN consortium/IBV(CSIC) | Antonio Rezusta L√≥pez et al |
| EPI_ISL_654418 | Spain/AR-IBV-98008133/2020 | 2020-10-13 | Europe/Spain/Aragon/Zaragoza | Servicio de Microbiolog√≠a, Hospital Miguel Servet, Zaragoza | SeqCOVID-SPAIN consortium/IBV(CSIC) | Antonio Rezusta L√≥pez et al |
| EPI_ISL_654419 | Spain/AR-IBV-98008142/2020 | 2020-10-13 | Europe/Spain/Aragon/Zaragoza | Servicio de Microbiolog√≠a, Hospital Miguel Servet, Zaragoza | SeqCOVID-SPAIN consortium/IBV(CSIC) | Antonio Rezusta L√≥pez et al |
| EPI_ISL_654420 | Spain/AR-IBV-98008160/2020 | 2020-10-14 | Europe/Spain/Aragon/Zaragoza | Servicio de Microbiolog√≠a, Hospital Miguel Servet, Zaragoza | SeqCOVID-SPAIN consortium/IBV(CSIC) | Antonio Rezusta L√≥pez et al |
| EPI_ISL_654421 | Spain/AR-IBV-98008011/2020 | 2020-09-30 | Europe/Spain/Aragon/Zaragoza | Servicio de Microbiolog√≠a, Hospital Miguel Servet, Zaragoza | SeqCOVID-SPAIN consortium/IBV(CSIC) | Antonio Rezusta L√≥pez et al |
| EPI_ISL_654424 | Spain/AR-IBV-98008066/2020 | 2020-09-30 | Europe/Spain/Aragon/Zaragoza | Servicio de Microbiolog√≠a, Hospital Miguel Servet, Zaragoza | SeqCOVID-SPAIN consortium/IBV(CSIC) | Antonio Rezusta L√≥pez et al |
| EPI_ISL_654425 | Spain/AR-IBV-98008080/2020 | 2020-09-30 | Europe/Spain/Aragon/Zaragoza | Servicio de Microbiolog√≠a, Hospital Miguel Servet, Zaragoza | SeqCOVID-SPAIN consortium/IBV(CSIC) | Antonio Rezusta L√≥pez et al |
| EPI_ISL_654427 | Spain/AR-IBV-98008123/2020 | 2020-10-13 | Europe/Spain/Aragon/Zaragoza | Servicio de Microbiolog√≠a, Hospital Miguel Servet, Zaragoza | SeqCOVID-SPAIN consortium/IBV(CSIC) | Antonio Rezusta L√≥pez et al |

| | | | | | | |
|---|---|---|---|---|---|---|
| EPI_ISL_654429 | Spain/AR-IBV-98008136/2020 | 2020-10-13 | Europe/Spain/Aragon/Zaragoza | Servicio de Microbiologv≠a, Hospital Miguel Servet, Zaragoza | SeqCOVID-SPAIN consortium/IBV(CSIC) | Antonio Rezusta Lv≥pez et al |
| EPI_ISL_654435 | Spain/AR-IBV-98008070/2020 | 2020-09-30 | Europe/Spain/Aragon/Zaragoza | Servicio de Microbiologv≠a, Hospital Miguel Servet, Zaragoza | SeqCOVID-SPAIN consortium/IBV(CSIC) | Antonio Rezusta Lv≥pez et al |
| EPI_ISL_654443 | Spain/AR-IBV-98008041/2020 | 2020-09-29 | Europe/Spain/Aragon/Zaragoza | Servicio de Microbiologv≠a, Hospital Miguel Servet, Zaragoza | SeqCOVID-SPAIN consortium/IBV(CSIC) | Antonio Rezusta Lv≥pez et al |
| EPI_ISL_654447 | Spain/AR-IBV-98008036/2020 | 2020-09-29 | Europe/Spain/Aragon/Zaragoza | Servicio de Microbiologv≠a, Hospital Miguel Servet, Zaragoza | SeqCOVID-SPAIN consortium/IBV(CSIC) | Antonio Rezusta Lv≥pez et al |
| EPI_ISL_654448 | Spain/AR-IBV-98008055/2020 | 2020-10-01 | Europe/Spain/Aragon/Zaragoza | Servicio de Microbiologv≠a, Hospital Miguel Servet, Zaragoza | SeqCOVID-SPAIN consortium/IBV(CSIC) | Antonio Rezusta Lv≥pez et al |
| EPI_ISL_654449 | Spain/AR-IBV-98008056/2020 | 2020-10-01 | Europe/Spain/Aragon/Zaragoza | Servicio de Microbiologv≠a, Hospital Miguel Servet, Zaragoza | SeqCOVID-SPAIN consortium/IBV(CSIC) | Antonio Rezusta Lv≥pez et al |
| EPI_ISL_654450 | Spain/AR-IBV-98008059/2020 | 2020-10-01 | Europe/Spain/Aragon/Zaragoza | Servicio de Microbiologv≠a, Hospital Miguel Servet, Zaragoza | SeqCOVID-SPAIN consortium/IBV(CSIC) | Antonio Rezusta Lv≥pez et al |
| EPI_ISL_654455 | Spain/AR-IBV-98008093/2020 | 2020-10-05 | Europe/Spain/Aragon/Zaragoza | Servicio de Microbiologv≠a, Hospital Miguel Servet, Zaragoza | SeqCOVID-SPAIN consortium/IBV(CSIC) | Antonio Rezusta Lv≥pez et al |
| EPI_ISL_654456 | Spain/AR-IBV-98008094/2020 | 2020-10-05 | Europe/Spain/Aragon/Zaragoza | Servicio de Microbiologv≠a, Hospital Miguel Servet, Zaragoza | SeqCOVID-SPAIN consortium/IBV(CSIC) | Antonio Rezusta Lv≥pez et al |
| EPI_ISL_654459 | Spain/AR-IBV-98008126/2020 | 2020-10-13 | Europe/Spain/Aragon/Zaragoza | Servicio de Microbiologv≠a, Hospital Miguel Servet, Zaragoza | SeqCOVID-SPAIN consortium/IBV(CSIC) | Antonio Rezusta Lv≥pez et al |
| EPI_ISL_654462 | Spain/AR-IBV-98008134/2020 | 2020-10-13 | Europe/Spain/Aragon/Zaragoza | Servicio de Microbiologv≠a, Hospital Miguel Servet, Zaragoza | SeqCOVID-SPAIN consortium/IBV(CSIC) | Antonio Rezusta Lv≥pez et al |
| EPI_ISL_654463 | Spain/AR-IBV-98008140/2020 | 2020-10-13 | Europe/Spain/Aragon/Zaragoza | Servicio de Microbiologv≠a, Hospital Miguel Servet, Zaragoza | SeqCOVID-SPAIN consortium/IBV(CSIC) | Antonio Rezusta Lv≥pez et al |
| EPI_ISL_654476 | Spain/VC-IBV-98004337/2020 | 2020-03-24 | Europe/Spain/Comunitat Valenciana/Ca | Servicio de Microbiologv≠a. Hospital General Universitario de Castellv≥n | SeqCOVID-SPAIN consortium/IBV(CSIC) | Rosario Moreno Muv±oz et al |

| | | | | | | |
|---|---|---|---|---|---|---|
| EPI_ISL_654479 | Spain/VC-IBV-98004349/2020 | 2020-03-25 | Europe/Spain/Comunitat Valenciana/Ca | Servicio de Microbiolog√≠a. Hospital General Universitario de Castell√≥n | SeqCOVID-SPAIN consortium/IBV(CSIC) | Rosario Moreno Mu√±oz et al |
| EPI_ISL_654483 | Spain/VC-IBV-98007297/2020 | 2020-09-10 | Europe/Spain/Comunitat Valenciana/Va | Servicio de Microbiolog√≠a. Hospital Cl√≠nico Universitario de Valencia | SeqCOVID-SPAIN consortium/IBV(CSIC) | David Navarro Ortega et al |
| EPI_ISL_654484 | Spain/VC-IBV-98007298/2020 | 2020-09-10 | Europe/Spain/Comunitat Valenciana/Va | Servicio de Microbiolog√≠a. Hospital Cl√≠nico Universitario de Valencia | SeqCOVID-SPAIN consortium/IBV(CSIC) | David Navarro Ortega et al |
| EPI_ISL_654486 | Spain/VC-IBV-98007308/2020 | 2020-09-14 | Europe/Spain/Comunitat Valenciana/Va | Servicio de Microbiolog√≠a. Hospital Cl√≠nico Universitario de Valencia | SeqCOVID-SPAIN consortium/IBV(CSIC) | David Navarro Ortega et al |
| EPI_ISL_654487 | Spain/VC-IBV-98007309/2020 | 2020-09-17 | Europe/Spain/Comunitat Valenciana/Va | Servicio de Microbiolog√≠a. Hospital Cl√≠nico Universitario de Valencia | SeqCOVID-SPAIN consortium/IBV(CSIC) | David Navarro Ortega et al |
| EPI_ISL_654488 | Spain/VC-IBV-98007319/2020 | 2020-09-17 | Europe/Spain/Comunitat Valenciana/Va | Servicio de Microbiolog√≠a. Hospital Cl√≠nico Universitario de Valencia | SeqCOVID-SPAIN consortium/IBV(CSIC) | David Navarro Ortega et al |
| EPI_ISL_654490 | Spain/VC-IBV-98007321/2020 | 2020-09-20 | Europe/Spain/Comunitat Valenciana/Va | Servicio de Microbiolog√≠a. Hospital Cl√≠nico Universitario de Valencia | SeqCOVID-SPAIN consortium/IBV(CSIC) | David Navarro Ortega et al |
| EPI_ISL_654492 | Spain/VC-IBV-98007323/2020 | 2020-09-20 | Europe/Spain/Comunitat Valenciana/Va | Servicio de Microbiolog√≠a. Hospital Cl√≠nico Universitario de Valencia | SeqCOVID-SPAIN consortium/IBV(CSIC) | David Navarro Ortega et al |
| EPI_ISL_654497 | Spain/VC-IBV-98008001/2020 | 2020-10-16 | Europe/Spain/Comunitat Valenciana/Sa | Servicio de Microbiolog√≠a. Hospital Cl√≠nico Universitario de Valencia | SeqCOVID-SPAIN consortium/IBV(CSIC) | David Navarro Ortega et al |
| EPI_ISL_654501 | Sweden/20-23816/2020 | 2020-10-19 | Europe/Sweden/Sweden/ | The Public Health Agency of Sweden | The Public Health Agency of Sweden | Anna-Malin Linde et al |
| EPI_ISL_654502 | Sweden/20-53268/2020 | 2020-10-27 | Europe/Sweden/Sweden/ | The Public Health Agency of Sweden | The Public Health Agency of Sweden | Anna-Malin Linde et al |
| EPI_ISL_654507 | Sweden/20-52706/2020 | 2020-06-14 | Europe/Sweden/Jonkoping/ | The Public Health Agency of Sweden | The Public Health Agency of Sweden | Anna-Malin Linde et al |
| EPI_ISL_654510 | Spain/CT-IBV-97007957/2020 | 2020-10-01 | Europe/Spain/Catalunya/Cerdanyola de | Servicio de Microbiolog√≠a, Laboratori Cl√≠nic Metropolitana Nord. Hospital Universitari Germans Trias i Pujol. Institut d‚ÄôInvestigaci√≥ en Ci√®ncies de la Salut Germans Trias i Pujol (IGTP) | SeqCOVID-SPAIN consortium/IBV(CSIC) | Elisa Martr√≥ et al |

| | | | | | | |
|---|---|---|---|---|---|---|
| EPI_ISL_654511 | Spain/CT-IBV-97007946/2020 | 2020-10-05 | Europe/Spain/Catalunya/Navas | Servicio de Microbiolog√≠a, Laboratori Cl√≠nic Metropolitana Nord. Hospital Universitari Germans Trias i Pujol. Institut d‚ÄôInvestigaci√≥ en Ci√®ncies de la Salut Germans Trias i Pujol (IGTP) | SeqCOVID-SPAIN consortium/IBV(CSIC) | Elisa Mart√≠ et al |
| EPI_ISL_654512 | Spain/CT-IBV-97007947/2020 | 2020-09-24 | Europe/Spain/Catalunya/Barcelona | Servicio de Microbiolog√≠a, Laboratori Cl√≠nic Metropolitana Nord. Hospital Universitari Germans Trias i Pujol. Institut d‚ÄôInvestigaci√≥ en Ci√®ncies de la Salut Germans Trias i Pujol (IGTP) | SeqCOVID-SPAIN consortium/IBV(CSIC) | Elisa Mart√≠ et al |
| EPI_ISL_654520 | Spain/CT-IBV-97007979/2020 | 2020-09-22 | Europe/Spain/Catalunya/Badalona | Servicio de Microbiolog√≠a, Laboratori Cl√≠nic Metropolitana Nord. Hospital Universitari Germans Trias i Pujol. Institut d‚ÄôInvestigaci√≥ en Ci√®ncies de la Salut Germans Trias i Pujol (IGTP) | SeqCOVID-SPAIN consortium/IBV(CSIC) | Elisa Mart√≠ et al |
| EPI_ISL_654521 | Spain/CT-IBV-97007980/2020 | 2020-09-30 | Europe/Spain/Catalunya/Badalona | Servicio de Microbiolog√≠a, Laboratori Cl√≠nic Metropolitana Nord. Hospital Universitari Germans Trias i Pujol. Institut d‚ÄôInvestigaci√≥ en Ci√®ncies de la Salut Germans Trias i Pujol (IGTP) | SeqCOVID-SPAIN consortium/IBV(CSIC) | Elisa Mart√≠ et al |
| EPI_ISL_654523 | Spain/CT-IBV-97007956/2020 | 2020-09-19 | Europe/Spain/Catalunya/Barcelona | Servicio de Microbiolog√≠a, Laboratori Cl√≠nic Metropolitana Nord. Hospital Universitari Germans Trias i Pujol. Institut d‚ÄôInvestigaci√≥ en Ci√®ncies de la Salut Germans Trias i Pujol (IGTP) | SeqCOVID-SPAIN consortium/IBV(CSIC) | Elisa Mart√≠ et al |
| EPI_ISL_654525 | Spain/CT-IBV-97007964/2020 | 2020-10-07 | Europe/Spain/Catalunya/Barcelona | Servicio de Microbiolog√≠a, Laboratori Cl√≠nic Metropolitana Nord. Hospital Universitari Germans Trias i Pujol. Institut d‚ÄôInvestigaci√≥ en Ci√®ncies de la Salut Germans Trias i Pujol (IGTP) | SeqCOVID-SPAIN consortium/IBV(CSIC) | Elisa Mart√≠ et al |
| EPI_ISL_654527 | Spain/CT-IBV-97007968/2020 | 2020-10-06 | Europe/Spain/Catalunya/Mataro | Servicio de Microbiolog√≠a, Laboratori Cl√≠nic Metropolitana Nord. Hospital Universitari Germans Trias i Pujol. Institut d‚ÄôInvestigaci√≥ en Ci√®ncies de la Salut Germans Trias i Pujol (IGTP) | SeqCOVID-SPAIN consortium/IBV(CSIC) | Elisa Mart√≠ et al |
| EPI_ISL_654528 | Spain/CT-IBV-97007981/2020 | 2020-10-05 | Europe/Spain/Catalunya/Mataro | Servicio de Microbiolog√≠a, Laboratori Cl√≠nic Metropolitana Nord. Hospital Universitari Germans Trias i Pujol. Institut d‚ÄôInvestigaci√≥ en Ci√®ncies de la Salut Germans Trias i Pujol (IGTP) | SeqCOVID-SPAIN consortium/IBV(CSIC) | Elisa Mart√≠ et al |

| | | | | | | |
|---|---|---|---|---|---|---|
| EPI_ISL_654529 | Spain/CT-IBV-97007955/2020 | 2020-10-07 | Europe/Spain/Catalunya/Moncada y Rei | Servicio de Microbiolog√≠a, Laboratori Cl√≠nic Metropolitana Nord. Hospital Universitari Germans Trias i Pujol. Institut d‚ÄôInvestigaci√≥ en Ci√®ncies de la Salut Germans Trias i Pujol (IGTP) | SeqCOVID-SPAIN consortium/IBV(CSIC) | Elisa Mart√≠ et al |
| EPI_ISL_654533 | Spain/CT-IBV-97007961/2020 | 2020-10-07 | Europe/Spain/Catalunya/Santa Coloma | Servicio de Microbiolog√≠a, Laboratori Cl√≠nic Metropolitana Nord. Hospital Universitari Germans Trias i Pujol. Institut d‚ÄôInvestigaci√≥ en Ci√®ncies de la Salut Germans Trias i Pujol (IGTP) | SeqCOVID-SPAIN consortium/IBV(CSIC) | Elisa Mart√≠ et al |
| EPI_ISL_654534 | Spain/CT-IBV-97007971/2020 | 2020-10-06 | Europe/Spain/Catalunya/Santa Coloma | Servicio de Microbiolog√≠a, Laboratori Cl√≠nic Metropolitana Nord. Hospital Universitari Germans Trias i Pujol. Institut d‚ÄôInvestigaci√≥ en Ci√®ncies de la Salut Germans Trias i Pujol (IGTP) | SeqCOVID-SPAIN consortium/IBV(CSIC) | Elisa Mart√≠ et al |
| EPI_ISL_654535 | Spain/CT-IBV-97007984/2020 | 2020-04-10 | Europe/Spain/Catalunya/Santa Coloma | Servicio de Microbiolog√≠a, Laboratori Cl√≠nic Metropolitana Nord. Hospital Universitari Germans Trias i Pujol. Institut d‚ÄôInvestigaci√≥ en Ci√®ncies de la Salut Germans Trias i Pujol (IGTP) | SeqCOVID-SPAIN consortium/IBV(CSIC) | Elisa Mart√≠ et al |
| EPI_ISL_654536 | Spain/CT-IBV-97007989/2020 | 2020-10-10 | Europe/Spain/Catalunya/Santa Coloma | Servicio de Microbiolog√≠a, Laboratori Cl√≠nic Metropolitana Nord. Hospital Universitari Germans Trias i Pujol. Institut d‚ÄôInvestigaci√≥ en Ci√®ncies de la Salut Germans Trias i Pujol (IGTP) | SeqCOVID-SPAIN consortium/IBV(CSIC) | Elisa Mart√≠ et al |
| EPI_ISL_654539 | Spain/PV-IBV-98008358/2020 | 2020-10-12 | Europe/Spain/Basque Country/San Seba | Servicio de Microbiolog√≠a. Hospital Universitario Donostia. OSI Donostialdea. √Årea de Enfermedades Infecciosas, Grupo de Infecci√≥n Respiratoria y Resistencia Antimicrobiana. Instituto de Investigaci√≥n Sanitaria Biodonostia | SeqCOVID-SPAIN consortium/IBV(CSIC) | Gustavo Cilla Eguiluz et al |
| EPI_ISL_654542 | Spain/PV-IBV-98008342/2020 | 2020-10-10 | Europe/Spain/Basque Country/San Seba | Servicio de Microbiolog√≠a. Hospital Universitario Donostia. OSI Donostialdea. √Årea de Enfermedades Infecciosas, Grupo de Infecci√≥n Respiratoria y Resistencia Antimicrobiana. Instituto de Investigaci√≥n Sanitaria Biodonostia | SeqCOVID-SPAIN consortium/IBV(CSIC) | Gustavo Cilla Eguiluz et al |

| | | | | | | |
|---|---|---|---|---|---|---|
| EPI_ISL_654543 | Spain/PV-IBV-98008291/2020 | 2020-10-04 | Europe/Spain/Basque Country/San Seba | Servicio de Microbiolog√≠a. Hospital Universitario Donostia. OSI Donostialdea. √Årea de Enfermedades Infecciosas, Grupo de Infecci√≥n Respiratoria y Resistencia Antimicrobiana. Instituto de Investigaci√≥n Sanitaria Biodonostia | SeqCOVID-SPAIN consortium/IBV(CSIC) | Gustavo Cilla Eguiluz et al |
| EPI_ISL_654546 | Spain/PV-IBV-98008329/2020 | 2020-10-09 | Europe/Spain/Basque Country/San Seba | Servicio de Microbiolog√≠a. Hospital Universitario Donostia. OSI Donostialdea. √Årea de Enfermedades Infecciosas, Grupo de Infecci√≥n Respiratoria y Resistencia Antimicrobiana. Instituto de Investigaci√≥n Sanitaria Biodonostia | SeqCOVID-SPAIN consortium/IBV(CSIC) | Gustavo Cilla Eguiluz et al |
| EPI_ISL_654553 | Spain/PV-IBV-98008265/2020 | 2020-10-01 | Europe/Spain/Basque Country/San Seba | Servicio de Microbiolog√≠a. Hospital Universitario Donostia. OSI Donostialdea. √Årea de Enfermedades Infecciosas, Grupo de Infecci√≥n Respiratoria y Resistencia Antimicrobiana. Instituto de Investigaci√≥n Sanitaria Biodonostia | SeqCOVID-SPAIN consortium/IBV(CSIC) | Gustavo Cilla Eguiluz et al |
| EPI_ISL_654554 | Spain/PV-IBV-98008266/2020 | 2020-10-01 | Europe/Spain/Basque Country/San Seba | Servicio de Microbiolog√≠a. Hospital Universitario Donostia. OSI Donostialdea. √Årea de Enfermedades Infecciosas, Grupo de Infecci√≥n Respiratoria y Resistencia Antimicrobiana. Instituto de Investigaci√≥n Sanitaria Biodonostia | SeqCOVID-SPAIN consortium/IBV(CSIC) | Gustavo Cilla Eguiluz et al |
| EPI_ISL_654556 | Spain/PV-IBV-98008274/2020 | 2020-10-02 | Europe/Spain/Basque Country/San Seba | Servicio de Microbiolog√≠a. Hospital Universitario Donostia. OSI Donostialdea. √Årea de Enfermedades Infecciosas, Grupo de Infecci√≥n Respiratoria y Resistencia Antimicrobiana. Instituto de Investigaci√≥n Sanitaria Biodonostia | SeqCOVID-SPAIN consortium/IBV(CSIC) | Gustavo Cilla Eguiluz et al |
| EPI_ISL_654566 | Spain/PV-IBV-98008323/2020 | 2020-10-08 | Europe/Spain/Basque Country/San Seba | Servicio de Microbiolog√≠a. Hospital Universitario Donostia. OSI Donostialdea. √Årea de Enfermedades Infecciosas, Grupo de Infecci√≥n Respiratoria y Resistencia Antimicrobiana. Instituto de Investigaci√≥n Sanitaria Biodonostia | SeqCOVID-SPAIN consortium/IBV(CSIC) | Gustavo Cilla Eguiluz et al |

| | | | | | | |
|---|---|---|---|---|---|---|
| EPI_ISL_654567 | Spain/PV-IBV-98008333/2020 | 2020-10-09 | Europe/Spain/Basque Country/San Seba | Servicio de Microbiolog√≠a. Hospital Universitario Donostia. OSI Donostialdea. √Årea de Enfermedades Infecciosas, Grupo de Infecci√≥n Respiratoria y Resistencia Antimicrobiana. Instituto de Investigaci√≥n Sanitaria Biodonostia | SeqCOVID-SPAIN consortium/IBV(CSIC) | Gustavo Cilla Eguiluz et al |
| EPI_ISL_654569 | Spain/PV-IBV-98008344/2020 | 2020-10-27 | Europe/Spain/Basque Country/San Seba | Servicio de Microbiolog√≠a. Hospital Universitario Donostia. OSI Donostialdea. √Årea de Enfermedades Infecciosas, Grupo de Infecci√≥n Respiratoria y Resistencia Antimicrobiana. Instituto de Investigaci√≥n Sanitaria Biodonostia | SeqCOVID-SPAIN consortium/IBV(CSIC) | Gustavo Cilla Eguiluz et al |
| EPI_ISL_654571 | Spain/PV-IBV-98008346/2020 | 2020-10-11 | Europe/Spain/Basque Country/San Seba | Servicio de Microbiolog√≠a. Hospital Universitario Donostia. OSI Donostialdea. √Årea de Enfermedades Infecciosas, Grupo de Infecci√≥n Respiratoria y Resistencia Antimicrobiana. Instituto de Investigaci√≥n Sanitaria Biodonostia | SeqCOVID-SPAIN consortium/IBV(CSIC) | Gustavo Cilla Eguiluz et al |
| EPI_ISL_654572 | Spain/PV-IBV-98008353/2020 | 2020-10-27 | Europe/Spain/Basque Country/San Seba | Servicio de Microbiolog√≠a. Hospital Universitario Donostia. OSI Donostialdea. √Årea de Enfermedades Infecciosas, Grupo de Infecci√≥n Respiratoria y Resistencia Antimicrobiana. Instituto de Investigaci√≥n Sanitaria Biodonostia | SeqCOVID-SPAIN consortium/IBV(CSIC) | Gustavo Cilla Eguiluz et al |
| EPI_ISL_654573 | Spain/PV-IBV-98008356/2020 | 2020-10-12 | Europe/Spain/Basque Country/San Seba | Servicio de Microbiolog√≠a. Hospital Universitario Donostia. OSI Donostialdea. √Årea de Enfermedades Infecciosas, Grupo de Infecci√≥n Respiratoria y Resistencia Antimicrobiana. Instituto de Investigaci√≥n Sanitaria Biodonostia | SeqCOVID-SPAIN consortium/IBV(CSIC) | Gustavo Cilla Eguiluz et al |
| EPI_ISL_654574 | Spain/PV-IBV-98008361/2020 | 2020-10-13 | Europe/Spain/Basque Country/San Seba | Servicio de Microbiolog√≠a. Hospital Universitario Donostia. OSI Donostialdea. √Årea de Enfermedades Infecciosas, Grupo de Infecci√≥n Respiratoria y Resistencia Antimicrobiana. Instituto de Investigaci√≥n Sanitaria Biodonostia | SeqCOVID-SPAIN consortium/IBV(CSIC) | Gustavo Cilla Eguiluz et al |

| | | | | | | |
|---|---|---|---|---|---|---|
| EPI_ISL_654576 | Spain/PV-IBV-98008364/2020 | 2020-10-13 | Europe/Spain/Basque Country/San Seba | Servicio de Microbiolog√≠a. Hospital Universitario Donostia. OSI Donostialdea. √Årea de Enfermedades Infecciosas, Grupo de Infecci√≥n Respiratoria y Resistencia Antimicrobiana. Instituto de Investigaci√≥n Sanitaria Biodonostia | SeqCOVID-SPAIN consortium/IBV(CSIC) | Gustavo Cilla Eguiluz et al |
| EPI_ISL_654577 | Spain/PV-IBV-98008369/2020 | 2020-10-14 | Europe/Spain/Basque Country/San Seba | Servicio de Microbiolog√≠a. Hospital Universitario Donostia. OSI Donostialdea. √Årea de Enfermedades Infecciosas, Grupo de Infecci√≥n Respiratoria y Resistencia Antimicrobiana. Instituto de Investigaci√≥n Sanitaria Biodonostia | SeqCOVID-SPAIN consortium/IBV(CSIC) | Gustavo Cilla Eguiluz et al |
| EPI_ISL_654578 | Spain/PV-IBV-98008371/2020 | 2020-10-14 | Europe/Spain/Basque Country/San Seba | Servicio de Microbiolog√≠a. Hospital Universitario Donostia. OSI Donostialdea. √Årea de Enfermedades Infecciosas, Grupo de Infecci√≥n Respiratoria y Resistencia Antimicrobiana. Instituto de Investigaci√≥n Sanitaria Biodonostia | SeqCOVID-SPAIN consortium/IBV(CSIC) | Gustavo Cilla Eguiluz et al |
| EPI_ISL_654580 | Spain/PV-IBV-98008379/2020 | 2020-10-15 | Europe/Spain/Basque Country/San Seba | Servicio de Microbiolog√≠a. Hospital Universitario Donostia. OSI Donostialdea. √Årea de Enfermedades Infecciosas, Grupo de Infecci√≥n Respiratoria y Resistencia Antimicrobiana. Instituto de Investigaci√≥n Sanitaria Biodonostia | SeqCOVID-SPAIN consortium/IBV(CSIC) | Gustavo Cilla Eguiluz et al |
| EPI_ISL_654584 | Spain/PV-IBV-98008389/2020 | 2020-10-16 | Europe/Spain/Basque Country/San Seba | Servicio de Microbiolog√≠a. Hospital Universitario Donostia. OSI Donostialdea. √Årea de Enfermedades Infecciosas, Grupo de Infecci√≥n Respiratoria y Resistencia Antimicrobiana. Instituto de Investigaci√≥n Sanitaria Biodonostia | SeqCOVID-SPAIN consortium/IBV(CSIC) | Gustavo Cilla Eguiluz et al |
| EPI_ISL_654585 | Spain/PV-IBV-98008391/2020 | 2020-10-16 | Europe/Spain/Basque Country/San Seba | Servicio de Microbiolog√≠a. Hospital Universitario Donostia. OSI Donostialdea. √Årea de Enfermedades Infecciosas, Grupo de Infecci√≥n Respiratoria y Resistencia Antimicrobiana. Instituto de Investigaci√≥n Sanitaria Biodonostia | SeqCOVID-SPAIN consortium/IBV(CSIC) | Gustavo Cilla Eguiluz et al |

| | | | | | | |
|---|---|---|---|---|---|---|
| EPI_ISL_654587 | Spain/PV-IBV-98008395/2020 | 2020-10-17 | Europe/Spain/Basque Country/San Seba | Servicio de Microbiolog√≠a. Hospital Universitario Donostia. OSI Donostialdea. √Årea de Enfermedades Infecciosas, Grupo de Infecci√≥n Respiratoria y Resistencia Antimicrobiana. Instituto de Investigaci√≥n Sanitaria Biodonostia | SeqCOVID-SPAIN consortium/IBV(CSIC) | Gustavo Cilla Eguiluz et al |
| EPI_ISL_654592 | Spain/PV-IBV-98008409/2020 | 2020-10-19 | Europe/Spain/Basque Country/San Seba | Servicio de Microbiolog√≠a. Hospital Universitario Donostia. OSI Donostialdea. √Årea de Enfermedades Infecciosas, Grupo de Infecci√≥n Respiratoria y Resistencia Antimicrobiana. Instituto de Investigaci√≥n Sanitaria Biodonostia | SeqCOVID-SPAIN consortium/IBV(CSIC) | Gustavo Cilla Eguiluz et al |
| EPI_ISL_654593 | Spain/PV-IBV-98008410/2020 | 2020-10-19 | Europe/Spain/Basque Country/San Seba | Servicio de Microbiolog√≠a. Hospital Universitario Donostia. OSI Donostialdea. √Årea de Enfermedades Infecciosas, Grupo de Infecci√≥n Respiratoria y Resistencia Antimicrobiana. Instituto de Investigaci√≥n Sanitaria Biodonostia | SeqCOVID-SPAIN consortium/IBV(CSIC) | Gustavo Cilla Eguiluz et al |
| EPI_ISL_654598 | Spain/PV-IBV-98008422/2020 | 2020-10-20 | Europe/Spain/Basque Country/San Seba | Servicio de Microbiolog√≠a. Hospital Universitario Donostia. OSI Donostialdea. √Årea de Enfermedades Infecciosas, Grupo de Infecci√≥n Respiratoria y Resistencia Antimicrobiana. Instituto de Investigaci√≥n Sanitaria Biodonostia | SeqCOVID-SPAIN consortium/IBV(CSIC) | Gustavo Cilla Eguiluz et al |
| EPI_ISL_654604 | Spain/PV-IBV-98008432/2020 | 2020-10-21 | Europe/Spain/Basque Country/San Seba | Servicio de Microbiolog√≠a. Hospital Universitario Donostia. OSI Donostialdea. √Årea de Enfermedades Infecciosas, Grupo de Infecci√≥n Respiratoria y Resistencia Antimicrobiana. Instituto de Investigaci√≥n Sanitaria Biodonostia | SeqCOVID-SPAIN consortium/IBV(CSIC) | Gustavo Cilla Eguiluz et al |
| EPI_ISL_654606 | Spain/PV-IBV-98008437/2020 | 2020-10-22 | Europe/Spain/Basque Country/San Seba | Servicio de Microbiolog√≠a. Hospital Universitario Donostia. OSI Donostialdea. √Årea de Enfermedades Infecciosas, Grupo de Infecci√≥n Respiratoria y Resistencia Antimicrobiana. Instituto de Investigaci√≥n Sanitaria Biodonostia | SeqCOVID-SPAIN consortium/IBV(CSIC) | Gustavo Cilla Eguiluz et al |

| | | | | | | |
|---|---|---|---|---|---|---|
| EPI_ISL_654607 | Spain/PV-IBV-98008439/2020 | 2020-10-22 | Europe/Spain/Basque Country/San Seba | Servicio de Microbiolog√≠a. Hospital Universitario Donostia. OSI Donostialdea. √Årea de Enfermedades Infecciosas, Grupo de Infecci√≥n Respiratoria y Resistencia Antimicrobiana. Instituto de Investigaci√≥n Sanitaria Biodonostia | SeqCOVID-SPAIN consortium/IBV(CSIC) | Gustavo Cilla Eguiluz et al |
| EPI_ISL_654609 | Spain/PV-IBV-98008443/2020 | 2020-10-23 | Europe/Spain/Basque Country/San Seba | Servicio de Microbiolog√≠a. Hospital Universitario Donostia. OSI Donostialdea. √Årea de Enfermedades Infecciosas, Grupo de Infecci√≥n Respiratoria y Resistencia Antimicrobiana. Instituto de Investigaci√≥n Sanitaria Biodonostia | SeqCOVID-SPAIN consortium/IBV(CSIC) | Gustavo Cilla Eguiluz et al |
| EPI_ISL_654610 | Spain/PV-IBV-98008447/2020 | 2020-10-23 | Europe/Spain/Basque Country/San Seba | Servicio de Microbiolog√≠a. Hospital Universitario Donostia. OSI Donostialdea. √Årea de Enfermedades Infecciosas, Grupo de Infecci√≥n Respiratoria y Resistencia Antimicrobiana. Instituto de Investigaci√≥n Sanitaria Biodonostia | SeqCOVID-SPAIN consortium/IBV(CSIC) | Gustavo Cilla Eguiluz et al |
| EPI_ISL_654615 | Spain/PV-IBV-98008461/2020 | 2020-10-25 | Europe/Spain/Basque Country/San Seba | Servicio de Microbiolog√≠a. Hospital Universitario Donostia. OSI Donostialdea. √Årea de Enfermedades Infecciosas, Grupo de Infecci√≥n Respiratoria y Resistencia Antimicrobiana. Instituto de Investigaci√≥n Sanitaria Biodonostia | SeqCOVID-SPAIN consortium/IBV(CSIC) | Gustavo Cilla Eguiluz et al |
| EPI_ISL_654617 | Spain/AS-IBV-98006622/2020 | 2020-03-21 | Europe/Spain/Asturias/Oviedo | Servicio de Microbiolog√≠a, Hospital Universitario Central de Asturias | SeqCOVID-SPAIN consortium/IBV(CSIC) | Cristi√°n Castell√≥ Abietar et al |
| EPI_ISL_654621 | Spain/AS-IBV-98006579/2020 | 2020-03-08 | Europe/Spain/Madrid/Madrid | Servicio de Microbiolog√≠a, Hospital Universitario Central de Asturias | SeqCOVID-SPAIN consortium/IBV(CSIC) | Cristi√°n Castell√≥ Abietar et al |
| EPI_ISL_654631 | Spain/AS-IBV-98006628/2020 | 2020-03-30 | Europe/Spain/Asturias/Gijon | Servicio de Microbiolog√≠a, Hospital Universitario Central de Asturias | SeqCOVID-SPAIN consortium/IBV(CSIC) | Cristi√°n Castell√≥ Abietar et al |
| EPI_ISL_654632 | Spain/AS-IBV-98006590/2020 | 2020-03-11 | Europe/Spain/Asturias/Grado | Servicio de Microbiolog√≠a, Hospital Universitario Central de Asturias | SeqCOVID-SPAIN consortium/IBV(CSIC) | Cristi√°n Castell√≥ Abietar et al |
| EPI_ISL_654638 | Spain/AS-IBV-98006592/2020 | 2020-03-12 | Europe/Spain/Asturias/Oviedo | Servicio de Microbiolog√≠a, Hospital Universitario Central de Asturias | SeqCOVID-SPAIN consortium/IBV(CSIC) | Cristi√°n Castell√≥ Abietar et al |

| | | | | | | |
|---|---|---|---|---|---|---|
| EPI_ISL_654640 | Spain/AS-IBV-98006595/2020 | 2020-03-12 | Europe/Spain/Asturias/Oviedo | Servicio de Microbiolog√≠a, Hospital Universitario Central de Asturias | SeqCOVID-SPAIN consortium/IBV(CSIC) | Cristi√°n Castell√≥ Abietar et al |
| EPI_ISL_654648 | Spain/AS-IBV-98006604/2020 | 2020-03-13 | Europe/Spain/Asturias/Oviedo | Servicio de Microbiolog√≠a, Hospital Universitario Central de Asturias | SeqCOVID-SPAIN consortium/IBV(CSIC) | Cristi√°n Castell√≥ Abietar et al |
| EPI_ISL_654654 | Spain/AS-IBV-98006611/2020 | 2020-03-13 | Europe/Spain/Asturias/Oviedo | Servicio de Microbiolog√≠a, Hospital Universitario Central de Asturias | SeqCOVID-SPAIN consortium/IBV(CSIC) | Cristi√°n Castell√≥ Abietar et al |
| EPI_ISL_654656 | Spain/AS-IBV-98006613/2020 | 2020-03-13 | Europe/Spain/Asturias/Oviedo | Servicio de Microbiolog√≠a, Hospital Universitario Central de Asturias | SeqCOVID-SPAIN consortium/IBV(CSIC) | Cristi√°n Castell√≥ Abietar et al |
| EPI_ISL_654657 | Spain/AS-IBV-98006616/2020 | 2020-03-14 | Europe/Spain/Asturias/Oviedo | Servicio de Microbiolog√≠a, Hospital Universitario Central de Asturias | SeqCOVID-SPAIN consortium/IBV(CSIC) | Cristi√°n Castell√≥ Abietar et al |
| EPI_ISL_654666 | Spain/AS-IBV-98006632/2020 | 2020-03-31 | Europe/Spain/Asturias/Oviedo | Servicio de Microbiolog√≠a, Hospital Universitario Central de Asturias | SeqCOVID-SPAIN consortium/IBV(CSIC) | Cristi√°n Castell√≥ Abietar et al |
| EPI_ISL_654696 | USA/MN-MDH-2015/2020 | 2020-09-28 | North America/USA/Minnesota/ | Essentia Health-St. Mary's Medical Center | Minnesota Department of Health, Public Health Laboratory | Matt Plumb et al |
| EPI_ISL_654700 | USA/MN-MDH-2019/2020 | 2020-10-30 | North America/USA/Minnesota/ | Minnesota Department of Health, Public Health Laboratory | Minnesota Department of Health, Public Health Laboratory | Matt Plumb et al |
| EPI_ISL_654718 | USA/MN-MDH-2037/2020 | 2020-11-03 | North America/USA/Minnesota/ | Minnesota Department of Health, Public Health Laboratory | Minnesota Department of Health, Public Health Laboratory | Matt Plumb et al |
| EPI_ISL_654720 | USA/MN-MDH-2039/2020 | 2020-11-03 | North America/USA/Minnesota/ | Minnesota Department of Health, Public Health Laboratory | Minnesota Department of Health, Public Health Laboratory | Matt Plumb et al |
| EPI_ISL_654723 | USA/MN-MDH-2042/2020 | 2020-11-03 | North America/USA/Minnesota/ | Minnesota Department of Health, Public Health Laboratory | Minnesota Department of Health, Public Health Laboratory | Matt Plumb et al |

| | | | | | | |
|---|---|---|---|---|---|---|
| EPI_ISL_654726 | USA/MN-MDH-2045/2020 | 2020-11-04 | North America/USA/Minnesota/ | Minnesota Department of Health, Public Health Laboratory | Minnesota Department of Health, Public Health Laboratory | Matt Plumb et al |
| EPI_ISL_654735 | USA/TX-DSHS-1236/2020 | 2020-05-01 | North America/USA/Texas/Lamar Count | Texas Department of State Health Services | Texas Department of State Health Services | Rashmi Tuladhar et al |
| EPI_ISL_654739 | USA/TX-DSHS-1394/2020 | 2020-05-17 | North America/USA/Texas/Titus County | Texas Department of State Health Services | Texas Department of State Health Services | Rashmi Tuladhar et al |
| EPI_ISL_654745 | USA/TX-DSHS-1370/2020 | 2020-05-09 | North America/USA/Texas/Titus County | Texas Department of State Health Services | Texas Department of State Health Services | Rashmi Tuladhar et al |
| EPI_ISL_654756 | USA/TX-DSHS-1372/2020 | 2020-05-11 | North America/USA/Texas/Gonzales Cou | Texas Department of State Health Services | Texas Department of State Health Services | Rashmi Tuladhar et al |
| EPI_ISL_654769 | USA/TX-DSHS-1364/2020 | 2020-05-01 | North America/USA/Texas/Williamson C | Texas Department of State Health Services | Texas Department of State Health Services | Rashmi Tuladhar et al |
| EPI_ISL_654770 | USA/TX-DSHS-1371/2020 | 2020-05-11 | North America/USA/Texas/Gonzales Cou | Texas Department of State Health Services | Texas Department of State Health Services | Rashmi Tuladhar et al |
| EPI_ISL_654774 | USA/TX-DSHS-1225/2020 | 2020-03-11 | North America/USA/Texas/Smith County | Texas Department of State Health Services | Texas Department of State Health Services | Rashmi Tuladhar et al |
| EPI_ISL_654818 | Ukraine/Chernivtsi_860/2020 | 2020-08-06 | Europe/Ukraine/Chernivtsi/ | Department of Respiratory and other Viral Infections of L.V.Gromashevsky Institute of Epidemiology & Infectious Diseases NAMS of Ukrain–µ | "Department of Respiratory and other Viral Infections of L.V.Gromashevsky Institute of Epidemiology & Infectious Diseases NAMS of Ukrain–µ, JSC ""Farmak""" | Alla Mironenko et al |

| | | | | | | |
|---|---|---|---|---|---|---|
| EPI_ISL_654819 | Ukraine/Chernivtsi_861/2020 | 2020-08-06 | Europe/Ukraine/Chernivtsi/ | Department of Respiratory and other Viral Infections of L.V.Gromashevsky Institute of Epidemiology & Infectious Diseases NAMS of Ukrain–µ | "Department of Respiratory and other Viral Infections of L.V.Gromashevsky Institute of Epidemiology & Infectious Diseases NAMS of Ukrain–µ, JSC ""Farmak""" | Alla Mironenko et al |
| EPI_ISL_654820 | Ukraine/Lutsk_824/2020 | 2020-08-04 | Europe/Ukraine/Volyn/Lutsk | Department of Respiratory and other Viral Infections of L.V.Gromashevsky Institute of Epidemiology & Infectious Diseases NAMS of Ukrain–µ | "Department of Respiratory and other Viral Infections of L.V.Gromashevsky Institute of Epidemiology & Infectious Diseases NAMS of Ukrain–µ, JSC ""Farmak""" | Alla Mironenko et al |
| EPI_ISL_654866 | Vietnam/PIHCM-14/2020 | 2020-01-31 | Asia/Vietnam/Ho Chi Minh City/ | Pasteur Institute in Ho Chi Minh city | Department of Microbiology and Immunology - Pasteur Institute in Ho Chi Minh city | Cao Minh Th·ʃØng et al |
| EPI_ISL_654895 | Sweden/20-22280/2020 | 2020-09-17 | Europe/Sweden/Skane/ | Klinisk mikrobiologi | The Public Health Agency of Sweden | Anna-Malin Linde et al |
| EPI_ISL_654898 | Sweden/20-22284/2020 | 2020-09-17 | Europe/Sweden/Skane/ | Klinisk mikrobiologi | The Public Health Agency of Sweden | Anna-Malin Linde et al |
| EPI_ISL_654911 | USA/TX-DSHS-1329/2020 | 2020-04-02 | North America/USA/Texas/Bexar County | Texas Department of State Health Services | Texas Department of State Health Services | Rashmi Tuladhar et al |
| EPI_ISL_654912 | USA/TX-DSHS-1330/2020 | 2020-04-02 | North America/USA/Texas/Bexar County | Texas Department of State Health Services | Texas Department of State Health Services | Rashmi Tuladhar et al |
| EPI_ISL_654926 | USA/TX-DSHS-1345/2020 | 2020-04-07 | North America/USA/Texas/Bexar County | Texas Department of State Health Services | Texas Department of State Health Services | Rashmi Tuladhar et al |

| | | | | | | |
|---|---|---|---|---|---|---|
| EPI_ISL_654933 | USA/TX-DSHS-1352/2020 | 2020-04-13 | North America/USA/Texas/Wilson Count | Texas Department of State Health Services | Texas Department of State Health Services | Rashmi Tuladhar et al |
| EPI_ISL_654946 | Sweden/20-52767/2020 | 2020-08-05 | Europe/Sweden/Kronoberg/ | Klinisk mikrobiologi | The Public Health Agency of Sweden | Anna-Malin Linde et al |
| EPI_ISL_655106 | England/QEUH-B68B28/2020 | 2020-11-11 | Europe/United Kingdom/England/ | Lighthouse Lab in Glasgow | Wellcome Sanger Institute for the COVID-19 Genomics UK (COG-UK) Consortium | Harper VanSteenhouse et al |
| EPI_ISL_655149 | England/QEUH-B6B9D0/2020 | 2020-11-11 | Europe/United Kingdom/England/ | Lighthouse Lab in Glasgow | Wellcome Sanger Institute for the COVID-19 Genomics UK (COG-UK) Consortium | Harper VanSteenhouse et al |
| EPI_ISL_655235 | England/QEUH-B6C975/2020 | 2020-11-11 | Europe/United Kingdom/England/ | Lighthouse Lab in Glasgow | Wellcome Sanger Institute for the COVID-19 Genomics UK (COG-UK) Consortium | Harper VanSteenhouse et al |
| EPI_ISL_655251 | England/QEUH-B67CEA/2020 | 2020-11-11 | Europe/United Kingdom/England/ | Lighthouse Lab in Glasgow | Wellcome Sanger Institute for the COVID-19 Genomics UK (COG-UK) Consortium | Harper VanSteenhouse et al |
| EPI_ISL_655374 | England/ALDP-B5B6EB/2020 | 2020-11-08 | Europe/United Kingdom/England/ | Lighthouse Lab in Alderley Park | Wellcome Sanger Institute for the COVID-19 Genomics UK (COG-UK) Consortium | Jacquelyn Wynn et al |
| EPI_ISL_655395 | England/ALDP-B5AF0F/2020 | 2020-11-09 | Europe/United Kingdom/England/ | Lighthouse Lab in Alderley Park | Wellcome Sanger Institute for the COVID-19 Genomics UK (COG-UK) Consortium | Jacquelyn Wynn et al |
| EPI_ISL_655648 | England/ALDP-B5BFA4/2020 | 2020-11-09 | Europe/United Kingdom/England/ | Lighthouse Lab in Alderley Park | Wellcome Sanger Institute for the COVID-19 Genomics UK (COG-UK) Consortium | Jacquelyn Wynn et al |

| | | | | | | |
|---|---|---|---|---|---|---|
| EPI_ISL_655964 | England/ALDP-B5C0DD/2020 | 2020-11-08 | Europe/United Kingdom/England/ | Lighthouse Lab in Alderley Park | Wellcome Sanger Institute for the COVID-19 Genomics UK (COG-UK) Consortium | Jacquelyn Wynn et al |
| EPI_ISL_656154 | England/ALDP-B472EE/2020 | 2020-11-07 | Europe/United Kingdom/England/ | Lighthouse Lab in Alderley Park | Wellcome Sanger Institute for the COVID-19 Genomics UK (COG-UK) Consortium | Jacquelyn Wynn et al |
| EPI_ISL_656286 | England/ALDP-B459FA/2020 | 2020-11-07 | Europe/United Kingdom/England/ | Lighthouse Lab in Alderley Park | Wellcome Sanger Institute for the COVID-19 Genomics UK (COG-UK) Consortium | Jacquelyn Wynn et al |
| EPI_ISL_656360 | England/ALDP-B62599/2020 | 2020-11-08 | Europe/United Kingdom/England/ | Lighthouse Lab in Alderley Park | Wellcome Sanger Institute for the COVID-19 Genomics UK (COG-UK) Consortium | Jacquelyn Wynn et al |
| EPI_ISL_656381 | England/ALDP-B6264B/2020 | 2020-11-08 | Europe/United Kingdom/England/ | Lighthouse Lab in Alderley Park | Wellcome Sanger Institute for the COVID-19 Genomics UK (COG-UK) Consortium | Jacquelyn Wynn et al |
| EPI_ISL_656404 | England/ALDP-B63404/2020 | 2020-11-09 | Europe/United Kingdom/England/ | Lighthouse Lab in Alderley Park | Wellcome Sanger Institute for the COVID-19 Genomics UK (COG-UK) Consortium | Jacquelyn Wynn et al |
| EPI_ISL_656674 | England/ALDP-B620D4/2020 | 2020-11-08 | Europe/United Kingdom/England/ | Lighthouse Lab in Alderley Park | Wellcome Sanger Institute for the COVID-19 Genomics UK (COG-UK) Consortium | Jacquelyn Wynn et al |
| EPI_ISL_656739 | England/CAMC-B53865/2020 | 2020-11-08 | Europe/United Kingdom/England/ | Lighthouse Lab in Cambridge | Wellcome Sanger Institute for the COVID-19 Genomics UK (COG-UK) Consortium | Rob Howes et al |

| | | | | | | |
|---|---|---|---|---|---|---|
| EPI_ISL_656966 | England/MILK-B56A6C/2020 | 2020-11-05 | Europe/United Kingdom/England/ | Lighthouse Lab in Milton Keynes | Wellcome Sanger Institute for the COVID-19 Genomics UK (COG-UK) Consortium | The Lighthouse Lab in Milton Keynes et al |
| EPI_ISL_657016 | England/MILK-B56B3C/2020 | 2020-11-06 | Europe/United Kingdom/England/ | Lighthouse Lab in Milton Keynes | Wellcome Sanger Institute for the COVID-19 Genomics UK (COG-UK) Consortium | The Lighthouse Lab in Milton Keynes et al |
| EPI_ISL_657053 | England/QEUH-B408C5/2020 | 2020-11-08 | Europe/United Kingdom/England/ | Lighthouse Lab in Glasgow | Wellcome Sanger Institute for the COVID-19 Genomics UK (COG-UK) Consortium | Harper VanSteenhouse et al |
| EPI_ISL_657157 | England/QEUH-B404FE/2020 | 2020-11-08 | Europe/United Kingdom/England/ | Lighthouse Lab in Glasgow | Wellcome Sanger Institute for the COVID-19 Genomics UK (COG-UK) Consortium | Harper VanSteenhouse et al |
| EPI_ISL_657212 | England/QEUH-B4331C/2020 | 2020-11-08 | Europe/United Kingdom/England/ | Lighthouse Lab in Glasgow | Wellcome Sanger Institute for the COVID-19 Genomics UK (COG-UK) Consortium | Harper VanSteenhouse et al |
| EPI_ISL_657220 | England/QEUH-B40027/2020 | 2020-11-08 | Europe/United Kingdom/England/ | Lighthouse Lab in Glasgow | Wellcome Sanger Institute for the COVID-19 Genomics UK (COG-UK) Consortium | Harper VanSteenhouse et al |
| EPI_ISL_657227 | England/QEUH-B40203/2020 | 2020-11-08 | Europe/United Kingdom/England/ | Lighthouse Lab in Glasgow | Wellcome Sanger Institute for the COVID-19 Genomics UK (COG-UK) Consortium | Harper VanSteenhouse et al |
| EPI_ISL_657334 | England/QEUH-B43604/2020 | 2020-11-08 | Europe/United Kingdom/England/ | Lighthouse Lab in Glasgow | Wellcome Sanger Institute for the COVID-19 Genomics UK (COG-UK) Consortium | Harper VanSteenhouse et al |

| | | | | | | |
|---|---|---|---|---|---|---|
| EPI_ISL_657429 | England/MILK-B55AF4/2020 | 2020-11-05 | Europe/United Kingdom/England/ | Lighthouse Lab in Milton Keynes | Wellcome Sanger Institute for the COVID-19 Genomics UK (COG-UK) Consortium | The Lighthouse Lab in Milton Keynes et al |
| EPI_ISL_657435 | England/CAMC-B527E1/2020 | 2020-11-08 | Europe/United Kingdom/England/ | Lighthouse Lab in Cambridge | Wellcome Sanger Institute for the COVID-19 Genomics UK (COG-UK) Consortium | Rob Howes et al |
| EPI_ISL_657450 | Spain/VC-IBV-98003588/2020 | 2020-03-31 | Europe/Spain/Comunitat Valenciana/Ca | Servicio de Microbiolog√≠a. Hospital General Universitario de Castell√≥n | SeqCOVID-SPAIN consortium/IBV(CSIC) | Rosario Moreno Mu√±oz et al |
| EPI_ISL_657460 | Spain/VC-IBV-98003619/2020 | 2020-03-17 | Europe/Spain/Comunitat Valenciana/Ca | Servicio de Microbiolog√≠a. Hospital General Universitario de Castell√≥n | SeqCOVID-SPAIN consortium/IBV(CSIC) | Rosario Moreno Mu√±oz et al |
| EPI_ISL_657466 | Spain/VC-IBV-98006456/2020 | 2020-06-27 | Europe/Spain/Comunitat Valenciana/Ca | Servicio de Microbiolog√≠a. Hospital General Universitario de Castell√≥n | SeqCOVID-SPAIN consortium/IBV(CSIC) | Rosario Moreno Mu√±oz et al |
| EPI_ISL_657618 | England/CAMC-B52D49/2020 | 2020-11-09 | Europe/United Kingdom/England/ | Lighthouse Lab in Cambridge | Wellcome Sanger Institute for the COVID-19 Genomics UK (COG-UK) Consortium | Rob Howes et al |
| EPI_ISL_657940 | England/MILK-B55195/2020 | 2020-11-06 | Europe/United Kingdom/England/ | Lighthouse Lab in Milton Keynes | Wellcome Sanger Institute for the COVID-19 Genomics UK (COG-UK) Consortium | The Lighthouse Lab in Milton Keynes et al |
| EPI_ISL_658059 | England/MILK-B54B2F/2020 | 2020-11-05 | Europe/United Kingdom/England/ | Lighthouse Lab in Milton Keynes | Wellcome Sanger Institute for the COVID-19 Genomics UK (COG-UK) Consortium | The Lighthouse Lab in Milton Keynes et al |
| EPI_ISL_658125 | England/CAMC-B5174C/2020 | 2020-11-08 | Europe/United Kingdom/England/ | Lighthouse Lab in Cambridge | Wellcome Sanger Institute for the COVID-19 Genomics UK (COG-UK) Consortium | Rob Howes et al |

| | | | | | | |
|---|---|---|---|---|---|---|
| EPI_ISL_658239 | England/CAMC-B5190A/2020 | 2020-11-08 | Europe/United Kingdom/England/ | Lighthouse Lab in Cambridge | Wellcome Sanger Institute for the COVID-19 Genomics UK (COG-UK) Consortium | Rob Howes et al |
| EPI_ISL_658302 | England/MILK-B5A497/2020 | 2020-11-06 | Europe/United Kingdom/England/ | Lighthouse Lab in Milton Keynes | Wellcome Sanger Institute for the COVID-19 Genomics UK (COG-UK) Consortium | The Lighthouse Lab in Milton Keynes et al |
| EPI_ISL_658579 | England/QEUH-B413E1/2020 | 2020-11-08 | Europe/United Kingdom/England/ | Lighthouse Lab in Glasgow | Wellcome Sanger Institute for the COVID-19 Genomics UK (COG-UK) Consortium | Harper VanSteenhouse et al |
| EPI_ISL_658691 | England/QEUH-B41439/2020 | 2020-11-08 | Europe/United Kingdom/England/ | Lighthouse Lab in Glasgow | Wellcome Sanger Institute for the COVID-19 Genomics UK (COG-UK) Consortium | Harper VanSteenhouse et al |
| EPI_ISL_658863 | Mexico/AGS-InDRE-78/2020 | 2020-11-02 | North America/Mexico/Aguascalientes/ | Instituto de Diagnostico y Referencia Epidemiologicos (INDRE) | Instituto de Diagnostico y Referencia Epidemiologicos (INDRE) | Gisela Barrera-Badillo et al |
| EPI_ISL_658868 | Mexico/AGS-InDRE-80/2020 | 2020-11-02 | North America/Mexico/Aguascalientes/ | Instituto de Diagnostico y Referencia Epidemiologicos (INDRE) | Instituto de Diagnostico y Referencia Epidemiologicos (INDRE) | Gisela Barrera-Badillo et al |
| EPI_ISL_658873 | Mexico/AGS-InDRE-82/2020 | 2020-11-02 | North America/Mexico/Aguascalientes/ | Instituto de Diagnostico y Referencia Epidemiologicos (INDRE) | Instituto de Diagnostico y Referencia Epidemiologicos (INDRE) | Gisela Barrera-Badillo et al |
| EPI_ISL_658875 | Mexico/AGS-InDRE-83/2020 | 2020-11-02 | North America/Mexico/Aguascalientes/ | Instituto de Diagnostico y Referencia Epidemiologicos (INDRE) | Instituto de Diagnostico y Referencia Epidemiologicos (INDRE) | Gisela Barrera-Badillo et al |

| | | | | | | |
|---|---|---|---|---|---|---|
| EPI_ISL_658878 | Mexico/QRO-InDRE-84/2020 | 2020-11-02 | North America/Mexico/Queretaro/ | Instituto de Diagnostico y Referencia Epidemiologicos (INDRE) | Instituto de Diagnostico y Referencia Epidemiologicos (INDRE) | Ernesto Ramirez-Gonzalez et al |
| EPI_ISL_658886 | Mexico/QRO-InDRE-87/2020 | 2020-11-03 | North America/Mexico/Queretaro/ | Instituto de Diagnostico y Referencia Epidemiologicos (INDRE) | Instituto de Diagnostico y Referencia Epidemiologicos (INDRE) | Ernesto Ramirez-Gonzalez et al |
| EPI_ISL_658888 | Mexico/QRO-InDRE-88/2020 | 2020-11-03 | North America/Mexico/Queretaro/ | Instituto de Diagnostico y Referencia Epidemiologicos (INDRE) | Instituto de Diagnostico y Referencia Epidemiologicos (INDRE) | Ernesto Ramirez-Gonzalez et al |
| EPI_ISL_658893 | Mexico/QRO-InDRE-90/2020 | 2020-11-03 | North America/Mexico/Queretaro/ | Instituto de Diagnostico y Referencia Epidemiologicos (INDRE) | Instituto de Diagnostico y Referencia Epidemiologicos (INDRE) | Ernesto Ramirez-Gonzalez et al |
| EPI_ISL_658896 | Mexico/GRO-InDRE-91/2020 | 2020-06-08 | North America/Mexico/Guerrero/ | Instituto de Diagnostico y Referencia Epidemiologicos (INDRE) | Instituto de Diagnostico y Referencia Epidemiologicos (INDRE) | Ernesto Ramirez-Gonzalez et al |
| EPI_ISL_658899 | Mexico/GRO-InDRE-92/2020 | 2020-07-18 | North America/Mexico/Guerrero/ | Instituto de Diagnostico y Referencia Epidemiologicos (INDRE) | Instituto de Diagnostico y Referencia Epidemiologicos (INDRE) | Gisela Barrera-Badillo et al |
| EPI_ISL_658901 | Mexico/GRO-InDRE-93/2020 | 2020-09-07 | North America/Mexico/Guerrero/ | Instituto de Diagnostico y Referencia Epidemiologicos (INDRE) | Instituto de Diagnostico y Referencia Epidemiologicos (INDRE) | Ernesto Ramirez-Gonzalez et al |
| EPI_ISL_658904 | Mexico/QROO-InDRE-53/2020 | 2020-04-10 | North America/Mexico/Quintana Roo/ | Instituto de Diagnostico y Referencia Epidemiologicos (INDRE) | Instituto de Diagnostico y Referencia Epidemiologicos (INDRE) | Ernesto Ramirez-Gonzalez et al |

| | | | | | | |
|---|---|---|---|---|---|---|
| EPI_ISL_658955 | England/MILK-B39531/2020 | 2020-11-02 | Europe/United Kingdom/England/ | Lighthouse Lab in Milton Keynes | Wellcome Sanger Institute for the COVID-19 Genomics UK (COG-UK) Consortium | The Lighthouse Lab in Milton Keynes et al |
| EPI_ISL_658989 | England/MILK-B38CB5/2020 | 2020-11-01 | Europe/United Kingdom/England/ | Lighthouse Lab in Milton Keynes | Wellcome Sanger Institute for the COVID-19 Genomics UK (COG-UK) Consortium | The Lighthouse Lab in Milton Keynes et al |
| EPI_ISL_658994 | England/MILK-B3A65B/2020 | 2020-11-02 | Europe/United Kingdom/England/ | Lighthouse Lab in Milton Keynes | Wellcome Sanger Institute for the COVID-19 Genomics UK (COG-UK) Consortium | The Lighthouse Lab in Milton Keynes et al |
| EPI_ISL_659043 | England/MILK-B384EA/2020 | 2020-11-01 | Europe/United Kingdom/England/ | Lighthouse Lab in Milton Keynes | Wellcome Sanger Institute for the COVID-19 Genomics UK (COG-UK) Consortium | The Lighthouse Lab in Milton Keynes et al |
| EPI_ISL_659108 | England/CAMC-B33ADE/2020 | 2020-11-05 | Europe/United Kingdom/England/ | Lighthouse Lab in Cambridge | Wellcome Sanger Institute for the COVID-19 Genomics UK (COG-UK) Consortium | Rob Howes et al |
| EPI_ISL_659323 | England/CAMC-B33519/2020 | 2020-11-05 | Europe/United Kingdom/England/ | Lighthouse Lab in Cambridge | Wellcome Sanger Institute for the COVID-19 Genomics UK (COG-UK) Consortium | Rob Howes et al |
| EPI_ISL_659436 | England/CAMC-B343D2/2020 | 2020-11-05 | Europe/United Kingdom/England/ | Lighthouse Lab in Cambridge | Wellcome Sanger Institute for the COVID-19 Genomics UK (COG-UK) Consortium | Rob Howes et al |
| EPI_ISL_659852 | England/QEUH-B3038B/2020 | 2020-11-04 | Europe/United Kingdom/England/ | Lighthouse Lab in Glasgow | Wellcome Sanger Institute for the COVID-19 Genomics UK (COG-UK) Consortium | Harper VanSteenhouse et al |

| | | | | | | |
|---|---|---|---|---|---|---|
| EPI_ISL_660002 | England/QEUH-B2EDBF/2020 | 2020-11-03 | Europe/United Kingdom/England/ | Lighthouse Lab in Glasgow | Wellcome Sanger Institute for the COVID-19 Genomics UK (COG-UK) Consortium | Harper VanSteenhouse et al |
| EPI_ISL_660068 | Mexico/SLP-InDRE-77/2020 | 2020-11-01 | North America/Mexico/San Luis Potosi/ | Instituto de Diagnostico y Referencia Epidemiologicos (INDRE) | Instituto de diagnV≥stico y Referencia Epidemiologicos (INDRE) Departamento de VirologV≠a | Gisela Barrera-Badillo et al |
| EPI_ISL_660069 | Ecuador/ZZL-3/2020 | 2020-07-20 | South America/Ecuador/Pichincha/Quito | Zurita & Zurita Laboratorios | Zurita & Zurita Laboratorios | Gabriela Sevillano Camilo Zurita-Salinas Karen Loaiza David Ortega-Paredes Jeannete Zurita et al |
| EPI_ISL_660070 | Ecuador/ZZL-4/2020 | 2020-11-26 | South America/Ecuador/Pichincha/Quito | Zurita & Zurita Laboratorios | Zurita & Zurita Laboratorios | Gabriela Sevillano Camilo Zurita-Salinas Karen Loaiza David Ortega-Paredes Jeannete Zurita et al |
| EPI_ISL_660075 | USA/MD-MDH-0374/2020 | 2020-10-26 | North America/USA/Maryland/ | MD PHL | MD PHL | Maryland Department of Health Laboratories Administration et al |
| EPI_ISL_660080 | USA/MD-MDH-0382/2020 | 2020-10-27 | North America/USA/Maryland/ | MD PHL | MD PHL | Maryland Department of Health Laboratories Administration et al |
| EPI_ISL_660086 | USA/MD-MDH-0392/2020 | 2020-11-02 | North America/USA/Maryland/ | MD PHL | MD PHL | Maryland Department of Health Laboratories Administration et al |
| EPI_ISL_660087 | USA/MD-MDH-0393/2020 | 2020-11-02 | North America/USA/Maryland/ | MD PHL | MD PHL | Maryland Department of Health Laboratories Administration et al |
| EPI_ISL_660098 | USA/MD-MDH-0405/2020 | 2020-11-04 | North America/USA/Maryland/ | MD PHL | MD PHL | Maryland Department of Health Laboratories Administration et al |
| EPI_ISL_660104 | USA/MD-MDH-0386/2020 | 2020-10-26 | North America/USA/Maryland/ | MD PHL | MD PHL | Maryland Department of Health Laboratories Administration et al |

| | | | | | | |
|---|---|---|---|---|---|---|
| EPI_ISL_660129 | SouthAfrica/Tygerberg_320_CC2/202 | 2020-10-09 | Africa/South Africa/Western Cape Provi | Ampath | National Health Laboratory Service (NHLS), Tygerberg | Susan Engelbrecht et al |
| EPI_ISL_660135 | SouthAfrica/Tygerberg_327_CC2/202 | 2020-10-09 | Africa/South Africa/Western Cape Provi | Lancet | National Health Laboratory Service (NHLS), Tygerberg | Susan Engelbrecht et al |
| EPI_ISL_660143 | SouthAfrica/Tygerberg_336_CC2/202 | 2020-10-09 | Africa/South Africa/Western Cape Provi | PathCare | National Health Laboratory Service (NHLS), Tygerberg | Susan Engelbrecht et al |
| EPI_ISL_660144 | SouthAfrica/Tygerberg_337_CC2/202 | 2020-10-09 | Africa/South Africa/Western Cape Provi | PathCare | National Health Laboratory Service (NHLS), Tygerberg | Susan Engelbrecht et al |
| EPI_ISL_660155 | SouthAfrica/Tygerberg_360_CC2/202 | 2020-10-13 | Africa/South Africa/Western Cape Provi | PathCare | National Health Laboratory Service (NHLS), Tygerberg | Susan Engelbrecht et al |
| EPI_ISL_660157 | SouthAfrica/Tygerberg_365_CC2/202 | 2020-10-16 | Africa/South Africa/Western Cape Provi | PathCare | National Health Laboratory Service (NHLS), Tygerberg | Susan Engelbrecht et al |
| EPI_ISL_660165 | SouthAfrica/KRISP-K004282/2020 | 2020-06-25 | Africa/South Africa/KwaZulu-Natal/eThe | NHLS-IALCH | KRISP, KZN Research Innovation and Sequencing Platform | Gazy I et al |
| EPI_ISL_660167 | SouthAfrica/KRISP-K004285/2020 | 2020-07-03 | Africa/South Africa/KwaZulu-Natal/eThe | NHLS-IALCH | KRISP, KZN Research Innovation and Sequencing Platform | Gazy I et al |
| EPI_ISL_660173 | SouthAfrica/KRISP-K004292/2020 | 2020-07-24 | Africa/South Africa/KwaZulu-Natal/eThe | NHLS-IALCH | KRISP, KZN Research Innovation and Sequencing Platform | Gazy I et al |

| | | | | | | |
|---|---|---|---|---|---|---|
| EPI_ISL_660174 | SouthAfrica/KRISP-K004293/2020 | 2020-07-24 | Africa/South Africa/KwaZulu-Natal/eThe | NHLS-IALCH | KRISP, KZN Research Innovation and Sequencing Platform | Gazy I et al |
| EPI_ISL_660175 | SouthAfrica/KRISP-K004294/2020 | 2020-07-27 | Africa/South Africa/KwaZulu-Natal/eThe | NHLS-IALCH | KRISP, KZN Research Innovation and Sequencing Platform | Gazy I et al |
| EPI_ISL_660178 | SouthAfrica/KRISP-K004298/2020 | 2020-08-14 | Africa/South Africa/KwaZulu-Natal/eThe | NHLS-IALCH | KRISP, KZN Research Innovation and Sequencing Platform | Gazy I et al |
| EPI_ISL_660180 | SouthAfrica/KRISP-K004300/2020 | 2020-08-17 | Africa/South Africa/KwaZulu-Natal/eThe | NHLS-IALCH | KRISP, KZN Research Innovation and Sequencing Platform | Gazy I et al |
| EPI_ISL_660181 | SouthAfrica/KRISP-K004302/2020 | 2020-08-18 | Africa/South Africa/KwaZulu-Natal/eThe | NHLS-IALCH | KRISP, KZN Research Innovation and Sequencing Platform | Gazy I et al |
| EPI_ISL_660182 | SouthAfrica/KRISP-K004303/2020 | 2020-08-19 | Africa/South Africa/KwaZulu-Natal/eThe | NHLS-IALCH | KRISP, KZN Research Innovation and Sequencing Platform | Gazy I et al |
| EPI_ISL_660183 | SouthAfrica/KRISP-K004304/2020 | 2020-08-31 | Africa/South Africa/KwaZulu-Natal/eThe | NHLS-IALCH | KRISP, KZN Research Innovation and Sequencing Platform | Gazy I et al |
| EPI_ISL_660199 | SouthAfrica/KRISP-K004323/2020 | 2020-10-25 | Africa/South Africa/KwaZulu-Natal/eThe | NHLS-IALCH | KRISP, KZN Research Innovation and Sequencing Platform | Giandhari J et al |

| | | | | | | |
|---|---|---|---|---|---|---|
| EPI_ISL_660203 | SouthAfrica/KRISP-K004327/2020 | 2020-10-26 | Africa/South Africa/KwaZulu-Natal/eThe | NHLS-IALCH | KRISP, KZN Research Innovation and Sequencing Platform | Giandhari J et al |
| EPI_ISL_660213 | SouthAfrica/KRISP-K004337/2020 | 2020-10-27 | Africa/South Africa/KwaZulu-Natal/Zulu | NHLS-IALCH | KRISP, KZN Research Innovation and Sequencing Platform | Giandhari J et al |
| EPI_ISL_660215 | SouthAfrica/KRISP-K004339/2020 | 2020-10-28 | Africa/South Africa/KwaZulu-Natal/Ugu | NHLS-IALCH | KRISP, KZN Research Innovation and Sequencing Platform | Giandhari J et al |
| EPI_ISL_660253 | SouthAfrica/KRISP-K004556/2020 | 2020-10-19 | Africa/South Africa/KwaZulu-Natal/Ama | NHLS-IALCH | KRISP, KZN Research Innovation and Sequencing Platform | Giandhari J et al |
| EPI_ISL_660256 | SouthAfrica/KRISP-K004723-CS1/2020 | 2020-11-18 | Africa/South Africa/KwaZulu-Natal/eThe | KRISP, KZN Research Innovation and Sequencing Platform | KRISP, KZN Research Innovation and Sequencing Platform | Giandhari J et al |
| EPI_ISL_660266 | Spain/CT-IBV-99009869/2020 | 2020-07-31 | Europe/Spain/Catalunya/Arenys de Mu | Servicio de Microbiolog√≠a, Laboratori Cl√≠nic Metropolitana Nord. Hospital Universitari Germans Trias i Pujol. Institut d‚ÄôInvestigaci√≥ en Ci√®ncies de la Salut Germans Trias i Pujol (IGTP) | SeqCOVID-SPAIN consortium/IBV(CSIC) | Elisa Mart√≠ et al |
| EPI_ISL_660272 | Spain/CT-IBV-99009875/2020 | 2020-08-01 | Europe/Spain/Catalunya/Badalona | Servicio de Microbiolog√≠a, Laboratori Cl√≠nic Metropolitana Nord. Hospital Universitari Germans Trias i Pujol. Institut d‚ÄôInvestigaci√≥ en Ci√®ncies de la Salut Germans Trias i Pujol (IGTP) | SeqCOVID-SPAIN consortium/IBV(CSIC) | Elisa Mart√≠ et al |
| EPI_ISL_660273 | Spain/CT-IBV-99009876/2020 | 2020-08-02 | Europe/Spain/Catalunya/Barcelona | Servicio de Microbiolog√≠a, Laboratori Cl√≠nic Metropolitana Nord. Hospital Universitari Germans Trias i Pujol. Institut d‚ÄôInvestigaci√≥ en Ci√®ncies de la Salut Germans Trias i Pujol (IGTP) | SeqCOVID-SPAIN consortium/IBV(CSIC) | Elisa Mart√≠ et al |

| | | | | | | |
|---|---|---|---|---|---|---|
| EPI_ISL_660276 | Spain/CT-IBV-99009879/2020 | 2020-07-31 | Europe/Spain/Aragon/Ayerbe | Servicio de Microbiolog√≠a, Laboratori Cl√≠nic Metropolitana Nord. Hospital Universitari Germans Trias i Pujol. Institut d,ÄôInvestigaci√≥ en Ci√®ncies de la Salut Germans Trias i Pujol (IGTP) | SeqCOVID-SPAIN consortium/IBV(CSIC) | Elisa Martr√≥ et al |
| EPI_ISL_660278 | Spain/CT-IBV-99009881/2020 | 2020-08-05 | Europe/Spain/Catalunya/La Llagosta | Servicio de Microbiolog√≠a, Laboratori Cl√≠nic Metropolitana Nord. Hospital Universitari Germans Trias i Pujol. Institut d,ÄôInvestigaci√≥ en Ci√®ncies de la Salut Germans Trias i Pujol (IGTP) | SeqCOVID-SPAIN consortium/IBV(CSIC) | Elisa Martr√≥ et al |
| EPI_ISL_660280 | Spain/CT-IBV-99009883/2020 | 2020-08-05 | Europe/Spain/Catalunya/Santa Coloma | Servicio de Microbiolog√≠a, Laboratori Cl√≠nic Metropolitana Nord. Hospital Universitari Germans Trias i Pujol. Institut d,ÄôInvestigaci√≥ en Ci√®ncies de la Salut Germans Trias i Pujol (IGTP) | SeqCOVID-SPAIN consortium/IBV(CSIC) | Elisa Martr√≥ et al |
| EPI_ISL_660281 | Spain/CT-IBV-99009884/2020 | 2020-08-05 | Europe/Spain/Catalunya/Llinars del Vall | Servicio de Microbiolog√≠a, Laboratori Cl√≠nic Metropolitana Nord. Hospital Universitari Germans Trias i Pujol. Institut d,ÄôInvestigaci√≥ en Ci√®ncies de la Salut Germans Trias i Pujol (IGTP) | SeqCOVID-SPAIN consortium/IBV(CSIC) | Elisa Martr√≥ et al |
| EPI_ISL_660282 | Spain/CT-IBV-99009885/2020 | 2020-08-05 | Europe/Spain/Catalunya/Granollers | Servicio de Microbiolog√≠a, Laboratori Cl√≠nic Metropolitana Nord. Hospital Universitari Germans Trias i Pujol. Institut d,ÄôInvestigaci√≥ en Ci√®ncies de la Salut Germans Trias i Pujol (IGTP) | SeqCOVID-SPAIN consortium/IBV(CSIC) | Elisa Martr√≥ et al |
| EPI_ISL_660285 | Spain/CT-IBV-99009888/2020 | 2020-08-06 | Europe/Spain/Catalunya/Bigues I Riells | Servicio de Microbiolog√≠a, Laboratori Cl√≠nic Metropolitana Nord. Hospital Universitari Germans Trias i Pujol. Institut d,ÄôInvestigaci√≥ en Ci√®ncies de la Salut Germans Trias i Pujol (IGTP) | SeqCOVID-SPAIN consortium/IBV(CSIC) | Elisa Martr√≥ et al |
| EPI_ISL_660287 | Spain/CT-IBV-99009890/2020 | 2020-08-06 | Europe/Spain/Catalunya/Santa Coloma | Servicio de Microbiolog√≠a, Laboratori Cl√≠nic Metropolitana Nord. Hospital Universitari Germans Trias i Pujol. Institut d,ÄôInvestigaci√≥ en Ci√®ncies de la Salut Germans Trias i Pujol (IGTP) | SeqCOVID-SPAIN consortium/IBV(CSIC) | Elisa Martr√≥ et al |
| EPI_ISL_660290 | Spain/CT-IBV-99009893/2020 | 2020-08-08 | Europe/Spain/Catalunya/L'Hospitalet de | Servicio de Microbiolog√≠a, Laboratori Cl√≠nic Metropolitana Nord. Hospital Universitari Germans Trias i Pujol. Institut d,ÄôInvestigaci√≥ en Ci√®ncies de la Salut Germans Trias i Pujol (IGTP) | SeqCOVID-SPAIN consortium/IBV(CSIC) | Elisa Martr√≥ et al |

| | | | | | | |
|---|---|---|---|---|---|---|
| EPI_ISL_660292 | Spain/CT-IBV-99009895/2020 | 2020-08-07 | Europe/Spain/Catalunya/Bellavista | Servicio de Microbiologv≠a, Laboratori Clv≠nic Metropolitana Nord. Hospital Universitari Germans Trias i Pujol. Institut d,ÄôInvestigaciv≥ en Civ®ncies de la Salut Germans Trias i Pujol (IGTP) | SeqCOVID-SPAIN consortium/IBV(CSIC) | Elisa Martrv≥ et al |
| EPI_ISL_660293 | Spain/CT-IBV-99009896/2020 | 2020-08-07 | Europe/Spain/Catalunya/Badalona | Servicio de Microbiologv≠a, Laboratori Clv≠nic Metropolitana Nord. Hospital Universitari Germans Trias i Pujol. Institut d,ÄôInvestigaciv≥ en Civ®ncies de la Salut Germans Trias i Pujol (IGTP) | SeqCOVID-SPAIN consortium/IBV(CSIC) | Elisa Martrv≥ et al |
| EPI_ISL_660294 | Spain/CT-IBV-99009897/2020 | 2020-08-07 | Europe/Spain/Catalunya/Lliv ßa de Vall | Servicio de Microbiologv≠a, Laboratori Clv≠nic Metropolitana Nord. Hospital Universitari Germans Trias i Pujol. Institut d,ÄôInvestigaciv≥ en Civ®ncies de la Salut Germans Trias i Pujol (IGTP) | SeqCOVID-SPAIN consortium/IBV(CSIC) | Elisa Martrv≥ et al |
| EPI_ISL_660295 | Spain/CT-IBV-99009898/2020 | 2020-08-07 | Europe/Spain/Catalunya/Mollet del Val | Servicio de Microbiologv≠a, Laboratori Clv≠nic Metropolitana Nord. Hospital Universitari Germans Trias i Pujol. Institut d,ÄôInvestigaciv≥ en Civ®ncies de la Salut Germans Trias i Pujol (IGTP) | SeqCOVID-SPAIN consortium/IBV(CSIC) | Elisa Martrv≥ et al |
| EPI_ISL_660298 | Spain/CT-IBV-99009901/2020 | 2020-08-08 | Europe/Spain/Catalunya/Mollet del Val | Servicio de Microbiologv≠a, Laboratori Clv≠nic Metropolitana Nord. Hospital Universitari Germans Trias i Pujol. Institut d,ÄôInvestigaciv≥ en Civ®ncies de la Salut Germans Trias i Pujol (IGTP) | SeqCOVID-SPAIN consortium/IBV(CSIC) | Elisa Martrv≥ et al |
| EPI_ISL_660300 | Spain/CT-IBV-99009903/2020 | 2020-08-08 | Europe/Spain/Catalunya/Cardedeu | Servicio de Microbiologv≠a, Laboratori Clv≠nic Metropolitana Nord. Hospital Universitari Germans Trias i Pujol. Institut d,ÄôInvestigaciv≥ en Civ®ncies de la Salut Germans Trias i Pujol (IGTP) | SeqCOVID-SPAIN consortium/IBV(CSIC) | Elisa Martrv≥ et al |
| EPI_ISL_660301 | Spain/CT-IBV-99009904/2020 | 2020-08-08 | Europe/Spain/Catalunya/Cardedeu | Servicio de Microbiologv≠a, Laboratori Clv≠nic Metropolitana Nord. Hospital Universitari Germans Trias i Pujol. Institut d,ÄôInvestigaciv≥ en Civ®ncies de la Salut Germans Trias i Pujol (IGTP) | SeqCOVID-SPAIN consortium/IBV(CSIC) | Elisa Martrv≥ et al |
| EPI_ISL_660303 | Spain/CT-IBV-99009906/2020 | 2020-08-08 | Europe/Spain/Catalunya/Cardedeu | Servicio de Microbiologv≠a, Laboratori Clv≠nic Metropolitana Nord. Hospital Universitari Germans Trias i Pujol. Institut d,ÄôInvestigaciv≥ en Civ®ncies de la Salut Germans Trias i Pujol (IGTP) | SeqCOVID-SPAIN consortium/IBV(CSIC) | Elisa Martrv≥ et al |

| | | | | | | |
|---|---|---|---|---|---|---|
| EPI_ISL_660304 | Spain/CT-IBV-99009907/2020 | 2020-09-04 | Europe/Spain/Catalunya/Barcelona | Servicio de Microbiolog√≠a, Laboratori Cl√≠nic Metropolitana Nord. Hospital Universitari Germans Trias i Pujol. Institut d‚ÄôInvestigaci√≥ en Ci√®ncies de la Salut Germans Trias i Pujol (IGTP) | SeqCOVID-SPAIN consortium/IBV(CSIC) | Elisa Mart√≠ et al |
| EPI_ISL_660305 | Spain/CT-IBV-99009908/2020 | 2020-08-09 | Europe/Spain/Catalunya/Mollet del Val | Servicio de Microbiolog√≠a, Laboratori Cl√≠nic Metropolitana Nord. Hospital Universitari Germans Trias i Pujol. Institut d‚ÄôInvestigaci√≥ en Ci√®ncies de la Salut Germans Trias i Pujol (IGTP) | SeqCOVID-SPAIN consortium/IBV(CSIC) | Elisa Mart√≠ et al |
| EPI_ISL_660313 | Spain/CT-IBV-99009916/2020 | 2020-08-11 | Europe/Spain/Catalunya/Teia | Servicio de Microbiolog√≠a, Laboratori Cl√≠nic Metropolitana Nord. Hospital Universitari Germans Trias i Pujol. Institut d‚ÄôInvestigaci√≥ en Ci√®ncies de la Salut Germans Trias i Pujol (IGTP) | SeqCOVID-SPAIN consortium/IBV(CSIC) | Elisa Mart√≠ et al |
| EPI_ISL_660315 | Spain/CT-IBV-99009918/2020 | 2020-08-11 | Europe/Spain/Catalunya/Badalona | Servicio de Microbiolog√≠a, Laboratori Cl√≠nic Metropolitana Nord. Hospital Universitari Germans Trias i Pujol. Institut d‚ÄôInvestigaci√≥ en Ci√®ncies de la Salut Germans Trias i Pujol (IGTP) | SeqCOVID-SPAIN consortium/IBV(CSIC) | Elisa Mart√≠ et al |
| EPI_ISL_660316 | Spain/CT-IBV-99009919/2020 | 2020-08-11 | Europe/Spain/Catalunya/Mollet del Val | Servicio de Microbiolog√≠a, Laboratori Cl√≠nic Metropolitana Nord. Hospital Universitari Germans Trias i Pujol. Institut d‚ÄôInvestigaci√≥ en Ci√®ncies de la Salut Germans Trias i Pujol (IGTP) | SeqCOVID-SPAIN consortium/IBV(CSIC) | Elisa Mart√≠ et al |
| EPI_ISL_660317 | Spain/CT-IBV-99009921/2020 | 2020-08-11 | Europe/Spain/Catalunya/Palau Solita i P | Servicio de Microbiolog√≠a, Laboratori Cl√≠nic Metropolitana Nord. Hospital Universitari Germans Trias i Pujol. Institut d‚ÄôInvestigaci√≥ en Ci√®ncies de la Salut Germans Trias i Pujol (IGTP) | SeqCOVID-SPAIN consortium/IBV(CSIC) | Elisa Mart√≠ et al |
| EPI_ISL_660319 | Spain/CT-IBV-99009923/2020 | 2020-08-12 | Europe/Spain/Catalunya/Sabadell | Servicio de Microbiolog√≠a, Laboratori Cl√≠nic Metropolitana Nord. Hospital Universitari Germans Trias i Pujol. Institut d‚ÄôInvestigaci√≥ en Ci√®ncies de la Salut Germans Trias i Pujol (IGTP) | SeqCOVID-SPAIN consortium/IBV(CSIC) | Elisa Mart√≠ et al |
| EPI_ISL_660325 | France/IDF-IPP9555/2020 | 2020-11-02 | Europe/France/Ile de France/Villeneuve | General practitioner | National Reference Center for Viruses of Respiratory Infections, Institut Pasteur, Paris | Marion Barbet et al |

| | | | | | | |
|---|---|---|---|---|---|---|
| EPI_ISL_660328 | France/ARA-SC323/2020 | 2020-03-23 | Europe/France/Auvergne-Rh√¥ne-Alpes/ | CHU de Saint-√âtienne H√¥pital Nord | CNR Virus des Infections Respiratoires - France SUD | Antonin Bal et al |
| EPI_ISL_660330 | France/ARA-SC326/2020 | 2020-03-24 | Europe/France/Auvergne-Rh√¥ne-Alpes/ | CHU de Saint-√âtienne H√¥pital Nord | CNR Virus des Infections Respiratoires - France SUD | Antonin Bal et al |
| EPI_ISL_660342 | France/ARA-SC349/2020 | 2020-04-11 | Europe/France/Auvergne-Rh√¥ne-Alpes/ | CHU de Saint-√âtienne H√¥pital Nord | CNR Virus des Infections Respiratoires - France SUD | Antonin Bal et al |
| EPI_ISL_660343 | France/ARA-SC352/2020 | 2020-04-15 | Europe/France/Auvergne-Rh√¥ne-Alpes/ | CHU de Saint-√âtienne H√¥pital Nord | CNR Virus des Infections Respiratoires - France SUD | Antonin Bal et al |
| EPI_ISL_660347 | France/ARA-SC362/2020 | 2020-03-07 | Europe/France/Auvergne-Rh√¥ne-Alpes/ | CHU de Saint-√âtienne H√¥pital Nord | CNR Virus des Infections Respiratoires - France SUD | Antonin Bal et al |
| EPI_ISL_660358 | France/ARA-SC386/2020 | 2020-03-17 | Europe/France/Auvergne-Rh√¥ne-Alpes/ | CHU Clermont-Ferrand | CNR Virus des Infections Respiratoires - France SUD | Antonin Bal et al |
| EPI_ISL_660360 | France/ARA-SC391/2020 | 2020-03-28 | Europe/France/Auvergne-Rh√¥ne-Alpes/ | CHU Clermont-Ferrand | CNR Virus des Infections Respiratoires - France SUD | Antonin Bal et al |
| EPI_ISL_660361 | France/ARA-SC396/2020 | 2020-04-05 | Europe/France/Auvergne-Rh√¥ne-Alpes/ | CHU Clermont-Ferrand | CNR Virus des Infections Respiratoires - France SUD | Antonin Bal et al |
| EPI_ISL_660362 | France/ARA-SC399/2020 | 2020-04-09 | Europe/France/Auvergne-Rh√¥ne-Alpes/ | CHU Clermont-Ferrand | CNR Virus des Infections Respiratoires - France SUD | Antonin Bal et al |
| EPI_ISL_660363 | France/ARA-SC400/2020 | 2020-04-14 | Europe/France/Auvergne-Rh√¥ne-Alpes/ | CHU Clermont-Ferrand | CNR Virus des Infections Respiratoires - France SUD | Antonin Bal et al |

| | | | | | | |
|---|---|---|---|---|---|---|
| EPI_ISL_660368 | France/ARA-SC409/2020 | 2020-05-02 | Europe/France/Auvergne-Rh√¥ne-Alpes/ | CHU Clermont-Ferrand | CNR Virus des Infections Respiratoires - France SUD | Antonin Bal et al |
| EPI_ISL_660371 | France/ARA-SC414/2020 | 2020-05-25 | Europe/France/Auvergne-Rh√¥ne-Alpes/ | CHU Clermont-Ferrand | CNR Virus des Infections Respiratoires - France SUD | Antonin Bal et al |
| EPI_ISL_660375 | France/OCC-SC422/2020 | 2020-04-11 | Europe/France/Occitanie/Toulouse | CHU Toulouse | CNR Virus des Infections Respiratoires - France SUD | Antonin Bal et al |
| EPI_ISL_660382 | Sweden/20-22134/2020 | 2020-04-04 | Europe/Sweden/Orebro/ | Orebro klinisk mikrobiologi | The Public Health Agency of Sweden | Anna-Malin Linde et al |
| EPI_ISL_660401 | Sweden/20-22306/2020 | 2020-04-02 | Europe/Sweden/Ostergotland/ | Klinsisk mikrobiologi Linkoping | The Public Health Agency of Sweden | Anna-Malin Linde et al |
| EPI_ISL_660406 | Sweden/20-22313/2020 | 2020-04-06 | Europe/Sweden/Ostergotland/ | Klinsisk mikrobiologi Linkoping | The Public Health Agency of Sweden | Anna-Malin Linde et al |
| EPI_ISL_660407 | Sweden/20-22315/2020 | 2020-04-07 | Europe/Sweden/Ostergotland/ | Klinsisk mikrobiologi Linkoping | The Public Health Agency of Sweden | Anna-Malin Linde et al |
| EPI_ISL_660409 | Sweden/20-22317/2020 | 2020-04-07 | Europe/Sweden/Ostergotland/ | Klinsisk mikrobiologi Linkoping | The Public Health Agency of Sweden | Anna-Malin Linde et al |
| EPI_ISL_660415 | Sweden/20-22561/2020 | 2020-06-22 | Europe/Sweden/Norrbotten/ | unknown | The Public Health Agency of Sweden | Anna-Malin Linde et al |
| EPI_ISL_660419 | Sweden/20-52655/2020 | 2020-05-22 | Europe/Sweden/Orebro/ | Klinisk mikrobiologi | The Public Health Agency of Sweden | Anna-Malin Linde et al |
| EPI_ISL_660420 | Sweden/20-52656/2020 | 2020-06-02 | Europe/Sweden/Orebro/ | Klinisk mikrobiologi | The Public Health Agency of Sweden | Anna-Malin Linde et al |
| EPI_ISL_660424 | Sweden/20-52944/2020 | 2020-09-23 | Europe/Sweden/Orebro/ | Orebro klinisk mikrobiologi | The Public Health Agency of Sweden | Anna-Malin Linde et al |
| EPI_ISL_660433 | Russia/MOS-CRIE-4906308/2020 | 2020-07-15 | Europe/Russia/Moscow Oblast/ | "Molecular diagnostic laboratory of Federal Budget Institution of Science ""Central Research Institute of Epidemiology"" of The Federal Service on Customers' Rights Protection and Human Well-being Surveillance" | Group of Genomics and Postgenomic Technologies of Central Research Institute of Epidemilology | Samoilov AE et al |

| | | | | | | |
|---|---|---|---|---|---|---|
| EPI_ISL_660435 | Russia/MOS-CRIE-5047186/2020 | 2020-07-16 | Europe/Russia/Moscow Oblast/ | "Molecular diagnostic laboratory of Federal Budget Institution of Science ""Central Research Institute of Epidemiology"" of The Federal Service on Customers' Rights Protection and Human Well-being Surveillance" | Group of Genomics and Postgenomic Technologies of Central Research Institute of Epidemilology | Samoilov AE et al |
| EPI_ISL_660436 | Russia/MOS-CRIE-5047186-2/2020 | 2020-07-16 | Europe/Russia/Moscow Oblast/ | "Molecular diagnostic laboratory of Federal Budget Institution of Science ""Central Research Institute of Epidemiology"" of The Federal Service on Customers' Rights Protection and Human Well-being Surveillance" | Group of Genomics and Postgenomic Technologies of Central Research Institute of Epidemilology | Samoilov AE et al |
| EPI_ISL_660437 | Russia/MOS-CRIE-7210452/2020 | 2020-08-24 | Europe/Russia/Moscow Oblast/ | "Molecular diagnostic laboratory of Federal Budget Institution of Science ""Central Research Institute of Epidemiology"" of The Federal Service on Customers' Rights Protection and Human Well-being Surveillance" | Group of Genomics and Postgenomic Technologies of Central Research Institute of Epidemilology | Samoilov AE et al |
| EPI_ISL_660438 | Russia/MOS-CRIE-7161283/2020 | 2020-08-24 | Europe/Russia/Moscow Oblast/ | "Molecular diagnostic laboratory of Federal Budget Institution of Science ""Central Research Institute of Epidemiology"" of The Federal Service on Customers' Rights Protection and Human Well-being Surveillance" | Group of Genomics and Postgenomic Technologies of Central Research Institute of Epidemilology | Samoilov AE et al |
| EPI_ISL_660452 | BurkinaFaso/2-035-1347/2020 | 2020-11-04 | Africa/Burkina Faso/Hauts-Bassins/Bobo | Laboratoire de Microbiologie CHU Souro Sanou | Centre Muraz | Abdoul-Salam Ouedraogo et al |
| EPI_ISL_660491 | BurkinaFaso/2342/2020 | 2020-09-09 | Africa/Burkina Faso/Hauts-Bassins/Bobo | Laboratoire de Microbiologie CHU Souro Sanou | Centre Muraz | Abdoul-Salam Ouedraogo et al |
| EPI_ISL_660504 | BurkinaFaso/3069/2020 | 2020-09-25 | Africa/Burkina Faso/Hauts-Bassins/Bobo | Laboratoire de Microbiologie CHU Souro Sanou | Centre Muraz | Abdoul-Salam Ouedraogo et al |
| EPI_ISL_660521 | BurkinaFaso/3535/2020 | 2020-10-09 | Africa/Burkina Faso/Hauts-Bassins/Bobo | Laboratoire de Microbiologie CHU Souro Sanou | Centre Muraz | Abdoul-Salam Ouedraogo et al |
| EPI_ISL_660529 | Ecuador/USFQ-228/2020 | 2020-10-01 | South America/Ecuador/Tungurahua/An | Institute of Microbiology, Universidad San Francisco de Quito | Institute of Microbiology, Universidad San Francisco de Quito | Sully M√°rquez et al |

| | | | | | | |
|---|---|---|---|---|---|---|
| EPI_ISL_660530 | Ecuador/USFQ-230/2020 | 2020-10-01 | South America/Ecuador/Tungurahua/An | Institute of Microbiology, Universidad San Francisco de Quito | Institute of Microbiology, Universidad San Francisco de Quito | Sully Mv°rquez et al |
| EPI_ISL_660531 | Ecuador/USFQ-231/2020 | 2020-10-01 | South America/Ecuador/Tungurahua/An | Institute of Microbiology, Universidad San Francisco de Quito | Institute of Microbiology, Universidad San Francisco de Quito | Sully Mv°rquez et al |
| EPI_ISL_660532 | Ecuador/USFQ-402/2020 | 2020-10-02 | South America/Ecuador/Pastaza/Puyo | Institute of Microbiology, Universidad San Francisco de Quito | Institute of Microbiology, Universidad San Francisco de Quito | Sully Mv°rquez et al |
| EPI_ISL_660533 | Ecuador/USFQ-485/2020 | 2020-10-05 | South America/Ecuador/Cotopaxi/Latacu | Institute of Microbiology, Universidad San Francisco de Quito | Institute of Microbiology, Universidad San Francisco de Quito | Sully Mv°rquez et al |
| EPI_ISL_660535 | Ecuador/USFQ-506/2020 | 2020-10-10 | South America/Ecuador/El Oro/Machala | Institute of Microbiology, Universidad San Francisco de Quito | Institute of Microbiology, Universidad San Francisco de Quito | Sully Mv°rquez et al |
| EPI_ISL_660536 | Ecuador/USFQ-509/2020 | 2020-10-10 | South America/Ecuador/El Oro/Machala | Institute of Microbiology, Universidad San Francisco de Quito | Institute of Microbiology, Universidad San Francisco de Quito | Sully Mv°rquez et al |
| EPI_ISL_660537 | Ecuador/USFQ-513/2020 | 2020-10-10 | South America/Ecuador/El Oro/Machala | Institute of Microbiology, Universidad San Francisco de Quito | Institute of Microbiology, Universidad San Francisco de Quito | Sully Mv°rquez et al |
| EPI_ISL_660539 | Ecuador/USFQ-520/2020 | 2020-10-02 | South America/Ecuador/Pastaza/Puyo | Institute of Microbiology, Universidad San Francisco de Quito | Institute of Microbiology, Universidad San Francisco de Quito | Sully Mv°rquez et al |
| EPI_ISL_660541 | Vietnam/38238/2020 | 2020-01-31 | Asia/Vietnam/Vinhphuc/ | National Influenza Center, National Institute of Hygiene and Epidemiology (NIHE) | National Influenza Center, National Institute of Hygiene and Epidemiology (NIHE) | Le Quynh Mai et al |

| | | | | | | |
|---|---|---|---|---|---|---|
| EPI_ISL_660543 | Taiwan/CGMH-CGU-31/2020 | 2020-07-31 | Asia/Taiwan/Taiwan/Taoyuan | Laboratory Medicine | Department of Laboratory Medicine, Lin-Kou Chang Gung Memorial Hospital, Taoyuan, Taiwan | Kuo-Chien Tsao et al |
| EPI_ISL_660553 | CzechRepublic/NRL-10233/2020 | 2020-10-21 | Europe/Czech Republic/Prague/ | The National Institute of Public Health | State Veterinary Institute Prague | Nagy et al |
| EPI_ISL_660554 | CzechRepublic/NRL-9962/2020 | 2020-09-24 | Europe/Czech Republic/South Moravian | The National Institute of Public Health | State Veterinary Institute Prague | Nagy et al |
| EPI_ISL_660555 | CzechRepublic/NRL-10281/2020 | 2020-10-22 | Europe/Czech Republic/Czech Republic/ | The National Institute of Public Health | State Veterinary Institute Prague | Nagy et al |
| EPI_ISL_660556 | CzechRepublic/NRL-10242/2020 | 2020-10-22 | Europe/Czech Republic/Central Bohemia | The National Institute of Public Health | State Veterinary Institute Prague | Nagy et al |
| EPI_ISL_660557 | CzechRepublic/NRL-10288/2020 | 2020-10-22 | Europe/Czech Republic/Prague/ | The National Institute of Public Health | State Veterinary Institute Prague | Nagy et al |
| EPI_ISL_660558 | CzechRepublic/NRL-9965/2020 | 2020-09-29 | Europe/Czech Republic/South Moravian | The National Institute of Public Health | State Veterinary Institute Prague | Nagy et al |
| EPI_ISL_660559 | CzechRepublic/NRL-9969/2020 | 2020-10-15 | Europe/Czech Republic/Prague/ | The National Institute of Public Health | State Veterinary Institute Prague | Nagy et al |
| EPI_ISL_660560 | CzechRepublic/NRL-9990/2020 | 2020-10-15 | Europe/Czech Republic/Prague/ | The National Institute of Public Health | State Veterinary Institute Prague | Nagy et al |
| EPI_ISL_660561 | CzechRepublic/NRL-10015/2020 | 2020-10-16 | Europe/Czech Republic/Prague/ | The National Institute of Public Health | State Veterinary Institute Prague | Nagy et al |
| EPI_ISL_660562 | CzechRepublic/NRL-10020/2020 | 2020-10-16 | Europe/Czech Republic/Prague/ | The National Institute of Public Health | State Veterinary Institute Prague | Nagy et al |
| EPI_ISL_660563 | CzechRepublic/NRL-10317/2020 | 2020-10-23 | Europe/Czech Republic/Prague/ | The National Institute of Public Health | State Veterinary Institute Prague | Nagy et al |
| EPI_ISL_660565 | CzechRepublic/NRL-10295/2020 | 2020-10-23 | Europe/Czech Republic/Southern Bohem | The National Institute of Public Health | State Veterinary Institute Prague | Nagy et al |
| EPI_ISL_660567 | CzechRepublic/NRL-10114/2020 | 2020-10-19 | Europe/Czech Republic/Prague/ | The National Institute of Public Health | State Veterinary Institute Prague | Nagy et al |
| EPI_ISL_660568 | CzechRepublic/NRL-10294/2020 | 2020-10-23 | Europe/Czech Republic/Southern Bohem | The National Institute of Public Health | State Veterinary Institute Prague | Nagy et al |
| EPI_ISL_660570 | CzechRepublic/NRL-10092/2020 | 2020-10-19 | Europe/Czech Republic/Prague/ | The National Institute of Public Health | State Veterinary Institute Prague | Nagy et al |
| EPI_ISL_660572 | CzechRepublic/NRL-10197/2020 | 2020-10-21 | Europe/Czech Republic/Prague/ | The National Institute of Public Health | State Veterinary Institute Prague | Nagy et al |

| | | | | | | |
|---|---|---|---|---|---|---|
| EPI_ISL_660573 | CzechRepublic/NRL-10207/2020 | 2020-10-21 | Europe/Czech Republic/Prague/ | The National Institute of Public Health | State Veterinary Institute Prague | Nagy et al |
| EPI_ISL_660574 | CzechRepublic/NRL-10336/2020 | 2020-10-23 | Europe/Czech Republic/Central Bohemia | The National Institute of Public Health | State Veterinary Institute Prague | Nagy et al |
| EPI_ISL_660578 | CzechRepublic/NRL-10269/2020 | 2020-10-22 | Europe/Czech Republic/Central Bohemia | The National Institute of Public Health | State Veterinary Institute Prague | Nagy et al |
| EPI_ISL_660579 | CzechRepublic/NRL-10056/2020 | 2020-10-16 | Europe/Czech Republic/Central Bohemia | The National Institute of Public Health | State Veterinary Institute Prague | Nagy et al |
| EPI_ISL_660580 | CzechRepublic/NRL-10227/2020 | 2020-10-21 | Europe/Czech Republic/Central Bohemia | The National Institute of Public Health | State Veterinary Institute Prague | Nagy et al |
| EPI_ISL_660582 | CzechRepublic/NRL-9978/2020 | 2020-10-15 | Europe/Czech Republic/Central Bohemia | The National Institute of Public Health | State Veterinary Institute Prague | Nagy et al |
| EPI_ISL_660583 | CzechRepublic/NRL-10208/2020 | 2020-10-21 | Europe/Czech Republic/Hradec Krv°lovv( | The National Institute of Public Health | State Veterinary Institute Prague | Nagy et al |
| EPI_ISL_660584 | CzechRepublic/NRL-10121/2020 | 2020-10-19 | Europe/Czech Republic/Prague/ | The National Institute of Public Health | State Veterinary Institute Prague | Nagy et al |
| EPI_ISL_660587 | CzechRepublic/NRL-10218/2020 | 2020-10-21 | Europe/Czech Republic/Prague/ | The National Institute of Public Health | State Veterinary Institute Prague | Nagy et al |
| EPI_ISL_660588 | CzechRepublic/NRL-10225/2020 | 2020-10-21 | Europe/Czech Republic/Prague/ | The National Institute of Public Health | State Veterinary Institute Prague | Nagy et al |
| EPI_ISL_660589 | CzechRepublic/NRL-9908/2020 | 2020-10-14 | Europe/Czech Republic/Prague/ | The National Institute of Public Health | State Veterinary Institute Prague | Nagy et al |
| EPI_ISL_660592 | CzechRepublic/NRL-10247/2020 | 2020-10-22 | Europe/Czech Republic/Prague/ | The National Institute of Public Health | State Veterinary Institute Prague | Nagy et al |
| EPI_ISL_660593 | CzechRepublic/NRL-10324/2020 | 2020-10-23 | Europe/Czech Republic/Prague/ | The National Institute of Public Health | State Veterinary Institute Prague | Nagy et al |
| EPI_ISL_660594 | CzechRepublic/NRL-10326/2020 | 2020-10-23 | Europe/Czech Republic/Prague/ | The National Institute of Public Health | State Veterinary Institute Prague | Nagy et al |
| EPI_ISL_660595 | CzechRepublic/NRL-10273/2020 | 2020-10-22 | Europe/Czech Republic/Prague/ | The National Institute of Public Health | State Veterinary Institute Prague | Nagy et al |
| EPI_ISL_660597 | CzechRepublic/NRL-9964/2020 | 2020-09-22 | Europe/Czech Republic/South Moravian | The National Institute of Public Health | State Veterinary Institute Prague | Nagy et al |
| EPI_ISL_660598 | CzechRepublic/NRL-10066/2020 | 2020-10-18 | Europe/Czech Republic/Czech Republic/ | The National Institute of Public Health | State Veterinary Institute Prague | Nagy et al |
| EPI_ISL_660648 | SouthAfrica/KRISP-K004632/2020 | 2020-11-04 | Africa/South Africa/KwaZulu-Natal/uMk | NHLS-IALCH | KRISP, KZN Research Innovation and Sequencing Platform | Giandhari J et al |

| | | | | | | |
|---|---|---|---|---|---|---|
| EPI_ISL_660670 | France/OCC-SC433/2020 | 2020-05-25 | Europe/France/Occitanie/Toulouse | CHU Toulouse | CNR Virus des Infections Respiratoires - France SUD | Antonin Bal et al |
| EPI_ISL_660672 | France/OCC-SC464/2020 | 2020-03-03 | Europe/France/Occitanie/Montpellier | CHU Montpellier | CNR Virus des Infections Respiratoires - France SUD | Antonin Bal et al |
| EPI_ISL_660674 | France/OCC-SC470/2020 | 2020-03-09 | Europe/France/Occitanie/Montpellier | CHU Montpellier | CNR Virus des Infections Respiratoires - France SUD | Antonin Bal et al |
| EPI_ISL_660680 | France/OCC-SC479/2020 | 2020-03-17 | Europe/France/Occitanie/Montpellier | CHU Montpellier | CNR Virus des Infections Respiratoires - France SUD | Antonin Bal et al |
| EPI_ISL_660681 | France/OCC-SC480/2020 | 2020-03-24 | Europe/France/Occitanie/Montpellier | CHU Montpellier | CNR Virus des Infections Respiratoires - France SUD | Antonin Bal et al |
| EPI_ISL_660682 | France/OCC-SC481/2020 | 2020-03-24 | Europe/France/Occitanie/Montpellier | CHU Montpellier | CNR Virus des Infections Respiratoires - France SUD | Antonin Bal et al |
| EPI_ISL_660685 | France/OCC-SC485/2020 | 2020-03-24 | Europe/France/Occitanie/Montpellier | CHU Montpellier | CNR Virus des Infections Respiratoires - France SUD | Antonin Bal et al |
| EPI_ISL_660688 | France/OCC-SC492/2020 | 2020-03-26 | Europe/France/Occitanie/Montpellier | CHU Montpellier | CNR Virus des Infections Respiratoires - France SUD | Antonin Bal et al |
| EPI_ISL_660690 | France/OCC-SC495/2020 | 2020-03-26 | Europe/France/Occitanie/Montpellier | CHU Montpellier | CNR Virus des Infections Respiratoires - France SUD | Antonin Bal et al |
| EPI_ISL_660691 | France/OCC-SC497/2020 | 2020-03-30 | Europe/France/Occitanie/Montpellier | CHU Montpellier | CNR Virus des Infections Respiratoires - France SUD | Antonin Bal et al |

| | | | | | | |
|---|---|---|---|---|---|---|
| EPI_ISL_660695 | France/OCC-SC501/2020 | 2020-04-06 | Europe/France/Occitanie/Montpellier | CHU Montpellier | CNR Virus des Infections Respiratoires - France SUD | Antonin Bal et al |
| EPI_ISL_660697 | France/OCC-SC505/2020 | 2020-04-22 | Europe/France/Occitanie/Montpellier | CHU Montpellier | CNR Virus des Infections Respiratoires - France SUD | Antonin Bal et al |
| EPI_ISL_660698 | France/OCC-SC506/2020 | 2020-04-22 | Europe/France/Occitanie/Montpellier | CHU Montpellier | CNR Virus des Infections Respiratoires - France SUD | Antonin Bal et al |
| EPI_ISL_660705 | France/OCC-SC517/2020 | 2020-04-29 | Europe/France/Occitanie/Montpellier | CHU Montpellier | CNR Virus des Infections Respiratoires - France SUD | Antonin Bal et al |
| EPI_ISL_660708 | France/OCC-SC521/2020 | 2020-05-11 | Europe/France/Occitanie/Montpellier | CHU Montpellier | CNR Virus des Infections Respiratoires - France SUD | Antonin Bal et al |
| EPI_ISL_660709 | France/OCC-SC522/2020 | 2020-05-11 | Europe/France/Occitanie/Montpellier | CHU Montpellier | CNR Virus des Infections Respiratoires - France SUD | Antonin Bal et al |
| EPI_ISL_660717 | France/OCC-SC541/2020 | 2020-03-30 | Europe/France/Occitanie/N√Æmes | CHU N√Æmes | CNR Virus des Infections Respiratoires - France SUD | Antonin Bal et al |
| EPI_ISL_660718 | France/OCC-SC542/2020 | 2020-03-30 | Europe/France/Occitanie/N√Æmes | CHU N√Æmes | CNR Virus des Infections Respiratoires - France SUD | Antonin Bal et al |
| EPI_ISL_660720 | France/OCC-SC548/2020 | 2020-04-03 | Europe/France/Occitanie/N√Æmes | CHU N√Æmes | CNR Virus des Infections Respiratoires - France SUD | Antonin Bal et al |
| EPI_ISL_660724 | France/OCC-SC555/2020 | 2020-04-04 | Europe/France/Occitanie/N√Æmes | CHU N√Æmes | CNR Virus des Infections Respiratoires - France SUD | Antonin Bal et al |

| | | | | | | |
|---|---|---|---|---|---|---|
| EPI_ISL_660728 | France/OCC-SC561/2020 | 2020-04-20 | Europe/France/Occitanie/N√Æmes | CHU N√Æmes | CNR Virus des Infections Respiratoires - France SUD | Antonin Bal et al |
| EPI_ISL_660730 | France/OCC-SC564/2020 | 2020-04-22 | Europe/France/Occitanie/N√Æmes | CHU N√Æmes | CNR Virus des Infections Respiratoires - France SUD | Antonin Bal et al |
| EPI_ISL_660731 | France/OCC-SC565/2020 | 2020-05-19 | Europe/France/Occitanie/N√Æmes | CHU N√Æmes | CNR Virus des Infections Respiratoires - France SUD | Antonin Bal et al |
| EPI_ISL_660739 | France/PAC-SC454/2020 | 2020-05-08 | Europe/France/Provence-Alpes-C√¥te d' | Unit√© des Virus √âmergents | CNR Virus des Infections Respiratoires - France SUD | Antonin Bal et al |
| EPI_ISL_660741 | France/PAC-SC459/2020 | 2020-05-14 | Europe/France/Provence-Alpes-C√¥te d' | Unit√© des Virus √âmergents | CNR Virus des Infections Respiratoires - France SUD | Antonin Bal et al |
| EPI_ISL_660769 | England/204491161/2020 | 2020-10-31 | Europe/United Kingdom/England/ | Respiratory Virus Unit, Microbiology Services Colindale, Public Health England | COVID-19 Genomics UK (COG-UK) Consortium | PHE Covid Sequencing Team et al |
| EPI_ISL_660796 | USA/WI-GMF-20481/2020 | 2020-09-15 | North America/USA/Wisconsin/Trempea | Gundersen Molecular Diagnostics Laboratory | Kabara Cancer Research Institute | Craig S. Richmond et al |
| EPI_ISL_660803 | USA/WI-GMF-20611/2020 | 2020-09-16 | North America/USA/Wisconsin/La Crosse | Gundersen Clinical Microbiology Laboratory | Kabara Cancer Research Institute | Craig S. Richmond et al |
| EPI_ISL_660804 | USA/WI-GMF-20612/2020 | 2020-09-16 | North America/USA/Wisconsin/La Crosse | Gundersen Clinical Microbiology Laboratory | Kabara Cancer Research Institute | Craig S. Richmond et al |
| EPI_ISL_660808 | USA/WI-GMF-20630/2020 | 2020-09-16 | North America/USA/Wisconsin/La Crosse | Gundersen Molecular Diagnostics Laboratory | Kabara Cancer Research Institute | Craig S. Richmond et al |
| EPI_ISL_660810 | USA/WI-GMF-20649/2020 | 2020-09-15 | North America/USA/Wisconsin/Vernon | Gundersen Molecular Diagnostics Laboratory | Kabara Cancer Research Institute | Craig S. Richmond et al |
| EPI_ISL_660812 | USA/WI-GMF-20745/2020 | 2020-09-17 | North America/USA/Wisconsin/Vernon | Gundersen Molecular Diagnostics Laboratory | Kabara Cancer Research Institute | Craig S. Richmond et al |
| EPI_ISL_660815 | USA/WI-GMF-20782/2020 | 2020-09-17 | North America/USA/Wisconsin/Trempea | Gundersen Molecular Diagnostics Laboratory | Kabara Cancer Research Institute | Craig S. Richmond et al |
| EPI_ISL_660818 | USA/WI-GMF-20859/2020 | 2020-09-17 | North America/USA/Wisconsin/La Crosse | Gundersen Molecular Diagnostics Laboratory | Kabara Cancer Research Institute | Craig S. Richmond et al |

| | | | | | | |
|---|---|---|---|---|---|---|
| EPI_ISL_660821 | USA/WI-GMF-20912/2020 | 2020-09-18 | North America/USA/Wisconsin/Monroe | Gundersen Molecular Diagnostics Laboratory | Kabara Cancer Research Institute | Craig S. Richmond et al |
| EPI_ISL_660822 | USA/WI-GMF-20969/2020 | 2020-09-18 | North America/USA/Wisconsin/Trempea | Gundersen Molecular Diagnostics Laboratory | Kabara Cancer Research Institute | Craig S. Richmond et al |
| EPI_ISL_660832 | USA/MN-GMF-21384/2020 | 2020-09-20 | North America/USA/Minnesota/Fillmore | Gundersen Molecular Diagnostics Laboratory | Kabara Cancer Research Institute | Craig S. Richmond et al |
| EPI_ISL_660838 | USA/WI-GMF-21746/2020 | 2020-09-21 | North America/USA/Wisconsin/Grant Co | Gundersen Molecular Diagnostics Laboratory | Kabara Cancer Research Institute | Craig S. Richmond et al |
| EPI_ISL_660841 | USA/MN-GMF-21836/2020 | 2020-09-22 | North America/USA/Minnesota/Housto | Gundersen Molecular Diagnostics Laboratory | Kabara Cancer Research Institute | Craig S. Richmond et al |
| EPI_ISL_660842 | USA/MN-GMF-21861/2020 | 2020-09-22 | North America/USA/Minnesota/Winona | Gundersen Molecular Diagnostics Laboratory | Kabara Cancer Research Institute | Craig S. Richmond et al |
| EPI_ISL_660847 | USA/WI-GMF-22070/2020 | 2020-09-23 | North America/USA/Wisconsin/Vernon | Gundersen Molecular Diagnostics Laboratory | Kabara Cancer Research Institute | Craig S. Richmond et al |
| EPI_ISL_660849 | USA/WI-GMF-22102/2020 | 2020-09-23 | North America/USA/Wisconsin/La Cross | Gundersen Molecular Diagnostics Laboratory | Kabara Cancer Research Institute | Craig S. Richmond et al |
| EPI_ISL_660851 | USA/WI-GMF-22117/2020 | 2020-09-23 | North America/USA/Wisconsin/La Cross | Gundersen Molecular Diagnostics Laboratory | Kabara Cancer Research Institute | Craig S. Richmond et al |
| EPI_ISL_660852 | USA/WI-GMF-22170/2020 | 2020-09-23 | North America/USA/Wisconsin/La Cross | Gundersen Molecular Diagnostics Laboratory | Kabara Cancer Research Institute | Craig S. Richmond et al |
| EPI_ISL_660856 | USA/MN-GMF-22344/2020 | 2020-09-25 | North America/USA/Minnesota/Housto | Gundersen Molecular Diagnostics Laboratory | Kabara Cancer Research Institute | Craig S. Richmond et al |
| EPI_ISL_660860 | USA/MN-GMF-22820/2020 | 2020-09-28 | North America/USA/Minnesota/Fillmore | Gundersen Molecular Diagnostics Laboratory | Kabara Cancer Research Institute | Craig S. Richmond et al |
| EPI_ISL_660864 | USA/WI-GMF-22921/2020 | 2020-09-28 | North America/USA/Wisconsin/La Cross | Gundersen Molecular Diagnostics Laboratory | Kabara Cancer Research Institute | Craig S. Richmond et al |
| EPI_ISL_660866 | USA/WI-GMF-23113/2020 | 2020-09-29 | North America/USA/Wisconsin/La Cross | Gundersen Molecular Diagnostics Laboratory | Kabara Cancer Research Institute | Craig S. Richmond et al |
| EPI_ISL_660879 | USA/IA-GMF-23647/2020 | 2020-10-02 | North America/USA/Iowa/Allamakee Co | Gundersen Clinical Microbiology Laboratory | Kabara Cancer Research Institute | Craig S. Richmond et al |
| EPI_ISL_660883 | USA/WI-GMF-23942/2020 | 2020-10-04 | North America/USA/Wisconsin/La Cross | Gundersen Molecular Diagnostics Laboratory | Kabara Cancer Research Institute | Craig S. Richmond et al |
| EPI_ISL_660888 | USA/WI-GMF-24039/2020 | 2020-10-04 | North America/USA/Wisconsin/La Cross | Gundersen Molecular Diagnostics Laboratory | Kabara Cancer Research Institute | Craig S. Richmond et al |
| EPI_ISL_660889 | USA/WI-GMF-24056/2020 | 2020-10-05 | North America/USA/Wisconsin/La Cross | Gundersen Molecular Diagnostics Laboratory | Kabara Cancer Research Institute | Craig S. Richmond et al |
| EPI_ISL_660910 | USA/WI-GMF-24758/2020 | 2020-10-08 | North America/USA/Wisconsin/La Cross | Gundersen Molecular Diagnostics Laboratory | Kabara Cancer Research Institute | Craig S. Richmond et al |
| EPI_ISL_660926 | USA/WI-GMF-25441/2020 | 2020-10-12 | North America/USA/Wisconsin/La Cross | Gundersen Molecular Diagnostics Laboratory | Kabara Cancer Research Institute | Craig S. Richmond et al |

| | | | | | | |
|---|---|---|---|---|---|---|
| EPI_ISL_660930 | USA/WI-GMF-25487/2020 | 2020-10-12 | North America/USA/Wisconsin/La Cross | Gundersen Molecular Diagnostics Laboratory | Kabara Cancer Research Institute | Craig S. Richmond et al |
| EPI_ISL_660936 | USA/WI-GMF-25857/2020 | 2020-10-14 | North America/USA/Wisconsin/La Cross | Gundersen Molecular Diagnostics Laboratory | Kabara Cancer Research Institute | Craig S. Richmond et al |
| EPI_ISL_660940 | USA/WI-GMF-25942/2020 | 2020-10-15 | North America/USA/Wisconsin/Juneau ( | Gundersen Molecular Diagnostics Laboratory | Kabara Cancer Research Institute | Craig S. Richmond et al |
| EPI_ISL_660941 | USA/WI-GMF-25948/2020 | 2020-10-15 | North America/USA/Wisconsin/Trempea | Gundersen Molecular Diagnostics Laboratory | Kabara Cancer Research Institute | Craig S. Richmond et al |
| EPI_ISL_660943 | USA/MN-GMF-26024/2020 | 2020-10-15 | North America/USA/Minnesota/Housto | Gundersen Molecular Diagnostics Laboratory | Kabara Cancer Research Institute | Craig S. Richmond et al |
| EPI_ISL_660952 | USA/WI-GMF-26493/2020 | 2020-10-18 | North America/USA/Wisconsin/La Cross | Gundersen Molecular Diagnostics Laboratory | Kabara Cancer Research Institute | Craig S. Richmond et al |
| EPI_ISL_660954 | USA/WI-GMF-26498/2020 | 2020-10-18 | North America/USA/Wisconsin/La Cross | Gundersen Molecular Diagnostics Laboratory | Kabara Cancer Research Institute | Craig S. Richmond et al |
| EPI_ISL_660958 | USA/WI-GMF-26667/2020 | 2020-10-19 | North America/USA/Wisconsin/La Cross | Gundersen Molecular Diagnostics Laboratory | Kabara Cancer Research Institute | Craig S. Richmond et al |
| EPI_ISL_660959 | USA/WI-GMF-26669/2020 | 2020-10-19 | North America/USA/Wisconsin/Vernon ( | Gundersen Molecular Diagnostics Laboratory | Kabara Cancer Research Institute | Craig S. Richmond et al |
| EPI_ISL_660971 | USA/WI-GMF-27181/2020 | 2020-10-22 | North America/USA/Wisconsin/Vernon ( | Gundersen Molecular Diagnostics Laboratory | Kabara Cancer Research Institute | Craig S. Richmond et al |
| EPI_ISL_660979 | USA/WI-GMF-27504/2020 | 2020-10-24 | North America/USA/Wisconsin/Monroe | Gundersen Molecular Diagnostics Laboratory | Kabara Cancer Research Institute | Craig S. Richmond et al |
| EPI_ISL_660996 | USA/WI-GMF-27942/2020 | 2020-10-27 | North America/USA/Wisconsin/Crawfor | Gundersen Molecular Diagnostics Laboratory | Kabara Cancer Research Institute | Craig S. Richmond et al |
| EPI_ISL_660999 | USA/WI-GMF-28004/2020 | 2020-10-27 | North America/USA/Wisconsin/Grant C | Gundersen Molecular Diagnostics Laboratory | Kabara Cancer Research Institute | Craig S. Richmond et al |
| EPI_ISL_661005 | USA/WI-GMF-28108/2020 | 2020-10-27 | North America/USA/Wisconsin/La Cross | Gundersen Molecular Diagnostics Laboratory | Kabara Cancer Research Institute | Craig S. Richmond et al |
| EPI_ISL_661007 | USA/WI-GMF-28119/2020 | 2020-10-27 | North America/USA/Wisconsin/La Cross | Gundersen Molecular Diagnostics Laboratory | Kabara Cancer Research Institute | Craig S. Richmond et al |
| EPI_ISL_661008 | USA/WI-GMF-28137/2020 | 2020-10-27 | North America/USA/Wisconsin/La Cross | Gundersen Molecular Diagnostics Laboratory | Kabara Cancer Research Institute | Craig S. Richmond et al |
| EPI_ISL_661016 | USA/WI-GMF-28395/2020 | 2020-10-28 | North America/USA/Wisconsin/La Cross | Gundersen Molecular Diagnostics Laboratory | Kabara Cancer Research Institute | Craig S. Richmond et al |
| EPI_ISL_661020 | USA/WI-GMF-28421/2020 | 2020-10-28 | North America/USA/Wisconsin/Vernon ( | Gundersen Molecular Diagnostics Laboratory | Kabara Cancer Research Institute | Craig S. Richmond et al |
| EPI_ISL_661028 | USA/WI-GMF-30231/2020 | 2020-10-30 | North America/USA/Wisconsin/La Cross | Gundersen Molecular Diagnostics Laboratory | Kabara Cancer Research Institute | Craig S. Richmond et al |
| EPI_ISL_661036 | USA/WI-GMF-30475/2020 | 2020-10-31 | North America/USA/Wisconsin/La Cross | Gundersen Molecular Diagnostics Laboratory | Kabara Cancer Research Institute | Craig S. Richmond et al |

| | | | | | | |
|---|---|---|---|---|---|---|
| EPI_ISL_661047 | USA/WI-GMF-30719/2020 | 2020-11-02 | North America/USA/Wisconsin/La Cross | Gundersen Molecular Diagnostics Laboratory | Kabara Cancer Research Institute | Craig S. Richmond et al |
| EPI_ISL_661060 | USA/MN-GMF-30871/2020 | 2020-11-02 | North America/USA/Minnesota/Winona | Gundersen Molecular Diagnostics Laboratory | Kabara Cancer Research Institute | Craig S. Richmond et al |
| EPI_ISL_661063 | USA/WI-GMF-30889/2020 | 2020-11-02 | North America/USA/Wisconsin/La Cross | Gundersen Molecular Diagnostics Laboratory | Kabara Cancer Research Institute | Craig S. Richmond et al |
| EPI_ISL_661065 | USA/WI-GMF-30903/2020 | 2020-11-02 | North America/USA/Wisconsin/Monroe | Gundersen Molecular Diagnostics Laboratory | Kabara Cancer Research Institute | Craig S. Richmond et al |
| EPI_ISL_661074 | USA/WI-GMF-31134/2020 | 2020-11-03 | North America/USA/Wisconsin/La Cross | Gundersen Molecular Diagnostics Laboratory | Kabara Cancer Research Institute | Craig S. Richmond et al |
| EPI_ISL_661076 | USA/MN-GMF-31153/2020 | 2020-11-03 | North America/USA/Minnesota/Winona | Gundersen Molecular Diagnostics Laboratory | Kabara Cancer Research Institute | Craig S. Richmond et al |
| EPI_ISL_661078 | USA/WI-GMF-31217/2020 | 2020-11-03 | North America/USA/Wisconsin/Buffalo C | Gundersen Molecular Diagnostics Laboratory | Kabara Cancer Research Institute | Craig S. Richmond et al |
| EPI_ISL_661090 | USA/WI-GMF-31430/2020 | 2020-11-04 | North America/USA/Wisconsin/La Cross | Gundersen Molecular Diagnostics Laboratory | Kabara Cancer Research Institute | Craig S. Richmond et al |
| EPI_ISL_661105 | USA/WI-GMF-31633/2020 | 2020-11-05 | North America/USA/Wisconsin/Trempea | Gundersen Molecular Diagnostics Laboratory | Kabara Cancer Research Institute | Craig S. Richmond et al |
| EPI_ISL_661109 | USA/WI-GMF-31666/2020 | 2020-11-05 | North America/USA/Wisconsin/La Cross | Gundersen Molecular Diagnostics Laboratory | Kabara Cancer Research Institute | Craig S. Richmond et al |
| EPI_ISL_661114 | USA/WI-GMF-31811/2020 | 2020-11-06 | North America/USA/Wisconsin/La Cross | Gundersen Molecular Diagnostics Laboratory | Kabara Cancer Research Institute | Craig S. Richmond et al |
| EPI_ISL_661118 | USA/WI-GMF-31893/2020 | 2020-11-06 | North America/USA/Wisconsin/La Cross | Gundersen Molecular Diagnostics Laboratory | Kabara Cancer Research Institute | Craig S. Richmond et al |
| EPI_ISL_661125 | USA/WI-GMF-32134/2020 | 2020-11-07 | North America/USA/Wisconsin/La Cross | Gundersen Molecular Diagnostics Laboratory | Kabara Cancer Research Institute | Craig S. Richmond et al |
| EPI_ISL_661131 | USA/WI-GMF-32237/2020 | 2020-11-08 | North America/USA/Wisconsin/La Cross | Gundersen Molecular Diagnostics Laboratory | Kabara Cancer Research Institute | Craig S. Richmond et al |
| EPI_ISL_661133 | USA/WI-GMF-32244/2020 | 2020-11-08 | North America/USA/Wisconsin/La Cross | Gundersen Molecular Diagnostics Laboratory | Kabara Cancer Research Institute | Craig S. Richmond et al |
| EPI_ISL_661138 | USA/WI-GMF-32355/2020 | 2020-11-08 | North America/USA/Wisconsin/Grant Co | Gundersen Molecular Diagnostics Laboratory | Kabara Cancer Research Institute | Craig S. Richmond et al |
| EPI_ISL_661151 | USA/WI-GMF-32511/2020 | 2020-11-09 | North America/USA/Wisconsin/La Cross | Gundersen Molecular Diagnostics Laboratory | Kabara Cancer Research Institute | Craig S. Richmond et al |
| EPI_ISL_661154 | USA/WI-GMF-32562/2020 | 2020-11-09 | North America/USA/Wisconsin/La Cross | Gundersen Molecular Diagnostics Laboratory | Kabara Cancer Research Institute | Craig S. Richmond et al |
| EPI_ISL_661160 | USA/MN-GMF-32638/2020 | 2020-11-09 | North America/USA/Minnesota/Winona | Gundersen Molecular Diagnostics Laboratory | Kabara Cancer Research Institute | Craig S. Richmond et al |
| EPI_ISL_661163 | USA/WI-GMF-32685/2020 | 2020-11-09 | North America/USA/Wisconsin/Vernon C | Gundersen Molecular Diagnostics Laboratory | Kabara Cancer Research Institute | Craig S. Richmond et al |

| | | | | | | |
|---|---|---|---|---|---|---|
| EPI_ISL_661165 | USA/WI-GMF-32806/2020 | 2020-11-10 | North America/USA/Wisconsin/La Crosse | Gundersen Molecular Diagnostics Laboratory | Kabara Cancer Research Institute | Craig S. Richmond et al |
| EPI_ISL_661173 | USA/WI-GMF-M3001/2020 | 2020-11-16 | North America/USA/Wisconsin/La Crosse | Gundersen Molecular Diagnostics Laboratory | Kabara Cancer Research Institute | Craig S. Richmond et al |
| EPI_ISL_661180 | Serbia/NoviSad-197031606/2020 | 2020-06-15 | Europe/Serbia/South Baƒçka District/No | Scientific Veterinary Institute Novi Sad | "Veterinary Specialized Institute ""Kraljevo"", Serbia" | Vidanovic et al |
| EPI_ISL_661184 | Serbia/Pancevo-210032506/2020 | 2020-06-24 | Europe/Serbia/South Banat District/Pan | Scientific Veterinary Institute Novi Sad | "Veterinary Specialized Institute ""Kraljevo"", Serbia" | Vidanovic et al |
| EPI_ISL_661189 | Serbia/Kikinda-000201303/2020 | 2020-03-12 | Europe/Serbia/North Banat District/Kiki | Scientific Veterinary Institute Novi Sad | "Veterinary Specialized Institute ""Kraljevo"", Serbia" | Vidanovic et al |
| EPI_ISL_661191 | Serbia/NoviSad-209542506/2020 | 2020-06-24 | Europe/Serbia/South Baƒçka District/No | Scientific Veterinary Institute Novi Sad | "Veterinary Specialized Institute ""Kraljevo"", Serbia" | Vidanovic et al |
| EPI_ISL_661194 | Serbia/NoviSad-283692307/2020 | 2020-07-22 | Europe/Serbia/South Baƒçka District/No | Scientific Veterinary Institute Novi Sad | "Veterinary Specialized Institute ""Kraljevo"", Serbia" | Vidanovic et al |
| EPI_ISL_661197 | Serbia/Vrsac-259991107/2020 | 2020-07-10 | Europe/Serbia/South Banat District/Vrs | Scientific Veterinary Institute Novi Sad | "Veterinary Specialized Institute ""Kraljevo"", Serbia" | Vidanovic et al |
| EPI_ISL_661222 | Belgium/ULG-10787/2020 | 2020-11-16 | Europe/Belgium/Li√®ge/ | Department of Clinical Microbiology | GIGA Medical Genomics | Keith Durkin et al |
| EPI_ISL_661232 | Belgium/ULG-10803/2020 | 2020-11-17 | Europe/Belgium/Li√®ge/ | Department of Clinical Microbiology | GIGA Medical Genomics | Keith Durkin et al |
| EPI_ISL_661241 | Belgium/ULG-10817/2020 | 2020-11-17 | Europe/Belgium/Li√®ge/ | Department of Clinical Microbiology | GIGA Medical Genomics | Keith Durkin et al |

| | | | | | | |
|---|---|---|---|---|---|---|
| EPI_ISL_661242 | Belgium/ULG-10819/2020 | 2020-11-18 | Europe/Belgium/Liv®ge/ | Department of Clinical Microbiology | GIGA Medical Genomics | Keith Durkin et al |
| EPI_ISL_661248 | Belgium/ULG-10830/2020 | 2020-11-13 | Europe/Belgium/Liv®ge/ | Department of Clinical Microbiology | GIGA Medical Genomics | Keith Durkin et al |
| EPI_ISL_661251 | Belgium/ULG-10834/2020 | 2020-11-13 | Europe/Belgium/Liv®ge/ | Department of Clinical Microbiology | GIGA Medical Genomics | Keith Durkin et al |
| EPI_ISL_661271 | Sichuan/SC-MY-001/2020 | 2020-03-10 | Asia/China/Sichuan/Chengdu | Research platform for Transfusion-transmitted Disease, Institute of Blood Transfusion, Chinese Academy of Medical Sciences | Research platform for Transfusion-transmitted Disease, Institute of Blood Transfusion, Chinese Academy of Medical Sciences | He et al |
| EPI_ISL_661274 | Sweden/20-09860/2020 | 2020-11-04 | Europe/Sweden/Skane/ | Klinisk mikrobiologi | The Public Health Agency of Sweden | Department of Microbiology et al |
| EPI_ISL_661276 | Sweden/20-09864/2020 | 2020-11-10 | Europe/Sweden/Skane/ | Klinisk mikrobiologi | The Public Health Agency of Sweden | Department of Microbiology et al |
| EPI_ISL_661283 | Sweden/20-52819/2020 | 2020-06-25 | Europe/Sweden/Gavleborgs Lan/ | Gavle klinisk mikrobiologi | The Public Health Agency of Sweden | Department of Microbiology et al |
| EPI_ISL_661285 | Sweden/20-53002/2020 | 2020-09-27 | Europe/Sweden/Ostergotland/ | Klinsisk mikrobiologi Linkoping | The Public Health Agency of Sweden | Department of Microbiology et al |
| EPI_ISL_661288 | Sweden/20-53180/2020 | 2020-10-21 | Europe/Sweden/Vastmanland/ | Klinisk mikrobiologi | The Public Health Agency of Sweden | Department of Microbiology et al |
| EPI_ISL_661289 | Sweden/20-53240/2020 | 2020-10-27 | Europe/Sweden/Ostergotland/ | Klinisk mikrobiologi | The Public Health Agency of Sweden | Department of Microbiology et al |
| EPI_ISL_661292 | Sweden/20-53277/2020 | 2020-10-30 | Europe/Sweden/Vasternorrland/ | Mikrobiologen | The Public Health Agency of Sweden | Department of Microbiology et al |
| EPI_ISL_661293 | Sweden/20-53288/2020 | 2020-10-30 | Europe/Sweden/Halland/ | Klinisk mikrobiologi | The Public Health Agency of Sweden | Department of Microbiology et al |
| EPI_ISL_661296 | Sweden/20-53292/2020 | 2020-10-26 | Europe/Sweden/Sormland/ | Unilabs | The Public Health Agency of Sweden | Department of Microbiology et al |
| EPI_ISL_661298 | Sweden/20-53295/2020 | 2020-10-26 | Europe/Sweden/Sormland/ | Unilabs | The Public Health Agency of Sweden | Department of Microbiology et al |
| EPI_ISL_661300 | Sweden/20-53380/2020 | 2020-11-02 | Europe/Sweden/Varmland/ | Laboratoriemedicin, Klinisk mikrobiologi | The Public Health Agency of Sweden | Department of Microbiology et al |
| EPI_ISL_661301 | Sweden/20-53401/2020 | 2020-11-04 | Europe/Sweden/Vastra Gotaland/ | Klinisk Mikrobiologi | The Public Health Agency of Sweden | Department of Microbiology et al |

| | | | | | | |
|---|---|---|---|---|---|---|
| EPI_ISL_661303 | Sweden/20-53492/2020 | 2020-11-13 | Europe/Sweden/Vastmanland/ | Klinisk mikrobiologi | The Public Health Agency of Sweden | Department of Microbiology et al |
| EPI_ISL_661304 | India/WB-IICB-031/2020 | 2020-10-29 | Asia/India/West Bengal/ | CSIR-Indian Institute of Chemical Biology, MEDICA Supercpecialty Hospital Kolkata | CSIR-Indian Institute of Chemical Biology, MEDICA Supercpecialty Hospital Kolkata | Sujay Krishna Maity et al |
| EPI_ISL_661305 | India/WB-IICB-032/2020 | 2020-10-29 | Asia/India/West Bengal/ | CSIR-Indian Institute of Chemical Biology, MEDICA Supercpecialty Hospital Kolkata | CSIR-Indian Institute of Chemical Biology, MEDICA Supercpecialty Hospital Kolkata | Sujay Krishna Maity et al |
| EPI_ISL_661306 | India/WB-IICB-033/2020 | 2020-10-29 | Asia/India/West Bengal/ | CSIR-Indian Institute of Chemical Biology, MEDICA Supercpecialty Hospital Kolkata | CSIR-Indian Institute of Chemical Biology, MEDICA Supercpecialty Hospital Kolkata | Sujay Krishna Maity et al |
| EPI_ISL_661307 | India/WB-IICB-034/2020 | 2020-10-29 | Asia/India/West Bengal/ | CSIR-Indian Institute of Chemical Biology, MEDICA Supercpecialty Hospital Kolkata | CSIR-Indian Institute of Chemical Biology, MEDICA Supercpecialty Hospital Kolkata | Sujay Krishna Maity et al |
| EPI_ISL_661308 | India/WB-IICB-035/2020 | 2020-10-29 | Asia/India/West Bengal/ | CSIR-Indian Institute of Chemical Biology, MEDICA Supercpecialty Hospital Kolkata | CSIR-Indian Institute of Chemical Biology, MEDICA Supercpecialty Hospital Kolkata | Sujay Krishna Maity et al |
| EPI_ISL_661309 | India/WB-IICB-036/2020 | 2020-10-29 | Asia/India/West Bengal/ | CSIR-Indian Institute of Chemical Biology, MEDICA Supercpecialty Hospital Kolkata | CSIR-Indian Institute of Chemical Biology, MEDICA Supercpecialty Hospital Kolkata | Sujay Krishna Maity et al |

| | | | | | | |
|---|---|---|---|---|---|---|
| EPI_ISL_661310 | India/WB-IICB-037/2020 | 2020-10-29 | Asia/India/West Bengal/ | CSIR-Indian Institute of Chemical Biology, MEDICA Supercpecialty Hospital Kolkata | CSIR-Indian Institute of Chemical Biology, MEDICA Supercpecialty Hospital Kolkata | Sujay Krishna Maity et al |
| EPI_ISL_661311 | India/WB-IICB-038/2020 | 2020-10-29 | Asia/India/West Bengal/ | CSIR-Indian Institute of Chemical Biology, MEDICA Supercpecialty Hospital Kolkata | CSIR-Indian Institute of Chemical Biology, MEDICA Supercpecialty Hospital Kolkata | Sujay Krishna Maity et al |
| EPI_ISL_661373 | England/QEUH-B6646B/2020 | 2020-11-11 | Europe/United Kingdom/England/ | Lighthouse Lab in Glasgow | Wellcome Sanger Institute for the COVID-19 Genomics UK (COG-UK) Consortium | Harper VanSteenhouse et al |
| EPI_ISL_661404 | England/QEUH-B679F2/2020 | 2020-11-11 | Europe/United Kingdom/England/ | Lighthouse Lab in Glasgow | Wellcome Sanger Institute for the COVID-19 Genomics UK (COG-UK) Consortium | Harper VanSteenhouse et al |
| EPI_ISL_661695 | England/CAMC-B78572/2020 | 2020-11-11 | Europe/United Kingdom/England/ | Lighthouse Lab in Cambridge | Wellcome Sanger Institute for the COVID-19 Genomics UK (COG-UK) Consortium | Rob Howes et al |
| EPI_ISL_661986 | England/ALDP-B746CE/2020 | 2020-11-12 | Europe/United Kingdom/England/ | Lighthouse Lab in Alderley Park | Wellcome Sanger Institute for the COVID-19 Genomics UK (COG-UK) Consortium | Jacquelyn Wynn et al |
| EPI_ISL_662008 | England/ALDP-B75618/2020 | 2020-11-12 | Europe/United Kingdom/England/ | Lighthouse Lab in Alderley Park | Wellcome Sanger Institute for the COVID-19 Genomics UK (COG-UK) Consortium | Jacquelyn Wynn et al |

| | | | | | | |
|---|---|---|---|---|---|---|
| EPI_ISL_662127 | England/QEUH-B68E3E/2020 | 2020-11-11 | Europe/United Kingdom/England/ | Lighthouse Lab in Glasgow | Wellcome Sanger Institute for the COVID-19 Genomics UK (COG-UK) Consortium | Harper VanSteenhouse et al |
| EPI_ISL_662306 | England/QEUH-B6693F/2020 | 2020-11-11 | Europe/United Kingdom/England/ | Lighthouse Lab in Glasgow | Wellcome Sanger Institute for the COVID-19 Genomics UK (COG-UK) Consortium | Harper VanSteenhouse et al |
| EPI_ISL_662526 | England/ALDP-B7353B/2020 | 2020-11-12 | Europe/United Kingdom/England/ | Lighthouse Lab in Alderley Park | Wellcome Sanger Institute for the COVID-19 Genomics UK (COG-UK) Consortium | Jacquelyn Wynn et al |
| EPI_ISL_662696 | England/QEUH-AF05CE/2020 | 2020-10-29 | Europe/United Kingdom/England/ | Lighthouse Lab in Glasgow | Wellcome Sanger Institute for the COVID-19 Genomics UK (COG-UK) Consortium | Harper VanSteenhouse et al |
| EPI_ISL_662797 | England/QEUH-AF06CB/2020 | 2020-10-29 | Europe/United Kingdom/England/ | Lighthouse Lab in Glasgow | Wellcome Sanger Institute for the COVID-19 Genomics UK (COG-UK) Consortium | Harper VanSteenhouse et al |
| EPI_ISL_662870 | England/QEUH-AF2878/2020 | 2020-10-30 | Europe/United Kingdom/England/ | Lighthouse Lab in Glasgow | Wellcome Sanger Institute for the COVID-19 Genomics UK (COG-UK) Consortium | Harper VanSteenhouse et al |
| EPI_ISL_663004 | England/QEUH-AF5022/2020 | 2020-10-30 | Europe/United Kingdom/England/ | Lighthouse Lab in Glasgow | Wellcome Sanger Institute for the COVID-19 Genomics UK (COG-UK) Consortium | Harper VanSteenhouse et al |
| EPI_ISL_663023 | England/QEUH-AF4CC5/2020 | 2020-10-30 | Europe/United Kingdom/England/ | Lighthouse Lab in Glasgow | Wellcome Sanger Institute for the COVID-19 Genomics UK (COG-UK) Consortium | Harper VanSteenhouse et al |

| | | | | | | |
|---|---|---|---|---|---|---|
| EPI_ISL_663057 | England/QEUH-AF5AE8/2020 | 2020-10-30 | Europe/United Kingdom/England/ | Lighthouse Lab in Glasgow | Wellcome Sanger Institute for the COVID-19 Genomics UK (COG-UK) Consortium | Harper VanSteenhouse et al |
| EPI_ISL_663113 | England/QEUH-AF5A7F/2020 | 2020-10-30 | Europe/United Kingdom/England/ | Lighthouse Lab in Glasgow | Wellcome Sanger Institute for the COVID-19 Genomics UK (COG-UK) Consortium | Harper VanSteenhouse et al |
| EPI_ISL_663212 | France/NAQ-SC262/2020 | 2020-03-23 | Europe/France/Nouvelle-Aquitaine/Limo | CHU de Limoges | CNR Virus des Infections Respiratoires - France SUD | Antonin Bal et al |
| EPI_ISL_663218 | France/NAQ-SC269/2020 | 2020-03-24 | Europe/France/Nouvelle-Aquitaine/Limo | CHU de Limoges | CNR Virus des Infections Respiratoires - France SUD | Antonin Bal et al |
| EPI_ISL_663221 | France/NAQ-SC272/2020 | 2020-03-25 | Europe/France/Nouvelle-Aquitaine/Limo | CHU de Limoges | CNR Virus des Infections Respiratoires - France SUD | Antonin Bal et al |
| EPI_ISL_663223 | France/NAQ-SC276/2020 | 2020-03-25 | Europe/France/Nouvelle-Aquitaine/Limo | CHU de Limoges | CNR Virus des Infections Respiratoires - France SUD | Antonin Bal et al |
| EPI_ISL_663226 | France/NAQ-SC287/2020 | 2020-04-10 | Europe/France/Nouvelle-Aquitaine/Limo | CHU de Limoges | CNR Virus des Infections Respiratoires - France SUD | Antonin Bal et al |
| EPI_ISL_663228 | France/NAQ-SC577/2020 | 2020-03-07 | Europe/France/Nouvelle-Aquitaine/Poit | CHU Poitiers | CNR Virus des Infections Respiratoires - France SUD | Antonin Bal et al |
| EPI_ISL_663233 | France/NAQ-SC582/2020 | 2020-03-09 | Europe/France/Nouvelle-Aquitaine/Poit | CHU Poitiers | CNR Virus des Infections Respiratoires - France SUD | Antonin Bal et al |

| | | | | | | |
|---|---|---|---|---|---|---|
| EPI_ISL_663236 | France/NAQ-SC585/2020 | 2020-05-13 | Europe/France/Nouvelle-Aquitaine/Poit | CHU Poitiers | CNR Virus des Infections Respiratoires - France SUD | Antonin Bal et al |
| EPI_ISL_663237 | France/NAQ-SC587/2020 | 2020-05-11 | Europe/France/Nouvelle-Aquitaine/Poit | CHU Poitiers | CNR Virus des Infections Respiratoires - France SUD | Antonin Bal et al |
| EPI_ISL_663239 | France/NAQ-SC592/2020 | 2020-05-01 | Europe/France/Nouvelle-Aquitaine/Poit | CHU Poitiers | CNR Virus des Infections Respiratoires - France SUD | Antonin Bal et al |
| EPI_ISL_663240 | France/NAQ-SC596/2020 | 2020-05-11 | Europe/France/Nouvelle-Aquitaine/Poit | CHU Poitiers | CNR Virus des Infections Respiratoires - France SUD | Antonin Bal et al |
| EPI_ISL_663242 | France/NAQ-SC598/2020 | 2020-06-20 | Europe/France/Nouvelle-Aquitaine/Poit | CHU Poitiers | CNR Virus des Infections Respiratoires - France SUD | Antonin Bal et al |
| EPI_ISL_663244 | France/OCC-SC567/2020 | 2020-05-20 | Europe/France/Occitanie/N√Æmes | CHU N√Æmes | CNR Virus des Infections Respiratoires - France SUD | Antonin Bal et al |
| EPI_ISL_663247 | France/OCC-SC570/2020 | 2020-05-26 | Europe/France/Occitanie/N√Æmes | CHU N√Æmes | CNR Virus des Infections Respiratoires - France SUD | Antonin Bal et al |
| EPI_ISL_663248 | France/OCC-SC571/2020 | 2020-05-26 | Europe/France/Occitanie/N√Æmes | CHU N√Æmes | CNR Virus des Infections Respiratoires - France SUD | Antonin Bal et al |
| EPI_ISL_663261 | France/PDL-SC615/2020 | 2020-03-30 | Europe/France/Pays de la Loire/Nantes | CHU Nantes | CNR Virus des Infections Respiratoires - France SUD | Antonin Bal et al |
| EPI_ISL_663263 | France/PDL-SC617/2020 | 2020-03-30 | Europe/France/Pays de la Loire/Nantes | CHU Nantes | CNR Virus des Infections Respiratoires - France SUD | Antonin Bal et al |

| | | | | | | |
|---|---|---|---|---|---|---|
| EPI_ISL_663274 | France/PDL-SC635/2020 | 2020-04-03 | Europe/France/Pays de la Loire/Nantes | CHU Nantes | CNR Virus des Infections Respiratoires - France SUD | Antonin Bal et al |
| EPI_ISL_663281 | France/PDL-SC643/2020 | 2020-04-05 | Europe/France/Pays de la Loire/Nantes | CHU Nantes | CNR Virus des Infections Respiratoires - France SUD | Antonin Bal et al |
| EPI_ISL_663387 | Australia/VIC16930/2020 | 2020-08-06 | Oceania/Australia/Victoria/ | Microbiological Diagnostic Unit - Public Health Laboratory (MDU-PHL) | MDU-PHL | Seemann T. et al |
| EPI_ISL_663756 | Australia/VIC17137/2020 | 2020-08-07 | Oceania/Australia/Victoria/ | Microbiological Diagnostic Unit - Public Health Laboratory (MDU-PHL) | MDU-PHL | Seemann T. et al |
| EPI_ISL_663781 | Australia/VIC17183/2020 | 2020-08-06 | Oceania/Australia/Victoria/ | Microbiological Diagnostic Unit - Public Health Laboratory (MDU-PHL) | MDU-PHL | Seemann T. et al |
| EPI_ISL_664105 | Norway/CS-03/2020 | 2020-04-13 | Europe/Norway/Viken/ | Dept. of Microbiology and Infection Control, Akershus University Hospital HF | Dept. of Microbiology and Infection Control, Akershus University Hospital HF | Hege Vangstein Aamot et al |
| EPI_ISL_664142 | England/LIVE-DBE7B4/2020 | 2020-10-29 | Europe/United Kingdom/England/ | Liverpool Clinical Laboratories | COVID-19 Genomics UK (COG-UK) Consortium | Sam Haldenby et al |
| EPI_ISL_664232 | England/LIVE-DBE7E1/2020 | 2020-10-27 | Europe/United Kingdom/England/ | Liverpool Clinical Laboratories | COVID-19 Genomics UK (COG-UK) Consortium | Sam Haldenby et al |
| EPI_ISL_664374 | England/ALDP-B1CC99/2020 | 2020-11-02 | Europe/United Kingdom/England/ | Centre for Enzyme Innovation, University of Portsmouth / Translational Research Laboratory, Portsmouth Hospitals NHS Trust | COVID-19 Genomics UK (COG-UK) Consortium | Angela Beckett et al |
| EPI_ISL_664592 | England/EXET-13A27F/2020 | 2020-10-31 | Europe/United Kingdom/England/ | University of Exeter | COVID-19 Genomics UK (COG-UK) Consortium | Ben Temperton et al |
| EPI_ISL_664677 | England/CAMB-1BAD76/2020 | 2020-11-12 | Europe/United Kingdom/England/ | Department of Pathology, University of Cambridge | COVID-19 Genomics UK (COG-UK) Consortium | Aminu S. Jahun et al |
| EPI_ISL_664715 | England/EXET-13AA68/2020 | 2020-11-07 | Europe/United Kingdom/England/ | University of Exeter | COVID-19 Genomics UK (COG-UK) Consortium | Ben Temperton et al |

| | | | | | | |
|---|---|---|---|---|---|---|
| EPI_ISL_665128 | England/LIVE-DBD15D/2020 | 2020-10-27 | Europe/United Kingdom/England/ | Liverpool Clinical Laboratories | COVID-19 Genomics UK (COG-UK) Consortium | Sam Haldenby et al |
| EPI_ISL_665179 | England/LOND-124F941/2020 | 2020-03-28 | Europe/United Kingdom/England/ | University College London Hospital | COVID-19 Genomics UK (COG-UK) Consortium | Judith Heaney et al |
| EPI_ISL_665249 | England/LOND-1250B56/2020 | 2020-11-03 | Europe/United Kingdom/England/ | University College London Hospital | COVID-19 Genomics UK (COG-UK) Consortium | Judith Heaney et al |
| EPI_ISL_665254 | Norway/CS-05/2020 | 2020-04-19 | Europe/Norway/Viken/ | Dept. of Microbiology and Infection Control, Akershus University Hospital HF | Dept. of Microbiology and Infection Control, Akershus University Hospital HF | Hege Vangstein Aamot et al |
| EPI_ISL_665255 | Norway/CS-06/2020 | 2020-04-27 | Europe/Norway/Viken/ | Dept. of Microbiology and Infection Control, Akershus University Hospital HF | Dept. of Microbiology and Infection Control, Akershus University Hospital HF | Hege Vangstein Aamot et al |
| EPI_ISL_665256 | Norway/CS-07/2020 | 2020-04-20 | Europe/Norway/Viken/ | Dept. of Microbiology and Infection Control, Akershus University Hospital HF | Dept. of Microbiology and Infection Control, Akershus University Hospital HF | Hege Vangstein Aamot et al |
| EPI_ISL_665512 | England/NOTT-11D227/2020 | 2020-11-21 | Europe/United Kingdom/England/ | Queens Medical Centre, Clinical Microbiology Department / DeepSeq Nottingham | COVID-19 Genomics UK (COG-UK) Consortium | Gemma Clark et al |
| EPI_ISL_665539 | Wales/PHWC-48C24E/2020 | 2020-11-09 | Europe/United Kingdom/Wales/ | Wales Specialist Virology Centre Sequencing lab: Pathogen Genomics Unit | COVID-19 Genomics UK (COG-UK) Consortium | Catherine Moore et al |
| EPI_ISL_665640 | Scotland/CVR5788/2020 | 2020-11-10 | Europe/United Kingdom/Scotland/ | West of Scotland Specialist Virology Centre, NHSGGC / MRC-University of Glasgow Centre for Virus Research | COVID-19 Genomics UK (COG-UK) Consortium | Ana da Silva Filipe et al |

| | | | | | | |
|---|---|---|---|---|---|---|
| EPI_ISL_666105 | Scotland/QEUH-AF1484/2020 | 2020-10-29 | Europe/United Kingdom/Scotland/ | Northumbria University / South Tees Hospitals NHS Foundation Trust / North Cumbria Integrated Care NHS Foundation Trust / North Tees and Hartlepool NHS Foundation Trust / Newcastle Hospitals NHS Foundation Trust | COVID-19 Genomics UK (COG-UK) Consortium | Darren L Smith et al |
| EPI_ISL_666293 | Wales/PHWC-48B889/2020 | 2020-11-05 | Europe/United Kingdom/Wales/ | Wales Specialist Virology Centre Sequencing lab: Pathogen Genomics Unit | COVID-19 Genomics UK (COG-UK) Consortium | Catherine Moore et al |
| EPI_ISL_666367 | Wales/PHWC-48BED2/2020 | 2020-11-01 | Europe/United Kingdom/Wales/ | Wales Specialist Virology Centre Sequencing lab: Pathogen Genomics Unit | COVID-19 Genomics UK (COG-UK) Consortium | Catherine Moore et al |
| EPI_ISL_666593 | Norway/CS-13/2020 | 2020-04-14 | Europe/Norway/Viken/ | Dept. of Microbiology and Infection Control, Akershus University Hospital HF | Dept. of Microbiology and Infection Control, Akershus University Hospital HF | Hege Vangstein Aamot et al |
| EPI_ISL_666601 | Norway/CS-21/2020 | 2020-03-12 | Europe/Norway/Viken/ | Dept. of Microbiology and Infection Control, Akershus University Hospital HF | Dept. of Microbiology and Infection Control, Akershus University Hospital HF | Hege Vangstein Aamot et al |
| EPI_ISL_666603 | Norway/CS-23/2020 | 2020-05-04 | Europe/Norway/Viken/ | Dept. of Microbiology and Infection Control, Akershus University Hospital HF | Dept. of Microbiology and Infection Control, Akershus University Hospital HF | Hege Vangstein Aamot et al |
| EPI_ISL_666605 | Norway/CS-25/2020 | 2020-03-24 | Europe/Norway/Viken/ | Dept. of Microbiology and Infection Control, Akershus University Hospital HF | Dept. of Microbiology and Infection Control, Akershus University Hospital HF | Hege Vangstein Aamot et al |

| | | | | | | |
|---|---|---|---|---|---|---|
| EPI_ISL_666606 | Norway/CS-26/2020 | 2020-04-06 | Europe/Norway/Viken/ | Dept. of Microbiology and Infection Control, Akershus University Hospital HF | Dept. of Microbiology and Infection Control, Akershus University Hospital HF | Hege Vangstein Aamot et al |
| EPI_ISL_666608 | Norway/CS-28/2020 | 2020-04-21 | Europe/Norway/Viken/ | Dept. of Microbiology and Infection Control, Akershus University Hospital HF | Dept. of Microbiology and Infection Control, Akershus University Hospital HF | Hege Vangstein Aamot et al |
| EPI_ISL_666610 | Norway/CS-29/2020 | 2020-04-24 | Europe/Norway/Viken/ | Dept. of Microbiology and Infection Control, Akershus University Hospital HF | Dept. of Microbiology and Infection Control, Akershus University Hospital HF | Hege Vangstein Aamot et al |
| EPI_ISL_666624 | Germany/NW-HHU-322/2020 | 2020-10-12 | Europe/Germany/North Rhine Westphal | ZOTZ KLIMAS MVZ D√ºsseldorf-Centrum GbR √úBAG f√ºr Labormedizin, Genetik, Zytologie, Pathologie | Center of Medical Microbiology, Virology, and Hospital Hygiene, University of Duesseldorf | Maximilian Damagnez et al |
| EPI_ISL_666625 | Germany/NW-HHU-324/2020 | 2020-10-12 | Europe/Germany/North Rhine Westphal | ZOTZ KLIMAS MVZ D√ºsseldorf-Centrum GbR √úBAG f√ºr Labormedizin, Genetik, Zytologie, Pathologie | Center of Medical Microbiology, Virology, and Hospital Hygiene, University of Duesseldorf | Maximilian Damagnez et al |
| EPI_ISL_666626 | Germany/NW-HHU-325/2020 | 2020-10-12 | Europe/Germany/North Rhine Westphal | ZOTZ KLIMAS MVZ D√ºsseldorf-Centrum GbR √úBAG f√ºr Labormedizin, Genetik, Zytologie, Pathologie | Center of Medical Microbiology, Virology, and Hospital Hygiene, University of Duesseldorf | Maximilian Damagnez et al |

| | | | | | | |
|---|---|---|---|---|---|---|
| EPI_ISL_666628 | Germany/NW-HHU-334/2020 | 2020-10-13 | Europe/Germany/North Rhine Westphal | ZOTZ KLIMAS MVZ D√ºsseldorf-Centrum GbR √úBAG f√ºr Labormedizin, Genetik, Zytologie, Pathologie | Center of Medical Microbiology, Virology, and Hospital Hygiene, University of Duesseldorf | Maximilian Damagnez et al |
| EPI_ISL_666633 | Germany/NW-HHU-346/2020 | 2020-09-04 | Europe/Germany/North Rhine Westphal | ZOTZ KLIMAS MVZ D√ºsseldorf-Centrum GbR √úBAG f√ºr Labormedizin, Genetik, Zytologie, Pathologie | Center of Medical Microbiology, Virology, and Hospital Hygiene, University of Duesseldorf | Maximilian Damagnez et al |
| EPI_ISL_666635 | Germany/NW-HHU-348/2020 | 2020-09-04 | Europe/Germany/North Rhine Westphal | ZOTZ KLIMAS MVZ D√ºsseldorf-Centrum GbR √úBAG f√ºr Labormedizin, Genetik, Zytologie, Pathologie | Center of Medical Microbiology, Virology, and Hospital Hygiene, University of Duesseldorf | Maximilian Damagnez et al |
| EPI_ISL_666639 | Germany/NW-HHU-352/2020 | 2020-09-10 | Europe/Germany/North Rhine Westphal | ZOTZ KLIMAS MVZ D√ºsseldorf-Centrum GbR √úBAG f√ºr Labormedizin, Genetik, Zytologie, Pathologie | Center of Medical Microbiology, Virology, and Hospital Hygiene, University of Duesseldorf | Maximilian Damagnez et al |
| EPI_ISL_666647 | Germany/NW-HHU-360/2020 | 2020-09-11 | Europe/Germany/North Rhine Westphal | ZOTZ KLIMAS MVZ D√ºsseldorf-Centrum GbR √úBAG f√ºr Labormedizin, Genetik, Zytologie, Pathologie | Center of Medical Microbiology, Virology, and Hospital Hygiene, University of Duesseldorf | Maximilian Damagnez et al |
| EPI_ISL_666648 | Germany/NW-HHU-361/2020 | 2020-09-11 | Europe/Germany/North Rhine Westphal | ZOTZ KLIMAS MVZ D√ºsseldorf-Centrum GbR √úBAG f√ºr Labormedizin, Genetik, Zytologie, Pathologie | Center of Medical Microbiology, Virology, and Hospital Hygiene, University of Duesseldorf | Maximilian Damagnez et al |
| EPI_ISL_666657 | France/ARA-SC029/2020 | 2020-03-23 | Europe/France/Auvergne-Rh√¥ne-Alpes/ | Laboratoire du Centre Hospitalier Annecy Genevois | CNR Virus des Infections Respiratoires - France SUD | Antonin Bal et al |

| | | | | | | |
|---|---|---|---|---|---|---|
| EPI_ISL_666659 | France/ARA-SC031/2020 | 2020-03-24 | Europe/France/Auvergne-Rhv¥ne-Alpes/ | Laboratoire du Centre Hospitalier Annecy Genevois | CNR Virus des Infections Respiratoires - France SUD | Antonin Bal et al |
| EPI_ISL_666664 | France/ARA-SC044/2020 | 2020-03-31 | Europe/France/Auvergne-Rhv¥ne-Alpes/ | Laboratoire du Centre Hospitalier Annecy Genevois | CNR Virus des Infections Respiratoires - France SUD | Antonin Bal et al |
| EPI_ISL_666675 | France/ARA-SC063/2020 | 2020-04-15 | Europe/France/Auvergne-Rhv¥ne-Alpes/ | Laboratoire du Centre Hospitalier Annecy Genevois | CNR Virus des Infections Respiratoires - France SUD | Antonin Bal et al |
| EPI_ISL_666678 | France/ARA-SC071/2020 | 2020-04-20 | Europe/France/Auvergne-Rhv¥ne-Alpes/ | Laboratoire du Centre Hospitalier Annecy Genevois | CNR Virus des Infections Respiratoires - France SUD | Antonin Bal et al |
| EPI_ISL_666686 | France/ARA-SC106/2020 | 2020-03-14 | Europe/France/Auvergne-Rhv¥ne-Alpes/ | Centre hospitalier Mv©tropole Savoie | CNR Virus des Infections Respiratoires - France SUD | Antonin Bal et al |
| EPI_ISL_666688 | Ireland/DL-NVRL-20G48995/2020 | 2020-04-06 | Europe/Ireland/Donegal/ | National Virus Reference Lab (NVRL) | Irish Coronavirus Sequencing Consortium-Teagasc Grange | Matthew McCabe et al |
| EPI_ISL_666708 | Ireland/DL-NVRL-20G45123/2020 | 2020-04-02 | Europe/Ireland/Donegal/ | National Virus Reference Lab (NVRL) | Irish Coronavirus Sequencing Consortium-Teagasc Grange | Matthew McCabe et al |
| EPI_ISL_666719 | France/NAQ-SC300/2020 | 2020-04-28 | Europe/France/Nouvelle-Aquitaine/Limo | CHU de Limoges | CNR Virus des Infections Respiratoires - France SUD | Antonin Bal et al |
| EPI_ISL_666720 | France/NAQ-SC301/2020 | 2020-04-27 | Europe/France/Nouvelle-Aquitaine/Limo | CHU de Limoges | CNR Virus des Infections Respiratoires - France SUD | Antonin Bal et al |
| EPI_ISL_666723 | France/PDL-SC656/2020 | 2020-04-24 | Europe/France/Pays de la Loire/Nantes | CHU Nantes | CNR Virus des Infections Respiratoires - France SUD | Antonin Bal et al |

| | | | | | | |
|---|---|---|---|---|---|---|
| EPI_ISL_666766 | England/C200044492/2020 | 2020-11-06 | Europe/United Kingdom/England/ | Respiratory Virus Unit, Microbiology Services Colindale, Public Health England | COVID-19 Genomics UK (COG-UK) Consortium | PHE Covid Sequencing Team et al |
| EPI_ISL_666811 | USA/MD-MDH-0444/2020 | 2020-11-10 | North America/USA/Maryland/ | Maryland Public Health Laboratory | Maryland Public Health Laboratory | Maryland Department of Health Laboratories Administration et al |
| EPI_ISL_666818 | USA/FL-BPHL-2196/2020 | 2020-07-27 | North America/USA/Florida/ | Florida Bureau of Public Health Laboratories | Florida Bureau of Public Health Laboratories | Sarah Schmedes et al |
| EPI_ISL_666828 | USA/FL-BPHL-2206/2020 | 2020-08-26 | North America/USA/Florida/ | Florida Bureau of Public Health Laboratories | Florida Bureau of Public Health Laboratories | Sarah Schmedes et al |
| EPI_ISL_666829 | USA/FL-BPHL-2207/2020 | 2020-08-29 | North America/USA/Florida/ | Florida Bureau of Public Health Laboratories | Florida Bureau of Public Health Laboratories | Sarah Schmedes et al |
| EPI_ISL_666842 | USA/FL-BPHL-2220/2020 | 2020-08-13 | North America/USA/Florida/ | Florida Bureau of Public Health Laboratories | Florida Bureau of Public Health Laboratories | Sarah Schmedes et al |
| EPI_ISL_666844 | USA/FL-BPHL-2222/2020 | 2020-08-31 | North America/USA/Florida/ | Florida Bureau of Public Health Laboratories | Florida Bureau of Public Health Laboratories | Sarah Schmedes et al |
| EPI_ISL_666846 | USA/FL-BPHL-2224/2020 | 2020-08-31 | North America/USA/Florida/ | Florida Bureau of Public Health Laboratories | Florida Bureau of Public Health Laboratories | Sarah Schmedes et al |
| EPI_ISL_666847 | USA/FL-BPHL-2225/2020 | 2020-08-31 | North America/USA/Florida/ | Florida Bureau of Public Health Laboratories | Florida Bureau of Public Health Laboratories | Sarah Schmedes et al |
| EPI_ISL_666848 | USA/FL-BPHL-2226/2020 | 2020-09-03 | North America/USA/Florida/ | Florida Bureau of Public Health Laboratories | Florida Bureau of Public Health Laboratories | Sarah Schmedes et al |
| EPI_ISL_666851 | USA/FL-BPHL-2229/2020 | 2020-09-08 | North America/USA/Florida/ | Florida Bureau of Public Health Laboratories | Florida Bureau of Public Health Laboratories | Sarah Schmedes et al |
| EPI_ISL_666853 | USA/FL-BPHL-2231/2020 | 2020-08-10 | North America/USA/Florida/ | Florida Bureau of Public Health Laboratories | Florida Bureau of Public Health Laboratories | Sarah Schmedes et al |
| EPI_ISL_666867 | USA/FL-BPHL-2245/2020 | 2020-09-04 | North America/USA/Florida/ | Florida Bureau of Public Health Laboratories | Florida Bureau of Public Health Laboratories | Sarah Schmedes et al |

| | | | | | | |
|---|---|---|---|---|---|---|
| EPI_ISL_666870 | France/NOR-UCN19/2020 | 2020-03-03 | Europe/France/Normandie/ | Laboratoire de Virologie, CHU de Caen, Normandie, France. | GRAM2.0, Universitv© de Caen Normandie | Meriadeg LE GOUIL et al |
| EPI_ISL_666872 | Belgium/ULG-10836/2020 | 2020-10-27 | Europe/Belgium/Liv®ge/ | Department of Clinical Microbiology | GIGA Medical Genomics | Keith Durkin et al |
| EPI_ISL_666874 | Belgium/ULG-10839/2020 | 2020-10-27 | Europe/Belgium/Liv®ge/ | Department of Clinical Microbiology | GIGA Medical Genomics | Keith Durkin et al |
| EPI_ISL_666877 | Belgium/ULG-10842/2020 | 2020-10-27 | Europe/Belgium/Liv®ge/ | Department of Clinical Microbiology | GIGA Medical Genomics | Keith Durkin et al |
| EPI_ISL_666879 | Belgium/ULG-10844/2020 | 2020-10-27 | Europe/Belgium/Liv®ge/ | Department of Clinical Microbiology | GIGA Medical Genomics | Keith Durkin et al |
| EPI_ISL_666882 | Belgium/ULG-10847/2020 | 2020-10-29 | Europe/Belgium/Liv®ge/ | Department of Clinical Microbiology | GIGA Medical Genomics | Keith Durkin et al |
| EPI_ISL_666883 | Belgium/ULG-10848/2020 | 2020-10-29 | Europe/Belgium/Liv®ge/ | Department of Clinical Microbiology | GIGA Medical Genomics | Keith Durkin et al |
| EPI_ISL_666899 | USA/MI-MDHHS-SC22233/2020 | 2020-11-03 | North America/USA/Michigan/ | Michigan Department of Health and Human Services, Bureau of Laboratories | Michigan Department of Health and Human Services, Bureau of Laboratories | Blankenship HM et al |
| EPI_ISL_666904 | USA/MI-MDHHS-SC22238/2020 | 2020-10-28 | North America/USA/Michigan/ | Michigan Department of Health and Human Services, Bureau of Laboratories | Michigan Department of Health and Human Services, Bureau of Laboratories | Blankenship HM et al |
| EPI_ISL_666911 | USA/MI-MDHHS-SC22245/2020 | 2020-11-05 | North America/USA/Michigan/ | Michigan Department of Health and Human Services, Bureau of Laboratories | Michigan Department of Health and Human Services, Bureau of Laboratories | Blankenship HM et al |
| EPI_ISL_666913 | USA/MI-MDHHS-SC22247/2020 | 2020-10-30 | North America/USA/Michigan/ | Michigan Department of Health and Human Services, Bureau of Laboratories | Michigan Department of Health and Human Services, Bureau of Laboratories | Blankenship HM et al |

| | | | | | | |
|---|---|---|---|---|---|---|
| EPI_ISL_666914 | USA/MI-MDHHS-SC22248/2020 | 2020-10-29 | North America/USA/Michigan/ | Michigan Department of Health and Human Services, Bureau of Laboratories | Michigan Department of Health and Human Services, Bureau of Laboratories | Blankenship HM et al |
| EPI_ISL_666916 | USA/MI-MDHHS-SC22250/2020 | 2020-10-29 | North America/USA/Michigan/ | Michigan Department of Health and Human Services, Bureau of Laboratories | Michigan Department of Health and Human Services, Bureau of Laboratories | Blankenship HM et al |
| EPI_ISL_666927 | USA/MI-MDHHS-SC22261/2020 | 2020-11-04 | North America/USA/Michigan/ | Michigan Department of Health and Human Services, Bureau of Laboratories | Michigan Department of Health and Human Services, Bureau of Laboratories | Blankenship HM et al |
| EPI_ISL_666930 | USA/MI-MDHHS-SC22264/2020 | 2020-11-07 | North America/USA/Michigan/ | Michigan Department of Health and Human Services, Bureau of Laboratories | Michigan Department of Health and Human Services, Bureau of Laboratories | Blankenship HM et al |
| EPI_ISL_666937 | USA/MI-MDHHS-SC22271/2020 | 2020-10-20 | North America/USA/Michigan/ | Michigan Department of Health and Human Services, Bureau of Laboratories | Michigan Department of Health and Human Services, Bureau of Laboratories | Blankenship HM et al |
| EPI_ISL_666939 | USA/MI-MDHHS-SC22273/2020 | 2020-10-28 | North America/USA/Michigan/ | Michigan Department of Health and Human Services, Bureau of Laboratories | Michigan Department of Health and Human Services, Bureau of Laboratories | Blankenship HM et al |

| | | | | | | |
|---|---|---|---|---|---|---|
| EPI_ISL_666954 | USA/MI-MDHHS-SC22288/2020 | 2020-11-02 | North America/USA/Michigan/ | Michigan Department of Health and Human Services, Bureau of Laboratories | Michigan Department of Health and Human Services, Bureau of Laboratories | Blankenship HM et al |
| EPI_ISL_666956 | USA/MI-MDHHS-SC22290/2020 | 2020-10-29 | North America/USA/Michigan/ | Michigan Department of Health and Human Services, Bureau of Laboratories | Michigan Department of Health and Human Services, Bureau of Laboratories | Blankenship HM et al |
| EPI_ISL_666958 | USA/MI-MDHHS-SC22292/2020 | 2020-10-30 | North America/USA/Michigan/ | Michigan Department of Health and Human Services, Bureau of Laboratories | Michigan Department of Health and Human Services, Bureau of Laboratories | Blankenship HM et al |
| EPI_ISL_666963 | USA/MI-MDHHS-SC22297/2020 | 2020-11-02 | North America/USA/Michigan/ | Michigan Department of Health and Human Services, Bureau of Laboratories | Michigan Department of Health and Human Services, Bureau of Laboratories | Blankenship HM et al |
| EPI_ISL_666965 | USA/CA-ALSR-4489/2020 | 2020-10-26 | North America/USA/California/San Dieg | San Diego County Public Health Laboratory | Andersen lab at Scripps Research | SEARCH Alliance San Diego with Tracy Basler et al |
| EPI_ISL_666968 | USA/CA-ALSR-4497/2020 | 2020-10-29 | North America/USA/California/San Dieg | San Diego County Public Health Laboratory | Andersen lab at Scripps Research | SEARCH Alliance San Diego with Tracy Basler et al |
| EPI_ISL_666993 | USA/CA-ALSR-4534/2020 | 2020-08-27 | North America/USA/California/San Dieg | San Diego County Public Health Laboratory | Andersen lab at Scripps Research | SEARCH Alliance San Diego with Tracy Basler et al |
| EPI_ISL_666995 | USA/CA-ALSR-4536/2020 | 2020-09-17 | North America/USA/California/San Dieg | San Diego County Public Health Laboratory | Andersen lab at Scripps Research | SEARCH Alliance San Diego with Tracy Basler et al |
| EPI_ISL_666996 | USA/CA-ALSR-4537/2020 | 2020-09-10 | North America/USA/California/San Dieg | San Diego County Public Health Laboratory | Andersen lab at Scripps Research | SEARCH Alliance San Diego with Tracy Basler et al |
| EPI_ISL_666997 | USA/CA-ALSR-4539/2020 | 2020-09-16 | North America/USA/California/San Dieg | San Diego County Public Health Laboratory | Andersen lab at Scripps Research | SEARCH Alliance San Diego with Tracy Basler et al |
| EPI_ISL_667004 | USA/CA-ALSR-4555/2020 | 2020-06-19 | North America/USA/California/San Dieg | San Diego County Public Health Laboratory | Andersen lab at Scripps Research | SEARCH Alliance San Diego with Tracy Basler et al |
| EPI_ISL_667005 | USA/CA-ALSR-4557/2020 | 2020-07-08 | North America/USA/California/San Dieg | San Diego County Public Health Laboratory | Andersen lab at Scripps Research | SEARCH Alliance San Diego with Tracy Basler et al |

| | | | | | | |
|---|---|---|---|---|---|---|
| EPI_ISL_667008 | USA/CA-ALSR-4562/2020 | 2020-07-08 | North America/USA/California/San Dieg | San Diego County Public Health Laboratory | Andersen lab at Scripps Research | SEARCH Alliance San Diego with Tracy Basler et al |
| EPI_ISL_667009 | USA/CA-ALSR-4564/2020 | 2020-07-08 | North America/USA/California/San Dieg | San Diego County Public Health Laboratory | Andersen lab at Scripps Research | SEARCH Alliance San Diego with Tracy Basler et al |
| EPI_ISL_667011 | USA/CA-ALSR-4567/2020 | 2020-07-07 | North America/USA/California/San Dieg | San Diego County Public Health Laboratory | Andersen lab at Scripps Research | SEARCH Alliance San Diego with Tracy Basler et al |
| EPI_ISL_667021 | USA/CA-ALSR-4585/2020 | 2020-07-11 | North America/USA/California/San Dieg | San Diego County Public Health Laboratory | Andersen lab at Scripps Research | SEARCH Alliance San Diego with Tracy Basler et al |
| EPI_ISL_667033 | USA/CA-ALSR-4604/2020 | 2020-07-16 | North America/USA/California/San Dieg | San Diego County Public Health Laboratory | Andersen lab at Scripps Research | SEARCH Alliance San Diego with Tracy Basler et al |
| EPI_ISL_667041 | USA/CA-ALSR-4619/2020 | 2020-07-08 | North America/USA/California/San Dieg | San Diego County Public Health Laboratory | Andersen lab at Scripps Research | SEARCH Alliance San Diego with Tracy Basler et al |
| EPI_ISL_667044 | USA/CA-ALSR-4624/2020 | 2020-10-22 | North America/USA/California/San Dieg | San Diego County Public Health Laboratory | Andersen lab at Scripps Research | SEARCH Alliance San Diego with Tracy Basler et al |
| EPI_ISL_667066 | USA/CA-ALSR-4666/2020 | 2020-11-11 | North America/USA/California/San Dieg | San Diego County Public Health Laboratory | Andersen lab at Scripps Research | SEARCH Alliance San Diego with Tracy Basler et al |
| EPI_ISL_667071 | USA/CA-ALSR-4675/2020 | 2020-11-10 | North America/USA/California/San Dieg | San Diego County Public Health Laboratory | Andersen lab at Scripps Research | SEARCH Alliance San Diego with Tracy Basler et al |
| EPI_ISL_667072 | USA/CA-ALSR-4676/2020 | 2020-11-10 | North America/USA/California/San Dieg | San Diego County Public Health Laboratory | Andersen lab at Scripps Research | SEARCH Alliance San Diego with Tracy Basler et al |
| EPI_ISL_667085 | USA/OR-OHSU-1886/2020 | 2020-07-28 | North America/USA/Oregon/Washingto | OHSU Lab Services Molecular Microbiology Lab | Oregon SARS-CoV-2 Genome Sequencing Center | Brendan L. O'Connell et al |
| EPI_ISL_667086 | USA/OR-OHSU-1972/2020 | 2020-07-30 | North America/USA/Oregon/Multnomal | OHSU Lab Services Molecular Microbiology Lab | Oregon SARS-CoV-2 Genome Sequencing Center | Brendan L. O'Connell et al |
| EPI_ISL_667089 | USA/OR-OHSU-2009/2020 | 2020-07-31 | North America/USA/Oregon/Washingto | OHSU Lab Services Molecular Microbiology Lab | Oregon SARS-CoV-2 Genome Sequencing Center | Brendan L. O'Connell et al |
| EPI_ISL_667091 | USA/OR-OHSU-2015/2020 | 2020-07-31 | North America/USA/Oregon/Clackamas | OHSU Lab Services Molecular Microbiology Lab | Oregon SARS-CoV-2 Genome Sequencing Center | Brendan L. O'Connell et al |
| EPI_ISL_667095 | USA/OR-OHSU-2027/2020 | 2020-07-31 | North America/USA/Oregon/Wasco Cou | OHSU Lab Services Molecular Microbiology Lab | Oregon SARS-CoV-2 Genome Sequencing Center | Brendan L. O'Connell et al |

| | | | | | | |
|---|---|---|---|---|---|---|
| EPI_ISL_667099 | USA/OR-OHSU-2035/2020 | 2020-08-01 | North America/USA/Oregon/Multnomal | OHSU Lab Services Molecular Microbiology Lab | Oregon SARS-CoV-2 Genome Sequencing Center | Brendan L. O'Connell et al |
| EPI_ISL_667100 | USA/OR-OHSU-2039/2020 | 2020-08-01 | North America/USA/Oregon/Multnomal | OHSU Lab Services Molecular Microbiology Lab | Oregon SARS-CoV-2 Genome Sequencing Center | Brendan L. O'Connell et al |
| EPI_ISL_667101 | USA/OR-OHSU-2043/2020 | 2020-08-01 | North America/USA/Oregon/Clackamas | OHSU Lab Services Molecular Microbiology Lab | Oregon SARS-CoV-2 Genome Sequencing Center | Brendan L. O'Connell et al |
| EPI_ISL_667102 | USA/WA-OHSU-2045/2020 | 2020-08-01 | North America/USA/Washington/Clark ( | OHSU Lab Services Molecular Microbiology Lab | Oregon SARS-CoV-2 Genome Sequencing Center | Brendan L. O'Connell et al |
| EPI_ISL_667106 | USA/OR-OHSU-2067/2020 | 2020-08-03 | North America/USA/Oregon/Multnomal | OHSU Lab Services Molecular Microbiology Lab | Oregon SARS-CoV-2 Genome Sequencing Center | Brendan L. O'Connell et al |
| EPI_ISL_667108 | USA/OR-OHSU-2071/2020 | 2020-08-03 | North America/USA/Oregon/Clackamas | OHSU Lab Services Molecular Microbiology Lab | Oregon SARS-CoV-2 Genome Sequencing Center | Brendan L. O'Connell et al |
| EPI_ISL_667113 | USA/OR-OHSU-2087/2020 | 2020-08-03 | North America/USA/Oregon/Wasco Cou | OHSU Lab Services Molecular Microbiology Lab | Oregon SARS-CoV-2 Genome Sequencing Center | Brendan L. O'Connell et al |
| EPI_ISL_667127 | USA/OR-OHSU-2145/2020 | 2020-08-05 | North America/USA/Oregon/Multnomal | OHSU Lab Services Molecular Microbiology Lab | Oregon SARS-CoV-2 Genome Sequencing Center | Brendan L. O'Connell et al |
| EPI_ISL_667147 | USA/OR-OHSU-2211/2020 | 2020-08-06 | North America/USA/Oregon/Washingto | OHSU Lab Services Molecular Microbiology Lab | Oregon SARS-CoV-2 Genome Sequencing Center | Brendan L. O'Connell et al |
| EPI_ISL_667149 | USA/OR-OHSU-2215/2020 | 2020-08-07 | North America/USA/Oregon/Multnomal | OHSU Lab Services Molecular Microbiology Lab | Oregon SARS-CoV-2 Genome Sequencing Center | Brendan L. O'Connell et al |

| | | | | | | |
|---|---|---|---|---|---|---|
| EPI_ISL_667150 | USA/OR-OHSU-2219/2020 | 2020-08-07 | North America/USA/Oregon/Washingto | OHSU Lab Services Molecular Microbiology Lab | Oregon SARS-CoV-2 Genome Sequencing Center | Brendan L. O'Connell et al |
| EPI_ISL_667154 | USA/OR-OHSU-2233/2020 | 2020-08-07 | North America/USA/Oregon/Washingto | OHSU Lab Services Molecular Microbiology Lab | Oregon SARS-CoV-2 Genome Sequencing Center | Brendan L. O'Connell et al |
| EPI_ISL_667156 | USA/OR-OHSU-2239/2020 | 2020-08-07 | North America/USA/Oregon/Multnomal | OHSU Lab Services Molecular Microbiology Lab | Oregon SARS-CoV-2 Genome Sequencing Center | Brendan L. O'Connell et al |
| EPI_ISL_667158 | USA/OR-OHSU-2249/2020 | 2020-08-07 | North America/USA/Oregon/Washingto | OHSU Lab Services Molecular Microbiology Lab | Oregon SARS-CoV-2 Genome Sequencing Center | Brendan L. O'Connell et al |
| EPI_ISL_667160 | USA/OR-OHSU-2253/2020 | 2020-08-08 | North America/USA/Oregon/Washingto | OHSU Lab Services Molecular Microbiology Lab | Oregon SARS-CoV-2 Genome Sequencing Center | Brendan L. O'Connell et al |
| EPI_ISL_667167 | USA/OR-OHSU-2275/2020 | 2020-08-08 | North America/USA/Oregon/Washingto | OHSU Lab Services Molecular Microbiology Lab | Oregon SARS-CoV-2 Genome Sequencing Center | Brendan L. O'Connell et al |
| EPI_ISL_667169 | USA/WA-OHSU-2287/2020 | 2020-08-10 | North America/USA/Washington/Clark ( | OHSU Lab Services Molecular Microbiology Lab | Oregon SARS-CoV-2 Genome Sequencing Center | Brendan L. O'Connell et al |
| EPI_ISL_667175 | USA/OR-OHSU-2301/2020 | 2020-08-10 | North America/USA/Oregon/Multnomal | OHSU Lab Services Molecular Microbiology Lab | Oregon SARS-CoV-2 Genome Sequencing Center | Brendan L. O'Connell et al |
| EPI_ISL_667176 | USA/OR-OHSU-2303/2020 | 2020-08-10 | North America/USA/Oregon/Washingto | OHSU Lab Services Molecular Microbiology Lab | Oregon SARS-CoV-2 Genome Sequencing Center | Brendan L. O'Connell et al |
| EPI_ISL_667180 | USA/OR-OHSU-2313/2020 | 2020-08-10 | North America/USA/Oregon/Multnomal | OHSU Lab Services Molecular Microbiology Lab | Oregon SARS-CoV-2 Genome Sequencing Center | Brendan L. O'Connell et al |

| | | | | | | |
|---|---|---|---|---|---|---|
| EPI_ISL_667185 | USA/OR-OHSU-2331/2020 | 2020-08-11 | North America/USA/Oregon/Multnomal | OHSU Lab Services Molecular Microbiology Lab | Oregon SARS-CoV-2 Genome Sequencing Center | Brendan L. O'Connell et al |
| EPI_ISL_667191 | USA/OR-OHSU-2353/2020 | 2020-08-12 | North America/USA/Oregon/Washingto | OHSU Lab Services Molecular Microbiology Lab | Oregon SARS-CoV-2 Genome Sequencing Center | Brendan L. O'Connell et al |
| EPI_ISL_667193 | USA/OR-OHSU-2365/2020 | 2020-08-12 | North America/USA/Oregon/Multnomal | OHSU Lab Services Molecular Microbiology Lab | Oregon SARS-CoV-2 Genome Sequencing Center | Brendan L. O'Connell et al |
| EPI_ISL_667197 | USA/OR-OHSU-2397/2020 | 2020-08-13 | North America/USA/Oregon/Clackamas | OHSU Lab Services Molecular Microbiology Lab | Oregon SARS-CoV-2 Genome Sequencing Center | Brendan L. O'Connell et al |
| EPI_ISL_667199 | USA/OR-OHSU-2409/2020 | 2020-08-13 | North America/USA/Oregon/Multnomal | OHSU Lab Services Molecular Microbiology Lab | Oregon SARS-CoV-2 Genome Sequencing Center | Brendan L. O'Connell et al |
| EPI_ISL_667201 | USA/OR-OHSU-2419/2020 | 2020-08-14 | North America/USA/Oregon/Washingto | OHSU Lab Services Molecular Microbiology Lab | Oregon SARS-CoV-2 Genome Sequencing Center | Brendan L. O'Connell et al |
| EPI_ISL_667208 | USA/OR-OHSU-2441/2020 | 2020-08-14 | North America/USA/Oregon/Washingto | OHSU Lab Services Molecular Microbiology Lab | Oregon SARS-CoV-2 Genome Sequencing Center | Brendan L. O'Connell et al |
| EPI_ISL_667218 | USA/OR-OHSU-2467/2020 | 2020-08-15 | North America/USA/Oregon/Washingto | OHSU Lab Services Molecular Microbiology Lab | Oregon SARS-CoV-2 Genome Sequencing Center | Brendan L. O'Connell et al |
| EPI_ISL_667224 | USA/OR-OHSU-2483/2020 | 2020-08-01 | North America/USA/Oregon/Yamhill Co | OHSU Lab Services Molecular Microbiology Lab | Oregon SARS-CoV-2 Genome Sequencing Center | Brendan L. O'Connell et al |
| EPI_ISL_667226 | USA/OR-OHSU-2489/2020 | 2020-08-17 | North America/USA/Oregon/Marion Co | OHSU Lab Services Molecular Microbiology Lab | Oregon SARS-CoV-2 Genome Sequencing Center | Brendan L. O'Connell et al |

| | | | | | | |
|---|---|---|---|---|---|---|
| EPI_ISL_667230 | USA/OR-OHSU-2503/2020 | 2020-10-26 | North America/USA/Oregon/ | OHSU Lab Services Molecular Microbiology Lab | Oregon SARS-CoV-2 Genome Sequencing Center | Brendan L. O'Connell et al |
| EPI_ISL_667231 | USA/OR-OHSU-2504/2020 | 2020-10-26 | North America/USA/Oregon/ | OHSU Lab Services Molecular Microbiology Lab | Oregon SARS-CoV-2 Genome Sequencing Center | Brendan L. O'Connell et al |
| EPI_ISL_667239 | USA/OR-OHSU-2527/2020 | 2020-08-18 | North America/USA/Oregon/Multnomal | OHSU Lab Services Molecular Microbiology Lab | Oregon SARS-CoV-2 Genome Sequencing Center | Brendan L. O'Connell et al |
| EPI_ISL_667240 | USA/OR-OHSU-2529/2020 | 2020-08-18 | North America/USA/Oregon/Multnomal | OHSU Lab Services Molecular Microbiology Lab | Oregon SARS-CoV-2 Genome Sequencing Center | Brendan L. O'Connell et al |
| EPI_ISL_667242 | USA/OR-OHSU-2537/2020 | 2020-08-19 | North America/USA/Oregon/Multnomal | OHSU Lab Services Molecular Microbiology Lab | Oregon SARS-CoV-2 Genome Sequencing Center | Brendan L. O'Connell et al |
| EPI_ISL_667244 | USA/OR-OHSU-2541/2020 | 2020-08-19 | North America/USA/Oregon/Multnomal | OHSU Lab Services Molecular Microbiology Lab | Oregon SARS-CoV-2 Genome Sequencing Center | Brendan L. O'Connell et al |
| EPI_ISL_667255 | USA/OR-OHSU-2575/2020 | 2020-08-20 | North America/USA/Oregon/Washingto | OHSU Lab Services Molecular Microbiology Lab | Oregon SARS-CoV-2 Genome Sequencing Center | Brendan L. O'Connell et al |
| EPI_ISL_667266 | USA/OR-OHSU-2623/2020 | 2020-08-22 | North America/USA/Oregon/Multnomal | OHSU Lab Services Molecular Microbiology Lab | Oregon SARS-CoV-2 Genome Sequencing Center | Brendan L. O'Connell et al |
| EPI_ISL_667267 | USA/OR-OHSU-2625/2020 | 2020-08-22 | North America/USA/Oregon/Washingto | OHSU Lab Services Molecular Microbiology Lab | Oregon SARS-CoV-2 Genome Sequencing Center | Brendan L. O'Connell et al |
| EPI_ISL_667268 | USA/OR-OHSU-2631/2020 | 2020-08-18 | North America/USA/Oregon/Marion Co | OHSU Lab Services Molecular Microbiology Lab | Oregon SARS-CoV-2 Genome Sequencing Center | Brendan L. O'Connell et al |

| | | | | | | |
|---|---|---|---|---|---|---|
| EPI_ISL_667269 | USA/OR-OHSU-2637/2020 | 2020-08-22 | North America/USA/Oregon/Multnomal | OHSU Lab Services Molecular Microbiology Lab | Oregon SARS-CoV-2 Genome Sequencing Center | Brendan L. O'Connell et al |
| EPI_ISL_667276 | USA/OR-OHSU-2661/2020 | 2020-08-24 | North America/USA/Oregon/Washingto | OHSU Lab Services Molecular Microbiology Lab | Oregon SARS-CoV-2 Genome Sequencing Center | Brendan L. O'Connell et al |
| EPI_ISL_667277 | USA/OR-OHSU-2667/2020 | 2020-08-24 | North America/USA/Oregon/Multnomal | OHSU Lab Services Molecular Microbiology Lab | Oregon SARS-CoV-2 Genome Sequencing Center | Brendan L. O'Connell et al |
| EPI_ISL_667278 | USA/OR-OHSU-2675/2020 | 2020-08-25 | North America/USA/Oregon/Multnomal | OHSU Lab Services Molecular Microbiology Lab | Oregon SARS-CoV-2 Genome Sequencing Center | Brendan L. O'Connell et al |
| EPI_ISL_667280 | USA/OR-OHSU-2681/2020 | 2020-08-25 | North America/USA/Oregon/Clackamas | OHSU Lab Services Molecular Microbiology Lab | Oregon SARS-CoV-2 Genome Sequencing Center | Brendan L. O'Connell et al |
| EPI_ISL_667286 | USA/OR-OHSU-2697/2020 | 2020-08-26 | North America/USA/Oregon/Washingto | OHSU Lab Services Molecular Microbiology Lab | Oregon SARS-CoV-2 Genome Sequencing Center | Brendan L. O'Connell et al |
| EPI_ISL_667289 | USA/OR-OHSU-2707/2020 | 2020-08-26 | North America/USA/Oregon/Multnomal | OHSU Lab Services Molecular Microbiology Lab | Oregon SARS-CoV-2 Genome Sequencing Center | Brendan L. O'Connell et al |
| EPI_ISL_667290 | USA/OR-OHSU-2709/2020 | 2020-08-26 | North America/USA/Oregon/Washingto | OHSU Lab Services Molecular Microbiology Lab | Oregon SARS-CoV-2 Genome Sequencing Center | Brendan L. O'Connell et al |
| EPI_ISL_667293 | USA/WA-OHSU-2719/2020 | 2020-08-27 | North America/USA/Washington/Clark ( | OHSU Lab Services Molecular Microbiology Lab | Oregon SARS-CoV-2 Genome Sequencing Center | Brendan L. O'Connell et al |
| EPI_ISL_667294 | USA/OR-OHSU-2723/2020 | 2020-08-27 | North America/USA/Oregon/Washingto | OHSU Lab Services Molecular Microbiology Lab | Oregon SARS-CoV-2 Genome Sequencing Center | Brendan L. O'Connell et al |

| | | | | | | |
|---|---|---|---|---|---|---|
| EPI_ISL_667297 | USA/OR-OHSU-2737/2020 | 2020-08-27 | North America/USA/Oregon/Multnomal | OHSU Lab Services Molecular Microbiology Lab | Oregon SARS-CoV-2 Genome Sequencing Center | Brendan L. O'Connell et al |
| EPI_ISL_667301 | USA/OR-OHSU-2753/2020 | 2020-08-28 | North America/USA/Oregon/Multnomal | OHSU Lab Services Molecular Microbiology Lab | Oregon SARS-CoV-2 Genome Sequencing Center | Brendan L. O'Connell et al |
| EPI_ISL_667305 | USA/OR-OHSU-2767/2020 | 2020-08-29 | North America/USA/Oregon/Washingto | OHSU Lab Services Molecular Microbiology Lab | Oregon SARS-CoV-2 Genome Sequencing Center | Brendan L. O'Connell et al |
| EPI_ISL_667311 | USA/OR-OHSU-2783/2020 | 2020-08-31 | North America/USA/Oregon/Washingto | OHSU Lab Services Molecular Microbiology Lab | Oregon SARS-CoV-2 Genome Sequencing Center | Brendan L. O'Connell et al |
| EPI_ISL_667313 | USA/OR-OHSU-2791/2020 | 2020-08-31 | North America/USA/Oregon/Washingto | OHSU Lab Services Molecular Microbiology Lab | Oregon SARS-CoV-2 Genome Sequencing Center | Brendan L. O'Connell et al |
| EPI_ISL_667315 | USA/OR-OHSU-2797/2020 | 2020-08-31 | North America/USA/Oregon/Washingto | OHSU Lab Services Molecular Microbiology Lab | Oregon SARS-CoV-2 Genome Sequencing Center | Brendan L. O'Connell et al |
| EPI_ISL_667318 | USA/OR-OHSU-2807/2020 | 2020-09-01 | North America/USA/Oregon/Multnomal | OHSU Lab Services Molecular Microbiology Lab | Oregon SARS-CoV-2 Genome Sequencing Center | Brendan L. O'Connell et al |
| EPI_ISL_667320 | USA/OR-OHSU-2811/2020 | 2020-09-01 | North America/USA/Oregon/Multnomal | OHSU Lab Services Molecular Microbiology Lab | Oregon SARS-CoV-2 Genome Sequencing Center | Brendan L. O'Connell et al |
| EPI_ISL_667323 | USA/OR-OHSU-2814/2020 | 2020-09-01 | North America/USA/Oregon/Wasco Cou | OHSU Lab Services Molecular Microbiology Lab | Oregon SARS-CoV-2 Genome Sequencing Center | Brendan L. O'Connell et al |
| EPI_ISL_667327 | USA/OR-OHSU-2819/2020 | 2020-09-02 | North America/USA/Oregon/Washingto | OHSU Lab Services Molecular Microbiology Lab | Oregon SARS-CoV-2 Genome Sequencing Center | Brendan L. O'Connell et al |

| | | | | | | |
|---|---|---|---|---|---|---|
| EPI_ISL_667328 | USA/OR-OHSU-2820/2020 | 2020-09-02 | North America/USA/Oregon/Washingto | OHSU Lab Services Molecular Microbiology Lab | Oregon SARS-CoV-2 Genome Sequencing Center | Brendan L. O'Connell et al |
| EPI_ISL_667331 | USA/OR-OHSU-2823/2020 | 2020-09-02 | North America/USA/Oregon/Washingto | OHSU Lab Services Molecular Microbiology Lab | Oregon SARS-CoV-2 Genome Sequencing Center | Brendan L. O'Connell et al |
| EPI_ISL_667332 | USA/OR-OHSU-2824/2020 | 2020-09-02 | North America/USA/Oregon/Washingto | OHSU Lab Services Molecular Microbiology Lab | Oregon SARS-CoV-2 Genome Sequencing Center | Brendan L. O'Connell et al |
| EPI_ISL_667333 | USA/OR-OHSU-2825/2020 | 2020-09-02 | North America/USA/Oregon/Washingto | OHSU Lab Services Molecular Microbiology Lab | Oregon SARS-CoV-2 Genome Sequencing Center | Brendan L. O'Connell et al |
| EPI_ISL_667336 | USA/OR-OHSU-2829/2020 | 2020-09-02 | North America/USA/Oregon/Multnomal | OHSU Lab Services Molecular Microbiology Lab | Oregon SARS-CoV-2 Genome Sequencing Center | Brendan L. O'Connell et al |
| EPI_ISL_667338 | USA/OR-OHSU-2831/2020 | 2020-09-02 | North America/USA/Oregon/Washingto | OHSU Lab Services Molecular Microbiology Lab | Oregon SARS-CoV-2 Genome Sequencing Center | Brendan L. O'Connell et al |
| EPI_ISL_667341 | USA/OR-OHSU-2835/2020 | 2020-09-02 | North America/USA/Oregon/Multnomal | OHSU Lab Services Molecular Microbiology Lab | Oregon SARS-CoV-2 Genome Sequencing Center | Brendan L. O'Connell et al |
| EPI_ISL_667343 | USA/OR-OHSU-2837/2020 | 2020-09-02 | North America/USA/Oregon/Multnomal | OHSU Lab Services Molecular Microbiology Lab | Oregon SARS-CoV-2 Genome Sequencing Center | Brendan L. O'Connell et al |
| EPI_ISL_667345 | USA/OR-OHSU-2840/2020 | 2020-09-02 | North America/USA/Oregon/Columbia C | OHSU Lab Services Molecular Microbiology Lab | Oregon SARS-CoV-2 Genome Sequencing Center | Brendan L. O'Connell et al |
| EPI_ISL_667351 | USA/OR-OHSU-2846/2020 | 2020-09-02 | North America/USA/Oregon/Wasco Cou | OHSU Lab Services Molecular Microbiology Lab | Oregon SARS-CoV-2 Genome Sequencing Center | Brendan L. O'Connell et al |

| | | | | | | |
|---|---|---|---|---|---|---|
| EPI_ISL_667352 | USA/OR-OHSU-2849/2020 | 2020-09-02 | North America/USA/Oregon/Washingto | OHSU Lab Services Molecular Microbiology Lab | Oregon SARS-CoV-2 Genome Sequencing Center | Brendan L. O'Connell et al |
| EPI_ISL_667354 | USA/OR-OHSU-2851/2020 | 2020-09-02 | North America/USA/Oregon/Marion Co | OHSU Lab Services Molecular Microbiology Lab | Oregon SARS-CoV-2 Genome Sequencing Center | Brendan L. O'Connell et al |
| EPI_ISL_667355 | USA/OR-OHSU-2852/2020 | 2020-09-02 | North America/USA/Oregon/Marion Co | OHSU Lab Services Molecular Microbiology Lab | Oregon SARS-CoV-2 Genome Sequencing Center | Brendan L. O'Connell et al |
| EPI_ISL_667357 | USA/OR-OHSU-2854/2020 | 2020-09-02 | North America/USA/Oregon/Wasco Cou | OHSU Lab Services Molecular Microbiology Lab | Oregon SARS-CoV-2 Genome Sequencing Center | Brendan L. O'Connell et al |
| EPI_ISL_667359 | USA/OR-OHSU-2856/2020 | 2020-09-02 | North America/USA/Oregon/Washingto | OHSU Lab Services Molecular Microbiology Lab | Oregon SARS-CoV-2 Genome Sequencing Center | Brendan L. O'Connell et al |
| EPI_ISL_667369 | USA/OR-OHSU-2866/2020 | 2020-09-04 | North America/USA/Oregon/Multnomal | OHSU Lab Services Molecular Microbiology Lab | Oregon SARS-CoV-2 Genome Sequencing Center | Brendan L. O'Connell et al |
| EPI_ISL_667374 | USA/OR-OHSU-2871/2020 | 2020-09-03 | North America/USA/Oregon/Multnomal | OHSU Lab Services Molecular Microbiology Lab | Oregon SARS-CoV-2 Genome Sequencing Center | Brendan L. O'Connell et al |
| EPI_ISL_667375 | USA/OR-OHSU-2872/2020 | 2020-09-03 | North America/USA/Oregon/Multnomal | OHSU Lab Services Molecular Microbiology Lab | Oregon SARS-CoV-2 Genome Sequencing Center | Brendan L. O'Connell et al |
| EPI_ISL_667376 | USA/OR-OHSU-2873/2020 | 2020-09-03 | North America/USA/Oregon/Multnomal | OHSU Lab Services Molecular Microbiology Lab | Oregon SARS-CoV-2 Genome Sequencing Center | Brendan L. O'Connell et al |
| EPI_ISL_667383 | USA/WA-OHSU-2882/2020 | 2020-09-04 | North America/USA/Washington/Clark ( | OHSU Lab Services Molecular Microbiology Lab | Oregon SARS-CoV-2 Genome Sequencing Center | Brendan L. O'Connell et al |

| | | | | | | |
|---|---|---|---|---|---|---|
| EPI_ISL_667388 | USA/OR-OHSU-2887/2020 | 2020-09-04 | North America/USA/Oregon/Washingto | OHSU Lab Services Molecular Microbiology Lab | Oregon SARS-CoV-2 Genome Sequencing Center | Brendan L. O'Connell et al |
| EPI_ISL_667422 | USA/OR-OHSU-2926/2020 | 2020-09-05 | North America/USA/Oregon/Washingto | OHSU Lab Services Molecular Microbiology Lab | Oregon SARS-CoV-2 Genome Sequencing Center | Brendan L. O'Connell et al |
| EPI_ISL_667426 | USA/OR-OHSU-2931/2020 | 2020-09-07 | North America/USA/Oregon/Multnomal | OHSU Lab Services Molecular Microbiology Lab | Oregon SARS-CoV-2 Genome Sequencing Center | Brendan L. O'Connell et al |
| EPI_ISL_667427 | USA/OR-OHSU-2943/2020 | 2020-09-01 | North America/USA/Oregon/Multnomal | OHSU Lab Services Molecular Microbiology Lab | Oregon SARS-CoV-2 Genome Sequencing Center | Brendan L. O'Connell et al |
| EPI_ISL_667436 | USA/OR-OHSU-2969/2020 | 2020-09-07 | North America/USA/Oregon/Yamhill Co | OHSU Lab Services Molecular Microbiology Lab | Oregon SARS-CoV-2 Genome Sequencing Center | Brendan L. O'Connell et al |
| EPI_ISL_667438 | USA/OR-OHSU-2971/2020 | 2020-09-07 | North America/USA/Oregon/Washingto | OHSU Lab Services Molecular Microbiology Lab | Oregon SARS-CoV-2 Genome Sequencing Center | Brendan L. O'Connell et al |
| EPI_ISL_667440 | USA/OR-OHSU-2975/2020 | 2020-09-09 | North America/USA/Oregon/Clackamas | OHSU Lab Services Molecular Microbiology Lab | Oregon SARS-CoV-2 Genome Sequencing Center | Brendan L. O'Connell et al |
| EPI_ISL_667445 | USA/OR-OHSU-2981/2020 | 2020-09-09 | North America/USA/Oregon/Wasco Cou | OHSU Lab Services Molecular Microbiology Lab | Oregon SARS-CoV-2 Genome Sequencing Center | Brendan L. O'Connell et al |
| EPI_ISL_667447 | USA/OR-OHSU-2983/2020 | 2020-09-09 | North America/USA/Oregon/Yamhill Co | OHSU Lab Services Molecular Microbiology Lab | Oregon SARS-CoV-2 Genome Sequencing Center | Brendan L. O'Connell et al |
| EPI_ISL_667452 | USA/OR-OHSU-2994/2020 | 2020-09-11 | North America/USA/Oregon/Multnomal | OHSU Lab Services Molecular Microbiology Lab | Oregon SARS-CoV-2 Genome Sequencing Center | Brendan L. O'Connell et al |

| | | | | | | |
|---|---|---|---|---|---|---|
| EPI_ISL_667457 | USA/OR-OHSU-3047/2020 | 2020-09-30 | North America/USA/Oregon/Washingto | OHSU Lab Services Molecular Microbiology Lab | Oregon SARS-CoV-2 Genome Sequencing Center | Brendan L. O'Connell et al |
| EPI_ISL_667458 | USA/OR-OHSU-3048/2020 | 2020-09-30 | North America/USA/Oregon/Washingto | OHSU Lab Services Molecular Microbiology Lab | Oregon SARS-CoV-2 Genome Sequencing Center | Brendan L. O'Connell et al |
| EPI_ISL_667462 | USA/OR-OHSU-3053/2020 | 2020-09-30 | North America/USA/Oregon/Washingto | OHSU Lab Services Molecular Microbiology Lab | Oregon SARS-CoV-2 Genome Sequencing Center | Brendan L. O'Connell et al |
| EPI_ISL_667482 | USA/OR-OHSU-3101/2020 | 2020-09-30 | North America/USA/Oregon/ | OHSU Lab Services Molecular Microbiology Lab | Oregon SARS-CoV-2 Genome Sequencing Center | Brendan L. O'Connell et al |
| EPI_ISL_667487 | USA/OR-OHSU-3115/2020 | 2020-10-02 | North America/USA/Oregon/Multnomal | OHSU Lab Services Molecular Microbiology Lab | Oregon SARS-CoV-2 Genome Sequencing Center | Brendan L. O'Connell et al |
| EPI_ISL_667491 | USA/OR-OHSU-3133/2020 | 2020-10-03 | North America/USA/Oregon/Multnomal | OHSU Lab Services Molecular Microbiology Lab | Oregon SARS-CoV-2 Genome Sequencing Center | Brendan L. O'Connell et al |
| EPI_ISL_667506 | USA/OR-OHSU-3561/2020 | 2020-10-21 | North America/USA/Oregon/ | OHSU Lab Services Molecular Microbiology Lab | Oregon SARS-CoV-2 Genome Sequencing Center | Brendan L. O'Connell et al |
| EPI_ISL_667526 | USA/OR-OHSU-4117/2020 | 2020-10-30 | North America/USA/Oregon/ | OHSU Lab Services Molecular Microbiology Lab | Oregon SARS-CoV-2 Genome Sequencing Center | Brendan L. O'Connell et al |
| EPI_ISL_667534 | USA/OR-OHSU-4292/2020 | 2020-11-03 | North America/USA/Oregon/ | OHSU Lab Services Molecular Microbiology Lab | Oregon SARS-CoV-2 Genome Sequencing Center | Brendan L. O'Connell et al |
| EPI_ISL_667535 | USA/OR-OHSU-4315/2020 | 2020-11-03 | North America/USA/Oregon/ | OHSU Lab Services Molecular Microbiology Lab | Oregon SARS-CoV-2 Genome Sequencing Center | Brendan L. O'Connell et al |

| | | | | | | |
|---|---|---|---|---|---|---|
| EPI_ISL_667537 | USA/OR-OHSU-4446/2020 | 2020-11-05 | North America/USA/Oregon/ | OHSU Lab Services Molecular Microbiology Lab | Oregon SARS-CoV-2 Genome Sequencing Center | Brendan L. O'Connell et al |
| EPI_ISL_667540 | USA/OR-OHSU-4349/2020 | 2020-11-03 | North America/USA/Oregon/ | OHSU Lab Services Molecular Microbiology Lab | Oregon SARS-CoV-2 Genome Sequencing Center | Brendan L. O'Connell et al |
| EPI_ISL_667545 | USA/OR-OHSU-4667/2020 | 2020-11-09 | North America/USA/Oregon/ | OHSU Lab Services Molecular Microbiology Lab | Oregon SARS-CoV-2 Genome Sequencing Center | Brendan L. O'Connell et al |
| EPI_ISL_667550 | USA/OR-OHSU-4350/2020 | 2020-11-03 | North America/USA/Oregon/ | OHSU Lab Services Molecular Microbiology Lab | Oregon SARS-CoV-2 Genome Sequencing Center | Brendan L. O'Connell et al |
| EPI_ISL_667551 | USA/OR-OHSU-2047/2020 | 2020-08-01 | North America/USA/Oregon/Multnomal | OHSU Lab Services Molecular Microbiology Lab | Oregon SARS-CoV-2 Genome Sequencing Center | Brendan L. O'Connell et al |
| EPI_ISL_667553 | USA/OR-OHSU-2157/2020 | 2020-08-05 | North America/USA/Oregon/Multnomal | OHSU Lab Services Molecular Microbiology Lab | Oregon SARS-CoV-2 Genome Sequencing Center | Brendan L. O'Connell et al |
| EPI_ISL_667556 | USA/OR-OHSU-2913/2020 | 2020-09-05 | North America/USA/Oregon/Washingto | OHSU Lab Services Molecular Microbiology Lab | Oregon SARS-CoV-2 Genome Sequencing Center | Brendan L. O'Connell et al |
| EPI_ISL_667566 | Japan/IC-0137/2020 | 20-Sep | Asia/Japan/Japan/ | Pathogen Genomics Center, National Institute of Infectious Diseases | Pathogen Genomics Center, National Institute of Infectious Diseases | Tsuyoshi Sekizuka et al |
| EPI_ISL_667577 | Japan/IC-0148/2020 | 20-Aug | Asia/Japan/Japan/ | Pathogen Genomics Center, National Institute of Infectious Diseases | Pathogen Genomics Center, National Institute of Infectious Diseases | Tsuyoshi Sekizuka et al |

| | | | | | | |
|---|---|---|---|---|---|---|
| EPI_ISL_667758 | Canada/UN-CRCHUM-PreFreeze-1/20 | 2020-09-04 | North America/Canada/Quebec/ | Microbiology, Infectious Diseases and Immunology, Centre de Recherche du Centre Hospitalier de l'Universite de Montreal | Microbiology, Infectious Diseases and Immunology, Centre de Recherche du Centre Hospitalier de l'Universite de Montreal | Benoit et al |
| EPI_ISL_667759 | Canada/UN-CRCHUM-PreFreeze-2/20 | 2020-09-04 | North America/Canada/Quebec/ | Microbiology, Infectious Diseases and Immunology, Centre de Recherche du Centre Hospitalier de l'Universite de Montreal | Microbiology, Infectious Diseases and Immunology, Centre de Recherche du Centre Hospitalier de l'Universite de Montreal | Benoit et al |
| EPI_ISL_667760 | Canada/UN-CRCHUM-PreFreeze-3/20 | 2020-09-04 | North America/Canada/Quebec/ | Microbiology, Infectious Diseases and Immunology, Centre de Recherche du Centre Hospitalier de l'Universite de Montreal | Microbiology, Infectious Diseases and Immunology, Centre de Recherche du Centre Hospitalier de l'Universite de Montreal | Benoit et al |
| EPI_ISL_667762 | Canada/UN-CRCHUM-PreFreeze-5/20 | 2020-09-04 | North America/Canada/Quebec/ | Microbiology, Infectious Diseases and Immunology, Centre de Recherche du Centre Hospitalier de l'Universite de Montreal | Microbiology, Infectious Diseases and Immunology, Centre de Recherche du Centre Hospitalier de l'Universite de Montreal | Benoit et al |

| | | | | | | |
|---|---|---|---|---|---|---|
| EPI_ISL_667763 | Canada/UN-CRCHUM-PreFreeze-6/20 | 2020-09-04 | North America/Canada/Quebec/ | Microbiology, Infectious Diseases and Immunology, Centre de Recherche du Centre Hospitalier de l'Universite de Montreal | Microbiology, Infectious Diseases and Immunology, Centre de Recherche du Centre Hospitalier de l'Universite de Montreal | Benoit et al |
| EPI_ISL_667766 | Canada/UN-CRCHUM-PreFreeze-9/20 | 2020-09-04 | North America/Canada/Quebec/ | Microbiology, Infectious Diseases and Immunology, Centre de Recherche du Centre Hospitalier de l'Universite de Montreal | Microbiology, Infectious Diseases and Immunology, Centre de Recherche du Centre Hospitalier de l'Universite de Montreal | Benoit et al |
| EPI_ISL_667767 | Canada/UN-CRCHUM-PreFreeze-10/2 | 2020-09-04 | North America/Canada/Quebec/ | Microbiology, Infectious Diseases and Immunology, Centre de Recherche du Centre Hospitalier de l'Universite de Montreal | Microbiology, Infectious Diseases and Immunology, Centre de Recherche du Centre Hospitalier de l'Universite de Montreal | Benoit et al |
| EPI_ISL_667768 | Canada/UN-CRCHUM-PostFreeze-1/2 | 2020-09-04 | North America/Canada/Quebec/ | Microbiology, Infectious Diseases and Immunology, Centre de Recherche du Centre Hospitalier de l'Universite de Montreal | Microbiology, Infectious Diseases and Immunology, Centre de Recherche du Centre Hospitalier de l'Universite de Montreal | Benoit et al |

| | | | | | | |
|---|---|---|---|---|---|---|
| EPI_ISL_667769 | Canada/UN-CRCHUM-PostFreeze-2/2( | 2020-09-04 | North America/Canada/Quebec/ | Microbiology, Infectious Diseases and Immunology, Centre de Recherche du Centre Hospitalier de l'Universite de Montreal | Microbiology, Infectious Diseases and Immunology, Centre de Recherche du Centre Hospitalier de l'Universite de Montreal | Benoit et al |
| EPI_ISL_667770 | Canada/UN-CRCHUM-PostFreeze-3/2( | 2020-09-04 | North America/Canada/Quebec/ | Microbiology, Infectious Diseases and Immunology, Centre de Recherche du Centre Hospitalier de l'Universite de Montreal | Microbiology, Infectious Diseases and Immunology, Centre de Recherche du Centre Hospitalier de l'Universite de Montreal | Benoit et al |
| EPI_ISL_667771 | Canada/UN-CRCHUM-PostFreeze-5/2( | 2020-09-04 | North America/Canada/Quebec/ | Microbiology, Infectious Diseases and Immunology, Centre de Recherche du Centre Hospitalier de l'Universite de Montreal | Microbiology, Infectious Diseases and Immunology, Centre de Recherche du Centre Hospitalier de l'Universite de Montreal | Benoit et al |
| EPI_ISL_667772 | Canada/UN-CRCHUM-PostFreeze-6/2( | 2020-09-04 | North America/Canada/Quebec/ | Microbiology, Infectious Diseases and Immunology, Centre de Recherche du Centre Hospitalier de l'Universite de Montreal | Microbiology, Infectious Diseases and Immunology, Centre de Recherche du Centre Hospitalier de l'Universite de Montreal | Benoit et al |

| | | | | | | |
|---|---|---|---|---|---|---|
| EPI_ISL_667774 | Canada/UN-CRCHUM-PostFreeze-9/2( | 2020-09-04 | North America/Canada/Quebec/ | Microbiology, Infectious Diseases and Immunology, Centre de Recherche du Centre Hospitalier de l'Universite de Montreal | Microbiology, Infectious Diseases and Immunology, Centre de Recherche du Centre Hospitalier de l'Universite de Montreal | Benoit et al |
| EPI_ISL_667775 | Canada/UN-CRCHUM-PostFreeze-10/: | 2020-09-04 | North America/Canada/Quebec/ | Microbiology, Infectious Diseases and Immunology, Centre de Recherche du Centre Hospitalier de l'Universite de Montreal | Microbiology, Infectious Diseases and Immunology, Centre de Recherche du Centre Hospitalier de l'Universite de Montreal | Benoit et al |
| EPI_ISL_667849 | England/QEUH-B7E961/2020 | 2020-11-14 | Europe/United Kingdom/England/ | Lighthouse Lab in Glasgow | Wellcome Sanger Institute for the COVID-19 Genomics UK (COG-UK) Consortium | Harper VanSteenhouse et al |
| EPI_ISL_667866 | England/QEUH-B7E600/2020 | 2020-11-14 | Europe/United Kingdom/England/ | Lighthouse Lab in Glasgow | Wellcome Sanger Institute for the COVID-19 Genomics UK (COG-UK) Consortium | Harper VanSteenhouse et al |
| EPI_ISL_667890 | England/CAMC-B79104/2020 | 2020-11-12 | Europe/United Kingdom/England/ | Lighthouse Lab in Cambridge | Wellcome Sanger Institute for the COVID-19 Genomics UK (COG-UK) Consortium | Rob Howes et al |
| EPI_ISL_668103 | England/CAMC-B78C13/2020 | 2020-11-12 | Europe/United Kingdom/England/ | Lighthouse Lab in Cambridge | Wellcome Sanger Institute for the COVID-19 Genomics UK (COG-UK) Consortium | Rob Howes et al |

| | | | | | | |
|---|---|---|---|---|---|---|
| EPI_ISL_668138 | England/CAMC-B77D99/2020 | 2020-11-12 | Europe/United Kingdom/England/ | Lighthouse Lab in Cambridge | Wellcome Sanger Institute for the COVID-19 Genomics UK (COG-UK) Consortium | Rob Howes et al |
| EPI_ISL_668422 | Norway/4891/2020 | 2020-10-22 | Europe/Norway/Nordland/ | Nordland Hospital - Bodo, Laboratory Department, Molecular Biology Unit | Norwegian Institute of Public Health, Department of Virology | Kathrine Stene-Johansen et al |
| EPI_ISL_668447 | SriLanka/C26818/2020 | 2020-11-03 | Asia/Sri Lanka/Colombo/ | Centre for Dengue Research, Department of Immunology and Molecular Medicine | Centre for Dengue Research, Department of Immunology and Molecular Medicine | Chandima Jeewandara et al |
| EPI_ISL_668450 | SriLanka/C28338/2020 | 2020-11-04 | Asia/Sri Lanka/Colombo/ | Centre for Dengue Research, Department of Immunology and Molecular Medicine | Centre for Dengue Research, Department of Immunology and Molecular Medicine | Chandima Jeewandara et al |
| EPI_ISL_668843 | Denmark/DCGC-9561/2020 | 2020-11-09 | Europe/Denmark/Hovedstaden/ | Department of Virus and Microbiological Special Diagnostics, Statens Serum Institut, Copenhagen, Denmark | Albertsen Lab, Department of Chemistry and Bioscience, Aalborg University, Denmark | Danish Covid-19 Genome Consortium et al |
| EPI_ISL_668933 | Denmark/DCGC-9369/2020 | 2020-11-16 | Europe/Denmark/Sjaelland/ | Department of Virus and Microbiological Special Diagnostics, Statens Serum Institut, Copenhagen, Denmark | Albertsen Lab, Department of Chemistry and Bioscience, Aalborg University, Denmark | Danish Covid-19 Genome Consortium et al |

| | | | | | | |
|---|---|---|---|---|---|---|
| EPI_ISL_669188 | Denmark/DCGC-9799/2020 | 2020-11-09 | Europe/Denmark/Hovedstaden/ | Department of Virus and Microbiological Special Diagnostics, Statens Serum Institut, Copenhagen, Denmark | Albertsen Lab, Department of Chemistry and Bioscience, Aalborg University, Denmark | Danish Covid-19 Genome Consortium et al |
| EPI_ISL_670701 | Denmark/DCGC-11987/2020 | 2020-09-07 | Europe/Denmark/Nordjylland/ | Department of Virus and Microbiological Special Diagnostics, Statens Serum Institut, Copenhagen, Denmark | Albertsen Lab, Department of Chemistry and Bioscience, Aalborg University, Denmark | Danish Covid-19 Genome Consortium et al |
| EPI_ISL_671268 | Denmark/DCGC-12021/2020 | 2020-03-09 | Europe/Denmark/Hovedstaden/ | Department of Virus and Microbiological Special Diagnostics, Statens Serum Institut, Copenhagen, Denmark | Albertsen Lab, Department of Chemistry and Bioscience, Aalborg University, Denmark | Danish Covid-19 Genome Consortium et al |
| EPI_ISL_671350 | Ireland/LK-NVRL-MPB53836/2020 | 2020-05-12 | Europe/Ireland/Limerick/ | National Virus Reference Laboratory | Irish Coronavirus Sequencing Consortium - Teagasc Moorepark | Calm Walsh et al |
| EPI_ISL_671364 | Ireland/D-NVRL-20Z04786/2020 | 2020-04-16 | Europe/Ireland/Dublin/ | National Virus Reference Laboratory | Irish Coronavirus Sequencing Consortium - Teagasc Moorepark | Calum Walsh et al |
| EPI_ISL_671371 | Ireland/WH-NVRL-20Z06323/2020 | 2020-04-17 | Europe/Ireland/Westmeath/ | National Virus Reference Laboratory | Irish Coronavirus Sequencing Consortium - Teagasc Moorepark | Calum Walsh et al |
| EPI_ISL_671375 | Ireland/KE-NVRL-20G27987/2020 | 2020-03-20 | Europe/Ireland/Kerry/ | National Virus Reference Laboratory | Irish Coronavirus Sequencing Consortium - Teagasc Moorepark | Calum Walsh et al |

| | | | | | | |
|---|---|---|---|---|---|---|
| EPI_ISL_671414 | Hungary/SRC-1014/2020 | 2020-09-02 | Europe/Hungary/Baranya County/ | National Laboratory of Virology, Szentv°gothai Research Centre | National Laboratory of Virology, Szentv°gothai Research Centre | Endre Gv°bor Tv≥th et al |
| EPI_ISL_671428 | Hungary/UD-56412/2020 | 2020-09-01 | Europe/Hungary/Szabolcs-Szatmv°r-Bere | University of Debrecen, Department of Medical Microbiology | National Laboratory of Virology, Szentv°gothai Research Centre | Endre Gv°bor Tv≥th et al |
| EPI_ISL_671431 | Hungary/UD-57410/2020 | 2020-09-02 | Europe/Hungary/Szabolcs-Szatmv°r-Bere | University of Debrecen, Department of Medical Microbiology | National Laboratory of Virology, Szentv°gothai Research Centre | Endre Gv°bor Tv≥th et al |
| EPI_ISL_671433 | Hungary/UD-57837/2020 | 2020-09-02 | Europe/Hungary/Hajdv∫-Bihar County/ | University of Debrecen, Department of Medical Microbiology | National Laboratory of Virology, Szentv°gothai Research Centre | Endre Gv°bor Tv≥th et al |
| EPI_ISL_671434 | Hungary/UD-57884/2020 | 2020-09-02 | Europe/Hungary/Hajdv∫-Bihar County/ | University of Debrecen, Department of Medical Microbiology | National Laboratory of Virology, Szentv°gothai Research Centre | Endre Gv°bor Tv≥th et al |
| EPI_ISL_671436 | Hungary/UD-57889/2020 | 2020-09-02 | Europe/Hungary/Hajdv∫-Bihar County/ | University of Debrecen, Department of Medical Microbiology | National Laboratory of Virology, Szentv°gothai Research Centre | Endre Gv°bor Tv≥th et al |
| EPI_ISL_671438 | Hungary/UD-57951/2020 | 2020-09-02 | Europe/Hungary/Gy≈ër-Moson-Sopron C | University of Debrecen, Department of Medical Microbiology | National Laboratory of Virology, Szentv°gothai Research Centre | Endre Gv°bor Tv≥th et al |
| EPI_ISL_671439 | Hungary/UD-57955/2020 | 2020-09-02 | Europe/Hungary/Hajdv∫-Bihar County/ | University of Debrecen, Department of Medical Microbiology | National Laboratory of Virology, Szentv°gothai Research Centre | Endre Gv°bor Tv≥th et al |

| | | | | | | |
|---|---|---|---|---|---|---|
| EPI_ISL_671440 | Hungary/UD-57958/2020 | 2020-09-02 | Europe/Hungary/Hajdv∫-Bihar County/ | University of Debrecen, Department of Medical Microbiology | National Laboratory of Virology, Szentv°gothai Research Centre | Endre Gv°bor Tv≥th et al |
| EPI_ISL_671442 | Hungary/UD-57972/2020 | 2020-09-02 | Europe/Hungary/Borsod-Abav∫j-Zemplv( | University of Debrecen, Department of Medical Microbiology | National Laboratory of Virology, Szentv°gothai Research Centre | Endre Gv°bor Tv≥th et al |
| EPI_ISL_671444 | Hungary/UD-57983/2020 | 2020-09-02 | Europe/Hungary/Borsod-Abav∫j-Zemplv( | University of Debrecen, Department of Medical Microbiology | National Laboratory of Virology, Szentv°gothai Research Centre | Endre Gv°bor Tv≥th et al |
| EPI_ISL_671445 | Hungary/UD-57985/2020 | 2020-09-02 | Europe/Hungary/Borsod-Abav∫j-Zemplv( | University of Debrecen, Department of Medical Microbiology | National Laboratory of Virology, Szentv°gothai Research Centre | Endre Gv°bor Tv≥th et al |
| EPI_ISL_671452 | Hungary/UD-58213/2020 | 2020-09-03 | Europe/Hungary/Szabolcs-Szatmv°r-Ber( | University of Debrecen, Department of Medical Microbiology | National Laboratory of Virology, Szentv°gothai Research Centre | Endre Gv°bor Tv≥th et al |
| EPI_ISL_671454 | Hungary/UD-58469/2020 | 2020-09-03 | Europe/Hungary/Hajdv∫-Bihar County/ | University of Debrecen, Department of Medical Microbiology | National Laboratory of Virology, Szentv°gothai Research Centre | Endre Gv°bor Tv≥th et al |
| EPI_ISL_671463 | Hungary/UD-90381/2020 | 2020-10-03 | Europe/Hungary/Veszprv©m County/ | University of Debrecen, Department of Medical Microbiology | National Laboratory of Virology, Szentv°gothai Research Centre | Endre Gv°bor Tv≥th et al |
| EPI_ISL_671466 | Hungary/UD-90395/2020 | 2020-10-04 | Europe/Hungary/Borsod-Abav∫j-Zemplv( | University of Debrecen, Department of Medical Microbiology | National Laboratory of Virology, Szentv°gothai Research Centre | Endre Gv°bor Tv≥th et al |

| | | | | | | |
|---|---|---|---|---|---|---|
| EPI_ISL_671471 | Hungary/UD-90469/2020 | 2020-10-04 | Europe/Hungary/Borsod-Aba√∫j-Zempl√ | University of Debrecen, Department of Medical Microbiology | National Laboratory of Virology, Szent√°gothai Research Centre | Endre G√°bor T√≥th et al |
| EPI_ISL_671476 | Hungary/UD-90625/2020 | 2020-10-04 | Europe/Hungary/Szabolcs-Szatm√°r-Ber | University of Debrecen, Department of Medical Microbiology | National Laboratory of Virology, Szent√°gothai Research Centre | Endre G√°bor T√≥th et al |
| EPI_ISL_671477 | Hungary/UD-90635/2020 | 2020-10-04 | Europe/Hungary/Szabolcs-Szatm√°r-Ber | University of Debrecen, Department of Medical Microbiology | National Laboratory of Virology, Szent√°gothai Research Centre | Endre G√°bor T√≥th et al |
| EPI_ISL_671486 | Hungary/UD-93823/2020 | 2020-10-08 | Europe/Hungary/Hajd√∫-Bihar County/ | University of Debrecen, Department of Medical Microbiology | National Laboratory of Virology, Szent√°gothai Research Centre | Endre G√°bor T√≥th et al |
| EPI_ISL_671487 | Hungary/UD-93827/2020 | 2020-10-08 | Europe/Hungary/Hajd√∫-Bihar County/ | University of Debrecen, Department of Medical Microbiology | National Laboratory of Virology, Szent√°gothai Research Centre | Endre G√°bor T√≥th et al |
| EPI_ISL_671492 | Hungary/UD-93998/2020 | 2020-10-08 | Europe/Hungary/Hajd√∫-Bihar County/ | University of Debrecen, Department of Medical Microbiology | National Laboratory of Virology, Szent√°gothai Research Centre | Endre G√°bor T√≥th et al |
| EPI_ISL_671496 | Canada/ON-PHL-20-02196/2020 | 20-Sep | North America/Canada/Ontario/ | Public Health Ontario Laboratory | Public Health Ontario Laboratory | Vanessa G Allen et al |
| EPI_ISL_671497 | Canada/ON-PHL-20-02197/2020 | 20-Sep | North America/Canada/Ontario/ | Public Health Ontario Laboratory | Public Health Ontario Laboratory | Vanessa G Allen et al |
| EPI_ISL_671498 | Canada/ON-PHL-20-02258/2020 | 20-Sep | North America/Canada/Ontario/ | Public Health Ontario Laboratory | Public Health Ontario Laboratory | Vanessa G Allen et al |

| | | | | | | |
|---|---|---|---|---|---|---|
| EPI_ISL_671499 | Canada/ON-PHL-20-02259/2020 | 20-Sep | North America/Canada/Ontario/ | Public Health Ontario Laboratory | Public Health Ontario Laboratory | Vanessa G Allen et al |
| EPI_ISL_671500 | Canada/ON-PHL-20-02222/2020 | 20-Sep | North America/Canada/Ontario/ | Public Health Ontario Laboratory | Public Health Ontario Laboratory | Vanessa G Allen et al |
| EPI_ISL_671501 | Canada/ON-PHL-20-02262/2020 | 20-Sep | North America/Canada/Ontario/ | Public Health Ontario Laboratory | Public Health Ontario Laboratory | Vanessa G Allen et al |
| EPI_ISL_671502 | Canada/ON-PHL-20-02227/2020 | 20-Sep | North America/Canada/Ontario/ | Public Health Ontario Laboratory | Public Health Ontario Laboratory | Vanessa G Allen et al |
| EPI_ISL_671503 | Canada/ON-PHL-20-02231/2020 | 20-Sep | North America/Canada/Ontario/ | Public Health Ontario Laboratory | Public Health Ontario Laboratory | Vanessa G Allen et al |
| EPI_ISL_671504 | Canada/ON-PHL-20-02233/2020 | 20-Sep | North America/Canada/Ontario/ | Public Health Ontario Laboratory | Public Health Ontario Laboratory | Vanessa G Allen et al |
| EPI_ISL_671505 | Canada/ON-PHL-20-02232/2020 | 20-Sep | North America/Canada/Ontario/ | Public Health Ontario Laboratory | Public Health Ontario Laboratory | Vanessa G Allen et al |
| EPI_ISL_671506 | Canada/ON-PHL-20-02234/2020 | 20-Sep | North America/Canada/Ontario/ | Public Health Ontario Laboratory | Public Health Ontario Laboratory | Vanessa G Allen et al |
| EPI_ISL_671507 | Canada/ON-PHL-20-02211/2020 | 20-Sep | North America/Canada/Ontario/ | Public Health Ontario Laboratory | Public Health Ontario Laboratory | Vanessa G Allen et al |
| EPI_ISL_671508 | Canada/ON-PHL-20-02209/2020 | 20-Sep | North America/Canada/Ontario/ | Public Health Ontario Laboratory | Public Health Ontario Laboratory | Vanessa G Allen et al |
| EPI_ISL_671509 | Canada/ON-PHL-20-02213/2020 | 20-Sep | North America/Canada/Ontario/ | Public Health Ontario Laboratory | Public Health Ontario Laboratory | Vanessa G Allen et al |
| EPI_ISL_671510 | Canada/ON-PHL-20-02239/2020 | 20-Sep | North America/Canada/Ontario/ | Public Health Ontario Laboratory | Public Health Ontario Laboratory | Vanessa G Allen et al |
| EPI_ISL_671511 | Canada/ON-PHL-20-02240/2020 | 20-Sep | North America/Canada/Ontario/ | Public Health Ontario Laboratory | Public Health Ontario Laboratory | Vanessa G Allen et al |

| | | | | | | |
|---|---|---|---|---|---|---|
| EPI_ISL_671512 | Canada/ON-PHL-20-02241/2020 | 20-Sep | North America/Canada/Ontario/ | Public Health Ontario Laboratory | Public Health Ontario Laboratory | Vanessa G Allen et al |
| EPI_ISL_671513 | Canada/ON-PHL-20-02243/2020 | 20-Sep | North America/Canada/Ontario/ | Public Health Ontario Laboratory | Public Health Ontario Laboratory | Vanessa G Allen et al |
| EPI_ISL_671514 | Canada/ON-PHL-20-02229/2020 | 20-Sep | North America/Canada/Ontario/ | Public Health Ontario Laboratory | Public Health Ontario Laboratory | Vanessa G Allen et al |
| EPI_ISL_671515 | Canada/ON-PHL-20-02230/2020 | 20-Sep | North America/Canada/Ontario/ | Public Health Ontario Laboratory | Public Health Ontario Laboratory | Vanessa G Allen et al |
| EPI_ISL_671516 | Canada/ON-PHL-20-02251/2020 | 20-Sep | North America/Canada/Ontario/ | Public Health Ontario Laboratory | Public Health Ontario Laboratory | Vanessa G Allen et al |
| EPI_ISL_671517 | Canada/ON-PHL-20-02252/2020 | 20-Sep | North America/Canada/Ontario/ | Public Health Ontario Laboratory | Public Health Ontario Laboratory | Vanessa G Allen et al |
| EPI_ISL_671519 | Canada/ON-PHL-20-02225/2020 | 20-Sep | North America/Canada/Ontario/ | Public Health Ontario Laboratory | Public Health Ontario Laboratory | Vanessa G Allen et al |
| EPI_ISL_671520 | Canada/ON-PHL-20-02208/2020 | 20-Sep | North America/Canada/Ontario/ | Public Health Ontario Laboratory | Public Health Ontario Laboratory | Vanessa G Allen et al |
| EPI_ISL_671521 | Canada/ON-PHL-20-02291/2020 | 20-Sep | North America/Canada/Ontario/ | Public Health Ontario Laboratory | Public Health Ontario Laboratory | Vanessa G Allen et al |
| EPI_ISL_671522 | Canada/ON-PHL-20-02292/2020 | 20-Sep | North America/Canada/Ontario/ | Public Health Ontario Laboratory | Public Health Ontario Laboratory | Vanessa G Allen et al |
| EPI_ISL_671523 | Canada/ON-PHL-20-02295/2020 | 20-Sep | North America/Canada/Ontario/ | Public Health Ontario Laboratory | Public Health Ontario Laboratory | Vanessa G Allen et al |
| EPI_ISL_671524 | Canada/ON-PHL-20-02298/2020 | 20-Sep | North America/Canada/Ontario/ | Public Health Ontario Laboratory | Public Health Ontario Laboratory | Vanessa G Allen et al |
| EPI_ISL_671525 | Canada/ON-PHL-20-02414/2020 | 20-Sep | North America/Canada/Ontario/ | Public Health Ontario Laboratory | Public Health Ontario Laboratory | Vanessa G Allen et al |

| | | | | | | |
|---|---|---|---|---|---|---|
| EPI_ISL_671526 | Canada/ON-PHL-20-02397/2020 | 20-Sep | North America/Canada/Ontario/ | Public Health Ontario Laboratory | Public Health Ontario Laboratory | Vanessa G Allen et al |
| EPI_ISL_671527 | Canada/ON-PHL-20-02398/2020 | 20-Sep | North America/Canada/Ontario/ | Public Health Ontario Laboratory | Public Health Ontario Laboratory | Vanessa G Allen et al |
| EPI_ISL_671528 | Canada/ON-PHL-20-02418/2020 | 20-Sep | North America/Canada/Ontario/ | Public Health Ontario Laboratory | Public Health Ontario Laboratory | Vanessa G Allen et al |
| EPI_ISL_671529 | Canada/ON-PHL-20-02417/2020 | 20-Sep | North America/Canada/Ontario/ | Public Health Ontario Laboratory | Public Health Ontario Laboratory | Vanessa G Allen et al |
| EPI_ISL_671530 | Canada/ON-PHL-20-02381/2020 | 20-Sep | North America/Canada/Ontario/ | Public Health Ontario Laboratory | Public Health Ontario Laboratory | Vanessa G Allen et al |
| EPI_ISL_671531 | Canada/ON-PHL-20-02419/2020 | 20-Sep | North America/Canada/Ontario/ | Public Health Ontario Laboratory | Public Health Ontario Laboratory | Vanessa G Allen et al |
| EPI_ISL_671532 | Canada/ON-PHL-20-02433/2020 | 20-Sep | North America/Canada/Ontario/ | Public Health Ontario Laboratory | Public Health Ontario Laboratory | Vanessa G Allen et al |
| EPI_ISL_671533 | Canada/ON-PHL-20-02400/2020 | 20-Sep | North America/Canada/Ontario/ | Public Health Ontario Laboratory | Public Health Ontario Laboratory | Vanessa G Allen et al |
| EPI_ISL_671534 | Canada/ON-PHL-20-02432/2020 | 20-Sep | North America/Canada/Ontario/ | Public Health Ontario Laboratory | Public Health Ontario Laboratory | Vanessa G Allen et al |
| EPI_ISL_671535 | Canada/ON-PHL-20-02401/2020 | 20-Sep | North America/Canada/Ontario/ | Public Health Ontario Laboratory | Public Health Ontario Laboratory | Vanessa G Allen et al |
| EPI_ISL_671536 | Canada/ON-PHL-20-02426/2020 | 20-Sep | North America/Canada/Ontario/ | Public Health Ontario Laboratory | Public Health Ontario Laboratory | Vanessa G Allen et al |
| EPI_ISL_671537 | Canada/ON-PHL-20-02394/2020 | 20-Sep | North America/Canada/Ontario/ | Public Health Ontario Laboratory | Public Health Ontario Laboratory | Vanessa G Allen et al |
| EPI_ISL_671538 | Canada/ON-PHL-20-02410/2020 | 20-Sep | North America/Canada/Ontario/ | Public Health Ontario Laboratory | Public Health Ontario Laboratory | Vanessa G Allen et al |

| | | | | | | |
|---|---|---|---|---|---|---|
| EPI_ISL_671539 | Canada/ON-PHL-20-02409/2020 | 20-Sep | North America/Canada/Ontario/ | Public Health Ontario Laboratory | Public Health Ontario Laboratory | Vanessa G Allen et al |
| EPI_ISL_671540 | Canada/ON-PHL-20-02412/2020 | 20-Sep | North America/Canada/Ontario/ | Public Health Ontario Laboratory | Public Health Ontario Laboratory | Vanessa G Allen et al |
| EPI_ISL_671541 | Canada/ON-PHL-20-02411/2020 | 20-Sep | North America/Canada/Ontario/ | Public Health Ontario Laboratory | Public Health Ontario Laboratory | Vanessa G Allen et al |
| EPI_ISL_671542 | Canada/ON-PHL-20-02449/2020 | 20-Sep | North America/Canada/Ontario/ | Public Health Ontario Laboratory | Public Health Ontario Laboratory | Vanessa G Allen et al |
| EPI_ISL_671543 | Canada/ON-PHL-20-02446/2020 | 20-Sep | North America/Canada/Ontario/ | Public Health Ontario Laboratory | Public Health Ontario Laboratory | Vanessa G Allen et al |
| EPI_ISL_671544 | Canada/ON-PHL-20-02445/2020 | 20-Sep | North America/Canada/Ontario/ | Public Health Ontario Laboratory | Public Health Ontario Laboratory | Vanessa G Allen et al |
| EPI_ISL_671545 | Canada/ON-PHL-20-02454/2020 | 20-Sep | North America/Canada/Ontario/ | Public Health Ontario Laboratory | Public Health Ontario Laboratory | Vanessa G Allen et al |
| EPI_ISL_671546 | Canada/ON-PHL-20-02299/2020 | 20-Sep | North America/Canada/Ontario/ | Public Health Ontario Laboratory | Public Health Ontario Laboratory | Vanessa G Allen et al |
| EPI_ISL_671547 | Canada/ON-PHL-20-02273/2020 | 20-Sep | North America/Canada/Ontario/ | Public Health Ontario Laboratory | Public Health Ontario Laboratory | Vanessa G Allen et al |
| EPI_ISL_671548 | Canada/ON-PHL-20-02297/2020 | 20-Sep | North America/Canada/Ontario/ | Public Health Ontario Laboratory | Public Health Ontario Laboratory | Vanessa G Allen et al |
| EPI_ISL_671549 | Canada/ON-PHL-20-02304/2020 | 20-Sep | North America/Canada/Ontario/ | Public Health Ontario Laboratory | Public Health Ontario Laboratory | Vanessa G Allen et al |
| EPI_ISL_671550 | Canada/ON-PHL-20-02306/2020 | 20-Sep | North America/Canada/Ontario/ | Public Health Ontario Laboratory | Public Health Ontario Laboratory | Vanessa G Allen et al |
| EPI_ISL_671551 | Canada/ON-PHL-20-02313/2020 | 20-Sep | North America/Canada/Ontario/ | Public Health Ontario Laboratory | Public Health Ontario Laboratory | Vanessa G Allen et al |

| | | | | | | |
|---|---|---|---|---|---|---|
| EPI_ISL_671552 | Canada/ON-PHL-20-02317/2020 | 20-Sep | North America/Canada/Ontario/ | Public Health Ontario Laboratory | Public Health Ontario Laboratory | Vanessa G Allen et al |
| EPI_ISL_671553 | Canada/ON-PHL-20-02318/2020 | 20-Sep | North America/Canada/Ontario/ | Public Health Ontario Laboratory | Public Health Ontario Laboratory | Vanessa G Allen et al |
| EPI_ISL_671554 | Canada/ON-PHL-20-02328/2020 | 20-Sep | North America/Canada/Ontario/ | Public Health Ontario Laboratory | Public Health Ontario Laboratory | Vanessa G Allen et al |
| EPI_ISL_671555 | Canada/ON-PHL-20-02329/2020 | 20-Sep | North America/Canada/Ontario/ | Public Health Ontario Laboratory | Public Health Ontario Laboratory | Vanessa G Allen et al |
| EPI_ISL_671556 | Canada/ON-PHL-20-02332/2020 | 20-Sep | North America/Canada/Ontario/ | Public Health Ontario Laboratory | Public Health Ontario Laboratory | Vanessa G Allen et al |
| EPI_ISL_671557 | Canada/ON-PHL-20-02300/2020 | 20-Sep | North America/Canada/Ontario/ | Public Health Ontario Laboratory | Public Health Ontario Laboratory | Vanessa G Allen et al |
| EPI_ISL_671558 | Canada/ON-PHL-20-02301/2020 | 20-Sep | North America/Canada/Ontario/ | Public Health Ontario Laboratory | Public Health Ontario Laboratory | Vanessa G Allen et al |
| EPI_ISL_671559 | Canada/ON-PHL-20-02271/2020 | 20-Sep | North America/Canada/Ontario/ | Public Health Ontario Laboratory | Public Health Ontario Laboratory | Vanessa G Allen et al |
| EPI_ISL_671560 | Canada/ON-PHL-20-02272/2020 | 20-Sep | North America/Canada/Ontario/ | Public Health Ontario Laboratory | Public Health Ontario Laboratory | Vanessa G Allen et al |
| EPI_ISL_671561 | Canada/ON-PHL-20-02290/2020 | 20-Sep | North America/Canada/Ontario/ | Public Health Ontario Laboratory | Public Health Ontario Laboratory | Vanessa G Allen et al |
| EPI_ISL_671562 | Canada/ON-PHL-20-02267/2020 | 20-Sep | North America/Canada/Ontario/ | Public Health Ontario Laboratory | Public Health Ontario Laboratory | Vanessa G Allen et al |
| EPI_ISL_671563 | Canada/ON-PHL-20-02268/2020 | 20-Sep | North America/Canada/Ontario/ | Public Health Ontario Laboratory | Public Health Ontario Laboratory | Vanessa G Allen et al |
| EPI_ISL_671564 | Canada/ON-PHL-20-02282/2020 | 20-Sep | North America/Canada/Ontario/ | Public Health Ontario Laboratory | Public Health Ontario Laboratory | Vanessa G Allen et al |

| | | | | | | |
|---|---|---|---|---|---|---|
| EPI_ISL_671565 | Canada/ON-PHL-20-02302/2020 | 20-Sep | North America/Canada/Ontario/ | Public Health Ontario Laboratory | Public Health Ontario Laboratory | Vanessa G Allen et al |
| EPI_ISL_671566 | Canada/ON-PHL-20-02334/2020 | 20-Sep | North America/Canada/Ontario/ | Public Health Ontario Laboratory | Public Health Ontario Laboratory | Vanessa G Allen et al |
| EPI_ISL_671567 | Canada/ON-PHL-20-02335/2020 | 20-Sep | North America/Canada/Ontario/ | Public Health Ontario Laboratory | Public Health Ontario Laboratory | Vanessa G Allen et al |
| EPI_ISL_671568 | Canada/ON-PHL-20-02485/2020 | 20-Sep | North America/Canada/Ontario/ | Public Health Ontario Laboratory | Public Health Ontario Laboratory | Vanessa G Allen et al |
| EPI_ISL_671569 | Canada/ON-PHL-20-02483/2020 | 20-Sep | North America/Canada/Ontario/ | Public Health Ontario Laboratory | Public Health Ontario Laboratory | Vanessa G Allen et al |
| EPI_ISL_671570 | Canada/ON-PHL-20-02455/2020 | 20-Sep | North America/Canada/Ontario/ | Public Health Ontario Laboratory | Public Health Ontario Laboratory | Vanessa G Allen et al |
| EPI_ISL_671571 | Canada/ON-PHL-20-02463/2020 | 20-Sep | North America/Canada/Ontario/ | Public Health Ontario Laboratory | Public Health Ontario Laboratory | Vanessa G Allen et al |
| EPI_ISL_671572 | Canada/ON-PHL-20-02470/2020 | 20-Sep | North America/Canada/Ontario/ | Public Health Ontario Laboratory | Public Health Ontario Laboratory | Vanessa G Allen et al |
| EPI_ISL_671573 | Canada/ON-PHL-20-02469/2020 | 20-Sep | North America/Canada/Ontario/ | Public Health Ontario Laboratory | Public Health Ontario Laboratory | Vanessa G Allen et al |
| EPI_ISL_671574 | Canada/ON-PHL-20-02468/2020 | 20-Sep | North America/Canada/Ontario/ | Public Health Ontario Laboratory | Public Health Ontario Laboratory | Vanessa G Allen et al |
| EPI_ISL_671575 | Canada/ON-PHL-20-02467/2020 | 20-Sep | North America/Canada/Ontario/ | Public Health Ontario Laboratory | Public Health Ontario Laboratory | Vanessa G Allen et al |
| EPI_ISL_671576 | Canada/ON-PHL-20-02466/2020 | 20-Sep | North America/Canada/Ontario/ | Public Health Ontario Laboratory | Public Health Ontario Laboratory | Vanessa G Allen et al |
| EPI_ISL_671577 | Canada/ON-PHL-20-02440/2020 | 20-Sep | North America/Canada/Ontario/ | Public Health Ontario Laboratory | Public Health Ontario Laboratory | Vanessa G Allen et al |

| | | | | | | |
|---|---|---|---|---|---|---|
| EPI_ISL_671578 | Canada/ON-PHL-20-02441/2020 | 20-Sep | North America/Canada/Ontario/ | Public Health Ontario Laboratory | Public Health Ontario Laboratory | Vanessa G Allen et al |
| EPI_ISL_671579 | Canada/ON-PHL-20-02459/2020 | 20-Sep | North America/Canada/Ontario/ | Public Health Ontario Laboratory | Public Health Ontario Laboratory | Vanessa G Allen et al |
| EPI_ISL_671580 | Canada/ON-PHL-20-02436/2020 | 20-Sep | North America/Canada/Ontario/ | Public Health Ontario Laboratory | Public Health Ontario Laboratory | Vanessa G Allen et al |
| EPI_ISL_671581 | Canada/ON-PHL-20-02437/2020 | 20-Sep | North America/Canada/Ontario/ | Public Health Ontario Laboratory | Public Health Ontario Laboratory | Vanessa G Allen et al |
| EPI_ISL_671582 | Canada/ON-PHL-20-02438/2020 | 20-Sep | North America/Canada/Ontario/ | Public Health Ontario Laboratory | Public Health Ontario Laboratory | Vanessa G Allen et al |
| EPI_ISL_671583 | Canada/ON-PHL-20-02480/2020 | 20-Sep | North America/Canada/Ontario/ | Public Health Ontario Laboratory | Public Health Ontario Laboratory | Vanessa G Allen et al |
| EPI_ISL_671584 | Canada/ON-PHL-20-02481/2020 | 20-Sep | North America/Canada/Ontario/ | Public Health Ontario Laboratory | Public Health Ontario Laboratory | Vanessa G Allen et al |
| EPI_ISL_671585 | Canada/ON-PHL-20-02479/2020 | 20-Sep | North America/Canada/Ontario/ | Public Health Ontario Laboratory | Public Health Ontario Laboratory | Vanessa G Allen et al |
| EPI_ISL_671586 | Canada/ON-PHL-20-02478/2020 | 20-Sep | North America/Canada/Ontario/ | Public Health Ontario Laboratory | Public Health Ontario Laboratory | Vanessa G Allen et al |
| EPI_ISL_671587 | Canada/ON-PHL-20-02475/2020 | 20-Sep | North America/Canada/Ontario/ | Public Health Ontario Laboratory | Public Health Ontario Laboratory | Vanessa G Allen et al |
| EPI_ISL_671588 | Canada/ON-PHL-20-02474/2020 | 20-Sep | North America/Canada/Ontario/ | Public Health Ontario Laboratory | Public Health Ontario Laboratory | Vanessa G Allen et al |
| EPI_ISL_671589 | Canada/ON-PHL-20-02493/2020 | 20-Sep | North America/Canada/Ontario/ | Public Health Ontario Laboratory | Public Health Ontario Laboratory | Vanessa G Allen et al |
| EPI_ISL_671590 | Canada/ON-PHL-20-02408/2020 | 20-Sep | North America/Canada/Ontario/ | Public Health Ontario Laboratory | Public Health Ontario Laboratory | Vanessa G Allen et al |

| | | | | | | |
|---|---|---|---|---|---|---|
| EPI_ISL_671591 | Canada/ON-PHL-20-02413/2020 | 20-Sep | North America/Canada/Ontario/ | Public Health Ontario Laboratory | Public Health Ontario Laboratory | Vanessa G Allen et al |
| EPI_ISL_671592 | Canada/ON-PHL-20-02422/2020 | 20-Sep | North America/Canada/Ontario/ | Public Health Ontario Laboratory | Public Health Ontario Laboratory | Vanessa G Allen et al |
| EPI_ISL_671593 | Canada/ON-PHL-20-02430/2020 | 20-Sep | North America/Canada/Ontario/ | Public Health Ontario Laboratory | Public Health Ontario Laboratory | Vanessa G Allen et al |
| EPI_ISL_671594 | Canada/ON-PHL-20-02424/2020 | 20-Sep | North America/Canada/Ontario/ | Public Health Ontario Laboratory | Public Health Ontario Laboratory | Vanessa G Allen et al |
| EPI_ISL_671595 | Canada/ON-PHL-20-02226/2020 | 20-Sep | North America/Canada/Ontario/ | Public Health Ontario Laboratory | Public Health Ontario Laboratory | Vanessa G Allen et al |
| EPI_ISL_671596 | Canada/ON-PHL-20-02406/2020 | 20-Sep | North America/Canada/Ontario/ | Public Health Ontario Laboratory | Public Health Ontario Laboratory | Vanessa G Allen et al |
| EPI_ISL_671597 | Canada/ON-PHL-20-02402/2020 | 20-Sep | North America/Canada/Ontario/ | Public Health Ontario Laboratory | Public Health Ontario Laboratory | Vanessa G Allen et al |
| EPI_ISL_671598 | Canada/ON-PHL-20-02457/2020 | 20-Sep | North America/Canada/Ontario/ | Public Health Ontario Laboratory | Public Health Ontario Laboratory | Vanessa G Allen et al |
| EPI_ISL_671599 | Canada/ON-PHL-20-02456/2020 | 20-Sep | North America/Canada/Ontario/ | Public Health Ontario Laboratory | Public Health Ontario Laboratory | Vanessa G Allen et al |
| EPI_ISL_671600 | Canada/ON-PHL-20-02344/2020 | 20-Sep | North America/Canada/Ontario/ | Public Health Ontario Laboratory | Public Health Ontario Laboratory | Vanessa G Allen et al |
| EPI_ISL_671601 | Canada/ON-PHL-20-02333/2020 | 20-Sep | North America/Canada/Ontario/ | Public Health Ontario Laboratory | Public Health Ontario Laboratory | Vanessa G Allen et al |
| EPI_ISL_671602 | Canada/ON-PHL-20-02452/2020 | 20-Sep | North America/Canada/Ontario/ | Public Health Ontario Laboratory | Public Health Ontario Laboratory | Vanessa G Allen et al |
| EPI_ISL_671603 | Canada/ON-PHL-20-02482/2020 | 20-Sep | North America/Canada/Ontario/ | Public Health Ontario Laboratory | Public Health Ontario Laboratory | Vanessa G Allen et al |

| | | | | | | |
|---|---|---|---|---|---|---|
| EPI_ISL_671604 | Canada/ON-PHL-20-02521/2020 | 20-Sep | North America/Canada/Ontario/ | Public Health Ontario Laboratory | Public Health Ontario Laboratory | Vanessa G Allen et al |
| EPI_ISL_671605 | Canada/ON-PHL-20-02074/2020 | 20-Sep | North America/Canada/Ontario/ | Public Health Ontario Laboratory | Public Health Ontario Laboratory | Vanessa G Allen et al |
| EPI_ISL_671606 | Canada/ON-PHL-20-02415/2020 | 20-Sep | North America/Canada/Ontario/ | Public Health Ontario Laboratory | Public Health Ontario Laboratory | Vanessa G Allen et al |
| EPI_ISL_671638 | USA/TX-DSHS-1412/2020 | 2020-06-09 | North America/USA/Texas/El Paso Coun | Texas Department of State Health Services | Texas Department of State Health Services | Rashmi Tuladhar et al |
| EPI_ISL_671639 | USA/TX-DSHS-1427/2020 | 2020-06-23 | North America/USA/Texas/Harris Count | Texas Department of State Health Services | Texas Department of State Health Services | Rashmi Tuladhar et al |
| EPI_ISL_671640 | USA/TX-DSHS-1410/2020 | 2020-06-09 | North America/USA/Texas/Dallas Count | Texas Department of State Health Services | Texas Department of State Health Services | Rashmi Tuladhar et al |
| EPI_ISL_671652 | Canada/ON-UHTC-0362/2020 | 2020-10-01 | North America/Canada/Ontario/ | Unity Health Toronto | Ontario Institute for Cancer Research | Ramzi Fattouh et al |
| EPI_ISL_671653 | Canada/ON-UHTC-0316/2020 | 2020-09-20 | North America/Canada/Ontario/ | Unity Health Toronto | Ontario Institute for Cancer Research | Ramzi Fattouh et al |
| EPI_ISL_671654 | Canada/ON-UHTC-0375/2020 | 2020-10-04 | North America/Canada/Ontario/ | Unity Health Toronto | Ontario Institute for Cancer Research | Ramzi Fattouh et al |
| EPI_ISL_671655 | Canada/ON-UHTC-0363/2020 | 2020-10-01 | North America/Canada/Ontario/ | Unity Health Toronto | Ontario Institute for Cancer Research | Ramzi Fattouh et al |
| EPI_ISL_671656 | Canada/ON-UHTC-0366/2020 | 2020-10-02 | North America/Canada/Ontario/ | Unity Health Toronto | Ontario Institute for Cancer Research | Ramzi Fattouh et al |
| EPI_ISL_671658 | Canada/ON-UHTC-0374/2020 | 2020-10-04 | North America/Canada/Ontario/ | Unity Health Toronto | Ontario Institute for Cancer Research | Ramzi Fattouh et al |
| EPI_ISL_671659 | Canada/ON-UHTC-0376/2020 | 2020-10-04 | North America/Canada/Ontario/ | Unity Health Toronto | Ontario Institute for Cancer Research | Ramzi Fattouh et al |

| | | | | | | |
|---|---|---|---|---|---|---|
| EPI_ISL_671660 | Canada/ON-UHTC-0369/2020 | 2020-10-01 | North America/Canada/Ontario/ | Unity Health Toronto | Ontario Institute for Cancer Research | Ramzi Fattouh et al |
| EPI_ISL_671661 | Canada/ON-UHTC-0359/2020 | 2020-10-01 | North America/Canada/Ontario/ | Unity Health Toronto | Ontario Institute for Cancer Research | Ramzi Fattouh et al |
| EPI_ISL_671662 | Canada/ON-UHTC-0370/2020 | 2020-10-01 | North America/Canada/Ontario/ | Unity Health Toronto | Ontario Institute for Cancer Research | Ramzi Fattouh et al |
| EPI_ISL_671663 | Canada/ON-UHTC-0360/2020 | 2020-09-30 | North America/Canada/Ontario/ | Unity Health Toronto | Ontario Institute for Cancer Research | Ramzi Fattouh et al |
| EPI_ISL_671664 | Canada/ON-UHTC-0371/2020 | 2020-10-01 | North America/Canada/Ontario/ | Unity Health Toronto | Ontario Institute for Cancer Research | Ramzi Fattouh et al |
| EPI_ISL_671665 | Canada/ON-UHTC-0373/2020 | 2020-10-03 | North America/Canada/Ontario/ | Unity Health Toronto | Ontario Institute for Cancer Research | Ramzi Fattouh et al |
| EPI_ISL_671666 | Canada/ON-UHTC-0365/2020 | 2020-10-02 | North America/Canada/Ontario/ | Unity Health Toronto | Ontario Institute for Cancer Research | Ramzi Fattouh et al |
| EPI_ISL_671667 | Canada/ON-UHTC-0377/2020 | 2020-10-04 | North America/Canada/Ontario/ | Unity Health Toronto | Ontario Institute for Cancer Research | Ramzi Fattouh et al |
| EPI_ISL_671668 | Canada/ON-UHTC-0364/2020 | 2020-10-01 | North America/Canada/Ontario/ | Unity Health Toronto | Ontario Institute for Cancer Research | Ramzi Fattouh et al |
| EPI_ISL_671669 | Canada/ON-UHTC-0367/2020 | 2020-10-02 | North America/Canada/Ontario/ | Unity Health Toronto | Ontario Institute for Cancer Research | Ramzi Fattouh et al |
| EPI_ISL_671670 | Canada/ON-UHTC-0361/2020 | 2020-09-30 | North America/Canada/Ontario/ | Unity Health Toronto | Ontario Institute for Cancer Research | Ramzi Fattouh et al |
| EPI_ISL_671671 | Canada/ON-UHTC-0368/2020 | 2020-10-01 | North America/Canada/Ontario/ | Unity Health Toronto | Ontario Institute for Cancer Research | Ramzi Fattouh et al |
| EPI_ISL_671672 | Canada/ON-UHTC-0315/2020 | 2020-09-20 | North America/Canada/Ontario/ | Unity Health Toronto | Ontario Institute for Cancer Research | Ramzi Fattouh et al |

| | | | | | | |
|---|---|---|---|---|---|---|
| EPI_ISL_671675 | USA/NY-NYCPHL-000972/2020 | 2020-09-16 | North America/USA/New York/New Yor | DOHMH Jamaica | New York City Public Health Laboratory | Jade Wang et al |
| EPI_ISL_671686 | USA/NY-NYCPHL-001103/2020 | 2020-10-18 | North America/USA/New York/New Yor | DOHMH Morrisania | New York City Public Health Laboratory | Jade Wang et al |
| EPI_ISL_671688 | USA/NY-NYCPHL-001105/2020 | 2020-10-18 | North America/USA/New York/New Yor | DOHMH Morrisania | New York City Public Health Laboratory | Jade Wang et al |
| EPI_ISL_671698 | USA/NY-NYCPHL-001115/2020 | 2020-10-20 | North America/USA/New York/New Yor | DOHMH Jamaica | New York City Public Health Laboratory | Jade Wang et al |
| EPI_ISL_671704 | USA/NY-NYCPHL-001121/2020 | 2020-10-22 | North America/USA/New York/New Yor | DOHMH Central Harlem | New York City Public Health Laboratory | Jade Wang et al |
| EPI_ISL_671705 | USA/NY-NYCPHL-001122/2020 | 2020-10-22 | North America/USA/New York/New Yor | DOHMH Corona | New York City Public Health Laboratory | Jade Wang et al |
| EPI_ISL_671714 | USA/NY-NYCPHL-001131/2020 | 2020-10-26 | North America/USA/New York/New Yor | DOHMH Jamaica | New York City Public Health Laboratory | Jade Wang et al |
| EPI_ISL_671717 | USA/NY-NYCPHL-001134/2020 | 2020-10-26 | North America/USA/New York/New Yor | DOHMH Corona | New York City Public Health Laboratory | Jade Wang et al |
| EPI_ISL_671723 | USA/NY-NYCPHL-001140/2020 | 2020-10-23 | North America/USA/New York/New Yor | DOHMH Jamaica | New York City Public Health Laboratory | Jade Wang et al |
| EPI_ISL_671732 | USA/NY-NYCPHL-001150/2020 | 2020-10-26 | North America/USA/New York/New Yor | DOHMH Corona | New York City Public Health Laboratory | Jade Wang et al |
| EPI_ISL_671739 | USA/NY-NYCPHL-001159/2020 | 2020-10-28 | North America/USA/New York/New Yor | DOHMH Jamaica | New York City Public Health Laboratory | Jade Wang et al |
| EPI_ISL_671747 | USA/NY-NYCPHL-001167/2020 | 2020-10-29 | North America/USA/New York/New Yor | DOHMH Corona | New York City Public Health Laboratory | Jade Wang et al |
| EPI_ISL_671748 | USA/NY-NYCPHL-001168/2020 | 2020-10-29 | North America/USA/New York/New Yor | DOHMH Riverside | New York City Public Health Laboratory | Jade Wang et al |

| | | | | | | |
|---|---|---|---|---|---|---|
| EPI_ISL_671751 | USA/NY-NYCPHL-001171/2020 | 2020-10-29 | North America/USA/New York/New Yor | DOHMH Morrisania | New York City Public Health Laboratory | Jade Wang et al |
| EPI_ISL_671761 | USA/NY-NYCPHL-001182/2020 | 2020-10-30 | North America/USA/New York/New Yor | DOHMH Corona | New York City Public Health Laboratory | Jade Wang et al |
| EPI_ISL_671803 | Spain/AR-IBV-97009524/2020 | 2020-10-05 | Europe/Spain/Aragon/Zaragoza | Hospital Cl√≠nico Universitario Lozano Blesa de Zaragoza (Espa√±a) | SeqCOVID-SPAIN consortium/IBV(CSIC) | Rafael Benito et al |
| EPI_ISL_671804 | Spain/AR-IBV-97009527/2020 | 2020-11-14 | Europe/Spain/Aragon/Zaragoza | Hospital Cl√≠nico Universitario Lozano Blesa de Zaragoza (Espa√±a) | SeqCOVID-SPAIN consortium/IBV(CSIC) | Rafael Benito et al |
| EPI_ISL_671806 | Spain/CT-IBV-97009125/2020 | 2020-08-23 | Europe/Spain/Catalunya/Barcelona | Hospital de la Santa Creu i Sant Pau. Servicio de Microbiolog√≠a | SeqCOVID-SPAIN consortium/IBV(CSIC) | Ferran Navarro et al |
| EPI_ISL_671815 | Spain/CT-IBV-97009141/2020 | 2020-09-01 | Europe/Spain/Catalunya/Barcelona | Hospital de la Santa Creu i Sant Pau. Servicio de Microbiolog√≠a | SeqCOVID-SPAIN consortium/IBV(CSIC) | Ferran Navarro et al |
| EPI_ISL_671822 | Spain/CT-IBV-97009156/2020 | 2020-09-07 | Europe/Spain/Catalunya/Barcelona | Hospital de la Santa Creu i Sant Pau. Servicio de Microbiolog√≠a | SeqCOVID-SPAIN consortium/IBV(CSIC) | Ferran Navarro et al |
| EPI_ISL_671830 | Spain/CT-IBV-97009179/2020 | 2020-09-15 | Europe/Spain/Catalunya/Barcelona | Hospital de la Santa Creu i Sant Pau. Servicio de Microbiolog√≠a | SeqCOVID-SPAIN consortium/IBV(CSIC) | Ferran Navarro et al |
| EPI_ISL_671835 | Spain/CT-IBV-97009529/2020 | 2020-10-27 | Europe/Spain/Catalunya/Badalona | Servicio de Microbiolog√≠a, Laboratori Cl√≠nic Metropolitana Nord. Hospital Universitari Germans Trias i Pujol. Institut d‚ÄôInvestigaci√≥ en Ci√®ncies de la Salut Germans Trias i Pujol (IGTP) | SeqCOVID-SPAIN consortium/IBV(CSIC) | Elisa Martr√≥ et al |
| EPI_ISL_671837 | Spain/CT-IBV-97009533/2020 | 2020-10-28 | Europe/Spain/Catalunya/Badalona | Servicio de Microbiolog√≠a, Laboratori Cl√≠nic Metropolitana Nord. Hospital Universitari Germans Trias i Pujol. Institut d‚ÄôInvestigaci√≥ en Ci√®ncies de la Salut Germans Trias i Pujol (IGTP) | SeqCOVID-SPAIN consortium/IBV(CSIC) | Elisa Martr√≥ et al |
| EPI_ISL_671838 | Spain/CT-IBV-97009534/2020 | 2020-10-28 | Europe/Spain/Catalunya/Sant Adria del | Servicio de Microbiolog√≠a, Laboratori Cl√≠nic Metropolitana Nord. Hospital Universitari Germans Trias i Pujol. Institut d‚ÄôInvestigaci√≥ en Ci√®ncies de la Salut Germans Trias i Pujol (IGTP) | SeqCOVID-SPAIN consortium/IBV(CSIC) | Elisa Martr√≥ et al |

| | | | | | | |
|---|---|---|---|---|---|---|
| EPI_ISL_671842 | Spain/CT-IBV-97009544/2020 | 2020-10-28 | Europe/Spain/Catalunya/Badalona | Servicio de Microbiolog√≠a, Laboratori Cl√≠nic Metropolitana Nord. Hospital Universitari Germans Trias i Pujol. Institut d‚ÄôInvestigaci√≥ en Ci√®ncies de la Salut Germans Trias i Pujol (IGTP) | SeqCOVID-SPAIN consortium/IBV(CSIC) | Elisa Mart√≠ et al |
| EPI_ISL_671843 | Spain/CT-IBV-97009546/2020 | 2020-10-28 | Europe/Spain/Catalunya/Santa Coloma | Servicio de Microbiolog√≠a, Laboratori Cl√≠nic Metropolitana Nord. Hospital Universitari Germans Trias i Pujol. Institut d‚ÄôInvestigaci√≥ en Ci√®ncies de la Salut Germans Trias i Pujol (IGTP) | SeqCOVID-SPAIN consortium/IBV(CSIC) | Elisa Mart√≠ et al |
| EPI_ISL_671854 | Spain/CT-IBV-97009557/2020 | 2020-07-18 | Europe/Spain/Catalunya/Mataro | Servicio de Microbiolog√≠a, Laboratori Cl√≠nic Metropolitana Nord. Hospital Universitari Germans Trias i Pujol. Institut d‚ÄôInvestigaci√≥ en Ci√®ncies de la Salut Germans Trias i Pujol (IGTP) | SeqCOVID-SPAIN consortium/IBV(CSIC) | Elisa Mart√≠ et al |
| EPI_ISL_671859 | Spain/CT-IBV-97009562/2020 | 2020-07-29 | Europe/Spain/Catalunya/Santa Coloma | Servicio de Microbiolog√≠a, Laboratori Cl√≠nic Metropolitana Nord. Hospital Universitari Germans Trias i Pujol. Institut d‚ÄôInvestigaci√≥ en Ci√®ncies de la Salut Germans Trias i Pujol (IGTP) | SeqCOVID-SPAIN consortium/IBV(CSIC) | Elisa Mart√≠ et al |
| EPI_ISL_671862 | Spain/CT-IBV-97009565/2020 | 2020-07-28 | Europe/Spain/Catalunya/Barcelona | Servicio de Microbiolog√≠a, Laboratori Cl√≠nic Metropolitana Nord. Hospital Universitari Germans Trias i Pujol. Institut d‚ÄôInvestigaci√≥ en Ci√®ncies de la Salut Germans Trias i Pujol (IGTP) | SeqCOVID-SPAIN consortium/IBV(CSIC) | Elisa Mart√≠ et al |
| EPI_ISL_671869 | Ireland/CO-NVRL-75IRL23254/2020 | 2020-10-06 | Europe/Ireland/Cork/ | National Virus Reference Laboratory | National Virus Reference Laboratory | Michael Carr et al |
| EPI_ISL_671901 | Ireland/D-NVRL-75IRL90324/2020 | 2020-10-14 | Europe/Ireland/Dublin/ | National Virus Reference Laboratory | National Virus Reference Laboratory | Michael Carr et al |
| EPI_ISL_671907 | Ireland/G-NVRL-76IRL13959/2020 | 2020-10-16 | Europe/Ireland/Galway/ | National Virus Reference Laboratory | National Virus Reference Laboratory | Michael Carr et al |
| EPI_ISL_671920 | Ireland/LK-NVRL-75IRL32488/2020 | 2020-10-07 | Europe/Ireland/Limerick/ | National Virus Reference Laboratory | National Virus Reference Laboratory | Michael Carr et al |
| EPI_ISL_671929 | Ireland/RN-NVRL-75IRL84062/2020 | 2020-10-13 | Europe/Ireland/Roscommon/ | National Virus Reference Laboratory | National Virus Reference Laboratory | Michael Carr et al |

| | | | | | | |
|---|---|---|---|---|---|---|
| EPI_ISL_671942 | France/OCC-95/2020 | 2020-09-08 | Europe/France/Occitanie/ | CHU Purpan - Laboratoire de Virologie - Institut F√©d√©ratif de Biologie | CHU Purpan - Laboratoire de Virologie - Institut F√©d√©ratif de Biologie | Latour J. et al |
| EPI_ISL_671944 | France/OCC-97/2020 | 2020-10-06 | Europe/France/Occitanie/ | CHU Purpan - Laboratoire de Virologie - Institut F√©d√©ratif de Biologie | CHU Purpan - Laboratoire de Virologie - Institut F√©d√©ratif de Biologie | Latour J. et al |
| EPI_ISL_671945 | France/OCC-98/2020 | 2020-10-05 | Europe/France/Occitanie/ | CHU Purpan - Laboratoire de Virologie - Institut F√©d√©ratif de Biologie | CHU Purpan - Laboratoire de Virologie - Institut F√©d√©ratif de Biologie | Latour J. et al |
| EPI_ISL_671947 | France/OCC-101/2020 | 2020-10-22 | Europe/France/Occitanie/ | CHU Purpan - Laboratoire de Virologie - Institut F√©d√©ratif de Biologie | CHU Purpan - Laboratoire de Virologie - Institut F√©d√©ratif de Biologie | Latour J. et al |
| EPI_ISL_671948 | France/OCC-102/2020 | 2020-10-22 | Europe/France/Occitanie/ | CHU Purpan - Laboratoire de Virologie - Institut F√©d√©ratif de Biologie | CHU Purpan - Laboratoire de Virologie - Institut F√©d√©ratif de Biologie | Latour J. et al |
| EPI_ISL_671949 | France/OCC-103/2020 | 2020-10-22 | Europe/France/Occitanie/ | CHU Purpan - Laboratoire de Virologie - Institut F√©d√©ratif de Biologie | CHU Purpan - Laboratoire de Virologie - Institut F√©d√©ratif de Biologie | Latour J. et al |
| EPI_ISL_671950 | France/OCC-104/2020 | 2020-10-22 | Europe/France/Occitanie/ | CHU Purpan - Laboratoire de Virologie - Institut F√©d√©ratif de Biologie | CHU Purpan - Laboratoire de Virologie - Institut F√©d√©ratif de Biologie | Latour J. et al |
| EPI_ISL_671952 | France/OCC-106/2020 | 2020-10-22 | Europe/France/Occitanie/ | CHU Purpan - Laboratoire de Virologie - Institut F√©d√©ratif de Biologie | CHU Purpan - Laboratoire de Virologie - Institut F√©d√©ratif de Biologie | Latour J. et al |

| | | | | | | |
|---|---|---|---|---|---|---|
| EPI_ISL_671953 | France/OCC-107/2020 | 2020-10-22 | Europe/France/Occitanie/ | CHU Purpan - Laboratoire de Virologie - Institut F√©d√©ratif de Biologie | CHU Purpan - Laboratoire de Virologie - Institut F√©d√©ratif de Biologie | Latour J. et al |
| EPI_ISL_671954 | France/OCC-108/2020 | 2020-10-22 | Europe/France/Occitanie/ | CHU Purpan - Laboratoire de Virologie - Institut F√©d√©ratif de Biologie | CHU Purpan - Laboratoire de Virologie - Institut F√©d√©ratif de Biologie | Latour J. et al |
| EPI_ISL_671956 | France/OCC-110/2020 | 2020-10-23 | Europe/France/Occitanie/ | CHU Purpan - Laboratoire de Virologie - Institut F√©d√©ratif de Biologie | CHU Purpan - Laboratoire de Virologie - Institut F√©d√©ratif de Biologie | Latour J. et al |
| EPI_ISL_671957 | France/OCC-111/2020 | 2020-10-22 | Europe/France/Occitanie/ | CHU Purpan - Laboratoire de Virologie - Institut F√©d√©ratif de Biologie | CHU Purpan - Laboratoire de Virologie - Institut F√©d√©ratif de Biologie | Latour J. et al |
| EPI_ISL_671958 | France/OCC-112/2020 | 2020-10-23 | Europe/France/Occitanie/ | CHU Purpan - Laboratoire de Virologie - Institut F√©d√©ratif de Biologie | CHU Purpan - Laboratoire de Virologie - Institut F√©d√©ratif de Biologie | Latour J. et al |
| EPI_ISL_671959 | France/OCC-113/2020 | 2020-10-22 | Europe/France/Occitanie/ | CHU Purpan - Laboratoire de Virologie - Institut F√©d√©ratif de Biologie | CHU Purpan - Laboratoire de Virologie - Institut F√©d√©ratif de Biologie | Latour J. et al |
| EPI_ISL_671960 | France/OCC-115/2020 | 2020-10-22 | Europe/France/Occitanie/ | CHU Purpan - Laboratoire de Virologie - Institut F√©d√©ratif de Biologie | CHU Purpan - Laboratoire de Virologie - Institut F√©d√©ratif de Biologie | Latour J. et al |
| EPI_ISL_671961 | France/OCC-116/2020 | 2020-10-22 | Europe/France/Occitanie/ | CHU Purpan - Laboratoire de Virologie - Institut F√©d√©ratif de Biologie | CHU Purpan - Laboratoire de Virologie - Institut F√©d√©ratif de Biologie | Latour J. et al |

| | | | | | | |
|---|---|---|---|---|---|---|
| EPI_ISL_671962 | France/OCC-117/2020 | 2020-10-22 | Europe/France/Occitanie/ | CHU Purpan - Laboratoire de Virologie - Institut F√©d√©ratif de Biologie | CHU Purpan - Laboratoire de Virologie - Institut F√©d√©ratif de Biologie | Latour J. et al |
| EPI_ISL_671963 | France/OCC-118/2020 | 2020-10-22 | Europe/France/Occitanie/ | CHU Purpan - Laboratoire de Virologie - Institut F√©d√©ratif de Biologie | CHU Purpan - Laboratoire de Virologie - Institut F√©d√©ratif de Biologie | Latour J. et al |
| EPI_ISL_671964 | France/OCC-119/2020 | 2020-09-12 | Europe/France/Occitanie/ | CHU Purpan - Laboratoire de Virologie - Institut F√©d√©ratif de Biologie | CHU Purpan - Laboratoire de Virologie - Institut F√©d√©ratif de Biologie | Latour J. et al |
| EPI_ISL_671965 | France/OCC-120/2020 | 2020-10-02 | Europe/France/Occitanie/ | CHU Purpan - Laboratoire de Virologie - Institut F√©d√©ratif de Biologie | CHU Purpan - Laboratoire de Virologie - Institut F√©d√©ratif de Biologie | Latour J. et al |
| EPI_ISL_671966 | France/OCC-121/2020 | 2020-10-13 | Europe/France/Occitanie/ | CHU Purpan - Laboratoire de Virologie - Institut F√©d√©ratif de Biologie | CHU Purpan - Laboratoire de Virologie - Institut F√©d√©ratif de Biologie | Latour J. et al |
| EPI_ISL_671967 | France/OCC-122/2020 | 2020-10-14 | Europe/France/Occitanie/ | CHU Purpan - Laboratoire de Virologie - Institut F√©d√©ratif de Biologie | CHU Purpan - Laboratoire de Virologie - Institut F√©d√©ratif de Biologie | Latour J. et al |
| EPI_ISL_671969 | France/OCC-124/2020 | 2020-10-22 | Europe/France/Occitanie/ | CHU Purpan - Laboratoire de Virologie - Institut F√©d√©ratif de Biologie | CHU Purpan - Laboratoire de Virologie - Institut F√©d√©ratif de Biologie | Latour J. et al |
| EPI_ISL_671971 | France/OCC-126/2020 | 2020-10-07 | Europe/France/Occitanie/ | CHU Purpan - Laboratoire de Virologie - Institut F√©d√©ratif de Biologie | CHU Purpan - Laboratoire de Virologie - Institut F√©d√©ratif de Biologie | Latour J. et al |

| | | | | | | |
|---|---|---|---|---|---|---|
| EPI_ISL_671972 | France/OCC-127/2020 | 2020-08-24 | Europe/France/Occitanie/ | CHU Purpan - Laboratoire de Virologie - Institut F√©d√©ratif de Biologie | CHU Purpan - Laboratoire de Virologie - Institut F√©d√©ratif de Biologie | Latour J. et al |
| EPI_ISL_671974 | ElSalvador/INS-01/2020 | 2020-09-16 | North America/El Salvador/San Salvado | Laboratorio de Virolog√≠a y Microbiolog√≠a Molecular, Depto. de Microbiolog√≠a, Facultad de Medicina, Universidad de El Salvador/INS-laboratorio de Ref. Ministerio de Salud | Laboratorio de Virolog√≠a y Microbiolog√≠a Molecular, Depto. de Microbiolog√≠a, Facultad de Medicina, Universidad de El Salvador/INS-laboratorio de Ref. Ministerio de Salud | Rivera NR et al |
| EPI_ISL_671975 | France/OCC-66/2020 | 2020-08-03 | Europe/France/Occitanie/ | CHU Purpan - Laboratoire de Virologie - Institut F√©d√©ratif de Biologie | CHU Purpan - Laboratoire de Virologie - Institut F√©d√©ratif de Biologie | Latour J. et al |
| EPI_ISL_672022 | USA/CA-CZB-13123/2020 | 2020-10-26 | North America/USA/California/Madera | Madera County Department of Public Health | Chan-Zuckerberg Biohub | CZB Cliahub Consortium et al |
| EPI_ISL_672026 | USA/CA-CZB-13481/2020 | 2020-10-23 | North America/USA/California/Madera | Madera County Department of Public Health | Chan-Zuckerberg Biohub | CZB Cliahub Consortium et al |
| EPI_ISL_672027 | USA/CA-CZB-9603/2020 | 2020-04-15 | North America/USA/California/Santa Cl | The Ashley Laboratory, Stanford University | Chan-Zuckerberg Biohub | CZB Cliahub Consortium et al |
| EPI_ISL_672029 | USA/CA-CZB-13409/2020 | 2020-09-02 | North America/USA/California/Alameda | Alameda County Public Health Lab | Chan-Zuckerberg Biohub | CZB Cliahub Consortium et al |
| EPI_ISL_672031 | USA/CA-CZB-11913/2020 | 2020-10-04 | North America/USA/California/Santa Cl | Santa Clara County Public Health Laboratory | Chan-Zuckerberg Biohub | CZB Cliahub Consortium et al |
| EPI_ISL_672036 | USA/CA-CZB-13460/2020 | 2020-09-09 | North America/USA/California/Alameda | Alameda County Public Health Lab | Chan-Zuckerberg Biohub | CZB Cliahub Consortium et al |
| EPI_ISL_672038 | USA/CA-CZB-13381/2020 | 2020-05-12 | North America/USA/California/Alameda | Alameda County Public Health Lab | Chan-Zuckerberg Biohub | CZB Cliahub Consortium et al |
| EPI_ISL_672053 | USA/CA-CZB-13082/2020 | 2020-05-07 | North America/USA/California/Alameda | Alameda County Public Health Lab | Chan-Zuckerberg Biohub | CZB Cliahub Consortium et al |

| | | | | | | |
|---|---|---|---|---|---|---|
| EPI_ISL_672056 | USA/CA-CZB-10003/2020 | 2020-07-09 | North America/USA/California/Santa Cl | The Ashley Laboratory, Stanford University | Chan-Zuckerberg Biohub | CZB Cliahub Consortium et al |
| EPI_ISL_672063 | USA/CA-CZB-10250/2020 | 2020-05-18 | North America/USA/California/Santa Cl | The Ashley Laboratory, Stanford University | Chan-Zuckerberg Biohub | CZB Cliahub Consortium et al |
| EPI_ISL_672065 | USA/CA-CZB-13012/2020 | 2020-10-26 | North America/USA/California/Santa Cl | Santa Clara County Public Health Laboratory | Chan-Zuckerberg Biohub | CZB Cliahub Consortium et al |
| EPI_ISL_672069 | USA/CA-CZB-12991/2020 | 2020-10-21 | North America/USA/California/Santa Cl | Santa Clara County Public Health Laboratory | Chan-Zuckerberg Biohub | CZB Cliahub Consortium et al |
| EPI_ISL_672072 | USA/CA-CZB-13300/2020 | 2020-10-31 | North America/USA/California/Santa Cl | Santa Clara County Public Health Laboratory | Chan-Zuckerberg Biohub | CZB Cliahub Consortium et al |
| EPI_ISL_672073 | USA/CA-CZB-13265/2020 | 2020-10-28 | North America/USA/California/Santa Cl | Santa Clara County Public Health Laboratory | Chan-Zuckerberg Biohub | CZB Cliahub Consortium et al |
| EPI_ISL_672075 | USA/CA-CZB-12941/2020 | 2020-10-18 | North America/USA/California/Orange C | Orange County Public Health Lab | Chan-Zuckerberg Biohub | CZB Cliahub Consortium et al |
| EPI_ISL_672077 | USA/CA-CZB-12959/2020 | 2020-07-21 | North America/USA/California/San Fran | San Francisco Public Health Laboratory | Chan-Zuckerberg Biohub | CZB Cliahub Consortium et al |
| EPI_ISL_672078 | USA/CA-CZB-13376/2020 | 2020-05-13 | North America/USA/California/Alameda | Alameda County Public Health Lab | Chan-Zuckerberg Biohub | CZB Cliahub Consortium et al |
| EPI_ISL_672086 | USA/CA-CZB-13385/2020 | 2020-08-31 | North America/USA/California/Alameda | Alameda County Public Health Lab | Chan-Zuckerberg Biohub | CZB Cliahub Consortium et al |
| EPI_ISL_672087 | USA/CA-CZB-5745/2020 | 2020-07-14 | North America/USA/California/San Luis | County of San Luis Obispo Public Health Laboratory | Chan-Zuckerberg Biohub | CZB Cliahub Consortium et al |
| EPI_ISL_672109 | USA/CA-CZB-12953/2020 | 2020-09-29 | North America/USA/California/Orange C | Orange County Public Health Lab | Chan-Zuckerberg Biohub | CZB Cliahub Consortium et al |
| EPI_ISL_672118 | USA/CA-CZB-9523/2020 | 2020-04-07 | North America/USA/California/Santa Cl | The Ashley Laboratory, Stanford University | Chan-Zuckerberg Biohub | CZB Cliahub Consortium et al |
| EPI_ISL_672120 | USA/CA-CZB-9527/2020 | 2020-04-07 | North America/USA/California/Santa Cl | The Ashley Laboratory, Stanford University | Chan-Zuckerberg Biohub | CZB Cliahub Consortium et al |
| EPI_ISL_672122 | USA/CA-CZB-9533/2020 | 2020-04-09 | North America/USA/California/Santa Cl | The Ashley Laboratory, Stanford University | Chan-Zuckerberg Biohub | CZB Cliahub Consortium et al |
| EPI_ISL_672133 | USA/CA-CZB-9563/2020 | 2020-04-16 | North America/USA/California/Santa Cl | The Ashley Laboratory, Stanford University | Chan-Zuckerberg Biohub | CZB Cliahub Consortium et al |
| EPI_ISL_672138 | USA/CA-CZB-9590/2020 | 2020-05-08 | North America/USA/California/Santa Cl | The Ashley Laboratory, Stanford University | Chan-Zuckerberg Biohub | CZB Cliahub Consortium et al |
| EPI_ISL_672148 | USA/CA-CZB-9648/2020 | 2020-04-27 | North America/USA/California/Santa Cl | The Ashley Laboratory, Stanford University | Chan-Zuckerberg Biohub | CZB Cliahub Consortium et al |
| EPI_ISL_672170 | USA/CA-CZB-9762/2020 | 2020-06-01 | North America/USA/California/Santa Cl | The Ashley Laboratory, Stanford University | Chan-Zuckerberg Biohub | CZB Cliahub Consortium et al |
| EPI_ISL_672171 | USA/CA-CZB-9768/2020 | 2020-06-06 | North America/USA/California/Santa Cl | The Ashley Laboratory, Stanford University | Chan-Zuckerberg Biohub | CZB Cliahub Consortium et al |

| | | | | | | |
|---|---|---|---|---|---|---|
| EPI_ISL_672179 | USA/CA-CZB-9792/2020 | 2020-06-04 | North America/USA/California/Santa Cl | The Ashley Laboratory, Stanford University | Chan-Zuckerberg Biohub | CZB Cliahub Consortium et al |
| EPI_ISL_672191 | USA/CA-CZB-10015/2020 | 2020-07-10 | North America/USA/California/Santa Cl | The Ashley Laboratory, Stanford University | Chan-Zuckerberg Biohub | CZB Cliahub Consortium et al |
| EPI_ISL_672202 | USA/CA-CZB-10065/2020 | 2020-06-20 | North America/USA/California/Santa Cl | The Ashley Laboratory, Stanford University | Chan-Zuckerberg Biohub | CZB Cliahub Consortium et al |
| EPI_ISL_672203 | USA/CA-CZB-10067/2020 | 2020-06-21 | North America/USA/California/Santa Cl | The Ashley Laboratory, Stanford University | Chan-Zuckerberg Biohub | CZB Cliahub Consortium et al |
| EPI_ISL_672210 | USA/CA-CZB-10091/2020 | 2020-06-21 | North America/USA/California/Santa Cl | The Ashley Laboratory, Stanford University | Chan-Zuckerberg Biohub | CZB Cliahub Consortium et al |
| EPI_ISL_672226 | USA/CA-CZB-10134/2020 | 2020-06-10 | North America/USA/California/Santa Cl | The Ashley Laboratory, Stanford University | Chan-Zuckerberg Biohub | CZB Cliahub Consortium et al |
| EPI_ISL_672238 | USA/CA-CZB-10161/2020 | 2020-06-22 | North America/USA/California/Santa Cl | The Ashley Laboratory, Stanford University | Chan-Zuckerberg Biohub | CZB Cliahub Consortium et al |
| EPI_ISL_672244 | USA/CA-CZB-10172/2020 | 2020-05-11 | North America/USA/California/Santa Cl | The Ashley Laboratory, Stanford University | Chan-Zuckerberg Biohub | CZB Cliahub Consortium et al |
| EPI_ISL_672246 | USA/CA-CZB-10178/2020 | 2020-05-13 | North America/USA/California/Santa Cl | The Ashley Laboratory, Stanford University | Chan-Zuckerberg Biohub | CZB Cliahub Consortium et al |
| EPI_ISL_672259 | USA/CA-CZB-10211/2020 | 2020-05-13 | North America/USA/California/Santa Cl | The Ashley Laboratory, Stanford University | Chan-Zuckerberg Biohub | CZB Cliahub Consortium et al |
| EPI_ISL_672262 | USA/CA-CZB-10220/2020 | 2020-05-11 | North America/USA/California/Santa Cl | The Ashley Laboratory, Stanford University | Chan-Zuckerberg Biohub | CZB Cliahub Consortium et al |
| EPI_ISL_672265 | USA/CA-CZB-10227/2020 | 2020-05-16 | North America/USA/California/Santa Cl | The Ashley Laboratory, Stanford University | Chan-Zuckerberg Biohub | CZB Cliahub Consortium et al |
| EPI_ISL_672269 | USA/CA-CZB-10236/2020 | 2020-05-17 | North America/USA/California/Santa Cl | The Ashley Laboratory, Stanford University | Chan-Zuckerberg Biohub | CZB Cliahub Consortium et al |
| EPI_ISL_672280 | USA/CA-CZB-11886/2020 | 2020-09-30 | North America/USA/California/Santa Cl | Santa Clara County Public Health Laboratory | Chan-Zuckerberg Biohub | CZB Cliahub Consortium et al |
| EPI_ISL_672286 | USA/CA-CZB-11901/2020 | 2020-10-03 | North America/USA/California/Santa Cl | Santa Clara County Public Health Laboratory | Chan-Zuckerberg Biohub | CZB Cliahub Consortium et al |
| EPI_ISL_672291 | USA/CA-CZB-11919/2020 | 2020-10-05 | North America/USA/California/Santa Cl | Santa Clara County Public Health Laboratory | Chan-Zuckerberg Biohub | CZB Cliahub Consortium et al |
| EPI_ISL_672297 | USA/CA-CZB-13000/2020 | 2020-10-21 | North America/USA/California/Santa Cl | Santa Clara County Public Health Laboratory | Chan-Zuckerberg Biohub | CZB Cliahub Consortium et al |
| EPI_ISL_672308 | USA/CA-CZB-13025/2020 | 2020-05-11 | North America/USA/California/Alameda | Alameda County Public Health Lab | Chan-Zuckerberg Biohub | CZB Cliahub Consortium et al |
| EPI_ISL_672312 | USA/CA-CZB-13029/2020 | 2020-05-11 | North America/USA/California/Alameda | Alameda County Public Health Lab | Chan-Zuckerberg Biohub | CZB Cliahub Consortium et al |
| EPI_ISL_672314 | USA/CA-CZB-13034/2020 | 2020-05-14 | North America/USA/California/Alameda | Alameda County Public Health Lab | Chan-Zuckerberg Biohub | CZB Cliahub Consortium et al |

| | | | | | | |
|---|---|---|---|---|---|---|
| EPI_ISL_672322 | USA/CA-CZB-13045/2020 | 2020-05-15 | North America/USA/California/Alameda | Alameda County Public Health Lab | Chan-Zuckerberg Biohub | CZB Cliahub Consortium et al |
| EPI_ISL_672324 | USA/CA-CZB-13047/2020 | 2020-05-15 | North America/USA/California/Alameda | Alameda County Public Health Lab | Chan-Zuckerberg Biohub | CZB Cliahub Consortium et al |
| EPI_ISL_672329 | USA/CA-CZB-13059/2020 | 2020-05-16 | North America/USA/California/Alameda | Alameda County Public Health Lab | Chan-Zuckerberg Biohub | CZB Cliahub Consortium et al |
| EPI_ISL_672335 | USA/CA-CZB-13066/2020 | 2020-05-16 | North America/USA/California/Alameda | Alameda County Public Health Lab | Chan-Zuckerberg Biohub | CZB Cliahub Consortium et al |
| EPI_ISL_672339 | USA/CA-CZB-13071/2020 | 2020-05-03 | North America/USA/California/Alameda | Alameda County Public Health Lab | Chan-Zuckerberg Biohub | CZB Cliahub Consortium et al |
| EPI_ISL_672348 | USA/CA-CZB-13094/2020 | 2020-05-09 | North America/USA/California/Alameda | Alameda County Public Health Lab | Chan-Zuckerberg Biohub | CZB Cliahub Consortium et al |
| EPI_ISL_672351 | USA/CA-CZB-13098/2020 | 2020-05-12 | North America/USA/California/Alameda | Alameda County Public Health Lab | Chan-Zuckerberg Biohub | CZB Cliahub Consortium et al |
| EPI_ISL_672357 | USA/CA-CZB-5762/2020 | 2020-07-12 | North America/USA/California/San Luis | County of San Luis Obispo Public Health Laboratory | Chan-Zuckerberg Biohub | CZB Cliahub Consortium et al |
| EPI_ISL_672358 | USA/CA-CZB-5757/2020 | 2020-07-17 | North America/USA/California/San Luis | County of San Luis Obispo Public Health Laboratory | Chan-Zuckerberg Biohub | CZB Cliahub Consortium et al |
| EPI_ISL_672366 | USA/CA-CZB-12944/2020 | 2020-10-20 | North America/USA/California/Orange C | Orange County Public Health Lab | Chan-Zuckerberg Biohub | CZB Cliahub Consortium et al |
| EPI_ISL_672367 | USA/CA-CZB-12945/2020 | 2020-10-19 | North America/USA/California/Orange C | Orange County Public Health Lab | Chan-Zuckerberg Biohub | CZB Cliahub Consortium et al |
| EPI_ISL_672370 | USA/CA-CZB-12948/2020 | 2020-10-20 | North America/USA/California/Orange C | Orange County Public Health Lab | Chan-Zuckerberg Biohub | CZB Cliahub Consortium et al |
| EPI_ISL_672371 | USA/CA-CZB-12950/2020 | 2020-10-22 | North America/USA/California/Orange C | Orange County Public Health Lab | Chan-Zuckerberg Biohub | CZB Cliahub Consortium et al |
| EPI_ISL_672375 | USA/CA-CZB-12957/2020 | 2020-07-21 | North America/USA/California/San Fran | San Francisco Public Health Laboratory | Chan-Zuckerberg Biohub | CZB Cliahub Consortium et al |
| EPI_ISL_672377 | USA/CA-CZB-12960/2020 | 2020-07-21 | North America/USA/California/San Fran | San Francisco Public Health Laboratory | Chan-Zuckerberg Biohub | CZB Cliahub Consortium et al |
| EPI_ISL_672380 | USA/CA-CZB-12965/2020 | 2020-07-27 | North America/USA/California/San Fran | San Francisco Public Health Laboratory | Chan-Zuckerberg Biohub | CZB Cliahub Consortium et al |
| EPI_ISL_672384 | USA/CA-CZB-12970/2020 | 2020-07-19 | North America/USA/California/San Fran | San Francisco Public Health Laboratory | Chan-Zuckerberg Biohub | CZB Cliahub Consortium et al |
| EPI_ISL_672386 | USA/CA-CZB-5845/2020 | 2020-07-22 | North America/USA/California/Marin Co | UCSF Clinical Microbiology Laboratory | Chan-Zuckerberg Biohub | CZB Cliahub Consortium et al |
| EPI_ISL_672404 | USA/CA-CZB-13132/2020 | 2020-10-26 | North America/USA/California/Madera | Madera County Department of Public Health | Chan-Zuckerberg Biohub | CZB Cliahub Consortium et al |
| EPI_ISL_672409 | USA/CA-CZB-13126/2020 | 2020-10-26 | North America/USA/California/Madera | Madera County Department of Public Health | Chan-Zuckerberg Biohub | CZB Cliahub Consortium et al |

| | | | | | | |
|---|---|---|---|---|---|---|
| EPI_ISL_672412 | USA/CA-CZB-13122/2020 | 2020-10-26 | North America/USA/California/Madera | Madera County Department of Public Health | Chan-Zuckerberg Biohub | CZB Cliahub Consortium et al |
| EPI_ISL_672419 | USA/CA-CZB-13115/2020 | 2020-10-27 | North America/USA/California/Madera | Madera County Department of Public Health | Chan-Zuckerberg Biohub | CZB Cliahub Consortium et al |
| EPI_ISL_672422 | USA/CA-CZB-13110/2020 | 2020-10-27 | North America/USA/California/Madera | Madera County Department of Public Health | Chan-Zuckerberg Biohub | CZB Cliahub Consortium et al |
| EPI_ISL_672436 | USA/CA-CZB-13312/2020 | 2020-11-02 | North America/USA/California/Santa Cl | Santa Clara County Public Health Laboratory | Chan-Zuckerberg Biohub | CZB Cliahub Consortium et al |
| EPI_ISL_672439 | USA/CA-CZB-13341/2020 | 2020-05-02 | North America/USA/California/Alameda | Alameda County Public Health Lab | Chan-Zuckerberg Biohub | CZB Cliahub Consortium et al |
| EPI_ISL_672454 | USA/CA-CZB-13384/2020 | 2020-08-31 | North America/USA/California/Alameda | Alameda County Public Health Lab | Chan-Zuckerberg Biohub | CZB Cliahub Consortium et al |
| EPI_ISL_672458 | USA/CA-CZB-13397/2020 | 2020-09-01 | North America/USA/California/Alameda | Alameda County Public Health Lab | Chan-Zuckerberg Biohub | CZB Cliahub Consortium et al |
| EPI_ISL_672465 | USA/CA-CZB-13418/2020 | 2020-09-03 | North America/USA/California/Alameda | Alameda County Public Health Lab | Chan-Zuckerberg Biohub | CZB Cliahub Consortium et al |
| EPI_ISL_672467 | USA/CA-CZB-13421/2020 | 2020-09-03 | North America/USA/California/Alameda | Alameda County Public Health Lab | Chan-Zuckerberg Biohub | CZB Cliahub Consortium et al |
| EPI_ISL_672470 | USA/CA-CZB-13444/2020 | 2020-09-04 | North America/USA/California/Alameda | Alameda County Public Health Lab | Chan-Zuckerberg Biohub | CZB Cliahub Consortium et al |
| EPI_ISL_672471 | USA/CA-CZB-13448/2020 | 2020-09-08 | North America/USA/California/Alameda | Alameda County Public Health Lab | Chan-Zuckerberg Biohub | CZB Cliahub Consortium et al |
| EPI_ISL_672474 | USA/CA-CZB-13454/2020 | 2020-09-09 | North America/USA/California/Alameda | Alameda County Public Health Lab | Chan-Zuckerberg Biohub | CZB Cliahub Consortium et al |
| EPI_ISL_672484 | USA/CA-CZB-13245/2020 | 2020-11-02 | North America/USA/California/Orange ( | Orange County Public Health Lab | Chan-Zuckerberg Biohub | CZB Cliahub Consortium et al |
| EPI_ISL_672489 | USA/CA-CZB-13461/2020 | 2020-09-10 | North America/USA/California/Contra C | Contra Costa County Public Health Lab | Chan-Zuckerberg Biohub | CZB Cliahub Consortium et al |
| EPI_ISL_672511 | USA/CA-CZB-13514/2020 | 2020-10-28 | North America/USA/California/Madera | Madera County Department of Public Health | Chan-Zuckerberg Biohub | CZB Cliahub Consortium et al |
| EPI_ISL_672523 | USA/CA-CZB-13487/2020 | 2020-10-24 | North America/USA/California/Madera | Madera County Department of Public Health | Chan-Zuckerberg Biohub | CZB Cliahub Consortium et al |
| EPI_ISL_672526 | USA/CA-CZB-13477/2020 | 2020-10-22 | North America/USA/California/Madera | Madera County Department of Public Health | Chan-Zuckerberg Biohub | CZB Cliahub Consortium et al |
| EPI_ISL_672528 | USA/CA-CZB-13475/2020 | 2020-10-22 | North America/USA/California/Madera | Madera County Department of Public Health | Chan-Zuckerberg Biohub | CZB Cliahub Consortium et al |
| EPI_ISL_672532 | USA/UT-UPHL-201204351/2020 | 2020-03-30 | North America/USA/Utah/ | Utah Public Health Laboratory | Utah Public Health Laboratory | Erin Young et al |
| EPI_ISL_672535 | USA/UT-UPHL-201214564/2020 | 2020-04-03 | North America/USA/Utah/ | Utah Public Health Laboratory | Utah Public Health Laboratory | Erin Young et al |

| | | | | | | |
|---|---|---|---|---|---|---|
| EPI_ISL_672540 | USA/UT-UPHL-201206488/2020 | 2020-04-01 | North America/USA/Utah/ | Utah Public Health Laboratory | Utah Public Health Laboratory | Erin Young et al |
| EPI_ISL_672559 | USA/UT-UPHL-201212641/2020 | 2020-04-01 | North America/USA/Utah/ | Utah Public Health Laboratory | Utah Public Health Laboratory | Erin Young et al |
| EPI_ISL_672562 | USA/UT-UPHL-201227098/2020 | 2020-04-06 | North America/USA/Utah/ | Utah Public Health Laboratory | Utah Public Health Laboratory | Erin Young et al |
| EPI_ISL_672672 | Brazil/MG-L2-MG07/2020 | 2020-04-01 | South America/Brazil/Minas Gerais/ | DB Diagnosticos do Brasil | Laborat√≥rio de Parasitologia M√©dica - Instituto de Medicina Tropical - Universidade de S√£o Paulo | Brazil-UK Centre for Arbovirus Discovery Diagnosis Genomics et al |
| EPI_ISL_672675 | Brazil/MG-L30-CD757/2020 | 2020-04-14 | South America/Brazil/Minas Gerais/ | DB Diagnosticos do Brasil | Laborat√≥rio de Parasitologia M√©dica - Instituto de Medicina Tropical - Universidade de S√£o Paulo | Brazil-UK Centre for Arbovirus Discovery Diagnosis Genomics et al |
| EPI_ISL_672677 | Brazil/MG-L30-CD785/2020 | 2020-04-17 | South America/Brazil/Minas Gerais/ | DB Diagnosticos do Brasil | Laborat√≥rio de Parasitologia M√©dica - Instituto de Medicina Tropical - Universidade de S√£o Paulo | Brazil-UK Centre for Arbovirus Discovery Diagnosis Genomics et al |
| EPI_ISL_672679 | Brazil/MG-L45-CD1172/2020 | 2020-05-18 | South America/Brazil/Minas Gerais/ | DB Diagnosticos do Brasil | Laborat√≥rio de Parasitologia M√©dica - Instituto de Medicina Tropical - Universidade de S√£o Paulo | Brazil-UK Centre for Arbovirus Discovery Diagnosis Genomics et al |
| EPI_ISL_672682 | Brazil/PE-L27-CD604/2020 | 2020-04-08 | South America/Brazil/Pernambuco/ | DB Diagnosticos do Brasil | Laborat√≥rio de Parasitologia M√©dica - Instituto de Medicina Tropical - Universidade de S√£o Paulo | Brazil-UK Centre for Arbovirus Discovery Diagnosis Genomics et al |

| | | | | | | |
|---|---|---|---|---|---|---|
| EPI_ISL_672702 | Brazil/SP-L25-CD627/2020 | 2020-04-07 | South America/Brazil/S√£o Paulo/ | Institute of Tropical Medicine at the University of S√£o Paulo (IMT-USP) | Laborat√≥rio de Parasitologia M√©dica - Instituto de Medicina Tropical - Universidade de S√£o Paulo | Brazil-UK Centre for Arbovirus Discovery Diagnosis Genomics et al |
| EPI_ISL_672706 | Brazil/SP-L25-CD638/2020 | 2020-04-08 | South America/Brazil/S√£o Paulo/ | Institute of Tropical Medicine at the University of S√£o Paulo (IMT-USP) | Laborat√≥rio de Parasitologia M√©dica - Instituto de Medicina Tropical - Universidade de S√£o Paulo | Brazil-UK Centre for Arbovirus Discovery Diagnosis Genomics et al |
| EPI_ISL_672709 | Brazil/SP-L25-CD643/2020 | 2020-04-09 | South America/Brazil/S√£o Paulo/ | Institute of Tropical Medicine at the University of S√£o Paulo (IMT-USP) | Laborat√≥rio de Parasitologia M√©dica - Instituto de Medicina Tropical - Universidade de S√£o Paulo | Brazil-UK Centre for Arbovirus Discovery Diagnosis Genomics et al |
| EPI_ISL_672715 | Brazil/SP-L25-CD661/2020 | 2020-04-14 | South America/Brazil/S√£o Paulo/ | Institute of Tropical Medicine at the University of S√£o Paulo (IMT-USP) | Laborat√≥rio de Parasitologia M√©dica - Instituto de Medicina Tropical - Universidade de S√£o Paulo | Brazil-UK Centre for Arbovirus Discovery Diagnosis Genomics et al |
| EPI_ISL_672718 | Brazil/SP-L25-CD666/2020 | 2020-04-14 | South America/Brazil/S√£o Paulo/ | Institute of Tropical Medicine at the University of S√£o Paulo (IMT-USP) | Laborat√≥rio de Parasitologia M√©dica - Instituto de Medicina Tropical - Universidade de S√£o Paulo | Brazil-UK Centre for Arbovirus Discovery Diagnosis Genomics et al |

| | | | | | | |
|---|---|---|---|---|---|---|
| EPI_ISL_672721 | Brazil/SP-L25-CD672/2020 | 2020-04-15 | South America/Brazil/S√£o Paulo/ | Institute of Tropical Medicine at the University of S√£o Paulo (IMT-USP) | Laborat√≥rio de Parasitologia M√©dica - Instituto de Medicina Tropical - Universidade de S√£o Paulo | Brazil-UK Centre for Arbovirus Discovery Diagnosis Genomics et al |
| EPI_ISL_672724 | Brazil/SP-L26-CD545/2020 | 2020-04-17 | South America/Brazil/S√£o Paulo/ | Hospital das Cl√≠nicas da Faculdade de Medicina da Universidade de S√£o Paulo (HC-FMUSP) | Laborat√≥rio de Parasitologia M√©dica - Instituto de Medicina Tropical - Universidade de S√£o Paulo | Brazil-UK Centre for Arbovirus Discovery Diagnosis Genomics et al |
| EPI_ISL_672725 | Brazil/SP-L26-CD547/2020 | 2020-04-17 | South America/Brazil/S√£o Paulo/ | Hospital das Cl√≠nicas da Faculdade de Medicina da Universidade de S√£o Paulo (HC-FMUSP) | Laborat√≥rio de Parasitologia M√©dica - Instituto de Medicina Tropical - Universidade de S√£o Paulo | Brazil-UK Centre for Arbovirus Discovery Diagnosis Genomics et al |
| EPI_ISL_672731 | Brazil/SP-L26-CD554/2020 | 2020-04-19 | South America/Brazil/S√£o Paulo/ | Hospital das Cl√≠nicas da Faculdade de Medicina da Universidade de S√£o Paulo (HC-FMUSP) | Laborat√≥rio de Parasitologia M√©dica - Instituto de Medicina Tropical - Universidade de S√£o Paulo | Brazil-UK Centre for Arbovirus Discovery Diagnosis Genomics et al |
| EPI_ISL_672733 | Brazil/SP-L26-CD556/2020 | 2020-04-20 | South America/Brazil/S√£o Paulo/ | Hospital das Cl√≠nicas da Faculdade de Medicina da Universidade de S√£o Paulo (HC-FMUSP) | Laborat√≥rio de Parasitologia M√©dica - Instituto de Medicina Tropical - Universidade de S√£o Paulo | Brazil-UK Centre for Arbovirus Discovery Diagnosis Genomics et al |

| | | | | | | |
|---|---|---|---|---|---|---|
| EPI_ISL_672743 | Brazil/SP-L26-CD572/2020 | 2020-04-23 | South America/Brazil/S√£o Paulo/ | Hospital das Cl√≠nicas da Faculdade de Medicina da Universidade de S√£o Paulo (HC-FMUSP) | Laborat√≥rio de Parasitologia M√©dica - Instituto de Medicina Tropical - Universidade de S√£o Paulo | Brazil-UK Centre for Arbovirus Discovery Diagnosis Genomics et al |
| EPI_ISL_672748 | Brazil/SP-L28-CD686/2020 | 2020-04-15 | South America/Brazil/S√£o Paulo/ | Institute of Tropical Medicine at the University of S√£o Paulo (IMT-USP) | Laborat√≥rio de Parasitologia M√©dica - Instituto de Medicina Tropical - Universidade de S√£o Paulo | Brazil-UK Centre for Arbovirus Discovery Diagnosis Genomics et al |
| EPI_ISL_672917 | England/CAMC-B09619/2020 | 2020-10-28 | Europe/United Kingdom/England/ | Lighthouse Lab in Cambridge | Wellcome Sanger Institute for the COVID-19 Genomics UK (COG-UK) Consortium | Rob Howes et al |
| EPI_ISL_673090 | England/CAMC-B09075/2020 | 2020-10-29 | Europe/United Kingdom/England/ | Lighthouse Lab in Cambridge | Wellcome Sanger Institute for the COVID-19 Genomics UK (COG-UK) Consortium | Rob Howes et al |
| EPI_ISL_673489 | England/CAMC-B99669/2020 | 2020-11-15 | Europe/United Kingdom/England/ | Lighthouse Lab in Cambridge | Wellcome Sanger Institute for the COVID-19 Genomics UK (COG-UK) Consortium | Rob Howes et al |
| EPI_ISL_673701 | England/CAMC-B98372/2020 | 2020-11-15 | Europe/United Kingdom/England/ | Lighthouse Lab in Cambridge | Wellcome Sanger Institute for the COVID-19 Genomics UK (COG-UK) Consortium | Rob Howes et al |
| EPI_ISL_673783 | England/CAMC-B98A9B/2020 | 2020-11-15 | Europe/United Kingdom/England/ | Lighthouse Lab in Cambridge | Wellcome Sanger Institute for the COVID-19 Genomics UK (COG-UK) Consortium | Rob Howes et al |

| | | | | | | |
|---|---|---|---|---|---|---|
| EPI_ISL_673944 | England/MILK-B8640B/2020 | 2020-11-13 | Europe/United Kingdom/England/ | Lighthouse Lab in Milton Keynes | Wellcome Sanger Institute for the COVID-19 Genomics UK (COG-UK) Consortium | The Lighthouse Lab in Milton Keynes et al |
| EPI_ISL_674162 | England/ALDP-B84A74/2020 | 2020-11-15 | Europe/United Kingdom/England/ | Lighthouse Lab in Alderley Park | Wellcome Sanger Institute for the COVID-19 Genomics UK (COG-UK) Consortium | Jacquelyn Wynn et al |
| EPI_ISL_674183 | England/MILK-B8C0D4/2020 | 2020-11-13 | Europe/United Kingdom/England/ | Lighthouse Lab in Milton Keynes | Wellcome Sanger Institute for the COVID-19 Genomics UK (COG-UK) Consortium | The Lighthouse Lab in Milton Keynes et al |
| EPI_ISL_674296 | England/ALDP-B83F67/2020 | 2020-11-14 | Europe/United Kingdom/England/ | Lighthouse Lab in Alderley Park | Wellcome Sanger Institute for the COVID-19 Genomics UK (COG-UK) Consortium | Jacquelyn Wynn et al |
| EPI_ISL_674338 | England/MILK-B8C168/2020 | 2020-11-13 | Europe/United Kingdom/England/ | Lighthouse Lab in Milton Keynes | Wellcome Sanger Institute for the COVID-19 Genomics UK (COG-UK) Consortium | The Lighthouse Lab in Milton Keynes et al |
| EPI_ISL_674394 | England/ALDP-B84CAB/2020 | 2020-11-15 | Europe/United Kingdom/England/ | Lighthouse Lab in Alderley Park | Wellcome Sanger Institute for the COVID-19 Genomics UK (COG-UK) Consortium | Jacquelyn Wynn et al |
| EPI_ISL_675098 | England/MILK-B9757D/2020 | 2020-11-13 | Europe/United Kingdom/England/ | Lighthouse Lab in Milton Keynes | Wellcome Sanger Institute for the COVID-19 Genomics UK (COG-UK) Consortium | The Lighthouse Lab in Milton Keynes et al |
| EPI_ISL_675641 | England/ALDP-B83019/2020 | 2020-11-14 | Europe/United Kingdom/England/ | Lighthouse Lab in Alderley Park | Wellcome Sanger Institute for the COVID-19 Genomics UK (COG-UK) Consortium | Jacquelyn Wynn et al |

| | | | | | | |
|---|---|---|---|---|---|---|
| EPI_ISL_675931 | England/MILK-B8C8AF/2020 | 2020-11-12 | Europe/United Kingdom/England/ | Lighthouse Lab in Milton Keynes | Wellcome Sanger Institute for the COVID-19 Genomics UK (COG-UK) Consortium | The Lighthouse Lab in Milton Keynes et al |
| EPI_ISL_676149 | England/MILK-B94DC3/2020 | 2020-11-14 | Europe/United Kingdom/England/ | Lighthouse Lab in Milton Keynes | Wellcome Sanger Institute for the COVID-19 Genomics UK (COG-UK) Consortium | The Lighthouse Lab in Milton Keynes et al |
| EPI_ISL_676489 | Sweden/20-09861/2020 | 2020-11-04 | Europe/Sweden/Skane/ | Klinisk mikrobiologi | The Public Health Agency of Sweden | Department of Microbiology et al |
| EPI_ISL_676490 | Sweden/20-09862/2020 | 2020-11-04 | Europe/Sweden/Skane/ | Klinisk mikrobiologi | The Public Health Agency of Sweden | Department of Microbiology et al |
| EPI_ISL_676492 | Sweden/20-52780/2020 | 2020-08-24 | Europe/Sweden/Vastmanland/ | Klinisk Mikrobiologi | The Public Health Agency of Sweden | Department of Microbiology et al |
| EPI_ISL_676501 | Sweden/20-53400/2020 | 2020-11-04 | Europe/Sweden/Vastra Gotaland/ | Klinisk Mikrobiologi | The Public Health Agency of Sweden | Department of Microbiology et al |
| EPI_ISL_676504 | Sweden/20-53525/2020 | 2020-11-22 | Europe/Sweden/Orebro/ | Klinisk mikrobiologi | The Public Health Agency of Sweden | Department of Microbiology et al |
| EPI_ISL_676505 | Sweden/20-53532/2020 | 2020-11-10 | Europe/Sweden/Jonkoping/ | Klinisk mikrobiologi | The Public Health Agency of Sweden | Department of Microbiology et al |
| EPI_ISL_676513 | Sweden/20-22131/2020 | 2020-08-25 | Europe/Sweden/Orebro/ | Orebro klinisk mikrobiologi | The Public Health Agency of Sweden | Department of Microbiology et al |
| EPI_ISL_676527 | Sweden/20-21508/2020 | 2020-07-10 | Europe/Sweden/Uppsala/ | Uppsala klinisk mikrobiologi | The Public Health Agency of Sweden | Department of Microbiology et al |
| EPI_ISL_676529 | Sweden/20-21511/2020 | 2020-07-13 | Europe/Sweden/Uppsala/ | Uppsala klinisk mikrobiologi | The Public Health Agency of Sweden | Department of Microbiology et al |
| EPI_ISL_676534 | Sweden/20-53067/2020 | 2020-10-02 | Europe/Sweden/Kalmar/ | Kalmar klinisk mikrobiologi | The Public Health Agency of Sweden | Department of Microbiology et al |
| EPI_ISL_676536 | Sweden/20-53325/2020 | 2020-10-26 | Europe/Sweden/Gavleborgs Lan/ | Laboratoriemedicin, Klinisk mikrobiologi | The Public Health Agency of Sweden | Department of Microbiology et al |
| EPI_ISL_676540 | Sweden/20-53382/2020 | 2020-11-03 | Europe/Sweden/Varmland/ | Laboratoriemedicin, Klinisk mikrobiologi | The Public Health Agency of Sweden | Department of Microbiology et al |
| EPI_ISL_676558 | France/ARA-26054/2020 | 2020-04-16 | Europe/France/Auvergne-Rhv¥ne-Alpes/ | CNR Virus des Infections Respiratoires - France SUD | CNR Virus des Infections Respiratoires - France SUD | Antonin Bal et al |

| | | | | | | |
|---|---|---|---|---|---|---|
| EPI_ISL_676568 | France/ARA-27161/2020 | 2020-04-18 | Europe/France/Auvergne-Rhv¥ne-Alpes/ | CNR Virus des Infections Respiratoires - France SUD | CNR Virus des Infections Respiratoires - France SUD | Antonin Bal et al |
| EPI_ISL_676572 | France/ARA-27314/2020 | 2020-04-19 | Europe/France/Auvergne-Rhv¥ne-Alpes/ | CNR Virus des Infections Respiratoires - France SUD | CNR Virus des Infections Respiratoires - France SUD | Antonin Bal et al |
| EPI_ISL_676573 | France/ARA-27377/2020 | 2020-04-18 | Europe/France/Auvergne-Rhv¥ne-Alpes/ | CNR Virus des Infections Respiratoires - France SUD | CNR Virus des Infections Respiratoires - France SUD | Antonin Bal et al |
| EPI_ISL_676575 | Serbia/SremskaMitrovica-01182120 | 2020-07-11 | Europe/Serbia/Srem District/Sremska M | Scientific Veterinary Institute Novi Sad | "Veterinary Specialized Institute ""Kraljevo"", Serbia" | Vidanovic et al |
| EPI_ISL_676576 | Serbia/Pancevo-007910104/2020 | 2020-03-31 | Europe/Serbia/South Banat District/Pan | Scientific Veterinary Institute Novi Sad | "Veterinary Specialized Institute ""Kraljevo"", Serbia" | Vidanovic et al |
| EPI_ISL_676579 | Serbia/SremskaMitrovica-01050090 | 2020-07-08 | Europe/Serbia/Srem District/Sremska M | Scientific Veterinary Institute Novi Sad | "Veterinary Specialized Institute ""Kraljevo"", Serbia" | Vidanovic et al |
| EPI_ISL_676581 | Serbia/SremskaMitrovica-02262090 | 2020-04-08 | Europe/Serbia/Srem District/Sremska M | Scientific Veterinary Institute Novi Sad | "Veterinary Specialized Institute ""Kraljevo"", Serbia" | Vidanovic et al |
| EPI_ISL_676583 | Serbia/NoviSad-212182506/2020 | 2020-06-24 | Europe/Serbia/South Baƒçka District/No | Scientific Veterinary Institute Novi Sad | "Veterinary Specialized Institute ""Kraljevo"", Serbia" | Vidanovic et al |

| | | | | | | |
|---|---|---|---|---|---|---|
| EPI_ISL_676585 | Serbia/Subotica-101360605/2020 | 2020-05-05 | Europe/Serbia/North Baſçka District/Sul | Scientific Veterinary Institute Novi Sad | "Veterinary Specialized Institute ""Kraljevo"", Serbia" | Vidanovic et al |
| EPI_ISL_676590 | Serbia/NoviSad-158472905/2020 | 2020-05-28 | Europe/Serbia/South Baſçka District/No | Scientific Veterinary Institute Novi Sad | "Veterinary Specialized Institute ""Kraljevo"", Serbia" | Vidanovic et al |
| EPI_ISL_676592 | Serbia/NoviSad-196221606/2020 | 2020-06-15 | Europe/Serbia/South Baſçka District/No | Scientific Veterinary Institute Novi Sad | "Veterinary Specialized Institute ""Kraljevo"", Serbia" | Vidanovic et al |
| EPI_ISL_676595 | Serbia/Vrsac-040471604/2020 | 2020-04-15 | Europe/Serbia/South Banat District/Vrs | Scientific Veterinary Institute Novi Sad | "Veterinary Specialized Institute ""Kraljevo"", Serbia" | Vidanovic et al |
| EPI_ISL_676598 | Serbia/Titel-059642304/2020 | 2020-04-22 | Europe/Serbia/South Baſçka District/Tit | Scientific Veterinary Institute Novi Sad | "Veterinary Specialized Institute ""Kraljevo"", Serbia" | Vidanovic et al |
| EPI_ISL_676599 | Serbia/Subotica-062172304/2020 | 2020-04-22 | Europe/Serbia/North Baſçka District/Sul | Scientific Veterinary Institute Novi Sad | "Veterinary Specialized Institute ""Kraljevo"", Serbia" | Vidanovic et al |
| EPI_ISL_676609 | USA/TX-DSHS-1470/2020 | 2020-06-22 | North America/USA/Texas/Hidalgo Cour | Texas Department of State Health Services | Texas Department of State Health Services | Rashmi Tuladhar et al |
| EPI_ISL_676613 | USA/TX-DSHS-1240/2020 | 2020-05-05 | North America/USA/Texas/Smith County | Texas Department of State Health Services | Texas Department of State Health Services | Rashmi Tuladhar et al |
| EPI_ISL_676614 | USA/TX-DSHS-1478/2020 | 2020-06-22 | North America/USA/Texas/Hidalgo Cour | Texas Department of State Health Services | Texas Department of State Health Services | Rashmi Tuladhar et al |

| | | | | | | |
|---|---|---|---|---|---|---|
| EPI_ISL_676617 | USA/TX-DSHS-1455/2020 | 2020-06-22 | North America/USA/Texas/Hidalgo Cour | Texas Department of State Health Services | Texas Department of State Health Services | Rashmi Tuladhar et al |
| EPI_ISL_676623 | USA/TX-DSHS-1464/2020 | 2020-06-22 | North America/USA/Texas/Hidalgo Cour | Texas Department of State Health Services | Texas Department of State Health Services | Rashmi Tuladhar et al |
| EPI_ISL_676625 | USA/TX-DSHS-1242/2020 | 2020-08-27 | North America/USA/Texas/Titus County | Texas Department of State Health Services | Texas Department of State Health Services | Rashmi Tuladhar et al |
| EPI_ISL_676635 | USA/TX-DSHS-1476/2020 | 2020-06-22 | North America/USA/Texas/Hidalgo Cour | Texas Department of State Health Services | Texas Department of State Health Services | Rashmi Tuladhar et al |
| EPI_ISL_676653 | USA/NY-Wadsworth-194562-01/2020 | 2020-08-07 | North America/USA/New York/Erie Cou | Wadsworth Center, New York State Department.of Health | Wadsworth Center, New York State Department.of Health | Kirsten St. George et al |
| EPI_ISL_676654 | USA/NY-Wadsworth-99944-01/2020 | 2020-05-14 | North America/USA/New York/Erie Cou | Wadsworth Center, New York State Department.of Health | Wadsworth Center, New York State Department.of Health | Kirsten St. George et al |
| EPI_ISL_676658 | USA/NY-Wadsworth-46277-01/2020 | 2020-04-13 | North America/USA/New York/Tioga Co | Wadsworth Center, New York State Department.of Health | Wadsworth Center, New York State Department.of Health | Kirsten St. George et al |
| EPI_ISL_676659 | USA/NY-Wadsworth-192881-01/2020 | 2020-07-02 | North America/USA/New York/Oneida C | Masonic Medical Research Institute | Wadsworth Center, New York State Department.of Health | Nathan Tucker et al |
| EPI_ISL_676663 | USA/NY-Wadsworth-192903-01/2020 | 2020-07-16 | North America/USA/New York/Oneida C | Masonic Medical Research Institute | Wadsworth Center, New York State Department.of Health | Nathan Tucker et al |

| | | | | | | |
|---|---|---|---|---|---|---|
| EPI_ISL_676673 | USA/NY-Wadsworth-226488-01/2020 | 2020-07-28 | North America/USA/New York/Oneida ( | Masonic Medical Research Institute | Wadsworth Center, New York State Department.of Health | Nathan Tucker et al |
| EPI_ISL_676676 | USA/NY-Wadsworth-176521-01/2020 | 2020-07-17 | North America/USA/New York/Jefferson | Wadsworth Center, New York State Department.of Health | Wadsworth Center, New York State Department.of Health | Kirsten St. George et al |
| EPI_ISL_676677 | USA/NY-Wadsworth-97859-01/2020 | 2020-05-13 | North America/USA/New York/Monroe | Wadsworth Center, New York State Department.of Health | Wadsworth Center, New York State Department.of Health | Kirsten St. George et al |
| EPI_ISL_676678 | USA/NY-Wadsworth-74733-01/2020 | 2020-05-01 | North America/USA/New York/Clinton ( | Wadsworth Center, New York State Department.of Health | Wadsworth Center, New York State Department.of Health | Kirsten St. George et al |
| EPI_ISL_676679 | USA/NY-Wadsworth-125725-01/2020 | 2020-05-30 | North America/USA/New York/Sullivan | Wadsworth Center, New York State Department.of Health | Wadsworth Center, New York State Department.of Health | Kirsten St. George et al |
| EPI_ISL_676683 | USA/NY-Wadsworth-84608-01/2020 | 2020-05-07 | North America/USA/New York/Monroe | Wadsworth Center, New York State Department.of Health | Wadsworth Center, New York State Department.of Health | Kirsten St. George et al |
| EPI_ISL_676684 | USA/NY-Wadsworth-57117-01/2020 | 2020-04-20 | North America/USA/New York/Westche | Wadsworth Center, New York State Department.of Health | Wadsworth Center, New York State Department.of Health | Kirsten St. George et al |
| EPI_ISL_676686 | USA/NY-Wadsworth-46244-01/2020 | 2020-04-11 | North America/USA/New York/Westche | Wadsworth Center, New York State Department.of Health | Wadsworth Center, New York State Department.of Health | Kirsten St. George et al |

| | | | | | | |
|---|---|---|---|---|---|---|
| EPI_ISL_676692 | USA/NY-Wadsworth-103460-01/2020 | 2020-05-15 | North America/USA/New York/Fulton C | Wadsworth Center, New York State Department.of Health | Wadsworth Center, New York State Department.of Health | Kirsten St. George et al |
| EPI_ISL_676701 | USA/NY-Wadsworth-142311-01/2020 | 2020-06-16 | North America/USA/New York/Onondag | Wadsworth Center, New York State Department.of Health | Wadsworth Center, New York State Department.of Health | Kirsten St. George et al |
| EPI_ISL_676704 | USA/NY-Wadsworth-19202-01/2020 | 2020-03-17 | North America/USA/New York/Montgo | Wadsworth Center, New York State Department.of Health | Wadsworth Center, New York State Department.of Health | Kirsten St. George et al |
| EPI_ISL_676712 | USA/NY-Wadsworth-103609-01/2020 | 2020-05-15 | North America/USA/New York/Columbi | Wadsworth Center, New York State Department.of Health | Wadsworth Center, New York State Department.of Health | Kirsten St. George et al |
| EPI_ISL_676713 | USA/NY-Wadsworth-192897-01/2020 | 2020-07-08 | North America/USA/New York/Oneida C | Masonic Medical Research Institute | Wadsworth Center, New York State Department.of Health | Nathan Tucker et al |
| EPI_ISL_676714 | USA/NY-Wadsworth-105920-01/2020 | 2020-05-17 | North America/USA/New York/Montgo | Wadsworth Center, New York State Department.of Health | Wadsworth Center, New York State Department.of Health | Kirsten St. George et al |
| EPI_ISL_676729 | USA/NY-Wadsworth-11205-01/2020 | 2020-03-06 | North America/USA/New York/Saratog | Wadsworth Center, New York State Department.of Health | Wadsworth Center, New York State Department.of Health | Kirsten St. George et al |
| EPI_ISL_676794 | USA/NY-Wadsworth-34564-01/2020 | 2020-03-31 | North America/USA/New York/Essex Co | Wadsworth Center, New York State Department.of Health | Wadsworth Center, New York State Department.of Health | Kirsten St. George et al |

| | | | | | | |
|---|---|---|---|---|---|---|
| EPI_ISL_676806 | USA/NY-Wadsworth-38889-01/2020 | 2020-04-05 | North America/USA/New York/Warren | Wadsworth Center, New York State Department.of Health | Wadsworth Center, New York State Department.of Health | Kirsten St. George et al |
| EPI_ISL_676824 | USA/NY-Wadsworth-41425-01/2020 | 2020-04-08 | North America/USA/New York/Westche | Wadsworth Center, New York State Department.of Health | Wadsworth Center, New York State Department.of Health | Kirsten St. George et al |
| EPI_ISL_676826 | USA/NY-Wadsworth-41429-01/2020 | 2020-04-08 | North America/USA/New York/Westche | Wadsworth Center, New York State Department.of Health | Wadsworth Center, New York State Department.of Health | Kirsten St. George et al |
| EPI_ISL_676827 | USA/NY-Wadsworth-41431-01/2020 | 2020-04-08 | North America/USA/New York/Westche | Wadsworth Center, New York State Department.of Health | Wadsworth Center, New York State Department.of Health | Kirsten St. George et al |
| EPI_ISL_676843 | USA/NY-Wadsworth-46181-01/2020 | 2020-04-13 | North America/USA/New York/Orange | Wadsworth Center, New York State Department.of Health | Wadsworth Center, New York State Department.of Health | Kirsten St. George et al |
| EPI_ISL_676845 | USA/NY-Wadsworth-46275-01/2020 | 2020-04-13 | North America/USA/New York/Tioga Co | Wadsworth Center, New York State Department.of Health | Wadsworth Center, New York State Department.of Health | Kirsten St. George et al |
| EPI_ISL_676852 | USA/NY-Wadsworth-46957-01/2020 | 2020-04-14 | North America/USA/New York/Westche | Wadsworth Center, New York State Department.of Health | Wadsworth Center, New York State Department.of Health | Kirsten St. George et al |
| EPI_ISL_676862 | USA/NY-Wadsworth-50111-01/2020 | 2020-04-16 | North America/USA/New York/Tioga Co | Wadsworth Center, New York State Department.of Health | Wadsworth Center, New York State Department.of Health | Kirsten St. George et al |

| | | | | | | |
|---|---|---|---|---|---|---|
| EPI_ISL_676873 | USA/NY-Wadsworth-65664-01/2020 | 2020-04-23 | North America/USA/New York/Fulton C | Wadsworth Center, New York State Department.of Health | Wadsworth Center, New York State Department.of Health | Kirsten St. George et al |
| EPI_ISL_676876 | USA/NY-Wadsworth-65679-01/2020 | 2020-04-23 | North America/USA/New York/Fulton C | Wadsworth Center, New York State Department.of Health | Wadsworth Center, New York State Department.of Health | Kirsten St. George et al |
| EPI_ISL_676889 | USA/NY-Wadsworth-72883-01/2020 | 2020-04-29 | North America/USA/New York/Columbia | Wadsworth Center, New York State Department.of Health | Wadsworth Center, New York State Department.of Health | Kirsten St. George et al |
| EPI_ISL_676893 | USA/NY-Wadsworth-75278-01/2020 | 2020-05-05 | North America/USA/New York/Cattarau | Wadsworth Center, New York State Department.of Health | Wadsworth Center, New York State Department.of Health | Kirsten St. George et al |
| EPI_ISL_676897 | USA/NY-Wadsworth-76313-01/2020 | 2020-05-01 | North America/USA/New York/Erie Cou | Wadsworth Center, New York State Department.of Health | Wadsworth Center, New York State Department.of Health | Kirsten St. George et al |
| EPI_ISL_676899 | USA/NY-Wadsworth-76680-01/2020 | 2020-05-02 | North America/USA/New York/Madison | Wadsworth Center, New York State Department.of Health | Wadsworth Center, New York State Department.of Health | Kirsten St. George et al |
| EPI_ISL_676904 | USA/NY-Wadsworth-77837-01/2020 | 2020-05-04 | North America/USA/New York/Oswego | Wadsworth Center, New York State Department.of Health | Wadsworth Center, New York State Department.of Health | Kirsten St. George et al |
| EPI_ISL_676907 | USA/NY-Wadsworth-79524-01/2020 | 2020-05-05 | North America/USA/New York/Putnam | Wadsworth Center, New York State Department.of Health | Wadsworth Center, New York State Department.of Health | Kirsten St. George et al |

| | | | | | | |
|---|---|---|---|---|---|---|
| EPI_ISL_676909 | USA/NY-Wadsworth-79894-01/2020 | 2020-05-05 | North America/USA/New York/Cattarau | Wadsworth Center, New York State Department.of Health | Wadsworth Center, New York State Department.of Health | Kirsten St. George et al |
| EPI_ISL_676910 | USA/NY-Wadsworth-79903-01/2020 | 2020-05-13 | North America/USA/New York/Cattarau | Wadsworth Center, New York State Department.of Health | Wadsworth Center, New York State Department.of Health | Kirsten St. George et al |
| EPI_ISL_676916 | USA/NY-Wadsworth-88095-01/2020 | 2020-05-09 | North America/USA/New York/Niagara | Wadsworth Center, New York State Department.of Health | Wadsworth Center, New York State Department.of Health | Kirsten St. George et al |
| EPI_ISL_676917 | USA/NY-Wadsworth-88316-01/2020 | 2020-05-09 | North America/USA/New York/Niagara | Wadsworth Center, New York State Department.of Health | Wadsworth Center, New York State Department.of Health | Kirsten St. George et al |
| EPI_ISL_676918 | USA/NY-Wadsworth-88325-01/2020 | 2020-05-09 | North America/USA/New York/Niagara | Wadsworth Center, New York State Department.of Health | Wadsworth Center, New York State Department.of Health | Kirsten St. George et al |
| EPI_ISL_676920 | USA/NY-Wadsworth-95277-01/2020 | 2020-05-12 | North America/USA/New York/Orange ( | Wadsworth Center, New York State Department.of Health | Wadsworth Center, New York State Department.of Health | Kirsten St. George et al |
| EPI_ISL_676921 | USA/NY-Wadsworth-95907-01/2020 | 2020-05-12 | North America/USA/New York/Monroe | Wadsworth Center, New York State Department.of Health | Wadsworth Center, New York State Department.of Health | Kirsten St. George et al |
| EPI_ISL_676927 | USA/NY-Wadsworth-97827-01/2020 | 2020-05-13 | North America/USA/New York/Monroe | Wadsworth Center, New York State Department.of Health | Wadsworth Center, New York State Department.of Health | Kirsten St. George et al |

| | | | | | | |
|---|---|---|---|---|---|---|
| EPI_ISL_676932 | USA/NY-Wadsworth-100184-01/2020 | 2020-05-13 | North America/USA/New York/Monroe | Wadsworth Center, New York State Department.of Health | Wadsworth Center, New York State Department.of Health | Kirsten St. George et al |
| EPI_ISL_676935 | USA/NY-Wadsworth-101230-01/2020 | 2020-05-14 | North America/USA/New York/Chenang | Wadsworth Center, New York State Department.of Health | Wadsworth Center, New York State Department.of Health | Kirsten St. George et al |
| EPI_ISL_676939 | USA/NY-Wadsworth-103456-01/2020 | 2020-05-15 | North America/USA/New York/Fulton C | Wadsworth Center, New York State Department.of Health | Wadsworth Center, New York State Department.of Health | Kirsten St. George et al |
| EPI_ISL_676949 | USA/NY-Wadsworth-109966-01/2020 | 2020-05-20 | North America/USA/New York/Broome | Wadsworth Center, New York State Department.of Health | Wadsworth Center, New York State Department.of Health | Kirsten St. George et al |
| EPI_ISL_676950 | USA/NY-Wadsworth-110122-01/2020 | 2020-05-20 | North America/USA/New York/Warren | Wadsworth Center, New York State Department.of Health | Wadsworth Center, New York State Department.of Health | Kirsten St. George et al |
| EPI_ISL_676952 | USA/NY-Wadsworth-114731-01/2020 | 2020-05-22 | North America/USA/New York/Delawar | Wadsworth Center, New York State Department.of Health | Wadsworth Center, New York State Department.of Health | Kirsten St. George et al |
| EPI_ISL_676954 | USA/NY-Wadsworth-116460-01/2020 | 2020-05-24 | North America/USA/New York/Chenang | Wadsworth Center, New York State Department.of Health | Wadsworth Center, New York State Department.of Health | Kirsten St. George et al |
| EPI_ISL_676955 | USA/NY-Wadsworth-116619-01/2020 | 2020-05-25 | North America/USA/New York/Columbia | Wadsworth Center, New York State Department.of Health | Wadsworth Center, New York State Department.of Health | Kirsten St. George et al |

| | | | | | | |
|---|---|---|---|---|---|---|
| EPI_ISL_676971 | USA/NY-Wadsworth-142766-01/2020 | 2020-04-10 | North America/USA/New York/Nassau C | Wadsworth Center, New York State Department.of Health | Wadsworth Center, New York State Department.of Health | Kirsten St. George et al |
| EPI_ISL_676979 | USA/NY-Wadsworth-143769-01/2020 | 2020-06-17 | North America/USA/New York/Schenect | Wadsworth Center, New York State Department.of Health | Wadsworth Center, New York State Department.of Health | Kirsten St. George et al |
| EPI_ISL_676987 | USA/NY-Wadsworth-159719-01/2020 | 2020-07-01 | North America/USA/New York/Cattarau | Wadsworth Center, New York State Department.of Health | Wadsworth Center, New York State Department.of Health | Kirsten St. George et al |
| EPI_ISL_676988 | USA/NY-Wadsworth-161631-01/2020 | 2020-07-06 | North America/USA/New York/Cattarau | Wadsworth Center, New York State Department.of Health | Wadsworth Center, New York State Department.of Health | Kirsten St. George et al |
| EPI_ISL_676993 | USA/NY-Wadsworth-170604-01/2020 | 2020-07-13 | North America/USA/New York/Cattarau | Wadsworth Center, New York State Department.of Health | Wadsworth Center, New York State Department.of Health | Kirsten St. George et al |
| EPI_ISL_676994 | USA/NY-Wadsworth-189267-01/2020 | 2020-07-31 | North America/USA/New York/Cattarau | Wadsworth Center, New York State Department.of Health | Wadsworth Center, New York State Department.of Health | Kirsten St. George et al |
| EPI_ISL_677005 | USA/NY-Wadsworth-192874-01/2020 | 2020-06-29 | North America/USA/New York/Oneida C | Masonic Medical Research Institute | Wadsworth Center, New York State Department.of Health | Nathan Tucker et al |
| EPI_ISL_677014 | USA/NY-Wadsworth-192885-01/2020 | 2020-07-03 | North America/USA/New York/Oneida C | Masonic Medical Research Institute | Wadsworth Center, New York State Department.of Health | Nathan Tucker et al |

| | | | | | | |
|---|---|---|---|---|---|---|
| EPI_ISL_677017 | USA/NY-Wadsworth-192889-01/2020 | 2020-07-04 | North America/USA/New York/Oneida ( | Masonic Medical Research Institute | Wadsworth Center, New York State Department.of Health | Nathan Tucker et al |
| EPI_ISL_677030 | USA/NY-Wadsworth-194545-01/2020 | 2020-08-07 | North America/USA/New York/Erie Cou | Wadsworth Center, New York State Department.of Health | Wadsworth Center, New York State Department.of Health | Kirsten St. George et al |
| EPI_ISL_677034 | USA/NY-Wadsworth-194570-01/2020 | 2020-08-07 | North America/USA/New York/Erie Cou | Wadsworth Center, New York State Department.of Health | Wadsworth Center, New York State Department.of Health | Kirsten St. George et al |
| EPI_ISL_677037 | USA/NY-Wadsworth-194588-01/2020 | 2020-08-07 | North America/USA/New York/Erie Cou | Wadsworth Center, New York State Department.of Health | Wadsworth Center, New York State Department.of Health | Kirsten St. George et al |
| EPI_ISL_677042 | USA/NY-Wadsworth-199731-01/2020 | 2020-08-17 | North America/USA/New York/Essex Co | Wadsworth Center, New York State Department.of Health | Wadsworth Center, New York State Department.of Health | Kirsten St. George et al |
| EPI_ISL_677045 | USA/NY-Wadsworth-199755-01/2020 | 2020-08-17 | North America/USA/New York/Essex Co | Wadsworth Center, New York State Department.of Health | Wadsworth Center, New York State Department.of Health | Kirsten St. George et al |
| EPI_ISL_677054 | USA/NY-Wadsworth-200219-01/2020 | 2020-06-24 | North America/USA/New York/Oneida ( | Masonic Medical Research Institute | Wadsworth Center, New York State Department.of Health | Nathan Tucker et al |
| EPI_ISL_677058 | USA/NY-Wadsworth-200223-01/2020 | 2020-06-28 | North America/USA/New York/Oneida ( | Masonic Medical Research Institute | Wadsworth Center, New York State Department.of Health | Nathan Tucker et al |

| | | | | | | |
|---|---|---|---|---|---|---|
| EPI_ISL_677066 | USA/NY-Wadsworth-200232-01/2020 | 2020-07-05 | North America/USA/New York/Oneida ( | Masonic Medical Research Institute | Wadsworth Center, New York State Department.of Health | Nathan Tucker et al |
| EPI_ISL_677067 | USA/NY-Wadsworth-200233-01/2020 | 2020-07-06 | North America/USA/New York/Oneida ( | Masonic Medical Research Institute | Wadsworth Center, New York State Department.of Health | Nathan Tucker et al |
| EPI_ISL_677070 | USA/NY-Wadsworth-200236-01/2020 | 2020-07-08 | North America/USA/New York/Oneida ( | Masonic Medical Research Institute | Wadsworth Center, New York State Department.of Health | Nathan Tucker et al |
| EPI_ISL_677076 | USA/NY-Wadsworth-200243-01/2020 | 2020-06-23 | North America/USA/New York/Oneida ( | Masonic Medical Research Institute | Wadsworth Center, New York State Department.of Health | Nathan Tucker et al |
| EPI_ISL_677077 | USA/NY-Wadsworth-201446-01/2020 | 2020-08-19 | North America/USA/New York/Essex Co | Wadsworth Center, New York State Department.of Health | Wadsworth Center, New York State Department.of Health | Kirsten St. George et al |
| EPI_ISL_677078 | USA/NY-Wadsworth-201485-01/2020 | 2020-08-19 | North America/USA/New York/Essex Co | Wadsworth Center, New York State Department.of Health | Wadsworth Center, New York State Department.of Health | Kirsten St. George et al |
| EPI_ISL_677082 | USA/NY-Wadsworth-203562-01/2020 | 2020-08-25 | North America/USA/New York/Essex Co | Wadsworth Center, New York State Department.of Health | Wadsworth Center, New York State Department.of Health | Kirsten St. George et al |
| EPI_ISL_677083 | USA/NY-Wadsworth-204625-01/2020 | 2020-08-26 | North America/USA/New York/Essex Co | Wadsworth Center, New York State Department.of Health | Wadsworth Center, New York State Department.of Health | Kirsten St. George et al |

| | | | | | | |
|---|---|---|---|---|---|---|
| EPI_ISL_677085 | USA/NY-Wadsworth-204627-01/2020 | 2020-08-26 | North America/USA/New York/Essex Co | Wadsworth Center, New York State Department.of Health | Wadsworth Center, New York State Department.of Health | Kirsten St. George et al |
| EPI_ISL_677087 | USA/NY-Wadsworth-204634-01/2020 | 2020-08-26 | North America/USA/New York/Essex Co | Wadsworth Center, New York State Department.of Health | Wadsworth Center, New York State Department.of Health | Kirsten St. George et al |
| EPI_ISL_677089 | USA/NY-Wadsworth-207202-01/2020 | 2020-09-01 | North America/USA/New York/Schenect | Wadsworth Center, New York State Department.of Health | Wadsworth Center, New York State Department.of Health | Kirsten St. George et al |
| EPI_ISL_677097 | USA/NY-Wadsworth-211750-01/2020 | 2020-09-04 | North America/USA/New York/Oneida ( | Wadsworth Center, New York State Department.of Health | Wadsworth Center, New York State Department.of Health | Kirsten St. George et al |
| EPI_ISL_677101 | USA/NY-Wadsworth-217641-01/2020 | 2020-09-15 | North America/USA/New York/Hamilton | Wadsworth Center, New York State Department.of Health | Wadsworth Center, New York State Department.of Health | Kirsten St. George et al |
| EPI_ISL_677104 | USA/NY-Wadsworth-223644-01/2020 | 2020-09-20 | North America/USA/New York/Otsego ( | Wadsworth Center, New York State Department.of Health | Wadsworth Center, New York State Department.of Health | Kirsten St. George et al |
| EPI_ISL_677106 | USA/NY-Wadsworth-224393-01/2020 | 2020-09-22 | North America/USA/New York/Niagara | Wadsworth Center, New York State Department.of Health | Wadsworth Center, New York State Department.of Health | Kirsten St. George et al |
| EPI_ISL_677108 | USA/NY-Wadsworth-224539-01/2020 | 2020-09-22 | North America/USA/New York/Rockland | Wadsworth Center, New York State Department.of Health | Wadsworth Center, New York State Department.of Health | Kirsten St. George et al |

| | | | | | | |
|---|---|---|---|---|---|---|
| EPI_ISL_677111 | USA/NY-Wadsworth-226476-01/2020 | 2020-08-22 | North America/USA/New York/Herkime | Masonic Medical Research Institute | Wadsworth Center, New York State Department.of Health | Nathan Tucker et al |
| EPI_ISL_677113 | USA/NY-Wadsworth-226481-01/2020 | 2020-08-04 | North America/USA/New York/Herkime | Masonic Medical Research Institute | Wadsworth Center, New York State Department.of Health | Nathan Tucker et al |
| EPI_ISL_677115 | USA/NY-Wadsworth-226483-01/2020 | 2020-07-28 | North America/USA/New York/Herkime | Masonic Medical Research Institute | Wadsworth Center, New York State Department.of Health | Nathan Tucker et al |
| EPI_ISL_677117 | USA/NY-Wadsworth-226485-01/2020 | 2020-08-12 | North America/USA/New York/Oneida ( | Masonic Medical Research Institute | Wadsworth Center, New York State Department.of Health | Nathan Tucker et al |
| EPI_ISL_677119 | USA/NY-Wadsworth-226487-01/2020 | 2020-08-23 | North America/USA/New York/Oneida ( | Masonic Medical Research Institute | Wadsworth Center, New York State Department.of Health | Nathan Tucker et al |
| EPI_ISL_677125 | USA/NY-Wadsworth-226515-01/2020 | 2020-08-18 | North America/USA/New York/Herkime | Masonic Medical Research Institute | Wadsworth Center, New York State Department.of Health | Nathan Tucker et al |
| EPI_ISL_677126 | USA/NY-Wadsworth-226516-01/2020 | 2020-08-20 | North America/USA/New York/Oneida ( | Masonic Medical Research Institute | Wadsworth Center, New York State Department.of Health | Nathan Tucker et al |
| EPI_ISL_677127 | USA/NY-Wadsworth-226517-01/2020 | 2020-08-03 | North America/USA/New York/Oneida ( | Masonic Medical Research Institute | Wadsworth Center, New York State Department.of Health | Nathan Tucker et al |

| | | | | | | |
|---|---|---|---|---|---|---|
| EPI_ISL_677128 | USA/NY-Wadsworth-226518-01/2020 | 2020-08-04 | North America/USA/New York/Oneida ( | Masonic Medical Research Institute | Wadsworth Center, New York State Department.of Health | Nathan Tucker et al |
| EPI_ISL_677130 | USA/NY-Wadsworth-226966-01/2020 | 2020-09-24 | North America/USA/New York/Washing | Wadsworth Center, New York State Department.of Health | Wadsworth Center, New York State Department.of Health | Kirsten St. George et al |
| EPI_ISL_677131 | USA/NY-Wadsworth-228415-01/2020 | 2020-09-25 | North America/USA/New York/Chemung | Wadsworth Center, New York State Department.of Health | Wadsworth Center, New York State Department.of Health | Kirsten St. George et al |
| EPI_ISL_677139 | USA/VA-DCLS-2101/2020 | 2020-10-21 | North America/USA/Virginia/ | Virginia Division of Consolidated Laboratory Services | Virginia Division of Consolidated Laboratory Services | Virginia DCLS et al |
| EPI_ISL_677145 | USA/VA-DCLS-2111/2020 | 2020-10-15 | North America/USA/Virginia/ | Virginia Division of Consolidated Laboratory Services | Virginia Division of Consolidated Laboratory Services | Virginia DCLS et al |
| EPI_ISL_677146 | USA/VA-DCLS-2112/2020 | 2020-10-19 | North America/USA/Virginia/ | Virginia Division of Consolidated Laboratory Services | Virginia Division of Consolidated Laboratory Services | Virginia DCLS et al |
| EPI_ISL_677153 | USA/VA-DCLS-2128/2020 | 2020-10-28 | North America/USA/Virginia/ | Virginia Division of Consolidated Laboratory Services | Virginia Division of Consolidated Laboratory Services | Virginia DCLS et al |
| EPI_ISL_677161 | USA/VA-DCLS-2142/2020 | 2020-10-28 | North America/USA/Virginia/ | Virginia Division of Consolidated Laboratory Services | Virginia Division of Consolidated Laboratory Services | Virginia DCLS et al |
| EPI_ISL_677168 | USA/VA-DCLS-2019/2020 | 2020-10-14 | North America/USA/Virginia/ | Virginia Division of Consolidated Laboratory Services | Virginia Division of Consolidated Laboratory Services | Virginia DCLS et al |

| | | | | | | |
|---|---|---|---|---|---|---|
| EPI_ISL_677172 | USA/VA-DCLS-2027/2020 | 2020-10-14 | North America/USA/Virginia/ | Virginia Division of Consolidated Laboratory Services | Virginia Division of Consolidated Laboratory Services | Virginia DCLS et al |
| EPI_ISL_677175 | USA/VA-DCLS-2031/2020 | 2020-10-14 | North America/USA/Virginia/ | Virginia Division of Consolidated Laboratory Services | Virginia Division of Consolidated Laboratory Services | Virginia DCLS et al |
| EPI_ISL_677179 | USA/VA-DCLS-2069/2020 | 2020-10-20 | North America/USA/Virginia/ | Virginia Division of Consolidated Laboratory Services | Virginia Division of Consolidated Laboratory Services | Virginia DCLS et al |
| EPI_ISL_677183 | USA/VA-DCLS-2075/2020 | 2020-10-20 | North America/USA/Virginia/ | Virginia Division of Consolidated Laboratory Services | Virginia Division of Consolidated Laboratory Services | Virginia DCLS et al |
| EPI_ISL_677184 | USA/VA-DCLS-2076/2020 | 2020-10-21 | North America/USA/Virginia/ | Virginia Division of Consolidated Laboratory Services | Virginia Division of Consolidated Laboratory Services | Virginia DCLS et al |
| EPI_ISL_677185 | USA/VA-DCLS-2078/2020 | 2020-10-21 | North America/USA/Virginia/ | Virginia Division of Consolidated Laboratory Services | Virginia Division of Consolidated Laboratory Services | Virginia DCLS et al |
| EPI_ISL_677189 | USA/VA-DCLS-2089/2020 | 2020-10-21 | North America/USA/Virginia/ | Virginia Division of Consolidated Laboratory Services | Virginia Division of Consolidated Laboratory Services | Virginia DCLS et al |
| EPI_ISL_677194 | USA/VA-DCLS-2165/2020 | 2020-11-06 | North America/USA/Virginia/ | Virginia Division of Consolidated Laboratory Services | Virginia Division of Consolidated Laboratory Services | Virginia DCLS et al |
| EPI_ISL_677205 | USA/VA-DCLS-2178/2020 | 2020-11-09 | North America/USA/Virginia/ | Virginia Division of Consolidated Laboratory Services | Virginia Division of Consolidated Laboratory Services | Virginia DCLS et al |
| EPI_ISL_677209 | USA/VA-DCLS-2183/2020 | 2020-11-09 | North America/USA/Virginia/ | Virginia Division of Consolidated Laboratory Services | Virginia Division of Consolidated Laboratory Services | Virginia DCLS et al |

| | | | | | | |
|---|---|---|---|---|---|---|
| EPI_ISL_677218 | USA/VA-DCLS-2046/2020 | 2020-10-26 | North America/USA/Virginia/ | Virginia Division of Consolidated Laboratory Services | Virginia Division of Consolidated Laboratory Services | Virginia DCLS et al |
| EPI_ISL_677226 | USA/VA-DCLS-2056/2020 | 2020-10-28 | North America/USA/Virginia/ | Virginia Division of Consolidated Laboratory Services | Virginia Division of Consolidated Laboratory Services | Virginia DCLS et al |
| EPI_ISL_677228 | USA/VA-DCLS-2058/2020 | 2020-10-28 | North America/USA/Virginia/ | Virginia Division of Consolidated Laboratory Services | Virginia Division of Consolidated Laboratory Services | Virginia DCLS et al |
| EPI_ISL_677231 | USA/VA-DCLS-2061/2020 | 2020-10-20 | North America/USA/Virginia/ | Virginia Division of Consolidated Laboratory Services | Virginia Division of Consolidated Laboratory Services | Virginia DCLS et al |
| EPI_ISL_677235 | USA/VA-DCLS-2115/2020 | 2020-10-24 | North America/USA/Virginia/ | Virginia Division of Consolidated Laboratory Services | Virginia Division of Consolidated Laboratory Services | Virginia DCLS et al |
| EPI_ISL_677239 | USA/VA-DCLS-2119/2020 | 2020-10-24 | North America/USA/Virginia/ | Virginia Division of Consolidated Laboratory Services | Virginia Division of Consolidated Laboratory Services | Virginia DCLS et al |
| EPI_ISL_677242 | USA/CO-CDPHE-2003120079/2020 | 2020-03-10 | North America/USA/Colorado/ | Colorado Department of Public Health and Environment | Colorado Department of Puplic Health and Environment | Laura Bankers et al |
| EPI_ISL_677243 | USA/CO-CDPHE-2009163590/2020 | 2020-09-16 | North America/USA/Colorado/ | Colorado Department of Public Health and Environment | Colorado Department of Puplic Health and Environment | Laura Bankers et al |
| EPI_ISL_677250 | USA/CO-CDPHE-2009260672/2020 | 2020-09-24 | North America/USA/Colorado/ | Colorado Department of Public Health and Environment | Colorado Department of Puplic Health and Environment | Laura Bankers et al |
| EPI_ISL_677262 | USA/CO-CDPHE-2009121107/2020 | 2020-09-11 | North America/USA/Colorado/ | Colorado Department of Public Health and Environment | Colorado Department of Puplic Health and Environment | Laura Bankers et al |

| | | | | | | |
|---|---|---|---|---|---|---|
| EPI_ISL_677263 | USA/CO-CDPHE-2010112609/2020 | 2020-10-08 | North America/USA/Colorado/ | Colorado Department of Public Health and Environment | Colorado Department of Puplic Health and Environment | Laura Bankers et al |
| EPI_ISL_677277 | USA/CO-CDPHE-2009163273/2020 | 2020-09-16 | North America/USA/Colorado/ | Colorado Department of Public Health and Environment | Colorado Department of Puplic Health and Environment | Laura Bankers et al |
| EPI_ISL_677291 | USA/CO-CDPHE-2005210130/2020 | 2020-05-20 | North America/USA/Colorado/ | Colorado Department of Public Health and Environment | Colorado Department of Puplic Health and Environment | Laura Bankers et al |
| EPI_ISL_677300 | USA/CO-CDPHE-2008161334/2020 | 2020-08-16 | North America/USA/Colorado/ | Colorado Department of Public Health and Environment | Colorado Department of Puplic Health and Environment | Laura Bankers et al |
| EPI_ISL_677301 | USA/CO-CDPHE-2008161388/2020 | 2020-08-16 | North America/USA/Colorado/ | Colorado Department of Public Health and Environment | Colorado Department of Puplic Health and Environment | Laura Bankers et al |
| EPI_ISL_677303 | USA/CO-CDPHE-2008170021/2020 | 2020-08-16 | North America/USA/Colorado/ | Colorado Department of Public Health and Environment | Colorado Department of Puplic Health and Environment | Laura Bankers et al |
| EPI_ISL_677305 | USA/CO-CDPHE-2008170097/2020 | 2020-08-16 | North America/USA/Colorado/ | Colorado Department of Public Health and Environment | Colorado Department of Puplic Health and Environment | Laura Bankers et al |
| EPI_ISL_677307 | USA/CO-CDPHE-2008170563/2020 | 2020-08-17 | North America/USA/Colorado/ | Colorado Department of Public Health and Environment | Colorado Department of Puplic Health and Environment | Laura Bankers et al |
| EPI_ISL_677308 | USA/CO-CDPHE-2008170936/2020 | 2020-08-17 | North America/USA/Colorado/ | Colorado Department of Public Health and Environment | Colorado Department of Puplic Health and Environment | Laura Bankers et al |
| EPI_ISL_677310 | USA/CO-CDPHE-2008181790/2020 | 2020-08-17 | North America/USA/Colorado/ | Colorado Department of Public Health and Environment | Colorado Department of Puplic Health and Environment | Laura Bankers et al |

| | | | | | | |
|---|---|---|---|---|---|---|
| EPI_ISL_677316 | USA/CO-CDPHE-2009151301/2020 | 2020-09-14 | North America/USA/Colorado/ | Colorado Department of Public Health and Environment | Colorado Department of Puplic Health and Environment | Laura Bankers et al |
| EPI_ISL_677317 | USA/CO-CDPHE-2009163495/2020 | 2020-09-16 | North America/USA/Colorado/ | Colorado Department of Public Health and Environment | Colorado Department of Puplic Health and Environment | Laura Bankers et al |
| EPI_ISL_677328 | USA/WI-UW-1919/2020 | 2020-10-28 | North America/USA/Wisconsin/Dane Co | University of Wisconsin-Madison AIDS Vaccine Research Laboratories | University of Wisconsin-Madison AIDS Vaccine Research Laboratories | Gage Moreno et al |
| EPI_ISL_677330 | USA/WI-UW-1921/2020 | 2020-10-30 | North America/USA/Wisconsin/Dane Co | University of Wisconsin-Madison AIDS Vaccine Research Laboratories | University of Wisconsin-Madison AIDS Vaccine Research Laboratories | Gage Moreno et al |
| EPI_ISL_677333 | USA/WI-UW-1924/2020 | 2020-10-30 | North America/USA/Wisconsin/Dane Co | University of Wisconsin-Madison AIDS Vaccine Research Laboratories | University of Wisconsin-Madison AIDS Vaccine Research Laboratories | Gage Moreno et al |
| EPI_ISL_677334 | USA/WI-UW-1925/2020 | 2020-10-30 | North America/USA/Wisconsin/Dane Co | University of Wisconsin-Madison AIDS Vaccine Research Laboratories | University of Wisconsin-Madison AIDS Vaccine Research Laboratories | Gage Moreno et al |
| EPI_ISL_677343 | USA/WI-UW-1934/2020 | 2020-09-17 | North America/USA/Wisconsin/Dane Co | University of Wisconsin-Madison AIDS Vaccine Research Laboratories | University of Wisconsin-Madison AIDS Vaccine Research Laboratories | Gage Moreno et al |
| EPI_ISL_677344 | USA/WI-UW-1935/2020 | 2020-09-27 | North America/USA/Wisconsin/Walwort | University of Wisconsin-Madison AIDS Vaccine Research Laboratories | University of Wisconsin-Madison AIDS Vaccine Research Laboratories | Gage Moreno et al |

| | | | | | | |
|---|---|---|---|---|---|---|
| EPI_ISL_677352 | USA/WI-UW-1944/2020 | 2020-07-18 | North America/USA/Wisconsin/Dane Co | University of Wisconsin-Madison AIDS Vaccine Research Laboratories | University of Wisconsin-Madison AIDS Vaccine Research Laboratories | Gage Moreno et al |
| EPI_ISL_677355 | USA/WI-UW-1947/2020 | 2020-09-29 | North America/USA/Wisconsin/Columbia | University of Wisconsin-Madison AIDS Vaccine Research Laboratories | University of Wisconsin-Madison AIDS Vaccine Research Laboratories | Gage Moreno et al |
| EPI_ISL_677359 | USA/WI-UW-1951/2020 | 2020-09-29 | North America/USA/Wisconsin/Dane Co | University of Wisconsin-Madison AIDS Vaccine Research Laboratories | University of Wisconsin-Madison AIDS Vaccine Research Laboratories | Gage Moreno et al |
| EPI_ISL_677364 | USA/IL-UW-1956/2020 | 2020-09-20 | North America/USA/Illinois/Winnebago | University of Wisconsin-Madison AIDS Vaccine Research Laboratories | University of Wisconsin-Madison AIDS Vaccine Research Laboratories | Gage Moreno et al |
| EPI_ISL_677369 | USA/IL-UW-1961/2020 | 2020-09-23 | North America/USA/Illinois/Winnebago | University of Wisconsin-Madison AIDS Vaccine Research Laboratories | University of Wisconsin-Madison AIDS Vaccine Research Laboratories | Gage Moreno et al |
| EPI_ISL_677377 | USA/WI-UW-1969/2020 | 2020-09-20 | North America/USA/Wisconsin/Dane Co | University of Wisconsin-Madison AIDS Vaccine Research Laboratories | University of Wisconsin-Madison AIDS Vaccine Research Laboratories | Gage Moreno et al |
| EPI_ISL_677378 | USA/WI-UW-1970/2020 | 2020-09-18 | North America/USA/Wisconsin/Dane Co | University of Wisconsin-Madison AIDS Vaccine Research Laboratories | University of Wisconsin-Madison AIDS Vaccine Research Laboratories | Gage Moreno et al |
| EPI_ISL_677381 | USA/WI-UW-1973/2020 | 2020-09-24 | North America/USA/Wisconsin/Green C | University of Wisconsin-Madison AIDS Vaccine Research Laboratories | University of Wisconsin-Madison AIDS Vaccine Research Laboratories | Gage Moreno et al |

| | | | | | | |
|---|---|---|---|---|---|---|
| EPI_ISL_677384 | USA/WI-UW-1976/2020 | 2020-09-29 | North America/USA/Wisconsin/Dane Co | University of Wisconsin-Madison AIDS Vaccine Research Laboratories | University of Wisconsin-Madison AIDS Vaccine Research Laboratories | Gage Moreno et al |
| EPI_ISL_677387 | USA/WI-UW-1979/2020 | 2020-09-21 | North America/USA/Wisconsin/Dane Co | University of Wisconsin-Madison AIDS Vaccine Research Laboratories | University of Wisconsin-Madison AIDS Vaccine Research Laboratories | Gage Moreno et al |
| EPI_ISL_677389 | USA/IL-UW-1981/2020 | 2020-09-23 | North America/USA/Illinois/Winnebago | University of Wisconsin-Madison AIDS Vaccine Research Laboratories | University of Wisconsin-Madison AIDS Vaccine Research Laboratories | Gage Moreno et al |
| EPI_ISL_677395 | USA/WI-UW-1987/2020 | 2020-09-25 | North America/USA/Wisconsin/Dane Co | University of Wisconsin-Madison AIDS Vaccine Research Laboratories | University of Wisconsin-Madison AIDS Vaccine Research Laboratories | Gage Moreno et al |
| EPI_ISL_677397 | USA/WI-UW-1989/2020 | 2020-09-26 | North America/USA/Wisconsin/Dane Co | University of Wisconsin-Madison AIDS Vaccine Research Laboratories | University of Wisconsin-Madison AIDS Vaccine Research Laboratories | Gage Moreno et al |
| EPI_ISL_677398 | USA/WI-UW-1990/2020 | 2020-09-22 | North America/USA/Wisconsin/Dane Co | University of Wisconsin-Madison AIDS Vaccine Research Laboratories | University of Wisconsin-Madison AIDS Vaccine Research Laboratories | Gage Moreno et al |
| EPI_ISL_677401 | USA/WI-UW-1993/2020 | 2020-09-25 | North America/USA/Wisconsin/Milwauk | University of Wisconsin-Madison AIDS Vaccine Research Laboratories | University of Wisconsin-Madison AIDS Vaccine Research Laboratories | Gage Moreno et al |
| EPI_ISL_677405 | USA/WI-UW-1997/2020 | 2020-09-25 | North America/USA/Wisconsin/Milwauk | University of Wisconsin-Madison AIDS Vaccine Research Laboratories | University of Wisconsin-Madison AIDS Vaccine Research Laboratories | Gage Moreno et al |

| | | | | | | |
|---|---|---|---|---|---|---|
| EPI_ISL_677407 | USA/WI-UW-1999/2020 | 2020-09-25 | North America/USA/Wisconsin/Milwauk | University of Wisconsin-Madison AIDS Vaccine Research Laboratories | University of Wisconsin-Madison AIDS Vaccine Research Laboratories | Gage Moreno et al |
| EPI_ISL_677408 | USA/WI-UW-2000/2020 | 2020-09-25 | North America/USA/Wisconsin/Milwauk | University of Wisconsin-Madison AIDS Vaccine Research Laboratories | University of Wisconsin-Madison AIDS Vaccine Research Laboratories | Gage Moreno et al |
| EPI_ISL_677410 | USA/WI-UW-2002/2020 | 2020-09-09 | North America/USA/Wisconsin/Milwauk | University of Wisconsin-Madison AIDS Vaccine Research Laboratories | University of Wisconsin-Madison AIDS Vaccine Research Laboratories | Gage Moreno et al |
| EPI_ISL_677417 | USA/WI-UW-2009/2020 | 2020-09-10 | North America/USA/Wisconsin/Milwauk | University of Wisconsin-Madison AIDS Vaccine Research Laboratories | University of Wisconsin-Madison AIDS Vaccine Research Laboratories | Gage Moreno et al |
| EPI_ISL_677418 | USA/WI-UW-2010/2020 | 2020-09-10 | North America/USA/Wisconsin/Milwauk | University of Wisconsin-Madison AIDS Vaccine Research Laboratories | University of Wisconsin-Madison AIDS Vaccine Research Laboratories | Gage Moreno et al |
| EPI_ISL_677425 | USA/IL-UW-2018/2020 | 2020-10-03 | North America/USA/Illinois/Ogle County | University of Wisconsin-Madison AIDS Vaccine Research Laboratories | University of Wisconsin-Madison AIDS Vaccine Research Laboratories | Gage Moreno et al |
| EPI_ISL_677426 | USA/WI-UW-2019/2020 | 2020-09-18 | North America/USA/Wisconsin/Dane Co | University of Wisconsin-Madison AIDS Vaccine Research Laboratories | University of Wisconsin-Madison AIDS Vaccine Research Laboratories | Gage Moreno et al |
| EPI_ISL_677430 | USA/WI-UW-2023/2020 | 2020-10-08 | North America/USA/Wisconsin/Dane Co | University of Wisconsin-Madison AIDS Vaccine Research Laboratories | University of Wisconsin-Madison AIDS Vaccine Research Laboratories | Gage Moreno et al |

| | | | | | | |
|---|---|---|---|---|---|---|
| EPI_ISL_677432 | USA/WI-UW-2025/2020 | 2020-09-26 | North America/USA/Wisconsin/Columbia | University of Wisconsin-Madison AIDS Vaccine Research Laboratories | University of Wisconsin-Madison AIDS Vaccine Research Laboratories | Gage Moreno et al |
| EPI_ISL_677434 | USA/WI-UW-2027/2020 | 2020-09-24 | North America/USA/Wisconsin/Dane Co | University of Wisconsin-Madison AIDS Vaccine Research Laboratories | University of Wisconsin-Madison AIDS Vaccine Research Laboratories | Gage Moreno et al |
| EPI_ISL_677439 | USA/IL-UW-2033/2020 | 2020-09-19 | North America/USA/Illinois/Winnebago | University of Wisconsin-Madison AIDS Vaccine Research Laboratories | University of Wisconsin-Madison AIDS Vaccine Research Laboratories | Gage Moreno et al |
| EPI_ISL_677445 | USA/WI-UW-2039/2020 | 2020-10-08 | North America/USA/Wisconsin/Dane Co | University of Wisconsin-Madison AIDS Vaccine Research Laboratories | University of Wisconsin-Madison AIDS Vaccine Research Laboratories | Gage Moreno et al |
| EPI_ISL_677453 | USA/WI-UW-2048/2020 | 2020-09-25 | North America/USA/Wisconsin/Dane Co | University of Wisconsin-Madison AIDS Vaccine Research Laboratories | University of Wisconsin-Madison AIDS Vaccine Research Laboratories | Gage Moreno et al |
| EPI_ISL_677461 | USA/WI-UW-2056/2020 | 2020-10-04 | North America/USA/Wisconsin/Dane Co | University of Wisconsin-Madison AIDS Vaccine Research Laboratories | University of Wisconsin-Madison AIDS Vaccine Research Laboratories | Gage Moreno et al |
| EPI_ISL_677488 | USA/WI-UW-2083/2020 | 2020-10-18 | North America/USA/Wisconsin/Dane Co | University of Wisconsin-Madison AIDS Vaccine Research Laboratories | University of Wisconsin-Madison AIDS Vaccine Research Laboratories | Gage Moreno et al |
| EPI_ISL_677490 | USA/IL-UW-2085/2020 | 2020-09-21 | North America/USA/Illinois/Winnebago | University of Wisconsin-Madison AIDS Vaccine Research Laboratories | University of Wisconsin-Madison AIDS Vaccine Research Laboratories | Gage Moreno et al |

| | | | | | | |
|---|---|---|---|---|---|---|
| EPI_ISL_677491 | USA/WI-UW-2086/2020 | 2020-09-23 | North America/USA/Wisconsin/Jefferson | University of Wisconsin-Madison AIDS Vaccine Research Laboratories | University of Wisconsin-Madison AIDS Vaccine Research Laboratories | Gage Moreno et al |
| EPI_ISL_677492 | USA/WI-UW-2087/2020 | 2020-09-18 | North America/USA/Wisconsin/Sauk Co | University of Wisconsin-Madison AIDS Vaccine Research Laboratories | University of Wisconsin-Madison AIDS Vaccine Research Laboratories | Gage Moreno et al |
| EPI_ISL_677493 | USA/WI-UW-2088/2020 | 2020-09-27 | North America/USA/Wisconsin/Dane Co | University of Wisconsin-Madison AIDS Vaccine Research Laboratories | University of Wisconsin-Madison AIDS Vaccine Research Laboratories | Gage Moreno et al |
| EPI_ISL_677496 | USA/WI-UW-2091/2020 | 2020-09-19 | North America/USA/Wisconsin/Columbia | University of Wisconsin-Madison AIDS Vaccine Research Laboratories | University of Wisconsin-Madison AIDS Vaccine Research Laboratories | Gage Moreno et al |
| EPI_ISL_677502 | USA/WI-UW-2097/2020 | 2020-10-27 | North America/USA/Wisconsin/Dane Co | University of Wisconsin-Madison AIDS Vaccine Research Laboratories | University of Wisconsin-Madison AIDS Vaccine Research Laboratories | Gage Moreno et al |
| EPI_ISL_677507 | USA/WI-UW-2102/2020 | 2020-11-09 | North America/USA/Wisconsin/Dane Co | University of Wisconsin-Madison AIDS Vaccine Research Laboratories | University of Wisconsin-Madison AIDS Vaccine Research Laboratories | Gage Moreno et al |
| EPI_ISL_677508 | USA/WI-UW-2103/2020 | 2020-11-09 | North America/USA/Wisconsin/Dane Co | University of Wisconsin-Madison AIDS Vaccine Research Laboratories | University of Wisconsin-Madison AIDS Vaccine Research Laboratories | Gage Moreno et al |
| EPI_ISL_677510 | USA/WI-UW-2105/2020 | 2020-09-29 | North America/USA/Wisconsin/Dane Co | University of Wisconsin-Madison AIDS Vaccine Research Laboratories | University of Wisconsin-Madison AIDS Vaccine Research Laboratories | Gage Moreno et al |

| | | | | | | |
|---|---|---|---|---|---|---|
| EPI_ISL_677514 | USA/WI-UW-2109/2020 | 2020-09-29 | North America/USA/Wisconsin/Dane Co | University of Wisconsin-Madison AIDS Vaccine Research Laboratories | University of Wisconsin-Madison AIDS Vaccine Research Laboratories | Gage Moreno et al |
| EPI_ISL_677515 | USA/WI-UW-2110/2020 | 2020-09-23 | North America/USA/Wisconsin/Dane Co | University of Wisconsin-Madison AIDS Vaccine Research Laboratories | University of Wisconsin-Madison AIDS Vaccine Research Laboratories | Gage Moreno et al |
| EPI_ISL_677517 | USA/IL-UW-2112/2020 | 2020-09-30 | North America/USA/Illinois/Winnebago | University of Wisconsin-Madison AIDS Vaccine Research Laboratories | University of Wisconsin-Madison AIDS Vaccine Research Laboratories | Gage Moreno et al |
| EPI_ISL_677518 | USA/WI-UW-2113/2020 | 2020-09-27 | North America/USA/Wisconsin/Dane Co | University of Wisconsin-Madison AIDS Vaccine Research Laboratories | University of Wisconsin-Madison AIDS Vaccine Research Laboratories | Gage Moreno et al |
| EPI_ISL_677525 | USA/WI-UW-2120/2020 | 2020-09-30 | North America/USA/Wisconsin/Dodge C | University of Wisconsin-Madison AIDS Vaccine Research Laboratories | University of Wisconsin-Madison AIDS Vaccine Research Laboratories | Gage Moreno et al |
| EPI_ISL_677536 | USA/WI-UW-2131/2020 | 2020-09-30 | North America/USA/Wisconsin/Dane Co | University of Wisconsin-Madison AIDS Vaccine Research Laboratories | University of Wisconsin-Madison AIDS Vaccine Research Laboratories | Gage Moreno et al |
| EPI_ISL_677538 | USA/WI-UW-2133/2020 | 2020-10-11 | North America/USA/Wisconsin/Dane Co | University of Wisconsin-Madison AIDS Vaccine Research Laboratories | University of Wisconsin-Madison AIDS Vaccine Research Laboratories | Gage Moreno et al |
| EPI_ISL_677539 | USA/WI-UW-2135/2020 | 2020-10-13 | North America/USA/Wisconsin/Dane Co | University of Wisconsin-Madison AIDS Vaccine Research Laboratories | University of Wisconsin-Madison AIDS Vaccine Research Laboratories | Gage Moreno et al |

| | | | | | | |
|---|---|---|---|---|---|---|
| EPI_ISL_677559 | USA/WI-UW-2155/2020 | 2020-10-15 | North America/USA/Wisconsin/Green Co | University of Wisconsin-Madison AIDS Vaccine Research Laboratories | University of Wisconsin-Madison AIDS Vaccine Research Laboratories | Gage Moreno et al |
| EPI_ISL_677570 | USA/WI-UW-2166/2020 | 2020-10-14 | North America/USA/Wisconsin/Columbia | University of Wisconsin-Madison AIDS Vaccine Research Laboratories | University of Wisconsin-Madison AIDS Vaccine Research Laboratories | Gage Moreno et al |
| EPI_ISL_677575 | USA/WI-UW-2171/2020 | 2020-10-15 | North America/USA/Wisconsin/Dane Co | University of Wisconsin-Madison AIDS Vaccine Research Laboratories | University of Wisconsin-Madison AIDS Vaccine Research Laboratories | Gage Moreno et al |
| EPI_ISL_677576 | USA/WI-UW-2172/2020 | 2020-10-15 | North America/USA/Wisconsin/Dane Co | University of Wisconsin-Madison AIDS Vaccine Research Laboratories | University of Wisconsin-Madison AIDS Vaccine Research Laboratories | Gage Moreno et al |
| EPI_ISL_677580 | USA/WI-UW-2176/2020 | 2020-10-28 | North America/USA/Wisconsin/Dane Co | University of Wisconsin-Madison AIDS Vaccine Research Laboratories | University of Wisconsin-Madison AIDS Vaccine Research Laboratories | Gage Moreno et al |
| EPI_ISL_677593 | USA/WI-UW-2190/2020 | 2020-10-25 | North America/USA/Wisconsin/Dane Co | University of Wisconsin-Madison AIDS Vaccine Research Laboratories | University of Wisconsin-Madison AIDS Vaccine Research Laboratories | Gage Moreno et al |
| EPI_ISL_677602 | USA/WI-UW-2199/2020 | 2020-10-24 | North America/USA/Wisconsin/Dane Co | University of Wisconsin-Madison AIDS Vaccine Research Laboratories | University of Wisconsin-Madison AIDS Vaccine Research Laboratories | Gage Moreno et al |
| EPI_ISL_677612 | USA/WI-UW-2209/2020 | 2020-10-26 | North America/USA/Wisconsin/Columbia | University of Wisconsin-Madison AIDS Vaccine Research Laboratories | University of Wisconsin-Madison AIDS Vaccine Research Laboratories | Gage Moreno et al |

| | | | | | | |
|---|---|---|---|---|---|---|
| EPI_ISL_677614 | USA/WI-UW-2211/2020 | 2020-10-25 | North America/USA/Wisconsin/Dane Co | University of Wisconsin-Madison AIDS Vaccine Research Laboratories | University of Wisconsin-Madison AIDS Vaccine Research Laboratories | Gage Moreno et al |
| EPI_ISL_677617 | USA/WI-UW-2214/2020 | 2020-10-26 | North America/USA/Wisconsin/Dane Co | University of Wisconsin-Madison AIDS Vaccine Research Laboratories | University of Wisconsin-Madison AIDS Vaccine Research Laboratories | Gage Moreno et al |
| EPI_ISL_677620 | USA/WI-UW-2217/2020 | 2020-09-26 | North America/USA/Wisconsin/Dane Co | University of Wisconsin-Madison AIDS Vaccine Research Laboratories | University of Wisconsin-Madison AIDS Vaccine Research Laboratories | Gage Moreno et al |
| EPI_ISL_677623 | USA/WI-UW-2220/2020 | 2020-09-26 | North America/USA/Wisconsin/Dane Co | University of Wisconsin-Madison AIDS Vaccine Research Laboratories | University of Wisconsin-Madison AIDS Vaccine Research Laboratories | Gage Moreno et al |
| EPI_ISL_677625 | USA/IL-UW-2222/2020 | 2020-09-28 | North America/USA/Illinois/Winnebago | University of Wisconsin-Madison AIDS Vaccine Research Laboratories | University of Wisconsin-Madison AIDS Vaccine Research Laboratories | Gage Moreno et al |
| EPI_ISL_677630 | USA/WI-UW-2227/2020 | 2020-10-02 | North America/USA/Wisconsin/Dane Co | University of Wisconsin-Madison AIDS Vaccine Research Laboratories | University of Wisconsin-Madison AIDS Vaccine Research Laboratories | Gage Moreno et al |
| EPI_ISL_677634 | Madagascar/VIRO-33698/2020 | 2020-09-16 | Africa/Madagascar/Analamanga/Antana | Virology Unit, Institut Pasteur de Madagascar | Virology Unit, Institut Pasteur de Madagascar | Christian Ranaivoson et al |
| EPI_ISL_677636 | Madagascar/VIRO-2728/2020 | 2020-03-26 | Africa/Madagascar/Toamasina/ | Virology Unit, Institut Pasteur de Madagascar | Virology Unit, Institut Pasteur de Madagascar | Christian Ranaivoson et al |
| EPI_ISL_677640 | USA/CO-CDPHE-2009031031/2020 | 2020-09-02 | North America/USA/Colorado/ | Colorado Department of Public Health and Environment | Colorado Department of Puplic Health and Environment | Laura Bankers et al |

| | | | | | | |
|---|---|---|---|---|---|---|
| EPI_ISL_677641 | USA/CO-CDPHE-2009042507/2020 | 2020-09-03 | North America/USA/Colorado/ | Colorado Department of Public Health and Environment | Colorado Department of Puplic Health and Environment | Laura Bankers et al |
| EPI_ISL_677642 | USA/CO-CDPHE-2009042555/2020 | 2020-09-03 | North America/USA/Colorado/ | Colorado Department of Public Health and Environment | Colorado Department of Puplic Health and Environment | Laura Bankers et al |
| EPI_ISL_677643 | USA/CO-CDPHE-2009090314/2020 | 2020-09-08 | North America/USA/Colorado/ | Colorado Department of Public Health and Environment | Colorado Department of Puplic Health and Environment | Laura Bankers et al |
| EPI_ISL_677659 | USA/WI-UW-2013/2020 | 2020-09-10 | North America/USA/Wisconsin/Milwauk | University of Wisconsin-Madison AIDS Vaccine Research Laboratories | University of Wisconsin-Madison AIDS Vaccine Research Laboratories | Gage Moreno et al |
| EPI_ISL_677668 | USA/NY-Wadsworth-176436-01/2020 | 2020-07-17 | North America/USA/New York/Jefferson | Wadsworth Center, New York State Department.of Health | Wadsworth Center, New York State Department.of Health | Kirsten St. George et al |
| EPI_ISL_677671 | USA/NY-Wadsworth-214818-01/2020 | 2020-09-10 | North America/USA/New York/Saratoga | Wadsworth Center, New York State Department.of Health | Wadsworth Center, New York State Department.of Health | Kirsten St. George et al |
| EPI_ISL_677672 | USA/NY-Wadsworth-226489-01/2020 | 2020-08-06 | North America/USA/New York/Oneida ( | Masonic Medical Research Institute | Wadsworth Center, New York State Department.of Health | Nathan Tucker et al |

| | | | | | | |
|---|---|---|---|---|---|---|
| EPI_ISL_677674 | NorthMacedonia/883/2020 | 2020-05-20 | Europe/North Macedonia/Pelagonia/Pri | General Hospital - Prilep | "Research Center for Genetic Engineering and Biotechnology ""Georgi D. Efremov,Äù , Macedonian Academy of Sciences and Arts" | RCGEB - MASA et al |
| EPI_ISL_677676 | NorthMacedonia/2227/2020 | 2020-06-03 | Europe/North Macedonia/Eastern Regio | Clinical Hospital - Shtip | "Research Center for Genetic Engineering and Biotechnology ""Georgi D. Efremov,Äù , Macedonian Academy of Sciences and Arts" | RCGEB - MASA et al |
| EPI_ISL_677688 | USA/VAN4606/2020 | 2020-05-11 | North America/USA/New York/New Yor | Vanda Pharmaceuticals Clinical Site | Vanda Pharmaceuticals | Vanda Pharmaceuticals et al |
| EPI_ISL_677695 | USA/VAN5674/2020 | 2020-09-03 | North America/USA/New York/New Yor | Vanda Pharmaceuticals Clinical Site | Vanda Pharmaceuticals | Vanda Pharmaceuticals et al |
| EPI_ISL_677696 | USA/VAN5074/2020 | 2020-09-14 | North America/USA/New York/New Yor | Vanda Pharmaceuticals Clinical Site | Vanda Pharmaceuticals | Vanda Pharmaceuticals et al |
| EPI_ISL_677699 | USA/VAN5434/2020 | 2020-09-16 | North America/USA/New York/New Yor | Vanda Pharmaceuticals Clinical Site | Vanda Pharmaceuticals | Vanda Pharmaceuticals et al |
| EPI_ISL_677708 | NorthMacedonia/4098/2020 | 2020-06-18 | Europe/North Macedonia/Skopje/ | Center for public health - Skopje | "Research Center for Genetic Engineering and Biotechnology ""Georgi D. Efremov,Äù , Macedonian Academy of Sciences and Arts" | RCGEB - MASA et al |

| | | | | | | |
|---|---|---|---|---|---|---|
| EPI_ISL_677715 | NorthMacedonia/6752/2020 | 2020-07-10 | Europe/North Macedonia/Pelagonia/Bit | Clinical Hospital - Bitola | "Research Center for Genetic Engineering and Biotechnology ""Georgi D. Efremov,Äù , Macedonian Academy of Sciences and Arts" | RCGEB - MASA et al |
| EPI_ISL_677717 | NorthMacedonia/7813/2020 | 2020-07-20 | Europe/North Macedonia/Eastern Regio | Clinical Hospital - Shtip | "Research Center for Genetic Engineering and Biotechnology ""Georgi D. Efremov,Äù , Macedonian Academy of Sciences and Arts" | RCGEB - MASA et al |
| EPI_ISL_677721 | NorthMacedonia/8881/2020 | 2020-07-30 | Europe/North Macedonia/Eastern Regio | Clinical Hospital - Shtip | "Research Center for Genetic Engineering and Biotechnology ""Georgi D. Efremov,Äù , Macedonian Academy of Sciences and Arts" | RCGEB - MASA et al |
| EPI_ISL_677722 | NorthMacedonia/9073/2020 | 2020-08-04 | Europe/North Macedonia/Southwestern | General Hospital - Ohrid | "Research Center for Genetic Engineering and Biotechnology ""Georgi D. Efremov,Äù , Macedonian Academy of Sciences and Arts" | RCGEB - MASA et al |

| | | | | | | |
|---|---|---|---|---|---|---|
| EPI_ISL_677723 | NorthMacedonia/9133/2020 | 2020-08-04 | Europe/North Macedonia/Skopje/ | Institute for Lung Diseases in Children - Skopje | "Research Center for Genetic Engineering and Biotechnology ""Georgi D. Efremov,Äù , Macedonian Academy of Sciences and Arts" | RCGEB - MASA et al |
| EPI_ISL_677725 | NorthMacedonia/9359/2020 | 2020-08-05 | Europe/North Macedonia/Southwesterr | General Hospital - Ohrid | "Research Center for Genetic Engineering and Biotechnology ""Georgi D. Efremov,Äù , Macedonian Academy of Sciences and Arts" | RCGEB - MASA et al |
| EPI_ISL_677740 | Hungary/US-32271w/2020 | 2020-09-01 | Europe/Hungary/B√°cs-Kiskun County/ | University of Szeged, Institute of Clinical Microbiology | National Laboratory of Virology, Szent√°gothai Research Centre | Endre G√°bor T√≥th et al |
| EPI_ISL_677741 | Hungary/US-32294w/2020 | 2020-09-01 | Europe/Hungary/Csongr√°d County/ | University of Szeged, Institute of Clinical Microbiology | National Laboratory of Virology, Szent√°gothai Research Centre | Endre G√°bor T√≥th et al |
| EPI_ISL_677747 | Hungary/US-32401w/2020 | 2020-09-02 | Europe/Hungary/Csongr√°d County/ | University of Szeged, Institute of Clinical Microbiology | National Laboratory of Virology, Szent√°gothai Research Centre | Endre G√°bor T√≥th et al |
| EPI_ISL_677752 | Hungary/US-32534w/2020 | 2020-09-02 | Europe/Hungary/Csongr√°d County/ | University of Szeged, Institute of Clinical Microbiology | National Laboratory of Virology, Szent√°gothai Research Centre | Endre G√°bor T√≥th et al |

| | | | | | | |
|---|---|---|---|---|---|---|
| EPI_ISL_677755 | Hungary/US-32545w/2020 | 2020-09-02 | Europe/Hungary/Csongr√°d County/ | University of Szeged, Institute of Clinical Microbiology | National Laboratory of Virology, Szent√°gothai Research Centre | Endre G√°bor T√≥th et al |
| EPI_ISL_677757 | Hungary/US-32566w/2020 | 2020-09-02 | Europe/Hungary/Csongr√°d County/ | University of Szeged, Institute of Clinical Microbiology | National Laboratory of Virology, Szent√°gothai Research Centre | Endre G√°bor T√≥th et al |
| EPI_ISL_677758 | Hungary/US-32570w/2020 | 2020-09-02 | Europe/Hungary/Csongr√°d County/ | University of Szeged, Institute of Clinical Microbiology | National Laboratory of Virology, Szent√°gothai Research Centre | Endre G√°bor T√≥th et al |
| EPI_ISL_677761 | Hungary/US-32620w/2020 | 2020-09-02 | Europe/Hungary/B√°cs-Kiskun County/ | University of Szeged, Institute of Clinical Microbiology | National Laboratory of Virology, Szent√°gothai Research Centre | Endre G√°bor T√≥th et al |
| EPI_ISL_677764 | Hungary/US-33735w/2020 | 2020-09-03 | Europe/Hungary/Csongr√°d County/ | University of Szeged, Institute of Clinical Microbiology | National Laboratory of Virology, Szent√°gothai Research Centre | Endre G√°bor T√≥th et al |
| EPI_ISL_677770 | Hungary/US-55383w/2020 | 2020-10-02 | Europe/Hungary/Csongr√°d County/ | University of Szeged, Institute of Clinical Microbiology | National Laboratory of Virology, Szent√°gothai Research Centre | Endre G√°bor T√≥th et al |
| EPI_ISL_677773 | Hungary/US-55491w/2020 | 2020-10-01 | Europe/Hungary/Csongr√°d County/ | University of Szeged, Institute of Clinical Microbiology | National Laboratory of Virology, Szent√°gothai Research Centre | Endre G√°bor T√≥th et al |
| EPI_ISL_677778 | Hungary/US-57395w/2020 | 2020-10-02 | Europe/Hungary/Csongr√°d County/ | University of Szeged, Institute of Clinical Microbiology | National Laboratory of Virology, Szent√°gothai Research Centre | Endre G√°bor T√≥th et al |

| | | | | | | |
|---|---|---|---|---|---|---|
| EPI_ISL_677785 | Hungary/US-57523w/2020 | 2020-10-02 | Europe/Hungary/B√°cs-Kiskun County/ | University of Szeged, Institute of Clinical Microbiology | National Laboratory of Virology, Szent√°gothai Research Centre | Endre G√°bor T√≥th et al |
| EPI_ISL_677788 | Hungary/US-57527w/2020 | 2020-10-02 | Europe/Hungary/B√°cs-Kiskun County/ | University of Szeged, Institute of Clinical Microbiology | National Laboratory of Virology, Szent√°gothai Research Centre | Endre G√°bor T√≥th et al |
| EPI_ISL_677793 | Hungary/US-57557w/2020 | 2020-10-02 | Europe/Hungary/B√°cs-Kiskun County/ | University of Szeged, Institute of Clinical Microbiology | National Laboratory of Virology, Szent√°gothai Research Centre | Endre G√°bor T√≥th et al |
| EPI_ISL_677794 | Hungary/US-57565w/2020 | 2020-10-02 | Europe/Hungary/B√©k√©s County/ | University of Szeged, Institute of Clinical Microbiology | National Laboratory of Virology, Szent√°gothai Research Centre | Endre G√°bor T√≥th et al |
| EPI_ISL_677795 | Hungary/US-57566w/2020 | 2020-10-01 | Europe/Hungary/B√©k√©s County/ | University of Szeged, Institute of Clinical Microbiology | National Laboratory of Virology, Szent√°gothai Research Centre | Endre G√°bor T√≥th et al |
| EPI_ISL_677796 | Hungary/US-57572w/2020 | 2020-10-02 | Europe/Hungary/B√©k√©s County/ | University of Szeged, Institute of Clinical Microbiology | National Laboratory of Virology, Szent√°gothai Research Centre | Endre G√°bor T√≥th et al |
| EPI_ISL_677799 | Hungary/US-57576w/2020 | 2020-10-01 | Europe/Hungary/B√©k√©s County/ | University of Szeged, Institute of Clinical Microbiology | National Laboratory of Virology, Szent√°gothai Research Centre | Endre G√°bor T√≥th et al |
| EPI_ISL_677809 | Hungary/US-60435w/2020 | 2020-10-02 | Europe/Hungary/B√©k√©s County/ | University of Szeged, Institute of Clinical Microbiology | National Laboratory of Virology, Szent√°gothai Research Centre | Endre G√°bor T√≥th et al |

| | | | | | | |
|---|---|---|---|---|---|---|
| EPI_ISL_677810 | Hungary/US-60505w/2020 | 2020-10-02 | Europe/Hungary/B√°cs-Kiskun County/ | University of Szeged, Institute of Clinical Microbiology | National Laboratory of Virology, Szent√°gothai Research Centre | Endre G√°bor T√≥th et al |
| EPI_ISL_677816 | Hungary/US-60592w/2020 | 2020-10-02 | Europe/Hungary/Csongr√°d County/ | University of Szeged, Institute of Clinical Microbiology | National Laboratory of Virology, Szent√°gothai Research Centre | Endre G√°bor T√≥th et al |
| EPI_ISL_677818 | Hungary/US-63527w/2020 | 2020-10-07 | Europe/Hungary/Csongr√°d County/ | University of Szeged, Institute of Clinical Microbiology | National Laboratory of Virology, Szent√°gothai Research Centre | Endre G√°bor T√≥th et al |
| EPI_ISL_677825 | USA/CA-IGI-0442/2020 | 2020-07-10 | North America/USA/California/Alameda | Innovative Genomics Institute, UC Berkeley | Innovative Genomics Institute, UC Berkeley | Stacia Wyman et al |
| EPI_ISL_677832 | USA/CA-IGI-0564/2020 | 2020-07-30 | North America/USA/California/Alameda | Innovative Genomics Institute, UC Berkeley | Innovative Genomics Institute, UC Berkeley | Stacia Wyman et al |
| EPI_ISL_677837 | USA/CA-IGI-0585/2020 | 2020-08-01 | North America/USA/California/Alameda | Innovative Genomics Institute, UC Berkeley | Innovative Genomics Institute, UC Berkeley | Stacia Wyman et al |
| EPI_ISL_677838 | USA/CA-IGI-0592/2020 | 2020-08-01 | North America/USA/California/Alameda | Innovative Genomics Institute, UC Berkeley | Innovative Genomics Institute, UC Berkeley | Stacia Wyman et al |
| EPI_ISL_677841 | USA/CA-IGI-0598/2020 | 2020-07-30 | North America/USA/California/Alameda | Innovative Genomics Institute, UC Berkeley | Innovative Genomics Institute, UC Berkeley | Stacia Wyman et al |
| EPI_ISL_677843 | USA/CA-IGI-0604/2020 | 2020-07-31 | North America/USA/California/Alameda | Innovative Genomics Institute, UC Berkeley | Innovative Genomics Institute, UC Berkeley | Stacia Wyman et al |

| | | | | | | |
|---|---|---|---|---|---|---|
| EPI_ISL_677844 | USA/CA-IGI-0608/2020 | 2020-08-01 | North America/USA/California/Alameda | Innovative Genomics Institute, UC Berkeley | Innovative Genomics Institute, UC Berkeley | Stacia Wyman et al |
| EPI_ISL_677852 | USA/CA-IGI-0625/2020 | 2020-08-03 | North America/USA/California/Alameda | Innovative Genomics Institute, UC Berkeley | Innovative Genomics Institute, UC Berkeley | Stacia Wyman et al |
| EPI_ISL_677855 | USA/CA-IGI-0639/2020 | 2020-08-03 | North America/USA/California/Alameda | Innovative Genomics Institute, UC Berkeley | Innovative Genomics Institute, UC Berkeley | Stacia Wyman et al |
| EPI_ISL_677858 | USA/CA-IGI-0647/2020 | 2020-07-29 | North America/USA/California/Alameda | Innovative Genomics Institute, UC Berkeley | Innovative Genomics Institute, UC Berkeley | Stacia Wyman et al |
| EPI_ISL_677862 | USA/CA-IGI-0656/2020 | 2020-08-01 | North America/USA/California/Alameda | Innovative Genomics Institute, UC Berkeley | Innovative Genomics Institute, UC Berkeley | Stacia Wyman et al |
| EPI_ISL_677865 | USA/CA-IGI-0672/2020 | 2020-08-01 | North America/USA/California/Alameda | Innovative Genomics Institute, UC Berkeley | Innovative Genomics Institute, UC Berkeley | Stacia Wyman et al |
| EPI_ISL_677870 | USA/CA-IGI-0687/2020 | 2020-07-31 | North America/USA/California/Alameda | Innovative Genomics Institute, UC Berkeley | Innovative Genomics Institute, UC Berkeley | Stacia Wyman et al |
| EPI_ISL_677872 | USA/CA-IGI-0691/2020 | 2020-08-07 | North America/USA/California/Alameda | Innovative Genomics Institute, UC Berkeley | Innovative Genomics Institute, UC Berkeley | Stacia Wyman et al |
| EPI_ISL_677884 | USA/CA-IGI-0741/2020 | 2020-07-30 | North America/USA/California/Alameda | Innovative Genomics Institute, UC Berkeley | Innovative Genomics Institute, UC Berkeley | Stacia Wyman et al |
| EPI_ISL_677886 | USA/CA-IGI-0765/2020 | 2020-07-31 | North America/USA/California/Alameda | Innovative Genomics Institute, UC Berkeley | Innovative Genomics Institute, UC Berkeley | Stacia Wyman et al |

| | | | | | | |
|---|---|---|---|---|---|---|
| EPI_ISL_677890 | USA/CA-IGI-0796/2020 | 2020-07-13 | North America/USA/California/Alameda | Innovative Genomics Institute, UC Berkeley | Innovative Genomics Institute, UC Berkeley | Stacia Wyman et al |
| EPI_ISL_677893 | USA/CA-IGI-0800/2020 | 2020-07-02 | North America/USA/California/Alameda | Innovative Genomics Institute, UC Berkeley | Innovative Genomics Institute, UC Berkeley | Stacia Wyman et al |
| EPI_ISL_677896 | USA/CA-IGI-0808/2020 | 2020-07-10 | North America/USA/California/Alameda | Innovative Genomics Institute, UC Berkeley | Innovative Genomics Institute, UC Berkeley | Stacia Wyman et al |
| EPI_ISL_677902 | USA/CA-IGI-0820/2020 | 2020-07-14 | North America/USA/California/Alameda | Innovative Genomics Institute, UC Berkeley | Innovative Genomics Institute, UC Berkeley | Stacia Wyman et al |
| EPI_ISL_677904 | USA/CA-IGI-0826/2020 | 2020-08-17 | North America/USA/California/Alameda | Innovative Genomics Institute, UC Berkeley | Innovative Genomics Institute, UC Berkeley | Stacia Wyman et al |
| EPI_ISL_677906 | USA/CA-IGI-0835/2020 | 2020-07-29 | North America/USA/California/Alameda | Innovative Genomics Institute, UC Berkeley | Innovative Genomics Institute, UC Berkeley | Stacia Wyman et al |
| EPI_ISL_677908 | SaudiArabia/KAUST-JEDDAH1330/20 | 2020-06-30 | Asia/Saudi Arabia/Jeddah/ | Pathogen Genomics Lab King Abdullah University of Science and Technology(KAUST) | Pathogen Genomics Lab King Abdullah University of Science and Technology(KAUST) | Sara Mfarrej et al |
| EPI_ISL_677910 | SaudiArabia/KAUST-JEDDAH1324/20 | 2020-06-30 | Asia/Saudi Arabia/Jeddah/ | Pathogen Genomics Lab King Abdullah University of Science and Technology(KAUST) | Pathogen Genomics Lab King Abdullah University of Science and Technology(KAUST) | Sara Mfarrej et al |

| | | | | | | |
|---|---|---|---|---|---|---|
| EPI_ISL_677911 | SaudiArabia/KAUST-JEDDAH1294/20 | 2020-06-30 | Asia/Saudi Arabia/Jeddah/ | Pathogen Genomics Lab King Abdullah University of Science and Technology(KAUST) | Pathogen Genomics Lab King Abdullah University of Science and Technology(KAUST) | Muhammad Shuaib et al |
| EPI_ISL_677912 | SaudiArabia/KAUST-JEDDAH1329/20 | 2020-06-30 | Asia/Saudi Arabia/Jeddah/ | Pathogen Genomics Lab King Abdullah University of Science and Technology(KAUST) | Pathogen Genomics Lab King Abdullah University of Science and Technology(KAUST) | Sara Mfarrej et al |
| EPI_ISL_677913 | SaudiArabia/KAUST-JEDDAH1326/20 | 2020-06-30 | Asia/Saudi Arabia/Jeddah/ | Pathogen Genomics Lab King Abdullah University of Science and Technology(KAUST) | Pathogen Genomics Lab King Abdullah University of Science and Technology(KAUST) | Sara Mfarrej et al |
| EPI_ISL_677914 | SaudiArabia/KAUST-JEDDAH1297/20 | 2020-06-30 | Asia/Saudi Arabia/Jeddah/ | Pathogen Genomics Lab King Abdullah University of Science and Technology(KAUST) | Pathogen Genomics Lab King Abdullah University of Science and Technology(KAUST) | Muhammad Shuaib et al |
| EPI_ISL_677917 | SaudiArabia/KAUST-JEDDAH1321/20 | 2020-06-30 | Asia/Saudi Arabia/Jeddah/ | Pathogen Genomics Lab King Abdullah University of Science and Technology(KAUST) | Pathogen Genomics Lab King Abdullah University of Science and Technology(KAUST) | Sara Mfarrej et al |

| | | | | | | |
|---|---|---|---|---|---|---|
| EPI_ISL_677918 | SaudiArabia/KAUST-JEDDAH1331/20 | 2020-06-30 | Asia/Saudi Arabia/Jeddah/ | Pathogen Genomics Lab King Abdullah University of Science and Technology(KAUST) | Pathogen Genomics Lab King Abdullah University of Science and Technology(KAUST) | Sara Mfarrej et al |
| EPI_ISL_677919 | SaudiArabia/KAUST-JEDDAH1334/20 | 2020-06-30 | Asia/Saudi Arabia/Jeddah/ | Pathogen Genomics Lab King Abdullah University of Science and Technology(KAUST) | Pathogen Genomics Lab King Abdullah University of Science and Technology(KAUST) | Sara Mfarrej et al |
| EPI_ISL_677920 | SaudiArabia/KAUST-JEDDAH1335/20 | 2020-06-30 | Asia/Saudi Arabia/Jeddah/ | Pathogen Genomics Lab King Abdullah University of Science and Technology(KAUST) | Pathogen Genomics Lab King Abdullah University of Science and Technology(KAUST) | Sara Mfarrej et al |
| EPI_ISL_677921 | SaudiArabia/KAUST-JEDDAH1320/20 | 2020-06-30 | Asia/Saudi Arabia/Jeddah/ | Pathogen Genomics Lab King Abdullah University of Science and Technology(KAUST) | Pathogen Genomics Lab King Abdullah University of Science and Technology(KAUST) | Sara Mfarrej et al |
| EPI_ISL_677923 | SaudiArabia/KAUST-JEDDAH1292/20 | 2020-06-30 | Asia/Saudi Arabia/Jeddah/ | Pathogen Genomics Lab King Abdullah University of Science and Technology(KAUST) | Pathogen Genomics Lab King Abdullah University of Science and Technology(KAUST) | Muhammad Shuaib et al |

| | | | | | | |
|---|---|---|---|---|---|---|
| EPI_ISL_677924 | SaudiArabia/KAUST-JEDDAH1295/20 | 2020-06-30 | Asia/Saudi Arabia/Jeddah/ | Pathogen Genomics Lab King Abdullah University of Science and Technology(KAUST) | Pathogen Genomics Lab King Abdullah University of Science and Technology(KAUST) | Muhammad Shuaib et al |
| EPI_ISL_677932 | SaudiArabia/KAUST-JEDDAH1325/20 | 2020-06-30 | Asia/Saudi Arabia/Jeddah/ | Pathogen Genomics Lab King Abdullah University of Science and Technology(KAUST) | Pathogen Genomics Lab King Abdullah University of Science and Technology(KAUST) | Sara Mfarrej et al |
| EPI_ISL_677933 | SaudiArabia/KAUST-JEDDAH1333/20 | 2020-06-30 | Asia/Saudi Arabia/Jeddah/ | Pathogen Genomics Lab King Abdullah University of Science and Technology(KAUST) | Pathogen Genomics Lab King Abdullah University of Science and Technology(KAUST) | Sara Mfarrej et al |
| EPI_ISL_677945 | SaudiArabia/KAUST-MADINAH683/2 | 2020-05-09 | Asia/Saudi Arabia/Madinah/ | Pathogen Genomics Lab King Abdullah University of Science and Technology(KAUST) | Pathogen Genomics Lab King Abdullah University of Science and Technology(KAUST) | Sara Mfarrej et al |
| EPI_ISL_677948 | SaudiArabia/KAUST-MADINAH1378/ | 2020-06-04 | Asia/Saudi Arabia/Madinah/ | Pathogen Genomics Lab King Abdullah University of Science and Technology(KAUST) | Pathogen Genomics Lab King Abdullah University of Science and Technology(KAUST) | Sara Mfarrej et al |

| | | | | | | |
|---|---|---|---|---|---|---|
| EPI_ISL_677949 | SaudiArabia/KAUST-MADINAH1364/ | 2020-06-04 | Asia/Saudi Arabia/Madinah/ | Pathogen Genomics Lab King Abdullah University of Science and Technology(KAUST) | Pathogen Genomics Lab King Abdullah University of Science and Technology(KAUST) | Muhammad Shuaib et al |
| EPI_ISL_677951 | SaudiArabia/KAUST-MADINAH1063/ | 2020-06-13 | Asia/Saudi Arabia/Madinah/ | Pathogen Genomics Lab King Abdullah University of Science and Technology(KAUST) | Pathogen Genomics Lab King Abdullah University of Science and Technology(KAUST) | Olga Douvropoulou et al |
| EPI_ISL_677952 | SaudiArabia/KAUST-MADINAH1282/ | 2020-06-25 | Asia/Saudi Arabia/Madinah/ | Pathogen Genomics Lab King Abdullah University of Science and Technology(KAUST) | Pathogen Genomics Lab King Abdullah University of Science and Technology(KAUST) | Sara Mfarrej et al |
| EPI_ISL_677953 | SaudiArabia/KAUST-MADINAH1479/ | 2020-06-19 | Asia/Saudi Arabia/Madinah/ | Pathogen Genomics Lab King Abdullah University of Science and Technology(KAUST) | Pathogen Genomics Lab King Abdullah University of Science and Technology(KAUST) | Sara Mfarrej et al |
| EPI_ISL_677954 | SaudiArabia/KAUST-MADINAH1370/ | 2020-06-04 | Asia/Saudi Arabia/Madinah/ | Pathogen Genomics Lab King Abdullah University of Science and Technology(KAUST) | Pathogen Genomics Lab King Abdullah University of Science and Technology(KAUST) | Muhammad Shuaib et al |

| | | | | | | |
|---|---|---|---|---|---|---|
| EPI_ISL_677956 | SaudiArabia/KAUST-MADINAH1490/ | 2020-06-19 | Asia/Saudi Arabia/Madinah/ | Pathogen Genomics Lab King Abdullah University of Science and Technology(KAUST) | Pathogen Genomics Lab King Abdullah University of Science and Technology(KAUST) | Sara Mfarrej et al |
| EPI_ISL_677959 | SaudiArabia/KAUST-MADINAH1371/ | 2020-06-11 | Asia/Saudi Arabia/Madinah/ | Pathogen Genomics Lab King Abdullah University of Science and Technology(KAUST) | Pathogen Genomics Lab King Abdullah University of Science and Technology(KAUST) | Sara Mfarrej et al |
| EPI_ISL_677961 | SaudiArabia/KAUST-MADINAH1380/ | 2020-06-04 | Asia/Saudi Arabia/Madinah/ | Pathogen Genomics Lab King Abdullah University of Science and Technology(KAUST) | Pathogen Genomics Lab King Abdullah University of Science and Technology(KAUST) | Sara Mfarrej et al |
| EPI_ISL_677965 | SaudiArabia/KAUST-MADINAH690/2 | 2020-05-09 | Asia/Saudi Arabia/Madinah/ | Pathogen Genomics Lab King Abdullah University of Science and Technology(KAUST) | Pathogen Genomics Lab King Abdullah University of Science and Technology(KAUST) | Sara Mfarrej et al |
| EPI_ISL_677969 | SaudiArabia/KAUST-MADINAH1482/ | 2020-06-19 | Asia/Saudi Arabia/Madinah/ | Pathogen Genomics Lab King Abdullah University of Science and Technology(KAUST) | Pathogen Genomics Lab King Abdullah University of Science and Technology(KAUST) | Sara Mfarrej et al |

| | | | | | | |
|---|---|---|---|---|---|---|
| EPI_ISL_677971 | SaudiArabia/KAUST-MADINAH1279/ | 2020-06-25 | Asia/Saudi Arabia/Madinah/ | Pathogen Genomics Lab King Abdullah University of Science and Technology(KAUST) | Pathogen Genomics Lab King Abdullah University of Science and Technology(KAUST) | Sara Mfarrej et al |
| EPI_ISL_677972 | SaudiArabia/KAUST-MADINAH1101/ | 2020-06-13 | Asia/Saudi Arabia/Madinah/ | Pathogen Genomics Lab King Abdullah University of Science and Technology(KAUST) | Pathogen Genomics Lab King Abdullah University of Science and Technology(KAUST) | Olga Douvropoulou et al |
| EPI_ISL_677974 | SaudiArabia/KAUST-MADINAH1489/ | 2020-06-19 | Asia/Saudi Arabia/Madinah/ | Pathogen Genomics Lab King Abdullah University of Science and Technology(KAUST) | Pathogen Genomics Lab King Abdullah University of Science and Technology(KAUST) | Raushan Nugmanova et al |
| EPI_ISL_677975 | SaudiArabia/KAUST-MADINAH1128/ | 2020-06-11 | Asia/Saudi Arabia/Madinah/ | Pathogen Genomics Lab King Abdullah University of Science and Technology(KAUST) | Pathogen Genomics Lab King Abdullah University of Science and Technology(KAUST) | Amit Kumar Subudhi et al |
| EPI_ISL_677977 | SaudiArabia/KAUST-MADINAH1117/ | 2020-06-11 | Asia/Saudi Arabia/Madinah/ | Pathogen Genomics Lab King Abdullah University of Science and Technology(KAUST) | Pathogen Genomics Lab King Abdullah University of Science and Technology(KAUST) | Amit Kumar Subudhi et al |

| | | | | | | |
|---|---|---|---|---|---|---|
| EPI_ISL_677979 | SaudiArabia/KAUST-MADINAH1119/ | 2020-06-11 | Asia/Saudi Arabia/Madinah/ | Pathogen Genomics Lab King Abdullah University of Science and Technology(KAUST) | Pathogen Genomics Lab King Abdullah University of Science and Technology(KAUST ) | Amit Kumar Subudhi et al |
| EPI_ISL_677980 | SaudiArabia/KAUST-MADINAH1120/ | 2020-06-11 | Asia/Saudi Arabia/Madinah/ | Pathogen Genomics Lab King Abdullah University of Science and Technology(KAUST) | Pathogen Genomics Lab King Abdullah University of Science and Technology(KAUST ) | Amit Kumar Subudhi et al |
| EPI_ISL_677983 | SaudiArabia/KAUST-MADINAH1123/ | 2020-06-11 | Asia/Saudi Arabia/Madinah/ | Pathogen Genomics Lab King Abdullah University of Science and Technology(KAUST) | Pathogen Genomics Lab King Abdullah University of Science and Technology(KAUST ) | Amit Kumar Subudhi et al |
| EPI_ISL_677988 | SaudiArabia/KAUST-MADINAH1130/ | 2020-06-13 | Asia/Saudi Arabia/Madinah/ | Pathogen Genomics Lab King Abdullah University of Science and Technology(KAUST) | Pathogen Genomics Lab King Abdullah University of Science and Technology(KAUST ) | Sara Mfarrej et al |
| EPI_ISL_677991 | SaudiArabia/KAUST-MADINAH1139/ | 2020-06-27 | Asia/Saudi Arabia/Madinah/ | Pathogen Genomics Lab King Abdullah University of Science and Technology(KAUST) | Pathogen Genomics Lab King Abdullah University of Science and Technology(KAUST ) | Olga Douvropoulou et al |

| | | | | | | |
|---|---|---|---|---|---|---|
| EPI_ISL_677992 | SaudiArabia/KAUST-MADINAH1140/ | 2020-06-27 | Asia/Saudi Arabia/Madinah/ | Pathogen Genomics Lab King Abdullah University of Science and Technology(KAUST) | Pathogen Genomics Lab King Abdullah University of Science and Technology(KAUST) | Raeece Naeem et al |
| EPI_ISL_677994 | SaudiArabia/KAUST-MADINAH1142/ | 2020-06-27 | Asia/Saudi Arabia/Madinah/ | Pathogen Genomics Lab King Abdullah University of Science and Technology(KAUST) | Pathogen Genomics Lab King Abdullah University of Science and Technology(KAUST) | Raeece Naeem et al |
| EPI_ISL_677995 | SaudiArabia/KAUST-MADINAH1143/ | 2020-06-27 | Asia/Saudi Arabia/Madinah/ | Pathogen Genomics Lab King Abdullah University of Science and Technology(KAUST) | Pathogen Genomics Lab King Abdullah University of Science and Technology(KAUST) | Raeece Naeem et al |
| EPI_ISL_677998 | SaudiArabia/KAUST-MADINAH1146/ | 2020-06-27 | Asia/Saudi Arabia/Madinah/ | Pathogen Genomics Lab King Abdullah University of Science and Technology(KAUST) | Pathogen Genomics Lab King Abdullah University of Science and Technology(KAUST) | Raeece Naeem et al |
| EPI_ISL_677999 | SaudiArabia/KAUST-MADINAH1147/ | 2020-06-27 | Asia/Saudi Arabia/Madinah/ | Pathogen Genomics Lab King Abdullah University of Science and Technology(KAUST) | Pathogen Genomics Lab King Abdullah University of Science and Technology(KAUST) | Raeece Naeem et al |

| | | | | | | |
|---|---|---|---|---|---|---|
| EPI_ISL_678000 | SaudiArabia/KAUST-MADINAH1148/ | 2020-06-27 | Asia/Saudi Arabia/Madinah/ | Pathogen Genomics Lab King Abdullah University of Science and Technology(KAUST) | Pathogen Genomics Lab King Abdullah University of Science and Technology(KAUST) | Raeece Naeem et al |
| EPI_ISL_678003 | SaudiArabia/KAUST-MADINAH1151/ | 2020-06-17 | Asia/Saudi Arabia/Madinah/ | Pathogen Genomics Lab King Abdullah University of Science and Technology(KAUST) | Pathogen Genomics Lab King Abdullah University of Science and Technology(KAUST) | Raeece Naeem et al |
| EPI_ISL_678009 | SaudiArabia/KAUST-MADINAH1284/ | 2020-06-25 | Asia/Saudi Arabia/Madinah/ | Pathogen Genomics Lab King Abdullah University of Science and Technology(KAUST) | Pathogen Genomics Lab King Abdullah University of Science and Technology(KAUST) | Sara Mfarrej et al |
| EPI_ISL_678017 | SaudiArabia/KAUST-MADINAH1375/ | 2020-06-04 | Asia/Saudi Arabia/Madinah/ | Pathogen Genomics Lab King Abdullah University of Science and Technology(KAUST) | Pathogen Genomics Lab King Abdullah University of Science and Technology(KAUST) | Muhammad Shuaib et al |
| EPI_ISL_678018 | SaudiArabia/KAUST-MADINAH1379/ | 2020-06-11 | Asia/Saudi Arabia/Madinah/ | Pathogen Genomics Lab King Abdullah University of Science and Technology(KAUST) | Pathogen Genomics Lab King Abdullah University of Science and Technology(KAUST) | Muhammad Shuaib et al |

| | | | | | | |
|---|---|---|---|---|---|---|
| EPI_ISL_678020 | SaudiArabia/KAUST-MADINAH1478/ | 2020-06-19 | Asia/Saudi Arabia/Madinah/ | Pathogen Genomics Lab King Abdullah University of Science and Technology(KAUST) | Pathogen Genomics Lab King Abdullah University of Science and Technology(KAUST ) | Raushan Nugmanova et al |
| EPI_ISL_678024 | SaudiArabia/KAUST-MADINAH1487/ | 2020-06-19 | Asia/Saudi Arabia/Madinah/ | Pathogen Genomics Lab King Abdullah University of Science and Technology(KAUST) | Pathogen Genomics Lab King Abdullah University of Science and Technology(KAUST ) | Raushan Nugmanova et al |
| EPI_ISL_678026 | SaudiArabia/KAUST-MADINAH210/2 | 2020-04-01 | Asia/Saudi Arabia/Madinah/ | Pathogen Genomics Lab King Abdullah University of Science and Technology(KAUST) | Pathogen Genomics Lab King Abdullah University of Science and Technology(KAUST ) | Sharif Hala et al |
| EPI_ISL_678034 | SaudiArabia/KAUST-MADINAH654/2 | 2020-05-08 | Asia/Saudi Arabia/Madinah/ | Pathogen Genomics Lab King Abdullah University of Science and Technology(KAUST) | Pathogen Genomics Lab King Abdullah University of Science and Technology(KAUST ) | Sara Mfarrej et al |
| EPI_ISL_678045 | SaudiArabia/KAUST-MAKKAH1513/2 | 2020-04-14 | Asia/Saudi Arabia/Makkah/ | Pathogen Genomics Lab King Abdullah University of Science and Technology(KAUST) | Pathogen Genomics Lab King Abdullah University of Science and Technology(KAUST ) | Amanda Ooi et al |

| | | | | | | |
|---|---|---|---|---|---|---|
| EPI_ISL_678049 | SaudiArabia/KAUST-MAKKAH1416/2 | 2020-06-28 | Asia/Saudi Arabia/Makkah/ | Pathogen Genomics Lab King Abdullah University of Science and Technology(KAUST) | Pathogen Genomics Lab King Abdullah University of Science and Technology(KAUST ) | Muhammad Shuaib et al |
| EPI_ISL_678064 | SaudiArabia/KAUST-MAKKAH1386/2 | 2020-06-23 | Asia/Saudi Arabia/Makkah/ | Pathogen Genomics Lab King Abdullah University of Science and Technology(KAUST) | Pathogen Genomics Lab King Abdullah University of Science and Technology(KAUST ) | Muhammad Shuaib et al |
| EPI_ISL_678066 | SaudiArabia/KAUST-MAKKAH223/20 | 2020-04-06 | Asia/Saudi Arabia/Makkah/ | Pathogen Genomics Lab King Abdullah University of Science and Technology(KAUST) | Pathogen Genomics Lab King Abdullah University of Science and Technology(KAUST ) | Fathia Ben Rached et al |
| EPI_ISL_678070 | SaudiArabia/KAUST-MAKKAH1413/2 | 2020-06-28 | Asia/Saudi Arabia/Makkah/ | Pathogen Genomics Lab King Abdullah University of Science and Technology(KAUST) | Pathogen Genomics Lab King Abdullah University of Science and Technology(KAUST ) | Muhammad Shuaib et al |
| EPI_ISL_678071 | SaudiArabia/KAUST-MAKKAH207/20 | 2020-04-06 | Asia/Saudi Arabia/Makkah/ | Pathogen Genomics Lab King Abdullah University of Science and Technology(KAUST) | Pathogen Genomics Lab King Abdullah University of Science and Technology(KAUST ) | Sharif Hala et al |

| | | | | | | |
|---|---|---|---|---|---|---|
| EPI_ISL_678074 | SaudiArabia/KAUST-MAKKAH191/20 | 2020-04-06 | Asia/Saudi Arabia/Makkah/ | Pathogen Genomics Lab King Abdullah University of Science and Technology(KAUST) | Pathogen Genomics Lab King Abdullah University of Science and Technology(KAUST) | Amit Kumar Subudhi et al |
| EPI_ISL_678077 | SaudiArabia/KAUST-MAKKAH199/20 | 2020-04-06 | Asia/Saudi Arabia/Makkah/ | Pathogen Genomics Lab King Abdullah University of Science and Technology(KAUST) | Pathogen Genomics Lab King Abdullah University of Science and Technology(KAUST) | Sharif Hala et al |
| EPI_ISL_678079 | SaudiArabia/KAUST-MAKKAH106/20 | 2020-03-24 | Asia/Saudi Arabia/Makkah/ | Pathogen Genomics Lab King Abdullah University of Science and Technology(KAUST) | Pathogen Genomics Lab King Abdullah University of Science and Technology(KAUST) | Sharif Hala et al |
| EPI_ISL_678084 | SaudiArabia/KAUST-MAKKAH1392/2 | 2020-06-23 | Asia/Saudi Arabia/Makkah/ | Pathogen Genomics Lab King Abdullah University of Science and Technology(KAUST) | Pathogen Genomics Lab King Abdullah University of Science and Technology(KAUST) | Muhammad Shuaib et al |
| EPI_ISL_678085 | SaudiArabia/KAUST-MAKKAH1395/2 | 2020-06-23 | Asia/Saudi Arabia/Makkah/ | Pathogen Genomics Lab King Abdullah University of Science and Technology(KAUST) | Pathogen Genomics Lab King Abdullah University of Science and Technology(KAUST) | Muhammad Shuaib et al |

| | | | | | | |
|---|---|---|---|---|---|---|
| EPI_ISL_678093 | SaudiArabia/KAUST-MAKKAH1421/2 | 2020-06-29 | Asia/Saudi Arabia/Makkah/ | Pathogen Genomics Lab King Abdullah University of Science and Technology(KAUST) | Pathogen Genomics Lab King Abdullah University of Science and Technology(KAUST) | Muhammad Shuaib et al |
| EPI_ISL_678094 | SaudiArabia/KAUST-MAKKAH1426/2 | 2020-07-02 | Asia/Saudi Arabia/Makkah/ | Pathogen Genomics Lab King Abdullah University of Science and Technology(KAUST) | Pathogen Genomics Lab King Abdullah University of Science and Technology(KAUST) | Olga Douvropoulou et al |
| EPI_ISL_678095 | SaudiArabia/KAUST-MAKKAH1427/2 | 2020-07-02 | Asia/Saudi Arabia/Makkah/ | Pathogen Genomics Lab King Abdullah University of Science and Technology(KAUST) | Pathogen Genomics Lab King Abdullah University of Science and Technology(KAUST) | Olga Douvropoulou et al |
| EPI_ISL_678110 | SaudiArabia/KAUST-MAKKAH1557/2 | 2020-04-14 | Asia/Saudi Arabia/Makkah/ | Pathogen Genomics Lab King Abdullah University of Science and Technology(KAUST) | Pathogen Genomics Lab King Abdullah University of Science and Technology(KAUST) | Amanda Ooi et al |
| EPI_ISL_678129 | SaudiArabia/KAUST-MAKKAH1623/2 | 2020-04-26 | Asia/Saudi Arabia/Makkah/ | Pathogen Genomics Lab King Abdullah University of Science and Technology(KAUST) | Pathogen Genomics Lab King Abdullah University of Science and Technology(KAUST) | Luke Esau et al |

| | | | | | | |
|---|---|---|---|---|---|---|
| EPI_ISL_678131 | SaudiArabia/KAUST-MAKKAH1630/2 | 2020-04-26 | Asia/Saudi Arabia/Makkah/ | Pathogen Genomics Lab King Abdullah University of Science and Technology(KAUST) | Pathogen Genomics Lab King Abdullah University of Science and Technology(KAUST) | Luke Esau et al |
| EPI_ISL_678134 | SaudiArabia/KAUST-MAKKAH1635/2 | 2020-04-26 | Asia/Saudi Arabia/Makkah/ | Pathogen Genomics Lab King Abdullah University of Science and Technology(KAUST) | Pathogen Genomics Lab King Abdullah University of Science and Technology(KAUST) | Luke Esau et al |
| EPI_ISL_678137 | SaudiArabia/KAUST-MAKKAH201/20 | 2020-04-06 | Asia/Saudi Arabia/Makkah/ | Pathogen Genomics Lab King Abdullah University of Science and Technology(KAUST) | Pathogen Genomics Lab King Abdullah University of Science and Technology(KAUST) | Sharif Hala et al |
| EPI_ISL_678145 | SaudiArabia/KAUST-MAKKAH819/20 | 2020-04-24 | Asia/Saudi Arabia/Makkah/ | Pathogen Genomics Lab King Abdullah University of Science and Technology(KAUST) | Pathogen Genomics Lab King Abdullah University of Science and Technology(KAUST) | Sara Mfarrej et al |
| EPI_ISL_678146 | SaudiArabia/KAUST-MAKKAH825/20 | 2020-04-24 | Asia/Saudi Arabia/Makkah/ | Pathogen Genomics Lab King Abdullah University of Science and Technology(KAUST) | Pathogen Genomics Lab King Abdullah University of Science and Technology(KAUST) | Sara Mfarrej et al |

| | | | | | | |
|---|---|---|---|---|---|---|
| EPI_ISL_678148 | SaudiArabia/KAUST-QATIF1355/2020 | 2020-06-15 | Asia/Saudi Arabia/Eastern Province/Qat | Pathogen Genomics Lab King Abdullah University of Science and Technology(KAUST) | Pathogen Genomics Lab King Abdullah University of Science and Technology(KAUST ) | Sara Mfarrej et al |
| EPI_ISL_678149 | SaudiArabia/KAUST-QATIF1451/2020 | 2020-06-27 | Asia/Saudi Arabia/Eastern Province/Qat | Pathogen Genomics Lab King Abdullah University of Science and Technology(KAUST) | Pathogen Genomics Lab King Abdullah University of Science and Technology(KAUST ) | Sara Mfarrej et al |
| EPI_ISL_678150 | SaudiArabia/KAUST-QATIF1447/2020 | 2020-06-27 | Asia/Saudi Arabia/Eastern Province/Qat | Pathogen Genomics Lab King Abdullah University of Science and Technology(KAUST) | Pathogen Genomics Lab King Abdullah University of Science and Technology(KAUST ) | Sara Mfarrej et al |
| EPI_ISL_678151 | SaudiArabia/KAUST-QATIF1441/2020 | 2020-06-27 | Asia/Saudi Arabia/Eastern Province/Qat | Pathogen Genomics Lab King Abdullah University of Science and Technology(KAUST) | Pathogen Genomics Lab King Abdullah University of Science and Technology(KAUST ) | Sara Mfarrej et al |
| EPI_ISL_678154 | SaudiArabia/KAUST-QATIF1353/2020 | 2020-06-20 | Asia/Saudi Arabia/Eastern Province/Qat | Pathogen Genomics Lab King Abdullah University of Science and Technology(KAUST) | Pathogen Genomics Lab King Abdullah University of Science and Technology(KAUST ) | Sara Mfarrej et al |

| | | | | | | |
|---|---|---|---|---|---|---|
| EPI_ISL_678155 | SaudiArabia/KAUST-QATIF1443/2020 | 2020-06-27 | Asia/Saudi Arabia/Eastern Province/Qat | Pathogen Genomics Lab King Abdullah University of Science and Technology(KAUST) | Pathogen Genomics Lab King Abdullah University of Science and Technology(KAUST) | Sara Mfarrej et al |
| EPI_ISL_678156 | SaudiArabia/KAUST-QATIF1448/2020 | 2020-06-27 | Asia/Saudi Arabia/Eastern Province/Qat | Pathogen Genomics Lab King Abdullah University of Science and Technology(KAUST) | Pathogen Genomics Lab King Abdullah University of Science and Technology(KAUST) | Sara Mfarrej et al |
| EPI_ISL_678158 | SaudiArabia/KAUST-QATIF1457/2020 | 2020-06-27 | Asia/Saudi Arabia/Eastern Province/Qat | Pathogen Genomics Lab King Abdullah University of Science and Technology(KAUST) | Pathogen Genomics Lab King Abdullah University of Science and Technology(KAUST) | Sara Mfarrej et al |
| EPI_ISL_678161 | SaudiArabia/KAUST-RIYADH1179/202 | 2020-06-05 | Asia/Saudi Arabia/Riyadh/ | Pathogen Genomics Lab King Abdullah University of Science and Technology(KAUST) | Pathogen Genomics Lab King Abdullah University of Science and Technology(KAUST) | Raeece Naeem et al |
| EPI_ISL_678162 | SaudiArabia/KAUST-RIYADH1285/202 | 2020-06-18 | Asia/Saudi Arabia/Riyadh/ | Pathogen Genomics Lab King Abdullah University of Science and Technology(KAUST) | Pathogen Genomics Lab King Abdullah University of Science and Technology(KAUST) | Sara Mfarrej et al |

| | | | | | | |
|---|---|---|---|---|---|---|
| EPI_ISL_678164 | SaudiArabia/KAUST-RIYADH1258/20 | 2020-07-29 | Asia/Saudi Arabia/Riyadh/ | Pathogen Genomics Lab King Abdullah University of Science and Technology(KAUST) | Pathogen Genomics Lab King Abdullah University of Science and Technology(KAUST) | Sara Mfarrej et al |
| EPI_ISL_678165 | SaudiArabia/KAUST-RIYADH1246/20 | 2020-07-29 | Asia/Saudi Arabia/Riyadh/ | Pathogen Genomics Lab King Abdullah University of Science and Technology(KAUST) | Pathogen Genomics Lab King Abdullah University of Science and Technology(KAUST) | Sara Mfarrej et al |
| EPI_ISL_678166 | SaudiArabia/KAUST-RIYADH1187/20 | 2020-06-15 | Asia/Saudi Arabia/Riyadh/ | Pathogen Genomics Lab King Abdullah University of Science and Technology(KAUST) | Pathogen Genomics Lab King Abdullah University of Science and Technology(KAUST) | Raeece Naeem et al |
| EPI_ISL_678167 | SaudiArabia/KAUST-RIYADH1155/20 | 2020-06-15 | Asia/Saudi Arabia/Riyadh/ | Pathogen Genomics Lab King Abdullah University of Science and Technology(KAUST) | Pathogen Genomics Lab King Abdullah University of Science and Technology(KAUST) | Olga Douvropoulou et al |
| EPI_ISL_678168 | SaudiArabia/KAUST-RIYADH1211/20 | 2020-06-05 | Asia/Saudi Arabia/Riyadh/ | Pathogen Genomics Lab King Abdullah University of Science and Technology(KAUST) | Pathogen Genomics Lab King Abdullah University of Science and Technology(KAUST) | Sara Mfarrej et al |

| | | | | | | |
|---|---|---|---|---|---|---|
| EPI_ISL_678170 | SaudiArabia/KAUST-RIYADH1231/202 | 2020-07-29 | Asia/Saudi Arabia/Riyadh/ | Pathogen Genomics Lab King Abdullah University of Science and Technology(KAUST) | Pathogen Genomics Lab King Abdullah University of Science and Technology(KAUST) | Muhammad Shuaib et al |
| EPI_ISL_678171 | SaudiArabia/KAUST-RIYADH1248/202 | 2020-08-11 | Asia/Saudi Arabia/Riyadh/ | Pathogen Genomics Lab King Abdullah University of Science and Technology(KAUST) | Pathogen Genomics Lab King Abdullah University of Science and Technology(KAUST) | Sara Mfarrej et al |
| EPI_ISL_678174 | SaudiArabia/KAUST-RIYADH1234/202 | 2020-07-29 | Asia/Saudi Arabia/Riyadh/ | Pathogen Genomics Lab King Abdullah University of Science and Technology(KAUST) | Pathogen Genomics Lab King Abdullah University of Science and Technology(KAUST) | Muhammad Shuaib et al |
| EPI_ISL_678175 | SaudiArabia/KAUST-RIYADH1166/202 | 2020-06-15 | Asia/Saudi Arabia/Riyadh/ | Pathogen Genomics Lab King Abdullah University of Science and Technology(KAUST) | Pathogen Genomics Lab King Abdullah University of Science and Technology(KAUST) | Raeece Naeem et al |
| EPI_ISL_678178 | SaudiArabia/KAUST-RIYADH1247/202 | 2020-07-29 | Asia/Saudi Arabia/Riyadh/ | Pathogen Genomics Lab King Abdullah University of Science and Technology(KAUST) | Pathogen Genomics Lab King Abdullah University of Science and Technology(KAUST) | Sara Mfarrej et al |

| | | | | | | |
|---|---|---|---|---|---|---|
| EPI_ISL_678179 | SaudiArabia/KAUST-RIYADH1255/202 | 2020-07-29 | Asia/Saudi Arabia/Riyadh/ | Pathogen Genomics Lab King Abdullah University of Science and Technology(KAUST) | Pathogen Genomics Lab King Abdullah University of Science and Technology(KAUST) | Sara Mfarrej et al |
| EPI_ISL_678180 | SaudiArabia/KAUST-RIYADH1446/202 | 2020-06-27 | Asia/Saudi Arabia/Riyadh/ | Pathogen Genomics Lab King Abdullah University of Science and Technology(KAUST) | Pathogen Genomics Lab King Abdullah University of Science and Technology(KAUST) | Olga Douvropoulou et al |
| EPI_ISL_678181 | SaudiArabia/KAUST-RIYADH1164/202 | 2020-06-05 | Asia/Saudi Arabia/Riyadh/ | Pathogen Genomics Lab King Abdullah University of Science and Technology(KAUST) | Pathogen Genomics Lab King Abdullah University of Science and Technology(KAUST) | Raeece Naeem et al |
| EPI_ISL_678182 | SaudiArabia/KAUST-RIYADH1256/202 | 2020-07-29 | Asia/Saudi Arabia/Riyadh/ | Pathogen Genomics Lab King Abdullah University of Science and Technology(KAUST) | Pathogen Genomics Lab King Abdullah University of Science and Technology(KAUST) | Sara Mfarrej et al |
| EPI_ISL_678183 | SaudiArabia/KAUST-RIYADH1193/202 | 2020-06-15 | Asia/Saudi Arabia/Riyadh/ | Pathogen Genomics Lab King Abdullah University of Science and Technology(KAUST) | Pathogen Genomics Lab King Abdullah University of Science and Technology(KAUST) | Sara Mfarrej et al |

| | | | | | | |
|---|---|---|---|---|---|---|
| EPI_ISL_678184 | SaudiArabia/KAUST-RIYADH1239/20[illegible] | 2020-07-29 | Asia/Saudi Arabia/Riyadh/ | Pathogen Genomics Lab King Abdullah University of Science and Technology(KAUST) | Pathogen Genomics Lab King Abdullah University of Science and Technology(KAUST) | Muhammad Shuaib et al |
| EPI_ISL_678189 | SaudiArabia/KAUST-RIYADH1181/20[illegible] | 2020-06-15 | Asia/Saudi Arabia/Riyadh/ | Pathogen Genomics Lab King Abdullah University of Science and Technology(KAUST) | Pathogen Genomics Lab King Abdullah University of Science and Technology(KAUST) | Raeece Naeem et al |
| EPI_ISL_678190 | SaudiArabia/KAUST-RIYADH1157/20[illegible] | 2020-06-15 | Asia/Saudi Arabia/Riyadh/ | Pathogen Genomics Lab King Abdullah University of Science and Technology(KAUST) | Pathogen Genomics Lab King Abdullah University of Science and Technology(KAUST) | Olga Douvropoulou et al |
| EPI_ISL_678192 | SaudiArabia/KAUST-RIYADH1171/20[illegible] | 2020-06-05 | Asia/Saudi Arabia/Riyadh/ | Pathogen Genomics Lab King Abdullah University of Science and Technology(KAUST) | Pathogen Genomics Lab King Abdullah University of Science and Technology(KAUST) | Raeece Naeem et al |
| EPI_ISL_678194 | SaudiArabia/KAUST-RIYADH1174/20[illegible] | 2020-06-05 | Asia/Saudi Arabia/Riyadh/ | Pathogen Genomics Lab King Abdullah University of Science and Technology(KAUST) | Pathogen Genomics Lab King Abdullah University of Science and Technology(KAUST) | Raeece Naeem et al |

| | | | | | | |
|---|---|---|---|---|---|---|
| EPI_ISL_678196 | SaudiArabia/KAUST-RIYADH1178/20[illegible] | 2020-06-05 | Asia/Saudi Arabia/Riyadh/ | Pathogen Genomics Lab King Abdullah University of Science and Technology(KAUST) | Pathogen Genomics Lab King Abdullah University of Science and Technology(KAUST) | Raeece Naeem et al |
| EPI_ISL_678199 | SaudiArabia/KAUST-RIYADH1192/20[illegible] | 2020-06-15 | Asia/Saudi Arabia/Riyadh/ | Pathogen Genomics Lab King Abdullah University of Science and Technology(KAUST) | Pathogen Genomics Lab King Abdullah University of Science and Technology(KAUST) | Raeece Naeem et al |
| EPI_ISL_678200 | SaudiArabia/KAUST-RIYADH1194/20[illegible] | 2020-06-15 | Asia/Saudi Arabia/Riyadh/ | Pathogen Genomics Lab King Abdullah University of Science and Technology(KAUST) | Pathogen Genomics Lab King Abdullah University of Science and Technology(KAUST) | Raushan Nugmanova et al |
| EPI_ISL_678201 | SaudiArabia/KAUST-RIYADH1195/20[illegible] | 2020-06-15 | Asia/Saudi Arabia/Riyadh/ | Pathogen Genomics Lab King Abdullah University of Science and Technology(KAUST) | Pathogen Genomics Lab King Abdullah University of Science and Technology(KAUST) | Raushan Nugmanova et al |
| EPI_ISL_678204 | SaudiArabia/KAUST-RIYADH1207/20[illegible] | 2020-06-15 | Asia/Saudi Arabia/Riyadh/ | Pathogen Genomics Lab King Abdullah University of Science and Technology(KAUST) | Pathogen Genomics Lab King Abdullah University of Science and Technology(KAUST) | Sara Mfarrej et al |

| | | | | | | |
|---|---|---|---|---|---|---|
| EPI_ISL_678206 | SaudiArabia/KAUST-RIYADH1214/20[illegible] | 2020-06-05 | Asia/Saudi Arabia/Riyadh/ | Pathogen Genomics Lab King Abdullah University of Science and Technology(KAUST) | Pathogen Genomics Lab King Abdullah University of Science and Technology(KAUST ) | Muhammad Shuaib et al |
| EPI_ISL_678207 | SaudiArabia/KAUST-RIYADH1215/20[illegible] | 2020-06-05 | Asia/Saudi Arabia/Riyadh/ | Pathogen Genomics Lab King Abdullah University of Science and Technology(KAUST) | Pathogen Genomics Lab King Abdullah University of Science and Technology(KAUST ) | Muhammad Shuaib et al |
| EPI_ISL_678208 | SaudiArabia/KAUST-RIYADH1216/20[illegible] | 2020-06-15 | Asia/Saudi Arabia/Riyadh/ | Pathogen Genomics Lab King Abdullah University of Science and Technology(KAUST) | Pathogen Genomics Lab King Abdullah University of Science and Technology(KAUST ) | Muhammad Shuaib et al |
| EPI_ISL_678210 | SaudiArabia/KAUST-RIYADH1220/20[illegible] | 2020-06-05 | Asia/Saudi Arabia/Riyadh/ | Pathogen Genomics Lab King Abdullah University of Science and Technology(KAUST) | Pathogen Genomics Lab King Abdullah University of Science and Technology(KAUST ) | Muhammad Shuaib et al |
| EPI_ISL_678211 | SaudiArabia/KAUST-RIYADH1222/20[illegible] | 2020-07-29 | Asia/Saudi Arabia/Riyadh/ | Pathogen Genomics Lab King Abdullah University of Science and Technology(KAUST) | Pathogen Genomics Lab King Abdullah University of Science and Technology(KAUST ) | Muhammad Shuaib et al |

| | | | | | | |
|---|---|---|---|---|---|---|
| EPI_ISL_678212 | SaudiArabia/KAUST-RIYADH1235/20[illegible] | 2020-08-11 | Asia/Saudi Arabia/Riyadh/ | Pathogen Genomics Lab King Abdullah University of Science and Technology(KAUST) | Pathogen Genomics Lab King Abdullah University of Science and Technology(KAUST) | Muhammad Shuaib et al |
| EPI_ISL_678213 | SaudiArabia/KAUST-RIYADH1241/20[illegible] | 2020-08-11 | Asia/Saudi Arabia/Riyadh/ | Pathogen Genomics Lab King Abdullah University of Science and Technology(KAUST) | Pathogen Genomics Lab King Abdullah University of Science and Technology(KAUST) | Muhammad Shuaib et al |
| EPI_ISL_678214 | SaudiArabia/KAUST-RIYADH1243/20[illegible] | 2020-07-29 | Asia/Saudi Arabia/Riyadh/ | Pathogen Genomics Lab King Abdullah University of Science and Technology(KAUST) | Pathogen Genomics Lab King Abdullah University of Science and Technology(KAUST) | Muhammad Shuaib et al |
| EPI_ISL_678215 | SaudiArabia/KAUST-RIYADH1249/20[illegible] | 2020-07-29 | Asia/Saudi Arabia/Riyadh/ | Pathogen Genomics Lab King Abdullah University of Science and Technology(KAUST) | Pathogen Genomics Lab King Abdullah University of Science and Technology(KAUST) | Sara Mfarrej et al |
| EPI_ISL_678216 | SaudiArabia/KAUST-RIYADH1250/20[illegible] | 2020-08-11 | Asia/Saudi Arabia/Riyadh/ | Pathogen Genomics Lab King Abdullah University of Science and Technology(KAUST) | Pathogen Genomics Lab King Abdullah University of Science and Technology(KAUST) | Sara Mfarrej et al |

| | | | | | | |
|---|---|---|---|---|---|---|
| EPI_ISL_678217 | SaudiArabia/KAUST-RIYADH1251/20 | 2020-08-11 | Asia/Saudi Arabia/Riyadh/ | Pathogen Genomics Lab King Abdullah University of Science and Technology(KAUST) | Pathogen Genomics Lab King Abdullah University of Science and Technology(KAUST ) | Sara Mfarrej et al |
| EPI_ISL_678218 | SaudiArabia/KAUST-RIYADH1254/20 | 2020-07-29 | Asia/Saudi Arabia/Riyadh/ | Pathogen Genomics Lab King Abdullah University of Science and Technology(KAUST) | Pathogen Genomics Lab King Abdullah University of Science and Technology(KAUST ) | Sara Mfarrej et al |
| EPI_ISL_678219 | SaudiArabia/KAUST-RIYADH1259/20 | 2020-07-29 | Asia/Saudi Arabia/Riyadh/ | Pathogen Genomics Lab King Abdullah University of Science and Technology(KAUST) | Pathogen Genomics Lab King Abdullah University of Science and Technology(KAUST ) | Sara Mfarrej et al |
| EPI_ISL_678220 | SaudiArabia/KAUST-RIYADH1261/20 | 2020-07-18 | Asia/Saudi Arabia/Riyadh/ | Pathogen Genomics Lab King Abdullah University of Science and Technology(KAUST) | Pathogen Genomics Lab King Abdullah University of Science and Technology(KAUST ) | Sara Mfarrej et al |
| EPI_ISL_678221 | SaudiArabia/KAUST-RIYADH1263/20 | 2020-07-18 | Asia/Saudi Arabia/Riyadh/ | Pathogen Genomics Lab King Abdullah University of Science and Technology(KAUST) | Pathogen Genomics Lab King Abdullah University of Science and Technology(KAUST ) | Sara Mfarrej et al |

| | | | | | | |
|---|---|---|---|---|---|---|
| EPI_ISL_678222 | SaudiArabia/KAUST-RIYADH1264/20 | 2020-07-18 | Asia/Saudi Arabia/Riyadh/ | Pathogen Genomics Lab King Abdullah University of Science and Technology(KAUST) | Pathogen Genomics Lab King Abdullah University of Science and Technology(KAUST) | Sara Mfarrej et al |
| EPI_ISL_678223 | SaudiArabia/KAUST-RIYADH1265/20 | 2020-07-18 | Asia/Saudi Arabia/Riyadh/ | Pathogen Genomics Lab King Abdullah University of Science and Technology(KAUST) | Pathogen Genomics Lab King Abdullah University of Science and Technology(KAUST) | Sara Mfarrej et al |
| EPI_ISL_678224 | SaudiArabia/KAUST-RIYADH1267/20 | 2020-07-18 | Asia/Saudi Arabia/Riyadh/ | Pathogen Genomics Lab King Abdullah University of Science and Technology(KAUST) | Pathogen Genomics Lab King Abdullah University of Science and Technology(KAUST) | Sara Mfarrej et al |
| EPI_ISL_678225 | SaudiArabia/KAUST-RIYADH1268/20 | 2020-07-18 | Asia/Saudi Arabia/Riyadh/ | Pathogen Genomics Lab King Abdullah University of Science and Technology(KAUST) | Pathogen Genomics Lab King Abdullah University of Science and Technology(KAUST) | Sara Mfarrej et al |
| EPI_ISL_678230 | SaudiArabia/KAUST-MAKKAH180/20 | 2020-04-06 | Asia/Saudi Arabia/Makkah/ | Pathogen Genomics Lab King Abdullah University of Science and Technology(KAUST) | Pathogen Genomics Lab King Abdullah University of Science and Technology(KAUST) | Amit Kumar Subudhi et al |

| | | | | | | |
|---|---|---|---|---|---|---|
| EPI_ISL_678234 | SaudiArabia/KAUST-MADINAH209/2 | 2020-04-01 | Asia/Saudi Arabia/Madinah/ | Pathogen Genomics Lab King Abdullah University of Science and Technology(KAUST) | Pathogen Genomics Lab King Abdullah University of Science and Technology(KAUST ) | Sharif Hala et al |
| EPI_ISL_678238 | SaudiArabia/KAUST-RIYADH1229/20 | 2020-08-11 | Asia/Saudi Arabia/Riyadh/ | Pathogen Genomics Lab King Abdullah University of Science and Technology(KAUST) | Pathogen Genomics Lab King Abdullah University of Science and Technology(KAUST ) | Muhammad Shuaib et al |
| EPI_ISL_678239 | SaudiArabia/KAUST-RIYADH1240/20 | 2020-07-29 | Asia/Saudi Arabia/Riyadh/ | Pathogen Genomics Lab King Abdullah University of Science and Technology(KAUST) | Pathogen Genomics Lab King Abdullah University of Science and Technology(KAUST ) | Muhammad Shuaib et al |
| EPI_ISL_678240 | SaudiArabia/KAUST-RIYADH1245/20 | 2020-07-29 | Asia/Saudi Arabia/Riyadh/ | Pathogen Genomics Lab King Abdullah University of Science and Technology(KAUST) | Pathogen Genomics Lab King Abdullah University of Science and Technology(KAUST ) | Sara Mfarrej et al |
| EPI_ISL_678243 | SaudiArabia/KAUST-JEDDAH1323/20 | 2020-06-30 | Asia/Saudi Arabia/Jeddah/ | Pathogen Genomics Lab King Abdullah University of Science and Technology(KAUST) | Pathogen Genomics Lab King Abdullah University of Science and Technology(KAUST ) | Sara Mfarrej et al |

| | | | | | | |
|---|---|---|---|---|---|---|
| EPI_ISL_678244 | SaudiArabia/KAUST-QATIF1354/2020 | 2020-06-15 | Asia/Saudi Arabia/Eastern Province/Qat | Pathogen Genomics Lab King Abdullah University of Science and Technology(KAUST) | Pathogen Genomics Lab King Abdullah University of Science and Technology(KAUST ) | Sara Mfarrej et al |
| EPI_ISL_678245 | SaudiArabia/KAUST-QATIF1472/2020 | 2020-06-27 | Asia/Saudi Arabia/Eastern Province/Qat | Pathogen Genomics Lab King Abdullah University of Science and Technology(KAUST) | Pathogen Genomics Lab King Abdullah University of Science and Technology(KAUST ) | Sara Mfarrej et al |
| EPI_ISL_678247 | SaudiArabia/KAUST-RIYADH1233/202 | 2020-08-11 | Asia/Saudi Arabia/Riyadh/ | Pathogen Genomics Lab King Abdullah University of Science and Technology (KAUST) | Pathogen Genomics Lab King Abdullah University of Science and Technology (KAUST) | Muhammad Shuaib et al |
| EPI_ISL_678250 | NorthMacedonia/9675/2020 | 2020-08-10 | Europe/North Macedonia/Eastern Regio | Clinical Hospital - Shtip | "Research Center for Genetic Engineering and Biotechnology ""Georgi D. Efremov,Äù , Macedonian Academy of Sciences and Arts" | RCGEB - MASA et al |
| EPI_ISL_678253 | NorthMacedonia/11567/2020 | 2020-09-07 | Europe/North Macedonia/Skopje/ | Institute for Lung Diseases in Children - Skopje | "Research Center for Genetic Engineering and Biotechnology ""Georgi D. Efremov,Äù , Macedonian Academy of Sciences and Arts" | RCGEB - MASA et al |

| | | | | | | |
|---|---|---|---|---|---|---|
| EPI_ISL_678259 | NorthMacedonia/11890/2020 | 2020-09-09 | Europe/North Macedonia/Eastern Regio | Clinical Hospital - Shtip | "Research Center for Genetic Engineering and Biotechnology ""Georgi D. Efremov,Äù , Macedonian Academy of Sciences and Arts" | RCGEB - MASA et al |
| EPI_ISL_678260 | NorthMacedonia/12026/2020 | 2020-09-11 | Europe/North Macedonia/Southwestern | General Hospital - Struga | "Research Center for Genetic Engineering and Biotechnology ""Georgi D. Efremov,Äù , Macedonian Academy of Sciences and Arts" | RCGEB - MASA et al |
| EPI_ISL_678265 | Bahrain/341016804_S1_L001/2020 | 2020-11-22 | Asia/Bahrain/Bahrain/ | Communicable Disease Laboratory, Public Health Directorate | Communicable Disease Laboratory, Public Health Directorate | Alwasti et al |
| EPI_ISL_678266 | Bahrain/341038078_S3_L001/2020 | 2020-11-21 | Asia/Bahrain/Bahrain/ | Communicable Disease Laboratory, Public Health Directorate | Communicable Disease Laboratory, Public Health Directorate | Alwasti et al |
| EPI_ISL_678269 | Bahrain/341063114_S18_L001/2020 | 2020-11-12 | Asia/Bahrain/Bahrain/ | Communicable Disease Laboratory, Public Health Directorate | Communicable Disease Laboratory, Public Health Directorate | Alwasti et al |
| EPI_ISL_678274 | Russia/SAR-RARI-3187/2020 | 2020-07-30 | Europe/Russia/Saratov/ | Mikrobiologie, RARI | Mikrobiologie, RARI | Krasnov et al |
| EPI_ISL_678275 | Russia/MOS-RARI-1054/2020 | 2020-03-18 | Europe/Russia/Moscow Oblast/ | Mikrobiologie, RARI | Mikrobiologie, RARI | Krasnov et al |
| EPI_ISL_678276 | Russia/SAM-RARI-1011/2020 | 2020-04-11 | Europe/Russia/Samara/ | Mikrobiologie, RARI | Mikrobiologie, RARI | Krasnov et al |
| EPI_ISL_678277 | Russia/SAR-RARI-3663/2020 | 2020-05-03 | Europe/Russia/Saratov/ | Mikrobiologie, RARI | Mikrobiologie, RARI | Krasnov et al |

| | | | | | | |
|---|---|---|---|---|---|---|
| EPI_ISL_678281 | Russia/SAR-RARI-3080/2020 | 2020-07-30 | Europe/Russia/Saratov/ | Mikrobiologie, RARI | Mikrobiologie, RARI | Krasnov et al |
| EPI_ISL_678282 | Russia/SAR-RARI-3656/2020 | 2020-07-31 | Europe/Russia/Saratov/ | Mikrobiologie, RARI | Mikrobiologie, RARI | Krasnov et al |
| EPI_ISL_678283 | Russia/SAR-RARI-3573/2020 | 2020-05-02 | Europe/Russia/Saratov/ | Mikrobiologie, RARI | Mikrobiologie, RARI | Krasnov et al |
| EPI_ISL_678284 | Russia/SAR-RARI-2272/2020 | 2020-07-28 | Europe/Russia/Saratov/ | Mikrobiologie, RARI | Mikrobiologie, RARI | Krasnov et al |
| EPI_ISL_678285 | Russia/SAR-RARI-3083/2020 | 2020-07-30 | Europe/Russia/Saratov/ | Mikrobiologie, RARI | Mikrobiologie, RARI | Krasnov et al |
| EPI_ISL_678286 | Russia/SAR-RARI-9895/2020 | 2020-07-22 | Europe/Russia/Saratov/ | Mikrobiologie, RARI | Mikrobiologie, RARI | Krasnov et al |
| EPI_ISL_678288 | Russia/PEN-RARI-953/2020 | 2020-04-08 | Europe/Russia/Penza/ | Mikrobiologie, RARI | Mikrobiologie, RARI | Krasnov et al |
| EPI_ISL_678289 | Russia/SAR-RARI-5473/2020 | 2020-08-11 | Europe/Russia/Saratov/ | Mikrobiologie, RARI | Mikrobiologie, RARI | Krasnov et al |
| EPI_ISL_678389 | Hungary/UD-94347/2020 | 2020-10-08 | Europe/Hungary/Borsod-Abav∫j-Zemplv | University of Debrecen, Department of Medical Microbiology | National Laboratory of Virology, Szentv°gothai Research Centre | Endre Gv°bor Tv≥th et al |
| EPI_ISL_678395 | Hungary/UD-94387/2020 | 2020-10-08 | Europe/Hungary/Borsod-Abav∫j-Zemplv | University of Debrecen, Department of Medical Microbiology | National Laboratory of Virology, Szentv°gothai Research Centre | Endre Gv°bor Tv≥th et al |
| EPI_ISL_678397 | Romania/ROSV-10281/2020 | 2020-04-25 | Europe/Romania/Suceava/ | Suceava County Emergency Hospital | """Stefan cel Mare"" University Metagenomics Lab" | Lobiuc Andrei et al |
| EPI_ISL_678410 | Belgium/ULG-10866/2020 | 2020-11-13 | Europe/Belgium/Liv®ge/ | Department of Clinical Microbiology | GIGA Medical Genomics | Keith Durkin et al |
| EPI_ISL_678417 | Belgium/ULG-10874/2020 | 2020-11-13 | Europe/Belgium/Liv®ge/ | Department of Clinical Microbiology | GIGA Medical Genomics | Keith Durkin et al |
| EPI_ISL_678420 | Belgium/ULG-10877/2020 | 2020-11-13 | Europe/Belgium/Liv®ge/ | Department of Clinical Microbiology | GIGA Medical Genomics | Keith Durkin et al |
| EPI_ISL_678425 | Belgium/ULG-10882/2020 | 2020-11-13 | Europe/Belgium/Liv®ge/ | Department of Clinical Microbiology | GIGA Medical Genomics | Keith Durkin et al |
| EPI_ISL_678428 | Belgium/ULG-10885/2020 | 2020-11-13 | Europe/Belgium/Liv®ge/ | Department of Clinical Microbiology | GIGA Medical Genomics | Keith Durkin et al |

| | | | | | | |
|---|---|---|---|---|---|---|
| EPI_ISL_678437 | Belgium/ULG-10895/2020 | 2020-11-14 | Europe/Belgium/Liv®ge/ | Department of Clinical Microbiology | GIGA Medical Genomics | Keith Durkin et al |
| EPI_ISL_678442 | Belgium/ULG-10900/2020 | 2020-11-14 | Europe/Belgium/Liv®ge/ | Department of Clinical Microbiology | GIGA Medical Genomics | Keith Durkin et al |
| EPI_ISL_678446 | Belgium/ULG-10904/2020 | 2020-11-14 | Europe/Belgium/Liv®ge/ | Department of Clinical Microbiology | GIGA Medical Genomics | Keith Durkin et al |
| EPI_ISL_678458 | Belgium/ULG-10917/2020 | 2020-11-24 | Europe/Belgium/Liv®ge/ | Department of Clinical Microbiology | GIGA Medical Genomics | Keith Durkin et al |
| EPI_ISL_678475 | Belgium/ULG-10936/2020 | 2020-11-24 | Europe/Belgium/Liv®ge/ | Department of Clinical Microbiology | GIGA Medical Genomics | Keith Durkin et al |
| EPI_ISL_678486 | Serbia/Sabac-000012108/2020 | 2020-08-20 | Europe/Serbia/Sabac/ | "Veterinary Specialized Institute ""Sabac"", Serbia" | "Veterinary Specialized Institute ""Kraljevo"", Serbia" | Vidanovic et al |
| EPI_ISL_678492 | Serbia/Sabac-000490807/2020 | 2020-07-07 | Europe/Serbia/Sabac/ | "Veterinary Specialized Institute ""Sabac"", Serbia" | "Veterinary Specialized Institute ""Kraljevo"", Serbia" | Vidanovic et al |
| EPI_ISL_678496 | France/ARA-12561/2020 | 2020-03-23 | Europe/France/Auvergne-Rhv¥ne-Alpes/ | CNR Virus des Infections Respiratoires - France SUD | CNR Virus des Infections Respiratoires - France SUD | Antonin Bal et al |
| EPI_ISL_678498 | France/ARA-15456/2020 | 2020-03-27 | Europe/France/Auvergne-Rhv¥ne-Alpes/ | CNR Virus des Infections Respiratoires - France SUD | CNR Virus des Infections Respiratoires - France SUD | Antonin Bal et al |
| EPI_ISL_678502 | France/ARA-18038/2020 | 2020-04-01 | Europe/France/Auvergne-Rhv¥ne-Alpes/ | CNR Virus des Infections Respiratoires - France SUD | CNR Virus des Infections Respiratoires - France SUD | Antonin Bal et al |
| EPI_ISL_678503 | France/ARA-18673/2020 | 2020-04-02 | Europe/France/Auvergne-Rhv¥ne-Alpes/ | CNR Virus des Infections Respiratoires - France SUD | CNR Virus des Infections Respiratoires - France SUD | Antonin Bal et al |
| EPI_ISL_678504 | France/ARA-193100/2020 | 2020-04-03 | Europe/France/Auvergne-Rhv¥ne-Alpes/ | CNR Virus des Infections Respiratoires - France SUD | CNR Virus des Infections Respiratoires - France SUD | Antonin Bal et al |

| | | | | | | |
|---|---|---|---|---|---|---|
| EPI_ISL_678507 | France/ARA-2804/2020 | 2020-04-20 | Europe/France/Auvergne-Rhv¥ne-Alpes/ | CNR Virus des Infections Respiratoires - France SUD | CNR Virus des Infections Respiratoires - France SUD | Antonin Bal et al |
| EPI_ISL_678508 | France/ARA-37861/2020 | 2020-05-15 | Europe/France/Auvergne-Rhv¥ne-Alpes/ | CNR Virus des Infections Respiratoires - France SUD | CNR Virus des Infections Respiratoires - France SUD | Antonin Bal et al |
| EPI_ISL_678514 | USA/UT-UPHL-201231865/2020 | 2020-04-05 | North America/USA/Utah/ | Utah Public Health Laboratory | Utah Public Health Laboratory | Erin Young et al |
| EPI_ISL_678516 | USA/UT-UPHL-201207200/2020 | 2020-04-05 | North America/USA/Utah/ | Utah Public Health Laboratory | Utah Public Health Laboratory | Erin Young et al |
| EPI_ISL_678534 | France/ARA-27774/2020 | 2020-04-20 | Europe/France/Auvergne-Rhv¥ne-Alpes/ | CNR Virus des Infections Respiratoires - France SUD | CNR Virus des Infections Respiratoires - France SUD | Antonin Bal et al |
| EPI_ISL_678535 | France/ARA-29004/2020 | 2020-04-22 | Europe/France/Auvergne-Rhv¥ne-Alpes/ | CNR Virus des Infections Respiratoires - France SUD | CNR Virus des Infections Respiratoires - France SUD | Antonin Bal et al |
| EPI_ISL_678538 | France/ARA-31289/2020 | 2020-04-27 | Europe/France/Auvergne-Rhv¥ne-Alpes/ | CNR Virus des Infections Respiratoires - France SUD | CNR Virus des Infections Respiratoires - France SUD | Antonin Bal et al |
| EPI_ISL_678541 | France/ARA-32910/2020 | 2020-04-30 | Europe/France/Auvergne-Rhv¥ne-Alpes/ | CNR Virus des Infections Respiratoires - France SUD | CNR Virus des Infections Respiratoires - France SUD | Antonin Bal et al |
| EPI_ISL_678542 | France/ARA-33366/2020 | 2020-05-03 | Europe/France/Auvergne-Rhv¥ne-Alpes/ | CNR Virus des Infections Respiratoires - France SUD | CNR Virus des Infections Respiratoires - France SUD | Antonin Bal et al |
| EPI_ISL_678545 | France/ARA-38660/2020 | 2020-05-18 | Europe/France/Auvergne-Rhv¥ne-Alpes/ | CNR Virus des Infections Respiratoires - France SUD | CNR Virus des Infections Respiratoires - France SUD | Antonin Bal et al |

| | | | | | | |
|---|---|---|---|---|---|---|
| EPI_ISL_678565 | SouthAfrica/KRISP-EC-K005276/2020 | 2020-11-19 | Africa/South Africa/Eastern Cape/Nelsoı | Netcare | KRISP, KZN Research Innovation and Sequencing Platform | Giandhari J et al |
| EPI_ISL_678606 | SouthAfrica/KRISP-K005313/2020 | 2020-10-29 | Africa/South Africa/KwaZulu-Natal/Ugu | NHLS-IALCH | KRISP, KZN Research Innovation and Sequencing Platform | Giandhari J et al |
| EPI_ISL_678699 | England/204580454/2020 | 2020-11-05 | Europe/United Kingdom/England/ | Respiratory Virus Unit, Microbiology Services Colindale, Public Health England | COVID-19 Genomics UK (COG-UK) Consortium | PHE Covid Sequencing Team et al |
| EPI_ISL_678760 | England/204790770/2020 | 2020-11-16 | Europe/United Kingdom/England/ | Respiratory Virus Unit, Microbiology Services Colindale, Public Health England | COVID-19 Genomics UK (COG-UK) Consortium | PHE Covid Sequencing Team et al |
| EPI_ISL_678848 | England/ALDP-B5D054/2020 | 2020-11-09 | Europe/United Kingdom/England/ | Department of Pathology, University of Cambridge | COVID-19 Genomics UK (COG-UK) Consortium | Aminu S. Jahun et al |
| EPI_ISL_678877 | England/ALDP-B5D25E/2020 | 2020-11-09 | Europe/United Kingdom/England/ | Department of Pathology, University of Cambridge | COVID-19 Genomics UK (COG-UK) Consortium | Aminu S. Jahun et al |
| EPI_ISL_679098 | England/BIRM-6A8FD/2020 | 2020-11-11 | Europe/United Kingdom/England/ | University of Birmingham | COVID-19 Genomics UK (COG-UK) Consortium | Institute of Microbiology et al |
| EPI_ISL_679108 | England/BIRM-6ACE2/2020 | 2020-11-14 | Europe/United Kingdom/England/ | University of Birmingham | COVID-19 Genomics UK (COG-UK) Consortium | Institute of Microbiology et al |
| EPI_ISL_679808 | England/MILK-B3BD73/2020 | 2020-11-01 | Europe/United Kingdom/England/ | Oxford Viromics, NDM, University of Oxford; Oxford University Hospitals; Basingstoke and North Hampshire Hospital | COVID-19 Genomics UK (COG-UK) Consortium | Tanya Golubchik et al |
| EPI_ISL_680209 | NorthernIreland/NIRE-109640/2020 | 2020-10-22 | Europe/United Kingdom/Northern Irelar | Regional Virus Laboratory, Belfast Health and Social Care Trust | COVID-19 Genomics UK (COG-UK) Consortium | Conall McCaughey et al |
| EPI_ISL_680273 | NorthernIreland/NIRE-233284/2020 | 2020-09-23 | Europe/United Kingdom/Northern Irelar | Regional Virus Laboratory, Belfast Health and Social Care Trust | COVID-19 Genomics UK (COG-UK) Consortium | Conall McCaughey et al |
| EPI_ISL_680330 | NorthernIreland/NIRE-233FC9/2020 | 2020-10-20 | Europe/United Kingdom/Northern Irelar | Regional Virus Laboratory, Belfast Health and Social Care Trust | COVID-19 Genomics UK (COG-UK) Consortium | Conall McCaughey et al |

| | | | | | | |
|---|---|---|---|---|---|---|
| EPI_ISL_680340 | NorthernIreland/NIRE-234089/2020 | 2020-10-12 | Europe/United Kingdom/Northern Irelar | Regional Virus Laboratory, Belfast Health and Social Care Trust | COVID-19 Genomics UK (COG-UK) Consortium | Conall McCaughey et al |
| EPI_ISL_680368 | NorthernIreland/NIRE-2342FC/2020 | 2020-10-15 | Europe/United Kingdom/Northern Irelar | Regional Virus Laboratory, Belfast Health and Social Care Trust | COVID-19 Genomics UK (COG-UK) Consortium | Conall McCaughey et al |
| EPI_ISL_680375 | NorthernIreland/NIRE-234371/2020 | 2020-10-15 | Europe/United Kingdom/Northern Irelar | Regional Virus Laboratory, Belfast Health and Social Care Trust | COVID-19 Genomics UK (COG-UK) Consortium | Conall McCaughey et al |
| EPI_ISL_680408 | NorthernIreland/NIRE-23458A/2020 | 2020-10-19 | Europe/United Kingdom/Northern Irelar | Regional Virus Laboratory, Belfast Health and Social Care Trust | COVID-19 Genomics UK (COG-UK) Consortium | Conall McCaughey et al |
| EPI_ISL_680497 | Scotland/EDB10196/2020 | 2020-09-17 | Europe/United Kingdom/Scotland/ | Virology Department, Royal Infirmary of Edinburgh, NHS Lothian / School of Biological Sciences, University of Edinburgh / Institute of Genetics and Molecular Medicine, University of Edinburgh | COVID-19 Genomics UK (COG-UK) Consortium | McHugh M et al |
| EPI_ISL_680644 | Wales/PHWC-48D571/2020 | 2020-11-12 | Europe/United Kingdom/Wales/ | Wales Specialist Virology Centre Sequencing lab: Pathogen Genomics Unit | COVID-19 Genomics UK (COG-UK) Consortium | Catherine Moore et al |
| EPI_ISL_681131 | Wales/PHWC-48F72E/2020 | 2020-11-10 | Europe/United Kingdom/Wales/ | Wales Specialist Virology Centre Sequencing lab: Pathogen Genomics Unit | COVID-19 Genomics UK (COG-UK) Consortium | Catherine Moore et al |
| EPI_ISL_681185 | Wales/PHWC-48FACB/2020 | 2020-11-10 | Europe/United Kingdom/Wales/ | Wales Specialist Virology Centre Sequencing lab: Pathogen Genomics Unit | COVID-19 Genomics UK (COG-UK) Consortium | Catherine Moore et al |
| EPI_ISL_681227 | Wales/PHWC-48FD95/2020 | 2020-11-11 | Europe/United Kingdom/Wales/ | Wales Specialist Virology Centre Sequencing lab: Pathogen Genomics Unit | COVID-19 Genomics UK (COG-UK) Consortium | Catherine Moore et al |
| EPI_ISL_681264 | Belgium/ULG-10634/2020 | 2020-10-26 | Europe/Belgium/Li√®ge/ | Department of Clinical Microbiology | GIGA Medical Genomics | Keith Durkin et al |
| EPI_ISL_681301 | Bahrain/920396988_S5_L001/2020 | 2020-09-11 | Asia/Bahrain/Bahrain/ | Communicable Disease Laboratory, Public Health Directorate | Communicable Disease Laboratory, Public Health Directorate | Alwasti et al |
| EPI_ISL_681320 | USA/WI-UW-1617/2020 | 2020-09-24 | North America/USA/Wisconsin/Dane Co | Pathology and Laboratory Medicine, UW-Madison | Pathology and Laboratory Medicine, UW-Madison | Moreno et al |

| | | | | | | |
|---|---|---|---|---|---|---|
| EPI_ISL_681419 | England/MILK-BAA911/2020 | 2020-11-18 | Europe/United Kingdom/England/ | Lighthouse Lab in Milton Keynes | Wellcome Sanger Institute for the COVID-19 Genomics UK (COG-UK) Consortium | The Lighthouse Lab in Milton Keynes et al |
| EPI_ISL_681653 | England/MILK-BAB066/2020 | 2020-11-18 | Europe/United Kingdom/England/ | Lighthouse Lab in Milton Keynes | Wellcome Sanger Institute for the COVID-19 Genomics UK (COG-UK) Consortium | The Lighthouse Lab in Milton Keynes et al |
| EPI_ISL_681683 | Chile/MA-CADIUMAG-17/2020 | 2020-05-04 | South America/Chile/Magallanes/Punta | Molecular Medicine Laboratory, University of Magallanes | Centro Asistencial Docente y de Investigacion, Universidad de Magallanes | Jorge Gonzv°lez et al |
| EPI_ISL_681684 | Chile/MA-CADIUMAG-38/2020 | 2020-05-28 | South America/Chile/Magallanes/Punta | Molecular Medicine Laboratory, University of Magallanes | Centro Asistencial Docente y de Investigacion, Universidad de Magallanes | Jorge Gonzv°lez et al |
| EPI_ISL_681685 | Chile/MA-CADIUMAG-37/2020 | 2020-06-17 | South America/Chile/Magallanes/Punta | Molecular Medicine Laboratory, University of Magallanes | Centro Asistencial Docente y de Investigacion, Universidad de Magallanes | Jorge Gonzv°lez et al |
| EPI_ISL_681686 | Chile/MA-CADIUMAG-18/2020 | 2020-06-18 | South America/Chile/Magallanes/Punta | Molecular Medicine Laboratory, University of Magallanes | Centro Asistencial Docente y de Investigacion, Universidad de Magallanes | Jorge Gonzv°lez et al |
| EPI_ISL_681687 | Chile/MA-CADIUMAG-22/2020 | 2020-06-20 | South America/Chile/Magallanes/Punta | Molecular Medicine Laboratory, University of Magallanes | Centro Asistencial Docente y de Investigacion, Universidad de Magallanes | Jorge Gonzv°lez et al |
| EPI_ISL_681688 | Chile/MA-CADIUMAG-23/2020 | 2020-06-20 | South America/Chile/Magallanes/Punta | Molecular Medicine Laboratory, University of Magallanes | Centro Asistencial Docente y de Investigacion, Universidad de Magallanes | Jorge Gonzv°lez et al |

| | | | | | | |
|---|---|---|---|---|---|---|
| EPI_ISL_681689 | Chile/MA-CADIUMAG-26/2020 | 2020-06-23 | South America/Chile/Magallanes/Puert | Molecular Medicine Laboratory, University of Magallanes | Centro Asistencial Docente y de Investigacion, Universidad de Magallanes | Jorge Gonzv°lez et al |
| EPI_ISL_681690 | Chile/MA-CADIUMAG-24/2020 | 2020-06-23 | South America/Chile/Magallanes/Puert | Molecular Medicine Laboratory, University of Magallanes | Centro Asistencial Docente y de Investigacion, Universidad de Magallanes | Jorge Gonzv°lez et al |
| EPI_ISL_681691 | Chile/MA-CADIUMAG-25/2020 | 2020-06-23 | South America/Chile/Magallanes/Puert | Molecular Medicine Laboratory, University of Magallanes | Centro Asistencial Docente y de Investigacion, Universidad de Magallanes | Jorge Gonzv°lez et al |
| EPI_ISL_681694 | Chile/MA-CADIUMAG-29/2020 | 2020-07-15 | South America/Chile/Magallanes/Punta | Molecular Medicine Laboratory, University of Magallanes | Centro Asistencial Docente y de Investigacion, Universidad de Magallanes | Jorge Gonzv°lez et al |
| EPI_ISL_681695 | Chile/MA-CADIUMAG-30/2020 | 2020-07-25 | South America/Chile/Magallanes/Punta | Molecular Medicine Laboratory, University of Magallanes | Centro Asistencial Docente y de Investigacion, Universidad de Magallanes | Jorge Gonzv°lez et al |
| EPI_ISL_681697 | Chile/MA-CADIUMAG-32/2020 | 2020-08-12 | South America/Chile/Magallanes/Punta | Molecular Medicine Laboratory, University of Magallanes | Centro Asistencial Docente y de Investigacion, Universidad de Magallanes | Jorge Gonzv°lez et al |
| EPI_ISL_681698 | Chile/MA-CADIUMAG-33/2020 | 2020-08-12 | South America/Chile/Magallanes/Porve | Molecular Medicine Laboratory, University of Magallanes | Centro Asistencial Docente y de Investigacion, Universidad de Magallanes | Jorge Gonzv°lez et al |
| EPI_ISL_681699 | Chile/MA-CADIUMAG-34/2020 | 2020-08-12 | South America/Chile/Magallanes/Porve | Molecular Medicine Laboratory, University of Magallanes | Centro Asistencial Docente y de Investigacion, Universidad de Magallanes | Jorge Gonzv°lez et al |

| | | | | | | |
|---|---|---|---|---|---|---|
| EPI_ISL_681702 | Ecuador/ZZ-SARS-1/2020 | 2020-03-23 | South America/Ecuador/Pichincha/Quito | Zurita & Zurita Laboratorios | Zurita & Zurita Laboratorios | Gabriela Sevillano et al |
| EPI_ISL_681703 | Ecuador/ZZ-SARS-2/2020 | 2020-04-30 | South America/Ecuador/Pichincha/Quito | Zurita & Zurita Laboratorios | Zurita & Zurita Laboratorios | Gabriela Sevillano et al |
| EPI_ISL_681705 | Ireland/CE-UHL101/2020 | 2020-05-21 | Europe/Ireland/Clare/ | University Hospital Limerick | Irish Coronavirus Sequencing Consortium - Teagasc Moorepark | Paul Cotter et al |
| EPI_ISL_681740 | Ireland/LK-UHL287/2020 | 2020-04-20 | Europe/Ireland/Limerick/Limerick City | University Hospital Limerick | Irish Coronavirus Sequencing Consortium - Teagasc Moorepark | Paul Cotter et al |
| EPI_ISL_681743 | Ireland/LK-UHL297/2020 | 2020-04-27 | Europe/Ireland/Limerick/ | University Hospital Limerick | Irish Coronavirus Sequencing Consortium - Teagasc Moorepark | Paul Cotter et al |
| EPI_ISL_681750 | Ireland/LK-UHL32/2020 | 2020-04-08 | Europe/Ireland/Limerick/Limerick City | University Hospital Limerick | Irish Coronavirus Sequencing Consortium - Teagasc Moorepark | Paul Cotter et al |
| EPI_ISL_681762 | Ireland/LK-UHL90/2020 | 2020-05-13 | Europe/Ireland/Limerick/Limerick City | University Hospital Limerick | Irish Coronavirus Sequencing Consortium - Teagasc Moorepark | Paul Cotter et al |
| EPI_ISL_681763 | Ireland/LK-UHL93/2020 | 2020-05-13 | Europe/Ireland/Limerick/Limerick City | University Hospital Limerick | Irish Coronavirus Sequencing Consortium - Teagasc Moorepark | Paul Cotter et al |

| | | | | | | |
|---|---|---|---|---|---|---|
| EPI_ISL_681782 | USA/GA-CDC-7364/2020 | 2020-10-22 | North America/USA/Georgia/ | GA Department of Public Health Laboratory | Pathogen Discovery, Respiratory Viruses Branch, Division of Viral Diseases, Centers for Disease Control and Prevention | Yan Li et al |
| EPI_ISL_681798 | USA/GA-CDC-7595/2020 | 2020-10-25 | North America/USA/Georgia/ | GA Department of Public Health Laboratory | Pathogen Discovery, Respiratory Viruses Branch, Division of Viral Diseases, Centers for Disease Control and Prevention | Yan Li et al |
| EPI_ISL_681814 | USA/GA-CDC-7799/2020 | 2020-10-27 | North America/USA/Georgia/ | GA Department of Public Health Laboratory | Pathogen Discovery, Respiratory Viruses Branch, Division of Viral Diseases, Centers for Disease Control and Prevention | Yan Li et al |
| EPI_ISL_681826 | USA/GA-CDC-7845/2020 | 2020-10-28 | North America/USA/Georgia/ | GA Department of Public Health Laboratory | Pathogen Discovery, Respiratory Viruses Branch, Division of Viral Diseases, Centers for Disease Control and Prevention | Yan Li et al |
| EPI_ISL_681845 | USA/TX-DSHS-1563/2020 | 2020-06-23 | North America/USA/Texas/Harris Count | Texas Department of State Health Services | Texas Department of State Health Services | Rashmi Tuladhar et al |

| | | | | | | |
|---|---|---|---|---|---|---|
| EPI_ISL_681846 | USA/TX-DSHS-1542/2020 | 2020-06-08 | North America/USA/Texas/El Paso Coun | Texas Department of State Health Services | Texas Department of State Health Services | Rashmi Tuladhar et al |
| EPI_ISL_681851 | USA/TX-DSHS-1538/2020 | 2020-06-08 | North America/USA/Texas/El Paso Coun | Texas Department of State Health Services | Texas Department of State Health Services | Rashmi Tuladhar et al |
| EPI_ISL_681854 | USA/TX-DSHS-1573/2020 | 2020-06-24 | North America/USA/Texas/Hidalgo Cour | Texas Department of State Health Services | Texas Department of State Health Services | Rashmi Tuladhar et al |
| EPI_ISL_681855 | USA/TX-DSHS-1571/2020 | 2020-06-24 | North America/USA/Texas/Hidalgo Cour | Texas Department of State Health Services | Texas Department of State Health Services | Rashmi Tuladhar et al |
| EPI_ISL_681856 | USA/TX-DSHS-1569/2020 | 2020-06-24 | North America/USA/Texas/Hidalgo Cour | Texas Department of State Health Services | Texas Department of State Health Services | Rashmi Tuladhar et al |
| EPI_ISL_681864 | USA/TX-DSHS-1566/2020 | 2020-06-23 | North America/USA/Texas/Harris Count | Texas Department of State Health Services | Texas Department of State Health Services | Rashmi Tuladhar et al |
| EPI_ISL_681919 | Ireland/D-NVRL-20G44042/2020 | 2020-03-28 | Europe/Ireland/Dublin/ | National Virus Reference Laboratory | Irish Coronavirus Sequencing Consortium - Teagasc Moorepark | Paul Cotter et al |
| EPI_ISL_681922 | Ireland/D-NVRL-20G41451/2020 | 2020-03-26 | Europe/Ireland/Dublin/ | National Virus Reference Laboratory | Irish Coronavirus Sequencing Consortium - Teagasc Moorepark | Paul Cotter et al |
| EPI_ISL_681924 | Ireland/WH-NVRL-20G49956/2020 | 2020-04-06 | Europe/Ireland/Westmeath/ | National Virus Reference Laboratory | Irish Coronavirus Sequencing Consortium - Teagasc Moorepark | Paul Cotter et al |
| EPI_ISL_681925 | Ireland/D-NVRL-20G43510/2020 | 2020-03-25 | Europe/Ireland/Dublin/ | National Virus Reference Laboratory | Irish Coronavirus Sequencing Consortium - Teagasc Moorepark | Paul Cotter et al |

| | | | | | | |
|---|---|---|---|---|---|---|
| EPI_ISL_681933 | USA/PA-MGEL-00445/2020 | 2020-07-25 | North America/USA/Pennsylvania/Alleg | UPMC Clinical Microbiology Laboratory | Microbial Genomic Epidemiology Laboratory, University of Pittsburgh | Mustapha M. Mustapha et al |
| EPI_ISL_681934 | USA/PA-MGEL-00446/2020 | 2020-07-24 | North America/USA/Pennsylvania/Alleg | UPMC Clinical Microbiology Laboratory | Microbial Genomic Epidemiology Laboratory, University of Pittsburgh | Mustapha M. Mustapha et al |
| EPI_ISL_681960 | USA/PA-MGEL-00204/2020 | 2020-04-10 | North America/USA/Pennsylvania/Alleg | UPMC Clinical Microbiology Laboratory | Microbial Genomic Epidemiology Laboratory, University of Pittsburgh | Mustapha M. Mustapha et al |
| EPI_ISL_681969 | USA/PA-MGEL-00221/2020 | 2020-04-13 | North America/USA/Pennsylvania/Alleg | UPMC Clinical Microbiology Laboratory | Microbial Genomic Epidemiology Laboratory, University of Pittsburgh | Mustapha M. Mustapha et al |
| EPI_ISL_681970 | USA/PA-MGEL-00223/2020 | 2020-04-13 | North America/USA/Pennsylvania/Alleg | UPMC Clinical Microbiology Laboratory | Microbial Genomic Epidemiology Laboratory, University of Pittsburgh | Mustapha M. Mustapha et al |
| EPI_ISL_681983 | USA/PA-MGEL-00246/2020 | 2020-04-19 | North America/USA/Pennsylvania/Alleg | UPMC Clinical Microbiology Laboratory | Microbial Genomic Epidemiology Laboratory, University of Pittsburgh | Mustapha M. Mustapha et al |

| | | | | | | |
|---|---|---|---|---|---|---|
| EPI_ISL_681984 | USA/PA-MGEL-00248/2020 | 2020-04-19 | North America/USA/Pennsylvania/Alleg | UPMC Clinical Microbiology Laboratory | Microbial Genomic Epidemiology Laboratory, University of Pittsburgh | Mustapha M. Mustapha et al |
| EPI_ISL_681987 | USA/PA-MGEL-00254/2020 | 2020-04-22 | North America/USA/Pennsylvania/Alleg | UPMC Clinical Microbiology Laboratory | Microbial Genomic Epidemiology Laboratory, University of Pittsburgh | Mustapha M. Mustapha et al |
| EPI_ISL_681998 | USA/PA-MGEL-00276/2020 | 2020-05-15 | North America/USA/Pennsylvania/Alleg | UPMC Clinical Microbiology Laboratory | Microbial Genomic Epidemiology Laboratory, University of Pittsburgh | Mustapha M. Mustapha et al |
| EPI_ISL_682009 | USA/PA-MGEL-00401/2020 | 2020-06-24 | North America/USA/Pennsylvania/Alleg | UPMC Clinical Microbiology Laboratory | Microbial Genomic Epidemiology Laboratory, University of Pittsburgh | Mustapha M. Mustapha et al |
| EPI_ISL_682010 | USA/PA-MGEL-00403/2020 | 2020-06-27 | North America/USA/Pennsylvania/Alleg | UPMC Clinical Microbiology Laboratory | Microbial Genomic Epidemiology Laboratory, University of Pittsburgh | Mustapha M. Mustapha et al |
| EPI_ISL_682016 | USA/PA-MGEL-00418/2020 | 2020-06-28 | North America/USA/Pennsylvania/Alleg | UPMC Clinical Microbiology Laboratory | Microbial Genomic Epidemiology Laboratory, University of Pittsburgh | Mustapha M. Mustapha et al |

| | | | | | | |
|---|---|---|---|---|---|---|
| EPI_ISL_682020 | USA/PA-MGEL-00439/2020 | 2020-07-04 | North America/USA/Pennsylvania/Alleg | UPMC Clinical Microbiology Laboratory | Microbial Genomic Epidemiology Laboratory, University of Pittsburgh | Mustapha M. Mustapha et al |
| EPI_ISL_682021 | USA/PA-MGEL-00440/2020 | 2020-07-11 | North America/USA/Pennsylvania/Alleg | UPMC Clinical Microbiology Laboratory | Microbial Genomic Epidemiology Laboratory, University of Pittsburgh | Mustapha M. Mustapha et al |
| EPI_ISL_682023 | USA/PA-MGEL-00473/2020 | 2020-08-13 | North America/USA/Pennsylvania/Alleg | UPMC Clinical Microbiology Laboratory | Microbial Genomic Epidemiology Laboratory, University of Pittsburgh | Mustapha M. Mustapha et al |
| EPI_ISL_682024 | USA/PA-MGEL-00475/2020 | 2020-08-13 | North America/USA/Pennsylvania/Alleg | UPMC Clinical Microbiology Laboratory | Microbial Genomic Epidemiology Laboratory, University of Pittsburgh | Mustapha M. Mustapha et al |
| EPI_ISL_682025 | USA/PA-MGEL-00486/2020 | 2020-07-16 | North America/USA/Pennsylvania/Alleg | UPMC Clinical Microbiology Laboratory | Microbial Genomic Epidemiology Laboratory, University of Pittsburgh | Mustapha M. Mustapha et al |
| EPI_ISL_682038 | USA/PA-MGEL-00710/2020 | 2020-09-10 | North America/USA/Pennsylvania/Alleg | UPMC Clinical Microbiology Laboratory | Microbial Genomic Epidemiology Laboratory, University of Pittsburgh | Mustapha M. Mustapha et al |

| | | | | | | |
|---|---|---|---|---|---|---|
| EPI_ISL_682043 | USA/PA-MGEL-00741/2020 | 2020-09-21 | North America/USA/Pennsylvania/Alleg | UPMC Clinical Microbiology Laboratory | Microbial Genomic Epidemiology Laboratory, University of Pittsburgh | Mustapha M. Mustapha et al |
| EPI_ISL_682046 | USA/PA-MGEL-00752/2020 | 2020-09-15 | North America/USA/Pennsylvania/Alleg | UPMC Clinical Microbiology Laboratory | Microbial Genomic Epidemiology Laboratory, University of Pittsburgh | Mustapha M. Mustapha et al |
| EPI_ISL_682048 | USA/PA-MGEL-00759/2020 | 2020-09-17 | North America/USA/Pennsylvania/Alleg | UPMC Clinical Microbiology Laboratory | Microbial Genomic Epidemiology Laboratory, University of Pittsburgh | Mustapha M. Mustapha et al |
| EPI_ISL_682049 | USA/PA-MGEL-00762/2020 | 2020-09-15 | North America/USA/Pennsylvania/Alleg | UPMC Clinical Microbiology Laboratory | Microbial Genomic Epidemiology Laboratory, University of Pittsburgh | Mustapha M. Mustapha et al |
| EPI_ISL_682050 | USA/PA-MGEL-00763/2020 | 2020-10-19 | North America/USA/Pennsylvania/Alleg | UPMC Clinical Microbiology Laboratory | Microbial Genomic Epidemiology Laboratory, University of Pittsburgh | Mustapha M. Mustapha et al |
| EPI_ISL_682051 | USA/PA-MGEL-00764/2020 | 2020-10-12 | North America/USA/Pennsylvania/Alleg | UPMC Clinical Microbiology Laboratory | Microbial Genomic Epidemiology Laboratory, University of Pittsburgh | Mustapha M. Mustapha et al |

| | | | | | | |
|---|---|---|---|---|---|---|
| EPI_ISL_682052 | USA/PA-MGEL-00765/2020 | 2020-10-19 | North America/USA/Pennsylvania/Alleg | UPMC Clinical Microbiology Laboratory | Microbial Genomic Epidemiology Laboratory, University of Pittsburgh | Mustapha M. Mustapha et al |
| EPI_ISL_682062 | USA/MD-MDH-0449/2020 | 2020-11-12 | North America/USA/Maryland/ | Maryland Public Health Laboratory | Maryland Public Health Laboratory | Maryland Department of Health Laboratories Administration et al |
| EPI_ISL_682071 | USA/MD-MDH-0460/2020 | 2020-11-11 | North America/USA/Maryland/ | Maryland Public Health Laboratory | Maryland Public Health Laboratory | Maryland Department of Health Laboratories Administration et al |
| EPI_ISL_682074 | USA/MD-MDH-0463/2020 | 2020-11-12 | North America/USA/Maryland/ | Maryland Public Health Laboratory | Maryland Public Health Laboratory | Maryland Department of Health Laboratories Administration et al |
| EPI_ISL_682078 | USA/MD-MDH-0468/2020 | 2020-11-11 | North America/USA/Maryland/ | Maryland Public Health Laboratory | Maryland Public Health Laboratory | Maryland Department of Health Laboratories Administration et al |
| EPI_ISL_682079 | USA/MI-UM-10036686754/2020 | 2020-10-29 | North America/USA/Michigan/ | University of Michigan Clinical Microbiology Laboratory | Lauring Lab, University of Michigan, Department of Microbiology and Immunology | Valesano et al |
| EPI_ISL_682087 | USA/MI-UM-10036682652/2020 | 2020-10-29 | North America/USA/Michigan/ | University of Michigan Clinical Microbiology Laboratory | Lauring Lab, University of Michigan, Department of Microbiology and Immunology | Valesano et al |
| EPI_ISL_682090 | USA/MI-UM-10036680667/2020 | 2020-10-29 | North America/USA/Michigan/ | University of Michigan Clinical Microbiology Laboratory | Lauring Lab, University of Michigan, Department of Microbiology and Immunology | Valesano et al |

| | | | | | | |
|---|---|---|---|---|---|---|
| EPI_ISL_682094 | USA/MI-UM-10036701487/2020 | 2020-10-30 | North America/USA/Michigan/ | University of Michigan Clinical Microbiology Laboratory | Lauring Lab, University of Michigan, Department of Microbiology and Immunology | Valesano et al |
| EPI_ISL_682099 | USA/MI-UM-10036686737/2020 | 2020-10-29 | North America/USA/Michigan/ | University of Michigan Clinical Microbiology Laboratory | Lauring Lab, University of Michigan, Department of Microbiology and Immunology | Valesano et al |
| EPI_ISL_682113 | USA/MI-UM-10036707756/2020 | 2020-10-30 | North America/USA/Michigan/ | University of Michigan Clinical Microbiology Laboratory | Lauring Lab, University of Michigan, Department of Microbiology and Immunology | Valesano et al |
| EPI_ISL_682123 | USA/MI-UM-10036704076/2020 | 2020-10-30 | North America/USA/Michigan/ | University of Michigan Clinical Microbiology Laboratory | Lauring Lab, University of Michigan, Department of Microbiology and Immunology | Valesano et al |
| EPI_ISL_682125 | USA/MI-UM-10036738721/2020 | 2020-11-02 | North America/USA/Michigan/ | University of Michigan Clinical Microbiology Laboratory | Lauring Lab, University of Michigan, Department of Microbiology and Immunology | Valesano et al |
| EPI_ISL_682138 | USA/MI-UM-10036711761/2020 | 2020-10-30 | North America/USA/Michigan/ | University of Michigan Clinical Microbiology Laboratory | Lauring Lab, University of Michigan, Department of Microbiology and Immunology | Valesano et al |

| | | | | | | |
|---|---|---|---|---|---|---|
| EPI_ISL_682144 | USA/MI-UM-10036721047/2020 | 2020-10-31 | North America/USA/Michigan/ | University of Michigan Clinical Microbiology Laboratory | Lauring Lab, University of Michigan, Department of Microbiology and Immunology | Valesano et al |
| EPI_ISL_682147 | USA/MI-UM-10036708302/2020 | 2020-10-30 | North America/USA/Michigan/ | University of Michigan Clinical Microbiology Laboratory | Lauring Lab, University of Michigan, Department of Microbiology and Immunology | Valesano et al |
| EPI_ISL_682153 | USA/MI-UM-10036710728/2020 | 2020-10-30 | North America/USA/Michigan/ | University of Michigan Clinical Microbiology Laboratory | Lauring Lab, University of Michigan, Department of Microbiology and Immunology | Valesano et al |
| EPI_ISL_682155 | USA/MI-UM-10036721859/2020 | 2020-10-31 | North America/USA/Michigan/ | University of Michigan Clinical Microbiology Laboratory | Lauring Lab, University of Michigan, Department of Microbiology and Immunology | Valesano et al |
| EPI_ISL_682167 | USA/MI-UM-10036742402/2020 | 2020-11-02 | North America/USA/Michigan/ | University of Michigan Clinical Microbiology Laboratory | Lauring Lab, University of Michigan, Department of Microbiology and Immunology | Valesano et al |
| EPI_ISL_682175 | USA/MI-UM-10036756404/2020 | 2020-11-03 | North America/USA/Michigan/ | University of Michigan Clinical Microbiology Laboratory | Lauring Lab, University of Michigan, Department of Microbiology and Immunology | Valesano et al |

| | | | | | | |
|---|---|---|---|---|---|---|
| EPI_ISL_682176 | USA/MI-UM-10036756586/2020 | 2020-11-03 | North America/USA/Michigan/ | University of Michigan Clinical Microbiology Laboratory | Lauring Lab, University of Michigan, Department of Microbiology and Immunology | Valesano et al |
| EPI_ISL_682191 | USA/MI-UM-10036743071/2020 | 2020-11-02 | North America/USA/Michigan/ | University of Michigan Clinical Microbiology Laboratory | Lauring Lab, University of Michigan, Department of Microbiology and Immunology | Valesano et al |
| EPI_ISL_682192 | USA/MI-UM-10036746277/2020 | 2020-11-02 | North America/USA/Michigan/ | University of Michigan Clinical Microbiology Laboratory | Lauring Lab, University of Michigan, Department of Microbiology and Immunology | Valesano et al |
| EPI_ISL_682193 | USA/MI-UM-10036755231/2020 | 2020-11-03 | North America/USA/Michigan/ | University of Michigan Clinical Microbiology Laboratory | Lauring Lab, University of Michigan, Department of Microbiology and Immunology | Valesano et al |
| EPI_ISL_682199 | USA/MI-UM-10036787296/2020 | 2020-11-04 | North America/USA/Michigan/ | University of Michigan Clinical Microbiology Laboratory | Lauring Lab, University of Michigan, Department of Microbiology and Immunology | Valesano et al |
| EPI_ISL_682200 | USA/MI-UM-10036767162/2020 | 2020-11-03 | North America/USA/Michigan/ | University of Michigan Clinical Microbiology Laboratory | Lauring Lab, University of Michigan, Department of Microbiology and Immunology | Valesano et al |

| | | | | | | |
|---|---|---|---|---|---|---|
| EPI_ISL_682201 | USA/MI-UM-10036754004/2020 | 2020-11-03 | North America/USA/Michigan/ | University of Michigan Clinical Microbiology Laboratory | Lauring Lab, University of Michigan, Department of Microbiology and Immunology | Valesano et al |
| EPI_ISL_682210 | USA/MI-UM-10036762912/2020 | 2020-11-03 | North America/USA/Michigan/ | University of Michigan Clinical Microbiology Laboratory | Lauring Lab, University of Michigan, Department of Microbiology and Immunology | Valesano et al |
| EPI_ISL_682211 | USA/MI-UM-10036758403/2020 | 2020-11-03 | North America/USA/Michigan/ | University of Michigan Clinical Microbiology Laboratory | Lauring Lab, University of Michigan, Department of Microbiology and Immunology | Valesano et al |
| EPI_ISL_682219 | USA/MI-UM-10036760166/2020 | 2020-11-03 | North America/USA/Michigan/ | University of Michigan Clinical Microbiology Laboratory | Lauring Lab, University of Michigan, Department of Microbiology and Immunology | Valesano et al |
| EPI_ISL_682223 | USA/MI-UM-10036762637/2020 | 2020-11-03 | North America/USA/Michigan/ | University of Michigan Clinical Microbiology Laboratory | Lauring Lab, University of Michigan, Department of Microbiology and Immunology | Valesano et al |
| EPI_ISL_682228 | USA/MI-UM-10036765841/2020 | 2020-11-03 | North America/USA/Michigan/ | University of Michigan Clinical Microbiology Laboratory | Lauring Lab, University of Michigan, Department of Microbiology and Immunology | Valesano et al |

| | | | | | | |
|---|---|---|---|---|---|---|
| EPI_ISL_682231 | USA/MI-UM-10036763086/2020 | 2020-11-03 | North America/USA/Michigan/ | University of Michigan Clinical Microbiology Laboratory | Lauring Lab, University of Michigan, Department of Microbiology and Immunology | Valesano et al |
| EPI_ISL_682238 | CostaRica/INC-0096/2020 | 2020-07-03 | North America/Costa Rica/Guanacaste/L | AREA DE SALUD LA CRUZ | Inciensa, Instituto Costarricense de Investigaci√≥n y Ense√±anza en Nutrici√≥n y Salud | Francisco Duarte et al |
| EPI_ISL_682241 | CostaRica/INC-0100/2020 | 2020-07-12 | North America/Costa Rica/Cartago CR/C | HOSPITAL SAN JUAN DE DIOS | Inciensa, Instituto Costarricense de Investigaci√≥n y Ense√±anza en Nutrici√≥n y Salud | Francisco Duarte et al |
| EPI_ISL_682242 | CostaRica/INC-0101/2020 | 2020-07-13 | North America/Costa Rica/Heredia/ | HOSPITAL SAN VICENTE DE PAUL | Inciensa, Instituto Costarricense de Investigaci√≥n y Ense√±anza en Nutrici√≥n y Salud | Francisco Duarte et al |
| EPI_ISL_682244 | CostaRica/INC-0103/2020 | 2020-07-15 | North America/Costa Rica/Alajuela/San | AREA DE SALUD LA CRUZ | Inciensa, Instituto Costarricense de Investigaci√≥n y Ense√±anza en Nutrici√≥n y Salud | Francisco Duarte et al |
| EPI_ISL_682245 | CostaRica/INC-0104/2020 | 2020-07-14 | North America/Costa Rica/San Jose/San | HOSPITAL SAN JUAN DE DIOS | Inciensa, Instituto Costarricense de Investigaci√≥n y Ense√±anza en Nutrici√≥n y Salud | Francisco Duarte et al |
| EPI_ISL_682248 | CostaRica/INC-0108/2020 | 2020-07-16 | North America/Costa Rica/Alajuela/Que | AREA DE SALUD CIUDAD QUESADA | Inciensa, Instituto Costarricense de Investigaci√≥n y Ense√±anza en Nutrici√≥n y Salud | Francisco Duarte et al |
| EPI_ISL_682262 | CostaRica/INC-0123/2020 | 2020-08-02 | North America/Costa Rica/San Jose/ | HOSPITAL SAN JUAN DE DIOS | Inciensa, Instituto Costarricense de Investigaci√≥n y Ense√±anza en Nutrici√≥n y Salud | Francisco Duarte et al |

| | | | | | | |
|---|---|---|---|---|---|---|
| EPI_ISL_682265 | CostaRica/INC-0126/2020 | 2020-08-07 | North America/Costa Rica/Alajuela/ | HOSPITAL UPALA | Inciensa, Instituto Costarricense de Investigaci√≥n y Ense√±anza en Nutrici√≥n y Salud | Francisco Duarte et al |
| EPI_ISL_682266 | CostaRica/INC-0127/2020 | 2020-08-06 | North America/Costa Rica/San Jose/Pica | HOSPITAL SAN JUAN DE DIOS | Inciensa, Instituto Costarricense de Investigaci√≥n y Ense√±anza en Nutrici√≥n y Salud | Francisco Duarte et al |
| EPI_ISL_682267 | CostaRica/INC-0128/2020 | 2020-08-08 | North America/Costa Rica/Limon/ | HOSPITAL SAN JUAN DE DIOS | Inciensa, Instituto Costarricense de Investigaci√≥n y Ense√±anza en Nutrici√≥n y Salud | Francisco Duarte et al |
| EPI_ISL_682268 | CostaRica/INC-0129/2020 | 2020-08-08 | North America/Costa Rica/San Jose/San | HOSPITAL SAN JUAN DE DIOS | Inciensa, Instituto Costarricense de Investigaci√≥n y Ense√±anza en Nutrici√≥n y Salud | Francisco Duarte et al |
| EPI_ISL_682269 | CostaRica/INC-0130/2020 | 2020-08-11 | North America/Costa Rica/San Jose/Los | HOSPITAL SAN JUAN DE DIOS | Inciensa, Instituto Costarricense de Investigaci√≥n y Ense√±anza en Nutrici√≥n y Salud | Francisco Duarte et al |
| EPI_ISL_682270 | CostaRica/INC-0131/2020 | 2020-08-10 | North America/Costa Rica/Cartago CR/L | HOSPITAL SAN JUAN DE DIOS | Inciensa, Instituto Costarricense de Investigaci√≥n y Ense√±anza en Nutrici√≥n y Salud | Francisco Duarte et al |
| EPI_ISL_682271 | CostaRica/INC-0132/2020 | 2020-08-10 | North America/Costa Rica/Cartago CR/S | HOSPITAL SAN JUAN DE DIOS | Inciensa, Instituto Costarricense de Investigaci√≥n y Ense√±anza en Nutrici√≥n y Salud | Francisco Duarte et al |
| EPI_ISL_682272 | CostaRica/INC-0133/2020 | 2020-08-13 | North America/Costa Rica/Cartago CR/Ll | HOSPITAL SAN JUAN DE DIOS | Inciensa, Instituto Costarricense de Investigaci√≥n y Ense√±anza en Nutrici√≥n y Salud | Francisco Duarte et al |

| | | | | | | |
|---|---|---|---|---|---|---|
| EPI_ISL_682273 | CostaRica/INC-0135/2020 | 2020-08-11 | North America/Costa Rica/Limon/ | HOSPITAL SAN JUAN DE DIOS | Inciensa, Instituto Costarricense de Investigaci√≥n y Ense√±anza en Nutrici√≥n y Salud | Francisco Duarte et al |
| EPI_ISL_682274 | CostaRica/INC-0136/2020 | 2020-08-13 | North America/Costa Rica/San Jose/ | HOSPITAL SAN JUAN DE DIOS | Inciensa, Instituto Costarricense de Investigaci√≥n y Ense√±anza en Nutrici√≥n y Salud | Francisco Duarte et al |
| EPI_ISL_682275 | CostaRica/INC-0137/2020 | 2020-08-17 | North America/Costa Rica/San Jose/Zap | LABORATORIOS LABIN | Inciensa, Instituto Costarricense de Investigaci√≥n y Ense√±anza en Nutrici√≥n y Salud | Francisco Duarte et al |
| EPI_ISL_682276 | CostaRica/INC-0138/2020 | 2020-08-17 | North America/Costa Rica/Heredia/San | LABORATORIOS LABIN | Inciensa, Instituto Costarricense de Investigaci√≥n y Ense√±anza en Nutrici√≥n y Salud | Francisco Duarte et al |
| EPI_ISL_682277 | CostaRica/INC-0139/2020 | 2020-08-17 | North America/Costa Rica/San Jose/San | LABORATORIOS LABIN | Inciensa, Instituto Costarricense de Investigaci√≥n y Ense√±anza en Nutrici√≥n y Salud | Francisco Duarte et al |
| EPI_ISL_682300 | Bahrain/340685804/2020 | 2020-09-14 | Asia/Bahrain/Bahrain/ | Communicable Disease Laboratory, Public Health Directorate | Communicable Disease Laboratory, Public Health Directorate | Alwasti et al |
| EPI_ISL_682303 | Bahrain/920407807/2020 | 2020-09-17 | Asia/Bahrain/Bahrain/ | Communicable Disease Laboratory, Public Health Directorate | Communicable Disease Laboratory, Public Health Directorate | Alwasti et al |
| EPI_ISL_682308 | Bahrain/920453107/2020 | 2020-10-11 | Asia/Bahrain/Bahrain/ | Communicable Disease Laboratory, Public Health Directorate | Communicable Disease Laboratory, Public Health Directorate | Alwasti et al |

| | | | | | | |
|---|---|---|---|---|---|---|
| EPI_ISL_682329 | SouthAfrica/UFS-VIRO-NGS-20/2020 | 2020-10-09 | Africa/South Africa/Free State/ | NHLS Universitas Academic | UFS Virology | PA Bester et al |
| EPI_ISL_682330 | SouthAfrica/UFS-VIRO-NGS-22/2020 | 2020-09-30 | Africa/South Africa/Free State/ | NHLS Universitas Academic | UFS Virology | PA Bester et al |
| EPI_ISL_682342 | SouthAfrica/UFS-VIRO-NGS-35/2020 | 2020-09-29 | Africa/South Africa/Free State/ | NHLS Universitas Academic | UFS Virology | PA Bester et al |
| EPI_ISL_683000 | Denmark/DCGC-12819/2020 | 2020-10-26 | Europe/Denmark/Midtjylland/ | Department of Virus and Microbiological Special Diagnostics, Statens Serum Institut, Copenhagen, Denmark | Albertsen Lab, Department of Chemistry and Bioscience, Aalborg University, Denmark | Danish Covid-19 Genome Consortium et al |
| EPI_ISL_683003 | Denmark/DCGC-12822/2020 | 2020-10-19 | Europe/Denmark/Nordjylland/ | Department of Virus and Microbiological Special Diagnostics, Statens Serum Institut, Copenhagen, Denmark | Albertsen Lab, Department of Chemistry and Bioscience, Aalborg University, Denmark | Danish Covid-19 Genome Consortium et al |
| EPI_ISL_683331 | France/ARA-SC011/2020 | 2020-03-02 | Europe/France/Auvergne-Rhv¥ne-Alpes/ | Laboratoire du Centre Hospitalier Annecy Genevois | CNR Virus des Infections Respiratoires - France SUD | Antonin Bal et al |
| EPI_ISL_683334 | France/ARA-100617/2020 | 2020-11-03 | Europe/France/Auvergne-Rhv¥ne-Alpes/ | CNR Virus des Infections Respiratoires - France SUD | CNR Virus des Infections Respiratoires - France SUD | Antonin Bal et al |
| EPI_ISL_683336 | France/ARA-101082/2020 | 2020-11-04 | Europe/France/Auvergne-Rhv¥ne-Alpes/ | CNR Virus des Infections Respiratoires - France SUD | CNR Virus des Infections Respiratoires - France SUD | Antonin Bal et al |
| EPI_ISL_683338 | France/ARA-101325/2020 | 2020-11-04 | Europe/France/Auvergne-Rhv¥ne-Alpes/ | CNR Virus des Infections Respiratoires - France SUD | CNR Virus des Infections Respiratoires - France SUD | Antonin Bal et al |
| EPI_ISL_683340 | France/ARA-102608/2020 | 2020-11-06 | Europe/France/Auvergne-Rhv¥ne-Alpes/ | CNR Virus des Infections Respiratoires - France SUD | CNR Virus des Infections Respiratoires - France SUD | Antonin Bal et al |

| | | | | | | |
|---|---|---|---|---|---|---|
| EPI_ISL_683342 | France/ARA-103182/2020 | 2020-11-06 | Europe/France/Auvergne-Rh√¥ne-Alpes/ | CNR Virus des Infections Respiratoires - France SUD | CNR Virus des Infections Respiratoires - France SUD | Antonin Bal et al |
| EPI_ISL_683343 | France/ARA-103382/2020 | 2020-11-09 | Europe/France/Auvergne-Rh√¥ne-Alpes/ | CNR Virus des Infections Respiratoires - France SUD | CNR Virus des Infections Respiratoires - France SUD | Antonin Bal et al |
| EPI_ISL_683345 | France/ARA-103906/2020 | 2020-11-09 | Europe/France/Auvergne-Rh√¥ne-Alpes/ | CNR Virus des Infections Respiratoires - France SUD | CNR Virus des Infections Respiratoires - France SUD | Antonin Bal et al |
| EPI_ISL_683348 | France/ARA-104202/2020 | 2020-11-09 | Europe/France/Auvergne-Rh√¥ne-Alpes/ | CNR Virus des Infections Respiratoires - France SUD | CNR Virus des Infections Respiratoires - France SUD | Antonin Bal et al |
| EPI_ISL_683349 | France/ARA-104314/2020 | 2020-11-09 | Europe/France/Auvergne-Rh√¥ne-Alpes/ | CNR Virus des Infections Respiratoires - France SUD | CNR Virus des Infections Respiratoires - France SUD | Antonin Bal et al |
| EPI_ISL_683350 | France/ARA-104350/2020 | 2020-11-09 | Europe/France/Auvergne-Rh√¥ne-Alpes/ | CNR Virus des Infections Respiratoires - France SUD | CNR Virus des Infections Respiratoires - France SUD | Antonin Bal et al |
| EPI_ISL_683351 | France/ARA-104306/2020 | 2020-11-09 | Europe/France/Auvergne-Rh√¥ne-Alpes/ | CNR Virus des Infections Respiratoires - France SUD | CNR Virus des Infections Respiratoires - France SUD | Antonin Bal et al |
| EPI_ISL_683353 | France/ARA-38548/2020 | 2020-05-18 | Europe/France/Auvergne-Rh√¥ne-Alpes/ | CNR Virus des Infections Respiratoires - France SUD | CNR Virus des Infections Respiratoires - France SUD | Antonin Bal et al |
| EPI_ISL_683358 | France/ARA-40331/2020 | 2020-05-24 | Europe/France/Auvergne-Rh√¥ne-Alpes/ | CNR Virus des Infections Respiratoires - France SUD | CNR Virus des Infections Respiratoires - France SUD | Antonin Bal et al |
| EPI_ISL_683359 | France/ARA-40765/2020 | 2020-05-25 | Europe/France/Auvergne-Rh√¥ne-Alpes/ | CNR Virus des Infections Respiratoires - France SUD | CNR Virus des Infections Respiratoires - France SUD | Antonin Bal et al |

| | | | | | | |
|---|---|---|---|---|---|---|
| EPI_ISL_683362 | France/ARA-41359/2020 | 2020-05-27 | Europe/France/Auvergne-Rh√¥ne-Alpes/ | CNR Virus des Infections Respiratoires - France SUD | CNR Virus des Infections Respiratoires - France SUD | Antonin Bal et al |
| EPI_ISL_683367 | France/ARA-59557/2020 | 2020-08-10 | Europe/France/Auvergne-Rh√¥ne-Alpes/ | CNR Virus des Infections Respiratoires - France SUD | CNR Virus des Infections Respiratoires - France SUD | Antonin Bal et al |
| EPI_ISL_683373 | France/ARA-60758/2020 | 2020-08-14 | Europe/France/Auvergne-Rh√¥ne-Alpes/ | CNR Virus des Infections Respiratoires - France SUD | CNR Virus des Infections Respiratoires - France SUD | Antonin Bal et al |
| EPI_ISL_683375 | France/ARA-62546/2020 | 2020-08-20 | Europe/France/Auvergne-Rh√¥ne-Alpes/ | CNR Virus des Infections Respiratoires - France SUD | CNR Virus des Infections Respiratoires - France SUD | Antonin Bal et al |
| EPI_ISL_683376 | France/ARA-62614/2020 | 2020-08-21 | Europe/France/Auvergne-Rh√¥ne-Alpes/ | CNR Virus des Infections Respiratoires - France SUD | CNR Virus des Infections Respiratoires - France SUD | Antonin Bal et al |
| EPI_ISL_683378 | France/ARA-63944/2020 | 2020-08-25 | Europe/France/Auvergne-Rh√¥ne-Alpes/ | CNR Virus des Infections Respiratoires - France SUD | CNR Virus des Infections Respiratoires - France SUD | Antonin Bal et al |
| EPI_ISL_683384 | France/ARA-66769/2020 | 2020-09-02 | Europe/France/Auvergne-Rh√¥ne-Alpes/ | CNR Virus des Infections Respiratoires - France SUD | CNR Virus des Infections Respiratoires - France SUD | Antonin Bal et al |
| EPI_ISL_683386 | France/ARA-67638/2020 | 2020-09-04 | Europe/France/Auvergne-Rh√¥ne-Alpes/ | CNR Virus des Infections Respiratoires - France SUD | CNR Virus des Infections Respiratoires - France SUD | Antonin Bal et al |
| EPI_ISL_683388 | France/ARA-67665/2020 | 2020-09-04 | Europe/France/Auvergne-Rh√¥ne-Alpes/ | CNR Virus des Infections Respiratoires - France SUD | CNR Virus des Infections Respiratoires - France SUD | Antonin Bal et al |
| EPI_ISL_683393 | France/ARA-68071/2020 | 2020-09-04 | Europe/France/Auvergne-Rh√¥ne-Alpes/ | CNR Virus des Infections Respiratoires - France SUD | CNR Virus des Infections Respiratoires - France SUD | Antonin Bal et al |

| | | | | | | |
|---|---|---|---|---|---|---|
| EPI_ISL_683394 | France/ARA-68072/2020 | 2020-09-04 | Europe/France/Auvergne-Rhv¥ne-Alpes/ | CNR Virus des Infections Respiratoires - France SUD | CNR Virus des Infections Respiratoires - France SUD | Antonin Bal et al |
| EPI_ISL_683395 | France/ARA-68073/2020 | 2020-09-04 | Europe/France/Auvergne-Rhv¥ne-Alpes/ | CNR Virus des Infections Respiratoires - France SUD | CNR Virus des Infections Respiratoires - France SUD | Antonin Bal et al |
| EPI_ISL_683396 | France/ARA-68082/2020 | 2020-09-04 | Europe/France/Auvergne-Rhv¥ne-Alpes/ | CNR Virus des Infections Respiratoires - France SUD | CNR Virus des Infections Respiratoires - France SUD | Antonin Bal et al |
| EPI_ISL_683399 | France/ARA-75260/2020 | 2020-09-19 | Europe/France/Auvergne-Rhv¥ne-Alpes/ | CNR Virus des Infections Respiratoires - France SUD | CNR Virus des Infections Respiratoires - France SUD | Antonin Bal et al |
| EPI_ISL_683400 | France/ARA-98691/2020 | 2020-11-01 | Europe/France/Auvergne-Rhv¥ne-Alpes/ | CNR Virus des Infections Respiratoires - France SUD | CNR Virus des Infections Respiratoires - France SUD | Antonin Bal et al |
| EPI_ISL_683402 | France/ARA-99504/2020 | 2020-11-02 | Europe/France/Auvergne-Rhv¥ne-Alpes/ | CNR Virus des Infections Respiratoires - France SUD | CNR Virus des Infections Respiratoires - France SUD | Antonin Bal et al |
| EPI_ISL_683421 | USA/TX-DSHS-1506/2020 | 2020-06-22 | North America/USA/Texas/Hidalgo Coun | Texas Department of State Health Services | Texas Department of State Health Services | Rashmi Tuladhar et al |
| EPI_ISL_683423 | USA/TX-DSHS-1508/2020 | 2020-06-22 | North America/USA/Texas/Hidalgo Coun | Texas Department of State Health Services | Texas Department of State Health Services | Rashmi Tuladhar et al |
| EPI_ISL_683427 | USA/TX-DSHS-1512/2020 | 2020-06-22 | North America/USA/Texas/Hidalgo Coun | Texas Department of State Health Services | Texas Department of State Health Services | Rashmi Tuladhar et al |
| EPI_ISL_683444 | USA/TX-DSHS-1530/2020 | 2020-06-23 | North America/USA/Texas/Harris Count | Texas Department of State Health Services | Texas Department of State Health Services | Rashmi Tuladhar et al |

| | | | | | | |
|---|---|---|---|---|---|---|
| EPI_ISL_683598 | Spain/CT-IBV-97010156/2020 | 2020-11-09 | Europe/Spain/Catalunya/Alella | Servicio de Microbiolog√≠a, Laboratori Cl√≠nic Metropolitana Nord. Hospital Universitari Germans Trias i Pujol. Institut d,ÄôInvestigaci√≥ en Ci√®ncies de la Salut Germans Trias i Pujol (IGTP) | SeqCOVID-SPAIN consortium/IBV(CSIC) | Elisa Martr√≥ et al |
| EPI_ISL_683602 | Spain/CT-IBV-97009573/2020 | 2020-09-17 | Europe/Spain/Catalunya/Badalona | Servicio de Microbiolog√≠a, Laboratori Cl√≠nic Metropolitana Nord. Hospital Universitari Germans Trias i Pujol. Institut d,ÄôInvestigaci√≥ en Ci√®ncies de la Salut Germans Trias i Pujol (IGTP) | SeqCOVID-SPAIN consortium/IBV(CSIC) | Elisa Martr√≥ et al |
| EPI_ISL_683603 | Spain/CT-IBV-97009575/2020 | 2020-09-16 | Europe/Spain/Catalunya/Badalona | Servicio de Microbiolog√≠a, Laboratori Cl√≠nic Metropolitana Nord. Hospital Universitari Germans Trias i Pujol. Institut d,ÄôInvestigaci√≥ en Ci√®ncies de la Salut Germans Trias i Pujol (IGTP) | SeqCOVID-SPAIN consortium/IBV(CSIC) | Elisa Martr√≥ et al |
| EPI_ISL_683605 | Spain/CT-IBV-97009585/2020 | 2020-08-17 | Europe/Spain/Catalunya/Badalona | Servicio de Microbiolog√≠a, Laboratori Cl√≠nic Metropolitana Nord. Hospital Universitari Germans Trias i Pujol. Institut d,ÄôInvestigaci√≥ en Ci√®ncies de la Salut Germans Trias i Pujol (IGTP) | SeqCOVID-SPAIN consortium/IBV(CSIC) | Elisa Martr√≥ et al |
| EPI_ISL_683607 | Spain/CT-IBV-97010109/2020 | 2020-11-10 | Europe/Spain/Catalunya/Badalona | Servicio de Microbiolog√≠a, Laboratori Cl√≠nic Metropolitana Nord. Hospital Universitari Germans Trias i Pujol. Institut d,ÄôInvestigaci√≥ en Ci√®ncies de la Salut Germans Trias i Pujol (IGTP) | SeqCOVID-SPAIN consortium/IBV(CSIC) | Elisa Martr√≥ et al |
| EPI_ISL_683609 | Spain/CT-IBV-97010121/2020 | 2020-11-16 | Europe/Spain/Catalunya/Badalona | Servicio de Microbiolog√≠a, Laboratori Cl√≠nic Metropolitana Nord. Hospital Universitari Germans Trias i Pujol. Institut d,ÄôInvestigaci√≥ en Ci√®ncies de la Salut Germans Trias i Pujol (IGTP) | SeqCOVID-SPAIN consortium/IBV(CSIC) | Elisa Martr√≥ et al |
| EPI_ISL_683610 | Spain/CT-IBV-97010132/2020 | 2020-11-14 | Europe/Spain/Catalunya/Badalona | Servicio de Microbiolog√≠a, Laboratori Cl√≠nic Metropolitana Nord. Hospital Universitari Germans Trias i Pujol. Institut d,ÄôInvestigaci√≥ en Ci√®ncies de la Salut Germans Trias i Pujol (IGTP) | SeqCOVID-SPAIN consortium/IBV(CSIC) | Elisa Martr√≥ et al |
| EPI_ISL_683611 | Spain/CT-IBV-97010137/2020 | 2020-11-03 | Europe/Spain/Catalunya/Badalona | Servicio de Microbiolog√≠a, Laboratori Cl√≠nic Metropolitana Nord. Hospital Universitari Germans Trias i Pujol. Institut d,ÄôInvestigaci√≥ en Ci√®ncies de la Salut Germans Trias i Pujol (IGTP) | SeqCOVID-SPAIN consortium/IBV(CSIC) | Elisa Martr√≥ et al |

| | | | | | | |
|---|---|---|---|---|---|---|
| EPI_ISL_683612 | Spain/CT-IBV-97010142/2020 | 2020-11-03 | Europe/Spain/Catalunya/Badalona | Servicio de Microbiolog√≠a, Laboratori Cl√≠nic Metropolitana Nord. Hospital Universitari Germans Trias i Pujol. Institut d‚ÄôInvestigaci√≥ en Ci√®ncies de la Salut Germans Trias i Pujol (IGTP) | SeqCOVID-SPAIN consortium/IBV(CSIC) | Elisa Martr√≥ et al |
| EPI_ISL_683615 | Spain/CT-IBV-97010146/2020 | 2020-11-03 | Europe/Spain/Catalunya/Badalona | Servicio de Microbiolog√≠a, Laboratori Cl√≠nic Metropolitana Nord. Hospital Universitari Germans Trias i Pujol. Institut d‚ÄôInvestigaci√≥ en Ci√®ncies de la Salut Germans Trias i Pujol (IGTP) | SeqCOVID-SPAIN consortium/IBV(CSIC) | Elisa Martr√≥ et al |
| EPI_ISL_683617 | Spain/CT-IBV-97010149/2020 | 2020-11-03 | Europe/Spain/Catalunya/Badalona | Servicio de Microbiolog√≠a, Laboratori Cl√≠nic Metropolitana Nord. Hospital Universitari Germans Trias i Pujol. Institut d‚ÄôInvestigaci√≥ en Ci√®ncies de la Salut Germans Trias i Pujol (IGTP) | SeqCOVID-SPAIN consortium/IBV(CSIC) | Elisa Martr√≥ et al |
| EPI_ISL_683619 | Spain/CT-IBV-97010153/2020 | 2020-11-06 | Europe/Spain/Catalunya/Badalona | Servicio de Microbiolog√≠a, Laboratori Cl√≠nic Metropolitana Nord. Hospital Universitari Germans Trias i Pujol. Institut d‚ÄôInvestigaci√≥ en Ci√®ncies de la Salut Germans Trias i Pujol (IGTP) | SeqCOVID-SPAIN consortium/IBV(CSIC) | Elisa Martr√≥ et al |
| EPI_ISL_683620 | Spain/CT-IBV-97010157/2020 | 2020-11-04 | Europe/Spain/Catalunya/Badalona | Servicio de Microbiolog√≠a, Laboratori Cl√≠nic Metropolitana Nord. Hospital Universitari Germans Trias i Pujol. Institut d‚ÄôInvestigaci√≥ en Ci√®ncies de la Salut Germans Trias i Pujol (IGTP) | SeqCOVID-SPAIN consortium/IBV(CSIC) | Elisa Martr√≥ et al |
| EPI_ISL_683621 | Spain/CT-IBV-97010161/2020 | 2020-11-09 | Europe/Spain/Catalunya/Badalona | Servicio de Microbiolog√≠a, Laboratori Cl√≠nic Metropolitana Nord. Hospital Universitari Germans Trias i Pujol. Institut d‚ÄôInvestigaci√≥ en Ci√®ncies de la Salut Germans Trias i Pujol (IGTP) | SeqCOVID-SPAIN consortium/IBV(CSIC) | Elisa Martr√≥ et al |
| EPI_ISL_683622 | Spain/CT-IBV-97010120/2020 | 2020-07-22 | Europe/Spain/Catalunya/Barcelona | Servicio de Microbiolog√≠a, Laboratori Cl√≠nic Metropolitana Nord. Hospital Universitari Germans Trias i Pujol. Institut d‚ÄôInvestigaci√≥ en Ci√®ncies de la Salut Germans Trias i Pujol (IGTP) | SeqCOVID-SPAIN consortium/IBV(CSIC) | Elisa Martr√≥ et al |
| EPI_ISL_683623 | Spain/CT-IBV-97010117/2020 | 2020-11-16 | Europe/Spain/Catalunya/Calella | Servicio de Microbiolog√≠a, Laboratori Cl√≠nic Metropolitana Nord. Hospital Universitari Germans Trias i Pujol. Institut d‚ÄôInvestigaci√≥ en Ci√®ncies de la Salut Germans Trias i Pujol (IGTP) | SeqCOVID-SPAIN consortium/IBV(CSIC) | Elisa Martr√≥ et al |

| | | | | | | |
|---|---|---|---|---|---|---|
| EPI_ISL_683625 | Spain/CT-IBV-97010126/2020 | 2020-11-11 | Europe/Spain/Catalunya/Cardedeu | Servicio de Microbiologv≠a, Laboratori Clv≠nic Metropolitana Nord. Hospital Universitari Germans Trias i Pujol. Institut d,ÄôInvestigaciv≥ en Civ®ncies de la Salut Germans Trias i Pujol (IGTP) | SeqCOVID-SPAIN consortium/IBV(CSIC) | Elisa Martrv≥ et al |
| EPI_ISL_683627 | Spain/CT-IBV-97010155/2020 | 2020-11-09 | Europe/Spain/Catalunya/El Masnou | Servicio de Microbiologv≠a, Laboratori Clv≠nic Metropolitana Nord. Hospital Universitari Germans Trias i Pujol. Institut d,ÄôInvestigaciv≥ en Civ®ncies de la Salut Germans Trias i Pujol (IGTP) | SeqCOVID-SPAIN consortium/IBV(CSIC) | Elisa Martrv≥ et al |
| EPI_ISL_683628 | Spain/CT-IBV-97009584/2020 | 2020-09-02 | Europe/Spain/Catalunya/Granollers | Servicio de Microbiologv≠a, Laboratori Clv≠nic Metropolitana Nord. Hospital Universitari Germans Trias i Pujol. Institut d,ÄôInvestigaciv≥ en Civ®ncies de la Salut Germans Trias i Pujol (IGTP) | SeqCOVID-SPAIN consortium/IBV(CSIC) | Elisa Martrv≥ et al |
| EPI_ISL_683631 | Spain/CT-IBV-97009587/2020 | 2020-08-19 | Europe/Spain/Catalunya/La Roca del Va | Servicio de Microbiologv≠a, Laboratori Clv≠nic Metropolitana Nord. Hospital Universitari Germans Trias i Pujol. Institut d,ÄôInvestigaciv≥ en Civ®ncies de la Salut Germans Trias i Pujol (IGTP) | SeqCOVID-SPAIN consortium/IBV(CSIC) | Elisa Martrv≥ et al |
| EPI_ISL_683637 | Spain/CT-IBV-97010113/2020 | 2020-11-12 | Europe/Spain/Catalunya/Mataro | Servicio de Microbiologv≠a, Laboratori Clv≠nic Metropolitana Nord. Hospital Universitari Germans Trias i Pujol. Institut d,ÄôInvestigaciv≥ en Civ®ncies de la Salut Germans Trias i Pujol (IGTP) | SeqCOVID-SPAIN consortium/IBV(CSIC) | Elisa Martrv≥ et al |
| EPI_ISL_683641 | Spain/CT-IBV-97010114/2020 | 2020-11-14 | Europe/Spain/Catalunya/Pineda de Mar | Servicio de Microbiologv≠a, Laboratori Clv≠nic Metropolitana Nord. Hospital Universitari Germans Trias i Pujol. Institut d,ÄôInvestigaciv≥ en Civ®ncies de la Salut Germans Trias i Pujol (IGTP) | SeqCOVID-SPAIN consortium/IBV(CSIC) | Elisa Martrv≥ et al |
| EPI_ISL_683642 | Spain/CT-IBV-97010124/2020 | 2020-11-13 | Europe/Spain/Catalunya/Premia de Mar | Servicio de Microbiologv≠a, Laboratori Clv≠nic Metropolitana Nord. Hospital Universitari Germans Trias i Pujol. Institut d,ÄôInvestigaciv≥ en Civ®ncies de la Salut Germans Trias i Pujol (IGTP) | SeqCOVID-SPAIN consortium/IBV(CSIC) | Elisa Martrv≥ et al |
| EPI_ISL_683643 | Spain/CT-IBV-97009578/2020 | 2020-08-21 | Europe/Spain/Catalunya/Ripollet | Servicio de Microbiologv≠a, Laboratori Clv≠nic Metropolitana Nord. Hospital Universitari Germans Trias i Pujol. Institut d,ÄôInvestigaciv≥ en Civ®ncies de la Salut Germans Trias i Pujol (IGTP) | SeqCOVID-SPAIN consortium/IBV(CSIC) | Elisa Martrv≥ et al |

| | | | | | | |
|---|---|---|---|---|---|---|
| EPI_ISL_683644 | Spain/CT-IBV-97009569/2020 | 2020-07-29 | Europe/Spain/Catalunya/Sabadell | Servicio de Microbiologv≠a, Laboratori Clv≠nic Metropolitana Nord. Hospital Universitari Germans Trias i Pujol. Institut d,ÄôInvestigaciv≥ en Civ®ncies de la Salut Germans Trias i Pujol (IGTP) | SeqCOVID-SPAIN consortium/IBV(CSIC) | Elisa Martrv≥ et al |
| EPI_ISL_683645 | Spain/CT-IBV-97009582/2020 | 2020-08-28 | Europe/Spain/Catalunya/Sabadell | Servicio de Microbiologv≠a, Laboratori Clv≠nic Metropolitana Nord. Hospital Universitari Germans Trias i Pujol. Institut d,ÄôInvestigaciv≥ en Civ®ncies de la Salut Germans Trias i Pujol (IGTP) | SeqCOVID-SPAIN consortium/IBV(CSIC) | Elisa Martrv≥ et al |
| EPI_ISL_683646 | Spain/CT-IBV-97010135/2020 | 2020-11-01 | Europe/Spain/Catalunya/Sant Adria del | Servicio de Microbiologv≠a, Laboratori Clv≠nic Metropolitana Nord. Hospital Universitari Germans Trias i Pujol. Institut d,ÄôInvestigaciv≥ en Civ®ncies de la Salut Germans Trias i Pujol (IGTP) | SeqCOVID-SPAIN consortium/IBV(CSIC) | Elisa Martrv≥ et al |
| EPI_ISL_683647 | Spain/CT-IBV-97010136/2020 | 2020-11-02 | Europe/Spain/Catalunya/Sant Adria del | Servicio de Microbiologv≠a, Laboratori Clv≠nic Metropolitana Nord. Hospital Universitari Germans Trias i Pujol. Institut d,ÄôInvestigaciv≥ en Civ®ncies de la Salut Germans Trias i Pujol (IGTP) | SeqCOVID-SPAIN consortium/IBV(CSIC) | Elisa Martrv≥ et al |
| EPI_ISL_683648 | Spain/CT-IBV-97010112/2020 | 2020-11-12 | Europe/Spain/Catalunya/Santa Coloma | Servicio de Microbiologv≠a, Laboratori Clv≠nic Metropolitana Nord. Hospital Universitari Germans Trias i Pujol. Institut d,ÄôInvestigaciv≥ en Civ®ncies de la Salut Germans Trias i Pujol (IGTP) | SeqCOVID-SPAIN consortium/IBV(CSIC) | Elisa Martrv≥ et al |
| EPI_ISL_683652 | Spain/CT-IBV-97009574/2020 | 2020-08-12 | Europe/Spain/Catalunya/Santa Coloma | Servicio de Microbiologv≠a, Laboratori Clv≠nic Metropolitana Nord. Hospital Universitari Germans Trias i Pujol. Institut d,ÄôInvestigaciv≥ en Civ®ncies de la Salut Germans Trias i Pujol (IGTP) | SeqCOVID-SPAIN consortium/IBV(CSIC) | Elisa Martrv≥ et al |
| EPI_ISL_683653 | Spain/CT-IBV-97009579/2020 | 2020-08-24 | Europe/Spain/Catalunya/Santa Coloma | Servicio de Microbiologv≠a, Laboratori Clv≠nic Metropolitana Nord. Hospital Universitari Germans Trias i Pujol. Institut d,ÄôInvestigaciv≥ en Civ®ncies de la Salut Germans Trias i Pujol (IGTP) | SeqCOVID-SPAIN consortium/IBV(CSIC) | Elisa Martrv≥ et al |
| EPI_ISL_683654 | Spain/CT-IBV-97009586/2020 | 2020-08-18 | Europe/Spain/Catalunya/Santa Coloma | Servicio de Microbiologv≠a, Laboratori Clv≠nic Metropolitana Nord. Hospital Universitari Germans Trias i Pujol. Institut d,ÄôInvestigaciv≥ en Civ®ncies de la Salut Germans Trias i Pujol (IGTP) | SeqCOVID-SPAIN consortium/IBV(CSIC) | Elisa Martrv≥ et al |

| | | | | | | |
|---|---|---|---|---|---|---|
| EPI_ISL_683655 | Spain/CT-IBV-97009576/2020 | 2020-08-17 | Europe/Spain/Catalunya/Tiana | Servicio de Microbiolog√≠a, Laboratori Cl√≠nic Metropolitana Nord. Hospital Universitari Germans Trias i Pujol. Institut d‚ÄôInvestigaci√≥ en Ci√®ncies de la Salut Germans Trias i Pujol (IGTP) | SeqCOVID-SPAIN consortium/IBV(CSIC) | Elisa Mart√≠ et al |
| EPI_ISL_683656 | Spain/CT-IBV-97010151/2020 | 2020-11-03 | Europe/Spain/Catalunya/Vilassar de Ma | Servicio de Microbiolog√≠a, Laboratori Cl√≠nic Metropolitana Nord. Hospital Universitari Germans Trias i Pujol. Institut d‚ÄôInvestigaci√≥ en Ci√®ncies de la Salut Germans Trias i Pujol (IGTP) | SeqCOVID-SPAIN consortium/IBV(CSIC) | Elisa Mart√≠ et al |
| EPI_ISL_683695 | USA/MN-MDH-2066/2020 | 2020-11-04 | North America/USA/Minnesota/ | Mayo Clinic & Mayo Clinic Laboratories | Minnesota Department of Health, Public Health Laboratory | Alexandra Lorentz et al |
| EPI_ISL_683696 | USA/MN-MDH-2067/2020 | 2020-10-30 | North America/USA/Minnesota/ | Essentia Health-St. Mary's Medical Center | Minnesota Department of Health, Public Health Laboratory | Alexandra Lorentz et al |
| EPI_ISL_683705 | USA/MN-MDH-2076/2020 | 2020-10-30 | North America/USA/Minnesota/ | Essentia Health-St. Mary's Medical Center | Minnesota Department of Health, Public Health Laboratory | Alexandra Lorentz et al |
| EPI_ISL_683707 | USA/MN-MDH-2078/2020 | 2020-10-31 | North America/USA/Minnesota/ | Essentia Health-St. Mary's Medical Center | Minnesota Department of Health, Public Health Laboratory | Alexandra Lorentz et al |
| EPI_ISL_683717 | USA/MN-MDH-2088/2020 | 2020-10-31 | North America/USA/Minnesota/ | Mayo Clinic & Mayo Clinic Laboratories | Minnesota Department of Health, Public Health Laboratory | Alexandra Lorentz et al |
| EPI_ISL_683721 | USA/MN-MDH-2092/2020 | 2020-10-29 | North America/USA/Minnesota/ | Mayo Clinic & Mayo Clinic Laboratories | Minnesota Department of Health, Public Health Laboratory | Alexandra Lorentz et al |
| EPI_ISL_683723 | USA/MN-MDH-2094/2020 | 2020-10-23 | North America/USA/Minnesota/ | Mayo Clinic & Mayo Clinic Laboratories | Minnesota Department of Health, Public Health Laboratory | Alexandra Lorentz et al |

| | | | | | | |
|---|---|---|---|---|---|---|
| EPI_ISL_683733 | USA/MN-MDH-2104/2020 | 2020-11-11 | North America/USA/Minnesota/ | Minnesota Department of Health, Public Health Laboratory | Minnesota Department of Health, Public Health Laboratory | Alexandra Lorentz et al |
| EPI_ISL_683739 | USA/MN-MDH-2110/2020 | 2020-11-03 | North America/USA/Minnesota/ | Minnesota Department of Health, Public Health Laboratory | Minnesota Department of Health, Public Health Laboratory | Alexandra Lorentz et al |
| EPI_ISL_683740 | USA/MN-MDH-2111/2020 | 2020-11-03 | North America/USA/Minnesota/ | Minnesota Department of Health, Public Health Laboratory | Minnesota Department of Health, Public Health Laboratory | Alexandra Lorentz et al |
| EPI_ISL_683742 | USA/MN-MDH-2113/2020 | 2020-11-03 | North America/USA/Minnesota/ | Minnesota Department of Health, Public Health Laboratory | Minnesota Department of Health, Public Health Laboratory | Alexandra Lorentz et al |
| EPI_ISL_683749 | USA/MN-MDH-2120/2020 | 2020-10-30 | North America/USA/Minnesota/ | Minnesota Department of Health, Public Health Laboratory | Minnesota Department of Health, Public Health Laboratory | Alexandra Lorentz et al |
| EPI_ISL_683750 | USA/MN-MDH-2121/2020 | 2020-10-30 | North America/USA/Minnesota/ | Minnesota Department of Health, Public Health Laboratory | Minnesota Department of Health, Public Health Laboratory | Alexandra Lorentz et al |
| EPI_ISL_683755 | USA/NY-NYCPHL-001403/2020 | 2020-11-18 | North America/USA/New York/New Yor | DOHMH PHL | New York City Public Health Laboratory | Jade Wang et al |
| EPI_ISL_683770 | USA/NY-NYCPHL-001472/2020 | 2020-11-27 | North America/USA/New York/New Yor | DOHMH Corona | New York City Public Health Laboratory | Jade Wang et al |
| EPI_ISL_683774 | USA/NY-NYCPHL-001199/2020 | 2020-11-02 | North America/USA/New York/New Yor | DOHMH PHL | New York City Public Health Laboratory | Jade Wang et al |
| EPI_ISL_683782 | USA/NY-NYCPHL-001406/2020 | 2020-11-18 | North America/USA/New York/New Yor | DOHMH Jamaica | New York City Public Health Laboratory | Jade Wang et al |
| EPI_ISL_683786 | USA/NY-NYCPHL-001414/2020 | 2020-11-19 | North America/USA/New York/New Yor | DOHMH Jamaica | New York City Public Health Laboratory | Jade Wang et al |

| | | | | | | |
|---|---|---|---|---|---|---|
| EPI_ISL_683793 | USA/NY-NYCPHL-001423/2020 | 2020-11-20 | North America/USA/New York/New Yor | DOHMH Corona | New York City Public Health Laboratory | Jade Wang et al |
| EPI_ISL_683800 | USA/NY-NYCPHL-001433/2020 | 2020-11-23 | North America/USA/New York/New Yor | DOHMH Corona | New York City Public Health Laboratory | Jade Wang et al |
| EPI_ISL_683810 | USA/NY-NYCPHL-001446/2020 | 2020-11-13 | North America/USA/New York/New Yor | DOHMH PHL | New York City Public Health Laboratory | Jade Wang et al |
| EPI_ISL_683813 | USA/NY-NYCPHL-001449/2020 | 2020-11-24 | North America/USA/New York/New Yor | DOHMH Central Harlem | New York City Public Health Laboratory | Jade Wang et al |
| EPI_ISL_683819 | USA/NY-NYCPHL-001457/2020 | 2020-11-25 | North America/USA/New York/New Yor | DOHMH Riverside | New York City Public Health Laboratory | Jade Wang et al |
| EPI_ISL_683829 | USA/NY-NYCPHL-001470/2020 | 2020-11-27 | North America/USA/New York/New Yor | DOHMH Central Harlem | New York City Public Health Laboratory | Jade Wang et al |
| EPI_ISL_683832 | USA/NY-NYCPHL-001476/2020 | 2020-11-27 | North America/USA/New York/New Yor | DOHMH Fort Greene | New York City Public Health Laboratory | Jade Wang et al |
| EPI_ISL_683835 | Mali/M00960/2020 | 2020-06-10 | Africa/Mali/Timbuktu/ | CICM | Malaria Research and Training Center (MRTC-Parasito) | Antoine Dara et al |
| EPI_ISL_683842 | USA/NY-NYCPHL-001385/2020 | 2020-11-17 | North America/USA/New York/New Yor | DOHMH Central Harlem | New York City Public Health Laboratory | Jade Wang et al |
| EPI_ISL_683846 | USA/NY-NYCPHL-001275/2020 | 2020-11-09 | North America/USA/New York/Westche | DOHMH PHL | New York City Public Health Laboratory | Jade Wang et al |
| EPI_ISL_683853 | USA/NY-NYCPHL-001261/2020 | 2020-11-07 | North America/USA/New York/New Yor | DOHMH Morrisania | New York City Public Health Laboratory | Jade Wang et al |
| EPI_ISL_683863 | USA/NY-NYCPHL-001298/2020 | 2020-11-09 | North America/USA/New York/New Yor | DOHMH Morrisania | New York City Public Health Laboratory | Jade Wang et al |
| EPI_ISL_683871 | USA/NY-NYCPHL-001379/2020 | 2020-11-17 | North America/USA/New York/New Yor | DOHMH PHL | New York City Public Health Laboratory | Jade Wang et al |

| | | | | | | |
|---|---|---|---|---|---|---|
| EPI_ISL_683876 | USA/NY-NYCPHL-001303/2020 | 2020-11-10 | North America/USA/New York/New Yor | DOHMH Corona | New York City Public Health Laboratory | Jade Wang et al |
| EPI_ISL_683880 | USA/NY-NYCPHL-001318/2020 | 2020-11-10 | North America/USA/New York/New Yor | DOHMH Morrisania | New York City Public Health Laboratory | Jade Wang et al |
| EPI_ISL_683885 | USA/NY-NYCPHL-001188/2020 | 2020-10-28 | North America/USA/New York/New Yor | DOHMH Jamaica | New York City Public Health Laboratory | Jade Wang et al |
| EPI_ISL_683891 | USA/NY-NYCPHL-001228/2020 | 2020-11-05 | North America/USA/New York/New Yor | DOHMH Crown Heights | New York City Public Health Laboratory | Jade Wang et al |
| EPI_ISL_683895 | USA/NY-NYCPHL-001256/2020 | 2020-11-06 | North America/USA/New York/New Yor | DOHMH Riverside | New York City Public Health Laboratory | Jade Wang et al |
| EPI_ISL_683905 | USA/NY-NYCPHL-001295/2020 | 2020-11-09 | North America/USA/New York/New Yor | DOHMH Morrisania | New York City Public Health Laboratory | Jade Wang et al |
| EPI_ISL_683908 | USA/NY-NYCPHL-001302/2020 | 2020-11-10 | North America/USA/New York/New Yor | DOHMH Corona | New York City Public Health Laboratory | Jade Wang et al |
| EPI_ISL_683913 | USA/NY-NYCPHL-001313/2020 | 2020-11-10 | North America/USA/New York/New Yor | DOHMH Fort Greene | New York City Public Health Laboratory | Jade Wang et al |
| EPI_ISL_683915 | USA/NY-NYCPHL-001315/2020 | 2020-11-10 | North America/USA/New York/New Yor | DOHMH Jamaica | New York City Public Health Laboratory | Jade Wang et al |
| EPI_ISL_683921 | USA/NY-NYCPHL-001327/2020 | 2020-11-12 | North America/USA/New York/New Yor | DOHMH Jamaica | New York City Public Health Laboratory | Jade Wang et al |
| EPI_ISL_683942 | USA/NY-NYCPHL-001238/2020 | 2020-11-06 | North America/USA/New York/New Yor | DOHMH Riverside | New York City Public Health Laboratory | Jade Wang et al |
| EPI_ISL_683944 | USA/NY-NYCPHL-001242/2020 | 2020-11-06 | North America/USA/New York/New Yor | DOHMH Jamaica | New York City Public Health Laboratory | Jade Wang et al |
| EPI_ISL_683949 | USA/NY-NYCPHL-001253/2020 | 2020-11-06 | North America/USA/New York/New Yor | DOHMH Riverside | New York City Public Health Laboratory | Jade Wang et al |

| | | | | | | |
|---|---|---|---|---|---|---|
| EPI_ISL_683959 | USA/NY-NYCPHL-001297/2020 | 2020-11-09 | North America/USA/New York/Westche | DOHMH Morrisania | New York City Public Health Laboratory | Jade Wang et al |
| EPI_ISL_683960 | USA/NY-NYCPHL-001301/2020 | 2020-11-10 | North America/USA/New York/New Yor | DOHMH Corona | New York City Public Health Laboratory | Jade Wang et al |
| EPI_ISL_683973 | USA/NY-NYCPHL-001338/2020 | 2020-11-13 | North America/USA/New York/New Yor | DOHMH Fort Greene | New York City Public Health Laboratory | Jade Wang et al |
| EPI_ISL_683981 | USA/NY-NYCPHL-001360/2020 | 2020-11-16 | North America/USA/New York/New Yor | DOHMH Chelsea | New York City Public Health Laboratory | Jade Wang et al |
| EPI_ISL_683987 | USA/NY-NYCPHL-001376/2020 | 2020-11-16 | North America/USA/New York/New Yor | DOHMH PHL | New York City Public Health Laboratory | Jade Wang et al |
| EPI_ISL_683993 | USA/NY-NYCPHL-001390/2020 | 2020-11-17 | North America/USA/New York/New Yor | DOHMH PHL | New York City Public Health Laboratory | Jade Wang et al |
| EPI_ISL_683994 | USA/NY-NYCPHL-001392/2020 | 2020-11-17 | North America/USA/New York/New Yor | DOHMH Riverside | New York City Public Health Laboratory | Jade Wang et al |
| EPI_ISL_684006 | USA/UT-UPHL-201201982/2020 | 2020-12-01 | North America/USA/Utah/ | Utah Public Health Laboratory | Utah Public Health Laboratory | Erin Young et al |
| EPI_ISL_684009 | USA/UT-UPHL-201207559/2020 | 2020-12-01 | North America/USA/Utah/ | Utah Public Health Laboratory | Utah Public Health Laboratory | Erin Young et al |
| EPI_ISL_684016 | USA/UT-UPHL-201218864/2020 | 2020-12-01 | North America/USA/Utah/ | Utah Public Health Laboratory | Utah Public Health Laboratory | Erin Young et al |
| EPI_ISL_684030 | Bahrain/341035032/2020 | 2020-11-15 | Asia/Bahrain/Bahrain/ | Communicable Disease Laboratory, Public Health Directorate | Communicable Disease Laboratory, Public Health Directorate | Alwasti et al |
| EPI_ISL_684031 | Bahrain/920509111/2020 | 2020-11-12 | Asia/Bahrain/Bahrain/ | Communicable Disease Laboratory, Public Health Directorate | Communicable Disease Laboratory, Public Health Directorate | Alwasti et al |
| EPI_ISL_684107 | France/ARA-29628/2020 | 2020-04-23 | Europe/France/Auvergne-Rhv¥ne-Alpes/ | CNR Virus des Infections Respiratoires - France SUD | CNR Virus des Infections Respiratoires - France SUD | Antonin Bal et al |

| | | | | | | |
|---|---|---|---|---|---|---|
| EPI_ISL_684110 | Japan/PG-0073/2020 | 20-Feb | Asia/Japan/Japan/ | Pathogen Genomics Center, National Institute of Infectious Diseases | Pathogen Genomics Center, National Institute of Infectious Diseases | Tsuyoshi Sekizuka et al |
| EPI_ISL_684111 | Japan/PG-0075/2020 | 20-Feb | Asia/Japan/Japan/ | Pathogen Genomics Center, National Institute of Infectious Diseases | Pathogen Genomics Center, National Institute of Infectious Diseases | Tsuyoshi Sekizuka et al |
| EPI_ISL_684112 | Japan/PG-0078/2020 | 20-Feb | Asia/Japan/Japan/ | Pathogen Genomics Center, National Institute of Infectious Diseases | Pathogen Genomics Center, National Institute of Infectious Diseases | Tsuyoshi Sekizuka et al |
| EPI_ISL_684113 | Japan/PG-0079/2020 | 20-Feb | Asia/Japan/Japan/ | Pathogen Genomics Center, National Institute of Infectious Diseases | Pathogen Genomics Center, National Institute of Infectious Diseases | Tsuyoshi Sekizuka et al |
| EPI_ISL_684127 | Japan/PG-0116/2020 | 20-Feb | Asia/Japan/Japan/ | Gunma Prefectural Institute of Public Health and Environmental Sciences | Pathogen Genomics Center, National Institute of Infectious Diseases | Tsuyoshi Sekizuka et al |
| EPI_ISL_684130 | Japan/PG-0142/2020 | 20-Feb | Asia/Japan/Japan/ | Pathogen Genomics Center, National Institute of Infectious Diseases | Pathogen Genomics Center, National Institute of Infectious Diseases | Tsuyoshi Sekizuka et al |
| EPI_ISL_684131 | Japan/PG-0154/2020 | 20-Mar | Asia/Japan/Japan/ | Pathogen Genomics Center, National Institute of Infectious Diseases | Pathogen Genomics Center, National Institute of Infectious Diseases | Tsuyoshi Sekizuka et al |
| EPI_ISL_684139 | Japan/PG-0199/2020 | 20-Feb | Asia/Japan/Japan/ | Pathogen Genomics Center, National Institute of Infectious Diseases | Pathogen Genomics Center, National Institute of Infectious Diseases | Tsuyoshi Sekizuka et al |

| | | | | | | |
|---|---|---|---|---|---|---|
| EPI_ISL_684160 | Japan/PG-0335/2020 | 20-Mar | Asia/Japan/Japan/ | Chiba Prefectural Institute of Public Health | Pathogen Genomics Center, National Institute of Infectious Diseases | Tsuyoshi Sekizuka et al |
| EPI_ISL_684163 | Japan/PG-0360/2020 | 20-Feb | Asia/Japan/Japan/ | Pathogen Genomics Center, National Institute of Infectious Diseases | Pathogen Genomics Center, National Institute of Infectious Diseases | Tsuyoshi Sekizuka et al |
| EPI_ISL_684164 | Japan/PG-0380/2020 | 20-Feb | Asia/Japan/Japan/ | Pathogen Genomics Center, National Institute of Infectious Diseases | Pathogen Genomics Center, National Institute of Infectious Diseases | Tsuyoshi Sekizuka et al |
| EPI_ISL_684237 | Japan/PG-0627/2020 | 20-Apr | Asia/Japan/Japan/ | Gunma Prefectural Institute of Public Health and Environmental Sciences | Pathogen Genomics Center, National Institute of Infectious Diseases | Tsuyoshi Sekizuka et al |
| EPI_ISL_684615 | Japan/PG-1005/2020 | 20-Feb | Asia/Japan/Japan/ | Tokyo Metropolitan Institute of Public Health | Pathogen Genomics Center, National Institute of Infectious Diseases | Tsuyoshi Sekizuka et al |
| EPI_ISL_684616 | Japan/PG-1006/2020 | 20-Feb | Asia/Japan/Japan/ | Tokyo Metropolitan Institute of Public Health | Pathogen Genomics Center, National Institute of Infectious Diseases | Tsuyoshi Sekizuka et al |
| EPI_ISL_684618 | Japan/PG-1008/2020 | 20-Mar | Asia/Japan/Japan/ | Tokyo Metropolitan Institute of Public Health | Pathogen Genomics Center, National Institute of Infectious Diseases | Tsuyoshi Sekizuka et al |
| EPI_ISL_684887 | Japan/PG-1278/2020 | 20-Mar | Asia/Japan/Japan/ | Saitama Prefectural Institute of Public Health | Pathogen Genomics Center, National Institute of Infectious Diseases | Tsuyoshi Sekizuka et al |

| | | | | | | |
|---|---|---|---|---|---|---|
| EPI_ISL_684926 | Japan/PG-1318/2020 | 20-Apr | Asia/Japan/Japan/ | Saitama Prefectural Institute of Public Health | Pathogen Genomics Center, National Institute of Infectious Diseases | Tsuyoshi Sekizuka et al |
| EPI_ISL_684929 | Japan/PG-1321/2020 | 20-Apr | Asia/Japan/Japan/ | Saitama Prefectural Institute of Public Health | Pathogen Genomics Center, National Institute of Infectious Diseases | Tsuyoshi Sekizuka et al |
| EPI_ISL_684961 | Japan/PG-1353/2020 | 20-Apr | Asia/Japan/Japan/ | Saitama Prefectural Institute of Public Health | Pathogen Genomics Center, National Institute of Infectious Diseases | Tsuyoshi Sekizuka et al |
| EPI_ISL_684969 | Japan/PG-1361/2020 | 20-Apr | Asia/Japan/Japan/ | Saitama Prefectural Institute of Public Health | Pathogen Genomics Center, National Institute of Infectious Diseases | Tsuyoshi Sekizuka et al |
| EPI_ISL_685061 | Japan/PG-1454/2020 | 20-Apr | Asia/Japan/Japan/ | Saitama Prefectural Institute of Public Health | Pathogen Genomics Center, National Institute of Infectious Diseases | Tsuyoshi Sekizuka et al |
| EPI_ISL_685079 | Japan/PG-1472/2020 | 20-Apr | Asia/Japan/Japan/ | Saitama Prefectural Institute of Public Health | Pathogen Genomics Center, National Institute of Infectious Diseases | Tsuyoshi Sekizuka et al |
| EPI_ISL_685143 | Japan/PG-1537/2020 | 20-Apr | Asia/Japan/Japan/ | Pathogen Genomics Center, National Institute of Infectious Diseases | Pathogen Genomics Center, National Institute of Infectious Diseases | Tsuyoshi Sekizuka et al |
| EPI_ISL_685264 | Japan/PG-1658/2020 | 20-Apr | Asia/Japan/Japan/ | Pathogen Genomics Center, National Institute of Infectious Diseases | Pathogen Genomics Center, National Institute of Infectious Diseases | Tsuyoshi Sekizuka et al |

| | | | | | | |
|---|---|---|---|---|---|---|
| EPI_ISL_685275 | Japan/PG-1669/2020 | 20-Apr | Asia/Japan/Japan/ | Pathogen Genomics Center, National Institute of Infectious Diseases | Pathogen Genomics Center, National Institute of Infectious Diseases | Tsuyoshi Sekizuka et al |
| EPI_ISL_685285 | Japan/PG-1679/2020 | 20-May | Asia/Japan/Japan/ | Pathogen Genomics Center, National Institute of Infectious Diseases | Pathogen Genomics Center, National Institute of Infectious Diseases | Tsuyoshi Sekizuka et al |
| EPI_ISL_685312 | Japan/PG-1706/2020 | 20-May | Asia/Japan/Japan/ | Pathogen Genomics Center, National Institute of Infectious Diseases | Pathogen Genomics Center, National Institute of Infectious Diseases | Tsuyoshi Sekizuka et al |
| EPI_ISL_685548 | Japan/PG-1942/2020 | 20-Apr | Asia/Japan/Japan/ | Pathogen Genomics Center, National Institute of Infectious Diseases | Pathogen Genomics Center, National Institute of Infectious Diseases | Tsuyoshi Sekizuka et al |
| EPI_ISL_685584 | Japan/PG-1978/2020 | 20-Feb | Asia/Japan/Japan/ | Pathogen Genomics Center, National Institute of Infectious Diseases | Pathogen Genomics Center, National Institute of Infectious Diseases | Tsuyoshi Sekizuka et al |
| EPI_ISL_685585 | Japan/PG-1979/2020 | 20-Feb | Asia/Japan/Japan/ | Pathogen Genomics Center, National Institute of Infectious Diseases | Pathogen Genomics Center, National Institute of Infectious Diseases | Tsuyoshi Sekizuka et al |
| EPI_ISL_685591 | Japan/PG-1985/2020 | 20-Apr | Asia/Japan/Japan/ | Pathogen Genomics Center, National Institute of Infectious Diseases | Pathogen Genomics Center, National Institute of Infectious Diseases | Tsuyoshi Sekizuka et al |
| EPI_ISL_685679 | Japan/PG-2074/2020 | 20-Apr | Asia/Japan/Japan/ | Tokyo Metropolitan Institute of Public Health | Pathogen Genomics Center, National Institute of Infectious Diseases | Tsuyoshi Sekizuka et al |

| | | | | | | |
|---|---|---|---|---|---|---|
| EPI_ISL_685688 | Japan/PG-2083/2020 | 20-Apr | Asia/Japan/Japan/ | Tokyo Metropolitan Institute of Public Health | Pathogen Genomics Center, National Institute of Infectious Diseases | Tsuyoshi Sekizuka et al |
| EPI_ISL_685751 | Japan/PG-2146/2020 | 20-Apr | Asia/Japan/Japan/ | Pathogen Genomics Center, National Institute of Infectious Diseases | Pathogen Genomics Center, National Institute of Infectious Diseases | Tsuyoshi Sekizuka et al |
| EPI_ISL_685780 | Japan/PG-2175/2020 | 20-Feb | Asia/Japan/Japan/ | Pathogen Genomics Center, National Institute of Infectious Diseases | Pathogen Genomics Center, National Institute of Infectious Diseases | Tsuyoshi Sekizuka et al |
| EPI_ISL_685911 | Japan/PG-2306/2020 | 20-May | Asia/Japan/Japan/ | Pathogen Genomics Center, National Institute of Infectious Diseases | Pathogen Genomics Center, National Institute of Infectious Diseases | Tsuyoshi Sekizuka et al |
| EPI_ISL_686079 | Japan/PG-2474/2020 | 20-May | Asia/Japan/Japan/ | Pathogen Genomics Center, National Institute of Infectious Diseases | Pathogen Genomics Center, National Institute of Infectious Diseases | Tsuyoshi Sekizuka et al |
| EPI_ISL_686140 | Japan/PG-8544/2020 | 20-Aug | Asia/Japan/Japan/ | Pathogen Genomics Center, National Institute of Infectious Diseases | Pathogen Genomics Center, National Institute of Infectious Diseases | Tsuyoshi Sekizuka et al |
| EPI_ISL_686226 | Japan/PG-8630/2020 | 20-Aug | Asia/Japan/Japan/ | Pathogen Genomics Center, National Institute of Infectious Diseases | Pathogen Genomics Center, National Institute of Infectious Diseases | Tsuyoshi Sekizuka et al |
| EPI_ISL_686241 | Japan/PG-8645/2020 | 20-Sep | Asia/Japan/Japan/ | Pathogen Genomics Center, National Institute of Infectious Diseases | Pathogen Genomics Center, National Institute of Infectious Diseases | Tsuyoshi Sekizuka et al |

| | | | | | | |
|---|---|---|---|---|---|---|
| EPI_ISL_686343 | Japan/PG-8747/2020 | 20-Sep | Asia/Japan/Japan/ | Pathogen Genomics Center, National Institute of Infectious Diseases | Pathogen Genomics Center, National Institute of Infectious Diseases | Tsuyoshi Sekizuka et al |
| EPI_ISL_686363 | Japan/PG-8767/2020 | 20-Sep | Asia/Japan/Japan/ | Pathogen Genomics Center, National Institute of Infectious Diseases | Pathogen Genomics Center, National Institute of Infectious Diseases | Tsuyoshi Sekizuka et al |
| EPI_ISL_686383 | Japan/PG-8795/2020 | 20-Aug | Asia/Japan/Japan/ | Pathogen Genomics Center, National Institute of Infectious Diseases | Pathogen Genomics Center, National Institute of Infectious Diseases | Tsuyoshi Sekizuka et al |
| EPI_ISL_686444 | Japan/PG-8898/2020 | 20-Aug | Asia/Japan/Japan/ | Pathogen Genomics Center, National Institute of Infectious Diseases | Pathogen Genomics Center, National Institute of Infectious Diseases | Tsuyoshi Sekizuka et al |
| EPI_ISL_686741 | Japan/PG-7639/2020 | 20-Aug | Asia/Japan/Japan/ | Chiba Prefectural Institute of Public Health | Pathogen Genomics Center, National Institute of Infectious Diseases | Tsuyoshi Sekizuka et al |
| EPI_ISL_686775 | Japan/PG-7673/2020 | 20-Aug | Asia/Japan/Japan/ | Chiba Prefectural Institute of Public Health | Pathogen Genomics Center, National Institute of Infectious Diseases | Tsuyoshi Sekizuka et al |
| EPI_ISL_687012 | Japan/PG-7911/2020 | 20-Aug | Asia/Japan/Japan/ | Pathogen Genomics Center, National Institute of Infectious Diseases | Pathogen Genomics Center, National Institute of Infectious Diseases | Tsuyoshi Sekizuka et al |
| EPI_ISL_687091 | Japan/PG-7990/2020 | 20-Aug | Asia/Japan/Japan/ | Pathogen Genomics Center, National Institute of Infectious Diseases | Pathogen Genomics Center, National Institute of Infectious Diseases | Tsuyoshi Sekizuka et al |

| | | | | | | |
|---|---|---|---|---|---|---|
| EPI_ISL_687147 | Japan/PG-8046/2020 | 20-Aug | Asia/Japan/Japan/ | Pathogen Genomics Center, National Institute of Infectious Diseases | Pathogen Genomics Center, National Institute of Infectious Diseases | Tsuyoshi Sekizuka et al |
| EPI_ISL_687218 | Japan/PG-8119/2020 | 20-Aug | Asia/Japan/Japan/ | Pathogen Genomics Center, National Institute of Infectious Diseases | Pathogen Genomics Center, National Institute of Infectious Diseases | Tsuyoshi Sekizuka et al |
| EPI_ISL_687254 | Japan/PG-8155/2020 | 20-Sep | Asia/Japan/Japan/ | Pathogen Genomics Center, National Institute of Infectious Diseases | Pathogen Genomics Center, National Institute of Infectious Diseases | Tsuyoshi Sekizuka et al |
| EPI_ISL_687334 | Japan/PG-8235/2020 | 20-Aug | Asia/Japan/Japan/ | Pathogen Genomics Center, National Institute of Infectious Diseases | Pathogen Genomics Center, National Institute of Infectious Diseases | Tsuyoshi Sekizuka et al |
| EPI_ISL_687362 | Japan/PG-8263/2020 | 20-Aug | Asia/Japan/Japan/ | Pathogen Genomics Center, National Institute of Infectious Diseases | Pathogen Genomics Center, National Institute of Infectious Diseases | Tsuyoshi Sekizuka et al |
| EPI_ISL_687542 | Japan/PG-8443/2020 | 20-Sep | Asia/Japan/Japan/ | Fukuoka Institute of Health and Environmental Sciences | Pathogen Genomics Center, National Institute of Infectious Diseases | Tsuyoshi Sekizuka et al |
| EPI_ISL_687680 | Japan/PG-6568/2020 | 20-Sep | Asia/Japan/Japan/ | Pathogen Genomics Center, National Institute of Infectious Diseases | Pathogen Genomics Center, National Institute of Infectious Diseases | Tsuyoshi Sekizuka et al |
| EPI_ISL_687723 | Japan/PG-6611/2020 | 20-Jul | Asia/Japan/Japan/ | Pathogen Genomics Center, National Institute of Infectious Diseases | Pathogen Genomics Center, National Institute of Infectious Diseases | Tsuyoshi Sekizuka et al |

| | | | | | | |
|---|---|---|---|---|---|---|
| EPI_ISL_687923 | Japan/PG-6814/2020 | 20-May | Asia/Japan/Japan/ | Pathogen Genomics Center, National Institute of Infectious Diseases | Pathogen Genomics Center, National Institute of Infectious Diseases | Tsuyoshi Sekizuka et al |
| EPI_ISL_687960 | Japan/PG-6852/2020 | 20-Sep | Asia/Japan/Japan/ | Pathogen Genomics Center, National Institute of Infectious Diseases | Pathogen Genomics Center, National Institute of Infectious Diseases | Tsuyoshi Sekizuka et al |
| EPI_ISL_688020 | Japan/PG-6912/2020 | 20-Sep | Asia/Japan/Japan/ | Pathogen Genomics Center, National Institute of Infectious Diseases | Pathogen Genomics Center, National Institute of Infectious Diseases | Tsuyoshi Sekizuka et al |
| EPI_ISL_688046 | Japan/PG-6941/2020 | 20-Sep | Asia/Japan/Japan/ | Pathogen Genomics Center, National Institute of Infectious Diseases | Pathogen Genomics Center, National Institute of Infectious Diseases | Tsuyoshi Sekizuka et al |
| EPI_ISL_688155 | Japan/PG-7050/2020 | 20-Aug | Asia/Japan/Japan/ | Pathogen Genomics Center, National Institute of Infectious Diseases | Pathogen Genomics Center, National Institute of Infectious Diseases | Tsuyoshi Sekizuka et al |
| EPI_ISL_688194 | Japan/PG-7089/2020 | 20-Aug | Asia/Japan/Japan/ | Pathogen Genomics Center, National Institute of Infectious Diseases | Pathogen Genomics Center, National Institute of Infectious Diseases | Tsuyoshi Sekizuka et al |
| EPI_ISL_688250 | Japan/PG-7145/2020 | 20-Aug | Asia/Japan/Japan/ | Pathogen Genomics Center, National Institute of Infectious Diseases | Pathogen Genomics Center, National Institute of Infectious Diseases | Tsuyoshi Sekizuka et al |
| EPI_ISL_688340 | Japan/PG-7236/2020 | 20-Aug | Asia/Japan/Japan/ | Saitama Prefectural Institute of Public Health | Pathogen Genomics Center, National Institute of Infectious Diseases | Tsuyoshi Sekizuka et al |

| | | | | | | |
|---|---|---|---|---|---|---|
| EPI_ISL_688343 | Japan/PG-7239/2020 | 20-Aug | Asia/Japan/Japan/ | Saitama Prefectural Institute of Public Health | Pathogen Genomics Center, National Institute of Infectious Diseases | Tsuyoshi Sekizuka et al |
| EPI_ISL_688344 | Japan/PG-7241/2020 | 20-Aug | Asia/Japan/Japan/ | Saitama Prefectural Institute of Public Health | Pathogen Genomics Center, National Institute of Infectious Diseases | Tsuyoshi Sekizuka et al |
| EPI_ISL_688627 | Japan/PG-5704/2020 | 20-Jul | Asia/Japan/Japan/ | Pathogen Genomics Center, National Institute of Infectious Diseases | Pathogen Genomics Center, National Institute of Infectious Diseases | Tsuyoshi Sekizuka et al |
| EPI_ISL_688753 | Japan/PG-5628/2020 | 20-Aug | Asia/Japan/Japan/ | Pathogen Genomics Center, National Institute of Infectious Diseases | Pathogen Genomics Center, National Institute of Infectious Diseases | Tsuyoshi Sekizuka et al |
| EPI_ISL_688779 | Japan/PG-5655/2020 | 20-Apr | Asia/Japan/Japan/ | Pathogen Genomics Center, National Institute of Infectious Diseases | Pathogen Genomics Center, National Institute of Infectious Diseases | Tsuyoshi Sekizuka et al |
| EPI_ISL_688809 | Japan/PG-5685/2020 | 20-Apr | Asia/Japan/Japan/ | Pathogen Genomics Center, National Institute of Infectious Diseases | Pathogen Genomics Center, National Institute of Infectious Diseases | Tsuyoshi Sekizuka et al |
| EPI_ISL_688829 | Japan/PG-5706/2020 | 20-Aug | Asia/Japan/Japan/ | Pathogen Genomics Center, National Institute of Infectious Diseases | Pathogen Genomics Center, National Institute of Infectious Diseases | Tsuyoshi Sekizuka et al |
| EPI_ISL_688960 | Japan/PG-5843/2020 | 20-Aug | Asia/Japan/Japan/ | Fukuoka Institute of Health and Environmental Sciences | Pathogen Genomics Center, National Institute of Infectious Diseases | Tsuyoshi Sekizuka et al |

| | | | | | | |
|---|---|---|---|---|---|---|
| EPI_ISL_689020 | Japan/PG-5905/2020 | 20-Aug | Asia/Japan/Japan/ | Pathogen Genomics Center, National Institute of Infectious Diseases | Pathogen Genomics Center, National Institute of Infectious Diseases | Tsuyoshi Sekizuka et al |
| EPI_ISL_689035 | Japan/PG-5920/2020 | 20-Aug | Asia/Japan/Japan/ | Pathogen Genomics Center, National Institute of Infectious Diseases | Pathogen Genomics Center, National Institute of Infectious Diseases | Tsuyoshi Sekizuka et al |
| EPI_ISL_689042 | Japan/PG-5927/2020 | 20-Aug | Asia/Japan/Japan/ | Pathogen Genomics Center, National Institute of Infectious Diseases | Pathogen Genomics Center, National Institute of Infectious Diseases | Tsuyoshi Sekizuka et al |
| EPI_ISL_689182 | Japan/PG-6070/2020 | 20-Jul | Asia/Japan/Japan/ | Pathogen Genomics Center, National Institute of Infectious Diseases | Pathogen Genomics Center, National Institute of Infectious Diseases | Tsuyoshi Sekizuka et al |
| EPI_ISL_689289 | Japan/PG-6177/2020 | 20-Aug | Asia/Japan/Japan/ | Pathogen Genomics Center, National Institute of Infectious Diseases | Pathogen Genomics Center, National Institute of Infectious Diseases | Tsuyoshi Sekizuka et al |
| EPI_ISL_689329 | Japan/PG-6217/2020 | 20-Aug | Asia/Japan/Japan/ | Pathogen Genomics Center, National Institute of Infectious Diseases | Pathogen Genomics Center, National Institute of Infectious Diseases | Tsuyoshi Sekizuka et al |
| EPI_ISL_689408 | Japan/PG-6297/2020 | 20-Jul | Asia/Japan/Japan/ | Pathogen Genomics Center, National Institute of Infectious Diseases | Pathogen Genomics Center, National Institute of Infectious Diseases | Tsuyoshi Sekizuka et al |
| EPI_ISL_689434 | Japan/PG-6323/2020 | 20-Aug | Asia/Japan/Japan/ | Pathogen Genomics Center, National Institute of Infectious Diseases | Pathogen Genomics Center, National Institute of Infectious Diseases | Tsuyoshi Sekizuka et al |

| | | | | | | |
|---|---|---|---|---|---|---|
| EPI_ISL_689528 | Japan/PG-6422/2020 | 20-Jul | Asia/Japan/Japan/ | Pathogen Genomics Center, National Institute of Infectious Diseases | Pathogen Genomics Center, National Institute of Infectious Diseases | Tsuyoshi Sekizuka et al |
| EPI_ISL_689578 | Japan/PG-6473/2020 | 20-Aug | Asia/Japan/Japan/ | Pathogen Genomics Center, National Institute of Infectious Diseases | Pathogen Genomics Center, National Institute of Infectious Diseases | Tsuyoshi Sekizuka et al |
| EPI_ISL_689581 | Japan/PG-6476/2020 | 20-Aug | Asia/Japan/Japan/ | Pathogen Genomics Center, National Institute of Infectious Diseases | Pathogen Genomics Center, National Institute of Infectious Diseases | Tsuyoshi Sekizuka et al |
| EPI_ISL_689603 | Japan/PG-6501/2020 | 20-Jul | Asia/Japan/Japan/ | Pathogen Genomics Center, National Institute of Infectious Diseases | Pathogen Genomics Center, National Institute of Infectious Diseases | Tsuyoshi Sekizuka et al |
| EPI_ISL_689949 | Japan/PG-4825/2020 | 20-Jul | Asia/Japan/Japan/ | Pathogen Genomics Center, National Institute of Infectious Diseases | Pathogen Genomics Center, National Institute of Infectious Diseases | Tsuyoshi Sekizuka et al |
| EPI_ISL_690105 | Japan/PG-4992/2020 | 20-Aug | Asia/Japan/Japan/ | Pathogen Genomics Center, National Institute of Infectious Diseases | Pathogen Genomics Center, National Institute of Infectious Diseases | Tsuyoshi Sekizuka et al |
| EPI_ISL_690150 | Japan/PG-5039/2020 | 20-Jul | Asia/Japan/Japan/ | Pathogen Genomics Center, National Institute of Infectious Diseases | Pathogen Genomics Center, National Institute of Infectious Diseases | Tsuyoshi Sekizuka et al |
| EPI_ISL_690236 | Japan/PG-5129/2020 | 20-Aug | Asia/Japan/Japan/ | Gunma Prefectural Institute of Public Health and Environmental Sciences | Pathogen Genomics Center, National Institute of Infectious Diseases | Tsuyoshi Sekizuka et al |

| | | | | | | |
|---|---|---|---|---|---|---|
| EPI_ISL_690261 | Japan/PG-5154/2020 | 20-Aug | Asia/Japan/Japan/ | Pathogen Genomics Center, National Institute of Infectious Diseases | Pathogen Genomics Center, National Institute of Infectious Diseases | Tsuyoshi Sekizuka et al |
| EPI_ISL_690321 | Japan/PG-5218/2020 | 20-Aug | Asia/Japan/Japan/ | Pathogen Genomics Center, National Institute of Infectious Diseases | Pathogen Genomics Center, National Institute of Infectious Diseases | Tsuyoshi Sekizuka et al |
| EPI_ISL_690570 | Japan/PG-5467/2020 | 20-Aug | Asia/Japan/Japan/ | Pathogen Genomics Center, National Institute of Infectious Diseases | Pathogen Genomics Center, National Institute of Infectious Diseases | Tsuyoshi Sekizuka et al |
| EPI_ISL_690624 | Japan/PG-3387/2020 | 20-May | Asia/Japan/Japan/ | Pathogen Genomics Center, National Institute of Infectious Diseases | Pathogen Genomics Center, National Institute of Infectious Diseases | Tsuyoshi Sekizuka et al |
| EPI_ISL_690672 | Japan/PG-2537/2020 | 20-May | Asia/Japan/Japan/ | Saitama Prefectural Institute of Public Health | Pathogen Genomics Center, National Institute of Infectious Diseases | Tsuyoshi Sekizuka et al |
| EPI_ISL_690745 | Japan/PG-2611/2020 | 20-Feb | Asia/Japan/Japan/ | Pathogen Genomics Center, National Institute of Infectious Diseases | Pathogen Genomics Center, National Institute of Infectious Diseases | Tsuyoshi Sekizuka et al |
| EPI_ISL_690756 | Japan/PG-2623/2020 | 20-Apr | Asia/Japan/Japan/ | Pathogen Genomics Center, National Institute of Infectious Diseases | Pathogen Genomics Center, National Institute of Infectious Diseases | Tsuyoshi Sekizuka et al |
| EPI_ISL_690968 | Japan/PG-2841/2020 | 20-Apr | Asia/Japan/Japan/ | Pathogen Genomics Center, National Institute of Infectious Diseases | Pathogen Genomics Center, National Institute of Infectious Diseases | Tsuyoshi Sekizuka et al |

| | | | | | | |
|---|---|---|---|---|---|---|
| EPI_ISL_690970 | Japan/PG-2843/2020 | 20-Apr | Asia/Japan/Japan/ | Pathogen Genomics Center, National Institute of Infectious Diseases | Pathogen Genomics Center, National Institute of Infectious Diseases | Tsuyoshi Sekizuka et al |
| EPI_ISL_691039 | Japan/PG-2913/2020 | 20-Apr | Asia/Japan/Japan/ | Pathogen Genomics Center, National Institute of Infectious Diseases | Pathogen Genomics Center, National Institute of Infectious Diseases | Tsuyoshi Sekizuka et al |
| EPI_ISL_691136 | Japan/PG-3012/2020 | 20-Apr | Asia/Japan/Japan/ | Pathogen Genomics Center, National Institute of Infectious Diseases | Pathogen Genomics Center, National Institute of Infectious Diseases | Tsuyoshi Sekizuka et al |
| EPI_ISL_691147 | Japan/PG-3023/2020 | 20-Apr | Asia/Japan/Japan/ | Pathogen Genomics Center, National Institute of Infectious Diseases | Pathogen Genomics Center, National Institute of Infectious Diseases | Tsuyoshi Sekizuka et al |
| EPI_ISL_691149 | Japan/PG-3026/2020 | 20-Apr | Asia/Japan/Japan/ | Pathogen Genomics Center, National Institute of Infectious Diseases | Pathogen Genomics Center, National Institute of Infectious Diseases | Tsuyoshi Sekizuka et al |
| EPI_ISL_691301 | Japan/PG-3181/2020 | 20-Apr | Asia/Japan/Japan/ | Pathogen Genomics Center, National Institute of Infectious Diseases | Pathogen Genomics Center, National Institute of Infectious Diseases | Tsuyoshi Sekizuka et al |
| EPI_ISL_691407 | Japan/PG-3292/2020 | 20-Apr | Asia/Japan/Japan/ | Pathogen Genomics Center, National Institute of Infectious Diseases | Pathogen Genomics Center, National Institute of Infectious Diseases | Tsuyoshi Sekizuka et al |
| EPI_ISL_691508 | Japan/PG-3394/2020 | 20-Apr | Asia/Japan/Japan/ | Pathogen Genomics Center, National Institute of Infectious Diseases | Pathogen Genomics Center, National Institute of Infectious Diseases | Tsuyoshi Sekizuka et al |

| | | | | | | |
|---|---|---|---|---|---|---|
| EPI_ISL_691608 | Japan/PG-3499/2020 | 20-Jul | Asia/Japan/Japan/ | Pathogen Genomics Center, National Institute of Infectious Diseases | Pathogen Genomics Center, National Institute of Infectious Diseases | Tsuyoshi Sekizuka et al |
| EPI_ISL_691612 | Spain/IB-IBV-99010822/2020 | 2020-08-03 | Europe/Spain/Balear Islands/Palma de M | Servicio de Microbiologv≠a, Hospital Universitario Son Espases | SeqCOVID-SPAIN consortium/IBV(CSIC) | Carla Lv≥pez-Causapv© et al |
| EPI_ISL_691613 | Spain/IB-IBV-99010830/2020 | 2020-08-13 | Europe/Spain/Balear Islands/Palma de M | Servicio de Microbiologv≠a, Hospital Universitario Son Espases | SeqCOVID-SPAIN consortium/IBV(CSIC) | Carla Lv≥pez-Causapv© et al |
| EPI_ISL_691614 | Spain/IB-IBV-99010842/2020 | 2020-09-09 | Europe/Spain/Balear Islands/Palma de M | Servicio de Microbiologv≠a, Hospital Universitario Son Espases | SeqCOVID-SPAIN consortium/IBV(CSIC) | Carla Lv≥pez-Causapv© et al |
| EPI_ISL_691615 | Spain/IB-IBV-99010784/2020 | 2020-10-15 | Europe/Spain/Balear Islands/Manacor | Servicio de Microbiologv≠a, Hospital Universitario Son Espases | SeqCOVID-SPAIN consortium/IBV(CSIC) | Carla Lv≥pez-Causapv© et al |
| EPI_ISL_691622 | Spain/IB-IBV-99010825/2020 | 2020-08-03 | Europe/Spain/Balear Islands/Palma de M | Servicio de Microbiologv≠a, Hospital Universitario Son Espases | SeqCOVID-SPAIN consortium/IBV(CSIC) | Carla Lv≥pez-Causapv© et al |
| EPI_ISL_691625 | Spain/IB-IBV-99010789/2020 | 2020-10-17 | Europe/Spain/Balear Islands/Ibiza | Servicio de Microbiologv≠a, Hospital Universitario Son Espases | SeqCOVID-SPAIN consortium/IBV(CSIC) | Carla Lv≥pez-Causapv© et al |
| EPI_ISL_691627 | Spain/IB-IBV-99010827/2020 | 2020-08-13 | Europe/Spain/Balear Islands/Palma de M | Servicio de Microbiologv≠a, Hospital Universitario Son Espases | SeqCOVID-SPAIN consortium/IBV(CSIC) | Carla Lv≥pez-Causapv© et al |
| EPI_ISL_691631 | Spain/IB-IBV-99010815/2020 | 2020-07-30 | Europe/Spain/Balear Islands/Palma de M | Servicio de Microbiologv≠a, Hospital Universitario Son Espases | SeqCOVID-SPAIN consortium/IBV(CSIC) | Carla Lv≥pez-Causapv© et al |
| EPI_ISL_691635 | Spain/IB-IBV-99010832/2020 | 2020-08-17 | Europe/Spain/Balear Islands/Palma de M | Servicio de Microbiologv≠a, Hospital Universitario Son Espases | SeqCOVID-SPAIN consortium/IBV(CSIC) | Carla Lv≥pez-Causapv© et al |
| EPI_ISL_691637 | Spain/IB-IBV-99010812/2020 | 2020-07-23 | Europe/Spain/Balear Islands/Can Pastilla | Servicio de Microbiologv≠a, Hospital Universitario Son Espases | SeqCOVID-SPAIN consortium/IBV(CSIC) | Carla Lv≥pez-Causapv© et al |
| EPI_ISL_691638 | Spain/IB-IBV-99010820/2020 | 2020-08-01 | Europe/Spain/Balear Islands/Palma de M | Servicio de Microbiologv≠a, Hospital Universitario Son Espases | SeqCOVID-SPAIN consortium/IBV(CSIC) | Carla Lv≥pez-Causapv© et al |
| EPI_ISL_691644 | Spain/IB-IBV-99010776/2020 | 2020-09-30 | Europe/Spain/Balear Islands/Ibiza | Servicio de Microbiologv≠a, Hospital Universitario Son Espases | SeqCOVID-SPAIN consortium/IBV(CSIC) | Carla Lv≥pez-Causapv© et al |

| | | | | | | |
|---|---|---|---|---|---|---|
| EPI_ISL_691646 | Spain/IB-IBV-99010844/2020 | 2020-09-05 | Europe/Spain/Balear Islands/Palma de M | Servicio de Microbiolog√≠a, Hospital Universitario Son Espases | SeqCOVID-SPAIN consortium/IBV(CSIC) | Carla L√≥pez-Causap√© et al |
| EPI_ISL_691647 | Spain/IB-IBV-99010828/2020 | 2020-08-13 | Europe/Spain/Balear Islands/Palma de M | Servicio de Microbiolog√≠a, Hospital Universitario Son Espases | SeqCOVID-SPAIN consortium/IBV(CSIC) | Carla L√≥pez-Causap√© et al |
| EPI_ISL_691648 | Spain/IB-IBV-99010843/2020 | 2020-09-09 | Europe/Spain/Balear Islands/Palma de M | Servicio de Microbiolog√≠a, Hospital Universitario Son Espases | SeqCOVID-SPAIN consortium/IBV(CSIC) | Carla L√≥pez-Causap√© et al |
| EPI_ISL_691649 | Spain/IB-IBV-99010834/2020 | 2020-08-16 | Europe/Spain/Balear Islands/Palma de M | Servicio de Microbiolog√≠a, Hospital Universitario Son Espases | SeqCOVID-SPAIN consortium/IBV(CSIC) | Carla L√≥pez-Causap√© et al |
| EPI_ISL_691650 | Spain/IB-IBV-99010833/2020 | 2020-08-12 | Europe/Spain/Balear Islands/Palma de M | Servicio de Microbiolog√≠a, Hospital Universitario Son Espases | SeqCOVID-SPAIN consortium/IBV(CSIC) | Carla L√≥pez-Causap√© et al |
| EPI_ISL_691653 | Spain/IB-IBV-99010824/2020 | 2020-08-03 | Europe/Spain/Balear Islands/Palma de M | Servicio de Microbiolog√≠a, Hospital Universitario Son Espases | SeqCOVID-SPAIN consortium/IBV(CSIC) | Carla L√≥pez-Causap√© et al |
| EPI_ISL_691654 | Spain/IB-IBV-99010837/2020 | 2020-08-20 | Europe/Spain/Balear Islands/Palma de M | Servicio de Microbiolog√≠a, Hospital Universitario Son Espases | SeqCOVID-SPAIN consortium/IBV(CSIC) | Carla L√≥pez-Causap√© et al |
| EPI_ISL_691658 | Spain/IB-IBV-99010821/2020 | 2020-08-02 | Europe/Spain/Balear Islands/Palma de M | Servicio de Microbiolog√≠a, Hospital Universitario Son Espases | SeqCOVID-SPAIN consortium/IBV(CSIC) | Carla L√≥pez-Causap√© et al |
| EPI_ISL_691668 | Spain/IB-IBV-99010775/2020 | 2020-09-30 | Europe/Spain/Balear Islands/Ibiza | Servicio de Microbiolog√≠a, Hospital Universitario Son Espases | SeqCOVID-SPAIN consortium/IBV(CSIC) | Carla L√≥pez-Causap√© et al |
| EPI_ISL_691669 | Spain/IB-IBV-99010777/2020 | 2020-10-06 | Europe/Spain/Balear Islands/Manacor | Servicio de Microbiolog√≠a, Hospital Universitario Son Espases | SeqCOVID-SPAIN consortium/IBV(CSIC) | Carla L√≥pez-Causap√© et al |
| EPI_ISL_691672 | Spain/IB-IBV-99010782/2020 | 2020-10-06 | Europe/Spain/Balear Islands/Campos | Servicio de Microbiolog√≠a, Hospital Universitario Son Espases | SeqCOVID-SPAIN consortium/IBV(CSIC) | Carla L√≥pez-Causap√© et al |
| EPI_ISL_691673 | Spain/IB-IBV-99010783/2020 | 2020-10-06 | Europe/Spain/Balear Islands/Vilafranca | Servicio de Microbiolog√≠a, Hospital Universitario Son Espases | SeqCOVID-SPAIN consortium/IBV(CSIC) | Carla L√≥pez-Causap√© et al |
| EPI_ISL_691678 | Spain/IB-IBV-99010804/2020 | 2020-09-30 | Europe/Spain/Balear Islands/Ibiza | Servicio de Microbiolog√≠a, Hospital Universitario Son Espases | SeqCOVID-SPAIN consortium/IBV(CSIC) | Carla L√≥pez-Causap√© et al |

| | | | | | | |
|---|---|---|---|---|---|---|
| EPI_ISL_691680 | Spain/IB-IBV-99010811/2020 | 2020-07-20 | Europe/Spain/Balear Islands/Can Pastilla | Servicio de Microbiolog√≠a, Hospital Universitario Son Espases | SeqCOVID-SPAIN consortium/IBV(CSIC) | Carla L√≥pez-Causap√© et al |
| EPI_ISL_691682 | Spain/CM-ISCIII-2010312/2020 | 2020-05-09 | Europe/Spain/Castilla la Mancha/ | Hospital General Universitario de Ciudad Real | Instituto de Salud Carlos III | Iglesias-Caballero et al |
| EPI_ISL_691684 | Spain/CM-ISCIII-2010740/2020 | 2020-05-13 | Europe/Spain/Castilla la Mancha/ | Complejo Hospitalario Universitario de Albacete | Instituto de Salud Carlos III | Iglesias-Caballero et al |
| EPI_ISL_691690 | Spain/CE-ISCIII-2016506/2020 | 2020-10-14 | Europe/Spain/Ceuta/ | Hospital Universitario de Ceuta | Instituto de Salud Carlos III | Iglesias-Caballero et al |
| EPI_ISL_691695 | Spain/CE-ISCIII-2016516/2020 | 2020-10-14 | Europe/Spain/Ceuta/ | Hospital Universitario de Ceuta | Instituto de Salud Carlos III | Iglesias-Caballero et al |
| EPI_ISL_691699 | Spain/CE-ISCIII-2016520/2020 | 2020-10-14 | Europe/Spain/Ceuta/ | Hospital Universitario de Ceuta | Instituto de Salud Carlos III | Iglesias-Caballero et al |
| EPI_ISL_691701 | Spain/MD-ISCIII-2014592/2020 | 2020-08-19 | Europe/Spain/Madrid/ | Hospital Cl√≠nico San Carlos | Instituto de Salud Carlos III | Iglesias-Caballero et al |
| EPI_ISL_691703 | Spain/MD-ISCIII-2014601/2020 | 2020-08-19 | Europe/Spain/Madrid/ | Hospital Cl√≠nico San Carlos | Instituto de Salud Carlos III | Iglesias-Caballero et al |
| EPI_ISL_691706 | Spain/MD-ISCIII-2014616/2020 | 2020-08-19 | Europe/Spain/Madrid/ | Hospital Cl√≠nico San Carlos | Instituto de Salud Carlos III | Iglesias-Caballero et al |
| EPI_ISL_691708 | Spain/MD-ISCIII-2014623/2020 | 2020-08-18 | Europe/Spain/Madrid/ | Hospital Cl√≠nico San Carlos | Instituto de Salud Carlos III | Iglesias-Caballero et al |
| EPI_ISL_691710 | Spain/MD-ISCIII-2014625/2020 | 2020-08-18 | Europe/Spain/Madrid/ | Hospital Cl√≠nico San Carlos | Instituto de Salud Carlos III | Iglesias-Caballero et al |
| EPI_ISL_691711 | Spain/MD-ISCIII-2014627/2020 | 2020-08-18 | Europe/Spain/Madrid/ | Hospital Cl√≠nico San Carlos | Instituto de Salud Carlos III | Iglesias-Caballero et al |
| EPI_ISL_691717 | Spain/MD-ISCIII-2014662/2020 | 2020-08-18 | Europe/Spain/Madrid/ | Hospital Cl√≠nico San Carlos | Instituto de Salud Carlos III | Iglesias-Caballero et al |
| EPI_ISL_691719 | Spain/MD-ISCIII-2014664/2020 | 2020-08-18 | Europe/Spain/Madrid/ | Hospital Cl√≠nico San Carlos | Instituto de Salud Carlos III | Iglesias-Caballero et al |
| EPI_ISL_691720 | Spain/MD-ISCIII-2014669/2020 | 2020-08-18 | Europe/Spain/Madrid/ | Hospital Cl√≠nico San Carlos | Instituto de Salud Carlos III | Iglesias-Caballero et al |
| EPI_ISL_691723 | Spain/MD-ISCIII-2014822/2020 | 2020-08-26 | Europe/Spain/Madrid/ | Hospital Cl√≠nico San Carlos | Instituto de Salud Carlos III | Iglesias-Caballero et al |
| EPI_ISL_691724 | Spain/MD-ISCIII-2014824/2020 | 2020-08-26 | Europe/Spain/Madrid/ | Hospital Cl√≠nico San Carlos | Instituto de Salud Carlos III | Iglesias-Caballero et al |
| EPI_ISL_691727 | Spain/MD-ISCIII-2014829/2020 | 2020-08-26 | Europe/Spain/Madrid/ | Hospital Cl√≠nico San Carlos | Instituto de Salud Carlos III | Iglesias-Caballero et al |
| EPI_ISL_691728 | Spain/MD-ISCIII-2014937/2020 | 2020-08-26 | Europe/Spain/Madrid/ | Hospital Cl√≠nico San Carlos | Instituto de Salud Carlos III | Iglesias-Caballero et al |

| | | | | | | |
|---|---|---|---|---|---|---|
| EPI_ISL_691729 | Spain/MD-ISCIII-2014833/2020 | 2020-09-01 | Europe/Spain/Madrid/ | Hospital Universitario Severo Ochoa | Instituto de Salud Carlos III | Iglesias-Caballero et al |
| EPI_ISL_691730 | Spain/MD-ISCIII-2015114/2020 | 2020-09-09 | Europe/Spain/Madrid/ | Hospital Universitario Severo Ochoa | Instituto de Salud Carlos III | Iglesias-Caballero et al |
| EPI_ISL_691732 | Spain/MD-ISCIII-2016128/2020 | 2020-10-09 | Europe/Spain/Madrid/ | Hospital de Madrid | Instituto de Salud Carlos III | Iglesias-Caballero et al |
| EPI_ISL_691893 | Japan/PG-3640/2020 | 20-Apr | Asia/Japan/Japan/ | Pathogen Genomics Center, National Institute of Infectious Diseases | Pathogen Genomics Center, National Institute of Infectious Diseases | Tsuyoshi Sekizuka et al |
| EPI_ISL_691928 | Japan/PG-3675/2020 | 20-Apr | Asia/Japan/Japan/ | Pathogen Genomics Center, National Institute of Infectious Diseases | Pathogen Genomics Center, National Institute of Infectious Diseases | Tsuyoshi Sekizuka et al |
| EPI_ISL_691994 | Japan/PG-3741/2020 | 20-Apr | Asia/Japan/Japan/ | Pathogen Genomics Center, National Institute of Infectious Diseases | Pathogen Genomics Center, National Institute of Infectious Diseases | Tsuyoshi Sekizuka et al |
| EPI_ISL_692151 | Japan/PG-3900/2020 | 20-Jul | Asia/Japan/Japan/ | Pathogen Genomics Center, National Institute of Infectious Diseases | Pathogen Genomics Center, National Institute of Infectious Diseases | Tsuyoshi Sekizuka et al |
| EPI_ISL_692260 | Japan/PG-4019/2020 | 20-Jul | Asia/Japan/Japan/ | Pathogen Genomics Center, National Institute of Infectious Diseases | Pathogen Genomics Center, National Institute of Infectious Diseases | Tsuyoshi Sekizuka et al |
| EPI_ISL_692337 | Japan/PG-4096/2020 | 20-Jul | Asia/Japan/Japan/ | Pathogen Genomics Center, National Institute of Infectious Diseases | Pathogen Genomics Center, National Institute of Infectious Diseases | Tsuyoshi Sekizuka et al |

| | | | | | | |
|---|---|---|---|---|---|---|
| EPI_ISL_692402 | Japan/PG-4163/2020 | 20-Jul | Asia/Japan/Japan/ | Pathogen Genomics Center, National Institute of Infectious Diseases | Pathogen Genomics Center, National Institute of Infectious Diseases | Tsuyoshi Sekizuka et al |
| EPI_ISL_692454 | Japan/PG-4216/2020 | 20-Jul | Asia/Japan/Japan/ | Fukuoka Institute of Health and Environmental Sciences | Pathogen Genomics Center, National Institute of Infectious Diseases | Tsuyoshi Sekizuka et al |
| EPI_ISL_692618 | Japan/PG-4391/2020 | 20-Jul | Asia/Japan/Japan/ | Pathogen Genomics Center, National Institute of Infectious Diseases | Pathogen Genomics Center, National Institute of Infectious Diseases | Tsuyoshi Sekizuka et al |
| EPI_ISL_692736 | France/ARA-61889/2020 | 2020-08-18 | Europe/France/Auvergne-Rh√¥ne-Alpes/ | CNR Virus des Infections Respiratoires - France SUD | CNR Virus des Infections Respiratoires - France SUD | Antonin Bal et al |
| EPI_ISL_692737 | France/ARA-66327/2020 | 2020-09-01 | Europe/France/Auvergne-Rh√¥ne-Alpes/ | CNR Virus des Infections Respiratoires - France SUD | CNR Virus des Infections Respiratoires - France SUD | Antonin Bal et al |
| EPI_ISL_692738 | France/ARA-66446/2020 | 2020-09-01 | Europe/France/Auvergne-Rh√¥ne-Alpes/ | CNR Virus des Infections Respiratoires - France SUD | CNR Virus des Infections Respiratoires - France SUD | Antonin Bal et al |
| EPI_ISL_692739 | France/ARA-67140/2020 | 2020-09-02 | Europe/France/Auvergne-Rh√¥ne-Alpes/ | CNR Virus des Infections Respiratoires - France SUD | CNR Virus des Infections Respiratoires - France SUD | Antonin Bal et al |
| EPI_ISL_692740 | France/ARA-67616/2020 | 2020-09-03 | Europe/France/Auvergne-Rh√¥ne-Alpes/ | CNR Virus des Infections Respiratoires - France SUD | CNR Virus des Infections Respiratoires - France SUD | Antonin Bal et al |
| EPI_ISL_692742 | France/ARA-67716/2020 | 2020-09-04 | Europe/France/Auvergne-Rh√¥ne-Alpes/ | CNR Virus des Infections Respiratoires - France SUD | CNR Virus des Infections Respiratoires - France SUD | Antonin Bal et al |

| | | | | | | |
|---|---|---|---|---|---|---|
| EPI_ISL_692743 | France/ARA-6773/2020 | 2020-09-04 | Europe/France/Auvergne-Rh√¥ne-Alpes/ | CNR Virus des Infections Respiratoires - France SUD | CNR Virus des Infections Respiratoires - France SUD | Antonin Bal et al |
| EPI_ISL_692744 | France/ARA-68122/2020 | 2020-09-05 | Europe/France/Auvergne-Rh√¥ne-Alpes/ | CNR Virus des Infections Respiratoires - France SUD | CNR Virus des Infections Respiratoires - France SUD | Antonin Bal et al |
| EPI_ISL_692745 | France/ARA-70123/2020 | 2020-09-09 | Europe/France/Auvergne-Rh√¥ne-Alpes/ | CNR Virus des Infections Respiratoires - France SUD | CNR Virus des Infections Respiratoires - France SUD | Antonin Bal et al |
| EPI_ISL_692747 | France/ARA-71440/2020 | 2020-09-11 | Europe/France/Auvergne-Rh√¥ne-Alpes/ | CNR Virus des Infections Respiratoires - France SUD | CNR Virus des Infections Respiratoires - France SUD | Antonin Bal et al |
| EPI_ISL_692749 | France/ARA-71532/2020 | 2020-09-12 | Europe/France/Auvergne-Rh√¥ne-Alpes/ | CNR Virus des Infections Respiratoires - France SUD | CNR Virus des Infections Respiratoires - France SUD | Antonin Bal et al |
| EPI_ISL_692750 | France/ARA-72111/2020 | 2020-09-14 | Europe/France/Auvergne-Rh√¥ne-Alpes/ | CNR Virus des Infections Respiratoires - France SUD | CNR Virus des Infections Respiratoires - France SUD | Antonin Bal et al |
| EPI_ISL_692751 | France/ARA-7212/2020 | 2020-09-14 | Europe/France/Auvergne-Rh√¥ne-Alpes/ | CNR Virus des Infections Respiratoires - France SUD | CNR Virus des Infections Respiratoires - France SUD | Antonin Bal et al |
| EPI_ISL_692753 | France/ARA-73624/2020 | 2020-09-16 | Europe/France/Auvergne-Rh√¥ne-Alpes/ | CNR Virus des Infections Respiratoires - France SUD | CNR Virus des Infections Respiratoires - France SUD | Antonin Bal et al |
| EPI_ISL_692754 | France/ARA-74140/2020 | 2020-09-17 | Europe/France/Auvergne-Rh√¥ne-Alpes/ | CNR Virus des Infections Respiratoires - France SUD | CNR Virus des Infections Respiratoires - France SUD | Antonin Bal et al |
| EPI_ISL_692755 | France/ARA-57804/2020 | 2020-08-03 | Europe/France/Auvergne-Rh√¥ne-Alpes/ | CNR Virus des Infections Respiratoires - France SUD | CNR Virus des Infections Respiratoires - France SUD | Antonin Bal et al |

| | | | | | | |
|---|---|---|---|---|---|---|
| EPI_ISL_692758 | France/ARA-62907/2020 | 2020-08-22 | Europe/France/Auvergne-Rhv¥ne-Alpes/ | CNR Virus des Infections Respiratoires - France SUD | CNR Virus des Infections Respiratoires - France SUD | Antonin Bal et al |
| EPI_ISL_692760 | France/ARA-64304/2020 | 2020-08-26 | Europe/France/Auvergne-Rhv¥ne-Alpes/ | CNR Virus des Infections Respiratoires - France SUD | CNR Virus des Infections Respiratoires - France SUD | Antonin Bal et al |
| EPI_ISL_692764 | France/ARA-67612/2020 | 2020-09-04 | Europe/France/Auvergne-Rhv¥ne-Alpes/ | CNR Virus des Infections Respiratoires - France SUD | CNR Virus des Infections Respiratoires - France SUD | Antonin Bal et al |
| EPI_ISL_692765 | France/ARA-67686/2020 | 2020-09-04 | Europe/France/Auvergne-Rhv¥ne-Alpes/ | CNR Virus des Infections Respiratoires - France SUD | CNR Virus des Infections Respiratoires - France SUD | Antonin Bal et al |
| EPI_ISL_692766 | France/ARA-67717/2020 | 2020-09-04 | Europe/France/Auvergne-Rhv¥ne-Alpes/ | CNR Virus des Infections Respiratoires - France SUD | CNR Virus des Infections Respiratoires - France SUD | Antonin Bal et al |
| EPI_ISL_692767 | France/ARA-6777/2020 | 2020-09-04 | Europe/France/Auvergne-Rhv¥ne-Alpes/ | CNR Virus des Infections Respiratoires - France SUD | CNR Virus des Infections Respiratoires - France SUD | Antonin Bal et al |
| EPI_ISL_692771 | USA/MA-MASPHL-01088/2020 | 2020-11-30 | North America/USA/Massachusetts/ | Massachusetts State Public Health Laboratory | Massachusetts State Public Health Laboratory | Andrew Lang et al |
| EPI_ISL_692778 | USA/MA-MASPHL-01035/2020 | 2020-11-17 | North America/USA/Massachusetts/ | Massachusetts State Public Health Laboratory | Massachusetts State Public Health Laboratory | Andrew Lang et al |
| EPI_ISL_692781 | USA/MA-MASPHL-01030/2020 | 2020-11-14 | North America/USA/Massachusetts/ | Massachusetts State Public Health Laboratory | Massachusetts State Public Health Laboratory | Andrew Lang et al |
| EPI_ISL_692784 | USA/MA-MASPHL-01027/2020 | 2020-11-05 | North America/USA/Massachusetts/ | Massachusetts State Public Health Laboratory | Massachusetts State Public Health Laboratory | Andrew Lang et al |
| EPI_ISL_692787 | USA/MA-MASPHL-01021/2020 | 2020-11-10 | North America/USA/Massachusetts/ | Massachusetts State Public Health Laboratory | Massachusetts State Public Health Laboratory | Andrew Lang et al |

| | | | | | | |
|---|---|---|---|---|---|---|
| EPI_ISL_692792 | USA/MA-MASPHL-01016/2020 | 2020-11-06 | North America/USA/Massachusetts/ | Massachusetts State Public Health Laboratory | Massachusetts State Public Health Laboratory | Andrew Lang et al |
| EPI_ISL_692799 | USA/MA-MASPHL-00993/2020 | 2020-11-08 | North America/USA/Massachusetts/ | Massachusetts State Public Health Laboratory | Massachusetts State Public Health Laboratory | Andrew Lang et al |
| EPI_ISL_692820 | USA/MA-MASPHL-00942/2020 | 2020-10-26 | North America/USA/Massachusetts/ | Massachusetts State Public Health Laboratory | Massachusetts State Public Health Laboratory | Andrew Lang et al |
| EPI_ISL_692825 | USA/MA-MASPHL-00928/2020 | 2020-05-28 | North America/USA/Massachusetts/ | Massachusetts State Public Health Laboratory | Massachusetts State Public Health Laboratory | Andrew Lang et al |
| EPI_ISL_692830 | USA/MA-MASPHL-00922/2020 | 2020-05-26 | North America/USA/Massachusetts/ | Massachusetts State Public Health Laboratory | Massachusetts State Public Health Laboratory | Andrew Lang et al |
| EPI_ISL_692832 | USA/MA-MASPHL-00918/2020 | 2020-04-28 | North America/USA/Massachusetts/ | Massachusetts State Public Health Laboratory | Massachusetts State Public Health Laboratory | Andrew Lang et al |
| EPI_ISL_692836 | USA/MA-MASPHL-00911/2020 | 2020-10-19 | North America/USA/Massachusetts/ | Massachusetts State Public Health Laboratory | Massachusetts State Public Health Laboratory | Andrew Lang et al |
| EPI_ISL_692843 | USA/MA-MASPHL-00903/2020 | 2020-10-19 | North America/USA/Massachusetts/ | Massachusetts State Public Health Laboratory | Massachusetts State Public Health Laboratory | Andrew Lang et al |
| EPI_ISL_692847 | USA/MA-MASPHL-00886/2020 | 2020-10-11 | North America/USA/Massachusetts/ | Massachusetts State Public Health Laboratory | Massachusetts State Public Health Laboratory | Andrew Lang et al |
| EPI_ISL_692853 | USA/MA-MASPHL-00879/2020 | 2020-10-16 | North America/USA/Massachusetts/ | Massachusetts State Public Health Laboratory | Massachusetts State Public Health Laboratory | Andrew Lang et al |
| EPI_ISL_692858 | USA/MA-MASPHL-00867/2020 | 2020-05-20 | North America/USA/Massachusetts/ | Massachusetts State Public Health Laboratory | Massachusetts State Public Health Laboratory | Andrew Lang et al |
| EPI_ISL_692869 | USA/MA-MASPHL-00834/2020 | 2020-10-14 | North America/USA/Massachusetts/ | Massachusetts State Public Health Laboratory | Massachusetts State Public Health Laboratory | Andrew Lang et al |
| EPI_ISL_692875 | USA/MA-MASPHL-00826/2020 | 2020-06-04 | North America/USA/Massachusetts/ | Massachusetts State Public Health Laboratory | Massachusetts State Public Health Laboratory | Andrew Lang et al |

| | | | | | | |
|---|---|---|---|---|---|---|
| EPI_ISL_692876 | USA/MA-MASPHL-00825/2020 | 2020-06-04 | North America/USA/Massachusetts/ | Massachusetts State Public Health Laboratory | Massachusetts State Public Health Laboratory | Andrew Lang et al |
| EPI_ISL_692887 | USA/MA-MASPHL-00813/2020 | 2020-05-26 | North America/USA/Massachusetts/ | Massachusetts State Public Health Laboratory | Massachusetts State Public Health Laboratory | Andrew Lang et al |
| EPI_ISL_692888 | USA/MA-MASPHL-00812/2020 | 2020-05-26 | North America/USA/Massachusetts/ | Massachusetts State Public Health Laboratory | Massachusetts State Public Health Laboratory | Andrew Lang et al |
| EPI_ISL_692891 | USA/MA-MASPHL-00809/2020 | 2020-05-26 | North America/USA/Massachusetts/ | Massachusetts State Public Health Laboratory | Massachusetts State Public Health Laboratory | Andrew Lang et al |
| EPI_ISL_692900 | USA/MA-MASPHL-00800/2020 | 2020-05-20 | North America/USA/Massachusetts/ | Massachusetts State Public Health Laboratory | Massachusetts State Public Health Laboratory | Andrew Lang et al |
| EPI_ISL_692909 | USA/MA-MASPHL-00789/2020 | 2020-09-27 | North America/USA/Massachusetts/ | Massachusetts State Public Health Laboratory | Massachusetts State Public Health Laboratory | Andrew Lang et al |
| EPI_ISL_692910 | USA/MA-MASPHL-00788/2020 | 2020-09-24 | North America/USA/Massachusetts/ | Massachusetts State Public Health Laboratory | Massachusetts State Public Health Laboratory | Andrew Lang et al |
| EPI_ISL_692916 | USA/MA-MASPHL-00780/2020 | 2020-09-25 | North America/USA/Massachusetts/ | Massachusetts State Public Health Laboratory | Massachusetts State Public Health Laboratory | Andrew Lang et al |
| EPI_ISL_692919 | USA/MA-MASPHL-00776/2020 | 2020-09-25 | North America/USA/Massachusetts/ | Massachusetts State Public Health Laboratory | Massachusetts State Public Health Laboratory | Andrew Lang et al |
| EPI_ISL_692920 | USA/MA-MASPHL-00775/2020 | 2020-09-25 | North America/USA/Massachusetts/ | Massachusetts State Public Health Laboratory | Massachusetts State Public Health Laboratory | Andrew Lang et al |
| EPI_ISL_692924 | USA/MA-MASPHL-00771/2020 | 2020-09-25 | North America/USA/Massachusetts/ | Massachusetts State Public Health Laboratory | Massachusetts State Public Health Laboratory | Andrew Lang et al |
| EPI_ISL_692926 | USA/MA-MASPHL-00748/2020 | 2020-09-21 | North America/USA/Massachusetts/ | Massachusetts State Public Health Laboratory | Massachusetts State Public Health Laboratory | Andrew Lang et al |
| EPI_ISL_692927 | USA/MA-MASPHL-00746/2020 | 2020-09-23 | North America/USA/Massachusetts/ | Massachusetts State Public Health Laboratory | Massachusetts State Public Health Laboratory | Andrew Lang et al |

| | | | | | | |
|---|---|---|---|---|---|---|
| EPI_ISL_692928 | USA/MA-MASPHL-00745/2020 | 2020-09-23 | North America/USA/Massachusetts/ | Massachusetts State Public Health Laboratory | Massachusetts State Public Health Laboratory | Andrew Lang et al |
| EPI_ISL_692931 | USA/MA-MASPHL-00742/2020 | 2020-09-23 | North America/USA/Massachusetts/ | Massachusetts State Public Health Laboratory | Massachusetts State Public Health Laboratory | Andrew Lang et al |
| EPI_ISL_692932 | USA/MA-MASPHL-00740/2020 | 2020-09-24 | North America/USA/Massachusetts/ | Massachusetts State Public Health Laboratory | Massachusetts State Public Health Laboratory | Andrew Lang et al |
| EPI_ISL_692933 | USA/MA-MASPHL-00739/2020 | 2020-09-24 | North America/USA/Massachusetts/ | Massachusetts State Public Health Laboratory | Massachusetts State Public Health Laboratory | Andrew Lang et al |
| EPI_ISL_692938 | USA/MA-MASPHL-00734/2020 | 2020-09-21 | North America/USA/Massachusetts/ | Massachusetts State Public Health Laboratory | Massachusetts State Public Health Laboratory | Andrew Lang et al |
| EPI_ISL_692942 | USA/MA-MASPHL-00730/2020 | 2020-09-22 | North America/USA/Massachusetts/ | Massachusetts State Public Health Laboratory | Massachusetts State Public Health Laboratory | Andrew Lang et al |
| EPI_ISL_692943 | USA/MA-MASPHL-00729/2020 | 2020-09-22 | North America/USA/Massachusetts/ | Massachusetts State Public Health Laboratory | Massachusetts State Public Health Laboratory | Andrew Lang et al |
| EPI_ISL_692946 | USA/MA-MASPHL-00726/2020 | 2020-09-24 | North America/USA/Massachusetts/ | Massachusetts State Public Health Laboratory | Massachusetts State Public Health Laboratory | Andrew Lang et al |
| EPI_ISL_692962 | USA/MA-MASPHL-00684/2020 | 2020-04-15 | North America/USA/Massachusetts/ | Massachusetts State Public Health Laboratory | Massachusetts State Public Health Laboratory | Andrew Lang et al |
| EPI_ISL_692973 | USA/MA-MASPHL-00662/2020 | 2020-05-18 | North America/USA/Massachusetts/ | Massachusetts State Public Health Laboratory | Massachusetts State Public Health Laboratory | Andrew Lang et al |
| EPI_ISL_692987 | USA/MA-MASPHL-00644/2020 | 2020-05-14 | North America/USA/Massachusetts/ | Massachusetts State Public Health Laboratory | Massachusetts State Public Health Laboratory | Andrew Lang et al |
| EPI_ISL_692996 | USA/MA-MASPHL-00632/2020 | 2020-05-12 | North America/USA/Massachusetts/ | Massachusetts State Public Health Laboratory | Massachusetts State Public Health Laboratory | Andrew Lang et al |
| EPI_ISL_693000 | USA/MA-MASPHL-00625/2020 | 2020-05-12 | North America/USA/Massachusetts/ | Massachusetts State Public Health Laboratory | Massachusetts State Public Health Laboratory | Andrew Lang et al |

| | | | | | | |
|---|---|---|---|---|---|---|
| EPI_ISL_693001 | USA/MA-MASPHL-00624/2020 | 2020-05-11 | North America/USA/Massachusetts/ | Massachusetts State Public Health Laboratory | Massachusetts State Public Health Laboratory | Andrew Lang et al |
| EPI_ISL_693012 | USA/MA-MASPHL-00608/2020 | 2020-05-11 | North America/USA/Massachusetts/ | Massachusetts State Public Health Laboratory | Massachusetts State Public Health Laboratory | Andrew Lang et al |
| EPI_ISL_693015 | USA/MA-MASPHL-00596/2020 | 2020-05-08 | North America/USA/Massachusetts/ | Massachusetts State Public Health Laboratory | Massachusetts State Public Health Laboratory | Andrew Lang et al |
| EPI_ISL_693018 | USA/MA-MASPHL-00591/2020 | 2020-05-08 | North America/USA/Massachusetts/ | Massachusetts State Public Health Laboratory | Massachusetts State Public Health Laboratory | Andrew Lang et al |
| EPI_ISL_693020 | USA/MA-MASPHL-00586/2020 | 2020-05-08 | North America/USA/Massachusetts/ | Massachusetts State Public Health Laboratory | Massachusetts State Public Health Laboratory | Andrew Lang et al |
| EPI_ISL_693022 | USA/MA-MASPHL-00582/2020 | 2020-05-08 | North America/USA/Massachusetts/ | Massachusetts State Public Health Laboratory | Massachusetts State Public Health Laboratory | Andrew Lang et al |
| EPI_ISL_693024 | USA/MA-MASPHL-00577/2020 | 2020-05-08 | North America/USA/Massachusetts/ | Massachusetts State Public Health Laboratory | Massachusetts State Public Health Laboratory | Andrew Lang et al |
| EPI_ISL_693031 | USA/MA-MASPHL-00556/2020 | 2020-05-08 | North America/USA/Massachusetts/ | Massachusetts State Public Health Laboratory | Massachusetts State Public Health Laboratory | Andrew Lang et al |
| EPI_ISL_693038 | USA/MA-MASPHL-00548/2020 | 2020-05-07 | North America/USA/Massachusetts/ | Massachusetts State Public Health Laboratory | Massachusetts State Public Health Laboratory | Andrew Lang et al |
| EPI_ISL_693050 | USA/MA-MASPHL-00528/2020 | 2020-05-05 | North America/USA/Massachusetts/ | Massachusetts State Public Health Laboratory | Massachusetts State Public Health Laboratory | Andrew Lang et al |
| EPI_ISL_693056 | USA/MA-MASPHL-00521/2020 | 2020-05-05 | North America/USA/Massachusetts/ | Massachusetts State Public Health Laboratory | Massachusetts State Public Health Laboratory | Andrew Lang et al |
| EPI_ISL_693062 | USA/MA-MASPHL-00512/2020 | 2020-05-03 | North America/USA/Massachusetts/ | Massachusetts State Public Health Laboratory | Massachusetts State Public Health Laboratory | Andrew Lang et al |
| EPI_ISL_693070 | USA/MA-MASPHL-00500/2020 | 2020-05-03 | North America/USA/Massachusetts/ | Massachusetts State Public Health Laboratory | Massachusetts State Public Health Laboratory | Andrew Lang et al |

| | | | | | | |
|---|---|---|---|---|---|---|
| EPI_ISL_693077 | USA/MA-MASPHL-00490/2020 | 2020-05-02 | North America/USA/Massachusetts/ | Massachusetts State Public Health Laboratory | Massachusetts State Public Health Laboratory | Andrew Lang et al |
| EPI_ISL_693081 | USA/MA-MASPHL-00486/2020 | 2020-05-01 | North America/USA/Massachusetts/ | Massachusetts State Public Health Laboratory | Massachusetts State Public Health Laboratory | Andrew Lang et al |
| EPI_ISL_693082 | USA/MA-MASPHL-00485/2020 | 2020-05-01 | North America/USA/Massachusetts/ | Massachusetts State Public Health Laboratory | Massachusetts State Public Health Laboratory | Andrew Lang et al |
| EPI_ISL_693083 | USA/MA-MASPHL-00484/2020 | 2020-05-01 | North America/USA/Massachusetts/ | Massachusetts State Public Health Laboratory | Massachusetts State Public Health Laboratory | Andrew Lang et al |
| EPI_ISL_693084 | USA/MA-MASPHL-00483/2020 | 2020-05-01 | North America/USA/Massachusetts/ | Massachusetts State Public Health Laboratory | Massachusetts State Public Health Laboratory | Andrew Lang et al |
| EPI_ISL_693093 | USA/MA-MASPHL-00470/2020 | 2020-04-30 | North America/USA/Massachusetts/ | Massachusetts State Public Health Laboratory | Massachusetts State Public Health Laboratory | Andrew Lang et al |
| EPI_ISL_693096 | USA/MA-MASPHL-00467/2020 | 2020-04-30 | North America/USA/Massachusetts/ | Massachusetts State Public Health Laboratory | Massachusetts State Public Health Laboratory | Andrew Lang et al |
| EPI_ISL_693119 | USA/MA-MASPHL-00438/2020 | 2020-04-28 | North America/USA/Massachusetts/ | Massachusetts State Public Health Laboratory | Massachusetts State Public Health Laboratory | Andrew Lang et al |
| EPI_ISL_693124 | USA/MA-MASPHL-00429/2020 | 2020-04-27 | North America/USA/Massachusetts/ | Massachusetts State Public Health Laboratory | Massachusetts State Public Health Laboratory | Andrew Lang et al |
| EPI_ISL_693135 | USA/MA-MASPHL-00416/2020 | 2020-04-27 | North America/USA/Massachusetts/ | Massachusetts State Public Health Laboratory | Massachusetts State Public Health Laboratory | Andrew Lang et al |
| EPI_ISL_693139 | USA/MA-MASPHL-00410/2020 | 2020-04-26 | North America/USA/Massachusetts/ | Massachusetts State Public Health Laboratory | Massachusetts State Public Health Laboratory | Andrew Lang et al |
| EPI_ISL_693142 | USA/MA-MASPHL-00406/2020 | 2020-04-25 | North America/USA/Massachusetts/ | Massachusetts State Public Health Laboratory | Massachusetts State Public Health Laboratory | Andrew Lang et al |
| EPI_ISL_693144 | USA/MA-MASPHL-00402/2020 | 2020-04-25 | North America/USA/Massachusetts/ | Massachusetts State Public Health Laboratory | Massachusetts State Public Health Laboratory | Andrew Lang et al |

| | | | | | | |
|---|---|---|---|---|---|---|
| EPI_ISL_693147 | USA/MA-MASPHL-00399/2020 | 2020-04-25 | North America/USA/Massachusetts/ | Massachusetts State Public Health Laboratory | Massachusetts State Public Health Laboratory | Andrew Lang et al |
| EPI_ISL_693153 | USA/MA-MASPHL-00392/2020 | 2020-05-12 | North America/USA/Massachusetts/ | Massachusetts State Public Health Laboratory | Massachusetts State Public Health Laboratory | Andrew Lang et al |
| EPI_ISL_693157 | USA/MA-MASPHL-00387/2020 | 2020-05-12 | North America/USA/Massachusetts/ | Massachusetts State Public Health Laboratory | Massachusetts State Public Health Laboratory | Andrew Lang et al |
| EPI_ISL_693171 | USA/MA-MASPHL-00362/2020 | 2020-05-20 | North America/USA/Massachusetts/ | Massachusetts State Public Health Laboratory | Massachusetts State Public Health Laboratory | Andrew Lang et al |
| EPI_ISL_693181 | USA/MA-MASPHL-00351/2020 | 2020-04-27 | North America/USA/Massachusetts/ | Massachusetts State Public Health Laboratory | Massachusetts State Public Health Laboratory | Andrew Lang et al |
| EPI_ISL_693190 | USA/MA-MASPHL-00327/2020 | 2020-04-28 | North America/USA/Massachusetts/ | Massachusetts State Public Health Laboratory | Massachusetts State Public Health Laboratory | Andrew Lang et al |
| EPI_ISL_693196 | Brazil/SP-243/2020 | 2020-05-02 | South America/Brazil/S√£o Paulo/S√£o | Hospital Santa Clara | Instituto Adolfo Lutz, Interdiciplinary Procedures Center, Strategic Laboratory | Claudio Tavares Sacchi et al |
| EPI_ISL_693198 | Brazil/SP-249/2020 | 2020-05-04 | South America/Brazil/S√£o Paulo/S√£o | Santa Casa de Misericordia de Sao Paulo - Hospital Central | Instituto Adolfo Lutz, Interdiciplinary Procedures Center, Strategic Laboratory | Claudio Tavares Sacchi et al |
| EPI_ISL_693199 | Brazil/SP-253/2020 | 2020-05-20 | South America/Brazil/S√£o Paulo/S√£o | Hospital do Servidor Publico Estadual Francisco Morato de Oliveira | Instituto Adolfo Lutz, Interdiciplinary Procedures Center, Strategic Laboratory | Claudio Tavares Sacchi et al |

| | | | | | | |
|---|---|---|---|---|---|---|
| EPI_ISL_693200 | Brazil/SP-571/2020 | 2020-04-29 | South America/Brazil/S√£o Paulo/Mairi | Hospital e Maternidade Mairipora | Instituto Adolfo Lutz, Interdiciplinary Procedures Center, Strategic Laboratory | Claudio Tavares Sacchi et al |
| EPI_ISL_693204 | Brazil/SP-352/2020 | 2020-06-22 | South America/Brazil/S√£o Paulo/Osasc | Pronto Socorro Dr. Conrado Cesarino Nuvolini | Instituto Adolfo Lutz, Interdiciplinary Procedures Center, Strategic Laboratory | Claudio Tavares Sacchi et al |
| EPI_ISL_693205 | Brazil/SP-577/2020 | 2020-06-22 | South America/Brazil/S√£o Paulo/Tarum | Hospital de Campanha Covid-19 Assis | Instituto Adolfo Lutz, Interdiciplinary Procedures Center, Strategic Laboratory | Claudio Tavares Sacchi et al |
| EPI_ISL_693206 | Brazil/SP-361/2020 | 2020-06-21 | South America/Brazil/S√£o Paulo/Campi | Hospital Municipal Mario Gatti | Instituto Adolfo Lutz, Interdiciplinary Procedures Center, Strategic Laboratory | Claudio Tavares Sacchi et al |
| EPI_ISL_693207 | Brazil/SP-366/2020 | 2020-06-24 | South America/Brazil/S√£o Paulo/Suma | Cs II Doutor Antonio Vicoso Moreira de Rezende | Instituto Adolfo Lutz, Interdiciplinary Procedures Center, Strategic Laboratory | Claudio Tavares Sacchi et al |
| EPI_ISL_693208 | Brazil/SP-400/2020 | 2020-07-01 | South America/Brazil/S√£o Paulo/Osasc | Hospital Municipal Antonio Giglio | Instituto Adolfo Lutz, Interdiciplinary Procedures Center, Strategic Laboratory | Claudio Tavares Sacchi et al |

| | | | | | | |
|---|---|---|---|---|---|---|
| EPI_ISL_693209 | Brazil/SP-401/2020 | 2020-07-01 | South America/Brazil/S√£o Paulo/Osasco | Hospital Municipal Antonio Giglio | Instituto Adolfo Lutz, Interdiciplinary Procedures Center, Strategic Laboratory | Claudio Tavares Sacchi et al |
| EPI_ISL_693210 | Brazil/SP-415/2020 | 2020-07-06 | South America/Brazil/S√£o Paulo/Osasco | Pronto-Socorro Dr. Osmar Mesquita | Instituto Adolfo Lutz, Interdiciplinary Procedures Center, Strategic Laboratory | Claudio Tavares Sacchi et al |
| EPI_ISL_693211 | Brazil/SP-416/2020 | 2020-07-06 | South America/Brazil/S√£o Paulo/Cunha | Santa Casa de Misericordia e Maternidade | Instituto Adolfo Lutz, Interdiciplinary Procedures Center, Strategic Laboratory | Claudio Tavares Sacchi et al |
| EPI_ISL_693212 | Brazil/SP-423/2020 | 2020-07-06 | South America/Brazil/S√£o Paulo/Braga | Santa Casa de Misericordia de Braganca Paulista | Instituto Adolfo Lutz, Interdiciplinary Procedures Center, Strategic Laboratory | Claudio Tavares Sacchi et al |
| EPI_ISL_693213 | Brazil/SP-390/2020 | 2020-06-18 | South America/Brazil/S√£o Paulo/Horto | Hospital E Maternidade Municipal Governador Mario Covas | Instituto Adolfo Lutz, Interdiciplinary Procedures Center, Strategic Laboratory | Claudio Tavares Sacchi et al |
| EPI_ISL_693214 | Brazil/SP-579/2020 | 2020-07-19 | South America/Brazil/S√£o Paulo/Sao S | Unidade de Pronto Atendimento Central de Caraguatatuba | Instituto Adolfo Lutz, Interdiciplinary Procedures Center, Strategic Laboratory | Claudio Tavares Sacchi et al |

| | | | | | | |
|---|---|---|---|---|---|---|
| EPI_ISL_693215 | Brazil/SP-589/2020 | 2020-07-06 | South America/Brazil/S√£o Paulo/Limeir | Secretaria Municipal de Sa√∫de de Iracemapolis | Instituto Adolfo Lutz, Interdiciplinary Procedures Center, Strategic Laboratory | Claudio Tavares Sacchi et al |
| EPI_ISL_693216 | Brazil/SP-590/2020 | 2020-07-05 | South America/Brazil/S√£o Paulo/Arara | Unidade de Vigil√¢ncia Epiudemiol√≥gica de Araras | Instituto Adolfo Lutz, Interdiciplinary Procedures Center, Strategic Laboratory | Claudio Tavares Sacchi et al |
| EPI_ISL_693217 | Brazil/SP-591/2020 | 2020-07-05 | South America/Brazil/S√£o Paulo/Arara | Unidade de Vigil√¢ncia Epiudemiol√≥gica de Araras | Instituto Adolfo Lutz, Interdiciplinary Procedures Center, Strategic Laboratory | Claudio Tavares Sacchi et al |
| EPI_ISL_693218 | Brazil/SP-594/2020 | 2020-06-16 | South America/Brazil/S√£o Paulo/Presic | Hospital Domingos Leonardo Ceravolo Presidente Prudente | Instituto Adolfo Lutz, Interdiciplinary Procedures Center, Strategic Laboratory | Claudio Tavares Sacchi et al |
| EPI_ISL_693219 | Brazil/SP-595/2020 | 2020-06-16 | South America/Brazil/S√£o Paulo/Taciba | Santa Casa da Miseric√≥rdia de Presidente Prudente | Instituto Adolfo Lutz, Interdiciplinary Procedures Center, Strategic Laboratory | Claudio Tavares Sacchi et al |
| EPI_ISL_693220 | Brazil/SP-596/2020 | 2020-07-01 | South America/Brazil/S√£o Paulo/Piracic | Laborat√≥rio Municipal de Piracicaba | Instituto Adolfo Lutz, Interdiciplinary Procedures Center, Strategic Laboratory | Claudio Tavares Sacchi et al |

| | | | | | | |
|---|---|---|---|---|---|---|
| EPI_ISL_693221 | Brazil/SP-597/2020 | 2020-06-16 | South America/Brazil/S√£o Paulo/Birigu | Secretaria Municipal de Sa√ude de Birigui | Instituto Adolfo Lutz, Interdiciplinary Procedures Center, Strategic Laboratory | Claudio Tavares Sacchi et al |
| EPI_ISL_693222 | Brazil/SP-598/2020 | 2020-06-16 | South America/Brazil/S√£o Paulo/Birigu | Secretaria Municipal de Sa√ude de Birigui | Instituto Adolfo Lutz, Interdiciplinary Procedures Center, Strategic Laboratory | Claudio Tavares Sacchi et al |
| EPI_ISL_693223 | Brazil/SP-643/2020 | 2020-07-02 | South America/Brazil/S√£o Paulo/Piraci | Laborat√≥rio Municipal de Piracicaba | Instituto Adolfo Lutz, Interdiciplinary Procedures Center, Strategic Laboratory | Claudio Tavares Sacchi et al |
| EPI_ISL_693224 | Brazil/SP-645/2020 | 2020-07-02 | South America/Brazil/S√£o Paulo/Piraci | Laborat√≥rio Municipal de Piracicaba | Instituto Adolfo Lutz, Interdiciplinary Procedures Center, Strategic Laboratory | Claudio Tavares Sacchi et al |
| EPI_ISL_693225 | Brazil/SP-652/2020 | 2020-07-06 | South America/Brazil/S√£o Paulo/Diade | Ubs Vila Rosa - Ol√≠mpia Gomes De Almeida | Instituto Adolfo Lutz, Interdiciplinary Procedures Center, Strategic Laboratory | Claudio Tavares Sacchi et al |
| EPI_ISL_693226 | Brazil/SP-653/2020 | 2020-06-16 | South America/Brazil/S√£o Paulo/Campi | Unidade de Pronto Atendimento Sao Jos√© | Instituto Adolfo Lutz, Interdiciplinary Procedures Center, Strategic Laboratory | Claudio Tavares Sacchi et al |

| | | | | | | |
|---|---|---|---|---|---|---|
| EPI_ISL_693227 | Brazil/SP-658/2020 | 2020-07-06 | South America/Brazil/S√£o Paulo/Sao B | UBS Vila Marchi | Instituto Adolfo Lutz, Interdiciplinary Procedures Center, Strategic Laboratory | Claudio Tavares Sacchi et al |
| EPI_ISL_693228 | Brazil/SP-661/2020 | 2020-06-22 | South America/Brazil/S√£o Paulo/Soroc | Secretaria Municipal de Sorocaba | Instituto Adolfo Lutz, Interdiciplinary Procedures Center, Strategic Laboratory | Claudio Tavares Sacchi et al |
| EPI_ISL_693229 | Brazil/SP-666/2020 | 2020-06-16 | South America/Brazil/S√£o Paulo/Guaru | Hospital 8 de Maio | Instituto Adolfo Lutz, Interdiciplinary Procedures Center, Strategic Laboratory | Claudio Tavares Sacchi et al |
| EPI_ISL_693230 | Brazil/SP-667/2020 | 2020-06-13 | South America/Brazil/S√£o Paulo/Cotia | Hospital e Pronto Socorro Portinari | Instituto Adolfo Lutz, Interdiciplinary Procedures Center, Strategic Laboratory | Claudio Tavares Sacchi et al |
| EPI_ISL_693231 | Brazil/SP-668/2020 | 2020-06-27 | South America/Brazil/S√£o Paulo/Jacare | Pronto Socorro Municipal de Santa Branca | Instituto Adolfo Lutz, Interdiciplinary Procedures Center, Strategic Laboratory | Claudio Tavares Sacchi et al |
| EPI_ISL_693232 | Brazil/SP-673/2020 | 2020-05-30 | South America/Brazil/S√£o Paulo/S√£o | Hospital e Pronto Socorro Portinari | Instituto Adolfo Lutz, Interdiciplinary Procedures Center, Strategic Laboratory | Claudio Tavares Sacchi et al |

| | | | | | | |
|---|---|---|---|---|---|---|
| EPI_ISL_693233 | Brazil/SP-678/2020 | 2020-06-12 | South America/Brazil/S√£o Paulo/Itapec | Hospital Santa Cruz | Instituto Adolfo Lutz, Interdiciplinary Procedures Center, Strategic Laboratory | Claudio Tavares Sacchi et al |
| EPI_ISL_693234 | Brazil/SP-688/2020 | 2020-06-30 | South America/Brazil/S√£o Paulo/Rio Gr | Upa Vereador Jose Da Rocha Goncalves | Instituto Adolfo Lutz, Interdiciplinary Procedures Center, Strategic Laboratory | Claudio Tavares Sacchi et al |
| EPI_ISL_693235 | Brazil/SP-689/2020 | 2020-06-30 | South America/Brazil/S√£o Paulo/Rio Gr | Casmi Centro Atendimento Saude da Mulher e Infancia | Instituto Adolfo Lutz, Interdiciplinary Procedures Center, Strategic Laboratory | Claudio Tavares Sacchi et al |
| EPI_ISL_693236 | Brazil/SP-692/2020 | 2020-07-02 | South America/Brazil/S√£o Paulo/S√£o | Hospital Santa Marcelina Sao Paulo | Instituto Adolfo Lutz, Interdiciplinary Procedures Center, Strategic Laboratory | Claudio Tavares Sacchi et al |
| EPI_ISL_693237 | Brazil/SP-694/2020 | 2020-07-01 | South America/Brazil/S√£o Paulo/Mogi | UPA Santa Isabel | Instituto Adolfo Lutz, Interdiciplinary Procedures Center, Strategic Laboratory | Claudio Tavares Sacchi et al |
| EPI_ISL_693238 | Brazil/SP-696/2020 | 2020-06-30 | South America/Brazil/S√£o Paulo/Santo | Secao Centro de Diagnostico Secedi | Instituto Adolfo Lutz, Interdiciplinary Procedures Center, Strategic Laboratory | Claudio Tavares Sacchi et al |

| | | | | | | |
|---|---|---|---|---|---|---|
| EPI_ISL_693239 | Brazil/SP-697/2020 | 2020-06-30 | South America/Brazil/S√£o Paulo/Santo | Secao Centro de Diagnostico Secedi | Instituto Adolfo Lutz, Interdiciplinary Procedures Center, Strategic Laboratory | Claudio Tavares Sacchi et al |
| EPI_ISL_693240 | Brazil/SP-698/2020 | 2020-06-30 | South America/Brazil/S√£o Paulo/Diade | Centro de Vigil√¢ncia a Saude de Diadema | Instituto Adolfo Lutz, Interdiciplinary Procedures Center, Strategic Laboratory | Claudio Tavares Sacchi et al |
| EPI_ISL_693241 | Brazil/SP-699/2020 | 2020-06-30 | South America/Brazil/S√£o Paulo/Ribeir | Hospital e Maternidade Sao Lucas | Instituto Adolfo Lutz, Interdiciplinary Procedures Center, Strategic Laboratory | Claudio Tavares Sacchi et al |
| EPI_ISL_693242 | Brazil/SP-702/2020 | 2020-06-30 | South America/Brazil/S√£o Paulo/Diade | Centro de Vigil√¢ncia a Saude de Diadema | Instituto Adolfo Lutz, Interdiciplinary Procedures Center, Strategic Laboratory | Claudio Tavares Sacchi et al |
| EPI_ISL_693243 | Brazil/SP-710/2020 | 2020-06-27 | South America/Brazil/S√£o Paulo/Piraci | Laborat√≥rio Municipal de Piracicaba | Instituto Adolfo Lutz, Interdiciplinary Procedures Center, Strategic Laboratory | Claudio Tavares Sacchi et al |
| EPI_ISL_693244 | Brazil/SP-716/2020 | 2020-07-02 | South America/Brazil/S√£o Paulo/Soroca | Centro M√©dico da Pol√≠cia Militar do Estado de Sao Paulo | Instituto Adolfo Lutz, Interdiciplinary Procedures Center, Strategic Laboratory | Claudio Tavares Sacchi et al |

| | | | | | | |
|---|---|---|---|---|---|---|
| EPI_ISL_693245 | Brazil/SP-719/2020 | 2020-07-02 | South America/Brazil/S√£o Paulo/Mogi | UPA Santa Isabel | Instituto Adolfo Lutz, Interdiciplinary Procedures Center, Strategic Laboratory | Claudio Tavares Sacchi et al |
| EPI_ISL_693246 | Brazil/SP-721/2020 | 2020-06-30 | South America/Brazil/S√£o Paulo/Rio Gr | Laboratorio Municipal de Rio Grande da Serra | Instituto Adolfo Lutz, Interdiciplinary Procedures Center, Strategic Laboratory | Claudio Tavares Sacchi et al |
| EPI_ISL_693247 | Brazil/SP-705/2020 | 2020-06-30 | South America/Brazil/S√£o Paulo/Santo | Secao Centro de Diagnostico Secedi | Instituto Adolfo Lutz, Interdiciplinary Procedures Center, Strategic Laboratory | Claudio Tavares Sacchi et al |
| EPI_ISL_693248 | Brazil/SP-735/2020 | 2020-07-06 | South America/Brazil/S√£o Paulo/Limeir | Centro Municipal de Epidemiologia e Imuniza√ß√µes | Instituto Adolfo Lutz, Interdiciplinary Procedures Center, Strategic Laboratory | Claudio Tavares Sacchi et al |
| EPI_ISL_693289 | Australia/QLD1292/2020 | 2020-03-23 | Oceania/Australia/Australia/ | unknown | Public Health Virology Laboratory, Forensic and Scientific Services (PHV-FSS) | Son Nguyen et al. et al |
| EPI_ISL_693290 | Australia/QLD1293/2020 | 2020-02-04 | Oceania/Australia/Australia/ | unknown | Public Health Virology Laboratory, Forensic and Scientific Services (PHV-FSS) | Son Nguyen et al. et al |
| EPI_ISL_693358 | Thailand/Bangkok-CONI-0280/2020 | 2020-03-14 | Asia/Thailand/Bangkok/ | Ramathibodi Hospital | COVID-19 Network Investigations (CONI) Alliance | Elizabeth Batty et al |

| | | | | | | |
|---|---|---|---|---|---|---|
| EPI_ISL_693360 | Thailand/Bangkok-CONI-0282/2020 | 2020-03-15 | Asia/Thailand/Bangkok/ | Ramathibodi Hospital | COVID-19 Network Investigations (CONI) Alliance | Elizabeth Batty et al |
| EPI_ISL_693362 | Thailand/Bangkok-CONI-0289/2020 | 2020-03-15 | Asia/Thailand/Bangkok/ | Ramathibodi Hospital | COVID-19 Network Investigations (CONI) Alliance | Elizabeth Batty et al |
| EPI_ISL_693386 | Thailand/Bangkok-CONI-0316/2020 | 2020-03-25 | Asia/Thailand/Bangkok/ | Ramathibodi Hospital | COVID-19 Network Investigations (CONI) Alliance | Elizabeth Batty et al |
| EPI_ISL_693387 | France/PAC02003/2020 | 2020-02-28 | Europe/France/Provence-Alpes-C√¥te d' | CHU de Nice - H√¥pital Archet 2 | CNR Virus des Infections Respiratoires - France SUD | Antonin Bal et al |
| EPI_ISL_693388 | France/PAC02004/2020 | 2020-02-29 | Europe/France/Provence-Alpes-C√¥te d' | CHU de Nice - H√¥pital Archet 2 | CNR Virus des Infections Respiratoires - France SUD | Antonin Bal et al |
| EPI_ISL_693390 | France/PAC02011/2020 | 2020-03-07 | Europe/France/Provence-Alpes-C√¥te d' | CHU de Nice - H√¥pital Archet 2 | CNR Virus des Infections Respiratoires - France SUD | Antonin Bal et al |
| EPI_ISL_693391 | France/ARA-SC088/2020 | 2020-03-05 | Europe/France/Auvergne-Rh√¥ne-Alpes/ | Centre hospitalier M√©tropole Savoie | CNR Virus des Infections Respiratoires - France SUD | Antonin Bal et al |
| EPI_ISL_693458 | England/204700071/2020 | 2020-11-15 | Europe/United Kingdom/England/ | Respiratory Virus Unit, Microbiology Services Colindale, Public Health England | COVID-19 Genomics UK (COG-UK) Consortium | PHE Covid Sequencing Team et al |
| EPI_ISL_693473 | PapuaNewGuinea/3/2020 | 2020-08-21 | Oceania/Papua New Guinea/Papua New | Central Public Health Laboratory | National Public Health Laboratory, National Centre for Infectious Diseases | Tze Minn Mak et al |

| | | | | | | |
|---|---|---|---|---|---|---|
| EPI_ISL_693477 | PapuaNewGuinea/7/2020 | 2020-08-29 | Oceania/Papua New Guinea/Papua New | Central Public Health Laboratory | National Public Health Laboratory, National Centre for Infectious Diseases | Tze Minn Mak et al |
| EPI_ISL_693488 | France/ARA-72249/2020 | 2020-09-14 | Europe/France/Auvergne-Rh√¥ne-Alpes/ | CNR Virus des Infections Respiratoires - France SUD | CNR Virus des Infections Respiratoires - France SUD | Antonin Bal et al |
| EPI_ISL_693489 | France/ARA-73557/2020 | 2020-09-16 | Europe/France/Auvergne-Rh√¥ne-Alpes/ | CNR Virus des Infections Respiratoires - France SUD | CNR Virus des Infections Respiratoires - France SUD | Antonin Bal et al |
| EPI_ISL_693492 | France/ARA-7096/2020 | 2020-09-11 | Europe/France/Auvergne-Rh√¥ne-Alpes/ | CNR Virus des Infections Respiratoires - France SUD | CNR Virus des Infections Respiratoires - France SUD | Antonin Bal et al |
| EPI_ISL_693494 | France/ARA-71481/2020 | 2020-09-12 | Europe/France/Auvergne-Rh√¥ne-Alpes/ | CNR Virus des Infections Respiratoires - France SUD | CNR Virus des Infections Respiratoires - France SUD | Antonin Bal et al |
| EPI_ISL_693496 | France/ARA-72054/2020 | 2020-09-14 | Europe/France/Auvergne-Rh√¥ne-Alpes/ | CNR Virus des Infections Respiratoires - France SUD | CNR Virus des Infections Respiratoires - France SUD | Antonin Bal et al |
| EPI_ISL_693497 | France/ARA-72179/2020 | 2020-09-14 | Europe/France/Auvergne-Rh√¥ne-Alpes/ | CNR Virus des Infections Respiratoires - France SUD | CNR Virus des Infections Respiratoires - France SUD | Antonin Bal et al |
| EPI_ISL_693498 | France/ARA-72266/2020 | 2020-09-14 | Europe/France/Auvergne-Rh√¥ne-Alpes/ | CNR Virus des Infections Respiratoires - France SUD | CNR Virus des Infections Respiratoires - France SUD | Antonin Bal et al |
| EPI_ISL_693499 | France/ARA-72824/2020 | 2020-09-15 | Europe/France/Auvergne-Rh√¥ne-Alpes/ | CNR Virus des Infections Respiratoires - France SUD | CNR Virus des Infections Respiratoires - France SUD | Antonin Bal et al |

| | | | | | | |
|---|---|---|---|---|---|---|
| EPI_ISL_693500 | France/ARA-74594/2020 | 2020-09-17 | Europe/France/Auvergne-Rh√¥ne-Alpes/ | CNR Virus des Infections Respiratoires - France SUD | CNR Virus des Infections Respiratoires - France SUD | Antonin Bal et al |
| EPI_ISL_693501 | France/ARA-75471/2020 | 2020-09-20 | Europe/France/Auvergne-Rh√¥ne-Alpes/ | CNR Virus des Infections Respiratoires - France SUD | CNR Virus des Infections Respiratoires - France SUD | Antonin Bal et al |
| EPI_ISL_693502 | France/ARA-75858/2020 | 2020-09-21 | Europe/France/Auvergne-Rh√¥ne-Alpes/ | CNR Virus des Infections Respiratoires - France SUD | CNR Virus des Infections Respiratoires - France SUD | Antonin Bal et al |
| EPI_ISL_693503 | France/ARA-76483/2020 | 2020-09-21 | Europe/France/Auvergne-Rh√¥ne-Alpes/ | CNR Virus des Infections Respiratoires - France SUD | CNR Virus des Infections Respiratoires - France SUD | Antonin Bal et al |
| EPI_ISL_693504 | France/ARA-76531/2020 | 2020-09-18 | Europe/France/Auvergne-Rh√¥ne-Alpes/ | CNR Virus des Infections Respiratoires - France SUD | CNR Virus des Infections Respiratoires - France SUD | Antonin Bal et al |
| EPI_ISL_693505 | France/ARA-76532/2020 | 2020-09-22 | Europe/France/Auvergne-Rh√¥ne-Alpes/ | CNR Virus des Infections Respiratoires - France SUD | CNR Virus des Infections Respiratoires - France SUD | Antonin Bal et al |
| EPI_ISL_693508 | France/ARA-77064/2020 | 2020-09-22 | Europe/France/Auvergne-Rh√¥ne-Alpes/ | CNR Virus des Infections Respiratoires - France SUD | CNR Virus des Infections Respiratoires - France SUD | Antonin Bal et al |
| EPI_ISL_693510 | France/ARA-77597/2020 | 2020-09-24 | Europe/France/Auvergne-Rh√¥ne-Alpes/ | CNR Virus des Infections Respiratoires - France SUD | CNR Virus des Infections Respiratoires - France SUD | Antonin Bal et al |
| EPI_ISL_693511 | France/ARA-77679/2020 | 2020-09-24 | Europe/France/Auvergne-Rh√¥ne-Alpes/ | CNR Virus des Infections Respiratoires - France SUD | CNR Virus des Infections Respiratoires - France SUD | Antonin Bal et al |
| EPI_ISL_693512 | France/ARA-78039/2020 | 2020-09-25 | Europe/France/Auvergne-Rh√¥ne-Alpes/ | CNR Virus des Infections Respiratoires - France SUD | CNR Virus des Infections Respiratoires - France SUD | Antonin Bal et al |

| | | | | | | |
|---|---|---|---|---|---|---|
| EPI_ISL_693513 | France/ARA-78043/2020 | 2020-09-24 | Europe/France/Auvergne-Rhv¥ne-Alpes/ | CNR Virus des Infections Respiratoires - France SUD | CNR Virus des Infections Respiratoires - France SUD | Antonin Bal et al |
| EPI_ISL_693514 | France/ARA-78085/2020 | 2020-09-24 | Europe/France/Auvergne-Rhv¥ne-Alpes/ | CNR Virus des Infections Respiratoires - France SUD | CNR Virus des Infections Respiratoires - France SUD | Antonin Bal et al |
| EPI_ISL_693523 | Portugal/PT1707/2020 | 2020-08-24 | Europe/Portugal/Portugal/ | Hospital Vila Franca de Xira | Instituto Nacional de Saude (INSA) | Borges et al et al |
| EPI_ISL_693527 | Portugal/PT1545/2020 | 2020-06-20 | Europe/Portugal/Portugal/ | Instituto Nacional de Saude (INSA) | Instituto Nacional de Saude (INSA) | Borges et al et al |
| EPI_ISL_693528 | Portugal/PT1539/2020 | 2020-06-04 | Europe/Portugal/Portugal/ | Instituto Nacional de Saude (INSA) | Instituto Nacional de Saude (INSA) | Borges et al et al |
| EPI_ISL_693529 | Portugal/PT1557/2020 | 2020-06-26 | Europe/Portugal/Portugal/ | Instituto Nacional de Saude (INSA) | Instituto Nacional de Saude (INSA) | Borges et al et al |
| EPI_ISL_693533 | Portugal/PT1580/2020 | 2020-07-01 | Europe/Portugal/Portugal/ | Instituto Nacional de Saude (INSA) | Instituto Nacional de Saude (INSA) | Borges et al et al |
| EPI_ISL_693534 | Portugal/PT1735/2020 | 2020-08-21 | Europe/Portugal/Portugal/ | Hospital Vila Franca de Xira | Instituto Nacional de Saude (INSA) | Borges et al et al |
| EPI_ISL_693535 | Portugal/PT1739/2020 | 2020-09-02 | Europe/Portugal/Portugal/ | Hospital Vila Franca de Xira | Instituto Nacional de Saude (INSA) | Borges et al et al |
| EPI_ISL_693542 | Portugal/PT1586/2020 | 2020-07-01 | Europe/Portugal/Portugal/ | Instituto Nacional de Saude (INSA) | Instituto Nacional de Saude (INSA) | Borges et al et al |
| EPI_ISL_693546 | Portugal/PT1527/2020 | 2020-05-11 | Europe/Portugal/Portugal/ | Instituto Nacional de Saude (INSA) | Instituto Nacional de Saude (INSA) | Borges et al et al |
| EPI_ISL_693549 | Portugal/PT1530/2020 | 2020-04-17 | Europe/Portugal/Portugal/ | Instituto Nacional de Saude (INSA) | Instituto Nacional de Saude (INSA) | Borges et al et al |
| EPI_ISL_693558 | Portugal/PT1544/2020 | 2020-06-21 | Europe/Portugal/Portugal/ | Instituto Nacional de Saude (INSA) | Instituto Nacional de Saude (INSA) | Borges et al et al |
| EPI_ISL_693565 | Portugal/PT1552/2020 | 2020-06-25 | Europe/Portugal/Portugal/ | Instituto Nacional de Saude (INSA) | Instituto Nacional de Saude (INSA) | Borges et al et al |
| EPI_ISL_693567 | Portugal/PT1554/2020 | 2020-06-26 | Europe/Portugal/Portugal/ | Instituto Nacional de Saude (INSA) | Instituto Nacional de Saude (INSA) | Borges et al et al |
| EPI_ISL_693568 | Portugal/PT1555/2020 | 2020-06-26 | Europe/Portugal/Portugal/ | Instituto Nacional de Saude (INSA) | Instituto Nacional de Saude (INSA) | Borges et al et al |
| EPI_ISL_693583 | Portugal/PT1575/2020 | 2020-06-29 | Europe/Portugal/Portugal/ | Instituto Nacional de Saude (INSA) | Instituto Nacional de Saude (INSA) | Borges et al et al |
| EPI_ISL_693593 | Portugal/PT1588/2020 | 2020-07-02 | Europe/Portugal/Portugal/ | Instituto Nacional de Saude (INSA) | Instituto Nacional de Saude (INSA) | Borges et al et al |

| | | | | | | |
|---|---|---|---|---|---|---|
| EPI_ISL_693596 | Portugal/PT1591/2020 | 2020-07-03 | Europe/Portugal/Portugal/ | Instituto Nacional de Saude (INSA) | Instituto Nacional de Saude (INSA) | Borges et al et al |
| EPI_ISL_693598 | Portugal/PT1593/2020 | 2020-07-03 | Europe/Portugal/Portugal/ | Instituto Nacional de Saude (INSA) | Instituto Nacional de Saude (INSA) | Borges et al et al |
| EPI_ISL_693601 | Portugal/PT1597/2020 | 2020-07-05 | Europe/Portugal/Portugal/ | Instituto Nacional de Saude (INSA) | Instituto Nacional de Saude (INSA) | Borges et al et al |
| EPI_ISL_693608 | Portugal/PT1605/2020 | 2020-07-07 | Europe/Portugal/Portugal/ | Instituto Nacional de Saude (INSA) | Instituto Nacional de Saude (INSA) | Borges et al et al |
| EPI_ISL_693616 | Portugal/PT1614/2020 | 2020-07-08 | Europe/Portugal/Portugal/ | Instituto Nacional de Saude (INSA) | Instituto Nacional de Saude (INSA) | Borges et al et al |
| EPI_ISL_693621 | Portugal/PT1620/2020 | 2020-07-09 | Europe/Portugal/Portugal/ | Instituto Nacional de Saude (INSA) | Instituto Nacional de Saude (INSA) | Borges et al et al |
| EPI_ISL_693623 | Portugal/PT1622/2020 | 2020-07-10 | Europe/Portugal/Portugal/ | Instituto Nacional de Saude (INSA) | Instituto Nacional de Saude (INSA) | Borges et al et al |
| EPI_ISL_693624 | Portugal/PT1623/2020 | 2020-07-10 | Europe/Portugal/Portugal/ | Instituto Nacional de Saude (INSA) | Instituto Nacional de Saude (INSA) | Borges et al et al |
| EPI_ISL_693627 | Portugal/PT1627/2020 | 2020-07-11 | Europe/Portugal/Portugal/ | Instituto Nacional de Saude (INSA) | Instituto Nacional de Saude (INSA) | Borges et al et al |
| EPI_ISL_693630 | Portugal/PT1701/2020 | 2020-08-24 | Europe/Portugal/Portugal/ | Hospital Vila Franca de Xira | Instituto Nacional de Saude (INSA) | Borges et al et al |
| EPI_ISL_693636 | Portugal/PT1708/2020 | 2020-08-24 | Europe/Portugal/Portugal/ | Hospital Vila Franca de Xira | Instituto Nacional de Saude (INSA) | Borges et al et al |
| EPI_ISL_693642 | Portugal/PT1715/2020 | 2020-08-24 | Europe/Portugal/Portugal/ | Hospital Vila Franca de Xira | Instituto Nacional de Saude (INSA) | Borges et al et al |
| EPI_ISL_693645 | Portugal/PT1718/2020 | 2020-08-24 | Europe/Portugal/Portugal/ | Hospital Vila Franca de Xira | Instituto Nacional de Saude (INSA) | Borges et al et al |
| EPI_ISL_693647 | Portugal/PT1720/2020 | 2020-08-24 | Europe/Portugal/Portugal/ | Hospital Vila Franca de Xira | Instituto Nacional de Saude (INSA) | Borges et al et al |
| EPI_ISL_693650 | Portugal/PT1724/2020 | 2020-08-24 | Europe/Portugal/Portugal/ | Hospital Vila Franca de Xira | Instituto Nacional de Saude (INSA) | Borges et al et al |
| EPI_ISL_693654 | Portugal/PT1728/2020 | 2020-08-24 | Europe/Portugal/Portugal/ | Hospital Vila Franca de Xira | Instituto Nacional de Saude (INSA) | Borges et al et al |
| EPI_ISL_693655 | Portugal/PT1731/2020 | 2020-08-24 | Europe/Portugal/Portugal/ | Hospital Vila Franca de Xira | Instituto Nacional de Saude (INSA) | Borges et al et al |
| EPI_ISL_693656 | Portugal/PT1733/2020 | 2020-08-24 | Europe/Portugal/Portugal/ | Hospital Vila Franca de Xira | Instituto Nacional de Saude (INSA) | Borges et al et al |
| EPI_ISL_693659 | USA/MO-WUSTL-C4/2020 | 2020-08-16 | North America/USA/Missouri/St. Louis I | Washington University in St. Louis | Washington University in St. Louis | David Wang et al |

| | | | | | | |
|---|---|---|---|---|---|---|
| EPI_ISL_693660 | CzechRepublic/NRL-10483/2020 | 2020-10-26 | Europe/Czech Republic/South Moravian | The National Institute of Public Health | State Veterinary Institute Prague | Nagy et al |
| EPI_ISL_693661 | CzechRepublic/NRL-10719/2020 | 2020-11-03 | Europe/Czech Republic/Central Bohemia | The National Institute of Public Health | State Veterinary Institute Prague | Nagy et al |
| EPI_ISL_693664 | CzechRepublic/NRL-10473/2020 | 2020-10-27 | Europe/Czech Republic/Hradec Krv°lovv | The National Institute of Public Health | State Veterinary Institute Prague | Nagy et al |
| EPI_ISL_693665 | CzechRepublic/NRL-10718/2020 | 2020-11-03 | Europe/Czech Republic/Prague/ | The National Institute of Public Health | State Veterinary Institute Prague | Nagy et al |
| EPI_ISL_693667 | CzechRepublic/NRL-10560/2020 | 2020-10-30 | Europe/Czech Republic/Prague/ | The National Institute of Public Health | State Veterinary Institute Prague | Nagy et al |
| EPI_ISL_693668 | CzechRepublic/NRL-10746/2020 | 2020-11-04 | Europe/Czech Republic/Prague/ | The National Institute of Public Health | State Veterinary Institute Prague | Nagy et al |
| EPI_ISL_693671 | CzechRepublic/NRL-10582/2020 | 2020-10-30 | Europe/Czech Republic/Moravian-Silesia | The National Institute of Public Health | State Veterinary Institute Prague | Nagy et al |
| EPI_ISL_693672 | CzechRepublic/NRL-10716/2020 | 2020-11-03 | Europe/Czech Republic/Prague/ | The National Institute of Public Health | State Veterinary Institute Prague | Nagy et al |
| EPI_ISL_693674 | CzechRepublic/NRL-10744/2020 | 2020-11-04 | Europe/Czech Republic/Central Bohemia | The National Institute of Public Health | State Veterinary Institute Prague | Nagy et al |
| EPI_ISL_693675 | CzechRepublic/NRL-10733/2020 | 2020-11-03 | Europe/Czech Republic/Czech Republic/ | The National Institute of Public Health | State Veterinary Institute Prague | Nagy et al |
| EPI_ISL_693676 | CzechRepublic/NRL-10565/2020 | 2020-10-30 | Europe/Czech Republic/Prague/ | The National Institute of Public Health | State Veterinary Institute Prague | Nagy et al |
| EPI_ISL_693677 | CzechRepublic/NRL-10739/2020 | 2020-11-03 | Europe/Czech Republic/Central Bohemia | The National Institute of Public Health | State Veterinary Institute Prague | Nagy et al |
| EPI_ISL_693678 | CzechRepublic/NRL-10712/2020 | 2020-11-03 | Europe/Czech Republic/Usti nad Labem/ | The National Institute of Public Health | State Veterinary Institute Prague | Nagy et al |
| EPI_ISL_693679 | CzechRepublic/NRL-10645/2020 | 2020-11-02 | Europe/Czech Republic/Prague/ | The National Institute of Public Health | State Veterinary Institute Prague | Nagy et al |
| EPI_ISL_693680 | CzechRepublic/NRL-10598/2020 | 2020-11-01 | Europe/Czech Republic/Prague/ | The National Institute of Public Health | State Veterinary Institute Prague | Nagy et al |
| EPI_ISL_693681 | CzechRepublic/NRL-10549/2020 | 2020-10-29 | Europe/Czech Republic/Prague/ | The National Institute of Public Health | State Veterinary Institute Prague | Nagy et al |
| EPI_ISL_693685 | CzechRepublic/NRL-10736/2020 | 2020-11-03 | Europe/Czech Republic/Prague/ | The National Institute of Public Health | State Veterinary Institute Prague | Nagy et al |
| EPI_ISL_693687 | CzechRepublic/NRL-10487/2020 | 2020-10-26 | Europe/Czech Republic/Prague/ | The National Institute of Public Health | State Veterinary Institute Prague | Nagy et al |
| EPI_ISL_693710 | USA/DE-DHSS-F982045/2020 | 2020-09-24 | North America/USA/Delaware/New Cas | Delaware Public Health Laboratory | Delaware Public Health Laboratory | Gregory Hovan et al |
| EPI_ISL_693718 | USA/DE-DHSS-F994362/2020 | 2020-09-16 | North America/USA/Delaware/New Cas | Delaware Public Health Laboratory | Delaware Public Health Laboratory | Gregory Hovan et al |

| | | | | | | |
|---|---|---|---|---|---|---|
| EPI_ISL_693719 | USA/DE-DHSS-F970597/2020 | 2020-06-12 | North America/USA/Delaware/New Cas | Delaware Public Health Laboratory | Delaware Public Health Laboratory | Gregory Hovan et al |
| EPI_ISL_693724 | USA/DE-DHSS-F995432/2020 | 2020-09-22 | North America/USA/Delaware/Kent Cou | Delaware Public Health Laboratory | Delaware Public Health Laboratory | Gregory Hovan et al |
| EPI_ISL_693727 | USA/DE-DHSS-F956491/2020 | 2020-05-27 | North America/USA/Delaware/Kent Cou | Delaware Public Health Laboratory | Delaware Public Health Laboratory | Gregory Hovan et al |
| EPI_ISL_693729 | USA/DE-DHSS-F980493/2020 | 2020-08-15 | North America/USA/Delaware/ | Delaware Public Health Laboratory | Delaware Public Health Laboratory | Gregory Hovan et al |
| EPI_ISL_693730 | USA/DE-DHSS-F956496/2020 | 2020-05-27 | North America/USA/Delaware/Kent Cou | Delaware Public Health Laboratory | Delaware Public Health Laboratory | Gregory Hovan et al |
| EPI_ISL_693733 | USA/DE-DHSS-F956499/2020 | 2020-05-27 | North America/USA/Delaware/Kent Cou | Delaware Public Health Laboratory | Delaware Public Health Laboratory | Gregory Hovan et al |
| EPI_ISL_693741 | USA/DE-DHSS-F957085/2020 | 2020-06-04 | North America/USA/Delaware/Kent Cou | Delaware Public Health Laboratory | Delaware Public Health Laboratory | Gregory Hovan et al |
| EPI_ISL_693745 | USA/DE-DHSS-F1003822/2020 | 2020-09-16 | North America/USA/Delaware/New Cas | Delaware Public Health Laboratory | Delaware Public Health Laboratory | Gregory Hovan et al |
| EPI_ISL_693746 | USA/DE-DHSS-F1007797/2020 | 2020-09-28 | North America/USA/Delaware/New Cas | Delaware Public Health Laboratory | Delaware Public Health Laboratory | Gregory Hovan et al |
| EPI_ISL_693751 | USA/DE-DHSS-F995385/2020 | 2020-09-21 | North America/USA/Delaware/New Cas | Delaware Public Health Laboratory | Delaware Public Health Laboratory | Gregory Hovan et al |
| EPI_ISL_693755 | USA/DE-DHSS-F970939/2020 | 2020-06-17 | North America/USA/Delaware/Sussex C | Delaware Public Health Laboratory | Delaware Public Health Laboratory | Gregory Hovan et al |
| EPI_ISL_693757 | USA/DE-DHSS-F995589/2020 | 2020-09-08 | North America/USA/Delaware/Sussex C | Delaware Public Health Laboratory | Delaware Public Health Laboratory | Gregory Hovan et al |
| EPI_ISL_693761 | France/BRE-IPP9949/2020 | 2020-09-09 | Europe/France/Bretagne/Rennes | Hospital | National Reference Center for Viruses of Respiratory Infections, Institut Pasteur, Paris | Marion Barbet et al |
| EPI_ISL_693764 | France/BRE-IPP9957/2020 | 2020-10-18 | Europe/France/Bretagne/Rennes | Hospital | National Reference Center for Viruses of Respiratory Infections, Institut Pasteur, Paris | Marion Barbet et al |

| | | | | | | |
|---|---|---|---|---|---|---|
| EPI_ISL_693765 | France/BRE-IPP9961/2020 | 2020-10-16 | Europe/France/Bretagne/Rennes | hospital | National Reference Center for Viruses of Respiratory Infections, Institut Pasteur, Paris | Marion Barbet et al |
| EPI_ISL_693766 | France/CVL-IPP9875/2020 | 2020-11-24 | Europe/France/Centre-Val de Loire/Crac | General practitioner | National Reference Center for Viruses of Respiratory Infections, Institut Pasteur, Paris | Marion Barbet et al |
| EPI_ISL_693767 | France/GES-IPP9562/2020 | 2020-11-03 | Europe/France/Grand Est/Brumath | General practitioner | National Reference Center for Viruses of Respiratory Infections, Institut Pasteur, Paris | Marion Barbet et al |
| EPI_ISL_693768 | France/IDF-IPP9592/2020 | 2020-10-31 | Europe/France/Ile de France/Egreville | General practitioner | National Reference Center for Viruses of Respiratory Infections, Institut Pasteur, Paris | Marion Barbet et al |
| EPI_ISL_693770 | France/IDF-IPP9725/2020 | 2020-11-16 | Europe/France/Ile de France/Paris | General practitioner | National Reference Center for Viruses of Respiratory Infections, Institut Pasteur, Paris | Marion Barbet et al |
| EPI_ISL_693771 | France/IDF-IPP9608/2020 | 2020-11-09 | Europe/France/Ile de France/Villeneuve | General practitioner | National Reference Center for Viruses of Respiratory Infections, Institut Pasteur, Paris | Marion Barbet et al |

| | | | | | | |
|---|---|---|---|---|---|---|
| EPI_ISL_693772 | France/HDF-IPP9631/2020 | 2020-11-09 | Europe/France/Hauts de France/Genech | General practitioner | National Reference Center for Viruses of Respiratory Infections, Institut Pasteur, Paris | Marion Barbet et al |
| EPI_ISL_693773 | France/HDF-IPP9591/2020 | 2020-10-23 | Europe/France/Hauts de France/Lille FR | General practitioner | National Reference Center for Viruses of Respiratory Infections, Institut Pasteur, Paris | Marion Barbet et al |
| EPI_ISL_693774 | France/HDF-IPP9877/2020 | 2020-11-25 | Europe/France/Hauts de France/Lille FR | General practitioner | National Reference Center for Viruses of Respiratory Infections, Institut Pasteur, Paris | Marion Barbet et al |
| EPI_ISL_693785 | Switzerland/BL-ETHZ-350011/2020 | 2020-11-03 | Europe/Switzerland/Basel-Land/ | Viollier AG | Department of Biosystems Science and Engineering, ETH Z√ºrich | Christian Beisel et al |
| EPI_ISL_693815 | Switzerland/ZH-ETHZ-350115/2020 | 2020-11-02 | Europe/Switzerland/Z√ºrich/ | Viollier AG | Department of Biosystems Science and Engineering, ETH Z√ºrich | Christian Beisel et al |
| EPI_ISL_693819 | Switzerland/LU-ETHZ-350120/2020 | 2020-11-02 | Europe/Switzerland/Lucerne/ | Viollier AG | Department of Biosystems Science and Engineering, ETH Z√ºrich | Christian Beisel et al |
| EPI_ISL_693831 | Switzerland/SO-ETHZ-350181/2020 | 2020-11-04 | Europe/Switzerland/Solothurn/ | Viollier AG | Department of Biosystems Science and Engineering, ETH Z√ºrich | Christian Beisel et al |

| | | | | | | |
|---|---|---|---|---|---|---|
| EPI_ISL_693834 | Switzerland/SO-ETHZ-350185/2020 | 2020-11-04 | Europe/Switzerland/Solothurn/ | Viollier AG | Department of Biosystems Science and Engineering, ETH Z√ºrich | Christian Beisel et al |
| EPI_ISL_693841 | Switzerland/VD-ETHZ-350192/2020 | 2020-11-04 | Europe/Switzerland/Vaud/ | Viollier AG | Department of Biosystems Science and Engineering, ETH Z√ºrich | Christian Beisel et al |
| EPI_ISL_693870 | Switzerland/BS-ETHZ-360010/2020 | 2020-11-12 | Europe/Switzerland/Basel-Stadt/ | Viollier AG | Department of Biosystems Science and Engineering, ETH Z√ºrich | Christian Beisel et al |
| EPI_ISL_693873 | Switzerland/BS-ETHZ-360014/2020 | 2020-11-12 | Europe/Switzerland/Basel-Stadt/ | Viollier AG | Department of Biosystems Science and Engineering, ETH Z√ºrich | Christian Beisel et al |
| EPI_ISL_693879 | Switzerland/BE-ETHZ-360023/2020 | 2020-11-10 | Europe/Switzerland/Bern/ | Viollier AG | Department of Biosystems Science and Engineering, ETH Z√ºrich | Christian Beisel et al |
| EPI_ISL_693897 | Switzerland/BE-ETHZ-360087/2020 | 2020-11-08 | Europe/Switzerland/Bern/ | Viollier AG | Department of Biosystems Science and Engineering, ETH Z√ºrich | Christian Beisel et al |
| EPI_ISL_693911 | Switzerland/ZH-ETHZ-360116/2020 | 2020-11-08 | Europe/Switzerland/Z√ºrich/ | Viollier AG | Department of Biosystems Science and Engineering, ETH Z√ºrich | Christian Beisel et al |
| EPI_ISL_693926 | Switzerland/BL-ETHZ-360154/2020 | 2020-11-06 | Europe/Switzerland/Basel-Land/ | Viollier AG | Department of Biosystems Science and Engineering, ETH Z√ºrich | Christian Beisel et al |

| | | | | | | |
|---|---|---|---|---|---|---|
| EPI_ISL_693980 | Switzerland/GE-ETHZ-370495/2020 | 2020-11-17 | Europe/Switzerland/Geneva/ | Viollier AG | Department of Biosystems Science and Engineering, ETH Z√ºrich | Christian Beisel et al |
| EPI_ISL_693990 | Switzerland/BE-ETHZ-370506/2020 | 2020-11-17 | Europe/Switzerland/Bern/ | Viollier AG | Department of Biosystems Science and Engineering, ETH Z√ºrich | Christian Beisel et al |
| EPI_ISL_694011 | USA/AZ-TG310526/2020 | 2020-05-15 | North America/USA/Arizona/ | TGen North | TGen North | Hayley D Yaglom et al |
| EPI_ISL_694020 | USA/AZ-TG324839/2020 | 2020-05-22 | North America/USA/Arizona/ | TGen North | TGen North | Hayley D Yaglom et al |
| EPI_ISL_694030 | USA/AZ-TG329217/2020 | 2020-05-27 | North America/USA/Arizona/ | TGen North | TGen North | Hayley D Yaglom et al |
| EPI_ISL_694052 | USA/AZ-TG271575/2020 | 2020-03-17 | North America/USA/Arizona/ | TGen North | TGen North | Jolene Bowers et al |
| EPI_ISL_694054 | USA/AZ-TG273328/2020 | 2020-04-02 | North America/USA/Arizona/ | AZ SPHL, Arizona Department of Health Services | TGen North | Jolene Bowers et al |
| EPI_ISL_694061 | USA/AZ-TG274709/2020 | 2020-04-06 | North America/USA/Arizona/ | AZ SPHL, Arizona Department of Health Services | TGen North | Jolene Bowers et al |
| EPI_ISL_694063 | USA/AZ-TG274721/2020 | 2020-04-06 | North America/USA/Arizona/ | AZ SPHL, Arizona Department of Health Services | TGen North | Jolene Bowers et al |
| EPI_ISL_694075 | USA/AZ-TG274769/2020 | 2020-04-08 | North America/USA/Arizona/ | AZ SPHL, Arizona Department of Health Services | TGen North | Jolene Bowers et al |
| EPI_ISL_694076 | USA/AZ-TG274773/2020 | 2020-04-08 | North America/USA/Arizona/ | AZ SPHL, Arizona Department of Health Services | TGen North | Jolene Bowers et al |
| EPI_ISL_694088 | USA/AZ-TG276136/2020 | 2020-04-09 | North America/USA/Arizona/ | AZ SPHL, Arizona Department of Health Services | TGen North | Jolene Bowers et al |
| EPI_ISL_694089 | USA/AZ-TG276138/2020 | 2020-04-09 | North America/USA/Arizona/ | AZ SPHL, Arizona Department of Health Services | TGen North | Jolene Bowers et al |
| EPI_ISL_694093 | USA/AZ-TG276152/2020 | 2020-04-09 | North America/USA/Arizona/ | AZ SPHL, Arizona Department of Health Services | TGen North | Jolene Bowers et al |
| EPI_ISL_694097 | USA/AZ-TG276177/2020 | 2020-04-10 | North America/USA/Arizona/ | AZ SPHL, Arizona Department of Health Services | TGen North | Jolene Bowers et al |
| EPI_ISL_694099 | USA/AZ-TG276181/2020 | 2020-04-10 | North America/USA/Arizona/ | AZ SPHL, Arizona Department of Health Services | TGen North | Jolene Bowers et al |
| EPI_ISL_694102 | USA/AZ-TG276458/2020 | 2020-04-11 | North America/USA/Arizona/ | AZ SPHL, Arizona Department of Health Services | TGen North | Jolene Bowers et al |
| EPI_ISL_694112 | USA/AZ-TG276577/2020 | 2020-04-15 | North America/USA/Arizona/ | AZ SPHL, Arizona Department of Health Services | TGen North | Jolene Bowers et al |

| | | | | | | |
|---|---|---|---|---|---|---|
| EPI_ISL_694116 | USA/AZ-TG276588/2020 | 2020-04-16 | North America/USA/Arizona/ | AZ SPHL, Arizona Department of Health Services | TGen North | Jolene Bowers et al |
| EPI_ISL_694118 | USA/AZ-TG276594/2020 | 2020-04-15 | North America/USA/Arizona/ | AZ SPHL, Arizona Department of Health Services | TGen North | Jolene Bowers et al |
| EPI_ISL_694129 | USA/AZ-TG276764/2020 | 2020-04-16 | North America/USA/Arizona/ | AZ SPHL, Arizona Department of Health Services | TGen North | Jolene Bowers et al |
| EPI_ISL_694135 | USA/AZ-TG276778/2020 | 2020-04-17 | North America/USA/Arizona/ | AZ SPHL, Arizona Department of Health Services | TGen North | Jolene Bowers et al |
| EPI_ISL_694157 | USA/AZ-TG280803/2020 | 2020-04-20 | North America/USA/Arizona/ | AZ SPHL, Arizona Department of Health Services | TGen North | Jolene Bowers et al |
| EPI_ISL_694160 | USA/AZ-TG280807/2020 | 2020-04-20 | North America/USA/Arizona/ | AZ SPHL, Arizona Department of Health Services | TGen North | Jolene Bowers et al |
| EPI_ISL_694161 | USA/AZ-TG280810/2020 | 2020-04-20 | North America/USA/Arizona/ | AZ SPHL, Arizona Department of Health Services | TGen North | Jolene Bowers et al |
| EPI_ISL_694167 | USA/AZ-TG280821/2020 | 2020-04-21 | North America/USA/Arizona/ | AZ SPHL, Arizona Department of Health Services | TGen North | Jolene Bowers et al |
| EPI_ISL_694173 | USA/AZ-TG280830/2020 | 2020-04-20 | North America/USA/Arizona/ | AZ SPHL, Arizona Department of Health Services | TGen North | Jolene Bowers et al |
| EPI_ISL_694180 | USA/AZ-TG280838/2020 | 2020-04-21 | North America/USA/Arizona/ | AZ SPHL, Arizona Department of Health Services | TGen North | Jolene Bowers et al |
| EPI_ISL_694186 | USA/AZ-TG280845/2020 | 2020-04-21 | North America/USA/Arizona/ | AZ SPHL, Arizona Department of Health Services | TGen North | Jolene Bowers et al |
| EPI_ISL_694202 | USA/AZ-TG280869/2020 | 2020-04-23 | North America/USA/Arizona/ | AZ SPHL, Arizona Department of Health Services | TGen North | Jolene Bowers et al |
| EPI_ISL_694203 | USA/AZ-TG280870/2020 | 2020-04-22 | North America/USA/Arizona/ | AZ SPHL, Arizona Department of Health Services | TGen North | Jolene Bowers et al |
| EPI_ISL_694206 | USA/AZ-TG280875/2020 | 2020-04-24 | North America/USA/Arizona/ | AZ SPHL, Arizona Department of Health Services | TGen North | Jolene Bowers et al |
| EPI_ISL_694209 | USA/AZ-TG280878/2020 | 2020-04-23 | North America/USA/Arizona/ | AZ SPHL, Arizona Department of Health Services | TGen North | Jolene Bowers et al |
| EPI_ISL_694223 | USA/AZ-TG281380/2020 | 2020-04-27 | North America/USA/Arizona/ | TGen North | TGen North | Jolene Bowers et al |
| EPI_ISL_694224 | USA/AZ-TG281576/2020 | 2020-04-30 | North America/USA/Arizona/ | TGen North | TGen North | Jolene Bowers et al |
| EPI_ISL_694226 | USA/AZ-TG283819/2020 | 2020-05-01 | North America/USA/Arizona/ | TGen North | TGen North | Jolene Bowers et al |
| EPI_ISL_694237 | USA/AZ-TG290705/2020 | 2020-05-05 | North America/USA/Arizona/ | TGen North | TGen North | Jolene Bowers et al |
| EPI_ISL_694252 | USA/AZ-TG305707/2020 | 2020-04-24 | North America/USA/Arizona/ | AZ SPHL, Arizona Department of Health Services | TGen North | Jolene Bowers et al |
| EPI_ISL_694253 | USA/AZ-TG305709/2020 | 2020-04-24 | North America/USA/Arizona/ | AZ SPHL, Arizona Department of Health Services | TGen North | Jolene Bowers et al |

| | | | | | | |
|---|---|---|---|---|---|---|
| EPI_ISL_694257 | USA/AZ-TG305724/2020 | 2020-04-25 | North America/USA/Arizona/ | AZ SPHL, Arizona Department of Health Services | TGen North | Jolene Bowers et al |
| EPI_ISL_694260 | USA/AZ-TG305737/2020 | 2020-04-27 | North America/USA/Arizona/ | AZ SPHL, Arizona Department of Health Services | TGen North | Jolene Bowers et al |
| EPI_ISL_694264 | USA/AZ-TG305749/2020 | 2020-04-27 | North America/USA/Arizona/ | AZ SPHL, Arizona Department of Health Services | TGen North | Jolene Bowers et al |
| EPI_ISL_694270 | USA/AZ-TG305766/2020 | 2020-04-24 | North America/USA/Arizona/ | AZ SPHL, Arizona Department of Health Services | TGen North | Jolene Bowers et al |
| EPI_ISL_694273 | USA/AZ-TG305772/2020 | 2020-04-27 | North America/USA/Arizona/ | AZ SPHL, Arizona Department of Health Services | TGen North | Jolene Bowers et al |
| EPI_ISL_694275 | USA/AZ-TG305783/2020 | 2020-04-27 | North America/USA/Arizona/ | AZ SPHL, Arizona Department of Health Services | TGen North | Jolene Bowers et al |
| EPI_ISL_694277 | USA/AZ-TG305789/2020 | 2020-04-28 | North America/USA/Arizona/ | AZ SPHL, Arizona Department of Health Services | TGen North | Jolene Bowers et al |
| EPI_ISL_694279 | USA/AZ-TG305796/2020 | 2020-04-27 | North America/USA/Arizona/ | AZ SPHL, Arizona Department of Health Services | TGen North | Jolene Bowers et al |
| EPI_ISL_694280 | USA/AZ-TG305800/2020 | 2020-04-28 | North America/USA/Arizona/ | AZ SPHL, Arizona Department of Health Services | TGen North | Jolene Bowers et al |
| EPI_ISL_694281 | USA/AZ-TG305802/2020 | 2020-04-28 | North America/USA/Arizona/ | AZ SPHL, Arizona Department of Health Services | TGen North | Jolene Bowers et al |
| EPI_ISL_694282 | USA/AZ-TG305807/2020 | 2020-04-28 | North America/USA/Arizona/ | AZ SPHL, Arizona Department of Health Services | TGen North | Jolene Bowers et al |
| EPI_ISL_694288 | USA/AZ-TG305888/2020 | 2020-04-29 | North America/USA/Arizona/ | AZ SPHL, Arizona Department of Health Services | TGen North | Jolene Bowers et al |
| EPI_ISL_694291 | USA/AZ-TG305899/2020 | 2020-04-29 | North America/USA/Arizona/ | AZ SPHL, Arizona Department of Health Services | TGen North | Jolene Bowers et al |
| EPI_ISL_694295 | USA/AZ-TG305910/2020 | 2020-04-29 | North America/USA/Arizona/ | AZ SPHL, Arizona Department of Health Services | TGen North | Jolene Bowers et al |
| EPI_ISL_694297 | USA/AZ-TG305914/2020 | 2020-04-29 | North America/USA/Arizona/ | AZ SPHL, Arizona Department of Health Services | TGen North | Jolene Bowers et al |
| EPI_ISL_694310 | USA/AZ-TG306001/2020 | 2020-04-30 | North America/USA/Arizona/ | AZ SPHL, Arizona Department of Health Services | TGen North | Jolene Bowers et al |
| EPI_ISL_694312 | USA/AZ-TG306005/2020 | 2020-04-30 | North America/USA/Arizona/ | AZ SPHL, Arizona Department of Health Services | TGen North | Jolene Bowers et al |
| EPI_ISL_694315 | USA/AZ-TG306016/2020 | 2020-04-30 | North America/USA/Arizona/ | AZ SPHL, Arizona Department of Health Services | TGen North | Jolene Bowers et al |
| EPI_ISL_694325 | USA/AZ-TG312663/2020 | 2020-05-16 | North America/USA/Arizona/ | TGen North | TGen North | Jolene Bowers et al |
| EPI_ISL_694330 | USA/AZ-TG318392/2020 | 2020-05-19 | North America/USA/Arizona/ | TGen North | TGen North | Jolene Bowers et al |
| EPI_ISL_694331 | USA/AZ-TG320609/2020 | 2020-05-16 | North America/USA/Arizona/ | TGen North | TGen North | Jolene Bowers et al |

| | | | | | | |
|---|---|---|---|---|---|---|
| EPI_ISL_694336 | USA/AZ-TG322725/2020 | 2020-05-20 | North America/USA/Arizona/ | TGen North | TGen North | Jolene Bowers et al |
| EPI_ISL_694337 | USA/AZ-TG322842/2020 | 2020-05-20 | North America/USA/Arizona/ | TGen North | TGen North | Jolene Bowers et al |
| EPI_ISL_694338 | USA/AZ-TG323137/2020 | 2020-05-21 | North America/USA/Arizona/ | TGen North | TGen North | Jolene Bowers et al |
| EPI_ISL_694354 | USA/AZ-TG332555/2020 | 2020-05-26 | North America/USA/Arizona/ | TGen North | TGen North | Jolene Bowers et al |
| EPI_ISL_694359 | USA/AZ-TG333630/2020 | 2020-05-27 | North America/USA/Arizona/ | TGen North | TGen North | Jolene Bowers et al |
| EPI_ISL_694360 | USA/AZ-TG333692/2020 | 2020-05-27 | North America/USA/Arizona/ | TGen North | TGen North | Jolene Bowers et al |
| EPI_ISL_694363 | USA/AZ-TG333885/2020 | 2020-05-28 | North America/USA/Arizona/ | TGen North | TGen North | Jolene Bowers et al |
| EPI_ISL_694369 | USA/AZ-TG336686/2020 | 2020-05-25 | North America/USA/Arizona/ | TGen North | TGen North | Jolene Bowers et al |
| EPI_ISL_694376 | USA/AZ-TG340339/2020 | 2020-05-25 | North America/USA/Arizona/ | TGen North | TGen North | Jolene Bowers et al |
| EPI_ISL_694380 | USA/AZ-TG341882/2020 | 2020-05-30 | North America/USA/Arizona/ | TGen North | TGen North | Jolene Bowers et al |
| EPI_ISL_694387 | USA/AZ-TG345029/2020 | 2020-06-01 | North America/USA/Arizona/ | TGen North | TGen North | Jolene Bowers et al |
| EPI_ISL_694398 | USA/AZ-TG348980/2020 | 2020-06-03 | North America/USA/Arizona/ | TGen North | TGen North | Jolene Bowers et al |
| EPI_ISL_694400 | USA/AZ-TG351477/2020 | 2020-06-03 | North America/USA/Arizona/ | TGen North | TGen North | Jolene Bowers et al |
| EPI_ISL_694423 | USA/AZ-TG352045/2020 | 2020-05-29 | North America/USA/Arizona/ | AZ SPHL, Arizona Department of Health Services | TGen North | Jolene Bowers et al |
| EPI_ISL_694424 | USA/AZ-TG352047/2020 | 2020-05-29 | North America/USA/Arizona/ | AZ SPHL, Arizona Department of Health Services | TGen North | Jolene Bowers et al |
| EPI_ISL_694430 | USA/AZ-TG352880/2020 | 2020-06-04 | North America/USA/Arizona/ | TGen North | TGen North | Jolene Bowers et al |
| EPI_ISL_694433 | USA/AZ-TG353013/2020 | 2020-06-04 | North America/USA/Arizona/ | TGen North | TGen North | Jolene Bowers et al |
| EPI_ISL_694436 | USA/AZ-TG354397/2020 | 2020-06-05 | North America/USA/Arizona/ | TGen North | TGen North | Jolene Bowers et al |
| EPI_ISL_694441 | USA/AZ-TG357125/2020 | 2020-06-08 | North America/USA/Arizona/ | TGen North | TGen North | Jolene Bowers et al |
| EPI_ISL_694445 | USA/AZ-TG359375/2020 | 2020-06-08 | North America/USA/Arizona/ | TGen North | TGen North | Jolene Bowers et al |
| EPI_ISL_694453 | USA/AZ-TG363238/2020 | 2020-06-10 | North America/USA/Arizona/ | TGen North | TGen North | Jolene Bowers et al |
| EPI_ISL_694463 | USA/AZ-TG366697/2020 | 2020-05-31 | North America/USA/Arizona/ | AZ SPHL, Arizona Department of Health Services | TGen North | Jolene Bowers et al |
| EPI_ISL_694467 | USA/AZ-TG366703/2020 | 2020-06-01 | North America/USA/Arizona/ | AZ SPHL, Arizona Department of Health Services | TGen North | Jolene Bowers et al |
| EPI_ISL_694469 | USA/AZ-TG366705/2020 | 2020-06-01 | North America/USA/Arizona/ | AZ SPHL, Arizona Department of Health Services | TGen North | Jolene Bowers et al |
| EPI_ISL_694470 | USA/AZ-TG366706/2020 | 2020-06-01 | North America/USA/Arizona/ | AZ SPHL, Arizona Department of Health Services | TGen North | Jolene Bowers et al |
| EPI_ISL_694476 | USA/AZ-TG366712/2020 | 2020-06-02 | North America/USA/Arizona/ | AZ SPHL, Arizona Department of Health Services | TGen North | Jolene Bowers et al |

| | | | | | | |
|---|---|---|---|---|---|---|
| EPI_ISL_694485 | USA/AZ-TG366721/2020 | 2020-06-02 | North America/USA/Arizona/ | AZ SPHL, Arizona Department of Health Services | TGen North | Jolene Bowers et al |
| EPI_ISL_694490 | USA/AZ-TG366731/2020 | 2020-06-02 | North America/USA/Arizona/ | AZ SPHL, Arizona Department of Health Services | TGen North | Jolene Bowers et al |
| EPI_ISL_694499 | USA/AZ-TG366743/2020 | 2020-06-02 | North America/USA/Arizona/ | AZ SPHL, Arizona Department of Health Services | TGen North | Jolene Bowers et al |
| EPI_ISL_694527 | USA/AZ-TG371267/2020 | 2020-05-15 | North America/USA/Arizona/ | AZ SPHL, Arizona Department of Health Services | TGen North | Jolene Bowers et al |
| EPI_ISL_694530 | USA/AZ-TG371271/2020 | 2020-05-15 | North America/USA/Arizona/ | AZ SPHL, Arizona Department of Health Services | TGen North | Jolene Bowers et al |
| EPI_ISL_694538 | USA/AZ-TG371279/2020 | 2020-05-16 | North America/USA/Arizona/ | AZ SPHL, Arizona Department of Health Services | TGen North | Jolene Bowers et al |
| EPI_ISL_694540 | USA/AZ-TG371281/2020 | 2020-05-16 | North America/USA/Arizona/ | AZ SPHL, Arizona Department of Health Services | TGen North | Jolene Bowers et al |
| EPI_ISL_694542 | USA/AZ-TG371283/2020 | 2020-05-16 | North America/USA/Arizona/ | AZ SPHL, Arizona Department of Health Services | TGen North | Jolene Bowers et al |
| EPI_ISL_694547 | USA/AZ-TG371288/2020 | 2020-05-19 | North America/USA/Arizona/ | AZ SPHL, Arizona Department of Health Services | TGen North | Jolene Bowers et al |
| EPI_ISL_694554 | USA/AZ-TG371298/2020 | 2020-05-19 | North America/USA/Arizona/ | AZ SPHL, Arizona Department of Health Services | TGen North | Jolene Bowers et al |
| EPI_ISL_694556 | USA/AZ-TG371301/2020 | 2020-05-19 | North America/USA/Arizona/ | AZ SPHL, Arizona Department of Health Services | TGen North | Jolene Bowers et al |
| EPI_ISL_694560 | USA/AZ-TG371307/2020 | 2020-05-18 | North America/USA/Arizona/ | AZ SPHL, Arizona Department of Health Services | TGen North | Jolene Bowers et al |
| EPI_ISL_694567 | USA/AZ-TG371315/2020 | 2020-05-20 | North America/USA/Arizona/ | AZ SPHL, Arizona Department of Health Services | TGen North | Jolene Bowers et al |
| EPI_ISL_694574 | USA/AZ-TG371324/2020 | 2020-05-21 | North America/USA/Arizona/ | AZ SPHL, Arizona Department of Health Services | TGen North | Jolene Bowers et al |
| EPI_ISL_694577 | USA/AZ-TG371333/2020 | 2020-05-22 | North America/USA/Arizona/ | AZ SPHL, Arizona Department of Health Services | TGen North | Jolene Bowers et al |
| EPI_ISL_694585 | USA/AZ-TG371341/2020 | 2020-05-22 | North America/USA/Arizona/ | AZ SPHL, Arizona Department of Health Services | TGen North | Jolene Bowers et al |
| EPI_ISL_694591 | USA/AZ-TG371349/2020 | 2020-05-22 | North America/USA/Arizona/ | AZ SPHL, Arizona Department of Health Services | TGen North | Jolene Bowers et al |
| EPI_ISL_694596 | USA/AZ-TG371355/2020 | 2020-05-22 | North America/USA/Arizona/ | AZ SPHL, Arizona Department of Health Services | TGen North | Jolene Bowers et al |
| EPI_ISL_694598 | USA/AZ-TG371441/2020 | 2020-05-29 | North America/USA/Arizona/ | AZ SPHL, Arizona Department of Health Services | TGen North | Jolene Bowers et al |
| EPI_ISL_694599 | USA/AZ-TG371442/2020 | 2020-05-29 | North America/USA/Arizona/ | AZ SPHL, Arizona Department of Health Services | TGen North | Jolene Bowers et al |

| | | | | | | |
|---|---|---|---|---|---|---|
| EPI_ISL_694606 | USA/AZ-TG375874/2020 | 2020-06-15 | North America/USA/Arizona/ | TGen North | TGen North | Jolene Bowers et al |
| EPI_ISL_694620 | USA/AZ-TG377256/2020 | 2020-06-15 | North America/USA/Arizona/ | TGen North | TGen North | Jolene Bowers et al |
| EPI_ISL_694623 | USA/AZ-TG377620/2020 | 2020-06-16 | North America/USA/Arizona/ | TGen North | TGen North | Jolene Bowers et al |
| EPI_ISL_694636 | USA/AZ-TG379342/2020 | 2020-06-16 | North America/USA/Arizona/ | TGen North | TGen North | Jolene Bowers et al |
| EPI_ISL_694638 | USA/AZ-TG379464/2020 | 2020-06-16 | North America/USA/Arizona/ | TGen North | TGen North | Jolene Bowers et al |
| EPI_ISL_694639 | USA/AZ-TG379485/2020 | 2020-06-16 | North America/USA/Arizona/ | TGen North | TGen North | Jolene Bowers et al |
| EPI_ISL_694640 | USA/AZ-TG379817/2020 | 2020-06-16 | North America/USA/Arizona/ | TGen North | TGen North | Jolene Bowers et al |
| EPI_ISL_694645 | USA/AZ-TG380033/2020 | 2020-06-16 | North America/USA/Arizona/ | TGen North | TGen North | Jolene Bowers et al |
| EPI_ISL_694653 | USA/AZ-TG381246/2020 | 2020-06-15 | North America/USA/Arizona/ | TGen North | TGen North | Jolene Bowers et al |
| EPI_ISL_694658 | USA/AZ-TG381367/2020 | 2020-06-15 | North America/USA/Arizona/ | TGen North | TGen North | Jolene Bowers et al |
| EPI_ISL_694661 | USA/AZ-TG381372/2020 | 2020-06-15 | North America/USA/Arizona/ | TGen North | TGen North | Jolene Bowers et al |
| EPI_ISL_694664 | USA/AZ-TG381594/2020 | 2020-06-15 | North America/USA/Arizona/ | TGen North | TGen North | Jolene Bowers et al |
| EPI_ISL_694667 | USA/AZ-TG381597/2020 | 2020-06-15 | North America/USA/Arizona/ | TGen North | TGen North | Jolene Bowers et al |
| EPI_ISL_694668 | USA/AZ-TG381598/2020 | 2020-06-15 | North America/USA/Arizona/ | TGen North | TGen North | Jolene Bowers et al |
| EPI_ISL_694669 | USA/AZ-TG381610/2020 | 2020-06-15 | North America/USA/Arizona/ | TGen North | TGen North | Jolene Bowers et al |
| EPI_ISL_694673 | USA/AZ-TG381616/2020 | 2020-06-15 | North America/USA/Arizona/ | TGen North | TGen North | Jolene Bowers et al |
| EPI_ISL_694676 | USA/AZ-TG382138/2020 | 2020-06-15 | North America/USA/Arizona/ | TGen North | TGen North | Jolene Bowers et al |
| EPI_ISL_694684 | USA/AZ-TG382290/2020 | 2020-06-15 | North America/USA/Arizona/ | TGen North | TGen North | Jolene Bowers et al |
| EPI_ISL_694687 | USA/AZ-TG383297/2020 | 2020-06-17 | North America/USA/Arizona/ | TGen North | TGen North | Jolene Bowers et al |
| EPI_ISL_694692 | USA/AZ-TG384418/2020 | 2020-06-17 | North America/USA/Arizona/ | TGen North | TGen North | Jolene Bowers et al |
| EPI_ISL_694694 | USA/AZ-TG384442/2020 | 2020-06-17 | North America/USA/Arizona/ | TGen North | TGen North | Jolene Bowers et al |
| EPI_ISL_694710 | USA/AZ-TG392982/2020 | 2020-06-18 | North America/USA/Arizona/ | TGen North | TGen North | Jolene Bowers et al |
| EPI_ISL_694711 | USA/AZ-TG393520/2020 | 2020-06-19 | North America/USA/Arizona/ | TGen North | TGen North | Jolene Bowers et al |
| EPI_ISL_694737 | USA/AZ-TG395893/2020 | 2020-06-05 | North America/USA/Arizona/ | AZ SPHL, Arizona Department of Health Services | TGen North | Jolene Bowers et al |
| EPI_ISL_694739 | USA/AZ-TG395895/2020 | 2020-06-05 | North America/USA/Arizona/ | AZ SPHL, Arizona Department of Health Services | TGen North | Jolene Bowers et al |
| EPI_ISL_694741 | USA/AZ-TG395897/2020 | 2020-06-05 | North America/USA/Arizona/ | AZ SPHL, Arizona Department of Health Services | TGen North | Jolene Bowers et al |
| EPI_ISL_694743 | USA/AZ-TG395899/2020 | 2020-06-05 | North America/USA/Arizona/ | AZ SPHL, Arizona Department of Health Services | TGen North | Jolene Bowers et al |
| EPI_ISL_694745 | USA/AZ-TG395901/2020 | 2020-06-08 | North America/USA/Arizona/ | AZ SPHL, Arizona Department of Health Services | TGen North | Jolene Bowers et al |

| | | | | | | |
|---|---|---|---|---|---|---|
| EPI_ISL_694759 | USA/AZ-TG395918/2020 | 2020-06-08 | North America/USA/Arizona/ | AZ SPHL, Arizona Department of Health Services | TGen North | Jolene Bowers et al |
| EPI_ISL_694765 | USA/AZ-TG395929/2020 | 2020-06-08 | North America/USA/Arizona/ | AZ SPHL, Arizona Department of Health Services | TGen North | Jolene Bowers et al |
| EPI_ISL_694769 | USA/AZ-TG395933/2020 | 2020-06-08 | North America/USA/Arizona/ | AZ SPHL, Arizona Department of Health Services | TGen North | Jolene Bowers et al |
| EPI_ISL_694780 | USA/AZ-TG395947/2020 | 2020-06-08 | North America/USA/Arizona/ | AZ SPHL, Arizona Department of Health Services | TGen North | Jolene Bowers et al |
| EPI_ISL_694782 | USA/AZ-TG395951/2020 | 2020-06-08 | North America/USA/Arizona/ | AZ SPHL, Arizona Department of Health Services | TGen North | Jolene Bowers et al |
| EPI_ISL_694785 | USA/AZ-TG395959/2020 | 2020-06-09 | North America/USA/Arizona/ | AZ SPHL, Arizona Department of Health Services | TGen North | Jolene Bowers et al |
| EPI_ISL_694802 | USA/AZ-TG396333/2020 | 2020-06-09 | North America/USA/Arizona/ | AZ SPHL, Arizona Department of Health Services | TGen North | Jolene Bowers et al |
| EPI_ISL_694809 | USA/AZ-TG396351/2020 | 2020-06-10 | North America/USA/Arizona/ | AZ SPHL, Arizona Department of Health Services | TGen North | Jolene Bowers et al |
| EPI_ISL_694812 | USA/AZ-TG396354/2020 | 2020-06-10 | North America/USA/Arizona/ | AZ SPHL, Arizona Department of Health Services | TGen North | Jolene Bowers et al |
| EPI_ISL_694817 | USA/AZ-TG396359/2020 | 2020-06-10 | North America/USA/Arizona/ | AZ SPHL, Arizona Department of Health Services | TGen North | Jolene Bowers et al |
| EPI_ISL_694818 | USA/AZ-TG396360/2020 | 2020-06-10 | North America/USA/Arizona/ | AZ SPHL, Arizona Department of Health Services | TGen North | Jolene Bowers et al |
| EPI_ISL_694822 | USA/AZ-TG396366/2020 | 2020-06-11 | North America/USA/Arizona/ | AZ SPHL, Arizona Department of Health Services | TGen North | Jolene Bowers et al |
| EPI_ISL_694838 | USA/AZ-TG396386/2020 | 2020-06-12 | North America/USA/Arizona/ | AZ SPHL, Arizona Department of Health Services | TGen North | Jolene Bowers et al |
| EPI_ISL_694839 | USA/AZ-TG396388/2020 | 2020-06-12 | North America/USA/Arizona/ | AZ SPHL, Arizona Department of Health Services | TGen North | Jolene Bowers et al |
| EPI_ISL_694841 | USA/AZ-TG396390/2020 | 2020-06-12 | North America/USA/Arizona/ | AZ SPHL, Arizona Department of Health Services | TGen North | Jolene Bowers et al |
| EPI_ISL_694848 | USA/AZ-TG398429/2020 | 2020-06-22 | North America/USA/Arizona/ | TGen North | TGen North | Jolene Bowers et al |
| EPI_ISL_694855 | USA/AZ-TG402010/2020 | 2020-06-23 | North America/USA/Arizona/ | TGen North | TGen North | Jolene Bowers et al |
| EPI_ISL_694858 | USA/AZ-TG402761/2020 | 2020-06-23 | North America/USA/Arizona/ | TGen North | TGen North | Jolene Bowers et al |
| EPI_ISL_694881 | USA/AZ-TG407779/2020 | 2020-06-25 | North America/USA/Arizona/ | TGen North | TGen North | Jolene Bowers et al |
| EPI_ISL_694885 | USA/AZ-TG408122/2020 | 2020-06-12 | North America/USA/Arizona/ | AZ SPHL, Arizona Department of Health Services | TGen North | Jolene Bowers et al |
| EPI_ISL_694887 | USA/AZ-TG408124/2020 | 2020-06-12 | North America/USA/Arizona/ | AZ SPHL, Arizona Department of Health Services | TGen North | Jolene Bowers et al |

| | | | | | | |
|---|---|---|---|---|---|---|
| EPI_ISL_694891 | USA/AZ-TG408128/2020 | 2020-06-12 | North America/USA/Arizona/ | AZ SPHL, Arizona Department of Health Services | TGen North | Jolene Bowers et al |
| EPI_ISL_694897 | USA/AZ-TG408140/2020 | 2020-06-14 | North America/USA/Arizona/ | AZ SPHL, Arizona Department of Health Services | TGen North | Jolene Bowers et al |
| EPI_ISL_694914 | USA/AZ-TG408162/2020 | 2020-06-15 | North America/USA/Arizona/ | AZ SPHL, Arizona Department of Health Services | TGen North | Jolene Bowers et al |
| EPI_ISL_694918 | USA/AZ-TG408167/2020 | 2020-06-16 | North America/USA/Arizona/ | AZ SPHL, Arizona Department of Health Services | TGen North | Jolene Bowers et al |
| EPI_ISL_694919 | USA/AZ-TG408168/2020 | 2020-06-16 | North America/USA/Arizona/ | AZ SPHL, Arizona Department of Health Services | TGen North | Jolene Bowers et al |
| EPI_ISL_694920 | USA/AZ-TG408169/2020 | 2020-06-16 | North America/USA/Arizona/ | AZ SPHL, Arizona Department of Health Services | TGen North | Jolene Bowers et al |
| EPI_ISL_694928 | USA/AZ-TG408188/2020 | 2020-06-16 | North America/USA/Arizona/ | AZ SPHL, Arizona Department of Health Services | TGen North | Jolene Bowers et al |
| EPI_ISL_694938 | USA/AZ-TG408200/2020 | 2020-06-17 | North America/USA/Arizona/ | AZ SPHL, Arizona Department of Health Services | TGen North | Jolene Bowers et al |
| EPI_ISL_694939 | USA/AZ-TG408201/2020 | 2020-06-16 | North America/USA/Arizona/ | AZ SPHL, Arizona Department of Health Services | TGen North | Jolene Bowers et al |
| EPI_ISL_694958 | USA/AZ-TG409875/2020 | 2020-06-24 | North America/USA/Arizona/ | TGen North | TGen North | Jolene Bowers et al |
| EPI_ISL_694972 | USA/AZ-TG411266/2020 | 2020-06-24 | North America/USA/Arizona/ | TGen North | TGen North | Jolene Bowers et al |
| EPI_ISL_694976 | USA/AZ-TG411559/2020 | 2020-06-25 | North America/USA/Arizona/ | TGen North | TGen North | Jolene Bowers et al |
| EPI_ISL_694982 | USA/AZ-TG412165/2020 | 2020-06-25 | North America/USA/Arizona/ | TGen North | TGen North | Jolene Bowers et al |
| EPI_ISL_694994 | USA/AZ-TG442565/2020 | 2020-06-25 | North America/USA/Arizona/ | TGen North | TGen North | Jolene Bowers et al |
| EPI_ISL_695004 | USA/AZ-TG445262/2020 | 2020-06-25 | North America/USA/Arizona/ | TGen North | TGen North | Jolene Bowers et al |
| EPI_ISL_695010 | USA/AZ-TG447453/2020 | 2020-06-26 | North America/USA/Arizona/ | TGen North | TGen North | Jolene Bowers et al |
| EPI_ISL_695012 | USA/AZ-TG447756/2020 | 2020-06-26 | North America/USA/Arizona/ | TGen North | TGen North | Jolene Bowers et al |
| EPI_ISL_695015 | USA/AZ-TG448276/2020 | 2020-06-26 | North America/USA/Arizona/ | TGen North | TGen North | Jolene Bowers et al |
| EPI_ISL_695017 | USA/AZ-TG448488/2020 | 2020-06-26 | North America/USA/Arizona/ | TGen North | TGen North | Jolene Bowers et al |
| EPI_ISL_695024 | USA/AZ-TG452852/2020 | 2020-06-29 | North America/USA/Arizona/ | TGen North | TGen North | Jolene Bowers et al |
| EPI_ISL_695043 | USA/AZ-TG456402/2020 | 2020-06-26 | North America/USA/Arizona/ | TGen North | TGen North | Jolene Bowers et al |
| EPI_ISL_695056 | USA/AZ-TG479329/2020 | 2020-07-08 | North America/USA/Arizona/ | TGen North | TGen North | Jolene Bowers et al |
| EPI_ISL_695067 | USA/AZ-TG480709/2020 | 2020-07-08 | North America/USA/Arizona/ | TGen North | TGen North | Jolene Bowers et al |
| EPI_ISL_695072 | USA/AZ-TG481283/2020 | 2020-07-07 | North America/USA/Arizona/ | TGen North | TGen North | Jolene Bowers et al |
| EPI_ISL_695078 | USA/AZ-TG485093/2020 | 2020-07-09 | North America/USA/Arizona/ | TGen North | TGen North | Jolene Bowers et al |
| EPI_ISL_695079 | USA/AZ-TG485788/2020 | 2020-07-09 | North America/USA/Arizona/ | TGen North | TGen North | Jolene Bowers et al |

| | | | | | | |
|---|---|---|---|---|---|---|
| EPI_ISL_695081 | USA/AZ-TG486040/2020 | 2020-07-09 | North America/USA/Arizona/ | TGen North | TGen North | Jolene Bowers et al |
| EPI_ISL_695084 | USA/AZ-TG486331/2020 | 2020-07-09 | North America/USA/Arizona/ | TGen North | TGen North | Jolene Bowers et al |
| EPI_ISL_695096 | USA/AZ-TG487161/2020 | 2020-07-09 | North America/USA/Arizona/ | TGen North | TGen North | Jolene Bowers et al |
| EPI_ISL_695097 | USA/AZ-TG489086/2020 | 2020-07-10 | North America/USA/Arizona/ | TGen North | TGen North | Jolene Bowers et al |
| EPI_ISL_695104 | USA/AZ-TG491469/2020 | 2020-06-17 | North America/USA/Arizona/ | AZ SPHL, Arizona Department of Health Services | TGen North | Jolene Bowers et al |
| EPI_ISL_695107 | USA/AZ-TG491472/2020 | 2020-06-17 | North America/USA/Arizona/ | AZ SPHL, Arizona Department of Health Services | TGen North | Jolene Bowers et al |
| EPI_ISL_695111 | USA/AZ-TG491478/2020 | 2020-06-18 | North America/USA/Arizona/ | AZ SPHL, Arizona Department of Health Services | TGen North | Jolene Bowers et al |
| EPI_ISL_695114 | USA/AZ-TG491482/2020 | 2020-06-18 | North America/USA/Arizona/ | AZ SPHL, Arizona Department of Health Services | TGen North | Jolene Bowers et al |
| EPI_ISL_695115 | USA/AZ-TG491483/2020 | 2020-06-18 | North America/USA/Arizona/ | AZ SPHL, Arizona Department of Health Services | TGen North | Jolene Bowers et al |
| EPI_ISL_695126 | USA/AZ-TG491497/2020 | 2020-06-18 | North America/USA/Arizona/ | AZ SPHL, Arizona Department of Health Services | TGen North | Jolene Bowers et al |
| EPI_ISL_695127 | USA/AZ-TG491498/2020 | 2020-06-18 | North America/USA/Arizona/ | AZ SPHL, Arizona Department of Health Services | TGen North | Jolene Bowers et al |
| EPI_ISL_695128 | USA/AZ-TG491499/2020 | 2020-06-18 | North America/USA/Arizona/ | AZ SPHL, Arizona Department of Health Services | TGen North | Jolene Bowers et al |
| EPI_ISL_695137 | USA/AZ-TG491513/2020 | 2020-06-19 | North America/USA/Arizona/ | AZ SPHL, Arizona Department of Health Services | TGen North | Jolene Bowers et al |
| EPI_ISL_695156 | USA/AZ-TG491540/2020 | 2020-06-21 | North America/USA/Arizona/ | AZ SPHL, Arizona Department of Health Services | TGen North | Jolene Bowers et al |
| EPI_ISL_695158 | USA/AZ-TG491542/2020 | 2020-06-21 | North America/USA/Arizona/ | AZ SPHL, Arizona Department of Health Services | TGen North | Jolene Bowers et al |
| EPI_ISL_695159 | USA/AZ-TG491543/2020 | 2020-06-21 | North America/USA/Arizona/ | AZ SPHL, Arizona Department of Health Services | TGen North | Jolene Bowers et al |
| EPI_ISL_695170 | USA/AZ-TG491557/2020 | 2020-06-22 | North America/USA/Arizona/ | AZ SPHL, Arizona Department of Health Services | TGen North | Jolene Bowers et al |
| EPI_ISL_695172 | USA/AZ-TG491559/2020 | 2020-06-22 | North America/USA/Arizona/ | AZ SPHL, Arizona Department of Health Services | TGen North | Jolene Bowers et al |
| EPI_ISL_695182 | USA/AZ-TG491590/2020 | 2020-06-21 | North America/USA/Arizona/ | AZ SPHL, Arizona Department of Health Services | TGen North | Jolene Bowers et al |
| EPI_ISL_695184 | USA/AZ-TG491596/2020 | 2020-06-22 | North America/USA/Arizona/ | AZ SPHL, Arizona Department of Health Services | TGen North | Jolene Bowers et al |
| EPI_ISL_695187 | USA/AZ-TG491601/2020 | 2020-06-23 | North America/USA/Arizona/ | AZ SPHL, Arizona Department of Health Services | TGen North | Jolene Bowers et al |

| | | | | | | |
|---|---|---|---|---|---|---|
| EPI_ISL_695190 | USA/AZ-TG491606/2020 | 2020-06-23 | North America/USA/Arizona/ | AZ SPHL, Arizona Department of Health Services | TGen North | Jolene Bowers et al |
| EPI_ISL_695192 | USA/AZ-TG491610/2020 | 2020-06-24 | North America/USA/Arizona/ | AZ SPHL, Arizona Department of Health Services | TGen North | Jolene Bowers et al |
| EPI_ISL_695197 | USA/AZ-TG491627/2020 | 2020-06-24 | North America/USA/Arizona/ | AZ SPHL, Arizona Department of Health Services | TGen North | Jolene Bowers et al |
| EPI_ISL_695209 | USA/AZ-TG491673/2020 | 2020-06-24 | North America/USA/Arizona/ | AZ SPHL, Arizona Department of Health Services | TGen North | Jolene Bowers et al |
| EPI_ISL_695214 | USA/AZ-TG491687/2020 | 2020-06-25 | North America/USA/Arizona/ | AZ SPHL, Arizona Department of Health Services | TGen North | Jolene Bowers et al |
| EPI_ISL_695224 | USA/AZ-TG491713/2020 | 2020-06-25 | North America/USA/Arizona/ | AZ SPHL, Arizona Department of Health Services | TGen North | Jolene Bowers et al |
| EPI_ISL_695232 | USA/AZ-TG493508/2020 | 2020-07-13 | North America/USA/Arizona/ | TGen North | TGen North | Jolene Bowers et al |
| EPI_ISL_695233 | USA/AZ-TG493511/2020 | 2020-07-13 | North America/USA/Arizona/ | TGen North | TGen North | Jolene Bowers et al |
| EPI_ISL_695249 | USA/AZ-TG494613/2020 | 2020-07-13 | North America/USA/Arizona/ | TGen North | TGen North | Jolene Bowers et al |
| EPI_ISL_695253 | USA/AZ-TG494844/2020 | 2020-07-10 | North America/USA/Arizona/ | TGen North | TGen North | Jolene Bowers et al |
| EPI_ISL_695256 | USA/AZ-TG494864/2020 | 2020-07-10 | North America/USA/Arizona/ | TGen North | TGen North | Jolene Bowers et al |
| EPI_ISL_695259 | USA/AZ-TG495423/2020 | 2020-07-12 | North America/USA/Arizona/ | TGen North | TGen North | Jolene Bowers et al |
| EPI_ISL_695265 | USA/AZ-TG497217/2020 | 2020-07-14 | North America/USA/Arizona/ | TGen North | TGen North | Jolene Bowers et al |
| EPI_ISL_695266 | USA/AZ-TG497941/2020 | 2020-07-14 | North America/USA/Arizona/ | TGen North | TGen North | Jolene Bowers et al |
| EPI_ISL_695267 | USA/AZ-TG498366/2020 | 2020-07-14 | North America/USA/Arizona/ | TGen North | TGen North | Jolene Bowers et al |
| EPI_ISL_695271 | USA/AZ-TG498783/2020 | 2020-07-14 | North America/USA/Arizona/ | TGen North | TGen North | Jolene Bowers et al |
| EPI_ISL_695272 | USA/AZ-TG498910/2020 | 2020-07-14 | North America/USA/Arizona/ | TGen North | TGen North | Jolene Bowers et al |
| EPI_ISL_695277 | USA/AZ-TG501918/2020 | 2020-07-15 | North America/USA/Arizona/ | TGen North | TGen North | Jolene Bowers et al |
| EPI_ISL_695286 | USA/AZ-TG502738/2020 | 2020-07-15 | North America/USA/Arizona/ | TGen North | TGen North | Jolene Bowers et al |
| EPI_ISL_695290 | USA/AZ-TG503851/2020 | 2020-07-15 | North America/USA/Arizona/ | TGen North | TGen North | Jolene Bowers et al |
| EPI_ISL_695291 | USA/AZ-TG505108/2020 | 2020-07-15 | North America/USA/Arizona/ | TGen North | TGen North | Jolene Bowers et al |
| EPI_ISL_695295 | USA/AZ-TG507252/2020 | 2020-07-16 | North America/USA/Arizona/ | TGen North | TGen North | Jolene Bowers et al |
| EPI_ISL_695300 | USA/AZ-TG507817/2020 | 2020-07-16 | North America/USA/Arizona/ | TGen North | TGen North | Jolene Bowers et al |
| EPI_ISL_695301 | USA/AZ-TG508113/2020 | 2020-07-16 | North America/USA/Arizona/ | TGen North | TGen North | Jolene Bowers et al |
| EPI_ISL_695303 | USA/AZ-TG508838/2020 | 2020-07-16 | North America/USA/Arizona/ | TGen North | TGen North | Jolene Bowers et al |
| EPI_ISL_695310 | USA/AZ-TG513826/2020 | 2020-07-17 | North America/USA/Arizona/ | TGen North | TGen North | Jolene Bowers et al |
| EPI_ISL_695321 | USA/AZ-TG517449/2020 | 2020-06-26 | North America/USA/Arizona/ | AZ SPHL, Arizona Department of Health Services | TGen North | Jolene Bowers et al |

| | | | | | | |
|---|---|---|---|---|---|---|
| EPI_ISL_695338 | USA/AZ-TG517510/2020 | 2020-06-30 | North America/USA/Arizona/ | AZ SPHL, Arizona Department of Health Services | TGen North | Jolene Bowers et al |
| EPI_ISL_695343 | USA/AZ-TG517518/2020 | 2020-06-29 | North America/USA/Arizona/ | AZ SPHL, Arizona Department of Health Services | TGen North | Jolene Bowers et al |
| EPI_ISL_695368 | USA/AZ-TG519189/2020 | 2020-07-01 | North America/USA/Arizona/ | AZ SPHL, Arizona Department of Health Services | TGen North | Jolene Bowers et al |
| EPI_ISL_695370 | USA/AZ-TG519200/2020 | 2020-07-01 | North America/USA/Arizona/ | AZ SPHL, Arizona Department of Health Services | TGen North | Jolene Bowers et al |
| EPI_ISL_695374 | USA/AZ-TG520830/2020 | 2020-07-20 | North America/USA/Arizona/ | TGen North | TGen North | Jolene Bowers et al |
| EPI_ISL_695379 | USA/AZ-TG521388/2020 | 2020-07-20 | North America/USA/Arizona/ | TGen North | TGen North | Jolene Bowers et al |
| EPI_ISL_695389 | USA/AZ-TG523563/2020 | 2020-07-16 | North America/USA/Arizona/ | TGen North | TGen North | Jolene Bowers et al |
| EPI_ISL_695393 | USA/AZ-TG532034/2020 | 2020-07-01 | North America/USA/Arizona/ | AZ SPHL, Arizona Department of Health Services | TGen North | Jolene Bowers et al |
| EPI_ISL_695400 | USA/AZ-TG532052/2020 | 2020-07-01 | North America/USA/Arizona/ | AZ SPHL, Arizona Department of Health Services | TGen North | Jolene Bowers et al |
| EPI_ISL_695402 | USA/AZ-TG532062/2020 | 2020-07-01 | North America/USA/Arizona/ | AZ SPHL, Arizona Department of Health Services | TGen North | Jolene Bowers et al |
| EPI_ISL_695403 | USA/AZ-TG532065/2020 | 2020-07-01 | North America/USA/Arizona/ | AZ SPHL, Arizona Department of Health Services | TGen North | Jolene Bowers et al |
| EPI_ISL_695407 | USA/AZ-TG532077/2020 | 2020-07-01 | North America/USA/Arizona/ | AZ SPHL, Arizona Department of Health Services | TGen North | Jolene Bowers et al |
| EPI_ISL_695408 | USA/AZ-TG532081/2020 | 2020-07-01 | North America/USA/Arizona/ | AZ SPHL, Arizona Department of Health Services | TGen North | Jolene Bowers et al |
| EPI_ISL_695417 | USA/AZ-TG532109/2020 | 2020-07-01 | North America/USA/Arizona/ | AZ SPHL, Arizona Department of Health Services | TGen North | Jolene Bowers et al |
| EPI_ISL_695418 | USA/AZ-TG532111/2020 | 2020-07-01 | North America/USA/Arizona/ | AZ SPHL, Arizona Department of Health Services | TGen North | Jolene Bowers et al |
| EPI_ISL_695420 | USA/AZ-TG532113/2020 | 2020-07-01 | North America/USA/Arizona/ | AZ SPHL, Arizona Department of Health Services | TGen North | Jolene Bowers et al |
| EPI_ISL_695422 | USA/AZ-TG532121/2020 | 2020-07-02 | North America/USA/Arizona/ | AZ SPHL, Arizona Department of Health Services | TGen North | Jolene Bowers et al |
| EPI_ISL_695424 | USA/AZ-TG532127/2020 | 2020-07-02 | North America/USA/Arizona/ | AZ SPHL, Arizona Department of Health Services | TGen North | Jolene Bowers et al |
| EPI_ISL_695431 | USA/AZ-TG547220/2020 | 2020-07-02 | North America/USA/Arizona/ | AZ SPHL, Arizona Department of Health Services | TGen North | Jolene Bowers et al |
| EPI_ISL_695434 | USA/AZ-TG547223/2020 | 2020-07-02 | North America/USA/Arizona/ | AZ SPHL, Arizona Department of Health Services | TGen North | Jolene Bowers et al |
| EPI_ISL_695437 | USA/AZ-TG547230/2020 | 2020-07-02 | North America/USA/Arizona/ | AZ SPHL, Arizona Department of Health Services | TGen North | Jolene Bowers et al |

| | | | | | | |
|---|---|---|---|---|---|---|
| EPI_ISL_695441 | USA/AZ-TG547242/2020 | 2020-07-02 | North America/USA/Arizona/ | AZ SPHL, Arizona Department of Health Services | TGen North | Jolene Bowers et al |
| EPI_ISL_695447 | USA/AZ-TG547251/2020 | 2020-07-03 | North America/USA/Arizona/ | AZ SPHL, Arizona Department of Health Services | TGen North | Jolene Bowers et al |
| EPI_ISL_695452 | USA/AZ-TG547256/2020 | 2020-07-03 | North America/USA/Arizona/ | AZ SPHL, Arizona Department of Health Services | TGen North | Jolene Bowers et al |
| EPI_ISL_695455 | USA/AZ-TG547261/2020 | 2020-07-04 | North America/USA/Arizona/ | AZ SPHL, Arizona Department of Health Services | TGen North | Jolene Bowers et al |
| EPI_ISL_695459 | USA/AZ-TG547275/2020 | 2020-07-04 | North America/USA/Arizona/ | AZ SPHL, Arizona Department of Health Services | TGen North | Jolene Bowers et al |
| EPI_ISL_695485 | USA/AZ-TG547335/2020 | 2020-07-06 | North America/USA/Arizona/ | AZ SPHL, Arizona Department of Health Services | TGen North | Jolene Bowers et al |
| EPI_ISL_695491 | USA/AZ-TG547343/2020 | 2020-07-06 | North America/USA/Arizona/ | AZ SPHL, Arizona Department of Health Services | TGen North | Jolene Bowers et al |
| EPI_ISL_695495 | USA/AZ-TG547347/2020 | 2020-07-06 | North America/USA/Arizona/ | AZ SPHL, Arizona Department of Health Services | TGen North | Jolene Bowers et al |
| EPI_ISL_695497 | USA/AZ-TG547351/2020 | 2020-07-06 | North America/USA/Arizona/ | AZ SPHL, Arizona Department of Health Services | TGen North | Jolene Bowers et al |
| EPI_ISL_695502 | USA/AZ-TG547357/2020 | 2020-07-07 | North America/USA/Arizona/ | AZ SPHL, Arizona Department of Health Services | TGen North | Jolene Bowers et al |
| EPI_ISL_695503 | USA/AZ-TG547358/2020 | 2020-07-07 | North America/USA/Arizona/ | AZ SPHL, Arizona Department of Health Services | TGen North | Jolene Bowers et al |
| EPI_ISL_695507 | USA/AZ-TG547364/2020 | 2020-07-07 | North America/USA/Arizona/ | AZ SPHL, Arizona Department of Health Services | TGen North | Jolene Bowers et al |
| EPI_ISL_695510 | USA/AZ-TG547372/2020 | 2020-07-07 | North America/USA/Arizona/ | AZ SPHL, Arizona Department of Health Services | TGen North | Jolene Bowers et al |
| EPI_ISL_695512 | USA/AZ-TG547375/2020 | 2020-07-07 | North America/USA/Arizona/ | AZ SPHL, Arizona Department of Health Services | TGen North | Jolene Bowers et al |
| EPI_ISL_695523 | USA/AZ-TG547955/2020 | 2020-07-27 | North America/USA/Arizona/ | TGen North | TGen North | Jolene Bowers et al |
| EPI_ISL_695525 | USA/AZ-TG548499/2020 | 2020-07-27 | North America/USA/Arizona/ | TGen North | TGen North | Jolene Bowers et al |
| EPI_ISL_695527 | USA/AZ-TG549110/2020 | 2020-07-27 | North America/USA/Arizona/ | TGen North | TGen North | Jolene Bowers et al |
| EPI_ISL_695538 | USA/AZ-TG554086/2020 | 2020-07-28 | North America/USA/Arizona/ | TGen North | TGen North | Jolene Bowers et al |
| EPI_ISL_695540 | USA/AZ-TG555472/2020 | 2020-07-28 | North America/USA/Arizona/ | TGen North | TGen North | Jolene Bowers et al |
| EPI_ISL_695549 | USA/AZ-TG558883/2020 | 2020-07-29 | North America/USA/Arizona/ | TGen North | TGen North | Jolene Bowers et al |
| EPI_ISL_695555 | USA/AZ-TG565575/2020 | 2020-07-31 | North America/USA/Arizona/ | TGen North | TGen North | Jolene Bowers et al |
| EPI_ISL_695556 | USA/AZ-TG565579/2020 | 2020-07-31 | North America/USA/Arizona/ | TGen North | TGen North | Jolene Bowers et al |
| EPI_ISL_695557 | USA/AZ-TG565817/2020 | 2020-07-29 | North America/USA/Arizona/ | TGen North | TGen North | Jolene Bowers et al |

| | | | | | | |
|---|---|---|---|---|---|---|
| EPI_ISL_695558 | USA/AZ-TG565842/2020 | 2020-07-31 | North America/USA/Arizona/ | TGen North | TGen North | Jolene Bowers et al |
| EPI_ISL_695564 | USA/AZ-TG569454/2020 | 2020-07-08 | North America/USA/Arizona/ | AZ SPHL, Arizona Department of Health Services | TGen North | Jolene Bowers et al |
| EPI_ISL_695568 | USA/AZ-TG569465/2020 | 2020-07-09 | North America/USA/Arizona/ | AZ SPHL, Arizona Department of Health Services | TGen North | Jolene Bowers et al |
| EPI_ISL_695570 | USA/AZ-TG569469/2020 | 2020-07-10 | North America/USA/Arizona/ | AZ SPHL, Arizona Department of Health Services | TGen North | Jolene Bowers et al |
| EPI_ISL_695571 | USA/AZ-TG569471/2020 | 2020-07-10 | North America/USA/Arizona/ | AZ SPHL, Arizona Department of Health Services | TGen North | Jolene Bowers et al |
| EPI_ISL_695577 | USA/AZ-TG569483/2020 | 2020-07-10 | North America/USA/Arizona/ | AZ SPHL, Arizona Department of Health Services | TGen North | Jolene Bowers et al |
| EPI_ISL_695579 | USA/AZ-TG569487/2020 | 2020-07-10 | North America/USA/Arizona/ | AZ SPHL, Arizona Department of Health Services | TGen North | Jolene Bowers et al |
| EPI_ISL_695587 | USA/AZ-TG569503/2020 | 2020-07-10 | North America/USA/Arizona/ | AZ SPHL, Arizona Department of Health Services | TGen North | Jolene Bowers et al |
| EPI_ISL_695588 | USA/AZ-TG569504/2020 | 2020-07-08 | North America/USA/Arizona/ | AZ SPHL, Arizona Department of Health Services | TGen North | Jolene Bowers et al |
| EPI_ISL_695590 | USA/AZ-TG569506/2020 | 2020-07-10 | North America/USA/Arizona/ | AZ SPHL, Arizona Department of Health Services | TGen North | Jolene Bowers et al |
| EPI_ISL_695592 | USA/AZ-TG569508/2020 | 2020-07-10 | North America/USA/Arizona/ | AZ SPHL, Arizona Department of Health Services | TGen North | Jolene Bowers et al |
| EPI_ISL_695600 | USA/AZ-TG569520/2020 | 2020-07-13 | North America/USA/Arizona/ | AZ SPHL, Arizona Department of Health Services | TGen North | Jolene Bowers et al |
| EPI_ISL_695601 | USA/AZ-TG569522/2020 | 2020-07-13 | North America/USA/Arizona/ | AZ SPHL, Arizona Department of Health Services | TGen North | Jolene Bowers et al |
| EPI_ISL_695604 | USA/AZ-TG569525/2020 | 2020-07-13 | North America/USA/Arizona/ | AZ SPHL, Arizona Department of Health Services | TGen North | Jolene Bowers et al |
| EPI_ISL_695605 | USA/AZ-TG569526/2020 | 2020-07-13 | North America/USA/Arizona/ | AZ SPHL, Arizona Department of Health Services | TGen North | Jolene Bowers et al |
| EPI_ISL_695615 | USA/AZ-TG571103/2020 | 2020-08-02 | North America/USA/Arizona/ | TGen North | TGen North | Jolene Bowers et al |
| EPI_ISL_695616 | USA/AZ-TG571107/2020 | 2020-08-03 | North America/USA/Arizona/ | TGen North | TGen North | Jolene Bowers et al |
| EPI_ISL_695626 | USA/AZ-TG579666/2020 | 2020-08-05 | North America/USA/Arizona/ | TGen North | TGen North | Jolene Bowers et al |
| EPI_ISL_695630 | USA/AZ-TG581075/2020 | 2020-08-05 | North America/USA/Arizona/ | TGen North | TGen North | Jolene Bowers et al |
| EPI_ISL_695637 | USA/AZ-TG594903/2020 | 2020-08-10 | North America/USA/Arizona/ | TGen North | TGen North | Jolene Bowers et al |
| EPI_ISL_695639 | USA/AZ-TG599215/2020 | 2020-08-12 | North America/USA/Arizona/ | TGen North | TGen North | Jolene Bowers et al |
| EPI_ISL_695640 | USA/AZ-TG600498/2020 | 2020-08-11 | North America/USA/Arizona/ | TGen North | TGen North | Jolene Bowers et al |
| EPI_ISL_695641 | USA/AZ-TG601033/2020 | 2020-08-11 | North America/USA/Arizona/ | TGen North | TGen North | Jolene Bowers et al |

| | | | | | | |
|---|---|---|---|---|---|---|
| EPI_ISL_695644 | USA/AZ-TG602415/2020 | 2020-07-14 | North America/USA/Arizona/ | AZ SPHL, Arizona Department of Health Services | TGen North | Jolene Bowers et al |
| EPI_ISL_695646 | USA/AZ-TG602442/2020 | 2020-07-15 | North America/USA/Arizona/ | AZ SPHL, Arizona Department of Health Services | TGen North | Jolene Bowers et al |
| EPI_ISL_695657 | USA/AZ-TG603454/2020 | 2020-08-12 | North America/USA/Arizona/ | TGen North | TGen North | Jolene Bowers et al |
| EPI_ISL_695659 | USA/AZ-TG610009/2020 | 2020-08-13 | North America/USA/Arizona/ | TGen North | TGen North | Jolene Bowers et al |
| EPI_ISL_695665 | USA/AZ-TG618007/2020 | 2020-07-17 | North America/USA/Arizona/ | AZ SPHL, Arizona Department of Health Services | TGen North | Jolene Bowers et al |
| EPI_ISL_695666 | USA/AZ-TG618010/2020 | 2020-07-17 | North America/USA/Arizona/ | AZ SPHL, Arizona Department of Health Services | TGen North | Jolene Bowers et al |
| EPI_ISL_695668 | USA/AZ-TG618013/2020 | 2020-07-17 | North America/USA/Arizona/ | AZ SPHL, Arizona Department of Health Services | TGen North | Jolene Bowers et al |
| EPI_ISL_695669 | USA/AZ-TG618014/2020 | 2020-07-17 | North America/USA/Arizona/ | AZ SPHL, Arizona Department of Health Services | TGen North | Jolene Bowers et al |
| EPI_ISL_695674 | USA/AZ-TG618027/2020 | 2020-07-18 | North America/USA/Arizona/ | AZ SPHL, Arizona Department of Health Services | TGen North | Jolene Bowers et al |
| EPI_ISL_695677 | USA/AZ-TG618033/2020 | 2020-07-20 | North America/USA/Arizona/ | AZ SPHL, Arizona Department of Health Services | TGen North | Jolene Bowers et al |
| EPI_ISL_695682 | USA/AZ-TG618038/2020 | 2020-07-20 | North America/USA/Arizona/ | AZ SPHL, Arizona Department of Health Services | TGen North | Jolene Bowers et al |
| EPI_ISL_695686 | USA/AZ-TG618044/2020 | 2020-07-20 | North America/USA/Arizona/ | AZ SPHL, Arizona Department of Health Services | TGen North | Jolene Bowers et al |
| EPI_ISL_695688 | USA/AZ-TG618046/2020 | 2020-07-20 | North America/USA/Arizona/ | AZ SPHL, Arizona Department of Health Services | TGen North | Jolene Bowers et al |
| EPI_ISL_695691 | USA/AZ-TG618049/2020 | 2020-07-20 | North America/USA/Arizona/ | AZ SPHL, Arizona Department of Health Services | TGen North | Jolene Bowers et al |
| EPI_ISL_695699 | USA/AZ-TG618074/2020 | 2020-07-22 | North America/USA/Arizona/ | AZ SPHL, Arizona Department of Health Services | TGen North | Jolene Bowers et al |
| EPI_ISL_695701 | USA/AZ-TG618079/2020 | 2020-07-23 | North America/USA/Arizona/ | AZ SPHL, Arizona Department of Health Services | TGen North | Jolene Bowers et al |
| EPI_ISL_695705 | USA/AZ-TG618660/2020 | 2020-08-17 | North America/USA/Arizona/ | TGen North | TGen North | Jolene Bowers et al |
| EPI_ISL_695707 | USA/AZ-TG619658/2020 | 2020-08-18 | North America/USA/Arizona/ | TGen North | TGen North | Jolene Bowers et al |
| EPI_ISL_695711 | USA/AZ-TG634236/2020 | 2020-08-20 | North America/USA/Arizona/ | TGen North | TGen North | Jolene Bowers et al |
| EPI_ISL_695712 | USA/AZ-TG637272/2020 | 2020-07-24 | North America/USA/Arizona/ | AZ SPHL, Arizona Department of Health Services | TGen North | Jolene Bowers et al |
| EPI_ISL_695713 | USA/AZ-TG637273/2020 | 2020-07-24 | North America/USA/Arizona/ | AZ SPHL, Arizona Department of Health Services | TGen North | Jolene Bowers et al |
| EPI_ISL_695720 | USA/AZ-TG637289/2020 | 2020-07-25 | North America/USA/Arizona/ | AZ SPHL, Arizona Department of Health Services | TGen North | Jolene Bowers et al |

| | | | | | | |
|---|---|---|---|---|---|---|
| EPI_ISL_695723 | USA/AZ-TG637296/2020 | 2020-07-27 | North America/USA/Arizona/ | AZ SPHL, Arizona Department of Health Services | TGen North | Jolene Bowers et al |
| EPI_ISL_695726 | USA/AZ-TG637302/2020 | 2020-07-28 | North America/USA/Arizona/ | AZ SPHL, Arizona Department of Health Services | TGen North | Jolene Bowers et al |
| EPI_ISL_695732 | USA/AZ-TG637311/2020 | 2020-07-29 | North America/USA/Arizona/ | AZ SPHL, Arizona Department of Health Services | TGen North | Jolene Bowers et al |
| EPI_ISL_695733 | USA/AZ-TG637312/2020 | 2020-07-29 | North America/USA/Arizona/ | AZ SPHL, Arizona Department of Health Services | TGen North | Jolene Bowers et al |
| EPI_ISL_695735 | USA/AZ-TG637314/2020 | 2020-07-29 | North America/USA/Arizona/ | AZ SPHL, Arizona Department of Health Services | TGen North | Jolene Bowers et al |
| EPI_ISL_695736 | USA/AZ-TG637315/2020 | 2020-07-30 | North America/USA/Arizona/ | AZ SPHL, Arizona Department of Health Services | TGen North | Jolene Bowers et al |
| EPI_ISL_695738 | USA/AZ-TG637325/2020 | 2020-07-31 | North America/USA/Arizona/ | AZ SPHL, Arizona Department of Health Services | TGen North | Jolene Bowers et al |
| EPI_ISL_695760 | USA/AZ-TG640787/2020 | 2020-04-27 | North America/USA/Arizona/ | Sonora Quest Laboratories, Laboratory Sciences of Arizona | TGen North | Jolene Bowers et al |
| EPI_ISL_695769 | USA/AZ-TG640804/2020 | 2020-04-27 | North America/USA/Arizona/ | Sonora Quest Laboratories, Laboratory Sciences of Arizona | TGen North | Jolene Bowers et al |
| EPI_ISL_695785 | USA/AZ-TG640835/2020 | 2020-04-27 | North America/USA/Arizona/ | Sonora Quest Laboratories, Laboratory Sciences of Arizona | TGen North | Jolene Bowers et al |
| EPI_ISL_695789 | USA/AZ-TG640844/2020 | 2020-04-27 | North America/USA/Arizona/ | Sonora Quest Laboratories, Laboratory Sciences of Arizona | TGen North | Jolene Bowers et al |
| EPI_ISL_695791 | USA/AZ-TG640846/2020 | 2020-04-27 | North America/USA/Arizona/ | Sonora Quest Laboratories, Laboratory Sciences of Arizona | TGen North | Jolene Bowers et al |
| EPI_ISL_695796 | USA/AZ-TG640853/2020 | 2020-04-27 | North America/USA/Arizona/ | Sonora Quest Laboratories, Laboratory Sciences of Arizona | TGen North | Jolene Bowers et al |
| EPI_ISL_695823 | USA/AZ-TG640950/2020 | 2020-04-10 | North America/USA/Arizona/ | Sonora Quest Laboratories, Laboratory Sciences of Arizona | TGen North | Jolene Bowers et al |
| EPI_ISL_695826 | USA/AZ-TG640962/2020 | 2020-04-27 | North America/USA/Arizona/ | Sonora Quest Laboratories, Laboratory Sciences of Arizona | TGen North | Jolene Bowers et al |
| EPI_ISL_695831 | USA/AZ-TG645813/2020 | 2020-08-04 | North America/USA/Arizona/ | AZ SPHL, Arizona Department of Health Services | TGen North | Jolene Bowers et al |
| EPI_ISL_695834 | USA/AZ-TG645821/2020 | 2020-08-07 | North America/USA/Arizona/ | AZ SPHL, Arizona Department of Health Services | TGen North | Jolene Bowers et al |
| EPI_ISL_695835 | USA/AZ-TG645822/2020 | 2020-08-07 | North America/USA/Arizona/ | AZ SPHL, Arizona Department of Health Services | TGen North | Jolene Bowers et al |
| EPI_ISL_695862 | USA/AZ-TG648623/2020 | 2020-04-27 | North America/USA/Arizona/ | Sonora Quest Laboratories, Laboratory Sciences of Arizona | TGen North | Jolene Bowers et al |
| EPI_ISL_695863 | USA/AZ-TG648625/2020 | 2020-04-27 | North America/USA/Arizona/ | Sonora Quest Laboratories, Laboratory Sciences of Arizona | TGen North | Jolene Bowers et al |

| | | | | | | |
|---|---|---|---|---|---|---|
| EPI_ISL_695873 | USA/AZ-TG648638/2020 | 2020-04-27 | North America/USA/Arizona/ | Sonora Quest Laboratories, Laboratory Sciences of Arizona | TGen North | Jolene Bowers et al |
| EPI_ISL_695882 | USA/AZ-TG648657/2020 | 2020-04-28 | North America/USA/Arizona/ | Sonora Quest Laboratories, Laboratory Sciences of Arizona | TGen North | Jolene Bowers et al |
| EPI_ISL_695886 | USA/AZ-TG648665/2020 | 2020-04-28 | North America/USA/Arizona/ | Sonora Quest Laboratories, Laboratory Sciences of Arizona | TGen North | Jolene Bowers et al |
| EPI_ISL_695904 | USA/AZ-TG648716/2020 | 2020-04-13 | North America/USA/Arizona/ | Sonora Quest Laboratories, Laboratory Sciences of Arizona | TGen North | Jolene Bowers et al |
| EPI_ISL_695906 | USA/AZ-TG648720/2020 | 2020-04-13 | North America/USA/Arizona/ | Sonora Quest Laboratories, Laboratory Sciences of Arizona | TGen North | Jolene Bowers et al |
| EPI_ISL_695907 | USA/AZ-TG648721/2020 | 2020-04-27 | North America/USA/Arizona/ | Sonora Quest Laboratories, Laboratory Sciences of Arizona | TGen North | Jolene Bowers et al |
| EPI_ISL_695914 | USA/AZ-TG648730/2020 | 2020-04-13 | North America/USA/Arizona/ | Sonora Quest Laboratories, Laboratory Sciences of Arizona | TGen North | Jolene Bowers et al |
| EPI_ISL_695915 | USA/AZ-TG648731/2020 | 2020-04-10 | North America/USA/Arizona/ | Sonora Quest Laboratories, Laboratory Sciences of Arizona | TGen North | Jolene Bowers et al |
| EPI_ISL_695917 | USA/AZ-TG648734/2020 | 2020-04-12 | North America/USA/Arizona/ | Sonora Quest Laboratories, Laboratory Sciences of Arizona | TGen North | Jolene Bowers et al |
| EPI_ISL_695918 | USA/AZ-TG648735/2020 | 2020-04-12 | North America/USA/Arizona/ | Sonora Quest Laboratories, Laboratory Sciences of Arizona | TGen North | Jolene Bowers et al |
| EPI_ISL_695923 | USA/AZ-TG648742/2020 | 2020-04-12 | North America/USA/Arizona/ | Sonora Quest Laboratories, Laboratory Sciences of Arizona | TGen North | Jolene Bowers et al |
| EPI_ISL_695925 | USA/AZ-TG648744/2020 | 2020-04-13 | North America/USA/Arizona/ | Sonora Quest Laboratories, Laboratory Sciences of Arizona | TGen North | Jolene Bowers et al |
| EPI_ISL_695928 | USA/AZ-TG648752/2020 | 2020-04-27 | North America/USA/Arizona/ | Sonora Quest Laboratories, Laboratory Sciences of Arizona | TGen North | Jolene Bowers et al |
| EPI_ISL_695947 | USA/AZ-TG654290/2020 | 2020-04-13 | North America/USA/Arizona/ | Sonora Quest Laboratories, Laboratory Sciences of Arizona | TGen North | Jolene Bowers et al |
| EPI_ISL_695954 | USA/AZ-TG654298/2020 | 2020-04-13 | North America/USA/Arizona/ | Sonora Quest Laboratories, Laboratory Sciences of Arizona | TGen North | Jolene Bowers et al |
| EPI_ISL_695961 | USA/AZ-TG654306/2020 | 2020-04-13 | North America/USA/Arizona/ | Sonora Quest Laboratories, Laboratory Sciences of Arizona | TGen North | Jolene Bowers et al |
| EPI_ISL_695965 | USA/AZ-TG654310/2020 | 2020-04-12 | North America/USA/Arizona/ | Sonora Quest Laboratories, Laboratory Sciences of Arizona | TGen North | Jolene Bowers et al |
| EPI_ISL_695970 | USA/AZ-TG654315/2020 | 2020-04-13 | North America/USA/Arizona/ | Sonora Quest Laboratories, Laboratory Sciences of Arizona | TGen North | Jolene Bowers et al |
| EPI_ISL_695983 | USA/AZ-TG654335/2020 | 2020-04-10 | North America/USA/Arizona/ | Sonora Quest Laboratories, Laboratory Sciences of Arizona | TGen North | Jolene Bowers et al |
| EPI_ISL_695984 | USA/AZ-TG654339/2020 | 2020-04-13 | North America/USA/Arizona/ | Sonora Quest Laboratories, Laboratory Sciences of Arizona | TGen North | Jolene Bowers et al |

| | | | | | | |
|---|---|---|---|---|---|---|
| EPI_ISL_695989 | USA/AZ-TG654347/2020 | 2020-04-13 | North America/USA/Arizona/ | Sonora Quest Laboratories, Laboratory Sciences of Arizona | TGen North | Jolene Bowers et al |
| EPI_ISL_696003 | USA/AZ-TG654366/2020 | 2020-04-13 | North America/USA/Arizona/ | Sonora Quest Laboratories, Laboratory Sciences of Arizona | TGen North | Jolene Bowers et al |
| EPI_ISL_696020 | USA/AZ-TG654730/2020 | 2020-04-29 | North America/USA/Arizona/ | Sonora Quest Laboratories, Laboratory Sciences of Arizona | TGen North | Jolene Bowers et al |
| EPI_ISL_696026 | USA/AZ-TG654772/2020 | 2020-04-03 | North America/USA/Arizona/ | Sonora Quest Laboratories, Laboratory Sciences of Arizona | TGen North | Jolene Bowers et al |
| EPI_ISL_696042 | USA/AZ-TG664237/2020 | 2020-04-27 | North America/USA/Arizona/ | Sonora Quest Laboratories, Laboratory Sciences of Arizona | TGen North | Jolene Bowers et al |
| EPI_ISL_696043 | USA/AZ-TG664239/2020 | 2020-04-28 | North America/USA/Arizona/ | Sonora Quest Laboratories, Laboratory Sciences of Arizona | TGen North | Jolene Bowers et al |
| EPI_ISL_696047 | USA/AZ-TG664243/2020 | 2020-04-28 | North America/USA/Arizona/ | Sonora Quest Laboratories, Laboratory Sciences of Arizona | TGen North | Jolene Bowers et al |
| EPI_ISL_696048 | USA/AZ-TG664244/2020 | 2020-04-28 | North America/USA/Arizona/ | Sonora Quest Laboratories, Laboratory Sciences of Arizona | TGen North | Jolene Bowers et al |
| EPI_ISL_696050 | USA/AZ-TG664247/2020 | 2020-04-28 | North America/USA/Arizona/ | Sonora Quest Laboratories, Laboratory Sciences of Arizona | TGen North | Jolene Bowers et al |
| EPI_ISL_696056 | USA/AZ-TG664254/2020 | 2020-04-28 | North America/USA/Arizona/ | Sonora Quest Laboratories, Laboratory Sciences of Arizona | TGen North | Jolene Bowers et al |
| EPI_ISL_696100 | USA/AZ-TG664309/2020 | 2020-04-28 | North America/USA/Arizona/ | Sonora Quest Laboratories, Laboratory Sciences of Arizona | TGen North | Jolene Bowers et al |
| EPI_ISL_696103 | USA/AZ-TG664470/2020 | 2020-04-20 | North America/USA/Arizona/ | Sonora Quest Laboratories, Laboratory Sciences of Arizona | TGen North | Jolene Bowers et al |
| EPI_ISL_696125 | USA/AZ-TG664501/2020 | 2020-04-18 | North America/USA/Arizona/ | Sonora Quest Laboratories, Laboratory Sciences of Arizona | TGen North | Jolene Bowers et al |
| EPI_ISL_696141 | USA/AZ-TG664524/2020 | 2020-04-20 | North America/USA/Arizona/ | Sonora Quest Laboratories, Laboratory Sciences of Arizona | TGen North | Jolene Bowers et al |
| EPI_ISL_696157 | USA/AZ-TG664540/2020 | 2020-05-02 | North America/USA/Arizona/ | Sonora Quest Laboratories, Laboratory Sciences of Arizona | TGen North | Jolene Bowers et al |
| EPI_ISL_696163 | USA/AZ-TG664546/2020 | 2020-04-20 | North America/USA/Arizona/ | Sonora Quest Laboratories, Laboratory Sciences of Arizona | TGen North | Jolene Bowers et al |
| EPI_ISL_696187 | USA/AZ-TG666097/2020 | 2020-05-02 | North America/USA/Arizona/ | Sonora Quest Laboratories, Laboratory Sciences of Arizona | TGen North | Jolene Bowers et al |
| EPI_ISL_696188 | USA/AZ-TG666098/2020 | 2020-05-02 | North America/USA/Arizona/ | Sonora Quest Laboratories, Laboratory Sciences of Arizona | TGen North | Jolene Bowers et al |
| EPI_ISL_696190 | USA/AZ-TG666100/2020 | 2020-05-01 | North America/USA/Arizona/ | Sonora Quest Laboratories, Laboratory Sciences of Arizona | TGen North | Jolene Bowers et al |
| EPI_ISL_696194 | USA/AZ-TG666104/2020 | 2020-05-01 | North America/USA/Arizona/ | Sonora Quest Laboratories, Laboratory Sciences of Arizona | TGen North | Jolene Bowers et al |

| | | | | | | |
|---|---|---|---|---|---|---|
| EPI_ISL_696196 | USA/AZ-TG666106/2020 | 2020-05-01 | North America/USA/Arizona/ | Sonora Quest Laboratories, Laboratory Sciences of Arizona | TGen North | Jolene Bowers et al |
| EPI_ISL_696197 | USA/AZ-TG666107/2020 | 2020-03-26 | North America/USA/Arizona/ | Sonora Quest Laboratories, Laboratory Sciences of Arizona | TGen North | Jolene Bowers et al |
| EPI_ISL_696206 | USA/AZ-TG666120/2020 | 2020-05-01 | North America/USA/Arizona/ | Sonora Quest Laboratories, Laboratory Sciences of Arizona | TGen North | Jolene Bowers et al |
| EPI_ISL_696217 | USA/AZ-TG666133/2020 | 2020-05-02 | North America/USA/Arizona/ | Sonora Quest Laboratories, Laboratory Sciences of Arizona | TGen North | Jolene Bowers et al |
| EPI_ISL_696233 | USA/AZ-TG666152/2020 | 2020-05-02 | North America/USA/Arizona/ | Sonora Quest Laboratories, Laboratory Sciences of Arizona | TGen North | Jolene Bowers et al |
| EPI_ISL_696236 | USA/AZ-TG666155/2020 | 2020-05-02 | North America/USA/Arizona/ | Sonora Quest Laboratories, Laboratory Sciences of Arizona | TGen North | Jolene Bowers et al |
| EPI_ISL_696237 | USA/AZ-TG666156/2020 | 2020-05-02 | North America/USA/Arizona/ | Sonora Quest Laboratories, Laboratory Sciences of Arizona | TGen North | Jolene Bowers et al |
| EPI_ISL_696243 | USA/AZ-TG666163/2020 | 2020-05-02 | North America/USA/Arizona/ | Sonora Quest Laboratories, Laboratory Sciences of Arizona | TGen North | Jolene Bowers et al |
| EPI_ISL_696266 | USA/AZ-TG668341/2020 | 2020-05-13 | North America/USA/Arizona/ | Sonora Quest Laboratories, Laboratory Sciences of Arizona | TGen North | Jolene Bowers et al |
| EPI_ISL_696270 | USA/AZ-TG668348/2020 | 2020-05-14 | North America/USA/Arizona/ | Sonora Quest Laboratories, Laboratory Sciences of Arizona | TGen North | Jolene Bowers et al |
| EPI_ISL_696271 | USA/AZ-TG668349/2020 | 2020-05-13 | North America/USA/Arizona/ | Sonora Quest Laboratories, Laboratory Sciences of Arizona | TGen North | Jolene Bowers et al |
| EPI_ISL_696274 | USA/AZ-TG668352/2020 | 2020-04-19 | North America/USA/Arizona/ | Sonora Quest Laboratories, Laboratory Sciences of Arizona | TGen North | Jolene Bowers et al |
| EPI_ISL_696277 | USA/AZ-TG668356/2020 | 2020-05-13 | North America/USA/Arizona/ | Sonora Quest Laboratories, Laboratory Sciences of Arizona | TGen North | Jolene Bowers et al |
| EPI_ISL_696283 | USA/AZ-TG668363/2020 | 2020-05-13 | North America/USA/Arizona/ | Sonora Quest Laboratories, Laboratory Sciences of Arizona | TGen North | Jolene Bowers et al |
| EPI_ISL_696294 | USA/AZ-TG668381/2020 | 2020-04-19 | North America/USA/Arizona/ | Sonora Quest Laboratories, Laboratory Sciences of Arizona | TGen North | Jolene Bowers et al |
| EPI_ISL_696295 | USA/AZ-TG668388/2020 | 2020-05-11 | North America/USA/Arizona/ | Sonora Quest Laboratories, Laboratory Sciences of Arizona | TGen North | Jolene Bowers et al |
| EPI_ISL_696300 | USA/AZ-TG668394/2020 | 2020-05-12 | North America/USA/Arizona/ | Sonora Quest Laboratories, Laboratory Sciences of Arizona | TGen North | Jolene Bowers et al |
| EPI_ISL_696306 | USA/AZ-TG668403/2020 | 2020-04-19 | North America/USA/Arizona/ | Sonora Quest Laboratories, Laboratory Sciences of Arizona | TGen North | Jolene Bowers et al |
| EPI_ISL_696312 | USA/AZ-TG668416/2020 | 2020-05-13 | North America/USA/Arizona/ | Sonora Quest Laboratories, Laboratory Sciences of Arizona | TGen North | Jolene Bowers et al |
| EPI_ISL_696317 | USA/AZ-TG670095/2020 | 2020-05-02 | North America/USA/Arizona/ | Sonora Quest Laboratories, Laboratory Sciences of Arizona | TGen North | Jolene Bowers et al |

| | | | | | | |
|---|---|---|---|---|---|---|
| EPI_ISL_696334 | USA/AZ-TG670118/2020 | 2020-05-02 | North America/USA/Arizona/ | Sonora Quest Laboratories, Laboratory Sciences of Arizona | TGen North | Jolene Bowers et al |
| EPI_ISL_696340 | USA/AZ-TG670127/2020 | 2020-05-02 | North America/USA/Arizona/ | Sonora Quest Laboratories, Laboratory Sciences of Arizona | TGen North | Jolene Bowers et al |
| EPI_ISL_696346 | USA/AZ-TG670138/2020 | 2020-05-02 | North America/USA/Arizona/ | Sonora Quest Laboratories, Laboratory Sciences of Arizona | TGen North | Jolene Bowers et al |
| EPI_ISL_696350 | USA/AZ-TG670145/2020 | 2020-04-01 | North America/USA/Arizona/ | Sonora Quest Laboratories, Laboratory Sciences of Arizona | TGen North | Jolene Bowers et al |
| EPI_ISL_696354 | USA/AZ-TG670151/2020 | 2020-04-30 | North America/USA/Arizona/ | Sonora Quest Laboratories, Laboratory Sciences of Arizona | TGen North | Jolene Bowers et al |
| EPI_ISL_696355 | USA/AZ-TG670152/2020 | 2020-05-01 | North America/USA/Arizona/ | Sonora Quest Laboratories, Laboratory Sciences of Arizona | TGen North | Jolene Bowers et al |
| EPI_ISL_696361 | USA/AZ-TG670161/2020 | 2020-04-30 | North America/USA/Arizona/ | Sonora Quest Laboratories, Laboratory Sciences of Arizona | TGen North | Jolene Bowers et al |
| EPI_ISL_696364 | USA/AZ-TG670165/2020 | 2020-04-04 | North America/USA/Arizona/ | Sonora Quest Laboratories, Laboratory Sciences of Arizona | TGen North | Jolene Bowers et al |
| EPI_ISL_696365 | USA/AZ-TG670167/2020 | 2020-04-30 | North America/USA/Arizona/ | Sonora Quest Laboratories, Laboratory Sciences of Arizona | TGen North | Jolene Bowers et al |
| EPI_ISL_696366 | USA/AZ-TG670168/2020 | 2020-04-30 | North America/USA/Arizona/ | Sonora Quest Laboratories, Laboratory Sciences of Arizona | TGen North | Jolene Bowers et al |
| EPI_ISL_696388 | USA/AZ-TG670196/2020 | 2020-04-29 | North America/USA/Arizona/ | Sonora Quest Laboratories, Laboratory Sciences of Arizona | TGen North | Jolene Bowers et al |
| EPI_ISL_696411 | USA/AZ-TG670229/2020 | 2020-05-01 | North America/USA/Arizona/ | Sonora Quest Laboratories, Laboratory Sciences of Arizona | TGen North | Jolene Bowers et al |
| EPI_ISL_696412 | USA/AZ-TG670230/2020 | 2020-05-01 | North America/USA/Arizona/ | Sonora Quest Laboratories, Laboratory Sciences of Arizona | TGen North | Jolene Bowers et al |
| EPI_ISL_696413 | USA/AZ-TG670231/2020 | 2020-04-30 | North America/USA/Arizona/ | Sonora Quest Laboratories, Laboratory Sciences of Arizona | TGen North | Jolene Bowers et al |
| EPI_ISL_696439 | USA/AZ-TG670263/2020 | 2020-04-30 | North America/USA/Arizona/ | Sonora Quest Laboratories, Laboratory Sciences of Arizona | TGen North | Jolene Bowers et al |
| EPI_ISL_696462 | SouthAfrica/NHLS-UCT-GS-1671-KRISI | 2020-11-19 | Africa/South Africa/Western Cape Provi | Sedgefield Clinic wc SGE & NHLS/UCT | KRISP, KZN Research Innovation and Sequencing Platform | Arash Iranzadeh et al |
| EPI_ISL_696465 | SouthAfrica/NHLS-UCT-GS-7010-KRISI | 2020-11-18 | Africa/South Africa/Western Cape Provi | Kwanokuthula CDC wc KWA & NHLS/UCT | KRISP, KZN Research Innovation and Sequencing Platform | Arash Iranzadeh et al |

| | | | | | | |
|---|---|---|---|---|---|---|
| EPI_ISL_696486 | SouthAfrica/NHLS-UCT-GS-7019-KRISI | 2020-11-20 | Africa/South Africa/Western Cape Provi | Hornlee Clinic wc HLC & NHLS/UCT | KRISP, KZN Research Innovation and Sequencing Platform | Arash Iranzadeh et al |
| EPI_ISL_696504 | SouthAfrica/NHLS-UCT-GS-7064-KRISI | 2020-11-25 | Africa/South Africa/Western Cape Provi | Pacaltsdorp Clinic wc PAC & NHLS/UCT | KRISP, KZN Research Innovation and Sequencing Platform | Arash Iranzadeh et al |
| EPI_ISL_696505 | SouthAfrica/NHLS-UCT-GS-7068-KRISI | 2020-11-25 | Africa/South Africa/Western Cape Provi | Khayelethu Clinic wc KLC & NHLS/UCT | KRISP, KZN Research Innovation and Sequencing Platform | Arash Iranzadeh et al |
| EPI_ISL_696521 | SouthAfrica/NHLS-UCT-GS-7034-KRISI | 2020-11-24 | Africa/South Africa/Western Cape Provi | Thembalethu CDC wc THC & NHLS/UC | KRISP, KZN Research Innovation and Sequencing Platform | Arash Iranzadeh et al |
| EPI_ISL_697784 | Ecuador/USFQ-185/2020 | 2020-08-12 | South America/Ecuador/Imbabura/Otav | Institute of Microbiology, Universidad San Francisco de Quito | Institute of Microbiology, Universidad San Francisco de Quito | Belv©n Prado-Vivar et al |
| EPI_ISL_697785 | Ecuador/USFQ-1188/2020 | 2020-08-12 | South America/Ecuador/Imbabura/Otav | Institute of Microbiology, Universidad San Francisco de Quito | Institute of Microbiology, Universidad San Francisco de Quito | Belv©n Prado-Vivar et al |
| EPI_ISL_697786 | Ecuador/USFQ-189/2020 | 2020-08-12 | South America/Ecuador/Imbabura/Otav | Institute of Microbiology, Universidad San Francisco de Quito | Institute of Microbiology, Universidad San Francisco de Quito | Belv©n Prado-Vivar et al |
| EPI_ISL_697789 | Ecuador/USFQ-344/2020 | 2020-09-02 | South America/Ecuador/Imbabura/Otav | Institute of Microbiology, Universidad San Francisco de Quito | Institute of Microbiology, Universidad San Francisco de Quito | Belv©n Prado-Vivar et al |
| EPI_ISL_697790 | Ecuador/USFQ-346/2020 | 2020-09-07 | South America/Ecuador/Imbabura/Otav | Institute of Microbiology, Universidad San Francisco de Quito | Institute of Microbiology, Universidad San Francisco de Quito | Belv©n Prado-Vivar et al |

| | | | | | | |
|---|---|---|---|---|---|---|
| EPI_ISL_697791 | Ecuador/USFQ-381/2020 | 2020-09-18 | South America/Ecuador/Imbabura/Otav | Institute of Microbiology, Universidad San Francisco de Quito | Institute of Microbiology, Universidad San Francisco de Quito | BelV©n Prado-Vivar et al |
| EPI_ISL_697792 | Ecuador/USFQ-405/2020 | 2020-09-02 | South America/Ecuador/Imbabura/San F | Institute of Microbiology, Universidad San Francisco de Quito | Institute of Microbiology, Universidad San Francisco de Quito | BelV©n Prado-Vivar et al |
| EPI_ISL_697793 | Ecuador/USFQ-433/2020 | 2020-09-07 | South America/Ecuador/Imbabura/Otav | Institute of Microbiology, Universidad San Francisco de Quito | Institute of Microbiology, Universidad San Francisco de Quito | BelV©n Prado-Vivar et al |
| EPI_ISL_697795 | Ecuador/USFQ-524/2020 | 2020-09-29 | South America/Ecuador/Morona Santiag | Institute of Microbiology, Universidad San Francisco de Quito | Institute of Microbiology, Universidad San Francisco de Quito | Andrea Macias et al |
| EPI_ISL_697796 | Ecuador/USFQ-527/2020 | 2020-05-26 | South America/Ecuador/Napo/Tena | Universidad Regional Amazonica IKIAM | Institute of Microbiology, Universidad San Francisco de Quito | Fabian Aguilar et al |
| EPI_ISL_697797 | Ecuador/USFQ-528/2020 | 2020-07-17 | South America/Ecuador/Guayas/Guayaq | Centro de Investigaciones, Universidad de Especialidades EspV≠ritu Santo | Institute of Microbiology, Universidad San Francisco de Quito | Derly Andrade et al |
| EPI_ISL_697798 | Ecuador/USFQ-531/2020 | 2020-11-30 | South America/Ecuador/Sucumbios/ | Institute of Microbiology, Universidad San Francisco de Quito | Institute of Microbiology, Universidad San Francisco de Quito | BelV©n Prado-Vivar et al |
| EPI_ISL_697799 | Ecuador/USFQ-533/2020 | 2020-11-30 | South America/Ecuador/Sucumbios/ | Institute of Microbiology, Universidad San Francisco de Quito | Institute of Microbiology, Universidad San Francisco de Quito | BelV©n Prado-Vivar et al |
| EPI_ISL_697800 | Ecuador/USFQ-539/2020 | 2020-11-30 | South America/Ecuador/Santa Elena/ | Institute of Microbiology, Universidad San Francisco de Quito | Institute of Microbiology, Universidad San Francisco de Quito | BelV©n Prado-Vivar et al |
| EPI_ISL_698079 | Canada/ON-UHTC_0398/2020 | 2020-11-24 | North America/Canada/Ontario/ | Unity Health Toronto | Ontario Institute for Cancer Research | Ramzi Fattouh et al |

| | | | | | | |
|---|---|---|---|---|---|---|
| EPI_ISL_698080 | Canada/ON-UHTC_0399/2020 | 2020-11-24 | North America/Canada/Ontario/ | Unity Health Toronto | Ontario Institute for Cancer Research | Ramzi Fattouh et al |
| EPI_ISL_698081 | Canada/ON-UHTC_0395/2020 | 2020-11-24 | North America/Canada/Ontario/ | Unity Health Toronto | Ontario Institute for Cancer Research | Ramzi Fattouh et al |
| EPI_ISL_698082 | Canada/ON-UHTC_0385/2020 | 2020-11-24 | North America/Canada/Ontario/ | Unity Health Toronto | Ontario Institute for Cancer Research | Ramzi Fattouh et al |
| EPI_ISL_698083 | Canada/ON-UHTC_0406/2020 | 2020-11-24 | North America/Canada/Ontario/ | Unity Health Toronto | Ontario Institute for Cancer Research | Ramzi Fattouh et al |
| EPI_ISL_698084 | Canada/ON-UHTC_0405/2020 | 2020-11-24 | North America/Canada/Ontario/ | Unity Health Toronto | Ontario Institute for Cancer Research | Ramzi Fattouh et al |
| EPI_ISL_698085 | Canada/ON-UHTC_0407/2020 | 2020-11-24 | North America/Canada/Ontario/ | Unity Health Toronto | Ontario Institute for Cancer Research | Ramzi Fattouh et al |
| EPI_ISL_698086 | Canada/ON-UHTC_0393/2020 | 2020-11-24 | North America/Canada/Ontario/ | Unity Health Toronto | Ontario Institute for Cancer Research | Ramzi Fattouh et al |
| EPI_ISL_698087 | Canada/ON-UHTC_0392/2020 | 2020-11-24 | North America/Canada/Ontario/ | Unity Health Toronto | Ontario Institute for Cancer Research | Ramzi Fattouh et al |
| EPI_ISL_698088 | Canada/ON-UHTC_0387/2020 | 2020-11-24 | North America/Canada/Ontario/ | Unity Health Toronto | Ontario Institute for Cancer Research | Ramzi Fattouh et al |
| EPI_ISL_698089 | Canada/ON-UHTC_0401/2020 | 2020-11-24 | North America/Canada/Ontario/ | Unity Health Toronto | Ontario Institute for Cancer Research | Ramzi Fattouh et al |
| EPI_ISL_698090 | Canada/ON-UHTC_0397/2020 | 2020-11-24 | North America/Canada/Ontario/ | Unity Health Toronto | Ontario Institute for Cancer Research | Ramzi Fattouh et al |
| EPI_ISL_698091 | Canada/ON-UHTC_0388/2020 | 2020-11-24 | North America/Canada/Ontario/ | Unity Health Toronto | Ontario Institute for Cancer Research | Ramzi Fattouh et al |
| EPI_ISL_698092 | Canada/ON-UHTC_0396/2020 | 2020-11-24 | North America/Canada/Ontario/ | Unity Health Toronto | Ontario Institute for Cancer Research | Ramzi Fattouh et al |

| | | | | | | |
|---|---|---|---|---|---|---|
| EPI_ISL_698093 | Canada/ON-UHTC_0409/2020 | 2020-11-24 | North America/Canada/Ontario/ | Unity Health Toronto | Ontario Institute for Cancer Research | Ramzi Fattouh et al |
| EPI_ISL_698094 | Canada/ON-UHTC_0382/2020 | 2020-11-24 | North America/Canada/Ontario/ | Unity Health Toronto | Ontario Institute for Cancer Research | Ramzi Fattouh et al |
| EPI_ISL_698095 | Canada/ON-UHTC_0389/2020 | 2020-11-24 | North America/Canada/Ontario/ | Unity Health Toronto | Ontario Institute for Cancer Research | Ramzi Fattouh et al |
| EPI_ISL_698096 | Canada/ON-UHTC_0391/2020 | 2020-11-24 | North America/Canada/Ontario/ | Unity Health Toronto | Ontario Institute for Cancer Research | Ramzi Fattouh et al |
| EPI_ISL_698097 | Canada/ON-UHTC_0400/2020 | 2020-11-24 | North America/Canada/Ontario/ | Unity Health Toronto | Ontario Institute for Cancer Research | Ramzi Fattouh et al |
| EPI_ISL_698098 | Canada/ON-UHTC_0394/2020 | 2020-11-24 | North America/Canada/Ontario/ | Unity Health Toronto | Ontario Institute for Cancer Research | Ramzi Fattouh et al |
| EPI_ISL_698099 | Canada/ON-UHTC_0384/2020 | 2020-11-24 | North America/Canada/Ontario/ | Unity Health Toronto | Ontario Institute for Cancer Research | Ramzi Fattouh et al |
| EPI_ISL_698100 | Canada/ON-UHTC_0390/2020 | 2020-11-24 | North America/Canada/Ontario/ | Unity Health Toronto | Ontario Institute for Cancer Research | Ramzi Fattouh et al |
| EPI_ISL_698101 | Canada/ON-UHTC_0383/2020 | 2020-11-24 | North America/Canada/Ontario/ | Unity Health Toronto | Ontario Institute for Cancer Research | Ramzi Fattouh et al |
| EPI_ISL_698102 | Canada/ON-UHTC_0386/2020 | 2020-11-24 | North America/Canada/Ontario/ | Unity Health Toronto | Ontario Institute for Cancer Research | Ramzi Fattouh et al |
| EPI_ISL_698103 | Canada/ON-UHTC_0381/2020 | 2020-11-24 | North America/Canada/Ontario/ | Unity Health Toronto | Ontario Institute for Cancer Research | Ramzi Fattouh et al |
| EPI_ISL_698104 | Canada/ON-UHTC_0402/2020 | 2020-11-24 | North America/Canada/Ontario/ | Unity Health Toronto | Ontario Institute for Cancer Research | Ramzi Fattouh et al |
| EPI_ISL_698117 | UnitedArabEmirates/1042/2020 | 2020-06-11 | Asia/United Arab Emirates/Abu Dhabi/ | Group 42 (G42) Healthcare, Abu Dhabi, United Arab Emirates; Department of Health, The United Arab Emirates | G42 Healthcare | Rong Liu et al |

| | | | | | | |
|---|---|---|---|---|---|---|
| EPI_ISL_698125 | UnitedArabEmirates/1050/2020 | 2020-06-11 | Asia/United Arab Emirates/Abu Dhabi/ | Group 42 (G42) Healthcare, Abu Dhabi, United Arab Emirates; Department of Health, The United Arab Emirates | G42 Healthcare | Rong Liu et al |
| EPI_ISL_698126 | UnitedArabEmirates/1051/2020 | 2020-06-11 | Asia/United Arab Emirates/Abu Dhabi/ | Group 42 (G42) Healthcare, Abu Dhabi, United Arab Emirates; Department of Health, The United Arab Emirates | G42 Healthcare | Rong Liu et al |
| EPI_ISL_698140 | UnitedArabEmirates/1065/2020 | 2020-06-11 | Asia/United Arab Emirates/Abu Dhabi/ | Group 42 (G42) Healthcare, Abu Dhabi, United Arab Emirates; Department of Health, The United Arab Emirates | G42 Healthcare | Rong Liu et al |
| EPI_ISL_698143 | UnitedArabEmirates/1068/2020 | 2020-06-11 | Asia/United Arab Emirates/Abu Dhabi/ | Group 42 (G42) Healthcare, Abu Dhabi, United Arab Emirates; Department of Health, The United Arab Emirates | G42 Healthcare | Rong Liu et al |
| EPI_ISL_698149 | UnitedArabEmirates/1075/2020 | 2020-06-12 | Asia/United Arab Emirates/Abu Dhabi/ | Group 42 (G42) Healthcare, Abu Dhabi, United Arab Emirates; Department of Health, The United Arab Emirates | G42 Healthcare | Rong Liu et al |
| EPI_ISL_698155 | UnitedArabEmirates/1081/2020 | 2020-05-15 | Asia/United Arab Emirates/Abu Dhabi/ | Group 42 (G42) Healthcare, Abu Dhabi, United Arab Emirates; Department of Health, The United Arab Emirates | G42 Healthcare | Rong Liu et al |
| EPI_ISL_698160 | UnitedArabEmirates/1086/2020 | 2020-05-16 | Asia/United Arab Emirates/Abu Dhabi/ | Group 42 (G42) Healthcare, Abu Dhabi, United Arab Emirates; Department of Health, The United Arab Emirates | G42 Healthcare | Rong Liu et al |
| EPI_ISL_698172 | UnitedArabEmirates/0134/2020 | 2020-05-18 | Asia/United Arab Emirates/Abu Dhabi/ | Group 42 (G42) Healthcare, Abu Dhabi, United Arab Emirates; Department of Health, The United Arab Emirates | G42 Healthcare | Rong Liu et al |
| EPI_ISL_698185 | UnitedArabEmirates/0491/2020 | 2020-05-18 | Asia/United Arab Emirates/Abu Dhabi/ | Group 42 (G42) Healthcare, Abu Dhabi, United Arab Emirates; Department of Health, The United Arab Emirates | G42 Healthcare | Rong Liu et al |
| EPI_ISL_698188 | UnitedArabEmirates/0527/2020 | 2020-05-18 | Asia/United Arab Emirates/Abu Dhabi/ | Group 42 (G42) Healthcare, Abu Dhabi, United Arab Emirates; Department of Health, The United Arab Emirates | G42 Healthcare | Rong Liu et al |
| EPI_ISL_698204 | UnitedArabEmirates/0252/2020 | 2020-05-17 | Asia/United Arab Emirates/Abu Dhabi/ | Group 42 (G42) Healthcare, Abu Dhabi, United Arab Emirates; Department of Health, The United Arab Emirates | G42 Healthcare | Rong Liu et al |
| EPI_ISL_698298 | UnitedArabEmirates/0108/2020 | 2020-05-15 | Asia/United Arab Emirates/Abu Dhabi/ | Group 42 (G42) Healthcare, Abu Dhabi, United Arab Emirates; Department of Health, The United Arab Emirates | G42 Healthcare | Rong Liu et al |
| EPI_ISL_698331 | UnitedArabEmirates/0144/2020 | 2020-05-18 | Asia/United Arab Emirates/Abu Dhabi/ | Group 42 (G42) Healthcare, Abu Dhabi, United Arab Emirates; Department of Health, The United Arab Emirates | G42 Healthcare | Rong Liu et al |

| | | | | | | |
|---|---|---|---|---|---|---|
| EPI_ISL_698367 | UnitedArabEmirates/0180/2020 | 2020-05-18 | Asia/United Arab Emirates/Abu Dhabi/ | Group 42 (G42) Healthcare, Abu Dhabi, United Arab Emirates; Department of Health, The United Arab Emirates | G42 Healthcare | Rong Liu et al |
| EPI_ISL_698379 | UnitedArabEmirates/0193/2020 | 2020-05-18 | Asia/United Arab Emirates/Abu Dhabi/ | Group 42 (G42) Healthcare, Abu Dhabi, United Arab Emirates; Department of Health, The United Arab Emirates | G42 Healthcare | Rong Liu et al |
| EPI_ISL_698389 | UnitedArabEmirates/0203/2020 | 2020-05-17 | Asia/United Arab Emirates/Abu Dhabi/ | Group 42 (G42) Healthcare, Abu Dhabi, United Arab Emirates; Department of Health, The United Arab Emirates | G42 Healthcare | Rong Liu et al |
| EPI_ISL_698395 | UnitedArabEmirates/0209/2020 | 2020-05-18 | Asia/United Arab Emirates/Abu Dhabi/ | Group 42 (G42) Healthcare, Abu Dhabi, United Arab Emirates; Department of Health, The United Arab Emirates | G42 Healthcare | Rong Liu et al |
| EPI_ISL_698440 | UnitedArabEmirates/0259/2020 | 2020-05-17 | Asia/United Arab Emirates/Abu Dhabi/ | Group 42 (G42) Healthcare, Abu Dhabi, United Arab Emirates; Department of Health, The United Arab Emirates | G42 Healthcare | Rong Liu et al |
| EPI_ISL_698469 | UnitedArabEmirates/0289/2020 | 2020-05-18 | Asia/United Arab Emirates/Abu Dhabi/ | Group 42 (G42) Healthcare, Abu Dhabi, United Arab Emirates; Department of Health, The United Arab Emirates | G42 Healthcare | Rong Liu et al |
| EPI_ISL_698513 | UnitedArabEmirates/0337/2020 | 2020-05-19 | Asia/United Arab Emirates/Abu Dhabi/ | Group 42 (G42) Healthcare, Abu Dhabi, United Arab Emirates; Department of Health, The United Arab Emirates | G42 Healthcare | Rong Liu et al |
| EPI_ISL_698523 | UnitedArabEmirates/0348/2020 | 2020-05-15 | Asia/United Arab Emirates/Abu Dhabi/ | Group 42 (G42) Healthcare, Abu Dhabi, United Arab Emirates; Department of Health, The United Arab Emirates | G42 Healthcare | Rong Liu et al |
| EPI_ISL_698560 | UnitedArabEmirates/0385/2020 | 2020-05-15 | Asia/United Arab Emirates/Abu Dhabi/ | Group 42 (G42) Healthcare, Abu Dhabi, United Arab Emirates; Department of Health, The United Arab Emirates | G42 Healthcare | Rong Liu et al |
| EPI_ISL_698574 | UnitedArabEmirates/0399/2020 | 2020-05-19 | Asia/United Arab Emirates/Abu Dhabi/ | Group 42 (G42) Healthcare, Abu Dhabi, United Arab Emirates; Department of Health, The United Arab Emirates | G42 Healthcare | Rong Liu et al |
| EPI_ISL_698588 | UnitedArabEmirates/0413/2020 | 2020-05-15 | Asia/United Arab Emirates/Abu Dhabi/ | Group 42 (G42) Healthcare, Abu Dhabi, United Arab Emirates; Department of Health, The United Arab Emirates | G42 Healthcare | Rong Liu et al |
| EPI_ISL_698589 | UnitedArabEmirates/0414/2020 | 2020-05-15 | Asia/United Arab Emirates/Abu Dhabi/ | Group 42 (G42) Healthcare, Abu Dhabi, United Arab Emirates; Department of Health, The United Arab Emirates | G42 Healthcare | Rong Liu et al |
| EPI_ISL_698615 | UnitedArabEmirates/0441/2020 | 2020-05-15 | Asia/United Arab Emirates/Abu Dhabi/ | Group 42 (G42) Healthcare, Abu Dhabi, United Arab Emirates; Department of Health, The United Arab Emirates | G42 Healthcare | Rong Liu et al |

| | | | | | | |
|---|---|---|---|---|---|---|
| EPI_ISL_698616 | UnitedArabEmirates/0442/2020 | 2020-05-17 | Asia/United Arab Emirates/Abu Dhabi/ | Group 42 (G42) Healthcare, Abu Dhabi, United Arab Emirates; Department of Health, The United Arab Emirates | G42 Healthcare | Rong Liu et al |
| EPI_ISL_698618 | UnitedArabEmirates/0445/2020 | 2020-05-18 | Asia/United Arab Emirates/Abu Dhabi/ | Group 42 (G42) Healthcare, Abu Dhabi, United Arab Emirates; Department of Health, The United Arab Emirates | G42 Healthcare | Rong Liu et al |
| EPI_ISL_698645 | UnitedArabEmirates/0476/2020 | 2020-05-18 | Asia/United Arab Emirates/Abu Dhabi/ | Group 42 (G42) Healthcare, Abu Dhabi, United Arab Emirates; Department of Health, The United Arab Emirates | G42 Healthcare | Rong Liu et al |
| EPI_ISL_698719 | UnitedArabEmirates/0558/2020 | 2020-05-19 | Asia/United Arab Emirates/Abu Dhabi/ | Group 42 (G42) Healthcare, Abu Dhabi, United Arab Emirates; Department of Health, The United Arab Emirates | G42 Healthcare | Rong Liu et al |
| EPI_ISL_698752 | UnitedArabEmirates/0594/2020 | 2020-05-15 | Asia/United Arab Emirates/Abu Dhabi/ | Group 42 (G42) Healthcare, Abu Dhabi, United Arab Emirates; Department of Health, The United Arab Emirates | G42 Healthcare | Rong Liu et al |
| EPI_ISL_698768 | UnitedArabEmirates/0612/2020 | 2020-05-15 | Asia/United Arab Emirates/Abu Dhabi/ | Group 42 (G42) Healthcare, Abu Dhabi, United Arab Emirates; Department of Health, The United Arab Emirates | G42 Healthcare | Rong Liu et al |
| EPI_ISL_698770 | UnitedArabEmirates/0614/2020 | 2020-05-15 | Asia/United Arab Emirates/Abu Dhabi/ | Group 42 (G42) Healthcare, Abu Dhabi, United Arab Emirates; Department of Health, The United Arab Emirates | G42 Healthcare | Rong Liu et al |
| EPI_ISL_698780 | UnitedArabEmirates/0624/2020 | 2020-05-15 | Asia/United Arab Emirates/Abu Dhabi/ | Group 42 (G42) Healthcare, Abu Dhabi, United Arab Emirates; Department of Health, The United Arab Emirates | G42 Healthcare | Rong Liu et al |
| EPI_ISL_698848 | UnitedArabEmirates/0697/2020 | 2020-05-15 | Asia/United Arab Emirates/Abu Dhabi/ | Group 42 (G42) Healthcare, Abu Dhabi, United Arab Emirates; Department of Health, The United Arab Emirates | G42 Healthcare | Rong Liu et al |
| EPI_ISL_698852 | UnitedArabEmirates/0701/2020 | 2020-05-15 | Asia/United Arab Emirates/Abu Dhabi/ | Group 42 (G42) Healthcare, Abu Dhabi, United Arab Emirates; Department of Health, The United Arab Emirates | G42 Healthcare | Rong Liu et al |
| EPI_ISL_698888 | UnitedArabEmirates/0739/2020 | 2020-05-15 | Asia/United Arab Emirates/Abu Dhabi/ | Group 42 (G42) Healthcare, Abu Dhabi, United Arab Emirates; Department of Health, The United Arab Emirates | G42 Healthcare | Rong Liu et al |
| EPI_ISL_698918 | UnitedArabEmirates/0775/2020 | 2020-05-19 | Asia/United Arab Emirates/Abu Dhabi/ | Group 42 (G42) Healthcare, Abu Dhabi, United Arab Emirates; Department of Health, The United Arab Emirates | G42 Healthcare | Rong Liu et al |
| EPI_ISL_698935 | UnitedArabEmirates/0795/2020 | 2020-05-19 | Asia/United Arab Emirates/Abu Dhabi/ | Group 42 (G42) Healthcare, Abu Dhabi, United Arab Emirates; Department of Health, The United Arab Emirates | G42 Healthcare | Rong Liu et al |

| | | | | | | |
|---|---|---|---|---|---|---|
| EPI_ISL_698941 | UnitedArabEmirates/0801/2020 | 2020-05-19 | Asia/United Arab Emirates/Abu Dhabi/ | Group 42 (G42) Healthcare, Abu Dhabi, United Arab Emirates; Department of Health, The United Arab Emirates | G42 Healthcare | Rong Liu et al |
| EPI_ISL_698975 | UnitedArabEmirates/0836/2020 | 2020-05-19 | Asia/United Arab Emirates/Abu Dhabi/ | Group 42 (G42) Healthcare, Abu Dhabi, United Arab Emirates; Department of Health, The United Arab Emirates | G42 Healthcare | Rong Liu et al |
| EPI_ISL_698996 | UnitedArabEmirates/0857/2020 | 2020-05-19 | Asia/United Arab Emirates/Abu Dhabi/ | Group 42 (G42) Healthcare, Abu Dhabi, United Arab Emirates; Department of Health, The United Arab Emirates | G42 Healthcare | Rong Liu et al |
| EPI_ISL_699006 | UnitedArabEmirates/0867/2020 | 2020-05-19 | Asia/United Arab Emirates/Abu Dhabi/ | Group 42 (G42) Healthcare, Abu Dhabi, United Arab Emirates; Department of Health, The United Arab Emirates | G42 Healthcare | Rong Liu et al |
| EPI_ISL_699042 | UnitedArabEmirates/0904/2020 | 2020-06-26 | Asia/United Arab Emirates/Abu Dhabi/ | Group 42 (G42) Healthcare, Abu Dhabi, United Arab Emirates; Department of Health, The United Arab Emirates | G42 Healthcare | Rong Liu et al |
| EPI_ISL_699086 | UnitedArabEmirates/0950/2020 | 2020-06-28 | Asia/United Arab Emirates/Abu Dhabi/ | Group 42 (G42) Healthcare, Abu Dhabi, United Arab Emirates; Department of Health, The United Arab Emirates | G42 Healthcare | Rong Liu et al |
| EPI_ISL_699102 | UnitedArabEmirates/0966/2020 | 2020-06-29 | Asia/United Arab Emirates/Abu Dhabi/ | Group 42 (G42) Healthcare, Abu Dhabi, United Arab Emirates; Department of Health, The United Arab Emirates | G42 Healthcare | Rong Liu et al |
| EPI_ISL_699104 | UnitedArabEmirates/0968/2020 | 2020-06-29 | Asia/United Arab Emirates/Abu Dhabi/ | Group 42 (G42) Healthcare, Abu Dhabi, United Arab Emirates; Department of Health, The United Arab Emirates | G42 Healthcare | Rong Liu et al |
| EPI_ISL_699112 | UnitedArabEmirates/0977/2020 | 2020-06-29 | Asia/United Arab Emirates/Abu Dhabi/ | Group 42 (G42) Healthcare, Abu Dhabi, United Arab Emirates; Department of Health, The United Arab Emirates | G42 Healthcare | Rong Liu et al |
| EPI_ISL_699114 | UnitedArabEmirates/0980/2020 | 2020-06-14 | Asia/United Arab Emirates/Abu Dhabi/ | Group 42 (G42) Healthcare, Abu Dhabi, United Arab Emirates; Department of Health, The United Arab Emirates | G42 Healthcare | Rong Liu et al |
| EPI_ISL_699118 | UnitedArabEmirates/0984/2020 | 2020-06-14 | Asia/United Arab Emirates/Abu Dhabi/ | Group 42 (G42) Healthcare, Abu Dhabi, United Arab Emirates; Department of Health, The United Arab Emirates | G42 Healthcare | Rong Liu et al |
| EPI_ISL_699127 | UnitedArabEmirates/0994/2020 | 2020-06-10 | Asia/United Arab Emirates/Abu Dhabi/ | Group 42 (G42) Healthcare, Abu Dhabi, United Arab Emirates; Department of Health, The United Arab Emirates | G42 Healthcare | Rong Liu et al |
| EPI_ISL_699132 | UnitedArabEmirates/1000/2020 | 2020-06-10 | Asia/United Arab Emirates/Abu Dhabi/ | Group 42 (G42) Healthcare, Abu Dhabi, United Arab Emirates; Department of Health, The United Arab Emirates | G42 Healthcare | Rong Liu et al |

| | | | | | | |
|---|---|---|---|---|---|---|
| EPI_ISL_699136 | UnitedArabEmirates/1004/2020 | 2020-06-10 | Asia/United Arab Emirates/Abu Dhabi/ | Group 42 (G42) Healthcare, Abu Dhabi, United Arab Emirates; Department of Health, The United Arab Emirates | G42 Healthcare | Rong Liu et al |
| EPI_ISL_699157 | UnitedArabEmirates/1025/2020 | 2020-06-11 | Asia/United Arab Emirates/Abu Dhabi/ | Group 42 (G42) Healthcare, Abu Dhabi, United Arab Emirates; Department of Health, The United Arab Emirates | G42 Healthcare | Rong Liu et al |
| EPI_ISL_699655 | Tunisia/61689/2020 | 2020-11-23 | Africa/Tunisia/Nabeul/ | 1-Laboratory of Microbiology, National Reference Lab, Charles Nicolle Hospital; 2-University of Tunis ElManar, Faculty of Medicine of Tunis, LR99ES09, Tunis, Tunisia | 1-Clinical and Experimental Pharmacology Lab, LR16SP02, National Center of Pharmacovigilance, University of Tunis El Manar, Tunis, Tunisia. 2-Neurodegenerative diseases and psychiatric troubles, LR18SP03, Razi Hospital, University of Tunis El Manar, Tunis, Tunisia. 3-Ministry of Health, National Observatory of New and Emerging Diseases, 1006, Tunis, Tunisia | Ilhem Boutiba-Ben Boubaker et al |
| EPI_ISL_699662 | India/MH-ACTREC-005/2020 | 2020-05-01 | Asia/India/Maharashtra/Thane | Hematopathology Laboratory, ACTREC, TMC | Hematopathology Laboratory, ACTREC, TMC | Hematopathology Laboratory et al |
| EPI_ISL_699663 | India/MH-ACTREC-006/2020 | 2020-05-02 | Asia/India/Maharashtra/Mumbai | Hematopathology Laboratory, ACTREC, TMC | Hematopathology Laboratory, ACTREC, TMC | Hematopathology Laboratory et al |
| EPI_ISL_699664 | India/MH-ACTREC-007/2020 | 2020-05-02 | Asia/India/Maharashtra/Raigad | Hematopathology Laboratory, ACTREC, TMC | Hematopathology Laboratory, ACTREC, TMC | Hematopathology Laboratory et al |

| | | | | | | |
|---|---|---|---|---|---|---|
| EPI_ISL_699668 | India/MH-ACTREC-011/2020 | 2020-05-02 | Asia/India/Maharashtra/Thane | Hematopathology Laboratory, ACTREC, TMC | Hematopathology Laboratory, ACTREC, TMC | Hematopathology Laboratory et al |
| EPI_ISL_699670 | India/MH-ACTREC-014/2020 | 2020-05-03 | Asia/India/Maharashtra/Raigad | Hematopathology Laboratory, ACTREC, TMC | Hematopathology Laboratory, ACTREC, TMC | Hematopathology Laboratory et al |
| EPI_ISL_699674 | India/MH-ACTREC-018/2020 | 2020-05-03 | Asia/India/Maharashtra/Raigad | Hematopathology Laboratory, ACTREC, TMC | Hematopathology Laboratory, ACTREC, TMC | Hematopathology Laboratory et al |
| EPI_ISL_699677 | India/MH-ACTREC-021/2020 | 2020-05-04 | Asia/India/Maharashtra/Raigad | Hematopathology Laboratory, ACTREC, TMC | Hematopathology Laboratory, ACTREC, TMC | Hematopathology Laboratory et al |
| EPI_ISL_699684 | India/MH-ACTREC-028/2020 | 2020-05-04 | Asia/India/Maharashtra/Raigad | Hematopathology Laboratory, ACTREC, TMC | Hematopathology Laboratory, ACTREC, TMC | Hematopathology Laboratory et al |
| EPI_ISL_699688 | India/MH-ACTREC-032/2020 | 2020-05-07 | Asia/India/Maharashtra/Raigad | Hematopathology Laboratory, ACTREC, TMC | Hematopathology Laboratory, ACTREC, TMC | Hematopathology Laboratory et al |
| EPI_ISL_699692 | India/MH-ACTREC-036/2020 | 2020-05-08 | Asia/India/Maharashtra/Raigad | Hematopathology Laboratory, ACTREC, TMC | Hematopathology Laboratory, ACTREC, TMC | Hematopathology Laboratory et al |
| EPI_ISL_699696 | India/MH-ACTREC-040/2020 | 2020-05-10 | Asia/India/Maharashtra/Mumbai | Hematopathology Laboratory, ACTREC, TMC | Hematopathology Laboratory, ACTREC, TMC | Hematopathology Laboratory et al |
| EPI_ISL_699701 | India/MH-ACTREC-045/2020 | 2020-05-12 | Asia/India/Maharashtra/Mumbai | Hematopathology Laboratory, ACTREC, TMC | Hematopathology Laboratory, ACTREC, TMC | Hematopathology Laboratory et al |
| EPI_ISL_699708 | India/MH-ACTREC-052/2020 | 2020-05-13 | Asia/India/Maharashtra/Raigad | Hematopathology Laboratory, ACTREC, TMC | Hematopathology Laboratory, ACTREC, TMC | Hematopathology Laboratory et al |
| EPI_ISL_699709 | India/MH-ACTREC-053/2020 | 2020-05-13 | Asia/India/Maharashtra/Mumbai | Hematopathology Laboratory, ACTREC, TMC | Hematopathology Laboratory, ACTREC, TMC | Hematopathology Laboratory et al |
| EPI_ISL_699712 | India/MH-ACTREC-056/2020 | 2020-05-14 | Asia/India/Maharashtra/Raigad | Hematopathology Laboratory, ACTREC, TMC | Hematopathology Laboratory, ACTREC, TMC | Hematopathology Laboratory et al |
| EPI_ISL_699717 | India/MH-ACTREC-061/2020 | 2020-05-16 | Asia/India/Maharashtra/Raigad | Hematopathology Laboratory, ACTREC, TMC | Hematopathology Laboratory, ACTREC, TMC | Hematopathology Laboratory et al |

| | | | | | | |
|---|---|---|---|---|---|---|
| EPI_ISL_699722 | India/MH-ACTREC-066/2020 | 2020-05-17 | Asia/India/Maharashtra/Raigad | Hematopathology Laboratory, ACTREC, TMC | Hematopathology Laboratory, ACTREC, TMC | Hematopathology Laboratory et al |
| EPI_ISL_699725 | India/MH-ACTREC-069/2020 | 2020-05-17 | Asia/India/Maharashtra/Raigad | Hematopathology Laboratory, ACTREC, TMC | Hematopathology Laboratory, ACTREC, TMC | Hematopathology Laboratory et al |
| EPI_ISL_699737 | India/MH-ACTREC-081/2020 | 2020-05-21 | Asia/India/Maharashtra/Mumbai | Hematopathology Laboratory, ACTREC, TMC | Hematopathology Laboratory, ACTREC, TMC | Hematopathology Laboratory et al |
| EPI_ISL_699739 | India/MH-ACTREC-083/2020 | 2020-05-21 | Asia/India/Maharashtra/Raigad | Hematopathology Laboratory, ACTREC, TMC | Hematopathology Laboratory, ACTREC, TMC | Hematopathology Laboratory et al |
| EPI_ISL_699740 | India/MH-ACTREC-084/2020 | 2020-05-21 | Asia/India/Maharashtra/Raigad | Hematopathology Laboratory, ACTREC, TMC | Hematopathology Laboratory, ACTREC, TMC | Hematopathology Laboratory et al |
| EPI_ISL_699742 | India/MH-ACTREC-086/2020 | 2020-05-21 | Asia/India/Maharashtra/Raigad | Hematopathology Laboratory, ACTREC, TMC | Hematopathology Laboratory, ACTREC, TMC | Hematopathology Laboratory et al |
| EPI_ISL_699743 | India/MH-ACTREC-087/2020 | 2020-05-21 | Asia/India/Maharashtra/Raigad | Hematopathology Laboratory, ACTREC, TMC | Hematopathology Laboratory, ACTREC, TMC | Hematopathology Laboratory et al |
| EPI_ISL_699750 | India/MH-ACTREC-094/2020 | 2020-05-22 | Asia/India/Maharashtra/Raigad | Hematopathology Laboratory, ACTREC, TMC | Hematopathology Laboratory, ACTREC, TMC | Hematopathology Laboratory et al |
| EPI_ISL_699754 | India/MH-ACTREC-098/2020 | 2020-05-23 | Asia/India/Maharashtra/Raigad | Hematopathology Laboratory, ACTREC, TMC | Hematopathology Laboratory, ACTREC, TMC | Hematopathology Laboratory et al |
| EPI_ISL_699764 | India/MH-ACTREC-108/2020 | 2020-05-24 | Asia/India/Maharashtra/Raigad | Hematopathology Laboratory, ACTREC, TMC | Hematopathology Laboratory, ACTREC, TMC | Hematopathology Laboratory et al |
| EPI_ISL_699766 | India/MH-ACTREC-110/2020 | 2020-05-24 | Asia/India/Maharashtra/Raigad | Hematopathology Laboratory, ACTREC, TMC | Hematopathology Laboratory, ACTREC, TMC | Hematopathology Laboratory et al |
| EPI_ISL_699778 | India/MH-ACTREC-122/2020 | 2020-05-25 | Asia/India/Maharashtra/Thane | Hematopathology Laboratory, ACTREC, TMC | Hematopathology Laboratory, ACTREC, TMC | Hematopathology Laboratory et al |
| EPI_ISL_699779 | India/MH-ACTREC-123/2020 | 2020-05-25 | Asia/India/Maharashtra/Thane | Hematopathology Laboratory, ACTREC, TMC | Hematopathology Laboratory, ACTREC, TMC | Hematopathology Laboratory et al |

| | | | | | | |
|---|---|---|---|---|---|---|
| EPI_ISL_699783 | India/MH-ACTREC-127/2020 | 2020-05-25 | Asia/India/Maharashtra/Mumbai | Hematopathology Laboratory, ACTREC, TMC | Hematopathology Laboratory, ACTREC, TMC | Hematopathology Laboratory et al |
| EPI_ISL_699784 | India/MH-ACTREC-128/2020 | 2020-05-26 | Asia/India/Maharashtra/Raigad | Hematopathology Laboratory, ACTREC, TMC | Hematopathology Laboratory, ACTREC, TMC | Hematopathology Laboratory et al |
| EPI_ISL_699795 | India/MH-ACTREC-139/2020 | 2020-05-28 | Asia/India/Maharashtra/Raigad | Hematopathology Laboratory, ACTREC, TMC | Hematopathology Laboratory, ACTREC, TMC | Hematopathology Laboratory et al |
| EPI_ISL_699807 | India/MH-ACTREC-151/2020 | 2020-06-02 | Asia/India/Maharashtra/Raigad | Hematopathology Laboratory, ACTREC, TMC | Hematopathology Laboratory, ACTREC, TMC | Hematopathology Laboratory et al |
| EPI_ISL_699811 | India/MH-ACTREC-155/2020 | 2020-06-02 | Asia/India/Maharashtra/Thane | Hematopathology Laboratory, ACTREC, TMC | Hematopathology Laboratory, ACTREC, TMC | Hematopathology Laboratory et al |
| EPI_ISL_699814 | India/MH-ACTREC-158/2020 | 2020-06-04 | Asia/India/Maharashtra/Raigad | Hematopathology Laboratory, ACTREC, TMC | Hematopathology Laboratory, ACTREC, TMC | Hematopathology Laboratory et al |
| EPI_ISL_699815 | India/MH-ACTREC-159/2020 | 2020-06-04 | Asia/India/Maharashtra/Raigad | Hematopathology Laboratory, ACTREC, TMC | Hematopathology Laboratory, ACTREC, TMC | Hematopathology Laboratory et al |
| EPI_ISL_699818 | India/MH-ACTREC-162/2020 | 2020-06-04 | Asia/India/Maharashtra/Raigad | Hematopathology Laboratory, ACTREC, TMC | Hematopathology Laboratory, ACTREC, TMC | Hematopathology Laboratory et al |
| EPI_ISL_699820 | India/MH-ACTREC-164/2020 | 2020-06-04 | Asia/India/Maharashtra/Raigad | Hematopathology Laboratory, ACTREC, TMC | Hematopathology Laboratory, ACTREC, TMC | Hematopathology Laboratory et al |
| EPI_ISL_699827 | India/MH-ACTREC-172/2020 | 2020-06-11 | Asia/India/Maharashtra/Raigad | Hematopathology Laboratory, ACTREC, TMC | Hematopathology Laboratory, ACTREC, TMC | Hematopathology Laboratory et al |
| EPI_ISL_699829 | India/MH-ACTREC-174/2020 | 2020-06-13 | Asia/India/Maharashtra/Mumbai Subur | Hematopathology Laboratory, ACTREC, TMC | Hematopathology Laboratory, ACTREC, TMC | Hematopathology Laboratory et al |
| EPI_ISL_699836 | India/MH-ACTREC-181/2020 | 2020-06-17 | Asia/India/Maharashtra/Raigad | Hematopathology Laboratory, ACTREC, TMC | Hematopathology Laboratory, ACTREC, TMC | Hematopathology Laboratory et al |
| EPI_ISL_699837 | India/MH-ACTREC-182/2020 | 2020-06-17 | Asia/India/Maharashtra/Raigad | Hematopathology Laboratory, ACTREC, TMC | Hematopathology Laboratory, ACTREC, TMC | Hematopathology Laboratory et al |

| | | | | | | |
|---|---|---|---|---|---|---|
| EPI_ISL_699839 | India/MH-ACTREC-184/2020 | 2020-06-17 | Asia/India/Maharashtra/Raigad | Hematopathology Laboratory, ACTREC, TMC | Hematopathology Laboratory, ACTREC, TMC | Hematopathology Laboratory et al |
| EPI_ISL_699845 | India/MH-ACTREC-191/2020 | 2020-06-22 | Asia/India/Maharashtra/Raigad | Hematopathology Laboratory, ACTREC, TMC | Hematopathology Laboratory, ACTREC, TMC | Hematopathology Laboratory et al |
| EPI_ISL_699850 | India/MH-ACTREC-196/2020 | 2020-06-22 | Asia/India/Maharashtra/Raigad | Hematopathology Laboratory, ACTREC, TMC | Hematopathology Laboratory, ACTREC, TMC | Hematopathology Laboratory et al |
| EPI_ISL_699854 | India/MH-ACTREC-200/2020 | 2020-06-25 | Asia/India/Maharashtra/Thane | Hematopathology Laboratory, ACTREC, TMC | Hematopathology Laboratory, ACTREC, TMC | Hematopathology Laboratory et al |
| EPI_ISL_699855 | India/MH-ACTREC-201/2020 | 2020-06-25 | Asia/India/Maharashtra/Raigad | Hematopathology Laboratory, ACTREC, TMC | Hematopathology Laboratory, ACTREC, TMC | Hematopathology Laboratory et al |
| EPI_ISL_699857 | India/MH-ACTREC-203/2020 | 2020-06-25 | Asia/India/Maharashtra/Raigad | Hematopathology Laboratory, ACTREC, TMC | Hematopathology Laboratory, ACTREC, TMC | Hematopathology Laboratory et al |
| EPI_ISL_699860 | India/MH-ACTREC-206/2020 | 2020-06-26 | Asia/India/Maharashtra/Raigad | Hematopathology Laboratory, ACTREC, TMC | Hematopathology Laboratory, ACTREC, TMC | Hematopathology Laboratory et al |
| EPI_ISL_699861 | India/MH-ACTREC-207/2020 | 2020-06-26 | Asia/India/Maharashtra/Raigad | Hematopathology Laboratory, ACTREC, TMC | Hematopathology Laboratory, ACTREC, TMC | Hematopathology Laboratory et al |
| EPI_ISL_699865 | India/MH-ACTREC-211/2020 | 2020-06-27 | Asia/India/Maharashtra/Raigad | Hematopathology Laboratory, ACTREC, TMC | Hematopathology Laboratory, ACTREC, TMC | Hematopathology Laboratory et al |
| EPI_ISL_699866 | India/MH-ACTREC-212/2020 | 2020-06-29 | Asia/India/Maharashtra/Mumbai | Hematopathology Laboratory, ACTREC, TMC | Hematopathology Laboratory, ACTREC, TMC | Hematopathology Laboratory et al |
| EPI_ISL_699867 | India/MH-ACTREC-213/2020 | 2020-06-29 | Asia/India/Maharashtra/Thane | Hematopathology Laboratory, ACTREC, TMC | Hematopathology Laboratory, ACTREC, TMC | Hematopathology Laboratory et al |
| EPI_ISL_699868 | India/MH-ACTREC-214/2020 | 2020-06-29 | Asia/India/Maharashtra/Raigad | Hematopathology Laboratory, ACTREC, TMC | Hematopathology Laboratory, ACTREC, TMC | Hematopathology Laboratory et al |
| EPI_ISL_699869 | India/MH-ACTREC-215/2020 | 2020-06-29 | Asia/India/Maharashtra/Raigad | Hematopathology Laboratory, ACTREC, TMC | Hematopathology Laboratory, ACTREC, TMC | Hematopathology Laboratory et al |

| | | | | | | |
|---|---|---|---|---|---|---|
| EPI_ISL_699870 | India/MH-ACTREC-216/2020 | 2020-06-29 | Asia/India/Maharashtra/Raigad | Hematopathology Laboratory, ACTREC, TMC | Hematopathology Laboratory, ACTREC, TMC | Hematopathology Laboratory et al |
| EPI_ISL_699873 | India/MH-ACTREC-219/2020 | 2020-06-29 | Asia/India/Maharashtra/Raigad | Hematopathology Laboratory, ACTREC, TMC | Hematopathology Laboratory, ACTREC, TMC | Hematopathology Laboratory et al |
| EPI_ISL_699875 | India/MH-ACTREC-221/2020 | 2020-06-30 | Asia/India/Maharashtra/Raigad | Hematopathology Laboratory, ACTREC, TMC | Hematopathology Laboratory, ACTREC, TMC | Hematopathology Laboratory et al |
| EPI_ISL_699876 | India/MH-ACTREC-222/2020 | 2020-06-30 | Asia/India/Maharashtra/Raigad | Hematopathology Laboratory, ACTREC, TMC | Hematopathology Laboratory, ACTREC, TMC | Hematopathology Laboratory et al |
| EPI_ISL_699878 | India/MH-ACTREC-224/2020 | 2020-07-01 | Asia/India/Maharashtra/Raigad | Hematopathology Laboratory, ACTREC, TMC | Hematopathology Laboratory, ACTREC, TMC | Hematopathology Laboratory et al |
| EPI_ISL_699881 | India/MH-ACTREC-227/2020 | 2020-07-02 | Asia/India/Maharashtra/Raigad | Hematopathology Laboratory, ACTREC, TMC | Hematopathology Laboratory, ACTREC, TMC | Hematopathology Laboratory et al |
| EPI_ISL_699882 | India/MH-ACTREC-228/2020 | 2020-07-02 | Asia/India/Maharashtra/Raigad | Hematopathology Laboratory, ACTREC, TMC | Hematopathology Laboratory, ACTREC, TMC | Hematopathology Laboratory et al |
| EPI_ISL_699884 | India/MH-ACTREC-231/2020 | 2020-07-02 | Asia/India/Maharashtra/Raigad | Hematopathology Laboratory, ACTREC, TMC | Hematopathology Laboratory, ACTREC, TMC | Hematopathology Laboratory et al |
| EPI_ISL_699885 | India/MH-ACTREC-232/2020 | 2020-07-03 | Asia/India/Maharashtra/Thane | Hematopathology Laboratory, ACTREC, TMC | Hematopathology Laboratory, ACTREC, TMC | Hematopathology Laboratory et al |
| EPI_ISL_699887 | India/MH-ACTREC-234/2020 | 2020-07-04 | Asia/India/Maharashtra/Thane | Hematopathology Laboratory, ACTREC, TMC | Hematopathology Laboratory, ACTREC, TMC | Hematopathology Laboratory et al |
| EPI_ISL_699888 | India/MH-ACTREC-235/2020 | 2020-07-04 | Asia/India/Maharashtra/Raigad | Hematopathology Laboratory, ACTREC, TMC | Hematopathology Laboratory, ACTREC, TMC | Hematopathology Laboratory et al |
| EPI_ISL_699889 | India/MH-ACTREC-236/2020 | 2020-07-05 | Asia/India/Maharashtra/Raigad | Hematopathology Laboratory, ACTREC, TMC | Hematopathology Laboratory, ACTREC, TMC | Hematopathology Laboratory et al |
| EPI_ISL_699890 | India/MH-ACTREC-237/2020 | 2020-07-06 | Asia/India/Maharashtra/Raigad | Hematopathology Laboratory, ACTREC, TMC | Hematopathology Laboratory, ACTREC, TMC | Hematopathology Laboratory et al |

| | | | | | | |
|---|---|---|---|---|---|---|
| EPI_ISL_699891 | India/MH-ACTREC-238/2020 | 2020-07-06 | Asia/India/Maharashtra/Raigad | Hematopathology Laboratory, ACTREC, TMC | Hematopathology Laboratory, ACTREC, TMC | Hematopathology Laboratory et al |
| EPI_ISL_699893 | India/MH-ACTREC-240/2020 | 2020-07-06 | Asia/India/Maharashtra/Raigad | Hematopathology Laboratory, ACTREC, TMC | Hematopathology Laboratory, ACTREC, TMC | Hematopathology Laboratory et al |
| EPI_ISL_699894 | India/MH-ACTREC-241/2020 | 2020-07-06 | Asia/India/Maharashtra/Raigad | Hematopathology Laboratory, ACTREC, TMC | Hematopathology Laboratory, ACTREC, TMC | Hematopathology Laboratory et al |
| EPI_ISL_699895 | India/MH-ACTREC-242/2020 | 2020-07-06 | Asia/India/Maharashtra/Raigad | Hematopathology Laboratory, ACTREC, TMC | Hematopathology Laboratory, ACTREC, TMC | Hematopathology Laboratory et al |
| EPI_ISL_699896 | India/MH-ACTREC-243/2020 | 2020-07-06 | Asia/India/Maharashtra/Raigad | Hematopathology Laboratory, ACTREC, TMC | Hematopathology Laboratory, ACTREC, TMC | Hematopathology Laboratory et al |
| EPI_ISL_699897 | India/MH-ACTREC-244/2020 | 2020-07-06 | Asia/India/Maharashtra/Raigad | Hematopathology Laboratory, ACTREC, TMC | Hematopathology Laboratory, ACTREC, TMC | Hematopathology Laboratory et al |
| EPI_ISL_699898 | India/MH-ACTREC-245/2020 | 2020-07-07 | Asia/India/Maharashtra/Raigad | Hematopathology Laboratory, ACTREC, TMC | Hematopathology Laboratory, ACTREC, TMC | Hematopathology Laboratory et al |
| EPI_ISL_699899 | India/MH-ACTREC-246/2020 | 2020-07-07 | Asia/India/Maharashtra/Raigad | Hematopathology Laboratory, ACTREC, TMC | Hematopathology Laboratory, ACTREC, TMC | Hematopathology Laboratory et al |
| EPI_ISL_699900 | India/MH-ACTREC-247/2020 | 2020-07-07 | Asia/India/Maharashtra/Raigad | Hematopathology Laboratory, ACTREC, TMC | Hematopathology Laboratory, ACTREC, TMC | Hematopathology Laboratory et al |
| EPI_ISL_699901 | India/MH-ACTREC-249/2020 | 2020-07-07 | Asia/India/Maharashtra/Raigad | Hematopathology Laboratory, ACTREC, TMC | Hematopathology Laboratory, ACTREC, TMC | Hematopathology Laboratory et al |
| EPI_ISL_699902 | India/MH-ACTREC-250/2020 | 2020-07-07 | Asia/India/Maharashtra/Raigad | Hematopathology Laboratory, ACTREC, TMC | Hematopathology Laboratory, ACTREC, TMC | Hematopathology Laboratory et al |
| EPI_ISL_699904 | India/MH-ACTREC-252/2020 | 2020-07-07 | Asia/India/Maharashtra/Raigad | Hematopathology Laboratory, ACTREC, TMC | Hematopathology Laboratory, ACTREC, TMC | Hematopathology Laboratory et al |
| EPI_ISL_699905 | India/MH-ACTREC-253/2020 | 2020-07-08 | Asia/India/Maharashtra/Raigad | Hematopathology Laboratory, ACTREC, TMC | Hematopathology Laboratory, ACTREC, TMC | Hematopathology Laboratory et al |

| | | | | | | |
|---|---|---|---|---|---|---|
| EPI_ISL_699906 | India/MH-ACTREC-254/2020 | 2020-07-08 | Asia/India/Maharashtra/Raigad | Hematopathology Laboratory, ACTREC, TMC | Hematopathology Laboratory, ACTREC, TMC | Hematopathology Laboratory et al |
| EPI_ISL_699907 | India/MH-ACTREC-255/2020 | 2020-07-08 | Asia/India/Maharashtra/Raigad | Hematopathology Laboratory, ACTREC, TMC | Hematopathology Laboratory, ACTREC, TMC | Hematopathology Laboratory et al |
| EPI_ISL_699908 | India/MH-ACTREC-256/2020 | 2020-07-08 | Asia/India/Maharashtra/Raigad | Hematopathology Laboratory, ACTREC, TMC | Hematopathology Laboratory, ACTREC, TMC | Hematopathology Laboratory et al |
| EPI_ISL_699909 | India/MH-ACTREC-257/2020 | 2020-07-08 | Asia/India/Maharashtra/Raigad | Hematopathology Laboratory, ACTREC, TMC | Hematopathology Laboratory, ACTREC, TMC | Hematopathology Laboratory et al |
| EPI_ISL_699910 | India/MH-ACTREC-258/2020 | 2020-07-08 | Asia/India/Maharashtra/Raigad | Hematopathology Laboratory, ACTREC, TMC | Hematopathology Laboratory, ACTREC, TMC | Hematopathology Laboratory et al |
| EPI_ISL_699911 | India/MH-ACTREC-259/2020 | 2020-07-08 | Asia/India/Maharashtra/Raigad | Hematopathology Laboratory, ACTREC, TMC | Hematopathology Laboratory, ACTREC, TMC | Hematopathology Laboratory et al |
| EPI_ISL_699912 | India/MH-ACTREC-260/2020 | 2020-07-08 | Asia/India/Maharashtra/Raigad | Hematopathology Laboratory, ACTREC, TMC | Hematopathology Laboratory, ACTREC, TMC | Hematopathology Laboratory et al |
| EPI_ISL_699913 | India/MH-ACTREC-261/2020 | 2020-07-08 | Asia/India/Maharashtra/Raigad | Hematopathology Laboratory, ACTREC, TMC | Hematopathology Laboratory, ACTREC, TMC | Hematopathology Laboratory et al |
| EPI_ISL_699914 | India/MH-ACTREC-262/2020 | 2020-07-08 | Asia/India/Maharashtra/Raigad | Hematopathology Laboratory, ACTREC, TMC | Hematopathology Laboratory, ACTREC, TMC | Hematopathology Laboratory et al |
| EPI_ISL_699915 | India/MH-ACTREC-263/2020 | 2020-07-08 | Asia/India/Maharashtra/Raigad | Hematopathology Laboratory, ACTREC, TMC | Hematopathology Laboratory, ACTREC, TMC | Hematopathology Laboratory et al |
| EPI_ISL_699916 | India/MH-ACTREC-264/2020 | 2020-07-08 | Asia/India/Maharashtra/Raigad | Hematopathology Laboratory, ACTREC, TMC | Hematopathology Laboratory, ACTREC, TMC | Hematopathology Laboratory et al |
| EPI_ISL_699917 | India/MH-ACTREC-265/2020 | 2020-07-08 | Asia/India/Maharashtra/Raigad | Hematopathology Laboratory, ACTREC, TMC | Hematopathology Laboratory, ACTREC, TMC | Hematopathology Laboratory et al |
| EPI_ISL_699918 | India/MH-ACTREC-266/2020 | 2020-07-08 | Asia/India/Maharashtra/Raigad | Hematopathology Laboratory, ACTREC, TMC | Hematopathology Laboratory, ACTREC, TMC | Hematopathology Laboratory et al |

| | | | | | | |
|---|---|---|---|---|---|---|
| EPI_ISL_699919 | India/MH-ACTREC-267/2020 | 2020-07-09 | Asia/India/Maharashtra/Thane | Hematopathology Laboratory, ACTREC, TMC | Hematopathology Laboratory, ACTREC, TMC | Hematopathology Laboratory et al |
| EPI_ISL_699920 | India/MH-ACTREC-268/2020 | 2020-07-09 | Asia/India/Maharashtra/Raigad | Hematopathology Laboratory, ACTREC, TMC | Hematopathology Laboratory, ACTREC, TMC | Hematopathology Laboratory et al |
| EPI_ISL_699921 | India/MH-ACTREC-269/2020 | 2020-07-09 | Asia/India/Maharashtra/Raigad | Hematopathology Laboratory, ACTREC, TMC | Hematopathology Laboratory, ACTREC, TMC | Hematopathology Laboratory et al |
| EPI_ISL_699922 | India/MH-ACTREC-270/2020 | 2020-07-09 | Asia/India/Maharashtra/Raigad | Hematopathology Laboratory, ACTREC, TMC | Hematopathology Laboratory, ACTREC, TMC | Hematopathology Laboratory et al |
| EPI_ISL_699923 | India/MH-ACTREC-271/2020 | 2020-07-10 | Asia/India/Maharashtra/Thane | Hematopathology Laboratory, ACTREC, TMC | Hematopathology Laboratory, ACTREC, TMC | Hematopathology Laboratory et al |
| EPI_ISL_699924 | India/MH-ACTREC-272/2020 | 2020-07-10 | Asia/India/Maharashtra/Thane | Hematopathology Laboratory, ACTREC, TMC | Hematopathology Laboratory, ACTREC, TMC | Hematopathology Laboratory et al |
| EPI_ISL_699925 | India/MH-ACTREC-273/2020 | 2020-07-10 | Asia/India/Maharashtra/Raigad | Hematopathology Laboratory, ACTREC, TMC | Hematopathology Laboratory, ACTREC, TMC | Hematopathology Laboratory et al |
| EPI_ISL_699926 | India/MH-ACTREC-274/2020 | 2020-07-10 | Asia/India/Maharashtra/Raigad | Hematopathology Laboratory, ACTREC, TMC | Hematopathology Laboratory, ACTREC, TMC | Hematopathology Laboratory et al |
| EPI_ISL_699927 | India/MH-ACTREC-275/2020 | 2020-07-10 | Asia/India/Maharashtra/Raigad | Hematopathology Laboratory, ACTREC, TMC | Hematopathology Laboratory, ACTREC, TMC | Hematopathology Laboratory et al |
| EPI_ISL_699928 | India/MH-ACTREC-276/2020 | 2020-07-10 | Asia/India/Maharashtra/Raigad | Hematopathology Laboratory, ACTREC, TMC | Hematopathology Laboratory, ACTREC, TMC | Hematopathology Laboratory et al |
| EPI_ISL_699929 | India/MH-ACTREC-277/2020 | 2020-07-10 | Asia/India/Maharashtra/Raigad | Hematopathology Laboratory, ACTREC, TMC | Hematopathology Laboratory, ACTREC, TMC | Hematopathology Laboratory et al |
| EPI_ISL_699930 | India/MH-ACTREC-278/2020 | 2020-07-11 | Asia/India/Maharashtra/Thane | Hematopathology Laboratory, ACTREC, TMC | Hematopathology Laboratory, ACTREC, TMC | Hematopathology Laboratory et al |
| EPI_ISL_699931 | India/MH-ACTREC-279/2020 | 2020-07-11 | Asia/India/Maharashtra/Thane | Hematopathology Laboratory, ACTREC, TMC | Hematopathology Laboratory, ACTREC, TMC | Hematopathology Laboratory et al |

| | | | | | | |
|---|---|---|---|---|---|---|
| EPI_ISL_699932 | India/MH-ACTREC-280/2020 | 2020-07-11 | Asia/India/Maharashtra/Raigad | Hematopathology Laboratory, ACTREC, TMC | Hematopathology Laboratory, ACTREC, TMC | Hematopathology Laboratory et al |
| EPI_ISL_699933 | India/MH-ACTREC-281/2020 | 2020-07-11 | Asia/India/Maharashtra/Raigad | Hematopathology Laboratory, ACTREC, TMC | Hematopathology Laboratory, ACTREC, TMC | Hematopathology Laboratory et al |
| EPI_ISL_699935 | India/MH-ACTREC-283/2020 | 2020-07-11 | Asia/India/Maharashtra/Mumbai | Hematopathology Laboratory, ACTREC, TMC | Hematopathology Laboratory, ACTREC, TMC | Hematopathology Laboratory et al |
| EPI_ISL_699936 | India/MH-ACTREC-284/2020 | 2020-07-12 | Asia/India/Maharashtra/Thane | Hematopathology Laboratory, ACTREC, TMC | Hematopathology Laboratory, ACTREC, TMC | Hematopathology Laboratory et al |
| EPI_ISL_699938 | India/MH-ACTREC-286/2020 | 2020-07-13 | Asia/India/Maharashtra/Vasai | Hematopathology Laboratory, ACTREC, TMC | Hematopathology Laboratory, ACTREC, TMC | Hematopathology Laboratory et al |
| EPI_ISL_699939 | India/MH-ACTREC-287/2020 | 2020-07-13 | Asia/India/Maharashtra/Raigad | Hematopathology Laboratory, ACTREC, TMC | Hematopathology Laboratory, ACTREC, TMC | Hematopathology Laboratory et al |
| EPI_ISL_699941 | India/MH-ACTREC-289/2020 | 2020-07-13 | Asia/India/Maharashtra/Raigad | Hematopathology Laboratory, ACTREC, TMC | Hematopathology Laboratory, ACTREC, TMC | Hematopathology Laboratory et al |
| EPI_ISL_699942 | India/MH-ACTREC-290/2020 | 2020-07-13 | Asia/India/Maharashtra/Raigad | Hematopathology Laboratory, ACTREC, TMC | Hematopathology Laboratory, ACTREC, TMC | Hematopathology Laboratory et al |
| EPI_ISL_699944 | India/MH-ACTREC-292/2020 | 2020-07-14 | Asia/India/Maharashtra/Raigad | Hematopathology Laboratory, ACTREC, TMC | Hematopathology Laboratory, ACTREC, TMC | Hematopathology Laboratory et al |
| EPI_ISL_699945 | India/MH-ACTREC-293/2020 | 2020-07-14 | Asia/India/Maharashtra/Raigad | Hematopathology Laboratory, ACTREC, TMC | Hematopathology Laboratory, ACTREC, TMC | Hematopathology Laboratory et al |
| EPI_ISL_699947 | India/MH-ACTREC-295/2020 | 2020-07-14 | Asia/India/Maharashtra/Thane | Hematopathology Laboratory, ACTREC, TMC | Hematopathology Laboratory, ACTREC, TMC | Hematopathology Laboratory et al |
| EPI_ISL_699948 | India/MH-ACTREC-296/2020 | 2020-07-14 | Asia/India/Maharashtra/Thane | Hematopathology Laboratory, ACTREC, TMC | Hematopathology Laboratory, ACTREC, TMC | Hematopathology Laboratory et al |
| EPI_ISL_699949 | India/MH-ACTREC-297/2020 | 2020-07-14 | Asia/India/Maharashtra/Raigad | Hematopathology Laboratory, ACTREC, TMC | Hematopathology Laboratory, ACTREC, TMC | Hematopathology Laboratory et al |

| | | | | | | |
|---|---|---|---|---|---|---|
| EPI_ISL_699950 | India/MH-ACTREC-298/2020 | 2020-07-14 | Asia/India/Maharashtra/Raigad | Hematopathology Laboratory, ACTREC, TMC | Hematopathology Laboratory, ACTREC, TMC | Hematopathology Laboratory et al |
| EPI_ISL_699951 | India/MH-ACTREC-299/2020 | 2020-07-14 | Asia/India/Maharashtra/Raigad | Hematopathology Laboratory, ACTREC, TMC | Hematopathology Laboratory, ACTREC, TMC | Hematopathology Laboratory et al |
| EPI_ISL_699952 | India/MH-ACTREC-301/2020 | 2020-07-14 | Asia/India/Maharashtra/Raigad | Hematopathology Laboratory, ACTREC, TMC | Hematopathology Laboratory, ACTREC, TMC | Hematopathology Laboratory et al |
| EPI_ISL_699953 | India/MH-ACTREC-302/2020 | 2020-07-15 | Asia/India/Maharashtra/Raigad | Hematopathology Laboratory, ACTREC, TMC | Hematopathology Laboratory, ACTREC, TMC | Hematopathology Laboratory et al |
| EPI_ISL_699954 | India/MH-ACTREC-303/2020 | 2020-07-15 | Asia/India/Maharashtra/Thane | Hematopathology Laboratory, ACTREC, TMC | Hematopathology Laboratory, ACTREC, TMC | Hematopathology Laboratory et al |
| EPI_ISL_699955 | India/MH-ACTREC-304/2020 | 2020-07-15 | Asia/India/Maharashtra/Raigad | Hematopathology Laboratory, ACTREC, TMC | Hematopathology Laboratory, ACTREC, TMC | Hematopathology Laboratory et al |
| EPI_ISL_699957 | India/MH-ACTREC-306/2020 | 2020-07-15 | Asia/India/Maharashtra/Raigad | Hematopathology Laboratory, ACTREC, TMC | Hematopathology Laboratory, ACTREC, TMC | Hematopathology Laboratory et al |
| EPI_ISL_699958 | India/MH-ACTREC-307/2020 | 2020-07-15 | Asia/India/Maharashtra/Raigad | Hematopathology Laboratory, ACTREC, TMC | Hematopathology Laboratory, ACTREC, TMC | Hematopathology Laboratory et al |
| EPI_ISL_699959 | India/MH-ACTREC-308/2020 | 2020-07-15 | Asia/India/Maharashtra/Raigad | Hematopathology Laboratory, ACTREC, TMC | Hematopathology Laboratory, ACTREC, TMC | Hematopathology Laboratory et al |
| EPI_ISL_699960 | India/MH-ACTREC-309/2020 | 2020-07-15 | Asia/India/Maharashtra/Raigad | Hematopathology Laboratory, ACTREC, TMC | Hematopathology Laboratory, ACTREC, TMC | Hematopathology Laboratory et al |
| EPI_ISL_699961 | India/MH-ACTREC-310/2020 | 2020-07-15 | Asia/India/Maharashtra/Raigad | Hematopathology Laboratory, ACTREC, TMC | Hematopathology Laboratory, ACTREC, TMC | Hematopathology Laboratory et al |
| EPI_ISL_699962 | India/MH-ACTREC-313/2020 | 2020-07-15 | Asia/India/Maharashtra/Raigad | Hematopathology Laboratory, ACTREC, TMC | Hematopathology Laboratory, ACTREC, TMC | Hematopathology Laboratory et al |
| EPI_ISL_699963 | India/MH-ACTREC-314/2020 | 2020-07-16 | Asia/India/Maharashtra/Raigad | Hematopathology Laboratory, ACTREC, TMC | Hematopathology Laboratory, ACTREC, TMC | Hematopathology Laboratory et al |

| | | | | | | |
|---|---|---|---|---|---|---|
| EPI_ISL_699964 | India/MH-ACTREC-315/2020 | 2020-07-16 | Asia/India/Maharashtra/Raigad | Hematopathology Laboratory, ACTREC, TMC | Hematopathology Laboratory, ACTREC, TMC | Hematopathology Laboratory et al |
| EPI_ISL_699966 | India/MH-ACTREC-317/2020 | 2020-07-16 | Asia/India/Maharashtra/Raigad | Hematopathology Laboratory, ACTREC, TMC | Hematopathology Laboratory, ACTREC, TMC | Hematopathology Laboratory et al |
| EPI_ISL_699968 | India/MH-ACTREC-319/2020 | 2020-07-16 | Asia/India/Maharashtra/Raigad | Hematopathology Laboratory, ACTREC, TMC | Hematopathology Laboratory, ACTREC, TMC | Hematopathology Laboratory et al |
| EPI_ISL_699969 | India/MH-ACTREC-320/2020 | 2020-07-16 | Asia/India/Maharashtra/Raigad | Hematopathology Laboratory, ACTREC, TMC | Hematopathology Laboratory, ACTREC, TMC | Hematopathology Laboratory et al |
| EPI_ISL_699970 | India/MH-ACTREC-321/2020 | 2020-07-16 | Asia/India/Maharashtra/Raigad | Hematopathology Laboratory, ACTREC, TMC | Hematopathology Laboratory, ACTREC, TMC | Hematopathology Laboratory et al |
| EPI_ISL_699971 | India/MH-ACTREC-322/2020 | 2020-07-17 | Asia/India/Maharashtra/Thane | Hematopathology Laboratory, ACTREC, TMC | Hematopathology Laboratory, ACTREC, TMC | Hematopathology Laboratory et al |
| EPI_ISL_699972 | India/MH-ACTREC-323/2020 | 2020-07-17 | Asia/India/Maharashtra/Raigad | Hematopathology Laboratory, ACTREC, TMC | Hematopathology Laboratory, ACTREC, TMC | Hematopathology Laboratory et al |
| EPI_ISL_699973 | India/MH-ACTREC-324/2020 | 2020-07-17 | Asia/India/Maharashtra/Raigad | Hematopathology Laboratory, ACTREC, TMC | Hematopathology Laboratory, ACTREC, TMC | Hematopathology Laboratory et al |
| EPI_ISL_699974 | India/MH-ACTREC-325/2020 | 2020-07-17 | Asia/India/Maharashtra/Raigad | Hematopathology Laboratory, ACTREC, TMC | Hematopathology Laboratory, ACTREC, TMC | Hematopathology Laboratory et al |
| EPI_ISL_699975 | India/MH-ACTREC-326/2020 | 2020-07-17 | Asia/India/Maharashtra/Raigad | Hematopathology Laboratory, ACTREC, TMC | Hematopathology Laboratory, ACTREC, TMC | Hematopathology Laboratory et al |
| EPI_ISL_699977 | India/MH-ACTREC-328/2020 | 2020-07-18 | Asia/India/Maharashtra/Raigad | Hematopathology Laboratory, ACTREC, TMC | Hematopathology Laboratory, ACTREC, TMC | Hematopathology Laboratory et al |
| EPI_ISL_699978 | India/MH-ACTREC-329/2020 | 2020-07-18 | Asia/India/Maharashtra/Raigad | Hematopathology Laboratory, ACTREC, TMC | Hematopathology Laboratory, ACTREC, TMC | Hematopathology Laboratory et al |
| EPI_ISL_699979 | India/MH-ACTREC-330/2020 | 2020-07-19 | Asia/India/Maharashtra/Thane | Hematopathology Laboratory, ACTREC, TMC | Hematopathology Laboratory, ACTREC, TMC | Hematopathology Laboratory et al |

| | | | | | | |
|---|---|---|---|---|---|---|
| EPI_ISL_699980 | India/MH-ACTREC-331/2020 | 2020-07-19 | Asia/India/Maharashtra/Raigad | Hematopathology Laboratory, ACTREC, TMC | Hematopathology Laboratory, ACTREC, TMC | Hematopathology Laboratory et al |
| EPI_ISL_699981 | India/MH-ACTREC-332/2020 | 2020-07-19 | Asia/India/Maharashtra/Raigad | Hematopathology Laboratory, ACTREC, TMC | Hematopathology Laboratory, ACTREC, TMC | Hematopathology Laboratory et al |
| EPI_ISL_699982 | India/MH-ACTREC-333/2020 | 2020-07-19 | Asia/India/Maharashtra/Thane | Hematopathology Laboratory, ACTREC, TMC | Hematopathology Laboratory, ACTREC, TMC | Hematopathology Laboratory et al |
| EPI_ISL_699983 | India/MH-ACTREC-334/2020 | 2020-07-19 | Asia/India/Maharashtra/Raigad | Hematopathology Laboratory, ACTREC, TMC | Hematopathology Laboratory, ACTREC, TMC | Hematopathology Laboratory et al |
| EPI_ISL_699984 | India/MH-ACTREC-335/2020 | 2020-07-19 | Asia/India/Maharashtra/Raigad | Hematopathology Laboratory, ACTREC, TMC | Hematopathology Laboratory, ACTREC, TMC | Hematopathology Laboratory et al |
| EPI_ISL_699985 | India/MH-ACTREC-336/2020 | 2020-07-19 | Asia/India/Maharashtra/Raigad | Hematopathology Laboratory, ACTREC, TMC | Hematopathology Laboratory, ACTREC, TMC | Hematopathology Laboratory et al |
| EPI_ISL_699986 | India/MH-ACTREC-337/2020 | 2020-07-19 | Asia/India/Maharashtra/Raigad | Hematopathology Laboratory, ACTREC, TMC | Hematopathology Laboratory, ACTREC, TMC | Hematopathology Laboratory et al |
| EPI_ISL_699987 | India/MH-ACTREC-338/2020 | 2020-07-20 | Asia/India/Maharashtra/Raigad | Hematopathology Laboratory, ACTREC, TMC | Hematopathology Laboratory, ACTREC, TMC | Hematopathology Laboratory et al |
| EPI_ISL_699988 | India/MH-ACTREC-339/2020 | 2020-07-20 | Asia/India/Maharashtra/Raigad | Hematopathology Laboratory, ACTREC, TMC | Hematopathology Laboratory, ACTREC, TMC | Hematopathology Laboratory et al |
| EPI_ISL_699989 | India/MH-ACTREC-340/2020 | 2020-07-20 | Asia/India/Maharashtra/Raigad | Hematopathology Laboratory, ACTREC, TMC | Hematopathology Laboratory, ACTREC, TMC | Hematopathology Laboratory et al |
| EPI_ISL_699990 | India/MH-ACTREC-341/2020 | 2020-07-20 | Asia/India/Maharashtra/Raigad | Hematopathology Laboratory, ACTREC, TMC | Hematopathology Laboratory, ACTREC, TMC | Hematopathology Laboratory et al |
| EPI_ISL_699991 | India/MH-ACTREC-342/2020 | 2020-07-20 | Asia/India/Maharashtra/Raigad | Hematopathology Laboratory, ACTREC, TMC | Hematopathology Laboratory, ACTREC, TMC | Hematopathology Laboratory et al |
| EPI_ISL_699992 | India/MH-ACTREC-343/2020 | 2020-07-20 | Asia/India/Maharashtra/Raigad | Hematopathology Laboratory, ACTREC, TMC | Hematopathology Laboratory, ACTREC, TMC | Hematopathology Laboratory et al |

| | | | | | | |
|---|---|---|---|---|---|---|
| EPI_ISL_699993 | India/MH-ACTREC-344/2020 | 2020-07-21 | Asia/India/Maharashtra/Raigad | Hematopathology Laboratory, ACTREC, TMC | Hematopathology Laboratory, ACTREC, TMC | Hematopathology Laboratory et al |
| EPI_ISL_699994 | India/MH-ACTREC-345/2020 | 2020-07-22 | Asia/India/Maharashtra/Raigad | Hematopathology Laboratory, ACTREC, TMC | Hematopathology Laboratory, ACTREC, TMC | Hematopathology Laboratory et al |
| EPI_ISL_699995 | India/MH-ACTREC-346/2020 | 2020-07-22 | Asia/India/Maharashtra/Mumbai | Hematopathology Laboratory, ACTREC, TMC | Hematopathology Laboratory, ACTREC, TMC | Hematopathology Laboratory et al |
| EPI_ISL_699997 | India/MH-ACTREC-348/2020 | 2020-07-22 | Asia/India/Maharashtra/Palghar | Hematopathology Laboratory, ACTREC, TMC | Hematopathology Laboratory, ACTREC, TMC | Hematopathology Laboratory et al |
| EPI_ISL_699998 | India/MH-ACTREC-349/2020 | 2020-07-22 | Asia/India/Maharashtra/Raigad | Hematopathology Laboratory, ACTREC, TMC | Hematopathology Laboratory, ACTREC, TMC | Hematopathology Laboratory et al |
| EPI_ISL_699999 | India/MH-ACTREC-350/2020 | 2020-07-22 | Asia/India/Maharashtra/Raigad | Hematopathology Laboratory, ACTREC, TMC | Hematopathology Laboratory, ACTREC, TMC | Hematopathology Laboratory et al |
| EPI_ISL_700000 | India/MH-ACTREC-351/2020 | 2020-07-22 | Asia/India/Maharashtra/Raigad | Hematopathology Laboratory, ACTREC, TMC | Hematopathology Laboratory, ACTREC, TMC | Hematopathology Laboratory et al |
| EPI_ISL_700002 | India/MH-ACTREC-353/2020 | 2020-07-22 | Asia/India/Maharashtra/Raigad | Hematopathology Laboratory, ACTREC, TMC | Hematopathology Laboratory, ACTREC, TMC | Hematopathology Laboratory et al |
| EPI_ISL_700003 | India/MH-ACTREC-354/2020 | 2020-07-22 | Asia/India/Maharashtra/Raigad | Hematopathology Laboratory, ACTREC, TMC | Hematopathology Laboratory, ACTREC, TMC | Hematopathology Laboratory et al |
| EPI_ISL_700004 | India/MH-ACTREC-355/2020 | 2020-07-22 | Asia/India/Maharashtra/Raigad | Hematopathology Laboratory, ACTREC, TMC | Hematopathology Laboratory, ACTREC, TMC | Hematopathology Laboratory et al |
| EPI_ISL_700005 | India/MH-ACTREC-356/2020 | 2020-07-23 | Asia/India/Maharashtra/Raigad | Hematopathology Laboratory, ACTREC, TMC | Hematopathology Laboratory, ACTREC, TMC | Hematopathology Laboratory et al |
| EPI_ISL_700006 | India/MH-ACTREC-357/2020 | 2020-07-23 | Asia/India/Maharashtra/Raigad | Hematopathology Laboratory, ACTREC, TMC | Hematopathology Laboratory, ACTREC, TMC | Hematopathology Laboratory et al |
| EPI_ISL_700007 | India/MH-ACTREC-358/2020 | 2020-07-23 | Asia/India/Maharashtra/Raigad | Hematopathology Laboratory, ACTREC, TMC | Hematopathology Laboratory, ACTREC, TMC | Hematopathology Laboratory et al |

| | | | | | | |
|---|---|---|---|---|---|---|
| EPI_ISL_700008 | India/MH-ACTREC-359/2020 | 2020-07-23 | Asia/India/Maharashtra/Raigad | Hematopathology Laboratory, ACTREC, TMC | Hematopathology Laboratory, ACTREC, TMC | Hematopathology Laboratory et al |
| EPI_ISL_700009 | India/MH-ACTREC-360/2020 | 2020-07-23 | Asia/India/Maharashtra/Raigad | Hematopathology Laboratory, ACTREC, TMC | Hematopathology Laboratory, ACTREC, TMC | Hematopathology Laboratory et al |
| EPI_ISL_700011 | India/MH-ACTREC-362/2020 | 2020-07-23 | Asia/India/Maharashtra/Raigad | Hematopathology Laboratory, ACTREC, TMC | Hematopathology Laboratory, ACTREC, TMC | Hematopathology Laboratory et al |
| EPI_ISL_700012 | India/MH-ACTREC-363/2020 | 2020-07-23 | Asia/India/Maharashtra/Raigad | Hematopathology Laboratory, ACTREC, TMC | Hematopathology Laboratory, ACTREC, TMC | Hematopathology Laboratory et al |
| EPI_ISL_700013 | India/MH-ACTREC-364/2020 | 2020-07-23 | Asia/India/Maharashtra/Raigad | Hematopathology Laboratory, ACTREC, TMC | Hematopathology Laboratory, ACTREC, TMC | Hematopathology Laboratory et al |
| EPI_ISL_700014 | India/MH-ACTREC-365/2020 | 2020-07-24 | Asia/India/Maharashtra/Thane | Hematopathology Laboratory, ACTREC, TMC | Hematopathology Laboratory, ACTREC, TMC | Hematopathology Laboratory et al |
| EPI_ISL_700015 | India/MH-ACTREC-366/2020 | 2020-07-24 | Asia/India/Maharashtra/Mumbai Subur | Hematopathology Laboratory, ACTREC, TMC | Hematopathology Laboratory, ACTREC, TMC | Hematopathology Laboratory et al |
| EPI_ISL_700017 | India/MH-ACTREC-368/2020 | 2020-07-24 | Asia/India/Maharashtra/Raigad | Hematopathology Laboratory, ACTREC, TMC | Hematopathology Laboratory, ACTREC, TMC | Hematopathology Laboratory et al |
| EPI_ISL_700018 | India/MH-ACTREC-369/2020 | 2020-07-24 | Asia/India/Maharashtra/Raigad | Hematopathology Laboratory, ACTREC, TMC | Hematopathology Laboratory, ACTREC, TMC | Hematopathology Laboratory et al |
| EPI_ISL_700019 | India/MH-ACTREC-370/2020 | 2020-07-24 | Asia/India/Maharashtra/Raigad | Hematopathology Laboratory, ACTREC, TMC | Hematopathology Laboratory, ACTREC, TMC | Hematopathology Laboratory et al |
| EPI_ISL_700020 | India/MH-ACTREC-371/2020 | 2020-07-24 | Asia/India/Maharashtra/Raigad | Hematopathology Laboratory, ACTREC, TMC | Hematopathology Laboratory, ACTREC, TMC | Hematopathology Laboratory et al |
| EPI_ISL_700021 | India/MH-ACTREC-372/2020 | 2020-07-24 | Asia/India/Maharashtra/Raigad | Hematopathology Laboratory, ACTREC, TMC | Hematopathology Laboratory, ACTREC, TMC | Hematopathology Laboratory et al |
| EPI_ISL_700022 | India/MH-ACTREC-373/2020 | 2020-07-24 | Asia/India/Maharashtra/Raigad | Hematopathology Laboratory, ACTREC, TMC | Hematopathology Laboratory, ACTREC, TMC | Hematopathology Laboratory et al |

| | | | | | | |
|---|---|---|---|---|---|---|
| EPI_ISL_700023 | India/MH-ACTREC-374/2020 | 2020-07-24 | Asia/India/Maharashtra/Raigad | Hematopathology Laboratory, ACTREC, TMC | Hematopathology Laboratory, ACTREC, TMC | Hematopathology Laboratory et al |
| EPI_ISL_700024 | India/MH-ACTREC-375/2020 | 2020-07-24 | Asia/India/Maharashtra/Raigad | Hematopathology Laboratory, ACTREC, TMC | Hematopathology Laboratory, ACTREC, TMC | Hematopathology Laboratory et al |
| EPI_ISL_700025 | India/MH-ACTREC-376/2020 | 2020-07-24 | Asia/India/Maharashtra/Raigad | Hematopathology Laboratory, ACTREC, TMC | Hematopathology Laboratory, ACTREC, TMC | Hematopathology Laboratory et al |
| EPI_ISL_700027 | India/MH-ACTREC-378/2020 | 2020-07-24 | Asia/India/Maharashtra/Raigad | Hematopathology Laboratory, ACTREC, TMC | Hematopathology Laboratory, ACTREC, TMC | Hematopathology Laboratory et al |
| EPI_ISL_700028 | India/MH-ACTREC-379/2020 | 2020-07-24 | Asia/India/Maharashtra/Raigad | Hematopathology Laboratory, ACTREC, TMC | Hematopathology Laboratory, ACTREC, TMC | Hematopathology Laboratory et al |
| EPI_ISL_700029 | India/MH-ACTREC-380/2020 | 2020-07-25 | Asia/India/Maharashtra/Raigad | Hematopathology Laboratory, ACTREC, TMC | Hematopathology Laboratory, ACTREC, TMC | Hematopathology Laboratory et al |
| EPI_ISL_700031 | India/MH-ACTREC-382/2020 | 2020-07-25 | Asia/India/Maharashtra/Raigad | Hematopathology Laboratory, ACTREC, TMC | Hematopathology Laboratory, ACTREC, TMC | Hematopathology Laboratory et al |
| EPI_ISL_700032 | India/MH-ACTREC-383/2020 | 2020-07-27 | Asia/India/Maharashtra/Mumbai | Hematopathology Laboratory, ACTREC, TMC | Hematopathology Laboratory, ACTREC, TMC | Hematopathology Laboratory et al |
| EPI_ISL_700033 | India/MH-ACTREC-384/2020 | 2020-07-27 | Asia/India/Maharashtra/Raigad | Hematopathology Laboratory, ACTREC, TMC | Hematopathology Laboratory, ACTREC, TMC | Hematopathology Laboratory et al |
| EPI_ISL_700034 | India/MH-ACTREC-385/2020 | 2020-07-27 | Asia/India/Maharashtra/Raigad | Hematopathology Laboratory, ACTREC, TMC | Hematopathology Laboratory, ACTREC, TMC | Hematopathology Laboratory et al |
| EPI_ISL_700035 | India/MH-ACTREC-386/2020 | 2020-07-27 | Asia/India/Maharashtra/Raigad | Hematopathology Laboratory, ACTREC, TMC | Hematopathology Laboratory, ACTREC, TMC | Hematopathology Laboratory et al |
| EPI_ISL_700036 | India/MH-ACTREC-387/2020 | 2020-07-28 | Asia/India/Maharashtra/Raigad | Hematopathology Laboratory, ACTREC, TMC | Hematopathology Laboratory, ACTREC, TMC | Hematopathology Laboratory et al |
| EPI_ISL_700037 | India/MH-ACTREC-388/2020 | 2020-07-28 | Asia/India/Maharashtra/Thane | Hematopathology Laboratory, ACTREC, TMC | Hematopathology Laboratory, ACTREC, TMC | Hematopathology Laboratory et al |

| | | | | | | |
|---|---|---|---|---|---|---|
| EPI_ISL_700038 | India/MH-ACTREC-389/2020 | 2020-07-28 | Asia/India/Maharashtra/Raigad | Hematopathology Laboratory, ACTREC, TMC | Hematopathology Laboratory, ACTREC, TMC | Hematopathology Laboratory et al |
| EPI_ISL_700039 | India/MH-ACTREC-390/2020 | 2020-07-29 | Asia/India/Maharashtra/Thane | Hematopathology Laboratory, ACTREC, TMC | Hematopathology Laboratory, ACTREC, TMC | Hematopathology Laboratory et al |
| EPI_ISL_700040 | India/MH-ACTREC-391/2020 | 2020-07-29 | Asia/India/Maharashtra/Thane | Hematopathology Laboratory, ACTREC, TMC | Hematopathology Laboratory, ACTREC, TMC | Hematopathology Laboratory et al |
| EPI_ISL_700041 | India/MH-ACTREC-392/2020 | 2020-07-29 | Asia/India/Maharashtra/Thane | Hematopathology Laboratory, ACTREC, TMC | Hematopathology Laboratory, ACTREC, TMC | Hematopathology Laboratory et al |
| EPI_ISL_700042 | India/MH-ACTREC-393/2020 | 2020-07-29 | Asia/India/Maharashtra/Mumbai | Hematopathology Laboratory, ACTREC, TMC | Hematopathology Laboratory, ACTREC, TMC | Hematopathology Laboratory et al |
| EPI_ISL_700043 | India/MH-ACTREC-394/2020 | 2020-07-29 | Asia/India/Maharashtra/Raigad | Hematopathology Laboratory, ACTREC, TMC | Hematopathology Laboratory, ACTREC, TMC | Hematopathology Laboratory et al |
| EPI_ISL_700044 | India/MH-ACTREC-395/2020 | 2020-07-29 | Asia/India/Maharashtra/Raigad | Hematopathology Laboratory, ACTREC, TMC | Hematopathology Laboratory, ACTREC, TMC | Hematopathology Laboratory et al |
| EPI_ISL_700045 | India/MH-ACTREC-396/2020 | 2020-07-29 | Asia/India/Maharashtra/Raigad | Hematopathology Laboratory, ACTREC, TMC | Hematopathology Laboratory, ACTREC, TMC | Hematopathology Laboratory et al |
| EPI_ISL_700046 | India/MH-ACTREC-397/2020 | 2020-07-29 | Asia/India/Maharashtra/Raigad | Hematopathology Laboratory, ACTREC, TMC | Hematopathology Laboratory, ACTREC, TMC | Hematopathology Laboratory et al |
| EPI_ISL_700047 | India/MH-ACTREC-398/2020 | 2020-07-29 | Asia/India/Maharashtra/Raigad | Hematopathology Laboratory, ACTREC, TMC | Hematopathology Laboratory, ACTREC, TMC | Hematopathology Laboratory et al |
| EPI_ISL_700048 | India/MH-ACTREC-399/2020 | 2020-07-29 | Asia/India/Maharashtra/Raigad | Hematopathology Laboratory, ACTREC, TMC | Hematopathology Laboratory, ACTREC, TMC | Hematopathology Laboratory et al |
| EPI_ISL_700049 | India/MH-ACTREC-400/2020 | 2020-07-29 | Asia/India/Maharashtra/Raigad | Hematopathology Laboratory, ACTREC, TMC | Hematopathology Laboratory, ACTREC, TMC | Hematopathology Laboratory et al |
| EPI_ISL_700050 | India/MH-ACTREC-401/2020 | 2020-07-29 | Asia/India/Maharashtra/Raigad | Hematopathology Laboratory, ACTREC, TMC | Hematopathology Laboratory, ACTREC, TMC | Hematopathology Laboratory et al |

| | | | | | | |
|---|---|---|---|---|---|---|
| EPI_ISL_700051 | India/MH-ACTREC-402/2020 | 2020-07-29 | Asia/India/Maharashtra/Raigad | Hematopathology Laboratory, ACTREC, TMC | Hematopathology Laboratory, ACTREC, TMC | Hematopathology Laboratory et al |
| EPI_ISL_700052 | India/MH-ACTREC-403/2020 | 2020-07-29 | Asia/India/Maharashtra/Raigad | Hematopathology Laboratory, ACTREC, TMC | Hematopathology Laboratory, ACTREC, TMC | Hematopathology Laboratory et al |
| EPI_ISL_700053 | India/MH-ACTREC-404/2020 | 2020-07-29 | Asia/India/Maharashtra/Raigad | Hematopathology Laboratory, ACTREC, TMC | Hematopathology Laboratory, ACTREC, TMC | Hematopathology Laboratory et al |
| EPI_ISL_700054 | India/MH-ACTREC-405/2020 | 2020-07-29 | Asia/India/Maharashtra/Raigad | Hematopathology Laboratory, ACTREC, TMC | Hematopathology Laboratory, ACTREC, TMC | Hematopathology Laboratory et al |
| EPI_ISL_700055 | India/MH-ACTREC-406/2020 | 2020-07-29 | Asia/India/Maharashtra/Raigad | Hematopathology Laboratory, ACTREC, TMC | Hematopathology Laboratory, ACTREC, TMC | Hematopathology Laboratory et al |
| EPI_ISL_700056 | India/MH-ACTREC-407/2020 | 2020-07-29 | Asia/India/Maharashtra/Raigad | Hematopathology Laboratory, ACTREC, TMC | Hematopathology Laboratory, ACTREC, TMC | Hematopathology Laboratory et al |
| EPI_ISL_700058 | India/MH-ACTREC-409/2020 | 2020-07-29 | Asia/India/Maharashtra/Raigad | Hematopathology Laboratory, ACTREC, TMC | Hematopathology Laboratory, ACTREC, TMC | Hematopathology Laboratory et al |
| EPI_ISL_700059 | India/MH-ACTREC-410/2020 | 2020-07-29 | Asia/India/Maharashtra/Raigad | Hematopathology Laboratory, ACTREC, TMC | Hematopathology Laboratory, ACTREC, TMC | Hematopathology Laboratory et al |
| EPI_ISL_700060 | India/MH-ACTREC-411/2020 | 2020-07-29 | Asia/India/Maharashtra/Raigad | Hematopathology Laboratory, ACTREC, TMC | Hematopathology Laboratory, ACTREC, TMC | Hematopathology Laboratory et al |
| EPI_ISL_700061 | India/MH-ACTREC-412/2020 | 2020-07-29 | Asia/India/Maharashtra/Raigad | Hematopathology Laboratory, ACTREC, TMC | Hematopathology Laboratory, ACTREC, TMC | Hematopathology Laboratory et al |
| EPI_ISL_700062 | India/MH-ACTREC-413/2020 | 2020-07-29 | Asia/India/Maharashtra/Raigad | Hematopathology Laboratory, ACTREC, TMC | Hematopathology Laboratory, ACTREC, TMC | Hematopathology Laboratory et al |
| EPI_ISL_700064 | India/MH-ACTREC-415/2020 | 2020-07-30 | Asia/India/Maharashtra/Raigad | Hematopathology Laboratory, ACTREC, TMC | Hematopathology Laboratory, ACTREC, TMC | Hematopathology Laboratory et al |
| EPI_ISL_700065 | India/MH-ACTREC-416/2020 | 2020-07-30 | Asia/India/Maharashtra/Raigad | Hematopathology Laboratory, ACTREC, TMC | Hematopathology Laboratory, ACTREC, TMC | Hematopathology Laboratory et al |

| | | | | | | |
|---|---|---|---|---|---|---|
| EPI_ISL_700067 | India/MH-ACTREC-418/2020 | 2020-07-30 | Asia/India/Maharashtra/Raigad | Hematopathology Laboratory, ACTREC, TMC | Hematopathology Laboratory, ACTREC, TMC | Hematopathology Laboratory et al |
| EPI_ISL_700068 | India/MH-ACTREC-419/2020 | 2020-07-30 | Asia/India/Maharashtra/Raigad | Hematopathology Laboratory, ACTREC, TMC | Hematopathology Laboratory, ACTREC, TMC | Hematopathology Laboratory et al |
| EPI_ISL_700069 | India/MH-ACTREC-420/2020 | 2020-07-30 | Asia/India/Maharashtra/Raigad | Hematopathology Laboratory, ACTREC, TMC | Hematopathology Laboratory, ACTREC, TMC | Hematopathology Laboratory et al |
| EPI_ISL_700070 | India/UP-ACTREC-421/2020 | 2020-07-31 | Asia/India/Uttar Pradesh/Gorakhpur | Hematopathology Laboratory, ACTREC, TMC | Hematopathology Laboratory, ACTREC, TMC | Hematopathology Laboratory et al |
| EPI_ISL_700071 | India/MH-ACTREC-422/2020 | 2020-07-31 | Asia/India/Maharashtra/Raigad | Hematopathology Laboratory, ACTREC, TMC | Hematopathology Laboratory, ACTREC, TMC | Hematopathology Laboratory et al |
| EPI_ISL_700072 | India/MH-ACTREC-423/2020 | 2020-07-31 | Asia/India/Maharashtra/Raigad | Hematopathology Laboratory, ACTREC, TMC | Hematopathology Laboratory, ACTREC, TMC | Hematopathology Laboratory et al |
| EPI_ISL_700073 | India/MH-ACTREC-424/2020 | 2020-07-31 | Asia/India/Maharashtra/Raigad | Hematopathology Laboratory, ACTREC, TMC | Hematopathology Laboratory, ACTREC, TMC | Hematopathology Laboratory et al |
| EPI_ISL_700074 | India/MH-ACTREC-425/2020 | 2020-07-31 | Asia/India/Maharashtra/Raigad | Hematopathology Laboratory, ACTREC, TMC | Hematopathology Laboratory, ACTREC, TMC | Hematopathology Laboratory et al |
| EPI_ISL_700075 | India/MH-ACTREC-426/2020 | 2020-07-31 | Asia/India/Maharashtra/Raigad | Hematopathology Laboratory, ACTREC, TMC | Hematopathology Laboratory, ACTREC, TMC | Hematopathology Laboratory et al |
| EPI_ISL_700076 | India/MH-ACTREC-427/2020 | 2020-07-31 | Asia/India/Maharashtra/Raigad | Hematopathology Laboratory, ACTREC, TMC | Hematopathology Laboratory, ACTREC, TMC | Hematopathology Laboratory et al |
| EPI_ISL_700077 | India/MH-ACTREC-428/2020 | 2020-07-31 | Asia/India/Maharashtra/Raigad | Hematopathology Laboratory, ACTREC, TMC | Hematopathology Laboratory, ACTREC, TMC | Hematopathology Laboratory et al |
| EPI_ISL_700078 | India/MH-ACTREC-429/2020 | 2020-07-31 | Asia/India/Maharashtra/Thane | Hematopathology Laboratory, ACTREC, TMC | Hematopathology Laboratory, ACTREC, TMC | Hematopathology Laboratory et al |
| EPI_ISL_700079 | India/MH-ACTREC-430/2020 | 2020-08-04 | Asia/India/Maharashtra/Raigad | Hematopathology Laboratory, ACTREC, TMC | Hematopathology Laboratory, ACTREC, TMC | Hematopathology Laboratory et al |

| | | | | | | |
|---|---|---|---|---|---|---|
| EPI_ISL_700080 | India/MH-ACTREC-431/2020 | 2020-08-04 | Asia/India/Maharashtra/Raigad | Hematopathology Laboratory, ACTREC, TMC | Hematopathology Laboratory, ACTREC, TMC | Hematopathology Laboratory et al |
| EPI_ISL_700081 | India/MH-ACTREC-432/2020 | 2020-08-04 | Asia/India/Maharashtra/Raigad | Hematopathology Laboratory, ACTREC, TMC | Hematopathology Laboratory, ACTREC, TMC | Hematopathology Laboratory et al |
| EPI_ISL_700082 | India/MH-ACTREC-433/2020 | 2020-08-04 | Asia/India/Maharashtra/Thane | Hematopathology Laboratory, ACTREC, TMC | Hematopathology Laboratory, ACTREC, TMC | Hematopathology Laboratory et al |
| EPI_ISL_700083 | India/MH-ACTREC-434/2020 | 2020-08-04 | Asia/India/Maharashtra/Raigad | Hematopathology Laboratory, ACTREC, TMC | Hematopathology Laboratory, ACTREC, TMC | Hematopathology Laboratory et al |
| EPI_ISL_700084 | India/MH-ACTREC-435/2020 | 2020-08-04 | Asia/India/Maharashtra/Raigad | Hematopathology Laboratory, ACTREC, TMC | Hematopathology Laboratory, ACTREC, TMC | Hematopathology Laboratory et al |
| EPI_ISL_700085 | India/MH-ACTREC-436/2020 | 2020-08-05 | Asia/India/Maharashtra/Mumbai | Hematopathology Laboratory, ACTREC, TMC | Hematopathology Laboratory, ACTREC, TMC | Hematopathology Laboratory et al |
| EPI_ISL_700086 | India/MH-ACTREC-437/2020 | 2020-08-05 | Asia/India/Maharashtra/Raigad | Hematopathology Laboratory, ACTREC, TMC | Hematopathology Laboratory, ACTREC, TMC | Hematopathology Laboratory et al |
| EPI_ISL_700087 | India/MH-ACTREC-438/2020 | 2020-08-05 | Asia/India/Maharashtra/Raigad | Hematopathology Laboratory, ACTREC, TMC | Hematopathology Laboratory, ACTREC, TMC | Hematopathology Laboratory et al |
| EPI_ISL_700088 | India/MH-ACTREC-439/2020 | 2020-08-05 | Asia/India/Maharashtra/Raigad | Hematopathology Laboratory, ACTREC, TMC | Hematopathology Laboratory, ACTREC, TMC | Hematopathology Laboratory et al |
| EPI_ISL_700089 | India/MH-ACTREC-440/2020 | 2020-08-05 | Asia/India/Maharashtra/Raigad | Hematopathology Laboratory, ACTREC, TMC | Hematopathology Laboratory, ACTREC, TMC | Hematopathology Laboratory et al |
| EPI_ISL_700090 | India/MH-ACTREC-441/2020 | 2020-08-05 | Asia/India/Maharashtra/Raigad | Hematopathology Laboratory, ACTREC, TMC | Hematopathology Laboratory, ACTREC, TMC | Hematopathology Laboratory et al |
| EPI_ISL_700091 | India/MH-ACTREC-442/2020 | 2020-08-05 | Asia/India/Maharashtra/Raigad | Hematopathology Laboratory, ACTREC, TMC | Hematopathology Laboratory, ACTREC, TMC | Hematopathology Laboratory et al |
| EPI_ISL_700092 | India/MH-ACTREC-444/2020 | 2020-08-05 | Asia/India/Maharashtra/Raigad | Hematopathology Laboratory, ACTREC, TMC | Hematopathology Laboratory, ACTREC, TMC | Hematopathology Laboratory et al |

| | | | | | | |
|---|---|---|---|---|---|---|
| EPI_ISL_700093 | India/MH-ACTREC-445/2020 | 2020-08-05 | Asia/India/Maharashtra/Raigad | Hematopathology Laboratory, ACTREC, TMC | Hematopathology Laboratory, ACTREC, TMC | Hematopathology Laboratory et al |
| EPI_ISL_700094 | India/MH-ACTREC-446/2020 | 2020-08-05 | Asia/India/Maharashtra/Thane | Hematopathology Laboratory, ACTREC, TMC | Hematopathology Laboratory, ACTREC, TMC | Hematopathology Laboratory et al |
| EPI_ISL_700095 | India/MH-ACTREC-447/2020 | 2020-08-07 | Asia/India/Maharashtra/Raigad | Hematopathology Laboratory, ACTREC, TMC | Hematopathology Laboratory, ACTREC, TMC | Hematopathology Laboratory et al |
| EPI_ISL_700097 | India/MH-ACTREC-449/2020 | 2020-08-07 | Asia/India/Maharashtra/Raigad | Hematopathology Laboratory, ACTREC, TMC | Hematopathology Laboratory, ACTREC, TMC | Hematopathology Laboratory et al |
| EPI_ISL_700098 | India/MH-ACTREC-450/2020 | 2020-08-07 | Asia/India/Maharashtra/Raigad | Hematopathology Laboratory, ACTREC, TMC | Hematopathology Laboratory, ACTREC, TMC | Hematopathology Laboratory et al |
| EPI_ISL_700099 | India/MH-ACTREC-451/2020 | 2020-08-07 | Asia/India/Maharashtra/Raigad | Hematopathology Laboratory, ACTREC, TMC | Hematopathology Laboratory, ACTREC, TMC | Hematopathology Laboratory et al |
| EPI_ISL_700100 | India/MH-ACTREC-452/2020 | 2020-08-08 | Asia/India/Maharashtra/Raigad | Hematopathology Laboratory, ACTREC, TMC | Hematopathology Laboratory, ACTREC, TMC | Hematopathology Laboratory et al |
| EPI_ISL_700101 | India/MH-ACTREC-453/2020 | 2020-08-08 | Asia/India/Maharashtra/Raigad | Hematopathology Laboratory, ACTREC, TMC | Hematopathology Laboratory, ACTREC, TMC | Hematopathology Laboratory et al |
| EPI_ISL_700102 | India/MH-ACTREC-454/2020 | 2020-08-09 | Asia/India/Maharashtra/Raigad | Hematopathology Laboratory, ACTREC, TMC | Hematopathology Laboratory, ACTREC, TMC | Hematopathology Laboratory et al |
| EPI_ISL_700103 | India/MH-ACTREC-455/2020 | 2020-08-09 | Asia/India/Maharashtra/Raigad | Hematopathology Laboratory, ACTREC, TMC | Hematopathology Laboratory, ACTREC, TMC | Hematopathology Laboratory et al |
| EPI_ISL_700104 | India/MH-ACTREC-456/2020 | 2020-08-09 | Asia/India/Maharashtra/Raigad | Hematopathology Laboratory, ACTREC, TMC | Hematopathology Laboratory, ACTREC, TMC | Hematopathology Laboratory et al |
| EPI_ISL_700105 | India/MH-ACTREC-457/2020 | 2020-08-09 | Asia/India/Maharashtra/Raigad | Hematopathology Laboratory, ACTREC, TMC | Hematopathology Laboratory, ACTREC, TMC | Hematopathology Laboratory et al |
| EPI_ISL_700106 | India/MH-ACTREC-458/2020 | 2020-08-09 | Asia/India/Maharashtra/Raigad | Hematopathology Laboratory, ACTREC, TMC | Hematopathology Laboratory, ACTREC, TMC | Hematopathology Laboratory et al |

| | | | | | | |
|---|---|---|---|---|---|---|
| EPI_ISL_700107 | India/MH-ACTREC-459/2020 | 2020-08-10 | Asia/India/Maharashtra/Raigad | Hematopathology Laboratory, ACTREC, TMC | Hematopathology Laboratory, ACTREC, TMC | Hematopathology Laboratory et al |
| EPI_ISL_700108 | India/MH-ACTREC-460/2020 | 2020-08-10 | Asia/India/Maharashtra/Raigad | Hematopathology Laboratory, ACTREC, TMC | Hematopathology Laboratory, ACTREC, TMC | Hematopathology Laboratory et al |
| EPI_ISL_700109 | India/MH-ACTREC-461/2020 | 2020-08-10 | Asia/India/Maharashtra/Thane | Hematopathology Laboratory, ACTREC, TMC | Hematopathology Laboratory, ACTREC, TMC | Hematopathology Laboratory et al |
| EPI_ISL_700110 | India/MH-ACTREC-462/2020 | 2020-08-10 | Asia/India/Maharashtra/Palghar | Hematopathology Laboratory, ACTREC, TMC | Hematopathology Laboratory, ACTREC, TMC | Hematopathology Laboratory et al |
| EPI_ISL_700111 | India/MH-ACTREC-463/2020 | 2020-08-10 | Asia/India/Maharashtra/Raigad | Hematopathology Laboratory, ACTREC, TMC | Hematopathology Laboratory, ACTREC, TMC | Hematopathology Laboratory et al |
| EPI_ISL_700112 | India/MH-ACTREC-464/2020 | 2020-08-10 | Asia/India/Maharashtra/Raigad | Hematopathology Laboratory, ACTREC, TMC | Hematopathology Laboratory, ACTREC, TMC | Hematopathology Laboratory et al |
| EPI_ISL_700113 | India/MH-ACTREC-465/2020 | 2020-08-10 | Asia/India/Maharashtra/Raigad | Hematopathology Laboratory, ACTREC, TMC | Hematopathology Laboratory, ACTREC, TMC | Hematopathology Laboratory et al |
| EPI_ISL_700114 | India/MH-ACTREC-466/2020 | 2020-08-10 | Asia/India/Maharashtra/Raigad | Hematopathology Laboratory, ACTREC, TMC | Hematopathology Laboratory, ACTREC, TMC | Hematopathology Laboratory et al |
| EPI_ISL_700115 | India/MH-ACTREC-467/2020 | 2020-08-10 | Asia/India/Maharashtra/Raigad | Hematopathology Laboratory, ACTREC, TMC | Hematopathology Laboratory, ACTREC, TMC | Hematopathology Laboratory et al |
| EPI_ISL_700116 | India/MH-ACTREC-468/2020 | 2020-08-10 | Asia/India/Maharashtra/Raigad | Hematopathology Laboratory, ACTREC, TMC | Hematopathology Laboratory, ACTREC, TMC | Hematopathology Laboratory et al |
| EPI_ISL_700117 | India/MH-ACTREC-469/2020 | 2020-08-11 | Asia/India/Maharashtra/Raigad | Hematopathology Laboratory, ACTREC, TMC | Hematopathology Laboratory, ACTREC, TMC | Hematopathology Laboratory et al |
| EPI_ISL_700118 | India/MH-ACTREC-470/2020 | 2020-08-11 | Asia/India/Maharashtra/Raigad | Hematopathology Laboratory, ACTREC, TMC | Hematopathology Laboratory, ACTREC, TMC | Hematopathology Laboratory et al |
| EPI_ISL_700119 | India/MH-ACTREC-471/2020 | 2020-08-11 | Asia/India/Maharashtra/Raigad | Hematopathology Laboratory, ACTREC, TMC | Hematopathology Laboratory, ACTREC, TMC | Hematopathology Laboratory et al |

| | | | | | | |
|---|---|---|---|---|---|---|
| EPI_ISL_700120 | India/MH-ACTREC-472/2020 | 2020-08-11 | Asia/India/Maharashtra/Raigad | Hematopathology Laboratory, ACTREC, TMC | Hematopathology Laboratory, ACTREC, TMC | Hematopathology Laboratory et al |
| EPI_ISL_700121 | India/MH-ACTREC-473/2020 | 2020-08-11 | Asia/India/Maharashtra/Raigad | Hematopathology Laboratory, ACTREC, TMC | Hematopathology Laboratory, ACTREC, TMC | Hematopathology Laboratory et al |
| EPI_ISL_700122 | India/MH-ACTREC-474/2020 | 2020-08-11 | Asia/India/Maharashtra/Raigad | Hematopathology Laboratory, ACTREC, TMC | Hematopathology Laboratory, ACTREC, TMC | Hematopathology Laboratory et al |
| EPI_ISL_700123 | India/MH-ACTREC-475/2020 | 2020-08-11 | Asia/India/Maharashtra/Raigad | Hematopathology Laboratory, ACTREC, TMC | Hematopathology Laboratory, ACTREC, TMC | Hematopathology Laboratory et al |
| EPI_ISL_700124 | India/MH-ACTREC-476/2020 | 2020-08-11 | Asia/India/Maharashtra/Raigad | Hematopathology Laboratory, ACTREC, TMC | Hematopathology Laboratory, ACTREC, TMC | Hematopathology Laboratory et al |
| EPI_ISL_700125 | India/MH-ACTREC-477/2020 | 2020-08-11 | Asia/India/Maharashtra/Mumbai | Hematopathology Laboratory, ACTREC, TMC | Hematopathology Laboratory, ACTREC, TMC | Hematopathology Laboratory et al |
| EPI_ISL_700126 | India/MH-ACTREC-478/2020 | 2020-08-11 | Asia/India/Maharashtra/Raigad | Hematopathology Laboratory, ACTREC, TMC | Hematopathology Laboratory, ACTREC, TMC | Hematopathology Laboratory et al |
| EPI_ISL_700127 | India/MH-ACTREC-479/2020 | 2020-08-11 | Asia/India/Maharashtra/Raigad | Hematopathology Laboratory, ACTREC, TMC | Hematopathology Laboratory, ACTREC, TMC | Hematopathology Laboratory et al |
| EPI_ISL_700128 | India/MH-ACTREC-480/2020 | 2020-08-12 | Asia/India/Maharashtra/Raigad | Hematopathology Laboratory, ACTREC, TMC | Hematopathology Laboratory, ACTREC, TMC | Hematopathology Laboratory et al |
| EPI_ISL_700129 | India/MH-ACTREC-481/2020 | 2020-08-13 | Asia/India/Maharashtra/Raigad | Hematopathology Laboratory, ACTREC, TMC | Hematopathology Laboratory, ACTREC, TMC | Hematopathology Laboratory et al |
| EPI_ISL_700131 | India/MH-ACTREC-483/2020 | 2020-08-13 | Asia/India/Maharashtra/Raigad | Hematopathology Laboratory, ACTREC, TMC | Hematopathology Laboratory, ACTREC, TMC | Hematopathology Laboratory et al |
| EPI_ISL_700132 | India/MH-ACTREC-484/2020 | 2020-08-13 | Asia/India/Maharashtra/Raigad | Hematopathology Laboratory, ACTREC, TMC | Hematopathology Laboratory, ACTREC, TMC | Hematopathology Laboratory et al |
| EPI_ISL_700133 | India/MH-ACTREC-485/2020 | 2020-08-13 | Asia/India/Maharashtra/Thane | Hematopathology Laboratory, ACTREC, TMC | Hematopathology Laboratory, ACTREC, TMC | Hematopathology Laboratory et al |

| | | | | | | |
|---|---|---|---|---|---|---|
| EPI_ISL_700135 | India/MH-ACTREC-487/2020 | 2020-08-13 | Asia/India/Maharashtra/Raigad | Hematopathology Laboratory, ACTREC, TMC | Hematopathology Laboratory, ACTREC, TMC | Hematopathology Laboratory et al |
| EPI_ISL_700136 | India/MH-ACTREC-488/2020 | 2020-08-13 | Asia/India/Maharashtra/Raigad | Hematopathology Laboratory, ACTREC, TMC | Hematopathology Laboratory, ACTREC, TMC | Hematopathology Laboratory et al |
| EPI_ISL_700137 | India/MH-ACTREC-489/2020 | 2020-08-13 | Asia/India/Maharashtra/Raigad | Hematopathology Laboratory, ACTREC, TMC | Hematopathology Laboratory, ACTREC, TMC | Hematopathology Laboratory et al |
| EPI_ISL_700138 | India/MH-ACTREC-490/2020 | 2020-08-13 | Asia/India/Maharashtra/Raigad | Hematopathology Laboratory, ACTREC, TMC | Hematopathology Laboratory, ACTREC, TMC | Hematopathology Laboratory et al |
| EPI_ISL_700139 | India/MH-ACTREC-491/2020 | 2020-08-14 | Asia/India/Maharashtra/Raigad | Hematopathology Laboratory, ACTREC, TMC | Hematopathology Laboratory, ACTREC, TMC | Hematopathology Laboratory et al |
| EPI_ISL_700141 | India/MH-ACTREC-493/2020 | 2020-08-14 | Asia/India/Maharashtra/Mumbai | Hematopathology Laboratory, ACTREC, TMC | Hematopathology Laboratory, ACTREC, TMC | Hematopathology Laboratory et al |
| EPI_ISL_700142 | India/MH-ACTREC-494/2020 | 2020-08-14 | Asia/India/Maharashtra/Raigad | Hematopathology Laboratory, ACTREC, TMC | Hematopathology Laboratory, ACTREC, TMC | Hematopathology Laboratory et al |
| EPI_ISL_700144 | India/MH-ACTREC-496/2020 | 2020-08-14 | Asia/India/Maharashtra/Raigad | Hematopathology Laboratory, ACTREC, TMC | Hematopathology Laboratory, ACTREC, TMC | Hematopathology Laboratory et al |
| EPI_ISL_700145 | India/MH-ACTREC-497/2020 | 2020-08-14 | Asia/India/Maharashtra/Raigad | Hematopathology Laboratory, ACTREC, TMC | Hematopathology Laboratory, ACTREC, TMC | Hematopathology Laboratory et al |
| EPI_ISL_700146 | India/MH-ACTREC-498/2020 | 2020-08-14 | Asia/India/Maharashtra/Raigad | Hematopathology Laboratory, ACTREC, TMC | Hematopathology Laboratory, ACTREC, TMC | Hematopathology Laboratory et al |
| EPI_ISL_700147 | India/MH-ACTREC-499/2020 | 2020-08-14 | Asia/India/Maharashtra/Raigad | Hematopathology Laboratory, ACTREC, TMC | Hematopathology Laboratory, ACTREC, TMC | Hematopathology Laboratory et al |
| EPI_ISL_700148 | India/MH-ACTREC-500/2020 | 2020-08-14 | Asia/India/Maharashtra/Raigad | Hematopathology Laboratory, ACTREC, TMC | Hematopathology Laboratory, ACTREC, TMC | Hematopathology Laboratory et al |
| EPI_ISL_700149 | India/MH-ACTREC-501/2020 | 2020-08-15 | Asia/India/Maharashtra/Raigad | Hematopathology Laboratory, ACTREC, TMC | Hematopathology Laboratory, ACTREC, TMC | Hematopathology Laboratory et al |

| | | | | | | |
|---|---|---|---|---|---|---|
| EPI_ISL_700150 | India/MH-ACTREC-502/2020 | 2020-08-15 | Asia/India/Maharashtra/Raigad | Hematopathology Laboratory, ACTREC, TMC | Hematopathology Laboratory, ACTREC, TMC | Hematopathology Laboratory et al |
| EPI_ISL_700151 | India/MH-ACTREC-503/2020 | 2020-08-15 | Asia/India/Maharashtra/Raigad | Hematopathology Laboratory, ACTREC, TMC | Hematopathology Laboratory, ACTREC, TMC | Hematopathology Laboratory et al |
| EPI_ISL_700152 | India/MH-ACTREC-504/2020 | 2020-08-17 | Asia/India/Maharashtra/Mumbai | Hematopathology Laboratory, ACTREC, TMC | Hematopathology Laboratory, ACTREC, TMC | Hematopathology Laboratory et al |
| EPI_ISL_700153 | India/MH-ACTREC-505/2020 | 2020-08-17 | Asia/India/Maharashtra/Palghar | Hematopathology Laboratory, ACTREC, TMC | Hematopathology Laboratory, ACTREC, TMC | Hematopathology Laboratory et al |
| EPI_ISL_700154 | India/MH-ACTREC-506/2020 | 2020-08-17 | Asia/India/Maharashtra/Raigad | Hematopathology Laboratory, ACTREC, TMC | Hematopathology Laboratory, ACTREC, TMC | Hematopathology Laboratory et al |
| EPI_ISL_700155 | India/MH-ACTREC-507/2020 | 2020-08-17 | Asia/India/Maharashtra/Raigad | Hematopathology Laboratory, ACTREC, TMC | Hematopathology Laboratory, ACTREC, TMC | Hematopathology Laboratory et al |
| EPI_ISL_700156 | India/MH-ACTREC-508/2020 | 2020-08-18 | Asia/India/Maharashtra/Thane | Hematopathology Laboratory, ACTREC, TMC | Hematopathology Laboratory, ACTREC, TMC | Hematopathology Laboratory et al |
| EPI_ISL_700158 | India/MH-ACTREC-510/2020 | 2020-08-19 | Asia/India/Maharashtra/Raigad | Hematopathology Laboratory, ACTREC, TMC | Hematopathology Laboratory, ACTREC, TMC | Hematopathology Laboratory et al |
| EPI_ISL_700159 | India/MH-ACTREC-511/2020 | 2020-08-19 | Asia/India/Maharashtra/Raigad | Hematopathology Laboratory, ACTREC, TMC | Hematopathology Laboratory, ACTREC, TMC | Hematopathology Laboratory et al |
| EPI_ISL_700160 | India/MH-ACTREC-513/2020 | 2020-08-19 | Asia/India/Maharashtra/Raigad | Hematopathology Laboratory, ACTREC, TMC | Hematopathology Laboratory, ACTREC, TMC | Hematopathology Laboratory et al |
| EPI_ISL_700161 | India/MH-ACTREC-514/2020 | 2020-08-19 | Asia/India/Maharashtra/Raigad | Hematopathology Laboratory, ACTREC, TMC | Hematopathology Laboratory, ACTREC, TMC | Hematopathology Laboratory et al |
| EPI_ISL_700162 | India/MH-ACTREC-515/2020 | 2020-08-19 | Asia/India/Maharashtra/Raigad | Hematopathology Laboratory, ACTREC, TMC | Hematopathology Laboratory, ACTREC, TMC | Hematopathology Laboratory et al |
| EPI_ISL_700163 | India/MH-ACTREC-516/2020 | 2020-08-19 | Asia/India/Maharashtra/Raigad | Hematopathology Laboratory, ACTREC, TMC | Hematopathology Laboratory, ACTREC, TMC | Hematopathology Laboratory et al |

| | | | | | | |
|---|---|---|---|---|---|---|
| EPI_ISL_700164 | India/MH-ACTREC-518/2020 | 2020-08-21 | Asia/India/Maharashtra/Raigad | Hematopathology Laboratory, ACTREC, TMC | Hematopathology Laboratory, ACTREC, TMC | Hematopathology Laboratory et al |
| EPI_ISL_700165 | India/MH-ACTREC-519/2020 | 2020-08-21 | Asia/India/Maharashtra/Raigad | Hematopathology Laboratory, ACTREC, TMC | Hematopathology Laboratory, ACTREC, TMC | Hematopathology Laboratory et al |
| EPI_ISL_700166 | India/MH-ACTREC-520/2020 | 2020-08-21 | Asia/India/Maharashtra/Navimumbai | Hematopathology Laboratory, ACTREC, TMC | Hematopathology Laboratory, ACTREC, TMC | Hematopathology Laboratory et al |
| EPI_ISL_700167 | India/MH-ACTREC-521/2020 | 2020-08-21 | Asia/India/Maharashtra/Raigad | Hematopathology Laboratory, ACTREC, TMC | Hematopathology Laboratory, ACTREC, TMC | Hematopathology Laboratory et al |
| EPI_ISL_700168 | India/MH-ACTREC-522/2020 | 2020-08-21 | Asia/India/Maharashtra/Raigad | Hematopathology Laboratory, ACTREC, TMC | Hematopathology Laboratory, ACTREC, TMC | Hematopathology Laboratory et al |
| EPI_ISL_700169 | India/MH-ACTREC-523/2020 | 2020-08-21 | Asia/India/Maharashtra/Raigad | Hematopathology Laboratory, ACTREC, TMC | Hematopathology Laboratory, ACTREC, TMC | Hematopathology Laboratory et al |
| EPI_ISL_700170 | India/MH-ACTREC-524/2020 | 2020-08-22 | Asia/India/Maharashtra/Raigad | Hematopathology Laboratory, ACTREC, TMC | Hematopathology Laboratory, ACTREC, TMC | Hematopathology Laboratory et al |
| EPI_ISL_700171 | India/MH-ACTREC-525/2020 | 2020-08-22 | Asia/India/Maharashtra/Raigad | Hematopathology Laboratory, ACTREC, TMC | Hematopathology Laboratory, ACTREC, TMC | Hematopathology Laboratory et al |
| EPI_ISL_700172 | India/MH-ACTREC-526/2020 | 2020-08-22 | Asia/India/Maharashtra/Mumbai | Hematopathology Laboratory, ACTREC, TMC | Hematopathology Laboratory, ACTREC, TMC | Hematopathology Laboratory et al |
| EPI_ISL_700173 | India/MH-ACTREC-527/2020 | 2020-08-22 | Asia/India/Maharashtra/Raigad | Hematopathology Laboratory, ACTREC, TMC | Hematopathology Laboratory, ACTREC, TMC | Hematopathology Laboratory et al |
| EPI_ISL_700174 | India/MH-ACTREC-528/2020 | 2020-08-24 | Asia/India/Maharashtra/Raigad | Hematopathology Laboratory, ACTREC, TMC | Hematopathology Laboratory, ACTREC, TMC | Hematopathology Laboratory et al |
| EPI_ISL_700175 | India/MH-ACTREC-529/2020 | 2020-08-25 | Asia/India/Maharashtra/Raigad | Hematopathology Laboratory, ACTREC, TMC | Hematopathology Laboratory, ACTREC, TMC | Hematopathology Laboratory et al |
| EPI_ISL_700176 | India/MH-ACTREC-530/2020 | 2020-08-25 | Asia/India/Maharashtra/Raigad | Hematopathology Laboratory, ACTREC, TMC | Hematopathology Laboratory, ACTREC, TMC | Hematopathology Laboratory et al |

| | | | | | | |
|---|---|---|---|---|---|---|
| EPI_ISL_700177 | India/MH-ACTREC-531/2020 | 2020-08-25 | Asia/India/Maharashtra/Raigad | Hematopathology Laboratory, ACTREC, TMC | Hematopathology Laboratory, ACTREC, TMC | Hematopathology Laboratory et al |
| EPI_ISL_700178 | India/MH-ACTREC-532/2020 | 2020-08-25 | Asia/India/Maharashtra/Raigad | Hematopathology Laboratory, ACTREC, TMC | Hematopathology Laboratory, ACTREC, TMC | Hematopathology Laboratory et al |
| EPI_ISL_700179 | India/MH-ACTREC-533/2020 | 2020-08-25 | Asia/India/Maharashtra/Raigad | Hematopathology Laboratory, ACTREC, TMC | Hematopathology Laboratory, ACTREC, TMC | Hematopathology Laboratory et al |
| EPI_ISL_700180 | India/MH-ACTREC-534/2020 | 2020-08-25 | Asia/India/Maharashtra/Raigad | Hematopathology Laboratory, ACTREC, TMC | Hematopathology Laboratory, ACTREC, TMC | Hematopathology Laboratory et al |
| EPI_ISL_700181 | India/MH-ACTREC-535/2020 | 2020-08-25 | Asia/India/Maharashtra/Raigad | Hematopathology Laboratory, ACTREC, TMC | Hematopathology Laboratory, ACTREC, TMC | Hematopathology Laboratory et al |
| EPI_ISL_700182 | India/MH-ACTREC-536/2020 | 2020-08-25 | Asia/India/Maharashtra/Raigad | Hematopathology Laboratory, ACTREC, TMC | Hematopathology Laboratory, ACTREC, TMC | Hematopathology Laboratory et al |
| EPI_ISL_700183 | India/MH-ACTREC-537/2020 | 2020-08-25 | Asia/India/Maharashtra/Mumbai | Hematopathology Laboratory, ACTREC, TMC | Hematopathology Laboratory, ACTREC, TMC | Hematopathology Laboratory et al |
| EPI_ISL_700184 | India/MH-ACTREC-538/2020 | 2020-08-25 | Asia/India/Maharashtra/Raigad | Hematopathology Laboratory, ACTREC, TMC | Hematopathology Laboratory, ACTREC, TMC | Hematopathology Laboratory et al |
| EPI_ISL_700185 | India/MH-ACTREC-539/2020 | 2020-08-25 | Asia/India/Maharashtra/Thane | Hematopathology Laboratory, ACTREC, TMC | Hematopathology Laboratory, ACTREC, TMC | Hematopathology Laboratory et al |
| EPI_ISL_700186 | India/MH-ACTREC-540/2020 | 2020-08-26 | Asia/India/Maharashtra/Mumbai | Hematopathology Laboratory, ACTREC, TMC | Hematopathology Laboratory, ACTREC, TMC | Hematopathology Laboratory et al |
| EPI_ISL_700187 | India/MH-ACTREC-541/2020 | 2020-08-26 | Asia/India/Maharashtra/Raigad | Hematopathology Laboratory, ACTREC, TMC | Hematopathology Laboratory, ACTREC, TMC | Hematopathology Laboratory et al |
| EPI_ISL_700188 | India/MH-ACTREC-542/2020 | 2020-08-26 | Asia/India/Maharashtra/Raigad | Hematopathology Laboratory, ACTREC, TMC | Hematopathology Laboratory, ACTREC, TMC | Hematopathology Laboratory et al |
| EPI_ISL_700189 | India/MH-ACTREC-543/2020 | 2020-08-26 | Asia/India/Maharashtra/Raigad | Hematopathology Laboratory, ACTREC, TMC | Hematopathology Laboratory, ACTREC, TMC | Hematopathology Laboratory et al |

| | | | | | | |
|---|---|---|---|---|---|---|
| EPI_ISL_700190 | India/MH-ACTREC-544/2020 | 2020-08-26 | Asia/India/Maharashtra/Raigad | Hematopathology Laboratory, ACTREC, TMC | Hematopathology Laboratory, ACTREC, TMC | Hematopathology Laboratory et al |
| EPI_ISL_700191 | India/MH-ACTREC-545/2020 | 2020-08-27 | Asia/India/Maharashtra/Raigad | Hematopathology Laboratory, ACTREC, TMC | Hematopathology Laboratory, ACTREC, TMC | Hematopathology Laboratory et al |
| EPI_ISL_700192 | India/MH-ACTREC-547/2020 | 2020-08-27 | Asia/India/Maharashtra/Raigad | Hematopathology Laboratory, ACTREC, TMC | Hematopathology Laboratory, ACTREC, TMC | Hematopathology Laboratory et al |
| EPI_ISL_700193 | India/MH-ACTREC-548/2020 | 2020-08-28 | Asia/India/Maharashtra/Raigad | Hematopathology Laboratory, ACTREC, TMC | Hematopathology Laboratory, ACTREC, TMC | Hematopathology Laboratory et al |
| EPI_ISL_700194 | India/MH-ACTREC-549/2020 | 2020-08-28 | Asia/India/Maharashtra/Raigad | Hematopathology Laboratory, ACTREC, TMC | Hematopathology Laboratory, ACTREC, TMC | Hematopathology Laboratory et al |
| EPI_ISL_700195 | India/MH-ACTREC-550/2020 | 2020-08-28 | Asia/India/Maharashtra/Raigad | Hematopathology Laboratory, ACTREC, TMC | Hematopathology Laboratory, ACTREC, TMC | Hematopathology Laboratory et al |
| EPI_ISL_700196 | India/MH-ACTREC-551/2020 | 2020-08-28 | Asia/India/Maharashtra/Raigad | Hematopathology Laboratory, ACTREC, TMC | Hematopathology Laboratory, ACTREC, TMC | Hematopathology Laboratory et al |
| EPI_ISL_700197 | India/MH-ACTREC-552/2020 | 2020-08-28 | Asia/India/Maharashtra/Raigad | Hematopathology Laboratory, ACTREC, TMC | Hematopathology Laboratory, ACTREC, TMC | Hematopathology Laboratory et al |
| EPI_ISL_700198 | India/MH-ACTREC-553/2020 | 2020-08-28 | Asia/India/Maharashtra/Raigad | Hematopathology Laboratory, ACTREC, TMC | Hematopathology Laboratory, ACTREC, TMC | Hematopathology Laboratory et al |
| EPI_ISL_700199 | India/MH-ACTREC-554/2020 | 2020-08-28 | Asia/India/Maharashtra/Pune | Hematopathology Laboratory, ACTREC, TMC | Hematopathology Laboratory, ACTREC, TMC | Hematopathology Laboratory et al |
| EPI_ISL_700200 | India/MH-ACTREC-555/2020 | 2020-08-28 | Asia/India/Maharashtra/Raigad | Hematopathology Laboratory, ACTREC, TMC | Hematopathology Laboratory, ACTREC, TMC | Hematopathology Laboratory et al |
| EPI_ISL_700201 | India/MH-ACTREC-556/2020 | 2020-08-28 | Asia/India/Maharashtra/Raigad | Hematopathology Laboratory, ACTREC, TMC | Hematopathology Laboratory, ACTREC, TMC | Hematopathology Laboratory et al |
| EPI_ISL_700202 | India/MH-ACTREC-557/2020 | 2020-08-29 | Asia/India/Maharashtra/Raigad | Hematopathology Laboratory, ACTREC, TMC | Hematopathology Laboratory, ACTREC, TMC | Hematopathology Laboratory et al |

| | | | | | | |
|---|---|---|---|---|---|---|
| EPI_ISL_700203 | India/MH-ACTREC-558/2020 | 2020-08-29 | Asia/India/Maharashtra/Mumbai | Hematopathology Laboratory, ACTREC, TMC | Hematopathology Laboratory, ACTREC, TMC | Hematopathology Laboratory et al |
| EPI_ISL_700204 | India/MH-ACTREC-559/2020 | 2020-08-29 | Asia/India/Maharashtra/Thane | Hematopathology Laboratory, ACTREC, TMC | Hematopathology Laboratory, ACTREC, TMC | Hematopathology Laboratory et al |
| EPI_ISL_700205 | India/MH-ACTREC-560/2020 | 2020-08-29 | Asia/India/Maharashtra/Raigad | Hematopathology Laboratory, ACTREC, TMC | Hematopathology Laboratory, ACTREC, TMC | Hematopathology Laboratory et al |
| EPI_ISL_700206 | India/MH-ACTREC-561/2020 | 2020-08-29 | Asia/India/Maharashtra/Raigad | Hematopathology Laboratory, ACTREC, TMC | Hematopathology Laboratory, ACTREC, TMC | Hematopathology Laboratory et al |
| EPI_ISL_700207 | India/MH-ACTREC-562/2020 | 2020-08-31 | Asia/India/Maharashtra/Thane | Hematopathology Laboratory, ACTREC, TMC | Hematopathology Laboratory, ACTREC, TMC | Hematopathology Laboratory et al |
| EPI_ISL_700208 | India/MH-ACTREC-563/2020 | 2020-08-31 | Asia/India/Maharashtra/Thane | Hematopathology Laboratory, ACTREC, TMC | Hematopathology Laboratory, ACTREC, TMC | Hematopathology Laboratory et al |
| EPI_ISL_700209 | India/MH-ACTREC-564/2020 | 2020-08-31 | Asia/India/Maharashtra/Raigad | Hematopathology Laboratory, ACTREC, TMC | Hematopathology Laboratory, ACTREC, TMC | Hematopathology Laboratory et al |
| EPI_ISL_700210 | India/MH-ACTREC-565/2020 | 2020-08-31 | Asia/India/Maharashtra/Raigad | Hematopathology Laboratory, ACTREC, TMC | Hematopathology Laboratory, ACTREC, TMC | Hematopathology Laboratory et al |
| EPI_ISL_700211 | India/MH-ACTREC-567/2020 | 2020-08-31 | Asia/India/Maharashtra/Raigad | Hematopathology Laboratory, ACTREC, TMC | Hematopathology Laboratory, ACTREC, TMC | Hematopathology Laboratory et al |
| EPI_ISL_700212 | India/MH-ACTREC-568/2020 | 2020-08-31 | Asia/India/Maharashtra/Raigad | Hematopathology Laboratory, ACTREC, TMC | Hematopathology Laboratory, ACTREC, TMC | Hematopathology Laboratory et al |
| EPI_ISL_700213 | India/MH-ACTREC-569/2020 | 2020-08-31 | Asia/India/Maharashtra/Raigad | Hematopathology Laboratory, ACTREC, TMC | Hematopathology Laboratory, ACTREC, TMC | Hematopathology Laboratory et al |
| EPI_ISL_700214 | India/MH-ACTREC-570/2020 | 2020-08-31 | Asia/India/Maharashtra/Raigad | Hematopathology Laboratory, ACTREC, TMC | Hematopathology Laboratory, ACTREC, TMC | Hematopathology Laboratory et al |
| EPI_ISL_700215 | India/MH-ACTREC-571/2020 | 2020-08-31 | Asia/India/Maharashtra/Thane | Hematopathology Laboratory, ACTREC, TMC | Hematopathology Laboratory, ACTREC, TMC | Hematopathology Laboratory et al |

| | | | | | | |
|---|---|---|---|---|---|---|
| EPI_ISL_700216 | India/MH-ACTREC-572/2020 | 2020-08-31 | Asia/India/Maharashtra/Thane | Hematopathology Laboratory, ACTREC, TMC | Hematopathology Laboratory, ACTREC, TMC | Hematopathology Laboratory et al |
| EPI_ISL_700217 | India/MH-ACTREC-573/2020 | 2020-08-31 | Asia/India/Maharashtra/Raigad | Hematopathology Laboratory, ACTREC, TMC | Hematopathology Laboratory, ACTREC, TMC | Hematopathology Laboratory et al |
| EPI_ISL_700218 | India/MH-ACTREC-574/2020 | 2020-09-01 | Asia/India/Maharashtra/Raigad | Hematopathology Laboratory, ACTREC, TMC | Hematopathology Laboratory, ACTREC, TMC | Hematopathology Laboratory et al |
| EPI_ISL_700219 | India/MH-ACTREC-575/2020 | 2020-09-01 | Asia/India/Maharashtra/Raigad | Hematopathology Laboratory, ACTREC, TMC | Hematopathology Laboratory, ACTREC, TMC | Hematopathology Laboratory et al |
| EPI_ISL_700220 | India/MH-ACTREC-576/2020 | 2020-09-02 | Asia/India/Maharashtra/Mumbai Subur | Hematopathology Laboratory, ACTREC, TMC | Hematopathology Laboratory, ACTREC, TMC | Hematopathology Laboratory et al |
| EPI_ISL_700221 | India/MH-ACTREC-578/2020 | 2020-09-02 | Asia/India/Maharashtra/Raigad | Hematopathology Laboratory, ACTREC, TMC | Hematopathology Laboratory, ACTREC, TMC | Hematopathology Laboratory et al |
| EPI_ISL_700222 | India/MH-ACTREC-580/2020 | 2020-09-02 | Asia/India/Maharashtra/Raigad | Hematopathology Laboratory, ACTREC, TMC | Hematopathology Laboratory, ACTREC, TMC | Hematopathology Laboratory et al |
| EPI_ISL_700223 | India/MH-ACTREC-582/2020 | 2020-09-02 | Asia/India/Maharashtra/Raigad | Hematopathology Laboratory, ACTREC, TMC | Hematopathology Laboratory, ACTREC, TMC | Hematopathology Laboratory et al |
| EPI_ISL_700224 | India/MH-ACTREC-585/2020 | 2020-09-02 | Asia/India/Maharashtra/Raigad | Hematopathology Laboratory, ACTREC, TMC | Hematopathology Laboratory, ACTREC, TMC | Hematopathology Laboratory et al |
| EPI_ISL_700225 | India/MH-ACTREC-586/2020 | 2020-09-03 | Asia/India/Maharashtra/Thane | Hematopathology Laboratory, ACTREC, TMC | Hematopathology Laboratory, ACTREC, TMC | Hematopathology Laboratory et al |
| EPI_ISL_700226 | India/MH-ACTREC-587/2020 | 2020-09-03 | Asia/India/Maharashtra/Thane | Hematopathology Laboratory, ACTREC, TMC | Hematopathology Laboratory, ACTREC, TMC | Hematopathology Laboratory et al |
| EPI_ISL_700227 | India/MH-ACTREC-588/2020 | 2020-09-04 | Asia/India/Maharashtra/Raigad | Hematopathology Laboratory, ACTREC, TMC | Hematopathology Laboratory, ACTREC, TMC | Hematopathology Laboratory et al |
| EPI_ISL_700228 | India/MH-ACTREC-590/2020 | 2020-09-04 | Asia/India/Maharashtra/Raigad | Hematopathology Laboratory, ACTREC, TMC | Hematopathology Laboratory, ACTREC, TMC | Hematopathology Laboratory et al |

| | | | | | | |
|---|---|---|---|---|---|---|
| EPI_ISL_700229 | India/MH-ACTREC-591/2020 | 2020-09-04 | Asia/India/Maharashtra/Raigad | Hematopathology Laboratory, ACTREC, TMC | Hematopathology Laboratory, ACTREC, TMC | Hematopathology Laboratory et al |
| EPI_ISL_700230 | India/MH-ACTREC-592/2020 | 2020-09-04 | Asia/India/Maharashtra/Raigad | Hematopathology Laboratory, ACTREC, TMC | Hematopathology Laboratory, ACTREC, TMC | Hematopathology Laboratory et al |
| EPI_ISL_700231 | India/MH-ACTREC-593/2020 | 2020-09-04 | Asia/India/Maharashtra/Thane | Hematopathology Laboratory, ACTREC, TMC | Hematopathology Laboratory, ACTREC, TMC | Hematopathology Laboratory et al |
| EPI_ISL_700232 | India/MH-ACTREC-594/2020 | 2020-09-04 | Asia/India/Maharashtra/Raigad | Hematopathology Laboratory, ACTREC, TMC | Hematopathology Laboratory, ACTREC, TMC | Hematopathology Laboratory et al |
| EPI_ISL_700233 | India/MH-ACTREC-595/2020 | 2020-09-05 | Asia/India/Maharashtra/Mumbai | Hematopathology Laboratory, ACTREC, TMC | Hematopathology Laboratory, ACTREC, TMC | Hematopathology Laboratory et al |
| EPI_ISL_700234 | India/MH-ACTREC-596/2020 | 2020-09-05 | Asia/India/Maharashtra/Raigad | Hematopathology Laboratory, ACTREC, TMC | Hematopathology Laboratory, ACTREC, TMC | Hematopathology Laboratory et al |
| EPI_ISL_700235 | India/MH-ACTREC-597/2020 | 2020-09-06 | Asia/India/Maharashtra/Raigad | Hematopathology Laboratory, ACTREC, TMC | Hematopathology Laboratory, ACTREC, TMC | Hematopathology Laboratory et al |
| EPI_ISL_700236 | India/MH-ACTREC-598/2020 | 2020-09-07 | Asia/India/Maharashtra/Raigad | Hematopathology Laboratory, ACTREC, TMC | Hematopathology Laboratory, ACTREC, TMC | Hematopathology Laboratory et al |
| EPI_ISL_700237 | India/MH-ACTREC-599/2020 | 2020-09-07 | Asia/India/Maharashtra/Raigad | Hematopathology Laboratory, ACTREC, TMC | Hematopathology Laboratory, ACTREC, TMC | Hematopathology Laboratory et al |
| EPI_ISL_700238 | India/MH-ACTREC-600/2020 | 2020-09-08 | Asia/India/Maharashtra/Raigad | Hematopathology Laboratory, ACTREC, TMC | Hematopathology Laboratory, ACTREC, TMC | Hematopathology Laboratory et al |
| EPI_ISL_700239 | India/MH-ACTREC-601/2020 | 2020-09-08 | Asia/India/Maharashtra/Raigad | Hematopathology Laboratory, ACTREC, TMC | Hematopathology Laboratory, ACTREC, TMC | Hematopathology Laboratory et al |
| EPI_ISL_700240 | India/MH-ACTREC-603/2020 | 2020-09-08 | Asia/India/Maharashtra/Raigad | Hematopathology Laboratory, ACTREC, TMC | Hematopathology Laboratory, ACTREC, TMC | Hematopathology Laboratory et al |
| EPI_ISL_700241 | India/MH-ACTREC-604/2020 | 2020-09-08 | Asia/India/Maharashtra/Mumbai | Hematopathology Laboratory, ACTREC, TMC | Hematopathology Laboratory, ACTREC, TMC | Hematopathology Laboratory et al |

| | | | | | | |
|---|---|---|---|---|---|---|
| EPI_ISL_700242 | India/MH-ACTREC-605/2020 | 2020-09-08 | Asia/India/Maharashtra/Raigad | Hematopathology Laboratory, ACTREC, TMC | Hematopathology Laboratory, ACTREC, TMC | Hematopathology Laboratory et al |
| EPI_ISL_700243 | India/MH-ACTREC-606/2020 | 2020-09-09 | Asia/India/Maharashtra/Raigad | Hematopathology Laboratory, ACTREC, TMC | Hematopathology Laboratory, ACTREC, TMC | Hematopathology Laboratory et al |
| EPI_ISL_700244 | India/MH-ACTREC-607/2020 | 2020-09-09 | Asia/India/Maharashtra/Raigad | Hematopathology Laboratory, ACTREC, TMC | Hematopathology Laboratory, ACTREC, TMC | Hematopathology Laboratory et al |
| EPI_ISL_700245 | India/MH-ACTREC-608/2020 | 2020-09-09 | Asia/India/Maharashtra/Raigad | Hematopathology Laboratory, ACTREC, TMC | Hematopathology Laboratory, ACTREC, TMC | Hematopathology Laboratory et al |
| EPI_ISL_700246 | India/MH-ACTREC-609/2020 | 2020-09-09 | Asia/India/Maharashtra/Raigad | Hematopathology Laboratory, ACTREC, TMC | Hematopathology Laboratory, ACTREC, TMC | Hematopathology Laboratory et al |
| EPI_ISL_700247 | India/MH-ACTREC-610/2020 | 2020-09-09 | Asia/India/Maharashtra/Raigad | Hematopathology Laboratory, ACTREC, TMC | Hematopathology Laboratory, ACTREC, TMC | Hematopathology Laboratory et al |
| EPI_ISL_700248 | India/MH-ACTREC-611/2020 | 2020-09-09 | Asia/India/Maharashtra/Raigad | Hematopathology Laboratory, ACTREC, TMC | Hematopathology Laboratory, ACTREC, TMC | Hematopathology Laboratory et al |
| EPI_ISL_700249 | India/MH-ACTREC-612/2020 | 2020-09-09 | Asia/India/Maharashtra/Raigad | Hematopathology Laboratory, ACTREC, TMC | Hematopathology Laboratory, ACTREC, TMC | Hematopathology Laboratory et al |
| EPI_ISL_700250 | India/MH-ACTREC-613/2020 | 2020-09-09 | Asia/India/Maharashtra/Raigad | Hematopathology Laboratory, ACTREC, TMC | Hematopathology Laboratory, ACTREC, TMC | Hematopathology Laboratory et al |
| EPI_ISL_700251 | India/MH-ACTREC-616/2020 | 2020-09-10 | Asia/India/Maharashtra/Mumbai | Hematopathology Laboratory, ACTREC, TMC | Hematopathology Laboratory, ACTREC, TMC | Hematopathology Laboratory et al |
| EPI_ISL_700252 | India/MH-ACTREC-617/2020 | 2020-09-10 | Asia/India/Maharashtra/Mumbai | Hematopathology Laboratory, ACTREC, TMC | Hematopathology Laboratory, ACTREC, TMC | Hematopathology Laboratory et al |
| EPI_ISL_700253 | India/MH-ACTREC-618/2020 | 2020-09-10 | Asia/India/Maharashtra/Raigad | Hematopathology Laboratory, ACTREC, TMC | Hematopathology Laboratory, ACTREC, TMC | Hematopathology Laboratory et al |
| EPI_ISL_700254 | India/MH-ACTREC-619/2020 | 2020-09-10 | Asia/India/Maharashtra/Mumbai | Hematopathology Laboratory, ACTREC, TMC | Hematopathology Laboratory, ACTREC, TMC | Hematopathology Laboratory et al |

| | | | | | | |
|---|---|---|---|---|---|---|
| EPI_ISL_700255 | India/MH-ACTREC-620/2020 | 2020-09-11 | Asia/India/Maharashtra/Raigad | Hematopathology Laboratory, ACTREC, TMC | Hematopathology Laboratory, ACTREC, TMC | Hematopathology Laboratory et al |
| EPI_ISL_700256 | India/MH-ACTREC-621/2020 | 2020-09-11 | Asia/India/Maharashtra/Raigad | Hematopathology Laboratory, ACTREC, TMC | Hematopathology Laboratory, ACTREC, TMC | Hematopathology Laboratory et al |
| EPI_ISL_700257 | India/MH-ACTREC-622/2020 | 2020-09-11 | Asia/India/Maharashtra/Raigad | Hematopathology Laboratory, ACTREC, TMC | Hematopathology Laboratory, ACTREC, TMC | Hematopathology Laboratory et al |
| EPI_ISL_700258 | India/MH-ACTREC-623/2020 | 2020-09-11 | Asia/India/Maharashtra/Raigad | Hematopathology Laboratory, ACTREC, TMC | Hematopathology Laboratory, ACTREC, TMC | Hematopathology Laboratory et al |
| EPI_ISL_700259 | India/MH-ACTREC-624/2020 | 2020-09-11 | Asia/India/Maharashtra/Raigad | Hematopathology Laboratory, ACTREC, TMC | Hematopathology Laboratory, ACTREC, TMC | Hematopathology Laboratory et al |
| EPI_ISL_700260 | India/MH-ACTREC-625/2020 | 2020-09-11 | Asia/India/Maharashtra/Raigad | Hematopathology Laboratory, ACTREC, TMC | Hematopathology Laboratory, ACTREC, TMC | Hematopathology Laboratory et al |
| EPI_ISL_700261 | India/MH-ACTREC-626/2020 | 2020-09-11 | Asia/India/Maharashtra/Raigad | Hematopathology Laboratory, ACTREC, TMC | Hematopathology Laboratory, ACTREC, TMC | Hematopathology Laboratory et al |
| EPI_ISL_700262 | India/MH-ACTREC-627/2020 | 2020-09-11 | Asia/India/Maharashtra/Raigad | Hematopathology Laboratory, ACTREC, TMC | Hematopathology Laboratory, ACTREC, TMC | Hematopathology Laboratory et al |
| EPI_ISL_700263 | India/MH-ACTREC-628/2020 | 2020-09-11 | Asia/India/Maharashtra/Raigad | Hematopathology Laboratory, ACTREC, TMC | Hematopathology Laboratory, ACTREC, TMC | Hematopathology Laboratory et al |
| EPI_ISL_700264 | India/MH-ACTREC-629/2020 | 2020-09-11 | Asia/India/Maharashtra/Thane | Hematopathology Laboratory, ACTREC, TMC | Hematopathology Laboratory, ACTREC, TMC | Hematopathology Laboratory et al |
| EPI_ISL_700265 | India/MH-ACTREC-632/2020 | 2020-09-12 | Asia/India/Maharashtra/Thane | Hematopathology Laboratory, ACTREC, TMC | Hematopathology Laboratory, ACTREC, TMC | Hematopathology Laboratory et al |
| EPI_ISL_700266 | India/MH-ACTREC-633/2020 | 2020-09-12 | Asia/India/Maharashtra/Thane | Hematopathology Laboratory, ACTREC, TMC | Hematopathology Laboratory, ACTREC, TMC | Hematopathology Laboratory et al |
| EPI_ISL_700267 | India/MH-ACTREC-634/2020 | 2020-09-12 | Asia/India/Maharashtra/Raigad | Hematopathology Laboratory, ACTREC, TMC | Hematopathology Laboratory, ACTREC, TMC | Hematopathology Laboratory et al |

| | | | | | | |
|---|---|---|---|---|---|---|
| EPI_ISL_700268 | India/MH-ACTREC-635/2020 | 2020-09-12 | Asia/India/Maharashtra/Raigad | Hematopathology Laboratory, ACTREC, TMC | Hematopathology Laboratory, ACTREC, TMC | Hematopathology Laboratory et al |
| EPI_ISL_700269 | India/MH-ACTREC-636/2020 | 2020-09-12 | Asia/India/Maharashtra/Raigad | Hematopathology Laboratory, ACTREC, TMC | Hematopathology Laboratory, ACTREC, TMC | Hematopathology Laboratory et al |
| EPI_ISL_700270 | India/MH-ACTREC-637/2020 | 2020-09-13 | Asia/India/Maharashtra/Raigad | Hematopathology Laboratory, ACTREC, TMC | Hematopathology Laboratory, ACTREC, TMC | Hematopathology Laboratory et al |
| EPI_ISL_700271 | India/MH-ACTREC-638/2020 | 2020-09-13 | Asia/India/Maharashtra/Thane | Hematopathology Laboratory, ACTREC, TMC | Hematopathology Laboratory, ACTREC, TMC | Hematopathology Laboratory et al |
| EPI_ISL_700272 | India/MH-ACTREC-639/2020 | 2020-09-13 | Asia/India/Maharashtra/Thane | Hematopathology Laboratory, ACTREC, TMC | Hematopathology Laboratory, ACTREC, TMC | Hematopathology Laboratory et al |
| EPI_ISL_700273 | India/MH-ACTREC-640/2020 | 2020-09-14 | Asia/India/Maharashtra/Raigad | Hematopathology Laboratory, ACTREC, TMC | Hematopathology Laboratory, ACTREC, TMC | Hematopathology Laboratory et al |
| EPI_ISL_700274 | India/MH-ACTREC-641/2020 | 2020-09-14 | Asia/India/Maharashtra/Raigad | Hematopathology Laboratory, ACTREC, TMC | Hematopathology Laboratory, ACTREC, TMC | Hematopathology Laboratory et al |
| EPI_ISL_700275 | India/MH-ACTREC-642/2020 | 2020-09-14 | Asia/India/Maharashtra/Raigad | Hematopathology Laboratory, ACTREC, TMC | Hematopathology Laboratory, ACTREC, TMC | Hematopathology Laboratory et al |
| EPI_ISL_700276 | India/MH-ACTREC-646/2020 | 2020-09-14 | Asia/India/Maharashtra/Raigad | Hematopathology Laboratory, ACTREC, TMC | Hematopathology Laboratory, ACTREC, TMC | Hematopathology Laboratory et al |
| EPI_ISL_700277 | India/MH-ACTREC-647/2020 | 2020-09-14 | Asia/India/Maharashtra/Raigad | Hematopathology Laboratory, ACTREC, TMC | Hematopathology Laboratory, ACTREC, TMC | Hematopathology Laboratory et al |
| EPI_ISL_700278 | India/MH-ACTREC-648/2020 | 2020-09-14 | Asia/India/Maharashtra/Raigad | Hematopathology Laboratory, ACTREC, TMC | Hematopathology Laboratory, ACTREC, TMC | Hematopathology Laboratory et al |
| EPI_ISL_700279 | India/MH-ACTREC-649/2020 | 2020-09-14 | Asia/India/Maharashtra/Raigad | Hematopathology Laboratory, ACTREC, TMC | Hematopathology Laboratory, ACTREC, TMC | Hematopathology Laboratory et al |
| EPI_ISL_700280 | India/MH-ACTREC-650/2020 | 2020-09-14 | Asia/India/Maharashtra/Thane | Hematopathology Laboratory, ACTREC, TMC | Hematopathology Laboratory, ACTREC, TMC | Hematopathology Laboratory et al |

| | | | | | | |
|---|---|---|---|---|---|---|
| EPI_ISL_700281 | India/MH-ACTREC-651/2020 | 2020-09-14 | Asia/India/Maharashtra/Raigad | Hematopathology Laboratory, ACTREC, TMC | Hematopathology Laboratory, ACTREC, TMC | Hematopathology Laboratory et al |
| EPI_ISL_700282 | India/MH-ACTREC-652/2020 | 2020-09-14 | Asia/India/Maharashtra/Raigad | Hematopathology Laboratory, ACTREC, TMC | Hematopathology Laboratory, ACTREC, TMC | Hematopathology Laboratory et al |
| EPI_ISL_700283 | India/MH-ACTREC-653/2020 | 2020-09-14 | Asia/India/Maharashtra/Raigad | Hematopathology Laboratory, ACTREC, TMC | Hematopathology Laboratory, ACTREC, TMC | Hematopathology Laboratory et al |
| EPI_ISL_700284 | India/MH-ACTREC-654/2020 | 2020-09-14 | Asia/India/Maharashtra/Raigad | Hematopathology Laboratory, ACTREC, TMC | Hematopathology Laboratory, ACTREC, TMC | Hematopathology Laboratory et al |
| EPI_ISL_700285 | India/MH-ACTREC-655/2020 | 2020-09-14 | Asia/India/Maharashtra/Raigad | Hematopathology Laboratory, ACTREC, TMC | Hematopathology Laboratory, ACTREC, TMC | Hematopathology Laboratory et al |
| EPI_ISL_700286 | India/MH-ACTREC-656/2020 | 2020-09-14 | Asia/India/Maharashtra/Raigad | Hematopathology Laboratory, ACTREC, TMC | Hematopathology Laboratory, ACTREC, TMC | Hematopathology Laboratory et al |
| EPI_ISL_700287 | India/MH-ACTREC-657/2020 | 2020-09-14 | Asia/India/Maharashtra/Raigad | Hematopathology Laboratory, ACTREC, TMC | Hematopathology Laboratory, ACTREC, TMC | Hematopathology Laboratory et al |
| EPI_ISL_700288 | India/MH-ACTREC-658/2020 | 2020-09-15 | Asia/India/Maharashtra/Raigad | Hematopathology Laboratory, ACTREC, TMC | Hematopathology Laboratory, ACTREC, TMC | Hematopathology Laboratory et al |
| EPI_ISL_700289 | India/MH-ACTREC-659/2020 | 2020-09-15 | Asia/India/Maharashtra/Mumbai | Hematopathology Laboratory, ACTREC, TMC | Hematopathology Laboratory, ACTREC, TMC | Hematopathology Laboratory et al |
| EPI_ISL_700290 | India/MH-ACTREC-660/2020 | 2020-09-15 | Asia/India/Maharashtra/Raigad | Hematopathology Laboratory, ACTREC, TMC | Hematopathology Laboratory, ACTREC, TMC | Hematopathology Laboratory et al |
| EPI_ISL_700291 | India/MH-ACTREC-662/2020 | 2020-09-15 | Asia/India/Maharashtra/Thane | Hematopathology Laboratory, ACTREC, TMC | Hematopathology Laboratory, ACTREC, TMC | Hematopathology Laboratory et al |
| EPI_ISL_700292 | India/MH-ACTREC-663/2020 | 2020-09-16 | Asia/India/Maharashtra/Raigad | Hematopathology Laboratory, ACTREC, TMC | Hematopathology Laboratory, ACTREC, TMC | Hematopathology Laboratory et al |
| EPI_ISL_700293 | India/MH-ACTREC-664/2020 | 2020-09-16 | Asia/India/Maharashtra/Raigad | Hematopathology Laboratory, ACTREC, TMC | Hematopathology Laboratory, ACTREC, TMC | Hematopathology Laboratory et al |

| | | | | | | |
|---|---|---|---|---|---|---|
| EPI_ISL_700294 | India/MH-ACTREC-668/2020 | 2020-09-16 | Asia/India/Maharashtra/Washim | Hematopathology Laboratory, ACTREC, TMC | Hematopathology Laboratory, ACTREC, TMC | Hematopathology Laboratory et al |
| EPI_ISL_700295 | India/MH-ACTREC-669/2020 | 2020-09-16 | Asia/India/Maharashtra/Thane | Hematopathology Laboratory, ACTREC, TMC | Hematopathology Laboratory, ACTREC, TMC | Hematopathology Laboratory et al |
| EPI_ISL_700296 | India/MH-ACTREC-670/2020 | 2020-09-18 | Asia/India/Maharashtra/Raigad | Hematopathology Laboratory, ACTREC, TMC | Hematopathology Laboratory, ACTREC, TMC | Hematopathology Laboratory et al |
| EPI_ISL_700297 | India/MH-ACTREC-671/2020 | 2020-09-18 | Asia/India/Maharashtra/Raigad | Hematopathology Laboratory, ACTREC, TMC | Hematopathology Laboratory, ACTREC, TMC | Hematopathology Laboratory et al |
| EPI_ISL_700298 | India/MH-ACTREC-672/2020 | 2020-09-18 | Asia/India/Maharashtra/Raigad | Hematopathology Laboratory, ACTREC, TMC | Hematopathology Laboratory, ACTREC, TMC | Hematopathology Laboratory et al |
| EPI_ISL_700299 | India/MH-ACTREC-673/2020 | 2020-09-18 | Asia/India/Maharashtra/Raigad | Hematopathology Laboratory, ACTREC, TMC | Hematopathology Laboratory, ACTREC, TMC | Hematopathology Laboratory et al |
| EPI_ISL_700300 | India/MH-ACTREC-675/2020 | 2020-09-18 | Asia/India/Maharashtra/Raigad | Hematopathology Laboratory, ACTREC, TMC | Hematopathology Laboratory, ACTREC, TMC | Hematopathology Laboratory et al |
| EPI_ISL_700301 | India/MH-ACTREC-676/2020 | 2020-09-18 | Asia/India/Maharashtra/Raigad | Hematopathology Laboratory, ACTREC, TMC | Hematopathology Laboratory, ACTREC, TMC | Hematopathology Laboratory et al |
| EPI_ISL_700302 | India/MH-ACTREC-677/2020 | 2020-09-18 | Asia/India/Maharashtra/Raigad | Hematopathology Laboratory, ACTREC, TMC | Hematopathology Laboratory, ACTREC, TMC | Hematopathology Laboratory et al |
| EPI_ISL_700303 | India/MH-ACTREC-678/2020 | 2020-09-18 | Asia/India/Maharashtra/Raigad | Hematopathology Laboratory, ACTREC, TMC | Hematopathology Laboratory, ACTREC, TMC | Hematopathology Laboratory et al |
| EPI_ISL_700304 | India/MH-ACTREC-679/2020 | 2020-09-19 | Asia/India/Maharashtra/Raigad | Hematopathology Laboratory, ACTREC, TMC | Hematopathology Laboratory, ACTREC, TMC | Hematopathology Laboratory et al |
| EPI_ISL_700305 | India/MH-ACTREC-680/2020 | 2020-09-19 | Asia/India/Maharashtra/Raigad | Hematopathology Laboratory, ACTREC, TMC | Hematopathology Laboratory, ACTREC, TMC | Hematopathology Laboratory et al |
| EPI_ISL_700306 | India/UP-ACTREC-681/2020 | 2020-09-19 | Asia/India/Uttar Pradesh/Gorakhpur | Hematopathology Laboratory, ACTREC, TMC | Hematopathology Laboratory, ACTREC, TMC | Hematopathology Laboratory et al |

| | | | | | | |
|---|---|---|---|---|---|---|
| EPI_ISL_700307 | India/MH-ACTREC-682/2020 | 2020-09-19 | Asia/India/Maharashtra/Raigad | Hematopathology Laboratory, ACTREC, TMC | Hematopathology Laboratory, ACTREC, TMC | Hematopathology Laboratory et al |
| EPI_ISL_700308 | India/MH-ACTREC-683/2020 | 2020-09-20 | Asia/India/Maharashtra/Raigad | Hematopathology Laboratory, ACTREC, TMC | Hematopathology Laboratory, ACTREC, TMC | Hematopathology Laboratory et al |
| EPI_ISL_700309 | India/MH-ACTREC-684/2020 | 2020-09-22 | Asia/India/Maharashtra/Raigad | Hematopathology Laboratory, ACTREC, TMC | Hematopathology Laboratory, ACTREC, TMC | Hematopathology Laboratory et al |
| EPI_ISL_700310 | India/MH-ACTREC-685/2020 | 2020-09-22 | Asia/India/Maharashtra/Hingoli | Hematopathology Laboratory, ACTREC, TMC | Hematopathology Laboratory, ACTREC, TMC | Hematopathology Laboratory et al |
| EPI_ISL_700311 | India/WB-ACTREC-686/2020 | 2020-09-23 | Asia/India/West Bengal/Murshidabad | Hematopathology Laboratory, ACTREC, TMC | Hematopathology Laboratory, ACTREC, TMC | Hematopathology Laboratory et al |
| EPI_ISL_700312 | India/MH-ACTREC-687/2020 | 2020-09-23 | Asia/India/Maharashtra/Thane | Hematopathology Laboratory, ACTREC, TMC | Hematopathology Laboratory, ACTREC, TMC | Hematopathology Laboratory et al |
| EPI_ISL_700313 | India/OR-ACTREC-688/2020 | 2020-09-24 | Asia/India/Odisha/Ganjam | Hematopathology Laboratory, ACTREC, TMC | Hematopathology Laboratory, ACTREC, TMC | Hematopathology Laboratory et al |
| EPI_ISL_700314 | India/MH-ACTREC-689/2020 | 2020-09-24 | Asia/India/Maharashtra/Thane | Hematopathology Laboratory, ACTREC, TMC | Hematopathology Laboratory, ACTREC, TMC | Hematopathology Laboratory et al |
| EPI_ISL_700315 | India/MH-ACTREC-690/2020 | 2020-09-24 | Asia/India/Maharashtra/Raigad | Hematopathology Laboratory, ACTREC, TMC | Hematopathology Laboratory, ACTREC, TMC | Hematopathology Laboratory et al |
| EPI_ISL_700316 | India/MH-ACTREC-691/2020 | 2020-09-24 | Asia/India/Maharashtra/Raigad | Hematopathology Laboratory, ACTREC, TMC | Hematopathology Laboratory, ACTREC, TMC | Hematopathology Laboratory et al |
| EPI_ISL_700317 | India/MH-ACTREC-692/2020 | 2020-09-24 | Asia/India/Maharashtra/Raigad | Hematopathology Laboratory, ACTREC, TMC | Hematopathology Laboratory, ACTREC, TMC | Hematopathology Laboratory et al |
| EPI_ISL_700318 | India/MH-ACTREC-693/2020 | 2020-09-24 | Asia/India/Maharashtra/Raigad | Hematopathology Laboratory, ACTREC, TMC | Hematopathology Laboratory, ACTREC, TMC | Hematopathology Laboratory et al |
| EPI_ISL_700319 | India/MH-ACTREC-694/2020 | 2020-09-25 | Asia/India/Maharashtra/Mumbai | Hematopathology Laboratory, ACTREC, TMC | Hematopathology Laboratory, ACTREC, TMC | Hematopathology Laboratory et al |

| | | | | | | |
|---|---|---|---|---|---|---|
| EPI_ISL_700320 | India/MH-ACTREC-695/2020 | 2020-09-25 | Asia/India/Maharashtra/Raigad | Hematopathology Laboratory, ACTREC, TMC | Hematopathology Laboratory, ACTREC, TMC | Hematopathology Laboratory et al |
| EPI_ISL_700321 | India/MH-ACTREC-696/2020 | 2020-09-25 | Asia/India/Maharashtra/Raigad | Hematopathology Laboratory, ACTREC, TMC | Hematopathology Laboratory, ACTREC, TMC | Hematopathology Laboratory et al |
| EPI_ISL_700322 | India/MH-ACTREC-697/2020 | 2020-09-25 | Asia/India/Maharashtra/Raigad | Hematopathology Laboratory, ACTREC, TMC | Hematopathology Laboratory, ACTREC, TMC | Hematopathology Laboratory et al |
| EPI_ISL_700323 | India/MH-ACTREC-698/2020 | 2020-09-25 | Asia/India/Maharashtra/Raigad | Hematopathology Laboratory, ACTREC, TMC | Hematopathology Laboratory, ACTREC, TMC | Hematopathology Laboratory et al |
| EPI_ISL_700324 | India/MH-ACTREC-699/2020 | 2020-09-25 | Asia/India/Maharashtra/Raigad | Hematopathology Laboratory, ACTREC, TMC | Hematopathology Laboratory, ACTREC, TMC | Hematopathology Laboratory et al |
| EPI_ISL_700325 | India/MH-ACTREC-700/2020 | 2020-09-25 | Asia/India/Maharashtra/Raigad | Hematopathology Laboratory, ACTREC, TMC | Hematopathology Laboratory, ACTREC, TMC | Hematopathology Laboratory et al |
| EPI_ISL_700327 | France/ARA-68001/2020 | 2020-09-04 | Europe/France/Auvergne-Rh√¥ne-Alpes/ | CNR Virus des Infections Respiratoires - France SUD | CNR Virus des Infections Respiratoires - France SUD | Antonin Bal et al |
| EPI_ISL_700328 | Bangladesh/CHRF-0085/2020 | 2020-09-16 | Asia/Bangladesh/Bangladesh/ | Child Health Research Foundation | Child Health Research Foundation | Senjuti Saha et al |
| EPI_ISL_700329 | Bangladesh/CHRF-0076/2020 | 2020-08-26 | Asia/Bangladesh/Bangladesh/ | Child Health Research Foundation | Child Health Research Foundation | Senjuti Saha et al |
| EPI_ISL_700330 | Bangladesh/CHRF-0075/2020 | 2020-08-23 | Asia/Bangladesh/Bangladesh/ | Child Health Research Foundation | Child Health Research Foundation | Senjuti Saha et al |
| EPI_ISL_700331 | Bangladesh/CHRF-0080/2020 | 2020-09-06 | Asia/Bangladesh/Bangladesh/ | Child Health Research Foundation | Child Health Research Foundation | Senjuti Saha et al |
| EPI_ISL_700332 | Bangladesh/CHRF-0082/2020 | 2020-09-12 | Asia/Bangladesh/Bangladesh/ | Child Health Research Foundation | Child Health Research Foundation | Senjuti Saha et al |
| EPI_ISL_700333 | Bangladesh/CHRF-0073/2020 | 2020-08-21 | Asia/Bangladesh/Bangladesh/ | Child Health Research Foundation | Child Health Research Foundation | Senjuti Saha et al |

| | | | | | | |
|---|---|---|---|---|---|---|
| EPI_ISL_700334 | Bangladesh/CHRF-0068/2020 | 2020-08-10 | Asia/Bangladesh/Bangladesh/ | Child Health Research Foundation | Child Health Research Foundation | Senjuti Saha et al |
| EPI_ISL_700335 | Bangladesh/CHRF-0078/2020 | 2020-08-31 | Asia/Bangladesh/Bangladesh/ | Child Health Research Foundation | Child Health Research Foundation | Senjuti Saha et al |
| EPI_ISL_700336 | Bangladesh/CHRF-0087/2020 | 2020-09-21 | Asia/Bangladesh/Bangladesh/ | Child Health Research Foundation | Child Health Research Foundation | Senjuti Saha et al |
| EPI_ISL_700337 | Bangladesh/CHRF-0071/2020 | 2020-08-20 | Asia/Bangladesh/Bangladesh/ | Child Health Research Foundation | Child Health Research Foundation | Senjuti Saha et al |
| EPI_ISL_700339 | Bangladesh/CHRF-0069/2020 | 2020-08-12 | Asia/Bangladesh/Bangladesh/ | Child Health Research Foundation | Child Health Research Foundation | Senjuti Saha et al |
| EPI_ISL_700340 | Bangladesh/CHRF-0070/2020 | 2020-08-15 | Asia/Bangladesh/Bangladesh/ | Child Health Research Foundation | Child Health Research Foundation | Senjuti Saha et al |
| EPI_ISL_700341 | Bangladesh/CHRF-0074/2020 | 2020-08-18 | Asia/Bangladesh/Bangladesh/ | Child Health Research Foundation | Child Health Research Foundation | Senjuti Saha et al |
| EPI_ISL_700342 | Bangladesh/CHRF-0079/2020 | 2020-09-01 | Asia/Bangladesh/Bangladesh/ | Child Health Research Foundation | Child Health Research Foundation | Senjuti Saha et al |
| EPI_ISL_700343 | Bangladesh/CHRF-0081/2020 | 2020-09-06 | Asia/Bangladesh/Bangladesh/ | Child Health Research Foundation | Child Health Research Foundation | Senjuti Saha et al |
| EPI_ISL_700344 | Bangladesh/CHRF-0083/2020 | 2020-09-13 | Asia/Bangladesh/Bangladesh/ | Child Health Research Foundation | Child Health Research Foundation | Senjuti Saha et al |
| EPI_ISL_700345 | Bangladesh/CHRF-0084/2020 | 2020-09-14 | Asia/Bangladesh/Bangladesh/ | Child Health Research Foundation | Child Health Research Foundation | Senjuti Saha et al |
| EPI_ISL_700346 | Bangladesh/CHRF-0086/2020 | 2020-09-17 | Asia/Bangladesh/Bangladesh/ | Child Health Research Foundation | Child Health Research Foundation | Senjuti Saha et al |
| EPI_ISL_700347 | Bangladesh/CHRF-0088/2020 | 2020-09-26 | Asia/Bangladesh/Bangladesh/ | Child Health Research Foundation | Child Health Research Foundation | Senjuti Saha et al |

| | | | | | | |
|---|---|---|---|---|---|---|
| EPI_ISL_700349 | France/ARA-SC149/2020 | 2020-03-09 | Europe/France/Auvergne-Rh√¥ne-Alpes/ | Laboratoire de virologie, CHU de Grenoble - CS 10217 - 38043 Grenoble cedex 10 | CNR Virus des Infections Respiratoires - France SUD | Antonin Bal et al |
| EPI_ISL_700352 | France/ARA-SC158/2020 | 2020-04-07 | Europe/France/Auvergne-Rh√¥ne-Alpes/ | Laboratoire de virologie, CHU de Grenoble - CS 10217 - 38043 Grenoble cedex 13 | CNR Virus des Infections Respiratoires - France SUD | Antonin Bal et al |
| EPI_ISL_700354 | France/ARA-SC166/2020 | 2020-04-11 | Europe/France/Auvergne-Rh√¥ne-Alpes/ | Laboratoire de virologie, CHU de Grenoble - CS 10217 - 38043 Grenoble cedex 15 | CNR Virus des Infections Respiratoires - France SUD | Antonin Bal et al |
| EPI_ISL_700355 | France/ARA-SC169/2020 | 2020-04-02 | Europe/France/Auvergne-Rh√¥ne-Alpes/ | Laboratoire de virologie, CHU de Grenoble - CS 10217 - 38043 Grenoble cedex 16 | CNR Virus des Infections Respiratoires - France SUD | Antonin Bal et al |
| EPI_ISL_700358 | France/ARA-SC174/2020 | 2020-04-06 | Europe/France/Auvergne-Rh√¥ne-Alpes/ | Laboratoire de virologie, CHU de Grenoble - CS 10217 - 38043 Grenoble cedex 19 | CNR Virus des Infections Respiratoires - France SUD | Antonin Bal et al |
| EPI_ISL_700364 | France/ARA-SC189/2020 | 2020-04-28 | Europe/France/Auvergne-Rh√¥ne-Alpes/ | Laboratoire de virologie, CHU de Grenoble - CS 10217 - 38043 Grenoble cedex 25 | CNR Virus des Infections Respiratoires - France SUD | Antonin Bal et al |
| EPI_ISL_700365 | France/ARA-SC190/2020 | 2020-04-28 | Europe/France/Auvergne-Rh√¥ne-Alpes/ | Laboratoire de virologie, CHU de Grenoble - CS 10217 - 38043 Grenoble cedex 26 | CNR Virus des Infections Respiratoires - France SUD | Antonin Bal et al |
| EPI_ISL_700370 | France/ARA-SC197/2020 | 2020-04-27 | Europe/France/Auvergne-Rh√¥ne-Alpes/ | Laboratoire de virologie, CHU de Grenoble - CS 10217 - 38043 Grenoble cedex 31 | CNR Virus des Infections Respiratoires - France SUD | Antonin Bal et al |
| EPI_ISL_700371 | France/PDL-N62441/2020 | 2020-03-16 | Europe/France/Pays de la Loire/Nantes | CHU Nantes | CNR Virus des Infections Respiratoires - France SUD | Antonin Bal et al |
| EPI_ISL_700375 | France/ARA-SC111/2020 | 2020-04-06 | Europe/France/Auvergne-Rh√¥ne-Alpes/ | Centre hospitalier M√©tropole Savoie | CNR Virus des Infections Respiratoires - France SUD | Antonin Bal et al |

| | | | | | | |
|---|---|---|---|---|---|---|
| EPI_ISL_700382 | France/ARA-SC118/2020 | 2020-04-13 | Europe/France/Auvergne-Rhv¥ne-Alpes/ | Centre hospitalier Mv©tropole Savoie | CNR Virus des Infections Respiratoires - France SUD | Antonin Bal et al |
| EPI_ISL_700383 | France/ARA-SC119/2020 | 2020-04-13 | Europe/France/Auvergne-Rhv¥ne-Alpes/ | Centre hospitalier Mv©tropole Savoie | CNR Virus des Infections Respiratoires - France SUD | Antonin Bal et al |
| EPI_ISL_700395 | France/ARA-SC131/2020 | 2020-04-20 | Europe/France/Auvergne-Rhv¥ne-Alpes/ | Centre hospitalier Mv©tropole Savoie | CNR Virus des Infections Respiratoires - France SUD | Antonin Bal et al |
| EPI_ISL_700397 | France/ARA-SC133/2020 | 2020-05-01 | Europe/France/Auvergne-Rhv¥ne-Alpes/ | Centre hospitalier Mv©tropole Savoie | CNR Virus des Infections Respiratoires - France SUD | Antonin Bal et al |
| EPI_ISL_700398 | France/ARA-SC134/2020 | 2020-05-09 | Europe/France/Auvergne-Rhv¥ne-Alpes/ | Centre hospitalier Mv©tropole Savoie | CNR Virus des Infections Respiratoires - France SUD | Antonin Bal et al |
| EPI_ISL_700402 | France/ARA-SC138/2020 | 2020-04-27 | Europe/France/Auvergne-Rhv¥ne-Alpes/ | Centre hospitalier Mv©tropole Savoie | CNR Virus des Infections Respiratoires - France SUD | Antonin Bal et al |
| EPI_ISL_700406 | France/ARA-SC142/2020 | 2020-05-01 | Europe/France/Auvergne-Rhv¥ne-Alpes/ | Centre hospitalier Mv©tropole Savoie | CNR Virus des Infections Respiratoires - France SUD | Antonin Bal et al |
| EPI_ISL_700421 | SouthAfrica/NHLS-UCT-GS-1493/2020 | 2020-08-11 | Africa/South Africa/Western Cape Provi | Victoria Hospital wc VHW | NHLS/UCT | Arash Iranzadeh et al |
| EPI_ISL_700423 | SouthAfrica/NHLS-UCT-GS-1379/2020 | 2020-11-13 | Africa/South Africa/Western Cape Provi | Khayelethu Clinic wc KLC | NHLS/UCT | Houriiyah Tegally et al |
| EPI_ISL_700426 | SouthAfrica/NHLS-UCT-GS-1332/2020 | 2020-11-04 | Africa/South Africa/Western Cape Provi | Knysna Hospital wc KNY | NHLS/UCT | Houriiyah Tegally et al |
| EPI_ISL_700427 | SouthAfrica/NHLS-UCT-GS-1386/2020 | 2020-11-12 | Africa/South Africa/Western Cape Provi | D'Almeida Clinic wc DAL | NHLS/UCT | Houriiyah Tegally et al |
| EPI_ISL_700435 | SouthAfrica/NHLS-UCT-GS-1025/2020 | 2020-10-26 | Africa/South Africa/Western Cape Provi | Pacaltsdorp Clinic wc PAC | NHLS/UCT | Houriiyah Tegally et al |
| EPI_ISL_700446 | SouthAfrica/NHLS-UCT-GS-1576/2020 | 2020-08-13 | Africa/South Africa/Western Cape Provi | 2 Military Hospital wc MAA | NHLS/UCT | Arash Iranzadeh et al |
| EPI_ISL_700447 | SouthAfrica/NHLS-UCT-GS-1525/2020 | 2020-08-12 | Africa/South Africa/Western Cape Provi | Oudtshoorn Hospital wc OUD | NHLS/UCT | Arash Iranzadeh et al |
| EPI_ISL_700463 | SouthAfrica/NHLS-UCT-GS-3258/2020 | 2020-04-29 | Africa/South Africa/Western Cape Provi | Hanover Park CHC wc HPH | NHLS/UCT | Arash Iranzadeh et al |
| EPI_ISL_700465 | SouthAfrica/NHLS-UCT-GS-1532/2020 | 2020-08-12 | Africa/South Africa/Western Cape Provi | Riversdale Clinic wc RAV | NHLS/UCT | Arash Iranzadeh et al |

| | | | | | | |
|---|---|---|---|---|---|---|
| EPI_ISL_700466 | SouthAfrica/NHLS-UCT-GS-2214/2020 | 2020-08-03 | Africa/South Africa/Western Cape Provi | Ladismith (Nissenville) Clinic wc LAF | NHLS/UCT | Arash Iranzadeh et al |
| EPI_ISL_700473 | SouthAfrica/NHLS-UCT-GP-5818/2020 | 2020-05-20 | Africa/South Africa/Western Cape Provi | Heideveld CDC wc HVP | NHLS/UCT | Arash Iranzadeh et al |
| EPI_ISL_700475 | SouthAfrica/NHLS-UCT-GS-1388/2020 | 2020-11-12 | Africa/South Africa/Western Cape Provi | Alma CDC wc AHC | NHLS/UCT | Houriiyah Tegally et al |
| EPI_ISL_700477 | SouthAfrica/NHLS-UCT-GP-5912/2020 | 2020-05-26 | Africa/South Africa/Western Cape Provi | Hanover Park CHC wc HPH | NHLS/UCT | Arash Iranzadeh et al |
| EPI_ISL_700479 | SouthAfrica/NHLS-UCT-GS-2232/2020 | 2020-08-06 | Africa/South Africa/Western Cape Provi | Mowbray Maternity Hospital wc MMH | NHLS/UCT | Arash Iranzadeh et al |
| EPI_ISL_700494 | SouthAfrica/NHLS-UCT-GP-5388/2020 | 2020-05-05 | Africa/South Africa/Western Cape Provi | Hanover Park CHC wc HPH | NHLS/UCT | Arash Iranzadeh et al |
| EPI_ISL_700495 | SouthAfrica/NHLS-UCT-GS-2217/2020 | 2020-08-06 | Africa/South Africa/Western Cape Provi | Kwanokuthula CDC wc KWA | NHLS/UCT | Arash Iranzadeh et al |
| EPI_ISL_700501 | SouthAfrica/NHLS-UCT-GS-1407/2020 | 2020-08-09 | Africa/South Africa/Western Cape Provi | Knysna Hospital wc KNY | NHLS/UCT | Arash Iranzadeh et al |
| EPI_ISL_700503 | SouthAfrica/NHLS-UCT-GP-5482/2020 | 2020-05-06 | Africa/South Africa/Western Cape Provi | Guguletu CHC wc GDH | NHLS/UCT | Arash Iranzadeh et al |
| EPI_ISL_700506 | SouthAfrica/NHLS-UCT-GS-1455/2020 | 2020-08-11 | Africa/South Africa/Western Cape Provi | Conville CDC wc CVC | NHLS/UCT | Arash Iranzadeh et al |
| EPI_ISL_700507 | SouthAfrica/NHLS-UCT-GS-1695/2020 | 2020-08-19 | Africa/South Africa/Western Cape Provi | Groote Schuur Hospital wc GSH | NHLS/UCT | Arash Iranzadeh et al |
| EPI_ISL_700508 | SouthAfrica/NHLS-UCT-GP-5326/2020 | 2020-05-04 | Africa/South Africa/Western Cape Provi | Heideveld CDC wc HVP | NHLS/UCT | Arash Iranzadeh et al |
| EPI_ISL_700509 | SouthAfrica/NHLS-UCT-GS-1582/2020 | 2020-08-14 | Africa/South Africa/Western Cape Provi | Knysna Hospital wc KNY | NHLS/UCT | Arash Iranzadeh et al |
| EPI_ISL_700513 | SouthAfrica/NHLS-UCT-GP-5557/2020 | 2020-05-08 | Africa/South Africa/Western Cape Provi | Dr Abdurahman CDC wc DAC | NHLS/UCT | Arash Iranzadeh et al |
| EPI_ISL_700514 | SouthAfrica/NHLS-UCT-GP-5312/2020 | 2020-05-03 | Africa/South Africa/Western Cape Provi | Guguletu CHC wc GDH | NHLS/UCT | Arash Iranzadeh et al |
| EPI_ISL_700516 | SouthAfrica/NHLS-UCT-GP-5671/2020 | 2020-05-14 | Africa/South Africa/Western Cape Provi | Hanover Park CHC wc HPH | NHLS/UCT | Arash Iranzadeh et al |
| EPI_ISL_700517 | SouthAfrica/NHLS-UCT-GP-5855/2020 | 2020-05-21 | Africa/South Africa/Western Cape Provi | Dr Abdurahman CDC wc DAC | NHLS/UCT | Arash Iranzadeh et al |
| EPI_ISL_700519 | SouthAfrica/NHLS-UCT-GS-0958/2020 | 2020-10-15 | Africa/South Africa/Western Cape Provi | Clinic-in-Asla | NHLS/UCT | Arash Iranzadeh et al |
| EPI_ISL_700521 | SouthAfrica/NHLS-UCT-GS-2211/2020 | 2020-08-05 | Africa/South Africa/Western Cape Provi | Alma CDC wc AHC | NHLS/UCT | Arash Iranzadeh et al |
| EPI_ISL_700527 | SouthAfrica/NHLS-UCT-GP-5020/2020 | 2020-04-15 | Africa/South Africa/Western Cape Provi | Vanguard CHC wc VGC | NHLS/UCT | Arash Iranzadeh et al |
| EPI_ISL_700528 | SouthAfrica/NHLS-UCT-GS-1971/2020 | 2020-08-26 | Africa/South Africa/Western Cape Provi | Plettenberg Bay Clinic wc PLC | NHLS/UCT | Arash Iranzadeh et al |
| EPI_ISL_700530 | SouthAfrica/NHLS-UCT-GP-5593/2020 | 2020-05-11 | Africa/South Africa/Western Cape Provi | Guguletu CHC wc GDH | NHLS/UCT | Arash Iranzadeh et al |
| EPI_ISL_700534 | SouthAfrica/NHLS-UCT-GS-2215/2020 | 2020-08-05 | Africa/South Africa/Western Cape Provi | Groote Schuur Hospital wc GSH | NHLS/UCT | Arash Iranzadeh et al |
| EPI_ISL_700535 | SouthAfrica/NHLS-UCT-GS-0684/2020 | 2020-10-23 | Africa/South Africa/Western Cape Provi | Kwanokuthula CDC wc KWA | NHLS/UCT | Houriiyah Tegally et al |
| EPI_ISL_700546 | SouthAfrica/NHLS-UCT-GS-2208/2020 | 2020-08-06 | Africa/South Africa/Western Cape Provi | Victoria Hospital wc VHW | NHLS/UCT | Arash Iranzadeh et al |
| EPI_ISL_700547 | SouthAfrica/NHLS-UCT-GP-5792/2020 | 2020-05-19 | Africa/South Africa/Western Cape Provi | Dr Abdurahman CDC wc DAC | NHLS/UCT | Arash Iranzadeh et al |
| EPI_ISL_700556 | SouthAfrica/NHLS-UCT-GS-0636/2020 | 2020-10-19 | Africa/South Africa/Western Cape Provi | Conville CDC wc CVC | NHLS/UCT | Arash Iranzadeh et al |
| EPI_ISL_700559 | SouthAfrica/NHLS-UCT-GS-2058/2020 | 2020-09-18 | Africa/South Africa/Western Cape Provi | Heideveld Emergency Centre | NHLS/UCT | Arash Iranzadeh et al |
| EPI_ISL_700561 | SouthAfrica/NHLS-UCT-GP-5689/2020 | 2020-05-15 | Africa/South Africa/Western Cape Provi | Hanover Park CHC wc HPH | NHLS/UCT | Arash Iranzadeh et al |
| EPI_ISL_700566 | SouthAfrica/NHLS-UCT-GP-5513/2020 | 2020-05-07 | Africa/South Africa/Western Cape Provi | Hanover Park CHC wc HPH | NHLS/UCT | Arash Iranzadeh et al |
| EPI_ISL_700574 | SouthAfrica/NHLS-UCT-GS-2980/2020 | 2020-05-10 | Africa/South Africa/Western Cape Provi | Heideveld Emergency Centre | NHLS/UCT | Arash Iranzadeh et al |

| | | | | | | |
|---|---|---|---|---|---|---|
| EPI_ISL_700577 | SouthAfrica/NHLS-UCT-GS-2284/2020 | 2020-08-08 | Africa/South Africa/Western Cape Provi | Groote Schuur Hospital wc GSH | NHLS/UCT | Arash Iranzadeh et al |
| EPI_ISL_700578 | SouthAfrica/NHLS-UCT-GS-2911/2020 | 2020-05-07 | Africa/South Africa/Western Cape Provi | Guguletu CHC wc GDH | NHLS/UCT | Arash Iranzadeh et al |
| EPI_ISL_700584 | SouthAfrica/NHLS-UCT-GP-5349/2020 | 2020-05-04 | Africa/South Africa/Western Cape Provi | Heideveld Emergency Centre | NHLS/UCT | Arash Iranzadeh et al |
| EPI_ISL_700588 | SouthAfrica/NHLS-UCT-GS-0619/2020 | 2020-10-22 | Africa/South Africa/Western Cape Provi | Sedgefield Clinic wc SGE | NHLS/UCT | Arash Iranzadeh et al |
| EPI_ISL_700592 | SouthAfrica/NHLS-UCT-GP-5840/2020 | 2020-05-21 | Africa/South Africa/Western Cape Provi | Dr Abdurahman CDC wc DAC | NHLS/UCT | Arash Iranzadeh et al |
| EPI_ISL_700593 | SouthAfrica/NHLS-UCT-GP-5890/2020 | 2020-05-24 | Africa/South Africa/Western Cape Provi | Hanover Park CHC wc HPH | NHLS/UCT | Arash Iranzadeh et al |
| EPI_ISL_700594 | SouthAfrica/NHLS-UCT-GS-2203/2020 | 2020-08-03 | Africa/South Africa/Western Cape Provi | Oudtshoorn Hospital wc OUD | NHLS/UCT | Arash Iranzadeh et al |
| EPI_ISL_700597 | SouthAfrica/NHLS-UCT-GS-1808/2020 | 2020-08-31 | Africa/South Africa/Western Cape Provi | Mitchells Plain Hospital wc MPH | NHLS/UCT | Arash Iranzadeh et al |
| EPI_ISL_700598 | SouthAfrica/NHLS-UCT-GS-2736/2020 | 2020-05-20 | Africa/South Africa/Western Cape Provi | Heideveld Emergency Centre | NHLS/UCT | Arash Iranzadeh et al |
| EPI_ISL_700701 | USA/TX-DSHS-1610/2020 | 2020-09-15 | North America/USA/Texas/Mclennan Co | Texas Department of State Health Services | Texas Department of State Health Services | Rashmi Tuladhar et al |
| EPI_ISL_700711 | USA/TX-DSHS-1627/2020 | 2020-10-22 | North America/USA/Texas/Travis Count | Texas Department of State Health Services | Texas Department of State Health Services | Rashmi Tuladhar et al |
| EPI_ISL_700715 | USA/TX-DSHS-1626/2020 | 2020-10-22 | North America/USA/Texas/Travis Count | Texas Department of State Health Services | Texas Department of State Health Services | Rashmi Tuladhar et al |
| EPI_ISL_700722 | USA/TX-DSHS-1600/2020 | 2020-08-24 | North America/USA/Texas/McCulloch Co | Texas Department of State Health Services | Texas Department of State Health Services | Rashmi Tuladhar et al |
| EPI_ISL_700725 | USA/TX-DSHS-1628/2020 | 2020-10-22 | North America/USA/Texas/Dallas Count | Texas Department of State Health Services | Texas Department of State Health Services | Rashmi Tuladhar et al |
| EPI_ISL_700739 | USA/TX-DSHS-1615/2020 | 2020-09-30 | North America/USA/Texas/Travis Count | Texas Department of State Health Services | Texas Department of State Health Services | Rashmi Tuladhar et al |
| EPI_ISL_701862 | England/ALDP-BBB4FE/2020 | 2020-11-21 | Europe/United Kingdom/England/ | Lighthouse Lab in Alderley Park | Wellcome Sanger Institute for the COVID-19 Genomics UK (COG-UK) Consortium | Jacquelyn Wynn et al |
| EPI_ISL_701979 | England/ALDP-BB53CA/2020 | 2020-11-20 | Europe/United Kingdom/England/ | Lighthouse Lab in Alderley Park | Wellcome Sanger Institute for the COVID-19 Genomics UK (COG-UK) Consortium | Jacquelyn Wynn et al |

| | | | | | | |
|---|---|---|---|---|---|---|
| EPI_ISL_701985 | England/ALDP-BB511B/2020 | 2020-11-21 | Europe/United Kingdom/England/ | Lighthouse Lab in Alderley Park | Wellcome Sanger Institute for the COVID-19 Genomics UK (COG-UK) Consortium | Jacquelyn Wynn et al |
| EPI_ISL_702132 | England/CAMC-BD22BB/2020 | 2020-11-28 | Europe/United Kingdom/England/ | Lighthouse Lab in Cambridge | Wellcome Sanger Institute for the COVID-19 Genomics UK (COG-UK) Consortium | Rob Howes et al |
| EPI_ISL_702150 | England/CAMC-BD46DB/2020 | 2020-11-27 | Europe/United Kingdom/England/ | Lighthouse Lab in Cambridge | Wellcome Sanger Institute for the COVID-19 Genomics UK (COG-UK) Consortium | Rob Howes et al |
| EPI_ISL_702188 | England/CAMC-BD1DF7/2020 | 2020-11-28 | Europe/United Kingdom/England/ | Lighthouse Lab in Cambridge | Wellcome Sanger Institute for the COVID-19 Genomics UK (COG-UK) Consortium | Rob Howes et al |
| EPI_ISL_702498 | England/LIVE-DC4E7A/2020 | 2020-11-11 | Europe/United Kingdom/England/ | Liverpool Clinical Laboratories | COVID-19 Genomics UK (COG-UK) Consortium | Sam Haldenby et al |
| EPI_ISL_702511 | England/ALDP-BBA671/2020 | 2020-11-20 | Europe/United Kingdom/England/ | Lighthouse Lab in Alderley Park | Wellcome Sanger Institute for the COVID-19 Genomics UK (COG-UK) Consortium | Jacquelyn Wynn et al |
| EPI_ISL_702531 | England/SHEF-C283F/2020 | 2020-11-27 | Europe/United Kingdom/England/ | Virology Department, Sheffield Teaching Hospitals NHS Foundation Trust/Department of Infection, Immunity and Cardiovascular Disease, The Medical School, University of Sheffield | COVID-19 Genomics UK (COG-UK) Consortium | Thushan de Silva et al |
| EPI_ISL_702704 | England/ALDP-BB2891/2020 | 2020-11-19 | Europe/United Kingdom/England/ | Lighthouse Lab in Alderley Park | Wellcome Sanger Institute for the COVID-19 Genomics UK (COG-UK) Consortium | Jacquelyn Wynn et al |
| EPI_ISL_702749 | England/NORW-F679F/2020 | 2020-12-01 | Europe/United Kingdom/England/ | Quadram Institute Bioscience | COVID-19 Genomics UK (COG-UK) Consortium | Dave J. Baker et al |

| | | | | | | |
|---|---|---|---|---|---|---|
| EPI_ISL_702924 | England/ALDP-BB9CBC/2020 | 2020-11-20 | Europe/United Kingdom/England/ | Lighthouse Lab in Alderley Park | Wellcome Sanger Institute for the COVID-19 Genomics UK (COG-UK) Consortium | Jacquelyn Wynn et al |
| EPI_ISL_703054 | England/ALDP-BB349C/2020 | 2020-11-18 | Europe/United Kingdom/England/ | Lighthouse Lab in Alderley Park | Wellcome Sanger Institute for the COVID-19 Genomics UK (COG-UK) Consortium | Jacquelyn Wynn et al |
| EPI_ISL_703586 | England/ALDP-BCA33B/2020 | 2020-11-23 | Europe/United Kingdom/England/ | Lighthouse Lab in Alderley Park | Wellcome Sanger Institute for the COVID-19 Genomics UK (COG-UK) Consortium | Jacquelyn Wynn et al |
| EPI_ISL_703703 | England/QEUH-BC400D/2020 | 2020-11-24 | Europe/United Kingdom/England/ | Lighthouse Lab in Glasgow | Wellcome Sanger Institute for the COVID-19 Genomics UK (COG-UK) Consortium | Harper VanSteenhouse et al |
| EPI_ISL_703767 | England/LOND-1264436/2020 | 2020-11-17 | Europe/United Kingdom/England/ | University College London, Great Ormond Street Hospital for Children NHS Foundation Trust, Imperial College Healthcare NHS Trust | COVID-19 Genomics UK (COG-UK) Consortium | Sergi Castellano et al |
| EPI_ISL_704058 | England/QEUH-BCBCE4/2020 | 2020-11-26 | Europe/United Kingdom/England/ | Lighthouse Lab in Glasgow | Wellcome Sanger Institute for the COVID-19 Genomics UK (COG-UK) Consortium | Harper VanSteenhouse et al |
| EPI_ISL_704228 | Scotland/CVR5978/2020 | 2020-11-20 | Europe/United Kingdom/Scotland/ | West of Scotland Specialist Virology Centre, NHSGGC / MRC-University of Glasgow Centre for Virus Research | COVID-19 Genomics UK (COG-UK) Consortium | Ana da Silva Filipe et al |
| EPI_ISL_704345 | England/QEUH-BCE2D0/2020 | 2020-11-26 | Europe/United Kingdom/England/ | Lighthouse Lab in Glasgow | Wellcome Sanger Institute for the COVID-19 Genomics UK (COG-UK) Consortium | Harper VanSteenhouse et al |

| | | | | | | |
|---|---|---|---|---|---|---|
| EPI_ISL_704437 | England/QEUH-BCD3FC/2020 | 2020-11-26 | Europe/United Kingdom/England/ | Lighthouse Lab in Glasgow | Wellcome Sanger Institute for the COVID-19 Genomics UK (COG-UK) Consortium | Harper VanSteenhouse et al |
| EPI_ISL_704441 | England/ALDP-B5D84D/2020 | 2020-11-08 | Europe/United Kingdom/England/ | Department of Pathology, University of Cambridge | COVID-19 Genomics UK (COG-UK) Consortium | Aminu S. Jahun et al |
| EPI_ISL_704458 | England/QEUH-BCBDB4/2020 | 2020-11-27 | Europe/United Kingdom/England/ | Lighthouse Lab in Glasgow | Wellcome Sanger Institute for the COVID-19 Genomics UK (COG-UK) Consortium | Harper VanSteenhouse et al |
| EPI_ISL_704471 | England/QEUH-BCC542/2020 | 2020-11-27 | Europe/United Kingdom/England/ | Lighthouse Lab in Glasgow | Wellcome Sanger Institute for the COVID-19 Genomics UK (COG-UK) Consortium | Harper VanSteenhouse et al |
| EPI_ISL_704503 | England/QEUH-BCE1D3/2020 | 2020-11-26 | Europe/United Kingdom/England/ | Lighthouse Lab in Glasgow | Wellcome Sanger Institute for the COVID-19 Genomics UK (COG-UK) Consortium | Harper VanSteenhouse et al |
| EPI_ISL_704547 | England/QEUH-BCE971/2020 | 2020-11-26 | Europe/United Kingdom/England/ | Lighthouse Lab in Glasgow | Wellcome Sanger Institute for the COVID-19 Genomics UK (COG-UK) Consortium | Harper VanSteenhouse et al |
| EPI_ISL_704650 | England/QEUH-BCE364/2020 | 2020-11-26 | Europe/United Kingdom/England/ | Lighthouse Lab in Glasgow | Wellcome Sanger Institute for the COVID-19 Genomics UK (COG-UK) Consortium | Harper VanSteenhouse et al |
| EPI_ISL_704756 | England/CAMC-BC0E70/2020 | 2020-11-22 | Europe/United Kingdom/England/ | Lighthouse Lab in Cambridge | Wellcome Sanger Institute for the COVID-19 Genomics UK (COG-UK) Consortium | Rob Howes et al |

| | | | | | | |
|---|---|---|---|---|---|---|
| EPI_ISL_704778 | England/CAMC-BC0001/2020 | 2020-11-22 | Europe/United Kingdom/England/ | Lighthouse Lab in Cambridge | Wellcome Sanger Institute for the COVID-19 Genomics UK (COG-UK) Consortium | Rob Howes et al |
| EPI_ISL_704918 | Wales/PHWC-490326/2020 | 2020-10-13 | Europe/United Kingdom/Wales/ | Wales Specialist Virology Centre Sequencing lab: Pathogen Genomics Unit | COVID-19 Genomics UK (COG-UK) Consortium | Catherine Moore et al |
| EPI_ISL_705175 | England/CAMC-BC0766/2020 | 2020-11-22 | Europe/United Kingdom/England/ | Lighthouse Lab in Cambridge | Wellcome Sanger Institute for the COVID-19 Genomics UK (COG-UK) Consortium | Rob Howes et al |
| EPI_ISL_705428 | England/MILK-B3C228/2020 | 2020-11-01 | Europe/United Kingdom/England/ | Oxford Viromics, NDM, University of Oxford; Oxford University Hospitals; Basingstoke and North Hampshire Hospital | COVID-19 Genomics UK (COG-UK) Consortium | Tanya Golubchik et al |
| EPI_ISL_705594 | England/CAMC-BBC509/2020 | 2020-11-18 | Europe/United Kingdom/England/ | Quadram Institute Bioscience | COVID-19 Genomics UK (COG-UK) Consortium | Dave J. Baker et al |
| EPI_ISL_705677 | Scotland/CVR5932/2020 | 2020-11-21 | Europe/United Kingdom/Scotland/ | West of Scotland Specialist Virology Centre, NHSGGC / MRC-University of Glasgow Centre for Virus Research | COVID-19 Genomics UK (COG-UK) Consortium | Ana da Silva Filipe et al |
| EPI_ISL_705698 | Scotland/CVR5961/2020 | 2020-11-24 | Europe/United Kingdom/Scotland/ | West of Scotland Specialist Virology Centre, NHSGGC / MRC-University of Glasgow Centre for Virus Research | COVID-19 Genomics UK (COG-UK) Consortium | Ana da Silva Filipe et al |
| EPI_ISL_705761 | Scotland/EDB10445/2020 | 2020-10-06 | Europe/United Kingdom/Scotland/ | Virology Department, Royal Infirmary of Edinburgh, NHS Lothian / School of Biological Sciences, University of Edinburgh / Institute of Genetics and Molecular Medicine, University of Edinburgh | COVID-19 Genomics UK (COG-UK) Consortium | McHugh M et al |
| EPI_ISL_705850 | England/LIVE-DC2E8B/2020 | 2020-11-25 | Europe/United Kingdom/England/ | Liverpool Clinical Laboratories | COVID-19 Genomics UK (COG-UK) Consortium | Sam Haldenby et al |
| EPI_ISL_706137 | England/MILK-B3C370/2020 | 2020-11-01 | Europe/United Kingdom/England/ | Oxford Viromics, NDM, University of Oxford; Oxford University Hospitals; Basingstoke and North Hampshire Hospital | COVID-19 Genomics UK (COG-UK) Consortium | Tanya Golubchik et al |
| EPI_ISL_706226 | England/MILK-B3D1A2/2020 | 2020-11-01 | Europe/United Kingdom/England/ | Oxford Viromics, NDM, University of Oxford; Oxford University Hospitals; Basingstoke and North Hampshire Hospital | COVID-19 Genomics UK (COG-UK) Consortium | Tanya Golubchik et al |

| | | | | | | |
|---|---|---|---|---|---|---|
| EPI_ISL_706303 | England/NORT-1B71109/2020 | 2020-11-14 | Europe/United Kingdom/England/ | Northumbria University / South Tees Hospitals NHS Foundation Trust / North Cumbria Integrated Care NHS Foundation Trust / North Tees and Hartlepool NHS Foundation Trust / Newcastle Hospitals NHS Foundation Trust | COVID-19 Genomics UK (COG-UK) Consortium | Darren L Smith et al |
| EPI_ISL_706462 | Wales/PHWC-490256/2020 | 2020-11-19 | Europe/United Kingdom/Wales/ | Wales Specialist Virology Centre Sequencing lab: Pathogen Genomics Unit | COVID-19 Genomics UK (COG-UK) Consortium | Catherine Moore et al |
| EPI_ISL_707692 | Indonesia/BT-SHSIU-01-3610/2020 | 2020-10-04 | Asia/Indonesia/Banten/ | Medical Research Center, Faculty of Medicine, Syarif Hidayatullah State Islamic University Jakarta | Medical Research Center, Faculty of Medicine, Syarif Hidayatullah State Islamic University Jakarta | Chris Adhiyanto et al |
| EPI_ISL_707695 | USA/CA-LACPHL-AB00003/2020 | 2020-10-05 | North America/USA/California/Los Ange | Los Angeles County Public Health Laboratory | Los Angeles County Public Health Laboratory | P. Hemarajata et al. et al |
| EPI_ISL_707696 | Indonesia/BT-SHSIU-01-4941/2020 | 2020-10-15 | Asia/Indonesia/Banten/ | Medical Research Center, Faculty of Medicine, Syarif Hidayatullah State Islamic University Jakarta | Medical Research Center, Faculty of Medicine, Syarif Hidayatullah State Islamic University Jakarta | Laifa Hendarmin et al |

| | | | | | | |
|---|---|---|---|---|---|---|
| EPI_ISL_707698 | Tunisia/61627/2020 | 2020-11-23 | Africa/Tunisia/Soliman/ | 1-Laboratory of Microbiology, National Reference Lab, Charles Nicolle Hospital; 2-University of Tunis ElManar, Faculty of Medicine of Tunis, LR99ES09, Tunis, Tunisia | 1-Clinical and Experimental Pharmacology Lab, LR16SP02, National Center of Pharmacovigilance, University of Tunis El Manar, Tunis, Tunisia. 2-Neurodegenerative diseases and psychiatric troubles, LR18SP03, Razi Hospital, University of Tunis El Manar, Tunis, Tunisia. 3-Ministry of Health, National Observatory of New and Emerging Diseases, 1006, Tunis, Tunisia | Ilhem Boutiba-Ben Boubaker et al |
| EPI_ISL_707723 | Belgium/ULG-10971/2020 | 2020-09-08 | Europe/Belgium/Liv®ge/ | Department of Clinical Microbiology | GIGA Medical Genomics | Keith Durkin et al |
| EPI_ISL_707741 | Belgium/ULG-10962/2020 | 2020-09-08 | Europe/Belgium/Liv®ge/ | Department of Clinical Microbiology | GIGA Medical Genomics | Keith Durkin et al |
| EPI_ISL_707754 | Belgium/ULG-10980/2020 | 2020-09-09 | Europe/Belgium/Liv®ge/ | Department of Clinical Microbiology | GIGA Medical Genomics | Keith Durkin et al |
| EPI_ISL_707761 | Belgium/ULG-10993/2020 | 2020-11-14 | Europe/Belgium/Liv®ge/ | Department of Clinical Microbiology | GIGA Medical Genomics | Keith Durkin et al |
| EPI_ISL_707763 | Belgium/ULG-10996/2020 | 2020-11-14 | Europe/Belgium/Liv®ge/ | Department of Clinical Microbiology | GIGA Medical Genomics | Keith Durkin et al |
| EPI_ISL_707764 | Belgium/ULG-10997/2020 | 2020-11-14 | Europe/Belgium/Liv®ge/ | Department of Clinical Microbiology | GIGA Medical Genomics | Keith Durkin et al |
| EPI_ISL_707767 | Belgium/ULG-11002/2020 | 2020-11-14 | Europe/Belgium/Liv®ge/ | Department of Clinical Microbiology | GIGA Medical Genomics | Keith Durkin et al |
| EPI_ISL_707769 | Belgium/ULG-11010/2020 | 2020-11-15 | Europe/Belgium/Liv®ge/ | Department of Clinical Microbiology | GIGA Medical Genomics | Keith Durkin et al |

| | | | | | | |
|---|---|---|---|---|---|---|
| EPI_ISL_707770 | Belgium/ULG-11011/2020 | 2020-11-15 | Europe/Belgium/Liv®ge/ | Department of Clinical Microbiology | GIGA Medical Genomics | Keith Durkin et al |
| EPI_ISL_707778 | Indonesia/BT-SHSIU-01-2566/2020 | 2020-09-30 | Asia/Indonesia/Banten/ | Medical Research Center, Faculty of Medicine, Syarif Hidayatullah State Islamic University Jakarta | Medical Research Center, Faculty of Medicine, Syarif Hidayatullah State Islamic University Jakarta | Erike A Suwarsono et al |
| EPI_ISL_707792 | Tunisia/6736/2020 | 2020-03-31 | Africa/Tunisia/Tunis/ | 1-Laboratory of Microbiology, National Reference Lab, Charles Nicolle Hospital; 2-University of Tunis ElManar, Faculty of Medicine of Tunis, LR99ES09, Tunis, Tunisia | 1-Clinical and Experimental Pharmacology Lab, LR16SP02, National Center of Pharmacovigilance, University of Tunis El Manar, Tunis, Tunisia. 2-Neurodegenerative diseases and psychiatric troubles, LR18SP03, Razi Hospital, University of Tunis El Manar, Tunis, Tunisia. 3-Ministry of Health, National Observatory of New and Emerging Diseases, 1006, Tunis, Tunisia | Ilhem Boutiba-Ben Boubaker et al |
| EPI_ISL_707819 | USA/MI-UM-10036764267/2020 | 2020-11-03 | North America/USA/Michigan/ | University of Michigan Clinical Microbiology Laboratory | Lauring Lab, University of Michigan, Department of Microbiology and Immunology | Valesano et al |

| | | | | | | |
|---|---|---|---|---|---|---|
| EPI_ISL_707823 | USA/MI-UM-10036784530/2020 | 2020-11-04 | North America/USA/Michigan/ | University of Michigan Clinical Microbiology Laboratory | Lauring Lab, University of Michigan, Department of Microbiology and Immunology | Valesano et al |
| EPI_ISL_707824 | USA/MI-UM-10036759411/2020 | 2020-11-03 | North America/USA/Michigan/ | University of Michigan Clinical Microbiology Laboratory | Lauring Lab, University of Michigan, Department of Microbiology and Immunology | Valesano et al |
| EPI_ISL_707834 | USA/MI-UM-10036799095/2020 | 2020-11-05 | North America/USA/Michigan/ | University of Michigan Clinical Microbiology Laboratory | Lauring Lab, University of Michigan, Department of Microbiology and Immunology | Valesano et al |
| EPI_ISL_707842 | USA/MI-UM-10036802563/2020 | 2020-11-05 | North America/USA/Michigan/ | University of Michigan Clinical Microbiology Laboratory | Lauring Lab, University of Michigan, Department of Microbiology and Immunology | Valesano et al |
| EPI_ISL_707845 | USA/MI-UM-10036782160/2020 | 2020-11-04 | North America/USA/Michigan/ | University of Michigan Clinical Microbiology Laboratory | Lauring Lab, University of Michigan, Department of Microbiology and Immunology | Valesano et al |
| EPI_ISL_707847 | USA/MI-UM-10036784170/2020 | 2020-11-04 | North America/USA/Michigan/ | University of Michigan Clinical Microbiology Laboratory | Lauring Lab, University of Michigan, Department of Microbiology and Immunology | Valesano et al |

| | | | | | | |
|---|---|---|---|---|---|---|
| EPI_ISL_707852 | USA/MI-UM-10036803122/2020 | 2020-11-05 | North America/USA/Michigan/ | University of Michigan Clinical Microbiology Laboratory | Lauring Lab, University of Michigan, Department of Microbiology and Immunology | Valesano et al |
| EPI_ISL_707856 | USA/MI-UM-10036797730/2020 | 2020-11-05 | North America/USA/Michigan/ | University of Michigan Clinical Microbiology Laboratory | Lauring Lab, University of Michigan, Department of Microbiology and Immunology | Valesano et al |
| EPI_ISL_707857 | USA/MI-UM-10036786992/2020 | 2020-11-04 | North America/USA/Michigan/ | University of Michigan Clinical Microbiology Laboratory | Lauring Lab, University of Michigan, Department of Microbiology and Immunology | Valesano et al |
| EPI_ISL_707865 | USA/MI-UM-10036789788/2020 | 2020-11-04 | North America/USA/Michigan/ | University of Michigan Clinical Microbiology Laboratory | Lauring Lab, University of Michigan, Department of Microbiology and Immunology | Valesano et al |
| EPI_ISL_707867 | USA/MI-UM-10036775247/2020 | 2020-11-04 | North America/USA/Michigan/ | University of Michigan Clinical Microbiology Laboratory | Lauring Lab, University of Michigan, Department of Microbiology and Immunology | Valesano et al |
| EPI_ISL_707868 | USA/MI-UM-10036782720/2020 | 2020-11-04 | North America/USA/Michigan/ | University of Michigan Clinical Microbiology Laboratory | Lauring Lab, University of Michigan, Department of Microbiology and Immunology | Valesano et al |

| | | | | | | |
|---|---|---|---|---|---|---|
| EPI_ISL_707873 | USA/MI-UM-10036782210/2020 | 2020-11-04 | North America/USA/Michigan/ | University of Michigan Clinical Microbiology Laboratory | Lauring Lab, University of Michigan, Department of Microbiology and Immunology | Valesano et al |
| EPI_ISL_707877 | USA/MI-UM-10036795528/2020 | 2020-11-05 | North America/USA/Michigan/ | University of Michigan Clinical Microbiology Laboratory | Lauring Lab, University of Michigan, Department of Microbiology and Immunology | Valesano et al |
| EPI_ISL_707879 | USA/MI-UM-10036784725/2020 | 2020-11-04 | North America/USA/Michigan/ | University of Michigan Clinical Microbiology Laboratory | Lauring Lab, University of Michigan, Department of Microbiology and Immunology | Valesano et al |
| EPI_ISL_707881 | USA/MI-UM-10036801511/2020 | 2020-11-05 | North America/USA/Michigan/ | University of Michigan Clinical Microbiology Laboratory | Lauring Lab, University of Michigan, Department of Microbiology and Immunology | Valesano et al |
| EPI_ISL_707884 | USA/CA-LACPHL-AC00002/2020 | 2020-08-18 | North America/USA/California/Los Ange | Los Angeles County Public Health Laboratory | Los Angeles County Public Health Laboratory | P. Hemarajata et al. et al |
| EPI_ISL_707885 | USA/CA-LACPHL-AC00004/2020 | 2020-08-18 | North America/USA/California/Los Ange | Los Angeles County Public Health Laboratory | Los Angeles County Public Health Laboratory | P. Hemarajata et al. et al |
| EPI_ISL_707888 | USA/CA-LACPHL-AC00007/2020 | 2020-08-18 | North America/USA/California/Los Ange | Los Angeles County Public Health Laboratory | Los Angeles County Public Health Laboratory | P. Hemarajata et al. et al |
| EPI_ISL_707890 | Indonesia/BT-SHSIU-01-3884/2020 | 2020-10-22 | Asia/Indonesia/Banten/ | Medical Research Center, Faculty of Medicine, Syarif Hidayatullah State Islamic University Jakarta | Medical Research Center, Faculty of Medicine, Syarif Hidayatullah State Islamic University Jakarta | Zeti Harriyati et al |

| | | | | | | |
|---|---|---|---|---|---|---|
| EPI_ISL_707903 | USA/CA-LACPHL-AE00001/2020 | 2020-06-05 | North America/USA/California/Los Ange | Los Angeles County Public Health Laboratory | Los Angeles County Public Health Laboratory | P. Hemarajata et al. et al |
| EPI_ISL_707905 | USA/CA-LACPHL-AE00004/2020 | 2020-06-16 | North America/USA/California/Los Ange | Los Angeles County Public Health Laboratory | Los Angeles County Public Health Laboratory | P. Hemarajata et al. et al |
| EPI_ISL_707907 | USA/CA-LACPHL-AE00009/2020 | 2020-07-01 | North America/USA/California/Los Ange | Los Angeles County Public Health Laboratory | Los Angeles County Public Health Laboratory | P. Hemarajata et al. et al |
| EPI_ISL_707908 | USA/CA-LACPHL-AE00011/2020 | 2020-07-01 | North America/USA/California/Los Ange | Los Angeles County Public Health Laboratory | Los Angeles County Public Health Laboratory | P. Hemarajata et al. et al |
| EPI_ISL_707911 | USA/CA-LACPHL-AE00014/2020 | 2020-07-02 | North America/USA/California/Los Ange | Los Angeles County Public Health Laboratory | Los Angeles County Public Health Laboratory | P. Hemarajata et al. et al |
| EPI_ISL_707916 | USA/CA-LACPHL-AE00019/2020 | 2020-07-03 | North America/USA/California/Los Ange | Los Angeles County Public Health Laboratory | Los Angeles County Public Health Laboratory | P. Hemarajata et al. et al |
| EPI_ISL_707925 | USA/CA-LACPHL-AE00028/2020 | 2020-07-08 | North America/USA/California/Los Ange | Los Angeles County Public Health Laboratory | Los Angeles County Public Health Laboratory | P. Hemarajata et al. et al |
| EPI_ISL_707932 | USA/CA-LACPHL-AE00030/2020 | 2020-07-09 | North America/USA/California/Los Ange | Los Angeles County Public Health Laboratory | Los Angeles County Public Health Laboratory | P. Hemarajata et al. et al |
| EPI_ISL_707937 | Turkey/DEN-PAU07/2020 | 2020-07-27 | Europe/Turkey/Denizli/ | Pamukkale University Hospital | Pamukkale University Department of Medical Genetics | Onur TOKGUN et al. et al |
| EPI_ISL_707938 | Turkey/DEN-PAU01/2020 | 2020-07-26 | Europe/Turkey/Denizli/ | Pamukkale University Hospital | Pamukkale University Department of Medical Genetics | Onur TOKGUN et al. et al |
| EPI_ISL_707939 | Turkey/DEN-PAU02/2020 | 2020-07-28 | Europe/Turkey/Denizli/ | Pamukkale University Hospital | Pamukkale University Department of Medical Genetics | Onur TOKGUN et al. et al |
| EPI_ISL_707943 | Germany/SL-SU-10426887/2020 | 2020-03-12 | Europe/Germany/Saarland/ | Virology, Universit√§tsklinikum des Saarlandes | Epigenetics, Saarland University | Kathrin Kattler et al |

| | | | | | | |
|---|---|---|---|---|---|---|
| EPI_ISL_707949 | Germany/SL-SU-10427450/2020 | 2020-03-15 | Europe/Germany/Saarland/ | Virology, Universit√§tsklinikum des Saarlandes | Epigenetics, Saarland University | Kathrin Kattler et al |
| EPI_ISL_707955 | Germany/SL-SU-10428507/2020 | 2020-03-21 | Europe/Germany/Saarland/ | Virology, Universit√§tsklinikum des Saarlandes | Epigenetics, Saarland University | Kathrin Kattler et al |
| EPI_ISL_707958 | Germany/SL-SU-10429377/2020 | 2020-03-25 | Europe/Germany/Saarland/ | Virology, Universit√§tsklinikum des Saarlandes | Epigenetics, Saarland University | Kathrin Kattler et al |
| EPI_ISL_707965 | Germany/SL-SU-10431371/2020 | 2020-04-02 | Europe/Germany/Saarland/ | Virology, Universit√§tsklinikum des Saarlandes | Epigenetics, Saarland University | Kathrin Kattler et al |
| EPI_ISL_707971 | Germany/SL-SU-10450152/2020 | 2020-10-11 | Europe/Germany/Saarland/ | Virology, Universit√§tsklinikum des Saarlandes | Epigenetics, Saarland University | Kathrin Kattler et al |
| EPI_ISL_707972 | Germany/SL-SU-10450307/2020 | 2020-10-12 | Europe/Germany/Saarland/ | Virology, Universit√§tsklinikum des Saarlandes | Epigenetics, Saarland University | Kathrin Kattler et al |
| EPI_ISL_707974 | Germany/SL-SU-10450495/2020 | 2020-10-12 | Europe/Germany/Saarland/ | Virology, Universit√§tsklinikum des Saarlandes | Epigenetics, Saarland University | Kathrin Kattler et al |
| EPI_ISL_707975 | Germany/SL-SU-10450502/2020 | 2020-10-12 | Europe/Germany/Saarland/ | Virology, Universit√§tsklinikum des Saarlandes | Epigenetics, Saarland University | Kathrin Kattler et al |
| EPI_ISL_707979 | Germany/SL-SU-10450618/2020 | 2020-10-13 | Europe/Germany/Saarland/ | Virology, Universit√§tsklinikum des Saarlandes | Epigenetics, Saarland University | Kathrin Kattler et al |
| EPI_ISL_707980 | Germany/SL-SU-10450646/2020 | 2020-10-13 | Europe/Germany/Saarland/ | Virology, Universit√§tsklinikum des Saarlandes | Epigenetics, Saarland University | Kathrin Kattler et al |
| EPI_ISL_707981 | Germany/SL-SU-10450660/2020 | 2020-10-13 | Europe/Germany/Saarland/ | Virology, Universit√§tsklinikum des Saarlandes | Epigenetics, Saarland University | Kathrin Kattler et al |
| EPI_ISL_707983 | Germany/SL-SU-10450682/2020 | 2020-10-13 | Europe/Germany/Saarland/ | Virology, Universit√§tsklinikum des Saarlandes | Epigenetics, Saarland University | Kathrin Kattler et al |
| EPI_ISL_707985 | Germany/SL-SU-10450978/2020 | 2020-10-14 | Europe/Germany/Saarland/ | Virology, Universit√§tsklinikum des Saarlandes | Epigenetics, Saarland University | Kathrin Kattler et al |

| | | | | | | |
|---|---|---|---|---|---|---|
| EPI_ISL_707990 | Germany/SL-SU-10451186/2020 | 2020-10-15 | Europe/Germany/Saarland/ | Virology, Universit√§tsklinikum des Saarlandes | Epigenetics, Saarland University | Kathrin Kattler et al |
| EPI_ISL_707991 | Germany/SL-SU-10451375/2020 | 2020-10-15 | Europe/Germany/Saarland/ | Virology, Universit√§tsklinikum des Saarlandes | Epigenetics, Saarland University | Kathrin Kattler et al |
| EPI_ISL_707994 | Germany/SL-SU-10451535/2020 | 2020-10-15 | Europe/Germany/Saarland/ | Virology, Universit√§tsklinikum des Saarlandes | Epigenetics, Saarland University | Kathrin Kattler et al |
| EPI_ISL_707995 | Germany/SL-SU-10451904/2020 | 2020-10-17 | Europe/Germany/Saarland/ | Virology, Universit√§tsklinikum des Saarlandes | Epigenetics, Saarland University | Kathrin Kattler et al |
| EPI_ISL_707998 | Germany/SL-SU-10452037/2020 | 2020-10-18 | Europe/Germany/Saarland/ | Virology, Universit√§tsklinikum des Saarlandes | Epigenetics, Saarland University | Kathrin Kattler et al |
| EPI_ISL_708000 | Germany/SL-SU-10452452/2020 | 2020-10-20 | Europe/Germany/Saarland/ | Virology, Universit√§tsklinikum des Saarlandes | Epigenetics, Saarland University | Kathrin Kattler et al |
| EPI_ISL_708001 | Germany/SL-SU-10452528/2020 | 2020-10-20 | Europe/Germany/Saarland/ | Virology, Universit√§tsklinikum des Saarlandes | Epigenetics, Saarland University | Kathrin Kattler et al |
| EPI_ISL_708004 | Germany/SL-SU-20181174/2020 | 2020-03-22 | Europe/Germany/Saarland/ | Virology, Universit√§tsklinikum des Saarlandes | Epigenetics, Saarland University | Kathrin Kattler et al |
| EPI_ISL_708010 | Germany/SL-SU-20238352/2020 | 2020-10-09 | Europe/Germany/Saarland/ | Virology, Universit√§tsklinikum des Saarlandes | Epigenetics, Saarland University | Kathrin Kattler et al |
| EPI_ISL_708011 | Germany/SL-SU-20238575/2020 | 2020-10-11 | Europe/Germany/Saarland/ | Virology, Universit√§tsklinikum des Saarlandes | Epigenetics, Saarland University | Kathrin Kattler et al |
| EPI_ISL_708015 | Germany/SL-SU-20242379/2020 | 2020-10-21 | Europe/Germany/Saarland/ | Virology, Universit√§tsklinikum des Saarlandes | Epigenetics, Saarland University | Kathrin Kattler et al |
| EPI_ISL_708017 | Germany/SL-SU-20183040/2020 | 2020-04-01 | Europe/Germany/Saarland/ | Virology, Universit√§tsklinikum des Saarlandes | Epigenetics, Saarland University | Kathrin Kattler et al |
| EPI_ISL_708019 | Germany/SL-SU-20186237/2020 | 2020-04-14 | Europe/Germany/Saarland/ | Virology, Universit√§tsklinikum des Saarlandes | Epigenetics, Saarland University | Kathrin Kattler et al |

| | | | | | | |
|---|---|---|---|---|---|---|
| EPI_ISL_708020 | Germany/SL-SU-20190128/2020 | 2020-04-28 | Europe/Germany/Saarland/ | Virology, Universit√§tsklinikum des Saarlandes | Epigenetics, Saarland University | Kathrin Kattler et al |
| EPI_ISL_708022 | Germany/SL-SU-OM002/2020 | 2020-06-19 | Europe/Germany/Saarland/ | Virology, Universit√§tsklinikum des Saarlandes | Epigenetics, Saarland University | Kathrin Kattler et al |
| EPI_ISL_708070 | Norway/5377/2020 | 2020-10-26 | Europe/Norway/Troms Og Finnmark/ | University Hospital of Northern Norway, Department for Microbiology and Infectious Disease Control | Norwegian Institute of Public Health, Department of Virology | Kathrine Stene-Johansen et al |
| EPI_ISL_708125 | Norway/5609/2020 | 2020-11-09 | Europe/Norway/Agder/ | Hospital of Southern Norway - Kristiansand, Department of Medical Microbiology | Norwegian Institute of Public Health, Department of Virology | Kathrine Stene-Johansen et al |
| EPI_ISL_708127 | Norway/5618/2020 | 2020-11-10 | Europe/Norway/Viken/ | Furst Medical Laboratory | Norwegian Institute of Public Health, Department of Virology | Kathrine Stene-Johansen et al |
| EPI_ISL_708138 | Norway/5657/2020 | 2020-11-17 | Europe/Norway/Viken/ | Medical Microbiology Unit, Department for Laboratory Medicine, Drammen Hospital, Vestre Viken Health Trust, | Norwegian Institute of Public Health, Department of Virology | Kathrine Stene-Johansen et al |
| EPI_ISL_708151 | Norway/5732/2020 | 2020-11-14 | Europe/Norway/Innlandet/ | Innlandet Hospital Trust, Division Lillehammer, Department for Medical Microbiology | Norwegian Institute of Public Health, Department of Virology | Kathrine Stene-Johansen et al |
| EPI_ISL_708184 | Turkey/DEN-PAU03/2020 | 2020-07-29 | Europe/Turkey/Denizli/ | Pamukkale University Hospital | Pamukkale University Department of Medical Genetics | Onur TOKGUN et al. et al |
| EPI_ISL_708185 | Turkey/DEN-PAU04/2020 | 2020-07-29 | Europe/Turkey/Denizli/ | Pamukkale University Hospital | Pamukkale University Department of Medical Genetics | Onur TOKGUN et al. et al |

| | | | | | | |
|---|---|---|---|---|---|---|
| EPI_ISL_708186 | Turkey/DEN-PAU08/2020 | 2020-08-26 | Europe/Turkey/Denizli/ | Pamukkale University Hospital | Pamukkale University Department of Medical Genetics | Onur TOKGUN et al. et al |
| EPI_ISL_708188 | Turkey/DEN-PAU10/2020 | 2020-08-27 | Europe/Turkey/Denizli/ | Pamukkale University Hospital | Pamukkale University Department of Medical Genetics | Onur TOKGUN et al. et al |
| EPI_ISL_708189 | Turkey/DEN-PAU11/2020 | 2020-08-26 | Europe/Turkey/Denizli/ | Pamukkale University Hospital | Pamukkale University Department of Medical Genetics | Onur TOKGUN et al. et al |
| EPI_ISL_708190 | Turkey/DEN-PAU12/2020 | 2020-07-30 | Europe/Turkey/Denizli/ | Pamukkale University Hospital | Pamukkale University Department of Medical Genetics | Onur TOKGUN et al. et al |
| EPI_ISL_708191 | Turkey/DEN-PAU13/2020 | 2020-07-30 | Europe/Turkey/Denizli/ | Pamukkale University Hospital | Pamukkale University Department of Medical Genetics | Onur TOKGUN et al. et al |
| EPI_ISL_708192 | Turkey/DEN-PAU15/2020 | 2020-08-28 | Europe/Turkey/Denizli/ | Pamukkale University Hospital | Pamukkale University Department of Medical Genetics | Onur TOKGUN et al. et al |
| EPI_ISL_708193 | Turkey/DEN-PAU17/2020 | 2020-08-26 | Europe/Turkey/Denizli/ | Pamukkale University Hospital | Pamukkale University Department of Medical Genetics | Onur TOKGUN et al. et al |
| EPI_ISL_708194 | Turkey/DEN-PAU19/2020 | 2020-08-25 | Europe/Turkey/Denizli/ | Pamukkale University Hospital | Pamukkale University Department of Medical Genetics | Onur TOKGUN et al. et al |
| EPI_ISL_708195 | Turkey/DEN-PAU14/2020 | 2020-08-26 | Europe/Turkey/Denizli/ | Pamukkale University Hospital | Pamukkale University Department of Medical Genetics | Onur TOKGUN et al. et al |

| | | | | | | |
|---|---|---|---|---|---|---|
| EPI_ISL_708207 | USA/MI-MDHHS-SC22504/2020 | 2020-11-16 | North America/USA/Michigan/ | Michigan Department of Health and Human Services, Bureau of Laboratories | Michigan Department of Health and Human Services, Bureau of Laboratories | Blankenship HM et al |
| EPI_ISL_708212 | USA/MI-MDHHS-SC22509/2020 | 2020-11-17 | North America/USA/Michigan/ | Michigan Department of Health and Human Services, Bureau of Laboratories | Michigan Department of Health and Human Services, Bureau of Laboratories | Blankenship HM et al |
| EPI_ISL_708213 | USA/MI-MDHHS-SC22510/2020 | 2020-11-18 | North America/USA/Michigan/ | Michigan Department of Health and Human Services, Bureau of Laboratories | Michigan Department of Health and Human Services, Bureau of Laboratories | Blankenship HM et al |
| EPI_ISL_708226 | USA/MI-MDHHS-SC22523/2020 | 2020-11-17 | North America/USA/Michigan/ | Michigan Department of Health and Human Services, Bureau of Laboratories | Michigan Department of Health and Human Services, Bureau of Laboratories | Blankenship HM et al |
| EPI_ISL_708229 | USA/MI-MDHHS-SC22526/2020 | 2020-11-18 | North America/USA/Michigan/ | Michigan Department of Health and Human Services, Bureau of Laboratories | Michigan Department of Health and Human Services, Bureau of Laboratories | Blankenship HM et al |
| EPI_ISL_708231 | USA/MI-MDHHS-SC22528/2020 | 2020-11-17 | North America/USA/Michigan/ | Michigan Department of Health and Human Services, Bureau of Laboratories | Michigan Department of Health and Human Services, Bureau of Laboratories | Blankenship HM et al |

| | | | | | | |
|---|---|---|---|---|---|---|
| EPI_ISL_708238 | USA/MI-MDHHS-SC22405/2020 | 2020-11-16 | North America/USA/Michigan/ | Michigan Department of Health and Human Services, Bureau of Laboratories | Michigan Department of Health and Human Services, Bureau of Laboratories | Blankenship HM et al |
| EPI_ISL_708240 | USA/MI-MDHHS-SC22407/2020 | 2020-11-18 | North America/USA/Michigan/ | Michigan Department of Health and Human Services, Bureau of Laboratories | Michigan Department of Health and Human Services, Bureau of Laboratories | Blankenship HM et al |
| EPI_ISL_708244 | USA/MI-MDHHS-SC22411/2020 | 2020-11-18 | North America/USA/Michigan/ | Michigan Department of Health and Human Services, Bureau of Laboratories | Michigan Department of Health and Human Services, Bureau of Laboratories | Blankenship HM et al |
| EPI_ISL_708254 | USA/MI-MDHHS-SC22421/2020 | 2020-11-14 | North America/USA/Michigan/ | Michigan Department of Health and Human Services, Bureau of Laboratories | Michigan Department of Health and Human Services, Bureau of Laboratories | Blankenship HM et al |
| EPI_ISL_708258 | USA/MI-MDHHS-SC22425/2020 | 2020-11-09 | North America/USA/Michigan/ | Michigan Department of Health and Human Services, Bureau of Laboratories | Michigan Department of Health and Human Services, Bureau of Laboratories | Blankenship HM et al |
| EPI_ISL_708270 | USA/MI-MDHHS-SC22437/2020 | 2020-11-18 | North America/USA/Michigan/ | Michigan Department of Health and Human Services, Bureau of Laboratories | Michigan Department of Health and Human Services, Bureau of Laboratories | Blankenship HM et al |

| | | | | | | |
|---|---|---|---|---|---|---|
| EPI_ISL_708272 | USA/MI-MDHHS-SC22439/2020 | 2020-11-18 | North America/USA/Michigan/ | Michigan Department of Health and Human Services, Bureau of Laboratories | Michigan Department of Health and Human Services, Bureau of Laboratories | Blankenship HM et al |
| EPI_ISL_708278 | USA/MI-MDHHS-SC22302/2020 | 2020-11-10 | North America/USA/Michigan/ | Michigan Department of Health and Human Services, Bureau of Laboratories | Michigan Department of Health and Human Services, Bureau of Laboratories | Blankenship HM et al |
| EPI_ISL_708283 | USA/MI-MDHHS-SC22307/2020 | 2020-11-08 | North America/USA/Michigan/ | Michigan Department of Health and Human Services, Bureau of Laboratories | Michigan Department of Health and Human Services, Bureau of Laboratories | Blankenship HM et al |
| EPI_ISL_708285 | USA/MI-MDHHS-SC22309/2020 | 2020-11-10 | North America/USA/Michigan/ | Michigan Department of Health and Human Services, Bureau of Laboratories | Michigan Department of Health and Human Services, Bureau of Laboratories | Blankenship HM et al |
| EPI_ISL_708290 | USA/MI-MDHHS-SC22314/2020 | 2020-11-10 | North America/USA/Michigan/ | Michigan Department of Health and Human Services, Bureau of Laboratories | Michigan Department of Health and Human Services, Bureau of Laboratories | Blankenship HM et al |
| EPI_ISL_708294 | USA/MI-MDHHS-SC22318/2020 | 2020-11-08 | North America/USA/Michigan/ | Michigan Department of Health and Human Services, Bureau of Laboratories | Michigan Department of Health and Human Services, Bureau of Laboratories | Blankenship HM et al |

| | | | | | | |
|---|---|---|---|---|---|---|
| EPI_ISL_708299 | USA/MI-MDHHS-SC22323/2020 | 2020-11-10 | North America/USA/Michigan/ | Michigan Department of Health and Human Services, Bureau of Laboratories | Michigan Department of Health and Human Services, Bureau of Laboratories | Blankenship HM et al |
| EPI_ISL_708302 | USA/MI-MDHHS-SC22326/2020 | 2020-11-10 | North America/USA/Michigan/ | Michigan Department of Health and Human Services, Bureau of Laboratories | Michigan Department of Health and Human Services, Bureau of Laboratories | Blankenship HM et al |
| EPI_ISL_708306 | Switzerland/GE-HUG-54280/2020 | 2020-10-25 | Europe/Switzerland/Geneva/ | University Hospitals of Geneva, Laboratory of Virology | University Hospitals of Geneva, Laboratory of Virology | Cordey Samuel et al |
| EPI_ISL_708312 | USA/MI-MDHHS-SC22333/2020 | 2020-11-05 | North America/USA/Michigan/ | Michigan Department of Health and Human Services, Bureau of Laboratories | Michigan Department of Health and Human Services, Bureau of Laboratories | Blankenship HM et al |
| EPI_ISL_708314 | USA/MI-MDHHS-SC22335/2020 | 2020-11-07 | North America/USA/Michigan/ | Michigan Department of Health and Human Services, Bureau of Laboratories | Michigan Department of Health and Human Services, Bureau of Laboratories | Blankenship HM et al |
| EPI_ISL_708324 | USA/MI-MDHHS-SC22344/2020 | 2020-11-04 | North America/USA/Michigan/ | Michigan Department of Health and Human Services, Bureau of Laboratories | Michigan Department of Health and Human Services, Bureau of Laboratories | Blankenship HM et al |
| EPI_ISL_708332 | USA/MI-MDHHS-SC22352/2020 | 2020-11-17 | North America/USA/Michigan/ | Michigan Department of Health and Human Services, Bureau of Laboratories | Michigan Department of Health and Human Services, Bureau of Laboratories | Blankenship HM et al |

| | | | | | | |
|---|---|---|---|---|---|---|
| EPI_ISL_708340 | USA/MI-MDHHS-SC22360/2020 | 2020-11-12 | North America/USA/Michigan/ | Michigan Department of Health and Human Services, Bureau of Laboratories | Michigan Department of Health and Human Services, Bureau of Laboratories | Blankenship HM et al |
| EPI_ISL_708341 | USA/MI-MDHHS-SC22361/2020 | 2020-10-31 | North America/USA/Michigan/ | Michigan Department of Health and Human Services, Bureau of Laboratories | Michigan Department of Health and Human Services, Bureau of Laboratories | Blankenship HM et al |
| EPI_ISL_708344 | USA/MI-MDHHS-SC22364/2020 | 2020-11-13 | North America/USA/Michigan/ | Michigan Department of Health and Human Services, Bureau of Laboratories | Michigan Department of Health and Human Services, Bureau of Laboratories | Blankenship HM et al |
| EPI_ISL_708347 | USA/MI-MDHHS-SC22367/2020 | 2020-11-02 | North America/USA/Michigan/ | Michigan Department of Health and Human Services, Bureau of Laboratories | Michigan Department of Health and Human Services, Bureau of Laboratories | Blankenship HM et al |
| EPI_ISL_708354 | USA/MI-MDHHS-SC22374/2020 | 2020-11-12 | North America/USA/Michigan/ | Michigan Department of Health and Human Services, Bureau of Laboratories | Michigan Department of Health and Human Services, Bureau of Laboratories | Blankenship HM et al |
| EPI_ISL_708360 | USA/MI-MDHHS-SC22380/2020 | 2020-11-09 | North America/USA/Michigan/ | Michigan Department of Health and Human Services, Bureau of Laboratories | Michigan Department of Health and Human Services, Bureau of Laboratories | Blankenship HM et al |

| | | | | | | |
|---|---|---|---|---|---|---|
| EPI_ISL_708361 | USA/MI-MDHHS-SC22381/2020 | 2020-11-12 | North America/USA/Michigan/ | Michigan Department of Health and Human Services, Bureau of Laboratories | Michigan Department of Health and Human Services, Bureau of Laboratories | Blankenship HM et al |
| EPI_ISL_708370 | USA/MI-MDHHS-SC22390/2020 | 2020-11-12 | North America/USA/Michigan/ | Michigan Department of Health and Human Services, Bureau of Laboratories | Michigan Department of Health and Human Services, Bureau of Laboratories | Blankenship HM et al |
| EPI_ISL_708376 | USA/MI-MDHHS-SC22396/2020 | 2020-11-05 | North America/USA/Michigan/ | Michigan Department of Health and Human Services, Bureau of Laboratories | Michigan Department of Health and Human Services, Bureau of Laboratories | Blankenship HM et al |
| EPI_ISL_708383 | USA/DE-DHSS-F1009945/2020 | 2020-10-15 | North America/USA/Delaware/New Cas | Delaware Public Health Lab | Delaware Public Health Lab | Gregory Hovan et al |
| EPI_ISL_708386 | USA/DE-DHSS-F980500/2020 | 2020-08-16 | North America/USA/Delaware/Kent Cou | Delaware Public Health Lab | Delaware Public Health Lab | Gregory Hovan et al |
| EPI_ISL_708388 | USA/DE-DHSS-F1009900/2020 | 2020-10-14 | North America/USA/Delaware/Kent Cou | Delaware Public Health Lab | Delaware Public Health Lab | Gregory Hovan et al |
| EPI_ISL_708392 | USA/DE-DHSS-F1016592/2020 | 2020-10-14 | North America/USA/Delaware/New Cas | Delaware Public Health Lab | Delaware Public Health Lab | Gregory Hovan et al |
| EPI_ISL_708401 | USA/DE-DHSS-F980693/2020 | 2020-08-19 | North America/USA/Delaware/New Cas | Delaware Public Health Lab | Delaware Public Health Lab | Gregory Hovan et al |
| EPI_ISL_708406 | USA/DE-DHSS-F995649/2020 | 2020-09-08 | North America/USA/Delaware/Sussex C | Delaware Public Health Lab | Delaware Public Health Lab | Gregory Hovan et al |
| EPI_ISL_708425 | USA/DE-DHSS-F981198/2020 | 2020-08-14 | North America/USA/Delaware/Kent Cou | Delaware Public Health Lab | Delaware Public Health Lab | Gregory Hovan et al |
| EPI_ISL_708434 | USA/DE-DHSS-F1001769/2020 | 2020-09-10 | North America/USA/Delaware/New Cas | Delaware Public Health Lab | Delaware Public Health Lab | Gregory Hovan et al |
| EPI_ISL_708435 | USA/DE-DHSS-F994223/2020 | 2020-09-14 | North America/USA/Delaware/Kent Cou | Delaware Public Health Lab | Delaware Public Health Lab | Gregory Hovan et al |
| EPI_ISL_708437 | USA/DE-DHSS-F1019096/2020 | 2020-10-28 | North America/USA/Delaware/Kent Cou | Delaware Public Health Lab | Delaware Public Health Lab | Gregory Hovan et al |
| EPI_ISL_708447 | USA/DE-DHSS-F1019092/2020 | 2020-10-28 | North America/USA/Delaware/Kent Cou | Delaware Public Health Lab | Delaware Public Health Lab | Gregory Hovan et al |

| | | | | | | |
|---|---|---|---|---|---|---|
| EPI_ISL_708448 | USA/DE-DHSS-F980816/2020 | 2020-08-24 | North America/USA/Delaware/Kent Cou | Delaware Public Health Lab | Delaware Public Health Lab | Gregory Hovan et al |
| EPI_ISL_708451 | USA/DE-DHSS-F1020823/2020 | 2020-10-29 | North America/USA/Delaware/Kent Cou | Delaware Public Health Lab | Delaware Public Health Lab | Gregory Hovan et al |
| EPI_ISL_708455 | Belgium/ULG-10986/2020 | 2020-10-25 | Europe/Belgium/Li√®ge/ | Department of Clinical Microbiology | GIGA Medical Genomics | Keith Durkin et al |
| EPI_ISL_708460 | USA/MN-MDH-2122/2020 | 2020-10-30 | North America/USA/Minnesota/ | Minnesota Department of Health, Public Health Laboratory | Minnesota Department of Health, Public Health Laboratory | Alexandra Lorentz et al |
| EPI_ISL_708490 | USA/MN-MDH-2152/2020 | 2020-11-25 | North America/USA/Minnesota/ | Minnesota Department of Health, Public Health Laboratory | Minnesota Department of Health, Public Health Laboratory | Alexandra Lorentz et al |
| EPI_ISL_708494 | USA/MN-MDH-2156/2020 | 2020-11-30 | North America/USA/Minnesota/ | Minnesota Department of Health, Public Health Laboratory | Minnesota Department of Health, Public Health Laboratory | Alexandra Lorentz et al |
| EPI_ISL_708505 | USA/CA-LACPHL-AD00005/2020 | 2020-05-05 | North America/USA/California/Los Ange | Los Angeles County Public Health Laboratory | Los Angeles County Public Health Laboratory | P. Hemarajata et al. et al |
| EPI_ISL_708506 | USA/CA-LACPHL-AD00006/2020 | 2020-04-17 | North America/USA/California/Los Ange | Los Angeles County Public Health Laboratory | Los Angeles County Public Health Laboratory | P. Hemarajata et al. et al |
| EPI_ISL_708529 | Brazil/SP-844R1/2020 | 2020-06-29 | South America/Brazil/S√£o Paulo/Ferna | Secret√°ria Municipal de Saude de Fernand√≥polis | Instituto Adolfo Lutz, Interdiciplinary Procedures Center, Strategic Laboratory | Claudio Tavares Sacchi et al |
| EPI_ISL_708530 | Brazil/SP-844R2/2020 | 2020-11-21 | South America/Brazil/S√£o Paulo/Ferna | Secretaria Municipal de Saude de Fernand√≥polis | Instituto Adolfo Lutz, Interdiciplinary Procedures Center, Strategic Laboratory | Claudio Tavares Sacchi et al |

| | | | | | | |
|---|---|---|---|---|---|---|
| EPI_ISL_708543 | USA/MI-MDHHS-SC22451/2020 | 2020-11-03 | North America/USA/Michigan/ | Michigan Department of Health and Human Services, Bureau of Laboratories | Michigan Department of Health and Human Services, Bureau of Laboratories | Blankenship HM et al |
| EPI_ISL_708546 | USA/MI-MDHHS-SC22453/2020 | 2020-11-10 | North America/USA/Michigan/ | Michigan Department of Health and Human Services, Bureau of Laboratories | Michigan Department of Health and Human Services, Bureau of Laboratories | Blankenship HM et al |
| EPI_ISL_708553 | USA/MI-MDHHS-SC22536/2020 | 2020-11-02 | North America/USA/Michigan/ | Michigan Department of Health and Human Services, Bureau of Laboratories | Michigan Department of Health and Human Services, Bureau of Laboratories | Blankenship HM et al |
| EPI_ISL_708561 | USA/MI-MDHHS-SC22538/2020 | 2020-11-02 | North America/USA/Michigan/ | Michigan Department of Health and Human Services, Bureau of Laboratories | Michigan Department of Health and Human Services, Bureau of Laboratories | Blankenship HM et al |
| EPI_ISL_708563 | USA/MI-MDHHS-SC22544/2020 | 2020-11-08 | North America/USA/Michigan/ | Michigan Department of Health and Human Services, Bureau of Laboratories | Michigan Department of Health and Human Services, Bureau of Laboratories | Blankenship HM et al |
| EPI_ISL_708569 | USA/MI-MDHHS-SC22467/2020 | 2020-11-03 | North America/USA/Michigan/ | Michigan Department of Health and Human Services, Bureau of Laboratories | Michigan Department of Health and Human Services, Bureau of Laboratories | Blankenship HM et al |

| | | | | | | |
|---|---|---|---|---|---|---|
| EPI_ISL_708577 | USA/MI-MDHHS-SC22476/2020 | 2020-10-29 | North America/USA/Michigan/ | Michigan Department of Health and Human Services, Bureau of Laboratories | Michigan Department of Health and Human Services, Bureau of Laboratories | Blankenship HM et al |
| EPI_ISL_708579 | USA/MI-MDHHS-SC22478/2020 | 2020-11-02 | North America/USA/Michigan/ | Michigan Department of Health and Human Services, Bureau of Laboratories | Michigan Department of Health and Human Services, Bureau of Laboratories | Blankenship HM et al |
| EPI_ISL_708587 | USA/MI-MDHHS-SC22486/2020 | 2020-11-08 | North America/USA/Michigan/ | Michigan Department of Health and Human Services, Bureau of Laboratories | Michigan Department of Health and Human Services, Bureau of Laboratories | Blankenship HM et al |
| EPI_ISL_708588 | USA/MI-MDHHS-SC22487/2020 | 2020-11-02 | North America/USA/Michigan/ | Michigan Department of Health and Human Services, Bureau of Laboratories | Michigan Department of Health and Human Services, Bureau of Laboratories | Blankenship HM et al |
| EPI_ISL_708590 | USA/MI-MDHHS-SC22489/2020 | 2020-11-02 | North America/USA/Michigan/ | Michigan Department of Health and Human Services, Bureau of Laboratories | Michigan Department of Health and Human Services, Bureau of Laboratories | Blankenship HM et al |
| EPI_ISL_708592 | USA/MI-MDHHS-SC22491/2020 | 2020-11-03 | North America/USA/Michigan/ | Michigan Department of Health and Human Services, Bureau of Laboratories | Michigan Department of Health and Human Services, Bureau of Laboratories | Blankenship HM et al |
| EPI_ISL_708609 | USA/TX-DSHS-1711/2020 | 2020-06-17 | North America/USA/Texas/El Paso Coun | Texas Department of State Health Services | Texas Department of State Health Services | Rashmi Tuladhar et al |

| | | | | | | |
|---|---|---|---|---|---|---|
| EPI_ISL_708615 | USA/TX-DSHS-1705/2020 | 2020-06-15 | North America/USA/Texas/Harris Count | Texas Department of State Health Services | Texas Department of State Health Services | Rashmi Tuladhar et al |
| EPI_ISL_708616 | USA/TX-DSHS-1714/2020 | 2020-06-20 | North America/USA/Texas/Harris Count | Texas Department of State Health Services | Texas Department of State Health Services | Rashmi Tuladhar et al |
| EPI_ISL_708618 | USA/TX-DSHS-1681/2020 | 2020-05-12 | North America/USA/Texas/El Paso Coun | Texas Department of State Health Services | Texas Department of State Health Services | Rashmi Tuladhar et al |
| EPI_ISL_708622 | USA/TX-DSHS-1684/2020 | 2020-05-14 | North America/USA/Texas/Harris Count | Texas Department of State Health Services | Texas Department of State Health Services | Rashmi Tuladhar et al |
| EPI_ISL_708635 | USA/TX-DSHS-1686/2020 | 2020-05-14 | North America/USA/Texas/Hidalgo Cour | Texas Department of State Health Services | Texas Department of State Health Services | Rashmi Tuladhar et al |
| EPI_ISL_708636 | USA/TX-DSHS-1725/2020 | 2020-06-19 | North America/USA/Texas/Harris Count | Texas Department of State Health Services | Texas Department of State Health Services | Rashmi Tuladhar et al |
| EPI_ISL_708652 | Indonesia/BT-SHSIU-01-4920/2020 | 2020-10-18 | Asia/Indonesia/Banten/ | Medical Research Center, Faculty of Medicine, Syarif Hidayatullah State Islamic University Jakarta | Medical Research Center, Faculty of Medicine, Syarif Hidayatullah State Islamic University Jakarta | Chris Adhiyanto et al |
| EPI_ISL_708724 | USA/CA-LACPHL-AE00032/2020 | 2020-10-15 | North America/USA/California/Los Ange | Los Angeles County Public Health Laboratory | Los Angeles County Public Health Laboratory | P. Hemarajata et al. et al |
| EPI_ISL_708728 | USA/CA-LACPHL-AE00036/2020 | 2020-09-01 | North America/USA/California/Los Ange | Los Angeles County Public Health Laboratory | Los Angeles County Public Health Laboratory | P. Hemarajata et al. et al |
| EPI_ISL_708734 | USA/CA-LACPHL-AE00042/2020 | 2020-07-17 | North America/USA/California/Los Ange | Los Angeles County Public Health Laboratory | Los Angeles County Public Health Laboratory | P. Hemarajata et al. et al |
| EPI_ISL_708815 | Thailand/Bangkok-14712/2020 | 2020-09-24 | Asia/Thailand/Bangkok/ | Urban Institute for Disease Prevention and Control | National Institute of Health, Department of Medical Sciences, Ministry of Public Health, Thailand | Pilailuk Okada et al |

| | | | | | | |
|---|---|---|---|---|---|---|
| EPI_ISL_708823 | Thailand/Bangkok-Tho07/2020 | 2020-10-31 | Asia/Thailand/Bangkok/ | Vibharam Hospital | National Institute of Health, Department of Medical Sciences, Ministry of Public Health, Thailand | Pilailuk Okada et al |
| EPI_ISL_708828 | UnitedArabEmirates/1092/2020 | 2020-05-15 | Asia/United Arab Emirates/Abu Dhabi/ | Group 42 (G42) Healthcare, Abu Dhabi, United Arab Emirates; Department of Health, The United Arab Emirates | G42 Healthcare | Rong Liu et al |
| EPI_ISL_708834 | UnitedArabEmirates/0758/2020 | 2020-05-15 | Asia/United Arab Emirates/Abu Dhabi/ | Group 42 (G42) Healthcare, Abu Dhabi, United Arab Emirates; Department of Health, The United Arab Emirates | G42 Healthcare | Rong Liu et al |
| EPI_ISL_708839 | Pakistan/NIBD-01-KHI/2020 | 2020-06-06 | Asia/Pakistan/Karachi/ | National Institute of Blood Diseases (NIBD), Molecular Biology Lab | Genomics Lab NIBD | Samina Naz Mukry et al |
| EPI_ISL_708840 | Pakistan/NIBD-02-KHI/2020 | 2020-06-10 | Asia/Pakistan/Karachi/ | National Institute of Blood Diseases (NIBD), Molecular Biology Lab | Genomics Lab NIBD | Samina Naz Mukry et al |
| EPI_ISL_708853 | England/ALDP-BE8EEB/2020 | 2020-12-03 | Europe/United Kingdom/England/ | Lighthouse Lab in Alderley Park | Wellcome Sanger Institute for the COVID-19 Genomics UK (COG-UK) Consortium | Jacquelyn Wynn et al |
| EPI_ISL_708905 | England/QEUH-BEB097/2020 | 2020-12-03 | Europe/United Kingdom/England/ | Lighthouse Lab in Glasgow | Wellcome Sanger Institute for the COVID-19 Genomics UK (COG-UK) Consortium | Harper VanSteenhouse et al |
| EPI_ISL_709044 | Pakistan/NIBD-03-KHI/2020 | 2020-06-10 | Asia/Pakistan/Karachi/ | National Institute of Blood Diseases (NIBD), Molecular Biology Lab | Genomics Lab NIBD | Samina Naz Mukry et al |
| EPI_ISL_709079 | England/QEUH-BF0A28/2020 | 2020-12-05 | Europe/United Kingdom/England/ | Lighthouse Lab in Glasgow | Wellcome Sanger Institute for the COVID-19 Genomics UK (COG-UK) Consortium | Harper VanSteenhouse et al |
| EPI_ISL_709495 | England/QEUH-C149B7/2020 | 2020-12-06 | Europe/United Kingdom/England/ | Lighthouse Lab in Glasgow | Wellcome Sanger Institute for the COVID-19 Genomics UK (COG-UK) Consortium | Harper VanSteenhouse et al |
| EPI_ISL_709542 | Pakistan/NIBD-04-KHI/2020 | 2020-10-15 | Asia/Pakistan/Karachi/ | National Institute of Blood Diseases (NIBD), Molecular Biology Lab | Genomics Lab NIBD | Samina Naz Mukry et al |

| | | | | | | |
|---|---|---|---|---|---|---|
| EPI_ISL_709830 | England/MILK-BE7013/2020 | 2020-11-29 | Europe/United Kingdom/England/ | Lighthouse Lab in Milton Keynes | Wellcome Sanger Institute for the COVID-19 Genomics UK (COG-UK) Consortium | The Lighthouse Lab in Milton Keynes et al |
| EPI_ISL_709895 | England/MILK-B94B14/2020 | 2020-11-13 | Europe/United Kingdom/England/ | Lighthouse Lab in Milton Keynes | Wellcome Sanger Institute for the COVID-19 Genomics UK (COG-UK) Consortium | The Lighthouse Lab in Milton Keynes et al |
| EPI_ISL_710035 | USA/MI-UM-10036783209/2020 | 2020-11-04 | North America/USA/Michigan/ | University of Michigan Clinical Microbiology Laboratory | Lauring Lab, University of Michigan, Department of Microbiology and Immunology | Valesano et al |
| EPI_ISL_710056 | USA/TX-DSHS-1679/2020 | 2020-06-24 | North America/USA/Texas/Hidalgo Cour | Texas Department of State Health Services | Texas Department of State Health Services | Anita Pokharel et al |
| EPI_ISL_710057 | USA/TX-DSHS-1639/2020 | 2020-06-24 | North America/USA/Texas/Hidalgo Cour | Texas Department of State Health Services | Texas Department of State Health Services | Anita Pokharel et al |
| EPI_ISL_710059 | USA/TX-DSHS-1673/2020 | 2020-06-24 | North America/USA/Texas/Hidalgo Cour | Texas Department of State Health Services | Texas Department of State Health Services | Anita Pokharel et al |
| EPI_ISL_710063 | USA/TX-DSHS-1672/2020 | 2020-06-24 | North America/USA/Texas/Hidalgo Cour | Texas Department of State Health Services | Texas Department of State Health Services | Anita Pokharel et al |
| EPI_ISL_710067 | USA/TX-DSHS-1644/2020 | 2020-06-24 | North America/USA/Texas/Hidalgo Cour | Texas Department of State Health Services | Texas Department of State Health Services | Anita Pokharel et al |
| EPI_ISL_710072 | USA/TX-DSHS-1652/2020 | 2020-06-24 | North America/USA/Texas/Hidalgo Cour | Texas Department of State Health Services | Texas Department of State Health Services | Anita Pokharel et al |
| EPI_ISL_710075 | USA/TX-DSHS-1646/2020 | 2020-06-24 | North America/USA/Texas/Hidalgo Cour | Texas Department of State Health Services | Texas Department of State Health Services | Anita Pokharel et al |
| EPI_ISL_710081 | USA/TX-DSHS-1676/2020 | 2020-06-24 | North America/USA/Texas/Hidalgo Cour | Texas Department of State Health Services | Texas Department of State Health Services | Anita Pokharel et al |

| | | | | | | |
|---|---|---|---|---|---|---|
| EPI_ISL_710097 | USA/MD-MDH-0483/2020 | 2020-11-12 | North America/USA/Maryland/ | Maryland Public Health Laboratory | Maryland Public Health Laboratory | Maryland Department of Health Laboratories Administration et al |
| EPI_ISL_710101 | USA/CA-LACPHL-AA00003/2020 | 2020-07-31 | North America/USA/California/Los Ange | Los Angeles County PHL | Los Angeles County PHL | P. Hemarajata et al. et al |
| EPI_ISL_710102 | USA/CA-LACPHL-AA00004/2020 | 2020-08-01 | North America/USA/California/Los Ange | Los Angeles County PHL | Los Angeles County PHL | P. Hemarajata et al. et al |
| EPI_ISL_710105 | USA/CA-LACPHL-AA00007/2020 | 2020-08-03 | North America/USA/California/Los Ange | Los Angeles County PHL | Los Angeles County PHL | P. Hemarajata et al. et al |
| EPI_ISL_710106 | USA/CA-LACPHL-AA00008/2020 | 2020-08-04 | North America/USA/California/Los Ange | Los Angeles County PHL | Los Angeles County PHL | P. Hemarajata et al. et al |
| EPI_ISL_710107 | USA/CA-LACPHL-AA00009/2020 | 2020-08-04 | North America/USA/California/Los Ange | Los Angeles County PHL | Los Angeles County PHL | P. Hemarajata et al. et al |
| EPI_ISL_710108 | USA/CA-LACPHL-AA00010/2020 | 2020-08-06 | North America/USA/California/Los Ange | Los Angeles County PHL | Los Angeles County PHL | P. Hemarajata et al. et al |
| EPI_ISL_710132 | USA/CO-CDPHE-2009163985/2020 | 2020-09-16 | North America/USA/Colorado/ | Colorado Department of Public Health and Environment | Colorado Department of Puplic Health and Environment | Laura Bankers et al |
| EPI_ISL_710135 | USA/CO-CDPHE-2004220716/2020 | 2020-04-22 | North America/USA/Colorado/ | Colorado Department of Public Health and Environment | Colorado Department of Puplic Health and Environment | Laura Bankers et al |
| EPI_ISL_710139 | USA/CO-CDPHE-2004230116/2020 | 2020-04-22 | North America/USA/Colorado/ | Colorado Department of Public Health and Environment | Colorado Department of Puplic Health and Environment | Laura Bankers et al |
| EPI_ISL_710141 | USA/CO-CDPHE-2008182422/2020 | 2020-08-18 | North America/USA/Colorado/ | Colorado Department of Public Health and Environment | Colorado Department of Puplic Health and Environment | Laura Bankers et al |
| EPI_ISL_710145 | USA/CO-CDPHE-2005220650/2020 | 2020-05-21 | North America/USA/Colorado/ | Colorado Department of Public Health and Environment | Colorado Department of Puplic Health and Environment | Laura Bankers et al |
| EPI_ISL_710155 | USA/CO-CDPHE-2010200214/2020 | 2020-10-19 | North America/USA/Colorado/ | Colorado Department of Public Health and Environment | Colorado Department of Puplic Health and Environment | Laura Bankers et al |

| | | | | | | |
|---|---|---|---|---|---|---|
| EPI_ISL_710156 | USA/CO-CDPHE-2005080929/2020 | 2020-05-08 | North America/USA/Colorado/ | Colorado Department of Public Health and Environment | Colorado Department of Puplic Health and Environment | Laura Bankers et al |
| EPI_ISL_710161 | USA/CO-CDPHE-2004220712/2020 | 2020-04-22 | North America/USA/Colorado/ | Colorado Department of Public Health and Environment | Colorado Department of Puplic Health and Environment | Laura Bankers et al |
| EPI_ISL_710163 | USA/CO-CDPHE-2008200804/2020 | 2020-08-19 | North America/USA/Colorado/ | Colorado Department of Public Health and Environment | Colorado Department of Puplic Health and Environment | Laura Bankers et al |
| EPI_ISL_710176 | USA/CO-CDPHE-2004220696/2020 | 2020-04-21 | North America/USA/Colorado/ | Colorado Department of Public Health and Environment | Colorado Department of Puplic Health and Environment | Laura Bankers et al |
| EPI_ISL_710178 | USA/CO-CDPHE-2004220694/2020 | 2020-04-21 | North America/USA/Colorado/ | Colorado Department of Public Health and Environment | Colorado Department of Puplic Health and Environment | Laura Bankers et al |
| EPI_ISL_710183 | USA/CO-CDPHE-2004220698/2020 | 2020-04-21 | North America/USA/Colorado/ | Colorado Department of Public Health and Environment | Colorado Department of Puplic Health and Environment | Laura Bankers et al |
| EPI_ISL_710191 | USA/CO-CDPHE-2004220761/2020 | 2020-04-21 | North America/USA/Colorado/ | Colorado Department of Public Health and Environment | Colorado Department of Puplic Health and Environment | Laura Bankers et al |
| EPI_ISL_710200 | USA/CO-CDPHE-2004220872/2020 | 2020-04-21 | North America/USA/Colorado/ | Colorado Department of Public Health and Environment | Colorado Department of Puplic Health and Environment | Laura Bankers et al |
| EPI_ISL_710201 | USA/CO-CDPHE-2004220878/2020 | 2020-04-21 | North America/USA/Colorado/ | Colorado Department of Public Health and Environment | Colorado Department of Puplic Health and Environment | Laura Bankers et al |
| EPI_ISL_710202 | USA/CO-CDPHE-2004230010/2020 | 2020-04-21 | North America/USA/Colorado/ | Colorado Department of Public Health and Environment | Colorado Department of Puplic Health and Environment | Laura Bankers et al |

| | | | | | | |
|---|---|---|---|---|---|---|
| EPI_ISL_710204 | USA/CO-CDPHE-2004230195/2020 | 2020-04-21 | North America/USA/Colorado/ | Colorado Department of Public Health and Environment | Colorado Department of Puplic Health and Environment | Laura Bankers et al |
| EPI_ISL_710212 | USA/CO-CDPHE-2005220143/2020 | 2020-05-21 | North America/USA/Colorado/ | Colorado Department of Public Health and Environment | Colorado Department of Puplic Health and Environment | Laura Bankers et al |
| EPI_ISL_710214 | USA/CO-CDPHE-2007230810/2020 | 2020-07-22 | North America/USA/Colorado/ | Colorado Department of Public Health and Environment | Colorado Department of Puplic Health and Environment | Laura Bankers et al |
| EPI_ISL_710220 | USA/CO-CDPHE-2008181078/2020 | 2020-08-17 | North America/USA/Colorado/ | Colorado Department of Public Health and Environment | Colorado Department of Puplic Health and Environment | Laura Bankers et al |
| EPI_ISL_710221 | USA/CO-CDPHE-2008181628/2020 | 2020-08-17 | North America/USA/Colorado/ | Colorado Department of Public Health and Environment | Colorado Department of Puplic Health and Environment | Laura Bankers et al |
| EPI_ISL_710222 | USA/CO-CDPHE-2009042396/2020 | 2020-09-03 | North America/USA/Colorado/ | Colorado Department of Public Health and Environment | Colorado Department of Puplic Health and Environment | Laura Bankers et al |
| EPI_ISL_710223 | USA/CO-CDPHE-2009053317/2020 | 2020-09-04 | North America/USA/Colorado/ | Colorado Department of Public Health and Environment | Colorado Department of Puplic Health and Environment | Laura Bankers et al |
| EPI_ISL_710224 | USA/CO-CDPHE-2009131675/2020 | 2020-09-12 | North America/USA/Colorado/ | Colorado Department of Public Health and Environment | Colorado Department of Puplic Health and Environment | Laura Bankers et al |
| EPI_ISL_710225 | USA/CO-CDPHE-2009131687/2020 | 2020-09-12 | North America/USA/Colorado/ | Colorado Department of Public Health and Environment | Colorado Department of Puplic Health and Environment | Laura Bankers et al |
| EPI_ISL_710226 | USA/CO-CDPHE-2009131689/2020 | 2020-09-12 | North America/USA/Colorado/ | Colorado Department of Public Health and Environment | Colorado Department of Puplic Health and Environment | Laura Bankers et al |

| | | | | | | |
|---|---|---|---|---|---|---|
| EPI_ISL_710233 | USA/CO-CDPHE-2009163956/2020 | 2020-09-16 | North America/USA/Colorado/ | Colorado Department of Public Health and Environment | Colorado Department of Puplic Health and Environment | Laura Bankers et al |
| EPI_ISL_710235 | USA/CO-CDPHE-2009164005/2020 | 2020-09-16 | North America/USA/Colorado/ | Colorado Department of Public Health and Environment | Colorado Department of Puplic Health and Environment | Laura Bankers et al |
| EPI_ISL_710248 | USA/CO-CDPHE-2005220217/2020 | 2020-05-21 | North America/USA/Colorado/ | Colorado Department of Public Health and Environment | Colorado Department of Puplic Health and Environment | Laura Bankers et al |
| EPI_ISL_710249 | USA/CO-CDPHE-2006020968/2020 | 2020-06-02 | North America/USA/Colorado/ | Colorado Department of Public Health and Environment | Colorado Department of Puplic Health and Environment | Laura Bankers et al |
| EPI_ISL_710250 | USA/CO-CDPHE-2008222966/2020 | 2020-08-22 | North America/USA/Colorado/ | Colorado Department of Public Health and Environment | Colorado Department of Puplic Health and Environment | Laura Bankers et al |
| EPI_ISL_710252 | USA/CO-CDPHE-2009170012/2020 | 2020-09-16 | North America/USA/Colorado/ | Colorado Department of Public Health and Environment | Colorado Department of Puplic Health and Environment | Laura Bankers et al |
| EPI_ISL_710265 | USA/CO-CDPHE-2009300877/2020 | 2020-09-28 | North America/USA/Colorado/ | Colorado Department of Public Health and Environment | Colorado Department of Puplic Health and Environment | Laura Bankers et al |
| EPI_ISL_710280 | USA/CO-CDPHE-2009170035/2020 | 2020-09-16 | North America/USA/Colorado/ | Colorado Department of Public Health and Environment | Colorado Department of Puplic Health and Environment | Laura Bankers et al |
| EPI_ISL_710297 | USA/CO-CDPHE-2005220680/2020 | 2020-05-21 | North America/USA/Colorado/ | Colorado Department of Public Health and Environment | Colorado Department of Puplic Health and Environment | Laura Bankers et al |
| EPI_ISL_710299 | USA/CO-CDPHE-2008282932/2020 | 2020-08-26 | North America/USA/Colorado/ | Colorado Department of Public Health and Environment | Colorado Department of Puplic Health and Environment | Laura Bankers et al |

| | | | | | | |
|---|---|---|---|---|---|---|
| EPI_ISL_710306 | USA/CO-CDPHE-2005220251/2020 | 2020-05-21 | North America/USA/Colorado/ | Colorado Department of Public Health and Environment | Colorado Department of Puplic Health and Environment | Laura Bankers et al |
| EPI_ISL_710315 | USA/CO-CDPHE-2005270733/2020 | 2020-05-26 | North America/USA/Colorado/ | Colorado Department of Public Health and Environment | Colorado Department of Puplic Health and Environment | Laura Bankers et al |
| EPI_ISL_710322 | USA/CO-CDPHE-2008081584/2020 | 2020-08-06 | North America/USA/Colorado/ | Colorado Department of Public Health and Environment | Colorado Department of Puplic Health and Environment | Laura Bankers et al |
| EPI_ISL_710325 | USA/CO-CDPHE-2008210131/2020 | 2020-08-20 | North America/USA/Colorado/ | Colorado Department of Public Health and Environment | Colorado Department of Puplic Health and Environment | Laura Bankers et al |
| EPI_ISL_710326 | USA/CO-CDPHE-2008222958/2020 | 2020-08-22 | North America/USA/Colorado/ | Colorado Department of Public Health and Environment | Colorado Department of Puplic Health and Environment | Laura Bankers et al |
| EPI_ISL_710332 | USA/CO-CDPHE-2009052131/2020 | 2020-09-03 | North America/USA/Colorado/ | Colorado Department of Public Health and Environment | Colorado Department of Puplic Health and Environment | Laura Bankers et al |
| EPI_ISL_710336 | USA/CO-CDPHE-2009164173/2020 | 2020-09-16 | North America/USA/Colorado/ | Colorado Department of Public Health and Environment | Colorado Department of Puplic Health and Environment | Laura Bankers et al |
| EPI_ISL_710338 | USA/CO-CDPHE-2009170073/2020 | 2020-09-16 | North America/USA/Colorado/ | Colorado Department of Public Health and Environment | Colorado Department of Puplic Health and Environment | Laura Bankers et al |
| EPI_ISL_710340 | USA/CO-CDPHE-2009170087/2020 | 2020-09-16 | North America/USA/Colorado/ | Colorado Department of Public Health and Environment | Colorado Department of Puplic Health and Environment | Laura Bankers et al |
| EPI_ISL_710343 | USA/CO-CDPHE-2009170327/2020 | 2020-09-16 | North America/USA/Colorado/ | Colorado Department of Public Health and Environment | Colorado Department of Puplic Health and Environment | Laura Bankers et al |

| | | | | | | |
|---|---|---|---|---|---|---|
| EPI_ISL_710350 | USA/CO-CDPHE-2010112598/2020 | 2020-10-08 | North America/USA/Colorado/ | Colorado Department of Public Health and Environment | Colorado Department of Puplic Health and Environment | Laura Bankers et al |
| EPI_ISL_710351 | USA/CO-CDPHE-2010120393/2020 | 2020-10-09 | North America/USA/Colorado/ | Colorado Department of Public Health and Environment | Colorado Department of Puplic Health and Environment | Laura Bankers et al |
| EPI_ISL_710361 | USA/CO-CDPHE-2010280189/2020 | 2020-10-26 | North America/USA/Colorado/ | Colorado Department of Public Health and Environment | Colorado Department of Puplic Health and Environment | Laura Bankers et al |
| EPI_ISL_710364 | USA/CO-CDPHE-2010280211/2020 | 2020-10-26 | North America/USA/Colorado/ | Colorado Department of Public Health and Environment | Colorado Department of Puplic Health and Environment | Laura Bankers et al |
| EPI_ISL_710370 | USA/CO-CDPHE-2011020263/2020 | 2020-11-01 | North America/USA/Colorado/ | Colorado Department of Public Health and Environment | Colorado Department of Puplic Health and Environment | Laura Bankers et al |
| EPI_ISL_710373 | USA/CO-CDPHE-2011020331/2020 | 2020-11-01 | North America/USA/Colorado/ | Colorado Department of Public Health and Environment | Colorado Department of Puplic Health and Environment | Laura Bankers et al |
| EPI_ISL_710376 | USA/TX-DSHS-1743/2020 | 2020-06-13 | North America/USA/Texas/Harris Count | Texas Department of State Health Services | Texas Department of State Health Services | Rashmi Tuladhar et al |
| EPI_ISL_710386 | USA/TX-DSHS-1763/2020 | 2020-06-21 | North America/USA/Texas/Hidalgo Cour | Texas Department of State Health Services | Texas Department of State Health Services | Rashmi Tuladhar et al |
| EPI_ISL_710392 | USA/TX-DSHS-1736/2020 | 2020-06-20 | North America/USA/Texas/Harris Count | Texas Department of State Health Services | Texas Department of State Health Services | Rashmi Tuladhar et al |
| EPI_ISL_710394 | USA/TX-DSHS-1738/2020 | 2020-06-20 | North America/USA/Texas/Harris Count | Texas Department of State Health Services | Texas Department of State Health Services | Rashmi Tuladhar et al |
| EPI_ISL_710396 | USA/TX-DSHS-1742/2020 | 2020-06-13 | North America/USA/Texas/Harris Count | Texas Department of State Health Services | Texas Department of State Health Services | Rashmi Tuladhar et al |

| | | | | | | |
|---|---|---|---|---|---|---|
| EPI_ISL_710398 | USA/TX-DSHS-1746/2020 | 2020-06-13 | North America/USA/Texas/Harris Count | Texas Department of State Health Services | Texas Department of State Health Services | Rashmi Tuladhar et al |
| EPI_ISL_710400 | USA/TX-DSHS-1748/2020 | 2020-06-13 | North America/USA/Texas/Harris Count | Texas Department of State Health Services | Texas Department of State Health Services | Rashmi Tuladhar et al |
| EPI_ISL_710409 | USA/TX-DSHS-1767/2020 | 2020-06-20 | North America/USA/Texas/Harris Count | Texas Department of State Health Services | Texas Department of State Health Services | Rashmi Tuladhar et al |
| EPI_ISL_710421 | USA/CA-LACPHL-AA00028/2020 | 2020-10-13 | North America/USA/California/Los Ange | Los Angeles County PHL | Los Angeles County PHL | P. Hemarajata et al. et al |
| EPI_ISL_710423 | USA/CA-LACPHL-AA00027/2020 | 2020-10-13 | North America/USA/California/Los Ange | Los Angeles County PHL | Los Angeles County PHL | P. Hemarajata et al. et al |
| EPI_ISL_710503 | Italy/SAR-ASSL/2020 | 2020-10-19 | Europe/Italy/Sardinia/ | "Laboratorio specialistico UOC Ematologia - Ospedale ""San Francesco"" - ATS-ASSL Nuoro" | "Laboratorio specialistico UOC Ematologia - Ospedale ""San Francesco"" - ATS-ASSL Nuoro" | Giovanna Piras et al |
| EPI_ISL_710532 | Tunisia/ADAGE-12622/2020 | 2020-09-20 | Africa/Tunisia/Monastir/ | H√¥pital Fattouma-Bourguiba de Monastir | Laboratoire des Proc√©d√©s de Criblage Mol√©culaire et Cellulaire-Centre de Biotechnologie de Sfax | Souissi et al |
| EPI_ISL_710534 | Tunisia/ADAGE-17873/2020 | 2020-10-14 | Africa/Tunisia/Monastir/ | H√¥pital Fattouma-Bourguiba de Monastir | Laboratoire des Proc√©d√©s de Criblage Mol√©culaire et Cellulaire-Centre de Biotechnologie de Sfax | Souissi et al |
| EPI_ISL_710540 | Tunisia/ADAGE-18212/2020 | 2020-10-16 | Africa/Tunisia/Monastir/ | H√¥pital Fattouma-Bourguiba de Monastir | Laboratoire des Proc√©d√©s de Criblage Mol√©culaire et Cellulaire-Centre de Biotechnologie de Sfax | Souissi et al |

| | | | | | | |
|---|---|---|---|---|---|---|
| EPI_ISL_710541 | Tunisia/ADAGE-18245/2020 | 2020-10-16 | Africa/Tunisia/Monastir/ | H√¥pital Fattouma-Bourguiba de Monastir | Laboratoire des Proc√©d√©s de Criblage Mol√©culaire et Cellulaire-Centre de Biotechnologie de Sfax | Souissi et al |
| EPI_ISL_710544 | Italy/LAZ-INMI-94/2020 | 2020-10-27 | Europe/Italy/Lazio/Rome | "National Institute for Infectious Diseases, INMI, ""L. Spallanzani"" IRCCS" | "National Institute for Infectious Diseases, INMI, ""L. Spallanzani"" IRCCS" | M Rueca et al |
| EPI_ISL_710545 | Italy/LAZ-INMI-95/2020 | 2020-10-27 | Europe/Italy/Lazio/Rome | "National Institute for Infectious Diseases, INMI, ""L. Spallanzani"" IRCCS" | "National Institute for Infectious Diseases, INMI, ""L. Spallanzani"" IRCCS" | F Messina et al |
| EPI_ISL_710549 | Croatia/Zagreb-12/2020 | 2020-08-24 | Europe/Croatia/Zagreb/ | University Hospital Dubrava | Ruƒëer Bo≈°kovic Institute; Forensic Science Centre Ivan Vuƒçetiƒá; University of Zagreb Faculty of Science | Robert Belu≈æiƒá et al |
| EPI_ISL_710556 | Croatia/Zagreb-4/2020 | 2020-08-30 | Europe/Croatia/Zagreb/ | University Hospital Dubrava | Ruƒëer Bo≈°kovic Institute; Forensic Science Centre Ivan Vuƒçetiƒá; University of Zagreb Faculty of Science | Robert Belu≈æiƒá et al |
| EPI_ISL_710558 | Croatia/Zagreb-9/2020 | 2020-09-06 | Europe/Croatia/Zagreb/ | University Hospital Dubrava | Ruƒëer Bo≈°kovic Institute; Forensic Science Centre Ivan Vuƒçetiƒá; University of Zagreb Faculty of Science | Robert Belu≈æiƒá et al |

| | | | | | | |
|---|---|---|---|---|---|---|
| EPI_ISL_710562 | Croatia/Zagreb-18/2020 | 2020-09-08 | Europe/Croatia/Zagreb/ | University Hospital Dubrava | Rufëer Bo≈°kovic Institute; Forensic Science Centre Ivan Vuƒçetiƒá; University of Zagreb Faculty of Science | Robert Belu≈æiƒá et al |
| EPI_ISL_710563 | Croatia/Zagreb-28/2020 | 2020-09-09 | Europe/Croatia/Zagreb/ | Sestre Milosrdnice University Hospital Center | Rufëer Bo≈°kovic Institute; Forensic Science Centre Ivan Vuƒçetiƒá; University of Zagreb Faculty of Science | Robert Belu≈æiƒá et al |
| EPI_ISL_710568 | Croatia/Zagreb-10/2020 | 2020-09-30 | Europe/Croatia/Zagreb/ | University Hospital Dubrava | Rufëer Bo≈°kovic Institute; Forensic Science Centre Ivan Vuƒçetiƒá; University of Zagreb Faculty of Science | Robert Belu≈æiƒá et al |
| EPI_ISL_710570 | Croatia/Zagreb-22/2020 | 2020-10-01 | Europe/Croatia/Zagreb/ | University Hospital Dubrava | Rufëer Bo≈°kovic Institute; Forensic Science Centre Ivan Vuƒçetiƒá; University of Zagreb Faculty of Science | Robert Belu≈æiƒá et al |
| EPI_ISL_710571 | Croatia/Zagreb-5/2020 | 2020-10-02 | Europe/Croatia/Zagreb/ | University Hospital Dubrava | Rufëer Bo≈°kovic Institute; Forensic Science Centre Ivan Vuƒçetiƒá; University of Zagreb Faculty of Science | Robert Belu≈æiƒá et al |

| | | | | | | |
|---|---|---|---|---|---|---|
| EPI_ISL_710572 | Croatia/Zagreb-8/2020 | 2020-10-04 | Europe/Croatia/Zagreb/ | University Hospital Dubrava | Ruƒëer Bo≈°kovic Institute; Forensic Science Centre Ivan Vuƒçetiƒá; University of Zagreb Faculty of Science | Robert Belu≈æiƒá et al |
| EPI_ISL_710573 | Croatia/Zagreb-26/2020 | 2020-10-04 | Europe/Croatia/Zagreb/ | University Hospital Dubrava | Ruƒëer Bo≈°kovic Institute; Forensic Science Centre Ivan Vuƒçetiƒá; University of Zagreb Faculty of Science | Robert Belu≈æiƒá et al |
| EPI_ISL_710574 | Croatia/Zagreb-1/2020 | 2020-10-05 | Europe/Croatia/Zagreb/ | University Hospital Dubrava | Ruƒëer Bo≈°kovic Institute; Forensic Science Centre Ivan Vuƒçetiƒá; University of Zagreb Faculty of Science | Robert Belu≈æiƒá et al |
| EPI_ISL_710575 | Tunisia/ADAGE-17786/2020 | 2020-10-13 | Africa/Tunisia/Monastir/ | H√¥pital Fattouma-Bourguiba de Monastir | Laboratoire des Proc√©d√©s de Criblage Mol√©culaire et Cellulaire-Centre de Biotechnologie de Sfax | Souissi et al |
| EPI_ISL_710577 | Sweden/20-24575/2020 | 2020-11-23 | Europe/Sweden/Ostergotland/ | Vardcentralen Brinken | The Public Health Agency of Sweden | Department of Microbiology et al |
| EPI_ISL_710579 | Sweden/20-23090/2020 | 2020-03-19 | Europe/Sweden/Uppsala/ | Uppsala klinisk mikrobiologi | The Public Health Agency of Sweden | Department of Microbiology et al |
| EPI_ISL_710581 | Sweden/20-53564/2020 | 2020-11-25 | Europe/Sweden/Ostergotland/ | The Public Health Agency of Sweden | The Public Health Agency of Sweden | Department of Microbiology et al |
| EPI_ISL_710582 | Sweden/20-53565/2020 | 2020-11-24 | Europe/Sweden/Ostergotland/ | The Public Health Agency of Sweden | The Public Health Agency of Sweden | Department of Microbiology et al |
| EPI_ISL_710585 | Sweden/20-24683/2020 | 2020-11-24 | Europe/Sweden/Dalarna/ | Orsa VC | The Public Health Agency of Sweden | Department of Microbiology et al |
| EPI_ISL_710586 | Sweden/20-24685/2020 | 2020-11-24 | Europe/Sweden/Dalarna/ | Orsa VC | The Public Health Agency of Sweden | Department of Microbiology et al |

| | | | | | | |
|---|---|---|---|---|---|---|
| EPI_ISL_710588 | Sweden/20-53552/2020 | 2020-11-25 | Europe/Sweden/Vastra Gotaland/ | Omtanken Grimmered | The Public Health Agency of Sweden | Department of Microbiology et al |
| EPI_ISL_710593 | Sweden/20-53306/2020 | 2020-11-02 | Europe/Sweden/Stockholm/ | MLF Care AB | The Public Health Agency of Sweden | Department of Microbiology et al |
| EPI_ISL_710599 | Sweden/20-53341/2020 | 2020-11-01 | Europe/Sweden/Halland/ | Klinisk mikrobiologi | The Public Health Agency of Sweden | Department of Microbiology et al |
| EPI_ISL_710602 | Sweden/20-53344/2020 | 2020-11-03 | Europe/Sweden/Halland/ | Klinisk mikrobiologi | The Public Health Agency of Sweden | Department of Microbiology et al |
| EPI_ISL_710606 | Sweden/20-53366/2020 | 2020-11-06 | Europe/Sweden/Skane/ | Halsomedicinskt Center | The Public Health Agency of Sweden | Department of Microbiology et al |
| EPI_ISL_710612 | Sweden/20-24688/2020 | 2020-11-23 | Europe/Sweden/Stockholm/ | Capio Bro VC | The Public Health Agency of Sweden | Department of Microbiology et al |
| EPI_ISL_710613 | Sweden/20-53337/2020 | 2020-11-04 | Europe/Sweden/Halland/ | Barnakuten | The Public Health Agency of Sweden | Department of Microbiology et al |
| EPI_ISL_710658 | Scotland/QEUH-C14CEB/2020 | 2020-12-06 | Europe/United Kingdom/Scotland/ | Lighthouse Lab in Glasgow | Wellcome Sanger Institute for the COVID-19 Genomics UK (COG-UK) Consortium | Harper VanSteenhouse et al |
| EPI_ISL_710690 | Scotland/QEUH-C1696A/2020 | 2020-12-06 | Europe/United Kingdom/Scotland/ | Lighthouse Lab in Glasgow | Wellcome Sanger Institute for the COVID-19 Genomics UK (COG-UK) Consortium | Harper VanSteenhouse et al |
| EPI_ISL_710692 | Scotland/QEUH-C148F6/2020 | 2020-12-06 | Europe/United Kingdom/Scotland/ | Lighthouse Lab in Glasgow | Wellcome Sanger Institute for the COVID-19 Genomics UK (COG-UK) Consortium | Harper VanSteenhouse et al |
| EPI_ISL_710706 | Wales/ALDP-BB4F12/2020 | 2020-11-20 | Europe/United Kingdom/Wales/ | Lighthouse Lab in Alderley Park | Wellcome Sanger Institute for the COVID-19 Genomics UK (COG-UK) Consortium | Jacquelyn Wynn et al |
| EPI_ISL_710800 | Scotland/QEUH-BCCD1D/2020 | 2020-11-26 | Europe/United Kingdom/Scotland/ | Lighthouse Lab in Glasgow | Wellcome Sanger Institute for the COVID-19 Genomics UK (COG-UK) Consortium | Harper VanSteenhouse et al |

| | | | | | | |
|---|---|---|---|---|---|---|
| EPI_ISL_710959 | Wales/ALDP-B1E2C2/2020 | 2020-11-01 | Europe/United Kingdom/Wales/ | Lighthouse Lab in Alderley Park | Wellcome Sanger Institute for the COVID-19 Genomics UK (COG-UK) Consortium | Jacquelyn Wynn et al |
| EPI_ISL_711011 | Wales/ALDP-B24AC1/2020 | 2020-11-04 | Europe/United Kingdom/Wales/ | Lighthouse Lab in Alderley Park | Wellcome Sanger Institute for the COVID-19 Genomics UK (COG-UK) Consortium | Jacquelyn Wynn et al |
| EPI_ISL_711057 | Tunisia/ADAGE-18503/2020 | 2020-10-19 | Africa/Tunisia/Monastir/ | H√¥pital Fattouma-Bourguiba de Monastir | Laboratoire des Proc√©d√©s de Criblage Mol√©culaire et Cellulaire-Centre de Biotechnologie de Sfax | Souissi et al |
| EPI_ISL_711798 | Denmark/DCGC-14498/2020 | 2020-11-30 | Europe/Denmark/Syddanmark/ | Department of Virus and Microbiological Special Diagnostics, Statens Serum Institut, Copenhagen, Denmark | Albertsen Lab, Department of Chemistry and Bioscience, Aalborg University, Denmark | Danish Covid-19 Genome Consortium et al |
| EPI_ISL_711996 | Denmark/DCGC-14805/2020 | 2020-11-23 | Europe/Denmark/Midtjylland/ | Department of Virus and Microbiological Special Diagnostics, Statens Serum Institut, Copenhagen, Denmark | Albertsen Lab, Department of Chemistry and Bioscience, Aalborg University, Denmark | Danish Covid-19 Genome Consortium et al |
| EPI_ISL_712060 | Tunisia/ADAGE-18505/2020 | 2020-10-19 | Africa/Tunisia/Monastir/ | H√¥pital Fattouma-Bourguiba de Monastir | Laboratoire des Proc√©d√©s de Criblage Mol√©culaire et Cellulaire-Centre de Biotechnologie de Sfax | Souissi et al |

| | | | | | | |
|---|---|---|---|---|---|---|
| EPI_ISL_712062 | Tunisia/ADAGE-24755/2020 | 2020-11-08 | Africa/Tunisia/Sfax/ | Laboratoire de Microbiologie- CHU Habib Bourguiba ,Äì Sfax adresse | Laboratoire des Procv©dv©s de Criblage Molv©culaire et Cellulaire-Centre de Biotechnologie de Sfax | Souissi et al |
| EPI_ISL_712065 | Tunisia/ADAGE-24776/2020 | 2020-11-08 | Africa/Tunisia/Sfax/ | Laboratoire de Microbiologie- CHU Habib Bourguiba ,Äì Sfax adresse | Laboratoire des Procv©dv©s de Criblage Molv©culaire et Cellulaire-Centre de Biotechnologie de Sfax | Souissi et al |
| EPI_ISL_712066 | Tunisia/ADAGE-24779/2020 | 2020-11-08 | Africa/Tunisia/Sfax/ | Laboratoire de Microbiologie- CHU Habib Bourguiba ,Äì Sfax adresse | Laboratoire des Procv©dv©s de Criblage Molv©culaire et Cellulaire-Centre de Biotechnologie de Sfax | Souissi et al |
| EPI_ISL_712068 | Tunisia/ADAGE-15425/2020 | 2020-09-21 | Africa/Tunisia/Sfax/ | Laboratoire de Microbiologie- CHU Habib Bourguiba ,Äì Sfax adresse | Laboratoire des Procv©dv©s de Criblage Molv©culaire et Cellulaire-Centre de Biotechnologie de Sfax | Souissi et al |
| EPI_ISL_712092 | SouthAfrica/N00358/2020 | 2020-09-28 | Africa/South Africa/Eastern Cape/Chris | East London NHLS Laboratory, Eastern Cape, South Africa | National Institute for Communicable Diseases of the National Health Laboratory Service | Mohale T et al |
| EPI_ISL_712568 | Tunisia/ADAGE-17149/2020 | 2020-09-29 | Africa/Tunisia/Sfax/ | Laboratoire de Microbiologie- CHU Habib Bourguiba ,Äì Sfax adresse | Laboratoire des Procv©dv©s de Criblage Molv©culaire et Cellulaire-Centre de Biotechnologie de Sfax | Souissi et al |

| | | | | | | |
|---|---|---|---|---|---|---|
| EPI_ISL_712599 | Denmark/DCGC-13126/2020 | 2020-11-30 | Europe/Denmark/Syddanmark/ | Department of Virus and Microbiological Special Diagnostics, Statens Serum Institut, Copenhagen, Denmark | Albertsen Lab, Department of Chemistry and Bioscience, Aalborg University, Denmark | Danish Covid-19 Genome Consortium et al |
| EPI_ISL_713697 | Denmark/DCGC-15212/2020 | 2020-11-23 | Europe/Denmark/Midtjylland/ | Department of Virus and Microbiological Special Diagnostics, Statens Serum Institut, Copenhagen, Denmark | Albertsen Lab, Department of Chemistry and Bioscience, Aalborg University, Denmark | Danish Covid-19 Genome Consortium et al |
| EPI_ISL_714031 | Denmark/DCGC-15805/2020 | 2020-11-23 | Europe/Denmark/Hovedstaden/ | Department of Virus and Microbiological Special Diagnostics, Statens Serum Institut, Copenhagen, Denmark | Albertsen Lab, Department of Chemistry and Bioscience, Aalborg University, Denmark | Danish Covid-19 Genome Consortium et al |
| EPI_ISL_714373 | Denmark/DCGC-16145/2020 | 2020-11-16 | Europe/Denmark/Midtjylland/ | Department of Virus and Microbiological Special Diagnostics, Statens Serum Institut, Copenhagen, Denmark | Albertsen Lab, Department of Chemistry and Bioscience, Aalborg University, Denmark | Danish Covid-19 Genome Consortium et al |
| EPI_ISL_717574 | Belgium/UGent-189/2020 | 2020-10-20 | Europe/Belgium/Gent/ | Lab voor klinische biologie | Onderzoeksgroep Virologie | Laurens Lambrechts et al |
| EPI_ISL_717577 | Belgium/UGent-226/2020 | 2020-11-15 | Europe/Belgium/Gent/ | Lab voor klinische biologie | Onderzoeksgroep Virologie | Nick Vereecke et al |
| EPI_ISL_717581 | Belgium/UGent-201/2020 | 2020-10-22 | Europe/Belgium/Gent/ | Lab voor klinische biologie | Onderzoeksgroep Virologie | Nick Vereecke et al |
| EPI_ISL_717591 | Belgium/UGent-207/2020 | 2020-10-22 | Europe/Belgium/Gent/ | Lab voor klinische biologie | Onderzoeksgroep Virologie | Laurens Lambrechts et al |
| EPI_ISL_717608 | Belgium/UGent-192/2020 | 2020-10-20 | Europe/Belgium/Gent/ | Lab voor klinische biologie | Onderzoeksgroep Virologie | Laurens Lambrechts et al |
| EPI_ISL_717612 | Belgium/UGent-211/2020 | 2020-10-22 | Europe/Belgium/Gent/ | Lab voor klinische biologie | Onderzoeksgroep Virologie | Laurens Lambrechts et al |

| | | | | | | |
|---|---|---|---|---|---|---|
| EPI_ISL_717613 | Belgium/UGent-212/2020 | 2020-10-23 | Europe/Belgium/Gent/ | Lab voor klinische biologie | Onderzoeksgroep Virologie | Laurens Lambrechts et al |
| EPI_ISL_717619 | Belgium/UGent-220/2020 | 2020-10-19 | Europe/Belgium/Gent/ | Lab voor klinische biologie | Onderzoeksgroep Virologie | Nick Vereecke et al |
| EPI_ISL_717623 | Belgium/UGent-230/2020 | 2020-11-12 | Europe/Belgium/Gent/ | Lab voor klinische biologie | Onderzoeksgroep Virologie | Laurens Lambrechts et al |
| EPI_ISL_717625 | Belgium/UGent-235/2020 | 2020-11-13 | Europe/Belgium/Gent/ | Lab voor klinische biologie | Onderzoeksgroep Virologie | Laurens Lambrechts et al |
| EPI_ISL_717626 | Belgium/UGent-240/2020 | 2020-11-08 | Europe/Belgium/Gent/ | Lab voor klinische biologie | Onderzoeksgroep Virologie | Nick Vereecke et al |
| EPI_ISL_717692 | Trinidad/TT6949/2020 | 2020-09-09 | South America/Trinidad and Tobago/Tri | Trinidad Public Health Laboratory | Carrington Lab, Department of PreClinical Sciences, Faculty of Medical Sciences, The University of the West Indies | Nikita S. D. Sahadeo et al |
| EPI_ISL_717695 | Trinidad/TT2238/2020 | 2020-08-26 | South America/Trinidad and Tobago/Tri | Trinidad Public Health Laboratory | Carrington Lab, Department of PreClinical Sciences, Faculty of Medical Sciences, The University of the West Indies | Nikita S. D. Sahadeo et al |
| EPI_ISL_717696 | Trinidad/TT7867/2020 | 2020-08-14 | South America/Trinidad and Tobago/Tri | Trinidad Public Health Laboratory | Carrington Lab, Department of PreClinical Sciences, Faculty of Medical Sciences, The University of the West Indies | Nikita S. D. Sahadeo et al |

| | | | | | | |
|---|---|---|---|---|---|---|
| EPI_ISL_717697 | Trinidad/TT4707/2020 | 2020-09-02 | South America/Trinidad and Tobago/Tri | Trinidad Public Health Laboratory | Carrington Lab, Department of PreClinical Sciences, Faculty of Medical Sciences, The University of the West Indies | Nikita S. D. Sahadeo et al |
| EPI_ISL_717700 | Trinidad/TT7923/2020 | 2020-09-14 | South America/Trinidad and Tobago/Tri | Trinidad Public Health Laboratory | Carrington Lab, Department of PreClinical Sciences, Faculty of Medical Sciences, The University of the West Indies | Nikita S. D. Sahadeo et al |
| EPI_ISL_717723 | Canada/ON-SLB3121/2020 | 2020-04-22 | North America/Canada/Ontario/ | Kingston Health Sciences Centre and Queen's University | Ontario Institute for Cancer Research | Prameet M. Sheth et al |
| EPI_ISL_717724 | Canada/ON-SLB3155/2020 | 2020-05-03 | North America/Canada/Ontario/ | Kingston Health Sciences Centre and Queen's University | Ontario Institute for Cancer Research | Prameet M. Sheth et al |
| EPI_ISL_717725 | Canada/ON-SLB3115/2020 | 2020-04-20 | North America/Canada/Ontario/ | Kingston Health Sciences Centre and Queen's University | Ontario Institute for Cancer Research | Prameet M. Sheth et al |
| EPI_ISL_717726 | Canada/ON-SLB3108/2020 | 2020-04-19 | North America/Canada/Ontario/ | Kingston Health Sciences Centre and Queen's University | Ontario Institute for Cancer Research | Prameet M. Sheth et al |
| EPI_ISL_717727 | Canada/ON-SLB3109/2020 | 2020-04-19 | North America/Canada/Ontario/ | Kingston Health Sciences Centre and Queen's University | Ontario Institute for Cancer Research | Prameet M. Sheth et al |
| EPI_ISL_717728 | Canada/ON-SLB3079/2020 | 2020-04-14 | North America/Canada/Ontario/ | Kingston Health Sciences Centre and Queen's University | Ontario Institute for Cancer Research | Prameet M. Sheth et al |
| EPI_ISL_717729 | Canada/ON-SLB3096/2020 | 2020-04-17 | North America/Canada/Ontario/ | Kingston Health Sciences Centre and Queen's University | Ontario Institute for Cancer Research | Prameet M. Sheth et al |
| EPI_ISL_717730 | Canada/ON-SLB3125/2020 | 2020-04-22 | North America/Canada/Ontario/ | Kingston Health Sciences Centre and Queen's University | Ontario Institute for Cancer Research | Prameet M. Sheth et al |

| | | | | | | |
|---|---|---|---|---|---|---|
| EPI_ISL_717731 | Canada/ON-SLB3167/2020 | 2020-05-08 | North America/Canada/Ontario/ | Kingston Health Sciences Centre and Queen's University | Ontario Institute for Cancer Research | Prameet M. Sheth et al |
| EPI_ISL_717732 | Canada/ON-SLB3153/2020 | 2020-05-02 | North America/Canada/Ontario/ | Kingston Health Sciences Centre and Queen's University | Ontario Institute for Cancer Research | Prameet M. Sheth et al |
| EPI_ISL_717733 | Canada/ON-SLB3139/2020 | 2020-04-25 | North America/Canada/Ontario/ | Kingston Health Sciences Centre and Queen's University | Ontario Institute for Cancer Research | Prameet M. Sheth et al |
| EPI_ISL_717734 | Canada/ON-SLB3242/2020 | 2020-11-24 | North America/Canada/Ontario/ | Kingston Health Sciences Centre and Queen's University | Ontario Institute for Cancer Research | Prameet M. Sheth et al |
| EPI_ISL_717735 | Canada/ON-SLB3101/2020 | 2020-04-18 | North America/Canada/Ontario/ | Kingston Health Sciences Centre and Queen's University | Ontario Institute for Cancer Research | Prameet M. Sheth et al |
| EPI_ISL_717736 | Canada/ON-SLB3119/2020 | 2020-04-21 | North America/Canada/Ontario/ | Kingston Health Sciences Centre and Queen's University | Ontario Institute for Cancer Research | Prameet M. Sheth et al |
| EPI_ISL_717737 | Canada/ON-SLB3243/2020 | 2020-11-24 | North America/Canada/Ontario/ | Kingston Health Sciences Centre and Queen's University | Ontario Institute for Cancer Research | Prameet M. Sheth et al |
| EPI_ISL_717738 | Canada/ON-SLB3140/2020 | 2020-04-25 | North America/Canada/Ontario/ | Kingston Health Sciences Centre and Queen's University | Ontario Institute for Cancer Research | Prameet M. Sheth et al |
| EPI_ISL_717739 | Canada/ON-SLB3144/2020 | 2020-04-27 | North America/Canada/Ontario/ | Kingston Health Sciences Centre and Queen's University | Ontario Institute for Cancer Research | Prameet M. Sheth et al |
| EPI_ISL_717740 | Canada/ON-SLB3099/2020 | 2020-04-17 | North America/Canada/Ontario/ | Kingston Health Sciences Centre and Queen's University | Ontario Institute for Cancer Research | Prameet M. Sheth et al |
| EPI_ISL_717741 | Canada/ON-SLB3244/2020 | 2020-11-24 | North America/Canada/Ontario/ | Kingston Health Sciences Centre and Queen's University | Ontario Institute for Cancer Research | Prameet M. Sheth et al |
| EPI_ISL_717742 | Canada/ON-SLB3105/2020 | 2020-04-18 | North America/Canada/Ontario/ | Kingston Health Sciences Centre and Queen's University | Ontario Institute for Cancer Research | Prameet M. Sheth et al |
| EPI_ISL_717743 | Canada/ON-SLB3132/2020 | 2020-04-23 | North America/Canada/Ontario/ | Kingston Health Sciences Centre and Queen's University | Ontario Institute for Cancer Research | Prameet M. Sheth et al |

| | | | | | | |
|---|---|---|---|---|---|---|
| EPI_ISL_717744 | Canada/ON-SLB3168/2020 | 2020-05-09 | North America/Canada/Ontario/ | Kingston Health Sciences Centre and Queen's University | Ontario Institute for Cancer Research | Prameet M. Sheth et al |
| EPI_ISL_717745 | Canada/ON-SLB3103/2020 | 2020-04-18 | North America/Canada/Ontario/ | Kingston Health Sciences Centre and Queen's University | Ontario Institute for Cancer Research | Prameet M. Sheth et al |
| EPI_ISL_717746 | Canada/ON-SLB3076/2020 | 2020-04-12 | North America/Canada/Ontario/ | Kingston Health Sciences Centre and Queen's University | Ontario Institute for Cancer Research | Prameet M. Sheth et al |
| EPI_ISL_717747 | Canada/ON-SLB3095/2020 | 2020-04-17 | North America/Canada/Ontario/ | Kingston Health Sciences Centre and Queen's University | Ontario Institute for Cancer Research | Prameet M. Sheth et al |
| EPI_ISL_717748 | Canada/ON-SLB3131/2020 | 2020-04-23 | North America/Canada/Ontario/ | Kingston Health Sciences Centre and Queen's University | Ontario Institute for Cancer Research | Prameet M. Sheth et al |
| EPI_ISL_717749 | Canada/ON-SLB3081/2020 | 2020-04-14 | North America/Canada/Ontario/ | Kingston Health Sciences Centre and Queen's University | Ontario Institute for Cancer Research | Prameet M. Sheth et al |
| EPI_ISL_717750 | Canada/ON-SLB3086/2020 | 2020-04-15 | North America/Canada/Ontario/ | Kingston Health Sciences Centre and Queen's University | Ontario Institute for Cancer Research | Prameet M. Sheth et al |
| EPI_ISL_717751 | Canada/ON-SLB3090/2020 | 2020-04-16 | North America/Canada/Ontario/ | Kingston Health Sciences Centre and Queen's University | Ontario Institute for Cancer Research | Prameet M. Sheth et al |
| EPI_ISL_717752 | Canada/ON-SLB3245/2020 | 2020-11-24 | North America/Canada/Ontario/ | Kingston Health Sciences Centre and Queen's University | Ontario Institute for Cancer Research | Prameet M. Sheth et al |
| EPI_ISL_717753 | Canada/ON-SLB3124/2020 | 2020-04-22 | North America/Canada/Ontario/ | Kingston Health Sciences Centre and Queen's University | Ontario Institute for Cancer Research | Prameet M. Sheth et al |
| EPI_ISL_717754 | Canada/ON-SLB3104/2020 | 2020-04-18 | North America/Canada/Ontario/ | Kingston Health Sciences Centre and Queen's University | Ontario Institute for Cancer Research | Prameet M. Sheth et al |
| EPI_ISL_717755 | Canada/ON-SLB3114/2020 | 2020-04-20 | North America/Canada/Ontario/ | Kingston Health Sciences Centre and Queen's University | Ontario Institute for Cancer Research | Prameet M. Sheth et al |
| EPI_ISL_717756 | Canada/ON-SLB3098/2020 | 2020-04-17 | North America/Canada/Ontario/ | Kingston Health Sciences Centre and Queen's University | Ontario Institute for Cancer Research | Prameet M. Sheth et al |

| | | | | | | |
|---|---|---|---|---|---|---|
| EPI_ISL_717757 | Canada/ON-SLB3110/2020 | 2020-04-19 | North America/Canada/Ontario/ | Kingston Health Sciences Centre and Queen's University | Ontario Institute for Cancer Research | Prameet M. Sheth et al |
| EPI_ISL_717758 | Canada/ON-SLB3141/2020 | 2020-04-26 | North America/Canada/Ontario/ | Kingston Health Sciences Centre and Queen's University | Ontario Institute for Cancer Research | Prameet M. Sheth et al |
| EPI_ISL_717759 | Canada/ON-SLB3106/2020 | 2020-04-18 | North America/Canada/Ontario/ | Kingston Health Sciences Centre and Queen's University | Ontario Institute for Cancer Research | Prameet M. Sheth et al |
| EPI_ISL_717760 | Canada/ON-SLB3082/2020 | 2020-04-14 | North America/Canada/Ontario/ | Kingston Health Sciences Centre and Queen's University | Ontario Institute for Cancer Research | Prameet M. Sheth et al |
| EPI_ISL_717761 | Canada/ON-SLB3147/2020 | 2020-04-28 | North America/Canada/Ontario/ | Kingston Health Sciences Centre and Queen's University | Ontario Institute for Cancer Research | Prameet M. Sheth et al |
| EPI_ISL_717762 | Canada/ON-SLB3078/2020 | 2020-04-14 | North America/Canada/Ontario/ | Kingston Health Sciences Centre and Queen's University | Ontario Institute for Cancer Research | Prameet M. Sheth et al |
| EPI_ISL_717763 | Canada/ON-SLB3148/2020 | 2020-04-28 | North America/Canada/Ontario/ | Kingston Health Sciences Centre and Queen's University | Ontario Institute for Cancer Research | Prameet M. Sheth et al |
| EPI_ISL_717764 | Canada/ON-SLB3080/2020 | 2020-04-14 | North America/Canada/Ontario/ | Kingston Health Sciences Centre and Queen's University | Ontario Institute for Cancer Research | Prameet M. Sheth et al |
| EPI_ISL_717774 | USA/WA-UW-28263/2020 | 2020-09-16 | North America/USA/Washington/ | UW Virology Lab | UW Virology Lab | Pavitra Roychoudhury et al |
| EPI_ISL_717776 | USA/WA-UW-29492/2020 | 2020-09-22 | North America/USA/Washington/ | UW Virology Lab | UW Virology Lab | Pavitra Roychoudhury et al |
| EPI_ISL_717784 | USA/WA-UW-9774/2020 | 2020-05-22 | North America/USA/Washington/ | UW Virology Lab | UW Virology Lab | Pavitra Roychoudhury et al |
| EPI_ISL_717785 | Brazil/RJ-00534/2020 | 2020-07-29 | South America/Brazil/Rio de Janeiro/Ita | LACEN RJ - Noel Nutels | Bioinformatics Laboratory / LNCC | Carolina M Voloch et al |
| EPI_ISL_717786 | Brazil/RJ-00406/2020 | 2020-07-28 | South America/Brazil/Rio de Janeiro/No | LACEN RJ - Noel Nutels | Bioinformatics Laboratory / LNCC | Carolina M Voloch et al |
| EPI_ISL_717787 | Brazil/RJ-00429/2020 | 2020-06-29 | South America/Brazil/Rio de Janeiro/Sa | LACEN RJ - Noel Nutels | Bioinformatics Laboratory / LNCC | Carolina M Voloch et al |
| EPI_ISL_717788 | Brazil/RJ-00510/2020 | 2020-07-27 | South America/Brazil/Rio de Janeiro/Sa | LACEN RJ - Noel Nutels | Bioinformatics Laboratory / LNCC | Carolina M Voloch et al |
| EPI_ISL_717789 | Brazil/RJ-00521/2020 | 2020-07-29 | South America/Brazil/Rio de Janeiro/No | LACEN RJ - Noel Nutels | Bioinformatics Laboratory / LNCC | Carolina M Voloch et al |
| EPI_ISL_717790 | Brazil/RJ-00513/2020 | 2020-07-28 | South America/Brazil/Rio de Janeiro/No | LACEN RJ - Noel Nutels | Bioinformatics Laboratory / LNCC | Carolina M Voloch et al |

| | | | | | | |
|---|---|---|---|---|---|---|
| EPI_ISL_717792 | Brazil/RJ-00523/2020 | 2020-07-29 | South America/Brazil/Rio de Janeiro/No | LACEN RJ - Noel Nutels | Bioinformatics Laboratory / LNCC | Carolina M Voloch et al |
| EPI_ISL_717793 | Brazil/RJ-00481/2020 | 2020-10-09 | South America/Brazil/Rio de Janeiro/Rio | Laboratorio de Virologia Molecular / UFRJ | Bioinformatics Laboratory / LNCC | Carolina M Voloch et al |
| EPI_ISL_717794 | Brazil/RJ-00430/2020 | 2020-07-29 | South America/Brazil/Rio de Janeiro/Ita | LACEN RJ - Noel Nutels | Bioinformatics Laboratory / LNCC | Carolina M Voloch et al |
| EPI_ISL_717797 | Brazil/RJ-00509/2020 | 2020-06-29 | South America/Brazil/Rio de Janeiro/Sa | LACEN RJ - Noel Nutels | Bioinformatics Laboratory / LNCC | Carolina M Voloch et al |
| EPI_ISL_717798 | Brazil/RJ-00431/2020 | 2020-07-28 | South America/Brazil/Rio de Janeiro/No | LACEN RJ - Noel Nutels | Bioinformatics Laboratory / LNCC | Carolina M Voloch et al |
| EPI_ISL_717799 | Brazil/RJ-00434/2020 | 2020-07-28 | South America/Brazil/Rio de Janeiro/No | LACEN RJ - Noel Nutels | Bioinformatics Laboratory / LNCC | Carolina M Voloch et al |
| EPI_ISL_717806 | Brazil/RJ-00402/2020 | 2020-09-03 | South America/Brazil/Rio de Janeiro/Rio | Laboratorio de Virologia Molecular / UFRJ | Bioinformatics Laboratory / LNCC | Carolina M Voloch et al |
| EPI_ISL_717807 | Brazil/RJ-00404/2020 | 2020-11-05 | South America/Brazil/Rio de Janeiro/Sv | Laboratorio de Virologia Molecular / UFRJ | Bioinformatics Laboratory / LNCC | Carolina M Voloch et al |
| EPI_ISL_717808 | Brazil/RJ-00405/2020 | 2020-11-03 | South America/Brazil/Rio de Janeiro/Rio | Laboratorio de Virologia Molecular / UFRJ | Bioinformatics Laboratory / LNCC | Carolina M Voloch et al |
| EPI_ISL_717809 | Brazil/RJ-00407/2020 | 2020-09-28 | South America/Brazil/Rio de Janeiro/Ma | LACEN Dr. Francisco Rimolo Neto | Bioinformatics Laboratory / LNCC | Carolina M Voloch et al |
| EPI_ISL_717811 | Brazil/RJ-00409/2020 | 2020-04-16 | South America/Brazil/Rio de Janeiro/Rio | Laboratorio de Virologia Molecular / UFRJ | Bioinformatics Laboratory / LNCC | Carolina M Voloch et al |
| EPI_ISL_717812 | Brazil/RJ-00410/2020 | 2020-06-01 | South America/Brazil/Rio de Janeiro/Rio | Laboratorio de Virologia Molecular / UFRJ | Bioinformatics Laboratory / LNCC | Carolina M Voloch et al |
| EPI_ISL_717813 | Brazil/RJ-00411/2020 | 2020-06-01 | South America/Brazil/Rio de Janeiro/Rio | Laboratorio de Virologia Molecular / UFRJ | Bioinformatics Laboratory / LNCC | Carolina M Voloch et al |
| EPI_ISL_717814 | Brazil/RJ-00412/2020 | 2020-09-03 | South America/Brazil/Rio de Janeiro/Nil | Laboratorio de Virologia Molecular / UFRJ | Bioinformatics Laboratory / LNCC | Carolina M Voloch et al |
| EPI_ISL_717815 | Brazil/RJ-00413/2020 | 2020-09-03 | South America/Brazil/Rio de Janeiro/Rio | Laboratorio de Virologia Molecular / UFRJ | Bioinformatics Laboratory / LNCC | Carolina M Voloch et al |
| EPI_ISL_717816 | Brazil/RJ-00414/2020 | 2020-09-03 | South America/Brazil/Rio de Janeiro/Rio | Laboratorio de Virologia Molecular / UFRJ | Bioinformatics Laboratory / LNCC | Carolina M Voloch et al |
| EPI_ISL_717817 | Brazil/RJ-00415/2020 | 2020-09-04 | South America/Brazil/Rio de Janeiro/Rio | Laboratorio de Virologia Molecular / UFRJ | Bioinformatics Laboratory / LNCC | Carolina M Voloch et al |
| EPI_ISL_717818 | Brazil/RJ-00416/2020 | 2020-10-08 | South America/Brazil/Rio de Janeiro/Rio | Laboratorio de Virologia Molecular / UFRJ | Bioinformatics Laboratory / LNCC | Carolina M Voloch et al |
| EPI_ISL_717819 | Brazil/RJ-00417/2020 | 2020-10-09 | South America/Brazil/Rio de Janeiro/Rio | Laboratorio de Virologia Molecular / UFRJ | Bioinformatics Laboratory / LNCC | Carolina M Voloch et al |
| EPI_ISL_717820 | Brazil/RJ-00418/2020 | 2020-10-26 | South America/Brazil/Rio de Janeiro/Du | Laboratorio de Virologia Molecular / UFRJ | Bioinformatics Laboratory / LNCC | Carolina M Voloch et al |

| | | | | | | |
|---|---|---|---|---|---|---|
| EPI_ISL_717821 | Brazil/RJ-00419/2020 | 2020-10-26 | South America/Brazil/Rio de Janeiro/Ric | Laboratorio de Virologia Molecular / UFRJ | Bioinformatics Laboratory / LNCC | Carolina M Voloch et al |
| EPI_ISL_717822 | Brazil/RJ-00420/2020 | 2020-10-26 | South America/Brazil/Rio de Janeiro/Du | Laboratorio de Virologia Molecular / UFRJ | Bioinformatics Laboratory / LNCC | Carolina M Voloch et al |
| EPI_ISL_717823 | Brazil/RJ-00421/2020 | 2020-10-29 | South America/Brazil/Rio de Janeiro/Ric | Laboratorio de Virologia Molecular / UFRJ | Bioinformatics Laboratory / LNCC | Carolina M Voloch et al |
| EPI_ISL_717824 | Brazil/RJ-00422/2020 | 2020-10-29 | South America/Brazil/Rio de Janeiro/Ric | Laboratorio de Virologia Molecular / UFRJ | Bioinformatics Laboratory / LNCC | Carolina M Voloch et al |
| EPI_ISL_717828 | Brazil/RJ-00426/2020 | 2020-11-04 | South America/Brazil/Rio de Janeiro/Ric | Laboratorio de Virologia Molecular / UFRJ | Bioinformatics Laboratory / LNCC | Carolina M Voloch et al |
| EPI_ISL_717829 | Brazil/RJ-00427/2020 | 2020-11-04 | South America/Brazil/Rio de Janeiro/Ric | Laboratorio de Virologia Molecular / UFRJ | Bioinformatics Laboratory / LNCC | Carolina M Voloch et al |
| EPI_ISL_717831 | Brazil/RJ-00432/2020 | 2020-10-30 | South America/Brazil/Rio de Janeiro/Sa | Laboratorio de Virologia Molecular / UFRJ | Bioinformatics Laboratory / LNCC | Carolina M Voloch et al |
| EPI_ISL_717832 | Brazil/RJ-00437/2020 | 2020-06-01 | South America/Brazil/Rio de Janeiro/Ma | LACEN Dr. Francisco Rimolo Neto | Bioinformatics Laboratory / LNCC | Carolina M Voloch et al |
| EPI_ISL_717833 | Brazil/RJ-00438/2020 | 2020-06-03 | South America/Brazil/Rio de Janeiro/Ma | LACEN Dr. Francisco Rimolo Neto | Bioinformatics Laboratory / LNCC | Carolina M Voloch et al |
| EPI_ISL_717834 | Brazil/RJ-00439/2020 | 2020-06-02 | South America/Brazil/Rio de Janeiro/Ma | LACEN Dr. Francisco Rimolo Neto | Bioinformatics Laboratory / LNCC | Carolina M Voloch et al |
| EPI_ISL_717835 | Brazil/RJ-00440/2020 | 2020-06-03 | South America/Brazil/Rio de Janeiro/Ma | LACEN Dr. Francisco Rimolo Neto | Bioinformatics Laboratory / LNCC | Carolina M Voloch et al |
| EPI_ISL_717836 | Brazil/RJ-00441/2020 | 2020-06-03 | South America/Brazil/Rio de Janeiro/Ma | LACEN Dr. Francisco Rimolo Neto | Bioinformatics Laboratory / LNCC | Carolina M Voloch et al |
| EPI_ISL_717838 | Brazil/RJ-00443/2020 | 2020-04-08 | South America/Brazil/Rio de Janeiro/Ric | Laboratorio de Virologia Molecular / UFRJ | Bioinformatics Laboratory / LNCC | Carolina M Voloch et al |
| EPI_ISL_717840 | Brazil/RJ-00445/2020 | 2020-04-13 | South America/Brazil/Rio de Janeiro/Ric | Laboratorio de Virologia Molecular / UFRJ | Bioinformatics Laboratory / LNCC | Carolina M Voloch et al |
| EPI_ISL_717841 | Brazil/RJ-00446/2020 | 2020-07-28 | South America/Brazil/Rio de Janeiro/Ma | LACEN Dr. Francisco Rimolo Neto | Bioinformatics Laboratory / LNCC | Carolina M Voloch et al |
| EPI_ISL_717845 | Brazil/RJ-00450/2020 | 2020-06-01 | South America/Brazil/Rio de Janeiro/No | Laboratorio de Virologia Molecular / UFRJ | Bioinformatics Laboratory / LNCC | Carolina M Voloch et al |
| EPI_ISL_717846 | Brazil/RJ-00451/2020 | 2020-06-01 | South America/Brazil/Rio de Janeiro/Ric | Laboratorio de Virologia Molecular / UFRJ | Bioinformatics Laboratory / LNCC | Carolina M Voloch et al |
| EPI_ISL_717847 | Brazil/RJ-00452/2020 | 2020-06-01 | South America/Brazil/Rio de Janeiro/Be | Laboratorio de Virologia Molecular / UFRJ | Bioinformatics Laboratory / LNCC | Carolina M Voloch et al |
| EPI_ISL_717848 | Brazil/RJ-00453/2020 | 2020-06-01 | South America/Brazil/Rio de Janeiro/Se | Laboratorio de Virologia Molecular / UFRJ | Bioinformatics Laboratory / LNCC | Carolina M Voloch et al |
| EPI_ISL_717849 | Brazil/RJ-00454/2020 | 2020-06-01 | South America/Brazil/Rio de Janeiro/Nil | Laboratorio de Virologia Molecular / UFRJ | Bioinformatics Laboratory / LNCC | Carolina M Voloch et al |

| | | | | | | |
|---|---|---|---|---|---|---|
| EPI_ISL_717850 | Brazil/RJ-00455/2020 | 2020-06-01 | South America/Brazil/Rio de Janeiro/Ita | Laboratorio de Virologia Molecular / UFRJ | Bioinformatics Laboratory / LNCC | Carolina M Voloch et al |
| EPI_ISL_717851 | Brazil/RJ-00456/2020 | 2020-06-01 | South America/Brazil/Rio de Janeiro/Ric | Laboratorio de Virologia Molecular / UFRJ | Bioinformatics Laboratory / LNCC | Carolina M Voloch et al |
| EPI_ISL_717852 | Brazil/RJ-00457/2020 | 2020-06-01 | South America/Brazil/Rio de Janeiro/Ric | Laboratorio de Virologia Molecular / UFRJ | Bioinformatics Laboratory / LNCC | Carolina M Voloch et al |
| EPI_ISL_717853 | Brazil/RJ-00458/2020 | 2020-07-01 | South America/Brazil/Rio de Janeiro/Ric | Laboratorio de Virologia Molecular / UFRJ | Bioinformatics Laboratory / LNCC | Carolina M Voloch et al |
| EPI_ISL_717854 | Brazil/RJ-00459/2020 | 2020-07-01 | South America/Brazil/Rio de Janeiro/Ric | Laboratorio de Virologia Molecular / UFRJ | Bioinformatics Laboratory / LNCC | Carolina M Voloch et al |
| EPI_ISL_717855 | Brazil/RJ-00460/2020 | 2020-07-01 | South America/Brazil/Rio de Janeiro/Ric | Laboratorio de Virologia Molecular / UFRJ | Bioinformatics Laboratory / LNCC | Carolina M Voloch et al |
| EPI_ISL_717856 | Brazil/RJ-00461/2020 | 2020-07-02 | South America/Brazil/Rio de Janeiro/Ric | Laboratorio de Virologia Molecular / UFRJ | Bioinformatics Laboratory / LNCC | Carolina M Voloch et al |
| EPI_ISL_717857 | Brazil/RJ-00462/2020 | 2020-07-02 | South America/Brazil/Rio de Janeiro/Ric | Laboratorio de Virologia Molecular / UFRJ | Bioinformatics Laboratory / LNCC | Carolina M Voloch et al |
| EPI_ISL_717858 | Brazil/RJ-00463/2020 | 2020-07-02 | South America/Brazil/Rio de Janeiro/Ric | Laboratorio de Virologia Molecular / UFRJ | Bioinformatics Laboratory / LNCC | Carolina M Voloch et al |
| EPI_ISL_717859 | Brazil/RJ-00464/2020 | 2020-07-03 | South America/Brazil/Rio de Janeiro/Ric | Laboratorio de Virologia Molecular / UFRJ | Bioinformatics Laboratory / LNCC | Carolina M Voloch et al |
| EPI_ISL_717860 | Brazil/RJ-00465/2020 | 2020-07-06 | South America/Brazil/Rio de Janeiro/Ric | Laboratorio de Virologia Molecular / UFRJ | Bioinformatics Laboratory / LNCC | Carolina M Voloch et al |
| EPI_ISL_717861 | Brazil/RJ-00466/2020 | 2020-07-08 | South America/Brazil/Rio de Janeiro/Ric | Laboratorio de Virologia Molecular / UFRJ | Bioinformatics Laboratory / LNCC | Carolina M Voloch et al |
| EPI_ISL_717862 | Brazil/RJ-00467/2020 | 2020-07-08 | South America/Brazil/Rio de Janeiro/Ric | Laboratorio de Virologia Molecular / UFRJ | Bioinformatics Laboratory / LNCC | Carolina M Voloch et al |
| EPI_ISL_717863 | Brazil/RJ-00468/2020 | 2020-08-03 | South America/Brazil/Rio de Janeiro/Sv | Laboratorio de Virologia Molecular / UFRJ | Bioinformatics Laboratory / LNCC | Carolina M Voloch et al |
| EPI_ISL_717864 | Brazil/RJ-00469/2020 | 2020-08-03 | South America/Brazil/Rio de Janeiro/Ric | Laboratorio de Virologia Molecular / UFRJ | Bioinformatics Laboratory / LNCC | Carolina M Voloch et al |
| EPI_ISL_717865 | Brazil/RJ-00470/2020 | 2020-08-03 | South America/Brazil/Rio de Janeiro/Gu | Laboratorio de Virologia Molecular / UFRJ | Bioinformatics Laboratory / LNCC | Carolina M Voloch et al |
| EPI_ISL_717866 | Brazil/RJ-00471/2020 | 2020-08-03 | South America/Brazil/Rio de Janeiro/Gu | Laboratorio de Virologia Molecular / UFRJ | Bioinformatics Laboratory / LNCC | Carolina M Voloch et al |
| EPI_ISL_717867 | Brazil/RJ-00472/2020 | 2020-08-04 | South America/Brazil/Rio de Janeiro/Ric | Laboratorio de Virologia Molecular / UFRJ | Bioinformatics Laboratory / LNCC | Carolina M Voloch et al |
| EPI_ISL_717868 | Brazil/RJ-00473/2020 | 2020-08-05 | South America/Brazil/Rio de Janeiro/Sv | Laboratorio de Virologia Molecular / UFRJ | Bioinformatics Laboratory / LNCC | Carolina M Voloch et al |
| EPI_ISL_717869 | Brazil/RJ-00474/2020 | 2020-09-03 | South America/Brazil/Rio de Janeiro/Ric | Laboratorio de Virologia Molecular / UFRJ | Bioinformatics Laboratory / LNCC | Carolina M Voloch et al |

| | | | | | | |
|---|---|---|---|---|---|---|
| EPI_ISL_717870 | Brazil/RJ-00475/2020 | 2020-09-04 | South America/Brazil/Rio de Janeiro/Rio | Laboratorio de Virologia Molecular / UFRJ | Bioinformatics Laboratory / LNCC | Carolina M Voloch et al |
| EPI_ISL_717871 | Brazil/RJ-00476/2020 | 2020-09-04 | South America/Brazil/Rio de Janeiro/Rio | Laboratorio de Virologia Molecular / UFRJ | Bioinformatics Laboratory / LNCC | Carolina M Voloch et al |
| EPI_ISL_717872 | Brazil/RJ-00477/2020 | 2020-09-04 | South America/Brazil/Rio de Janeiro/Rio | Laboratorio de Virologia Molecular / UFRJ | Bioinformatics Laboratory / LNCC | Carolina M Voloch et al |
| EPI_ISL_717873 | Brazil/RJ-00478/2020 | 2020-09-08 | South America/Brazil/Rio de Janeiro/Rio | Laboratorio de Virologia Molecular / UFRJ | Bioinformatics Laboratory / LNCC | Carolina M Voloch et al |
| EPI_ISL_717874 | Brazil/RJ-00479/2020 | 2020-10-09 | South America/Brazil/Rio de Janeiro/Sv | Laboratorio de Virologia Molecular / UFRJ | Bioinformatics Laboratory / LNCC | Carolina M Voloch et al |
| EPI_ISL_717875 | Brazil/RJ-00480/2020 | 2020-10-09 | South America/Brazil/Rio de Janeiro/Du | Laboratorio de Virologia Molecular / UFRJ | Bioinformatics Laboratory / LNCC | Carolina M Voloch et al |
| EPI_ISL_717876 | Brazil/RJ-00482/2020 | 2020-10-09 | South America/Brazil/Rio de Janeiro/Rio | Laboratorio de Virologia Molecular / UFRJ | Bioinformatics Laboratory / LNCC | Carolina M Voloch et al |
| EPI_ISL_717877 | Brazil/RJ-00483/2020 | 2020-10-26 | South America/Brazil/Rio de Janeiro/Rio | Laboratorio de Virologia Molecular / UFRJ | Bioinformatics Laboratory / LNCC | Carolina M Voloch et al |
| EPI_ISL_717878 | Brazil/RJ-00484/2020 | 2020-10-26 | South America/Brazil/Rio de Janeiro/Rio | Laboratorio de Virologia Molecular / UFRJ | Bioinformatics Laboratory / LNCC | Carolina M Voloch et al |
| EPI_ISL_717879 | Brazil/RJ-00485/2020 | 2020-10-26 | South America/Brazil/Rio de Janeiro/Rio | Laboratorio de Virologia Molecular / UFRJ | Bioinformatics Laboratory / LNCC | Carolina M Voloch et al |
| EPI_ISL_717880 | Brazil/RJ-00486/2020 | 2020-10-26 | South America/Brazil/Rio de Janeiro/Nit | Laboratorio de Virologia Molecular / UFRJ | Bioinformatics Laboratory / LNCC | Carolina M Voloch et al |
| EPI_ISL_717881 | Brazil/RJ-00487/2020 | 2020-10-26 | South America/Brazil/Rio de Janeiro/Sv | Laboratorio de Virologia Molecular / UFRJ | Bioinformatics Laboratory / LNCC | Carolina M Voloch et al |
| EPI_ISL_717882 | Brazil/RJ-00488/2020 | 2020-10-26 | South America/Brazil/Rio de Janeiro/Rio | Laboratorio de Virologia Molecular / UFRJ | Bioinformatics Laboratory / LNCC | Carolina M Voloch et al |
| EPI_ISL_717883 | Brazil/RJ-00489/2020 | 2020-10-27 | South America/Brazil/Rio de Janeiro/Rio | Laboratorio de Virologia Molecular / UFRJ | Bioinformatics Laboratory / LNCC | Carolina M Voloch et al |
| EPI_ISL_717884 | Brazil/RJ-00490/2020 | 2020-10-27 | South America/Brazil/Rio de Janeiro/Sv | Laboratorio de Virologia Molecular / UFRJ | Bioinformatics Laboratory / LNCC | Carolina M Voloch et al |
| EPI_ISL_717885 | Brazil/RJ-00491/2020 | 2020-10-27 | South America/Brazil/Rio de Janeiro/Rio | Laboratorio de Virologia Molecular / UFRJ | Bioinformatics Laboratory / LNCC | Carolina M Voloch et al |
| EPI_ISL_717886 | Brazil/RJ-00492/2020 | 2020-10-27 | South America/Brazil/Rio de Janeiro/Rio | Laboratorio de Virologia Molecular / UFRJ | Bioinformatics Laboratory / LNCC | Carolina M Voloch et al |
| EPI_ISL_717887 | Brazil/RJ-00493/2020 | 2020-10-29 | South America/Brazil/Rio de Janeiro/Rio | Laboratorio de Virologia Molecular / UFRJ | Bioinformatics Laboratory / LNCC | Carolina M Voloch et al |
| EPI_ISL_717888 | Brazil/RJ-00494/2020 | 2020-10-29 | South America/Brazil/Rio de Janeiro/Rio | Laboratorio de Virologia Molecular / UFRJ | Bioinformatics Laboratory / LNCC | Carolina M Voloch et al |
| EPI_ISL_717889 | Brazil/RJ-00495/2020 | 2020-10-30 | South America/Brazil/Rio de Janeiro/Rio | Laboratorio de Virologia Molecular / UFRJ | Bioinformatics Laboratory / LNCC | Carolina M Voloch et al |

| | | | | | | |
|---|---|---|---|---|---|---|
| EPI_ISL_717891 | Brazil/RJ-00497/2020 | 2020-11-03 | South America/Brazil/Rio de Janeiro/Ric | Laboratorio de Virologia Molecular / UFRJ | Bioinformatics Laboratory / LNCC | Carolina M Voloch et al |
| EPI_ISL_717894 | Brazil/RJ-00500/2020 | 2020-11-04 | South America/Brazil/Rio de Janeiro/Ita | Laboratorio de Virologia Molecular / UFRJ | Bioinformatics Laboratory / LNCC | Carolina M Voloch et al |
| EPI_ISL_717895 | Brazil/RJ-00501/2020 | 2020-11-04 | South America/Brazil/Rio de Janeiro/Ric | Laboratorio de Virologia Molecular / UFRJ | Bioinformatics Laboratory / LNCC | Carolina M Voloch et al |
| EPI_ISL_717897 | Brazil/RJ-00503/2020 | 2020-11-04 | South America/Brazil/Rio de Janeiro/Ric | Laboratorio de Virologia Molecular / UFRJ | Bioinformatics Laboratory / LNCC | Carolina M Voloch et al |
| EPI_ISL_717899 | Brazil/RJ-00505/2020 | 2020-07-27 | South America/Brazil/Rio de Janeiro/Ric | LACEN RJ - Noel Nutels | Bioinformatics Laboratory / LNCC | Carolina M Voloch et al |
| EPI_ISL_717900 | Brazil/RJ-00507/2020 | 2020-07-27 | South America/Brazil/Rio de Janeiro/Ric | LACEN RJ - Noel Nutels | Bioinformatics Laboratory / LNCC | Carolina M Voloch et al |
| EPI_ISL_717901 | Brazil/RJ-00508/2020 | 2020-07-30 | South America/Brazil/Rio de Janeiro/Ric | LACEN RJ - Noel Nutels | Bioinformatics Laboratory / LNCC | Carolina M Voloch et al |
| EPI_ISL_717902 | Brazil/RJ-00511/2020 | 2020-07-27 | South America/Brazil/Rio de Janeiro/Sa | LACEN RJ - Noel Nutels | Bioinformatics Laboratory / LNCC | Carolina M Voloch et al |
| EPI_ISL_717903 | Brazil/RJ-00512/2020 | 2020-07-28 | South America/Brazil/Rio de Janeiro/Ma | LACEN RJ - Noel Nutels | Bioinformatics Laboratory / LNCC | Carolina M Voloch et al |
| EPI_ISL_717904 | Brazil/RJ-00515/2020 | 2020-07-28 | South America/Brazil/Rio de Janeiro/No | LACEN RJ - Noel Nutels | Bioinformatics Laboratory / LNCC | Carolina M Voloch et al |
| EPI_ISL_717905 | Brazil/RJ-00516/2020 | 2020-07-28 | South America/Brazil/Rio de Janeiro/Pe | LACEN RJ - Noel Nutels | Bioinformatics Laboratory / LNCC | Carolina M Voloch et al |
| EPI_ISL_717906 | Brazil/RJ-00517/2020 | 2020-07-29 | South America/Brazil/Rio de Janeiro/Pe | LACEN RJ - Noel Nutels | Bioinformatics Laboratory / LNCC | Carolina M Voloch et al |
| EPI_ISL_717907 | Brazil/RJ-00518/2020 | 2020-07-18 | South America/Brazil/Rio de Janeiro/Tre | LACEN RJ - Noel Nutels | Bioinformatics Laboratory / LNCC | Carolina M Voloch et al |
| EPI_ISL_717908 | Brazil/RJ-00520/2020 | 2020-08-01 | South America/Brazil/Rio de Janeiro/Tre | LACEN RJ - Noel Nutels | Bioinformatics Laboratory / LNCC | Carolina M Voloch et al |
| EPI_ISL_717909 | Brazil/RJ-00522/2020 | 2020-07-29 | South America/Brazil/Rio de Janeiro/No | LACEN RJ - Noel Nutels | Bioinformatics Laboratory / LNCC | Carolina M Voloch et al |
| EPI_ISL_717910 | Brazil/RJ-00524/2020 | 2020-05-31 | South America/Brazil/Rio de Janeiro/Ma | LACEN Dr. Francisco Rimolo Neto | Bioinformatics Laboratory / LNCC | Carolina M Voloch et al |
| EPI_ISL_717911 | Brazil/RJ-00525/2020 | 2020-06-01 | South America/Brazil/Rio de Janeiro/Ma | LACEN Dr. Francisco Rimolo Neto | Bioinformatics Laboratory / LNCC | Carolina M Voloch et al |
| EPI_ISL_717912 | Brazil/RJ-00526/2020 | 2020-06-02 | South America/Brazil/Rio de Janeiro/Ma | LACEN Dr. Francisco Rimolo Neto | Bioinformatics Laboratory / LNCC | Carolina M Voloch et al |
| EPI_ISL_717913 | Brazil/RJ-00527/2020 | 2020-07-29 | South America/Brazil/Rio de Janeiro/Ma | LACEN Dr. Francisco Rimolo Neto | Bioinformatics Laboratory / LNCC | Carolina M Voloch et al |
| EPI_ISL_717914 | Brazil/RJ-00528/2020 | 2020-07-29 | South America/Brazil/Rio de Janeiro/Ma | LACEN Dr. Francisco Rimolo Neto | Bioinformatics Laboratory / LNCC | Carolina M Voloch et al |

| | | | | | | |
|---|---|---|---|---|---|---|
| EPI_ISL_717915 | Brazil/RJ-00529/2020 | 2020-07-29 | South America/Brazil/Rio de Janeiro/Ma | LACEN Dr. Francisco Rimolo Neto | Bioinformatics Laboratory / LNCC | Carolina M Voloch et al |
| EPI_ISL_717916 | Brazil/RJ-00530/2020 | 2020-07-29 | South America/Brazil/Rio de Janeiro/Ma | LACEN Dr. Francisco Rimolo Neto | Bioinformatics Laboratory / LNCC | Carolina M Voloch et al |
| EPI_ISL_717917 | Brazil/RJ-00531/2020 | 2020-07-29 | South America/Brazil/Rio de Janeiro/Ma | LACEN Dr. Francisco Rimolo Neto | Bioinformatics Laboratory / LNCC | Carolina M Voloch et al |
| EPI_ISL_717918 | Brazil/RJ-00532/2020 | 2020-06-02 | South America/Brazil/Rio de Janeiro/Ma | LACEN Dr. Francisco Rimolo Neto | Bioinformatics Laboratory / LNCC | Carolina M Voloch et al |
| EPI_ISL_717919 | Brazil/RJ-00533/2020 | 2020-06-02 | South America/Brazil/Rio de Janeiro/Ma | LACEN Dr. Francisco Rimolo Neto | Bioinformatics Laboratory / LNCC | Carolina M Voloch et al |
| EPI_ISL_717920 | Brazil/RJ-00535/2020 | 2020-10-29 | South America/Brazil/Rio de Janeiro/Ric | Laboratorio de Virologia Molecular / UFRJ | Bioinformatics Laboratory / LNCC | Carolina M Voloch et al |
| EPI_ISL_717921 | Brazil/RJ-00536/2020 | 2020-10-26 | South America/Brazil/Rio de Janeiro/Ca | Laboratorio de Virologia Molecular / UFRJ | Bioinformatics Laboratory / LNCC | Carolina M Voloch et al |
| EPI_ISL_717922 | Brazil/RJ-00537/2020 | 2020-10-29 | South America/Brazil/Rio de Janeiro/Ric | Laboratorio de Virologia Molecular / UFRJ | Bioinformatics Laboratory / LNCC | Carolina M Voloch et al |
| EPI_ISL_717924 | Brazil/RJ-00540/2020 | 2020-10-29 | South America/Brazil/Rio de Janeiro/Ric | Laboratorio de Virologia Molecular / UFRJ | Bioinformatics Laboratory / LNCC | Carolina M Voloch et al |
| EPI_ISL_717925 | Brazil/RJ-00541/2020 | 2020-10-09 | South America/Brazil/Rio de Janeiro/Ric | Laboratorio de Virologia Molecular / UFRJ | Bioinformatics Laboratory / LNCC | Carolina M Voloch et al |
| EPI_ISL_717926 | Brazil/RJ-00542/2020 | 2020-10-26 | South America/Brazil/Rio de Janeiro/Du | Laboratorio de Virologia Molecular / UFRJ | Bioinformatics Laboratory / LNCC | Carolina M Voloch et al |
| EPI_ISL_717927 | Brazil/RJ-00543/2020 | 2020-10-27 | South America/Brazil/Rio de Janeiro/Ric | Laboratorio de Virologia Molecular / UFRJ | Bioinformatics Laboratory / LNCC | Carolina M Voloch et al |
| EPI_ISL_717928 | Brazil/RJ-00544/2020 | 2020-10-27 | South America/Brazil/Rio de Janeiro/Ric | Laboratorio de Virologia Molecular / UFRJ | Bioinformatics Laboratory / LNCC | Carolina M Voloch et al |
| EPI_ISL_717929 | Brazil/RJ-00545/2020 | 2020-10-27 | South America/Brazil/Rio de Janeiro/Du | Laboratorio de Virologia Molecular / UFRJ | Bioinformatics Laboratory / LNCC | Carolina M Voloch et al |
| EPI_ISL_717930 | Brazil/RJ-00546/2020 | 2020-10-27 | South America/Brazil/Rio de Janeiro/Ric | Laboratorio de Virologia Molecular / UFRJ | Bioinformatics Laboratory / LNCC | Carolina M Voloch et al |
| EPI_ISL_717931 | Brazil/RJ-00547/2020 | 2020-10-27 | South America/Brazil/Rio de Janeiro/Ric | Laboratorio de Virologia Molecular / UFRJ | Bioinformatics Laboratory / LNCC | Carolina M Voloch et al |
| EPI_ISL_717932 | Brazil/RJ-00548/2020 | 2020-10-29 | South America/Brazil/Rio de Janeiro/Ric | Laboratorio de Virologia Molecular / UFRJ | Bioinformatics Laboratory / LNCC | Carolina M Voloch et al |
| EPI_ISL_717933 | Brazil/RJ-00549/2020 | 2020-10-29 | South America/Brazil/Rio de Janeiro/Ric | Laboratorio de Virologia Molecular / UFRJ | Bioinformatics Laboratory / LNCC | Carolina M Voloch et al |
| EPI_ISL_717934 | Brazil/RJ-00550/2020 | 2020-10-29 | South America/Brazil/Rio de Janeiro/Ric | Laboratorio de Virologia Molecular / UFRJ | Bioinformatics Laboratory / LNCC | Carolina M Voloch et al |
| EPI_ISL_717935 | Brazil/RJ-00551/2020 | 2020-10-29 | South America/Brazil/Rio de Janeiro/Ric | Laboratorio de Virologia Molecular / UFRJ | Bioinformatics Laboratory / LNCC | Carolina M Voloch et al |

| | | | | | | |
|---|---|---|---|---|---|---|
| EPI_ISL_717936 | Brazil/RJ-00552/2020 | 2020-10-29 | South America/Brazil/Rio de Janeiro/Rio | Laboratorio de Virologia Molecular / UFRJ | Bioinformatics Laboratory / LNCC | Carolina M Voloch et al |
| EPI_ISL_717937 | Brazil/RJ-00553/2020 | 2020-10-29 | South America/Brazil/Rio de Janeiro/Du | Laboratorio de Virologia Molecular / UFRJ | Bioinformatics Laboratory / LNCC | Carolina M Voloch et al |
| EPI_ISL_717938 | Brazil/RJ-00554/2020 | 2020-10-29 | South America/Brazil/Rio de Janeiro/Nit | Laboratorio de Virologia Molecular / UFRJ | Bioinformatics Laboratory / LNCC | Carolina M Voloch et al |
| EPI_ISL_717939 | Brazil/RJ-00555/2020 | 2020-10-29 | South America/Brazil/Rio de Janeiro/Rio | Laboratorio de Virologia Molecular / UFRJ | Bioinformatics Laboratory / LNCC | Carolina M Voloch et al |
| EPI_ISL_717940 | Brazil/RJ-00556/2020 | 2020-10-30 | South America/Brazil/Rio de Janeiro/Nit | Laboratorio de Virologia Molecular / UFRJ | Bioinformatics Laboratory / LNCC | Carolina M Voloch et al |
| EPI_ISL_717941 | Brazil/RJ-00557/2020 | 2020-10-30 | South America/Brazil/Rio de Janeiro/Rio | Laboratorio de Virologia Molecular / UFRJ | Bioinformatics Laboratory / LNCC | Carolina M Voloch et al |
| EPI_ISL_717942 | Brazil/RJ-00558/2020 | 2020-11-03 | South America/Brazil/Rio de Janeiro/Rio | Laboratorio de Virologia Molecular / UFRJ | Bioinformatics Laboratory / LNCC | Carolina M Voloch et al |
| EPI_ISL_717943 | Brazil/RJ-00559/2020 | 2020-11-03 | South America/Brazil/Rio de Janeiro/Rio | Laboratorio de Virologia Molecular / UFRJ | Bioinformatics Laboratory / LNCC | Carolina M Voloch et al |
| EPI_ISL_717945 | Brazil/RJ-00561/2020 | 2020-11-03 | South America/Brazil/Rio de Janeiro/Rio | Laboratorio de Virologia Molecular / UFRJ | Bioinformatics Laboratory / LNCC | Carolina M Voloch et al |
| EPI_ISL_717947 | Brazil/RJ-00563/2020 | 2020-11-03 | South America/Brazil/Rio de Janeiro/Rio | Laboratorio de Virologia Molecular / UFRJ | Bioinformatics Laboratory / LNCC | Carolina M Voloch et al |
| EPI_ISL_717948 | Brazil/RJ-00564/2020 | 2020-11-03 | South America/Brazil/Rio de Janeiro/Rio | Laboratorio de Virologia Molecular / UFRJ | Bioinformatics Laboratory / LNCC | Carolina M Voloch et al |
| EPI_ISL_717950 | Brazil/RJ-00566/2020 | 2020-11-03 | South America/Brazil/Rio de Janeiro/Rio | Laboratorio de Virologia Molecular / UFRJ | Bioinformatics Laboratory / LNCC | Carolina M Voloch et al |
| EPI_ISL_717951 | Brazil/RJ-00567/2020 | 2020-11-04 | South America/Brazil/Rio de Janeiro/Rio | Laboratorio de Virologia Molecular / UFRJ | Bioinformatics Laboratory / LNCC | Carolina M Voloch et al |
| EPI_ISL_717952 | Brazil/RJ-00568/2020 | 2020-11-04 | South America/Brazil/Rio de Janeiro/Rio | Laboratorio de Virologia Molecular / UFRJ | Bioinformatics Laboratory / LNCC | Carolina M Voloch et al |
| EPI_ISL_717956 | Brazil/RJ-00572/2020 | 2020-11-05 | South America/Brazil/Rio de Janeiro/Rio | Laboratorio de Virologia Molecular / UFRJ | Bioinformatics Laboratory / LNCC | Carolina M Voloch et al |
| EPI_ISL_717957 | Brazil/RJ-00573/2020 | 2020-11-05 | South America/Brazil/Rio de Janeiro/Rio | Laboratorio de Virologia Molecular / UFRJ | Bioinformatics Laboratory / LNCC | Carolina M Voloch et al |
| EPI_ISL_717958 | Brazil/RJ-00574/2020 | 2020-07-28 | South America/Brazil/Rio de Janeiro/Ma | LACEN Dr. Francisco Rimolo Neto | Bioinformatics Laboratory / LNCC | Carolina M Voloch et al |
| EPI_ISL_717959 | Brazil/RJ-00575/2020 | 2020-08-03 | South America/Brazil/Rio de Janeiro/Rio | Laboratorio de Virologia Molecular / UFRJ | Bioinformatics Laboratory / LNCC | Carolina M Voloch et al |
| EPI_ISL_717960 | Brazil/RJ-00576/2020 | 2020-10-09 | South America/Brazil/Rio de Janeiro/Rio | Laboratorio de Virologia Molecular / UFRJ | Bioinformatics Laboratory / LNCC | Carolina M Voloch et al |
| EPI_ISL_717962 | Brazil/RJ-00578/2020 | 2020-07-28 | South America/Brazil/Rio de Janeiro/Ma | LACEN RJ - Noel Nutels | Bioinformatics Laboratory / LNCC | Carolina M Voloch et al |

| | | | | | | |
|---|---|---|---|---|---|---|
| EPI_ISL_717963 | Brazil/RJ-00579/2020 | 2020-07-29 | South America/Brazil/Rio de Janeiro/Ma | LACEN Dr. Francisco Rimolo Neto | Bioinformatics Laboratory / LNCC | Carolina M Voloch et al |
| EPI_ISL_717964 | Brazil/RJ-00580/2020 | 2020-07-29 | South America/Brazil/Rio de Janeiro/Ma | LACEN Dr. Francisco Rimolo Neto | Bioinformatics Laboratory / LNCC | Carolina M Voloch et al |
| EPI_ISL_717965 | Russia/MOS-CRIE-13449451/2020 | 2020-11-07 | Europe/Russia/Moscow Oblast/ | "Molecular diagnostic laboratory of Federal Budget Institution of Science ""Central Research Institute of Epidemiology"" of The Federal Service on Customers' Rights Protection and Human Well-being Surveillance" | Group of Genomics and Postgenomic Technologies of Central Research Institute of Epidemilology | Samoilov AE et al |
| EPI_ISL_717966 | Russia/MOS-CRIE-13455107/2020 | 2020-11-07 | Europe/Russia/Moscow Oblast/ | "Molecular diagnostic laboratory of Federal Budget Institution of Science ""Central Research Institute of Epidemiology"" of The Federal Service on Customers' Rights Protection and Human Well-being Surveillance" | Group of Genomics and Postgenomic Technologies of Central Research Institute of Epidemilology | Samoilov AE et al |
| EPI_ISL_717967 | Russia/MOS-CRIE-13294236/2020 | 2020-11-06 | Europe/Russia/Moscow Oblast/ | "Molecular diagnostic laboratory of Federal Budget Institution of Science ""Central Research Institute of Epidemiology"" of The Federal Service on Customers' Rights Protection and Human Well-being Surveillance" | Group of Genomics and Postgenomic Technologies of Central Research Institute of Epidemilology | Samoilov AE et al |
| EPI_ISL_717969 | Russia/MOS-CRIE-13454908/2020 | 2020-11-07 | Europe/Russia/Moscow Oblast/ | "Molecular diagnostic laboratory of Federal Budget Institution of Science ""Central Research Institute of Epidemiology"" of The Federal Service on Customers' Rights Protection and Human Well-being Surveillance" | Group of Genomics and Postgenomic Technologies of Central Research Institute of Epidemilology | Samoilov AE et al |
| EPI_ISL_717970 | Russia/MOS-CRIE-13379339/2020 | 2020-11-06 | Europe/Russia/Moscow Oblast/ | "Molecular diagnostic laboratory of Federal Budget Institution of Science ""Central Research Institute of Epidemiology"" of The Federal Service on Customers' Rights Protection and Human Well-being Surveillance" | Group of Genomics and Postgenomic Technologies of Central Research Institute of Epidemilology | Samoilov AE et al |

| | | | | | | |
|---|---|---|---|---|---|---|
| EPI_ISL_717971 | Russia/MOS-CRIE-13383171/2020 | 2020-11-06 | Europe/Russia/Moscow Oblast/ | "Molecular diagnostic laboratory of Federal Budget Institution of Science ""Central Research Institute of Epidemiology"" of The Federal Service on Customers' Rights Protection and Human Well-being Surveillance" | Group of Genomics and Postgenomic Technologies of Central Research Institute of Epidemilology | Samoilov AE et al |
| EPI_ISL_717972 | Russia/MOS-CRIE-13312464/2020 | 2020-11-06 | Europe/Russia/Moscow Oblast/ | "Molecular diagnostic laboratory of Federal Budget Institution of Science ""Central Research Institute of Epidemiology"" of The Federal Service on Customers' Rights Protection and Human Well-being Surveillance" | Group of Genomics and Postgenomic Technologies of Central Research Institute of Epidemilology | Samoilov AE et al |
| EPI_ISL_717973 | Russia/MOS-CRIE-13385510/2020 | 2020-11-06 | Europe/Russia/Moscow Oblast/ | "Molecular diagnostic laboratory of Federal Budget Institution of Science ""Central Research Institute of Epidemiology"" of The Federal Service on Customers' Rights Protection and Human Well-being Surveillance" | Group of Genomics and Postgenomic Technologies of Central Research Institute of Epidemilology | Samoilov AE et al |
| EPI_ISL_717974 | Russia/MOS-CRIE-13386579/2020 | 2020-11-06 | Europe/Russia/Moscow Oblast/ | "Molecular diagnostic laboratory of Federal Budget Institution of Science ""Central Research Institute of Epidemiology"" of The Federal Service on Customers' Rights Protection and Human Well-being Surveillance" | Group of Genomics and Postgenomic Technologies of Central Research Institute of Epidemilology | Samoilov AE et al |
| EPI_ISL_717975 | Slovakia/UKBA-320/2020 | 2020-09-16 | Europe/Slovakia/Bratislava/ | Institute of Virology, Biomedical Research Center of the Slovak Academy of Sciences, Bratislava | Faculty of Natural Sciences, Comenius University, Bratislava | Bro≈àa Brejovv° et al |
| EPI_ISL_717982 | Belgium/UGent-301/2020 | 2020-11-30 | Europe/Belgium/Gent/ | Lab voor klinische biologie | Onderzoeksgroep Virologie | Nick Vereecke et al |
| EPI_ISL_717991 | Belgium/UGent-251/2020 | 2020-10-26 | Europe/Belgium/Gent/ | Lab voor klinische biologie | Onderzoeksgroep Virologie | Laurens Lambrechts et al |
| EPI_ISL_717997 | Belgium/UGent-257/2020 | 2020-11-04 | Europe/Belgium/Gent/ | Lab voor klinische biologie | Onderzoeksgroep Virologie | Laurens Lambrechts et al |
| EPI_ISL_718003 | Belgium/UGent-265/2020 | 2020-11-04 | Europe/Belgium/Gent/ | Lab voor klinische biologie | Onderzoeksgroep Virologie | Nick Vereecke et al |

| | | | | | | |
|---|---|---|---|---|---|---|
| EPI_ISL_718008 | Belgium/UGent-271/2020 | 2020-11-19 | Europe/Belgium/Gent/ | Lab voor klinische biologie | Onderzoeksgroep Virologie | Nick Vereecke et al |
| EPI_ISL_718010 | Belgium/UGent-274/2020 | 2020-11-16 | Europe/Belgium/Gent/ | Lab voor klinische biologie | Onderzoeksgroep Virologie | Laurens Lambrechts et al |
| EPI_ISL_718012 | Belgium/UGent-276/2020 | 2020-11-16 | Europe/Belgium/Gent/ | Lab voor klinische biologie | Onderzoeksgroep Virologie | Laurens Lambrechts et al |
| EPI_ISL_718015 | Belgium/UGent-281/2020 | 2020-11-26 | Europe/Belgium/Gent/ | Lab voor klinische biologie | Onderzoeksgroep Virologie | Laurens Lambrechts et al |
| EPI_ISL_718018 | Belgium/UGent-288/2020 | 2020-11-19 | Europe/Belgium/Gent/ | Lab voor klinische biologie | Onderzoeksgroep Virologie | Nick Vereecke et al |
| EPI_ISL_718028 | Germany/NW-HHU-331/2020 | 2020-10-12 | Europe/Germany/North Rhine Westphal | ZOTZ KLIMAS MVZ D√ºsseldorf-Centrum GbR √úBAG f√ºr Labormedizin, Genetik, Zytologie, Pathologie | Center of Medical Microbiology, Virology, and Hospital Hygiene, University of Duesseldorf | Maximilian Damagnez et al |
| EPI_ISL_718029 | Germany/NW-HHU-337/2020 | 2020-10-13 | Europe/Germany/North Rhine Westphal | ZOTZ KLIMAS MVZ D√ºsseldorf-Centrum GbR √úBAG f√ºr Labormedizin, Genetik, Zytologie, Pathologie | Center of Medical Microbiology, Virology, and Hospital Hygiene, University of Duesseldorf | Maximilian Damagnez et al |
| EPI_ISL_718030 | Germany/NW-HHU-328/2020 | 2020-10-12 | Europe/Germany/North Rhine Westphal | ZOTZ KLIMAS MVZ D√ºsseldorf-Centrum GbR √úBAG f√ºr Labormedizin, Genetik, Zytologie, Pathologie | Center of Medical Microbiology, Virology, and Hospital Hygiene, University of Duesseldorf | Maximilian Damagnez et al |
| EPI_ISL_718033 | Germany/NW-HHU-332/2020 | 2020-10-12 | Europe/Germany/North Rhine Westphal | ZOTZ KLIMAS MVZ D√ºsseldorf-Centrum GbR √úBAG f√ºr Labormedizin, Genetik, Zytologie, Pathologie | Center of Medical Microbiology, Virology, and Hospital Hygiene, University of Duesseldorf | Maximilian Damagnez et al |
| EPI_ISL_718035 | Germany/NW-HHU-335/2020 | 2020-10-13 | Europe/Germany/North Rhine Westphal | ZOTZ KLIMAS MVZ D√ºsseldorf-Centrum GbR √úBAG f√ºr Labormedizin, Genetik, Zytologie, Pathologie | Center of Medical Microbiology, Virology, and Hospital Hygiene, University of Duesseldorf | Maximilian Damagnez et al |

| | | | | | | |
|---|---|---|---|---|---|---|
| EPI_ISL_718036 | Germany/NW-HHU-336/2020 | 2020-10-13 | Europe/Germany/North Rhine Westphal | ZOTZ KLIMAS MVZ D√ºsseldorf-Centrum GbR √úBAG f√ºr Labormedizin, Genetik, Zytologie, Pathologie | Center of Medical Microbiology, Virology, and Hospital Hygiene, University of Duesseldorf | Maximilian Damagnez et al |
| EPI_ISL_718041 | Germany/NW-HHU-377/2020 | 2020-11-26 | Europe/Germany/North Rhine Westphal | ZOTZ KLIMAS MVZ D√ºsseldorf-Centrum GbR √úBAG f√ºr Labormedizin, Genetik, Zytologie, Pathologie | Center of Medical Microbiology, Virology, and Hospital Hygiene, University of Duesseldorf | Maximilian Damagnez et al |
| EPI_ISL_718042 | Germany/NW-HHU-383/2020 | 2020-11-26 | Europe/Germany/North Rhine Westphal | ZOTZ KLIMAS MVZ D√ºsseldorf-Centrum GbR √úBAG f√ºr Labormedizin, Genetik, Zytologie, Pathologie | Center of Medical Microbiology, Virology, and Hospital Hygiene, University of Duesseldorf | Maximilian Damagnez et al |
| EPI_ISL_718043 | Germany/NW-HHU-375/2020 | 2020-11-26 | Europe/Germany/North Rhine Westphal | ZOTZ KLIMAS MVZ D√ºsseldorf-Centrum GbR √úBAG f√ºr Labormedizin, Genetik, Zytologie, Pathologie | Center of Medical Microbiology, Virology, and Hospital Hygiene, University of Duesseldorf | Maximilian Damagnez et al |
| EPI_ISL_718045 | Germany/NW-HHU-366/2020 | 2020-11-26 | Europe/Germany/North Rhine Westphal | ZOTZ KLIMAS MVZ D√ºsseldorf-Centrum GbR √úBAG f√ºr Labormedizin, Genetik, Zytologie, Pathologie | Center of Medical Microbiology, Virology, and Hospital Hygiene, University of Duesseldorf | Maximilian Damagnez et al |
| EPI_ISL_718046 | Germany/NW-HHU-367/2020 | 2020-11-26 | Europe/Germany/North Rhine Westphal | ZOTZ KLIMAS MVZ D√ºsseldorf-Centrum GbR √úBAG f√ºr Labormedizin, Genetik, Zytologie, Pathologie | Center of Medical Microbiology, Virology, and Hospital Hygiene, University of Duesseldorf | Maximilian Damagnez et al |

| | | | | | | |
|---|---|---|---|---|---|---|
| EPI_ISL_718047 | Germany/NW-HHU-379/2020 | 2020-11-26 | Europe/Germany/North Rhine Westphal | ZOTZ KLIMAS MVZ D√ºsseldorf-Centrum GbR √úBAG f√ºr Labormedizin, Genetik, Zytologie, Pathologie | Center of Medical Microbiology, Virology, and Hospital Hygiene, University of Duesseldorf | Maximilian Damagnez et al |
| EPI_ISL_718048 | Germany/NW-HHU-380/2020 | 2020-11-26 | Europe/Germany/North Rhine Westphal | ZOTZ KLIMAS MVZ D√ºsseldorf-Centrum GbR √úBAG f√ºr Labormedizin, Genetik, Zytologie, Pathologie | Center of Medical Microbiology, Virology, and Hospital Hygiene, University of Duesseldorf | Maximilian Damagnez et al |
| EPI_ISL_718049 | Germany/NW-HHU-368/2020 | 2020-11-26 | Europe/Germany/North Rhine Westphal | ZOTZ KLIMAS MVZ D√ºsseldorf-Centrum GbR √úBAG f√ºr Labormedizin, Genetik, Zytologie, Pathologie | Center of Medical Microbiology, Virology, and Hospital Hygiene, University of Duesseldorf | Maximilian Damagnez et al |
| EPI_ISL_718054 | Germany/NW-HHU-370/2020 | 2020-11-26 | Europe/Germany/North Rhine Westphal | ZOTZ KLIMAS MVZ D√ºsseldorf-Centrum GbR √úBAG f√ºr Labormedizin, Genetik, Zytologie, Pathologie | Center of Medical Microbiology, Virology, and Hospital Hygiene, University of Duesseldorf | Maximilian Damagnez et al |
| EPI_ISL_718055 | Germany/NW-HHU-372/2020 | 2020-11-26 | Europe/Germany/North Rhine Westphal | ZOTZ KLIMAS MVZ D√ºsseldorf-Centrum GbR √úBAG f√ºr Labormedizin, Genetik, Zytologie, Pathologie | Center of Medical Microbiology, Virology, and Hospital Hygiene, University of Duesseldorf | Maximilian Damagnez et al |
| EPI_ISL_718056 | Germany/NW-HHU-373/2020 | 2020-11-26 | Europe/Germany/North Rhine Westphal | ZOTZ KLIMAS MVZ D√ºsseldorf-Centrum GbR √úBAG f√ºr Labormedizin, Genetik, Zytologie, Pathologie | Center of Medical Microbiology, Virology, and Hospital Hygiene, University of Duesseldorf | Maximilian Damagnez et al |

| | | | | | | |
|---|---|---|---|---|---|---|
| EPI_ISL_718057 | Germany/NW-HHU-374/2020 | 2020-11-26 | Europe/Germany/North Rhine Westphal | ZOTZ KLIMAS MVZ Dv°sseldorf-Centrum GbR √úBAG fv°r Labormedizin, Genetik, Zytologie, Pathologie | Center of Medical Microbiology, Virology, and Hospital Hygiene, University of Duesseldorf | Maximilian Damagnez et al |
| EPI_ISL_718060 | Germany/NW-HHU-393/2020 | 2020-11-01 | Europe/Germany/North Rhine Westphal | ZOTZ KLIMAS MVZ Dv°sseldorf-Centrum GbR √úBAG fv°r Labormedizin, Genetik, Zytologie, Pathologie | Center of Medical Microbiology, Virology, and Hospital Hygiene, University of Duesseldorf | Maximilian Damagnez et al |
| EPI_ISL_718061 | Germany/NW-HHU-400/2020 | 2020-11-01 | Europe/Germany/North Rhine Westphal | ZOTZ KLIMAS MVZ Dv°sseldorf-Centrum GbR √úBAG fv°r Labormedizin, Genetik, Zytologie, Pathologie | Center of Medical Microbiology, Virology, and Hospital Hygiene, University of Duesseldorf | Maximilian Damagnez et al |
| EPI_ISL_718065 | Germany/NW-HHU-397/2020 | 2020-11-01 | Europe/Germany/North Rhine Westphal | ZOTZ KLIMAS MVZ Dv°sseldorf-Centrum GbR √úBAG fv°r Labormedizin, Genetik, Zytologie, Pathologie | Center of Medical Microbiology, Virology, and Hospital Hygiene, University of Duesseldorf | Maximilian Damagnez et al |
| EPI_ISL_718066 | Germany/NW-HHU-402/2020 | 2020-11-01 | Europe/Germany/North Rhine Westphal | ZOTZ KLIMAS MVZ Dv°sseldorf-Centrum GbR √úBAG fv°r Labormedizin, Genetik, Zytologie, Pathologie | Center of Medical Microbiology, Virology, and Hospital Hygiene, University of Duesseldorf | Maximilian Damagnez et al |
| EPI_ISL_718067 | Germany/NW-HHU-403/2020 | 2020-11-01 | Europe/Germany/North Rhine Westphal | ZOTZ KLIMAS MVZ Dv°sseldorf-Centrum GbR √úBAG fv°r Labormedizin, Genetik, Zytologie, Pathologie | Center of Medical Microbiology, Virology, and Hospital Hygiene, University of Duesseldorf | Maximilian Damagnez et al |

| | | | | | | |
|---|---|---|---|---|---|---|
| EPI_ISL_718068 | Germany/NW-HHU-386/2020 | 2020-11-01 | Europe/Germany/North Rhine Westphal | ZOTZ KLIMAS MVZ Dv∫sseldorf-Centrum GbR √úBAG fv∫r Labormedizin, Genetik, Zytologie, Pathologie | Center of Medical Microbiology, Virology, and Hospital Hygiene, University of Duesseldorf | Maximilian Damagnez et al |
| EPI_ISL_718069 | Germany/NW-HHU-387/2020 | 2020-11-01 | Europe/Germany/North Rhine Westphal | ZOTZ KLIMAS MVZ Dv∫sseldorf-Centrum GbR √úBAG fv∫r Labormedizin, Genetik, Zytologie, Pathologie | Center of Medical Microbiology, Virology, and Hospital Hygiene, University of Duesseldorf | Maximilian Damagnez et al |
| EPI_ISL_718077 | Germany/NW-HHU-398/2020 | 2020-11-01 | Europe/Germany/North Rhine Westphal | ZOTZ KLIMAS MVZ Dv∫sseldorf-Centrum GbR √úBAG fv∫r Labormedizin, Genetik, Zytologie, Pathologie | Center of Medical Microbiology, Virology, and Hospital Hygiene, University of Duesseldorf | Maximilian Damagnez et al |
| EPI_ISL_718079 | Germany/NW-HHU-401/2020 | 2020-11-01 | Europe/Germany/North Rhine Westphal | ZOTZ KLIMAS MVZ Dv∫sseldorf-Centrum GbR √úBAG fv∫r Labormedizin, Genetik, Zytologie, Pathologie | Center of Medical Microbiology, Virology, and Hospital Hygiene, University of Duesseldorf | Maximilian Damagnez et al |
| EPI_ISL_718081 | Germany/NW-HHU-405/2020 | 2020-11-01 | Europe/Germany/North Rhine Westphal | ZOTZ KLIMAS MVZ Dv∫sseldorf-Centrum GbR √úBAG fv∫r Labormedizin, Genetik, Zytologie, Pathologie | Center of Medical Microbiology, Virology, and Hospital Hygiene, University of Duesseldorf | Maximilian Damagnez et al |
| EPI_ISL_718082 | Germany/NW-HHU-406/2020 | 2020-11-01 | Europe/Germany/North Rhine Westphal | ZOTZ KLIMAS MVZ Dv∫sseldorf-Centrum GbR √úBAG fv∫r Labormedizin, Genetik, Zytologie, Pathologie | Center of Medical Microbiology, Virology, and Hospital Hygiene, University of Duesseldorf | Maximilian Damagnez et al |

| | | | | | | |
|---|---|---|---|---|---|---|
| EPI_ISL_718083 | Germany/NW-HHU-409/2020 | 2020-11-01 | Europe/Germany/North Rhine Westphal | ZOTZ KLIMAS MVZ D√ºsseldorf-Centrum GbR √úBAG f√ºr Labormedizin, Genetik, Zytologie, Pathologie | Center of Medical Microbiology, Virology, and Hospital Hygiene, University of Duesseldorf | Maximilian Damagnez et al |
| EPI_ISL_718084 | Germany/NW-HHU-410/2020 | 2020-11-11 | Europe/Germany/North Rhine Westphal | ZOTZ KLIMAS MVZ D√ºsseldorf-Centrum GbR √úBAG f√ºr Labormedizin, Genetik, Zytologie, Pathologie | Center of Medical Microbiology, Virology, and Hospital Hygiene, University of Duesseldorf | Maximilian Damagnez et al |
| EPI_ISL_718087 | Germany/NW-HHU-413/2020 | 2020-11-11 | Europe/Germany/North Rhine Westphal | ZOTZ KLIMAS MVZ D√ºsseldorf-Centrum GbR √úBAG f√ºr Labormedizin, Genetik, Zytologie, Pathologie | Center of Medical Microbiology, Virology, and Hospital Hygiene, University of Duesseldorf | Maximilian Damagnez et al |
| EPI_ISL_718088 | Germany/NW-HHU-414/2020 | 2020-11-11 | Europe/Germany/North Rhine Westphal | ZOTZ KLIMAS MVZ D√ºsseldorf-Centrum GbR √úBAG f√ºr Labormedizin, Genetik, Zytologie, Pathologie | Center of Medical Microbiology, Virology, and Hospital Hygiene, University of Duesseldorf | Maximilian Damagnez et al |
| EPI_ISL_718090 | Germany/NW-HHU-416/2020 | 2020-11-11 | Europe/Germany/North Rhine Westphal | ZOTZ KLIMAS MVZ D√ºsseldorf-Centrum GbR √úBAG f√ºr Labormedizin, Genetik, Zytologie, Pathologie | Center of Medical Microbiology, Virology, and Hospital Hygiene, University of Duesseldorf | Maximilian Damagnez et al |
| EPI_ISL_718091 | Germany/NW-HHU-417/2020 | 2020-11-11 | Europe/Germany/North Rhine Westphal | ZOTZ KLIMAS MVZ D√ºsseldorf-Centrum GbR √úBAG f√ºr Labormedizin, Genetik, Zytologie, Pathologie | Center of Medical Microbiology, Virology, and Hospital Hygiene, University of Duesseldorf | Maximilian Damagnez et al |

| | | | | | | |
|---|---|---|---|---|---|---|
| EPI_ISL_718092 | Germany/NW-HHU-418/2020 | 2020-11-11 | Europe/Germany/North Rhine Westphal | ZOTZ KLIMAS MVZ D√ºsseldorf-Centrum GbR √úBAG f√ºr Labormedizin, Genetik, Zytologie, Pathologie | Center of Medical Microbiology, Virology, and Hospital Hygiene, University of Duesseldorf | Maximilian Damagnez et al |
| EPI_ISL_718094 | Germany/NW-HHU-420/2020 | 2020-11-11 | Europe/Germany/North Rhine Westphal | ZOTZ KLIMAS MVZ D√ºsseldorf-Centrum GbR √úBAG f√ºr Labormedizin, Genetik, Zytologie, Pathologie | Center of Medical Microbiology, Virology, and Hospital Hygiene, University of Duesseldorf | Maximilian Damagnez et al |
| EPI_ISL_718096 | Germany/NW-HHU-422/2020 | 2020-11-13 | Europe/Germany/North Rhine Westphal | ZOTZ KLIMAS MVZ D√ºsseldorf-Centrum GbR √úBAG f√ºr Labormedizin, Genetik, Zytologie, Pathologie | Center of Medical Microbiology, Virology, and Hospital Hygiene, University of Duesseldorf | Maximilian Damagnez et al |
| EPI_ISL_718097 | Germany/NW-HHU-423/2020 | 2020-11-13 | Europe/Germany/North Rhine Westphal | ZOTZ KLIMAS MVZ D√ºsseldorf-Centrum GbR √úBAG f√ºr Labormedizin, Genetik, Zytologie, Pathologie | Center of Medical Microbiology, Virology, and Hospital Hygiene, University of Duesseldorf | Maximilian Damagnez et al |
| EPI_ISL_718100 | Germany/NW-HHU-426/2020 | 2020-11-13 | Europe/Germany/North Rhine Westphal | ZOTZ KLIMAS MVZ D√ºsseldorf-Centrum GbR √úBAG f√ºr Labormedizin, Genetik, Zytologie, Pathologie | Center of Medical Microbiology, Virology, and Hospital Hygiene, University of Duesseldorf | Maximilian Damagnez et al |
| EPI_ISL_718103 | Germany/NW-HHU-429/2020 | 2020-11-13 | Europe/Germany/North Rhine Westphal | ZOTZ KLIMAS MVZ D√ºsseldorf-Centrum GbR √úBAG f√ºr Labormedizin, Genetik, Zytologie, Pathologie | Center of Medical Microbiology, Virology, and Hospital Hygiene, University of Duesseldorf | Maximilian Damagnez et al |
| EPI_ISL_718105 | Canada/ON-SLB2861/2020 | 2020-04-07 | North America/Canada/Ontario/ | Kingston Health Sciences Centre and Queen's University | Ontario Institute for Cancer Research | Prameet M. Sheth et al |

| | | | | | | |
|---|---|---|---|---|---|---|
| EPI_ISL_718133 | Malaysia/UNIMAS-M3851/2020 | 2020-05-11 | Asia/Malaysia/Sarawak/ | Ministry of Health Hospitals | Institute of Health and Community Medicine | David Perera et al |
| EPI_ISL_718135 | Malaysia/UNIMAS-C121/2020 | 2020-05-29 | Asia/Malaysia/Sarawak/ | Ministry of Health Hospitals | Institute of Health and Community Medicine | David Perera et al |
| EPI_ISL_718145 | Malaysia/UNIMAS-M0710/2020 | 2020-04-13 | Asia/Malaysia/Sarawak/ | Ministry of Health Hospitals | Institute of Health and Community Medicine | David Perera et al |
| EPI_ISL_718166 | Malaysia/UNIMAS-M3482/2020 | 2020-05-10 | Asia/Malaysia/Sarawak/ | Ministry of Health Hospitals | Institute of Health and Community Medicine | David Perera et al |
| EPI_ISL_718167 | Malaysia/UNIMAS-B066/2020 | 2020-11-01 | Asia/Malaysia/Sarawak/ | Borneo Medical Centre | Institute of Health and Community Medicine | David Perera et al |
| EPI_ISL_718169 | Malaysia/UNIMAS-15723/2020 | 2020-10-28 | Asia/Malaysia/Sarawak/ | Ministry of Health Hospitals | Institute of Health and Community Medicine | David Perera et al |
| EPI_ISL_718172 | Malaysia/UNIMAS-15790/2020 | 2020-11-18 | Asia/Malaysia/Sarawak/ | Ministry of Health Hospitals | Institute of Health and Community Medicine | David Perera et al |
| EPI_ISL_718176 | Malaysia/UNIMAS-B074/2020 | 2020-11-08 | Asia/Malaysia/Sarawak/ | Borneo Medical Centre | Institute of Health and Community Medicine | David Perera et al |
| EPI_ISL_718184 | Malaysia/UNIMAS-15765/2020 | 2020-11-09 | Asia/Malaysia/Sarawak/ | Ministry of Health Hospitals | Institute of Health and Community Medicine | David Perera et al |
| EPI_ISL_718186 | Malaysia/UNIMAS-15764/2020 | 2020-11-13 | Asia/Malaysia/Sarawak/ | Ministry of Health Hospitals | Institute of Health and Community Medicine | David Perera et al |
| EPI_ISL_718187 | Malaysia/UNIMAS-15780/2020 | 2020-11-13 | Asia/Malaysia/Sarawak/ | Ministry of Health Hospitals | Institute of Health and Community Medicine | David Perera et al |
| EPI_ISL_718189 | Malaysia/UNIMAS-15763/2020 | 2020-11-11 | Asia/Malaysia/Sarawak/ | Ministry of Health Hospitals | Institute of Health and Community Medicine | David Perera et al |
| EPI_ISL_718191 | Malaysia/UNIMAS-B057/2020 | 2020-11-02 | Asia/Malaysia/Sarawak/ | Borneo Medical Centre | Institute of Health and Community Medicine | David Perera et al |

| | | | | | | |
|---|---|---|---|---|---|---|
| EPI_ISL_718194 | Malaysia/UNIMAS-15777/2020 | 2020-11-08 | Asia/Malaysia/Sarawak/ | Ministry of Health Hospitals | Institute of Health and Community Medicine | David Perera et al |
| EPI_ISL_718198 | Malaysia/UNIMAS-B037/2020 | 2020-10-26 | Asia/Malaysia/Sarawak/ | Borneo Medical Centre | Institute of Health and Community Medicine | David Perera et al |
| EPI_ISL_718199 | Malaysia/UNIMAS-15766/2020 | 2020-11-09 | Asia/Malaysia/Sarawak/ | Ministry of Health Hospitals | Institute of Health and Community Medicine | David Perera et al |
| EPI_ISL_718201 | Malaysia/UNIMAS-15731/2020 | 2020-11-06 | Asia/Malaysia/Sarawak/ | Ministry of Health Hospitals | Institute of Health and Community Medicine | David Perera et al |
| EPI_ISL_718204 | Malaysia/UNIMAS-B021/2020 | 2020-10-25 | Asia/Malaysia/Sarawak/ | Borneo Medical Centre | Institute of Health and Community Medicine | David Perera et al |
| EPI_ISL_718207 | Malaysia/UNIMAS-15736/2020 | 2020-11-04 | Asia/Malaysia/Sarawak/ | Ministry of Health Hospitals | Institute of Health and Community Medicine | David Perera et al |
| EPI_ISL_718208 | Malaysia/UNIMAS-15712/2020 | 2020-10-30 | Asia/Malaysia/Sarawak/ | Ministry of Health Hospitals | Institute of Health and Community Medicine | David Perera et al |
| EPI_ISL_718212 | Malaysia/UNIMAS-15683/2020 | 2020-10-22 | Asia/Malaysia/Sarawak/ | Ministry of Health Hospitals | Institute of Health and Community Medicine | David Perera et al |
| EPI_ISL_718226 | France/BRE-IPP9966/2020 | 2020-10-31 | Europe/France/Bretagne/Rennes | Hospital | National Reference Center for Viruses of Respiratory Infections, Institut Pasteur, Paris | Marion Barbet et al |
| EPI_ISL_718228 | France/BRE-IPP9969/2020 | 2020-10-27 | Europe/France/Bretagne/Rennes | Hospital | National Reference Center for Viruses of Respiratory Infections, Institut Pasteur, Paris | Marion Barbet et al |

| | | | | | | |
|---|---|---|---|---|---|---|
| EPI_ISL_718229 | France/BRE-IPP9970/2020 | 2020-09-14 | Europe/France/Bretagne/Rennes | Hospital | National Reference Center for Viruses of Respiratory Infections, Institut Pasteur, Paris | Marion Barbet et al |
| EPI_ISL_718230 | France/BRE-IPP9971/2020 | 2020-09-26 | Europe/France/Bretagne/Rennes | Hospital | National Reference Center for Viruses of Respiratory Infections, Institut Pasteur, Paris | Marion Barbet et al |
| EPI_ISL_718231 | France/BRE-IPP9972/2020 | 2020-10-24 | Europe/France/Bretagne/Rennes | Hospital | National Reference Center for Viruses of Respiratory Infections, Institut Pasteur, Paris | Marion Barbet et al |
| EPI_ISL_718232 | France/BRE-IPP9973/2020 | 2020-10-24 | Europe/France/Bretagne/Rennes | Hospital | National Reference Center for Viruses of Respiratory Infections, Institut Pasteur, Paris | Marion Barbet et al |
| EPI_ISL_718234 | France/BRE-IPP9976/2020 | 2020-10-25 | Europe/France/Bretagne/Rennes | Hospital | National Reference Center for Viruses of Respiratory Infections, Institut Pasteur, Paris | Marion Barbet et al |
| EPI_ISL_718239 | France/BRE-IPP9982/2020 | 2020-10-23 | Europe/France/Bretagne/Rennes | Hospital | National Reference Center for Viruses of Respiratory Infections, Institut Pasteur, Paris | Marion Barbet et al |

| | | | | | | |
|---|---|---|---|---|---|---|
| EPI_ISL_718241 | France/BRE-IPP9984/2020 | 2020-10-03 | Europe/France/Bretagne/Rennes | Hospital | National Reference Center for Viruses of Respiratory Infections, Institut Pasteur, Paris | Marion Barbet et al |
| EPI_ISL_718242 | France/BRE-IPP9986/2020 | 2020-10-17 | Europe/France/Bretagne/Rennes | Hospital | National Reference Center for Viruses of Respiratory Infections, Institut Pasteur, Paris | Marion Barbet et al |
| EPI_ISL_718244 | France/BRE-IPP9989/2020 | 2020-10-10 | Europe/France/Bretagne/Rennes | Hospital | National Reference Center for Viruses of Respiratory Infections, Institut Pasteur, Paris | Marion Barbet et al |
| EPI_ISL_718246 | France/BRE-IPP9992/2020 | 2020-10-10 | Europe/France/Bretagne/Rennes | Hospital | National Reference Center for Viruses of Respiratory Infections, Institut Pasteur, Paris | Marion Barbet et al |
| EPI_ISL_718247 | France/BRE-IPP9994/2020 | 2020-10-30 | Europe/France/Bretagne/Rennes | Hospital | National Reference Center for Viruses of Respiratory Infections, Institut Pasteur, Paris | Marion Barbet et al |
| EPI_ISL_718248 | France/BRE-IPP9995/2020 | 2020-11-02 | Europe/France/Bretagne/Rennes | Hospital | National Reference Center for Viruses of Respiratory Infections, Institut Pasteur, Paris | Marion Barbet et al |

| | | | | | | |
|---|---|---|---|---|---|---|
| EPI_ISL_718249 | France/BRE-IPP9996/2020 | 2020-10-27 | Europe/France/Bretagne/Rennes | Hospital | National Reference Center for Viruses of Respiratory Infections, Institut Pasteur, Paris | Marion Barbet et al |
| EPI_ISL_718250 | Slovakia/UKBA-402/2020 | 2020-11-22 | Europe/Slovakia/Bratislava/ | Institute of Virology, Biomedical Research Center of the Slovak Academy of Sciences, Bratislava | Faculty of Natural Sciences, Comenius University, Bratislava | Kristv≠na Bor≈°ov√° et al |
| EPI_ISL_718255 | Slovakia/UKBA-407/2020 | 2020-11-30 | Europe/Slovakia/Bratislava/ | Institute of Virology, Biomedical Research Center of the Slovak Academy of Sciences, Bratislava | Faculty of Natural Sciences, Comenius University, Bratislava | Bro≈àa Brejov√° et al |
| EPI_ISL_718256 | Slovakia/UKBA-408/2020 | 2020-11-30 | Europe/Slovakia/Bratislava/ | Institute of Virology, Biomedical Research Center of the Slovak Academy of Sciences, Bratislava | Faculty of Natural Sciences, Comenius University, Bratislava | Bro≈àa Brejov√° et al |
| EPI_ISL_718258 | Slovakia/UKBA-410/2020 | 2020-11-19 | Europe/Slovakia/Nitra/ | Institute of Virology, Biomedical Research Center of the Slovak Academy of Sciences, Bratislava | Faculty of Natural Sciences, Comenius University, Bratislava | Vikt√≥ria Hodorov√° et al |
| EPI_ISL_718259 | Slovakia/UKBA-411/2020 | 2020-11-13 | Europe/Slovakia/Bratislava/ | Institute of Virology, Biomedical Research Center of the Slovak Academy of Sciences, Bratislava | Faculty of Natural Sciences, Comenius University, Bratislava | Bro≈àa Brejov√° et al |
| EPI_ISL_718263 | Italy/LAZ-INMI-98/2020 | 2020-11-04 | Europe/Italy/Lazio/Rome | "National Institute for Infectious Diseases, INMI, ""L. Spallanzani"" IRCCS" | "National Institute for Infectious Diseases, INMI, ""L. Spallanzani"" IRCCS" | E Giombini et al |

| | | | | | | |
|---|---|---|---|---|---|---|
| EPI_ISL_718264 | Italy/LAZ-INMI-99/2020 | 2020-11-03 | Europe/Italy/Lazio/Rome | "National Institute for Infectious Diseases, INMI, ""L. Spallanzani"" IRCCS" | "National Institute for Infectious Diseases, INMI, ""L. Spallanzani"" IRCCS" | F Messina et al |
| EPI_ISL_718265 | Italy/LAZ-INMI-100/2020 | 2020-11-03 | Europe/Italy/Lazio/Rome | "National Institute for Infectious Diseases, INMI, ""L. Spallanzani"" IRCCS" | "National Institute for Infectious Diseases, INMI, ""L. Spallanzani"" IRCCS" | B Bartolini et al |
| EPI_ISL_718273 | Japan/FHU-CS24-0221/2020 | 2020-02-21 | Asia/Japan/Aichi/ | Fujita Health University Okazaki Medical Center | Department of Virology and Parasitology, Fujita Health University School of Medicine | Takayuki Murata et al |
| EPI_ISL_718280 | Malaysia/IMR-WI132/2020 | 2020-12-19 | Asia/Malaysia/Malaysia/ | Institute for Medical Research, Infectious Disease Research Centre, National Institutes of Health, Ministry of Health Malaysia | Institute for Medical Research, Infectious Disease Research Centre, National Institutes of Health, Ministry of Health Malaysia | Suppiah J et al |
| EPI_ISL_718296 | Malaysia/IMR-CV139158/2020 | 2020-09-01 | Asia/Malaysia/Malaysia/ | Institute for Medical Research, Infectious Disease Research Centre, National Institutes of Health, Ministry of Health Malaysia | Institute for Medical Research, Infectious Disease Research Centre, National Institutes of Health, Ministry of Health Malaysia | Suppiah J et al |

| | | | | | | |
|---|---|---|---|---|---|---|
| EPI_ISL_718314 | Malaysia/IMR-WI310/2020 | 2020-11-01 | Asia/Malaysia/Malaysia/ | Institute for Medical Research, Infectious Disease Research Centre, National Institutes of Health, Ministry of Health Malaysia | Institute for Medical Research, Infectious Disease Research Centre, National Institutes of Health, Ministry of Health Malaysia | Suppiah J et al |
| EPI_ISL_718406 | England/CAMC-C3FDC9/2020 | 2020-12-10 | Europe/United Kingdom/England/ | Lighthouse Lab in Cambridge | Wellcome Sanger Institute for the COVID-19 Genomics UK (COG-UK) Consortium | Rob Howes et al |
| EPI_ISL_718811 | England/ALDP-C481A3/2020 | 2020-12-12 | Europe/United Kingdom/England/ | Lighthouse Lab in Alderley Park | Wellcome Sanger Institute for the COVID-19 Genomics UK (COG-UK) Consortium | Jacquelyn Wynn et al |
| EPI_ISL_719062 | England/CAMC-C4376A/2020 | 2020-12-10 | Europe/United Kingdom/England/ | Lighthouse Lab in Cambridge | Wellcome Sanger Institute for the COVID-19 Genomics UK (COG-UK) Consortium | Rob Howes et al |
| EPI_ISL_719200 | England/CAMC-C3E4D4/2020 | 2020-12-10 | Europe/United Kingdom/England/ | Lighthouse Lab in Cambridge | Wellcome Sanger Institute for the COVID-19 Genomics UK (COG-UK) Consortium | Rob Howes et al |
| EPI_ISL_719210 | England/CAMC-C42385/2020 | 2020-12-10 | Europe/United Kingdom/England/ | Lighthouse Lab in Cambridge | Wellcome Sanger Institute for the COVID-19 Genomics UK (COG-UK) Consortium | Rob Howes et al |
| EPI_ISL_719213 | England/CAMC-C3E51D/2020 | 2020-12-10 | Europe/United Kingdom/England/ | Lighthouse Lab in Cambridge | Wellcome Sanger Institute for the COVID-19 Genomics UK (COG-UK) Consortium | Rob Howes et al |

| | | | | | | |
|---|---|---|---|---|---|---|
| EPI_ISL_719221 | England/CAMC-C43858/2020 | 2020-12-10 | Europe/United Kingdom/England/ | Lighthouse Lab in Cambridge | Wellcome Sanger Institute for the COVID-19 Genomics UK (COG-UK) Consortium | Rob Howes et al |
| EPI_ISL_719393 | England/CAMC-C3FBB0/2020 | 2020-12-10 | Europe/United Kingdom/England/ | Lighthouse Lab in Cambridge | Wellcome Sanger Institute for the COVID-19 Genomics UK (COG-UK) Consortium | Rob Howes et al |
| EPI_ISL_719546 | England/CAMC-C3FCAE/2020 | 2020-12-10 | Europe/United Kingdom/England/ | Lighthouse Lab in Cambridge | Wellcome Sanger Institute for the COVID-19 Genomics UK (COG-UK) Consortium | Rob Howes et al |
| EPI_ISL_719623 | England/QEUH-BA076C/2020 | 2020-11-18 | Europe/United Kingdom/England/ | Lighthouse Lab in Glasgow | Wellcome Sanger Institute for the COVID-19 Genomics UK (COG-UK) Consortium | Harper VanSteenhouse et al |
| EPI_ISL_719648 | England/QEUH-BA06E7/2020 | 2020-11-18 | Europe/United Kingdom/England/ | Lighthouse Lab in Glasgow | Wellcome Sanger Institute for the COVID-19 Genomics UK (COG-UK) Consortium | Harper VanSteenhouse et al |
| EPI_ISL_719840 | England/QEUH-BA2797/2020 | 2020-11-18 | Europe/United Kingdom/England/ | Lighthouse Lab in Glasgow | Wellcome Sanger Institute for the COVID-19 Genomics UK (COG-UK) Consortium | Harper VanSteenhouse et al |
| EPI_ISL_719946 | England/QEUH-BA1FDC/2020 | 2020-11-18 | Europe/United Kingdom/England/ | Lighthouse Lab in Glasgow | Wellcome Sanger Institute for the COVID-19 Genomics UK (COG-UK) Consortium | Harper VanSteenhouse et al |
| EPI_ISL_719951 | England/QEUH-BA1B8D/2020 | 2020-11-18 | Europe/United Kingdom/England/ | Lighthouse Lab in Glasgow | Wellcome Sanger Institute for the COVID-19 Genomics UK (COG-UK) Consortium | Harper VanSteenhouse et al |

| | | | | | | |
|---|---|---|---|---|---|---|
| EPI_ISL_720061 | England/MILK-BB156E/2020 | 2020-11-17 | Europe/United Kingdom/England/ | Lighthouse Lab in Milton Keynes | Wellcome Sanger Institute for the COVID-19 Genomics UK (COG-UK) Consortium | The Lighthouse Lab in Milton Keynes et al |
| EPI_ISL_720117 | England/MILK-BB12D0/2020 | 2020-11-17 | Europe/United Kingdom/England/ | Lighthouse Lab in Milton Keynes | Wellcome Sanger Institute for the COVID-19 Genomics UK (COG-UK) Consortium | The Lighthouse Lab in Milton Keynes et al |
| EPI_ISL_720223 | England/MILK-BB01C5/2020 | 2020-11-18 | Europe/United Kingdom/England/ | Lighthouse Lab in Milton Keynes | Wellcome Sanger Institute for the COVID-19 Genomics UK (COG-UK) Consortium | The Lighthouse Lab in Milton Keynes et al |
| EPI_ISL_720466 | England/MILK-BAB8E6/2020 | 2020-11-17 | Europe/United Kingdom/England/ | Lighthouse Lab in Milton Keynes | Wellcome Sanger Institute for the COVID-19 Genomics UK (COG-UK) Consortium | The Lighthouse Lab in Milton Keynes et al |
| EPI_ISL_720520 | England/MILK-BABF4B/2020 | 2020-11-18 | Europe/United Kingdom/England/ | Lighthouse Lab in Milton Keynes | Wellcome Sanger Institute for the COVID-19 Genomics UK (COG-UK) Consortium | The Lighthouse Lab in Milton Keynes et al |
| EPI_ISL_720595 | England/MILK-BB19BD/2020 | 2020-11-18 | Europe/United Kingdom/England/ | Lighthouse Lab in Milton Keynes | Wellcome Sanger Institute for the COVID-19 Genomics UK (COG-UK) Consortium | The Lighthouse Lab in Milton Keynes et al |
| EPI_ISL_720619 | England/MILK-BAFCF5/2020 | 2020-11-17 | Europe/United Kingdom/England/ | Lighthouse Lab in Milton Keynes | Wellcome Sanger Institute for the COVID-19 Genomics UK (COG-UK) Consortium | The Lighthouse Lab in Milton Keynes et al |
| EPI_ISL_720667 | England/MILK-BABF3C/2020 | 2020-11-18 | Europe/United Kingdom/England/ | Lighthouse Lab in Milton Keynes | Wellcome Sanger Institute for the COVID-19 Genomics UK (COG-UK) Consortium | The Lighthouse Lab in Milton Keynes et al |

| | | | | | | |
|---|---|---|---|---|---|---|
| EPI_ISL_720904 | England/QEUH-C01443/2020 | 2020-12-04 | Europe/United Kingdom/England/ | Lighthouse Lab in Glasgow | Wellcome Sanger Institute for the COVID-19 Genomics UK (COG-UK) Consortium | Harper VanSteenhouse et al |
| EPI_ISL_721083 | England/QEUH-BA323B/2020 | 2020-11-18 | Europe/United Kingdom/England/ | Lighthouse Lab in Glasgow | Wellcome Sanger Institute for the COVID-19 Genomics UK (COG-UK) Consortium | Harper VanSteenhouse et al |
| EPI_ISL_721095 | England/QEUH-B9E145/2020 | 2020-11-17 | Europe/United Kingdom/England/ | Lighthouse Lab in Glasgow | Wellcome Sanger Institute for the COVID-19 Genomics UK (COG-UK) Consortium | Harper VanSteenhouse et al |
| EPI_ISL_721178 | England/QEUH-BA4285/2020 | 2020-11-18 | Europe/United Kingdom/England/ | Lighthouse Lab in Glasgow | Wellcome Sanger Institute for the COVID-19 Genomics UK (COG-UK) Consortium | Harper VanSteenhouse et al |
| EPI_ISL_721327 | England/ALDP-BC7D5E/2020 | 2020-11-23 | Europe/United Kingdom/England/ | Lighthouse Lab in Alderley Park | Wellcome Sanger Institute for the COVID-19 Genomics UK (COG-UK) Consortium | Jacquelyn Wynn et al |
| EPI_ISL_721502 | England/ALDP-BC7B63/2020 | 2020-11-23 | Europe/United Kingdom/England/ | Lighthouse Lab in Alderley Park | Wellcome Sanger Institute for the COVID-19 Genomics UK (COG-UK) Consortium | Jacquelyn Wynn et al |
| EPI_ISL_721529 | England/MILK-BE6EDE/2020 | 2020-11-28 | Europe/United Kingdom/England/ | Lighthouse Lab in Milton Keynes | Wellcome Sanger Institute for the COVID-19 Genomics UK (COG-UK) Consortium | The Lighthouse Lab in Milton Keynes et al |
| EPI_ISL_721545 | England/QEUH-AAF2D6/2020 | 2020-10-21 | Europe/United Kingdom/England/ | Lighthouse Lab in Glasgow | Wellcome Sanger Institute for the COVID-19 Genomics UK (COG-UK) Consortium | Harper VanSteenhouse et al |

| | | | | | | |
|---|---|---|---|---|---|---|
| EPI_ISL_721559 | England/QEUH-A9F28E/2020 | 2020-10-19 | Europe/United Kingdom/England/ | Lighthouse Lab in Glasgow | Wellcome Sanger Institute for the COVID-19 Genomics UK (COG-UK) Consortium | Harper VanSteenhouse et al |
| EPI_ISL_721624 | Italy/LAZ-INMI-101/2020 | 2020-11-17 | Europe/Italy/Lazio/Rome | "National Institute for Infectious Diseases, INMI, ""L. Spallanzani"" IRCCS" | "National Institute for Infectious Diseases, INMI, ""L. Spallanzani"" IRCCS" | C.E.M Gruber et al |
| EPI_ISL_721625 | Italy/LAZ-INMI-102/2020 | 2020-11-17 | Europe/Italy/Lazio/Rome | "National Institute for Infectious Diseases, INMI, ""L. Spallanzani"" IRCCS" | "National Institute for Infectious Diseases, INMI, ""L. Spallanzani"" IRCCS" | B Bartolini et al |
| EPI_ISL_721626 | Italy/LAZ-INMI-103/2020 | 2020-11-20 | Europe/Italy/Lazio/Rome | "National Institute for Infectious Diseases, INMI, ""L. Spallanzani"" IRCCS" | "National Institute for Infectious Diseases, INMI, ""L. Spallanzani"" IRCCS" | M Rueca et al |
| EPI_ISL_721628 | Italy/LAZ-INMI-105/2020 | 2020-11-17 | Europe/Italy/Lazio/Rome | "National Institute for Infectious Diseases, INMI, ""L. Spallanzani"" IRCCS" | "National Institute for Infectious Diseases, INMI, ""L. Spallanzani"" IRCCS" | E Giombini et al |
| EPI_ISL_721629 | India/MH-AFCO-14752/2020 | 2020-07-22 | Asia/India/Maharashtra/ | Armed Forces Medical College | National Centre For Cell Science | Dhiraj Paul et al |
| EPI_ISL_721630 | India/MH-AFCO-14778/2020 | 2020-07-22 | Asia/India/Maharashtra/ | Armed Forces Medical College | National Centre For Cell Science | Dhiraj Paul et al |
| EPI_ISL_721631 | India/MH-AFCO-15230/2020 | 2020-07-25 | Asia/India/Maharashtra/ | Armed Forces Medical College | National Centre For Cell Science | Dhiraj Paul et al |
| EPI_ISL_721632 | India/MH-AFCO-15275/2020 | 2020-07-25 | Asia/India/Maharashtra/ | Armed Forces Medical College | National Centre For Cell Science | Dhiraj Paul et al |
| EPI_ISL_721633 | India/MH-AFCO-15381/2020 | 2020-07-26 | Asia/India/Maharashtra/ | Armed Forces Medical College | National Centre For Cell Science | Dhiraj Paul et al |
| EPI_ISL_721635 | India/MH-AFCO-15531/2020 | 2020-07-27 | Asia/India/Maharashtra/ | Armed Forces Medical College | National Centre For Cell Science | Dhiraj Paul et al |

| | | | | | | |
|---|---|---|---|---|---|---|
| EPI_ISL_721636 | India/MH-AFCO-15533/2020 | 2020-07-27 | Asia/India/Maharashtra/ | Armed Forces Medical College | National Centre For Cell Science | Dhiraj Paul et al |
| EPI_ISL_721637 | India/MH-AFCO-8108/2020 | 2020-06-16 | Asia/India/Maharashtra/ | Armed Forces Medical College | National Centre For Cell Science | Dhiraj Paul et al |
| EPI_ISL_721638 | India/MH-AFCO-8189/2020 | 2020-06-17 | Asia/India/Maharashtra/ | Armed Forces Medical College | National Centre For Cell Science | Dhiraj Paul et al |
| EPI_ISL_721639 | India/MH-AFCO-8677/2020 | 2020-06-19 | Asia/India/Maharashtra/ | Armed Forces Medical College | National Centre For Cell Science | Dhiraj Paul et al |
| EPI_ISL_721643 | India/MH-AFCO-9320/2020 | 2020-06-23 | Asia/India/Maharashtra/ | Armed Forces Medical College | National Centre For Cell Science | Dhiraj Paul et al |
| EPI_ISL_721646 | India/MH-NCCS-NR-10609/2020 | 2020-07-10 | Asia/India/Maharashtra/ | National Centre For Cell Science | National Centre For Cell Science | Dhiraj Paul et al |
| EPI_ISL_721647 | India/MH-NCCS-NR-10951/2020 | 2020-07-11 | Asia/India/Maharashtra/ | National Centre For Cell Science | National Centre For Cell Science | Dhiraj Paul et al |
| EPI_ISL_721648 | India/MH-NCCS-NR-11448/2020 | 2020-07-13 | Asia/India/Maharashtra/ | National Centre For Cell Science | National Centre For Cell Science | Dhiraj Paul et al |
| EPI_ISL_721650 | India/MH-NCCS-NR-11843/2020 | 2020-07-15 | Asia/India/Maharashtra/ | National Centre For Cell Science | National Centre For Cell Science | Dhiraj Paul et al |
| EPI_ISL_721653 | India/MH-NCCS-NR-13749/2020 | 2020-07-22 | Asia/India/Maharashtra/ | National Centre For Cell Science | National Centre For Cell Science | Dhiraj Paul et al |
| EPI_ISL_721655 | India/MH-NCCS-NR-14607/2020 | 2020-07-26 | Asia/India/Maharashtra/ | National Centre For Cell Science | National Centre For Cell Science | Dhiraj Paul et al |
| EPI_ISL_721656 | India/MH-NCCS-NR-15788/2020 | 2020-07-30 | Asia/India/Maharashtra/ | National Centre For Cell Science | National Centre For Cell Science | Dhiraj Paul et al |
| EPI_ISL_721659 | India/MH-NCCS-NR-6668/2020 | 2020-06-19 | Asia/India/Maharashtra/ | National Centre For Cell Science | National Centre For Cell Science | Dhiraj Paul et al |
| EPI_ISL_721661 | India/MH-NCCS-NR-7614/2020 | 2020-06-26 | Asia/India/Maharashtra/ | National Centre For Cell Science | National Centre For Cell Science | Dhiraj Paul et al |
| EPI_ISL_721663 | India/MH-NCCS-NR-7636/2020 | 2020-06-26 | Asia/India/Maharashtra/ | National Centre For Cell Science | National Centre For Cell Science | Dhiraj Paul et al |
| EPI_ISL_721664 | India/MH-NCCS-NR-8720/2020 | 2020-07-01 | Asia/India/Maharashtra/ | National Centre For Cell Science | National Centre For Cell Science | Dhiraj Paul et al |
| EPI_ISL_721665 | India/MH-NCCS-NR-8737/2020 | 2020-07-01 | Asia/India/Maharashtra/ | National Centre For Cell Science | National Centre For Cell Science | Dhiraj Paul et al |
| EPI_ISL_721667 | India/MH-NCCS-NR-9234/2020 | 2020-06-03 | Asia/India/Maharashtra/ | National Centre For Cell Science | National Centre For Cell Science | Dhiraj Paul et al |

| | | | | | | |
|---|---|---|---|---|---|---|
| EPI_ISL_721679 | Switzerland/BE-ETHZ-380384/2020 | 2020-11-23 | Europe/Switzerland/Bern/ | Viollier AG | Department of Biosystems Science and Engineering, ETH Z√ºrich | Christian Beisel et al |
| EPI_ISL_721728 | Switzerland/AG-ETHZ-380323/2020 | 2020-11-26 | Europe/Switzerland/Aargau/ | Viollier AG | Department of Biosystems Science and Engineering, ETH Z√ºrich | Christian Beisel et al |
| EPI_ISL_721732 | Switzerland/AG-ETHZ-380337/2020 | 2020-11-26 | Europe/Switzerland/Aargau/ | Viollier AG | Department of Biosystems Science and Engineering, ETH Z√ºrich | Christian Beisel et al |
| EPI_ISL_721750 | Switzerland/BE-ETHZ-340115/2020 | 2020-10-26 | Europe/Switzerland/Bern/ | Viollier AG | Department of Biosystems Science and Engineering, ETH Z√ºrich | Christian Beisel et al |
| EPI_ISL_721770 | Switzerland/BE-ETHZ-380402/2020 | 2020-11-23 | Europe/Switzerland/Bern/ | Viollier AG | Department of Biosystems Science and Engineering, ETH Z√ºrich | Christian Beisel et al |
| EPI_ISL_721782 | Switzerland/BL-ETHZ-380273/2020 | 2020-11-22 | Europe/Switzerland/Basel-Land/ | Viollier AG | Department of Biosystems Science and Engineering, ETH Z√ºrich | Christian Beisel et al |
| EPI_ISL_721787 | Switzerland/BL-ETHZ-380283/2020 | 2020-11-25 | Europe/Switzerland/Basel-Land/ | Viollier AG | Department of Biosystems Science and Engineering, ETH Z√ºrich | Christian Beisel et al |
| EPI_ISL_721810 | Switzerland/GE-ETHZ-340103/2020 | 2020-10-26 | Europe/Switzerland/Geneva/ | Viollier AG | Department of Biosystems Science and Engineering, ETH Z√ºrich | Christian Beisel et al |

| | | | | | | |
|---|---|---|---|---|---|---|
| EPI_ISL_721815 | Switzerland/GR-ETHZ-340106/2020 | 2020-10-26 | Europe/Switzerland/Graubvºnden/ | Viollier AG | Department of Biosystems Science and Engineering, ETH Zv°rich | Christian Beisel et al |
| EPI_ISL_721818 | Switzerland/GR-ETHZ-390381/2020 | 2020-12-01 | Europe/Switzerland/Graubv°nden/ | Viollier AG | Department of Biosystems Science and Engineering, ETH Zv°rich | Christian Beisel et al |
| EPI_ISL_721833 | Switzerland/NE-ETHZ-390320/2020 | 2020-11-29 | Europe/Switzerland/Neuchv¢tel/ | Viollier AG | Department of Biosystems Science and Engineering, ETH Zv°rich | Christian Beisel et al |
| EPI_ISL_721838 | Switzerland/NE-ETHZ-390327/2020 | 2020-11-29 | Europe/Switzerland/Neuchv¢tel/ | Viollier AG | Department of Biosystems Science and Engineering, ETH Zv°rich | Christian Beisel et al |
| EPI_ISL_721842 | Switzerland/NE-ETHZ-390332/2020 | 2020-11-30 | Europe/Switzerland/Neuchv¢tel/ | Viollier AG | Department of Biosystems Science and Engineering, ETH Zv°rich | Christian Beisel et al |
| EPI_ISL_721883 | Switzerland/SO-ETHZ-380301/2020 | 2020-11-25 | Europe/Switzerland/Solothurn/ | Viollier AG | Department of Biosystems Science and Engineering, ETH Zv°rich | Christian Beisel et al |
| EPI_ISL_721896 | Switzerland/SO-ETHZ-390342/2020 | 2020-11-30 | Europe/Switzerland/Solothurn/ | Viollier AG | Department of Biosystems Science and Engineering, ETH Zv°rich | Christian Beisel et al |
| EPI_ISL_721906 | Switzerland/TG-ETHZ-390379/2020 | 2020-12-01 | Europe/Switzerland/Thurgau/ | Viollier AG | Department of Biosystems Science and Engineering, ETH Zv°rich | Christian Beisel et al |

| | | | | | | |
|---|---|---|---|---|---|---|
| EPI_ISL_721917 | Switzerland/VD-ETHZ-390324/2020 | 2020-11-29 | Europe/Switzerland/Vaud/ | Viollier AG | Department of Biosystems Science and Engineering, ETH Zv∫rich | Christian Beisel et al |
| EPI_ISL_721957 | Switzerland/ZH-ETHZ-380381/2020 | 2020-11-23 | Europe/Switzerland/Zv∫rich/ | Viollier AG | Department of Biosystems Science and Engineering, ETH Zv∫rich | Christian Beisel et al |
| EPI_ISL_721961 | Switzerland/ZH-ETHZ-390218/2020 | 2020-11-27 | Europe/Switzerland/Zv∫rich/ | Viollier AG | Department of Biosystems Science and Engineering, ETH Zv∫rich | Christian Beisel et al |
| EPI_ISL_721988 | Brazil/SP-L39-CD1045/2020 | 2020-05-13 | South America/Brazil/S√£o Paulo/ | Hospital das Cl√≠nicas Universidade de S√£o Paulo Medical School | Laborat√≥rio de Parasitologia M√©dica - Instituto de Medicina Tropical - Universidade de S√£o Paulo | Brazil-UK Centre for Arbovirus Discovery Diagnosis Genomics et al |
| EPI_ISL_721990 | Brazil/SP-L39-CD1028/2020 | 2020-05-12 | South America/Brazil/S√£o Paulo/ | Hospital das Cl√≠nicas Universidade de S√£o Paulo Medical School | Laborat√≥rio de Parasitologia M√©dica - Instituto de Medicina Tropical - Universidade de S√£o Paulo | Brazil-UK Centre for Arbovirus Discovery Diagnosis Genomics et al |
| EPI_ISL_721991 | Brazil/SP-L39-CD1031/2020 | 2020-05-12 | South America/Brazil/S√£o Paulo/ | Hospital das Cl√≠nicas Universidade de S√£o Paulo Medical School | Laborat√≥rio de Parasitologia M√©dica - Instituto de Medicina Tropical - Universidade de S√£o Paulo | Brazil-UK Centre for Arbovirus Discovery Diagnosis Genomics et al |

| | | | | | | |
|---|---|---|---|---|---|---|
| EPI_ISL_721994 | Brazil/SP-L36-CD934/2020 | 2020-04-30 | South America/Brazil/S√£o Paulo/ | Hospital das Cl√≠nicas Universidade de S√£o Paulo Medical School | Laborat√≥rio de Parasitologia M√©dica - Instituto de Medicina Tropical - Universidade de S√£o Paulo | Brazil-UK Centre for Arbovirus Discovery Diagnosis Genomics et al |
| EPI_ISL_721995 | Brazil/SP-L38-CD1014/2020 | 2020-05-10 | South America/Brazil/S√£o Paulo/ | Hospital das Cl√≠nicas Universidade de S√£o Paulo Medical School | Laborat√≥rio de Parasitologia M√©dica - Instituto de Medicina Tropical - Universidade de S√£o Paulo | Brazil-UK Centre for Arbovirus Discovery Diagnosis Genomics et al |
| EPI_ISL_721997 | Brazil/SP-L35-CD892/2020 | 2020-04-25 | South America/Brazil/S√£o Paulo/ | Hospital das Cl√≠nicas Universidade de S√£o Paulo Medical School | Laborat√≥rio de Parasitologia M√©dica - Instituto de Medicina Tropical - Universidade de S√£o Paulo | Brazil-UK Centre for Arbovirus Discovery Diagnosis Genomics et al |
| EPI_ISL_722006 | Brazil/SP-L38-CD1011/2020 | 2020-05-10 | South America/Brazil/S√£o Paulo/ | Hospital das Cl√≠nicas Universidade de S√£o Paulo Medical School | Laborat√≥rio de Parasitologia M√©dica - Instituto de Medicina Tropical - Universidade de S√£o Paulo | Brazil-UK Centre for Arbovirus Discovery Diagnosis Genomics et al |
| EPI_ISL_722007 | Brazil/SP-L39-CD1027/2020 | 2020-05-12 | South America/Brazil/S√£o Paulo/ | Hospital das Cl√≠nicas Universidade de S√£o Paulo Medical School | Laborat√≥rio de Parasitologia M√©dica - Instituto de Medicina Tropical - Universidade de S√£o Paulo | Brazil-UK Centre for Arbovirus Discovery Diagnosis Genomics et al |

| | | | | | | |
|---|---|---|---|---|---|---|
| EPI_ISL_722009 | Brazil/SP-L33-CD856/2020 | 2020-04-22 | South America/Brazil/S√£o Paulo/ | Hospital das Cl√≠nicas Universidade de S√£o Paulo Medical School | Laborat√≥rio de Parasitologia M√©dica - Instituto de Medicina Tropical - Universidade de S√£o Paulo | Brazil-UK Centre for Arbovirus Discovery Diagnosis Genomics et al |
| EPI_ISL_722010 | Brazil/SP-L39-CD1032/2020 | 2020-05-12 | South America/Brazil/S√£o Paulo/ | Hospital das Cl√≠nicas Universidade de S√£o Paulo Medical School | Laborat√≥rio de Parasitologia M√©dica - Instituto de Medicina Tropical - Universidade de S√£o Paulo | Brazil-UK Centre for Arbovirus Discovery Diagnosis Genomics et al |
| EPI_ISL_722011 | Brazil/SP-L39-CD1036/2020 | 2020-05-12 | South America/Brazil/S√£o Paulo/ | Hospital das Cl√≠nicas Universidade de S√£o Paulo Medical School | Laborat√≥rio de Parasitologia M√©dica - Instituto de Medicina Tropical - Universidade de S√£o Paulo | Brazil-UK Centre for Arbovirus Discovery Diagnosis Genomics et al |
| EPI_ISL_722014 | Brazil/SP-L33-CD852/2020 | 2020-04-17 | South America/Brazil/S√£o Paulo/ | Hospital das Cl√≠nicas Universidade de S√£o Paulo Medical School | Laborat√≥rio de Parasitologia M√©dica - Instituto de Medicina Tropical - Universidade de S√£o Paulo | Brazil-UK Centre for Arbovirus Discovery Diagnosis Genomics et al |
| EPI_ISL_722017 | Brazil/SP-L39-CD1052/2020 | 2020-05-14 | South America/Brazil/S√£o Paulo/ | Hospital das Cl√≠nicas Universidade de S√£o Paulo Medical School | Laborat√≥rio de Parasitologia M√©dica - Instituto de Medicina Tropical - Universidade de S√£o Paulo | Brazil-UK Centre for Arbovirus Discovery Diagnosis Genomics et al |

| | | | | | | |
|---|---|---|---|---|---|---|
| EPI_ISL_722022 | Brazil/SP-L37-CD965/2020 | 2020-05-04 | South America/Brazil/S√£o Paulo/ | Hospital das Cl√≠nicas Universidade de S√£o Paulo Medical School | Laborat√≥rio de Parasitologia M√©dica - Instituto de Medicina Tropical - Universidade de S√£o Paulo | Brazil-UK Centre for Arbovirus Discovery Diagnosis Genomics et al |
| EPI_ISL_722023 | Brazil/SP-L31-CD798/2020 | 2020-04-03 | South America/Brazil/S√£o Paulo/ | Hospital das Cl√≠nicas Universidade de S√£o Paulo Medical School | Laborat√≥rio de Parasitologia M√©dica - Instituto de Medicina Tropical - Universidade de S√£o Paulo | Brazil-UK Centre for Arbovirus Discovery Diagnosis Genomics et al |
| EPI_ISL_722026 | Brazil/SP-L33-CD869/2020 | 2020-04-22 | South America/Brazil/S√£o Paulo/ | Hospital das Cl√≠nicas Universidade de S√£o Paulo Medical School | Laborat√≥rio de Parasitologia M√©dica - Instituto de Medicina Tropical - Universidade de S√£o Paulo | Brazil-UK Centre for Arbovirus Discovery Diagnosis Genomics et al |
| EPI_ISL_722032 | Brazil/SP-L32-CD836/2020 | 2020-04-13 | South America/Brazil/S√£o Paulo/ | Hospital das Cl√≠nicas Universidade de S√£o Paulo Medical School | Laborat√≥rio de Parasitologia M√©dica - Instituto de Medicina Tropical - Universidade de S√£o Paulo | Brazil-UK Centre for Arbovirus Discovery Diagnosis Genomics et al |
| EPI_ISL_722040 | Brazil/SP-L36-CD952/2020 | 2020-05-02 | South America/Brazil/S√£o Paulo/ | Hospital das Cl√≠nicas Universidade de S√£o Paulo Medical School | Laborat√≥rio de Parasitologia M√©dica - Instituto de Medicina Tropical - Universidade de S√£o Paulo | Brazil-UK Centre for Arbovirus Discovery Diagnosis Genomics et al |

| | | | | | | |
|---|---|---|---|---|---|---|
| EPI_ISL_722044 | Brazil/SP-L32-CD837/2020 | 2020-04-24 | South America/Brazil/S√£o Paulo/ | Hospital das Cl√≠nicas Universidade de S√£o Paulo Medical School | Laborat√≥rio de Parasitologia M√©dica - Instituto de Medicina Tropical - Universidade de S√£o Paulo | Brazil-UK Centre for Arbovirus Discovery Diagnosis Genomics et al |
| EPI_ISL_722046 | Brazil/SP-L34-CD873/2020 | 2020-04-23 | South America/Brazil/S√£o Paulo/ | Hospital das Cl√≠nicas Universidade de S√£o Paulo Medical School | Laborat√≥rio de Parasitologia M√©dica - Instituto de Medicina Tropical - Universidade de S√£o Paulo | Brazil-UK Centre for Arbovirus Discovery Diagnosis Genomics et al |
| EPI_ISL_722049 | Brazil/SP-L34-CD883/2020 | 2020-04-23 | South America/Brazil/S√£o Paulo/ | Hospital das Cl√≠nicas Universidade de S√£o Paulo Medical School | Laborat√≥rio de Parasitologia M√©dica - Instituto de Medicina Tropical - Universidade de S√£o Paulo | Brazil-UK Centre for Arbovirus Discovery Diagnosis Genomics et al |
| EPI_ISL_722055 | Brazil/SP-L32-CD820/2020 | 2020-04-08 | South America/Brazil/S√£o Paulo/ | Hospital das Cl√≠nicas Universidade de S√£o Paulo Medical School | Laborat√≥rio de Parasitologia M√©dica - Instituto de Medicina Tropical - Universidade de S√£o Paulo | Brazil-UK Centre for Arbovirus Discovery Diagnosis Genomics et al |
| EPI_ISL_722058 | Brazil/SP-L39-CD1035/2020 | 2020-05-12 | South America/Brazil/S√£o Paulo/ | Hospital das Cl√≠nicas Universidade de S√£o Paulo Medical School | Laborat√≥rio de Parasitologia M√©dica - Instituto de Medicina Tropical - Universidade de S√£o Paulo | Brazil-UK Centre for Arbovirus Discovery Diagnosis Genomics et al |

| | | | | | | |
|---|---|---|---|---|---|---|
| EPI_ISL_722059 | Brazil/SP-L32-CD827/2020 | 2020-04-09 | South America/Brazil/S√£o Paulo/ | Hospital das Cl√≠nicas Universidade de S√£o Paulo Medical School | Laborat√≥rio de Parasitologia M√©dica - Instituto de Medicina Tropical - Universidade de S√£o Paulo | Brazil-UK Centre for Arbovirus Discovery Diagnosis Genomics et al |
| EPI_ISL_722062 | Brazil/SP-L36-CD945/2020 | 2020-05-01 | South America/Brazil/S√£o Paulo/ | Hospital das Cl√≠nicas Universidade de S√£o Paulo Medical School | Laborat√≥rio de Parasitologia M√©dica - Instituto de Medicina Tropical - Universidade de S√£o Paulo | Brazil-UK Centre for Arbovirus Discovery Diagnosis Genomics et al |
| EPI_ISL_722066 | Brazil/SP-L39-CD1044/2020 | 2020-05-13 | South America/Brazil/S√£o Paulo/ | Hospital das Cl√≠nicas Universidade de S√£o Paulo Medical School | Laborat√≥rio de Parasitologia M√©dica - Instituto de Medicina Tropical - Universidade de S√£o Paulo | Brazil-UK Centre for Arbovirus Discovery Diagnosis Genomics et al |
| EPI_ISL_722070 | Brazil/SP-L39-CD1033/2020 | 2020-05-12 | South America/Brazil/S√£o Paulo/ | Hospital das Cl√≠nicas Universidade de S√£o Paulo Medical School | Laborat√≥rio de Parasitologia M√©dica - Instituto de Medicina Tropical - Universidade de S√£o Paulo | Brazil-UK Centre for Arbovirus Discovery Diagnosis Genomics et al |
| EPI_ISL_722075 | Brazil/SP-L32-CD833/2020 | 2020-04-09 | South America/Brazil/S√£o Paulo/ | Hospital das Cl√≠nicas Universidade de S√£o Paulo Medical School | Laborat√≥rio de Parasitologia M√©dica - Instituto de Medicina Tropical - Universidade de S√£o Paulo | Brazil-UK Centre for Arbovirus Discovery Diagnosis Genomics et al |

| | | | | | | |
|---|---|---|---|---|---|---|
| EPI_ISL_722077 | Brazil/SP-L37-CD990/2020 | 2020-05-05 | South America/Brazil/S√£o Paulo/ | Hospital das Cl√≠nicas Universidade de S√£o Paulo Medical School | Laborat√≥rio de Parasitologia M√©dica - Instituto de Medicina Tropical - Universidade de S√£o Paulo | Brazil-UK Centre for Arbovirus Discovery Diagnosis Genomics et al |
| EPI_ISL_722086 | Brazil/SP-L36-CD942/2020 | 2020-05-01 | South America/Brazil/S√£o Paulo/ | Hospital das Cl√≠nicas Universidade de S√£o Paulo Medical School | Laborat√≥rio de Parasitologia M√©dica - Instituto de Medicina Tropical - Universidade de S√£o Paulo | Brazil-UK Centre for Arbovirus Discovery Diagnosis Genomics et al |
| EPI_ISL_722107 | Brazil/SP-L36-CD943/2020 | 2020-05-01 | South America/Brazil/S√£o Paulo/ | Hospital das Cl√≠nicas Universidade de S√£o Paulo Medical School | Laborat√≥rio de Parasitologia M√©dica - Instituto de Medicina Tropical - Universidade de S√£o Paulo | Brazil-UK Centre for Arbovirus Discovery Diagnosis Genomics et al |
| EPI_ISL_722114 | Brazil/SP-L33-CD854/2020 | 2020-04-17 | South America/Brazil/S√£o Paulo/ | Hospital das Cl√≠nicas Universidade de S√£o Paulo Medical School | Laborat√≥rio de Parasitologia M√©dica - Instituto de Medicina Tropical - Universidade de S√£o Paulo | Brazil-UK Centre for Arbovirus Discovery Diagnosis Genomics et al |
| EPI_ISL_722117 | Brazil/SP-L36-CD954/2020 | 2020-05-02 | South America/Brazil/S√£o Paulo/ | Hospital das Cl√≠nicas Universidade de S√£o Paulo Medical School | Laborat√≥rio de Parasitologia M√©dica - Instituto de Medicina Tropical - Universidade de S√£o Paulo | Brazil-UK Centre for Arbovirus Discovery Diagnosis Genomics et al |

| | | | | | | |
|---|---|---|---|---|---|---|
| EPI_ISL_722119 | Brazil/SP-L31-CD800/2020 | 2020-04-03 | South America/Brazil/S√£o Paulo/ | Hospital das Cl√≠nicas Universidade de S√£o Paulo Medical School | Laborat√≥rio de Parasitologia M√©dica - Instituto de Medicina Tropical - Universidade de S√£o Paulo | Brazil-UK Centre for Arbovirus Discovery Diagnosis Genomics et al |
| EPI_ISL_722121 | Brazil/SP-L37-CD992/2020 | 2020-05-05 | South America/Brazil/S√£o Paulo/ | Hospital das Cl√≠nicas Universidade de S√£o Paulo Medical School | Laborat√≥rio de Parasitologia M√©dica - Instituto de Medicina Tropical - Universidade de S√£o Paulo | Brazil-UK Centre for Arbovirus Discovery Diagnosis Genomics et al |
| EPI_ISL_722125 | Brazil/SP-L36-CD924/2020 | 2020-04-28 | South America/Brazil/S√£o Paulo/ | Hospital das Cl√≠nicas Universidade de S√£o Paulo Medical School | Laborat√≥rio de Parasitologia M√©dica - Instituto de Medicina Tropical - Universidade de S√£o Paulo | Brazil-UK Centre for Arbovirus Discovery Diagnosis Genomics et al |
| EPI_ISL_722126 | Brazil/SP-L36-CD926/2020 | 2020-04-29 | South America/Brazil/S√£o Paulo/ | Hospital das Cl√≠nicas Universidade de S√£o Paulo Medical School | Laborat√≥rio de Parasitologia M√©dica - Instituto de Medicina Tropical - Universidade de S√£o Paulo | Brazil-UK Centre for Arbovirus Discovery Diagnosis Genomics et al |
| EPI_ISL_722131 | Brazil/SP-L28-CD675/2020 | 2020-04-15 | South America/Brazil/S√£o Paulo/ | Instituto de Medicina Tropical Universidade de S√£o Paulo | Laborat√≥rio de Parasitologia M√©dica - Instituto de Medicina Tropical - Universidade de S√£o Paulo | Brazil-UK Centre for Arbovirus Discovery Diagnosis Genomics et al |

| | | | | | | |
|---|---|---|---|---|---|---|
| EPI_ISL_722132 | Brazil/SP-L28-CD674/2020 | 2020-04-15 | South America/Brazil/S√£o Paulo/ | Instituto de Medicina Tropical Universidade de S√£o Paulo | Laborat√≥rio de Parasitologia M√©dica - Instituto de Medicina Tropical - Universidade de S√£o Paulo | Brazil-UK Centre for Arbovirus Discovery Diagnosis Genomics et al |
| EPI_ISL_722140 | Brazil/MA-L28-CD618/2020 | 2020-04-09 | South America/Brazil/Maranh√£o/ | DB Diagnosticos do Brasil | Laborat√≥rio de Parasitologia M√©dica - Instituto de Medicina Tropical - Universidade de S√£o Paulo | Brazil-UK Centre for Arbovirus Discovery Diagnosis Genomics et al |
| EPI_ISL_722142 | Brazil/PR-L29-CD717/2020 | 2020-04-11 | South America/Brazil/Paran√°/ | DB Diagnosticos do Brasil | Laborat√≥rio de Parasitologia M√©dica - Instituto de Medicina Tropical - Universidade de S√£o Paulo | Brazil-UK Centre for Arbovirus Discovery Diagnosis Genomics et al |
| EPI_ISL_722152 | Brazil/MA-L29-CD734/2020 | 2020-04-13 | South America/Brazil/Maranh√£o/ | DB Diagnosticos do Brasil | Laborat√≥rio de Parasitologia M√©dica - Instituto de Medicina Tropical - Universidade de S√£o Paulo | Brazil-UK Centre for Arbovirus Discovery Diagnosis Genomics et al |
| EPI_ISL_722164 | Brazil/PE-L29-CD729/2020 | 2020-04-12 | South America/Brazil/Pernambuco/ | DB Diagnosticos do Brasil | Laborat√≥rio de Parasitologia M√©dica - Instituto de Medicina Tropical - Universidade de S√£o Paulo | Brazil-UK Centre for Arbovirus Discovery Diagnosis Genomics et al |

| | | | | | | |
|---|---|---|---|---|---|---|
| EPI_ISL_722165 | Brazil/PA-L30-CD779/2020 | 2020-04-16 | South America/Brazil/Para/ | DB Diagnosticos do Brasil | Laboratv≥rio de Parasitologia Mv©dica - Instituto de Medicina Tropical - Universidade de Sv£o Paulo | Brazil-UK Centre for Arbovirus Discovery Diagnosis Genomics et al |
| EPI_ISL_722177 | India/MH-NCCS-NR-14432/2020 | 2020-07-25 | Asia/India/Maharashtra/ | National Centre For Cell Science | National Centre For Cell Science | Dhiraj Paul et al |
| EPI_ISL_722178 | India/MH-NCCS-NR-8740/2020 | 2020-07-01 | Asia/India/Maharashtra/ | National Centre For Cell Science | National Centre For Cell Science | Dhiraj Paul et al |
| EPI_ISL_722180 | India/MH-NCCS-NR-12775/2020 | 2020-07-18 | Asia/India/Maharashtra/ | National Centre For Cell Science | National Centre For Cell Science | Dhiraj Paul et al |
| EPI_ISL_722181 | India/MH-NCCS-NR-8031/2020 | 2020-06-28 | Asia/India/Maharashtra/ | National Centre For Cell Science | National Centre For Cell Science | Dhiraj Paul et al |
| EPI_ISL_722182 | India/MH-NCCS-NR-6984/2020 | 2020-06-21 | Asia/India/Maharashtra/ | National Centre For Cell Science | National Centre For Cell Science | Dhiraj Paul et al |
| EPI_ISL_722186 | India/MH-NCCS-NR-7779/2020 | 2020-06-26 | Asia/India/Maharashtra/ | National Centre For Cell Science | National Centre For Cell Science | Dhiraj Paul et al |
| EPI_ISL_722190 | India/MH-AFCO-8225/2020 | 2020-06-17 | Asia/India/Maharashtra/ | Armed Forces Medical College | National Centre For Cell Science | Dhiraj Paul et al |
| EPI_ISL_722191 | India/MH-AFCO-8699/2020 | 2020-06-19 | Asia/India/Maharashtra/ | Armed Forces Medical College | National Centre For Cell Science | Dhiraj Paul et al |
| EPI_ISL_722193 | India/MH-AFCO-9124/2020 | 2020-06-22 | Asia/India/Maharashtra/ | Armed Forces Medical College | National Centre For Cell Science | Dhiraj Paul et al |
| EPI_ISL_722195 | India/MH-AFCO-9304/2020 | 2020-06-23 | Asia/India/Maharashtra/ | Armed Forces Medical College | National Centre For Cell Science | Dhiraj Paul et al |
| EPI_ISL_722198 | India/MH-AFCO-15278/2020 | 2020-07-25 | Asia/India/Maharashtra/ | Armed Forces Medical College | National Centre For Cell Science | Dhiraj Paul et al |
| EPI_ISL_722201 | BosniaandHerzegovina/AGC-MOS-00 | 2020-11-20 | Europe/Bosnia and Herzegovina/Mostar | Vitalis Mostar | Alea Genetic Center | Salihefendic L. et al |
| EPI_ISL_722209 | BosniaandHerzegovina/AGC-SAR-002 | 2020-11-20 | Europe/Bosnia and Herzegovina/Sarajev | Dom Zdravlja Sarajevo | Alea Genetic Center | Salihefendic L. et al |
| EPI_ISL_722211 | Spain/VC-IBV-98004476/2020 | 2020-03-24 | Europe/Spain/Comunitat Valenciana/Ca | Servicio de Microbiologv≠a. Hospital General Universitario de Castellv≥n | SeqCOVID-SPAIN consortium/IBV(CSIC) | Rosario Moreno Muv±oz et al |
| EPI_ISL_722218 | Spain/AR-IBV-98010710/2020 | 2020-10-07 | Europe/Spain/Aragon/Zaragoza | Servicio de Microbiologv≠a, Hospital Miguel Servet, Zaragoza | SeqCOVID-SPAIN consortium/IBV(CSIC) | Antonio Rezusta Lv≥pez et al |

| | | | | | | |
|---|---|---|---|---|---|---|
| EPI_ISL_722219 | Spain/AR-IBV-98010607/2020 | 2020-10-02 | Europe/Spain/Aragon/Zaragoza | Servicio de Microbiolog√≠a, Hospital Miguel Servet, Zaragoza | SeqCOVID-SPAIN consortium/IBV(CSIC) | Antonio Rezusta L√≥pez et al |
| EPI_ISL_722220 | Spain/AR-IBV-98010717/2020 | 2020-10-07 | Europe/Spain/Aragon/Zaragoza | Servicio de Microbiolog√≠a, Hospital Miguel Servet, Zaragoza | SeqCOVID-SPAIN consortium/IBV(CSIC) | Antonio Rezusta L√≥pez et al |
| EPI_ISL_722222 | Spain/AR-IBV-98010584/2020 | 2020-10-05 | Europe/Spain/Aragon/Zaragoza | Servicio de Microbiolog√≠a, Hospital Miguel Servet, Zaragoza | SeqCOVID-SPAIN consortium/IBV(CSIC) | Antonio Rezusta L√≥pez et al |
| EPI_ISL_722224 | Spain/AR-IBV-98010617/2020 | 2020-09-30 | Europe/Spain/Aragon/Zaragoza | Servicio de Microbiolog√≠a, Hospital Miguel Servet, Zaragoza | SeqCOVID-SPAIN consortium/IBV(CSIC) | Antonio Rezusta L√≥pez et al |
| EPI_ISL_722227 | Spain/AR-IBV-98010626/2020 | 2020-09-30 | Europe/Spain/Aragon/Zaragoza | Servicio de Microbiolog√≠a, Hospital Miguel Servet, Zaragoza | SeqCOVID-SPAIN consortium/IBV(CSIC) | Antonio Rezusta L√≥pez et al |
| EPI_ISL_722228 | Spain/AR-IBV-98010592/2020 | 2020-10-03 | Europe/Spain/Aragon/Zaragoza | Servicio de Microbiolog√≠a, Hospital Miguel Servet, Zaragoza | SeqCOVID-SPAIN consortium/IBV(CSIC) | Antonio Rezusta L√≥pez et al |
| EPI_ISL_722239 | Spain/VC-IBV-98004495/2020 | 2020-03-26 | Europe/Spain/Comunitat Valenciana/Ca | Servicio de Microbiolog√≠a. Hospital General Universitario de Castell√≥n | SeqCOVID-SPAIN consortium/IBV(CSIC) | Rosario Moreno Mu√±oz et al |
| EPI_ISL_722242 | Spain/AR-IBV-98010577/2020 | 2020-11-11 | Europe/Spain/Aragon/Zaragoza | Servicio de Microbiolog√≠a, Hospital Miguel Servet, Zaragoza | SeqCOVID-SPAIN consortium/IBV(CSIC) | Antonio Rezusta L√≥pez et al |
| EPI_ISL_722243 | Spain/AR-IBV-98010579/2020 | 2020-11-11 | Europe/Spain/Aragon/Zaragoza | Servicio de Microbiolog√≠a, Hospital Miguel Servet, Zaragoza | SeqCOVID-SPAIN consortium/IBV(CSIC) | Antonio Rezusta L√≥pez et al |
| EPI_ISL_722246 | Spain/AR-IBV-98010585/2020 | 2020-10-05 | Europe/Spain/Aragon/Zaragoza | Servicio de Microbiolog√≠a, Hospital Miguel Servet, Zaragoza | SeqCOVID-SPAIN consortium/IBV(CSIC) | Antonio Rezusta L√≥pez et al |
| EPI_ISL_722251 | Spain/AR-IBV-98010599/2020 | 2020-10-02 | Europe/Spain/Aragon/Zaragoza | Servicio de Microbiolog√≠a, Hospital Miguel Servet, Zaragoza | SeqCOVID-SPAIN consortium/IBV(CSIC) | Antonio Rezusta L√≥pez et al |
| EPI_ISL_722254 | Spain/AR-IBV-98010608/2020 | 2020-10-02 | Europe/Spain/Aragon/Zaragoza | Servicio de Microbiolog√≠a, Hospital Miguel Servet, Zaragoza | SeqCOVID-SPAIN consortium/IBV(CSIC) | Antonio Rezusta L√≥pez et al |
| EPI_ISL_722256 | Spain/AR-IBV-98010614/2020 | 2020-10-01 | Europe/Spain/Aragon/Zaragoza | Servicio de Microbiolog√≠a, Hospital Miguel Servet, Zaragoza | SeqCOVID-SPAIN consortium/IBV(CSIC) | Antonio Rezusta L√≥pez et al |

| | | | | | | |
|---|---|---|---|---|---|---|
| EPI_ISL_722257 | Spain/AR-IBV-98010615/2020 | 2020-09-30 | Europe/Spain/Aragon/Zaragoza | Servicio de Microbiolog√≠a, Hospital Miguel Servet, Zaragoza | SeqCOVID-SPAIN consortium/IBV(CSIC) | Antonio Rezusta L√≥pez et al |
| EPI_ISL_722258 | Spain/AR-IBV-98010619/2020 | 2020-11-10 | Europe/Spain/Aragon/Zaragoza | Servicio de Microbiolog√≠a, Hospital Miguel Servet, Zaragoza | SeqCOVID-SPAIN consortium/IBV(CSIC) | Antonio Rezusta L√≥pez et al |
| EPI_ISL_722259 | Spain/AR-IBV-98010620/2020 | 2020-11-10 | Europe/Spain/Aragon/Zaragoza | Servicio de Microbiolog√≠a, Hospital Miguel Servet, Zaragoza | SeqCOVID-SPAIN consortium/IBV(CSIC) | Antonio Rezusta L√≥pez et al |
| EPI_ISL_722260 | Spain/AR-IBV-98010621/2020 | 2020-09-30 | Europe/Spain/Aragon/Zaragoza | Servicio de Microbiolog√≠a, Hospital Miguel Servet, Zaragoza | SeqCOVID-SPAIN consortium/IBV(CSIC) | Antonio Rezusta L√≥pez et al |
| EPI_ISL_722261 | Spain/AR-IBV-98010622/2020 | 2020-10-01 | Europe/Spain/Aragon/Zaragoza | Servicio de Microbiolog√≠a, Hospital Miguel Servet, Zaragoza | SeqCOVID-SPAIN consortium/IBV(CSIC) | Antonio Rezusta L√≥pez et al |
| EPI_ISL_722263 | Spain/AR-IBV-98010634/2020 | 2020-11-11 | Europe/Spain/Aragon/Zaragoza | Servicio de Microbiolog√≠a, Hospital Miguel Servet, Zaragoza | SeqCOVID-SPAIN consortium/IBV(CSIC) | Antonio Rezusta L√≥pez et al |
| EPI_ISL_722264 | Spain/AR-IBV-98010640/2020 | 2020-10-06 | Europe/Spain/Aragon/Zaragoza | Servicio de Microbiolog√≠a, Hospital Miguel Servet, Zaragoza | SeqCOVID-SPAIN consortium/IBV(CSIC) | Antonio Rezusta L√≥pez et al |
| EPI_ISL_722272 | Spain/AR-IBV-98010737/2020 | 2020-11-12 | Europe/Spain/Aragon/Zaragoza | Servicio de Microbiolog√≠a, Hospital Miguel Servet, Zaragoza | SeqCOVID-SPAIN consortium/IBV(CSIC) | Antonio Rezusta L√≥pez et al |
| EPI_ISL_722275 | Netherlands/ZH-EMC-954/2020 | 2020-11-02 | Europe/Netherlands/South Holland/ | Dutch COVID-19 response team | Erasmus Medical Center | Bas Oude Munnink et al |
| EPI_ISL_722278 | Netherlands/NB-EMC-373/2020 | 2020-09-28 | Europe/Netherlands/North Brabant/ | Dutch COVID-19 response team | Erasmus Medical Center | Bas Oude Munnink et al |
| EPI_ISL_722287 | Netherlands/NB-EMC-347/2020 | 2020-10-25 | Europe/Netherlands/North Brabant/ | Dutch COVID-19 response team | Erasmus Medical Center | Bas Oude Munnink et al |
| EPI_ISL_722291 | Netherlands/NB-EMC-416/2020 | 2020-11-09 | Europe/Netherlands/North Brabant/ | Dutch COVID-19 response team | Erasmus Medical Center | Bas Oude Munnink et al |
| EPI_ISL_722298 | Netherlands/ZH-EMC-963/2020 | 2020-10-15 | Europe/Netherlands/South Holland/ | Dutch COVID-19 response team | Erasmus Medical Center | Bas Oude Munnink et al |
| EPI_ISL_722304 | Netherlands/ZH-EMC-1020/2020 | 2020-11-09 | Europe/Netherlands/South Holland/ | Dutch COVID-19 response team | Erasmus Medical Center | Bas Oude Munnink et al |
| EPI_ISL_722308 | Netherlands/ZH-EMC-944/2020 | 2020-10-19 | Europe/Netherlands/South Holland/ | Dutch COVID-19 response team | Erasmus Medical Center | Bas Oude Munnink et al |
| EPI_ISL_722310 | Netherlands/NB-EMC-388/2020 | 2020-09-30 | Europe/Netherlands/North Brabant/ | Dutch COVID-19 response team | Erasmus Medical Center | Bas Oude Munnink et al |

| | | | | | | |
|---|---|---|---|---|---|---|
| EPI_ISL_722319 | Netherlands/ZH-EMC-1040/2020 | 2020-09-22 | Europe/Netherlands/South Holland/ | Dutch COVID-19 response team | Erasmus Medical Center | Bas Oude Munnink et al |
| EPI_ISL_722323 | Netherlands/GE-EMC-396/2020 | 2020-11-09 | Europe/Netherlands/Gelderland/ | Dutch COVID-19 response team | Erasmus Medical Center | Bas Oude Munnink et al |
| EPI_ISL_722324 | Netherlands/ZH-EMC-928/2020 | 2020-10-27 | Europe/Netherlands/South Holland/ | Dutch COVID-19 response team | Erasmus Medical Center | Bas Oude Munnink et al |
| EPI_ISL_722331 | Netherlands/UT-EMC-186/2020 | 2020-11-16 | Europe/Netherlands/Utrecht/ | Dutch COVID-19 response team | Erasmus Medical Center | Bas Oude Munnink et al |
| EPI_ISL_722352 | Netherlands/NB-EMC-448/2020 | 2020-11-28 | Europe/Netherlands/North Brabant/ | Dutch COVID-19 response team | Erasmus Medical Center | Bas Oude Munnink et al |
| EPI_ISL_722366 | Netherlands/ZH-EMC-950/2020 | 2020-10-13 | Europe/Netherlands/South Holland/ | Dutch COVID-19 response team | Erasmus Medical Center | Bas Oude Munnink et al |
| EPI_ISL_722378 | Netherlands/NB-EMC-431/2020 | 2020-11-07 | Europe/Netherlands/North Brabant/ | Dutch COVID-19 response team | Erasmus Medical Center | Bas Oude Munnink et al |
| EPI_ISL_722403 | Netherlands/NB-EMC-369/2020 | 2020-09-30 | Europe/Netherlands/North Brabant/ | Dutch COVID-19 response team | Erasmus Medical Center | Bas Oude Munnink et al |
| EPI_ISL_722404 | Netherlands/NB-EMC-333/2020 | 2020-10-23 | Europe/Netherlands/North Brabant/ | Dutch COVID-19 response team | Erasmus Medical Center | Bas Oude Munnink et al |
| EPI_ISL_722406 | Netherlands/ZH-EMC-1035/2020 | 2020-11-23 | Europe/Netherlands/South Holland/ | Dutch COVID-19 response team | Erasmus Medical Center | Bas Oude Munnink et al |
| EPI_ISL_722412 | Netherlands/LI-EMC-101/2020 | 2020-10-30 | Europe/Netherlands/Limburg/ | Dutch COVID-19 response team | Erasmus Medical Center | Bas Oude Munnink et al |
| EPI_ISL_722436 | Netherlands/OV-EMC-97/2020 | 2020-09-09 | Europe/Netherlands/Overijssel/ | Dutch COVID-19 response team | Erasmus Medical Center | Bas Oude Munnink et al |
| EPI_ISL_722439 | Netherlands/LI-EMC-127/2020 | 2020-10-14 | Europe/Netherlands/Limburg/ | Dutch COVID-19 response team | Erasmus Medical Center | Bas Oude Munnink et al |
| EPI_ISL_722443 | Netherlands/GE-EMC-419/2020 | 2020-10-24 | Europe/Netherlands/Gelderland/ | Dutch COVID-19 response team | Erasmus Medical Center | Bas Oude Munnink et al |
| EPI_ISL_722444 | Netherlands/GE-EMC-384/2020 | 2020-10-29 | Europe/Netherlands/Gelderland/ | Dutch COVID-19 response team | Erasmus Medical Center | Bas Oude Munnink et al |
| EPI_ISL_722493 | Netherlands/ZH-EMC-1067/2020 | 2020-10-01 | Europe/Netherlands/South Holland/ | Dutch COVID-19 response team | Erasmus Medical Center | Bas Oude Munnink et al |
| EPI_ISL_722496 | Netherlands/NB-EMC-324/2020 | 2020-10-10 | Europe/Netherlands/North Brabant/ | Dutch COVID-19 response team | Erasmus Medical Center | Bas Oude Munnink et al |
| EPI_ISL_722502 | Netherlands/DR-EMC-51/2020 | 2020-10-14 | Europe/Netherlands/Drenthe/ | Dutch COVID-19 response team | Erasmus Medical Center | Bas Oude Munnink et al |
| EPI_ISL_722503 | Netherlands/ZH-EMC-1002/2020 | 2020-10-14 | Europe/Netherlands/South Holland/ | Dutch COVID-19 response team | Erasmus Medical Center | Bas Oude Munnink et al |
| EPI_ISL_722504 | Netherlands/ZH-EMC-1073/2020 | 2020-10-14 | Europe/Netherlands/South Holland/ | Dutch COVID-19 response team | Erasmus Medical Center | Bas Oude Munnink et al |

| | | | | | | |
|---|---|---|---|---|---|---|
| EPI_ISL_722505 | Netherlands/ZH-EMC-1074/2020 | 2020-10-14 | Europe/Netherlands/South Holland/ | Dutch COVID-19 response team | Erasmus Medical Center | Bas Oude Munnink et al |
| EPI_ISL_722506 | Netherlands/ZH-EMC-1075/2020 | 2020-10-14 | Europe/Netherlands/South Holland/ | Dutch COVID-19 response team | Erasmus Medical Center | Bas Oude Munnink et al |
| EPI_ISL_722507 | Netherlands/ZH-EMC-931/2020 | 2020-10-15 | Europe/Netherlands/South Holland/ | Dutch COVID-19 response team | Erasmus Medical Center | Bas Oude Munnink et al |
| EPI_ISL_722510 | Netherlands/ZH-EMC-961/2020 | 2020-10-17 | Europe/Netherlands/South Holland/ | Dutch COVID-19 response team | Erasmus Medical Center | Bas Oude Munnink et al |
| EPI_ISL_722511 | Netherlands/GE-EMC-397/2020 | 2020-10-18 | Europe/Netherlands/Gelderland/ | Dutch COVID-19 response team | Erasmus Medical Center | Bas Oude Munnink et al |
| EPI_ISL_722514 | Netherlands/UT-EMC-190/2020 | 2020-10-19 | Europe/Netherlands/Utrecht/ | Dutch COVID-19 response team | Erasmus Medical Center | Bas Oude Munnink et al |
| EPI_ISL_722516 | Netherlands/ZH-EMC-979/2020 | 2020-10-19 | Europe/Netherlands/South Holland/ | Dutch COVID-19 response team | Erasmus Medical Center | Bas Oude Munnink et al |
| EPI_ISL_722519 | Netherlands/ZH-EMC-923/2020 | 2020-10-20 | Europe/Netherlands/South Holland/ | Dutch COVID-19 response team | Erasmus Medical Center | Bas Oude Munnink et al |
| EPI_ISL_722521 | Netherlands/ZH-EMC-933/2020 | 2020-10-20 | Europe/Netherlands/South Holland/ | Dutch COVID-19 response team | Erasmus Medical Center | Bas Oude Munnink et al |
| EPI_ISL_722525 | Netherlands/NB-EMC-345/2020 | 2020-10-21 | Europe/Netherlands/North Brabant/ | Dutch COVID-19 response team | Erasmus Medical Center | Bas Oude Munnink et al |
| EPI_ISL_722526 | Netherlands/NB-EMC-346/2020 | 2020-10-21 | Europe/Netherlands/North Brabant/ | Dutch COVID-19 response team | Erasmus Medical Center | Bas Oude Munnink et al |
| EPI_ISL_722529 | Netherlands/NB-EMC-471/2020 | 2020-10-22 | Europe/Netherlands/North Brabant/ | Dutch COVID-19 response team | Erasmus Medical Center | Bas Oude Munnink et al |
| EPI_ISL_722533 | Netherlands/NB-EMC-352/2020 | 2020-10-22 | Europe/Netherlands/North Brabant/ | Dutch COVID-19 response team | Erasmus Medical Center | Bas Oude Munnink et al |
| EPI_ISL_722534 | Netherlands/UT-EMC-192/2020 | 2020-10-22 | Europe/Netherlands/Utrecht/ | Dutch COVID-19 response team | Erasmus Medical Center | Bas Oude Munnink et al |
| EPI_ISL_722537 | Netherlands/GE-EMC-382/2020 | 2020-10-23 | Europe/Netherlands/Gelderland/ | Dutch COVID-19 response team | Erasmus Medical Center | Bas Oude Munnink et al |
| EPI_ISL_722538 | Netherlands/NB-EMC-332/2020 | 2020-10-23 | Europe/Netherlands/North Brabant/ | Dutch COVID-19 response team | Erasmus Medical Center | Bas Oude Munnink et al |
| EPI_ISL_722540 | Netherlands/NB-EMC-335/2020 | 2020-10-23 | Europe/Netherlands/North Brabant/ | Dutch COVID-19 response team | Erasmus Medical Center | Bas Oude Munnink et al |
| EPI_ISL_722546 | Netherlands/GE-EMC-403/2020 | 2020-10-25 | Europe/Netherlands/Gelderland/ | Dutch COVID-19 response team | Erasmus Medical Center | Bas Oude Munnink et al |
| EPI_ISL_722547 | Netherlands/ZH-EMC-1049/2020 | 2020-10-25 | Europe/Netherlands/South Holland/ | Dutch COVID-19 response team | Erasmus Medical Center | Bas Oude Munnink et al |
| EPI_ISL_722551 | Netherlands/NB-EMC-337/2020 | 2020-10-26 | Europe/Netherlands/North Brabant/ | Dutch COVID-19 response team | Erasmus Medical Center | Bas Oude Munnink et al |

| | | | | | | |
|---|---|---|---|---|---|---|
| EPI_ISL_722552 | Netherlands/NB-EMC-338/2020 | 2020-10-26 | Europe/Netherlands/North Brabant/ | Dutch COVID-19 response team | Erasmus Medical Center | Bas Oude Munnink et al |
| EPI_ISL_722553 | Netherlands/NB-EMC-339/2020 | 2020-10-26 | Europe/Netherlands/North Brabant/ | Dutch COVID-19 response team | Erasmus Medical Center | Bas Oude Munnink et al |
| EPI_ISL_722554 | Netherlands/ZH-EMC-926/2020 | 2020-10-26 | Europe/Netherlands/South Holland/ | Dutch COVID-19 response team | Erasmus Medical Center | Bas Oude Munnink et al |
| EPI_ISL_722558 | Netherlands/UT-EMC-177/2020 | 2020-10-27 | Europe/Netherlands/Utrecht/ | Dutch COVID-19 response team | Erasmus Medical Center | Bas Oude Munnink et al |
| EPI_ISL_722559 | Netherlands/GE-EMC-357/2020 | 2020-10-28 | Europe/Netherlands/Gelderland/ | Dutch COVID-19 response team | Erasmus Medical Center | Bas Oude Munnink et al |
| EPI_ISL_722561 | Netherlands/NB-EMC-365/2020 | 2020-10-28 | Europe/Netherlands/North Brabant/ | Dutch COVID-19 response team | Erasmus Medical Center | Bas Oude Munnink et al |
| EPI_ISL_722567 | Netherlands/NB-EMC-348/2020 | 2020-10-29 | Europe/Netherlands/North Brabant/ | Dutch COVID-19 response team | Erasmus Medical Center | Bas Oude Munnink et al |
| EPI_ISL_722570 | Netherlands/GE-EMC-369/2020 | 2020-10-03 | Europe/Netherlands/Gelderland/ | Dutch COVID-19 response team | Erasmus Medical Center | Bas Oude Munnink et al |
| EPI_ISL_722574 | Netherlands/LI-EMC-104/2020 | 2020-10-30 | Europe/Netherlands/Limburg/ | Dutch COVID-19 response team | Erasmus Medical Center | Bas Oude Munnink et al |
| EPI_ISL_722575 | Netherlands/LI-EMC-105/2020 | 2020-10-30 | Europe/Netherlands/Limburg/ | Dutch COVID-19 response team | Erasmus Medical Center | Bas Oude Munnink et al |
| EPI_ISL_722579 | Netherlands/LI-EMC-111/2020 | 2020-10-30 | Europe/Netherlands/Limburg/ | Dutch COVID-19 response team | Erasmus Medical Center | Bas Oude Munnink et al |
| EPI_ISL_722582 | Netherlands/LI-EMC-108/2020 | 2020-10-31 | Europe/Netherlands/Limburg/ | Dutch COVID-19 response team | Erasmus Medical Center | Bas Oude Munnink et al |
| EPI_ISL_722584 | Netherlands/NB-EMC-349/2020 | 2020-10-31 | Europe/Netherlands/North Brabant/ | Dutch COVID-19 response team | Erasmus Medical Center | Bas Oude Munnink et al |
| EPI_ISL_722592 | Netherlands/NB-EMC-387/2020 | 2020-10-05 | Europe/Netherlands/North Brabant/ | Dutch COVID-19 response team | Erasmus Medical Center | Bas Oude Munnink et al |
| EPI_ISL_722616 | Netherlands/NB-EMC-424/2020 | 2020-11-11 | Europe/Netherlands/North Brabant/ | Dutch COVID-19 response team | Erasmus Medical Center | Bas Oude Munnink et al |
| EPI_ISL_722629 | Netherlands/GE-EMC-367/2020 | 2020-11-13 | Europe/Netherlands/Gelderland/ | Dutch COVID-19 response team | Erasmus Medical Center | Bas Oude Munnink et al |
| EPI_ISL_722634 | Netherlands/ZH-EMC-1018/2020 | 2020-11-13 | Europe/Netherlands/South Holland/ | Dutch COVID-19 response team | Erasmus Medical Center | Bas Oude Munnink et al |
| EPI_ISL_722665 | Netherlands/ZH-EMC-975/2020 | 2020-11-17 | Europe/Netherlands/South Holland/ | Dutch COVID-19 response team | Erasmus Medical Center | Bas Oude Munnink et al |
| EPI_ISL_722667 | Netherlands/GE-EMC-407/2020 | 2020-11-18 | Europe/Netherlands/Gelderland/ | Dutch COVID-19 response team | Erasmus Medical Center | Bas Oude Munnink et al |
| EPI_ISL_722677 | Netherlands/ZH-EMC-1004/2020 | 2020-11-19 | Europe/Netherlands/South Holland/ | Dutch COVID-19 response team | Erasmus Medical Center | Bas Oude Munnink et al |

| | | | | | | |
|---|---|---|---|---|---|---|
| EPI_ISL_722696 | Netherlands/ZH-EMC-1044/2020 | 2020-11-27 | Europe/Netherlands/South Holland/ | Dutch COVID-19 response team | Erasmus Medical Center | Bas Oude Munnink et al |
| EPI_ISL_722702 | Netherlands/ZH-EMC-907/2020 | 2020-11-03 | Europe/Netherlands/South Holland/ | Dutch COVID-19 response team | Erasmus Medical Center | Bas Oude Munnink et al |
| EPI_ISL_722717 | Netherlands/GE-EMC-361/2020 | 2020-11-04 | Europe/Netherlands/Gelderland/ | Dutch COVID-19 response team | Erasmus Medical Center | Bas Oude Munnink et al |
| EPI_ISL_722727 | Netherlands/LI-EMC-113/2020 | 2020-11-05 | Europe/Netherlands/Limburg/ | Dutch COVID-19 response team | Erasmus Medical Center | Bas Oude Munnink et al |
| EPI_ISL_722804 | Netherlands/NB-EMC-455/2020 | 2020-07-15 | Europe/Netherlands/North Brabant/ | Dutch COVID-19 response team | Erasmus Medical Center | Bas Oude Munnink et al |
| EPI_ISL_722821 | Netherlands/OV-EMC-100/2020 | 2020-09-16 | Europe/Netherlands/Overijssel/ | Dutch COVID-19 response team | Erasmus Medical Center | Bas Oude Munnink et al |
| EPI_ISL_722825 | Netherlands/OV-EMC-105/2020 | 2020-09-19 | Europe/Netherlands/Overijssel/ | Dutch COVID-19 response team | Erasmus Medical Center | Bas Oude Munnink et al |
| EPI_ISL_722826 | Netherlands/ZH-EMC-991/2020 | 2020-09-19 | Europe/Netherlands/South Holland/ | Dutch COVID-19 response team | Erasmus Medical Center | Bas Oude Munnink et al |
| EPI_ISL_722828 | Netherlands/ZH-EMC-960/2020 | 2020-09-24 | Europe/Netherlands/South Holland/ | Dutch COVID-19 response team | Erasmus Medical Center | Bas Oude Munnink et al |
| EPI_ISL_722842 | Netherlands/OV-EMC-85/2020 | 2020-09-04 | Europe/Netherlands/Overijssel/ | Dutch COVID-19 response team | Erasmus Medical Center | Bas Oude Munnink et al |
| EPI_ISL_722854 | Italy/APU-IZSPB-199APT/2020 | 2020-08-28 | Europe/Italy/Apulia/Bari | "I.R.C.C.S. ""S. De Bellis"" - Ente Ospedaliero" | Istituto Zooprofilattico Sperimentale della Puglia e della Basilicata | Parisi A. et al |
| EPI_ISL_722856 | Italy/APU-IZSPB-153APT/2020 | 2020-07-18 | Europe/Italy/Apulia/Brindisi | Dipartimento di Scienze Biomediche e Oncologia Umana - Azienda Ospedaliero Universitaria Consorziale Policlinico | Istituto Zooprofilattico Sperimentale della Puglia e della Basilicata | Parisi A. et al |
| EPI_ISL_722864 | Italy/APU-IZSPB-186PT/2020 | 2020-08-28 | Europe/Italy/Apulia/Bari | "I.R.C.C.S. ""S. De Bellis"" - Ente Ospedaliero" | Istituto Zooprofilattico Sperimentale della Puglia e della Basilicata | Parisi A. et al |
| EPI_ISL_722868 | Italy/APU-IZSPB-197PT/2020 | 2020-08-28 | Europe/Italy/Apulia/Bari | "I.R.C.C.S. ""S. De Bellis"" - Ente Ospedaliero" | Istituto Zooprofilattico Sperimentale della Puglia e della Basilicata | Parisi A. et al |

| | | | | | | |
|---|---|---|---|---|---|---|
| EPI_ISL_722871 | Italy/APU-IZSPB-203PT/2020 | 2020-08-28 | Europe/Italy/Apulia/Bari | "I.R.C.C.S. ""S. De Bellis"" - Ente Ospedaliero" | Istituto Zooprofilattico Sperimentale della Puglia e della Basilicata | Parisi A. et al |
| EPI_ISL_722873 | Italy/BAS-IZSPB-338PT/2020 | 2020-10-27 | Europe/Italy/Basilicata/Matera | Istituto Zooprofilattico Sperimentale della Puglia e della Basilicata | Istituto Zooprofilattico Sperimentale della Puglia e della Basilicata | Parisi A. et al |
| EPI_ISL_722875 | Italy/BAS-IZSPB-341PT/2020 | 2020-10-28 | Europe/Italy/Basilicata/Matera | Istituto Zooprofilattico Sperimentale della Puglia e della Basilicata | Istituto Zooprofilattico Sperimentale della Puglia e della Basilicata | Parisi A. et al |
| EPI_ISL_722876 | Italy/BAS-IZSPB-351PT/2020 | 2020-10-28 | Europe/Italy/Basilicata/Matera | Istituto Zooprofilattico Sperimentale della Puglia e della Basilicata | Istituto Zooprofilattico Sperimentale della Puglia e della Basilicata | Parisi A. et al |
| EPI_ISL_722878 | Italy/BAS-IZSPB-364PT/2020 | 2020-10-15 | Europe/Italy/Basilicata/Matera | Istituto Zooprofilattico Sperimentale della Puglia e della Basilicata | Istituto Zooprofilattico Sperimentale della Puglia e della Basilicata | Parisi A. et al |
| EPI_ISL_722879 | Italy/BAS-IZSPB-368PT/2020 | 2020-10-16 | Europe/Italy/Basilicata/Potenza | Istituto Zooprofilattico Sperimentale della Puglia e della Basilicata | Istituto Zooprofilattico Sperimentale della Puglia e della Basilicata | Parisi A. et al |
| EPI_ISL_722880 | Italy/BAS-IZSPB-373PT/2020 | 2020-10-16 | Europe/Italy/Basilicata/Matera | Istituto Zooprofilattico Sperimentale della Puglia e della Basilicata | Istituto Zooprofilattico Sperimentale della Puglia e della Basilicata | Parisi A. et al |
| EPI_ISL_722881 | Italy/BAS-IZSPB-374PT/2020 | 2020-10-16 | Europe/Italy/Basilicata/Matera | Istituto Zooprofilattico Sperimentale della Puglia e della Basilicata | Istituto Zooprofilattico Sperimentale della Puglia e della Basilicata | Parisi A. et al |

| | | | | | | |
|---|---|---|---|---|---|---|
| EPI_ISL_722884 | Italy/BAS-IZSPB-383PT/2020 | 2020-10-25 | Europe/Italy/Basilicata/Matera | Istituto Zooprofilattico Sperimentale della Puglia e della Basilicata | Istituto Zooprofilattico Sperimentale della Puglia e della Basilicata | Parisi A. et al |
| EPI_ISL_722885 | Italy/BAS-IZSPB-384PT/2020 | 2020-10-20 | Europe/Italy/Basilicata/Matera | Istituto Zooprofilattico Sperimentale della Puglia e della Basilicata | Istituto Zooprofilattico Sperimentale della Puglia e della Basilicata | Parisi A. et al |
| EPI_ISL_722886 | Italy/BAS-IZSPB-385PT/2020 | 2020-10-20 | Europe/Italy/Basilicata/Potenza | Istituto Zooprofilattico Sperimentale della Puglia e della Basilicata | Istituto Zooprofilattico Sperimentale della Puglia e della Basilicata | Parisi A. et al |
| EPI_ISL_722888 | Italy/BAS-IZSPB-389PT/2020 | 2020-10-25 | Europe/Italy/Basilicata/Potenza | Istituto Zooprofilattico Sperimentale della Puglia e della Basilicata | Istituto Zooprofilattico Sperimentale della Puglia e della Basilicata | Parisi A. et al |
| EPI_ISL_722889 | Italy/BAS-IZSPB-390PT/2020 | 2020-10-25 | Europe/Italy/Basilicata/Matera | Istituto Zooprofilattico Sperimentale della Puglia e della Basilicata | Istituto Zooprofilattico Sperimentale della Puglia e della Basilicata | Parisi A. et al |
| EPI_ISL_722891 | Italy/BAS-IZSPB-398PT/2020 | 2020-10-25 | Europe/Italy/Basilicata/Matera | Istituto Zooprofilattico Sperimentale della Puglia e della Basilicata | Istituto Zooprofilattico Sperimentale della Puglia e della Basilicata | Parisi A. et al |
| EPI_ISL_722893 | Italy/APU-IZSPB-332PT/2020 | 2020-10-10 | Europe/Italy/Apulia/Foggia | Istituto Zooprofilattico Sperimentale della Puglia e della Basilicata | Istituto Zooprofilattico Sperimentale della Puglia e della Basilicata | Parisi A. et al |
| EPI_ISL_722894 | Italy/APU-IZSPB-333PT/2020 | 2020-11-05 | Europe/Italy/Apulia/Foggia | Istituto Zooprofilattico Sperimentale della Puglia e della Basilicata | Istituto Zooprofilattico Sperimentale della Puglia e della Basilicata | Parisi A. et al |

| | | | | | | |
|---|---|---|---|---|---|---|
| EPI_ISL_722895 | Italy/APU-IZSPB-187PT/2020 | 2020-08-28 | Europe/Italy/Apulia/Bari | "I.R.C.C.S. ""S. De Bellis"" - Ente Ospedaliero" | Istituto Zooprofilattico Sperimentale della Puglia e della Basilicata | Parisi A. et al |
| EPI_ISL_722898 | Italy/APU-IZSPB-180PT/2020 | 2020-08-28 | Europe/Italy/Apulia/Bari | "I.R.C.C.S. ""S. De Bellis"" - Ente Ospedaliero" | Istituto Zooprofilattico Sperimentale della Puglia e della Basilicata | Parisi A. et al |
| EPI_ISL_722903 | Italy/BAS-IZSPB-358PT/2020 | 2020-10-29 | Europe/Italy/Basilicata/Potenza | Istituto Zooprofilattico Sperimentale della Puglia e della Basilicata | Istituto Zooprofilattico Sperimentale della Puglia e della Basilicata | Parisi A. et al |
| EPI_ISL_722904 | Italy/BAS-IZSPB-370PT/2020 | 2020-10-16 | Europe/Italy/Basilicata/Potenza | Istituto Zooprofilattico Sperimentale della Puglia e della Basilicata | Istituto Zooprofilattico Sperimentale della Puglia e della Basilicata | Parisi A. et al |
| EPI_ISL_722905 | Italy/BAS-IZSPB-372PT/2020 | 2020-10-28 | Europe/Italy/Basilicata/Matera | Istituto Zooprofilattico Sperimentale della Puglia e della Basilicata | Istituto Zooprofilattico Sperimentale della Puglia e della Basilicata | Parisi A. et al |
| EPI_ISL_722909 | Italy/APU-IZSPB-334PT/2020 | 2020-11-04 | Europe/Italy/Apulia/Foggia | Istituto Zooprofilattico Sperimentale della Puglia e della Basilicata | Istituto Zooprofilattico Sperimentale della Puglia e della Basilicata | Parisi A. et al |
| EPI_ISL_722911 | Italy/APU-IZSPB-331PT/2020 | 2020-11-05 | Europe/Italy/Apulia/Foggia | Istituto Zooprofilattico Sperimentale della Puglia e della Basilicata | Istituto Zooprofilattico Sperimentale della Puglia e della Basilicata | Parisi A. et al |
| EPI_ISL_722920 | Italy/BAS-IZSPB-386PT/2020 | 2020-10-25 | Europe/Italy/Basilicata/Matera | Istituto Zooprofilattico Sperimentale della Puglia e della Basilicata | Istituto Zooprofilattico Sperimentale della Puglia e della Basilicata | Parisi A. et al |

| | | | | | | |
|---|---|---|---|---|---|---|
| EPI_ISL_722924 | Italy/BAS-IZSPB-395PT/2020 | 2020-10-21 | Europe/Italy/Basilicata/Matera | Istituto Zooprofilattico Sperimentale della Puglia e della Basilicata | Istituto Zooprofilattico Sperimentale della Puglia e della Basilicata | Parisi A. et al |
| EPI_ISL_722925 | Italy/BAS-IZSPB-397PT/2020 | 2020-10-21 | Europe/Italy/Basilicata/Matera | Istituto Zooprofilattico Sperimentale della Puglia e della Basilicata | Istituto Zooprofilattico Sperimentale della Puglia e della Basilicata | Parisi A. et al |
| EPI_ISL_722936 | Belgium/ULG-11017/2020 | 2020-11-28 | Europe/Belgium/Liv®ge/ | Department of Clinical Microbiology | GIGA Medical Genomics | Keith Durkin et al |
| EPI_ISL_722937 | Belgium/ULG-11018/2020 | 2020-11-28 | Europe/Belgium/Liv®ge/ | Department of Clinical Microbiology | GIGA Medical Genomics | Keith Durkin et al |
| EPI_ISL_722943 | Belgium/ULG-11025/2020 | 2020-11-29 | Europe/Belgium/Liv®ge/ | Department of Clinical Microbiology | GIGA Medical Genomics | Keith Durkin et al |
| EPI_ISL_722950 | Belgium/ULG-11033/2020 | 2020-11-30 | Europe/Belgium/Liv®ge/ | Department of Clinical Microbiology | GIGA Medical Genomics | Keith Durkin et al |
| EPI_ISL_722953 | Belgium/ULG-11039/2020 | 2020-11-30 | Europe/Belgium/Liv®ge/ | Department of Clinical Microbiology | GIGA Medical Genomics | Keith Durkin et al |
| EPI_ISL_722955 | Belgium/ULG-11042/2020 | 2020-11-30 | Europe/Belgium/Liv®ge/ | Department of Clinical Microbiology | GIGA Medical Genomics | Keith Durkin et al |
| EPI_ISL_722956 | Belgium/ULG-11043/2020 | 2020-11-30 | Europe/Belgium/Liv®ge/ | Department of Clinical Microbiology | GIGA Medical Genomics | Keith Durkin et al |
| EPI_ISL_722957 | Belgium/ULG-11044/2020 | 2020-11-30 | Europe/Belgium/Liv®ge/ | Department of Clinical Microbiology | GIGA Medical Genomics | Keith Durkin et al |
| EPI_ISL_723001 | England/205080610/2020 | 2020-12-09 | Europe/United Kingdom/England/ | Respiratory Virus Unit, National Infection Service, Public Health England | COVID-19 Genomics UK (COG-UK) Consortium | PHE Covid Sequencing Team et al |
| EPI_ISL_723048 | India/MH-ACTREC-229/2020 | 2020-07-02 | Asia/India/Maharashtra/Raigad | Hematopathology Laboratory, ACTREC, TMC | Hematopathology Laboratory, ACTREC, TMC | Hematopathology Laboratory et al |
| EPI_ISL_723049 | India/MH-ACTREC-248/2020 | 2020-07-07 | Asia/India/Maharashtra/Thane | Hematopathology Laboratory, ACTREC, TMC | Hematopathology Laboratory, ACTREC, TMC | Hematopathology Laboratory et al |
| EPI_ISL_723050 | India/MH-ACTREC-300/2020 | 2020-07-14 | Asia/India/Maharashtra/Raigad | Hematopathology Laboratory, ACTREC, TMC | Hematopathology Laboratory, ACTREC, TMC | Hematopathology Laboratory et al |

| | | | | | | |
|---|---|---|---|---|---|---|
| EPI_ISL_723051 | India/MH-ACTREC-311/2020 | 2020-07-15 | Asia/India/Maharashtra/Raigad | Hematopathology Laboratory, ACTREC, TMC | Hematopathology Laboratory, ACTREC, TMC | Hematopathology Laboratory et al |
| EPI_ISL_723052 | India/MH-ACTREC-312/2020 | 2020-07-15 | Asia/India/Maharashtra/Raigad | Hematopathology Laboratory, ACTREC, TMC | Hematopathology Laboratory, ACTREC, TMC | Hematopathology Laboratory et al |
| EPI_ISL_723053 | India/MH-ACTREC-443/2020 | 2020-08-05 | Asia/India/Maharashtra/Raigad | Hematopathology Laboratory, ACTREC, TMC | Hematopathology Laboratory, ACTREC, TMC | Hematopathology Laboratory et al |
| EPI_ISL_723054 | India/MH-ACTREC-512/2020 | 2020-08-19 | Asia/India/Maharashtra/Thane | Hematopathology Laboratory, ACTREC, TMC | Hematopathology Laboratory, ACTREC, TMC | Hematopathology Laboratory et al |
| EPI_ISL_723055 | India/MH-ACTREC-517/2020 | 2020-08-20 | Asia/India/Maharashtra/Thane | Hematopathology Laboratory, ACTREC, TMC | Hematopathology Laboratory, ACTREC, TMC | Hematopathology Laboratory et al |
| EPI_ISL_723057 | India/MH-ACTREC-566/2020 | 2020-08-31 | Asia/India/Maharashtra/Raigad | Hematopathology Laboratory, ACTREC, TMC | Hematopathology Laboratory, ACTREC, TMC | Hematopathology Laboratory et al |
| EPI_ISL_723058 | India/MH-ACTREC-577/2020 | 2020-09-02 | Asia/India/Maharashtra/Raigad | Hematopathology Laboratory, ACTREC, TMC | Hematopathology Laboratory, ACTREC, TMC | Hematopathology Laboratory et al |
| EPI_ISL_723059 | India/MH-ACTREC-579/2020 | 2020-09-02 | Asia/India/Maharashtra/Raigad | Hematopathology Laboratory, ACTREC, TMC | Hematopathology Laboratory, ACTREC, TMC | Hematopathology Laboratory et al |
| EPI_ISL_723060 | India/MH-ACTREC-581/2020 | 2020-09-02 | Asia/India/Maharashtra/Raigad | Hematopathology Laboratory, ACTREC, TMC | Hematopathology Laboratory, ACTREC, TMC | Hematopathology Laboratory et al |
| EPI_ISL_723061 | India/MH-ACTREC-583/2020 | 2020-09-02 | Asia/India/Maharashtra/Raigad | Hematopathology Laboratory, ACTREC, TMC | Hematopathology Laboratory, ACTREC, TMC | Hematopathology Laboratory et al |
| EPI_ISL_723062 | India/MH-ACTREC-584/2020 | 2020-09-02 | Asia/India/Maharashtra/Raigad | Hematopathology Laboratory, ACTREC, TMC | Hematopathology Laboratory, ACTREC, TMC | Hematopathology Laboratory et al |
| EPI_ISL_723063 | India/MH-ACTREC-589/2020 | 2020-09-04 | Asia/India/Maharashtra/Raigad | Hematopathology Laboratory, ACTREC, TMC | Hematopathology Laboratory, ACTREC, TMC | Hematopathology Laboratory et al |
| EPI_ISL_723064 | India/MH-ACTREC-602/2020 | 2020-09-08 | Asia/India/Maharashtra/Thane | Hematopathology Laboratory, ACTREC, TMC | Hematopathology Laboratory, ACTREC, TMC | Hematopathology Laboratory et al |

| | | | | | | |
|---|---|---|---|---|---|---|
| EPI_ISL_723065 | India/MH-ACTREC-614/2020 | 2020-09-09 | Asia/India/Maharashtra/Raigad | Hematopathology Laboratory, ACTREC, TMC | Hematopathology Laboratory, ACTREC, TMC | Hematopathology Laboratory et al |
| EPI_ISL_723066 | India/MH-ACTREC-615/2020 | 2020-09-09 | Asia/India/Maharashtra/Raigad | Hematopathology Laboratory, ACTREC, TMC | Hematopathology Laboratory, ACTREC, TMC | Hematopathology Laboratory et al |
| EPI_ISL_723067 | India/MH-ACTREC-630/2020 | 2020-09-11 | Asia/India/Maharashtra/Raigad | Hematopathology Laboratory, ACTREC, TMC | Hematopathology Laboratory, ACTREC, TMC | Hematopathology Laboratory et al |
| EPI_ISL_723068 | India/MH-ACTREC-631/2020 | 2020-09-11 | Asia/India/Maharashtra/Raigad | Hematopathology Laboratory, ACTREC, TMC | Hematopathology Laboratory, ACTREC, TMC | Hematopathology Laboratory et al |
| EPI_ISL_723069 | India/MH-ACTREC-643/2020 | 2020-09-14 | Asia/India/Maharashtra/Mumbai Subur | Hematopathology Laboratory, ACTREC, TMC | Hematopathology Laboratory, ACTREC, TMC | Hematopathology Laboratory et al |
| EPI_ISL_723070 | India/MH-ACTREC-644/2020 | 2020-09-14 | Asia/India/Maharashtra/Raigad | Hematopathology Laboratory, ACTREC, TMC | Hematopathology Laboratory, ACTREC, TMC | Hematopathology Laboratory et al |
| EPI_ISL_723071 | India/MH-ACTREC-645/2020 | 2020-09-14 | Asia/India/Maharashtra/Raigad | Hematopathology Laboratory, ACTREC, TMC | Hematopathology Laboratory, ACTREC, TMC | Hematopathology Laboratory et al |
| EPI_ISL_723072 | India/MH-ACTREC-661/2020 | 2020-09-15 | Asia/India/Maharashtra/Raigad | Hematopathology Laboratory, ACTREC, TMC | Hematopathology Laboratory, ACTREC, TMC | Hematopathology Laboratory et al |
| EPI_ISL_723073 | India/MH-ACTREC-665/2020 | 2020-09-16 | Asia/India/Maharashtra/Raigad | Hematopathology Laboratory, ACTREC, TMC | Hematopathology Laboratory, ACTREC, TMC | Hematopathology Laboratory et al |
| EPI_ISL_723074 | India/MH-ACTREC-666/2020 | 2020-09-16 | Asia/India/Maharashtra/Amravati | Hematopathology Laboratory, ACTREC, TMC | Hematopathology Laboratory, ACTREC, TMC | Hematopathology Laboratory et al |
| EPI_ISL_723075 | India/MH-ACTREC-667/2020 | 2020-09-16 | Asia/India/Maharashtra/Washim | Hematopathology Laboratory, ACTREC, TMC | Hematopathology Laboratory, ACTREC, TMC | Hematopathology Laboratory et al |
| EPI_ISL_723076 | India/MH-ACTREC-674/2020 | 2020-09-18 | Asia/India/Maharashtra/Raigad | Hematopathology Laboratory, ACTREC, TMC | Hematopathology Laboratory, ACTREC, TMC | Hematopathology Laboratory et al |
| EPI_ISL_723081 | Germany/BW-UKT-a092/2020 | 2020-04-16 | Europe/Germany/Baden-Wuerttemberg | Institute of Medical Genetics and Applied Genomics | Institute of Medical Genetics and Applied Genomics | Caspar Gross et al |

| | | | | | | |
|---|---|---|---|---|---|---|
| EPI_ISL_723082 | Germany/BW-UKT-a025/2020 | 2020-05-13 | Europe/Germany/Baden-Wuerttemberg | Institute of Medical Genetics and Applied Genomics | Institute of Medical Genetics and Applied Genomics | Caspar Gross et al |
| EPI_ISL_723087 | Germany/BW-UKT-a032/2020 | 2020-03-22 | Europe/Germany/Baden-Wuerttemberg | Institute of Medical Genetics and Applied Genomics | Institute of Medical Genetics and Applied Genomics | Caspar Gross et al |
| EPI_ISL_723093 | Germany/BW-UKT-a002/2020 | 2020-03-23 | Europe/Germany/Baden-Wuerttemberg | Institute of Medical Genetics and Applied Genomics | Institute of Medical Genetics and Applied Genomics | Caspar Gross et al |
| EPI_ISL_723103 | Germany/BW-UKT-a046/2020 | 2020-03-26 | Europe/Germany/Baden-Wuerttemberg | Institute of Medical Genetics and Applied Genomics | Institute of Medical Genetics and Applied Genomics | Caspar Gross et al |
| EPI_ISL_723106 | Germany/BW-UKT-a014/2020 | 2020-03-31 | Europe/Germany/Baden-Wuerttemberg | Institute of Medical Genetics and Applied Genomics | Institute of Medical Genetics and Applied Genomics | Caspar Gross et al |
| EPI_ISL_723108 | Germany/BW-UKT-a016/2020 | 2020-04-02 | Europe/Germany/Baden-Wuerttemberg | Institute of Medical Genetics and Applied Genomics | Institute of Medical Genetics and Applied Genomics | Caspar Gross et al |
| EPI_ISL_723109 | Germany/BW-UKT-a117/2020 | 2020-04-02 | Europe/Germany/Baden-Wuerttemberg | Institute of Medical Genetics and Applied Genomics | Institute of Medical Genetics and Applied Genomics | Caspar Gross et al |
| EPI_ISL_723110 | Germany/BW-UKT-a017/2020 | 2020-04-03 | Europe/Germany/Baden-Wuerttemberg | Institute of Medical Genetics and Applied Genomics | Institute of Medical Genetics and Applied Genomics | Caspar Gross et al |
| EPI_ISL_723122 | USA/MN-MDH-2171/2020 | 2020-11-15 | North America/USA/Minnesota/ | Minnesota Department of Health, Public Health Laboratory | Minnesota Department of Health, Public Health Laboratory | Alexandra Lorentz et al |
| EPI_ISL_723134 | USA/MN-MDH-2183/2020 | 2020-11-29 | North America/USA/Minnesota/ | Minnesota Department of Health, Public Health Laboratory | Minnesota Department of Health, Public Health Laboratory | Alexandra Lorentz et al |

| | | | | | | |
|---|---|---|---|---|---|---|
| EPI_ISL_723139 | USA/MN-MDH-2188/2020 | 2020-12-07 | North America/USA/Minnesota/ | Minnesota Department of Health, Public Health Laboratory | Minnesota Department of Health, Public Health Laboratory | Alexandra Lorentz et al |
| EPI_ISL_723140 | USA/MN-MDH-2189/2020 | 2020-12-07 | North America/USA/Minnesota/ | Minnesota Department of Health, Public Health Laboratory | Minnesota Department of Health, Public Health Laboratory | Alexandra Lorentz et al |
| EPI_ISL_723197 | Netherlands/NB-RIVM-20274/2020 | 2020-11-26 | Europe/Netherlands/North Brabant/ | Dutch COVID-19 response team | National Institute for Public Health and the Environment (RIVM) | Adam Meijer et al |
| EPI_ISL_723205 | Netherlands/NH-RIVM-20006/2020 | 2020-11-21 | Europe/Netherlands/North Holland/ | Dutch COVID-19 response team | National Institute for Public Health and the Environment (RIVM) | Adam Meijer et al |
| EPI_ISL_723208 | Netherlands/NH-RIVM-20009/2020 | 2020-11-21 | Europe/Netherlands/North Holland/ | Dutch COVID-19 response team | National Institute for Public Health and the Environment (RIVM) | Adam Meijer et al |
| EPI_ISL_723228 | Netherlands/LI-RIVM-20031/2020 | 2020-11-26 | Europe/Netherlands/Limburg/ | Dutch COVID-19 response team | National Institute for Public Health and the Environment (RIVM) | Adam Meijer et al |
| EPI_ISL_723232 | Netherlands/GE-RIVM-20045/2020 | 2020-11-09 | Europe/Netherlands/Gelderland/ | Dutch COVID-19 response team | National Institute for Public Health and the Environment (RIVM) | Adam Meijer et al |
| EPI_ISL_723240 | Netherlands/UT-RIVM-20056/2020 | 2020-11-13 | Europe/Netherlands/Utrecht/ | Dutch COVID-19 response team | National Institute for Public Health and the Environment (RIVM) | Adam Meijer et al |

| | | | | | | |
|---|---|---|---|---|---|---|
| EPI_ISL_723242 | Netherlands/NH-RIVM-20058/2020 | 2020-11-28 | Europe/Netherlands/North Holland/ | Dutch COVID-19 response team | National Institute for Public Health and the Environment (RIVM) | Adam Meijer et al |
| EPI_ISL_723261 | Netherlands/NH-RIVM-20082/2020 | 2020-11-13 | Europe/Netherlands/North Holland/ | Dutch COVID-19 response team | National Institute for Public Health and the Environment (RIVM) | Adam Meijer et al |
| EPI_ISL_723271 | Netherlands/OV-RIVM-20093/2020 | 2020-11-11 | Europe/Netherlands/Overijssel/ | Dutch COVID-19 response team | National Institute for Public Health and the Environment (RIVM) | Adam Meijer et al |
| EPI_ISL_723293 | Netherlands/OV-RIVM-20118/2020 | 2020-11-18 | Europe/Netherlands/Overijssel/ | Dutch COVID-19 response team | National Institute for Public Health and the Environment (RIVM) | Adam Meijer et al |
| EPI_ISL_723305 | Netherlands/NH-RIVM-20134/2020 | 2020-11-14 | Europe/Netherlands/North Holland/ | Dutch COVID-19 response team | National Institute for Public Health and the Environment (RIVM) | Adam Meijer et al |
| EPI_ISL_723308 | Netherlands/UT-RIVM-20137/2020 | 2020-11-24 | Europe/Netherlands/Utrecht/ | Dutch COVID-19 response team | National Institute for Public Health and the Environment (RIVM) | Adam Meijer et al |
| EPI_ISL_723317 | Netherlands/FR-RIVM-20147/2020 | 2020-11-23 | Europe/Netherlands/Friesland/ | Dutch COVID-19 response team | National Institute for Public Health and the Environment (RIVM) | Adam Meijer et al |
| EPI_ISL_723325 | Netherlands/OV-RIVM-20158/2020 | 2020-11-17 | Europe/Netherlands/Overijssel/ | Dutch COVID-19 response team | National Institute for Public Health and the Environment (RIVM) | Adam Meijer et al |

| | | | | | | |
|---|---|---|---|---|---|---|
| EPI_ISL_723329 | Netherlands/UT-RIVM-20162/2020 | 2020-11-29 | Europe/Netherlands/Utrecht/ | Dutch COVID-19 response team | National Institute for Public Health and the Environment (RIVM) | Adam Meijer et al |
| EPI_ISL_723338 | Netherlands/UT-RIVM-20177/2020 | 2020-11-30 | Europe/Netherlands/Utrecht/ | Dutch COVID-19 response team | National Institute for Public Health and the Environment (RIVM) | Adam Meijer et al |
| EPI_ISL_723361 | Netherlands/GE-RIVM-20208/2020 | 2020-11-24 | Europe/Netherlands/Gelderland/ | Dutch COVID-19 response team | National Institute for Public Health and the Environment (RIVM) | Adam Meijer et al |
| EPI_ISL_723386 | Netherlands/LI-RIVM-20237/2020 | 2020-11-30 | Europe/Netherlands/Limburg/ | Dutch COVID-19 response team | National Institute for Public Health and the Environment (RIVM) | Adam Meijer et al |
| EPI_ISL_723411 | Netherlands/NB-RIVM-20268/2020 | 2020-11-17 | Europe/Netherlands/North Brabant/ | Dutch COVID-19 response team | National Institute for Public Health and the Environment (RIVM) | Adam Meijer et al |
| EPI_ISL_723422 | Netherlands/NB-RIVM-20282/2020 | 2020-11-28 | Europe/Netherlands/North Brabant/ | Dutch COVID-19 response team | National Institute for Public Health and the Environment (RIVM) | Adam Meijer et al |
| EPI_ISL_723471 | USA/VA-DCLS-2295/2020 | 2020-10-08 | North America/USA/Virginia/ | Virginia DCLS | Virginia DCLS | Virginia DCLS et al |
| EPI_ISL_723477 | USA/VA-DCLS-2396/2020 | 2020-11-24 | North America/USA/Virginia/ | Virginia DCLS | Virginia DCLS | Virginia DCLS et al |
| EPI_ISL_723480 | USA/VA-DCLS-2414/2020 | 2020-11-23 | North America/USA/Virginia/ | Virginia DCLS | Virginia DCLS | Virginia DCLS et al |
| EPI_ISL_723485 | USA/VA-DCLS-2421/2020 | 2020-11-23 | North America/USA/Virginia/ | Virginia DCLS | Virginia DCLS | Virginia DCLS et al |
| EPI_ISL_723488 | USA/VA-DCLS-2424/2020 | 2020-11-23 | North America/USA/Virginia/ | Virginia DCLS | Virginia DCLS | Virginia DCLS et al |
| EPI_ISL_723495 | USA/VA-DCLS-2192/2020 | 2020-11-12 | North America/USA/Virginia/ | Virginia Division of Consolidated Laboratory Services (DCLS) | Virginia Division of Consolidated Laboratory Services (DCLS) | Virginia DCLS et al |

| | | | | | | |
|---|---|---|---|---|---|---|
| EPI_ISL_723497 | USA/VA-DCLS-2250/2020 | 2020-11-03 | North America/USA/Virginia/ | Virginia Division of Consolidated Laboratory Services (DCLS) | Virginia Division of Consolidated Laboratory Services (DCLS) | Virginia DCLS et al |
| EPI_ISL_723498 | USA/VA-DCLS-2251/2020 | 2020-11-03 | North America/USA/Virginia/ | Virginia Division of Consolidated Laboratory Services (DCLS) | Virginia Division of Consolidated Laboratory Services (DCLS) | Virginia DCLS et al |
| EPI_ISL_723499 | USA/VA-DCLS-2252/2020 | 2020-11-04 | North America/USA/Virginia/ | Virginia Division of Consolidated Laboratory Services (DCLS) | Virginia Division of Consolidated Laboratory Services (DCLS) | Virginia DCLS et al |
| EPI_ISL_723511 | USA/VA-DCLS-2267/2020 | 2020-11-13 | North America/USA/Virginia/ | Virginia Division of Consolidated Laboratory Services (DCLS) | Virginia Division of Consolidated Laboratory Services (DCLS) | Virginia DCLS et al |
| EPI_ISL_723572 | England/ALDP-B5DBF9/2020 | 2020-11-10 | Europe/United Kingdom/England/ | Northumbria University / South Tees Hospitals NHS Foundation Trust / North Cumbria Integrated Care NHS Foundation Trust / North Tees and Hartlepool NHS Foundation Trust / Newcastle Hospitals NHS Foundation Trust | COVID-19 Genomics UK (COG-UK) Consortium | Darren L Smith et al |
| EPI_ISL_723927 | England/CAMB-A97848/2020 | 2020-12-11 | Europe/United Kingdom/England/ | Department of Pathology, University of Cambridge | COVID-19 Genomics UK (COG-UK) Consortium | Aminu S. Jahun et al |
| EPI_ISL_723949 | England/CAMC-B7AC20/2020 | 2020-11-12 | Europe/United Kingdom/England/ | Oxford Viromics, NDM, University of Oxford; Oxford University Hospitals; Basingstoke and North Hampshire Hospital | COVID-19 Genomics UK (COG-UK) Consortium | Tanya Golubchik et al |
| EPI_ISL_723984 | England/CAMC-B7B120/2020 | 2020-11-12 | Europe/United Kingdom/England/ | Oxford Viromics, NDM, University of Oxford; Oxford University Hospitals; Basingstoke and North Hampshire Hospital | COVID-19 Genomics UK (COG-UK) Consortium | Tanya Golubchik et al |
| EPI_ISL_724188 | England/CAMC-BBC92B/2020 | 2020-11-17 | Europe/United Kingdom/England/ | Quadram Institute Bioscience | COVID-19 Genomics UK (COG-UK) Consortium | Dave J. Baker et al |
| EPI_ISL_724294 | England/EXET-13DE3C/2020 | 2020-12-03 | Europe/United Kingdom/England/ | University of Exeter | COVID-19 Genomics UK (COG-UK) Consortium | Ben Temperton et al |
| EPI_ISL_724485 | England/LIVE-DCC06D/2020 | 2020-11-24 | Europe/United Kingdom/England/ | Liverpool Clinical Laboratories | COVID-19 Genomics UK (COG-UK) Consortium | Sam Haldenby et al |

| | | | | | | |
|---|---|---|---|---|---|---|
| EPI_ISL_724501 | England/LIVE-DCC382/2020 | 2020-11-26 | Europe/United Kingdom/England/ | Liverpool Clinical Laboratories | COVID-19 Genomics UK (COG-UK) Consortium | Sam Haldenby et al |
| EPI_ISL_724710 | England/LOND-DADAE/2020 | 2020-11-08 | Europe/United Kingdom/England/ | University College London, Great Ormond Street Hospital for Children NHS Foundation Trust, Imperial College Healthcare NHS Trust | COVID-19 Genomics UK (COG-UK) Consortium | Sergi Castellano et al |
| EPI_ISL_724974 | England/NORT-1B87328/2020 | 2020-10-29 | Europe/United Kingdom/England/ | Northumbria University / South Tees Hospitals NHS Foundation Trust / North Cumbria Integrated Care NHS Foundation Trust / North Tees and Hartlepool NHS Foundation Trust / Newcastle Hospitals NHS Foundation Trust | COVID-19 Genomics UK (COG-UK) Consortium | Darren L Smith et al |
| EPI_ISL_725042 | England/NORW-21F707/2020 | 2020-12-07 | Europe/United Kingdom/England/ | Quadram Institute Bioscience | COVID-19 Genomics UK (COG-UK) Consortium | Dave J. Baker et al |
| EPI_ISL_725093 | England/NORW-220DB6/2020 | 2020-12-12 | Europe/United Kingdom/England/ | Quadram Institute Bioscience | COVID-19 Genomics UK (COG-UK) Consortium | Dave J. Baker et al |
| EPI_ISL_725195 | England/NORW-F4148/2020 | 2020-11-16 | Europe/United Kingdom/England/ | Quadram Institute Bioscience | COVID-19 Genomics UK (COG-UK) Consortium | Dave J. Baker et al |
| EPI_ISL_725210 | England/NORW-F4360/2020 | 2020-11-20 | Europe/United Kingdom/England/ | Quadram Institute Bioscience | COVID-19 Genomics UK (COG-UK) Consortium | Dave J. Baker et al |
| EPI_ISL_725233 | England/NORW-F5226/2020 | 2020-11-17 | Europe/United Kingdom/England/ | Quadram Institute Bioscience | COVID-19 Genomics UK (COG-UK) Consortium | Dave J. Baker et al |
| EPI_ISL_725279 | England/NORW-F596D/2020 | 2020-11-04 | Europe/United Kingdom/England/ | Quadram Institute Bioscience | COVID-19 Genomics UK (COG-UK) Consortium | Dave J. Baker et al |
| EPI_ISL_725285 | England/NORW-F59E5/2020 | 2020-11-13 | Europe/United Kingdom/England/ | Quadram Institute Bioscience | COVID-19 Genomics UK (COG-UK) Consortium | Dave J. Baker et al |
| EPI_ISL_725446 | England/NORW-F9E6A/2020 | 2020-12-09 | Europe/United Kingdom/England/ | Quadram Institute Bioscience | COVID-19 Genomics UK (COG-UK) Consortium | Dave J. Baker et al |
| EPI_ISL_725707 | Wales/PHWC-492834/2020 | 2020-11-29 | Europe/United Kingdom/Wales/ | Wales Specialist Virology Centre Sequencing lab: Pathogen Genomics Unit | COVID-19 Genomics UK (COG-UK) Consortium | Catherine Moore et al |

| | | | | | | |
|---|---|---|---|---|---|---|
| EPI_ISL_726343 | Wales/PHWC-495400/2020 | 2020-11-22 | Europe/United Kingdom/Wales/ | Wales Specialist Virology Centre Sequencing lab: Pathogen Genomics Unit | COVID-19 Genomics UK (COG-UK) Consortium | Catherine Moore et al |
| EPI_ISL_726795 | Wales/PHWC-4971F8/2020 | 2020-11-26 | Europe/United Kingdom/Wales/ | Wales Specialist Virology Centre Sequencing lab: Pathogen Genomics Unit | COVID-19 Genomics UK (COG-UK) Consortium | Catherine Moore et al |
| EPI_ISL_727064 | Wales/PHWC-498397/2020 | 2020-12-01 | Europe/United Kingdom/Wales/ | Wales Specialist Virology Centre Sequencing lab: Pathogen Genomics Unit | COVID-19 Genomics UK (COG-UK) Consortium | Catherine Moore et al |
| EPI_ISL_727256 | Wales/PHWC-4990EA/2020 | 2020-12-03 | Europe/United Kingdom/Wales/ | Wales Specialist Virology Centre Sequencing lab: Pathogen Genomics Unit | COVID-19 Genomics UK (COG-UK) Consortium | Catherine Moore et al |
| EPI_ISL_727269 | Wales/PHWC-4991C9/2020 | 2020-12-02 | Europe/United Kingdom/Wales/ | Wales Specialist Virology Centre Sequencing lab: Pathogen Genomics Unit | COVID-19 Genomics UK (COG-UK) Consortium | Catherine Moore et al |
| EPI_ISL_727462 | Wales/PHWC-499ED1/2020 | 2020-11-24 | Europe/United Kingdom/Wales/ | Wales Specialist Virology Centre Sequencing lab: Pathogen Genomics Unit | COVID-19 Genomics UK (COG-UK) Consortium | Catherine Moore et al |
| EPI_ISL_727549 | Wales/PHWC-49A474/2020 | 2020-12-02 | Europe/United Kingdom/Wales/ | Wales Specialist Virology Centre Sequencing lab: Pathogen Genomics Unit | COVID-19 Genomics UK (COG-UK) Consortium | Catherine Moore et al |
| EPI_ISL_727969 | England/SHEF-10D4503/2020 | 2020-11-26 | Europe/United Kingdom/England/ | Virology Department, Sheffield Teaching Hospitals NHS Foundation Trust/Department of Infection, Immunity and Cardiovascular Disease, The Medical School, University of Sheffield | COVID-19 Genomics UK (COG-UK) Consortium | Thushan de Silva et al |
| EPI_ISL_727998 | Japan/FHU-CS27-0221/2020 | 2020-02-21 | Asia/Japan/Aichi/ | Fujita Health University Okazaki Medical Center | Department of Virology and Parasitology, Fujita Health University School of Medicine | Takayuki Murata et al |
| EPI_ISL_727999 | Japan/FHU-CS29-0221/2020 | 2020-02-21 | Asia/Japan/Aichi/ | Fujita Health University Okazaki Medical Center | Department of Virology and Parasitology, Fujita Health University School of Medicine | Takayuki Murata et al |

| | | | | | | |
|---|---|---|---|---|---|---|
| EPI_ISL_728000 | Japan/FHU-CS56-0221/2020 | 2020-02-21 | Asia/Japan/Aichi/ | Fujita Health University Okazaki Medical Center | Department of Virology and Parasitology, Fujita Health University School of Medicin | Takayuki Murata et al |
| EPI_ISL_728006 | USA/WI-UW-2236/2020 | 2020-11-16 | North America/USA/Wisconsin/Dane Co | University of Wisconsin-Madison AIDS Vaccine Research Laboratories | University of Wisconsin-Madison AIDS Vaccine Research Laboratories | Gage Moreno et al |
| EPI_ISL_728009 | USA/WI-UW-2240/2020 | 2020-11-11 | North America/USA/Wisconsin/Dane Co | University of Wisconsin-Madison AIDS Vaccine Research Laboratories | University of Wisconsin-Madison AIDS Vaccine Research Laboratories | Gage Moreno et al |
| EPI_ISL_728041 | USA/WI-UW-2273/2020 | 2020-10-25 | North America/USA/Wisconsin/Dane Co | University of Wisconsin-Madison AIDS Vaccine Research Laboratories | University of Wisconsin-Madison AIDS Vaccine Research Laboratories | Gage Moreno et al |
| EPI_ISL_728042 | USA/WI-UW-2275/2020 | 2020-11-05 | North America/USA/Wisconsin/Dane Co | University of Wisconsin-Madison AIDS Vaccine Research Laboratories | University of Wisconsin-Madison AIDS Vaccine Research Laboratories | Gage Moreno et al |
| EPI_ISL_728056 | USA/WI-UW-2290/2020 | 2020-11-12 | North America/USA/Wisconsin/Marquet | University of Wisconsin-Madison AIDS Vaccine Research Laboratories | University of Wisconsin-Madison AIDS Vaccine Research Laboratories | Gage Moreno et al |
| EPI_ISL_728072 | USA/WI-UW-2306/2020 | 2020-11-26 | North America/USA/Wisconsin/Dane Co | University of Wisconsin-Madison AIDS Vaccine Research Laboratories | University of Wisconsin-Madison AIDS Vaccine Research Laboratories | Gage Moreno et al |
| EPI_ISL_728074 | USA/WI-UW-2308/2020 | 2020-11-26 | North America/USA/Wisconsin/Dane Co | University of Wisconsin-Madison AIDS Vaccine Research Laboratories | University of Wisconsin-Madison AIDS Vaccine Research Laboratories | Gage Moreno et al |

| | | | | | | |
|---|---|---|---|---|---|---|
| EPI_ISL_728078 | USA/WI-UW-2312/2020 | 2020-11-10 | North America/USA/Wisconsin/Dane Co | University of Wisconsin-Madison AIDS Vaccine Research Laboratories | University of Wisconsin-Madison AIDS Vaccine Research Laboratories | Gage Moreno et al |
| EPI_ISL_728090 | USA/WI-UW-2324/2020 | 2020-11-21 | North America/USA/Wisconsin/Dane Co | University of Wisconsin-Madison AIDS Vaccine Research Laboratories | University of Wisconsin-Madison AIDS Vaccine Research Laboratories | Gage Moreno et al |
| EPI_ISL_728100 | USA/WI-UW-2334/2020 | 2020-11-17 | North America/USA/Wisconsin/Dane Co | University of Wisconsin-Madison AIDS Vaccine Research Laboratories | University of Wisconsin-Madison AIDS Vaccine Research Laboratories | Gage Moreno et al |
| EPI_ISL_728102 | USA/WI-UW-2336/2020 | 2020-11-19 | North America/USA/Wisconsin/Dane Co | University of Wisconsin-Madison AIDS Vaccine Research Laboratories | University of Wisconsin-Madison AIDS Vaccine Research Laboratories | Gage Moreno et al |
| EPI_ISL_728103 | USA/WI-UW-2337/2020 | 2020-11-13 | North America/USA/Wisconsin/Dane Co | University of Wisconsin-Madison AIDS Vaccine Research Laboratories | University of Wisconsin-Madison AIDS Vaccine Research Laboratories | Gage Moreno et al |
| EPI_ISL_728111 | USA/WI-UW-2345/2020 | 2020-11-13 | North America/USA/Wisconsin/Dane Co | University of Wisconsin-Madison AIDS Vaccine Research Laboratories | University of Wisconsin-Madison AIDS Vaccine Research Laboratories | Gage Moreno et al |
| EPI_ISL_728131 | USA/WI-UW-2365/2020 | 2020-11-13 | North America/USA/Wisconsin/Dane Co | University of Wisconsin-Madison AIDS Vaccine Research Laboratories | University of Wisconsin-Madison AIDS Vaccine Research Laboratories | Gage Moreno et al |
| EPI_ISL_728133 | USA/WI-UW-2367/2020 | 2020-11-16 | North America/USA/Wisconsin/Dane Co | University of Wisconsin-Madison AIDS Vaccine Research Laboratories | University of Wisconsin-Madison AIDS Vaccine Research Laboratories | Gage Moreno et al |

| | | | | | | |
|---|---|---|---|---|---|---|
| EPI_ISL_728139 | USA/WI-UW-2373/2020 | 2020-11-02 | North America/USA/Wisconsin/Green C | University of Wisconsin-Madison AIDS Vaccine Research Laboratories | University of Wisconsin-Madison AIDS Vaccine Research Laboratories | Gage Moreno et al |
| EPI_ISL_728144 | USA/WI-UW-2379/2020 | 2020-11-01 | North America/USA/Wisconsin/Dane Co | University of Wisconsin-Madison AIDS Vaccine Research Laboratories | University of Wisconsin-Madison AIDS Vaccine Research Laboratories | Gage Moreno et al |
| EPI_ISL_728147 | USA/WI-UW-2382/2020 | 2020-10-29 | North America/USA/Wisconsin/Dane Co | University of Wisconsin-Madison AIDS Vaccine Research Laboratories | University of Wisconsin-Madison AIDS Vaccine Research Laboratories | Gage Moreno et al |
| EPI_ISL_728152 | USA/WI-UW-2387/2020 | 2020-10-29 | North America/USA/Wisconsin/Dane Co | University of Wisconsin-Madison AIDS Vaccine Research Laboratories | University of Wisconsin-Madison AIDS Vaccine Research Laboratories | Gage Moreno et al |
| EPI_ISL_728154 | Japan/FHU-CS77-0222/2020 | 2020-02-22 | Asia/Japan/Aichi/ | Fujita Health University Okazaki Medical Center | Department of Virology and Parasitology, Fujita Health University School of Medicine | Takayuki Murata et al |
| EPI_ISL_728155 | Japan/FHU-CS84-0222/2020 | 2020-02-22 | Asia/Japan/Aichi/ | Fujita Health University Okazaki Medical Center | Department of Virology and Parasitology, Fujita Health University School of Medicine | Takayuki Murata et al |
| EPI_ISL_728156 | Japan/FHU-CS108-0222/2020 | 2020-02-22 | Asia/Japan/Aichi/ | Fujita Health University Okazaki Medical Center | Department of Virology and Parasitology, Fujita Health University School of Medicine | Takayuki Murata et al |

| | | | | | | |
|---|---|---|---|---|---|---|
| EPI_ISL_728159 | Japan/FHU-CS108-0224/2020 | 2020-02-24 | Asia/Japan/Aichi/ | Fujita Health University Okazaki Medical Center | Department of Virology and Parasitology, Fujita Health University School of Medicine | Takayuki Murata et al |
| EPI_ISL_728160 | Malaysia/IMR-WC194899/2020 | 2020-10-02 | Asia/Malaysia/Malaysia/ | Institute for Medical Research, Infectious Disease Research Centre, National Institutes of Health, Ministry of Health Malaysia | Institute for Medical Research, Infectious Disease Research Centre, National Institutes of Health, Ministry of Health Malaysia | Suppiah J et al |
| EPI_ISL_728178 | Malaysia/IMR-WC257003/2020 | 2020-11-06 | Asia/Malaysia/Malaysia/ | Institute for Medical Research, Infectious Disease Research Centre, National Institutes of Health, Ministry of Health Malaysia | Institute for Medical Research, Infectious Disease Research Centre, National Institutes of Health, Ministry of Health Malaysia | Suppiah J et al |
| EPI_ISL_728206 | Japan/FHU-CS24-0227/2020 | 2020-02-27 | Asia/Japan/Aichi/ | Fujita Health University Okazaki Medical Center | Fujita health University, Department of Microbiology | Takayuki Murata et al |
| EPI_ISL_728207 | Japan/FHU-CS56-0224/2020 | 2020-02-24 | Asia/Japan/Aichi/ | Fujita Health University Okazaki Medical Center | Fujita health University, Department of Microbiology | Takayuki Murata et al |
| EPI_ISL_728208 | Japan/FHU-CS84-0228/2020 | 2020-02-28 | Asia/Japan/Aichi/ | Fujita Health University Okazaki Medical Center | Fujita health University, Department of Microbiology | Takayuki Murata et al |

| | | | | | | |
|---|---|---|---|---|---|---|
| EPI_ISL_728210 | Malaysia/IMR-WC258707/2020 | 2020-11-06 | Asia/Malaysia/Malaysia/ | Institute for Medical Research, Infectious Disease Research Centre, National Institutes of Health, Ministry of Health Malaysia | Institute for Medical Research, Infectious Disease Research Centre, National Institutes of Health, Ministry of Health Malaysia | Suppiah J et al |
| EPI_ISL_728212 | Malaysia/IMR-WC268317/2020 | 2020-11-11 | Asia/Malaysia/Malaysia/ | Institute for Medical Research, Infectious Disease Research Centre, National Institutes of Health, Ministry of Health Malaysia | Institute for Medical Research, Infectious Disease Research Centre, National Institutes of Health, Ministry of Health Malaysia | Suppiah J et al |
| EPI_ISL_728224 | Morocco/RMPS-20/2020 | 2020-04-01 | Africa/Morocco/Morocco/ | LNR National Reference Laboratory, Mohammed VI University of Health Sciences | Medical Biotechnology Laboratory, Rabat Medical and Pharmacy School, Mohammed The Vth University in Rabat | Souad KARTTI et al |
| EPI_ISL_728243 | Malaysia/IMR-WC268619/2020 | 2020-11-11 | Asia/Malaysia/Malaysia/ | Institute for Medical Research, Infectious Disease Research Centre, National Institutes of Health, Ministry of Health Malaysia | Institute for Medical Research, Infectious Disease Research Centre, National Institutes of Health, Ministry of Health Malaysia | Suppiah J et al |

| | | | | | | |
|---|---|---|---|---|---|---|
| EPI_ISL_728250 | Malaysia/IMR-WC277457/2020 | 2020-11-16 | Asia/Malaysia/Malaysia/ | Institute for Medical Research, Infectious Disease Research Centre, National Institutes of Health, Ministry of Health Malaysia | Institute for Medical Research, Infectious Disease Research Centre, National Institutes of Health, Ministry of Health Malaysia | Suppiah J et al |
| EPI_ISL_728251 | Malaysia/IMR-WC280729/2020 | 2020-11-18 | Asia/Malaysia/Malaysia/ | Institute for Medical Research, Infectious Disease Research Centre, National Institutes of Health, Ministry of Health Malaysia | Institute for Medical Research, Infectious Disease Research Centre, National Institutes of Health, Ministry of Health Malaysia | Suppiah J et al |
| EPI_ISL_728252 | Malaysia/IMR-WC280757/2020 | 2020-11-18 | Asia/Malaysia/Malaysia/ | Institute for Medical Research, Infectious Disease Research Centre, National Institutes of Health, Ministry of Health Malaysia | Institute for Medical Research, Infectious Disease Research Centre, National Institutes of Health, Ministry of Health Malaysia | Suppiah J et al |
| EPI_ISL_728255 | India/MH-BJMC-22259/2020 | 2020-07-18 | Asia/India/Maharashtra/ | B.J. Govt. Medical College | National Centre For Cell Science | Dhiraj Paul et al |
| EPI_ISL_728258 | India/MH-BJMC-13585/2020 | 2020-06-29 | Asia/India/Maharashtra/ | B.J. Govt. Medical College | National Centre For Cell Science | Dhiraj Paul et al |
| EPI_ISL_728259 | India/MH-BJMC-8677/2020 | 2020-06-02 | Asia/India/Maharashtra/ | B.J. Govt. Medical College | National Centre For Cell Science | Dhiraj Paul et al |
| EPI_ISL_728260 | India/MH-BJMC-9070/2020 | 2020-06-04 | Asia/India/Maharashtra/ | B.J. Govt. Medical College | National Centre For Cell Science | Dhiraj Paul et al |
| EPI_ISL_728262 | India/MH-BJMC-9065/2020 | 2020-06-04 | Asia/India/Maharashtra/ | B.J. Govt. Medical College | National Centre For Cell Science | Dhiraj Paul et al |
| EPI_ISL_728265 | India/MH-BJMC-13327/2020 | 2020-06-23 | Asia/India/Maharashtra/ | B.J. Govt. Medical College | National Centre For Cell Science | Dhiraj Paul et al |
| EPI_ISL_728267 | India/MH-BJMC-11034/2020 | 2020-06-13 | Asia/India/Maharashtra/ | B.J. Govt. Medical College | National Centre For Cell Science | Dhiraj Paul et al |

| | | | | | | |
|---|---|---|---|---|---|---|
| EPI_ISL_728269 | India/MH-BJMC-22738/2020 | 2020-07-18 | Asia/India/Maharashtra/ | B.J. Govt. Medical College | National Centre For Cell Science | Dhiraj Paul et al |
| EPI_ISL_728270 | India/MH-BJMC-11052/2020 | 2020-06-13 | Asia/India/Maharashtra/ | B.J. Govt. Medical College | National Centre For Cell Science | Dhiraj Paul et al |
| EPI_ISL_728271 | India/MH-BJMC-19749/2020 | 2020-07-11 | Asia/India/Maharashtra/ | B.J. Govt. Medical College | National Centre For Cell Science | Dhiraj Paul et al |
| EPI_ISL_728282 | Italy/LAZ-INMI-109/2020 | 2020-12-01 | Europe/Italy/Lazio/Rome | "National Institute for Infectious Diseases, INMI, ""L. Spallanzani"" IRCCS" | "National Institute for Infectious Diseases, INMI, ""L. Spallanzani"" IRCCS" | M. Rueca et al |
| EPI_ISL_728284 | Italy/LAZ-INMI-111/2020 | 2020-12-01 | Europe/Italy/Lazio/Rome | "National Institute for Infectious Diseases, INMI, ""L. Spallanzani"" IRCCS" | "National Institute for Infectious Diseases, INMI, ""L. Spallanzani"" IRCCS" | M Rueca et al |
| EPI_ISL_728286 | Italy/LAZ-INMI-113/2020 | 2020-11-26 | Europe/Italy/Lazio/Rome | "National Institute for Infectious Diseases, INMI, ""L. Spallanzani"" IRCCS" | "National Institute for Infectious Diseases, INMI, ""L. Spallanzani"" IRCCS" | E Giombini et al |
| EPI_ISL_728287 | Italy/LAZ-INMI-114/2020 | 2020-11-30 | Europe/Italy/Lazio/Rome | "National Institute for Infectious Diseases, INMI, ""L. Spallanzani"" IRCCS" | "National Institute for Infectious Diseases, INMI, ""L. Spallanzani"" IRCCS" | C.E.M Gruber et al |
| EPI_ISL_728293 | Germany/TH-IIMK-CaSe-4/2020 | 2020-04-03 | Europe/Germany/Thuringia/ | Jena University Hospital, Institute for Infectious Diseases and Infection Control | Institute of infectious medicine & hospital hygiene, CaSe-Group | Spott et al |
| EPI_ISL_728304 | France/ARA-105635/2020 | 2020-11-12 | Europe/France/Auvergne-Rh√¥ne-Alpes/ | CNR Virus des Infections Respiratoires - France SUD | CNR Virus des Infections Respiratoires - France SUD | Antonin Bal et al |

| | | | | | | |
|---|---|---|---|---|---|---|
| EPI_ISL_728305 | France/ARA-105681/2020 | 2020-11-12 | Europe/France/Auvergne-Rhv¥ne-Alpes/ | CNR Virus des Infections Respiratoires - France SUD | CNR Virus des Infections Respiratoires - France SUD | Antonin Bal et al |
| EPI_ISL_728307 | France/ARA-105851/2020 | 2020-11-12 | Europe/France/Auvergne-Rhv¥ne-Alpes/ | CNR Virus des Infections Respiratoires - France SUD | CNR Virus des Infections Respiratoires - France SUD | Antonin Bal et al |
| EPI_ISL_728308 | France/ARA-106294/2020 | 2020-11-13 | Europe/France/Auvergne-Rhv¥ne-Alpes/ | CNR Virus des Infections Respiratoires - France SUD | CNR Virus des Infections Respiratoires - France SUD | Antonin Bal et al |
| EPI_ISL_728310 | France/ARA-107339/2020 | 2020-11-15 | Europe/France/Auvergne-Rhv¥ne-Alpes/ | CNR Virus des Infections Respiratoires - France SUD | CNR Virus des Infections Respiratoires - France SUD | Antonin Bal et al |
| EPI_ISL_728311 | France/ARA-107421/2020 | 2020-11-16 | Europe/France/Auvergne-Rhv¥ne-Alpes/ | CNR Virus des Infections Respiratoires - France SUD | CNR Virus des Infections Respiratoires - France SUD | Antonin Bal et al |
| EPI_ISL_728312 | France/ARA-107506/2020 | 2020-11-16 | Europe/France/Auvergne-Rhv¥ne-Alpes/ | CNR Virus des Infections Respiratoires - France SUD | CNR Virus des Infections Respiratoires - France SUD | Antonin Bal et al |
| EPI_ISL_728313 | France/ARA-107560/2020 | 2020-11-16 | Europe/France/Auvergne-Rhv¥ne-Alpes/ | CNR Virus des Infections Respiratoires - France SUD | CNR Virus des Infections Respiratoires - France SUD | Antonin Bal et al |
| EPI_ISL_728315 | France/ARA-107604/2020 | 2020-11-16 | Europe/France/Auvergne-Rhv¥ne-Alpes/ | CNR Virus des Infections Respiratoires - France SUD | CNR Virus des Infections Respiratoires - France SUD | Antonin Bal et al |
| EPI_ISL_728316 | France/ARA-107712/2020 | 2020-11-16 | Europe/France/Auvergne-Rhv¥ne-Alpes/ | CNR Virus des Infections Respiratoires - France SUD | CNR Virus des Infections Respiratoires - France SUD | Antonin Bal et al |
| EPI_ISL_728317 | France/ARA-107994/2020 | 2020-11-17 | Europe/France/Auvergne-Rhv¥ne-Alpes/ | CNR Virus des Infections Respiratoires - France SUD | CNR Virus des Infections Respiratoires - France SUD | Antonin Bal et al |

| | | | | | | |
|---|---|---|---|---|---|---|
| EPI_ISL_728318 | France/ARA-108014/2020 | 2020-11-17 | Europe/France/Auvergne-Rhv¥ne-Alpes/ | CNR Virus des Infections Respiratoires - France SUD | CNR Virus des Infections Respiratoires - France SUD | Antonin Bal et al |
| EPI_ISL_728319 | France/ARA-108039/2020 | 2020-11-17 | Europe/France/Auvergne-Rhv¥ne-Alpes/ | CNR Virus des Infections Respiratoires - France SUD | CNR Virus des Infections Respiratoires - France SUD | Antonin Bal et al |
| EPI_ISL_728320 | France/ARA-108102/2020 | 2020-11-17 | Europe/France/Auvergne-Rhv¥ne-Alpes/ | CNR Virus des Infections Respiratoires - France SUD | CNR Virus des Infections Respiratoires - France SUD | Antonin Bal et al |
| EPI_ISL_728324 | Germany/TH-IIMK-CaSe-12/2020 | 2020-11-30 | Europe/Germany/Thuringia/ | Jena University Hospital, Institute for Infectious Diseases and Infection Control | Institute of infectious medicine & hospital hygiene, CaSe-Group | Spott et al |
| EPI_ISL_728325 | India/MH-BJMC-8674/2020 | 2020-06-02 | Asia/India/Maharashtra/ | B.J. Govt. Medical College | National Centre For Cell Science | Dhiraj Paul et al |
| EPI_ISL_728327 | India/MH-BJMC-21565/2020 | 2020-07-16 | Asia/India/Maharashtra/ | B.J. Govt. Medical College | National Centre For Cell Science | Dhiraj Paul et al |
| EPI_ISL_728328 | India/MH-BJMC-8827/2020 | 2020-06-03 | Asia/India/Maharashtra/ | B.J. Govt. Medical College | National Centre For Cell Science | Dhiraj Paul et al |
| EPI_ISL_728330 | India/MH-BJMC-6666/2020 | 2020-06-21 | Asia/India/Maharashtra/ | B.J. Govt. Medical College | National Centre For Cell Science | Dhiraj Paul et al |
| EPI_ISL_728331 | Germany/TH-IIMK-CaSe-13/2020 | 2020-12-01 | Europe/Germany/Thuringia/ | Jena University Hospital, Institute for Infectious Diseases and Infection Control | Institute of infectious medicine & hospital hygiene, CaSe-Group | Spott et al |
| EPI_ISL_728335 | Germany/TH-IIMK-CaSe-15/2020 | 2020-12-03 | Europe/Germany/Thuringia/ | Jena University Hospital, Institute for Infectious Diseases and Infection Control | Institute of infectious medicine & hospital hygiene, CaSe-Group | Spott et al |
| EPI_ISL_728337 | Germany/TH-IIMK-CaSe-16/2020 | 2020-12-06 | Europe/Germany/Thuringia/ | Jena University Hospital, Institute for Infectious Diseases and Infection Control | Institute of infectious medicine & hospital hygiene, CaSe-Group | Spott et al |

| | | | | | | |
|---|---|---|---|---|---|---|
| EPI_ISL_728338 | Germany/TH-IIMK-CaSe-17/2020 | 2020-12-06 | Europe/Germany/Thuringia/ | Jena University Hospital, Institute for Infectious Diseases and Infection Control | Institute of infectious medicine & hospital hygiene, CaSe-Group | Spott et al |
| EPI_ISL_728362 | Spain/CT-HUVH-GEST1-3/2020 | 2020-04-21 | Europe/Spain/Catalunya/ | Hospital Universitari Vall d'Hebron (HUVH) - Vall d'Hebron Research Institute (VHIR) | Hospital Universitari Vall d'Hebron - Vall d'Hebron Research Institute (VHIR) | Maria Pi√±ana et al |
| EPI_ISL_728364 | Spain/CT-HUVH-GEST1-2/2020 | 2020-04-21 | Europe/Spain/Catalunya/ | Hospital Universitari Vall d'Hebron (HUVH) - Vall d'Hebron Research Institute (VHIR) | Hospital Universitari Vall d'Hebron (HUVH) - Vall d'Hebron Research Institute (VHIR) | Maria Pi√±ana et al |
| EPI_ISL_728372 | USA/MI-UM-10036774731/2020 | 2020-11-04 | North America/USA/Michigan/ | University of Michigan Clinical Microbiology Laboratory | Lauring Lab, University of Michigan, Department of Microbiology and Immunology | Valesano et al |
| EPI_ISL_728378 | USA/MI-UM-10036821132/2020 | 2020-11-06 | North America/USA/Michigan/ | University of Michigan Clinical Microbiology Laboratory | Lauring Lab, University of Michigan, Department of Microbiology and Immunology | Valesano et al |
| EPI_ISL_728381 | USA/MI-UM-10036802807/2020 | 2020-11-05 | North America/USA/Michigan/ | University of Michigan Clinical Microbiology Laboratory | Lauring Lab, University of Michigan, Department of Microbiology and Immunology | Valesano et al |
| EPI_ISL_728396 | USA/MI-UM-10036819118/2020 | 2020-11-06 | North America/USA/Michigan/ | University of Michigan Clinical Microbiology Laboratory | Lauring Lab, University of Michigan, Department of Microbiology and Immunology | Valesano et al |

| | | | | | | |
|---|---|---|---|---|---|---|
| EPI_ISL_728399 | USA/MI-UM-10036821294/2020 | 2020-11-06 | North America/USA/Michigan/ | University of Michigan Clinical Microbiology Laboratory | Lauring Lab, University of Michigan, Department of Microbiology and Immunology | Valesano et al |
| EPI_ISL_728408 | USA/MI-UM-10036823312/2020 | 2020-11-06 | North America/USA/Michigan/ | University of Michigan Clinical Microbiology Laboratory | Lauring Lab, University of Michigan, Department of Microbiology and Immunology | Valesano et al |
| EPI_ISL_728409 | USA/MI-UM-10036820004/2020 | 2020-11-06 | North America/USA/Michigan/ | University of Michigan Clinical Microbiology Laboratory | Lauring Lab, University of Michigan, Department of Microbiology and Immunology | Valesano et al |
| EPI_ISL_728416 | USA/MI-UM-10036814157/2020 | 2020-11-06 | North America/USA/Michigan/ | University of Michigan Clinical Microbiology Laboratory | Lauring Lab, University of Michigan, Department of Microbiology and Immunology | Valesano et al |
| EPI_ISL_728440 | USA/MI-UM-10036833206/2020 | 2020-11-07 | North America/USA/Michigan/ | University of Michigan Clinical Microbiology Laboratory | Lauring Lab, University of Michigan, Department of Microbiology and Immunology | Valesano et al |
| EPI_ISL_728447 | USA/MI-UM-10036850527/2020 | 2020-11-09 | North America/USA/Michigan/ | University of Michigan Clinical Microbiology Laboratory | Lauring Lab, University of Michigan, Department of Microbiology and Immunology | Valesano et al |

| | | | | | | |
|---|---|---|---|---|---|---|
| EPI_ISL_728450 | USA/MI-UM-10036850459/2020 | 2020-11-09 | North America/USA/Michigan/ | University of Michigan Clinical Microbiology Laboratory | Lauring Lab, University of Michigan, Department of Microbiology and Immunology | Valesano et al |
| EPI_ISL_728451 | USA/MI-UM-10036847408/2020 | 2020-11-09 | North America/USA/Michigan/ | University of Michigan Clinical Microbiology Laboratory | Lauring Lab, University of Michigan, Department of Microbiology and Immunology | Valesano et al |
| EPI_ISL_728461 | USA/MI-UM-10036835773/2020 | 2020-11-07 | North America/USA/Michigan/ | University of Michigan Clinical Microbiology Laboratory | Lauring Lab, University of Michigan, Department of Microbiology and Immunology | Valesano et al |
| EPI_ISL_728462 | USA/MI-UM-10036832225/2020 | 2020-11-07 | North America/USA/Michigan/ | University of Michigan Clinical Microbiology Laboratory | Lauring Lab, University of Michigan, Department of Microbiology and Immunology | Valesano et al |
| EPI_ISL_728466 | USA/MI-UM-10036835677/2020 | 2020-11-07 | North America/USA/Michigan/ | University of Michigan Clinical Microbiology Laboratory | Lauring Lab, University of Michigan, Department of Microbiology and Immunology | Valesano et al |
| EPI_ISL_728471 | USA/MI-UM-10036835605/2020 | 2020-11-07 | North America/USA/Michigan/ | University of Michigan Clinical Microbiology Laboratory | Lauring Lab, University of Michigan, Department of Microbiology and Immunology | Valesano et al |

| | | | | | | |
|---|---|---|---|---|---|---|
| EPI_ISL_728472 | USA/MI-UM-10036834187/2020 | 2020-11-07 | North America/USA/Michigan/ | University of Michigan Clinical Microbiology Laboratory | Lauring Lab, University of Michigan, Department of Microbiology and Immunology | Valesano et al |
| EPI_ISL_728490 | USA/MI-UM-10036857965/2020 | 2020-11-09 | North America/USA/Michigan/ | University of Michigan Clinical Microbiology Laboratory | Lauring Lab, University of Michigan, Department of Microbiology and Immunology | Valesano et al |
| EPI_ISL_728500 | USA/MI-UM-10036858924/2020 | 2020-11-09 | North America/USA/Michigan/ | University of Michigan Clinical Microbiology Laboratory | Lauring Lab, University of Michigan, Department of Microbiology and Immunology | Valesano et al |
| EPI_ISL_728501 | USA/MI-UM-10036850953/2020 | 2020-11-09 | North America/USA/Michigan/ | University of Michigan Clinical Microbiology Laboratory | Lauring Lab, University of Michigan, Department of Microbiology and Immunology | Valesano et al |
| EPI_ISL_728514 | USA/MI-UM-10036861170/2020 | 2020-11-09 | North America/USA/Michigan/ | University of Michigan Clinical Microbiology Laboratory | Lauring Lab, University of Michigan, Department of Microbiology and Immunology | Valesano et al |
| EPI_ISL_728516 | France/ARA-107987/2020 | 2020-11-17 | Europe/France/Auvergne-Rhv¥ne-Alpes/ | CNR Virus des Infections Respiratoires - France SUD | CNR Virus des Infections Respiratoires - France SUD | Antonin Bal et al |
| EPI_ISL_728518 | France/ARA-108484/2020 | 2020-11-17 | Europe/France/Auvergne-Rhv¥ne-Alpes/ | CNR Virus des Infections Respiratoires - France SUD | CNR Virus des Infections Respiratoires - France SUD | Antonin Bal et al |

| | | | | | | |
|---|---|---|---|---|---|---|
| EPI_ISL_728520 | France/ARA-108658/2020 | 2020-11-18 | Europe/France/Auvergne-Rh√¥ne-Alpes/ | CNR Virus des Infections Respiratoires - France SUD | CNR Virus des Infections Respiratoires - France SUD | Antonin Bal et al |
| EPI_ISL_728521 | France/ARA-108709/2020 | 2020-11-18 | Europe/France/Auvergne-Rh√¥ne-Alpes/ | CNR Virus des Infections Respiratoires - France SUD | CNR Virus des Infections Respiratoires - France SUD | Antonin Bal et al |
| EPI_ISL_728522 | France/ARA-108748/2020 | 2020-11-18 | Europe/France/Auvergne-Rh√¥ne-Alpes/ | CNR Virus des Infections Respiratoires - France SUD | CNR Virus des Infections Respiratoires - France SUD | Antonin Bal et al |
| EPI_ISL_728523 | France/ARA-108867/2020 | 2020-11-18 | Europe/France/Auvergne-Rh√¥ne-Alpes/ | CNR Virus des Infections Respiratoires - France SUD | CNR Virus des Infections Respiratoires - France SUD | Antonin Bal et al |
| EPI_ISL_728527 | France/ARA-109213/2020 | 2020-11-19 | Europe/France/Auvergne-Rh√¥ne-Alpes/ | CNR Virus des Infections Respiratoires - France SUD | CNR Virus des Infections Respiratoires - France SUD | Antonin Bal et al |
| EPI_ISL_728528 | France/ARA-109322/2020 | 2020-11-19 | Europe/France/Auvergne-Rh√¥ne-Alpes/ | CNR Virus des Infections Respiratoires - France SUD | CNR Virus des Infections Respiratoires - France SUD | Antonin Bal et al |
| EPI_ISL_728530 | France/ARA-109346/2020 | 2020-11-19 | Europe/France/Auvergne-Rh√¥ne-Alpes/ | CNR Virus des Infections Respiratoires - France SUD | CNR Virus des Infections Respiratoires - France SUD | Antonin Bal et al |
| EPI_ISL_728531 | France/ARA-109382/2020 | 2020-11-19 | Europe/France/Auvergne-Rh√¥ne-Alpes/ | CNR Virus des Infections Respiratoires - France SUD | CNR Virus des Infections Respiratoires - France SUD | Antonin Bal et al |
| EPI_ISL_728532 | France/ARA-109528/2020 | 2020-11-19 | Europe/France/Auvergne-Rh√¥ne-Alpes/ | CNR Virus des Infections Respiratoires - France SUD | CNR Virus des Infections Respiratoires - France SUD | Antonin Bal et al |
| EPI_ISL_728533 | France/ARA-109714/2020 | 2020-11-19 | Europe/France/Auvergne-Rh√¥ne-Alpes/ | CNR Virus des Infections Respiratoires - France SUD | CNR Virus des Infections Respiratoires - France SUD | Antonin Bal et al |

| | | | | | | |
|---|---|---|---|---|---|---|
| EPI_ISL_728534 | France/ARA-109965/2020 | 2020-11-20 | Europe/France/Auvergne-Rh√¥ne-Alpes/ | CNR Virus des Infections Respiratoires - France SUD | CNR Virus des Infections Respiratoires - France SUD | Antonin Bal et al |
| EPI_ISL_728535 | France/ARA-109970/2020 | 2020-11-20 | Europe/France/Auvergne-Rh√¥ne-Alpes/ | CNR Virus des Infections Respiratoires - France SUD | CNR Virus des Infections Respiratoires - France SUD | Antonin Bal et al |
| EPI_ISL_728537 | France/ARA-110022/2020 | 2020-11-20 | Europe/France/Auvergne-Rh√¥ne-Alpes/ | CNR Virus des Infections Respiratoires - France SUD | CNR Virus des Infections Respiratoires - France SUD | Antonin Bal et al |
| EPI_ISL_728539 | France/ARA-110538/2020 | 2020-11-21 | Europe/France/Auvergne-Rh√¥ne-Alpes/ | CNR Virus des Infections Respiratoires - France SUD | CNR Virus des Infections Respiratoires - France SUD | Antonin Bal et al |
| EPI_ISL_728540 | France/ARA-110872/2020 | 2020-11-23 | Europe/France/Auvergne-Rh√¥ne-Alpes/ | CNR Virus des Infections Respiratoires - France SUD | CNR Virus des Infections Respiratoires - France SUD | Antonin Bal et al |
| EPI_ISL_728542 | France/ARA-110995/2020 | 2020-11-23 | Europe/France/Auvergne-Rh√¥ne-Alpes/ | CNR Virus des Infections Respiratoires - France SUD | CNR Virus des Infections Respiratoires - France SUD | Antonin Bal et al |
| EPI_ISL_728543 | France/ARA-111014/2020 | 2020-11-23 | Europe/France/Auvergne-Rh√¥ne-Alpes/ | CNR Virus des Infections Respiratoires - France SUD | CNR Virus des Infections Respiratoires - France SUD | Antonin Bal et al |
| EPI_ISL_728545 | France/ARA-111219/2020 | 2020-11-23 | Europe/France/Auvergne-Rh√¥ne-Alpes/ | CNR Virus des Infections Respiratoires - France SUD | CNR Virus des Infections Respiratoires - France SUD | Antonin Bal et al |
| EPI_ISL_728547 | France/ARA-111501/2020 | 2020-11-24 | Europe/France/Auvergne-Rh√¥ne-Alpes/ | CNR Virus des Infections Respiratoires - France SUD | CNR Virus des Infections Respiratoires - France SUD | Antonin Bal et al |
| EPI_ISL_728550 | France/ARA-111638/2020 | 2020-11-24 | Europe/France/Auvergne-Rh√¥ne-Alpes/ | CNR Virus des Infections Respiratoires - France SUD | CNR Virus des Infections Respiratoires - France SUD | Antonin Bal et al |

| | | | | | | |
|---|---|---|---|---|---|---|
| EPI_ISL_728551 | France/ARA-111718/2020 | 2020-11-24 | Europe/France/Auvergne-Rhv¥ne-Alpes/ | CNR Virus des Infections Respiratoires - France SUD | CNR Virus des Infections Respiratoires - France SUD | Antonin Bal et al |
| EPI_ISL_728600 | Netherlands/UT-RIVM-20380/2020 | 2020-12-07 | Europe/Netherlands/Utrecht/ | Dutch COVID-19 response team | National Institute for Public Health and the Environment (RIVM) | Adam Meijer et al |
| EPI_ISL_728643 | Netherlands/DR-RIVM-20423/2020 | 2020-11-30 | Europe/Netherlands/Drenthe/ | Dutch COVID-19 response team | National Institute for Public Health and the Environment (RIVM) | Adam Meijer et al |
| EPI_ISL_728647 | Netherlands/GR-RIVM-20427/2020 | 2020-12-01 | Europe/Netherlands/Groningen/ | Dutch COVID-19 response team | National Institute for Public Health and the Environment (RIVM) | Adam Meijer et al |
| EPI_ISL_728655 | Netherlands/NH-RIVM-20436/2020 | 2020-12-09 | Europe/Netherlands/North Holland/ | Dutch COVID-19 response team | National Institute for Public Health and the Environment (RIVM) | Adam Meijer et al |
| EPI_ISL_728689 | Netherlands/GE-RIVM-20474/2020 | 2020-12-03 | Europe/Netherlands/Gelderland/ | Dutch COVID-19 response team | National Institute for Public Health and the Environment (RIVM) | Adam Meijer et al |
| EPI_ISL_728704 | Netherlands/ZH-RIVM-20489/2020 | 2020-10-26 | Europe/Netherlands/South Holland/ | Dutch COVID-19 response team | National Institute for Public Health and the Environment (RIVM) | Adam Meijer et al |
| EPI_ISL_728705 | Netherlands/ZH-RIVM-20490/2020 | 2020-10-26 | Europe/Netherlands/South Holland/ | Dutch COVID-19 response team | National Institute for Public Health and the Environment (RIVM) | Adam Meijer et al |

| | | | | | | |
|---|---|---|---|---|---|---|
| EPI_ISL_728710 | Netherlands/OV-RIVM-20495/2020 | 2020-11-21 | Europe/Netherlands/Overijssel/ | Dutch COVID-19 response team | National Institute for Public Health and the Environment (RIVM) | Adam Meijer et al |
| EPI_ISL_728728 | Netherlands/OV-RIVM-20513/2020 | 2020-11-11 | Europe/Netherlands/Overijssel/ | Dutch COVID-19 response team | National Institute for Public Health and the Environment (RIVM) | Adam Meijer et al |
| EPI_ISL_728733 | Netherlands/OV-RIVM-20518/2020 | 2020-11-27 | Europe/Netherlands/Overijssel/ | Dutch COVID-19 response team | National Institute for Public Health and the Environment (RIVM) | Adam Meijer et al |
| EPI_ISL_728746 | Netherlands/NH-RIVM-20531/2020 | 2020-12-08 | Europe/Netherlands/North Holland/ | Dutch COVID-19 response team | National Institute for Public Health and the Environment (RIVM) | Adam Meijer et al |
| EPI_ISL_728756 | Netherlands/OV-RIVM-20541/2020 | 2020-10-06 | Europe/Netherlands/Overijssel/ | Dutch COVID-19 response team | National Institute for Public Health and the Environment (RIVM) | Adam Meijer et al |
| EPI_ISL_728781 | Switzerland/ZH-ETHZ-350339/2020 | 2020-11-03 | Europe/Switzerland/Zv∫rich/ | Viollier AG | Department of Biosystems Science and Engineering, ETH Zv∫rich | Chaoran Chen et al |
| EPI_ISL_728795 | Switzerland/BL-ETHZ-390075/2020 | 2020-11-29 | Europe/Switzerland/Basel-Land/ | Viollier AG | Department of Biosystems Science and Engineering, ETH Zv∫rich | Chaoran Chen et al |
| EPI_ISL_728799 | Switzerland/NE-ETHZ-350425/2020 | 2020-11-04 | Europe/Switzerland/Neuchv¢tel/ | Viollier AG | Department of Biosystems Science and Engineering, ETH Zv∫rich | Chaoran Chen et al |

| | | | | | | |
|---|---|---|---|---|---|---|
| EPI_ISL_728805 | Switzerland/ZH-ETHZ-350275/2020 | 2020-10-30 | Europe/Switzerland/Zvºrich/ | Viollier AG | Department of Biosystems Science and Engineering, ETH Zvºrich | Chaoran Chen et al |
| EPI_ISL_728810 | Switzerland/NE-ETHZ-350455/2020 | 2020-11-03 | Europe/Switzerland/Neuchv¢tel/ | Viollier AG | Department of Biosystems Science and Engineering, ETH Zvºrich | Chaoran Chen et al |
| EPI_ISL_728838 | Switzerland/BE-ETHZ-360293/2020 | 2020-11-06 | Europe/Switzerland/Bern/ | Viollier AG | Department of Biosystems Science and Engineering, ETH Zvºrich | Chaoran Chen et al |
| EPI_ISL_728844 | Switzerland/BL-ETHZ-350326/2020 | 2020-11-03 | Europe/Switzerland/Basel-Land/ | Viollier AG | Department of Biosystems Science and Engineering, ETH Zvºrich | Chaoran Chen et al |
| EPI_ISL_728857 | Switzerland/BE-ETHZ-360292/2020 | 2020-11-06 | Europe/Switzerland/Bern/ | Viollier AG | Department of Biosystems Science and Engineering, ETH Zvºrich | Chaoran Chen et al |
| EPI_ISL_728915 | Switzerland/GR-ETHZ-350337/2020 | 2020-11-03 | Europe/Switzerland/Graubvºnden/ | Viollier AG | Department of Biosystems Science and Engineering, ETH Zvºrich | Chaoran Chen et al |
| EPI_ISL_729069 | Switzerland/ZH-ETHZ-390422/2020 | 2020-11-04 | Europe/Switzerland/Zvºrich/ | Viollier AG | Department of Biosystems Science and Engineering, ETH Zvºrich | Christian Beisel et al |
| EPI_ISL_729123 | Switzerland/BE-ETHZ-400071/2020 | 2020-12-06 | Europe/Switzerland/Bern/ | Viollier AG | Department of Biosystems Science and Engineering, ETH Zvºrich | Chaoran Chen et al |

| | | | | | | |
|---|---|---|---|---|---|---|
| EPI_ISL_729125 | Switzerland/BE-ETHZ-400074/2020 | 2020-12-06 | Europe/Switzerland/Bern/ | Viollier AG | Department of Biosystems Science and Engineering, ETH Zvºrich | Chaoran Chen et al |
| EPI_ISL_729179 | Switzerland/BE-ETHZ-400349/2020 | 2020-12-08 | Europe/Switzerland/Bern/ | Viollier AG | Department of Biosystems Science and Engineering, ETH Zvºrich | Christian Beisel et al |
| EPI_ISL_729196 | Switzerland/BE-ETHZ-350336/2020 | 2020-11-03 | Europe/Switzerland/Bern/ | Viollier AG | Department of Biosystems Science and Engineering, ETH Zvºrich | Chaoran Chen et al |
| EPI_ISL_729197 | Switzerland/BL-ETHZ-350342/2020 | 2020-11-02 | Europe/Switzerland/Basel-Land/ | Viollier AG | Department of Biosystems Science and Engineering, ETH Zvºrich | Chaoran Chen et al |
| EPI_ISL_729198 | Switzerland/AG-ETHZ-350351/2020 | 2020-11-02 | Europe/Switzerland/Aargau/ | Viollier AG | Department of Biosystems Science and Engineering, ETH Zvºrich | Chaoran Chen et al |
| EPI_ISL_729205 | Switzerland/NE-ETHZ-350457/2020 | 2020-11-03 | Europe/Switzerland/Neuchv¢tel/ | Viollier AG | Department of Biosystems Science and Engineering, ETH Zvºrich | Chaoran Chen et al |
| EPI_ISL_729208 | Switzerland/NE-ETHZ-350463/2020 | 2020-11-03 | Europe/Switzerland/Neuchv¢tel/ | Viollier AG | Department of Biosystems Science and Engineering, ETH Zvºrich | Chaoran Chen et al |
| EPI_ISL_729211 | Switzerland/BE-ETHZ-360343/2020 | 2020-11-06 | Europe/Switzerland/Bern/ | Viollier AG | Department of Biosystems Science and Engineering, ETH Zvºrich | Chaoran Chen et al |

| | | | | | | |
|---|---|---|---|---|---|---|
| EPI_ISL_729255 | Switzerland/SO-ETHZ-360209/2020 | 2020-11-09 | Europe/Switzerland/Solothurn/ | Viollier AG | Department of Biosystems Science and Engineering, ETH Z√ºrich | Chaoran Chen et al |
| EPI_ISL_729258 | Switzerland/NE-ETHZ-350441/2020 | 2020-11-04 | Europe/Switzerland/Neuch√¢tel/ | Viollier AG | Department of Biosystems Science and Engineering, ETH Z√ºrich | Chaoran Chen et al |
| EPI_ISL_729259 | Canada/ON-S1841/2020 | 2020-11-24 | North America/Canada/Ontario/Toronto | Toronto Invasive Bacterial Diseases Network | McMaster University | Allison McGeer et al |
| EPI_ISL_729260 | Canada/ON-S1798/2020 | 2020-11-24 | North America/Canada/Ontario/Toronto | Toronto Invasive Bacterial Diseases Network | McMaster University | Allison McGeer et al |
| EPI_ISL_729261 | Canada/ON-S1843/2020 | 2020-11-24 | North America/Canada/Ontario/Toronto | Toronto Invasive Bacterial Diseases Network | McMaster University | Allison McGeer et al |
| EPI_ISL_729262 | Canada/ON-S1853/2020 | 2020-11-24 | North America/Canada/Ontario/Toronto | Toronto Invasive Bacterial Diseases Network | McMaster University | Allison McGeer et al |
| EPI_ISL_729263 | Canada/ON-S1808/2020 | 2020-11-24 | North America/Canada/Ontario/Toronto | Toronto Invasive Bacterial Diseases Network | McMaster University | Allison McGeer et al |
| EPI_ISL_729264 | Canada/ON-S1849/2020 | 2020-11-24 | North America/Canada/Ontario/Toronto | Toronto Invasive Bacterial Diseases Network | McMaster University | Allison McGeer et al |
| EPI_ISL_729265 | Canada/ON-S1913/2020 | 2020-11-24 | North America/Canada/Ontario/Toronto | Toronto Invasive Bacterial Diseases Network | McMaster University | Allison McGeer et al |
| EPI_ISL_729266 | Canada/ON-S1923/2020 | 2020-11-24 | North America/Canada/Ontario/Toronto | Toronto Invasive Bacterial Diseases Network | McMaster University | Allison McGeer et al |
| EPI_ISL_729267 | Canada/ON-S1894/2020 | 2020-11-24 | North America/Canada/Ontario/Toronto | Toronto Invasive Bacterial Diseases Network | McMaster University | Allison McGeer et al |
| EPI_ISL_729268 | Canada/ON-S1814/2020 | 2020-11-24 | North America/Canada/Ontario/Toronto | Toronto Invasive Bacterial Diseases Network | McMaster University | Allison McGeer et al |
| EPI_ISL_729269 | Canada/ON-S1924/2020 | 2020-11-24 | North America/Canada/Ontario/Toronto | Toronto Invasive Bacterial Diseases Network | McMaster University | Allison McGeer et al |
| EPI_ISL_729270 | Canada/ON-S1909/2020 | 2020-11-24 | North America/Canada/Ontario/Toronto | Toronto Invasive Bacterial Diseases Network | McMaster University | Allison McGeer et al |
| EPI_ISL_729271 | Canada/ON-S1799/2020 | 2020-11-24 | North America/Canada/Ontario/Toronto | Toronto Invasive Bacterial Diseases Network | McMaster University | Allison McGeer et al |
| EPI_ISL_729272 | Canada/ON-S1800/2020 | 2020-11-24 | North America/Canada/Ontario/Toronto | Toronto Invasive Bacterial Diseases Network | McMaster University | Allison McGeer et al |
| EPI_ISL_729273 | Canada/ON-S1801/2020 | 2020-11-24 | North America/Canada/Ontario/Toronto | Toronto Invasive Bacterial Diseases Network | McMaster University | Allison McGeer et al |

| | | | | | | |
|---|---|---|---|---|---|---|
| EPI_ISL_729274 | Canada/ON-S1802/2020 | 2020-11-24 | North America/Canada/Ontario/Toronto | Toronto Invasive Bacterial Diseases Network | McMaster University | Allison McGeer et al |
| EPI_ISL_729275 | Canada/ON-S1803/2020 | 2020-11-24 | North America/Canada/Ontario/Toronto | Toronto Invasive Bacterial Diseases Network | McMaster University | Allison McGeer et al |
| EPI_ISL_729276 | Canada/ON-S1804/2020 | 2020-11-24 | North America/Canada/Ontario/Toronto | Toronto Invasive Bacterial Diseases Network | McMaster University | Allison McGeer et al |
| EPI_ISL_729277 | Canada/ON-S1805/2020 | 2020-11-24 | North America/Canada/Ontario/Toronto | Toronto Invasive Bacterial Diseases Network | McMaster University | Allison McGeer et al |
| EPI_ISL_729278 | Canada/ON-S1806/2020 | 2020-11-24 | North America/Canada/Ontario/Toronto | Toronto Invasive Bacterial Diseases Network | McMaster University | Allison McGeer et al |
| EPI_ISL_729279 | Canada/ON-S1807/2020 | 2020-11-24 | North America/Canada/Ontario/Toronto | Toronto Invasive Bacterial Diseases Network | McMaster University | Allison McGeer et al |
| EPI_ISL_729280 | Canada/ON-S1811/2020 | 2020-11-24 | North America/Canada/Ontario/Toronto | Toronto Invasive Bacterial Diseases Network | McMaster University | Allison McGeer et al |
| EPI_ISL_729281 | Canada/ON-S1812/2020 | 2020-11-24 | North America/Canada/Ontario/Toronto | Toronto Invasive Bacterial Diseases Network | McMaster University | Allison McGeer et al |
| EPI_ISL_729282 | Canada/ON-S1813/2020 | 2020-11-24 | North America/Canada/Ontario/Toronto | Toronto Invasive Bacterial Diseases Network | McMaster University | Allison McGeer et al |
| EPI_ISL_729283 | Canada/ON-S1815/2020 | 2020-11-24 | North America/Canada/Ontario/Toronto | Toronto Invasive Bacterial Diseases Network | McMaster University | Allison McGeer et al |
| EPI_ISL_729284 | Canada/ON-S1820/2020 | 2020-11-24 | North America/Canada/Ontario/Toronto | Toronto Invasive Bacterial Diseases Network | McMaster University | Allison McGeer et al |
| EPI_ISL_729285 | Canada/ON-S1821/2020 | 2020-11-24 | North America/Canada/Ontario/Toronto | Toronto Invasive Bacterial Diseases Network | McMaster University | Allison McGeer et al |
| EPI_ISL_729286 | Canada/ON-S1822/2020 | 2020-11-24 | North America/Canada/Ontario/Toronto | Toronto Invasive Bacterial Diseases Network | McMaster University | Allison McGeer et al |
| EPI_ISL_729287 | Canada/ON-S1823/2020 | 2020-11-24 | North America/Canada/Ontario/Toronto | Toronto Invasive Bacterial Diseases Network | McMaster University | Allison McGeer et al |
| EPI_ISL_729288 | Canada/ON-S1824/2020 | 2020-11-24 | North America/Canada/Ontario/Toronto | Toronto Invasive Bacterial Diseases Network | McMaster University | Allison McGeer et al |
| EPI_ISL_729289 | Canada/ON-S1825/2020 | 2020-11-24 | North America/Canada/Ontario/Toronto | Toronto Invasive Bacterial Diseases Network | McMaster University | Allison McGeer et al |
| EPI_ISL_729290 | Canada/ON-S1826/2020 | 2020-11-24 | North America/Canada/Ontario/Toronto | Toronto Invasive Bacterial Diseases Network | McMaster University | Allison McGeer et al |
| EPI_ISL_729291 | Canada/ON-S1827/2020 | 2020-11-24 | North America/Canada/Ontario/Toronto | Toronto Invasive Bacterial Diseases Network | McMaster University | Allison McGeer et al |
| EPI_ISL_729292 | Canada/ON-S1828/2020 | 2020-11-24 | North America/Canada/Ontario/Toronto | Toronto Invasive Bacterial Diseases Network | McMaster University | Allison McGeer et al |
| EPI_ISL_729293 | Canada/ON-S1829/2020 | 2020-11-24 | North America/Canada/Ontario/Toronto | Toronto Invasive Bacterial Diseases Network | McMaster University | Allison McGeer et al |

| | | | | | | |
|---|---|---|---|---|---|---|
| EPI_ISL_729294 | Canada/ON-S1830/2020 | 2020-11-24 | North America/Canada/Ontario/Toronto | Toronto Invasive Bacterial Diseases Network | McMaster University | Allison McGeer et al |
| EPI_ISL_729295 | Canada/ON-S1831/2020 | 2020-11-24 | North America/Canada/Ontario/Toronto | Toronto Invasive Bacterial Diseases Network | McMaster University | Allison McGeer et al |
| EPI_ISL_729296 | Canada/ON-S1832/2020 | 2020-11-24 | North America/Canada/Ontario/Toronto | Toronto Invasive Bacterial Diseases Network | McMaster University | Allison McGeer et al |
| EPI_ISL_729297 | Canada/ON-S1833/2020 | 2020-11-24 | North America/Canada/Ontario/Toronto | Toronto Invasive Bacterial Diseases Network | McMaster University | Allison McGeer et al |
| EPI_ISL_729298 | Canada/ON-S1834/2020 | 2020-11-24 | North America/Canada/Ontario/Toronto | Toronto Invasive Bacterial Diseases Network | McMaster University | Allison McGeer et al |
| EPI_ISL_729299 | Canada/ON-S1835/2020 | 2020-11-24 | North America/Canada/Ontario/Toronto | Toronto Invasive Bacterial Diseases Network | McMaster University | Allison McGeer et al |
| EPI_ISL_729300 | Canada/ON-S1836/2020 | 2020-11-24 | North America/Canada/Ontario/Toronto | Toronto Invasive Bacterial Diseases Network | McMaster University | Allison McGeer et al |
| EPI_ISL_729301 | Canada/ON-S1837/2020 | 2020-11-24 | North America/Canada/Ontario/Toronto | Toronto Invasive Bacterial Diseases Network | McMaster University | Allison McGeer et al |
| EPI_ISL_729302 | Canada/ON-S1838/2020 | 2020-11-24 | North America/Canada/Ontario/Toronto | Toronto Invasive Bacterial Diseases Network | McMaster University | Allison McGeer et al |
| EPI_ISL_729303 | Canada/ON-S1840/2020 | 2020-11-24 | North America/Canada/Ontario/Toronto | Toronto Invasive Bacterial Diseases Network | McMaster University | Allison McGeer et al |
| EPI_ISL_729304 | Canada/ON-S1842/2020 | 2020-11-24 | North America/Canada/Ontario/Toronto | Toronto Invasive Bacterial Diseases Network | McMaster University | Allison McGeer et al |
| EPI_ISL_729305 | Canada/ON-S1844/2020 | 2020-11-24 | North America/Canada/Ontario/Toronto | Toronto Invasive Bacterial Diseases Network | McMaster University | Allison McGeer et al |
| EPI_ISL_729306 | Canada/ON-S1846/2020 | 2020-11-24 | North America/Canada/Ontario/Toronto | Toronto Invasive Bacterial Diseases Network | McMaster University | Allison McGeer et al |
| EPI_ISL_729307 | Canada/ON-S1847/2020 | 2020-11-24 | North America/Canada/Ontario/Toronto | Toronto Invasive Bacterial Diseases Network | McMaster University | Allison McGeer et al |
| EPI_ISL_729308 | Canada/ON-S1850/2020 | 2020-11-24 | North America/Canada/Ontario/Toronto | Toronto Invasive Bacterial Diseases Network | McMaster University | Allison McGeer et al |
| EPI_ISL_729309 | Canada/ON-S1851/2020 | 2020-11-24 | North America/Canada/Ontario/Toronto | Toronto Invasive Bacterial Diseases Network | McMaster University | Allison McGeer et al |
| EPI_ISL_729310 | Canada/ON-S1852/2020 | 2020-11-24 | North America/Canada/Ontario/Toronto | Toronto Invasive Bacterial Diseases Network | McMaster University | Allison McGeer et al |
| EPI_ISL_729311 | Canada/ON-S1886/2020 | 2020-11-24 | North America/Canada/Ontario/Toronto | Toronto Invasive Bacterial Diseases Network | McMaster University | Allison McGeer et al |
| EPI_ISL_729312 | Canada/ON-S1887/2020 | 2020-11-24 | North America/Canada/Ontario/Toronto | Toronto Invasive Bacterial Diseases Network | McMaster University | Allison McGeer et al |
| EPI_ISL_729313 | Canada/ON-S1888/2020 | 2020-11-24 | North America/Canada/Ontario/Toronto | Toronto Invasive Bacterial Diseases Network | McMaster University | Allison McGeer et al |

| | | | | | | |
|---|---|---|---|---|---|---|
| EPI_ISL_729314 | Canada/ON-S1890/2020 | 2020-11-24 | North America/Canada/Ontario/Toronto | Toronto Invasive Bacterial Diseases Network | McMaster University | Allison McGeer et al |
| EPI_ISL_729315 | Canada/ON-S1892/2020 | 2020-11-24 | North America/Canada/Ontario/Toronto | Toronto Invasive Bacterial Diseases Network | McMaster University | Allison McGeer et al |
| EPI_ISL_729316 | Canada/ON-S1893/2020 | 2020-11-24 | North America/Canada/Ontario/Toronto | Toronto Invasive Bacterial Diseases Network | McMaster University | Allison McGeer et al |
| EPI_ISL_729317 | Canada/ON-S1900/2020 | 2020-11-24 | North America/Canada/Ontario/Toronto | Toronto Invasive Bacterial Diseases Network | McMaster University | Allison McGeer et al |
| EPI_ISL_729318 | Canada/ON-S1901/2020 | 2020-11-24 | North America/Canada/Ontario/Toronto | Toronto Invasive Bacterial Diseases Network | McMaster University | Allison McGeer et al |
| EPI_ISL_729319 | Canada/ON-S1903/2020 | 2020-11-24 | North America/Canada/Ontario/Toronto | Toronto Invasive Bacterial Diseases Network | McMaster University | Allison McGeer et al |
| EPI_ISL_729320 | Canada/ON-S1904/2020 | 2020-11-24 | North America/Canada/Ontario/Toronto | Toronto Invasive Bacterial Diseases Network | McMaster University | Allison McGeer et al |
| EPI_ISL_729321 | Canada/ON-S1905/2020 | 2020-11-24 | North America/Canada/Ontario/Toronto | Toronto Invasive Bacterial Diseases Network | McMaster University | Allison McGeer et al |
| EPI_ISL_729322 | Canada/ON-S1906/2020 | 2020-11-24 | North America/Canada/Ontario/Toronto | Toronto Invasive Bacterial Diseases Network | McMaster University | Allison McGeer et al |
| EPI_ISL_729323 | Canada/ON-S1908/2020 | 2020-11-24 | North America/Canada/Ontario/Toronto | Toronto Invasive Bacterial Diseases Network | McMaster University | Allison McGeer et al |
| EPI_ISL_729324 | Canada/ON-S1912/2020 | 2020-11-24 | North America/Canada/Ontario/Toronto | Toronto Invasive Bacterial Diseases Network | McMaster University | Allison McGeer et al |
| EPI_ISL_729325 | Canada/ON-S1915/2020 | 2020-11-24 | North America/Canada/Ontario/Toronto | Toronto Invasive Bacterial Diseases Network | McMaster University | Allison McGeer et al |
| EPI_ISL_729326 | Canada/ON-S1917/2020 | 2020-11-24 | North America/Canada/Ontario/Toronto | Toronto Invasive Bacterial Diseases Network | McMaster University | Allison McGeer et al |
| EPI_ISL_729327 | Canada/ON-S1918/2020 | 2020-11-24 | North America/Canada/Ontario/Toronto | Toronto Invasive Bacterial Diseases Network | McMaster University | Allison McGeer et al |
| EPI_ISL_729328 | Canada/ON-S1920/2020 | 2020-11-24 | North America/Canada/Ontario/Toronto | Toronto Invasive Bacterial Diseases Network | McMaster University | Allison McGeer et al |
| EPI_ISL_729329 | Canada/ON-S1921/2020 | 2020-11-24 | North America/Canada/Ontario/Toronto | Toronto Invasive Bacterial Diseases Network | McMaster University | Allison McGeer et al |
| EPI_ISL_729330 | Canada/ON-S1922/2020 | 2020-11-24 | North America/Canada/Ontario/Toronto | Toronto Invasive Bacterial Diseases Network | McMaster University | Allison McGeer et al |
| EPI_ISL_729331 | Canada/ON-S1925/2020 | 2020-11-24 | North America/Canada/Ontario/Toronto | Toronto Invasive Bacterial Diseases Network | McMaster University | Allison McGeer et al |
| EPI_ISL_729332 | Canada/ON-S1930/2020 | 2020-11-24 | North America/Canada/Ontario/Toronto | Toronto Invasive Bacterial Diseases Network | McMaster University | Allison McGeer et al |
| EPI_ISL_729333 | Canada/ON-S1931/2020 | 2020-11-24 | North America/Canada/Ontario/Toronto | Toronto Invasive Bacterial Diseases Network | McMaster University | Allison McGeer et al |

| | | | | | | |
|---|---|---|---|---|---|---|
| EPI_ISL_729334 | Canada/ON-S1932/2020 | 2020-11-24 | North America/Canada/Ontario/Toronto | Toronto Invasive Bacterial Diseases Network | McMaster University | Allison McGeer et al |
| EPI_ISL_729335 | Canada/ON-S1934/2020 | 2020-11-24 | North America/Canada/Ontario/Toronto | Toronto Invasive Bacterial Diseases Network | McMaster University | Allison McGeer et al |
| EPI_ISL_729338 | Germany/SH-ChVir20742/2020 | 2020-11-17 | Europe/Germany/Schleswig-Holstein/ | A. Krumbholz, Labor Dr. Krause und Kollegen MVZ GmbH, Kiel | Charit√© Universit√§tsmedizin Berlin, Institut f√ºr Virologie | Victor M Corman et al |
| EPI_ISL_729341 | Germany/SH-ChVir9983/2020 | 2020-11-02 | Europe/Germany/Schleswig-Holstein/ | A. Krumbholz, Labor Dr. Krause und Kollegen MVZ GmbH, Kiel | Charit√© Universit√§tsmedizin Berlin, Institut f√ºr Virologie | Victor M Corman et al |
| EPI_ISL_729345 | Germany/SH-ChVir20029/2020 | 2020-11-06 | Europe/Germany/Schleswig-Holstein/ | A. Krumbholz, Labor Dr. Krause und Kollegen MVZ GmbH, Kiel | Charit√© Universit√§tsmedizin Berlin, Institut f√ºr Virologie | Victor M Corman et al |
| EPI_ISL_729348 | Germany/BE-ChVir8986/2020 | 2020-09-21 | Europe/Germany/Berlin/ | Charit√© Universit√§tsmedizin Berlin, Institut f√ºr Virologie/Labor Berlin | Charit√© Universit√§tsmedizin Berlin, Institut f√ºr Virologie | Victor M Corman et al |
| EPI_ISL_729357 | Germany/BE-ChVir9039/2020 | 2020-09-21 | Europe/Germany/Berlin/ | Charit√© Universit√§tsmedizin Berlin, Institut f√ºr Virologie/Labor Berlin | Charit√© Universit√§tsmedizin Berlin, Institut f√ºr Virologie | Victor M Corman et al |
| EPI_ISL_729359 | Germany/SH-ChVir9115/2020 | 2020-09-14 | Europe/Germany/Schleswig-Holstein/ | A. Krumbholz, Labor Dr. Krause und Kollegen MVZ GmbH, Kiel | Charit√© Universit√§tsmedizin Berlin, Institut f√ºr Virologie | Victor M Corman et al |
| EPI_ISL_729362 | Germany/SH-ChVir9125/2020 | 2020-09-21 | Europe/Germany/Schleswig-Holstein/ | A. Krumbholz, Labor Dr. Krause und Kollegen MVZ GmbH, Kiel | Charit√© Universit√§tsmedizin Berlin, Institut f√ºr Virologie | Victor M Corman et al |
| EPI_ISL_729363 | Germany/SH-ChVir9635/2020 | 2020-10-16 | Europe/Germany/Schleswig-Holstein/ | A. Krumbholz, Labor Dr. Krause und Kollegen MVZ GmbH, Kiel | Charit√© Universit√§tsmedizin Berlin, Institut f√ºr Virologie | Victor M Corman et al |
| EPI_ISL_729367 | Germany/ChVir9639/2020 | 2020-10-16 | Europe/Germany/Germany/ | A. Krumbholz, Labor Dr. Krause und Kollegen MVZ GmbH, Kiel | Charit√© Universit√§tsmedizin Berlin, Institut f√ºr Virologie | Victor M Corman et al |

| | | | | | | |
|---|---|---|---|---|---|---|
| EPI_ISL_729371 | Germany/SH-ChVir20741/2020 | 2020-11-17 | Europe/Germany/Schleswig-Holstein/ | A. Krumbholz, Labor Dr. Krause und Kollegen MVZ GmbH, Kiel | Charit√© Universit√§tsmedizin Berlin, Institut f√ºr Virologie | Victor M Corman et al |
| EPI_ISL_729372 | Germany/SH-ChVir20045/2020 | 2020-11-06 | Europe/Germany/Schleswig-Holstein/ | A. Krumbholz, Labor Dr. Krause und Kollegen MVZ GmbH, Kiel | Charit√© Universit√§tsmedizin Berlin, Institut f√ºr Virologie | Victor M Corman et al |
| EPI_ISL_729375 | Germany/SH-ChVir9119/2020 | 2020-09-16 | Europe/Germany/Schleswig-Holstein/ | A. Krumbholz, Labor Dr. Krause und Kollegen MVZ GmbH, Kiel | Charit√© Universit√§tsmedizin Berlin, Institut f√ºr Virologie | Victor M Corman et al |
| EPI_ISL_729377 | Germany/BE-ChVir8992/2020 | 2020-09-21 | Europe/Germany/Berlin/ | Charit√© Universit√§tsmedizin Berlin, Institut f√ºr Virologie/Labor Berlin | Charit√© Universit√§tsmedizin Berlin, Institut f√ºr Virologie | Victor M Corman et al |
| EPI_ISL_729381 | Germany/SH-ChVir3194/2020 | 2020-03-22 | Europe/Germany/Schleswig-Holstein/ | A. Krumbholz, Labor Dr. Krause und Kollegen MVZ GmbH, Kiel | Charit√© Universit√§tsmedizin Berlin, Institut f√ºr Virologie | Victor M Corman et al |
| EPI_ISL_729385 | Germany/SH-ChVir20767/2020 | 2020-11-23 | Europe/Germany/Schleswig-Holstein/ | A. Krumbholz, Labor Dr. Krause und Kollegen MVZ GmbH, Kiel | Charit√© Universit√§tsmedizin Berlin, Institut f√ºr Virologie | Victor M Corman et al |
| EPI_ISL_729391 | Germany/SH-ChVir20039/2020 | 2020-11-06 | Europe/Germany/Schleswig-Holstein/ | A. Krumbholz, Labor Dr. Krause und Kollegen MVZ GmbH, Kiel | Charit√© Universit√§tsmedizin Berlin, Institut f√ºr Virologie | Victor M Corman et al |
| EPI_ISL_729392 | Germany/SH-ChVir20799/2020 | 2020-12-02 | Europe/Germany/Schleswig-Holstein/ | A. Krumbholz, Labor Dr. Krause und Kollegen MVZ GmbH, Kiel | Charit√© Universit√§tsmedizin Berlin, Institut f√ºr Virologie | Victor M Corman et al |
| EPI_ISL_729397 | Germany/SH-ChVir3221/2020 | 2020-03-23 | Europe/Germany/Schleswig-Holstein/ | A. Krumbholz, Labor Dr. Krause und Kollegen MVZ GmbH, Kiel | Charit√© Universit√§tsmedizin Berlin, Institut f√ºr Virologie | Victor M Corman et al |
| EPI_ISL_729401 | Germany/SH-ChVir9980/2020 | 2020-11-02 | Europe/Germany/Schleswig-Holstein/ | A. Krumbholz, Labor Dr. Krause und Kollegen MVZ GmbH, Kiel | Charit√© Universit√§tsmedizin Berlin, Institut f√ºr Virologie | Victor M Corman et al |

| | | | | | | |
|---|---|---|---|---|---|---|
| EPI_ISL_729409 | Germany/BE-ChVir9031/2020 | 2020-09-20 | Europe/Germany/Berlin/ | Charit√© Universit√§tsmedizin Berlin, Institut f√ºr Virologie/Labor Berlin | Charit√© Universit√§tsmedizin Berlin, Institut f√ºr Virologie | Victor M Corman et al |
| EPI_ISL_729414 | Germany/SH-ChVir9979/2020 | 2020-11-02 | Europe/Germany/Schleswig-Holstein/ | A. Krumbholz, Labor Dr. Krause und Kollegen MVZ GmbH, Kiel | Charit√© Universit√§tsmedizin Berlin, Institut f√ºr Virologie | Victor M Corman et al |
| EPI_ISL_729417 | Germany/SH-ChVir20054/2020 | 2020-11-06 | Europe/Germany/Schleswig-Holstein/ | A. Krumbholz, Labor Dr. Krause und Kollegen MVZ GmbH, Kiel | Charit√© Universit√§tsmedizin Berlin, Institut f√ºr Virologie | Victor M Corman et al |
| EPI_ISL_729420 | Germany/SH-ChVir20770/2020 | 2020-11-26 | Europe/Germany/Schleswig-Holstein/ | A. Krumbholz, Labor Dr. Krause und Kollegen MVZ GmbH, Kiel | Charit√© Universit√§tsmedizin Berlin, Institut f√ºr Virologie | Victor M Corman et al |
| EPI_ISL_729421 | Germany/SH-ChVir9290/2020 | 2020-10-07 | Europe/Germany/Schleswig-Holstein/ | A. Krumbholz, Labor Dr. Krause und Kollegen MVZ GmbH, Kiel | Charit√© Universit√§tsmedizin Berlin, Institut f√ºr Virologie | Victor M Corman et al |
| EPI_ISL_729423 | Germany/SH-ChVir9633/2020 | 2020-10-16 | Europe/Germany/Schleswig-Holstein/ | A. Krumbholz, Labor Dr. Krause und Kollegen MVZ GmbH, Kiel | Charit√© Universit√§tsmedizin Berlin, Institut f√ºr Virologie | Victor M Corman et al |
| EPI_ISL_729424 | Germany/SH-ChVir9933/2020 | 2020-10-31 | Europe/Germany/Schleswig-Holstein/ | A. Krumbholz, Labor Dr. Krause und Kollegen MVZ GmbH, Kiel | Charit√© Universit√§tsmedizin Berlin, Institut f√ºr Virologie | Victor M Corman et al |
| EPI_ISL_729427 | Germany/SH-ChVir9944/2020 | 2020-11-02 | Europe/Germany/Schleswig-Holstein/ | A. Krumbholz, Labor Dr. Krause und Kollegen MVZ GmbH, Kiel | Charit√© Universit√§tsmedizin Berlin, Institut f√ºr Virologie | Victor M Corman et al |
| EPI_ISL_729429 | Germany/SH-ChVir9953/2020 | 2020-11-03 | Europe/Germany/Schleswig-Holstein/ | A. Krumbholz, Labor Dr. Krause und Kollegen MVZ GmbH, Kiel | Charit√© Universit√§tsmedizin Berlin, Institut f√ºr Virologie | Victor M Corman et al |
| EPI_ISL_729433 | Germany/SH-ChVir20009/2020 | 2020-11-04 | Europe/Germany/Schleswig-Holstein/ | A. Krumbholz, Labor Dr. Krause und Kollegen MVZ GmbH, Kiel | Charit√© Universit√§tsmedizin Berlin, Institut f√ºr Virologie | Victor M Corman et al |

| | | | | | | |
|---|---|---|---|---|---|---|
| EPI_ISL_729434 | Germany/SH-ChVir20013/2020 | 2020-11-04 | Europe/Germany/Schleswig-Holstein/ | A. Krumbholz, Labor Dr. Krause und Kollegen MVZ GmbH, Kiel | Charit√© Universit√§tsmedizin Berlin, Institut f√ºr Virologie | Victor M Corman et al |
| EPI_ISL_729437 | Germany/SH-ChVir20027/2020 | 2020-11-05 | Europe/Germany/Schleswig-Holstein/ | A. Krumbholz, Labor Dr. Krause und Kollegen MVZ GmbH, Kiel | Charit√© Universit√§tsmedizin Berlin, Institut f√ºr Virologie | Victor M Corman et al |
| EPI_ISL_729438 | Germany/HH-ChVir20032/2020 | 2020-11-06 | Europe/Germany/Hamburg/ | A. Krumbholz, Labor Dr. Krause und Kollegen MVZ GmbH, Kiel | Charit√© Universit√§tsmedizin Berlin, Institut f√ºr Virologie | Victor M Corman et al |
| EPI_ISL_729439 | Germany/SH-ChVir20033/2020 | 2020-11-06 | Europe/Germany/Schleswig-Holstein/ | A. Krumbholz, Labor Dr. Krause und Kollegen MVZ GmbH, Kiel | Charit√© Universit√§tsmedizin Berlin, Institut f√ºr Virologie | Victor M Corman et al |
| EPI_ISL_729440 | Germany/SH-ChVir20041/2020 | 2020-11-06 | Europe/Germany/Schleswig-Holstein/ | A. Krumbholz, Labor Dr. Krause und Kollegen MVZ GmbH, Kiel | Charit√© Universit√§tsmedizin Berlin, Institut f√ºr Virologie | Victor M Corman et al |
| EPI_ISL_729443 | Germany/SH-ChVir20309/2020 | 2020-11-16 | Europe/Germany/Schleswig-Holstein/ | A. Krumbholz, Labor Dr. Krause und Kollegen MVZ GmbH, Kiel | Charit√© Universit√§tsmedizin Berlin, Institut f√ºr Virologie | Victor M Corman et al |
| EPI_ISL_729445 | Germany/SH-ChVir20747/2020 | 2020-11-19 | Europe/Germany/Schleswig-Holstein/ | A. Krumbholz, Labor Dr. Krause und Kollegen MVZ GmbH, Kiel | Charit√© Universit√§tsmedizin Berlin, Institut f√ºr Virologie | Victor M Corman et al |
| EPI_ISL_729446 | Germany/SH-ChVir20749/2020 | 2020-11-19 | Europe/Germany/Schleswig-Holstein/ | A. Krumbholz, Labor Dr. Krause und Kollegen MVZ GmbH, Kiel | Charit√© Universit√§tsmedizin Berlin, Institut f√ºr Virologie | Victor M Corman et al |
| EPI_ISL_729448 | Germany/SH-ChVir20774/2020 | 2020-11-26 | Europe/Germany/Schleswig-Holstein/ | A. Krumbholz, Labor Dr. Krause und Kollegen MVZ GmbH, Kiel | Charit√© Universit√§tsmedizin Berlin, Institut f√ºr Virologie | Victor M Corman et al |
| EPI_ISL_729449 | Germany/SH-ChVir20791/2020 | 2020-12-02 | Europe/Germany/Schleswig-Holstein/ | A. Krumbholz, Labor Dr. Krause und Kollegen MVZ GmbH, Kiel | Charit√© Universit√§tsmedizin Berlin, Institut f√ºr Virologie | Victor M Corman et al |

| | | | | | | |
|---|---|---|---|---|---|---|
| EPI_ISL_729452 | Germany/SH-ChVir20831/2020 | 2020-11-30 | Europe/Germany/Schleswig-Holstein/ | A. Krumbholz, Labor Dr. Krause und Kollegen MVZ GmbH, Kiel | Charitv© Universitv§tsmedizin Berlin, Institut fvºr Virologie | Victor M Corman et al |
| EPI_ISL_729462 | Germany/SH-ChVir9943/2020 | 2020-11-03 | Europe/Germany/Schleswig-Holstein/ | A. Krumbholz, Labor Dr. Krause und Kollegen MVZ GmbH, Kiel | Charitv© Universitv§tsmedizin Berlin, Institut fvºr Virologie | Victor M Corman et al |
| EPI_ISL_729463 | Germany/SH-ChVir20810/2020 | 2020-12-01 | Europe/Germany/Schleswig-Holstein/ | A. Krumbholz, Labor Dr. Krause und Kollegen MVZ GmbH, Kiel | Charitv© Universitv§tsmedizin Berlin, Institut fvºr Virologie | Victor M Corman et al |
| EPI_ISL_729464 | Germany/SH-ChVir20773/2020 | 2020-11-26 | Europe/Germany/Schleswig-Holstein/ | A. Krumbholz, Labor Dr. Krause und Kollegen MVZ GmbH, Kiel | Charitv© Universitv§tsmedizin Berlin, Institut fvºr Virologie | Victor M Corman et al |
| EPI_ISL_729465 | Germany/SH-ChVir3210/2020 | 2020-03-22 | Europe/Germany/Schleswig-Holstein/ | A. Krumbholz, Labor Dr. Krause und Kollegen MVZ GmbH, Kiel | Charitv© Universitv§tsmedizin Berlin, Institut fvºr Virologie | Victor M Corman et al |
| EPI_ISL_729471 | Germany/SH-ChVir9982/2020 | 2020-11-02 | Europe/Germany/Schleswig-Holstein/ | A. Krumbholz, Labor Dr. Krause und Kollegen MVZ GmbH, Kiel | Charitv© Universitv§tsmedizin Berlin, Institut fvºr Virologie | Victor M Corman et al |
| EPI_ISL_729473 | Germany/BW-ChVir1600/2020 | 2020-02-26 | Europe/Germany/Baden-Wuerttemberg | Charitv© Universitv§tsmedizin Berlin, Institut fvºr Virologie/Labor Berlin | Charitv© Universitv§tsmedizin Berlin, Institut fvºr Virologie | Victor M Corman et al |
| EPI_ISL_729475 | Germany/BW-ChVir1611/2020 | 2020-02-26 | Europe/Germany/Baden-Wuerttemberg | Charitv© Universitv§tsmedizin Berlin, Institut fvºr Virologie/Labor Berlin | Charitv© Universitv§tsmedizin Berlin, Institut fvºr Virologie | Victor M Corman et al |
| EPI_ISL_729478 | Germany/NW-ChVir1683/2020 | 2020-02-28 | Europe/Germany/North Rhine Westphal | Charitv© Universitv§tsmedizin Berlin, Institut fvºr Virologie/Labor Berlin | Charitv© Universitv§tsmedizin Berlin, Institut fvºr Virologie | Victor M Corman et al |
| EPI_ISL_729479 | Germany/BE-ChVir1685/2020 | 2020-03-02 | Europe/Germany/Berlin/ | Charitv© Universitv§tsmedizin Berlin, Institut fvºr Virologie/Labor Berlin | Charitv© Universitv§tsmedizin Berlin, Institut fvºr Virologie | Victor M Corman et al |

| | | | | | | |
|---|---|---|---|---|---|---|
| EPI_ISL_729481 | Germany/BE-ChVir1716/2020 | 2020-03-04 | Europe/Germany/Berlin/ | Charit√© Universit√§tsmedizin Berlin, Institut f√ºr Virologie/Labor Berlin | Charit√© Universit√§tsmedizin Berlin, Institut f√ºr Virologie | Victor M Corman et al |
| EPI_ISL_729484 | Germany/BW-ChVir1902/2020 | 2020-03-05 | Europe/Germany/Baden-Wuerttemberg | Charit√© Universit√§tsmedizin Berlin, Institut f√ºr Virologie/Labor Berlin | Charit√© Universit√§tsmedizin Berlin, Institut f√ºr Virologie | Victor M Corman et al |
| EPI_ISL_729488 | Germany/BW-ChVir1946/2020 | 2020-03-05 | Europe/Germany/Baden-Wuerttemberg | Charit√© Universit√§tsmedizin Berlin, Institut f√ºr Virologie/Labor Berlin | Charit√© Universit√§tsmedizin Berlin, Institut f√ºr Virologie | Victor M Corman et al |
| EPI_ISL_729490 | Germany/BE-ChVir1962/2020 | 2020-03-07 | Europe/Germany/Berlin/ | Charit√© Universit√§tsmedizin Berlin, Institut f√ºr Virologie/Labor Berlin | Charit√© Universit√§tsmedizin Berlin, Institut f√ºr Virologie | Victor M Corman et al |
| EPI_ISL_729495 | Germany/BY-ChVir1981/2020 | 2020-03-06 | Europe/Germany/Bavaria/ | Charit√© Universit√§tsmedizin Berlin, Institut f√ºr Virologie/Labor Berlin | Charit√© Universit√§tsmedizin Berlin, Institut f√ºr Virologie | Victor M Corman et al |
| EPI_ISL_729497 | Germany/ChVir1994/2020 | 2020-03-08 | Europe/Germany/Germany/ | Charit√© Universit√§tsmedizin Berlin, Institut f√ºr Virologie/Labor Berlin | Charit√© Universit√§tsmedizin Berlin, Institut f√ºr Virologie | Victor M Corman et al |
| EPI_ISL_729501 | Germany/NI-ChVir2098/2020 | 2020-03-11 | Europe/Germany/Niedersachsen/ | A. Krumbholz, Labor Dr. Krause und Kollegen MVZ GmbH, Kiel | Charit√© Universit√§tsmedizin Berlin, Institut f√ºr Virologie | Victor M Corman et al |
| EPI_ISL_729505 | Germany/SH-ChVir2109/2020 | 2020-03-13 | Europe/Germany/Schleswig-Holstein/ | A. Krumbholz, Labor Dr. Krause und Kollegen MVZ GmbH, Kiel | Charit√© Universit√§tsmedizin Berlin, Institut f√ºr Virologie | Victor M Corman et al |
| EPI_ISL_729507 | Germany/SH-ChVir2114/2020 | 2020-03-15 | Europe/Germany/Schleswig-Holstein/ | A. Krumbholz, Labor Dr. Krause und Kollegen MVZ GmbH, Kiel | Charit√© Universit√§tsmedizin Berlin, Institut f√ºr Virologie | Victor M Corman et al |
| EPI_ISL_729516 | Germany/SH-ChVir3034/2020 | 2020-03-17 | Europe/Germany/Schleswig-Holstein/ | A. Krumbholz, Labor Dr. Krause und Kollegen MVZ GmbH, Kiel | Charit√© Universit√§tsmedizin Berlin, Institut f√ºr Virologie | Victor M Corman et al |

| | | | | | | |
|---|---|---|---|---|---|---|
| EPI_ISL_729522 | Germany/SH-ChVir3052/2020 | 2020-03-20 | Europe/Germany/Schleswig-Holstein/ | A. Krumbholz, Labor Dr. Krause und Kollegen MVZ GmbH, Kiel | Charitv© Universitv§tsmedizin Berlin, Institut fvºr Virologie | Victor M Corman et al |
| EPI_ISL_729538 | Germany/SH-ChVir3192/2020 | 2020-03-22 | Europe/Germany/Schleswig-Holstein/ | A. Krumbholz, Labor Dr. Krause und Kollegen MVZ GmbH, Kiel | Charitv© Universitv§tsmedizin Berlin, Institut fvºr Virologie | Victor M Corman et al |
| EPI_ISL_729546 | Germany/SH-ChVir3218/2020 | 2020-03-23 | Europe/Germany/Schleswig-Holstein/ | A. Krumbholz, Labor Dr. Krause und Kollegen MVZ GmbH, Kiel | Charitv© Universitv§tsmedizin Berlin, Institut fvºr Virologie | Victor M Corman et al |
| EPI_ISL_729550 | Germany/SH-ChVir4733/2020 | 2020-04-08 | Europe/Germany/Schleswig-Holstein/ | A. Krumbholz, Labor Dr. Krause und Kollegen MVZ GmbH, Kiel | Charitv© Universitv§tsmedizin Berlin, Institut fvºr Virologie | Victor M Corman et al |
| EPI_ISL_729581 | Germany/SH-ChVir8326/2020 | 2020-07-28 | Europe/Germany/Schleswig-Holstein/ | A. Krumbholz, Labor Dr. Krause und Kollegen MVZ GmbH, Kiel | Charitv© Universitv§tsmedizin Berlin, Institut fvºr Virologie | Victor M Corman et al |
| EPI_ISL_729589 | Germany/SH-ChVir8595/2020 | 2020-08-19 | Europe/Germany/Schleswig-Holstein/ | A. Krumbholz, Labor Dr. Krause und Kollegen MVZ GmbH, Kiel | Charitv© Universitv§tsmedizin Berlin, Institut fvºr Virologie | Victor M Corman et al |
| EPI_ISL_729590 | Germany/SH-ChVir8596/2020 | 2020-08-19 | Europe/Germany/Schleswig-Holstein/ | A. Krumbholz, Labor Dr. Krause und Kollegen MVZ GmbH, Kiel | Charitv© Universitv§tsmedizin Berlin, Institut fvºr Virologie | Victor M Corman et al |
| EPI_ISL_729591 | Germany/SH-ChVir8599/2020 | 2020-08-19 | Europe/Germany/Schleswig-Holstein/ | A. Krumbholz, Labor Dr. Krause und Kollegen MVZ GmbH, Kiel | Charitv© Universitv§tsmedizin Berlin, Institut fvºr Virologie | Victor M Corman et al |
| EPI_ISL_729592 | Germany/SH-ChVir8604/2020 | 2020-08-22 | Europe/Germany/Schleswig-Holstein/ | A. Krumbholz, Labor Dr. Krause und Kollegen MVZ GmbH, Kiel | Charitv© Universitv§tsmedizin Berlin, Institut fvºr Virologie | Victor M Corman et al |
| EPI_ISL_729600 | Germany/BE-ChVir8991/2020 | 2020-09-21 | Europe/Germany/Berlin/ | Charitv© Universitv§tsmedizin Berlin, Institut fvºr Virologie/Labor Berlin | Charitv© Universitv§tsmedizin Berlin, Institut fvºr Virologie | Victor M Corman et al |

| | | | | | | |
|---|---|---|---|---|---|---|
| EPI_ISL_729601 | Germany/BE-ChVir9004/2020 | 2020-09-22 | Europe/Germany/Berlin/ | Charit√© Universit√§tsmedizin Berlin, Institut f√ºr Virologie/Labor Berlin | Charit√© Universit√§tsmedizin Berlin, Institut f√ºr Virologie | Victor M Corman et al |
| EPI_ISL_729602 | Germany/BE-ChVir9006/2020 | 2020-09-22 | Europe/Germany/Berlin/ | Charit√© Universit√§tsmedizin Berlin, Institut f√ºr Virologie/Labor Berlin | Charit√© Universit√§tsmedizin Berlin, Institut f√ºr Virologie | Victor M Corman et al |
| EPI_ISL_729605 | Germany/BE-ChVir9029/2020 | 2020-09-20 | Europe/Germany/Berlin/ | Charit√© Universit√§tsmedizin Berlin, Institut f√ºr Virologie/Labor Berlin | Charit√© Universit√§tsmedizin Berlin, Institut f√ºr Virologie | Victor M Corman et al |
| EPI_ISL_729607 | Germany/BE-ChVir9034/2020 | 2020-09-20 | Europe/Germany/Berlin/ | Charit√© Universit√§tsmedizin Berlin, Institut f√ºr Virologie/Labor Berlin | Charit√© Universit√§tsmedizin Berlin, Institut f√ºr Virologie | Victor M Corman et al |
| EPI_ISL_729610 | Germany/SH-ChVir9131/2020 | 2020-09-28 | Europe/Germany/Schleswig-Holstein/ | A. Krumbholz, Labor Dr. Krause und Kollegen MVZ GmbH, Kiel | Charit√© Universit√§tsmedizin Berlin, Institut f√ºr Virologie | Victor M Corman et al |
| EPI_ISL_729611 | Germany/SH-ChVir9277/2020 | 2020-09-28 | Europe/Germany/Schleswig-Holstein/ | A. Krumbholz, Labor Dr. Krause und Kollegen MVZ GmbH, Kiel | Charit√© Universit√§tsmedizin Berlin, Institut f√ºr Virologie | Victor M Corman et al |
| EPI_ISL_729616 | Germany/SH-ChVir9275/2020 | 2020-09-28 | Europe/Germany/Schleswig-Holstein/ | A. Krumbholz, Labor Dr. Krause und Kollegen MVZ GmbH, Kiel | Charit√© Universit√§tsmedizin Berlin, Institut f√ºr Virologie | Victor M Corman et al |
| EPI_ISL_729618 | Germany/SH-ChVir9287/2020 | 2020-10-03 | Europe/Germany/Schleswig-Holstein/ | A. Krumbholz, Labor Dr. Krause und Kollegen MVZ GmbH, Kiel | Charit√© Universit√§tsmedizin Berlin, Institut f√ºr Virologie | Victor M Corman et al |
| EPI_ISL_729619 | Germany/SH-ChVir9636/2020 | 2020-10-16 | Europe/Germany/Schleswig-Holstein/ | A. Krumbholz, Labor Dr. Krause und Kollegen MVZ GmbH, Kiel | Charit√© Universit√§tsmedizin Berlin, Institut f√ºr Virologie | Victor M Corman et al |
| EPI_ISL_729622 | Germany/SH-ChVir9946/2020 | 2020-11-02 | Europe/Germany/Schleswig-Holstein/ | A. Krumbholz, Labor Dr. Krause und Kollegen MVZ GmbH, Kiel | Charit√© Universit√§tsmedizin Berlin, Institut f√ºr Virologie | Victor M Corman et al |

| | | | | | | |
|---|---|---|---|---|---|---|
| EPI_ISL_729624 | Germany/SH-ChVir9995/2020 | 2020-11-04 | Europe/Germany/Schleswig-Holstein/ | A. Krumbholz, Labor Dr. Krause und Kollegen MVZ GmbH, Kiel | Charitv© Universitv§tsmedizin Berlin, Institut fvºr Virologie | Victor M Corman et al |
| EPI_ISL_729625 | Germany/SH-ChVir20001/2020 | 2020-11-04 | Europe/Germany/Schleswig-Holstein/ | A. Krumbholz, Labor Dr. Krause und Kollegen MVZ GmbH, Kiel | Charitv© Universitv§tsmedizin Berlin, Institut fvºr Virologie | Victor M Corman et al |
| EPI_ISL_729627 | Germany/SH-ChVir20005/2020 | 2020-11-04 | Europe/Germany/Schleswig-Holstein/ | A. Krumbholz, Labor Dr. Krause und Kollegen MVZ GmbH, Kiel | Charitv© Universitv§tsmedizin Berlin, Institut fvºr Virologie | Victor M Corman et al |
| EPI_ISL_729628 | Germany/NW-ChVir20006/2020 | 2020-11-04 | Europe/Germany/North Rhine Westphal | A. Krumbholz, Labor Dr. Krause und Kollegen MVZ GmbH, Kiel | Charitv© Universitv§tsmedizin Berlin, Institut fvºr Virologie | Victor M Corman et al |
| EPI_ISL_729629 | Germany/SH-ChVir20012/2020 | 2020-11-04 | Europe/Germany/Schleswig-Holstein/ | A. Krumbholz, Labor Dr. Krause und Kollegen MVZ GmbH, Kiel | Charitv© Universitv§tsmedizin Berlin, Institut fvºr Virologie | Victor M Corman et al |
| EPI_ISL_729630 | Germany/SH-ChVir20031/2020 | 2020-11-06 | Europe/Germany/Schleswig-Holstein/ | A. Krumbholz, Labor Dr. Krause und Kollegen MVZ GmbH, Kiel | Charitv© Universitv§tsmedizin Berlin, Institut fvºr Virologie | Victor M Corman et al |
| EPI_ISL_729631 | Germany/SH-ChVir20036/2020 | 2020-11-06 | Europe/Germany/Schleswig-Holstein/ | A. Krumbholz, Labor Dr. Krause und Kollegen MVZ GmbH, Kiel | Charitv© Universitv§tsmedizin Berlin, Institut fvºr Virologie | Victor M Corman et al |
| EPI_ISL_729633 | Germany/SH-ChVir20043/2020 | 2020-11-06 | Europe/Germany/Schleswig-Holstein/ | A. Krumbholz, Labor Dr. Krause und Kollegen MVZ GmbH, Kiel | Charitv© Universitv§tsmedizin Berlin, Institut fvºr Virologie | Victor M Corman et al |
| EPI_ISL_729634 | Germany/SH-ChVir20049/2020 | 2020-11-06 | Europe/Germany/Schleswig-Holstein/ | A. Krumbholz, Labor Dr. Krause und Kollegen MVZ GmbH, Kiel | Charitv© Universitv§tsmedizin Berlin, Institut fvºr Virologie | Victor M Corman et al |
| EPI_ISL_729635 | Germany/SH-ChVir20296/2020 | 2020-11-13 | Europe/Germany/Schleswig-Holstein/ | A. Krumbholz, Labor Dr. Krause und Kollegen MVZ GmbH, Kiel | Charitv© Universitv§tsmedizin Berlin, Institut fvºr Virologie | Victor M Corman et al |

| | | | | | | |
|---|---|---|---|---|---|---|
| EPI_ISL_729636 | Germany/SH-ChVir20297/2020 | 2020-11-13 | Europe/Germany/Schleswig-Holstein/ | A. Krumbholz, Labor Dr. Krause und Kollegen MVZ GmbH, Kiel | Charit√© Universit√§tsmedizin Berlin, Institut f√ºr Virologie | Victor M Corman et al |
| EPI_ISL_729638 | Germany/SH-ChVir20300/2020 | 2020-11-14 | Europe/Germany/Schleswig-Holstein/ | A. Krumbholz, Labor Dr. Krause und Kollegen MVZ GmbH, Kiel | Charit√© Universit√§tsmedizin Berlin, Institut f√ºr Virologie | Victor M Corman et al |
| EPI_ISL_729640 | Germany/SH-ChVir20302/2020 | 2020-11-14 | Europe/Germany/Schleswig-Holstein/ | A. Krumbholz, Labor Dr. Krause und Kollegen MVZ GmbH, Kiel | Charit√© Universit√§tsmedizin Berlin, Institut f√ºr Virologie | Victor M Corman et al |
| EPI_ISL_729641 | Germany/ChVir20304/2020 | 2020-11-14 | Europe/Germany/Germany/ | A. Krumbholz, Labor Dr. Krause und Kollegen MVZ GmbH, Kiel | Charit√© Universit√§tsmedizin Berlin, Institut f√ºr Virologie | Victor M Corman et al |
| EPI_ISL_729643 | Germany/SH-ChVir20306/2020 | 2020-11-15 | Europe/Germany/Schleswig-Holstein/ | A. Krumbholz, Labor Dr. Krause und Kollegen MVZ GmbH, Kiel | Charit√© Universit√§tsmedizin Berlin, Institut f√ºr Virologie | Victor M Corman et al |
| EPI_ISL_729645 | Germany/SH-ChVir20308/2020 | 2020-11-16 | Europe/Germany/Schleswig-Holstein/ | A. Krumbholz, Labor Dr. Krause und Kollegen MVZ GmbH, Kiel | Charit√© Universit√§tsmedizin Berlin, Institut f√ºr Virologie | Victor M Corman et al |
| EPI_ISL_729646 | Germany/ChVir20311/2020 | 2020-11-16 | Europe/Germany/Germany/ | A. Krumbholz, Labor Dr. Krause und Kollegen MVZ GmbH, Kiel | Charit√© Universit√§tsmedizin Berlin, Institut f√ºr Virologie | Victor M Corman et al |
| EPI_ISL_729648 | Germany/SH-ChVir20313/2020 | 2020-11-16 | Europe/Germany/Schleswig-Holstein/ | A. Krumbholz, Labor Dr. Krause und Kollegen MVZ GmbH, Kiel | Charit√© Universit√§tsmedizin Berlin, Institut f√ºr Virologie | Victor M Corman et al |
| EPI_ISL_729649 | Germany/ChVir20314/2020 | 2020-11-16 | Europe/Germany/Germany/ | A. Krumbholz, Labor Dr. Krause und Kollegen MVZ GmbH, Kiel | Charit√© Universit√§tsmedizin Berlin, Institut f√ºr Virologie | Victor M Corman et al |
| EPI_ISL_729654 | Germany/SH-ChVir20292/2020 | 2020-11-12 | Europe/Germany/Schleswig-Holstein/ | A. Krumbholz, Labor Dr. Krause und Kollegen MVZ GmbH, Kiel | Charit√© Universit√§tsmedizin Berlin, Institut f√ºr Virologie | Victor M Corman et al |

| | | | | | | |
|---|---|---|---|---|---|---|
| EPI_ISL_729656 | Germany/SH-ChVir20295/2020 | 2020-11-12 | Europe/Germany/Schleswig-Holstein/ | A. Krumbholz, Labor Dr. Krause und Kollegen MVZ GmbH, Kiel | Charitv© Universitv§tsmedizin Berlin, Institut fvºr Virologie | Victor M Corman et al |
| EPI_ISL_729657 | Germany/SH-ChVir9978/2020 | 2020-11-02 | Europe/Germany/Schleswig-Holstein/ | A. Krumbholz, Labor Dr. Krause und Kollegen MVZ GmbH, Kiel | Charitv© Universitv§tsmedizin Berlin, Institut fvºr Virologie | Victor M Corman et al |
| EPI_ISL_729658 | Germany/SH-ChVir9994/2020 | 2020-11-04 | Europe/Germany/Schleswig-Holstein/ | A. Krumbholz, Labor Dr. Krause und Kollegen MVZ GmbH, Kiel | Charitv© Universitv§tsmedizin Berlin, Institut fvºr Virologie | Victor M Corman et al |
| EPI_ISL_729659 | Germany/SH-ChVir20028/2020 | 2020-11-06 | Europe/Germany/Schleswig-Holstein/ | A. Krumbholz, Labor Dr. Krause und Kollegen MVZ GmbH, Kiel | Charitv© Universitv§tsmedizin Berlin, Institut fvºr Virologie | Victor M Corman et al |
| EPI_ISL_729660 | Germany/SH-ChVir9989/2020 | 2020-11-03 | Europe/Germany/Schleswig-Holstein/ | A. Krumbholz, Labor Dr. Krause und Kollegen MVZ GmbH, Kiel | Charitv© Universitv§tsmedizin Berlin, Institut fvºr Virologie | Victor M Corman et al |
| EPI_ISL_729663 | Germany/SH-ChVir20048/2020 | 2020-11-06 | Europe/Germany/Schleswig-Holstein/ | A. Krumbholz, Labor Dr. Krause und Kollegen MVZ GmbH, Kiel | Charitv© Universitv§tsmedizin Berlin, Institut fvºr Virologie | Victor M Corman et al |
| EPI_ISL_729665 | Germany/SH-ChVir9991/2020 | 2020-11-04 | Europe/Germany/Schleswig-Holstein/ | A. Krumbholz, Labor Dr. Krause und Kollegen MVZ GmbH, Kiel | Charitv© Universitv§tsmedizin Berlin, Institut fvºr Virologie | Victor M Corman et al |
| EPI_ISL_729667 | Germany/SH-ChVir9999/2020 | 2020-11-04 | Europe/Germany/Schleswig-Holstein/ | A. Krumbholz, Labor Dr. Krause und Kollegen MVZ GmbH, Kiel | Charitv© Universitv§tsmedizin Berlin, Institut fvºr Virologie | Victor M Corman et al |
| EPI_ISL_729669 | Germany/SH-ChVir20000/2020 | 2020-11-04 | Europe/Germany/Schleswig-Holstein/ | A. Krumbholz, Labor Dr. Krause und Kollegen MVZ GmbH, Kiel | Charitv© Universitv§tsmedizin Berlin, Institut fvºr Virologie | Victor M Corman et al |
| EPI_ISL_729671 | Germany/SH-ChVir20728/2020 | 2020-11-18 | Europe/Germany/Schleswig-Holstein/ | A. Krumbholz, Labor Dr. Krause und Kollegen MVZ GmbH, Kiel | Charitv© Universitv§tsmedizin Berlin, Institut fvºr Virologie | Victor M Corman et al |

| | | | | | | |
|---|---|---|---|---|---|---|
| EPI_ISL_729673 | Germany/BY-ChVir20731/2020 | 2020-11-18 | Europe/Germany/Bavaria/ | A. Krumbholz, Labor Dr. Krause und Kollegen MVZ GmbH, Kiel | Charit√© Universit√§tsmedizin Berlin, Institut f√ºr Virologie | Victor M Corman et al |
| EPI_ISL_729677 | Germany/SH-ChVir20737/2020 | 2020-11-17 | Europe/Germany/Schleswig-Holstein/ | A. Krumbholz, Labor Dr. Krause und Kollegen MVZ GmbH, Kiel | Charit√© Universit√§tsmedizin Berlin, Institut f√ºr Virologie | Victor M Corman et al |
| EPI_ISL_729678 | Germany/SH-ChVir20739/2020 | 2020-11-17 | Europe/Germany/Schleswig-Holstein/ | A. Krumbholz, Labor Dr. Krause und Kollegen MVZ GmbH, Kiel | Charit√© Universit√§tsmedizin Berlin, Institut f√ºr Virologie | Victor M Corman et al |
| EPI_ISL_729679 | Germany/SH-ChVir20740/2020 | 2020-11-17 | Europe/Germany/Schleswig-Holstein/ | A. Krumbholz, Labor Dr. Krause und Kollegen MVZ GmbH, Kiel | Charit√© Universit√§tsmedizin Berlin, Institut f√ºr Virologie | Victor M Corman et al |
| EPI_ISL_729680 | Germany/SH-ChVir20743/2020 | 2020-11-19 | Europe/Germany/Schleswig-Holstein/ | A. Krumbholz, Labor Dr. Krause und Kollegen MVZ GmbH, Kiel | Charit√© Universit√§tsmedizin Berlin, Institut f√ºr Virologie | Victor M Corman et al |
| EPI_ISL_729684 | Germany/SH-ChVir20758/2020 | 2020-11-23 | Europe/Germany/Schleswig-Holstein/ | A. Krumbholz, Labor Dr. Krause und Kollegen MVZ GmbH, Kiel | Charit√© Universit√§tsmedizin Berlin, Institut f√ºr Virologie | Victor M Corman et al |
| EPI_ISL_729686 | Germany/SH-ChVir20775/2020 | 2020-11-26 | Europe/Germany/Schleswig-Holstein/ | A. Krumbholz, Labor Dr. Krause und Kollegen MVZ GmbH, Kiel | Charit√© Universit√§tsmedizin Berlin, Institut f√ºr Virologie | Victor M Corman et al |
| EPI_ISL_729690 | Germany/SH-ChVir20780/2020 | 2020-11-27 | Europe/Germany/Schleswig-Holstein/ | A. Krumbholz, Labor Dr. Krause und Kollegen MVZ GmbH, Kiel | Charit√© Universit√§tsmedizin Berlin, Institut f√ºr Virologie | Victor M Corman et al |
| EPI_ISL_729692 | Germany/SH-ChVir20782/2020 | 2020-11-28 | Europe/Germany/Schleswig-Holstein/ | A. Krumbholz, Labor Dr. Krause und Kollegen MVZ GmbH, Kiel | Charit√© Universit√§tsmedizin Berlin, Institut f√ºr Virologie | Victor M Corman et al |
| EPI_ISL_729694 | Germany/SH-ChVir20784/2020 | 2020-11-28 | Europe/Germany/Schleswig-Holstein/ | A. Krumbholz, Labor Dr. Krause und Kollegen MVZ GmbH, Kiel | Charit√© Universit√§tsmedizin Berlin, Institut f√ºr Virologie | Victor M Corman et al |

| | | | | | | |
|---|---|---|---|---|---|---|
| EPI_ISL_729695 | Germany/SH-ChVir20785/2020 | 2020-11-28 | Europe/Germany/Schleswig-Holstein/ | A. Krumbholz, Labor Dr. Krause und Kollegen MVZ GmbH, Kiel | Charit√© Universit√§tsmedizin Berlin, Institut f√ºr Virologie | Victor M Corman et al |
| EPI_ISL_729702 | Germany/SH-ChVir20794/2020 | 2020-12-02 | Europe/Germany/Schleswig-Holstein/ | A. Krumbholz, Labor Dr. Krause und Kollegen MVZ GmbH, Kiel | Charit√© Universit√§tsmedizin Berlin, Institut f√ºr Virologie | Victor M Corman et al |
| EPI_ISL_729704 | Germany/SH-ChVir20796/2020 | 2020-11-29 | Europe/Germany/Schleswig-Holstein/ | A. Krumbholz, Labor Dr. Krause und Kollegen MVZ GmbH, Kiel | Charit√© Universit√§tsmedizin Berlin, Institut f√ºr Virologie | Victor M Corman et al |
| EPI_ISL_729706 | Germany/SH-ChVir20798/2020 | 2020-12-02 | Europe/Germany/Schleswig-Holstein/ | A. Krumbholz, Labor Dr. Krause und Kollegen MVZ GmbH, Kiel | Charit√© Universit√§tsmedizin Berlin, Institut f√ºr Virologie | Victor M Corman et al |
| EPI_ISL_729709 | Germany/SH-ChVir20803/2020 | 2020-12-02 | Europe/Germany/Schleswig-Holstein/ | A. Krumbholz, Labor Dr. Krause und Kollegen MVZ GmbH, Kiel | Charit√© Universit√§tsmedizin Berlin, Institut f√ºr Virologie | Victor M Corman et al |
| EPI_ISL_729710 | Germany/SH-ChVir20807/2020 | 2020-12-01 | Europe/Germany/Schleswig-Holstein/ | A. Krumbholz, Labor Dr. Krause und Kollegen MVZ GmbH, Kiel | Charit√© Universit√§tsmedizin Berlin, Institut f√ºr Virologie | Victor M Corman et al |
| EPI_ISL_729712 | Germany/SH-ChVir20811/2020 | 2020-12-01 | Europe/Germany/Schleswig-Holstein/ | A. Krumbholz, Labor Dr. Krause und Kollegen MVZ GmbH, Kiel | Charit√© Universit√§tsmedizin Berlin, Institut f√ºr Virologie | Victor M Corman et al |
| EPI_ISL_729720 | Germany/SH-ChVir20823/2020 | 2020-11-29 | Europe/Germany/Schleswig-Holstein/ | A. Krumbholz, Labor Dr. Krause und Kollegen MVZ GmbH, Kiel | Charit√© Universit√§tsmedizin Berlin, Institut f√ºr Virologie | Victor M Corman et al |
| EPI_ISL_729722 | Germany/SH-ChVir20825/2020 | 2020-11-30 | Europe/Germany/Schleswig-Holstein/ | A. Krumbholz, Labor Dr. Krause und Kollegen MVZ GmbH, Kiel | Charit√© Universit√§tsmedizin Berlin, Institut f√ºr Virologie | Victor M Corman et al |
| EPI_ISL_729723 | Germany/SH-ChVir20841/2020 | 2020-11-29 | Europe/Germany/Schleswig-Holstein/ | A. Krumbholz, Labor Dr. Krause und Kollegen MVZ GmbH, Kiel | Charit√© Universit√§tsmedizin Berlin, Institut f√ºr Virologie | Victor M Corman et al |

| | | | | | | |
|---|---|---|---|---|---|---|
| EPI_ISL_729725 | Germany/SH-ChVir20846/2020 | 2020-12-03 | Europe/Germany/Schleswig-Holstein/ | A. Krumbholz, Labor Dr. Krause und Kollegen MVZ GmbH, Kiel | Charit√© Universit√§tsmedizin Berlin, Institut f√ºr Virologie | Victor M Corman et al |
| EPI_ISL_729726 | Germany/SH-ChVir20847/2020 | 2020-12-03 | Europe/Germany/Schleswig-Holstein/ | A. Krumbholz, Labor Dr. Krause und Kollegen MVZ GmbH, Kiel | Charit√© Universit√§tsmedizin Berlin, Institut f√ºr Virologie | Victor M Corman et al |
| EPI_ISL_729728 | Germany/SH-ChVir20826/2020 | 2020-11-30 | Europe/Germany/Schleswig-Holstein/ | A. Krumbholz, Labor Dr. Krause und Kollegen MVZ GmbH, Kiel | Charit√© Universit√§tsmedizin Berlin, Institut f√ºr Virologie | Victor M Corman et al |
| EPI_ISL_729730 | Germany/SH-ChVir20828/2020 | 2020-11-30 | Europe/Germany/Schleswig-Holstein/ | A. Krumbholz, Labor Dr. Krause und Kollegen MVZ GmbH, Kiel | Charit√© Universit√§tsmedizin Berlin, Institut f√ºr Virologie | Victor M Corman et al |
| EPI_ISL_729731 | Germany/SH-ChVir20829/2020 | 2020-11-30 | Europe/Germany/Schleswig-Holstein/ | A. Krumbholz, Labor Dr. Krause und Kollegen MVZ GmbH, Kiel | Charit√© Universit√§tsmedizin Berlin, Institut f√ºr Virologie | Victor M Corman et al |
| EPI_ISL_729732 | Germany/SH-ChVir20833/2020 | 2020-11-30 | Europe/Germany/Schleswig-Holstein/ | A. Krumbholz, Labor Dr. Krause und Kollegen MVZ GmbH, Kiel | Charit√© Universit√§tsmedizin Berlin, Institut f√ºr Virologie | Victor M Corman et al |
| EPI_ISL_729733 | Germany/SH-ChVir20835/2020 | 2020-11-30 | Europe/Germany/Schleswig-Holstein/ | A. Krumbholz, Labor Dr. Krause und Kollegen MVZ GmbH, Kiel | Charit√© Universit√§tsmedizin Berlin, Institut f√ºr Virologie | Victor M Corman et al |
| EPI_ISL_729734 | Germany/SH-ChVir20836/2020 | 2020-11-29 | Europe/Germany/Schleswig-Holstein/ | A. Krumbholz, Labor Dr. Krause und Kollegen MVZ GmbH, Kiel | Charit√© Universit√§tsmedizin Berlin, Institut f√ºr Virologie | Victor M Corman et al |
| EPI_ISL_729745 | USA/CT-Yale-467/2020 | 2020-09-12 | North America/USA/Connecticut/ | Connecticut Department of Health | Grubaugh Lab - Yale School of Public Health | Joseph Fauver et al |
| EPI_ISL_729754 | USA/CT-Yale-453/2020 | 2020-09-12 | North America/USA/Connecticut/ | Connecticut Department of Health | Grubaugh Lab - Yale School of Public Health | Joseph Fauver et al |

| | | | | | | |
|---|---|---|---|---|---|---|
| EPI_ISL_729755 | USA/CT-Yale-484/2020 | 2020-09-18 | North America/USA/Connecticut/ | Connecticut Department of Health | Grubaugh Lab - Yale School of Public Health | Joseph Fauver et al |
| EPI_ISL_729765 | USA/CT-Yale-480/2020 | 2020-09-18 | North America/USA/Connecticut/ | Connecticut Department of Health | Grubaugh Lab - Yale School of Public Health | Joseph Fauver et al |
| EPI_ISL_729778 | USA/CT-Yale-479/2020 | 2020-09-18 | North America/USA/Connecticut/ | Connecticut Department of Health | Grubaugh Lab - Yale School of Public Health | Joseph Fauver et al |
| EPI_ISL_729794 | Brazil/RS-6195/2020 | 2020-05-11 | South America/Brazil/Rio Grande do Sul | Laboratorio Central de Saude Publica do Estado do Rio Grande do Sul (LACEN-RS) | Laboratory of Respiratory Viruses and Measles, Oswaldo Cruz Institute, FIOCRUZ | Paola Resende et al |
| EPI_ISL_729795 | Brazil/RS-6219/2020 | 2020-05-27 | South America/Brazil/Rio Grande do Sul | Laboratorio Central de Saude Publica do Estado do Rio Grande do Sul (LACEN-RS) | Laboratory of Respiratory Viruses and Measles, Oswaldo Cruz Institute, FIOCRUZ | Paola Resende et al |
| EPI_ISL_729796 | Brazil/RS-6220/2020 | 2020-05-27 | South America/Brazil/Rio Grande do Sul | Laboratorio Central de Saude Publica do Estado do Rio Grande do Sul (LACEN-RS) | Laboratory of Respiratory Viruses and Measles, Oswaldo Cruz Institute, FIOCRUZ | Paola Resende et al |
| EPI_ISL_729797 | Brazil/RS-6180/2020 | 2020-05-06 | South America/Brazil/Rio Grande do Sul | Laboratorio Central de Saude Publica do Estado do Rio Grande do Sul (LACEN-RS) | Laboratory of Respiratory Viruses and Measles, Oswaldo Cruz Institute, FIOCRUZ | Paola Resende et al |
| EPI_ISL_729798 | Brazil/RS-6187/2020 | 2020-05-09 | South America/Brazil/Rio Grande do Sul | Laboratorio Central de Saude Publica do Estado do Rio Grande do Sul (LACEN-RS) | Laboratory of Respiratory Viruses and Measles, Oswaldo Cruz Institute, FIOCRUZ | Paola Resende et al |

| | | | | | | |
|---|---|---|---|---|---|---|
| EPI_ISL_729799 | Brazil/RS-15290/2020 | 2020-08-15 | South America/Brazil/Rio Grande do Sul | Laboratorio Central de Saude Publica do Estado do Rio Grande do Sul (LACEN-RS) | Laboratory of Respiratory Viruses and Measles, Oswaldo Cruz Institute, FIOCRUZ | Paola Resende et al |
| EPI_ISL_729800 | Brazil/RS-6228/2020 | 2020-05-28 | South America/Brazil/Rio Grande do Sul | Laboratorio Central de Saude Publica do Estado do Rio Grande do Sul (LACEN-RS) | Laboratory of Respiratory Viruses and Measles, Oswaldo Cruz Institute, FIOCRUZ | Paola Resende et al |
| EPI_ISL_729801 | Brazil/RS-15270/2020 | 2020-07-13 | South America/Brazil/Rio Grande do Sul | Laboratorio Central de Saude Publica do Estado do Rio Grande do Sul (LACEN-RS) | Laboratory of Respiratory Viruses and Measles, Oswaldo Cruz Institute, FIOCRUZ | Paola Resende et al |
| EPI_ISL_729802 | Brazil/RS-15283/2020 | 2020-08-09 | South America/Brazil/Rio Grande do Sul | Laboratorio Central de Saude Publica do Estado do Rio Grande do Sul (LACEN-RS) | Laboratory of Respiratory Viruses and Measles, Oswaldo Cruz Institute, FIOCRUZ | Paola Resende et al |
| EPI_ISL_729803 | Brazil/RS-15286/2020 | 2020-08-14 | South America/Brazil/Rio Grande do Sul | Laboratorio Central de Saude Publica do Estado do Rio Grande do Sul (LACEN-RS) | Laboratory of Respiratory Viruses and Measles, Oswaldo Cruz Institute, FIOCRUZ | Paola Resende et al |
| EPI_ISL_729804 | Brazil/RS-2525/2020 | 2020-03-09 | South America/Brazil/Rio Grande do Sul | Laboratorio Central de Saude Publica do Estado do Rio Grande do Sul (LACEN-RS) | Laboratory of Respiratory Viruses and Measles, Oswaldo Cruz Institute, FIOCRUZ | Paola Resende et al |

| | | | | | | |
|---|---|---|---|---|---|---|
| EPI_ISL_729817 | Brazil/RS-2565/2020 | 2020-04-03 | South America/Brazil/Rio Grande do Sul | Laboratorio Central de Saude Publica do Estado do Rio Grande do Sul (LACEN-RS) | Laboratory of Respiratory Viruses and Measles, Oswaldo Cruz Institute, FIOCRUZ | Paola Resende et al |
| EPI_ISL_729820 | Brazil/RS-6177/2020 | 2020-05-06 | South America/Brazil/Rio Grande do Sul | Laboratorio Central de Saude Publica do Estado do Rio Grande do Sul (LACEN-RS) | Laboratory of Respiratory Viruses and Measles, Oswaldo Cruz Institute, FIOCRUZ | Paola Resende et al |
| EPI_ISL_729821 | Brazil/RS-6179/2020 | 2020-05-06 | South America/Brazil/Rio Grande do Sul | Laboratorio Central de Saude Publica do Estado do Rio Grande do Sul (LACEN-RS) | Laboratory of Respiratory Viruses and Measles, Oswaldo Cruz Institute, FIOCRUZ | Paola Resende et al |
| EPI_ISL_729822 | Brazil/RS-6183/2020 | 2020-05-06 | South America/Brazil/Rio Grande do Sul | Laboratorio Central de Saude Publica do Estado do Rio Grande do Sul (LACEN-RS) | Laboratory of Respiratory Viruses and Measles, Oswaldo Cruz Institute, FIOCRUZ | Paola Resende et al |
| EPI_ISL_729823 | Brazil/RS-6184/2020 | 2020-05-08 | South America/Brazil/Rio Grande do Sul | Laboratorio Central de Saude Publica do Estado do Rio Grande do Sul (LACEN-RS) | Laboratory of Respiratory Viruses and Measles, Oswaldo Cruz Institute, FIOCRUZ | Paola Resende et al |
| EPI_ISL_729824 | Brazil/RS-6188/2020 | 2020-05-10 | South America/Brazil/Rio Grande do Sul | Laboratorio Central de Saude Publica do Estado do Rio Grande do Sul (LACEN-RS) | Laboratory of Respiratory Viruses and Measles, Oswaldo Cruz Institute, FIOCRUZ | Paola Resende et al |

| | | | | | | |
|---|---|---|---|---|---|---|
| EPI_ISL_729825 | Brazil/RS-6189/2020 | 2020-05-10 | South America/Brazil/Rio Grande do Sul | Laboratorio Central de Saude Publica do Estado do Rio Grande do Sul (LACEN-RS) | Laboratory of Respiratory Viruses and Measles, Oswaldo Cruz Institute, FIOCRUZ | Paola Resende et al |
| EPI_ISL_729826 | Brazil/RS-6190/2020 | 2020-05-11 | South America/Brazil/Rio Grande do Sul | Laboratorio Central de Saude Publica do Estado do Rio Grande do Sul (LACEN-RS) | Laboratory of Respiratory Viruses and Measles, Oswaldo Cruz Institute, FIOCRUZ | Paola Resende et al |
| EPI_ISL_729827 | Brazil/RS-6192/2020 | 2020-05-12 | South America/Brazil/Rio Grande do Sul | Laboratorio Central de Saude Publica do Estado do Rio Grande do Sul (LACEN-RS) | Laboratory of Respiratory Viruses and Measles, Oswaldo Cruz Institute, FIOCRUZ | Paola Resende et al |
| EPI_ISL_729828 | Brazil/RS-6196/2020 | 2020-05-12 | South America/Brazil/Rio Grande do Sul | Laboratorio Central de Saude Publica do Estado do Rio Grande do Sul (LACEN-RS) | Laboratory of Respiratory Viruses and Measles, Oswaldo Cruz Institute, FIOCRUZ | Paola Resende et al |
| EPI_ISL_729829 | Brazil/RS-6197/2020 | 2020-05-13 | South America/Brazil/Rio Grande do Sul | Laboratorio Central de Saude Publica do Estado do Rio Grande do Sul (LACEN-RS) | Laboratory of Respiratory Viruses and Measles, Oswaldo Cruz Institute, FIOCRUZ | Paola Resende et al |
| EPI_ISL_729830 | Brazil/RS-6198/2020 | 2020-05-14 | South America/Brazil/Rio Grande do Sul | Laboratorio Central de Saude Publica do Estado do Rio Grande do Sul (LACEN-RS) | Laboratory of Respiratory Viruses and Measles, Oswaldo Cruz Institute, FIOCRUZ | Paola Resende et al |

| | | | | | | |
|---|---|---|---|---|---|---|
| EPI_ISL_729831 | Brazil/RS-6203/2020 | 2020-05-14 | South America/Brazil/Rio Grande do Sul | Laboratorio Central de Saude Publica do Estado do Rio Grande do Sul (LACEN-RS) | Laboratory of Respiratory Viruses and Measles, Oswaldo Cruz Institute, FIOCRUZ | Paola Resende et al |
| EPI_ISL_729832 | Brazil/RS-6205/2020 | 2020-05-18 | South America/Brazil/Rio Grande do Sul | Laboratorio Central de Saude Publica do Estado do Rio Grande do Sul (LACEN-RS) | Laboratory of Respiratory Viruses and Measles, Oswaldo Cruz Institute, FIOCRUZ | Paola Resende et al |
| EPI_ISL_729833 | Brazil/RS-6208/2020 | 2020-05-19 | South America/Brazil/Rio Grande do Sul | Laboratorio Central de Saude Publica do Estado do Rio Grande do Sul (LACEN-RS) | Laboratory of Respiratory Viruses and Measles, Oswaldo Cruz Institute, FIOCRUZ | Paola Resende et al |
| EPI_ISL_729835 | Brazil/RS-6215/2020 | 2020-05-20 | South America/Brazil/Rio Grande do Sul | Laboratorio Central de Saude Publica do Estado do Rio Grande do Sul (LACEN-RS) | Laboratory of Respiratory Viruses and Measles, Oswaldo Cruz Institute, FIOCRUZ | Paola Resende et al |
| EPI_ISL_729836 | Brazil/RS-6218/2020 | 2020-05-27 | South America/Brazil/Rio Grande do Sul | Laboratorio Central de Saude Publica do Estado do Rio Grande do Sul (LACEN-RS) | Laboratory of Respiratory Viruses and Measles, Oswaldo Cruz Institute, FIOCRUZ | Paola Resende et al |
| EPI_ISL_729837 | Brazil/RS-6222/2020 | 2020-05-28 | South America/Brazil/Rio Grande do Sul | Laboratorio Central de Saude Publica do Estado do Rio Grande do Sul (LACEN-RS) | Laboratory of Respiratory Viruses and Measles, Oswaldo Cruz Institute, FIOCRUZ | Paola Resende et al |

| | | | | | | |
|---|---|---|---|---|---|---|
| EPI_ISL_729838 | Brazil/RS-6227/2020 | 2020-05-29 | South America/Brazil/Rio Grande do Sul | Laboratorio Central de Saude Publica do Estado do Rio Grande do Sul (LACEN-RS) | Laboratory of Respiratory Viruses and Measles, Oswaldo Cruz Institute, FIOCRUZ | Paola Resende et al |
| EPI_ISL_729839 | Brazil/RS-6231/2020 | 2020-05-29 | South America/Brazil/Rio Grande do Sul | Laboratorio Central de Saude Publica do Estado do Rio Grande do Sul (LACEN-RS) | Laboratory of Respiratory Viruses and Measles, Oswaldo Cruz Institute, FIOCRUZ | Paola Resende et al |
| EPI_ISL_729840 | Brazil/RS-6232/2020 | 2020-05-28 | South America/Brazil/Rio Grande do Sul | Laboratorio Central de Saude Publica do Estado do Rio Grande do Sul (LACEN-RS) | Laboratory of Respiratory Viruses and Measles, Oswaldo Cruz Institute, FIOCRUZ | Paola Resende et al |
| EPI_ISL_729841 | Brazil/RS-6240/2020 | 2020-06-01 | South America/Brazil/Rio Grande do Sul | Laboratorio Central de Saude Publica do Estado do Rio Grande do Sul (LACEN-RS) | Laboratory of Respiratory Viruses and Measles, Oswaldo Cruz Institute, FIOCRUZ | Paola Resende et al |
| EPI_ISL_729842 | Brazil/RS-6241/2020 | 2020-06-01 | South America/Brazil/Rio Grande do Sul | Laboratorio Central de Saude Publica do Estado do Rio Grande do Sul (LACEN-RS) | Laboratory of Respiratory Viruses and Measles, Oswaldo Cruz Institute, FIOCRUZ | Paola Resende et al |
| EPI_ISL_729843 | Brazil/RS-6242/2020 | 2020-05-31 | South America/Brazil/Rio Grande do Sul | Laboratorio Central de Saude Publica do Estado do Rio Grande do Sul (LACEN-RS) | Laboratory of Respiratory Viruses and Measles, Oswaldo Cruz Institute, FIOCRUZ | Paola Resende et al |

| | | | | | | |
|---|---|---|---|---|---|---|
| EPI_ISL_729844 | Brazil/RS-15273/2020 | 2020-07-17 | South America/Brazil/Rio Grande do Sul | Laboratorio Central de Saude Publica do Estado do Rio Grande do Sul (LACEN-RS) | Laboratory of Respiratory Viruses and Measles, Oswaldo Cruz Institute, FIOCRUZ | Paola Resende et al |
| EPI_ISL_729845 | Brazil/RS-15279/2020 | 2020-07-28 | South America/Brazil/Rio Grande do Sul | Laboratorio Central de Saude Publica do Estado do Rio Grande do Sul (LACEN-RS) | Laboratory of Respiratory Viruses and Measles, Oswaldo Cruz Institute, FIOCRUZ | Paola Resende et al |
| EPI_ISL_729846 | Brazil/RS-6226/2020 | 2020-05-29 | South America/Brazil/Rio Grande do Sul | Laboratorio Central de Saude Publica do Estado do Rio Grande do Sul (LACEN-RS) | Laboratory of Respiratory Viruses and Measles, Oswaldo Cruz Institute, FIOCRUZ | Paola Resende et al |
| EPI_ISL_729847 | Brazil/RS-6243/2020 | 2020-06-02 | South America/Brazil/Rio Grande do Sul | Laboratorio Central de Saude Publica do Estado do Rio Grande do Sul (LACEN-RS) | Laboratory of Respiratory Viruses and Measles, Oswaldo Cruz Institute, FIOCRUZ | Paola Resende et al |
| EPI_ISL_729848 | Brazil/RS-15278/2020 | 2020-07-27 | South America/Brazil/Rio Grande do Sul | Laboratorio Central de Saude Publica do Estado do Rio Grande do Sul (LACEN-RS) | Laboratory of Respiratory Viruses and Measles, Oswaldo Cruz Institute, FIOCRUZ | Paola Resende et al |
| EPI_ISL_729849 | Brazil/RS-15288/2020 | 2020-08-12 | South America/Brazil/Rio Grande do Sul | Laboratorio Central de Saude Publica do Estado do Rio Grande do Sul (LACEN-RS) | Laboratory of Respiratory Viruses and Measles, Oswaldo Cruz Institute, FIOCRUZ | Paola Resende et al |

| | | | | | | |
|---|---|---|---|---|---|---|
| EPI_ISL_729850 | Brazil/RS-15274/2020 | 2020-07-17 | South America/Brazil/Rio Grande do Sul | Laboratorio Central de Saude Publica do Estado do Rio Grande do Sul (LACEN-RS) | Laboratory of Respiratory Viruses and Measles, Oswaldo Cruz Institute, FIOCRUZ | Paola Resende et al |
| EPI_ISL_729851 | Brazil/RS-15280/2020 | 2020-07-28 | South America/Brazil/Rio Grande do Sul | Laboratorio Central de Saude Publica do Estado do Rio Grande do Sul (LACEN-RS) | Laboratory of Respiratory Viruses and Measles, Oswaldo Cruz Institute, FIOCRUZ | Paola Resende et al |
| EPI_ISL_729852 | Brazil/RS-15281/2020 | 2020-08-07 | South America/Brazil/Rio Grande do Sul | Laboratorio Central de Saude Publica do Estado do Rio Grande do Sul (LACEN-RS) | Laboratory of Respiratory Viruses and Measles, Oswaldo Cruz Institute, FIOCRUZ | Paola Resende et al |
| EPI_ISL_729853 | Brazil/RS-15284/2020 | 2020-08-12 | South America/Brazil/Rio Grande do Sul | Laboratorio Central de Saude Publica do Estado do Rio Grande do Sul (LACEN-RS) | Laboratory of Respiratory Viruses and Measles, Oswaldo Cruz Institute, FIOCRUZ | Paola Resende et al |
| EPI_ISL_729854 | Brazil/RS-15289/2020 | 2020-08-14 | South America/Brazil/Rio Grande do Sul | Laboratorio Central de Saude Publica do Estado do Rio Grande do Sul (LACEN-RS) | Laboratory of Respiratory Viruses and Measles, Oswaldo Cruz Institute, FIOCRUZ | Paola Resende et al |
| EPI_ISL_729856 | Brazil/RS-15275/2020 | 2020-07-18 | South America/Brazil/Rio Grande do Sul | Laboratorio Central de Saude Publica do Estado do Rio Grande do Sul (LACEN-RS) | Laboratory of Respiratory Viruses and Measles, Oswaldo Cruz Institute, FIOCRUZ | Paola Resende et al |

| | | | | | | |
|---|---|---|---|---|---|---|
| EPI_ISL_729857 | Brazil/RS-15276/2020 | 2020-07-20 | South America/Brazil/Rio Grande do Sul | Laboratorio Central de Saude Publica do Estado do Rio Grande do Sul (LACEN-RS) | Laboratory of Respiratory Viruses and Measles, Oswaldo Cruz Institute, FIOCRUZ | Paola Resende et al |
| EPI_ISL_729858 | Brazil/RS-15282/2020 | 2020-08-06 | South America/Brazil/Rio Grande do Sul | Laboratorio Central de Saude Publica do Estado do Rio Grande do Sul (LACEN-RS) | Laboratory of Respiratory Viruses and Measles, Oswaldo Cruz Institute, FIOCRUZ | Paola Resende et al |
| EPI_ISL_729859 | Brazil/RS-15285/2020 | 2020-08-14 | South America/Brazil/Rio Grande do Sul | Laboratorio Central de Saude Publica do Estado do Rio Grande do Sul (LACEN-RS) | Laboratory of Respiratory Viruses and Measles, Oswaldo Cruz Institute, FIOCRUZ | Paola Resende et al |
| EPI_ISL_729860 | Brazil/RS-15287/2020 | 2020-08-13 | South America/Brazil/Rio Grande do Sul | Laboratorio Central de Saude Publica do Estado do Rio Grande do Sul (LACEN-RS) | Laboratory of Respiratory Viruses and Measles, Oswaldo Cruz Institute, FIOCRUZ | Paola Resende et al |
| EPI_ISL_729861 | Brazil/RS-15292/2020 | 2020-08-16 | South America/Brazil/Rio Grande do Sul | Laboratorio Central de Saude Publica do Estado do Rio Grande do Sul (LACEN-RS) | Laboratory of Respiratory Viruses and Measles, Oswaldo Cruz Institute, FIOCRUZ | Paola Resende et al |
| EPI_ISL_729866 | Peru/LIM-UPCH-0203/2020 | 2020-10-15 | South America/Peru/Lima/Lima | Laboratorio de Referencia Nacional de Virus Respiratorios, Instituto Nacional de Salud Peru | Laboratorio de Genv≥mica Microbiana, Universidad Peruana Cayetano Heredia | Pablo Tsukayama et al |

| | | | | | | |
|---|---|---|---|---|---|---|
| EPI_ISL_729868 | Peru/LIM-UPCH-0238/2020 | 2020-10-15 | South America/Peru/Lima/Lima | Laboratorio de Referencia Nacional de Virus Respiratorios, Instituto Nacional de Salud Peru | Laboratorio de Gen√≥mica Microbiana, Universidad Peruana Cayetano Heredia | Pablo Tsukayama et al |
| EPI_ISL_729869 | Peru/AMA-UPCH-0255/2020 | 2020-09-21 | South America/Peru/Amazonas PE/Bong | Instituto de Medicina Tropical, Universidad Nacional Toribio Rodr√≠guez de Mendoza de Amazonas | Laboratorio de Gen√≥mica Microbiana, Universidad Peruana Cayetano Heredia | Pablo Tsukayama et al |
| EPI_ISL_729870 | Peru/AMA-UPCH-0269/2020 | 2020-10-01 | South America/Peru/Amazonas PE/Bong | Instituto de Medicina Tropical, Universidad Nacional Toribio Rodr√≠guez de Mendoza de Amazonas | Laboratorio de Gen√≥mica Microbiana, Universidad Peruana Cayetano Heredia | Pablo Tsukayama et al |
| EPI_ISL_729871 | Peru/AMA-UPCH-0277/2020 | 2020-09-16 | South America/Peru/Amazonas PE/Chac | Instituto de Medicina Tropical, Universidad Nacional Toribio Rodr√≠guez de Mendoza de Amazonas | Laboratorio de Gen√≥mica Microbiana, Universidad Peruana Cayetano Heredia | Pablo Tsukayama et al |
| EPI_ISL_729873 | Peru/LIM-UPCH-0165/2020 | 2020-10-14 | South America/Peru/Lima/Lima | Laboratorio de Referencia Nacional de Virus Respiratorios, Instituto Nacional de Salud Peru | Laboratorio de Gen√≥mica Microbiana, Universidad Peruana Cayetano Heredia | Pablo Tsukayama et al |
| EPI_ISL_729874 | Peru/LIM-UPCH-0167/2020 | 2020-10-14 | South America/Peru/Lima/Lima | Laboratorio de Referencia Nacional de Virus Respiratorios, Instituto Nacional de Salud Peru | Laboratorio de Gen√≥mica Microbiana, Universidad Peruana Cayetano Heredia | Pablo Tsukayama et al |

| | | | | | | |
|---|---|---|---|---|---|---|
| EPI_ISL_729876 | Peru/LIM-UPCH-0169/2020 | 2020-10-12 | South America/Peru/Lima/Lima | Laboratorio de Referencia Nacional de Virus Respiratorios, Instituto Nacional de Salud Peru | Laboratorio de Genv≥mica Microbiana, Universidad Peruana Cayetano Heredia | Pablo Tsukayama et al |
| EPI_ISL_729877 | Peru/LIM-UPCH-0170/2020 | 2020-10-14 | South America/Peru/Lima/Lima | Laboratorio de Referencia Nacional de Virus Respiratorios, Instituto Nacional de Salud Peru | Laboratorio de Genv≥mica Microbiana, Universidad Peruana Cayetano Heredia | Pablo Tsukayama et al |
| EPI_ISL_729879 | Peru/LIM-UPCH-0173/2020 | 2020-10-13 | South America/Peru/Lima/Lima | Laboratorio de Referencia Nacional de Virus Respiratorios, Instituto Nacional de Salud Peru | Laboratorio de Genv≥mica Microbiana, Universidad Peruana Cayetano Heredia | Pablo Tsukayama et al |
| EPI_ISL_729881 | Peru/LIM-UPCH-0176/2020 | 2020-10-14 | South America/Peru/Lima/Lima | Laboratorio de Referencia Nacional de Virus Respiratorios, Instituto Nacional de Salud Peru | Laboratorio de Genv≥mica Microbiana, Universidad Peruana Cayetano Heredia | Pablo Tsukayama et al |
| EPI_ISL_729882 | Peru/LIM-UPCH-0179/2020 | 2020-10-13 | South America/Peru/Lima/Lima | Laboratorio de Referencia Nacional de Virus Respiratorios, Instituto Nacional de Salud Peru | Laboratorio de Genv≥mica Microbiana, Universidad Peruana Cayetano Heredia | Pablo Tsukayama et al |
| EPI_ISL_729883 | Peru/LIM-UPCH-0182/2020 | 2020-10-13 | South America/Peru/Lima/Lima | Laboratorio de Referencia Nacional de Virus Respiratorios, Instituto Nacional de Salud Peru | Laboratorio de Genv≥mica Microbiana, Universidad Peruana Cayetano Heredia | Pablo Tsukayama et al |

| | | | | | | |
|---|---|---|---|---|---|---|
| EPI_ISL_729884 | Peru/LIM-UPCH-0184/2020 | 2020-10-13 | South America/Peru/Lima/Lima | Laboratorio de Referencia Nacional de Virus Respiratorios, Instituto Nacional de Salud Peru | Laboratorio de Genv≥mica Microbiana, Universidad Peruana Cayetano Heredia | Pablo Tsukayama et al |
| EPI_ISL_729885 | Peru/LIM-UPCH-0187/2020 | 2020-10-13 | South America/Peru/Lima/Lima | Laboratorio de Referencia Nacional de Virus Respiratorios, Instituto Nacional de Salud Peru | Laboratorio de Genv≥mica Microbiana, Universidad Peruana Cayetano Heredia | Pablo Tsukayama et al |
| EPI_ISL_729886 | Peru/LIM-UPCH-0189/2020 | 2020-10-14 | South America/Peru/Lima/Lima | Laboratorio de Referencia Nacional de Virus Respiratorios, Instituto Nacional de Salud Peru | Laboratorio de Genv≥mica Microbiana, Universidad Peruana Cayetano Heredia | Pablo Tsukayama et al |
| EPI_ISL_729887 | Peru/LIM-UPCH-0191/2020 | 2020-10-15 | South America/Peru/Lima/Lima | Laboratorio de Referencia Nacional de Virus Respiratorios, Instituto Nacional de Salud Peru | Laboratorio de Genv≥mica Microbiana, Universidad Peruana Cayetano Heredia | Pablo Tsukayama et al |
| EPI_ISL_729891 | Peru/LIM-UPCH-0197/2020 | 2020-10-14 | South America/Peru/Lima/Lima | Laboratorio de Referencia Nacional de Virus Respiratorios, Instituto Nacional de Salud Peru | Laboratorio de Genv≥mica Microbiana, Universidad Peruana Cayetano Heredia | Pablo Tsukayama et al |
| EPI_ISL_729892 | Peru/LIM-UPCH-0198/2020 | 2020-10-16 | South America/Peru/Lima/Lima | Laboratorio de Referencia Nacional de Virus Respiratorios, Instituto Nacional de Salud Peru | Laboratorio de Genv≥mica Microbiana, Universidad Peruana Cayetano Heredia | Pablo Tsukayama et al |

| | | | | | | |
|---|---|---|---|---|---|---|
| EPI_ISL_729893 | Peru/LIM-UPCH-0200/2020 | 2020-10-15 | South America/Peru/Lima/Lima | Laboratorio de Referencia Nacional de Virus Respiratorios, Instituto Nacional de Salud Peru | Laboratorio de Genv≥mica Microbiana, Universidad Peruana Cayetano Heredia | Pablo Tsukayama et al |
| EPI_ISL_729894 | Peru/LIM-UPCH-0202/2020 | 2020-10-15 | South America/Peru/Lima/Lima | Laboratorio de Referencia Nacional de Virus Respiratorios, Instituto Nacional de Salud Peru | Laboratorio de Genv≥mica Microbiana, Universidad Peruana Cayetano Heredia | Pablo Tsukayama et al |
| EPI_ISL_729895 | Peru/LIM-UPCH-0204/2020 | 2020-10-15 | South America/Peru/Lima/Lima | Laboratorio de Referencia Nacional de Virus Respiratorios, Instituto Nacional de Salud Peru | Laboratorio de Genv≥mica Microbiana, Universidad Peruana Cayetano Heredia | Pablo Tsukayama et al |
| EPI_ISL_729896 | Peru/LIM-UPCH-0205/2020 | 2020-10-15 | South America/Peru/Lima/Lima | Laboratorio de Referencia Nacional de Virus Respiratorios, Instituto Nacional de Salud Peru | Laboratorio de Genv≥mica Microbiana, Universidad Peruana Cayetano Heredia | Pablo Tsukayama et al |
| EPI_ISL_729897 | Peru/LIM-UPCH-0207/2020 | 2020-10-16 | South America/Peru/Lima/Lima | Laboratorio de Referencia Nacional de Virus Respiratorios, Instituto Nacional de Salud Peru | Laboratorio de Genv≥mica Microbiana, Universidad Peruana Cayetano Heredia | Pablo Tsukayama et al |
| EPI_ISL_729898 | Peru/LIM-UPCH-0236/2020 | 2020-10-14 | South America/Peru/Lima/Lima | Laboratorio de Referencia Nacional de Virus Respiratorios, Instituto Nacional de Salud Peru | Laboratorio de Genv≥mica Microbiana, Universidad Peruana Cayetano Heredia | Pablo Tsukayama et al |

| | | | | | | |
|---|---|---|---|---|---|---|
| EPI_ISL_729899 | Peru/LIM-UPCH-0240/2020 | 2020-10-15 | South America/Peru/Lima/Lima | Laboratorio de Referencia Nacional de Virus Respiratorios, Instituto Nacional de Salud Peru | Laboratorio de Gen√≥mica Microbiana, Universidad Peruana Cayetano Heredia | Pablo Tsukayama et al |
| EPI_ISL_729901 | Peru/LIM-UPCH-0243/2020 | 2020-10-14 | South America/Peru/Lima/Lima | Laboratorio de Referencia Nacional de Virus Respiratorios, Instituto Nacional de Salud Peru | Laboratorio de Gen√≥mica Microbiana, Universidad Peruana Cayetano Heredia | Pablo Tsukayama et al |
| EPI_ISL_729902 | Peru/AMA-UPCH-0256/2020 | 2020-08-29 | South America/Peru/Amazonas PE/Bong | Instituto de Medicina Tropical, Universidad Nacional Toribio Rodr√≠guez de Mendoza de Amazonas | Laboratorio de Gen√≥mica Microbiana, Universidad Peruana Cayetano Heredia | Pablo Tsukayama et al |
| EPI_ISL_729903 | Peru/AMA-UPCH-0257/2020 | 2020-09-28 | South America/Peru/Amazonas PE/Chac | Instituto de Medicina Tropical, Universidad Nacional Toribio Rodr√≠guez de Mendoza de Amazonas | Laboratorio de Gen√≥mica Microbiana, Universidad Peruana Cayetano Heredia | Pablo Tsukayama et al |
| EPI_ISL_729904 | Peru/AMA-UPCH-0258/2020 | 2020-09-08 | South America/Peru/Amazonas PE/Bong | Instituto de Medicina Tropical, Universidad Nacional Toribio Rodr√≠guez de Mendoza de Amazonas | Laboratorio de Gen√≥mica Microbiana, Universidad Peruana Cayetano Heredia | Pablo Tsukayama et al |
| EPI_ISL_729905 | Peru/AMA-UPCH-0259/2020 | 2020-09-08 | South America/Peru/Amazonas PE/Chac | Instituto de Medicina Tropical, Universidad Nacional Toribio Rodr√≠guez de Mendoza de Amazonas | Laboratorio de Gen√≥mica Microbiana, Universidad Peruana Cayetano Heredia | Pablo Tsukayama et al |

| | | | | | | |
|---|---|---|---|---|---|---|
| EPI_ISL_729906 | Peru/AMA-UPCH-0260/2020 | 2020-09-17 | South America/Peru/Amazonas PE/Chac | Instituto de Medicina Tropical, Universidad Nacional Toribio Rodr√≠guez de Mendoza de Amazonas | Laboratorio de Gen√≥mica Microbiana, Universidad Peruana Cayetano Heredia | Pablo Tsukayama et al |
| EPI_ISL_729907 | Peru/AMA-UPCH-0261/2020 | 2020-09-28 | South America/Peru/Amazonas PE/Chac | Instituto de Medicina Tropical, Universidad Nacional Toribio Rodr√≠guez de Mendoza de Amazonas | Laboratorio de Gen√≥mica Microbiana, Universidad Peruana Cayetano Heredia | Pablo Tsukayama et al |
| EPI_ISL_729908 | Peru/AMA-UPCH-0262/2020 | 2020-09-21 | South America/Peru/Amazonas PE/Chac | Instituto de Medicina Tropical, Universidad Nacional Toribio Rodr√≠guez de Mendoza de Amazonas | Laboratorio de Gen√≥mica Microbiana, Universidad Peruana Cayetano Heredia | Pablo Tsukayama et al |
| EPI_ISL_729909 | Peru/AMA-UPCH-0263/2020 | 2020-09-21 | South America/Peru/Amazonas PE/Bong | Instituto de Medicina Tropical, Universidad Nacional Toribio Rodr√≠guez de Mendoza de Amazonas | Laboratorio de Gen√≥mica Microbiana, Universidad Peruana Cayetano Heredia | Pablo Tsukayama et al |
| EPI_ISL_729910 | Peru/AMA-UPCH-0264/2020 | 2020-08-20 | South America/Peru/Amazonas PE/Chac | Instituto de Medicina Tropical, Universidad Nacional Toribio Rodr√≠guez de Mendoza de Amazonas | Laboratorio de Gen√≥mica Microbiana, Universidad Peruana Cayetano Heredia | Pablo Tsukayama et al |
| EPI_ISL_729911 | Peru/AMA-UPCH-0265/2020 | 2020-09-21 | South America/Peru/Amazonas PE/Chac | Instituto de Medicina Tropical, Universidad Nacional Toribio Rodr√≠guez de Mendoza de Amazonas | Laboratorio de Gen√≥mica Microbiana, Universidad Peruana Cayetano Heredia | Pablo Tsukayama et al |

| | | | | | | |
|---|---|---|---|---|---|---|
| EPI_ISL_729912 | Peru/AMA-UPCH-0266/2020 | 2020-08-26 | South America/Peru/Amazonas PE/Chac | Instituto de Medicina Tropical, Universidad Nacional Toribio Rodr√≠guez de Mendoza de Amazonas | Laboratorio de Gen√≥mica Microbiana, Universidad Peruana Cayetano Heredia | Pablo Tsukayama et al |
| EPI_ISL_729913 | Peru/AMA-UPCH-0267/2020 | 2020-08-18 | South America/Peru/Amazonas PE/Chac | Instituto de Medicina Tropical, Universidad Nacional Toribio Rodr√≠guez de Mendoza de Amazonas | Laboratorio de Gen√≥mica Microbiana, Universidad Peruana Cayetano Heredia | Pablo Tsukayama et al |
| EPI_ISL_729914 | Peru/AMA-UPCH-0268/2020 | 2020-08-17 | South America/Peru/Amazonas PE/Chac | Instituto de Medicina Tropical, Universidad Nacional Toribio Rodr√≠guez de Mendoza de Amazonas | Laboratorio de Gen√≥mica Microbiana, Universidad Peruana Cayetano Heredia | Pablo Tsukayama et al |
| EPI_ISL_729915 | Peru/AMA-UPCH-0270/2020 | 2020-09-21 | South America/Peru/Amazonas PE/Bong | Instituto de Medicina Tropical, Universidad Nacional Toribio Rodr√≠guez de Mendoza de Amazonas | Laboratorio de Gen√≥mica Microbiana, Universidad Peruana Cayetano Heredia | Pablo Tsukayama et al |
| EPI_ISL_729916 | Peru/AMA-UPCH-0272/2020 | 2020-09-21 | South America/Peru/Amazonas PE/Luya | Instituto de Medicina Tropical, Universidad Nacional Toribio Rodr√≠guez de Mendoza de Amazonas | Laboratorio de Gen√≥mica Microbiana, Universidad Peruana Cayetano Heredia | Pablo Tsukayama et al |
| EPI_ISL_729917 | Peru/AMA-UPCH-0273/2020 | 2020-08-11 | South America/Peru/Amazonas PE/Chac | Instituto de Medicina Tropical, Universidad Nacional Toribio Rodr√≠guez de Mendoza de Amazonas | Laboratorio de Gen√≥mica Microbiana, Universidad Peruana Cayetano Heredia | Pablo Tsukayama et al |

| | | | | | | |
|---|---|---|---|---|---|---|
| EPI_ISL_729918 | Peru/AMA-UPCH-0274/2020 | 2020-08-11 | South America/Peru/Amazonas PE/Chac | Instituto de Medicina Tropical, Universidad Nacional Toribio Rodr√≠guez de Mendoza de Amazonas | Laboratorio de Gen√≥mica Microbiana, Universidad Peruana Cayetano Heredia | Pablo Tsukayama et al |
| EPI_ISL_729919 | Peru/AMA-UPCH-0275/2020 | 2020-08-26 | South America/Peru/Amazonas PE/Chac | Instituto de Medicina Tropical, Universidad Nacional Toribio Rodr√≠guez de Mendoza de Amazonas | Laboratorio de Gen√≥mica Microbiana, Universidad Peruana Cayetano Heredia | Pablo Tsukayama et al |
| EPI_ISL_729928 | Nigeria/BE-CV243/2020 | 2020-07-05 | Africa/Nigeria/Benue State/ | Nigeria Centre for Disease Control (NCDC) | African Centre of Excellence for Genomics of Infectious Diseases (ACEGID), Redeemer's University, Ede, Osun State, Nigeria | Oluniyi P.E. et al et al |
| EPI_ISL_729929 | Nigeria/NI-CV244/2020 | 2020-07-02 | Africa/Nigeria/Niger State/ | Nigeria Centre for Disease Control (NCDC) | African Centre of Excellence for Genomics of Infectious Diseases (ACEGID), Redeemer's University, Ede, Osun State, Nigeria | Oluniyi P.E. et al et al |

| | | | | | | |
|---|---|---|---|---|---|---|
| EPI_ISL_729931 | Nigeria/FC-CV246/2020 | 2020-07-07 | Africa/Nigeria/Abuja/ | Nigeria Centre for Disease Control (NCDC) | African Centre of Excellence for Genomics of Infectious Diseases (ACEGID), Redeemer's University, Ede, Osun State, Nigeria | Oluniyi P.E. et al et al |
| EPI_ISL_729932 | Nigeria/FC-CV247/2020 | 2020-07-08 | Africa/Nigeria/Abuja/ | Nigeria Centre for Disease Control (NCDC) | African Centre of Excellence for Genomics of Infectious Diseases (ACEGID), Redeemer's University, Ede, Osun State, Nigeria | Oluniyi P.E. et al et al |
| EPI_ISL_729933 | Nigeria/FC-CV248/2020 | 2020-07-09 | Africa/Nigeria/Abuja/ | Nigeria Centre for Disease Control (NCDC) | African Centre of Excellence for Genomics of Infectious Diseases (ACEGID), Redeemer's University, Ede, Osun State, Nigeria | Oluniyi P.E. et al et al |
| EPI_ISL_729945 | Nigeria/LA-CV261/2020 | 2020-05-14 | Africa/Nigeria/Lagos/ | Nigeria Centre for Disease Control (NCDC) | African Centre of Excellence for Genomics of Infectious Diseases (ACEGID), Redeemer's University, Ede, Osun State, Nigeria | Oluniyi P.E. et al et al |

| | | | | | | |
|---|---|---|---|---|---|---|
| EPI_ISL_729953 | Nigeria/AK-CV269/2020 | 2020-06-29 | Africa/Nigeria/Akwa-Ibom/ | Nigeria Centre for Disease Control (NCDC) | African Centre of Excellence for Genomics of Infectious Diseases (ACEGID), Redeemer's University, Ede, Osun State, Nigeria | Oluniyi P.E. et al et al |
| EPI_ISL_729955 | Nigeria/AK-CV271/2020 | 2020-07-01 | Africa/Nigeria/Akwa-Ibom/ | Nigeria Centre for Disease Control (NCDC) | African Centre of Excellence for Genomics of Infectious Diseases (ACEGID), Redeemer's University, Ede, Osun State, Nigeria | Oluniyi P.E. et al et al |
| EPI_ISL_729956 | Nigeria/AK-CV272/2020 | 2020-07-12 | Africa/Nigeria/Akwa-Ibom/ | Nigeria Centre for Disease Control (NCDC) | African Centre of Excellence for Genomics of Infectious Diseases (ACEGID), Redeemer's University, Ede, Osun State, Nigeria | Oluniyi P.E. et al et al |
| EPI_ISL_729958 | Nigeria/AK-CV274/2020 | 2020-07-25 | Africa/Nigeria/Akwa-Ibom/ | Nigeria Centre for Disease Control (NCDC) | African Centre of Excellence for Genomics of Infectious Diseases (ACEGID), Redeemer's University, Ede, Osun State, Nigeria | Oluniyi P.E. et al et al |

| | | | | | | |
|---|---|---|---|---|---|---|
| EPI_ISL_729967 | Nigeria/OS-CV286/2020 | 2020-06-23 | Africa/Nigeria/Osun State/ | Nigeria Centre for Disease Control (NCDC) | African Centre of Excellence for Genomics of Infectious Diseases (ACEGID), Redeemer's University, Ede, Osun State, Nigeria | Oluniyi P.E. et al et al |
| EPI_ISL_729968 | Nigeria/OS-CV287/2020 | 2020-06-27 | Africa/Nigeria/Osun State/ | Nigeria Centre for Disease Control (NCDC) | African Centre of Excellence for Genomics of Infectious Diseases (ACEGID), Redeemer's University, Ede, Osun State, Nigeria | Oluniyi P.E. et al et al |
| EPI_ISL_729969 | Nigeria/OS-CV288/2020 | 2020-06-30 | Africa/Nigeria/Osun State/ | Nigeria Centre for Disease Control (NCDC) | African Centre of Excellence for Genomics of Infectious Diseases (ACEGID), Redeemer's University, Ede, Osun State, Nigeria | Oluniyi P.E. et al et al |
| EPI_ISL_729974 | Nigeria/OS-CV295/2020 | 2020-07-08 | Africa/Nigeria/Osun State/ | Nigeria Centre for Disease Control (NCDC) | African Centre of Excellence for Genomics of Infectious Diseases (ACEGID), Redeemer's University, Ede, Osun State, Nigeria | Oluniyi P.E. et al et al |

| | | | | | | |
|---|---|---|---|---|---|---|
| EPI_ISL_729975 | Nigeria/OS-CV296/2020 | 2020-08-03 | Africa/Nigeria/Osun State/ | Nigeria Centre for Disease Control (NCDC) | African Centre of Excellence for Genomics of Infectious Diseases (ACEGID), Redeemer's University, Ede, Osun State, Nigeria | Oluniyi P.E. et al et al |
| EPI_ISL_729976 | Nigeria/OS-CV297/2020 | 2020-08-04 | Africa/Nigeria/Osun State/ | Nigeria Centre for Disease Control (NCDC) | African Centre of Excellence for Genomics of Infectious Diseases (ACEGID), Redeemer's University, Ede, Osun State, Nigeria | Oluniyi P.E. et al et al |
| EPI_ISL_729977 | Nigeria/OS-CV299/2020 | 2020-08-07 | Africa/Nigeria/Osun State/ | Nigeria Centre for Disease Control (NCDC) | African Centre of Excellence for Genomics of Infectious Diseases (ACEGID), Redeemer's University, Ede, Osun State, Nigeria | Oluniyi P.E. et al et al |
| EPI_ISL_729978 | Nigeria/OS-CV300/2020 | 2020-08-31 | Africa/Nigeria/Osun State/ | Nigeria Centre for Disease Control (NCDC) | African Centre of Excellence for Genomics of Infectious Diseases (ACEGID), Redeemer's University, Ede, Osun State, Nigeria | Oluniyi P.E. et al et al |

| | | | | | | |
|---|---|---|---|---|---|---|
| EPI_ISL_729979 | Nigeria/OS-CV301/2020 | 2020-09-01 | Africa/Nigeria/Osun State/ | Nigeria Centre for Disease Control (NCDC) | African Centre of Excellence for Genomics of Infectious Diseases (ACEGID), Redeemer's University, Ede, Osun State, Nigeria | Oluniyi P.E. et al et al |
| EPI_ISL_729980 | Nigeria/OS-CV302/2020 | 2020-09-03 | Africa/Nigeria/Osun State/ | Nigeria Centre for Disease Control (NCDC) | African Centre of Excellence for Genomics of Infectious Diseases (ACEGID), Redeemer's University, Ede, Osun State, Nigeria | Oluniyi P.E. et al et al |
| EPI_ISL_729981 | Nigeria/OS-CV303/2020 | 2020-08-17 | Africa/Nigeria/Osun State/ | Nigeria Centre for Disease Control (NCDC) | African Centre of Excellence for Genomics of Infectious Diseases (ACEGID), Redeemer's University, Ede, Osun State, Nigeria | Oluniyi P.E. et al et al |
| EPI_ISL_729982 | Nigeria/OS-CV304/2020 | 2020-10-05 | Africa/Nigeria/Osun State/ | Nigeria Centre for Disease Control (NCDC) | African Centre of Excellence for Genomics of Infectious Diseases (ACEGID), Redeemer's University, Ede, Osun State, Nigeria | Oluniyi P.E. et al et al |

| | | | | | | |
|---|---|---|---|---|---|---|
| EPI_ISL_729986 | Nigeria/AK-CV308/2020 | 2020-06-12 | Africa/Nigeria/Akwa-Ibom/ | Nigeria Centre for Disease Control (NCDC) | African Centre of Excellence for Genomics of Infectious Diseases (ACEGID), Redeemer's University, Ede, Osun State, Nigeria | Oluniyi P.E. et al et al |
| EPI_ISL_729987 | Nigeria/AK-CV313/2020 | 2020-06-27 | Africa/Nigeria/Akwa-Ibom/ | Nigeria Centre for Disease Control (NCDC) | African Centre of Excellence for Genomics of Infectious Diseases (ACEGID), Redeemer's University, Ede, Osun State, Nigeria | Oluniyi P.E. et al et al |
| EPI_ISL_729988 | Nigeria/AK-CV314/2020 | 2020-06-29 | Africa/Nigeria/Akwa-Ibom/ | Nigeria Centre for Disease Control (NCDC) | African Centre of Excellence for Genomics of Infectious Diseases (ACEGID), Redeemer's University, Ede, Osun State, Nigeria | Oluniyi P.E. et al et al |
| EPI_ISL_729989 | Nigeria/AK-CV317/2020 | 2020-07-04 | Africa/Nigeria/Akwa-Ibom/ | Nigeria Centre for Disease Control (NCDC) | African Centre of Excellence for Genomics of Infectious Diseases (ACEGID), Redeemer's University, Ede, Osun State, Nigeria | Oluniyi P.E. et al et al |

| | | | | | | |
|---|---|---|---|---|---|---|
| EPI_ISL_729990 | Nigeria/AK-CV319/2020 | 2020-07-17 | Africa/Nigeria/Akwa-Ibom/ | Nigeria Centre for Disease Control (NCDC) | African Centre of Excellence for Genomics of Infectious Diseases (ACEGID), Redeemer's University, Ede, Osun State, Nigeria | Oluniyi P.E. et al et al |
| EPI_ISL_729993 | Nigeria/AK-CV322/2020 | 2020-07-20 | Africa/Nigeria/Akwa-Ibom/ | Nigeria Centre for Disease Control (NCDC) | African Centre of Excellence for Genomics of Infectious Diseases (ACEGID), Redeemer's University, Ede, Osun State, Nigeria | Oluniyi P.E. et al et al |
| EPI_ISL_729994 | Nigeria/AK-CV324/2020 | 2020-07-27 | Africa/Nigeria/Akwa-Ibom/ | Nigeria Centre for Disease Control (NCDC) | African Centre of Excellence for Genomics of Infectious Diseases (ACEGID), Redeemer's University, Ede, Osun State, Nigeria | Oluniyi P.E. et al et al |
| EPI_ISL_730011 | Nigeria/BO-CV346/2020 | 2020-05-16 | Africa/Nigeria/Borno State/ | Nigeria Centre for Disease Control (NCDC) | African Centre of Excellence for Genomics of Infectious Diseases (ACEGID), Redeemer's University, Ede, Osun State, Nigeria | Oluniyi P.E. et al et al |

| | | | | | | |
|---|---|---|---|---|---|---|
| EPI_ISL_730014 | Nigeria/BO-CV349/2020 | 2020-05-19 | Africa/Nigeria/Borno State/ | Nigeria Centre for Disease Control (NCDC) | African Centre of Excellence for Genomics of Infectious Diseases (ACEGID), Redeemer's University, Ede, Osun State, Nigeria | Oluniyi P.E. et al et al |
| EPI_ISL_730015 | Nigeria/BO-CV350/2020 | 2020-05-23 | Africa/Nigeria/Borno State/ | Nigeria Centre for Disease Control (NCDC) | African Centre of Excellence for Genomics of Infectious Diseases (ACEGID), Redeemer's University, Ede, Osun State, Nigeria | Oluniyi P.E. et al et al |
| EPI_ISL_730016 | Nigeria/NA-CV351/2020 | 2020-05-31 | Africa/Nigeria/Nasarawa State/ | Nigeria Centre for Disease Control (NCDC) | African Centre of Excellence for Genomics of Infectious Diseases (ACEGID), Redeemer's University, Ede, Osun State, Nigeria | Oluniyi P.E. et al et al |
| EPI_ISL_730017 | Nigeria/BE-CV352/2020 | 2020-06-01 | Africa/Nigeria/Benue State/ | Nigeria Centre for Disease Control (NCDC) | African Centre of Excellence for Genomics of Infectious Diseases (ACEGID), Redeemer's University, Ede, Osun State, Nigeria | Oluniyi P.E. et al et al |

| | | | | | | |
|---|---|---|---|---|---|---|
| EPI_ISL_730019 | Nigeria/FC-CV354/2020 | 2020-06-24 | Africa/Nigeria/Abuja/ | Nigeria Centre for Disease Control (NCDC) | African Centre of Excellence for Genomics of Infectious Diseases (ACEGID), Redeemer's University, Ede, Osun State, Nigeria | Oluniyi P.E. et al et al |
| EPI_ISL_730020 | Nigeria/FC-CV355/2020 | 2020-06-26 | Africa/Nigeria/Abuja/ | Nigeria Centre for Disease Control (NCDC) | African Centre of Excellence for Genomics of Infectious Diseases (ACEGID), Redeemer's University, Ede, Osun State, Nigeria | Oluniyi P.E. et al et al |
| EPI_ISL_730022 | Nigeria/FC-CV357/2020 | 2020-06-26 | Africa/Nigeria/Abuja/ | Nigeria Centre for Disease Control (NCDC) | African Centre of Excellence for Genomics of Infectious Diseases (ACEGID), Redeemer's University, Ede, Osun State, Nigeria | Oluniyi P.E. et al et al |
| EPI_ISL_730033 | Nigeria/LA-CV374/2020 | 2020-05-16 | Africa/Nigeria/Lagos/ | Nigeria Centre for Disease Control (NCDC) | African Centre of Excellence for Genomics of Infectious Diseases (ACEGID), Redeemer's University, Ede, Osun State, Nigeria | Oluniyi P.E. et al et al |

| | | | | | | |
|---|---|---|---|---|---|---|
| EPI_ISL_730040 | Nigeria/OG-CV387/2020 | 2020-05-29 | Africa/Nigeria/Ogun State/ | Nigeria Centre for Disease Control (NCDC) | African Centre of Excellence for Genomics of Infectious Diseases (ACEGID), Redeemer's University, Ede, Osun State, Nigeria | Oluniyi P.E. et al et al |
| EPI_ISL_730058 | USA/CT-Yale-574/2020 | 2020-11-02 | North America/USA/Connecticut/ | Connecticut Department of Health | Grubaugh Lab - Yale School of Public Health | Joseph Fauver et al |
| EPI_ISL_730062 | USA/CT-Yale-598/2020 | 2020-05-01 | North America/USA/Connecticut/ | Yale Clinical Virology Laboratory | Grubaugh Lab - Yale School of Public Health | Joseph Fauver et al |
| EPI_ISL_730079 | USA/CT-Yale-624/2020 | 2020-06-03 | North America/USA/Connecticut/ | Yale Clinical Virology Laboratory | Grubaugh Lab - Yale School of Public Health | Joseph Fauver et al |
| EPI_ISL_730082 | USA/CT-Yale-628/2020 | 2020-07-10 | North America/USA/Connecticut/ | Yale Clinical Virology Laboratory | Grubaugh Lab - Yale School of Public Health | Joseph Fauver et al |
| EPI_ISL_730086 | USA/CA-ALSR-4693/2020 | 2020-11-11 | North America/USA/California/San Dieg | San Diego County Public Health Laboratory | Andersen lab at Scripps Research | SEARCH Alliance San Diego with Tracy Basler et al |
| EPI_ISL_730087 | USA/CA-ALSR-4694/2020 | 2020-10-07 | North America/USA/California/San Dieg | San Diego County Public Health Laboratory | Andersen lab at Scripps Research | SEARCH Alliance San Diego with Tracy Basler et al |
| EPI_ISL_730097 | USA/CA-ALSR-4710/2020 | 2020-09-29 | North America/USA/California/San Dieg | San Diego County Public Health Laboratory | Andersen lab at Scripps Research | SEARCH Alliance San Diego with Tracy Basler et al |
| EPI_ISL_730100 | USA/CA-ALSR-4715/2020 | 2020-10-29 | North America/USA/California/San Dieg | San Diego County Public Health Laboratory | Andersen lab at Scripps Research | SEARCH Alliance San Diego with Tracy Basler et al |
| EPI_ISL_730101 | USA/CA-ALSR-4716/2020 | 2020-09-21 | North America/USA/California/San Dieg | San Diego County Public Health Laboratory | Andersen lab at Scripps Research | SEARCH Alliance San Diego with Tracy Basler et al |
| EPI_ISL_730102 | USA/CA-ALSR-4718/2020 | 2020-10-09 | North America/USA/California/San Dieg | San Diego County Public Health Laboratory | Andersen lab at Scripps Research | SEARCH Alliance San Diego with Tracy Basler et al |
| EPI_ISL_730105 | USA/CA-ALSR-4722/2020 | 2020-09-29 | North America/USA/California/San Dieg | San Diego County Public Health Laboratory | Andersen lab at Scripps Research | SEARCH Alliance San Diego with Tracy Basler et al |
| EPI_ISL_730109 | USA/CA-ALSR-4726/2020 | 2020-09-21 | North America/USA/California/San Dieg | San Diego County Public Health Laboratory | Andersen lab at Scripps Research | SEARCH Alliance San Diego with Tracy Basler et al |
| EPI_ISL_730111 | USA/CA-ALSR-4728/2020 | 2020-10-30 | North America/USA/California/San Dieg | San Diego County Public Health Laboratory | Andersen lab at Scripps Research | SEARCH Alliance San Diego with Tracy Basler et al |

| | | | | | | |
|---|---|---|---|---|---|---|
| EPI_ISL_730112 | USA/CA-ALSR-4730/2020 | 2020-10-28 | North America/USA/California/Orange ( | San Diego County Public Health Laboratory | Andersen lab at Scripps Research | SEARCH Alliance San Diego with Tracy Basler et al |
| EPI_ISL_730115 | USA/CA-ALSR-4736/2020 | 2020-09-17 | North America/USA/California/San Dieg | San Diego County Public Health Laboratory | Andersen lab at Scripps Research | SEARCH Alliance San Diego with Tracy Basler et al |
| EPI_ISL_730117 | USA/CA-ALSR-4739/2020 | 2020-10-30 | North America/USA/California/San Dieg | San Diego County Public Health Laboratory | Andersen lab at Scripps Research | SEARCH Alliance San Diego with Tracy Basler et al |
| EPI_ISL_730122 | USA/CA-ALSR-4751/2020 | 2020-10-06 | North America/USA/California/San Dieg | San Diego County Public Health Laboratory | Andersen lab at Scripps Research | SEARCH Alliance San Diego with Tracy Basler et al |
| EPI_ISL_730127 | Mexico/BCN-ALSR-4762/2020 | 2020-08-24 | North America/Mexico/Baja California/T | Sharp HealthCare Laboratory | Andersen lab at Scripps Research | SEARCH Alliance San Diego with Aaron Harding et al |
| EPI_ISL_730128 | Mexico/BCN-ALSR-4766/2020 | 2020-08-24 | North America/Mexico/Baja California/T | Sharp HealthCare Laboratory | Andersen lab at Scripps Research | SEARCH Alliance San Diego with Aaron Harding et al |
| EPI_ISL_730129 | Mexico/BCN-ALSR-4768/2020 | 2020-08-25 | North America/Mexico/Baja California/T | Sharp HealthCare Laboratory | Andersen lab at Scripps Research | SEARCH Alliance San Diego with Aaron Harding et al |
| EPI_ISL_730130 | Mexico/BCN-ALSR-4770/2020 | 2020-08-24 | North America/Mexico/Baja California/T | Sharp HealthCare Laboratory | Andersen lab at Scripps Research | SEARCH Alliance San Diego with Aaron Harding et al |
| EPI_ISL_730131 | Mexico/BCN-ALSR-4771/2020 | 2020-08-22 | North America/Mexico/Baja California/T | Sharp HealthCare Laboratory | Andersen lab at Scripps Research | SEARCH Alliance San Diego with Aaron Harding et al |
| EPI_ISL_730132 | Mexico/BCN-ALSR-4772/2020 | 2020-08-25 | North America/Mexico/Baja California/E | Sharp HealthCare Laboratory | Andersen lab at Scripps Research | SEARCH Alliance San Diego with Aaron Harding et al |
| EPI_ISL_730133 | Mexico/BCN-ALSR-4774/2020 | 2020-08-25 | North America/Mexico/Baja California/R | Sharp HealthCare Laboratory | Andersen lab at Scripps Research | SEARCH Alliance San Diego with Aaron Harding et al |
| EPI_ISL_730134 | Mexico/BCN-ALSR-4775/2020 | 2020-08-25 | North America/Mexico/Baja California/T | Sharp HealthCare Laboratory | Andersen lab at Scripps Research | SEARCH Alliance San Diego with Aaron Harding et al |
| EPI_ISL_730135 | Mexico/BCN-ALSR-4777/2020 | 2020-08-25 | North America/Mexico/Baja California/T | Sharp HealthCare Laboratory | Andersen lab at Scripps Research | SEARCH Alliance San Diego with Aaron Harding et al |
| EPI_ISL_730136 | Mexico/BCN-ALSR-4778/2020 | 2020-08-22 | North America/Mexico/Baja California/T | Sharp HealthCare Laboratory | Andersen lab at Scripps Research | SEARCH Alliance San Diego with Aaron Harding et al |
| EPI_ISL_730140 | USA/CA-ALSR-4787/2020 | 2020-05-05 | North America/USA/California/San Dieg | Scripps Medical Laboratory | Andersen lab at Scripps Research | SEARCH Alliance San Diego with Michael Quigley et al |
| EPI_ISL_730143 | USA/CA-ALSR-4791/2020 | 2020-05-01 | North America/USA/California/San Dieg | Scripps Medical Laboratory | Andersen lab at Scripps Research | SEARCH Alliance San Diego with Michael Quigley et al |
| EPI_ISL_730150 | USA/CA-ALSR-4800/2020 | 2020-05-02 | North America/USA/California/San Dieg | Scripps Medical Laboratory | Andersen lab at Scripps Research | SEARCH Alliance San Diego with Michael Quigley et al |
| EPI_ISL_730153 | USA/CA-ALSR-4805/2020 | 2020-04-01 | North America/USA/California/San Dieg | Scripps Medical Laboratory | Andersen lab at Scripps Research | SEARCH Alliance San Diego with Michael Quigley et al |
| EPI_ISL_730160 | USA/CA-ALSR-4820/2020 | 2020-04-03 | North America/USA/California/San Dieg | Scripps Medical Laboratory | Andersen lab at Scripps Research | SEARCH Alliance San Diego with Michael Quigley et al |
| EPI_ISL_730162 | USA/CA-ALSR-4822/2020 | 2020-11-04 | North America/USA/California/San Dieg | San Diego County Public Health Laboratory | Andersen lab at Scripps Research | SEARCH Alliance San Diego with Tracy Basler et al |

| | | | | | | |
|---|---|---|---|---|---|---|
| EPI_ISL_730165 | USA/CA-ALSR-4828/2020 | 2020-10-28 | North America/USA/California/San Dieg | San Diego County Public Health Laboratory | Andersen lab at Scripps Research | SEARCH Alliance San Diego with Tracy Basler et al |
| EPI_ISL_730167 | USA/CA-ALSR-4831/2020 | 2020-11-04 | North America/USA/California/San Dieg | San Diego County Public Health Laboratory | Andersen lab at Scripps Research | SEARCH Alliance San Diego with Tracy Basler et al |
| EPI_ISL_730171 | USA/CA-ALSR-4839/2020 | 2020-11-04 | North America/USA/California/San Dieg | San Diego County Public Health Laboratory | Andersen lab at Scripps Research | SEARCH Alliance San Diego with Tracy Basler et al |
| EPI_ISL_730181 | USA/CA-ALSR-4861/2020 | 2020-09-09 | North America/USA/California/San Dieg | San Diego County Public Health Laboratory | Andersen lab at Scripps Research | SEARCH Alliance San Diego with Tracy Basler et al |
| EPI_ISL_730184 | USA/CA-ALSR-4864/2020 | 2020-08-20 | North America/USA/California/San Dieg | San Diego County Public Health Laboratory | Andersen lab at Scripps Research | SEARCH Alliance San Diego with Tracy Basler et al |
| EPI_ISL_730190 | USA/CA-ALSR-4871/2020 | 2020-11-02 | North America/USA/California/San Dieg | San Diego County Public Health Laboratory | Andersen lab at Scripps Research | SEARCH Alliance San Diego with Tracy Basler et al |
| EPI_ISL_730195 | USA/CA-ALSR-4879/2020 | 2020-11-09 | North America/USA/California/San Dieg | San Diego County Public Health Laboratory | Andersen lab at Scripps Research | SEARCH Alliance San Diego with Tracy Basler et al |
| EPI_ISL_730197 | Mexico/BCN-ALSR-4888/2020 | 2020-07-03 | North America/Mexico/Baja California/T | Genomica Lab Molecular, M,àö¬©xico | Andersen lab at Scripps Research | SEARCH Alliance San Diego with Jonathan Gonzalez Garcia et al |
| EPI_ISL_730198 | Mexico/BCN-ALSR-4893/2020 | 2020-07-04 | North America/Mexico/Baja California/T | Genomica Lab Molecular, M,àö¬©xico | Andersen lab at Scripps Research | SEARCH Alliance San Diego with Jonathan Gonzalez Garcia et al |
| EPI_ISL_730199 | Mexico/BCN-ALSR-4894/2020 | 2020-07-04 | North America/Mexico/Baja California/T | Genomica Lab Molecular, M,àö¬©xico | Andersen lab at Scripps Research | SEARCH Alliance San Diego with Jonathan Gonzalez Garcia et al |
| EPI_ISL_730200 | Mexico/BCN-ALSR-4896/2020 | 2020-07-07 | North America/Mexico/Baja California/T | Genomica Lab Molecular, M,àö¬©xico | Andersen lab at Scripps Research | SEARCH Alliance San Diego with Jonathan Gonzalez Garcia et al |
| EPI_ISL_730201 | Mexico/BCN-ALSR-4898/2020 | 2020-07-08 | North America/Mexico/Baja California/T | Genomica Lab Molecular, M,àö¬©xico | Andersen lab at Scripps Research | SEARCH Alliance San Diego with Jonathan Gonzalez Garcia et al |
| EPI_ISL_730202 | Mexico/BCN-ALSR-4899/2020 | 2020-07-08 | North America/Mexico/Baja California/T | Genomica Lab Molecular, M,àö¬©xico | Andersen lab at Scripps Research | SEARCH Alliance San Diego with Jonathan Gonzalez Garcia et al |
| EPI_ISL_730203 | Mexico/BCN-ALSR-4900/2020 | 2020-07-08 | North America/Mexico/Baja California/T | Genomica Lab Molecular, M,àö¬©xico | Andersen lab at Scripps Research | SEARCH Alliance San Diego with Jonathan Gonzalez Garcia et al |
| EPI_ISL_730204 | Mexico/BCN-ALSR-4902/2020 | 2020-07-08 | North America/Mexico/Baja California/T | Genomica Lab Molecular, M,àö¬©xico | Andersen lab at Scripps Research | SEARCH Alliance San Diego with Jonathan Gonzalez Garcia et al |
| EPI_ISL_730205 | Mexico/BCN-ALSR-4903/2020 | 2020-07-08 | North America/Mexico/Baja California/T | Genomica Lab Molecular, M,àö¬©xico | Andersen lab at Scripps Research | SEARCH Alliance San Diego with Jonathan Gonzalez Garcia et al |

| | | | | | | |
|---|---|---|---|---|---|---|
| EPI_ISL_730206 | Mexico/BCN-ALSR-4904/2020 | 2020-07-11 | North America/Mexico/Baja California/T | Genomica Lab Molecular, M,àö¬©xico | Andersen lab at Scripps Research | SEARCH Alliance San Diego with Jonathan Gonzalez Garcia et al |
| EPI_ISL_730208 | Mexico/BCN-ALSR-4910/2020 | 2020-07-17 | North America/Mexico/Baja California/T | Genomica Lab Molecular, M,àö¬©xico | Andersen lab at Scripps Research | SEARCH Alliance San Diego with Jonathan Gonzalez Garcia et al |
| EPI_ISL_730209 | Mexico/BCN-ALSR-4917/2020 | 2020-08-03 | North America/Mexico/Baja California/T | Genomica Lab Molecular, M,àö¬©xico | Andersen lab at Scripps Research | SEARCH Alliance San Diego with Jonathan Gonzalez Garcia et al |
| EPI_ISL_730210 | Mexico/BCN-ALSR-4918/2020 | 2020-08-08 | North America/Mexico/Baja California/T | Genomica Lab Molecular, M,àö¬©xico | Andersen lab at Scripps Research | SEARCH Alliance San Diego with Jonathan Gonzalez Garcia et al |
| EPI_ISL_730211 | Mexico/BCN-ALSR-4920/2020 | 2020-08-09 | North America/Mexico/Baja California/T | Genomica Lab Molecular, M,àö¬©xico | Andersen lab at Scripps Research | SEARCH Alliance San Diego with Jonathan Gonzalez Garcia et al |
| EPI_ISL_730212 | Mexico/BCN-ALSR-4922/2020 | 2020-08-13 | North America/Mexico/Baja California/T | Genomica Lab Molecular, M,àö¬©xico | Andersen lab at Scripps Research | SEARCH Alliance San Diego with Jonathan Gonzalez Garcia et al |
| EPI_ISL_730213 | Mexico/BCN-ALSR-4923/2020 | 2020-08-13 | North America/Mexico/Baja California/T | Genomica Lab Molecular, M,àö¬©xico | Andersen lab at Scripps Research | SEARCH Alliance San Diego with Jonathan Gonzalez Garcia et al |
| EPI_ISL_730214 | Mexico/BCN-ALSR-4924/2020 | 2020-08-20 | North America/Mexico/Baja California/T | Genomica Lab Molecular, M,àö¬©xico | Andersen lab at Scripps Research | SEARCH Alliance San Diego with Jonathan Gonzalez Garcia et al |
| EPI_ISL_730215 | Mexico/BCN-ALSR-4926/2020 | 2020-08-25 | North America/Mexico/Baja California/T | Genomica Lab Molecular, M,àö¬©xico | Andersen lab at Scripps Research | SEARCH Alliance San Diego with Jonathan Gonzalez Garcia et al |
| EPI_ISL_730216 | Mexico/BCN-ALSR-4927/2020 | 2020-08-25 | North America/Mexico/Baja California/T | Genomica Lab Molecular, M,àö¬©xico | Andersen lab at Scripps Research | SEARCH Alliance San Diego with Jonathan Gonzalez Garcia et al |
| EPI_ISL_730217 | Mexico/BCN-ALSR-4928/2020 | 2020-08-25 | North America/Mexico/Baja California/T | Genomica Lab Molecular, M,àö¬©xico | Andersen lab at Scripps Research | SEARCH Alliance San Diego with Jonathan Gonzalez Garcia et al |
| EPI_ISL_730218 | Mexico/BCN-ALSR-4929/2020 | 2020-08-25 | North America/Mexico/Baja California/T | Genomica Lab Molecular, M,àö¬©xico | Andersen lab at Scripps Research | SEARCH Alliance San Diego with Jonathan Gonzalez Garcia et al |
| EPI_ISL_730219 | Mexico/BCN-ALSR-4934/2020 | 2020-08-26 | North America/Mexico/Baja California/T | Genomica Lab Molecular, M,àö¬©xico | Andersen lab at Scripps Research | SEARCH Alliance San Diego with Jonathan Gonzalez Garcia et al |

| | | | | | | |
|---|---|---|---|---|---|---|
| EPI_ISL_730220 | Mexico/BCN-ALSR-4935/2020 | 2020-08-26 | North America/Mexico/Baja California/T | Genomica Lab Molecular, M‚àö¬©xico | Andersen lab at Scripps Research | SEARCH Alliance San Diego with Jonathan Gonzalez Garcia et al |
| EPI_ISL_730221 | Mexico/BCN-ALSR-4937/2020 | 2020-08-31 | North America/Mexico/Baja California/T | Genomica Lab Molecular, M‚àö¬©xico | Andersen lab at Scripps Research | SEARCH Alliance San Diego with Jonathan Gonzalez Garcia et al |
| EPI_ISL_730222 | Mexico/BCN-ALSR-4942/2020 | 2020-08-02 | North America/Mexico/Baja California/T | Genomica Lab Molecular, M‚àö¬©xico | Andersen lab at Scripps Research | SEARCH Alliance San Diego with Jonathan Gonzalez Garcia et al |
| EPI_ISL_730223 | Mexico/BCN-ALSR-4943/2020 | 2020-08-02 | North America/Mexico/Baja California/T | Genomica Lab Molecular, M‚àö¬©xico | Andersen lab at Scripps Research | SEARCH Alliance San Diego with Jonathan Gonzalez Garcia et al |
| EPI_ISL_730224 | Mexico/BCN-ALSR-4946/2020 | 2020-08-02 | North America/Mexico/Baja California/T | Genomica Lab Molecular, M‚àö¬©xico | Andersen lab at Scripps Research | SEARCH Alliance San Diego with Jonathan Gonzalez Garcia et al |
| EPI_ISL_730225 | Mexico/BCN-ALSR-4947/2020 | 2020-08-02 | North America/Mexico/Baja California/T | Genomica Lab Molecular, M‚àö¬©xico | Andersen lab at Scripps Research | SEARCH Alliance San Diego with Jonathan Gonzalez Garcia et al |
| EPI_ISL_730226 | Mexico/BCN-ALSR-4951/2020 | 2020-08-02 | North America/Mexico/Baja California/T | Genomica Lab Molecular, M‚àö¬©xico | Andersen lab at Scripps Research | SEARCH Alliance San Diego with Jonathan Gonzalez Garcia et al |
| EPI_ISL_730227 | Mexico/BCN-ALSR-4952/2020 | 2020-08-02 | North America/Mexico/Baja California/T | Genomica Lab Molecular, M‚àö¬©xico | Andersen lab at Scripps Research | SEARCH Alliance San Diego with Jonathan Gonzalez Garcia et al |
| EPI_ISL_730228 | Mexico/BCN-ALSR-4953/2020 | 2020-08-02 | North America/Mexico/Baja California/T | Genomica Lab Molecular, M‚àö¬©xico | Andersen lab at Scripps Research | SEARCH Alliance San Diego with Jonathan Gonzalez Garcia et al |
| EPI_ISL_730229 | Jordan/AM-ALSR-4955/2020 | 2020-09-12 | Asia/Jordan/Amman/ | Biolab Diagnostic Laboratories | Andersen lab at Scripps Research | Issa Abu-Dayyeh et al |
| EPI_ISL_730237 | Jordan/AM-ALSR-4964/2020 | 2020-10-03 | Asia/Jordan/Amman/ | Biolab Diagnostic Laboratories | Andersen lab at Scripps Research | Issa Abu-Dayyeh et al |
| EPI_ISL_730243 | Jordan/AM-ALSR-4970/2020 | 2020-10-03 | Asia/Jordan/Amman/ | Biolab Diagnostic Laboratories | Andersen lab at Scripps Research | Issa Abu-Dayyeh et al |
| EPI_ISL_730252 | Jordan/AM-ALSR-4979/2020 | 2020-10-06 | Asia/Jordan/Amman/ | Biolab Diagnostic Laboratories | Andersen lab at Scripps Research | Issa Abu-Dayyeh et al |
| EPI_ISL_730253 | Jordan/AM-ALSR-4980/2020 | 2020-10-07 | Asia/Jordan/Amman/ | Biolab Diagnostic Laboratories | Andersen lab at Scripps Research | Issa Abu-Dayyeh et al |
| EPI_ISL_730278 | Jordan/AM-ALSR-5005/2020 | 2020-10-10 | Asia/Jordan/Amman/ | Biolab Diagnostic Laboratories | Andersen lab at Scripps Research | Issa Abu-Dayyeh et al |
| EPI_ISL_730283 | Jordan/AM-ALSR-5010/2020 | 2020-10-06 | Asia/Jordan/Amman/ | Biolab Diagnostic Laboratories | Andersen lab at Scripps Research | Issa Abu-Dayyeh et al |

| | | | | | | |
|---|---|---|---|---|---|---|
| EPI_ISL_730296 | USA/CA-ALSR-5025/2020 | 2020-12-05 | North America/USA/California/San Dieg | San Diego County Public Health Laboratory | Andersen lab at Scripps Research | SEARCH Alliance San Diego with Tracy Basler et al |
| EPI_ISL_730297 | USA/CA-ALSR-5026/2020 | 2020-11-15 | North America/USA/California/San Dieg | San Diego County Public Health Laboratory | Andersen lab at Scripps Research | SEARCH Alliance San Diego with Tracy Basler et al |
| EPI_ISL_730298 | USA/CA-ALSR-5027/2020 | 2020-11-12 | North America/USA/California/San Dieg | San Diego County Public Health Laboratory | Andersen lab at Scripps Research | SEARCH Alliance San Diego with Tracy Basler et al |
| EPI_ISL_730306 | USA/CA-ALSR-5035/2020 | 2020-11-15 | North America/USA/California/San Dieg | San Diego County Public Health Laboratory | Andersen lab at Scripps Research | SEARCH Alliance San Diego with Tracy Basler et al |
| EPI_ISL_730314 | USA/CA-ALSR-5047/2020 | 2020-12-06 | North America/USA/California/San Dieg | San Diego County Public Health Laboratory | Andersen lab at Scripps Research | SEARCH Alliance San Diego with Tracy Basler et al |
| EPI_ISL_730315 | USA/CA-ALSR-5048/2020 | 2020-12-05 | North America/USA/California/San Dieg | San Diego County Public Health Laboratory | Andersen lab at Scripps Research | SEARCH Alliance San Diego with Tracy Basler et al |
| EPI_ISL_730318 | USA/CA-ALSR-5051/2020 | 2020-09-21 | North America/USA/California/San Dieg | San Diego County Public Health Laboratory | Andersen lab at Scripps Research | SEARCH Alliance San Diego with Tracy Basler et al |
| EPI_ISL_730333 | USA/CA-ALSR-5072/2020 | 2020-09-21 | North America/USA/California/San Dieg | San Diego County Public Health Laboratory | Andersen lab at Scripps Research | SEARCH Alliance San Diego with Tracy Basler et al |
| EPI_ISL_730337 | USA/CA-ALSR-5077/2020 | 2020-09-21 | North America/USA/California/San Dieg | San Diego County Public Health Laboratory | Andersen lab at Scripps Research | SEARCH Alliance San Diego with Tracy Basler et al |
| EPI_ISL_730344 | USA/CA-ALSR-5089/2020 | 2020-05-24 | North America/USA/California/San Dieg | San Diego County Public Health Laboratory | Andersen lab at Scripps Research | SEARCH Alliance San Diego with Tracy Basler et al |
| EPI_ISL_730353 | USA/CA-ALSR-5104/2020 | 2020-12-03 | North America/USA/California/San Dieg | San Diego County Public Health Laboratory | Andersen lab at Scripps Research | SEARCH Alliance San Diego with Tracy Basler et al |
| EPI_ISL_730356 | USA/CA-ALSR-5109/2020 | 2020-11-12 | North America/USA/California/San Dieg | San Diego County Public Health Laboratory | Andersen lab at Scripps Research | SEARCH Alliance San Diego with Tracy Basler et al |
| EPI_ISL_730358 | USA/CA-ALSR-5111/2020 | 2020-12-03 | North America/USA/California/San Dieg | San Diego County Public Health Laboratory | Andersen lab at Scripps Research | SEARCH Alliance San Diego with Tracy Basler et al |
| EPI_ISL_730361 | USA/CA-ALSR-5115/2020 | 2020-12-03 | North America/USA/California/Kern Cou | San Diego County Public Health Laboratory | Andersen lab at Scripps Research | SEARCH Alliance San Diego with Tracy Basler et al |
| EPI_ISL_730363 | USA/CA-ALSR-5118/2020 | 2020-12-03 | North America/USA/California/San Dieg | San Diego County Public Health Laboratory | Andersen lab at Scripps Research | SEARCH Alliance San Diego with Tracy Basler et al |
| EPI_ISL_730366 | USA/CA-ALSR-5123/2020 | 2020-12-03 | North America/USA/California/San Dieg | San Diego County Public Health Laboratory | Andersen lab at Scripps Research | SEARCH Alliance San Diego with Tracy Basler et al |
| EPI_ISL_730381 | Jordan/AM-ALSR-5142/2020 | 2020-10-15 | Asia/Jordan/Amman/ | Biolab Diagnostic Laboratories | Andersen lab at Scripps Research | Issa Abu-Dayyeh et al |
| EPI_ISL_730382 | Jordan/AM-ALSR-5143/2020 | 2020-10-12 | Asia/Jordan/Amman/ | Biolab Diagnostic Laboratories | Andersen lab at Scripps Research | Issa Abu-Dayyeh et al |
| EPI_ISL_730387 | Jordan/AM-ALSR-5148/2020 | 2020-10-16 | Asia/Jordan/Amman/ | Biolab Diagnostic Laboratories | Andersen lab at Scripps Research | Issa Abu-Dayyeh et al |
| EPI_ISL_730388 | Jordan/AM-ALSR-5149/2020 | 2020-10-18 | Asia/Jordan/Amman/ | Biolab Diagnostic Laboratories | Andersen lab at Scripps Research | Issa Abu-Dayyeh et al |

| | | | | | | |
|---|---|---|---|---|---|---|
| EPI_ISL_730395 | Jordan/AM-ALSR-5156/2020 | 2020-10-15 | Asia/Jordan/Amman/ | Biolab Diagnostic Laboratories | Andersen lab at Scripps Research | Issa Abu-Dayyeh et al |
| EPI_ISL_730402 | Jordan/AM-ALSR-5163/2020 | 2020-10-16 | Asia/Jordan/Amman/ | Biolab Diagnostic Laboratories | Andersen lab at Scripps Research | Issa Abu-Dayyeh et al |
| EPI_ISL_730403 | Jordan/AM-ALSR-5164/2020 | 2020-10-20 | Asia/Jordan/Amman/ | Biolab Diagnostic Laboratories | Andersen lab at Scripps Research | Issa Abu-Dayyeh et al |
| EPI_ISL_730405 | Jordan/AM-ALSR-5166/2020 | 2020-10-15 | Asia/Jordan/Amman/ | Biolab Diagnostic Laboratories | Andersen lab at Scripps Research | Issa Abu-Dayyeh et al |
| EPI_ISL_730407 | Jordan/AM-ALSR-5168/2020 | 2020-10-16 | Asia/Jordan/Amman/ | Biolab Diagnostic Laboratories | Andersen lab at Scripps Research | Issa Abu-Dayyeh et al |
| EPI_ISL_730408 | Jordan/AM-ALSR-5169/2020 | 2020-10-16 | Asia/Jordan/Amman/ | Biolab Diagnostic Laboratories | Andersen lab at Scripps Research | Issa Abu-Dayyeh et al |
| EPI_ISL_730412 | Jordan/AM-ALSR-5173/2020 | 2020-10-12 | Asia/Jordan/Amman/ | Biolab Diagnostic Laboratories | Andersen lab at Scripps Research | Issa Abu-Dayyeh et al |
| EPI_ISL_730414 | Jordan/AM-ALSR-5175/2020 | 2020-10-16 | Asia/Jordan/Amman/ | Biolab Diagnostic Laboratories | Andersen lab at Scripps Research | Issa Abu-Dayyeh et al |
| EPI_ISL_730419 | Jordan/AM-ALSR-5180/2020 | 2020-10-12 | Asia/Jordan/Amman/ | Biolab Diagnostic Laboratories | Andersen lab at Scripps Research | Issa Abu-Dayyeh et al |
| EPI_ISL_730423 | Jordan/AM-ALSR-5184/2020 | 2020-10-15 | Asia/Jordan/Amman/ | Biolab Diagnostic Laboratories | Andersen lab at Scripps Research | Issa Abu-Dayyeh et al |
| EPI_ISL_730453 | Jordan/IR-ALSR-5215/2020 | 2020-10-18 | Asia/Jordan/Irbid/ | Biolab Diagnostic Laboratories | Andersen lab at Scripps Research | Issa Abu-Dayyeh et al |
| EPI_ISL_730458 | Jordan/IR-ALSR-5220/2020 | 2020-10-20 | Asia/Jordan/Irbid/ | Biolab Diagnostic Laboratories | Andersen lab at Scripps Research | Issa Abu-Dayyeh et al |
| EPI_ISL_730470 | Jordan/IR-ALSR-5232/2020 | 2020-10-18 | Asia/Jordan/Irbid/ | Biolab Diagnostic Laboratories | Andersen lab at Scripps Research | Issa Abu-Dayyeh et al |
| EPI_ISL_730471 | Jordan/IR-ALSR-5233/2020 | 2020-10-19 | Asia/Jordan/Irbid/ | Biolab Diagnostic Laboratories | Andersen lab at Scripps Research | Issa Abu-Dayyeh et al |
| EPI_ISL_730475 | Jordan/IR-ALSR-5237/2020 | 2020-10-07 | Asia/Jordan/Irbid/ | Biolab Diagnostic Laboratories | Andersen lab at Scripps Research | Issa Abu-Dayyeh et al |
| EPI_ISL_730484 | Jordan/AQ-ALSR-5246/2020 | 2020-10-05 | Asia/Jordan/Aqaba/ | Biolab Diagnostic Laboratories | Andersen lab at Scripps Research | Issa Abu-Dayyeh et al |
| EPI_ISL_730490 | Jordan/AQ-ALSR-5252/2020 | 2020-10-15 | Asia/Jordan/Aqaba/ | Biolab Diagnostic Laboratories | Andersen lab at Scripps Research | Issa Abu-Dayyeh et al |
| EPI_ISL_730496 | Jordan/AQ-ALSR-5258/2020 | 2020-10-17 | Asia/Jordan/Aqaba/ | Biolab Diagnostic Laboratories | Andersen lab at Scripps Research | Issa Abu-Dayyeh et al |
| EPI_ISL_730501 | Jordan/AQ-ALSR-5263/2020 | 2020-10-12 | Asia/Jordan/Aqaba/ | Biolab Diagnostic Laboratories | Andersen lab at Scripps Research | Issa Abu-Dayyeh et al |
| EPI_ISL_730510 | Jordan/AQ-ALSR-5272/2020 | 2020-10-06 | Asia/Jordan/Aqaba/ | Biolab Diagnostic Laboratories | Andersen lab at Scripps Research | Issa Abu-Dayyeh et al |

| | | | | | | |
|---|---|---|---|---|---|---|
| EPI_ISL_730517 | Jordan/IR-ALSR-5279/2020 | 2020-11-03 | Asia/Jordan/Irbid/ | Biolab Diagnostic Laboratories | Andersen lab at Scripps Research | Issa Abu-Dayyeh et al |
| EPI_ISL_730519 | Jordan/AM-ALSR-5281/2020 | 2020-11-04 | Asia/Jordan/Amman/ | Biolab Diagnostic Laboratories | Andersen lab at Scripps Research | Issa Abu-Dayyeh et al |
| EPI_ISL_730526 | Jordan/AM-ALSR-5289/2020 | 2020-11-06 | Asia/Jordan/Amman/ | Biolab Diagnostic Laboratories | Andersen lab at Scripps Research | Issa Abu-Dayyeh et al |
| EPI_ISL_730528 | Jordan/AM-ALSR-5291/2020 | 2020-11-02 | Asia/Jordan/Amman/ | Biolab Diagnostic Laboratories | Andersen lab at Scripps Research | Issa Abu-Dayyeh et al |
| EPI_ISL_730529 | Jordan/AM-ALSR-5292/2020 | 2020-11-04 | Asia/Jordan/Amman/ | Biolab Diagnostic Laboratories | Andersen lab at Scripps Research | Issa Abu-Dayyeh et al |
| EPI_ISL_730530 | Jordan/AM-ALSR-5293/2020 | 2020-11-04 | Asia/Jordan/Amman/ | Biolab Diagnostic Laboratories | Andersen lab at Scripps Research | Issa Abu-Dayyeh et al |
| EPI_ISL_730531 | Jordan/AM-ALSR-5294/2020 | 2020-11-06 | Asia/Jordan/Amman/ | Biolab Diagnostic Laboratories | Andersen lab at Scripps Research | Issa Abu-Dayyeh et al |
| EPI_ISL_730533 | Jordan/AZ-ALSR-5297/2020 | 2020-11-01 | Asia/Jordan/Zarqa/ | Biolab Diagnostic Laboratories | Andersen lab at Scripps Research | Issa Abu-Dayyeh et al |
| EPI_ISL_730534 | Jordan/AM-ALSR-5298/2020 | 2020-11-04 | Asia/Jordan/Amman/ | Biolab Diagnostic Laboratories | Andersen lab at Scripps Research | Issa Abu-Dayyeh et al |
| EPI_ISL_730535 | Jordan/AM-ALSR-5299/2020 | 2020-11-06 | Asia/Jordan/Amman/ | Biolab Diagnostic Laboratories | Andersen lab at Scripps Research | Issa Abu-Dayyeh et al |
| EPI_ISL_730536 | Jordan/AM-ALSR-5300/2020 | 2020-11-06 | Asia/Jordan/Amman/ | Biolab Diagnostic Laboratories | Andersen lab at Scripps Research | Issa Abu-Dayyeh et al |
| EPI_ISL_730538 | Jordan/IR-ALSR-5302/2020 | 2020-11-03 | Asia/Jordan/Irbid/ | Biolab Diagnostic Laboratories | Andersen lab at Scripps Research | Issa Abu-Dayyeh et al |
| EPI_ISL_730539 | Jordan/AM-ALSR-5303/2020 | 2020-11-04 | Asia/Jordan/Amman/ | Biolab Diagnostic Laboratories | Andersen lab at Scripps Research | Issa Abu-Dayyeh et al |
| EPI_ISL_730541 | Jordan/AM-ALSR-5305/2020 | 2020-11-07 | Asia/Jordan/Amman/ | Biolab Diagnostic Laboratories | Andersen lab at Scripps Research | Issa Abu-Dayyeh et al |
| EPI_ISL_730543 | Jordan/AM-ALSR-5307/2020 | 2020-11-02 | Asia/Jordan/Amman/ | Biolab Diagnostic Laboratories | Andersen lab at Scripps Research | Issa Abu-Dayyeh et al |
| EPI_ISL_730544 | Jordan/AM-ALSR-5308/2020 | 2020-11-02 | Asia/Jordan/Amman/ | Biolab Diagnostic Laboratories | Andersen lab at Scripps Research | Issa Abu-Dayyeh et al |
| EPI_ISL_730546 | Jordan/AM-ALSR-5310/2020 | 2020-11-07 | Asia/Jordan/Amman/ | Biolab Diagnostic Laboratories | Andersen lab at Scripps Research | Issa Abu-Dayyeh et al |
| EPI_ISL_730548 | Jordan/AM-ALSR-5312/2020 | 2020-11-02 | Asia/Jordan/Amman/ | Biolab Diagnostic Laboratories | Andersen lab at Scripps Research | Issa Abu-Dayyeh et al |
| EPI_ISL_730552 | Jordan/AM-ALSR-5316/2020 | 2020-11-06 | Asia/Jordan/Amman/ | Biolab Diagnostic Laboratories | Andersen lab at Scripps Research | Issa Abu-Dayyeh et al |
| EPI_ISL_730553 | Jordan/AM-ALSR-5317/2020 | 2020-11-07 | Asia/Jordan/Amman/ | Biolab Diagnostic Laboratories | Andersen lab at Scripps Research | Issa Abu-Dayyeh et al |

| | | | | | | |
|---|---|---|---|---|---|---|
| EPI_ISL_730555 | Jordan/AM-ALSR-5319/2020 | 2020-11-03 | Asia/Jordan/Amman/ | Biolab Diagnostic Laboratories | Andersen lab at Scripps Research | Issa Abu-Dayyeh et al |
| EPI_ISL_730557 | Jordan/AM-ALSR-5321/2020 | 2020-11-04 | Asia/Jordan/Amman/ | Biolab Diagnostic Laboratories | Andersen lab at Scripps Research | Issa Abu-Dayyeh et al |
| EPI_ISL_730558 | Jordan/AM-ALSR-5322/2020 | 2020-11-06 | Asia/Jordan/Amman/ | Biolab Diagnostic Laboratories | Andersen lab at Scripps Research | Issa Abu-Dayyeh et al |
| EPI_ISL_730559 | Jordan/AM-ALSR-5323/2020 | 2020-11-07 | Asia/Jordan/Amman/ | Biolab Diagnostic Laboratories | Andersen lab at Scripps Research | Issa Abu-Dayyeh et al |
| EPI_ISL_730560 | Jordan/AM-ALSR-5324/2020 | 2020-11-03 | Asia/Jordan/Amman/ | Biolab Diagnostic Laboratories | Andersen lab at Scripps Research | Issa Abu-Dayyeh et al |
| EPI_ISL_730561 | Jordan/AM-ALSR-5325/2020 | 2020-11-04 | Asia/Jordan/Amman/ | Biolab Diagnostic Laboratories | Andersen lab at Scripps Research | Issa Abu-Dayyeh et al |
| EPI_ISL_730562 | Jordan/AM-ALSR-5326/2020 | 2020-11-03 | Asia/Jordan/Amman/ | Biolab Diagnostic Laboratories | Andersen lab at Scripps Research | Issa Abu-Dayyeh et al |
| EPI_ISL_730563 | Jordan/AM-ALSR-5327/2020 | 2020-11-04 | Asia/Jordan/Amman/ | Biolab Diagnostic Laboratories | Andersen lab at Scripps Research | Issa Abu-Dayyeh et al |
| EPI_ISL_730567 | Turkey/GUMV_36208/2020 | 2020-09-30 | Europe/Turkey/Ankara/ | Gazi University Faculty of Medicine, Medical Virology Laboratory | Gazi University Faculty of Medicine, Medical Virology Laboratory | Erdem ≈ûahin et al |
| EPI_ISL_730568 | Turkey/Ankara_GUMV_25210/2020 | 2020-09-01 | Europe/Turkey/Ankara/ | Gazi University Faculty of Medicine, Medical Virology Laboratory | Gazi University Faculty of Medicine, Medical Virology Laboratory | Erdem ≈ûahin et al |
| EPI_ISL_730569 | Turkey/Ankara_GUMV_31394/2020 | 2020-09-14 | Europe/Turkey/Ankara/ | Gazi University Faculty of Medicine, Medical Virology Laboratory | Gazi University Faculty of Medicine, Medical Virology Laboratory | Erdem ≈ûahin et al |
| EPI_ISL_730570 | Turkey/Ankara_GUMV_37230/2020 | 2020-10-04 | Europe/Turkey/Ankara/ | Gazi University Faculty of Medicine, Medical Virology Laboratory | Gazi University Faculty of Medicine, Medical Virology Laboratory | Erdem ≈ûahin et al |

| | | | | | | |
|---|---|---|---|---|---|---|
| EPI_ISL_730571 | Turkey/Ankara_GUMV_22790/2020 | 2020-08-26 | Europe/Turkey/Ankara/ | Gazi University Faculty of Medicine, Medical Virology Laboratory | Gazi University Faculty of Medicine, Medical Virology Laboratory | Erdem ≈ûahin et al |
| EPI_ISL_730572 | Turkey/Ankara_GUMV_22731/2020 | 2020-08-26 | Europe/Turkey/Ankara/ | Gazi University Faculty of Medicine, Medical Virology Laboratory | Gazi University Faculty of Medicine, Medical Virology Laboratory | Erdem ≈ûahin et al |
| EPI_ISL_730573 | Turkey/Ankara_GUMV_28458/2020 | 2020-09-07 | Europe/Turkey/Ankara/ | Gazi University Faculty of Medicine, Medical Virology Laboratory | Gazi University Faculty of Medicine, Medical Virology Laboratory | Erdem ≈ûahin et al |
| EPI_ISL_730574 | Turkey/Ankara_GUMV_18121/2020 | 2020-08-11 | Europe/Turkey/Ankara/ | Gazi University Faculty of Medicine, Medical Virology Laboratory | Gazi University Faculty of Medicine, Medical Virology Laboratory | Erdem ≈ûahin et al |
| EPI_ISL_730575 | Turkey/Ankara_33508/2020 | 2020-09-20 | Europe/Turkey/Ankara/ | Gazi University Faculty of Medicine, Medical Virology Laboratory | Gazi University Faculty of Medicine, Medical Virology Laboratory | Erdem ≈ûahin et al |
| EPI_ISL_730576 | Turkey/Ankara_GUMV_10711/2020 | 2020-06-25 | Europe/Turkey/Ankara/ | Gazi University Faculty of Medicine, Medical Virology Laboratory | Gazi University Faculty of Medicine, Medical Virology Laboratory | Erdem ≈ûahin et al |
| EPI_ISL_730577 | Turkey/Ankara_GUMV_42541/2020 | 2020-10-24 | Europe/Turkey/Ankara/ | Gazi University Faculty of Medicine, Medical Virology Laboratory | Gazi University Faculty of Medicine, Medical Virology Laboratory | Erdem ≈ûahin et al |
| EPI_ISL_730620 | Turkey/Ankara_GUMV_41954/2020 | 2020-10-22 | Europe/Turkey/Ankara/ | Gazi University Faculty of Medicine, Medical Virology Laboratory | Gazi University Faculty of Medicine, Medical Virology Laboratory | Erdem ≈ûahin et al |

| | | | | | | |
|---|---|---|---|---|---|---|
| EPI_ISL_730641 | France/ARA-104482/2020 | 2020-11-09 | Europe/France/Auvergne-Rhv¥ne-Alpes/ | CNR Virus des Infections Respiratoires - France SUD | CNR Virus des Infections Respiratoires - France SUD | Antonin Bal et al |
| EPI_ISL_730642 | France/ARA-104489/2020 | 2020-11-09 | Europe/France/Auvergne-Rhv¥ne-Alpes/ | CNR Virus des Infections Respiratoires - France SUD | CNR Virus des Infections Respiratoires - France SUD | Antonin Bal et al |
| EPI_ISL_730643 | France/ARA-104519/2020 | 2020-11-10 | Europe/France/Auvergne-Rhv¥ne-Alpes/ | CNR Virus des Infections Respiratoires - France SUD | CNR Virus des Infections Respiratoires - France SUD | Antonin Bal et al |
| EPI_ISL_730645 | France/ARA-104821/2020 | 2020-11-10 | Europe/France/Auvergne-Rhv¥ne-Alpes/ | CNR Virus des Infections Respiratoires - France SUD | CNR Virus des Infections Respiratoires - France SUD | Antonin Bal et al |
| EPI_ISL_730647 | France/ARA-104824/2020 | 2020-11-10 | Europe/France/Auvergne-Rhv¥ne-Alpes/ | CNR Virus des Infections Respiratoires - France SUD | CNR Virus des Infections Respiratoires - France SUD | Antonin Bal et al |
| EPI_ISL_730649 | France/ARA-104848/2020 | 2020-11-10 | Europe/France/Auvergne-Rhv¥ne-Alpes/ | CNR Virus des Infections Respiratoires - France SUD | CNR Virus des Infections Respiratoires - France SUD | Antonin Bal et al |
| EPI_ISL_730650 | France/ARA-105054/2020 | 2020-11-11 | Europe/France/Auvergne-Rhv¥ne-Alpes/ | CNR Virus des Infections Respiratoires - France SUD | CNR Virus des Infections Respiratoires - France SUD | Antonin Bal et al |
| EPI_ISL_730653 | Italy/TE.237004/2020 | 2020-11-04 | Europe/Italy/Apulia/ | University of Bari, Valenzano, Italy | "Istituto Zooprofilattico Sperimentale dell'Abruzzo e del Molise ""G. Caporale""." | N. Decaro et al |
| EPI_ISL_730706 | England/ALDP-C39F7E/2020 | 2020-12-09 | Europe/United Kingdom/England/ | Lighthouse Lab in Alderley Park | Wellcome Sanger Institute for the COVID-19 Genomics UK (COG-UK) Consortium | Jacquelyn Wynn et al |

| | | | | | | |
|---|---|---|---|---|---|---|
| EPI_ISL_730712 | England/ALDP-C3DED7/2020 | 2020-12-10 | Europe/United Kingdom/England/ | Lighthouse Lab in Alderley Park | Wellcome Sanger Institute for the COVID-19 Genomics UK (COG-UK) Consortium | Jacquelyn Wynn et al |
| EPI_ISL_730730 | England/ALDP-C3A9D9/2020 | 2020-12-08 | Europe/United Kingdom/England/ | Lighthouse Lab in Alderley Park | Wellcome Sanger Institute for the COVID-19 Genomics UK (COG-UK) Consortium | Jacquelyn Wynn et al |
| EPI_ISL_730750 | England/ALDP-C3D88E/2020 | 2020-12-10 | Europe/United Kingdom/England/ | Lighthouse Lab in Alderley Park | Wellcome Sanger Institute for the COVID-19 Genomics UK (COG-UK) Consortium | Jacquelyn Wynn et al |
| EPI_ISL_730809 | England/QEUH-C3226D/2020 | 2020-12-11 | Europe/United Kingdom/England/ | Lighthouse Lab in Glasgow | Wellcome Sanger Institute for the COVID-19 Genomics UK (COG-UK) Consortium | Harper VanSteenhouse et al |
| EPI_ISL_730812 | England/ALDP-C3B64A/2020 | 2020-12-09 | Europe/United Kingdom/England/ | Lighthouse Lab in Alderley Park | Wellcome Sanger Institute for the COVID-19 Genomics UK (COG-UK) Consortium | Jacquelyn Wynn et al |
| EPI_ISL_730954 | England/ALDP-C3C421/2020 | 2020-12-10 | Europe/United Kingdom/England/ | Lighthouse Lab in Alderley Park | Wellcome Sanger Institute for the COVID-19 Genomics UK (COG-UK) Consortium | Jacquelyn Wynn et al |
| EPI_ISL_731226 | England/ALDP-C3D059/2020 | 2020-12-10 | Europe/United Kingdom/England/ | Lighthouse Lab in Alderley Park | Wellcome Sanger Institute for the COVID-19 Genomics UK (COG-UK) Consortium | Jacquelyn Wynn et al |
| EPI_ISL_731501 | England/QEUH-C30A0D/2020 | 2020-12-10 | Europe/United Kingdom/England/ | Lighthouse Lab in Glasgow | Wellcome Sanger Institute for the COVID-19 Genomics UK (COG-UK) Consortium | Harper VanSteenhouse et al |

| | | | | | | |
|---|---|---|---|---|---|---|
| EPI_ISL_731505 | England/QEUH-C30B46/2020 | 2020-12-11 | Europe/United Kingdom/England/ | Lighthouse Lab in Glasgow | Wellcome Sanger Institute for the COVID-19 Genomics UK (COG-UK) Consortium | Harper VanSteenhouse et al |
| EPI_ISL_731583 | England/ALDP-C3BFB8/2020 | 2020-12-10 | Europe/United Kingdom/England/ | Lighthouse Lab in Alderley Park | Wellcome Sanger Institute for the COVID-19 Genomics UK (COG-UK) Consortium | Jacquelyn Wynn et al |
| EPI_ISL_731592 | England/ALDP-C3B273/2020 | 2020-12-09 | Europe/United Kingdom/England/ | Lighthouse Lab in Alderley Park | Wellcome Sanger Institute for the COVID-19 Genomics UK (COG-UK) Consortium | Jacquelyn Wynn et al |
| EPI_ISL_731899 | Portugal/PT1815/2020 | 2020-11-13 | Europe/Portugal/Portugal/ | Instituto Nacional de Saude (INSA) | Instituto Nacional de Saude (INSA) | Borges et al et al |
| EPI_ISL_731906 | Portugal/PT1932/2020 | 2020-11-15 | Europe/Portugal/Portugal/ | Instituto Nacional de Saude (INSA) and Instituto Gulbenkian de Ciencia (IGC) | Instituto Nacional de Saude (INSA) and Instituto Gulbenkian de Ciencia (IGC) | Borges et al et al |
| EPI_ISL_731920 | Portugal/PT1645/2020 | 2020-11-17 | Europe/Portugal/Portugal/ | Instituto Nacional de Saude (INSA) | Instituto Nacional de Saude (INSA) | Borges et al et al |
| EPI_ISL_731922 | Portugal/PT1881/2020 | 2020-11-11 | Europe/Portugal/Portugal/ | Instituto Nacional de Saude (INSA) | Instituto Nacional de Saude (INSA) | Borges et al et al |
| EPI_ISL_731924 | Portugal/PT1914/2020 | 2020-11-13 | Europe/Portugal/Portugal/ | Instituto Nacional de Saude (INSA) and Instituto Gulbenkian de Ciencia (IGC) | Instituto Nacional de Saude (INSA) and Instituto Gulbenkian de Ciencia (IGC) | Borges et al et al |
| EPI_ISL_731928 | Portugal/PT2022/2020 | 2020-11-19 | Europe/Portugal/Portugal/ | Instituto Nacional de Saude (INSA) and Instituto Gulbenkian de Ciencia (IGC) | Instituto Nacional de Saude (INSA) and Instituto Gulbenkian de Ciencia (IGC) | Borges et al et al |
| EPI_ISL_731945 | Portugal/PT1635/2020 | 2020-05-28 | Europe/Portugal/Portugal/ | Instituto Nacional de Saude (INSA) | Instituto Nacional de Saude (INSA) | Borges et al et al |
| EPI_ISL_731947 | Portugal/PT1637/2020 | 2020-05-13 | Europe/Portugal/Portugal/ | Instituto Nacional de Saude (INSA) | Instituto Nacional de Saude (INSA) | Borges et al et al |

| | | | | | | |
|---|---|---|---|---|---|---|
| EPI_ISL_731959 | Portugal/PT1657/2020 | 2020-11-09 | Europe/Portugal/Portugal/ | Instituto Nacional de Saude (INSA) | Instituto Nacional de Saude (INSA) | Borges et al et al |
| EPI_ISL_731964 | Portugal/PT1663/2020 | 2020-11-12 | Europe/Portugal/Portugal/ | Instituto Nacional de Saude (INSA) | Instituto Nacional de Saude (INSA) | Borges et al et al |
| EPI_ISL_731977 | Portugal/PT1740/2020 | 2020-11-09 | Europe/Portugal/Portugal/ | Instituto Nacional de Saude (INSA) | Instituto Nacional de Saude (INSA) | Borges et al et al |
| EPI_ISL_732003 | Portugal/PT1772/2020 | 2020-11-09 | Europe/Portugal/Portugal/ | Instituto Nacional de Saude (INSA) | Instituto Nacional de Saude (INSA) | Borges et al et al |
| EPI_ISL_732034 | Portugal/PT1809/2020 | 2020-11-11 | Europe/Portugal/Portugal/ | Instituto Nacional de Saude (INSA) | Instituto Nacional de Saude (INSA) | Borges et al et al |
| EPI_ISL_732052 | Portugal/PT1831/2020 | 2020-11-15 | Europe/Portugal/Portugal/ | Instituto Nacional de Saude (INSA) | Instituto Nacional de Saude (INSA) | Borges et al et al |
| EPI_ISL_732055 | Portugal/PT1834/2020 | 2020-11-09 | Europe/Portugal/Portugal/ | Instituto Nacional de Saude (INSA) | Instituto Nacional de Saude (INSA) | Borges et al et al |
| EPI_ISL_732069 | Portugal/PT1849/2020 | 2020-11-12 | Europe/Portugal/Portugal/ | Instituto Nacional de Saude (INSA) | Instituto Nacional de Saude (INSA) | Borges et al et al |
| EPI_ISL_732075 | Portugal/PT1856/2020 | 2020-11-13 | Europe/Portugal/Portugal/ | Instituto Nacional de Saude (INSA) | Instituto Nacional de Saude (INSA) | Borges et al et al |
| EPI_ISL_732081 | Portugal/PT1862/2020 | 2020-11-09 | Europe/Portugal/Portugal/ | Instituto Nacional de Saude (INSA) | Instituto Nacional de Saude (INSA) | Borges et al et al |
| EPI_ISL_732083 | Portugal/PT1864/2020 | 2020-11-10 | Europe/Portugal/Portugal/ | Instituto Nacional de Saude (INSA) | Instituto Nacional de Saude (INSA) | Borges et al et al |
| EPI_ISL_732087 | Portugal/PT1868/2020 | 2020-11-10 | Europe/Portugal/Portugal/ | Instituto Nacional de Saude (INSA) | Instituto Nacional de Saude (INSA) | Borges et al et al |
| EPI_ISL_732088 | Portugal/PT1869/2020 | 2020-11-10 | Europe/Portugal/Portugal/ | Instituto Nacional de Saude (INSA) | Instituto Nacional de Saude (INSA) | Borges et al et al |
| EPI_ISL_732096 | Portugal/PT1879/2020 | 2020-11-12 | Europe/Portugal/Portugal/ | Instituto Nacional de Saude (INSA) | Instituto Nacional de Saude (INSA) | Borges et al et al |
| EPI_ISL_732101 | Portugal/PT1886/2020 | 2020-11-12 | Europe/Portugal/Portugal/ | Instituto Nacional de Saude (INSA) | Instituto Nacional de Saude (INSA) | Borges et al et al |
| EPI_ISL_732104 | Portugal/PT1889/2020 | 2020-11-12 | Europe/Portugal/Portugal/ | Instituto Nacional de Saude (INSA) | Instituto Nacional de Saude (INSA) | Borges et al et al |
| EPI_ISL_732114 | Portugal/PT1900/2020 | 2020-12-02 | Europe/Portugal/Portugal/ | Instituto Nacional de Saude (INSA) | Instituto Nacional de Saude (INSA) | Borges et al et al |
| EPI_ISL_732118 | Portugal/PT1904/2020 | 2020-11-09 | Europe/Portugal/Portugal/ | Instituto Nacional de Saude (INSA) and Instituto Gulbenkian de Ciencia (IGC) | Instituto Nacional de Saude (INSA) and Instituto Gulbenkian de Ciencia (IGC) | Borges et al et al |

| | | | | | | |
|---|---|---|---|---|---|---|
| EPI_ISL_732120 | Portugal/PT1907/2020 | 2020-11-11 | Europe/Portugal/Portugal/ | Instituto Nacional de Saude (INSA) and Instituto Gulbenkian de Ciencia (IGC) | Instituto Nacional de Saude (INSA) and Instituto Gulbenkian de Ciencia (IGC) | Borges et al et al |
| EPI_ISL_732135 | Portugal/PT1923/2020 | 2020-11-14 | Europe/Portugal/Portugal/ | Instituto Nacional de Saude (INSA) and Instituto Gulbenkian de Ciencia (IGC) | Instituto Nacional de Saude (INSA) and Instituto Gulbenkian de Ciencia (IGC) | Borges et al et al |
| EPI_ISL_732138 | Portugal/PT1927/2020 | 2020-11-15 | Europe/Portugal/Portugal/ | Instituto Nacional de Saude (INSA) and Instituto Gulbenkian de Ciencia (IGC) | Instituto Nacional de Saude (INSA) and Instituto Gulbenkian de Ciencia (IGC) | Borges et al et al |
| EPI_ISL_732140 | Portugal/PT1929/2020 | 2020-11-15 | Europe/Portugal/Portugal/ | Instituto Nacional de Saude (INSA) and Instituto Gulbenkian de Ciencia (IGC) | Instituto Nacional de Saude (INSA) and Instituto Gulbenkian de Ciencia (IGC) | Borges et al et al |
| EPI_ISL_732142 | Portugal/PT1931/2020 | 2020-11-15 | Europe/Portugal/Portugal/ | Instituto Nacional de Saude (INSA) and Instituto Gulbenkian de Ciencia (IGC) | Instituto Nacional de Saude (INSA) and Instituto Gulbenkian de Ciencia (IGC) | Borges et al et al |
| EPI_ISL_732148 | Portugal/PT1681/2020 | 2020-11-09 | Europe/Portugal/Portugal/ | Instituto Nacional de Saude (INSA) and Instituto Gulbenkian de Ciencia (IGC) | Instituto Nacional de Saude (INSA) and Instituto Gulbenkian de Ciencia (IGC) | Borges et al et al |
| EPI_ISL_732189 | Portugal/PT1964/2020 | 2020-11-24 | Europe/Portugal/Portugal/ | Instituto Nacional de Saude (INSA) and Instituto Gulbenkian de Ciencia (IGC) | Instituto Nacional de Saude (INSA) and Instituto Gulbenkian de Ciencia (IGC) | Borges et al et al |
| EPI_ISL_732190 | Portugal/PT1965/2020 | 2020-11-24 | Europe/Portugal/Portugal/ | Instituto Nacional de Saude (INSA) and Instituto Gulbenkian de Ciencia (IGC) | Instituto Nacional de Saude (INSA) and Instituto Gulbenkian de Ciencia (IGC) | Borges et al et al |

| | | | | | | |
|---|---|---|---|---|---|---|
| EPI_ISL_732194 | Portugal/PT1969/2020 | 2020-11-24 | Europe/Portugal/Portugal/ | Instituto Nacional de Saude (INSA) and Instituto Gulbenkian de Ciencia (IGC) | Instituto Nacional de Saude (INSA) and Instituto Gulbenkian de Ciencia (IGC) | Borges et al et al |
| EPI_ISL_732197 | Portugal/PT1973/2020 | 2020-11-09 | Europe/Portugal/Portugal/ | Instituto Nacional de Saude (INSA) and Instituto Gulbenkian de Ciencia (IGC) | Instituto Nacional de Saude (INSA) and Instituto Gulbenkian de Ciencia (IGC) | Borges et al et al |
| EPI_ISL_732199 | Portugal/PT1976/2020 | 2020-11-09 | Europe/Portugal/Portugal/ | Instituto Nacional de Saude (INSA) and Instituto Gulbenkian de Ciencia (IGC) | Instituto Nacional de Saude (INSA) and Instituto Gulbenkian de Ciencia (IGC) | Borges et al et al |
| EPI_ISL_732210 | Portugal/PT1987/2020 | 2020-11-11 | Europe/Portugal/Portugal/ | Instituto Nacional de Saude (INSA) and Instituto Gulbenkian de Ciencia (IGC) | Instituto Nacional de Saude (INSA) and Instituto Gulbenkian de Ciencia (IGC) | Borges et al et al |
| EPI_ISL_732211 | Portugal/PT1988/2020 | 2020-11-11 | Europe/Portugal/Portugal/ | Instituto Nacional de Saude (INSA) and Instituto Gulbenkian de Ciencia (IGC) | Instituto Nacional de Saude (INSA) and Instituto Gulbenkian de Ciencia (IGC) | Borges et al et al |
| EPI_ISL_732214 | Portugal/PT1992/2020 | 2020-11-13 | Europe/Portugal/Portugal/ | Instituto Nacional de Saude (INSA) and Instituto Gulbenkian de Ciencia (IGC) | Instituto Nacional de Saude (INSA) and Instituto Gulbenkian de Ciencia (IGC) | Borges et al et al |
| EPI_ISL_732215 | Portugal/PT1993/2020 | 2020-11-14 | Europe/Portugal/Portugal/ | Instituto Nacional de Saude (INSA) and Instituto Gulbenkian de Ciencia (IGC) | Instituto Nacional de Saude (INSA) and Instituto Gulbenkian de Ciencia (IGC) | Borges et al et al |
| EPI_ISL_732224 | Portugal/PT2003/2020 | 2020-11-12 | Europe/Portugal/Portugal/ | Instituto Nacional de Saude (INSA) and Instituto Gulbenkian de Ciencia (IGC) | Instituto Nacional de Saude (INSA) and Instituto Gulbenkian de Ciencia (IGC) | Borges et al et al |

| | | | | | | |
|---|---|---|---|---|---|---|
| EPI_ISL_732245 | Portugal/PT2029/2020 | 2020-11-23 | Europe/Portugal/Portugal/ | Instituto Nacional de Saude (INSA) and Instituto Gulbenkian de Ciencia (IGC) | Instituto Nacional de Saude (INSA) and Instituto Gulbenkian de Ciencia (IGC) | Borges et al et al |
| EPI_ISL_732251 | Portugal/PT2035/2020 | 2020-11-09 | Europe/Portugal/Portugal/ | Instituto Nacional de Saude (INSA) and Instituto Gulbenkian de Ciencia (IGC) | Instituto Nacional de Saude (INSA) and Instituto Gulbenkian de Ciencia (IGC) | Borges et al et al |
| EPI_ISL_732271 | Portugal/PT2057/2020 | 2020-11-09 | Europe/Portugal/Portugal/ | Instituto Nacional de Saude (INSA) and Instituto Gulbenkian de Ciencia (IGC) | Instituto Nacional de Saude (INSA) and Instituto Gulbenkian de Ciencia (IGC) | Borges et al et al |
| EPI_ISL_732280 | Portugal/PT2066/2020 | 2020-11-11 | Europe/Portugal/Portugal/ | Instituto Nacional de Saude (INSA) and Instituto Gulbenkian de Ciencia (IGC) | Instituto Nacional de Saude (INSA) and Instituto Gulbenkian de Ciencia (IGC) | Borges et al et al |
| EPI_ISL_732282 | Portugal/PT2068/2020 | 2020-11-11 | Europe/Portugal/Portugal/ | Instituto Nacional de Saude (INSA) and Instituto Gulbenkian de Ciencia (IGC) | Instituto Nacional de Saude (INSA) and Instituto Gulbenkian de Ciencia (IGC) | Borges et al et al |
| EPI_ISL_732388 | Ireland/D-NVRL-79IRL01712/2020 | 2020-11-28 | Europe/Ireland/Dublin/ | National Virus Reference Laboratory | National Virus Reference Laboratory | Michael Carr et al |
| EPI_ISL_732389 | Ireland/D-NVRL-20ILR10836/2020 | 2020-11-15 | Europe/Ireland/Dublin/ | National Virus Reference Laboratory | National Virus Reference Laboratory | Michael Carr et al |
| EPI_ISL_732392 | Ireland/D-NVRL-20ILR10860/2020 | 2020-11-23 | Europe/Ireland/Dublin/ | National Virus Reference Laboratory | National Virus Reference Laboratory | Michael Carr et al |
| EPI_ISL_732395 | Ireland/LH-NVRL-20ILR08024/2020 | 2020-11-10 | Europe/Ireland/Louth/ | National Virus Reference Laboratory | National Virus Reference Laboratory | Michael Carr et al |
| EPI_ISL_732398 | Ireland/D-NVRL-20IRL76603/2020 | 2020-11-04 | Europe/Ireland/Dublin/ | National Virus Reference Laboratory | National Virus Reference Laboratory | Michael Carr et al |

| | | | | | | |
|---|---|---|---|---|---|---|
| EPI_ISL_732405 | Ireland/D-NVRL-20ILR10849/2020 | 2020-11-24 | Europe/Ireland/Dublin/ | National Virus Reference Laboratory | National Virus Reference Laboratory | Michael Carr et al |
| EPI_ISL_732412 | Ireland/LH-NVRL-20ILR08013/2020 | 2020-11-10 | Europe/Ireland/Louth/ | National Virus Reference Laboratory | National Virus Reference Laboratory | Michael Carr et al |
| EPI_ISL_732461 | Ireland/SO-NVRL-76IRL31587/2020 | 2020-10-18 | Europe/Ireland/Sligo/ | National Virus Reference Laboratory | National Virus Reference Laboratory | Michael Carr et al |
| EPI_ISL_732481 | Ireland/KE-NVRL-76IRL26703/2020 | 2020-10-18 | Europe/Ireland/Kildare/ | National Virus Reference Laboratory | National Virus Reference Laboratory | Michael Carr et al |
| EPI_ISL_732492 | Ireland/KK-NVRL-76IRL28885/2020 | 2020-10-18 | Europe/Ireland/Kilkenny/ | National Virus Reference Laboratory | National Virus Reference Laboratory | Michael Carr et al |
| EPI_ISL_732529 | Germany/BY-IMB11051/2020 | 2020-09-24 | Europe/Germany/Bavaria/Munich | Bundeswehr Institute of Microbiology | Bundeswehr Institute of Microbiology | Markus Antwerpen et al |
| EPI_ISL_732530 | Germany/BY-IMB00331/2020 | 2020-02-03 | Europe/Germany/Bavaria/Munich | Bundeswehr Institute of Microbiology | Bundeswehr Institute of Microbiology | Markus Antwerpen et al |
| EPI_ISL_732532 | Germany/BY-IMB09442/2020 | 2020-08-11 | Europe/Germany/Bavaria/Munich | Bundeswehr Institute of Microbiology | Bundeswehr Institute of Microbiology | Elham Khatamzas et al |
| EPI_ISL_732536 | Germany/BY-IMB07990/2020 | 2020-06-29 | Europe/Germany/Bavaria/Munich | Bundeswehr Institute of Microbiology | Bundeswehr Institute of Microbiology | Elham Khatamzas et al |
| EPI_ISL_732541 | Germany/BY-IMB00234/2020 | 2020-02-01 | Europe/Germany/Bavaria/Munich | Bundeswehr Institute of Microbiology | Bundeswehr Institute of Microbiology | Markus Antwerpen et al |
| EPI_ISL_732547 | Germany/BY-IMB02460/2020 | 2020-03-19 | Europe/Germany/Bavaria/Munich | Bundeswehr Institute of Microbiology | Bundeswehr Institute of Microbiology | Markus Antwerpen et al |
| EPI_ISL_732549 | Germany/BY-IMB02858/2020 | 2020-03-26 | Europe/Germany/Bavaria/Munich | Bundeswehr Institute of Microbiology | Bundeswehr Institute of Microbiology | Markus Antwerpen et al |
| EPI_ISL_732554 | Germany/BY-IMB04157/2020 | 2020-04-14 | Europe/Germany/Bavaria/Munich | Bundeswehr Institute of Microbiology | Bundeswehr Institute of Microbiology | Markus Antwerpen et al |

| | | | | | | |
|---|---|---|---|---|---|---|
| EPI_ISL_732557 | Germany/BY-IMB06128/2020 | 2020-05-13 | Europe/Germany/Bavaria/Munich | Bundeswehr Institute of Microbiology | Bundeswehr Institute of Microbiology | Elham Khatamzas et al |
| EPI_ISL_732560 | Germany/BY-IMB06494/2020 | 2020-05-26 | Europe/Germany/Bavaria/Munich | Bundeswehr Institute of Microbiology | Bundeswehr Institute of Microbiology | Elham Khatamzas et al |
| EPI_ISL_732626 | Finland/S10CF2/2020 | 2020-10-30 | Europe/Finland/Uusimaa/ | Department of Virology and Immunology, University of Helsinki and Helsinki University Hospital, Huslab Finland | Department of Virology, Faculty of Medicine, University of Helsinki, Helsinki, Finland | Teemu Smura et al |
| EPI_ISL_732636 | Finland/S1CD10/2020 | 2020-10-30 | Europe/Finland/Uusimaa/ | Department of Virology and Immunology, University of Helsinki and Helsinki University Hospital, Huslab Finland | Department of Virology, Faculty of Medicine, University of Helsinki, Helsinki, Finland | Teemu Smura et al |
| EPI_ISL_732658 | Germany/BY-IMB11525/2020 | 2020-10-07 | Europe/Germany/Bavaria/Munich | Bundeswehr Institute of Microbiology | Bundeswehr Institute of Microbiology | Elham Khatamzas et al |
| EPI_ISL_732661 | Russia/MOS-CRIE-13384051/2020 | 2020-11-07 | Europe/Russia/Moscow Oblast/ | "Molecular diagnostic laboratory of Federal Budget Institution of Science ""Central Research Institute of Epidemiology"" of The Federal Service on Customers' Rights Protection and Human Well-being Surveillance" | Group of Genomics and Postgenomic Technologies of Central Research Institute of Epidemilology | Samoilov AE et al |
| EPI_ISL_732662 | Russia/MOS-CRIE-13364310/2020 | 2020-11-06 | Europe/Russia/Moscow Oblast/ | "Molecular diagnostic laboratory of Federal Budget Institution of Science ""Central Research Institute of Epidemiology"" of The Federal Service on Customers' Rights Protection and Human Well-being Surveillance" | Group of Genomics and Postgenomic Technologies of Central Research Institute of Epidemilology | Samoilov AE et al |

| | | | | | | |
|---|---|---|---|---|---|---|
| EPI_ISL_732663 | Russia/MOS-CRIE-13385391/2020 | 2020-11-06 | Europe/Russia/Moscow Oblast/ | "Molecular diagnostic laboratory of Federal Budget Institution of Science ""Central Research Institute of Epidemiology"" of The Federal Service on Customers' Rights Protection and Human Well-being Surveillance" | Group of Genomics and Postgenomic Technologies of Central Research Institute of Epidemilology | Samoilov AE et al |
| EPI_ISL_732665 | Russia/MOS-CRIE-13381937/2020 | 2020-11-06 | Europe/Russia/Moscow Oblast/ | "Molecular diagnostic laboratory of Federal Budget Institution of Science ""Central Research Institute of Epidemiology"" of The Federal Service on Customers' Rights Protection and Human Well-being Surveillance" | Group of Genomics and Postgenomic Technologies of Central Research Institute of Epidemilology | Samoilov AE et al |
| EPI_ISL_732666 | Russia/MOS-CRIE-13363841/2020 | 2020-11-06 | Europe/Russia/Moscow Oblast/ | "Molecular diagnostic laboratory of Federal Budget Institution of Science ""Central Research Institute of Epidemiology"" of The Federal Service on Customers' Rights Protection and Human Well-being Surveillance" | Group of Genomics and Postgenomic Technologies of Central Research Institute of Epidemilology | Samoilov AE et al |
| EPI_ISL_732668 | Russia/MOS-CRIE-13383434/2020 | 2020-11-06 | Europe/Russia/Moscow Oblast/ | "Molecular diagnostic laboratory of Federal Budget Institution of Science ""Central Research Institute of Epidemiology"" of The Federal Service on Customers' Rights Protection and Human Well-being Surveillance" | Group of Genomics and Postgenomic Technologies of Central Research Institute of Epidemilology | Samoilov AE et al |
| EPI_ISL_732670 | Russia/MOS-CRIE-13385401/2020 | 2020-11-07 | Europe/Russia/Moscow Oblast/ | "Molecular diagnostic laboratory of Federal Budget Institution of Science ""Central Research Institute of Epidemiology"" of The Federal Service on Customers' Rights Protection and Human Well-being Surveillance" | Group of Genomics and Postgenomic Technologies of Central Research Institute of Epidemilology | Samoilov AE et al |

| | | | | | | |
|---|---|---|---|---|---|---|
| EPI_ISL_732671 | Russia/MOS-CRIE-13431175/2020 | 2020-11-06 | Europe/Russia/Moscow Oblast/ | "Molecular diagnostic laboratory of Federal Budget Institution of Science ""Central Research Institute of Epidemiology"" of The Federal Service on Customers' Rights Protection and Human Well-being Surveillance" | Group of Genomics and Postgenomic Technologies of Central Research Institute of Epidemilology | Samoilov AE et al |
| EPI_ISL_732672 | Russia/MOS-CRIE-13385721/2020 | 2020-11-06 | Europe/Russia/Moscow Oblast/ | "Molecular diagnostic laboratory of Federal Budget Institution of Science ""Central Research Institute of Epidemiology"" of The Federal Service on Customers' Rights Protection and Human Well-being Surveillance" | Group of Genomics and Postgenomic Technologies of Central Research Institute of Epidemilology | Samoilov AE et al |
| EPI_ISL_732673 | Russia/MOS-CRIE-13365319/2020 | 2020-11-06 | Europe/Russia/Moscow Oblast/ | "Molecular diagnostic laboratory of Federal Budget Institution of Science ""Central Research Institute of Epidemiology"" of The Federal Service on Customers' Rights Protection and Human Well-being Surveillance" | Group of Genomics and Postgenomic Technologies of Central Research Institute of Epidemilology | Samoilov AE et al |
| EPI_ISL_732675 | Russia/MOS-CRIE-13444338/2020 | 2020-11-06 | Europe/Russia/Moscow Oblast/ | "Molecular diagnostic laboratory of Federal Budget Institution of Science ""Central Research Institute of Epidemiology"" of The Federal Service on Customers' Rights Protection and Human Well-being Surveillance" | Group of Genomics and Postgenomic Technologies of Central Research Institute of Epidemilology | Samoilov AE et al |
| EPI_ISL_732676 | Russia/MOS-CRIE-13298544/2020 | 2020-11-06 | Europe/Russia/Moscow Oblast/ | "Molecular diagnostic laboratory of Federal Budget Institution of Science ""Central Research Institute of Epidemiology"" of The Federal Service on Customers' Rights Protection and Human Well-being Surveillance" | Group of Genomics and Postgenomic Technologies of Central Research Institute of Epidemilology | Samoilov AE et al |
| EPI_ISL_732677 | France/ARA-100319/2020 | 2020-11-03 | Europe/France/Auvergne-Rh√¥ne-Alpes/ | CNR Virus des Infections Respiratoires - France SUD | CNR Virus des Infections Respiratoires - France SUD | Antonin Bal et al |

| | | | | | | |
|---|---|---|---|---|---|---|
| EPI_ISL_732678 | France/ARA-100644/2020 | 2020-11-03 | Europe/France/Auvergne-Rhv¥ne-Alpes/ | CNR Virus des Infections Respiratoires - France SUD | CNR Virus des Infections Respiratoires - France SUD | Antonin Bal et al |
| EPI_ISL_732679 | France/ARA-101085/2020 | 2020-11-04 | Europe/France/Auvergne-Rhv¥ne-Alpes/ | CNR Virus des Infections Respiratoires - France SUD | CNR Virus des Infections Respiratoires - France SUD | Antonin Bal et al |
| EPI_ISL_732681 | France/ARA-102363/2020 | 2020-11-05 | Europe/France/Auvergne-Rhv¥ne-Alpes/ | CNR Virus des Infections Respiratoires - France SUD | CNR Virus des Infections Respiratoires - France SUD | Antonin Bal et al |
| EPI_ISL_732684 | France/ARA-106011/2020 | 2020-11-12 | Europe/France/Auvergne-Rhv¥ne-Alpes/ | CNR Virus des Infections Respiratoires - France SUD | CNR Virus des Infections Respiratoires - France SUD | Antonin Bal et al |
| EPI_ISL_732685 | France/ARA-106089/2020 | 2020-11-12 | Europe/France/Auvergne-Rhv¥ne-Alpes/ | CNR Virus des Infections Respiratoires - France SUD | CNR Virus des Infections Respiratoires - France SUD | Antonin Bal et al |
| EPI_ISL_732686 | France/ARA-107483/2020 | 2020-11-16 | Europe/France/Auvergne-Rhv¥ne-Alpes/ | CNR Virus des Infections Respiratoires - France SUD | CNR Virus des Infections Respiratoires - France SUD | Antonin Bal et al |
| EPI_ISL_732687 | France/ARA-108320/2020 | 2020-11-17 | Europe/France/Auvergne-Rhv¥ne-Alpes/ | CNR Virus des Infections Respiratoires - France SUD | CNR Virus des Infections Respiratoires - France SUD | Antonin Bal et al |
| EPI_ISL_732689 | France/ARA-110756/2020 | 2020-11-22 | Europe/France/Auvergne-Rhv¥ne-Alpes/ | CNR Virus des Infections Respiratoires - France SUD | CNR Virus des Infections Respiratoires - France SUD | Antonin Bal et al |
| EPI_ISL_732690 | France/ARA-111424/2020 | 2020-11-23 | Europe/France/Auvergne-Rhv¥ne-Alpes/ | CNR Virus des Infections Respiratoires - France SUD | CNR Virus des Infections Respiratoires - France SUD | Antonin Bal et al |
| EPI_ISL_732691 | France/ARA-111572/2020 | 2020-11-24 | Europe/France/Auvergne-Rhv¥ne-Alpes/ | CNR Virus des Infections Respiratoires - France SUD | CNR Virus des Infections Respiratoires - France SUD | Antonin Bal et al |

| | | | | | | |
|---|---|---|---|---|---|---|
| EPI_ISL_732692 | France/ARA-111841/2020 | 2020-11-24 | Europe/France/Auvergne-Rh√¥ne-Alpes/ | CNR Virus des Infections Respiratoires - France SUD | CNR Virus des Infections Respiratoires - France SUD | Antonin Bal et al |
| EPI_ISL_732693 | France/ARA-75541/2020 | 2020-09-20 | Europe/France/Auvergne-Rh√¥ne-Alpes/ | CNR Virus des Infections Respiratoires - France SUD | CNR Virus des Infections Respiratoires - France SUD | Antonin Bal et al |
| EPI_ISL_732695 | France/ARA-83616/2020 | 2020-09-26 | Europe/France/Auvergne-Rh√¥ne-Alpes/ | CNR Virus des Infections Respiratoires - France SUD | CNR Virus des Infections Respiratoires - France SUD | Antonin Bal et al |
| EPI_ISL_732696 | France/ARA-86532/2020 | 2020-10-13 | Europe/France/Auvergne-Rh√¥ne-Alpes/ | CNR Virus des Infections Respiratoires - France SUD | CNR Virus des Infections Respiratoires - France SUD | Antonin Bal et al |
| EPI_ISL_732697 | France/ARA-86645/2020 | 2020-10-13 | Europe/France/Auvergne-Rh√¥ne-Alpes/ | CNR Virus des Infections Respiratoires - France SUD | CNR Virus des Infections Respiratoires - France SUD | Antonin Bal et al |
| EPI_ISL_732698 | France/ARA-87533/2020 | 2020-10-14 | Europe/France/Auvergne-Rh√¥ne-Alpes/ | CNR Virus des Infections Respiratoires - France SUD | CNR Virus des Infections Respiratoires - France SUD | Antonin Bal et al |
| EPI_ISL_732699 | France/ARA-95401/2020 | 2020-10-27 | Europe/France/Auvergne-Rh√¥ne-Alpes/ | CNR Virus des Infections Respiratoires - France SUD | CNR Virus des Infections Respiratoires - France SUD | Antonin Bal et al |
| EPI_ISL_732701 | France/ARA-96089/2020 | 2020-10-28 | Europe/France/Auvergne-Rh√¥ne-Alpes/ | CNR Virus des Infections Respiratoires - France SUD | CNR Virus des Infections Respiratoires - France SUD | Antonin Bal et al |
| EPI_ISL_732702 | France/ARA-96707/2020 | 2020-10-29 | Europe/France/Auvergne-Rh√¥ne-Alpes/ | CNR Virus des Infections Respiratoires - France SUD | CNR Virus des Infections Respiratoires - France SUD | Antonin Bal et al |
| EPI_ISL_732704 | USA/NM-DOH-2020314707/2020 | 2020-08-27 | North America/USA/New Mexico/ | New Mexico Department of Health Scientific Laboratory | New Mexico Department of Health Scientific Laboratory | Ellie Johnson et al |

| | | | | | | |
|---|---|---|---|---|---|---|
| EPI_ISL_732707 | USA/NM-DOH-2020315222/2020 | 2020-08-27 | North America/USA/New Mexico/ | New Mexico Department of Health Scientific Laboratory | New Mexico Department of Health Scientific Laboratory | Ellie Johnson et al |
| EPI_ISL_732708 | USA/NM-DOH-2020315376/2020 | 2020-08-27 | North America/USA/New Mexico/ | New Mexico Department of Health Scientific Laboratory | New Mexico Department of Health Scientific Laboratory | Ellie Johnson et al |
| EPI_ISL_732709 | USA/NM-DOH-2020315490/2020 | 2020-08-27 | North America/USA/New Mexico/ | New Mexico Department of Health Scientific Laboratory | New Mexico Department of Health Scientific Laboratory | Ellie Johnson et al |
| EPI_ISL_732712 | USA/NM-DOH-2020315536/2020 | 2020-08-27 | North America/USA/New Mexico/ | New Mexico Department of Health Scientific Laboratory | New Mexico Department of Health Scientific Laboratory | Ellie Johnson et al |
| EPI_ISL_732722 | USA/NM-DOH-2020316601/2020 | 2020-08-27 | North America/USA/New Mexico/ | New Mexico Department of Health Scientific Laboratory | New Mexico Department of Health Scientific Laboratory | Ellie Johnson et al |
| EPI_ISL_732726 | USA/NM-DOH-2020317191/2020 | 2020-08-26 | North America/USA/New Mexico/ | New Mexico Department of Health Scientific Laboratory | New Mexico Department of Health Scientific Laboratory | Ellie Johnson et al |
| EPI_ISL_732732 | USA/NM-DOH-2020318368/2020 | 2020-08-27 | North America/USA/New Mexico/ | New Mexico Department of Health Scientific Laboratory | New Mexico Department of Health Scientific Laboratory | Ellie Johnson et al |
| EPI_ISL_732734 | USA/NM-DOH-2020318505/2020 | 2020-08-29 | North America/USA/New Mexico/ | New Mexico Department of Health Scientific Laboratory | New Mexico Department of Health Scientific Laboratory | Ellie Johnson et al |
| EPI_ISL_732736 | USA/NM-DOH-2020318654/2020 | 2020-08-28 | North America/USA/New Mexico/ | New Mexico Department of Health Scientific Laboratory | New Mexico Department of Health Scientific Laboratory | Ellie Johnson et al |
| EPI_ISL_732737 | USA/NM-DOH-2020318667/2020 | 2020-08-28 | North America/USA/New Mexico/ | New Mexico Department of Health Scientific Laboratory | New Mexico Department of Health Scientific Laboratory | Ellie Johnson et al |

| | | | | | | |
|---|---|---|---|---|---|---|
| EPI_ISL_732739 | USA/NM-DOH-2020318741/2020 | 2020-08-28 | North America/USA/New Mexico/ | New Mexico Department of Health Scientific Laboratory | New Mexico Department of Health Scientific Laboratory | Ellie Johnson et al |
| EPI_ISL_732740 | USA/NM-DOH-2020318805/2020 | 2020-08-27 | North America/USA/New Mexico/ | New Mexico Department of Health Scientific Laboratory | New Mexico Department of Health Scientific Laboratory | Ellie Johnson et al |
| EPI_ISL_732742 | USA/NM-DOH-2020318937/2020 | 2020-08-27 | North America/USA/New Mexico/ | New Mexico Department of Health Scientific Laboratory | New Mexico Department of Health Scientific Laboratory | Ellie Johnson et al |
| EPI_ISL_732743 | USA/NM-DOH-2020318939/2020 | 2020-08-27 | North America/USA/New Mexico/ | New Mexico Department of Health Scientific Laboratory | New Mexico Department of Health Scientific Laboratory | Ellie Johnson et al |
| EPI_ISL_732747 | USA/NM-DOH-2020319240/2020 | 2020-08-28 | North America/USA/New Mexico/ | New Mexico Department of Health Scientific Laboratory | New Mexico Department of Health Scientific Laboratory | Ellie Johnson et al |
| EPI_ISL_732749 | Spain/RI-IBV-99010903/2020 | 2020-04-20 | Europe/Spain/La Rioja/Santo Domingo d | Centro de Investigaci√≥n Biom√©dica de La Rioja - Hospital San Pedro Logro√±o | SeqCOVID-SPAIN consortium/IBV(CSIC) | Mar√≠a de Toro et al |
| EPI_ISL_732753 | Spain/RI-IBV-99010907/2020 | 2020-11-12 | Europe/Spain/La Rioja/Santo Domingo d | Centro de Investigaci√≥n Biom√©dica de La Rioja - Hospital San Pedro Logro√±o | SeqCOVID-SPAIN consortium/IBV(CSIC) | Mar√≠a de Toro et al |
| EPI_ISL_732756 | Spain/RI-IBV-99010911/2020 | 2020-11-13 | Europe/Spain/La Rioja/Logrono | Centro de Investigaci√≥n Biom√©dica de La Rioja - Hospital San Pedro Logro√±o | SeqCOVID-SPAIN consortium/IBV(CSIC) | Mar√≠a de Toro et al |
| EPI_ISL_732757 | Spain/RI-IBV-99010912/2020 | 2020-11-04 | Europe/Spain/La Rioja/Logrono | Centro de Investigaci√≥n Biom√©dica de La Rioja - Hospital San Pedro Logro√±o | SeqCOVID-SPAIN consortium/IBV(CSIC) | Mar√≠a de Toro et al |
| EPI_ISL_732764 | Spain/RI-IBV-99010925/2020 | 2020-03-26 | Europe/Spain/La Rioja/Albelda de Iregua | Centro de Investigaci√≥n Biom√©dica de La Rioja - Hospital San Pedro Logro√±o | SeqCOVID-SPAIN consortium/IBV(CSIC) | Mar√≠a de Toro et al |
| EPI_ISL_732772 | Spain/RI-IBV-99010934/2020 | 2020-11-11 | Europe/Spain/La Rioja/Tudelilla | Centro de Investigaci√≥n Biom√©dica de La Rioja - Hospital San Pedro Logro√±o | SeqCOVID-SPAIN consortium/IBV(CSIC) | Mar√≠a de Toro et al |

| | | | | | | |
|---|---|---|---|---|---|---|
| EPI_ISL_732776 | Spain/RI-IBV-99010941/2020 | 2020-07-17 | Europe/Spain/La Rioja/Logrono | Centro de Investigaci√≥n Biom√©dica de La Rioja - Hospital San Pedro Logro√±o | SeqCOVID-SPAIN consortium/IBV(CSIC) | Mar√≠a de Toro et al |
| EPI_ISL_732780 | Spain/RI-IBV-99010949/2020 | 2020-08-15 | Europe/Spain/La Rioja/Logrono | Centro de Investigaci√≥n Biom√©dica de La Rioja - Hospital San Pedro Logro√±o | SeqCOVID-SPAIN consortium/IBV(CSIC) | Mar√≠a de Toro et al |
| EPI_ISL_732782 | Spain/RI-IBV-99010951/2020 | 2020-08-25 | Europe/Spain/La Rioja/Logrono | Centro de Investigaci√≥n Biom√©dica de La Rioja - Hospital San Pedro Logro√±o | SeqCOVID-SPAIN consortium/IBV(CSIC) | Mar√≠a de Toro et al |
| EPI_ISL_732786 | Spain/RI-IBV-99010958/2020 | 2020-10-27 | Europe/Spain/La Rioja/Logrono | Centro de Investigaci√≥n Biom√©dica de La Rioja - Hospital San Pedro Logro√±o | SeqCOVID-SPAIN consortium/IBV(CSIC) | Mar√≠a de Toro et al |
| EPI_ISL_732787 | Spain/RI-IBV-99010959/2020 | 2020-10-31 | Europe/Spain/La Rioja/Logrono | Centro de Investigaci√≥n Biom√©dica de La Rioja - Hospital San Pedro Logro√±o | SeqCOVID-SPAIN consortium/IBV(CSIC) | Mar√≠a de Toro et al |
| EPI_ISL_732789 | Spain/RI-IBV-99010961/2020 | 2020-11-10 | Europe/Spain/La Rioja/Logrono | Centro de Investigaci√≥n Biom√©dica de La Rioja - Hospital San Pedro Logro√±o | SeqCOVID-SPAIN consortium/IBV(CSIC) | Mar√≠a de Toro et al |
| EPI_ISL_732794 | Spain/RI-IBV-99010966/2020 | 2020-09-30 | Europe/Spain/La Rioja/Calahorra | Centro de Investigaci√≥n Biom√©dica de La Rioja - Hospital San Pedro Logro√±o | SeqCOVID-SPAIN consortium/IBV(CSIC) | Mar√≠a de Toro et al |
| EPI_ISL_732795 | Spain/RI-IBV-99010967/2020 | 2020-10-21 | Europe/Spain/La Rioja/Calahorra | Centro de Investigaci√≥n Biom√©dica de La Rioja - Hospital San Pedro Logro√±o | SeqCOVID-SPAIN consortium/IBV(CSIC) | Mar√≠a de Toro et al |
| EPI_ISL_732798 | Spain/RI-IBV-99010970/2020 | 2020-11-09 | Europe/Spain/La Rioja/Rincon de Soto | Centro de Investigaci√≥n Biom√©dica de La Rioja - Hospital San Pedro Logro√±o | SeqCOVID-SPAIN consortium/IBV(CSIC) | Mar√≠a de Toro et al |
| EPI_ISL_732799 | Spain/RI-IBV-99010971/2020 | 2020-11-07 | Europe/Spain/La Rioja/Calahorra | Centro de Investigaci√≥n Biom√©dica de La Rioja - Hospital San Pedro Logro√±o | SeqCOVID-SPAIN consortium/IBV(CSIC) | Mar√≠a de Toro et al |
| EPI_ISL_732800 | Spain/RI-IBV-99010972/2020 | 2020-10-09 | Europe/Spain/La Rioja/Alfaro | Centro de Investigaci√≥n Biom√©dica de La Rioja - Hospital San Pedro Logro√±o | SeqCOVID-SPAIN consortium/IBV(CSIC) | Mar√≠a de Toro et al |
| EPI_ISL_732801 | Spain/RI-IBV-99010973/2020 | 2020-09-23 | Europe/Spain/La Rioja/Logrono | Centro de Investigaci√≥n Biom√©dica de La Rioja - Hospital San Pedro Logro√±o | SeqCOVID-SPAIN consortium/IBV(CSIC) | Mar√≠a de Toro et al |
| EPI_ISL_732804 | Spain/RI-IBV-99010976/2020 | 2020-10-01 | Europe/Spain/La Rioja/Calahorra | Centro de Investigaci√≥n Biom√©dica de La Rioja - Hospital San Pedro Logro√±o | SeqCOVID-SPAIN consortium/IBV(CSIC) | Mar√≠a de Toro et al |

| | | | | | | |
|---|---|---|---|---|---|---|
| EPI_ISL_732810 | Spain/RI-IBV-99010987/2020 | 2020-08-28 | Europe/Spain/La Rioja/Logrono | Centro de Investigaci√≥n Biom√©dica de La Rioja - Hospital San Pedro Logro√±o | SeqCOVID-SPAIN consortium/IBV(CSIC) | Mar√≠a de Toro et al |
| EPI_ISL_732813 | Spain/RI-IBV-99010990/2020 | 2020-10-18 | Europe/Spain/La Rioja/Logrono | Centro de Investigaci√≥n Biom√©dica de La Rioja - Hospital San Pedro Logro√±o | SeqCOVID-SPAIN consortium/IBV(CSIC) | Mar√≠a de Toro et al |
| EPI_ISL_732814 | Spain/RI-IBV-99010991/2020 | 2020-10-18 | Europe/Spain/La Rioja/Logrono | Centro de Investigaci√≥n Biom√©dica de La Rioja - Hospital San Pedro Logro√±o | SeqCOVID-SPAIN consortium/IBV(CSIC) | Mar√≠a de Toro et al |
| EPI_ISL_732815 | Spain/RI-IBV-99010992/2020 | 2020-09-10 | Europe/Spain/La Rioja/Logrono | Centro de Investigaci√≥n Biom√©dica de La Rioja - Hospital San Pedro Logro√±o | SeqCOVID-SPAIN consortium/IBV(CSIC) | Mar√≠a de Toro et al |
| EPI_ISL_732817 | Spain/RI-IBV-99010994/2020 | 2020-10-22 | Europe/Spain/La Rioja/Logrono | Centro de Investigaci√≥n Biom√©dica de La Rioja - Hospital San Pedro Logro√±o | SeqCOVID-SPAIN consortium/IBV(CSIC) | Mar√≠a de Toro et al |
| EPI_ISL_732818 | Spain/RI-IBV-99010995/2020 | 2020-10-14 | Europe/Spain/La Rioja/Aleson | Centro de Investigaci√≥n Biom√©dica de La Rioja - Hospital San Pedro Logro√±o | SeqCOVID-SPAIN consortium/IBV(CSIC) | Mar√≠a de Toro et al |
| EPI_ISL_732821 | USA/NE-002-17/2020 | 2020-09-26 | North America/USA/Nebraska/ | Nebraska Public Health Laboratory | UNMC COVID-19 Response Team | UNMC COVID-19 Response Team et al |
| EPI_ISL_732825 | USA/NE-006-17/2020 | 2020-09-26 | North America/USA/Nebraska/ | Nebraska Public Health Laboratory | UNMC COVID-19 Response Team | UNMC COVID-19 Response Team et al |
| EPI_ISL_732971 | Russia/SPE-RII-30769V/2020 | 2020-08-20 | Europe/Russia/Saint-Petersburg/ | WHO National Influenza Centre Russian Federation | WHO National Influenza Centre Russian Federation | Andrey Komissarov et al |
| EPI_ISL_732972 | Russia/SPE-RII-29894S/2020 | 2020-08-02 | Europe/Russia/Saint-Petersburg/ | WHO National Influenza Centre Russian Federation | WHO National Influenza Centre Russian Federation | Andrey Komissarov et al |
| EPI_ISL_732974 | Russia/KR-RII-12884S/2020 | 2020-11-18 | Europe/Russia/Karelia/Derevyanka | WHO National Influenza Centre Russian Federation | WHO National Influenza Centre Russian Federation | Andrey Komissarov et al |
| EPI_ISL_732975 | Russia/KR-RII-12754S/2020 | 2020-11-16 | Europe/Russia/Karelia/Podporozhye | WHO National Influenza Centre Russian Federation | WHO National Influenza Centre Russian Federation | Andrey Komissarov et al |

| | | | | | | |
|---|---|---|---|---|---|---|
| EPI_ISL_732976 | Russia/KR-RII-12753S/2020 | 2020-11-16 | Europe/Russia/Karelia/Podporozhye | WHO National Influenza Centre Russian Federation | WHO National Influenza Centre Russian Federation | Andrey Komissarov et al |
| EPI_ISL_732980 | Russia/KR-RII-12221S/2020 | 2020-11-16 | Europe/Russia/Karelia/Povenets | WHO National Influenza Centre Russian Federation | WHO National Influenza Centre Russian Federation | Andrey Komissarov et al |
| EPI_ISL_732983 | Russia/KR-RII-12210S/2020 | 2020-11-17 | Europe/Russia/Karelia/Kazhma | WHO National Influenza Centre Russian Federation | WHO National Influenza Centre Russian Federation | Andrey Komissarov et al |
| EPI_ISL_732985 | Russia/KR-RII-12198S/2020 | 2020-11-18 | Europe/Russia/Karelia/Shuya (Karelia) | WHO National Influenza Centre Russian Federation | WHO National Influenza Centre Russian Federation | Andrey Komissarov et al |
| EPI_ISL_732987 | Russia/SPE-RII-10016S/2020 | 2020-11-16 | Europe/Russia/Saint-Petersburg/ | WHO National Influenza Centre Russian Federation | WHO National Influenza Centre Russian Federation | Andrey Komissarov et al |
| EPI_ISL_732988 | Russia/SPE-RII-9960S/2020 | 2020-11-16 | Europe/Russia/Saint-Petersburg/ | WHO National Influenza Centre Russian Federation | WHO National Influenza Centre Russian Federation | Andrey Komissarov et al |
| EPI_ISL_732989 | Russia/SPE-RII-6981S/2020 | 2020-11-06 | Europe/Russia/Saint-Petersburg/ | WHO National Influenza Centre Russian Federation | WHO National Influenza Centre Russian Federation | Andrey Komissarov et al |
| EPI_ISL_732990 | Russia/SVE-RII-MH183S/2020 | 2020-07-08 | Europe/Russia/Sverdlovsk/Verkhnyaya | UMMC-Health | WHO National Influenza Centre Russian Federation | Andrey Komissarov et al |
| EPI_ISL_732991 | Russia/SVE-RII-MH186S/2020 | 2020-07-08 | Europe/Russia/Sverdlovsk/Verkhnyaya | UMMC-Health | WHO National Influenza Centre Russian Federation | Andrey Komissarov et al |
| EPI_ISL_732992 | Russia/SVE-RII-MH189S/2020 | 2020-07-08 | Europe/Russia/Sverdlovsk/Yekaterinbur | UMMC-Health | WHO National Influenza Centre Russian Federation | Andrey Komissarov et al |

| | | | | | | |
|---|---|---|---|---|---|---|
| EPI_ISL_732994 | Russia/SVE-RII-MH212S/2020 | 2020-07-23 | Europe/Russia/Sverdlovsk/Yekaterinbur | UMMC-Health | WHO National Influenza Centre Russian Federation | Andrey Komissarov et al |
| EPI_ISL_732995 | Russia/SVE-RII-MH216S/2020 | 2020-07-25 | Europe/Russia/Sverdlovsk/Yekaterinbur | UMMC-Health | WHO National Influenza Centre Russian Federation | Andrey Komissarov et al |
| EPI_ISL_732997 | Russia/SVE-RII-MH222S/2020 | 2020-07-29 | Europe/Russia/Sverdlovsk/Yekaterinbur | UMMC-Health | WHO National Influenza Centre Russian Federation | Andrey Komissarov et al |
| EPI_ISL_732998 | Russia/SVE-RII-MH223S/2020 | 2020-07-29 | Europe/Russia/Sverdlovsk/Yekaterinbur | UMMC-Health | WHO National Influenza Centre Russian Federation | Andrey Komissarov et al |
| EPI_ISL_733000 | Russia/SVE-RII-MH230S/2020 | 2020-08-08 | Europe/Russia/Sverdlovsk/Yekaterinbur | UMMC-Health | WHO National Influenza Centre Russian Federation | Andrey Komissarov et al |
| EPI_ISL_733001 | Russia/SVE-RII-MH238S/2020 | 2020-08-08 | Europe/Russia/Sverdlovsk/Yekaterinbur | UMMC-Health | WHO National Influenza Centre Russian Federation | Andrey Komissarov et al |
| EPI_ISL_733002 | Russia/SVE-RII-MH239S/2020 | 2020-08-08 | Europe/Russia/Sverdlovsk/Yekaterinbur | UMMC-Health | WHO National Influenza Centre Russian Federation | Andrey Komissarov et al |
| EPI_ISL_733003 | Russia/SVE-RII-MH245S/2020 | 2020-08-08 | Europe/Russia/Sverdlovsk/Yekaterinbur | UMMC-Health | WHO National Influenza Centre Russian Federation | Andrey Komissarov et al |
| EPI_ISL_733004 | Russia/SVE-RII-MH248S/2020 | 2020-08-10 | Europe/Russia/Sverdlovsk/Yekaterinbur | UMMC-Health | WHO National Influenza Centre Russian Federation | Andrey Komissarov et al |
| EPI_ISL_733005 | Russia/PRI-RII-MH745S/2020 | 2020-08-07 | Europe/Russia/Primorsky/Vladivostok | WHO National Influenza Centre Russian Federation | WHO National Influenza Centre Russian Federation | Andrey Komissarov et al |

| | | | | | | |
|---|---|---|---|---|---|---|
| EPI_ISL_733006 | Russia/PRI-RII-MH750S/2020 | 2020-08-08 | Europe/Russia/Primorsky/Vladivostok | WHO National Influenza Centre Russian Federation | WHO National Influenza Centre Russian Federation | Andrey Komissarov et al |
| EPI_ISL_733007 | Russia/PRI-RII-MH755S/2020 | 2020-08-09 | Europe/Russia/Primorsky/Vladivostok | WHO National Influenza Centre Russian Federation | WHO National Influenza Centre Russian Federation | Andrey Komissarov et al |
| EPI_ISL_733008 | Russia/PRI-RII-MH757S/2020 | 2020-08-10 | Europe/Russia/Primorsky/Vladivostok | WHO National Influenza Centre Russian Federation | WHO National Influenza Centre Russian Federation | Andrey Komissarov et al |
| EPI_ISL_733009 | Russia/PRI-RII-MH760S/2020 | 2020-08-10 | Europe/Russia/Primorsky/Vladivostok | WHO National Influenza Centre Russian Federation | WHO National Influenza Centre Russian Federation | Andrey Komissarov et al |
| EPI_ISL_733010 | Russia/PRI-RII-MH762S/2020 | 2020-08-11 | Europe/Russia/Primorsky/Vladivostok | WHO National Influenza Centre Russian Federation | WHO National Influenza Centre Russian Federation | Andrey Komissarov et al |
| EPI_ISL_733011 | Russia/PRI-RII-MH765S/2020 | 2020-08-11 | Europe/Russia/Primorsky/Vladivostok | WHO National Influenza Centre Russian Federation | WHO National Influenza Centre Russian Federation | Andrey Komissarov et al |
| EPI_ISL_733012 | Russia/PRI-RII-MH766S/2020 | 2020-08-11 | Europe/Russia/Primorsky/Vladivostok | WHO National Influenza Centre Russian Federation | WHO National Influenza Centre Russian Federation | Andrey Komissarov et al |
| EPI_ISL_733013 | Russia/PRI-RII-MH768S/2020 | 2020-08-11 | Europe/Russia/Primorsky/Vladivostok | WHO National Influenza Centre Russian Federation | WHO National Influenza Centre Russian Federation | Andrey Komissarov et al |
| EPI_ISL_733014 | Russia/PRI-RII-MH770S/2020 | 2020-08-11 | Europe/Russia/Primorsky/Vladivostok | WHO National Influenza Centre Russian Federation | WHO National Influenza Centre Russian Federation | Andrey Komissarov et al |
| EPI_ISL_733015 | Russia/PRI-RII-MH795S/2020 | 2020-07-26 | Europe/Russia/Primorsky/Ussuriysk | WHO National Influenza Centre Russian Federation | WHO National Influenza Centre Russian Federation | Andrey Komissarov et al |

| | | | | | | |
|---|---|---|---|---|---|---|
| EPI_ISL_733017 | Russia/PRI-RII-MH817S/2020 | 2020-08-07 | Europe/Russia/Primorsky/Ussuriysk | WHO National Influenza Centre Russian Federation | WHO National Influenza Centre Russian Federation | Andrey Komissarov et al |
| EPI_ISL_733018 | Russia/PRI-RII-MH829S/2020 | 2020-08-07 | Europe/Russia/Primorsky/Ussuriysk | WHO National Influenza Centre Russian Federation | WHO National Influenza Centre Russian Federation | Andrey Komissarov et al |
| EPI_ISL_733019 | Russia/PRI-RII-MH892S/2020 | 2020-08-02 | Europe/Russia/Primorsky/Vladivostok | WHO National Influenza Centre Russian Federation | WHO National Influenza Centre Russian Federation | Andrey Komissarov et al |
| EPI_ISL_733020 | Russia/PRI-RII-MH905S/2020 | 2020-08-12 | Europe/Russia/Primorsky/Vladivostok | WHO National Influenza Centre Russian Federation | WHO National Influenza Centre Russian Federation | Andrey Komissarov et al |
| EPI_ISL_733022 | Russia/PRI-RII-MH931S/2020 | 2020-08-12 | Europe/Russia/Primorsky/Vladivostok | WHO National Influenza Centre Russian Federation | WHO National Influenza Centre Russian Federation | Andrey Komissarov et al |
| EPI_ISL_733025 | Russia/SVE-RII-MH2834S/2020 | 2020-09-24 | Europe/Russia/Sverdlovsk/Revda | HELIX LLC | WHO National Influenza Centre Russian Federation | Andrey Komissarov et al |
| EPI_ISL_733028 | Russia/SVE-RII-MH2841S/2020 | 2020-09-26 | Europe/Russia/Sverdlovsk/Svobodny (Sv | HELIX LLC | WHO National Influenza Centre Russian Federation | Andrey Komissarov et al |
| EPI_ISL_733031 | Russia/SVE-RII-MH2844S/2020 | 2020-09-26 | Europe/Russia/Sverdlovsk/Svobodny (Sv | HELIX LLC | WHO National Influenza Centre Russian Federation | Andrey Komissarov et al |
| EPI_ISL_733032 | Russia/SVE-RII-MH2845S/2020 | 2020-09-24 | Europe/Russia/Sverdlovsk/Yekaterinbur | HELIX LLC | WHO National Influenza Centre Russian Federation | Andrey Komissarov et al |
| EPI_ISL_733034 | Russia/SVE-RII-MH2850S/2020 | 2020-09-24 | Europe/Russia/Sverdlovsk/Yekaterinbur | HELIX LLC | WHO National Influenza Centre Russian Federation | Andrey Komissarov et al |

| | | | | | | |
|---|---|---|---|---|---|---|
| EPI_ISL_733035 | Russia/SVE-RII-MH2851S/2020 | 2020-09-21 | Europe/Russia/Sverdlovsk/Yekaterinbur | HELIX LLC | WHO National Influenza Centre Russian Federation | Andrey Komissarov et al |
| EPI_ISL_733038 | Russia/TAM-RII-MH2856S/2020 | 2020-09-26 | Europe/Russia/Tambov Region/Morshan | HELIX LLC | WHO National Influenza Centre Russian Federation | Andrey Komissarov et al |
| EPI_ISL_733039 | Russia/TAM-RII-MH2857S/2020 | 2020-09-20 | Europe/Russia/Tambov Region/Tambov | HELIX LLC | WHO National Influenza Centre Russian Federation | Andrey Komissarov et al |
| EPI_ISL_733046 | Russia/MOW-RII-MH2874S/2020 | 2020-09-17 | Europe/Russia/Moscow Oblast/Moscow | HELIX LLC | WHO National Influenza Centre Russian Federation | Andrey Komissarov et al |
| EPI_ISL_733047 | Russia/MOW-RII-MH2876S/2020 | 2020-09-27 | Europe/Russia/Moscow Oblast/Moscow | HELIX LLC | WHO National Influenza Centre Russian Federation | Andrey Komissarov et al |
| EPI_ISL_733048 | Russia/MOW-RII-MH2878S/2020 | 2020-09-21 | Europe/Russia/Moscow Oblast/Moscow | HELIX LLC | WHO National Influenza Centre Russian Federation | Andrey Komissarov et al |
| EPI_ISL_733051 | Russia/MOW-RII-MH2881S/2020 | 2020-09-25 | Europe/Russia/Moscow Oblast/Moscow | HELIX LLC | WHO National Influenza Centre Russian Federation | Andrey Komissarov et al |
| EPI_ISL_733052 | Russia/MOW-RII-MH2882S/2020 | 2020-09-27 | Europe/Russia/Moscow Oblast/Moscow | HELIX LLC | WHO National Influenza Centre Russian Federation | Andrey Komissarov et al |
| EPI_ISL_733053 | Russia/MOW-RII-MH2884S/2020 | 2020-09-23 | Europe/Russia/Moscow Oblast/Moscow | HELIX LLC | WHO National Influenza Centre Russian Federation | Andrey Komissarov et al |
| EPI_ISL_733054 | Russia/MOW-RII-MH2885S/2020 | 2020-09-23 | Europe/Russia/Moscow Oblast/Moscow | HELIX LLC | WHO National Influenza Centre Russian Federation | Andrey Komissarov et al |

| | | | | | | |
|---|---|---|---|---|---|---|
| EPI_ISL_733056 | Russia/MOW-RII-MH2888S/2020 | 2020-09-25 | Europe/Russia/Moscow Oblast/Moscow | HELIX LLC | WHO National Influenza Centre Russian Federation | Andrey Komissarov et al |
| EPI_ISL_733057 | Russia/MOW-RII-MH2890S/2020 | 2020-09-22 | Europe/Russia/Moscow Oblast/Moscow | HELIX LLC | WHO National Influenza Centre Russian Federation | Andrey Komissarov et al |
| EPI_ISL_733060 | Russia/MOW-RII-MH2899S/2020 | 2020-09-25 | Europe/Russia/Moscow Oblast/Moscow | HELIX LLC | WHO National Influenza Centre Russian Federation | Andrey Komissarov et al |
| EPI_ISL_733068 | Russia/MOW-RII-MH2914S/2020 | 2020-09-28 | Europe/Russia/Moscow Oblast/Moscow | HELIX LLC | WHO National Influenza Centre Russian Federation | Andrey Komissarov et al |
| EPI_ISL_733074 | Russia/MOW-RII-MH2929S/2020 | 2020-09-27 | Europe/Russia/Moscow Oblast/Moscow | HELIX LLC | WHO National Influenza Centre Russian Federation | Andrey Komissarov et al |
| EPI_ISL_733077 | Crimea/RII-MH2936S/2020 | 2020-09-21 | Europe/Ukraine/Crimea/Simpheropol | HELIX LLC | WHO National Influenza Centre Russian Federation | Andrey Komissarov et al |
| EPI_ISL_733078 | Crimea/RII-MH2937S/2020 | 2020-09-21 | Europe/Ukraine/Crimea/Alushta | HELIX LLC | WHO National Influenza Centre Russian Federation | Andrey Komissarov et al |
| EPI_ISL_733081 | Russia/SVE-RII-MH2960S/2020 | 2020-09-30 | Europe/Russia/Sverdlovsk/Nizhny Tagil | HELIX LLC | WHO National Influenza Centre Russian Federation | Andrey Komissarov et al |
| EPI_ISL_733082 | Russia/SVE-RII-MH2962S/2020 | 2020-10-01 | Europe/Russia/Sverdlovsk/Yekaterinbur | HELIX LLC | WHO National Influenza Centre Russian Federation | Andrey Komissarov et al |
| EPI_ISL_733084 | Russia/SVE-RII-MH2964S/2020 | 2020-10-02 | Europe/Russia/Sverdlovsk/Yekaterinbur | HELIX LLC | WHO National Influenza Centre Russian Federation | Andrey Komissarov et al |

| | | | | | | |
|---|---|---|---|---|---|---|
| EPI_ISL_733085 | Russia/SVE-RII-MH2968S/2020 | 2020-10-03 | Europe/Russia/Sverdlovsk/Svobodny (Sv | HELIX LLC | WHO National Influenza Centre Russian Federation | Andrey Komissarov et al |
| EPI_ISL_733086 | Russia/SVE-RII-MH2973S/2020 | 2020-10-02 | Europe/Russia/Sverdlovsk/Kamensk-Ura | HELIX LLC | WHO National Influenza Centre Russian Federation | Andrey Komissarov et al |
| EPI_ISL_733087 | Russia/SVE-RII-MH2978S/2020 | 2020-10-03 | Europe/Russia/Sverdlovsk/Yekaterinbur | HELIX LLC | WHO National Influenza Centre Russian Federation | Andrey Komissarov et al |
| EPI_ISL_733088 | Russia/SVE-RII-MH3109S/2020 | 2020-10-03 | Europe/Russia/Sverdlovsk/Yekaterinbur | HELIX LLC | WHO National Influenza Centre Russian Federation | Andrey Komissarov et al |
| EPI_ISL_733089 | Russia/SVE-RII-MH3110S/2020 | 2020-10-03 | Europe/Russia/Sverdlovsk/Serov | HELIX LLC | WHO National Influenza Centre Russian Federation | Andrey Komissarov et al |
| EPI_ISL_733090 | Russia/SVE-RII-MH3111S/2020 | 2020-10-03 | Europe/Russia/Sverdlovsk/Yekaterinbur | HELIX LLC | WHO National Influenza Centre Russian Federation | Andrey Komissarov et al |
| EPI_ISL_733092 | Russia/TYU-RII-MH3116S/2020 | 2020-10-03 | Europe/Russia/Tyumen Region/Tyumen | HELIX LLC | WHO National Influenza Centre Russian Federation | Andrey Komissarov et al |
| EPI_ISL_733093 | Russia/LEN-RII-MH3118S/2020 | 2020-09-30 | Europe/Russia/Leningrad Oblast/Tosno | HELIX LLC | WHO National Influenza Centre Russian Federation | Andrey Komissarov et al |
| EPI_ISL_733094 | Russia/LEN-RII-MH3119S/2020 | 2020-10-01 | Europe/Russia/Leningrad Oblast/Gatchir | HELIX LLC | WHO National Influenza Centre Russian Federation | Andrey Komissarov et al |
| EPI_ISL_733095 | Russia/LEN-RII-MH3120S/2020 | 2020-09-30 | Europe/Russia/Leningrad Oblast/Kirovsk | HELIX LLC | WHO National Influenza Centre Russian Federation | Andrey Komissarov et al |

| | | | | | | |
|---|---|---|---|---|---|---|
| EPI_ISL_733096 | Russia/LEN-RII-MH3121S/2020 | 2020-10-01 | Europe/Russia/Leningrad Oblast/Vsevol | HELIX LLC | WHO National Influenza Centre Russian Federation | Andrey Komissarov et al |
| EPI_ISL_733097 | Russia/LEN-RII-MH3122S/2020 | 2020-09-29 | Europe/Russia/Leningrad Oblast/Gatchi | HELIX LLC | WHO National Influenza Centre Russian Federation | Andrey Komissarov et al |
| EPI_ISL_733098 | Russia/KDA-RII-MH3351S/2020 | 2020-10-07 | Europe/Russia/Krasnodar/Anapa | HELIX LLC | WHO National Influenza Centre Russian Federation | Andrey Komissarov et al |
| EPI_ISL_733100 | Russia/MOW-RII-MH3353S/2020 | 2020-10-07 | Europe/Russia/Moscow Oblast/Moscow | HELIX LLC | WHO National Influenza Centre Russian Federation | Andrey Komissarov et al |
| EPI_ISL_733101 | Russia/KDA-RII-MH3354S/2020 | 2020-10-07 | Europe/Russia/Krasnodar/Anapa | HELIX LLC | WHO National Influenza Centre Russian Federation | Andrey Komissarov et al |
| EPI_ISL_733102 | Russia/SE-RII-MH3355S/2020 | 2020-10-07 | Europe/Russia/Republic of North Ossetia | HELIX LLC | WHO National Influenza Centre Russian Federation | Andrey Komissarov et al |
| EPI_ISL_733103 | Russia/SE-RII-MH3356S/2020 | 2020-10-07 | Europe/Russia/Republic of North Ossetia | HELIX LLC | WHO National Influenza Centre Russian Federation | Andrey Komissarov et al |
| EPI_ISL_733104 | Russia/KLU-RII-MH3358S/2020 | 2020-10-06 | Europe/Russia/Kaluga/Tovarkovo | HELIX LLC | WHO National Influenza Centre Russian Federation | Andrey Komissarov et al |
| EPI_ISL_733106 | Russia/RYA-RII-MH3361S/2020 | 2020-10-07 | Europe/Russia/Ryazan/Ryazan | HELIX LLC | WHO National Influenza Centre Russian Federation | Andrey Komissarov et al |
| EPI_ISL_733107 | Russia/STA-RII-MH3363S/2020 | 2020-10-07 | Europe/Russia/Stavropol/Yessentuki | HELIX LLC | WHO National Influenza Centre Russian Federation | Andrey Komissarov et al |

| | | | | | | |
|---|---|---|---|---|---|---|
| EPI_ISL_733109 | Russia/IVA-RII-MH3366S/2020 | 2020-10-07 | Europe/Russia/Ivanovo/Ivanovo | HELIX LLC | WHO National Influenza Centre Russian Federation | Andrey Komissarov et al |
| EPI_ISL_733110 | Russia/ROS-RII-MH3369S/2020 | 2020-10-06 | Europe/Russia/Rostov/Kamensk-Shakhti | HELIX LLC | WHO National Influenza Centre Russian Federation | Andrey Komissarov et al |
| EPI_ISL_733112 | Russia/TUL-RII-MH3371S/2020 | 2020-10-07 | Europe/Russia/Tula Region/Novomoskov | HELIX LLC | WHO National Influenza Centre Russian Federation | Andrey Komissarov et al |
| EPI_ISL_733113 | Russia/TUL-RII-MH3372S/2020 | 2020-10-07 | Europe/Russia/Tula Region/Novomoskov | HELIX LLC | WHO National Influenza Centre Russian Federation | Andrey Komissarov et al |
| EPI_ISL_733114 | Russia/STA-RII-MH3373S/2020 | 2020-10-07 | Europe/Russia/Stavropol/Yessentuki | HELIX LLC | WHO National Influenza Centre Russian Federation | Andrey Komissarov et al |
| EPI_ISL_733117 | Russia/ROS-RII-MH3377S/2020 | 2020-10-07 | Europe/Russia/Rostov/Zernograd | HELIX LLC | WHO National Influenza Centre Russian Federation | Andrey Komissarov et al |
| EPI_ISL_733118 | Russia/STA-RII-MH3378S/2020 | 2020-10-07 | Europe/Russia/Stavropol/Stavropol | HELIX LLC | WHO National Influenza Centre Russian Federation | Andrey Komissarov et al |
| EPI_ISL_733120 | Russia/KC-RII-MH3381S/2020 | 2020-10-06 | Europe/Russia/Karachay-Cherkess/Zelen | HELIX LLC | WHO National Influenza Centre Russian Federation | Andrey Komissarov et al |
| EPI_ISL_733121 | Russia/YAR-RII-MH3382S/2020 | 2020-10-07 | Europe/Russia/Yaroslavl/Yaroslavl | HELIX LLC | WHO National Influenza Centre Russian Federation | Andrey Komissarov et al |
| EPI_ISL_733122 | Russia/TAM-RII-MH3383S/2020 | 2020-10-07 | Europe/Russia/Tambov Region/Tambov | HELIX LLC | WHO National Influenza Centre Russian Federation | Andrey Komissarov et al |

| | | | | | | |
|---|---|---|---|---|---|---|
| EPI_ISL_733123 | Russia/VGG-RII-MH3388S/2020 | 2020-10-06 | Europe/Russia/Volgograd Region/Volgog | HELIX LLC | WHO National Influenza Centre Russian Federation | Andrey Komissarov et al |
| EPI_ISL_733124 | Russia/KDA-RII-MH3389S/2020 | 2020-10-07 | Europe/Russia/Krasnodar/Anapa | HELIX LLC | WHO National Influenza Centre Russian Federation | Andrey Komissarov et al |
| EPI_ISL_733125 | Russia/KO-RII-MH3391S/2020 | 2020-10-07 | Europe/Russia/Komi/Ukhta | HELIX LLC | WHO National Influenza Centre Russian Federation | Andrey Komissarov et al |
| EPI_ISL_733126 | Russia/MOS-RII-MH3393S/2020 | 2020-10-07 | Europe/Russia/Moscow Oblast/Sergiyev | HELIX LLC | WHO National Influenza Centre Russian Federation | Andrey Komissarov et al |
| EPI_ISL_733127 | Russia/MOW-RII-MH3394S/2020 | 2020-10-07 | Europe/Russia/Moscow Oblast/Moscow | HELIX LLC | WHO National Influenza Centre Russian Federation | Andrey Komissarov et al |
| EPI_ISL_733128 | Russia/MOW-RII-MH3395S/2020 | 2020-10-07 | Europe/Russia/Moscow Oblast/Moscow | HELIX LLC | WHO National Influenza Centre Russian Federation | Andrey Komissarov et al |
| EPI_ISL_733129 | Russia/MOS-RII-MH3397S/2020 | 2020-10-07 | Europe/Russia/Moscow Oblast/Kalininet | HELIX LLC | WHO National Influenza Centre Russian Federation | Andrey Komissarov et al |
| EPI_ISL_733130 | Russia/MOW-RII-MH3398S/2020 | 2020-10-07 | Europe/Russia/Moscow Oblast/Moscow | HELIX LLC | WHO National Influenza Centre Russian Federation | Andrey Komissarov et al |
| EPI_ISL_733131 | Russia/MOW-RII-MH3400S/2020 | 2020-10-07 | Europe/Russia/Moscow Oblast/Moscow | HELIX LLC | WHO National Influenza Centre Russian Federation | Andrey Komissarov et al |
| EPI_ISL_733132 | Russia/MOW-RII-MH3401S/2020 | 2020-10-07 | Europe/Russia/Moscow Oblast/Moscow | HELIX LLC | WHO National Influenza Centre Russian Federation | Andrey Komissarov et al |

| | | | | | | |
|---|---|---|---|---|---|---|
| EPI_ISL_733133 | Russia/TUL-RII-MH3405S/2020 | 2020-10-07 | Europe/Russia/Tula Region/Novomoskov | HELIX LLC | WHO National Influenza Centre Russian Federation | Andrey Komissarov et al |
| EPI_ISL_733134 | Russia/MOS-RII-MH3406S/2020 | 2020-10-07 | Europe/Russia/Moscow Oblast/Naro-For | HELIX LLC | WHO National Influenza Centre Russian Federation | Andrey Komissarov et al |
| EPI_ISL_733135 | Russia/STA-RII-MH3407S/2020 | 2020-10-07 | Europe/Russia/Stavropol/Stavropol | HELIX LLC | WHO National Influenza Centre Russian Federation | Andrey Komissarov et al |
| EPI_ISL_733136 | Russia/BRY-RII-MH3408S/2020 | 2020-10-07 | Europe/Russia/Bryansk Oblast/Bryansk | HELIX LLC | WHO National Influenza Centre Russian Federation | Andrey Komissarov et al |
| EPI_ISL_733137 | Russia/KO-RII-MH3409S/2020 | 2020-10-06 | Europe/Russia/Komi/Ukhta | HELIX LLC | WHO National Influenza Centre Russian Federation | Andrey Komissarov et al |
| EPI_ISL_733138 | Russia/KHM-RII-MH3413S/2020 | 2020-10-06 | Europe/Russia/Khanty-Mansi/Surgut | HELIX LLC | WHO National Influenza Centre Russian Federation | Andrey Komissarov et al |
| EPI_ISL_733139 | Russia/VOR-RII-MH3414S/2020 | 2020-10-07 | Europe/Russia/Voronezh/Voronezh | HELIX LLC | WHO National Influenza Centre Russian Federation | Andrey Komissarov et al |
| EPI_ISL_733140 | Russia/KHM-RII-MH3415S/2020 | 2020-10-06 | Europe/Russia/Khanty-Mansi/Surgut | HELIX LLC | WHO National Influenza Centre Russian Federation | Andrey Komissarov et al |
| EPI_ISL_733141 | Russia/VOR-RII-MH3416S/2020 | 2020-10-07 | Europe/Russia/Voronezh/Voronezh | HELIX LLC | WHO National Influenza Centre Russian Federation | Andrey Komissarov et al |
| EPI_ISL_733142 | Russia/KLU-RII-MH3421S/2020 | 2020-10-06 | Europe/Russia/Kaluga/Tovarkovo | HELIX LLC | WHO National Influenza Centre Russian Federation | Andrey Komissarov et al |

| | | | | | | |
|---|---|---|---|---|---|---|
| EPI_ISL_733143 | Russia/VLA-RII-MH3422S/2020 | 2020-10-07 | Europe/Russia/Vladimir Region/Vladimi | HELIX LLC | WHO National Influenza Centre Russian Federation | Andrey Komissarov et al |
| EPI_ISL_733144 | Russia/KLU-RII-MH3423S/2020 | 2020-10-06 | Europe/Russia/Kaluga/Tovarkovo | HELIX LLC | WHO National Influenza Centre Russian Federation | Andrey Komissarov et al |
| EPI_ISL_733146 | Russia/ROS-RII-MH3425S/2020 | 2020-10-06 | Europe/Russia/Rostov/Kamensk-Shakhti | HELIX LLC | WHO National Influenza Centre Russian Federation | Andrey Komissarov et al |
| EPI_ISL_733147 | Russia/ORE-RII-MH3429S/2020 | 2020-10-07 | Europe/Russia/Orenburg/Orenburg | HELIX LLC | WHO National Influenza Centre Russian Federation | Andrey Komissarov et al |
| EPI_ISL_733149 | Russia/MOW-RII-MH3431S/2020 | 2020-10-07 | Europe/Russia/Moscow Oblast/Moscow | HELIX LLC | WHO National Influenza Centre Russian Federation | Andrey Komissarov et al |
| EPI_ISL_733150 | Russia/KO-RII-MH3432S/2020 | 2020-10-06 | Europe/Russia/Komi/Sosnogorsk | HELIX LLC | WHO National Influenza Centre Russian Federation | Andrey Komissarov et al |
| EPI_ISL_733151 | Crimea/RII-MH3437S/2020 | 2020-10-07 | Europe/Ukraine/Crimea/Simpheropol | HELIX LLC | WHO National Influenza Centre Russian Federation | Andrey Komissarov et al |
| EPI_ISL_733152 | Russia/TUL-RII-MH3439S/2020 | 2020-10-07 | Europe/Russia/Tula Region/Novomoskov | HELIX LLC | WHO National Influenza Centre Russian Federation | Andrey Komissarov et al |
| EPI_ISL_733153 | Russia/ORE-RII-MH3441S/2020 | 2020-10-07 | Europe/Russia/Orenburg/Orenburg | HELIX LLC | WHO National Influenza Centre Russian Federation | Andrey Komissarov et al |
| EPI_ISL_733154 | Russia/MO-RII-MH3571S/2020 | 2020-10-13 | Europe/Russia/Republic of Mordovia/Sa | WHO National Influenza Centre Russian Federation | WHO National Influenza Centre Russian Federation | Andrey Komissarov et al |

| | | | | | | |
|---|---|---|---|---|---|---|
| EPI_ISL_733155 | Russia/MO-RII-MH3582S/2020 | 2020-10-13 | Europe/Russia/Republic of Mordovia/Sa | WHO National Influenza Centre Russian Federation | WHO National Influenza Centre Russian Federation | Andrey Komissarov et al |
| EPI_ISL_733156 | Russia/MOW-RII-MH4070S/2020 | 2020-06-09 | Europe/Russia/Moscow Oblast/Moscow | Pathogenic Microorganisms Variability Laboratory | WHO National Influenza Centre Russian Federation | Andrey Komissarov et al |
| EPI_ISL_733158 | Russia/MOW-RII-MH4074S/2020 | 2020-06-06 | Europe/Russia/Moscow Oblast/Moscow | Pathogenic Microorganisms Variability Laboratory | WHO National Influenza Centre Russian Federation | Andrey Komissarov et al |
| EPI_ISL_733160 | Russia/MOW-RII-MH4077S/2020 | 2020-06-18 | Europe/Russia/Moscow Oblast/Moscow | Pathogenic Microorganisms Variability Laboratory | WHO National Influenza Centre Russian Federation | Andrey Komissarov et al |
| EPI_ISL_733162 | Russia/MOW-RII-MH4085S/2020 | 2020-06-01 | Europe/Russia/Moscow Oblast/Moscow | Pathogenic Microorganisms Variability Laboratory | WHO National Influenza Centre Russian Federation | Andrey Komissarov et al |
| EPI_ISL_733163 | Russia/MOW-RII-MH4105S/2020 | 2020-05-12 | Europe/Russia/Moscow Oblast/Moscow | Pathogenic Microorganisms Variability Laboratory | WHO National Influenza Centre Russian Federation | Andrey Komissarov et al |
| EPI_ISL_733164 | Russia/AST-RII-MH4118S/2020 | 2020-08-04 | Europe/Russia/Astrakhan Region/Astrak | Pathogenic Microorganisms Variability Laboratory | WHO National Influenza Centre Russian Federation | Andrey Komissarov et al |
| EPI_ISL_733165 | Russia/AST-RII-MH4119S/2020 | 2020-08-05 | Europe/Russia/Astrakhan Region/Astrak | Pathogenic Microorganisms Variability Laboratory | WHO National Influenza Centre Russian Federation | Andrey Komissarov et al |
| EPI_ISL_733166 | Russia/AST-RII-MH4121S/2020 | 2020-08-05 | Europe/Russia/Astrakhan Region/Astrak | Pathogenic Microorganisms Variability Laboratory | WHO National Influenza Centre Russian Federation | Andrey Komissarov et al |
| EPI_ISL_733167 | Russia/AST-RII-MH4130S/2020 | 2020-08-04 | Europe/Russia/Astrakhan Region/Astrak | Pathogenic Microorganisms Variability Laboratory | WHO National Influenza Centre Russian Federation | Andrey Komissarov et al |

| | | | | | | |
|---|---|---|---|---|---|---|
| EPI_ISL_733168 | Russia/AST-RII-MH4131S/2020 | 2020-08-04 | Europe/Russia/Astrakhan Region/Astrak | Pathogenic Microorganisms Variability Laboratory | WHO National Influenza Centre Russian Federation | Andrey Komissarov et al |
| EPI_ISL_733169 | Russia/AST-RII-MH4135S/2020 | 2020-08-04 | Europe/Russia/Astrakhan Region/Astrak | Pathogenic Microorganisms Variability Laboratory | WHO National Influenza Centre Russian Federation | Andrey Komissarov et al |
| EPI_ISL_733174 | Russia/AST-RII-MH4150S/2020 | 2020-08-13 | Europe/Russia/Astrakhan Region/Astrak | Pathogenic Microorganisms Variability Laboratory | WHO National Influenza Centre Russian Federation | Andrey Komissarov et al |
| EPI_ISL_733176 | Russia/AST-RII-MH4152S/2020 | 2020-08-11 | Europe/Russia/Astrakhan Region/Astrak | Pathogenic Microorganisms Variability Laboratory | WHO National Influenza Centre Russian Federation | Andrey Komissarov et al |
| EPI_ISL_733177 | Russia/AST-RII-MH4153S/2020 | 2020-08-18 | Europe/Russia/Astrakhan Region/Astrak | Pathogenic Microorganisms Variability Laboratory | WHO National Influenza Centre Russian Federation | Andrey Komissarov et al |
| EPI_ISL_733178 | Russia/AST-RII-MH4155S/2020 | 2020-08-18 | Europe/Russia/Astrakhan Region/Astrak | Pathogenic Microorganisms Variability Laboratory | WHO National Influenza Centre Russian Federation | Andrey Komissarov et al |
| EPI_ISL_733179 | Russia/AST-RII-MH4156S/2020 | 2020-08-18 | Europe/Russia/Astrakhan Region/Astrak | Pathogenic Microorganisms Variability Laboratory | WHO National Influenza Centre Russian Federation | Andrey Komissarov et al |
| EPI_ISL_733180 | Russia/AST-RII-MH4157S/2020 | 2020-08-18 | Europe/Russia/Astrakhan Region/Astrak | Pathogenic Microorganisms Variability Laboratory | WHO National Influenza Centre Russian Federation | Andrey Komissarov et al |
| EPI_ISL_733181 | Russia/AST-RII-MH4159S/2020 | 2020-08-05 | Europe/Russia/Astrakhan Region/Astrak | Pathogenic Microorganisms Variability Laboratory | WHO National Influenza Centre Russian Federation | Andrey Komissarov et al |
| EPI_ISL_733182 | Russia/AST-RII-MH4160S/2020 | 2020-08-05 | Europe/Russia/Astrakhan Region/Astrak | Pathogenic Microorganisms Variability Laboratory | WHO National Influenza Centre Russian Federation | Andrey Komissarov et al |

| | | | | | | |
|---|---|---|---|---|---|---|
| EPI_ISL_733183 | Russia/ORE-RII-MH4312S/2020 | 2020-09-08 | Europe/Russia/Orenburg/Orenburg | WHO National Influenza Centre Russian Federation | WHO National Influenza Centre Russian Federation | Andrey Komissarov et al |
| EPI_ISL_733184 | Russia/ORE-RII-MH4423S/2020 | 2020-10-29 | Europe/Russia/Orenburg/Orenburg | WHO National Influenza Centre Russian Federation | WHO National Influenza Centre Russian Federation | Andrey Komissarov et al |
| EPI_ISL_733188 | Russia/AST-RII-MH4439S/2020 | 2020-08-15 | Europe/Russia/Astrakhan Region/Astrak | Pathogenic Microorganisms Variability Laboratory | WHO National Influenza Centre Russian Federation | Andrey Komissarov et al |
| EPI_ISL_733189 | Russia/AST-RII-MH4442S/2020 | 2020-08-11 | Europe/Russia/Astrakhan Region/Astrak | Pathogenic Microorganisms Variability Laboratory | WHO National Influenza Centre Russian Federation | Andrey Komissarov et al |
| EPI_ISL_733190 | Russia/AST-RII-MH4444S/2020 | 2020-10-01 | Europe/Russia/Astrakhan Region/Astrak | Pathogenic Microorganisms Variability Laboratory | WHO National Influenza Centre Russian Federation | Andrey Komissarov et al |
| EPI_ISL_733191 | Russia/AST-RII-MH4447S/2020 | 2020-09-09 | Europe/Russia/Astrakhan Region/Astrak | Pathogenic Microorganisms Variability Laboratory | WHO National Influenza Centre Russian Federation | Andrey Komissarov et al |
| EPI_ISL_733193 | Russia/AST-RII-MH4450S/2020 | 2020-09-04 | Europe/Russia/Astrakhan Region/Astrak | Pathogenic Microorganisms Variability Laboratory | WHO National Influenza Centre Russian Federation | Andrey Komissarov et al |
| EPI_ISL_733194 | Russia/AST-RII-MH4453S/2020 | 2020-09-14 | Europe/Russia/Astrakhan Region/Astrak | Pathogenic Microorganisms Variability Laboratory | WHO National Influenza Centre Russian Federation | Andrey Komissarov et al |
| EPI_ISL_733195 | Russia/AST-RII-MH4454S/2020 | 2020-08-15 | Europe/Russia/Astrakhan Region/Astrak | Pathogenic Microorganisms Variability Laboratory | WHO National Influenza Centre Russian Federation | Andrey Komissarov et al |
| EPI_ISL_733196 | Russia/AST-RII-MH4455S/2020 | 2020-08-11 | Europe/Russia/Astrakhan Region/Astrak | Pathogenic Microorganisms Variability Laboratory | WHO National Influenza Centre Russian Federation | Andrey Komissarov et al |

| | | | | | | |
|---|---|---|---|---|---|---|
| EPI_ISL_733197 | Russia/AST-RII-MH4456S/2020 | 2020-08-15 | Europe/Russia/Astrakhan Region/Astrak | Pathogenic Microorganisms Variability Laboratory | WHO National Influenza Centre Russian Federation | Andrey Komissarov et al |
| EPI_ISL_733198 | Russia/AST-RII-MH4457S/2020 | 2020-08-02 | Europe/Russia/Astrakhan Region/Astrak | Pathogenic Microorganisms Variability Laboratory | WHO National Influenza Centre Russian Federation | Andrey Komissarov et al |
| EPI_ISL_733200 | Russia/AST-RII-MH4459S/2020 | 2020-08-15 | Europe/Russia/Astrakhan Region/Astrak | Pathogenic Microorganisms Variability Laboratory | WHO National Influenza Centre Russian Federation | Andrey Komissarov et al |
| EPI_ISL_733201 | Russia/AST-RII-MH4460S/2020 | 2020-08-15 | Europe/Russia/Astrakhan Region/Astrak | Pathogenic Microorganisms Variability Laboratory | WHO National Influenza Centre Russian Federation | Andrey Komissarov et al |
| EPI_ISL_733202 | Russia/AST-RII-MH4462S/2020 | 2020-08-13 | Europe/Russia/Astrakhan Region/Astrak | Pathogenic Microorganisms Variability Laboratory | WHO National Influenza Centre Russian Federation | Andrey Komissarov et al |
| EPI_ISL_733204 | Russia/AST-RII-MH4466S/2020 | 2020-09-10 | Europe/Russia/Astrakhan Region/Astrak | Pathogenic Microorganisms Variability Laboratory | WHO National Influenza Centre Russian Federation | Andrey Komissarov et al |
| EPI_ISL_733205 | Russia/AST-RII-MH4468S/2020 | 2020-09-23 | Europe/Russia/Astrakhan Region/Astrak | Pathogenic Microorganisms Variability Laboratory | WHO National Influenza Centre Russian Federation | Andrey Komissarov et al |
| EPI_ISL_733207 | Russia/AST-RII-MH4474S/2020 | 2020-09-23 | Europe/Russia/Astrakhan Region/Astrak | Pathogenic Microorganisms Variability Laboratory | WHO National Influenza Centre Russian Federation | Andrey Komissarov et al |
| EPI_ISL_733210 | Russia/AST-RII-MH4490S/2020 | 2020-08-06 | Europe/Russia/Astrakhan Region/Astrak | Pathogenic Microorganisms Variability Laboratory | WHO National Influenza Centre Russian Federation | Andrey Komissarov et al |
| EPI_ISL_733211 | Russia/AST-RII-MH4494S/2020 | 2020-08-06 | Europe/Russia/Astrakhan Region/Astrak | Pathogenic Microorganisms Variability Laboratory | WHO National Influenza Centre Russian Federation | Andrey Komissarov et al |

| | | | | | | |
|---|---|---|---|---|---|---|
| EPI_ISL_733215 | Russia/AST-RII-MH4571S/2020 | 2020-08-12 | Europe/Russia/Astrakhan Region/Astrak | Pathogenic Microorganisms Variability Laboratory | WHO National Influenza Centre Russian Federation | Andrey Komissarov et al |
| EPI_ISL_733216 | Russia/AST-RII-MH4585S/2020 | 2020-09-21 | Europe/Russia/Astrakhan Region/Voloda | Pathogenic Microorganisms Variability Laboratory | WHO National Influenza Centre Russian Federation | Andrey Komissarov et al |
| EPI_ISL_733218 | Russia/AST-RII-MH4592S/2020 | 2020-09-21 | Europe/Russia/Astrakhan Region/Voloda | Pathogenic Microorganisms Variability Laboratory | WHO National Influenza Centre Russian Federation | Andrey Komissarov et al |
| EPI_ISL_733222 | Russia/AST-RII-MH4609S/2020 | 2020-08-25 | Europe/Russia/Astrakhan Region/Astrak | Pathogenic Microorganisms Variability Laboratory | WHO National Influenza Centre Russian Federation | Andrey Komissarov et al |
| EPI_ISL_733224 | Russia/SVE-RII-MH4660S/2020 | 2020-08-25 | Europe/Russia/Sverdlovsk/Yekaterinbur | UMMC-Health | WHO National Influenza Centre Russian Federation | Andrey Komissarov et al |
| EPI_ISL_733225 | Russia/SVE-RII-MH4662S/2020 | 2020-08-29 | Europe/Russia/Sverdlovsk/Yekaterinbur | UMMC-Health | WHO National Influenza Centre Russian Federation | Andrey Komissarov et al |
| EPI_ISL_733226 | Russia/SVE-RII-MH4663S/2020 | 2020-08-29 | Europe/Russia/Sverdlovsk/Yekaterinbur | UMMC-Health | WHO National Influenza Centre Russian Federation | Andrey Komissarov et al |
| EPI_ISL_733227 | Russia/SVE-RII-MH4666S/2020 | 2020-08-31 | Europe/Russia/Sverdlovsk/Yekaterinbur | UMMC-Health | WHO National Influenza Centre Russian Federation | Andrey Komissarov et al |
| EPI_ISL_733229 | Russia/SVE-RII-MH4669S/2020 | 2020-09-08 | Europe/Russia/Sverdlovsk/Yekaterinbur | UMMC-Health | WHO National Influenza Centre Russian Federation | Andrey Komissarov et al |
| EPI_ISL_733231 | Russia/SVE-RII-MH4673S/2020 | 2020-09-18 | Europe/Russia/Sverdlovsk/Yekaterinbur | UMMC-Health | WHO National Influenza Centre Russian Federation | Andrey Komissarov et al |

| | | | | | | |
|---|---|---|---|---|---|---|
| EPI_ISL_733233 | Russia/SVE-RII-MH4681S/2020 | 2020-09-25 | Europe/Russia/Sverdlovsk/Yekaterinbur | UMMC-Health | WHO National Influenza Centre Russian Federation | Andrey Komissarov et al |
| EPI_ISL_733235 | Russia/SVE-RII-MH4684S/2020 | 2020-09-26 | Europe/Russia/Sverdlovsk/Yekaterinbur | UMMC-Health | WHO National Influenza Centre Russian Federation | Andrey Komissarov et al |
| EPI_ISL_733236 | Russia/SVE-RII-MH4685S/2020 | 2020-09-30 | Europe/Russia/Sverdlovsk/Yekaterinbur | UMMC-Health | WHO National Influenza Centre Russian Federation | Andrey Komissarov et al |
| EPI_ISL_733239 | Russia/SVE-RII-MH4694S/2020 | 2020-10-22 | Europe/Russia/Sverdlovsk/Yekaterinbur | UMMC-Health | WHO National Influenza Centre Russian Federation | Andrey Komissarov et al |
| EPI_ISL_733240 | Russia/SVE-RII-MH4700S/2020 | 2020-10-19 | Europe/Russia/Sverdlovsk/Yekaterinbur | UMMC-Health | WHO National Influenza Centre Russian Federation | Andrey Komissarov et al |
| EPI_ISL_733241 | Russia/SAR-RII-MH4702S/2020 | 2020-11-11 | Europe/Russia/Saratov/Balakovo | WHO National Influenza Centre Russian Federation | WHO National Influenza Centre Russian Federation | Andrey Komissarov et al |
| EPI_ISL_733243 | Russia/KDA-RII-MH4710S/2020 | 2020-10-05 | Europe/Russia/Krasnodar/Krasnodar | WHO National Influenza Centre Russian Federation | WHO National Influenza Centre Russian Federation | Andrey Komissarov et al |
| EPI_ISL_733244 | Russia/KDA-RII-MH4712S/2020 | 2020-10-05 | Europe/Russia/Krasnodar/Krasnodar | WHO National Influenza Centre Russian Federation | WHO National Influenza Centre Russian Federation | Andrey Komissarov et al |
| EPI_ISL_733245 | Russia/KDA-RII-MH4714S/2020 | 2020-10-06 | Europe/Russia/Krasnodar/Krasnodar | WHO National Influenza Centre Russian Federation | WHO National Influenza Centre Russian Federation | Andrey Komissarov et al |
| EPI_ISL_733246 | Russia/KDA-RII-MH4721S/2020 | 2020-10-05 | Europe/Russia/Krasnodar/Krasnodar | WHO National Influenza Centre Russian Federation | WHO National Influenza Centre Russian Federation | Andrey Komissarov et al |

| | | | | | | |
|---|---|---|---|---|---|---|
| EPI_ISL_733247 | Russia/KDA-RII-MH4727S/2020 | 2020-10-11 | Europe/Russia/Krasnodar/Krasnodar | WHO National Influenza Centre Russian Federation | WHO National Influenza Centre Russian Federation | Andrey Komissarov et al |
| EPI_ISL_733248 | Russia/KDA-RII-MH4730S/2020 | 2020-10-14 | Europe/Russia/Krasnodar/Krasnodar | WHO National Influenza Centre Russian Federation | WHO National Influenza Centre Russian Federation | Andrey Komissarov et al |
| EPI_ISL_733249 | Russia/KDA-RII-MH4731S/2020 | 2020-10-14 | Europe/Russia/Krasnodar/Krasnodar | WHO National Influenza Centre Russian Federation | WHO National Influenza Centre Russian Federation | Andrey Komissarov et al |
| EPI_ISL_733250 | Russia/KDA-RII-MH4734S/2020 | 2020-10-15 | Europe/Russia/Krasnodar/Krasnodar | WHO National Influenza Centre Russian Federation | WHO National Influenza Centre Russian Federation | Andrey Komissarov et al |
| EPI_ISL_733251 | Russia/KDA-RII-MH4736S/2020 | 2020-10-16 | Europe/Russia/Krasnodar/Krasnodar | WHO National Influenza Centre Russian Federation | WHO National Influenza Centre Russian Federation | Andrey Komissarov et al |
| EPI_ISL_733252 | Russia/KDA-RII-MH4740S/2020 | 2020-10-17 | Europe/Russia/Krasnodar/Krasnodar | WHO National Influenza Centre Russian Federation | WHO National Influenza Centre Russian Federation | Andrey Komissarov et al |
| EPI_ISL_733254 | Russia/KDA-RII-MH4744S/2020 | 2020-10-20 | Europe/Russia/Krasnodar/Krasnodar | WHO National Influenza Centre Russian Federation | WHO National Influenza Centre Russian Federation | Andrey Komissarov et al |
| EPI_ISL_733256 | Russia/KDA-RII-MH4747S/2020 | 2020-10-21 | Europe/Russia/Krasnodar/Krasnodar | WHO National Influenza Centre Russian Federation | WHO National Influenza Centre Russian Federation | Andrey Komissarov et al |
| EPI_ISL_733260 | Russia/KDA-RII-MH4785S/2020 | 2020-11-06 | Europe/Russia/Krasnodar/Krasnodar | WHO National Influenza Centre Russian Federation | WHO National Influenza Centre Russian Federation | Andrey Komissarov et al |
| EPI_ISL_733261 | Russia/KDA-RII-MH4793S/2020 | 2020-09-26 | Europe/Russia/Krasnodar/Krasnodar | WHO National Influenza Centre Russian Federation | WHO National Influenza Centre Russian Federation | Andrey Komissarov et al |

| | | | | | | |
|---|---|---|---|---|---|---|
| EPI_ISL_733262 | Russia/KDA-RII-MH4797S/2020 | 2020-09-28 | Europe/Russia/Krasnodar/Krasnodar | WHO National Influenza Centre Russian Federation | WHO National Influenza Centre Russian Federation | Andrey Komissarov et al |
| EPI_ISL_733263 | Russia/KDA-RII-MH4806S/2020 | 2020-09-30 | Europe/Russia/Krasnodar/Krasnodar | WHO National Influenza Centre Russian Federation | WHO National Influenza Centre Russian Federation | Andrey Komissarov et al |
| EPI_ISL_733264 | Russia/KDA-RII-MH4815S/2020 | 2020-10-02 | Europe/Russia/Krasnodar/Krasnodar | WHO National Influenza Centre Russian Federation | WHO National Influenza Centre Russian Federation | Andrey Komissarov et al |
| EPI_ISL_733265 | Russia/KDA-RII-MH4816S/2020 | 2020-10-02 | Europe/Russia/Krasnodar/Krasnodar | WHO National Influenza Centre Russian Federation | WHO National Influenza Centre Russian Federation | Andrey Komissarov et al |
| EPI_ISL_733267 | Russia/KDA-RII-MH4821S/2020 | 2020-10-03 | Europe/Russia/Krasnodar/Krasnodar | WHO National Influenza Centre Russian Federation | WHO National Influenza Centre Russian Federation | Andrey Komissarov et al |
| EPI_ISL_733271 | Russia/KDA-RII-MH4845S/2020 | 2020-10-12 | Europe/Russia/Krasnodar/Krasnodar | WHO National Influenza Centre Russian Federation | WHO National Influenza Centre Russian Federation | Andrey Komissarov et al |
| EPI_ISL_733272 | Russia/KDA-RII-MH4846S/2020 | 2020-10-13 | Europe/Russia/Krasnodar/Krasnodar | WHO National Influenza Centre Russian Federation | WHO National Influenza Centre Russian Federation | Andrey Komissarov et al |
| EPI_ISL_733273 | Russia/KDA-RII-MH4849S/2020 | 2020-10-15 | Europe/Russia/Krasnodar/Krasnodar | WHO National Influenza Centre Russian Federation | WHO National Influenza Centre Russian Federation | Andrey Komissarov et al |
| EPI_ISL_733275 | Russia/KDA-RII-MH4855S/2020 | 2020-10-18 | Europe/Russia/Krasnodar/Krasnodar | WHO National Influenza Centre Russian Federation | WHO National Influenza Centre Russian Federation | Andrey Komissarov et al |
| EPI_ISL_733277 | Russia/KDA-RII-MH4868S/2020 | 2020-10-21 | Europe/Russia/Krasnodar/Krasnodar | WHO National Influenza Centre Russian Federation | WHO National Influenza Centre Russian Federation | Andrey Komissarov et al |

| | | | | | | |
|---|---|---|---|---|---|---|
| EPI_ISL_733278 | Russia/KDA-RII-MH4871S/2020 | 2020-10-21 | Europe/Russia/Krasnodar/Krasnodar | WHO National Influenza Centre Russian Federation | WHO National Influenza Centre Russian Federation | Andrey Komissarov et al |
| EPI_ISL_733280 | Russia/KDA-RII-MH4887S/2020 | 2020-10-25 | Europe/Russia/Krasnodar/Krasnodar | WHO National Influenza Centre Russian Federation | WHO National Influenza Centre Russian Federation | Andrey Komissarov et al |
| EPI_ISL_733282 | Russia/KDA-RII-MH4901S/2020 | 2020-10-14 | Europe/Russia/Krasnodar/Armavir | WHO National Influenza Centre Russian Federation | WHO National Influenza Centre Russian Federation | Andrey Komissarov et al |
| EPI_ISL_733283 | Russia/KDA-RII-MH4912S/2020 | 2020-10-15 | Europe/Russia/Krasnodar/Armavir | WHO National Influenza Centre Russian Federation | WHO National Influenza Centre Russian Federation | Andrey Komissarov et al |
| EPI_ISL_733285 | Russia/KDA-RII-MH4919S/2020 | 2020-10-17 | Europe/Russia/Krasnodar/Armavir | WHO National Influenza Centre Russian Federation | WHO National Influenza Centre Russian Federation | Andrey Komissarov et al |
| EPI_ISL_733287 | Russia/KDA-RII-MH4928S/2020 | 2020-10-14 | Europe/Russia/Krasnodar/Armavir | WHO National Influenza Centre Russian Federation | WHO National Influenza Centre Russian Federation | Andrey Komissarov et al |
| EPI_ISL_733289 | Russia/KDA-RII-MH4936S/2020 | 2020-10-14 | Europe/Russia/Krasnodar/Armavir | WHO National Influenza Centre Russian Federation | WHO National Influenza Centre Russian Federation | Andrey Komissarov et al |
| EPI_ISL_733290 | Russia/KDA-RII-MH4938S/2020 | 2020-10-14 | Europe/Russia/Krasnodar/Armavir | WHO National Influenza Centre Russian Federation | WHO National Influenza Centre Russian Federation | Andrey Komissarov et al |
| EPI_ISL_733293 | Russia/KDA-RII-MH5029S/2020 | 2020-10-05 | Europe/Russia/Krasnodar/Novorossiysk | WHO National Influenza Centre Russian Federation | WHO National Influenza Centre Russian Federation | Andrey Komissarov et al |
| EPI_ISL_733295 | Russia/KDA-RII-MH5090S/2020 | 2020-11-05 | Europe/Russia/Krasnodar/Novorossiysk | WHO National Influenza Centre Russian Federation | WHO National Influenza Centre Russian Federation | Andrey Komissarov et al |

| | | | | | | |
|---|---|---|---|---|---|---|
| EPI_ISL_733297 | Russia/MUR-RII-MH5220S/2020 | 2020-09-29 | Europe/Russia/Murmansk/Murmansk | WHO National Influenza Centre Russian Federation | WHO National Influenza Centre Russian Federation | Andrey Komissarov et al |
| EPI_ISL_733301 | Russia/PSK-RII-MH5593S/2020 | 2020-11-09 | Europe/Russia/Pskov Oblast/Velikiye Lu | HELIX LLC | WHO National Influenza Centre Russian Federation | Andrey Komissarov et al |
| EPI_ISL_733302 | Russia/LEN-RII-MH5595S/2020 | 2020-11-09 | Europe/Russia/Leningrad Oblast/Kirovsk | HELIX LLC | WHO National Influenza Centre Russian Federation | Andrey Komissarov et al |
| EPI_ISL_733303 | Russia/PSK-RII-MH5604S/2020 | 2020-11-09 | Europe/Russia/Pskov Oblast/Velikiye Lu | HELIX LLC | WHO National Influenza Centre Russian Federation | Andrey Komissarov et al |
| EPI_ISL_733304 | Russia/LEN-RII-MH5616S/2020 | 2020-11-09 | Europe/Russia/Leningrad Oblast/Gatchin | HELIX LLC | WHO National Influenza Centre Russian Federation | Andrey Komissarov et al |
| EPI_ISL_733305 | Russia/PSK-RII-MH5619S/2020 | 2020-11-09 | Europe/Russia/Pskov Oblast/Velikiye Lu | HELIX LLC | WHO National Influenza Centre Russian Federation | Andrey Komissarov et al |
| EPI_ISL_733307 | Russia/NGR-RII-MH5625S/2020 | 2020-11-09 | Europe/Russia/Novgorod/Veliky Novgor | HELIX LLC | WHO National Influenza Centre Russian Federation | Andrey Komissarov et al |
| EPI_ISL_733308 | Russia/PSK-RII-MH5633S/2020 | 2020-11-09 | Europe/Russia/Pskov Oblast/Velikiye Lu | HELIX LLC | WHO National Influenza Centre Russian Federation | Andrey Komissarov et al |
| EPI_ISL_733309 | Russia/LEN-RII-MH5654S/2020 | 2020-11-09 | Europe/Russia/Leningrad Oblast/Yanino- | HELIX LLC | WHO National Influenza Centre Russian Federation | Andrey Komissarov et al |
| EPI_ISL_733310 | Russia/PSK-RII-MH5658S/2020 | 2020-11-09 | Europe/Russia/Pskov Oblast/Velikiye Lu | HELIX LLC | WHO National Influenza Centre Russian Federation | Andrey Komissarov et al |

| | | | | | | |
|---|---|---|---|---|---|---|
| EPI_ISL_733311 | Russia/PSK-RII-MH5661S/2020 | 2020-11-13 | Europe/Russia/Pskov Oblast/Velikiye Lu | HELIX LLC | WHO National Influenza Centre Russian Federation | Andrey Komissarov et al |
| EPI_ISL_733312 | Russia/PSK-RII-MH5666S/2020 | 2020-11-14 | Europe/Russia/Pskov Oblast/Velikiye Lu | HELIX LLC | WHO National Influenza Centre Russian Federation | Andrey Komissarov et al |
| EPI_ISL_733314 | Russia/PSK-RII-MH5736S/2020 | 2020-11-09 | Europe/Russia/Pskov Oblast/Borovichi (I | HELIX LLC | WHO National Influenza Centre Russian Federation | Andrey Komissarov et al |
| EPI_ISL_733317 | Russia/LEN-RII-MH5773S/2020 | 2020-11-09 | Europe/Russia/Leningrad Oblast/Volkho | HELIX LLC | WHO National Influenza Centre Russian Federation | Andrey Komissarov et al |
| EPI_ISL_733318 | Russia/SPE-RII-MH5887S/2020 | 2020-11-05 | Europe/Russia/Saint-Petersburg/Saint-P | HELIX LLC | WHO National Influenza Centre Russian Federation | Andrey Komissarov et al |
| EPI_ISL_733320 | Russia/SPE-RII-MH5889S/2020 | 2020-11-06 | Europe/Russia/Saint-Petersburg/Saint-P | HELIX LLC | WHO National Influenza Centre Russian Federation | Andrey Komissarov et al |
| EPI_ISL_733321 | Russia/SPE-RII-MH5892S/2020 | 2020-11-05 | Europe/Russia/Saint-Petersburg/Saint-P | HELIX LLC | WHO National Influenza Centre Russian Federation | Andrey Komissarov et al |
| EPI_ISL_733323 | Russia/SPE-RII-MH5894S/2020 | 2020-11-06 | Europe/Russia/Saint-Petersburg/Saint-P | HELIX LLC | WHO National Influenza Centre Russian Federation | Andrey Komissarov et al |
| EPI_ISL_733325 | Russia/SPE-RII-MH5897S/2020 | 2020-11-05 | Europe/Russia/Saint-Petersburg/Saint-P | HELIX LLC | WHO National Influenza Centre Russian Federation | Andrey Komissarov et al |
| EPI_ISL_733331 | Russia/LEN-RII-MH5948S/2020 | 2020-11-11 | Europe/Russia/Leningrad Oblast/Volkho | HELIX LLC | WHO National Influenza Centre Russian Federation | Andrey Komissarov et al |

| | | | | | | |
|---|---|---|---|---|---|---|
| EPI_ISL_733332 | Russia/PSK-RII-MH5952S/2020 | 2020-11-10 | Europe/Russia/Pskov Oblast/Borovichi (F | HELIX LLC | WHO National Influenza Centre Russian Federation | Andrey Komissarov et al |
| EPI_ISL_733333 | Russia/NGR-RII-MH5963S/2020 | 2020-11-14 | Europe/Russia/Novgorod/Borovichi (Nov | HELIX LLC | WHO National Influenza Centre Russian Federation | Andrey Komissarov et al |
| EPI_ISL_733334 | Russia/MUR-RII-MH6034S/2020 | 2020-11-11 | Europe/Russia/Murmansk/Murmansk | HELIX LLC | WHO National Influenza Centre Russian Federation | Andrey Komissarov et al |
| EPI_ISL_733338 | Russia/MUR-RII-MH6068S/2020 | 2020-11-11 | Europe/Russia/Murmansk/Murmansk | HELIX LLC | WHO National Influenza Centre Russian Federation | Andrey Komissarov et al |
| EPI_ISL_733340 | Russia/MUR-RII-MH6076S/2020 | 2020-11-13 | Europe/Russia/Murmansk/Severomorsk | HELIX LLC | WHO National Influenza Centre Russian Federation | Andrey Komissarov et al |
| EPI_ISL_733342 | Russia/PSK-RII-MH6081S/2020 | 2020-11-14 | Europe/Russia/Pskov Oblast/Velikiye Lu | HELIX LLC | WHO National Influenza Centre Russian Federation | Andrey Komissarov et al |
| EPI_ISL_733343 | Russia/PSK-RII-MH6083S/2020 | 2020-11-14 | Europe/Russia/Pskov Oblast/Velikiye Lu | HELIX LLC | WHO National Influenza Centre Russian Federation | Andrey Komissarov et al |
| EPI_ISL_733344 | Russia/PSK-RII-MH6086S/2020 | 2020-11-14 | Europe/Russia/Pskov Oblast/Velikiye Lu | HELIX LLC | WHO National Influenza Centre Russian Federation | Andrey Komissarov et al |
| EPI_ISL_733347 | Russia/PSK-RII-MH6102S/2020 | 2020-11-13 | Europe/Russia/Pskov Oblast/Velikiye Lu | HELIX LLC | WHO National Influenza Centre Russian Federation | Andrey Komissarov et al |
| EPI_ISL_733350 | Russia/NGR-RII-MH6127S/2020 | 2020-11-06 | Europe/Russia/Novgorod/Veliky Novgor | HELIX LLC | WHO National Influenza Centre Russian Federation | Andrey Komissarov et al |

| | | | | | | |
|---|---|---|---|---|---|---|
| EPI_ISL_733351 | Russia/LEN-RII-MH6145S/2020 | 2020-11-06 | Europe/Russia/Leningrad Oblast/Volkho | HELIX LLC | WHO National Influenza Centre Russian Federation | Andrey Komissarov et al |
| EPI_ISL_733352 | Russia/NGR-RII-MH6147S/2020 | 2020-11-06 | Europe/Russia/Novgorod/Borovichi (Nov | HELIX LLC | WHO National Influenza Centre Russian Federation | Andrey Komissarov et al |
| EPI_ISL_733353 | Russia/PSK-RII-MH6156S/2020 | 2020-11-14 | Europe/Russia/Pskov Oblast/Velikiye Lu | HELIX LLC | WHO National Influenza Centre Russian Federation | Andrey Komissarov et al |
| EPI_ISL_733354 | Russia/PSK-RII-MH6161S/2020 | 2020-11-14 | Europe/Russia/Pskov Oblast/Velikiye Lu | HELIX LLC | WHO National Influenza Centre Russian Federation | Andrey Komissarov et al |
| EPI_ISL_733355 | Russia/MUR-RII-MH6168S/2020 | 2020-11-13 | Europe/Russia/Murmansk/Severomorsk | HELIX LLC | WHO National Influenza Centre Russian Federation | Andrey Komissarov et al |
| EPI_ISL_733357 | Russia/KR-RII-MH6176S/2020 | 2020-11-13 | Europe/Russia/Karelia/Petrozavodsk | HELIX LLC | WHO National Influenza Centre Russian Federation | Andrey Komissarov et al |
| EPI_ISL_733359 | Russia/KR-RII-MH6188S/2020 | 2020-11-13 | Europe/Russia/Karelia/Petrozavodsk | HELIX LLC | WHO National Influenza Centre Russian Federation | Andrey Komissarov et al |
| EPI_ISL_733361 | Russia/PSK-RII-MH6191S/2020 | 2020-11-13 | Europe/Russia/Pskov Oblast/Borovichi (F | HELIX LLC | WHO National Influenza Centre Russian Federation | Andrey Komissarov et al |
| EPI_ISL_733366 | Russia/MUR-RII-MH6271S/2020 | 2020-11-12 | Europe/Russia/Murmansk/Murmansk | HELIX LLC | WHO National Influenza Centre Russian Federation | Andrey Komissarov et al |
| EPI_ISL_733368 | Russia/PSK-RII-MH6287S/2020 | 2020-11-12 | Europe/Russia/Pskov Oblast/Velikiye Lu | HELIX LLC | WHO National Influenza Centre Russian Federation | Andrey Komissarov et al |

| | | | | | | |
|---|---|---|---|---|---|---|
| EPI_ISL_733369 | Russia/NGR-RII-MH6288S/2020 | 2020-11-12 | Europe/Russia/Novgorod/Veliky Novgor | HELIX LLC | WHO National Influenza Centre Russian Federation | Andrey Komissarov et al |
| EPI_ISL_733370 | Russia/PSK-RII-MH6290S/2020 | 2020-11-09 | Europe/Russia/Pskov Oblast/Velikiye Lu | HELIX LLC | WHO National Influenza Centre Russian Federation | Andrey Komissarov et al |
| EPI_ISL_733372 | Russia/PSK-RII-MH6292S/2020 | 2020-11-10 | Europe/Russia/Pskov Oblast/Velikiye Lu | HELIX LLC | WHO National Influenza Centre Russian Federation | Andrey Komissarov et al |
| EPI_ISL_733373 | Russia/NGR-RII-MH6295S/2020 | 2020-11-12 | Europe/Russia/Novgorod/Veliky Novgor | HELIX LLC | WHO National Influenza Centre Russian Federation | Andrey Komissarov et al |
| EPI_ISL_733376 | Russia/NGR-RII-MH6306S/2020 | 2020-11-12 | Europe/Russia/Novgorod/Veliky Novgor | HELIX LLC | WHO National Influenza Centre Russian Federation | Andrey Komissarov et al |
| EPI_ISL_733378 | Russia/PSK-RII-MH6314S/2020 | 2020-11-12 | Europe/Russia/Pskov Oblast/Velikiye Lu | HELIX LLC | WHO National Influenza Centre Russian Federation | Andrey Komissarov et al |
| EPI_ISL_733379 | Russia/PSK-RII-MH6316S/2020 | 2020-11-12 | Europe/Russia/Pskov Oblast/Velikiye Lu | HELIX LLC | WHO National Influenza Centre Russian Federation | Andrey Komissarov et al |
| EPI_ISL_733380 | Russia/PSK-RII-MH6320S/2020 | 2020-11-12 | Europe/Russia/Pskov Oblast/Velikiye Lu | HELIX LLC | WHO National Influenza Centre Russian Federation | Andrey Komissarov et al |
| EPI_ISL_733381 | Russia/NGR-RII-MH6321S/2020 | 2020-11-12 | Europe/Russia/Novgorod/Veliky Novgor | HELIX LLC | WHO National Influenza Centre Russian Federation | Andrey Komissarov et al |
| EPI_ISL_733384 | Russia/MUR-RII-MH6337S/2020 | 2020-11-11 | Europe/Russia/Murmansk/Severomorsk | HELIX LLC | WHO National Influenza Centre Russian Federation | Andrey Komissarov et al |

| | | | | | | |
|---|---|---|---|---|---|---|
| EPI_ISL_733385 | Russia/NGR-RII-MH6339S/2020 | 2020-11-10 | Europe/Russia/Novgorod/Veliky Novgor | HELIX LLC | WHO National Influenza Centre Russian Federation | Andrey Komissarov et al |
| EPI_ISL_733387 | Russia/MUR-RII-MH6348S/2020 | 2020-11-10 | Europe/Russia/Murmansk/Murmansk | HELIX LLC | WHO National Influenza Centre Russian Federation | Andrey Komissarov et al |
| EPI_ISL_733392 | Russia/NVS-RII-MH1159V/2020 | 2020-08-04 | Europe/Russia/Novosibirsk/Novosibirsk | WHO National Influenza Centre Russian Federation | WHO National Influenza Centre Russian Federation | Andrey Komissarov et al |
| EPI_ISL_733394 | Russia/CE-RII-MH2523V/2020 | 2020-09-26 | Europe/Russia/Chechen Republic/Grozny | WHO National Influenza Centre Russian Federation | WHO National Influenza Centre Russian Federation | Andrey Komissarov et al |
| EPI_ISL_733395 | Russia/CE-RII-MH2636V/2020 | 2020-09-11 | Europe/Russia/Chechen Republic/Grozny | WHO National Influenza Centre Russian Federation | WHO National Influenza Centre Russian Federation | Andrey Komissarov et al |
| EPI_ISL_733397 | Russia/NGR-RII-MH4539S/2020 | 2020-10-29 | Europe/Russia/Novgorod/Veliky Novgor | HELIX LLC | WHO National Influenza Centre Russian Federation | Andrey Komissarov et al |
| EPI_ISL_733399 | Russia/SPE-RII-MH6849S/2020 | 2020-11-13 | Europe/Russia/Saint-Petersburg/Saint-P | HELIX LLC | WHO National Influenza Centre Russian Federation | Andrey Komissarov et al |
| EPI_ISL_733400 | Russia/NGR-RII-MH6870S/2020 | 2020-11-05 | Europe/Russia/Novgorod/Borovichi (Nov | HELIX LLC | WHO National Influenza Centre Russian Federation | Andrey Komissarov et al |
| EPI_ISL_733405 | Russia/SVE-RII-MH8982S/2020 | 2020-10-01 | Europe/Russia/Sverdlovsk/Yekaterinbur | UMMC-Health | WHO National Influenza Centre Russian Federation | Andrey Komissarov et al |
| EPI_ISL_733406 | Russia/SVE-RII-MH9000S/2020 | 2020-10-01 | Europe/Russia/Sverdlovsk/Dvurechensk | UMMC-Health | WHO National Influenza Centre Russian Federation | Andrey Komissarov et al |

| | | | | | | |
|---|---|---|---|---|---|---|
| EPI_ISL_733407 | Russia/SVE-RII-MH9083S/2020 | 2020-10-01 | Europe/Russia/Sverdlovsk/Bobrovsky | UMMC-Health | WHO National Influenza Centre Russian Federation | Andrey Komissarov et al |
| EPI_ISL_733411 | Russia/SVE-RII-MH9112S/2020 | 2020-10-01 | Europe/Russia/Sverdlovsk/Verkhnyaya | UMMC-Health | WHO National Influenza Centre Russian Federation | Andrey Komissarov et al |
| EPI_ISL_733412 | Russia/SVE-RII-MH9113S/2020 | 2020-10-01 | Europe/Russia/Sverdlovsk/Verkhnyaya | UMMC-Health | WHO National Influenza Centre Russian Federation | Andrey Komissarov et al |
| EPI_ISL_733414 | Russia/SVE-RII-MH9206S/2020 | 2020-10-03 | Europe/Russia/Sverdlovsk/Yekaterinbur | UMMC-Health | WHO National Influenza Centre Russian Federation | Andrey Komissarov et al |
| EPI_ISL_733415 | Russia/SVE-RII-MH9209S/2020 | 2020-10-03 | Europe/Russia/Sverdlovsk/Nizhny Tagil | UMMC-Health | WHO National Influenza Centre Russian Federation | Andrey Komissarov et al |
| EPI_ISL_733416 | Russia/SVE-RII-MH9221S/2020 | 2020-10-04 | Europe/Russia/Sverdlovsk/Nizhny Tagil | UMMC-Health | WHO National Influenza Centre Russian Federation | Andrey Komissarov et al |
| EPI_ISL_733417 | Russia/MOS-RII-MH9243S/2020 | 2020-10-14 | Europe/Russia/Moscow Oblast/Korolyov | HELIX LLC | WHO National Influenza Centre Russian Federation | Andrey Komissarov et al |
| EPI_ISL_733420 | Russia/MOS-RII-MH9250S/2020 | 2020-10-11 | Europe/Russia/Moscow Oblast/Lyuberts | HELIX LLC | WHO National Influenza Centre Russian Federation | Andrey Komissarov et al |
| EPI_ISL_733423 | Russia/MOS-RII-MH9264S/2020 | 2020-10-14 | Europe/Russia/Moscow Oblast/Lyuberts | HELIX LLC | WHO National Influenza Centre Russian Federation | Andrey Komissarov et al |
| EPI_ISL_733424 | Russia/KHM-RII-MH9266S/2020 | 2020-10-10 | Europe/Russia/Khanty-Mansi/Nizhnevar | HELIX LLC | WHO National Influenza Centre Russian Federation | Andrey Komissarov et al |

| | | | | | | |
|---|---|---|---|---|---|---|
| EPI_ISL_733425 | Russia/MOS-RII-MH9267S/2020 | 2020-10-11 | Europe/Russia/Moscow Oblast/Korolyov | HELIX LLC | WHO National Influenza Centre Russian Federation | Andrey Komissarov et al |
| EPI_ISL_733426 | Russia/IVA-RII-MH9268S/2020 | 2020-10-14 | Europe/Russia/Ivanovo/Ivanovo | HELIX LLC | WHO National Influenza Centre Russian Federation | Andrey Komissarov et al |
| EPI_ISL_733428 | Russia/MOS-RII-MH9271S/2020 | 2020-10-14 | Europe/Russia/Moscow Oblast/Lyuberts | HELIX LLC | WHO National Influenza Centre Russian Federation | Andrey Komissarov et al |
| EPI_ISL_733429 | Russia/MOS-RII-MH9279S/2020 | 2020-10-15 | Europe/Russia/Moscow Oblast/Lapino | HELIX LLC | WHO National Influenza Centre Russian Federation | Andrey Komissarov et al |
| EPI_ISL_733430 | Russia/MOS-RII-MH9280S/2020 | 2020-10-11 | Europe/Russia/Moscow Oblast/Kubinka | HELIX LLC | WHO National Influenza Centre Russian Federation | Andrey Komissarov et al |
| EPI_ISL_733433 | Crimea/RII-MH9286S/2020 | 2020-10-13 | Europe/Ukraine/Crimea/Yevpatoriya | HELIX LLC | WHO National Influenza Centre Russian Federation | Andrey Komissarov et al |
| EPI_ISL_733436 | Russia/KDA-RII-MH9294S/2020 | 2020-10-11 | Europe/Russia/Krasnodar/Anapa | HELIX LLC | WHO National Influenza Centre Russian Federation | Andrey Komissarov et al |
| EPI_ISL_733439 | Russia/MOS-RII-MH9309S/2020 | 2020-10-11 | Europe/Russia/Moscow Oblast/Lapino | HELIX LLC | WHO National Influenza Centre Russian Federation | Andrey Komissarov et al |
| EPI_ISL_733440 | Russia/RYA-RII-MH9318S/2020 | 2020-10-11 | Europe/Russia/Ryazan/Ryazan | HELIX LLC | WHO National Influenza Centre Russian Federation | Andrey Komissarov et al |
| EPI_ISL_733441 | Russia/RYA-RII-MH9326S/2020 | 2020-10-14 | Europe/Russia/Ryazan/Ryazan | HELIX LLC | WHO National Influenza Centre Russian Federation | Andrey Komissarov et al |

| | | | | | | |
|---|---|---|---|---|---|---|
| EPI_ISL_733442 | Russia/ORL-RII-MH9331S/2020 | 2020-10-12 | Europe/Russia/Oryol/Oryol | HELIX LLC | WHO National Influenza Centre Russian Federation | Andrey Komissarov et al |
| EPI_ISL_733443 | Russia/ORE-RII-MH9335S/2020 | 2020-10-13 | Europe/Russia/Orenburg/Orsk | HELIX LLC | WHO National Influenza Centre Russian Federation | Andrey Komissarov et al |
| EPI_ISL_733445 | Russia/MOS-RII-MH9340S/2020 | 2020-10-11 | Europe/Russia/Moscow Oblast/Zvenigor | HELIX LLC | WHO National Influenza Centre Russian Federation | Andrey Komissarov et al |
| EPI_ISL_733446 | Russia/MOS-RII-MH9341S/2020 | 2020-10-14 | Europe/Russia/Moscow Oblast/Naro-For | HELIX LLC | WHO National Influenza Centre Russian Federation | Andrey Komissarov et al |
| EPI_ISL_733451 | Russia/MOS-RII-MH9358S/2020 | 2020-10-14 | Europe/Russia/Moscow Oblast/Elektrost | HELIX LLC | WHO National Influenza Centre Russian Federation | Andrey Komissarov et al |
| EPI_ISL_733452 | Crimea/RII-MH9362S/2020 | 2020-10-14 | Europe/Ukraine/Crimea/Novofedorovka | HELIX LLC | WHO National Influenza Centre Russian Federation | Andrey Komissarov et al |
| EPI_ISL_733454 | Russia/RYA-RII-MH9366S/2020 | 2020-10-11 | Europe/Russia/Ryazan/Ryazan | HELIX LLC | WHO National Influenza Centre Russian Federation | Andrey Komissarov et al |
| EPI_ISL_733455 | Russia/MOS-RII-MH9369S/2020 | 2020-10-14 | Europe/Russia/Moscow Oblast/Pushkino | HELIX LLC | WHO National Influenza Centre Russian Federation | Andrey Komissarov et al |
| EPI_ISL_733456 | Russia/MOS-RII-MH9370S/2020 | 2020-10-14 | Europe/Russia/Moscow Oblast/Naro-For | HELIX LLC | WHO National Influenza Centre Russian Federation | Andrey Komissarov et al |
| EPI_ISL_733457 | Russia/MOW-RII-MH9374S/2020 | 2020-10-11 | Europe/Russia/Moscow Oblast/Moscow | HELIX LLC | WHO National Influenza Centre Russian Federation | Andrey Komissarov et al |

| | | | | | | |
|---|---|---|---|---|---|---|
| EPI_ISL_733458 | Russia/VGG-RII-MH9375S/2020 | 2020-10-10 | Europe/Russia/Volgograd Region/Volgog | HELIX LLC | WHO National Influenza Centre Russian Federation | Andrey Komissarov et al |
| EPI_ISL_733462 | Russia/AMU-RII-MH10154S/2020 | 2020-11-20 | Europe/Russia/Amur Region/Yekaterino | WHO National Influenza Centre Russian Federation | WHO National Influenza Centre Russian Federation | Andrey Komissarov et al |
| EPI_ISL_733463 | Russia/AMU-RII-MH10159S/2020 | 2020-11-23 | Europe/Russia/Amur Region/Blagovesho | WHO National Influenza Centre Russian Federation | WHO National Influenza Centre Russian Federation | Andrey Komissarov et al |
| EPI_ISL_733464 | Russia/AMU-RII-MH10160S/2020 | 2020-11-19 | Europe/Russia/Amur Region/Blagovesho | WHO National Influenza Centre Russian Federation | WHO National Influenza Centre Russian Federation | Andrey Komissarov et al |
| EPI_ISL_733465 | Russia/AMU-RII-MH10166S/2020 | 2020-11-23 | Europe/Russia/Amur Region/Blagovesho | WHO National Influenza Centre Russian Federation | WHO National Influenza Centre Russian Federation | Andrey Komissarov et al |
| EPI_ISL_733473 | Russia/AMU-RII-MH10205S/2020 | 2020-11-25 | Europe/Russia/Amur Region/Progress | WHO National Influenza Centre Russian Federation | WHO National Influenza Centre Russian Federation | Andrey Komissarov et al |
| EPI_ISL_733476 | Russia/AMU-RII-MH10219S/2020 | 2020-11-19 | Europe/Russia/Amur Region/Arkhara | WHO National Influenza Centre Russian Federation | WHO National Influenza Centre Russian Federation | Andrey Komissarov et al |
| EPI_ISL_733477 | Russia/AMU-RII-MH10223S/2020 | 2020-11-18 | Europe/Russia/Amur Region/Blagovesho | WHO National Influenza Centre Russian Federation | WHO National Influenza Centre Russian Federation | Andrey Komissarov et al |
| EPI_ISL_733482 | Russia/OMS-ORINFI-MH10267S/2020 | 2020-04-19 | Europe/Russia/Omsk/Omsk | Omsk Research Institute of Natural Focal Infections | WHO National Influenza Centre Russian Federation | Andrey Komissarov et al |
| EPI_ISL_733484 | Russia/OMS-ORINFI-MH10275S/2020 | 2020-11-02 | Europe/Russia/Omsk/Omsk | Omsk Research Institute of Natural Focal Infections | WHO National Influenza Centre Russian Federation | Andrey Komissarov et al |

| | | | | | | |
|---|---|---|---|---|---|---|
| EPI_ISL_733485 | Russia/OMS-ORINFI-MH10280S/2020 | 2020-11-06 | Europe/Russia/Omsk/Tara | Omsk Research Institute of Natural Focal Infections | WHO National Influenza Centre Russian Federation | Andrey Komissarov et al |
| EPI_ISL_733489 | Russia/OMS-ORINFI-MH10285M/2020 | 2020-11-12 | Europe/Russia/Omsk/Tara | Omsk Research Institute of Natural Focal Infections | WHO National Influenza Centre Russian Federation | Andrey Komissarov et al |
| EPI_ISL_733490 | Russia/OMS-ORINFI-MH10288M/2020 | 2020-11-12 | Europe/Russia/Omsk/Tara | Omsk Research Institute of Natural Focal Infections | WHO National Influenza Centre Russian Federation | Andrey Komissarov et al |
| EPI_ISL_733493 | Russia/OMS-ORINFI-MH10294S/2020 | 2020-11-17 | Europe/Russia/Omsk/Tara | Omsk Research Institute of Natural Focal Infections | WHO National Influenza Centre Russian Federation | Andrey Komissarov et al |
| EPI_ISL_733495 | Russia/KR-RII-12146S/2020 | 2020-11-17 | Europe/Russia/Karelia/Petrozavodsk | WHO National Influenza Centre Russian Federation | WHO National Influenza Centre Russian Federation | Andrey Komissarov et al |

| | | | | | | |
|---|---|---|---|---|---|---|
| EPI_ISL_733499 | Tunisia/63686/2020 | 2020-12-01 | Africa/Tunisia/Nabeul/ | 1-Laboratory of Microbiology, National Reference Lab, Charles Nicolle Hospital; 2-University of Tunis ElManar, Faculty of Medicine of Tunis, LR99ES09, Tunis, Tunisia | 1-Clinical and Experimental Pharmacology Lab, LR16SP02, National Center of Pharmacovigilance, University of Tunis El Manar, Tunis, Tunisia. 2-Neurodegenerative diseases and psychiatric troubles, LR18SP03, Razi Hospital, University of Tunis El Manar, Tunis, Tunisia. 3-Ministry of Health, National Observatory of New and Emerging Diseases, 1006, Tunis, Tunisia | Ilhem Boutiba-Ben Boubaker et al |
| EPI_ISL_733502 | Germany/NW-HHU-439/2020 | 2020-10-25 | Europe/Germany/North Rhine Westphal | ZOTZ KLIMAS MVZ Dvºsseldorf-Centrum GbR √úBAG fvºr Labormedizin, Genetik, Zytologie, Pathologie | Center of Medical Microbiology, Virology, and Hospital Hygiene, University of Duesseldorf | Maximilian Damagnez et al |
| EPI_ISL_733503 | Germany/NW-HHU-447/2020 | 2020-10-26 | Europe/Germany/North Rhine Westphal | ZOTZ KLIMAS MVZ Dvºsseldorf-Centrum GbR √úBAG fvºr Labormedizin, Genetik, Zytologie, Pathologie | Center of Medical Microbiology, Virology, and Hospital Hygiene, University of Duesseldorf | Maximilian Damagnez et al |

| | | | | | | |
|---|---|---|---|---|---|---|
| EPI_ISL_733504 | Germany/NW-HHU-448/2020 | 2020-10-26 | Europe/Germany/North Rhine Westphal | ZOTZ KLIMAS MVZ D√ºsseldorf-Centrum GbR √úBAG f√ºr Labormedizin, Genetik, Zytologie, Pathologie | Center of Medical Microbiology, Virology, and Hospital Hygiene, University of Duesseldorf | Maximilian Damagnez et al |
| EPI_ISL_733506 | Germany/NW-HHU-431/2020 | 2020-10-25 | Europe/Germany/North Rhine Westphal | ZOTZ KLIMAS MVZ D√ºsseldorf-Centrum GbR √úBAG f√ºr Labormedizin, Genetik, Zytologie, Pathologie | Center of Medical Microbiology, Virology, and Hospital Hygiene, University of Duesseldorf | Maximilian Damagnez et al |
| EPI_ISL_733508 | Germany/NW-HHU-433/2020 | 2020-10-25 | Europe/Germany/North Rhine Westphal | ZOTZ KLIMAS MVZ D√ºsseldorf-Centrum GbR √úBAG f√ºr Labormedizin, Genetik, Zytologie, Pathologie | Center of Medical Microbiology, Virology, and Hospital Hygiene, University of Duesseldorf | Maximilian Damagnez et al |
| EPI_ISL_733512 | Germany/NW-HHU-437/2020 | 2020-10-25 | Europe/Germany/North Rhine Westphal | ZOTZ KLIMAS MVZ D√ºsseldorf-Centrum GbR √úBAG f√ºr Labormedizin, Genetik, Zytologie, Pathologie | Center of Medical Microbiology, Virology, and Hospital Hygiene, University of Duesseldorf | Maximilian Damagnez et al |
| EPI_ISL_733518 | Germany/NW-HHU-445/2020 | 2020-10-25 | Europe/Germany/North Rhine Westphal | ZOTZ KLIMAS MVZ D√ºsseldorf-Centrum GbR √úBAG f√ºr Labormedizin, Genetik, Zytologie, Pathologie | Center of Medical Microbiology, Virology, and Hospital Hygiene, University of Duesseldorf | Maximilian Damagnez et al |
| EPI_ISL_733520 | Germany/NW-HHU-449/2020 | 2020-10-26 | Europe/Germany/North Rhine Westphal | ZOTZ KLIMAS MVZ D√ºsseldorf-Centrum GbR √úBAG f√ºr Labormedizin, Genetik, Zytologie, Pathologie | Center of Medical Microbiology, Virology, and Hospital Hygiene, University of Duesseldorf | Maximilian Damagnez et al |
| EPI_ISL_733898 | England/CAMC-C2D362/2020 | 2020-12-07 | Europe/United Kingdom/England/ | Lighthouse Lab in Cambridge | Wellcome Sanger Institute for the COVID-19 Genomics UK (COG-UK) Consortium | Rob Howes et al |

| | | | | | | |
|---|---|---|---|---|---|---|
| EPI_ISL_734008 | England/QEUH-C337D6/2020 | 2020-12-10 | Europe/United Kingdom/England/ | Lighthouse Lab in Glasgow | Wellcome Sanger Institute for the COVID-19 Genomics UK (COG-UK) Consortium | Harper VanSteenhouse et al |
| EPI_ISL_734168 | France/BRE-IPP9985/2020 | 2020-10-10 | Europe/France/Bretagne/Rennes | CHRU Pontchaillou - Laboratoire de Virologie | National Reference Center for Viruses of Respiratory Infections, Institut Pasteur, Paris | Marion Barbet et al |
| EPI_ISL_734169 | France/BRE-IPP9997/2020 | 2020-11-05 | Europe/France/Bretagne/Rennes | CHRU Pontchaillou - Laboratoire de Virologie | National Reference Center for Viruses of Respiratory Infections, Institut Pasteur, Paris | Marion Barbet et al |
| EPI_ISL_734173 | USA/VA-DCLS-2402/2020 | 2020-11-16 | North America/USA/Virginia/ | Virginia Division of Consolidated Laboratory Services (DCLS) | Virginia Division of Consolidated Laboratory Services (DCLS) | Virginia DCLS et al |
| EPI_ISL_734175 | USA/VA-DCLS-2404/2020 | 2020-11-16 | North America/USA/Virginia/ | Virginia Division of Consolidated Laboratory Services (DCLS) | Virginia Division of Consolidated Laboratory Services (DCLS) | Virginia DCLS et al |
| EPI_ISL_734177 | USA/VA-DCLS-2406/2020 | 2020-11-20 | North America/USA/Virginia/ | Virginia Division of Consolidated Laboratory Services (DCLS) | Virginia Division of Consolidated Laboratory Services (DCLS) | Virginia DCLS et al |
| EPI_ISL_734178 | USA/VA-DCLS-2407/2020 | 2020-11-20 | North America/USA/Virginia/ | Virginia Division of Consolidated Laboratory Services (DCLS) | Virginia Division of Consolidated Laboratory Services (DCLS) | Virginia DCLS et al |
| EPI_ISL_734182 | USA/VA-DCLS-2411/2020 | 2020-11-18 | North America/USA/Virginia/ | Virginia Division of Consolidated Laboratory Services (DCLS) | Virginia Division of Consolidated Laboratory Services (DCLS) | Virginia DCLS et al |

| | | | | | | |
|---|---|---|---|---|---|---|
| EPI_ISL_734186 | USA/VA-DCLS-2437/2020 | 2020-11-17 | North America/USA/Virginia/ | Virginia Division of Consolidated Laboratory Services (DCLS) | Virginia Division of Consolidated Laboratory Services (DCLS) | Virginia DCLS et al |
| EPI_ISL_734187 | USA/VA-DCLS-2438/2020 | 2020-11-17 | North America/USA/Virginia/ | Virginia Division of Consolidated Laboratory Services (DCLS) | Virginia Division of Consolidated Laboratory Services (DCLS) | Virginia DCLS et al |
| EPI_ISL_734191 | USA/VA-DCLS-2442/2020 | 2020-11-17 | North America/USA/Virginia/ | Virginia Division of Consolidated Laboratory Services (DCLS) | Virginia Division of Consolidated Laboratory Services (DCLS) | Virginia DCLS et al |
| EPI_ISL_734193 | USA/VA-DCLS-2444/2020 | 2020-11-20 | North America/USA/Virginia/ | Virginia Division of Consolidated Laboratory Services (DCLS) | Virginia Division of Consolidated Laboratory Services (DCLS) | Virginia DCLS et al |
| EPI_ISL_734195 | USA/VA-DCLS-2448/2020 | 2020-11-20 | North America/USA/Virginia/ | Virginia Division of Consolidated Laboratory Services (DCLS) | Virginia Division of Consolidated Laboratory Services (DCLS) | Virginia DCLS et al |
| EPI_ISL_734202 | USA/VA-DCLS-2457/2020 | 2020-11-18 | North America/USA/Virginia/ | Virginia Division of Consolidated Laboratory Services (DCLS) | Virginia Division of Consolidated Laboratory Services (DCLS) | Virginia DCLS et al |
| EPI_ISL_734208 | USA/VA-DCLS-2222/2020 | 2020-11-03 | North America/USA/Virginia/ | Virginia Division of Consolidated Laboratory Services (DCLS) | Virginia Division of Consolidated Laboratory Services (DCLS) | Virginia DCLS et al |
| EPI_ISL_734214 | USA/VA-DCLS-2234/2020 | 2020-11-03 | North America/USA/Virginia/ | Virginia Division of Consolidated Laboratory Services (DCLS) | Virginia Division of Consolidated Laboratory Services (DCLS) | Virginia DCLS et al |
| EPI_ISL_734234 | USA/VA-DCLS-2147/2020 | 2020-10-27 | North America/USA/Virginia/ | Virginia Division of Consolidated Laboratory Services (DCLS) | Virginia Division of Consolidated Laboratory Services (DCLS) | Virginia DCLS et al |
| EPI_ISL_734240 | USA/VA-DCLS-2158/2020 | 2020-10-28 | North America/USA/Virginia/ | Virginia Division of Consolidated Laboratory Services (DCLS) | Virginia Division of Consolidated Laboratory Services (DCLS) | Virginia DCLS et al |

| | | | | | | |
|---|---|---|---|---|---|---|
| EPI_ISL_734245 | USA/VA-DCLS-2164/2020 | 2020-11-01 | North America/USA/Virginia/ | Virginia Division of Consolidated Laboratory Services (DCLS) | Virginia Division of Consolidated Laboratory Services (DCLS) | Virginia DCLS et al |
| EPI_ISL_734247 | USA/VA-DCLS-2194/2020 | 2020-11-04 | North America/USA/Virginia/ | Virginia Division of Consolidated Laboratory Services (DCLS) | Virginia Division of Consolidated Laboratory Services (DCLS) | Virginia DCLS et al |
| EPI_ISL_734253 | USA/VA-DCLS-2537/2020 | 2020-11-14 | North America/USA/Virginia/ | Virginia Division of Consolidated Laboratory Services (DCLS) | Virginia Division of Consolidated Laboratory Services (DCLS) | Virginia DCLS et al |
| EPI_ISL_734266 | USA/VA-DCLS-2316/2020 | 2020-10-22 | North America/USA/Virginia/ | Virginia Division of Consolidated Laboratory Services (DCLS) | Virginia Division of Consolidated Laboratory Services (DCLS) | Virginia DCLS et al |
| EPI_ISL_734270 | USA/VA-DCLS-2322/2020 | 2020-10-27 | North America/USA/Virginia/ | Virginia Division of Consolidated Laboratory Services (DCLS) | Virginia Division of Consolidated Laboratory Services (DCLS) | Virginia DCLS et al |
| EPI_ISL_734273 | USA/VA-DCLS-2325/2020 | 2020-10-27 | North America/USA/Virginia/ | Virginia Division of Consolidated Laboratory Services (DCLS) | Virginia Division of Consolidated Laboratory Services (DCLS) | Virginia DCLS et al |
| EPI_ISL_734279 | USA/VA-DCLS-2333/2020 | 2020-11-09 | North America/USA/Virginia/ | Virginia Division of Consolidated Laboratory Services (DCLS) | Virginia Division of Consolidated Laboratory Services (DCLS) | Virginia DCLS et al |
| EPI_ISL_734283 | USA/VA-DCLS-2508/2020 | 2020-11-16 | North America/USA/Virginia/ | Virginia Division of Consolidated Laboratory Services (DCLS) | Virginia Division of Consolidated Laboratory Services (DCLS) | Virginia DCLS et al |
| EPI_ISL_734284 | USA/VA-DCLS-2509/2020 | 2020-11-16 | North America/USA/Virginia/ | Virginia Division of Consolidated Laboratory Services (DCLS) | Virginia Division of Consolidated Laboratory Services (DCLS) | Virginia DCLS et al |

| | | | | | | |
|---|---|---|---|---|---|---|
| EPI_ISL_734287 | USA/NY-Wadsworth-210263-01/2020 | 2020-09-03 | North America/USA/New York/Orleans | Wadsworth Center, New York State Department.of Health | Wadsworth Center, New York State Department.of Health | Kirsten St. George et al |
| EPI_ISL_734292 | USA/NY-Wadsworth-269599-01/2020 | 2020-11-10 | North America/USA/New York/Wyomin | Wadsworth Center, New York State Department.of Health | Wadsworth Center, New York State Department.of Health | Kirsten St. George et al |
| EPI_ISL_734295 | USA/NY-Wadsworth-269593-01/2020 | 2020-11-10 | North America/USA/New York/Wyomin | Wadsworth Center, New York State Department.of Health | Wadsworth Center, New York State Department.of Health | Kirsten St. George et al |
| EPI_ISL_734299 | USA/NY-Wadsworth-251003-01/2020 | 2020-10-21 | North America/USA/New York/Chemun | Wadsworth Center, New York State Department.of Health | Wadsworth Center, New York State Department.of Health | Kirsten St. George et al |
| EPI_ISL_734300 | USA/NY-Wadsworth-251080-01/2020 | 2020-10-21 | North America/USA/New York/Chemun | Wadsworth Center, New York State Department.of Health | Wadsworth Center, New York State Department.of Health | Kirsten St. George et al |
| EPI_ISL_734309 | USA/NY-Wadsworth-233280-01/2020 | 2020-10-01 | North America/USA/New York/Greene | Wadsworth Center, New York State Department.of Health | Wadsworth Center, New York State Department.of Health | Kirsten St. George et al |
| EPI_ISL_734320 | USA/NY-Wadsworth-237076-01/2020 | 2020-10-05 | North America/USA/New York/Greene | Wadsworth Center, New York State Department.of Health | Wadsworth Center, New York State Department.of Health | Kirsten St. George et al |
| EPI_ISL_734323 | USA/NY-Wadsworth-237088-01/2020 | 2020-10-05 | North America/USA/New York/Greene | Wadsworth Center, New York State Department.of Health | Wadsworth Center, New York State Department.of Health | Kirsten St. George et al |

| | | | | | | |
|---|---|---|---|---|---|---|
| EPI_ISL_734325 | USA/NY-Wadsworth-237093-01/2020 | 2020-10-05 | North America/USA/New York/Greene ( | Wadsworth Center, New York State Department.of Health | Wadsworth Center, New York State Department.of Health | Kirsten St. George et al |
| EPI_ISL_734329 | USA/NY-Wadsworth-241177-01/2020 | 2020-10-08 | North America/USA/New York/Albany ( | Wadsworth Center, New York State Department.of Health | Wadsworth Center, New York State Department.of Health | Kirsten St. George et al |
| EPI_ISL_734332 | USA/NY-Wadsworth-241579-01/2020 | 2020-10-09 | North America/USA/New York/Steuben | Wadsworth Center, New York State Department.of Health | Wadsworth Center, New York State Department.of Health | Kirsten St. George et al |
| EPI_ISL_734339 | USA/NY-Wadsworth-242694-01/2020 | 2020-10-11 | North America/USA/New York/Greene ( | Wadsworth Center, New York State Department.of Health | Wadsworth Center, New York State Department.of Health | Kirsten St. George et al |
| EPI_ISL_734345 | USA/NY-Wadsworth-242752-01/2020 | 2020-10-11 | North America/USA/New York/Greene ( | Wadsworth Center, New York State Department.of Health | Wadsworth Center, New York State Department.of Health | Kirsten St. George et al |
| EPI_ISL_734355 | USA/NY-Wadsworth-247518-01/2020 | 2020-10-17 | North America/USA/New York/Clinton ( | Wadsworth Center, New York State Department.of Health | Wadsworth Center, New York State Department.of Health | Kirsten St. George et al |
| EPI_ISL_734356 | USA/NY-Wadsworth-247525-01/2020 | 2020-10-17 | North America/USA/New York/Clinton ( | Wadsworth Center, New York State Department.of Health | Wadsworth Center, New York State Department.of Health | Kirsten St. George et al |
| EPI_ISL_734363 | USA/NY-Wadsworth-247867-01/2020 | 2020-10-17 | North America/USA/New York/Chemung | Wadsworth Center, New York State Department.of Health | Wadsworth Center, New York State Department.of Health | Kirsten St. George et al |

| | | | | | | |
|---|---|---|---|---|---|---|
| EPI_ISL_734365 | USA/NY-Wadsworth-248011-01/2020 | 2020-10-18 | North America/USA/New York/Chemung | Wadsworth Center, New York State Department.of Health | Wadsworth Center, New York State Department.of Health | Kirsten St. George et al |
| EPI_ISL_734369 | USA/NY-Wadsworth-248541-01/2020 | 2020-10-19 | North America/USA/New York/Greene ( | Wadsworth Center, New York State Department.of Health | Wadsworth Center, New York State Department.of Health | Kirsten St. George et al |
| EPI_ISL_734374 | USA/NY-Wadsworth-248789-01/2020 | 2020-10-19 | North America/USA/New York/Chemung | Wadsworth Center, New York State Department.of Health | Wadsworth Center, New York State Department.of Health | Kirsten St. George et al |
| EPI_ISL_734381 | USA/NY-Wadsworth-251287-01/2020 | 2020-10-20 | North America/USA/New York/Cayuga ( | Wadsworth Center, New York State Department.of Health | Wadsworth Center, New York State Department.of Health | Kirsten St. George et al |
| EPI_ISL_734383 | USA/NY-Wadsworth-251307-01/2020 | 2020-10-20 | North America/USA/New York/Cayuga ( | Wadsworth Center, New York State Department.of Health | Wadsworth Center, New York State Department.of Health | Kirsten St. George et al |
| EPI_ISL_734384 | USA/NY-Wadsworth-251308-01/2020 | 2020-10-20 | North America/USA/New York/Cayuga ( | Wadsworth Center, New York State Department.of Health | Wadsworth Center, New York State Department.of Health | Kirsten St. George et al |
| EPI_ISL_734388 | USA/NY-Wadsworth-251942-01/2020 | 2020-10-22 | North America/USA/New York/Greene ( | Wadsworth Center, New York State Department.of Health | Wadsworth Center, New York State Department.of Health | Kirsten St. George et al |
| EPI_ISL_734397 | USA/NY-Wadsworth-251975-01/2020 | 2020-10-22 | North America/USA/New York/Greene ( | Wadsworth Center, New York State Department.of Health | Wadsworth Center, New York State Department.of Health | Kirsten St. George et al |

| | | | | | | |
|---|---|---|---|---|---|---|
| EPI_ISL_734430 | USA/NY-Wadsworth-269609-01/2020 | 2020-11-10 | North America/USA/New York/Wyomin | Wadsworth Center, New York State Department.of Health | Wadsworth Center, New York State Department.of Health | Kirsten St. George et al |
| EPI_ISL_734432 | USA/NY-Wadsworth-269616-01/2020 | 2020-11-10 | North America/USA/New York/Wyomin | Wadsworth Center, New York State Department.of Health | Wadsworth Center, New York State Department.of Health | Kirsten St. George et al |
| EPI_ISL_734434 | USA/NY-Wadsworth-269621-01/2020 | 2020-11-10 | North America/USA/New York/Wyomin | Wadsworth Center, New York State Department.of Health | Wadsworth Center, New York State Department.of Health | Kirsten St. George et al |
| EPI_ISL_734439 | USA/NY-Wadsworth-269636-01/2020 | 2020-11-10 | North America/USA/New York/Wyomin | Wadsworth Center, New York State Department.of Health | Wadsworth Center, New York State Department.of Health | Kirsten St. George et al |
| EPI_ISL_734452 | USA/NY-Wadsworth-281382-01/2020 | 2020-10-15 | North America/USA/New York/Oneida ( | Masonic Medical Research Institute | Wadsworth Center, New York State Department.of Health | Kirsten St. George et al |
| EPI_ISL_734457 | USA/NY-Wadsworth-281388-01/2020 | 2020-10-20 | North America/USA/New York/Oneida ( | Masonic Medical Research Institute | Wadsworth Center, New York State Department.of Health | Kirsten St. George et al |
| EPI_ISL_734471 | USA/NY-Wadsworth-281405-01/2020 | 2020-11-03 | North America/USA/New York/Oneida ( | Masonic Medical Research Institute | Wadsworth Center, New York State Department.of Health | Kirsten St. George et al |
| EPI_ISL_734473 | USA/NY-Wadsworth-281407-01/2020 | 2020-11-03 | North America/USA/New York/Oneida ( | Masonic Medical Research Institute | Wadsworth Center, New York State Department.of Health | Kirsten St. George et al |

| | | | | | | |
|---|---|---|---|---|---|---|
| EPI_ISL_734474 | USA/NY-Wadsworth-281408-01/2020 | 2020-11-03 | North America/USA/New York/Oneida C | Masonic Medical Research Institute | Wadsworth Center, New York State Department.of Health | Kirsten St. George et al |
| EPI_ISL_734476 | USA/NY-Wadsworth-281411-01/2020 | 2020-11-05 | North America/USA/New York/Oneida C | Masonic Medical Research Institute | Wadsworth Center, New York State Department.of Health | Kirsten St. George et al |
| EPI_ISL_734477 | USA/NY-Wadsworth-281412-01/2020 | 2020-11-05 | North America/USA/New York/Onondag | Masonic Medical Research Institute | Wadsworth Center, New York State Department.of Health | Kirsten St. George et al |
| EPI_ISL_734484 | Belgium/rega-10191290/2020 | 2020-10-19 | Europe/Belgium/Turnhout/Merksplas | UZ Leuven, National Reference Laboratory for Coronaviruses, Laboratory Medicine, Leuven, Belgium | KU Leuven, Rega Institute, Clinical and Epidemiological Virology | Tony Wawina-Bokalanga et al |
| EPI_ISL_734497 | Belgium/rega-10021225/2020 | 2020-10-02 | Europe/Belgium/Leuven/Winksele | UZ Leuven, National Reference Laboratory for Coronaviruses, Laboratory Medicine, Leuven, Belgium | KU Leuven, Rega Institute, Clinical and Epidemiological Virology | Tony Wawina-Bokalanga et al |
| EPI_ISL_734498 | Belgium/rega-10191273/2020 | 2020-10-19 | Europe/Belgium/Leuven/Bost | UZ Leuven, National Reference Laboratory for Coronaviruses, Laboratory Medicine, Leuven, Belgium | KU Leuven, Rega Institute, Clinical and Epidemiological Virology | Tony Wawina-Bokalanga et al |
| EPI_ISL_734509 | Belgium/rega-0319860/2020 | 2020-03-19 | Europe/Belgium/Brussels/Auderghem | UZ Leuven, National Reference Laboratory for Coronaviruses, Laboratory Medicine, Leuven, Belgium | KU Leuven, Rega Institute, Clinical and Epidemiological Virology | Tony Wawina-Bokalanga et al |
| EPI_ISL_734514 | Belgium/rega-0322864/2020 | 2020-03-22 | Europe/Belgium/Nivelles/Chaumont-Gist | UZ Leuven, National Reference Laboratory for Coronaviruses, Laboratory Medicine, Leuven, Belgium | KU Leuven, Rega Institute, Clinical and Epidemiological Virology | Tony Wawina-Bokalanga et al |

| | | | | | | |
|---|---|---|---|---|---|---|
| EPI_ISL_734527 | Belgium/rega-0331877/2020 | 2020-03-31 | Europe/Belgium/Namur/Ligny | UZ Leuven, National Reference Laboratory for Coronaviruses, Laboratory Medicine, Leuven, Belgium | KU Leuven, Rega Institute, Clinical and Epidemiological Virology | Tony Wawina-Bokalanga et al |
| EPI_ISL_734532 | Belgium/rega-0403882/2020 | 2020-04-03 | Europe/Belgium/Nivelles/Beauvechain | UZ Leuven, National Reference Laboratory for Coronaviruses, Laboratory Medicine, Leuven, Belgium | KU Leuven, Rega Institute, Clinical and Epidemiological Virology | Tony Wawina-Bokalanga et al |
| EPI_ISL_734551 | Belgium/rega-0409561/2020 | 2020-04-09 | Europe/Belgium/Soignies/Braine-le-Com | UZ Leuven, National Reference Laboratory for Coronaviruses, Laboratory Medicine, Leuven, Belgium | KU Leuven, Rega Institute, Clinical and Epidemiological Virology | Tony Wawina-Bokalanga et al |
| EPI_ISL_734555 | Belgium/rega-0409565/2020 | 2020-04-09 | Europe/Belgium/Leuven/Tienen | UZ Leuven, National Reference Laboratory for Coronaviruses, Laboratory Medicine, Leuven, Belgium | KU Leuven, Rega Institute, Clinical and Epidemiological Virology | Tony Wawina-Bokalanga et al |
| EPI_ISL_734558 | Belgium/rega-0409568/2020 | 2020-04-09 | Europe/Belgium/Leuven/Tienen | UZ Leuven, National Reference Laboratory for Coronaviruses, Laboratory Medicine, Leuven, Belgium | KU Leuven, Rega Institute, Clinical and Epidemiological Virology | Tony Wawina-Bokalanga et al |
| EPI_ISL_734559 | Belgium/rega-0409569/2020 | 2020-04-09 | Europe/Belgium/Dinant/Maffe | UZ Leuven, National Reference Laboratory for Coronaviruses, Laboratory Medicine, Leuven, Belgium | KU Leuven, Rega Institute, Clinical and Epidemiological Virology | Tony Wawina-Bokalanga et al |
| EPI_ISL_734569 | Belgium/rega-0413577/2020 | 2020-04-13 | Europe/Belgium/Leuven/Tienen | UZ Leuven, National Reference Laboratory for Coronaviruses, Laboratory Medicine, Leuven, Belgium | KU Leuven, Rega Institute, Clinical and Epidemiological Virology | Tony Wawina-Bokalanga et al |
| EPI_ISL_734570 | Belgium/rega-0413578/2020 | 2020-04-13 | Europe/Belgium/Antwerpen/Boom | UZ Leuven, National Reference Laboratory for Coronaviruses, Laboratory Medicine, Leuven, Belgium | KU Leuven, Rega Institute, Clinical and Epidemiological Virology | Tony Wawina-Bokalanga et al |

| | | | | | | |
|---|---|---|---|---|---|---|
| EPI_ISL_734571 | Belgium/rega-0413579/2020 | 2020-04-13 | Europe/Belgium/Leuven/Aarschot | UZ Leuven, National Reference Laboratory for Coronaviruses, Laboratory Medicine, Leuven, Belgium | KU Leuven, Rega Institute, Clinical and Epidemiological Virology | Tony Wawina-Bokalanga et al |
| EPI_ISL_734574 | Belgium/rega-0414491/2020 | 2020-04-14 | Europe/Belgium/Brussels/Forest | UZ Leuven, National Reference Laboratory for Coronaviruses, Laboratory Medicine, Leuven, Belgium | KU Leuven, Rega Institute, Clinical and Epidemiological Virology | Tony Wawina-Bokalanga et al |
| EPI_ISL_734577 | Belgium/rega-0414494/2020 | 2020-04-14 | Europe/Belgium/Ath/ | UZ Leuven, National Reference Laboratory for Coronaviruses, Laboratory Medicine, Leuven, Belgium | KU Leuven, Rega Institute, Clinical and Epidemiological Virology | Tony Wawina-Bokalanga et al |
| EPI_ISL_734586 | Belgium/rega-0415905/2020 | 2020-04-15 | Europe/Belgium/Nivelles/Waterloo | UZ Leuven, National Reference Laboratory for Coronaviruses, Laboratory Medicine, Leuven, Belgium | KU Leuven, Rega Institute, Clinical and Epidemiological Virology | Tony Wawina-Bokalanga et al |
| EPI_ISL_734589 | Belgium/rega-0416497/2020 | 2020-04-16 | Europe/Belgium/Antwerpen/Niel | UZ Leuven, National Reference Laboratory for Coronaviruses, Laboratory Medicine, Leuven, Belgium | KU Leuven, Rega Institute, Clinical and Epidemiological Virology | Tony Wawina-Bokalanga et al |
| EPI_ISL_734592 | Belgium/rega-0416500/2020 | 2020-04-16 | Europe/Belgium/Leuven/Huldenberg | UZ Leuven, National Reference Laboratory for Coronaviruses, Laboratory Medicine, Leuven, Belgium | KU Leuven, Rega Institute, Clinical and Epidemiological Virology | Tony Wawina-Bokalanga et al |
| EPI_ISL_734599 | Belgium/rega-0417507/2020 | 2020-04-17 | Europe/Belgium/Hasselt/Sint-Truiden | UZ Leuven, National Reference Laboratory for Coronaviruses, Laboratory Medicine, Leuven, Belgium | KU Leuven, Rega Institute, Clinical and Epidemiological Virology | Tony Wawina-Bokalanga et al |
| EPI_ISL_734604 | Belgium/rega-0417512/2020 | 2020-04-17 | Europe/Belgium/Gent/ | UZ Leuven, National Reference Laboratory for Coronaviruses, Laboratory Medicine, Leuven, Belgium | KU Leuven, Rega Institute, Clinical and Epidemiological Virology | Tony Wawina-Bokalanga et al |

| | | | | | | |
|---|---|---|---|---|---|---|
| EPI_ISL_734609 | Belgium/rega-0417517/2020 | 2020-04-17 | Europe/Belgium/Leuven/Tervuren | UZ Leuven, National Reference Laboratory for Coronaviruses, Laboratory Medicine, Leuven, Belgium | KU Leuven, Rega Institute, Clinical and Epidemiological Virology | Tony Wawina-Bokalanga et al |
| EPI_ISL_734613 | Belgium/rega-0417521/2020 | 2020-04-17 | Europe/Belgium/Leuven/Kortenberg | UZ Leuven, National Reference Laboratory for Coronaviruses, Laboratory Medicine, Leuven, Belgium | KU Leuven, Rega Institute, Clinical and Epidemiological Virology | Tony Wawina-Bokalanga et al |
| EPI_ISL_734615 | Belgium/rega-0417524/2020 | 2020-04-17 | Europe/Belgium/Tournai-Mouscron/Mou | UZ Leuven, National Reference Laboratory for Coronaviruses, Laboratory Medicine, Leuven, Belgium | KU Leuven, Rega Institute, Clinical and Epidemiological Virology | Tony Wawina-Bokalanga et al |
| EPI_ISL_734616 | Belgium/rega-0417525/2020 | 2020-04-17 | Europe/Belgium/Tournai-Mouscron/Mou | UZ Leuven, National Reference Laboratory for Coronaviruses, Laboratory Medicine, Leuven, Belgium | KU Leuven, Rega Institute, Clinical and Epidemiological Virology | Tony Wawina-Bokalanga et al |
| EPI_ISL_734619 | Belgium/rega-0417908/2020 | 2020-04-17 | Europe/Belgium/Nivelles/Mont-Saint-Gu | UZ Leuven, National Reference Laboratory for Coronaviruses, Laboratory Medicine, Leuven, Belgium | KU Leuven, Rega Institute, Clinical and Epidemiological Virology | Tony Wawina-Bokalanga et al |
| EPI_ISL_734622 | Belgium/rega-0418529/2020 | 2020-04-18 | Europe/Belgium/Hasselt/Paal | UZ Leuven, National Reference Laboratory for Coronaviruses, Laboratory Medicine, Leuven, Belgium | KU Leuven, Rega Institute, Clinical and Epidemiological Virology | Tony Wawina-Bokalanga et al |
| EPI_ISL_734626 | Belgium/rega-0419533/2020 | 2020-04-19 | Europe/Belgium/Leuven/Tienen | UZ Leuven, National Reference Laboratory for Coronaviruses, Laboratory Medicine, Leuven, Belgium | KU Leuven, Rega Institute, Clinical and Epidemiological Virology | Tony Wawina-Bokalanga et al |
| EPI_ISL_734644 | Belgium/rega-0423597/2020 | 2020-04-23 | Europe/Belgium/Brussels/Anderlecht | UZ Leuven, National Reference Laboratory for Coronaviruses, Laboratory Medicine, Leuven, Belgium | KU Leuven, Rega Institute, Clinical and Epidemiological Virology | Tony Wawina-Bokalanga et al |

| | | | | | | |
|---|---|---|---|---|---|---|
| EPI_ISL_734669 | Belgium/rega-0427658/2020 | 2020-04-27 | Europe/Belgium/Tournai-Mouscron/Brur | UZ Leuven, National Reference Laboratory for Coronaviruses, Laboratory Medicine, Leuven, Belgium | KU Leuven, Rega Institute, Clinical and Epidemiological Virology | Tony Wawina-Bokalanga et al |
| EPI_ISL_734675 | Belgium/rega-0429622/2020 | 2020-04-29 | Europe/Belgium/Hasselt/Ham | UZ Leuven, National Reference Laboratory for Coronaviruses, Laboratory Medicine, Leuven, Belgium | KU Leuven, Rega Institute, Clinical and Epidemiological Virology | Tony Wawina-Bokalanga et al |
| EPI_ISL_734678 | Belgium/rega-0429625/2020 | 2020-04-29 | Europe/Belgium/Leuven/ | UZ Leuven, National Reference Laboratory for Coronaviruses, Laboratory Medicine, Leuven, Belgium | KU Leuven, Rega Institute, Clinical and Epidemiological Virology | Tony Wawina-Bokalanga et al |
| EPI_ISL_734686 | Belgium/rega-0429911/2020 | 2020-04-29 | Europe/Belgium/Namur/Jemeppe-sur-Sa | UZ Leuven, National Reference Laboratory for Coronaviruses, Laboratory Medicine, Leuven, Belgium | KU Leuven, Rega Institute, Clinical and Epidemiological Virology | Tony Wawina-Bokalanga et al |
| EPI_ISL_734687 | Belgium/rega-0430630/2020 | 2020-04-30 | Europe/Belgium/Leuven/ | UZ Leuven, National Reference Laboratory for Coronaviruses, Laboratory Medicine, Leuven, Belgium | KU Leuven, Rega Institute, Clinical and Epidemiological Virology | Tony Wawina-Bokalanga et al |
| EPI_ISL_734688 | Belgium/rega-0430631/2020 | 2020-04-30 | Europe/Belgium/Leuven/Averbode | UZ Leuven, National Reference Laboratory for Coronaviruses, Laboratory Medicine, Leuven, Belgium | KU Leuven, Rega Institute, Clinical and Epidemiological Virology | Tony Wawina-Bokalanga et al |
| EPI_ISL_734690 | Belgium/rega-0501633/2020 | 2020-05-01 | Europe/Belgium/Tournai-Mouscron/Mou | UZ Leuven, National Reference Laboratory for Coronaviruses, Laboratory Medicine, Leuven, Belgium | KU Leuven, Rega Institute, Clinical and Epidemiological Virology | Tony Wawina-Bokalanga et al |
| EPI_ISL_734694 | Belgium/rega-0501666/2020 | 2020-05-01 | Europe/Belgium/Tournai-Mouscron/Her | UZ Leuven, National Reference Laboratory for Coronaviruses, Laboratory Medicine, Leuven, Belgium | KU Leuven, Rega Institute, Clinical and Epidemiological Virology | Tony Wawina-Bokalanga et al |

| | | | | | | |
|---|---|---|---|---|---|---|
| EPI_ISL_734696 | Belgium/rega-0502638/2020 | 2020-05-02 | Europe/Belgium/Ath/Quevaucamps | UZ Leuven, National Reference Laboratory for Coronaviruses, Laboratory Medicine, Leuven, Belgium | KU Leuven, Rega Institute, Clinical and Epidemiological Virology | Tony Wawina-Bokalanga et al |
| EPI_ISL_734709 | Belgium/rega-0507539/2020 | 2020-05-07 | Europe/Belgium/Nivelles/Waterloo | UZ Leuven, National Reference Laboratory for Coronaviruses, Laboratory Medicine, Leuven, Belgium | KU Leuven, Rega Institute, Clinical and Epidemiological Virology | Tony Wawina-Bokalanga et al |
| EPI_ISL_734715 | Belgium/rega-0508546/2020 | 2020-05-08 | Europe/Belgium/Leuven/Wilsele | UZ Leuven, National Reference Laboratory for Coronaviruses, Laboratory Medicine, Leuven, Belgium | KU Leuven, Rega Institute, Clinical and Epidemiological Virology | Tony Wawina-Bokalanga et al |
| EPI_ISL_734723 | Belgium/rega-0509652/2020 | 2020-05-09 | Europe/Belgium/Tournai-Mouscron/Gau | UZ Leuven, National Reference Laboratory for Coronaviruses, Laboratory Medicine, Leuven, Belgium | KU Leuven, Rega Institute, Clinical and Epidemiological Virology | Tony Wawina-Bokalanga et al |
| EPI_ISL_734726 | Belgium/rega-0512550/2020 | 2020-05-12 | Europe/Belgium/Dendermonde/Zele | UZ Leuven, National Reference Laboratory for Coronaviruses, Laboratory Medicine, Leuven, Belgium | KU Leuven, Rega Institute, Clinical and Epidemiological Virology | Tony Wawina-Bokalanga et al |
| EPI_ISL_734730 | Belgium/rega-0513668/2020 | 2020-05-13 | Europe/Belgium/Leuven/Wilsele | UZ Leuven, National Reference Laboratory for Coronaviruses, Laboratory Medicine, Leuven, Belgium | KU Leuven, Rega Institute, Clinical and Epidemiological Virology | Tony Wawina-Bokalanga et al |
| EPI_ISL_734731 | Belgium/rega-0514551/2020 | 2020-05-14 | Europe/Belgium/Leuven/Bekkevoort | UZ Leuven, National Reference Laboratory for Coronaviruses, Laboratory Medicine, Leuven, Belgium | KU Leuven, Rega Institute, Clinical and Epidemiological Virology | Tony Wawina-Bokalanga et al |
| EPI_ISL_734738 | Belgium/rega-0517554/2020 | 2020-05-17 | Europe/Belgium/Turnhout/Westerlo | UZ Leuven, National Reference Laboratory for Coronaviruses, Laboratory Medicine, Leuven, Belgium | KU Leuven, Rega Institute, Clinical and Epidemiological Virology | Tony Wawina-Bokalanga et al |

| | | | | | | |
|---|---|---|---|---|---|---|
| EPI_ISL_734744 | Belgium/rega-0526920/2020 | 2020-05-26 | Europe/Belgium/Leuven/ | UZ Leuven, National Reference Laboratory for Coronaviruses, Laboratory Medicine, Leuven, Belgium | KU Leuven, Rega Institute, Clinical and Epidemiological Virology | Tony Wawina-Bokalanga et al |
| EPI_ISL_734745 | Belgium/rega-0528690/2020 | 2020-05-28 | Europe/Belgium/Hasselt/Stevoort | UZ Leuven, National Reference Laboratory for Coronaviruses, Laboratory Medicine, Leuven, Belgium | KU Leuven, Rega Institute, Clinical and Epidemiological Virology | Tony Wawina-Bokalanga et al |
| EPI_ISL_734753 | Belgium/rega-0615692/2020 | 2020-06-15 | Europe/Belgium/Leuven/ | UZ Leuven, National Reference Laboratory for Coronaviruses, Laboratory Medicine, Leuven, Belgium | KU Leuven, Rega Institute, Clinical and Epidemiological Virology | Tony Wawina-Bokalanga et al |
| EPI_ISL_734754 | Belgium/rega-0615693/2020 | 2020-06-15 | Europe/Belgium/Leuven/Tervuren | UZ Leuven, National Reference Laboratory for Coronaviruses, Laboratory Medicine, Leuven, Belgium | KU Leuven, Rega Institute, Clinical and Epidemiological Virology | Tony Wawina-Bokalanga et al |
| EPI_ISL_734757 | Belgium/rega-0616695/2020 | 2020-06-16 | Europe/Belgium/Leuven/Kortenberg | UZ Leuven, National Reference Laboratory for Coronaviruses, Laboratory Medicine, Leuven, Belgium | KU Leuven, Rega Institute, Clinical and Epidemiological Virology | Tony Wawina-Bokalanga et al |
| EPI_ISL_734762 | Belgium/rega-0616835/2020 | 2020-06-16 | Europe/Belgium/Kortrijk/Harelbeke | UZ Leuven, National Reference Laboratory for Coronaviruses, Laboratory Medicine, Leuven, Belgium | KU Leuven, Rega Institute, Clinical and Epidemiological Virology | Tony Wawina-Bokalanga et al |
| EPI_ISL_734774 | Belgium/rega-0620851/2020 | 2020-06-20 | Europe/Belgium/Antwerpen/ | UZ Leuven, National Reference Laboratory for Coronaviruses, Laboratory Medicine, Leuven, Belgium | KU Leuven, Rega Institute, Clinical and Epidemiological Virology | Tony Wawina-Bokalanga et al |
| EPI_ISL_734818 | Belgium/rega-0722814/2020 | 2020-07-22 | Europe/Belgium/Turnhout/Laakdal | UZ Leuven, National Reference Laboratory for Coronaviruses, Laboratory Medicine, Leuven, Belgium | KU Leuven, Rega Institute, Clinical and Epidemiological Virology | Tony Wawina-Bokalanga et al |

| | | | | | | |
|---|---|---|---|---|---|---|
| EPI_ISL_734821 | Belgium/rega-0722817/2020 | 2020-07-22 | Europe/Belgium/Halle-Vilvoorde/Sint-G | UZ Leuven, National Reference Laboratory for Coronaviruses, Laboratory Medicine, Leuven, Belgium | KU Leuven, Rega Institute, Clinical and Epidemiological Virology | Tony Wawina-Bokalanga et al |
| EPI_ISL_734836 | Belgium/rega-0731789/2020 | 2020-07-31 | Europe/Belgium/Antwerpen/ | UZ Leuven, National Reference Laboratory for Coronaviruses, Laboratory Medicine, Leuven, Belgium | KU Leuven, Rega Institute, Clinical and Epidemiological Virology | Tony Wawina-Bokalanga et al |
| EPI_ISL_734846 | Belgium/rega-0801769/2020 | 2020-08-01 | Europe/Belgium/Antwerpen/Merksem | UZ Leuven, National Reference Laboratory for Coronaviruses, Laboratory Medicine, Leuven, Belgium | KU Leuven, Rega Institute, Clinical and Epidemiological Virology | Tony Wawina-Bokalanga et al |
| EPI_ISL_734847 | Belgium/rega-0801770/2020 | 2020-08-01 | Europe/Belgium/Brussels/Evere | UZ Leuven, National Reference Laboratory for Coronaviruses, Laboratory Medicine, Leuven, Belgium | KU Leuven, Rega Institute, Clinical and Epidemiological Virology | Tony Wawina-Bokalanga et al |
| EPI_ISL_734848 | Belgium/rega-0801771/2020 | 2020-08-01 | Europe/Belgium/Brussels/Schaerbeek | UZ Leuven, National Reference Laboratory for Coronaviruses, Laboratory Medicine, Leuven, Belgium | KU Leuven, Rega Institute, Clinical and Epidemiological Virology | Tony Wawina-Bokalanga et al |
| EPI_ISL_734856 | Belgium/rega-0801793/2020 | 2020-08-01 | Europe/Belgium/Sint-Niklaas/ | UZ Leuven, National Reference Laboratory for Coronaviruses, Laboratory Medicine, Leuven, Belgium | KU Leuven, Rega Institute, Clinical and Epidemiological Virology | Tony Wawina-Bokalanga et al |
| EPI_ISL_734861 | Belgium/rega-0801798/2020 | 2020-08-01 | Europe/Belgium/Antwerpen/Ranst | UZ Leuven, National Reference Laboratory for Coronaviruses, Laboratory Medicine, Leuven, Belgium | KU Leuven, Rega Institute, Clinical and Epidemiological Virology | Tony Wawina-Bokalanga et al |
| EPI_ISL_734900 | Belgium/rega-0803757/2020 | 2020-08-03 | Europe/Belgium/Antwerpen/Merksem | UZ Leuven, National Reference Laboratory for Coronaviruses, Laboratory Medicine, Leuven, Belgium | KU Leuven, Rega Institute, Clinical and Epidemiological Virology | Tony Wawina-Bokalanga et al |

| | | | | | | |
|---|---|---|---|---|---|---|
| EPI_ISL_734909 | Belgium/rega-0804729/2020 | 2020-08-04 | Europe/Belgium/Antwerpen/Deurne | UZ Leuven, National Reference Laboratory for Coronaviruses, Laboratory Medicine, Leuven, Belgium | KU Leuven, Rega Institute, Clinical and Epidemiological Virology | Tony Wawina-Bokalanga et al |
| EPI_ISL_734917 | Belgium/rega-0908933/2020 | 2020-09-08 | Europe/Belgium/Leuven/Wilsele | UZ Leuven, National Reference Laboratory for Coronaviruses, Laboratory Medicine, Leuven, Belgium | KU Leuven, Rega Institute, Clinical and Epidemiological Virology | Tony Wawina-Bokalanga et al |
| EPI_ISL_734921 | Belgium/rega-0909932/2020 | 2020-09-09 | Europe/Belgium/Leuven/Bierbeek | UZ Leuven, National Reference Laboratory for Coronaviruses, Laboratory Medicine, Leuven, Belgium | KU Leuven, Rega Institute, Clinical and Epidemiological Virology | Tony Wawina-Bokalanga et al |
| EPI_ISL_734934 | Belgium/rega-0914947/2020 | 2020-09-14 | Europe/Belgium/Halle-Vilvoorde/Zavent | UZ Leuven, National Reference Laboratory for Coronaviruses, Laboratory Medicine, Leuven, Belgium | KU Leuven, Rega Institute, Clinical and Epidemiological Virology | Tony Wawina-Bokalanga et al |
| EPI_ISL_734936 | Belgium/rega-09151223/2020 | 2020-09-15 | Europe/Belgium/Leuven/Tervuren | UZ Leuven, National Reference Laboratory for Coronaviruses, Laboratory Medicine, Leuven, Belgium | KU Leuven, Rega Institute, Clinical and Epidemiological Virology | Tony Wawina-Bokalanga et al |
| EPI_ISL_734944 | Belgium/rega-0915954/2020 | 2020-09-15 | Europe/Belgium/Turnhout/Geel | UZ Leuven, National Reference Laboratory for Coronaviruses, Laboratory Medicine, Leuven, Belgium | KU Leuven, Rega Institute, Clinical and Epidemiological Virology | Tony Wawina-Bokalanga et al |
| EPI_ISL_734947 | Belgium/rega-0915957/2020 | 2020-09-15 | Europe/Belgium/Halle-Vilvoorde/Overijs | UZ Leuven, National Reference Laboratory for Coronaviruses, Laboratory Medicine, Leuven, Belgium | KU Leuven, Rega Institute, Clinical and Epidemiological Virology | Tony Wawina-Bokalanga et al |
| EPI_ISL_734953 | Belgium/rega-0916960/2020 | 2020-09-16 | Europe/Belgium/Leuven/Kessel-Lo | UZ Leuven, National Reference Laboratory for Coronaviruses, Laboratory Medicine, Leuven, Belgium | KU Leuven, Rega Institute, Clinical and Epidemiological Virology | Tony Wawina-Bokalanga et al |

| | | | | | | |
|---|---|---|---|---|---|---|
| EPI_ISL_734961 | Belgium/rega-0916971/2020 | 2020-09-16 | Europe/Belgium/Leuven/Begijnendijk | UZ Leuven, National Reference Laboratory for Coronaviruses, Laboratory Medicine, Leuven, Belgium | KU Leuven, Rega Institute, Clinical and Epidemiological Virology | Tony Wawina-Bokalanga et al |
| EPI_ISL_734965 | Belgium/rega-0916975/2020 | 2020-09-16 | Europe/Belgium/Halle-Vilvoorde/Steenc | UZ Leuven, National Reference Laboratory for Coronaviruses, Laboratory Medicine, Leuven, Belgium | KU Leuven, Rega Institute, Clinical and Epidemiological Virology | Tony Wawina-Bokalanga et al |
| EPI_ISL_734967 | Belgium/rega-0916977/2020 | 2020-09-16 | Europe/Belgium/Halle-Vilvoorde/Hoeila | UZ Leuven, National Reference Laboratory for Coronaviruses, Laboratory Medicine, Leuven, Belgium | KU Leuven, Rega Institute, Clinical and Epidemiological Virology | Tony Wawina-Bokalanga et al |
| EPI_ISL_734969 | Belgium/rega-0916979/2020 | 2020-09-16 | Europe/Belgium/Leuven/Wilsele | UZ Leuven, National Reference Laboratory for Coronaviruses, Laboratory Medicine, Leuven, Belgium | KU Leuven, Rega Institute, Clinical and Epidemiological Virology | Tony Wawina-Bokalanga et al |
| EPI_ISL_734973 | Belgium/rega-09171048/2020 | 2020-09-17 | Europe/Belgium/Leuven/Tervuren | UZ Leuven, National Reference Laboratory for Coronaviruses, Laboratory Medicine, Leuven, Belgium | KU Leuven, Rega Institute, Clinical and Epidemiological Virology | Tony Wawina-Bokalanga et al |
| EPI_ISL_734976 | Belgium/rega-0917984/2020 | 2020-09-17 | Europe/Belgium/Halle-Vilvoorde/Steenc | UZ Leuven, National Reference Laboratory for Coronaviruses, Laboratory Medicine, Leuven, Belgium | KU Leuven, Rega Institute, Clinical and Epidemiological Virology | Tony Wawina-Bokalanga et al |
| EPI_ISL_734992 | Belgium/rega-09181049/2020 | 2020-09-18 | Europe/Belgium/Leuven/Bost | UZ Leuven, National Reference Laboratory for Coronaviruses, Laboratory Medicine, Leuven, Belgium | KU Leuven, Rega Institute, Clinical and Epidemiological Virology | Tony Wawina-Bokalanga et al |
| EPI_ISL_734997 | Belgium/rega-09181055/2020 | 2020-09-18 | Europe/Belgium/Halle-Vilvoorde/Steenc | UZ Leuven, National Reference Laboratory for Coronaviruses, Laboratory Medicine, Leuven, Belgium | KU Leuven, Rega Institute, Clinical and Epidemiological Virology | Tony Wawina-Bokalanga et al |

| | | | | | | |
|---|---|---|---|---|---|---|
| EPI_ISL_735010 | Belgium/rega-09221012/2020 | 2020-09-22 | Europe/Belgium/Leuven/Heverlee | UZ Leuven, National Reference Laboratory for Coronaviruses, Laboratory Medicine, Leuven, Belgium | KU Leuven, Rega Institute, Clinical and Epidemiological Virology | Tony Wawina-Bokalanga et al |
| EPI_ISL_735012 | Belgium/rega-09221014/2020 | 2020-09-22 | Europe/Belgium/Leuven/Haacht | UZ Leuven, National Reference Laboratory for Coronaviruses, Laboratory Medicine, Leuven, Belgium | KU Leuven, Rega Institute, Clinical and Epidemiological Virology | Tony Wawina-Bokalanga et al |
| EPI_ISL_735015 | Belgium/rega-09221017/2020 | 2020-09-22 | Europe/Belgium/Leuven/Bost | UZ Leuven, National Reference Laboratory for Coronaviruses, Laboratory Medicine, Leuven, Belgium | KU Leuven, Rega Institute, Clinical and Epidemiological Virology | Tony Wawina-Bokalanga et al |
| EPI_ISL_735016 | Belgium/rega-09221018/2020 | 2020-09-22 | Europe/Belgium/Leuven/Bost | UZ Leuven, National Reference Laboratory for Coronaviruses, Laboratory Medicine, Leuven, Belgium | KU Leuven, Rega Institute, Clinical and Epidemiological Virology | Tony Wawina-Bokalanga et al |
| EPI_ISL_735020 | Belgium/rega-09221022/2020 | 2020-09-22 | Europe/Belgium/Leuven/Bertem | UZ Leuven, National Reference Laboratory for Coronaviruses, Laboratory Medicine, Leuven, Belgium | KU Leuven, Rega Institute, Clinical and Epidemiological Virology | Tony Wawina-Bokalanga et al |
| EPI_ISL_735035 | Belgium/rega-09231248/2020 | 2020-09-23 | Europe/Belgium/Leuven/Kessel-Lo | UZ Leuven, National Reference Laboratory for Coronaviruses, Laboratory Medicine, Leuven, Belgium | KU Leuven, Rega Institute, Clinical and Epidemiological Virology | Tony Wawina-Bokalanga et al |
| EPI_ISL_735046 | Belgium/rega-10031028/2020 | 2020-10-03 | Europe/Belgium/Leuven/Heverlee | UZ Leuven, National Reference Laboratory for Coronaviruses, Laboratory Medicine, Leuven, Belgium | KU Leuven, Rega Institute, Clinical and Epidemiological Virology | Tony Wawina-Bokalanga et al |
| EPI_ISL_735049 | Belgium/rega-10061009/2020 | 2020-10-06 | Europe/Belgium/Halle-Vilvoorde/Overijs | UZ Leuven, National Reference Laboratory for Coronaviruses, Laboratory Medicine, Leuven, Belgium | KU Leuven, Rega Institute, Clinical and Epidemiological Virology | Tony Wawina-Bokalanga et al |

| | | | | | | |
|---|---|---|---|---|---|---|
| EPI_ISL_735050 | Belgium/rega-10061010/2020 | 2020-10-06 | Europe/Belgium/Halle-Vilvoorde/Overijs | UZ Leuven, National Reference Laboratory for Coronaviruses, Laboratory Medicine, Leuven, Belgium | KU Leuven, Rega Institute, Clinical and Epidemiological Virology | Tony Wawina-Bokalanga et al |
| EPI_ISL_735056 | Belgium/rega-10061035/2020 | 2020-10-06 | Europe/Belgium/Halle-Vilvoorde/Hoeila | UZ Leuven, National Reference Laboratory for Coronaviruses, Laboratory Medicine, Leuven, Belgium | KU Leuven, Rega Institute, Clinical and Epidemiological Virology | Tony Wawina-Bokalanga et al |
| EPI_ISL_735059 | Belgium/rega-10061039/2020 | 2020-10-06 | Europe/Belgium/Leuven/ | UZ Leuven, National Reference Laboratory for Coronaviruses, Laboratory Medicine, Leuven, Belgium | KU Leuven, Rega Institute, Clinical and Epidemiological Virology | Tony Wawina-Bokalanga et al |
| EPI_ISL_735060 | Belgium/rega-10061040/2020 | 2020-10-06 | Europe/Belgium/Leuven/Haasrode | UZ Leuven, National Reference Laboratory for Coronaviruses, Laboratory Medicine, Leuven, Belgium | KU Leuven, Rega Institute, Clinical and Epidemiological Virology | Tony Wawina-Bokalanga et al |
| EPI_ISL_735061 | Belgium/rega-10061041/2020 | 2020-10-06 | Europe/Belgium/Halle-Vilvoorde/Overijs | UZ Leuven, National Reference Laboratory for Coronaviruses, Laboratory Medicine, Leuven, Belgium | KU Leuven, Rega Institute, Clinical and Epidemiological Virology | Tony Wawina-Bokalanga et al |
| EPI_ISL_735065 | Belgium/rega-10061045/2020 | 2020-10-06 | Europe/Belgium/Leuven/ | UZ Leuven, National Reference Laboratory for Coronaviruses, Laboratory Medicine, Leuven, Belgium | KU Leuven, Rega Institute, Clinical and Epidemiological Virology | Tony Wawina-Bokalanga et al |
| EPI_ISL_735066 | Belgium/rega-10061046/2020 | 2020-10-06 | Europe/Belgium/Halle-Vilvoorde/Steen | UZ Leuven, National Reference Laboratory for Coronaviruses, Laboratory Medicine, Leuven, Belgium | KU Leuven, Rega Institute, Clinical and Epidemiological Virology | Tony Wawina-Bokalanga et al |
| EPI_ISL_735067 | Belgium/rega-10061193/2020 | 2020-10-06 | Europe/Belgium/Leuven/Bost | UZ Leuven, National Reference Laboratory for Coronaviruses, Laboratory Medicine, Leuven, Belgium | KU Leuven, Rega Institute, Clinical and Epidemiological Virology | Tony Wawina-Bokalanga et al |

| | | | | | | |
|---|---|---|---|---|---|---|
| EPI_ISL_735068 | Belgium/rega-10061194/2020 | 2020-10-06 | Europe/Belgium/Leuven/Bost | UZ Leuven, National Reference Laboratory for Coronaviruses, Laboratory Medicine, Leuven, Belgium | KU Leuven, Rega Institute, Clinical and Epidemiological Virology | Tony Wawina-Bokalanga et al |
| EPI_ISL_735069 | Belgium/rega-10061195/2020 | 2020-10-06 | Europe/Belgium/Halle-Vilvoorde/Wezen | UZ Leuven, National Reference Laboratory for Coronaviruses, Laboratory Medicine, Leuven, Belgium | KU Leuven, Rega Institute, Clinical and Epidemiological Virology | Tony Wawina-Bokalanga et al |
| EPI_ISL_735071 | Belgium/rega-10061197/2020 | 2020-10-06 | Europe/Belgium/Leuven/Tervuren | UZ Leuven, National Reference Laboratory for Coronaviruses, Laboratory Medicine, Leuven, Belgium | KU Leuven, Rega Institute, Clinical and Epidemiological Virology | Tony Wawina-Bokalanga et al |
| EPI_ISL_735072 | Belgium/rega-10061198/2020 | 2020-10-06 | Europe/Belgium/Leuven/Winksele | UZ Leuven, National Reference Laboratory for Coronaviruses, Laboratory Medicine, Leuven, Belgium | KU Leuven, Rega Institute, Clinical and Epidemiological Virology | Tony Wawina-Bokalanga et al |
| EPI_ISL_735074 | Belgium/rega-10061255/2020 | 2020-10-06 | Europe/Belgium/Halle-Vilvoorde/Overijs | UZ Leuven, National Reference Laboratory for Coronaviruses, Laboratory Medicine, Leuven, Belgium | KU Leuven, Rega Institute, Clinical and Epidemiological Virology | Tony Wawina-Bokalanga et al |
| EPI_ISL_735078 | Belgium/rega-10071243/2020 | 2020-10-07 | Europe/Belgium/Halle-Vilvoorde/Hoeila | UZ Leuven, National Reference Laboratory for Coronaviruses, Laboratory Medicine, Leuven, Belgium | KU Leuven, Rega Institute, Clinical and Epidemiological Virology | Tony Wawina-Bokalanga et al |
| EPI_ISL_735079 | Belgium/rega-10071249/2020 | 2020-10-07 | Europe/Belgium/Leuven/Heverlee | UZ Leuven, National Reference Laboratory for Coronaviruses, Laboratory Medicine, Leuven, Belgium | KU Leuven, Rega Institute, Clinical and Epidemiological Virology | Tony Wawina-Bokalanga et al |
| EPI_ISL_735082 | Belgium/rega-10081220/2020 | 2020-10-08 | Europe/Belgium/Turnhout/Kasterlee | UZ Leuven, National Reference Laboratory for Coronaviruses, Laboratory Medicine, Leuven, Belgium | KU Leuven, Rega Institute, Clinical and Epidemiological Virology | Tony Wawina-Bokalanga et al |

| | | | | | | |
|---|---|---|---|---|---|---|
| EPI_ISL_735088 | Belgium/rega-10081239/2020 | 2020-10-08 | Europe/Belgium/Brussels/Woluwe-Saint | UZ Leuven, National Reference Laboratory for Coronaviruses, Laboratory Medicine, Leuven, Belgium | KU Leuven, Rega Institute, Clinical and Epidemiological Virology | Tony Wawina-Bokalanga et al |
| EPI_ISL_735090 | Belgium/rega-10081245/2020 | 2020-10-08 | Europe/Belgium/Halle-Vilvoorde/Hoeila | UZ Leuven, National Reference Laboratory for Coronaviruses, Laboratory Medicine, Leuven, Belgium | KU Leuven, Rega Institute, Clinical and Epidemiological Virology | Tony Wawina-Bokalanga et al |
| EPI_ISL_735094 | Belgium/rega-10081257/2020 | 2020-10-08 | Europe/Belgium/Turnhout/Oud-Turnhou | UZ Leuven, National Reference Laboratory for Coronaviruses, Laboratory Medicine, Leuven, Belgium | KU Leuven, Rega Institute, Clinical and Epidemiological Virology | Tony Wawina-Bokalanga et al |
| EPI_ISL_735111 | Belgium/rega-10181071/2020 | 2020-10-18 | Europe/Belgium/Halle-Vilvoorde/Overijs | UZ Leuven, National Reference Laboratory for Coronaviruses, Laboratory Medicine, Leuven, Belgium | KU Leuven, Rega Institute, Clinical and Epidemiological Virology | Tony Wawina-Bokalanga et al |
| EPI_ISL_735116 | Belgium/rega-10181076/2020 | 2020-10-18 | Europe/Belgium/Leuven/ | UZ Leuven, National Reference Laboratory for Coronaviruses, Laboratory Medicine, Leuven, Belgium | KU Leuven, Rega Institute, Clinical and Epidemiological Virology | Tony Wawina-Bokalanga et al |
| EPI_ISL_735118 | Belgium/rega-10181078/2020 | 2020-10-16 | Europe/Belgium/Leuven/Tervuren | UZ Leuven, National Reference Laboratory for Coronaviruses, Laboratory Medicine, Leuven, Belgium | KU Leuven, Rega Institute, Clinical and Epidemiological Virology | Tony Wawina-Bokalanga et al |
| EPI_ISL_735124 | Belgium/rega-10181084/2020 | 2020-10-18 | Europe/Belgium/Leuven/Winksele | UZ Leuven, National Reference Laboratory for Coronaviruses, Laboratory Medicine, Leuven, Belgium | KU Leuven, Rega Institute, Clinical and Epidemiological Virology | Tony Wawina-Bokalanga et al |
| EPI_ISL_735125 | Belgium/rega-10181085/2020 | 2020-10-18 | Europe/Belgium/Leuven/Kessel-Lo | UZ Leuven, National Reference Laboratory for Coronaviruses, Laboratory Medicine, Leuven, Belgium | KU Leuven, Rega Institute, Clinical and Epidemiological Virology | Tony Wawina-Bokalanga et al |

| | | | | | | |
|---|---|---|---|---|---|---|
| EPI_ISL_735127 | Belgium/rega-10181087/2020 | 2020-10-18 | Europe/Belgium/Leuven/Kessel-Lo | UZ Leuven, National Reference Laboratory for Coronaviruses, Laboratory Medicine, Leuven, Belgium | KU Leuven, Rega Institute, Clinical and Epidemiological Virology | Tony Wawina-Bokalanga et al |
| EPI_ISL_735134 | Belgium/rega-10181113/2020 | 2020-10-18 | Europe/Belgium/Leuven/Kessel-Lo | UZ Leuven, National Reference Laboratory for Coronaviruses, Laboratory Medicine, Leuven, Belgium | KU Leuven, Rega Institute, Clinical and Epidemiological Virology | Tony Wawina-Bokalanga et al |
| EPI_ISL_735136 | Belgium/rega-10181164/2020 | 2020-10-18 | Europe/Belgium/Leuven/ | UZ Leuven, National Reference Laboratory for Coronaviruses, Laboratory Medicine, Leuven, Belgium | KU Leuven, Rega Institute, Clinical and Epidemiological Virology | Tony Wawina-Bokalanga et al |
| EPI_ISL_735147 | Belgium/rega-10191100/2020 | 2020-10-19 | Europe/Belgium/Leuven/Bunsbeek | UZ Leuven, National Reference Laboratory for Coronaviruses, Laboratory Medicine, Leuven, Belgium | KU Leuven, Rega Institute, Clinical and Epidemiological Virology | Tony Wawina-Bokalanga et al |
| EPI_ISL_735148 | Belgium/rega-10191101/2020 | 2020-10-19 | Europe/Belgium/Antwerpen/ | UZ Leuven, National Reference Laboratory for Coronaviruses, Laboratory Medicine, Leuven, Belgium | KU Leuven, Rega Institute, Clinical and Epidemiological Virology | Tony Wawina-Bokalanga et al |
| EPI_ISL_735149 | Belgium/rega-10191102/2020 | 2020-10-19 | Europe/Belgium/Leuven/Tervuren | UZ Leuven, National Reference Laboratory for Coronaviruses, Laboratory Medicine, Leuven, Belgium | KU Leuven, Rega Institute, Clinical and Epidemiological Virology | Tony Wawina-Bokalanga et al |
| EPI_ISL_735152 | Belgium/rega-10191109/2020 | 2020-10-19 | Europe/Belgium/Leuven/Bost | UZ Leuven, National Reference Laboratory for Coronaviruses, Laboratory Medicine, Leuven, Belgium | KU Leuven, Rega Institute, Clinical and Epidemiological Virology | Tony Wawina-Bokalanga et al |
| EPI_ISL_735154 | Belgium/rega-10191123/2020 | 2020-10-19 | Europe/Belgium/Turnhout/Rijkevorsel | UZ Leuven, National Reference Laboratory for Coronaviruses, Laboratory Medicine, Leuven, Belgium | KU Leuven, Rega Institute, Clinical and Epidemiological Virology | Tony Wawina-Bokalanga et al |

| | | | | | | |
|---|---|---|---|---|---|---|
| EPI_ISL_735156 | Belgium/rega-10191125/2020 | 2020-10-19 | Europe/Belgium/Hasselt/Nieuwerkerken | UZ Leuven, National Reference Laboratory for Coronaviruses, Laboratory Medicine, Leuven, Belgium | KU Leuven, Rega Institute, Clinical and Epidemiological Virology | Tony Wawina-Bokalanga et al |
| EPI_ISL_735159 | Belgium/rega-10191128/2020 | 2020-10-19 | Europe/Belgium/Liv®ge/Ans | UZ Leuven, National Reference Laboratory for Coronaviruses, Laboratory Medicine, Leuven, Belgium | KU Leuven, Rega Institute, Clinical and Epidemiological Virology | Tony Wawina-Bokalanga et al |
| EPI_ISL_735161 | Belgium/rega-10191130/2020 | 2020-10-16 | Europe/Belgium/Halle-Vilvoorde/Hoeila | UZ Leuven, National Reference Laboratory for Coronaviruses, Laboratory Medicine, Leuven, Belgium | KU Leuven, Rega Institute, Clinical and Epidemiological Virology | Tony Wawina-Bokalanga et al |
| EPI_ISL_735164 | Belgium/rega-10191228/2020 | 2020-10-19 | Europe/Belgium/Mechelen/Schriek | UZ Leuven, National Reference Laboratory for Coronaviruses, Laboratory Medicine, Leuven, Belgium | KU Leuven, Rega Institute, Clinical and Epidemiological Virology | Tony Wawina-Bokalanga et al |
| EPI_ISL_735170 | Belgium/rega-10191259/2020 | 2020-10-19 | Europe/Belgium/Halle-Vilvoorde/Kraain | UZ Leuven, National Reference Laboratory for Coronaviruses, Laboratory Medicine, Leuven, Belgium | KU Leuven, Rega Institute, Clinical and Epidemiological Virology | Tony Wawina-Bokalanga et al |
| EPI_ISL_735172 | Belgium/rega-10191262/2020 | 2020-10-19 | Europe/Belgium/Leuven/Bost | UZ Leuven, National Reference Laboratory for Coronaviruses, Laboratory Medicine, Leuven, Belgium | KU Leuven, Rega Institute, Clinical and Epidemiological Virology | Tony Wawina-Bokalanga et al |
| EPI_ISL_735174 | Belgium/rega-10191264/2020 | 2020-10-19 | Europe/Belgium/Leuven/Kessel-Lo | UZ Leuven, National Reference Laboratory for Coronaviruses, Laboratory Medicine, Leuven, Belgium | KU Leuven, Rega Institute, Clinical and Epidemiological Virology | Tony Wawina-Bokalanga et al |
| EPI_ISL_735182 | Belgium/rega-10191272/2020 | 2020-10-19 | Europe/Belgium/Halle-Vilvoorde/Overijs | UZ Leuven, National Reference Laboratory for Coronaviruses, Laboratory Medicine, Leuven, Belgium | KU Leuven, Rega Institute, Clinical and Epidemiological Virology | Tony Wawina-Bokalanga et al |

| | | | | | | |
|---|---|---|---|---|---|---|
| EPI_ISL_735183 | Belgium/rega-10191274/2020 | 2020-10-19 | Europe/Belgium/Leuven/Haacht | UZ Leuven, National Reference Laboratory for Coronaviruses, Laboratory Medicine, Leuven, Belgium | KU Leuven, Rega Institute, Clinical and Epidemiological Virology | Tony Wawina-Bokalanga et al |
| EPI_ISL_735185 | Belgium/rega-10191276/2020 | 2020-10-19 | Europe/Belgium/Leuven/Winksele | UZ Leuven, National Reference Laboratory for Coronaviruses, Laboratory Medicine, Leuven, Belgium | KU Leuven, Rega Institute, Clinical and Epidemiological Virology | Tony Wawina-Bokalanga et al |
| EPI_ISL_735194 | Belgium/rega-10191288/2020 | 2020-10-19 | Europe/Belgium/Halle-Vilvoorde/Overijs | UZ Leuven, National Reference Laboratory for Coronaviruses, Laboratory Medicine, Leuven, Belgium | KU Leuven, Rega Institute, Clinical and Epidemiological Virology | Tony Wawina-Bokalanga et al |
| EPI_ISL_735195 | Belgium/rega-10191289/2020 | 2020-10-19 | Europe/Belgium/Halle-Vilvoorde/Nosseg | UZ Leuven, National Reference Laboratory for Coronaviruses, Laboratory Medicine, Leuven, Belgium | KU Leuven, Rega Institute, Clinical and Epidemiological Virology | Tony Wawina-Bokalanga et al |
| EPI_ISL_735201 | Belgium/rega-10191297/2020 | 2020-10-19 | Europe/Belgium/Leuven/Bost | UZ Leuven, National Reference Laboratory for Coronaviruses, Laboratory Medicine, Leuven, Belgium | KU Leuven, Rega Institute, Clinical and Epidemiological Virology | Tony Wawina-Bokalanga et al |
| EPI_ISL_735204 | Belgium/rega-10191301/2020 | 2020-10-19 | Europe/Belgium/Leuven/Kortenberg | UZ Leuven, National Reference Laboratory for Coronaviruses, Laboratory Medicine, Leuven, Belgium | KU Leuven, Rega Institute, Clinical and Epidemiological Virology | Tony Wawina-Bokalanga et al |
| EPI_ISL_735205 | Belgium/rega-10191302/2020 | 2020-10-19 | Europe/Belgium/Leuven/Winksele | UZ Leuven, National Reference Laboratory for Coronaviruses, Laboratory Medicine, Leuven, Belgium | KU Leuven, Rega Institute, Clinical and Epidemiological Virology | Tony Wawina-Bokalanga et al |
| EPI_ISL_735207 | Belgium/rega-10191304/2020 | 2020-10-19 | Europe/Belgium/Leuven/Blanden | UZ Leuven, National Reference Laboratory for Coronaviruses, Laboratory Medicine, Leuven, Belgium | KU Leuven, Rega Institute, Clinical and Epidemiological Virology | Tony Wawina-Bokalanga et al |

| | | | | | | |
|---|---|---|---|---|---|---|
| EPI_ISL_735208 | Belgium/rega-10191305/2020 | 2020-10-19 | Europe/Belgium/Leuven/Heverlee | UZ Leuven, National Reference Laboratory for Coronaviruses, Laboratory Medicine, Leuven, Belgium | KU Leuven, Rega Institute, Clinical and Epidemiological Virology | Tony Wawina-Bokalanga et al |
| EPI_ISL_735211 | Belgium/rega-10201105/2020 | 2020-10-20 | Europe/Belgium/Halle-Vilvoorde/Grimb | UZ Leuven, National Reference Laboratory for Coronaviruses, Laboratory Medicine, Leuven, Belgium | KU Leuven, Rega Institute, Clinical and Epidemiological Virology | Tony Wawina-Bokalanga et al |
| EPI_ISL_735218 | Belgium/rega-11031207/2020 | 2020-11-03 | Europe/Belgium/Tielt/Meulebeke | UZ Leuven, National Reference Laboratory for Coronaviruses, Laboratory Medicine, Leuven, Belgium | KU Leuven, Rega Institute, Clinical and Epidemiological Virology | Tony Wawina-Bokalanga et al |
| EPI_ISL_735220 | Belgium/rega-11031209/2020 | 2020-11-03 | Europe/Belgium/Antwerpen/ | UZ Leuven, National Reference Laboratory for Coronaviruses, Laboratory Medicine, Leuven, Belgium | KU Leuven, Rega Institute, Clinical and Epidemiological Virology | Tony Wawina-Bokalanga et al |
| EPI_ISL_735225 | Belgium/rega-11271172/2020 | 2020-11-27 | Europe/Belgium/Tongeren/Bilzen | UZ Leuven, National Reference Laboratory for Coronaviruses, Laboratory Medicine, Leuven, Belgium | KU Leuven, Rega Institute, Clinical and Epidemiological Virology | Tony Wawina-Bokalanga et al |
| EPI_ISL_735226 | Belgium/rega-11271173/2020 | 2020-11-27 | Europe/Belgium/Tongeren/Bilzen | UZ Leuven, National Reference Laboratory for Coronaviruses, Laboratory Medicine, Leuven, Belgium | KU Leuven, Rega Institute, Clinical and Epidemiological Virology | Tony Wawina-Bokalanga et al |
| EPI_ISL_735228 | Belgium/rega-11271175/2020 | 2020-11-27 | Europe/Belgium/Tongeren/Bilzen | UZ Leuven, National Reference Laboratory for Coronaviruses, Laboratory Medicine, Leuven, Belgium | KU Leuven, Rega Institute, Clinical and Epidemiological Virology | Tony Wawina-Bokalanga et al |
| EPI_ISL_735233 | Belgium/rega-11271180/2020 | 2020-11-27 | Europe/Belgium/Tongeren/Bilzen | UZ Leuven, National Reference Laboratory for Coronaviruses, Laboratory Medicine, Leuven, Belgium | KU Leuven, Rega Institute, Clinical and Epidemiological Virology | Tony Wawina-Bokalanga et al |

| | | | | | | |
|---|---|---|---|---|---|---|
| EPI_ISL_735235 | Belgium/rega-11271182/2020 | 2020-11-27 | Europe/Belgium/Tongeren/ | UZ Leuven, National Reference Laboratory for Coronaviruses, Laboratory Medicine, Leuven, Belgium | KU Leuven, Rega Institute, Clinical and Epidemiological Virology | Tony Wawina-Bokalanga et al |
| EPI_ISL_735236 | Belgium/rega-11301199/2020 | 2020-11-30 | Europe/Belgium/Eeklo/Assenede | UZ Leuven, National Reference Laboratory for Coronaviruses, Laboratory Medicine, Leuven, Belgium | KU Leuven, Rega Institute, Clinical and Epidemiological Virology | Tony Wawina-Bokalanga et al |
| EPI_ISL_735241 | France/HDF-rega-0413575/2020 | 2020-04-13 | Europe/France/Hauts de France/Lyz-Les- | UZ Leuven, National Reference Laboratory for Coronaviruses, Laboratory Medicine, Leuven, Belgium | KU Leuven, Rega Institute, Clinical and Epidemiological Virology | Tony Wawina-Bokalanga et al |
| EPI_ISL_735242 | France/HDF-rega-0414581/2020 | 2020-04-14 | Europe/France/Hauts de France/Tourcoi | UZ Leuven, National Reference Laboratory for Coronaviruses, Laboratory Medicine, Leuven, Belgium | KU Leuven, Rega Institute, Clinical and Epidemiological Virology | Tony Wawina-Bokalanga et al |
| EPI_ISL_735245 | France/IDF-rega-0508543/2020 | 2020-05-08 | Europe/France/Ile de France/Groslay | UZ Leuven, National Reference Laboratory for Coronaviruses, Laboratory Medicine, Leuven, Belgium | KU Leuven, Rega Institute, Clinical and Epidemiological Virology | Tony Wawina-Bokalanga et al |
| EPI_ISL_735259 | Turkey/IST-GLAB-005/2020 | 2020-04-13 | Europe/Turkey/Istanbul/ | Genomic Laboratory (GLAB) (Conjoint lab of Health Directorate of Istanbul and Istanbul Technical University) | Genomic Laboratory (GLAB), Istanbul Technical University | Ilker Karacan et al |
| EPI_ISL_735260 | Turkey/IST-GLAB-010/2020 | 2020-04-15 | Europe/Turkey/Istanbul/ | Genomic Laboratory (GLAB) (Conjoint lab of Health Directorate of Istanbul and Istanbul Technical University) | Genomic Laboratory (GLAB), Istanbul Technical University | Ilker Karacan et al |
| EPI_ISL_735261 | Turkey/IST-GLAB-026/2020 | 2020-04-17 | Europe/Turkey/Istanbul/ | Genomic Laboratory (GLAB) (Conjoint lab of Health Directorate of Istanbul and Istanbul Technical University) | Genomic Laboratory (GLAB), Istanbul Technical University | Ilker Karacan et al |

| | | | | | | |
|---|---|---|---|---|---|---|
| EPI_ISL_735262 | Turkey/IST-GLAB-028/2020 | 2020-04-17 | Europe/Turkey/Istanbul/ | Genomic Laboratory (GLAB) (Conjoint lab of Health Directorate of Istanbul and Istanbul Technical University) | Genomic Laboratory (GLAB), Istanbul Technical University | Ilker Karacan et al |
| EPI_ISL_735263 | Turkey/IST-GLAB-056/2020 | 2020-04-22 | Europe/Turkey/Istanbul/ | Genomic Laboratory (GLAB) (Conjoint lab of Health Directorate of Istanbul and Istanbul Technical University) | Genomic Laboratory (GLAB), Istanbul Technical University | Ilker Karacan et al |
| EPI_ISL_735264 | Turkey/IST-GLAB-060/2020 | 2020-04-22 | Europe/Turkey/Istanbul/ | Genomic Laboratory (GLAB) (Conjoint lab of Health Directorate of Istanbul and Istanbul Technical University) | Genomic Laboratory (GLAB), Istanbul Technical University | Ilker Karacan et al |
| EPI_ISL_735265 | Turkey/IST-GLAB-061/2020 | 2020-04-22 | Europe/Turkey/Istanbul/ | Genomic Laboratory (GLAB) (Conjoint lab of Health Directorate of Istanbul and Istanbul Technical University) | Genomic Laboratory (GLAB), Istanbul Technical University | Ilker Karacan et al |
| EPI_ISL_735267 | Turkey/IST-GLAB-119/2020 | 2020-04-29 | Europe/Turkey/Istanbul/ | Genomic Laboratory (GLAB) (Conjoint lab of Health Directorate of Istanbul and Istanbul Technical University) | Genomic Laboratory (GLAB), Istanbul Technical University | Ilker Karacan et al |
| EPI_ISL_735268 | Turkey/IST-GLAB-120/2020 | 2020-04-30 | Europe/Turkey/Istanbul/ | Genomic Laboratory (GLAB) (Conjoint lab of Health Directorate of Istanbul and Istanbul Technical University) | Genomic Laboratory (GLAB), Istanbul Technical University | Ilker Karacan et al |
| EPI_ISL_735270 | Turkey/IST-GLAB-122/2020 | 2020-04-29 | Europe/Turkey/Istanbul/ | Genomic Laboratory (GLAB) (Conjoint lab of Health Directorate of Istanbul and Istanbul Technical University) | Genomic Laboratory (GLAB), Istanbul Technical University | Ilker Karacan et al |
| EPI_ISL_735271 | Turkey/IST-GLAB-123/2020 | 2020-04-29 | Europe/Turkey/Istanbul/ | Genomic Laboratory (GLAB) (Conjoint lab of Health Directorate of Istanbul and Istanbul Technical University) | Genomic Laboratory (GLAB), Istanbul Technical University | Ilker Karacan et al |

| | | | | | | |
|---|---|---|---|---|---|---|
| EPI_ISL_735272 | Turkey/IST-GLAB-124/2020 | 2020-04-29 | Europe/Turkey/Istanbul/ | Genomic Laboratory (GLAB) (Conjoint lab of Health Directorate of Istanbul and Istanbul Technical University) | Genomic Laboratory (GLAB), Istanbul Technical University | Ilker Karacan et al |
| EPI_ISL_735273 | Turkey/IST-GLAB-127/2020 | 2020-04-30 | Europe/Turkey/Istanbul/ | Genomic Laboratory (GLAB) (Conjoint lab of Health Directorate of Istanbul and Istanbul Technical University) | Genomic Laboratory (GLAB), Istanbul Technical University | Ilker Karacan et al |
| EPI_ISL_735274 | Turkey/IST-GLAB-128/2020 | 2020-04-29 | Europe/Turkey/Istanbul/ | Genomic Laboratory (GLAB) (Conjoint lab of Health Directorate of Istanbul and Istanbul Technical University) | Genomic Laboratory (GLAB), Istanbul Technical University | Ilker Karacan et al |
| EPI_ISL_735275 | Turkey/IST-GLAB-129/2020 | 2020-04-29 | Europe/Turkey/Istanbul/ | Genomic Laboratory (GLAB) (Conjoint lab of Health Directorate of Istanbul and Istanbul Technical University) | Genomic Laboratory (GLAB), Istanbul Technical University | Ilker Karacan et al |
| EPI_ISL_735277 | Turkey/IST-GLAB-140/2020 | 2020-04-30 | Europe/Turkey/Istanbul/ | Genomic Laboratory (GLAB) (Conjoint lab of Health Directorate of Istanbul and Istanbul Technical University) | Genomic Laboratory (GLAB), Istanbul Technical University | Ilker Karacan et al |
| EPI_ISL_735278 | Turkey/IST-GLAB-141/2020 | 2020-04-30 | Europe/Turkey/Istanbul/ | Genomic Laboratory (GLAB) (Conjoint lab of Health Directorate of Istanbul and Istanbul Technical University) | Genomic Laboratory (GLAB), Istanbul Technical University | Ilker Karacan et al |
| EPI_ISL_735279 | Turkey/IST-GLAB-142/2020 | 2020-04-30 | Europe/Turkey/Istanbul/ | Genomic Laboratory (GLAB) (Conjoint lab of Health Directorate of Istanbul and Istanbul Technical University) | Genomic Laboratory (GLAB), Istanbul Technical University | Ilker Karacan et al |
| EPI_ISL_735280 | Turkey/IST-GLAB-143/2020 | 2020-04-30 | Europe/Turkey/Istanbul/ | Genomic Laboratory (GLAB) (Conjoint lab of Health Directorate of Istanbul and Istanbul Technical University) | Genomic Laboratory (GLAB), Istanbul Technical University | Ilker Karacan et al |

| | | | | | | |
|---|---|---|---|---|---|---|
| EPI_ISL_735281 | Turkey/IST-GLAB-145/2020 | 2020-04-21 | Europe/Turkey/Istanbul/ | Genomic Laboratory (GLAB) (Conjoint lab of Health Directorate of Istanbul and Istanbul Technical University) | Genomic Laboratory (GLAB), Istanbul Technical University | Ilker Karacan et al |
| EPI_ISL_735282 | Turkey/IST-GLAB-146/2020 | 2020-05-03 | Europe/Turkey/Istanbul/ | Genomic Laboratory (GLAB) (Conjoint lab of Health Directorate of Istanbul and Istanbul Technical University) | Genomic Laboratory (GLAB), Istanbul Technical University | Ilker Karacan et al |
| EPI_ISL_735283 | Turkey/IST-GLAB-148/2020 | 2020-05-02 | Europe/Turkey/Istanbul/ | Genomic Laboratory (GLAB) (Conjoint lab of Health Directorate of Istanbul and Istanbul Technical University) | Genomic Laboratory (GLAB), Istanbul Technical University | Ilker Karacan et al |
| EPI_ISL_735285 | Turkey/IST-GLAB-160/2020 | 2020-05-03 | Europe/Turkey/Istanbul/ | Genomic Laboratory (GLAB) (Conjoint lab of Health Directorate of Istanbul and Istanbul Technical University) | Genomic Laboratory (GLAB), Istanbul Technical University | Ilker Karacan et al |
| EPI_ISL_735286 | Turkey/IST-GLAB-172/2020 | 2020-05-04 | Europe/Turkey/Istanbul/ | Genomic Laboratory (GLAB) (Conjoint lab of Health Directorate of Istanbul and Istanbul Technical University) | Genomic Laboratory (GLAB), Istanbul Technical University | Ilker Karacan et al |
| EPI_ISL_735287 | Turkey/IST-GLAB-186/2020 | 2020-05-06 | Europe/Turkey/Istanbul/ | Genomic Laboratory (GLAB) (Conjoint lab of Health Directorate of Istanbul and Istanbul Technical University) | Genomic Laboratory (GLAB), Istanbul Technical University | Ilker Karacan et al |
| EPI_ISL_735288 | Turkey/IST-GLAB-218/2020 | 2020-05-09 | Europe/Turkey/Istanbul/ | Genomic Laboratory (GLAB) (Conjoint lab of Health Directorate of Istanbul and Istanbul Technical University) | Genomic Laboratory (GLAB), Istanbul Technical University | Ilker Karacan et al |
| EPI_ISL_735290 | Turkey/IST-GLAB-222/2020 | 2020-05-13 | Europe/Turkey/Istanbul/ | Genomic Laboratory (GLAB) (Conjoint lab of Health Directorate of Istanbul and Istanbul Technical University) | Genomic Laboratory (GLAB), Istanbul Technical University | Ilker Karacan et al |

| | | | | | | |
|---|---|---|---|---|---|---|
| EPI_ISL_735291 | Turkey/IST-GLAB-223/2020 | 2020-05-13 | Europe/Turkey/Istanbul/ | Genomic Laboratory (GLAB) (Conjoint lab of Health Directorate of Istanbul and Istanbul Technical University) | Genomic Laboratory (GLAB), Istanbul Technical University | Ilker Karacan et al |
| EPI_ISL_735294 | Turkey/IST-GLAB-227/2020 | 2020-05-14 | Europe/Turkey/Istanbul/ | Genomic Laboratory (GLAB) (Conjoint lab of Health Directorate of Istanbul and Istanbul Technical University) | Genomic Laboratory (GLAB), Istanbul Technical University | Ilker Karacan et al |
| EPI_ISL_735295 | Turkey/IST-GLAB-228/2020 | 2020-05-14 | Europe/Turkey/Istanbul/ | Genomic Laboratory (GLAB) (Conjoint lab of Health Directorate of Istanbul and Istanbul Technical University) | Genomic Laboratory (GLAB), Istanbul Technical University | Ilker Karacan et al |
| EPI_ISL_735296 | Turkey/IST-GLAB-229/2020 | 2020-05-15 | Europe/Turkey/Istanbul/ | Genomic Laboratory (GLAB) (Conjoint lab of Health Directorate of Istanbul and Istanbul Technical University) | Genomic Laboratory (GLAB), Istanbul Technical University | Ilker Karacan et al |
| EPI_ISL_735299 | Turkey/IST-GLAB-232/2020 | 2020-05-15 | Europe/Turkey/Istanbul/ | Genomic Laboratory (GLAB) (Conjoint lab of Health Directorate of Istanbul and Istanbul Technical University) | Genomic Laboratory (GLAB), Istanbul Technical University | Ilker Karacan et al |
| EPI_ISL_735304 | Turkey/IST-GLAB-237/2020 | 2020-05-15 | Europe/Turkey/Istanbul/ | Genomic Laboratory (GLAB) (Conjoint lab of Health Directorate of Istanbul and Istanbul Technical University) | Genomic Laboratory (GLAB), Istanbul Technical University | Ilker Karacan et al |
| EPI_ISL_735305 | Turkey/IST-GLAB-238/2020 | 2020-05-15 | Europe/Turkey/Istanbul/ | Genomic Laboratory (GLAB) (Conjoint lab of Health Directorate of Istanbul and Istanbul Technical University) | Genomic Laboratory (GLAB), Istanbul Technical University | Ilker Karacan et al |
| EPI_ISL_735308 | Turkey/IST-GLAB-241/2020 | 2020-05-16 | Europe/Turkey/Istanbul/ | Genomic Laboratory (GLAB) (Conjoint lab of Health Directorate of Istanbul and Istanbul Technical University) | Genomic Laboratory (GLAB), Istanbul Technical University | Ilker Karacan et al |

| | | | | | | |
|---|---|---|---|---|---|---|
| EPI_ISL_735310 | Turkey/IST-GLAB-244/2020 | 2020-05-16 | Europe/Turkey/Istanbul/ | Genomic Laboratory (GLAB) (Conjoint lab of Health Directorate of Istanbul and Istanbul Technical University) | Genomic Laboratory (GLAB), Istanbul Technical University | Ilker Karacan et al |
| EPI_ISL_735311 | Turkey/IST-GLAB-245/2020 | 2020-05-16 | Europe/Turkey/Istanbul/ | Genomic Laboratory (GLAB) (Conjoint lab of Health Directorate of Istanbul and Istanbul Technical University) | Genomic Laboratory (GLAB), Istanbul Technical University | Ilker Karacan et al |
| EPI_ISL_735312 | Turkey/IST-GLAB-246/2020 | 2020-05-17 | Europe/Turkey/Istanbul/ | Genomic Laboratory (GLAB) (Conjoint lab of Health Directorate of Istanbul and Istanbul Technical University) | Genomic Laboratory (GLAB), Istanbul Technical University | Ilker Karacan et al |
| EPI_ISL_735314 | Turkey/IST-GLAB-248/2020 | 2020-05-18 | Europe/Turkey/Istanbul/ | Genomic Laboratory (GLAB) (Conjoint lab of Health Directorate of Istanbul and Istanbul Technical University) | Genomic Laboratory (GLAB), Istanbul Technical University | Ilker Karacan et al |
| EPI_ISL_735316 | Turkey/IST-GLAB-250/2020 | 2020-05-19 | Europe/Turkey/Istanbul/ | Genomic Laboratory (GLAB) (Conjoint lab of Health Directorate of Istanbul and Istanbul Technical University) | Genomic Laboratory (GLAB), Istanbul Technical University | Ilker Karacan et al |
| EPI_ISL_735317 | Turkey/IST-GLAB-252/2020 | 2020-05-19 | Europe/Turkey/Istanbul/ | Genomic Laboratory (GLAB) (Conjoint lab of Health Directorate of Istanbul and Istanbul Technical University) | Genomic Laboratory (GLAB), Istanbul Technical University | Ilker Karacan et al |
| EPI_ISL_735318 | Turkey/IST-GLAB-253/2020 | 2020-05-19 | Europe/Turkey/Istanbul/ | Genomic Laboratory (GLAB) (Conjoint lab of Health Directorate of Istanbul and Istanbul Technical University) | Genomic Laboratory (GLAB), Istanbul Technical University | Ilker Karacan et al |
| EPI_ISL_735319 | Turkey/IST-GLAB-254/2020 | 2020-05-19 | Europe/Turkey/Istanbul/ | Genomic Laboratory (GLAB) (Conjoint lab of Health Directorate of Istanbul and Istanbul Technical University) | Genomic Laboratory (GLAB), Istanbul Technical University | Ilker Karacan et al |

| | | | | | | |
|---|---|---|---|---|---|---|
| EPI_ISL_735320 | Turkey/IST-GLAB-255/2020 | 2020-05-19 | Europe/Turkey/Istanbul/ | Genomic Laboratory (GLAB) (Conjoint lab of Health Directorate of Istanbul and Istanbul Technical University) | Genomic Laboratory (GLAB), Istanbul Technical University | Ilker Karacan et al |
| EPI_ISL_735321 | Turkey/IST-GLAB-258/2020 | 2020-05-19 | Europe/Turkey/Istanbul/ | Genomic Laboratory (GLAB) (Conjoint lab of Health Directorate of Istanbul and Istanbul Technical University) | Genomic Laboratory (GLAB), Istanbul Technical University | Ilker Karacan et al |
| EPI_ISL_735322 | Turkey/IST-GLAB-261/2020 | 2020-05-19 | Europe/Turkey/Istanbul/ | Genomic Laboratory (GLAB) (Conjoint lab of Health Directorate of Istanbul and Istanbul Technical University) | Genomic Laboratory (GLAB), Istanbul Technical University | Ilker Karacan et al |
| EPI_ISL_735323 | Turkey/IST-GLAB-263/2020 | 2020-05-19 | Europe/Turkey/Istanbul/ | Genomic Laboratory (GLAB) (Conjoint lab of Health Directorate of Istanbul and Istanbul Technical University) | Genomic Laboratory (GLAB), Istanbul Technical University | Ilker Karacan et al |
| EPI_ISL_735328 | Turkey/IST-GLAB-273/2020 | 2020-05-21 | Europe/Turkey/Istanbul/ | Genomic Laboratory (GLAB) (Conjoint lab of Health Directorate of Istanbul and Istanbul Technical University) | Genomic Laboratory (GLAB), Istanbul Technical University | Ilker Karacan et al |
| EPI_ISL_735329 | Turkey/IST-GLAB-274/2020 | 2020-05-21 | Europe/Turkey/Istanbul/ | Genomic Laboratory (GLAB) (Conjoint lab of Health Directorate of Istanbul and Istanbul Technical University) | Genomic Laboratory (GLAB), Istanbul Technical University | Ilker Karacan et al |
| EPI_ISL_735337 | Turkey/IST-GLAB-293/2020 | 2020-05-23 | Europe/Turkey/Istanbul/ | Genomic Laboratory (GLAB) (Conjoint lab of Health Directorate of Istanbul and Istanbul Technical University) | Genomic Laboratory (GLAB), Istanbul Technical University | Ilker Karacan et al |
| EPI_ISL_735338 | Turkey/IST-GLAB-295/2020 | 2020-05-23 | Europe/Turkey/Istanbul/ | Genomic Laboratory (GLAB) (Conjoint lab of Health Directorate of Istanbul and Istanbul Technical University) | Genomic Laboratory (GLAB), Istanbul Technical University | Ilker Karacan et al |

| | | | | | | |
|---|---|---|---|---|---|---|
| EPI_ISL_735341 | Turkey/IST-GLAB-301/2020 | 2020-05-25 | Europe/Turkey/Istanbul/ | Genomic Laboratory (GLAB) (Conjoint lab of Health Directorate of Istanbul and Istanbul Technical University) | Genomic Laboratory (GLAB), Istanbul Technical University | Ilker Karacan et al |
| EPI_ISL_735343 | Turkey/IST-GLAB-303/2020 | 2020-05-28 | Europe/Turkey/Istanbul/ | Genomic Laboratory (GLAB) (Conjoint lab of Health Directorate of Istanbul and Istanbul Technical University) | Genomic Laboratory (GLAB), Istanbul Technical University | Ilker Karacan et al |
| EPI_ISL_735344 | Turkey/IST-GLAB-304/2020 | 2020-05-28 | Europe/Turkey/Istanbul/ | Genomic Laboratory (GLAB) (Conjoint lab of Health Directorate of Istanbul and Istanbul Technical University) | Genomic Laboratory (GLAB), Istanbul Technical University | Ilker Karacan et al |
| EPI_ISL_735346 | Turkey/IST-GLAB-312/2020 | 2020-06-19 | Europe/Turkey/Istanbul/ | Genomic Laboratory (GLAB) (Conjoint lab of Health Directorate of Istanbul and Istanbul Technical University) | Genomic Laboratory (GLAB), Istanbul Technical University | Ilker Karacan et al |
| EPI_ISL_735347 | Turkey/IST-GLAB-313/2020 | 2020-06-19 | Europe/Turkey/Istanbul/ | Genomic Laboratory (GLAB) (Conjoint lab of Health Directorate of Istanbul and Istanbul Technical University) | Genomic Laboratory (GLAB), Istanbul Technical University | Ilker Karacan et al |
| EPI_ISL_735349 | Turkey/IST-GLAB-320/2020 | 2020-06-19 | Europe/Turkey/Istanbul/ | Genomic Laboratory (GLAB) (Conjoint lab of Health Directorate of Istanbul and Istanbul Technical University) | Genomic Laboratory (GLAB), Istanbul Technical University | Ilker Karacan et al |
| EPI_ISL_735350 | Turkey/IST-GLAB-323/2020 | 2020-06-23 | Europe/Turkey/Istanbul/ | Genomic Laboratory (GLAB) (Conjoint lab of Health Directorate of Istanbul and Istanbul Technical University) | Genomic Laboratory (GLAB), Istanbul Technical University | Ilker Karacan et al |
| EPI_ISL_735351 | Turkey/IST-GLAB-325/2020 | 2020-06-23 | Europe/Turkey/Istanbul/ | Genomic Laboratory (GLAB) (Conjoint lab of Health Directorate of Istanbul and Istanbul Technical University) | Genomic Laboratory (GLAB), Istanbul Technical University | Ilker Karacan et al |

| | | | | | | |
|---|---|---|---|---|---|---|
| EPI_ISL_735352 | Turkey/IST-GLAB-335/2020 | 2020-06-23 | Europe/Turkey/Istanbul/ | Genomic Laboratory (GLAB) (Conjoint lab of Health Directorate of Istanbul and Istanbul Technical University) | Genomic Laboratory (GLAB), Istanbul Technical University | Ilker Karacan et al |
| EPI_ISL_735353 | Turkey/IST-GLAB-337/2020 | 2020-06-23 | Europe/Turkey/Istanbul/ | Genomic Laboratory (GLAB) (Conjoint lab of Health Directorate of Istanbul and Istanbul Technical University) | Genomic Laboratory (GLAB), Istanbul Technical University | Ilker Karacan et al |
| EPI_ISL_735354 | Turkey/IST-GLAB-338/2020 | 2020-06-23 | Europe/Turkey/Istanbul/ | Genomic Laboratory (GLAB) (Conjoint lab of Health Directorate of Istanbul and Istanbul Technical University) | Genomic Laboratory (GLAB), Istanbul Technical University | Ilker Karacan et al |
| EPI_ISL_735355 | Turkey/IST-GLAB-340/2020 | 2020-06-24 | Europe/Turkey/Istanbul/ | Genomic Laboratory (GLAB) (Conjoint lab of Health Directorate of Istanbul and Istanbul Technical University) | Genomic Laboratory (GLAB), Istanbul Technical University | Ilker Karacan et al |
| EPI_ISL_735357 | Turkey/IST-GLAB-342/2020 | 2020-06-24 | Europe/Turkey/Istanbul/ | Genomic Laboratory (GLAB) (Conjoint lab of Health Directorate of Istanbul and Istanbul Technical University) | Genomic Laboratory (GLAB), Istanbul Technical University | Ilker Karacan et al |
| EPI_ISL_735359 | Turkey/IST-GLAB-347/2020 | 2020-06-24 | Europe/Turkey/Istanbul/ | Genomic Laboratory (GLAB) (Conjoint lab of Health Directorate of Istanbul and Istanbul Technical University) | Genomic Laboratory (GLAB), Istanbul Technical University | Ilker Karacan et al |
| EPI_ISL_735360 | Turkey/IST-GLAB-348/2020 | 2020-06-24 | Europe/Turkey/Istanbul/ | Genomic Laboratory (GLAB) (Conjoint lab of Health Directorate of Istanbul and Istanbul Technical University) | Genomic Laboratory (GLAB), Istanbul Technical University | Ilker Karacan et al |
| EPI_ISL_735361 | Turkey/IST-GLAB-349/2020 | 2020-06-24 | Europe/Turkey/Istanbul/ | Genomic Laboratory (GLAB) (Conjoint lab of Health Directorate of Istanbul and Istanbul Technical University) | Genomic Laboratory (GLAB), Istanbul Technical University | Ilker Karacan et al |

| | | | | | | |
|---|---|---|---|---|---|---|
| EPI_ISL_735362 | Turkey/IST-GLAB-350/2020 | 2020-06-24 | Europe/Turkey/Istanbul/ | Genomic Laboratory (GLAB) (Conjoint lab of Health Directorate of Istanbul and Istanbul Technical University) | Genomic Laboratory (GLAB), Istanbul Technical University | Ilker Karacan et al |
| EPI_ISL_735363 | Turkey/IST-GLAB-351/2020 | 2020-06-24 | Europe/Turkey/Istanbul/ | Genomic Laboratory (GLAB) (Conjoint lab of Health Directorate of Istanbul and Istanbul Technical University) | Genomic Laboratory (GLAB), Istanbul Technical University | Ilker Karacan et al |
| EPI_ISL_735364 | Turkey/IST-GLAB-352/2020 | 2020-06-24 | Europe/Turkey/Istanbul/ | Genomic Laboratory (GLAB) (Conjoint lab of Health Directorate of Istanbul and Istanbul Technical University) | Genomic Laboratory (GLAB), Istanbul Technical University | Ilker Karacan et al |
| EPI_ISL_735365 | Turkey/IST-GLAB-353/2020 | 2020-06-24 | Europe/Turkey/Istanbul/ | Genomic Laboratory (GLAB) (Conjoint lab of Health Directorate of Istanbul and Istanbul Technical University) | Genomic Laboratory (GLAB), Istanbul Technical University | Ilker Karacan et al |
| EPI_ISL_735366 | Turkey/IST-GLAB-A1/2020 | 2020-06-01 | Europe/Turkey/Istanbul/ | Genomic Laboratory (GLAB) (Conjoint lab of Health Directorate of Istanbul and Istanbul Technical University) | Genomic Laboratory (GLAB), Istanbul Technical University | Ilker Karacan et al |
| EPI_ISL_735367 | Turkey/IST-GLAB-A10/2020 | 2020-06-01 | Europe/Turkey/Istanbul/ | Genomic Laboratory (GLAB) (Conjoint lab of Health Directorate of Istanbul and Istanbul Technical University) | Genomic Laboratory (GLAB), Istanbul Technical University | Ilker Karacan et al |
| EPI_ISL_735369 | Turkey/IST-GLAB-A4/2020 | 2020-06-01 | Europe/Turkey/Istanbul/ | Genomic Laboratory (GLAB) (Conjoint lab of Health Directorate of Istanbul and Istanbul Technical University) | Genomic Laboratory (GLAB), Istanbul Technical University | Ilker Karacan et al |
| EPI_ISL_735370 | Turkey/IST-GLAB-A5/2020 | 2020-06-01 | Europe/Turkey/Istanbul/ | Genomic Laboratory (GLAB) (Conjoint lab of Health Directorate of Istanbul and Istanbul Technical University) | Genomic Laboratory (GLAB), Istanbul Technical University | Ilker Karacan et al |

| | | | | | | |
|---|---|---|---|---|---|---|
| EPI_ISL_735371 | Turkey/IST-GLAB-A6/2020 | 2020-06-01 | Europe/Turkey/Istanbul/ | Genomic Laboratory (GLAB) (Conjoint lab of Health Directorate of Istanbul and Istanbul Technical University) | Genomic Laboratory (GLAB), Istanbul Technical University | Ilker Karacan et al |
| EPI_ISL_735372 | Turkey/IST-GLAB-A7/2020 | 2020-06-01 | Europe/Turkey/Istanbul/ | Genomic Laboratory (GLAB) (Conjoint lab of Health Directorate of Istanbul and Istanbul Technical University) | Genomic Laboratory (GLAB), Istanbul Technical University | Ilker Karacan et al |
| EPI_ISL_735373 | Turkey/IST-GLAB-A9/2020 | 2020-06-01 | Europe/Turkey/Istanbul/ | Genomic Laboratory (GLAB) (Conjoint lab of Health Directorate of Istanbul and Istanbul Technical University) | Genomic Laboratory (GLAB), Istanbul Technical University | Ilker Karacan et al |
| EPI_ISL_735375 | USA/CA-LACPHL-AE00044/2020 | 2020-07-09 | North America/USA/California/Los Ange | Los Angeles County Public Health Laboratory | Los Angeles County Public Health Laboratory | P. Hemarajata et al. et al |
| EPI_ISL_735379 | USA/CA-LACPHL-AE00050/2020 | 2020-07-13 | North America/USA/California/Los Ange | Los Angeles County Public Health Laboratory | Los Angeles County Public Health Laboratory | P. Hemarajata et al. et al |
| EPI_ISL_735381 | USA/CA-LACPHL-AE00052/2020 | 2020-07-15 | North America/USA/California/Los Ange | Los Angeles County Public Health Laboratory | Los Angeles County Public Health Laboratory | P. Hemarajata et al. et al |
| EPI_ISL_735391 | France/CVL-SC719/2020 | 2020-12-21 | Europe/France/Centre-Val de Loire/Tour | CHU Tours | CNR Virus des Infections Respiratoires - France SUD | Antonin Bal et al |
| EPI_ISL_735396 | Brazil/SP-585/2020 | 2020-07-05 | South America/Brazil/S√£o Paulo/Taboa | Hospital de Camplanha COVID 19 SER | Instituto Adolfo Lutz, Interdiciplinary Procedures Center, Strategic Laboratory | Claudio Tavares Sacchi et al |
| EPI_ISL_735397 | Brazil/SP-586/2020 | 2020-07-09 | South America/Brazil/S√£o Paulo/Horto | Unidade Respirat√≥ria Nova Hortolandia | Instituto Adolfo Lutz, Interdiciplinary Procedures Center, Strategic Laboratory | Claudio Tavares Sacchi et al |

| | | | | | | |
|---|---|---|---|---|---|---|
| EPI_ISL_735398 | Brazil/SP-737/2020 | 2020-07-09 | South America/Brazil/S√£o Paulo/S√£o [illegible] | Laboratorio Fleury | Instituto Adolfo Lutz, Interdiciplinary Procedures Center, Strategic Laboratory | Claudio Tavares Sacchi et al |
| EPI_ISL_735400 | Brazil/SP-739/2020 | 2020-07-09 | South America/Brazil/S√£o Paulo/Santo | Instituto Adolfo Lutz - Regional de Santos | Instituto Adolfo Lutz, Interdiciplinary Procedures Center, Strategic Laboratory | Claudio Tavares Sacchi et al |
| EPI_ISL_735401 | Brazil/SP-740/2020 | 2020-07-04 | South America/Brazil/S√£o Paulo/Rio Cl | Instituto Adolfo Lutz - Regional de Rio Claro | Instituto Adolfo Lutz, Interdiciplinary Procedures Center, Strategic Laboratory | Claudio Tavares Sacchi et al |
| EPI_ISL_735402 | Brazil/SP-742/2020 | 2020-07-04 | South America/Brazil/S√£o Paulo/Rio Cl | Instituto Adolfo Lutz - Regional de Rio Claro | Instituto Adolfo Lutz, Interdiciplinary Procedures Center, Strategic Laboratory | Claudio Tavares Sacchi et al |
| EPI_ISL_735403 | Brazil/SP-743/2020 | 2020-07-04 | South America/Brazil/S√£o Paulo/Rio Cl | Instituto Adolfo Lutz - Regional de Rio Claro | Instituto Adolfo Lutz, Interdiciplinary Procedures Center, Strategic Laboratory | Claudio Tavares Sacchi et al |
| EPI_ISL_735404 | Brazil/SP-744/2020 | 2020-07-04 | South America/Brazil/S√£o Paulo/Rio Cl | Instituto Adolfo Lutz - Regional de Rio Claro | Instituto Adolfo Lutz, Interdiciplinary Procedures Center, Strategic Laboratory | Claudio Tavares Sacchi et al |

| | | | | | | |
|---|---|---|---|---|---|---|
| EPI_ISL_735405 | Brazil/SP-745/2020 | 2020-06-16 | South America/Brazil/S√£o Paulo/Birigu | Secretaria Minucipal de Saude de Birigui | Instituto Adolfo Lutz, Interdiciplinary Procedures Center, Strategic Laboratory | Claudio Tavares Sacchi et al |
| EPI_ISL_735406 | Brazil/SP-748/2020 | 2020-07-08 | South America/Brazil/S√£o Paulo/Santa | Unidade de Pronto Atendimento UPA I Sta Isabel | Instituto Adolfo Lutz, Interdiciplinary Procedures Center, Strategic Laboratory | Claudio Tavares Sacchi et al |
| EPI_ISL_735407 | Brazil/SP-749/2020 | 2020-06-26 | South America/Brazil/S√£o Paulo/Marili | Santa Casa de Marilia | Instituto Adolfo Lutz, Interdiciplinary Procedures Center, Strategic Laboratory | Claudio Tavares Sacchi et al |
| EPI_ISL_735408 | Brazil/SP-750/2020 | 2020-06-25 | South America/Brazil/S√£o Paulo/Jandir | COVID 19 Centro de Combate ao Coronavirus CCC Jandira | Instituto Adolfo Lutz, Interdiciplinary Procedures Center, Strategic Laboratory | Claudio Tavares Sacchi et al |
| EPI_ISL_735409 | Brazil/SP-751/2020 | 2020-06-17 | South America/Brazil/S√£o Paulo/Campi | Unidade de Pronto Atendimento Carlos Lourenco | Instituto Adolfo Lutz, Interdiciplinary Procedures Center, Strategic Laboratory | Claudio Tavares Sacchi et al |
| EPI_ISL_735410 | Brazil/SP-754/2020 | 2020-07-03 | South America/Brazil/S√£o Paulo/Rio Cl | Instituto Adolfo Lutz - Regional de Rio Claro | Instituto Adolfo Lutz, Interdiciplinary Procedures Center, Strategic Laboratory | Claudio Tavares Sacchi et al |

| | | | | | | |
|---|---|---|---|---|---|---|
| EPI_ISL_735411 | Brazil/SP-756/2020 | 2020-07-06 | South America/Brazil/S√£o Paulo/Diade | Centro de Vigilancia a Saude de Diadema | Instituto Adolfo Lutz, Interdiciplinary Procedures Center, Strategic Laboratory | Claudio Tavares Sacchi et al |
| EPI_ISL_735412 | Brazil/SP-758/2020 | 2020-06-15 | South America/Brazil/S√£o Paulo/S√£o | Hospital e Pronto Socorro Portinari | Instituto Adolfo Lutz, Interdiciplinary Procedures Center, Strategic Laboratory | Claudio Tavares Sacchi et al |
| EPI_ISL_735413 | Brazil/SP-759/2020 | 2020-06-17 | South America/Brazil/S√£o Paulo/S√£o | Militello Centro de Diagnosticos e Biopesquisa Clinica | Instituto Adolfo Lutz, Interdiciplinary Procedures Center, Strategic Laboratory | Claudio Tavares Sacchi et al |
| EPI_ISL_735414 | Brazil/SP-761/2020 | 2020-06-26 | South America/Brazil/S√£o Paulo/Mong | Unidade de Pronto Atendimento de Agenor de Campos | Instituto Adolfo Lutz, Interdiciplinary Procedures Center, Strategic Laboratory | Claudio Tavares Sacchi et al |
| EPI_ISL_735415 | Brazil/SP-762/2020 | 2020-06-25 | South America/Brazil/S√£o Paulo/Mong | Unidade de Pronto Atendimento de Agenor de Campos | Instituto Adolfo Lutz, Interdiciplinary Procedures Center, Strategic Laboratory | Claudio Tavares Sacchi et al |
| EPI_ISL_735416 | Brazil/SP-764/2020 | 2020-06-26 | South America/Brazil/S√£o Paulo/Moco | Centro de Saude II Dr Jose Paione Mococa | Instituto Adolfo Lutz, Interdiciplinary Procedures Center, Strategic Laboratory | Claudio Tavares Sacchi et al |

| | | | | | | |
|---|---|---|---|---|---|---|
| EPI_ISL_735417 | Brazil/SP-765/2020 | 2020-06-26 | South America/Brazil/S√£o Paulo/Mong | Unidade de Pronto Atendimento de Agenor de Campos | Instituto Adolfo Lutz, Interdiciplinary Procedures Center, Strategic Laboratory | Claudio Tavares Sacchi et al |
| EPI_ISL_735418 | Brazil/SP-766/2020 | 2020-06-27 | South America/Brazil/S√£o Paulo/Tauba | Hospital Regional do Vale do Paraiba | Instituto Adolfo Lutz, Interdiciplinary Procedures Center, Strategic Laboratory | Claudio Tavares Sacchi et al |
| EPI_ISL_735419 | Brazil/SP-770/2020 | 2020-07-01 | South America/Brazil/S√£o Paulo/Juquia | UBS Alvarenga | Instituto Adolfo Lutz, Interdiciplinary Procedures Center, Strategic Laboratory | Claudio Tavares Sacchi et al |
| EPI_ISL_735420 | Brazil/SP-772/2020 | 2020-07-01 | South America/Brazil/S√£o Paulo/Sao B | UBS Riacho Grande | Instituto Adolfo Lutz, Interdiciplinary Procedures Center, Strategic Laboratory | Claudio Tavares Sacchi et al |
| EPI_ISL_735421 | Brazil/SP-773/2020 | 2020-07-01 | South America/Brazil/S√£o Paulo/Sao B | UBS Sta Terezinha | Instituto Adolfo Lutz, Interdiciplinary Procedures Center, Strategic Laboratory | Claudio Tavares Sacchi et al |
| EPI_ISL_735422 | Brazil/SP-774/2020 | 2020-06-20 | South America/Brazil/S√£o Paulo/Sao B | UBS Dematchi | Instituto Adolfo Lutz, Interdiciplinary Procedures Center, Strategic Laboratory | Claudio Tavares Sacchi et al |

| | | | | | | |
|---|---|---|---|---|---|---|
| EPI_ISL_735423 | Brazil/SP-783/2020 | 2020-07-01 | South America/Brazil/S√£o Paulo/Diade | Centro de Vigilancia a Saude de Diadema | Instituto Adolfo Lutz, Interdiciplinary Procedures Center, Strategic Laboratory | Claudio Tavares Sacchi et al |
| EPI_ISL_735424 | Brazil/SP-784/2020 | 2020-07-01 | South America/Brazil/S√£o Paulo/Diade | Centro de Vigilancia a Saude de Diadema | Instituto Adolfo Lutz, Interdiciplinary Procedures Center, Strategic Laboratory | Claudio Tavares Sacchi et al |
| EPI_ISL_735425 | Brazil/SP-785/2020 | 2020-06-20 | South America/Brazil/S√£o Paulo/Ribeir | Hospital e Maternidade Sao Lucas | Instituto Adolfo Lutz, Interdiciplinary Procedures Center, Strategic Laboratory | Claudio Tavares Sacchi et al |
| EPI_ISL_735426 | Brazil/SP-786/2020 | 2020-07-01 | South America/Brazil/S√£o Paulo/Diade | Centro de Vigilancia a Saude de Diadema | Instituto Adolfo Lutz, Interdiciplinary Procedures Center, Strategic Laboratory | Claudio Tavares Sacchi et al |
| EPI_ISL_735427 | Brazil/SP-789/2020 | 2020-07-01 | South America/Brazil/S√£o Paulo/Santo | Instituto Adolfo Lutz - Regional de Santos | Instituto Adolfo Lutz, Interdiciplinary Procedures Center, Strategic Laboratory | Claudio Tavares Sacchi et al |
| EPI_ISL_735428 | Brazil/SP-791/2020 | 2020-07-02 | South America/Brazil/S√£o Paulo/S√£o | Hospital Nipo Brasileiro | Instituto Adolfo Lutz, Interdiciplinary Procedures Center, Strategic Laboratory | Claudio Tavares Sacchi et al |

| | | | | | | |
|---|---|---|---|---|---|---|
| EPI_ISL_735429 | Brazil/SP-792/2020 | 2020-07-01 | South America/Brazil/S√£o Paulo/S√£o | Hospital Nipo Brasileiro | Instituto Adolfo Lutz, Interdiciplinary Procedures Center, Strategic Laboratory | Claudio Tavares Sacchi et al |
| EPI_ISL_735430 | Brazil/SP-793/2020 | 2020-07-01 | South America/Brazil/S√£o Paulo/Santo | Instituto Adolfo Lutz - Regional de Santos | Instituto Adolfo Lutz, Interdiciplinary Procedures Center, Strategic Laboratory | Claudio Tavares Sacchi et al |
| EPI_ISL_735431 | Brazil/SP-794/2020 | 2020-07-02 | South America/Brazil/S√£o Paulo/S√£o | Hospital Nipo Brasileiro | Instituto Adolfo Lutz, Interdiciplinary Procedures Center, Strategic Laboratory | Claudio Tavares Sacchi et al |
| EPI_ISL_735432 | Brazil/SP-795/2020 | 2020-07-02 | South America/Brazil/S√£o Paulo/S√£o | Hospital Nipo Brasileiro | Instituto Adolfo Lutz, Interdiciplinary Procedures Center, Strategic Laboratory | Claudio Tavares Sacchi et al |
| EPI_ISL_735433 | Brazil/SP-796/2020 | 2020-07-01 | South America/Brazil/S√£o Paulo/Cajati | Posto de Atendimento Saude Cidade Pasc Cajati | Instituto Adolfo Lutz, Interdiciplinary Procedures Center, Strategic Laboratory | Claudio Tavares Sacchi et al |
| EPI_ISL_735435 | Turkey/IST-GLAB-294/2020 | 2020-05-23 | Europe/Turkey/Istanbul/ | Genomic Laboratory (GLAB) (Conjoint lab of Health Directorate of Istanbul and Istanbul Technical University) | Genomic Laboratory (GLAB), Istanbul Technical University | Ilker Karacan et al |
| EPI_ISL_735437 | Rwanda/NRLNAT1003/2020 | 2020-08-30 | Africa/Rwanda/Rwanda/ | Nucleic Acid Testing - Rwanda National Reference Laboratory | GIGA Medical Genomics | Yvan Butera et al |
| EPI_ISL_735445 | Rwanda/NRLNAT1005/2020 | 2020-10-22 | Africa/Rwanda/Kigali/ | Rwanda National Reference Laboratory | GIGA Medical Genomics | Yvan Butera et al |
| EPI_ISL_735451 | USA/WA-UW-29724/2020 | 2020-09-28 | North America/USA/Washington/ | UW Virology Lab | UW Virology Lab | Pavitra Roychoudhury et al |

| | | | | | | |
|---|---|---|---|---|---|---|
| EPI_ISL_735458 | USA/WA-UW-30946/2020 | 2020-10-10 | North America/USA/Washington/ | UW Virology Lab | UW Virology Lab | Pavitra Roychoudhury et al |
| EPI_ISL_735469 | USA/WA-UW-31239/2020 | 2020-10-13 | North America/USA/Washington/ | UW Virology Lab | UW Virology Lab | Pavitra Roychoudhury et al |
| EPI_ISL_735472 | USA/WA-UW-31382/2020 | 2020-10-10 | North America/USA/Washington/ | UW Virology Lab | UW Virology Lab | Pavitra Roychoudhury et al |
| EPI_ISL_735474 | USA/WA-UW-31404/2020 | 2020-10-10 | North America/USA/Washington/ | UW Virology Lab | UW Virology Lab | Pavitra Roychoudhury et al |
| EPI_ISL_735489 | USA/WA-UW-31637/2020 | 2020-10-18 | North America/USA/Washington/ | UW Virology Lab | UW Virology Lab | Pavitra Roychoudhury et al |
| EPI_ISL_735490 | Bangladesh/CU-CTG-02/2020 | 2020-11-26 | Asia/Bangladesh/Chattogram/ | Rangamati General Hospital RT-PCR lab, | Central Biological Research Laboratory and Department of Biochemistry and Molecular Biology | Robiul Hasan Bhuiyan et al |
| EPI_ISL_735492 | Bangladesh/CU-CTG-04/2020 | 2020-10-16 | Asia/Bangladesh/Chattogram/ | Kumilla Medical College | Central Biological Research Laboratory and Department of Biochemistry and Molecular Biology | Robiul Hasan Bhuiyan et al |
| EPI_ISL_735493 | Bangladesh/CU-CTG-09/2020 | 2020-11-22 | Asia/Bangladesh/Chattogram/Cox's Baza | Cox's Bazar Medicila College | Central Biological Research Laboratory and Department of Biochemistry and Molecular Biology Central Biological Research Laboratory and Department of Biochemistry and Molecular Biology | Robiul Hasan Bhuiyan et al |
| EPI_ISL_735494 | Bangladesh/CU-CTG-13/2020 | 2020-11-24 | Asia/Bangladesh/Chattogram/Bandarba | Cox's Bazar Medical College | Central Biological Research Laboratory and Department of Biochemistry and Molecular Biology | Md. Imranul Hoq et al |

| | | | | | | |
|---|---|---|---|---|---|---|
| EPI_ISL_735496 | Bangladesh/CU-CTG-24/2020 | 2020-11-23 | Asia/Bangladesh/Chattogram/Khagracha | Chattogram Veterinary and Animal Sciences University | Central Biological Research Laboratory and Department of Biochemistry and Molecular Biology | Md. Imranul Hoq et al |
| EPI_ISL_735497 | Bangladesh/CU-CTG-32/2020 | 2020-11-21 | Asia/Bangladesh/Chattogram/ | Chattogram Veterinary and Animal Sciences University | Central Biological Research Laboratory and Department of Biochemistry and Molecular Biology | Mohammad Omar Faruque et al |
| EPI_ISL_735501 | Bangladesh/CU-CTG-71/2020 | 2020-08-23 | Asia/Bangladesh/Chattogram/Feni | Abdul Malek Ukil Medical College, Noakhali | Central Biological Research Laboratory and Department of Biochemistry and Molecular Biology | H. M. Abdullah Al Masud et al |
| EPI_ISL_735502 | Bangladesh/CU-CTG-76/2020 | 2020-08-26 | Asia/Bangladesh/Chattogram/Noakhali | Abdul Malek Ukil Medical College, Noakhali | Central Biological Research Laboratory and Department of Biochemistry and Molecular Biology | H. M. Abdullah Al Masud et al |
| EPI_ISL_735532 | England/MILK-C61739/2020 | 2020-12-13 | Europe/United Kingdom/England/ | Lighthouse Lab in Milton Keynes | Wellcome Sanger Institute for the COVID-19 Genomics UK (COG-UK) Consortium | The Lighthouse Lab in Milton Keynes et al |
| EPI_ISL_735536 | England/MILK-C61818/2020 | 2020-12-13 | Europe/United Kingdom/England/ | Lighthouse Lab in Milton Keynes | Wellcome Sanger Institute for the COVID-19 Genomics UK (COG-UK) Consortium | The Lighthouse Lab in Milton Keynes et al |
| EPI_ISL_735548 | England/MILK-C61308/2020 | 2020-12-13 | Europe/United Kingdom/England/ | Lighthouse Lab in Milton Keynes | Wellcome Sanger Institute for the COVID-19 Genomics UK (COG-UK) Consortium | The Lighthouse Lab in Milton Keynes et al |

| | | | | | | |
|---|---|---|---|---|---|---|
| EPI_ISL_735692 | England/MILK-C6389E/2020 | 2020-12-13 | Europe/United Kingdom/England/ | Lighthouse Lab in Milton Keynes | Wellcome Sanger Institute for the COVID-19 Genomics UK (COG-UK) Consortium | The Lighthouse Lab in Milton Keynes et al |
| EPI_ISL_735902 | England/ALDP-C585FF/2020 | 2020-12-14 | Europe/United Kingdom/England/ | Lighthouse Lab in Alderley Park | Wellcome Sanger Institute for the COVID-19 Genomics UK (COG-UK) Consortium | Jacquelyn Wynn et al |
| EPI_ISL_736163 | England/ALDP-C58322/2020 | 2020-12-13 | Europe/United Kingdom/England/ | Lighthouse Lab in Alderley Park | Wellcome Sanger Institute for the COVID-19 Genomics UK (COG-UK) Consortium | Jacquelyn Wynn et al |
| EPI_ISL_736220 | England/QEUH-C579D6/2020 | 2020-12-12 | Europe/United Kingdom/England/ | Lighthouse Lab in Glasgow | Wellcome Sanger Institute for the COVID-19 Genomics UK (COG-UK) Consortium | Harper VanSteenhouse et al |
| EPI_ISL_736325 | England/QEUH-C542B0/2020 | 2020-12-11 | Europe/United Kingdom/England/ | Lighthouse Lab in Glasgow | Wellcome Sanger Institute for the COVID-19 Genomics UK (COG-UK) Consortium | Harper VanSteenhouse et al |
| EPI_ISL_736491 | England/QEUH-C4BD33/2020 | 2020-12-14 | Europe/United Kingdom/England/ | Lighthouse Lab in Glasgow | Wellcome Sanger Institute for the COVID-19 Genomics UK (COG-UK) Consortium | Harper VanSteenhouse et al |
| EPI_ISL_736700 | England/QEUH-C4BE7C/2020 | 2020-12-14 | Europe/United Kingdom/England/ | Lighthouse Lab in Glasgow | Wellcome Sanger Institute for the COVID-19 Genomics UK (COG-UK) Consortium | Harper VanSteenhouse et al |
| EPI_ISL_736703 | England/MILK-C504EB/2020 | 2020-12-11 | Europe/United Kingdom/England/ | Lighthouse Lab in Milton Keynes | Wellcome Sanger Institute for the COVID-19 Genomics UK (COG-UK) Consortium | The Lighthouse Lab in Milton Keynes et al |

| | | | | | | |
|---|---|---|---|---|---|---|
| EPI_ISL_736779 | Italy/CAM-TIGEM-24/2020 | 2020-10-26 | Europe/Italy/Campania/ | Istituto Zooprofilattico Sperimentale del Mezzogiorno | TIGEM | Patrizia Annunziata et al |
| EPI_ISL_736783 | Italy/CAM-TIGEM-178/2020 | 2020-12-21 | Europe/Italy/Campania/ | Istituto Zooprofilattico Sperimentale del Mezzogiorno | TIGEM | Patrizia Annunziata et al |
| EPI_ISL_736788 | Italy/CAM-TIGEM-17/2020 | 2020-10-26 | Europe/Italy/Campania/ | Istituto Zooprofilattico Sperimentale del Mezzogiorno | TIGEM | Patrizia Annunziata et al |
| EPI_ISL_736790 | Italy/CAM-TIGEM-104/2020 | 2020-10-26 | Europe/Italy/Campania/ | Istituto Zooprofilattico Sperimentale del Mezzogiorno | TIGEM | Patrizia Annunziata et al |
| EPI_ISL_736791 | Italy/CAM-TIGEM-1/2020 | 2020-10-26 | Europe/Italy/Campania/ | Istituto Zooprofilattico Sperimentale del Mezzogiorno | TIGEM | Patrizia Annunziata et al |
| EPI_ISL_736792 | Italy/CAM-TIGEM-2/2020 | 2020-10-26 | Europe/Italy/Campania/ | Istituto Zooprofilattico Sperimentale del Mezzogiorno | TIGEM | Patrizia Annunziata et al |
| EPI_ISL_736794 | Italy/CAM-TIGEM-4/2020 | 2020-10-26 | Europe/Italy/Campania/ | Istituto Zooprofilattico Sperimentale del Mezzogiorno | TIGEM | Patrizia Annunziata et al |
| EPI_ISL_736797 | Italy/CAM-TIGEM-8/2020 | 2020-10-26 | Europe/Italy/Campania/ | Istituto Zooprofilattico Sperimentale del Mezzogiorno | TIGEM | Patrizia Annunziata et al |
| EPI_ISL_736798 | Italy/CAM-TIGEM-9/2020 | 2020-10-26 | Europe/Italy/Campania/ | Istituto Zooprofilattico Sperimentale del Mezzogiorno | TIGEM | Patrizia Annunziata et al |
| EPI_ISL_736799 | Italy/CAM-TIGEM-11/2020 | 2020-10-26 | Europe/Italy/Campania/ | Istituto Zooprofilattico Sperimentale del Mezzogiorno | TIGEM | Patrizia Annunziata et al |
| EPI_ISL_736800 | Italy/CAM-TIGEM-13/2020 | 2020-10-26 | Europe/Italy/Campania/ | Istituto Zooprofilattico Sperimentale del Mezzogiorno | TIGEM | Patrizia Annunziata et al |
| EPI_ISL_736801 | Italy/CAM-TIGEM-14/2020 | 2020-10-26 | Europe/Italy/Campania/ | Istituto Zooprofilattico Sperimentale del Mezzogiorno | TIGEM | Patrizia Annunziata et al |
| EPI_ISL_736803 | Italy/CAM-TIGEM-16/2020 | 2020-10-26 | Europe/Italy/Campania/ | Istituto Zooprofilattico Sperimentale del Mezzogiorno | TIGEM | Patrizia Annunziata et al |
| EPI_ISL_736805 | Italy/CAM-TIGEM-26/2020 | 2020-10-26 | Europe/Italy/Campania/ | Istituto Zooprofilattico Sperimentale del Mezzogiorno | TIGEM | Patrizia Annunziata et al |
| EPI_ISL_736807 | Italy/CAM-TIGEM-31/2020 | 2020-10-26 | Europe/Italy/Campania/ | Istituto Zooprofilattico Sperimentale del Mezzogiorno | TIGEM | Patrizia Annunziata et al |
| EPI_ISL_736808 | Italy/CAM-TIGEM-32/2020 | 2020-10-26 | Europe/Italy/Campania/ | Istituto Zooprofilattico Sperimentale del Mezzogiorno | TIGEM | Patrizia Annunziata et al |
| EPI_ISL_736810 | Italy/CAM-TIGEM-34/2020 | 2020-10-26 | Europe/Italy/Campania/ | Istituto Zooprofilattico Sperimentale del Mezzogiorno | TIGEM | Patrizia Annunziata et al |
| EPI_ISL_736813 | Italy/CAM-TIGEM-37/2020 | 2020-10-26 | Europe/Italy/Campania/ | Istituto Zooprofilattico Sperimentale del Mezzogiorno | TIGEM | Patrizia Annunziata et al |
| EPI_ISL_736814 | Italy/CAM-TIGEM-38/2020 | 2020-10-26 | Europe/Italy/Campania/ | Istituto Zooprofilattico Sperimentale del Mezzogiorno | TIGEM | Patrizia Annunziata et al |
| EPI_ISL_736817 | Italy/CAM-TIGEM-42/2020 | 2020-10-26 | Europe/Italy/Campania/ | Istituto Zooprofilattico Sperimentale del Mezzogiorno | TIGEM | Patrizia Annunziata et al |

| | | | | | | |
|---|---|---|---|---|---|---|
| EPI_ISL_736819 | Italy/CAM-TIGEM-47/2020 | 2020-10-26 | Europe/Italy/Campania/ | Istituto Zooprofilattico Sperimentale del Mezzogiorno | TIGEM | Patrizia Annunziata et al |
| EPI_ISL_736820 | Italy/CAM-TIGEM-48/2020 | 2020-10-26 | Europe/Italy/Campania/ | Istituto Zooprofilattico Sperimentale del Mezzogiorno | TIGEM | Patrizia Annunziata et al |
| EPI_ISL_736821 | Italy/CAM-TIGEM-49/2020 | 2020-10-26 | Europe/Italy/Campania/ | Istituto Zooprofilattico Sperimentale del Mezzogiorno | TIGEM | Patrizia Annunziata et al |
| EPI_ISL_736822 | Italy/CAM-TIGEM-50/2020 | 2020-10-26 | Europe/Italy/Campania/ | Istituto Zooprofilattico Sperimentale del Mezzogiorno | TIGEM | Patrizia Annunziata et al |
| EPI_ISL_736823 | Italy/CAM-TIGEM-53/2020 | 2020-10-26 | Europe/Italy/Campania/ | Istituto Zooprofilattico Sperimentale del Mezzogiorno | TIGEM | Patrizia Annunziata et al |
| EPI_ISL_736826 | Italy/CAM-TIGEM-58/2020 | 2020-10-26 | Europe/Italy/Campania/ | Istituto Zooprofilattico Sperimentale del Mezzogiorno | TIGEM | Patrizia Annunziata et al |
| EPI_ISL_736828 | Italy/CAM-TIGEM-60/2020 | 2020-10-26 | Europe/Italy/Campania/ | Istituto Zooprofilattico Sperimentale del Mezzogiorno | TIGEM | Patrizia Annunziata et al |
| EPI_ISL_736829 | Italy/CAM-TIGEM-61/2020 | 2020-10-26 | Europe/Italy/Campania/ | Istituto Zooprofilattico Sperimentale del Mezzogiorno | TIGEM | Patrizia Annunziata et al |
| EPI_ISL_736830 | Italy/CAM-TIGEM-62/2020 | 2020-10-26 | Europe/Italy/Campania/ | Istituto Zooprofilattico Sperimentale del Mezzogiorno | TIGEM | Patrizia Annunziata et al |
| EPI_ISL_736832 | Italy/CAM-TIGEM-64/2020 | 2020-10-26 | Europe/Italy/Campania/ | Istituto Zooprofilattico Sperimentale del Mezzogiorno | TIGEM | Patrizia Annunziata et al |
| EPI_ISL_736833 | Italy/CAM-TIGEM-70/2020 | 2020-10-26 | Europe/Italy/Campania/ | Istituto Zooprofilattico Sperimentale del Mezzogiorno | TIGEM | Patrizia Annunziata et al |
| EPI_ISL_736834 | Italy/CAM-TIGEM-71/2020 | 2020-10-26 | Europe/Italy/Campania/ | Istituto Zooprofilattico Sperimentale del Mezzogiorno | TIGEM | Patrizia Annunziata et al |
| EPI_ISL_736837 | Italy/CAM-TIGEM-80/2020 | 2020-10-26 | Europe/Italy/Campania/ | Istituto Zooprofilattico Sperimentale del Mezzogiorno | TIGEM | Patrizia Annunziata et al |
| EPI_ISL_736840 | Italy/CAM-TIGEM-94/2020 | 2020-10-26 | Europe/Italy/Campania/ | Istituto Zooprofilattico Sperimentale del Mezzogiorno | TIGEM | Patrizia Annunziata et al |
| EPI_ISL_736842 | Italy/CAM-TIGEM-97/2020 | 2020-10-26 | Europe/Italy/Campania/ | Istituto Zooprofilattico Sperimentale del Mezzogiorno | TIGEM | Patrizia Annunziata et al |
| EPI_ISL_736846 | Italy/CAM-TIGEM-105/2020 | 2020-10-26 | Europe/Italy/Campania/ | Istituto Zooprofilattico Sperimentale del Mezzogiorno | TIGEM | Patrizia Annunziata et al |
| EPI_ISL_736847 | Italy/CAM-TIGEM-107/2020 | 2020-10-26 | Europe/Italy/Campania/ | Istituto Zooprofilattico Sperimentale del Mezzogiorno | TIGEM | Patrizia Annunziata et al |
| EPI_ISL_736849 | Italy/CAM-TIGEM-111/2020 | 2020-10-26 | Europe/Italy/Campania/ | Istituto Zooprofilattico Sperimentale del Mezzogiorno | TIGEM | Patrizia Annunziata et al |
| EPI_ISL_736850 | Italy/CAM-TIGEM-112/2020 | 2020-10-26 | Europe/Italy/Campania/ | Istituto Zooprofilattico Sperimentale del Mezzogiorno | TIGEM | Patrizia Annunziata et al |
| EPI_ISL_736855 | Italy/CAM-TIGEM-121/2020 | 2020-10-26 | Europe/Italy/Campania/ | Istituto Zooprofilattico Sperimentale del Mezzogiorno | TIGEM | Patrizia Annunziata et al |

| | | | | | | |
|---|---|---|---|---|---|---|
| EPI_ISL_736856 | Italy/CAM-TIGEM-122/2020 | 2020-10-26 | Europe/Italy/Campania/ | Istituto Zooprofilattico Sperimentale del Mezzogiorno | TIGEM | Patrizia Annunziata et al |
| EPI_ISL_736858 | Italy/CAM-TIGEM-124/2020 | 2020-10-26 | Europe/Italy/Campania/ | Istituto Zooprofilattico Sperimentale del Mezzogiorno | TIGEM | Patrizia Annunziata et al |
| EPI_ISL_736859 | Italy/CAM-TIGEM-126/2020 | 2020-10-26 | Europe/Italy/Campania/ | Istituto Zooprofilattico Sperimentale del Mezzogiorno | TIGEM | Patrizia Annunziata et al |
| EPI_ISL_736860 | Italy/CAM-TIGEM-127/2020 | 2020-10-26 | Europe/Italy/Campania/ | Istituto Zooprofilattico Sperimentale del Mezzogiorno | TIGEM | Patrizia Annunziata et al |
| EPI_ISL_736861 | Italy/CAM-TIGEM-130/2020 | 2020-10-26 | Europe/Italy/Campania/ | Istituto Zooprofilattico Sperimentale del Mezzogiorno | TIGEM | Patrizia Annunziata et al |
| EPI_ISL_736864 | Italy/CAM-TIGEM-135/2020 | 2020-10-26 | Europe/Italy/Campania/ | Istituto Zooprofilattico Sperimentale del Mezzogiorno | TIGEM | Patrizia Annunziata et al |
| EPI_ISL_736866 | Italy/CAM-TIGEM-138/2020 | 2020-10-26 | Europe/Italy/Campania/ | Istituto Zooprofilattico Sperimentale del Mezzogiorno | TIGEM | Patrizia Annunziata et al |
| EPI_ISL_736869 | Italy/CAM-TIGEM-142/2020 | 2020-10-26 | Europe/Italy/Campania/ | Istituto Zooprofilattico Sperimentale del Mezzogiorno | TIGEM | Patrizia Annunziata et al |
| EPI_ISL_736870 | Italy/CAM-TIGEM-144/2020 | 2020-10-26 | Europe/Italy/Campania/ | Istituto Zooprofilattico Sperimentale del Mezzogiorno | TIGEM | Patrizia Annunziata et al |
| EPI_ISL_736875 | Italy/CAM-TIGEM-151/2020 | 2020-10-26 | Europe/Italy/Campania/ | Istituto Zooprofilattico Sperimentale del Mezzogiorno | TIGEM | Patrizia Annunziata et al |
| EPI_ISL_736877 | Italy/CAM-TIGEM-155/2020 | 2020-10-26 | Europe/Italy/Campania/ | Istituto Zooprofilattico Sperimentale del Mezzogiorno | TIGEM | Patrizia Annunziata et al |
| EPI_ISL_736879 | Italy/CAM-TIGEM-158/2020 | 2020-10-26 | Europe/Italy/Campania/ | Istituto Zooprofilattico Sperimentale del Mezzogiorno | TIGEM | Patrizia Annunziata et al |
| EPI_ISL_736882 | Italy/CAM-TIGEM-161/2020 | 2020-10-26 | Europe/Italy/Campania/ | Istituto Zooprofilattico Sperimentale del Mezzogiorno | TIGEM | Patrizia Annunziata et al |
| EPI_ISL_736883 | Italy/CAM-TIGEM-162/2020 | 2020-10-26 | Europe/Italy/Campania/ | Istituto Zooprofilattico Sperimentale del Mezzogiorno | TIGEM | Patrizia Annunziata et al |
| EPI_ISL_736887 | Italy/CAM-TIGEM-169/2020 | 2020-10-26 | Europe/Italy/Campania/ | Istituto Zooprofilattico Sperimentale del Mezzogiorno | TIGEM | Patrizia Annunziata et al |
| EPI_ISL_736927 | SouthAfrica/KRISP-MDSH920916/202 | 2020-12-07 | Africa/South Africa/KwaZulu-Natal/eThe | MDS | KRISP, KZN Research Innovation and Sequencing Platform | Giandhari J et al |
| EPI_ISL_736929 | SouthAfrica/KRISP-WC-SA04352428/2 | 2020-11-24 | Africa/South Africa/Western Cape Provi | NHLS-UCT | KRISP, KZN Research Innovation and Sequencing Platform | Arash Iranzadeh et al |

| | | | | | | |
|---|---|---|---|---|---|---|
| EPI_ISL_736968 | SouthAfrica/KRISP-MDSH920888/202 | 2020-12-06 | Africa/South Africa/KwaZulu-Natal/eThe | MDS | KRISP, KZN Research Innovation and Sequencing Platform | Giandhari J et al |
| EPI_ISL_736970 | SouthAfrica/KRISP-EC-MDSH924795/ | 2020-12-07 | Africa/South Africa/Eastern Cape/Sarah | NHLS-IALCH | KRISP, KZN Research Innovation and Sequencing Platform | Giandhari J et al |
| EPI_ISL_736975 | SouthAfrica/KRISP-EC-MDSH925058/ | 2020-12-08 | Africa/South Africa/Eastern Cape/Sarah | NHLS-IALCH | KRISP, KZN Research Innovation and Sequencing Platform | Giandhari J et al |
| EPI_ISL_736985 | SouthAfrica/KRISP-CH01110222/2020 | 2020-11-25 | Africa/South Africa/KwaZulu-Natal/Ugu | NHLS-IALCH | KRISP, KZN Research Innovation and Sequencing Platform | Giandhari J et al |
| EPI_ISL_736986 | SouthAfrica/KRISP-CH01110224/2020 | 2020-11-25 | Africa/South Africa/KwaZulu-Natal/Ugu | NHLS-IALCH | KRISP, KZN Research Innovation and Sequencing Platform | Giandhari J et al |
| EPI_ISL_736998 | CzechRepublic/NRL_10348/2020 | 2020-10-23 | Europe/Czech Republic/Prague/ | The National Institute of Public Health | State Veterinary Institute Prague | Nagy et al |
| EPI_ISL_737001 | CzechRepublic/NRL_10386/2020 | 2020-10-25 | Europe/Czech Republic/Prague/ | The National Institute of Public Health | State Veterinary Institute Prague | Nagy et al |
| EPI_ISL_737002 | CzechRepublic/NRL_10398/2020 | 2020-10-26 | Europe/Czech Republic/Prague/ | The National Institute of Public Health | State Veterinary Institute Prague | Nagy et al |
| EPI_ISL_737003 | CzechRepublic/NRL_10405/2020 | 2020-10-26 | Europe/Czech Republic/Central Bohemia | The National Institute of Public Health | State Veterinary Institute Prague | Nagy et al |
| EPI_ISL_737004 | CzechRepublic/NRL_10406/2020 | 2020-10-26 | Europe/Czech Republic/Central Bohemia | The National Institute of Public Health | State Veterinary Institute Prague | Nagy et al |
| EPI_ISL_737005 | CzechRepublic/NRL_10410/2020 | 2020-10-26 | Europe/Czech Republic/Central Bohemia | The National Institute of Public Health | State Veterinary Institute Prague | Nagy et al |
| EPI_ISL_737006 | CzechRepublic/NRL_10411/2020 | 2020-10-26 | Europe/Czech Republic/Central Bohemia | The National Institute of Public Health | State Veterinary Institute Prague | Nagy et al |

| | | | | | | |
|---|---|---|---|---|---|---|
| EPI_ISL_737007 | CzechRepublic/NRL_10416/2020 | 2020-10-26 | Europe/Czech Republic/Prague/ | The National Institute of Public Health | State Veterinary Institute Prague | Nagy et al |
| EPI_ISL_737008 | CzechRepublic/NRL_10417/2020 | 2020-10-26 | Europe/Czech Republic/Central Bohemia | The National Institute of Public Health | State Veterinary Institute Prague | Nagy et al |
| EPI_ISL_737009 | CzechRepublic/NRL_10418/2020 | 2020-10-26 | Europe/Czech Republic/Central Bohemia | The National Institute of Public Health | State Veterinary Institute Prague | Nagy et al |
| EPI_ISL_737011 | CzechRepublic/NRL_10420/2020 | 2020-10-26 | Europe/Czech Republic/Prague/ | The National Institute of Public Health | State Veterinary Institute Prague | Nagy et al |
| EPI_ISL_737012 | CzechRepublic/NRL_10421/2020 | 2020-10-26 | Europe/Czech Republic/Prague/ | The National Institute of Public Health | State Veterinary Institute Prague | Nagy et al |
| EPI_ISL_737013 | CzechRepublic/NRL_10472/2020 | 2020-10-27 | Europe/Czech Republic/Prague/ | The National Institute of Public Health | State Veterinary Institute Prague | Nagy et al |
| EPI_ISL_737014 | CzechRepublic/NRL_10749/2020 | 2020-11-02 | Europe/Czech Republic/Usti nad Labem/ | The National Institute of Public Health | State Veterinary Institute Prague | Nagy et al |
| EPI_ISL_737015 | CzechRepublic/NRL_10750/2020 | 2020-11-02 | Europe/Czech Republic/Usti nad Labem/ | The National Institute of Public Health | State Veterinary Institute Prague | Nagy et al |
| EPI_ISL_737016 | CzechRepublic/NRL_10804/2020 | 2020-11-01 | Europe/Czech Republic/South Moravian | The National Institute of Public Health | State Veterinary Institute Prague | Nagy et al |
| EPI_ISL_737017 | CzechRepublic/NRL_10875/2020 | 2020-11-04 | Europe/Czech Republic/Central Bohemia | The National Institute of Public Health | State Veterinary Institute Prague | Nagy et al |
| EPI_ISL_737018 | CzechRepublic/NRL_10882/2020 | 2020-11-06 | Europe/Czech Republic/Prague/ | The National Institute of Public Health | State Veterinary Institute Prague | Nagy et al |
| EPI_ISL_737019 | CzechRepublic/NRL_10908/2020 | 2020-11-05 | Europe/Czech Republic/Central Bohemia | The National Institute of Public Health | State Veterinary Institute Prague | Nagy et al |
| EPI_ISL_737020 | CzechRepublic/NRL_10961/2020 | 2020-11-09 | Europe/Czech Republic/Olomouc Region/ | The National Institute of Public Health | State Veterinary Institute Prague | Nagy et al |
| EPI_ISL_737021 | CzechRepublic/NRL_10980/2020 | 2020-11-09 | Europe/Czech Republic/Prague/ | The National Institute of Public Health | State Veterinary Institute Prague | Nagy et al |
| EPI_ISL_737022 | CzechRepublic/NRL_10885/2020 | 2020-11-04 | Europe/Czech Republic/Prague/ | The National Institute of Public Health | State Veterinary Institute Prague | Nagy et al |
| EPI_ISL_737023 | CzechRepublic/NRL_10888/2020 | 2020-11-04 | Europe/Czech Republic/Prague/ | The National Institute of Public Health | State Veterinary Institute Prague | Nagy et al |
| EPI_ISL_737025 | CzechRepublic/NRL_10985/2020 | 2020-11-06 | Europe/Czech Republic/Central Bohemia | The National Institute of Public Health | State Veterinary Institute Prague | Nagy et al |
| EPI_ISL_737026 | CzechRepublic/NRL_11016/2020 | 2020-11-08 | Europe/Czech Republic/Prague/ | The National Institute of Public Health | State Veterinary Institute Prague | Nagy et al |
| EPI_ISL_737027 | CzechRepublic/NRL_11017/2020 | 2020-11-09 | Europe/Czech Republic/Central Bohemia | The National Institute of Public Health | State Veterinary Institute Prague | Nagy et al |
| EPI_ISL_737028 | CzechRepublic/NRL_10571/2020 | 2020-10-30 | Europe/Czech Republic/Prague/ | The National Institute of Public Health | State Veterinary Institute Prague | Nagy et al |

| | | | | | | |
|---|---|---|---|---|---|---|
| EPI_ISL_737029 | CzechRepublic/NRL_10533/2020 | 2020-10-29 | Europe/Czech Republic/Prague/ | The National Institute of Public Health | State Veterinary Institute Prague | Nagy et al |
| EPI_ISL_737030 | CzechRepublic/NRL_10569/2020 | 2020-10-30 | Europe/Czech Republic/Prague/ | The National Institute of Public Health | State Veterinary Institute Prague | Nagy et al |
| EPI_ISL_737031 | CzechRepublic/NRL_10593/2020 | 2020-10-30 | Europe/Czech Republic/Prague/ | The National Institute of Public Health | State Veterinary Institute Prague | Nagy et al |
| EPI_ISL_737033 | CzechRepublic/NRL_10490/2020 | 2020-10-27 | Europe/Czech Republic/Prague/ | The National Institute of Public Health | State Veterinary Institute Prague | Nagy et al |
| EPI_ISL_737085 | USA/WA-UW-32751/2020 | 2020-10-19 | North America/USA/Washington/ | UW Virology Lab | UW Virology Lab | Pavitra Roychoudhury et al |
| EPI_ISL_737096 | USA/WA-UW-32904/2020 | 2020-10-17 | North America/USA/Washington/ | UW Virology Lab | UW Virology Lab | Pavitra Roychoudhury et al |
| EPI_ISL_737098 | USA/WA-UW-32915/2020 | 2020-10-17 | North America/USA/Washington/ | UW Virology Lab | UW Virology Lab | Pavitra Roychoudhury et al |
| EPI_ISL_737099 | USA/WA-UW-32920/2020 | 2020-10-17 | North America/USA/Washington/ | UW Virology Lab | UW Virology Lab | Pavitra Roychoudhury et al |
| EPI_ISL_737110 | USA/MI-MDHHS-SC22555/2020 | 2020-11-10 | North America/USA/Michigan/ | Michigan Department of Health and Human Services, Bureau of Laboratories | Michigan Department of Health and Human Services, Bureau of Laboratories | Blankenship HM et al |
| EPI_ISL_737111 | USA/MI-MDHHS-SC22633/2020 | 2020-12-01 | North America/USA/Michigan/ | Michigan Department of Health and Human Services, Bureau of Laboratories | Michigan Department of Health and Human Services, Bureau of Laboratories | Blankenship HM et al |
| EPI_ISL_737114 | USA/MI-MDHHS-SC22634/2020 | 2020-11-30 | North America/USA/Michigan/ | Michigan Department of Health and Human Services, Bureau of Laboratories | Michigan Department of Health and Human Services, Bureau of Laboratories | Blankenship HM et al |
| EPI_ISL_737116 | USA/MI-MDHHS-SC22635/2020 | 2020-12-03 | North America/USA/Michigan/ | Michigan Department of Health and Human Services, Bureau of Laboratories | Michigan Department of Health and Human Services, Bureau of Laboratories | Blankenship HM et al |

| | | | | | | |
|---|---|---|---|---|---|---|
| EPI_ISL_737119 | USA/MI-MDHHS-SC22557/2020 | 2020-11-10 | North America/USA/Michigan/ | Michigan Department of Health and Human Services, Bureau of Laboratories | Michigan Department of Health and Human Services, Bureau of Laboratories | Blankenship HM et al |
| EPI_ISL_737140 | USA/MI-MDHHS-SC22570/2020 | 2020-11-03 | North America/USA/Michigan/ | Michigan Department of Health and Human Services, Bureau of Laboratories | Michigan Department of Health and Human Services, Bureau of Laboratories | Blankenship HM et al |
| EPI_ISL_737148 | USA/MI-MDHHS-SC22578/2020 | 2020-11-23 | North America/USA/Michigan/ | Michigan Department of Health and Human Services, Bureau of Laboratories | Michigan Department of Health and Human Services, Bureau of Laboratories | Blankenship HM et al |
| EPI_ISL_737151 | USA/MI-MDHHS-SC22549/2020 | 2020-11-23 | North America/USA/Michigan/ | Michigan Department of Health and Human Services, Bureau of Laboratories | Michigan Department of Health and Human Services, Bureau of Laboratories | Blankenship HM et al |
| EPI_ISL_737154 | USA/MI-MDHHS-SC22580/2020 | 2020-11-18 | North America/USA/Michigan/ | Michigan Department of Health and Human Services, Bureau of Laboratories | Michigan Department of Health and Human Services, Bureau of Laboratories | Blankenship HM et al |
| EPI_ISL_737159 | USA/MI-MDHHS-SC22586/2020 | 2020-11-24 | North America/USA/Michigan/ | Michigan Department of Health and Human Services, Bureau of Laboratories | Michigan Department of Health and Human Services, Bureau of Laboratories | Blankenship HM et al |

| | | | | | | |
|---|---|---|---|---|---|---|
| EPI_ISL_737180 | USA/MI-MDHHS-SC22607/2020 | 2020-11-19 | North America/USA/Michigan/ | Michigan Department of Health and Human Services, Bureau of Laboratories | Michigan Department of Health and Human Services, Bureau of Laboratories | Blankenship HM et al |
| EPI_ISL_737183 | USA/MI-MDHHS-SC22610/2020 | 2020-11-23 | North America/USA/Michigan/ | Michigan Department of Health and Human Services, Bureau of Laboratories | Michigan Department of Health and Human Services, Bureau of Laboratories | Blankenship HM et al |
| EPI_ISL_737187 | USA/MI-MDHHS-SC22614/2020 | 2020-11-09 | North America/USA/Michigan/ | Michigan Department of Health and Human Services, Bureau of Laboratories | Michigan Department of Health and Human Services, Bureau of Laboratories | Blankenship HM et al |
| EPI_ISL_737192 | USA/MI-MDHHS-SC22619/2020 | 2020-11-10 | North America/USA/Michigan/ | Michigan Department of Health and Human Services, Bureau of Laboratories | Michigan Department of Health and Human Services, Bureau of Laboratories | Blankenship HM et al |
| EPI_ISL_737193 | USA/MI-MDHHS-SC22620/2020 | 2020-11-19 | North America/USA/Michigan/ | Michigan Department of Health and Human Services, Bureau of Laboratories | Michigan Department of Health and Human Services, Bureau of Laboratories | Blankenship HM et al |
| EPI_ISL_737196 | USA/MI-MDHHS-SC22623/2020 | 2020-11-23 | North America/USA/Michigan/ | Michigan Department of Health and Human Services, Bureau of Laboratories | Michigan Department of Health and Human Services, Bureau of Laboratories | Blankenship HM et al |

| | | | | | | |
|---|---|---|---|---|---|---|
| EPI_ISL_737210 | Finland/13ES23H9/2020 | 2020-08-13 | Europe/Finland/Uusimaa/ | Department of Virology and Immunology, University of Helsinki and Helsinki University Hospital, Huslab Finland | Department of Virology, Faculty of Medicine, University of Helsinki, Helsinki, Finland | Teemu Smura et al |
| EPI_ISL_737236 | USA/CA-LACPHL-AF00001/2020 | 2020-11-26 | North America/USA/California/Los Ange | Los Angeles County PHL | Los Angeles County PHL | P. Hemarajata et al. et al |
| EPI_ISL_737237 | USA/CA-LACPHL-AF00002/2020 | 2020-12-18 | North America/USA/California/Los Ange | Los Angeles County PHL | Los Angeles County PHL | P. Hemarajata et al. et al |
| EPI_ISL_737244 | USA/CA-LACPHL-AF00009/2020 | 2020-12-01 | North America/USA/California/Los Ange | Los Angeles County PHL | Los Angeles County PHL | P. Hemarajata et al. et al |
| EPI_ISL_737246 | USA/CA-LACPHL-AF00011/2020 | 2020-12-15 | North America/USA/California/Los Ange | Los Angeles County PHL | Los Angeles County PHL | P. Hemarajata et al. et al |
| EPI_ISL_737252 | USA/CA-LACPHL-AF00017/2020 | 2020-12-14 | North America/USA/California/Los Ange | Los Angeles County PHL | Los Angeles County PHL | P. Hemarajata et al. et al |
| EPI_ISL_737253 | USA/CA-LACPHL-AF00018/2020 | 2020-12-15 | North America/USA/California/Los Ange | Los Angeles County PHL | Los Angeles County PHL | P. Hemarajata et al. et al |
| EPI_ISL_737257 | USA/CA-LACPHL-AF00022/2020 | 2020-12-16 | North America/USA/California/Los Ange | Los Angeles County PHL | Los Angeles County PHL | P. Hemarajata et al. et al |
| EPI_ISL_737261 | USA/CA-LACPHL-AF00026/2020 | 2020-12-16 | North America/USA/California/Los Ange | Los Angeles County PHL | Los Angeles County PHL | P. Hemarajata et al. et al |
| EPI_ISL_737269 | USA/MI-MDHHS-SC22655/2020 | 2020-11-16 | North America/USA/Michigan/ | Michigan Department of Health and Human Services, Bureau of Laboratories | Michigan Department of Health and Human Services, Bureau of Laboratories | Blankenship HM et al |
| EPI_ISL_737274 | USA/MI-MDHHS-SC22660/2020 | 2020-11-13 | North America/USA/Michigan/ | Michigan Department of Health and Human Services, Bureau of Laboratories | Michigan Department of Health and Human Services, Bureau of Laboratories | Blankenship HM et al |
| EPI_ISL_737280 | USA/MI-MDHHS-SC22666/2020 | 2020-11-12 | North America/USA/Michigan/ | Michigan Department of Health and Human Services, Bureau of Laboratories | Michigan Department of Health and Human Services, Bureau of Laboratories | Blankenship HM et al |

| | | | | | | |
|---|---|---|---|---|---|---|
| EPI_ISL_737286 | USA/MI-MDHHS-SC22672/2020 | 2020-11-23 | North America/USA/Michigan/ | Michigan Department of Health and Human Services, Bureau of Laboratories | Michigan Department of Health and Human Services, Bureau of Laboratories | Blankenship HM et al |
| EPI_ISL_737287 | USA/MI-MDHHS-SC22673/2020 | 2020-11-18 | North America/USA/Michigan/ | Michigan Department of Health and Human Services, Bureau of Laboratories | Michigan Department of Health and Human Services, Bureau of Laboratories | Blankenship HM et al |
| EPI_ISL_737289 | USA/MI-MDHHS-SC22675/2020 | 2020-11-27 | North America/USA/Michigan/ | Michigan Department of Health and Human Services, Bureau of Laboratories | Michigan Department of Health and Human Services, Bureau of Laboratories | Blankenship HM et al |
| EPI_ISL_737293 | USA/MI-MDHHS-SC22679/2020 | 2020-11-26 | North America/USA/Michigan/ | Michigan Department of Health and Human Services, Bureau of Laboratories | Michigan Department of Health and Human Services, Bureau of Laboratories | Blankenship HM et al |
| EPI_ISL_737295 | USA/MI-MDHHS-SC22681/2020 | 2020-11-24 | North America/USA/Michigan/ | Michigan Department of Health and Human Services, Bureau of Laboratories | Michigan Department of Health and Human Services, Bureau of Laboratories | Blankenship HM et al |
| EPI_ISL_737298 | USA/MI-MDHHS-SC22684/2020 | 2020-11-09 | North America/USA/Michigan/ | Michigan Department of Health and Human Services, Bureau of Laboratories | Michigan Department of Health and Human Services, Bureau of Laboratories | Blankenship HM et al |

| | | | | | | |
|---|---|---|---|---|---|---|
| EPI_ISL_737301 | USA/MI-MDHHS-SC22687/2020 | 2020-12-07 | North America/USA/Michigan/ | Michigan Department of Health and Human Services, Bureau of Laboratories | Michigan Department of Health and Human Services, Bureau of Laboratories | Blankenship HM et al |
| EPI_ISL_737308 | USA/MI-MDHHS-SC22694/2020 | 2020-11-26 | North America/USA/Michigan/ | Michigan Department of Health and Human Services, Bureau of Laboratories | Michigan Department of Health and Human Services, Bureau of Laboratories | Blankenship HM et al |
| EPI_ISL_737430 | Switzerland/BL-ETHZ-410150/2020 | 2020-12-15 | Europe/Switzerland/Basel-Land/ | Viollier AG | Department of Biosystems Science and Engineering, ETH Z√ºrich | Chaoran Chen et al |
| EPI_ISL_737484 | Switzerland/AG-ETHZ-410157/2020 | 2020-12-17 | Europe/Switzerland/Aargau/ | Viollier AG | Department of Biosystems Science and Engineering, ETH Z√ºrich | Chaoran Chen et al |
| EPI_ISL_737487 | Switzerland/SO-ETHZ-410745/2020 | 2020-12-12 | Europe/Switzerland/Solothurn/ | Viollier AG | Department of Biosystems Science and Engineering, ETH Z√ºrich | Chaoran Chen et al |
| EPI_ISL_737521 | Switzerland/BE-ETHZ-410881/2020 | 2020-12-14 | Europe/Switzerland/Bern/ | Viollier AG | Department of Biosystems Science and Engineering, ETH Z√ºrich | Chaoran Chen et al |
| EPI_ISL_737528 | Switzerland/SO-ETHZ-410741/2020 | 2020-12-12 | Europe/Switzerland/Solothurn/ | Viollier AG | Department of Biosystems Science and Engineering, ETH Z√ºrich | Chaoran Chen et al |

| | | | | | | |
|---|---|---|---|---|---|---|
| EPI_ISL_737553 | Switzerland/BE-ETHZ-410568/2020 | 2020-11-26 | Europe/Switzerland/Bern/ | Viollier AG | Department of Biosystems Science and Engineering, ETH Zv∫rich | Chaoran Chen et al |
| EPI_ISL_737599 | Switzerland/BE-ETHZ-410244/2020 | 2020-11-27 | Europe/Switzerland/Bern/ | Viollier AG | Department of Biosystems Science and Engineering, ETH Zv∫rich | Chaoran Chen et al |
| EPI_ISL_737688 | Switzerland/BE-ETHZ-320581/2020 | 2020-10-12 | Europe/Switzerland/Bern/ | Viollier AG | Department of Biosystems Science and Engineering, ETH Zv∫rich | Chaoran Chen et al |
| EPI_ISL_737719 | Switzerland/BE-ETHZ-410206/2020 | 2020-12-13 | Europe/Switzerland/Bern/ | Viollier AG | Department of Biosystems Science and Engineering, ETH Zv∫rich | Chaoran Chen et al |
| EPI_ISL_737731 | Switzerland/BE-ETHZ-410294/2020 | 2020-12-15 | Europe/Switzerland/Bern/ | Viollier AG | Department of Biosystems Science and Engineering, ETH Zv∫rich | Chaoran Chen et al |
| EPI_ISL_737748 | Switzerland/BE-ETHZ-410918/2020 | 2020-11-30 | Europe/Switzerland/Bern/ | Viollier AG | Department of Biosystems Science and Engineering, ETH Zv∫rich | Chaoran Chen et al |
| EPI_ISL_737777 | Switzerland/SH-ETHZ-410536/2020 | 2020-11-26 | Europe/Switzerland/Schaffhausen/ | Viollier AG | Department of Biosystems Science and Engineering, ETH Zv∫rich | Chaoran Chen et al |
| EPI_ISL_737811 | Switzerland/SO-ETHZ-410956/2020 | 2020-11-27 | Europe/Switzerland/Solothurn/ | Viollier AG | Department of Biosystems Science and Engineering, ETH Zv∫rich | Chaoran Chen et al |

| | | | | | | |
|---|---|---|---|---|---|---|
| EPI_ISL_737839 | Switzerland/VS-ETHZ-360051/2020 | 2020-11-09 | Europe/Switzerland/Valais/ | Viollier AG | Department of Biosystems Science and Engineering, ETH Zv∫rich | Chaoran Chen et al |
| EPI_ISL_737848 | Switzerland/VS-ETHZ-410248/2020 | 2020-11-27 | Europe/Switzerland/Valais/ | Viollier AG | Department of Biosystems Science and Engineering, ETH Zv∫rich | Chaoran Chen et al |
| EPI_ISL_737850 | Switzerland/VS-ETHZ-410378/2020 | 2020-11-26 | Europe/Switzerland/Valais/ | Viollier AG | Department of Biosystems Science and Engineering, ETH Zv∫rich | Chaoran Chen et al |
| EPI_ISL_737853 | Switzerland/VS-ETHZ-410436/2020 | 2020-11-24 | Europe/Switzerland/Valais/ | Viollier AG | Department of Biosystems Science and Engineering, ETH Zv∫rich | Chaoran Chen et al |
| EPI_ISL_737895 | Switzerland/VD-ETHZ-320522/2020 | 2020-10-13 | Europe/Switzerland/Vaud/ | Viollier AG | Department of Biosystems Science and Engineering, ETH Zv∫rich | Chaoran Chen et al |
| EPI_ISL_737910 | Switzerland/ZH-ETHZ-410025/2020 | 2020-12-16 | Europe/Switzerland/Zv∫rich/ | Viollier AG | Department of Biosystems Science and Engineering, ETH Zv∫rich | Chaoran Chen et al |
| EPI_ISL_737920 | Switzerland/ZH-ETHZ-410529/2020 | 2020-11-26 | Europe/Switzerland/Zv∫rich/ | Viollier AG | Department of Biosystems Science and Engineering, ETH Zv∫rich | Chaoran Chen et al |
| EPI_ISL_737945 | Uganda/UG100/2020 | 2020-08-14 | Africa/Uganda/Uganda/ | Uganda Central Public Health Lab and Uganda Virus Research Institute | MRC/UVRI & LSHTM Uganda Research Unit | Matthew Cotten et al |
| EPI_ISL_737998 | Uganda/UG087/2020 | 2020-08-25 | Africa/Uganda/Uganda/ | Uganda Central Public Health Lab and Uganda Virus Research Institute | MRC/UVRI & LSHTM Uganda Research Unit | Matthew Cotten et al |

| | | | | | | |
|---|---|---|---|---|---|---|
| EPI_ISL_738017 | Uganda/UG110/2020 | 2020-08-11 | Africa/Uganda/Uganda/ | Uganda Central Public Health Lab and Uganda Virus Research Institute | MRC/UVRI & LSHTM Uganda Research Unit | Matthew Cotten et al |
| EPI_ISL_738026 | Uganda/UG119/2020 | 2020-09-01 | Africa/Uganda/Uganda/ | Uganda Central Public Health Lab and Uganda Virus Research Institute | MRC/UVRI & LSHTM Uganda Research Unit | Matthew Cotten et al |
| EPI_ISL_738029 | Uganda/UG122/2020 | 2020-09-01 | Africa/Uganda/Uganda/ | Uganda Central Public Health Lab and Uganda Virus Research Institute | MRC/UVRI & LSHTM Uganda Research Unit | Matthew Cotten et al |
| EPI_ISL_738031 | Uganda/UG124/2020 | 2020-09-01 | Africa/Uganda/Uganda/ | Uganda Central Public Health Lab and Uganda Virus Research Institute | MRC/UVRI & LSHTM Uganda Research Unit | Matthew Cotten et al |
| EPI_ISL_738033 | Uganda/UG126/2020 | 2020-09-01 | Africa/Uganda/Uganda/ | Uganda Central Public Health Lab and Uganda Virus Research Institute | MRC/UVRI & LSHTM Uganda Research Unit | Matthew Cotten et al |
| EPI_ISL_738034 | Uganda/UG127/2020 | 2020-09-01 | Africa/Uganda/Uganda/ | Uganda Central Public Health Lab and Uganda Virus Research Institute | MRC/UVRI & LSHTM Uganda Research Unit | Matthew Cotten et al |
| EPI_ISL_738039 | Uganda/UG132/2020 | 2020-10-01 | Africa/Uganda/Uganda/ | Uganda Central Public Health Lab and Uganda Virus Research Institute | MRC/UVRI & LSHTM Uganda Research Unit | Matthew Cotten et al |
| EPI_ISL_738084 | HongKong/HKPU31_2801/2020 | 2020-02-08 | Asia/Hong Kong/Hong Kong/ | Department of Health Technology and Informatics, Hong Kong Polytechnic University of Hong Kong | Department of Health Technology and Informatics, Hong Kong Polytechnic University of Hong Kong | Leung et al |
| EPI_ISL_738092 | Portugal/PT2092/2020 | 2020-12-20 | Europe/Portugal/Portugal/ | Instituto Nacional de Saude (INSA) | Instituto Nacional de Saude (INSA) | Borges et al et al |
| EPI_ISL_738093 | Portugal/PT2095/2020 | 2020-12-19 | Europe/Portugal/Portugal/ | Instituto Nacional de Saude (INSA) | Instituto Nacional de Saude (INSA) | Borges et al et al |
| EPI_ISL_738099 | Portugal/PT2099/2020 | 2020-12-18 | Europe/Portugal/Portugal/ | Instituto Nacional de Saude (INSA) | Instituto Nacional de Saude (INSA) | Borges et al et al |
| EPI_ISL_738105 | Portugal/PT2106/2020 | 2020-12-17 | Europe/Portugal/Portugal/ | Instituto Nacional de Saude (INSA) | Instituto Nacional de Saude (INSA) | Borges et al et al |
| EPI_ISL_738108 | Portugal/PT2110/2020 | 2020-12-07 | Europe/Portugal/Portugal/ | Instituto Nacional de Saude (INSA) | Instituto Nacional de Saude (INSA) | Borges et al et al |
| EPI_ISL_738112 | Portugal/PT2109/2020 | 2020-12-08 | Europe/Portugal/Portugal/ | Instituto Nacional de Saude (INSA) | Instituto Nacional de Saude (INSA) | Borges et al et al |

| | | | | | | |
|---|---|---|---|---|---|---|
| EPI_ISL_738113 | Portugal/PT2111/2020 | 2020-12-05 | Europe/Portugal/Portugal/ | Instituto Nacional de Saude (INSA) | Instituto Nacional de Saude (INSA) | Borges et al et al |
| EPI_ISL_738119 | Portugal/PT2087/2020 | 2020-11-08 | Europe/Portugal/Portugal/ | Instituto Nacional de Saude (INSA) | Instituto Nacional de Saude (INSA) | Borges et al et al |
| EPI_ISL_738129 | Italy/CAM-IZSM-88763-ORS/2020 | 2020-10-22 | Europe/Italy/Campania/ | IZSM-U.O.C. Virologia | Istituto Zooprofilattico Sperimentale del Mezzogiorno | Maurizio Viscardi et al |
| EPI_ISL_738139 | SouthKorea/KDCA0001/2020 | 2020-12-22 | Asia/South Korea/South Korea/ | Division of Emerging Infectious Diseases, Bureau of Infectious Diseases Diagnosis Control, Korea Disease Control and Prevention Agency | Division of Emerging Infectious Diseases, Bureau of Infectious Diseases Diagnosis Control, Korea Disease Control and Prevention Agency | Jeong-Min Kim et al |
| EPI_ISL_738141 | SouthKorea/KDCA0002/2020 | 2020-12-22 | Asia/South Korea/South Korea/ | Division of Emerging Infectious Diseases, Bureau of Infectious Diseases Diagnosis Control, Korea Disease Control and Prevention Agency | Division of Emerging Infectious Diseases, Bureau of Infectious Diseases Diagnosis Control, Korea Disease Control and Prevention Agency | Jeong-Min Kim et al |
| EPI_ISL_738142 | SouthKorea/KDCA0003/2020 | 2020-12-22 | Asia/South Korea/South Korea/ | Division of Emerging Infectious Diseases, Bureau of Infectious Diseases Diagnosis Control, Korea Disease Control and Prevention Agency | Division of Emerging Infectious Diseases, Bureau of Infectious Diseases Diagnosis Control, Korea Disease Control and Prevention Agency | Jeong-Min Kim et al |

| | | | | | | |
|---|---|---|---|---|---|---|
| EPI_ISL_738144 | Italy/APU-POLBA04/2020 | 2020-12-19 | Europe/Italy/Apulia/ | University of Bari Biomedical Sciences and Human Oncology | University of Bari Biomedical Sciences and Human Oncology | Maria Chironna et al |
| EPI_ISL_738147 | Italy/TUS-C44/2020 | 2020-02-25 | Europe/Italy/Tuscany/Florence | Microbiology and Virology Unit, Florence Careggi University Hospital | Microbiology and Virology Unit, Florence Careggi University Hospital | Vincenzo Di Pilato et al |
| EPI_ISL_738154 | USA/TX-DSHS-1815/2020 | 2020-06-22 | North America/USA/Texas/Hidalgo Coun | Texas Department of State Health Services | Texas Department of State Health Services | Anita Pokharel et al |
| EPI_ISL_738162 | USA/TX-DSHS-1787/2020 | 2020-06-13 | North America/USA/Texas/Harris Count | Texas Department of State Health Services | Texas Department of State Health Services | Anita Pokharel et al |
| EPI_ISL_738165 | USA/TX-DSHS-1790/2020 | 2020-06-13 | North America/USA/Texas/Harris Count | Texas Department of State Health Services | Texas Department of State Health Services | Anita Pokharel et al |
| EPI_ISL_738179 | USA/TX-DSHS-1809/2020 | 2020-06-15 | North America/USA/Texas/Harris Count | Texas Department of State Health Services | Texas Department of State Health Services | Anita Pokharel et al |
| EPI_ISL_738186 | USA/TX-DSHS-1819/2020 | 2020-06-22 | North America/USA/Texas/Hidalgo Coun | Texas Department of State Health Services | Texas Department of State Health Services | Anita Pokharel et al |
| EPI_ISL_738189 | USA/TX-DSHS-1822/2020 | 2020-06-22 | North America/USA/Texas/Hidalgo Coun | Texas Department of State Health Services | Texas Department of State Health Services | Anita Pokharel et al |
| EPI_ISL_738194 | Italy/TUS-C198/2020 | 2020-02-24 | Europe/Italy/Tuscany/ | Microbiology and Virology Unit, Florence Careggi University Hospital | Microbiology and Virology Unit, Florence Careggi University Hospital | Vincenzo Di Pilato et al |
| EPI_ISL_738201 | Belgium/rega-03041161/2020 | 2020-03-04 | Europe/Belgium/Nivelles/Ottignies | UZ Leuven, National Reference Laboratory for Coronaviruses, Laboratory Medicine, Leuven, Belgium | KU Leuven, Rega Institute, Clinical and Epidemiological Virology | Tony Wawina-Bokalanga et al |

| | | | | | | |
|---|---|---|---|---|---|---|
| EPI_ISL_738212 | Belgium/rega-0414677/2020 | 2020-04-14 | Europe/Belgium/Brussels/Anderlecht | UZ Leuven, National Reference Laboratory for Coronaviruses, Laboratory Medicine, Leuven, Belgium | KU Leuven, Rega Institute, Clinical and Epidemiological Virology | Tony Wawina-Bokalanga et al |
| EPI_ISL_738216 | Belgium/rega-0512675/2020 | 2020-05-12 | Europe/Belgium/Leuven/Oud-Heverlee | UZ Leuven, National Reference Laboratory for Coronaviruses, Laboratory Medicine, Leuven, Belgium | KU Leuven, Rega Institute, Clinical and Epidemiological Virology | Tony Wawina-Bokalanga et al |
| EPI_ISL_738237 | Belgium/rega-09221149/2020 | 2020-09-22 | Europe/Belgium/Brussels/Haren | UZ Leuven, National Reference Laboratory for Coronaviruses, Laboratory Medicine, Leuven, Belgium | KU Leuven, Rega Institute, Clinical and Epidemiological Virology | Tony Wawina-Bokalanga et al |
| EPI_ISL_738241 | France/HDF-rega-0416679/2020 | 2020-04-16 | Europe/France/Hauts de France/Bousbe | UZ Leuven, National Reference Laboratory for Coronaviruses, Laboratory Medicine, Leuven, Belgium | KU Leuven, Rega Institute, Clinical and Epidemiological Virology | Tony Wawina-Bokalanga et al |
| EPI_ISL_738243 | Italy/TUS-C266/2020 | 2020-03-03 | Europe/Italy/Tuscany/Florence | Microbiology and Virology Unit, Florence Careggi University Hospital | Microbiology and Virology Unit, Florence Careggi University Hospital | Vincenzo Di Pilato et al |
| EPI_ISL_738252 | USA/MA-Yale-446/2020 | 2020-04-15 | North America/USA/Massachusetts/ | Connecticut Veterans' Affairs Hospital | Grubaugh Lab - Yale School of Public Health | Joseph Fauver et al |
| EPI_ISL_738254 | USA/MA-Yale-442/2020 | 2020-04-14 | North America/USA/Massachusetts/ | Connecticut Veterans' Affairs Hospital | Grubaugh Lab - Yale School of Public Health | Joseph Fauver et al |
| EPI_ISL_738261 | USA/MA-Yale-435/2020 | 2020-04-09 | North America/USA/Massachusetts/ | Connecticut Veterans' Affairs Hospital | Grubaugh Lab - Yale School of Public Health | Joseph Fauver et al |
| EPI_ISL_738275 | USA/MA-Yale-419/2020 | 2020-04-24 | North America/USA/Massachusetts/ | Connecticut Veterans' Affairs Hospital | Grubaugh Lab - Yale School of Public Health | Joseph Fauver et al |
| EPI_ISL_738280 | USA/MA-Yale-414/2020 | 2020-04-20 | North America/USA/Massachusetts/ | Connecticut Veterans' Affairs Hospital | Grubaugh Lab - Yale School of Public Health | Joseph Fauver et al |

| | | | | | | |
|---|---|---|---|---|---|---|
| EPI_ISL_738290 | USA/CT-Yale-585/2020 | 2020-04-05 | North America/USA/Connecticut/ | Yale Pathology Lab | Grubaugh Lab - Yale School of Public Health | Joseph Fauver et al |
| EPI_ISL_738293 | USA/CT-Yale-582/2020 | 2020-05-27 | North America/USA/Connecticut/ | Yale Pathology Lab | Grubaugh Lab - Yale School of Public Health | Joseph Fauver et al |
| EPI_ISL_738305 | USA/CT-Yale-403/2020 | 2020-03-31 | North America/USA/Connecticut/ | Connecticut Veterans' Affairs Hospital | Grubaugh Lab - Yale School of Public Health | Joseph Fauver et al |
| EPI_ISL_738310 | USA/CT-Yale-397/2020 | 2020-05-05 | North America/USA/Connecticut/ | Connecticut Veterans' Affairs Hospital | Grubaugh Lab - Yale School of Public Health | Joseph Fauver et al |
| EPI_ISL_738317 | Romania/USAFSAM-S556/2020 | 2020-11-28 | Europe/Romania/Romania/ | Landstuhl Regional Medical Center | United States Air Force School of Aerospace Medicine | Anthony Fries et al |
| EPI_ISL_738318 | Romania/USAFSAM-S553/2020 | 2020-11-28 | Europe/Romania/Romania/ | Landstuhl Regional Medical Center | United States Air Force School of Aerospace Medicine | Anthony Fries et al |
| EPI_ISL_738319 | Romania/USAFSAM-S539/2020 | 2020-11-28 | Europe/Romania/Romania/ | Landstuhl Regional Medical Center | United States Air Force School of Aerospace Medicine | Anthony Fries et al |
| EPI_ISL_738320 | Romania/USAFSAM-S574/2020 | 2020-11-28 | Europe/Romania/Romania/ | Landstuhl Regional Medical Center | United States Air Force School of Aerospace Medicine | Anthony Fries et al |
| EPI_ISL_738321 | Romania/USAFSAM-S577/2020 | 2020-11-28 | Europe/Romania/Romania/ | Landstuhl Regional Medical Center | United States Air Force School of Aerospace Medicine | Anthony Fries et al |
| EPI_ISL_738322 | Romania/USAFSAM-S578/2020 | 2020-11-28 | Europe/Romania/Romania/ | Landstuhl Regional Medical Center | United States Air Force School of Aerospace Medicine | Anthony Fries et al |
| EPI_ISL_738325 | Romania/USAFSAM-S566/2020 | 2020-11-28 | Europe/Romania/Romania/ | Landstuhl Regional Medical Center | United States Air Force School of Aerospace Medicine | Anthony Fries et al |

| | | | | | | |
|---|---|---|---|---|---|---|
| EPI_ISL_738326 | Romania/USAFSAM-S542/2020 | 2020-11-28 | Europe/Romania/Romania/ | Landstuhl Regional Medical Center | United States Air Force School of Aerospace Medicine | Anthony Fries et al |
| EPI_ISL_738327 | Romania/USAFSAM-S543/2020 | 2020-11-28 | Europe/Romania/Romania/ | Landstuhl Regional Medical Center | United States Air Force School of Aerospace Medicine | Anthony Fries et al |
| EPI_ISL_738329 | Romania/USAFSAM-S546/2020 | 2020-11-28 | Europe/Romania/Romania/ | Landstuhl Regional Medical Center | United States Air Force School of Aerospace Medicine | Anthony Fries et al |
| EPI_ISL_738330 | Romania/USAFSAM-S547/2020 | 2020-11-28 | Europe/Romania/Romania/ | Landstuhl Regional Medical Center | United States Air Force School of Aerospace Medicine | Anthony Fries et al |
| EPI_ISL_738331 | Romania/USAFSAM-S548/2020 | 2020-11-28 | Europe/Romania/Romania/ | Landstuhl Regional Medical Center | United States Air Force School of Aerospace Medicine | Anthony Fries et al |
| EPI_ISL_738332 | Romania/USAFSAM-S549/2020 | 2020-11-28 | Europe/Romania/Romania/ | Landstuhl Regional Medical Center | United States Air Force School of Aerospace Medicine | Anthony Fries et al |
| EPI_ISL_738333 | Romania/USAFSAM-S550/2020 | 2020-11-28 | Europe/Romania/Romania/ | Landstuhl Regional Medical Center | United States Air Force School of Aerospace Medicine | Anthony Fries et al |
| EPI_ISL_738335 | Romania/USAFSAM-S554/2020 | 2020-11-28 | Europe/Romania/Romania/ | Landstuhl Regional Medical Center | United States Air Force School of Aerospace Medicine | Anthony Fries et al |
| EPI_ISL_738336 | Romania/USAFSAM-S555/2020 | 2020-11-28 | Europe/Romania/Romania/ | Landstuhl Regional Medical Center | United States Air Force School of Aerospace Medicine | Anthony Fries et al |
| EPI_ISL_738337 | Romania/USAFSAM-S557/2020 | 2020-11-28 | Europe/Romania/Romania/ | Landstuhl Regional Medical Center | United States Air Force School of Aerospace Medicine | Anthony Fries et al |

| | | | | | | |
|---|---|---|---|---|---|---|
| EPI_ISL_738338 | Romania/USAFSAM-S558/2020 | 2020-11-28 | Europe/Romania/Romania/ | Landstuhl Regional Medical Center | United States Air Force School of Aerospace Medicine | Anthony Fries et al |
| EPI_ISL_738339 | Romania/USAFSAM-S559/2020 | 2020-11-28 | Europe/Romania/Romania/ | Landstuhl Regional Medical Center | United States Air Force School of Aerospace Medicine | Anthony Fries et al |
| EPI_ISL_738340 | Romania/USAFSAM-S562/2020 | 2020-11-28 | Europe/Romania/Romania/ | Landstuhl Regional Medical Center | United States Air Force School of Aerospace Medicine | Anthony Fries et al |
| EPI_ISL_738341 | Romania/USAFSAM-S564/2020 | 2020-11-28 | Europe/Romania/Romania/ | Landstuhl Regional Medical Center | United States Air Force School of Aerospace Medicine | Anthony Fries et al |
| EPI_ISL_738342 | Romania/USAFSAM-S565/2020 | 2020-11-28 | Europe/Romania/Romania/ | Landstuhl Regional Medical Center | United States Air Force School of Aerospace Medicine | Anthony Fries et al |
| EPI_ISL_738343 | Romania/USAFSAM-S569/2020 | 2020-11-28 | Europe/Romania/Romania/ | Landstuhl Regional Medical Center | United States Air Force School of Aerospace Medicine | Anthony Fries et al |
| EPI_ISL_738344 | Romania/USAFSAM-S570/2020 | 2020-11-28 | Europe/Romania/Romania/ | Landstuhl Regional Medical Center | United States Air Force School of Aerospace Medicine | Anthony Fries et al |
| EPI_ISL_738345 | Romania/USAFSAM-S571/2020 | 2020-11-28 | Europe/Romania/Romania/ | Landstuhl Regional Medical Center | United States Air Force School of Aerospace Medicine | Anthony Fries et al |
| EPI_ISL_738347 | Romania/USAFSAM-S573/2020 | 2020-11-28 | Europe/Romania/Romania/ | Landstuhl Regional Medical Center | United States Air Force School of Aerospace Medicine | Anthony Fries et al |
| EPI_ISL_738348 | Romania/USAFSAM-S575/2020 | 2020-11-28 | Europe/Romania/Romania/ | Landstuhl Regional Medical Center | United States Air Force School of Aerospace Medicine | Anthony Fries et al |

| | | | | | | |
|---|---|---|---|---|---|---|
| EPI_ISL_738349 | Romania/USAFSAM-S576/2020 | 2020-11-28 | Europe/Romania/Romania/ | Landstuhl Regional Medical Center | United States Air Force School of Aerospace Medicine | Anthony Fries et al |
| EPI_ISL_738358 | USA/NH-UPHL-2012218768/2020 | 2020-12-18 | North America/USA/New Hampshire/ | Utah Public Health Laboratory | Utah Public Health Laboratory | Erin Young et al |
| EPI_ISL_738367 | Belgium/rega-12111313/2020 | 2020-12-11 | Europe/Belgium/Turnhout/Mol | UZ Leuven, National Reference Laboratory for Coronaviruses, Laboratory Medicine, Leuven, Belgium | KU Leuven, Rega Institute, Clinical and Epidemiological Virology | Tony Wawina-Bokalanga et al |
| EPI_ISL_738391 | Belgium/rega-12111338/2020 | 2020-12-11 | Europe/Belgium/Turnhout/Mol | UZ Leuven, National Reference Laboratory for Coronaviruses, Laboratory Medicine, Leuven, Belgium | KU Leuven, Rega Institute, Clinical and Epidemiological Virology | Tony Wawina-Bokalanga et al |
| EPI_ISL_738421 | Belgium/rega-12111368/2020 | 2020-12-11 | Europe/Belgium/Turnhout/Mol | UZ Leuven, National Reference Laboratory for Coronaviruses, Laboratory Medicine, Leuven, Belgium | KU Leuven, Rega Institute, Clinical and Epidemiological Virology | Tony Wawina-Bokalanga et al |
| EPI_ISL_738436 | Belgium/rega-10141384/2020 | 2020-10-14 | Europe/Belgium/Halle-Vilvoorde/Wezen | UZ Leuven, National Reference Laboratory for Coronaviruses, Laboratory Medicine, Leuven, Belgium | KU Leuven, Rega Institute, Clinical and Epidemiological Virology | Tony Wawina-Bokalanga et al |
| EPI_ISL_738453 | Belgium/rega-10141401/2020 | 2020-10-14 | Europe/Belgium/Leuven/Bertem | UZ Leuven, National Reference Laboratory for Coronaviruses, Laboratory Medicine, Leuven, Belgium | KU Leuven, Rega Institute, Clinical and Epidemiological Virology | Tony Wawina-Bokalanga et al |
| EPI_ISL_738470 | Belgium/rega-10131418/2020 | 2020-10-13 | Europe/Belgium/Halle-Vilvoorde/Kraain | UZ Leuven, National Reference Laboratory for Coronaviruses, Laboratory Medicine, Leuven, Belgium | KU Leuven, Rega Institute, Clinical and Epidemiological Virology | Tony Wawina-Bokalanga et al |

| | | | | | | |
|---|---|---|---|---|---|---|
| EPI_ISL_738496 | Belgium/rega-12111444/2020 | 2020-12-11 | Europe/Belgium/Turnhout/Arendonk | UZ Leuven, National Reference Laboratory for Coronaviruses, Laboratory Medicine, Leuven, Belgium | KU Leuven, Rega Institute, Clinical and Epidemiological Virology | Tony Wawina-Bokalanga et al |
| EPI_ISL_738508 | USA/CA-CZB-15112/2020 | 2020-11-22 | North America/USA/California/Santa Cl | Santa Clara County Public Health Laboratory | Chan-Zuckerberg Biohub | CZB Cliahub Consortium et al |
| EPI_ISL_738518 | USA/CA-CZB-15451/2020 | 2020-12-03 | North America/USA/California/Orange C | Orange County Public Health Lab | Chan-Zuckerberg Biohub | CZB Cliahub Consortium et al |
| EPI_ISL_738523 | USA/CA-CZB-14277/2020 | 2020-08-27 | North America/USA/California/Alameda | Alameda County Public Health Lab | Chan-Zuckerberg Biohub | CZB Cliahub Consortium et al |
| EPI_ISL_738537 | USA/CA-CZB-14320/2020 | 2020-08-28 | North America/USA/California/Alameda | Alameda County Public Health Lab | Chan-Zuckerberg Biohub | CZB Cliahub Consortium et al |
| EPI_ISL_738548 | USA/CA-CZB-15479/2020 | 2020-12-04 | North America/USA/California/Orange C | Orange County Public Health Lab | Chan-Zuckerberg Biohub | CZB Cliahub Consortium et al |
| EPI_ISL_738550 | USA/CA-CZB-15973/2020 | 2020-11-30 | North America/USA/California/Santa Cl | Santa Clara County Public Health Laboratory | Chan-Zuckerberg Biohub | CZB Cliahub Consortium et al |
| EPI_ISL_738554 | USA/CA-CZB-13763/2020 | 2020-09-17 | North America/USA/California/Alameda | Alameda County Public Health Lab | Chan-Zuckerberg Biohub | CZB Cliahub Consortium et al |
| EPI_ISL_738555 | USA/CA-CZB-15940/2020 | 2020-12-02 | North America/USA/California/Santa Cl | Santa Clara County Public Health Laboratory | Chan-Zuckerberg Biohub | CZB Cliahub Consortium et al |
| EPI_ISL_738561 | USA/CA-CZB-13855/2020 | 2020-08-14 | North America/USA/California/Alameda | Alameda County Public Health Lab | Chan-Zuckerberg Biohub | CZB Cliahub Consortium et al |
| EPI_ISL_738566 | USA/CA-CZB-13664/2020 | 2020-11-04 | North America/USA/California/Santa Cl | Santa Clara County Public Health Laboratory | Chan-Zuckerberg Biohub | CZB Cliahub Consortium et al |
| EPI_ISL_738570 | USA/CA-CZB-15419/2020 | 2020-12-02 | North America/USA/California/Orange C | Orange County Public Health Lab | Chan-Zuckerberg Biohub | CZB Cliahub Consortium et al |
| EPI_ISL_738578 | USA/CA-CZB-14564/2020 | 2020-11-14 | North America/USA/California/Santa Cl | Santa Clara County Public Health Laboratory | Chan-Zuckerberg Biohub | CZB Cliahub Consortium et al |
| EPI_ISL_738584 | USA/CA-CZB-14365/2020 | 2020-07-09 | North America/USA/California/Alameda | Alameda County Public Health Lab | Chan-Zuckerberg Biohub | CZB Cliahub Consortium et al |
| EPI_ISL_738587 | USA/CA-CZB-14108/2020 | 2020-05-25 | North America/USA/California/Alameda | Alameda County Public Health Lab | Chan-Zuckerberg Biohub | CZB Cliahub Consortium et al |
| EPI_ISL_738592 | USA/CA-CZB-14299/2020 | 2020-08-24 | North America/USA/California/Alameda | Alameda County Public Health Lab | Chan-Zuckerberg Biohub | CZB Cliahub Consortium et al |
| EPI_ISL_738593 | USA/CA-CZB-15327/2020 | 2020-11-25 | North America/USA/California/Santa Cl | Santa Clara County Public Health Laboratory | Chan-Zuckerberg Biohub | CZB Cliahub Consortium et al |
| EPI_ISL_738594 | USA/CA-CZB-14681/2020 | 2020-11-17 | North America/USA/California/Orange C | Orange County Public Health Lab | Chan-Zuckerberg Biohub | CZB Cliahub Consortium et al |

| | | | | | | |
|---|---|---|---|---|---|---|
| EPI_ISL_738595 | USA/CA-CZB-15253/2020 | 2020-11-24 | North America/USA/California/Santa Cl | Santa Clara County Public Health Laboratory | Chan-Zuckerberg Biohub | CZB Cliahub Consortium et al |
| EPI_ISL_738598 | USA/CA-CZB-15219/2020 | 2020-11-27 | North America/USA/California/Santa Cl | Santa Clara County Public Health Laboratory | Chan-Zuckerberg Biohub | CZB Cliahub Consortium et al |
| EPI_ISL_738601 | USA/CA-CZB-15007/2020 | 2020-11-18 | North America/USA/California/Santa Cl | Santa Clara County Public Health Laboratory | Chan-Zuckerberg Biohub | CZB Cliahub Consortium et al |
| EPI_ISL_738622 | USA/CA-CZB-14353/2020 | 2020-07-15 | North America/USA/California/Alameda | Alameda County Public Health Lab | Chan-Zuckerberg Biohub | CZB Cliahub Consortium et al |
| EPI_ISL_738631 | USA/CA-CZB-15146/2020 | 2020-11-23 | North America/USA/California/Santa Cl | Santa Clara County Public Health Laboratory | Chan-Zuckerberg Biohub | CZB Cliahub Consortium et al |
| EPI_ISL_738640 | USA/CA-CZB-15408/2020 | 2020-12-02 | North America/USA/California/Orange ( | Orange County Public Health Lab | Chan-Zuckerberg Biohub | CZB Cliahub Consortium et al |
| EPI_ISL_738641 | USA/CA-CZB-15417/2020 | 2020-12-02 | North America/USA/California/Orange ( | Orange County Public Health Lab | Chan-Zuckerberg Biohub | CZB Cliahub Consortium et al |
| EPI_ISL_738646 | USA/CA-CZB-15436/2020 | 2020-12-02 | North America/USA/California/Orange ( | Orange County Public Health Lab | Chan-Zuckerberg Biohub | CZB Cliahub Consortium et al |
| EPI_ISL_738653 | USA/CA-CZB-15173/2020 | 2020-11-23 | North America/USA/California/Santa Cl | Santa Clara County Public Health Laboratory | Chan-Zuckerberg Biohub | CZB Cliahub Consortium et al |
| EPI_ISL_738659 | USA/CA-CZB-14381/2020 | 2020-07-07 | North America/USA/California/Alameda | Alameda County Public Health Lab | Chan-Zuckerberg Biohub | CZB Cliahub Consortium et al |
| EPI_ISL_738665 | USA/CA-CZB-14123/2020 | 2020-05-22 | North America/USA/California/Alameda | Alameda County Public Health Lab | Chan-Zuckerberg Biohub | CZB Cliahub Consortium et al |
| EPI_ISL_738672 | USA/CA-CZB-14536/2020 | 2020-11-11 | North America/USA/California/Santa Cl | Santa Clara County Public Health Laboratory | Chan-Zuckerberg Biohub | CZB Cliahub Consortium et al |
| EPI_ISL_738674 | USA/CA-CZB-13822/2020 | 2020-08-18 | North America/USA/California/Alameda | Alameda County Public Health Lab | Chan-Zuckerberg Biohub | CZB Cliahub Consortium et al |
| EPI_ISL_738688 | USA/CA-CZB-15454/2020 | 2020-12-04 | North America/USA/California/Orange ( | Orange County Public Health Lab | Chan-Zuckerberg Biohub | CZB Cliahub Consortium et al |
| EPI_ISL_738689 | USA/CA-CZB-15125/2020 | 2020-11-23 | North America/USA/California/Santa Cl | Santa Clara County Public Health Laboratory | Chan-Zuckerberg Biohub | CZB Cliahub Consortium et al |
| EPI_ISL_738693 | USA/CA-CZB-14635/2020 | 2020-11-16 | North America/USA/California/Orange ( | Orange County Public Health Lab | Chan-Zuckerberg Biohub | CZB Cliahub Consortium et al |
| EPI_ISL_738704 | USA/CA-CZB-15459/2020 | 2020-12-04 | North America/USA/California/Orange ( | Orange County Public Health Lab | Chan-Zuckerberg Biohub | CZB Cliahub Consortium et al |
| EPI_ISL_738714 | USA/CA-CZB-16171/2020 | 2020-11-25 | North America/USA/California/San Fran | Renegade | Chan-Zuckerberg Biohub | CZB Cliahub Consortium et al |
| EPI_ISL_738716 | USA/CA-CZB-13661/2020 | 2020-11-04 | North America/USA/California/Santa Cl | Santa Clara County Public Health Laboratory | Chan-Zuckerberg Biohub | CZB Cliahub Consortium et al |
| EPI_ISL_738727 | USA/CA-CZB-16172/2020 | 2020-11-25 | North America/USA/California/San Fran | Renegade | Chan-Zuckerberg Biohub | CZB Cliahub Consortium et al |

| | | | | | | |
|---|---|---|---|---|---|---|
| EPI_ISL_738728 | USA/CA-CZB-14541/2020 | 2020-11-12 | North America/USA/California/Santa Cl | Santa Clara County Public Health Laboratory | Chan-Zuckerberg Biohub | CZB Cliahub Consortium et al |
| EPI_ISL_738739 | USA/CA-CZB-14943/2020 | 2020-11-29 | North America/USA/California/Orange C | Orange County Public Health Lab | Chan-Zuckerberg Biohub | CZB Cliahub Consortium et al |
| EPI_ISL_738740 | USA/CA-CZB-14944/2020 | 2020-11-29 | North America/USA/California/Orange C | Orange County Public Health Lab | Chan-Zuckerberg Biohub | CZB Cliahub Consortium et al |
| EPI_ISL_738744 | USA/CA-CZB-13816/2020 | 2020-08-17 | North America/USA/California/Alameda | Alameda County Public Health Lab | Chan-Zuckerberg Biohub | CZB Cliahub Consortium et al |
| EPI_ISL_738745 | USA/CA-CZB-13738/2020 | 2020-09-09 | North America/USA/California/Alameda | Alameda County Public Health Lab | Chan-Zuckerberg Biohub | CZB Cliahub Consortium et al |
| EPI_ISL_738757 | USA/CA-CZB-14210/2020 | 2020-11-10 | North America/USA/California/Orange C | Orange County Public Health Lab | Chan-Zuckerberg Biohub | CZB Cliahub Consortium et al |
| EPI_ISL_738769 | USA/CA-CZB-15008/2020 | 2020-11-18 | North America/USA/California/Santa Cl | Santa Clara County Public Health Laboratory | Chan-Zuckerberg Biohub | CZB Cliahub Consortium et al |
| EPI_ISL_738773 | USA/CA-CZB-14113/2020 | 2020-05-25 | North America/USA/California/Alameda | Alameda County Public Health Lab | Chan-Zuckerberg Biohub | CZB Cliahub Consortium et al |
| EPI_ISL_738778 | USA/CA-CZB-14716/2020 | 2020-09-23 | North America/USA/California/Humbold | Humboldt County Public Health Laboratory | Chan-Zuckerberg Biohub | CZB Cliahub Consortium et al |
| EPI_ISL_738783 | USA/CA-CZB-13833/2020 | 2020-08-17 | North America/USA/California/Alameda | Alameda County Public Health Lab | Chan-Zuckerberg Biohub | CZB Cliahub Consortium et al |
| EPI_ISL_738787 | USA/CA-CZB-15455/2020 | 2020-12-04 | North America/USA/California/Orange C | Orange County Public Health Lab | Chan-Zuckerberg Biohub | CZB Cliahub Consortium et al |
| EPI_ISL_738788 | USA/CA-CZB-14134/2020 | 2020-05-23 | North America/USA/California/Alameda | Alameda County Public Health Lab | Chan-Zuckerberg Biohub | CZB Cliahub Consortium et al |
| EPI_ISL_738792 | USA/CA-CZB-13774/2020 | 2020-09-21 | North America/USA/California/Alameda | Alameda County Public Health Lab | Chan-Zuckerberg Biohub | CZB Cliahub Consortium et al |
| EPI_ISL_738797 | USA/CA-CZB-14139/2020 | 2020-09-15 | North America/USA/California/Alameda | Alameda County Public Health Lab | Chan-Zuckerberg Biohub | CZB Cliahub Consortium et al |
| EPI_ISL_738803 | USA/CA-CZB-15135/2020 | 2020-11-23 | North America/USA/California/Santa Cl | Santa Clara County Public Health Laboratory | Chan-Zuckerberg Biohub | CZB Cliahub Consortium et al |
| EPI_ISL_738805 | USA/CA-CZB-15958/2020 | 2020-11-30 | North America/USA/California/Santa Cl | Santa Clara County Public Health Laboratory | Chan-Zuckerberg Biohub | CZB Cliahub Consortium et al |
| EPI_ISL_738812 | USA/CA-CZB-15167/2020 | 2020-11-23 | North America/USA/California/Santa Cl | Santa Clara County Public Health Laboratory | Chan-Zuckerberg Biohub | CZB Cliahub Consortium et al |
| EPI_ISL_738818 | USA/CA-CZB-14654/2020 | 2020-11-16 | North America/USA/California/Orange C | Orange County Public Health Lab | Chan-Zuckerberg Biohub | CZB Cliahub Consortium et al |
| EPI_ISL_738820 | USA/CA-CZB-11254/2020 | 2020-04-24 | North America/USA/California/ | Napa-Solano-Yolo- Marin County (NSYM) Public Health Laboratories | Chan-Zuckerberg Biohub | CZB Cliahub Consortium et al |
| EPI_ISL_738824 | USA/CA-CZB-14645/2020 | 2020-11-16 | North America/USA/California/Orange C | Orange County Public Health Lab | Chan-Zuckerberg Biohub | CZB Cliahub Consortium et al |

| | | | | | | |
|---|---|---|---|---|---|---|
| EPI_ISL_738825 | USA/CA-CZB-13906/2020 | 2020-08-08 | North America/USA/California/Alameda | Alameda County Public Health Lab | Chan-Zuckerberg Biohub | CZB Cliahub Consortium et al |
| EPI_ISL_738826 | USA/CA-CZB-14887/2020 | 2020-11-24 | North America/USA/California/Orange ( | Orange County Public Health Lab | Chan-Zuckerberg Biohub | CZB Cliahub Consortium et al |
| EPI_ISL_738829 | USA/CA-CZB-15469/2020 | 2020-12-06 | North America/USA/California/Orange ( | Orange County Public Health Lab | Chan-Zuckerberg Biohub | CZB Cliahub Consortium et al |
| EPI_ISL_738835 | USA/CA-CZB-13844/2020 | 2020-08-17 | North America/USA/California/Alameda | Alameda County Public Health Lab | Chan-Zuckerberg Biohub | CZB Cliahub Consortium et al |
| EPI_ISL_738836 | USA/CA-CZB-14606/2020 | 2020-10-29 | North America/USA/California/ | Napa-Solano-Yolo- Marin County (NSYM) Public Health Laboratories | Chan-Zuckerberg Biohub | CZB Cliahub Consortium et al |
| EPI_ISL_738838 | USA/CA-CZB-13946/2020 | 2020-06-22 | North America/USA/California/Alameda | Alameda County Public Health Lab | Chan-Zuckerberg Biohub | CZB Cliahub Consortium et al |
| EPI_ISL_738845 | USA/CA-CZB-14674/2020 | 2020-11-16 | North America/USA/California/Orange ( | Orange County Public Health Lab | Chan-Zuckerberg Biohub | CZB Cliahub Consortium et al |
| EPI_ISL_738859 | USA/CA-CZB-15274/2020 | 2020-11-28 | North America/USA/California/Santa Cl | Santa Clara County Public Health Laboratory | Chan-Zuckerberg Biohub | CZB Cliahub Consortium et al |
| EPI_ISL_738860 | USA/CA-CZB-14642/2020 | 2020-11-16 | North America/USA/California/Orange ( | Orange County Public Health Lab | Chan-Zuckerberg Biohub | CZB Cliahub Consortium et al |
| EPI_ISL_738861 | USA/CA-CZB-14563/2020 | 2020-11-14 | North America/USA/California/Santa Cl | Santa Clara County Public Health Laboratory | Chan-Zuckerberg Biohub | CZB Cliahub Consortium et al |
| EPI_ISL_738875 | USA/CA-CZB-14823/2020 | 2020-11-18 | North America/USA/California/Humbold | Humboldt County Public Health Laboratory | Chan-Zuckerberg Biohub | CZB Cliahub Consortium et al |
| EPI_ISL_738879 | USA/CA-CZB-14748/2020 | 2020-10-23 | North America/USA/California/Humbold | Humboldt County Public Health Laboratory | Chan-Zuckerberg Biohub | CZB Cliahub Consortium et al |
| EPI_ISL_738881 | USA/CA-CZB-14034/2020 | 2020-07-08 | North America/USA/California/Alameda | Alameda County Public Health Lab | Chan-Zuckerberg Biohub | CZB Cliahub Consortium et al |
| EPI_ISL_738890 | USA/CA-CZB-15482/2020 | 2020-09-04 | North America/USA/California/San Luis | County of San Luis Obispo Public Health Laboratory | Chan-Zuckerberg Biohub | CZB Cliahub Consortium et al |
| EPI_ISL_738897 | USA/CA-CZB-11250/2020 | 2020-05-02 | North America/USA/California/ | Napa-Solano-Yolo- Marin County (NSYM) Public Health Laboratories | Chan-Zuckerberg Biohub | CZB Cliahub Consortium et al |
| EPI_ISL_738901 | USA/CA-CZB-16066/2020 | 2020-11-29 | North America/USA/California/San Fran | Renegade | Chan-Zuckerberg Biohub | CZB Cliahub Consortium et al |
| EPI_ISL_738903 | USA/CA-CZB-15069/2020 | 2020-11-19 | North America/USA/California/Santa Cl | Santa Clara County Public Health Laboratory | Chan-Zuckerberg Biohub | CZB Cliahub Consortium et al |
| EPI_ISL_738912 | USA/CA-CZB-14778/2020 | 2020-11-10 | North America/USA/California/Humbold | Humboldt County Public Health Laboratory | Chan-Zuckerberg Biohub | CZB Cliahub Consortium et al |
| EPI_ISL_738914 | USA/CA-CZB-13997/2020 | 2020-06-12 | North America/USA/California/Alameda | Alameda County Public Health Lab | Chan-Zuckerberg Biohub | CZB Cliahub Consortium et al |
| EPI_ISL_738917 | USA/CA-CZB-14815/2020 | 2020-11-18 | North America/USA/California/Humbold | Humboldt County Public Health Laboratory | Chan-Zuckerberg Biohub | CZB Cliahub Consortium et al |

| | | | | | | |
|---|---|---|---|---|---|---|
| EPI_ISL_738923 | USA/CA-CZB-14791/2020 | 2020-11-12 | North America/USA/California/Humbold | Humboldt County Public Health Laboratory | Chan-Zuckerberg Biohub | CZB Cliahub Consortium et al |
| EPI_ISL_738939 | USA/CA-CZB-15443/2020 | 2020-12-03 | North America/USA/California/Orange ( | Orange County Public Health Lab | Chan-Zuckerberg Biohub | CZB Cliahub Consortium et al |
| EPI_ISL_738950 | USA/CA-CZB-14103/2020 | 2020-05-23 | North America/USA/California/Alameda | Alameda County Public Health Lab | Chan-Zuckerberg Biohub | CZB Cliahub Consortium et al |
| EPI_ISL_738962 | USA/CA-CZB-14553/2020 | 2020-11-14 | North America/USA/California/Santa Cl | Santa Clara County Public Health Laboratory | Chan-Zuckerberg Biohub | CZB Cliahub Consortium et al |
| EPI_ISL_738963 | USA/CA-CZB-13789/2020 | 2020-09-23 | North America/USA/California/Alameda | Alameda County Public Health Lab | Chan-Zuckerberg Biohub | CZB Cliahub Consortium et al |
| EPI_ISL_738964 | USA/CA-CZB-14152/2020 | 2020-10-15 | North America/USA/California/Madera | Madera County Department of Public Health | Chan-Zuckerberg Biohub | CZB Cliahub Consortium et al |
| EPI_ISL_738966 | USA/CA-CZB-14650/2020 | 2020-11-16 | North America/USA/California/Orange ( | Orange County Public Health Lab | Chan-Zuckerberg Biohub | CZB Cliahub Consortium et al |
| EPI_ISL_738972 | USA/CA-CZB-14528/2020 | 2020-11-11 | North America/USA/California/Santa Cl | Santa Clara County Public Health Laboratory | Chan-Zuckerberg Biohub | CZB Cliahub Consortium et al |
| EPI_ISL_738974 | USA/CA-CZB-13897/2020 | 2020-08-10 | North America/USA/California/Alameda | Alameda County Public Health Lab | Chan-Zuckerberg Biohub | CZB Cliahub Consortium et al |
| EPI_ISL_738976 | USA/CA-CZB-13948/2020 | 2020-06-22 | North America/USA/California/Alameda | Alameda County Public Health Lab | Chan-Zuckerberg Biohub | CZB Cliahub Consortium et al |
| EPI_ISL_738987 | USA/CA-CZB-15155/2020 | 2020-11-23 | North America/USA/California/Santa Cl | Santa Clara County Public Health Laboratory | Chan-Zuckerberg Biohub | CZB Cliahub Consortium et al |
| EPI_ISL_738988 | USA/CA-CZB-14734/2020 | 2020-10-09 | North America/USA/California/Humbold | Humboldt County Public Health Laboratory | Chan-Zuckerberg Biohub | CZB Cliahub Consortium et al |
| EPI_ISL_738990 | USA/CA-CZB-15491/2020 | 2020-09-17 | North America/USA/California/San Luis | County of San Luis Obispo Public Health Laboratory | Chan-Zuckerberg Biohub | CZB Cliahub Consortium et al |
| EPI_ISL_738991 | USA/CA-CZB-14878/2020 | 2020-11-21 | North America/USA/California/Orange ( | Orange County Public Health Lab | Chan-Zuckerberg Biohub | CZB Cliahub Consortium et al |
| EPI_ISL_738999 | USA/CA-CZB-14849/2020 | 2020-11-20 | North America/USA/California/Humbold | Humboldt County Public Health Laboratory | Chan-Zuckerberg Biohub | CZB Cliahub Consortium et al |
| EPI_ISL_739003 | USA/CA-CZB-14118/2020 | 2020-05-24 | North America/USA/California/Alameda | Alameda County Public Health Lab | Chan-Zuckerberg Biohub | CZB Cliahub Consortium et al |
| EPI_ISL_739009 | USA/CA-CZB-15959/2020 | 2020-11-25 | North America/USA/California/Santa Cl | Santa Clara County Public Health Laboratory | Chan-Zuckerberg Biohub | CZB Cliahub Consortium et al |
| EPI_ISL_739017 | USA/CA-CZB-15494/2020 | 2020-09-22 | North America/USA/California/San Luis | County of San Luis Obispo Public Health Laboratory | Chan-Zuckerberg Biohub | CZB Cliahub Consortium et al |
| EPI_ISL_739019 | USA/CA-CZB-13681/2020 | 2020-11-04 | North America/USA/California/Santa Cl | Santa Clara County Public Health Laboratory | Chan-Zuckerberg Biohub | CZB Cliahub Consortium et al |
| EPI_ISL_739020 | USA/CA-CZB-15325/2020 | 2020-11-25 | North America/USA/California/Santa Cl | Santa Clara County Public Health Laboratory | Chan-Zuckerberg Biohub | CZB Cliahub Consortium et al |

| | | | | | | |
|---|---|---|---|---|---|---|
| EPI_ISL_739048 | USA/CA-CZB-14686/2020 | 2020-11-16 | North America/USA/California/Orange ( | Orange County Public Health Lab | Chan-Zuckerberg Biohub | CZB Cliahub Consortium et al |
| EPI_ISL_739052 | USA/CA-CZB-14760/2020 | 2020-11-04 | North America/USA/California/Humbold | Humboldt County Public Health Laboratory | Chan-Zuckerberg Biohub | CZB Cliahub Consortium et al |
| EPI_ISL_739065 | USA/CA-CZB-15188/2020 | 2020-11-23 | North America/USA/California/Santa Cl | Santa Clara County Public Health Laboratory | Chan-Zuckerberg Biohub | CZB Cliahub Consortium et al |
| EPI_ISL_739067 | USA/CA-CZB-15322/2020 | 2020-11-25 | North America/USA/California/Santa Cl | Santa Clara County Public Health Laboratory | Chan-Zuckerberg Biohub | CZB Cliahub Consortium et al |
| EPI_ISL_739069 | USA/CA-CZB-14934/2020 | 2020-11-28 | North America/USA/California/Orange ( | Orange County Public Health Lab | Chan-Zuckerberg Biohub | CZB Cliahub Consortium et al |
| EPI_ISL_739073 | USA/CA-CZB-13838/2020 | 2020-08-12 | North America/USA/California/Alameda | Alameda County Public Health Lab | Chan-Zuckerberg Biohub | CZB Cliahub Consortium et al |
| EPI_ISL_739075 | USA/CA-CZB-13970/2020 | 2020-06-18 | North America/USA/California/Alameda | Alameda County Public Health Lab | Chan-Zuckerberg Biohub | CZB Cliahub Consortium et al |
| EPI_ISL_739077 | USA/CA-CZB-14363/2020 | 2020-07-09 | North America/USA/California/Alameda | Alameda County Public Health Lab | Chan-Zuckerberg Biohub | CZB Cliahub Consortium et al |
| EPI_ISL_739085 | USA/CA-CZB-15105/2020 | 2020-11-20 | North America/USA/California/Santa Cl | Santa Clara County Public Health Laboratory | Chan-Zuckerberg Biohub | CZB Cliahub Consortium et al |
| EPI_ISL_739086 | USA/CA-CZB-14724/2020 | 2020-09-30 | North America/USA/California/Humbold | Humboldt County Public Health Laboratory | Chan-Zuckerberg Biohub | CZB Cliahub Consortium et al |
| EPI_ISL_739092 | USA/CA-CZB-14679/2020 | 2020-11-17 | North America/USA/California/Orange ( | Orange County Public Health Lab | Chan-Zuckerberg Biohub | CZB Cliahub Consortium et al |
| EPI_ISL_739096 | USA/CA-CZB-13890/2020 | 2020-08-10 | North America/USA/California/Alameda | Alameda County Public Health Lab | Chan-Zuckerberg Biohub | CZB Cliahub Consortium et al |
| EPI_ISL_739100 | USA/CA-CZB-14016/2020 | 2020-06-07 | North America/USA/California/Alameda | Alameda County Public Health Lab | Chan-Zuckerberg Biohub | CZB Cliahub Consortium et al |
| EPI_ISL_739102 | USA/CA-CZB-15495/2020 | 2020-06-15 | North America/USA/California/San Luis | County of San Luis Obispo Public Health Laboratory | Chan-Zuckerberg Biohub | CZB Cliahub Consortium et al |
| EPI_ISL_739115 | USA/CA-CZB-13807/2020 | 2020-08-17 | North America/USA/California/Alameda | Alameda County Public Health Lab | Chan-Zuckerberg Biohub | CZB Cliahub Consortium et al |
| EPI_ISL_739116 | USA/CA-CZB-15426/2020 | 2020-12-02 | North America/USA/California/Orange ( | Orange County Public Health Lab | Chan-Zuckerberg Biohub | CZB Cliahub Consortium et al |
| EPI_ISL_739132 | USA/CA-CZB-14946/2020 | 2020-11-29 | North America/USA/California/Orange ( | Orange County Public Health Lab | Chan-Zuckerberg Biohub | CZB Cliahub Consortium et al |
| EPI_ISL_739133 | USA/CA-CZB-14153/2020 | 2020-10-15 | North America/USA/California/Madera | Madera County Department of Public Health | Chan-Zuckerberg Biohub | CZB Cliahub Consortium et al |
| EPI_ISL_739151 | USA/CA-CZB-14200/2020 | 2020-11-09 | North America/USA/California/Orange ( | Orange County Public Health Lab | Chan-Zuckerberg Biohub | CZB Cliahub Consortium et al |
| EPI_ISL_739155 | USA/CA-CZB-14853/2020 | 2020-11-22 | North America/USA/California/Humbold | Humboldt County Public Health Laboratory | Chan-Zuckerberg Biohub | CZB Cliahub Consortium et al |

| | | | | | | |
|---|---|---|---|---|---|---|
| EPI_ISL_739156 | USA/CA-CZB-14209/2020 | 2020-11-10 | North America/USA/California/Orange ( | Orange County Public Health Lab | Chan-Zuckerberg Biohub | CZB Cliahub Consortium et al |
| EPI_ISL_739158 | USA/CA-CZB-13864/2020 | 2020-08-14 | North America/USA/California/Alameda | Alameda County Public Health Lab | Chan-Zuckerberg Biohub | CZB Cliahub Consortium et al |
| EPI_ISL_739163 | USA/CA-CZB-13885/2020 | 2020-08-11 | North America/USA/California/Alameda | Alameda County Public Health Lab | Chan-Zuckerberg Biohub | CZB Cliahub Consortium et al |
| EPI_ISL_739173 | USA/CA-CZB-13742/2020 | 2020-09-10 | North America/USA/California/Alameda | Alameda County Public Health Lab | Chan-Zuckerberg Biohub | CZB Cliahub Consortium et al |
| EPI_ISL_739174 | USA/CA-CZB-14129/2020 | 2020-05-27 | North America/USA/California/Alameda | Alameda County Public Health Lab | Chan-Zuckerberg Biohub | CZB Cliahub Consortium et al |
| EPI_ISL_739175 | USA/CA-CZB-14226/2020 | 2020-11-13 | North America/USA/California/Orange ( | Orange County Public Health Lab | Chan-Zuckerberg Biohub | CZB Cliahub Consortium et al |
| EPI_ISL_739177 | USA/CA-CZB-14741/2020 | 2020-10-16 | North America/USA/California/Humbold | Humboldt County Public Health Laboratory | Chan-Zuckerberg Biohub | CZB Cliahub Consortium et al |
| EPI_ISL_739179 | USA/CA-CZB-14306/2020 | 2020-08-24 | North America/USA/California/Alameda | Alameda County Public Health Lab | Chan-Zuckerberg Biohub | CZB Cliahub Consortium et al |
| EPI_ISL_739180 | USA/CA-CZB-15190/2020 | 2020-11-23 | North America/USA/California/Santa Cl | Santa Clara County Public Health Laboratory | Chan-Zuckerberg Biohub | CZB Cliahub Consortium et al |
| EPI_ISL_739186 | USA/CA-CZB-15968/2020 | 2020-11-30 | North America/USA/California/Santa Cl | Santa Clara County Public Health Laboratory | Chan-Zuckerberg Biohub | CZB Cliahub Consortium et al |
| EPI_ISL_739193 | USA/CA-CZB-14713/2020 | 2020-10-23 | North America/USA/California/Tulare C | Tulare County Public Health Lab | Chan-Zuckerberg Biohub | CZB Cliahub Consortium et al |
| EPI_ISL_739197 | USA/CA-CZB-14154/2020 | 2020-10-15 | North America/USA/California/Madera | Madera County Department of Public Health | Chan-Zuckerberg Biohub | CZB Cliahub Consortium et al |
| EPI_ISL_739199 | USA/CA-CZB-14230/2020 | 2020-11-13 | North America/USA/California/Orange ( | Orange County Public Health Lab | Chan-Zuckerberg Biohub | CZB Cliahub Consortium et al |
| EPI_ISL_739202 | USA/CA-CZB-13779/2020 | 2020-09-22 | North America/USA/California/Alameda | Alameda County Public Health Lab | Chan-Zuckerberg Biohub | CZB Cliahub Consortium et al |
| EPI_ISL_739203 | USA/CA-CZB-14590/2020 | 2020-10-08 | North America/USA/California/Marin C | UCSF Clinical Microbiology Laboratory | Chan-Zuckerberg Biohub | CZB Cliahub Consortium et al |
| EPI_ISL_739210 | USA/CA-CZB-14799/2020 | 2020-11-16 | North America/USA/California/Humbold | Humboldt County Public Health Laboratory | Chan-Zuckerberg Biohub | CZB Cliahub Consortium et al |
| EPI_ISL_739218 | USA/CA-CZB-14259/2020 | 2020-08-25 | North America/USA/California/Alameda | Alameda County Public Health Lab | Chan-Zuckerberg Biohub | CZB Cliahub Consortium et al |
| EPI_ISL_739219 | USA/CA-CZB-15178/2020 | 2020-11-23 | North America/USA/California/Santa Cl | Santa Clara County Public Health Laboratory | Chan-Zuckerberg Biohub | CZB Cliahub Consortium et al |
| EPI_ISL_739226 | USA/CA-CZB-13917/2020 | 2020-08-06 | North America/USA/California/Alameda | Alameda County Public Health Lab | Chan-Zuckerberg Biohub | CZB Cliahub Consortium et al |
| EPI_ISL_739234 | USA/CA-CZB-15170/2020 | 2020-11-23 | North America/USA/California/Santa Cl | Santa Clara County Public Health Laboratory | Chan-Zuckerberg Biohub | CZB Cliahub Consortium et al |

| | | | | | | |
|---|---|---|---|---|---|---|
| EPI_ISL_739236 | USA/CA-CZB-14387/2020 | 2020-06-29 | North America/USA/California/Alameda | Alameda County Public Health Lab | Chan-Zuckerberg Biohub | CZB Cliahub Consortium et al |
| EPI_ISL_739238 | USA/CA-CZB-14636/2020 | 2020-11-16 | North America/USA/California/Orange ( | Orange County Public Health Lab | Chan-Zuckerberg Biohub | CZB Cliahub Consortium et al |
| EPI_ISL_739241 | USA/CA-CZB-14417/2020 | 2020-07-08 | North America/USA/California/Alameda | Alameda County Public Health Lab | Chan-Zuckerberg Biohub | CZB Cliahub Consortium et al |
| EPI_ISL_739248 | USA/CA-CZB-15321/2020 | 2020-11-25 | North America/USA/California/Santa Cl | Santa Clara County Public Health Laboratory | Chan-Zuckerberg Biohub | CZB Cliahub Consortium et al |
| EPI_ISL_739249 | USA/CA-CZB-15347/2020 | 2020-05-13 | North America/USA/California/Santa Cl | Santa Clara County Public Health Laboratory | Chan-Zuckerberg Biohub | CZB Cliahub Consortium et al |
| EPI_ISL_739262 | USA/CA-CZB-13927/2020 | 2020-08-05 | North America/USA/California/Alameda | Alameda County Public Health Lab | Chan-Zuckerberg Biohub | CZB Cliahub Consortium et al |
| EPI_ISL_739264 | USA/CA-CZB-14722/2020 | 2020-09-28 | North America/USA/California/Humbolc | Humboldt County Public Health Laboratory | Chan-Zuckerberg Biohub | CZB Cliahub Consortium et al |
| EPI_ISL_739265 | USA/CA-CZB-15472/2020 | 2020-12-05 | North America/USA/California/Orange ( | Orange County Public Health Lab | Chan-Zuckerberg Biohub | CZB Cliahub Consortium et al |
| EPI_ISL_739268 | USA/CA-CZB-14137/2020 | 2020-05-25 | North America/USA/California/Alameda | Alameda County Public Health Lab | Chan-Zuckerberg Biohub | CZB Cliahub Consortium et al |
| EPI_ISL_739273 | USA/CA-CZB-15295/2020 | 2020-11-20 | North America/USA/California/Santa Cl | Santa Clara County Public Health Laboratory | Chan-Zuckerberg Biohub | CZB Cliahub Consortium et al |
| EPI_ISL_739276 | USA/CA-CZB-14240/2020 | 2020-11-17 | North America/USA/California/Orange ( | Orange County Public Health Lab | Chan-Zuckerberg Biohub | CZB Cliahub Consortium et al |
| EPI_ISL_739280 | USA/CA-CZB-13823/2020 | 2020-08-17 | North America/USA/California/Alameda | Alameda County Public Health Lab | Chan-Zuckerberg Biohub | CZB Cliahub Consortium et al |
| EPI_ISL_739283 | USA/CA-CZB-13772/2020 | 2020-09-20 | North America/USA/California/Alameda | Alameda County Public Health Lab | Chan-Zuckerberg Biohub | CZB Cliahub Consortium et al |
| EPI_ISL_739285 | USA/CA-CZB-13793/2020 | 2020-09-24 | North America/USA/California/Alameda | Alameda County Public Health Lab | Chan-Zuckerberg Biohub | CZB Cliahub Consortium et al |
| EPI_ISL_739299 | USA/CA-CZB-14841/2020 | 2020-11-20 | North America/USA/California/Humbolc | Humboldt County Public Health Laboratory | Chan-Zuckerberg Biohub | CZB Cliahub Consortium et al |
| EPI_ISL_739302 | USA/CA-CZB-13891/2020 | 2020-08-10 | North America/USA/California/Alameda | Alameda County Public Health Lab | Chan-Zuckerberg Biohub | CZB Cliahub Consortium et al |
| EPI_ISL_739321 | USA/CA-CZB-13702/2020 | 2020-11-04 | North America/USA/California/Santa Cl | Santa Clara County Public Health Laboratory | Chan-Zuckerberg Biohub | CZB Cliahub Consortium et al |
| EPI_ISL_739324 | USA/CA-CZB-14383/2020 | 2020-07-08 | North America/USA/California/Alameda | Alameda County Public Health Lab | Chan-Zuckerberg Biohub | CZB Cliahub Consortium et al |
| EPI_ISL_739326 | USA/CA-CZB-13856/2020 | 2020-08-12 | North America/USA/California/Alameda | Alameda County Public Health Lab | Chan-Zuckerberg Biohub | CZB Cliahub Consortium et al |
| EPI_ISL_739329 | USA/CA-CZB-14953/2020 | 2020-06-06 | North America/USA/California/Montere | Monterey County Public Health Lab | Chan-Zuckerberg Biohub | CZB Cliahub Consortium et al |

| | | | | | | |
|---|---|---|---|---|---|---|
| EPI_ISL_739332 | USA/CA-CZB-14583/2020 | 2020-11-17 | North America/USA/California/Santa Cl | Santa Clara County Public Health Laboratory | Chan-Zuckerberg Biohub | CZB Cliahub Consortium et al |
| EPI_ISL_739336 | USA/CA-CZB-14948/2020 | 2020-09-18 | North America/USA/California/Montere | Monterey County Public Health Lab | Chan-Zuckerberg Biohub | CZB Cliahub Consortium et al |
| EPI_ISL_739340 | USA/CA-CZB-14394/2020 | 2020-07-02 | North America/USA/California/Alameda | Alameda County Public Health Lab | Chan-Zuckerberg Biohub | CZB Cliahub Consortium et al |
| EPI_ISL_739342 | USA/CA-CZB-15475/2020 | 2020-12-05 | North America/USA/California/Orange ( | Orange County Public Health Lab | Chan-Zuckerberg Biohub | CZB Cliahub Consortium et al |
| EPI_ISL_739343 | USA/CA-CZB-15059/2020 | 2020-11-18 | North America/USA/California/Santa Cl | Santa Clara County Public Health Laboratory | Chan-Zuckerberg Biohub | CZB Cliahub Consortium et al |
| EPI_ISL_739346 | USA/CA-CZB-14587/2020 | 2020-11-13 | North America/USA/California/Santa Cl | Santa Clara County Public Health Laboratory | Chan-Zuckerberg Biohub | CZB Cliahub Consortium et al |
| EPI_ISL_739347 | USA/CA-CZB-14892/2020 | 2020-11-25 | North America/USA/California/Orange ( | Orange County Public Health Lab | Chan-Zuckerberg Biohub | CZB Cliahub Consortium et al |
| EPI_ISL_739356 | USA/CA-CZB-13868/2020 | 2020-08-19 | North America/USA/California/Alameda | Alameda County Public Health Lab | Chan-Zuckerberg Biohub | CZB Cliahub Consortium et al |
| EPI_ISL_739360 | USA/CA-CZB-15099/2020 | 2020-11-20 | North America/USA/California/Santa Cl | Santa Clara County Public Health Laboratory | Chan-Zuckerberg Biohub | CZB Cliahub Consortium et al |
| EPI_ISL_739365 | USA/CA-CZB-15067/2020 | 2020-11-17 | North America/USA/California/Santa Cl | Santa Clara County Public Health Laboratory | Chan-Zuckerberg Biohub | CZB Cliahub Consortium et al |
| EPI_ISL_739369 | USA/CA-CZB-13716/2020 | 2020-11-05 | North America/USA/California/Orange ( | Orange County Public Health Lab | Chan-Zuckerberg Biohub | CZB Cliahub Consortium et al |
| EPI_ISL_739376 | USA/CA-CZB-15023/2020 | 2020-11-18 | North America/USA/California/Santa Cl | Santa Clara County Public Health Laboratory | Chan-Zuckerberg Biohub | CZB Cliahub Consortium et al |
| EPI_ISL_739377 | USA/CA-CZB-15092/2020 | 2020-11-20 | North America/USA/California/Santa Cl | Santa Clara County Public Health Laboratory | Chan-Zuckerberg Biohub | CZB Cliahub Consortium et al |
| EPI_ISL_739379 | USA/CA-CZB-14239/2020 | 2020-11-16 | North America/USA/California/Orange ( | Orange County Public Health Lab | Chan-Zuckerberg Biohub | CZB Cliahub Consortium et al |
| EPI_ISL_739384 | USA/CA-CZB-13725/2020 | 2020-11-09 | North America/USA/California/Orange ( | Orange County Public Health Lab | Chan-Zuckerberg Biohub | CZB Cliahub Consortium et al |
| EPI_ISL_739385 | USA/CA-CZB-14010/2020 | 2020-06-08 | North America/USA/California/Alameda | Alameda County Public Health Lab | Chan-Zuckerberg Biohub | CZB Cliahub Consortium et al |
| EPI_ISL_739386 | USA/CA-CZB-14225/2020 | 2020-11-13 | North America/USA/California/Orange ( | Orange County Public Health Lab | Chan-Zuckerberg Biohub | CZB Cliahub Consortium et al |
| EPI_ISL_739387 | USA/CA-CZB-13870/2020 | 2020-08-17 | North America/USA/California/Alameda | Alameda County Public Health Lab | Chan-Zuckerberg Biohub | CZB Cliahub Consortium et al |
| EPI_ISL_739389 | USA/CA-CZB-14797/2020 | 2020-11-16 | North America/USA/California/Humbold | Humboldt County Public Health Laboratory | Chan-Zuckerberg Biohub | CZB Cliahub Consortium et al |
| EPI_ISL_739392 | USA/CA-CZB-15328/2020 | 2020-11-25 | North America/USA/California/Santa Cl | Santa Clara County Public Health Laboratory | Chan-Zuckerberg Biohub | CZB Cliahub Consortium et al |

| | | | | | | |
|---|---|---|---|---|---|---|
| EPI_ISL_739393 | USA/CA-CZB-14160/2020 | 2020-11-04 | North America/USA/California/Madera | Madera County Department of Public Health | Chan-Zuckerberg Biohub | CZB Cliahub Consortium et al |
| EPI_ISL_739404 | USA/CA-CZB-15055/2020 | 2020-11-18 | North America/USA/California/Santa Cl | Santa Clara County Public Health Laboratory | Chan-Zuckerberg Biohub | CZB Cliahub Consortium et al |
| EPI_ISL_739406 | USA/CA-CZB-13802/2020 | 2020-08-12 | North America/USA/California/Alameda | Alameda County Public Health Lab | Chan-Zuckerberg Biohub | CZB Cliahub Consortium et al |
| EPI_ISL_739416 | USA/CA-CZB-15156/2020 | 2020-11-23 | North America/USA/California/Santa Cl | Santa Clara County Public Health Laboratory | Chan-Zuckerberg Biohub | CZB Cliahub Consortium et al |
| EPI_ISL_739424 | USA/CA-CZB-14112/2020 | 2020-05-25 | North America/USA/California/Alameda | Alameda County Public Health Lab | Chan-Zuckerberg Biohub | CZB Cliahub Consortium et al |
| EPI_ISL_739427 | USA/CA-CZB-14243/2020 | 2020-08-27 | North America/USA/California/Alameda | Alameda County Public Health Lab | Chan-Zuckerberg Biohub | CZB Cliahub Consortium et al |
| EPI_ISL_739436 | USA/CA-CZB-14806/2020 | 2020-11-17 | North America/USA/California/Humbold | Humboldt County Public Health Laboratory | Chan-Zuckerberg Biohub | CZB Cliahub Consortium et al |
| EPI_ISL_739448 | USA/CA-CZB-13806/2020 | 2020-08-17 | North America/USA/California/Alameda | Alameda County Public Health Lab | Chan-Zuckerberg Biohub | CZB Cliahub Consortium et al |
| EPI_ISL_739450 | USA/CA-CZB-14907/2020 | 2020-11-25 | North America/USA/California/Orange ( | Orange County Public Health Lab | Chan-Zuckerberg Biohub | CZB Cliahub Consortium et al |
| EPI_ISL_739453 | USA/CA-CZB-14578/2020 | 2020-11-17 | North America/USA/California/Santa Cl | Santa Clara County Public Health Laboratory | Chan-Zuckerberg Biohub | CZB Cliahub Consortium et al |
| EPI_ISL_739455 | USA/CA-CZB-14928/2020 | 2020-11-27 | North America/USA/California/Orange ( | Orange County Public Health Lab | Chan-Zuckerberg Biohub | CZB Cliahub Consortium et al |
| EPI_ISL_739456 | USA/CA-CZB-15238/2020 | 2020-11-24 | North America/USA/California/Santa Cl | Santa Clara County Public Health Laboratory | Chan-Zuckerberg Biohub | CZB Cliahub Consortium et al |
| EPI_ISL_739459 | USA/CA-CZB-14390/2020 | 2020-07-01 | North America/USA/California/Alameda | Alameda County Public Health Lab | Chan-Zuckerberg Biohub | CZB Cliahub Consortium et al |
| EPI_ISL_739465 | USA/CA-CZB-14667/2020 | 2020-11-16 | North America/USA/California/Orange ( | Orange County Public Health Lab | Chan-Zuckerberg Biohub | CZB Cliahub Consortium et al |
| EPI_ISL_739466 | USA/CA-CZB-16075/2020 | 2020-11-29 | North America/USA/California/San Fran | Renegade | Chan-Zuckerberg Biohub | CZB Cliahub Consortium et al |
| EPI_ISL_739469 | USA/CA-CZB-11271/2020 | 2020-09-11 | North America/USA/California/ | Napa-Solano-Yolo- Marin County (NSYM) Public Health Laboratories | Chan-Zuckerberg Biohub | CZB Cliahub Consortium et al |
| EPI_ISL_739470 | USA/CA-CZB-13865/2020 | 2020-08-12 | North America/USA/California/Alameda | Alameda County Public Health Lab | Chan-Zuckerberg Biohub | CZB Cliahub Consortium et al |
| EPI_ISL_739472 | USA/CA-CZB-13867/2020 | 2020-08-20 | North America/USA/California/Alameda | Alameda County Public Health Lab | Chan-Zuckerberg Biohub | CZB Cliahub Consortium et al |
| EPI_ISL_739473 | USA/CA-CZB-14254/2020 | 2020-08-27 | North America/USA/California/Alameda | Alameda County Public Health Lab | Chan-Zuckerberg Biohub | CZB Cliahub Consortium et al |
| EPI_ISL_739475 | USA/CA-CZB-14830/2020 | 2020-11-18 | North America/USA/California/Humbold | Humboldt County Public Health Laboratory | Chan-Zuckerberg Biohub | CZB Cliahub Consortium et al |

| | | | | | | |
|---|---|---|---|---|---|---|
| EPI_ISL_739487 | USA/CA-CZB-15971/2020 | 2020-11-30 | North America/USA/California/Santa Cl | Santa Clara County Public Health Laboratory | Chan-Zuckerberg Biohub | CZB Cliahub Consortium et al |
| EPI_ISL_739490 | USA/CA-CZB-13715/2020 | 2020-11-05 | North America/USA/California/Orange C | Orange County Public Health Lab | Chan-Zuckerberg Biohub | CZB Cliahub Consortium et al |
| EPI_ISL_739493 | USA/CA-CZB-14695/2020 | 2020-11-17 | North America/USA/California/Orange C | Orange County Public Health Lab | Chan-Zuckerberg Biohub | CZB Cliahub Consortium et al |
| EPI_ISL_739497 | USA/CA-CZB-14902/2020 | 2020-11-25 | North America/USA/California/Orange C | Orange County Public Health Lab | Chan-Zuckerberg Biohub | CZB Cliahub Consortium et al |
| EPI_ISL_739499 | USA/CA-CZB-14856/2020 | 2020-11-23 | North America/USA/California/Humbold | Humboldt County Public Health Laboratory | Chan-Zuckerberg Biohub | CZB Cliahub Consortium et al |
| EPI_ISL_739505 | USA/CA-CZB-15933/2020 | 2020-12-02 | North America/USA/California/Santa Cl | Santa Clara County Public Health Laboratory | Chan-Zuckerberg Biohub | CZB Cliahub Consortium et al |
| EPI_ISL_739527 | USA/CA-CZB-14775/2020 | 2020-11-09 | North America/USA/California/Humbold | Humboldt County Public Health Laboratory | Chan-Zuckerberg Biohub | CZB Cliahub Consortium et al |
| EPI_ISL_739533 | USA/CA-CZB-14737/2020 | 2020-10-12 | North America/USA/California/Humbold | Humboldt County Public Health Laboratory | Chan-Zuckerberg Biohub | CZB Cliahub Consortium et al |
| EPI_ISL_739536 | USA/CA-CZB-14511/2020 | 2020-11-11 | North America/USA/California/Santa Cl | Santa Clara County Public Health Laboratory | Chan-Zuckerberg Biohub | CZB Cliahub Consortium et al |
| EPI_ISL_739540 | USA/CA-CZB-13718/2020 | 2020-11-06 | North America/USA/California/Orange C | Orange County Public Health Lab | Chan-Zuckerberg Biohub | CZB Cliahub Consortium et al |
| EPI_ISL_739543 | USA/CA-CZB-15028/2020 | 2020-11-18 | North America/USA/California/Santa Cl | Santa Clara County Public Health Laboratory | Chan-Zuckerberg Biohub | CZB Cliahub Consortium et al |
| EPI_ISL_739550 | USA/CA-CZB-15230/2020 | 2020-11-24 | North America/USA/California/Santa Cl | Santa Clara County Public Health Laboratory | Chan-Zuckerberg Biohub | CZB Cliahub Consortium et al |
| EPI_ISL_739555 | USA/CA-CZB-13836/2020 | 2020-08-15 | North America/USA/California/Alameda | Alameda County Public Health Lab | Chan-Zuckerberg Biohub | CZB Cliahub Consortium et al |
| EPI_ISL_739557 | USA/CA-CZB-11273/2020 | 2020-09-11 | North America/USA/California/ | Napa-Solano-Yolo- Marin County (NSYM) Public Health Laboratories | Chan-Zuckerberg Biohub | CZB Cliahub Consortium et al |
| EPI_ISL_739571 | USA/CA-CZB-14729/2020 | 2020-10-05 | North America/USA/California/Humbold | Humboldt County Public Health Laboratory | Chan-Zuckerberg Biohub | CZB Cliahub Consortium et al |
| EPI_ISL_739577 | USA/CA-CZB-14938/2020 | 2020-11-28 | North America/USA/California/Orange C | Orange County Public Health Lab | Chan-Zuckerberg Biohub | CZB Cliahub Consortium et al |
| EPI_ISL_739583 | USA/CA-CZB-13743/2020 | 2020-09-10 | North America/USA/California/Alameda | Alameda County Public Health Lab | Chan-Zuckerberg Biohub | CZB Cliahub Consortium et al |
| EPI_ISL_739586 | USA/CA-CZB-15465/2020 | 2020-12-05 | North America/USA/California/Orange C | Orange County Public Health Lab | Chan-Zuckerberg Biohub | CZB Cliahub Consortium et al |
| EPI_ISL_739587 | USA/CA-CZB-14056/2020 | 2020-06-28 | North America/USA/California/Alameda | Alameda County Public Health Lab | Chan-Zuckerberg Biohub | CZB Cliahub Consortium et al |
| EPI_ISL_739588 | USA/CA-CZB-15488/2020 | 2020-09-15 | North America/USA/California/San Luis | County of San Luis Obispo Public Health Laboratory | Chan-Zuckerberg Biohub | CZB Cliahub Consortium et al |

| | | | | | | |
|---|---|---|---|---|---|---|
| EPI_ISL_739592 | USA/CA-CZB-15467/2020 | 2020-12-05 | North America/USA/California/Orange ( | Orange County Public Health Lab | Chan-Zuckerberg Biohub | CZB Cliahub Consortium et al |
| EPI_ISL_739602 | USA/CA-CZB-15294/2020 | 2020-11-20 | North America/USA/California/Santa Cl | Santa Clara County Public Health Laboratory | Chan-Zuckerberg Biohub | CZB Cliahub Consortium et al |
| EPI_ISL_739609 | USA/CA-CZB-15438/2020 | 2020-12-01 | North America/USA/California/Orange ( | Orange County Public Health Lab | Chan-Zuckerberg Biohub | CZB Cliahub Consortium et al |
| EPI_ISL_739614 | USA/CA-CZB-14675/2020 | 2020-11-16 | North America/USA/California/Orange ( | Orange County Public Health Lab | Chan-Zuckerberg Biohub | CZB Cliahub Consortium et al |
| EPI_ISL_739619 | USA/CA-CZB-14066/2020 | 2020-06-24 | North America/USA/California/Alameda | Alameda County Public Health Lab | Chan-Zuckerberg Biohub | CZB Cliahub Consortium et al |
| EPI_ISL_739622 | USA/CA-CZB-14020/2020 | 2020-07-12 | North America/USA/California/Alameda | Alameda County Public Health Lab | Chan-Zuckerberg Biohub | CZB Cliahub Consortium et al |
| EPI_ISL_739623 | USA/CA-CZB-13744/2020 | 2020-09-11 | North America/USA/California/Alameda | Alameda County Public Health Lab | Chan-Zuckerberg Biohub | CZB Cliahub Consortium et al |
| EPI_ISL_739626 | USA/CA-CZB-15169/2020 | 2020-11-23 | North America/USA/California/Santa Cl | Santa Clara County Public Health Laboratory | Chan-Zuckerberg Biohub | CZB Cliahub Consortium et al |
| EPI_ISL_739630 | USA/CA-CZB-16059/2020 | 2020-11-29 | North America/USA/California/San Fran | Renegade | Chan-Zuckerberg Biohub | CZB Cliahub Consortium et al |
| EPI_ISL_739632 | USA/CA-CZB-14594/2020 | 2020-10-14 | North America/USA/California/Marin Co | UCSF Clinical Microbiology Laboratory | Chan-Zuckerberg Biohub | CZB Cliahub Consortium et al |
| EPI_ISL_739633 | USA/CA-CZB-14600/2020 | 2020-10-17 | North America/USA/California/Marin Co | UCSF Clinical Microbiology Laboratory | Chan-Zuckerberg Biohub | CZB Cliahub Consortium et al |
| EPI_ISL_739640 | USA/CA-CZB-14132/2020 | 2020-05-25 | North America/USA/California/Alameda | Alameda County Public Health Lab | Chan-Zuckerberg Biohub | CZB Cliahub Consortium et al |
| EPI_ISL_739650 | USA/CA-CZB-15957/2020 | 2020-11-30 | North America/USA/California/Santa Cl | Santa Clara County Public Health Laboratory | Chan-Zuckerberg Biohub | CZB Cliahub Consortium et al |
| EPI_ISL_739662 | Canada/BC-BCCDC-9557/2020 | 20-Dec | North America/Canada/British Columbia | BCCDC Public Health Laboratory | BCCDC Public Health Laboratory | Prystajecky Natalie et al |
| EPI_ISL_739663 | Colombia/SAN-INS-0904/2020 | 2020-10-31 | South America/Colombia/Santander/Bu | Instituto Nacional de Salud, Bogot√°, Colombia | Instituto Nacional de Salud, Bogot√°, Colombia | Katherine Laiton-Donato et al |
| EPI_ISL_739671 | Colombia/DC-INS-4283/2020 | 2020-11-06 | South America/Colombia/Bogota/ | Instituto Nacional de Salud, Bogot√°, Colombia | Instituto Nacional de Salud, Bogot√°, Colombia | Katherine Laiton-Donato et al |
| EPI_ISL_739672 | Colombia/CAQ-INS-003003/2020 | 2020-11-20 | South America/Colombia/Caqueta/ | Instituto Nacional de Salud, Bogot√°, Colombia | Instituto Nacional de Salud, Bogot√°, Colombia | Katherine Laiton-Donato et al |
| EPI_ISL_739673 | Colombia/DC-INS-4301/2020 | 2020-11-09 | South America/Colombia/Bogota/ | Instituto Nacional de Salud, Bogot√°, Colombia | Instituto Nacional de Salud, Bogot√°, Colombia | Katherine Laiton-Donato et al |

| | | | | | | |
|---|---|---|---|---|---|---|
| EPI_ISL_739675 | Colombia/ANT-INS-103236/2020 | 2020-10-20 | South America/Colombia/Antioquia/Me | Instituto Nacional de Salud, Bogotv°, Colombia | Instituto Nacional de Salud, Bogotv°, Colombia | Katherine Laiton-Donato et al |
| EPI_ISL_739680 | Colombia/ANT-INS-103235/2020 | 2020-11-13 | South America/Colombia/Antioquia/Me | Instituto Nacional de Salud, Bogotv°, Colombia | Instituto Nacional de Salud, Bogotv°, Colombia | Katherine Laiton-Donato et al |
| EPI_ISL_739697 | Luxembourg/LNS0022046/2020 | 2020-11-21 | Europe/Luxembourg/Luxembourg/ | Laboratoire national de santv©, Microbiology, Virology | Laboratoire national de santv©, Microbiology, Microbial Genomics Platform | Anke Wienecke-Baldacchino et al |
| EPI_ISL_739718 | Luxembourg/LNS0121983/2020 | 2020-10-30 | Europe/Luxembourg/Luxembourg/ | Laboratoire national de santv©, Microbiology, Virology | Laboratoire national de santv©, Microbiology, Microbial Genomics Platform | Anke Wienecke-Baldacchino et al |
| EPI_ISL_740032 | Luxembourg/LNS1987832/2020 | 2020-07-13 | Europe/Luxembourg/Luxembourg/ | Laboratoire national de santv©, Microbiology, Virology | Laboratoire national de santv©, Microbiology, Microbial Genomics Platform | Anke Wienecke-Baldacchino et al |
| EPI_ISL_740045 | Luxembourg/LNS2038651/2020 | 2020-11-12 | Europe/Luxembourg/Luxembourg/ | Laboratoire national de santv©, Microbiology, Virology | Laboratoire national de santv©, Microbiology, Microbial Genomics Platform | Anke Wienecke-Baldacchino et al |
| EPI_ISL_740296 | Luxembourg/LNS3355829/2020 | 2020-10-23 | Europe/Luxembourg/Luxembourg/ | Laboratoire national de santv©, Microbiology, Virology | Laboratoire national de santv©, Microbiology, Microbial Genomics Platform | Anke Wienecke-Baldacchino et al |

| | | | | | | |
|---|---|---|---|---|---|---|
| EPI_ISL_740409 | Luxembourg/LNS4048104/2020 | 2020-10-20 | Europe/Luxembourg/Luxembourg/ | Laboratoire national de sant√©, Microbiology, Virology | Laboratoire national de sant√©, Microbiology, Microbial Genomics Platform | Anke Wienecke-Baldacchino et al |
| EPI_ISL_740548 | Canada/BC-BCCDC-6390/2020 | 20-Oct | North America/Canada/British Columbia | BCCDC Public Health Laboratory | BCCDC Public Health Laboratory | Prystajecky Natalie et al |
| EPI_ISL_740550 | Canada/BC-BCCDC-6392/2020 | 20-Oct | North America/Canada/British Columbia | BCCDC Public Health Laboratory | BCCDC Public Health Laboratory | Prystajecky Natalie et al |
| EPI_ISL_740551 | Canada/BC-BCCDC-6394/2020 | 20-Oct | North America/Canada/British Columbia | BCCDC Public Health Laboratory | BCCDC Public Health Laboratory | Prystajecky Natalie et al |
| EPI_ISL_740552 | Canada/BC-BCCDC-6395/2020 | 20-Oct | North America/Canada/British Columbia | BCCDC Public Health Laboratory | BCCDC Public Health Laboratory | Prystajecky Natalie et al |
| EPI_ISL_740553 | Canada/BC-BCCDC-6396/2020 | 20-Oct | North America/Canada/British Columbia | BCCDC Public Health Laboratory | BCCDC Public Health Laboratory | Prystajecky Natalie et al |
| EPI_ISL_740554 | Canada/BC-BCCDC-6398/2020 | 20-Oct | North America/Canada/British Columbia | BCCDC Public Health Laboratory | BCCDC Public Health Laboratory | Prystajecky Natalie et al |
| EPI_ISL_740555 | Canada/BC-BCCDC-6399/2020 | 20-Oct | North America/Canada/British Columbia | BCCDC Public Health Laboratory | BCCDC Public Health Laboratory | Prystajecky Natalie et al |
| EPI_ISL_740557 | Canada/BC-BCCDC-6401/2020 | 20-Oct | North America/Canada/British Columbia | BCCDC Public Health Laboratory | BCCDC Public Health Laboratory | Prystajecky Natalie et al |
| EPI_ISL_740558 | Canada/BC-BCCDC-6403/2020 | 20-Oct | North America/Canada/British Columbia | BCCDC Public Health Laboratory | BCCDC Public Health Laboratory | Prystajecky Natalie et al |
| EPI_ISL_740559 | Canada/BC-BCCDC-6404/2020 | 20-Oct | North America/Canada/British Columbia | BCCDC Public Health Laboratory | BCCDC Public Health Laboratory | Prystajecky Natalie et al |
| EPI_ISL_740560 | Canada/BC-BCCDC-6405/2020 | 20-Oct | North America/Canada/British Columbia | BCCDC Public Health Laboratory | BCCDC Public Health Laboratory | Prystajecky Natalie et al |
| EPI_ISL_740561 | Canada/BC-BCCDC-6406/2020 | 20-Oct | North America/Canada/British Columbia | BCCDC Public Health Laboratory | BCCDC Public Health Laboratory | Prystajecky Natalie et al |
| EPI_ISL_740562 | Canada/BC-BCCDC-6407/2020 | 20-Oct | North America/Canada/British Columbia | BCCDC Public Health Laboratory | BCCDC Public Health Laboratory | Prystajecky Natalie et al |
| EPI_ISL_740563 | Canada/BC-BCCDC-6408/2020 | 20-Oct | North America/Canada/British Columbia | BCCDC Public Health Laboratory | BCCDC Public Health Laboratory | Prystajecky Natalie et al |
| EPI_ISL_740564 | Canada/BC-BCCDC-6409/2020 | 20-Oct | North America/Canada/British Columbia | BCCDC Public Health Laboratory | BCCDC Public Health Laboratory | Prystajecky Natalie et al |
| EPI_ISL_740565 | Canada/BC-BCCDC-6410/2020 | 20-Oct | North America/Canada/British Columbia | BCCDC Public Health Laboratory | BCCDC Public Health Laboratory | Prystajecky Natalie et al |

| | | | | | | |
|---|---|---|---|---|---|---|
| EPI_ISL_740566 | Canada/BC-BCCDC-6411/2020 | 20-Oct | North America/Canada/British Columbia | BCCDC Public Health Laboratory | BCCDC Public Health Laboratory | Prystajecky Natalie et al |
| EPI_ISL_740567 | Canada/BC-BCCDC-6412/2020 | 20-Oct | North America/Canada/British Columbia | BCCDC Public Health Laboratory | BCCDC Public Health Laboratory | Prystajecky Natalie et al |
| EPI_ISL_740568 | Canada/BC-BCCDC-6413/2020 | 20-Oct | North America/Canada/British Columbia | BCCDC Public Health Laboratory | BCCDC Public Health Laboratory | Prystajecky Natalie et al |
| EPI_ISL_740569 | Canada/BC-BCCDC-6414/2020 | 20-Oct | North America/Canada/British Columbia | BCCDC Public Health Laboratory | BCCDC Public Health Laboratory | Prystajecky Natalie et al |
| EPI_ISL_740570 | Canada/BC-BCCDC-6416/2020 | 20-Oct | North America/Canada/British Columbia | BCCDC Public Health Laboratory | BCCDC Public Health Laboratory | Prystajecky Natalie et al |
| EPI_ISL_740571 | Canada/BC-BCCDC-6417/2020 | 20-Oct | North America/Canada/British Columbia | BCCDC Public Health Laboratory | BCCDC Public Health Laboratory | Prystajecky Natalie et al |
| EPI_ISL_740572 | Canada/BC-BCCDC-6418/2020 | 20-Oct | North America/Canada/British Columbia | BCCDC Public Health Laboratory | BCCDC Public Health Laboratory | Prystajecky Natalie et al |
| EPI_ISL_740573 | Canada/BC-BCCDC-6419/2020 | 20-Oct | North America/Canada/British Columbia | BCCDC Public Health Laboratory | BCCDC Public Health Laboratory | Prystajecky Natalie et al |
| EPI_ISL_740574 | Canada/BC-BCCDC-6420/2020 | 20-Oct | North America/Canada/British Columbia | BCCDC Public Health Laboratory | BCCDC Public Health Laboratory | Prystajecky Natalie et al |
| EPI_ISL_740575 | Canada/BC-BCCDC-6421/2020 | 20-Oct | North America/Canada/British Columbia | BCCDC Public Health Laboratory | BCCDC Public Health Laboratory | Prystajecky Natalie et al |
| EPI_ISL_740576 | Canada/BC-BCCDC-6422/2020 | 20-Oct | North America/Canada/British Columbia | BCCDC Public Health Laboratory | BCCDC Public Health Laboratory | Prystajecky Natalie et al |
| EPI_ISL_740577 | Canada/BC-BCCDC-6423/2020 | 20-Oct | North America/Canada/British Columbia | BCCDC Public Health Laboratory | BCCDC Public Health Laboratory | Prystajecky Natalie et al |
| EPI_ISL_740578 | Canada/BC-BCCDC-6425/2020 | 20-Oct | North America/Canada/British Columbia | BCCDC Public Health Laboratory | BCCDC Public Health Laboratory | Prystajecky Natalie et al |
| EPI_ISL_740579 | Canada/BC-BCCDC-6426/2020 | 20-Oct | North America/Canada/British Columbia | BCCDC Public Health Laboratory | BCCDC Public Health Laboratory | Prystajecky Natalie et al |
| EPI_ISL_740580 | Canada/BC-BCCDC-6427/2020 | 20-Oct | North America/Canada/British Columbia | BCCDC Public Health Laboratory | BCCDC Public Health Laboratory | Prystajecky Natalie et al |
| EPI_ISL_740581 | Canada/BC-BCCDC-6428/2020 | 20-Oct | North America/Canada/British Columbia | BCCDC Public Health Laboratory | BCCDC Public Health Laboratory | Prystajecky Natalie et al |
| EPI_ISL_740582 | Canada/BC-BCCDC-6429/2020 | 20-Oct | North America/Canada/British Columbia | BCCDC Public Health Laboratory | BCCDC Public Health Laboratory | Prystajecky Natalie et al |
| EPI_ISL_740583 | Canada/BC-BCCDC-6430/2020 | 20-Oct | North America/Canada/British Columbia | BCCDC Public Health Laboratory | BCCDC Public Health Laboratory | Prystajecky Natalie et al |
| EPI_ISL_740584 | Canada/BC-BCCDC-6431/2020 | 20-Oct | North America/Canada/British Columbia | BCCDC Public Health Laboratory | BCCDC Public Health Laboratory | Prystajecky Natalie et al |
| EPI_ISL_740585 | Canada/BC-BCCDC-6432/2020 | 20-Oct | North America/Canada/British Columbia | BCCDC Public Health Laboratory | BCCDC Public Health Laboratory | Prystajecky Natalie et al |

| | | | | | | |
|---|---|---|---|---|---|---|
| EPI_ISL_740586 | Canada/BC-BCCDC-6433/2020 | 20-Oct | North America/Canada/British Columbia | BCCDC Public Health Laboratory | BCCDC Public Health Laboratory | Prystajecky Natalie et al |
| EPI_ISL_740587 | Canada/BC-BCCDC-6435/2020 | 20-Oct | North America/Canada/British Columbia | BCCDC Public Health Laboratory | BCCDC Public Health Laboratory | Prystajecky Natalie et al |
| EPI_ISL_740588 | Canada/BC-BCCDC-6436/2020 | 20-Oct | North America/Canada/British Columbia | BCCDC Public Health Laboratory | BCCDC Public Health Laboratory | Prystajecky Natalie et al |
| EPI_ISL_740589 | Canada/BC-BCCDC-6437/2020 | 20-Oct | North America/Canada/British Columbia | BCCDC Public Health Laboratory | BCCDC Public Health Laboratory | Prystajecky Natalie et al |
| EPI_ISL_740590 | Canada/BC-BCCDC-6438/2020 | 20-Oct | North America/Canada/British Columbia | BCCDC Public Health Laboratory | BCCDC Public Health Laboratory | Prystajecky Natalie et al |
| EPI_ISL_740591 | Canada/BC-BCCDC-6439/2020 | 20-Oct | North America/Canada/British Columbia | BCCDC Public Health Laboratory | BCCDC Public Health Laboratory | Prystajecky Natalie et al |
| EPI_ISL_740592 | Canada/BC-BCCDC-6440/2020 | 20-Oct | North America/Canada/British Columbia | BCCDC Public Health Laboratory | BCCDC Public Health Laboratory | Prystajecky Natalie et al |
| EPI_ISL_740593 | Canada/BC-BCCDC-6443/2020 | 20-Oct | North America/Canada/British Columbia | BCCDC Public Health Laboratory | BCCDC Public Health Laboratory | Prystajecky Natalie et al |
| EPI_ISL_740594 | Canada/BC-BCCDC-6444/2020 | 20-Oct | North America/Canada/British Columbia | BCCDC Public Health Laboratory | BCCDC Public Health Laboratory | Prystajecky Natalie et al |
| EPI_ISL_740595 | Canada/BC-BCCDC-6445/2020 | 20-Oct | North America/Canada/British Columbia | BCCDC Public Health Laboratory | BCCDC Public Health Laboratory | Prystajecky Natalie et al |
| EPI_ISL_740596 | Canada/BC-BCCDC-6446/2020 | 20-Oct | North America/Canada/British Columbia | BCCDC Public Health Laboratory | BCCDC Public Health Laboratory | Prystajecky Natalie et al |
| EPI_ISL_740597 | Canada/BC-BCCDC-6449/2020 | 20-Oct | North America/Canada/British Columbia | BCCDC Public Health Laboratory | BCCDC Public Health Laboratory | Prystajecky Natalie et al |
| EPI_ISL_740598 | Canada/BC-BCCDC-6450/2020 | 20-Oct | North America/Canada/British Columbia | BCCDC Public Health Laboratory | BCCDC Public Health Laboratory | Prystajecky Natalie et al |
| EPI_ISL_740599 | Canada/BC-BCCDC-6451/2020 | 20-Oct | North America/Canada/British Columbia | BCCDC Public Health Laboratory | BCCDC Public Health Laboratory | Prystajecky Natalie et al |
| EPI_ISL_740600 | Canada/BC-BCCDC-6452/2020 | 20-Oct | North America/Canada/British Columbia | BCCDC Public Health Laboratory | BCCDC Public Health Laboratory | Prystajecky Natalie et al |
| EPI_ISL_740601 | Canada/BC-BCCDC-6453/2020 | 20-Oct | North America/Canada/British Columbia | BCCDC Public Health Laboratory | BCCDC Public Health Laboratory | Prystajecky Natalie et al |
| EPI_ISL_740602 | Canada/BC-BCCDC-6454/2020 | 20-Oct | North America/Canada/British Columbia | BCCDC Public Health Laboratory | BCCDC Public Health Laboratory | Prystajecky Natalie et al |
| EPI_ISL_740603 | Canada/BC-BCCDC-6455/2020 | 20-Oct | North America/Canada/British Columbia | BCCDC Public Health Laboratory | BCCDC Public Health Laboratory | Prystajecky Natalie et al |
| EPI_ISL_740604 | Canada/BC-BCCDC-6456/2020 | 20-Oct | North America/Canada/British Columbia | BCCDC Public Health Laboratory | BCCDC Public Health Laboratory | Prystajecky Natalie et al |
| EPI_ISL_740605 | Canada/BC-BCCDC-6457/2020 | 20-Oct | North America/Canada/British Columbia | BCCDC Public Health Laboratory | BCCDC Public Health Laboratory | Prystajecky Natalie et al |

| | | | | | | |
|---|---|---|---|---|---|---|
| EPI_ISL_740606 | Canada/BC-BCCDC-6458/2020 | 20-Oct | North America/Canada/British Columbia | BCCDC Public Health Laboratory | BCCDC Public Health Laboratory | Prystajecky Natalie et al |
| EPI_ISL_740607 | Canada/BC-BCCDC-6460/2020 | 20-Oct | North America/Canada/British Columbia | BCCDC Public Health Laboratory | BCCDC Public Health Laboratory | Prystajecky Natalie et al |
| EPI_ISL_740608 | Canada/BC-BCCDC-6463/2020 | 20-Oct | North America/Canada/British Columbia | BCCDC Public Health Laboratory | BCCDC Public Health Laboratory | Prystajecky Natalie et al |
| EPI_ISL_740609 | Canada/BC-BCCDC-6464/2020 | 20-Oct | North America/Canada/British Columbia | BCCDC Public Health Laboratory | BCCDC Public Health Laboratory | Prystajecky Natalie et al |
| EPI_ISL_740610 | Canada/BC-BCCDC-6465/2020 | 20-Oct | North America/Canada/British Columbia | BCCDC Public Health Laboratory | BCCDC Public Health Laboratory | Prystajecky Natalie et al |
| EPI_ISL_740611 | Canada/BC-BCCDC-6466/2020 | 20-Oct | North America/Canada/British Columbia | BCCDC Public Health Laboratory | BCCDC Public Health Laboratory | Prystajecky Natalie et al |
| EPI_ISL_740612 | Canada/BC-BCCDC-6467/2020 | 20-Oct | North America/Canada/British Columbia | BCCDC Public Health Laboratory | BCCDC Public Health Laboratory | Prystajecky Natalie et al |
| EPI_ISL_740614 | Canada/BC-BCCDC-6469/2020 | 20-Oct | North America/Canada/British Columbia | BCCDC Public Health Laboratory | BCCDC Public Health Laboratory | Prystajecky Natalie et al |
| EPI_ISL_740615 | Canada/BC-BCCDC-6470/2020 | 20-Oct | North America/Canada/British Columbia | BCCDC Public Health Laboratory | BCCDC Public Health Laboratory | Prystajecky Natalie et al |
| EPI_ISL_740616 | Canada/BC-BCCDC-6471/2020 | 20-Oct | North America/Canada/British Columbia | BCCDC Public Health Laboratory | BCCDC Public Health Laboratory | Prystajecky Natalie et al |
| EPI_ISL_740617 | Canada/BC-BCCDC-6472/2020 | 20-Oct | North America/Canada/British Columbia | BCCDC Public Health Laboratory | BCCDC Public Health Laboratory | Prystajecky Natalie et al |
| EPI_ISL_740618 | Canada/BC-BCCDC-6473/2020 | 20-Oct | North America/Canada/British Columbia | BCCDC Public Health Laboratory | BCCDC Public Health Laboratory | Prystajecky Natalie et al |
| EPI_ISL_740619 | Canada/BC-BCCDC-6474/2020 | 20-Oct | North America/Canada/British Columbia | BCCDC Public Health Laboratory | BCCDC Public Health Laboratory | Prystajecky Natalie et al |
| EPI_ISL_740620 | Canada/BC-BCCDC-6479/2020 | 20-Oct | North America/Canada/British Columbia | BCCDC Public Health Laboratory | BCCDC Public Health Laboratory | Prystajecky Natalie et al |
| EPI_ISL_740621 | Canada/BC-BCCDC-6481/2020 | 20-Oct | North America/Canada/British Columbia | BCCDC Public Health Laboratory | BCCDC Public Health Laboratory | Prystajecky Natalie et al |
| EPI_ISL_740622 | Canada/BC-BCCDC-6482/2020 | 20-Oct | North America/Canada/British Columbia | BCCDC Public Health Laboratory | BCCDC Public Health Laboratory | Prystajecky Natalie et al |
| EPI_ISL_740623 | Canada/BC-BCCDC-7230/2020 | 20-Nov | North America/Canada/British Columbia | BCCDC Public Health Laboratory | BCCDC Public Health Laboratory | Prystajecky Natalie et al |
| EPI_ISL_740624 | Canada/BC-BCCDC-7232/2020 | 20-Nov | North America/Canada/British Columbia | BCCDC Public Health Laboratory | BCCDC Public Health Laboratory | Prystajecky Natalie et al |
| EPI_ISL_740625 | Canada/BC-BCCDC-7233/2020 | 20-Nov | North America/Canada/British Columbia | BCCDC Public Health Laboratory | BCCDC Public Health Laboratory | Prystajecky Natalie et al |
| EPI_ISL_740626 | Canada/BC-BCCDC-7234/2020 | 20-Nov | North America/Canada/British Columbia | BCCDC Public Health Laboratory | BCCDC Public Health Laboratory | Prystajecky Natalie et al |

| | | | | | | |
|---|---|---|---|---|---|---|
| EPI_ISL_740627 | Canada/BC-BCCDC-7235/2020 | 20-Nov | North America/Canada/British Columbia | BCCDC Public Health Laboratory | BCCDC Public Health Laboratory | Prystajecky Natalie et al |
| EPI_ISL_740628 | Canada/BC-BCCDC-7236/2020 | 20-Nov | North America/Canada/British Columbia | BCCDC Public Health Laboratory | BCCDC Public Health Laboratory | Prystajecky Natalie et al |
| EPI_ISL_740629 | Canada/BC-BCCDC-7237/2020 | 20-Nov | North America/Canada/British Columbia | BCCDC Public Health Laboratory | BCCDC Public Health Laboratory | Prystajecky Natalie et al |
| EPI_ISL_740630 | Canada/BC-BCCDC-7238/2020 | 20-Nov | North America/Canada/British Columbia | BCCDC Public Health Laboratory | BCCDC Public Health Laboratory | Prystajecky Natalie et al |
| EPI_ISL_740631 | Canada/BC-BCCDC-7239/2020 | 20-Nov | North America/Canada/British Columbia | BCCDC Public Health Laboratory | BCCDC Public Health Laboratory | Prystajecky Natalie et al |
| EPI_ISL_740632 | Canada/BC-BCCDC-7241/2020 | 20-Nov | North America/Canada/British Columbia | BCCDC Public Health Laboratory | BCCDC Public Health Laboratory | Prystajecky Natalie et al |
| EPI_ISL_740633 | Canada/BC-BCCDC-7242/2020 | 20-Nov | North America/Canada/British Columbia | BCCDC Public Health Laboratory | BCCDC Public Health Laboratory | Prystajecky Natalie et al |
| EPI_ISL_740634 | Canada/BC-BCCDC-7243/2020 | 20-Nov | North America/Canada/British Columbia | BCCDC Public Health Laboratory | BCCDC Public Health Laboratory | Prystajecky Natalie et al |
| EPI_ISL_740635 | Canada/BC-BCCDC-7245/2020 | 20-Nov | North America/Canada/British Columbia | BCCDC Public Health Laboratory | BCCDC Public Health Laboratory | Prystajecky Natalie et al |
| EPI_ISL_740636 | Canada/BC-BCCDC-7247/2020 | 20-Nov | North America/Canada/British Columbia | BCCDC Public Health Laboratory | BCCDC Public Health Laboratory | Prystajecky Natalie et al |
| EPI_ISL_740637 | Canada/BC-BCCDC-7248/2020 | 20-Nov | North America/Canada/British Columbia | BCCDC Public Health Laboratory | BCCDC Public Health Laboratory | Prystajecky Natalie et al |
| EPI_ISL_740638 | Canada/BC-BCCDC-7249/2020 | 20-Nov | North America/Canada/British Columbia | BCCDC Public Health Laboratory | BCCDC Public Health Laboratory | Prystajecky Natalie et al |
| EPI_ISL_740639 | Canada/BC-BCCDC-7250/2020 | 20-Nov | North America/Canada/British Columbia | BCCDC Public Health Laboratory | BCCDC Public Health Laboratory | Prystajecky Natalie et al |
| EPI_ISL_740640 | Canada/BC-BCCDC-7252/2020 | 20-Nov | North America/Canada/British Columbia | BCCDC Public Health Laboratory | BCCDC Public Health Laboratory | Prystajecky Natalie et al |
| EPI_ISL_740641 | Canada/BC-BCCDC-7253/2020 | 20-Nov | North America/Canada/British Columbia | BCCDC Public Health Laboratory | BCCDC Public Health Laboratory | Prystajecky Natalie et al |
| EPI_ISL_740642 | Canada/BC-BCCDC-7256/2020 | 20-Nov | North America/Canada/British Columbia | BCCDC Public Health Laboratory | BCCDC Public Health Laboratory | Prystajecky Natalie et al |
| EPI_ISL_740643 | Canada/BC-BCCDC-7258/2020 | 20-Nov | North America/Canada/British Columbia | BCCDC Public Health Laboratory | BCCDC Public Health Laboratory | Prystajecky Natalie et al |
| EPI_ISL_740644 | Canada/BC-BCCDC-7259/2020 | 20-Nov | North America/Canada/British Columbia | BCCDC Public Health Laboratory | BCCDC Public Health Laboratory | Prystajecky Natalie et al |
| EPI_ISL_740645 | Canada/BC-BCCDC-7262/2020 | 20-Nov | North America/Canada/British Columbia | BCCDC Public Health Laboratory | BCCDC Public Health Laboratory | Prystajecky Natalie et al |
| EPI_ISL_740646 | Canada/BC-BCCDC-7264/2020 | 20-Nov | North America/Canada/British Columbia | BCCDC Public Health Laboratory | BCCDC Public Health Laboratory | Prystajecky Natalie et al |

| | | | | | | |
|---|---|---|---|---|---|---|
| EPI_ISL_740647 | Canada/BC-BCCDC-7265/2020 | 20-Nov | North America/Canada/British Columbia | BCCDC Public Health Laboratory | BCCDC Public Health Laboratory | Prystajecky Natalie et al |
| EPI_ISL_740648 | Canada/BC-BCCDC-7266/2020 | 20-Nov | North America/Canada/British Columbia | BCCDC Public Health Laboratory | BCCDC Public Health Laboratory | Prystajecky Natalie et al |
| EPI_ISL_740649 | Canada/BC-BCCDC-7268/2020 | 20-Nov | North America/Canada/British Columbia | BCCDC Public Health Laboratory | BCCDC Public Health Laboratory | Prystajecky Natalie et al |
| EPI_ISL_740650 | Canada/BC-BCCDC-7269/2020 | 20-Nov | North America/Canada/British Columbia | BCCDC Public Health Laboratory | BCCDC Public Health Laboratory | Prystajecky Natalie et al |
| EPI_ISL_740651 | Canada/BC-BCCDC-7270/2020 | 20-Nov | North America/Canada/British Columbia | BCCDC Public Health Laboratory | BCCDC Public Health Laboratory | Prystajecky Natalie et al |
| EPI_ISL_740652 | Canada/BC-BCCDC-7273/2020 | 20-Nov | North America/Canada/British Columbia | BCCDC Public Health Laboratory | BCCDC Public Health Laboratory | Prystajecky Natalie et al |
| EPI_ISL_740653 | Canada/BC-BCCDC-7274/2020 | 20-Nov | North America/Canada/British Columbia | BCCDC Public Health Laboratory | BCCDC Public Health Laboratory | Prystajecky Natalie et al |
| EPI_ISL_740654 | Canada/BC-BCCDC-7280/2020 | 20-Nov | North America/Canada/British Columbia | BCCDC Public Health Laboratory | BCCDC Public Health Laboratory | Prystajecky Natalie et al |
| EPI_ISL_740655 | Canada/BC-BCCDC-7282/2020 | 20-Oct | North America/Canada/British Columbia | BCCDC Public Health Laboratory | BCCDC Public Health Laboratory | Prystajecky Natalie et al |
| EPI_ISL_740656 | Canada/BC-BCCDC-7283/2020 | 20-Nov | North America/Canada/British Columbia | BCCDC Public Health Laboratory | BCCDC Public Health Laboratory | Prystajecky Natalie et al |
| EPI_ISL_740657 | Canada/BC-BCCDC-7286/2020 | 20-Nov | North America/Canada/British Columbia | BCCDC Public Health Laboratory | BCCDC Public Health Laboratory | Prystajecky Natalie et al |
| EPI_ISL_740658 | Canada/BC-BCCDC-7287/2020 | 20-Nov | North America/Canada/British Columbia | BCCDC Public Health Laboratory | BCCDC Public Health Laboratory | Prystajecky Natalie et al |
| EPI_ISL_740659 | Canada/BC-BCCDC-7288/2020 | 20-Nov | North America/Canada/British Columbia | BCCDC Public Health Laboratory | BCCDC Public Health Laboratory | Prystajecky Natalie et al |
| EPI_ISL_740660 | Canada/BC-BCCDC-7289/2020 | 20-Nov | North America/Canada/British Columbia | BCCDC Public Health Laboratory | BCCDC Public Health Laboratory | Prystajecky Natalie et al |
| EPI_ISL_740661 | Canada/BC-BCCDC-7290/2020 | 20-Nov | North America/Canada/British Columbia | BCCDC Public Health Laboratory | BCCDC Public Health Laboratory | Prystajecky Natalie et al |
| EPI_ISL_740665 | Canada/BC-BCCDC-7296/2020 | 20-Nov | North America/Canada/British Columbia | BCCDC Public Health Laboratory | BCCDC Public Health Laboratory | Prystajecky Natalie et al |
| EPI_ISL_740668 | Canada/BC-BCCDC-7299/2020 | 20-Nov | North America/Canada/British Columbia | BCCDC Public Health Laboratory | BCCDC Public Health Laboratory | Prystajecky Natalie et al |
| EPI_ISL_740669 | Canada/BC-BCCDC-7300/2020 | 20-Nov | North America/Canada/British Columbia | BCCDC Public Health Laboratory | BCCDC Public Health Laboratory | Prystajecky Natalie et al |
| EPI_ISL_740670 | Canada/BC-BCCDC-7301/2020 | 20-Nov | North America/Canada/British Columbia | BCCDC Public Health Laboratory | BCCDC Public Health Laboratory | Prystajecky Natalie et al |
| EPI_ISL_740677 | Canada/BC-BCCDC-7311/2020 | 20-Oct | North America/Canada/British Columbia | BCCDC Public Health Laboratory | BCCDC Public Health Laboratory | Prystajecky Natalie et al |

| | | | | | | |
|---|---|---|---|---|---|---|
| EPI_ISL_740678 | Canada/BC-BCCDC-7314/2020 | 20-Nov | North America/Canada/British Columbia | BCCDC Public Health Laboratory | BCCDC Public Health Laboratory | Prystajecky Natalie et al |
| EPI_ISL_740679 | Canada/BC-BCCDC-7315/2020 | 20-Nov | North America/Canada/British Columbia | BCCDC Public Health Laboratory | BCCDC Public Health Laboratory | Prystajecky Natalie et al |
| EPI_ISL_740680 | Canada/BC-BCCDC-7316/2020 | 20-Nov | North America/Canada/British Columbia | BCCDC Public Health Laboratory | BCCDC Public Health Laboratory | Prystajecky Natalie et al |
| EPI_ISL_740681 | Canada/BC-BCCDC-7318/2020 | 20-Oct | North America/Canada/British Columbia | BCCDC Public Health Laboratory | BCCDC Public Health Laboratory | Prystajecky Natalie et al |
| EPI_ISL_740682 | Canada/BC-BCCDC-7319/2020 | 20-Oct | North America/Canada/British Columbia | BCCDC Public Health Laboratory | BCCDC Public Health Laboratory | Prystajecky Natalie et al |
| EPI_ISL_740683 | Canada/BC-BCCDC-7320/2020 | 20-Nov | North America/Canada/British Columbia | BCCDC Public Health Laboratory | BCCDC Public Health Laboratory | Prystajecky Natalie et al |
| EPI_ISL_740684 | Canada/BC-BCCDC-7321/2020 | 20-Oct | North America/Canada/British Columbia | BCCDC Public Health Laboratory | BCCDC Public Health Laboratory | Prystajecky Natalie et al |
| EPI_ISL_740685 | Canada/BC-BCCDC-7510/2020 | 20-Nov | North America/Canada/British Columbia | BCCDC Public Health Laboratory | BCCDC Public Health Laboratory | Prystajecky Natalie et al |
| EPI_ISL_740686 | Canada/BC-BCCDC-7515/2020 | 20-Nov | North America/Canada/British Columbia | BCCDC Public Health Laboratory | BCCDC Public Health Laboratory | Prystajecky Natalie et al |
| EPI_ISL_740687 | Canada/BC-BCCDC-7517/2020 | 20-Nov | North America/Canada/British Columbia | BCCDC Public Health Laboratory | BCCDC Public Health Laboratory | Prystajecky Natalie et al |
| EPI_ISL_740688 | Canada/BC-BCCDC-7518/2020 | 20-Nov | North America/Canada/British Columbia | BCCDC Public Health Laboratory | BCCDC Public Health Laboratory | Prystajecky Natalie et al |
| EPI_ISL_740689 | Canada/BC-BCCDC-7520/2020 | 20-Nov | North America/Canada/British Columbia | BCCDC Public Health Laboratory | BCCDC Public Health Laboratory | Prystajecky Natalie et al |
| EPI_ISL_740690 | Canada/BC-BCCDC-7521/2020 | 20-Nov | North America/Canada/British Columbia | BCCDC Public Health Laboratory | BCCDC Public Health Laboratory | Prystajecky Natalie et al |
| EPI_ISL_740691 | Canada/BC-BCCDC-7522/2020 | 20-Nov | North America/Canada/British Columbia | BCCDC Public Health Laboratory | BCCDC Public Health Laboratory | Prystajecky Natalie et al |
| EPI_ISL_740692 | Canada/BC-BCCDC-7523/2020 | 20-Nov | North America/Canada/British Columbia | BCCDC Public Health Laboratory | BCCDC Public Health Laboratory | Prystajecky Natalie et al |
| EPI_ISL_740693 | Canada/BC-BCCDC-7524/2020 | 20-Nov | North America/Canada/British Columbia | BCCDC Public Health Laboratory | BCCDC Public Health Laboratory | Prystajecky Natalie et al |
| EPI_ISL_740694 | Canada/BC-BCCDC-7525/2020 | 20-Nov | North America/Canada/British Columbia | BCCDC Public Health Laboratory | BCCDC Public Health Laboratory | Prystajecky Natalie et al |
| EPI_ISL_740695 | Canada/BC-BCCDC-7527/2020 | 20-Nov | North America/Canada/British Columbia | BCCDC Public Health Laboratory | BCCDC Public Health Laboratory | Prystajecky Natalie et al |
| EPI_ISL_740696 | Canada/BC-BCCDC-7528/2020 | 20-Nov | North America/Canada/British Columbia | BCCDC Public Health Laboratory | BCCDC Public Health Laboratory | Prystajecky Natalie et al |
| EPI_ISL_740697 | Canada/BC-BCCDC-7529/2020 | 20-Nov | North America/Canada/British Columbia | BCCDC Public Health Laboratory | BCCDC Public Health Laboratory | Prystajecky Natalie et al |

| | | | | | | |
|---|---|---|---|---|---|---|
| EPI_ISL_740698 | Canada/BC-BCCDC-7530/2020 | 20-Nov | North America/Canada/British Columbia | BCCDC Public Health Laboratory | BCCDC Public Health Laboratory | Prystajecky Natalie et al |
| EPI_ISL_740700 | Canada/BC-BCCDC-7532/2020 | 20-Nov | North America/Canada/British Columbia | BCCDC Public Health Laboratory | BCCDC Public Health Laboratory | Prystajecky Natalie et al |
| EPI_ISL_740701 | Canada/BC-BCCDC-7534/2020 | 20-Nov | North America/Canada/British Columbia | BCCDC Public Health Laboratory | BCCDC Public Health Laboratory | Prystajecky Natalie et al |
| EPI_ISL_740702 | Canada/BC-BCCDC-7535/2020 | 20-Nov | North America/Canada/British Columbia | BCCDC Public Health Laboratory | BCCDC Public Health Laboratory | Prystajecky Natalie et al |
| EPI_ISL_740703 | Canada/BC-BCCDC-7536/2020 | 20-Nov | North America/Canada/British Columbia | BCCDC Public Health Laboratory | BCCDC Public Health Laboratory | Prystajecky Natalie et al |
| EPI_ISL_740704 | Canada/BC-BCCDC-7537/2020 | 20-Nov | North America/Canada/British Columbia | BCCDC Public Health Laboratory | BCCDC Public Health Laboratory | Prystajecky Natalie et al |
| EPI_ISL_740705 | Canada/BC-BCCDC-7538/2020 | 20-Nov | North America/Canada/British Columbia | BCCDC Public Health Laboratory | BCCDC Public Health Laboratory | Prystajecky Natalie et al |
| EPI_ISL_740706 | Canada/BC-BCCDC-7540/2020 | 20-Nov | North America/Canada/British Columbia | BCCDC Public Health Laboratory | BCCDC Public Health Laboratory | Prystajecky Natalie et al |
| EPI_ISL_740707 | Canada/BC-BCCDC-7544/2020 | 20-Nov | North America/Canada/British Columbia | BCCDC Public Health Laboratory | BCCDC Public Health Laboratory | Prystajecky Natalie et al |
| EPI_ISL_740708 | Canada/BC-BCCDC-7545/2020 | 20-Nov | North America/Canada/British Columbia | BCCDC Public Health Laboratory | BCCDC Public Health Laboratory | Prystajecky Natalie et al |
| EPI_ISL_740709 | Canada/BC-BCCDC-7546/2020 | 20-Nov | North America/Canada/British Columbia | BCCDC Public Health Laboratory | BCCDC Public Health Laboratory | Prystajecky Natalie et al |
| EPI_ISL_740710 | Canada/BC-BCCDC-7547/2020 | 20-Nov | North America/Canada/British Columbia | BCCDC Public Health Laboratory | BCCDC Public Health Laboratory | Prystajecky Natalie et al |
| EPI_ISL_740711 | Canada/BC-BCCDC-7548/2020 | 20-Nov | North America/Canada/British Columbia | BCCDC Public Health Laboratory | BCCDC Public Health Laboratory | Prystajecky Natalie et al |
| EPI_ISL_740712 | Canada/BC-BCCDC-7549/2020 | 20-Nov | North America/Canada/British Columbia | BCCDC Public Health Laboratory | BCCDC Public Health Laboratory | Prystajecky Natalie et al |
| EPI_ISL_740713 | Canada/BC-BCCDC-7550/2020 | 20-Nov | North America/Canada/British Columbia | BCCDC Public Health Laboratory | BCCDC Public Health Laboratory | Prystajecky Natalie et al |
| EPI_ISL_740714 | Canada/BC-BCCDC-7551/2020 | 20-Nov | North America/Canada/British Columbia | BCCDC Public Health Laboratory | BCCDC Public Health Laboratory | Prystajecky Natalie et al |
| EPI_ISL_740715 | Canada/BC-BCCDC-7552/2020 | 20-Nov | North America/Canada/British Columbia | BCCDC Public Health Laboratory | BCCDC Public Health Laboratory | Prystajecky Natalie et al |
| EPI_ISL_740716 | Canada/BC-BCCDC-7553/2020 | 20-Nov | North America/Canada/British Columbia | BCCDC Public Health Laboratory | BCCDC Public Health Laboratory | Prystajecky Natalie et al |
| EPI_ISL_740717 | Canada/BC-BCCDC-7555/2020 | 20-Nov | North America/Canada/British Columbia | BCCDC Public Health Laboratory | BCCDC Public Health Laboratory | Prystajecky Natalie et al |
| EPI_ISL_740718 | Canada/BC-BCCDC-7556/2020 | 20-Nov | North America/Canada/British Columbia | BCCDC Public Health Laboratory | BCCDC Public Health Laboratory | Prystajecky Natalie et al |

| | | | | | | |
|---|---|---|---|---|---|---|
| EPI_ISL_740719 | Canada/BC-BCCDC-7557/2020 | 20-Nov | North America/Canada/British Columbia | BCCDC Public Health Laboratory | BCCDC Public Health Laboratory | Prystajecky Natalie et al |
| EPI_ISL_740720 | Canada/BC-BCCDC-7558/2020 | 20-Nov | North America/Canada/British Columbia | BCCDC Public Health Laboratory | BCCDC Public Health Laboratory | Prystajecky Natalie et al |
| EPI_ISL_740721 | Canada/BC-BCCDC-7559/2020 | 20-Nov | North America/Canada/British Columbia | BCCDC Public Health Laboratory | BCCDC Public Health Laboratory | Prystajecky Natalie et al |
| EPI_ISL_740723 | Canada/BC-BCCDC-7562/2020 | 20-Nov | North America/Canada/British Columbia | BCCDC Public Health Laboratory | BCCDC Public Health Laboratory | Prystajecky Natalie et al |
| EPI_ISL_740724 | Canada/BC-BCCDC-7563/2020 | 20-Nov | North America/Canada/British Columbia | BCCDC Public Health Laboratory | BCCDC Public Health Laboratory | Prystajecky Natalie et al |
| EPI_ISL_740725 | Canada/BC-BCCDC-7564/2020 | 20-Nov | North America/Canada/British Columbia | BCCDC Public Health Laboratory | BCCDC Public Health Laboratory | Prystajecky Natalie et al |
| EPI_ISL_740726 | Canada/BC-BCCDC-7565/2020 | 20-Nov | North America/Canada/British Columbia | BCCDC Public Health Laboratory | BCCDC Public Health Laboratory | Prystajecky Natalie et al |
| EPI_ISL_740727 | Canada/BC-BCCDC-7566/2020 | 20-Nov | North America/Canada/British Columbia | BCCDC Public Health Laboratory | BCCDC Public Health Laboratory | Prystajecky Natalie et al |
| EPI_ISL_740728 | Canada/BC-BCCDC-7569/2020 | 20-Nov | North America/Canada/British Columbia | BCCDC Public Health Laboratory | BCCDC Public Health Laboratory | Prystajecky Natalie et al |
| EPI_ISL_740729 | Canada/BC-BCCDC-7574/2020 | 20-Nov | North America/Canada/British Columbia | BCCDC Public Health Laboratory | BCCDC Public Health Laboratory | Prystajecky Natalie et al |
| EPI_ISL_740730 | Canada/BC-BCCDC-7575/2020 | 20-Nov | North America/Canada/British Columbia | BCCDC Public Health Laboratory | BCCDC Public Health Laboratory | Prystajecky Natalie et al |
| EPI_ISL_740731 | Canada/BC-BCCDC-7576/2020 | 20-Nov | North America/Canada/British Columbia | BCCDC Public Health Laboratory | BCCDC Public Health Laboratory | Prystajecky Natalie et al |
| EPI_ISL_740732 | Canada/BC-BCCDC-7580/2020 | 20-Nov | North America/Canada/British Columbia | BCCDC Public Health Laboratory | BCCDC Public Health Laboratory | Prystajecky Natalie et al |
| EPI_ISL_740735 | Canada/BC-BCCDC-7587/2020 | 20-Nov | North America/Canada/British Columbia | BCCDC Public Health Laboratory | BCCDC Public Health Laboratory | Prystajecky Natalie et al |
| EPI_ISL_740736 | Canada/BC-BCCDC-7588/2020 | 20-Nov | North America/Canada/British Columbia | BCCDC Public Health Laboratory | BCCDC Public Health Laboratory | Prystajecky Natalie et al |
| EPI_ISL_740737 | Canada/BC-BCCDC-7590/2020 | 20-Nov | North America/Canada/British Columbia | BCCDC Public Health Laboratory | BCCDC Public Health Laboratory | Prystajecky Natalie et al |
| EPI_ISL_740738 | Canada/BC-BCCDC-7591/2020 | 20-Nov | North America/Canada/British Columbia | BCCDC Public Health Laboratory | BCCDC Public Health Laboratory | Prystajecky Natalie et al |
| EPI_ISL_740739 | Canada/BC-BCCDC-7592/2020 | 20-Nov | North America/Canada/British Columbia | BCCDC Public Health Laboratory | BCCDC Public Health Laboratory | Prystajecky Natalie et al |
| EPI_ISL_740740 | Canada/BC-BCCDC-7593/2020 | 20-Nov | North America/Canada/British Columbia | BCCDC Public Health Laboratory | BCCDC Public Health Laboratory | Prystajecky Natalie et al |
| EPI_ISL_740741 | Canada/BC-BCCDC-7594/2020 | 20-Nov | North America/Canada/British Columbia | BCCDC Public Health Laboratory | BCCDC Public Health Laboratory | Prystajecky Natalie et al |

| | | | | | | |
|---|---|---|---|---|---|---|
| EPI_ISL_740742 | Canada/BC-BCCDC-7595/2020 | 20-Nov | North America/Canada/British Columbia | BCCDC Public Health Laboratory | BCCDC Public Health Laboratory | Prystajecky Natalie et al |
| EPI_ISL_740743 | Canada/BC-BCCDC-7596/2020 | 20-Nov | North America/Canada/British Columbia | BCCDC Public Health Laboratory | BCCDC Public Health Laboratory | Prystajecky Natalie et al |
| EPI_ISL_740744 | Canada/BC-BCCDC-7599/2020 | 20-Nov | North America/Canada/British Columbia | BCCDC Public Health Laboratory | BCCDC Public Health Laboratory | Prystajecky Natalie et al |
| EPI_ISL_740745 | Canada/BC-BCCDC-7600/2020 | 20-Nov | North America/Canada/British Columbia | BCCDC Public Health Laboratory | BCCDC Public Health Laboratory | Prystajecky Natalie et al |
| EPI_ISL_740746 | Canada/BC-BCCDC-7601/2020 | 20-Nov | North America/Canada/British Columbia | BCCDC Public Health Laboratory | BCCDC Public Health Laboratory | Prystajecky Natalie et al |
| EPI_ISL_740747 | Canada/BC-BCCDC-7602/2020 | 20-Nov | North America/Canada/British Columbia | BCCDC Public Health Laboratory | BCCDC Public Health Laboratory | Prystajecky Natalie et al |
| EPI_ISL_740748 | Canada/BC-BCCDC-7603/2020 | 20-Nov | North America/Canada/British Columbia | BCCDC Public Health Laboratory | BCCDC Public Health Laboratory | Prystajecky Natalie et al |
| EPI_ISL_740749 | Canada/BC-BCCDC-8356/2020 | 20-Nov | North America/Canada/British Columbia | BCCDC Public Health Laboratory | BCCDC Public Health Laboratory | Prystajecky Natalie et al |
| EPI_ISL_740750 | Canada/BC-BCCDC-8358/2020 | 20-Nov | North America/Canada/British Columbia | BCCDC Public Health Laboratory | BCCDC Public Health Laboratory | Prystajecky Natalie et al |
| EPI_ISL_740751 | Canada/BC-BCCDC-8360/2020 | 20-Nov | North America/Canada/British Columbia | BCCDC Public Health Laboratory | BCCDC Public Health Laboratory | Prystajecky Natalie et al |
| EPI_ISL_740752 | Canada/BC-BCCDC-8362/2020 | 20-Nov | North America/Canada/British Columbia | BCCDC Public Health Laboratory | BCCDC Public Health Laboratory | Prystajecky Natalie et al |
| EPI_ISL_740753 | Canada/BC-BCCDC-8363/2020 | 20-Nov | North America/Canada/British Columbia | BCCDC Public Health Laboratory | BCCDC Public Health Laboratory | Prystajecky Natalie et al |
| EPI_ISL_740754 | Canada/BC-BCCDC-8364/2020 | 20-Nov | North America/Canada/British Columbia | BCCDC Public Health Laboratory | BCCDC Public Health Laboratory | Prystajecky Natalie et al |
| EPI_ISL_740755 | Canada/BC-BCCDC-8366/2020 | 20-Nov | North America/Canada/British Columbia | BCCDC Public Health Laboratory | BCCDC Public Health Laboratory | Prystajecky Natalie et al |
| EPI_ISL_740756 | Canada/BC-BCCDC-8367/2020 | 20-Nov | North America/Canada/British Columbia | BCCDC Public Health Laboratory | BCCDC Public Health Laboratory | Prystajecky Natalie et al |
| EPI_ISL_740757 | Canada/BC-BCCDC-8368/2020 | 20-Nov | North America/Canada/British Columbia | BCCDC Public Health Laboratory | BCCDC Public Health Laboratory | Prystajecky Natalie et al |
| EPI_ISL_740758 | Canada/BC-BCCDC-8369/2020 | 20-Nov | North America/Canada/British Columbia | BCCDC Public Health Laboratory | BCCDC Public Health Laboratory | Prystajecky Natalie et al |
| EPI_ISL_740759 | Canada/BC-BCCDC-8370/2020 | 20-Nov | North America/Canada/British Columbia | BCCDC Public Health Laboratory | BCCDC Public Health Laboratory | Prystajecky Natalie et al |
| EPI_ISL_740760 | Canada/BC-BCCDC-8371/2020 | 20-Nov | North America/Canada/British Columbia | BCCDC Public Health Laboratory | BCCDC Public Health Laboratory | Prystajecky Natalie et al |
| EPI_ISL_740761 | Canada/BC-BCCDC-8375/2020 | 20-Nov | North America/Canada/British Columbia | BCCDC Public Health Laboratory | BCCDC Public Health Laboratory | Prystajecky Natalie et al |

| | | | | | | |
|---|---|---|---|---|---|---|
| EPI_ISL_740762 | Canada/BC-BCCDC-8377/2020 | 20-Nov | North America/Canada/British Columbia | BCCDC Public Health Laboratory | BCCDC Public Health Laboratory | Prystajecky Natalie et al |
| EPI_ISL_740763 | Canada/BC-BCCDC-8379/2020 | 20-Nov | North America/Canada/British Columbia | BCCDC Public Health Laboratory | BCCDC Public Health Laboratory | Prystajecky Natalie et al |
| EPI_ISL_740764 | Canada/BC-BCCDC-8380/2020 | 20-Nov | North America/Canada/British Columbia | BCCDC Public Health Laboratory | BCCDC Public Health Laboratory | Prystajecky Natalie et al |
| EPI_ISL_740765 | Canada/BC-BCCDC-8381/2020 | 20-Nov | North America/Canada/British Columbia | BCCDC Public Health Laboratory | BCCDC Public Health Laboratory | Prystajecky Natalie et al |
| EPI_ISL_740766 | Canada/BC-BCCDC-8384/2020 | 20-Nov | North America/Canada/British Columbia | BCCDC Public Health Laboratory | BCCDC Public Health Laboratory | Prystajecky Natalie et al |
| EPI_ISL_740767 | Canada/BC-BCCDC-8387/2020 | 20-Oct | North America/Canada/British Columbia | BCCDC Public Health Laboratory | BCCDC Public Health Laboratory | Prystajecky Natalie et al |
| EPI_ISL_740768 | Canada/BC-BCCDC-8388/2020 | 20-Nov | North America/Canada/British Columbia | BCCDC Public Health Laboratory | BCCDC Public Health Laboratory | Prystajecky Natalie et al |
| EPI_ISL_740769 | Canada/BC-BCCDC-8392/2020 | 20-Nov | North America/Canada/British Columbia | BCCDC Public Health Laboratory | BCCDC Public Health Laboratory | Prystajecky Natalie et al |
| EPI_ISL_740770 | Canada/BC-BCCDC-8393/2020 | 20-Nov | North America/Canada/British Columbia | BCCDC Public Health Laboratory | BCCDC Public Health Laboratory | Prystajecky Natalie et al |
| EPI_ISL_740771 | Canada/BC-BCCDC-8394/2020 | 20-Nov | North America/Canada/British Columbia | BCCDC Public Health Laboratory | BCCDC Public Health Laboratory | Prystajecky Natalie et al |
| EPI_ISL_740773 | Canada/BC-BCCDC-8397/2020 | 20-Nov | North America/Canada/British Columbia | BCCDC Public Health Laboratory | BCCDC Public Health Laboratory | Prystajecky Natalie et al |
| EPI_ISL_740774 | Canada/BC-BCCDC-8398/2020 | 20-Nov | North America/Canada/British Columbia | BCCDC Public Health Laboratory | BCCDC Public Health Laboratory | Prystajecky Natalie et al |
| EPI_ISL_740775 | Canada/BC-BCCDC-8399/2020 | 20-Nov | North America/Canada/British Columbia | BCCDC Public Health Laboratory | BCCDC Public Health Laboratory | Prystajecky Natalie et al |
| EPI_ISL_740776 | Canada/BC-BCCDC-8401/2020 | 20-Nov | North America/Canada/British Columbia | BCCDC Public Health Laboratory | BCCDC Public Health Laboratory | Prystajecky Natalie et al |
| EPI_ISL_740777 | Canada/BC-BCCDC-8405/2020 | 20-Oct | North America/Canada/British Columbia | BCCDC Public Health Laboratory | BCCDC Public Health Laboratory | Prystajecky Natalie et al |
| EPI_ISL_740778 | Canada/BC-BCCDC-8406/2020 | 20-Oct | North America/Canada/British Columbia | BCCDC Public Health Laboratory | BCCDC Public Health Laboratory | Prystajecky Natalie et al |
| EPI_ISL_740779 | Canada/BC-BCCDC-8407/2020 | 20-Oct | North America/Canada/British Columbia | BCCDC Public Health Laboratory | BCCDC Public Health Laboratory | Prystajecky Natalie et al |
| EPI_ISL_740780 | Canada/BC-BCCDC-8408/2020 | 20-Oct | North America/Canada/British Columbia | BCCDC Public Health Laboratory | BCCDC Public Health Laboratory | Prystajecky Natalie et al |
| EPI_ISL_740781 | Canada/BC-BCCDC-8409/2020 | 20-Oct | North America/Canada/British Columbia | BCCDC Public Health Laboratory | BCCDC Public Health Laboratory | Prystajecky Natalie et al |
| EPI_ISL_740782 | Canada/BC-BCCDC-8410/2020 | 20-Nov | North America/Canada/British Columbia | BCCDC Public Health Laboratory | BCCDC Public Health Laboratory | Prystajecky Natalie et al |

| | | | | | | |
|---|---|---|---|---|---|---|
| EPI_ISL_740783 | Canada/BC-BCCDC-8411/2020 | 20-Nov | North America/Canada/British Columbia | BCCDC Public Health Laboratory | BCCDC Public Health Laboratory | Prystajecky Natalie et al |
| EPI_ISL_740784 | Canada/BC-BCCDC-8412/2020 | 20-Oct | North America/Canada/British Columbia | BCCDC Public Health Laboratory | BCCDC Public Health Laboratory | Prystajecky Natalie et al |
| EPI_ISL_740785 | Canada/BC-BCCDC-8413/2020 | 20-Oct | North America/Canada/British Columbia | BCCDC Public Health Laboratory | BCCDC Public Health Laboratory | Prystajecky Natalie et al |
| EPI_ISL_740786 | Canada/BC-BCCDC-8416/2020 | 20-Oct | North America/Canada/British Columbia | BCCDC Public Health Laboratory | BCCDC Public Health Laboratory | Prystajecky Natalie et al |
| EPI_ISL_740787 | Canada/BC-BCCDC-8417/2020 | 20-Oct | North America/Canada/British Columbia | BCCDC Public Health Laboratory | BCCDC Public Health Laboratory | Prystajecky Natalie et al |
| EPI_ISL_740788 | Canada/BC-BCCDC-8418/2020 | 20-Oct | North America/Canada/British Columbia | BCCDC Public Health Laboratory | BCCDC Public Health Laboratory | Prystajecky Natalie et al |
| EPI_ISL_740789 | Canada/BC-BCCDC-8419/2020 | 20-Oct | North America/Canada/British Columbia | BCCDC Public Health Laboratory | BCCDC Public Health Laboratory | Prystajecky Natalie et al |
| EPI_ISL_740790 | Canada/BC-BCCDC-8420/2020 | 20-Nov | North America/Canada/British Columbia | BCCDC Public Health Laboratory | BCCDC Public Health Laboratory | Prystajecky Natalie et al |
| EPI_ISL_740791 | Canada/BC-BCCDC-8422/2020 | 20-Nov | North America/Canada/British Columbia | BCCDC Public Health Laboratory | BCCDC Public Health Laboratory | Prystajecky Natalie et al |
| EPI_ISL_740792 | Canada/BC-BCCDC-8425/2020 | 20-Nov | North America/Canada/British Columbia | BCCDC Public Health Laboratory | BCCDC Public Health Laboratory | Prystajecky Natalie et al |
| EPI_ISL_740793 | Canada/BC-BCCDC-8426/2020 | 20-Nov | North America/Canada/British Columbia | BCCDC Public Health Laboratory | BCCDC Public Health Laboratory | Prystajecky Natalie et al |
| EPI_ISL_740794 | Canada/BC-BCCDC-8428/2020 | 20-Nov | North America/Canada/British Columbia | BCCDC Public Health Laboratory | BCCDC Public Health Laboratory | Prystajecky Natalie et al |
| EPI_ISL_740795 | Canada/BC-BCCDC-8429/2020 | 20-Nov | North America/Canada/British Columbia | BCCDC Public Health Laboratory | BCCDC Public Health Laboratory | Prystajecky Natalie et al |
| EPI_ISL_740796 | Canada/BC-BCCDC-8430/2020 | 20-Nov | North America/Canada/British Columbia | BCCDC Public Health Laboratory | BCCDC Public Health Laboratory | Prystajecky Natalie et al |
| EPI_ISL_740797 | Canada/BC-BCCDC-8431/2020 | 20-Nov | North America/Canada/British Columbia | BCCDC Public Health Laboratory | BCCDC Public Health Laboratory | Prystajecky Natalie et al |
| EPI_ISL_740799 | Canada/BC-BCCDC-8433/2020 | 20-Nov | North America/Canada/British Columbia | BCCDC Public Health Laboratory | BCCDC Public Health Laboratory | Prystajecky Natalie et al |
| EPI_ISL_740800 | Canada/BC-BCCDC-8435/2020 | 20-Nov | North America/Canada/British Columbia | BCCDC Public Health Laboratory | BCCDC Public Health Laboratory | Prystajecky Natalie et al |
| EPI_ISL_740801 | Canada/BC-BCCDC-8437/2020 | 20-Nov | North America/Canada/British Columbia | BCCDC Public Health Laboratory | BCCDC Public Health Laboratory | Prystajecky Natalie et al |
| EPI_ISL_740802 | Canada/BC-BCCDC-8440/2020 | 20-Nov | North America/Canada/British Columbia | BCCDC Public Health Laboratory | BCCDC Public Health Laboratory | Prystajecky Natalie et al |
| EPI_ISL_740803 | Canada/BC-BCCDC-8441/2020 | 20-Nov | North America/Canada/British Columbia | BCCDC Public Health Laboratory | BCCDC Public Health Laboratory | Prystajecky Natalie et al |

| | | | | | | |
|---|---|---|---|---|---|---|
| EPI_ISL_740804 | Canada/BC-BCCDC-8443/2020 | 20-Nov | North America/Canada/British Columbia | BCCDC Public Health Laboratory | BCCDC Public Health Laboratory | Prystajecky Natalie et al |
| EPI_ISL_740805 | Canada/BC-BCCDC-8444/2020 | 20-Nov | North America/Canada/British Columbia | BCCDC Public Health Laboratory | BCCDC Public Health Laboratory | Prystajecky Natalie et al |
| EPI_ISL_740806 | Canada/BC-BCCDC-8447/2020 | 20-Nov | North America/Canada/British Columbia | BCCDC Public Health Laboratory | BCCDC Public Health Laboratory | Prystajecky Natalie et al |
| EPI_ISL_740807 | Canada/BC-BCCDC-8449/2020 | 20-Nov | North America/Canada/British Columbia | BCCDC Public Health Laboratory | BCCDC Public Health Laboratory | Prystajecky Natalie et al |
| EPI_ISL_740808 | Canada/BC-BCCDC-8544/2020 | 20-Nov | North America/Canada/British Columbia | BCCDC Public Health Laboratory | BCCDC Public Health Laboratory | Prystajecky Natalie et al |
| EPI_ISL_740809 | Canada/BC-BCCDC-8545/2020 | 20-Nov | North America/Canada/British Columbia | BCCDC Public Health Laboratory | BCCDC Public Health Laboratory | Prystajecky Natalie et al |
| EPI_ISL_740810 | Canada/BC-BCCDC-8546/2020 | 20-Nov | North America/Canada/British Columbia | BCCDC Public Health Laboratory | BCCDC Public Health Laboratory | Prystajecky Natalie et al |
| EPI_ISL_740811 | Canada/BC-BCCDC-8547/2020 | 20-Nov | North America/Canada/British Columbia | BCCDC Public Health Laboratory | BCCDC Public Health Laboratory | Prystajecky Natalie et al |
| EPI_ISL_740812 | Canada/BC-BCCDC-8548/2020 | 20-Nov | North America/Canada/British Columbia | BCCDC Public Health Laboratory | BCCDC Public Health Laboratory | Prystajecky Natalie et al |
| EPI_ISL_740813 | Canada/BC-BCCDC-8549/2020 | 20-Nov | North America/Canada/British Columbia | BCCDC Public Health Laboratory | BCCDC Public Health Laboratory | Prystajecky Natalie et al |
| EPI_ISL_740815 | Canada/BC-BCCDC-8555/2020 | 20-Nov | North America/Canada/British Columbia | BCCDC Public Health Laboratory | BCCDC Public Health Laboratory | Prystajecky Natalie et al |
| EPI_ISL_740816 | Canada/BC-BCCDC-8557/2020 | 20-Nov | North America/Canada/British Columbia | BCCDC Public Health Laboratory | BCCDC Public Health Laboratory | Prystajecky Natalie et al |
| EPI_ISL_740817 | Canada/BC-BCCDC-8558/2020 | 20-Nov | North America/Canada/British Columbia | BCCDC Public Health Laboratory | BCCDC Public Health Laboratory | Prystajecky Natalie et al |
| EPI_ISL_740818 | Canada/BC-BCCDC-8559/2020 | 20-Nov | North America/Canada/British Columbia | BCCDC Public Health Laboratory | BCCDC Public Health Laboratory | Prystajecky Natalie et al |
| EPI_ISL_740819 | Canada/BC-BCCDC-8560/2020 | 20-Nov | North America/Canada/British Columbia | BCCDC Public Health Laboratory | BCCDC Public Health Laboratory | Prystajecky Natalie et al |
| EPI_ISL_740820 | Canada/BC-BCCDC-8561/2020 | 20-Nov | North America/Canada/British Columbia | BCCDC Public Health Laboratory | BCCDC Public Health Laboratory | Prystajecky Natalie et al |
| EPI_ISL_740821 | Canada/BC-BCCDC-8562/2020 | 20-Nov | North America/Canada/British Columbia | BCCDC Public Health Laboratory | BCCDC Public Health Laboratory | Prystajecky Natalie et al |
| EPI_ISL_740822 | Canada/BC-BCCDC-8563/2020 | 20-Nov | North America/Canada/British Columbia | BCCDC Public Health Laboratory | BCCDC Public Health Laboratory | Prystajecky Natalie et al |
| EPI_ISL_740823 | Canada/BC-BCCDC-8564/2020 | 20-Nov | North America/Canada/British Columbia | BCCDC Public Health Laboratory | BCCDC Public Health Laboratory | Prystajecky Natalie et al |
| EPI_ISL_740824 | Canada/BC-BCCDC-8565/2020 | 20-Nov | North America/Canada/British Columbia | BCCDC Public Health Laboratory | BCCDC Public Health Laboratory | Prystajecky Natalie et al |

| | | | | | | |
|---|---|---|---|---|---|---|
| EPI_ISL_740825 | Canada/BC-BCCDC-8566/2020 | 20-Nov | North America/Canada/British Columbia | BCCDC Public Health Laboratory | BCCDC Public Health Laboratory | Prystajecky Natalie et al |
| EPI_ISL_740826 | Canada/BC-BCCDC-8576/2020 | 20-Nov | North America/Canada/British Columbia | BCCDC Public Health Laboratory | BCCDC Public Health Laboratory | Prystajecky Natalie et al |
| EPI_ISL_740827 | Canada/BC-BCCDC-8577/2020 | 20-Nov | North America/Canada/British Columbia | BCCDC Public Health Laboratory | BCCDC Public Health Laboratory | Prystajecky Natalie et al |
| EPI_ISL_740828 | Canada/BC-BCCDC-8578/2020 | 20-Nov | North America/Canada/British Columbia | BCCDC Public Health Laboratory | BCCDC Public Health Laboratory | Prystajecky Natalie et al |
| EPI_ISL_740830 | Canada/BC-BCCDC-8581/2020 | 20-Nov | North America/Canada/British Columbia | BCCDC Public Health Laboratory | BCCDC Public Health Laboratory | Prystajecky Natalie et al |
| EPI_ISL_740831 | Canada/BC-BCCDC-8584/2020 | 20-Nov | North America/Canada/British Columbia | BCCDC Public Health Laboratory | BCCDC Public Health Laboratory | Prystajecky Natalie et al |
| EPI_ISL_740832 | Canada/BC-BCCDC-8585/2020 | 20-Nov | North America/Canada/British Columbia | BCCDC Public Health Laboratory | BCCDC Public Health Laboratory | Prystajecky Natalie et al |
| EPI_ISL_740833 | Canada/BC-BCCDC-8586/2020 | 20-Nov | North America/Canada/British Columbia | BCCDC Public Health Laboratory | BCCDC Public Health Laboratory | Prystajecky Natalie et al |
| EPI_ISL_740834 | Canada/BC-BCCDC-8587/2020 | 20-Nov | North America/Canada/British Columbia | BCCDC Public Health Laboratory | BCCDC Public Health Laboratory | Prystajecky Natalie et al |
| EPI_ISL_740835 | Canada/BC-BCCDC-8588/2020 | 20-Nov | North America/Canada/British Columbia | BCCDC Public Health Laboratory | BCCDC Public Health Laboratory | Prystajecky Natalie et al |
| EPI_ISL_740836 | Canada/BC-BCCDC-8589/2020 | 20-Nov | North America/Canada/British Columbia | BCCDC Public Health Laboratory | BCCDC Public Health Laboratory | Prystajecky Natalie et al |
| EPI_ISL_740837 | Canada/BC-BCCDC-8590/2020 | 20-Nov | North America/Canada/British Columbia | BCCDC Public Health Laboratory | BCCDC Public Health Laboratory | Prystajecky Natalie et al |
| EPI_ISL_740838 | Canada/BC-BCCDC-8591/2020 | 20-Nov | North America/Canada/British Columbia | BCCDC Public Health Laboratory | BCCDC Public Health Laboratory | Prystajecky Natalie et al |
| EPI_ISL_740840 | Canada/BC-BCCDC-8593/2020 | 20-Nov | North America/Canada/British Columbia | BCCDC Public Health Laboratory | BCCDC Public Health Laboratory | Prystajecky Natalie et al |
| EPI_ISL_740841 | Canada/BC-BCCDC-8595/2020 | 20-Nov | North America/Canada/British Columbia | BCCDC Public Health Laboratory | BCCDC Public Health Laboratory | Prystajecky Natalie et al |
| EPI_ISL_740842 | Canada/BC-BCCDC-8597/2020 | 20-Nov | North America/Canada/British Columbia | BCCDC Public Health Laboratory | BCCDC Public Health Laboratory | Prystajecky Natalie et al |
| EPI_ISL_740843 | Canada/BC-BCCDC-8598/2020 | 20-Nov | North America/Canada/British Columbia | BCCDC Public Health Laboratory | BCCDC Public Health Laboratory | Prystajecky Natalie et al |
| EPI_ISL_740844 | Canada/BC-BCCDC-8600/2020 | 20-Nov | North America/Canada/British Columbia | BCCDC Public Health Laboratory | BCCDC Public Health Laboratory | Prystajecky Natalie et al |
| EPI_ISL_740845 | Canada/BC-BCCDC-8601/2020 | 20-Nov | North America/Canada/British Columbia | BCCDC Public Health Laboratory | BCCDC Public Health Laboratory | Prystajecky Natalie et al |
| EPI_ISL_740846 | Canada/BC-BCCDC-8603/2020 | 20-Nov | North America/Canada/British Columbia | BCCDC Public Health Laboratory | BCCDC Public Health Laboratory | Prystajecky Natalie et al |

| | | | | | | |
|---|---|---|---|---|---|---|
| EPI_ISL_740847 | Canada/BC-BCCDC-8604/2020 | 20-Nov | North America/Canada/British Columbia | BCCDC Public Health Laboratory | BCCDC Public Health Laboratory | Prystajecky Natalie et al |
| EPI_ISL_740849 | Canada/BC-BCCDC-8607/2020 | 20-Nov | North America/Canada/British Columbia | BCCDC Public Health Laboratory | BCCDC Public Health Laboratory | Prystajecky Natalie et al |
| EPI_ISL_740850 | Canada/BC-BCCDC-8608/2020 | 20-Nov | North America/Canada/British Columbia | BCCDC Public Health Laboratory | BCCDC Public Health Laboratory | Prystajecky Natalie et al |
| EPI_ISL_740851 | Canada/BC-BCCDC-8612/2020 | 20-Nov | North America/Canada/British Columbia | BCCDC Public Health Laboratory | BCCDC Public Health Laboratory | Prystajecky Natalie et al |
| EPI_ISL_740853 | Canada/BC-BCCDC-8616/2020 | 20-Nov | North America/Canada/British Columbia | BCCDC Public Health Laboratory | BCCDC Public Health Laboratory | Prystajecky Natalie et al |
| EPI_ISL_740854 | Canada/BC-BCCDC-8618/2020 | 20-Nov | North America/Canada/British Columbia | BCCDC Public Health Laboratory | BCCDC Public Health Laboratory | Prystajecky Natalie et al |
| EPI_ISL_740855 | Canada/BC-BCCDC-8619/2020 | 20-Nov | North America/Canada/British Columbia | BCCDC Public Health Laboratory | BCCDC Public Health Laboratory | Prystajecky Natalie et al |
| EPI_ISL_740856 | Canada/BC-BCCDC-8621/2020 | 20-Nov | North America/Canada/British Columbia | BCCDC Public Health Laboratory | BCCDC Public Health Laboratory | Prystajecky Natalie et al |
| EPI_ISL_740857 | Canada/BC-BCCDC-8625/2020 | 20-Nov | North America/Canada/British Columbia | BCCDC Public Health Laboratory | BCCDC Public Health Laboratory | Prystajecky Natalie et al |
| EPI_ISL_740858 | Canada/BC-BCCDC-8627/2020 | 20-Nov | North America/Canada/British Columbia | BCCDC Public Health Laboratory | BCCDC Public Health Laboratory | Prystajecky Natalie et al |
| EPI_ISL_740859 | Canada/BC-BCCDC-8633/2020 | 20-Nov | North America/Canada/British Columbia | BCCDC Public Health Laboratory | BCCDC Public Health Laboratory | Prystajecky Natalie et al |
| EPI_ISL_740860 | Canada/BC-BCCDC-8635/2020 | 20-Nov | North America/Canada/British Columbia | BCCDC Public Health Laboratory | BCCDC Public Health Laboratory | Prystajecky Natalie et al |
| EPI_ISL_740861 | Canada/BC-BCCDC-8636/2020 | 20-Nov | North America/Canada/British Columbia | BCCDC Public Health Laboratory | BCCDC Public Health Laboratory | Prystajecky Natalie et al |
| EPI_ISL_740862 | Canada/BC-BCCDC-8637/2020 | 20-Nov | North America/Canada/British Columbia | BCCDC Public Health Laboratory | BCCDC Public Health Laboratory | Prystajecky Natalie et al |
| EPI_ISL_740942 | England/ALDP-B75BDD/2020 | 2020-11-12 | Europe/United Kingdom/England/ | Oxford Viromics, NDM, University of Oxford; Oxford University Hospitals; Basingstoke and North Hampshire Hospital | COVID-19 Genomics UK (COG-UK) Consortium | Tanya Golubchik et al |
| EPI_ISL_740943 | England/ALDP-B75BEC/2020 | 2020-11-12 | Europe/United Kingdom/England/ | Oxford Viromics, NDM, University of Oxford; Oxford University Hospitals; Basingstoke and North Hampshire Hospital | COVID-19 Genomics UK (COG-UK) Consortium | Tanya Golubchik et al |
| EPI_ISL_741425 | England/NORT-1B63225/2020 | 2020-12-01 | Europe/United Kingdom/England/ | Northumbria University / South Tees Hospitals NHS Foundation Trust / North Cumbria Integrated Care NHS Foundation Trust / North Tees and Hartlepool NHS Foundation Trust / Newcastle Hospitals NHS Foundation Trust | COVID-19 Genomics UK (COG-UK) Consortium | Darren L Smith et al |

| | | | | | | |
|---|---|---|---|---|---|---|
| EPI_ISL_741536 | England/NORW-222E39/2020 | 2020-12-19 | Europe/United Kingdom/England/ | Quadram Institute Bioscience | COVID-19 Genomics UK (COG-UK) Consortium | Dave J. Baker et al |
| EPI_ISL_741838 | England/QEUH-B7CFF8/2020 | 2020-11-14 | Europe/United Kingdom/England/ | Oxford Viromics, NDM, University of Oxford; Oxford University Hospitals; Basingstoke and North Hampshire Hospital | COVID-19 Genomics UK (COG-UK) Consortium | Tanya Golubchik et al |
| EPI_ISL_741919 | England/QEUH-B7DD39/2020 | 2020-11-14 | Europe/United Kingdom/England/ | Oxford Viromics, NDM, University of Oxford; Oxford University Hospitals; Basingstoke and North Hampshire Hospital | COVID-19 Genomics UK (COG-UK) Consortium | Tanya Golubchik et al |
| EPI_ISL_742083 | England/SHEF-10D5FF5/2020 | 2020-12-19 | Europe/United Kingdom/England/ | Virology Department, Sheffield Teaching Hospitals NHS Foundation Trust/Department of Infection, Immunity and Cardiovascular Disease, The Medical School, University of Sheffield | COVID-19 Genomics UK (COG-UK) Consortium | Thushan de Silva et al |
| EPI_ISL_742147 | NorthernIreland/NIRE-2357FC/2020 | 2020-11-05 | Europe/United Kingdom/Northern Irelar | Regional Virus Laboratory, Belfast Health and Social Care Trust | COVID-19 Genomics UK (COG-UK) Consortium | Conall McCaughey et al |
| EPI_ISL_742642 | Wales/PHWC-49CABC/2020 | 2020-12-04 | Europe/United Kingdom/Wales/ | Wales Specialist Virology Centre Sequencing lab: Pathogen Genomics Unit | COVID-19 Genomics UK (COG-UK) Consortium | Catherine Moore et al |
| EPI_ISL_742644 | Wales/PHWC-49CAE9/2020 | 2020-12-04 | Europe/United Kingdom/Wales/ | Wales Specialist Virology Centre Sequencing lab: Pathogen Genomics Unit | COVID-19 Genomics UK (COG-UK) Consortium | Catherine Moore et al |
| EPI_ISL_742688 | Wales/PHWC-49CDB3/2020 | 2020-12-09 | Europe/United Kingdom/Wales/ | Wales Specialist Virology Centre Sequencing lab: Pathogen Genomics Unit | COVID-19 Genomics UK (COG-UK) Consortium | Catherine Moore et al |
| EPI_ISL_742774 | Wales/PHWC-49D3B0/2020 | 2020-12-10 | Europe/United Kingdom/Wales/ | Wales Specialist Virology Centre Sequencing lab: Pathogen Genomics Unit | COVID-19 Genomics UK (COG-UK) Consortium | Catherine Moore et al |
| EPI_ISL_742980 | Wales/PHWC-49E2B2/2020 | 2020-12-07 | Europe/United Kingdom/Wales/ | Wales Specialist Virology Centre Sequencing lab: Pathogen Genomics Unit | COVID-19 Genomics UK (COG-UK) Consortium | Catherine Moore et al |
| EPI_ISL_742991 | Wales/PHWC-49E364/2020 | 2020-12-07 | Europe/United Kingdom/Wales/ | Wales Specialist Virology Centre Sequencing lab: Pathogen Genomics Unit | COVID-19 Genomics UK (COG-UK) Consortium | Catherine Moore et al |
| EPI_ISL_743229 | Wales/PHWC-49F512/2020 | 2020-12-07 | Europe/United Kingdom/Wales/ | Wales Specialist Virology Centre Sequencing lab: Pathogen Genomics Unit | COVID-19 Genomics UK (COG-UK) Consortium | Catherine Moore et al |
| EPI_ISL_743274 | Wales/PHWC-49F819/2020 | 2020-12-08 | Europe/United Kingdom/Wales/ | Wales Specialist Virology Centre Sequencing lab: Pathogen Genomics Unit | COVID-19 Genomics UK (COG-UK) Consortium | Catherine Moore et al |

| | | | | | | |
|---|---|---|---|---|---|---|
| EPI_ISL_743386 | Wales/PHWC-4A003B/2020 | 2020-11-30 | Europe/United Kingdom/Wales/ | Wales Specialist Virology Centre Sequencing lab: Pathogen Genomics Unit | COVID-19 Genomics UK (COG-UK) Consortium | Catherine Moore et al |
| EPI_ISL_743736 | Wales/PHWC-4A187E/2020 | 2020-12-10 | Europe/United Kingdom/Wales/ | Wales Specialist Virology Centre Sequencing lab: Pathogen Genomics Unit | COVID-19 Genomics UK (COG-UK) Consortium | Catherine Moore et al |
| EPI_ISL_744163 | Luxembourg/LNS4933943/2020 | 2020-10-22 | Europe/Luxembourg/Luxembourg/ | Laboratoire national de santv©, Microbiology, Virology | Laboratoire national de santv©, Microbiology, Microbial Genomics Platform | Anke Wienecke-Baldacchino et al |
| EPI_ISL_744165 | Luxembourg/LNS4969344/2020 | 2020-09-23 | Europe/Luxembourg/Luxembourg/ | Laboratoire national de santv©, Microbiology, Virology | Laboratoire national de santv©, Microbiology, Microbial Genomics Platform | Anke Wienecke-Baldacchino et al |
| EPI_ISL_744171 | Luxembourg/LNS4993901/2020 | 2020-11-04 | Europe/Luxembourg/Luxembourg/ | Laboratoire national de santv©, Microbiology, Virology | Laboratoire national de santv©, Microbiology, Microbial Genomics Platform | Anke Wienecke-Baldacchino et al |
| EPI_ISL_744296 | Luxembourg/LNS5766113/2020 | 2020-10-19 | Europe/Luxembourg/Luxembourg/ | Laboratoire national de santv©, Microbiology, Virology | Laboratoire national de santv©, Microbiology, Microbial Genomics Platform | Anke Wienecke-Baldacchino et al |
| EPI_ISL_744367 | Luxembourg/LNS6133647/2020 | 2020-06-29 | Europe/Luxembourg/Luxembourg/ | Laboratoire national de santv©, Microbiology, Virology | Laboratoire national de santv©, Microbiology, Microbial Genomics Platform | Anke Wienecke-Baldacchino et al |

| | | | | | | |
|---|---|---|---|---|---|---|
| EPI_ISL_744436 | Luxembourg/LNS6593260/2020 | 2020-10-29 | Europe/Luxembourg/Luxembourg/ | Laboratoire national de santv©, Microbiology, Virology | Laboratoire national de santv©, Microbiology, Microbial Genomics Platform | Anke Wienecke-Baldacchino et al |
| EPI_ISL_744439 | Luxembourg/LNS6600836/2020 | 2020-10-24 | Europe/Luxembourg/Luxembourg/ | Laboratoire national de santv©, Microbiology, Virology | Laboratoire national de santv©, Microbiology, Microbial Genomics Platform | Anke Wienecke-Baldacchino et al |
| EPI_ISL_744496 | Luxembourg/LNS7026792/2020 | 2020-09-21 | Europe/Luxembourg/Luxembourg/ | Laboratoire national de santv©, Microbiology, Virology | Laboratoire national de santv©, Microbiology, Microbial Genomics Platform | Anke Wienecke-Baldacchino et al |
| EPI_ISL_744584 | Luxembourg/LNS7532467/2020 | 2020-10-25 | Europe/Luxembourg/Luxembourg/ | Laboratoire national de santv©, Microbiology, Virology | Laboratoire national de santv©, Microbiology, Microbial Genomics Platform | Anke Wienecke-Baldacchino et al |
| EPI_ISL_744732 | Luxembourg/LNS8387253/2020 | 2020-11-02 | Europe/Luxembourg/Luxembourg/ | Laboratoire national de santv©, Microbiology, Virology | Laboratoire national de santv©, Microbiology, Microbial Genomics Platform | Anke Wienecke-Baldacchino et al |

| | | | | | | |
|---|---|---|---|---|---|---|
| EPI_ISL_744835 | Luxembourg/LNS9007346/2020 | 2020-11-06 | Europe/Luxembourg/Luxembourg/ | Laboratoire national de santÃ©, Microbiology, Virology | Laboratoire national de santÃ©, Microbiology, Microbial Genomics Platform | Anke Wienecke-Baldacchino et al |
| EPI_ISL_744897 | Luxembourg/LNS9385975/2020 | 2020-10-24 | Europe/Luxembourg/Luxembourg/ | Laboratoire national de santÃ©, Microbiology, Virology | Laboratoire national de santÃ©, Microbiology, Microbial Genomics Platform | Anke Wienecke-Baldacchino et al |
| EPI_ISL_744983 | Luxembourg/LNS9789059/2020 | 2020-11-17 | Europe/Luxembourg/Luxembourg/ | Laboratoire national de santÃ©, Microbiology, Virology | Laboratoire national de santÃ©, Microbiology, Microbial Genomics Platform | Anke Wienecke-Baldacchino et al |
| EPI_ISL_745033 | Indonesia/JB-WJHL-ITB-L18952/2020 | 2020-10-04 | Asia/Indonesia/West Java/ | West Java Health Laboratory | West Java Health Laboratory | Azzania Fibriani et al |
| EPI_ISL_745069 | Israel/CVL-46324/2020 | 2020-12-13 | Asia/Israel/Tel Aviv District/ | Israel Central Virology laboratory | Israel Central Virology laboratory | Neta Zuckerman et al |
| EPI_ISL_745126 | SouthAfrica/Tygerberg-386/2020 | 2020-12-01 | Africa/South Africa/Northern Cape/Nam | Okiep CHC | National Health Laboratory Service (NHLS), Tygerberg | Susan Engelbrecht et al |
| EPI_ISL_745130 | SouthAfrica/Tygerberg-405/2020 | 2020-12-04 | Africa/South Africa/Western Cape Provi | Langebaan Clinic wc LBC | National Health Laboratory Service (NHLS), Tygerberg | Susan Engelbrecht et al |
| EPI_ISL_745132 | SouthAfrica/Tygerberg-393/2020 | 2020-12-03 | Africa/South Africa/Western Cape Provi | Dr Ivan Toms Clinic wc IVT | National Health Laboratory Service (NHLS), Tygerberg | Susan Engelbrecht et al |

| | | | | | | |
|---|---|---|---|---|---|---|
| EPI_ISL_745133 | SouthAfrica/Tygerberg-447/2020 | 2020-12-07 | Africa/South Africa/Western Cape Provi | Kraaifontein CHC wc KFP | National Health Laboratory Service (NHLS), Tygerberg | Susan Engelbrecht et al |
| EPI_ISL_745136 | SouthAfrica/Tygerberg-445/2020 | 2020-12-07 | Africa/South Africa/Western Cape Provi | Kraaifontein CHC wc KFP | National Health Laboratory Service (NHLS), Tygerberg | Susan Engelbrecht et al |
| EPI_ISL_745144 | SouthAfrica/Tygerberg-404/2020 | 2020-12-04 | Africa/South Africa/Northern Cape/Nam | Joe Slovo Hospital | National Health Laboratory Service (NHLS), Tygerberg | Susan Engelbrecht et al |
| EPI_ISL_745148 | SouthAfrica/Tygerberg-451/2020 | 2020-12-07 | Africa/South Africa/Western Cape Provi | Tygerberg Hospital wc TBH | National Health Laboratory Service (NHLS), Tygerberg | Susan Engelbrecht et al |
| EPI_ISL_745152 | SouthAfrica/Tygerberg-383/2020 | 2020-12-01 | Africa/South Africa/Eastern Cape/Nelso | Laetitia Bam CHC | National Health Laboratory Service (NHLS), Tygerberg | Susan Engelbrecht et al |
| EPI_ISL_745154 | SouthAfrica/Tygerberg-388/2020 | 2020-12-02 | Africa/South Africa/Western Cape Provi | Dr Ivan Toms Clinic wc IVT | National Health Laboratory Service (NHLS), Tygerberg | Susan Engelbrecht et al |
| EPI_ISL_745155 | SouthAfrica/Tygerberg-390/2020 | 2020-12-02 | Africa/South Africa/Western Cape Provi | Langebaan Clinic wc LBC | National Health Laboratory Service (NHLS), Tygerberg | Susan Engelbrecht et al |
| EPI_ISL_745156 | SouthAfrica/Tygerberg-392/2020 | 2020-12-02 | Africa/South Africa/Western Cape Provi | SAS Saldanha | National Health Laboratory Service (NHLS), Tygerberg | Susan Engelbrecht et al |
| EPI_ISL_745163 | SouthAfrica/Tygerberg-412/2020 | 2020-12-05 | Africa/South Africa/Western Cape Provi | Khayelitsha (Site B) CHC wc KHC | National Health Laboratory Service (NHLS), Tygerberg | Susan Engelbrecht et al |
| EPI_ISL_745176 | SouthAfrica/Tygerberg-440/2020 | 2020-12-07 | Africa/South Africa/Western Cape Provi | Eerste River Hospital wc ERH | National Health Laboratory Service (NHLS), Tygerberg | Susan Engelbrecht et al |

| | | | | | | |
|---|---|---|---|---|---|---|
| EPI_ISL_745182 | SouthAfrica/Tygerberg-456/2020 | 2020-12-07 | Africa/South Africa/Western Cape Provi | Tygerberg Hospital wc TBH | National Health Laboratory Service (NHLS), Tygerberg | Susan Engelbrecht et al |
| EPI_ISL_745183 | SouthAfrica/Tygerberg-457/2020 | 2020-12-07 | Africa/South Africa/Western Cape Provi | Tygerberg Hospital wc TBH | National Health Laboratory Service (NHLS), Tygerberg | Susan Engelbrecht et al |
| EPI_ISL_745187 | SouthAfrica/Tygerberg-395/2020 | 2020-12-03 | Africa/South Africa/Northern Cape/Nam | Port Nolloth Hospital | National Health Laboratory Service (NHLS), Tygerberg | Susan Engelbrecht et al |
| EPI_ISL_745193 | Italy/APU-IZSPB-399PT/2020 | 2020-12-21 | Europe/Italy/Apulia/Bari | Ospedale Di Venere | Istituto Zooprofilattico Sperimentale della Puglia e della Basilicata | Parisi A. et al |
| EPI_ISL_745194 | India/TG-CCMB-BB560/2020 | 2020-12-25 | Asia/India/Telangana/ | Osmania Medical College | CSIR-Centre for Cellular and Molecular Biology | Dr.V.Sudha Rani et al |
| EPI_ISL_745196 | India/TG-CCMB-BB565/2020 | 2020-12-25 | Asia/India/Telangana/ | Osmania Medical College | CSIR-Centre for Cellular and Molecular Biology | Dr.V.Sudha Rani et al |
| EPI_ISL_745197 | India/TG-CCMB-BB566/2020 | 2020-12-25 | Asia/India/Telangana/ | Kakatiya Medical College | CSIR-Centre for Cellular and Molecular Biology | Dr.V.Sudha Rani et al |
| EPI_ISL_745198 | India/TG-CCMB-BB568/2020 | 2020-12-25 | Asia/India/Telangana/ | ESI Hospital | CSIR-Centre for Cellular and Molecular Biology | Dr.V.Sudha Rani et al |
| EPI_ISL_745199 | India/AP-CCMB-BB567/2020 | 2020-12-25 | Asia/India/Andhra Pradesh/ | Rangaraya Medical College | CSIR-Centre for Cellular and Molecular Biology | Dr.V.Sudha Rani et al |
| EPI_ISL_745200 | India/TG-CCMB-BB647/2020 | 2020-12-25 | Asia/India/Telangana/ | Nizams Institute of Medical Sciences | CSIR-Centre for Cellular and Molecular Biology | Dr.V.Sudha Rani et al |
| EPI_ISL_745201 | India/AP-CCMB-BB648/2020 | 2020-12-25 | Asia/India/Andhra Pradesh/ | Guntur General Hospital | CSIR-Centre for Cellular and Molecular Biology | Dr.V.Sudha Rani et al |
| EPI_ISL_745216 | Belgium/UGent-330/2020 | 2020-12-11 | Europe/Belgium/Gent/ | Lab voor klinische biologie | Onderzoeksgroep Virologie | Nick Vereecke et al |

| | | | | | | |
|---|---|---|---|---|---|---|
| EPI_ISL_745223 | Russia/MOS-CRIE-7182855/2020 | 2020-08-24 | Europe/Russia/Moscow Oblast/ | "Molecular diagnostic laboratory of Federal Budget Institution of Science ""Central Research Institute of Epidemiology"" of The Federal Service on Customers' Rights Protection and Human Well-being Surveillance" | Group of Genomics and Postgenomic Technologies of Central Research Institute of Epidemilology | Samoilov AE et al |
| EPI_ISL_745224 | Russia/MOS-CRIE-13476745/2020 | 2020-11-07 | Europe/Russia/Moscow Oblast/ | "Molecular diagnostic laboratory of Federal Budget Institution of Science ""Central Research Institute of Epidemiology"" of The Federal Service on Customers' Rights Protection and Human Well-being Surveillance" | Group of Genomics and Postgenomic Technologies of Central Research Institute of Epidemilology | Samoilov AE et al |
| EPI_ISL_745226 | Russia/MOS-CRIE-13380492/2020 | 2020-11-06 | Europe/Russia/Moscow Oblast/ | "Molecular diagnostic laboratory of Federal Budget Institution of Science ""Central Research Institute of Epidemiology"" of The Federal Service on Customers' Rights Protection and Human Well-being Surveillance" | Group of Genomics and Postgenomic Technologies of Central Research Institute of Epidemilology | Samoilov AE et al |
| EPI_ISL_745228 | Russia/MOS-CRIE-13352861/2020 | 2020-11-06 | Europe/Russia/Moscow Oblast/ | "Molecular diagnostic laboratory of Federal Budget Institution of Science ""Central Research Institute of Epidemiology"" of The Federal Service on Customers' Rights Protection and Human Well-being Surveillance" | Group of Genomics and Postgenomic Technologies of Central Research Institute of Epidemilology | Samoilov AE et al |
| EPI_ISL_745229 | Russia/MOS-CRIE-13677408/2020 | 2020-11-09 | Europe/Russia/Moscow Oblast/ | "Molecular diagnostic laboratory of Federal Budget Institution of Science ""Central Research Institute of Epidemiology"" of The Federal Service on Customers' Rights Protection and Human Well-being Surveillance" | Group of Genomics and Postgenomic Technologies of Central Research Institute of Epidemilology | Samoilov AE et al |

| | | | | | | |
|---|---|---|---|---|---|---|
| EPI_ISL_745230 | Russia/MOS-CRIE-13659370/2020 | 2020-11-09 | Europe/Russia/Moscow Oblast/ | "Molecular diagnostic laboratory of Federal Budget Institution of Science ""Central Research Institute of Epidemiology"" of The Federal Service on Customers' Rights Protection and Human Well-being Surveillance" | Group of Genomics and Postgenomic Technologies of Central Research Institute of Epidemilology | Samoilov AE et al |
| EPI_ISL_745231 | Russia/MOS-CRIE-13661344/2020 | 2020-11-09 | Europe/Russia/Moscow Oblast/ | "Molecular diagnostic laboratory of Federal Budget Institution of Science ""Central Research Institute of Epidemiology"" of The Federal Service on Customers' Rights Protection and Human Well-being Surveillance" | Group of Genomics and Postgenomic Technologies of Central Research Institute of Epidemilology | Samoilov AE et al |
| EPI_ISL_745232 | Russia/MOS-CRIE-13594312/2020 | 2020-11-09 | Europe/Russia/Moscow Oblast/ | "Molecular diagnostic laboratory of Federal Budget Institution of Science ""Central Research Institute of Epidemiology"" of The Federal Service on Customers' Rights Protection and Human Well-being Surveillance" | Group of Genomics and Postgenomic Technologies of Central Research Institute of Epidemilology | Samoilov AE et al |
| EPI_ISL_745233 | Russia/MOS-CRIE-13370222/2020 | 2020-11-06 | Europe/Russia/Moscow Oblast/ | "Molecular diagnostic laboratory of Federal Budget Institution of Science ""Central Research Institute of Epidemiology"" of The Federal Service on Customers' Rights Protection and Human Well-being Surveillance" | Group of Genomics and Postgenomic Technologies of Central Research Institute of Epidemilology | Samoilov AE et al |
| EPI_ISL_745234 | Russia/MOS-CRIE-13384497/2020 | 2020-11-06 | Europe/Russia/Moscow Oblast/ | "Molecular diagnostic laboratory of Federal Budget Institution of Science ""Central Research Institute of Epidemiology"" of The Federal Service on Customers' Rights Protection and Human Well-being Surveillance" | Group of Genomics and Postgenomic Technologies of Central Research Institute of Epidemilology | Samoilov AE et al |

| | | | | | | |
|---|---|---|---|---|---|---|
| EPI_ISL_745235 | Russia/MOS-CRIE-13372214/2020 | 2020-11-06 | Europe/Russia/Moscow Oblast/ | "Molecular diagnostic laboratory of Federal Budget Institution of Science ""Central Research Institute of Epidemiology"" of The Federal Service on Customers' Rights Protection and Human Well-being Surveillance" | Group of Genomics and Postgenomic Technologies of Central Research Institute of Epidemilology | Samoilov AE et al |
| EPI_ISL_745238 | Russia/MOS-CRIE-13381035/2020 | 2020-11-06 | Europe/Russia/Moscow Oblast/ | "Molecular diagnostic laboratory of Federal Budget Institution of Science ""Central Research Institute of Epidemiology"" of The Federal Service on Customers' Rights Protection and Human Well-being Surveillance" | Group of Genomics and Postgenomic Technologies of Central Research Institute of Epidemilology | Samoilov AE et al |
| EPI_ISL_745239 | Russia/MOS-CRIE-13380178/2020 | 2020-11-06 | Europe/Russia/Moscow Oblast/ | "Molecular diagnostic laboratory of Federal Budget Institution of Science ""Central Research Institute of Epidemiology"" of The Federal Service on Customers' Rights Protection and Human Well-being Surveillance" | Group of Genomics and Postgenomic Technologies of Central Research Institute of Epidemilology | Samoilov AE et al |
| EPI_ISL_745240 | Russia/MOS-CRIE-13351661/2020 | 2020-11-06 | Europe/Russia/Moscow Oblast/ | "Molecular diagnostic laboratory of Federal Budget Institution of Science ""Central Research Institute of Epidemiology"" of The Federal Service on Customers' Rights Protection and Human Well-being Surveillance" | Group of Genomics and Postgenomic Technologies of Central Research Institute of Epidemilology | Samoilov AE et al |
| EPI_ISL_745242 | Russia/MOS-CRIE-13389205/2020 | 2020-11-07 | Europe/Russia/Moscow Oblast/ | "Molecular diagnostic laboratory of Federal Budget Institution of Science ""Central Research Institute of Epidemiology"" of The Federal Service on Customers' Rights Protection and Human Well-being Surveillance" | Group of Genomics and Postgenomic Technologies of Central Research Institute of Epidemilology | Samoilov AE et al |

| | | | | | | |
|---|---|---|---|---|---|---|
| EPI_ISL_745243 | Russia/MOS-CRIE-13472261/2020 | 2020-11-07 | Europe/Russia/Moscow Oblast/ | "Molecular diagnostic laboratory of Federal Budget Institution of Science ""Central Research Institute of Epidemiology"" of The Federal Service on Customers' Rights Protection and Human Well-being Surveillance" | Group of Genomics and Postgenomic Technologies of Central Research Institute of Epidemilology | Samoilov AE et al |
| EPI_ISL_745245 | Russia/MOS-CRIE-13678204/2020 | 2020-11-09 | Europe/Russia/Moscow Oblast/ | "Molecular diagnostic laboratory of Federal Budget Institution of Science ""Central Research Institute of Epidemiology"" of The Federal Service on Customers' Rights Protection and Human Well-being Surveillance" | Group of Genomics and Postgenomic Technologies of Central Research Institute of Epidemilology | Samoilov AE et al |
| EPI_ISL_745248 | Russia/MOS-CRIE-13675465/2020 | 2020-11-09 | Europe/Russia/Moscow Oblast/ | "Molecular diagnostic laboratory of Federal Budget Institution of Science ""Central Research Institute of Epidemiology"" of The Federal Service on Customers' Rights Protection and Human Well-being Surveillance" | Group of Genomics and Postgenomic Technologies of Central Research Institute of Epidemilology | Samoilov AE et al |
| EPI_ISL_745249 | Russia/MOS-CRIE-13681164/2020 | 2020-11-09 | Europe/Russia/Moscow Oblast/ | "Molecular diagnostic laboratory of Federal Budget Institution of Science ""Central Research Institute of Epidemiology"" of The Federal Service on Customers' Rights Protection and Human Well-being Surveillance" | Group of Genomics and Postgenomic Technologies of Central Research Institute of Epidemilology | Samoilov AE et al |
| EPI_ISL_745250 | Russia/MOS-CRIE-13668879/2020 | 2020-11-09 | Europe/Russia/Moscow Oblast/ | "Molecular diagnostic laboratory of Federal Budget Institution of Science ""Central Research Institute of Epidemiology"" of The Federal Service on Customers' Rights Protection and Human Well-being Surveillance" | Group of Genomics and Postgenomic Technologies of Central Research Institute of Epidemilology | Samoilov AE et al |

| | | | | | | |
|---|---|---|---|---|---|---|
| EPI_ISL_745251 | Russia/MOS-CRIE-13679342/2020 | 2020-11-09 | Europe/Russia/Moscow Oblast/ | "Molecular diagnostic laboratory of Federal Budget Institution of Science ""Central Research Institute of Epidemiology"" of The Federal Service on Customers' Rights Protection and Human Well-being Surveillance" | Group of Genomics and Postgenomic Technologies of Central Research Institute of Epidemilology | Samoilov AE et al |
| EPI_ISL_745254 | Russia/MOS-CRIE-13642350/2020 | 2020-11-09 | Europe/Russia/Moscow Oblast/ | "Molecular diagnostic laboratory of Federal Budget Institution of Science ""Central Research Institute of Epidemiology"" of The Federal Service on Customers' Rights Protection and Human Well-being Surveillance" | Group of Genomics and Postgenomic Technologies of Central Research Institute of Epidemilology | Samoilov AE et al |
| EPI_ISL_745255 | Russia/MOS-CRIE-13670631/2020 | 2020-11-09 | Europe/Russia/Moscow Oblast/ | "Molecular diagnostic laboratory of Federal Budget Institution of Science ""Central Research Institute of Epidemiology"" of The Federal Service on Customers' Rights Protection and Human Well-being Surveillance" | Group of Genomics and Postgenomic Technologies of Central Research Institute of Epidemilology | Samoilov AE et al |
| EPI_ISL_745256 | Russia/MOS-CRIE-13678675/2020 | 2020-11-09 | Europe/Russia/Moscow Oblast/ | "Molecular diagnostic laboratory of Federal Budget Institution of Science ""Central Research Institute of Epidemiology"" of The Federal Service on Customers' Rights Protection and Human Well-being Surveillance" | Group of Genomics and Postgenomic Technologies of Central Research Institute of Epidemilology | Samoilov AE et al |
| EPI_ISL_745258 | Russia/MOS-CRIE-13678038/2020 | 2020-11-09 | Europe/Russia/Moscow Oblast/ | "Molecular diagnostic laboratory of Federal Budget Institution of Science ""Central Research Institute of Epidemiology"" of The Federal Service on Customers' Rights Protection and Human Well-being Surveillance" | Group of Genomics and Postgenomic Technologies of Central Research Institute of Epidemilology | Samoilov AE et al |

| | | | | | | |
|---|---|---|---|---|---|---|
| EPI_ISL_745259 | Russia/MOS-CRIE-7172114/2020 | 2020-08-24 | Europe/Russia/Moscow Oblast/ | "Molecular diagnostic laboratory of Federal Budget Institution of Science ""Central Research Institute of Epidemiology"" of The Federal Service on Customers' Rights Protection and Human Well-being Surveillance" | Group of Genomics and Postgenomic Technologies of Central Research Institute of Epidemilology | Samoilov AE et al |
| EPI_ISL_745260 | Canada/QC-L00314539/2020 | 2020-12-15 | North America/Canada/Quebec/ | CHUM-Site Glen-LAB Microbiologie | Laboratoire de santV© publique du QuV©bec | Sandrine Moreira et al |
| EPI_ISL_745264 | USA/TX-DSHS-1873/2020 | 2020-06-24 | North America/USA/Texas/Hidalgo Cour | Texas Department of State Health Services | Texas Department of State Health Services | Rashmi Tuladhar et al |
| EPI_ISL_745265 | USA/TX-DSHS-1835/2020 | 2020-06-23 | North America/USA/Texas/Harris Count | Texas Department of State Health Services | Texas Department of State Health Services | Rashmi Tuladhar et al |
| EPI_ISL_745267 | USA/TX-DSHS-1872/2020 | 2020-06-24 | North America/USA/Texas/Hidalgo Cour | Texas Department of State Health Services | Texas Department of State Health Services | Rashmi Tuladhar et al |
| EPI_ISL_745269 | USA/TX-DSHS-1849/2020 | 2020-06-23 | North America/USA/Texas/Harris Count | Texas Department of State Health Services | Texas Department of State Health Services | Rashmi Tuladhar et al |
| EPI_ISL_745283 | USA/TX-DSHS-1839/2020 | 2020-06-23 | North America/USA/Texas/Harris Count | Texas Department of State Health Services | Texas Department of State Health Services | Rashmi Tuladhar et al |
| EPI_ISL_745287 | USA/TX-DSHS-1848/2020 | 2020-06-22 | North America/USA/Texas/Harris Count | Texas Department of State Health Services | Texas Department of State Health Services | Rashmi Tuladhar et al |
| EPI_ISL_745297 | USA/TX-DSHS-1862/2020 | 2020-06-24 | North America/USA/Texas/Hidalgo Cour | Texas Department of State Health Services | Texas Department of State Health Services | Rashmi Tuladhar et al |
| EPI_ISL_745298 | USA/TX-DSHS-1863/2020 | 2020-06-24 | North America/USA/Texas/Hidalgo Cour | Texas Department of State Health Services | Texas Department of State Health Services | Rashmi Tuladhar et al |
| EPI_ISL_745303 | USA/TX-DSHS-1870/2020 | 2020-06-24 | North America/USA/Texas/Hidalgo Cour | Texas Department of State Health Services | Texas Department of State Health Services | Rashmi Tuladhar et al |
| EPI_ISL_745312 | France/ARA-SC713/2020 | 2020-10-24 | Europe/France/Auvergne-RhV¥ne-Alpes/ | CHU Clermont-Ferrand | CNR Virus des Infections Respiratoires - France SUD | Antonin Bal et al |

| | | | | | | |
|---|---|---|---|---|---|---|
| EPI_ISL_745313 | France/ARA-SC714/2020 | 2020-11-02 | Europe/France/Auvergne-Rh√¥ne-Alpes/ | CHU Clermont-Ferrand | CNR Virus des Infections Respiratoires - France SUD | Antonin Bal et al |
| EPI_ISL_745317 | France/ARA-113925/2020 | 2020-11-30 | Europe/France/Auvergne-Rh√¥ne-Alpes/ | CNR Virus des Infections Respiratoires - France SUD | CNR Virus des Infections Respiratoires - France SUD | Antonin Bal et al |
| EPI_ISL_745322 | France/ARA-122089/2020 | 2020-12-19 | Europe/France/Auvergne-Rh√¥ne-Alpes/ | CNR Virus des Infections Respiratoires - France SUD | CNR Virus des Infections Respiratoires - France SUD | Antonin Bal et al |
| EPI_ISL_745326 | France/ARA-114092/2020 | 2020-11-30 | Europe/France/Auvergne-Rh√¥ne-Alpes/ | CNR Virus des Infections Respiratoires - France SUD | CNR Virus des Infections Respiratoires - France SUD | Antonin Bal et al |
| EPI_ISL_745328 | France/ARA-114632/2020 | 2020-12-01 | Europe/France/Auvergne-Rh√¥ne-Alpes/ | CNR Virus des Infections Respiratoires - France SUD | CNR Virus des Infections Respiratoires - France SUD | Antonin Bal et al |
| EPI_ISL_745331 | France/ARA-118288/2020 | 2020-12-10 | Europe/France/Auvergne-Rh√¥ne-Alpes/ | CNR Virus des Infections Respiratoires - France SUD | CNR Virus des Infections Respiratoires - France SUD | Antonin Bal et al |
| EPI_ISL_745335 | France/ARA-SC715/2020 | 2020-11-20 | Europe/France/Auvergne-Rh√¥ne-Alpes/ | CHU Clermont-Ferrand | CNR Virus des Infections Respiratoires - France SUD | Antonin Bal et al |
| EPI_ISL_745336 | France/ARA-SC718/2020 | 2020-12-01 | Europe/France/Auvergne-Rh√¥ne-Alpes/ | CHU Clermont-Ferrand | CNR Virus des Infections Respiratoires - France SUD | Antonin Bal et al |
| EPI_ISL_745337 | France/ARA-112613/2020 | 2020-11-26 | Europe/France/Auvergne-Rh√¥ne-Alpes/ | CNR Virus des Infections Respiratoires - France SUD | CNR Virus des Infections Respiratoires - France SUD | Antonin Bal et al |
| EPI_ISL_745338 | France/ARA-112755/2020 | 2020-11-26 | Europe/France/Auvergne-Rh√¥ne-Alpes/ | CNR Virus des Infections Respiratoires - France SUD | CNR Virus des Infections Respiratoires - France SUD | Antonin Bal et al |

| | | | | | | |
|---|---|---|---|---|---|---|
| EPI_ISL_745341 | France/ARA-113019/2020 | 2020-11-27 | Europe/France/Auvergne-Rh√¥ne-Alpes/ | CNR Virus des Infections Respiratoires - France SUD | CNR Virus des Infections Respiratoires - France SUD | Antonin Bal et al |
| EPI_ISL_745342 | France/ARA-113354/2020 | 2020-11-27 | Europe/France/Auvergne-Rh√¥ne-Alpes/ | CNR Virus des Infections Respiratoires - France SUD | CNR Virus des Infections Respiratoires - France SUD | Antonin Bal et al |
| EPI_ISL_745344 | France/ARA-113416/2020 | 2020-11-28 | Europe/France/Auvergne-Rh√¥ne-Alpes/ | CNR Virus des Infections Respiratoires - France SUD | CNR Virus des Infections Respiratoires - France SUD | Antonin Bal et al |
| EPI_ISL_745345 | France/ARA-113487/2020 | 2020-11-28 | Europe/France/Auvergne-Rh√¥ne-Alpes/ | CNR Virus des Infections Respiratoires - France SUD | CNR Virus des Infections Respiratoires - France SUD | Antonin Bal et al |
| EPI_ISL_745346 | France/ARA-113814/2020 | 2020-11-30 | Europe/France/Auvergne-Rh√¥ne-Alpes/ | CNR Virus des Infections Respiratoires - France SUD | CNR Virus des Infections Respiratoires - France SUD | Antonin Bal et al |
| EPI_ISL_745348 | France/ARA-114176/2020 | 2020-11-30 | Europe/France/Auvergne-Rh√¥ne-Alpes/ | CNR Virus des Infections Respiratoires - France SUD | CNR Virus des Infections Respiratoires - France SUD | Antonin Bal et al |
| EPI_ISL_745349 | France/ARA-114292/2020 | 2020-11-30 | Europe/France/Auvergne-Rh√¥ne-Alpes/ | CNR Virus des Infections Respiratoires - France SUD | CNR Virus des Infections Respiratoires - France SUD | Antonin Bal et al |
| EPI_ISL_745351 | France/ARA-114442/2020 | 2020-12-01 | Europe/France/Auvergne-Rh√¥ne-Alpes/ | CNR Virus des Infections Respiratoires - France SUD | CNR Virus des Infections Respiratoires - France SUD | Antonin Bal et al |
| EPI_ISL_745357 | France/ARA-115468/2020 | 2020-12-03 | Europe/France/Auvergne-Rh√¥ne-Alpes/ | CNR Virus des Infections Respiratoires - France SUD | CNR Virus des Infections Respiratoires - France SUD | Antonin Bal et al |
| EPI_ISL_745360 | France/ARA-115793/2020 | 2020-12-04 | Europe/France/Auvergne-Rh√¥ne-Alpes/ | CNR Virus des Infections Respiratoires - France SUD | CNR Virus des Infections Respiratoires - France SUD | Antonin Bal et al |

| | | | | | | |
|---|---|---|---|---|---|---|
| EPI_ISL_745361 | France/ARA-115811/2020 | 2020-12-03 | Europe/France/Auvergne-Rh√¥ne-Alpes/ | CNR Virus des Infections Respiratoires - France SUD | CNR Virus des Infections Respiratoires - France SUD | Antonin Bal et al |
| EPI_ISL_745364 | France/ARA-116443/2020 | 2020-12-06 | Europe/France/Auvergne-Rh√¥ne-Alpes/ | CNR Virus des Infections Respiratoires - France SUD | CNR Virus des Infections Respiratoires - France SUD | Antonin Bal et al |
| EPI_ISL_745369 | France/ARA-116701/2020 | 2020-12-07 | Europe/France/Auvergne-Rh√¥ne-Alpes/ | CNR Virus des Infections Respiratoires - France SUD | CNR Virus des Infections Respiratoires - France SUD | Antonin Bal et al |
| EPI_ISL_745376 | France/ARA-117764/2020 | 2020-12-09 | Europe/France/Auvergne-Rh√¥ne-Alpes/ | CNR Virus des Infections Respiratoires - France SUD | CNR Virus des Infections Respiratoires - France SUD | Antonin Bal et al |
| EPI_ISL_745386 | France/ARA-119333/2020 | 2020-12-13 | Europe/France/Auvergne-Rh√¥ne-Alpes/ | CNR Virus des Infections Respiratoires - France SUD | CNR Virus des Infections Respiratoires - France SUD | Antonin Bal et al |
| EPI_ISL_745388 | France/ARA-119460/2020 | 2020-12-13 | Europe/France/Auvergne-Rh√¥ne-Alpes/ | CNR Virus des Infections Respiratoires - France SUD | CNR Virus des Infections Respiratoires - France SUD | Antonin Bal et al |
| EPI_ISL_745391 | France/ARA-121169/2020 | 2020-12-17 | Europe/France/Auvergne-Rh√¥ne-Alpes/ | CNR Virus des Infections Respiratoires - France SUD | CNR Virus des Infections Respiratoires - France SUD | Antonin Bal et al |
| EPI_ISL_745395 | France/ARA-57377/2020 | 2020-08-02 | Europe/France/Auvergne-Rh√¥ne-Alpes/ | CNR Virus des Infections Respiratoires - France SUD | CNR Virus des Infections Respiratoires - France SUD | Antonin Bal et al |
| EPI_ISL_745396 | France/ARA-113522/2020 | 2020-11-28 | Europe/France/Auvergne-Rh√¥ne-Alpes/ | CNR Virus des Infections Respiratoires - France SUD | CNR Virus des Infections Respiratoires - France SUD | Antonin Bal et al |
| EPI_ISL_745414 | USA/NY-NYCPHL-001480/2020 | 2020-11-27 | North America/USA/New York/New Yor | DOHMH Jamaica | New York City Public Health Laboratory | Jade Wang et al |

| | | | | | | |
|---|---|---|---|---|---|---|
| EPI_ISL_745415 | USA/NY-NYCPHL-001483/2020 | 2020-11-27 | North America/USA/New York/New Yor | DOHMH Jamaica | New York City Public Health Laboratory | Jade Wang et al |
| EPI_ISL_745423 | USA/NY-NYCPHL-001496/2020 | 2020-11-30 | North America/USA/New York/New Yor | DOHMH Corona | New York City Public Health Laboratory | Jade Wang et al |
| EPI_ISL_745428 | USA/NY-NYCPHL-001507/2020 | 2020-11-30 | North America/USA/New York/New Yor | DOHMH Riverside | New York City Public Health Laboratory | Jade Wang et al |
| EPI_ISL_745440 | USA/NY-NYCPHL-001525/2020 | 2020-11-30 | North America/USA/New York/New Yor | DOHMH Jamaica | New York City Public Health Laboratory | Jade Wang et al |
| EPI_ISL_745456 | USA/NY-NYCPHL-001539/2020 | 2020-12-01 | North America/USA/New York/New Yor | DOHMH PHL | New York City Public Health Laboratory | Jade Wang et al |
| EPI_ISL_745460 | USA/NY-NYCPHL-001544/2020 | 2020-12-02 | North America/USA/New York/New Yor | DOHMH Corona | New York City Public Health Laboratory | Jade Wang et al |
| EPI_ISL_745463 | USA/NY-NYCPHL-001548/2020 | 2020-12-02 | North America/USA/New York/New Yor | DOHMH Fort Greene | New York City Public Health Laboratory | Jade Wang et al |
| EPI_ISL_745467 | USA/NY-NYCPHL-001558/2020 | 2020-12-02 | North America/USA/New York/New Yor | DOHMH Corona | New York City Public Health Laboratory | Jade Wang et al |
| EPI_ISL_745474 | USA/UT-UPHL-2012864166/2020 | 2020-10-25 | North America/USA/Utah/ | Ginkgo Bioworks Clinical Laboratory | Utah Public Health Laboratory | Erin L. Young et al |
| EPI_ISL_745476 | USA/UT-UPHL-2012681747/2020 | 2020-08-17 | North America/USA/Utah/ | Ginkgo Bioworks Clinical Laboratory | Utah Public Health Laboratory | Erin L. Young et al |
| EPI_ISL_745477 | USA/UT-UPHL-2012650899/2020 | 2020-08-03 | North America/USA/Utah/ | Ginkgo Bioworks Clinical Laboratory | Utah Public Health Laboratory | Erin L. Young et al |
| EPI_ISL_745481 | USA/UT-UPHL-2012285603/2020 | 2020-08-17 | North America/USA/Utah/ | Ginkgo Bioworks Clinical Laboratory | Utah Public Health Laboratory | Erin L. Young et al |
| EPI_ISL_745482 | USA/UT-UPHL-2012453443/2020 | 2020-08-08 | North America/USA/Utah/ | Ginkgo Bioworks Clinical Laboratory | Utah Public Health Laboratory | Erin L. Young et al |
| EPI_ISL_745484 | USA/UT-UPHL-2012367107/2020 | 2020-07-27 | North America/USA/Utah/ | Ginkgo Bioworks Clinical Laboratory | Utah Public Health Laboratory | Erin L. Young et al |
| EPI_ISL_745486 | USA/UT-UPHL-2012781796/2020 | 2020-07-25 | North America/USA/Utah/ | Ginkgo Bioworks Clinical Laboratory | Utah Public Health Laboratory | Erin L. Young et al |
| EPI_ISL_745490 | USA/UT-UPHL-2012367591/2020 | 2020-08-03 | North America/USA/Utah/ | Ginkgo Bioworks Clinical Laboratory | Utah Public Health Laboratory | Erin L. Young et al |

| | | | | | | |
|---|---|---|---|---|---|---|
| EPI_ISL_745492 | USA/UT-UPHL-2012757008/2020 | 2020-07-27 | North America/USA/Utah/ | Ginkgo Bioworks Clinical Laboratory | Utah Public Health Laboratory | Erin L. Young et al |
| EPI_ISL_745495 | USA/UT-UPHL-2012924903/2020 | 2020-08-12 | North America/USA/Utah/ | Ginkgo Bioworks Clinical Laboratory | Utah Public Health Laboratory | Erin L. Young et al |
| EPI_ISL_745496 | USA/UT-UPHL-2012167424/2020 | 2020-08-03 | North America/USA/Utah/ | Ginkgo Bioworks Clinical Laboratory | Utah Public Health Laboratory | Erin L. Young et al |
| EPI_ISL_745497 | USA/UT-UPHL-2012930871/2020 | 2020-08-11 | North America/USA/Utah/ | Ginkgo Bioworks Clinical Laboratory | Utah Public Health Laboratory | Erin L. Young et al |
| EPI_ISL_745499 | USA/UT-UPHL-2012190379/2020 | 2020-08-18 | North America/USA/Utah/ | Ginkgo Bioworks Clinical Laboratory | Utah Public Health Laboratory | Erin L. Young et al |
| EPI_ISL_745500 | USA/UT-UPHL-2012659055/2020 | 2020-08-17 | North America/USA/Utah/ | Ginkgo Bioworks Clinical Laboratory | Utah Public Health Laboratory | Erin L. Young et al |
| EPI_ISL_745501 | USA/UT-UPHL-2012898524/2020 | 2020-07-26 | North America/USA/Utah/ | Ginkgo Bioworks Clinical Laboratory | Utah Public Health Laboratory | Erin L. Young et al |
| EPI_ISL_745505 | USA/UT-UPHL-2012819912/2020 | 2020-08-17 | North America/USA/Utah/ | Ginkgo Bioworks Clinical Laboratory | Utah Public Health Laboratory | Erin L. Young et al |
| EPI_ISL_745510 | USA/UT-UPHL-2012446164/2020 | 2020-07-25 | North America/USA/Utah/ | Ginkgo Bioworks Clinical Laboratory | Utah Public Health Laboratory | Erin L. Young et al |
| EPI_ISL_745518 | USA/UT-UPHL-2012105008/2020 | 2020-08-10 | North America/USA/Utah/ | Ginkgo Bioworks Clinical Laboratory | Utah Public Health Laboratory | Erin L. Young et al |
| EPI_ISL_745529 | USA/UT-UPHL-2012641846/2020 | 2020-08-15 | North America/USA/Utah/ | Ginkgo Bioworks Clinical Laboratory | Utah Public Health Laboratory | Erin L. Young et al |
| EPI_ISL_745534 | USA/UT-UPHL-2012885221/2020 | 2020-08-03 | North America/USA/Utah/ | Ginkgo Bioworks Clinical Laboratory | Utah Public Health Laboratory | Erin L. Young et al |
| EPI_ISL_745536 | USA/UT-UPHL-2012797881/2020 | 2020-08-06 | North America/USA/Utah/ | Ginkgo Bioworks Clinical Laboratory | Utah Public Health Laboratory | Erin L. Young et al |
| EPI_ISL_745542 | USA/UT-UPHL-2012619179/2020 | 2020-10-27 | North America/USA/Utah/ | Ginkgo Bioworks Clinical Laboratory | Utah Public Health Laboratory | Erin L. Young et al |
| EPI_ISL_745545 | USA/UT-UPHL-2012520384/2020 | 2020-10-23 | North America/USA/Utah/ | Ginkgo Bioworks Clinical Laboratory | Utah Public Health Laboratory | Erin L. Young et al |
| EPI_ISL_745547 | USA/UT-UPHL-2012764192/2020 | 2020-07-25 | North America/USA/Utah/ | Ginkgo Bioworks Clinical Laboratory | Utah Public Health Laboratory | Erin L. Young et al |
| EPI_ISL_745551 | USA/UT-UPHL-2012335532/2020 | 2020-08-11 | North America/USA/Utah/ | Ginkgo Bioworks Clinical Laboratory | Utah Public Health Laboratory | Erin L. Young et al |
| EPI_ISL_745557 | USA/UT-UPHL-2012377203/2020 | 2020-08-10 | North America/USA/Utah/ | Ginkgo Bioworks Clinical Laboratory | Utah Public Health Laboratory | Erin L. Young et al |
| EPI_ISL_745566 | USA/UT-UPHL-2012967715/2020 | 2020-08-03 | North America/USA/Utah/ | Ginkgo Bioworks Clinical Laboratory | Utah Public Health Laboratory | Erin L. Young et al |
| EPI_ISL_745567 | USA/UT-UPHL-2012437075/2020 | 2020-08-17 | North America/USA/Utah/ | Ginkgo Bioworks Clinical Laboratory | Utah Public Health Laboratory | Erin L. Young et al |

| | | | | | | |
|---|---|---|---|---|---|---|
| EPI_ISL_745576 | USA/UT-UPHL-2012053525/2020 | 2020-08-15 | North America/USA/Utah/ | Ginkgo Bioworks Clinical Laboratory | Utah Public Health Laboratory | Erin L. Young et al |
| EPI_ISL_745585 | USA/UT-UPHL-2012627192/2020 | 2020-08-14 | North America/USA/Utah/ | Ginkgo Bioworks Clinical Laboratory | Utah Public Health Laboratory | Erin L. Young et al |
| EPI_ISL_745587 | USA/UT-UPHL-2012069870/2020 | 2020-08-16 | North America/USA/Utah/ | Ginkgo Bioworks Clinical Laboratory | Utah Public Health Laboratory | Erin L. Young et al |
| EPI_ISL_745588 | USA/UT-UPHL-2012234499/2020 | 2020-08-03 | North America/USA/Utah/ | Ginkgo Bioworks Clinical Laboratory | Utah Public Health Laboratory | Erin L. Young et al |
| EPI_ISL_745591 | USA/UT-UPHL-2012902224/2020 | 2020-07-26 | North America/USA/Utah/ | Ginkgo Bioworks Clinical Laboratory | Utah Public Health Laboratory | Erin L. Young et al |
| EPI_ISL_745597 | USA/UT-UPHL-2012311714/2020 | 2020-08-08 | North America/USA/Utah/ | Ginkgo Bioworks Clinical Laboratory | Utah Public Health Laboratory | Erin L. Young et al |
| EPI_ISL_745602 | USA/UT-UPHL-2012290626/2020 | 2020-08-03 | North America/USA/Utah/ | Ginkgo Bioworks Clinical Laboratory | Utah Public Health Laboratory | Erin L. Young et al |
| EPI_ISL_745618 | USA/UT-UPHL-2012046697/2020 | 2020-08-08 | North America/USA/Utah/ | Ginkgo Bioworks Clinical Laboratory | Utah Public Health Laboratory | Erin L. Young et al |
| EPI_ISL_745621 | USA/UT-UPHL-2012729310/2020 | 2020-07-23 | North America/USA/Utah/ | Ginkgo Bioworks Clinical Laboratory | Utah Public Health Laboratory | Erin L. Young et al |
| EPI_ISL_745623 | USA/UT-UPHL-2012502940/2020 | 2020-10-02 | North America/USA/Utah/ | Ginkgo Bioworks Clinical Laboratory | Utah Public Health Laboratory | Erin L. Young et al |
| EPI_ISL_745631 | USA/UT-UPHL-2012907458/2020 | 2020-08-17 | North America/USA/Utah/ | Ginkgo Bioworks Clinical Laboratory | Utah Public Health Laboratory | Erin L. Young et al |
| EPI_ISL_745641 | USA/UT-UPHL-2012548250/2020 | 2020-08-11 | North America/USA/Utah/ | Ginkgo Bioworks Clinical Laboratory | Utah Public Health Laboratory | Erin L. Young et al |
| EPI_ISL_745646 | USA/UT-UPHL-2012546380/2020 | 2020-10-23 | North America/USA/Utah/ | Ginkgo Bioworks Clinical Laboratory | Utah Public Health Laboratory | Erin L. Young et al |
| EPI_ISL_745651 | USA/UT-UPHL-2012361831/2020 | 2020-08-09 | North America/USA/Utah/ | Ginkgo Bioworks Clinical Laboratory | Utah Public Health Laboratory | Erin L. Young et al |
| EPI_ISL_745658 | USA/UT-UPHL-2012255611/2020 | 2020-10-26 | North America/USA/Utah/ | Ginkgo Bioworks Clinical Laboratory | Utah Public Health Laboratory | Erin L. Young et al |
| EPI_ISL_745659 | USA/UT-UPHL-2012332988/2020 | 2020-07-26 | North America/USA/Utah/ | Ginkgo Bioworks Clinical Laboratory | Utah Public Health Laboratory | Erin L. Young et al |
| EPI_ISL_745663 | USA/UT-UPHL-2012692663/2020 | 2020-08-11 | North America/USA/Utah/ | Ginkgo Bioworks Clinical Laboratory | Utah Public Health Laboratory | Erin L. Young et al |
| EPI_ISL_745667 | USA/UT-UPHL-2012855927/2020 | 2020-08-13 | North America/USA/Utah/ | Ginkgo Bioworks Clinical Laboratory | Utah Public Health Laboratory | Erin L. Young et al |
| EPI_ISL_745673 | USA/UT-UPHL-2012590314/2020 | 2020-07-27 | North America/USA/Utah/ | Ginkgo Bioworks Clinical Laboratory | Utah Public Health Laboratory | Erin L. Young et al |
| EPI_ISL_745677 | USA/UT-UPHL-2012647991/2020 | 2020-07-27 | North America/USA/Utah/ | Ginkgo Bioworks Clinical Laboratory | Utah Public Health Laboratory | Erin L. Young et al |

| | | | | | | |
|---|---|---|---|---|---|---|
| EPI_ISL_745681 | USA/UT-UPHL-2012408644/2020 | 2020-07-26 | North America/USA/Utah/ | Ginkgo Bioworks Clinical Laboratory | Utah Public Health Laboratory | Erin L. Young et al |
| EPI_ISL_745686 | USA/UT-UPHL-2012756486/2020 | 2020-07-26 | North America/USA/Utah/ | Ginkgo Bioworks Clinical Laboratory | Utah Public Health Laboratory | Erin L. Young et al |
| EPI_ISL_745690 | USA/UT-UPHL-2012885396/2020 | 2020-07-27 | North America/USA/Utah/ | Ginkgo Bioworks Clinical Laboratory | Utah Public Health Laboratory | Erin L. Young et al |
| EPI_ISL_745695 | USA/UT-UPHL-2012227413/2020 | 2020-07-27 | North America/USA/Utah/ | Ginkgo Bioworks Clinical Laboratory | Utah Public Health Laboratory | Erin L. Young et al |
| EPI_ISL_745696 | USA/UT-UPHL-2012145786/2020 | 2020-07-27 | North America/USA/Utah/ | Ginkgo Bioworks Clinical Laboratory | Utah Public Health Laboratory | Erin L. Young et al |
| EPI_ISL_745701 | USA/UT-UPHL-2012657838/2020 | 2020-07-27 | North America/USA/Utah/ | Ginkgo Bioworks Clinical Laboratory | Utah Public Health Laboratory | Erin L. Young et al |
| EPI_ISL_745702 | USA/UT-UPHL-2012689482/2020 | 2020-07-25 | North America/USA/Utah/ | Ginkgo Bioworks Clinical Laboratory | Utah Public Health Laboratory | Erin L. Young et al |
| EPI_ISL_745706 | USA/UT-UPHL-2012403517/2020 | 2020-07-25 | North America/USA/Utah/ | Ginkgo Bioworks Clinical Laboratory | Utah Public Health Laboratory | Erin L. Young et al |
| EPI_ISL_745707 | USA/UT-UPHL-2012343311/2020 | 2020-07-27 | North America/USA/Utah/ | Ginkgo Bioworks Clinical Laboratory | Utah Public Health Laboratory | Erin L. Young et al |
| EPI_ISL_745708 | USA/UT-UPHL-2012769332/2020 | 2020-07-26 | North America/USA/Utah/ | Ginkgo Bioworks Clinical Laboratory | Utah Public Health Laboratory | Erin L. Young et al |
| EPI_ISL_745711 | USA/UT-UPHL-2012907859/2020 | 2020-07-27 | North America/USA/Utah/ | Ginkgo Bioworks Clinical Laboratory | Utah Public Health Laboratory | Erin L. Young et al |
| EPI_ISL_745715 | USA/UT-UPHL-2012529719/2020 | 2020-07-27 | North America/USA/Utah/ | Ginkgo Bioworks Clinical Laboratory | Utah Public Health Laboratory | Erin L. Young et al |
| EPI_ISL_745721 | USA/UT-UPHL-2012379451/2020 | 2020-08-03 | North America/USA/Utah/ | Ginkgo Bioworks Clinical Laboratory | Utah Public Health Laboratory | Erin L. Young et al |
| EPI_ISL_745722 | USA/UT-UPHL-2012652261/2020 | 2020-08-03 | North America/USA/Utah/ | Ginkgo Bioworks Clinical Laboratory | Utah Public Health Laboratory | Erin L. Young et al |
| EPI_ISL_745723 | USA/UT-UPHL-2012620376/2020 | 2020-08-03 | North America/USA/Utah/ | Ginkgo Bioworks Clinical Laboratory | Utah Public Health Laboratory | Erin L. Young et al |
| EPI_ISL_745725 | USA/UT-UPHL-2012061461/2020 | 2020-08-03 | North America/USA/Utah/ | Ginkgo Bioworks Clinical Laboratory | Utah Public Health Laboratory | Erin L. Young et al |
| EPI_ISL_745727 | USA/UT-UPHL-2012322333/2020 | 2020-08-03 | North America/USA/Utah/ | Ginkgo Bioworks Clinical Laboratory | Utah Public Health Laboratory | Erin L. Young et al |
| EPI_ISL_745732 | USA/UT-UPHL-2012067509/2020 | 2020-08-02 | North America/USA/Utah/ | Ginkgo Bioworks Clinical Laboratory | Utah Public Health Laboratory | Erin L. Young et al |
| EPI_ISL_745738 | USA/UT-UPHL-2012772719/2020 | 2020-08-03 | North America/USA/Utah/ | Ginkgo Bioworks Clinical Laboratory | Utah Public Health Laboratory | Erin L. Young et al |
| EPI_ISL_745742 | USA/UT-UPHL-2012807295/2020 | 2020-08-03 | North America/USA/Utah/ | Ginkgo Bioworks Clinical Laboratory | Utah Public Health Laboratory | Erin L. Young et al |

| | | | | | | |
|---|---|---|---|---|---|---|
| EPI_ISL_745744 | USA/UT-UPHL-2012874225/2020 | 2020-08-03 | North America/USA/Utah/ | Ginkgo Bioworks Clinical Laboratory | Utah Public Health Laboratory | Erin L. Young et al |
| EPI_ISL_745752 | USA/UT-UPHL-2012079566/2020 | 2020-08-03 | North America/USA/Utah/ | Ginkgo Bioworks Clinical Laboratory | Utah Public Health Laboratory | Erin L. Young et al |
| EPI_ISL_745754 | USA/UT-UPHL-2012296348/2020 | 2020-08-03 | North America/USA/Utah/ | Ginkgo Bioworks Clinical Laboratory | Utah Public Health Laboratory | Erin L. Young et al |
| EPI_ISL_745763 | USA/UT-UPHL-2012979596/2020 | 2020-08-03 | North America/USA/Utah/ | Ginkgo Bioworks Clinical Laboratory | Utah Public Health Laboratory | Erin L. Young et al |
| EPI_ISL_745768 | USA/UT-UPHL-2012828260/2020 | 2020-08-03 | North America/USA/Utah/ | Ginkgo Bioworks Clinical Laboratory | Utah Public Health Laboratory | Erin L. Young et al |
| EPI_ISL_745771 | USA/UT-UPHL-2012403274/2020 | 2020-08-09 | North America/USA/Utah/ | Ginkgo Bioworks Clinical Laboratory | Utah Public Health Laboratory | Erin L. Young et al |
| EPI_ISL_745774 | USA/UT-UPHL-2012491321/2020 | 2020-08-09 | North America/USA/Utah/ | Ginkgo Bioworks Clinical Laboratory | Utah Public Health Laboratory | Erin L. Young et al |
| EPI_ISL_745778 | USA/UT-UPHL-2012614422/2020 | 2020-08-09 | North America/USA/Utah/ | Ginkgo Bioworks Clinical Laboratory | Utah Public Health Laboratory | Erin L. Young et al |
| EPI_ISL_745781 | USA/UT-UPHL-2012018896/2020 | 2020-08-09 | North America/USA/Utah/ | Ginkgo Bioworks Clinical Laboratory | Utah Public Health Laboratory | Erin L. Young et al |
| EPI_ISL_745784 | USA/UT-UPHL-2012118130/2020 | 2020-08-08 | North America/USA/Utah/ | Ginkgo Bioworks Clinical Laboratory | Utah Public Health Laboratory | Erin L. Young et al |
| EPI_ISL_745789 | USA/UT-UPHL-2012030607/2020 | 2020-08-09 | North America/USA/Utah/ | Ginkgo Bioworks Clinical Laboratory | Utah Public Health Laboratory | Erin L. Young et al |
| EPI_ISL_745790 | USA/UT-UPHL-2012811862/2020 | 2020-08-08 | North America/USA/Utah/ | Ginkgo Bioworks Clinical Laboratory | Utah Public Health Laboratory | Erin L. Young et al |
| EPI_ISL_745791 | USA/UT-UPHL-2012022396/2020 | 2020-08-09 | North America/USA/Utah/ | Ginkgo Bioworks Clinical Laboratory | Utah Public Health Laboratory | Erin L. Young et al |
| EPI_ISL_745793 | USA/UT-UPHL-2012102790/2020 | 2020-08-08 | North America/USA/Utah/ | Ginkgo Bioworks Clinical Laboratory | Utah Public Health Laboratory | Erin L. Young et al |
| EPI_ISL_745794 | USA/UT-UPHL-2012077077/2020 | 2020-08-08 | North America/USA/Utah/ | Ginkgo Bioworks Clinical Laboratory | Utah Public Health Laboratory | Erin L. Young et al |
| EPI_ISL_745802 | USA/UT-UPHL-2012942531/2020 | 2020-08-09 | North America/USA/Utah/ | Ginkgo Bioworks Clinical Laboratory | Utah Public Health Laboratory | Erin L. Young et al |
| EPI_ISL_745803 | USA/UT-UPHL-2012842708/2020 | 2020-08-08 | North America/USA/Utah/ | Ginkgo Bioworks Clinical Laboratory | Utah Public Health Laboratory | Erin L. Young et al |
| EPI_ISL_745806 | USA/UT-UPHL-2012606373/2020 | 2020-08-08 | North America/USA/Utah/ | Ginkgo Bioworks Clinical Laboratory | Utah Public Health Laboratory | Erin L. Young et al |
| EPI_ISL_745810 | USA/UT-UPHL-2012274873/2020 | 2020-08-08 | North America/USA/Utah/ | Ginkgo Bioworks Clinical Laboratory | Utah Public Health Laboratory | Erin L. Young et al |
| EPI_ISL_745811 | USA/UT-UPHL-2012905899/2020 | 2020-08-08 | North America/USA/Utah/ | Ginkgo Bioworks Clinical Laboratory | Utah Public Health Laboratory | Erin L. Young et al |

| | | | | | | |
|---|---|---|---|---|---|---|
| EPI_ISL_745813 | USA/UT-UPHL-2012502791/2020 | 2020-08-08 | North America/USA/Utah/ | Ginkgo Bioworks Clinical Laboratory | Utah Public Health Laboratory | Erin L. Young et al |
| EPI_ISL_745814 | USA/UT-UPHL-2012321043/2020 | 2020-08-09 | North America/USA/Utah/ | Ginkgo Bioworks Clinical Laboratory | Utah Public Health Laboratory | Erin L. Young et al |
| EPI_ISL_745822 | USA/UT-UPHL-2012742622/2020 | 2020-08-10 | North America/USA/Utah/ | Ginkgo Bioworks Clinical Laboratory | Utah Public Health Laboratory | Erin L. Young et al |
| EPI_ISL_745824 | USA/UT-UPHL-2012156788/2020 | 2020-08-10 | North America/USA/Utah/ | Ginkgo Bioworks Clinical Laboratory | Utah Public Health Laboratory | Erin L. Young et al |
| EPI_ISL_745825 | USA/UT-UPHL-2012789048/2020 | 2020-08-10 | North America/USA/Utah/ | Ginkgo Bioworks Clinical Laboratory | Utah Public Health Laboratory | Erin L. Young et al |
| EPI_ISL_745830 | USA/UT-UPHL-2012581328/2020 | 2020-08-10 | North America/USA/Utah/ | Ginkgo Bioworks Clinical Laboratory | Utah Public Health Laboratory | Erin L. Young et al |
| EPI_ISL_745832 | USA/UT-UPHL-2012063197/2020 | 2020-08-10 | North America/USA/Utah/ | Ginkgo Bioworks Clinical Laboratory | Utah Public Health Laboratory | Erin L. Young et al |
| EPI_ISL_745834 | USA/UT-UPHL-2012487788/2020 | 2020-08-10 | North America/USA/Utah/ | Ginkgo Bioworks Clinical Laboratory | Utah Public Health Laboratory | Erin L. Young et al |
| EPI_ISL_745835 | USA/UT-UPHL-2012890972/2020 | 2020-08-09 | North America/USA/Utah/ | Ginkgo Bioworks Clinical Laboratory | Utah Public Health Laboratory | Erin L. Young et al |
| EPI_ISL_745845 | USA/UT-UPHL-2012380154/2020 | 2020-08-10 | North America/USA/Utah/ | Ginkgo Bioworks Clinical Laboratory | Utah Public Health Laboratory | Erin L. Young et al |
| EPI_ISL_745848 | USA/UT-UPHL-2012753144/2020 | 2020-08-09 | North America/USA/Utah/ | Ginkgo Bioworks Clinical Laboratory | Utah Public Health Laboratory | Erin L. Young et al |
| EPI_ISL_745855 | USA/UT-UPHL-2012766698/2020 | 2020-08-10 | North America/USA/Utah/ | Ginkgo Bioworks Clinical Laboratory | Utah Public Health Laboratory | Erin L. Young et al |
| EPI_ISL_745857 | USA/UT-UPHL-2012846734/2020 | 2020-08-09 | North America/USA/Utah/ | Ginkgo Bioworks Clinical Laboratory | Utah Public Health Laboratory | Erin L. Young et al |
| EPI_ISL_745859 | USA/UT-UPHL-2012191494/2020 | 2020-08-10 | North America/USA/Utah/ | Ginkgo Bioworks Clinical Laboratory | Utah Public Health Laboratory | Erin L. Young et al |
| EPI_ISL_745861 | USA/UT-UPHL-2012514756/2020 | 2020-08-10 | North America/USA/Utah/ | Ginkgo Bioworks Clinical Laboratory | Utah Public Health Laboratory | Erin L. Young et al |
| EPI_ISL_745863 | USA/UT-UPHL-2012724093/2020 | 2020-08-10 | North America/USA/Utah/ | Ginkgo Bioworks Clinical Laboratory | Utah Public Health Laboratory | Erin L. Young et al |
| EPI_ISL_745879 | USA/UT-UPHL-2012972043/2020 | 2020-08-13 | North America/USA/Utah/ | Ginkgo Bioworks Clinical Laboratory | Utah Public Health Laboratory | Erin L. Young et al |
| EPI_ISL_745880 | USA/UT-UPHL-2012338415/2020 | 2020-08-13 | North America/USA/Utah/ | Ginkgo Bioworks Clinical Laboratory | Utah Public Health Laboratory | Erin L. Young et al |
| EPI_ISL_745886 | USA/UT-UPHL-2012380818/2020 | 2020-08-12 | North America/USA/Utah/ | Ginkgo Bioworks Clinical Laboratory | Utah Public Health Laboratory | Erin L. Young et al |
| EPI_ISL_745889 | USA/UT-UPHL-2012543390/2020 | 2020-08-12 | North America/USA/Utah/ | Ginkgo Bioworks Clinical Laboratory | Utah Public Health Laboratory | Erin L. Young et al |

| | | | | | | |
|---|---|---|---|---|---|---|
| EPI_ISL_745890 | USA/UT-UPHL-2012442845/2020 | 2020-08-12 | North America/USA/Utah/ | Ginkgo Bioworks Clinical Laboratory | Utah Public Health Laboratory | Erin L. Young et al |
| EPI_ISL_745892 | USA/UT-UPHL-2012337841/2020 | 2020-08-10 | North America/USA/Utah/ | Ginkgo Bioworks Clinical Laboratory | Utah Public Health Laboratory | Erin L. Young et al |
| EPI_ISL_745893 | USA/UT-UPHL-2012000183/2020 | 2020-08-13 | North America/USA/Utah/ | Ginkgo Bioworks Clinical Laboratory | Utah Public Health Laboratory | Erin L. Young et al |
| EPI_ISL_745899 | USA/UT-UPHL-2012218419/2020 | 2020-08-12 | North America/USA/Utah/ | Ginkgo Bioworks Clinical Laboratory | Utah Public Health Laboratory | Erin L. Young et al |
| EPI_ISL_745905 | USA/UT-UPHL-2012247385/2020 | 2020-08-12 | North America/USA/Utah/ | Ginkgo Bioworks Clinical Laboratory | Utah Public Health Laboratory | Erin L. Young et al |
| EPI_ISL_745907 | USA/UT-UPHL-2012286179/2020 | 2020-08-10 | North America/USA/Utah/ | Ginkgo Bioworks Clinical Laboratory | Utah Public Health Laboratory | Erin L. Young et al |
| EPI_ISL_745909 | USA/UT-UPHL-2012841191/2020 | 2020-08-13 | North America/USA/Utah/ | Ginkgo Bioworks Clinical Laboratory | Utah Public Health Laboratory | Erin L. Young et al |
| EPI_ISL_745910 | USA/UT-UPHL-2012989138/2020 | 2020-08-13 | North America/USA/Utah/ | Ginkgo Bioworks Clinical Laboratory | Utah Public Health Laboratory | Erin L. Young et al |
| EPI_ISL_745914 | USA/UT-UPHL-2012955404/2020 | 2020-08-10 | North America/USA/Utah/ | Ginkgo Bioworks Clinical Laboratory | Utah Public Health Laboratory | Erin L. Young et al |
| EPI_ISL_745920 | USA/UT-UPHL-2012047870/2020 | 2020-08-12 | North America/USA/Utah/ | Ginkgo Bioworks Clinical Laboratory | Utah Public Health Laboratory | Erin L. Young et al |
| EPI_ISL_745924 | USA/UT-UPHL-2012233218/2020 | 2020-08-13 | North America/USA/Utah/ | Ginkgo Bioworks Clinical Laboratory | Utah Public Health Laboratory | Erin L. Young et al |
| EPI_ISL_745925 | USA/UT-UPHL-2012640082/2020 | 2020-08-12 | North America/USA/Utah/ | Ginkgo Bioworks Clinical Laboratory | Utah Public Health Laboratory | Erin L. Young et al |
| EPI_ISL_745926 | USA/UT-UPHL-2012036591/2020 | 2020-08-12 | North America/USA/Utah/ | Ginkgo Bioworks Clinical Laboratory | Utah Public Health Laboratory | Erin L. Young et al |
| EPI_ISL_745929 | USA/UT-UPHL-2012750625/2020 | 2020-08-11 | North America/USA/Utah/ | Ginkgo Bioworks Clinical Laboratory | Utah Public Health Laboratory | Erin L. Young et al |
| EPI_ISL_745931 | USA/UT-UPHL-2012242806/2020 | 2020-08-12 | North America/USA/Utah/ | Ginkgo Bioworks Clinical Laboratory | Utah Public Health Laboratory | Erin L. Young et al |
| EPI_ISL_745942 | USA/UT-UPHL-2012334615/2020 | 2020-08-13 | North America/USA/Utah/ | Ginkgo Bioworks Clinical Laboratory | Utah Public Health Laboratory | Erin L. Young et al |
| EPI_ISL_745946 | USA/UT-UPHL-2012773576/2020 | 2020-08-11 | North America/USA/Utah/ | Ginkgo Bioworks Clinical Laboratory | Utah Public Health Laboratory | Erin L. Young et al |
| EPI_ISL_745950 | USA/UT-UPHL-2012505034/2020 | 2020-08-12 | North America/USA/Utah/ | Ginkgo Bioworks Clinical Laboratory | Utah Public Health Laboratory | Erin L. Young et al |
| EPI_ISL_745951 | USA/UT-UPHL-2012071943/2020 | 2020-08-15 | North America/USA/Utah/ | Ginkgo Bioworks Clinical Laboratory | Utah Public Health Laboratory | Erin L. Young et al |
| EPI_ISL_745954 | USA/UT-UPHL-2012082051/2020 | 2020-08-11 | North America/USA/Utah/ | Ginkgo Bioworks Clinical Laboratory | Utah Public Health Laboratory | Erin L. Young et al |

| | | | | | | |
|---|---|---|---|---|---|---|
| EPI_ISL_745958 | USA/UT-UPHL-2012634496/2020 | 2020-08-10 | North America/USA/Utah/ | Ginkgo Bioworks Clinical Laboratory | Utah Public Health Laboratory | Erin L. Young et al |
| EPI_ISL_745962 | USA/UT-UPHL-2012370469/2020 | 2020-08-11 | North America/USA/Utah/ | Ginkgo Bioworks Clinical Laboratory | Utah Public Health Laboratory | Erin L. Young et al |
| EPI_ISL_745965 | USA/UT-UPHL-2012259833/2020 | 2020-08-12 | North America/USA/Utah/ | Ginkgo Bioworks Clinical Laboratory | Utah Public Health Laboratory | Erin L. Young et al |
| EPI_ISL_745968 | USA/UT-UPHL-2012205637/2020 | 2020-08-11 | North America/USA/Utah/ | Ginkgo Bioworks Clinical Laboratory | Utah Public Health Laboratory | Erin L. Young et al |
| EPI_ISL_745973 | USA/UT-UPHL-2012133597/2020 | 2020-08-11 | North America/USA/Utah/ | Ginkgo Bioworks Clinical Laboratory | Utah Public Health Laboratory | Erin L. Young et al |
| EPI_ISL_745975 | USA/UT-UPHL-2012400155/2020 | 2020-08-11 | North America/USA/Utah/ | Ginkgo Bioworks Clinical Laboratory | Utah Public Health Laboratory | Erin L. Young et al |
| EPI_ISL_745980 | USA/UT-UPHL-2012725139/2020 | 2020-08-08 | North America/USA/Utah/ | Ginkgo Bioworks Clinical Laboratory | Utah Public Health Laboratory | Erin L. Young et al |
| EPI_ISL_745982 | USA/UT-UPHL-2012212287/2020 | 2020-09-22 | North America/USA/Utah/ | Ginkgo Bioworks Clinical Laboratory | Utah Public Health Laboratory | Erin L. Young et al |
| EPI_ISL_745983 | USA/UT-UPHL-2012798103/2020 | 2020-08-04 | North America/USA/Utah/ | Ginkgo Bioworks Clinical Laboratory | Utah Public Health Laboratory | Erin L. Young et al |
| EPI_ISL_745988 | USA/UT-UPHL-2012432100/2020 | 2020-08-13 | North America/USA/Utah/ | Ginkgo Bioworks Clinical Laboratory | Utah Public Health Laboratory | Erin L. Young et al |
| EPI_ISL_745991 | USA/UT-UPHL-2012498286/2020 | 2020-08-06 | North America/USA/Utah/ | Ginkgo Bioworks Clinical Laboratory | Utah Public Health Laboratory | Erin L. Young et al |
| EPI_ISL_746001 | USA/UT-UPHL-2012780542/2020 | 2020-08-11 | North America/USA/Utah/ | Ginkgo Bioworks Clinical Laboratory | Utah Public Health Laboratory | Erin L. Young et al |
| EPI_ISL_746002 | USA/UT-UPHL-2012968336/2020 | 2020-08-13 | North America/USA/Utah/ | Ginkgo Bioworks Clinical Laboratory | Utah Public Health Laboratory | Erin L. Young et al |
| EPI_ISL_746008 | USA/UT-UPHL-2012006545/2020 | 2020-09-23 | North America/USA/Utah/ | Ginkgo Bioworks Clinical Laboratory | Utah Public Health Laboratory | Erin L. Young et al |
| EPI_ISL_746009 | USA/UT-UPHL-2012543435/2020 | 2020-08-13 | North America/USA/Utah/ | Ginkgo Bioworks Clinical Laboratory | Utah Public Health Laboratory | Erin L. Young et al |
| EPI_ISL_746010 | USA/UT-UPHL-2012395754/2020 | 2020-08-13 | North America/USA/Utah/ | Ginkgo Bioworks Clinical Laboratory | Utah Public Health Laboratory | Erin L. Young et al |
| EPI_ISL_746018 | USA/UT-UPHL-2012829129/2020 | 2020-08-06 | North America/USA/Utah/ | Ginkgo Bioworks Clinical Laboratory | Utah Public Health Laboratory | Erin L. Young et al |
| EPI_ISL_746020 | USA/UT-UPHL-2012716871/2020 | 2020-08-17 | North America/USA/Utah/ | Ginkgo Bioworks Clinical Laboratory | Utah Public Health Laboratory | Erin L. Young et al |
| EPI_ISL_746025 | USA/UT-UPHL-2012698910/2020 | 2020-08-04 | North America/USA/Utah/ | Ginkgo Bioworks Clinical Laboratory | Utah Public Health Laboratory | Erin L. Young et al |
| EPI_ISL_746028 | USA/UT-UPHL-2012049713/2020 | 2020-08-08 | North America/USA/Utah/ | Ginkgo Bioworks Clinical Laboratory | Utah Public Health Laboratory | Erin L. Young et al |

| | | | | | | |
|---|---|---|---|---|---|---|
| EPI_ISL_746030 | USA/UT-UPHL-2012481939/2020 | 2020-08-13 | North America/USA/Utah/ | Ginkgo Bioworks Clinical Laboratory | Utah Public Health Laboratory | Erin L. Young et al |
| EPI_ISL_746031 | USA/UT-UPHL-2012198597/2020 | 2020-08-04 | North America/USA/Utah/ | Ginkgo Bioworks Clinical Laboratory | Utah Public Health Laboratory | Erin L. Young et al |
| EPI_ISL_746037 | USA/UT-UPHL-2012036481/2020 | 2020-08-17 | North America/USA/Utah/ | Ginkgo Bioworks Clinical Laboratory | Utah Public Health Laboratory | Erin L. Young et al |
| EPI_ISL_746047 | USA/UT-UPHL-2012392041/2020 | 2020-08-05 | North America/USA/Utah/ | Ginkgo Bioworks Clinical Laboratory | Utah Public Health Laboratory | Erin L. Young et al |
| EPI_ISL_746049 | USA/UT-UPHL-2012859839/2020 | 2020-08-08 | North America/USA/Utah/ | Ginkgo Bioworks Clinical Laboratory | Utah Public Health Laboratory | Erin L. Young et al |
| EPI_ISL_746050 | USA/UT-UPHL-2012081770/2020 | 2020-07-31 | North America/USA/Utah/ | Ginkgo Bioworks Clinical Laboratory | Utah Public Health Laboratory | Erin L. Young et al |
| EPI_ISL_746054 | USA/UT-UPHL-2012762917/2020 | 2020-08-17 | North America/USA/Utah/ | Ginkgo Bioworks Clinical Laboratory | Utah Public Health Laboratory | Erin L. Young et al |
| EPI_ISL_746063 | USA/UT-UPHL-2012913551/2020 | 2020-08-06 | North America/USA/Utah/ | Ginkgo Bioworks Clinical Laboratory | Utah Public Health Laboratory | Erin L. Young et al |
| EPI_ISL_746067 | USA/UT-UPHL-2012580181/2020 | 2020-08-17 | North America/USA/Utah/ | Ginkgo Bioworks Clinical Laboratory | Utah Public Health Laboratory | Erin L. Young et al |
| EPI_ISL_746068 | USA/UT-UPHL-2012944794/2020 | 2020-08-17 | North America/USA/Utah/ | Ginkgo Bioworks Clinical Laboratory | Utah Public Health Laboratory | Erin L. Young et al |
| EPI_ISL_746069 | USA/UT-UPHL-2012787592/2020 | 2020-08-05 | North America/USA/Utah/ | Ginkgo Bioworks Clinical Laboratory | Utah Public Health Laboratory | Erin L. Young et al |
| EPI_ISL_746070 | USA/UT-UPHL-2012080934/2020 | 2020-08-04 | North America/USA/Utah/ | Ginkgo Bioworks Clinical Laboratory | Utah Public Health Laboratory | Erin L. Young et al |
| EPI_ISL_746079 | USA/UT-UPHL-2012372658/2020 | 2020-08-01 | North America/USA/Utah/ | Ginkgo Bioworks Clinical Laboratory | Utah Public Health Laboratory | Erin L. Young et al |
| EPI_ISL_746090 | USA/UT-UPHL-2012776941/2020 | 2020-08-17 | North America/USA/Utah/ | Ginkgo Bioworks Clinical Laboratory | Utah Public Health Laboratory | Erin L. Young et al |
| EPI_ISL_746092 | USA/UT-UPHL-2012734844/2020 | 2020-08-17 | North America/USA/Utah/ | Ginkgo Bioworks Clinical Laboratory | Utah Public Health Laboratory | Erin L. Young et al |
| EPI_ISL_746093 | USA/UT-UPHL-2012159552/2020 | 2020-08-06 | North America/USA/Utah/ | Ginkgo Bioworks Clinical Laboratory | Utah Public Health Laboratory | Erin L. Young et al |
| EPI_ISL_746102 | USA/UT-UPHL-2012266977/2020 | 2020-10-02 | North America/USA/Utah/ | Ginkgo Bioworks Clinical Laboratory | Utah Public Health Laboratory | Erin L. Young et al |
| EPI_ISL_746111 | USA/UT-UPHL-2012332844/2020 | 2020-08-14 | North America/USA/Utah/ | Ginkgo Bioworks Clinical Laboratory | Utah Public Health Laboratory | Erin L. Young et al |
| EPI_ISL_746117 | USA/UT-UPHL-2012256876/2020 | 2020-10-02 | North America/USA/Utah/ | Ginkgo Bioworks Clinical Laboratory | Utah Public Health Laboratory | Erin L. Young et al |
| EPI_ISL_746119 | USA/UT-UPHL-2012780804/2020 | 2020-08-14 | North America/USA/Utah/ | Ginkgo Bioworks Clinical Laboratory | Utah Public Health Laboratory | Erin L. Young et al |

| | | | | | | |
|---|---|---|---|---|---|---|
| EPI_ISL_746120 | USA/UT-UPHL-2012738554/2020 | 2020-08-18 | North America/USA/Utah/ | Ginkgo Bioworks Clinical Laboratory | Utah Public Health Laboratory | Erin L. Young et al |
| EPI_ISL_746128 | USA/UT-UPHL-2012106527/2020 | 2020-09-05 | North America/USA/Utah/ | Ginkgo Bioworks Clinical Laboratory | Utah Public Health Laboratory | Erin L. Young et al |
| EPI_ISL_746131 | USA/UT-UPHL-2012854117/2020 | 2020-10-02 | North America/USA/Utah/ | Ginkgo Bioworks Clinical Laboratory | Utah Public Health Laboratory | Erin L. Young et al |
| EPI_ISL_746135 | USA/UT-UPHL-2012533179/2020 | 2020-09-07 | North America/USA/Utah/ | Ginkgo Bioworks Clinical Laboratory | Utah Public Health Laboratory | Erin L. Young et al |
| EPI_ISL_746137 | USA/UT-UPHL-2012536667/2020 | 2020-09-09 | North America/USA/Utah/ | Ginkgo Bioworks Clinical Laboratory | Utah Public Health Laboratory | Erin L. Young et al |
| EPI_ISL_746145 | USA/UT-UPHL-2012975721/2020 | 2020-08-15 | North America/USA/Utah/ | Ginkgo Bioworks Clinical Laboratory | Utah Public Health Laboratory | Erin L. Young et al |
| EPI_ISL_746146 | USA/UT-UPHL-2012517618/2020 | 2020-08-18 | North America/USA/Utah/ | Ginkgo Bioworks Clinical Laboratory | Utah Public Health Laboratory | Erin L. Young et al |
| EPI_ISL_746150 | USA/UT-UPHL-2012564151/2020 | 2020-08-18 | North America/USA/Utah/ | Ginkgo Bioworks Clinical Laboratory | Utah Public Health Laboratory | Erin L. Young et al |
| EPI_ISL_746151 | USA/UT-UPHL-2012045630/2020 | 2020-08-18 | North America/USA/Utah/ | Ginkgo Bioworks Clinical Laboratory | Utah Public Health Laboratory | Erin L. Young et al |
| EPI_ISL_746152 | USA/UT-UPHL-2012607685/2020 | 2020-08-14 | North America/USA/Utah/ | Ginkgo Bioworks Clinical Laboratory | Utah Public Health Laboratory | Erin L. Young et al |
| EPI_ISL_746153 | USA/UT-UPHL-2012468965/2020 | 2020-08-17 | North America/USA/Utah/ | Ginkgo Bioworks Clinical Laboratory | Utah Public Health Laboratory | Erin L. Young et al |
| EPI_ISL_746155 | USA/UT-UPHL-2012040579/2020 | 2020-08-14 | North America/USA/Utah/ | Ginkgo Bioworks Clinical Laboratory | Utah Public Health Laboratory | Erin L. Young et al |
| EPI_ISL_746158 | USA/UT-UPHL-2012214216/2020 | 2020-08-13 | North America/USA/Utah/ | Ginkgo Bioworks Clinical Laboratory | Utah Public Health Laboratory | Erin L. Young et al |
| EPI_ISL_746169 | USA/UT-UPHL-2012130118/2020 | 2020-08-17 | North America/USA/Utah/ | Ginkgo Bioworks Clinical Laboratory | Utah Public Health Laboratory | Erin L. Young et al |
| EPI_ISL_746179 | USA/UT-UPHL-2012086479/2020 | 2020-08-14 | North America/USA/Utah/ | Ginkgo Bioworks Clinical Laboratory | Utah Public Health Laboratory | Erin L. Young et al |
| EPI_ISL_746183 | USA/UT-UPHL-2012444066/2020 | 2020-08-13 | North America/USA/Utah/ | Ginkgo Bioworks Clinical Laboratory | Utah Public Health Laboratory | Erin L. Young et al |
| EPI_ISL_746191 | USA/UT-UPHL-2012038375/2020 | 2020-08-17 | North America/USA/Utah/ | Ginkgo Bioworks Clinical Laboratory | Utah Public Health Laboratory | Erin L. Young et al |
| EPI_ISL_746198 | USA/UT-UPHL-2012432369/2020 | 2020-08-17 | North America/USA/Utah/ | Ginkgo Bioworks Clinical Laboratory | Utah Public Health Laboratory | Erin L. Young et al |
| EPI_ISL_746199 | USA/UT-UPHL-2012580849/2020 | 2020-08-17 | North America/USA/Utah/ | Ginkgo Bioworks Clinical Laboratory | Utah Public Health Laboratory | Erin L. Young et al |
| EPI_ISL_746202 | USA/UT-UPHL-2012525389/2020 | 2020-08-14 | North America/USA/Utah/ | Ginkgo Bioworks Clinical Laboratory | Utah Public Health Laboratory | Erin L. Young et al |

| | | | | | | |
|---|---|---|---|---|---|---|
| EPI_ISL_746203 | USA/UT-UPHL-2012218264/2020 | 2020-08-17 | North America/USA/Utah/ | Ginkgo Bioworks Clinical Laboratory | Utah Public Health Laboratory | Erin L. Young et al |
| EPI_ISL_746211 | USA/UT-UPHL-2012244887/2020 | 2020-10-08 | North America/USA/Utah/ | Ginkgo Bioworks Clinical Laboratory | Utah Public Health Laboratory | Erin L. Young et al |
| EPI_ISL_746216 | USA/UT-UPHL-2012255039/2020 | 2020-10-19 | North America/USA/Utah/ | Ginkgo Bioworks Clinical Laboratory | Utah Public Health Laboratory | Erin L. Young et al |
| EPI_ISL_746217 | USA/UT-UPHL-2012089430/2020 | 2020-10-21 | North America/USA/Utah/ | Ginkgo Bioworks Clinical Laboratory | Utah Public Health Laboratory | Erin L. Young et al |
| EPI_ISL_746219 | USA/UT-UPHL-2012125874/2020 | 2020-10-22 | North America/USA/Utah/ | Ginkgo Bioworks Clinical Laboratory | Utah Public Health Laboratory | Erin L. Young et al |
| EPI_ISL_746228 | USA/UT-UPHL-2012973122/2020 | 2020-10-26 | North America/USA/Utah/ | Ginkgo Bioworks Clinical Laboratory | Utah Public Health Laboratory | Erin L. Young et al |
| EPI_ISL_746233 | USA/UT-UPHL-2012876879/2020 | 2020-10-27 | North America/USA/Utah/ | Ginkgo Bioworks Clinical Laboratory | Utah Public Health Laboratory | Erin L. Young et al |
| EPI_ISL_746235 | USA/UT-UPHL-2012427069/2020 | 2020-10-26 | North America/USA/Utah/ | Ginkgo Bioworks Clinical Laboratory | Utah Public Health Laboratory | Erin L. Young et al |
| EPI_ISL_746263 | USA/UT-UPHL-2012434776/2020 | 2020-10-12 | North America/USA/Utah/ | Ginkgo Bioworks Clinical Laboratory | Utah Public Health Laboratory | Erin L. Young et al |
| EPI_ISL_746264 | USA/UT-UPHL-2012956729/2020 | 2020-10-12 | North America/USA/Utah/ | Ginkgo Bioworks Clinical Laboratory | Utah Public Health Laboratory | Erin L. Young et al |
| EPI_ISL_746270 | USA/UT-UPHL-2012682075/2020 | 2020-08-16 | North America/USA/Utah/ | Ginkgo Bioworks Clinical Laboratory | Utah Public Health Laboratory | Erin L. Young et al |
| EPI_ISL_746271 | USA/UT-UPHL-2012130640/2020 | 2020-10-19 | North America/USA/Utah/ | Ginkgo Bioworks Clinical Laboratory | Utah Public Health Laboratory | Erin L. Young et al |
| EPI_ISL_746276 | USA/UT-UPHL-2012852183/2020 | 2020-08-16 | North America/USA/Utah/ | Ginkgo Bioworks Clinical Laboratory | Utah Public Health Laboratory | Erin L. Young et al |
| EPI_ISL_746282 | USA/UT-UPHL-2012414451/2020 | 2020-10-24 | North America/USA/Utah/ | Ginkgo Bioworks Clinical Laboratory | Utah Public Health Laboratory | Erin L. Young et al |
| EPI_ISL_746283 | USA/UT-UPHL-2012036570/2020 | 2020-08-15 | North America/USA/Utah/ | Ginkgo Bioworks Clinical Laboratory | Utah Public Health Laboratory | Erin L. Young et al |
| EPI_ISL_746284 | USA/UT-UPHL-2012776444/2020 | 2020-10-23 | North America/USA/Utah/ | Ginkgo Bioworks Clinical Laboratory | Utah Public Health Laboratory | Erin L. Young et al |
| EPI_ISL_746287 | USA/UT-UPHL-2012739315/2020 | 2020-08-15 | North America/USA/Utah/ | Ginkgo Bioworks Clinical Laboratory | Utah Public Health Laboratory | Erin L. Young et al |
| EPI_ISL_746290 | USA/UT-UPHL-2012148746/2020 | 2020-08-15 | North America/USA/Utah/ | Ginkgo Bioworks Clinical Laboratory | Utah Public Health Laboratory | Erin L. Young et al |
| EPI_ISL_746293 | USA/UT-UPHL-2012133022/2020 | 2020-08-16 | North America/USA/Utah/ | Ginkgo Bioworks Clinical Laboratory | Utah Public Health Laboratory | Erin L. Young et al |
| EPI_ISL_746297 | USA/UT-UPHL-2012190312/2020 | 2020-08-15 | North America/USA/Utah/ | Ginkgo Bioworks Clinical Laboratory | Utah Public Health Laboratory | Erin L. Young et al |

| | | | | | | |
|---|---|---|---|---|---|---|
| EPI_ISL_746304 | USA/UT-UPHL-2012098996/2020 | 2020-08-15 | North America/USA/Utah/ | Ginkgo Bioworks Clinical Laboratory | Utah Public Health Laboratory | Erin L. Young et al |
| EPI_ISL_746319 | Bangladesh/JUST-GC55-034/2020 | 2020-06-26 | Asia/Bangladesh/Khulna/Satkhira | Genome Center | Genome Center | Md. Shazid Hasan et al |
| EPI_ISL_746334 | USA/UT-UPHL-2012005997/2020 | 2020-06-17 | North America/USA/Utah/ | Utah Public Health Laboratory | Utah Public Health Laboratory | Erin Young et al |
| EPI_ISL_746338 | USA/UT-UPHL-2012441822/2020 | 2020-06-18 | North America/USA/Utah/ | Utah Public Health Laboratory | Utah Public Health Laboratory | Erin Young et al |
| EPI_ISL_746350 | USA/UT-UPHL-2012884106/2020 | 2020-06-16 | North America/USA/Utah/ | Utah Public Health Laboratory | Utah Public Health Laboratory | Erin Young et al |
| EPI_ISL_746364 | USA/UT-UPHL-2012191112/2020 | 2020-06-18 | North America/USA/Utah/ | Utah Public Health Laboratory | Utah Public Health Laboratory | Erin Young et al |
| EPI_ISL_746378 | USA/UT-UPHL-2012681522/2020 | 2020-06-18 | North America/USA/Utah/ | Utah Public Health Laboratory | Utah Public Health Laboratory | Erin Young et al |
| EPI_ISL_746382 | USA/UT-UPHL-2012790266/2020 | 2020-06-18 | North America/USA/Utah/ | Utah Public Health Laboratory | Utah Public Health Laboratory | Erin Young et al |
| EPI_ISL_746387 | USA/UT-UPHL-2012984468/2020 | 2020-06-16 | North America/USA/Utah/ | Utah Public Health Laboratory | Utah Public Health Laboratory | Erin Young et al |
| EPI_ISL_746398 | USA/UT-UPHL-2012691035/2020 | 2020-11-30 | North America/USA/Utah/ | Utah Public Health Laboratory | Utah Public Health Laboratory | Erin Young et al |
| EPI_ISL_746403 | USA/UT-UPHL-2012789877/2020 | 2020-11-30 | North America/USA/Utah/ | Utah Public Health Laboratory | Utah Public Health Laboratory | Erin Young et al |
| EPI_ISL_746408 | USA/UT-UPHL-2012773212/2020 | 2020-11-28 | North America/USA/Utah/ | Utah Public Health Laboratory | Utah Public Health Laboratory | Erin Young et al |
| EPI_ISL_746411 | USA/UT-UPHL-2012681356/2020 | 2020-06-18 | North America/USA/Utah/ | Utah Public Health Laboratory | Utah Public Health Laboratory | Erin Young et al |
| EPI_ISL_746424 | USA/UT-UPHL-2012438158/2020 | 2020-06-18 | North America/USA/Utah/ | Utah Public Health Laboratory | Utah Public Health Laboratory | Erin Young et al |
| EPI_ISL_746426 | USA/UT-UPHL-2012127660/2020 | 2020-11-30 | North America/USA/Utah/ | Utah Public Health Laboratory | Utah Public Health Laboratory | Erin Young et al |
| EPI_ISL_746434 | USA/UT-UPHL-2012222800/2020 | 2020-11-30 | North America/USA/Utah/ | Utah Public Health Laboratory | Utah Public Health Laboratory | Erin Young et al |
| EPI_ISL_746438 | USA/UT-UPHL-2012275312/2020 | 2020-06-19 | North America/USA/Utah/ | Utah Public Health Laboratory | Utah Public Health Laboratory | Erin Young et al |
| EPI_ISL_746449 | USA/UT-UPHL-2012582762/2020 | 2020-06-18 | North America/USA/Utah/ | Utah Public Health Laboratory | Utah Public Health Laboratory | Erin Young et al |
| EPI_ISL_746458 | USA/UT-UPHL-2012683062/2020 | 2020-06-19 | North America/USA/Utah/ | Utah Public Health Laboratory | Utah Public Health Laboratory | Erin Young et al |
| EPI_ISL_746462 | USA/UT-UPHL-2012743524/2020 | 2020-06-19 | North America/USA/Utah/ | Utah Public Health Laboratory | Utah Public Health Laboratory | Erin Young et al |

| | | | | | | |
|---|---|---|---|---|---|---|
| EPI_ISL_746467 | USA/UT-UPHL-2012796558/2020 | 2020-11-29 | North America/USA/Utah/ | Utah Public Health Laboratory | Utah Public Health Laboratory | Erin Young et al |
| EPI_ISL_746468 | USA/UT-UPHL-2012825995/2020 | 2020-06-19 | North America/USA/Utah/ | Utah Public Health Laboratory | Utah Public Health Laboratory | Erin Young et al |
| EPI_ISL_746477 | USA/UT-UPHL-2012964498/2020 | 2020-06-18 | North America/USA/Utah/ | Utah Public Health Laboratory | Utah Public Health Laboratory | Erin Young et al |
| EPI_ISL_746480 | Chile/AR-192894/2020 | 2020-09-29 | South America/Chile/Araucan√≠a/Temu | Genetica Molecular and Subdepartamento de Virologia ISP Chile | Instituto de Salud Publica de Chile | Javier Tognarelli et al |
| EPI_ISL_746481 | Chile/AR-179001/2020 | 2020-09-15 | South America/Chile/Araucan√≠a/Temu | Genetica Molecular and Subdepartamento de Virologia ISP Chile | Instituto de Salud Publica de Chile | Javier Tognarelli et al |
| EPI_ISL_746482 | Chile/RM-82489/2020 | 2020-05-29 | South America/Chile/Region Metropolit | Genetica Molecular and Subdepartamento de Virologia ISP Chile | Instituto de Salud Publica de Chile | Javier Tognarelli et al |
| EPI_ISL_746484 | Chile/RM-88166-TC1/2020 | 2020-06-03 | South America/Chile/Region Metropolit | Genetica Molecular and Subdepartamento de Virologia ISP Chile | Instituto de Salud Publica de Chile | Javier Tognarelli et al |
| EPI_ISL_746485 | Chile/RM-131478/2020 | 2020-07-22 | South America/Chile/Region Metropolit | Genetica Molecular and Subdepartamento de Virologia ISP Chile | Instituto de Salud Publica de Chile | Javier Tognarelli et al |
| EPI_ISL_746487 | Chile/RM-155625/2020 | 2020-08-19 | South America/Chile/Region Metropolit | Genetica Molecular and Subdepartamento de Virologia ISP Chile | Instituto de Salud Publica de Chile | Javier Tognarelli et al |
| EPI_ISL_746488 | Chile/RM-155645/2020 | 2020-08-19 | South America/Chile/Region Metropolit | Genetica Molecular and Subdepartamento de Virologia ISP Chile | Instituto de Salud Publica de Chile | Javier Tognarelli et al |
| EPI_ISL_746489 | Chile/RM-72436/2020 | 2020-05-23 | South America/Chile/Region Metropolit | Genetica Molecular and Subdepartamento de Virologia ISP Chile | Instituto de Salud Publica de Chile | Javier Tognarelli et al |
| EPI_ISL_746493 | Chile/RM-82462/2020 | 2020-05-30 | South America/Chile/Region Metropolit | Genetica Molecular and Subdepartamento de Virologia ISP Chile | Instituto de Salud Publica de Chile | Javier Tognarelli et al |
| EPI_ISL_746499 | Chile/AR-172172/2020 | 2020-09-08 | South America/Chile/Araucan√≠a/Temu | Genetica Molecular and Subdepartamento de Virologia ISP Chile | Instituto de Salud Publica de Chile | Javier Tognarelli et al |
| EPI_ISL_746500 | Chile/AR-178998/2020 | 2020-09-15 | South America/Chile/Araucan√≠a/Temu | Genetica Molecular and Subdepartamento de Virologia ISP Chile | Instituto de Salud Publica de Chile | Javier Tognarelli et al |
| EPI_ISL_746501 | Chile/AR-192896/2020 | 2020-09-29 | South America/Chile/Araucan√≠a/Temu | Genetica Molecular and Subdepartamento de Virologia ISP Chile | Instituto de Salud Publica de Chile | Javier Tognarelli et al |
| EPI_ISL_746504 | Chile/AR-208476/2020 | 2020-10-16 | South America/Chile/Araucan√≠a/Temu | Genetica Molecular and Subdepartamento de Virologia ISP Chile | Instituto de Salud Publica de Chile | Javier Tognarelli et al |
| EPI_ISL_746505 | Chile/AR-208477/2020 | 2020-10-16 | South America/Chile/Araucan√≠a/Temu | Genetica Molecular and Subdepartamento de Virologia ISP Chile | Instituto de Salud Publica de Chile | Javier Tognarelli et al |
| EPI_ISL_746506 | Chile/BI-109143/2020 | 2020-06-24 | South America/Chile/Region Metropolit | Genetica Molecular and Subdepartamento de Virologia ISP Chile | Instituto de Salud Publica de Chile | Javier Tognarelli et al |
| EPI_ISL_746510 | Chile/BI-185934/2020 | 2020-09-23 | South America/Chile/Biob√≠o/Concepcio | Genetica Molecular and Subdepartamento de Virologia ISP Chile | Instituto de Salud Publica de Chile | Javier Tognarelli et al |
| EPI_ISL_746512 | Chile/BI-194034/2020 | 2020-09-30 | South America/Chile/Biob√≠o/Concepcio | Genetica Molecular and Subdepartamento de Virologia ISP Chile | Instituto de Salud Publica de Chile | Javier Tognarelli et al |

| | | | | | | |
|---|---|---|---|---|---|---|
| EPI_ISL_746513 | Chile/BI-199845/2020 | 2020-10-07 | South America/Chile/Biobv≠o/Concepcio | Genetica Molecular and Subdepartamento de Virologia ISP Chile | Instituto de Salud Publica de Chile | Javier Tognarelli et al |
| EPI_ISL_746515 | Chile/BI-199849/2020 | 2020-10-07 | South America/Chile/Biobv≠o/Concepcio | Genetica Molecular and Subdepartamento de Virologia ISP Chile | Instituto de Salud Publica de Chile | Javier Tognarelli et al |
| EPI_ISL_746519 | Chile/BI-206420/2020 | 2020-10-15 | South America/Chile/Biobv≠o/Concepcio | Genetica Molecular and Subdepartamento de Virologia ISP Chile | Instituto de Salud Publica de Chile | Javier Tognarelli et al |
| EPI_ISL_746521 | Chile/MA-194107-B/2020 | 2020-09-27 | South America/Chile/Magallanes/Punta | Genetica Molecular and Subdepartamento de Virologia ISP Chile | Instituto de Salud Publica de Chile | Javier Tognarelli et al |
| EPI_ISL_746522 | Chile/MA-194108-B/2020 | 2020-09-27 | South America/Chile/Magallanes/Punta | Genetica Molecular and Subdepartamento de Virologia ISP Chile | Instituto de Salud Publica de Chile | Javier Tognarelli et al |
| EPI_ISL_746523 | Chile/MA-194109-B/2020 | 2020-09-27 | South America/Chile/Magallanes/Punta | Genetica Molecular and Subdepartamento de Virologia ISP Chile | Instituto de Salud Publica de Chile | Javier Tognarelli et al |
| EPI_ISL_746525 | Chile/MA-194112-B/2020 | 2020-09-27 | South America/Chile/Magallanes/Punta | Genetica Molecular and Subdepartamento de Virologia ISP Chile | Instituto de Salud Publica de Chile | Javier Tognarelli et al |
| EPI_ISL_746526 | Chile/MA-194113-B/2020 | 2020-09-27 | South America/Chile/Magallanes/Punta | Genetica Molecular and Subdepartamento de Virologia ISP Chile | Instituto de Salud Publica de Chile | Javier Tognarelli et al |
| EPI_ISL_746527 | Chile/MA-194114-B/2020 | 2020-09-27 | South America/Chile/Magallanes/Punta | Genetica Molecular and Subdepartamento de Virologia ISP Chile | Instituto de Salud Publica de Chile | Javier Tognarelli et al |
| EPI_ISL_746529 | Chile/MA-194116-B/2020 | 2020-09-27 | South America/Chile/Magallanes/Punta | Genetica Molecular and Subdepartamento de Virologia ISP Chile | Instituto de Salud Publica de Chile | Javier Tognarelli et al |
| EPI_ISL_746530 | Chile/MA-201640/2020 | 2020-10-08 | South America/Chile/Magallanes/Punta | Genetica Molecular and Subdepartamento de Virologia ISP Chile | Instituto de Salud Publica de Chile | Javier Tognarelli et al |
| EPI_ISL_746534 | Chile/MA-212212/2020 | 2020-10-22 | South America/Chile/Magallanes/Punta | Genetica Molecular and Subdepartamento de Virologia ISP Chile | Instituto de Salud Publica de Chile | Javier Tognarelli et al |
| EPI_ISL_746535 | Chile/MA-212213/2020 | 2020-10-22 | South America/Chile/Magallanes/Punta | Genetica Molecular and Subdepartamento de Virologia ISP Chile | Instituto de Salud Publica de Chile | Javier Tognarelli et al |
| EPI_ISL_746537 | Chile/NB-175522/2020 | 2020-09-11 | South America/Chile/Vëuble/Chillan | Genetica Molecular and Subdepartamento de Virologia ISP Chile | Instituto de Salud Publica de Chile | Javier Tognarelli et al |
| EPI_ISL_746545 | Chile/RM-108589/2020 | 2020-06-23 | South America/Chile/Region Metropolit | Genetica Molecular and Subdepartamento de Virologia ISP Chile | Instituto de Salud Publica de Chile | Javier Tognarelli et al |
| EPI_ISL_746546 | Chile/RM-108734/2020 | 2020-06-23 | South America/Chile/Region Metropolit | Genetica Molecular and Subdepartamento de Virologia ISP Chile | Instituto de Salud Publica de Chile | Javier Tognarelli et al |
| EPI_ISL_746547 | Chile/RM-108736/2020 | 2020-06-23 | South America/Chile/Region Metropolit | Genetica Molecular and Subdepartamento de Virologia ISP Chile | Instituto de Salud Publica de Chile | Javier Tognarelli et al |
| EPI_ISL_746548 | Chile/RM-108996/2020 | 2020-06-23 | South America/Chile/Region Metropolit | Genetica Molecular and Subdepartamento de Virologia ISP Chile | Instituto de Salud Publica de Chile | Javier Tognarelli et al |
| EPI_ISL_746549 | Chile/RM-108997/2020 | 2020-06-23 | South America/Chile/Region Metropolit | Genetica Molecular and Subdepartamento de Virologia ISP Chile | Instituto de Salud Publica de Chile | Javier Tognarelli et al |
| EPI_ISL_746550 | Chile/RM-108999/2020 | 2020-06-23 | South America/Chile/Region Metropolit | Genetica Molecular and Subdepartamento de Virologia ISP Chile | Instituto de Salud Publica de Chile | Javier Tognarelli et al |

| | | | | | | |
|---|---|---|---|---|---|---|
| EPI_ISL_746551 | Chile/RM-109139/2020 | 2020-06-24 | South America/Chile/Region Metropolit | Genetica Molecular and Subdepartamento de Virologia ISP Chile | Instituto de Salud Publica de Chile | Javier Tognarelli et al |
| EPI_ISL_746552 | Chile/RM-109226/2020 | 2020-06-23 | South America/Chile/Region Metropolit | Genetica Molecular and Subdepartamento de Virologia ISP Chile | Instituto de Salud Publica de Chile | Javier Tognarelli et al |
| EPI_ISL_746553 | Chile/RM-109231/2020 | 2020-06-23 | South America/Chile/Region Metropolit | Genetica Molecular and Subdepartamento de Virologia ISP Chile | Instituto de Salud Publica de Chile | Javier Tognarelli et al |
| EPI_ISL_746554 | Chile/RM-109455/2020 | 2020-06-23 | South America/Chile/Region Metropolit | Genetica Molecular and Subdepartamento de Virologia ISP Chile | Instituto de Salud Publica de Chile | Javier Tognarelli et al |
| EPI_ISL_746555 | Chile/RM-109521/2020 | 2020-06-24 | South America/Chile/Region Metropolit | Genetica Molecular and Subdepartamento de Virologia ISP Chile | Instituto de Salud Publica de Chile | Javier Tognarelli et al |
| EPI_ISL_746556 | Chile/RM-109567/2020 | 2020-06-24 | South America/Chile/Region Metropolit | Genetica Molecular and Subdepartamento de Virologia ISP Chile | Instituto de Salud Publica de Chile | Javier Tognarelli et al |
| EPI_ISL_746557 | Chile/RM-109578/2020 | 2020-06-24 | South America/Chile/Region Metropolit | Genetica Molecular and Subdepartamento de Virologia ISP Chile | Instituto de Salud Publica de Chile | Javier Tognarelli et al |
| EPI_ISL_746558 | Chile/RM-109586/2020 | 2020-06-24 | South America/Chile/Region Metropolit | Genetica Molecular and Subdepartamento de Virologia ISP Chile | Instituto de Salud Publica de Chile | Javier Tognarelli et al |
| EPI_ISL_746559 | Chile/RM-109587/2020 | 2020-06-24 | South America/Chile/Region Metropolit | Genetica Molecular and Subdepartamento de Virologia ISP Chile | Instituto de Salud Publica de Chile | Javier Tognarelli et al |
| EPI_ISL_746560 | Chile/RM-109600/2020 | 2020-06-24 | South America/Chile/Region Metropolit | Genetica Molecular and Subdepartamento de Virologia ISP Chile | Instituto de Salud Publica de Chile | Javier Tognarelli et al |
| EPI_ISL_746561 | Chile/RM-109602/2020 | 2020-06-24 | South America/Chile/Region Metropolit | Genetica Molecular and Subdepartamento de Virologia ISP Chile | Instituto de Salud Publica de Chile | Javier Tognarelli et al |
| EPI_ISL_746562 | Chile/RM-110507/2020 | 2020-06-25 | South America/Chile/Region Metropolit | Genetica Molecular and Subdepartamento de Virologia ISP Chile | Instituto de Salud Publica de Chile | Javier Tognarelli et al |
| EPI_ISL_746563 | Chile/RM-110508/2020 | 2020-06-25 | South America/Chile/Region Metropolit | Genetica Molecular and Subdepartamento de Virologia ISP Chile | Instituto de Salud Publica de Chile | Javier Tognarelli et al |
| EPI_ISL_746564 | Chile/RM-110525/2020 | 2020-06-25 | South America/Chile/Region Metropolit | Genetica Molecular and Subdepartamento de Virologia ISP Chile | Instituto de Salud Publica de Chile | Javier Tognarelli et al |
| EPI_ISL_746565 | Chile/RM-110750/2020 | 2020-06-25 | South America/Chile/Region Metropolit | Genetica Molecular and Subdepartamento de Virologia ISP Chile | Instituto de Salud Publica de Chile | Javier Tognarelli et al |
| EPI_ISL_746566 | Chile/RM-110802/2020 | 2020-06-25 | South America/Chile/Region Metropolit | Genetica Molecular and Subdepartamento de Virologia ISP Chile | Instituto de Salud Publica de Chile | Javier Tognarelli et al |
| EPI_ISL_746567 | Chile/RM-110804/2020 | 2020-06-25 | South America/Chile/Region Metropolit | Genetica Molecular and Subdepartamento de Virologia ISP Chile | Instituto de Salud Publica de Chile | Javier Tognarelli et al |
| EPI_ISL_746568 | Chile/RM-110815/2020 | 2020-06-25 | South America/Chile/Region Metropolit | Genetica Molecular and Subdepartamento de Virologia ISP Chile | Instituto de Salud Publica de Chile | Javier Tognarelli et al |
| EPI_ISL_746569 | Chile/RM-110874/2020 | 2020-06-26 | South America/Chile/Region Metropolit | Genetica Molecular and Subdepartamento de Virologia ISP Chile | Instituto de Salud Publica de Chile | Javier Tognarelli et al |
| EPI_ISL_746570 | Chile/RM-110881/2020 | 2020-06-26 | South America/Chile/Region Metropolit | Genetica Molecular and Subdepartamento de Virologia ISP Chile | Instituto de Salud Publica de Chile | Javier Tognarelli et al |

| | | | | | | |
|---|---|---|---|---|---|---|
| EPI_ISL_746571 | Chile/RM-110884/2020 | 2020-06-26 | South America/Chile/Region Metropolit | Genetica Molecular and Subdepartamento de Virologia ISP Chile | Instituto de Salud Publica de Chile | Javier Tognarelli et al |
| EPI_ISL_746572 | Chile/RM-110889/2020 | 2020-06-26 | South America/Chile/Region Metropolit | Genetica Molecular and Subdepartamento de Virologia ISP Chile | Instituto de Salud Publica de Chile | Javier Tognarelli et al |
| EPI_ISL_746573 | Chile/RM-110918/2020 | 2020-06-26 | South America/Chile/Region Metropolit | Genetica Molecular and Subdepartamento de Virologia ISP Chile | Instituto de Salud Publica de Chile | Javier Tognarelli et al |
| EPI_ISL_746574 | Chile/RM-110919/2020 | 2020-06-26 | South America/Chile/Region Metropolit | Genetica Molecular and Subdepartamento de Virologia ISP Chile | Instituto de Salud Publica de Chile | Javier Tognarelli et al |
| EPI_ISL_746575 | Chile/RM-110962/2020 | 2020-06-25 | South America/Chile/Region Metropolit | Genetica Molecular and Subdepartamento de Virologia ISP Chile | Instituto de Salud Publica de Chile | Javier Tognarelli et al |
| EPI_ISL_746576 | Chile/RM-110966/2020 | 2020-06-25 | South America/Chile/Region Metropolit | Genetica Molecular and Subdepartamento de Virologia ISP Chile | Instituto de Salud Publica de Chile | Javier Tognarelli et al |
| EPI_ISL_746578 | Chile/RM-110981/2020 | 2020-06-25 | South America/Chile/Region Metropolit | Genetica Molecular and Subdepartamento de Virologia ISP Chile | Instituto de Salud Publica de Chile | Javier Tognarelli et al |
| EPI_ISL_746579 | Chile/RM-110983/2020 | 2020-06-25 | South America/Chile/Region Metropolit | Genetica Molecular and Subdepartamento de Virologia ISP Chile | Instituto de Salud Publica de Chile | Javier Tognarelli et al |
| EPI_ISL_746580 | Chile/RM-110987/2020 | 2020-06-26 | South America/Chile/Region Metropolit | Genetica Molecular and Subdepartamento de Virologia ISP Chile | Instituto de Salud Publica de Chile | Javier Tognarelli et al |
| EPI_ISL_746581 | Chile/RM-111206/2020 | 2020-06-26 | South America/Chile/Region Metropolit | Genetica Molecular and Subdepartamento de Virologia ISP Chile | Instituto de Salud Publica de Chile | Javier Tognarelli et al |
| EPI_ISL_746582 | Chile/RM-111211/2020 | 2020-06-26 | South America/Chile/Region Metropolit | Genetica Molecular and Subdepartamento de Virologia ISP Chile | Instituto de Salud Publica de Chile | Javier Tognarelli et al |
| EPI_ISL_746583 | Chile/RM-111216/2020 | 2020-06-25 | South America/Chile/Region Metropolit | Genetica Molecular and Subdepartamento de Virologia ISP Chile | Instituto de Salud Publica de Chile | Javier Tognarelli et al |
| EPI_ISL_746584 | Chile/RM-112466/2020 | 2020-06-28 | South America/Chile/Region Metropolit | Genetica Molecular and Subdepartamento de Virologia ISP Chile | Instituto de Salud Publica de Chile | Javier Tognarelli et al |
| EPI_ISL_746585 | Chile/RM-112467/2020 | 2020-06-28 | South America/Chile/Region Metropolit | Genetica Molecular and Subdepartamento de Virologia ISP Chile | Instituto de Salud Publica de Chile | Javier Tognarelli et al |
| EPI_ISL_746586 | Chile/RM-112634/2020 | 2020-06-28 | South America/Chile/Region Metropolit | Genetica Molecular and Subdepartamento de Virologia ISP Chile | Instituto de Salud Publica de Chile | Javier Tognarelli et al |
| EPI_ISL_746587 | Chile/RM-112635/2020 | 2020-06-28 | South America/Chile/Region Metropolit | Genetica Molecular and Subdepartamento de Virologia ISP Chile | Instituto de Salud Publica de Chile | Javier Tognarelli et al |
| EPI_ISL_746588 | Chile/RM-112637/2020 | 2020-06-28 | South America/Chile/Region Metropolit | Genetica Molecular and Subdepartamento de Virologia ISP Chile | Instituto de Salud Publica de Chile | Javier Tognarelli et al |
| EPI_ISL_746589 | Chile/RM-112760/2020 | 2020-06-28 | South America/Chile/Region Metropolit | Genetica Molecular and Subdepartamento de Virologia ISP Chile | Instituto de Salud Publica de Chile | Javier Tognarelli et al |
| EPI_ISL_746590 | Chile/RM-112763/2020 | 2020-06-28 | South America/Chile/Region Metropolit | Genetica Molecular and Subdepartamento de Virologia ISP Chile | Instituto de Salud Publica de Chile | Javier Tognarelli et al |
| EPI_ISL_746591 | Chile/RM-112769/2020 | 2020-06-28 | South America/Chile/Region Metropolit | Genetica Molecular and Subdepartamento de Virologia ISP Chile | Instituto de Salud Publica de Chile | Javier Tognarelli et al |

| | | | | | | |
|---|---|---|---|---|---|---|
| EPI_ISL_746592 | Chile/RM-112773/2020 | 2020-06-28 | South America/Chile/Region Metropolit | Genetica Molecular and Subdepartamento de Virologia ISP Chile | Instituto de Salud Publica de Chile | Javier Tognarelli et al |
| EPI_ISL_746593 | Chile/RM-112777/2020 | 2020-06-28 | South America/Chile/Region Metropolit | Genetica Molecular and Subdepartamento de Virologia ISP Chile | Instituto de Salud Publica de Chile | Javier Tognarelli et al |
| EPI_ISL_746595 | Chile/RM-126534/2020 | 2020-07-16 | South America/Chile/Region Metropolit | Genetica Molecular and Subdepartamento de Virologia ISP Chile | Instituto de Salud Publica de Chile | Javier Tognarelli et al |
| EPI_ISL_746596 | Chile/RM-126787/2020 | 2020-07-17 | South America/Chile/Region Metropolit | Genetica Molecular and Subdepartamento de Virologia ISP Chile | Instituto de Salud Publica de Chile | Javier Tognarelli et al |
| EPI_ISL_746597 | Chile/RM-127204/2020 | 2020-07-17 | South America/Chile/Region Metropolit | Genetica Molecular and Subdepartamento de Virologia ISP Chile | Instituto de Salud Publica de Chile | Javier Tognarelli et al |
| EPI_ISL_746601 | Chile/RM-127562/2020 | 2020-07-17 | South America/Chile/Region Metropolit | Genetica Molecular and Subdepartamento de Virologia ISP Chile | Instituto de Salud Publica de Chile | Javier Tognarelli et al |
| EPI_ISL_746603 | Chile/RM-127568/2020 | 2020-07-18 | South America/Chile/Region Metropolit | Genetica Molecular and Subdepartamento de Virologia ISP Chile | Instituto de Salud Publica de Chile | Javier Tognarelli et al |
| EPI_ISL_746605 | Chile/RM-127602/2020 | 2020-07-17 | South America/Chile/Region Metropolit | Genetica Molecular and Subdepartamento de Virologia ISP Chile | Instituto de Salud Publica de Chile | Javier Tognarelli et al |
| EPI_ISL_746606 | Chile/RM-127633/2020 | 2020-07-18 | South America/Chile/Region Metropolit | Genetica Molecular and Subdepartamento de Virologia ISP Chile | Instituto de Salud Publica de Chile | Javier Tognarelli et al |
| EPI_ISL_746607 | Chile/RM-127636/2020 | 2020-07-17 | South America/Chile/Region Metropolit | Genetica Molecular and Subdepartamento de Virologia ISP Chile | Instituto de Salud Publica de Chile | Javier Tognarelli et al |
| EPI_ISL_746608 | Chile/RM-127644/2020 | 2020-07-17 | South America/Chile/Region Metropolit | Genetica Molecular and Subdepartamento de Virologia ISP Chile | Instituto de Salud Publica de Chile | Javier Tognarelli et al |
| EPI_ISL_746609 | Chile/RM-127666/2020 | 2020-07-18 | South America/Chile/Region Metropolit | Genetica Molecular and Subdepartamento de Virologia ISP Chile | Instituto de Salud Publica de Chile | Javier Tognarelli et al |
| EPI_ISL_746610 | Chile/RM-127822/2020 | 2020-07-18 | South America/Chile/Region Metropolit | Genetica Molecular and Subdepartamento de Virologia ISP Chile | Instituto de Salud Publica de Chile | Javier Tognarelli et al |
| EPI_ISL_746612 | Chile/RM-127976/2020 | 2020-07-18 | South America/Chile/Region Metropolit | Genetica Molecular and Subdepartamento de Virologia ISP Chile | Instituto de Salud Publica de Chile | Javier Tognarelli et al |
| EPI_ISL_746613 | Chile/RM-128039/2020 | 2020-07-18 | South America/Chile/Region Metropolit | Genetica Molecular and Subdepartamento de Virologia ISP Chile | Instituto de Salud Publica de Chile | Javier Tognarelli et al |
| EPI_ISL_746615 | Chile/RM-128084/2020 | 2020-07-18 | South America/Chile/Region Metropolit | Genetica Molecular and Subdepartamento de Virologia ISP Chile | Instituto de Salud Publica de Chile | Javier Tognarelli et al |
| EPI_ISL_746616 | Chile/RM-128176/2020 | 2020-07-18 | South America/Chile/Region Metropolit | Genetica Molecular and Subdepartamento de Virologia ISP Chile | Instituto de Salud Publica de Chile | Javier Tognarelli et al |
| EPI_ISL_746617 | Chile/RM-128225/2020 | 2020-07-18 | South America/Chile/Region Metropolit | Genetica Molecular and Subdepartamento de Virologia ISP Chile | Instituto de Salud Publica de Chile | Javier Tognarelli et al |
| EPI_ISL_746618 | Chile/RM-128269/2020 | 2020-07-19 | South America/Chile/Region Metropolit | Genetica Molecular and Subdepartamento de Virologia ISP Chile | Instituto de Salud Publica de Chile | Javier Tognarelli et al |
| EPI_ISL_746619 | Chile/RM-128531/2020 | 2020-07-19 | South America/Chile/Region Metropolit | Genetica Molecular and Subdepartamento de Virologia ISP Chile | Instituto de Salud Publica de Chile | Javier Tognarelli et al |

| | | | | | | |
|---|---|---|---|---|---|---|
| EPI_ISL_746620 | Chile/RM-130072/2020 | 2020-07-21 | South America/Chile/Region Metropolit | Genetica Molecular and Subdepartamento de Virologia ISP Chile | Instituto de Salud Publica de Chile | Javier Tognarelli et al |
| EPI_ISL_746621 | Chile/RM-130073/2020 | 2020-07-21 | South America/Chile/Region Metropolit | Genetica Molecular and Subdepartamento de Virologia ISP Chile | Instituto de Salud Publica de Chile | Javier Tognarelli et al |
| EPI_ISL_746622 | Chile/RM-130081/2020 | 2020-07-21 | South America/Chile/Region Metropolit | Genetica Molecular and Subdepartamento de Virologia ISP Chile | Instituto de Salud Publica de Chile | Javier Tognarelli et al |
| EPI_ISL_746623 | Chile/RM-130924/2020 | 2020-07-22 | South America/Chile/Region Metropolit | Genetica Molecular and Subdepartamento de Virologia ISP Chile | Instituto de Salud Publica de Chile | Javier Tognarelli et al |
| EPI_ISL_746625 | Chile/RM-131381/2020 | 2020-07-23 | South America/Chile/Region Metropolit | Genetica Molecular and Subdepartamento de Virologia ISP Chile | Instituto de Salud Publica de Chile | Javier Tognarelli et al |
| EPI_ISL_746626 | Chile/RM-131430/2020 | 2020-07-23 | South America/Chile/Region Metropolit | Genetica Molecular and Subdepartamento de Virologia ISP Chile | Instituto de Salud Publica de Chile | Javier Tognarelli et al |
| EPI_ISL_746628 | Chile/RM-132398/2020 | 2020-07-24 | South America/Chile/Region Metropolit | Genetica Molecular and Subdepartamento de Virologia ISP Chile | Instituto de Salud Publica de Chile | Javier Tognarelli et al |
| EPI_ISL_746629 | Chile/RM-133091/2020 | 2020-07-24 | South America/Chile/Region Metropolit | Genetica Molecular and Subdepartamento de Virologia ISP Chile | Instituto de Salud Publica de Chile | Javier Tognarelli et al |
| EPI_ISL_746632 | Chile/RM-133191/2020 | 2020-07-25 | South America/Chile/Region Metropolit | Genetica Molecular and Subdepartamento de Virologia ISP Chile | Instituto de Salud Publica de Chile | Javier Tognarelli et al |
| EPI_ISL_746634 | Chile/RM-133318/2020 | 2020-07-25 | South America/Chile/Region Metropolit | Genetica Molecular and Subdepartamento de Virologia ISP Chile | Instituto de Salud Publica de Chile | Javier Tognarelli et al |
| EPI_ISL_746635 | Chile/RM-133547/2020 | 2020-07-25 | South America/Chile/Region Metropolit | Genetica Molecular and Subdepartamento de Virologia ISP Chile | Instituto de Salud Publica de Chile | Javier Tognarelli et al |
| EPI_ISL_746636 | Chile/RM-133558/2020 | 2020-07-26 | South America/Chile/Region Metropolit | Genetica Molecular and Subdepartamento de Virologia ISP Chile | Instituto de Salud Publica de Chile | Javier Tognarelli et al |
| EPI_ISL_746637 | Chile/RM-133562/2020 | 2020-07-26 | South America/Chile/Region Metropolit | Genetica Molecular and Subdepartamento de Virologia ISP Chile | Instituto de Salud Publica de Chile | Javier Tognarelli et al |
| EPI_ISL_746638 | Chile/RM-133625/2020 | 2020-07-25 | South America/Chile/Region Metropolit | Genetica Molecular and Subdepartamento de Virologia ISP Chile | Instituto de Salud Publica de Chile | Javier Tognarelli et al |
| EPI_ISL_746640 | Chile/RM-133746/2020 | 2020-07-26 | South America/Chile/Region Metropolit | Genetica Molecular and Subdepartamento de Virologia ISP Chile | Instituto de Salud Publica de Chile | Javier Tognarelli et al |
| EPI_ISL_746643 | Chile/RM-134067/2020 | 2020-07-27 | South America/Chile/Region Metropolit | Genetica Molecular and Subdepartamento de Virologia ISP Chile | Instituto de Salud Publica de Chile | Javier Tognarelli et al |
| EPI_ISL_746644 | Chile/RM-145979/2020 | 2020-08-09 | South America/Chile/Region Metropolit | Genetica Molecular and Subdepartamento de Virologia ISP Chile | Instituto de Salud Publica de Chile | Javier Tognarelli et al |
| EPI_ISL_746645 | Chile/RM-146431/2020 | 2020-08-10 | South America/Chile/Region Metropolit | Genetica Molecular and Subdepartamento de Virologia ISP Chile | Instituto de Salud Publica de Chile | Javier Tognarelli et al |
| EPI_ISL_746647 | Chile/RM-150435/2020 | 2020-08-13 | South America/Chile/Region Metropolit | Genetica Molecular and Subdepartamento de Virologia ISP Chile | Instituto de Salud Publica de Chile | Javier Tognarelli et al |
| EPI_ISL_746650 | Chile/RM-153333/2020 | 2020-08-17 | South America/Chile/Region Metropolit | Genetica Molecular and Subdepartamento de Virologia ISP Chile | Instituto de Salud Publica de Chile | Javier Tognarelli et al |

| | | | | | | |
|---|---|---|---|---|---|---|
| EPI_ISL_746651 | Chile/RM-153360/2020 | 2020-08-17 | South America/Chile/Region Metropolit | Genetica Molecular and Subdepartamento de Virologia ISP Chile | Instituto de Salud Publica de Chile | Javier Tognarelli et al |
| EPI_ISL_746652 | Chile/RM-153390/2020 | 2020-08-17 | South America/Chile/Region Metropolit | Genetica Molecular and Subdepartamento de Virologia ISP Chile | Instituto de Salud Publica de Chile | Javier Tognarelli et al |
| EPI_ISL_746653 | Chile/RM-153914/2020 | 2020-08-18 | South America/Chile/Region Metropolit | Genetica Molecular and Subdepartamento de Virologia ISP Chile | Instituto de Salud Publica de Chile | Javier Tognarelli et al |
| EPI_ISL_746654 | Chile/RM-153915/2020 | 2020-08-18 | South America/Chile/Region Metropolit | Genetica Molecular and Subdepartamento de Virologia ISP Chile | Instituto de Salud Publica de Chile | Javier Tognarelli et al |
| EPI_ISL_746655 | Chile/RM-154196/2020 | 2020-08-18 | South America/Chile/Region Metropolit | Genetica Molecular and Subdepartamento de Virologia ISP Chile | Instituto de Salud Publica de Chile | Javier Tognarelli et al |
| EPI_ISL_746656 | Chile/RM-154362/2020 | 2020-08-18 | South America/Chile/Region Metropolit | Genetica Molecular and Subdepartamento de Virologia ISP Chile | Instituto de Salud Publica de Chile | Javier Tognarelli et al |
| EPI_ISL_746657 | Chile/RM-154655/2020 | 2020-08-18 | South America/Chile/Region Metropolit | Genetica Molecular and Subdepartamento de Virologia ISP Chile | Instituto de Salud Publica de Chile | Javier Tognarelli et al |
| EPI_ISL_746659 | Chile/RM-155380/2020 | 2020-08-19 | South America/Chile/Region Metropolit | Genetica Molecular and Subdepartamento de Virologia ISP Chile | Instituto de Salud Publica de Chile | Javier Tognarelli et al |
| EPI_ISL_746660 | Chile/RM-155487/2020 | 2020-08-18 | South America/Chile/Region Metropolit | Genetica Molecular and Subdepartamento de Virologia ISP Chile | Instituto de Salud Publica de Chile | Javier Tognarelli et al |
| EPI_ISL_746661 | Chile/RM-155633/2020 | 2020-08-19 | South America/Chile/Region Metropolit | Genetica Molecular and Subdepartamento de Virologia ISP Chile | Instituto de Salud Publica de Chile | Javier Tognarelli et al |
| EPI_ISL_746662 | Chile/RM-155699/2020 | 2020-08-19 | South America/Chile/Region Metropolit | Genetica Molecular and Subdepartamento de Virologia ISP Chile | Instituto de Salud Publica de Chile | Javier Tognarelli et al |
| EPI_ISL_746663 | Chile/RM-155723/2020 | 2020-08-19 | South America/Chile/Region Metropolit | Genetica Molecular and Subdepartamento de Virologia ISP Chile | Instituto de Salud Publica de Chile | Javier Tognarelli et al |
| EPI_ISL_746665 | Chile/RM-155990/2020 | 2020-08-19 | South America/Chile/Region Metropolit | Genetica Molecular and Subdepartamento de Virologia ISP Chile | Instituto de Salud Publica de Chile | Javier Tognarelli et al |
| EPI_ISL_746666 | Chile/RM-156030/2020 | 2020-08-20 | South America/Chile/Region Metropolit | Genetica Molecular and Subdepartamento de Virologia ISP Chile | Instituto de Salud Publica de Chile | Javier Tognarelli et al |
| EPI_ISL_746667 | Chile/RM-156453/2020 | 2020-08-20 | South America/Chile/Region Metropolit | Genetica Molecular and Subdepartamento de Virologia ISP Chile | Instituto de Salud Publica de Chile | Javier Tognarelli et al |
| EPI_ISL_746669 | Chile/RM-156709/2020 | 2020-08-21 | South America/Chile/Region Metropolit | Genetica Molecular and Subdepartamento de Virologia ISP Chile | Instituto de Salud Publica de Chile | Javier Tognarelli et al |
| EPI_ISL_746673 | Chile/RM-157307/2020 | 2020-08-21 | South America/Chile/Region Metropolit | Genetica Molecular and Subdepartamento de Virologia ISP Chile | Instituto de Salud Publica de Chile | Javier Tognarelli et al |
| EPI_ISL_746674 | Chile/RM-157308/2020 | 2020-08-20 | South America/Chile/Region Metropolit | Genetica Molecular and Subdepartamento de Virologia ISP Chile | Instituto de Salud Publica de Chile | Javier Tognarelli et al |
| EPI_ISL_746676 | Chile/RM-157618/2020 | 2020-08-21 | South America/Chile/Region Metropolit | Genetica Molecular and Subdepartamento de Virologia ISP Chile | Instituto de Salud Publica de Chile | Javier Tognarelli et al |
| EPI_ISL_746677 | Chile/RM-157632/2020 | 2020-08-21 | South America/Chile/Region Metropolit | Genetica Molecular and Subdepartamento de Virologia ISP Chile | Instituto de Salud Publica de Chile | Javier Tognarelli et al |

| | | | | | | |
|---|---|---|---|---|---|---|
| EPI_ISL_746678 | Chile/RM-157643/2020 | 2020-08-21 | South America/Chile/Region Metropolit | Genetica Molecular and Subdepartamento de Virologia ISP Chile | Instituto de Salud Publica de Chile | Javier Tognarelli et al |
| EPI_ISL_746679 | Chile/RM-157658/2020 | 2020-08-21 | South America/Chile/Region Metropolit | Genetica Molecular and Subdepartamento de Virologia ISP Chile | Instituto de Salud Publica de Chile | Javier Tognarelli et al |
| EPI_ISL_746680 | Chile/RM-157770/2020 | 2020-08-21 | South America/Chile/Region Metropolit | Genetica Molecular and Subdepartamento de Virologia ISP Chile | Instituto de Salud Publica de Chile | Javier Tognarelli et al |
| EPI_ISL_746682 | Chile/RM-157801/2020 | 2020-08-22 | South America/Chile/Region Metropolit | Genetica Molecular and Subdepartamento de Virologia ISP Chile | Instituto de Salud Publica de Chile | Javier Tognarelli et al |
| EPI_ISL_746683 | Chile/RM-157806/2020 | 2020-08-22 | South America/Chile/Region Metropolit | Genetica Molecular and Subdepartamento de Virologia ISP Chile | Instituto de Salud Publica de Chile | Javier Tognarelli et al |
| EPI_ISL_746685 | Chile/RM-158079/2020 | 2020-08-21 | South America/Chile/Region Metropolit | Genetica Molecular and Subdepartamento de Virologia ISP Chile | Instituto de Salud Publica de Chile | Javier Tognarelli et al |
| EPI_ISL_746686 | Chile/RM-158209/2020 | 2020-08-22 | South America/Chile/Region Metropolit | Genetica Molecular and Subdepartamento de Virologia ISP Chile | Instituto de Salud Publica de Chile | Javier Tognarelli et al |
| EPI_ISL_746687 | Chile/RM-158239/2020 | 2020-08-22 | South America/Chile/Region Metropolit | Genetica Molecular and Subdepartamento de Virologia ISP Chile | Instituto de Salud Publica de Chile | Javier Tognarelli et al |
| EPI_ISL_746689 | Chile/RM-163995/2020 | 2020-08-29 | South America/Chile/Region Metropolit | Genetica Molecular and Subdepartamento de Virologia ISP Chile | Instituto de Salud Publica de Chile | Javier Tognarelli et al |
| EPI_ISL_746690 | Chile/RM-164001/2020 | 2020-08-29 | South America/Chile/Region Metropolit | Genetica Molecular and Subdepartamento de Virologia ISP Chile | Instituto de Salud Publica de Chile | Javier Tognarelli et al |
| EPI_ISL_746691 | Chile/RM-164078/2020 | 2020-08-29 | South America/Chile/Region Metropolit | Genetica Molecular and Subdepartamento de Virologia ISP Chile | Instituto de Salud Publica de Chile | Javier Tognarelli et al |
| EPI_ISL_746692 | Chile/RM-164508/2020 | 2020-08-30 | South America/Chile/Region Metropolit | Genetica Molecular and Subdepartamento de Virologia ISP Chile | Instituto de Salud Publica de Chile | Javier Tognarelli et al |
| EPI_ISL_746694 | Chile/RM-164804/2020 | 2020-08-31 | South America/Chile/Region Metropolit | Genetica Molecular and Subdepartamento de Virologia ISP Chile | Instituto de Salud Publica de Chile | Javier Tognarelli et al |
| EPI_ISL_746697 | Chile/RM-165843/2020 | 2020-08-31 | South America/Chile/Region Metropolit | Genetica Molecular and Subdepartamento de Virologia ISP Chile | Instituto de Salud Publica de Chile | Javier Tognarelli et al |
| EPI_ISL_746698 | Chile/RM-166412/2020 | 2020-09-01 | South America/Chile/Region Metropolit | Genetica Molecular and Subdepartamento de Virologia ISP Chile | Instituto de Salud Publica de Chile | Javier Tognarelli et al |
| EPI_ISL_746701 | Chile/RM-167590/2020 | 2020-09-01 | South America/Chile/Region Metropolit | Genetica Molecular and Subdepartamento de Virologia ISP Chile | Instituto de Salud Publica de Chile | Javier Tognarelli et al |
| EPI_ISL_746702 | Chile/RM-168269/2020 | 2020-09-03 | South America/Chile/Region Metropolit | Genetica Molecular and Subdepartamento de Virologia ISP Chile | Instituto de Salud Publica de Chile | Javier Tognarelli et al |
| EPI_ISL_746706 | Chile/RM-169155/2020 | 2020-09-03 | South America/Chile/Region Metropolit | Genetica Molecular and Subdepartamento de Virologia ISP Chile | Instituto de Salud Publica de Chile | Javier Tognarelli et al |
| EPI_ISL_746709 | Chile/RM-170043/2020 | 2020-09-04 | South America/Chile/Region Metropolit | Genetica Molecular and Subdepartamento de Virologia ISP Chile | Instituto de Salud Publica de Chile | Javier Tognarelli et al |
| EPI_ISL_746711 | Chile/RM-170137/2020 | 2020-09-04 | South America/Chile/Region Metropolit | Genetica Molecular and Subdepartamento de Virologia ISP Chile | Instituto de Salud Publica de Chile | Javier Tognarelli et al |

| | | | | | | |
|---|---|---|---|---|---|---|
| EPI_ISL_746712 | Chile/RM-170148/2020 | 2020-09-04 | South America/Chile/Region Metropolit | Genetica Molecular and Subdepartamento de Virologia ISP Chile | Instituto de Salud Publica de Chile | Javier Tognarelli et al |
| EPI_ISL_746716 | Chile/RM-170368/2020 | 2020-09-05 | South America/Chile/Region Metropolit | Genetica Molecular and Subdepartamento de Virologia ISP Chile | Instituto de Salud Publica de Chile | Javier Tognarelli et al |
| EPI_ISL_746717 | Chile/RM-170370/2020 | 2020-09-05 | South America/Chile/Region Metropolit | Genetica Molecular and Subdepartamento de Virologia ISP Chile | Instituto de Salud Publica de Chile | Javier Tognarelli et al |
| EPI_ISL_746718 | Chile/RM-170468/2020 | 2020-09-04 | South America/Chile/Region Metropolit | Genetica Molecular and Subdepartamento de Virologia ISP Chile | Instituto de Salud Publica de Chile | Javier Tognarelli et al |
| EPI_ISL_746720 | Chile/RM-170589/2020 | 2020-09-05 | South America/Chile/Region Metropolit | Genetica Molecular and Subdepartamento de Virologia ISP Chile | Instituto de Salud Publica de Chile | Javier Tognarelli et al |
| EPI_ISL_746721 | Chile/RM-170707/2020 | 2020-09-05 | South America/Chile/Region Metropolit | Genetica Molecular and Subdepartamento de Virologia ISP Chile | Instituto de Salud Publica de Chile | Javier Tognarelli et al |
| EPI_ISL_746722 | Chile/RM-170921/2020 | 2020-09-06 | South America/Chile/Region Metropolit | Genetica Molecular and Subdepartamento de Virologia ISP Chile | Instituto de Salud Publica de Chile | Javier Tognarelli et al |
| EPI_ISL_746724 | Chile/RM-170990/2020 | 2020-09-07 | South America/Chile/Region Metropolit | Genetica Molecular and Subdepartamento de Virologia ISP Chile | Instituto de Salud Publica de Chile | Javier Tognarelli et al |
| EPI_ISL_746725 | Chile/RM-171011/2020 | 2020-09-06 | South America/Chile/Region Metropolit | Genetica Molecular and Subdepartamento de Virologia ISP Chile | Instituto de Salud Publica de Chile | Javier Tognarelli et al |
| EPI_ISL_746726 | Chile/RM-171015/2020 | 2020-09-07 | South America/Chile/Region Metropolit | Genetica Molecular and Subdepartamento de Virologia ISP Chile | Instituto de Salud Publica de Chile | Javier Tognarelli et al |
| EPI_ISL_746731 | Chile/RM-183871/2020 | 2020-09-21 | South America/Chile/Region Metropolit | Genetica Molecular and Subdepartamento de Virologia ISP Chile | Instituto de Salud Publica de Chile | Javier Tognarelli et al |
| EPI_ISL_746732 | Chile/RM-184415/2020 | 2020-09-21 | South America/Chile/Region Metropolit | Genetica Molecular and Subdepartamento de Virologia ISP Chile | Instituto de Salud Publica de Chile | Javier Tognarelli et al |
| EPI_ISL_746733 | Chile/RM-185039/2020 | 2020-09-21 | South America/Chile/Region Metropolit | Genetica Molecular and Subdepartamento de Virologia ISP Chile | Instituto de Salud Publica de Chile | Javier Tognarelli et al |
| EPI_ISL_746734 | Chile/RM-185819/2020 | 2020-09-22 | South America/Chile/Region Metropolit | Genetica Molecular and Subdepartamento de Virologia ISP Chile | Instituto de Salud Publica de Chile | Javier Tognarelli et al |
| EPI_ISL_746735 | Chile/RM-185867/2020 | 2020-09-22 | South America/Chile/Region Metropolit | Genetica Molecular and Subdepartamento de Virologia ISP Chile | Instituto de Salud Publica de Chile | Javier Tognarelli et al |
| EPI_ISL_746736 | Chile/RM-187626/2020 | 2020-09-23 | South America/Chile/Region Metropolit | Genetica Molecular and Subdepartamento de Virologia ISP Chile | Instituto de Salud Publica de Chile | Javier Tognarelli et al |
| EPI_ISL_746737 | Chile/RM-187968/2020 | 2020-09-23 | South America/Chile/Region Metropolit | Genetica Molecular and Subdepartamento de Virologia ISP Chile | Instituto de Salud Publica de Chile | Javier Tognarelli et al |
| EPI_ISL_746739 | Chile/RM-188963/2020 | 2020-09-24 | South America/Chile/Region Metropolit | Genetica Molecular and Subdepartamento de Virologia ISP Chile | Instituto de Salud Publica de Chile | Javier Tognarelli et al |
| EPI_ISL_746740 | Chile/RM-189170/2020 | 2020-09-24 | South America/Chile/Region Metropolit | Genetica Molecular and Subdepartamento de Virologia ISP Chile | Instituto de Salud Publica de Chile | Javier Tognarelli et al |
| EPI_ISL_746742 | Chile/RM-190290/2020 | 2020-09-25 | South America/Chile/Region Metropolit | Genetica Molecular and Subdepartamento de Virologia ISP Chile | Instituto de Salud Publica de Chile | Javier Tognarelli et al |

| | | | | | | |
|---|---|---|---|---|---|---|
| EPI_ISL_746743 | Chile/RM-190581/2020 | 2020-09-25 | South America/Chile/Region Metropolit | Genetica Molecular and Subdepartamento de Virologia ISP Chile | Instituto de Salud Publica de Chile | Javier Tognarelli et al |
| EPI_ISL_746744 | Chile/RM-190882/2020 | 2020-09-25 | South America/Chile/Region Metropolit | Genetica Molecular and Subdepartamento de Virologia ISP Chile | Instituto de Salud Publica de Chile | Javier Tognarelli et al |
| EPI_ISL_746745 | Chile/RM-191185/2020 | 2020-09-26 | South America/Chile/Region Metropolit | Genetica Molecular and Subdepartamento de Virologia ISP Chile | Instituto de Salud Publica de Chile | Javier Tognarelli et al |
| EPI_ISL_746746 | Chile/RM-191226/2020 | 2020-09-26 | South America/Chile/Region Metropolit | Genetica Molecular and Subdepartamento de Virologia ISP Chile | Instituto de Salud Publica de Chile | Javier Tognarelli et al |
| EPI_ISL_746747 | Chile/RM-191308/2020 | 2020-09-26 | South America/Chile/Region Metropolit | Genetica Molecular and Subdepartamento de Virologia ISP Chile | Instituto de Salud Publica de Chile | Javier Tognarelli et al |
| EPI_ISL_746748 | Chile/RM-191591/2020 | 2020-09-26 | South America/Chile/Region Metropolit | Genetica Molecular and Subdepartamento de Virologia ISP Chile | Instituto de Salud Publica de Chile | Javier Tognarelli et al |
| EPI_ISL_746749 | Chile/RM-191594/2020 | 2020-09-27 | South America/Chile/Region Metropolit | Genetica Molecular and Subdepartamento de Virologia ISP Chile | Instituto de Salud Publica de Chile | Javier Tognarelli et al |
| EPI_ISL_746751 | Chile/RM-191972/2020 | 2020-09-28 | South America/Chile/Region Metropolit | Genetica Molecular and Subdepartamento de Virologia ISP Chile | Instituto de Salud Publica de Chile | Javier Tognarelli et al |
| EPI_ISL_746755 | Chile/RM-193870/2020 | 2020-09-29 | South America/Chile/Region Metropolit | Genetica Molecular and Subdepartamento de Virologia ISP Chile | Instituto de Salud Publica de Chile | Javier Tognarelli et al |
| EPI_ISL_746756 | Chile/RM-194896/2020 | 2020-09-30 | South America/Chile/Region Metropolit | Genetica Molecular and Subdepartamento de Virologia ISP Chile | Instituto de Salud Publica de Chile | Javier Tognarelli et al |
| EPI_ISL_746759 | Chile/RM-203937/2020 | 2020-10-10 | South America/Chile/Region Metropolit | Genetica Molecular and Subdepartamento de Virologia ISP Chile | Instituto de Salud Publica de Chile | Javier Tognarelli et al |
| EPI_ISL_746762 | Chile/RM-204484/2020 | 2020-10-12 | South America/Chile/Region Metropolit | Genetica Molecular and Subdepartamento de Virologia ISP Chile | Instituto de Salud Publica de Chile | Javier Tognarelli et al |
| EPI_ISL_746764 | Chile/RM-204591/2020 | 2020-10-12 | South America/Chile/Region Metropolit | Genetica Molecular and Subdepartamento de Virologia ISP Chile | Instituto de Salud Publica de Chile | Javier Tognarelli et al |
| EPI_ISL_746767 | Chile/RM-207231/2020 | 2020-10-14 | South America/Chile/Region Metropolit | Genetica Molecular and Subdepartamento de Virologia ISP Chile | Instituto de Salud Publica de Chile | Javier Tognarelli et al |
| EPI_ISL_746768 | Chile/RM-207382/2020 | 2020-10-15 | South America/Chile/Region Metropolit | Genetica Molecular and Subdepartamento de Virologia ISP Chile | Instituto de Salud Publica de Chile | Javier Tognarelli et al |
| EPI_ISL_746769 | Chile/RM-207982/2020 | 2020-10-15 | South America/Chile/Region Metropolit | Genetica Molecular and Subdepartamento de Virologia ISP Chile | Instituto de Salud Publica de Chile | Javier Tognarelli et al |
| EPI_ISL_746774 | Chile/RM-72426/2020 | 2020-05-22 | South America/Chile/Region Metropolit | Genetica Molecular and Subdepartamento de Virologia ISP Chile | Instituto de Salud Publica de Chile | Javier Tognarelli et al |
| EPI_ISL_746775 | Chile/RM-72428/2020 | 2020-05-22 | South America/Chile/Region Metropolit | Genetica Molecular and Subdepartamento de Virologia ISP Chile | Instituto de Salud Publica de Chile | Javier Tognarelli et al |
| EPI_ISL_746777 | Chile/RM-72434/2020 | 2020-05-23 | South America/Chile/Region Metropolit | Genetica Molecular and Subdepartamento de Virologia ISP Chile | Instituto de Salud Publica de Chile | Javier Tognarelli et al |
| EPI_ISL_746779 | Chile/RM-72817/2020 | 2020-05-22 | South America/Chile/Region Metropolit | Genetica Molecular and Subdepartamento de Virologia ISP Chile | Instituto de Salud Publica de Chile | Javier Tognarelli et al |

| | | | | | | |
|---|---|---|---|---|---|---|
| EPI_ISL_746780 | Chile/RM-72820/2020 | 2020-05-22 | South America/Chile/Region Metropolit | Genetica Molecular and Subdepartamento de Virologia ISP Chile | Instituto de Salud Publica de Chile | Javier Tognarelli et al |
| EPI_ISL_746781 | Chile/RM-72821/2020 | 2020-05-23 | South America/Chile/Region Metropolit | Genetica Molecular and Subdepartamento de Virologia ISP Chile | Instituto de Salud Publica de Chile | Javier Tognarelli et al |
| EPI_ISL_746782 | Chile/RM-72843/2020 | 2020-05-22 | South America/Chile/Region Metropolit | Genetica Molecular and Subdepartamento de Virologia ISP Chile | Instituto de Salud Publica de Chile | Javier Tognarelli et al |
| EPI_ISL_746783 | Chile/RM-72846/2020 | 2020-05-22 | South America/Chile/Region Metropolit | Genetica Molecular and Subdepartamento de Virologia ISP Chile | Instituto de Salud Publica de Chile | Javier Tognarelli et al |
| EPI_ISL_746784 | Chile/RM-72850/2020 | 2020-05-22 | South America/Chile/Region Metropolit | Genetica Molecular and Subdepartamento de Virologia ISP Chile | Instituto de Salud Publica de Chile | Javier Tognarelli et al |
| EPI_ISL_746785 | Chile/RM-72851/2020 | 2020-05-20 | South America/Chile/Region Metropolit | Genetica Molecular and Subdepartamento de Virologia ISP Chile | Instituto de Salud Publica de Chile | Javier Tognarelli et al |
| EPI_ISL_746787 | Chile/RM-73219/2020 | 2020-05-22 | South America/Chile/Region Metropolit | Genetica Molecular and Subdepartamento de Virologia ISP Chile | Instituto de Salud Publica de Chile | Javier Tognarelli et al |
| EPI_ISL_746791 | Chile/RM-73255/2020 | 2020-05-22 | South America/Chile/Region Metropolit | Genetica Molecular and Subdepartamento de Virologia ISP Chile | Instituto de Salud Publica de Chile | Javier Tognarelli et al |
| EPI_ISL_746792 | Chile/RM-82478/2020 | 2020-05-29 | South America/Chile/Region Metropolit | Genetica Molecular and Subdepartamento de Virologia ISP Chile | Instituto de Salud Publica de Chile | Javier Tognarelli et al |
| EPI_ISL_746794 | Chile/RM-82495/2020 | 2020-05-29 | South America/Chile/Region Metropolit | Genetica Molecular and Subdepartamento de Virologia ISP Chile | Instituto de Salud Publica de Chile | Javier Tognarelli et al |
| EPI_ISL_746795 | Chile/RM-83539/2020 | 2020-05-30 | South America/Chile/Region Metropolit | Genetica Molecular and Subdepartamento de Virologia ISP Chile | Instituto de Salud Publica de Chile | Javier Tognarelli et al |
| EPI_ISL_746796 | Chile/RM-83540/2020 | 2020-05-31 | South America/Chile/Region Metropolit | Genetica Molecular and Subdepartamento de Virologia ISP Chile | Instituto de Salud Publica de Chile | Javier Tognarelli et al |
| EPI_ISL_746797 | Chile/RM-83548/2020 | 2020-05-30 | South America/Chile/Region Metropolit | Genetica Molecular and Subdepartamento de Virologia ISP Chile | Instituto de Salud Publica de Chile | Javier Tognarelli et al |
| EPI_ISL_746798 | Chile/RM-83553/2020 | 2020-05-30 | South America/Chile/Region Metropolit | Genetica Molecular and Subdepartamento de Virologia ISP Chile | Instituto de Salud Publica de Chile | Javier Tognarelli et al |
| EPI_ISL_746799 | Chile/RM-83568/2020 | 2020-05-31 | South America/Chile/Region Metropolit | Genetica Molecular and Subdepartamento de Virologia ISP Chile | Instituto de Salud Publica de Chile | Javier Tognarelli et al |
| EPI_ISL_746800 | Chile/RM-83613/2020 | 2020-05-30 | South America/Chile/Region Metropolit | Genetica Molecular and Subdepartamento de Virologia ISP Chile | Instituto de Salud Publica de Chile | Javier Tognarelli et al |
| EPI_ISL_746801 | Chile/RM-84213/2020 | 2020-05-31 | South America/Chile/Region Metropolit | Genetica Molecular and Subdepartamento de Virologia ISP Chile | Instituto de Salud Publica de Chile | Javier Tognarelli et al |
| EPI_ISL_746802 | Chile/RM-84393/2020 | 2020-05-31 | South America/Chile/Region Metropolit | Genetica Molecular and Subdepartamento de Virologia ISP Chile | Instituto de Salud Publica de Chile | Javier Tognarelli et al |
| EPI_ISL_746804 | Chile/RM-84400/2020 | 2020-05-31 | South America/Chile/Region Metropolit | Genetica Molecular and Subdepartamento de Virologia ISP Chile | Instituto de Salud Publica de Chile | Javier Tognarelli et al |
| EPI_ISL_746805 | Chile/RM-84488/2020 | 2020-05-31 | South America/Chile/Region Metropolit | Genetica Molecular and Subdepartamento de Virologia ISP Chile | Instituto de Salud Publica de Chile | Javier Tognarelli et al |

| | | | | | | |
|---|---|---|---|---|---|---|
| EPI_ISL_746807 | Chile/RM-84569/2020 | 2020-05-31 | South America/Chile/Region Metropolit | Genetica Molecular and Subdepartamento de Virologia ISP Chile | Instituto de Salud Publica de Chile | Javier Tognarelli et al |
| EPI_ISL_746808 | Chile/RM-84641/2020 | 2020-05-31 | South America/Chile/Region Metropolit | Genetica Molecular and Subdepartamento de Virologia ISP Chile | Instituto de Salud Publica de Chile | Javier Tognarelli et al |
| EPI_ISL_746809 | Chile/RM-84921/2020 | 2020-05-31 | South America/Chile/Region Metropolit | Genetica Molecular and Subdepartamento de Virologia ISP Chile | Instituto de Salud Publica de Chile | Javier Tognarelli et al |
| EPI_ISL_746810 | Chile/RM-84953/2020 | 2020-05-31 | South America/Chile/Region Metropolit | Genetica Molecular and Subdepartamento de Virologia ISP Chile | Instituto de Salud Publica de Chile | Javier Tognarelli et al |
| EPI_ISL_746811 | Chile/RM-85089/2020 | 2020-06-01 | South America/Chile/Region Metropolit | Genetica Molecular and Subdepartamento de Virologia ISP Chile | Instituto de Salud Publica de Chile | Javier Tognarelli et al |
| EPI_ISL_746812 | Chile/RM-85307/2020 | 2020-06-01 | South America/Chile/Region Metropolit | Genetica Molecular and Subdepartamento de Virologia ISP Chile | Instituto de Salud Publica de Chile | Javier Tognarelli et al |
| EPI_ISL_746813 | Chile/RM-85457/2020 | 2020-05-31 | South America/Chile/Region Metropolit | Genetica Molecular and Subdepartamento de Virologia ISP Chile | Instituto de Salud Publica de Chile | Javier Tognarelli et al |
| EPI_ISL_746814 | Chile/RM-85695/2020 | 2020-05-31 | South America/Chile/Region Metropolit | Genetica Molecular and Subdepartamento de Virologia ISP Chile | Instituto de Salud Publica de Chile | Javier Tognarelli et al |
| EPI_ISL_746815 | Chile/RM-86748-TC1/2020 | 2020-06-01 | South America/Chile/Region Metropolit | Genetica Molecular and Subdepartamento de Virologia ISP Chile | Instituto de Salud Publica de Chile | Javier Tognarelli et al |
| EPI_ISL_746816 | Chile/RM-87391-TC1/2020 | 2020-06-02 | South America/Chile/Region Metropolit | Genetica Molecular and Subdepartamento de Virologia ISP Chile | Instituto de Salud Publica de Chile | Javier Tognarelli et al |
| EPI_ISL_746817 | Chile/RM-87410-TC1/2020 | 2020-06-01 | South America/Chile/Region Metropolit | Genetica Molecular and Subdepartamento de Virologia ISP Chile | Instituto de Salud Publica de Chile | Javier Tognarelli et al |
| EPI_ISL_746818 | Chile/RM-87464-TC1/2020 | 2020-06-02 | South America/Chile/Region Metropolit | Genetica Molecular and Subdepartamento de Virologia ISP Chile | Instituto de Salud Publica de Chile | Javier Tognarelli et al |
| EPI_ISL_746819 | Chile/RM-87471-TC1/2020 | 2020-06-02 | South America/Chile/Region Metropolit | Genetica Molecular and Subdepartamento de Virologia ISP Chile | Instituto de Salud Publica de Chile | Javier Tognarelli et al |
| EPI_ISL_746820 | Chile/RM-87477-TC1/2020 | 2020-06-02 | South America/Chile/Region Metropolit | Genetica Molecular and Subdepartamento de Virologia ISP Chile | Instituto de Salud Publica de Chile | Javier Tognarelli et al |
| EPI_ISL_746821 | Chile/RM-87636-TC1/2020 | 2020-06-02 | South America/Chile/Region Metropolit | Genetica Molecular and Subdepartamento de Virologia ISP Chile | Instituto de Salud Publica de Chile | Javier Tognarelli et al |
| EPI_ISL_746822 | Chile/RM-87637-TC1/2020 | 2020-06-03 | South America/Chile/Region Metropolit | Genetica Molecular and Subdepartamento de Virologia ISP Chile | Instituto de Salud Publica de Chile | Javier Tognarelli et al |
| EPI_ISL_746823 | Chile/RM-88024-TC1/2020 | 2020-06-02 | South America/Chile/Region Metropolit | Genetica Molecular and Subdepartamento de Virologia ISP Chile | Instituto de Salud Publica de Chile | Javier Tognarelli et al |
| EPI_ISL_746824 | Chile/RM-88139-TC1/2020 | 2020-06-03 | South America/Chile/Region Metropolit | Genetica Molecular and Subdepartamento de Virologia ISP Chile | Instituto de Salud Publica de Chile | Javier Tognarelli et al |
| EPI_ISL_746825 | Chile/RM-88162-TC1/2020 | 2020-06-02 | South America/Chile/Region Metropolit | Genetica Molecular and Subdepartamento de Virologia ISP Chile | Instituto de Salud Publica de Chile | Javier Tognarelli et al |

| | | | | | | |
|---|---|---|---|---|---|---|
| EPI_ISL_746833 | Italy/LAZ-INMI-126/2020 | 2020-12-18 | Europe/Italy/Lazio/ | "National Institute for Infectious Diseases, INMI, ""L. Spallanzani"" IRCCS" | "National Institute for Infectious Diseases, INMI, ""L. Spallanzani"" IRCCS" | F Messina et al |
| EPI_ISL_746877 | USA/UT-UPHL-2012637301/2020 | 2020-11-30 | North America/USA/Utah/ | Utah Public Health Laboratory | Utah Public Health Laboratory | Erin Young et al |
| EPI_ISL_746880 | USA/UT-UPHL-2012480272/2020 | 2020-12-03 | North America/USA/Utah/ | Utah Public Health Laboratory | Utah Public Health Laboratory | Erin Young et al |
| EPI_ISL_746887 | USA/UT-UPHL-2012326830/2020 | 2020-12-03 | North America/USA/Utah/ | Utah Public Health Laboratory | Utah Public Health Laboratory | Erin Young et al |
| EPI_ISL_746901 | USA/UT-UPHL-2012312267/2020 | 2020-11-30 | North America/USA/Utah/ | Utah Public Health Laboratory | Utah Public Health Laboratory | Erin Young et al |
| EPI_ISL_746916 | USA/UT-UPHL-2012091937/2020 | 2020-11-30 | North America/USA/Utah/ | Utah Public Health Laboratory | Utah Public Health Laboratory | Erin Young et al |
| EPI_ISL_746919 | USA/UT-UPHL-2012036707/2020 | 2020-11-29 | North America/USA/Utah/ | Utah Public Health Laboratory | Utah Public Health Laboratory | Erin Young et al |
| EPI_ISL_746922 | USA/UT-UPHL-2012422031/2020 | 2020-11-29 | North America/USA/Utah/ | Utah Public Health Laboratory | Utah Public Health Laboratory | Erin Young et al |
| EPI_ISL_746931 | USA/UT-UPHL-2012020835/2020 | 2020-11-30 | North America/USA/Utah/ | Utah Public Health Laboratory | Utah Public Health Laboratory | Erin Young et al |
| EPI_ISL_746955 | USA/UT-UPHL-2012089416/2020 | 2020-11-27 | North America/USA/Utah/ | Utah Public Health Laboratory | Utah Public Health Laboratory | Erin Young et al |
| EPI_ISL_746956 | USA/UT-UPHL-2012172138/2020 | 2020-11-30 | North America/USA/Utah/ | Utah Public Health Laboratory | Utah Public Health Laboratory | Erin Young et al |
| EPI_ISL_746960 | USA/UT-UPHL-2012189554/2020 | 2020-11-30 | North America/USA/Utah/ | Utah Public Health Laboratory | Utah Public Health Laboratory | Erin Young et al |
| EPI_ISL_746962 | USA/UT-UPHL-2012146142/2020 | 2020-12-01 | North America/USA/Utah/ | Utah Public Health Laboratory | Utah Public Health Laboratory | Erin Young et al |
| EPI_ISL_746963 | USA/UT-UPHL-2012463625/2020 | 2020-11-20 | North America/USA/Utah/ | Utah Public Health Laboratory | Utah Public Health Laboratory | Erin Young et al |
| EPI_ISL_746973 | USA/UT-UPHL-2012811501/2020 | 2020-11-30 | North America/USA/Utah/ | Utah Public Health Laboratory | Utah Public Health Laboratory | Erin Young et al |
| EPI_ISL_746974 | USA/UT-UPHL-2012618844/2020 | 2020-11-23 | North America/USA/Utah/ | Utah Public Health Laboratory | Utah Public Health Laboratory | Erin Young et al |
| EPI_ISL_746975 | USA/UT-UPHL-2012124802/2020 | 2020-12-01 | North America/USA/Utah/ | Utah Public Health Laboratory | Utah Public Health Laboratory | Erin Young et al |
| EPI_ISL_746977 | USA/UT-UPHL-2012642336/2020 | 2020-11-30 | North America/USA/Utah/ | Utah Public Health Laboratory | Utah Public Health Laboratory | Erin Young et al |

| | | | | | | |
|---|---|---|---|---|---|---|
| EPI_ISL_746989 | USA/UT-UPHL-2012982319/2020 | 2020-11-30 | North America/USA/Utah/ | Utah Public Health Laboratory | Utah Public Health Laboratory | Erin Young et al |
| EPI_ISL_746993 | USA/UT-UPHL-2012802557/2020 | 2020-12-02 | North America/USA/Utah/ | Utah Public Health Laboratory | Utah Public Health Laboratory | Erin Young et al |
| EPI_ISL_746996 | USA/UT-UPHL-2012209911/2020 | 2020-11-18 | North America/USA/Utah/ | Utah Public Health Laboratory | Utah Public Health Laboratory | Erin Young et al |
| EPI_ISL_747001 | USA/UT-UPHL-2012637932/2020 | 2020-11-24 | North America/USA/Utah/ | Utah Public Health Laboratory | Utah Public Health Laboratory | Erin Young et al |
| EPI_ISL_747023 | USA/UT-UPHL-2012583894/2020 | 2020-12-03 | North America/USA/Utah/ | Utah Public Health Laboratory | Utah Public Health Laboratory | Erin Young et al |
| EPI_ISL_747041 | USA/NY-CDC-2-3693655/2020 | 2020-10-05 | North America/USA/New York/ | Respiratory Viruses Branch, Centers for Disease Control and Prevention | Respiratory Viruses Branch, Centers for Disease Control and Prevention | Queen et al |
| EPI_ISL_747044 | USA/AL-CDC-2-3693782/2020 | 2020-11-04 | North America/USA/Alabama/ | Respiratory Viruses Branch, Centers for Disease Control and Prevention | Respiratory Viruses Branch, Centers for Disease Control and Prevention | Queen et al |
| EPI_ISL_747046 | USA/AL-CDC-2-3693696/2020 | 2020-11-04 | North America/USA/Alabama/ | Respiratory Viruses Branch, Centers for Disease Control and Prevention | Respiratory Viruses Branch, Centers for Disease Control and Prevention | Queen et al |
| EPI_ISL_747052 | USA/PA-CDC-2-3693665/2020 | 2020-11-07 | North America/USA/Pennsylvania/ | Respiratory Viruses Branch, Centers for Disease Control and Prevention | Respiratory Viruses Branch, Centers for Disease Control and Prevention | Queen et al |
| EPI_ISL_747065 | USA/AR-CDC-2-3693682/2020 | 2020-10-28 | North America/USA/Arkansas/ | Respiratory Viruses Branch, Centers for Disease Control and Prevention | Respiratory Viruses Branch, Centers for Disease Control and Prevention | Queen et al |
| EPI_ISL_747067 | USA/AR-CDC-2-3693691/2020 | 2020-10-30 | North America/USA/Arkansas/ | Respiratory Viruses Branch, Centers for Disease Control and Prevention | Respiratory Viruses Branch, Centers for Disease Control and Prevention | Queen et al |

| | | | | | | |
|---|---|---|---|---|---|---|
| EPI_ISL_747069 | USA/LA-CDC-2-3693793/2020 | 2020-11-04 | North America/USA/Louisiana/ | Respiratory Viruses Branch, Centers for Disease Control and Prevention | Respiratory Viruses Branch, Centers for Disease Control and Prevention | Queen et al |
| EPI_ISL_747074 | USA/UT-CDC-2-3693760/2020 | 2020-11-05 | North America/USA/Utah/ | Respiratory Viruses Branch, Centers for Disease Control and Prevention | Respiratory Viruses Branch, Centers for Disease Control and Prevention | Queen et al |
| EPI_ISL_747076 | USA/UT-CDC-2-3693656/2020 | 2020-11-05 | North America/USA/Utah/ | Respiratory Viruses Branch, Centers for Disease Control and Prevention | Respiratory Viruses Branch, Centers for Disease Control and Prevention | Queen et al |
| EPI_ISL_747080 | USA/WI-CDC-2-3693678/2020 | 2020-11-07 | North America/USA/Wisconsin/ | Respiratory Viruses Branch, Centers for Disease Control and Prevention | Respiratory Viruses Branch, Centers for Disease Control and Prevention | Queen et al |
| EPI_ISL_747085 | USA/IN-CDC-2-3693679/2020 | 2020-11-07 | North America/USA/Indiana/ | Respiratory Viruses Branch, Centers for Disease Control and Prevention | Respiratory Viruses Branch, Centers for Disease Control and Prevention | Queen et al |
| EPI_ISL_747090 | USA/KY-CDC-2-3693781/2020 | 2020-11-04 | North America/USA/Kentucky/ | Respiratory Viruses Branch, Centers for Disease Control and Prevention | Respiratory Viruses Branch, Centers for Disease Control and Prevention | Queen et al |
| EPI_ISL_747091 | USA/KY-CDC-2-3693686/2020 | 2020-11-04 | North America/USA/Kentucky/ | Respiratory Viruses Branch, Centers for Disease Control and Prevention | Respiratory Viruses Branch, Centers for Disease Control and Prevention | Queen et al |
| EPI_ISL_747108 | USA/NJ-CDC-2-3693666/2020 | 2020-10-31 | North America/USA/New Jersey/ | Respiratory Viruses Branch, Centers for Disease Control and Prevention | Respiratory Viruses Branch, Centers for Disease Control and Prevention | Queen et al |

| | | | | | | |
|---|---|---|---|---|---|---|
| EPI_ISL_747119 | USA/NC-CDC-2-3693555/2020 | 2020-10-30 | North America/USA/North Carolina/ | Respiratory Viruses Branch, Centers for Disease Control and Prevention | Respiratory Viruses Branch, Centers for Disease Control and Prevention | Queen et al |
| EPI_ISL_747134 | USA/CT-CDC-2-3693591/2020 | 2020-11-05 | North America/USA/Connecticut/ | Respiratory Viruses Branch, Centers for Disease Control and Prevention | Respiratory Viruses Branch, Centers for Disease Control and Prevention | Queen et al |
| EPI_ISL_747137 | USA/KS-CDC-2-3693574/2020 | 2020-11-03 | North America/USA/Kansas/ | Respiratory Viruses Branch, Centers for Disease Control and Prevention | Respiratory Viruses Branch, Centers for Disease Control and Prevention | Queen et al |
| EPI_ISL_747141 | USA/MO-CDC-2-3693556/2020 | 2020-11-02 | North America/USA/Missouri/ | Respiratory Viruses Branch, Centers for Disease Control and Prevention | Respiratory Viruses Branch, Centers for Disease Control and Prevention | Queen et al |
| EPI_ISL_747144 | USA/MO-CDC-2-3693577/2020 | 2020-11-04 | North America/USA/Missouri/ | Respiratory Viruses Branch, Centers for Disease Control and Prevention | Respiratory Viruses Branch, Centers for Disease Control and Prevention | Queen et al |
| EPI_ISL_747148 | USA/TX-CDC-2-3693572/2020 | 2020-10-27 | North America/USA/Texas/ | Respiratory Viruses Branch, Centers for Disease Control and Prevention | Respiratory Viruses Branch, Centers for Disease Control and Prevention | Queen et al |
| EPI_ISL_747150 | USA/TX-CDC-2-3693478/2020 | 2020-11-04 | North America/USA/Texas/ | Respiratory Viruses Branch, Centers for Disease Control and Prevention | Respiratory Viruses Branch, Centers for Disease Control and Prevention | Queen et al |
| EPI_ISL_747152 | USA/NV-CDC-2-3693465/2020 | 2020-11-07 | North America/USA/Nevada/ | Respiratory Viruses Branch, Centers for Disease Control and Prevention | Respiratory Viruses Branch, Centers for Disease Control and Prevention | Queen et al |

| | | | | | | |
|---|---|---|---|---|---|---|
| EPI_ISL_747163 | USA/OK-CDC-2-3693462/2020 | 2020-11-17 | North America/USA/Oklahoma/ | Respiratory Viruses Branch, Centers for Disease Control and Prevention | Respiratory Viruses Branch, Centers for Disease Control and Prevention | Queen et al |
| EPI_ISL_747170 | USA/OR-CDC-2-3693481/2020 | 2020-11-23 | North America/USA/Oregon/ | Respiratory Viruses Branch, Centers for Disease Control and Prevention | Respiratory Viruses Branch, Centers for Disease Control and Prevention | Queen et al |
| EPI_ISL_747174 | USA/NC-CDC-2-3693498/2020 | 2020-11-09 | North America/USA/North Carolina/ | Respiratory Viruses Branch, Centers for Disease Control and Prevention | Respiratory Viruses Branch, Centers for Disease Control and Prevention | Queen et al |
| EPI_ISL_747184 | USA/TN-CDC-2-3693597/2020 | 2020-11-25 | North America/USA/Tennessee/ | Respiratory Viruses Branch, Centers for Disease Control and Prevention | Respiratory Viruses Branch, Centers for Disease Control and Prevention | Queen et al |
| EPI_ISL_747189 | USA/OR-CDC-2-3693562/2020 | 2020-11-15 | North America/USA/Oregon/ | Respiratory Viruses Branch, Centers for Disease Control and Prevention | Respiratory Viruses Branch, Centers for Disease Control and Prevention | Queen et al |
| EPI_ISL_747192 | USA/OR-CDC-2-3693600/2020 | 2020-11-14 | North America/USA/Oregon/ | Respiratory Viruses Branch, Centers for Disease Control and Prevention | Respiratory Viruses Branch, Centers for Disease Control and Prevention | Queen et al |
| EPI_ISL_747201 | USA/WA-UW-45072/2020 | 2020-12-08 | North America/USA/Washington/ | UW Virology Lab | UW Virology Lab | Pavitra Roychoudhury et al |
| EPI_ISL_747206 | USA/WA-UW-45088/2020 | 2020-12-08 | North America/USA/Washington/ | UW Virology Lab | UW Virology Lab | Pavitra Roychoudhury et al |
| EPI_ISL_747225 | USA/WA-UW-45122/2020 | 2020-12-08 | North America/USA/Washington/ | UW Virology Lab | UW Virology Lab | Pavitra Roychoudhury et al |
| EPI_ISL_747229 | USA/WA-UW-45145/2020 | 2020-12-08 | North America/USA/Washington/ | UW Virology Lab | UW Virology Lab | Pavitra Roychoudhury et al |
| EPI_ISL_747232 | USA/WA-UW-45190/2020 | 2020-12-07 | North America/USA/Washington/ | UW Virology Lab | UW Virology Lab | Pavitra Roychoudhury et al |
| EPI_ISL_747233 | Indonesia/JK-NIHRD-IST002/2020 | 2020-09-18 | Asia/Indonesia/Jakarta/ | National Institute of Health Research and Development | National Institute of Health Research and Development | Subangkit et al |

| | | | | | | |
|---|---|---|---|---|---|---|
| EPI_ISL_747235 | Indonesia/JB-WJHL-ITB-A02050/2020 | 2020-10-21 | Asia/Indonesia/West Java/ | Sukabumi Health Laboratory | West Java Health Laboratory; School of Life Sciences and Technology, Institut Teknologi Bandung | Azzania Fibriani et al |
| EPI_ISL_747236 | Indonesia/JB-WJHL-ITB-A03267/2020 | 2020-10-23 | Asia/Indonesia/West Java/ | Sukabumi Public Health | West Java Health Laboratory; School of Life Sciences and Technology, Institut Teknologi Bandung | Azzania Fibriani et al |
| EPI_ISL_747237 | Indonesia/BB-NIHRD-PME9185/2020 | 2020-10-01 | Asia/Indonesia/Kepulauan Bangka Belitu | National Institute of Health Research and Development | National Institute of Health Research and Development | Subangkit et al |
| EPI_ISL_747238 | Indonesia/JB-WJHL-ITB-R104416/2020 | 2020-09-23 | Asia/Indonesia/West Java/ | Bogor Public Health | West Java Health Laboratory; School of Life Sciences and Technology, Institut Teknologi Bandung | Azzania Fibriani et al |
| EPI_ISL_747239 | Indonesia/JB-WJHL-ITB-R109969/2020 | 2020-10-02 | Asia/Indonesia/West Java/ | Kuningan Publlic Health Office | West Java Health Laboratory; School of Life Sciences and Technology, Institut Teknologi Bandung | Azzania Fibriani et al |
| EPI_ISL_747240 | Indonesia/JB-WJHL-ITB-R119840/2020 | 2020-10-14 | Asia/Indonesia/West Java/ | Al Islam Hospital | West Java Health Laboratory; School of Life Sciences and Technology, Institut Teknologi Bandung | Azzania Fibriani et al |

| | | | | | | |
|---|---|---|---|---|---|---|
| EPI_ISL_747241 | Indonesia/JB-WJHL-ITB-R120975/2020 | 2020-10-15 | Asia/Indonesia/West Java/ | Subang Public Health Office | West Java Health Laboratory; School of Life Sciences and Technology, Institut Teknologi Bandung | Azzania Fibriani et al |
| EPI_ISL_747242 | Mexico/BCN-CICESE-BC2/2020 | 2020-08-28 | North America/Mexico/Baja California/ | Pathogen Lab (BSL3), Biomedical Innovation Department, Applied and Experimental Biology Division, Scientific Research Center and High Education from Ensenada (CICESE) | Pathogen Laboratory (BSL3), Biomedical Innovation Department, Experimental and Applied Biology Division, Scientific Research Center and High Education from Ensenada (CICESE) | Cervantes-Luevano K et al |
| EPI_ISL_747243 | Mexico/BCN-CICESE-BC5/2020 | 2020-09-21 | North America/Mexico/Baja California/ | General Hospital Laboratory | Pathogen Laboratory (BSL3), Biomedical Innovation Department, Experimental and Applied Biology Division, Scientific Research Center and High Education from Ensenada (CICESE) | Cervantes-Luevano K. et al |
| EPI_ISL_747244 | India/KA-NIMH-SEQ-8/2020 | 2020-12-22 | Asia/India/Karnataka/ | BBMP Urban PHC | Department of Neurovirology, National Institute of Mental Health and Neurosciences (NIMHANS) | Chitra Pattabiraman et al |

| | | | | | | |
|---|---|---|---|---|---|---|
| EPI_ISL_747257 | SouthKorea/KDCA0016/2020 | 2020-08-26 | Asia/South Korea/South Korea/ | Division of Emerging Infectious Diseases, Bureau of Infectious Diseases Diagnosis Control, Korea Disease Control and Prevention Agency | Division of Emerging Infectious Diseases, Bureau of Infectious Diseases Diagnosis Control, Korea Disease Control and Prevention Agency | Ae Kyung Park et al |
| EPI_ISL_747261 | SouthKorea/KDCA0020/2020 | 2020-09-04 | Asia/South Korea/South Korea/ | Division of Emerging Infectious Diseases, Bureau of Infectious Diseases Diagnosis Control, Korea Disease Control and Prevention Agency | Division of Emerging Infectious Diseases, Bureau of Infectious Diseases Diagnosis Control, Korea Disease Control and Prevention Agency | Ae Kyung Park et al |
| EPI_ISL_747283 | Bangladesh/DNAS-Jakir/2020 | 2020-12-06 | Asia/Bangladesh/Dhaka/ | DNA SOLUTION LTD. | DNA SOLUTION LTD. | Md. Imran Khan et al |
| EPI_ISL_747287 | SouthKorea/KDCA0046/2020 | 2020-09-24 | Asia/South Korea/South Korea/ | Division of Emerging Infectious Diseases, Bureau of Infectious Diseases Diagnosis Control, Korea Disease Control and Prevention Agency | Division of Emerging Infectious Diseases, Bureau of Infectious Diseases Diagnosis Control, Korea Disease Control and Prevention Agency | Ae Kyung Park et al |

| | | | | | | |
|---|---|---|---|---|---|---|
| EPI_ISL_747323 | SouthKorea/KDCA0083/2020 | 2020-09-24 | Asia/South Korea/South Korea/ | Division of Emerging Infectious Diseases, Bureau of Infectious Diseases Diagnosis Control, Korea Disease Control and Prevention Agency | Division of Emerging Infectious Diseases, Bureau of Infectious Diseases Diagnosis Control, Korea Disease Control and Prevention Agency | Ae Kyung Park et al |
| EPI_ISL_747327 | SouthKorea/KDCA0088/2020 | 2020-10-02 | Asia/South Korea/South Korea/ | Division of Emerging Infectious Diseases, Bureau of Infectious Diseases Diagnosis Control, Korea Disease Control and Prevention Agency | Division of Emerging Infectious Diseases, Bureau of Infectious Diseases Diagnosis Control, Korea Disease Control and Prevention Agency | Ae Kyung Park et al |
| EPI_ISL_747343 | SouthKorea/KDCA0107/2020 | 2020-09-25 | Asia/South Korea/South Korea/ | Division of Emerging Infectious Diseases, Bureau of Infectious Diseases Diagnosis Control, Korea Disease Control and Prevention Agency | Division of Emerging Infectious Diseases, Bureau of Infectious Diseases Diagnosis Control, Korea Disease Control and Prevention Agency | Ae Kyung Park et al |

| | | | | | | |
|---|---|---|---|---|---|---|
| EPI_ISL_747349 | SouthKorea/KDCA0114/2020 | 2020-09-29 | Asia/South Korea/South Korea/ | Division of Emerging Infectious Diseases, Bureau of Infectious Diseases Diagnosis Control, Korea Disease Control and Prevention Agency | Division of Emerging Infectious Diseases, Bureau of Infectious Diseases Diagnosis Control, Korea Disease Control and Prevention Agency | Ae Kyung Park et al |
| EPI_ISL_747353 | SouthKorea/KDCA0118/2020 | 2020-09-30 | Asia/South Korea/South Korea/ | Division of Emerging Infectious Diseases, Bureau of Infectious Diseases Diagnosis Control, Korea Disease Control and Prevention Agency | Division of Emerging Infectious Diseases, Bureau of Infectious Diseases Diagnosis Control, Korea Disease Control and Prevention Agency | Ae Kyung Park et al |
| EPI_ISL_747371 | SouthKorea/KDCA0136/2020 | 2020-10-13 | Asia/South Korea/South Korea/ | Division of Emerging Infectious Diseases, Bureau of Infectious Diseases Diagnosis Control, Korea Disease Control and Prevention Agency | Division of Emerging Infectious Diseases, Bureau of Infectious Diseases Diagnosis Control, Korea Disease Control and Prevention Agency | Ae Kyung Park et al |

| | | | | | | |
|---|---|---|---|---|---|---|
| EPI_ISL_747425 | SouthKorea/KDCA0189/2020 | 2020-11-22 | Asia/South Korea/South Korea/ | Division of Emerging Infectious Diseases, Bureau of Infectious Diseases Diagnosis Control, Korea Disease Control and Prevention Agency | Division of Emerging Infectious Diseases, Bureau of Infectious Diseases Diagnosis Control, Korea Disease Control and Prevention Agency | Ae Kyung Park et al |
| EPI_ISL_747459 | Italy/VEN-IZSVe-20RS2160-29_VR/20 | 2020-11-26 | Europe/Italy/Veneto/Verona IT | Ospedale San Bonifacio | Istituto Zooprofilattico Sperimentale delle Venezie | Adelaide Milani et al |
| EPI_ISL_747460 | Italy/VEN-IZSVe-20RS2160-30_VR/20 | 2020-11-26 | Europe/Italy/Veneto/Verona IT | Ospedale San Bonifacio | Istituto Zooprofilattico Sperimentale delle Venezie | Adelaide Milani et al |
| EPI_ISL_747461 | Italy/VEN-IZSVe-20RS2160-31_VR/20 | 2020-11-26 | Europe/Italy/Veneto/Verona IT | Ospedale San Bonifacio | Istituto Zooprofilattico Sperimentale delle Venezie | Adelaide Milani et al |
| EPI_ISL_747462 | Italy/VEN-IZSVe-20RS2160-32_VR/20 | 2020-11-26 | Europe/Italy/Veneto/Verona IT | Ospedale San Bonifacio | Istituto Zooprofilattico Sperimentale delle Venezie | Adelaide Milani et al |
| EPI_ISL_747463 | Italy/VEN-IZSVe-20RS2160-33_VR/20 | 2020-11-26 | Europe/Italy/Veneto/Verona IT | Ospedale San Bonifacio | Istituto Zooprofilattico Sperimentale delle Venezie | Adelaide Milani et al |
| EPI_ISL_747464 | Italy/VEN-IZSVe-20RS2160-35_VR/20 | 2020-11-30 | Europe/Italy/Veneto/Verona IT | Ospedale San Bonifacio | Istituto Zooprofilattico Sperimentale delle Venezie | Adelaide Milani et al |
| EPI_ISL_747465 | Italy/VEN-IZSVe-20RS2161-11_VR/20 | 2020-11-04 | Europe/Italy/Veneto/Verona IT | Ospedale Mater Salutis | Istituto Zooprofilattico Sperimentale delle Venezie | Adelaide Milani et al |

| | | | | | | |
|---|---|---|---|---|---|---|
| EPI_ISL_747466 | Italy/VEN-IZSVe-20RS2161-13_VR/20 | 2020-11-09 | Europe/Italy/Veneto/Verona IT | Ospedale Mater Salutis | Istituto Zooprofilattico Sperimentale delle Venezie | Adelaide Milani et al |
| EPI_ISL_747467 | Italy/VEN-IZSVe-20RS2161-14_VR/20 | 2020-11-09 | Europe/Italy/Veneto/Verona IT | Ospedale Mater Salutis | Istituto Zooprofilattico Sperimentale delle Venezie | Adelaide Milani et al |
| EPI_ISL_747468 | Italy/VEN-IZSVe-20RS2161-17_VR/20 | 2020-11-15 | Europe/Italy/Veneto/Verona IT | Ospedale Mater Salutis | Istituto Zooprofilattico Sperimentale delle Venezie | Adelaide Milani et al |
| EPI_ISL_747470 | Italy/VEN-IZSVe-20RS2161-20_VR/20 | 2020-11-14 | Europe/Italy/Veneto/Verona IT | Ospedale Mater Salutis | Istituto Zooprofilattico Sperimentale delle Venezie | Adelaide Milani et al |
| EPI_ISL_747471 | Italy/VEN-IZSVe-20RS2163-1_VR/202 | 2020-11-20 | Europe/Italy/Veneto/Verona IT | ULSS9 Distretto di Bussolengo | Istituto Zooprofilattico Sperimentale delle Venezie | Adelaide Milani et al |
| EPI_ISL_747472 | Italy/VEN-IZSVe-20RS2163-2_VR/202 | 2020-11-20 | Europe/Italy/Veneto/Verona IT | ULSS9 Distretto di Bussolengo | Istituto Zooprofilattico Sperimentale delle Venezie | Adelaide Milani et al |
| EPI_ISL_747473 | Italy/VEN-IZSVe-20RS2163-3_VR/202 | 2020-11-21 | Europe/Italy/Veneto/Verona IT | ULSS9 Distretto di Bussolengo | Istituto Zooprofilattico Sperimentale delle Venezie | Adelaide Milani et al |
| EPI_ISL_747475 | Italy/VEN-IZSVe-20RS2163-6_VR/202 | 2020-11-23 | Europe/Italy/Veneto/Verona IT | ULSS9 Distretto di Bussolengo | Istituto Zooprofilattico Sperimentale delle Venezie | Adelaide Milani et al |
| EPI_ISL_747476 | Italy/VEN-IZSVe-20RS2163-7_VR/202 | 2020-11-24 | Europe/Italy/Veneto/Verona IT | ULSS9 Distretto di Bussolengo | Istituto Zooprofilattico Sperimentale delle Venezie | Adelaide Milani et al |
| EPI_ISL_747478 | Italy/VEN-IZSVe-20RS2165-3_RO/202 | 2020-11-24 | Europe/Italy/Veneto/Rovigo | ULSS 5 Polesana | Istituto Zooprofilattico Sperimentale delle Venezie | Adelaide Milani et al |

| | | | | | | |
|---|---|---|---|---|---|---|
| EPI_ISL_747479 | Italy/VEN-IZSVe-20RS2165-5_RO/202 | 2020-11-24 | Europe/Italy/Veneto/Rovigo | ULSS 5 Polesana | Istituto Zooprofilattico Sperimentale delle Venezie | Adelaide Milani et al |
| EPI_ISL_747480 | Italy/VEN-IZSVe-20RS2165-6_RO/202 | 2020-11-25 | Europe/Italy/Veneto/Rovigo | ULSS 5 Polesana | Istituto Zooprofilattico Sperimentale delle Venezie | Adelaide Milani et al |
| EPI_ISL_747483 | Italy/VEN-IZSVe-20RS2160-34_VR/20 | 2020-11-30 | Europe/Italy/Veneto/Verona IT | Ospedale San Bonifacio | Istituto Zooprofilattico Sperimentale delle Venezie | Adelaide Milani et al |
| EPI_ISL_747484 | Italy/VEN-IZSVe-20RS2161-12_VR/20 | 2020-11-10 | Europe/Italy/Veneto/Verona IT | Ospedale Mater Salutis | Istituto Zooprofilattico Sperimentale delle Venezie | Adelaide Milani et al |
| EPI_ISL_747486 | Italy/VEN-IZSVe-20RS2165-1_RO/202 | 2020-11-24 | Europe/Italy/Veneto/Rovigo | ULSS 5 Polesana | Istituto Zooprofilattico Sperimentale delle Venezie | Adelaide Milani et al |
| EPI_ISL_747487 | Italy/VEN-IZSVe-20RS2165-2_RO/202 | 2020-11-24 | Europe/Italy/Veneto/Rovigo | ULSS 5 Polesana | Istituto Zooprofilattico Sperimentale delle Venezie | Adelaide Milani et al |
| EPI_ISL_747488 | Italy/VEN-IZSVe-20RS2165-4_RO/202 | 2020-11-24 | Europe/Italy/Veneto/Rovigo | ULSS 5 Polesana | Istituto Zooprofilattico Sperimentale delle Venezie | Adelaide Milani et al |
| EPI_ISL_747489 | Italy/VEN-IZSVe-20RS2165-7_RO/202 | 2020-11-25 | Europe/Italy/Veneto/Rovigo | ULSS 5 Polesana | Istituto Zooprofilattico Sperimentale delle Venezie | Adelaide Milani et al |
| EPI_ISL_747502 | England/205082500/2020 | 2020-12-11 | Europe/United Kingdom/England/ | Respiratory Virus Unit, National Infection Service, Public Health England | COVID-19 Genomics UK (COG-UK) Consortium | PHE Covid Sequencing Team et al |

| | | | | | | |
|---|---|---|---|---|---|---|
| EPI_ISL_747627 | Denmark/DCGC-20260/2020 | 2020-11-30 | Europe/Denmark/Syddanmark/ | Department of Virus and Microbiological Special Diagnostics, Statens Serum Institut, Copenhagen, Denmark | Albertsen Lab, Department of Chemistry and Bioscience, Aalborg University, Denmark | Danish Covid-19 Genome Consortium et al |
| EPI_ISL_747976 | Denmark/DCGC-20090/2020 | 2020-12-07 | Europe/Denmark/Hovedstaden/ | Department of Virus and Microbiological Special Diagnostics, Statens Serum Institut, Copenhagen, Denmark | Albertsen Lab, Department of Chemistry and Bioscience, Aalborg University, Denmark | Danish Covid-19 Genome Consortium et al |
| EPI_ISL_747993 | Denmark/DCGC-20117/2020 | 2020-12-07 | Europe/Denmark/Hovedstaden/ | Department of Virus and Microbiological Special Diagnostics, Statens Serum Institut, Copenhagen, Denmark | Albertsen Lab, Department of Chemistry and Bioscience, Aalborg University, Denmark | Danish Covid-19 Genome Consortium et al |
| EPI_ISL_748031 | Denmark/DCGC-20179/2020 | 2020-11-30 | Europe/Denmark/Sjaelland/ | Department of Virus and Microbiological Special Diagnostics, Statens Serum Institut, Copenhagen, Denmark | Albertsen Lab, Department of Chemistry and Bioscience, Aalborg University, Denmark | Danish Covid-19 Genome Consortium et al |
| EPI_ISL_748411 | Denmark/DCGC-19046/2020 | 2020-12-07 | Europe/Denmark/Midtjylland/ | Department of Virus and Microbiological Special Diagnostics, Statens Serum Institut, Copenhagen, Denmark | Albertsen Lab, Department of Chemistry and Bioscience, Aalborg University, Denmark | Danish Covid-19 Genome Consortium et al |

| | | | | | | |
|---|---|---|---|---|---|---|
| EPI_ISL_748746 | Denmark/DCGC-19084/2020 | 2020-12-07 | Europe/Denmark/Syddanmark/ | Department of Virus and Microbiological Special Diagnostics, Statens Serum Institut, Copenhagen, Denmark | Albertsen Lab, Department of Chemistry and Bioscience, Aalborg University, Denmark | Danish Covid-19 Genome Consortium et al |
| EPI_ISL_748969 | Denmark/DCGC-19470/2020 | 2020-12-07 | Europe/Denmark/Syddanmark/ | Department of Virus and Microbiological Special Diagnostics, Statens Serum Institut, Copenhagen, Denmark | Albertsen Lab, Department of Chemistry and Bioscience, Aalborg University, Denmark | Danish Covid-19 Genome Consortium et al |
| EPI_ISL_749042 | Denmark/DCGC-19583/2020 | 2020-12-07 | Europe/Denmark/Hovedstaden/ | Department of Virus and Microbiological Special Diagnostics, Statens Serum Institut, Copenhagen, Denmark | Albertsen Lab, Department of Chemistry and Bioscience, Aalborg University, Denmark | Danish Covid-19 Genome Consortium et al |
| EPI_ISL_751125 | Denmark/DCGC-17652/2020 | 2020-12-14 | Europe/Denmark/Syddanmark/ | Department of Virus and Microbiological Special Diagnostics, Statens Serum Institut, Copenhagen, Denmark | Albertsen Lab, Department of Chemistry and Bioscience, Aalborg University, Denmark | Danish Covid-19 Genome Consortium et al |
| EPI_ISL_751205 | SaudiArabia/KAUST-MAKKAH734/20 | 2020-04-17 | Asia/Saudi Arabia/Makkah/ | Pathogen Genomics Lab King Abdullah University of Science and Technology(KAUST) | Pathogen Genomics Lab King Abdullah University of Science and Technology(KAUST) | Fathia Ben Rached et al |

| | | | | | | |
|---|---|---|---|---|---|---|
| EPI_ISL_751208 | SaudiArabia/KAUST-JEDDAH832/202 | 2020-05-24 | Asia/Saudi Arabia/Jeddah/ | Pathogen Genomics Lab King Abdullah University of Science and Technology(KAUST) | Pathogen Genomics Lab King Abdullah University of Science and Technology(KAUST) | Sharif Hala et al |
| EPI_ISL_751209 | SaudiArabia/KAUST-MADINAH1657/ | 2020-08-23 | Asia/Saudi Arabia/Madinah/ | Pathogen Genomics Lab King Abdullah University of Science and Technology(KAUST) | Pathogen Genomics Lab King Abdullah University of Science and Technology(KAUST) | Sara Mfarrej et al |
| EPI_ISL_751214 | SaudiArabia/KAUST-MADINAH1498/ | 2020-07-17 | Asia/Saudi Arabia/Madinah/ | Pathogen Genomics Lab King Abdullah University of Science and Technology(KAUST) | Pathogen Genomics Lab King Abdullah University of Science and Technology(KAUST) | Sara Mfarrej et al |
| EPI_ISL_751216 | SaudiArabia/KAUST-MADINAH1659/ | 2020-08-23 | Asia/Saudi Arabia/Madinah/ | Pathogen Genomics Lab King Abdullah University of Science and Technology(KAUST) | Pathogen Genomics Lab King Abdullah University of Science and Technology(KAUST) | Sara Mfarrej et al |
| EPI_ISL_751217 | SaudiArabia/KAUST-MADINAH1671/ | 2020-08-23 | Asia/Saudi Arabia/Madinah/ | Pathogen Genomics Lab King Abdullah University of Science and Technology(KAUST) | Pathogen Genomics Lab King Abdullah University of Science and Technology(KAUST) | Sara Mfarrej et al |

| | | | | | | |
|---|---|---|---|---|---|---|
| EPI_ISL_751219 | SaudiArabia/KAUST-JEDDAH881/202 | 2020-05-24 | Asia/Saudi Arabia/Jeddah/ | Pathogen Genomics Lab King Abdullah University of Science and Technology(KAUST) | Pathogen Genomics Lab King Abdullah University of Science and Technology(KAUST ) | Fathia Ben Rached et al |
| EPI_ISL_751220 | SaudiArabia/KAUST-JEDDAH885/202 | 2020-05-24 | Asia/Saudi Arabia/Jeddah/ | Pathogen Genomics Lab King Abdullah University of Science and Technology(KAUST) | Pathogen Genomics Lab King Abdullah University of Science and Technology(KAUST ) | Fathia Ben Rached et al |
| EPI_ISL_751222 | SaudiArabia/KAUST-RIYADH1173/20 | 2020-06-15 | Asia/Saudi Arabia/Riyadh/ | Pathogen Genomics Lab King Abdullah University of Science and Technology(KAUST) | Pathogen Genomics Lab King Abdullah University of Science and Technology(KAUST ) | Raeece Naeem et al |
| EPI_ISL_751225 | SaudiArabia/KAUST-MADINAH1669/ | 2020-08-23 | Asia/Saudi Arabia/Madinah/ | Pathogen Genomics Lab King Abdullah University of Science and Technology(KAUST) | Pathogen Genomics Lab King Abdullah University of Science and Technology(KAUST ) | Raushan Nugmanova et al |
| EPI_ISL_751226 | SaudiArabia/KAUST-MADINAH1648/ | 2020-07-17 | Asia/Saudi Arabia/Madinah/ | Pathogen Genomics Lab King Abdullah University of Science and Technology(KAUST) | Pathogen Genomics Lab King Abdullah University of Science and Technology(KAUST ) | Sara Mfarrej et al |

| | | | | | | |
|---|---|---|---|---|---|---|
| EPI_ISL_751227 | SaudiArabia/KAUST-MADINAH1677/ | 2020-08-23 | Asia/Saudi Arabia/Madinah/ | Pathogen Genomics Lab King Abdullah University of Science and Technology(KAUST) | Pathogen Genomics Lab King Abdullah University of Science and Technology(KAUST) | Sara Mfarrej et al |
| EPI_ISL_751228 | SaudiArabia/KAUST-MADINAH1497/ | 2020-07-17 | Asia/Saudi Arabia/Madinah/ | Pathogen Genomics Lab King Abdullah University of Science and Technology(KAUST) | Pathogen Genomics Lab King Abdullah University of Science and Technology(KAUST) | Sara Mfarrej et al |
| EPI_ISL_751230 | SaudiArabia/KAUST-MADINAH1661/ | 2020-08-23 | Asia/Saudi Arabia/Madinah/ | Pathogen Genomics Lab King Abdullah University of Science and Technology(KAUST) | Pathogen Genomics Lab King Abdullah University of Science and Technology(KAUST) | Sara Mfarrej et al |
| EPI_ISL_751231 | SaudiArabia/KAUST-MADINAH1689/ | 2020-08-23 | Asia/Saudi Arabia/Madinah/ | Pathogen Genomics Lab King Abdullah University of Science and Technology(KAUST) | Pathogen Genomics Lab King Abdullah University of Science and Technology(KAUST) | Sara Mfarrej et al |
| EPI_ISL_751232 | SaudiArabia/KAUST-MADINAH1674/ | 2020-08-23 | Asia/Saudi Arabia/Madinah/ | Pathogen Genomics Lab King Abdullah University of Science and Technology(KAUST) | Pathogen Genomics Lab King Abdullah University of Science and Technology(KAUST) | Sara Mfarrej et al |

| | | | | | | |
|---|---|---|---|---|---|---|
| EPI_ISL_751233 | SaudiArabia/KAUST-MADINAH1667/ | 2020-08-23 | Asia/Saudi Arabia/Madinah/ | Pathogen Genomics Lab King Abdullah University of Science and Technology(KAUST) | Pathogen Genomics Lab King Abdullah University of Science and Technology(KAUST) | Olga Douvropoulou et al |
| EPI_ISL_751234 | SaudiArabia/KAUST-MADINAH1651/ | 2020-07-17 | Asia/Saudi Arabia/Madinah/ | Pathogen Genomics Lab King Abdullah University of Science and Technology(KAUST) | Pathogen Genomics Lab King Abdullah University of Science and Technology(KAUST) | Sara Mfarrej et al |
| EPI_ISL_751235 | SaudiArabia/KAUST-MADINAH1655/ | 2020-08-23 | Asia/Saudi Arabia/Madinah/ | Pathogen Genomics Lab King Abdullah University of Science and Technology(KAUST) | Pathogen Genomics Lab King Abdullah University of Science and Technology(KAUST) | Sara Mfarrej et al |
| EPI_ISL_751320 | Italy/VEN-UniVR-119/2020 | 2020-03-29 | Europe/Italy/Veneto/Verona IT | IRCCS Sacro Cuore Don Calabria Hospital, Department of Infectious, Tropical Diseases & Microbiology | University of Verona, Department of Biotechnology | Antonio Mori et al |
| EPI_ISL_751321 | Italy/VEN-UniVR-130/2020 | 2020-04-01 | Europe/Italy/Veneto/Verona IT | IRCCS Sacro Cuore Don Calabria Hospital, Department of Infectious, Tropical Diseases & Microbiology | University of Verona, Department of Biotechnology | Antonio Mori et al |
| EPI_ISL_751322 | Italy/VEN-UniVR-151/2020 | 2020-04-13 | Europe/Italy/Veneto/Verona IT | IRCCS Sacro Cuore Don Calabria Hospital, Department of Infectious, Tropical Diseases & Microbiology | University of Verona, Department of Biotechnology | Antonio Mori et al |
| EPI_ISL_751332 | Italy/VEN-UniVR-32/2020 | 2020-08-20 | Europe/Italy/Veneto/Verona IT | IRCCS Sacro Cuore Don Calabria Hospital, Department of Infectious, Tropical Diseases & Microbiology | University of Verona, Department of Biotechnology | Antonio Mori et al |
| EPI_ISL_751333 | Italy/VEN-UniVR-24/2020 | 2020-04-30 | Europe/Italy/Veneto/Verona IT | IRCCS Sacro Cuore Don Calabria Hospital, Department of Infectious, Tropical Diseases & Microbiology | University of Verona, Department of Biotechnology | Antonio Mori et al |

| | | | | | | |
|---|---|---|---|---|---|---|
| EPI_ISL_751336 | Italy/VEN-UniVR-13/2020 | 2020-04-20 | Europe/Italy/Veneto/Verona IT | IRCCS Sacro Cuore Don Calabria Hospital, Department of Infectious, Tropical Diseases & Microbiology | University of Verona, Department of Biotechnology | Antonio Mori et al |
| EPI_ISL_751338 | Italy/VEN-UniVR-17/2020 | 2020-04-20 | Europe/Italy/Veneto/Verona IT | IRCCS Sacro Cuore Don Calabria Hospital, Department of Infectious, Tropical Diseases & Microbiology | University of Verona, Department of Biotechnology | Antonio Mori et al |
| EPI_ISL_751343 | Italy/VEN-UniVR-22/2020 | 2020-08-13 | Europe/Italy/Veneto/Verona IT | IRCCS Sacro Cuore Don Calabria Hospital, Department of Infectious, Tropical Diseases & Microbiology | University of Verona, Department of Biotechnology | Antonio Mori et al |
| EPI_ISL_751347 | Italy/VEN-UniVR-35/2020 | 2020-03-07 | Europe/Italy/Veneto/Verona IT | IRCCS Sacro Cuore Don Calabria Hospital, Department of Infectious, Tropical Diseases & Microbiology | University of Verona, Department of Biotechnology | Antonio Mori et al |
| EPI_ISL_751351 | Italy/VEN-UniVR-39/2020 | 2020-03-10 | Europe/Italy/Veneto/Verona IT | IRCCS Sacro Cuore Don Calabria Hospital, Department of Infectious, Tropical Diseases & Microbiology | University of Verona, Department of Biotechnology | Antonio Mori et al |
| EPI_ISL_751352 | Italy/VEN-UniVR-40/2020 | 2020-03-10 | Europe/Italy/Veneto/Verona IT | IRCCS Sacro Cuore Don Calabria Hospital, Department of Infectious, Tropical Diseases & Microbiology | University of Verona, Department of Biotechnology | Antonio Mori et al |
| EPI_ISL_751355 | Italy/VEN-UniVR-44/2020 | 2020-03-12 | Europe/Italy/Veneto/Verona IT | IRCCS Sacro Cuore Don Calabria Hospital, Department of Infectious, Tropical Diseases & Microbiology | University of Verona, Department of Biotechnology | Antonio Mori et al |
| EPI_ISL_751361 | Italy/VEN-UniVR-51/2020 | 2020-03-12 | Europe/Italy/Veneto/Verona IT | IRCCS Sacro Cuore Don Calabria Hospital, Department of Infectious, Tropical Diseases & Microbiology | University of Verona, Department of Biotechnology | Antonio Mori et al |
| EPI_ISL_751362 | Italy/VEN-UniVR-52/2020 | 2020-03-12 | Europe/Italy/Veneto/Verona IT | IRCCS Sacro Cuore Don Calabria Hospital, Department of Infectious, Tropical Diseases & Microbiology | University of Verona, Department of Biotechnology | Antonio Mori et al |
| EPI_ISL_751363 | Italy/VEN-UniVR-53/2020 | 2020-03-12 | Europe/Italy/Veneto/Verona IT | IRCCS Sacro Cuore Don Calabria Hospital, Department of Infectious, Tropical Diseases & Microbiology | University of Verona, Department of Biotechnology | Antonio Mori et al |

| | | | | | | |
|---|---|---|---|---|---|---|
| EPI_ISL_751364 | Italy/VEN-UniVR-54/2020 | 2020-03-12 | Europe/Italy/Veneto/Verona IT | IRCCS Sacro Cuore Don Calabria Hospital, Department of Infectious, Tropical Diseases & Microbiology | University of Verona, Department of Biotechnology | Antonio Mori et al |
| EPI_ISL_751365 | Italy/VEN-UniVR-55/2020 | 2020-03-13 | Europe/Italy/Veneto/Verona IT | IRCCS Sacro Cuore Don Calabria Hospital, Department of Infectious, Tropical Diseases & Microbiology | University of Verona, Department of Biotechnology | Antonio Mori et al |
| EPI_ISL_751366 | Italy/VEN-UniVR-56/2020 | 2020-03-13 | Europe/Italy/Veneto/Verona IT | IRCCS Sacro Cuore Don Calabria Hospital, Department of Infectious, Tropical Diseases & Microbiology | University of Verona, Department of Biotechnology | Antonio Mori et al |
| EPI_ISL_751367 | Italy/VEN-UniVR-57/2020 | 2020-03-13 | Europe/Italy/Veneto/Verona IT | IRCCS Sacro Cuore Don Calabria Hospital, Department of Infectious, Tropical Diseases & Microbiology | University of Verona, Department of Biotechnology | Antonio Mori et al |
| EPI_ISL_751369 | Italy/VEN-UniVR-59/2020 | 2020-03-13 | Europe/Italy/Veneto/Verona IT | IRCCS Sacro Cuore Don Calabria Hospital, Department of Infectious, Tropical Diseases & Microbiology | University of Verona, Department of Biotechnology | Antonio Mori et al |
| EPI_ISL_751372 | Italy/VEN-UniVR-62/2020 | 2020-03-13 | Europe/Italy/Veneto/Verona IT | IRCCS Sacro Cuore Don Calabria Hospital, Department of Infectious, Tropical Diseases & Microbiology | University of Verona, Department of Biotechnology | Antonio Mori et al |
| EPI_ISL_751374 | Italy/VEN-UniVR-65/2020 | 2020-03-13 | Europe/Italy/Veneto/Verona IT | IRCCS Sacro Cuore Don Calabria Hospital, Department of Infectious, Tropical Diseases & Microbiology | University of Verona, Department of Biotechnology | Antonio Mori et al |
| EPI_ISL_751375 | Italy/VEN-UniVR-66/2020 | 2020-03-14 | Europe/Italy/Veneto/Verona IT | IRCCS Sacro Cuore Don Calabria Hospital, Department of Infectious, Tropical Diseases & Microbiology | University of Verona, Department of Biotechnology | Antonio Mori et al |
| EPI_ISL_751378 | Italy/VEN-UniVR-69/2020 | 2020-03-14 | Europe/Italy/Veneto/Verona IT | IRCCS Sacro Cuore Don Calabria Hospital, Department of Infectious, Tropical Diseases & Microbiology | University of Verona, Department of Biotechnology | Antonio Mori et al |
| EPI_ISL_751380 | Italy/VEN-UniVR-71/2020 | 2020-03-16 | Europe/Italy/Veneto/Verona IT | IRCCS Sacro Cuore Don Calabria Hospital, Department of Infectious, Tropical Diseases & Microbiology | University of Verona, Department of Biotechnology | Antonio Mori et al |

| | | | | | | |
|---|---|---|---|---|---|---|
| EPI_ISL_751382 | Italy/VEN-UniVR-73/2020 | 2020-03-19 | Europe/Italy/Veneto/Verona IT | IRCCS Sacro Cuore Don Calabria Hospital, Department of Infectious, Tropical Diseases & Microbiology | University of Verona, Department of Biotechnology | Antonio Mori et al |
| EPI_ISL_751383 | Italy/VEN-UniVR-74/2020 | 2020-03-20 | Europe/Italy/Veneto/Verona IT | IRCCS Sacro Cuore Don Calabria Hospital, Department of Infectious, Tropical Diseases & Microbiology | University of Verona, Department of Biotechnology | Antonio Mori et al |
| EPI_ISL_751384 | Italy/VEN-UniVR-75/2020 | 2020-03-20 | Europe/Italy/Veneto/Verona IT | IRCCS Sacro Cuore Don Calabria Hospital, Department of Infectious, Tropical Diseases & Microbiology | University of Verona, Department of Biotechnology | Antonio Mori et al |
| EPI_ISL_751395 | Italy/VEN-UniVR-87/2020 | 2020-03-23 | Europe/Italy/Veneto/Verona IT | IRCCS Sacro Cuore Don Calabria Hospital, Department of Infectious, Tropical Diseases & Microbiology | University of Verona, Department of Biotechnology | Antonio Mori et al |
| EPI_ISL_751397 | Italy/VEN-UniVR-89/2020 | 2020-03-24 | Europe/Italy/Veneto/Verona IT | IRCCS Sacro Cuore Don Calabria Hospital, Department of Infectious, Tropical Diseases & Microbiology | University of Verona, Department of Biotechnology | Antonio Mori et al |
| EPI_ISL_751400 | Italy/VEN-UniVR-92/2020 | 2020-03-24 | Europe/Italy/Veneto/Verona IT | IRCCS Sacro Cuore Don Calabria Hospital, Department of Infectious, Tropical Diseases & Microbiology | University of Verona, Department of Biotechnology | Antonio Mori et al |
| EPI_ISL_751402 | Italy/VEN-UniVR-94/2020 | 2020-03-25 | Europe/Italy/Veneto/Verona IT | IRCCS Sacro Cuore Don Calabria Hospital, Department of Infectious, Tropical Diseases & Microbiology | University of Verona, Department of Biotechnology | Antonio Mori et al |
| EPI_ISL_751403 | Italy/VEN-UniVR-95/2020 | 2020-03-25 | Europe/Italy/Veneto/Verona IT | IRCCS Sacro Cuore Don Calabria Hospital, Department of Infectious, Tropical Diseases & Microbiology | University of Verona, Department of Biotechnology | Antonio Mori et al |
| EPI_ISL_751407 | Italy/VEN-UniVR-101/2020 | 2020-03-25 | Europe/Italy/Veneto/Verona IT | IRCCS Sacro Cuore Don Calabria Hospital, Department of Infectious, Tropical Diseases & Microbiology | University of Verona, Department of Biotechnology | Antonio Mori et al |
| EPI_ISL_751410 | Italy/VEN-UniVR-105/2020 | 2020-03-26 | Europe/Italy/Veneto/Verona IT | IRCCS Sacro Cuore Don Calabria Hospital, Department of Infectious, Tropical Diseases & Microbiology | University of Verona, Department of Biotechnology | Antonio Mori et al |

| | | | | | | |
|---|---|---|---|---|---|---|
| EPI_ISL_751411 | Italy/VEN-UniVR-106/2020 | 2020-03-26 | Europe/Italy/Veneto/Verona IT | IRCCS Sacro Cuore Don Calabria Hospital, Department of Infectious, Tropical Diseases & Microbiology | University of Verona, Department of Biotechnology | Antonio Mori et al |
| EPI_ISL_751413 | Italy/VEN-UniVR-109/2020 | 2020-03-27 | Europe/Italy/Veneto/Verona IT | IRCCS Sacro Cuore Don Calabria Hospital, Department of Infectious, Tropical Diseases & Microbiology | University of Verona, Department of Biotechnology | Antonio Mori et al |
| EPI_ISL_751414 | Italy/VEN-UniVR-111/2020 | 2020-03-28 | Europe/Italy/Veneto/Verona IT | IRCCS Sacro Cuore Don Calabria Hospital, Department of Infectious, Tropical Diseases & Microbiology | University of Verona, Department of Biotechnology | Antonio Mori et al |
| EPI_ISL_751415 | Italy/VEN-UniVR-112/2020 | 2020-03-28 | Europe/Italy/Veneto/Verona IT | IRCCS Sacro Cuore Don Calabria Hospital, Department of Infectious, Tropical Diseases & Microbiology | University of Verona, Department of Biotechnology | Antonio Mori et al |
| EPI_ISL_751416 | Italy/VEN-UniVR-113/2020 | 2020-03-28 | Europe/Italy/Veneto/Verona IT | IRCCS Sacro Cuore Don Calabria Hospital, Department of Infectious, Tropical Diseases & Microbiology | University of Verona, Department of Biotechnology | Antonio Mori et al |
| EPI_ISL_751422 | Italy/VEN-UniVR-122/2020 | 2020-03-30 | Europe/Italy/Veneto/Verona IT | IRCCS Sacro Cuore Don Calabria Hospital, Department of Infectious, Tropical Diseases & Microbiology | University of Verona, Department of Biotechnology | Antonio Mori et al |
| EPI_ISL_751423 | Italy/VEN-UniVR-123/2020 | 2020-03-30 | Europe/Italy/Veneto/Verona IT | IRCCS Sacro Cuore Don Calabria Hospital, Department of Infectious, Tropical Diseases & Microbiology | University of Verona, Department of Biotechnology | Antonio Mori et al |
| EPI_ISL_751425 | Italy/VEN-UniVR-125/2020 | 2020-03-30 | Europe/Italy/Veneto/Verona IT | IRCCS Sacro Cuore Don Calabria Hospital, Department of Infectious, Tropical Diseases & Microbiology | University of Verona, Department of Biotechnology | Antonio Mori et al |
| EPI_ISL_751426 | Italy/VEN-UniVR-126/2020 | 2020-03-30 | Europe/Italy/Veneto/Verona IT | IRCCS Sacro Cuore Don Calabria Hospital, Department of Infectious, Tropical Diseases & Microbiology | University of Verona, Department of Biotechnology | Antonio Mori et al |
| EPI_ISL_751430 | Italy/VEN-UniVR-133/2020 | 2020-04-02 | Europe/Italy/Veneto/Verona IT | IRCCS Sacro Cuore Don Calabria Hospital, Department of Infectious, Tropical Diseases & Microbiology | University of Verona, Department of Biotechnology | Antonio Mori et al |

| | | | | | | |
|---|---|---|---|---|---|---|
| EPI_ISL_751431 | Italy/VEN-UniVR-134/2020 | 2020-04-02 | Europe/Italy/Veneto/Verona IT | IRCCS Sacro Cuore Don Calabria Hospital, Department of Infectious, Tropical Diseases & Microbiology | University of Verona, Department of Biotechnology | Antonio Mori et al |
| EPI_ISL_751432 | Italy/VEN-UniVR-135/2020 | 2020-04-02 | Europe/Italy/Veneto/Verona IT | IRCCS Sacro Cuore Don Calabria Hospital, Department of Infectious, Tropical Diseases & Microbiology | University of Verona, Department of Biotechnology | Antonio Mori et al |
| EPI_ISL_751435 | Italy/VEN-UniVR-138/2020 | 2020-04-04 | Europe/Italy/Veneto/Verona IT | IRCCS Sacro Cuore Don Calabria Hospital, Department of Infectious, Tropical Diseases & Microbiology | University of Verona, Department of Biotechnology | Antonio Mori et al |
| EPI_ISL_751436 | Italy/VEN-UniVR-139/2020 | 2020-04-04 | Europe/Italy/Veneto/Verona IT | IRCCS Sacro Cuore Don Calabria Hospital, Department of Infectious, Tropical Diseases & Microbiology | University of Verona, Department of Biotechnology | Antonio Mori et al |
| EPI_ISL_751438 | Italy/VEN-UniVR-141/2020 | 2020-04-05 | Europe/Italy/Veneto/Verona IT | IRCCS Sacro Cuore Don Calabria Hospital, Department of Infectious, Tropical Diseases & Microbiology | University of Verona, Department of Biotechnology | Antonio Mori et al |
| EPI_ISL_751439 | Italy/VEN-UniVR-142/2020 | 2020-04-05 | Europe/Italy/Veneto/Verona IT | IRCCS Sacro Cuore Don Calabria Hospital, Department of Infectious, Tropical Diseases & Microbiology | University of Verona, Department of Biotechnology | Antonio Mori et al |
| EPI_ISL_751440 | Italy/VEN-UniVR-143/2020 | 2020-04-06 | Europe/Italy/Veneto/Verona IT | IRCCS Sacro Cuore Don Calabria Hospital, Department of Infectious, Tropical Diseases & Microbiology | University of Verona, Department of Biotechnology | Antonio Mori et al |
| EPI_ISL_751444 | Italy/VEN-UniVR-147/2020 | 2020-04-09 | Europe/Italy/Veneto/Verona IT | IRCCS Sacro Cuore Don Calabria Hospital, Department of Infectious, Tropical Diseases & Microbiology | University of Verona, Department of Biotechnology | Antonio Mori et al |
| EPI_ISL_751448 | France/OCC-128/2020 | 2020-10-30 | Europe/France/Occitanie/ | CHU Purpan - Laboratoire de Virologie - Institut F√©d√©ratif de Biologie | CHU Purpan - Laboratoire de Virologie - Institut F√©d√©ratif de Biologie | Latour J. et al |

| | | | | | | |
|---|---|---|---|---|---|---|
| EPI_ISL_751449 | France/OCC-129/2020 | 2020-11-09 | Europe/France/Occitanie/ | CHU Purpan - Laboratoire de Virologie - Institut F√©d√©ratif de Biologie | CHU Purpan - Laboratoire de Virologie - Institut F√©d√©ratif de Biologie | Latour J. et al |
| EPI_ISL_751450 | France/OCC-130/2020 | 2020-11-02 | Europe/France/Occitanie/ | CHU Purpan - Laboratoire de Virologie - Institut F√©d√©ratif de Biologie | CHU Purpan - Laboratoire de Virologie - Institut F√©d√©ratif de Biologie | Latour J. et al |
| EPI_ISL_751451 | France/OCC-131/2020 | 2020-11-02 | Europe/France/Occitanie/ | CHU Purpan - Laboratoire de Virologie - Institut F√©d√©ratif de Biologie | CHU Purpan - Laboratoire de Virologie - Institut F√©d√©ratif de Biologie | Latour J. et al |
| EPI_ISL_751452 | France/OCC-132/2020 | 2020-11-02 | Europe/France/Occitanie/ | CHU Purpan - Laboratoire de Virologie - Institut F√©d√©ratif de Biologie | CHU Purpan - Laboratoire de Virologie - Institut F√©d√©ratif de Biologie | Latour J. et al |
| EPI_ISL_751453 | France/OCC-133/2020 | 2020-11-02 | Europe/France/Occitanie/ | CHU Purpan - Laboratoire de Virologie - Institut F√©d√©ratif de Biologie | CHU Purpan - Laboratoire de Virologie - Institut F√©d√©ratif de Biologie | Latour J. et al |
| EPI_ISL_751454 | France/OCC-134/2020 | 2020-11-18 | Europe/France/Occitanie/ | CHU Purpan - Laboratoire de Virologie - Institut F√©d√©ratif de Biologie | CHU Purpan - Laboratoire de Virologie - Institut F√©d√©ratif de Biologie | Latour J. et al |
| EPI_ISL_751455 | France/OCC-135/2020 | 2020-11-16 | Europe/France/Occitanie/ | CHU Purpan - Laboratoire de Virologie - Institut F√©d√©ratif de Biologie | CHU Purpan - Laboratoire de Virologie - Institut F√©d√©ratif de Biologie | Latour J. et al |
| EPI_ISL_751456 | France/OCC-136/2020 | 2020-11-16 | Europe/France/Occitanie/ | CHU Purpan - Laboratoire de Virologie - Institut F√©d√©ratif de Biologie | CHU Purpan - Laboratoire de Virologie - Institut F√©d√©ratif de Biologie | Latour J. et al |

| | | | | | | |
|---|---|---|---|---|---|---|
| EPI_ISL_751458 | France/OCC-138/2020 | 2020-11-23 | Europe/France/Occitanie/ | CHU Purpan - Laboratoire de Virologie - Institut F√©d√©ratif de Biologie | CHU Purpan - Laboratoire de Virologie - Institut F√©d√©ratif de Biologie | Latour J. et al |
| EPI_ISL_751459 | France/OCC-139/2020 | 2020-11-23 | Europe/France/Occitanie/ | CHU Purpan - Laboratoire de Virologie - Institut F√©d√©ratif de Biologie | CHU Purpan - Laboratoire de Virologie - Institut F√©d√©ratif de Biologie | Latour J. et al |
| EPI_ISL_751460 | France/OCC-140/2020 | 2020-11-23 | Europe/France/Occitanie/ | CHU Purpan - Laboratoire de Virologie - Institut F√©d√©ratif de Biologie | CHU Purpan - Laboratoire de Virologie - Institut F√©d√©ratif de Biologie | Latour J. et al |
| EPI_ISL_751463 | France/OCC-143/2020 | 2020-11-18 | Europe/France/Occitanie/ | CHU Purpan - Laboratoire de Virologie - Institut F√©d√©ratif de Biologie | CHU Purpan - Laboratoire de Virologie - Institut F√©d√©ratif de Biologie | Latour J. et al |
| EPI_ISL_751464 | France/OCC-144/2020 | 2020-11-20 | Europe/France/Occitanie/ | CHU Purpan - Laboratoire de Virologie - Institut F√©d√©ratif de Biologie | CHU Purpan - Laboratoire de Virologie - Institut F√©d√©ratif de Biologie | Latour J. et al |
| EPI_ISL_751465 | France/OCC-145/2020 | 2020-11-16 | Europe/France/Occitanie/ | CHU Purpan - Laboratoire de Virologie - Institut F√©d√©ratif de Biologie | CHU Purpan - Laboratoire de Virologie - Institut F√©d√©ratif de Biologie | Latour J. et al |
| EPI_ISL_751466 | France/OCC-146/2020 | 2020-11-13 | Europe/France/Occitanie/ | CHU Purpan - Laboratoire de Virologie - Institut F√©d√©ratif de Biologie | CHU Purpan - Laboratoire de Virologie - Institut F√©d√©ratif de Biologie | Latour J. et al |
| EPI_ISL_751467 | France/OCC-147/2020 | 2020-11-12 | Europe/France/Occitanie/ | CHU Purpan - Laboratoire de Virologie - Institut F√©d√©ratif de Biologie | CHU Purpan - Laboratoire de Virologie - Institut F√©d√©ratif de Biologie | Latour J. et al |

| | | | | | | |
|---|---|---|---|---|---|---|
| EPI_ISL_751468 | France/OCC-148/2020 | 2020-11-12 | Europe/France/Occitanie/ | CHU Purpan - Laboratoire de Virologie - Institut F√©d√©ratif de Biologie | CHU Purpan - Laboratoire de Virologie - Institut F√©d√©ratif de Biologie | Latour J. et al |
| EPI_ISL_751469 | France/OCC-149/2020 | 2020-11-14 | Europe/France/Occitanie/ | CHU Purpan - Laboratoire de Virologie - Institut F√©d√©ratif de Biologie | CHU Purpan - Laboratoire de Virologie - Institut F√©d√©ratif de Biologie | Latour J. et al |
| EPI_ISL_751471 | France/OCC-151/2020 | 2020-11-15 | Europe/France/Occitanie/ | CHU Purpan - Laboratoire de Virologie - Institut F√©d√©ratif de Biologie | CHU Purpan - Laboratoire de Virologie - Institut F√©d√©ratif de Biologie | Latour J. et al |
| EPI_ISL_751473 | France/OCC-153/2020 | 2020-11-21 | Europe/France/Occitanie/ | CHU Purpan - Laboratoire de Virologie - Institut F√©d√©ratif de Biologie | CHU Purpan - Laboratoire de Virologie - Institut F√©d√©ratif de Biologie | Latour J. et al |
| EPI_ISL_751474 | France/OCC-154/2020 | 2020-11-18 | Europe/France/Occitanie/ | CHU Purpan - Laboratoire de Virologie - Institut F√©d√©ratif de Biologie | CHU Purpan - Laboratoire de Virologie - Institut F√©d√©ratif de Biologie | Latour J. et al |
| EPI_ISL_751475 | France/OCC-155/2020 | 2020-11-18 | Europe/France/Occitanie/ | CHU Purpan - Laboratoire de Virologie - Institut F√©d√©ratif de Biologie | CHU Purpan - Laboratoire de Virologie - Institut F√©d√©ratif de Biologie | Latour J. et al |
| EPI_ISL_751477 | France/OCC-158/2020 | 2020-10-13 | Europe/France/Occitanie/ | CHU Purpan - Laboratoire de Virologie - Institut F√©d√©ratif de Biologie | CHU Purpan - Laboratoire de Virologie - Institut F√©d√©ratif de Biologie | Latour J. et al |
| EPI_ISL_751479 | France/OCC-160/2020 | 2020-10-23 | Europe/France/Occitanie/ | CHU Purpan - Laboratoire de Virologie - Institut F√©d√©ratif de Biologie | CHU Purpan - Laboratoire de Virologie - Institut F√©d√©ratif de Biologie | Latour J. et al |

| | | | | | | |
|---|---|---|---|---|---|---|
| EPI_ISL_751480 | France/OCC-161/2020 | 2020-10-09 | Europe/France/Occitanie/ | CHU Purpan - Laboratoire de Virologie - Institut F√©d√©ratif de Biologie | CHU Purpan - Laboratoire de Virologie - Institut F√©d√©ratif de Biologie | Latour J. et al |
| EPI_ISL_751481 | France/OCC-162/2020 | 2020-10-23 | Europe/France/Occitanie/ | CHU Purpan - Laboratoire de Virologie - Institut F√©d√©ratif de Biologie | CHU Purpan - Laboratoire de Virologie - Institut F√©d√©ratif de Biologie | Latour J. et al |
| EPI_ISL_751483 | France/OCC-164/2020 | 2020-10-29 | Europe/France/Occitanie/ | CHU Purpan - Laboratoire de Virologie - Institut F√©d√©ratif de Biologie | CHU Purpan - Laboratoire de Virologie - Institut F√©d√©ratif de Biologie | Latour J. et al |
| EPI_ISL_751484 | France/OCC-165/2020 | 2020-10-29 | Europe/France/Occitanie/ | CHU Purpan - Laboratoire de Virologie - Institut F√©d√©ratif de Biologie | CHU Purpan - Laboratoire de Virologie - Institut F√©d√©ratif de Biologie | Latour J. et al |
| EPI_ISL_751485 | France/OCC-166/2020 | 2020-10-29 | Europe/France/Occitanie/ | CHU Purpan - Laboratoire de Virologie - Institut F√©d√©ratif de Biologie | CHU Purpan - Laboratoire de Virologie - Institut F√©d√©ratif de Biologie | Latour J. et al |
| EPI_ISL_751487 | France/OCC-168/2020 | 2020-10-16 | Europe/France/Occitanie/ | CHU Purpan - Laboratoire de Virologie - Institut F√©d√©ratif de Biologie | CHU Purpan - Laboratoire de Virologie - Institut F√©d√©ratif de Biologie | Latour J. et al |
| EPI_ISL_751489 | France/OCC-170/2020 | 2020-10-24 | Europe/France/Occitanie/ | CHU Purpan - Laboratoire de Virologie - Institut F√©d√©ratif de Biologie | CHU Purpan - Laboratoire de Virologie - Institut F√©d√©ratif de Biologie | Latour J. et al |
| EPI_ISL_751490 | France/OCC-171/2020 | 2020-10-30 | Europe/France/Occitanie/ | CHU Purpan - Laboratoire de Virologie - Institut F√©d√©ratif de Biologie | CHU Purpan - Laboratoire de Virologie - Institut F√©d√©ratif de Biologie | Latour J. et al |

| | | | | | | |
|---|---|---|---|---|---|---|
| EPI_ISL_751491 | France/OCC-172/2020 | 2020-11-10 | Europe/France/Occitanie/ | CHU Purpan - Laboratoire de Virologie - Institut F√©d√©ratif de Biologie | CHU Purpan - Laboratoire de Virologie - Institut F√©d√©ratif de Biologie | Latour J. et al |
| EPI_ISL_751492 | France/OCC-173/2020 | 2020-11-10 | Europe/France/Occitanie/ | CHU Purpan - Laboratoire de Virologie - Institut F√©d√©ratif de Biologie | CHU Purpan - Laboratoire de Virologie - Institut F√©d√©ratif de Biologie | Latour J. et al |
| EPI_ISL_751495 | France/OCC-177/2020 | 2020-10-05 | Europe/France/Occitanie/ | CHU Purpan - Laboratoire de Virologie - Institut F√©d√©ratif de Biologie | CHU Purpan - Laboratoire de Virologie - Institut F√©d√©ratif de Biologie | Latour J. et al |
| EPI_ISL_751496 | France/OCC-178/2020 | 2020-10-09 | Europe/France/Occitanie/ | CHU Purpan - Laboratoire de Virologie - Institut F√©d√©ratif de Biologie | CHU Purpan - Laboratoire de Virologie - Institut F√©d√©ratif de Biologie | Latour J. et al |
| EPI_ISL_751497 | France/OCC-179/2020 | 2020-11-14 | Europe/France/Occitanie/ | CHU Purpan - Laboratoire de Virologie - Institut F√©d√©ratif de Biologie | CHU Purpan - Laboratoire de Virologie - Institut F√©d√©ratif de Biologie | Latour J. et al |
| EPI_ISL_751498 | France/OCC-180/2020 | 2020-11-02 | Europe/France/Occitanie/ | CHU Purpan - Laboratoire de Virologie - Institut F√©d√©ratif de Biologie | CHU Purpan - Laboratoire de Virologie - Institut F√©d√©ratif de Biologie | Latour J. et al |
| EPI_ISL_751499 | France/OCC-181/2020 | 2020-10-01 | Europe/France/Occitanie/ | CHU Purpan - Laboratoire de Virologie - Institut F√©d√©ratif de Biologie | CHU Purpan - Laboratoire de Virologie - Institut F√©d√©ratif de Biologie | Latour J. et al |

| | | | | | | |
|---|---|---|---|---|---|---|
| EPI_ISL_751500 | SaudiArabia/KAUST-MADINAH1496/ | 2020-07-17 | Asia/Saudi Arabia/Madinah/ | Pathogen Genomics Lab King Abdullah University of Science and Technology(KAUST) | Pathogen Genomics Lab King Abdullah University of Science and Technology(KAUST) | Sara Mfarrej et al |
| EPI_ISL_751501 | SaudiArabia/KAUST-MADINAH1688/ | 2020-08-23 | Asia/Saudi Arabia/Madinah/ | Pathogen Genomics Lab King Abdullah University of Science and Technology(KAUST) | Pathogen Genomics Lab King Abdullah University of Science and Technology(KAUST) | Sara Mfarrej et al |
| EPI_ISL_751504 | USA/TX-DSHS-1893/2020 | 2020-07-08 | North America/USA/Texas/Harris Count | Texas Department of State Health Services | Texas Department of State Health Services | Rashmi Tuladhar et al |
| EPI_ISL_751506 | USA/TX-DSHS-1895/2020 | 2020-07-08 | North America/USA/Texas/Harris Count | Texas Department of State Health Services | Texas Department of State Health Services | Rashmi Tuladhar et al |
| EPI_ISL_751510 | USA/TX-DSHS-1908/2020 | 2020-07-11 | North America/USA/Texas/Dallas Count | Texas Department of State Health Services | Texas Department of State Health Services | Rashmi Tuladhar et al |
| EPI_ISL_751513 | USA/TX-DSHS-1894/2020 | 2020-07-08 | North America/USA/Texas/Harris Count | Texas Department of State Health Services | Texas Department of State Health Services | Rashmi Tuladhar et al |
| EPI_ISL_751527 | USA/TX-DSHS-1915/2020 | 2020-07-11 | North America/USA/Texas/Dallas Count | Texas Department of State Health Services | Texas Department of State Health Services | Rashmi Tuladhar et al |
| EPI_ISL_751547 | USA/DE-CDC-2-3714057/2020 | 2020-10-22 | North America/USA/Delaware/ | DE Public Health Laboratory | Genomics and Discovery, Respiratory Viruses Branch, Division of Viral Diseases, Centers for Disease Control and Prevention | Krista Queen et al |

| | | | | | | |
|---|---|---|---|---|---|---|
| EPI_ISL_751558 | USA/NH-CDC-2-3714170/2020 | 2020-11-16 | North America/USA/New Hampshire/ | NH Dept. of Health and Human Services Public Health Labs | Genomics and Discovery, Respiratory Viruses Branch, Division of Viral Diseases, Centers for Disease Control and Prevention | Krista Queen et al |
| EPI_ISL_751567 | USA/ID-CDC-2-3714453/2020 | 2020-11-15 | North America/USA/Idaho/ | ID Bureau of Laboratories | Genomics and Discovery, Respiratory Viruses Branch, Division of Viral Diseases, Centers for Disease Control and Prevention | Krista Queen et al |
| EPI_ISL_751574 | USA/AR-CDC-2-3713957/2020 | 2020-09-02 | North America/USA/Arkansas/ | AR Dept. of Health-Public Health Lab | Genomics and Discovery, Respiratory Viruses Branch, Division of Viral Diseases, Centers for Disease Control and Prevention | Krista Queen et al |
| EPI_ISL_751582 | USA/NH-CDC-2-3714179/2020 | 2020-11-06 | North America/USA/New Hampshire/ | NH Dept. of Health and Human Services Public Health Labs | Genomics and Discovery, Respiratory Viruses Branch, Division of Viral Diseases, Centers for Disease Control and Prevention | Krista Queen et al |

| | | | | | | |
|---|---|---|---|---|---|---|
| EPI_ISL_751588 | USA/VA-CDC-2-3714379/2020 | 2020-11-18 | North America/USA/Virginia/ | VA-Division of Consolidated Laboratory Services | Genomics and Discovery, Respiratory Viruses Branch, Division of Viral Diseases, Centers for Disease Control and Prevention | Krista Queen et al |
| EPI_ISL_751590 | USA/ID-CDC-2-3714455/2020 | 2020-11-18 | North America/USA/Idaho/ | ID Bureau of Laboratories | Genomics and Discovery, Respiratory Viruses Branch, Division of Viral Diseases, Centers for Disease Control and Prevention | Krista Queen et al |
| EPI_ISL_751592 | USA/AK-CDC-2-3714493/2020 | 2020-11-17 | North America/USA/Alaska/ | AK State Public Health Lab, State Health Department | Genomics and Discovery, Respiratory Viruses Branch, Division of Viral Diseases, Centers for Disease Control and Prevention | Krista Queen et al |
| EPI_ISL_751594 | USA/KY-CDC-2-3714554/2020 | 2020-11-27 | North America/USA/Kentucky/ | KY State Public Health Lab | Genomics and Discovery, Respiratory Viruses Branch, Division of Viral Diseases, Centers for Disease Control and Prevention | Krista Queen et al |

| | | | | | | |
|---|---|---|---|---|---|---|
| EPI_ISL_751597 | USA/AK-CDC-2-3714579/2020 | 2020-11-15 | North America/USA/Alaska/ | AK State Public Health Lab, State Health Department | Genomics and Discovery, Respiratory Viruses Branch, Division of Viral Diseases, Centers for Disease Control and Prevention | Krista Queen et al |
| EPI_ISL_751599 | USA/IN-CDC-2-3713959/2020 | 2020-12-04 | North America/USA/Indiana/ | IN State Department of Health Laboratory Services | Genomics and Discovery, Respiratory Viruses Branch, Division of Viral Diseases, Centers for Disease Control and Prevention | Krista Queen et al |
| EPI_ISL_751604 | USA/MD-CDC-2-3713976/2020 | 2020-12-03 | North America/USA/Maryland/ | MD DOH Laboratories Administration | Genomics and Discovery, Respiratory Viruses Branch, Division of Viral Diseases, Centers for Disease Control and Prevention | Krista Queen et al |
| EPI_ISL_751605 | USA/DE-CDC-2-3713979/2020 | 2020-10-28 | North America/USA/Delaware/ | DE Public Health Laboratory | Genomics and Discovery, Respiratory Viruses Branch, Division of Viral Diseases, Centers for Disease Control and Prevention | Krista Queen et al |

| | | | | | | |
|---|---|---|---|---|---|---|
| EPI_ISL_751606 | USA/MD-CDC-2-3713984/2020 | 2020-11-22 | North America/USA/Maryland/ | MD DOH Laboratories Administration | Genomics and Discovery, Respiratory Viruses Branch, Division of Viral Diseases, Centers for Disease Control and Prevention | Krista Queen et al |
| EPI_ISL_751619 | USA/WY-CDC-2-3714478/2020 | 2020-12-05 | North America/USA/Wyoming/ | WY Public Health Laboratory | Genomics and Discovery, Respiratory Viruses Branch, Division of Viral Diseases, Centers for Disease Control and Prevention | Krista Queen et al |
| EPI_ISL_751621 | USA/DE-CDC-2-3713992/2020 | 2020-10-28 | North America/USA/Delaware/ | DE Public Health Laboratory | Genomics and Discovery, Respiratory Viruses Branch, Division of Viral Diseases, Centers for Disease Control and Prevention | Krista Queen et al |
| EPI_ISL_751628 | USA/MD-CDC-2-3714081/2020 | 2020-12-03 | North America/USA/Maryland/ | MD DOH Laboratories Administration | Genomics and Discovery, Respiratory Viruses Branch, Division of Viral Diseases, Centers for Disease Control and Prevention | Krista Queen et al |

| | | | | | | |
|---|---|---|---|---|---|---|
| EPI_ISL_751631 | USA/DE-CDC-2-3714090/2020 | 2020-10-27 | North America/USA/Delaware/ | DE Public Health Laboratory | Genomics and Discovery, Respiratory Viruses Branch, Division of Viral Diseases, Centers for Disease Control and Prevention | Krista Queen et al |
| EPI_ISL_751641 | USA/MN-CDC-2-3714282/2020 | 2020-11-18 | North America/USA/Minnesota/ | MN PHL Division, Minnesota Department of Health | Genomics and Discovery, Respiratory Viruses Branch, Division of Viral Diseases, Centers for Disease Control and Prevention | Krista Queen et al |
| EPI_ISL_751643 | USA/WY-CDC-2-3714354/2020 | 2020-12-05 | North America/USA/Wyoming/ | WY Public Health Laboratory | Genomics and Discovery, Respiratory Viruses Branch, Division of Viral Diseases, Centers for Disease Control and Prevention | Krista Queen et al |
| EPI_ISL_751649 | USA/CT-CDC-2-3713968/2020 | 2020-11-20 | North America/USA/Connecticut/ | CT-Dr. Katherine A. Kelley State Public Health Lab | Genomics and Discovery, Respiratory Viruses Branch, Division of Viral Diseases, Centers for Disease Control and Prevention | Krista Queen et al |

| | | | | | | |
|---|---|---|---|---|---|---|
| EPI_ISL_751652 | USA/AR-CDC-2-3713986/2020 | 2020-09-02 | North America/USA/Arkansas/ | AR Dept. of Health-Public Health Lab | Genomics and Discovery, Respiratory Viruses Branch, Division of Viral Diseases, Centers for Disease Control and Prevention | Krista Queen et al |
| EPI_ISL_751654 | USA/MN-CDC-2-3714089/2020 | 2020-11-09 | North America/USA/Minnesota/ | MN PHL Division, Minnesota Department of Health | Genomics and Discovery, Respiratory Viruses Branch, Division of Viral Diseases, Centers for Disease Control and Prevention | Krista Queen et al |
| EPI_ISL_751668 | USA/AL-CDC-2-3714551/2020 | 2020-11-04 | North America/USA/Alabama/ | AL Dept. of Public Health Bureau of Clinical Laboratories | Genomics and Discovery, Respiratory Viruses Branch, Division of Viral Diseases, Centers for Disease Control and Prevention | Krista Queen et al |
| EPI_ISL_751670 | USA/KY-CDC-2-3714580/2020 | 2020-12-03 | North America/USA/Kentucky/ | KY State Public Health Lab | Genomics and Discovery, Respiratory Viruses Branch, Division of Viral Diseases, Centers for Disease Control and Prevention | Krista Queen et al |

| | | | | | | |
|---|---|---|---|---|---|---|
| EPI_ISL_751671 | USA/RI-CDC-2-3714588/2020 | 2020-12-03 | North America/USA/Rhode Island/ | RI State Health Laboratories | Genomics and Discovery, Respiratory Viruses Branch, Division of Viral Diseases, Centers for Disease Control and Prevention | Krista Queen et al |
| EPI_ISL_751685 | USA/VA-CDC-2-3714292/2020 | 2020-11-09 | North America/USA/Virginia/ | VA-Division of Consolidated Laboratory Services | Genomics and Discovery, Respiratory Viruses Branch, Division of Viral Diseases, Centers for Disease Control and Prevention | Krista Queen et al |
| EPI_ISL_751687 | USA/IA-CDC-2-3714371/2020 | 2020-11-29 | North America/USA/Iowa/ | IA State Hygienic Laboratory | Genomics and Discovery, Respiratory Viruses Branch, Division of Viral Diseases, Centers for Disease Control and Prevention | Krista Queen et al |
| EPI_ISL_751691 | USA/WY-CDC-2-3714476/2020 | 2020-12-04 | North America/USA/Wyoming/ | WY Public Health Laboratory | Genomics and Discovery, Respiratory Viruses Branch, Division of Viral Diseases, Centers for Disease Control and Prevention | Krista Queen et al |

| | | | | | | |
|---|---|---|---|---|---|---|
| EPI_ISL_751694 | USA/AK-CDC-2-3714492/2020 | 2020-11-16 | North America/USA/Alaska/ | AK State Public Health Lab, State Health Department | Genomics and Discovery, Respiratory Viruses Branch, Division of Viral Diseases, Centers for Disease Control and Prevention | Krista Queen et al |
| EPI_ISL_751697 | USA/AR-CDC-2-3713953/2020 | 2020-09-01 | North America/USA/Arkansas/ | AR Dept. of Health-Public Health Lab | Genomics and Discovery, Respiratory Viruses Branch, Division of Viral Diseases, Centers for Disease Control and Prevention | Krista Queen et al |
| EPI_ISL_751698 | USA/AR-CDC-2-3713965/2020 | 2020-09-02 | North America/USA/Arkansas/ | AR Dept. of Health-Public Health Lab | Genomics and Discovery, Respiratory Viruses Branch, Division of Viral Diseases, Centers for Disease Control and Prevention | Krista Queen et al |
| EPI_ISL_751699 | USA/AR-CDC-2-3713967/2020 | 2020-09-02 | North America/USA/Arkansas/ | AR Dept. of Health-Public Health Lab | Genomics and Discovery, Respiratory Viruses Branch, Division of Viral Diseases, Centers for Disease Control and Prevention | Krista Queen et al |

| | | | | | | |
|---|---|---|---|---|---|---|
| EPI_ISL_751709 | USA/NJ-CDC-2-3714156/2020 | 2020-11-21 | North America/USA/New Jersey/ | NJ Public Health and Environmental Laboratories | Genomics and Discovery, Respiratory Viruses Branch, Division of Viral Diseases, Centers for Disease Control and Prevention | Krista Queen et al |
| EPI_ISL_751718 | USA/SC-CDC-2-3714351/2020 | 2020-12-02 | North America/USA/South Carolina/ | SC Dept of Health and Env. Control-Bureau of Laboratories | Genomics and Discovery, Respiratory Viruses Branch, Division of Viral Diseases, Centers for Disease Control and Prevention | Krista Queen et al |
| EPI_ISL_751719 | USA/ID-CDC-2-3714381/2020 | 2020-11-10 | North America/USA/Idaho/ | ID Bureau of Laboratories | Genomics and Discovery, Respiratory Viruses Branch, Division of Viral Diseases, Centers for Disease Control and Prevention | Krista Queen et al |
| EPI_ISL_751724 | USA/KS-CDC-2-3713958/2020 | 2020-11-02 | North America/USA/Kansas/ | KS Health and Environmental Laboratories | Genomics and Discovery, Respiratory Viruses Branch, Division of Viral Diseases, Centers for Disease Control and Prevention | Krista Queen et al |

| | | | | | | |
|---|---|---|---|---|---|---|
| EPI_ISL_751726 | USA/DE-CDC-2-3713996/2020 | 2020-10-21 | North America/USA/Delaware/ | DE Public Health Laboratory | Genomics and Discovery, Respiratory Viruses Branch, Division of Viral Diseases, Centers for Disease Control and Prevention | Krista Queen et al |
| EPI_ISL_751746 | USA/WV-CDC-2-3714397/2020 | 2020-12-01 | North America/USA/West Virginia/ | WVDHHR - Office of Laboratory Services | Genomics and Discovery, Respiratory Viruses Branch, Division of Viral Diseases, Centers for Disease Control and Prevention | Krista Queen et al |
| EPI_ISL_751754 | USA/KY-CDC-2-3714582/2020 | 2020-12-03 | North America/USA/Kentucky/ | KY State Public Health Lab | Genomics and Discovery, Respiratory Viruses Branch, Division of Viral Diseases, Centers for Disease Control and Prevention | Krista Queen et al |
| EPI_ISL_751760 | USA/AR-CDC-2-3713966/2020 | 2020-09-02 | North America/USA/Arkansas/ | AR Dept. of Health-Public Health Lab | Genomics and Discovery, Respiratory Viruses Branch, Division of Viral Diseases, Centers for Disease Control and Prevention | Krista Queen et al |

| | | | | | | |
|---|---|---|---|---|---|---|
| EPI_ISL_751772 | USA/HI-CDC-2-3714093/2020 | 2020-11-30 | North America/USA/Hawaii/ | HI Dept. of Health, State Laboratories Division | Genomics and Discovery, Respiratory Viruses Branch, Division of Viral Diseases, Centers for Disease Control and Prevention | Krista Queen et al |
| EPI_ISL_751782 | USA/ID-CDC-2-3714366/2020 | 2020-11-18 | North America/USA/Idaho/ | ID Bureau of Laboratories | Genomics and Discovery, Respiratory Viruses Branch, Division of Viral Diseases, Centers for Disease Control and Prevention | Krista Queen et al |
| EPI_ISL_751792 | USA/AL-CDC-2-3714557/2020 | 2020-11-04 | North America/USA/Alabama/ | AL Dept. of Public Health Bureau of Clinical Laboratories | Genomics and Discovery, Respiratory Viruses Branch, Division of Viral Diseases, Centers for Disease Control and Prevention | Krista Queen et al |
| EPI_ISL_751795 | USA/RI-CDC-2-3714578/2020 | 2020-12-04 | North America/USA/Rhode Island/ | RI State Health Laboratories | Genomics and Discovery, Respiratory Viruses Branch, Division of Viral Diseases, Centers for Disease Control and Prevention | Krista Queen et al |
| EPI_ISL_751796 | Canada/ON-PHL-20-04108/2020 | 20-Dec | North America/Canada/Ontario/ | Public Health Ontario Laboratory | Public Health Ontario Laboratory | Vanessa G Allen et al |

| | | | | | | |
|---|---|---|---|---|---|---|
| EPI_ISL_751797 | Canada/ON-PHL-20-04109/2020 | 20-Dec | North America/Canada/Ontario/ | Public Health Ontario Laboratory | Public Health Ontario Laboratory | Vanessa G Allen et al |
| EPI_ISL_751798 | Canada/ON-PHL-20-04116/2020 | 20-Dec | North America/Canada/Ontario/ | Public Health Ontario Laboratory | Public Health Ontario Laboratory | Vanessa G Allen et al |
| EPI_ISL_751804 | USA/NM-DOH-2020617125/2020 | 2020-12-12 | North America/USA/New Mexico/ | New Mexico Department of Health Scientific Laboratory | New Mexico Department of Health Scientific Laboratory | D'eldra Malone et al |
| EPI_ISL_751807 | USA/NM-DOH-2020617002/2020 | 2020-12-12 | North America/USA/New Mexico/ | New Mexico Department of Health Scientific Laboratory | New Mexico Department of Health Scientific Laboratory | D'eldra Malone et al |
| EPI_ISL_751810 | USA/NM-DOH-2020616233/2020 | 2020-12-12 | North America/USA/New Mexico/ | New Mexico Department of Health Scientific Laboratory | New Mexico Department of Health Scientific Laboratory | D'eldra Malone et al |
| EPI_ISL_751813 | USA/NM-DOH-2020616318/2020 | 2020-12-12 | North America/USA/New Mexico/ | New Mexico Department of Health Scientific Laboratory | New Mexico Department of Health Scientific Laboratory | D'eldra Malone et al |
| EPI_ISL_751816 | USA/NM-DOH-2020616323/2020 | 2020-12-12 | North America/USA/New Mexico/ | New Mexico Department of Health Scientific Laboratory | New Mexico Department of Health Scientific Laboratory | D'eldra Malone et al |
| EPI_ISL_751827 | USA/NM-DOH-2020616689/2020 | 2020-12-12 | North America/USA/New Mexico/ | New Mexico Department of Health Scientific Laboratory | New Mexico Department of Health Scientific Laboratory | D'eldra Malone et al |
| EPI_ISL_751828 | USA/NM-DOH-2020616696/2020 | 2020-12-12 | North America/USA/New Mexico/ | New Mexico Department of Health Scientific Laboratory | New Mexico Department of Health Scientific Laboratory | D'eldra Malone et al |
| EPI_ISL_751837 | USA/NM-DOH-2020617022/2020 | 2020-12-12 | North America/USA/New Mexico/ | New Mexico Department of Health Scientific Laboratory | New Mexico Department of Health Scientific Laboratory | D'eldra Malone et al |

| | | | | | | |
|---|---|---|---|---|---|---|
| EPI_ISL_751839 | USA/NM-DOH-2020617051/2020 | 2020-12-12 | North America/USA/New Mexico/ | New Mexico Department of Health Scientific Laboratory | New Mexico Department of Health Scientific Laboratory | D'eldra Malone et al |
| EPI_ISL_751840 | USA/NM-DOH-2020617070/2020 | 2020-12-12 | North America/USA/New Mexico/ | New Mexico Department of Health Scientific Laboratory | New Mexico Department of Health Scientific Laboratory | D'eldra Malone et al |
| EPI_ISL_751843 | USA/NM-DOH-2020617085/2020 | 2020-12-12 | North America/USA/New Mexico/ | New Mexico Department of Health Scientific Laboratory | New Mexico Department of Health Scientific Laboratory | D'eldra Malone et al |
| EPI_ISL_751845 | USA/NM-DOH-2020617173/2020 | 2020-12-11 | North America/USA/New Mexico/ | New Mexico Department of Health Scientific Laboratory | New Mexico Department of Health Scientific Laboratory | D'eldra Malone et al |
| EPI_ISL_752522 | Ireland/MH-NVRL-77IRL72666/2020 | 2020-11-05 | Europe/Ireland/Meath/ | National Virus Reference Laboratory | National Virus Reference Laboratory | Michael Carr et al |
| EPI_ISL_752567 | Ireland/G-NVRL-78IRL74237/2020 | 2020-11-23 | Europe/Ireland/Galway/ | National Virus Reference Laboratory | National Virus Reference Laboratory | Michael Carr et al |
| EPI_ISL_752613 | USA/HI-H200019/2020 | 2020-04-20 | North America/USA/Hawaii/Hawaii Cou | State Laboratories Division, Hawaii State Department of Health | State Laboratories Division, Hawaii State Department of Health | Pamela O'Brien et al |
| EPI_ISL_752616 | USA/HI-H200030/2020 | 2020-03-19 | North America/USA/Hawaii/Honolulu C | State Laboratories Division, Hawaii State Department of Health | State Laboratories Division, Hawaii State Department of Health | Pamela O'Brien et al |
| EPI_ISL_752623 | USA/HI-H200039/2020 | 2020-04-15 | North America/USA/Hawaii/Maui Count | State Laboratories Division, Hawaii State Department of Health | State Laboratories Division, Hawaii State Department of Health | Pamela O'Brien et al |

| | | | | | | |
|---|---|---|---|---|---|---|
| EPI_ISL_752655 | USA/HI-H200088/2020 | 2020-06-18 | North America/USA/Hawaii/Honolulu C | State Laboratories Division, Hawaii State Department of Health | State Laboratories Division, Hawaii State Department of Health | Pamela O'Brien et al |
| EPI_ISL_752658 | USA/HI-H200091/2020 | 2020-06-21 | North America/USA/Hawaii/ | State Laboratories Division, Hawaii State Department of Health | State Laboratories Division, Hawaii State Department of Health | Pamela O'Brien et al |
| EPI_ISL_752660 | USA/HI-H200093/2020 | 2020-03-28 | North America/USA/Hawaii/ | State Laboratories Division, Hawaii State Department of Health | State Laboratories Division, Hawaii State Department of Health | Pamela O'Brien et al |
| EPI_ISL_752663 | USA/HI-H200099/2020 | 2020-05-10 | North America/USA/Hawaii/ | State Laboratories Division, Hawaii State Department of Health | State Laboratories Division, Hawaii State Department of Health | Pamela O'Brien et al |
| EPI_ISL_752664 | USA/HI-H200100/2020 | 2020-06-19 | North America/USA/Hawaii/Kauai Coun | State Laboratories Division, Hawaii State Department of Health | State Laboratories Division, Hawaii State Department of Health | Pamela O'Brien et al |
| EPI_ISL_752665 | USA/HI-H200101/2020 | 2020-06-17 | North America/USA/Hawaii/Kauai Coun | State Laboratories Division, Hawaii State Department of Health | State Laboratories Division, Hawaii State Department of Health | Pamela O'Brien et al |
| EPI_ISL_752679 | USA/HI-H200116/2020 | 2020-06-26 | North America/USA/Hawaii/Honolulu C | State Laboratories Division, Hawaii State Department of Health | State Laboratories Division, Hawaii State Department of Health | Pamela O'Brien et al |
| EPI_ISL_752684 | USA/HI-H200121/2020 | 2020-06-12 | North America/USA/Hawaii/Honolulu C | State Laboratories Division, Hawaii State Department of Health | State Laboratories Division, Hawaii State Department of Health | Pamela O'Brien et al |

| | | | | | | |
|---|---|---|---|---|---|---|
| EPI_ISL_752696 | USA/HI-H200133/2020 | 2020-07-09 | North America/USA/Hawaii/ | State Laboratories Division, Hawaii State Department of Health | State Laboratories Division, Hawaii State Department of Health | Pamela O'Brien et al |
| EPI_ISL_752705 | USA/HI-H200142/2020 | 2020-06-28 | North America/USA/Hawaii/Honolulu C | State Laboratories Division, Hawaii State Department of Health | State Laboratories Division, Hawaii State Department of Health | Pamela O'Brien et al |
| EPI_ISL_752709 | USA/HI-H200146/2020 | 2020-07-13 | North America/USA/Hawaii/ | State Laboratories Division, Hawaii State Department of Health | State Laboratories Division, Hawaii State Department of Health | Pamela O'Brien et al |
| EPI_ISL_752713 | USA/HI-H200150/2020 | 2020-07-18 | North America/USA/Hawaii/Honolulu C | State Laboratories Division, Hawaii State Department of Health | State Laboratories Division, Hawaii State Department of Health | Pamela O'Brien et al |
| EPI_ISL_752722 | USA/HI-H200160/2020 | 2020-07-24 | North America/USA/Hawaii/Honolulu C | State Laboratories Division, Hawaii State Department of Health | State Laboratories Division, Hawaii State Department of Health | Pamela O'Brien et al |
| EPI_ISL_752724 | USA/HI-H200163/2020 | 2020-07-24 | North America/USA/Hawaii/Honolulu C | State Laboratories Division, Hawaii State Department of Health | State Laboratories Division, Hawaii State Department of Health | Pamela O'Brien et al |
| EPI_ISL_752725 | USA/HI-H200164/2020 | 2020-07-24 | North America/USA/Hawaii/Honolulu C | State Laboratories Division, Hawaii State Department of Health | State Laboratories Division, Hawaii State Department of Health | Pamela O'Brien et al |
| EPI_ISL_752732 | USA/HI-H200171/2020 | 2020-07-27 | North America/USA/Hawaii/ | State Laboratories Division, Hawaii State Department of Health | State Laboratories Division, Hawaii State Department of Health | Pamela O'Brien et al |

| | | | | | | |
|---|---|---|---|---|---|---|
| EPI_ISL_752740 | USA/HI-H200180/2020 | 2020-07-29 | North America/USA/Hawaii/Honolulu C | State Laboratories Division, Hawaii State Department of Health | State Laboratories Division, Hawaii State Department of Health | Pamela O'Brien et al |
| EPI_ISL_752742 | USA/HI-H200182/2020 | 2020-07-30 | North America/USA/Hawaii/Honolulu C | State Laboratories Division, Hawaii State Department of Health | State Laboratories Division, Hawaii State Department of Health | Pamela O'Brien et al |
| EPI_ISL_752745 | USA/HI-H200185/2020 | 2020-07-30 | North America/USA/Hawaii/Honolulu C | State Laboratories Division, Hawaii State Department of Health | State Laboratories Division, Hawaii State Department of Health | Pamela O'Brien et al |
| EPI_ISL_752746 | USA/HI-H200187/2020 | 2020-07-31 | North America/USA/Hawaii/Honolulu C | State Laboratories Division, Hawaii State Department of Health | State Laboratories Division, Hawaii State Department of Health | Pamela O'Brien et al |
| EPI_ISL_752749 | USA/HI-H200190/2020 | 2020-08-09 | North America/USA/Hawaii/Honolulu C | State Laboratories Division, Hawaii State Department of Health | State Laboratories Division, Hawaii State Department of Health | Pamela O'Brien et al |
| EPI_ISL_752758 | USA/HI-H200199/2020 | 2020-08-10 | North America/USA/Hawaii/Honolulu C | State Laboratories Division, Hawaii State Department of Health | State Laboratories Division, Hawaii State Department of Health | Pamela O'Brien et al |
| EPI_ISL_752767 | USA/HI-H200208/2020 | 2020-08-12 | North America/USA/Hawaii/Honolulu C | State Laboratories Division, Hawaii State Department of Health | State Laboratories Division, Hawaii State Department of Health | Pamela O'Brien et al |
| EPI_ISL_752775 | USA/HI-H200216/2020 | 2020-08-10 | North America/USA/Hawaii/Honolulu C | State Laboratories Division, Hawaii State Department of Health | State Laboratories Division, Hawaii State Department of Health | Pamela O'Brien et al |

| | | | | | | |
|---|---|---|---|---|---|---|
| EPI_ISL_752778 | USA/HI-H200219/2020 | 2020-08-10 | North America/USA/Hawaii/Honolulu C | State Laboratories Division, Hawaii State Department of Health | State Laboratories Division, Hawaii State Department of Health | Pamela O'Brien et al |
| EPI_ISL_752781 | USA/HI-H200222/2020 | 2020-08-11 | North America/USA/Hawaii/Honolulu C | State Laboratories Division, Hawaii State Department of Health | State Laboratories Division, Hawaii State Department of Health | Pamela O'Brien et al |
| EPI_ISL_752785 | USA/HI-H200226/2020 | 2020-08-11 | North America/USA/Hawaii/Honolulu C | State Laboratories Division, Hawaii State Department of Health | State Laboratories Division, Hawaii State Department of Health | Pamela O'Brien et al |
| EPI_ISL_752786 | USA/HI-H200227/2020 | 2020-08-11 | North America/USA/Hawaii/Honolulu C | State Laboratories Division, Hawaii State Department of Health | State Laboratories Division, Hawaii State Department of Health | Pamela O'Brien et al |
| EPI_ISL_752787 | USA/HI-H200228/2020 | 2020-08-11 | North America/USA/Hawaii/Honolulu C | State Laboratories Division, Hawaii State Department of Health | State Laboratories Division, Hawaii State Department of Health | Pamela O'Brien et al |
| EPI_ISL_752790 | USA/HI-H200231/2020 | 2020-08-11 | North America/USA/Hawaii/Honolulu C | State Laboratories Division, Hawaii State Department of Health | State Laboratories Division, Hawaii State Department of Health | Pamela O'Brien et al |
| EPI_ISL_752812 | USA/HI-H200253/2020 | 2020-08-18 | North America/USA/Hawaii/Honolulu C | State Laboratories Division, Hawaii State Department of Health | State Laboratories Division, Hawaii State Department of Health | Pamela O'Brien et al |
| EPI_ISL_752814 | USA/HI-H200255/2020 | 2020-08-19 | North America/USA/Hawaii/Honolulu C | State Laboratories Division, Hawaii State Department of Health | State Laboratories Division, Hawaii State Department of Health | Pamela O'Brien et al |

| | | | | | | |
|---|---|---|---|---|---|---|
| EPI_ISL_752818 | USA/HI-H200259/2020 | 2020-08-25 | North America/USA/Hawaii/Honolulu C | State Laboratories Division, Hawaii State Department of Health | State Laboratories Division, Hawaii State Department of Health | Pamela O'Brien et al |
| EPI_ISL_752825 | USA/HI-H200266/2020 | 2020-08-31 | North America/USA/Hawaii/Honolulu C | State Laboratories Division, Hawaii State Department of Health | State Laboratories Division, Hawaii State Department of Health | Pamela O'Brien et al |
| EPI_ISL_752840 | USA/HI-H200281/2020 | 2020-09-09 | North America/USA/Hawaii/Honolulu C | State Laboratories Division, Hawaii State Department of Health | State Laboratories Division, Hawaii State Department of Health | Pamela O'Brien et al |
| EPI_ISL_752859 | USA/HI-H200300/2020 | 2020-08-11 | North America/USA/Hawaii/Honolulu C | State Laboratories Division, Hawaii State Department of Health | State Laboratories Division, Hawaii State Department of Health | Pamela O'Brien et al |
| EPI_ISL_752864 | USA/HI-H200305/2020 | 2020-08-12 | North America/USA/Hawaii/Honolulu C | State Laboratories Division, Hawaii State Department of Health | State Laboratories Division, Hawaii State Department of Health | Pamela O'Brien et al |
| EPI_ISL_752870 | USA/HI-H200311/2020 | 2020-08-12 | North America/USA/Hawaii/Honolulu C | State Laboratories Division, Hawaii State Department of Health | State Laboratories Division, Hawaii State Department of Health | Pamela O'Brien et al |
| EPI_ISL_752872 | USA/HI-H200313/2020 | 2020-08-16 | North America/USA/Hawaii/Honolulu C | State Laboratories Division, Hawaii State Department of Health | State Laboratories Division, Hawaii State Department of Health | Pamela O'Brien et al |
| EPI_ISL_752880 | USA/HI-H200322/2020 | 2020-08-17 | North America/USA/Hawaii/Honolulu C | State Laboratories Division, Hawaii State Department of Health | State Laboratories Division, Hawaii State Department of Health | Pamela O'Brien et al |

| | | | | | | |
|---|---|---|---|---|---|---|
| EPI_ISL_752885 | USA/HI-H200327/2020 | 2020-08-20 | North America/USA/Hawaii/Honolulu C | State Laboratories Division, Hawaii State Department of Health | State Laboratories Division, Hawaii State Department of Health | Pamela O'Brien et al |
| EPI_ISL_752888 | USA/HI-H200330/2020 | 2020-08-22 | North America/USA/Hawaii/Honolulu C | State Laboratories Division, Hawaii State Department of Health | State Laboratories Division, Hawaii State Department of Health | Pamela O'Brien et al |
| EPI_ISL_752890 | USA/HI-H200332/2020 | 2020-08-25 | North America/USA/Hawaii/Honolulu C | State Laboratories Division, Hawaii State Department of Health | State Laboratories Division, Hawaii State Department of Health | Pamela O'Brien et al |
| EPI_ISL_752893 | USA/HI-H200335/2020 | 2020-08-15 | North America/USA/Hawaii/Honolulu C | State Laboratories Division, Hawaii State Department of Health | State Laboratories Division, Hawaii State Department of Health | Pamela O'Brien et al |
| EPI_ISL_752895 | USA/HI-H200337/2020 | 2020-08-16 | North America/USA/Hawaii/Honolulu C | State Laboratories Division, Hawaii State Department of Health | State Laboratories Division, Hawaii State Department of Health | Pamela O'Brien et al |
| EPI_ISL_752898 | USA/HI-H200342/2020 | 2020-08-27 | North America/USA/Hawaii/Honolulu C | State Laboratories Division, Hawaii State Department of Health | State Laboratories Division, Hawaii State Department of Health | Pamela O'Brien et al |
| EPI_ISL_752901 | USA/HI-H200345/2020 | 2020-08-31 | North America/USA/Hawaii/Honolulu C | State Laboratories Division, Hawaii State Department of Health | State Laboratories Division, Hawaii State Department of Health | Pamela O'Brien et al |
| EPI_ISL_752902 | USA/HI-H200346/2020 | 2020-09-08 | North America/USA/Hawaii/Honolulu C | State Laboratories Division, Hawaii State Department of Health | State Laboratories Division, Hawaii State Department of Health | Pamela O'Brien et al |

| | | | | | | |
|---|---|---|---|---|---|---|
| EPI_ISL_752906 | USA/HI-H200350/2020 | 2020-09-15 | North America/USA/Hawaii/Honolulu C | State Laboratories Division, Hawaii State Department of Health | State Laboratories Division, Hawaii State Department of Health | Pamela O'Brien et al |
| EPI_ISL_752908 | USA/HI-H200352/2020 | 2020-09-16 | North America/USA/Hawaii/Honolulu C | State Laboratories Division, Hawaii State Department of Health | State Laboratories Division, Hawaii State Department of Health | Pamela O'Brien et al |
| EPI_ISL_752909 | USA/HI-H200353/2020 | 2020-09-17 | North America/USA/Hawaii/Honolulu C | State Laboratories Division, Hawaii State Department of Health | State Laboratories Division, Hawaii State Department of Health | Pamela O'Brien et al |
| EPI_ISL_752917 | USA/HI-H200361/2020 | 2020-09-28 | North America/USA/Hawaii/Honolulu C | State Laboratories Division, Hawaii State Department of Health | State Laboratories Division, Hawaii State Department of Health | Pamela O'Brien et al |
| EPI_ISL_752921 | USA/HI-H200365/2020 | 2020-09-29 | North America/USA/Hawaii/Honolulu C | State Laboratories Division, Hawaii State Department of Health | State Laboratories Division, Hawaii State Department of Health | Pamela O'Brien et al |
| EPI_ISL_752926 | USA/HI-H200377/2020 | 2020-09-19 | North America/USA/Hawaii/Hawaii Cou | State Laboratories Division, Hawaii State Department of Health | State Laboratories Division, Hawaii State Department of Health | Pamela O'Brien et al |
| EPI_ISL_752927 | USA/HI-H200379/2020 | 2020-09-12 | North America/USA/Hawaii/Hawaii Cou | State Laboratories Division, Hawaii State Department of Health | State Laboratories Division, Hawaii State Department of Health | Pamela O'Brien et al |
| EPI_ISL_752928 | USA/HI-H200381/2020 | 2020-09-18 | North America/USA/Hawaii/Honolulu C | State Laboratories Division, Hawaii State Department of Health | State Laboratories Division, Hawaii State Department of Health | Pamela O'Brien et al |

| | | | | | | |
|---|---|---|---|---|---|---|
| EPI_ISL_752933 | USA/HI-H200386/2020 | 2020-09-25 | North America/USA/Hawaii/Honolulu C | State Laboratories Division, Hawaii State Department of Health | State Laboratories Division, Hawaii State Department of Health | Pamela O'Brien et al |
| EPI_ISL_752936 | USA/HI-H200389/2020 | 2020-09-29 | North America/USA/Hawaii/Honolulu C | State Laboratories Division, Hawaii State Department of Health | State Laboratories Division, Hawaii State Department of Health | Pamela O'Brien et al |
| EPI_ISL_752938 | USA/HI-H200391/2020 | 2020-09-29 | North America/USA/Hawaii/Honolulu C | State Laboratories Division, Hawaii State Department of Health | State Laboratories Division, Hawaii State Department of Health | Pamela O'Brien et al |
| EPI_ISL_752941 | USA/HI-H200395/2020 | 2020-09-09 | North America/USA/Hawaii/Hawaii Cou | State Laboratories Division, Hawaii State Department of Health | State Laboratories Division, Hawaii State Department of Health | Pamela O'Brien et al |
| EPI_ISL_752942 | USA/HI-H200396/2020 | 2020-09-09 | North America/USA/Hawaii/Hawaii Cou | State Laboratories Division, Hawaii State Department of Health | State Laboratories Division, Hawaii State Department of Health | Pamela O'Brien et al |
| EPI_ISL_752943 | USA/HI-H200398/2020 | 2020-09-09 | North America/USA/Hawaii/Hawaii Cou | State Laboratories Division, Hawaii State Department of Health | State Laboratories Division, Hawaii State Department of Health | Pamela O'Brien et al |
| EPI_ISL_752945 | USA/HI-H200400/2020 | 2020-09-09 | North America/USA/Hawaii/Hawaii Cou | State Laboratories Division, Hawaii State Department of Health | State Laboratories Division, Hawaii State Department of Health | Pamela O'Brien et al |
| EPI_ISL_752952 | USA/HI-H200407/2020 | 2020-07-31 | North America/USA/Hawaii/Honolulu C | State Laboratories Division, Hawaii State Department of Health | State Laboratories Division, Hawaii State Department of Health | Pamela O'Brien et al |

| | | | | | | |
|---|---|---|---|---|---|---|
| EPI_ISL_752956 | USA/HI-H200412/2020 | 2020-08-06 | North America/USA/Hawaii/Honolulu C | State Laboratories Division, Hawaii State Department of Health | State Laboratories Division, Hawaii State Department of Health | Pamela O'Brien et al |
| EPI_ISL_752957 | USA/HI-H200413/2020 | 2020-08-07 | North America/USA/Hawaii/Honolulu C | State Laboratories Division, Hawaii State Department of Health | State Laboratories Division, Hawaii State Department of Health | Pamela O'Brien et al |
| EPI_ISL_752960 | USA/HI-H200416/2020 | 2020-08-10 | North America/USA/Hawaii/Honolulu C | State Laboratories Division, Hawaii State Department of Health | State Laboratories Division, Hawaii State Department of Health | Pamela O'Brien et al |
| EPI_ISL_752961 | USA/HI-H200417/2020 | 2020-08-10 | North America/USA/Hawaii/Honolulu C | State Laboratories Division, Hawaii State Department of Health | State Laboratories Division, Hawaii State Department of Health | Pamela O'Brien et al |
| EPI_ISL_752962 | USA/HI-H200418/2020 | 2020-08-11 | North America/USA/Hawaii/Honolulu C | State Laboratories Division, Hawaii State Department of Health | State Laboratories Division, Hawaii State Department of Health | Pamela O'Brien et al |
| EPI_ISL_752969 | USA/HI-H200425/2020 | 2020-09-23 | North America/USA/Hawaii/Honolulu C | State Laboratories Division, Hawaii State Department of Health | State Laboratories Division, Hawaii State Department of Health | Pamela O'Brien et al |
| EPI_ISL_752971 | USA/HI-H200427/2020 | 2020-09-24 | North America/USA/Hawaii/Honolulu C | State Laboratories Division, Hawaii State Department of Health | State Laboratories Division, Hawaii State Department of Health | Pamela O'Brien et al |
| EPI_ISL_752974 | USA/HI-H200430/2020 | 2020-09-30 | North America/USA/Hawaii/Hawaii Cou | State Laboratories Division, Hawaii State Department of Health | State Laboratories Division, Hawaii State Department of Health | Pamela O'Brien et al |

| | | | | | | |
|---|---|---|---|---|---|---|
| EPI_ISL_752976 | USA/HI-H200433/2020 | 2020-10-01 | North America/USA/Hawaii/Hawaii Cou | State Laboratories Division, Hawaii State Department of Health | State Laboratories Division, Hawaii State Department of Health | Pamela O'Brien et al |
| EPI_ISL_752977 | USA/HI-H200434/2020 | 2020-10-01 | North America/USA/Hawaii/Hawaii Cou | State Laboratories Division, Hawaii State Department of Health | State Laboratories Division, Hawaii State Department of Health | Pamela O'Brien et al |
| EPI_ISL_752979 | USA/HI-H200436/2020 | 2020-10-03 | North America/USA/Hawaii/Hawaii Cou | State Laboratories Division, Hawaii State Department of Health | State Laboratories Division, Hawaii State Department of Health | Pamela O'Brien et al |
| EPI_ISL_752984 | USA/HI-H200441/2020 | 2020-10-07 | North America/USA/Hawaii/Hawaii Cou | State Laboratories Division, Hawaii State Department of Health | State Laboratories Division, Hawaii State Department of Health | Pamela O'Brien et al |
| EPI_ISL_752989 | USA/HI-H200446/2020 | 2020-10-08 | North America/USA/Hawaii/Hawaii Cou | State Laboratories Division, Hawaii State Department of Health | State Laboratories Division, Hawaii State Department of Health | Pamela O'Brien et al |
| EPI_ISL_752992 | USA/HI-H200449/2020 | 2020-10-09 | North America/USA/Hawaii/Hawaii Cou | State Laboratories Division, Hawaii State Department of Health | State Laboratories Division, Hawaii State Department of Health | Pamela O'Brien et al |
| EPI_ISL_752999 | USA/HI-H200456/2020 | 2020-09-15 | North America/USA/Hawaii/Honolulu C | State Laboratories Division, Hawaii State Department of Health | State Laboratories Division, Hawaii State Department of Health | Pamela O'Brien et al |
| EPI_ISL_753000 | USA/HI-H200457/2020 | 2020-09-15 | North America/USA/Hawaii/Honolulu C | State Laboratories Division, Hawaii State Department of Health | State Laboratories Division, Hawaii State Department of Health | Pamela O'Brien et al |

| | | | | | | |
|---|---|---|---|---|---|---|
| EPI_ISL_753004 | USA/HI-H200461/2020 | 2020-10-08 | North America/USA/Hawaii/Honolulu C | State Laboratories Division, Hawaii State Department of Health | State Laboratories Division, Hawaii State Department of Health | Pamela O'Brien et al |
| EPI_ISL_753009 | USA/HI-H200466/2020 | 2020-10-03 | North America/USA/Hawaii/Hawaii Cou | State Laboratories Division, Hawaii State Department of Health | State Laboratories Division, Hawaii State Department of Health | Pamela O'Brien et al |
| EPI_ISL_753027 | USA/HI-H200486/2020 | 2020-09-24 | North America/USA/Hawaii/Honolulu C | State Laboratories Division, Hawaii State Department of Health | State Laboratories Division, Hawaii State Department of Health | Pamela O'Brien et al |
| EPI_ISL_753030 | USA/HI-H200490/2020 | 2020-09-30 | North America/USA/Hawaii/Honolulu C | State Laboratories Division, Hawaii State Department of Health | State Laboratories Division, Hawaii State Department of Health | Pamela O'Brien et al |
| EPI_ISL_753035 | USA/HI-H200496/2020 | 2020-10-09 | North America/USA/Hawaii/Honolulu C | State Laboratories Division, Hawaii State Department of Health | State Laboratories Division, Hawaii State Department of Health | Pamela O'Brien et al |
| EPI_ISL_753042 | USA/HI-H200503/2020 | 2020-08-02 | North America/USA/Hawaii/Honolulu C | State Laboratories Division, Hawaii State Department of Health | State Laboratories Division, Hawaii State Department of Health | Pamela O'Brien et al |
| EPI_ISL_753043 | USA/HI-H200504/2020 | 2020-08-02 | North America/USA/Hawaii/Honolulu C | State Laboratories Division, Hawaii State Department of Health | State Laboratories Division, Hawaii State Department of Health | Pamela O'Brien et al |
| EPI_ISL_753044 | USA/HI-H200505/2020 | 2020-08-11 | North America/USA/Hawaii/ | State Laboratories Division, Hawaii State Department of Health | State Laboratories Division, Hawaii State Department of Health | Pamela O'Brien et al |

| | | | | | | |
|---|---|---|---|---|---|---|
| EPI_ISL_753051 | USA/HI-H200515/2020 | 2020-08-18 | North America/USA/Hawaii/Honolulu C | State Laboratories Division, Hawaii State Department of Health | State Laboratories Division, Hawaii State Department of Health | Pamela O'Brien et al |
| EPI_ISL_753057 | USA/HI-H200523/2020 | 2020-11-02 | North America/USA/Hawaii/Honolulu C | State Laboratories Division, Hawaii State Department of Health | State Laboratories Division, Hawaii State Department of Health | Pamela O'Brien et al |
| EPI_ISL_753058 | USA/HI-H200525/2020 | 2020-11-02 | North America/USA/Hawaii/Honolulu C | State Laboratories Division, Hawaii State Department of Health | State Laboratories Division, Hawaii State Department of Health | Pamela O'Brien et al |
| EPI_ISL_753060 | USA/HI-H200527/2020 | 2020-11-02 | North America/USA/Hawaii/Honolulu C | State Laboratories Division, Hawaii State Department of Health | State Laboratories Division, Hawaii State Department of Health | Pamela O'Brien et al |
| EPI_ISL_753061 | USA/HI-H200528/2020 | 2020-11-02 | North America/USA/Hawaii/Honolulu C | State Laboratories Division, Hawaii State Department of Health | State Laboratories Division, Hawaii State Department of Health | Pamela O'Brien et al |
| EPI_ISL_753062 | USA/HI-H200529/2020 | 2020-11-03 | North America/USA/Hawaii/Honolulu C | State Laboratories Division, Hawaii State Department of Health | State Laboratories Division, Hawaii State Department of Health | Pamela O'Brien et al |
| EPI_ISL_753063 | USA/HI-H200530/2020 | 2020-10-24 | North America/USA/Hawaii/Maui Count | State Laboratories Division, Hawaii State Department of Health | State Laboratories Division, Hawaii State Department of Health | Pamela O'Brien et al |
| EPI_ISL_753074 | USA/HI-H200543/2020 | 2020-11-05 | North America/USA/Hawaii/Honolulu C | State Laboratories Division, Hawaii State Department of Health | State Laboratories Division, Hawaii State Department of Health | Pamela O'Brien et al |

| | | | | | | |
|---|---|---|---|---|---|---|
| EPI_ISL_753092 | USA/HI-H200564/2020 | 2020-10-24 | North America/USA/Hawaii/Maui Count | State Laboratories Division, Hawaii State Department of Health | State Laboratories Division, Hawaii State Department of Health | Pamela O'Brien et al |
| EPI_ISL_753101 | USA/HI-H200574/2020 | 2020-09-21 | North America/USA/Hawaii/Honolulu C | State Laboratories Division, Hawaii State Department of Health | State Laboratories Division, Hawaii State Department of Health | Pamela O'Brien et al |
| EPI_ISL_753104 | USA/HI-H200577/2020 | 2020-09-24 | North America/USA/Hawaii/Honolulu C | State Laboratories Division, Hawaii State Department of Health | State Laboratories Division, Hawaii State Department of Health | Pamela O'Brien et al |
| EPI_ISL_753107 | USA/HI-H200580/2020 | 2020-10-15 | North America/USA/Hawaii/Honolulu C | State Laboratories Division, Hawaii State Department of Health | State Laboratories Division, Hawaii State Department of Health | Pamela O'Brien et al |
| EPI_ISL_753109 | USA/HI-H200583/2020 | 2020-10-29 | North America/USA/Hawaii/Honolulu C | State Laboratories Division, Hawaii State Department of Health | State Laboratories Division, Hawaii State Department of Health | Pamela O'Brien et al |
| EPI_ISL_753112 | USA/HI-H200586/2020 | 2020-11-05 | North America/USA/Hawaii/Honolulu C | State Laboratories Division, Hawaii State Department of Health | State Laboratories Division, Hawaii State Department of Health | Pamela O'Brien et al |
| EPI_ISL_753118 | USA/HI-H200592/2020 | 2020-11-21 | North America/USA/Hawaii/Honolulu C | State Laboratories Division, Hawaii State Department of Health | State Laboratories Division, Hawaii State Department of Health | Pamela O'Brien et al |
| EPI_ISL_753120 | USA/HI-H200594/2020 | 2020-11-23 | North America/USA/Hawaii/Honolulu C | State Laboratories Division, Hawaii State Department of Health | State Laboratories Division, Hawaii State Department of Health | Pamela O'Brien et al |

| | | | | | | |
|---|---|---|---|---|---|---|
| EPI_ISL_753138 | USA/HI-H200612/2020 | 2020-11-23 | North America/USA/Hawaii/Honolulu C | State Laboratories Division, Hawaii State Department of Health | State Laboratories Division, Hawaii State Department of Health | Pamela O'Brien et al |
| EPI_ISL_753139 | USA/HI-H200613/2020 | 2020-11-23 | North America/USA/Hawaii/Honolulu C | State Laboratories Division, Hawaii State Department of Health | State Laboratories Division, Hawaii State Department of Health | Pamela O'Brien et al |
| EPI_ISL_753144 | USA/HI-H200618/2020 | 2020-11-23 | North America/USA/Hawaii/Honolulu C | State Laboratories Division, Hawaii State Department of Health | State Laboratories Division, Hawaii State Department of Health | Pamela O'Brien et al |
| EPI_ISL_753148 | USA/HI-H200622/2020 | 2020-09-24 | North America/USA/Hawaii/Honolulu C | State Laboratories Division, Hawaii State Department of Health | State Laboratories Division, Hawaii State Department of Health | Pamela O'Brien et al |
| EPI_ISL_753153 | USA/HI-H200628/2020 | 2020-11-13 | North America/USA/Hawaii/Honolulu C | State Laboratories Division, Hawaii State Department of Health | State Laboratories Division, Hawaii State Department of Health | Pamela O'Brien et al |
| EPI_ISL_753154 | USA/HI-H200629/2020 | 2020-11-23 | North America/USA/Hawaii/Honolulu C | State Laboratories Division, Hawaii State Department of Health | State Laboratories Division, Hawaii State Department of Health | Pamela O'Brien et al |
| EPI_ISL_753162 | USA/HI-H200638/2020 | 2020-11-23 | North America/USA/Hawaii/Honolulu C | State Laboratories Division, Hawaii State Department of Health | State Laboratories Division, Hawaii State Department of Health | Pamela O'Brien et al |
| EPI_ISL_753163 | USA/HI-H200639/2020 | 2020-11-23 | North America/USA/Hawaii/Honolulu C | State Laboratories Division, Hawaii State Department of Health | State Laboratories Division, Hawaii State Department of Health | Pamela O'Brien et al |

| | | | | | | |
|---|---|---|---|---|---|---|
| EPI_ISL_753166 | USA/HI-H200642/2020 | 2020-11-23 | North America/USA/Hawaii/Honolulu C | State Laboratories Division, Hawaii State Department of Health | State Laboratories Division, Hawaii State Department of Health | Pamela O'Brien et al |
| EPI_ISL_753168 | USA/HI-H200644/2020 | 2020-11-23 | North America/USA/Hawaii/Honolulu C | State Laboratories Division, Hawaii State Department of Health | State Laboratories Division, Hawaii State Department of Health | Pamela O'Brien et al |
| EPI_ISL_753173 | USA/HI-H200649/2020 | 2020-11-23 | North America/USA/Hawaii/Kauai Coun | State Laboratories Division, Hawaii State Department of Health | State Laboratories Division, Hawaii State Department of Health | Pamela O'Brien et al |
| EPI_ISL_753175 | USA/HI-H200652/2020 | 2020-11-25 | North America/USA/Hawaii/Honolulu C | State Laboratories Division, Hawaii State Department of Health | State Laboratories Division, Hawaii State Department of Health | Pamela O'Brien et al |
| EPI_ISL_753177 | USA/HI-H200654/2020 | 2020-11-30 | North America/USA/Hawaii/Honolulu C | State Laboratories Division, Hawaii State Department of Health | State Laboratories Division, Hawaii State Department of Health | Pamela O'Brien et al |
| EPI_ISL_753184 | USA/HI-H200661/2020 | 2020-12-01 | North America/USA/Hawaii/Honolulu C | State Laboratories Division, Hawaii State Department of Health | State Laboratories Division, Hawaii State Department of Health | Pamela O'Brien et al |
| EPI_ISL_753189 | USA/HI-H200666/2020 | 2020-12-01 | North America/USA/Hawaii/Honolulu C | State Laboratories Division, Hawaii State Department of Health | State Laboratories Division, Hawaii State Department of Health | Pamela O'Brien et al |
| EPI_ISL_753195 | USA/HI-H200672/2020 | 2020-09-21 | North America/USA/Hawaii/Honolulu C | State Laboratories Division, Hawaii State Department of Health | State Laboratories Division, Hawaii State Department of Health | Pamela O'Brien et al |

| | | | | | | |
|---|---|---|---|---|---|---|
| EPI_ISL_753199 | USA/HI-H200676/2020 | 2020-11-25 | North America/USA/Hawaii/Honolulu C | State Laboratories Division, Hawaii State Department of Health | State Laboratories Division, Hawaii State Department of Health | Pamela O'Brien et al |
| EPI_ISL_753200 | USA/HI-H200677/2020 | 2020-11-30 | North America/USA/Hawaii/Honolulu C | State Laboratories Division, Hawaii State Department of Health | State Laboratories Division, Hawaii State Department of Health | Pamela O'Brien et al |
| EPI_ISL_753203 | USA/HI-H200680/2020 | 2020-12-01 | North America/USA/Hawaii/Honolulu C | State Laboratories Division, Hawaii State Department of Health | State Laboratories Division, Hawaii State Department of Health | Pamela O'Brien et al |
| EPI_ISL_753204 | USA/HI-H200681/2020 | 2020-12-01 | North America/USA/Hawaii/Honolulu C | State Laboratories Division, Hawaii State Department of Health | State Laboratories Division, Hawaii State Department of Health | Pamela O'Brien et al |
| EPI_ISL_753213 | USA/HI-H200691/2020 | 2020-12-01 | North America/USA/Hawaii/Honolulu C | State Laboratories Division, Hawaii State Department of Health | State Laboratories Division, Hawaii State Department of Health | Pamela O'Brien et al |
| EPI_ISL_753215 | USA/HI-H200693/2020 | 2020-09-21 | North America/USA/Hawaii/Honolulu C | State Laboratories Division, Hawaii State Department of Health | State Laboratories Division, Hawaii State Department of Health | Pamela O'Brien et al |
| EPI_ISL_753219 | USA/HI-H200698/2020 | 2020-12-01 | North America/USA/Hawaii/Honolulu C | State Laboratories Division, Hawaii State Department of Health | State Laboratories Division, Hawaii State Department of Health | Pamela O'Brien et al |
| EPI_ISL_753226 | USA/HI-H200705/2020 | 2020-12-09 | North America/USA/Hawaii/Honolulu C | State Laboratories Division, Hawaii State Department of Health | State Laboratories Division, Hawaii State Department of Health | Pamela O'Brien et al |

| | | | | | | |
|---|---|---|---|---|---|---|
| EPI_ISL_753235 | USA/HI-H200714/2020 | 2020-12-09 | North America/USA/Hawaii/Honolulu C | State Laboratories Division, Hawaii State Department of Health | State Laboratories Division, Hawaii State Department of Health | Pamela O'Brien et al |
| EPI_ISL_753244 | USA/HI-H200723/2020 | 2020-12-09 | North America/USA/Hawaii/Honolulu C | State Laboratories Division, Hawaii State Department of Health | State Laboratories Division, Hawaii State Department of Health | Pamela O'Brien et al |
| EPI_ISL_753246 | USA/HI-H200725/2020 | 2020-12-09 | North America/USA/Hawaii/Honolulu C | State Laboratories Division, Hawaii State Department of Health | State Laboratories Division, Hawaii State Department of Health | Pamela O'Brien et al |
| EPI_ISL_753248 | USA/HI-H200727/2020 | 2020-12-09 | North America/USA/Hawaii/Honolulu C | State Laboratories Division, Hawaii State Department of Health | State Laboratories Division, Hawaii State Department of Health | Pamela O'Brien et al |
| EPI_ISL_753258 | USA/HI-H200737/2020 | 2020-12-09 | North America/USA/Hawaii/Honolulu C | State Laboratories Division, Hawaii State Department of Health | State Laboratories Division, Hawaii State Department of Health | Pamela O'Brien et al |
| EPI_ISL_753272 | USA/CA-CHLA-PLM26509492/2020 | 2020-11-03 | North America/USA/California/Los Ange | Clinical virology Laboratory, Children's Hospital Los Angeles | Center for Personalized Medicine, Children's Hospital Los Angeles | Gai et al et al |
| EPI_ISL_753285 | USA/CA-CHLA-PLM01113476/2020 | 2020-11-03 | North America/USA/California/Los Ange | Clinical virology Laboratory, Children's Hospital Los Angeles | Center for Personalized Medicine, Children's Hospital Los Angeles | Gai et al et al |

| | | | | | | |
|---|---|---|---|---|---|---|
| EPI_ISL_753294 | USA/CA-CHLA-PLM28873007/2020 | 2020-11-09 | North America/USA/California/Los Ange | Clinical virology Laboratory, Children's Hospital Los Angeles | Center for Personalized Medicine, Children's Hospital Los Angeles | Gai et al et al |
| EPI_ISL_753299 | USA/CA-CHLA-PLM65167224/2020 | 2020-11-09 | North America/USA/California/Los Ange | Clinical virology Laboratory, Children's Hospital Los Angeles | Center for Personalized Medicine, Children's Hospital Los Angeles | Gai et al et al |
| EPI_ISL_753311 | USA/CA-CHLA-PLM87213925/2020 | 2020-11-13 | North America/USA/California/Los Ange | Clinical virology Laboratory, Children's Hospital Los Angeles | Center for Personalized Medicine, Children's Hospital Los Angeles | Gai et al et al |
| EPI_ISL_753314 | USA/CA-CHLA-PLM32086413/2020 | 2020-11-13 | North America/USA/California/Los Ange | Clinical virology Laboratory, Children's Hospital Los Angeles | Center for Personalized Medicine, Children's Hospital Los Angeles | Gai et al et al |
| EPI_ISL_753316 | USA/CA-CHLA-PLM94151982/2020 | 2020-11-13 | North America/USA/California/Los Ange | Clinical virology Laboratory, Children's Hospital Los Angeles | Center for Personalized Medicine, Children's Hospital Los Angeles | Gai et al et al |
| EPI_ISL_753319 | USA/CA-CHLA-PLM02775277/2020 | 2020-11-13 | North America/USA/California/Los Ange | Clinical virology Laboratory, Children's Hospital Los Angeles | Center for Personalized Medicine, Children's Hospital Los Angeles | Gai et al et al |

| | | | | | | |
|---|---|---|---|---|---|---|
| EPI_ISL_753325 | USA/CA-CHLA-PLM47014149/2020 | 2020-11-07 | North America/USA/California/Los Ange | Clinical virology Laboratory, Children's Hospital Los Angeles | Center for Personalized Medicine, Children's Hospital Los Angeles | Gai et al et al |
| EPI_ISL_753327 | USA/CA-CHLA-PLM14752712/2020 | 2020-11-09 | North America/USA/California/Los Ange | Clinical virology Laboratory, Children's Hospital Los Angeles | Center for Personalized Medicine, Children's Hospital Los Angeles | Gai et al et al |
| EPI_ISL_753328 | USA/CA-CHLA-PLM67956098/2020 | 2020-11-09 | North America/USA/California/Los Ange | Clinical virology Laboratory, Children's Hospital Los Angeles | Center for Personalized Medicine, Children's Hospital Los Angeles | Gai et al et al |
| EPI_ISL_753332 | USA/CA-CHLA-PLM70845114/2020 | 2020-11-15 | North America/USA/California/Los Ange | Clinical virology Laboratory, Children's Hospital Los Angeles | Center for Personalized Medicine, Children's Hospital Los Angeles | Gai et al et al |
| EPI_ISL_753337 | USA/CA-CHLA-PLM50981298/2020 | 2020-11-15 | North America/USA/California/Los Ange | Clinical virology Laboratory, Children's Hospital Los Angeles | Center for Personalized Medicine, Children's Hospital Los Angeles | Gai et al et al |
| EPI_ISL_753341 | USA/CA-CHLA-PLM54818837/2020 | 2020-11-15 | North America/USA/California/Los Ange | Clinical virology Laboratory, Children's Hospital Los Angeles | Center for Personalized Medicine, Children's Hospital Los Angeles | Gai et al et al |

| | | | | | | |
|---|---|---|---|---|---|---|
| EPI_ISL_753347 | USA/CA-CHLA-PLM07026043/2020 | 2020-11-16 | North America/USA/California/Los Ange | Clinical virology Laboratory, Children's Hospital Los Angeles | Center for Personalized Medicine, Children's Hospital Los Angeles | Gai et al et al |
| EPI_ISL_753363 | USA/CA-CHLA-PLM04975012/2020 | 2020-11-17 | North America/USA/California/Los Ange | Clinical virology Laboratory, Children's Hospital Los Angeles | Center for Personalized Medicine, Children's Hospital Los Angeles | Gai et al et al |
| EPI_ISL_753367 | USA/CA-CHLA-PLM53174364/2020 | 2020-11-18 | North America/USA/California/Los Ange | Clinical virology Laboratory, Children's Hospital Los Angeles | Center for Personalized Medicine, Children's Hospital Los Angeles | Gai et al et al |
| EPI_ISL_753378 | USA/CA-CHLA-PLM76320099/2020 | 2020-11-18 | North America/USA/California/Los Ange | Clinical virology Laboratory, Children's Hospital Los Angeles | Center for Personalized Medicine, Children's Hospital Los Angeles | Gai et al et al |
| EPI_ISL_753381 | USA/CA-CHLA-PLM41302615/2020 | 2020-11-19 | North America/USA/California/Los Ange | Clinical virology Laboratory, Children's Hospital Los Angeles | Center for Personalized Medicine, Children's Hospital Los Angeles | Gai et al et al |
| EPI_ISL_753384 | USA/CA-CHLA-PLM47954122/2020 | 2020-11-19 | North America/USA/California/Los Ange | Clinical virology Laboratory, Children's Hospital Los Angeles | Center for Personalized Medicine, Children's Hospital Los Angeles | Gai et al et al |

| | | | | | | |
|---|---|---|---|---|---|---|
| EPI_ISL_753392 | USA/CA-CHLA-PLM35584098/2020 | 2020-11-19 | North America/USA/California/Los Ange | Clinical virology Laboratory, Children's Hospital Los Angeles | Center for Personalized Medicine, Children's Hospital Los Angeles | Gai et al et al |
| EPI_ISL_753393 | USA/CA-CHLA-PLM41214732/2020 | 2020-11-20 | North America/USA/California/Los Ange | Clinical virology Laboratory, Children's Hospital Los Angeles | Center for Personalized Medicine, Children's Hospital Los Angeles | Gai et al et al |
| EPI_ISL_753402 | USA/CA-CHLA-PLM32737996/2020 | 2020-11-21 | North America/USA/California/Los Ange | Clinical virology Laboratory, Children's Hospital Los Angeles | Center for Personalized Medicine, Children's Hospital Los Angeles | Gai et al et al |
| EPI_ISL_753408 | USA/CA-CHLA-PLM38887875/2020 | 2020-11-18 | North America/USA/California/Los Ange | Clinical virology Laboratory, Children's Hospital Los Angeles | Center for Personalized Medicine, Children's Hospital Los Angeles | Gai et al et al |
| EPI_ISL_753410 | USA/CA-CHLA-PLM57057181/2020 | 2020-11-22 | North America/USA/California/Los Ange | Clinical virology Laboratory, Children's Hospital Los Angeles | Center for Personalized Medicine, Children's Hospital Los Angeles | Gai et al et al |
| EPI_ISL_753411 | USA/CA-CHLA-PLM78059394/2020 | 2020-11-21 | North America/USA/California/Los Ange | Clinical virology Laboratory, Children's Hospital Los Angeles | Center for Personalized Medicine, Children's Hospital Los Angeles | Gai et al et al |

| | | | | | | |
|---|---|---|---|---|---|---|
| EPI_ISL_753417 | USA/CA-CHLA-PLM14088876/2020 | 2020-11-30 | North America/USA/California/Los Ange | Clinical virology Laboratory, Children's Hospital Los Angeles | Center for Personalized Medicine, Children's Hospital Los Angeles | Gai et al et al |
| EPI_ISL_753422 | USA/CA-CHLA-PLM36879580/2020 | 2020-11-30 | North America/USA/California/Los Ange | Clinical virology Laboratory, Children's Hospital Los Angeles | Center for Personalized Medicine, Children's Hospital Los Angeles | Gai et al et al |
| EPI_ISL_753423 | USA/CA-CHLA-PLM04548495/2020 | 2020-11-30 | North America/USA/California/Los Ange | Clinical virology Laboratory, Children's Hospital Los Angeles | Center for Personalized Medicine, Children's Hospital Los Angeles | Gai et al et al |
| EPI_ISL_753424 | USA/CA-CHLA-PLM51044374/2020 | 2020-11-30 | North America/USA/California/Los Ange | Clinical virology Laboratory, Children's Hospital Los Angeles | Center for Personalized Medicine, Children's Hospital Los Angeles | Gai et al et al |
| EPI_ISL_753428 | USA/CA-CHLA-PLM49123428/2020 | 2020-11-30 | North America/USA/California/Los Ange | Clinical virology Laboratory, Children's Hospital Los Angeles | Center for Personalized Medicine, Children's Hospital Los Angeles | Gai et al et al |
| EPI_ISL_753441 | USA/CA-CHLA-PLM47567508/2020 | 2020-11-30 | North America/USA/California/Los Ange | Clinical virology Laboratory, Children's Hospital Los Angeles | Center for Personalized Medicine, Children's Hospital Los Angeles | Gai et al et al |

| | | | | | | |
|---|---|---|---|---|---|---|
| EPI_ISL_753448 | USA/CA-CHLA-PLM02905782/2020 | 2020-11-30 | North America/USA/California/Los Ange | Clinical virology Laboratory, Children's Hospital Los Angeles | Center for Personalized Medicine, Children's Hospital Los Angeles | Gai et al et al |
| EPI_ISL_753469 | USA/CA-CHLA-PLM48900225/2020 | 2020-11-30 | North America/USA/California/Los Ange | Clinical virology Laboratory, Children's Hospital Los Angeles | Center for Personalized Medicine, Children's Hospital Los Angeles | Gai et al et al |
| EPI_ISL_753477 | USA/CA-CHLA-PLM38859034/2020 | 2020-11-30 | North America/USA/California/Los Ange | Clinical virology Laboratory, Children's Hospital Los Angeles | Center for Personalized Medicine, Children's Hospital Los Angeles | Gai et al et al |
| EPI_ISL_753479 | USA/CA-CHLA-PLM02129348/2020 | 2020-11-30 | North America/USA/California/Los Ange | Clinical virology Laboratory, Children's Hospital Los Angeles | Center for Personalized Medicine, Children's Hospital Los Angeles | Gai et al et al |
| EPI_ISL_753482 | USA/CA-CHLA-PLM53657240/2020 | 2020-11-30 | North America/USA/California/Los Ange | Clinical virology Laboratory, Children's Hospital Los Angeles | Center for Personalized Medicine, Children's Hospital Los Angeles | Gai et al et al |
| EPI_ISL_753485 | USA/CA-CHLA-PLM42821873/2020 | 2020-11-30 | North America/USA/California/Los Ange | Clinical virology Laboratory, Children's Hospital Los Angeles | Center for Personalized Medicine, Children's Hospital Los Angeles | Gai et al et al |

| | | | | | | |
|---|---|---|---|---|---|---|
| EPI_ISL_753489 | USA/CA-CHLA-PLM48892933/2020 | 2020-11-30 | North America/USA/California/Los Ange | Clinical virology Laboratory, Children's Hospital Los Angeles | Center for Personalized Medicine, Children's Hospital Los Angeles | Gai et al et al |
| EPI_ISL_753493 | USA/CA-CHLA-PLM92698286/2020 | 2020-11-30 | North America/USA/California/Los Ange | Clinical virology Laboratory, Children's Hospital Los Angeles | Center for Personalized Medicine, Children's Hospital Los Angeles | Gai et al et al |
| EPI_ISL_753499 | USA/CA-CHLA-PLM16398520/2020 | 2020-11-30 | North America/USA/California/Los Ange | Clinical virology Laboratory, Children's Hospital Los Angeles | Center for Personalized Medicine, Children's Hospital Los Angeles | Gai et al et al |
| EPI_ISL_753510 | USA/CA-CHLA-PLM57308775/2020 | 2020-11-30 | North America/USA/California/Los Ange | Clinical virology Laboratory, Children's Hospital Los Angeles | Center for Personalized Medicine, Children's Hospital Los Angeles | Gai et al et al |
| EPI_ISL_753512 | USA/CA-CHLA-PLM57076996/2020 | 2020-11-30 | North America/USA/California/Los Ange | Clinical virology Laboratory, Children's Hospital Los Angeles | Center for Personalized Medicine, Children's Hospital Los Angeles | Gai et al et al |
| EPI_ISL_753521 | USA/CA-CHLA-PLM38778784/2020 | 2020-11-30 | North America/USA/California/Los Ange | Clinical virology Laboratory, Children's Hospital Los Angeles | Center for Personalized Medicine, Children's Hospital Los Angeles | Gai et al et al |

| | | | | | | |
|---|---|---|---|---|---|---|
| EPI_ISL_753523 | USA/CA-CHLA-PLM59547698/2020 | 2020-11-30 | North America/USA/California/Los Ange | Clinical virology Laboratory, Children's Hospital Los Angeles | Center for Personalized Medicine, Children's Hospital Los Angeles | Gai et al et al |
| EPI_ISL_753530 | USA/CA-CHLA-PLM59996756/2020 | 2020-11-28 | North America/USA/California/Los Ange | Clinical virology Laboratory, Children's Hospital Los Angeles | Center for Personalized Medicine, Children's Hospital Los Angeles | Gai et al et al |
| EPI_ISL_753535 | USA/CA-CHLA-PLM71343358/2020 | 2020-11-30 | North America/USA/California/Los Ange | Clinical virology Laboratory, Children's Hospital Los Angeles | Center for Personalized Medicine, Children's Hospital Los Angeles | Gai et al et al |
| EPI_ISL_753537 | USA/CA-CHLA-PLM07635073/2020 | 2020-11-30 | North America/USA/California/Los Ange | Clinical virology Laboratory, Children's Hospital Los Angeles | Center for Personalized Medicine, Children's Hospital Los Angeles | Gai et al et al |
| EPI_ISL_753542 | USA/CA-CHLA-PLM78910262/2020 | 2020-11-30 | North America/USA/California/Los Ange | Clinical virology Laboratory, Children's Hospital Los Angeles | Center for Personalized Medicine, Children's Hospital Los Angeles | Gai et al et al |
| EPI_ISL_753545 | USA/CA-CHLA-PLM62737694/2020 | 2020-11-30 | North America/USA/California/Los Ange | Clinical virology Laboratory, Children's Hospital Los Angeles | Center for Personalized Medicine, Children's Hospital Los Angeles | Gai et al et al |

| | | | | | | |
|---|---|---|---|---|---|---|
| EPI_ISL_753559 | USA/CA-CHLA-PLM99693185/2020 | 2020-12-01 | North America/USA/California/Los Ange | Clinical virology Laboratory, Children's Hospital Los Angeles | Center for Personalized Medicine, Children's Hospital Los Angeles | Gai et al et al |
| EPI_ISL_753562 | USA/CA-CHLA-PLM87172348/2020 | 2020-12-01 | North America/USA/California/Los Ange | Clinical virology Laboratory, Children's Hospital Los Angeles | Center for Personalized Medicine, Children's Hospital Los Angeles | Gai et al et al |
| EPI_ISL_753563 | USA/CA-CHLA-PLM41746412/2020 | 2020-12-01 | North America/USA/California/Los Ange | Clinical virology Laboratory, Children's Hospital Los Angeles | Center for Personalized Medicine, Children's Hospital Los Angeles | Gai et al et al |
| EPI_ISL_753573 | USA/CA-CHLA-PLM48917501/2020 | 2020-12-01 | North America/USA/California/Los Ange | Clinical virology Laboratory, Children's Hospital Los Angeles | Center for Personalized Medicine, Children's Hospital Los Angeles | Gai et al et al |
| EPI_ISL_753579 | USA/CA-CHLA-PLM50354444/2020 | 2020-12-02 | North America/USA/California/Los Ange | Clinical virology Laboratory, Children's Hospital Los Angeles | Center for Personalized Medicine, Children's Hospital Los Angeles | Gai et al et al |
| EPI_ISL_753587 | USA/CA-CHLA-PLM53624288/2020 | 2020-12-02 | North America/USA/California/Los Ange | Clinical virology Laboratory, Children's Hospital Los Angeles | Center for Personalized Medicine, Children's Hospital Los Angeles | Gai et al et al |

| | | | | | | |
|---|---|---|---|---|---|---|
| EPI_ISL_753590 | USA/CA-CHLA-PLM04375107/2020 | 2020-12-02 | North America/USA/California/Los Ange | Clinical virology Laboratory, Children's Hospital Los Angeles | Center for Personalized Medicine, Children's Hospital Los Angeles | Gai et al et al |
| EPI_ISL_753607 | USA/CA-CHLA-PLM77894582/2020 | 2020-12-03 | North America/USA/California/Los Ange | Clinical virology Laboratory, Children's Hospital Los Angeles | Center for Personalized Medicine, Children's Hospital Los Angeles | Gai et al et al |
| EPI_ISL_753617 | USA/CA-CHLA-PLM21825921/2020 | 2020-12-04 | North America/USA/California/Los Ange | Clinical virology Laboratory, Children's Hospital Los Angeles | Center for Personalized Medicine, Children's Hospital Los Angeles | Gai et al et al |
| EPI_ISL_753619 | USA/CA-CHLA-PLM62569795/2020 | 2020-12-04 | North America/USA/California/Los Ange | Clinical virology Laboratory, Children's Hospital Los Angeles | Center for Personalized Medicine, Children's Hospital Los Angeles | Gai et al et al |
| EPI_ISL_753623 | USA/CA-CHLA-PLM67203110/2020 | 2020-12-04 | North America/USA/California/Los Ange | Clinical virology Laboratory, Children's Hospital Los Angeles | Center for Personalized Medicine, Children's Hospital Los Angeles | Gai et al et al |
| EPI_ISL_753637 | USA/CA-CHLA-PLM02726594/2020 | 2020-12-04 | North America/USA/California/Los Ange | Clinical virology Laboratory, Children's Hospital Los Angeles | Center for Personalized Medicine, Children's Hospital Los Angeles | Gai et al et al |

| | | | | | | |
|---|---|---|---|---|---|---|
| EPI_ISL_753645 | USA/CA-CHLA-PLM40483831/2020 | 2020-11-29 | North America/USA/California/Los Ange | Clinical virology Laboratory, Children's Hospital Los Angeles | Center for Personalized Medicine, Children's Hospital Los Angeles | Gai et al et al |
| EPI_ISL_753650 | USA/CA-CHLA-PLM97776917/2020 | 2020-11-29 | North America/USA/California/Los Ange | Clinical virology Laboratory, Children's Hospital Los Angeles | Center for Personalized Medicine, Children's Hospital Los Angeles | Gai et al et al |
| EPI_ISL_753651 | USA/CA-CHLA-PLM51328819/2020 | 2020-11-29 | North America/USA/California/Los Ange | Clinical virology Laboratory, Children's Hospital Los Angeles | Center for Personalized Medicine, Children's Hospital Los Angeles | Gai et al et al |
| EPI_ISL_753663 | USA/CA-CHLA-PLM42701908/2020 | 2020-12-10 | North America/USA/California/Los Ange | Clinical virology Laboratory, Children's Hospital Los Angeles | Center for Personalized Medicine, Children's Hospital Los Angeles | Gai et al et al |
| EPI_ISL_753668 | USA/CA-CHLA-PLM80644511/2020 | 2020-12-07 | North America/USA/California/Los Ange | Clinical virology Laboratory, Children's Hospital Los Angeles | Center for Personalized Medicine, Children's Hospital Los Angeles | Gai et al et al |
| EPI_ISL_753670 | USA/CA-CHLA-PLM41118038/2020 | 2020-12-07 | North America/USA/California/Los Ange | Clinical virology Laboratory, Children's Hospital Los Angeles | Center for Personalized Medicine, Children's Hospital Los Angeles | Gai et al et al |

| | | | | | | |
|---|---|---|---|---|---|---|
| EPI_ISL_753681 | USA/CA-CHLA-PLM99533378/2020 | 2020-12-17 | North America/USA/California/Los Ange | Clinical virology Laboratory, Children's Hospital Los Angeles | Center for Personalized Medicine, Children's Hospital Los Angeles | Gai et al et al |
| EPI_ISL_753686 | USA/CA-CHLA-PLM50798548/2020 | 2020-12-17 | North America/USA/California/Los Ange | Clinical virology Laboratory, Children's Hospital Los Angeles | Center for Personalized Medicine, Children's Hospital Los Angeles | Gai et al et al |
| EPI_ISL_753697 | Bangladesh/SUST-GEB-Sunamganj-1/ | 2020-12-02 | Asia/Bangladesh/Sylhet/Sunamganj | Specialized Lab for COVID-19 Detection, Department of Genetic Engineering and Biotechnology | Specialized Lab for COVID-19 Detection, Department of Genetic Engineering and Biotechnology | Shamsul H. Prodhan et al |
| EPI_ISL_753699 | Indonesia/KS-NIHRD-PME4901/2020 | 2020-08-04 | Asia/Indonesia/South Kalimantan/ | RSUD Ulin Banjarmasin Kalimantan Selatan | National Institute of Health Research and Development | Subangkit et al |
| EPI_ISL_753708 | Germany/BE-ChVir-W1699-8236/202 | 2020-07-29 | Europe/Germany/Berlin/ | Charit√© Universit√§tsmedizin Berlin, Institut f√ºr Virologie/Labor Berlin | Charit√© Universit√§tsmedizin Berlin, Institut f√ºr Virologie | Victor M Corman et al |
| EPI_ISL_753710 | Germany/BE-ChVir-D2048-8293/2020 | 2020-10-06 | Europe/Germany/Berlin/ | Charit√© Universit√§tsmedizin Berlin, Institut f√ºr Virologie/Labor Berlin | Charit√© Universit√§tsmedizin Berlin, Institut f√ºr Virologie | Victor M Corman et al |
| EPI_ISL_753714 | Germany/BE-ChVir-W2212-8208/202 | 2020-11-18 | Europe/Germany/Berlin/ | Charit√© Universit√§tsmedizin Berlin, Institut f√ºr Virologie/Labor Berlin | Charit√© Universit√§tsmedizin Berlin, Institut f√ºr Virologie | Victor M Corman et al |
| EPI_ISL_753716 | Germany/BE-ChVir-D2049-8234/2020 | 2020-10-05 | Europe/Germany/Berlin/ | Charit√© Universit√§tsmedizin Berlin, Institut f√ºr Virologie/Labor Berlin | Charit√© Universit√§tsmedizin Berlin, Institut f√ºr Virologie | Victor M Corman et al |

| | | | | | | |
|---|---|---|---|---|---|---|
| EPI_ISL_753719 | Germany/BB-ChVir-D2047-5009/2020 | 2020-10-15 | Europe/Germany/Brandenburg/ | Charit√© Universit√§tsmedizin Berlin, Institut f√ºr Virologie/Labor Berlin | Charit√© Universit√§tsmedizin Berlin, Institut f√ºr Virologie | Victor M Corman et al |
| EPI_ISL_753731 | Germany/BE-ChVir-D1937-8281/2020 | 2020-09-25 | Europe/Germany/Berlin/ | Charit√© Universit√§tsmedizin Berlin, Institut f√ºr Virologie/Labor Berlin | Charit√© Universit√§tsmedizin Berlin, Institut f√ºr Virologie | Victor M Corman et al |
| EPI_ISL_753734 | Germany/BE-ChVir-W1730-6753/2020 | 2020-08-08 | Europe/Germany/Berlin/ | Charit√© Universit√§tsmedizin Berlin, Institut f√ºr Virologie/Labor Berlin | Charit√© Universit√§tsmedizin Berlin, Institut f√ºr Virologie | Victor M Corman et al |
| EPI_ISL_753746 | Germany/BE-ChVir-W1317-7453/2020 | 2020-04-26 | Europe/Germany/Berlin/ | Charit√© Universit√§tsmedizin Berlin, Institut f√ºr Virologie/Labor Berlin | Charit√© Universit√§tsmedizin Berlin, Institut f√ºr Virologie | Victor M Corman et al |
| EPI_ISL_753761 | Germany/BE-ChVir-W1341-4391/2020 | 2020-04-10 | Europe/Germany/Berlin/ | Charit√© Universit√§tsmedizin Berlin, Institut f√ºr Virologie/Labor Berlin | Charit√© Universit√§tsmedizin Berlin, Institut f√ºr Virologie | Victor M Corman et al |
| EPI_ISL_753764 | Germany/BE-ChVir-W1532-5774/2020 | 2020-04-05 | Europe/Germany/Berlin/ | Charit√© Universit√§tsmedizin Berlin, Institut f√ºr Virologie/Labor Berlin | Charit√© Universit√§tsmedizin Berlin, Institut f√ºr Virologie | Victor M Corman et al |
| EPI_ISL_753767 | Germany/un-ChVir-W1318-5622/2020 | 2020-04-29 | Europe/Germany/Germany/ | Charit√© Universit√§tsmedizin Berlin, Institut f√ºr Virologie/Labor Berlin | Charit√© Universit√§tsmedizin Berlin, Institut f√ºr Virologie | Victor M Corman et al |
| EPI_ISL_753770 | Germany/BB-ChVir-D932-5620/2020 | 2020-03-14 | Europe/Germany/Brandenburg/ | Charit√© Universit√§tsmedizin Berlin, Institut f√ºr Virologie/Labor Berlin | Charit√© Universit√§tsmedizin Berlin, Institut f√ºr Virologie | Victor M Corman et al |
| EPI_ISL_753771 | Germany/BB-ChVir-W1549-7455/2020 | 2020-06-17 | Europe/Germany/Brandenburg/ | Charit√© Universit√§tsmedizin Berlin, Institut f√ºr Virologie/Labor Berlin | Charit√© Universit√§tsmedizin Berlin, Institut f√ºr Virologie | Victor M Corman et al |
| EPI_ISL_753772 | Germany/BE-ChVir-W1321-4373/2020 | 2020-05-04 | Europe/Germany/Berlin/ | Charit√© Universit√§tsmedizin Berlin, Institut f√ºr Virologie/Labor Berlin | Charit√© Universit√§tsmedizin Berlin, Institut f√ºr Virologie | Victor M Corman et al |

| | | | | | | |
|---|---|---|---|---|---|---|
| EPI_ISL_753773 | Germany/BE-ChVir-D1987-4459/2020 | 2020-10-02 | Europe/Germany/Berlin/ | Charit√© Universit√§tsmedizin Berlin, Institut f√ºr Virologie/Labor Berlin | Charit√© Universit√§tsmedizin Berlin, Institut f√ºr Virologie | Victor M Corman et al |
| EPI_ISL_753777 | Germany/BE-ChVir-D1944-8293/2020 | 2020-10-01 | Europe/Germany/Berlin/ | Charit√© Universit√§tsmedizin Berlin, Institut f√ºr Virologie/Labor Berlin | Charit√© Universit√§tsmedizin Berlin, Institut f√ºr Virologie | Victor M Corman et al |
| EPI_ISL_753779 | Germany/BE-ChVir-D1951-5060/2020 | 2020-10-10 | Europe/Germany/Berlin/ | Charit√© Universit√§tsmedizin Berlin, Institut f√ºr Virologie/Labor Berlin | Charit√© Universit√§tsmedizin Berlin, Institut f√ºr Virologie | Victor M Corman et al |
| EPI_ISL_753785 | Germany/BE-ChVir-D1936-8209/2020 | 2020-09-24 | Europe/Germany/Berlin/ | Charit√© Universit√§tsmedizin Berlin, Institut f√ºr Virologie/Labor Berlin | Charit√© Universit√§tsmedizin Berlin, Institut f√ºr Virologie | Victor M Corman et al |
| EPI_ISL_753788 | Germany/BE-ChVir-D1946-8235/2020 | 2020-10-01 | Europe/Germany/Berlin/ | Charit√© Universit√§tsmedizin Berlin, Institut f√ºr Virologie/Labor Berlin | Charit√© Universit√§tsmedizin Berlin, Institut f√ºr Virologie | Victor M Corman et al |
| EPI_ISL_753792 | Germany/BE-ChVir-D2014-8229/2020 | 2020-10-14 | Europe/Germany/Berlin/ | Charit√© Universit√§tsmedizin Berlin, Institut f√ºr Virologie/Labor Berlin | Charit√© Universit√§tsmedizin Berlin, Institut f√ºr Virologie | Victor M Corman et al |
| EPI_ISL_753795 | Germany/BE-ChVir-W2118-4404/202 | 2020-10-12 | Europe/Germany/Berlin/ | Charit√© Universit√§tsmedizin Berlin, Institut f√ºr Virologie/Labor Berlin | Charit√© Universit√§tsmedizin Berlin, Institut f√ºr Virologie | Victor M Corman et al |
| EPI_ISL_753796 | Germany/NW-ChVir-D2005-8224/202 | 2020-10-19 | Europe/Germany/North Rhine Westphal | Charit√© Universit√§tsmedizin Berlin, Institut f√ºr Virologie/Labor Berlin | Charit√© Universit√§tsmedizin Berlin, Institut f√ºr Virologie | Victor M Corman et al |
| EPI_ISL_753797 | Germany/SN-ChVir-W1313-4308/202 | 2020-04-16 | Europe/Germany/Saxony/ | Charit√© Universit√§tsmedizin Berlin, Institut f√ºr Virologie/Labor Berlin | Charit√© Universit√§tsmedizin Berlin, Institut f√ºr Virologie | Victor M Corman et al |
| EPI_ISL_753798 | Germany/SN-ChVir-W1314-4321/202 | 2020-04-17 | Europe/Germany/Saxony/ | Charit√© Universit√§tsmedizin Berlin, Institut f√ºr Virologie/Labor Berlin | Charit√© Universit√§tsmedizin Berlin, Institut f√ºr Virologie | Victor M Corman et al |

| | | | | | | |
|---|---|---|---|---|---|---|
| EPI_ISL_753801 | Germany/HB-ChVir-D1974-8282/2020 | 2020-10-05 | Europe/Germany/Bremen/ | Charit√© Universit√§tsmedizin Berlin, Institut f√ºr Virologie/Labor Berlin | Charit√© Universit√§tsmedizin Berlin, Institut f√ºr Virologie | Victor M Corman et al |
| EPI_ISL_753808 | Germany/BE-ChVir-W1447-8299/2020 | 2020-06-23 | Europe/Germany/Berlin/ | Charit√© Universit√§tsmedizin Berlin, Institut f√ºr Virologie/Labor Berlin | Charit√© Universit√§tsmedizin Berlin, Institut f√ºr Virologie | Victor M Corman et al |
| EPI_ISL_753815 | Germany/BE-ChVir-D1972-8263/2020 | 2020-10-05 | Europe/Germany/Berlin/ | Charit√© Universit√§tsmedizin Berlin, Institut f√ºr Virologie/Labor Berlin | Charit√© Universit√§tsmedizin Berlin, Institut f√ºr Virologie | Victor M Corman et al |
| EPI_ISL_753816 | Germany/BE-ChVir-D1977-5942/2020 | 2020-10-08 | Europe/Germany/Berlin/ | Charit√© Universit√§tsmedizin Berlin, Institut f√ºr Virologie/Labor Berlin | Charit√© Universit√§tsmedizin Berlin, Institut f√ºr Virologie | Victor M Corman et al |
| EPI_ISL_753818 | Germany/BE-ChVir-D1979-4481/2020 | 2020-10-09 | Europe/Germany/Berlin/ | Charit√© Universit√§tsmedizin Berlin, Institut f√ºr Virologie/Labor Berlin | Charit√© Universit√§tsmedizin Berlin, Institut f√ºr Virologie | Victor M Corman et al |
| EPI_ISL_753823 | Germany/BE-ChVir-W1319-2159/2020 | 2020-05-04 | Europe/Germany/Berlin/ | Charit√© Universit√§tsmedizin Berlin, Institut f√ºr Virologie/Labor Berlin | Charit√© Universit√§tsmedizin Berlin, Institut f√ºr Virologie | Victor M Corman et al |
| EPI_ISL_753824 | Germany/BE-ChVir-W1323-5723/2020 | 2020-05-11 | Europe/Germany/Berlin/ | Charit√© Universit√§tsmedizin Berlin, Institut f√ºr Virologie/Labor Berlin | Charit√© Universit√§tsmedizin Berlin, Institut f√ºr Virologie | Victor M Corman et al |
| EPI_ISL_753829 | Germany/BE-ChVir-W2207-8292/2020 | 2020-11-22 | Europe/Germany/Berlin/ | Charit√© Universit√§tsmedizin Berlin, Institut f√ºr Virologie/Labor Berlin | Charit√© Universit√§tsmedizin Berlin, Institut f√ºr Virologie | Victor M Corman et al |
| EPI_ISL_753833 | Germany/BE-ChVir-W2219-4467/2020 | 2020-11-16 | Europe/Germany/Berlin/ | Charit√© Universit√§tsmedizin Berlin, Institut f√ºr Virologie/Labor Berlin | Charit√© Universit√§tsmedizin Berlin, Institut f√ºr Virologie | Victor M Corman et al |
| EPI_ISL_753843 | Germany/BB-ChVir-W1041-5644/2020 | 2020-03-25 | Europe/Germany/Brandenburg/ | Charit√© Universit√§tsmedizin Berlin, Institut f√ºr Virologie/Labor Berlin | Charit√© Universit√§tsmedizin Berlin, Institut f√ºr Virologie | Victor M Corman et al |

| | | | | | | |
|---|---|---|---|---|---|---|
| EPI_ISL_753847 | Germany/BB-ChVir-W1066-7201/202( | 2020-04-01 | Europe/Germany/Brandenburg/ | Charit√© Universit√§tsmedizin Berlin, Institut f√ºr Virologie/Labor Berlin | Charit√© Universit√§tsmedizin Berlin, Institut f√ºr Virologie | Victor M Corman et al |
| EPI_ISL_753858 | Germany/BB-ChVir-W1646-6734/202( | 2020-08-05 | Europe/Germany/Brandenburg/ | Charit√© Universit√§tsmedizin Berlin, Institut f√ºr Virologie/Labor Berlin | Charit√© Universit√§tsmedizin Berlin, Institut f√ºr Virologie | Victor M Corman et al |
| EPI_ISL_753859 | Germany/BB-ChVir-W2128-8294/202( | 2020-10-09 | Europe/Germany/Brandenburg/ | Charit√© Universit√§tsmedizin Berlin, Institut f√ºr Virologie/Labor Berlin | Charit√© Universit√§tsmedizin Berlin, Institut f√ºr Virologie | Victor M Corman et al |
| EPI_ISL_753861 | Germany/BE-ChVir-D1913-7445/2020 | 2020-10-03 | Europe/Germany/Berlin/ | Charit√© Universit√§tsmedizin Berlin, Institut f√ºr Virologie/Labor Berlin | Charit√© Universit√§tsmedizin Berlin, Institut f√ºr Virologie | Victor M Corman et al |
| EPI_ISL_753863 | Germany/BE-ChVir-D1916-8268/2020 | 2020-10-06 | Europe/Germany/Berlin/ | Charit√© Universit√§tsmedizin Berlin, Institut f√ºr Virologie/Labor Berlin | Charit√© Universit√§tsmedizin Berlin, Institut f√ºr Virologie | Victor M Corman et al |
| EPI_ISL_753865 | Germany/BE-ChVir-D1919-4432/2020 | 2020-10-04 | Europe/Germany/Berlin/ | Charit√© Universit√§tsmedizin Berlin, Institut f√ºr Virologie/Labor Berlin | Charit√© Universit√§tsmedizin Berlin, Institut f√ºr Virologie | Victor M Corman et al |
| EPI_ISL_753868 | Germany/BE-ChVir-D1923-8213/2020 | 2020-10-05 | Europe/Germany/Berlin/ | Charit√© Universit√§tsmedizin Berlin, Institut f√ºr Virologie/Labor Berlin | Charit√© Universit√§tsmedizin Berlin, Institut f√ºr Virologie | Victor M Corman et al |
| EPI_ISL_753869 | Germany/BE-ChVir-D1924-8211/2020 | 2020-10-09 | Europe/Germany/Berlin/ | Charit√© Universit√§tsmedizin Berlin, Institut f√ºr Virologie/Labor Berlin | Charit√© Universit√§tsmedizin Berlin, Institut f√ºr Virologie | Victor M Corman et al |
| EPI_ISL_753871 | Germany/BE-ChVir-D1927-7462/2020 | 2020-10-09 | Europe/Germany/Berlin/ | Charit√© Universit√§tsmedizin Berlin, Institut f√ºr Virologie/Labor Berlin | Charit√© Universit√§tsmedizin Berlin, Institut f√ºr Virologie | Victor M Corman et al |
| EPI_ISL_753872 | Germany/BE-ChVir-D1928-7459/2020 | 2020-10-09 | Europe/Germany/Berlin/ | Charit√© Universit√§tsmedizin Berlin, Institut f√ºr Virologie/Labor Berlin | Charit√© Universit√§tsmedizin Berlin, Institut f√ºr Virologie | Victor M Corman et al |

| | | | | | | |
|---|---|---|---|---|---|---|
| EPI_ISL_753873 | Germany/BE-ChVir-D1930-8296/2020 | 2020-10-10 | Europe/Germany/Berlin/ | Charit√© Universit√§tsmedizin Berlin, Institut f√ºr Virologie/Labor Berlin | Charit√© Universit√§tsmedizin Berlin, Institut f√ºr Virologie | Victor M Corman et al |
| EPI_ISL_753874 | Germany/BE-ChVir-D1932-8298/2020 | 2020-10-08 | Europe/Germany/Berlin/ | Charit√© Universit√§tsmedizin Berlin, Institut f√ºr Virologie/Labor Berlin | Charit√© Universit√§tsmedizin Berlin, Institut f√ºr Virologie | Victor M Corman et al |
| EPI_ISL_753875 | Germany/BE-ChVir-D1935-5099/2020 | 2020-10-10 | Europe/Germany/Berlin/ | Charit√© Universit√§tsmedizin Berlin, Institut f√ºr Virologie/Labor Berlin | Charit√© Universit√§tsmedizin Berlin, Institut f√ºr Virologie | Victor M Corman et al |
| EPI_ISL_753879 | Germany/BE-ChVir-D1943-5032/2020 | 2020-10-01 | Europe/Germany/Berlin/ | Charit√© Universit√§tsmedizin Berlin, Institut f√ºr Virologie/Labor Berlin | Charit√© Universit√§tsmedizin Berlin, Institut f√ºr Virologie | Victor M Corman et al |
| EPI_ISL_753883 | Germany/BE-ChVir-D1953-7492/2020 | 2020-10-10 | Europe/Germany/Berlin/ | Charit√© Universit√§tsmedizin Berlin, Institut f√ºr Virologie/Labor Berlin | Charit√© Universit√§tsmedizin Berlin, Institut f√ºr Virologie | Victor M Corman et al |
| EPI_ISL_753885 | Germany/BE-ChVir-D1957-4437/2020 | 2020-09-26 | Europe/Germany/Berlin/ | Charit√© Universit√§tsmedizin Berlin, Institut f√ºr Virologie/Labor Berlin | Charit√© Universit√§tsmedizin Berlin, Institut f√ºr Virologie | Victor M Corman et al |
| EPI_ISL_753886 | Germany/BE-ChVir-D1958-8276/2020 | 2020-09-28 | Europe/Germany/Berlin/ | Charit√© Universit√§tsmedizin Berlin, Institut f√ºr Virologie/Labor Berlin | Charit√© Universit√§tsmedizin Berlin, Institut f√ºr Virologie | Victor M Corman et al |
| EPI_ISL_753888 | Germany/BE-ChVir-D1996-4483/2020 | 2020-10-13 | Europe/Germany/Berlin/ | Charit√© Universit√§tsmedizin Berlin, Institut f√ºr Virologie/Labor Berlin | Charit√© Universit√§tsmedizin Berlin, Institut f√ºr Virologie | Victor M Corman et al |
| EPI_ISL_753889 | Germany/BE-ChVir-D1997-8209/2020 | 2020-10-05 | Europe/Germany/Berlin/ | Charit√© Universit√§tsmedizin Berlin, Institut f√ºr Virologie/Labor Berlin | Charit√© Universit√§tsmedizin Berlin, Institut f√ºr Virologie | Victor M Corman et al |
| EPI_ISL_753892 | Germany/BE-ChVir-D2000-8271/2020 | 2020-10-15 | Europe/Germany/Berlin/ | Charit√© Universit√§tsmedizin Berlin, Institut f√ºr Virologie/Labor Berlin | Charit√© Universit√§tsmedizin Berlin, Institut f√ºr Virologie | Victor M Corman et al |

| | | | | | | |
|---|---|---|---|---|---|---|
| EPI_ISL_753893 | Germany/BE-ChVir-D2003-5974/2020 | 2020-10-15 | Europe/Germany/Berlin/ | Charit√© Universit√§tsmedizin Berlin, Institut f√ºr Virologie/Labor Berlin | Charit√© Universit√§tsmedizin Berlin, Institut f√ºr Virologie | Victor M Corman et al |
| EPI_ISL_753898 | Germany/BE-ChVir-D2010-8268/2020 | 2020-10-15 | Europe/Germany/Berlin/ | Charit√© Universit√§tsmedizin Berlin, Institut f√ºr Virologie/Labor Berlin | Charit√© Universit√§tsmedizin Berlin, Institut f√ºr Virologie | Victor M Corman et al |
| EPI_ISL_753901 | Germany/BE-ChVir-D2016-5933/2020 | 2020-10-18 | Europe/Germany/Berlin/ | Charit√© Universit√§tsmedizin Berlin, Institut f√ºr Virologie/Labor Berlin | Charit√© Universit√§tsmedizin Berlin, Institut f√ºr Virologie | Victor M Corman et al |
| EPI_ISL_753902 | Germany/BE-ChVir-D2017-8214/2020 | 2020-10-19 | Europe/Germany/Berlin/ | Charit√© Universit√§tsmedizin Berlin, Institut f√ºr Virologie/Labor Berlin | Charit√© Universit√§tsmedizin Berlin, Institut f√ºr Virologie | Victor M Corman et al |
| EPI_ISL_753903 | Germany/BE-ChVir-D2020-4418/2020 | 2020-10-11 | Europe/Germany/Berlin/ | Charit√© Universit√§tsmedizin Berlin, Institut f√ºr Virologie/Labor Berlin | Charit√© Universit√§tsmedizin Berlin, Institut f√ºr Virologie | Victor M Corman et al |
| EPI_ISL_753904 | Germany/BE-ChVir-D2021-4462/2020 | 2020-10-10 | Europe/Germany/Berlin/ | Charit√© Universit√§tsmedizin Berlin, Institut f√ºr Virologie/Labor Berlin | Charit√© Universit√§tsmedizin Berlin, Institut f√ºr Virologie | Victor M Corman et al |
| EPI_ISL_753911 | Germany/BE-ChVir-D2033-5902/2020 | 2020-10-06 | Europe/Germany/Berlin/ | Charit√© Universit√§tsmedizin Berlin, Institut f√ºr Virologie/Labor Berlin | Charit√© Universit√§tsmedizin Berlin, Institut f√ºr Virologie | Victor M Corman et al |
| EPI_ISL_753913 | Germany/BE-ChVir-D2035-8263/2020 | 2020-10-09 | Europe/Germany/Berlin/ | Charit√© Universit√§tsmedizin Berlin, Institut f√ºr Virologie/Labor Berlin | Charit√© Universit√§tsmedizin Berlin, Institut f√ºr Virologie | Victor M Corman et al |
| EPI_ISL_753914 | Germany/BE-ChVir-D2036-8292/2020 | 2020-10-08 | Europe/Germany/Berlin/ | Charit√© Universit√§tsmedizin Berlin, Institut f√ºr Virologie/Labor Berlin | Charit√© Universit√§tsmedizin Berlin, Institut f√ºr Virologie | Victor M Corman et al |
| EPI_ISL_753922 | Germany/BE-ChVir-D909-4341/2020 | 2020-03-16 | Europe/Germany/Berlin/ | Charit√© Universit√§tsmedizin Berlin, Institut f√ºr Virologie/Labor Berlin | Charit√© Universit√§tsmedizin Berlin, Institut f√ºr Virologie | Victor M Corman et al |

| | | | | | | |
|---|---|---|---|---|---|---|
| EPI_ISL_753925 | Germany/BE-ChVir-W1045-5646/202( | 2020-03-26 | Europe/Germany/Berlin/ | Charit√© Universit√§tsmedizin Berlin, Institut f√ºr Virologie/Labor Berlin | Charit√© Universit√§tsmedizin Berlin, Institut f√ºr Virologie | Victor M Corman et al |
| EPI_ISL_753926 | Germany/BE-ChVir-W1048-4346/202( | 2020-03-27 | Europe/Germany/Berlin/ | Charit√© Universit√§tsmedizin Berlin, Institut f√ºr Virologie/Labor Berlin | Charit√© Universit√§tsmedizin Berlin, Institut f√ºr Virologie | Victor M Corman et al |
| EPI_ISL_753928 | Germany/BE-ChVir-W1060-4393/202( | 2020-03-30 | Europe/Germany/Berlin/ | Charit√© Universit√§tsmedizin Berlin, Institut f√ºr Virologie/Labor Berlin | Charit√© Universit√§tsmedizin Berlin, Institut f√ºr Virologie | Victor M Corman et al |
| EPI_ISL_753930 | Germany/BE-ChVir-W1072-4344/202( | 2020-04-02 | Europe/Germany/Berlin/ | Charit√© Universit√§tsmedizin Berlin, Institut f√ºr Virologie/Labor Berlin | Charit√© Universit√§tsmedizin Berlin, Institut f√ºr Virologie | Victor M Corman et al |
| EPI_ISL_753943 | Germany/BE-ChVir-W1149-8118/202( | 2020-04-09 | Europe/Germany/Berlin/ | Charit√© Universit√§tsmedizin Berlin, Institut f√ºr Virologie/Labor Berlin | Charit√© Universit√§tsmedizin Berlin, Institut f√ºr Virologie | Victor M Corman et al |
| EPI_ISL_753947 | Germany/BE-ChVir-W1282-4384/202( | 2020-04-17 | Europe/Germany/Berlin/ | Charit√© Universit√§tsmedizin Berlin, Institut f√ºr Virologie/Labor Berlin | Charit√© Universit√§tsmedizin Berlin, Institut f√ºr Virologie | Victor M Corman et al |
| EPI_ISL_753949 | Germany/BE-ChVir-W1291-8183/202( | 2020-04-21 | Europe/Germany/Berlin/ | Charit√© Universit√§tsmedizin Berlin, Institut f√ºr Virologie/Labor Berlin | Charit√© Universit√§tsmedizin Berlin, Institut f√ºr Virologie | Victor M Corman et al |
| EPI_ISL_753951 | Germany/BE-ChVir-W1299-4302/202( | 2020-04-28 | Europe/Germany/Berlin/ | Charit√© Universit√§tsmedizin Berlin, Institut f√ºr Virologie/Labor Berlin | Charit√© Universit√§tsmedizin Berlin, Institut f√ºr Virologie | Victor M Corman et al |
| EPI_ISL_753952 | Germany/BE-ChVir-W1303-8106/202( | 2020-04-30 | Europe/Germany/Berlin/ | Charit√© Universit√§tsmedizin Berlin, Institut f√ºr Virologie/Labor Berlin | Charit√© Universit√§tsmedizin Berlin, Institut f√ºr Virologie | Victor M Corman et al |
| EPI_ISL_753953 | Germany/BE-ChVir-W1309-8155/202( | 2020-05-08 | Europe/Germany/Berlin/ | Charit√© Universit√§tsmedizin Berlin, Institut f√ºr Virologie/Labor Berlin | Charit√© Universit√§tsmedizin Berlin, Institut f√ºr Virologie | Victor M Corman et al |

| | | | | | | |
|---|---|---|---|---|---|---|
| EPI_ISL_753970 | Germany/BE-ChVir-W1445-6284/202( | 2020-06-29 | Europe/Germany/Berlin/ | Charit√© Universit√§tsmedizin Berlin, Institut f√ºr Virologie/Labor Berlin | Charit√© Universit√§tsmedizin Berlin, Institut f√ºr Virologie | Victor M Corman et al |
| EPI_ISL_753979 | Germany/BE-ChVir-W1645-6781/202( | 2020-08-05 | Europe/Germany/Berlin/ | Charit√© Universit√§tsmedizin Berlin, Institut f√ºr Virologie/Labor Berlin | Charit√© Universit√§tsmedizin Berlin, Institut f√ºr Virologie | Victor M Corman et al |
| EPI_ISL_753981 | Germany/BE-ChVir-W1649-7416/202( | 2020-07-16 | Europe/Germany/Berlin/ | Charit√© Universit√§tsmedizin Berlin, Institut f√ºr Virologie/Labor Berlin | Charit√© Universit√§tsmedizin Berlin, Institut f√ºr Virologie | Victor M Corman et al |
| EPI_ISL_753985 | Germany/BE-ChVir-W1690-8217/202( | 2020-07-17 | Europe/Germany/Berlin/ | Charit√© Universit√§tsmedizin Berlin, Institut f√ºr Virologie/Labor Berlin | Charit√© Universit√§tsmedizin Berlin, Institut f√ºr Virologie | Victor M Corman et al |
| EPI_ISL_753990 | Germany/BE-ChVir-W1695-8202/202( | 2020-07-01 | Europe/Germany/Berlin/ | Charit√© Universit√§tsmedizin Berlin, Institut f√ºr Virologie/Labor Berlin | Charit√© Universit√§tsmedizin Berlin, Institut f√ºr Virologie | Victor M Corman et al |
| EPI_ISL_753997 | Germany/BE-ChVir-W1706-8239/202( | 2020-08-15 | Europe/Germany/Berlin/ | Charit√© Universit√§tsmedizin Berlin, Institut f√ºr Virologie/Labor Berlin | Charit√© Universit√§tsmedizin Berlin, Institut f√ºr Virologie | Victor M Corman et al |
| EPI_ISL_753998 | Germany/BE-ChVir-W1708-6787/202( | 2020-08-05 | Europe/Germany/Berlin/ | Charit√© Universit√§tsmedizin Berlin, Institut f√ºr Virologie/Labor Berlin | Charit√© Universit√§tsmedizin Berlin, Institut f√ºr Virologie | Victor M Corman et al |
| EPI_ISL_754016 | Germany/BE-ChVir-W1731-2742/202( | 2020-08-07 | Europe/Germany/Berlin/ | Charit√© Universit√§tsmedizin Berlin, Institut f√ºr Virologie/Labor Berlin | Charit√© Universit√§tsmedizin Berlin, Institut f√ºr Virologie | Victor M Corman et al |
| EPI_ISL_754018 | Germany/BE-ChVir-W2122-5977/202( | 2020-10-15 | Europe/Germany/Berlin/ | Charit√© Universit√§tsmedizin Berlin, Institut f√ºr Virologie/Labor Berlin | Charit√© Universit√§tsmedizin Berlin, Institut f√ºr Virologie | Victor M Corman et al |
| EPI_ISL_754021 | Germany/BE-ChVir-W2125-4401/202( | 2020-09-27 | Europe/Germany/Berlin/ | Charit√© Universit√§tsmedizin Berlin, Institut f√ºr Virologie/Labor Berlin | Charit√© Universit√§tsmedizin Berlin, Institut f√ºr Virologie | Victor M Corman et al |

| | | | | | | |
|---|---|---|---|---|---|---|
| EPI_ISL_754022 | Germany/BW-ChVir-W1424-8276/202 | 2020-06-28 | Europe/Germany/Baden-Wuerttemberg | Charit√© Universit√§tsmedizin Berlin, Institut f√ºr Virologie/Labor Berlin | Charit√© Universit√§tsmedizin Berlin, Institut f√ºr Virologie | Victor M Corman et al |
| EPI_ISL_754032 | Germany/BY-ChVir-W1126-5740/2020 | 2020-03-27 | Europe/Germany/Bavaria/ | Charit√© Universit√§tsmedizin Berlin, Institut f√ºr Virologie/Labor Berlin | Charit√© Universit√§tsmedizin Berlin, Institut f√ºr Virologie | Victor M Corman et al |
| EPI_ISL_754042 | Germany/HE-ChVir-W1339-8077/202 | 2020-05-18 | Europe/Germany/Hesse/ | Charit√© Universit√§tsmedizin Berlin, Institut f√ºr Virologie/Labor Berlin | Charit√© Universit√§tsmedizin Berlin, Institut f√ºr Virologie | Victor M Corman et al |
| EPI_ISL_754045 | Germany/NI-ChVir-W1707-6795/2020 | 2020-08-07 | Europe/Germany/Niedersachsen/ | Charit√© Universit√§tsmedizin Berlin, Institut f√ºr Virologie/Labor Berlin | Charit√© Universit√§tsmedizin Berlin, Institut f√ºr Virologie | Victor M Corman et al |
| EPI_ISL_754047 | Germany/NW-ChVir-W1049-5662/20 | 2020-03-26 | Europe/Germany/North Rhine Westphal | Charit√© Universit√§tsmedizin Berlin, Institut f√ºr Virologie/Labor Berlin | Charit√© Universit√§tsmedizin Berlin, Institut f√ºr Virologie | Victor M Corman et al |
| EPI_ISL_754051 | Germany/SN-ChVir-W1157-4330/202 | 2020-04-06 | Europe/Germany/Saxony/ | Charit√© Universit√§tsmedizin Berlin, Institut f√ºr Virologie/Labor Berlin | Charit√© Universit√§tsmedizin Berlin, Institut f√ºr Virologie | Victor M Corman et al |
| EPI_ISL_754052 | Germany/SN-ChVir-W1275-4371/202 | 2020-04-15 | Europe/Germany/Saxony/ | Charit√© Universit√§tsmedizin Berlin, Institut f√ºr Virologie/Labor Berlin | Charit√© Universit√§tsmedizin Berlin, Institut f√ºr Virologie | Victor M Corman et al |
| EPI_ISL_754053 | Germany/SN-ChVir-W1276-4387/202 | 2020-04-15 | Europe/Germany/Saxony/ | Charit√© Universit√§tsmedizin Berlin, Institut f√ºr Virologie/Labor Berlin | Charit√© Universit√§tsmedizin Berlin, Institut f√ºr Virologie | Victor M Corman et al |
| EPI_ISL_754055 | Germany/BE-ChVir-D697-8137/2020 | 2020-03-05 | Europe/Germany/Berlin/ | Charit√© Universit√§tsmedizin Berlin, Institut f√ºr Virologie/Labor Berlin | Charit√© Universit√§tsmedizin Berlin, Institut f√ºr Virologie | Victor M Corman et al |
| EPI_ISL_754057 | Germany/BE-ChVir-D751-5650/2020 | 2020-03-13 | Europe/Germany/Berlin/ | Charit√© Universit√§tsmedizin Berlin, Institut f√ºr Virologie/Labor Berlin | Charit√© Universit√§tsmedizin Berlin, Institut f√ºr Virologie | Victor M Corman et al |

| | | | | | | |
|---|---|---|---|---|---|---|
| EPI_ISL_754058 | Germany/BE-ChVir-D754-8111/2020 | 2020-03-16 | Europe/Germany/Berlin/ | Charit√© Universit√§tsmedizin Berlin, Institut f√ºr Virologie/Labor Berlin | Charit√© Universit√§tsmedizin Berlin, Institut f√ºr Virologie | Victor M Corman et al |
| EPI_ISL_754059 | Bangladesh/SUST-GEB-Sunamganj-4/2 | 2020-12-02 | Asia/Bangladesh/Sylhet/Sunamganj | Specialized Lab for COVID-19 Detection, Department of Genetic Engineering and Biotechnology | Specialized Lab for COVID-19 Detection, Department of Genetic Engineering and Biotechnology | Md. Nazmul Hasan et al |
| EPI_ISL_754064 | Nepal/617/2020 | 2020-08-10 | Asia/Nepal/Kavre/ | Kathmandu University | Nepal Health Research Council | Pradip Gyanwali et al |
| EPI_ISL_754068 | Nepal/2097/2020 | 2020-07-30 | Asia/Nepal/Kathmandu/ | Nepal Korea Friendship Municipality Hospital | Nepal Health Research Council | Pradip Gyanwali et al |
| EPI_ISL_754072 | Nepal/459A/2020 | 2020-08-03 | Asia/Nepal/Baglung/ | National Public Health Laboratory | Nepal Health Research Council | Pradip Gyanwali et al |
| EPI_ISL_754073 | Nepal/70N/2020 | 2020-08-03 | Asia/Nepal/Rautahat/ | National Public Health Laboratory | Nepal Health Research Council | Pradip Gyanwali et al |
| EPI_ISL_754103 | Singapore/1494/2020 | 2020-12-21 | Asia/Singapore/Singapore/ | National Public Health Laboratory, National Centre for Infectious Diseases | National Public Health Laboratory, National Centre for Infectious Diseases | Tze Minn Mak et al |
| EPI_ISL_754112 | USA/CA-LACPHL-AF00029/2020 | 2020-12-20 | North America/USA/California/Los Ange | UCLA Clinical Micro Lab | Los Angeles County PHL | P. Hemarajata et al. et al |
| EPI_ISL_754119 | USA/CA-LACPHL-AF00036/2020 | 2020-12-20 | North America/USA/California/Los Ange | UCLA Clinical Micro Lab | Los Angeles County PHL | P. Hemarajata et al. et al |
| EPI_ISL_754124 | USA/CA-LACPHL-AF00049/2020 | 2020-12-19 | North America/USA/California/Los Ange | USC Clinical Lab | Los Angeles County PHL | P. Hemarajata et al. et al |
| EPI_ISL_754131 | USA/CA-LACPHL-AF00056/2020 | 2020-12-17 | North America/USA/California/Los Ange | Los Angeles County PHL | Los Angeles County PHL | P. Hemarajata et al. et al |
| EPI_ISL_754136 | France/OCC-182/2020 | 2020-09-07 | Europe/France/Occitanie/ | CHU Purpan - Laboratoire de Virologie - Institut F√©d√©ratif de Biologie | CHU Purpan - Laboratoire de Virologie - Institut F√©d√©ratif de Biologie | Latour J. et al |

| | | | | | | |
|---|---|---|---|---|---|---|
| EPI_ISL_754137 | France/OCC-183/2020 | 2020-10-27 | Europe/France/Occitanie/ | CHU Purpan - Laboratoire de Virologie - Institut F√©d√©ratif de Biologie | CHU Purpan - Laboratoire de Virologie - Institut F√©d√©ratif de Biologie | Latour J. et al |
| EPI_ISL_754138 | France/OCC-184/2020 | 2020-10-11 | Europe/France/Occitanie/ | CHU Purpan - Laboratoire de Virologie - Institut F√©d√©ratif de Biologie | CHU Purpan - Laboratoire de Virologie - Institut F√©d√©ratif de Biologie | Latour J. et al |
| EPI_ISL_754144 | France/BRE-IPP10831/2020 | 2020-11-20 | Europe/France/Bretagne/Quimper | Laboratoire de Microbiologie | National Reference Center for Viruses of Respiratory Infections, Institut Pasteur, Paris | Marion Barbet et al |
| EPI_ISL_754145 | France/BRE-IPP10838/2020 | 2020-11-25 | Europe/France/Bretagne/Quimper | Laboratoire de Microbiologie | National Reference Center for Viruses of Respiratory Infections, Institut Pasteur, Paris | Marion Barbet et al |
| EPI_ISL_754148 | France/BRE-IPP10856/2020 | 2020-12-02 | Europe/France/Bretagne/Quimper | Laboratoire de Microbiologie | National Reference Center for Viruses of Respiratory Infections, Institut Pasteur, Paris | Marion Barbet et al |
| EPI_ISL_754152 | France/BRE-IPP10833/2020 | 2020-11-23 | Europe/France/Bretagne/Quimper | Laboratoire de Microbiologie | National Reference Center for Viruses of Respiratory Infections, Institut Pasteur, Paris | Marion Barbet et al |
| EPI_ISL_754153 | France/BRE-IPP10835/2020 | 2020-11-23 | Europe/France/Bretagne/Quimper | Laboratoire de Microbiologie | National Reference Center for Viruses of Respiratory Infections, Institut Pasteur, Paris | Marion Barbet et al |

| | | | | | | |
|---|---|---|---|---|---|---|
| EPI_ISL_754154 | France/BRE-IPP10841/2020 | 2020-11-20 | Europe/France/Bretagne/Quimper | Laboratoire de Microbiologie | National Reference Center for Viruses of Respiratory Infections, Institut Pasteur, Paris | Marion Barbet et al |
| EPI_ISL_754155 | France/BRE-IPP10843/2020 | 2020-11-26 | Europe/France/Bretagne/Quimper | Laboratoire de Microbiologie | National Reference Center for Viruses of Respiratory Infections, Institut Pasteur, Paris | Marion Barbet et al |
| EPI_ISL_754157 | France/BRE-IPP10853/2020 | 2020-11-30 | Europe/France/Bretagne/Quimper | Laboratoire de Microbiologie | National Reference Center for Viruses of Respiratory Infections, Institut Pasteur, Paris | Marion Barbet et al |
| EPI_ISL_754161 | France/BRE-IPP10865/2020 | 2020-11-18 | Europe/France/Bretagne/Quimper | Laboratoire de Microbiologie | National Reference Center for Viruses of Respiratory Infections, Institut Pasteur, Paris | Marion Barbet et al |
| EPI_ISL_754169 | France/BRE-IPP10836/2020 | 2020-11-15 | Europe/France/Bretagne/Quimper | Laboratoire de Microbiologie | National Reference Center for Viruses of Respiratory Infections, Institut Pasteur, Paris | Marion Barbet et al |
| EPI_ISL_754173 | Bangladesh/SUST-GEB-Habiganj-3/20 | 2020-11-25 | Asia/Bangladesh/Sylhet/Habiganj | Specialized Lab for COVID-19 Detection, Department of Genetic Engineering and Biotechnology | Specialized Lab for COVID-19 Detection, Department of Genetic Engineering and Biotechnology | Md. Nazmul Hasan et al |
| EPI_ISL_754174 | Germany/BW-ChVir21528/2020 | 2020-12-21 | Europe/Germany/Baden-Wuerttemberg | Charitv© Universitv§tsmedizin Berlin, Institut fvºr Virologie | Charitv© Universitv§tsmedizin Berlin, Institut fvºr Virologie | Victor M Corman et al |

| | | | | | | |
|---|---|---|---|---|---|---|
| EPI_ISL_754175 | Germany/NI-ChVir21542/2020 | 2020-12-20 | Europe/Germany/Niedersachsen/ | Charit√© Universit√§tsmedizin Berlin, Institut f√ºr Virologie | Charit√© Universit√§tsmedizin Berlin, Institut f√ºr Virologie | Victor M Corman et al |
| EPI_ISL_754177 | Bangladesh/SUST-GEB-Habiganj-4/20 | 2020-11-25 | Asia/Bangladesh/Sylhet/Habiganj | Specialized Lab for COVID-19 Detection, Department of Genetic Engineering and Biotechnology | Specialized Lab for COVID-19 Detection, Department of Genetic Engineering and Biotechnology | Nurnabi Azad Jewel et al |
| EPI_ISL_754179 | Bangladesh/SUST-GEB-Habiganj-6/20 | 2020-11-12 | Asia/Bangladesh/Sylhet/Habiganj | Specialized Lab for COVID-19 Detection, Department of Genetic Engineering and Biotechnology | Specialized Lab for COVID-19 Detection, Department of Genetic Engineering and Biotechnology | Hammadul Hoque et al |
| EPI_ISL_754181 | Georgia/Tb-97937/2020 | 2020-09-23 | Asia/Georgia/Tbilisi/ | Department for Virology, Molecular Biology and Genome Research, R. G. Lugar Center for Public Health Research, National Center for Disease Control and Public Health (NCDC) of Georgia. | Department for Virology, Molecular Biology and Genome Research, R. G. Lugar Center for Public Health Research, National Center for Disease Control and Public Health (NCDC) of Georgia. | Meri Pantsulaia et al |
| EPI_ISL_754186 | Germany/BE-ChVir-W2121-4431/2020 | 2020-10-15 | Europe/Germany/Berlin/ | Charit√© Universit√§tsmedizin Berlin, Institut f√ºr Virologie/Labor Berlin | Charit√© Universit√§tsmedizin Berlin, Institut f√ºr Virologie | Victor M Corman et al |
| EPI_ISL_754189 | Germany/BE-ChVir-D1947-5915/2020 | 2020-10-04 | Europe/Germany/Berlin/ | Charit√© Universit√§tsmedizin Berlin, Institut f√ºr Virologie/Labor Berlin | Charit√© Universit√§tsmedizin Berlin, Institut f√ºr Virologie | Victor M Corman et al |

| | | | | | | |
|---|---|---|---|---|---|---|
| EPI_ISL_754190 | Germany/BE-ChVir-W2120-5976/202[illegible] | 2020-10-15 | Europe/Germany/Berlin/ | Charit√© Universit√§tsmedizin Berlin, Institut f√ºr Virologie/Labor Berlin | Charit√© Universit√§tsmedizin Berlin, Institut f√ºr Virologie | Victor M Corman et al |
| EPI_ISL_754195 | Georgia/Tb-72720/2020 | 2020-09-02 | Asia/Georgia/Tbilisi/ | Department for Virology, Molecular Biology and Genome Research, R. G. Lugar Center for Public Health Research, National Center for Disease Control and Public Health (NCDC) of Georgia. | Department for Virology, Molecular Biology and Genome Research, R. G. Lugar Center for Public Health Research, National Center for Disease Control and Public Health (NCDC) of Georgia. | Gvantsa Brachveli et al |
| EPI_ISL_754196 | Russia/MOS-CRIE-13606547/2020 | 2020-11-09 | Europe/Russia/Moscow Oblast/ | "Molecular diagnostic laboratory of Federal Budget Institution of Science ""Central Research Institute of Epidemiology"" of The Federal Service on Customers' Rights Protection and Human Well-being Surveillance" | Group of Genomics and Postgenomic Technologies of Central Research Institute of Epidemilology | Samoilov AE et al |
| EPI_ISL_754197 | Russia/MOS-CRIE-13604992/2020 | 2020-11-09 | Europe/Russia/Moscow Oblast/ | "Molecular diagnostic laboratory of Federal Budget Institution of Science ""Central Research Institute of Epidemiology"" of The Federal Service on Customers' Rights Protection and Human Well-being Surveillance" | Group of Genomics and Postgenomic Technologies of Central Research Institute of Epidemilology | Samoilov AE et al |
| EPI_ISL_754198 | Russia/MOS-CRIE-13604226/2020 | 2020-11-09 | Europe/Russia/Moscow Oblast/ | "Molecular diagnostic laboratory of Federal Budget Institution of Science ""Central Research Institute of Epidemiology"" of The Federal Service on Customers' Rights Protection and Human Well-being Surveillance" | Group of Genomics and Postgenomic Technologies of Central Research Institute of Epidemilology | Samoilov AE et al |

| | | | | | | |
|---|---|---|---|---|---|---|
| EPI_ISL_754199 | Russia/MOS-CRIE-13642264/2020 | 2020-11-09 | Europe/Russia/Moscow Oblast/ | "Molecular diagnostic laboratory of Federal Budget Institution of Science ""Central Research Institute of Epidemiology"" of The Federal Service on Customers' Rights Protection and Human Well-being Surveillance" | Group of Genomics and Postgenomic Technologies of Central Research Institute of Epidemilology | Samoilov AE et al |
| EPI_ISL_754200 | Russia/MOS-CRIE-13637504/2020 | 2020-11-09 | Europe/Russia/Moscow Oblast/ | "Molecular diagnostic laboratory of Federal Budget Institution of Science ""Central Research Institute of Epidemiology"" of The Federal Service on Customers' Rights Protection and Human Well-being Surveillance" | Group of Genomics and Postgenomic Technologies of Central Research Institute of Epidemilology | Samoilov AE et al |
| EPI_ISL_754203 | Russia/MOS-CRIE-13652490/2020 | 2020-11-09 | Europe/Russia/Moscow Oblast/ | "Molecular diagnostic laboratory of Federal Budget Institution of Science ""Central Research Institute of Epidemiology"" of The Federal Service on Customers' Rights Protection and Human Well-being Surveillance" | Group of Genomics and Postgenomic Technologies of Central Research Institute of Epidemilology | Samoilov AE et al |
| EPI_ISL_754204 | Russia/MOS-CRIE-13651164/2020 | 2020-11-09 | Europe/Russia/Moscow Oblast/ | "Molecular diagnostic laboratory of Federal Budget Institution of Science ""Central Research Institute of Epidemiology"" of The Federal Service on Customers' Rights Protection and Human Well-being Surveillance" | Group of Genomics and Postgenomic Technologies of Central Research Institute of Epidemilology | Samoilov AE et al |
| EPI_ISL_754205 | Russia/MOS-CRIE-13675724/2020 | 2020-11-09 | Europe/Russia/Moscow Oblast/ | "Molecular diagnostic laboratory of Federal Budget Institution of Science ""Central Research Institute of Epidemiology"" of The Federal Service on Customers' Rights Protection and Human Well-being Surveillance" | Group of Genomics and Postgenomic Technologies of Central Research Institute of Epidemilology | Samoilov AE et al |

| | | | | | | |
|---|---|---|---|---|---|---|
| EPI_ISL_754206 | Russia/MOS-CRIE-13674129/2020 | 2020-11-09 | Europe/Russia/Moscow Oblast/ | "Molecular diagnostic laboratory of Federal Budget Institution of Science ""Central Research Institute of Epidemiology"" of The Federal Service on Customers' Rights Protection and Human Well-being Surveillance" | Group of Genomics and Postgenomic Technologies of Central Research Institute of Epidemilology | Samoilov AE et al |
| EPI_ISL_754207 | Russia/MOS-CRIE-13669866/2020 | 2020-11-09 | Europe/Russia/Moscow Oblast/ | "Molecular diagnostic laboratory of Federal Budget Institution of Science ""Central Research Institute of Epidemiology"" of The Federal Service on Customers' Rights Protection and Human Well-being Surveillance" | Group of Genomics and Postgenomic Technologies of Central Research Institute of Epidemilology | Samoilov AE et al |
| EPI_ISL_754208 | Russia/MOS-CRIE-13663504/2020 | 2020-11-09 | Europe/Russia/Moscow Oblast/ | "Molecular diagnostic laboratory of Federal Budget Institution of Science ""Central Research Institute of Epidemiology"" of The Federal Service on Customers' Rights Protection and Human Well-being Surveillance" | Group of Genomics and Postgenomic Technologies of Central Research Institute of Epidemilology | Samoilov AE et al |
| EPI_ISL_754210 | Russia/MOS-CRIE-13666589/2020 | 2020-11-09 | Europe/Russia/Moscow Oblast/ | "Molecular diagnostic laboratory of Federal Budget Institution of Science ""Central Research Institute of Epidemiology"" of The Federal Service on Customers' Rights Protection and Human Well-being Surveillance" | Group of Genomics and Postgenomic Technologies of Central Research Institute of Epidemilology | Samoilov AE et al |
| EPI_ISL_754211 | Russia/MOS-CRIE-13652759/2020 | 2020-11-09 | Europe/Russia/Moscow Oblast/ | "Molecular diagnostic laboratory of Federal Budget Institution of Science ""Central Research Institute of Epidemiology"" of The Federal Service on Customers' Rights Protection and Human Well-being Surveillance" | Group of Genomics and Postgenomic Technologies of Central Research Institute of Epidemilology | Samoilov AE et al |

| | | | | | | |
|---|---|---|---|---|---|---|
| EPI_ISL_754212 | Russia/MOS-CRIE-13670319/2020 | 2020-11-09 | Europe/Russia/Moscow Oblast/ | "Molecular diagnostic laboratory of Federal Budget Institution of Science ""Central Research Institute of Epidemiology"" of The Federal Service on Customers' Rights Protection and Human Well-being Surveillance" | Group of Genomics and Postgenomic Technologies of Central Research Institute of Epidemilology | Samoilov AE et al |
| EPI_ISL_754214 | Russia/MOS-CRIE-13674490/2020 | 2020-11-09 | Europe/Russia/Moscow Oblast/ | "Molecular diagnostic laboratory of Federal Budget Institution of Science ""Central Research Institute of Epidemiology"" of The Federal Service on Customers' Rights Protection and Human Well-being Surveillance" | Group of Genomics and Postgenomic Technologies of Central Research Institute of Epidemilology | Samoilov AE et al |
| EPI_ISL_754216 | USA/WY-WPHL-20094982/2020 | 2020-12-12 | North America/USA/Wyoming/ | Wyoming Public Health Laboratory | Wyoming Public Health Laboratory | Noah Hull et al |
| EPI_ISL_754229 | Belarus/MI-RII-MH11337S/2020 | 2020-04-24 | Europe/Belarus/Minsk Region/Uzda | The Republican Research and Practical Center for Epidemiology and Microbiology (RRPCEM) | WHO National Influenza Centre Russian Federation | Elena Gasich et al |
| EPI_ISL_754231 | Belarus/MI-RII-MH11353S/2020 | 2020-07-28 | Europe/Belarus/Minsk Region/Smalyavi | The Republican Research and Practical Center for Epidemiology and Microbiology (RRPCEM) | WHO National Influenza Centre Russian Federation | Elena Gasich et al |
| EPI_ISL_754233 | Belarus/MI-RII-MH11355S/2020 | 2020-11-23 | Europe/Belarus/Minsk Region/Kolodishc | The Republican Research and Practical Center for Epidemiology and Microbiology (RRPCEM) | WHO National Influenza Centre Russian Federation | Elena Gasich et al |
| EPI_ISL_754234 | Belarus/VI-RII-MH11358S/2020 | 2020-10-22 | Europe/Belarus/Vitebsk Region/Dubrow | The Republican Research and Practical Center for Epidemiology and Microbiology (RRPCEM) | WHO National Influenza Centre Russian Federation | Elena Gasich et al |
| EPI_ISL_754238 | Romania/Bucuresti_407958/2020 | 2020-12-29 | Europe/Romania/Romania/ | Laboratory for Respiratory Viruses, Cantacuzino National Military-Medical Institute for Research and Development | Cantacuzino Institute Virology | Luiza Ustea et al |

| | | | | | | |
|---|---|---|---|---|---|---|
| EPI_ISL_754239 | Indonesia/JB-TFRIC19-R46646/2020 | 2020-07-20 | Asia/Indonesia/West Java/Bandung | Santosa Hospital Bandung Central | """School of Life Sciences and Technology & School of Pharmacy-Institut Teknologi Bandung; Molecular Genetics Laboratory-Faculty of Medicine-Universitas Padjadjaran; Laboratorium Kesehatan Provinsi Jawa Barat""" | Marselina Irasonia Tan et al |
| EPI_ISL_754241 | Indonesia/JB-TFRIC19-R50090/2020 | 2020-07-24 | Asia/Indonesia/West Java/Tasikmalaya | Dinkes Tasikmalaya | """School of Life Sciences and Technology & School of Pharmacy-Institut Teknologi Bandung; Molecular Genetics Laboratory-Faculty of Medicine-Universitas Padjadjaran; Laboratorium Kesehatan Provinsi Jawa Barat""" | Husna Nugrahapraja et al |
| EPI_ISL_754338 | England/204890704/2020 | 2020-11-28 | Europe/United Kingdom/England/ | Respiratory Virus Unit, National Infection Service, Public Health England | COVID-19 Genomics UK (COG-UK) Consortium | PHE Covid Sequencing Team et al |
| EPI_ISL_754389 | USA/WY-WPHL-20156911/2020 | 2020-12-02 | North America/USA/Wyoming/ | Wyoming Public Health Laboratory | Wyoming Public Health Laboratory | Noah Hull et al |

| | | | | | | |
|---|---|---|---|---|---|---|
| EPI_ISL_754391 | Chile/RM-126796/2020 | 2020-07-17 | South America/Chile/Region Metropolit | Genetica Molecular and Subdepartamento de Virologia ISP Chile | Insituto de Salud Publica de Chile | Javier Tognarelli et al |
| EPI_ISL_754392 | Chile/RM-170923/2020 | 2020-09-06 | South America/Chile/Region Metropolit | Genetica Molecular and Subdepartamento de Virologia ISP Chile | Insituto de Salud Publica de Chile | Javier Tognarelli et al |
| EPI_ISL_754393 | Chile/RM-191288/2020 | 2020-09-26 | South America/Chile/Region Metropolit | Genetica Molecular and Subdepartamento de Virologia ISP Chile | Insituto de Salud Publica de Chile | Javier Tognarelli et al |
| EPI_ISL_754410 | USA/NY-Wadsworth-280624-01/2020 | 2020-11-30 | North America/USA/New York/Washing | Wadsworth Center, New York State Department.of Health | Wadsworth Center, New York State Department.of Health | Kirsten St. George et al |
| EPI_ISL_754411 | USA/NY-Wadsworth-281613-01/2020 | 2020-11-30 | North America/USA/New York/Montgo | Wadsworth Center, New York State Department.of Health | Wadsworth Center, New York State Department.of Health | Kirsten St. George et al |
| EPI_ISL_754414 | USA/NY-Wadsworth-281984-01/2020 | 2020-12-01 | North America/USA/New York/Albany ( | Wadsworth Center, New York State Department.of Health | Wadsworth Center, New York State Department.of Health | Kirsten St. George et al |
| EPI_ISL_754418 | USA/NY-Wadsworth-281619-01/2020 | 2020-12-01 | North America/USA/New York/Warren | Wadsworth Center, New York State Department.of Health | Wadsworth Center, New York State Department.of Health | Kirsten St. George et al |
| EPI_ISL_754421 | USA/NY-Wadsworth-281667-01/2020 | 2020-11-30 | North America/USA/New York/Cattarau | Wadsworth Center, New York State Department.of Health | Wadsworth Center, New York State Department.of Health | Kirsten St. George et al |
| EPI_ISL_754426 | USA/NY-Wadsworth-281872-01/2020 | 2020-12-01 | North America/USA/New York/Wayne ( | Wadsworth Center, New York State Department.of Health | Wadsworth Center, New York State Department.of Health | Kirsten St. George et al |

| | | | | | | |
|---|---|---|---|---|---|---|
| EPI_ISL_754429 | USA/NY-Wadsworth-282596-01/2020 | 2020-12-02 | North America/USA/New York/Greene ( | Wadsworth Center, New York State Department.of Health | Wadsworth Center, New York State Department.of Health | Kirsten St. George et al |
| EPI_ISL_754449 | USA/NY-Wadsworth-283604-01/2020 | 2020-12-07 | North America/USA/New York/Albany ( | Wadsworth Center, New York State Department.of Health | Wadsworth Center, New York State Department.of Health | Kirsten St. George et al |
| EPI_ISL_754455 | USA/NY-Wadsworth-284730-01/2020 | 2020-12-08 | North America/USA/New York/Columbia | Wadsworth Center, New York State Department.of Health | Wadsworth Center, New York State Department.of Health | Kirsten St. George et al |
| EPI_ISL_754456 | USA/NY-Wadsworth-284731-01/2020 | 2020-12-08 | North America/USA/New York/Columbia | Wadsworth Center, New York State Department.of Health | Wadsworth Center, New York State Department.of Health | Kirsten St. George et al |
| EPI_ISL_754462 | USA/NY-Wadsworth-285193-01/2020 | 2020-12-07 | North America/USA/New York/Oswego | Wadsworth Center, New York State Department.of Health | Wadsworth Center, New York State Department.of Health | Kirsten St. George et al |
| EPI_ISL_754488 | USA/NY-Wadsworth-280011-01/2020 | 2020-11-22 | North America/USA/New York/Wyomin | Wadsworth Center, New York State Department.of Health | Wadsworth Center, New York State Department.of Health | Kirsten St. George et al |
| EPI_ISL_754489 | USA/NY-Wadsworth-280303-01/2020 | 2020-11-26 | North America/USA/New York/Albany ( | Wadsworth Center, New York State Department.of Health | Wadsworth Center, New York State Department.of Health | Kirsten St. George et al |
| EPI_ISL_754494 | USA/NY-Wadsworth-280101-01/2020 | 2020-11-25 | North America/USA/New York/Oneida ( | Wadsworth Center, New York State Department.of Health | Wadsworth Center, New York State Department.of Health | Kirsten St. George et al |

| | | | | | | |
|---|---|---|---|---|---|---|
| EPI_ISL_754499 | USA/NY-Wadsworth-280499-01/2020 | 2020-11-29 | North America/USA/New York/Saratoga | Wadsworth Center, New York State Department.of Health | Wadsworth Center, New York State Department.of Health | Kirsten St. George et al |
| EPI_ISL_754500 | USA/NY-Wadsworth-280462-01/2020 | 2020-11-29 | North America/USA/New York/Saratoga | Wadsworth Center, New York State Department.of Health | Wadsworth Center, New York State Department.of Health | Kirsten St. George et al |
| EPI_ISL_754513 | USA/NY-Wadsworth-263735-01/2020 | 2020-11-04 | North America/USA/New York/Greene ( | Wadsworth Center, New York State Department.of Health | Wadsworth Center, New York State Department.of Health | Kirsten St. George et al |
| EPI_ISL_754517 | USA/NY-Wadsworth-264206-01/2020 | 2020-11-05 | North America/USA/New York/Greene ( | Wadsworth Center, New York State Department.of Health | Wadsworth Center, New York State Department.of Health | Kirsten St. George et al |
| EPI_ISL_754548 | USA/NY-Wadsworth-285721-01/2020 | 2020-12-09 | North America/USA/New York/Greene ( | Wadsworth Center, New York State Department.of Health | Wadsworth Center, New York State Department.of Health | Kirsten St. George et al |
| EPI_ISL_754550 | USA/NY-Wadsworth-286011-01/2020 | 2020-12-10 | North America/USA/New York/Albany ( | Wadsworth Center, New York State Department.of Health | Wadsworth Center, New York State Department.of Health | Kirsten St. George et al |
| EPI_ISL_754554 | USA/NY-Wadsworth-286021-01/2020 | 2020-12-10 | North America/USA/New York/Albany ( | Wadsworth Center, New York State Department.of Health | Wadsworth Center, New York State Department.of Health | Kirsten St. George et al |
| EPI_ISL_754571 | USA/NY-Wadsworth-286312-01/2020 | 2020-12-10 | North America/USA/New York/Fulton C | Wadsworth Center, New York State Department.of Health | Wadsworth Center, New York State Department.of Health | Kirsten St. George et al |

| | | | | | | |
|---|---|---|---|---|---|---|
| EPI_ISL_754574 | USA/NY-Wadsworth-286381-01/2020 | 2020-12-11 | North America/USA/New York/Albany C | Wadsworth Center, New York State Department.of Health | Wadsworth Center, New York State Department.of Health | Kirsten St. George et al |
| EPI_ISL_754579 | USA/NY-Wadsworth-275442-01/2020 | 2020-11-16 | North America/USA/New York/Hamilto | Wadsworth Center, New York State Department.of Health | Wadsworth Center, New York State Department.of Health | Kirsten St. George et al |
| EPI_ISL_754586 | USA/NY-Wadsworth-277125-01/2020 | 2020-11-19 | North America/USA/New York/Warren | Wadsworth Center, New York State Department.of Health | Wadsworth Center, New York State Department.of Health | Kirsten St. George et al |
| EPI_ISL_754589 | USA/NY-Wadsworth-277731-01/2020 | 2020-11-20 | North America/USA/New York/Warren | Wadsworth Center, New York State Department.of Health | Wadsworth Center, New York State Department.of Health | Kirsten St. George et al |
| EPI_ISL_754593 | USA/NY-Wadsworth-278047-01/2020 | 2020-11-22 | North America/USA/New York/Wyomin | Wadsworth Center, New York State Department.of Health | Wadsworth Center, New York State Department.of Health | Kirsten St. George et al |
| EPI_ISL_754595 | USA/NY-Wadsworth-278036-01/2020 | 2020-11-22 | North America/USA/New York/Wyomin | Wadsworth Center, New York State Department.of Health | Wadsworth Center, New York State Department.of Health | Kirsten St. George et al |
| EPI_ISL_754596 | USA/NY-Wadsworth-278037-01/2020 | 2020-11-22 | North America/USA/New York/Wyomin | Wadsworth Center, New York State Department.of Health | Wadsworth Center, New York State Department.of Health | Kirsten St. George et al |
| EPI_ISL_754601 | USA/NY-Wadsworth-278033-01/2020 | 2020-11-22 | North America/USA/New York/Wyomin | Wadsworth Center, New York State Department.of Health | Wadsworth Center, New York State Department.of Health | Kirsten St. George et al |

| | | | | | | |
|---|---|---|---|---|---|---|
| EPI_ISL_754611 | USA/NY-Wadsworth-278032-01/2020 | 2020-11-22 | North America/USA/New York/Wyomin | Wadsworth Center, New York State Department.of Health | Wadsworth Center, New York State Department.of Health | Kirsten St. George et al |
| EPI_ISL_754613 | USA/NY-Wadsworth-278021-01/2020 | 2020-11-22 | North America/USA/New York/Wyomin | Wadsworth Center, New York State Department.of Health | Wadsworth Center, New York State Department.of Health | Kirsten St. George et al |
| EPI_ISL_754614 | USA/NY-Wadsworth-278024-01/2020 | 2020-11-22 | North America/USA/New York/Wyomin | Wadsworth Center, New York State Department.of Health | Wadsworth Center, New York State Department.of Health | Kirsten St. George et al |
| EPI_ISL_754615 | USA/NY-Wadsworth-277985-01/2020 | 2020-11-22 | North America/USA/New York/Wyomin | Wadsworth Center, New York State Department.of Health | Wadsworth Center, New York State Department.of Health | Kirsten St. George et al |
| EPI_ISL_754618 | USA/NY-Wadsworth-278398-01/2020 | 2020-11-20 | North America/USA/New York/Erie Cou | Wadsworth Center, New York State Department.of Health | Wadsworth Center, New York State Department.of Health | Kirsten St. George et al |
| EPI_ISL_754625 | USA/WI-UW-2403/2020 | 2020-12-03 | North America/USA/Wisconsin/Dane Co | University of Wisconsin-Madison AIDS Vaccine Research Laboratories | University of Wisconsin-Madison AIDS Vaccine Research Laboratories | Gage Moreno et al |
| EPI_ISL_754626 | USA/WI-UW-2390/2020 | 2020-10-30 | North America/USA/Wisconsin/Columbia | University of Wisconsin-Madison AIDS Vaccine Research Laboratories | University of Wisconsin-Madison AIDS Vaccine Research Laboratories | Gage Moreno et al |
| EPI_ISL_754627 | USA/WI-UW-2391/2020 | 2020-10-30 | North America/USA/Wisconsin/Dane Co | University of Wisconsin-Madison AIDS Vaccine Research Laboratories | University of Wisconsin-Madison AIDS Vaccine Research Laboratories | Gage Moreno et al |

| | | | | | | |
|---|---|---|---|---|---|---|
| EPI_ISL_754630 | USA/WI-UW-2426/2020 | 2020-12-11 | North America/USA/Wisconsin/Dane Co | University of Wisconsin-Madison AIDS Vaccine Research Laboratories | University of Wisconsin-Madison AIDS Vaccine Research Laboratories | Gage Moreno et al |
| EPI_ISL_754631 | USA/WI-UW-2427/2020 | 2020-12-11 | North America/USA/Wisconsin/Columbia | University of Wisconsin-Madison AIDS Vaccine Research Laboratories | University of Wisconsin-Madison AIDS Vaccine Research Laboratories | Gage Moreno et al |
| EPI_ISL_754639 | USA/WI-UW-2432/2020 | 2020-12-14 | North America/USA/Wisconsin/Dane Co | University of Wisconsin-Madison AIDS Vaccine Research Laboratories | University of Wisconsin-Madison AIDS Vaccine Research Laboratories | Gage Moreno et al |
| EPI_ISL_754648 | USA/WI-UW-2392/2020 | 2020-12-16 | North America/USA/Wisconsin/Milwauk | University of Wisconsin-Madison AIDS Vaccine Research Laboratories | University of Wisconsin-Madison AIDS Vaccine Research Laboratories | Gage Moreno et al |
| EPI_ISL_754652 | USA/WI-UW-2440/2020 | 2020-12-16 | North America/USA/Wisconsin/Dane Co | University of Wisconsin-Madison AIDS Vaccine Research Laboratories | University of Wisconsin-Madison AIDS Vaccine Research Laboratories | Gage Moreno et al |
| EPI_ISL_754660 | USA/WI-UW-2405/2020 | 2020-12-04 | North America/USA/Wisconsin/Dane Co | University of Wisconsin-Madison AIDS Vaccine Research Laboratories | University of Wisconsin-Madison AIDS Vaccine Research Laboratories | Gage Moreno et al |
| EPI_ISL_754689 | USA/NY-Wadsworth-248115-01/2020 | 2020-10-18 | North America/USA/New York/Chemung | Wadsworth Center, New York State Department.of Health | Wadsworth Center, New York State Department.of Health | Kirsten St. George et al |
| EPI_ISL_754692 | USA/NY-Wadsworth-251060-01/2020 | 2020-10-21 | North America/USA/New York/Chemung | Wadsworth Center, New York State Department.of Health | Wadsworth Center, New York State Department.of Health | Kirsten St. George et al |

| | | | | | | |
|---|---|---|---|---|---|---|
| EPI_ISL_754702 | USA/NY-Wadsworth-248111-01/2020 | 2020-10-18 | North America/USA/New York/Chemung | Wadsworth Center, New York State Department.of Health | Wadsworth Center, New York State Department.of Health | Kirsten St. George et al |
| EPI_ISL_754705 | USA/NY-Wadsworth-247823-01/2020 | 2020-10-17 | North America/USA/New York/Chemung | Wadsworth Center, New York State Department.of Health | Wadsworth Center, New York State Department.of Health | Kirsten St. George et al |
| EPI_ISL_754718 | USA/NY-Wadsworth-248646-01/2020 | 2020-10-19 | North America/USA/New York/Chemung | Wadsworth Center, New York State Department.of Health | Wadsworth Center, New York State Department.of Health | Kirsten St. George et al |
| EPI_ISL_754730 | USA/NY-Wadsworth-249754-01/2020 | 2020-10-20 | North America/USA/New York/Chemung | Wadsworth Center, New York State Department.of Health | Wadsworth Center, New York State Department.of Health | Kirsten St. George et al |
| EPI_ISL_754735 | USA/NY-Wadsworth-249770-01/2020 | 2020-10-20 | North America/USA/New York/Chemung | Wadsworth Center, New York State Department.of Health | Wadsworth Center, New York State Department.of Health | Kirsten St. George et al |
| EPI_ISL_754737 | USA/NY-Wadsworth-249779-01/2020 | 2020-10-20 | North America/USA/New York/Chemung | Wadsworth Center, New York State Department.of Health | Wadsworth Center, New York State Department.of Health | Kirsten St. George et al |
| EPI_ISL_754738 | USA/NY-Wadsworth-249783-01/2020 | 2020-10-20 | North America/USA/New York/Chemung | Wadsworth Center, New York State Department.of Health | Wadsworth Center, New York State Department.of Health | Kirsten St. George et al |
| EPI_ISL_754740 | USA/NY-Wadsworth-249890-01/2020 | 2020-10-20 | North America/USA/New York/Chemung | Wadsworth Center, New York State Department.of Health | Wadsworth Center, New York State Department.of Health | Kirsten St. George et al |

| | | | | | | |
|---|---|---|---|---|---|---|
| EPI_ISL_754742 | USA/NY-Wadsworth-249933-01/2020 | 2020-10-20 | North America/USA/New York/Chemung | Wadsworth Center, New York State Department.of Health | Wadsworth Center, New York State Department.of Health | Kirsten St. George et al |
| EPI_ISL_754744 | USA/NY-Wadsworth-251941-01/2020 | 2020-10-22 | North America/USA/New York/Greene ( | Wadsworth Center, New York State Department.of Health | Wadsworth Center, New York State Department.of Health | Kirsten St. George et al |
| EPI_ISL_754745 | USA/NY-Wadsworth-251938-01/2020 | 2020-10-22 | North America/USA/New York/Greene ( | Wadsworth Center, New York State Department.of Health | Wadsworth Center, New York State Department.of Health | Kirsten St. George et al |
| EPI_ISL_754759 | USA/GA-EHC-043Q/2020 | 2020-03-15 | North America/USA/Georgia/ | Emory Molecular Diagnostics Laboratory, Emory Healthcare | Piantadosi Lab, Emory Department of Pathology | Ahmed Babiker et al |
| EPI_ISL_754763 | USA/GA-EHC-053A/2020 | 2020-03-16 | North America/USA/Georgia/ | Emory Molecular Diagnostics Laboratory, Emory Healthcare | Piantadosi Lab, Emory Department of Pathology | Ahmed Babiker et al |
| EPI_ISL_754769 | USA/GA-EHC-139K/2020 | 2020-04-06 | North America/USA/Georgia/ | Emory Molecular Diagnostics Laboratory, Emory Healthcare | Piantadosi Lab, Emory Department of Pathology | Ahmed Babiker et al |
| EPI_ISL_754772 | USA/GA-EHC-417C/2020 | 2020-11-30 | North America/USA/Georgia/ | Emory Molecular Diagnostics Laboratory, Emory Healthcare | Piantadosi Lab, Emory Department of Pathology | Ahmed Babiker et al |
| EPI_ISL_754777 | USA/GA-EHC-060H/2020 | 2020-03-11 | North America/USA/Georgia/ | Emory Molecular Diagnostics Laboratory, Emory Healthcare | Piantadosi Lab, Emory Department of Pathology | Ahmed Babiker et al |
| EPI_ISL_754784 | USA/NY-Wadsworth-252190-01/2020 | 2020-10-22 | North America/USA/New York/Chemung | Wadsworth Center, New York State Department.of Health | Wadsworth Center, New York State Department.of Health | Kirsten St. George et al |

| | | | | | | |
|---|---|---|---|---|---|---|
| EPI_ISL_754792 | USA/NY-Wadsworth-251959-01/2020 | 2020-10-22 | North America/USA/New York/Greene ( | Wadsworth Center, New York State Department.of Health | Wadsworth Center, New York State Department.of Health | Kirsten St. George et al |
| EPI_ISL_754794 | USA/NY-Wadsworth-252159-01/2020 | 2020-10-22 | North America/USA/New York/Chemung | Wadsworth Center, New York State Department.of Health | Wadsworth Center, New York State Department.of Health | Kirsten St. George et al |
| EPI_ISL_754795 | USA/NY-Wadsworth-251971-01/2020 | 2020-10-22 | North America/USA/New York/Greene ( | Wadsworth Center, New York State Department.of Health | Wadsworth Center, New York State Department.of Health | Kirsten St. George et al |
| EPI_ISL_754806 | USA/NY-Wadsworth-251043-01/2020 | 2020-10-21 | North America/USA/New York/Chemung | Wadsworth Center, New York State Department.of Health | Wadsworth Center, New York State Department.of Health | Kirsten St. George et al |
| EPI_ISL_754830 | USA/NY-Wadsworth-250969-01/2020 | 2020-10-21 | North America/USA/New York/Chemung | Wadsworth Center, New York State Department.of Health | Wadsworth Center, New York State Department.of Health | Kirsten St. George et al |
| EPI_ISL_754840 | USA/NY-Wadsworth-252077-01/2020 | 2020-10-22 | North America/USA/New York/Chemung | Wadsworth Center, New York State Department.of Health | Wadsworth Center, New York State Department.of Health | Kirsten St. George et al |
| EPI_ISL_754843 | France/GES-IPP10958/2020 | 2020-12-17 | Europe/France/Grand Est/Reims | Laboratoire de Virologie H√¥pital Robert Debr√© | National Reference Center for Viruses of Respiratory Infections, Institut Pasteur, Paris | Marion Barbet et al |

| | | | | | | |
|---|---|---|---|---|---|---|
| EPI_ISL_754846 | France/GES-IPP10971/2020 | 2020-12-18 | Europe/France/Grand Est/Reims | Laboratoire de Virologie H√¥pital Robert Debr√© | National Reference Center for Viruses of Respiratory Infections, Institut Pasteur, Paris | Marion Barbet et al |
| EPI_ISL_754847 | France/BFC-IPP10929/2020 | 2020-12-21 | Europe/France/Bourgogne-France-Comt | Korian Villa d'Azon | National Reference Center for Viruses of Respiratory Infections, Institut Pasteur, Paris | Marion Barbet et al |
| EPI_ISL_754856 | France/BRE-IPP11310/2020 | 2020-09-16 | Europe/France/Bretagne/Brest | CHU - H√¥pital Cavale Blanche - Labo. de Virologie | National Reference Center for Viruses of Respiratory Infections, Institut Pasteur, Paris | Marion Barbet et al |
| EPI_ISL_754857 | USA/CA-IGI-0376/2020 | 2020-07-22 | North America/USA/California/Alameda | Innovative Genomics Institute, UC Berkeley | Innovative Genomics Institute, UC Berkeley | Stacia Wyman et al |
| EPI_ISL_754866 | USA/CA-IGI-0325/2020 | 2020-07-22 | North America/USA/California/Contra C | Innovative Genomics Institute, UC Berkeley | Innovative Genomics Institute, UC Berkeley | Stacia Wyman et al |
| EPI_ISL_754872 | USA/CA-IGI-0343/2020 | 2020-07-20 | North America/USA/California/Alameda | Innovative Genomics Institute, UC Berkeley | Innovative Genomics Institute, UC Berkeley | Stacia Wyman et al |
| EPI_ISL_754873 | USA/CA-IGI-0349/2020 | 2020-07-21 | North America/USA/California/Alameda | Innovative Genomics Institute, UC Berkeley | Innovative Genomics Institute, UC Berkeley | Stacia Wyman et al |
| EPI_ISL_754874 | USA/CA-IGI-0350/2020 | 2020-07-17 | North America/USA/California/Contra C | Innovative Genomics Institute, UC Berkeley | Innovative Genomics Institute, UC Berkeley | Stacia Wyman et al |

| | | | | | | |
|---|---|---|---|---|---|---|
| EPI_ISL_754879 | USA/CA-IGI-0386/2020 | 2020-07-14 | North America/USA/California/Alameda | Innovative Genomics Institute, UC Berkeley | Innovative Genomics Institute, UC Berkeley | Stacia Wyman et al |
| EPI_ISL_754881 | USA/CA-IGI-0391/2020 | 2020-07-09 | North America/USA/California/Alameda | Innovative Genomics Institute, UC Berkeley | Innovative Genomics Institute, UC Berkeley | Stacia Wyman et al |
| EPI_ISL_754889 | USA/CA-IGI-0407/2020 | 2020-07-08 | North America/USA/California/Alameda | Innovative Genomics Institute, UC Berkeley | Innovative Genomics Institute, UC Berkeley | Stacia Wyman et al |
| EPI_ISL_754894 | USA/CA-IGI-0415/2020 | 2020-07-09 | North America/USA/California/Contra C | Innovative Genomics Institute, UC Berkeley | Innovative Genomics Institute, UC Berkeley | Stacia Wyman et al |
| EPI_ISL_754899 | USA/CA-IGI-0431/2020 | 2020-07-10 | North America/USA/California/Alameda | Innovative Genomics Institute, UC Berkeley | Innovative Genomics Institute, UC Berkeley | Stacia Wyman et al |
| EPI_ISL_754902 | Poland/Masovia-14965/2020 | 2020-09-27 | Europe/Poland/Masovia/ | Laboratory Diagnostics and Clinical Immunology of Developmental Age, Medical University of Warsaw | genXone SA, Research & Development Laboratory; The Faculty of Mathematics, Informatics and Mechanics of the University of Warsaw | Maciej Sykulski et al |
| EPI_ISL_754904 | Poland/Masovia-12865/2020 | 2020-08-28 | Europe/Poland/Masovia/ | Laboratory Diagnostics and Clinical Immunology of Developmental Age, Medical University of Warsaw | genXone SA, Research & Development Laboratory; The Faculty of Mathematics, Informatics and Mechanics of the University of Warsaw | Maciej Sykulski et al |

| | | | | | | |
|---|---|---|---|---|---|---|
| EPI_ISL_754905 | Poland/Masovia-13529/2020 | 2020-09-06 | Europe/Poland/Masovia/ | Laboratory Diagnostics and Clinical Immunology of Developmental Age, Medical University of Warsaw | genXone SA, Research & Development Laboratory; The Faculty of Mathematics, Informatics and Mechanics of the University of Warsaw | Maciej Sykulski et al |
| EPI_ISL_754906 | Poland/Masovia-12866/2020 | 2020-08-28 | Europe/Poland/Masovia/ | Laboratory Diagnostics and Clinical Immunology of Developmental Age, Medical University of Warsaw | genXone SA, Research & Development Laboratory; The Faculty of Mathematics, Informatics and Mechanics of the University of Warsaw | Maciej Sykulski et al |
| EPI_ISL_754907 | Poland/Masovia-13450/2020 | 2020-09-04 | Europe/Poland/Masovia/ | Laboratory Diagnostics and Clinical Immunology of Developmental Age, Medical University of Warsaw | genXone SA, Research & Development Laboratory; The Faculty of Mathematics, Informatics and Mechanics of the University of Warsaw | Maciej Sykulski et al |
| EPI_ISL_754909 | Poland/Masovia-14512/2020 | 2020-09-20 | Europe/Poland/Masovia/ | Laboratory Diagnostics and Clinical Immunology of Developmental Age, Medical University of Warsaw | genXone SA, Research & Development Laboratory; The Faculty of Mathematics, Informatics and Mechanics of the University of Warsaw | Maciej Sykulski et al |

| | | | | | | |
|---|---|---|---|---|---|---|
| EPI_ISL_754910 | Poland/Masovia-14790/2020 | 2020-09-24 | Europe/Poland/Masovia/ | Laboratory Diagnostics and Clinical Immunology of Developmental Age, Medical University of Warsaw | genXone SA, Research & Development Laboratory; The Faculty of Mathematics, Informatics and Mechanics of the University of Warsaw | Maciej Sykulski et al |
| EPI_ISL_754911 | Poland/Masovia-4249/2020 | 2020-05-26 | Europe/Poland/Masovia/ | Laboratory Diagnostics and Clinical Immunology of Developmental Age, Medical University of Warsaw | genXone SA, Research & Development Laboratory; The Faculty of Mathematics, Informatics and Mechanics of the University of Warsaw | Maciej Sykulski et al |
| EPI_ISL_754912 | Poland/Masovia-14376/2020 | 2020-09-17 | Europe/Poland/Masovia/ | Laboratory Diagnostics and Clinical Immunology of Developmental Age, Medical University of Warsaw | genXone SA, Research & Development Laboratory; The Faculty of Mathematics, Informatics and Mechanics of the University of Warsaw | Maciej Sykulski et al |
| EPI_ISL_754913 | Poland/Masovia-13802/2020 | 2020-09-09 | Europe/Poland/Masovia/ | Laboratory Diagnostics and Clinical Immunology of Developmental Age, Medical University of Warsaw | genXone SA, Research & Development Laboratory; The Faculty of Mathematics, Informatics and Mechanics of the University of Warsaw | Maciej Sykulski et al |

| | | | | | | |
|---|---|---|---|---|---|---|
| EPI_ISL_754917 | USA/CA-CDPH180/2020 | 2020-09-28 | North America/USA/California/Imperial | California Department of Public Health | California Department of Public Health | CDPH IDLB COVIDNet et al |
| EPI_ISL_754919 | USA/CA-CDPH136/2020 | 2020-08-11 | North America/USA/California/Inyo Cou | California Department of Public Health | California Department of Public Health | CDPH IDLB COVIDNet et al |
| EPI_ISL_754922 | USA/CA-CDPH115/2020 | 2020-09-18 | North America/USA/California/Fresno C | California Department of Public Health | California Department of Public Health | CDPH IDLB COVIDNet et al |
| EPI_ISL_754925 | USA/CA-CDPH109/2020 | 2020-09-17 | North America/USA/California/Kings Co | California Department of Public Health | California Department of Public Health | CDPH IDLB COVIDNet et al |
| EPI_ISL_754928 | USA/CA-CDPH119/2020 | 2020-09-20 | North America/USA/California/Kern Cou | California Department of Public Health | California Department of Public Health | CDPH IDLB COVIDNet et al |
| EPI_ISL_754932 | USA/CA-CDPH074/2020 | 2020-07-09 | North America/USA/California/Marin Co | California Department of Public Health | California Department of Public Health | CDPH IDLB COVIDNet et al |
| EPI_ISL_754933 | USA/CA-CDPH133/2020 | 2020-07-08 | North America/USA/California/Colusa C | California Department of Public Health | California Department of Public Health | CDPH IDLB COVIDNet et al |
| EPI_ISL_754934 | USA/CA-CDPH153/2020 | 2020-09-28 | North America/USA/California/Stanislau | California Department of Public Health | California Department of Public Health | CDPH IDLB COVIDNet et al |
| EPI_ISL_754936 | USA/CA-CDPH103/2020 | 2020-07-28 | North America/USA/California/ | California Department of Public Health | California Department of Public Health | CDPH IDLB COVIDNet et al |
| EPI_ISL_754938 | USA/CA-CDPH087/2020 | 2020-08-05 | North America/USA/California/Butte Co | California Department of Public Health | California Department of Public Health | CDPH IDLB COVIDNet et al |
| EPI_ISL_754940 | USA/CA-CDPH122/2020 | 2020-09-21 | North America/USA/California/Kern Cou | California Department of Public Health | California Department of Public Health | CDPH IDLB COVIDNet et al |
| EPI_ISL_754952 | USA/CA-CDPH156/2020 | 2020-09-30 | North America/USA/California/San Joac | California Department of Public Health | California Department of Public Health | CDPH IDLB COVIDNet et al |
| EPI_ISL_754954 | USA/CA-CDPH114/2020 | 2020-09-18 | North America/USA/California/Madera | California Department of Public Health | California Department of Public Health | CDPH IDLB COVIDNet et al |

| | | | | | | |
|---|---|---|---|---|---|---|
| EPI_ISL_754961 | USA/CA-CDPH075/2020 | 2020-07-20 | North America/USA/California/Sonoma | California Department of Public Health | California Department of Public Health | CDPH IDLB COVIDNet et al |
| EPI_ISL_754965 | USA/CA-CDPH232/2020 | 2020-11-14 | North America/USA/California/Alameda | California Department of Public Health | California Department of Public Health | CDPH IDLB COVIDNet et al |
| EPI_ISL_754967 | USA/CA-CDPH218/2020 | 2020-11-15 | North America/USA/California/Alameda | California Department of Public Health | California Department of Public Health | CDPH IDLB COVIDNet et al |
| EPI_ISL_754968 | USA/CA-CDPH083/2020 | 2020-07-20 | North America/USA/California/Mono Co | California Department of Public Health | California Department of Public Health | CDPH IDLB COVIDNet et al |
| EPI_ISL_754969 | USA/CA-CDPH071/2020 | 2020-07-01 | North America/USA/California/Marin Co | California Department of Public Health | California Department of Public Health | CDPH IDLB COVIDNet et al |
| EPI_ISL_754970 | USA/CA-CDPH129/2020 | 2020-06-18 | North America/USA/California/Marin Co | California Department of Public Health | California Department of Public Health | CDPH IDLB COVIDNet et al |
| EPI_ISL_754975 | USA/CA-CDPH141/2020 | 2020-09-25 | North America/USA/California/Tulare Co | California Department of Public Health | California Department of Public Health | CDPH IDLB COVIDNet et al |
| EPI_ISL_754976 | USA/CA-CDPH161/2020 | 2020-06-24 | North America/USA/California/Stanislau | California Department of Public Health | California Department of Public Health | CDPH IDLB COVIDNet et al |
| EPI_ISL_754978 | USA/CA-CDPH095/2020 | 2020-08-17 | North America/USA/California/Marin Co | California Department of Public Health | California Department of Public Health | CDPH IDLB COVIDNet et al |
| EPI_ISL_754979 | USA/CA-CDPH146/2020 | 2020-09-26 | North America/USA/California/Stanislau | California Department of Public Health | California Department of Public Health | CDPH IDLB COVIDNet et al |
| EPI_ISL_754995 | USA/CA-CDPH147/2020 | 2020-09-26 | North America/USA/California/Stanislau | California Department of Public Health | California Department of Public Health | CDPH IDLB COVIDNet et al |
| EPI_ISL_754997 | USA/CA-CDPH247/2020 | 2020-11-15 | North America/USA/California/Alameda | California Department of Public Health | California Department of Public Health | CDPH IDLB COVIDNet et al |
| EPI_ISL_754999 | USA/CA-CDPH102/2020 | 2020-08-17 | North America/USA/California/Butte Co | California Department of Public Health | California Department of Public Health | CDPH IDLB COVIDNet et al |

| | | | | | | |
|---|---|---|---|---|---|---|
| EPI_ISL_755001 | USA/CA-CDPH099/2020 | 2020-08-17 | North America/USA/California/Marin Co | California Department of Public Health | California Department of Public Health | CDPH IDLB COVIDNet et al |
| EPI_ISL_755012 | USA/CA-CDPH137/2020 | 2020-08-11 | North America/USA/California/Inyo Cou | California Department of Public Health | California Department of Public Health | CDPH IDLB COVIDNet et al |
| EPI_ISL_755013 | USA/CA-CDPH219/2020 | 2020-11-15 | North America/USA/California/Alameda | California Department of Public Health | California Department of Public Health | CDPH IDLB COVIDNet et al |
| EPI_ISL_755015 | USA/CA-CDPH294/2020 | 2020-12-04 | North America/USA/California/Solano C | California Department of Public Health | California Department of Public Health | CDPH IDLB COVIDNet et al |
| EPI_ISL_755017 | USA/CA-CDPH128/2020 | 2020-09-21 | North America/USA/California/Kern Cou | California Department of Public Health | California Department of Public Health | CDPH IDLB COVIDNet et al |
| EPI_ISL_755019 | USA/CA-CDPH163/2020 | 2020-07-06 | North America/USA/California/San Dieg | California Department of Public Health | California Department of Public Health | CDPH IDLB COVIDNet et al |
| EPI_ISL_755032 | USA/CA-CDPH159/2020 | 2020-09-28 | North America/USA/California/ | California Department of Public Health | California Department of Public Health | CDPH IDLB COVIDNet et al |
| EPI_ISL_755033 | USA/CA-CDPH045/2020 | 2020-05-19 | North America/USA/California/Los Ange | California Department of Public Health | California Department of Public Health | CDPH IDLB COVIDNet et al |
| EPI_ISL_755044 | USA/CA-CDPH084/2020 | 2020-07-28 | North America/USA/California/ | California Department of Public Health | California Department of Public Health | CDPH IDLB COVIDNet et al |
| EPI_ISL_755045 | USA/CA-CDPH085/2020 | 2020-07-27 | North America/USA/California/Butte Co | California Department of Public Health | California Department of Public Health | CDPH IDLB COVIDNet et al |
| EPI_ISL_755047 | USA/CA-CDPH088/2020 | 2020-08-05 | North America/USA/California/Butte Co | California Department of Public Health | California Department of Public Health | CDPH IDLB COVIDNet et al |
| EPI_ISL_755048 | USA/CA-CDPH093/2020 | 2020-08-17 | North America/USA/California/Marin Co | California Department of Public Health | California Department of Public Health | CDPH IDLB COVIDNet et al |
| EPI_ISL_755052 | USA/CA-CDPH112/2020 | 2020-09-18 | North America/USA/California/Madera | California Department of Public Health | California Department of Public Health | CDPH IDLB COVIDNet et al |

| | | | | | | |
|---|---|---|---|---|---|---|
| EPI_ISL_755055 | USA/CA-CDPH131/2020 | 2020-07-06 | North America/USA/California/Sacrame | California Department of Public Health | California Department of Public Health | CDPH IDLB COVIDNet et al |
| EPI_ISL_755060 | USA/CA-CDPH160/2020 | 2020-06-24 | North America/USA/California/ | California Department of Public Health | California Department of Public Health | CDPH IDLB COVIDNet et al |
| EPI_ISL_755075 | USA/CA-CDPH223/2020 | 2020-11-15 | North America/USA/California/Alameda | California Department of Public Health | California Department of Public Health | CDPH IDLB COVIDNet et al |
| EPI_ISL_755082 | USA/CA-CDPH236/2020 | 2020-11-15 | North America/USA/California/Alameda | California Department of Public Health | California Department of Public Health | CDPH IDLB COVIDNet et al |
| EPI_ISL_755083 | USA/CA-CDPH237/2020 | 2020-11-15 | North America/USA/California/Alameda | California Department of Public Health | California Department of Public Health | CDPH IDLB COVIDNet et al |
| EPI_ISL_755085 | USA/CA-CDPH240/2020 | 2020-11-15 | North America/USA/California/Alameda | California Department of Public Health | California Department of Public Health | CDPH IDLB COVIDNet et al |
| EPI_ISL_755091 | USA/CA-CDPH255/2020 | 2020-11-14 | North America/USA/California/Alameda | California Department of Public Health | California Department of Public Health | CDPH IDLB COVIDNet et al |
| EPI_ISL_755100 | USA/CA-CDPH273/2020 | 2020-11-15 | North America/USA/California/Alameda | California Department of Public Health | California Department of Public Health | CDPH IDLB COVIDNet et al |
| EPI_ISL_755108 | USA/CA-CDPH283/2020 | 2020-11-14 | North America/USA/California/Alameda | California Department of Public Health | California Department of Public Health | CDPH IDLB COVIDNet et al |
| EPI_ISL_755112 | USA/CA-CDPH293/2020 | 2020-12-04 | North America/USA/California/Solano C | California Department of Public Health | California Department of Public Health | CDPH IDLB COVIDNet et al |
| EPI_ISL_755115 | USA/CA-SEARCH-5567/2020 | 2020-11-29 | North America/USA/California/San Dieg | San Diego County Public Health Laboratory | Andersen lab at Scripps Research | SEARCH Alliance San Diego with Tracy Basler et al |
| EPI_ISL_755123 | Jordan/SEARCH-5485/2020 | 2020-09-13 | Asia/Jordan/Amman/ | Biolab Diagnostic Laboratories | Andersen lab at Scripps Research | Issa Abu-Dayyeh et al |
| EPI_ISL_755133 | USA/CA-SEARCH-5345/2020 | 2020-12-09 | North America/USA/California/San Dieg | UCSD EXCITE lab | Andersen lab at Scripps Research | SEARCH Alliance San Diego et al |
| EPI_ISL_755151 | USA/CA-SEARCH-5332/2020 | 2020-12-07 | North America/USA/California/San Dieg | UCSD EXCITE lab | Andersen lab at Scripps Research | SEARCH Alliance San Diego et al |
| EPI_ISL_755155 | USA/CA-SEARCH-5337/2020 | 2020-12-09 | North America/USA/California/San Dieg | UCSD EXCITE lab | Andersen lab at Scripps Research | SEARCH Alliance San Diego et al |

| | | | | | | |
|---|---|---|---|---|---|---|
| EPI_ISL_755158 | USA/CA-SEARCH-5341/2020 | 2020-12-08 | North America/USA/California/San Dieg | UCSD EXCITE lab | Andersen lab at Scripps Research | SEARCH Alliance San Diego et al |
| EPI_ISL_755168 | USA/CA-SEARCH-5353/2020 | 2020-12-10 | North America/USA/California/San Dieg | UCSD EXCITE lab | Andersen lab at Scripps Research | SEARCH Alliance San Diego et al |
| EPI_ISL_755173 | USA/CA-SEARCH-5361/2020 | 2020-12-15 | North America/USA/California/San Dieg | UCSD EXCITE lab | Andersen lab at Scripps Research | SEARCH Alliance San Diego et al |
| EPI_ISL_755175 | USA/CA-SEARCH-5363/2020 | 2020-12-14 | North America/USA/California/San Dieg | UCSD EXCITE lab | Andersen lab at Scripps Research | SEARCH Alliance San Diego et al |
| EPI_ISL_755176 | USA/CA-SEARCH-5364/2020 | 2020-12-14 | North America/USA/California/San Dieg | UCSD EXCITE lab | Andersen lab at Scripps Research | SEARCH Alliance San Diego et al |
| EPI_ISL_755179 | USA/CA-SEARCH-5367/2020 | 2020-12-15 | North America/USA/California/San Dieg | UCSD EXCITE lab | Andersen lab at Scripps Research | SEARCH Alliance San Diego et al |
| EPI_ISL_755180 | USA/CA-SEARCH-5368/2020 | 2020-12-15 | North America/USA/California/San Dieg | UCSD EXCITE lab | Andersen lab at Scripps Research | SEARCH Alliance San Diego et al |
| EPI_ISL_755190 | USA/CA-SEARCH-5378/2020 | 2020-12-04 | North America/USA/California/San Dieg | UCSD EXCITE lab | Andersen lab at Scripps Research | SEARCH Alliance San Diego et al |
| EPI_ISL_755191 | USA/CA-SEARCH-5379/2020 | 2020-12-04 | North America/USA/California/San Dieg | UCSD EXCITE lab | Andersen lab at Scripps Research | SEARCH Alliance San Diego et al |
| EPI_ISL_755196 | USA/CA-SEARCH-5384/2020 | 2020-12-05 | North America/USA/California/San Dieg | UCSD EXCITE lab | Andersen lab at Scripps Research | SEARCH Alliance San Diego et al |
| EPI_ISL_755200 | USA/CA-SEARCH-5389/2020 | 2020-12-01 | North America/USA/California/San Dieg | UCSD EXCITE lab | Andersen lab at Scripps Research | SEARCH Alliance San Diego et al |
| EPI_ISL_755205 | USA/CA-SEARCH-5394/2020 | 2020-12-02 | North America/USA/California/San Dieg | UCSD EXCITE lab | Andersen lab at Scripps Research | SEARCH Alliance San Diego et al |
| EPI_ISL_755206 | USA/CA-SEARCH-5395/2020 | 2020-12-03 | North America/USA/California/San Dieg | UCSD EXCITE lab | Andersen lab at Scripps Research | SEARCH Alliance San Diego et al |
| EPI_ISL_755212 | Jordan/SEARCH-5401/2020 | 2020-10-07 | Asia/Jordan/Aqaba/ | Biolab Diagnostic Laboratories | Andersen lab at Scripps Research | Issa Abu-Dayyeh et al |
| EPI_ISL_755213 | Jordan/SEARCH-5402/2020 | 2020-11-07 | Asia/Jordan/Amman/ | Biolab Diagnostic Laboratories | Andersen lab at Scripps Research | Issa Abu-Dayyeh et al |
| EPI_ISL_755218 | USA/CA-SEARCH-5407/2020 | 2020-12-01 | North America/USA/California/San Dieg | UCSD EXCITE lab | Andersen lab at Scripps Research | SEARCH Alliance San Diego et al |
| EPI_ISL_755222 | USA/CA-SEARCH-5412/2020 | 2020-11-25 | North America/USA/California/San Dieg | UCSD EXCITE lab | Andersen lab at Scripps Research | SEARCH Alliance San Diego et al |
| EPI_ISL_755223 | USA/CA-SEARCH-5413/2020 | 2020-11-28 | North America/USA/California/San Dieg | UCSD EXCITE lab | Andersen lab at Scripps Research | SEARCH Alliance San Diego et al |
| EPI_ISL_755225 | USA/CA-SEARCH-5415/2020 | 2020-11-29 | North America/USA/California/San Dieg | UCSD EXCITE lab | Andersen lab at Scripps Research | SEARCH Alliance San Diego et al |
| EPI_ISL_755239 | Jordan/SEARCH-5432/2020 | 2020-09-07 | Asia/Jordan/Amman/ | Biolab Diagnostic Laboratories | Andersen lab at Scripps Research | Issa Abu-Dayyeh et al |

| | | | | | | |
|---|---|---|---|---|---|---|
| EPI_ISL_755241 | Jordan/SEARCH-5445/2020 | 2020-09-09 | Asia/Jordan/Amman/ | Biolab Diagnostic Laboratories | Andersen lab at Scripps Research | Issa Abu-Dayyeh et al |
| EPI_ISL_755242 | Jordan/SEARCH-5446/2020 | 2020-09-13 | Asia/Jordan/Amman/ | Biolab Diagnostic Laboratories | Andersen lab at Scripps Research | Issa Abu-Dayyeh et al |
| EPI_ISL_755244 | Jordan/SEARCH-5453/2020 | 2020-09-13 | Asia/Jordan/Amman/ | Biolab Diagnostic Laboratories | Andersen lab at Scripps Research | Issa Abu-Dayyeh et al |
| EPI_ISL_755245 | Jordan/SEARCH-5457/2020 | 2020-09-07 | Asia/Jordan/Amman/ | Biolab Diagnostic Laboratories | Andersen lab at Scripps Research | Issa Abu-Dayyeh et al |
| EPI_ISL_755248 | Jordan/SEARCH-5461/2020 | 2020-09-15 | Asia/Jordan/Amman/ | Biolab Diagnostic Laboratories | Andersen lab at Scripps Research | Issa Abu-Dayyeh et al |
| EPI_ISL_755249 | Jordan/SEARCH-5462/2020 | 2020-09-07 | Asia/Jordan/Amman/ | Biolab Diagnostic Laboratories | Andersen lab at Scripps Research | Issa Abu-Dayyeh et al |
| EPI_ISL_755250 | Jordan/SEARCH-5463/2020 | 2020-09-08 | Asia/Jordan/Amman/ | Biolab Diagnostic Laboratories | Andersen lab at Scripps Research | Issa Abu-Dayyeh et al |
| EPI_ISL_755256 | Jordan/SEARCH-5476/2020 | 2020-09-08 | Asia/Jordan/Amman/ | Biolab Diagnostic Laboratories | Andersen lab at Scripps Research | Issa Abu-Dayyeh et al |
| EPI_ISL_755261 | Jordan/SEARCH-5484/2020 | 2020-09-08 | Asia/Jordan/Amman/ | Biolab Diagnostic Laboratories | Andersen lab at Scripps Research | Issa Abu-Dayyeh et al |
| EPI_ISL_755263 | Jordan/SEARCH-5490/2020 | 2020-09-07 | Asia/Jordan/Aqaba/ | Biolab Diagnostic Laboratories | Andersen lab at Scripps Research | Issa Abu-Dayyeh et al |
| EPI_ISL_755264 | Jordan/SEARCH-5497/2020 | 2020-09-07 | Asia/Jordan/Amman/ | Biolab Diagnostic Laboratories | Andersen lab at Scripps Research | Issa Abu-Dayyeh et al |
| EPI_ISL_755267 | Jordan/SEARCH-5500/2020 | 2020-09-14 | Asia/Jordan/Amman/ | Biolab Diagnostic Laboratories | Andersen lab at Scripps Research | Issa Abu-Dayyeh et al |
| EPI_ISL_755272 | USA/CA-SEARCH-5511/2020 | 2020-11-23 | North America/USA/California/San Dieg | San Diego County Public Health Laboratory | Andersen lab at Scripps Research | SEARCH Alliance San Diego with Tracy Basler et al |
| EPI_ISL_755274 | USA/CA-SEARCH-5515/2020 | 2020-11-17 | North America/USA/California/San Dieg | San Diego County Public Health Laboratory | Andersen lab at Scripps Research | SEARCH Alliance San Diego with Tracy Basler et al |
| EPI_ISL_755285 | USA/CA-SEARCH-5544/2020 | 2020-11-30 | North America/USA/California/San Dieg | San Diego County Public Health Laboratory | Andersen lab at Scripps Research | SEARCH Alliance San Diego with Tracy Basler et al |
| EPI_ISL_755299 | USA/CA-SEARCH-5570/2020 | 2020-11-22 | North America/USA/California/San Dieg | San Diego County Public Health Laboratory | Andersen lab at Scripps Research | SEARCH Alliance San Diego with Tracy Basler et al |
| EPI_ISL_755302 | Chile/LI-4/2020 | 2020-08-20 | South America/Chile/O'Higgins/San Frai | Laboratorio de Bioingenier√≠a, Instituto de Ciencias de la Ingenier√≠a, Universidad de O¬¥Higgins | Center for Mathematical Modeling and Center for Genome Regulation. Santiago, Chile | M. Latorre et al |

| | | | | | | |
|---|---|---|---|---|---|---|
| EPI_ISL_755303 | Chile/LI-1-4/2020 | 2020-08-03 | South America/Chile/O'Higgins/Lolol | Laboratorio de Bioingenier√≠a, Instituto de Ciencias de la Ingenier√≠a, Universidad de O¬¥Higgins | Center for Mathematical Modeling and Center for Genome Regulation. Santiago, Chile | M. Latorre et al |
| EPI_ISL_755311 | USA/ME-HETL-J0107/2020 | 2020-08-27 | North America/USA/Maine/ | Maine Health and Environmental Testing Laboratory | Tewhey Lab, The Jackson Laboratory | Matluk et al |
| EPI_ISL_755320 | USA/ME-HETL-J0118/2020 | 2020-09-01 | North America/USA/Maine/ | Maine Health and Environmental Testing Laboratory | Tewhey Lab, The Jackson Laboratory | Matluk et al |
| EPI_ISL_755323 | USA/ME-HETL-J0124/2020 | 2020-09-03 | North America/USA/Maine/ | Maine Health and Environmental Testing Laboratory | Tewhey Lab, The Jackson Laboratory | Matluk et al |
| EPI_ISL_755328 | USA/ME-HETL-J0131/2020 | 2020-09-08 | North America/USA/Maine/ | Maine Health and Environmental Testing Laboratory | Tewhey Lab, The Jackson Laboratory | Matluk et al |
| EPI_ISL_755329 | USA/ME-HETL-J0132/2020 | 2020-09-08 | North America/USA/Maine/ | Maine Health and Environmental Testing Laboratory | Tewhey Lab, The Jackson Laboratory | Matluk et al |
| EPI_ISL_755331 | USA/ME-HETL-J0136/2020 | 2020-09-09 | North America/USA/Maine/ | Maine Health and Environmental Testing Laboratory | Tewhey Lab, The Jackson Laboratory | Matluk et al |
| EPI_ISL_755332 | USA/ME-HETL-J0137/2020 | 2020-09-08 | North America/USA/Maine/ | Maine Health and Environmental Testing Laboratory | Tewhey Lab, The Jackson Laboratory | Matluk et al |
| EPI_ISL_755334 | USA/ME-HETL-J0140/2020 | 2020-09-10 | North America/USA/Maine/ | Maine Health and Environmental Testing Laboratory | Tewhey Lab, The Jackson Laboratory | Matluk et al |
| EPI_ISL_755340 | USA/ME-HETL-J0150/2020 | 2020-09-11 | North America/USA/Maine/ | Maine Health and Environmental Testing Laboratory | Tewhey Lab, The Jackson Laboratory | Matluk et al |
| EPI_ISL_755345 | USA/ME-HETL-J0162/2020 | 2020-09-14 | North America/USA/Maine/ | Maine Health and Environmental Testing Laboratory | Tewhey Lab, The Jackson Laboratory | Matluk et al |
| EPI_ISL_755349 | USA/ME-HETL-J0167/2020 | 2020-09-15 | North America/USA/Maine/ | Maine Health and Environmental Testing Laboratory | Tewhey Lab, The Jackson Laboratory | Matluk et al |

| | | | | | | |
|---|---|---|---|---|---|---|
| EPI_ISL_755350 | USA/ME-HETL-J0168/2020 | 2020-09-14 | North America/USA/Maine/ | Maine Health and Environmental Testing Laboratory | Tewhey Lab, The Jackson Laboratory | Matluk et al |
| EPI_ISL_755353 | USA/ME-HETL-J0172/2020 | 2020-09-15 | North America/USA/Maine/ | Maine Health and Environmental Testing Laboratory | Tewhey Lab, The Jackson Laboratory | Matluk et al |
| EPI_ISL_755355 | USA/ME-HETL-J0174/2020 | 2020-09-15 | North America/USA/Maine/ | Maine Health and Environmental Testing Laboratory | Tewhey Lab, The Jackson Laboratory | Matluk et al |
| EPI_ISL_755356 | USA/ME-HETL-J0175/2020 | 2020-09-15 | North America/USA/Maine/ | Maine Health and Environmental Testing Laboratory | Tewhey Lab, The Jackson Laboratory | Matluk et al |
| EPI_ISL_755359 | USA/ME-HETL-J0179/2020 | 2020-09-16 | North America/USA/Maine/ | Maine Health and Environmental Testing Laboratory | Tewhey Lab, The Jackson Laboratory | Matluk et al |
| EPI_ISL_755370 | USA/ME-HETL-J0199/2020 | 2020-09-18 | North America/USA/Maine/ | Maine Health and Environmental Testing Laboratory | Tewhey Lab, The Jackson Laboratory | Matluk et al |
| EPI_ISL_755374 | USA/ME-HETL-J0210/2020 | 2020-09-22 | North America/USA/Maine/ | Maine Health and Environmental Testing Laboratory | Tewhey Lab, The Jackson Laboratory | Matluk et al |
| EPI_ISL_755376 | USA/ME-HETL-J0213/2020 | 2020-09-22 | North America/USA/Maine/ | Maine Health and Environmental Testing Laboratory | Tewhey Lab, The Jackson Laboratory | Matluk et al |
| EPI_ISL_755379 | USA/ME-HETL-J0216/2020 | 2020-09-23 | North America/USA/Maine/ | Maine Health and Environmental Testing Laboratory | Tewhey Lab, The Jackson Laboratory | Matluk et al |
| EPI_ISL_755402 | USA/ME-HETL-J0247/2020 | 2020-09-28 | North America/USA/Maine/ | Maine Health and Environmental Testing Laboratory | Tewhey Lab, The Jackson Laboratory | Matluk et al |
| EPI_ISL_755404 | USA/ME-HETL-J0251/2020 | 2020-09-28 | North America/USA/Maine/ | Maine Health and Environmental Testing Laboratory | Tewhey Lab, The Jackson Laboratory | Matluk et al |
| EPI_ISL_755408 | USA/ME-HETL-J0260/2020 | 2020-09-29 | North America/USA/Maine/ | Maine Health and Environmental Testing Laboratory | Tewhey Lab, The Jackson Laboratory | Matluk et al |
| EPI_ISL_755409 | USA/ME-HETL-J0261/2020 | 2020-09-29 | North America/USA/Maine/ | Maine Health and Environmental Testing Laboratory | Tewhey Lab, The Jackson Laboratory | Matluk et al |

| | | | | | | |
|---|---|---|---|---|---|---|
| EPI_ISL_755412 | USA/ME-HETL-J0266/2020 | 2020-09-29 | North America/USA/Maine/ | Maine Health and Environmental Testing Laboratory | Tewhey Lab, The Jackson Laboratory | Matluk et al |
| EPI_ISL_755415 | USA/ME-HETL-J0269/2020 | 2020-09-29 | North America/USA/Maine/ | Maine Health and Environmental Testing Laboratory | Tewhey Lab, The Jackson Laboratory | Matluk et al |
| EPI_ISL_755421 | USA/ME-HETL-J0275/2020 | 2020-10-03 | North America/USA/Maine/ | Maine Health and Environmental Testing Laboratory | Tewhey Lab, The Jackson Laboratory | Matluk et al |
| EPI_ISL_755422 | USA/ME-HETL-J0276/2020 | 2020-10-04 | North America/USA/Maine/ | Maine Health and Environmental Testing Laboratory | Tewhey Lab, The Jackson Laboratory | Matluk et al |
| EPI_ISL_755423 | USA/ME-HETL-J0277/2020 | 2020-10-04 | North America/USA/Maine/ | Maine Health and Environmental Testing Laboratory | Tewhey Lab, The Jackson Laboratory | Matluk et al |
| EPI_ISL_755427 | USA/ME-HETL-J0284/2020 | 2020-10-05 | North America/USA/Maine/ | Maine Health and Environmental Testing Laboratory | Tewhey Lab, The Jackson Laboratory | Matluk et al |
| EPI_ISL_755435 | USA/ME-HETL-J0296/2020 | 2020-11-30 | North America/USA/Maine/ | Maine Health and Environmental Testing Laboratory | Tewhey Lab, The Jackson Laboratory | Matluk et al |
| EPI_ISL_755438 | USA/ME-HETL-J0300/2020 | 2020-11-30 | North America/USA/Maine/ | Maine Health and Environmental Testing Laboratory | Tewhey Lab, The Jackson Laboratory | Matluk et al |
| EPI_ISL_755439 | USA/ME-HETL-J0301/2020 | 2020-11-30 | North America/USA/Maine/ | Maine Health and Environmental Testing Laboratory | Tewhey Lab, The Jackson Laboratory | Matluk et al |
| EPI_ISL_755452 | USA/ME-HETL-J0323/2020 | 2020-11-30 | North America/USA/Maine/ | Maine Health and Environmental Testing Laboratory | Tewhey Lab, The Jackson Laboratory | Matluk et al |
| EPI_ISL_755457 | USA/ME-HETL-J0329/2020 | 2020-11-24 | North America/USA/Maine/ | Maine Health and Environmental Testing Laboratory | Tewhey Lab, The Jackson Laboratory | Matluk et al |
| EPI_ISL_755460 | USA/ME-HETL-J0332/2020 | 2020-11-24 | North America/USA/Maine/ | Maine Health and Environmental Testing Laboratory | Tewhey Lab, The Jackson Laboratory | Matluk et al |
| EPI_ISL_755466 | USA/ME-HETL-J0338/2020 | 2020-11-24 | North America/USA/Maine/ | Maine Health and Environmental Testing Laboratory | Tewhey Lab, The Jackson Laboratory | Matluk et al |

| | | | | | | |
|---|---|---|---|---|---|---|
| EPI_ISL_755481 | USA/ME-HETL-J0358/2020 | 2020-11-24 | North America/USA/Maine/ | Maine Health and Environmental Testing Laboratory | Tewhey Lab, The Jackson Laboratory | Matluk et al |
| EPI_ISL_755484 | USA/ME-HETL-J0361/2020 | 2020-11-30 | North America/USA/Maine/ | Maine Health and Environmental Testing Laboratory | Tewhey Lab, The Jackson Laboratory | Matluk et al |
| EPI_ISL_755508 | USA/ME-HETL-J0397/2020 | 2020-12-15 | North America/USA/Maine/ | Maine Health and Environmental Testing Laboratory | Tewhey Lab, The Jackson Laboratory | Matluk et al |
| EPI_ISL_755511 | USA/ME-HETL-J0402/2020 | 2020-12-16 | North America/USA/Maine/ | Maine Health and Environmental Testing Laboratory | Tewhey Lab, The Jackson Laboratory | Matluk et al |
| EPI_ISL_755520 | USA/ME-HETL-J0414/2020 | 2020-12-18 | North America/USA/Maine/ | Maine Health and Environmental Testing Laboratory | Tewhey Lab, The Jackson Laboratory | Matluk et al |
| EPI_ISL_755545 | USA/ME-HETL-J0447/2020 | 2020-12-17 | North America/USA/Maine/ | Maine Health and Environmental Testing Laboratory | Tewhey Lab, The Jackson Laboratory | Matluk et al |
| EPI_ISL_755546 | USA/ME-HETL-J0448/2020 | 2020-12-17 | North America/USA/Maine/ | Maine Health and Environmental Testing Laboratory | Tewhey Lab, The Jackson Laboratory | Matluk et al |
| EPI_ISL_755549 | USA/ME-HETL-J0453/2020 | 2020-12-17 | North America/USA/Maine/ | Maine Health and Environmental Testing Laboratory | Tewhey Lab, The Jackson Laboratory | Matluk et al |
| EPI_ISL_755550 | USA/ME-HETL-J0457/2020 | 2020-12-03 | North America/USA/Maine/ | Maine Health and Environmental Testing Laboratory | Tewhey Lab, The Jackson Laboratory | Matluk et al |
| EPI_ISL_755556 | USA/ME-HETL-J0464/2020 | 2020-12-15 | North America/USA/Maine/ | Maine Health and Environmental Testing Laboratory | Tewhey Lab, The Jackson Laboratory | Matluk et al |
| EPI_ISL_755558 | USA/ME-HETL-J0467/2020 | 2020-12-14 | North America/USA/Maine/ | Maine Health and Environmental Testing Laboratory | Tewhey Lab, The Jackson Laboratory | Matluk et al |
| EPI_ISL_755560 | USA/ME-HETL-J0469/2020 | 2020-12-16 | North America/USA/Maine/ | Maine Health and Environmental Testing Laboratory | Tewhey Lab, The Jackson Laboratory | Matluk et al |
| EPI_ISL_755562 | USA/ME-HETL-J0471/2020 | 2020-12-11 | North America/USA/Maine/ | Maine Health and Environmental Testing Laboratory | Tewhey Lab, The Jackson Laboratory | Matluk et al |

| | | | | | | |
|---|---|---|---|---|---|---|
| EPI_ISL_755567 | USA/ME-HETL-J0480/2020 | 2020-12-15 | North America/USA/Maine/ | Maine Health and Environmental Testing Laboratory | Tewhey Lab, The Jackson Laboratory | Matluk et al |
| EPI_ISL_755598 | USA/FL-CDC-STM-P004/2020 | 2020-12-19 | North America/USA/Florida/ | Helix/Illumina | Genomics and Discovery, Respiratory Viruses Branch, Division of Viral Diseases, Centers for Disease Control and Prevention | Peter W. Cook et al |
| EPI_ISL_755606 | USA/FL-CDC-STM-P014/2020 | 2020-12-19 | North America/USA/Florida/ | Helix/Illumina | Genomics and Discovery, Respiratory Viruses Branch, Division of Viral Diseases, Centers for Disease Control and Prevention | Peter W. Cook et al |
| EPI_ISL_755611 | USA/MA-CDC-STM-P021/2020 | 2020-12-19 | North America/USA/Massachusetts/ | Helix/Illumina | Genomics and Discovery, Respiratory Viruses Branch, Division of Viral Diseases, Centers for Disease Control and Prevention | Peter W. Cook et al |
| EPI_ISL_755616 | USA/IN-CDC-STM-P053/2020 | 2020-12-18 | North America/USA/Indiana/ | Helix/Illumina | Genomics and Discovery, Respiratory Viruses Branch, Division of Viral Diseases, Centers for Disease Control and Prevention | Peter W. Cook et al |

| | | | | | | |
|---|---|---|---|---|---|---|
| EPI_ISL_755639 | Germany/BY-IMB14592/2020 | 2020-12-20 | Europe/Germany/Bavaria/Munich | Bundeswehr Institute of Microbiology | Bundeswehr Institute of Microbiology | Malena Bestehorn-Willmann et al |
| EPI_ISL_755640 | Brazil/SP-874/2020 | 2020-12-03 | South America/Brazil/S√£o Paulo/S√£o I | Instituto Adolfo Lutz - Central | Instituto Adolfo Lutz, Interdiciplinary Procedures Center, Strategic Laboratory | Claudio Tavares Sacchi et al |
| EPI_ISL_755641 | Brazil/SP-875/2020 | 2020-12-01 | South America/Brazil/S√£o Paulo/Sao B | Instituto Adolfo Lutz - Regional de Santo Andre | Instituto Adolfo Lutz, Interdiciplinary Procedures Center, Strategic Laboratory | Claudio Tavares Sacchi et al |
| EPI_ISL_755642 | Brazil/SP-876/2020 | 2020-12-05 | South America/Brazil/S√£o Paulo/Mogi | Instituto Adolfo Lutz - Central | Instituto Adolfo Lutz, Interdiciplinary Procedures Center, Strategic Laboratory | Claudio Tavares Sacchi et al |
| EPI_ISL_755643 | Brazil/SP-877/2020 | 2020-12-07 | South America/Brazil/S√£o Paulo/Mogi | Instituto Adolfo Lutz - Central | Instituto Adolfo Lutz, Interdiciplinary Procedures Center, Strategic Laboratory | Claudio Tavares Sacchi et al |
| EPI_ISL_755644 | Brazil/SP-878/2020 | 2020-12-08 | South America/Brazil/S√£o Paulo/Embu | Lab LOC - Itapecerica da Serra | Instituto Adolfo Lutz, Interdiciplinary Procedures Center, Strategic Laboratory | Claudio Tavares Sacchi et al |
| EPI_ISL_755645 | Brazil/SP-879/2020 | 2020-12-08 | South America/Brazil/S√£o Paulo/Taboa | Lab LOC - Itapecerica da Serra | Instituto Adolfo Lutz, Interdiciplinary Procedures Center, Strategic Laboratory | Claudio Tavares Sacchi et al |

| | | | | | | |
|---|---|---|---|---|---|---|
| EPI_ISL_755646 | Brazil/SP-880/2020 | 2020-12-07 | South America/Brazil/S√£o Paulo/Sao B | Instituto Adolfo Lutz - Regional de Santo Andre | Instituto Adolfo Lutz, Interdiciplinary Procedures Center, Strategic Laboratory | Claudio Tavares Sacchi et al |
| EPI_ISL_755647 | Brazil/SP-881/2020 | 2020-12-08 | South America/Brazil/S√£o Paulo/Sao B | Instituto Adolfo Lutz - Regional de Santo Andre | Instituto Adolfo Lutz, Interdiciplinary Procedures Center, Strategic Laboratory | Claudio Tavares Sacchi et al |
| EPI_ISL_755648 | Brazil/SP-882/2020 | 2020-12-08 | South America/Brazil/S√£o Paulo/Carag | Instituto Adolfo Lutz - Regional de Taubate | Instituto Adolfo Lutz, Interdiciplinary Procedures Center, Strategic Laboratory | Claudio Tavares Sacchi et al |
| EPI_ISL_755649 | Brazil/SP-883/2020 | 2020-12-06 | South America/Brazil/S√£o Paulo/Sao B | Instituto Adolfo Lutz - Regional de Santo Andre | Instituto Adolfo Lutz, Interdiciplinary Procedures Center, Strategic Laboratory | Claudio Tavares Sacchi et al |
| EPI_ISL_755650 | Brazil/SP-884/2020 | 2020-12-08 | South America/Brazil/S√£o Paulo/Carag | Instituto Adolfo Lutz - Regional de Taubate | Instituto Adolfo Lutz, Interdiciplinary Procedures Center, Strategic Laboratory | Claudio Tavares Sacchi et al |
| EPI_ISL_755651 | Brazil/SP-885/2020 | 2020-12-10 | South America/Brazil/S√£o Paulo/Poa | Instituto Adolfo Lutz - Central | Instituto Adolfo Lutz, Interdiciplinary Procedures Center, Strategic Laboratory | Claudio Tavares Sacchi et al |

| | | | | | | |
|---|---|---|---|---|---|---|
| EPI_ISL_755652 | Brazil/SP-886/2020 | 2020-12-09 | South America/Brazil/S√£o Paulo/Embu | Lab LOC - Itapecerica da Serra | Instituto Adolfo Lutz, Interdiciplinary Procedures Center, Strategic Laboratory | Claudio Tavares Sacchi et al |
| EPI_ISL_755653 | Brazil/SP-887/2020 | 2020-12-12 | South America/Brazil/S√£o Paulo/Guara | Instituto Adolfo Lutz - Central | Instituto Adolfo Lutz, Interdiciplinary Procedures Center, Strategic Laboratory | Claudio Tavares Sacchi et al |
| EPI_ISL_755654 | Brazil/SP-888/2020 | 2020-12-11 | South America/Brazil/S√£o Paulo/S√£o | Instituto Adolfo Lutz - Central | Instituto Adolfo Lutz, Interdiciplinary Procedures Center, Strategic Laboratory | Claudio Tavares Sacchi et al |
| EPI_ISL_755655 | Brazil/SP-889/2020 | 2020-12-11 | South America/Brazil/S√£o Paulo/Indaia | Instituto Adolfo Lutz - Regional de Campinas | Instituto Adolfo Lutz, Interdiciplinary Procedures Center, Strategic Laboratory | Claudio Tavares Sacchi et al |
| EPI_ISL_755656 | Canada/ON-S1935/2020 | 2020-11-24 | North America/Canada/Ontario/Toronto | Toronto Invasive Bacterial Diseases Network | McMaster University | Allison McGeer et al |
| EPI_ISL_755657 | Canada/ON-S1936/2020 | 2020-11-24 | North America/Canada/Ontario/Toronto | Toronto Invasive Bacterial Diseases Network | McMaster University | Allison McGeer et al |
| EPI_ISL_755658 | Canada/ON-S1937/2020 | 2020-11-24 | North America/Canada/Ontario/Toronto | Toronto Invasive Bacterial Diseases Network | McMaster University | Allison McGeer et al |
| EPI_ISL_755659 | Canada/ON-S1938/2020 | 2020-11-24 | North America/Canada/Ontario/Toronto | Toronto Invasive Bacterial Diseases Network | McMaster University | Allison McGeer et al |
| EPI_ISL_755660 | Canada/ON-S1939/2020 | 2020-11-24 | North America/Canada/Ontario/Toronto | Toronto Invasive Bacterial Diseases Network | McMaster University | Allison McGeer et al |
| EPI_ISL_755661 | Canada/ON-S1941/2020 | 2020-11-24 | North America/Canada/Ontario/Toronto | Toronto Invasive Bacterial Diseases Network | McMaster University | Allison McGeer et al |
| EPI_ISL_755662 | Canada/ON-S1942/2020 | 2020-11-24 | North America/Canada/Ontario/Toronto | Toronto Invasive Bacterial Diseases Network | McMaster University | Allison McGeer et al |
| EPI_ISL_755663 | Canada/ON-S1943/2020 | 2020-11-24 | North America/Canada/Ontario/Toronto | Toronto Invasive Bacterial Diseases Network | McMaster University | Allison McGeer et al |

| | | | | | | |
|---|---|---|---|---|---|---|
| EPI_ISL_755664 | Canada/ON-S1944/2020 | 2020-11-24 | North America/Canada/Ontario/Toronto | Toronto Invasive Bacterial Diseases Network | McMaster University | Allison McGeer et al |
| EPI_ISL_755665 | Canada/ON-S1945/2020 | 2020-11-24 | North America/Canada/Ontario/Toronto | Toronto Invasive Bacterial Diseases Network | McMaster University | Allison McGeer et al |
| EPI_ISL_755666 | Canada/ON-S1946/2020 | 2020-11-24 | North America/Canada/Ontario/Toronto | Toronto Invasive Bacterial Diseases Network | McMaster University | Allison McGeer et al |
| EPI_ISL_755667 | Canada/ON-S1947/2020 | 2020-11-24 | North America/Canada/Ontario/Toronto | Toronto Invasive Bacterial Diseases Network | McMaster University | Allison McGeer et al |
| EPI_ISL_755668 | Canada/ON-S1948/2020 | 2020-11-24 | North America/Canada/Ontario/Toronto | Toronto Invasive Bacterial Diseases Network | McMaster University | Allison McGeer et al |
| EPI_ISL_755669 | Canada/ON-S1949/2020 | 2020-11-24 | North America/Canada/Ontario/Toronto | Toronto Invasive Bacterial Diseases Network | McMaster University | Allison McGeer et al |
| EPI_ISL_755670 | Canada/ON-S1950/2020 | 2020-11-24 | North America/Canada/Ontario/Toronto | Toronto Invasive Bacterial Diseases Network | McMaster University | Allison McGeer et al |
| EPI_ISL_755671 | Canada/ON-S1951/2020 | 2020-11-24 | North America/Canada/Ontario/Toronto | Toronto Invasive Bacterial Diseases Network | McMaster University | Allison McGeer et al |
| EPI_ISL_755672 | Canada/ON-S1952/2020 | 2020-11-24 | North America/Canada/Ontario/Toronto | Toronto Invasive Bacterial Diseases Network | McMaster University | Allison McGeer et al |
| EPI_ISL_755673 | Canada/ON-S1953/2020 | 2020-11-24 | North America/Canada/Ontario/Toronto | Toronto Invasive Bacterial Diseases Network | McMaster University | Allison McGeer et al |
| EPI_ISL_755674 | Canada/ON-S1954/2020 | 2020-11-24 | North America/Canada/Ontario/Toronto | Toronto Invasive Bacterial Diseases Network | McMaster University | Allison McGeer et al |
| EPI_ISL_755675 | Canada/ON-S1955/2020 | 2020-11-24 | North America/Canada/Ontario/Toronto | Toronto Invasive Bacterial Diseases Network | McMaster University | Allison McGeer et al |
| EPI_ISL_755676 | Canada/ON-S1957/2020 | 2020-11-24 | North America/Canada/Ontario/Toronto | Toronto Invasive Bacterial Diseases Network | McMaster University | Allison McGeer et al |
| EPI_ISL_755677 | Canada/ON-S1958/2020 | 2020-11-24 | North America/Canada/Ontario/Toronto | Toronto Invasive Bacterial Diseases Network | McMaster University | Allison McGeer et al |
| EPI_ISL_755678 | Canada/ON-S1962/2020 | 2020-11-24 | North America/Canada/Ontario/Toronto | Toronto Invasive Bacterial Diseases Network | McMaster University | Allison McGeer et al |
| EPI_ISL_755679 | Canada/ON-S1963/2020 | 2020-11-24 | North America/Canada/Ontario/Toronto | Toronto Invasive Bacterial Diseases Network | McMaster University | Allison McGeer et al |
| EPI_ISL_755680 | Canada/ON-S1964/2020 | 2020-11-24 | North America/Canada/Ontario/Toronto | Toronto Invasive Bacterial Diseases Network | McMaster University | Allison McGeer et al |
| EPI_ISL_755681 | Canada/ON-S1965/2020 | 2020-11-24 | North America/Canada/Ontario/Toronto | Toronto Invasive Bacterial Diseases Network | McMaster University | Allison McGeer et al |
| EPI_ISL_755682 | Canada/ON-S1966/2020 | 2020-11-24 | North America/Canada/Ontario/Toronto | Toronto Invasive Bacterial Diseases Network | McMaster University | Allison McGeer et al |
| EPI_ISL_755683 | Canada/ON-S1967/2020 | 2020-11-24 | North America/Canada/Ontario/Toronto | Toronto Invasive Bacterial Diseases Network | McMaster University | Allison McGeer et al |

| | | | | | | |
|---|---|---|---|---|---|---|
| EPI_ISL_755684 | Canada/ON-S1968/2020 | 2020-11-24 | North America/Canada/Ontario/Toronto | Toronto Invasive Bacterial Diseases Network | McMaster University | Allison McGeer et al |
| EPI_ISL_755685 | Canada/ON-S1971/2020 | 2020-11-24 | North America/Canada/Ontario/Toronto | Toronto Invasive Bacterial Diseases Network | McMaster University | Allison McGeer et al |
| EPI_ISL_755686 | Canada/ON-S1972/2020 | 2020-11-24 | North America/Canada/Ontario/Toronto | Toronto Invasive Bacterial Diseases Network | McMaster University | Allison McGeer et al |
| EPI_ISL_755687 | Canada/ON-S1973/2020 | 2020-11-24 | North America/Canada/Ontario/Toronto | Toronto Invasive Bacterial Diseases Network | McMaster University | Allison McGeer et al |
| EPI_ISL_755688 | Canada/ON-S1974/2020 | 2020-11-24 | North America/Canada/Ontario/Toronto | Toronto Invasive Bacterial Diseases Network | McMaster University | Allison McGeer et al |
| EPI_ISL_755689 | Canada/ON-S1975/2020 | 2020-11-24 | North America/Canada/Ontario/Toronto | Toronto Invasive Bacterial Diseases Network | McMaster University | Allison McGeer et al |
| EPI_ISL_755690 | Canada/ON-S1976/2020 | 2020-11-24 | North America/Canada/Ontario/Toronto | Toronto Invasive Bacterial Diseases Network | McMaster University | Allison McGeer et al |
| EPI_ISL_755691 | Canada/ON-S1977/2020 | 2020-11-24 | North America/Canada/Ontario/Toronto | Toronto Invasive Bacterial Diseases Network | McMaster University | Allison McGeer et al |
| EPI_ISL_755692 | Canada/ON-S1978/2020 | 2020-11-24 | North America/Canada/Ontario/Toronto | Toronto Invasive Bacterial Diseases Network | McMaster University | Allison McGeer et al |
| EPI_ISL_755693 | Canada/ON-S1979/2020 | 2020-11-24 | North America/Canada/Ontario/Toronto | Toronto Invasive Bacterial Diseases Network | McMaster University | Allison McGeer et al |
| EPI_ISL_755694 | Canada/ON-S1981/2020 | 2020-11-24 | North America/Canada/Ontario/Toronto | Toronto Invasive Bacterial Diseases Network | McMaster University | Allison McGeer et al |
| EPI_ISL_755695 | Canada/ON-S1982/2020 | 2020-11-24 | North America/Canada/Ontario/Toronto | Toronto Invasive Bacterial Diseases Network | McMaster University | Allison McGeer et al |
| EPI_ISL_755696 | Canada/ON-S1983/2020 | 2020-11-24 | North America/Canada/Ontario/Toronto | Toronto Invasive Bacterial Diseases Network | McMaster University | Allison McGeer et al |
| EPI_ISL_755697 | Canada/ON-S1984/2020 | 2020-11-24 | North America/Canada/Ontario/Toronto | Toronto Invasive Bacterial Diseases Network | McMaster University | Allison McGeer et al |
| EPI_ISL_755698 | Canada/ON-S1985/2020 | 2020-11-24 | North America/Canada/Ontario/Toronto | Toronto Invasive Bacterial Diseases Network | McMaster University | Allison McGeer et al |
| EPI_ISL_755699 | Canada/ON-S1986/2020 | 2020-11-24 | North America/Canada/Ontario/Toronto | Toronto Invasive Bacterial Diseases Network | McMaster University | Allison McGeer et al |
| EPI_ISL_755700 | Canada/ON-S1987/2020 | 2020-11-24 | North America/Canada/Ontario/Toronto | Toronto Invasive Bacterial Diseases Network | McMaster University | Allison McGeer et al |
| EPI_ISL_755701 | Canada/ON-S1988/2020 | 2020-11-24 | North America/Canada/Ontario/Toronto | Toronto Invasive Bacterial Diseases Network | McMaster University | Allison McGeer et al |
| EPI_ISL_755702 | Canada/ON-S1991/2020 | 2020-11-24 | North America/Canada/Ontario/Toronto | Toronto Invasive Bacterial Diseases Network | McMaster University | Allison McGeer et al |
| EPI_ISL_755703 | Canada/ON-S1992/2020 | 2020-11-24 | North America/Canada/Ontario/Toronto | Toronto Invasive Bacterial Diseases Network | McMaster University | Allison McGeer et al |

| | | | | | | |
|---|---|---|---|---|---|---|
| EPI_ISL_755704 | Canada/ON-S1994/2020 | 2020-11-24 | North America/Canada/Ontario/Toronto | Toronto Invasive Bacterial Diseases Network | McMaster University | Allison McGeer et al |
| EPI_ISL_755705 | Canada/ON-S1995/2020 | 2020-11-24 | North America/Canada/Ontario/Toronto | Toronto Invasive Bacterial Diseases Network | McMaster University | Allison McGeer et al |
| EPI_ISL_755706 | Canada/ON-S1996/2020 | 2020-11-24 | North America/Canada/Ontario/Toronto | Toronto Invasive Bacterial Diseases Network | McMaster University | Allison McGeer et al |
| EPI_ISL_755707 | Canada/ON-S1997/2020 | 2020-11-24 | North America/Canada/Ontario/Toronto | Toronto Invasive Bacterial Diseases Network | McMaster University | Allison McGeer et al |
| EPI_ISL_755708 | Canada/ON-S2000/2020 | 2020-11-24 | North America/Canada/Ontario/Toronto | Toronto Invasive Bacterial Diseases Network | McMaster University | Allison McGeer et al |
| EPI_ISL_755709 | Canada/ON-S2004/2020 | 2020-11-24 | North America/Canada/Ontario/Toronto | Toronto Invasive Bacterial Diseases Network | McMaster University | Allison McGeer et al |
| EPI_ISL_755710 | Canada/ON-S2005/2020 | 2020-11-24 | North America/Canada/Ontario/Toronto | Toronto Invasive Bacterial Diseases Network | McMaster University | Allison McGeer et al |
| EPI_ISL_755711 | Canada/ON-S2006/2020 | 2020-11-24 | North America/Canada/Ontario/Toronto | Toronto Invasive Bacterial Diseases Network | McMaster University | Allison McGeer et al |
| EPI_ISL_755712 | Canada/ON-A1/2020 | 2020-11-24 | North America/Canada/Ontario/Brampt | Toronto Invasive Bacterial Diseases Network | McMaster University | David Richardson et al |
| EPI_ISL_755713 | Canada/ON-A10/2020 | 2020-11-24 | North America/Canada/Ontario/Brampt | Toronto Invasive Bacterial Diseases Network | McMaster University | David Richardson et al |
| EPI_ISL_755714 | Canada/ON-A11/2020 | 2020-11-24 | North America/Canada/Ontario/Brampt | Toronto Invasive Bacterial Diseases Network | McMaster University | David Richardson et al |
| EPI_ISL_755715 | Canada/ON-A12/2020 | 2020-11-24 | North America/Canada/Ontario/Brampt | Toronto Invasive Bacterial Diseases Network | McMaster University | David Richardson et al |
| EPI_ISL_755716 | Canada/ON-A13/2020 | 2020-11-24 | North America/Canada/Ontario/Brampt | Toronto Invasive Bacterial Diseases Network | McMaster University | David Richardson et al |
| EPI_ISL_755717 | Canada/ON-A15/2020 | 2020-11-24 | North America/Canada/Ontario/Brampt | Toronto Invasive Bacterial Diseases Network | McMaster University | David Richardson et al |
| EPI_ISL_755718 | Canada/ON-A16/2020 | 2020-11-24 | North America/Canada/Ontario/Brampt | Toronto Invasive Bacterial Diseases Network | McMaster University | David Richardson et al |
| EPI_ISL_755719 | Canada/ON-A19/2020 | 2020-11-24 | North America/Canada/Ontario/Brampt | Toronto Invasive Bacterial Diseases Network | McMaster University | David Richardson et al |
| EPI_ISL_755720 | Canada/ON-A2/2020 | 2020-11-24 | North America/Canada/Ontario/Brampt | Toronto Invasive Bacterial Diseases Network | McMaster University | David Richardson et al |
| EPI_ISL_755721 | Canada/ON-A21/2020 | 2020-11-24 | North America/Canada/Ontario/Brampt | Toronto Invasive Bacterial Diseases Network | McMaster University | David Richardson et al |
| EPI_ISL_755722 | Canada/ON-A23/2020 | 2020-11-24 | North America/Canada/Ontario/Brampt | Toronto Invasive Bacterial Diseases Network | McMaster University | David Richardson et al |
| EPI_ISL_755723 | Canada/ON-A25/2020 | 2020-11-24 | North America/Canada/Ontario/Brampt | Toronto Invasive Bacterial Diseases Network | McMaster University | David Richardson et al |

| | | | | | | |
|---|---|---|---|---|---|---|
| EPI_ISL_755724 | Canada/ON-A26/2020 | 2020-11-24 | North America/Canada/Ontario/Brampt | Toronto Invasive Bacterial Diseases Network | McMaster University | David Richardson et al |
| EPI_ISL_755725 | Canada/ON-A27/2020 | 2020-11-24 | North America/Canada/Ontario/Brampt | Toronto Invasive Bacterial Diseases Network | McMaster University | David Richardson et al |
| EPI_ISL_755726 | Canada/ON-A28/2020 | 2020-11-24 | North America/Canada/Ontario/Brampt | Toronto Invasive Bacterial Diseases Network | McMaster University | David Richardson et al |
| EPI_ISL_755727 | Canada/ON-A3/2020 | 2020-11-24 | North America/Canada/Ontario/Brampt | Toronto Invasive Bacterial Diseases Network | McMaster University | David Richardson et al |
| EPI_ISL_755728 | Canada/ON-A30/2020 | 2020-11-24 | North America/Canada/Ontario/Brampt | Toronto Invasive Bacterial Diseases Network | McMaster University | David Richardson et al |
| EPI_ISL_755729 | Canada/ON-A31/2020 | 2020-11-24 | North America/Canada/Ontario/Brampt | Toronto Invasive Bacterial Diseases Network | McMaster University | David Richardson et al |
| EPI_ISL_755730 | Canada/ON-A32/2020 | 2020-11-24 | North America/Canada/Ontario/Brampt | Toronto Invasive Bacterial Diseases Network | McMaster University | David Richardson et al |
| EPI_ISL_755731 | Canada/ON-A4/2020 | 2020-11-24 | North America/Canada/Ontario/Brampt | Toronto Invasive Bacterial Diseases Network | McMaster University | David Richardson et al |
| EPI_ISL_755732 | Canada/ON-A5/2020 | 2020-11-24 | North America/Canada/Ontario/Brampt | Toronto Invasive Bacterial Diseases Network | McMaster University | David Richardson et al |
| EPI_ISL_755733 | Canada/ON-A6/2020 | 2020-11-24 | North America/Canada/Ontario/Brampt | Toronto Invasive Bacterial Diseases Network | McMaster University | David Richardson et al |
| EPI_ISL_755734 | Canada/ON-A7/2020 | 2020-11-24 | North America/Canada/Ontario/Brampt | Toronto Invasive Bacterial Diseases Network | McMaster University | David Richardson et al |
| EPI_ISL_755735 | Canada/ON-A8/2020 | 2020-11-24 | North America/Canada/Ontario/Brampt | Toronto Invasive Bacterial Diseases Network | McMaster University | David Richardson et al |
| EPI_ISL_755736 | Canada/ON-A9/2020 | 2020-11-24 | North America/Canada/Ontario/Brampt | Toronto Invasive Bacterial Diseases Network | McMaster University | David Richardson et al |
| EPI_ISL_755737 | Canada/ON-CVD00072/2020 | 2020-11-24 | North America/Canada/Ontario/Toronto | Toronto Invasive Bacterial Diseases Network | McMaster University | Allison McGeer et al |
| EPI_ISL_755738 | Canada/ON-CVD00183/2020 | 2020-11-24 | North America/Canada/Ontario/Toronto | Toronto Invasive Bacterial Diseases Network | McMaster University | Allison McGeer et al |
| EPI_ISL_755739 | Canada/ON-CVD01417/2020 | 2020-11-24 | North America/Canada/Ontario/Toronto | Toronto Invasive Bacterial Diseases Network | McMaster University | Allison McGeer et al |
| EPI_ISL_755740 | Canada/ON-S269/2020 | 2020-03-27 | North America/Canada/Ontario/Toronto | Toronto Invasive Bacterial Diseases Network | McMaster University | Allison McGeer et al |
| EPI_ISL_755741 | Canada/ON-S832/2020 | 2020-04-11 | North America/Canada/Ontario/Toronto | Toronto Invasive Bacterial Diseases Network | McMaster University | Allison McGeer et al |
| EPI_ISL_755742 | Canada/ON-SB-01/2020 | 2020-10-21 | North America/Canada/Ontario/Toronto | Toronto Invasive Bacterial Diseases Network | McMaster University | Allison McGeer et al |
| EPI_ISL_755743 | Canada/ON-SB-02/2020 | 2020-10-21 | North America/Canada/Ontario/Toronto | Toronto Invasive Bacterial Diseases Network | McMaster University | Allison McGeer et al |

| | | | | | | |
|---|---|---|---|---|---|---|
| EPI_ISL_755744 | Canada/ON-SB-04/2020 | 2020-10-21 | North America/Canada/Ontario/Toronto | Toronto Invasive Bacterial Diseases Network | McMaster University | Allison McGeer et al |
| EPI_ISL_755745 | Canada/ON-SB-06/2020 | 2020-10-14 | North America/Canada/Ontario/Toronto | Toronto Invasive Bacterial Diseases Network | McMaster University | Allison McGeer et al |
| EPI_ISL_755746 | Canada/ON-SB-07/2020 | 2020-10-01 | North America/Canada/Ontario/Toronto | Toronto Invasive Bacterial Diseases Network | McMaster University | Allison McGeer et al |
| EPI_ISL_755747 | Canada/ON-SB-08/2020 | 2020-09-23 | North America/Canada/Ontario/Toronto | Toronto Invasive Bacterial Diseases Network | McMaster University | Allison McGeer et al |
| EPI_ISL_755748 | Canada/ON-SB-09/2020 | 2020-10-23 | North America/Canada/Ontario/Toronto | Toronto Invasive Bacterial Diseases Network | McMaster University | Allison McGeer et al |
| EPI_ISL_755749 | Canada/ON-SB-10/2020 | 2020-10-21 | North America/Canada/Ontario/Toronto | Toronto Invasive Bacterial Diseases Network | McMaster University | Allison McGeer et al |
| EPI_ISL_755750 | Canada/ON-SB-11/2020 | 2020-10-24 | North America/Canada/Ontario/Toronto | Toronto Invasive Bacterial Diseases Network | McMaster University | Allison McGeer et al |
| EPI_ISL_755751 | Canada/ON-SB-12/2020 | 2020-10-24 | North America/Canada/Ontario/Toronto | Toronto Invasive Bacterial Diseases Network | McMaster University | Allison McGeer et al |
| EPI_ISL_755752 | Canada/ON-S133/2020 | 2020-03-23 | North America/Canada/Ontario/Toronto | Toronto Invasive Bacterial Diseases Network | McMaster University | Allison McGeer et al |
| EPI_ISL_755753 | Canada/ON-S134/2020 | 2020-03-23 | North America/Canada/Ontario/Toronto | Toronto Invasive Bacterial Diseases Network | McMaster University | Allison McGeer et al |
| EPI_ISL_755754 | Canada/ON-S158/2020 | 2020-03-24 | North America/Canada/Ontario/Toronto | Toronto Invasive Bacterial Diseases Network | McMaster University | Allison McGeer et al |
| EPI_ISL_755755 | Canada/ON-S1628/2020 | 2020-09-14 | North America/Canada/Ontario/Toronto | Toronto Invasive Bacterial Diseases Network | McMaster University | Allison McGeer et al |
| EPI_ISL_755756 | Canada/ON-S1629/2020 | 2020-09-14 | North America/Canada/Ontario/Toronto | Toronto Invasive Bacterial Diseases Network | McMaster University | Allison McGeer et al |
| EPI_ISL_755757 | Canada/ON-S1636/2020 | 2020-09-15 | North America/Canada/Ontario/Toronto | Toronto Invasive Bacterial Diseases Network | McMaster University | Allison McGeer et al |
| EPI_ISL_755758 | Canada/ON-S1637/2020 | 2020-09-15 | North America/Canada/Ontario/Toronto | Toronto Invasive Bacterial Diseases Network | McMaster University | Allison McGeer et al |
| EPI_ISL_755759 | Canada/ON-S1638/2020 | 2020-09-15 | North America/Canada/Ontario/Toronto | Toronto Invasive Bacterial Diseases Network | McMaster University | Allison McGeer et al |
| EPI_ISL_755760 | Canada/ON-S165/2020 | 2020-03-24 | North America/Canada/Ontario/Toronto | Toronto Invasive Bacterial Diseases Network | McMaster University | Allison McGeer et al |
| EPI_ISL_755761 | Canada/ON-S1760/2020 | 2020-11-09 | North America/Canada/Ontario/Toronto | Toronto Invasive Bacterial Diseases Network | McMaster University | Allison McGeer et al |
| EPI_ISL_755762 | Canada/ON-S1761/2020 | 2020-11-09 | North America/Canada/Ontario/Toronto | Toronto Invasive Bacterial Diseases Network | McMaster University | Allison McGeer et al |
| EPI_ISL_755763 | Canada/ON-S1762/2020 | 2020-11-09 | North America/Canada/Ontario/Toronto | Toronto Invasive Bacterial Diseases Network | McMaster University | Allison McGeer et al |

| | | | | | | |
|---|---|---|---|---|---|---|
| EPI_ISL_755764 | Canada/ON-S1763/2020 | 2020-11-08 | North America/Canada/Ontario/Toronto | Toronto Invasive Bacterial Diseases Network | McMaster University | Allison McGeer et al |
| EPI_ISL_755765 | Canada/ON-S1764/2020 | 2020-11-08 | North America/Canada/Ontario/Toronto | Toronto Invasive Bacterial Diseases Network | McMaster University | Allison McGeer et al |
| EPI_ISL_755766 | Canada/ON-S1765/2020 | 2020-11-08 | North America/Canada/Ontario/Toronto | Toronto Invasive Bacterial Diseases Network | McMaster University | Allison McGeer et al |
| EPI_ISL_755767 | Canada/ON-S1766/2020 | 2020-11-08 | North America/Canada/Ontario/Toronto | Toronto Invasive Bacterial Diseases Network | McMaster University | Allison McGeer et al |
| EPI_ISL_755768 | Canada/ON-S1767/2020 | 2020-11-08 | North America/Canada/Ontario/Toronto | Toronto Invasive Bacterial Diseases Network | McMaster University | Allison McGeer et al |
| EPI_ISL_755769 | Canada/ON-S1768/2020 | 2020-11-08 | North America/Canada/Ontario/Toronto | Toronto Invasive Bacterial Diseases Network | McMaster University | Allison McGeer et al |
| EPI_ISL_755770 | Canada/ON-S1769/2020 | 2020-11-08 | North America/Canada/Ontario/Toronto | Toronto Invasive Bacterial Diseases Network | McMaster University | Allison McGeer et al |
| EPI_ISL_755771 | Canada/ON-S1770/2020 | 2020-11-08 | North America/Canada/Ontario/Toronto | Toronto Invasive Bacterial Diseases Network | McMaster University | Allison McGeer et al |
| EPI_ISL_755772 | Canada/ON-S1771/2020 | 2020-11-08 | North America/Canada/Ontario/Toronto | Toronto Invasive Bacterial Diseases Network | McMaster University | Allison McGeer et al |
| EPI_ISL_755773 | Canada/ON-S1772/2020 | 2020-11-08 | North America/Canada/Ontario/Toronto | Toronto Invasive Bacterial Diseases Network | McMaster University | Allison McGeer et al |
| EPI_ISL_755774 | Canada/ON-S1774/2020 | 2020-11-08 | North America/Canada/Ontario/Toronto | Toronto Invasive Bacterial Diseases Network | McMaster University | Allison McGeer et al |
| EPI_ISL_755775 | Canada/ON-S1775/2020 | 2020-11-08 | North America/Canada/Ontario/Toronto | Toronto Invasive Bacterial Diseases Network | McMaster University | Allison McGeer et al |
| EPI_ISL_755776 | Canada/ON-S1778/2020 | 2020-11-08 | North America/Canada/Ontario/Toronto | Toronto Invasive Bacterial Diseases Network | McMaster University | Allison McGeer et al |
| EPI_ISL_755777 | Canada/ON-S1779/2020 | 2020-11-08 | North America/Canada/Ontario/Toronto | Toronto Invasive Bacterial Diseases Network | McMaster University | Allison McGeer et al |
| EPI_ISL_755778 | Canada/ON-S1780/2020 | 2020-11-08 | North America/Canada/Ontario/Toronto | Toronto Invasive Bacterial Diseases Network | McMaster University | Allison McGeer et al |
| EPI_ISL_755779 | Canada/ON-S1781/2020 | 2020-11-08 | North America/Canada/Ontario/Toronto | Toronto Invasive Bacterial Diseases Network | McMaster University | Allison McGeer et al |
| EPI_ISL_755780 | Canada/ON-S1782/2020 | 2020-11-08 | North America/Canada/Ontario/Toronto | Toronto Invasive Bacterial Diseases Network | McMaster University | Allison McGeer et al |
| EPI_ISL_755781 | Canada/ON-S1783/2020 | 2020-11-08 | North America/Canada/Ontario/Toronto | Toronto Invasive Bacterial Diseases Network | McMaster University | Allison McGeer et al |
| EPI_ISL_755782 | Canada/ON-S1785/2020 | 2020-11-15 | North America/Canada/Ontario/Toronto | Toronto Invasive Bacterial Diseases Network | McMaster University | Allison McGeer et al |
| EPI_ISL_755783 | Canada/ON-S1786/2020 | 2020-11-15 | North America/Canada/Ontario/Toronto | Toronto Invasive Bacterial Diseases Network | McMaster University | Allison McGeer et al |

| | | | | | | |
|---|---|---|---|---|---|---|
| EPI_ISL_755784 | Canada/ON-S1787/2020 | 2020-11-15 | North America/Canada/Ontario/Toronto | Toronto Invasive Bacterial Diseases Network | McMaster University | Allison McGeer et al |
| EPI_ISL_755786 | Canada/ON-S1790/2020 | 2020-11-15 | North America/Canada/Ontario/Toronto | Toronto Invasive Bacterial Diseases Network | McMaster University | Allison McGeer et al |
| EPI_ISL_755787 | Canada/ON-S1792/2020 | 2020-11-15 | North America/Canada/Ontario/Toronto | Toronto Invasive Bacterial Diseases Network | McMaster University | Allison McGeer et al |
| EPI_ISL_755788 | Canada/ON-S1793/2020 | 2020-11-15 | North America/Canada/Ontario/Toronto | Toronto Invasive Bacterial Diseases Network | McMaster University | Allison McGeer et al |
| EPI_ISL_755789 | Canada/ON-S1794/2020 | 2020-11-15 | North America/Canada/Ontario/Toronto | Toronto Invasive Bacterial Diseases Network | McMaster University | Allison McGeer et al |
| EPI_ISL_755790 | Canada/ON-S1795/2020 | 2020-11-15 | North America/Canada/Ontario/Toronto | Toronto Invasive Bacterial Diseases Network | McMaster University | Allison McGeer et al |
| EPI_ISL_755791 | Canada/ON-S1796/2020 | 2020-11-15 | North America/Canada/Ontario/Toronto | Toronto Invasive Bacterial Diseases Network | McMaster University | Allison McGeer et al |
| EPI_ISL_755792 | Canada/ON-S1797/2020 | 2020-11-15 | North America/Canada/Ontario/Toronto | Toronto Invasive Bacterial Diseases Network | McMaster University | Allison McGeer et al |
| EPI_ISL_755793 | Canada/ON-S187/2020 | 2020-03-25 | North America/Canada/Ontario/Toronto | Toronto Invasive Bacterial Diseases Network | McMaster University | Allison McGeer et al |
| EPI_ISL_755794 | Canada/ON-S19/2020 | 2020-03-16 | North America/Canada/Ontario/Toronto | Toronto Invasive Bacterial Diseases Network | McMaster University | Allison McGeer et al |
| EPI_ISL_755795 | Canada/ON-S191/2020 | 2020-03-25 | North America/Canada/Ontario/Toronto | Toronto Invasive Bacterial Diseases Network | McMaster University | Allison McGeer et al |
| EPI_ISL_755796 | Canada/ON-S193/2020 | 2020-03-25 | North America/Canada/Ontario/Toronto | Toronto Invasive Bacterial Diseases Network | McMaster University | Allison McGeer et al |
| EPI_ISL_755797 | Canada/ON-S194/2020 | 2020-03-27 | North America/Canada/Ontario/Toronto | Toronto Invasive Bacterial Diseases Network | McMaster University | Allison McGeer et al |
| EPI_ISL_755798 | Canada/ON-S195/2020 | 2020-03-25 | North America/Canada/Ontario/Toronto | Toronto Invasive Bacterial Diseases Network | McMaster University | Allison McGeer et al |
| EPI_ISL_755799 | Canada/ON-S196/2020 | 2020-03-26 | North America/Canada/Ontario/Toronto | Toronto Invasive Bacterial Diseases Network | McMaster University | Allison McGeer et al |
| EPI_ISL_755800 | Canada/ON-S198/2020 | 2020-03-27 | North America/Canada/Ontario/Toronto | Toronto Invasive Bacterial Diseases Network | McMaster University | Allison McGeer et al |
| EPI_ISL_755801 | Canada/ON-S199/2020 | 2020-03-26 | North America/Canada/Ontario/Toronto | Toronto Invasive Bacterial Diseases Network | McMaster University | Allison McGeer et al |
| EPI_ISL_755802 | Canada/ON-S2016/2020 | 2020-11-28 | North America/Canada/Ontario/Toronto | Toronto Invasive Bacterial Diseases Network | McMaster University | Allison McGeer et al |
| EPI_ISL_755803 | Canada/ON-S2017/2020 | 2020-12-01 | North America/Canada/Ontario/Toronto | Toronto Invasive Bacterial Diseases Network | McMaster University | Allison McGeer et al |
| EPI_ISL_755804 | Canada/ON-S2018/2020 | 2020-12-17 | North America/Canada/Ontario/Toronto | Toronto Invasive Bacterial Diseases Network | McMaster University | Allison McGeer et al |

| | | | | | | |
|---|---|---|---|---|---|---|
| EPI_ISL_755805 | Canada/ON-S2019/2020 | 2020-11-16 | North America/Canada/Ontario/Toronto | Toronto Invasive Bacterial Diseases Network | McMaster University | Allison McGeer et al |
| EPI_ISL_755806 | Canada/ON-S2020/2020 | 2020-11-16 | North America/Canada/Ontario/Toronto | Toronto Invasive Bacterial Diseases Network | McMaster University | Allison McGeer et al |
| EPI_ISL_755807 | Canada/ON-S2021/2020 | 2020-11-16 | North America/Canada/Ontario/Toronto | Toronto Invasive Bacterial Diseases Network | McMaster University | Allison McGeer et al |
| EPI_ISL_755808 | Canada/ON-S2022/2020 | 2020-09-07 | North America/Canada/Ontario/Toronto | Toronto Invasive Bacterial Diseases Network | McMaster University | Allison McGeer et al |
| EPI_ISL_755809 | Canada/ON-S2023/2020 | 2020-09-12 | North America/Canada/Ontario/Toronto | Toronto Invasive Bacterial Diseases Network | McMaster University | Allison McGeer et al |
| EPI_ISL_755810 | Canada/ON-S2025/2020 | 2020-10-10 | North America/Canada/Ontario/Toronto | Toronto Invasive Bacterial Diseases Network | McMaster University | Allison McGeer et al |
| EPI_ISL_755811 | Canada/ON-S2027/2020 | 2020-10-13 | North America/Canada/Ontario/Toronto | Toronto Invasive Bacterial Diseases Network | McMaster University | Allison McGeer et al |
| EPI_ISL_755812 | Canada/ON-S2031/2020 | 2020-12-21 | North America/Canada/Ontario/Toronto | Toronto Invasive Bacterial Diseases Network | McMaster University | Allison McGeer et al |
| EPI_ISL_755813 | Canada/ON-S2032/2020 | 2020-12-21 | North America/Canada/Ontario/Toronto | Toronto Invasive Bacterial Diseases Network | McMaster University | Allison McGeer et al |
| EPI_ISL_755814 | Canada/ON-S204/2020 | 2020-03-26 | North America/Canada/Ontario/Toronto | Toronto Invasive Bacterial Diseases Network | McMaster University | Allison McGeer et al |
| EPI_ISL_755815 | Canada/ON-S207/2020 | 2020-03-25 | North America/Canada/Ontario/Toronto | Toronto Invasive Bacterial Diseases Network | McMaster University | Allison McGeer et al |
| EPI_ISL_755816 | Canada/ON-S208/2020 | 2020-03-25 | North America/Canada/Ontario/Toronto | Toronto Invasive Bacterial Diseases Network | McMaster University | Allison McGeer et al |
| EPI_ISL_755817 | Canada/ON-S209/2020 | 2020-03-26 | North America/Canada/Ontario/Toronto | Toronto Invasive Bacterial Diseases Network | McMaster University | Allison McGeer et al |
| EPI_ISL_755818 | Canada/ON-S210/2020 | 2020-03-26 | North America/Canada/Ontario/Toronto | Toronto Invasive Bacterial Diseases Network | McMaster University | Allison McGeer et al |
| EPI_ISL_755819 | Canada/ON-S211/2020 | 2020-03-25 | North America/Canada/Ontario/Toronto | Toronto Invasive Bacterial Diseases Network | McMaster University | Allison McGeer et al |
| EPI_ISL_755820 | Canada/ON-S214/2020 | 2020-03-27 | North America/Canada/Ontario/Toronto | Toronto Invasive Bacterial Diseases Network | McMaster University | Allison McGeer et al |
| EPI_ISL_755821 | Canada/ON-S216/2020 | 2020-03-26 | North America/Canada/Ontario/Toronto | Toronto Invasive Bacterial Diseases Network | McMaster University | Allison McGeer et al |
| EPI_ISL_755822 | Canada/ON-S217/2020 | 2020-03-26 | North America/Canada/Ontario/Toronto | Toronto Invasive Bacterial Diseases Network | McMaster University | Allison McGeer et al |
| EPI_ISL_755823 | Canada/ON-S219/2020 | 2020-03-26 | North America/Canada/Ontario/Toronto | Toronto Invasive Bacterial Diseases Network | McMaster University | Allison McGeer et al |
| EPI_ISL_755824 | Canada/ON-S221/2020 | 2020-03-26 | North America/Canada/Ontario/Toronto | Toronto Invasive Bacterial Diseases Network | McMaster University | Allison McGeer et al |

| | | | | | | |
|---|---|---|---|---|---|---|
| EPI_ISL_755825 | Canada/ON-S225/2020 | 2020-03-26 | North America/Canada/Ontario/Toronto | Toronto Invasive Bacterial Diseases Network | McMaster University | Allison McGeer et al |
| EPI_ISL_755826 | Canada/ON-S231/2020 | 2020-03-27 | North America/Canada/Ontario/Toronto | Toronto Invasive Bacterial Diseases Network | McMaster University | Allison McGeer et al |
| EPI_ISL_755827 | Canada/ON-S237/2020 | 2020-03-26 | North America/Canada/Ontario/Toronto | Toronto Invasive Bacterial Diseases Network | McMaster University | Allison McGeer et al |
| EPI_ISL_755828 | Canada/ON-S242/2020 | 2020-03-27 | North America/Canada/Ontario/Toronto | Toronto Invasive Bacterial Diseases Network | McMaster University | Allison McGeer et al |
| EPI_ISL_755829 | Canada/ON-S243/2020 | 2020-03-27 | North America/Canada/Ontario/Toronto | Toronto Invasive Bacterial Diseases Network | McMaster University | Allison McGeer et al |
| EPI_ISL_755830 | Canada/ON-S244/2020 | 2020-03-27 | North America/Canada/Ontario/Toronto | Toronto Invasive Bacterial Diseases Network | McMaster University | Allison McGeer et al |
| EPI_ISL_755831 | Canada/ON-S246/2020 | 2020-03-27 | North America/Canada/Ontario/Toronto | Toronto Invasive Bacterial Diseases Network | McMaster University | Allison McGeer et al |
| EPI_ISL_755832 | Canada/ON-S247/2020 | 2020-03-27 | North America/Canada/Ontario/Toronto | Toronto Invasive Bacterial Diseases Network | McMaster University | Allison McGeer et al |
| EPI_ISL_755833 | Canada/ON-S248/2020 | 2020-03-27 | North America/Canada/Ontario/Toronto | Toronto Invasive Bacterial Diseases Network | McMaster University | Allison McGeer et al |
| EPI_ISL_755834 | Canada/ON-S250/2020 | 2020-03-27 | North America/Canada/Ontario/Toronto | Toronto Invasive Bacterial Diseases Network | McMaster University | Allison McGeer et al |
| EPI_ISL_755835 | Canada/ON-S251/2020 | 2020-03-27 | North America/Canada/Ontario/Toronto | Toronto Invasive Bacterial Diseases Network | McMaster University | Allison McGeer et al |
| EPI_ISL_755836 | Canada/ON-S252/2020 | 2020-03-27 | North America/Canada/Ontario/Toronto | Toronto Invasive Bacterial Diseases Network | McMaster University | Allison McGeer et al |
| EPI_ISL_755837 | Canada/ON-S258/2020 | 2020-03-27 | North America/Canada/Ontario/Toronto | Toronto Invasive Bacterial Diseases Network | McMaster University | Allison McGeer et al |
| EPI_ISL_755838 | Canada/ON-S259/2020 | 2020-03-27 | North America/Canada/Ontario/Toronto | Toronto Invasive Bacterial Diseases Network | McMaster University | Allison McGeer et al |
| EPI_ISL_755839 | Canada/ON-S260/2020 | 2020-03-27 | North America/Canada/Ontario/Toronto | Toronto Invasive Bacterial Diseases Network | McMaster University | Allison McGeer et al |
| EPI_ISL_755840 | Canada/ON-S261/2020 | 2020-03-28 | North America/Canada/Ontario/Toronto | Toronto Invasive Bacterial Diseases Network | McMaster University | Allison McGeer et al |
| EPI_ISL_755841 | Canada/ON-S265/2020 | 2020-03-27 | North America/Canada/Ontario/Toronto | Toronto Invasive Bacterial Diseases Network | McMaster University | Allison McGeer et al |
| EPI_ISL_755842 | Canada/ON-S267/2020 | 2020-03-27 | North America/Canada/Ontario/Toronto | Toronto Invasive Bacterial Diseases Network | McMaster University | Allison McGeer et al |
| EPI_ISL_755843 | Canada/ON-S268/2020 | 2020-03-27 | North America/Canada/Ontario/Toronto | Toronto Invasive Bacterial Diseases Network | McMaster University | Allison McGeer et al |
| EPI_ISL_755844 | Canada/ON-S271/2020 | 2020-03-28 | North America/Canada/Ontario/Toronto | Toronto Invasive Bacterial Diseases Network | McMaster University | Allison McGeer et al |

| | | | | | | |
|---|---|---|---|---|---|---|
| EPI_ISL_755845 | Canada/ON-S272/2020 | 2020-03-28 | North America/Canada/Ontario/Toronto | Toronto Invasive Bacterial Diseases Network | McMaster University | Allison McGeer et al |
| EPI_ISL_755846 | Canada/ON-S273/2020 | 2020-03-27 | North America/Canada/Ontario/Toronto | Toronto Invasive Bacterial Diseases Network | McMaster University | Allison McGeer et al |
| EPI_ISL_755847 | Canada/ON-S274/2020 | 2020-03-28 | North America/Canada/Ontario/Toronto | Toronto Invasive Bacterial Diseases Network | McMaster University | Allison McGeer et al |
| EPI_ISL_755848 | Canada/ON-S275/2020 | 2020-03-28 | North America/Canada/Ontario/Toronto | Toronto Invasive Bacterial Diseases Network | McMaster University | Allison McGeer et al |
| EPI_ISL_755849 | Canada/ON-S276/2020 | 2020-03-28 | North America/Canada/Ontario/Toronto | Toronto Invasive Bacterial Diseases Network | McMaster University | Allison McGeer et al |
| EPI_ISL_755850 | Canada/ON-S279/2020 | 2020-03-28 | North America/Canada/Ontario/Toronto | Toronto Invasive Bacterial Diseases Network | McMaster University | Allison McGeer et al |
| EPI_ISL_755851 | Canada/ON-S282/2020 | 2020-03-28 | North America/Canada/Ontario/Toronto | Toronto Invasive Bacterial Diseases Network | McMaster University | Allison McGeer et al |
| EPI_ISL_755852 | Canada/ON-S285/2020 | 2020-03-28 | North America/Canada/Ontario/Toronto | Toronto Invasive Bacterial Diseases Network | McMaster University | Allison McGeer et al |
| EPI_ISL_755853 | Canada/ON-S286/2020 | 2020-03-28 | North America/Canada/Ontario/Toronto | Toronto Invasive Bacterial Diseases Network | McMaster University | Allison McGeer et al |
| EPI_ISL_755854 | Canada/ON-S287/2020 | 2020-03-28 | North America/Canada/Ontario/Toronto | Toronto Invasive Bacterial Diseases Network | McMaster University | Allison McGeer et al |
| EPI_ISL_755855 | Canada/ON-S291/2020 | 2020-03-28 | North America/Canada/Ontario/Toronto | Toronto Invasive Bacterial Diseases Network | McMaster University | Allison McGeer et al |
| EPI_ISL_755856 | Canada/ON-S293/2020 | 2020-03-28 | North America/Canada/Ontario/Toronto | Toronto Invasive Bacterial Diseases Network | McMaster University | Allison McGeer et al |
| EPI_ISL_755857 | Canada/ON-S294/2020 | 2020-03-28 | North America/Canada/Ontario/Toronto | Toronto Invasive Bacterial Diseases Network | McMaster University | Allison McGeer et al |
| EPI_ISL_755858 | Canada/ON-S298/2020 | 2020-03-28 | North America/Canada/Ontario/Toronto | Toronto Invasive Bacterial Diseases Network | McMaster University | Allison McGeer et al |
| EPI_ISL_755859 | Canada/ON-S300/2020 | 2020-03-28 | North America/Canada/Ontario/Toronto | Toronto Invasive Bacterial Diseases Network | McMaster University | Allison McGeer et al |
| EPI_ISL_755860 | Canada/ON-S304/2020 | 2020-03-29 | North America/Canada/Ontario/Toronto | Toronto Invasive Bacterial Diseases Network | McMaster University | Allison McGeer et al |
| EPI_ISL_755861 | Canada/ON-S317/2020 | 2020-03-28 | North America/Canada/Ontario/Toronto | Toronto Invasive Bacterial Diseases Network | McMaster University | Allison McGeer et al |
| EPI_ISL_755862 | Canada/ON-S321/2020 | 2020-03-30 | North America/Canada/Ontario/Toronto | Toronto Invasive Bacterial Diseases Network | McMaster University | Allison McGeer et al |
| EPI_ISL_755863 | Canada/ON-S329/2020 | 2020-03-29 | North America/Canada/Ontario/Toronto | Toronto Invasive Bacterial Diseases Network | McMaster University | Allison McGeer et al |
| EPI_ISL_755864 | Canada/ON-S331/2020 | 2020-03-28 | North America/Canada/Ontario/Toronto | Toronto Invasive Bacterial Diseases Network | McMaster University | Allison McGeer et al |

| | | | | | | |
|---|---|---|---|---|---|---|
| EPI_ISL_755865 | Canada/ON-S334/2020 | 2020-03-29 | North America/Canada/Ontario/Toronto | Toronto Invasive Bacterial Diseases Network | McMaster University | Allison McGeer et al |
| EPI_ISL_755866 | Canada/ON-S335/2020 | 2020-03-29 | North America/Canada/Ontario/Toronto | Toronto Invasive Bacterial Diseases Network | McMaster University | Allison McGeer et al |
| EPI_ISL_755867 | Canada/ON-S339/2020 | 2020-03-29 | North America/Canada/Ontario/Toronto | Toronto Invasive Bacterial Diseases Network | McMaster University | Allison McGeer et al |
| EPI_ISL_755868 | Canada/ON-S341/2020 | 2020-03-29 | North America/Canada/Ontario/Toronto | Toronto Invasive Bacterial Diseases Network | McMaster University | Allison McGeer et al |
| EPI_ISL_755869 | Canada/ON-S343/2020 | 2020-03-29 | North America/Canada/Ontario/Toronto | Toronto Invasive Bacterial Diseases Network | McMaster University | Allison McGeer et al |
| EPI_ISL_755870 | Canada/ON-S346/2020 | 2020-03-29 | North America/Canada/Ontario/Toronto | Toronto Invasive Bacterial Diseases Network | McMaster University | Allison McGeer et al |
| EPI_ISL_755871 | Canada/ON-S351/2020 | 2020-03-29 | North America/Canada/Ontario/Toronto | Toronto Invasive Bacterial Diseases Network | McMaster University | Allison McGeer et al |
| EPI_ISL_755872 | Canada/ON-S356/2020 | 2020-03-29 | North America/Canada/Ontario/Toronto | Toronto Invasive Bacterial Diseases Network | McMaster University | Allison McGeer et al |
| EPI_ISL_755873 | Canada/ON-S369/2020 | 2020-03-30 | North America/Canada/Ontario/Toronto | Toronto Invasive Bacterial Diseases Network | McMaster University | Allison McGeer et al |
| EPI_ISL_755874 | Canada/ON-S370/2020 | 2020-03-30 | North America/Canada/Ontario/Toronto | Toronto Invasive Bacterial Diseases Network | McMaster University | Allison McGeer et al |
| EPI_ISL_755875 | Canada/ON-S380/2020 | 2020-03-30 | North America/Canada/Ontario/Toronto | Toronto Invasive Bacterial Diseases Network | McMaster University | Allison McGeer et al |
| EPI_ISL_755876 | Canada/ON-S381/2020 | 2020-03-30 | North America/Canada/Ontario/Toronto | Toronto Invasive Bacterial Diseases Network | McMaster University | Allison McGeer et al |
| EPI_ISL_755877 | Canada/ON-S385/2020 | 2020-03-30 | North America/Canada/Ontario/Toronto | Toronto Invasive Bacterial Diseases Network | McMaster University | Allison McGeer et al |
| EPI_ISL_755878 | Canada/ON-S394/2020 | 2020-03-31 | North America/Canada/Ontario/Toronto | Toronto Invasive Bacterial Diseases Network | McMaster University | Allison McGeer et al |
| EPI_ISL_755879 | Canada/ON-S395/2020 | 2020-03-31 | North America/Canada/Ontario/Toronto | Toronto Invasive Bacterial Diseases Network | McMaster University | Allison McGeer et al |
| EPI_ISL_755880 | Canada/ON-S397/2020 | 2020-03-31 | North America/Canada/Ontario/Toronto | Toronto Invasive Bacterial Diseases Network | McMaster University | Allison McGeer et al |
| EPI_ISL_755881 | Canada/ON-S59/2020 | 2020-03-18 | North America/Canada/Ontario/Toronto | Toronto Invasive Bacterial Diseases Network | McMaster University | Allison McGeer et al |
| EPI_ISL_755883 | Canada/ON-S1494/2020 | 2020-08-28 | North America/Canada/Ontario/Toronto | Toronto Invasive Bacterial Diseases Network | McMaster University | Allison McGeer et al |
| EPI_ISL_755884 | Canada/ON-S1495/2020 | 2020-08-28 | North America/Canada/Ontario/Toronto | Toronto Invasive Bacterial Diseases Network | McMaster University | Allison McGeer et al |
| EPI_ISL_755885 | Canada/ON-S1496/2020 | 2020-08-28 | North America/Canada/Ontario/Toronto | Toronto Invasive Bacterial Diseases Network | McMaster University | Allison McGeer et al |

| | | | | | | |
|---|---|---|---|---|---|---|
| EPI_ISL_755886 | Canada/ON-S1497/2020 | 2020-08-28 | North America/Canada/Ontario/Toronto | Toronto Invasive Bacterial Diseases Network | McMaster University | Allison McGeer et al |
| EPI_ISL_755887 | Canada/ON-S1498/2020 | 2020-08-29 | North America/Canada/Ontario/Toronto | Toronto Invasive Bacterial Diseases Network | McMaster University | Allison McGeer et al |
| EPI_ISL_755888 | Canada/ON-S1499/2020 | 2020-08-29 | North America/Canada/Ontario/Toronto | Toronto Invasive Bacterial Diseases Network | McMaster University | Allison McGeer et al |
| EPI_ISL_755889 | Canada/ON-S1500/2020 | 2020-08-30 | North America/Canada/Ontario/Toronto | Toronto Invasive Bacterial Diseases Network | McMaster University | Allison McGeer et al |
| EPI_ISL_755890 | Canada/ON-S1501/2020 | 2020-08-30 | North America/Canada/Ontario/Toronto | Toronto Invasive Bacterial Diseases Network | McMaster University | Allison McGeer et al |
| EPI_ISL_755891 | Canada/ON-S1503/2020 | 2020-08-30 | North America/Canada/Ontario/Toronto | Toronto Invasive Bacterial Diseases Network | McMaster University | Allison McGeer et al |
| EPI_ISL_755892 | Canada/ON-S1504/2020 | 2020-08-30 | North America/Canada/Ontario/Toronto | Toronto Invasive Bacterial Diseases Network | McMaster University | Allison McGeer et al |
| EPI_ISL_755893 | Canada/ON-S1508/2020 | 2020-08-30 | North America/Canada/Ontario/Toronto | Toronto Invasive Bacterial Diseases Network | McMaster University | Allison McGeer et al |
| EPI_ISL_755894 | Canada/ON-S1511/2020 | 2020-08-30 | North America/Canada/Ontario/Toronto | Toronto Invasive Bacterial Diseases Network | McMaster University | Allison McGeer et al |
| EPI_ISL_755895 | Canada/ON-S1512/2020 | 2020-08-30 | North America/Canada/Ontario/Toronto | Toronto Invasive Bacterial Diseases Network | McMaster University | Allison McGeer et al |
| EPI_ISL_755896 | Canada/ON-S1513/2020 | 2020-08-30 | North America/Canada/Ontario/Toronto | Toronto Invasive Bacterial Diseases Network | McMaster University | Allison McGeer et al |
| EPI_ISL_755897 | Canada/ON-S1514/2020 | 2020-08-30 | North America/Canada/Ontario/Toronto | Toronto Invasive Bacterial Diseases Network | McMaster University | Allison McGeer et al |
| EPI_ISL_755898 | Canada/ON-S1515/2020 | 2020-08-31 | North America/Canada/Ontario/Toronto | Toronto Invasive Bacterial Diseases Network | McMaster University | Allison McGeer et al |
| EPI_ISL_755899 | Canada/ON-S1517/2020 | 2020-08-30 | North America/Canada/Ontario/Toronto | Toronto Invasive Bacterial Diseases Network | McMaster University | Allison McGeer et al |
| EPI_ISL_755900 | Canada/ON-S1519/2020 | 2020-08-31 | North America/Canada/Ontario/Toronto | Toronto Invasive Bacterial Diseases Network | McMaster University | Allison McGeer et al |
| EPI_ISL_755901 | Canada/ON-S1520/2020 | 2020-08-31 | North America/Canada/Ontario/Toronto | Toronto Invasive Bacterial Diseases Network | McMaster University | Allison McGeer et al |
| EPI_ISL_755902 | Canada/ON-S1523/2020 | 2020-08-31 | North America/Canada/Ontario/Toronto | Toronto Invasive Bacterial Diseases Network | McMaster University | Allison McGeer et al |
| EPI_ISL_755903 | Canada/ON-S1524/2020 | 2020-08-30 | North America/Canada/Ontario/Toronto | Toronto Invasive Bacterial Diseases Network | McMaster University | Allison McGeer et al |
| EPI_ISL_755904 | Canada/ON-S1528/2020 | 2020-08-31 | North America/Canada/Ontario/Toronto | Toronto Invasive Bacterial Diseases Network | McMaster University | Allison McGeer et al |
| EPI_ISL_755905 | Canada/ON-S1529/2020 | 2020-08-31 | North America/Canada/Ontario/Toronto | Toronto Invasive Bacterial Diseases Network | McMaster University | Allison McGeer et al |

| | | | | | | |
|---|---|---|---|---|---|---|
| EPI_ISL_755906 | Canada/ON-S1533/2020 | 2020-09-01 | North America/Canada/Ontario/Toronto | Toronto Invasive Bacterial Diseases Network | McMaster University | Allison McGeer et al |
| EPI_ISL_755907 | Canada/ON-S1534/2020 | 2020-09-01 | North America/Canada/Ontario/Toronto | Toronto Invasive Bacterial Diseases Network | McMaster University | Allison McGeer et al |
| EPI_ISL_755908 | Canada/ON-S1535/2020 | 2020-09-01 | North America/Canada/Ontario/Toronto | Toronto Invasive Bacterial Diseases Network | McMaster University | Allison McGeer et al |
| EPI_ISL_755909 | Canada/ON-S1536/2020 | 2020-09-01 | North America/Canada/Ontario/Toronto | Toronto Invasive Bacterial Diseases Network | McMaster University | Allison McGeer et al |
| EPI_ISL_755910 | Canada/ON-S1537/2020 | 2020-09-01 | North America/Canada/Ontario/Toronto | Toronto Invasive Bacterial Diseases Network | McMaster University | Allison McGeer et al |
| EPI_ISL_755913 | Canada/ON-S1553/2020 | 2020-09-04 | North America/Canada/Ontario/Toronto | Toronto Invasive Bacterial Diseases Network | McMaster University | Allison McGeer et al |
| EPI_ISL_755914 | Canada/ON-S1555/2020 | 2020-09-04 | North America/Canada/Ontario/Toronto | Toronto Invasive Bacterial Diseases Network | McMaster University | Allison McGeer et al |
| EPI_ISL_755915 | Canada/ON-S1556/2020 | 2020-09-04 | North America/Canada/Ontario/Toronto | Toronto Invasive Bacterial Diseases Network | McMaster University | Allison McGeer et al |
| EPI_ISL_755916 | Canada/ON-S1558/2020 | 2020-09-03 | North America/Canada/Ontario/Toronto | Toronto Invasive Bacterial Diseases Network | McMaster University | Allison McGeer et al |
| EPI_ISL_755917 | Canada/ON-S1559/2020 | 2020-09-04 | North America/Canada/Ontario/Toronto | Toronto Invasive Bacterial Diseases Network | McMaster University | Allison McGeer et al |
| EPI_ISL_755919 | Canada/ON-S1564/2020 | 2020-09-04 | North America/Canada/Ontario/Toronto | Toronto Invasive Bacterial Diseases Network | McMaster University | Allison McGeer et al |
| EPI_ISL_755920 | Canada/ON-S1565/2020 | 2020-09-02 | North America/Canada/Ontario/Toronto | Toronto Invasive Bacterial Diseases Network | McMaster University | Allison McGeer et al |
| EPI_ISL_755921 | Canada/ON-S1568/2020 | 2020-09-02 | North America/Canada/Ontario/Toronto | Toronto Invasive Bacterial Diseases Network | McMaster University | Allison McGeer et al |
| EPI_ISL_755922 | Canada/ON-S1569/2020 | 2020-09-02 | North America/Canada/Ontario/Toronto | Toronto Invasive Bacterial Diseases Network | McMaster University | Allison McGeer et al |
| EPI_ISL_755923 | Canada/ON-S1574/2020 | 2020-09-01 | North America/Canada/Ontario/Toronto | Toronto Invasive Bacterial Diseases Network | McMaster University | Allison McGeer et al |
| EPI_ISL_755924 | Canada/ON-S1576/2020 | 2020-09-01 | North America/Canada/Ontario/Toronto | Toronto Invasive Bacterial Diseases Network | McMaster University | Allison McGeer et al |
| EPI_ISL_755926 | Canada/ON-S1581/2020 | 2020-09-04 | North America/Canada/Ontario/Toronto | Toronto Invasive Bacterial Diseases Network | McMaster University | Allison McGeer et al |
| EPI_ISL_755927 | Canada/ON-S1582/2020 | 2020-09-04 | North America/Canada/Ontario/Toronto | Toronto Invasive Bacterial Diseases Network | McMaster University | Allison McGeer et al |
| EPI_ISL_755928 | Canada/ON-S1584/2020 | 2020-09-04 | North America/Canada/Ontario/Toronto | Toronto Invasive Bacterial Diseases Network | McMaster University | Allison McGeer et al |
| EPI_ISL_755929 | Canada/ON-S1585/2020 | 2020-09-03 | North America/Canada/Ontario/Toronto | Toronto Invasive Bacterial Diseases Network | McMaster University | Allison McGeer et al |

| | | | | | | |
|---|---|---|---|---|---|---|
| EPI_ISL_755930 | Canada/ON-S1589/2020 | 2020-09-03 | North America/Canada/Ontario/Toronto | Toronto Invasive Bacterial Diseases Network | McMaster University | Allison McGeer et al |
| EPI_ISL_755931 | Canada/ON-S1590/2020 | 2020-09-04 | North America/Canada/Ontario/Toronto | Toronto Invasive Bacterial Diseases Network | McMaster University | Allison McGeer et al |
| EPI_ISL_755932 | Canada/ON-S1591/2020 | 2020-09-04 | North America/Canada/Ontario/Toronto | Toronto Invasive Bacterial Diseases Network | McMaster University | Allison McGeer et al |
| EPI_ISL_755933 | Canada/ON-S1592/2020 | 2020-09-04 | North America/Canada/Ontario/Toronto | Toronto Invasive Bacterial Diseases Network | McMaster University | Allison McGeer et al |
| EPI_ISL_755934 | Canada/ON-S1593/2020 | 2020-09-04 | North America/Canada/Ontario/Toronto | Toronto Invasive Bacterial Diseases Network | McMaster University | Allison McGeer et al |
| EPI_ISL_755935 | Canada/ON-S1594/2020 | 2020-09-04 | North America/Canada/Ontario/Toronto | Toronto Invasive Bacterial Diseases Network | McMaster University | Allison McGeer et al |
| EPI_ISL_755936 | Canada/ON-S1595/2020 | 2020-09-04 | North America/Canada/Ontario/Toronto | Toronto Invasive Bacterial Diseases Network | McMaster University | Allison McGeer et al |
| EPI_ISL_755937 | Canada/ON-S1597/2020 | 2020-09-04 | North America/Canada/Ontario/Toronto | Toronto Invasive Bacterial Diseases Network | McMaster University | Allison McGeer et al |
| EPI_ISL_755938 | Canada/ON-S1598/2020 | 2020-09-04 | North America/Canada/Ontario/Toronto | Toronto Invasive Bacterial Diseases Network | McMaster University | Allison McGeer et al |
| EPI_ISL_755939 | Canada/ON-S1599/2020 | 2020-09-04 | North America/Canada/Ontario/Toronto | Toronto Invasive Bacterial Diseases Network | McMaster University | Allison McGeer et al |
| EPI_ISL_755961 | Finland/9LS7BE11/2020 | 2020-10-09 | Europe/Finland/Uusimaa/ | Department of Virology and Immunology, University of Helsinki and Helsinki University Hospital, Huslab Finland | Department of Virology, Faculty of Medicine, University of Helsinki, Helsinki, Finland | Teemu Smura et al |
| EPI_ISL_755997 | Finland/4LS19A5/2020 | 2020-08-08 | Europe/Finland/Uusimaa/ | Department of Virology and Immunology, University of Helsinki and Helsinki University Hospital, Huslab Finland | Department of Virology, Faculty of Medicine, University of Helsinki, Helsinki, Finland | Teemu Smura et al |
| EPI_ISL_756113 | Finland/2LS12G2/2020 | 2020-10-02 | Europe/Finland/Uusimaa/ | Department of Virology and Immunology, University of Helsinki and Helsinki University Hospital, Huslab Finland | Department of Virology, Faculty of Medicine, University of Helsinki, Helsinki, Finland | Teemu Smura et al |
| EPI_ISL_756184 | USA/WY-WPHL-20157383/2020 | 2020-12-02 | North America/USA/Wyoming/ | Wyoming Public Health Laboratory | Wyoming Public Health Laboratory | Noah Hull et al |

| | | | | | | |
|---|---|---|---|---|---|---|
| EPI_ISL_756187 | USA/WY-WPHL-20157593/2020 | 2020-12-03 | North America/USA/Wyoming/ | Wyoming Public Health Laboratory | Wyoming Public Health Laboratory | Noah Hull et al |
| EPI_ISL_756201 | USA/WA-UW-46903/2020 | 2020-12-12 | North America/USA/Washington/ | UW Virology Lab | UW Virology Lab | Pavitra Roychoudhury et al |
| EPI_ISL_756204 | USA/WA-UW-46910/2020 | 2020-12-07 | North America/USA/Washington/ | UW Virology Lab | UW Virology Lab | Pavitra Roychoudhury et al |
| EPI_ISL_756209 | USA/WA-UW-46945/2020 | 2020-12-13 | North America/USA/Washington/ | UW Virology Lab | UW Virology Lab | Pavitra Roychoudhury et al |
| EPI_ISL_756217 | USA/WA-UW-46999/2020 | 2020-12-12 | North America/USA/Washington/ | UW Virology Lab | UW Virology Lab | Pavitra Roychoudhury et al |
| EPI_ISL_756218 | USA/WA-UW-47030/2020 | 2020-12-14 | North America/USA/Washington/ | UW Virology Lab | UW Virology Lab | Pavitra Roychoudhury et al |
| EPI_ISL_756220 | USA/WA-UW-47034/2020 | 2020-12-14 | North America/USA/Washington/ | UW Virology Lab | UW Virology Lab | Pavitra Roychoudhury et al |
| EPI_ISL_756232 | USA/WA-UW-47132/2020 | 2020-12-15 | North America/USA/Washington/ | UW Virology Lab | UW Virology Lab | Pavitra Roychoudhury et al |
| EPI_ISL_756240 | USA/WA-UW-47151/2020 | 2020-12-17 | North America/USA/Washington/ | UW Virology Lab | UW Virology Lab | Pavitra Roychoudhury et al |
| EPI_ISL_756241 | USA/WY-WPHL-20157765/2020 | 2020-12-03 | North America/USA/Wyoming/ | Wyoming Public Health Laboratory | Wyoming Public Health Laboratory | Noah Hull et al |
| EPI_ISL_756243 | USA/WY-WPHL-20158237/2020 | 2020-12-03 | North America/USA/Wyoming/ | Wyoming Public Health Laboratory | Wyoming Public Health Laboratory | Noah Hull et al |
| EPI_ISL_756253 | USA/WY-WPHL-20158485/2020 | 2020-12-04 | North America/USA/Wyoming/ | Wyoming Public Health Laboratory | Wyoming Public Health Laboratory | Noah Hull et al |
| EPI_ISL_756262 | USA/WY-WPHL-20170197/2020 | 2020-12-23 | North America/USA/Wyoming/ | Wyoming Public Health Laboratory | Wyoming Public Health Laboratory | Noah Hull et al |
| EPI_ISL_756268 | USA/WY-WPHL-20170351/2020 | 2020-12-24 | North America/USA/Wyoming/ | Wyoming Public Health Laboratory | Wyoming Public Health Laboratory | Noah Hull et al |
| EPI_ISL_756271 | Spain/MD-20510356/2020 | 2020-12-22 | Europe/Spain/Madrid/ | Hospital General Universitario Gregorio Marav√±√≥n | Hospital General Universitario Gregorio Marav√±√≥n | Sergio Buenestado Serrano et al |
| EPI_ISL_756275 | USA/HI-H200397/2020 | 2020-09-07 | North America/USA/Hawaii/Hawaii Cou | State Laboratories Division, Hawaii State Department of Health | State Laboratories Division, Hawaii State Department of Health | Pamela O'Brien et al |
| EPI_ISL_756277 | USA/HI-H200581/2020 | 2020-10-29 | North America/USA/Hawaii/Honolulu C | State Laboratories Division, Hawaii State Department of Health | State Laboratories Division, Hawaii State Department of Health | Pamela O'Brien et al |

| | | | | | | |
|---|---|---|---|---|---|---|
| EPI_ISL_756293 | Brazil/BA-1/2020 | 2020-06-01 | South America/Brazil/Bahia/ | Center for Biotechnology and Cell Therapy, S√£o Rafael Hospital, Salvador, Brazil | Center for Biotechnology and Cell Therapy, S√£o Rafael Hospital, Salvador, Brazil | Carolina Kymie Vasques Nonaka et al |
| EPI_ISL_756294 | Brazil/BA-2/2020 | 2020-10-26 | South America/Brazil/Bahia/ | Center for Biotechnology and Cell Therapy, S√£o Rafael Hospital, Salvador, Brazil | Center for Biotechnology and Cell Therapy, S√£o Rafael Hospital, Salvador, Brazil | Carolina Kymie Vasques Nonaka et al |
| EPI_ISL_756323 | USA/CA-IGI-0421/2020 | 2020-07-09 | North America/USA/California/Alameda | Innovative Genomics Institute, UC Berkeley | Innovative Genomics Institute, UC Berkeley | Stacia Wyman et al |
| EPI_ISL_756326 | USA/CA-IGI-0428/2020 | 2020-07-14 | North America/USA/California/Alameda | Innovative Genomics Institute, UC Berkeley | Innovative Genomics Institute, UC Berkeley | Stacia Wyman et al |
| EPI_ISL_756333 | USA/CA-IGI-0555/2020 | 2020-07-24 | North America/USA/California/Alameda | Innovative Genomics Institute, UC Berkeley | Innovative Genomics Institute, UC Berkeley | Stacia Wyman et al |
| EPI_ISL_756363 | Trinidad/TT6851/2020 | 2020-09-09 | South America/Trinidad and Tobago/Tri | Trinidad Public Health Laboratory | Carrington Lab, Department of PreClinical Sciences, Faculty of Medical Sciences, The University of the West Indies | Nikita S. D. Sahadeo et al |
| EPI_ISL_756376 | Indonesia/JB-NIHRDC-0056536/2020 | 2020-09-16 | Asia/Indonesia/West Java/ | National Institute of Health Research and Development | National Institute of Health Research and Development | Ramadhany et al |
| EPI_ISL_756401 | Indonesia/SU-NIHRDC-0045870/2020 | 2020-08-28 | Asia/Indonesia/North Sumatra/ | National Institute of Health Research and Development | National Institute of Health Research and Development | Adam et al |

| | | | | | | |
|---|---|---|---|---|---|---|
| EPI_ISL_756547 | England/ALDP-C8B599/2020 | 2020-12-16 | Europe/United Kingdom/England/ | Lighthouse Lab in Alderley Park | Wellcome Sanger Institute for the COVID-19 Genomics UK (COG-UK) Consortium | Jacquelyn Wynn et al |
| EPI_ISL_756972 | England/MILK-C7A9EC/2020 | 2020-12-15 | Europe/United Kingdom/England/ | Lighthouse Lab in Milton Keynes | Wellcome Sanger Institute for the COVID-19 Genomics UK (COG-UK) Consortium | The Lighthouse Lab in Milton Keynes et al |
| EPI_ISL_756982 | England/QEUH-C7D12D/2020 | 2020-12-16 | Europe/United Kingdom/England/ | Lighthouse Lab in Glasgow | Wellcome Sanger Institute for the COVID-19 Genomics UK (COG-UK) Consortium | Harper VanSteenhouse et al |
| EPI_ISL_756984 | England/MILK-C7B24A/2020 | 2020-12-15 | Europe/United Kingdom/England/ | Lighthouse Lab in Milton Keynes | Wellcome Sanger Institute for the COVID-19 Genomics UK (COG-UK) Consortium | The Lighthouse Lab in Milton Keynes et al |
| EPI_ISL_756985 | England/MILK-C78483/2020 | 2020-12-15 | Europe/United Kingdom/England/ | Lighthouse Lab in Milton Keynes | Wellcome Sanger Institute for the COVID-19 Genomics UK (COG-UK) Consortium | The Lighthouse Lab in Milton Keynes et al |
| EPI_ISL_757009 | England/QEUH-C7FFB8/2020 | 2020-12-16 | Europe/United Kingdom/England/ | Lighthouse Lab in Glasgow | Wellcome Sanger Institute for the COVID-19 Genomics UK (COG-UK) Consortium | Harper VanSteenhouse et al |
| EPI_ISL_757094 | Scotland/QEUH-C4C181/2020 | 2020-12-14 | Europe/United Kingdom/Scotland/ | Lighthouse Lab in Glasgow | Wellcome Sanger Institute for the COVID-19 Genomics UK (COG-UK) Consortium | Harper VanSteenhouse et al |
| EPI_ISL_757110 | Scotland/QEUH-C4C163/2020 | 2020-12-14 | Europe/United Kingdom/Scotland/ | Lighthouse Lab in Glasgow | Wellcome Sanger Institute for the COVID-19 Genomics UK (COG-UK) Consortium | Harper VanSteenhouse et al |

| | | | | | | |
|---|---|---|---|---|---|---|
| EPI_ISL_757126 | Scotland/QEUH-C30F86/2020 | 2020-12-11 | Europe/United Kingdom/Scotland/ | Lighthouse Lab in Glasgow | Wellcome Sanger Institute for the COVID-19 Genomics UK (COG-UK) Consortium | Harper VanSteenhouse et al |
| EPI_ISL_757285 | Pakistan/MU-NIH-K14/2020 | 2020-12-25 | Asia/Pakistan/Rawalpindi/ | Department of Virology, Public Health Laboratories Division | Department of Virology, Public Health Laboratories Division | Massab Umair et al |
| EPI_ISL_757286 | Pakistan/MU-NIH-K53/2020 | 2020-12-28 | Asia/Pakistan/Sindh/ | Department of Virology, Public Health Laboratories Division | Department of Virology, Public Health Laboratories Division | Massab Umair et al |
| EPI_ISL_757287 | Indonesia/AC-NIHRD-C0057926/2020 | 2020-09-15 | Asia/Indonesia/Aceh/ | National Institute of Health Research and Development | National Institute of Health Research and Development | Rukminiati et al |
| EPI_ISL_757368 | Finland/20MS4A4/2020 | 2020-03-20 | Europe/Finland/Uusimaa/ | Department of Virology and Immunology, University of Helsinki and Helsinki University Hospital, Huslab Finland | Department of Virology, Faculty of Medicine, University of Helsinki, Helsinki, Finland | Teemu Smura et al |
| EPI_ISL_757880 | Denmark/DCGC-21570/2020 | 2020-12-21 | Europe/Denmark/Hovedstaden/ | Department of Virus and Microbiological Special Diagnostics, Statens Serum Institut, Copenhagen, Denmark | Albertsen Lab, Department of Chemistry and Bioscience, Aalborg University, Denmark | Danish Covid-19 Genome Consortium et al |
| EPI_ISL_758681 | Denmark/DCGC-22330/2020 | 2020-12-14 | Europe/Denmark/Hovedstaden/ | Department of Virus and Microbiological Special Diagnostics, Statens Serum Institut, Copenhagen, Denmark | Albertsen Lab, Department of Chemistry and Bioscience, Aalborg University, Denmark | Danish Covid-19 Genome Consortium et al |

| | | | | | | |
|---|---|---|---|---|---|---|
| EPI_ISL_759490 | Denmark/DCGC-20984/2020 | 2020-12-21 | Europe/Denmark/Hovedstaden/ | Department of Virus and Microbiological Special Diagnostics, Statens Serum Institut, Copenhagen, Denmark | Albertsen Lab, Department of Chemistry and Bioscience, Aalborg University, Denmark | Danish Covid-19 Genome Consortium et al |
| EPI_ISL_759701 | Indonesia/JT-UNS-18_39701/2020 | 2020-10-14 | Asia/Indonesia/Central Java/ | RS UNS, Surakarta | UNS/RS-UNS, Faculty of Medicine | Betty Suryawati et al |
| EPI_ISL_759702 | Indonesia/JT-UNS-7-27136/2020 | 2020-09-10 | Asia/Indonesia/Central Java/ | RS UNS Surakarta | Universitas Sebelas Maret (UNS), Rumah sakit Universitas Sebelas Maret (RS-UNS) Surakarta, National Institute of Health Research and Development, Indonesian Ministry of Health | Betty Suryawati et al |
| EPI_ISL_759703 | Indonesia/JT-UNS-27-41398/2020 | 2020-10-19 | Asia/Indonesia/Central Java/ | RSUD Bung Karno | Universitas Sebelas Maret (UNS), Rumah sakit Universitas Sebelas Maret (RS-UNS) Surakarta, National Institute of Health Research and Development, Indonesian Ministry of Health | Betty Suryawati et al |
| EPI_ISL_759719 | Portugal/PT2128/2020 | 2020-12-23 | Europe/Portugal/Portugal/ | Instituto Nacional de Saude (INSA) | Instituto Nacional de Saude (INSA) | Borges et al et al |
| EPI_ISL_759720 | Portugal/PT2129/2020 | 2020-12-23 | Europe/Portugal/Portugal/ | Instituto Nacional de Saude (INSA) | Instituto Nacional de Saude (INSA) | Borges et al et al |
| EPI_ISL_759722 | Portugal/PT2131/2020 | 2020-11-19 | Europe/Portugal/Portugal/ | Instituto Nacional de Saude (INSA) | Instituto Nacional de Saude (INSA) | Borges et al et al |

| | | | | | | |
|---|---|---|---|---|---|---|
| EPI_ISL_759723 | Portugal/PT2132/2020 | 2020-12-28 | Europe/Portugal/Portugal/ | Instituto Nacional de Saude (INSA) | Instituto Nacional de Saude (INSA) | Borges et al et al |
| EPI_ISL_759731 | USA/WI-UW-2456/2020 | 2020-12-21 | North America/USA/Wisconsin/Dane Co | University of Wisconsin-Madison AIDS Vaccine Research Laboratories | University of Wisconsin-Madison AIDS Vaccine Research Laboratories | Gage Moreno et al |
| EPI_ISL_759739 | USA/WI-UW-2464/2020 | 2020-12-18 | North America/USA/Wisconsin/Dane Co | University of Wisconsin-Madison AIDS Vaccine Research Laboratories | University of Wisconsin-Madison AIDS Vaccine Research Laboratories | Gage Moreno et al |
| EPI_ISL_759741 | USA/WI-UW-2466/2020 | 2020-12-28 | North America/USA/Wisconsin/Dane Co | University of Wisconsin-Madison AIDS Vaccine Research Laboratories | University of Wisconsin-Madison AIDS Vaccine Research Laboratories | Gage Moreno et al |
| EPI_ISL_759749 | USA/WI-UW-2475/2020 | 2020-12-28 | North America/USA/Wisconsin/Dane Co | University of Wisconsin-Madison AIDS Vaccine Research Laboratories | University of Wisconsin-Madison AIDS Vaccine Research Laboratories | Gage Moreno et al |
| EPI_ISL_759750 | USA/WI-UW-2476/2020 | 2020-12-28 | North America/USA/Wisconsin/Dane Co | University of Wisconsin-Madison AIDS Vaccine Research Laboratories | University of Wisconsin-Madison AIDS Vaccine Research Laboratories | Gage Moreno et al |
| EPI_ISL_759752 | USA/WI-UW-2478/2020 | 2020-12-24 | North America/USA/Wisconsin/Dane Co | University of Wisconsin-Madison AIDS Vaccine Research Laboratories | University of Wisconsin-Madison AIDS Vaccine Research Laboratories | Gage Moreno et al |
| EPI_ISL_759755 | USA/WI-UW-2481/2020 | 2020-12-25 | North America/USA/Wisconsin/Dane Co | University of Wisconsin-Madison AIDS Vaccine Research Laboratories | University of Wisconsin-Madison AIDS Vaccine Research Laboratories | Gage Moreno et al |

| | | | | | | |
|---|---|---|---|---|---|---|
| EPI_ISL_759757 | USA/WI-UW-2483/2020 | 2020-12-26 | North America/USA/Wisconsin/Dane Co | University of Wisconsin-Madison AIDS Vaccine Research Laboratories | University of Wisconsin-Madison AIDS Vaccine Research Laboratories | Gage Moreno et al |
| EPI_ISL_759842 | Indonesia/JT-UNS-26-41182/2020 | 2020-10-19 | Asia/Indonesia/Central Java/ | Rumah Sakit Universitas Sebelas Maret (RS-UNS) | Universitas Sebelas Maret (UNS); Rumah Sakit Universitas Sebelas Maret (RS-UNS), Surakarta; National Institute of Health Research and Development, Indonesian Ministry of Health, Jakarta. | Betty Suryawati et al |
| EPI_ISL_759862 | USA/UN-PB-Wuhan-Hu-1-3/2020 | 2020-04-04 | North America/USA/USA/ | Molecular, ProgenaBiome | Molecular, ProgenaBiome | Papoutsis et al |
| EPI_ISL_759864 | USA/UN-PB-Wuhan-Hu-1-5/2020 | 2020-04-05 | North America/USA/USA/ | Molecular, ProgenaBiome | Molecular, ProgenaBiome | Papoutsis et al |
| EPI_ISL_759901 | Finland/25LS10CE11/2020 | 2020-10-25 | Europe/Finland/Uusimaa/ | Department of Virology and Immunology, University of Helsinki and Helsinki University Hospital, Huslab Finland | Department of Virology, Faculty of Medicine, University of Helsinki, Helsinki, Finland | Teemu Smura et al |
| EPI_ISL_759958 | Indonesia/JI-ITDua-5748NTv/2020 | 2020-04-29 | Asia/Indonesia/East Java/Surabaya | Universitas Airlangga Hospital | Institute of Tropical Disease, Universitas Airlangga | Maria I Lusida et al |
| EPI_ISL_759962 | Indonesia/JI-ITDua-4134NTv/2020 | 2020-04-17 | Asia/Indonesia/East Java/Surabaya | RSUD Dr. Soetomo | Institute of Tropical Disease, Universitas Airlangga | Krisnoadi Rahardjo et al |
| EPI_ISL_759963 | Indonesia/JI-ITDua-12323Nvv/2020 | 2020-06-09 | Asia/Indonesia/East Java/Sidoarjo | RSUD Sidoarjo | Institute of Tropical Disease, Universitas Airlangga | Aldise M Nastri et al |

| | | | | | | |
|---|---|---|---|---|---|---|
| EPI_ISL_759967 | Indonesia/JI-ITDua-1609Nv/2020 | 2020-04-01 | Asia/Indonesia/East Java/Tulungagung | RSUD Dr. Iskak Tulungagung | Institute of Tropical Disease, Universitas Airlangga | Rima R Prasetya et al |
| EPI_ISL_759968 | Indonesia/JI-ITDua-28257NT/2020 | 2020-10-02 | Asia/Indonesia/East Java/Surabaya | RSI Darus Syifa' | Institute of Tropical Disease, Universitas Airlangga | Maria I Lusida et al |
| EPI_ISL_760008 | SouthKorea/KDCA0243/2020 | 2020-08-24 | Asia/South Korea/South Korea/ | Division of Emerging Infectious Diseases, Bureau of Infectious Diseases Diagnosis Control, Korea Disease Control and Prevention Agency | Division of Emerging Infectious Diseases, Bureau of Infectious Diseases Diagnosis Control, Korea Disease Control and Prevention Agency | Ae Kyung Park et al |
| EPI_ISL_760076 | SouthKorea/KDCA0282/2020 | 2020-10-24 | Asia/South Korea/South Korea/ | Division of Emerging Infectious Diseases, Bureau of Infectious Diseases Diagnosis Control, Korea Disease Control and Prevention Agency | Division of Emerging Infectious Diseases, Bureau of Infectious Diseases Diagnosis Control, Korea Disease Control and Prevention Agency | Ae Kyung Park et al |
| EPI_ISL_760162 | SouthKorea/KDCA0378/2020 | 2020-11-04 | Asia/South Korea/South Korea/ | Division of Emerging Infectious Diseases, Bureau of Infectious Diseases Diagnosis Control, Korea Disease Control and Prevention Agency | Division of Emerging Infectious Diseases, Bureau of Infectious Diseases Diagnosis Control, Korea Disease Control and Prevention Agency | Ae Kyung Park et al |

| | | | | | | |
|---|---|---|---|---|---|---|
| EPI_ISL_760196 | SouthKorea/KDCA0412/2020 | 2020-11-12 | Asia/South Korea/South Korea/ | Division of Emerging Infectious Diseases, Bureau of Infectious Diseases Diagnosis Control, Korea Disease Control and Prevention Agency | Division of Emerging Infectious Diseases, Bureau of Infectious Diseases Diagnosis Control, Korea Disease Control and Prevention Agency | Ae Kyung Park et al |
| EPI_ISL_760209 | SouthKorea/KDCA0425/2020 | 2020-11-14 | Asia/South Korea/South Korea/ | Division of Emerging Infectious Diseases, Bureau of Infectious Diseases Diagnosis Control, Korea Disease Control and Prevention Agency | Division of Emerging Infectious Diseases, Bureau of Infectious Diseases Diagnosis Control, Korea Disease Control and Prevention Agency | Ae Kyung Park et al |
| EPI_ISL_760229 | SouthKorea/KDCA0445/2020 | 2020-11-14 | Asia/South Korea/South Korea/ | Division of Emerging Infectious Diseases, Bureau of Infectious Diseases Diagnosis Control, Korea Disease Control and Prevention Agency | Division of Emerging Infectious Diseases, Bureau of Infectious Diseases Diagnosis Control, Korea Disease Control and Prevention Agency | Ae Kyung Park et al |
| EPI_ISL_760603 | England/ALDP-CBE885/2020 | 2020-12-20 | Europe/United Kingdom/England/ | Lighthouse Lab in Alderley Park | Wellcome Sanger Institute for the COVID-19 Genomics UK (COG-UK) Consortium | Jacquelyn Wynn et al |

| | | | | | | |
|---|---|---|---|---|---|---|
| EPI_ISL_761460 | England/MILK-CA6325/2020 | 2020-12-18 | Europe/United Kingdom/England/ | Lighthouse Lab in Milton Keynes | Wellcome Sanger Institute for the COVID-19 Genomics UK (COG-UK) Consortium | The Lighthouse Lab in Milton Keynes et al |
| EPI_ISL_761513 | England/MILK-CA63AD/2020 | 2020-12-18 | Europe/United Kingdom/England/ | Lighthouse Lab in Milton Keynes | Wellcome Sanger Institute for the COVID-19 Genomics UK (COG-UK) Consortium | The Lighthouse Lab in Milton Keynes et al |
| EPI_ISL_761585 | England/MILK-CA64C8/2020 | 2020-12-16 | Europe/United Kingdom/England/ | Lighthouse Lab in Milton Keynes | Wellcome Sanger Institute for the COVID-19 Genomics UK (COG-UK) Consortium | The Lighthouse Lab in Milton Keynes et al |
| EPI_ISL_761807 | England/ALDP-CB69B7/2020 | 2020-12-18 | Europe/United Kingdom/England/ | Lighthouse Lab in Alderley Park | Wellcome Sanger Institute for the COVID-19 Genomics UK (COG-UK) Consortium | Jacquelyn Wynn et al |
| EPI_ISL_761822 | England/ALDP-CB503B/2020 | 2020-12-18 | Europe/United Kingdom/England/ | Lighthouse Lab in Alderley Park | Wellcome Sanger Institute for the COVID-19 Genomics UK (COG-UK) Consortium | Jacquelyn Wynn et al |
| EPI_ISL_762138 | England/CAMC-CA0A35/2020 | 2020-12-18 | Europe/United Kingdom/England/ | Lighthouse Lab in Cambridge | Wellcome Sanger Institute for the COVID-19 Genomics UK (COG-UK) Consortium | Rob Howes et al |
| EPI_ISL_762441 | England/MILK-CA9B2A/2020 | 2020-12-17 | Europe/United Kingdom/England/ | Lighthouse Lab in Milton Keynes | Wellcome Sanger Institute for the COVID-19 Genomics UK (COG-UK) Consortium | The Lighthouse Lab in Milton Keynes et al |
| EPI_ISL_762633 | England/QEUH-C7C540/2020 | 2020-12-16 | Europe/United Kingdom/England/ | Lighthouse Lab in Glasgow | Wellcome Sanger Institute for the COVID-19 Genomics UK (COG-UK) Consortium | Harper VanSteenhouse et al |

| | | | | | | |
|---|---|---|---|---|---|---|
| EPI_ISL_763001 | Germany/TH-RKI-NP-0062/2020 | 2020-12-01 | Europe/Germany/Thuringia/ | Unit 17: Influenza & Other Respiratory Viruses, German National Influenza Center | Project group Epidemiology of Highly Pathogenic Microorganisms, Robert Koch-Institute | Ariane Dvºx et al |
| EPI_ISL_763013 | Germany/SN-RKI-NP-0063/2020 | 2020-12-15 | Europe/Germany/Saxony/ | Unit 17: Influenza & Other Respiratory Viruses, German National Influenza Center | Project group Epidemiology of Highly Pathogenic Microorganisms, Robert Koch-Institute | Ariane Dvºx et al |
| EPI_ISL_763018 | Germany/SN-RKI-NP-0028/2020 | 2020-12-08 | Europe/Germany/Saxony/ | Unit 17: Influenza & Other Respiratory Viruses, German National Influenza Center | Project group Epidemiology of Highly Pathogenic Microorganisms, Robert Koch-Institute | Ariane Dvºx et al |
| EPI_ISL_763019 | Germany/SN-RKI-NP-0024/2020 | 2020-12-07 | Europe/Germany/Saxony/ | Unit 17: Influenza & Other Respiratory Viruses, German National Influenza Center | Project group Epidemiology of Highly Pathogenic Microorganisms, Robert Koch-Institute | Ariane Dvºx et al |
| EPI_ISL_763021 | Germany/SN-RKI-NP-0013/2020 | 2020-12-03 | Europe/Germany/Saxony/ | Unit 17: Influenza & Other Respiratory Viruses, German National Influenza Center | Project group Epidemiology of Highly Pathogenic Microorganisms, Robert Koch-Institute | Ariane Dvºx et al |
| EPI_ISL_763036 | Germany/BY-RKI-NP-0060/2020 | 2020-12-14 | Europe/Germany/Bavaria/ | Unit 17: Influenza & Other Respiratory Viruses, German National Influenza Center | Project group Epidemiology of Highly Pathogenic Microorganisms, Robert Koch-Institute | Ariane Dvºx et al |

| | | | | | | |
|---|---|---|---|---|---|---|
| EPI_ISL_763038 | Germany/BW-RKI-NP-0075/2020 | 2020-12-21 | Europe/Germany/Baden-Wuerttemberg | Unit 17: Influenza & Other Respiratory Viruses, German National Influenza Center | Project group Epidemiology of Highly Pathogenic Microorganisms, Robert Koch-Institute | Ariane Dvºx et al |
| EPI_ISL_763039 | Germany/BW-RKI-NP-0071/2020 | 2020-12-14 | Europe/Germany/Baden-Wuerttemberg | Unit 17: Influenza & Other Respiratory Viruses, German National Influenza Center | Project group Epidemiology of Highly Pathogenic Microorganisms, Robert Koch-Institute | Ariane Dvºx et al |
| EPI_ISL_763043 | Germany/BW-RKI-NP-0040/2020 | 2020-12-09 | Europe/Germany/Baden-Wuerttemberg | Unit 17: Influenza & Other Respiratory Viruses, German National Influenza Center | Project group Epidemiology of Highly Pathogenic Microorganisms, Robert Koch-Institute | Ariane Dvºx et al |
| EPI_ISL_763046 | Germany/BW-RKI-NP-0023/2020 | 2020-12-07 | Europe/Germany/Baden-Wuerttemberg | Unit 17: Influenza & Other Respiratory Viruses, German National Influenza Center | Project group Epidemiology of Highly Pathogenic Microorganisms, Robert Koch-Institute | Ariane Dvºx et al |
| EPI_ISL_763051 | Germany/BW-RKI-NP-0002/2020 | 2020-11-23 | Europe/Germany/Baden-Wuerttemberg | Unit 17: Influenza & Other Respiratory Viruses, German National Influenza Center | Project group Epidemiology of Highly Pathogenic Microorganisms, Robert Koch-Institute | Ariane Dvºx et al |
| EPI_ISL_763052 | Germany/BB-RKI-NP-0077/2020 | 2020-12-21 | Europe/Germany/Brandenburg/ | Unit 17: Influenza & Other Respiratory Viruses, German National Influenza Center | Project group Epidemiology of Highly Pathogenic Microorganisms, Robert Koch-Institute | Ariane Dvºx et al |

| | | | | | | |
|---|---|---|---|---|---|---|
| EPI_ISL_763053 | Germany/BB-RKI-NP-0068/2020 | 2020-12-16 | Europe/Germany/Brandenburg/ | Unit 17: Influenza & Other Respiratory Viruses, German National Influenza Center | Project group Epidemiology of Highly Pathogenic Microorganisms, Robert Koch-Institute | Ariane Dvºx et al |
| EPI_ISL_763054 | Germany/BB-RKI-NP-0067/2020 | 2020-12-17 | Europe/Germany/Brandenburg/ | Unit 17: Influenza & Other Respiratory Viruses, German National Influenza Center | Project group Epidemiology of Highly Pathogenic Microorganisms, Robert Koch-Institute | Ariane Dvºx et al |
| EPI_ISL_763058 | Germany/BB-RKI-NP-0055/2020 | 2020-12-14 | Europe/Germany/Brandenburg/ | Unit 17: Influenza & Other Respiratory Viruses, German National Influenza Center | Project group Epidemiology of Highly Pathogenic Microorganisms, Robert Koch-Institute | Ariane Dvºx et al |
| EPI_ISL_763062 | Georgia/Tb-171516/2020 | 2020-12-21 | Asia/Georgia/Tbilisi/ | Department for Virology, Molecular Biology and Genome Research, R. G. Lugar Center for Public Health Research, National Center for Disease Control and Public Health (NCDC) of Georgia. | Department for Virology, Molecular Biology and Genome Research, R. G. Lugar Center for Public Health Research, National Center for Disease Control and Public Health (NCDC) of Georgia. | Giorgi Tomashvili et al |

| | | | | | | |
|---|---|---|---|---|---|---|
| EPI_ISL_763063 | Indonesia/JT-UNS-13-38697/2020 | 2020-10-12 | Asia/Indonesia/Central Java/ | RS Panti Waluyo, Surakarta | Universitas Sebelas Maret (UNS); Rumah Sakit UNS (RS-UNS); National Institute of Health Research and Development, Indonesian Ministry of Health. | Betty Suryawati et al |
| EPI_ISL_763064 | Indonesia/JT-UNS-9-27310/2020 | 2020-09-10 | Asia/Indonesia/Central Java/ | RS Kasih Ibu, Surakarta | Universitas Sebelas Maret (UNS); Rumah Sakit UNS (RS-UNS); National Institute of Health Research and Development, Indonesian Ministry of Health. | Betty Suryawati et al |

| | | | | | | |
|---|---|---|---|---|---|---|
| EPI_ISL_763065 | Tunisia/6895/2020 | 2020-04-01 | Africa/Tunisia/Tunis/ | 1-Laboratory of Microbiology, National Reference Lab, Charles Nicolle Hospital; 2-University of Tunis ElManar, Faculty of Medicine of Tunis, LR99ES09, Tunis, Tunisia | 1-Clinical and Experimental Pharmacology Lab, LR16SP02, National Center of Pharmacovigilance, University of Tunis El Manar, Tunis, Tunisia. 2-Neurodegenerative diseases and psychiatric troubles, LR18SP03, Razi Hospital, University of Tunis El Manar, Tunis, Tunisia. 3-Ministry of Health, National Observatory of New and Emerging Diseases, 1006, Tunis, Tunisia | Ilhem Boutiba-Ben Boubaker et al |
| EPI_ISL_763066 | Indonesia/JT-UNS-12-37578/2020 | 2020-10-09 | Asia/Indonesia/Central Java/ | Puskesmas Grogol, Sukoharjo | Universitas Sebelas Maret (UNS); Rumah Sakit UNS (RS-UNS); National Institute of Health Research and Development, Indonesian Ministry of Health. | Yulia Sari et al |

| | | | | | | |
|---|---|---|---|---|---|---|
| EPI_ISL_763067 | Tunisia/7899/2020 | 2020-04-06 | Africa/Tunisia/Tunis/ | 1-Laboratory of Microbiology, National Reference Lab, Charles Nicolle Hospital; 2-University of Tunis ElManar, Faculty of Medicine of Tunis, LR99ES09, Tunis, Tunisia | 1-Clinical and Experimental Pharmacology Lab, LR16SP02, National Center of Pharmacovigilance, University of Tunis El Manar, Tunis, Tunisia. 2-Neurodegenerative diseases and psychiatric troubles, LR18SP03, Razi Hospital, University of Tunis El Manar, Tunis, Tunisia. 3-Ministry of Health, National Observatory of New and Emerging Diseases, 1006, Tunis, Tunisia | Ilhem Boutiba-Ben Boubaker et al |
| EPI_ISL_763069 | Italy/UMB-IZSUM-48682-1/2020 | 2020-10-29 | Europe/Italy/Umbria/ | Istituto Zooprofilattico Sperimentale dell' Umbria e delle Marche -Togo Rosati | Istituto Superiore di Sanitv† | Massimo Biagetti et al |
| EPI_ISL_763071 | Italy/UMB-IZSUM-48682-9/2020 | 2020-10-29 | Europe/Italy/Umbria/ | Istituto Zooprofilattico Sperimentale dell' Umbria e delle Marche -Togo Rosati | Istituto Superiore di Sanitv† | Massimo Biagetti et al |
| EPI_ISL_763073 | Italy/UMB-IZSUM-48682-18/2020 | 2020-10-29 | Europe/Italy/Umbria/ | Istituto Zooprofilattico Sperimentale dell' Umbria e delle Marche -Togo Rosati | Istituto Superiore di Sanitv† | Massimo Biagetti et al |
| EPI_ISL_763074 | Brazil/SP-900B/2020 | 2020-12-21 | South America/Brazil/Sv£o Paulo/Sv£o | Diagnosticos da America - DASA | Instituto Adolfo Lutz, Interdiciplinary Procedures Center, Strategic Laboratory | Claudio Tavares Sacchi et al |

| | | | | | | |
|---|---|---|---|---|---|---|
| EPI_ISL_763075 | Brazil/SP-901B/2020 | 2020-12-22 | South America/Brazil/S√£o Paulo/S√£o | Diagnosticos da America - DASA | Instituto Adolfo Lutz, Interdiciplinary Procedures Center, Strategic Laboratory | Claudio Tavares Sacchi et al |
| EPI_ISL_763078 | Italy/VEN-IZSVe-20RS2272-36_PD/20 | 2020-12-15 | Europe/Italy/Veneto/Padova | Microbiologia e Virologia | Istituto Zooprofilattico Sperimentale delle Venezie | Adelaide Milani et al |
| EPI_ISL_763082 | Italy/VEN-IZSVe-20RS2272-40_PD/20 | 2020-12-15 | Europe/Italy/Veneto/Padova | Microbiologia e Virologia | Istituto Zooprofilattico Sperimentale delle Venezie | Adelaide Milani et al |
| EPI_ISL_763084 | Italy/VEN-IZSVe-20RS2272-42_PD/20 | 2020-12-20 | Europe/Italy/Veneto/Padova | Microbiologia e Virologia | Istituto Zooprofilattico Sperimentale delle Venezie | Adelaide Milani et al |
| EPI_ISL_763088 | Germany/TH-IIMK-CaSe-23/2020 | 2020-03-24 | Europe/Germany/Thuringia/ | Jena University Hospital, Institute for Infectious Diseases and Infection Control | Institute of infectious medicine & hospital hygiene, CaSe-Group | Spott et al |
| EPI_ISL_763090 | Germany/TH-IIMK-CaSe-25/2020 | 2020-03-25 | Europe/Germany/Thuringia/ | Jena University Hospital, Institute for Infectious Diseases and Infection Control | Institute of infectious medicine & hospital hygiene, CaSe-Group | Spott et al |
| EPI_ISL_763092 | Germany/TH-IIMK-CaSe-27/2020 | 2020-04-01 | Europe/Germany/Thuringia/ | Jena University Hospital, Institute for Infectious Diseases and Infection Control | Institute of infectious medicine & hospital hygiene, CaSe-Group | Spott et al |
| EPI_ISL_763093 | Germany/TH-IIMK-CaSe-28/2020 | 2020-04-03 | Europe/Germany/Thuringia/ | Jena University Hospital, Institute for Infectious Diseases and Infection Control | Institute of infectious medicine & hospital hygiene, CaSe-Group | Spott et al |
| EPI_ISL_763094 | Italy/UMB-IZSUM-48682-12/2020 | 2020-10-29 | Europe/Italy/Umbria/ | Istituto Zooprofilattico Sperimentale dell' Umbria e delle Marche -Togo Rosati | Istituto Superiore di Sanit√† | Massimo Biagetti et al |

| | | | | | | |
|---|---|---|---|---|---|---|
| EPI_ISL_763095 | Italy/UMB-IZSUM-48682-17/2020 | 2020-10-19 | Europe/Italy/Umbria/ | Istituto Zooprofilattico Sperimentale dell' Umbria e delle Marche -Togo Rosati | Istituto Superiore di Sanitv† | Massimo Biagetti et al |
| EPI_ISL_763096 | Italy/UMB-IZSUM-48682-15/2020 | 2020-10-29 | Europe/Italy/Umbria/ | Istituto Zooprofilattico Sperimentale dell' Umbria e delle Marche -Togo Rosati | Istituto Superiore di Sanitv† | Massimo Biagetti et al |
| EPI_ISL_763097 | Italy/UMB-IZSUM-48682-20/2020 | 2020-10-29 | Europe/Italy/Umbria/ | Istituto Zooprofilattico Sperimentale dell' Umbria e delle Marche -Togo Rosati | Istituto Superiore di Sanitv† | Massimo Biagetti et al |
| EPI_ISL_763098 | Italy/UMB-IZSUM-48682-10/2020 | 2020-10-29 | Europe/Italy/Umbria/ | Istituto Zooprofilattico Sperimentale dell' Umbria e delle Marche -Togo Rosati | Istituto Superiore di Sanitv† | Massimo Biagetti et al |
| EPI_ISL_763099 | Belgium/un-EMC-7/2020 | 2020-11-25 | Europe/Belgium/Belgium/ | Dutch COVID-19 response team | Erasmus Medical Center | Bas Oude Munnink et al |
| EPI_ISL_763111 | Netherlands/LI-EMC-143/2020 | 2020-11-18 | Europe/Netherlands/Limburg/ | Dutch COVID-19 response team | Erasmus Medical Center | Bas Oude Munnink et al |
| EPI_ISL_763117 | Netherlands/LI-EMC-149/2020 | 2020-11-21 | Europe/Netherlands/Limburg/ | Dutch COVID-19 response team | Erasmus Medical Center | Bas Oude Munnink et al |
| EPI_ISL_763125 | Netherlands/LI-EMC-157/2020 | 2020-11-27 | Europe/Netherlands/Limburg/ | Dutch COVID-19 response team | Erasmus Medical Center | Bas Oude Munnink et al |
| EPI_ISL_763126 | Netherlands/LI-EMC-158/2020 | 2020-12-08 | Europe/Netherlands/Limburg/ | Dutch COVID-19 response team | Erasmus Medical Center | Bas Oude Munnink et al |
| EPI_ISL_763150 | Netherlands/OV-EMC-110/2020 | 2020-11-07 | Europe/Netherlands/Overijssel/ | Dutch COVID-19 response team | Erasmus Medical Center | Bas Oude Munnink et al |
| EPI_ISL_763157 | Netherlands/OV-EMC-117/2020 | 2020-11-07 | Europe/Netherlands/Overijssel/ | Dutch COVID-19 response team | Erasmus Medical Center | Bas Oude Munnink et al |
| EPI_ISL_763158 | Netherlands/OV-EMC-118/2020 | 2020-11-07 | Europe/Netherlands/Overijssel/ | Dutch COVID-19 response team | Erasmus Medical Center | Bas Oude Munnink et al |
| EPI_ISL_763167 | Netherlands/OV-EMC-128/2020 | 2020-11-22 | Europe/Netherlands/Overijssel/ | Dutch COVID-19 response team | Erasmus Medical Center | Bas Oude Munnink et al |
| EPI_ISL_763201 | Netherlands/ZH-EMC-1122/2020 | 2020-12-15 | Europe/Netherlands/South Holland/ | Dutch COVID-19 response team | Erasmus Medical Center | Bas Oude Munnink et al |
| EPI_ISL_763219 | Netherlands/ZH-EMC-1142/2020 | 2020-12-19 | Europe/Netherlands/South Holland/ | Dutch COVID-19 response team | Erasmus Medical Center | Bas Oude Munnink et al |
| EPI_ISL_763236 | Netherlands/ZH-EMC-1161/2020 | 2020-11-23 | Europe/Netherlands/South Holland/ | Dutch COVID-19 response team | Erasmus Medical Center | Bas Oude Munnink et al |
| EPI_ISL_763265 | Netherlands/ZH-EMC-1191/2020 | 2020-12-20 | Europe/Netherlands/South Holland/ | Dutch COVID-19 response team | Erasmus Medical Center | Bas Oude Munnink et al |
| EPI_ISL_763303 | Netherlands/ZH-EMC-1234/2020 | 2020-12-23 | Europe/Netherlands/South Holland/ | Dutch COVID-19 response team | Erasmus Medical Center | Bas Oude Munnink et al |
| EPI_ISL_763306 | Netherlands/ZH-EMC-1237/2020 | 2020-09-17 | Europe/Netherlands/South Holland/ | Dutch COVID-19 response team | Erasmus Medical Center | Bas Oude Munnink et al |
| EPI_ISL_763314 | Netherlands/ZH-EMC-1245/2020 | 2020-12-23 | Europe/Netherlands/South Holland/ | Dutch COVID-19 response team | Erasmus Medical Center | Bas Oude Munnink et al |

| | | | | | | |
|---|---|---|---|---|---|---|
| EPI_ISL_763320 | Italy/UMB-IZSUM-48682-19/2020 | 2020-10-29 | Europe/Italy/Umbria/ | Istituto Zooprofilattico Sperimentale dell' Umbria e delle Marche -Togo Rosati | Istituto Superiore di Sanitv† | Massimo Biagetti et al |
| EPI_ISL_763324 | Italy/UMB-IZSUM-48682-6/2020 | 2020-10-29 | Europe/Italy/Umbria/ | Istituto Zooprofilattico Sperimentale dell' Umbria e delle Marche -Togo Rosati | Istituto Superiore di Sanitv† | Massimo Biagetti et al |
| EPI_ISL_763326 | Italy/UMB-IZSUM-48682-23/2020 | 2020-10-29 | Europe/Italy/Umbria/ | Istituto Zooprofilattico Sperimentale dell' Umbria e delle Marche -Togo Rosati | Istituto Superiore di Sanitv† | Massimo Biagetti et al |
| EPI_ISL_763328 | Italy/UMB-IZSUM-48682-21/2020 | 2020-10-29 | Europe/Italy/Umbria/ | Istituto Zooprofilattico Sperimentale dell' Umbria e delle Marche -Togo Rosati | Istituto Superiore di Sanitv† | Massimo Biagetti et al |
| EPI_ISL_763330 | Italy/UMB-IZSUM-48682-2/2020 | 2020-10-29 | Europe/Italy/Umbria/ | Istituto Zooprofilattico Sperimentale dell' Umbria e delle Marche -Togo Rosati | Istituto Superiore di Sanitv† | Massimo Biagetti et al |
| EPI_ISL_763334 | USA/FL-BPHL-2251/2020 | 2020-11-24 | North America/USA/Florida/ | Florida Bureau of Public Health Laboratories | Florida Bureau of Public Health Laboratories | Sarah Schmedes et al |
| EPI_ISL_763338 | USA/FL-BPHL-2255/2020 | 2020-11-18 | North America/USA/Florida/ | Florida Bureau of Public Health Laboratories | Florida Bureau of Public Health Laboratories | Sarah Schmedes et al |
| EPI_ISL_763339 | USA/FL-BPHL-2256/2020 | 2020-11-17 | North America/USA/Florida/ | Florida Bureau of Public Health Laboratories | Florida Bureau of Public Health Laboratories | Sarah Schmedes et al |
| EPI_ISL_763345 | USA/FL-BPHL-2262/2020 | 2020-12-01 | North America/USA/Florida/ | Florida Bureau of Public Health Laboratories | Florida Bureau of Public Health Laboratories | Sarah Schmedes et al |
| EPI_ISL_763411 | Wales/PHWC-4A6A55/2020 | 2020-12-17 | Europe/United Kingdom/Wales/ | Wales Specialist Virology Centre Sequencing lab: Pathogen Genomics Unit | COVID-19 Genomics UK (COG-UK) Consortium | Catherine Moore et al |
| EPI_ISL_763446 | NorthernIreland/NIRE-234D73/2020 | 2020-10-18 | Europe/United Kingdom/Northern Irelar | Regional Virus Laboratory, Belfast Health and Social Care Trust | COVID-19 Genomics UK (COG-UK) Consortium | Conall McCaughey et al |
| EPI_ISL_763487 | Wales/PHWC-4A3B28/2020 | 2020-12-13 | Europe/United Kingdom/Wales/ | Wales Specialist Virology Centre Sequencing lab: Pathogen Genomics Unit | COVID-19 Genomics UK (COG-UK) Consortium | Catherine Moore et al |
| EPI_ISL_763553 | England/EXET-13ED2F/2020 | 2020-12-14 | Europe/United Kingdom/England/ | University of Exeter | COVID-19 Genomics UK (COG-UK) Consortium | Ben Temperton et al |
| EPI_ISL_763561 | England/EXET-13EA28/2020 | 2020-12-12 | Europe/United Kingdom/England/ | University of Exeter | COVID-19 Genomics UK (COG-UK) Consortium | Ben Temperton et al |
| EPI_ISL_763974 | England/OXON-F415C9/2020 | 2020-12-15 | Europe/United Kingdom/England/ | Oxford Viromics, NDM, University of Oxford; Oxford University Hospitals; Basingstoke and North Hampshire Hospital | COVID-19 Genomics UK (COG-UK) Consortium | Tanya Golubchik et al |

| | | | | | | |
|---|---|---|---|---|---|---|
| EPI_ISL_764187 | NorthernIreland/NIRE-2349E8/2020 | 2020-10-23 | Europe/United Kingdom/Northern Irelar | Regional Virus Laboratory, Belfast Health and Social Care Trust | COVID-19 Genomics UK (COG-UK) Consortium | Conall McCaughey et al |
| EPI_ISL_764263 | NorthernIreland/NIRE-235361/2020 | 2020-10-19 | Europe/United Kingdom/Northern Irelar | Regional Virus Laboratory, Belfast Health and Social Care Trust | COVID-19 Genomics UK (COG-UK) Consortium | Conall McCaughey et al |
| EPI_ISL_764432 | NorthernIreland/NIRE-235413/2020 | 2020-11-03 | Europe/United Kingdom/Northern Irelar | Regional Virus Laboratory, Belfast Health and Social Care Trust | COVID-19 Genomics UK (COG-UK) Consortium | Conall McCaughey et al |
| EPI_ISL_764438 | NorthernIreland/NIRE-2354E6/2020 | 2020-11-13 | Europe/United Kingdom/Northern Irelar | Regional Virus Laboratory, Belfast Health and Social Care Trust | COVID-19 Genomics UK (COG-UK) Consortium | Conall McCaughey et al |
| EPI_ISL_764518 | England/NOTT-121865/2020 | 2020-12-23 | Europe/United Kingdom/England/ | Queens Medical Centre, Clinical Microbiology Department / DeepSeq Nottingham | COVID-19 Genomics UK (COG-UK) Consortium | Gemma Clark et al |
| EPI_ISL_764571 | Scotland/OXON-F4157E/2020 | 2020-12-16 | Europe/United Kingdom/Scotland/ | Oxford Viromics, NDM, University of Oxford; Oxford University Hospitals; Basingstoke and North Hampshire Hospital | COVID-19 Genomics UK (COG-UK) Consortium | Tanya Golubchik et al |
| EPI_ISL_764944 | Wales/PHWC-4A59B3/2020 | 2020-12-15 | Europe/United Kingdom/Wales/ | Wales Specialist Virology Centre Sequencing lab: Pathogen Genomics Unit | COVID-19 Genomics UK (COG-UK) Consortium | Catherine Moore et al |
| EPI_ISL_765018 | Wales/PHWC-4A628A/2020 | 2020-12-16 | Europe/United Kingdom/Wales/ | Wales Specialist Virology Centre Sequencing lab: Pathogen Genomics Unit | COVID-19 Genomics UK (COG-UK) Consortium | Catherine Moore et al |
| EPI_ISL_765211 | Portugal/PT1666/2020 | 2020-11-12 | Europe/Portugal/Portugal/ | Instituto Nacional de Saude (INSA) | Instituto Nacional de Saude (INSA) | Borges et al et al |
| EPI_ISL_765212 | Portugal/PT1752/2020 | 2020-11-11 | Europe/Portugal/Portugal/ | Instituto Nacional de Saude (INSA) | Instituto Nacional de Saude (INSA) | Borges et al et al |
| EPI_ISL_765218 | Portugal/PT2038/2020 | 2020-11-09 | Europe/Portugal/Portugal/ | Instituto Nacional de Saude (INSA) and Instituto Gulbenkian de Ciencia (IGC) | Instituto Nacional de Saude (INSA) and Instituto Gulbenkian de Ciencia (IGC) | Borges et al et al |
| EPI_ISL_765220 | Portugal/PT1568/2020 | 2020-06-28 | Europe/Portugal/Portugal/ | Instituto Nacional de Saude (INSA) | Instituto Nacional de Saude (INSA) | Borges et al et al |
| EPI_ISL_765480 | USA/NY-Wadsworth-213972-01/2020 | 2020-09-09 | North America/USA/New York/Chemung | Wadsworth Center, New York State Department.of Health | Wadsworth Center, New York State Department.of Health | Kirsten St. George et al |

| | | | | | | |
|---|---|---|---|---|---|---|
| EPI_ISL_765484 | USA/NY-Wadsworth-213973-01/2020 | 2020-09-09 | North America/USA/New York/Chemung | Wadsworth Center, New York State Department.of Health | Wadsworth Center, New York State Department.of Health | Kirsten St. George et al |
| EPI_ISL_765489 | USA/NY-Wadsworth-290352-01/2020 | 2020-12-15 | North America/USA/New York/ | MONTEFIORE MEDICAL CENTER LABORATORIES | Wadsworth Center, New York State Department.of Health | Kirsten St. George et al |
| EPI_ISL_765498 | USA/NY-Wadsworth-213917-01/2020 | 2020-09-08 | North America/USA/New York/Otsego C | Wadsworth Center, New York State Department.of Health | Wadsworth Center, New York State Department.of Health | Kirsten St. George et al |
| EPI_ISL_765500 | USA/NY-Wadsworth-213737-01/2020 | 2020-09-09 | North America/USA/New York/Warren | Wadsworth Center, New York State Department.of Health | Wadsworth Center, New York State Department.of Health | Kirsten St. George et al |
| EPI_ISL_765508 | USA/NY-Wadsworth-220236-01/2020 | 2020-09-17 | North America/USA/New York/Saratoga | Wadsworth Center, New York State Department.of Health | Wadsworth Center, New York State Department.of Health | Kirsten St. George et al |
| EPI_ISL_765509 | USA/NY-Wadsworth-222417-01/2020 | 2020-09-19 | North America/USA/New York/Saratoga | Wadsworth Center, New York State Department.of Health | Wadsworth Center, New York State Department.of Health | Kirsten St. George et al |
| EPI_ISL_765511 | USA/NY-Wadsworth-223805-01/2020 | 2020-09-21 | North America/USA/New York/Chemung | Wadsworth Center, New York State Department.of Health | Wadsworth Center, New York State Department.of Health | Kirsten St. George et al |
| EPI_ISL_765512 | USA/NY-Wadsworth-223775-01/2020 | 2020-09-21 | North America/USA/New York/Chemung | Wadsworth Center, New York State Department.of Health | Wadsworth Center, New York State Department.of Health | Kirsten St. George et al |

| | | | | | | |
|---|---|---|---|---|---|---|
| EPI_ISL_765519 | USA/NY-Wadsworth-290318-01/2020 | 2020-12-14 | North America/USA/New York/Saratoga | SARATOGA HOSPITAL LABORATORY | Wadsworth Center, New York State Department.of Health | Kirsten St. George et al |
| EPI_ISL_765523 | USA/NY-Wadsworth-290324-01/2020 | 2020-12-15 | North America/USA/New York/Saratoga | SARATOGA HOSPITAL LABORATORY | Wadsworth Center, New York State Department.of Health | Kirsten St. George et al |
| EPI_ISL_765527 | USA/NY-Wadsworth-290328-01/2020 | 2020-12-21 | North America/USA/New York/ | SARATOGA HOSPITAL LABORATORY | Wadsworth Center, New York State Department.of Health | Kirsten St. George et al |
| EPI_ISL_765534 | USA/NY-Wadsworth-290342-01/2020 | 2020-12-18 | North America/USA/New York/Bronx | MONTEFIORE MEDICAL CENTER LABORATORIES | Wadsworth Center, New York State Department.of Health | Kirsten St. George et al |
| EPI_ISL_765535 | USA/NY-Wadsworth-290345-01/2020 | 2020-12-18 | North America/USA/New York/Bronx | MONTEFIORE MEDICAL CENTER LABORATORIES | Wadsworth Center, New York State Department.of Health | Kirsten St. George et al |
| EPI_ISL_765544 | USA/NY-Wadsworth-290361-01/2020 | 2020-12-16 | North America/USA/New York/Bronx | MONTEFIORE MEDICAL CENTER LABORATORIES | Wadsworth Center, New York State Department.of Health | Kirsten St. George et al |
| EPI_ISL_765550 | USA/NY-Wadsworth-290369-01/2020 | 2020-12-18 | North America/USA/New York/Bronx | MONTEFIORE MEDICAL CENTER LABORATORIES | Wadsworth Center, New York State Department.of Health | Kirsten St. George et al |
| EPI_ISL_765553 | USA/NY-Wadsworth-290374-01/2020 | 2020-12-18 | North America/USA/New York/Bronx | MONTEFIORE MEDICAL CENTER LABORATORIES | Wadsworth Center, New York State Department.of Health | Kirsten St. George et al |

| | | | | | | |
|---|---|---|---|---|---|---|
| EPI_ISL_765556 | USA/NY-Wadsworth-290377-01/2020 | 2020-12-16 | North America/USA/New York/Bronx | MONTEFIORE MEDICAL CENTER LABORATORIES | Wadsworth Center, New York State Department.of Health | Kirsten St. George et al |
| EPI_ISL_765578 | USA/CO-Broad-CMU_00114/2020 | 2020-11-10 | North America/USA/Colorado/ | Colorado Mesa University | Infectious Disease Program, Broad Institute of Harvard and MIT | Lemieux et al |
| EPI_ISL_765583 | USA/MA-MGH-02662/2020 | 2020-09-23 | North America/USA/Massachusetts/ | Brigham and Womens Hospital | Infectious Disease Program, Broad Institute of Harvard and MIT | Lemieux et al |
| EPI_ISL_765587 | USA/MA-MGH-02694/2020 | 2020-09-14 | North America/USA/Massachusetts/ | Brigham and Womens Hospital | Infectious Disease Program, Broad Institute of Harvard and MIT | Lemieux et al |
| EPI_ISL_765589 | USA/MA-MGH-02696/2020 | 2020-09-27 | North America/USA/Massachusetts/ | Brigham and Womens Hospital | Infectious Disease Program, Broad Institute of Harvard and MIT | Lemieux et al |
| EPI_ISL_765592 | USA/MA-MGH-02703/2020 | 2020-09-25 | North America/USA/Massachusetts/ | Brigham and Womens Hospital | Infectious Disease Program, Broad Institute of Harvard and MIT | Lemieux et al |
| EPI_ISL_765599 | USA/MA-MGH-02721/2020 | 2020-09-25 | North America/USA/Massachusetts/ | Brigham and Womens Hospital | Infectious Disease Program, Broad Institute of Harvard and MIT | Lemieux et al |
| EPI_ISL_765605 | USA/MA-MGH-02732/2020 | 2020-09-30 | North America/USA/Massachusetts/ | Brigham and Womens Hospital | Infectious Disease Program, Broad Institute of Harvard and MIT | Lemieux et al |
| EPI_ISL_765607 | USA/MA-MGH-02734/2020 | 2020-09-29 | North America/USA/Massachusetts/ | Brigham and Womens Hospital | Infectious Disease Program, Broad Institute of Harvard and MIT | Lemieux et al |

| | | | | | | |
|---|---|---|---|---|---|---|
| EPI_ISL_765609 | USA/MA-MGH-02739/2020 | 2020-09-29 | North America/USA/Massachusetts/ | Brigham and Womens Hospital | Infectious Disease Program, Broad Institute of Harvard and MIT | Lemieux et al |
| EPI_ISL_765615 | USA/MA-MGH-02750/2020 | 2020-09-25 | North America/USA/Massachusetts/ | Massachusetts General Hospital | Infectious Disease Program, Broad Institute of Harvard and MIT | Lemieux et al |
| EPI_ISL_765616 | USA/MA-MGH-02751/2020 | 2020-09-23 | North America/USA/Massachusetts/ | Massachusetts General Hospital | Infectious Disease Program, Broad Institute of Harvard and MIT | Lemieux et al |
| EPI_ISL_765619 | USA/MA-MGH-02757/2020 | 2020-09-23 | North America/USA/Massachusetts/ | Massachusetts General Hospital | Infectious Disease Program, Broad Institute of Harvard and MIT | Lemieux et al |
| EPI_ISL_765626 | USA/MA-MGH-02769/2020 | 2020-09-24 | North America/USA/Massachusetts/ | Massachusetts General Hospital | Infectious Disease Program, Broad Institute of Harvard and MIT | Lemieux et al |
| EPI_ISL_765628 | USA/MA-MGH-02776/2020 | 2020-09-23 | North America/USA/Massachusetts/ | Massachusetts General Hospital | Infectious Disease Program, Broad Institute of Harvard and MIT | Lemieux et al |
| EPI_ISL_765633 | USA/MA-MGH-02784/2020 | 2020-09-20 | North America/USA/Massachusetts/ | Massachusetts General Hospital | Infectious Disease Program, Broad Institute of Harvard and MIT | Lemieux et al |
| EPI_ISL_765635 | USA/MA-MGH-02786/2020 | 2020-09-23 | North America/USA/Massachusetts/ | Massachusetts General Hospital | Infectious Disease Program, Broad Institute of Harvard and MIT | Lemieux et al |
| EPI_ISL_765636 | USA/MA-MGH-02788/2020 | 2020-09-23 | North America/USA/Massachusetts/ | Massachusetts General Hospital | Infectious Disease Program, Broad Institute of Harvard and MIT | Lemieux et al |
| EPI_ISL_765639 | USA/MA-MGH-02804/2020 | 2020-09-24 | North America/USA/Massachusetts/ | Massachusetts General Hospital | Infectious Disease Program, Broad Institute of Harvard and MIT | Lemieux et al |

| | | | | | | |
|---|---|---|---|---|---|---|
| EPI_ISL_765641 | USA/MA-MGH-02808/2020 | 2020-09-25 | North America/USA/Massachusetts/ | Massachusetts General Hospital | Infectious Disease Program, Broad Institute of Harvard and MIT | Lemieux et al |
| EPI_ISL_765646 | USA/MA-MGH-02815/2020 | 2020-09-24 | North America/USA/Massachusetts/ | Massachusetts General Hospital | Infectious Disease Program, Broad Institute of Harvard and MIT | Lemieux et al |
| EPI_ISL_765650 | USA/MA-MGH-02825/2020 | 2020-10-13 | North America/USA/Massachusetts/ | Massachusetts General Hospital | Infectious Disease Program, Broad Institute of Harvard and MIT | Lemieux et al |
| EPI_ISL_765663 | USA/MA-MGH-02951/2020 | 2020-07-06 | North America/USA/Massachusetts/ | Massachusetts General Hospital | Infectious Disease Program, Broad Institute of Harvard and MIT | Lemieux et al |
| EPI_ISL_765667 | USA/MA-MGH-02973/2020 | 2020-07-10 | North America/USA/Massachusetts/ | Massachusetts General Hospital | Infectious Disease Program, Broad Institute of Harvard and MIT | Lemieux et al |
| EPI_ISL_765668 | USA/MA-MGH-02975/2020 | 2020-07-10 | North America/USA/Massachusetts/ | Massachusetts General Hospital | Infectious Disease Program, Broad Institute of Harvard and MIT | Lemieux et al |
| EPI_ISL_765669 | USA/MA-MGH-02978/2020 | 2020-07-10 | North America/USA/Massachusetts/ | Massachusetts General Hospital | Infectious Disease Program, Broad Institute of Harvard and MIT | Lemieux et al |
| EPI_ISL_765672 | USA/MA-MGH-02990/2020 | 2020-07-14 | North America/USA/Massachusetts/ | Massachusetts General Hospital | Infectious Disease Program, Broad Institute of Harvard and MIT | Lemieux et al |
| EPI_ISL_765678 | USA/MA-MGH-03006/2020 | 2020-07-19 | North America/USA/Massachusetts/ | Massachusetts General Hospital | Infectious Disease Program, Broad Institute of Harvard and MIT | Lemieux et al |
| EPI_ISL_765679 | USA/MA-MGH-03007/2020 | 2020-07-20 | North America/USA/Massachusetts/ | Massachusetts General Hospital | Infectious Disease Program, Broad Institute of Harvard and MIT | Lemieux et al |

| | | | | | | |
|---|---|---|---|---|---|---|
| EPI_ISL_765686 | USA/MA-MGH-03031/2020 | 2020-07-25 | North America/USA/Massachusetts/ | Massachusetts General Hospital | Infectious Disease Program, Broad Institute of Harvard and MIT | Lemieux et al |
| EPI_ISL_765689 | USA/MA-MGH-03040/2020 | 2020-08-01 | North America/USA/Massachusetts/ | Massachusetts General Hospital | Infectious Disease Program, Broad Institute of Harvard and MIT | Lemieux et al |
| EPI_ISL_765691 | USA/MA-MGH-03043/2020 | 2020-08-01 | North America/USA/Massachusetts/ | Massachusetts General Hospital | Infectious Disease Program, Broad Institute of Harvard and MIT | Lemieux et al |
| EPI_ISL_765693 | USA/MA-MGH-03048/2020 | 2020-08-03 | North America/USA/Massachusetts/ | Massachusetts General Hospital | Infectious Disease Program, Broad Institute of Harvard and MIT | Lemieux et al |
| EPI_ISL_765702 | USA/MA-MGH-03083/2020 | 2020-08-14 | North America/USA/Massachusetts/ | Massachusetts General Hospital | Infectious Disease Program, Broad Institute of Harvard and MIT | Lemieux et al |
| EPI_ISL_765704 | USA/MA-MGH-03101/2020 | 2020-08-31 | North America/USA/Massachusetts/ | Massachusetts General Hospital | Infectious Disease Program, Broad Institute of Harvard and MIT | Lemieux et al |
| EPI_ISL_765708 | USA/MA-MGH-03116/2020 | 2020-08-14 | North America/USA/Massachusetts/ | Massachusetts General Hospital | Infectious Disease Program, Broad Institute of Harvard and MIT | Lemieux et al |
| EPI_ISL_765711 | USA/MA-MGH-03124/2020 | 2020-11-15 | North America/USA/Massachusetts/ | Massachusetts General Hospital | Infectious Disease Program, Broad Institute of Harvard and MIT | Lemieux et al |
| EPI_ISL_765715 | USA/MA-MGH-03141/2020 | 2020-09-05 | North America/USA/Massachusetts/ | Massachusetts General Hospital | Infectious Disease Program, Broad Institute of Harvard and MIT | Lemieux et al |
| EPI_ISL_765716 | USA/MA-MGH-03144/2020 | 2020-09-06 | North America/USA/Massachusetts/ | Massachusetts General Hospital | Infectious Disease Program, Broad Institute of Harvard and MIT | Lemieux et al |

| | | | | | | |
|---|---|---|---|---|---|---|
| EPI_ISL_765719 | USA/MA-MGH-03147/2020 | 2020-09-07 | North America/USA/Massachusetts/ | Massachusetts General Hospital | Infectious Disease Program, Broad Institute of Harvard and MIT | Lemieux et al |
| EPI_ISL_765720 | USA/MA-MGH-03149/2020 | 2020-09-08 | North America/USA/Massachusetts/ | Massachusetts General Hospital | Infectious Disease Program, Broad Institute of Harvard and MIT | Lemieux et al |
| EPI_ISL_765724 | USA/MA-MGH-03161/2020 | 2020-08-28 | North America/USA/Massachusetts/ | Massachusetts General Hospital | Infectious Disease Program, Broad Institute of Harvard and MIT | Lemieux et al |
| EPI_ISL_765725 | USA/MA-MGH-03162/2020 | 2020-08-30 | North America/USA/Massachusetts/ | Massachusetts General Hospital | Infectious Disease Program, Broad Institute of Harvard and MIT | Lemieux et al |
| EPI_ISL_765726 | USA/MA-MGH-03165/2020 | 2020-09-11 | North America/USA/Massachusetts/ | Massachusetts General Hospital | Infectious Disease Program, Broad Institute of Harvard and MIT | Lemieux et al |
| EPI_ISL_765727 | USA/MA-MGH-03167/2020 | 2020-08-29 | North America/USA/Massachusetts/ | Massachusetts General Hospital | Infectious Disease Program, Broad Institute of Harvard and MIT | Lemieux et al |
| EPI_ISL_765728 | USA/MA-MGH-03168/2020 | 2020-09-11 | North America/USA/Massachusetts/ | Massachusetts General Hospital | Infectious Disease Program, Broad Institute of Harvard and MIT | Lemieux et al |
| EPI_ISL_765729 | USA/MA-MGH-03172/2020 | 2020-09-24 | North America/USA/Massachusetts/ | Massachusetts General Hospital | Infectious Disease Program, Broad Institute of Harvard and MIT | Lemieux et al |
| EPI_ISL_765732 | USA/MA-MGH-03175/2020 | 2020-09-24 | North America/USA/Massachusetts/ | Massachusetts General Hospital | Infectious Disease Program, Broad Institute of Harvard and MIT | Lemieux et al |
| EPI_ISL_765733 | USA/MA-MGH-03176/2020 | 2020-09-24 | North America/USA/Massachusetts/ | Massachusetts General Hospital | Infectious Disease Program, Broad Institute of Harvard and MIT | Lemieux et al |

| | | | | | | |
|---|---|---|---|---|---|---|
| EPI_ISL_765734 | USA/MA-MGH-03177/2020 | 2020-09-24 | North America/USA/Massachusetts/ | Massachusetts General Hospital | Infectious Disease Program, Broad Institute of Harvard and MIT | Lemieux et al |
| EPI_ISL_765735 | USA/MA-MGH-03178/2020 | 2020-09-25 | North America/USA/Massachusetts/ | Massachusetts General Hospital | Infectious Disease Program, Broad Institute of Harvard and MIT | Lemieux et al |
| EPI_ISL_765739 | USA/MA-MGH-03184/2020 | 2020-09-26 | North America/USA/Massachusetts/ | Massachusetts General Hospital | Infectious Disease Program, Broad Institute of Harvard and MIT | Lemieux et al |
| EPI_ISL_765742 | USA/MA-MGH-03191/2020 | 2020-09-28 | North America/USA/Massachusetts/ | Massachusetts General Hospital | Infectious Disease Program, Broad Institute of Harvard and MIT | Lemieux et al |
| EPI_ISL_765753 | USA/MA-MGH-03234/2020 | 2020-10-09 | North America/USA/Massachusetts/ | Massachusetts General Hospital | Infectious Disease Program, Broad Institute of Harvard and MIT | Lemieux et al |
| EPI_ISL_765755 | USA/MA-MGH-03242/2020 | 2020-10-12 | North America/USA/Massachusetts/ | Massachusetts General Hospital | Infectious Disease Program, Broad Institute of Harvard and MIT | Lemieux et al |
| EPI_ISL_765757 | USA/MA-MGH-03247/2020 | 2020-10-14 | North America/USA/Massachusetts/ | Massachusetts General Hospital | Infectious Disease Program, Broad Institute of Harvard and MIT | Lemieux et al |
| EPI_ISL_765788 | USA/MA-MGH-03330/2020 | 2020-11-02 | North America/USA/Massachusetts/ | Massachusetts General Hospital | Infectious Disease Program, Broad Institute of Harvard and MIT | Lemieux et al |
| EPI_ISL_765791 | USA/MA-MGH-03336/2020 | 2020-11-03 | North America/USA/Massachusetts/ | Massachusetts General Hospital | Infectious Disease Program, Broad Institute of Harvard and MIT | Lemieux et al |
| EPI_ISL_765792 | USA/MA-MGH-03337/2020 | 2020-11-04 | North America/USA/Massachusetts/ | Massachusetts General Hospital | Infectious Disease Program, Broad Institute of Harvard and MIT | Lemieux et al |

| | | | | | | |
|---|---|---|---|---|---|---|
| EPI_ISL_765793 | USA/MA-MGH-03339/2020 | 2020-11-04 | North America/USA/Massachusetts/ | Massachusetts General Hospital | Infectious Disease Program, Broad Institute of Harvard and MIT | Lemieux et al |
| EPI_ISL_765799 | USA/MA-MGH-03350/2020 | 2020-11-05 | North America/USA/Massachusetts/ | Massachusetts General Hospital | Infectious Disease Program, Broad Institute of Harvard and MIT | Lemieux et al |
| EPI_ISL_765812 | USA/MA-MGH-03386/2020 | 2020-11-09 | North America/USA/Massachusetts/ | Massachusetts General Hospital | Infectious Disease Program, Broad Institute of Harvard and MIT | Lemieux et al |
| EPI_ISL_765821 | USA/MA-MGH-03403/2020 | 2020-12-07 | North America/USA/Massachusetts/ | North Shore Medical Center | Infectious Disease Program, Broad Institute of Harvard and MIT | Lemieux et al |
| EPI_ISL_765822 | USA/MA-MGH-03405/2020 | 2020-12-07 | North America/USA/Massachusetts/ | North Shore Medical Center | Infectious Disease Program, Broad Institute of Harvard and MIT | Lemieux et al |
| EPI_ISL_765823 | USA/MA-MGH-03407/2020 | 2020-12-07 | North America/USA/Massachusetts/ | North Shore Medical Center | Infectious Disease Program, Broad Institute of Harvard and MIT | Lemieux et al |
| EPI_ISL_765828 | USA/MA-MGH-03418/2020 | 2020-11-11 | North America/USA/Massachusetts/ | Massachusetts General Hospital | Infectious Disease Program, Broad Institute of Harvard and MIT | Lemieux et al |
| EPI_ISL_765833 | USA/MA-MGH-03428/2020 | 2020-11-12 | North America/USA/Massachusetts/ | Massachusetts General Hospital | Infectious Disease Program, Broad Institute of Harvard and MIT | Lemieux et al |
| EPI_ISL_765855 | USA/MA-MGH-03479/2020 | 2020-11-20 | North America/USA/Massachusetts/ | Massachusetts General Hospital | Infectious Disease Program, Broad Institute of Harvard and MIT | Lemieux et al |
| EPI_ISL_765856 | USA/MA-MGH-03483/2020 | 2020-11-20 | North America/USA/Massachusetts/ | Massachusetts General Hospital | Infectious Disease Program, Broad Institute of Harvard and MIT | Lemieux et al |

| | | | | | | |
|---|---|---|---|---|---|---|
| EPI_ISL_765862 | USA/MA-MGH-03496/2020 | 2020-11-23 | North America/USA/Massachusetts/ | Massachusetts General Hospital | Infectious Disease Program, Broad Institute of Harvard and MIT | Lemieux et al |
| EPI_ISL_765865 | USA/MA-MGH-03506/2020 | 2020-11-24 | North America/USA/Massachusetts/ | Massachusetts General Hospital | Infectious Disease Program, Broad Institute of Harvard and MIT | Lemieux et al |
| EPI_ISL_765868 | USA/MA-MGH-03517/2020 | 2020-11-25 | North America/USA/Massachusetts/ | Massachusetts General Hospital | Infectious Disease Program, Broad Institute of Harvard and MIT | Lemieux et al |
| EPI_ISL_765870 | USA/MA-MGH-03522/2020 | 2020-11-25 | North America/USA/Massachusetts/ | Massachusetts General Hospital | Infectious Disease Program, Broad Institute of Harvard and MIT | Lemieux et al |
| EPI_ISL_765874 | USA/MA-MGH-03528/2020 | 2020-11-27 | North America/USA/Massachusetts/ | Massachusetts General Hospital | Infectious Disease Program, Broad Institute of Harvard and MIT | Lemieux et al |
| EPI_ISL_765882 | USA/MA-MGH-03555/2020 | 2020-11-30 | North America/USA/Massachusetts/ | Massachusetts General Hospital | Infectious Disease Program, Broad Institute of Harvard and MIT | Lemieux et al |
| EPI_ISL_765884 | USA/MA-MGH-03561/2020 | 2020-11-30 | North America/USA/Massachusetts/ | Massachusetts General Hospital | Infectious Disease Program, Broad Institute of Harvard and MIT | Lemieux et al |
| EPI_ISL_765885 | USA/MA-MGH-03563/2020 | 2020-11-30 | North America/USA/Massachusetts/ | Massachusetts General Hospital | Infectious Disease Program, Broad Institute of Harvard and MIT | Lemieux et al |
| EPI_ISL_765898 | USA/TX-DSHS-1922/2020 | 2020-06-20 | North America/USA/Texas/Harris Count | TXDSHS | TXDSHS | Rashmi Tuladhar et al |
| EPI_ISL_765902 | USA/TX-DSHS-1928/2020 | 2020-06-21 | North America/USA/Texas/Hidalgo Cour | TXDSHS | TXDSHS | Rashmi Tuladhar et al |
| EPI_ISL_765913 | USA/TX-DSHS-1939/2020 | 2020-06-21 | North America/USA/Texas/Hidalgo Cour | TXDSHS | TXDSHS | Rashmi Tuladhar et al |
| EPI_ISL_765925 | USA/TX-DSHS-1934/2020 | 2020-06-21 | North America/USA/Texas/Hidalgo Cour | TXDSHS | TXDSHS | Rashmi Tuladhar et al |
| EPI_ISL_765941 | USA/TX-DSHS-1920/2020 | 2020-06-20 | North America/USA/Texas/Harris Count | TXDSHS | TXDSHS | Rashmi Tuladhar et al |

| | | | | | | |
|---|---|---|---|---|---|---|
| EPI_ISL_765942 | USA/AZ-WL-CVG00126/2020 | 2020-10-31 | North America/USA/Arizona/Pima Coun | Worobey Lab, Department of Ecology and Evolutionary Biology, University of Arizona | Worobey Lab, Department of Ecology and Evolutionary Biology, University of Arizona | Brendan Larsen et al |
| EPI_ISL_765943 | USA/AZ-WL-CVG00007/2020 | 2020-10-05 | North America/USA/Arizona/Pima Coun | Worobey Lab, Department of Ecology and Evolutionary Biology, University of Arizona | Worobey Lab, Department of Ecology and Evolutionary Biology, University of Arizona | Brendan Larsen et al |
| EPI_ISL_765945 | USA/AZ-WL-9900192/2020 | 2020-09-28 | North America/USA/Arizona/Pima Coun | Worobey Lab, Department of Ecology and Evolutionary Biology, University of Arizona | Worobey Lab, Department of Ecology and Evolutionary Biology, University of Arizona | Brendan Larsen et al |
| EPI_ISL_765946 | USA/AZ-WL-9900204/2020 | 2020-09-29 | North America/USA/Arizona/Pima Coun | Worobey Lab, Department of Ecology and Evolutionary Biology, University of Arizona | Worobey Lab, Department of Ecology and Evolutionary Biology, University of Arizona | Brendan Larsen et al |
| EPI_ISL_765947 | USA/AZ-WL-9900234/2020 | 2020-09-30 | North America/USA/Arizona/Pima Coun | Worobey Lab, Department of Ecology and Evolutionary Biology, University of Arizona | Worobey Lab, Department of Ecology and Evolutionary Biology, University of Arizona | Brendan Larsen et al |

| | | | | | | |
|---|---|---|---|---|---|---|
| EPI_ISL_765955 | USA/AZ-WL-CVG00023/2020 | 2020-10-05 | North America/USA/Arizona/Pima Coun | Worobey Lab, Department of Ecology and Evolutionary Biology, University of Arizona | Worobey Lab, Department of Ecology and Evolutionary Biology, University of Arizona | Brendan Larsen et al |
| EPI_ISL_765964 | USA/WY-WPHL-20160842/2020 | 2020-12-07 | North America/USA/Wyoming/ | Wyoming Public Health Laboratory | Wyoming Public Health Laboratory | Noah Hull et al |
| EPI_ISL_765972 | USA/WY-WPHL-20161708/2020 | 2020-12-08 | North America/USA/Wyoming/ | Wyoming Public Health Laboratory | Wyoming Public Health Laboratory | Noah Hull et al |
| EPI_ISL_765976 | USA/WY-WPHL-20162212/2020 | 2020-12-09 | North America/USA/Wyoming/ | Wyoming Public Health Laboratory | Wyoming Public Health Laboratory | Noah Hull et al |
| EPI_ISL_765986 | USA/MA-MGH-03584/2020 | 2020-12-15 | North America/USA/Massachusetts/ | Massachusetts General Hospital | Infectious Disease Program, Broad Institute of Harvard and MIT | Lemieux et al |
| EPI_ISL_765989 | USA/CA-LACPHL-AF00041/2020 | 2020-12-20 | North America/USA/California/Los Ange | UCLA Clinical Micro Lab | Los Angeles County PHL | P. Hemarajata et al. et al |
| EPI_ISL_765994 | USA/CA-LACPHL-AE00055/2020 | 2020-07-07 | North America/USA/California/Los Ange | USC Clinical Lab | Los Angeles County PHL | P. Hemarajata et al. et al |
| EPI_ISL_766002 | USA/CA-LACPHL-AF00065/2020 | 2020-12-21 | North America/USA/California/Los Ange | Los Angeles County PHL | Los Angeles County PHL | P. Hemarajata et al. et al |
| EPI_ISL_766012 | USA/CA-LACPHL-AF00075/2020 | 2020-12-21 | North America/USA/California/Los Ange | Los Angeles County PHL | Los Angeles County PHL | P. Hemarajata et al. et al |
| EPI_ISL_766028 | Germany/NI-IOV-60201584/2020 | 2020-11-30 | Europe/Germany/Niedersachsen/Hanno | Hannover Medical School, Institute of Virology | Hannover Medical School, Institute of Virology | Lars Steinbrvºck et al |
| EPI_ISL_766033 | Indonesia/NT-EIJK14/2020 | 2020-03-28 | Asia/Indonesia/East Nusa Tenggara/Kup | RSUD Prof. DR. W. Z. Johannes Kupang | Eijkman Institute for Molecular Biology, Ministry of Research and Technology/National Agency for Research and Innovation | Frilasita A Yudhaputri et al |

| | | | | | | |
|---|---|---|---|---|---|---|
| EPI_ISL_766037 | Indonesia/JK-EIJK22/2020 | 2020-04-13 | Asia/Indonesia/Jakarta/ | Puskesmas Palmerah | Eijkman Institute for Molecular Biology, Ministry of Research and Technology/National Agency for Research and Innovation | Frilasita A Yudhaputri et al |
| EPI_ISL_766040 | Indonesia/JK-EIJK28/2020 | 2020-04-17 | Asia/Indonesia/Jakarta/ | RS Tingkat II Moh. Ridwan Meuraksa | Eijkman Institute for Molecular Biology, Ministry of Research and Technology/National Agency for Research and Innovation | Frilasita A Yudhaputri et al |
| EPI_ISL_766044 | Indonesia/JK-EIJK33/2020 | 2020-04-23 | Asia/Indonesia/Jakarta/ | RSUP Fatmawati | Eijkman Institute for Molecular Biology, Ministry of Research and Technology/National Agency for Research and Innovation | Frilasita A Yudhaputri et al |
| EPI_ISL_766046 | Indonesia/JK-EIJK37/2020 | 2020-05-10 | Asia/Indonesia/Jakarta/ | RSUP Fatmawati | Eijkman Institute for Molecular Biology, Ministry of Research and Technology/National Agency for Research and Innovation | Frilasita A Yudhaputri et al |
| EPI_ISL_766047 | Indonesia/JK-EIJK44/2020 | 2020-06-04 | Asia/Indonesia/Jakarta/ | RS Kramat 128 | Eijkman Institute for Molecular Biology, Ministry of Research and Technology/National Agency for Research and Innovation | Frilasita A Yudhaputri et al |

| | | | | | | |
|---|---|---|---|---|---|---|
| EPI_ISL_766093 | England/205181915/2020 | 2020-12-17 | Europe/United Kingdom/England/ | Respiratory Virus Unit, National Infection Service, Public Health England | COVID-19 Genomics UK (COG-UK) Consortium | PHE Covid Sequencing Team et al |
| EPI_ISL_766199 | England/205190979/2020 | 2020-12-20 | Europe/United Kingdom/England/ | Respiratory Virus Unit, National Infection Service, Public Health England | COVID-19 Genomics UK (COG-UK) Consortium | PHE Covid Sequencing Team et al |
| EPI_ISL_766221 | England/205360507/2020 | 2020-12-30 | Europe/United Kingdom/England/ | Respiratory Virus Unit, National Infection Service, Public Health England | COVID-19 Genomics UK (COG-UK) Consortium | PHE Covid Sequencing Team et al |
| EPI_ISL_766570 | Oman/89477/2020 | 2020-12-22 | Asia/Oman/Muscat/ | Oman-National Influenza Center | Oman-National Influenza Center | Samiha Al-Kharusi et al |
| EPI_ISL_766571 | Italy/VEN-IZSVe-20RS2155-23_VI/20 | 2020-11-25 | Europe/Italy/Veneto/Vicenza | ULSS 7 Pedemontana - Distretto 1 | Istituto Zooprofilattico Sperimentale delle Venezie | Adelaide Milani et al |
| EPI_ISL_766572 | Italy/VEN-IZSVe-20RS2155-22_VI/20 | 2020-11-24 | Europe/Italy/Veneto/Vicenza | ULSS 7 Pedemontana - Distretto 1 | Istituto Zooprofilattico Sperimentale delle Venezie | Adelaide Milani et al |
| EPI_ISL_766574 | Italy/VEN-IZSVe-20RS2293-1_VI/202 | 2020-12-03 | Europe/Italy/Veneto/Vicenza | ULSS 8 Berica | Istituto Zooprofilattico Sperimentale delle Venezie | Adelaide Milani et al |
| EPI_ISL_766576 | Italy/VEN-IZSVe-20RS2293-4_VI/202 | 2020-12-02 | Europe/Italy/Veneto/Vicenza | ULSS 8 Berica | Istituto Zooprofilattico Sperimentale delle Venezie | Adelaide Milani et al |
| EPI_ISL_766579 | Italy/VEN-IZSVe-20RS2293-7_VI/202 | 2020-12-21 | Europe/Italy/Veneto/Vicenza | ULSS 8 Berica | Istituto Zooprofilattico Sperimentale delle Venezie | Adelaide Milani et al |
| EPI_ISL_766580 | Italy/VEN-IZSVe-20RS2299-1_TV/202 | 2020-12-19 | Europe/Italy/Veneto/Treviso | ULSS 2 Treviso | Istituto Zooprofilattico Sperimentale delle Venezie | Adelaide Milani et al |
| EPI_ISL_766589 | Switzerland/VD-UZH-IMV143/2020 | 2020-12-22 | Europe/Switzerland/Vaud/ | Dr. Boubaker Karim laboratory | Institute of Medical Virology, University of Zurich | Stefan Schmutz et al |

| | | | | | | |
|---|---|---|---|---|---|---|
| EPI_ISL_766599 | Sweden/20-33511/2020 | 2020-12-23 | Europe/Sweden/Dalarna/ | The Public Health Agency of Sweden | The Public Health Agency of Sweden | Department of Microbiology et al |
| EPI_ISL_766606 | Sweden/20-33523/2020 | 2020-12-23 | Europe/Sweden/Dalarna/ | The Public Health Agency of Sweden | The Public Health Agency of Sweden | Department of Microbiology et al |
| EPI_ISL_766614 | Sweden/20-53563/2020 | 2020-11-25 | Europe/Sweden/Ostergotland/ | unknown | The Public Health Agency of Sweden | Department of Microbiology et al |
| EPI_ISL_766615 | Sweden/20-53603/2020 | 2020-10-30 | Europe/Sweden/Vastra Gotaland/ | Klinisk mikrobiologi | The Public Health Agency of Sweden | Department of Microbiology et al |
| EPI_ISL_766616 | Sweden/20-53606/2020 | 2020-10-26 | Europe/Sweden/Vastra Gotaland/ | Klinisk mikrobiologi | The Public Health Agency of Sweden | Department of Microbiology et al |
| EPI_ISL_766618 | Sweden/20-53612/2020 | 2020-11-21 | Europe/Sweden/Vastra Gotaland/ | Klinisk mikrobiologi | The Public Health Agency of Sweden | Department of Microbiology et al |
| EPI_ISL_766619 | Sweden/20-53619/2020 | 2020-10-20 | Europe/Sweden/Vastra Gotaland/ | Klinisk mikrobiologi | The Public Health Agency of Sweden | Department of Microbiology et al |
| EPI_ISL_766620 | Sweden/20-53620/2020 | 2020-11-20 | Europe/Sweden/Vastra Gotaland/ | Klinisk mikrobiologi | The Public Health Agency of Sweden | Department of Microbiology et al |
| EPI_ISL_766623 | Sweden/20-53638/2020 | 2020-11-02 | Europe/Sweden/Vastra Gotaland/ | Klinisk mikrobiologi | The Public Health Agency of Sweden | Department of Microbiology et al |
| EPI_ISL_766624 | Sweden/20-53640/2020 | 2020-11-02 | Europe/Sweden/Vastra Gotaland/ | Klinisk mikrobiologi | The Public Health Agency of Sweden | Department of Microbiology et al |
| EPI_ISL_766628 | Sweden/20-52903/2020 | 2020-09-15 | Europe/Sweden/Stockholm/ | Klinisk mikrobiologi | The Public Health Agency of Sweden | Department of Microbiology et al |
| EPI_ISL_766633 | Sweden/20-53761/2020 | 2020-12-17 | Europe/Sweden/Ostergotland/ | Klinisk mikrobiologi | The Public Health Agency of Sweden | Department of Microbiology et al |
| EPI_ISL_766636 | Sweden/20-53766/2020 | 2020-12-20 | Europe/Sweden/Ostergotland/ | Klinisk mikrobiologi | The Public Health Agency of Sweden | Department of Microbiology et al |
| EPI_ISL_766642 | Sweden/20-24578/2020 | 2020-11-23 | Europe/Sweden/Ostergotland/ | Vardcentralen Brinken | The Public Health Agency of Sweden | Department of Microbiology et al |
| EPI_ISL_766643 | Sweden/20-24736/2020 | 2020-11-25 | Europe/Sweden/Ostergotland/ | Vardcentralen Brinken | The Public Health Agency of Sweden | Department of Microbiology et al |
| EPI_ISL_766670 | USA/TX-DSHS-2054/2020 | 2020-12-03 | North America/USA/Texas/Williamson C | Texas Department of State Health Services | Texas Department of State Health Services | Rashmi Tuladhar et al |
| EPI_ISL_766685 | USA/TX-DSHS-2037/2020 | 2020-11-04 | North America/USA/Texas/Collin County | Texas Department of State Health Services | Texas Department of State Health Services | Rashmi Tuladhar et al |
| EPI_ISL_766687 | USA/TX-DSHS-2028/2020 | 2020-07-23 | North America/USA/Texas/Travis Count | Texas Department of State Health Services | Texas Department of State Health Services | Rashmi Tuladhar et al |

| | | | | | | |
|---|---|---|---|---|---|---|
| EPI_ISL_766689 | USA/TX-DSHS-2050/2020 | 2020-12-03 | North America/USA/Texas/Williamson C | Texas Department of State Health Services | Texas Department of State Health Services | Rashmi Tuladhar et al |
| EPI_ISL_766704 | Sweden/20-33508/2020 | 2020-12-23 | Europe/Sweden/Dalarna/ | The Public Health Agency of Sweden | The Public Health Agency of Sweden | Department of Microbiology et al |
| EPI_ISL_766717 | Sweden/20-13170/2020 | 2020-12-25 | Europe/Sweden/Stockholm/ | A05 Biomedicum | The Public Health Agency of Sweden | Department of Microbiology et al |
| EPI_ISL_766730 | Sweden/20-53925/2020 | 2020-12-22 | Europe/Sweden/Stockholm/ | Klinisk mikrobiologi | The Public Health Agency of Sweden | Department of Microbiology et al |
| EPI_ISL_766735 | USA/TX-DSHS-1978/2020 | 2020-06-14 | North America/USA/Texas/Cameron Cou | Texas Department of State Health Services | Texas Department of State Health Services | Rashmi Tuladhar et al |
| EPI_ISL_766761 | USA/TX-DSHS-1975/2020 | 2020-06-14 | North America/USA/Texas/Cameron Cou | Texas Department of State Health Services | Texas Department of State Health Services | Rashmi Tuladhar et al |
| EPI_ISL_766862 | Algeria/G42452-8847/2020 | 2020-06-24 | Africa/Algeria/Bouira/ | NIC Viral Respiratory Unit - Institut Pasteur of Algeria | National Reference Center for Viruses of Respiratory Infections, Institut Pasteur, Paris | M√©lanie Albert et al |
| EPI_ISL_766863 | Algeria/G37318-8849/2020 | 2020-06-15 | Africa/Algeria/Tipaza/ | NIC Viral Respiratory Unit - Institut Pasteur of Algeria | National Reference Center for Viruses of Respiratory Infections, Institut Pasteur, Paris | M√©lanie Albert et al |
| EPI_ISL_766864 | Algeria/G35155-8850/2020 | 2020-07-15 | Africa/Algeria/Bouira/ | NIC Viral Respiratory Unit - Institut Pasteur of Algeria | National Reference Center for Viruses of Respiratory Infections, Institut Pasteur, Paris | M√©lanie Albert et al |
| EPI_ISL_766866 | Algeria/G38882-8853/2020 | 2020-06-20 | Africa/Algeria/Bouira/ | NIC Viral Respiratory Unit - Institut Pasteur of Algeria | National Reference Center for Viruses of Respiratory Infections, Institut Pasteur, Paris | M√©lanie Albert et al |

| | | | | | | |
|---|---|---|---|---|---|---|
| EPI_ISL_766867 | Algeria/G37138-8854/2020 | 2020-06-18 | Africa/Algeria/Alger/ | NIC Viral Respiratory Unit - Institut Pasteur of Algeria | National Reference Center for Viruses of Respiratory Infections, Institut Pasteur, Paris | M√©lanie Albert et al |
| EPI_ISL_766871 | Algeria/G38599-8859/2020 | 2020-06-20 | Africa/Algeria/Laghouat/ | NIC Viral Respiratory Unit - Institut Pasteur of Algeria | National Reference Center for Viruses of Respiratory Infections, Institut Pasteur, Paris | M√©lanie Albert et al |
| EPI_ISL_766873 | Algeria/G40009-8862/2020 | 2020-06-22 | Africa/Algeria/Bordj-Bou-Arreridj/ | NIC Viral Respiratory Unit - Institut Pasteur of Algeria | National Reference Center for Viruses of Respiratory Infections, Institut Pasteur, Paris | M√©lanie Albert et al |
| EPI_ISL_766875 | Algeria/G37308-8864/2020 | 2020-06-17 | Africa/Algeria/Ouargla/ | NIC Viral Respiratory Unit - Institut Pasteur of Algeria | National Reference Center for Viruses of Respiratory Infections, Institut Pasteur, Paris | M√©lanie Albert et al |
| EPI_ISL_766894 | USA/NM-DOH-2020632229/2020 | 2020-12-20 | North America/USA/New Mexico/ | New Mexico Department of Health Scientific Laboratory | New Mexico Department of Health Scientific Laboratory | D'eldra Malone et al |
| EPI_ISL_766901 | USA/NM-DOH-2020631558/2020 | 2020-12-19 | North America/USA/New Mexico/ | New Mexico Department of Health Scientific Laboratory | New Mexico Department of Health Scientific Laboratory | D'eldra Malone et al |
| EPI_ISL_766902 | USA/NM-DOH-2020631590/2020 | 2020-12-19 | North America/USA/New Mexico/ | New Mexico Department of Health Scientific Laboratory | New Mexico Department of Health Scientific Laboratory | D'eldra Malone et al |
| EPI_ISL_766903 | USA/NM-DOH-2020631598/2020 | 2020-12-19 | North America/USA/New Mexico/ | New Mexico Department of Health Scientific Laboratory | New Mexico Department of Health Scientific Laboratory | D'eldra Malone et al |

| | | | | | | |
|---|---|---|---|---|---|---|
| EPI_ISL_766904 | USA/NM-DOH-2020631623/2020 | 2020-12-19 | North America/USA/New Mexico/ | New Mexico Department of Health Scientific Laboratory | New Mexico Department of Health Scientific Laboratory | D'eldra Malone et al |
| EPI_ISL_766910 | USA/NM-DOH-2020631840/2020 | 2020-12-18 | North America/USA/New Mexico/ | New Mexico Department of Health Scientific Laboratory | New Mexico Department of Health Scientific Laboratory | D'eldra Malone et al |
| EPI_ISL_766920 | USA/NM-DOH-2020632249/2020 | 2020-12-20 | North America/USA/New Mexico/ | New Mexico Department of Health Scientific Laboratory | New Mexico Department of Health Scientific Laboratory | D'eldra Malone et al |
| EPI_ISL_766976 | USA/DE-DHSS-F1018850/2020 | 2020-11-22 | North America/USA/Delaware/Kent Cou | Delaware Public Health Laboratory | Delaware Public Health Laboratory | Gregory Hovan et al |
| EPI_ISL_766987 | USA/DE-DHSS-FLW00407812/2020 | 2020-12-02 | North America/USA/Delaware/Kent Cou | Delaware Public Health Laboratory | Delaware Public Health Laboratory | Gregory Hovan et al |
| EPI_ISL_766992 | USA/DE-DHSS-FLW00407322/2020 | 2020-12-02 | North America/USA/Delaware/Kent Cou | Delaware Public Health Laboratory | Delaware Public Health Laboratory | Gregory Hovan et al |
| EPI_ISL_766995 | USA/DE-DHSS-F1006556/2020 | 2020-11-12 | North America/USA/Delaware/New Cas | Delaware Public Health Laboratory | Delaware Public Health Laboratory | Gregory Hovan et al |
| EPI_ISL_766998 | USA/DE-DHSS-F986562/2020 | 2020-11-09 | North America/USA/Delaware/Kent Cou | Delaware Public Health Laboratory | Delaware Public Health Laboratory | Gregory Hovan et al |
| EPI_ISL_767007 | USA/DE-DHSS-F987154/2020 | 2020-11-30 | North America/USA/Delaware/New Cas | Delaware Public Health Laboratory | Delaware Public Health Laboratory | Gregory Hovan et al |
| EPI_ISL_767010 | USA/DE-DHSS-FLW00407861/2020 | 2020-12-02 | North America/USA/Delaware/New Cas | Delaware Public Health Laboratory | Delaware Public Health Laboratory | Gregory Hovan et al |
| EPI_ISL_767044 | USA/NM-DOH-2020642971/2020 | 2020-12-27 | North America/USA/New Mexico/ | New Mexico Department of Health Scientific Laboratory | New Mexico Department of Health Scientific Laboratory | D'eldra Malone et al |
| EPI_ISL_767045 | USA/NM-DOH-2020640998/2020 | 2020-12-26 | North America/USA/New Mexico/ | New Mexico Department of Health Scientific Laboratory | New Mexico Department of Health Scientific Laboratory | D'eldra Malone et al |
| EPI_ISL_767052 | USA/NM-DOH-2020641432/2020 | 2020-12-26 | North America/USA/New Mexico/ | New Mexico Department of Health Scientific Laboratory | New Mexico Department of Health Scientific Laboratory | D'eldra Malone et al |

| | | | | | | |
|---|---|---|---|---|---|---|
| EPI_ISL_767058 | USA/NM-DOH-2020641094/2020 | 2020-12-26 | North America/USA/New Mexico/ | New Mexico Department of Health Scientific Laboratory | New Mexico Department of Health Scientific Laboratory | D'eldra Malone et al |
| EPI_ISL_767065 | USA/NM-DOH-2020642119/2020 | 2020-12-27 | North America/USA/New Mexico/ | New Mexico Department of Health Scientific Laboratory | New Mexico Department of Health Scientific Laboratory | D'eldra Malone et al |
| EPI_ISL_767072 | USA/NM-DOH-2020642540/2020 | 2020-12-26 | North America/USA/New Mexico/ | New Mexico Department of Health Scientific Laboratory | New Mexico Department of Health Scientific Laboratory | D'eldra Malone et al |
| EPI_ISL_767073 | USA/NM-DOH-2020642973/2020 | 2020-12-27 | North America/USA/New Mexico/ | New Mexico Department of Health Scientific Laboratory | New Mexico Department of Health Scientific Laboratory | D'eldra Malone et al |
| EPI_ISL_767074 | USA/NM-DOH-2020642974/2020 | 2020-12-28 | North America/USA/New Mexico/ | New Mexico Department of Health Scientific Laboratory | New Mexico Department of Health Scientific Laboratory | D'eldra Malone et al |
| EPI_ISL_767080 | USA/NM-DOH-2020643043/2020 | 2020-12-28 | North America/USA/New Mexico/ | New Mexico Department of Health Scientific Laboratory | New Mexico Department of Health Scientific Laboratory | D'eldra Malone et al |
| EPI_ISL_767142 | Wales/ALDP-C47F6E/2020 | 2020-12-12 | Europe/United Kingdom/Wales/ | Lighthouse Lab in Alderley Park | Wellcome Sanger Institute for the COVID-19 Genomics UK (COG-UK) Consortium | Jacquelyn Wynn et al |
| EPI_ISL_767352 | USA/MI-MDHHS-SC22703/2020 | 2020-12-10 | North America/USA/Michigan/ | Michigan Department of Health and Human Services, Bureau of Laboratories | Michigan Department of Health and Human Services, Bureau of Laboratories | Blankenship HM et al |

| | | | | | | |
|---|---|---|---|---|---|---|
| EPI_ISL_767360 | USA/MI-MDHHS-SC22711/2020 | 2020-11-14 | North America/USA/Michigan/ | Michigan Department of Health and Human Services, Bureau of Laboratories | Michigan Department of Health and Human Services, Bureau of Laboratories | Blankenship HM et al |
| EPI_ISL_767417 | USA/NY-Wadsworth-290737-01/2020 | 2020-12-21 | North America/USA/New York/Monroe | URMC LABS | Wadsworth Center, New York State Department.of Health | Kirsten St. George et al |
| EPI_ISL_767428 | USA/NY-Wadsworth-225796-01/2020 | 2020-09-23 | North America/USA/New York/Chemung | Wadsworth Center, New York State Department.of Health | Wadsworth Center, New York State Department.of Health | Kirsten St. George et al |
| EPI_ISL_767431 | USA/NY-Wadsworth-226213-01/2020 | 2020-09-23 | North America/USA/New York/Chemung | Wadsworth Center, New York State Department.of Health | Wadsworth Center, New York State Department.of Health | Kirsten St. George et al |
| EPI_ISL_767432 | USA/NY-Wadsworth-226222-01/2020 | 2020-09-23 | North America/USA/New York/Chemung | Wadsworth Center, New York State Department.of Health | Wadsworth Center, New York State Department.of Health | Kirsten St. George et al |
| EPI_ISL_767435 | USA/NY-Wadsworth-226232-01/2020 | 2020-09-23 | North America/USA/New York/Chemung | Wadsworth Center, New York State Department.of Health | Wadsworth Center, New York State Department.of Health | Kirsten St. George et al |
| EPI_ISL_767441 | USA/NY-Wadsworth-228324-01/2020 | 2020-09-25 | North America/USA/New York/Steuben | Wadsworth Center, New York State Department.of Health | Wadsworth Center, New York State Department.of Health | Kirsten St. George et al |
| EPI_ISL_767442 | USA/NY-Wadsworth-228345-01/2020 | 2020-09-25 | North America/USA/New York/Steuben | Wadsworth Center, New York State Department.of Health | Wadsworth Center, New York State Department.of Health | Kirsten St. George et al |

| | | | | | | |
|---|---|---|---|---|---|---|
| EPI_ISL_767449 | USA/NY-Wadsworth-229230-01/2020 | 2020-09-26 | North America/USA/New York/Steuben | Wadsworth Center, New York State Department.of Health | Wadsworth Center, New York State Department.of Health | Kirsten St. George et al |
| EPI_ISL_767451 | USA/NY-Wadsworth-286666-01/2020 | 2020-12-14 | North America/USA/New York/Warren | Wadsworth Center, New York State Department.of Health | Wadsworth Center, New York State Department.of Health | Kirsten St. George et al |
| EPI_ISL_767453 | USA/NY-Wadsworth-286676-01/2020 | 2020-12-14 | North America/USA/New York/Montgor | Wadsworth Center, New York State Department.of Health | Wadsworth Center, New York State Department.of Health | Kirsten St. George et al |
| EPI_ISL_767454 | USA/NY-Wadsworth-287341-01/2020 | 2020-12-15 | North America/USA/New York/Albany ( | Wadsworth Center, New York State Department.of Health | Wadsworth Center, New York State Department.of Health | Kirsten St. George et al |
| EPI_ISL_767460 | USA/NY-Wadsworth-287417-01/2020 | 2020-12-15 | North America/USA/New York/Columbia | Wadsworth Center, New York State Department.of Health | Wadsworth Center, New York State Department.of Health | Kirsten St. George et al |
| EPI_ISL_767462 | USA/NY-Wadsworth-287495-01/2020 | 2020-12-15 | North America/USA/New York/Albany ( | Wadsworth Center, New York State Department.of Health | Wadsworth Center, New York State Department.of Health | Kirsten St. George et al |
| EPI_ISL_767471 | USA/NY-Wadsworth-287929-01/2020 | 2020-12-15 | North America/USA/New York/Albany ( | Wadsworth Center, New York State Department.of Health | Wadsworth Center, New York State Department.of Health | Kirsten St. George et al |
| EPI_ISL_767474 | USA/NY-Wadsworth-288186-01/2020 | 2020-12-15 | North America/USA/New York/Cattarau | Wadsworth Center, New York State Department.of Health | Wadsworth Center, New York State Department.of Health | Kirsten St. George et al |

| | | | | | | |
|---|---|---|---|---|---|---|
| EPI_ISL_767475 | USA/NY-Wadsworth-288188-01/2020 | 2020-12-15 | North America/USA/New York/Cattarau | Wadsworth Center, New York State Department.of Health | Wadsworth Center, New York State Department.of Health | Kirsten St. George et al |
| EPI_ISL_767484 | USA/NY-Wadsworth-288783-01/2020 | 2020-12-18 | North America/USA/New York/Albany C | Wadsworth Center, New York State Department.of Health | Wadsworth Center, New York State Department.of Health | Kirsten St. George et al |
| EPI_ISL_767487 | USA/NY-Wadsworth-288824-01/2020 | 2020-12-18 | North America/USA/New York/Albany C | Wadsworth Center, New York State Department.of Health | Wadsworth Center, New York State Department.of Health | Kirsten St. George et al |
| EPI_ISL_767499 | USA/NY-Wadsworth-289161-01/2020 | 2020-12-21 | North America/USA/New York/Albany C | Wadsworth Center, New York State Department.of Health | Wadsworth Center, New York State Department.of Health | Kirsten St. George et al |
| EPI_ISL_767501 | USA/NY-Wadsworth-289459-01/2020 | 2020-12-21 | North America/USA/New York/Broome | Wadsworth Center, New York State Department.of Health | Wadsworth Center, New York State Department.of Health | Kirsten St. George et al |
| EPI_ISL_767502 | USA/NY-Wadsworth-289753-01/2020 | 2020-12-21 | North America/USA/New York/Saratoga | Wadsworth Center, New York State Department.of Health | Wadsworth Center, New York State Department.of Health | Kirsten St. George et al |
| EPI_ISL_767515 | USA/NY-Wadsworth-289892-01/2020 | 2020-12-22 | North America/USA/New York/Cattarau | Wadsworth Center, New York State Department.of Health | Wadsworth Center, New York State Department.of Health | Kirsten St. George et al |
| EPI_ISL_767516 | USA/NY-Wadsworth-289895-01/2020 | 2020-12-22 | North America/USA/New York/Cattarau | Wadsworth Center, New York State Department.of Health | Wadsworth Center, New York State Department.of Health | Kirsten St. George et al |

| | | | | | | |
|---|---|---|---|---|---|---|
| EPI_ISL_767522 | USA/NY-Wadsworth-289958-01/2020 | 2020-12-21 | North America/USA/New York/Hamilton | Wadsworth Center, New York State Department.of Health | Wadsworth Center, New York State Department.of Health | Kirsten St. George et al |
| EPI_ISL_767541 | USA/NY-Wadsworth-290708-01/2020 | 2020-12-21 | North America/USA/New York/Ontario | URMC LABS | Wadsworth Center, New York State Department.of Health | Kirsten St. George et al |
| EPI_ISL_767543 | USA/NY-Wadsworth-290710-01/2020 | 2020-12-20 | North America/USA/New York/Monroe | URMC LABS | Wadsworth Center, New York State Department.of Health | Kirsten St. George et al |
| EPI_ISL_767544 | USA/NY-Wadsworth-290711-01/2020 | 2020-12-20 | North America/USA/New York/Chemung | URMC LABS | Wadsworth Center, New York State Department.of Health | Kirsten St. George et al |
| EPI_ISL_767551 | USA/NY-Wadsworth-290723-01/2020 | 2020-12-20 | North America/USA/New York/Monroe | URMC LABS | Wadsworth Center, New York State Department.of Health | Kirsten St. George et al |
| EPI_ISL_767552 | USA/NY-Wadsworth-290725-01/2020 | 2020-12-20 | North America/USA/New York/Ontario | URMC LABS | Wadsworth Center, New York State Department.of Health | Kirsten St. George et al |
| EPI_ISL_767564 | USA/NY-Wadsworth-290742-01/2020 | 2020-12-21 | North America/USA/New York/Allegany | URMC LABS | Wadsworth Center, New York State Department.of Health | Kirsten St. George et al |
| EPI_ISL_767570 | USA/NY-Wadsworth-290751-01/2020 | 2020-12-21 | North America/USA/New York/Steuben | URMC LABS | Wadsworth Center, New York State Department.of Health | Kirsten St. George et al |

| | | | | | | |
|---|---|---|---|---|---|---|
| EPI_ISL_767577 | USA/NY-Wadsworth-290760-01/2020 | 2020-12-21 | North America/USA/New York/Monroe | URMC LABS | Wadsworth Center, New York State Department.of Health | Kirsten St. George et al |
| EPI_ISL_767579 | USA/NY-Wadsworth-290762-01/2020 | 2020-12-21 | North America/USA/New York/Monroe | URMC LABS | Wadsworth Center, New York State Department.of Health | Kirsten St. George et al |
| EPI_ISL_767583 | USA/NY-Wadsworth-292244-01/2020 | 2020-12-01 | North America/USA/New York/Kings Co | MEMORIAL SLOAN KETTERING CANCER CENTER | Wadsworth Center, New York State Department.of Health | Kirsten St. George et al |
| EPI_ISL_767586 | USA/NY-Wadsworth-291993-01/2020 | 2020-12-20 | North America/USA/New York/Westche | WHITE PLAINS HOSPITAL CENTER LABORATORY | Wadsworth Center, New York State Department.of Health | Kirsten St. George et al |
| EPI_ISL_767597 | USA/NY-Wadsworth-292287-01/2020 | 2020-12-02 | North America/USA/New York/Suffolk C | MEMORIAL SLOAN KETTERING CANCER CENTER | Wadsworth Center, New York State Department.of Health | Kirsten St. George et al |
| EPI_ISL_767599 | USA/NY-Wadsworth-291670-01/2020 | 2020-12-22 | North America/USA/New York/Saratoga | SARATOGA HOSPITAL LABORATORY | Wadsworth Center, New York State Department.of Health | Kirsten St. George et al |
| EPI_ISL_767607 | USA/NY-Wadsworth-291743-01/2020 | 2020-12-25 | North America/USA/New York/Saratoga | SARATOGA HOSPITAL LABORATORY | Wadsworth Center, New York State Department.of Health | Kirsten St. George et al |
| EPI_ISL_767630 | USA/NY-Wadsworth-292248-01/2020 | 2020-12-01 | North America/USA/New York/Suffolk C | MEMORIAL SLOAN KETTERING CANCER CENTER | Wadsworth Center, New York State Department.of Health | Kirsten St. George et al |

| | | | | | | |
|---|---|---|---|---|---|---|
| EPI_ISL_767677 | USA/NY-Wadsworth-292400-01/2020 | 2020-12-21 | North America/USA/New York/Westche | BIO-REFERENCE LABORATORIES | Wadsworth Center, New York State Department.of Health | Kirsten St. George et al |
| EPI_ISL_767683 | USA/NY-Wadsworth-292445-01/2020 | 2020-12-04 | North America/USA/New York/Richmon | MEMORIAL SLOAN KETTERING CANCER CENTER | Wadsworth Center, New York State Department.of Health | Kirsten St. George et al |
| EPI_ISL_767684 | USA/NY-Wadsworth-292446-01/2020 | 2020-12-04 | North America/USA/New York/Kings Co | MEMORIAL SLOAN KETTERING CANCER CENTER | Wadsworth Center, New York State Department.of Health | Kirsten St. George et al |
| EPI_ISL_767687 | USA/NY-Wadsworth-292470-01/2020 | 2020-12-06 | North America/USA/New York/Suffolk C | MEMORIAL SLOAN KETTERING CANCER CENTER | Wadsworth Center, New York State Department.of Health | Kirsten St. George et al |
| EPI_ISL_767694 | USA/NY-Wadsworth-292649-01/2020 | 2020-12-07 | North America/USA/New York/Suffolk C | MEMORIAL SLOAN KETTERING CANCER CENTER | Wadsworth Center, New York State Department.of Health | Kirsten St. George et al |
| EPI_ISL_767697 | USA/NY-Wadsworth-292655-01/2020 | 2020-12-07 | North America/USA/New York/Westche | MEMORIAL SLOAN KETTERING CANCER CENTER | Wadsworth Center, New York State Department.of Health | Kirsten St. George et al |
| EPI_ISL_767699 | USA/NY-Wadsworth-292657-01/2020 | 2020-12-07 | North America/USA/New York/New Yor | MEMORIAL SLOAN KETTERING CANCER CENTER | Wadsworth Center, New York State Department.of Health | Kirsten St. George et al |
| EPI_ISL_767714 | USA/NY-Wadsworth-292733-01/2020 | 2020-12-06 | North America/USA/New York/Queens | MEMORIAL SLOAN KETTERING CANCER CENTER | Wadsworth Center, New York State Department.of Health | Kirsten St. George et al |

| | | | | | | |
|---|---|---|---|---|---|---|
| EPI_ISL_767725 | Ireland/WW-NVRL-MPA51513/2020 | 2020-04-11 | Europe/Ireland/Wicklow/ | National Virus Reference Laboratory | Irish Coronavirus Sequencing Consortium - Teagasc Moorepark | Calum Walsh et al |
| EPI_ISL_767727 | Ireland/D-NVRL-MPA89258/2020 | 2020-04-08 | Europe/Ireland/Dublin/ | National Virus Reference Laboratory | Irish Coronavirus Sequencing Consortium - Teagasc Moorepark | Calum Walsh et al |
| EPI_ISL_767737 | Ireland/CO-NVRL-MPZ33926/2020 | 2020-04-11 | Europe/Ireland/Cork/ | National Virus Reference Laboratory | Irish Coronavirus Sequencing Consortium - Teagasc Moorepark | Calum Walsh et al |
| EPI_ISL_767738 | Ireland/CW-NVRL-MPA64650/2020 | 2020-04-11 | Europe/Ireland/Carlow/ | National Virus Reference Laboratory | Irish Coronavirus Sequencing Consortium - Teagasc Moorepark | Calum Walsh et al |
| EPI_ISL_767757 | Ireland/D-NVRL-MPG23468/2020 | 2020-04-11 | Europe/Ireland/Dublin/ | National Virus Reference Laboratory | Irish Coronavirus Sequencing Consortium - Teagasc Moorepark | Calum Walsh et al |
| EPI_ISL_767763 | Ireland/D-NVRL-MPW29674/2020 | 2020-04-07 | Europe/Ireland/Dublin/ | National Virus Reference Laboratory | Irish Coronavirus Sequencing Consortium - Teagasc Moorepark | Calum Walsh et al |
| EPI_ISL_767782 | Ireland/D-NVRL-MPL38042/2020 | 2020-04-11 | Europe/Ireland/Dublin/ | National Virus Reference Laboratory | Irish Coronavirus Sequencing Consortium - Teagasc Moorepark | Calum Walsh et al |
| EPI_ISL_767804 | Ireland/KK-NVRL-MPQ56853/2020 | 2020-04-10 | Europe/Ireland/Kilkenny/ | National Virus Reference Laboratory | Irish Coronavirus Sequencing Consortium - Teagasc Moorepark | Calum Walsh et al |

| | | | | | | |
|---|---|---|---|---|---|---|
| EPI_ISL_767811 | Ireland/LK-NVRL-MPW54586/2020 | 2020-05-20 | Europe/Ireland/Limerick/ | National Virus Reference Laboratory | Irish Coronavirus Sequencing Consortium - Teagasc Moorepark | Calum Walsh et al |
| EPI_ISL_767816 | Ireland/MH-NVRL-MPK48258/2020 | 2020-05-20 | Europe/Ireland/Meath/ | National Virus Reference Laboratory | Irish Coronavirus Sequencing Consortium - Teagasc Moorepark | Calum Walsh et al |
| EPI_ISL_767942 | Switzerland/AG-ETHZ-420310/2020 | 2020-12-20 | Europe/Switzerland/Aargau/ | Viollier AG | Department of Biosystems Science and Engineering, ETH Zvºrich | Chaoran Chen et al |
| EPI_ISL_768056 | Switzerland/BL-ETHZ-420207/2020 | 2020-12-19 | Europe/Switzerland/Basel-Land/ | Viollier AG | Department of Biosystems Science and Engineering, ETH Zvºrich | Chaoran Chen et al |
| EPI_ISL_768163 | Switzerland/SH-ETHZ-420382/2020 | 2020-12-21 | Europe/Switzerland/Schaffhausen/ | Viollier AG | Department of Biosystems Science and Engineering, ETH Zvºrich | Chaoran Chen et al |
| EPI_ISL_768248 | Switzerland/ZG-ETHZ-420048/2020 | 2020-12-18 | Europe/Switzerland/Zug/ | Viollier AG | Department of Biosystems Science and Engineering, ETH Zvºrich | Chaoran Chen et al |
| EPI_ISL_768253 | Switzerland/ZH-ETHZ-420172/2020 | 2020-12-18 | Europe/Switzerland/Zvºrich/ | Viollier AG | Department of Biosystems Science and Engineering, ETH Zvºrich | Chaoran Chen et al |
| EPI_ISL_768325 | USA/LA-EVTL1055/2020 | 2020-11-25 | North America/USA/Louisiana/Webster | LSUHS Emerging Viral Threat Laboratory | Microbial Genome Sequencing Center | Jeremy P. Kamil et al |
| EPI_ISL_768336 | USA/LA-EVTL1066/2020 | 2020-11-27 | North America/USA/Louisiana/Sabine Pa | LSUHS Emerging Viral Threat Laboratory | Microbial Genome Sequencing Center | Jeremy P. Kamil et al |

| | | | | | | |
|---|---|---|---|---|---|---|
| EPI_ISL_768339 | USA/LA-EVTL1069/2020 | 2020-11-27 | North America/USA/Louisiana/Sabine Pa | LSUHS Emerging Viral Threat Laboratory | Microbial Genome Sequencing Center | Jeremy P. Kamil et al |
| EPI_ISL_768352 | USA/LA-EVTL1082/2020 | 2020-11-28 | North America/USA/Louisiana/Webster | LSUHS Emerging Viral Threat Laboratory | Microbial Genome Sequencing Center | Jeremy P. Kamil et al |
| EPI_ISL_768354 | USA/LA-EVTL1084/2020 | 2020-11-23 | North America/USA/Louisiana/Webster | LSUHS Emerging Viral Threat Laboratory | Microbial Genome Sequencing Center | Jeremy P. Kamil et al |
| EPI_ISL_768362 | USA/LA-EVTL1092/2020 | 2020-12-01 | North America/USA/Louisiana/Bossier P | LSUHS Emerging Viral Threat Laboratory | Microbial Genome Sequencing Center | Jeremy P. Kamil et al |
| EPI_ISL_768384 | USA/LA-EVTL1114/2020 | 2020-12-04 | North America/USA/Louisiana/Caddo Pa | LSUHS Emerging Viral Threat Laboratory | Microbial Genome Sequencing Center | Jeremy P. Kamil et al |
| EPI_ISL_768389 | USA/LA-EVTL1119/2020 | 2020-12-04 | North America/USA/Louisiana/Bossier P | LSUHS Emerging Viral Threat Laboratory | Microbial Genome Sequencing Center | Jeremy P. Kamil et al |
| EPI_ISL_768390 | USA/LA-EVTL1120/2020 | 2020-12-04 | North America/USA/Louisiana/Bossier P | LSUHS Emerging Viral Threat Laboratory | Microbial Genome Sequencing Center | Jeremy P. Kamil et al |
| EPI_ISL_768391 | USA/LA-EVTL1121/2020 | 2020-12-04 | North America/USA/Louisiana/Caddo Pa | LSUHS Emerging Viral Threat Laboratory | Microbial Genome Sequencing Center | Jeremy P. Kamil et al |
| EPI_ISL_768404 | USA/LA-EVTL1135/2020 | 2020-12-07 | North America/USA/Louisiana/Caddo Pa | LSUHS Emerging Viral Threat Laboratory | Microbial Genome Sequencing Center | Jeremy P. Kamil et al |
| EPI_ISL_768406 | USA/LA-EVTL1137/2020 | 2020-12-07 | North America/USA/Louisiana/Caddo Pa | LSUHS Emerging Viral Threat Laboratory | Microbial Genome Sequencing Center | Jeremy P. Kamil et al |
| EPI_ISL_768412 | USA/LA-EVTL1143/2020 | 2020-12-08 | North America/USA/Louisiana/Caddo Pa | LSUHS Emerging Viral Threat Laboratory | Microbial Genome Sequencing Center | Jeremy P. Kamil et al |
| EPI_ISL_768422 | USA/LA-EVTL1153/2020 | 2020-12-10 | North America/USA/Louisiana/Bossier P | LSUHS Emerging Viral Threat Laboratory | Microbial Genome Sequencing Center | Jeremy P. Kamil et al |
| EPI_ISL_768425 | USA/LA-EVTL1156/2020 | 2020-12-10 | North America/USA/Louisiana/Webster | LSUHS Emerging Viral Threat Laboratory | Microbial Genome Sequencing Center | Jeremy P. Kamil et al |

| | | | | | | |
|---|---|---|---|---|---|---|
| EPI_ISL_768428 | USA/LA-EVTL1159/2020 | 2020-12-10 | North America/USA/Louisiana/Webster | LSUHS Emerging Viral Threat Laboratory | Microbial Genome Sequencing Center | Jeremy P. Kamil et al |
| EPI_ISL_768436 | USA/LA-EVTL1167/2020 | 2020-12-10 | North America/USA/Louisiana/Caddo Pa | LSUHS Emerging Viral Threat Laboratory | Microbial Genome Sequencing Center | Jeremy P. Kamil et al |
| EPI_ISL_768438 | USA/LA-EVTL1169/2020 | 2020-12-10 | North America/USA/Louisiana/Caddo Pa | LSUHS Emerging Viral Threat Laboratory | Microbial Genome Sequencing Center | Jeremy P. Kamil et al |
| EPI_ISL_768440 | USA/LA-EVTL1171/2020 | 2020-12-11 | North America/USA/Louisiana/Bienville | LSUHS Emerging Viral Threat Laboratory | Microbial Genome Sequencing Center | Jeremy P. Kamil et al |
| EPI_ISL_768446 | USA/LA-EVTL1177/2020 | 2020-12-11 | North America/USA/Louisiana/Bossier P | LSUHS Emerging Viral Threat Laboratory | Microbial Genome Sequencing Center | Jeremy P. Kamil et al |
| EPI_ISL_768450 | USA/LA-EVTL1181/2020 | 2020-12-11 | North America/USA/Louisiana/Bossier P | LSUHS Emerging Viral Threat Laboratory | Microbial Genome Sequencing Center | Jeremy P. Kamil et al |
| EPI_ISL_768458 | USA/LA-EVTL1189/2020 | 2020-12-11 | North America/USA/Louisiana/Bienville | LSUHS Emerging Viral Threat Laboratory | Microbial Genome Sequencing Center | Jeremy P. Kamil et al |
| EPI_ISL_768468 | USA/LA-EVTL1199/2020 | 2020-12-11 | North America/USA/Louisiana/Caddo Pa | LSUHS Emerging Viral Threat Laboratory | Microbial Genome Sequencing Center | Jeremy P. Kamil et al |
| EPI_ISL_768475 | USA/LA-EVTL1206/2020 | 2020-12-14 | North America/USA/Louisiana/DeSoto P | LSUHS Emerging Viral Threat Laboratory | Microbial Genome Sequencing Center | Jeremy P. Kamil et al |
| EPI_ISL_768484 | USA/LA-EVTL1215/2020 | 2020-12-15 | North America/USA/Louisiana/Caddo Pa | LSUHS Emerging Viral Threat Laboratory | Microbial Genome Sequencing Center | Jeremy P. Kamil et al |
| EPI_ISL_768494 | USA/LA-EVTL1225/2020 | 2020-12-15 | North America/USA/Louisiana/Caddo Pa | LSUHS Emerging Viral Threat Laboratory | Microbial Genome Sequencing Center | Jeremy P. Kamil et al |
| EPI_ISL_768498 | USA/LA-EVTL1229/2020 | 2020-12-15 | North America/USA/Louisiana/Sabine Pa | LSUHS Emerging Viral Threat Laboratory | Microbial Genome Sequencing Center | Jennifer L. Carroll et al |
| EPI_ISL_768504 | USA/LA-EVTL1235/2020 | 2020-12-17 | North America/USA/Louisiana/Claiborne | LSUHS Emerging Viral Threat Laboratory | Microbial Genome Sequencing Center | Jennifer L. Carroll et al |

| | | | | | | |
|---|---|---|---|---|---|---|
| EPI_ISL_768537 | Thailand/SamutSakorn_1184-19/202 | 2020-12-19 | Asia/Thailand/Samut Sakhon/ | Regional Medical Sciences Center 5 Samut Songkhram | National Institute of Health, Department of Medical Sciences, Ministry of Public Health, Thailand | Pilailuk Okada et al |
| EPI_ISL_768688 | Japan/IC-0479/2020 | 20-Dec | Asia/Japan/Japan/ | Pathogen Genomics Center, National Institute of Infectious Diseases | Pathogen Genomics Center, National Institute of Infectious Diseases | Tsuyoshi Sekizuka et al |
| EPI_ISL_768709 | Japan/IC-0500/2020 | 20-Dec | Asia/Japan/Japan/ | Pathogen Genomics Center, National Institute of Infectious Diseases | Pathogen Genomics Center, National Institute of Infectious Diseases | Tsuyoshi Sekizuka et al |
| EPI_ISL_768726 | Germany/TH-IIMK-CaSe-31/2020 | 2020-11-02 | Europe/Germany/Thuringia/ | Jena University Hospital, Institute for Infectious Diseases and Infection Control | Institute of infectious medicine & hospital hygiene, CaSe-Group | Spott et al |
| EPI_ISL_768727 | Germany/TH-IIMK-CaSe-32/2020 | 2020-11-16 | Europe/Germany/Thuringia/ | Jena University Hospital, Institute for Infectious Diseases and Infection Control | Institute of infectious medicine & hospital hygiene, CaSe-Group | Spott et al |
| EPI_ISL_768728 | Bangladesh/CHRF-0032/2020 | 2020-06-09 | Asia/Bangladesh/Dhaka/ | Child Health Research Foundation | Child Health Research Foundation | Senjuti Saha et al |
| EPI_ISL_768742 | Bangladesh/CHRF-0077/2020 | 2020-08-29 | Asia/Bangladesh/Dhaka/ | Child Health Research Foundation | Child Health Research Foundation | Senjuti Saha et al |
| EPI_ISL_768763 | Ireland/D-NVRL-AIIDV0432v2-2/2020 | 2020-05-18 | Europe/Ireland/Dublin/ | AIID | Irish Coronavirus Sequencing Consortium - National Virus Reference Laboratory | Michael Carr et al |

| | | | | | | |
|---|---|---|---|---|---|---|
| EPI_ISL_768780 | Ireland/D-NVRL-AIIDV1239v1/2020 | 2020-10-20 | Europe/Ireland/Dublin/ | AIID | Irish Coronavirus Sequencing Consortium - National Virus Reference Laboratory | Michael Carr et al |
| EPI_ISL_768799 | Lebanon/LAU-uk3/2020 | 2020-12-21 | Asia/Lebanon/Beirut/ | MoPH (Lebanese ministry of public health) | Microbial Genomics lab LAU | Hamad Hassan et al |
| EPI_ISL_768823 | France/NAQ-SC768/2020 | 2020-12-24 | Europe/France/Nouvelle-Aquitaine/Borc | CHU Bordeaux | CNR Virus des Infections Respiratoires - France SUD | Antonin Bal et al |
| EPI_ISL_768824 | France/NAQ-SC785/2020 | 2020-12-28 | Europe/France/Nouvelle-Aquitaine/La B | Laboratoire BIOLAB33 | CNR Virus des Infections Respiratoires - France SUD | Antonin Bal et al |
| EPI_ISL_768827 | France/PAC-NRC2929/2020 | 2020-12-22 | Europe/France/Provence-Alpes-CV¥te d' | CNR Virus des Infections Respiratoires - France SUD | CNR Virus des Infections Respiratoires - France SUD | Antonin Bal et al |
| EPI_ISL_768828 | France/PAC-NRC2933/2020 | 2020-12-22 | Europe/France/Provence-Alpes-CV¥te d' | CNR Virus des Infections Respiratoires - France SUD | CNR Virus des Infections Respiratoires - France SUD | Antonin Bal et al |
| EPI_ISL_768840 | Morocco/FMP-41/2020 | 2020-12-17 | Africa/Morocco/Rabat/ | Laboratoire Biolife | Laboratoire de Biotechnologie | Mouna Ouadghiri et al |
| EPI_ISL_769158 | England/CAMC-C738AA/2020 | 2020-12-15 | Europe/United Kingdom/England/ | Lighthouse Lab in Cambridge | Wellcome Sanger Institute for the COVID-19 Genomics UK (COG-UK) Consortium | Rob Howes et al |
| EPI_ISL_769471 | England/QEUH-C5BF94/2020 | 2020-12-14 | Europe/United Kingdom/England/ | Lighthouse Lab in Glasgow | Wellcome Sanger Institute for the COVID-19 Genomics UK (COG-UK) Consortium | Harper VanSteenhouse et al |

| | | | | | | |
|---|---|---|---|---|---|---|
| EPI_ISL_769670 | England/MILK-C4FE96/2020 | 2020-12-11 | Europe/United Kingdom/England/ | Lighthouse Lab in Milton Keynes | Wellcome Sanger Institute for the COVID-19 Genomics UK (COG-UK) Consortium | The Lighthouse Lab in Milton Keynes et al |
| EPI_ISL_769707 | England/MILK-C51866/2020 | 2020-12-11 | Europe/United Kingdom/England/ | Lighthouse Lab in Milton Keynes | Wellcome Sanger Institute for the COVID-19 Genomics UK (COG-UK) Consortium | The Lighthouse Lab in Milton Keynes et al |
| EPI_ISL_769812 | England/MILK-C5156F/2020 | 2020-12-11 | Europe/United Kingdom/England/ | Lighthouse Lab in Milton Keynes | Wellcome Sanger Institute for the COVID-19 Genomics UK (COG-UK) Consortium | The Lighthouse Lab in Milton Keynes et al |
| EPI_ISL_769863 | Morocco/FMP-42/2020 | 2020-12-14 | Africa/Morocco/Casablanca/ | Laboratoire Biolife | Laboratoire de Biotechnologie | Mouna Ouadghiri et al |
| EPI_ISL_769881 | USA/WY-WPHL-20063101/2020 | 2020-08-17 | North America/USA/Wyoming/ | Wyoming Public Health Laboratory | Wyoming Public Health Laboratory | Noah Hull et al |
| EPI_ISL_769885 | USA/WY-WPHL-20062989/2020 | 2020-08-18 | North America/USA/Wyoming/ | Wyoming Public Health Laboratory | Wyoming Public Health Laboratory | Noah Hull et al |
| EPI_ISL_769887 | USA/WY-WPHL-20062881/2020 | 2020-08-17 | North America/USA/Wyoming/ | Wyoming Public Health Laboratory | Wyoming Public Health Laboratory | Noah Hull et al |
| EPI_ISL_769888 | USA/WY-WPHL-20062878/2020 | 2020-08-17 | North America/USA/Wyoming/ | Wyoming Public Health Laboratory | Wyoming Public Health Laboratory | Noah Hull et al |
| EPI_ISL_769889 | USA/WY-WPHL-20062877/2020 | 2020-08-17 | North America/USA/Wyoming/ | Wyoming Public Health Laboratory | Wyoming Public Health Laboratory | Noah Hull et al |
| EPI_ISL_769890 | USA/WY-WPHL-20062765/2020 | 2020-08-17 | North America/USA/Wyoming/ | Wyoming Public Health Laboratory | Wyoming Public Health Laboratory | Noah Hull et al |
| EPI_ISL_769896 | USA/WY-WPHL-20062180/2020 | 2020-08-17 | North America/USA/Wyoming/ | Wyoming Public Health Laboratory | Wyoming Public Health Laboratory | Noah Hull et al |
| EPI_ISL_769904 | USA/WY-WPHL-20061820/2020 | 2020-08-14 | North America/USA/Wyoming/ | Wyoming Public Health Laboratory | Wyoming Public Health Laboratory | Noah Hull et al |
| EPI_ISL_769909 | USA/WY-WPHL-20044665/2020 | 2020-07-16 | North America/USA/Wyoming/ | Wyoming Public Health Laboratory | Wyoming Public Health Laboratory | Noah Hull et al |
| EPI_ISL_769913 | USA/WY-WPHL-20044542/2020 | 2020-07-16 | North America/USA/Wyoming/ | Wyoming Public Health Laboratory | Wyoming Public Health Laboratory | Noah Hull et al |

| | | | | | | |
|---|---|---|---|---|---|---|
| EPI_ISL_769920 | USA/NY-Wadsworth-292759-01/2020 | 2020-12-18 | North America/USA/New York/Albany ( | Albany Medical Center Hospital Clinical Laboratories | Wadsworth Center, New York State Department.of Health | Kirsten St. George et al |
| EPI_ISL_769921 | USA/NY-Wadsworth-292810-01/2020 | 2020-12-26 | North America/USA/New York/Albany ( | Albany Medical Center Hospital Clinical Laboratories | Wadsworth Center, New York State Department.of Health | Kirsten St. George et al |
| EPI_ISL_769922 | USA/NY-Wadsworth-292738-01/2020 | 2020-12-21 | North America/USA/New York/Greene ( | Albany Medical Center Hospital Clinical Laboratories | Wadsworth Center, New York State Department.of Health | Kirsten St. George et al |
| EPI_ISL_769923 | USA/NY-Wadsworth-292744-01/2020 | 2020-12-21 | North America/USA/New York/Columbia | Albany Medical Center Hospital Clinical Laboratories | Wadsworth Center, New York State Department.of Health | Kirsten St. George et al |
| EPI_ISL_769926 | USA/NY-Wadsworth-292748-01/2020 | 2020-12-20 | North America/USA/New York/Dutchess | Albany Medical Center Hospital Clinical Laboratories | Wadsworth Center, New York State Department.of Health | Kirsten St. George et al |
| EPI_ISL_769938 | USA/NY-Wadsworth-292762-01/2020 | 2020-12-26 | North America/USA/New York/Columbia | Albany Medical Center Hospital Clinical Laboratories | Wadsworth Center, New York State Department.of Health | Kirsten St. George et al |
| EPI_ISL_769943 | USA/NY-Wadsworth-292771-01/2020 | 2020-12-26 | North America/USA/New York/Columbia | Albany Medical Center Hospital Clinical Laboratories | Wadsworth Center, New York State Department.of Health | Kirsten St. George et al |
| EPI_ISL_769945 | USA/NY-Wadsworth-292774-01/2020 | 2020-12-26 | North America/USA/New York/Columbia | Albany Medical Center Hospital Clinical Laboratories | Wadsworth Center, New York State Department.of Health | Kirsten St. George et al |

| | | | | | | |
|---|---|---|---|---|---|---|
| EPI_ISL_769987 | CostaRica/INC-0162/2020 | 2020-10-13 | North America/Costa Rica/Heredia/San | Laboratorio Clinico Labin | Inciensa, Instituto Costarricense de Investigaci√≥n y Ense√±anza en Nutrici√≥n y Salud | Francisco Duarte et al |
| EPI_ISL_769990 | CostaRica/INC-0144/2020 | 2020-09-08 | North America/Costa Rica/Alajuela/Gre | Hospital Dr. Rafael A. Calderon Guardia | Inciensa, Instituto Costarricense de Investigaci√≥n y Ense√±anza en Nutrici√≥n y Salud | Francisco Duarte et al |
| EPI_ISL_769991 | CostaRica/INC-0148/2020 | 2020-09-15 | North America/Costa Rica/Alajuela/Alaj | Laboratorio Clinico Labin | Inciensa, Instituto Costarricense de Investigaci√≥n y Ense√±anza en Nutrici√≥n y Salud | Francisco Duarte et al |
| EPI_ISL_769992 | CostaRica/INC-0155/2020 | 2020-09-30 | North America/Costa Rica/Alajuela/San | Hospital Metropolitano | Inciensa, Instituto Costarricense de Investigaci√≥n y Ense√±anza en Nutrici√≥n y Salud | Francisco Duarte et al |
| EPI_ISL_769993 | CostaRica/INC-0143/2020 | 2020-09-05 | North America/Costa Rica/Cartago CR/ | Area De Salud El Guarco | Inciensa, Instituto Costarricense de Investigaci√≥n y Ense√±anza en Nutrici√≥n y Salud | Francisco Duarte et al |
| EPI_ISL_769994 | CostaRica/INC-0146/2020 | 2020-09-10 | North America/Costa Rica/Cartago CR/C | Area De Salud Catedral Noreste | Inciensa, Instituto Costarricense de Investigaci√≥n y Ense√±anza en Nutrici√≥n y Salud | Francisco Duarte et al |
| EPI_ISL_769995 | CostaRica/INC-0154/2020 | 2020-09-30 | North America/Costa Rica/Cartago CR/E | Hospital Metropolitano | Inciensa, Instituto Costarricense de Investigaci√≥n y Ense√±anza en Nutrici√≥n y Salud | Francisco Duarte et al |
| EPI_ISL_769996 | CostaRica/INC-0163/2020 | 2020-10-14 | North America/Costa Rica/Cartago CR/C | Area De Salud San Juan-San Diego-Concepcion 2 | Inciensa, Instituto Costarricense de Investigaci√≥n y Ense√±anza en Nutrici√≥n y Salud | Francisco Duarte et al |

| | | | | | | |
|---|---|---|---|---|---|---|
| EPI_ISL_770000 | CostaRica/INC-0158/2020 | 2020-10-05 | North America/Costa Rica/Guanacaste/L | Area De Salud La Cruz | Inciensa, Instituto Costarricense de Investigaci√≥n y Ense√±anza en Nutrici√≥n y Salud | Francisco Duarte et al |
| EPI_ISL_770001 | CostaRica/INC-0160/2020 | 2020-10-07 | North America/Costa Rica/Guanacaste/L | Area De Salud La Cruz | Inciensa, Instituto Costarricense de Investigaci√≥n y Ense√±anza en Nutrici√≥n y Salud | Francisco Duarte et al |
| EPI_ISL_770002 | CostaRica/INC-0161/2020 | 2020-10-09 | North America/Costa Rica/Guanacaste/L | Area De Salud La Cruz | Inciensa, Instituto Costarricense de Investigaci√≥n y Ense√±anza en Nutrici√≥n y Salud | Francisco Duarte et al |
| EPI_ISL_770003 | CostaRica/INC-0173/2020 | 2020-11-04 | North America/Costa Rica/Guanacaste/L | Area De Salud La Cruz | Inciensa, Instituto Costarricense de Investigaci√≥n y Ense√±anza en Nutrici√≥n y Salud | Francisco Duarte et al |
| EPI_ISL_770004 | CostaRica/INC-0159/2020 | 2020-10-06 | North America/Costa Rica/Heredia/San | Clinica Biblica | Inciensa, Instituto Costarricense de Investigaci√≥n y Ense√±anza en Nutrici√≥n y Salud | Francisco Duarte et al |
| EPI_ISL_770005 | CostaRica/INC-0185/2020 | 2020-11-25 | North America/Costa Rica/Heredia/Para | Area De Salud Moravia | Inciensa, Instituto Costarricense de Investigaci√≥n y Ense√±anza en Nutrici√≥n y Salud | Francisco Duarte et al |
| EPI_ISL_770006 | CostaRica/INC-0142/2020 | 2020-09-05 | North America/Costa Rica/Puntarenas/P | Hle - Asociacion Hogar De Ancianos De Palmar Sur De Osa | Inciensa, Instituto Costarricense de Investigaci√≥n y Ense√±anza en Nutrici√≥n y Salud | Francisco Duarte et al |
| EPI_ISL_770007 | CostaRica/INC-0157/2020 | 2020-09-30 | North America/Costa Rica/Puntarenas/P | Area De Salud San Rafael | Inciensa, Instituto Costarricense de Investigaci√≥n y Ense√±anza en Nutrici√≥n y Salud | Francisco Duarte et al |

| | | | | | | |
|---|---|---|---|---|---|---|
| EPI_ISL_770008 | CostaRica/INC-0168/2020 | 2020-10-21 | North America/Costa Rica/Puntarenas/L | Area De Salud Corredores | Inciensa, Instituto Costarricense de Investigaci√≥n y Ense√±anza en Nutrici√≥n y Salud | Francisco Duarte et al |
| EPI_ISL_770009 | CostaRica/INC-0140/2020 | 2020-07-21 | North America/Costa Rica/San Jose/San | Area De Salud Escazu (Coopesana) | Inciensa, Instituto Costarricense de Investigaci√≥n y Ense√±anza en Nutrici√≥n y Salud | Francisco Duarte et al |
| EPI_ISL_770010 | CostaRica/INC-0141/2020 | 2020-09-03 | North America/Costa Rica/San Jose/Pav | Area De Salud Escazu (Coopesana) | Inciensa, Instituto Costarricense de Investigaci√≥n y Ense√±anza en Nutrici√≥n y Salud | Francisco Duarte et al |
| EPI_ISL_770011 | CostaRica/INC-0145/2020 | 2020-09-07 | North America/Costa Rica/San Jose/Pat | Area De Salud Coronado | Inciensa, Instituto Costarricense de Investigaci√≥n y Ense√±anza en Nutrici√≥n y Salud | Francisco Duarte et al |
| EPI_ISL_770012 | CostaRica/INC-0149/2020 | 2020-09-19 | North America/Costa Rica/San Jose/Pat | Area De Salud Coronado | Inciensa, Instituto Costarricense de Investigaci√≥n y Ense√±anza en Nutrici√≥n y Salud | Francisco Duarte et al |
| EPI_ISL_770013 | CostaRica/INC-0150/2020 | 2020-09-22 | North America/Costa Rica/San Jose/Pav | Hle - Asilos De Ancianos | Inciensa, Instituto Costarricense de Investigaci√≥n y Ense√±anza en Nutrici√≥n y Salud | Francisco Duarte et al |
| EPI_ISL_770014 | CostaRica/INC-0151/2020 | 2020-09-23 | North America/Costa Rica/San Jose/Tirr | Area De Salud Curridabat 2 | Inciensa, Instituto Costarricense de Investigaci√≥n y Ense√±anza en Nutrici√≥n y Salud | Francisco Duarte et al |
| EPI_ISL_770015 | CostaRica/INC-0164/2020 | 2020-10-16 | North America/Costa Rica/San Jose/San | Area De Salud San Francisco-San Antonio (Coopesana) | Inciensa, Instituto Costarricense de Investigaci√≥n y Ense√±anza en Nutrici√≥n y Salud | Francisco Duarte et al |

| | | | | | | |
|---|---|---|---|---|---|---|
| EPI_ISL_770016 | CostaRica/INC-0166/2020 | 2020-10-19 | North America/Costa Rica/San Jose/Puri | Area De Salud Goicoechea 2 - Clinica Dr. Jimenez Nuv±ez | Inciensa, Instituto Costarricense de Investigaciv≥n y Ensev±anza en Nutriciv≥n y Salud | Francisco Duarte et al |
| EPI_ISL_770017 | CostaRica/INC-0167/2020 | 2020-10-21 | North America/Costa Rica/San Jose/Pere | Area De Salud Perez Zeledon | Inciensa, Instituto Costarricense de Investigaciv≥n y Ensev±anza en Nutriciv≥n y Salud | Francisco Duarte et al |
| EPI_ISL_770018 | CostaRica/INC-0169/2020 | 2020-10-27 | North America/Costa Rica/San Jose/Llor | Laboratorio Cenahce | Inciensa, Instituto Costarricense de Investigaciv≥n y Ensev±anza en Nutriciv≥n y Salud | Francisco Duarte et al |
| EPI_ISL_770023 | CostaRica/INC-0184/2020 | 2020-11-24 | North America/Costa Rica/San Jose/San | Area De Salud San Francisco-San Antonio (Coopesana) | Inciensa, Instituto Costarricense de Investigaciv≥n y Ensev±anza en Nutriciv≥n y Salud | Francisco Duarte et al |
| EPI_ISL_770026 | CostaRica/INC-0180/2020 | 2020-11-17 | North America/Costa Rica/Cartago CR/S | Area De Salud Catedral Noreste | Inciensa, Instituto Costarricense de Investigaciv≥n y Ensev±anza en Nutriciv≥n y Salud | Francisco Duarte et al |
| EPI_ISL_770027 | CostaRica/INC-0156/2020 | 2020-10-01 | North America/Costa Rica/Heredia/Mer | Area De Salud La Cruz | Inciensa, Instituto Costarricense de Investigaciv≥n y Ensev±anza en Nutriciv≥n y Salud | Francisco Duarte et al |
| EPI_ISL_770028 | CostaRica/INC-0147/2020 | 2020-09-17 | North America/Costa Rica/Limon/Limon | Hospital Dr. Raul Blanco Cervantes | Inciensa, Instituto Costarricense de Investigaciv≥n y Ensev±anza en Nutriciv≥n y Salud | Francisco Duarte et al |
| EPI_ISL_770030 | CostaRica/INC-0175/2020 | 2020-11-05 | North America/Costa Rica/Puntarenas/B | Area De Salud Buenos Aires | Inciensa, Instituto Costarricense de Investigaciv≥n y Ensev±anza en Nutriciv≥n y Salud | Francisco Duarte et al |

| | | | | | | |
|---|---|---|---|---|---|---|
| EPI_ISL_770031 | CostaRica/INC-0152/2020 | 2020-09-26 | North America/Costa Rica/San Jose/San | Hle-Asociacion De Atencion Integral Del Anciano San Cayetano | Inciensa, Instituto Costarricense de Investigaci√≥n y Ense√±anza en Nutrici√≥n y Salud | Francisco Duarte et al |
| EPI_ISL_770032 | CostaRica/INC-0153/2020 | 2020-09-30 | North America/Costa Rica/San Jose/San | Hle-Asociacion De Atencion Integral Del Anciano San Cayetano | Inciensa, Instituto Costarricense de Investigaci√≥n y Ense√±anza en Nutrici√≥n y Salud | Francisco Duarte et al |
| EPI_ISL_770049 | Latvia/214/2020 | 2020-09-13 | Europe/Latvia/Latvia/ | CentrƒÅlƒÅ Laboratorija | Latvian Biomedical Research and Study Centre | Ivars Silamiƒ∑elis et al |
| EPI_ISL_770072 | USA/WY-WPHL-20063191/2020 | 2020-08-18 | North America/USA/Wyoming/ | Wyoming Public Health Laboratory | Wyoming Public Health Laboratory | Noah Hull et al |
| EPI_ISL_770073 | USA/WY-WPHL-20063192/2020 | 2020-08-18 | North America/USA/Wyoming/ | Wyoming Public Health Laboratory | Wyoming Public Health Laboratory | Noah Hull et al |
| EPI_ISL_770074 | USA/WY-WPHL-20063197/2020 | 2020-08-17 | North America/USA/Wyoming/ | Wyoming Public Health Laboratory | Wyoming Public Health Laboratory | Noah Hull et al |
| EPI_ISL_770078 | USA/WY-WPHL-20063394/2020 | 2020-08-19 | North America/USA/Wyoming/ | Wyoming Public Health Laboratory | Wyoming Public Health Laboratory | Noah Hull et al |
| EPI_ISL_770080 | USA/WY-WPHL-20094746/2020 | 2020-10-11 | North America/USA/Wyoming/ | Wyoming Public Health Laboratory | Wyoming Public Health Laboratory | Noah Hull et al |
| EPI_ISL_770081 | USA/WY-WPHL-20095100/2020 | 2020-10-12 | North America/USA/Wyoming/ | Wyoming Public Health Laboratory | Wyoming Public Health Laboratory | Noah Hull et al |
| EPI_ISL_770086 | USA/WY-WPHL-20148235/2020 | 2020-11-23 | North America/USA/Wyoming/ | Wyoming Public Health Laboratory | Wyoming Public Health Laboratory | Noah Hull et al |
| EPI_ISL_770103 | USA/WY-WPHL-20149786/2020 | 2020-11-24 | North America/USA/Wyoming/ | Wyoming Public Health Laboratory | Wyoming Public Health Laboratory | Noah Hull et al |
| EPI_ISL_770106 | USA/WY-WPHL-20150313/2020 | 2020-11-24 | North America/USA/Wyoming/ | Wyoming Public Health Laboratory | Wyoming Public Health Laboratory | Noah Hull et al |
| EPI_ISL_770107 | USA/WY-WPHL-20150375/2020 | 2020-11-24 | North America/USA/Wyoming/ | Wyoming Public Health Laboratory | Wyoming Public Health Laboratory | Noah Hull et al |
| EPI_ISL_770121 | USA/WY-WPHL-20063090/2020 | 2020-08-17 | North America/USA/Wyoming/ | Wyoming Public Health Laboratory | Wyoming Public Health Laboratory | Noah Hull et al |
| EPI_ISL_770123 | USA/WY-WPHL-20062503/2020 | 2020-08-17 | North America/USA/Wyoming/ | Wyoming Public Health Laboratory | Wyoming Public Health Laboratory | Noah Hull et al |
| EPI_ISL_770453 | USA/WY-WPHL-20171805/2020 | 2020-12-29 | North America/USA/Wyoming/ | Wyoming Public Health Laboratory | Wyoming Public Health Laboratory | Noah Hull et al |

| | | | | | | |
|---|---|---|---|---|---|---|
| EPI_ISL_770457 | USA/WY-WPHL-20171966/2020 | 2020-12-29 | North America/USA/Wyoming/ | Wyoming Public Health Laboratory | Wyoming Public Health Laboratory | Noah Hull et al |
| EPI_ISL_770478 | USA/WY-WPHL-20172451/2020 | 2020-12-30 | North America/USA/Wyoming/ | Wyoming Public Health Laboratory | Wyoming Public Health Laboratory | Noah Hull et al |
| EPI_ISL_770480 | USA/WY-WPHL-20172245/2020 | 2020-12-30 | North America/USA/Wyoming/ | Wyoming Public Health Laboratory | Wyoming Public Health Laboratory | Noah Hull et al |
| EPI_ISL_770485 | USA/WY-WPHL-20172449/2020 | 2020-12-30 | North America/USA/Wyoming/ | Wyoming Public Health Laboratory | Wyoming Public Health Laboratory | Noah Hull et al |
| EPI_ISL_770500 | Lithuania/MR-LUHS-Eilnr234964/202 | 2020-12-17 | Europe/Lithuania/Marijampoles Apskrit | Lithuanian University of Health Sciences Hospital, Department of Laboratory Medicine | Lithuanian University of Health Sciences, Molecular cardiology lab. | Lukas Zemaitis et al |
| EPI_ISL_770524 | Lithuania/MR-LUHS-Eilnr238208/202 | 2020-12-22 | Europe/Lithuania/Kauno Apskritis/ | Lithuanian University of Health Sciences Hospital, Department of Laboratory Medicine | Lithuanian University of Health Sciences, Molecular cardiology lab. | Lukas Zemaitis et al |
| EPI_ISL_770532 | Lithuania/MR-LUHS-Eilnr238418/202 | 2020-12-22 | Europe/Lithuania/Kauno Apskritis/ | Lithuanian University of Health Sciences Hospital, Department of Laboratory Medicine | Lithuanian University of Health Sciences, Molecular cardiology lab. | Lukas Zemaitis et al |
| EPI_ISL_770551 | Brazil/RS-00651/2020 | 2020-11-25 | South America/Brazil/Rio Grande do Sul | Laboratv≥rio de Microbiologia Molecular - Universidade FEEVALE | Bioinformatics Laboratory / LNCC | Felipe Benites et al |
| EPI_ISL_770553 | Brazil/RS-00628/2020 | 2020-11-24 | South America/Brazil/Rio Grande do Sul | Laboratv≥rio de Microbiologia Molecular - Universidade FEEVALE | Bioinformatics Laboratory / LNCC | Felipe Benites et al |
| EPI_ISL_770554 | Brazil/RS-00682/2020 | 2020-11-27 | South America/Brazil/Rio Grande do Sul | Laboratv≥rio de Microbiologia Molecular - Universidade FEEVALE | Bioinformatics Laboratory / LNCC | Felipe Benites et al |
| EPI_ISL_770555 | Brazil/RS-00603/2020 | 2020-11-23 | South America/Brazil/Rio Grande do Sul | Laboratv≥rio de Microbiologia Molecular - Universidade FEEVALE | Bioinformatics Laboratory / LNCC | Felipe Benites et al |
| EPI_ISL_770557 | Brazil/RS-00605/2020 | 2020-11-24 | South America/Brazil/Rio Grande do Sul | Laboratv≥rio de Microbiologia Molecular - Universidade FEEVALE | Bioinformatics Laboratory / LNCC | Felipe Benites et al |
| EPI_ISL_770558 | Brazil/RS-00606/2020 | 2020-11-24 | South America/Brazil/Rio Grande do Sul | Laboratv≥rio de Microbiologia Molecular - Universidade FEEVALE | Bioinformatics Laboratory / LNCC | Felipe Benites et al |
| EPI_ISL_770560 | Brazil/RS-00608/2020 | 2020-11-24 | South America/Brazil/Rio Grande do Sul | Laboratv≥rio de Microbiologia Molecular - Universidade FEEVALE | Bioinformatics Laboratory / LNCC | Felipe Benites et al |
| EPI_ISL_770561 | Brazil/RS-00609/2020 | 2020-11-24 | South America/Brazil/Rio Grande do Sul | Laboratv≥rio de Microbiologia Molecular - Universidade FEEVALE | Bioinformatics Laboratory / LNCC | Felipe Benites et al |

| | | | | | | |
|---|---|---|---|---|---|---|
| EPI_ISL_770562 | Brazil/RS-00610/2020 | 2020-11-24 | South America/Brazil/Rio Grande do Sul | Laboratv≥rio de Microbiologia Molecular - Universidade FEEVALE | Bioinformatics Laboratory / LNCC | Felipe Benites et al |
| EPI_ISL_770563 | Brazil/RS-00611/2020 | 2020-11-24 | South America/Brazil/Rio Grande do Sul | Laboratv≥rio de Microbiologia Molecular - Universidade FEEVALE | Bioinformatics Laboratory / LNCC | Felipe Benites et al |
| EPI_ISL_770564 | Brazil/RS-00612/2020 | 2020-11-24 | South America/Brazil/Rio Grande do Sul | Laboratv≥rio de Microbiologia Molecular - Universidade FEEVALE | Bioinformatics Laboratory / LNCC | Felipe Benites et al |
| EPI_ISL_770566 | Brazil/RS-00614/2020 | 2020-11-24 | South America/Brazil/Rio Grande do Sul | Laboratv≥rio de Microbiologia Molecular - Universidade FEEVALE | Bioinformatics Laboratory / LNCC | Felipe Benites et al |
| EPI_ISL_770567 | Brazil/RS-00615/2020 | 2020-11-25 | South America/Brazil/Rio Grande do Sul | Laboratv≥rio de Microbiologia Molecular - Universidade FEEVALE | Bioinformatics Laboratory / LNCC | Felipe Benites et al |
| EPI_ISL_770569 | Brazil/RS-00617/2020 | 2020-11-25 | South America/Brazil/Rio Grande do Sul | Laboratv≥rio de Microbiologia Molecular - Universidade FEEVALE | Bioinformatics Laboratory / LNCC | Felipe Benites et al |
| EPI_ISL_770571 | Brazil/RS-00619/2020 | 2020-11-25 | South America/Brazil/Rio Grande do Sul | Laboratv≥rio de Microbiologia Molecular - Universidade FEEVALE | Bioinformatics Laboratory / LNCC | Felipe Benites et al |
| EPI_ISL_770572 | Brazil/RS-00620/2020 | 2020-11-24 | South America/Brazil/Rio Grande do Sul | Laboratv≥rio de Microbiologia Molecular - Universidade FEEVALE | Bioinformatics Laboratory / LNCC | Felipe Benites et al |
| EPI_ISL_770573 | Brazil/RS-00622/2020 | 2020-11-23 | South America/Brazil/Rio Grande do Sul | Laboratv≥rio de Microbiologia Molecular - Universidade FEEVALE | Bioinformatics Laboratory / LNCC | Felipe Benites et al |
| EPI_ISL_770574 | Brazil/RS-00623/2020 | 2020-11-23 | South America/Brazil/Rio Grande do Sul | Laboratv≥rio de Microbiologia Molecular - Universidade FEEVALE | Bioinformatics Laboratory / LNCC | Felipe Benites et al |
| EPI_ISL_770575 | Brazil/RS-00625/2020 | 2020-11-23 | South America/Brazil/Rio Grande do Sul | Laboratv≥rio de Microbiologia Molecular - Universidade FEEVALE | Bioinformatics Laboratory / LNCC | Felipe Benites et al |
| EPI_ISL_770577 | Brazil/RS-00627/2020 | 2020-11-24 | South America/Brazil/Rio Grande do Sul | Laboratv≥rio de Microbiologia Molecular - Universidade FEEVALE | Bioinformatics Laboratory / LNCC | Felipe Benites et al |
| EPI_ISL_770578 | Brazil/RS-00629/2020 | 2020-11-24 | South America/Brazil/Rio Grande do Sul | Laboratv≥rio de Microbiologia Molecular - Universidade FEEVALE | Bioinformatics Laboratory / LNCC | Felipe Benites et al |
| EPI_ISL_770580 | Brazil/RS-00632/2020 | 2020-11-25 | South America/Brazil/Rio Grande do Sul | Laboratv≥rio de Microbiologia Molecular - Universidade FEEVALE | Bioinformatics Laboratory / LNCC | Felipe Benites et al |
| EPI_ISL_770581 | Brazil/RS-00633/2020 | 2020-11-24 | South America/Brazil/Rio Grande do Sul | Laboratv≥rio de Microbiologia Molecular - Universidade FEEVALE | Bioinformatics Laboratory / LNCC | Felipe Benites et al |
| EPI_ISL_770583 | Brazil/RS-00635/2020 | 2020-11-25 | South America/Brazil/Rio Grande do Sul | Laboratv≥rio de Microbiologia Molecular - Universidade FEEVALE | Bioinformatics Laboratory / LNCC | Felipe Benites et al |
| EPI_ISL_770585 | Brazil/RS-00637/2020 | 2020-11-24 | South America/Brazil/Rio Grande do Sul | Laboratv≥rio de Microbiologia Molecular - Universidade FEEVALE | Bioinformatics Laboratory / LNCC | Felipe Benites et al |
| EPI_ISL_770586 | Brazil/RS-00638/2020 | 2020-11-25 | South America/Brazil/Rio Grande do Sul | Laboratv≥rio de Microbiologia Molecular - Universidade FEEVALE | Bioinformatics Laboratory / LNCC | Felipe Benites et al |
| EPI_ISL_770587 | Brazil/RS-00639/2020 | 2020-11-25 | South America/Brazil/Rio Grande do Sul | Laboratv≥rio de Microbiologia Molecular - Universidade FEEVALE | Bioinformatics Laboratory / LNCC | Felipe Benites et al |
| EPI_ISL_770588 | Brazil/RS-00640/2020 | 2020-11-24 | South America/Brazil/Rio Grande do Sul | Laboratv≥rio de Microbiologia Molecular - Universidade FEEVALE | Bioinformatics Laboratory / LNCC | Felipe Benites et al |

| | | | | | | |
|---|---|---|---|---|---|---|
| EPI_ISL_770590 | Brazil/RS-00643/2020 | 2020-11-25 | South America/Brazil/Rio Grande do Sul | Laboratv≥rio de Microbiologia Molecular - Universidade FEEVALE | Bioinformatics Laboratory / LNCC | Felipe Benites et al |
| EPI_ISL_770591 | Brazil/RS-00645/2020 | 2020-11-25 | South America/Brazil/Rio Grande do Sul | Laboratv≥rio de Microbiologia Molecular - Universidade FEEVALE | Bioinformatics Laboratory / LNCC | Felipe Benites et al |
| EPI_ISL_770592 | Brazil/RS-00646/2020 | 2020-11-25 | South America/Brazil/Rio Grande do Sul | Laboratv≥rio de Microbiologia Molecular - Universidade FEEVALE | Bioinformatics Laboratory / LNCC | Felipe Benites et al |
| EPI_ISL_770593 | Brazil/RS-00647/2020 | 2020-11-25 | South America/Brazil/Rio Grande do Sul | Laboratv≥rio de Microbiologia Molecular - Universidade FEEVALE | Bioinformatics Laboratory / LNCC | Felipe Benites et al |
| EPI_ISL_770595 | Brazil/RS-00649/2020 | 2020-11-25 | South America/Brazil/Rio Grande do Sul | Laboratv≥rio de Microbiologia Molecular - Universidade FEEVALE | Bioinformatics Laboratory / LNCC | Felipe Benites et al |
| EPI_ISL_770597 | Brazil/RS-00652/2020 | 2020-11-25 | South America/Brazil/Rio Grande do Sul | Laboratv≥rio de Microbiologia Molecular - Universidade FEEVALE | Bioinformatics Laboratory / LNCC | Felipe Benites et al |
| EPI_ISL_770598 | Brazil/RS-00653/2020 | 2020-11-24 | South America/Brazil/Rio Grande do Sul | Laboratv≥rio de Microbiologia Molecular - Universidade FEEVALE | Bioinformatics Laboratory / LNCC | Felipe Benites et al |
| EPI_ISL_770599 | Brazil/RS-00654/2020 | 2020-11-25 | South America/Brazil/Rio Grande do Sul | Laboratv≥rio de Microbiologia Molecular - Universidade FEEVALE | Bioinformatics Laboratory / LNCC | Felipe Benites et al |
| EPI_ISL_770600 | Brazil/RS-00656/2020 | 2020-11-26 | South America/Brazil/Rio Grande do Sul | Laboratv≥rio de Microbiologia Molecular - Universidade FEEVALE | Bioinformatics Laboratory / LNCC | Felipe Benites et al |
| EPI_ISL_770601 | Brazil/RS-00657/2020 | 2020-11-25 | South America/Brazil/Rio Grande do Sul | Laboratv≥rio de Microbiologia Molecular - Universidade FEEVALE | Bioinformatics Laboratory / LNCC | Felipe Benites et al |
| EPI_ISL_770602 | Brazil/RS-00659/2020 | 2020-11-26 | South America/Brazil/Rio Grande do Sul | Laboratv≥rio de Microbiologia Molecular - Universidade FEEVALE | Bioinformatics Laboratory / LNCC | Felipe Benites et al |
| EPI_ISL_770603 | Brazil/RS-00660/2020 | 2020-11-26 | South America/Brazil/Rio Grande do Sul | Laboratv≥rio de Microbiologia Molecular - Universidade FEEVALE | Bioinformatics Laboratory / LNCC | Felipe Benites et al |
| EPI_ISL_770605 | Brazil/RS-00662/2020 | 2020-11-26 | South America/Brazil/Rio Grande do Sul | Laboratv≥rio de Microbiologia Molecular - Universidade FEEVALE | Bioinformatics Laboratory / LNCC | Felipe Benites et al |
| EPI_ISL_770606 | Brazil/RS-00663/2020 | 2020-11-26 | South America/Brazil/Rio Grande do Sul | Laboratv≥rio de Microbiologia Molecular - Universidade FEEVALE | Bioinformatics Laboratory / LNCC | Felipe Benites et al |
| EPI_ISL_770607 | Brazil/RS-00665/2020 | 2020-11-26 | South America/Brazil/Rio Grande do Sul | Laboratv≥rio de Microbiologia Molecular - Universidade FEEVALE | Bioinformatics Laboratory / LNCC | Felipe Benites et al |
| EPI_ISL_770608 | Brazil/RS-00666/2020 | 2020-11-26 | South America/Brazil/Rio Grande do Sul | Laboratv≥rio de Microbiologia Molecular - Universidade FEEVALE | Bioinformatics Laboratory / LNCC | Felipe Benites et al |
| EPI_ISL_770609 | Brazil/RS-00667/2020 | 2020-11-26 | South America/Brazil/Rio Grande do Sul | Laboratv≥rio de Microbiologia Molecular - Universidade FEEVALE | Bioinformatics Laboratory / LNCC | Felipe Benites et al |
| EPI_ISL_770610 | Brazil/RS-00668/2020 | 2020-11-26 | South America/Brazil/Rio Grande do Sul | Laboratv≥rio de Microbiologia Molecular - Universidade FEEVALE | Bioinformatics Laboratory / LNCC | Felipe Benites et al |
| EPI_ISL_770612 | Brazil/RS-00670/2020 | 2020-11-26 | South America/Brazil/Rio Grande do Sul | Laboratv≥rio de Microbiologia Molecular - Universidade FEEVALE | Bioinformatics Laboratory / LNCC | Felipe Benites et al |
| EPI_ISL_770614 | Brazil/RS-00673/2020 | 2020-11-27 | South America/Brazil/Rio Grande do Sul | Laboratv≥rio de Microbiologia Molecular - Universidade FEEVALE | Bioinformatics Laboratory / LNCC | Felipe Benites et al |

| | | | | | | |
|---|---|---|---|---|---|---|
| EPI_ISL_770615 | Brazil/RS-00674/2020 | 2020-11-27 | South America/Brazil/Rio Grande do Sul | Laboratv≥rio de Microbiologia Molecular - Universidade FEEVALE | Bioinformatics Laboratory / LNCC | Felipe Benites et al |
| EPI_ISL_770618 | Brazil/RS-00679/2020 | 2020-11-27 | South America/Brazil/Rio Grande do Sul | Laboratv≥rio de Microbiologia Molecular - Universidade FEEVALE | Bioinformatics Laboratory / LNCC | Felipe Benites et al |
| EPI_ISL_770619 | Brazil/RS-00680/2020 | 2020-11-27 | South America/Brazil/Rio Grande do Sul | Laboratv≥rio de Microbiologia Molecular - Universidade FEEVALE | Bioinformatics Laboratory / LNCC | Felipe Benites et al |
| EPI_ISL_770620 | Brazil/RS-00681/2020 | 2020-11-27 | South America/Brazil/Rio Grande do Sul | Laboratv≥rio de Microbiologia Molecular - Universidade FEEVALE | Bioinformatics Laboratory / LNCC | Felipe Benites et al |
| EPI_ISL_770621 | Brazil/RS-00683/2020 | 2020-11-27 | South America/Brazil/Rio Grande do Sul | Laboratv≥rio de Microbiologia Molecular - Universidade FEEVALE | Bioinformatics Laboratory / LNCC | Felipe Benites et al |
| EPI_ISL_770622 | Brazil/RS-00684/2020 | 2020-11-27 | South America/Brazil/Rio Grande do Sul | Laboratv≥rio de Microbiologia Molecular - Universidade FEEVALE | Bioinformatics Laboratory / LNCC | Felipe Benites et al |
| EPI_ISL_770625 | Brazil/RS-00689/2020 | 2020-11-27 | South America/Brazil/Rio Grande do Sul | Laboratv≥rio de Microbiologia Molecular - Universidade FEEVALE | Bioinformatics Laboratory / LNCC | Felipe Benites et al |
| EPI_ISL_770626 | Brazil/RS-00690/2020 | 2020-11-27 | South America/Brazil/Rio Grande do Sul | Laboratv≥rio de Microbiologia Molecular - Universidade FEEVALE | Bioinformatics Laboratory / LNCC | Felipe Benites et al |
| EPI_ISL_770628 | Brazil/RS-00692/2020 | 2020-11-28 | South America/Brazil/Rio Grande do Sul | Laboratv≥rio de Microbiologia Molecular - Universidade FEEVALE | Bioinformatics Laboratory / LNCC | Felipe Benites et al |
| EPI_ISL_770629 | Brazil/RS-00693/2020 | 2020-11-27 | South America/Brazil/Rio Grande do Sul | Laboratv≥rio de Microbiologia Molecular - Universidade FEEVALE | Bioinformatics Laboratory / LNCC | Felipe Benites et al |
| EPI_ISL_770633 | Russia/KYA-RII-MH10304S/2020 | 2020-11-26 | Europe/Russia/Krasnoyarsk/Krasnoyarsk | WHO National Influenza Centre Russian Federation | WHO National Influenza Centre Russian Federation | Andrey Komissarov et al |
| EPI_ISL_770635 | Russia/KYA-RII-MH10349S/2020 | 2020-11-19 | Europe/Russia/Krasnoyarsk/Krasnoyarsk | WHO National Influenza Centre Russian Federation | WHO National Influenza Centre Russian Federation | Andrey Komissarov et al |
| EPI_ISL_770647 | Russia/SVE-RII-MH8648S/2020 | 2020-09-29 | Europe/Russia/Sverdlovsk/Nizhny Tagil | UMMC-Health | WHO National Influenza Centre Russian Federation | Andrey Komissarov et al |
| EPI_ISL_770651 | Russia/SVE-RII-MH8658S/2020 | 2020-09-28 | Europe/Russia/Sverdlovsk/Sysert | UMMC-Health | WHO National Influenza Centre Russian Federation | Andrey Komissarov et al |
| EPI_ISL_770652 | Russia/SVE-RII-MH8647S/2020 | 2020-09-29 | Europe/Russia/Sverdlovsk/Nizhny Tagil | UMMC-Health | WHO National Influenza Centre Russian Federation | Andrey Komissarov et al |

| | | | | | | |
|---|---|---|---|---|---|---|
| EPI_ISL_770655 | Russia/SVE-RII-MH8699S/2020 | 2020-09-29 | Europe/Russia/Sverdlovsk/Verkhnyaya | UMMC-Health | WHO National Influenza Centre Russian Federation | Andrey Komissarov et al |
| EPI_ISL_770656 | Russia/SVE-RII-MH8945S/2020 | 2020-10-03 | Europe/Russia/Sverdlovsk/ | UMMC-Health | WHO National Influenza Centre Russian Federation | Andrey Komissarov et al |
| EPI_ISL_770658 | Russia/KYA-RII-MH10338S/2020 | 2020-11-20 | Europe/Russia/Krasnoyarsk/Krasnoyarsk | WHO National Influenza Centre Russian Federation | WHO National Influenza Centre Russian Federation | Andrey Komissarov et al |
| EPI_ISL_770664 | Russia/SVE-RII-MH8932S/2020 | 2020-10-02 | Europe/Russia/Sverdlovsk/Verkhnyaya | UMMC-Health | WHO National Influenza Centre Russian Federation | Andrey Komissarov et al |
| EPI_ISL_770667 | Russia/SVE-RII-MH8732S/2020 | 2020-09-29 | Europe/Russia/Sverdlovsk/Yekaterinbur | UMMC-Health | WHO National Influenza Centre Russian Federation | Andrey Komissarov et al |
| EPI_ISL_770670 | Russia/MOW-RII-MH4088S/2020 | 2020-06-09 | Europe/Russia/Moscow Oblast/Moscow | Pathogenic Microorganisms Variability Laboratory | WHO National Influenza Centre Russian Federation | Andrey Komissarov et al |
| EPI_ISL_770673 | Russia/KYA-RII-MH10306S/2020 | 2020-11-23 | Europe/Russia/Krasnoyarsk/Krasnoyarsk | WHO National Influenza Centre Russian Federation | WHO National Influenza Centre Russian Federation | Andrey Komissarov et al |
| EPI_ISL_770674 | Russia/KYA-RII-MH10309S/2020 | 2020-11-24 | Europe/Russia/Krasnoyarsk/Krasnoyarsk | WHO National Influenza Centre Russian Federation | WHO National Influenza Centre Russian Federation | Andrey Komissarov et al |
| EPI_ISL_770689 | Germany/NW-HHU-455/2020 | 2020-12-11 | Europe/Germany/North Rhine Westphal | ZOTZ KLIMAS MVZ Dvºsseldorf-Centrum GbR √úBAG fvºr Labormedizin, Genetik, Zytologie, Pathologie | Center of Medical Microbiology, Virology, and Hospital Hygiene, University of Duesseldorf | Maximilian Damagnez et al |

| | | | | | | |
|---|---|---|---|---|---|---|
| EPI_ISL_770693 | Germany/NW-HHU-459/2020 | 2020-12-11 | Europe/Germany/North Rhine Westphal | ZOTZ KLIMAS MVZ D√ºsseldorf-Centrum GbR √úBAG f√ºr Labormedizin, Genetik, Zytologie, Pathologie | Center of Medical Microbiology, Virology, and Hospital Hygiene, University of Duesseldorf | Maximilian Damagnez et al |
| EPI_ISL_770695 | Germany/NW-HHU-461/2020 | 2020-12-11 | Europe/Germany/North Rhine Westphal | ZOTZ KLIMAS MVZ D√ºsseldorf-Centrum GbR √úBAG f√ºr Labormedizin, Genetik, Zytologie, Pathologie | Center of Medical Microbiology, Virology, and Hospital Hygiene, University of Duesseldorf | Maximilian Damagnez et al |
| EPI_ISL_770699 | Germany/NW-HHU-465/2020 | 2020-12-11 | Europe/Germany/North Rhine Westphal | ZOTZ KLIMAS MVZ D√ºsseldorf-Centrum GbR √úBAG f√ºr Labormedizin, Genetik, Zytologie, Pathologie | Center of Medical Microbiology, Virology, and Hospital Hygiene, University of Duesseldorf | Maximilian Damagnez et al |
| EPI_ISL_770704 | Germany/NW-HHU-470/2020 | 2020-12-11 | Europe/Germany/North Rhine Westphal | ZOTZ KLIMAS MVZ D√ºsseldorf-Centrum GbR √úBAG f√ºr Labormedizin, Genetik, Zytologie, Pathologie | Center of Medical Microbiology, Virology, and Hospital Hygiene, University of Duesseldorf | Maximilian Damagnez et al |
| EPI_ISL_770705 | Germany/NW-HHU-471/2020 | 2020-12-11 | Europe/Germany/North Rhine Westphal | ZOTZ KLIMAS MVZ D√ºsseldorf-Centrum GbR √úBAG f√ºr Labormedizin, Genetik, Zytologie, Pathologie | Center of Medical Microbiology, Virology, and Hospital Hygiene, University of Duesseldorf | Maximilian Damagnez et al |
| EPI_ISL_770711 | Germany/NW-HHU-477/2020 | 2020-12-11 | Europe/Germany/North Rhine Westphal | ZOTZ KLIMAS MVZ D√ºsseldorf-Centrum GbR √úBAG f√ºr Labormedizin, Genetik, Zytologie, Pathologie | Center of Medical Microbiology, Virology, and Hospital Hygiene, University of Duesseldorf | Maximilian Damagnez et al |

| | | | | | | |
|---|---|---|---|---|---|---|
| EPI_ISL_770713 | Germany/NW-HHU-479/2020 | 2020-12-06 | Europe/Germany/North Rhine Westphal | ZOTZ KLIMAS MVZ D√ºsseldorf-Centrum GbR √úBAG f√ºr Labormedizin, Genetik, Zytologie, Pathologie | Center of Medical Microbiology, Virology, and Hospital Hygiene, University of Duesseldorf | Maximilian Damagnez et al |
| EPI_ISL_770717 | Germany/NW-HHU-483/2020 | 2020-11-30 | Europe/Germany/North Rhine Westphal | ZOTZ KLIMAS MVZ D√ºsseldorf-Centrum GbR √úBAG f√ºr Labormedizin, Genetik, Zytologie, Pathologie | Center of Medical Microbiology, Virology, and Hospital Hygiene, University of Duesseldorf | Maximilian Damagnez et al |
| EPI_ISL_770718 | Germany/NW-HHU-484/2020 | 2020-11-30 | Europe/Germany/North Rhine Westphal | ZOTZ KLIMAS MVZ D√ºsseldorf-Centrum GbR √úBAG f√ºr Labormedizin, Genetik, Zytologie, Pathologie | Center of Medical Microbiology, Virology, and Hospital Hygiene, University of Duesseldorf | Maximilian Damagnez et al |
| EPI_ISL_770719 | Germany/NW-HHU-485/2020 | 2020-12-01 | Europe/Germany/North Rhine Westphal | ZOTZ KLIMAS MVZ D√ºsseldorf-Centrum GbR √úBAG f√ºr Labormedizin, Genetik, Zytologie, Pathologie | Center of Medical Microbiology, Virology, and Hospital Hygiene, University of Duesseldorf | Maximilian Damagnez et al |
| EPI_ISL_770722 | Germany/NW-HHU-506/2020 | 2020-09-17 | Europe/Germany/North Rhine Westphal | ZOTZ KLIMAS MVZ D√ºsseldorf-Centrum GbR √úBAG f√ºr Labormedizin, Genetik, Zytologie, Pathologie | Center of Medical Microbiology, Virology, and Hospital Hygiene, University of Duesseldorf | Maximilian Damagnez et al |
| EPI_ISL_770723 | Germany/NW-HHU-507/2020 | 2020-09-17 | Europe/Germany/North Rhine Westphal | ZOTZ KLIMAS MVZ D√ºsseldorf-Centrum GbR √úBAG f√ºr Labormedizin, Genetik, Zytologie, Pathologie | Center of Medical Microbiology, Virology, and Hospital Hygiene, University of Duesseldorf | Maximilian Damagnez et al |

| | | | | | | |
|---|---|---|---|---|---|---|
| EPI_ISL_770725 | Germany/NW-HHU-509/2020 | 2020-09-18 | Europe/Germany/North Rhine Westphal | ZOTZ KLIMAS MVZ D√ºsseldorf-Centrum GbR √úBAG f√ºr Labormedizin, Genetik, Zytologie, Pathologie | Center of Medical Microbiology, Virology, and Hospital Hygiene, University of Duesseldorf | Maximilian Damagnez et al |
| EPI_ISL_770726 | Germany/NW-HHU-510/2020 | 2020-09-18 | Europe/Germany/North Rhine Westphal | ZOTZ KLIMAS MVZ D√ºsseldorf-Centrum GbR √úBAG f√ºr Labormedizin, Genetik, Zytologie, Pathologie | Center of Medical Microbiology, Virology, and Hospital Hygiene, University of Duesseldorf | Maximilian Damagnez et al |
| EPI_ISL_770728 | Germany/NW-HHU-512/2020 | 2020-09-19 | Europe/Germany/North Rhine Westphal | ZOTZ KLIMAS MVZ D√ºsseldorf-Centrum GbR √úBAG f√ºr Labormedizin, Genetik, Zytologie, Pathologie | Center of Medical Microbiology, Virology, and Hospital Hygiene, University of Duesseldorf | Maximilian Damagnez et al |
| EPI_ISL_770731 | Germany/NW-HHU-317/2020 | 2020-10-06 | Europe/Germany/North Rhine Westphal | ZOTZ KLIMAS MVZ D√ºsseldorf-Centrum GbR √úBAG f√ºr Labormedizin, Genetik, Zytologie, Pathologie | Center of Medical Microbiology, Virology, and Hospital Hygiene, University of Duesseldorf | Maximilian Damagnez et al |
| EPI_ISL_770732 | Germany/NW-HHU-318/2020 | 2020-10-06 | Europe/Germany/North Rhine Westphal | ZOTZ KLIMAS MVZ D√ºsseldorf-Centrum GbR √úBAG f√ºr Labormedizin, Genetik, Zytologie, Pathologie | Center of Medical Microbiology, Virology, and Hospital Hygiene, University of Duesseldorf | Maximilian Damagnez et al |
| EPI_ISL_770738 | Germany/NW-HHU-489/2020 | 2020-12-03 | Europe/Germany/North Rhine Westphal | ZOTZ KLIMAS MVZ D√ºsseldorf-Centrum GbR √úBAG f√ºr Labormedizin, Genetik, Zytologie, Pathologie | Center of Medical Microbiology, Virology, and Hospital Hygiene, University of Duesseldorf | Maximilian Damagnez et al |

| | | | | | | |
|---|---|---|---|---|---|---|
| EPI_ISL_770741 | Germany/NW-HHU-492/2020 | 2020-12-03 | Europe/Germany/North Rhine Westphal | ZOTZ KLIMAS MVZ D√ºsseldorf-Centrum GbR √úBAG f√ºr Labormedizin, Genetik, Zytologie, Pathologie | Center of Medical Microbiology, Virology, and Hospital Hygiene, University of Duesseldorf | Maximilian Damagnez et al |
| EPI_ISL_770747 | Germany/NW-HHU-499/2020 | 2020-12-03 | Europe/Germany/North Rhine Westphal | ZOTZ KLIMAS MVZ D√ºsseldorf-Centrum GbR √úBAG f√ºr Labormedizin, Genetik, Zytologie, Pathologie | Center of Medical Microbiology, Virology, and Hospital Hygiene, University of Duesseldorf | Maximilian Damagnez et al |
| EPI_ISL_770748 | Germany/NW-HHU-500/2020 | 2020-12-03 | Europe/Germany/North Rhine Westphal | ZOTZ KLIMAS MVZ D√ºsseldorf-Centrum GbR √úBAG f√ºr Labormedizin, Genetik, Zytologie, Pathologie | Center of Medical Microbiology, Virology, and Hospital Hygiene, University of Duesseldorf | Maximilian Damagnez et al |
| EPI_ISL_770750 | Germany/NW-HHU-502/2020 | 2020-12-03 | Europe/Germany/North Rhine Westphal | ZOTZ KLIMAS MVZ D√ºsseldorf-Centrum GbR √úBAG f√ºr Labormedizin, Genetik, Zytologie, Pathologie | Center of Medical Microbiology, Virology, and Hospital Hygiene, University of Duesseldorf | Maximilian Damagnez et al |
| EPI_ISL_770751 | Germany/NW-HHU-503/2020 | 2020-12-03 | Europe/Germany/North Rhine Westphal | ZOTZ KLIMAS MVZ D√ºsseldorf-Centrum GbR √úBAG f√ºr Labormedizin, Genetik, Zytologie, Pathologie | Center of Medical Microbiology, Virology, and Hospital Hygiene, University of Duesseldorf | Maximilian Damagnez et al |
| EPI_ISL_770752 | Germany/NW-HHU-277/2020 | 2020-09-28 | Europe/Germany/North Rhine Westphal | ZOTZ KLIMAS MVZ D√ºsseldorf-Centrum GbR √úBAG f√ºr Labormedizin, Genetik, Zytologie, Pathologie | Center of Medical Microbiology, Virology, and Hospital Hygiene, University of Duesseldorf | Maximilian Damagnez et al |

| | | | | | | |
|---|---|---|---|---|---|---|
| EPI_ISL_770755 | Germany/NW-HHU-280/2020 | 2020-09-28 | Europe/Germany/North Rhine Westphal | ZOTZ KLIMAS MVZ Dv∫sseldorf-Centrum GbR √úBAG fv∫r Labormedizin, Genetik, Zytologie, Pathologie | Center of Medical Microbiology, Virology, and Hospital Hygiene, University of Duesseldorf | Maximilian Damagnez et al |
| EPI_ISL_770759 | Germany/NW-HHU-284/2020 | 2020-09-29 | Europe/Germany/North Rhine Westphal | ZOTZ KLIMAS MVZ Dv∫sseldorf-Centrum GbR √úBAG fv∫r Labormedizin, Genetik, Zytologie, Pathologie | Center of Medical Microbiology, Virology, and Hospital Hygiene, University of Duesseldorf | Maximilian Damagnez et al |
| EPI_ISL_770769 | Germany/NW-HHU-294/2020 | 2020-10-01 | Europe/Germany/North Rhine Westphal | ZOTZ KLIMAS MVZ Dv∫sseldorf-Centrum GbR √úBAG fv∫r Labormedizin, Genetik, Zytologie, Pathologie | Center of Medical Microbiology, Virology, and Hospital Hygiene, University of Duesseldorf | Maximilian Damagnez et al |
| EPI_ISL_770780 | USA/MN-MDH-2206/2020 | 2020-11-16 | North America/USA/Minnesota/ | Minnesota Department of Health, Public Health Laboratory | Minnesota Department of Health, Public Health Laboratory | Alexandra Lorentz et al |
| EPI_ISL_770791 | USA/MN-MDH-2217/2020 | 2020-12-07 | North America/USA/Minnesota/ | Minnesota Department of Health, Public Health Laboratory | Minnesota Department of Health, Public Health Laboratory | Alexandra Lorentz et al |
| EPI_ISL_770801 | USA/MN-MDH-2227/2020 | 2020-12-05 | North America/USA/Minnesota/ | Vault Health | Minnesota Department of Health, Public Health Laboratory | Alexandra Lorentz et al |
| EPI_ISL_770822 | Canada/ON-E69b/2020 | 2020-04-03 | North America/Canada/Ontario/Ottawa | Eastern Ontario Regional Laboratory Association | McMaster University | Leanne Mortimer et al |
| EPI_ISL_770823 | Canada/ON-E70b/2020 | 2020-03-28 | North America/Canada/Ontario/Ottawa | Eastern Ontario Regional Laboratory Association | McMaster University | Leanne Mortimer et al |
| EPI_ISL_770824 | Canada/ON-E71b/2020 | 2020-04-03 | North America/Canada/Ontario/Ottawa | Eastern Ontario Regional Laboratory Association | McMaster University | Leanne Mortimer et al |
| EPI_ISL_770825 | Canada/ON-E72b/2020 | 2020-04-03 | North America/Canada/Ontario/Ottawa | Eastern Ontario Regional Laboratory Association | McMaster University | Leanne Mortimer et al |
| EPI_ISL_770826 | Canada/ON-E74b/2020 | 2020-10-10 | North America/Canada/Ontario/Ottawa | Eastern Ontario Regional Laboratory Association | McMaster University | Leanne Mortimer et al |

| | | | | | | |
|---|---|---|---|---|---|---|
| EPI_ISL_770827 | Canada/ON-E76b/2020 | 2020-05-06 | North America/Canada/Ontario/Ottawa | Eastern Ontario Regional Laboratory Association | McMaster University | Leanne Mortimer et al |
| EPI_ISL_770828 | Canada/ON-S162-KN/2020 | 2020-03-24 | North America/Canada/Ontario/Toronto | Toronto Invasive Bacterial Diseases Network | McMaster University | Allison McGeer et al |
| EPI_ISL_770829 | Canada/ON-S461-KN/2020 | 2020-04-01 | North America/Canada/Ontario/Toronto | Toronto Invasive Bacterial Diseases Network | McMaster University | Allison McGeer et al |
| EPI_ISL_770830 | Canada/ON-S1453/2020 | 20-Aug | North America/Canada/Ontario/Toronto | Toronto Invasive Bacterial Diseases Network | McMaster University | Allison McGeer et al |
| EPI_ISL_770831 | Canada/ON-S1458/2020 | 20-Aug | North America/Canada/Ontario/Toronto | Toronto Invasive Bacterial Diseases Network | McMaster University | Allison McGeer et al |
| EPI_ISL_770832 | Canada/ON-S1459/2020 | 20-Aug | North America/Canada/Ontario/Toronto | Toronto Invasive Bacterial Diseases Network | McMaster University | Allison McGeer et al |
| EPI_ISL_770833 | Canada/ON-S1460/2020 | 20-Aug | North America/Canada/Ontario/Toronto | Toronto Invasive Bacterial Diseases Network | McMaster University | Allison McGeer et al |
| EPI_ISL_770834 | Canada/ON-S1461/2020 | 20-Aug | North America/Canada/Ontario/Toronto | Toronto Invasive Bacterial Diseases Network | McMaster University | Allison McGeer et al |
| EPI_ISL_770835 | Canada/ON-S1462/2020 | 20-Aug | North America/Canada/Ontario/Toronto | Toronto Invasive Bacterial Diseases Network | McMaster University | Allison McGeer et al |
| EPI_ISL_770836 | Canada/ON-S1463/2020 | 20-Aug | North America/Canada/Ontario/Toronto | Toronto Invasive Bacterial Diseases Network | McMaster University | Allison McGeer et al |
| EPI_ISL_770837 | Canada/ON-S1464/2020 | 20-Aug | North America/Canada/Ontario/Toronto | Toronto Invasive Bacterial Diseases Network | McMaster University | Allison McGeer et al |
| EPI_ISL_770838 | Canada/ON-S1467/2020 | 20-Aug | North America/Canada/Ontario/Toronto | Toronto Invasive Bacterial Diseases Network | McMaster University | Allison McGeer et al |
| EPI_ISL_770839 | Canada/ON-S1486/2020 | 20-Aug | North America/Canada/Ontario/Toronto | Toronto Invasive Bacterial Diseases Network | McMaster University | Allison McGeer et al |
| EPI_ISL_770840 | Canada/ON-S1487/2020 | 20-Aug | North America/Canada/Ontario/Toronto | Toronto Invasive Bacterial Diseases Network | McMaster University | Allison McGeer et al |
| EPI_ISL_770841 | Canada/ON-S1489/2020 | 20-Aug | North America/Canada/Ontario/Toronto | Toronto Invasive Bacterial Diseases Network | McMaster University | Allison McGeer et al |
| EPI_ISL_770842 | Canada/ON-S1490/2020 | 20-Aug | North America/Canada/Ontario/Toronto | Toronto Invasive Bacterial Diseases Network | McMaster University | Allison McGeer et al |
| EPI_ISL_770843 | Canada/ON-S1491/2020 | 20-Aug | North America/Canada/Ontario/Toronto | Toronto Invasive Bacterial Diseases Network | McMaster University | Allison McGeer et al |
| EPI_ISL_770844 | Canada/ON-S1493/2020 | 20-Aug | North America/Canada/Ontario/Toronto | Toronto Invasive Bacterial Diseases Network | McMaster University | Allison McGeer et al |
| EPI_ISL_771155 | USA/CO-CDPHE-2011210355/2020 | 2020-11-19 | North America/USA/Colorado/ | Colorado Department of Public Health and Environment | Colorado Department of Puplic Health and Environment | Laura Bankers et al |

| | | | | | | |
|---|---|---|---|---|---|---|
| EPI_ISL_771161 | USA/CO-CDPHE-2011193762/2020 | 2020-11-17 | North America/USA/Colorado/ | Colorado Department of Public Health and Environment | Colorado Department of Puplic Health and Environment | Laura Bankers et al |
| EPI_ISL_771172 | USA/CO-CDPHE-2100038277/2020 | 2020-11-17 | North America/USA/Colorado/ | Colorado Department of Public Health and Environment | Colorado Department of Puplic Health and Environment | Laura Bankers et al |
| EPI_ISL_771183 | USA/CO-CDPHE-2100008733/2020 | 2020-11-20 | North America/USA/Colorado/ | Colorado Department of Public Health and Environment | Colorado Department of Puplic Health and Environment | Laura Bankers et al |
| EPI_ISL_771189 | USA/CO-CDPHE-2100013054/2020 | 2020-11-19 | North America/USA/Colorado/ | Colorado Department of Public Health and Environment | Colorado Department of Puplic Health and Environment | Laura Bankers et al |
| EPI_ISL_771217 | USA/CO-CDPHE-2100036208/2020 | 2020-11-24 | North America/USA/Colorado/ | Colorado Department of Public Health and Environment | Colorado Department of Puplic Health and Environment | Laura Bankers et al |
| EPI_ISL_771227 | USA/CO-CDPHE-2005060754/2020 | 2020-05-05 | North America/USA/Colorado/ | Colorado Department of Public Health and Environment | Colorado Department of Puplic Health and Environment | Laura Bankers et al |
| EPI_ISL_771229 | USA/CO-CDPHE-2007082647/2020 | 2020-07-08 | North America/USA/Colorado/ | Colorado Department of Public Health and Environment | Colorado Department of Puplic Health and Environment | Laura Bankers et al |
| EPI_ISL_771231 | USA/CO-CDPHE-2007152397/2020 | 2020-07-15 | North America/USA/Colorado/ | Colorado Department of Public Health and Environment | Colorado Department of Puplic Health and Environment | Laura Bankers et al |
| EPI_ISL_771245 | USA/CO-CDPHE-2011103380/2020 | 2020-11-06 | North America/USA/Colorado/ | Colorado Department of Public Health and Environment | Colorado Department of Puplic Health and Environment | Laura Bankers et al |
| EPI_ISL_771246 | USA/CO-CDPHE-2011113930/2020 | 2020-11-11 | North America/USA/Colorado/ | Colorado Department of Public Health and Environment | Colorado Department of Puplic Health and Environment | Laura Bankers et al |

| | | | | | | |
|---|---|---|---|---|---|---|
| EPI_ISL_771247 | USA/CO-CDPHE-2011162142/2020 | 2020-11-16 | North America/USA/Colorado/ | Colorado Department of Public Health and Environment | Colorado Department of Puplic Health and Environment | Laura Bankers et al |
| EPI_ISL_771252 | USA/CO-CDPHE-2011193717/2020 | 2020-11-18 | North America/USA/Colorado/ | Colorado Department of Public Health and Environment | Colorado Department of Puplic Health and Environment | Laura Bankers et al |
| EPI_ISL_771257 | USA/CO-CDPHE-2011200477/2020 | 2020-11-17 | North America/USA/Colorado/ | Colorado Department of Public Health and Environment | Colorado Department of Puplic Health and Environment | Laura Bankers et al |
| EPI_ISL_771260 | USA/CO-CDPHE-2011200553/2020 | 2020-11-19 | North America/USA/Colorado/ | Colorado Department of Public Health and Environment | Colorado Department of Puplic Health and Environment | Laura Bankers et al |
| EPI_ISL_771261 | USA/CO-CDPHE-2011200641/2020 | 2020-11-17 | North America/USA/Colorado/ | Colorado Department of Public Health and Environment | Colorado Department of Puplic Health and Environment | Laura Bankers et al |
| EPI_ISL_771262 | USA/CO-CDPHE-2011201244/2020 | 2020-11-19 | North America/USA/Colorado/ | Colorado Department of Public Health and Environment | Colorado Department of Puplic Health and Environment | Laura Bankers et al |
| EPI_ISL_771291 | USA/CO-CDPHE-2100008783/2020 | 2020-11-20 | North America/USA/Colorado/ | Colorado Department of Public Health and Environment | Colorado Department of Puplic Health and Environment | Laura Bankers et al |
| EPI_ISL_771293 | USA/CO-CDPHE-2100010210/2020 | 2020-11-19 | North America/USA/Colorado/ | Colorado Department of Public Health and Environment | Colorado Department of Puplic Health and Environment | Laura Bankers et al |
| EPI_ISL_771295 | USA/CO-CDPHE-2100010222/2020 | 2020-11-19 | North America/USA/Colorado/ | Colorado Department of Public Health and Environment | Colorado Department of Puplic Health and Environment | Laura Bankers et al |
| EPI_ISL_771296 | USA/CO-CDPHE-2100010692/2020 | 2020-11-19 | North America/USA/Colorado/ | Colorado Department of Public Health and Environment | Colorado Department of Puplic Health and Environment | Laura Bankers et al |

| | | | | | | |
|---|---|---|---|---|---|---|
| EPI_ISL_771298 | USA/CO-CDPHE-2100012987/2020 | 2020-11-19 | North America/USA/Colorado/ | Colorado Department of Public Health and Environment | Colorado Department of Puplic Health and Environment | Laura Bankers et al |
| EPI_ISL_771310 | USA/CO-CDPHE-2100015096/2020 | 2020-11-21 | North America/USA/Colorado/ | Colorado Department of Public Health and Environment | Colorado Department of Puplic Health and Environment | Laura Bankers et al |
| EPI_ISL_771312 | USA/CO-CDPHE-2100015182/2020 | 2020-11-21 | North America/USA/Colorado/ | Colorado Department of Public Health and Environment | Colorado Department of Puplic Health and Environment | Laura Bankers et al |
| EPI_ISL_771316 | USA/CO-CDPHE-2100015827/2020 | 2020-11-20 | North America/USA/Colorado/ | Colorado Department of Public Health and Environment | Colorado Department of Puplic Health and Environment | Laura Bankers et al |
| EPI_ISL_771317 | USA/CO-CDPHE-2100015900/2020 | 2020-11-20 | North America/USA/Colorado/ | Colorado Department of Public Health and Environment | Colorado Department of Puplic Health and Environment | Laura Bankers et al |
| EPI_ISL_771333 | USA/CO-CDPHE-2100023678/2020 | 2020-11-23 | North America/USA/Colorado/ | Colorado Department of Public Health and Environment | Colorado Department of Puplic Health and Environment | Laura Bankers et al |
| EPI_ISL_771338 | USA/CO-CDPHE-2100028879/2020 | 2020-11-24 | North America/USA/Colorado/ | Colorado Department of Public Health and Environment | Colorado Department of Puplic Health and Environment | Laura Bankers et al |
| EPI_ISL_771339 | USA/CO-CDPHE-2100030859/2020 | 2020-11-25 | North America/USA/Colorado/ | Colorado Department of Public Health and Environment | Colorado Department of Puplic Health and Environment | Laura Bankers et al |
| EPI_ISL_771344 | USA/CO-CDPHE-2100036501/2020 | 2020-11-24 | North America/USA/Colorado/ | Colorado Department of Public Health and Environment | Colorado Department of Puplic Health and Environment | Laura Bankers et al |
| EPI_ISL_771347 | USA/CO-CDPHE-2100037717/2020 | 2020-11-25 | North America/USA/Colorado/ | Colorado Department of Public Health and Environment | Colorado Department of Puplic Health and Environment | Laura Bankers et al |

| | | | | | | |
|---|---|---|---|---|---|---|
| EPI_ISL_771353 | USA/CO-CDPHE-2100041649/2020 | 2020-11-27 | North America/USA/Colorado/ | Colorado Department of Public Health and Environment | Colorado Department of Puplic Health and Environment | Laura Bankers et al |
| EPI_ISL_771358 | USA/CO-CDPHE-2100042764/2020 | 2020-11-27 | North America/USA/Colorado/ | Colorado Department of Public Health and Environment | Colorado Department of Puplic Health and Environment | Laura Bankers et al |
| EPI_ISL_771365 | USA/WA-S2920/2020 | 2020-07-24 | North America/USA/Washington/Pacific | Washington State Department of Health | Seattle Flu Study | Deborah A. Nickerson et al |
| EPI_ISL_774875 | Bangladesh/DNAS_DRICM_CPH_isl_5 | 2020-04-29 | Asia/Bangladesh/Dhaka/ | Designated Reference Institute for Chemical Measurements (DRICM) | DNA SOLUTION LTD. | Md. Imran Khan et al |
| EPI_ISL_774883 | Bangladesh/DNAS_DRCIM_CPH_isl_5 | 2020-04-29 | Asia/Bangladesh/Dhaka/ | Designated Reference Institute for Chemical Measurements (DRICM) | DNA SOLUTION LTD. | Md. Imran Khan et al |
| EPI_ISL_774884 | Bangladesh/DNAS_DRCIM_CPH_isl_5 | 2020-04-29 | Asia/Bangladesh/Dhaka/ | Designated Reference Institute for Chemical Measurements (DRICM) | DNA SOLUTION LTD. | Md. Imran Khan et al |
| EPI_ISL_774892 | Bangladesh/DNAS_DRCIM_CPH_isl_4 | 2020-04-29 | Asia/Bangladesh/Dhaka/ | Designated Reference Institute for Chemical Measurements (DRICM) | DNA SOLUTION LTD. | Md. Imran Khan et al |
| EPI_ISL_774901 | Bangladesh/DNAS_DRCIM_isl_75/20 | 2020-04-29 | Asia/Bangladesh/Dhaka/ | Designated Reference Institute for Chemical Measurements (DRICM) | DNA SOLUTION LTD. | Md. Imran Khan et al |
| EPI_ISL_774906 | Bangladesh/DNAS_DRCIM_isl_6841/ | 2020-04-29 | Asia/Bangladesh/Dhaka/ | Designated Reference Institute for Chemical Measurements (DRICM) | DNA SOLUTION LTD. | Md. Imran Khan et al |
| EPI_ISL_774911 | Bangladesh/DNAS_DRCIM_isl_17/20 | 2020-04-29 | Asia/Bangladesh/Dhaka/ | Designated Reference Institute for Chemical Measurements (DRICM) | DNA SOLUTION LTD. | Md. Imran Khan et al |
| EPI_ISL_774915 | Bangladesh/DNAS_DRCIM_isl_BRB4/ | 2020-07-03 | Asia/Bangladesh/Dhaka/ | Designated Reference Institute for Chemical Measurements (DRICM) | DNA SOLUTION LTD. | Md. Imran Khan et al |
| EPI_ISL_774917 | Bangladesh/DNAS_DRCIM_isl_P2-98/ | 2020-07-02 | Asia/Bangladesh/Dhaka/ | Designated Reference Institute for Chemical Measurements (DRICM) | DNA SOLUTION LTD. | Md. Imran Khan et al |
| EPI_ISL_774919 | Bangladesh/DNAS_DRCIM_isl_SM-01 | 2020-07-04 | Asia/Bangladesh/Dhaka/ | Designated Reference Institute for Chemical Measurements (DRICM) | DNA SOLUTION LTD. | Md. Imran Khan et al |
| EPI_ISL_774920 | Bangladesh/DNAS_DRCIM_isl_R-88/2 | 2020-07-02 | Asia/Bangladesh/Dhaka/ | Designated Reference Institute for Chemical Measurements (DRICM) | DNA SOLUTION LTD. | Md. Imran Khan et al |
| EPI_ISL_774921 | Bangladesh/DNAS_isl_P2-104/2020 | 2020-07-02 | Asia/Bangladesh/Dhaka/ | Designated Reference Institute for Chemical Measurements (DRICM) | DNA SOLUTION LTD. | Md. Imran Khan et al |
| EPI_ISL_774922 | Bangladesh/DNAS_DRCIM_isl_Amal/ | 2020-07-02 | Asia/Bangladesh/Dhaka/ | Designated Reference Institute for Chemical Measurements (DRICM) | DNA SOLUTION LTD. | Md. Imran Khan et al |
| EPI_ISL_774923 | Bangladesh/DNAS_DRCIM_isl_MH-0 | 2020-07-02 | Asia/Bangladesh/Dhaka/ | Designated Reference Institute for Chemical Measurements (DRICM) | DNA SOLUTION LTD. | Md. Imran Khan et al |
| EPI_ISL_774927 | Bangladesh/DNAS_DRCIM_isl_P2-10 | 2020-07-02 | Asia/Bangladesh/Dhaka/ | Designated Reference Institute for Chemical Measurements (DRICM) | DNA SOLUTION LTD. | Md. Imran Khan et al |

| | | | | | | |
|---|---|---|---|---|---|---|
| EPI_ISL_774929 | Bangladesh/DNAS_DRCIM_isl_P2-107 | 2020-07-02 | Asia/Bangladesh/Dhaka/ | Designated Reference Institute for Chemical Measurements (DRICM) | DNA SOLUTION LTD. | Md. Imran Khan et al |
| EPI_ISL_774932 | Bangladesh/DNAS_DRCIM_isl_R-36/2 | 2020-06-30 | Asia/Bangladesh/Dhaka/ | Designated Reference Institute for Chemical Measurements (DRICM) | DNA SOLUTION LTD. | Md. Imran Khan et al |
| EPI_ISL_774936 | Bangladesh/DNAS_DRCIM_isl_Sik-bip | 2020-06-30 | Asia/Bangladesh/Dhaka/ | Designated Reference Institute for Chemical Measurements (DRICM) | DNA SOLUTION LTD. | Md. Imran Khan et al |
| EPI_ISL_774938 | Bangladesh/DNAS_DRCIM_isl_SIBL-7/ | 2020-06-30 | Asia/Bangladesh/Dhaka/ | Designated Reference Institute for Chemical Measurements (DRICM) | DNA SOLUTION LTD. | Md. Imran Khan et al |
| EPI_ISL_774941 | Bangladesh/DNAS_DRCIM_isl_IH-03/ | 2020-06-30 | Asia/Bangladesh/Dhaka/ | Designated Reference Institute for Chemical Measurements (DRICM) | DNA SOLUTION LTD. | Md. Imran Khan et al |
| EPI_ISL_774942 | Bangladesh/DNAS_DRCIM_isl_KHL-01 | 2020-07-05 | Asia/Bangladesh/Dhaka/ | Designated Reference Institute for Chemical Measurements (DRICM) | DNA SOLUTION LTD. | Md. Imran Khan et al |
| EPI_ISL_774943 | Bangladesh/DNAS_DRCIM_isl_HH_13 | 2020-06-30 | Asia/Bangladesh/Dhaka/ | Designated Reference Institute for Chemical Measurements (DRICM) | DNA SOLUTION LTD. | Md. Imran Khan et al |
| EPI_ISL_774947 | Bangladesh/DNAS_DRCIM_isl_R-101/ | 2020-07-04 | Asia/Bangladesh/Dhaka/ | Designated Reference Institute for Chemical Measurements (DRICM) | DNA SOLUTION LTD. | Md. Imran Khan et al |
| EPI_ISL_774948 | Bangladesh/DNAS_DRCIM_isl_Sik-bip | 2020-06-30 | Asia/Bangladesh/Dhaka/ | Designated Reference Institute for Chemical Measurements (DRICM) | DNA SOLUTION LTD. | Md. Imran Khan et al |
| EPI_ISL_774949 | Bangladesh/DNAS_DRCIM_isl_HH-12 | 2020-06-30 | Asia/Bangladesh/Dhaka/ | Designated Reference Institute for Chemical Measurements (DRICM) | DNA SOLUTION LTD. | Md. Imran Khan et al |
| EPI_ISL_774950 | Bangladesh/DNAS_DRCIM_isl_GL-01/ | 2020-07-04 | Asia/Bangladesh/Dhaka/ | Designated Reference Institute for Chemical Measurements (DRICM) | DNA SOLUTION LTD. | Md. Imran Khan et al |
| EPI_ISL_774951 | Bangladesh/DNAS_DRCIM_isl_HH-11 | 2020-06-30 | Asia/Bangladesh/Dhaka/ | Designated Reference Institute for Chemical Measurements (DRICM) | DNA SOLUTION LTD. | Md. Imran Khan et al |
| EPI_ISL_774953 | Bangladesh/DNAS_DRCIM_isl_SIBL-0 | 2020-06-28 | Asia/Bangladesh/Dhaka/ | Designated Reference Institute for Chemical Measurements (DRICM) | DNA SOLUTION LTD. | Md. Imran Khan et al |
| EPI_ISL_774960 | Bangladesh/DNAS_DRCIM_isl_SIBL-0 | 2020-06-28 | Asia/Bangladesh/Dhaka/ | Designated Reference Institute for Chemical Measurements (DRICM) | DNA SOLUTION LTD. | Md. Imran Khan et al |
| EPI_ISL_774962 | Bangladesh/DNAS_DRCIM_isl_BRB-3/ | 2020-06-30 | Asia/Bangladesh/Dhaka/ | Designated Reference Institute for Chemical Measurements (DRICM) | DNA SOLUTION LTD. | Md. Imran Khan et al |
| EPI_ISL_774963 | Bangladesh/DNAS_DRCIM_isl_MMC- | 2020-06-28 | Asia/Bangladesh/Dhaka/ | Designated Reference Institute for Chemical Measurements (DRICM) | DNA SOLUTION LTD. | Md. Imran Khan et al |
| EPI_ISL_774966 | Bangladesh/DNAS_DRCIM_isl_R-97/2 | 2020-07-04 | Asia/Bangladesh/Dhaka/ | Designated Reference Institute for Chemical Measurements (DRICM) | DNA SOLUTION LTD. | Md. Imran Khan et al |
| EPI_ISL_774967 | Bangladesh/DNAS_DRICM_isl_19183 | 2020-06-28 | Asia/Bangladesh/Dhaka/ | Designated Reference Institute for Chemical Measurements (DRICM) | DNA SOLUTION LTD. | Md. Imran Khan et al |
| EPI_ISL_774968 | Bangladesh/DNAS_DRICM_isl_19189 | 2020-06-28 | Asia/Bangladesh/Dhaka/ | Designated Reference Institute for Chemical Measurements (DRICM) | DNA SOLUTION LTD. | Md. Imran Khan et al |
| EPI_ISL_774969 | Bangladesh/DNAS_DRICM_isl_ICHRI_ | 2020-06-28 | Asia/Bangladesh/Dhaka/ | Designated Reference Institute for Chemical Measurements (DRICM) | DNA SOLUTION LTD. | Md. Imran Khan et al |

| | | | | | | |
|---|---|---|---|---|---|---|
| EPI_ISL_774971 | Bangladesh/DNAS_DRICM_isl_MH-32 | 2020-07-11 | Asia/Bangladesh/Dhaka/ | Designated Reference Institute for Chemical Measurements (DRICM) | DNA SOLUTION LTD. | Md. Imran Khan et al |
| EPI_ISL_774972 | Bangladesh/DNAS_DRICM_isl_SIBL-5/ | 2020-07-11 | Asia/Bangladesh/Dhaka/ | Designated Reference Institute for Chemical Measurements (DRICM) | DNA SOLUTION LTD. | Md. Imran Khan et al |
| EPI_ISL_774973 | Bangladesh/DNAS_DRICM_isl_MH-22 | 2020-07-11 | Asia/Bangladesh/Dhaka/ | Designated Reference Institute for Chemical Measurements (DRICM) | DNA SOLUTION LTD. | Md. Imran Khan et al |
| EPI_ISL_774976 | Bangladesh/GRMDRC/2020 | 2020-12-19 | Asia/Bangladesh/Dhaka/Dhamrai | Gonoshasthya-RNA Molecular Diagnostic and Research Center | Gonoshasthya-RNA Molecular Diagnostic and Research Center | Nihad Adnan et al |
| EPI_ISL_774977 | Bangladesh/DNAS_DRICM_isl_19205 | 2020-06-28 | Asia/Bangladesh/Dhaka/ | Designated Reference Institute for Chemical Measurements (DRICM) | DNA SOLUTION LTD. | Md. Imran Khan et al |
| EPI_ISL_774978 | Bangladesh/DNAS_DRICM_isl_B-4/20 | 2020-07-12 | Asia/Bangladesh/Dhaka/ | Designated Reference Institute for Chemical Measurements (DRICM) | DNA SOLUTION LTD. | Md. Imran Khan et al |
| EPI_ISL_774980 | Bangladesh/DNAS_DRICM_isl_B-2/20 | 2020-07-12 | Asia/Bangladesh/Dhaka/ | Designated Reference Institute for Chemical Measurements (DRICM) | DNA SOLUTION LTD. | Md. Imran Khan et al |
| EPI_ISL_774982 | Bangladesh/DNAS_DRICM_isl_MH-09 | 2020-07-12 | Asia/Bangladesh/Dhaka/ | Designated Reference Institute for Chemical Measurements (DRICM) | DNA SOLUTION LTD. | Md. Imran Khan et al |
| EPI_ISL_774985 | Bangladesh/DNAS_DRICM_isl_Iffat/2 | 2020-07-10 | Asia/Bangladesh/Dhaka/ | Designated Reference Institute for Chemical Measurements (DRICM) | DNA SOLUTION LTD. | Md. Imran Khan et al |
| EPI_ISL_774986 | Bangladesh/DNAS_DRICM_isl_IH-2/2 | 2020-07-12 | Asia/Bangladesh/Dhaka/ | Designated Reference Institute for Chemical Measurements (DRICM) | DNA SOLUTION LTD. | Md. Imran Khan et al |
| EPI_ISL_774988 | Bangladesh/DNAS_DRICM_isl_18927 | 2020-06-27 | Asia/Bangladesh/Dhaka/ | Designated Reference Institute for Chemical Measurements (DRICM) | DNA SOLUTION LTD. | Md. Imran Khan et al |
| EPI_ISL_774989 | Bangladesh/DNAS_DRICM_isl_HH-08 | 2020-07-11 | Asia/Bangladesh/Dhaka/ | Designated Reference Institute for Chemical Measurements (DRICM) | DNA SOLUTION LTD. | Md. Imran Khan et al |
| EPI_ISL_774990 | Bangladesh/DNAS_DRICM_isl_MM-5 | 2020-06-29 | Asia/Bangladesh/Dhaka/ | Designated Reference Institute for Chemical Measurements (DRICM) | DNA SOLUTION LTD. | Md. Imran Khan et al |
| EPI_ISL_774995 | Bangladesh/DNAS_DRICM_isl_Enaye | 2020-07-10 | Asia/Bangladesh/Dhaka/ | Designated Reference Institute for Chemical Measurements (DRICM) | DNA SOLUTION LTD. | Md. Imran Khan et al |
| EPI_ISL_774997 | Bangladesh/DNAS_DRICM_isl_OPS_3 | 2020-06-28 | Asia/Bangladesh/Dhaka/ | Designated Reference Institute for Chemical Measurements (DRICM) | DNA SOLUTION LTD. | Md. Imran Khan et al |
| EPI_ISL_774998 | Bangladesh/DNAS_DRICM_isl_Ismail/ | 2020-06-28 | Asia/Bangladesh/Dhaka/ | Designated Reference Institute for Chemical Measurements (DRICM) | DNA SOLUTION LTD. | Md. Imran Khan et al |
| EPI_ISL_775001 | Bangladesh/DNAS_DRICM_isl_UMCH | 2020-06-28 | Asia/Bangladesh/Dhaka/ | Designated Reference Institute for Chemical Measurements (DRICM) | DNA SOLUTION LTD. | Md. Imran Khan et al |
| EPI_ISL_775002 | Bangladesh/DNAS_DRICM_isl_18974 | 2020-06-27 | Asia/Bangladesh/Dhaka/ | Designated Reference Institute for Chemical Measurements (DRICM) | DNA SOLUTION LTD. | Md. Imran Khan et al |
| EPI_ISL_775005 | Bangladesh/DNAS_DRICM_isl_R-6/20 | 2020-07-17 | Asia/Bangladesh/Dhaka/ | Designated Reference Institute for Chemical Measurements (DRICM) | DNA SOLUTION LTD. | Md. Imran Khan et al |

| | | | | | | |
|---|---|---|---|---|---|---|
| EPI_ISL_775008 | Bangladesh/DNAS_DRICM_isl_MH-25 | 2020-06-28 | Asia/Bangladesh/Dhaka/ | Designated Reference Institute for Chemical Measurements (DRICM) | DNA SOLUTION LTD. | Md. Imran Khan et al |
| EPI_ISL_775013 | Bangladesh/DNAS_DRICM_isl_S-1/20 | 2020-07-22 | Asia/Bangladesh/Dhaka/ | Designated Reference Institute for Chemical Measurements (DRICM) | DNA SOLUTION LTD. | Md. Imran Khan et al |
| EPI_ISL_775014 | Bangladesh/DNAS_DRICM_isl_SM-14 | 2020-07-20 | Asia/Bangladesh/Dhaka/ | Designated Reference Institute for Chemical Measurements (DRICM) | DNA SOLUTION LTD. | Md. Imran Khan et al |
| EPI_ISL_775016 | Bangladesh/DNAS_DRICM_isl_SM-12 | 2020-07-20 | Asia/Bangladesh/Dhaka/ | Designated Reference Institute for Chemical Measurements (DRICM) | DNA SOLUTION LTD. | Md. Imran Khan et al |
| EPI_ISL_775017 | Bangladesh/DNAS_DRICM_isl_Sharm | 2020-07-21 | Asia/Bangladesh/Dhaka/ | Designated Reference Institute for Chemical Measurements (DRICM) | DNA SOLUTION LTD. | Md. Imran Khan et al |
| EPI_ISL_775021 | Canada/ON-PHL-20-03810/2020 | 20-Nov | North America/Canada/Ontario/ | Public Health Ontario Laboratory | Public Health Ontario Laboratory | Vanessa G Allen et al |
| EPI_ISL_775022 | Canada/ON-PHL-20-03811/2020 | 20-Nov | North America/Canada/Ontario/ | Public Health Ontario Laboratory | Public Health Ontario Laboratory | Vanessa G Allen et al |
| EPI_ISL_775023 | Canada/ON-PHL-20-03812/2020 | 20-Nov | North America/Canada/Ontario/ | Public Health Ontario Laboratory | Public Health Ontario Laboratory | Vanessa G Allen et al |
| EPI_ISL_775024 | Canada/ON-PHL-20-03813/2020 | 20-Nov | North America/Canada/Ontario/ | Public Health Ontario Laboratory | Public Health Ontario Laboratory | Vanessa G Allen et al |
| EPI_ISL_775025 | Canada/ON-PHL-20-03814/2020 | 20-Nov | North America/Canada/Ontario/ | Public Health Ontario Laboratory | Public Health Ontario Laboratory | Vanessa G Allen et al |
| EPI_ISL_775026 | Canada/ON-PHL-20-03815/2020 | 20-Nov | North America/Canada/Ontario/ | Public Health Ontario Laboratory | Public Health Ontario Laboratory | Vanessa G Allen et al |
| EPI_ISL_775027 | Canada/ON-PHL-20-03816/2020 | 20-Nov | North America/Canada/Ontario/ | Public Health Ontario Laboratory | Public Health Ontario Laboratory | Vanessa G Allen et al |
| EPI_ISL_775028 | Canada/ON-PHL-20-03817/2020 | 20-Nov | North America/Canada/Ontario/ | Public Health Ontario Laboratory | Public Health Ontario Laboratory | Vanessa G Allen et al |
| EPI_ISL_775029 | Canada/ON-PHL-20-03819/2020 | 20-Nov | North America/Canada/Ontario/ | Public Health Ontario Laboratory | Public Health Ontario Laboratory | Vanessa G Allen et al |
| EPI_ISL_775030 | Canada/ON-PHL-20-03820/2020 | 20-Nov | North America/Canada/Ontario/ | Public Health Ontario Laboratory | Public Health Ontario Laboratory | Vanessa G Allen et al |

| | | | | | | |
|---|---|---|---|---|---|---|
| EPI_ISL_775031 | Canada/ON-PHL-20-03821/2020 | 20-Nov | North America/Canada/Ontario/ | Public Health Ontario Laboratory | Public Health Ontario Laboratory | Vanessa G Allen et al |
| EPI_ISL_775032 | Canada/ON-PHL-20-03823/2020 | 20-Nov | North America/Canada/Ontario/ | Public Health Ontario Laboratory | Public Health Ontario Laboratory | Vanessa G Allen et al |
| EPI_ISL_775033 | Canada/ON-PHL-20-03833/2020 | 20-Nov | North America/Canada/Ontario/ | Public Health Ontario Laboratory | Public Health Ontario Laboratory | Vanessa G Allen et al |
| EPI_ISL_775034 | Canada/ON-PHL-20-03834/2020 | 20-Nov | North America/Canada/Ontario/ | Public Health Ontario Laboratory | Public Health Ontario Laboratory | Vanessa G Allen et al |
| EPI_ISL_775035 | Canada/ON-PHL-20-03751/2020 | 20-Nov | North America/Canada/Ontario/ | Public Health Ontario Laboratory | Public Health Ontario Laboratory | Vanessa G Allen et al |
| EPI_ISL_775036 | Canada/ON-PHL-20-03752/2020 | 20-Nov | North America/Canada/Ontario/ | Public Health Ontario Laboratory | Public Health Ontario Laboratory | Vanessa G Allen et al |
| EPI_ISL_775037 | Canada/ON-PHL-20-03754/2020 | 20-Nov | North America/Canada/Ontario/ | Public Health Ontario Laboratory | Public Health Ontario Laboratory | Vanessa G Allen et al |
| EPI_ISL_775038 | Canada/ON-PHL-20-03755/2020 | 20-Nov | North America/Canada/Ontario/ | Public Health Ontario Laboratory | Public Health Ontario Laboratory | Vanessa G Allen et al |
| EPI_ISL_775039 | Canada/ON-PHL-20-03756/2020 | 20-Nov | North America/Canada/Ontario/ | Public Health Ontario Laboratory | Public Health Ontario Laboratory | Vanessa G Allen et al |
| EPI_ISL_775040 | Canada/ON-PHL-20-03757/2020 | 20-Nov | North America/Canada/Ontario/ | Public Health Ontario Laboratory | Public Health Ontario Laboratory | Vanessa G Allen et al |
| EPI_ISL_775041 | Canada/ON-PHL-20-03758/2020 | 20-Nov | North America/Canada/Ontario/ | Public Health Ontario Laboratory | Public Health Ontario Laboratory | Vanessa G Allen et al |
| EPI_ISL_775042 | Canada/ON-PHL-20-03759/2020 | 20-Nov | North America/Canada/Ontario/ | Public Health Ontario Laboratory | Public Health Ontario Laboratory | Vanessa G Allen et al |
| EPI_ISL_775043 | Canada/ON-PHL-20-03782/2020 | 20-Nov | North America/Canada/Ontario/ | Public Health Ontario Laboratory | Public Health Ontario Laboratory | Vanessa G Allen et al |

| | | | | | | |
|---|---|---|---|---|---|---|
| EPI_ISL_775044 | Canada/ON-PHL-20-03783/2020 | 20-Nov | North America/Canada/Ontario/ | Public Health Ontario Laboratory | Public Health Ontario Laboratory | Vanessa G Allen et al |
| EPI_ISL_775045 | Canada/ON-PHL-20-03784/2020 | 20-Nov | North America/Canada/Ontario/ | Public Health Ontario Laboratory | Public Health Ontario Laboratory | Vanessa G Allen et al |
| EPI_ISL_775046 | Canada/ON-PHL-20-03785/2020 | 20-Nov | North America/Canada/Ontario/ | Public Health Ontario Laboratory | Public Health Ontario Laboratory | Vanessa G Allen et al |
| EPI_ISL_775047 | Canada/ON-PHL-20-03786/2020 | 20-Nov | North America/Canada/Ontario/ | Public Health Ontario Laboratory | Public Health Ontario Laboratory | Vanessa G Allen et al |
| EPI_ISL_775048 | Canada/ON-PHL-20-03787/2020 | 20-Nov | North America/Canada/Ontario/ | Public Health Ontario Laboratory | Public Health Ontario Laboratory | Vanessa G Allen et al |
| EPI_ISL_775049 | Canada/ON-PHL-20-03788/2020 | 20-Nov | North America/Canada/Ontario/ | Public Health Ontario Laboratory | Public Health Ontario Laboratory | Vanessa G Allen et al |
| EPI_ISL_775050 | Canada/ON-PHL-20-03789/2020 | 20-Nov | North America/Canada/Ontario/ | Public Health Ontario Laboratory | Public Health Ontario Laboratory | Vanessa G Allen et al |
| EPI_ISL_775051 | Canada/ON-PHL-20-03790/2020 | 20-Nov | North America/Canada/Ontario/ | Public Health Ontario Laboratory | Public Health Ontario Laboratory | Vanessa G Allen et al |
| EPI_ISL_775052 | Canada/ON-PHL-20-03791/2020 | 20-Nov | North America/Canada/Ontario/ | Public Health Ontario Laboratory | Public Health Ontario Laboratory | Vanessa G Allen et al |
| EPI_ISL_775053 | Canada/ON-PHL-20-03792/2020 | 20-Nov | North America/Canada/Ontario/ | Public Health Ontario Laboratory | Public Health Ontario Laboratory | Vanessa G Allen et al |
| EPI_ISL_775054 | Canada/ON-PHL-20-03793/2020 | 20-Nov | North America/Canada/Ontario/ | Public Health Ontario Laboratory | Public Health Ontario Laboratory | Vanessa G Allen et al |
| EPI_ISL_775055 | Canada/ON-PHL-20-03794/2020 | 20-Nov | North America/Canada/Ontario/ | Public Health Ontario Laboratory | Public Health Ontario Laboratory | Vanessa G Allen et al |
| EPI_ISL_775056 | Canada/ON-PHL-20-03795/2020 | 20-Nov | North America/Canada/Ontario/ | Public Health Ontario Laboratory | Public Health Ontario Laboratory | Vanessa G Allen et al |

| | | | | | | |
|---|---|---|---|---|---|---|
| EPI_ISL_775057 | Canada/ON-PHL-20-03796/2020 | 20-Nov | North America/Canada/Ontario/ | Public Health Ontario Laboratory | Public Health Ontario Laboratory | Vanessa G Allen et al |
| EPI_ISL_775058 | Canada/ON-PHL-20-03797/2020 | 20-Nov | North America/Canada/Ontario/ | Public Health Ontario Laboratory | Public Health Ontario Laboratory | Vanessa G Allen et al |
| EPI_ISL_775059 | Canada/ON-PHL-20-03798/2020 | 20-Nov | North America/Canada/Ontario/ | Public Health Ontario Laboratory | Public Health Ontario Laboratory | Vanessa G Allen et al |
| EPI_ISL_775060 | Canada/ON-PHL-20-03799/2020 | 20-Nov | North America/Canada/Ontario/ | Public Health Ontario Laboratory | Public Health Ontario Laboratory | Vanessa G Allen et al |
| EPI_ISL_775061 | Canada/ON-PHL-20-03800/2020 | 20-Nov | North America/Canada/Ontario/ | Public Health Ontario Laboratory | Public Health Ontario Laboratory | Vanessa G Allen et al |
| EPI_ISL_775062 | Canada/ON-PHL-20-03801/2020 | 20-Nov | North America/Canada/Ontario/ | Public Health Ontario Laboratory | Public Health Ontario Laboratory | Vanessa G Allen et al |
| EPI_ISL_775063 | Canada/ON-PHL-20-03802/2020 | 20-Nov | North America/Canada/Ontario/ | Public Health Ontario Laboratory | Public Health Ontario Laboratory | Vanessa G Allen et al |
| EPI_ISL_775064 | Canada/ON-PHL-20-03803/2020 | 20-Nov | North America/Canada/Ontario/ | Public Health Ontario Laboratory | Public Health Ontario Laboratory | Vanessa G Allen et al |
| EPI_ISL_775065 | Canada/ON-PHL-20-03804/2020 | 20-Nov | North America/Canada/Ontario/ | Public Health Ontario Laboratory | Public Health Ontario Laboratory | Vanessa G Allen et al |
| EPI_ISL_775066 | Canada/ON-PHL-20-03805/2020 | 20-Nov | North America/Canada/Ontario/ | Public Health Ontario Laboratory | Public Health Ontario Laboratory | Vanessa G Allen et al |
| EPI_ISL_775067 | Canada/ON-PHL-20-03806/2020 | 20-Nov | North America/Canada/Ontario/ | Public Health Ontario Laboratory | Public Health Ontario Laboratory | Vanessa G Allen et al |
| EPI_ISL_775068 | Canada/ON-PHL-20-03807/2020 | 20-Nov | North America/Canada/Ontario/ | Public Health Ontario Laboratory | Public Health Ontario Laboratory | Vanessa G Allen et al |
| EPI_ISL_775069 | Canada/ON-PHL-20-03808/2020 | 20-Nov | North America/Canada/Ontario/ | Public Health Ontario Laboratory | Public Health Ontario Laboratory | Vanessa G Allen et al |

| | | | | | | |
|---|---|---|---|---|---|---|
| EPI_ISL_775070 | Canada/ON-PHL-20-03760/2020 | 20-Nov | North America/Canada/Ontario/ | Public Health Ontario Laboratory | Public Health Ontario Laboratory | Vanessa G Allen et al |
| EPI_ISL_775071 | Canada/ON-PHL-20-03761/2020 | 20-Nov | North America/Canada/Ontario/ | Public Health Ontario Laboratory | Public Health Ontario Laboratory | Vanessa G Allen et al |
| EPI_ISL_775072 | Canada/ON-PHL-20-03762/2020 | 20-Nov | North America/Canada/Ontario/ | Public Health Ontario Laboratory | Public Health Ontario Laboratory | Vanessa G Allen et al |
| EPI_ISL_775073 | Canada/ON-PHL-20-03763/2020 | 20-Nov | North America/Canada/Ontario/ | Public Health Ontario Laboratory | Public Health Ontario Laboratory | Vanessa G Allen et al |
| EPI_ISL_775074 | Canada/ON-PHL-20-03764/2020 | 20-Nov | North America/Canada/Ontario/ | Public Health Ontario Laboratory | Public Health Ontario Laboratory | Vanessa G Allen et al |
| EPI_ISL_775075 | Canada/ON-PHL-20-03765/2020 | 20-Nov | North America/Canada/Ontario/ | Public Health Ontario Laboratory | Public Health Ontario Laboratory | Vanessa G Allen et al |
| EPI_ISL_775076 | Canada/ON-PHL-20-03766/2020 | 20-Nov | North America/Canada/Ontario/ | Public Health Ontario Laboratory | Public Health Ontario Laboratory | Vanessa G Allen et al |
| EPI_ISL_775077 | Canada/ON-PHL-20-03767/2020 | 20-Nov | North America/Canada/Ontario/ | Public Health Ontario Laboratory | Public Health Ontario Laboratory | Vanessa G Allen et al |
| EPI_ISL_775078 | Canada/ON-PHL-20-03768/2020 | 20-Nov | North America/Canada/Ontario/ | Public Health Ontario Laboratory | Public Health Ontario Laboratory | Vanessa G Allen et al |
| EPI_ISL_775079 | Canada/ON-PHL-20-03770/2020 | 20-Nov | North America/Canada/Ontario/ | Public Health Ontario Laboratory | Public Health Ontario Laboratory | Vanessa G Allen et al |
| EPI_ISL_775080 | Canada/ON-PHL-20-03771/2020 | 20-Nov | North America/Canada/Ontario/ | Public Health Ontario Laboratory | Public Health Ontario Laboratory | Vanessa G Allen et al |
| EPI_ISL_775081 | Canada/ON-PHL-20-03772/2020 | 20-Nov | North America/Canada/Ontario/ | Public Health Ontario Laboratory | Public Health Ontario Laboratory | Vanessa G Allen et al |
| EPI_ISL_775082 | Canada/ON-PHL-20-03773/2020 | 20-Nov | North America/Canada/Ontario/ | Public Health Ontario Laboratory | Public Health Ontario Laboratory | Vanessa G Allen et al |

| | | | | | | |
|---|---|---|---|---|---|---|
| EPI_ISL_775083 | Canada/ON-PHL-20-03774/2020 | 20-Nov | North America/Canada/Ontario/ | Public Health Ontario Laboratory | Public Health Ontario Laboratory | Vanessa G Allen et al |
| EPI_ISL_775084 | Canada/ON-PHL-20-03775/2020 | 20-Nov | North America/Canada/Ontario/ | Public Health Ontario Laboratory | Public Health Ontario Laboratory | Vanessa G Allen et al |
| EPI_ISL_775085 | Canada/ON-PHL-20-03776/2020 | 20-Nov | North America/Canada/Ontario/ | Public Health Ontario Laboratory | Public Health Ontario Laboratory | Vanessa G Allen et al |
| EPI_ISL_775086 | Canada/ON-PHL-20-03777/2020 | 20-Nov | North America/Canada/Ontario/ | Public Health Ontario Laboratory | Public Health Ontario Laboratory | Vanessa G Allen et al |
| EPI_ISL_775087 | Canada/ON-PHL-20-03779/2020 | 20-Nov | North America/Canada/Ontario/ | Public Health Ontario Laboratory | Public Health Ontario Laboratory | Vanessa G Allen et al |
| EPI_ISL_775089 | Canada/ON-PHL-20-03781/2020 | 20-Nov | North America/Canada/Ontario/ | Public Health Ontario Laboratory | Public Health Ontario Laboratory | Vanessa G Allen et al |
| EPI_ISL_775090 | Canada/ON-PHL-20-03651/2020 | 20-Nov | North America/Canada/Ontario/ | Public Health Ontario Laboratory | Public Health Ontario Laboratory | Vanessa G Allen et al |
| EPI_ISL_775091 | Canada/ON-PHL-20-03652/2020 | 20-Nov | North America/Canada/Ontario/ | Public Health Ontario Laboratory | Public Health Ontario Laboratory | Vanessa G Allen et al |
| EPI_ISL_775092 | Canada/ON-PHL-20-03653/2020 | 20-Nov | North America/Canada/Ontario/ | Public Health Ontario Laboratory | Public Health Ontario Laboratory | Vanessa G Allen et al |
| EPI_ISL_775093 | Canada/ON-PHL-20-03654/2020 | 20-Nov | North America/Canada/Ontario/ | Public Health Ontario Laboratory | Public Health Ontario Laboratory | Vanessa G Allen et al |
| EPI_ISL_775094 | Canada/ON-PHL-20-03655/2020 | 20-Nov | North America/Canada/Ontario/ | Public Health Ontario Laboratory | Public Health Ontario Laboratory | Vanessa G Allen et al |
| EPI_ISL_775095 | Canada/ON-PHL-20-03656/2020 | 20-Nov | North America/Canada/Ontario/ | Public Health Ontario Laboratory | Public Health Ontario Laboratory | Vanessa G Allen et al |
| EPI_ISL_775096 | Canada/ON-PHL-20-03658/2020 | 20-Nov | North America/Canada/Ontario/ | Public Health Ontario Laboratory | Public Health Ontario Laboratory | Vanessa G Allen et al |

| | | | | | | |
|---|---|---|---|---|---|---|
| EPI_ISL_775097 | Canada/ON-PHL-20-03659/2020 | 20-Nov | North America/Canada/Ontario/ | Public Health Ontario Laboratory | Public Health Ontario Laboratory | Vanessa G Allen et al |
| EPI_ISL_775098 | Canada/ON-PHL-20-03660/2020 | 20-Nov | North America/Canada/Ontario/ | Public Health Ontario Laboratory | Public Health Ontario Laboratory | Vanessa G Allen et al |
| EPI_ISL_775099 | Canada/ON-PHL-20-03661/2020 | 20-Nov | North America/Canada/Ontario/ | Public Health Ontario Laboratory | Public Health Ontario Laboratory | Vanessa G Allen et al |
| EPI_ISL_775100 | Canada/ON-PHL-20-03662/2020 | 20-Nov | North America/Canada/Ontario/ | Public Health Ontario Laboratory | Public Health Ontario Laboratory | Vanessa G Allen et al |
| EPI_ISL_775101 | Canada/ON-PHL-20-03663/2020 | 20-Nov | North America/Canada/Ontario/ | Public Health Ontario Laboratory | Public Health Ontario Laboratory | Vanessa G Allen et al |
| EPI_ISL_775102 | Canada/ON-PHL-20-03664/2020 | 20-Nov | North America/Canada/Ontario/ | Public Health Ontario Laboratory | Public Health Ontario Laboratory | Vanessa G Allen et al |
| EPI_ISL_775103 | Canada/ON-PHL-20-03665/2020 | 20-Nov | North America/Canada/Ontario/ | Public Health Ontario Laboratory | Public Health Ontario Laboratory | Vanessa G Allen et al |
| EPI_ISL_775104 | Canada/ON-PHL-20-03666/2020 | 20-Nov | North America/Canada/Ontario/ | Public Health Ontario Laboratory | Public Health Ontario Laboratory | Vanessa G Allen et al |
| EPI_ISL_775105 | Canada/ON-PHL-20-03745/2020 | 20-Nov | North America/Canada/Ontario/ | Public Health Ontario Laboratory | Public Health Ontario Laboratory | Vanessa G Allen et al |
| EPI_ISL_775106 | Canada/ON-PHL-20-03747/2020 | 20-Nov | North America/Canada/Ontario/ | Public Health Ontario Laboratory | Public Health Ontario Laboratory | Vanessa G Allen et al |
| EPI_ISL_775107 | Canada/ON-PHL-20-03748/2020 | 20-Nov | North America/Canada/Ontario/ | Public Health Ontario Laboratory | Public Health Ontario Laboratory | Vanessa G Allen et al |
| EPI_ISL_775108 | Canada/ON-PHL-20-03749/2020 | 20-Nov | North America/Canada/Ontario/ | Public Health Ontario Laboratory | Public Health Ontario Laboratory | Vanessa G Allen et al |
| EPI_ISL_775109 | Canada/ON-PHL-20-03750/2020 | 20-Nov | North America/Canada/Ontario/ | Public Health Ontario Laboratory | Public Health Ontario Laboratory | Vanessa G Allen et al |

| | | | | | | |
|---|---|---|---|---|---|---|
| EPI_ISL_775110 | Canada/ON-PHL-20-03602/2020 | 20-Nov | North America/Canada/Ontario/ | Public Health Ontario Laboratory | Public Health Ontario Laboratory | Vanessa G Allen et al |
| EPI_ISL_775111 | Canada/ON-PHL-20-03603/2020 | 20-Nov | North America/Canada/Ontario/ | Public Health Ontario Laboratory | Public Health Ontario Laboratory | Vanessa G Allen et al |
| EPI_ISL_775112 | Canada/ON-PHL-20-03604/2020 | 20-Nov | North America/Canada/Ontario/ | Public Health Ontario Laboratory | Public Health Ontario Laboratory | Vanessa G Allen et al |
| EPI_ISL_775113 | Canada/ON-PHL-20-03605/2020 | 20-Nov | North America/Canada/Ontario/ | Public Health Ontario Laboratory | Public Health Ontario Laboratory | Vanessa G Allen et al |
| EPI_ISL_775114 | Canada/ON-PHL-20-03606/2020 | 20-Nov | North America/Canada/Ontario/ | Public Health Ontario Laboratory | Public Health Ontario Laboratory | Vanessa G Allen et al |
| EPI_ISL_775115 | Canada/ON-PHL-20-03607/2020 | 20-Nov | North America/Canada/Ontario/ | Public Health Ontario Laboratory | Public Health Ontario Laboratory | Vanessa G Allen et al |
| EPI_ISL_775116 | Canada/ON-PHL-20-03608/2020 | 20-Nov | North America/Canada/Ontario/ | Public Health Ontario Laboratory | Public Health Ontario Laboratory | Vanessa G Allen et al |
| EPI_ISL_775117 | Canada/ON-PHL-20-03609/2020 | 20-Nov | North America/Canada/Ontario/ | Public Health Ontario Laboratory | Public Health Ontario Laboratory | Vanessa G Allen et al |
| EPI_ISL_775118 | Canada/ON-PHL-20-03610/2020 | 20-Nov | North America/Canada/Ontario/ | Public Health Ontario Laboratory | Public Health Ontario Laboratory | Vanessa G Allen et al |
| EPI_ISL_775119 | Canada/ON-PHL-20-03611/2020 | 20-Nov | North America/Canada/Ontario/ | Public Health Ontario Laboratory | Public Health Ontario Laboratory | Vanessa G Allen et al |
| EPI_ISL_775120 | Canada/ON-PHL-20-03612/2020 | 20-Nov | North America/Canada/Ontario/ | Public Health Ontario Laboratory | Public Health Ontario Laboratory | Vanessa G Allen et al |
| EPI_ISL_775121 | Canada/ON-PHL-20-03613/2020 | 20-Nov | North America/Canada/Ontario/ | Public Health Ontario Laboratory | Public Health Ontario Laboratory | Vanessa G Allen et al |
| EPI_ISL_775122 | Canada/ON-PHL-20-03615/2020 | 20-Nov | North America/Canada/Ontario/ | Public Health Ontario Laboratory | Public Health Ontario Laboratory | Vanessa G Allen et al |

| | | | | | | |
|---|---|---|---|---|---|---|
| EPI_ISL_775123 | Canada/ON-PHL-20-03616/2020 | 20-Nov | North America/Canada/Ontario/ | Public Health Ontario Laboratory | Public Health Ontario Laboratory | Vanessa G Allen et al |
| EPI_ISL_775124 | Canada/ON-PHL-20-03617/2020 | 20-Nov | North America/Canada/Ontario/ | Public Health Ontario Laboratory | Public Health Ontario Laboratory | Vanessa G Allen et al |
| EPI_ISL_775125 | Canada/ON-PHL-20-03618/2020 | 20-Nov | North America/Canada/Ontario/ | Public Health Ontario Laboratory | Public Health Ontario Laboratory | Vanessa G Allen et al |
| EPI_ISL_775126 | Canada/ON-PHL-20-03619/2020 | 20-Nov | North America/Canada/Ontario/ | Public Health Ontario Laboratory | Public Health Ontario Laboratory | Vanessa G Allen et al |
| EPI_ISL_775128 | Canada/ON-PHL-20-03621/2020 | 20-Nov | North America/Canada/Ontario/ | Public Health Ontario Laboratory | Public Health Ontario Laboratory | Vanessa G Allen et al |
| EPI_ISL_775129 | Canada/ON-PHL-20-03622/2020 | 20-Nov | North America/Canada/Ontario/ | Public Health Ontario Laboratory | Public Health Ontario Laboratory | Vanessa G Allen et al |
| EPI_ISL_775130 | Canada/ON-PHL-20-03623/2020 | 20-Nov | North America/Canada/Ontario/ | Public Health Ontario Laboratory | Public Health Ontario Laboratory | Vanessa G Allen et al |
| EPI_ISL_775131 | Canada/ON-PHL-20-03625/2020 | 20-Nov | North America/Canada/Ontario/ | Public Health Ontario Laboratory | Public Health Ontario Laboratory | Vanessa G Allen et al |
| EPI_ISL_775132 | Canada/ON-PHL-20-03626/2020 | 20-Nov | North America/Canada/Ontario/ | Public Health Ontario Laboratory | Public Health Ontario Laboratory | Vanessa G Allen et al |
| EPI_ISL_775133 | Canada/ON-PHL-20-03627/2020 | 20-Nov | North America/Canada/Ontario/ | Public Health Ontario Laboratory | Public Health Ontario Laboratory | Vanessa G Allen et al |
| EPI_ISL_775134 | Canada/ON-PHL-20-03628/2020 | 20-Nov | North America/Canada/Ontario/ | Public Health Ontario Laboratory | Public Health Ontario Laboratory | Vanessa G Allen et al |
| EPI_ISL_775135 | Canada/ON-PHL-20-03629/2020 | 20-Nov | North America/Canada/Ontario/ | Public Health Ontario Laboratory | Public Health Ontario Laboratory | Vanessa G Allen et al |
| EPI_ISL_775136 | Canada/ON-PHL-20-03632/2020 | 20-Nov | North America/Canada/Ontario/ | Public Health Ontario Laboratory | Public Health Ontario Laboratory | Vanessa G Allen et al |

| | | | | | | |
|---|---|---|---|---|---|---|
| EPI_ISL_775137 | Canada/ON-PHL-20-03633/2020 | 20-Nov | North America/Canada/Ontario/ | Public Health Ontario Laboratory | Public Health Ontario Laboratory | Vanessa G Allen et al |
| EPI_ISL_775138 | Canada/ON-PHL-20-03634/2020 | 20-Nov | North America/Canada/Ontario/ | Public Health Ontario Laboratory | Public Health Ontario Laboratory | Vanessa G Allen et al |
| EPI_ISL_775139 | Canada/ON-PHL-20-03635/2020 | 20-Nov | North America/Canada/Ontario/ | Public Health Ontario Laboratory | Public Health Ontario Laboratory | Vanessa G Allen et al |
| EPI_ISL_775140 | Canada/ON-PHL-20-03636/2020 | 20-Nov | North America/Canada/Ontario/ | Public Health Ontario Laboratory | Public Health Ontario Laboratory | Vanessa G Allen et al |
| EPI_ISL_775141 | Canada/ON-PHL-20-03637/2020 | 20-Nov | North America/Canada/Ontario/ | Public Health Ontario Laboratory | Public Health Ontario Laboratory | Vanessa G Allen et al |
| EPI_ISL_775142 | Canada/ON-PHL-20-03638/2020 | 20-Nov | North America/Canada/Ontario/ | Public Health Ontario Laboratory | Public Health Ontario Laboratory | Vanessa G Allen et al |
| EPI_ISL_775143 | Canada/ON-PHL-20-03639/2020 | 20-Nov | North America/Canada/Ontario/ | Public Health Ontario Laboratory | Public Health Ontario Laboratory | Vanessa G Allen et al |
| EPI_ISL_775144 | Canada/ON-PHL-20-03640/2020 | 20-Nov | North America/Canada/Ontario/ | Public Health Ontario Laboratory | Public Health Ontario Laboratory | Vanessa G Allen et al |
| EPI_ISL_775145 | Canada/ON-PHL-20-03641/2020 | 20-Nov | North America/Canada/Ontario/ | Public Health Ontario Laboratory | Public Health Ontario Laboratory | Vanessa G Allen et al |
| EPI_ISL_775146 | Canada/ON-PHL-20-03642/2020 | 20-Nov | North America/Canada/Ontario/ | Public Health Ontario Laboratory | Public Health Ontario Laboratory | Vanessa G Allen et al |
| EPI_ISL_775147 | Canada/ON-PHL-20-03643/2020 | 20-Nov | North America/Canada/Ontario/ | Public Health Ontario Laboratory | Public Health Ontario Laboratory | Vanessa G Allen et al |
| EPI_ISL_775148 | Canada/ON-PHL-20-03644/2020 | 20-Nov | North America/Canada/Ontario/ | Public Health Ontario Laboratory | Public Health Ontario Laboratory | Vanessa G Allen et al |
| EPI_ISL_775149 | Canada/ON-PHL-20-03645/2020 | 20-Nov | North America/Canada/Ontario/ | Public Health Ontario Laboratory | Public Health Ontario Laboratory | Vanessa G Allen et al |

| | | | | | | |
|---|---|---|---|---|---|---|
| EPI_ISL_775150 | Canada/ON-PHL-20-03646/2020 | 20-Nov | North America/Canada/Ontario/ | Public Health Ontario Laboratory | Public Health Ontario Laboratory | Vanessa G Allen et al |
| EPI_ISL_775151 | Canada/ON-PHL-20-03647/2020 | 20-Nov | North America/Canada/Ontario/ | Public Health Ontario Laboratory | Public Health Ontario Laboratory | Vanessa G Allen et al |
| EPI_ISL_775152 | Canada/ON-PHL-20-03649/2020 | 20-Nov | North America/Canada/Ontario/ | Public Health Ontario Laboratory | Public Health Ontario Laboratory | Vanessa G Allen et al |
| EPI_ISL_775153 | Canada/ON-PHL-20-03667/2020 | 20-Nov | North America/Canada/Ontario/ | Public Health Ontario Laboratory | Public Health Ontario Laboratory | Vanessa G Allen et al |
| EPI_ISL_775154 | Canada/ON-PHL-20-03668/2020 | 20-Nov | North America/Canada/Ontario/ | Public Health Ontario Laboratory | Public Health Ontario Laboratory | Vanessa G Allen et al |
| EPI_ISL_775155 | Canada/ON-PHL-20-03669/2020 | 20-Nov | North America/Canada/Ontario/ | Public Health Ontario Laboratory | Public Health Ontario Laboratory | Vanessa G Allen et al |
| EPI_ISL_775156 | Canada/ON-PHL-20-03670/2020 | 20-Nov | North America/Canada/Ontario/ | Public Health Ontario Laboratory | Public Health Ontario Laboratory | Vanessa G Allen et al |
| EPI_ISL_775157 | Canada/ON-PHL-20-03671/2020 | 20-Nov | North America/Canada/Ontario/ | Public Health Ontario Laboratory | Public Health Ontario Laboratory | Vanessa G Allen et al |
| EPI_ISL_775158 | Canada/ON-PHL-20-03672/2020 | 20-Nov | North America/Canada/Ontario/ | Public Health Ontario Laboratory | Public Health Ontario Laboratory | Vanessa G Allen et al |
| EPI_ISL_775159 | Canada/ON-PHL-20-03675/2020 | 20-Nov | North America/Canada/Ontario/ | Public Health Ontario Laboratory | Public Health Ontario Laboratory | Vanessa G Allen et al |
| EPI_ISL_775160 | Canada/ON-PHL-20-03676/2020 | 20-Nov | North America/Canada/Ontario/ | Public Health Ontario Laboratory | Public Health Ontario Laboratory | Vanessa G Allen et al |
| EPI_ISL_775161 | Canada/ON-PHL-20-03677/2020 | 20-Nov | North America/Canada/Ontario/ | Public Health Ontario Laboratory | Public Health Ontario Laboratory | Vanessa G Allen et al |
| EPI_ISL_775162 | Canada/ON-PHL-20-03678/2020 | 20-Nov | North America/Canada/Ontario/ | Public Health Ontario Laboratory | Public Health Ontario Laboratory | Vanessa G Allen et al |

| | | | | | | |
|---|---|---|---|---|---|---|
| EPI_ISL_775163 | Canada/ON-PHL-20-03679/2020 | 20-Nov | North America/Canada/Ontario/ | Public Health Ontario Laboratory | Public Health Ontario Laboratory | Vanessa G Allen et al |
| EPI_ISL_775164 | Canada/ON-PHL-20-03680/2020 | 20-Nov | North America/Canada/Ontario/ | Public Health Ontario Laboratory | Public Health Ontario Laboratory | Vanessa G Allen et al |
| EPI_ISL_775165 | Canada/ON-PHL-20-03681/2020 | 20-Nov | North America/Canada/Ontario/ | Public Health Ontario Laboratory | Public Health Ontario Laboratory | Vanessa G Allen et al |
| EPI_ISL_775166 | Canada/ON-PHL-20-03683/2020 | 20-Nov | North America/Canada/Ontario/ | Public Health Ontario Laboratory | Public Health Ontario Laboratory | Vanessa G Allen et al |
| EPI_ISL_775167 | Canada/ON-PHL-20-03684/2020 | 20-Nov | North America/Canada/Ontario/ | Public Health Ontario Laboratory | Public Health Ontario Laboratory | Vanessa G Allen et al |
| EPI_ISL_775168 | Canada/ON-PHL-20-03685/2020 | 20-Nov | North America/Canada/Ontario/ | Public Health Ontario Laboratory | Public Health Ontario Laboratory | Vanessa G Allen et al |
| EPI_ISL_775169 | Canada/ON-PHL-20-03686/2020 | 20-Nov | North America/Canada/Ontario/ | Public Health Ontario Laboratory | Public Health Ontario Laboratory | Vanessa G Allen et al |
| EPI_ISL_775170 | Canada/ON-PHL-20-03687/2020 | 20-Nov | North America/Canada/Ontario/ | Public Health Ontario Laboratory | Public Health Ontario Laboratory | Vanessa G Allen et al |
| EPI_ISL_775171 | Canada/ON-PHL-20-03688/2020 | 20-Nov | North America/Canada/Ontario/ | Public Health Ontario Laboratory | Public Health Ontario Laboratory | Vanessa G Allen et al |
| EPI_ISL_775172 | Canada/ON-PHL-20-03689/2020 | 20-Nov | North America/Canada/Ontario/ | Public Health Ontario Laboratory | Public Health Ontario Laboratory | Vanessa G Allen et al |
| EPI_ISL_775173 | Canada/ON-PHL-20-03690/2020 | 20-Nov | North America/Canada/Ontario/ | Public Health Ontario Laboratory | Public Health Ontario Laboratory | Vanessa G Allen et al |
| EPI_ISL_775174 | Canada/ON-PHL-20-03691/2020 | 20-Nov | North America/Canada/Ontario/ | Public Health Ontario Laboratory | Public Health Ontario Laboratory | Vanessa G Allen et al |
| EPI_ISL_775175 | Canada/ON-PHL-20-03692/2020 | 20-Nov | North America/Canada/Ontario/ | Public Health Ontario Laboratory | Public Health Ontario Laboratory | Vanessa G Allen et al |

| | | | | | | |
|---|---|---|---|---|---|---|
| EPI_ISL_775176 | Canada/ON-PHL-20-03693/2020 | 20-Nov | North America/Canada/Ontario/ | Public Health Ontario Laboratory | Public Health Ontario Laboratory | Vanessa G Allen et al |
| EPI_ISL_775177 | Canada/ON-PHL-20-03694/2020 | 20-Nov | North America/Canada/Ontario/ | Public Health Ontario Laboratory | Public Health Ontario Laboratory | Vanessa G Allen et al |
| EPI_ISL_775178 | Canada/ON-PHL-20-03695/2020 | 20-Nov | North America/Canada/Ontario/ | Public Health Ontario Laboratory | Public Health Ontario Laboratory | Vanessa G Allen et al |
| EPI_ISL_775179 | Canada/ON-PHL-20-03696/2020 | 20-Nov | North America/Canada/Ontario/ | Public Health Ontario Laboratory | Public Health Ontario Laboratory | Vanessa G Allen et al |
| EPI_ISL_775180 | Canada/ON-PHL-20-03697/2020 | 20-Nov | North America/Canada/Ontario/ | Public Health Ontario Laboratory | Public Health Ontario Laboratory | Vanessa G Allen et al |
| EPI_ISL_775181 | Canada/ON-PHL-20-03698/2020 | 20-Nov | North America/Canada/Ontario/ | Public Health Ontario Laboratory | Public Health Ontario Laboratory | Vanessa G Allen et al |
| EPI_ISL_775182 | Canada/ON-PHL-20-03699/2020 | 20-Nov | North America/Canada/Ontario/ | Public Health Ontario Laboratory | Public Health Ontario Laboratory | Vanessa G Allen et al |
| EPI_ISL_775183 | Canada/ON-PHL-20-03700/2020 | 20-Nov | North America/Canada/Ontario/ | Public Health Ontario Laboratory | Public Health Ontario Laboratory | Vanessa G Allen et al |
| EPI_ISL_775184 | Canada/ON-PHL-20-03701/2020 | 20-Nov | North America/Canada/Ontario/ | Public Health Ontario Laboratory | Public Health Ontario Laboratory | Vanessa G Allen et al |
| EPI_ISL_775185 | Canada/ON-PHL-20-03702/2020 | 20-Nov | North America/Canada/Ontario/ | Public Health Ontario Laboratory | Public Health Ontario Laboratory | Vanessa G Allen et al |
| EPI_ISL_775186 | Canada/ON-PHL-20-03703/2020 | 20-Nov | North America/Canada/Ontario/ | Public Health Ontario Laboratory | Public Health Ontario Laboratory | Vanessa G Allen et al |
| EPI_ISL_775187 | Canada/ON-PHL-20-03704/2020 | 20-Nov | North America/Canada/Ontario/ | Public Health Ontario Laboratory | Public Health Ontario Laboratory | Vanessa G Allen et al |
| EPI_ISL_775188 | Canada/ON-PHL-20-03705/2020 | 20-Nov | North America/Canada/Ontario/ | Public Health Ontario Laboratory | Public Health Ontario Laboratory | Vanessa G Allen et al |

| | | | | | | |
|---|---|---|---|---|---|---|
| EPI_ISL_775189 | Canada/ON-PHL-20-03707/2020 | 20-Nov | North America/Canada/Ontario/ | Public Health Ontario Laboratory | Public Health Ontario Laboratory | Vanessa G Allen et al |
| EPI_ISL_775190 | Canada/ON-PHL-20-03708/2020 | 20-Nov | North America/Canada/Ontario/ | Public Health Ontario Laboratory | Public Health Ontario Laboratory | Vanessa G Allen et al |
| EPI_ISL_775191 | Canada/ON-PHL-20-03709/2020 | 20-Nov | North America/Canada/Ontario/ | Public Health Ontario Laboratory | Public Health Ontario Laboratory | Vanessa G Allen et al |
| EPI_ISL_775192 | Canada/ON-PHL-20-03710/2020 | 20-Nov | North America/Canada/Ontario/ | Public Health Ontario Laboratory | Public Health Ontario Laboratory | Vanessa G Allen et al |
| EPI_ISL_775193 | Canada/ON-PHL-20-03711/2020 | 20-Nov | North America/Canada/Ontario/ | Public Health Ontario Laboratory | Public Health Ontario Laboratory | Vanessa G Allen et al |
| EPI_ISL_775194 | Canada/ON-PHL-20-03712/2020 | 20-Nov | North America/Canada/Ontario/ | Public Health Ontario Laboratory | Public Health Ontario Laboratory | Vanessa G Allen et al |
| EPI_ISL_775195 | Canada/ON-PHL-20-03713/2020 | 20-Nov | North America/Canada/Ontario/ | Public Health Ontario Laboratory | Public Health Ontario Laboratory | Vanessa G Allen et al |
| EPI_ISL_775196 | Canada/ON-PHL-20-03714/2020 | 20-Nov | North America/Canada/Ontario/ | Public Health Ontario Laboratory | Public Health Ontario Laboratory | Vanessa G Allen et al |
| EPI_ISL_775197 | Canada/ON-PHL-20-03715/2020 | 20-Nov | North America/Canada/Ontario/ | Public Health Ontario Laboratory | Public Health Ontario Laboratory | Vanessa G Allen et al |
| EPI_ISL_775198 | Canada/ON-PHL-20-03716/2020 | 20-Nov | North America/Canada/Ontario/ | Public Health Ontario Laboratory | Public Health Ontario Laboratory | Vanessa G Allen et al |
| EPI_ISL_775199 | Canada/ON-PHL-20-03718/2020 | 20-Nov | North America/Canada/Ontario/ | Public Health Ontario Laboratory | Public Health Ontario Laboratory | Vanessa G Allen et al |
| EPI_ISL_775200 | Canada/ON-PHL-20-03719/2020 | 20-Nov | North America/Canada/Ontario/ | Public Health Ontario Laboratory | Public Health Ontario Laboratory | Vanessa G Allen et al |
| EPI_ISL_775201 | Canada/ON-PHL-20-03720/2020 | 20-Nov | North America/Canada/Ontario/ | Public Health Ontario Laboratory | Public Health Ontario Laboratory | Vanessa G Allen et al |

| | | | | | | |
|---|---|---|---|---|---|---|
| EPI_ISL_775202 | Canada/ON-PHL-20-03721/2020 | 20-Nov | North America/Canada/Ontario/ | Public Health Ontario Laboratory | Public Health Ontario Laboratory | Vanessa G Allen et al |
| EPI_ISL_775203 | Canada/ON-PHL-20-03723/2020 | 20-Nov | North America/Canada/Ontario/ | Public Health Ontario Laboratory | Public Health Ontario Laboratory | Vanessa G Allen et al |
| EPI_ISL_775204 | Canada/ON-PHL-20-03724/2020 | 20-Nov | North America/Canada/Ontario/ | Public Health Ontario Laboratory | Public Health Ontario Laboratory | Vanessa G Allen et al |
| EPI_ISL_775205 | Canada/ON-PHL-20-03732/2020 | 20-Nov | North America/Canada/Ontario/ | Public Health Ontario Laboratory | Public Health Ontario Laboratory | Vanessa G Allen et al |
| EPI_ISL_775206 | Canada/ON-PHL-20-03733/2020 | 20-Nov | North America/Canada/Ontario/ | Public Health Ontario Laboratory | Public Health Ontario Laboratory | Vanessa G Allen et al |
| EPI_ISL_775207 | Canada/ON-PHL-20-03737/2020 | 20-Nov | North America/Canada/Ontario/ | Public Health Ontario Laboratory | Public Health Ontario Laboratory | Vanessa G Allen et al |
| EPI_ISL_775208 | Canada/ON-PHL-20-03740/2020 | 20-Nov | North America/Canada/Ontario/ | Public Health Ontario Laboratory | Public Health Ontario Laboratory | Vanessa G Allen et al |
| EPI_ISL_775209 | Canada/ON-PHL-20-03741/2020 | 20-Nov | North America/Canada/Ontario/ | Public Health Ontario Laboratory | Public Health Ontario Laboratory | Vanessa G Allen et al |
| EPI_ISL_775210 | Canada/ON-PHL-20-03742/2020 | 20-Nov | North America/Canada/Ontario/ | Public Health Ontario Laboratory | Public Health Ontario Laboratory | Vanessa G Allen et al |
| EPI_ISL_775211 | Canada/ON-PHL-20-03743/2020 | 20-Nov | North America/Canada/Ontario/ | Public Health Ontario Laboratory | Public Health Ontario Laboratory | Vanessa G Allen et al |
| EPI_ISL_775212 | Canada/ON-PHL-20-03744/2020 | 20-Nov | North America/Canada/Ontario/ | Public Health Ontario Laboratory | Public Health Ontario Laboratory | Vanessa G Allen et al |
| EPI_ISL_775214 | Bangladesh/GRMDRC-5/2020 | 2020-11-14 | Asia/Bangladesh/Dhaka/ | Gonoshasthya-RNA Molecular Research Center | Gonoshasthya-RNA Molecular Research Center | Mohd. Raeed Jamiruddin et al |
| EPI_ISL_775217 | Bangladesh/GRMDRC-8/2020 | 2020-09-05 | Asia/Bangladesh/Dhaka/ | Gonoshasthya-RNA Molecular Research Center | Gonoshasthya-RNA Molecular Research Center | Nihad Adnan et al |

| | | | | | | |
|---|---|---|---|---|---|---|
| EPI_ISL_775218 | Bangladesh/GRMDRC-9/2020 | 2020-12-15 | Asia/Bangladesh/Dhaka/ | Gonoshasthya-RNA Molecular Research Center | Gonoshasthya-RNA Molecular Research Center | Mohd. Raeed Jamiruddin et al |
| EPI_ISL_775219 | Morocco/FMP-43/2020 | 2020-12-14 | Africa/Morocco/Temara/ | Laboratoire Biolife | Laboratoire de Biotechnologie | Mouna Ouadghiri et al |
| EPI_ISL_775220 | Morocco/FMP-44/2020 | 2020-12-15 | Africa/Morocco/Temara/ | Laboratoire Biolife | Laboratoire de Biotechnologie | Mouna Ouadghiri et al |
| EPI_ISL_775222 | Morocco/FMP-46/2020 | 2020-12-16 | Africa/Morocco/SalV©/ | Laboratoire Biolife | Laboratoire de Biotechnologie | Mouna Ouadghiri et al |
| EPI_ISL_775225 | Morocco/FMP-49/2020 | 2020-12-14 | Africa/Morocco/Temara/ | Laboratoire Biolife | Laboratoire de Biotechnologie | Mouna Ouadghiri et al |
| EPI_ISL_775239 | Italy/CAM-INTPas-14/2020 | 2020-04-02 | Europe/Italy/Campania/ | INT Fondazione Pascale | INT Fondazione Pascale | INT Fondazione Pascale et al |
| EPI_ISL_775240 | Italy/CAM-INTPas-15/2020 | 2020-04-03 | Europe/Italy/Campania/ | INT Fondazione Pascale | INT Fondazione Pascale | INT Fondazione Pascale et al |
| EPI_ISL_775256 | Morocco/FMP-57/2020 | 2020-12-21 | Africa/Morocco/Temara/ | Laboratoire Biolife | Laboratoire de Biotechnologie | Mouna Ouadghiri et al |
| EPI_ISL_775257 | Italy/CAM-INTPas-19/2020 | 2020-03-30 | Europe/Italy/Campania/ | INT Fondazione Pascale | INT Fondazione Pascale | INT Fondazione Pascale et al |
| EPI_ISL_775258 | Morocco/FMP-58/2020 | 2020-12-21 | Africa/Morocco/Temara/ | Laboratoire Biolife | Laboratoire de Biotechnologie | Mouna Ouadghiri et al |
| EPI_ISL_775263 | Morocco/FMP-61/2020 | 2020-12-21 | Africa/Morocco/Rabat/ | Laboratoire Biolife | Laboratoire de Biotechnologie | Mouna Ouadghiri et al |
| EPI_ISL_775268 | Slovenia/MB4/2020 | 2020-12-15 | Europe/Slovenia/Slovenia/ | National Laboratory for Health, Environment and Food | National Laboratory for Health, Environment and Food | Aleksander Mahnic et al |
| EPI_ISL_775394 | Norway/6572/2020 | 2020-12-06 | Europe/Norway/Viken/ | Medical Microbiology Unit, Department for Laboratory Medicine, Drammen Hospital, Vestre Viken Health Trust, | Norwegian Institute of Public Health, Department of Virology | Kathrine Stene-Johansen et al |
| EPI_ISL_775398 | Norway/6593/2020 | 2020-11-23 | Europe/Norway/Nordland/ | Nordland Hospital - Bodo, Laboratory Department, Molecular Biology Unit | Norwegian Institute of Public Health, Department of Virology | Kathrine Stene-Johansen et al |

| | | | | | | |
|---|---|---|---|---|---|---|
| EPI_ISL_775524 | Norway/7104/2020 | 2020-12-21 | Europe/Norway/Viken/ | Furst Medical Laboratory | Norwegian Institute of Public Health, Department of Virology | Kathrine Stene-Johansen et al |
| EPI_ISL_775532 | Sweden/20-09927/2020 | 2020-10-27 | Europe/Sweden/Stockholm/ | Klinisk mikrobiologi, Viruslab | The Public Health Agency of Sweden | Department of Microbiology et al |
| EPI_ISL_775544 | Sweden/20-53721/2020 | 2020-12-07 | Europe/Sweden/Stockholm/ | Klinisk mikrobiologi, Viruslab | The Public Health Agency of Sweden | Department of Microbiology et al |
| EPI_ISL_775545 | Sweden/20-53724/2020 | 2020-12-08 | Europe/Sweden/Stockholm/ | Klinisk mikrobiologi, Viruslab | The Public Health Agency of Sweden | Department of Microbiology et al |
| EPI_ISL_775546 | Sweden/20-53777/2020 | 2020-12-18 | Europe/Sweden/Vastmanland/ | Klinisk Mikrobiologi | The Public Health Agency of Sweden | Department of Microbiology et al |
| EPI_ISL_775549 | Sweden/20-13018/2020 | 2020-12-09 | Europe/Sweden/Stockholm/ | Klinisk mikrobiologi | The Public Health Agency of Sweden | Department of Microbiology et al |
| EPI_ISL_775552 | Sweden/20-13038/2020 | 2020-12-29 | Europe/Sweden/Stockholm/ | ABC | The Public Health Agency of Sweden | Department of Microbiology et al |
| EPI_ISL_775556 | Sweden/20-13049/2020 | 2020-12-29 | Europe/Sweden/Stockholm/ | ABC | The Public Health Agency of Sweden | Department of Microbiology et al |
| EPI_ISL_775563 | Sweden/20-13101/2020 | 2020-12-29 | Europe/Sweden/Stockholm/ | ABC | The Public Health Agency of Sweden | Department of Microbiology et al |
| EPI_ISL_775580 | Sweden/20-13146/2020 | 2020-12-23 | Europe/Sweden/Stockholm/ | ABC | The Public Health Agency of Sweden | Department of Microbiology et al |
| EPI_ISL_775588 | Sweden/20-53843/2020 | 2020-12-14 | Europe/Sweden/Ostergotland/ | Klinisk mikrobiologi | The Public Health Agency of Sweden | Department of Microbiology et al |
| EPI_ISL_775589 | Sweden/20-53844/2020 | 2020-12-21 | Europe/Sweden/Vastra Gotaland/ | Laboratorium for klinisk mikrobiologi | The Public Health Agency of Sweden | Department of Microbiology et al |
| EPI_ISL_775591 | Sweden/20-53852/2020 | 2020-12-21 | Europe/Sweden/Vastra Gotaland/ | Klinisk mikrobiologi | The Public Health Agency of Sweden | Department of Microbiology et al |
| EPI_ISL_775594 | Indonesia/LA-NIHRD-PME8719/2020 | 2020-09-18 | Asia/Indonesia/Lampung/ | Balitvet Lampung | National Institute of Health Research and Development | Nugraha et al |
| EPI_ISL_775596 | Indonesia/BA-NIHRD-PME5833/2020 | 2020-08-01 | Asia/Indonesia/Bali/ | Lab. Biologi Molekular FKIK Unwar, Bali | National Institute of Health Research and Development | Ikawati et al |

| | | | | | | |
|---|---|---|---|---|---|---|
| EPI_ISL_775597 | Indonesia/PA-NIHRD-PME8510/2020 | 2020-09-26 | Asia/Indonesia/West Papua/ | RS Pertamina Sorong | National Institute of Health Research and Development | Puspa et al |
| EPI_ISL_775598 | Indonesia/NB-NIHRD-PME1393/2020 | 2020-06-01 | Asia/Indonesia/West Nusa Tenggara/ | RSUD Kota Mataram | National Institute of Health Research and Development | Puspa et al |
| EPI_ISL_775599 | Indonesia/SS-NIHRD-PME1692/2020 | 2020-06-09 | Asia/Indonesia/South Sulawesi/ | Balai Besar Veteriner Maros | National Institute of Health Research and Development | Subangkit et al |
| EPI_ISL_775600 | USA/OH-ODH-SC35063/2020 | 2020-04-23 | North America/USA/Ohio/ | Ohio Department of Health Laboratory | Ohio Department of Health Laboratory | Holmes et al |
| EPI_ISL_775602 | USA/OH-ODH-SC35065/2020 | 2020-04-23 | North America/USA/Ohio/ | Ohio Department of Health Laboratory | Ohio Department of Health Laboratory | Holmes et al |
| EPI_ISL_775605 | USA/OH-ODH-SC35076/2020 | 2020-04-23 | North America/USA/Ohio/ | Ohio Department of Health Laboratory | Ohio Department of Health Laboratory | Holmes et al |
| EPI_ISL_775606 | USA/OH-ODH-SC35077/2020 | 2020-04-23 | North America/USA/Ohio/ | Ohio Department of Health Laboratory | Ohio Department of Health Laboratory | Holmes et al |
| EPI_ISL_775607 | Germany/HH-hpi-p2373/2020 | 2020-03-24 | Europe/Germany/Hamburg/ | University Medical Center Hamburg Eppendorf | Heinrich Pette Institute, Leibniz Institute for Experimental Virology | Alexis Robitaille et al |
| EPI_ISL_775608 | Germany/HH-hpi-p1910/2020 | 2020-03-24 | Europe/Germany/Hamburg/ | University Medical Center Hamburg Eppendorf | Heinrich Pette Institute, Leibniz Institute for Experimental Virology | Alexis Robitaille et al |
| EPI_ISL_775611 | Germany/HH-hpi-p2315/2020 | 2020-03-23 | Europe/Germany/Hamburg/ | University Medical Center Hamburg Eppendorf | Heinrich Pette Institute, Leibniz Institute for Experimental Virology | Alexis Robitaille et al |

| | | | | | | |
|---|---|---|---|---|---|---|
| EPI_ISL_775622 | Germany/HH-hpi-p1604/2020 | 2020-03-16 | Europe/Germany/Hamburg/ | University Medical Center Hamburg Eppendorf | Heinrich Pette Institute, Leibniz Institute for Experimental Virology | Alexis Robitaille et al |
| EPI_ISL_775624 | Germany/HH-hpi-p1346/2020 | 2020-03-26 | Europe/Germany/Hamburg/ | University Medical Center Hamburg Eppendorf | Heinrich Pette Institute, Leibniz Institute for Experimental Virology | Alexis Robitaille et al |
| EPI_ISL_775627 | Germany/HH-hpi-p1348/2020 | 2020-03-26 | Europe/Germany/Hamburg/ | University Medical Center Hamburg Eppendorf | Heinrich Pette Institute, Leibniz Institute for Experimental Virology | Alexis Robitaille et al |
| EPI_ISL_775631 | Germany/HH-hpi-p1977/2020 | 2020-03-26 | Europe/Germany/Hamburg/ | University Medical Center Hamburg Eppendorf | Heinrich Pette Institute, Leibniz Institute for Experimental Virology | Alexis Robitaille et al |
| EPI_ISL_775637 | Germany/HH-hpi-p1557/2020 | 2020-03-15 | Europe/Germany/Hamburg/ | University Medical Center Hamburg Eppendorf | Heinrich Pette Institute, Leibniz Institute for Experimental Virology | Alexis Robitaille et al |
| EPI_ISL_775639 | Germany/HH-hpi-p1654/2020 | 2020-03-17 | Europe/Germany/Hamburg/ | University Medical Center Hamburg Eppendorf | Heinrich Pette Institute, Leibniz Institute for Experimental Virology | Alexis Robitaille et al |
| EPI_ISL_775643 | Germany/HH-hpi-p2033/2020 | 2020-03-21 | Europe/Germany/Hamburg/ | University Medical Center Hamburg Eppendorf | Heinrich Pette Institute, Leibniz Institute for Experimental Virology | Alexis Robitaille et al |
| EPI_ISL_775645 | Germany/HH-hpi-p1564/2020 | 2020-03-15 | Europe/Germany/Hamburg/ | University Medical Center Hamburg Eppendorf | Heinrich Pette Institute, Leibniz Institute for Experimental Virology | Alexis Robitaille et al |

| | | | | | | |
|---|---|---|---|---|---|---|
| EPI_ISL_775652 | Germany/HH-hpi-p2389/2020 | 2020-06-11 | Europe/Germany/Hamburg/ | University Medical Center Hamburg Eppendorf | Heinrich Pette Institute, Leibniz Institute for Experimental Virology | Alexis Robitaille et al |
| EPI_ISL_775655 | Germany/HH-hpi-p2399/2020 | 2020-04-29 | Europe/Germany/Hamburg/ | University Medical Center Hamburg Eppendorf | Heinrich Pette Institute, Leibniz Institute for Experimental Virology | Alexis Robitaille et al |
| EPI_ISL_775659 | Germany/HH-hpi-p1660/2020 | 2020-03-18 | Europe/Germany/Hamburg/ | University Medical Center Hamburg Eppendorf | Heinrich Pette Institute, Leibniz Institute for Experimental Virology | Alexis Robitaille et al |
| EPI_ISL_775661 | Germany/HH-hpi-p2086/2020 | 2020-03-20 | Europe/Germany/Hamburg/ | University Medical Center Hamburg Eppendorf | Heinrich Pette Institute, Leibniz Institute for Experimental Virology | Alexis Robitaille et al |
| EPI_ISL_775667 | Germany/HH-hpi-p1336/2020 | 2020-03-24 | Europe/Germany/Hamburg/ | University Medical Center Hamburg Eppendorf | Heinrich Pette Institute, Leibniz Institute for Experimental Virology | Alexis Robitaille et al |
| EPI_ISL_775669 | Germany/HH-hpi-p2050/2020 | 2020-03-21 | Europe/Germany/Hamburg/ | University Medical Center Hamburg Eppendorf | Heinrich Pette Institute, Leibniz Institute for Experimental Virology | Alexis Robitaille et al |
| EPI_ISL_775677 | Germany/HH-hpi-p1676/2020 | 2020-03-18 | Europe/Germany/Hamburg/ | University Medical Center Hamburg Eppendorf | Heinrich Pette Institute, Leibniz Institute for Experimental Virology | Alexis Robitaille et al |
| EPI_ISL_775682 | Germany/HH-hpi-p2311/2020 | 2020-03-15 | Europe/Germany/Hamburg/ | University Medical Center Hamburg Eppendorf | Heinrich Pette Institute, Leibniz Institute for Experimental Virology | Alexis Robitaille et al |

| | | | | | | |
|---|---|---|---|---|---|---|
| EPI_ISL_775683 | Germany/HH-hpi-p2062/2020 | 2020-03-21 | Europe/Germany/Hamburg/ | University Medical Center Hamburg Eppendorf | Heinrich Pette Institute, Leibniz Institute for Experimental Virology | Alexis Robitaille et al |
| EPI_ISL_775685 | Germany/HH-hpi-p1841/2020 | 2020-03-22 | Europe/Germany/Hamburg/ | University Medical Center Hamburg Eppendorf | Heinrich Pette Institute, Leibniz Institute for Experimental Virology | Alexis Robitaille et al |
| EPI_ISL_775688 | Germany/HH-hpi-p1723/2020 | 2020-03-15 | Europe/Germany/Hamburg/ | University Medical Center Hamburg Eppendorf | Heinrich Pette Institute, Leibniz Institute for Experimental Virology | Alexis Robitaille et al |
| EPI_ISL_775692 | Germany/HH-hpi-p2121/2020 | 2020-03-22 | Europe/Germany/Hamburg/ | University Medical Center Hamburg Eppendorf | Heinrich Pette Institute, Leibniz Institute for Experimental Virology | Alexis Robitaille et al |
| EPI_ISL_775695 | Germany/HH-hpi-p1931/2020 | 2020-03-26 | Europe/Germany/Hamburg/ | University Medical Center Hamburg Eppendorf | Heinrich Pette Institute, Leibniz Institute for Experimental Virology | Alexis Robitaille et al |
| EPI_ISL_775696 | Germany/HH-hpi-p2409/2020 | 2020-04-25 | Europe/Germany/Hamburg/ | University Medical Center Hamburg Eppendorf | Heinrich Pette Institute, Leibniz Institute for Experimental Virology | Alexis Robitaille et al |
| EPI_ISL_775699 | Germany/HH-hpi-p2310/2020 | 2020-03-18 | Europe/Germany/Hamburg/ | University Medical Center Hamburg Eppendorf | Heinrich Pette Institute, Leibniz Institute for Experimental Virology | Alexis Robitaille et al |
| EPI_ISL_775700 | Germany/HH-hpi-p1440/2020 | 2020-03-04 | Europe/Germany/Hamburg/ | University Medical Center Hamburg Eppendorf | Heinrich Pette Institute, Leibniz Institute for Experimental Virology | Alexis Robitaille et al |

| | | | | | | |
|---|---|---|---|---|---|---|
| EPI_ISL_775710 | Germany/HH-hpi-p1927/2020 | 2020-03-25 | Europe/Germany/Hamburg/ | University Medical Center Hamburg Eppendorf | Heinrich Pette Institute, Leibniz Institute for Experimental Virology | Alexis Robitaille et al |
| EPI_ISL_775711 | Germany/HH-hpi-p2323/2020 | 2020-03-21 | Europe/Germany/Hamburg/ | University Medical Center Hamburg Eppendorf | Heinrich Pette Institute, Leibniz Institute for Experimental Virology | Alexis Robitaille et al |
| EPI_ISL_775713 | Germany/HH-hpi-p2143/2020 | 2020-03-22 | Europe/Germany/Hamburg/ | University Medical Center Hamburg Eppendorf | Heinrich Pette Institute, Leibniz Institute for Experimental Virology | Alexis Robitaille et al |
| EPI_ISL_775715 | Germany/HH-hpi-p1683/2020 | 2020-03-15 | Europe/Germany/Hamburg/ | University Medical Center Hamburg Eppendorf | Heinrich Pette Institute, Leibniz Institute for Experimental Virology | Alexis Robitaille et al |
| EPI_ISL_775716 | Germany/HH-hpi-p2363/2020 | 2020-03-15 | Europe/Germany/Hamburg/ | University Medical Center Hamburg Eppendorf | Heinrich Pette Institute, Leibniz Institute for Experimental Virology | Alexis Robitaille et al |
| EPI_ISL_775723 | Germany/HH-hpi-p2415/2020 | 2020-04-21 | Europe/Germany/Hamburg/ | University Medical Center Hamburg Eppendorf | Heinrich Pette Institute, Leibniz Institute for Experimental Virology | Alexis Robitaille et al |
| EPI_ISL_775726 | Germany/HH-hpi-p1801/2020 | 2020-03-19 | Europe/Germany/Hamburg/ | University Medical Center Hamburg Eppendorf | Heinrich Pette Institute, Leibniz Institute for Experimental Virology | Alexis Robitaille et al |
| EPI_ISL_775729 | Germany/HH-hpi-p1878/2020 | 2020-03-24 | Europe/Germany/Hamburg/ | University Medical Center Hamburg Eppendorf | Heinrich Pette Institute, Leibniz Institute for Experimental Virology | Alexis Robitaille et al |

| | | | | | | |
|---|---|---|---|---|---|---|
| EPI_ISL_775734 | Germany/HH-hpi-p2072/2020 | 2020-03-20 | Europe/Germany/Hamburg/ | University Medical Center Hamburg Eppendorf | Heinrich Pette Institute, Leibniz Institute for Experimental Virology | Alexis Robitaille et al |
| EPI_ISL_775738 | Germany/HH-hpi-p2326/2020 | 2020-03-21 | Europe/Germany/Hamburg/ | University Medical Center Hamburg Eppendorf | Heinrich Pette Institute, Leibniz Institute for Experimental Virology | Alexis Robitaille et al |
| EPI_ISL_775743 | Germany/HH-hpi-p1595/2020 | 2020-03-16 | Europe/Germany/Hamburg/ | University Medical Center Hamburg Eppendorf | Heinrich Pette Institute, Leibniz Institute for Experimental Virology | Alexis Robitaille et al |
| EPI_ISL_775745 | Germany/HH-hpi-p2301/2020 | 2020-03-12 | Europe/Germany/Hamburg/ | University Medical Center Hamburg Eppendorf | Heinrich Pette Institute, Leibniz Institute for Experimental Virology | Alexis Robitaille et al |
| EPI_ISL_775746 | Germany/HH-hpi-p2377/2020 | 2020-05-15 | Europe/Germany/Hamburg/ | University Medical Center Hamburg Eppendorf | Heinrich Pette Institute, Leibniz Institute for Experimental Virology | Alexis Robitaille et al |
| EPI_ISL_775755 | Germany/HH-hpi-p2083/2020 | 2020-03-20 | Europe/Germany/Hamburg/ | University Medical Center Hamburg Eppendorf | Heinrich Pette Institute, Leibniz Institute for Experimental Virology | Alexis Robitaille et al |
| EPI_ISL_775756 | Germany/HH-hpi-p1909/2020 | 2020-03-24 | Europe/Germany/Hamburg/ | University Medical Center Hamburg Eppendorf | Heinrich Pette Institute, Leibniz Institute for Experimental Virology | Alexis Robitaille et al |
| EPI_ISL_775764 | Germany/HH-hpi-p1634/2020 | 2020-03-17 | Europe/Germany/Hamburg/ | University Medical Center Hamburg Eppendorf | Heinrich Pette Institute, Leibniz Institute for Experimental Virology | Alexis Robitaille et al |

| | | | | | | |
|---|---|---|---|---|---|---|
| EPI_ISL_775780 | Germany/HH-hpi-p1284/2020 | 2020-03-16 | Europe/Germany/Hamburg/ | University Medical Center Hamburg Eppendorf | Heinrich Pette Institute, Leibniz Institute for Experimental Virology | Alexis Robitaille et al |
| EPI_ISL_775795 | Germany/HH-hpi-p1349/2020 | 2020-03-26 | Europe/Germany/Hamburg/ | University Medical Center Hamburg Eppendorf | Heinrich Pette Institute, Leibniz Institute for Experimental Virology | Alexis Robitaille et al |
| EPI_ISL_775797 | Germany/HH-hpi-p2038/2020 | 2020-03-20 | Europe/Germany/Hamburg/ | University Medical Center Hamburg Eppendorf | Heinrich Pette Institute, Leibniz Institute for Experimental Virology | Alexis Robitaille et al |
| EPI_ISL_775806 | Germany/HH-hpi-p1849/2020 | 2020-03-22 | Europe/Germany/Hamburg/ | University Medical Center Hamburg Eppendorf | Heinrich Pette Institute, Leibniz Institute for Experimental Virology | Alexis Robitaille et al |
| EPI_ISL_775809 | Germany/HH-hpi-p1667/2020 | 2020-03-18 | Europe/Germany/Hamburg/ | University Medical Center Hamburg Eppendorf | Heinrich Pette Institute, Leibniz Institute for Experimental Virology | Alexis Robitaille et al |
| EPI_ISL_775816 | Germany/HH-hpi-p1354/2020 | 2020-03-27 | Europe/Germany/Hamburg/ | University Medical Center Hamburg Eppendorf | Heinrich Pette Institute, Leibniz Institute for Experimental Virology | Alexis Robitaille et al |
| EPI_ISL_775817 | Germany/HH-hpi-p2085/2020 | 2020-03-20 | Europe/Germany/Hamburg/ | University Medical Center Hamburg Eppendorf | Heinrich Pette Institute, Leibniz Institute for Experimental Virology | Alexis Robitaille et al |
| EPI_ISL_775826 | Germany/HH-hpi-p1898/2020 | 2020-03-24 | Europe/Germany/Hamburg/ | University Medical Center Hamburg Eppendorf | Heinrich Pette Institute, Leibniz Institute for Experimental Virology | Alexis Robitaille et al |

| | | | | | | |
|---|---|---|---|---|---|---|
| EPI_ISL_775830 | Germany/HH-hpi-p1798/2020 | 2020-03-19 | Europe/Germany/Hamburg/ | University Medical Center Hamburg Eppendorf | Heinrich Pette Institute, Leibniz Institute for Experimental Virology | Alexis Robitaille et al |
| EPI_ISL_775835 | Germany/HH-hpi-p1532/2020 | 2020-03-15 | Europe/Germany/Hamburg/ | University Medical Center Hamburg Eppendorf | Heinrich Pette Institute, Leibniz Institute for Experimental Virology | Alexis Robitaille et al |
| EPI_ISL_775916 | Germany/HH-hpi-p1792/2020 | 2020-03-19 | Europe/Germany/Hamburg/ | University Medical Center Hamburg Eppendorf | Heinrich Pette Institute, Leibniz Institute for Experimental Virology | Alexis Robitaille et al |
| EPI_ISL_775929 | Germany/HH-hpi-p1633/2020 | 2020-03-17 | Europe/Germany/Hamburg/ | University Medical Center Hamburg Eppendorf | Heinrich Pette Institute, Leibniz Institute for Experimental Virology | Alexis Robitaille et al |
| EPI_ISL_775932 | Germany/HH-hpi-p1859/2020 | 2020-03-23 | Europe/Germany/Hamburg/ | University Medical Center Hamburg Eppendorf | Heinrich Pette Institute, Leibniz Institute for Experimental Virology | Alexis Robitaille et al |
| EPI_ISL_775943 | Germany/HH-hpi-p1416/2020 | 2020-03-03 | Europe/Germany/Hamburg/ | University Medical Center Hamburg Eppendorf | Heinrich Pette Institute, Leibniz Institute for Experimental Virology | Alexis Robitaille et al |
| EPI_ISL_775944 | Germany/HH-hpi-p1418/2020 | 2020-03-10 | Europe/Germany/Hamburg/ | University Medical Center Hamburg Eppendorf | Heinrich Pette Institute, Leibniz Institute for Experimental Virology | Alexis Robitaille et al |
| EPI_ISL_775947 | Germany/HH-hpi-p1423/2020 | 2020-03-10 | Europe/Germany/Hamburg/ | University Medical Center Hamburg Eppendorf | Heinrich Pette Institute, Leibniz Institute for Experimental Virology | Alexis Robitaille et al |

| | | | | | | |
|---|---|---|---|---|---|---|
| EPI_ISL_775950 | Germany/HH-hpi-p1429/2020 | 2020-03-09 | Europe/Germany/Hamburg/ | University Medical Center Hamburg Eppendorf | Heinrich Pette Institute, Leibniz Institute for Experimental Virology | Alexis Robitaille et al |
| EPI_ISL_775952 | Germany/HH-hpi-p1431/2020 | 2020-03-09 | Europe/Germany/Hamburg/ | University Medical Center Hamburg Eppendorf | Heinrich Pette Institute, Leibniz Institute for Experimental Virology | Alexis Robitaille et al |
| EPI_ISL_775960 | Germany/HH-hpi-p1441/2020 | 2020-03-05 | Europe/Germany/Hamburg/ | University Medical Center Hamburg Eppendorf | Heinrich Pette Institute, Leibniz Institute for Experimental Virology | Alexis Robitaille et al |
| EPI_ISL_775963 | Germany/HH-hpi-p1444/2020 | 2020-03-11 | Europe/Germany/Hamburg/ | University Medical Center Hamburg Eppendorf | Heinrich Pette Institute, Leibniz Institute for Experimental Virology | Alexis Robitaille et al |
| EPI_ISL_775973 | Germany/HH-hpi-p1460/2020 | 2020-03-10 | Europe/Germany/Hamburg/ | University Medical Center Hamburg Eppendorf | Heinrich Pette Institute, Leibniz Institute for Experimental Virology | Alexis Robitaille et al |
| EPI_ISL_775975 | Germany/HH-hpi-p1462/2020 | 2020-03-12 | Europe/Germany/Hamburg/ | University Medical Center Hamburg Eppendorf | Heinrich Pette Institute, Leibniz Institute for Experimental Virology | Alexis Robitaille et al |
| EPI_ISL_775977 | Germany/HH-hpi-p1464/2020 | 2020-03-11 | Europe/Germany/Hamburg/ | University Medical Center Hamburg Eppendorf | Heinrich Pette Institute, Leibniz Institute for Experimental Virology | Alexis Robitaille et al |
| EPI_ISL_775979 | Germany/HH-hpi-p1467/2020 | 2020-03-11 | Europe/Germany/Hamburg/ | University Medical Center Hamburg Eppendorf | Heinrich Pette Institute, Leibniz Institute for Experimental Virology | Alexis Robitaille et al |

| | | | | | | |
|---|---|---|---|---|---|---|
| EPI_ISL_775980 | Germany/HH-hpi-p1468/2020 | 2020-03-10 | Europe/Germany/Hamburg/ | University Medical Center Hamburg Eppendorf | Heinrich Pette Institute, Leibniz Institute for Experimental Virology | Alexis Robitaille et al |
| EPI_ISL_775984 | Germany/HH-hpi-p1472/2020 | 2020-03-13 | Europe/Germany/Hamburg/ | University Medical Center Hamburg Eppendorf | Heinrich Pette Institute, Leibniz Institute for Experimental Virology | Alexis Robitaille et al |
| EPI_ISL_775991 | Germany/HH-hpi-p1480/2020 | 2020-03-14 | Europe/Germany/Hamburg/ | University Medical Center Hamburg Eppendorf | Heinrich Pette Institute, Leibniz Institute for Experimental Virology | Alexis Robitaille et al |
| EPI_ISL_775992 | Germany/HH-hpi-p1481/2020 | 2020-03-11 | Europe/Germany/Hamburg/ | University Medical Center Hamburg Eppendorf | Heinrich Pette Institute, Leibniz Institute for Experimental Virology | Alexis Robitaille et al |
| EPI_ISL_775995 | Germany/HH-hpi-p1485/2020 | 2020-03-13 | Europe/Germany/Hamburg/ | University Medical Center Hamburg Eppendorf | Heinrich Pette Institute, Leibniz Institute for Experimental Virology | Alexis Robitaille et al |
| EPI_ISL_775999 | Germany/HH-hpi-p1491/2020 | 2020-03-11 | Europe/Germany/Hamburg/ | University Medical Center Hamburg Eppendorf | Heinrich Pette Institute, Leibniz Institute for Experimental Virology | Alexis Robitaille et al |
| EPI_ISL_776009 | Germany/HH-hpi-p1503/2020 | 2020-03-14 | Europe/Germany/Hamburg/ | University Medical Center Hamburg Eppendorf | Heinrich Pette Institute, Leibniz Institute for Experimental Virology | Alexis Robitaille et al |
| EPI_ISL_776041 | Germany/HH-hpi-p1559/2020 | 2020-03-15 | Europe/Germany/Hamburg/ | University Medical Center Hamburg Eppendorf | Heinrich Pette Institute, Leibniz Institute for Experimental Virology | Alexis Robitaille et al |

| | | | | | | |
|---|---|---|---|---|---|---|
| EPI_ISL_776042 | Germany/HH-hpi-p1560/2020 | 2020-03-15 | Europe/Germany/Hamburg/ | University Medical Center Hamburg Eppendorf | Heinrich Pette Institute, Leibniz Institute for Experimental Virology | Alexis Robitaille et al |
| EPI_ISL_776045 | Germany/HH-hpi-p1565/2020 | 2020-03-15 | Europe/Germany/Hamburg/ | University Medical Center Hamburg Eppendorf | Heinrich Pette Institute, Leibniz Institute for Experimental Virology | Alexis Robitaille et al |
| EPI_ISL_776051 | Germany/HH-hpi-p1572/2020 | 2020-03-15 | Europe/Germany/Hamburg/ | University Medical Center Hamburg Eppendorf | Heinrich Pette Institute, Leibniz Institute for Experimental Virology | Alexis Robitaille et al |
| EPI_ISL_776052 | Germany/HH-hpi-p1574/2020 | 2020-03-15 | Europe/Germany/Hamburg/ | University Medical Center Hamburg Eppendorf | Heinrich Pette Institute, Leibniz Institute for Experimental Virology | Alexis Robitaille et al |
| EPI_ISL_776053 | Germany/HH-hpi-p1575/2020 | 2020-03-16 | Europe/Germany/Hamburg/ | University Medical Center Hamburg Eppendorf | Heinrich Pette Institute, Leibniz Institute for Experimental Virology | Alexis Robitaille et al |
| EPI_ISL_776056 | Germany/HH-hpi-p1581/2020 | 2020-03-16 | Europe/Germany/Hamburg/ | University Medical Center Hamburg Eppendorf | Heinrich Pette Institute, Leibniz Institute for Experimental Virology | Alexis Robitaille et al |
| EPI_ISL_776059 | Germany/HH-hpi-p1584/2020 | 2020-03-16 | Europe/Germany/Hamburg/ | University Medical Center Hamburg Eppendorf | Heinrich Pette Institute, Leibniz Institute for Experimental Virology | Alexis Robitaille et al |
| EPI_ISL_776063 | Germany/HH-hpi-p1591/2020 | 2020-03-16 | Europe/Germany/Hamburg/ | University Medical Center Hamburg Eppendorf | Heinrich Pette Institute, Leibniz Institute for Experimental Virology | Alexis Robitaille et al |

| | | | | | | |
|---|---|---|---|---|---|---|
| EPI_ISL_776065 | Germany/HH-hpi-p1598/2020 | 2020-03-16 | Europe/Germany/Hamburg/ | University Medical Center Hamburg Eppendorf | Heinrich Pette Institute, Leibniz Institute for Experimental Virology | Alexis Robitaille et al |
| EPI_ISL_776071 | Germany/HH-hpi-p1617/2020 | 2020-03-17 | Europe/Germany/Hamburg/ | University Medical Center Hamburg Eppendorf | Heinrich Pette Institute, Leibniz Institute for Experimental Virology | Alexis Robitaille et al |
| EPI_ISL_776076 | Germany/HH-hpi-p1626/2020 | 2020-03-15 | Europe/Germany/Hamburg/ | University Medical Center Hamburg Eppendorf | Heinrich Pette Institute, Leibniz Institute for Experimental Virology | Alexis Robitaille et al |
| EPI_ISL_776080 | Germany/HH-hpi-p1643/2020 | 2020-03-17 | Europe/Germany/Hamburg/ | University Medical Center Hamburg Eppendorf | Heinrich Pette Institute, Leibniz Institute for Experimental Virology | Alexis Robitaille et al |
| EPI_ISL_776082 | Germany/HH-hpi-p1648/2020 | 2020-03-17 | Europe/Germany/Hamburg/ | University Medical Center Hamburg Eppendorf | Heinrich Pette Institute, Leibniz Institute for Experimental Virology | Alexis Robitaille et al |
| EPI_ISL_776085 | Germany/HH-hpi-p1659/2020 | 2020-03-18 | Europe/Germany/Hamburg/ | University Medical Center Hamburg Eppendorf | Heinrich Pette Institute, Leibniz Institute for Experimental Virology | Alexis Robitaille et al |
| EPI_ISL_776090 | Germany/HH-hpi-p1670/2020 | 2020-03-18 | Europe/Germany/Hamburg/ | University Medical Center Hamburg Eppendorf | Heinrich Pette Institute, Leibniz Institute for Experimental Virology | Alexis Robitaille et al |
| EPI_ISL_776102 | Germany/HH-hpi-p1719/2020 | 2020-03-15 | Europe/Germany/Hamburg/ | University Medical Center Hamburg Eppendorf | Heinrich Pette Institute, Leibniz Institute for Experimental Virology | Alexis Robitaille et al |

| | | | | | | |
|---|---|---|---|---|---|---|
| EPI_ISL_776104 | Germany/HH-hpi-p1724/2020 | 2020-03-15 | Europe/Germany/Hamburg/ | University Medical Center Hamburg Eppendorf | Heinrich Pette Institute, Leibniz Institute for Experimental Virology | Alexis Robitaille et al |
| EPI_ISL_776111 | Germany/HH-hpi-p1731/2020 | 2020-03-15 | Europe/Germany/Hamburg/ | University Medical Center Hamburg Eppendorf | Heinrich Pette Institute, Leibniz Institute for Experimental Virology | Alexis Robitaille et al |
| EPI_ISL_776112 | Germany/HH-hpi-p1732/2020 | 2020-03-14 | Europe/Germany/Hamburg/ | University Medical Center Hamburg Eppendorf | Heinrich Pette Institute, Leibniz Institute for Experimental Virology | Alexis Robitaille et al |
| EPI_ISL_776119 | Germany/HH-hpi-p1740/2020 | 2020-03-12 | Europe/Germany/Hamburg/ | University Medical Center Hamburg Eppendorf | Heinrich Pette Institute, Leibniz Institute for Experimental Virology | Alexis Robitaille et al |
| EPI_ISL_776145 | Germany/HH-hpi-p1771/2020 | 2020-03-19 | Europe/Germany/Hamburg/ | University Medical Center Hamburg Eppendorf | Heinrich Pette Institute, Leibniz Institute for Experimental Virology | Alexis Robitaille et al |
| EPI_ISL_776146 | Germany/HH-hpi-p1772/2020 | 2020-03-19 | Europe/Germany/Hamburg/ | University Medical Center Hamburg Eppendorf | Heinrich Pette Institute, Leibniz Institute for Experimental Virology | Alexis Robitaille et al |
| EPI_ISL_776149 | Germany/HH-hpi-p1776/2020 | 2020-03-19 | Europe/Germany/Hamburg/ | University Medical Center Hamburg Eppendorf | Heinrich Pette Institute, Leibniz Institute for Experimental Virology | Alexis Robitaille et al |
| EPI_ISL_776158 | Germany/HH-hpi-p1787/2020 | 2020-03-19 | Europe/Germany/Hamburg/ | University Medical Center Hamburg Eppendorf | Heinrich Pette Institute, Leibniz Institute for Experimental Virology | Alexis Robitaille et al |

| | | | | | | |
|---|---|---|---|---|---|---|
| EPI_ISL_776164 | Germany/HH-hpi-p1795/2020 | 2020-03-19 | Europe/Germany/Hamburg/ | University Medical Center Hamburg Eppendorf | Heinrich Pette Institute, Leibniz Institute for Experimental Virology | Alexis Robitaille et al |
| EPI_ISL_776177 | Germany/HH-hpi-p1821/2020 | 2020-03-22 | Europe/Germany/Hamburg/ | University Medical Center Hamburg Eppendorf | Heinrich Pette Institute, Leibniz Institute for Experimental Virology | Alexis Robitaille et al |
| EPI_ISL_776178 | Germany/HH-hpi-p1822/2020 | 2020-03-22 | Europe/Germany/Hamburg/ | University Medical Center Hamburg Eppendorf | Heinrich Pette Institute, Leibniz Institute for Experimental Virology | Alexis Robitaille et al |
| EPI_ISL_776181 | Germany/HH-hpi-p1827/2020 | 2020-03-22 | Europe/Germany/Hamburg/ | University Medical Center Hamburg Eppendorf | Heinrich Pette Institute, Leibniz Institute for Experimental Virology | Alexis Robitaille et al |
| EPI_ISL_776183 | Germany/HH-hpi-p1829/2020 | 2020-03-22 | Europe/Germany/Hamburg/ | University Medical Center Hamburg Eppendorf | Heinrich Pette Institute, Leibniz Institute for Experimental Virology | Alexis Robitaille et al |
| EPI_ISL_776185 | Germany/HH-hpi-p1832/2020 | 2020-03-22 | Europe/Germany/Hamburg/ | University Medical Center Hamburg Eppendorf | Heinrich Pette Institute, Leibniz Institute for Experimental Virology | Alexis Robitaille et al |
| EPI_ISL_776186 | Germany/HH-hpi-p1833/2020 | 2020-03-22 | Europe/Germany/Hamburg/ | University Medical Center Hamburg Eppendorf | Heinrich Pette Institute, Leibniz Institute for Experimental Virology | Alexis Robitaille et al |
| EPI_ISL_776192 | Germany/HH-hpi-p1839/2020 | 2020-03-22 | Europe/Germany/Hamburg/ | University Medical Center Hamburg Eppendorf | Heinrich Pette Institute, Leibniz Institute for Experimental Virology | Alexis Robitaille et al |

| | | | | | | |
|---|---|---|---|---|---|---|
| EPI_ISL_776213 | Germany/HH-hpi-p1867/2020 | 2020-03-23 | Europe/Germany/Hamburg/ | University Medical Center Hamburg Eppendorf | Heinrich Pette Institute, Leibniz Institute for Experimental Virology | Alexis Robitaille et al |
| EPI_ISL_776215 | Germany/HH-hpi-p1869/2020 | 2020-03-23 | Europe/Germany/Hamburg/ | University Medical Center Hamburg Eppendorf | Heinrich Pette Institute, Leibniz Institute for Experimental Virology | Alexis Robitaille et al |
| EPI_ISL_776216 | Germany/HH-hpi-p1871/2020 | 2020-03-23 | Europe/Germany/Hamburg/ | University Medical Center Hamburg Eppendorf | Heinrich Pette Institute, Leibniz Institute for Experimental Virology | Alexis Robitaille et al |
| EPI_ISL_776220 | Germany/HH-hpi-p1875/2020 | 2020-03-23 | Europe/Germany/Hamburg/ | University Medical Center Hamburg Eppendorf | Heinrich Pette Institute, Leibniz Institute for Experimental Virology | Alexis Robitaille et al |
| EPI_ISL_776223 | Germany/HH-hpi-p1879/2020 | 2020-03-24 | Europe/Germany/Hamburg/ | University Medical Center Hamburg Eppendorf | Heinrich Pette Institute, Leibniz Institute for Experimental Virology | Alexis Robitaille et al |
| EPI_ISL_776225 | Germany/HH-hpi-p1881/2020 | 2020-03-24 | Europe/Germany/Hamburg/ | University Medical Center Hamburg Eppendorf | Heinrich Pette Institute, Leibniz Institute for Experimental Virology | Alexis Robitaille et al |
| EPI_ISL_776226 | Germany/HH-hpi-p1882/2020 | 2020-03-24 | Europe/Germany/Hamburg/ | University Medical Center Hamburg Eppendorf | Heinrich Pette Institute, Leibniz Institute for Experimental Virology | Alexis Robitaille et al |
| EPI_ISL_776231 | Germany/HH-hpi-p1888/2020 | 2020-03-24 | Europe/Germany/Hamburg/ | University Medical Center Hamburg Eppendorf | Heinrich Pette Institute, Leibniz Institute for Experimental Virology | Alexis Robitaille et al |

| | | | | | | |
|---|---|---|---|---|---|---|
| EPI_ISL_776232 | Germany/HH-hpi-p1891/2020 | 2020-03-24 | Europe/Germany/Hamburg/ | University Medical Center Hamburg Eppendorf | Heinrich Pette Institute, Leibniz Institute for Experimental Virology | Alexis Robitaille et al |
| EPI_ISL_776234 | Germany/HH-hpi-p1894/2020 | 2020-03-24 | Europe/Germany/Hamburg/ | University Medical Center Hamburg Eppendorf | Heinrich Pette Institute, Leibniz Institute for Experimental Virology | Alexis Robitaille et al |
| EPI_ISL_776246 | Germany/HH-hpi-p1915/2020 | 2020-03-25 | Europe/Germany/Hamburg/ | University Medical Center Hamburg Eppendorf | Heinrich Pette Institute, Leibniz Institute for Experimental Virology | Alexis Robitaille et al |
| EPI_ISL_776250 | Germany/HH-hpi-p1919/2020 | 2020-03-25 | Europe/Germany/Hamburg/ | University Medical Center Hamburg Eppendorf | Heinrich Pette Institute, Leibniz Institute for Experimental Virology | Alexis Robitaille et al |
| EPI_ISL_776252 | Germany/HH-hpi-p1921/2020 | 2020-03-25 | Europe/Germany/Hamburg/ | University Medical Center Hamburg Eppendorf | Heinrich Pette Institute, Leibniz Institute for Experimental Virology | Alexis Robitaille et al |
| EPI_ISL_776262 | Germany/HH-hpi-p1934/2020 | 2020-03-26 | Europe/Germany/Hamburg/ | University Medical Center Hamburg Eppendorf | Heinrich Pette Institute, Leibniz Institute for Experimental Virology | Alexis Robitaille et al |
| EPI_ISL_776264 | Germany/HH-hpi-p1936/2020 | 2020-03-26 | Europe/Germany/Hamburg/ | University Medical Center Hamburg Eppendorf | Heinrich Pette Institute, Leibniz Institute for Experimental Virology | Alexis Robitaille et al |
| EPI_ISL_776266 | Germany/HH-hpi-p1938/2020 | 2020-03-26 | Europe/Germany/Hamburg/ | University Medical Center Hamburg Eppendorf | Heinrich Pette Institute, Leibniz Institute for Experimental Virology | Alexis Robitaille et al |

| | | | | | | |
|---|---|---|---|---|---|---|
| EPI_ISL_776268 | Germany/HH-hpi-p1941/2020 | 2020-03-27 | Europe/Germany/Hamburg/ | University Medical Center Hamburg Eppendorf | Heinrich Pette Institute, Leibniz Institute for Experimental Virology | Alexis Robitaille et al |
| EPI_ISL_776269 | Germany/HH-hpi-p1942/2020 | 2020-03-27 | Europe/Germany/Hamburg/ | University Medical Center Hamburg Eppendorf | Heinrich Pette Institute, Leibniz Institute for Experimental Virology | Alexis Robitaille et al |
| EPI_ISL_776272 | Germany/HH-hpi-p1945/2020 | 2020-03-27 | Europe/Germany/Hamburg/ | University Medical Center Hamburg Eppendorf | Heinrich Pette Institute, Leibniz Institute for Experimental Virology | Alexis Robitaille et al |
| EPI_ISL_776277 | Germany/HH-hpi-p1950/2020 | 2020-03-27 | Europe/Germany/Hamburg/ | University Medical Center Hamburg Eppendorf | Heinrich Pette Institute, Leibniz Institute for Experimental Virology | Alexis Robitaille et al |
| EPI_ISL_776279 | Germany/HH-hpi-p1952/2020 | 2020-03-24 | Europe/Germany/Hamburg/ | University Medical Center Hamburg Eppendorf | Heinrich Pette Institute, Leibniz Institute for Experimental Virology | Alexis Robitaille et al |
| EPI_ISL_776280 | Germany/HH-hpi-p1953/2020 | 2020-03-24 | Europe/Germany/Hamburg/ | University Medical Center Hamburg Eppendorf | Heinrich Pette Institute, Leibniz Institute for Experimental Virology | Alexis Robitaille et al |
| EPI_ISL_776284 | Germany/HH-hpi-p1959/2020 | 2020-03-25 | Europe/Germany/Hamburg/ | University Medical Center Hamburg Eppendorf | Heinrich Pette Institute, Leibniz Institute for Experimental Virology | Alexis Robitaille et al |
| EPI_ISL_776285 | Germany/HH-hpi-p1960/2020 | 2020-03-25 | Europe/Germany/Hamburg/ | University Medical Center Hamburg Eppendorf | Heinrich Pette Institute, Leibniz Institute for Experimental Virology | Alexis Robitaille et al |

| | | | | | | |
|---|---|---|---|---|---|---|
| EPI_ISL_776287 | Germany/HH-hpi-p1962/2020 | 2020-03-25 | Europe/Germany/Hamburg/ | University Medical Center Hamburg Eppendorf | Heinrich Pette Institute, Leibniz Institute for Experimental Virology | Alexis Robitaille et al |
| EPI_ISL_776288 | Germany/HH-hpi-p1965/2020 | 2020-03-25 | Europe/Germany/Hamburg/ | University Medical Center Hamburg Eppendorf | Heinrich Pette Institute, Leibniz Institute for Experimental Virology | Alexis Robitaille et al |
| EPI_ISL_776295 | Germany/HH-hpi-p1974/2020 | 2020-03-26 | Europe/Germany/Hamburg/ | University Medical Center Hamburg Eppendorf | Heinrich Pette Institute, Leibniz Institute for Experimental Virology | Alexis Robitaille et al |
| EPI_ISL_776304 | Germany/HH-hpi-p1986/2020 | 2020-03-26 | Europe/Germany/Hamburg/ | University Medical Center Hamburg Eppendorf | Heinrich Pette Institute, Leibniz Institute for Experimental Virology | Alexis Robitaille et al |
| EPI_ISL_776306 | Germany/HH-hpi-p1988/2020 | 2020-03-25 | Europe/Germany/Hamburg/ | University Medical Center Hamburg Eppendorf | Heinrich Pette Institute, Leibniz Institute for Experimental Virology | Alexis Robitaille et al |
| EPI_ISL_776311 | Germany/HH-hpi-p1994/2020 | 2020-03-27 | Europe/Germany/Hamburg/ | University Medical Center Hamburg Eppendorf | Heinrich Pette Institute, Leibniz Institute for Experimental Virology | Alexis Robitaille et al |
| EPI_ISL_776320 | Germany/HH-hpi-p2012/2020 | 2020-03-21 | Europe/Germany/Hamburg/ | University Medical Center Hamburg Eppendorf | Heinrich Pette Institute, Leibniz Institute for Experimental Virology | Alexis Robitaille et al |
| EPI_ISL_776324 | Germany/HH-hpi-p2018/2020 | 2020-03-21 | Europe/Germany/Hamburg/ | University Medical Center Hamburg Eppendorf | Heinrich Pette Institute, Leibniz Institute for Experimental Virology | Alexis Robitaille et al |

| | | | | | | |
|---|---|---|---|---|---|---|
| EPI_ISL_776330 | Germany/HH-hpi-p2028/2020 | 2020-03-21 | Europe/Germany/Hamburg/ | University Medical Center Hamburg Eppendorf | Heinrich Pette Institute, Leibniz Institute for Experimental Virology | Alexis Robitaille et al |
| EPI_ISL_776335 | Germany/HH-hpi-p2043/2020 | 2020-03-20 | Europe/Germany/Hamburg/ | University Medical Center Hamburg Eppendorf | Heinrich Pette Institute, Leibniz Institute for Experimental Virology | Alexis Robitaille et al |
| EPI_ISL_776344 | Germany/HH-hpi-p2060/2020 | 2020-03-21 | Europe/Germany/Hamburg/ | University Medical Center Hamburg Eppendorf | Heinrich Pette Institute, Leibniz Institute for Experimental Virology | Alexis Robitaille et al |
| EPI_ISL_776346 | Germany/HH-hpi-p2063/2020 | 2020-03-21 | Europe/Germany/Hamburg/ | University Medical Center Hamburg Eppendorf | Heinrich Pette Institute, Leibniz Institute for Experimental Virology | Alexis Robitaille et al |
| EPI_ISL_776349 | Germany/HH-hpi-p2080/2020 | 2020-03-20 | Europe/Germany/Hamburg/ | University Medical Center Hamburg Eppendorf | Heinrich Pette Institute, Leibniz Institute for Experimental Virology | Alexis Robitaille et al |
| EPI_ISL_776353 | Germany/HH-hpi-p2107/2020 | 2020-03-22 | Europe/Germany/Hamburg/ | University Medical Center Hamburg Eppendorf | Heinrich Pette Institute, Leibniz Institute for Experimental Virology | Alexis Robitaille et al |
| EPI_ISL_776357 | Germany/HH-hpi-p2113/2020 | 2020-03-22 | Europe/Germany/Hamburg/ | University Medical Center Hamburg Eppendorf | Heinrich Pette Institute, Leibniz Institute for Experimental Virology | Alexis Robitaille et al |
| EPI_ISL_776359 | Germany/HH-hpi-p2115/2020 | 2020-03-22 | Europe/Germany/Hamburg/ | University Medical Center Hamburg Eppendorf | Heinrich Pette Institute, Leibniz Institute for Experimental Virology | Alexis Robitaille et al |

| | | | | | | |
|---|---|---|---|---|---|---|
| EPI_ISL_776360 | Germany/HH-hpi-p2116/2020 | 2020-03-22 | Europe/Germany/Hamburg/ | University Medical Center Hamburg Eppendorf | Heinrich Pette Institute, Leibniz Institute for Experimental Virology | Alexis Robitaille et al |
| EPI_ISL_776361 | Germany/HH-hpi-p2118/2020 | 2020-03-22 | Europe/Germany/Hamburg/ | University Medical Center Hamburg Eppendorf | Heinrich Pette Institute, Leibniz Institute for Experimental Virology | Alexis Robitaille et al |
| EPI_ISL_776363 | Germany/HH-hpi-p2120/2020 | 2020-03-22 | Europe/Germany/Hamburg/ | University Medical Center Hamburg Eppendorf | Heinrich Pette Institute, Leibniz Institute for Experimental Virology | Alexis Robitaille et al |
| EPI_ISL_776365 | Germany/HH-hpi-p2125/2020 | 2020-03-22 | Europe/Germany/Hamburg/ | University Medical Center Hamburg Eppendorf | Heinrich Pette Institute, Leibniz Institute for Experimental Virology | Alexis Robitaille et al |
| EPI_ISL_776368 | Germany/HH-hpi-p2128/2020 | 2020-03-22 | Europe/Germany/Hamburg/ | University Medical Center Hamburg Eppendorf | Heinrich Pette Institute, Leibniz Institute for Experimental Virology | Alexis Robitaille et al |
| EPI_ISL_776373 | Germany/HH-hpi-p2135/2020 | 2020-03-21 | Europe/Germany/Hamburg/ | University Medical Center Hamburg Eppendorf | Heinrich Pette Institute, Leibniz Institute for Experimental Virology | Alexis Robitaille et al |
| EPI_ISL_776377 | Germany/HH-hpi-p2145/2020 | 2020-03-27 | Europe/Germany/Hamburg/ | University Medical Center Hamburg Eppendorf | Heinrich Pette Institute, Leibniz Institute for Experimental Virology | Alexis Robitaille et al |
| EPI_ISL_776380 | Germany/HH-hpi-p2148/2020 | 2020-03-28 | Europe/Germany/Hamburg/ | University Medical Center Hamburg Eppendorf | Heinrich Pette Institute, Leibniz Institute for Experimental Virology | Alexis Robitaille et al |

| | | | | | | |
|---|---|---|---|---|---|---|
| EPI_ISL_776382 | Germany/HH-hpi-p2150/2020 | 2020-03-27 | Europe/Germany/Hamburg/ | University Medical Center Hamburg Eppendorf | Heinrich Pette Institute, Leibniz Institute for Experimental Virology | Alexis Robitaille et al |
| EPI_ISL_776390 | Germany/HH-hpi-p2158/2020 | 2020-03-28 | Europe/Germany/Hamburg/ | University Medical Center Hamburg Eppendorf | Heinrich Pette Institute, Leibniz Institute for Experimental Virology | Alexis Robitaille et al |
| EPI_ISL_776394 | Germany/HH-hpi-p2164/2020 | 2020-03-28 | Europe/Germany/Hamburg/ | University Medical Center Hamburg Eppendorf | Heinrich Pette Institute, Leibniz Institute for Experimental Virology | Alexis Robitaille et al |
| EPI_ISL_776405 | Germany/HH-hpi-p2175/2020 | 2020-03-21 | Europe/Germany/Hamburg/ | University Medical Center Hamburg Eppendorf | Heinrich Pette Institute, Leibniz Institute for Experimental Virology | Alexis Robitaille et al |
| EPI_ISL_776406 | Germany/HH-hpi-p2177/2020 | 2020-03-28 | Europe/Germany/Hamburg/ | University Medical Center Hamburg Eppendorf | Heinrich Pette Institute, Leibniz Institute for Experimental Virology | Alexis Robitaille et al |
| EPI_ISL_776418 | Germany/HH-hpi-p2190/2020 | 2020-03-18 | Europe/Germany/Hamburg/ | University Medical Center Hamburg Eppendorf | Heinrich Pette Institute, Leibniz Institute for Experimental Virology | Alexis Robitaille et al |
| EPI_ISL_776419 | Germany/HH-hpi-p2191/2020 | 2020-03-21 | Europe/Germany/Hamburg/ | University Medical Center Hamburg Eppendorf | Heinrich Pette Institute, Leibniz Institute for Experimental Virology | Alexis Robitaille et al |
| EPI_ISL_776420 | Germany/HH-hpi-p2192/2020 | 2020-03-21 | Europe/Germany/Hamburg/ | University Medical Center Hamburg Eppendorf | Heinrich Pette Institute, Leibniz Institute for Experimental Virology | Alexis Robitaille et al |

| | | | | | | |
|---|---|---|---|---|---|---|
| EPI_ISL_776427 | Germany/HH-hpi-p2199/2020 | 2020-03-21 | Europe/Germany/Hamburg/ | University Medical Center Hamburg Eppendorf | Heinrich Pette Institute, Leibniz Institute for Experimental Virology | Alexis Robitaille et al |
| EPI_ISL_776429 | Germany/HH-hpi-p2296/2020 | 2020-03-12 | Europe/Germany/Hamburg/ | University Medical Center Hamburg Eppendorf | Heinrich Pette Institute, Leibniz Institute for Experimental Virology | Alexis Robitaille et al |
| EPI_ISL_776445 | Germany/HH-hpi-p2333/2020 | 2020-03-28 | Europe/Germany/Hamburg/ | University Medical Center Hamburg Eppendorf | Heinrich Pette Institute, Leibniz Institute for Experimental Virology | Alexis Robitaille et al |
| EPI_ISL_776449 | Germany/HH-hpi-p2354/2020 | 2020-03-14 | Europe/Germany/Hamburg/ | University Medical Center Hamburg Eppendorf | Heinrich Pette Institute, Leibniz Institute for Experimental Virology | Alexis Robitaille et al |
| EPI_ISL_776454 | Germany/HH-hpi-p2367/2020 | 2020-03-25 | Europe/Germany/Hamburg/ | University Medical Center Hamburg Eppendorf | Heinrich Pette Institute, Leibniz Institute for Experimental Virology | Alexis Robitaille et al |
| EPI_ISL_776455 | Germany/HH-hpi-p2376/2020 | 2020-05-13 | Europe/Germany/Hamburg/ | University Medical Center Hamburg Eppendorf | Heinrich Pette Institute, Leibniz Institute for Experimental Virology | Alexis Robitaille et al |
| EPI_ISL_776458 | Germany/HH-hpi-p2380/2020 | 2020-06-04 | Europe/Germany/Hamburg/ | University Medical Center Hamburg Eppendorf | Heinrich Pette Institute, Leibniz Institute for Experimental Virology | Alexis Robitaille et al |
| EPI_ISL_776472 | Germany/HH-hpi-p2397/2020 | 2020-04-24 | Europe/Germany/Hamburg/ | University Medical Center Hamburg Eppendorf | Heinrich Pette Institute, Leibniz Institute for Experimental Virology | Alexis Robitaille et al |

| | | | | | | |
|---|---|---|---|---|---|---|
| EPI_ISL_776480 | Germany/HH-hpi-p2407/2020 | 2020-04-25 | Europe/Germany/Hamburg/ | University Medical Center Hamburg Eppendorf | Heinrich Pette Institute, Leibniz Institute for Experimental Virology | Alexis Robitaille et al |
| EPI_ISL_776483 | Germany/HH-hpi-p2412/2020 | 2020-04-25 | Europe/Germany/Hamburg/ | University Medical Center Hamburg Eppendorf | Heinrich Pette Institute, Leibniz Institute for Experimental Virology | Alexis Robitaille et al |
| EPI_ISL_776484 | Germany/HH-hpi-p2413/2020 | 2020-04-29 | Europe/Germany/Hamburg/ | University Medical Center Hamburg Eppendorf | Heinrich Pette Institute, Leibniz Institute for Experimental Virology | Alexis Robitaille et al |
| EPI_ISL_776488 | Germany/HH-hpi-p2419/2020 | 2020-04-20 | Europe/Germany/Hamburg/ | University Medical Center Hamburg Eppendorf | Heinrich Pette Institute, Leibniz Institute for Experimental Virology | Alexis Robitaille et al |
| EPI_ISL_776492 | Germany/HH-hpi-p2423/2020 | 2020-04-26 | Europe/Germany/Hamburg/ | University Medical Center Hamburg Eppendorf | Heinrich Pette Institute, Leibniz Institute for Experimental Virology | Alexis Robitaille et al |
| EPI_ISL_776497 | Germany/HH-hpi-p2428/2020 | 2020-05-20 | Europe/Germany/Hamburg/ | University Medical Center Hamburg Eppendorf | Heinrich Pette Institute, Leibniz Institute for Experimental Virology | Alexis Robitaille et al |
| EPI_ISL_776499 | Germany/HH-hpi-p2430/2020 | 2020-04-20 | Europe/Germany/Hamburg/ | University Medical Center Hamburg Eppendorf | Heinrich Pette Institute, Leibniz Institute for Experimental Virology | Alexis Robitaille et al |
| EPI_ISL_776500 | Germany/HH-hpi-p2431/2020 | 2020-04-21 | Europe/Germany/Hamburg/ | University Medical Center Hamburg Eppendorf | Heinrich Pette Institute, Leibniz Institute for Experimental Virology | Alexis Robitaille et al |

| | | | | | | |
|---|---|---|---|---|---|---|
| EPI_ISL_776508 | Germany/HH-hpi-p2443/2020 | 2020-04-19 | Europe/Germany/Hamburg/ | University Medical Center Hamburg Eppendorf | Heinrich Pette Institute, Leibniz Institute for Experimental Virology | Alexis Robitaille et al |
| EPI_ISL_776512 | Germany/HH-hpi-p2450/2020 | 2020-04-19 | Europe/Germany/Hamburg/ | University Medical Center Hamburg Eppendorf | Heinrich Pette Institute, Leibniz Institute for Experimental Virology | Alexis Robitaille et al |
| EPI_ISL_776526 | Germany/HH-hpi-p2467/2020 | 2020-04-28 | Europe/Germany/Hamburg/ | University Medical Center Hamburg Eppendorf | Heinrich Pette Institute, Leibniz Institute for Experimental Virology | Alexis Robitaille et al |
| EPI_ISL_776527 | Germany/HH-hpi-p2468/2020 | 2020-04-26 | Europe/Germany/Hamburg/ | University Medical Center Hamburg Eppendorf | Heinrich Pette Institute, Leibniz Institute for Experimental Virology | Alexis Robitaille et al |
| EPI_ISL_776529 | Germany/HH-hpi-p2471/2020 | 2020-06-19 | Europe/Germany/Hamburg/ | University Medical Center Hamburg Eppendorf | Heinrich Pette Institute, Leibniz Institute for Experimental Virology | Alexis Robitaille et al |
| EPI_ISL_776548 | Germany/NI-hpi-p1458/2020 | 2020-03-11 | Europe/Germany/Niedersachsen/ | University Medical Center Hamburg Eppendorf | Heinrich Pette Institute, Leibniz Institute for Experimental Virology | Alexis Robitaille et al |
| EPI_ISL_776552 | Germany/NI-hpi-p1746/2020 | 2020-03-14 | Europe/Germany/Niedersachsen/ | University Medical Center Hamburg Eppendorf | Heinrich Pette Institute, Leibniz Institute for Experimental Virology | Alexis Robitaille et al |
| EPI_ISL_776560 | Germany/HH-hpi-p1364/2020 | 2020-03-28 | Europe/Germany/Hamburg/ | University Medical Center Hamburg Eppendorf | Heinrich Pette Institute, Leibniz Institute for Experimental Virology | Alexis Robitaille et al |

| | | | | | | |
|---|---|---|---|---|---|---|
| EPI_ISL_776564 | Germany/HH-hpi-p1939/2020 | 2020-03-25 | Europe/Germany/Hamburg/ | University Medical Center Hamburg Eppendorf | Heinrich Pette Institute, Leibniz Institute for Experimental Virology | Alexis Robitaille et al |
| EPI_ISL_776565 | Germany/SH-hpi-p1907/2020 | 2020-03-24 | Europe/Germany/Schleswig-Holstein/ | University Medical Center Hamburg Eppendorf | Heinrich Pette Institute, Leibniz Institute for Experimental Virology | Alexis Robitaille et al |
| EPI_ISL_776566 | Germany/SH-hpi-p2305/2020 | 2020-03-12 | Europe/Germany/Schleswig-Holstein/ | University Medical Center Hamburg Eppendorf | Heinrich Pette Institute, Leibniz Institute for Experimental Virology | Alexis Robitaille et al |
| EPI_ISL_776575 | Israel/CVL-94-ngs/2020 | 2020-12-27 | Asia/Israel/Israel/ | Israel Central Virology laboratory | Israel Central Virology laboratory | Neta Zuckerman et al |
| EPI_ISL_776631 | Israel/CVL-40-ngs/2020 | 2020-12-24 | Asia/Israel/Israel/ | Israel Central Virology laboratory | Israel Central Virology laboratory | Neta Zuckerman et al |
| EPI_ISL_776657 | USA/WA-UW-5501/2020 | 2020-04-30 | North America/USA/Washington/ | UW Virology Lab | UW Virology Lab | Pavitra Roychoudhury et al |
| EPI_ISL_776660 | USA/WA-UW-9453/2020 | 2020-05-19 | North America/USA/Washington/ | UW Virology Lab | UW Virology Lab | Pavitra Roychoudhury et al |
| EPI_ISL_776662 | USA/WA-UW-47334/2020 | 2020-12-14 | North America/USA/Washington/ | UW Virology Lab | UW Virology Lab | Pavitra Roychoudhury et al |
| EPI_ISL_776664 | USA/WA-UW-13887/2020 | 2020-07-06 | North America/USA/Washington/ | UW Virology Lab | UW Virology Lab | Pavitra Roychoudhury et al |
| EPI_ISL_776682 | USA/WA-UW-14623/2020 | 2020-07-12 | North America/USA/Washington/ | UW Virology Lab | UW Virology Lab | Pavitra Roychoudhury et al |
| EPI_ISL_776698 | USA/WA-UW-47265/2020 | 2020-12-07 | North America/USA/Washington/ | UW Virology Lab | UW Virology Lab | Pavitra Roychoudhury et al |
| EPI_ISL_776708 | USA/WA-UW-47337/2020 | 2020-12-12 | North America/USA/Washington/ | UW Virology Lab | UW Virology Lab | Pavitra Roychoudhury et al |
| EPI_ISL_776710 | USA/WA-UW-47361/2020 | 2020-12-14 | North America/USA/Washington/ | UW Virology Lab | UW Virology Lab | Pavitra Roychoudhury et al |
| EPI_ISL_776714 | USA/WA-UW-47379/2020 | 2020-12-14 | North America/USA/Washington/ | UW Virology Lab | UW Virology Lab | Pavitra Roychoudhury et al |
| EPI_ISL_776724 | USA/WA-UW-47418/2020 | 2020-12-11 | North America/USA/Washington/ | UW Virology Lab | UW Virology Lab | Pavitra Roychoudhury et al |
| EPI_ISL_776744 | USA/WA-UW-47422/2020 | 2020-12-11 | North America/USA/Washington/ | UW Virology Lab | UW Virology Lab | Pavitra Roychoudhury et al |
| EPI_ISL_776747 | USA/WA-UW-8508/2020 | 2020-05-04 | North America/USA/Washington/ | UW Virology Lab | UW Virology Lab | Pavitra Roychoudhury et al |

| | | | | | | |
|---|---|---|---|---|---|---|
| EPI_ISL_776750 | Brazil/SP-815/2020 | 2020-04-17 | South America/Brazil/S√£o Paulo/S√£o | Instituto Adolfo Lutz - Central | Instituto Adolfo Lutz, Interdiciplinary Procedures Center, Strategic Laboratory | Claudio Tavares Sacchi et al |
| EPI_ISL_776751 | Brazil/SP-816/2020 | 2020-03-24 | South America/Brazil/S√£o Paulo/S√£o | Instituto Adolfo Lutz - Central | Instituto Adolfo Lutz, Interdiciplinary Procedures Center, Strategic Laboratory | Claudio Tavares Sacchi et al |
| EPI_ISL_776764 | Brazil/SP-776/2020 | 2020-07-01 | South America/Brazil/S√£o Paulo/Rio Gr | Instituto Adolfo Lutz - Regional de Santo Andre | Instituto Adolfo Lutz, Interdiciplinary Procedures Center, Strategic Laboratory | Claudio Tavares Sacchi et al |
| EPI_ISL_776765 | Brazil/SP-777/2020 | 2020-07-01 | South America/Brazil/S√£o Paulo/Rio Gr | Instituto Adolfo Lutz - Regional de Santo Andre | Instituto Adolfo Lutz, Interdiciplinary Procedures Center, Strategic Laboratory | Claudio Tavares Sacchi et al |
| EPI_ISL_776767 | Brazil/SP-871/2020 | 2020-11-27 | South America/Brazil/S√£o Paulo/Candi | Instituto Adolfo Lutz - Regional de Marilia | Instituto Adolfo Lutz, Interdiciplinary Procedures Center, Strategic Laboratory | Claudio Tavares Sacchi et al |
| EPI_ISL_776768 | Brazil/SP-872/2020 | 2020-12-01 | South America/Brazil/S√£o Paulo/Ara√ß | Instituto Adolfo Lutz - Regional de Aracatuba | Instituto Adolfo Lutz, Interdiciplinary Procedures Center, Strategic Laboratory | Claudio Tavares Sacchi et al |

| | | | | | | |
|---|---|---|---|---|---|---|
| EPI_ISL_776769 | Brazil/SP-873/2020 | 2020-11-30 | South America/Brazil/S√£o Paulo/Sao B | Instituto Adolfo Lutz - Regional de Santo Andre | Instituto Adolfo Lutz, Interdiciplinary Procedures Center, Strategic Laboratory | Claudio Tavares Sacchi et al |
| EPI_ISL_776841 | Israel/CVL-128-ngs/2020 | 2020-12-27 | Asia/Israel/Israel/ | Israel Central Virology laboratory | Israel Central Virology laboratory | Neta Zuckerman et al |
| EPI_ISL_776852 | Israel/CVL-203-ngs/2020 | 2020-12-27 | Asia/Israel/Israel/ | Israel Central Virology laboratory | Israel Central Virology laboratory | Neta Zuckerman et al |
| EPI_ISL_776872 | USA/OH-ODH-SC35535/2020 | 2020-04-26 | North America/USA/Ohio/ | Ohio Department of Health Laboratory | Ohio Department of Health Laboratory | Holmes et al |
| EPI_ISL_776877 | Spain/M-20166261/2020 | 2020-05-28 | Europe/Spain/Madrid/ | Hospital General Universitario Gregorio Marav√±√≥n | Hospital General Universitario Gregorio Marav√±√≥n | Sergio Buenestado Serrano et al |
| EPI_ISL_776879 | Italy/CAM-TIGEM-196/2020 | 2020-10-26 | Europe/Italy/Campania/ | Istituto Zooprofilattico Sperimentale del Mezzogiorno | TIGEM | Patrizia Annunziata et al |
| EPI_ISL_776880 | Italy/CAM-TIGEM-202/2020 | 2020-10-26 | Europe/Italy/Campania/ | Istituto Zooprofilattico Sperimentale del Mezzogiorno | TIGEM | Patrizia Annunziata et al |
| EPI_ISL_776882 | Italy/CAM-TIGEM-195/2020 | 2020-10-26 | Europe/Italy/Campania/ | Istituto Zooprofilattico Sperimentale del Mezzogiorno | TIGEM | Patrizia Annunziata et al |
| EPI_ISL_776884 | Italy/CAM-TIGEM-194/2020 | 2020-10-26 | Europe/Italy/Campania/ | Istituto Zooprofilattico Sperimentale del Mezzogiorno | TIGEM | Patrizia Annunziata et al |
| EPI_ISL_776885 | Italy/CAM-TIGEM-219/2020 | 2020-10-26 | Europe/Italy/Campania/ | Istituto Zooprofilattico Sperimentale del Mezzogiorno | TIGEM | Patrizia Annunziata et al |
| EPI_ISL_776886 | Italy/CAM-TIGEM-207/2020 | 2020-10-26 | Europe/Italy/Campania/ | Istituto Zooprofilattico Sperimentale del Mezzogiorno | TIGEM | Patrizia Annunziata et al |
| EPI_ISL_776887 | Italy/CAM-TIGEM-230/2020 | 2020-10-26 | Europe/Italy/Campania/ | Istituto Zooprofilattico Sperimentale del Mezzogiorno | TIGEM | Patrizia Annunziata et al |
| EPI_ISL_776888 | Italy/CAM-TIGEM-208/2020 | 2020-10-26 | Europe/Italy/Campania/ | Istituto Zooprofilattico Sperimentale del Mezzogiorno | TIGEM | Patrizia Annunziata et al |
| EPI_ISL_776891 | Italy/CAM-TIGEM-216/2020 | 2020-10-26 | Europe/Italy/Campania/ | Istituto Zooprofilattico Sperimentale del Mezzogiorno | TIGEM | Patrizia Annunziata et al |
| EPI_ISL_776893 | Italy/CAM-TIGEM-233/2020 | 2020-10-26 | Europe/Italy/Campania/ | Istituto Zooprofilattico Sperimentale del Mezzogiorno | TIGEM | Patrizia Annunziata et al |

| | | | | | | |
|---|---|---|---|---|---|---|
| EPI_ISL_776896 | Italy/CAM-TIGEM-247/2020 | 2020-10-26 | Europe/Italy/Campania/ | Istituto Zooprofilattico Sperimentale del Mezzogiorno | TIGEM | Patrizia Annunziata et al |
| EPI_ISL_776898 | Italy/CAM-TIGEM-239/2020 | 2020-10-26 | Europe/Italy/Campania/ | Istituto Zooprofilattico Sperimentale del Mezzogiorno | TIGEM | Patrizia Annunziata et al |
| EPI_ISL_776900 | Italy/CAM-TIGEM-245/2020 | 2020-10-26 | Europe/Italy/Campania/ | Istituto Zooprofilattico Sperimentale del Mezzogiorno | TIGEM | Patrizia Annunziata et al |
| EPI_ISL_776903 | Italy/CAM-TIGEM-226/2020 | 2020-10-26 | Europe/Italy/Campania/ | Istituto Zooprofilattico Sperimentale del Mezzogiorno | TIGEM | Patrizia Annunziata et al |
| EPI_ISL_776908 | Italy/CAM-TIGEM-227/2020 | 2020-10-26 | Europe/Italy/Campania/ | Istituto Zooprofilattico Sperimentale del Mezzogiorno | TIGEM | Patrizia Annunziata et al |
| EPI_ISL_776910 | Italy/CAM-TIGEM-197/2020 | 2020-10-26 | Europe/Italy/Campania/ | Istituto Zooprofilattico Sperimentale del Mezzogiorno | TIGEM | Patrizia Annunziata et al |
| EPI_ISL_776912 | Italy/CAM-TIGEM-243/2020 | 2020-10-26 | Europe/Italy/Campania/ | Istituto Zooprofilattico Sperimentale del Mezzogiorno | TIGEM | Patrizia Annunziata et al |
| EPI_ISL_776913 | Italy/CAM-TIGEM-235/2020 | 2020-10-26 | Europe/Italy/Campania/ | Istituto Zooprofilattico Sperimentale del Mezzogiorno | TIGEM | Patrizia Annunziata et al |
| EPI_ISL_776914 | Italy/CAM-TIGEM-249/2020 | 2020-10-26 | Europe/Italy/Campania/ | Istituto Zooprofilattico Sperimentale del Mezzogiorno | TIGEM | Patrizia Annunziata et al |
| EPI_ISL_776915 | Italy/CAM-TIGEM-198/2020 | 2020-10-26 | Europe/Italy/Campania/ | Istituto Zooprofilattico Sperimentale del Mezzogiorno | TIGEM | Patrizia Annunziata et al |
| EPI_ISL_776916 | Italy/CAM-TIGEM-212/2020 | 2020-10-26 | Europe/Italy/Campania/ | Istituto Zooprofilattico Sperimentale del Mezzogiorno | TIGEM | Patrizia Annunziata et al |
| EPI_ISL_776917 | Italy/CAM-TIGEM-215/2020 | 2020-10-26 | Europe/Italy/Campania/ | Istituto Zooprofilattico Sperimentale del Mezzogiorno | TIGEM | Patrizia Annunziata et al |
| EPI_ISL_776918 | Italy/CAM-TIGEM-214/2020 | 2020-10-26 | Europe/Italy/Campania/ | Istituto Zooprofilattico Sperimentale del Mezzogiorno | TIGEM | Patrizia Annunziata et al |
| EPI_ISL_776920 | Italy/CAM-TIGEM-201/2020 | 2020-10-26 | Europe/Italy/Campania/ | Istituto Zooprofilattico Sperimentale del Mezzogiorno | TIGEM | Patrizia Annunziata et al |
| EPI_ISL_776924 | Italy/CAM-TIGEM-263/2020 | 2020-10-26 | Europe/Italy/Campania/ | Istituto Zooprofilattico Sperimentale del Mezzogiorno | TIGEM | Patrizia Annunziata et al |
| EPI_ISL_776932 | Italy/CAM-TIGEM-284/2020 | 2020-10-26 | Europe/Italy/Campania/ | Istituto Zooprofilattico Sperimentale del Mezzogiorno | TIGEM | Patrizia Annunziata et al |
| EPI_ISL_776934 | Italy/CAM-TIGEM-281/2020 | 2020-10-26 | Europe/Italy/Campania/ | Istituto Zooprofilattico Sperimentale del Mezzogiorno | TIGEM | Patrizia Annunziata et al |
| EPI_ISL_776935 | Italy/CAM-TIGEM-286/2020 | 2020-10-26 | Europe/Italy/Campania/ | Istituto Zooprofilattico Sperimentale del Mezzogiorno | TIGEM | Patrizia Annunziata et al |
| EPI_ISL_776938 | Italy/CAM-TIGEM-258/2020 | 2020-10-26 | Europe/Italy/Campania/ | Istituto Zooprofilattico Sperimentale del Mezzogiorno | TIGEM | Patrizia Annunziata et al |
| EPI_ISL_776941 | Italy/CAM-TIGEM-255/2020 | 2020-10-26 | Europe/Italy/Campania/ | Istituto Zooprofilattico Sperimentale del Mezzogiorno | TIGEM | Patrizia Annunziata et al |

| | | | | | | |
|---|---|---|---|---|---|---|
| EPI_ISL_776942 | Italy/CAM-TIGEM-278/2020 | 2020-10-26 | Europe/Italy/Campania/ | Istituto Zooprofilattico Sperimentale del Mezzogiorno | TIGEM | Patrizia Annunziata et al |
| EPI_ISL_776945 | Italy/CAM-TIGEM-257/2020 | 2020-10-26 | Europe/Italy/Campania/ | Istituto Zooprofilattico Sperimentale del Mezzogiorno | TIGEM | Patrizia Annunziata et al |
| EPI_ISL_776947 | Italy/CAM-TIGEM-260/2020 | 2020-10-26 | Europe/Italy/Campania/ | Istituto Zooprofilattico Sperimentale del Mezzogiorno | TIGEM | Patrizia Annunziata et al |
| EPI_ISL_776948 | Italy/CAM-TIGEM-256/2020 | 2020-10-26 | Europe/Italy/Campania/ | Istituto Zooprofilattico Sperimentale del Mezzogiorno | TIGEM | Patrizia Annunziata et al |
| EPI_ISL_776949 | Italy/CAM-TIGEM-270/2020 | 2020-10-26 | Europe/Italy/Campania/ | Istituto Zooprofilattico Sperimentale del Mezzogiorno | TIGEM | Patrizia Annunziata et al |
| EPI_ISL_776953 | Italy/CAM-TIGEM-276/2020 | 2020-10-26 | Europe/Italy/Campania/ | Istituto Zooprofilattico Sperimentale del Mezzogiorno | TIGEM | Patrizia Annunziata et al |
| EPI_ISL_776954 | Italy/CAM-TIGEM-297/2020 | 2020-10-26 | Europe/Italy/Campania/ | Istituto Zooprofilattico Sperimentale del Mezzogiorno | TIGEM | Patrizia Annunziata et al |
| EPI_ISL_776955 | Italy/CAM-TIGEM-277/2020 | 2020-10-26 | Europe/Italy/Campania/ | Istituto Zooprofilattico Sperimentale del Mezzogiorno | TIGEM | Patrizia Annunziata et al |
| EPI_ISL_776956 | Italy/CAM-TIGEM-262/2020 | 2020-10-26 | Europe/Italy/Campania/ | Istituto Zooprofilattico Sperimentale del Mezzogiorno | TIGEM | Patrizia Annunziata et al |
| EPI_ISL_776957 | Italy/CAM-TIGEM-275/2020 | 2020-10-26 | Europe/Italy/Campania/ | Istituto Zooprofilattico Sperimentale del Mezzogiorno | TIGEM | Patrizia Annunziata et al |
| EPI_ISL_776962 | Italy/CAM-TIGEM-381/2020 | 2020-09-29 | Europe/Italy/Campania/ | Istituto Zooprofilattico Sperimentale del Mezzogiorno | TIGEM | Patrizia Annunziata et al |
| EPI_ISL_776963 | Italy/CAM-TIGEM-363/2020 | 2020-09-29 | Europe/Italy/Campania/ | Istituto Zooprofilattico Sperimentale del Mezzogiorno | TIGEM | Patrizia Annunziata et al |
| EPI_ISL_776971 | Italy/CAM-TIGEM-324/2020 | 2020-09-02 | Europe/Italy/Campania/ | Istituto Zooprofilattico Sperimentale del Mezzogiorno | TIGEM | Patrizia Annunziata et al |
| EPI_ISL_776972 | Italy/CAM-TIGEM-351/2020 | 2020-09-02 | Europe/Italy/Campania/ | Istituto Zooprofilattico Sperimentale del Mezzogiorno | TIGEM | Patrizia Annunziata et al |
| EPI_ISL_776974 | Italy/CAM-TIGEM-341/2020 | 2020-09-03 | Europe/Italy/Campania/ | Istituto Zooprofilattico Sperimentale del Mezzogiorno | TIGEM | Patrizia Annunziata et al |
| EPI_ISL_776978 | Italy/CAM-TIGEM-300/2020 | 2020-09-07 | Europe/Italy/Campania/ | Istituto Zooprofilattico Sperimentale del Mezzogiorno | TIGEM | Patrizia Annunziata et al |
| EPI_ISL_776988 | Italy/CAM-TIGEM-354/2020 | 2020-09-10 | Europe/Italy/Campania/ | Istituto Zooprofilattico Sperimentale del Mezzogiorno | TIGEM | Patrizia Annunziata et al |
| EPI_ISL_776993 | Italy/CAM-TIGEM-360/2020 | 2020-09-14 | Europe/Italy/Campania/ | Istituto Zooprofilattico Sperimentale del Mezzogiorno | TIGEM | Patrizia Annunziata et al |
| EPI_ISL_776995 | Italy/CAM-TIGEM-379/2020 | 2020-09-15 | Europe/Italy/Campania/ | Istituto Zooprofilattico Sperimentale del Mezzogiorno | TIGEM | Patrizia Annunziata et al |
| EPI_ISL_776997 | Italy/CAM-TIGEM-365/2020 | 2020-09-20 | Europe/Italy/Campania/ | Istituto Zooprofilattico Sperimentale del Mezzogiorno | TIGEM | Patrizia Annunziata et al |

| | | | | | | |
|---|---|---|---|---|---|---|
| EPI_ISL_777002 | Italy/CAM-TIGEM-371/2020 | 2020-09-20 | Europe/Italy/Campania/ | Istituto Zooprofilattico Sperimentale del Mezzogiorno | TIGEM | Patrizia Annunziata et al |
| EPI_ISL_777003 | Italy/CAM-TIGEM-382/2020 | 2020-09-20 | Europe/Italy/Campania/ | Istituto Zooprofilattico Sperimentale del Mezzogiorno | TIGEM | Patrizia Annunziata et al |
| EPI_ISL_777004 | Italy/CAM-TIGEM-370/2020 | 2020-09-20 | Europe/Italy/Campania/ | Istituto Zooprofilattico Sperimentale del Mezzogiorno | TIGEM | Patrizia Annunziata et al |
| EPI_ISL_777007 | Italy/CAM-TIGEM-383/2020 | 2020-09-20 | Europe/Italy/Campania/ | Istituto Zooprofilattico Sperimentale del Mezzogiorno | TIGEM | Patrizia Annunziata et al |
| EPI_ISL_777008 | Italy/CAM-TIGEM-308/2020 | 2020-09-21 | Europe/Italy/Campania/ | Istituto Zooprofilattico Sperimentale del Mezzogiorno | TIGEM | Patrizia Annunziata et al |
| EPI_ISL_777235 | England/QEUH-C93965/2020 | 2020-12-17 | Europe/United Kingdom/England/ | Lighthouse Lab in Glasgow | Wellcome Sanger Institute for the COVID-19 Genomics UK (COG-UK) Consortium | Harper VanSteenhouse et al |
| EPI_ISL_777584 | England/ALDP-C8DE41/2020 | 2020-12-15 | Europe/United Kingdom/England/ | Lighthouse Lab in Alderley Park | Wellcome Sanger Institute for the COVID-19 Genomics UK (COG-UK) Consortium | Jacquelyn Wynn et al |
| EPI_ISL_777681 | England/CAMC-CB7CDB/2020 | 2020-12-19 | Europe/United Kingdom/England/ | Lighthouse Lab in Cambridge | Wellcome Sanger Institute for the COVID-19 Genomics UK (COG-UK) Consortium | Rob Howes et al |
| EPI_ISL_777684 | England/CAMC-CB87D9/2020 | 2020-12-19 | Europe/United Kingdom/England/ | Lighthouse Lab in Cambridge | Wellcome Sanger Institute for the COVID-19 Genomics UK (COG-UK) Consortium | Rob Howes et al |
| EPI_ISL_777732 | England/CAMC-CB8A85/2020 | 2020-12-18 | Europe/United Kingdom/England/ | Lighthouse Lab in Cambridge | Wellcome Sanger Institute for the COVID-19 Genomics UK (COG-UK) Consortium | Rob Howes et al |
| EPI_ISL_777851 | England/CAMC-CB7D32/2020 | 2020-12-18 | Europe/United Kingdom/England/ | Lighthouse Lab in Cambridge | Wellcome Sanger Institute for the COVID-19 Genomics UK (COG-UK) Consortium | Rob Howes et al |

| | | | | | | |
|---|---|---|---|---|---|---|
| EPI_ISL_777881 | England/CAMC-CB6B93/2020 | 2020-12-19 | Europe/United Kingdom/England/ | Lighthouse Lab in Cambridge | Wellcome Sanger Institute for the COVID-19 Genomics UK (COG-UK) Consortium | Rob Howes et al |
| EPI_ISL_778061 | England/CAMC-CB8EB6/2020 | 2020-12-18 | Europe/United Kingdom/England/ | Lighthouse Lab in Cambridge | Wellcome Sanger Institute for the COVID-19 Genomics UK (COG-UK) Consortium | Rob Howes et al |
| EPI_ISL_778137 | England/CAMC-CB8F77/2020 | 2020-12-18 | Europe/United Kingdom/England/ | Lighthouse Lab in Cambridge | Wellcome Sanger Institute for the COVID-19 Genomics UK (COG-UK) Consortium | Rob Howes et al |
| EPI_ISL_778174 | England/CAMC-CB8EF2/2020 | 2020-12-18 | Europe/United Kingdom/England/ | Lighthouse Lab in Cambridge | Wellcome Sanger Institute for the COVID-19 Genomics UK (COG-UK) Consortium | Rob Howes et al |
| EPI_ISL_778643 | England/MILK-C51927/2020 | 2020-12-11 | Europe/United Kingdom/England/ | Lighthouse Lab in Milton Keynes | Wellcome Sanger Institute for the COVID-19 Genomics UK (COG-UK) Consortium | The Lighthouse Lab in Milton Keynes et al |
| EPI_ISL_778646 | Italy/CAM-TIGEM-589/2020 | 2020-04-12 | Europe/Italy/Campania/ | Istituto Zooprofilattico Sperimentale del Mezzogiorno | TIGEM | Patrizia Annunziata et al |
| EPI_ISL_778647 | Italy/CAM-TIGEM-636/2020 | 2020-03-29 | Europe/Italy/Campania/ | Istituto Zooprofilattico Sperimentale del Mezzogiorno | TIGEM | Patrizia Annunziata et al |
| EPI_ISL_778648 | Italy/CAM-TIGEM-600/2020 | 2020-04-03 | Europe/Italy/Campania/ | Istituto Zooprofilattico Sperimentale del Mezzogiorno | TIGEM | Patrizia Annunziata et al |
| EPI_ISL_778649 | Italy/CAM-TIGEM-601/2020 | 2020-04-03 | Europe/Italy/Campania/ | Istituto Zooprofilattico Sperimentale del Mezzogiorno | TIGEM | Patrizia Annunziata et al |
| EPI_ISL_778650 | Italy/CAM-TIGEM-652/2020 | 2020-03-29 | Europe/Italy/Campania/ | Istituto Zooprofilattico Sperimentale del Mezzogiorno | TIGEM | Patrizia Annunziata et al |
| EPI_ISL_778651 | Italy/CAM-TIGEM-654/2020 | 2020-03-29 | Europe/Italy/Campania/ | Istituto Zooprofilattico Sperimentale del Mezzogiorno | TIGEM | Patrizia Annunziata et al |
| EPI_ISL_778655 | Italy/CAM-TIGEM-613/2020 | 2020-04-08 | Europe/Italy/Campania/ | Istituto Zooprofilattico Sperimentale del Mezzogiorno | TIGEM | Patrizia Annunziata et al |

| | | | | | | |
|---|---|---|---|---|---|---|
| EPI_ISL_778659 | Italy/CAM-TIGEM-581/2020 | 2020-04-13 | Europe/Italy/Campania/ | Istituto Zooprofilattico Sperimentale del Mezzogiorno | TIGEM | Patrizia Annunziata et al |
| EPI_ISL_778662 | Italy/CAM-TIGEM-595/2020 | 2020-04-13 | Europe/Italy/Campania/ | Istituto Zooprofilattico Sperimentale del Mezzogiorno | TIGEM | Patrizia Annunziata et al |
| EPI_ISL_778666 | Italy/CAM-TIGEM-588/2020 | 2020-04-14 | Europe/Italy/Campania/ | Istituto Zooprofilattico Sperimentale del Mezzogiorno | TIGEM | Patrizia Annunziata et al |
| EPI_ISL_778671 | Italy/CAM-TIGEM-608/2020 | 2020-04-18 | Europe/Italy/Campania/ | Istituto Zooprofilattico Sperimentale del Mezzogiorno | TIGEM | Patrizia Annunziata et al |
| EPI_ISL_778672 | Italy/CAM-TIGEM-605/2020 | 2020-04-18 | Europe/Italy/Campania/ | Istituto Zooprofilattico Sperimentale del Mezzogiorno | TIGEM | Patrizia Annunziata et al |
| EPI_ISL_778674 | Italy/CAM-TIGEM-610/2020 | 2020-04-20 | Europe/Italy/Campania/ | Istituto Zooprofilattico Sperimentale del Mezzogiorno | TIGEM | Patrizia Annunziata et al |
| EPI_ISL_778676 | Italy/CAM-TIGEM-618/2020 | 2020-04-21 | Europe/Italy/Campania/ | Istituto Zooprofilattico Sperimentale del Mezzogiorno | TIGEM | Patrizia Annunziata et al |
| EPI_ISL_778680 | Italy/CAM-TIGEM-623/2020 | 2020-04-21 | Europe/Italy/Campania/ | Istituto Zooprofilattico Sperimentale del Mezzogiorno | TIGEM | Patrizia Annunziata et al |
| EPI_ISL_778681 | Italy/CAM-TIGEM-616/2020 | 2020-04-21 | Europe/Italy/Campania/ | Istituto Zooprofilattico Sperimentale del Mezzogiorno | TIGEM | Patrizia Annunziata et al |
| EPI_ISL_778684 | Italy/CAM-TIGEM-631/2020 | 2020-04-27 | Europe/Italy/Campania/ | Istituto Zooprofilattico Sperimentale del Mezzogiorno | TIGEM | Patrizia Annunziata et al |
| EPI_ISL_778693 | Italy/CAM-TIGEM-389/2020 | 2020-09-29 | Europe/Italy/Campania/ | Istituto Zooprofilattico Sperimentale del Mezzogiorno | TIGEM | Patrizia Annunziata et al |
| EPI_ISL_778696 | Italy/CAM-TIGEM-564/2020 | 2020-04-03 | Europe/Italy/Campania/ | Istituto Zooprofilattico Sperimentale del Mezzogiorno | TIGEM | Patrizia Annunziata et al |
| EPI_ISL_778698 | Italy/CAM-TIGEM-566/2020 | 2020-04-03 | Europe/Italy/Campania/ | Istituto Zooprofilattico Sperimentale del Mezzogiorno | TIGEM | Patrizia Annunziata et al |
| EPI_ISL_778701 | Italy/CAM-TIGEM-571/2020 | 2020-04-03 | Europe/Italy/Campania/ | Istituto Zooprofilattico Sperimentale del Mezzogiorno | TIGEM | Patrizia Annunziata et al |
| EPI_ISL_778702 | Italy/CAM-TIGEM-572/2020 | 2020-04-03 | Europe/Italy/Campania/ | Istituto Zooprofilattico Sperimentale del Mezzogiorno | TIGEM | Patrizia Annunziata et al |
| EPI_ISL_778707 | Italy/CAM-TIGEM-400/2020 | 2020-09-15 | Europe/Italy/Campania/ | Istituto Zooprofilattico Sperimentale del Mezzogiorno | TIGEM | Patrizia Annunziata et al |
| EPI_ISL_778708 | Italy/CAM-TIGEM-412/2020 | 2020-09-15 | Europe/Italy/Campania/ | Istituto Zooprofilattico Sperimentale del Mezzogiorno | TIGEM | Patrizia Annunziata et al |
| EPI_ISL_778711 | Italy/CAM-TIGEM-414/2020 | 2020-09-15 | Europe/Italy/Campania/ | Istituto Zooprofilattico Sperimentale del Mezzogiorno | TIGEM | Patrizia Annunziata et al |
| EPI_ISL_778714 | Italy/CAM-TIGEM-419/2020 | 2020-09-15 | Europe/Italy/Campania/ | Istituto Zooprofilattico Sperimentale del Mezzogiorno | TIGEM | Patrizia Annunziata et al |
| EPI_ISL_778717 | Italy/CAM-TIGEM-408/2020 | 2020-09-15 | Europe/Italy/Campania/ | Istituto Zooprofilattico Sperimentale del Mezzogiorno | TIGEM | Patrizia Annunziata et al |

| | | | | | | |
|---|---|---|---|---|---|---|
| EPI_ISL_778722 | Italy/CAM-TIGEM-561/2020 | 2020-04-09 | Europe/Italy/Campania/ | Istituto Zooprofilattico Sperimentale del Mezzogiorno | TIGEM | Patrizia Annunziata et al |
| EPI_ISL_778728 | Italy/CAM-TIGEM-575/2020 | 2020-04-11 | Europe/Italy/Campania/ | Istituto Zooprofilattico Sperimentale del Mezzogiorno | TIGEM | Patrizia Annunziata et al |
| EPI_ISL_778729 | Italy/CAM-TIGEM-545/2020 | 2020-04-13 | Europe/Italy/Campania/ | Istituto Zooprofilattico Sperimentale del Mezzogiorno | TIGEM | Patrizia Annunziata et al |
| EPI_ISL_778730 | Italy/CAM-TIGEM-552/2020 | 2020-04-13 | Europe/Italy/Campania/ | Istituto Zooprofilattico Sperimentale del Mezzogiorno | TIGEM | Patrizia Annunziata et al |
| EPI_ISL_778733 | Italy/CAM-TIGEM-555/2020 | 2020-04-13 | Europe/Italy/Campania/ | Istituto Zooprofilattico Sperimentale del Mezzogiorno | TIGEM | Patrizia Annunziata et al |
| EPI_ISL_778734 | Italy/CAM-TIGEM-547/2020 | 2020-04-13 | Europe/Italy/Campania/ | Istituto Zooprofilattico Sperimentale del Mezzogiorno | TIGEM | Patrizia Annunziata et al |
| EPI_ISL_778735 | Italy/CAM-TIGEM-558/2020 | 2020-04-20 | Europe/Italy/Campania/ | Istituto Zooprofilattico Sperimentale del Mezzogiorno | TIGEM | Patrizia Annunziata et al |
| EPI_ISL_778736 | Italy/CAM-TIGEM-562/2020 | 2020-04-29 | Europe/Italy/Campania/ | Istituto Zooprofilattico Sperimentale del Mezzogiorno | TIGEM | Patrizia Annunziata et al |
| EPI_ISL_778758 | Italy/CAM-TIGEM-439/2020 | 2020-06-30 | Europe/Italy/Campania/ | Istituto Zooprofilattico Sperimentale del Mezzogiorno | TIGEM | Patrizia Annunziata et al |
| EPI_ISL_778777 | Italy/CAM-TIGEM-469/2020 | 2020-07-02 | Europe/Italy/Campania/ | Istituto Zooprofilattico Sperimentale del Mezzogiorno | TIGEM | Patrizia Annunziata et al |
| EPI_ISL_778782 | Italy/CAM-TIGEM-478/2020 | 2020-07-02 | Europe/Italy/Campania/ | Istituto Zooprofilattico Sperimentale del Mezzogiorno | TIGEM | Patrizia Annunziata et al |
| EPI_ISL_778785 | Italy/CAM-TIGEM-483/2020 | 2020-06-28 | Europe/Italy/Campania/ | Istituto Zooprofilattico Sperimentale del Mezzogiorno | TIGEM | Patrizia Annunziata et al |
| EPI_ISL_778805 | Italy/CAM-TIGEM-507/2020 | 2020-06-29 | Europe/Italy/Campania/ | Istituto Zooprofilattico Sperimentale del Mezzogiorno | TIGEM | Patrizia Annunziata et al |
| EPI_ISL_778843 | Argentina/C97133/2020 | 2020-11-20 | South America/Argentina/Argentina/ | Servicio Virosis Respiratorias-Departamento Virolog√≠a-INEI | Instituto Nacional Enfermedades Infecciosas C.G.Malbran | Baumeister E. et al |
| EPI_ISL_778845 | Canada/ON-PHL-20-03614/2020 | 20-Nov | North America/Canada/Ontario/ | Public Health Ontario Laboratory | Public Health Ontario Laboratory | Vanessa G Allen et al |
| EPI_ISL_778846 | Canada/ON-PHL-20-03650/2020 | 20-Nov | North America/Canada/Ontario/ | Public Health Ontario Laboratory | Public Health Ontario Laboratory | Vanessa G Allen et al |
| EPI_ISL_778848 | Canada/ON-PHL-20-03673/2020 | 20-Nov | North America/Canada/Ontario/ | Public Health Ontario Laboratory | Public Health Ontario Laboratory | Vanessa G Allen et al |

| | | | | | | |
|---|---|---|---|---|---|---|
| EPI_ISL_778849 | Canada/ON-PHL-20-03674/2020 | 20-Nov | North America/Canada/Ontario/ | Public Health Ontario Laboratory | Public Health Ontario Laboratory | Vanessa G Allen et al |
| EPI_ISL_778850 | Canada/ON-PHL-20-03725/2020 | 20-Nov | North America/Canada/Ontario/ | Public Health Ontario Laboratory | Public Health Ontario Laboratory | Vanessa G Allen et al |
| EPI_ISL_778851 | Canada/ON-PHL-20-03729/2020 | 20-Nov | North America/Canada/Ontario/ | Public Health Ontario Laboratory | Public Health Ontario Laboratory | Vanessa G Allen et al |
| EPI_ISL_778852 | Canada/ON-PHL-20-03753/2020 | 20-Nov | North America/Canada/Ontario/ | Public Health Ontario Laboratory | Public Health Ontario Laboratory | Vanessa G Allen et al |
| EPI_ISL_778853 | Canada/ON-PHL-20-03778/2020 | 20-Nov | North America/Canada/Ontario/ | Public Health Ontario Laboratory | Public Health Ontario Laboratory | Vanessa G Allen et al |
| EPI_ISL_778854 | Canada/ON-PHL-20-03822/2020 | 20-Nov | North America/Canada/Ontario/ | Public Health Ontario Laboratory | Public Health Ontario Laboratory | Vanessa G Allen et al |
| EPI_ISL_778858 | USA/OH-ODH-SC35119/2020 | 2020-04-23 | North America/USA/Ohio/ | Ohio Department of Health Laboratory | Ohio Department of Health Laboratory | Holmes et al |
| EPI_ISL_778872 | USA/MD-MDH-0487/2020 | 2020-11-30 | North America/USA/Maryland/ | Maryland Public Health Laboratory | Maryland Public Health Laboratory | Maryland Department of Health Laboratories Administration et al |
| EPI_ISL_778875 | USA/MD-MDH-0494/2020 | 2020-12-02 | North America/USA/Maryland/ | Maryland Public Health Laboratory | Maryland Public Health Laboratory | Maryland Department of Health Laboratories Administration et al |
| EPI_ISL_778876 | USA/MD-MDH-0495/2020 | 2020-12-02 | North America/USA/Maryland/ | Maryland Public Health Laboratory | Maryland Public Health Laboratory | Maryland Department of Health Laboratories Administration et al |
| EPI_ISL_778880 | USA/MD-MDH-0499/2020 | 2020-12-23 | North America/USA/Maryland/ | Maryland Public Health Laboratory | Maryland Public Health Laboratory | Maryland Department of Health Laboratories Administration et al |
| EPI_ISL_778894 | USA/MD-MDH-0513/2020 | 2020-11-30 | North America/USA/Maryland/ | Maryland Public Health Laboratory | Maryland Public Health Laboratory | Maryland Department of Health Laboratories Administration et al |
| EPI_ISL_778897 | USA/MD-MDH-0517/2020 | 2020-11-30 | North America/USA/Maryland/ | Maryland Public Health Laboratory | Maryland Public Health Laboratory | Maryland Department of Health Laboratories Administration et al |

| | | | | | | |
|---|---|---|---|---|---|---|
| EPI_ISL_778912 | USA/MD-MDH-0536/2020 | 2020-11-30 | North America/USA/Maryland/ | Maryland Public Health Laboratory | Maryland Public Health Laboratory | Maryland Department of Health Laboratories Administration et al |
| EPI_ISL_778919 | USA/LA-EVTL1253/2020 | 2020-12-04 | North America/USA/Louisiana/Bienville | LSUHS Emerging Viral Threat Laboratory | Microbial Genome Sequencing Center | Jeremy P. Kamil et al |
| EPI_ISL_778923 | USA/LA-EVTL1257/2020 | 2020-12-04 | North America/USA/Louisiana/Bienville | LSUHS Emerging Viral Threat Laboratory | Microbial Genome Sequencing Center | Jeremy P. Kamil et al |
| EPI_ISL_778927 | USA/LA-EVTL1261/2020 | 2020-12-04 | North America/USA/Louisiana/Bienville | LSUHS Emerging Viral Threat Laboratory | Microbial Genome Sequencing Center | Jeremy P. Kamil et al |
| EPI_ISL_778929 | USA/LA-EVTL1263/2020 | 2020-12-04 | North America/USA/Louisiana/Bienville | LSUHS Emerging Viral Threat Laboratory | Microbial Genome Sequencing Center | Jeremy P. Kamil et al |
| EPI_ISL_778936 | USA/LA-EVTL1270/2020 | 2020-12-04 | North America/USA/Louisiana/Bienville | LSUHS Emerging Viral Threat Laboratory | Microbial Genome Sequencing Center | Jeremy P. Kamil et al |
| EPI_ISL_778940 | USA/LA-EVTL1274/2020 | 2020-12-04 | North America/USA/Louisiana/Bienville | LSUHS Emerging Viral Threat Laboratory | Microbial Genome Sequencing Center | Jeremy P. Kamil et al |
| EPI_ISL_778948 | USA/LA-EVTL1282/2020 | 2020-12-10 | North America/USA/Louisiana/Caddo Pa | LSUHS Emerging Viral Threat Laboratory | Microbial Genome Sequencing Center | Jeremy P. Kamil et al |
| EPI_ISL_778951 | USA/LA-EVTL1285/2020 | 2020-12-10 | North America/USA/Louisiana/Caddo Pa | LSUHS Emerging Viral Threat Laboratory | Microbial Genome Sequencing Center | Jeremy P. Kamil et al |
| EPI_ISL_778961 | USA/LA-EVTL1295/2020 | 2020-12-11 | North America/USA/Louisiana/Bossier P | LSUHS Emerging Viral Threat Laboratory | Microbial Genome Sequencing Center | Jeremy P. Kamil et al |
| EPI_ISL_778971 | USA/LA-EVTL1305/2020 | 2020-12-11 | North America/USA/Louisiana/Bossier P | LSUHS Emerging Viral Threat Laboratory | Microbial Genome Sequencing Center | Jeremy P. Kamil et al |
| EPI_ISL_778975 | USA/LA-EVTL1309/2020 | 2020-12-11 | North America/USA/Louisiana/Bossier P | LSUHS Emerging Viral Threat Laboratory | Microbial Genome Sequencing Center | Jeremy P. Kamil et al |
| EPI_ISL_778985 | USA/LA-EVTL1319/2020 | 2020-12-11 | North America/USA/Louisiana/Bossier P | LSUHS Emerging Viral Threat Laboratory | Microbial Genome Sequencing Center | Jeremy P. Kamil et al |

| | | | | | | |
|---|---|---|---|---|---|---|
| EPI_ISL_779003 | USA/LA-EVTL1337/2020 | 2020-12-11 | North America/USA/Louisiana/Bossier P | LSUHS Emerging Viral Threat Laboratory | Microbial Genome Sequencing Center | Jeremy P. Kamil et al |
| EPI_ISL_779004 | USA/LA-EVTL1338/2020 | 2020-12-11 | North America/USA/Louisiana/Bossier P | LSUHS Emerging Viral Threat Laboratory | Microbial Genome Sequencing Center | Jeremy P. Kamil et al |
| EPI_ISL_779009 | USA/LA-EVTL1343/2020 | 2020-12-11 | North America/USA/Louisiana/Bossier P | LSUHS Emerging Viral Threat Laboratory | Microbial Genome Sequencing Center | Jeremy P. Kamil et al |
| EPI_ISL_779015 | USA/LA-EVTL1349/2020 | 2020-12-11 | North America/USA/Louisiana/Bossier P | LSUHS Emerging Viral Threat Laboratory | Microbial Genome Sequencing Center | Jeremy P. Kamil et al |
| EPI_ISL_779019 | USA/LA-EVTL1353/2020 | 2020-12-11 | North America/USA/Louisiana/Bossier P | LSUHS Emerging Viral Threat Laboratory | Microbial Genome Sequencing Center | Jeremy P. Kamil et al |
| EPI_ISL_779028 | USA/LA-EVTL1362/2020 | 2020-12-11 | North America/USA/Louisiana/Bossier P | LSUHS Emerging Viral Threat Laboratory | Microbial Genome Sequencing Center | Jeremy P. Kamil et al |
| EPI_ISL_779036 | USA/LA-EVTL1370/2020 | 2020-12-11 | North America/USA/Louisiana/Bossier P | LSUHS Emerging Viral Threat Laboratory | Microbial Genome Sequencing Center | Jeremy P. Kamil et al |
| EPI_ISL_779044 | USA/LA-EVTL1378/2020 | 2020-12-11 | North America/USA/Louisiana/Bossier P | LSUHS Emerging Viral Threat Laboratory | Microbial Genome Sequencing Center | Jeremy P. Kamil et al |
| EPI_ISL_779052 | USA/LA-EVTL1386/2020 | 2020-12-11 | North America/USA/Louisiana/Bossier P | LSUHS Emerging Viral Threat Laboratory | Microbial Genome Sequencing Center | Jeremy P. Kamil et al |
| EPI_ISL_779058 | USA/LA-EVTL1392/2020 | 2020-12-11 | North America/USA/Louisiana/Bossier P | LSUHS Emerging Viral Threat Laboratory | Microbial Genome Sequencing Center | Jeremy P. Kamil et al |
| EPI_ISL_779063 | USA/LA-EVTL1397/2020 | 2020-12-11 | North America/USA/Louisiana/Bossier P | LSUHS Emerging Viral Threat Laboratory | Microbial Genome Sequencing Center | Jeremy P. Kamil et al |
| EPI_ISL_779071 | USA/LA-EVTL1405/2020 | 2020-12-11 | North America/USA/Louisiana/Bossier P | LSUHS Emerging Viral Threat Laboratory | Microbial Genome Sequencing Center | Jeremy P. Kamil et al |
| EPI_ISL_779073 | USA/LA-EVTL1407/2020 | 2020-12-11 | North America/USA/Louisiana/Bossier P | LSUHS Emerging Viral Threat Laboratory | Microbial Genome Sequencing Center | Jeremy P. Kamil et al |

| | | | | | | |
|---|---|---|---|---|---|---|
| EPI_ISL_779079 | USA/LA-EVTL1413/2020 | 2020-12-11 | North America/USA/Louisiana/Bossier P | LSUHS Emerging Viral Threat Laboratory | Microbial Genome Sequencing Center | Jeremy P. Kamil et al |
| EPI_ISL_779080 | USA/LA-EVTL1414/2020 | 2020-12-11 | North America/USA/Louisiana/Bossier P | LSUHS Emerging Viral Threat Laboratory | Microbial Genome Sequencing Center | Jeremy P. Kamil et al |
| EPI_ISL_779082 | USA/LA-EVTL1416/2020 | 2020-12-14 | North America/USA/Louisiana/Bossier P | LSUHS Emerging Viral Threat Laboratory | Microbial Genome Sequencing Center | Jeremy P. Kamil et al |
| EPI_ISL_779083 | USA/LA-EVTL1417/2020 | 2020-12-14 | North America/USA/Louisiana/Caddo Pa | LSUHS Emerging Viral Threat Laboratory | Microbial Genome Sequencing Center | Jeremy P. Kamil et al |
| EPI_ISL_779106 | USA/LA-EVTL1440/2020 | 2020-12-15 | North America/USA/Louisiana/Bossier P | LSUHS Emerging Viral Threat Laboratory | Microbial Genome Sequencing Center | Jennifer L. Carroll et al |
| EPI_ISL_779109 | USA/LA-EVTL1443/2020 | 2020-12-17 | North America/USA/Louisiana/Bossier P | LSUHS Emerging Viral Threat Laboratory | Microbial Genome Sequencing Center | Jennifer L. Carroll et al |
| EPI_ISL_779110 | USA/LA-EVTL1444/2020 | 2020-12-17 | North America/USA/Louisiana/Bossier P | LSUHS Emerging Viral Threat Laboratory | Microbial Genome Sequencing Center | Jennifer L. Carroll et al |
| EPI_ISL_779120 | USA/LA-EVTL1454/2020 | 2020-12-17 | North America/USA/Louisiana/Caddo Pa | LSUHS Emerging Viral Threat Laboratory | Microbial Genome Sequencing Center | Maarten Van Diest et al |
| EPI_ISL_779136 | USA/WY-WPHL-20152870/2020 | 2020-11-29 | North America/USA/Wyoming/ | Wyoming Public Health Laboratory | Wyoming Public Health Laboratory | Noah Hull et al |
| EPI_ISL_779155 | Brazil/RS-00601/2020 | 2020-11-23 | South America/Brazil/Rio Grande do Sul | Laboratv≥rio de Microbiologia Molecular - Universidade FEEVALE | Bioinformatics Laboratory / LNCC | Felipe Benites et al |
| EPI_ISL_779156 | Brazil/RS-00602/2020 | 2020-11-23 | South America/Brazil/Rio Grande do Sul | Laboratv≥rio de Microbiologia Molecular - Universidade FEEVALE | Bioinformatics Laboratory / LNCC | Felipe Benites et al |
| EPI_ISL_779159 | Brazil/RS-00631/2020 | 2020-11-25 | South America/Brazil/Rio Grande do Sul | Laboratv≥rio de Microbiologia Molecular - Universidade FEEVALE | Bioinformatics Laboratory / LNCC | Felipe Benites et al |
| EPI_ISL_779163 | Brazil/RS-00664/2020 | 2020-11-26 | South America/Brazil/Rio Grande do Sul | Laboratv≥rio de Microbiologia Molecular - Universidade FEEVALE | Bioinformatics Laboratory / LNCC | Felipe Benites et al |
| EPI_ISL_779165 | Brazil/RS-00677/2020 | 2020-11-27 | South America/Brazil/Rio Grande do Sul | Laboratv≥rio de Microbiologia Molecular - Universidade FEEVALE | Bioinformatics Laboratory / LNCC | Felipe Benites et al |
| EPI_ISL_779166 | Brazil/RS-00678/2020 | 2020-11-27 | South America/Brazil/Rio Grande do Sul | Laboratv≥rio de Microbiologia Molecular - Universidade FEEVALE | Bioinformatics Laboratory / LNCC | Felipe Benites et al |
| EPI_ISL_779167 | Brazil/RS-00687/2020 | 2020-11-27 | South America/Brazil/Rio Grande do Sul | Laboratv≥rio de Microbiologia Molecular - Universidade FEEVALE | Bioinformatics Laboratory / LNCC | Felipe Benites et al |

| | | | | | | |
|---|---|---|---|---|---|---|
| EPI_ISL_779168 | Brazil/RS-00688/2020 | 2020-11-27 | South America/Brazil/Rio Grande do Sul | Laboratv≥rio de Microbiologia Molecular - Universidade FEEVALE | Bioinformatics Laboratory / LNCC | Felipe Benites et al |
| EPI_ISL_779173 | Mexico/NLE-UANL-006/2020 | 2020-05-18 | North America/Mexico/Nuevo Leon/ | Laboratorio de Infectologv≠a, Servicio de Infectologv≠a, Hospital Universitario Dr. Josv© Eleuterio Gonzv°lez - Universidad Autv≥noma de Nuevo Lev≥n | Laboratorio de Infectologv≠a Molecular, Departamento de Bioquv≠mica y Medicina Molecular,Facultad de Medicina - Universidad Autv≥noma de Nuevo Lev≥n | Kame A. Galv°n-Huerta et al |
| EPI_ISL_779174 | Mexico/NLE-UANL-007/2020 | 2020-05-19 | North America/Mexico/Nuevo Leon/ | Laboratorio de Infectologv≠a, Servicio de Infectologv≠a, Hospital Universitario Dr. Josv© Eleuterio Gonzv°lez - Universidad Autv≥noma de Nuevo Lev≥n | Laboratorio de Infectologv≠a Molecular, Departamento de Bioquv≠mica y Medicina Molecular,Facultad de Medicina - Universidad Autv≥noma de Nuevo Lev≥n | Kame A. Galv°n-Huerta et al |
| EPI_ISL_779176 | Mexico/NLE-UANL-009/2020 | 2020-05-19 | North America/Mexico/Nuevo Leon/ | Laboratorio de Infectologv≠a, Servicio de Infectologv≠a, Hospital Universitario Dr. Josv© Eleuterio Gonzv°lez - Universidad Autv≥noma de Nuevo Lev≥n | Laboratorio de Infectologv≠a Molecular, Departamento de Bioquv≠mica y Medicina Molecular,Facultad de Medicina - Universidad Autv≥noma de Nuevo Lev≥n | Kame A. Galv°n-Huerta et al |

| | | | | | | |
|---|---|---|---|---|---|---|
| EPI_ISL_779177 | Mexico/NLE-UANL-012/2020 | 2020-05-23 | North America/Mexico/Nuevo Leon/ | Laboratorio de Infectolog√≠a, Servicio de Infectolog√≠a, Hospital Universitario Dr. Jos√© Eleuterio Gonz√°lez - Universidad Aut√≥noma de Nuevo Le√≥n | Laboratorio de Infectolog√≠a Molecular, Departamento de Bioqu√≠mica y Medicina Molecular,Facultad de Medicina - Universidad Aut√≥noma de Nuevo Le√≥n | Kame A. Gal√°n-Huerta et al |
| EPI_ISL_779178 | Mexico/NLE-UANL-013/2020 | 2020-05-23 | North America/Mexico/Nuevo Leon/ | Laboratorio de Infectolog√≠a, Servicio de Infectolog√≠a, Hospital Universitario Dr. Jos√© Eleuterio Gonz√°lez - Universidad Aut√≥noma de Nuevo Le√≥n | Laboratorio de Infectolog√≠a Molecular, Departamento de Bioqu√≠mica y Medicina Molecular,Facultad de Medicina - Universidad Aut√≥noma de Nuevo Le√≥n | Kame A. Gal√°n-Huerta et al |
| EPI_ISL_779179 | Mexico/NLE-UANL-014/2020 | 2020-06-17 | North America/Mexico/Nuevo Leon/ | Laboratorio de Infectolog√≠a, Servicio de Infectolog√≠a, Hospital Universitario Dr. Jos√© Eleuterio Gonz√°lez - Universidad Aut√≥noma de Nuevo Le√≥n | Laboratorio de Infectolog√≠a Molecular, Departamento de Bioqu√≠mica y Medicina Molecular,Facultad de Medicina - Universidad Aut√≥noma de Nuevo Le√≥n | Kame A. Gal√°n-Huerta et al |

| | | | | | | |
|---|---|---|---|---|---|---|
| EPI_ISL_779180 | Mexico/NLE-UANL-015/2020 | 2020-06-17 | North America/Mexico/Nuevo Leon/ | Laboratorio de Infectologv≠a, Servicio de Infectologv≠a, Hospital Universitario Dr. Josv© Eleuterio Gonzv°lez - Universidad Autv≥noma de Nuevo Lev≥n | Laboratorio de Infectologv≠a Molecular, Departamento de Bioquv≠mica y Medicina Molecular,Facultad de Medicina - Universidad Autv≥noma de Nuevo Lev≥n | Kame A. Galv°n-Huerta et al |
| EPI_ISL_779181 | Mexico/NLE-UANL-017/2020 | 2020-06-17 | North America/Mexico/Nuevo Leon/ | Laboratorio de Infectologv≠a, Servicio de Infectologv≠a, Hospital Universitario Dr. Josv© Eleuterio Gonzv°lez - Universidad Autv≥noma de Nuevo Lev≥n | Laboratorio de Infectologv≠a Molecular, Departamento de Bioquv≠mica y Medicina Molecular,Facultad de Medicina - Universidad Autv≥noma de Nuevo Lev≥n | Kame A. Galv°n-Huerta et al |
| EPI_ISL_779182 | Mexico/NLE-UANL-018/2020 | 2020-07-05 | North America/Mexico/Nuevo Leon/ | Laboratorio de Infectologv≠a, Servicio de Infectologv≠a, Hospital Universitario Dr. Josv© Eleuterio Gonzv°lez - Universidad Autv≥noma de Nuevo Lev≥n | Laboratorio de Infectologv≠a Molecular, Departamento de Bioquv≠mica y Medicina Molecular,Facultad de Medicina - Universidad Autv≥noma de Nuevo Lev≥n | Kame A. Galv°n-Huerta et al |

| | | | | | | |
|---|---|---|---|---|---|---|
| EPI_ISL_779184 | Mexico/NLE-UANL-020/2020 | 2020-07-05 | North America/Mexico/Nuevo Leon/ | Laboratorio de Infectologv≠a, Servicio de Infectologv≠a, Hospital Universitario Dr. Josv© Eleuterio Gonzv°lez - Universidad Autv≥noma de Nuevo Lev≥n | Laboratorio de Infectologv≠a Molecular, Departamento de Bioquv≠mica y Medicina Molecular,Facultad de Medicina - Universidad Autv≥noma de Nuevo Lev≥n | Kame A. Galv°n-Huerta et al |
| EPI_ISL_779185 | Mexico/NLE-UANL-022/2020 | 2020-11-19 | North America/Mexico/Nuevo Leon/ | Laboratorio Estatal de Salud Pv∫blica de Nuevo Lev≥n | Laboratorio de Infectologv≠a Molecular, Departamento de Bioquv≠mica y Medicina Molecular,Facultad de Medicina - Universidad Autv≥noma de Nuevo Lev≥n | Kame A. Galv°n-Huerta et al |
| EPI_ISL_779187 | Mexico/NLE-UANL-024/2020 | 2020-12-19 | North America/Mexico/Nuevo Leon/ | Laboratorio Estatal de Salud Pv∫blica de Nuevo Lev≥n | Laboratorio de Infectologv≠a Molecular, Departamento de Bioquv≠mica y Medicina Molecular,Facultad de Medicina - Universidad Autv≥noma de Nuevo Lev≥n | Kame A. Galv°n-Huerta et al |

| | | | | | | |
|---|---|---|---|---|---|---|
| EPI_ISL_779188 | Mexico/NLE-UANL-026/2020 | 2020-11-17 | North America/Mexico/Nuevo Leon/ | Laboratorio Estatal de Salud P√∫blica de Nuevo Le√≥n | Laboratorio de Infectolog√≠a Molecular, Departamento de Bioqu√≠mica y Medicina Molecular,Facultad de Medicina - Universidad Aut√≥noma de Nuevo Le√≥n | Kame A. Gal√°n-Huerta et al |
| EPI_ISL_779191 | Mexico/NLE-UANL-030/2020 | 2020-11-14 | North America/Mexico/Nuevo Leon/ | Laboratorio Estatal de Salud P√∫blica de Nuevo Le√≥n | Laboratorio de Infectolog√≠a Molecular, Departamento de Bioqu√≠mica y Medicina Molecular,Facultad de Medicina - Universidad Aut√≥noma de Nuevo Le√≥n | Kame A. Gal√°n-Huerta et al |
| EPI_ISL_779193 | Mexico/NLE-UANL-032/2020 | 2020-12-04 | North America/Mexico/Nuevo Leon/ | Laboratorio Estatal de Salud P√∫blica de Nuevo Le√≥n | Laboratorio de Infectolog√≠a Molecular, Departamento de Bioqu√≠mica y Medicina Molecular,Facultad de Medicina - Universidad Aut√≥noma de Nuevo Le√≥n | Kame A. Gal√°n-Huerta et al |

| | | | | | | |
|---|---|---|---|---|---|---|
| EPI_ISL_779194 | Mexico/NLE-UANL-033/2020 | 2020-12-12 | North America/Mexico/Nuevo Leon/ | Laboratorio Estatal de Salud P√∫blica de Nuevo Le√≥n | Laboratorio de Infectolog√≠a Molecular, Departamento de Bioqu√≠mica y Medicina Molecular,Facultad de Medicina - Universidad Aut√≥noma de Nuevo Le√≥n | Kame A. Gal√°n-Huerta et al |
| EPI_ISL_779195 | Mexico/NLE-UANL-034/2020 | 2020-12-05 | North America/Mexico/Nuevo Leon/ | Laboratorio Estatal de Salud P√∫blica de Nuevo Le√≥n | Laboratorio de Infectolog√≠a Molecular, Departamento de Bioqu√≠mica y Medicina Molecular,Facultad de Medicina - Universidad Aut√≥noma de Nuevo Le√≥n | Kame A. Gal√°n-Huerta et al |
| EPI_ISL_779196 | Mexico/NLE-UANL-035/2020 | 2020-11-16 | North America/Mexico/Nuevo Leon/ | Laboratorio Estatal de Salud P√∫blica de Nuevo Le√≥n | Laboratorio de Infectolog√≠a Molecular, Departamento de Bioqu√≠mica y Medicina Molecular,Facultad de Medicina - Universidad Aut√≥noma de Nuevo Le√≥n | Kame A. Gal√°n-Huerta et al |

| | | | | | | |
|---|---|---|---|---|---|---|
| EPI_ISL_779197 | Mexico/NLE-UANL-036/2020 | 2020-12-05 | North America/Mexico/Nuevo Leon/ | Laboratorio Estatal de Salud P√∫blica de Nuevo Le√≥n | Laboratorio de Infectolog√≠a Molecular, Departamento de Bioqu√≠mica y Medicina Molecular,Facultad de Medicina - Universidad Aut√≥noma de Nuevo Le√≥n | Kame A. Gal√°n-Huerta et al |
| EPI_ISL_779199 | Japan/IC-0516/2021 | 21-Jan | Asia/Japan/Japan/ | Pathogen Genomics Center, National Institute of Infectious Diseases | Pathogen Genomics Center, National Institute of Infectious Diseases | Tsuyoshi Sekizuka et al |
| EPI_ISL_779200 | Japan/IC-0517/2021 | 21-Jan | Asia/Japan/Japan/ | Pathogen Genomics Center, National Institute of Infectious Diseases | Pathogen Genomics Center, National Institute of Infectious Diseases | Tsuyoshi Sekizuka et al |
| EPI_ISL_779214 | Japan/IC-0531/2020 | 20-Dec | Asia/Japan/Japan/ | Pathogen Genomics Center, National Institute of Infectious Diseases | Pathogen Genomics Center, National Institute of Infectious Diseases | Tsuyoshi Sekizuka et al |
| EPI_ISL_779219 | Japan/IC-0536/2021 | 21-Jan | Asia/Japan/Japan/ | Pathogen Genomics Center, National Institute of Infectious Diseases | Pathogen Genomics Center, National Institute of Infectious Diseases | Tsuyoshi Sekizuka et al |
| EPI_ISL_779239 | Japan/IC-0556/2021 | 21-Jan | Asia/Japan/Japan/ | Pathogen Genomics Center, National Institute of Infectious Diseases | Pathogen Genomics Center, National Institute of Infectious Diseases | Tsuyoshi Sekizuka et al |
| EPI_ISL_779254 | France/UN-FRA-WGS157/2020 | 2020-03-06 | Europe/France/France/ | Virology, CHU Pitie Salpetriere Charles Foix | Virology, CHU Pitie Salpetriere Charles Foix | Marot et al |

| | | | | | | |
|---|---|---|---|---|---|---|
| EPI_ISL_779255 | Pakistan/JRCGR-KHI05/2020 | 2020-05-20 | Asia/Pakistan/Karachi/ | Jamil-ur-Rahman Center for Genome Research, Dr. Panjwani Center for Molecular Medicine and Drug Research | Jamil-ur-Rahman Center for Genome Research, Dr. Panjwani Center for Molecular Medicine and Drug Research | Shakeel et al |
| EPI_ISL_779256 | Pakistan/JRCGR-KHI06/2020 | 2020-05-20 | Asia/Pakistan/Karachi/ | Jamil-ur-Rahman Center for Genome Research, Dr. Panjwani Center for Molecular Medicine and Drug Research | Jamil-ur-Rahman Center for Genome Research, Dr. Panjwani Center for Molecular Medicine and Drug Research | Shakeel et al |
| EPI_ISL_779257 | Pakistan/JRCGR-KHI22/2020 | 2020-05-03 | Asia/Pakistan/Karachi/ | Jamil-ur-Rahman Center for Genome Research, Dr. Panjwani Center for Molecular Medicine and Drug Research | Jamil-ur-Rahman Center for Genome Research, Dr. Panjwani Center for Molecular Medicine and Drug Research | Shakeel et al |
| EPI_ISL_779258 | Pakistan/JRCGR-KHI07/2020 | 2020-05-20 | Asia/Pakistan/Karachi/ | Jamil-ur-Rahman Center for Genome Research, Dr. Panjwani Center for Molecular Medicine and Drug Research | Jamil-ur-Rahman Center for Genome Research, Dr. Panjwani Center for Molecular Medicine and Drug Research | Shakeel et al |
| EPI_ISL_779259 | Pakistan/JRCGR-KHI25/2020 | 2020-06-02 | Asia/Pakistan/Karachi/ | Jamil-ur-Rahman Center for Genome Research, Dr. Panjwani Center for Molecular Medicine and Drug Research | Jamil-ur-Rahman Center for Genome Research, Dr. Panjwani Center for Molecular Medicine and Drug Research | Shakeel et al |

| | | | | | | |
|---|---|---|---|---|---|---|
| EPI_ISL_779260 | Pakistan/JRCGR-KHI26/2020 | 2020-06-02 | Asia/Pakistan/Karachi/ | Jamil-ur-Rahman Center for Genome Research, Dr. Panjwani Center for Molecular Medicine and Drug Research | Jamil-ur-Rahman Center for Genome Research, Dr. Panjwani Center for Molecular Medicine and Drug Research | Shakeel et al |
| EPI_ISL_779261 | Pakistan/JRCGR-KHI27/2020 | 2020-06-02 | Asia/Pakistan/Karachi/ | Jamil-ur-Rahman Center for Genome Research, Dr. Panjwani Center for Molecular Medicine and Drug Research | Jamil-ur-Rahman Center for Genome Research, Dr. Panjwani Center for Molecular Medicine and Drug Research | Shakeel et al |
| EPI_ISL_779262 | Pakistan/JRCGR-KHI28/2020 | 2020-06-02 | Asia/Pakistan/Karachi/ | Jamil-ur-Rahman Center for Genome Research, Dr. Panjwani Center for Molecular Medicine and Drug Research | Jamil-ur-Rahman Center for Genome Research, Dr. Panjwani Center for Molecular Medicine and Drug Research | Shakeel et al |
| EPI_ISL_779263 | Pakistan/JRCGR-KHI46/2020 | 2020-05-03 | Asia/Pakistan/Karachi/ | Jamil-ur-Rahman Center for Genome Research, Dr. Panjwani Center for Molecular Medicine and Drug Research | Jamil-ur-Rahman Center for Genome Research, Dr. Panjwani Center for Molecular Medicine and Drug Research | Shakeel et al |
| EPI_ISL_779264 | Pakistan/JRCGR-KHI04/2020 | 2020-06-03 | Asia/Pakistan/Karachi/ | Jamil-ur-Rahman Center for Genome Research, Dr. Panjwani Center for Molecular Medicine and Drug Research | Jamil-ur-Rahman Center for Genome Research, Dr. Panjwani Center for Molecular Medicine and Drug Research | Shakeel et al |

| | | | | | | |
|---|---|---|---|---|---|---|
| EPI_ISL_779265 | Pakistan/JRCGR-KHI29/2020 | 2020-06-02 | Asia/Pakistan/Karachi/ | Jamil-ur-Rahman Center for Genome Research, Dr. Panjwani Center for Molecular Medicine and Drug Research | Jamil-ur-Rahman Center for Genome Research, Dr. Panjwani Center for Molecular Medicine and Drug Research | Shakeel et al |
| EPI_ISL_779266 | Pakistan/JRCGR-KHI30/2020 | 2020-06-02 | Asia/Pakistan/Karachi/ | Jamil-ur-Rahman Center for Genome Research, Dr. Panjwani Center for Molecular Medicine and Drug Research | Jamil-ur-Rahman Center for Genome Research, Dr. Panjwani Center for Molecular Medicine and Drug Research | Shakeel et al |
| EPI_ISL_779267 | Pakistan/JRCGR-KHI40/2020 | 2020-06-02 | Asia/Pakistan/Karachi/ | Jamil-ur-Rahman Center for Genome Research, Dr. Panjwani Center for Molecular Medicine and Drug Research | Jamil-ur-Rahman Center for Genome Research, Dr. Panjwani Center for Molecular Medicine and Drug Research | Shakeel et al |
| EPI_ISL_779268 | Pakistan/JRCGR-KHI03/2020 | 2020-06-03 | Asia/Pakistan/Karachi/ | Jamil-ur-Rahman Center for Genome Research, Dr. Panjwani Center for Molecular Medicine and Drug Research | Jamil-ur-Rahman Center for Genome Research, Dr. Panjwani Center for Molecular Medicine and Drug Research | Shakeel et al |
| EPI_ISL_779269 | Pakistan/JRCGR-KHI02/2020 | 2020-06-04 | Asia/Pakistan/Karachi/ | Jamil-ur-Rahman Center for Genome Research, Dr. Panjwani Center for Molecular Medicine and Drug Research | Jamil-ur-Rahman Center for Genome Research, Dr. Panjwani Center for Molecular Medicine and Drug Research | Shakeel et al |

| | | | | | | |
|---|---|---|---|---|---|---|
| EPI_ISL_779270 | Pakistan/JRCGR-KHI10/2020 | 2020-06-14 | Asia/Pakistan/Karachi/ | Jamil-ur-Rahman Center for Genome Research, Dr. Panjwani Center for Molecular Medicine and Drug Research | Jamil-ur-Rahman Center for Genome Research, Dr. Panjwani Center for Molecular Medicine and Drug Research | Shakeel et al |
| EPI_ISL_779271 | Pakistan/JRCGR-KHI12/2020 | 2020-06-14 | Asia/Pakistan/Karachi/ | Jamil-ur-Rahman Center for Genome Research, Dr. Panjwani Center for Molecular Medicine and Drug Research | Jamil-ur-Rahman Center for Genome Research, Dr. Panjwani Center for Molecular Medicine and Drug Research | Shakeel et al |
| EPI_ISL_779272 | Pakistan/JRCGR-KHI18/2020 | 2020-07-02 | Asia/Pakistan/Karachi/ | Jamil-ur-Rahman Center for Genome Research, Dr. Panjwani Center for Molecular Medicine and Drug Research | Jamil-ur-Rahman Center for Genome Research, Dr. Panjwani Center for Molecular Medicine and Drug Research | Shakeel et al |
| EPI_ISL_779273 | Pakistan/JRCGR-KHI36/2020 | 2020-08-10 | Asia/Pakistan/Karachi/ | Jamil-ur-Rahman Center for Genome Research, Dr. Panjwani Center for Molecular Medicine and Drug Research | Jamil-ur-Rahman Center for Genome Research, Dr. Panjwani Center for Molecular Medicine and Drug Research | Shakeel et al |
| EPI_ISL_779274 | Pakistan/JRCGR-KHI43/2020 | 2020-07-02 | Asia/Pakistan/Karachi/ | Jamil-ur-Rahman Center for Genome Research, Dr. Panjwani Center for Molecular Medicine and Drug Research | Jamil-ur-Rahman Center for Genome Research, Dr. Panjwani Center for Molecular Medicine and Drug Research | Shakeel et al |

| | | | | | | |
|---|---|---|---|---|---|---|
| EPI_ISL_779275 | Pakistan/JRCGR-KHI15/2020 | 2020-08-10 | Asia/Pakistan/Karachi/ | Jamil-ur-Rahman Center for Genome Research, Dr. Panjwani Center for Molecular Medicine and Drug Research | Jamil-ur-Rahman Center for Genome Research, Dr. Panjwani Center for Molecular Medicine and Drug Research | Shakeel et al |
| EPI_ISL_779276 | Pakistan/JRCGR-KHI20/2020 | 2020-07-02 | Asia/Pakistan/Karachi/ | Jamil-ur-Rahman Center for Genome Research, Dr. Panjwani Center for Molecular Medicine and Drug Research | Jamil-ur-Rahman Center for Genome Research, Dr. Panjwani Center for Molecular Medicine and Drug Research | Shakeel et al |
| EPI_ISL_779277 | Pakistan/JRCGR-KHI35/2020 | 2020-08-10 | Asia/Pakistan/Karachi/ | Jamil-ur-Rahman Center for Genome Research, Dr. Panjwani Center for Molecular Medicine and Drug Research | Jamil-ur-Rahman Center for Genome Research, Dr. Panjwani Center for Molecular Medicine and Drug Research | Shakeel et al |
| EPI_ISL_779278 | Pakistan/JRCGR-KHI09/2020 | 2020-06-14 | Asia/Pakistan/Karachi/ | Jamil-ur-Rahman Center for Genome Research, Dr. Panjwani Center for Molecular Medicine and Drug Research | Jamil-ur-Rahman Center for Genome Research, Dr. Panjwani Center for Molecular Medicine and Drug Research | Shakeel et al |
| EPI_ISL_779279 | Pakistan/JRCGR-KHI11/2020 | 2020-06-14 | Asia/Pakistan/Karachi/ | Jamil-ur-Rahman Center for Genome Research, Dr. Panjwani Center for Molecular Medicine and Drug Research | Jamil-ur-Rahman Center for Genome Research, Dr. Panjwani Center for Molecular Medicine and Drug Research | Shakeel et al |

| | | | | | | |
|---|---|---|---|---|---|---|
| EPI_ISL_779280 | Pakistan/JRCGR-KHI32/2020 | 2020-06-14 | Asia/Pakistan/Karachi/ | Jamil-ur-Rahman Center for Genome Research, Dr. Panjwani Center for Molecular Medicine and Drug Research | Jamil-ur-Rahman Center for Genome Research, Dr. Panjwani Center for Molecular Medicine and Drug Research | Shakeel et al |
| EPI_ISL_779281 | Pakistan/JRCGR-KHI48/2020 | 2020-08-04 | Asia/Pakistan/Karachi/ | Jamil-ur-Rahman Center for Genome Research, Dr. Panjwani Center for Molecular Medicine and Drug Research | Jamil-ur-Rahman Center for Genome Research, Dr. Panjwani Center for Molecular Medicine and Drug Research | Shakeel et al |
| EPI_ISL_779282 | Pakistan/JRCGR-KHI45/2020 | 2020-08-10 | Asia/Pakistan/Karachi/ | Jamil-ur-Rahman Center for Genome Research, Dr. Panjwani Center for Molecular Medicine and Drug Research | Jamil-ur-Rahman Center for Genome Research, Dr. Panjwani Center for Molecular Medicine and Drug Research | Shakeel et al |
| EPI_ISL_779283 | Pakistan/JRCGR-KHI37/2020 | 2020-08-10 | Asia/Pakistan/Karachi/ | Jamil-ur-Rahman Center for Genome Research, Dr. Panjwani Center for Molecular Medicine and Drug Research | Jamil-ur-Rahman Center for Genome Research, Dr. Panjwani Center for Molecular Medicine and Drug Research | Shakeel et al |
| EPI_ISL_779284 | Pakistan/JRCGR-KHI13/2020 | 2020-08-10 | Asia/Pakistan/Karachi/ | Jamil-ur-Rahman Center for Genome Research, Dr. Panjwani Center for Molecular Medicine and Drug Research | Jamil-ur-Rahman Center for Genome Research, Dr. Panjwani Center for Molecular Medicine and Drug Research | Shakeel et al |

| | | | | | | |
|---|---|---|---|---|---|---|
| EPI_ISL_779285 | Pakistan/JRCGR-KHI47/2020 | 2020-08-10 | Asia/Pakistan/Karachi/ | Jamil-ur-Rahman Center for Genome Research, Dr. Panjwani Center for Molecular Medicine and Drug Research | Jamil-ur-Rahman Center for Genome Research, Dr. Panjwani Center for Molecular Medicine and Drug Research | Shakeel et al |
| EPI_ISL_779287 | Pakistan/JRCGR-KHI41/2020 | 2020-08-10 | Asia/Pakistan/Karachi/ | Jamil-ur-Rahman Center for Genome Research, Dr. Panjwani Center for Molecular Medicine and Drug Research | Jamil-ur-Rahman Center for Genome Research, Dr. Panjwani Center for Molecular Medicine and Drug Research | Shakeel et al |
| EPI_ISL_779288 | Pakistan/JRCGR-KHI33/2020 | 2020-06-14 | Asia/Pakistan/Karachi/ | Jamil-ur-Rahman Center for Genome Research, Dr. Panjwani Center for Molecular Medicine and Drug Research | Jamil-ur-Rahman Center for Genome Research, Dr. Panjwani Center for Molecular Medicine and Drug Research | Shakeel et al |
| EPI_ISL_779289 | Pakistan/JRCGR-KHI42/2020 | 2020-07-02 | Asia/Pakistan/Karachi/ | Jamil-ur-Rahman Center for Genome Research, Dr. Panjwani Center for Molecular Medicine and Drug Research | Jamil-ur-Rahman Center for Genome Research, Dr. Panjwani Center for Molecular Medicine and Drug Research | Shakeel et al |
| EPI_ISL_779290 | Pakistan/JRCGR-KHI17/2020 | 2020-07-02 | Asia/Pakistan/Karachi/ | Jamil-ur-Rahman Center for Genome Research, Dr. Panjwani Center for Molecular Medicine and Drug Research | Jamil-ur-Rahman Center for Genome Research, Dr. Panjwani Center for Molecular Medicine and Drug Research | Shakeel et al |

| | | | | | | |
|---|---|---|---|---|---|---|
| EPI_ISL_779299 | USA/UT-UPHL-2012250113/2020 | 2020-11-30 | North America/USA/Utah/ | Utah Public Health Laboratory, Utah Public Health Laboratory Infectious Disease submission group | Utah Public Health Laboratory, Utah Public Health Laboratory Infectious Disease submission group | Gallagher et al |
| EPI_ISL_779303 | USA/UT-UPHL-2012336575/2020 | 2020-11-30 | North America/USA/Utah/ | Utah Public Health Laboratory, Utah Public Health Laboratory Infectious Disease submission group | Utah Public Health Laboratory, Utah Public Health Laboratory Infectious Disease submission group | Gallagher et al |
| EPI_ISL_779308 | USA/UT-UPHL-2012500518/2020 | 2020-11-30 | North America/USA/Utah/ | Utah Public Health Laboratory, Utah Public Health Laboratory Infectious Disease submission group | Utah Public Health Laboratory, Utah Public Health Laboratory Infectious Disease submission group | Gallagher et al |
| EPI_ISL_779309 | USA/UT-UPHL-2012528694/2020 | 2020-11-30 | North America/USA/Utah/ | Utah Public Health Laboratory, Utah Public Health Laboratory Infectious Disease submission group | Utah Public Health Laboratory, Utah Public Health Laboratory Infectious Disease submission group | Gallagher et al |
| EPI_ISL_779319 | USA/WI-UW-2487/2020 | 2020-12-30 | North America/USA/Wisconsin/Dane Co | University of Wisconsin-Madison AIDS Vaccine Research Laboratories | University of Wisconsin-Madison AIDS Vaccine Research Laboratories | Gage Moreno et al |
| EPI_ISL_779326 | USA/WI-UW-2494/2020 | 2020-12-30 | North America/USA/Wisconsin/Dane Co | University of Wisconsin-Madison AIDS Vaccine Research Laboratories | University of Wisconsin-Madison AIDS Vaccine Research Laboratories | Gage Moreno et al |

| | | | | | | |
|---|---|---|---|---|---|---|
| EPI_ISL_779333 | USA/WI-UW-2501/2020 | 2020-12-30 | North America/USA/Wisconsin/Dane Co | University of Wisconsin-Madison AIDS Vaccine Research Laboratories | University of Wisconsin-Madison AIDS Vaccine Research Laboratories | Gage Moreno et al |
| EPI_ISL_779338 | USA/WI-UW-2506/2020 | 2020-12-30 | North America/USA/Wisconsin/Dane Co | University of Wisconsin-Madison AIDS Vaccine Research Laboratories | University of Wisconsin-Madison AIDS Vaccine Research Laboratories | Gage Moreno et al |
| EPI_ISL_779341 | USA/WI-UW-2509/2020 | 2020-12-30 | North America/USA/Wisconsin/Dane Co | University of Wisconsin-Madison AIDS Vaccine Research Laboratories | University of Wisconsin-Madison AIDS Vaccine Research Laboratories | Gage Moreno et al |
| EPI_ISL_779353 | USA/WI-UW-2522/2020 | 2020-12-30 | North America/USA/Wisconsin/Dane Co | University of Wisconsin-Madison AIDS Vaccine Research Laboratories | University of Wisconsin-Madison AIDS Vaccine Research Laboratories | Gage Moreno et al |
| EPI_ISL_779354 | USA/WI-UW-2523/2020 | 2020-12-30 | North America/USA/Wisconsin/Dane Co | University of Wisconsin-Madison AIDS Vaccine Research Laboratories | University of Wisconsin-Madison AIDS Vaccine Research Laboratories | Gage Moreno et al |
| EPI_ISL_779366 | USA/WI-UW-2535/2020 | 2020-12-30 | North America/USA/Wisconsin/Dane Co | University of Wisconsin-Madison AIDS Vaccine Research Laboratories | University of Wisconsin-Madison AIDS Vaccine Research Laboratories | Gage Moreno et al |
| EPI_ISL_779382 | USA/WI-UW-2553/2020 | 2020-12-29 | North America/USA/Wisconsin/Dane Co | University of Wisconsin-Madison AIDS Vaccine Research Laboratories | University of Wisconsin-Madison AIDS Vaccine Research Laboratories | Gage Moreno et al |
| EPI_ISL_779509 | Australia/VIC17860/2020 | 2020-08-30 | Oceania/Australia/Victoria/ | Microbiological Diagnostic Unit - Public Health Laboratory (MDU-PHL) | MDU-PHL | Seemann T. et al |

| | | | | | | |
|---|---|---|---|---|---|---|
| EPI_ISL_779651 | Slovakia/UKBA-501/2020 | 2020-12-28 | Europe/Slovakia/Pavlovce nad Uhom/ | Institute of Virology, Biomedical Research Center of the Slovak Academy of Sciences, Bratislava | Faculty of Natural Sciences, Comenius University, Bratislava | Viktv≥ria ƒåabanovv° et al |
| EPI_ISL_779652 | Slovakia/UKBA-502/2020 | 2020-12-28 | Europe/Slovakia/Pavlovce nad Uhom/ | Institute of Virology, Biomedical Research Center of the Slovak Academy of Sciences, Bratislava | Faculty of Natural Sciences, Comenius University, Bratislava | Kristv≠na Bor≈°ovv° et al |
| EPI_ISL_779655 | Slovakia/UKBA-505/2020 | 2020-12-28 | Europe/Slovakia/Pavlovce nad Uhom/ | Institute of Virology, Biomedical Research Center of the Slovak Academy of Sciences, Bratislava | Faculty of Natural Sciences, Comenius University, Bratislava | Viktv≥ria ƒåabanovv° et al |
| EPI_ISL_779658 | Slovakia/UKBA-508/2020 | 2020-12-28 | Europe/Slovakia/Pavlovce nad Uhom/ | Institute of Virology, Biomedical Research Center of the Slovak Academy of Sciences, Bratislava | Faculty of Natural Sciences, Comenius University, Bratislava | Bro≈àa Brejovv° et al |
| EPI_ISL_779659 | Slovakia/UKBA-509/2020 | 2020-12-28 | Europe/Slovakia/Pavlovce nad Uhom/ | Institute of Virology, Biomedical Research Center of the Slovak Academy of Sciences, Bratislava | Faculty of Natural Sciences, Comenius University, Bratislava | Bro≈àa Brejovv° et al |
| EPI_ISL_779661 | Japan/IC-0575/2021 | 21-Jan | Asia/Japan/Japan/ | Pathogen Genomics Center, National Institute of Infectious Diseases | Pathogen Genomics Center, National Institute of Infectious Diseases | Tsuyoshi Sekizuka et al |
| EPI_ISL_779664 | Japan/IC-0578/2021 | 21-Jan | Asia/Japan/Japan/ | Pathogen Genomics Center, National Institute of Infectious Diseases | Pathogen Genomics Center, National Institute of Infectious Diseases | Tsuyoshi Sekizuka et al |
| EPI_ISL_779687 | Japan/IC-0601/2021 | 21-Jan | Asia/Japan/Japan/ | Pathogen Genomics Center, National Institute of Infectious Diseases | Pathogen Genomics Center, National Institute of Infectious Diseases | Tsuyoshi Sekizuka et al |

| | | | | | | |
|---|---|---|---|---|---|---|
| EPI_ISL_779690 | Japan/IC-0604/2021 | 21-Jan | Asia/Japan/Japan/ | Pathogen Genomics Center, National Institute of Infectious Diseases | Pathogen Genomics Center, National Institute of Infectious Diseases | Tsuyoshi Sekizuka et al |
| EPI_ISL_779694 | Japan/IC-0608/2021 | 21-Jan | Asia/Japan/Japan/ | Pathogen Genomics Center, National Institute of Infectious Diseases | Pathogen Genomics Center, National Institute of Infectious Diseases | Tsuyoshi Sekizuka et al |
| EPI_ISL_779696 | Japan/IC-0610/2021 | 21-Jan | Asia/Japan/Japan/ | Pathogen Genomics Center, National Institute of Infectious Diseases | Pathogen Genomics Center, National Institute of Infectious Diseases | Tsuyoshi Sekizuka et al |
| EPI_ISL_779703 | India/MH-CMP-19/2020 | 2020-08-24 | Asia/India/Maharashtra/Mumbai | The Foundation for Medical Research | The Foundation for Medical Research | Ayan Mandal et al |
| EPI_ISL_779704 | Italy/LOM-UniMI05/2020 | 2020-02-24 | Europe/Italy/Lombardy/ | Laboratory of Infectious Diseases, Department of Biomedical and Clinical Sciences L. Sacco, University of Milan | Laboratory of Infectious Diseases, Department of Biomedical and Clinical Sciences L. Sacco, University of Milan | Alessia Lai et al |
| EPI_ISL_779705 | India/MH-CMP-30/2020 | 2020-09-09 | Asia/India/Maharashtra/Mumbai | The Foundation for Medical Research | The Foundation for Medical Research | Ayan Mandal et al |
| EPI_ISL_779708 | India/MH-CMP-34/2020 | 2020-09-09 | Asia/India/Maharashtra/Mumbai | The Foundation for Medical Research | The Foundation for Medical Research | Ayan Mandal et al |
| EPI_ISL_779709 | Italy/LOM-UniMI07/2020 | 2020-02-24 | Europe/Italy/Lombardy/ | Laboratory of Infectious Diseases, Department of Biomedical and Clinical Sciences L. Sacco, University of Milan | Laboratory of Infectious Diseases, Department of Biomedical and Clinical Sciences L. Sacco, University of Milan | Alessia Lai et al |

| | | | | | | |
|---|---|---|---|---|---|---|
| EPI_ISL_779710 | India/MH-CMP-35/2020 | 2020-09-09 | Asia/India/Maharashtra/Mumbai | The Foundation for Medical Research | The Foundation for Medical Research | Ayan Mandal et al |
| EPI_ISL_779712 | Italy/LOM-UniMI08/2020 | 2020-02-24 | Europe/Italy/Lombardy/ | Laboratory of Infectious Diseases, Department of Biomedical and Clinical Sciences L. Sacco, University of Milan | Laboratory of Infectious Diseases, Department of Biomedical and Clinical Sciences L. Sacco, University of Milan | Alessia Lai et al |
| EPI_ISL_779713 | Italy/LOM-UniMI09/2020 | 2020-02-24 | Europe/Italy/Lombardy/ | Laboratory of Infectious Diseases, Department of Biomedical and Clinical Sciences L. Sacco, University of Milan | Laboratory of Infectious Diseases, Department of Biomedical and Clinical Sciences L. Sacco, University of Milan | Alessia Lai et al |
| EPI_ISL_779720 | Spain/MD-IBV-97010036/2020 | 2020-03-10 | Europe/Spain/Madrid/Madrid | IdISSC/Hospital Cl√≠nico San Carlos de Madrid,Äã | SeqCOVID-SPAIN consortium/IBV(CSIC) | Alberto Delgado-Iribarren et al |
| EPI_ISL_779728 | Spain/MD-IBV-97010046/2020 | 2020-03-10 | Europe/Spain/Madrid/Mostoles | IdISSC/Hospital Cl√≠nico San Carlos de Madrid,Äã | SeqCOVID-SPAIN consortium/IBV(CSIC) | Alberto Delgado-Iribarren et al |
| EPI_ISL_779733 | Spain/AR-IBV-97010750/2020 | 2020-10-03 | Europe/Spain/Aragon/Zaragoza | Servicio de Microbiolog√≠a, Hospital Miguel Servet, Zaragoza | SeqCOVID-SPAIN consortium/IBV(CSIC) | Antonio Rezusta L√≥pez et al |
| EPI_ISL_779734 | Spain/AR-IBV-97005114/2020 | 2020-04-26 | Europe/Spain/Aragon/Zaragoza | Servicio de Microbiolog√≠a, Hospital Miguel Servet, Zaragoza | SeqCOVID-SPAIN consortium/IBV(CSIC) | Antonio Rezusta L√≥pez et al |
| EPI_ISL_779745 | Spain/MD-IBV-97010025/2020 | 2020-03-24 | Europe/Spain/Madrid/Madrid | IdISSC/Hospital Cl√≠nico San Carlos de Madrid,Äã | SeqCOVID-SPAIN consortium/IBV(CSIC) | Alberto Delgado-Iribarren et al |
| EPI_ISL_779753 | Spain/AR-IBV-97010742/2020 | 2020-10-02 | Europe/Spain/Aragon/Zaragoza | Servicio de Microbiolog√≠a, Hospital Miguel Servet, Zaragoza | SeqCOVID-SPAIN consortium/IBV(CSIC) | Antonio Rezusta L√≥pez et al |
| EPI_ISL_779755 | Spain/MD-IBV-97010058/2020 | 2020-03-12 | Europe/Spain/Madrid/Madrid | IdISSC/Hospital Cl√≠nico San Carlos de Madrid,Äã | SeqCOVID-SPAIN consortium/IBV(CSIC) | Alberto Delgado-Iribarren et al |

| | | | | | | |
|---|---|---|---|---|---|---|
| EPI_ISL_779774 | Spain/AR-IBV-97010739/2020 | 2020-10-03 | Europe/Spain/Aragon/Zaragoza | Servicio de Microbiolog√≠a, Hospital Miguel Servet, Zaragoza | SeqCOVID-SPAIN consortium/IBV(CSIC) | Antonio Rezusta L√≥pez et al |
| EPI_ISL_779777 | Spain/AR-IBV-97010744/2020 | 2020-10-02 | Europe/Spain/Aragon/Zaragoza | Servicio de Microbiolog√≠a, Hospital Miguel Servet, Zaragoza | SeqCOVID-SPAIN consortium/IBV(CSIC) | Antonio Rezusta L√≥pez et al |
| EPI_ISL_779778 | Spain/AR-IBV-97010745/2020 | 2020-10-02 | Europe/Spain/Aragon/Zaragoza | Servicio de Microbiolog√≠a, Hospital Miguel Servet, Zaragoza | SeqCOVID-SPAIN consortium/IBV(CSIC) | Antonio Rezusta L√≥pez et al |
| EPI_ISL_779779 | Spain/AR-IBV-97010746/2020 | 2020-10-03 | Europe/Spain/Aragon/Zaragoza | Servicio de Microbiolog√≠a, Hospital Miguel Servet, Zaragoza | SeqCOVID-SPAIN consortium/IBV(CSIC) | Antonio Rezusta L√≥pez et al |
| EPI_ISL_779781 | France/ARA-SC739/2020 | 2020-11-17 | Europe/France/Auvergne-Rh√¥ne-Alpes/ | Laboratoire de virologie, CHU de Grenoble | CNR Virus des Infections Respiratoires - France SUD | Antonin Bal et al |
| EPI_ISL_779782 | France/ARA-SC741/2020 | 2020-11-17 | Europe/France/Auvergne-Rh√¥ne-Alpes/ | Laboratoire de virologie, CHU de Grenoble | CNR Virus des Infections Respiratoires - France SUD | Antonin Bal et al |
| EPI_ISL_779783 | France/ARA-124622/2020 | 2020-12-24 | Europe/France/Auvergne-Rh√¥ne-Alpes/ | CNR Virus des Infections Respiratoires - France SUD | CNR Virus des Infections Respiratoires - France SUD | Antonin Bal et al |
| EPI_ISL_779798 | France/ARA-123003/2020 | 2020-12-21 | Europe/France/Auvergne-Rh√¥ne-Alpes/ | CNR Virus des Infections Respiratoires - France SUD | CNR Virus des Infections Respiratoires - France SUD | Antonin Bal et al |
| EPI_ISL_779799 | France/ARA-123089/2020 | 2020-12-21 | Europe/France/Auvergne-Rh√¥ne-Alpes/ | CNR Virus des Infections Respiratoires - France SUD | CNR Virus des Infections Respiratoires - France SUD | Antonin Bal et al |
| EPI_ISL_779800 | France/ARA-123093/2020 | 2020-12-21 | Europe/France/Auvergne-Rh√¥ne-Alpes/ | CNR Virus des Infections Respiratoires - France SUD | CNR Virus des Infections Respiratoires - France SUD | Antonin Bal et al |
| EPI_ISL_779804 | France/ARA-123946/2020 | 2020-12-23 | Europe/France/Auvergne-Rh√¥ne-Alpes/ | CNR Virus des Infections Respiratoires - France SUD | CNR Virus des Infections Respiratoires - France SUD | Antonin Bal et al |

| | | | | | | |
|---|---|---|---|---|---|---|
| EPI_ISL_779805 | France/ARA-124345/2020 | 2020-12-24 | Europe/France/Auvergne-Rhv¥ne-Alpes/ | CNR Virus des Infections Respiratoires - France SUD | CNR Virus des Infections Respiratoires - France SUD | Antonin Bal et al |
| EPI_ISL_779807 | France/ARA-124840/2020 | 2020-12-25 | Europe/France/Auvergne-Rhv¥ne-Alpes/ | CNR Virus des Infections Respiratoires - France SUD | CNR Virus des Infections Respiratoires - France SUD | Antonin Bal et al |
| EPI_ISL_779809 | France/ARA-125033/2020 | 2020-12-26 | Europe/France/Auvergne-Rhv¥ne-Alpes/ | CNR Virus des Infections Respiratoires - France SUD | CNR Virus des Infections Respiratoires - France SUD | Antonin Bal et al |
| EPI_ISL_779817 | France/ARA-SC723/2020 | 2020-11-15 | Europe/France/Auvergne-Rhv¥ne-Alpes/ | Laboratoire de virologie, CHU de Grenoble | CNR Virus des Infections Respiratoires - France SUD | Antonin Bal et al |
| EPI_ISL_779818 | France/ARA-SC724/2020 | 2020-11-15 | Europe/France/Auvergne-Rhv¥ne-Alpes/ | Laboratoire de virologie, CHU de Grenoble | CNR Virus des Infections Respiratoires - France SUD | Antonin Bal et al |
| EPI_ISL_779820 | France/ARA-SC726/2020 | 2020-11-15 | Europe/France/Auvergne-Rhv¥ne-Alpes/ | Laboratoire de virologie, CHU de Grenoble | CNR Virus des Infections Respiratoires - France SUD | Antonin Bal et al |
| EPI_ISL_779821 | France/ARA-SC727/2020 | 2020-11-16 | Europe/France/Auvergne-Rhv¥ne-Alpes/ | Laboratoire de virologie, CHU de Grenoble | CNR Virus des Infections Respiratoires - France SUD | Antonin Bal et al |
| EPI_ISL_779822 | France/ARA-SC728/2020 | 2020-11-16 | Europe/France/Auvergne-Rhv¥ne-Alpes/ | Laboratoire de virologie, CHU de Grenoble | CNR Virus des Infections Respiratoires - France SUD | Antonin Bal et al |
| EPI_ISL_779826 | France/ARA-SC732/2020 | 2020-11-16 | Europe/France/Auvergne-Rhv¥ne-Alpes/ | Laboratoire de virologie, CHU de Grenoble | CNR Virus des Infections Respiratoires - France SUD | Antonin Bal et al |
| EPI_ISL_779827 | France/ARA-SC733/2020 | 2020-11-16 | Europe/France/Auvergne-Rhv¥ne-Alpes/ | Laboratoire de virologie, CHU de Grenoble | CNR Virus des Infections Respiratoires - France SUD | Antonin Bal et al |

| | | | | | | |
|---|---|---|---|---|---|---|
| EPI_ISL_779828 | France/ARA-SC734/2020 | 2020-11-17 | Europe/France/Auvergne-Rhv¥ne-Alpes/ | Laboratoire de virologie, CHU de Grenoble | CNR Virus des Infections Respiratoires - France SUD | Antonin Bal et al |
| EPI_ISL_779829 | France/ARA-SC735/2020 | 2020-11-17 | Europe/France/Auvergne-Rhv¥ne-Alpes/ | Laboratoire de virologie, CHU de Grenoble | CNR Virus des Infections Respiratoires - France SUD | Antonin Bal et al |
| EPI_ISL_779830 | France/ARA-SC736/2020 | 2020-11-17 | Europe/France/Auvergne-Rhv¥ne-Alpes/ | Laboratoire de virologie, CHU de Grenoble | CNR Virus des Infections Respiratoires - France SUD | Antonin Bal et al |
| EPI_ISL_779835 | France/ARA-SC743/2020 | 2020-11-17 | Europe/France/Auvergne-Rhv¥ne-Alpes/ | Laboratoire de virologie, CHU de Grenoble | CNR Virus des Infections Respiratoires - France SUD | Antonin Bal et al |
| EPI_ISL_779837 | France/NAQ-SC769/2020 | 2020-12-09 | Europe/France/Nouvelle-Aquitaine/Bord | CHU Bordeaux | CNR Virus des Infections Respiratoires - France SUD | Antonin Bal et al |
| EPI_ISL_779838 | France/NAQ-SC770/2020 | 2020-11-24 | Europe/France/Nouvelle-Aquitaine/Bord | CHU Bordeaux | CNR Virus des Infections Respiratoires - France SUD | Antonin Bal et al |
| EPI_ISL_779839 | France/NAQ-SC771/2020 | 2020-12-17 | Europe/France/Nouvelle-Aquitaine/Bord | CHU Bordeaux | CNR Virus des Infections Respiratoires - France SUD | Antonin Bal et al |
| EPI_ISL_779841 | France/NAQ-SC778/2020 | 2020-12-18 | Europe/France/Nouvelle-Aquitaine/Bord | CHU Bordeaux | CNR Virus des Infections Respiratoires - France SUD | Antonin Bal et al |
| EPI_ISL_779848 | France/PAC-SC782/2020 | 2020-12-29 | Europe/France/Provence-Alpes-Cv¥te d' | Laboratoire BIOMED 05 | CNR Virus des Infections Respiratoires - France SUD | Antonin Bal et al |
| EPI_ISL_779857 | Spain/IB-IBV-99011606/2020 | 2020-12-11 | Europe/Spain/Balear Islands/Ibiza | Servicio de Microbiologv≠a, Hospital Universitario Son Espases | SeqCOVID-SPAIN consortium/IBV(CSIC) | Carla Lv≥pez-Causapv© et al |

| | | | | | | |
|---|---|---|---|---|---|---|
| EPI_ISL_779858 | Spain/IB-IBV-99011571/2020 | 2020-12-14 | Europe/Spain/Balear Islands/Menorca | Servicio de Microbiolog√≠a, Hospital Universitario Son Espases | SeqCOVID-SPAIN consortium/IBV(CSIC) | Carla L√≥pez-Causap√© et al |
| EPI_ISL_779863 | Spain/IB-IBV-99011590/2020 | 2020-12-05 | Europe/Spain/Balear Islands/Pollenca | Servicio de Microbiolog√≠a, Hospital Universitario Son Espases | SeqCOVID-SPAIN consortium/IBV(CSIC) | Carla L√≥pez-Causap√© et al |
| EPI_ISL_779866 | Spain/IB-IBV-99011526/2020 | 2020-12-18 | Europe/Spain/Balear Islands/Manacor | Servicio de Microbiolog√≠a, Hospital Universitario Son Espases | SeqCOVID-SPAIN consortium/IBV(CSIC) | Carla L√≥pez-Causap√© et al |
| EPI_ISL_779879 | Spain/IB-IBV-99011521/2020 | 2020-12-05 | Europe/Spain/Balear Islands/Inca | Servicio de Microbiolog√≠a, Hospital Universitario Son Espases | SeqCOVID-SPAIN consortium/IBV(CSIC) | Carla L√≥pez-Causap√© et al |
| EPI_ISL_779890 | Spain/IB-IBV-99011546/2020 | 2020-12-13 | Europe/Spain/Balear Islands/Manacor | Servicio de Microbiolog√≠a, Hospital Universitario Son Espases | SeqCOVID-SPAIN consortium/IBV(CSIC) | Carla L√≥pez-Causap√© et al |
| EPI_ISL_779892 | Spain/IB-IBV-99011550/2020 | 2020-12-21 | Europe/Spain/Balear Islands/Menorca | Servicio de Microbiolog√≠a, Hospital Universitario Son Espases | SeqCOVID-SPAIN consortium/IBV(CSIC) | Carla L√≥pez-Causap√© et al |
| EPI_ISL_779901 | Spain/IB-IBV-99011568/2020 | 2020-12-13 | Europe/Spain/Balear Islands/Palma de M | Servicio de Microbiolog√≠a, Hospital Universitario Son Espases | SeqCOVID-SPAIN consortium/IBV(CSIC) | Carla L√≥pez-Causap√© et al |
| EPI_ISL_779907 | Spain/IB-IBV-99011584/2020 | 2020-12-15 | Europe/Spain/Balear Islands/Palma de M | Servicio de Microbiolog√≠a, Hospital Universitario Son Espases | SeqCOVID-SPAIN consortium/IBV(CSIC) | Carla L√≥pez-Causap√© et al |
| EPI_ISL_779914 | Spain/IB-IBV-99011597/2020 | 2020-12-23 | Europe/Spain/Balear Islands/Ibiza | Servicio de Microbiolog√≠a, Hospital Universitario Son Espases | SeqCOVID-SPAIN consortium/IBV(CSIC) | Carla L√≥pez-Causap√© et al |
| EPI_ISL_779916 | Spain/IB-IBV-99011599/2020 | 2020-12-02 | Europe/Spain/Balear Islands/Palma de M | Servicio de Microbiolog√≠a, Hospital Universitario Son Espases | SeqCOVID-SPAIN consortium/IBV(CSIC) | Carla L√≥pez-Causap√© et al |
| EPI_ISL_779924 | Germany/NW-HHU-530/2020 | 2020-12-15 | Europe/Germany/North Rhine Westphal | Center of Medical Microbiology, Virology, and Hospital Hygiene, University of Duesseldorf | Center of Medical Microbiology, Virology, and Hospital Hygiene, University of Duesseldorf | Maximilian Damagnez et al |

| | | | | | | |
|---|---|---|---|---|---|---|
| EPI_ISL_779926 | Germany/NW-HHU-518/2020 | 2020-06-25 | Europe/Germany/North Rhine Westphal | Center of Medical Microbiology, Virology, and Hospital Hygiene, University of Duesseldorf | Center of Medical Microbiology, Virology, and Hospital Hygiene, University of Duesseldorf | Maximilian Damagnez et al |
| EPI_ISL_779927 | Germany/NW-HHU-537/2020 | 2020-12-15 | Europe/Germany/North Rhine Westphal | Center of Medical Microbiology, Virology, and Hospital Hygiene, University of Duesseldorf | Center of Medical Microbiology, Virology, and Hospital Hygiene, University of Duesseldorf | Maximilian Damagnez et al |
| EPI_ISL_779931 | Germany/NW-HHU-545/2020 | 2020-12-16 | Europe/Germany/North Rhine Westphal | Center of Medical Microbiology, Virology, and Hospital Hygiene, University of Duesseldorf | Center of Medical Microbiology, Virology, and Hospital Hygiene, University of Duesseldorf | Maximilian Damagnez et al |
| EPI_ISL_779935 | Germany/NW-HHU-524/2020 | 2020-10-22 | Europe/Germany/North Rhine Westphal | Center of Medical Microbiology, Virology, and Hospital Hygiene, University of Duesseldorf | Center of Medical Microbiology, Virology, and Hospital Hygiene, University of Duesseldorf | Maximilian Damagnez et al |
| EPI_ISL_779939 | Germany/NW-HHU-528/2020 | 2020-12-14 | Europe/Germany/North Rhine Westphal | Center of Medical Microbiology, Virology, and Hospital Hygiene, University of Duesseldorf | Center of Medical Microbiology, Virology, and Hospital Hygiene, University of Duesseldorf | Maximilian Damagnez et al |
| EPI_ISL_779941 | Germany/NW-HHU-531/2020 | 2020-12-15 | Europe/Germany/North Rhine Westphal | Center of Medical Microbiology, Virology, and Hospital Hygiene, University of Duesseldorf | Center of Medical Microbiology, Virology, and Hospital Hygiene, University of Duesseldorf | Maximilian Damagnez et al |

| | | | | | | |
|---|---|---|---|---|---|---|
| EPI_ISL_779942 | Germany/NW-HHU-532/2020 | 2020-12-15 | Europe/Germany/North Rhine Westphal | Center of Medical Microbiology, Virology, and Hospital Hygiene, University of Duesseldorf | Center of Medical Microbiology, Virology, and Hospital Hygiene, University of Duesseldorf | Maximilian Damagnez et al |
| EPI_ISL_779943 | Germany/NW-HHU-533/2020 | 2020-12-15 | Europe/Germany/North Rhine Westphal | Center of Medical Microbiology, Virology, and Hospital Hygiene, University of Duesseldorf | Center of Medical Microbiology, Virology, and Hospital Hygiene, University of Duesseldorf | Maximilian Damagnez et al |
| EPI_ISL_779945 | Germany/NW-HHU-535/2020 | 2020-12-15 | Europe/Germany/North Rhine Westphal | Center of Medical Microbiology, Virology, and Hospital Hygiene, University of Duesseldorf | Center of Medical Microbiology, Virology, and Hospital Hygiene, University of Duesseldorf | Maximilian Damagnez et al |
| EPI_ISL_779957 | Spain/MD-IBV-99012327/2020 | 2020-11-20 | Europe/Spain/Madrid/Madrid | Hospital General Universitario Gregorio Marav√±√≥n | SeqCOVID-SPAIN consortium/IBV(CSIC) | Dar√≠o Garc√≠a de Viedma et al |
| EPI_ISL_779958 | Spain/MD-IBV-99012335/2020 | 2020-11-20 | Europe/Spain/Madrid/Madrid | Hospital General Universitario Gregorio Marav√±√≥n | SeqCOVID-SPAIN consortium/IBV(CSIC) | Dar√≠o Garc√≠a de Viedma et al |
| EPI_ISL_779960 | Spain/MD-IBV-99012357/2020 | 2020-11-22 | Europe/Spain/Madrid/Madrid | Hospital General Universitario Gregorio Marav√±√≥n | SeqCOVID-SPAIN consortium/IBV(CSIC) | Dar√≠o Garc√≠a de Viedma et al |
| EPI_ISL_779962 | Spain/MD-IBV-99012347/2020 | 2020-11-20 | Europe/Spain/Madrid/Madrid | Hospital General Universitario Gregorio Marav√±√≥n | SeqCOVID-SPAIN consortium/IBV(CSIC) | Dar√≠o Garc√≠a de Viedma et al |
| EPI_ISL_779963 | Spain/MD-IBV-99012371/2020 | 2020-11-18 | Europe/Spain/Madrid/Madrid | Hospital General Universitario Gregorio Marav√±√≥n | SeqCOVID-SPAIN consortium/IBV(CSIC) | Dar√≠o Garc√≠a de Viedma et al |
| EPI_ISL_779964 | Spain/MD-IBV-99012340/2020 | 2020-11-20 | Europe/Spain/Madrid/Madrid | Hospital General Universitario Gregorio Marav√±√≥n | SeqCOVID-SPAIN consortium/IBV(CSIC) | Dar√≠o Garc√≠a de Viedma et al |
| EPI_ISL_779965 | Spain/MD-IBV-99012325/2020 | 2020-11-20 | Europe/Spain/Madrid/Madrid | Hospital General Universitario Gregorio Marav√±√≥n | SeqCOVID-SPAIN consortium/IBV(CSIC) | Dar√≠o Garc√≠a de Viedma et al |

| | | | | | | |
|---|---|---|---|---|---|---|
| EPI_ISL_779967 | Spain/MD-IBV-99012336/2020 | 2020-11-20 | Europe/Spain/Madrid/Madrid | Hospital General Universitario Gregorio Marav√±√≥n | SeqCOVID-SPAIN consortium/IBV(CSIC) | Dar√≠o Garc√≠a de Viedma et al |
| EPI_ISL_779975 | Spain/MD-IBV-99012315/2020 | 2020-11-17 | Europe/Spain/Madrid/Madrid | Hospital General Universitario Gregorio Marav√±√≥n | SeqCOVID-SPAIN consortium/IBV(CSIC) | Dar√≠o Garc√≠a de Viedma et al |
| EPI_ISL_779979 | Spain/MD-IBV-99012324/2020 | 2020-11-20 | Europe/Spain/Madrid/Madrid | Hospital General Universitario Gregorio Marav√±√≥n | SeqCOVID-SPAIN consortium/IBV(CSIC) | Dar√≠o Garc√≠a de Viedma et al |
| EPI_ISL_779980 | Spain/MD-IBV-99012326/2020 | 2020-11-20 | Europe/Spain/Madrid/Madrid | Hospital General Universitario Gregorio Marav√±√≥n | SeqCOVID-SPAIN consortium/IBV(CSIC) | Dar√≠o Garc√≠a de Viedma et al |
| EPI_ISL_779982 | Spain/MD-IBV-99012329/2020 | 2020-11-20 | Europe/Spain/Madrid/Madrid | Hospital General Universitario Gregorio Marav√±√≥n | SeqCOVID-SPAIN consortium/IBV(CSIC) | Dar√≠o Garc√≠a de Viedma et al |
| EPI_ISL_779983 | Spain/MD-IBV-99012330/2020 | 2020-11-20 | Europe/Spain/Madrid/Madrid | Hospital General Universitario Gregorio Marav√±√≥n | SeqCOVID-SPAIN consortium/IBV(CSIC) | Dar√≠o Garc√≠a de Viedma et al |
| EPI_ISL_779985 | Spain/MD-IBV-99012332/2020 | 2020-11-20 | Europe/Spain/Madrid/Madrid | Hospital General Universitario Gregorio Marav√±√≥n | SeqCOVID-SPAIN consortium/IBV(CSIC) | Dar√≠o Garc√≠a de Viedma et al |
| EPI_ISL_779986 | Spain/MD-IBV-99012333/2020 | 2020-11-20 | Europe/Spain/Madrid/Madrid | Hospital General Universitario Gregorio Marav√±√≥n | SeqCOVID-SPAIN consortium/IBV(CSIC) | Dar√≠o Garc√≠a de Viedma et al |
| EPI_ISL_779989 | Spain/MD-IBV-99012339/2020 | 2020-11-20 | Europe/Spain/Madrid/Madrid | Hospital General Universitario Gregorio Marav√±√≥n | SeqCOVID-SPAIN consortium/IBV(CSIC) | Dar√≠o Garc√≠a de Viedma et al |
| EPI_ISL_779993 | Spain/MD-IBV-99012344/2020 | 2020-11-20 | Europe/Spain/Madrid/Madrid | Hospital General Universitario Gregorio Marav√±√≥n | SeqCOVID-SPAIN consortium/IBV(CSIC) | Dar√≠o Garc√≠a de Viedma et al |
| EPI_ISL_779994 | Spain/MD-IBV-99012345/2020 | 2020-11-20 | Europe/Spain/Madrid/Madrid | Hospital General Universitario Gregorio Marav√±√≥n | SeqCOVID-SPAIN consortium/IBV(CSIC) | Dar√≠o Garc√≠a de Viedma et al |
| EPI_ISL_779996 | Spain/MD-IBV-99012348/2020 | 2020-11-20 | Europe/Spain/Madrid/Madrid | Hospital General Universitario Gregorio Marav√±√≥n | SeqCOVID-SPAIN consortium/IBV(CSIC) | Dar√≠o Garc√≠a de Viedma et al |
| EPI_ISL_779997 | Spain/MD-IBV-99012350/2020 | 2020-11-22 | Europe/Spain/Madrid/Madrid | Hospital General Universitario Gregorio Marav√±√≥n | SeqCOVID-SPAIN consortium/IBV(CSIC) | Dar√≠o Garc√≠a de Viedma et al |

| | | | | | | |
|---|---|---|---|---|---|---|
| EPI_ISL_780000 | Spain/MD-IBV-99012353/2020 | 2020-11-22 | Europe/Spain/Madrid/Madrid | Hospital General Universitario Gregorio Marav√±√≥n | SeqCOVID-SPAIN consortium/IBV(CSIC) | Dar√≠o Garc√≠a de Viedma et al |
| EPI_ISL_780001 | Spain/MD-IBV-99012355/2020 | 2020-11-22 | Europe/Spain/Madrid/Madrid | Hospital General Universitario Gregorio Marav√±√≥n | SeqCOVID-SPAIN consortium/IBV(CSIC) | Dar√≠o Garc√≠a de Viedma et al |
| EPI_ISL_780002 | Spain/MD-IBV-99012358/2020 | 2020-11-22 | Europe/Spain/Madrid/Madrid | Hospital General Universitario Gregorio Marav√±√≥n | SeqCOVID-SPAIN consortium/IBV(CSIC) | Dar√≠o Garc√≠a de Viedma et al |
| EPI_ISL_780004 | Spain/MD-IBV-99012360/2020 | 2020-11-22 | Europe/Spain/Madrid/Madrid | Hospital General Universitario Gregorio Marav√±√≥n | SeqCOVID-SPAIN consortium/IBV(CSIC) | Dar√≠o Garc√≠a de Viedma et al |
| EPI_ISL_780005 | Spain/MD-IBV-99012361/2020 | 2020-11-22 | Europe/Spain/Madrid/Madrid | Hospital General Universitario Gregorio Marav√±√≥n | SeqCOVID-SPAIN consortium/IBV(CSIC) | Dar√≠o Garc√≠a de Viedma et al |
| EPI_ISL_780006 | Spain/MD-IBV-99012363/2020 | 2020-11-22 | Europe/Spain/Madrid/Madrid | Hospital General Universitario Gregorio Marav√±√≥n | SeqCOVID-SPAIN consortium/IBV(CSIC) | Dar√≠o Garc√≠a de Viedma et al |
| EPI_ISL_780007 | Spain/MD-IBV-99012364/2020 | 2020-11-22 | Europe/Spain/Madrid/Madrid | Hospital General Universitario Gregorio Marav√±√≥n | SeqCOVID-SPAIN consortium/IBV(CSIC) | Dar√≠o Garc√≠a de Viedma et al |
| EPI_ISL_780008 | Spain/MD-IBV-99012365/2020 | 2020-11-22 | Europe/Spain/Madrid/Madrid | Hospital General Universitario Gregorio Marav√±√≥n | SeqCOVID-SPAIN consortium/IBV(CSIC) | Dar√≠o Garc√≠a de Viedma et al |
| EPI_ISL_780009 | Spain/MD-IBV-99012368/2020 | 2020-11-21 | Europe/Spain/Madrid/Madrid | Hospital General Universitario Gregorio Marav√±√≥n | SeqCOVID-SPAIN consortium/IBV(CSIC) | Dar√≠o Garc√≠a de Viedma et al |
| EPI_ISL_780010 | Spain/MD-IBV-99012369/2020 | 2020-11-21 | Europe/Spain/Madrid/Madrid | Hospital General Universitario Gregorio Marav√±√≥n | SeqCOVID-SPAIN consortium/IBV(CSIC) | Dar√≠o Garc√≠a de Viedma et al |
| EPI_ISL_780011 | Spain/MD-IBV-99012370/2020 | 2020-11-21 | Europe/Spain/Madrid/Madrid | Hospital General Universitario Gregorio Marav√±√≥n | SeqCOVID-SPAIN consortium/IBV(CSIC) | Dar√≠o Garc√≠a de Viedma et al |
| EPI_ISL_780016 | Spain/MD-IBV-99012378/2020 | 2020-11-18 | Europe/Spain/Madrid/Madrid | Hospital General Universitario Gregorio Marav√±√≥n | SeqCOVID-SPAIN consortium/IBV(CSIC) | Dar√≠o Garc√≠a de Viedma et al |
| EPI_ISL_780017 | Spain/MD-IBV-99012380/2020 | 2020-11-18 | Europe/Spain/Madrid/Madrid | Hospital General Universitario Gregorio Marav√±√≥n | SeqCOVID-SPAIN consortium/IBV(CSIC) | Dar√≠o Garc√≠a de Viedma et al |

| | | | | | | |
|---|---|---|---|---|---|---|
| EPI_ISL_780018 | Spain/MD-IBV-99012383/2020 | 2020-11-18 | Europe/Spain/Madrid/Madrid | Hospital General Universitario Gregorio Marav±V≥n | SeqCOVID-SPAIN consortium/IBV(CSIC) | Dar√≠o Garc√≠a de Viedma et al |
| EPI_ISL_780020 | Spain/MD-IBV-99012385/2020 | 2020-11-18 | Europe/Spain/Madrid/Madrid | Hospital General Universitario Gregorio Marav±V≥n | SeqCOVID-SPAIN consortium/IBV(CSIC) | Dar√≠o Garc√≠a de Viedma et al |
| EPI_ISL_780021 | Spain/MD-IBV-99012387/2020 | 2020-10-29 | Europe/Spain/Madrid/Madrid | Hospital General Universitario Gregorio Marav±V≥n | SeqCOVID-SPAIN consortium/IBV(CSIC) | Dar√≠o Garc√≠a de Viedma et al |
| EPI_ISL_780022 | Spain/MD-IBV-99012388/2020 | 2020-10-31 | Europe/Spain/Madrid/Madrid | Hospital General Universitario Gregorio Marav±V≥n | SeqCOVID-SPAIN consortium/IBV(CSIC) | Dar√≠o Garc√≠a de Viedma et al |
| EPI_ISL_780024 | Spain/MD-IBV-99012390/2020 | 2020-11-05 | Europe/Spain/Madrid/Madrid | Hospital General Universitario Gregorio Marav±V≥n | SeqCOVID-SPAIN consortium/IBV(CSIC) | Dar√≠o Garc√≠a de Viedma et al |
| EPI_ISL_780027 | Spain/MD-IBV-99012394/2020 | 2020-11-14 | Europe/Spain/Madrid/Madrid | Hospital General Universitario Gregorio Marav±V≥n | SeqCOVID-SPAIN consortium/IBV(CSIC) | Dar√≠o Garc√≠a de Viedma et al |
| EPI_ISL_780028 | Spain/MD-IBV-99012395/2020 | 2020-10-12 | Europe/Spain/Madrid/Madrid | Hospital General Universitario Gregorio Marav±V≥n | SeqCOVID-SPAIN consortium/IBV(CSIC) | Dar√≠o Garc√≠a de Viedma et al |
| EPI_ISL_780029 | Spain/MD-IBV-99012451/2020 | 2020-11-19 | Europe/Spain/Madrid/Madrid | Hospital General Universitario Gregorio Marav±V≥n | SeqCOVID-SPAIN consortium/IBV(CSIC) | Dar√≠o Garc√≠a de Viedma et al |
| EPI_ISL_780032 | Spain/MD-IBV-99012416/2020 | 2020-11-17 | Europe/Spain/Madrid/Madrid | Hospital General Universitario Gregorio Marav±V≥n | SeqCOVID-SPAIN consortium/IBV(CSIC) | Dar√≠o Garc√≠a de Viedma et al |
| EPI_ISL_780033 | Spain/MD-IBV-99012422/2020 | 2020-11-23 | Europe/Spain/Madrid/Madrid | Hospital General Universitario Gregorio Marav±V≥n | SeqCOVID-SPAIN consortium/IBV(CSIC) | Dar√≠o Garc√≠a de Viedma et al |
| EPI_ISL_780035 | Spain/MD-IBV-99012415/2020 | 2020-11-24 | Europe/Spain/Madrid/Madrid | Hospital General Universitario Gregorio Marav±V≥n | SeqCOVID-SPAIN consortium/IBV(CSIC) | Dar√≠o Garc√≠a de Viedma et al |
| EPI_ISL_780037 | Spain/MD-IBV-99012428/2020 | 2020-11-29 | Europe/Spain/Madrid/Madrid | Hospital General Universitario Gregorio Marav±V≥n | SeqCOVID-SPAIN consortium/IBV(CSIC) | Dar√≠o Garc√≠a de Viedma et al |
| EPI_ISL_780039 | Spain/MD-IBV-99012419/2020 | 2020-11-17 | Europe/Spain/Madrid/Madrid | Hospital General Universitario Gregorio Marav±V≥n | SeqCOVID-SPAIN consortium/IBV(CSIC) | Dar√≠o Garc√≠a de Viedma et al |

| | | | | | | |
|---|---|---|---|---|---|---|
| EPI_ISL_780040 | Spain/MD-IBV-99012482/2020 | 2020-11-24 | Europe/Spain/Madrid/Madrid | Hospital General Universitario Gregorio Marav√±√≥n | SeqCOVID-SPAIN consortium/IBV(CSIC) | Dar√≠o Garc√≠a de Viedma et al |
| EPI_ISL_780041 | Spain/MD-IBV-99012399/2020 | 2020-11-12 | Europe/Spain/Madrid/Madrid | Hospital General Universitario Gregorio Marav√±√≥n | SeqCOVID-SPAIN consortium/IBV(CSIC) | Dar√≠o Garc√≠a de Viedma et al |
| EPI_ISL_780042 | Spain/MD-IBV-99012455/2020 | 2020-11-19 | Europe/Spain/Madrid/Madrid | Hospital General Universitario Gregorio Marav√±√≥n | SeqCOVID-SPAIN consortium/IBV(CSIC) | Dar√≠o Garc√≠a de Viedma et al |
| EPI_ISL_780044 | Spain/MD-IBV-99012423/2020 | 2020-11-24 | Europe/Spain/Madrid/Madrid | Hospital General Universitario Gregorio Marav√±√≥n | SeqCOVID-SPAIN consortium/IBV(CSIC) | Dar√≠o Garc√≠a de Viedma et al |
| EPI_ISL_780045 | Spain/MD-IBV-99012417/2020 | 2020-11-17 | Europe/Spain/Madrid/Madrid | Hospital General Universitario Gregorio Marav√±√≥n | SeqCOVID-SPAIN consortium/IBV(CSIC) | Dar√≠o Garc√≠a de Viedma et al |
| EPI_ISL_780047 | Spain/MD-IBV-99012414/2020 | 2020-11-14 | Europe/Spain/Madrid/Madrid | Hospital General Universitario Gregorio Marav√±√≥n | SeqCOVID-SPAIN consortium/IBV(CSIC) | Dar√≠o Garc√≠a de Viedma et al |
| EPI_ISL_780048 | Spain/MD-IBV-99012436/2020 | 2020-11-02 | Europe/Spain/Madrid/Madrid | Hospital General Universitario Gregorio Marav√±√≥n | SeqCOVID-SPAIN consortium/IBV(CSIC) | Dar√≠o Garc√≠a de Viedma et al |
| EPI_ISL_780054 | Spain/MD-IBV-99012481/2020 | 2020-11-24 | Europe/Spain/Madrid/Madrid | Hospital General Universitario Gregorio Marav√±√≥n | SeqCOVID-SPAIN consortium/IBV(CSIC) | Dar√≠o Garc√≠a de Viedma et al |
| EPI_ISL_780055 | Spain/MD-IBV-99012420/2020 | 2020-11-16 | Europe/Spain/Madrid/Madrid | Hospital General Universitario Gregorio Marav√±√≥n | SeqCOVID-SPAIN consortium/IBV(CSIC) | Dar√≠o Garc√≠a de Viedma et al |
| EPI_ISL_780058 | Spain/MD-IBV-99012400/2020 | 2020-11-04 | Europe/Spain/Madrid/Madrid | Hospital General Universitario Gregorio Marav√±√≥n | SeqCOVID-SPAIN consortium/IBV(CSIC) | Dar√≠o Garc√≠a de Viedma et al |
| EPI_ISL_780059 | Spain/MD-IBV-99012401/2020 | 2020-11-05 | Europe/Spain/Madrid/Madrid | Hospital General Universitario Gregorio Marav√±√≥n | SeqCOVID-SPAIN consortium/IBV(CSIC) | Dar√≠o Garc√≠a de Viedma et al |
| EPI_ISL_780062 | Spain/MD-IBV-99012407/2020 | 2020-11-06 | Europe/Spain/Madrid/Madrid | Hospital General Universitario Gregorio Marav√±√≥n | SeqCOVID-SPAIN consortium/IBV(CSIC) | Dar√≠o Garc√≠a de Viedma et al |
| EPI_ISL_780063 | Spain/MD-IBV-99012408/2020 | 2020-11-18 | Europe/Spain/Madrid/Madrid | Hospital General Universitario Gregorio Marav√±√≥n | SeqCOVID-SPAIN consortium/IBV(CSIC) | Dar√≠o Garc√≠a de Viedma et al |

| | | | | | | |
|---|---|---|---|---|---|---|
| EPI_ISL_780064 | Spain/MD-IBV-99012410/2020 | 2020-11-11 | Europe/Spain/Madrid/Madrid | Hospital General Universitario Gregorio Marav√±√≥n | SeqCOVID-SPAIN consortium/IBV(CSIC) | Dar√≠o Garc√≠a de Viedma et al |
| EPI_ISL_780065 | Spain/MD-IBV-99012411/2020 | 2020-11-16 | Europe/Spain/Madrid/Madrid | Hospital General Universitario Gregorio Marav√±√≥n | SeqCOVID-SPAIN consortium/IBV(CSIC) | Dar√≠o Garc√≠a de Viedma et al |
| EPI_ISL_780067 | Spain/MD-IBV-99012418/2020 | 2020-11-17 | Europe/Spain/Madrid/Madrid | Hospital General Universitario Gregorio Marav√±√≥n | SeqCOVID-SPAIN consortium/IBV(CSIC) | Dar√≠o Garc√≠a de Viedma et al |
| EPI_ISL_780068 | Spain/MD-IBV-99012425/2020 | 2020-10-05 | Europe/Spain/Madrid/Madrid | Hospital General Universitario Gregorio Marav√±√≥n | SeqCOVID-SPAIN consortium/IBV(CSIC) | Dar√≠o Garc√≠a de Viedma et al |
| EPI_ISL_780070 | Spain/MD-IBV-99012429/2020 | 2020-11-27 | Europe/Spain/Madrid/Madrid | Hospital General Universitario Gregorio Marav√±√≥n | SeqCOVID-SPAIN consortium/IBV(CSIC) | Dar√≠o Garc√≠a de Viedma et al |
| EPI_ISL_780072 | Spain/MD-IBV-99012431/2020 | 2020-11-26 | Europe/Spain/Madrid/Madrid | Hospital General Universitario Gregorio Marav√±√≥n | SeqCOVID-SPAIN consortium/IBV(CSIC) | Dar√≠o Garc√≠a de Viedma et al |
| EPI_ISL_780074 | Spain/MD-IBV-99012437/2020 | 2020-11-01 | Europe/Spain/Madrid/Madrid | Hospital General Universitario Gregorio Marav√±√≥n | SeqCOVID-SPAIN consortium/IBV(CSIC) | Dar√≠o Garc√≠a de Viedma et al |
| EPI_ISL_780077 | Spain/MD-IBV-99012442/2020 | 2020-11-27 | Europe/Spain/Madrid/Madrid | Hospital General Universitario Gregorio Marav√±√≥n | SeqCOVID-SPAIN consortium/IBV(CSIC) | Dar√≠o Garc√≠a de Viedma et al |
| EPI_ISL_780079 | Spain/MD-IBV-99012444/2020 | 2020-11-19 | Europe/Spain/Madrid/Madrid | Hospital General Universitario Gregorio Marav√±√≥n | SeqCOVID-SPAIN consortium/IBV(CSIC) | Dar√≠o Garc√≠a de Viedma et al |
| EPI_ISL_780080 | Spain/MD-IBV-99012445/2020 | 2020-11-19 | Europe/Spain/Madrid/Madrid | Hospital General Universitario Gregorio Marav√±√≥n | SeqCOVID-SPAIN consortium/IBV(CSIC) | Dar√≠o Garc√≠a de Viedma et al |
| EPI_ISL_780083 | Spain/MD-IBV-99012453/2020 | 2020-11-19 | Europe/Spain/Madrid/Madrid | Hospital General Universitario Gregorio Marav√±√≥n | SeqCOVID-SPAIN consortium/IBV(CSIC) | Dar√≠o Garc√≠a de Viedma et al |
| EPI_ISL_780084 | Spain/MD-IBV-99012454/2020 | 2020-11-19 | Europe/Spain/Madrid/Madrid | Hospital General Universitario Gregorio Marav√±√≥n | SeqCOVID-SPAIN consortium/IBV(CSIC) | Dar√≠o Garc√≠a de Viedma et al |
| EPI_ISL_780085 | Spain/MD-IBV-99012456/2020 | 2020-11-19 | Europe/Spain/Madrid/Madrid | Hospital General Universitario Gregorio Marav√±√≥n | SeqCOVID-SPAIN consortium/IBV(CSIC) | Dar√≠o Garc√≠a de Viedma et al |

| | | | | | | |
|---|---|---|---|---|---|---|
| EPI_ISL_780086 | Spain/MD-IBV-99012457/2020 | 2020-11-19 | Europe/Spain/Madrid/Madrid | Hospital General Universitario Gregorio Marav±v≥n | SeqCOVID-SPAIN consortium/IBV(CSIC) | Darv≠o Garcv≠a de Viedma et al |
| EPI_ISL_780089 | Spain/MD-IBV-99012464/2020 | 2020-11-19 | Europe/Spain/Madrid/Madrid | Hospital General Universitario Gregorio Marav±v≥n | SeqCOVID-SPAIN consortium/IBV(CSIC) | Darv≠o Garcv≠a de Viedma et al |
| EPI_ISL_780090 | Spain/MD-IBV-99012466/2020 | 2020-11-14 | Europe/Spain/Madrid/Madrid | Hospital General Universitario Gregorio Marav±v≥n | SeqCOVID-SPAIN consortium/IBV(CSIC) | Darv≠o Garcv≠a de Viedma et al |
| EPI_ISL_780091 | Spain/MD-IBV-99012467/2020 | 2020-11-14 | Europe/Spain/Madrid/Madrid | Hospital General Universitario Gregorio Marav±v≥n | SeqCOVID-SPAIN consortium/IBV(CSIC) | Darv≠o Garcv≠a de Viedma et al |
| EPI_ISL_780093 | Spain/MD-IBV-99012469/2020 | 2020-11-14 | Europe/Spain/Madrid/Madrid | Hospital General Universitario Gregorio Marav±v≥n | SeqCOVID-SPAIN consortium/IBV(CSIC) | Darv≠o Garcv≠a de Viedma et al |
| EPI_ISL_780095 | Spain/MD-IBV-99012480/2020 | 2020-11-24 | Europe/Spain/Madrid/Madrid | Hospital General Universitario Gregorio Marav±v≥n | SeqCOVID-SPAIN consortium/IBV(CSIC) | Darv≠o Garcv≠a de Viedma et al |
| EPI_ISL_780104 | USA/TX-HMH-MCoV-9584/2020 | 2020-07-05 | North America/USA/Texas/Houston | Houston Methodist Hospital | Houston Methodist Hospital | S. Wesley Long et al |
| EPI_ISL_780111 | USA/TX-HMH-MCoV-9944/2020 | 2020-07-06 | North America/USA/Texas/Houston | Houston Methodist Hospital | Houston Methodist Hospital | S. Wesley Long et al |
| EPI_ISL_780112 | USA/TX-HMH-MCoV-9997/2020 | 2020-07-06 | North America/USA/Texas/Houston | Houston Methodist Hospital | Houston Methodist Hospital | S. Wesley Long et al |
| EPI_ISL_780113 | USA/TX-HMH-MCoV-9981/2020 | 2020-07-06 | North America/USA/Texas/Houston | Houston Methodist Hospital | Houston Methodist Hospital | S. Wesley Long et al |
| EPI_ISL_780119 | USA/TX-HMH-MCoV-9658/2020 | 2020-07-04 | North America/USA/Texas/Houston | Houston Methodist Hospital | Houston Methodist Hospital | S. Wesley Long et al |
| EPI_ISL_780123 | USA/TX-HMH-MCoV-9909/2020 | 2020-07-06 | North America/USA/Texas/Houston | Houston Methodist Hospital | Houston Methodist Hospital | S. Wesley Long et al |
| EPI_ISL_780124 | USA/TX-HMH-MCoV-9717/2020 | 2020-07-04 | North America/USA/Texas/Houston | Houston Methodist Hospital | Houston Methodist Hospital | S. Wesley Long et al |

| | | | | | | |
|---|---|---|---|---|---|---|
| EPI_ISL_780145 | USA/TX-HMH-MCoV-9556/2020 | 2020-07-03 | North America/USA/Texas/Houston | Houston Methodist Hospital | Houston Methodist Hospital | S. Wesley Long et al |
| EPI_ISL_780153 | USA/TX-HMH-MCoV-9550/2020 | 2020-07-05 | North America/USA/Texas/Houston | Houston Methodist Hospital | Houston Methodist Hospital | S. Wesley Long et al |
| EPI_ISL_780159 | USA/TX-HMH-MCoV-9562/2020 | 2020-07-02 | North America/USA/Texas/Houston | Houston Methodist Hospital | Houston Methodist Hospital | S. Wesley Long et al |
| EPI_ISL_780166 | USA/TX-HMH-MCoV-9578/2020 | 2020-07-04 | North America/USA/Texas/Houston | Houston Methodist Hospital | Houston Methodist Hospital | S. Wesley Long et al |
| EPI_ISL_780172 | USA/TX-HMH-MCoV-9590/2020 | 2020-07-02 | North America/USA/Texas/Houston | Houston Methodist Hospital | Houston Methodist Hospital | S. Wesley Long et al |
| EPI_ISL_780174 | USA/TX-HMH-MCoV-9598/2020 | 2020-07-04 | North America/USA/Texas/Houston | Houston Methodist Hospital | Houston Methodist Hospital | S. Wesley Long et al |
| EPI_ISL_780176 | USA/TX-HMH-MCoV-9600/2020 | 2020-07-04 | North America/USA/Texas/Houston | Houston Methodist Hospital | Houston Methodist Hospital | S. Wesley Long et al |
| EPI_ISL_780177 | USA/TX-HMH-MCoV-9601/2020 | 2020-07-07 | North America/USA/Texas/Houston | Houston Methodist Hospital | Houston Methodist Hospital | S. Wesley Long et al |
| EPI_ISL_780179 | USA/TX-HMH-MCoV-9603/2020 | 2020-07-03 | North America/USA/Texas/Houston | Houston Methodist Hospital | Houston Methodist Hospital | S. Wesley Long et al |
| EPI_ISL_780180 | USA/TX-HMH-MCoV-9605/2020 | 2020-07-04 | North America/USA/Texas/Houston | Houston Methodist Hospital | Houston Methodist Hospital | S. Wesley Long et al |
| EPI_ISL_780182 | USA/TX-HMH-MCoV-9608/2020 | 2020-07-04 | North America/USA/Texas/Houston | Houston Methodist Hospital | Houston Methodist Hospital | S. Wesley Long et al |
| EPI_ISL_780184 | USA/TX-HMH-MCoV-9616/2020 | 2020-07-04 | North America/USA/Texas/Houston | Houston Methodist Hospital | Houston Methodist Hospital | S. Wesley Long et al |
| EPI_ISL_780190 | USA/TX-HMH-MCoV-9627/2020 | 2020-07-04 | North America/USA/Texas/Houston | Houston Methodist Hospital | Houston Methodist Hospital | S. Wesley Long et al |

| | | | | | | |
|---|---|---|---|---|---|---|
| EPI_ISL_780193 | USA/TX-HMH-MCoV-9632/2020 | 2020-07-04 | North America/USA/Texas/Houston | Houston Methodist Hospital | Houston Methodist Hospital | S. Wesley Long et al |
| EPI_ISL_780198 | USA/TX-HMH-MCoV-9643/2020 | 2020-07-03 | North America/USA/Texas/Houston | Houston Methodist Hospital | Houston Methodist Hospital | S. Wesley Long et al |
| EPI_ISL_780200 | USA/TX-HMH-MCoV-9646/2020 | 2020-07-04 | North America/USA/Texas/Houston | Houston Methodist Hospital | Houston Methodist Hospital | S. Wesley Long et al |
| EPI_ISL_780202 | USA/TX-HMH-MCoV-9648/2020 | 2020-07-04 | North America/USA/Texas/Houston | Houston Methodist Hospital | Houston Methodist Hospital | S. Wesley Long et al |
| EPI_ISL_780205 | USA/TX-HMH-MCoV-9653/2020 | 2020-07-04 | North America/USA/Texas/Houston | Houston Methodist Hospital | Houston Methodist Hospital | S. Wesley Long et al |
| EPI_ISL_780216 | USA/TX-HMH-MCoV-9674/2020 | 2020-07-02 | North America/USA/Texas/Houston | Houston Methodist Hospital | Houston Methodist Hospital | S. Wesley Long et al |
| EPI_ISL_780219 | USA/TX-HMH-MCoV-9677/2020 | 2020-07-05 | North America/USA/Texas/Houston | Houston Methodist Hospital | Houston Methodist Hospital | S. Wesley Long et al |
| EPI_ISL_780230 | USA/TX-HMH-MCoV-9700/2020 | 2020-07-07 | North America/USA/Texas/Houston | Houston Methodist Hospital | Houston Methodist Hospital | S. Wesley Long et al |
| EPI_ISL_780232 | USA/TX-HMH-MCoV-9702/2020 | 2020-07-03 | North America/USA/Texas/Houston | Houston Methodist Hospital | Houston Methodist Hospital | S. Wesley Long et al |
| EPI_ISL_780237 | USA/TX-HMH-MCoV-9712/2020 | 2020-07-07 | North America/USA/Texas/Houston | Houston Methodist Hospital | Houston Methodist Hospital | S. Wesley Long et al |
| EPI_ISL_780240 | USA/TX-HMH-MCoV-9718/2020 | 2020-07-06 | North America/USA/Texas/Houston | Houston Methodist Hospital | Houston Methodist Hospital | S. Wesley Long et al |
| EPI_ISL_780241 | USA/TX-HMH-MCoV-9720/2020 | 2020-07-06 | North America/USA/Texas/Houston | Houston Methodist Hospital | Houston Methodist Hospital | S. Wesley Long et al |
| EPI_ISL_780242 | USA/TX-HMH-MCoV-9723/2020 | 2020-07-07 | North America/USA/Texas/Houston | Houston Methodist Hospital | Houston Methodist Hospital | S. Wesley Long et al |

| | | | | | | |
|---|---|---|---|---|---|---|
| EPI_ISL_780244 | USA/TX-HMH-MCoV-9725/2020 | 2020-06-29 | North America/USA/Texas/Houston | Houston Methodist Hospital | Houston Methodist Hospital | S. Wesley Long et al |
| EPI_ISL_780247 | USA/TX-HMH-MCoV-9734/2020 | 2020-07-06 | North America/USA/Texas/Houston | Houston Methodist Hospital | Houston Methodist Hospital | S. Wesley Long et al |
| EPI_ISL_780252 | USA/TX-HMH-MCoV-9743/2020 | 2020-07-06 | North America/USA/Texas/Houston | Houston Methodist Hospital | Houston Methodist Hospital | S. Wesley Long et al |
| EPI_ISL_780260 | USA/TX-HMH-MCoV-9763/2020 | 2020-07-04 | North America/USA/Texas/Houston | Houston Methodist Hospital | Houston Methodist Hospital | S. Wesley Long et al |
| EPI_ISL_780262 | USA/TX-HMH-MCoV-9767/2020 | 2020-07-06 | North America/USA/Texas/Houston | Houston Methodist Hospital | Houston Methodist Hospital | S. Wesley Long et al |
| EPI_ISL_780263 | USA/TX-HMH-MCoV-9768/2020 | 2020-07-06 | North America/USA/Texas/Houston | Houston Methodist Hospital | Houston Methodist Hospital | S. Wesley Long et al |
| EPI_ISL_780266 | USA/TX-HMH-MCoV-9771/2020 | 2020-06-19 | North America/USA/Texas/Houston | Houston Methodist Hospital | Houston Methodist Hospital | S. Wesley Long et al |
| EPI_ISL_780272 | USA/TX-HMH-MCoV-9778/2020 | 2020-07-06 | North America/USA/Texas/Houston | Houston Methodist Hospital | Houston Methodist Hospital | S. Wesley Long et al |
| EPI_ISL_780280 | USA/TX-HMH-MCoV-9795/2020 | 2020-07-04 | North America/USA/Texas/Houston | Houston Methodist Hospital | Houston Methodist Hospital | S. Wesley Long et al |
| EPI_ISL_780284 | USA/TX-HMH-MCoV-9804/2020 | 2020-07-05 | North America/USA/Texas/Houston | Houston Methodist Hospital | Houston Methodist Hospital | S. Wesley Long et al |
| EPI_ISL_780289 | USA/TX-HMH-MCoV-9814/2020 | 2020-07-06 | North America/USA/Texas/Houston | Houston Methodist Hospital | Houston Methodist Hospital | S. Wesley Long et al |
| EPI_ISL_780290 | USA/TX-HMH-MCoV-9815/2020 | 2020-07-06 | North America/USA/Texas/Houston | Houston Methodist Hospital | Houston Methodist Hospital | S. Wesley Long et al |
| EPI_ISL_780291 | USA/TX-HMH-MCoV-9816/2020 | 2020-07-06 | North America/USA/Texas/Houston | Houston Methodist Hospital | Houston Methodist Hospital | S. Wesley Long et al |

| | | | | | | |
|---|---|---|---|---|---|---|
| EPI_ISL_780294 | USA/TX-HMH-MCoV-9823/2020 | 2020-07-06 | North America/USA/Texas/Houston | Houston Methodist Hospital | Houston Methodist Hospital | S. Wesley Long et al |
| EPI_ISL_780305 | USA/TX-HMH-MCoV-9853/2020 | 2020-07-06 | North America/USA/Texas/Houston | Houston Methodist Hospital | Houston Methodist Hospital | S. Wesley Long et al |
| EPI_ISL_780307 | USA/TX-HMH-MCoV-9856/2020 | 2020-07-06 | North America/USA/Texas/Houston | Houston Methodist Hospital | Houston Methodist Hospital | S. Wesley Long et al |
| EPI_ISL_780320 | USA/TX-HMH-MCoV-9880/2020 | 2020-07-06 | North America/USA/Texas/Houston | Houston Methodist Hospital | Houston Methodist Hospital | S. Wesley Long et al |
| EPI_ISL_780328 | USA/TX-HMH-MCoV-9895/2020 | 2020-07-06 | North America/USA/Texas/Houston | Houston Methodist Hospital | Houston Methodist Hospital | S. Wesley Long et al |
| EPI_ISL_780333 | USA/TX-HMH-MCoV-9903/2020 | 2020-07-06 | North America/USA/Texas/Houston | Houston Methodist Hospital | Houston Methodist Hospital | S. Wesley Long et al |
| EPI_ISL_780335 | USA/TX-HMH-MCoV-9906/2020 | 2020-07-06 | North America/USA/Texas/Houston | Houston Methodist Hospital | Houston Methodist Hospital | S. Wesley Long et al |
| EPI_ISL_780343 | USA/TX-HMH-MCoV-9924/2020 | 2020-07-06 | North America/USA/Texas/Houston | Houston Methodist Hospital | Houston Methodist Hospital | S. Wesley Long et al |
| EPI_ISL_780353 | USA/TX-HMH-MCoV-9954/2020 | 2020-07-06 | North America/USA/Texas/Houston | Houston Methodist Hospital | Houston Methodist Hospital | S. Wesley Long et al |
| EPI_ISL_780355 | USA/TX-HMH-MCoV-9961/2020 | 2020-07-06 | North America/USA/Texas/Houston | Houston Methodist Hospital | Houston Methodist Hospital | S. Wesley Long et al |
| EPI_ISL_780356 | USA/TX-HMH-MCoV-9963/2020 | 2020-07-06 | North America/USA/Texas/Houston | Houston Methodist Hospital | Houston Methodist Hospital | S. Wesley Long et al |
| EPI_ISL_780361 | USA/TX-HMH-MCoV-9973/2020 | 2020-07-06 | North America/USA/Texas/Houston | Houston Methodist Hospital | Houston Methodist Hospital | S. Wesley Long et al |
| EPI_ISL_780364 | USA/TX-HMH-MCoV-9977/2020 | 2020-07-06 | North America/USA/Texas/Houston | Houston Methodist Hospital | Houston Methodist Hospital | S. Wesley Long et al |

| | | | | | | |
|---|---|---|---|---|---|---|
| EPI_ISL_780365 | USA/TX-HMH-MCoV-9982/2020 | 2020-07-06 | North America/USA/Texas/Houston | Houston Methodist Hospital | Houston Methodist Hospital | S. Wesley Long et al |
| EPI_ISL_780369 | USA/TX-HMH-MCoV-9989/2020 | 2020-07-06 | North America/USA/Texas/Houston | Houston Methodist Hospital | Houston Methodist Hospital | S. Wesley Long et al |
| EPI_ISL_780372 | USA/TX-HMH-MCoV-9992/2020 | 2020-07-06 | North America/USA/Texas/Houston | Houston Methodist Hospital | Houston Methodist Hospital | S. Wesley Long et al |
| EPI_ISL_780381 | Italy/LOM-UniMI14/2020 | 2020-03-17 | Europe/Italy/Lombardy/ | Laboratory of Infectious Diseases, Department of Biomedical and Clinical Sciences L. Sacco, University of Milan | Laboratory of Infectious Diseases, Department of Biomedical and Clinical Sciences L. Sacco, University of Milan | Alessia Lai et al |
| EPI_ISL_780414 | Spain/MD-IBV-99012489/2020 | 2020-10-29 | Europe/Spain/Madrid/Madrid | Hospital General Universitario Gregorio Marav√±√≥n | SeqCOVID-SPAIN consortium/IBV(CSIC) | Dar√≠o Garc√≠a de Viedma et al |
| EPI_ISL_780418 | France/HDF-CERBA-0160-202011/202 | 2020-06-03 | Europe/France/Hauts de France/ | CERBA LAB | CERBA LAB | Roquebert B et al |
| EPI_ISL_780430 | USA/TX-HMH-MCoV-8515/2020 | 2020-07-01 | North America/USA/Texas/Houston | Houston Methodist Hospital | Houston Methodist Hospital | S. Wesley Long et al |
| EPI_ISL_780431 | USA/TX-HMH-MCoV-9169/2020 | 2020-07-02 | North America/USA/Texas/Houston | Houston Methodist Hospital | Houston Methodist Hospital | S. Wesley Long et al |
| EPI_ISL_780440 | USA/TX-HMH-MCoV-9487/2020 | 2020-07-03 | North America/USA/Texas/Houston | Houston Methodist Hospital | Houston Methodist Hospital | S. Wesley Long et al |
| EPI_ISL_780442 | USA/TX-HMH-MCoV-8546/2020 | 2020-07-01 | North America/USA/Texas/Houston | Houston Methodist Hospital | Houston Methodist Hospital | S. Wesley Long et al |
| EPI_ISL_780448 | USA/TX-HMH-MCoV-9521/2020 | 2020-07-03 | North America/USA/Texas/Houston | Houston Methodist Hospital | Houston Methodist Hospital | S. Wesley Long et al |
| EPI_ISL_780450 | USA/TX-HMH-MCoV-9215/2020 | 2020-07-02 | North America/USA/Texas/Houston | Houston Methodist Hospital | Houston Methodist Hospital | S. Wesley Long et al |

| | | | | | | |
|---|---|---|---|---|---|---|
| EPI_ISL_780451 | USA/TX-HMH-MCoV-8648/2020 | 2020-07-01 | North America/USA/Texas/Houston | Houston Methodist Hospital | Houston Methodist Hospital | S. Wesley Long et al |
| EPI_ISL_780452 | USA/TX-HMH-MCoV-9051/2020 | 2020-07-01 | North America/USA/Texas/Houston | Houston Methodist Hospital | Houston Methodist Hospital | S. Wesley Long et al |
| EPI_ISL_780453 | USA/TX-HMH-MCoV-9132/2020 | 2020-07-03 | North America/USA/Texas/Houston | Houston Methodist Hospital | Houston Methodist Hospital | S. Wesley Long et al |
| EPI_ISL_780455 | USA/TX-HMH-MCoV-8602/2020 | 2020-07-01 | North America/USA/Texas/Houston | Houston Methodist Hospital | Houston Methodist Hospital | S. Wesley Long et al |
| EPI_ISL_780456 | USA/TX-HMH-MCoV-9470/2020 | 2020-07-03 | North America/USA/Texas/Houston | Houston Methodist Hospital | Houston Methodist Hospital | S. Wesley Long et al |
| EPI_ISL_780458 | USA/TX-HMH-MCoV-8509/2020 | 2020-07-01 | North America/USA/Texas/Houston | Houston Methodist Hospital | Houston Methodist Hospital | S. Wesley Long et al |
| EPI_ISL_780459 | USA/TX-HMH-MCoV-8545/2020 | 2020-07-01 | North America/USA/Texas/Houston | Houston Methodist Hospital | Houston Methodist Hospital | S. Wesley Long et al |
| EPI_ISL_780460 | USA/TX-HMH-MCoV-8644/2020 | 2020-07-01 | North America/USA/Texas/Houston | Houston Methodist Hospital | Houston Methodist Hospital | S. Wesley Long et al |
| EPI_ISL_780462 | USA/TX-HMH-MCoV-8496/2020 | 2020-07-01 | North America/USA/Texas/Houston | Houston Methodist Hospital | Houston Methodist Hospital | S. Wesley Long et al |
| EPI_ISL_780467 | USA/TX-HMH-MCoV-9415/2020 | 2020-07-03 | North America/USA/Texas/Houston | Houston Methodist Hospital | Houston Methodist Hospital | S. Wesley Long et al |
| EPI_ISL_780468 | USA/TX-HMH-MCoV-9137/2020 | 2020-07-02 | North America/USA/Texas/Houston | Houston Methodist Hospital | Houston Methodist Hospital | S. Wesley Long et al |
| EPI_ISL_780469 | USA/TX-HMH-MCoV-9221/2020 | 2020-07-02 | North America/USA/Texas/Houston | Houston Methodist Hospital | Houston Methodist Hospital | S. Wesley Long et al |
| EPI_ISL_780473 | USA/TX-HMH-MCoV-8565/2020 | 2020-07-01 | North America/USA/Texas/Houston | Houston Methodist Hospital | Houston Methodist Hospital | S. Wesley Long et al |

| | | | | | | |
|---|---|---|---|---|---|---|
| EPI_ISL_780475 | USA/TX-HMH-MCoV-8582/2020 | 2020-07-01 | North America/USA/Texas/Houston | Houston Methodist Hospital | Houston Methodist Hospital | S. Wesley Long et al |
| EPI_ISL_780480 | USA/TX-HMH-MCoV-9365/2020 | 2020-07-03 | North America/USA/Texas/Houston | Houston Methodist Hospital | Houston Methodist Hospital | S. Wesley Long et al |
| EPI_ISL_780494 | USA/TX-HMH-MCoV-8517/2020 | 2020-07-01 | North America/USA/Texas/Houston | Houston Methodist Hospital | Houston Methodist Hospital | S. Wesley Long et al |
| EPI_ISL_780498 | USA/TX-HMH-MCoV-8781/2020 | 2020-07-01 | North America/USA/Texas/Houston | Houston Methodist Hospital | Houston Methodist Hospital | S. Wesley Long et al |
| EPI_ISL_780501 | USA/TX-HMH-MCoV-8997/2020 | 2020-07-02 | North America/USA/Texas/Houston | Houston Methodist Hospital | Houston Methodist Hospital | S. Wesley Long et al |
| EPI_ISL_780504 | USA/TX-HMH-MCoV-9309/2020 | 2020-07-03 | North America/USA/Texas/Houston | Houston Methodist Hospital | Houston Methodist Hospital | S. Wesley Long et al |
| EPI_ISL_780506 | USA/TX-HMH-MCoV-8973/2020 | 2020-07-02 | North America/USA/Texas/Houston | Houston Methodist Hospital | Houston Methodist Hospital | S. Wesley Long et al |
| EPI_ISL_780510 | USA/TX-HMH-MCoV-9213/2020 | 2020-07-02 | North America/USA/Texas/Houston | Houston Methodist Hospital | Houston Methodist Hospital | S. Wesley Long et al |
| EPI_ISL_780518 | USA/TX-HMH-MCoV-9227/2020 | 2020-07-02 | North America/USA/Texas/Houston | Houston Methodist Hospital | Houston Methodist Hospital | S. Wesley Long et al |
| EPI_ISL_780523 | USA/TX-HMH-MCoV-8541/2020 | 2020-07-01 | North America/USA/Texas/Houston | Houston Methodist Hospital | Houston Methodist Hospital | S. Wesley Long et al |
| EPI_ISL_780526 | USA/TX-HMH-MCoV-9500/2020 | 2020-07-03 | North America/USA/Texas/Houston | Houston Methodist Hospital | Houston Methodist Hospital | S. Wesley Long et al |
| EPI_ISL_780527 | USA/TX-HMH-MCoV-9494/2020 | 2020-07-03 | North America/USA/Texas/Houston | Houston Methodist Hospital | Houston Methodist Hospital | S. Wesley Long et al |
| EPI_ISL_780531 | USA/TX-HMH-MCoV-9117/2020 | 2020-07-02 | North America/USA/Texas/Houston | Houston Methodist Hospital | Houston Methodist Hospital | S. Wesley Long et al |

| | | | | | | |
|---|---|---|---|---|---|---|
| EPI_ISL_780533 | USA/TX-HMH-MCoV-8584/2020 | 2020-07-01 | North America/USA/Texas/Houston | Houston Methodist Hospital | Houston Methodist Hospital | S. Wesley Long et al |
| EPI_ISL_780536 | USA/TX-HMH-MCoV-8604/2020 | 2020-07-01 | North America/USA/Texas/Houston | Houston Methodist Hospital | Houston Methodist Hospital | S. Wesley Long et al |
| EPI_ISL_780546 | USA/TX-HMH-MCoV-9264/2020 | 2020-07-03 | North America/USA/Texas/Houston | Houston Methodist Hospital | Houston Methodist Hospital | S. Wesley Long et al |
| EPI_ISL_780555 | USA/TX-HMH-MCoV-8450/2020 | 2020-07-02 | North America/USA/Texas/Houston | Houston Methodist Hospital | Houston Methodist Hospital | S. Wesley Long et al |
| EPI_ISL_780560 | USA/TX-HMH-MCoV-9239/2020 | 2020-07-03 | North America/USA/Texas/Houston | Houston Methodist Hospital | Houston Methodist Hospital | S. Wesley Long et al |
| EPI_ISL_780573 | USA/TX-HMH-MCoV-8461/2020 | 2020-07-01 | North America/USA/Texas/Houston | Houston Methodist Hospital | Houston Methodist Hospital | S. Wesley Long et al |
| EPI_ISL_780574 | USA/TX-HMH-MCoV-8462/2020 | 2020-07-01 | North America/USA/Texas/Houston | Houston Methodist Hospital | Houston Methodist Hospital | S. Wesley Long et al |
| EPI_ISL_780578 | USA/TX-HMH-MCoV-8467/2020 | 2020-07-01 | North America/USA/Texas/Houston | Houston Methodist Hospital | Houston Methodist Hospital | S. Wesley Long et al |
| EPI_ISL_780584 | USA/TX-HMH-MCoV-8479/2020 | 2020-07-01 | North America/USA/Texas/Houston | Houston Methodist Hospital | Houston Methodist Hospital | S. Wesley Long et al |
| EPI_ISL_780585 | USA/TX-HMH-MCoV-8480/2020 | 2020-07-01 | North America/USA/Texas/Houston | Houston Methodist Hospital | Houston Methodist Hospital | S. Wesley Long et al |
| EPI_ISL_780589 | USA/TX-HMH-MCoV-8490/2020 | 2020-07-01 | North America/USA/Texas/Houston | Houston Methodist Hospital | Houston Methodist Hospital | S. Wesley Long et al |
| EPI_ISL_780592 | USA/TX-HMH-MCoV-8495/2020 | 2020-07-01 | North America/USA/Texas/Houston | Houston Methodist Hospital | Houston Methodist Hospital | S. Wesley Long et al |
| EPI_ISL_780597 | USA/TX-HMH-MCoV-8504/2020 | 2020-07-01 | North America/USA/Texas/Houston | Houston Methodist Hospital | Houston Methodist Hospital | S. Wesley Long et al |

| | | | | | | |
|---|---|---|---|---|---|---|
| EPI_ISL_780600 | USA/TX-HMH-MCoV-8513/2020 | 2020-07-01 | North America/USA/Texas/Houston | Houston Methodist Hospital | Houston Methodist Hospital | S. Wesley Long et al |
| EPI_ISL_780623 | USA/TX-HMH-MCoV-8551/2020 | 2020-07-01 | North America/USA/Texas/Houston | Houston Methodist Hospital | Houston Methodist Hospital | S. Wesley Long et al |
| EPI_ISL_780625 | USA/TX-HMH-MCoV-8556/2020 | 2020-07-01 | North America/USA/Texas/Houston | Houston Methodist Hospital | Houston Methodist Hospital | S. Wesley Long et al |
| EPI_ISL_780626 | USA/TX-HMH-MCoV-8558/2020 | 2020-07-02 | North America/USA/Texas/Houston | Houston Methodist Hospital | Houston Methodist Hospital | S. Wesley Long et al |
| EPI_ISL_780636 | USA/TX-HMH-MCoV-8587/2020 | 2020-07-01 | North America/USA/Texas/Houston | Houston Methodist Hospital | Houston Methodist Hospital | S. Wesley Long et al |
| EPI_ISL_780646 | USA/TX-HMH-MCoV-8605/2020 | 2020-07-01 | North America/USA/Texas/Houston | Houston Methodist Hospital | Houston Methodist Hospital | S. Wesley Long et al |
| EPI_ISL_780650 | USA/TX-HMH-MCoV-8611/2020 | 2020-07-01 | North America/USA/Texas/Houston | Houston Methodist Hospital | Houston Methodist Hospital | S. Wesley Long et al |
| EPI_ISL_780651 | USA/TX-HMH-MCoV-8613/2020 | 2020-07-01 | North America/USA/Texas/Houston | Houston Methodist Hospital | Houston Methodist Hospital | S. Wesley Long et al |
| EPI_ISL_780660 | USA/TX-HMH-MCoV-8626/2020 | 2020-07-01 | North America/USA/Texas/Houston | Houston Methodist Hospital | Houston Methodist Hospital | S. Wesley Long et al |
| EPI_ISL_780662 | USA/TX-HMH-MCoV-8631/2020 | 2020-07-01 | North America/USA/Texas/Houston | Houston Methodist Hospital | Houston Methodist Hospital | S. Wesley Long et al |
| EPI_ISL_780663 | USA/TX-HMH-MCoV-8633/2020 | 2020-07-01 | North America/USA/Texas/Houston | Houston Methodist Hospital | Houston Methodist Hospital | S. Wesley Long et al |
| EPI_ISL_780666 | USA/TX-HMH-MCoV-8637/2020 | 2020-07-01 | North America/USA/Texas/Houston | Houston Methodist Hospital | Houston Methodist Hospital | S. Wesley Long et al |
| EPI_ISL_780667 | USA/TX-HMH-MCoV-8638/2020 | 2020-07-01 | North America/USA/Texas/Houston | Houston Methodist Hospital | Houston Methodist Hospital | S. Wesley Long et al |

| | | | | | | |
|---|---|---|---|---|---|---|
| EPI_ISL_780672 | USA/TX-HMH-MCoV-8642/2020 | 2020-07-01 | North America/USA/Texas/Houston | Houston Methodist Hospital | Houston Methodist Hospital | S. Wesley Long et al |
| EPI_ISL_780680 | USA/TX-HMH-MCoV-8654/2020 | 2020-07-01 | North America/USA/Texas/Houston | Houston Methodist Hospital | Houston Methodist Hospital | S. Wesley Long et al |
| EPI_ISL_780691 | USA/TX-HMH-MCoV-8673/2020 | 2020-07-01 | North America/USA/Texas/Houston | Houston Methodist Hospital | Houston Methodist Hospital | S. Wesley Long et al |
| EPI_ISL_780701 | USA/TX-HMH-MCoV-8688/2020 | 2020-07-02 | North America/USA/Texas/Houston | Houston Methodist Hospital | Houston Methodist Hospital | S. Wesley Long et al |
| EPI_ISL_780705 | USA/TX-HMH-MCoV-8693/2020 | 2020-07-01 | North America/USA/Texas/Houston | Houston Methodist Hospital | Houston Methodist Hospital | S. Wesley Long et al |
| EPI_ISL_780707 | USA/TX-HMH-MCoV-8696/2020 | 2020-07-01 | North America/USA/Texas/Houston | Houston Methodist Hospital | Houston Methodist Hospital | S. Wesley Long et al |
| EPI_ISL_780711 | USA/TX-HMH-MCoV-8707/2020 | 2020-07-01 | North America/USA/Texas/Houston | Houston Methodist Hospital | Houston Methodist Hospital | S. Wesley Long et al |
| EPI_ISL_780713 | USA/TX-HMH-MCoV-8710/2020 | 2020-07-02 | North America/USA/Texas/Houston | Houston Methodist Hospital | Houston Methodist Hospital | S. Wesley Long et al |
| EPI_ISL_780717 | USA/TX-HMH-MCoV-8717/2020 | 2020-07-01 | North America/USA/Texas/Houston | Houston Methodist Hospital | Houston Methodist Hospital | S. Wesley Long et al |
| EPI_ISL_780721 | USA/TX-HMH-MCoV-8726/2020 | 2020-07-01 | North America/USA/Texas/Houston | Houston Methodist Hospital | Houston Methodist Hospital | S. Wesley Long et al |
| EPI_ISL_780726 | USA/TX-HMH-MCoV-8737/2020 | 2020-07-01 | North America/USA/Texas/Houston | Houston Methodist Hospital | Houston Methodist Hospital | S. Wesley Long et al |
| EPI_ISL_780729 | USA/TX-HMH-MCoV-8742/2020 | 2020-07-01 | North America/USA/Texas/Houston | Houston Methodist Hospital | Houston Methodist Hospital | S. Wesley Long et al |
| EPI_ISL_780737 | USA/TX-HMH-MCoV-8758/2020 | 2020-07-01 | North America/USA/Texas/Houston | Houston Methodist Hospital | Houston Methodist Hospital | S. Wesley Long et al |

| | | | | | | |
|---|---|---|---|---|---|---|
| EPI_ISL_780741 | USA/TX-HMH-MCoV-8766/2020 | 2020-07-01 | North America/USA/Texas/Houston | Houston Methodist Hospital | Houston Methodist Hospital | S. Wesley Long et al |
| EPI_ISL_780742 | USA/TX-HMH-MCoV-8767/2020 | 2020-07-01 | North America/USA/Texas/Houston | Houston Methodist Hospital | Houston Methodist Hospital | S. Wesley Long et al |
| EPI_ISL_780744 | USA/TX-HMH-MCoV-8770/2020 | 2020-07-01 | North America/USA/Texas/Houston | Houston Methodist Hospital | Houston Methodist Hospital | S. Wesley Long et al |
| EPI_ISL_780745 | USA/TX-HMH-MCoV-8773/2020 | 2020-07-02 | North America/USA/Texas/Houston | Houston Methodist Hospital | Houston Methodist Hospital | S. Wesley Long et al |
| EPI_ISL_780749 | USA/TX-HMH-MCoV-8783/2020 | 2020-07-01 | North America/USA/Texas/Houston | Houston Methodist Hospital | Houston Methodist Hospital | S. Wesley Long et al |
| EPI_ISL_780751 | USA/TX-HMH-MCoV-8785/2020 | 2020-07-02 | North America/USA/Texas/Houston | Houston Methodist Hospital | Houston Methodist Hospital | S. Wesley Long et al |
| EPI_ISL_780753 | USA/TX-HMH-MCoV-8787/2020 | 2020-07-01 | North America/USA/Texas/Houston | Houston Methodist Hospital | Houston Methodist Hospital | S. Wesley Long et al |
| EPI_ISL_780756 | USA/TX-HMH-MCoV-8790/2020 | 2020-07-01 | North America/USA/Texas/Houston | Houston Methodist Hospital | Houston Methodist Hospital | S. Wesley Long et al |
| EPI_ISL_780758 | USA/TX-HMH-MCoV-8792/2020 | 2020-07-01 | North America/USA/Texas/Houston | Houston Methodist Hospital | Houston Methodist Hospital | S. Wesley Long et al |
| EPI_ISL_780764 | USA/TX-HMH-MCoV-8800/2020 | 2020-07-01 | North America/USA/Texas/Houston | Houston Methodist Hospital | Houston Methodist Hospital | S. Wesley Long et al |
| EPI_ISL_780766 | USA/TX-HMH-MCoV-8803/2020 | 2020-07-01 | North America/USA/Texas/Houston | Houston Methodist Hospital | Houston Methodist Hospital | S. Wesley Long et al |
| EPI_ISL_780769 | USA/TX-HMH-MCoV-8807/2020 | 2020-07-01 | North America/USA/Texas/Houston | Houston Methodist Hospital | Houston Methodist Hospital | S. Wesley Long et al |
| EPI_ISL_780772 | USA/TX-HMH-MCoV-8814/2020 | 2020-07-01 | North America/USA/Texas/Houston | Houston Methodist Hospital | Houston Methodist Hospital | S. Wesley Long et al |

| | | | | | | |
|---|---|---|---|---|---|---|
| EPI_ISL_780776 | USA/TX-HMH-MCoV-8823/2020 | 2020-07-01 | North America/USA/Texas/Houston | Houston Methodist Hospital | Houston Methodist Hospital | S. Wesley Long et al |
| EPI_ISL_780777 | USA/TX-HMH-MCoV-8824/2020 | 2020-07-01 | North America/USA/Texas/Houston | Houston Methodist Hospital | Houston Methodist Hospital | S. Wesley Long et al |
| EPI_ISL_780785 | USA/TX-HMH-MCoV-8837/2020 | 2020-07-01 | North America/USA/Texas/Houston | Houston Methodist Hospital | Houston Methodist Hospital | S. Wesley Long et al |
| EPI_ISL_780787 | USA/TX-HMH-MCoV-8839/2020 | 2020-07-01 | North America/USA/Texas/Houston | Houston Methodist Hospital | Houston Methodist Hospital | S. Wesley Long et al |
| EPI_ISL_780788 | USA/TX-HMH-MCoV-8840/2020 | 2020-07-01 | North America/USA/Texas/Houston | Houston Methodist Hospital | Houston Methodist Hospital | S. Wesley Long et al |
| EPI_ISL_780792 | USA/TX-HMH-MCoV-8846/2020 | 2020-07-02 | North America/USA/Texas/Houston | Houston Methodist Hospital | Houston Methodist Hospital | S. Wesley Long et al |
| EPI_ISL_780796 | USA/TX-HMH-MCoV-8852/2020 | 2020-07-02 | North America/USA/Texas/Houston | Houston Methodist Hospital | Houston Methodist Hospital | S. Wesley Long et al |
| EPI_ISL_780802 | USA/TX-HMH-MCoV-8865/2020 | 2020-07-02 | North America/USA/Texas/Houston | Houston Methodist Hospital | Houston Methodist Hospital | S. Wesley Long et al |
| EPI_ISL_780804 | USA/TX-HMH-MCoV-8876/2020 | 2020-07-02 | North America/USA/Texas/Houston | Houston Methodist Hospital | Houston Methodist Hospital | S. Wesley Long et al |
| EPI_ISL_780808 | USA/TX-HMH-MCoV-8881/2020 | 2020-07-02 | North America/USA/Texas/Houston | Houston Methodist Hospital | Houston Methodist Hospital | S. Wesley Long et al |
| EPI_ISL_780812 | USA/TX-HMH-MCoV-8888/2020 | 2020-07-02 | North America/USA/Texas/Houston | Houston Methodist Hospital | Houston Methodist Hospital | S. Wesley Long et al |
| EPI_ISL_780815 | USA/TX-HMH-MCoV-8894/2020 | 2020-07-02 | North America/USA/Texas/Houston | Houston Methodist Hospital | Houston Methodist Hospital | S. Wesley Long et al |
| EPI_ISL_780818 | USA/TX-HMH-MCoV-8898/2020 | 2020-07-02 | North America/USA/Texas/Houston | Houston Methodist Hospital | Houston Methodist Hospital | S. Wesley Long et al |

| | | | | | | |
|---|---|---|---|---|---|---|
| EPI_ISL_780821 | USA/TX-HMH-MCoV-8903/2020 | 2020-07-02 | North America/USA/Texas/Houston | Houston Methodist Hospital | Houston Methodist Hospital | S. Wesley Long et al |
| EPI_ISL_780829 | USA/TX-HMH-MCoV-8916/2020 | 2020-07-02 | North America/USA/Texas/Houston | Houston Methodist Hospital | Houston Methodist Hospital | S. Wesley Long et al |
| EPI_ISL_780834 | USA/TX-HMH-MCoV-8923/2020 | 2020-07-02 | North America/USA/Texas/Houston | Houston Methodist Hospital | Houston Methodist Hospital | S. Wesley Long et al |
| EPI_ISL_780837 | USA/TX-HMH-MCoV-8929/2020 | 2020-07-01 | North America/USA/Texas/Houston | Houston Methodist Hospital | Houston Methodist Hospital | S. Wesley Long et al |
| EPI_ISL_780840 | USA/TX-HMH-MCoV-8932/2020 | 2020-07-02 | North America/USA/Texas/Houston | Houston Methodist Hospital | Houston Methodist Hospital | S. Wesley Long et al |
| EPI_ISL_780842 | USA/TX-HMH-MCoV-8934/2020 | 2020-07-02 | North America/USA/Texas/Houston | Houston Methodist Hospital | Houston Methodist Hospital | S. Wesley Long et al |
| EPI_ISL_780844 | USA/TX-HMH-MCoV-8936/2020 | 2020-07-02 | North America/USA/Texas/Houston | Houston Methodist Hospital | Houston Methodist Hospital | S. Wesley Long et al |
| EPI_ISL_780845 | USA/TX-HMH-MCoV-8938/2020 | 2020-07-02 | North America/USA/Texas/Houston | Houston Methodist Hospital | Houston Methodist Hospital | S. Wesley Long et al |
| EPI_ISL_780847 | USA/TX-HMH-MCoV-8941/2020 | 2020-07-02 | North America/USA/Texas/Houston | Houston Methodist Hospital | Houston Methodist Hospital | S. Wesley Long et al |
| EPI_ISL_780848 | USA/TX-HMH-MCoV-8942/2020 | 2020-07-02 | North America/USA/Texas/Houston | Houston Methodist Hospital | Houston Methodist Hospital | S. Wesley Long et al |
| EPI_ISL_780852 | USA/TX-HMH-MCoV-8946/2020 | 2020-07-02 | North America/USA/Texas/Houston | Houston Methodist Hospital | Houston Methodist Hospital | S. Wesley Long et al |
| EPI_ISL_780854 | USA/TX-HMH-MCoV-8953/2020 | 2020-07-02 | North America/USA/Texas/Houston | Houston Methodist Hospital | Houston Methodist Hospital | S. Wesley Long et al |
| EPI_ISL_780855 | USA/TX-HMH-MCoV-8955/2020 | 2020-07-02 | North America/USA/Texas/Houston | Houston Methodist Hospital | Houston Methodist Hospital | S. Wesley Long et al |

| | | | | | | |
|---|---|---|---|---|---|---|
| EPI_ISL_780857 | USA/TX-HMH-MCoV-8957/2020 | 2020-07-02 | North America/USA/Texas/Houston | Houston Methodist Hospital | Houston Methodist Hospital | S. Wesley Long et al |
| EPI_ISL_780862 | USA/TX-HMH-MCoV-8962/2020 | 2020-07-02 | North America/USA/Texas/Houston | Houston Methodist Hospital | Houston Methodist Hospital | S. Wesley Long et al |
| EPI_ISL_780866 | USA/TX-HMH-MCoV-8970/2020 | 2020-07-02 | North America/USA/Texas/Houston | Houston Methodist Hospital | Houston Methodist Hospital | S. Wesley Long et al |
| EPI_ISL_780871 | USA/TX-HMH-MCoV-8977/2020 | 2020-07-02 | North America/USA/Texas/Houston | Houston Methodist Hospital | Houston Methodist Hospital | S. Wesley Long et al |
| EPI_ISL_780872 | USA/TX-HMH-MCoV-8979/2020 | 2020-07-02 | North America/USA/Texas/Houston | Houston Methodist Hospital | Houston Methodist Hospital | S. Wesley Long et al |
| EPI_ISL_780879 | USA/TX-HMH-MCoV-8993/2020 | 2020-07-01 | North America/USA/Texas/Houston | Houston Methodist Hospital | Houston Methodist Hospital | S. Wesley Long et al |
| EPI_ISL_780887 | USA/TX-HMH-MCoV-9003/2020 | 2020-07-02 | North America/USA/Texas/Houston | Houston Methodist Hospital | Houston Methodist Hospital | S. Wesley Long et al |
| EPI_ISL_780890 | USA/TX-HMH-MCoV-9006/2020 | 2020-07-02 | North America/USA/Texas/Houston | Houston Methodist Hospital | Houston Methodist Hospital | S. Wesley Long et al |
| EPI_ISL_780897 | USA/TX-HMH-MCoV-9014/2020 | 2020-07-02 | North America/USA/Texas/Houston | Houston Methodist Hospital | Houston Methodist Hospital | S. Wesley Long et al |
| EPI_ISL_780904 | USA/TX-HMH-MCoV-9028/2020 | 2020-07-02 | North America/USA/Texas/Houston | Houston Methodist Hospital | Houston Methodist Hospital | S. Wesley Long et al |
| EPI_ISL_780907 | USA/TX-HMH-MCoV-9031/2020 | 2020-07-02 | North America/USA/Texas/Houston | Houston Methodist Hospital | Houston Methodist Hospital | S. Wesley Long et al |
| EPI_ISL_780912 | USA/TX-HMH-MCoV-9037/2020 | 2020-07-02 | North America/USA/Texas/Houston | Houston Methodist Hospital | Houston Methodist Hospital | S. Wesley Long et al |
| EPI_ISL_780914 | USA/TX-HMH-MCoV-9041/2020 | 2020-07-03 | North America/USA/Texas/Houston | Houston Methodist Hospital | Houston Methodist Hospital | S. Wesley Long et al |

| | | | | | | |
|---|---|---|---|---|---|---|
| EPI_ISL_780925 | USA/TX-HMH-MCoV-9061/2020 | 2020-07-02 | North America/USA/Texas/Houston | Houston Methodist Hospital | Houston Methodist Hospital | S. Wesley Long et al |
| EPI_ISL_780928 | USA/TX-HMH-MCoV-9072/2020 | 2020-07-02 | North America/USA/Texas/Houston | Houston Methodist Hospital | Houston Methodist Hospital | S. Wesley Long et al |
| EPI_ISL_780931 | USA/TX-HMH-MCoV-9078/2020 | 2020-07-02 | North America/USA/Texas/Houston | Houston Methodist Hospital | Houston Methodist Hospital | S. Wesley Long et al |
| EPI_ISL_780932 | USA/TX-HMH-MCoV-9080/2020 | 2020-07-02 | North America/USA/Texas/Houston | Houston Methodist Hospital | Houston Methodist Hospital | S. Wesley Long et al |
| EPI_ISL_780935 | USA/TX-HMH-MCoV-9085/2020 | 2020-07-02 | North America/USA/Texas/Houston | Houston Methodist Hospital | Houston Methodist Hospital | S. Wesley Long et al |
| EPI_ISL_780939 | USA/TX-HMH-MCoV-9094/2020 | 2020-07-02 | North America/USA/Texas/Houston | Houston Methodist Hospital | Houston Methodist Hospital | S. Wesley Long et al |
| EPI_ISL_780944 | USA/TX-HMH-MCoV-9103/2020 | 2020-07-02 | North America/USA/Texas/Houston | Houston Methodist Hospital | Houston Methodist Hospital | S. Wesley Long et al |
| EPI_ISL_780945 | USA/TX-HMH-MCoV-9104/2020 | 2020-07-03 | North America/USA/Texas/Houston | Houston Methodist Hospital | Houston Methodist Hospital | S. Wesley Long et al |
| EPI_ISL_780948 | USA/TX-HMH-MCoV-9115/2020 | 2020-07-02 | North America/USA/Texas/Houston | Houston Methodist Hospital | Houston Methodist Hospital | S. Wesley Long et al |
| EPI_ISL_780949 | USA/TX-HMH-MCoV-9116/2020 | 2020-07-03 | North America/USA/Texas/Houston | Houston Methodist Hospital | Houston Methodist Hospital | S. Wesley Long et al |
| EPI_ISL_780953 | USA/TX-HMH-MCoV-9123/2020 | 2020-07-02 | North America/USA/Texas/Houston | Houston Methodist Hospital | Houston Methodist Hospital | S. Wesley Long et al |
| EPI_ISL_780956 | USA/TX-HMH-MCoV-9126/2020 | 2020-07-02 | North America/USA/Texas/Houston | Houston Methodist Hospital | Houston Methodist Hospital | S. Wesley Long et al |
| EPI_ISL_780958 | USA/TX-HMH-MCoV-9128/2020 | 2020-07-02 | North America/USA/Texas/Houston | Houston Methodist Hospital | Houston Methodist Hospital | S. Wesley Long et al |

| | | | | | | |
|---|---|---|---|---|---|---|
| EPI_ISL_780965 | USA/TX-HMH-MCoV-9140/2020 | 2020-07-02 | North America/USA/Texas/Houston | Houston Methodist Hospital | Houston Methodist Hospital | S. Wesley Long et al |
| EPI_ISL_780967 | USA/TX-HMH-MCoV-9143/2020 | 2020-07-02 | North America/USA/Texas/Houston | Houston Methodist Hospital | Houston Methodist Hospital | S. Wesley Long et al |
| EPI_ISL_780982 | USA/TX-HMH-MCoV-9176/2020 | 2020-07-02 | North America/USA/Texas/Houston | Houston Methodist Hospital | Houston Methodist Hospital | S. Wesley Long et al |
| EPI_ISL_780986 | USA/TX-HMH-MCoV-9183/2020 | 2020-07-02 | North America/USA/Texas/Houston | Houston Methodist Hospital | Houston Methodist Hospital | S. Wesley Long et al |
| EPI_ISL_780987 | USA/TX-HMH-MCoV-9184/2020 | 2020-07-02 | North America/USA/Texas/Houston | Houston Methodist Hospital | Houston Methodist Hospital | S. Wesley Long et al |
| EPI_ISL_780992 | USA/TX-HMH-MCoV-9195/2020 | 2020-07-02 | North America/USA/Texas/Houston | Houston Methodist Hospital | Houston Methodist Hospital | S. Wesley Long et al |
| EPI_ISL_780996 | USA/TX-HMH-MCoV-9200/2020 | 2020-07-02 | North America/USA/Texas/Houston | Houston Methodist Hospital | Houston Methodist Hospital | S. Wesley Long et al |
| EPI_ISL_781007 | USA/TX-HMH-MCoV-9225/2020 | 2020-07-02 | North America/USA/Texas/Houston | Houston Methodist Hospital | Houston Methodist Hospital | S. Wesley Long et al |
| EPI_ISL_781014 | USA/TX-HMH-MCoV-9236/2020 | 2020-07-03 | North America/USA/Texas/Houston | Houston Methodist Hospital | Houston Methodist Hospital | S. Wesley Long et al |
| EPI_ISL_781019 | USA/TX-HMH-MCoV-9256/2020 | 2020-07-03 | North America/USA/Texas/Houston | Houston Methodist Hospital | Houston Methodist Hospital | S. Wesley Long et al |
| EPI_ISL_781021 | USA/TX-HMH-MCoV-9258/2020 | 2020-07-02 | North America/USA/Texas/Houston | Houston Methodist Hospital | Houston Methodist Hospital | S. Wesley Long et al |
| EPI_ISL_781024 | USA/TX-HMH-MCoV-9268/2020 | 2020-07-04 | North America/USA/Texas/Houston | Houston Methodist Hospital | Houston Methodist Hospital | S. Wesley Long et al |
| EPI_ISL_781025 | USA/TX-HMH-MCoV-9271/2020 | 2020-07-03 | North America/USA/Texas/Houston | Houston Methodist Hospital | Houston Methodist Hospital | S. Wesley Long et al |

| | | | | | | |
|---|---|---|---|---|---|---|
| EPI_ISL_781026 | USA/TX-HMH-MCoV-9273/2020 | 2020-07-03 | North America/USA/Texas/Houston | Houston Methodist Hospital | Houston Methodist Hospital | S. Wesley Long et al |
| EPI_ISL_781029 | USA/TX-HMH-MCoV-9277/2020 | 2020-07-03 | North America/USA/Texas/Houston | Houston Methodist Hospital | Houston Methodist Hospital | S. Wesley Long et al |
| EPI_ISL_781038 | USA/TX-HMH-MCoV-9287/2020 | 2020-07-03 | North America/USA/Texas/Houston | Houston Methodist Hospital | Houston Methodist Hospital | S. Wesley Long et al |
| EPI_ISL_781039 | USA/TX-HMH-MCoV-9288/2020 | 2020-07-03 | North America/USA/Texas/Houston | Houston Methodist Hospital | Houston Methodist Hospital | S. Wesley Long et al |
| EPI_ISL_781048 | USA/TX-HMH-MCoV-9303/2020 | 2020-07-03 | North America/USA/Texas/Houston | Houston Methodist Hospital | Houston Methodist Hospital | S. Wesley Long et al |
| EPI_ISL_781051 | USA/TX-HMH-MCoV-9308/2020 | 2020-07-03 | North America/USA/Texas/Houston | Houston Methodist Hospital | Houston Methodist Hospital | S. Wesley Long et al |
| EPI_ISL_781056 | USA/TX-HMH-MCoV-9315/2020 | 2020-07-03 | North America/USA/Texas/Houston | Houston Methodist Hospital | Houston Methodist Hospital | S. Wesley Long et al |
| EPI_ISL_781059 | USA/TX-HMH-MCoV-9318/2020 | 2020-07-03 | North America/USA/Texas/Houston | Houston Methodist Hospital | Houston Methodist Hospital | S. Wesley Long et al |
| EPI_ISL_781061 | USA/TX-HMH-MCoV-9322/2020 | 2020-07-03 | North America/USA/Texas/Houston | Houston Methodist Hospital | Houston Methodist Hospital | S. Wesley Long et al |
| EPI_ISL_781069 | USA/TX-HMH-MCoV-9342/2020 | 2020-07-03 | North America/USA/Texas/Houston | Houston Methodist Hospital | Houston Methodist Hospital | S. Wesley Long et al |
| EPI_ISL_781076 | USA/TX-HMH-MCoV-9358/2020 | 2020-07-03 | North America/USA/Texas/Houston | Houston Methodist Hospital | Houston Methodist Hospital | S. Wesley Long et al |
| EPI_ISL_781083 | USA/TX-HMH-MCoV-9370/2020 | 2020-07-03 | North America/USA/Texas/Houston | Houston Methodist Hospital | Houston Methodist Hospital | S. Wesley Long et al |
| EPI_ISL_781088 | USA/TX-HMH-MCoV-9383/2020 | 2020-07-03 | North America/USA/Texas/Houston | Houston Methodist Hospital | Houston Methodist Hospital | S. Wesley Long et al |

| | | | | | | |
|---|---|---|---|---|---|---|
| EPI_ISL_781097 | USA/TX-HMH-MCoV-9399/2020 | 2020-07-02 | North America/USA/Texas/Houston | Houston Methodist Hospital | Houston Methodist Hospital | S. Wesley Long et al |
| EPI_ISL_781100 | USA/TX-HMH-MCoV-9402/2020 | 2020-07-03 | North America/USA/Texas/Houston | Houston Methodist Hospital | Houston Methodist Hospital | S. Wesley Long et al |
| EPI_ISL_781101 | USA/TX-HMH-MCoV-9403/2020 | 2020-07-03 | North America/USA/Texas/Houston | Houston Methodist Hospital | Houston Methodist Hospital | S. Wesley Long et al |
| EPI_ISL_781105 | USA/TX-HMH-MCoV-9409/2020 | 2020-07-03 | North America/USA/Texas/Houston | Houston Methodist Hospital | Houston Methodist Hospital | S. Wesley Long et al |
| EPI_ISL_781108 | USA/TX-HMH-MCoV-9414/2020 | 2020-07-03 | North America/USA/Texas/Houston | Houston Methodist Hospital | Houston Methodist Hospital | S. Wesley Long et al |
| EPI_ISL_781109 | USA/TX-HMH-MCoV-9416/2020 | 2020-07-03 | North America/USA/Texas/Houston | Houston Methodist Hospital | Houston Methodist Hospital | S. Wesley Long et al |
| EPI_ISL_781115 | USA/TX-HMH-MCoV-9426/2020 | 2020-07-03 | North America/USA/Texas/Houston | Houston Methodist Hospital | Houston Methodist Hospital | S. Wesley Long et al |
| EPI_ISL_781118 | USA/TX-HMH-MCoV-9430/2020 | 2020-07-03 | North America/USA/Texas/Houston | Houston Methodist Hospital | Houston Methodist Hospital | S. Wesley Long et al |
| EPI_ISL_781119 | USA/TX-HMH-MCoV-9431/2020 | 2020-07-03 | North America/USA/Texas/Houston | Houston Methodist Hospital | Houston Methodist Hospital | S. Wesley Long et al |
| EPI_ISL_781123 | USA/TX-HMH-MCoV-9441/2020 | 2020-07-03 | North America/USA/Texas/Houston | Houston Methodist Hospital | Houston Methodist Hospital | S. Wesley Long et al |
| EPI_ISL_781134 | USA/TX-HMH-MCoV-9461/2020 | 2020-07-03 | North America/USA/Texas/Houston | Houston Methodist Hospital | Houston Methodist Hospital | S. Wesley Long et al |
| EPI_ISL_781136 | USA/TX-HMH-MCoV-9463/2020 | 2020-07-03 | North America/USA/Texas/Houston | Houston Methodist Hospital | Houston Methodist Hospital | S. Wesley Long et al |
| EPI_ISL_781139 | USA/TX-HMH-MCoV-9466/2020 | 2020-07-03 | North America/USA/Texas/Houston | Houston Methodist Hospital | Houston Methodist Hospital | S. Wesley Long et al |

| | | | | | | |
|---|---|---|---|---|---|---|
| EPI_ISL_781149 | USA/TX-HMH-MCoV-9485/2020 | 2020-07-03 | North America/USA/Texas/Houston | Houston Methodist Hospital | Houston Methodist Hospital | S. Wesley Long et al |
| EPI_ISL_781150 | USA/TX-HMH-MCoV-9486/2020 | 2020-07-03 | North America/USA/Texas/Houston | Houston Methodist Hospital | Houston Methodist Hospital | S. Wesley Long et al |
| EPI_ISL_781152 | USA/TX-HMH-MCoV-9492/2020 | 2020-07-03 | North America/USA/Texas/Houston | Houston Methodist Hospital | Houston Methodist Hospital | S. Wesley Long et al |
| EPI_ISL_781153 | USA/TX-HMH-MCoV-9501/2020 | 2020-07-03 | North America/USA/Texas/Houston | Houston Methodist Hospital | Houston Methodist Hospital | S. Wesley Long et al |
| EPI_ISL_781154 | USA/TX-HMH-MCoV-9502/2020 | 2020-07-03 | North America/USA/Texas/Houston | Houston Methodist Hospital | Houston Methodist Hospital | S. Wesley Long et al |
| EPI_ISL_781155 | USA/TX-HMH-MCoV-9504/2020 | 2020-07-03 | North America/USA/Texas/Houston | Houston Methodist Hospital | Houston Methodist Hospital | S. Wesley Long et al |
| EPI_ISL_781157 | USA/TX-HMH-MCoV-9511/2020 | 2020-07-03 | North America/USA/Texas/Houston | Houston Methodist Hospital | Houston Methodist Hospital | S. Wesley Long et al |
| EPI_ISL_781165 | USA/TX-HMH-MCoV-9526/2020 | 2020-07-05 | North America/USA/Texas/Houston | Houston Methodist Hospital | Houston Methodist Hospital | S. Wesley Long et al |
| EPI_ISL_781167 | USA/TX-HMH-MCoV-9529/2020 | 2020-07-04 | North America/USA/Texas/Houston | Houston Methodist Hospital | Houston Methodist Hospital | S. Wesley Long et al |
| EPI_ISL_781172 | USA/TX-HMH-MCoV-9538/2020 | 2020-07-04 | North America/USA/Texas/Houston | Houston Methodist Hospital | Houston Methodist Hospital | S. Wesley Long et al |
| EPI_ISL_781173 | USA/TX-HMH-MCoV-9540/2020 | 2020-07-03 | North America/USA/Texas/Houston | Houston Methodist Hospital | Houston Methodist Hospital | S. Wesley Long et al |
| EPI_ISL_781175 | USA/TX-HMH-MCoV-9542/2020 | 2020-07-04 | North America/USA/Texas/Houston | Houston Methodist Hospital | Houston Methodist Hospital | S. Wesley Long et al |
| EPI_ISL_781176 | USA/TX-HMH-MCoV-9544/2020 | 2020-07-03 | North America/USA/Texas/Houston | Houston Methodist Hospital | Houston Methodist Hospital | S. Wesley Long et al |

| | | | | | | |
|---|---|---|---|---|---|---|
| EPI_ISL_781177 | USA/TX-HMH-MCoV-9546/2020 | 2020-06-22 | North America/USA/Texas/Houston | Houston Methodist Hospital | Houston Methodist Hospital | S. Wesley Long et al |
| EPI_ISL_781181 | England/CAMC-CF9A3D/2020 | 2020-12-19 | Europe/United Kingdom/England/ | Lighthouse Lab in Cambridge | Wellcome Sanger Institute for the COVID-19 Genomics UK (COG-UK) Consortium | Rob Howes et al |
| EPI_ISL_781272 | England/MILK-D09CF7/2020 | 2020-12-21 | Europe/United Kingdom/England/ | Lighthouse Lab in Milton Keynes | Wellcome Sanger Institute for the COVID-19 Genomics UK (COG-UK) Consortium | The Lighthouse Lab in Milton Keynes et al |
| EPI_ISL_782139 | England/ALDP-CF2973/2020 | 2020-12-17 | Europe/United Kingdom/England/ | Lighthouse Lab in Alderley Park | Wellcome Sanger Institute for the COVID-19 Genomics UK (COG-UK) Consortium | Jacquelyn Wynn et al |
| EPI_ISL_782328 | England/ALDP-CEC2DB/2020 | 2020-12-21 | Europe/United Kingdom/England/ | Lighthouse Lab in Alderley Park | Wellcome Sanger Institute for the COVID-19 Genomics UK (COG-UK) Consortium | Jacquelyn Wynn et al |
| EPI_ISL_782370 | England/QEUH-CCCF52/2020 | 2020-12-23 | Europe/United Kingdom/England/ | Lighthouse Lab in Glasgow | Wellcome Sanger Institute for the COVID-19 Genomics UK (COG-UK) Consortium | Harper VanSteenhouse et al |
| EPI_ISL_783020 | England/MILK-C3086D/2020 | 2020-12-07 | Europe/United Kingdom/England/ | Lighthouse Lab in Milton Keynes | Wellcome Sanger Institute for the COVID-19 Genomics UK (COG-UK) Consortium | The Lighthouse Lab in Milton Keynes et al |
| EPI_ISL_783078 | England/MILK-BAF8F1/2020 | 2020-11-17 | Europe/United Kingdom/England/ | Lighthouse Lab in Milton Keynes | Wellcome Sanger Institute for the COVID-19 Genomics UK (COG-UK) Consortium | The Lighthouse Lab in Milton Keynes et al |

| | | | | | | |
|---|---|---|---|---|---|---|
| EPI_ISL_783168 | England/MILK-BAA234/2020 | 2020-11-17 | Europe/United Kingdom/England/ | Lighthouse Lab in Milton Keynes | Wellcome Sanger Institute for the COVID-19 Genomics UK (COG-UK) Consortium | The Lighthouse Lab in Milton Keynes et al |
| EPI_ISL_783208 | England/MILK-BAC78E/2020 | 2020-11-18 | Europe/United Kingdom/England/ | Lighthouse Lab in Milton Keynes | Wellcome Sanger Institute for the COVID-19 Genomics UK (COG-UK) Consortium | The Lighthouse Lab in Milton Keynes et al |
| EPI_ISL_783243 | England/MILK-BAA2DA/2020 | 2020-11-17 | Europe/United Kingdom/England/ | Lighthouse Lab in Milton Keynes | Wellcome Sanger Institute for the COVID-19 Genomics UK (COG-UK) Consortium | The Lighthouse Lab in Milton Keynes et al |
| EPI_ISL_783497 | England/CAMC-B77BCC/2020 | 2020-11-12 | Europe/United Kingdom/England/ | Lighthouse Lab in Cambridge | Wellcome Sanger Institute for the COVID-19 Genomics UK (COG-UK) Consortium | Rob Howes et al |
| EPI_ISL_783499 | England/CAMC-B786BB/2020 | 2020-11-12 | Europe/United Kingdom/England/ | Lighthouse Lab in Cambridge | Wellcome Sanger Institute for the COVID-19 Genomics UK (COG-UK) Consortium | Rob Howes et al |
| EPI_ISL_783541 | USA/TX-HMH-MCoV-8368/2020 | 2020-06-29 | North America/USA/Texas/Houston | Houston Methodist Hospital | Houston Methodist Hospital | S. Wesley Long et al |
| EPI_ISL_783543 | USA/TX-HMH-MCoV-19518/2020 | 2020-11-30 | North America/USA/Texas/Houston | Houston Methodist Hospital | Houston Methodist Hospital | S. Wesley Long et al |
| EPI_ISL_783549 | USA/TX-HMH-MCoV-19645/2020 | 2020-11-16 | North America/USA/Texas/Houston | Houston Methodist Hospital | Houston Methodist Hospital | S. Wesley Long et al |
| EPI_ISL_783552 | USA/TX-HMH-MCoV-19230/2020 | 2020-11-23 | North America/USA/Texas/Houston | Houston Methodist Hospital | Houston Methodist Hospital | S. Wesley Long et al |
| EPI_ISL_783554 | USA/TX-HMH-MCoV-8245/2020 | 2020-07-07 | North America/USA/Texas/Houston | Houston Methodist Hospital | Houston Methodist Hospital | S. Wesley Long et al |

| | | | | | | |
|---|---|---|---|---|---|---|
| EPI_ISL_783562 | USA/TX-HMH-MCoV-19800/2020 | 2020-11-26 | North America/USA/Texas/Houston | Houston Methodist Hospital | Houston Methodist Hospital | S. Wesley Long et al |
| EPI_ISL_783564 | USA/TX-HMH-MCoV-19803/2020 | 2020-11-25 | North America/USA/Texas/Houston | Houston Methodist Hospital | Houston Methodist Hospital | S. Wesley Long et al |
| EPI_ISL_783573 | USA/TX-HMH-MCoV-8354/2020 | 2020-07-07 | North America/USA/Texas/Houston | Houston Methodist Hospital | Houston Methodist Hospital | S. Wesley Long et al |
| EPI_ISL_783575 | USA/TX-HMH-MCoV-8109/2020 | 2020-06-30 | North America/USA/Texas/Houston | Houston Methodist Hospital | Houston Methodist Hospital | S. Wesley Long et al |
| EPI_ISL_783584 | USA/TX-HMH-MCoV-8265/2020 | 2020-07-07 | North America/USA/Texas/Houston | Houston Methodist Hospital | Houston Methodist Hospital | S. Wesley Long et al |
| EPI_ISL_783587 | USA/TX-HMH-MCoV-8327/2020 | 2020-07-07 | North America/USA/Texas/Houston | Houston Methodist Hospital | Houston Methodist Hospital | S. Wesley Long et al |
| EPI_ISL_783590 | USA/TX-HMH-MCoV-8136/2020 | 2020-06-29 | North America/USA/Texas/Houston | Houston Methodist Hospital | Houston Methodist Hospital | S. Wesley Long et al |
| EPI_ISL_783594 | USA/TX-HMH-MCoV-19041/2020 | 2020-11-23 | North America/USA/Texas/Houston | Houston Methodist Hospital | Houston Methodist Hospital | S. Wesley Long et al |
| EPI_ISL_783595 | USA/TX-HMH-MCoV-19042/2020 | 2020-11-23 | North America/USA/Texas/Houston | Houston Methodist Hospital | Houston Methodist Hospital | S. Wesley Long et al |
| EPI_ISL_783599 | USA/TX-HMH-MCoV-19048/2020 | 2020-11-25 | North America/USA/Texas/Houston | Houston Methodist Hospital | Houston Methodist Hospital | S. Wesley Long et al |
| EPI_ISL_783603 | USA/TX-HMH-MCoV-19054/2020 | 2020-11-23 | North America/USA/Texas/Houston | Houston Methodist Hospital | Houston Methodist Hospital | S. Wesley Long et al |
| EPI_ISL_783618 | USA/TX-HMH-MCoV-19087/2020 | 2020-11-25 | North America/USA/Texas/Houston | Houston Methodist Hospital | Houston Methodist Hospital | S. Wesley Long et al |
| EPI_ISL_783620 | USA/TX-HMH-MCoV-19090/2020 | 2020-11-24 | North America/USA/Texas/Houston | Houston Methodist Hospital | Houston Methodist Hospital | S. Wesley Long et al |

| | | | | | | |
|---|---|---|---|---|---|---|
| EPI_ISL_783623 | USA/TX-HMH-MCoV-19097/2020 | 2020-11-23 | North America/USA/Texas/Houston | Houston Methodist Hospital | Houston Methodist Hospital | S. Wesley Long et al |
| EPI_ISL_783633 | USA/TX-HMH-MCoV-19113/2020 | 2020-11-23 | North America/USA/Texas/Houston | Houston Methodist Hospital | Houston Methodist Hospital | S. Wesley Long et al |
| EPI_ISL_783637 | USA/TX-HMH-MCoV-19119/2020 | 2020-11-23 | North America/USA/Texas/Houston | Houston Methodist Hospital | Houston Methodist Hospital | S. Wesley Long et al |
| EPI_ISL_783640 | USA/TX-HMH-MCoV-19123/2020 | 2020-11-23 | North America/USA/Texas/Houston | Houston Methodist Hospital | Houston Methodist Hospital | S. Wesley Long et al |
| EPI_ISL_783641 | USA/TX-HMH-MCoV-19124/2020 | 2020-11-24 | North America/USA/Texas/Houston | Houston Methodist Hospital | Houston Methodist Hospital | S. Wesley Long et al |
| EPI_ISL_783658 | USA/TX-HMH-MCoV-19153/2020 | 2020-11-23 | North America/USA/Texas/Houston | Houston Methodist Hospital | Houston Methodist Hospital | S. Wesley Long et al |
| EPI_ISL_783659 | USA/TX-HMH-MCoV-19154/2020 | 2020-11-23 | North America/USA/Texas/Houston | Houston Methodist Hospital | Houston Methodist Hospital | S. Wesley Long et al |
| EPI_ISL_783669 | USA/TX-HMH-MCoV-19173/2020 | 2020-11-24 | North America/USA/Texas/Houston | Houston Methodist Hospital | Houston Methodist Hospital | S. Wesley Long et al |
| EPI_ISL_783678 | USA/TX-HMH-MCoV-19188/2020 | 2020-11-23 | North America/USA/Texas/Houston | Houston Methodist Hospital | Houston Methodist Hospital | S. Wesley Long et al |
| EPI_ISL_783687 | USA/TX-HMH-MCoV-19202/2020 | 2020-11-28 | North America/USA/Texas/Houston | Houston Methodist Hospital | Houston Methodist Hospital | S. Wesley Long et al |
| EPI_ISL_783691 | USA/TX-HMH-MCoV-19206/2020 | 2020-11-24 | North America/USA/Texas/Houston | Houston Methodist Hospital | Houston Methodist Hospital | S. Wesley Long et al |
| EPI_ISL_783693 | USA/TX-HMH-MCoV-19212/2020 | 2020-11-27 | North America/USA/Texas/Houston | Houston Methodist Hospital | Houston Methodist Hospital | S. Wesley Long et al |
| EPI_ISL_783711 | USA/TX-HMH-MCoV-19239/2020 | 2020-11-23 | North America/USA/Texas/Houston | Houston Methodist Hospital | Houston Methodist Hospital | S. Wesley Long et al |

| | | | | | | |
|---|---|---|---|---|---|---|
| EPI_ISL_783719 | USA/TX-HMH-MCoV-19251/2020 | 2020-11-24 | North America/USA/Texas/Houston | Houston Methodist Hospital | Houston Methodist Hospital | S. Wesley Long et al |
| EPI_ISL_783725 | USA/TX-HMH-MCoV-19265/2020 | 2020-11-23 | North America/USA/Texas/Houston | Houston Methodist Hospital | Houston Methodist Hospital | S. Wesley Long et al |
| EPI_ISL_783726 | USA/TX-HMH-MCoV-19267/2020 | 2020-11-24 | North America/USA/Texas/Houston | Houston Methodist Hospital | Houston Methodist Hospital | S. Wesley Long et al |
| EPI_ISL_783728 | USA/TX-HMH-MCoV-19269/2020 | 2020-11-27 | North America/USA/Texas/Houston | Houston Methodist Hospital | Houston Methodist Hospital | S. Wesley Long et al |
| EPI_ISL_783732 | USA/TX-HMH-MCoV-19273/2020 | 2020-11-27 | North America/USA/Texas/Houston | Houston Methodist Hospital | Houston Methodist Hospital | S. Wesley Long et al |
| EPI_ISL_783749 | USA/TX-HMH-MCoV-19303/2020 | 2020-11-27 | North America/USA/Texas/Houston | Houston Methodist Hospital | Houston Methodist Hospital | S. Wesley Long et al |
| EPI_ISL_783757 | USA/TX-HMH-MCoV-19318/2020 | 2020-11-28 | North America/USA/Texas/Houston | Houston Methodist Hospital | Houston Methodist Hospital | S. Wesley Long et al |
| EPI_ISL_783772 | USA/TX-HMH-MCoV-19337/2020 | 2020-11-27 | North America/USA/Texas/Houston | Houston Methodist Hospital | Houston Methodist Hospital | S. Wesley Long et al |
| EPI_ISL_783776 | USA/TX-HMH-MCoV-19343/2020 | 2020-11-29 | North America/USA/Texas/Houston | Houston Methodist Hospital | Houston Methodist Hospital | S. Wesley Long et al |
| EPI_ISL_783777 | USA/TX-HMH-MCoV-19344/2020 | 2020-11-29 | North America/USA/Texas/Houston | Houston Methodist Hospital | Houston Methodist Hospital | S. Wesley Long et al |
| EPI_ISL_783781 | USA/TX-HMH-MCoV-19349/2020 | 2020-11-28 | North America/USA/Texas/Houston | Houston Methodist Hospital | Houston Methodist Hospital | S. Wesley Long et al |
| EPI_ISL_783783 | USA/TX-HMH-MCoV-19352/2020 | 2020-11-28 | North America/USA/Texas/Houston | Houston Methodist Hospital | Houston Methodist Hospital | S. Wesley Long et al |
| EPI_ISL_783785 | USA/TX-HMH-MCoV-19354/2020 | 2020-11-27 | North America/USA/Texas/Houston | Houston Methodist Hospital | Houston Methodist Hospital | S. Wesley Long et al |

| | | | | | | |
|---|---|---|---|---|---|---|
| EPI_ISL_783787 | USA/TX-HMH-MCoV-19356/2020 | 2020-11-29 | North America/USA/Texas/Houston | Houston Methodist Hospital | Houston Methodist Hospital | S. Wesley Long et al |
| EPI_ISL_783794 | USA/TX-HMH-MCoV-19365/2020 | 2020-11-28 | North America/USA/Texas/Houston | Houston Methodist Hospital | Houston Methodist Hospital | S. Wesley Long et al |
| EPI_ISL_783800 | USA/TX-HMH-MCoV-19371/2020 | 2020-11-28 | North America/USA/Texas/Houston | Houston Methodist Hospital | Houston Methodist Hospital | S. Wesley Long et al |
| EPI_ISL_783801 | USA/TX-HMH-MCoV-19372/2020 | 2020-11-27 | North America/USA/Texas/Houston | Houston Methodist Hospital | Houston Methodist Hospital | S. Wesley Long et al |
| EPI_ISL_783819 | USA/TX-HMH-MCoV-19403/2020 | 2020-11-30 | North America/USA/Texas/Houston | Houston Methodist Hospital | Houston Methodist Hospital | S. Wesley Long et al |
| EPI_ISL_783827 | USA/TX-HMH-MCoV-19415/2020 | 2020-11-30 | North America/USA/Texas/Houston | Houston Methodist Hospital | Houston Methodist Hospital | S. Wesley Long et al |
| EPI_ISL_783837 | USA/TX-HMH-MCoV-19431/2020 | 2020-11-30 | North America/USA/Texas/Houston | Houston Methodist Hospital | Houston Methodist Hospital | S. Wesley Long et al |
| EPI_ISL_783838 | USA/TX-HMH-MCoV-19432/2020 | 2020-11-30 | North America/USA/Texas/Houston | Houston Methodist Hospital | Houston Methodist Hospital | S. Wesley Long et al |
| EPI_ISL_783845 | USA/TX-HMH-MCoV-19447/2020 | 2020-11-29 | North America/USA/Texas/Houston | Houston Methodist Hospital | Houston Methodist Hospital | S. Wesley Long et al |
| EPI_ISL_783847 | USA/TX-HMH-MCoV-19451/2020 | 2020-11-30 | North America/USA/Texas/Houston | Houston Methodist Hospital | Houston Methodist Hospital | S. Wesley Long et al |
| EPI_ISL_783848 | USA/TX-HMH-MCoV-19452/2020 | 2020-11-30 | North America/USA/Texas/Houston | Houston Methodist Hospital | Houston Methodist Hospital | S. Wesley Long et al |
| EPI_ISL_783865 | USA/TX-HMH-MCoV-19480/2020 | 2020-11-30 | North America/USA/Texas/Houston | Houston Methodist Hospital | Houston Methodist Hospital | S. Wesley Long et al |
| EPI_ISL_783866 | USA/TX-HMH-MCoV-19482/2020 | 2020-11-30 | North America/USA/Texas/Houston | Houston Methodist Hospital | Houston Methodist Hospital | S. Wesley Long et al |

| | | | | | | |
|---|---|---|---|---|---|---|
| EPI_ISL_783871 | USA/TX-HMH-MCoV-19487/2020 | 2020-11-17 | North America/USA/Texas/Houston | Houston Methodist Hospital | Houston Methodist Hospital | S. Wesley Long et al |
| EPI_ISL_783874 | USA/TX-HMH-MCoV-19490/2020 | 2020-11-17 | North America/USA/Texas/Houston | Houston Methodist Hospital | Houston Methodist Hospital | S. Wesley Long et al |
| EPI_ISL_783880 | USA/TX-HMH-MCoV-19497/2020 | 2020-11-17 | North America/USA/Texas/Houston | Houston Methodist Hospital | Houston Methodist Hospital | S. Wesley Long et al |
| EPI_ISL_783881 | USA/TX-HMH-MCoV-19500/2020 | 2020-11-16 | North America/USA/Texas/Houston | Houston Methodist Hospital | Houston Methodist Hospital | S. Wesley Long et al |
| EPI_ISL_783890 | USA/TX-HMH-MCoV-19510/2020 | 2020-11-17 | North America/USA/Texas/Houston | Houston Methodist Hospital | Houston Methodist Hospital | S. Wesley Long et al |
| EPI_ISL_783895 | USA/TX-HMH-MCoV-19516/2020 | 2020-11-30 | North America/USA/Texas/Houston | Houston Methodist Hospital | Houston Methodist Hospital | S. Wesley Long et al |
| EPI_ISL_783903 | USA/TX-HMH-MCoV-19526/2020 | 2020-11-17 | North America/USA/Texas/Houston | Houston Methodist Hospital | Houston Methodist Hospital | S. Wesley Long et al |
| EPI_ISL_783908 | USA/TX-HMH-MCoV-19532/2020 | 2020-11-17 | North America/USA/Texas/Houston | Houston Methodist Hospital | Houston Methodist Hospital | S. Wesley Long et al |
| EPI_ISL_783916 | USA/TX-HMH-MCoV-19541/2020 | 2020-11-30 | North America/USA/Texas/Houston | Houston Methodist Hospital | Houston Methodist Hospital | S. Wesley Long et al |
| EPI_ISL_783925 | USA/TX-HMH-MCoV-19553/2020 | 2020-11-30 | North America/USA/Texas/Houston | Houston Methodist Hospital | Houston Methodist Hospital | S. Wesley Long et al |
| EPI_ISL_783930 | USA/TX-HMH-MCoV-19558/2020 | 2020-11-17 | North America/USA/Texas/Houston | Houston Methodist Hospital | Houston Methodist Hospital | S. Wesley Long et al |
| EPI_ISL_783934 | USA/TX-HMH-MCoV-19562/2020 | 2020-11-24 | North America/USA/Texas/Houston | Houston Methodist Hospital | Houston Methodist Hospital | S. Wesley Long et al |
| EPI_ISL_783939 | USA/TX-HMH-MCoV-19567/2020 | 2020-11-30 | North America/USA/Texas/Houston | Houston Methodist Hospital | Houston Methodist Hospital | S. Wesley Long et al |

| | | | | | | |
|---|---|---|---|---|---|---|
| EPI_ISL_783940 | USA/TX-HMH-MCoV-19568/2020 | 2020-11-17 | North America/USA/Texas/Houston | Houston Methodist Hospital | Houston Methodist Hospital | S. Wesley Long et al |
| EPI_ISL_783960 | USA/TX-HMH-MCoV-19589/2020 | 2020-11-21 | North America/USA/Texas/Houston | Houston Methodist Hospital | Houston Methodist Hospital | S. Wesley Long et al |
| EPI_ISL_783970 | USA/TX-HMH-MCoV-19600/2020 | 2020-11-24 | North America/USA/Texas/Houston | Houston Methodist Hospital | Houston Methodist Hospital | S. Wesley Long et al |
| EPI_ISL_783980 | USA/TX-HMH-MCoV-19611/2020 | 2020-11-28 | North America/USA/Texas/Houston | Houston Methodist Hospital | Houston Methodist Hospital | S. Wesley Long et al |
| EPI_ISL_783981 | USA/TX-HMH-MCoV-19613/2020 | 2020-11-19 | North America/USA/Texas/Houston | Houston Methodist Hospital | Houston Methodist Hospital | S. Wesley Long et al |
| EPI_ISL_783986 | USA/TX-HMH-MCoV-19619/2020 | 2020-11-25 | North America/USA/Texas/Houston | Houston Methodist Hospital | Houston Methodist Hospital | S. Wesley Long et al |
| EPI_ISL_783988 | USA/TX-HMH-MCoV-19622/2020 | 2020-11-22 | North America/USA/Texas/Houston | Houston Methodist Hospital | Houston Methodist Hospital | S. Wesley Long et al |
| EPI_ISL_783991 | USA/TX-HMH-MCoV-19625/2020 | 2020-11-16 | North America/USA/Texas/Houston | Houston Methodist Hospital | Houston Methodist Hospital | S. Wesley Long et al |
| EPI_ISL_783992 | USA/TX-HMH-MCoV-19627/2020 | 2020-11-16 | North America/USA/Texas/Houston | Houston Methodist Hospital | Houston Methodist Hospital | S. Wesley Long et al |
| EPI_ISL_784021 | USA/TX-HMH-MCoV-19662/2020 | 2020-11-29 | North America/USA/Texas/Houston | Houston Methodist Hospital | Houston Methodist Hospital | S. Wesley Long et al |
| EPI_ISL_784026 | USA/TX-HMH-MCoV-19667/2020 | 2020-11-19 | North America/USA/Texas/Houston | Houston Methodist Hospital | Houston Methodist Hospital | S. Wesley Long et al |
| EPI_ISL_784043 | USA/TX-HMH-MCoV-19689/2020 | 2020-11-28 | North America/USA/Texas/Houston | Houston Methodist Hospital | Houston Methodist Hospital | S. Wesley Long et al |
| EPI_ISL_784047 | USA/TX-HMH-MCoV-19693/2020 | 2020-11-30 | North America/USA/Texas/Houston | Houston Methodist Hospital | Houston Methodist Hospital | S. Wesley Long et al |

| | | | | | | |
|---|---|---|---|---|---|---|
| EPI_ISL_784049 | USA/TX-HMH-MCoV-19695/2020 | 2020-11-25 | North America/USA/Texas/Houston | Houston Methodist Hospital | Houston Methodist Hospital | S. Wesley Long et al |
| EPI_ISL_784051 | USA/TX-HMH-MCoV-19697/2020 | 2020-11-23 | North America/USA/Texas/Houston | Houston Methodist Hospital | Houston Methodist Hospital | S. Wesley Long et al |
| EPI_ISL_784058 | USA/TX-HMH-MCoV-19706/2020 | 2020-11-21 | North America/USA/Texas/Houston | Houston Methodist Hospital | Houston Methodist Hospital | S. Wesley Long et al |
| EPI_ISL_784075 | USA/TX-HMH-MCoV-19727/2020 | 2020-11-22 | North America/USA/Texas/Houston | Houston Methodist Hospital | Houston Methodist Hospital | S. Wesley Long et al |
| EPI_ISL_784080 | USA/TX-HMH-MCoV-19732/2020 | 2020-11-25 | North America/USA/Texas/Houston | Houston Methodist Hospital | Houston Methodist Hospital | S. Wesley Long et al |
| EPI_ISL_784081 | USA/TX-HMH-MCoV-19735/2020 | 2020-11-19 | North America/USA/Texas/Houston | Houston Methodist Hospital | Houston Methodist Hospital | S. Wesley Long et al |
| EPI_ISL_784092 | USA/TX-HMH-MCoV-19748/2020 | 2020-11-19 | North America/USA/Texas/Houston | Houston Methodist Hospital | Houston Methodist Hospital | S. Wesley Long et al |
| EPI_ISL_784099 | USA/TX-HMH-MCoV-19756/2020 | 2020-11-16 | North America/USA/Texas/Houston | Houston Methodist Hospital | Houston Methodist Hospital | S. Wesley Long et al |
| EPI_ISL_784100 | USA/TX-HMH-MCoV-19757/2020 | 2020-11-25 | North America/USA/Texas/Houston | Houston Methodist Hospital | Houston Methodist Hospital | S. Wesley Long et al |
| EPI_ISL_784101 | USA/TX-HMH-MCoV-19758/2020 | 2020-11-21 | North America/USA/Texas/Houston | Houston Methodist Hospital | Houston Methodist Hospital | S. Wesley Long et al |
| EPI_ISL_784109 | USA/TX-HMH-MCoV-19767/2020 | 2020-11-24 | North America/USA/Texas/Houston | Houston Methodist Hospital | Houston Methodist Hospital | S. Wesley Long et al |
| EPI_ISL_784120 | USA/TX-HMH-MCoV-19781/2020 | 2020-11-28 | North America/USA/Texas/Houston | Houston Methodist Hospital | Houston Methodist Hospital | S. Wesley Long et al |
| EPI_ISL_784150 | USA/TX-HMH-MCoV-19822/2020 | 2020-11-18 | North America/USA/Texas/Houston | Houston Methodist Hospital | Houston Methodist Hospital | S. Wesley Long et al |

| | | | | | | |
|---|---|---|---|---|---|---|
| EPI_ISL_784154 | USA/TX-HMH-MCoV-19826/2020 | 2020-11-16 | North America/USA/Texas/Houston | Houston Methodist Hospital | Houston Methodist Hospital | S. Wesley Long et al |
| EPI_ISL_784158 | USA/TX-HMH-MCoV-19830/2020 | 2020-11-26 | North America/USA/Texas/Houston | Houston Methodist Hospital | Houston Methodist Hospital | S. Wesley Long et al |
| EPI_ISL_784173 | USA/TX-HMH-MCoV-19851/2020 | 2020-11-23 | North America/USA/Texas/Houston | Houston Methodist Hospital | Houston Methodist Hospital | S. Wesley Long et al |
| EPI_ISL_784177 | USA/TX-HMH-MCoV-19855/2020 | 2020-11-28 | North America/USA/Texas/Houston | Houston Methodist Hospital | Houston Methodist Hospital | S. Wesley Long et al |
| EPI_ISL_784184 | USA/TX-HMH-MCoV-8079/2020 | 2020-06-30 | North America/USA/Texas/Houston | Houston Methodist Hospital | Houston Methodist Hospital | S. Wesley Long et al |
| EPI_ISL_784186 | USA/TX-HMH-MCoV-8083/2020 | 2020-06-29 | North America/USA/Texas/Houston | Houston Methodist Hospital | Houston Methodist Hospital | S. Wesley Long et al |
| EPI_ISL_784189 | USA/TX-HMH-MCoV-8089/2020 | 2020-07-07 | North America/USA/Texas/Houston | Houston Methodist Hospital | Houston Methodist Hospital | S. Wesley Long et al |
| EPI_ISL_784193 | USA/TX-HMH-MCoV-8094/2020 | 2020-06-29 | North America/USA/Texas/Houston | Houston Methodist Hospital | Houston Methodist Hospital | S. Wesley Long et al |
| EPI_ISL_784196 | USA/TX-HMH-MCoV-8100/2020 | 2020-06-29 | North America/USA/Texas/Houston | Houston Methodist Hospital | Houston Methodist Hospital | S. Wesley Long et al |
| EPI_ISL_784199 | USA/TX-HMH-MCoV-8104/2020 | 2020-06-30 | North America/USA/Texas/Houston | Houston Methodist Hospital | Houston Methodist Hospital | S. Wesley Long et al |
| EPI_ISL_784205 | USA/TX-HMH-MCoV-8113/2020 | 2020-06-30 | North America/USA/Texas/Houston | Houston Methodist Hospital | Houston Methodist Hospital | S. Wesley Long et al |
| EPI_ISL_784223 | USA/TX-HMH-MCoV-8133/2020 | 2020-06-30 | North America/USA/Texas/Houston | Houston Methodist Hospital | Houston Methodist Hospital | S. Wesley Long et al |
| EPI_ISL_784226 | USA/TX-HMH-MCoV-8139/2020 | 2020-06-30 | North America/USA/Texas/Houston | Houston Methodist Hospital | Houston Methodist Hospital | S. Wesley Long et al |

| | | | | | | |
|---|---|---|---|---|---|---|
| EPI_ISL_784231 | USA/TX-HMH-MCoV-8144/2020 | 2020-06-30 | North America/USA/Texas/Houston | Houston Methodist Hospital | Houston Methodist Hospital | S. Wesley Long et al |
| EPI_ISL_784234 | USA/TX-HMH-MCoV-8151/2020 | 2020-06-30 | North America/USA/Texas/Houston | Houston Methodist Hospital | Houston Methodist Hospital | S. Wesley Long et al |
| EPI_ISL_784242 | USA/TX-HMH-MCoV-8166/2020 | 2020-06-30 | North America/USA/Texas/Houston | Houston Methodist Hospital | Houston Methodist Hospital | S. Wesley Long et al |
| EPI_ISL_784247 | USA/TX-HMH-MCoV-8176/2020 | 2020-07-07 | North America/USA/Texas/Houston | Houston Methodist Hospital | Houston Methodist Hospital | S. Wesley Long et al |
| EPI_ISL_784263 | USA/TX-HMH-MCoV-8204/2020 | 2020-07-07 | North America/USA/Texas/Houston | Houston Methodist Hospital | Houston Methodist Hospital | S. Wesley Long et al |
| EPI_ISL_784268 | USA/TX-HMH-MCoV-8209/2020 | 2020-07-07 | North America/USA/Texas/Houston | Houston Methodist Hospital | Houston Methodist Hospital | S. Wesley Long et al |
| EPI_ISL_784273 | USA/TX-HMH-MCoV-8220/2020 | 2020-07-07 | North America/USA/Texas/Houston | Houston Methodist Hospital | Houston Methodist Hospital | S. Wesley Long et al |
| EPI_ISL_784279 | USA/TX-HMH-MCoV-8231/2020 | 2020-07-07 | North America/USA/Texas/Houston | Houston Methodist Hospital | Houston Methodist Hospital | S. Wesley Long et al |
| EPI_ISL_784283 | USA/TX-HMH-MCoV-8238/2020 | 2020-07-07 | North America/USA/Texas/Houston | Houston Methodist Hospital | Houston Methodist Hospital | S. Wesley Long et al |
| EPI_ISL_784287 | USA/TX-HMH-MCoV-8244/2020 | 2020-07-07 | North America/USA/Texas/Houston | Houston Methodist Hospital | Houston Methodist Hospital | S. Wesley Long et al |
| EPI_ISL_784288 | USA/TX-HMH-MCoV-8246/2020 | 2020-07-07 | North America/USA/Texas/Houston | Houston Methodist Hospital | Houston Methodist Hospital | S. Wesley Long et al |
| EPI_ISL_784292 | USA/TX-HMH-MCoV-8252/2020 | 2020-07-07 | North America/USA/Texas/Houston | Houston Methodist Hospital | Houston Methodist Hospital | S. Wesley Long et al |
| EPI_ISL_784293 | USA/TX-HMH-MCoV-8253/2020 | 2020-07-07 | North America/USA/Texas/Houston | Houston Methodist Hospital | Houston Methodist Hospital | S. Wesley Long et al |

| | | | | | | |
|---|---|---|---|---|---|---|
| EPI_ISL_784295 | USA/TX-HMH-MCoV-8256/2020 | 2020-07-07 | North America/USA/Texas/Houston | Houston Methodist Hospital | Houston Methodist Hospital | S. Wesley Long et al |
| EPI_ISL_784298 | USA/TX-HMH-MCoV-8262/2020 | 2020-07-07 | North America/USA/Texas/Houston | Houston Methodist Hospital | Houston Methodist Hospital | S. Wesley Long et al |
| EPI_ISL_784301 | USA/TX-HMH-MCoV-8271/2020 | 2020-07-07 | North America/USA/Texas/Houston | Houston Methodist Hospital | Houston Methodist Hospital | S. Wesley Long et al |
| EPI_ISL_784302 | USA/TX-HMH-MCoV-8272/2020 | 2020-07-05 | North America/USA/Texas/Houston | Houston Methodist Hospital | Houston Methodist Hospital | S. Wesley Long et al |
| EPI_ISL_784316 | USA/TX-HMH-MCoV-8299/2020 | 2020-07-08 | North America/USA/Texas/Houston | Houston Methodist Hospital | Houston Methodist Hospital | S. Wesley Long et al |
| EPI_ISL_784317 | USA/TX-HMH-MCoV-8300/2020 | 2020-07-07 | North America/USA/Texas/Houston | Houston Methodist Hospital | Houston Methodist Hospital | S. Wesley Long et al |
| EPI_ISL_784321 | USA/TX-HMH-MCoV-8309/2020 | 2020-07-07 | North America/USA/Texas/Houston | Houston Methodist Hospital | Houston Methodist Hospital | S. Wesley Long et al |
| EPI_ISL_784329 | USA/TX-HMH-MCoV-8320/2020 | 2020-07-07 | North America/USA/Texas/Houston | Houston Methodist Hospital | Houston Methodist Hospital | S. Wesley Long et al |
| EPI_ISL_784334 | USA/TX-HMH-MCoV-8326/2020 | 2020-07-08 | North America/USA/Texas/Houston | Houston Methodist Hospital | Houston Methodist Hospital | S. Wesley Long et al |
| EPI_ISL_784342 | USA/TX-HMH-MCoV-8341/2020 | 2020-07-07 | North America/USA/Texas/Houston | Houston Methodist Hospital | Houston Methodist Hospital | S. Wesley Long et al |
| EPI_ISL_784354 | USA/TX-HMH-MCoV-8364/2020 | 2020-07-07 | North America/USA/Texas/Houston | Houston Methodist Hospital | Houston Methodist Hospital | S. Wesley Long et al |
| EPI_ISL_784360 | USA/TX-HMH-MCoV-8380/2020 | 2020-06-23 | North America/USA/Texas/Houston | Houston Methodist Hospital | Houston Methodist Hospital | S. Wesley Long et al |
| EPI_ISL_784380 | USA/TX-HMH-MCoV-8412/2020 | 2020-06-26 | North America/USA/Texas/Houston | Houston Methodist Hospital | Houston Methodist Hospital | S. Wesley Long et al |

| | | | | | | |
|---|---|---|---|---|---|---|
| EPI_ISL_784381 | USA/TX-HMH-MCoV-8413/2020 | 2020-07-01 | North America/USA/Texas/Houston | Houston Methodist Hospital | Houston Methodist Hospital | S. Wesley Long et al |
| EPI_ISL_784383 | USA/TX-HMH-MCoV-8416/2020 | 2020-07-01 | North America/USA/Texas/Houston | Houston Methodist Hospital | Houston Methodist Hospital | S. Wesley Long et al |
| EPI_ISL_784386 | USA/TX-HMH-MCoV-8420/2020 | 2020-07-01 | North America/USA/Texas/Houston | Houston Methodist Hospital | Houston Methodist Hospital | S. Wesley Long et al |
| EPI_ISL_784409 | USA/TX-HMH-MCoV-18061/2020 | 2020-11-13 | North America/USA/Texas/Houston | Houston Methodist Hospital | Houston Methodist Hospital | S. Wesley Long et al |
| EPI_ISL_784411 | USA/TX-HMH-MCoV-18063/2020 | 2020-11-16 | North America/USA/Texas/Houston | Houston Methodist Hospital | Houston Methodist Hospital | S. Wesley Long et al |
| EPI_ISL_784413 | USA/TX-HMH-MCoV-18065/2020 | 2020-11-16 | North America/USA/Texas/Houston | Houston Methodist Hospital | Houston Methodist Hospital | S. Wesley Long et al |
| EPI_ISL_784419 | USA/TX-HMH-MCoV-18071/2020 | 2020-11-12 | North America/USA/Texas/Houston | Houston Methodist Hospital | Houston Methodist Hospital | S. Wesley Long et al |
| EPI_ISL_784427 | USA/TX-HMH-MCoV-18079/2020 | 2020-11-15 | North America/USA/Texas/Houston | Houston Methodist Hospital | Houston Methodist Hospital | S. Wesley Long et al |
| EPI_ISL_784438 | USA/TX-HMH-MCoV-18090/2020 | 2020-11-16 | North America/USA/Texas/Houston | Houston Methodist Hospital | Houston Methodist Hospital | S. Wesley Long et al |
| EPI_ISL_784441 | USA/TX-HMH-MCoV-18093/2020 | 2020-11-14 | North America/USA/Texas/Houston | Houston Methodist Hospital | Houston Methodist Hospital | S. Wesley Long et al |
| EPI_ISL_784448 | USA/TX-HMH-MCoV-18100/2020 | 2020-11-16 | North America/USA/Texas/Houston | Houston Methodist Hospital | Houston Methodist Hospital | S. Wesley Long et al |
| EPI_ISL_784451 | USA/TX-HMH-MCoV-18103/2020 | 2020-11-12 | North America/USA/Texas/Houston | Houston Methodist Hospital | Houston Methodist Hospital | S. Wesley Long et al |
| EPI_ISL_784453 | USA/TX-HMH-MCoV-18105/2020 | 2020-11-12 | North America/USA/Texas/Houston | Houston Methodist Hospital | Houston Methodist Hospital | S. Wesley Long et al |

| | | | | | | |
|---|---|---|---|---|---|---|
| EPI_ISL_784454 | USA/TX-HMH-MCoV-18106/2020 | 2020-11-17 | North America/USA/Texas/Houston | Houston Methodist Hospital | Houston Methodist Hospital | S. Wesley Long et al |
| EPI_ISL_784460 | USA/TX-HMH-MCoV-18112/2020 | 2020-11-16 | North America/USA/Texas/Houston | Houston Methodist Hospital | Houston Methodist Hospital | S. Wesley Long et al |
| EPI_ISL_784462 | USA/TX-HMH-MCoV-18114/2020 | 2020-11-16 | North America/USA/Texas/Houston | Houston Methodist Hospital | Houston Methodist Hospital | S. Wesley Long et al |
| EPI_ISL_784463 | USA/TX-HMH-MCoV-18115/2020 | 2020-11-13 | North America/USA/Texas/Houston | Houston Methodist Hospital | Houston Methodist Hospital | S. Wesley Long et al |
| EPI_ISL_784464 | USA/TX-HMH-MCoV-18116/2020 | 2020-11-14 | North America/USA/Texas/Houston | Houston Methodist Hospital | Houston Methodist Hospital | S. Wesley Long et al |
| EPI_ISL_784469 | USA/TX-HMH-MCoV-18121/2020 | 2020-11-17 | North America/USA/Texas/Houston | Houston Methodist Hospital | Houston Methodist Hospital | S. Wesley Long et al |
| EPI_ISL_784470 | USA/TX-HMH-MCoV-18122/2020 | 2020-11-17 | North America/USA/Texas/Houston | Houston Methodist Hospital | Houston Methodist Hospital | S. Wesley Long et al |
| EPI_ISL_784485 | USA/TX-HMH-MCoV-18137/2020 | 2020-11-11 | North America/USA/Texas/Houston | Houston Methodist Hospital | Houston Methodist Hospital | S. Wesley Long et al |
| EPI_ISL_784498 | USA/TX-HMH-MCoV-18150/2020 | 2020-11-14 | North America/USA/Texas/Houston | Houston Methodist Hospital | Houston Methodist Hospital | S. Wesley Long et al |
| EPI_ISL_784503 | USA/TX-HMH-MCoV-18155/2020 | 2020-11-14 | North America/USA/Texas/Houston | Houston Methodist Hospital | Houston Methodist Hospital | S. Wesley Long et al |
| EPI_ISL_784504 | USA/TX-HMH-MCoV-18156/2020 | 2020-11-14 | North America/USA/Texas/Houston | Houston Methodist Hospital | Houston Methodist Hospital | S. Wesley Long et al |
| EPI_ISL_784505 | USA/TX-HMH-MCoV-18157/2020 | 2020-11-12 | North America/USA/Texas/Houston | Houston Methodist Hospital | Houston Methodist Hospital | S. Wesley Long et al |
| EPI_ISL_784511 | USA/TX-HMH-MCoV-18163/2020 | 2020-11-13 | North America/USA/Texas/Houston | Houston Methodist Hospital | Houston Methodist Hospital | S. Wesley Long et al |

| | | | | | | |
|---|---|---|---|---|---|---|
| EPI_ISL_784513 | USA/TX-HMH-MCoV-18165/2020 | 2020-11-14 | North America/USA/Texas/Houston | Houston Methodist Hospital | Houston Methodist Hospital | S. Wesley Long et al |
| EPI_ISL_784514 | USA/TX-HMH-MCoV-18166/2020 | 2020-11-17 | North America/USA/Texas/Houston | Houston Methodist Hospital | Houston Methodist Hospital | S. Wesley Long et al |
| EPI_ISL_784520 | USA/TX-HMH-MCoV-18172/2020 | 2020-11-15 | North America/USA/Texas/Houston | Houston Methodist Hospital | Houston Methodist Hospital | S. Wesley Long et al |
| EPI_ISL_784530 | USA/TX-HMH-MCoV-18182/2020 | 2020-11-14 | North America/USA/Texas/Houston | Houston Methodist Hospital | Houston Methodist Hospital | S. Wesley Long et al |
| EPI_ISL_784532 | USA/TX-HMH-MCoV-18184/2020 | 2020-11-12 | North America/USA/Texas/Houston | Houston Methodist Hospital | Houston Methodist Hospital | S. Wesley Long et al |
| EPI_ISL_784542 | USA/TX-HMH-MCoV-18194/2020 | 2020-11-15 | North America/USA/Texas/Houston | Houston Methodist Hospital | Houston Methodist Hospital | S. Wesley Long et al |
| EPI_ISL_784545 | USA/TX-HMH-MCoV-18197/2020 | 2020-11-15 | North America/USA/Texas/Houston | Houston Methodist Hospital | Houston Methodist Hospital | S. Wesley Long et al |
| EPI_ISL_784548 | USA/TX-HMH-MCoV-18200/2020 | 2020-11-13 | North America/USA/Texas/Houston | Houston Methodist Hospital | Houston Methodist Hospital | S. Wesley Long et al |
| EPI_ISL_784555 | USA/TX-HMH-MCoV-18207/2020 | 2020-11-11 | North America/USA/Texas/Houston | Houston Methodist Hospital | Houston Methodist Hospital | S. Wesley Long et al |
| EPI_ISL_784556 | USA/TX-HMH-MCoV-18208/2020 | 2020-11-12 | North America/USA/Texas/Houston | Houston Methodist Hospital | Houston Methodist Hospital | S. Wesley Long et al |
| EPI_ISL_784558 | USA/TX-HMH-MCoV-18210/2020 | 2020-11-13 | North America/USA/Texas/Houston | Houston Methodist Hospital | Houston Methodist Hospital | S. Wesley Long et al |
| EPI_ISL_784563 | USA/TX-HMH-MCoV-18215/2020 | 2020-11-13 | North America/USA/Texas/Houston | Houston Methodist Hospital | Houston Methodist Hospital | S. Wesley Long et al |
| EPI_ISL_784568 | USA/TX-HMH-MCoV-18220/2020 | 2020-11-17 | North America/USA/Texas/Houston | Houston Methodist Hospital | Houston Methodist Hospital | S. Wesley Long et al |

| | | | | | | |
|---|---|---|---|---|---|---|
| EPI_ISL_784572 | USA/TX-HMH-MCoV-18224/2020 | 2020-11-16 | North America/USA/Texas/Houston | Houston Methodist Hospital | Houston Methodist Hospital | S. Wesley Long et al |
| EPI_ISL_784576 | USA/TX-HMH-MCoV-18228/2020 | 2020-11-12 | North America/USA/Texas/Houston | Houston Methodist Hospital | Houston Methodist Hospital | S. Wesley Long et al |
| EPI_ISL_784581 | USA/TX-HMH-MCoV-18233/2020 | 2020-11-14 | North America/USA/Texas/Houston | Houston Methodist Hospital | Houston Methodist Hospital | S. Wesley Long et al |
| EPI_ISL_784586 | USA/TX-HMH-MCoV-18238/2020 | 2020-11-17 | North America/USA/Texas/Houston | Houston Methodist Hospital | Houston Methodist Hospital | S. Wesley Long et al |
| EPI_ISL_784589 | USA/TX-HMH-MCoV-18241/2020 | 2020-11-17 | North America/USA/Texas/Houston | Houston Methodist Hospital | Houston Methodist Hospital | S. Wesley Long et al |
| EPI_ISL_784593 | USA/TX-HMH-MCoV-18245/2020 | 2020-11-17 | North America/USA/Texas/Houston | Houston Methodist Hospital | Houston Methodist Hospital | S. Wesley Long et al |
| EPI_ISL_784594 | USA/TX-HMH-MCoV-18246/2020 | 2020-11-17 | North America/USA/Texas/Houston | Houston Methodist Hospital | Houston Methodist Hospital | S. Wesley Long et al |
| EPI_ISL_784600 | USA/TX-HMH-MCoV-18252/2020 | 2020-11-17 | North America/USA/Texas/Houston | Houston Methodist Hospital | Houston Methodist Hospital | S. Wesley Long et al |
| EPI_ISL_784603 | USA/TX-HMH-MCoV-18255/2020 | 2020-11-17 | North America/USA/Texas/Houston | Houston Methodist Hospital | Houston Methodist Hospital | S. Wesley Long et al |
| EPI_ISL_784609 | USA/TX-HMH-MCoV-18261/2020 | 2020-11-17 | North America/USA/Texas/Houston | Houston Methodist Hospital | Houston Methodist Hospital | S. Wesley Long et al |
| EPI_ISL_784614 | USA/TX-HMH-MCoV-18266/2020 | 2020-11-17 | North America/USA/Texas/Houston | Houston Methodist Hospital | Houston Methodist Hospital | S. Wesley Long et al |
| EPI_ISL_784621 | USA/TX-HMH-MCoV-18273/2020 | 2020-11-17 | North America/USA/Texas/Houston | Houston Methodist Hospital | Houston Methodist Hospital | S. Wesley Long et al |
| EPI_ISL_784655 | USA/TX-HMH-MCoV-18307/2020 | 2020-11-17 | North America/USA/Texas/Houston | Houston Methodist Hospital | Houston Methodist Hospital | S. Wesley Long et al |

| | | | | | | |
|---|---|---|---|---|---|---|
| EPI_ISL_784657 | USA/TX-HMH-MCoV-18309/2020 | 2020-11-17 | North America/USA/Texas/Houston | Houston Methodist Hospital | Houston Methodist Hospital | S. Wesley Long et al |
| EPI_ISL_784660 | USA/TX-HMH-MCoV-18312/2020 | 2020-11-17 | North America/USA/Texas/Houston | Houston Methodist Hospital | Houston Methodist Hospital | S. Wesley Long et al |
| EPI_ISL_784661 | USA/TX-HMH-MCoV-18313/2020 | 2020-11-17 | North America/USA/Texas/Houston | Houston Methodist Hospital | Houston Methodist Hospital | S. Wesley Long et al |
| EPI_ISL_784666 | USA/TX-HMH-MCoV-18318/2020 | 2020-11-17 | North America/USA/Texas/Houston | Houston Methodist Hospital | Houston Methodist Hospital | S. Wesley Long et al |
| EPI_ISL_784669 | USA/TX-HMH-MCoV-18321/2020 | 2020-11-17 | North America/USA/Texas/Houston | Houston Methodist Hospital | Houston Methodist Hospital | S. Wesley Long et al |
| EPI_ISL_784672 | USA/TX-HMH-MCoV-18324/2020 | 2020-11-17 | North America/USA/Texas/Houston | Houston Methodist Hospital | Houston Methodist Hospital | S. Wesley Long et al |
| EPI_ISL_784680 | USA/TX-HMH-MCoV-18332/2020 | 2020-11-18 | North America/USA/Texas/Houston | Houston Methodist Hospital | Houston Methodist Hospital | S. Wesley Long et al |
| EPI_ISL_784700 | USA/TX-HMH-MCoV-18352/2020 | 2020-11-18 | North America/USA/Texas/Houston | Houston Methodist Hospital | Houston Methodist Hospital | S. Wesley Long et al |
| EPI_ISL_784707 | USA/TX-HMH-MCoV-18359/2020 | 2020-11-20 | North America/USA/Texas/Houston | Houston Methodist Hospital | Houston Methodist Hospital | S. Wesley Long et al |
| EPI_ISL_784708 | USA/TX-HMH-MCoV-18360/2020 | 2020-11-18 | North America/USA/Texas/Houston | Houston Methodist Hospital | Houston Methodist Hospital | S. Wesley Long et al |
| EPI_ISL_784715 | USA/TX-HMH-MCoV-18367/2020 | 2020-11-18 | North America/USA/Texas/Houston | Houston Methodist Hospital | Houston Methodist Hospital | S. Wesley Long et al |
| EPI_ISL_784729 | USA/TX-HMH-MCoV-18381/2020 | 2020-11-18 | North America/USA/Texas/Houston | Houston Methodist Hospital | Houston Methodist Hospital | S. Wesley Long et al |
| EPI_ISL_784733 | USA/TX-HMH-MCoV-18385/2020 | 2020-11-18 | North America/USA/Texas/Houston | Houston Methodist Hospital | Houston Methodist Hospital | S. Wesley Long et al |

| | | | | | | |
|---|---|---|---|---|---|---|
| EPI_ISL_784742 | USA/TX-HMH-MCoV-18394/2020 | 2020-11-18 | North America/USA/Texas/Houston | Houston Methodist Hospital | Houston Methodist Hospital | S. Wesley Long et al |
| EPI_ISL_784745 | USA/TX-HMH-MCoV-18397/2020 | 2020-11-19 | North America/USA/Texas/Houston | Houston Methodist Hospital | Houston Methodist Hospital | S. Wesley Long et al |
| EPI_ISL_784752 | USA/TX-HMH-MCoV-18404/2020 | 2020-11-18 | North America/USA/Texas/Houston | Houston Methodist Hospital | Houston Methodist Hospital | S. Wesley Long et al |
| EPI_ISL_784760 | USA/TX-HMH-MCoV-18412/2020 | 2020-11-18 | North America/USA/Texas/Houston | Houston Methodist Hospital | Houston Methodist Hospital | S. Wesley Long et al |
| EPI_ISL_784763 | USA/TX-HMH-MCoV-18415/2020 | 2020-11-18 | North America/USA/Texas/Houston | Houston Methodist Hospital | Houston Methodist Hospital | S. Wesley Long et al |
| EPI_ISL_784768 | USA/TX-HMH-MCoV-18420/2020 | 2020-11-18 | North America/USA/Texas/Houston | Houston Methodist Hospital | Houston Methodist Hospital | S. Wesley Long et al |
| EPI_ISL_784773 | USA/TX-HMH-MCoV-18425/2020 | 2020-11-20 | North America/USA/Texas/Houston | Houston Methodist Hospital | Houston Methodist Hospital | S. Wesley Long et al |
| EPI_ISL_784779 | USA/TX-HMH-MCoV-18431/2020 | 2020-11-20 | North America/USA/Texas/Houston | Houston Methodist Hospital | Houston Methodist Hospital | S. Wesley Long et al |
| EPI_ISL_784784 | USA/TX-HMH-MCoV-18436/2020 | 2020-11-18 | North America/USA/Texas/Houston | Houston Methodist Hospital | Houston Methodist Hospital | S. Wesley Long et al |
| EPI_ISL_784785 | USA/TX-HMH-MCoV-18437/2020 | 2020-11-18 | North America/USA/Texas/Houston | Houston Methodist Hospital | Houston Methodist Hospital | S. Wesley Long et al |
| EPI_ISL_784786 | USA/TX-HMH-MCoV-18438/2020 | 2020-11-20 | North America/USA/Texas/Houston | Houston Methodist Hospital | Houston Methodist Hospital | S. Wesley Long et al |
| EPI_ISL_784790 | USA/TX-HMH-MCoV-18442/2020 | 2020-11-20 | North America/USA/Texas/Houston | Houston Methodist Hospital | Houston Methodist Hospital | S. Wesley Long et al |
| EPI_ISL_784791 | USA/TX-HMH-MCoV-18443/2020 | 2020-11-19 | North America/USA/Texas/Houston | Houston Methodist Hospital | Houston Methodist Hospital | S. Wesley Long et al |

| | | | | | | |
|---|---|---|---|---|---|---|
| EPI_ISL_784792 | USA/TX-HMH-MCoV-18444/2020 | 2020-11-19 | North America/USA/Texas/Houston | Houston Methodist Hospital | Houston Methodist Hospital | S. Wesley Long et al |
| EPI_ISL_784797 | USA/TX-HMH-MCoV-18449/2020 | 2020-11-18 | North America/USA/Texas/Houston | Houston Methodist Hospital | Houston Methodist Hospital | S. Wesley Long et al |
| EPI_ISL_784807 | USA/TX-HMH-MCoV-18459/2020 | 2020-11-18 | North America/USA/Texas/Houston | Houston Methodist Hospital | Houston Methodist Hospital | S. Wesley Long et al |
| EPI_ISL_784810 | USA/TX-HMH-MCoV-18462/2020 | 2020-11-18 | North America/USA/Texas/Houston | Houston Methodist Hospital | Houston Methodist Hospital | S. Wesley Long et al |
| EPI_ISL_784813 | USA/TX-HMH-MCoV-18465/2020 | 2020-11-20 | North America/USA/Texas/Houston | Houston Methodist Hospital | Houston Methodist Hospital | S. Wesley Long et al |
| EPI_ISL_784814 | USA/TX-HMH-MCoV-18466/2020 | 2020-11-20 | North America/USA/Texas/Houston | Houston Methodist Hospital | Houston Methodist Hospital | S. Wesley Long et al |
| EPI_ISL_784818 | USA/TX-HMH-MCoV-18470/2020 | 2020-11-20 | North America/USA/Texas/Houston | Houston Methodist Hospital | Houston Methodist Hospital | S. Wesley Long et al |
| EPI_ISL_784831 | USA/TX-HMH-MCoV-18483/2020 | 2020-11-18 | North America/USA/Texas/Houston | Houston Methodist Hospital | Houston Methodist Hospital | S. Wesley Long et al |
| EPI_ISL_784833 | USA/TX-HMH-MCoV-18485/2020 | 2020-11-18 | North America/USA/Texas/Houston | Houston Methodist Hospital | Houston Methodist Hospital | S. Wesley Long et al |
| EPI_ISL_784834 | USA/TX-HMH-MCoV-18486/2020 | 2020-11-18 | North America/USA/Texas/Houston | Houston Methodist Hospital | Houston Methodist Hospital | S. Wesley Long et al |
| EPI_ISL_784838 | USA/TX-HMH-MCoV-18490/2020 | 2020-11-19 | North America/USA/Texas/Houston | Houston Methodist Hospital | Houston Methodist Hospital | S. Wesley Long et al |
| EPI_ISL_784850 | USA/TX-HMH-MCoV-18502/2020 | 2020-11-19 | North America/USA/Texas/Houston | Houston Methodist Hospital | Houston Methodist Hospital | S. Wesley Long et al |
| EPI_ISL_784854 | USA/TX-HMH-MCoV-18506/2020 | 2020-11-20 | North America/USA/Texas/Houston | Houston Methodist Hospital | Houston Methodist Hospital | S. Wesley Long et al |

| | | | | | | |
|---|---|---|---|---|---|---|
| EPI_ISL_784856 | USA/TX-HMH-MCoV-18508/2020 | 2020-11-18 | North America/USA/Texas/Houston | Houston Methodist Hospital | Houston Methodist Hospital | S. Wesley Long et al |
| EPI_ISL_784858 | USA/TX-HMH-MCoV-18510/2020 | 2020-11-18 | North America/USA/Texas/Houston | Houston Methodist Hospital | Houston Methodist Hospital | S. Wesley Long et al |
| EPI_ISL_784864 | USA/TX-HMH-MCoV-18516/2020 | 2020-11-19 | North America/USA/Texas/Houston | Houston Methodist Hospital | Houston Methodist Hospital | S. Wesley Long et al |
| EPI_ISL_784884 | USA/TX-HMH-MCoV-18536/2020 | 2020-11-20 | North America/USA/Texas/Houston | Houston Methodist Hospital | Houston Methodist Hospital | S. Wesley Long et al |
| EPI_ISL_784899 | USA/TX-HMH-MCoV-18551/2020 | 2020-11-19 | North America/USA/Texas/Houston | Houston Methodist Hospital | Houston Methodist Hospital | S. Wesley Long et al |
| EPI_ISL_784900 | USA/TX-HMH-MCoV-18552/2020 | 2020-11-20 | North America/USA/Texas/Houston | Houston Methodist Hospital | Houston Methodist Hospital | S. Wesley Long et al |
| EPI_ISL_784901 | USA/TX-HMH-MCoV-18553/2020 | 2020-11-19 | North America/USA/Texas/Houston | Houston Methodist Hospital | Houston Methodist Hospital | S. Wesley Long et al |
| EPI_ISL_784909 | USA/TX-HMH-MCoV-18561/2020 | 2020-11-20 | North America/USA/Texas/Houston | Houston Methodist Hospital | Houston Methodist Hospital | S. Wesley Long et al |
| EPI_ISL_784912 | USA/TX-HMH-MCoV-18564/2020 | 2020-11-19 | North America/USA/Texas/Houston | Houston Methodist Hospital | Houston Methodist Hospital | S. Wesley Long et al |
| EPI_ISL_784922 | USA/TX-HMH-MCoV-18574/2020 | 2020-11-20 | North America/USA/Texas/Houston | Houston Methodist Hospital | Houston Methodist Hospital | S. Wesley Long et al |
| EPI_ISL_784925 | USA/TX-HMH-MCoV-18577/2020 | 2020-11-19 | North America/USA/Texas/Houston | Houston Methodist Hospital | Houston Methodist Hospital | S. Wesley Long et al |
| EPI_ISL_784935 | USA/TX-HMH-MCoV-18587/2020 | 2020-11-20 | North America/USA/Texas/Houston | Houston Methodist Hospital | Houston Methodist Hospital | S. Wesley Long et al |
| EPI_ISL_784936 | USA/TX-HMH-MCoV-18588/2020 | 2020-11-19 | North America/USA/Texas/Houston | Houston Methodist Hospital | Houston Methodist Hospital | S. Wesley Long et al |

| | | | | | | |
|---|---|---|---|---|---|---|
| EPI_ISL_784946 | USA/TX-HMH-MCoV-18598/2020 | 2020-11-19 | North America/USA/Texas/Houston | Houston Methodist Hospital | Houston Methodist Hospital | S. Wesley Long et al |
| EPI_ISL_784947 | USA/TX-HMH-MCoV-18599/2020 | 2020-11-19 | North America/USA/Texas/Houston | Houston Methodist Hospital | Houston Methodist Hospital | S. Wesley Long et al |
| EPI_ISL_784952 | USA/TX-HMH-MCoV-18604/2020 | 2020-11-20 | North America/USA/Texas/Houston | Houston Methodist Hospital | Houston Methodist Hospital | S. Wesley Long et al |
| EPI_ISL_784958 | USA/TX-HMH-MCoV-18610/2020 | 2020-11-19 | North America/USA/Texas/Houston | Houston Methodist Hospital | Houston Methodist Hospital | S. Wesley Long et al |
| EPI_ISL_784962 | USA/TX-HMH-MCoV-18614/2020 | 2020-11-19 | North America/USA/Texas/Houston | Houston Methodist Hospital | Houston Methodist Hospital | S. Wesley Long et al |
| EPI_ISL_784973 | USA/TX-HMH-MCoV-18625/2020 | 2020-11-22 | North America/USA/Texas/Houston | Houston Methodist Hospital | Houston Methodist Hospital | S. Wesley Long et al |
| EPI_ISL_784974 | USA/TX-HMH-MCoV-18626/2020 | 2020-11-20 | North America/USA/Texas/Houston | Houston Methodist Hospital | Houston Methodist Hospital | S. Wesley Long et al |
| EPI_ISL_784978 | USA/TX-HMH-MCoV-18630/2020 | 2020-11-20 | North America/USA/Texas/Houston | Houston Methodist Hospital | Houston Methodist Hospital | S. Wesley Long et al |
| EPI_ISL_784983 | USA/TX-HMH-MCoV-18635/2020 | 2020-11-20 | North America/USA/Texas/Houston | Houston Methodist Hospital | Houston Methodist Hospital | S. Wesley Long et al |
| EPI_ISL_784988 | USA/TX-HMH-MCoV-18640/2020 | 2020-11-20 | North America/USA/Texas/Houston | Houston Methodist Hospital | Houston Methodist Hospital | S. Wesley Long et al |
| EPI_ISL_784990 | USA/TX-HMH-MCoV-18642/2020 | 2020-11-20 | North America/USA/Texas/Houston | Houston Methodist Hospital | Houston Methodist Hospital | S. Wesley Long et al |
| EPI_ISL_784997 | USA/TX-HMH-MCoV-18649/2020 | 2020-11-23 | North America/USA/Texas/Houston | Houston Methodist Hospital | Houston Methodist Hospital | S. Wesley Long et al |
| EPI_ISL_785003 | USA/TX-HMH-MCoV-18655/2020 | 2020-11-20 | North America/USA/Texas/Houston | Houston Methodist Hospital | Houston Methodist Hospital | S. Wesley Long et al |

| | | | | | | |
|---|---|---|---|---|---|---|
| EPI_ISL_785006 | USA/TX-HMH-MCoV-18658/2020 | 2020-11-21 | North America/USA/Texas/Houston | Houston Methodist Hospital | Houston Methodist Hospital | S. Wesley Long et al |
| EPI_ISL_785007 | USA/TX-HMH-MCoV-18659/2020 | 2020-11-17 | North America/USA/Texas/Houston | Houston Methodist Hospital | Houston Methodist Hospital | S. Wesley Long et al |
| EPI_ISL_785010 | USA/TX-HMH-MCoV-18662/2020 | 2020-11-21 | North America/USA/Texas/Houston | Houston Methodist Hospital | Houston Methodist Hospital | S. Wesley Long et al |
| EPI_ISL_785012 | USA/TX-HMH-MCoV-18664/2020 | 2020-11-19 | North America/USA/Texas/Houston | Houston Methodist Hospital | Houston Methodist Hospital | S. Wesley Long et al |
| EPI_ISL_785013 | USA/TX-HMH-MCoV-18665/2020 | 2020-11-19 | North America/USA/Texas/Houston | Houston Methodist Hospital | Houston Methodist Hospital | S. Wesley Long et al |
| EPI_ISL_785014 | USA/TX-HMH-MCoV-18666/2020 | 2020-11-20 | North America/USA/Texas/Houston | Houston Methodist Hospital | Houston Methodist Hospital | S. Wesley Long et al |
| EPI_ISL_785018 | USA/TX-HMH-MCoV-18670/2020 | 2020-11-21 | North America/USA/Texas/Houston | Houston Methodist Hospital | Houston Methodist Hospital | S. Wesley Long et al |
| EPI_ISL_785030 | USA/TX-HMH-MCoV-18682/2020 | 2020-11-21 | North America/USA/Texas/Houston | Houston Methodist Hospital | Houston Methodist Hospital | S. Wesley Long et al |
| EPI_ISL_785033 | USA/TX-HMH-MCoV-18685/2020 | 2020-11-21 | North America/USA/Texas/Houston | Houston Methodist Hospital | Houston Methodist Hospital | S. Wesley Long et al |
| EPI_ISL_785038 | USA/TX-HMH-MCoV-18690/2020 | 2020-11-19 | North America/USA/Texas/Houston | Houston Methodist Hospital | Houston Methodist Hospital | S. Wesley Long et al |
| EPI_ISL_785040 | USA/TX-HMH-MCoV-18692/2020 | 2020-11-21 | North America/USA/Texas/Houston | Houston Methodist Hospital | Houston Methodist Hospital | S. Wesley Long et al |
| EPI_ISL_785049 | USA/TX-HMH-MCoV-18701/2020 | 2020-11-19 | North America/USA/Texas/Houston | Houston Methodist Hospital | Houston Methodist Hospital | S. Wesley Long et al |
| EPI_ISL_785068 | USA/TX-HMH-MCoV-18721/2020 | 2020-11-23 | North America/USA/Texas/Houston | Houston Methodist Hospital | Houston Methodist Hospital | S. Wesley Long et al |

| | | | | | | |
|---|---|---|---|---|---|---|
| EPI_ISL_785076 | USA/TX-HMH-MCoV-18731/2020 | 2020-11-24 | North America/USA/Texas/Houston | Houston Methodist Hospital | Houston Methodist Hospital | S. Wesley Long et al |
| EPI_ISL_785080 | USA/TX-HMH-MCoV-18738/2020 | 2020-11-21 | North America/USA/Texas/Houston | Houston Methodist Hospital | Houston Methodist Hospital | S. Wesley Long et al |
| EPI_ISL_785083 | USA/TX-HMH-MCoV-18742/2020 | 2020-11-22 | North America/USA/Texas/Houston | Houston Methodist Hospital | Houston Methodist Hospital | S. Wesley Long et al |
| EPI_ISL_785094 | USA/TX-HMH-MCoV-18757/2020 | 2020-11-23 | North America/USA/Texas/Houston | Houston Methodist Hospital | Houston Methodist Hospital | S. Wesley Long et al |
| EPI_ISL_785095 | USA/TX-HMH-MCoV-18758/2020 | 2020-11-23 | North America/USA/Texas/Houston | Houston Methodist Hospital | Houston Methodist Hospital | S. Wesley Long et al |
| EPI_ISL_785100 | USA/TX-HMH-MCoV-18768/2020 | 2020-11-23 | North America/USA/Texas/Houston | Houston Methodist Hospital | Houston Methodist Hospital | S. Wesley Long et al |
| EPI_ISL_785103 | USA/TX-HMH-MCoV-18771/2020 | 2020-11-22 | North America/USA/Texas/Houston | Houston Methodist Hospital | Houston Methodist Hospital | S. Wesley Long et al |
| EPI_ISL_785105 | USA/TX-HMH-MCoV-18773/2020 | 2020-11-21 | North America/USA/Texas/Houston | Houston Methodist Hospital | Houston Methodist Hospital | S. Wesley Long et al |
| EPI_ISL_785117 | USA/TX-HMH-MCoV-18787/2020 | 2020-11-21 | North America/USA/Texas/Houston | Houston Methodist Hospital | Houston Methodist Hospital | S. Wesley Long et al |
| EPI_ISL_785122 | USA/TX-HMH-MCoV-18795/2020 | 2020-11-17 | North America/USA/Texas/Houston | Houston Methodist Hospital | Houston Methodist Hospital | S. Wesley Long et al |
| EPI_ISL_785128 | USA/TX-HMH-MCoV-18801/2020 | 2020-11-21 | North America/USA/Texas/Houston | Houston Methodist Hospital | Houston Methodist Hospital | S. Wesley Long et al |
| EPI_ISL_785134 | USA/TX-HMH-MCoV-18807/2020 | 2020-11-22 | North America/USA/Texas/Houston | Houston Methodist Hospital | Houston Methodist Hospital | S. Wesley Long et al |
| EPI_ISL_785142 | USA/TX-HMH-MCoV-18816/2020 | 2020-11-26 | North America/USA/Texas/Houston | Houston Methodist Hospital | Houston Methodist Hospital | S. Wesley Long et al |

| | | | | | | |
|---|---|---|---|---|---|---|
| EPI_ISL_785145 | USA/TX-HMH-MCoV-18821/2020 | 2020-11-24 | North America/USA/Texas/Houston | Houston Methodist Hospital | Houston Methodist Hospital | S. Wesley Long et al |
| EPI_ISL_785149 | USA/TX-HMH-MCoV-18828/2020 | 2020-11-25 | North America/USA/Texas/Houston | Houston Methodist Hospital | Houston Methodist Hospital | S. Wesley Long et al |
| EPI_ISL_785153 | USA/TX-HMH-MCoV-18833/2020 | 2020-11-24 | North America/USA/Texas/Houston | Houston Methodist Hospital | Houston Methodist Hospital | S. Wesley Long et al |
| EPI_ISL_785154 | USA/TX-HMH-MCoV-18834/2020 | 2020-11-24 | North America/USA/Texas/Houston | Houston Methodist Hospital | Houston Methodist Hospital | S. Wesley Long et al |
| EPI_ISL_785155 | USA/TX-HMH-MCoV-18835/2020 | 2020-11-23 | North America/USA/Texas/Houston | Houston Methodist Hospital | Houston Methodist Hospital | S. Wesley Long et al |
| EPI_ISL_785156 | USA/TX-HMH-MCoV-18837/2020 | 2020-11-25 | North America/USA/Texas/Houston | Houston Methodist Hospital | Houston Methodist Hospital | S. Wesley Long et al |
| EPI_ISL_785161 | USA/TX-HMH-MCoV-18842/2020 | 2020-11-25 | North America/USA/Texas/Houston | Houston Methodist Hospital | Houston Methodist Hospital | S. Wesley Long et al |
| EPI_ISL_785162 | USA/TX-HMH-MCoV-18843/2020 | 2020-11-23 | North America/USA/Texas/Houston | Houston Methodist Hospital | Houston Methodist Hospital | S. Wesley Long et al |
| EPI_ISL_785163 | USA/TX-HMH-MCoV-18844/2020 | 2020-11-24 | North America/USA/Texas/Houston | Houston Methodist Hospital | Houston Methodist Hospital | S. Wesley Long et al |
| EPI_ISL_785166 | USA/TX-HMH-MCoV-18849/2020 | 2020-11-26 | North America/USA/Texas/Houston | Houston Methodist Hospital | Houston Methodist Hospital | S. Wesley Long et al |
| EPI_ISL_785182 | USA/TX-HMH-MCoV-18866/2020 | 2020-11-26 | North America/USA/Texas/Houston | Houston Methodist Hospital | Houston Methodist Hospital | S. Wesley Long et al |
| EPI_ISL_785183 | USA/TX-HMH-MCoV-18867/2020 | 2020-11-24 | North America/USA/Texas/Houston | Houston Methodist Hospital | Houston Methodist Hospital | S. Wesley Long et al |
| EPI_ISL_785194 | USA/TX-HMH-MCoV-18881/2020 | 2020-11-23 | North America/USA/Texas/Houston | Houston Methodist Hospital | Houston Methodist Hospital | S. Wesley Long et al |

| | | | | | | |
|---|---|---|---|---|---|---|
| EPI_ISL_785203 | USA/TX-HMH-MCoV-18895/2020 | 2020-11-25 | North America/USA/Texas/Houston | Houston Methodist Hospital | Houston Methodist Hospital | S. Wesley Long et al |
| EPI_ISL_785206 | USA/TX-HMH-MCoV-18898/2020 | 2020-11-25 | North America/USA/Texas/Houston | Houston Methodist Hospital | Houston Methodist Hospital | S. Wesley Long et al |
| EPI_ISL_785213 | USA/TX-HMH-MCoV-18908/2020 | 2020-11-25 | North America/USA/Texas/Houston | Houston Methodist Hospital | Houston Methodist Hospital | S. Wesley Long et al |
| EPI_ISL_785231 | USA/TX-HMH-MCoV-18935/2020 | 2020-11-25 | North America/USA/Texas/Houston | Houston Methodist Hospital | Houston Methodist Hospital | S. Wesley Long et al |
| EPI_ISL_785234 | USA/TX-HMH-MCoV-18938/2020 | 2020-11-26 | North America/USA/Texas/Houston | Houston Methodist Hospital | Houston Methodist Hospital | S. Wesley Long et al |
| EPI_ISL_785235 | USA/TX-HMH-MCoV-18939/2020 | 2020-11-25 | North America/USA/Texas/Houston | Houston Methodist Hospital | Houston Methodist Hospital | S. Wesley Long et al |
| EPI_ISL_785245 | USA/TX-HMH-MCoV-18953/2020 | 2020-11-26 | North America/USA/Texas/Houston | Houston Methodist Hospital | Houston Methodist Hospital | S. Wesley Long et al |
| EPI_ISL_785248 | USA/TX-HMH-MCoV-18960/2020 | 2020-11-27 | North America/USA/Texas/Houston | Houston Methodist Hospital | Houston Methodist Hospital | S. Wesley Long et al |
| EPI_ISL_785251 | USA/TX-HMH-MCoV-18965/2020 | 2020-11-25 | North America/USA/Texas/Houston | Houston Methodist Hospital | Houston Methodist Hospital | S. Wesley Long et al |
| EPI_ISL_785256 | USA/TX-HMH-MCoV-18973/2020 | 2020-11-25 | North America/USA/Texas/Houston | Houston Methodist Hospital | Houston Methodist Hospital | S. Wesley Long et al |
| EPI_ISL_785262 | USA/TX-HMH-MCoV-18980/2020 | 2020-11-26 | North America/USA/Texas/Houston | Houston Methodist Hospital | Houston Methodist Hospital | S. Wesley Long et al |
| EPI_ISL_785266 | USA/TX-HMH-MCoV-18985/2020 | 2020-11-25 | North America/USA/Texas/Houston | Houston Methodist Hospital | Houston Methodist Hospital | S. Wesley Long et al |
| EPI_ISL_785271 | USA/TX-HMH-MCoV-18996/2020 | 2020-11-27 | North America/USA/Texas/Houston | Houston Methodist Hospital | Houston Methodist Hospital | S. Wesley Long et al |

| | | | | | | |
|---|---|---|---|---|---|---|
| EPI_ISL_785275 | USA/TX-HMH-MCoV-19001/2020 | 2020-11-25 | North America/USA/Texas/Houston | Houston Methodist Hospital | Houston Methodist Hospital | S. Wesley Long et al |
| EPI_ISL_785276 | USA/TX-HMH-MCoV-19003/2020 | 2020-11-25 | North America/USA/Texas/Houston | Houston Methodist Hospital | Houston Methodist Hospital | S. Wesley Long et al |
| EPI_ISL_785277 | USA/TX-HMH-MCoV-19004/2020 | 2020-11-25 | North America/USA/Texas/Houston | Houston Methodist Hospital | Houston Methodist Hospital | S. Wesley Long et al |
| EPI_ISL_785280 | USA/TX-HMH-MCoV-19010/2020 | 2020-11-23 | North America/USA/Texas/Houston | Houston Methodist Hospital | Houston Methodist Hospital | S. Wesley Long et al |
| EPI_ISL_785281 | USA/TX-HMH-MCoV-19011/2020 | 2020-11-26 | North America/USA/Texas/Houston | Houston Methodist Hospital | Houston Methodist Hospital | S. Wesley Long et al |
| EPI_ISL_785289 | USA/TX-HMH-MCoV-19023/2020 | 2020-11-25 | North America/USA/Texas/Houston | Houston Methodist Hospital | Houston Methodist Hospital | S. Wesley Long et al |
| EPI_ISL_785297 | USA/TX-HMH-MCoV-19040/2020 | 2020-11-25 | North America/USA/Texas/Houston | Houston Methodist Hospital | Houston Methodist Hospital | S. Wesley Long et al |
| EPI_ISL_785299 | USA/TX-HMH-MCoV-15615/2020 | 2020-10-23 | North America/USA/Texas/Houston | Houston Methodist Hospital | Houston Methodist Hospital | S. Wesley Long et al |
| EPI_ISL_785300 | USA/TX-HMH-MCoV-15616/2020 | 2020-10-26 | North America/USA/Texas/Houston | Houston Methodist Hospital | Houston Methodist Hospital | S. Wesley Long et al |
| EPI_ISL_785302 | USA/TX-HMH-MCoV-15618/2020 | 2020-10-26 | North America/USA/Texas/Houston | Houston Methodist Hospital | Houston Methodist Hospital | S. Wesley Long et al |
| EPI_ISL_785305 | USA/TX-HMH-MCoV-15621/2020 | 2020-10-26 | North America/USA/Texas/Houston | Houston Methodist Hospital | Houston Methodist Hospital | S. Wesley Long et al |
| EPI_ISL_785333 | USA/TX-HMH-MCoV-15649/2020 | 2020-07-09 | North America/USA/Texas/Houston | Houston Methodist Hospital | Houston Methodist Hospital | S. Wesley Long et al |
| EPI_ISL_785337 | USA/TX-HMH-MCoV-15653/2020 | 2020-07-09 | North America/USA/Texas/Houston | Houston Methodist Hospital | Houston Methodist Hospital | S. Wesley Long et al |

| | | | | | | |
|---|---|---|---|---|---|---|
| EPI_ISL_785340 | USA/TX-HMH-MCoV-15656/2020 | 2020-07-09 | North America/USA/Texas/Houston | Houston Methodist Hospital | Houston Methodist Hospital | S. Wesley Long et al |
| EPI_ISL_785341 | USA/TX-HMH-MCoV-15657/2020 | 2020-07-09 | North America/USA/Texas/Houston | Houston Methodist Hospital | Houston Methodist Hospital | S. Wesley Long et al |
| EPI_ISL_785343 | USA/TX-HMH-MCoV-15659/2020 | 2020-07-09 | North America/USA/Texas/Houston | Houston Methodist Hospital | Houston Methodist Hospital | S. Wesley Long et al |
| EPI_ISL_785349 | USA/TX-HMH-MCoV-15665/2020 | 2020-07-09 | North America/USA/Texas/Houston | Houston Methodist Hospital | Houston Methodist Hospital | S. Wesley Long et al |
| EPI_ISL_785351 | USA/TX-HMH-MCoV-15667/2020 | 2020-07-09 | North America/USA/Texas/Houston | Houston Methodist Hospital | Houston Methodist Hospital | S. Wesley Long et al |
| EPI_ISL_785354 | USA/TX-HMH-MCoV-15670/2020 | 2020-07-09 | North America/USA/Texas/Houston | Houston Methodist Hospital | Houston Methodist Hospital | S. Wesley Long et al |
| EPI_ISL_785357 | USA/TX-HMH-MCoV-15673/2020 | 2020-07-09 | North America/USA/Texas/Houston | Houston Methodist Hospital | Houston Methodist Hospital | S. Wesley Long et al |
| EPI_ISL_785365 | USA/TX-HMH-MCoV-15681/2020 | 2020-07-09 | North America/USA/Texas/Houston | Houston Methodist Hospital | Houston Methodist Hospital | S. Wesley Long et al |
| EPI_ISL_785366 | USA/TX-HMH-MCoV-15682/2020 | 2020-07-09 | North America/USA/Texas/Houston | Houston Methodist Hospital | Houston Methodist Hospital | S. Wesley Long et al |
| EPI_ISL_785368 | USA/TX-HMH-MCoV-15684/2020 | 2020-07-09 | North America/USA/Texas/Houston | Houston Methodist Hospital | Houston Methodist Hospital | S. Wesley Long et al |
| EPI_ISL_785374 | USA/TX-HMH-MCoV-15690/2020 | 2020-07-09 | North America/USA/Texas/Houston | Houston Methodist Hospital | Houston Methodist Hospital | S. Wesley Long et al |
| EPI_ISL_785375 | USA/TX-HMH-MCoV-15691/2020 | 2020-07-09 | North America/USA/Texas/Houston | Houston Methodist Hospital | Houston Methodist Hospital | S. Wesley Long et al |
| EPI_ISL_785380 | USA/TX-HMH-MCoV-15696/2020 | 2020-07-09 | North America/USA/Texas/Houston | Houston Methodist Hospital | Houston Methodist Hospital | S. Wesley Long et al |

| | | | | | | |
|---|---|---|---|---|---|---|
| EPI_ISL_785382 | USA/TX-HMH-MCoV-15698/2020 | 2020-07-09 | North America/USA/Texas/Houston | Houston Methodist Hospital | Houston Methodist Hospital | S. Wesley Long et al |
| EPI_ISL_785384 | USA/TX-HMH-MCoV-15700/2020 | 2020-07-10 | North America/USA/Texas/Houston | Houston Methodist Hospital | Houston Methodist Hospital | S. Wesley Long et al |
| EPI_ISL_785385 | USA/TX-HMH-MCoV-15701/2020 | 2020-07-09 | North America/USA/Texas/Houston | Houston Methodist Hospital | Houston Methodist Hospital | S. Wesley Long et al |
| EPI_ISL_785386 | USA/TX-HMH-MCoV-15702/2020 | 2020-07-09 | North America/USA/Texas/Houston | Houston Methodist Hospital | Houston Methodist Hospital | S. Wesley Long et al |
| EPI_ISL_785388 | USA/TX-HMH-MCoV-15704/2020 | 2020-07-09 | North America/USA/Texas/Houston | Houston Methodist Hospital | Houston Methodist Hospital | S. Wesley Long et al |
| EPI_ISL_785394 | USA/TX-HMH-MCoV-15710/2020 | 2020-07-09 | North America/USA/Texas/Houston | Houston Methodist Hospital | Houston Methodist Hospital | S. Wesley Long et al |
| EPI_ISL_785403 | USA/TX-HMH-MCoV-15719/2020 | 2020-07-09 | North America/USA/Texas/Houston | Houston Methodist Hospital | Houston Methodist Hospital | S. Wesley Long et al |
| EPI_ISL_785405 | USA/TX-HMH-MCoV-15721/2020 | 2020-07-09 | North America/USA/Texas/Houston | Houston Methodist Hospital | Houston Methodist Hospital | S. Wesley Long et al |
| EPI_ISL_785411 | USA/TX-HMH-MCoV-15727/2020 | 2020-07-09 | North America/USA/Texas/Houston | Houston Methodist Hospital | Houston Methodist Hospital | S. Wesley Long et al |
| EPI_ISL_785415 | USA/TX-HMH-MCoV-15731/2020 | 2020-07-09 | North America/USA/Texas/Houston | Houston Methodist Hospital | Houston Methodist Hospital | S. Wesley Long et al |
| EPI_ISL_785417 | USA/TX-HMH-MCoV-15733/2020 | 2020-07-09 | North America/USA/Texas/Houston | Houston Methodist Hospital | Houston Methodist Hospital | S. Wesley Long et al |
| EPI_ISL_785419 | USA/TX-HMH-MCoV-15735/2020 | 2020-07-09 | North America/USA/Texas/Houston | Houston Methodist Hospital | Houston Methodist Hospital | S. Wesley Long et al |
| EPI_ISL_785423 | USA/TX-HMH-MCoV-15739/2020 | 2020-07-09 | North America/USA/Texas/Houston | Houston Methodist Hospital | Houston Methodist Hospital | S. Wesley Long et al |

| | | | | | | |
|---|---|---|---|---|---|---|
| EPI_ISL_785428 | USA/TX-HMH-MCoV-15744/2020 | 2020-07-10 | North America/USA/Texas/Houston | Houston Methodist Hospital | Houston Methodist Hospital | S. Wesley Long et al |
| EPI_ISL_785431 | USA/TX-HMH-MCoV-15747/2020 | 2020-07-09 | North America/USA/Texas/Houston | Houston Methodist Hospital | Houston Methodist Hospital | S. Wesley Long et al |
| EPI_ISL_785433 | USA/TX-HMH-MCoV-15749/2020 | 2020-07-09 | North America/USA/Texas/Houston | Houston Methodist Hospital | Houston Methodist Hospital | S. Wesley Long et al |
| EPI_ISL_785439 | USA/TX-HMH-MCoV-15755/2020 | 2020-07-10 | North America/USA/Texas/Houston | Houston Methodist Hospital | Houston Methodist Hospital | S. Wesley Long et al |
| EPI_ISL_785443 | USA/TX-HMH-MCoV-15759/2020 | 2020-07-09 | North America/USA/Texas/Houston | Houston Methodist Hospital | Houston Methodist Hospital | S. Wesley Long et al |
| EPI_ISL_785452 | USA/TX-HMH-MCoV-15768/2020 | 2020-07-10 | North America/USA/Texas/Houston | Houston Methodist Hospital | Houston Methodist Hospital | S. Wesley Long et al |
| EPI_ISL_785453 | USA/TX-HMH-MCoV-15769/2020 | 2020-07-10 | North America/USA/Texas/Houston | Houston Methodist Hospital | Houston Methodist Hospital | S. Wesley Long et al |
| EPI_ISL_785457 | USA/TX-HMH-MCoV-15773/2020 | 2020-07-09 | North America/USA/Texas/Houston | Houston Methodist Hospital | Houston Methodist Hospital | S. Wesley Long et al |
| EPI_ISL_785463 | USA/TX-HMH-MCoV-15779/2020 | 2020-07-10 | North America/USA/Texas/Houston | Houston Methodist Hospital | Houston Methodist Hospital | S. Wesley Long et al |
| EPI_ISL_785465 | USA/TX-HMH-MCoV-15781/2020 | 2020-07-10 | North America/USA/Texas/Houston | Houston Methodist Hospital | Houston Methodist Hospital | S. Wesley Long et al |
| EPI_ISL_785466 | USA/TX-HMH-MCoV-15782/2020 | 2020-07-10 | North America/USA/Texas/Houston | Houston Methodist Hospital | Houston Methodist Hospital | S. Wesley Long et al |
| EPI_ISL_785467 | USA/TX-HMH-MCoV-15783/2020 | 2020-07-09 | North America/USA/Texas/Houston | Houston Methodist Hospital | Houston Methodist Hospital | S. Wesley Long et al |
| EPI_ISL_785468 | USA/TX-HMH-MCoV-15784/2020 | 2020-07-09 | North America/USA/Texas/Houston | Houston Methodist Hospital | Houston Methodist Hospital | S. Wesley Long et al |

| | | | | | | |
|---|---|---|---|---|---|---|
| EPI_ISL_785471 | USA/TX-HMH-MCoV-15787/2020 | 2020-07-09 | North America/USA/Texas/Houston | Houston Methodist Hospital | Houston Methodist Hospital | S. Wesley Long et al |
| EPI_ISL_785472 | USA/TX-HMH-MCoV-15788/2020 | 2020-07-09 | North America/USA/Texas/Houston | Houston Methodist Hospital | Houston Methodist Hospital | S. Wesley Long et al |
| EPI_ISL_785474 | USA/TX-HMH-MCoV-15790/2020 | 2020-07-10 | North America/USA/Texas/Houston | Houston Methodist Hospital | Houston Methodist Hospital | S. Wesley Long et al |
| EPI_ISL_785480 | USA/TX-HMH-MCoV-15796/2020 | 2020-07-09 | North America/USA/Texas/Houston | Houston Methodist Hospital | Houston Methodist Hospital | S. Wesley Long et al |
| EPI_ISL_785488 | USA/TX-HMH-MCoV-15804/2020 | 2020-07-10 | North America/USA/Texas/Houston | Houston Methodist Hospital | Houston Methodist Hospital | S. Wesley Long et al |
| EPI_ISL_785489 | USA/TX-HMH-MCoV-15805/2020 | 2020-07-10 | North America/USA/Texas/Houston | Houston Methodist Hospital | Houston Methodist Hospital | S. Wesley Long et al |
| EPI_ISL_785490 | USA/TX-HMH-MCoV-15806/2020 | 2020-07-10 | North America/USA/Texas/Houston | Houston Methodist Hospital | Houston Methodist Hospital | S. Wesley Long et al |
| EPI_ISL_785495 | USA/TX-HMH-MCoV-15811/2020 | 2020-07-09 | North America/USA/Texas/Houston | Houston Methodist Hospital | Houston Methodist Hospital | S. Wesley Long et al |
| EPI_ISL_785499 | USA/TX-HMH-MCoV-15815/2020 | 2020-07-05 | North America/USA/Texas/Houston | Houston Methodist Hospital | Houston Methodist Hospital | S. Wesley Long et al |
| EPI_ISL_785500 | USA/TX-HMH-MCoV-15816/2020 | 2020-07-10 | North America/USA/Texas/Houston | Houston Methodist Hospital | Houston Methodist Hospital | S. Wesley Long et al |
| EPI_ISL_785501 | USA/TX-HMH-MCoV-15817/2020 | 2020-07-10 | North America/USA/Texas/Houston | Houston Methodist Hospital | Houston Methodist Hospital | S. Wesley Long et al |
| EPI_ISL_785504 | USA/TX-HMH-MCoV-15820/2020 | 2020-07-09 | North America/USA/Texas/Houston | Houston Methodist Hospital | Houston Methodist Hospital | S. Wesley Long et al |
| EPI_ISL_785511 | USA/TX-HMH-MCoV-15827/2020 | 2020-07-10 | North America/USA/Texas/Houston | Houston Methodist Hospital | Houston Methodist Hospital | S. Wesley Long et al |

| | | | | | | |
|---|---|---|---|---|---|---|
| EPI_ISL_785520 | USA/TX-HMH-MCoV-15836/2020 | 2020-07-10 | North America/USA/Texas/Houston | Houston Methodist Hospital | Houston Methodist Hospital | S. Wesley Long et al |
| EPI_ISL_785521 | USA/TX-HMH-MCoV-15837/2020 | 2020-07-10 | North America/USA/Texas/Houston | Houston Methodist Hospital | Houston Methodist Hospital | S. Wesley Long et al |
| EPI_ISL_785524 | USA/TX-HMH-MCoV-15840/2020 | 2020-07-10 | North America/USA/Texas/Houston | Houston Methodist Hospital | Houston Methodist Hospital | S. Wesley Long et al |
| EPI_ISL_785530 | USA/TX-HMH-MCoV-15846/2020 | 2020-07-10 | North America/USA/Texas/Houston | Houston Methodist Hospital | Houston Methodist Hospital | S. Wesley Long et al |
| EPI_ISL_785533 | USA/TX-HMH-MCoV-15849/2020 | 2020-07-10 | North America/USA/Texas/Houston | Houston Methodist Hospital | Houston Methodist Hospital | S. Wesley Long et al |
| EPI_ISL_785540 | USA/TX-HMH-MCoV-15856/2020 | 2020-07-10 | North America/USA/Texas/Houston | Houston Methodist Hospital | Houston Methodist Hospital | S. Wesley Long et al |
| EPI_ISL_785544 | USA/TX-HMH-MCoV-15860/2020 | 2020-07-10 | North America/USA/Texas/Houston | Houston Methodist Hospital | Houston Methodist Hospital | S. Wesley Long et al |
| EPI_ISL_785548 | USA/TX-HMH-MCoV-15864/2020 | 2020-07-10 | North America/USA/Texas/Houston | Houston Methodist Hospital | Houston Methodist Hospital | S. Wesley Long et al |
| EPI_ISL_785552 | USA/TX-HMH-MCoV-15868/2020 | 2020-07-10 | North America/USA/Texas/Houston | Houston Methodist Hospital | Houston Methodist Hospital | S. Wesley Long et al |
| EPI_ISL_785553 | USA/TX-HMH-MCoV-15869/2020 | 2020-07-10 | North America/USA/Texas/Houston | Houston Methodist Hospital | Houston Methodist Hospital | S. Wesley Long et al |
| EPI_ISL_785556 | USA/TX-HMH-MCoV-15872/2020 | 2020-07-10 | North America/USA/Texas/Houston | Houston Methodist Hospital | Houston Methodist Hospital | S. Wesley Long et al |
| EPI_ISL_785562 | USA/TX-HMH-MCoV-15878/2020 | 2020-07-10 | North America/USA/Texas/Houston | Houston Methodist Hospital | Houston Methodist Hospital | S. Wesley Long et al |
| EPI_ISL_785570 | USA/TX-HMH-MCoV-15886/2020 | 2020-07-10 | North America/USA/Texas/Houston | Houston Methodist Hospital | Houston Methodist Hospital | S. Wesley Long et al |

| | | | | | | |
|---|---|---|---|---|---|---|
| EPI_ISL_785579 | USA/TX-HMH-MCoV-15895/2020 | 2020-07-10 | North America/USA/Texas/Houston | Houston Methodist Hospital | Houston Methodist Hospital | S. Wesley Long et al |
| EPI_ISL_785584 | USA/TX-HMH-MCoV-15900/2020 | 2020-07-10 | North America/USA/Texas/Houston | Houston Methodist Hospital | Houston Methodist Hospital | S. Wesley Long et al |
| EPI_ISL_785585 | USA/TX-HMH-MCoV-15901/2020 | 2020-07-10 | North America/USA/Texas/Houston | Houston Methodist Hospital | Houston Methodist Hospital | S. Wesley Long et al |
| EPI_ISL_785587 | USA/TX-HMH-MCoV-15903/2020 | 2020-07-10 | North America/USA/Texas/Houston | Houston Methodist Hospital | Houston Methodist Hospital | S. Wesley Long et al |
| EPI_ISL_785588 | USA/TX-HMH-MCoV-15904/2020 | 2020-07-10 | North America/USA/Texas/Houston | Houston Methodist Hospital | Houston Methodist Hospital | S. Wesley Long et al |
| EPI_ISL_785590 | USA/TX-HMH-MCoV-15906/2020 | 2020-07-10 | North America/USA/Texas/Houston | Houston Methodist Hospital | Houston Methodist Hospital | S. Wesley Long et al |
| EPI_ISL_785598 | USA/TX-HMH-MCoV-15914/2020 | 2020-07-10 | North America/USA/Texas/Houston | Houston Methodist Hospital | Houston Methodist Hospital | S. Wesley Long et al |
| EPI_ISL_785606 | USA/TX-HMH-MCoV-15922/2020 | 2020-07-10 | North America/USA/Texas/Houston | Houston Methodist Hospital | Houston Methodist Hospital | S. Wesley Long et al |
| EPI_ISL_785614 | USA/TX-HMH-MCoV-15930/2020 | 2020-07-10 | North America/USA/Texas/Houston | Houston Methodist Hospital | Houston Methodist Hospital | S. Wesley Long et al |
| EPI_ISL_785619 | USA/TX-HMH-MCoV-15935/2020 | 2020-07-10 | North America/USA/Texas/Houston | Houston Methodist Hospital | Houston Methodist Hospital | S. Wesley Long et al |
| EPI_ISL_785620 | USA/TX-HMH-MCoV-15936/2020 | 2020-07-10 | North America/USA/Texas/Houston | Houston Methodist Hospital | Houston Methodist Hospital | S. Wesley Long et al |
| EPI_ISL_785628 | USA/TX-HMH-MCoV-15944/2020 | 2020-07-10 | North America/USA/Texas/Houston | Houston Methodist Hospital | Houston Methodist Hospital | S. Wesley Long et al |
| EPI_ISL_785632 | USA/TX-HMH-MCoV-15948/2020 | 2020-07-10 | North America/USA/Texas/Houston | Houston Methodist Hospital | Houston Methodist Hospital | S. Wesley Long et al |

| | | | | | | |
|---|---|---|---|---|---|---|
| EPI_ISL_785633 | USA/TX-HMH-MCoV-15949/2020 | 2020-07-10 | North America/USA/Texas/Houston | Houston Methodist Hospital | Houston Methodist Hospital | S. Wesley Long et al |
| EPI_ISL_785636 | USA/TX-HMH-MCoV-15952/2020 | 2020-07-10 | North America/USA/Texas/Houston | Houston Methodist Hospital | Houston Methodist Hospital | S. Wesley Long et al |
| EPI_ISL_785638 | USA/TX-HMH-MCoV-15954/2020 | 2020-07-10 | North America/USA/Texas/Houston | Houston Methodist Hospital | Houston Methodist Hospital | S. Wesley Long et al |
| EPI_ISL_785643 | USA/TX-HMH-MCoV-15959/2020 | 2020-07-10 | North America/USA/Texas/Houston | Houston Methodist Hospital | Houston Methodist Hospital | S. Wesley Long et al |
| EPI_ISL_785650 | USA/TX-HMH-MCoV-15966/2020 | 2020-07-10 | North America/USA/Texas/Houston | Houston Methodist Hospital | Houston Methodist Hospital | S. Wesley Long et al |
| EPI_ISL_785652 | USA/TX-HMH-MCoV-15968/2020 | 2020-07-10 | North America/USA/Texas/Houston | Houston Methodist Hospital | Houston Methodist Hospital | S. Wesley Long et al |
| EPI_ISL_785654 | USA/TX-HMH-MCoV-15970/2020 | 2020-07-10 | North America/USA/Texas/Houston | Houston Methodist Hospital | Houston Methodist Hospital | S. Wesley Long et al |
| EPI_ISL_785657 | USA/TX-HMH-MCoV-15973/2020 | 2020-07-10 | North America/USA/Texas/Houston | Houston Methodist Hospital | Houston Methodist Hospital | S. Wesley Long et al |
| EPI_ISL_785659 | USA/TX-HMH-MCoV-15975/2020 | 2020-07-10 | North America/USA/Texas/Houston | Houston Methodist Hospital | Houston Methodist Hospital | S. Wesley Long et al |
| EPI_ISL_785661 | USA/TX-HMH-MCoV-15977/2020 | 2020-07-10 | North America/USA/Texas/Houston | Houston Methodist Hospital | Houston Methodist Hospital | S. Wesley Long et al |
| EPI_ISL_785667 | USA/TX-HMH-MCoV-15983/2020 | 2020-07-10 | North America/USA/Texas/Houston | Houston Methodist Hospital | Houston Methodist Hospital | S. Wesley Long et al |
| EPI_ISL_785669 | USA/TX-HMH-MCoV-15985/2020 | 2020-07-10 | North America/USA/Texas/Houston | Houston Methodist Hospital | Houston Methodist Hospital | S. Wesley Long et al |
| EPI_ISL_785670 | USA/TX-HMH-MCoV-15986/2020 | 2020-07-10 | North America/USA/Texas/Houston | Houston Methodist Hospital | Houston Methodist Hospital | S. Wesley Long et al |

| | | | | | | |
|---|---|---|---|---|---|---|
| EPI_ISL_785675 | USA/TX-HMH-MCoV-15991/2020 | 2020-07-10 | North America/USA/Texas/Houston | Houston Methodist Hospital | Houston Methodist Hospital | S. Wesley Long et al |
| EPI_ISL_785679 | USA/TX-HMH-MCoV-15995/2020 | 2020-07-10 | North America/USA/Texas/Houston | Houston Methodist Hospital | Houston Methodist Hospital | S. Wesley Long et al |
| EPI_ISL_785683 | USA/TX-HMH-MCoV-15999/2020 | 2020-07-10 | North America/USA/Texas/Houston | Houston Methodist Hospital | Houston Methodist Hospital | S. Wesley Long et al |
| EPI_ISL_785691 | USA/TX-HMH-MCoV-16007/2020 | 2020-07-10 | North America/USA/Texas/Houston | Houston Methodist Hospital | Houston Methodist Hospital | S. Wesley Long et al |
| EPI_ISL_785697 | USA/TX-HMH-MCoV-16013/2020 | 2020-07-10 | North America/USA/Texas/Houston | Houston Methodist Hospital | Houston Methodist Hospital | S. Wesley Long et al |
| EPI_ISL_785708 | USA/TX-HMH-MCoV-16024/2020 | 2020-10-29 | North America/USA/Texas/Houston | Houston Methodist Hospital | Houston Methodist Hospital | S. Wesley Long et al |
| EPI_ISL_785715 | USA/TX-HMH-MCoV-16031/2020 | 2020-10-27 | North America/USA/Texas/Houston | Houston Methodist Hospital | Houston Methodist Hospital | S. Wesley Long et al |
| EPI_ISL_785718 | USA/TX-HMH-MCoV-16034/2020 | 2020-10-15 | North America/USA/Texas/Houston | Houston Methodist Hospital | Houston Methodist Hospital | S. Wesley Long et al |
| EPI_ISL_785720 | USA/TX-HMH-MCoV-16036/2020 | 2020-10-27 | North America/USA/Texas/Houston | Houston Methodist Hospital | Houston Methodist Hospital | S. Wesley Long et al |
| EPI_ISL_785734 | USA/TX-HMH-MCoV-16050/2020 | 2020-10-27 | North America/USA/Texas/Houston | Houston Methodist Hospital | Houston Methodist Hospital | S. Wesley Long et al |
| EPI_ISL_785742 | USA/TX-HMH-MCoV-16058/2020 | 2020-10-28 | North America/USA/Texas/Houston | Houston Methodist Hospital | Houston Methodist Hospital | S. Wesley Long et al |
| EPI_ISL_785752 | USA/TX-HMH-MCoV-16068/2020 | 2020-10-27 | North America/USA/Texas/Houston | Houston Methodist Hospital | Houston Methodist Hospital | S. Wesley Long et al |
| EPI_ISL_785753 | USA/TX-HMH-MCoV-16069/2020 | 2020-10-27 | North America/USA/Texas/Houston | Houston Methodist Hospital | Houston Methodist Hospital | S. Wesley Long et al |

| | | | | | | |
|---|---|---|---|---|---|---|
| EPI_ISL_785759 | USA/TX-HMH-MCoV-16075/2020 | 2020-10-27 | North America/USA/Texas/Houston | Houston Methodist Hospital | Houston Methodist Hospital | S. Wesley Long et al |
| EPI_ISL_785766 | USA/TX-HMH-MCoV-16082/2020 | 2020-10-28 | North America/USA/Texas/Houston | Houston Methodist Hospital | Houston Methodist Hospital | S. Wesley Long et al |
| EPI_ISL_785790 | USA/TX-HMH-MCoV-16106/2020 | 2020-10-28 | North America/USA/Texas/Houston | Houston Methodist Hospital | Houston Methodist Hospital | S. Wesley Long et al |
| EPI_ISL_785792 | USA/TX-HMH-MCoV-16108/2020 | 2020-10-27 | North America/USA/Texas/Houston | Houston Methodist Hospital | Houston Methodist Hospital | S. Wesley Long et al |
| EPI_ISL_785794 | USA/TX-HMH-MCoV-16110/2020 | 2020-10-27 | North America/USA/Texas/Houston | Houston Methodist Hospital | Houston Methodist Hospital | S. Wesley Long et al |
| EPI_ISL_785795 | USA/TX-HMH-MCoV-16111/2020 | 2020-10-27 | North America/USA/Texas/Houston | Houston Methodist Hospital | Houston Methodist Hospital | S. Wesley Long et al |
| EPI_ISL_785800 | USA/TX-HMH-MCoV-16116/2020 | 2020-10-27 | North America/USA/Texas/Houston | Houston Methodist Hospital | Houston Methodist Hospital | S. Wesley Long et al |
| EPI_ISL_785803 | USA/TX-HMH-MCoV-16119/2020 | 2020-10-29 | North America/USA/Texas/Houston | Houston Methodist Hospital | Houston Methodist Hospital | S. Wesley Long et al |
| EPI_ISL_785814 | USA/TX-HMH-MCoV-16130/2020 | 2020-10-29 | North America/USA/Texas/Houston | Houston Methodist Hospital | Houston Methodist Hospital | S. Wesley Long et al |
| EPI_ISL_785823 | USA/TX-HMH-MCoV-16139/2020 | 2020-10-26 | North America/USA/Texas/Houston | Houston Methodist Hospital | Houston Methodist Hospital | S. Wesley Long et al |
| EPI_ISL_785827 | USA/TX-HMH-MCoV-16143/2020 | 2020-10-28 | North America/USA/Texas/Houston | Houston Methodist Hospital | Houston Methodist Hospital | S. Wesley Long et al |
| EPI_ISL_785831 | USA/TX-HMH-MCoV-16147/2020 | 2020-10-28 | North America/USA/Texas/Houston | Houston Methodist Hospital | Houston Methodist Hospital | S. Wesley Long et al |
| EPI_ISL_785846 | USA/TX-HMH-MCoV-16162/2020 | 2020-10-30 | North America/USA/Texas/Houston | Houston Methodist Hospital | Houston Methodist Hospital | S. Wesley Long et al |

| | | | | | | |
|---|---|---|---|---|---|---|
| EPI_ISL_785850 | USA/TX-HMH-MCoV-16166/2020 | 2020-10-29 | North America/USA/Texas/Houston | Houston Methodist Hospital | Houston Methodist Hospital | S. Wesley Long et al |
| EPI_ISL_785856 | USA/TX-HMH-MCoV-16172/2020 | 2020-10-27 | North America/USA/Texas/Houston | Houston Methodist Hospital | Houston Methodist Hospital | S. Wesley Long et al |
| EPI_ISL_785879 | USA/TX-HMH-MCoV-16195/2020 | 2020-10-28 | North America/USA/Texas/Houston | Houston Methodist Hospital | Houston Methodist Hospital | S. Wesley Long et al |
| EPI_ISL_785887 | USA/TX-HMH-MCoV-16203/2020 | 2020-10-29 | North America/USA/Texas/Houston | Houston Methodist Hospital | Houston Methodist Hospital | S. Wesley Long et al |
| EPI_ISL_785892 | USA/TX-HMH-MCoV-16208/2020 | 2020-10-26 | North America/USA/Texas/Houston | Houston Methodist Hospital | Houston Methodist Hospital | S. Wesley Long et al |
| EPI_ISL_785894 | USA/TX-HMH-MCoV-16210/2020 | 2020-10-30 | North America/USA/Texas/Houston | Houston Methodist Hospital | Houston Methodist Hospital | S. Wesley Long et al |
| EPI_ISL_785901 | USA/TX-HMH-MCoV-16217/2020 | 2020-10-30 | North America/USA/Texas/Houston | Houston Methodist Hospital | Houston Methodist Hospital | S. Wesley Long et al |
| EPI_ISL_785903 | USA/TX-HMH-MCoV-16219/2020 | 2020-10-30 | North America/USA/Texas/Houston | Houston Methodist Hospital | Houston Methodist Hospital | S. Wesley Long et al |
| EPI_ISL_785918 | USA/TX-HMH-MCoV-16234/2020 | 2020-10-31 | North America/USA/Texas/Houston | Houston Methodist Hospital | Houston Methodist Hospital | S. Wesley Long et al |
| EPI_ISL_785924 | USA/TX-HMH-MCoV-16240/2020 | 2020-10-30 | North America/USA/Texas/Houston | Houston Methodist Hospital | Houston Methodist Hospital | S. Wesley Long et al |
| EPI_ISL_785926 | USA/TX-HMH-MCoV-16242/2020 | 2020-10-30 | North America/USA/Texas/Houston | Houston Methodist Hospital | Houston Methodist Hospital | S. Wesley Long et al |
| EPI_ISL_785927 | USA/TX-HMH-MCoV-16243/2020 | 2020-10-30 | North America/USA/Texas/Houston | Houston Methodist Hospital | Houston Methodist Hospital | S. Wesley Long et al |
| EPI_ISL_785929 | USA/TX-HMH-MCoV-16245/2020 | 2020-10-31 | North America/USA/Texas/Houston | Houston Methodist Hospital | Houston Methodist Hospital | S. Wesley Long et al |

| | | | | | | |
|---|---|---|---|---|---|---|
| EPI_ISL_785933 | USA/TX-HMH-MCoV-16249/2020 | 2020-10-28 | North America/USA/Texas/Houston | Houston Methodist Hospital | Houston Methodist Hospital | S. Wesley Long et al |
| EPI_ISL_785948 | USA/TX-HMH-MCoV-16264/2020 | 2020-10-30 | North America/USA/Texas/Houston | Houston Methodist Hospital | Houston Methodist Hospital | S. Wesley Long et al |
| EPI_ISL_785950 | USA/TX-HMH-MCoV-16266/2020 | 2020-10-30 | North America/USA/Texas/Houston | Houston Methodist Hospital | Houston Methodist Hospital | S. Wesley Long et al |
| EPI_ISL_785957 | USA/TX-HMH-MCoV-16273/2020 | 2020-10-27 | North America/USA/Texas/Houston | Houston Methodist Hospital | Houston Methodist Hospital | S. Wesley Long et al |
| EPI_ISL_785970 | USA/TX-HMH-MCoV-16286/2020 | 2020-10-26 | North America/USA/Texas/Houston | Houston Methodist Hospital | Houston Methodist Hospital | S. Wesley Long et al |
| EPI_ISL_785975 | USA/TX-HMH-MCoV-16291/2020 | 2020-10-30 | North America/USA/Texas/Houston | Houston Methodist Hospital | Houston Methodist Hospital | S. Wesley Long et al |
| EPI_ISL_785977 | USA/TX-HMH-MCoV-16293/2020 | 2020-10-31 | North America/USA/Texas/Houston | Houston Methodist Hospital | Houston Methodist Hospital | S. Wesley Long et al |
| EPI_ISL_786005 | USA/TX-HMH-MCoV-16321/2020 | 2020-11-02 | North America/USA/Texas/Houston | Houston Methodist Hospital | Houston Methodist Hospital | S. Wesley Long et al |
| EPI_ISL_786011 | USA/TX-HMH-MCoV-16327/2020 | 2020-11-01 | North America/USA/Texas/Houston | Houston Methodist Hospital | Houston Methodist Hospital | S. Wesley Long et al |
| EPI_ISL_786017 | USA/TX-HMH-MCoV-16333/2020 | 2020-11-02 | North America/USA/Texas/Houston | Houston Methodist Hospital | Houston Methodist Hospital | S. Wesley Long et al |
| EPI_ISL_786019 | USA/TX-HMH-MCoV-16335/2020 | 2020-10-26 | North America/USA/Texas/Houston | Houston Methodist Hospital | Houston Methodist Hospital | S. Wesley Long et al |
| EPI_ISL_786025 | USA/TX-HMH-MCoV-16341/2020 | 2020-11-02 | North America/USA/Texas/Houston | Houston Methodist Hospital | Houston Methodist Hospital | S. Wesley Long et al |
| EPI_ISL_786047 | USA/TX-HMH-MCoV-16363/2020 | 2020-11-01 | North America/USA/Texas/Houston | Houston Methodist Hospital | Houston Methodist Hospital | S. Wesley Long et al |

| | | | | | | |
|---|---|---|---|---|---|---|
| EPI_ISL_786051 | USA/TX-HMH-MCoV-16367/2020 | 2020-11-02 | North America/USA/Texas/Houston | Houston Methodist Hospital | Houston Methodist Hospital | S. Wesley Long et al |
| EPI_ISL_786052 | USA/TX-HMH-MCoV-16368/2020 | 2020-11-02 | North America/USA/Texas/Houston | Houston Methodist Hospital | Houston Methodist Hospital | S. Wesley Long et al |
| EPI_ISL_786061 | USA/TX-HMH-MCoV-16377/2020 | 2020-11-02 | North America/USA/Texas/Houston | Houston Methodist Hospital | Houston Methodist Hospital | S. Wesley Long et al |
| EPI_ISL_786063 | USA/TX-HMH-MCoV-16379/2020 | 2020-11-02 | North America/USA/Texas/Houston | Houston Methodist Hospital | Houston Methodist Hospital | S. Wesley Long et al |
| EPI_ISL_786066 | USA/TX-HMH-MCoV-16382/2020 | 2020-11-02 | North America/USA/Texas/Houston | Houston Methodist Hospital | Houston Methodist Hospital | S. Wesley Long et al |
| EPI_ISL_786071 | USA/TX-HMH-MCoV-16387/2020 | 2020-11-01 | North America/USA/Texas/Houston | Houston Methodist Hospital | Houston Methodist Hospital | S. Wesley Long et al |
| EPI_ISL_786076 | USA/TX-HMH-MCoV-16392/2020 | 2020-11-02 | North America/USA/Texas/Houston | Houston Methodist Hospital | Houston Methodist Hospital | S. Wesley Long et al |
| EPI_ISL_786099 | USA/TX-HMH-MCoV-17951/2020 | 2020-11-16 | North America/USA/Texas/Houston | Houston Methodist Hospital | Houston Methodist Hospital | S. Wesley Long et al |
| EPI_ISL_786107 | USA/TX-HMH-MCoV-17959/2020 | 2020-11-16 | North America/USA/Texas/Houston | Houston Methodist Hospital | Houston Methodist Hospital | S. Wesley Long et al |
| EPI_ISL_786118 | USA/TX-HMH-MCoV-17970/2020 | 2020-11-16 | North America/USA/Texas/Houston | Houston Methodist Hospital | Houston Methodist Hospital | S. Wesley Long et al |
| EPI_ISL_786123 | USA/TX-HMH-MCoV-17975/2020 | 2020-11-16 | North America/USA/Texas/Houston | Houston Methodist Hospital | Houston Methodist Hospital | S. Wesley Long et al |
| EPI_ISL_786131 | USA/TX-HMH-MCoV-17983/2020 | 2020-11-16 | North America/USA/Texas/Houston | Houston Methodist Hospital | Houston Methodist Hospital | S. Wesley Long et al |
| EPI_ISL_786139 | USA/TX-HMH-MCoV-17991/2020 | 2020-11-16 | North America/USA/Texas/Houston | Houston Methodist Hospital | Houston Methodist Hospital | S. Wesley Long et al |

| | | | | | | |
|---|---|---|---|---|---|---|
| EPI_ISL_786151 | USA/TX-HMH-MCoV-18003/2020 | 2020-11-16 | North America/USA/Texas/Houston | Houston Methodist Hospital | Houston Methodist Hospital | S. Wesley Long et al |
| EPI_ISL_786156 | USA/TX-HMH-MCoV-18008/2020 | 2020-11-16 | North America/USA/Texas/Houston | Houston Methodist Hospital | Houston Methodist Hospital | S. Wesley Long et al |
| EPI_ISL_786176 | USA/TX-HMH-MCoV-18028/2020 | 2020-11-16 | North America/USA/Texas/Houston | Houston Methodist Hospital | Houston Methodist Hospital | S. Wesley Long et al |
| EPI_ISL_786201 | USA/TX-HMH-MCoV-14716/2020 | 2020-10-01 | North America/USA/Texas/Houston | Houston Methodist Hospital | Houston Methodist Hospital | S. Wesley Long et al |
| EPI_ISL_786207 | USA/TX-HMH-MCoV-14722/2020 | 2020-10-12 | North America/USA/Texas/Houston | Houston Methodist Hospital | Houston Methodist Hospital | S. Wesley Long et al |
| EPI_ISL_786208 | USA/TX-HMH-MCoV-14723/2020 | 2020-10-12 | North America/USA/Texas/Houston | Houston Methodist Hospital | Houston Methodist Hospital | S. Wesley Long et al |
| EPI_ISL_786212 | USA/TX-HMH-MCoV-14727/2020 | 2020-10-01 | North America/USA/Texas/Houston | Houston Methodist Hospital | Houston Methodist Hospital | S. Wesley Long et al |
| EPI_ISL_786220 | USA/TX-HMH-MCoV-14735/2020 | 2020-10-12 | North America/USA/Texas/Houston | Houston Methodist Hospital | Houston Methodist Hospital | S. Wesley Long et al |
| EPI_ISL_786222 | USA/TX-HMH-MCoV-14737/2020 | 2020-10-12 | North America/USA/Texas/Houston | Houston Methodist Hospital | Houston Methodist Hospital | S. Wesley Long et al |
| EPI_ISL_786225 | USA/TX-HMH-MCoV-14740/2020 | 2020-10-01 | North America/USA/Texas/Houston | Houston Methodist Hospital | Houston Methodist Hospital | S. Wesley Long et al |
| EPI_ISL_786229 | USA/TX-HMH-MCoV-14744/2020 | 2020-10-11 | North America/USA/Texas/Houston | Houston Methodist Hospital | Houston Methodist Hospital | S. Wesley Long et al |
| EPI_ISL_786230 | USA/TX-HMH-MCoV-14745/2020 | 2020-10-12 | North America/USA/Texas/Houston | Houston Methodist Hospital | Houston Methodist Hospital | S. Wesley Long et al |
| EPI_ISL_786239 | USA/TX-HMH-MCoV-14754/2020 | 2020-10-01 | North America/USA/Texas/Houston | Houston Methodist Hospital | Houston Methodist Hospital | S. Wesley Long et al |

| | | | | | | |
|---|---|---|---|---|---|---|
| EPI_ISL_786240 | USA/TX-HMH-MCoV-14755/2020 | 2020-10-08 | North America/USA/Texas/Houston | Houston Methodist Hospital | Houston Methodist Hospital | S. Wesley Long et al |
| EPI_ISL_786247 | USA/TX-HMH-MCoV-14762/2020 | 2020-10-12 | North America/USA/Texas/Houston | Houston Methodist Hospital | Houston Methodist Hospital | S. Wesley Long et al |
| EPI_ISL_786250 | USA/TX-HMH-MCoV-14765/2020 | 2020-10-01 | North America/USA/Texas/Houston | Houston Methodist Hospital | Houston Methodist Hospital | S. Wesley Long et al |
| EPI_ISL_786269 | USA/TX-HMH-MCoV-14784/2020 | 2020-10-13 | North America/USA/Texas/Houston | Houston Methodist Hospital | Houston Methodist Hospital | S. Wesley Long et al |
| EPI_ISL_786271 | USA/TX-HMH-MCoV-14786/2020 | 2020-10-13 | North America/USA/Texas/Houston | Houston Methodist Hospital | Houston Methodist Hospital | S. Wesley Long et al |
| EPI_ISL_786275 | USA/TX-HMH-MCoV-14790/2020 | 2020-10-12 | North America/USA/Texas/Houston | Houston Methodist Hospital | Houston Methodist Hospital | S. Wesley Long et al |
| EPI_ISL_786276 | USA/TX-HMH-MCoV-14791/2020 | 2020-10-12 | North America/USA/Texas/Houston | Houston Methodist Hospital | Houston Methodist Hospital | S. Wesley Long et al |
| EPI_ISL_786292 | USA/TX-HMH-MCoV-14807/2020 | 2020-10-13 | North America/USA/Texas/Houston | Houston Methodist Hospital | Houston Methodist Hospital | S. Wesley Long et al |
| EPI_ISL_786294 | USA/TX-HMH-MCoV-14809/2020 | 2020-10-13 | North America/USA/Texas/Houston | Houston Methodist Hospital | Houston Methodist Hospital | S. Wesley Long et al |
| EPI_ISL_786298 | USA/TX-HMH-MCoV-14814/2020 | 2020-10-12 | North America/USA/Texas/Houston | Houston Methodist Hospital | Houston Methodist Hospital | S. Wesley Long et al |
| EPI_ISL_786300 | USA/TX-HMH-MCoV-14816/2020 | 2020-10-13 | North America/USA/Texas/Houston | Houston Methodist Hospital | Houston Methodist Hospital | S. Wesley Long et al |
| EPI_ISL_786302 | USA/TX-HMH-MCoV-14818/2020 | 2020-10-13 | North America/USA/Texas/Houston | Houston Methodist Hospital | Houston Methodist Hospital | S. Wesley Long et al |
| EPI_ISL_786303 | USA/TX-HMH-MCoV-14819/2020 | 2020-10-13 | North America/USA/Texas/Houston | Houston Methodist Hospital | Houston Methodist Hospital | S. Wesley Long et al |

| | | | | | | |
|---|---|---|---|---|---|---|
| EPI_ISL_786315 | USA/TX-HMH-MCoV-14831/2020 | 2020-10-13 | North America/USA/Texas/Houston | Houston Methodist Hospital | Houston Methodist Hospital | S. Wesley Long et al |
| EPI_ISL_786320 | USA/TX-HMH-MCoV-14836/2020 | 2020-10-12 | North America/USA/Texas/Houston | Houston Methodist Hospital | Houston Methodist Hospital | S. Wesley Long et al |
| EPI_ISL_786322 | USA/TX-HMH-MCoV-14838/2020 | 2020-10-12 | North America/USA/Texas/Houston | Houston Methodist Hospital | Houston Methodist Hospital | S. Wesley Long et al |
| EPI_ISL_786324 | USA/TX-HMH-MCoV-14840/2020 | 2020-10-13 | North America/USA/Texas/Houston | Houston Methodist Hospital | Houston Methodist Hospital | S. Wesley Long et al |
| EPI_ISL_786326 | USA/TX-HMH-MCoV-14842/2020 | 2020-10-13 | North America/USA/Texas/Houston | Houston Methodist Hospital | Houston Methodist Hospital | S. Wesley Long et al |
| EPI_ISL_786338 | USA/TX-HMH-MCoV-14854/2020 | 2020-10-13 | North America/USA/Texas/Houston | Houston Methodist Hospital | Houston Methodist Hospital | S. Wesley Long et al |
| EPI_ISL_786343 | USA/TX-HMH-MCoV-14859/2020 | 2020-10-12 | North America/USA/Texas/Houston | Houston Methodist Hospital | Houston Methodist Hospital | S. Wesley Long et al |
| EPI_ISL_786345 | USA/TX-HMH-MCoV-14861/2020 | 2020-10-12 | North America/USA/Texas/Houston | Houston Methodist Hospital | Houston Methodist Hospital | S. Wesley Long et al |
| EPI_ISL_786346 | USA/TX-HMH-MCoV-14862/2020 | 2020-10-12 | North America/USA/Texas/Houston | Houston Methodist Hospital | Houston Methodist Hospital | S. Wesley Long et al |
| EPI_ISL_786347 | USA/TX-HMH-MCoV-14863/2020 | 2020-10-13 | North America/USA/Texas/Houston | Houston Methodist Hospital | Houston Methodist Hospital | S. Wesley Long et al |
| EPI_ISL_786349 | USA/TX-HMH-MCoV-14865/2020 | 2020-10-12 | North America/USA/Texas/Houston | Houston Methodist Hospital | Houston Methodist Hospital | S. Wesley Long et al |
| EPI_ISL_786353 | USA/TX-HMH-MCoV-14869/2020 | 2020-10-13 | North America/USA/Texas/Houston | Houston Methodist Hospital | Houston Methodist Hospital | S. Wesley Long et al |
| EPI_ISL_786354 | USA/TX-HMH-MCoV-14870/2020 | 2020-10-13 | North America/USA/Texas/Houston | Houston Methodist Hospital | Houston Methodist Hospital | S. Wesley Long et al |

| | | | | | | |
|---|---|---|---|---|---|---|
| EPI_ISL_786356 | USA/TX-HMH-MCoV-14872/2020 | 2020-10-13 | North America/USA/Texas/Houston | Houston Methodist Hospital | Houston Methodist Hospital | S. Wesley Long et al |
| EPI_ISL_786377 | USA/TX-HMH-MCoV-14893/2020 | 2020-10-15 | North America/USA/Texas/Houston | Houston Methodist Hospital | Houston Methodist Hospital | S. Wesley Long et al |
| EPI_ISL_786379 | USA/TX-HMH-MCoV-14895/2020 | 2020-10-13 | North America/USA/Texas/Houston | Houston Methodist Hospital | Houston Methodist Hospital | S. Wesley Long et al |
| EPI_ISL_786381 | USA/TX-HMH-MCoV-14897/2020 | 2020-10-14 | North America/USA/Texas/Houston | Houston Methodist Hospital | Houston Methodist Hospital | S. Wesley Long et al |
| EPI_ISL_786391 | USA/TX-HMH-MCoV-14907/2020 | 2020-10-12 | North America/USA/Texas/Houston | Houston Methodist Hospital | Houston Methodist Hospital | S. Wesley Long et al |
| EPI_ISL_786398 | USA/TX-HMH-MCoV-14914/2020 | 2020-07-05 | North America/USA/Texas/Houston | Houston Methodist Hospital | Houston Methodist Hospital | S. Wesley Long et al |
| EPI_ISL_786405 | USA/TX-HMH-MCoV-14921/2020 | 2020-10-15 | North America/USA/Texas/Houston | Houston Methodist Hospital | Houston Methodist Hospital | S. Wesley Long et al |
| EPI_ISL_786422 | USA/TX-HMH-MCoV-14938/2020 | 2020-10-15 | North America/USA/Texas/Houston | Houston Methodist Hospital | Houston Methodist Hospital | S. Wesley Long et al |
| EPI_ISL_786423 | USA/TX-HMH-MCoV-14939/2020 | 2020-10-15 | North America/USA/Texas/Houston | Houston Methodist Hospital | Houston Methodist Hospital | S. Wesley Long et al |
| EPI_ISL_786427 | USA/TX-HMH-MCoV-14943/2020 | 2020-10-13 | North America/USA/Texas/Houston | Houston Methodist Hospital | Houston Methodist Hospital | S. Wesley Long et al |
| EPI_ISL_786438 | USA/TX-HMH-MCoV-14954/2020 | 2020-10-15 | North America/USA/Texas/Houston | Houston Methodist Hospital | Houston Methodist Hospital | S. Wesley Long et al |
| EPI_ISL_786445 | USA/TX-HMH-MCoV-14961/2020 | 2020-10-14 | North America/USA/Texas/Houston | Houston Methodist Hospital | Houston Methodist Hospital | S. Wesley Long et al |
| EPI_ISL_786451 | USA/TX-HMH-MCoV-14967/2020 | 2020-10-15 | North America/USA/Texas/Houston | Houston Methodist Hospital | Houston Methodist Hospital | S. Wesley Long et al |

| | | | | | | |
|---|---|---|---|---|---|---|
| EPI_ISL_786453 | USA/TX-HMH-MCoV-14969/2020 | 2020-10-15 | North America/USA/Texas/Houston | Houston Methodist Hospital | Houston Methodist Hospital | S. Wesley Long et al |
| EPI_ISL_786458 | USA/TX-HMH-MCoV-14974/2020 | 2020-10-16 | North America/USA/Texas/Houston | Houston Methodist Hospital | Houston Methodist Hospital | S. Wesley Long et al |
| EPI_ISL_786462 | USA/TX-HMH-MCoV-14978/2020 | 2020-10-16 | North America/USA/Texas/Houston | Houston Methodist Hospital | Houston Methodist Hospital | S. Wesley Long et al |
| EPI_ISL_786465 | USA/TX-HMH-MCoV-14981/2020 | 2020-10-15 | North America/USA/Texas/Houston | Houston Methodist Hospital | Houston Methodist Hospital | S. Wesley Long et al |
| EPI_ISL_786478 | USA/TX-HMH-MCoV-14994/2020 | 2020-10-15 | North America/USA/Texas/Houston | Houston Methodist Hospital | Houston Methodist Hospital | S. Wesley Long et al |
| EPI_ISL_786483 | USA/TX-HMH-MCoV-14999/2020 | 2020-10-16 | North America/USA/Texas/Houston | Houston Methodist Hospital | Houston Methodist Hospital | S. Wesley Long et al |
| EPI_ISL_786489 | USA/TX-HMH-MCoV-15005/2020 | 2020-10-15 | North America/USA/Texas/Houston | Houston Methodist Hospital | Houston Methodist Hospital | S. Wesley Long et al |
| EPI_ISL_786499 | USA/TX-HMH-MCoV-15015/2020 | 2020-10-15 | North America/USA/Texas/Houston | Houston Methodist Hospital | Houston Methodist Hospital | S. Wesley Long et al |
| EPI_ISL_786504 | USA/TX-HMH-MCoV-15020/2020 | 2020-10-16 | North America/USA/Texas/Houston | Houston Methodist Hospital | Houston Methodist Hospital | S. Wesley Long et al |
| EPI_ISL_786522 | USA/TX-HMH-MCoV-15038/2020 | 2020-10-16 | North America/USA/Texas/Houston | Houston Methodist Hospital | Houston Methodist Hospital | S. Wesley Long et al |
| EPI_ISL_786531 | USA/TX-HMH-MCoV-15047/2020 | 2020-10-16 | North America/USA/Texas/Houston | Houston Methodist Hospital | Houston Methodist Hospital | S. Wesley Long et al |
| EPI_ISL_786538 | USA/TX-HMH-MCoV-15054/2020 | 2020-10-16 | North America/USA/Texas/Houston | Houston Methodist Hospital | Houston Methodist Hospital | S. Wesley Long et al |
| EPI_ISL_786545 | USA/TX-HMH-MCoV-15061/2020 | 2020-10-16 | North America/USA/Texas/Houston | Houston Methodist Hospital | Houston Methodist Hospital | S. Wesley Long et al |

| | | | | | | |
|---|---|---|---|---|---|---|
| EPI_ISL_786549 | USA/TX-HMH-MCoV-15065/2020 | 2020-10-16 | North America/USA/Texas/Houston | Houston Methodist Hospital | Houston Methodist Hospital | S. Wesley Long et al |
| EPI_ISL_786564 | USA/TX-HMH-MCoV-15080/2020 | 2020-10-17 | North America/USA/Texas/Houston | Houston Methodist Hospital | Houston Methodist Hospital | S. Wesley Long et al |
| EPI_ISL_786566 | USA/TX-HMH-MCoV-15082/2020 | 2020-10-17 | North America/USA/Texas/Houston | Houston Methodist Hospital | Houston Methodist Hospital | S. Wesley Long et al |
| EPI_ISL_786577 | USA/TX-HMH-MCoV-15093/2020 | 2020-10-17 | North America/USA/Texas/Houston | Houston Methodist Hospital | Houston Methodist Hospital | S. Wesley Long et al |
| EPI_ISL_786578 | USA/TX-HMH-MCoV-15094/2020 | 2020-10-17 | North America/USA/Texas/Houston | Houston Methodist Hospital | Houston Methodist Hospital | S. Wesley Long et al |
| EPI_ISL_786586 | USA/TX-HMH-MCoV-15102/2020 | 2020-10-16 | North America/USA/Texas/Houston | Houston Methodist Hospital | Houston Methodist Hospital | S. Wesley Long et al |
| EPI_ISL_786593 | USA/TX-HMH-MCoV-15109/2020 | 2020-10-21 | North America/USA/Texas/Houston | Houston Methodist Hospital | Houston Methodist Hospital | S. Wesley Long et al |
| EPI_ISL_786599 | USA/TX-HMH-MCoV-15115/2020 | 2020-10-17 | North America/USA/Texas/Houston | Houston Methodist Hospital | Houston Methodist Hospital | S. Wesley Long et al |
| EPI_ISL_786611 | USA/TX-HMH-MCoV-15127/2020 | 2020-10-17 | North America/USA/Texas/Houston | Houston Methodist Hospital | Houston Methodist Hospital | S. Wesley Long et al |
| EPI_ISL_786617 | USA/TX-HMH-MCoV-15133/2020 | 2020-10-19 | North America/USA/Texas/Houston | Houston Methodist Hospital | Houston Methodist Hospital | S. Wesley Long et al |
| EPI_ISL_786623 | USA/TX-HMH-MCoV-15139/2020 | 2020-10-20 | North America/USA/Texas/Houston | Houston Methodist Hospital | Houston Methodist Hospital | S. Wesley Long et al |
| EPI_ISL_786629 | USA/TX-HMH-MCoV-15145/2020 | 2020-10-19 | North America/USA/Texas/Houston | Houston Methodist Hospital | Houston Methodist Hospital | S. Wesley Long et al |
| EPI_ISL_786645 | USA/TX-HMH-MCoV-15161/2020 | 2020-10-20 | North America/USA/Texas/Houston | Houston Methodist Hospital | Houston Methodist Hospital | S. Wesley Long et al |

| | | | | | | |
|---|---|---|---|---|---|---|
| EPI_ISL_786647 | USA/TX-HMH-MCoV-15163/2020 | 2020-10-19 | North America/USA/Texas/Houston | Houston Methodist Hospital | Houston Methodist Hospital | S. Wesley Long et al |
| EPI_ISL_786651 | USA/TX-HMH-MCoV-15167/2020 | 2020-10-21 | North America/USA/Texas/Houston | Houston Methodist Hospital | Houston Methodist Hospital | S. Wesley Long et al |
| EPI_ISL_786661 | USA/TX-HMH-MCoV-15177/2020 | 2020-10-20 | North America/USA/Texas/Houston | Houston Methodist Hospital | Houston Methodist Hospital | S. Wesley Long et al |
| EPI_ISL_786665 | USA/TX-HMH-MCoV-15181/2020 | 2020-10-19 | North America/USA/Texas/Houston | Houston Methodist Hospital | Houston Methodist Hospital | S. Wesley Long et al |
| EPI_ISL_786666 | USA/TX-HMH-MCoV-15182/2020 | 2020-10-19 | North America/USA/Texas/Houston | Houston Methodist Hospital | Houston Methodist Hospital | S. Wesley Long et al |
| EPI_ISL_786677 | USA/TX-HMH-MCoV-15193/2020 | 2020-10-19 | North America/USA/Texas/Houston | Houston Methodist Hospital | Houston Methodist Hospital | S. Wesley Long et al |
| EPI_ISL_786681 | USA/TX-HMH-MCoV-15197/2020 | 2020-10-21 | North America/USA/Texas/Houston | Houston Methodist Hospital | Houston Methodist Hospital | S. Wesley Long et al |
| EPI_ISL_786683 | USA/TX-HMH-MCoV-15199/2020 | 2020-10-21 | North America/USA/Texas/Houston | Houston Methodist Hospital | Houston Methodist Hospital | S. Wesley Long et al |
| EPI_ISL_786684 | USA/TX-HMH-MCoV-15200/2020 | 2020-10-20 | North America/USA/Texas/Houston | Houston Methodist Hospital | Houston Methodist Hospital | S. Wesley Long et al |
| EPI_ISL_786686 | USA/TX-HMH-MCoV-15202/2020 | 2020-10-20 | North America/USA/Texas/Houston | Houston Methodist Hospital | Houston Methodist Hospital | S. Wesley Long et al |
| EPI_ISL_786688 | USA/TX-HMH-MCoV-15204/2020 | 2020-10-19 | North America/USA/Texas/Houston | Houston Methodist Hospital | Houston Methodist Hospital | S. Wesley Long et al |
| EPI_ISL_786701 | USA/TX-HMH-MCoV-15217/2020 | 2020-10-19 | North America/USA/Texas/Houston | Houston Methodist Hospital | Houston Methodist Hospital | S. Wesley Long et al |
| EPI_ISL_786703 | USA/TX-HMH-MCoV-15219/2020 | 2020-10-20 | North America/USA/Texas/Houston | Houston Methodist Hospital | Houston Methodist Hospital | S. Wesley Long et al |

| | | | | | | |
|---|---|---|---|---|---|---|
| EPI_ISL_786705 | USA/TX-HMH-MCoV-15221/2020 | 2020-10-20 | North America/USA/Texas/Houston | Houston Methodist Hospital | Houston Methodist Hospital | S. Wesley Long et al |
| EPI_ISL_786712 | USA/TX-HMH-MCoV-15228/2020 | 2020-10-19 | North America/USA/Texas/Houston | Houston Methodist Hospital | Houston Methodist Hospital | S. Wesley Long et al |
| EPI_ISL_786713 | USA/TX-HMH-MCoV-15229/2020 | 2020-10-19 | North America/USA/Texas/Houston | Houston Methodist Hospital | Houston Methodist Hospital | S. Wesley Long et al |
| EPI_ISL_786716 | USA/TX-HMH-MCoV-15232/2020 | 2020-10-20 | North America/USA/Texas/Houston | Houston Methodist Hospital | Houston Methodist Hospital | S. Wesley Long et al |
| EPI_ISL_786719 | USA/TX-HMH-MCoV-15235/2020 | 2020-10-20 | North America/USA/Texas/Houston | Houston Methodist Hospital | Houston Methodist Hospital | S. Wesley Long et al |
| EPI_ISL_786724 | USA/TX-HMH-MCoV-15240/2020 | 2020-10-19 | North America/USA/Texas/Houston | Houston Methodist Hospital | Houston Methodist Hospital | S. Wesley Long et al |
| EPI_ISL_786726 | USA/TX-HMH-MCoV-15242/2020 | 2020-10-19 | North America/USA/Texas/Houston | Houston Methodist Hospital | Houston Methodist Hospital | S. Wesley Long et al |
| EPI_ISL_786727 | USA/TX-HMH-MCoV-15243/2020 | 2020-10-19 | North America/USA/Texas/Houston | Houston Methodist Hospital | Houston Methodist Hospital | S. Wesley Long et al |
| EPI_ISL_786730 | USA/TX-HMH-MCoV-15246/2020 | 2020-10-20 | North America/USA/Texas/Houston | Houston Methodist Hospital | Houston Methodist Hospital | S. Wesley Long et al |
| EPI_ISL_786747 | USA/TX-HMH-MCoV-15263/2020 | 2020-10-19 | North America/USA/Texas/Houston | Houston Methodist Hospital | Houston Methodist Hospital | S. Wesley Long et al |
| EPI_ISL_786761 | USA/TX-HMH-MCoV-15277/2020 | 2020-10-22 | North America/USA/Texas/Houston | Houston Methodist Hospital | Houston Methodist Hospital | S. Wesley Long et al |
| EPI_ISL_786771 | USA/TX-HMH-MCoV-15287/2020 | 2020-10-22 | North America/USA/Texas/Houston | Houston Methodist Hospital | Houston Methodist Hospital | S. Wesley Long et al |
| EPI_ISL_786779 | USA/TX-HMH-MCoV-15295/2020 | 2020-10-19 | North America/USA/Texas/Houston | Houston Methodist Hospital | Houston Methodist Hospital | S. Wesley Long et al |

| | | | | | | |
|---|---|---|---|---|---|---|
| EPI_ISL_786785 | USA/TX-HMH-MCoV-15301/2020 | 2020-10-21 | North America/USA/Texas/Houston | Houston Methodist Hospital | Houston Methodist Hospital | S. Wesley Long et al |
| EPI_ISL_786787 | USA/TX-HMH-MCoV-15303/2020 | 2020-10-19 | North America/USA/Texas/Houston | Houston Methodist Hospital | Houston Methodist Hospital | S. Wesley Long et al |
| EPI_ISL_786800 | USA/TX-HMH-MCoV-15316/2020 | 2020-10-19 | North America/USA/Texas/Houston | Houston Methodist Hospital | Houston Methodist Hospital | S. Wesley Long et al |
| EPI_ISL_786804 | USA/TX-HMH-MCoV-15320/2020 | 2020-10-19 | North America/USA/Texas/Houston | Houston Methodist Hospital | Houston Methodist Hospital | S. Wesley Long et al |
| EPI_ISL_786810 | USA/TX-HMH-MCoV-15326/2020 | 2020-10-19 | North America/USA/Texas/Houston | Houston Methodist Hospital | Houston Methodist Hospital | S. Wesley Long et al |
| EPI_ISL_786817 | USA/TX-HMH-MCoV-15333/2020 | 2020-10-21 | North America/USA/Texas/Houston | Houston Methodist Hospital | Houston Methodist Hospital | S. Wesley Long et al |
| EPI_ISL_786818 | USA/TX-HMH-MCoV-15334/2020 | 2020-10-21 | North America/USA/Texas/Houston | Houston Methodist Hospital | Houston Methodist Hospital | S. Wesley Long et al |
| EPI_ISL_786824 | USA/TX-HMH-MCoV-15340/2020 | 2020-10-19 | North America/USA/Texas/Houston | Houston Methodist Hospital | Houston Methodist Hospital | S. Wesley Long et al |
| EPI_ISL_786830 | USA/TX-HMH-MCoV-15346/2020 | 2020-10-21 | North America/USA/Texas/Houston | Houston Methodist Hospital | Houston Methodist Hospital | S. Wesley Long et al |
| EPI_ISL_786841 | USA/TX-HMH-MCoV-15357/2020 | 2020-10-21 | North America/USA/Texas/Houston | Houston Methodist Hospital | Houston Methodist Hospital | S. Wesley Long et al |
| EPI_ISL_786855 | USA/TX-HMH-MCoV-15371/2020 | 2020-10-21 | North America/USA/Texas/Houston | Houston Methodist Hospital | Houston Methodist Hospital | S. Wesley Long et al |
| EPI_ISL_786861 | USA/TX-HMH-MCoV-15377/2020 | 2020-10-21 | North America/USA/Texas/Houston | Houston Methodist Hospital | Houston Methodist Hospital | S. Wesley Long et al |
| EPI_ISL_786869 | USA/TX-HMH-MCoV-15385/2020 | 2020-10-21 | North America/USA/Texas/Houston | Houston Methodist Hospital | Houston Methodist Hospital | S. Wesley Long et al |

| | | | | | | |
|---|---|---|---|---|---|---|
| EPI_ISL_786872 | USA/TX-HMH-MCoV-15388/2020 | 2020-10-22 | North America/USA/Texas/Houston | Houston Methodist Hospital | Houston Methodist Hospital | S. Wesley Long et al |
| EPI_ISL_786880 | USA/TX-HMH-MCoV-15396/2020 | 2020-10-22 | North America/USA/Texas/Houston | Houston Methodist Hospital | Houston Methodist Hospital | S. Wesley Long et al |
| EPI_ISL_786886 | USA/TX-HMH-MCoV-15402/2020 | 2020-10-22 | North America/USA/Texas/Houston | Houston Methodist Hospital | Houston Methodist Hospital | S. Wesley Long et al |
| EPI_ISL_786889 | USA/TX-HMH-MCoV-15405/2020 | 2020-10-21 | North America/USA/Texas/Houston | Houston Methodist Hospital | Houston Methodist Hospital | S. Wesley Long et al |
| EPI_ISL_786895 | USA/TX-HMH-MCoV-15411/2020 | 2020-10-21 | North America/USA/Texas/Houston | Houston Methodist Hospital | Houston Methodist Hospital | S. Wesley Long et al |
| EPI_ISL_786899 | USA/TX-HMH-MCoV-15415/2020 | 2020-10-20 | North America/USA/Texas/Houston | Houston Methodist Hospital | Houston Methodist Hospital | S. Wesley Long et al |
| EPI_ISL_786904 | USA/TX-HMH-MCoV-15420/2020 | 2020-10-22 | North America/USA/Texas/Houston | Houston Methodist Hospital | Houston Methodist Hospital | S. Wesley Long et al |
| EPI_ISL_786905 | USA/TX-HMH-MCoV-15421/2020 | 2020-10-21 | North America/USA/Texas/Houston | Houston Methodist Hospital | Houston Methodist Hospital | S. Wesley Long et al |
| EPI_ISL_786910 | USA/TX-HMH-MCoV-15426/2020 | 2020-10-21 | North America/USA/Texas/Houston | Houston Methodist Hospital | Houston Methodist Hospital | S. Wesley Long et al |
| EPI_ISL_786912 | USA/TX-HMH-MCoV-15428/2020 | 2020-10-22 | North America/USA/Texas/Houston | Houston Methodist Hospital | Houston Methodist Hospital | S. Wesley Long et al |
| EPI_ISL_786917 | USA/TX-HMH-MCoV-15433/2020 | 2020-10-22 | North America/USA/Texas/Houston | Houston Methodist Hospital | Houston Methodist Hospital | S. Wesley Long et al |
| EPI_ISL_786943 | USA/TX-HMH-MCoV-15459/2020 | 2020-10-23 | North America/USA/Texas/Houston | Houston Methodist Hospital | Houston Methodist Hospital | S. Wesley Long et al |
| EPI_ISL_786945 | USA/TX-HMH-MCoV-15461/2020 | 2020-10-23 | North America/USA/Texas/Houston | Houston Methodist Hospital | Houston Methodist Hospital | S. Wesley Long et al |

| | | | | | | |
|---|---|---|---|---|---|---|
| EPI_ISL_786953 | USA/TX-HMH-MCoV-15469/2020 | 2020-10-23 | North America/USA/Texas/Houston | Houston Methodist Hospital | Houston Methodist Hospital | S. Wesley Long et al |
| EPI_ISL_786956 | USA/TX-HMH-MCoV-15472/2020 | 2020-10-23 | North America/USA/Texas/Houston | Houston Methodist Hospital | Houston Methodist Hospital | S. Wesley Long et al |
| EPI_ISL_786961 | USA/TX-HMH-MCoV-15477/2020 | 2020-10-23 | North America/USA/Texas/Houston | Houston Methodist Hospital | Houston Methodist Hospital | S. Wesley Long et al |
| EPI_ISL_786967 | USA/TX-HMH-MCoV-15483/2020 | 2020-10-25 | North America/USA/Texas/Houston | Houston Methodist Hospital | Houston Methodist Hospital | S. Wesley Long et al |
| EPI_ISL_786968 | USA/TX-HMH-MCoV-15484/2020 | 2020-10-24 | North America/USA/Texas/Houston | Houston Methodist Hospital | Houston Methodist Hospital | S. Wesley Long et al |
| EPI_ISL_786975 | USA/TX-HMH-MCoV-15491/2020 | 2020-10-24 | North America/USA/Texas/Houston | Houston Methodist Hospital | Houston Methodist Hospital | S. Wesley Long et al |
| EPI_ISL_786977 | USA/TX-HMH-MCoV-15493/2020 | 2020-10-23 | North America/USA/Texas/Houston | Houston Methodist Hospital | Houston Methodist Hospital | S. Wesley Long et al |
| EPI_ISL_786994 | USA/TX-HMH-MCoV-15510/2020 | 2020-10-23 | North America/USA/Texas/Houston | Houston Methodist Hospital | Houston Methodist Hospital | S. Wesley Long et al |
| EPI_ISL_787003 | USA/TX-HMH-MCoV-15519/2020 | 2020-10-24 | North America/USA/Texas/Houston | Houston Methodist Hospital | Houston Methodist Hospital | S. Wesley Long et al |
| EPI_ISL_787008 | USA/TX-HMH-MCoV-15524/2020 | 2020-10-23 | North America/USA/Texas/Houston | Houston Methodist Hospital | Houston Methodist Hospital | S. Wesley Long et al |
| EPI_ISL_787012 | USA/TX-HMH-MCoV-15528/2020 | 2020-10-23 | North America/USA/Texas/Houston | Houston Methodist Hospital | Houston Methodist Hospital | S. Wesley Long et al |
| EPI_ISL_787017 | USA/TX-HMH-MCoV-15533/2020 | 2020-10-23 | North America/USA/Texas/Houston | Houston Methodist Hospital | Houston Methodist Hospital | S. Wesley Long et al |
| EPI_ISL_787023 | USA/TX-HMH-MCoV-15539/2020 | 2020-10-22 | North America/USA/Texas/Houston | Houston Methodist Hospital | Houston Methodist Hospital | S. Wesley Long et al |

| | | | | | | |
|---|---|---|---|---|---|---|
| EPI_ISL_787025 | USA/TX-HMH-MCoV-15541/2020 | 2020-10-24 | North America/USA/Texas/Houston | Houston Methodist Hospital | Houston Methodist Hospital | S. Wesley Long et al |
| EPI_ISL_787046 | USA/TX-HMH-MCoV-15562/2020 | 2020-10-26 | North America/USA/Texas/Houston | Houston Methodist Hospital | Houston Methodist Hospital | S. Wesley Long et al |
| EPI_ISL_787047 | USA/TX-HMH-MCoV-15563/2020 | 2020-10-26 | North America/USA/Texas/Houston | Houston Methodist Hospital | Houston Methodist Hospital | S. Wesley Long et al |
| EPI_ISL_787048 | USA/TX-HMH-MCoV-15564/2020 | 2020-10-26 | North America/USA/Texas/Houston | Houston Methodist Hospital | Houston Methodist Hospital | S. Wesley Long et al |
| EPI_ISL_787050 | USA/TX-HMH-MCoV-15566/2020 | 2020-10-23 | North America/USA/Texas/Houston | Houston Methodist Hospital | Houston Methodist Hospital | S. Wesley Long et al |
| EPI_ISL_787053 | USA/TX-HMH-MCoV-15569/2020 | 2020-10-26 | North America/USA/Texas/Houston | Houston Methodist Hospital | Houston Methodist Hospital | S. Wesley Long et al |
| EPI_ISL_787070 | USA/TX-HMH-MCoV-15586/2020 | 2020-10-26 | North America/USA/Texas/Houston | Houston Methodist Hospital | Houston Methodist Hospital | S. Wesley Long et al |
| EPI_ISL_787074 | USA/TX-HMH-MCoV-15590/2020 | 2020-10-24 | North America/USA/Texas/Houston | Houston Methodist Hospital | Houston Methodist Hospital | S. Wesley Long et al |
| EPI_ISL_787079 | USA/TX-HMH-MCoV-15595/2020 | 2020-10-26 | North America/USA/Texas/Houston | Houston Methodist Hospital | Houston Methodist Hospital | S. Wesley Long et al |
| EPI_ISL_787083 | USA/TX-HMH-MCoV-15599/2020 | 2020-10-26 | North America/USA/Texas/Houston | Houston Methodist Hospital | Houston Methodist Hospital | S. Wesley Long et al |
| EPI_ISL_787088 | USA/TX-HMH-MCoV-15604/2020 | 2020-10-26 | North America/USA/Texas/Houston | Houston Methodist Hospital | Houston Methodist Hospital | S. Wesley Long et al |
| EPI_ISL_787090 | USA/TX-HMH-MCoV-15606/2020 | 2020-10-26 | North America/USA/Texas/Houston | Houston Methodist Hospital | Houston Methodist Hospital | S. Wesley Long et al |
| EPI_ISL_787096 | USA/TX-HMH-MCoV-15612/2020 | 2020-10-26 | North America/USA/Texas/Houston | Houston Methodist Hospital | Houston Methodist Hospital | S. Wesley Long et al |

| | | | | | | |
|---|---|---|---|---|---|---|
| EPI_ISL_787099 | USA/TX-HMH-MCoV-13806/2020 | 2020-08-18 | North America/USA/Texas/Houston | Houston Methodist Hospital | Houston Methodist Hospital | S. Wesley Long et al |
| EPI_ISL_787100 | USA/TX-HMH-MCoV-13807/2020 | 2020-08-17 | North America/USA/Texas/Houston | Houston Methodist Hospital | Houston Methodist Hospital | S. Wesley Long et al |
| EPI_ISL_787105 | USA/TX-HMH-MCoV-13812/2020 | 2020-08-17 | North America/USA/Texas/Houston | Houston Methodist Hospital | Houston Methodist Hospital | S. Wesley Long et al |
| EPI_ISL_787107 | USA/TX-HMH-MCoV-13814/2020 | 2020-08-17 | North America/USA/Texas/Houston | Houston Methodist Hospital | Houston Methodist Hospital | S. Wesley Long et al |
| EPI_ISL_787110 | USA/TX-HMH-MCoV-13817/2020 | 2020-08-17 | North America/USA/Texas/Houston | Houston Methodist Hospital | Houston Methodist Hospital | S. Wesley Long et al |
| EPI_ISL_787115 | USA/TX-HMH-MCoV-13822/2020 | 2020-08-17 | North America/USA/Texas/Houston | Houston Methodist Hospital | Houston Methodist Hospital | S. Wesley Long et al |
| EPI_ISL_787123 | USA/TX-HMH-MCoV-13830/2020 | 2020-08-17 | North America/USA/Texas/Houston | Houston Methodist Hospital | Houston Methodist Hospital | S. Wesley Long et al |
| EPI_ISL_787125 | USA/TX-HMH-MCoV-13832/2020 | 2020-08-17 | North America/USA/Texas/Houston | Houston Methodist Hospital | Houston Methodist Hospital | S. Wesley Long et al |
| EPI_ISL_787126 | USA/TX-HMH-MCoV-13833/2020 | 2020-08-17 | North America/USA/Texas/Houston | Houston Methodist Hospital | Houston Methodist Hospital | S. Wesley Long et al |
| EPI_ISL_787127 | USA/TX-HMH-MCoV-13834/2020 | 2020-08-17 | North America/USA/Texas/Houston | Houston Methodist Hospital | Houston Methodist Hospital | S. Wesley Long et al |
| EPI_ISL_787131 | USA/TX-HMH-MCoV-13838/2020 | 2020-08-16 | North America/USA/Texas/Houston | Houston Methodist Hospital | Houston Methodist Hospital | S. Wesley Long et al |
| EPI_ISL_787133 | USA/TX-HMH-MCoV-13840/2020 | 2020-08-17 | North America/USA/Texas/Houston | Houston Methodist Hospital | Houston Methodist Hospital | S. Wesley Long et al |
| EPI_ISL_787134 | USA/TX-HMH-MCoV-13841/2020 | 2020-08-17 | North America/USA/Texas/Houston | Houston Methodist Hospital | Houston Methodist Hospital | S. Wesley Long et al |

| | | | | | | |
|---|---|---|---|---|---|---|
| EPI_ISL_787145 | USA/TX-HMH-MCoV-13852/2020 | 2020-08-17 | North America/USA/Texas/Houston | Houston Methodist Hospital | Houston Methodist Hospital | S. Wesley Long et al |
| EPI_ISL_787152 | USA/TX-HMH-MCoV-13859/2020 | 2020-08-17 | North America/USA/Texas/Houston | Houston Methodist Hospital | Houston Methodist Hospital | S. Wesley Long et al |
| EPI_ISL_787160 | USA/TX-HMH-MCoV-13867/2020 | 2020-08-17 | North America/USA/Texas/Houston | Houston Methodist Hospital | Houston Methodist Hospital | S. Wesley Long et al |
| EPI_ISL_787161 | USA/TX-HMH-MCoV-13868/2020 | 2020-08-17 | North America/USA/Texas/Houston | Houston Methodist Hospital | Houston Methodist Hospital | S. Wesley Long et al |
| EPI_ISL_787165 | USA/TX-HMH-MCoV-13872/2020 | 2020-08-17 | North America/USA/Texas/Houston | Houston Methodist Hospital | Houston Methodist Hospital | S. Wesley Long et al |
| EPI_ISL_787168 | USA/TX-HMH-MCoV-13875/2020 | 2020-08-17 | North America/USA/Texas/Houston | Houston Methodist Hospital | Houston Methodist Hospital | S. Wesley Long et al |
| EPI_ISL_787169 | USA/TX-HMH-MCoV-13876/2020 | 2020-08-17 | North America/USA/Texas/Houston | Houston Methodist Hospital | Houston Methodist Hospital | S. Wesley Long et al |
| EPI_ISL_787170 | USA/TX-HMH-MCoV-13877/2020 | 2020-08-17 | North America/USA/Texas/Houston | Houston Methodist Hospital | Houston Methodist Hospital | S. Wesley Long et al |
| EPI_ISL_787173 | USA/TX-HMH-MCoV-13880/2020 | 2020-08-17 | North America/USA/Texas/Houston | Houston Methodist Hospital | Houston Methodist Hospital | S. Wesley Long et al |
| EPI_ISL_787178 | USA/TX-HMH-MCoV-13886/2020 | 2020-08-16 | North America/USA/Texas/Houston | Houston Methodist Hospital | Houston Methodist Hospital | S. Wesley Long et al |
| EPI_ISL_787179 | USA/TX-HMH-MCoV-13887/2020 | 2020-08-17 | North America/USA/Texas/Houston | Houston Methodist Hospital | Houston Methodist Hospital | S. Wesley Long et al |
| EPI_ISL_787183 | USA/TX-HMH-MCoV-13891/2020 | 2020-08-17 | North America/USA/Texas/Houston | Houston Methodist Hospital | Houston Methodist Hospital | S. Wesley Long et al |
| EPI_ISL_787185 | USA/TX-HMH-MCoV-13893/2020 | 2020-08-17 | North America/USA/Texas/Houston | Houston Methodist Hospital | Houston Methodist Hospital | S. Wesley Long et al |

| | | | | | | |
|---|---|---|---|---|---|---|
| EPI_ISL_787188 | USA/TX-HMH-MCoV-13896/2020 | 2020-08-17 | North America/USA/Texas/Houston | Houston Methodist Hospital | Houston Methodist Hospital | S. Wesley Long et al |
| EPI_ISL_787189 | USA/TX-HMH-MCoV-13897/2020 | 2020-08-16 | North America/USA/Texas/Houston | Houston Methodist Hospital | Houston Methodist Hospital | S. Wesley Long et al |
| EPI_ISL_787190 | USA/TX-HMH-MCoV-13898/2020 | 2020-08-16 | North America/USA/Texas/Houston | Houston Methodist Hospital | Houston Methodist Hospital | S. Wesley Long et al |
| EPI_ISL_787199 | USA/TX-HMH-MCoV-13907/2020 | 2020-08-17 | North America/USA/Texas/Houston | Houston Methodist Hospital | Houston Methodist Hospital | S. Wesley Long et al |
| EPI_ISL_787202 | USA/TX-HMH-MCoV-13910/2020 | 2020-08-16 | North America/USA/Texas/Houston | Houston Methodist Hospital | Houston Methodist Hospital | S. Wesley Long et al |
| EPI_ISL_787206 | USA/TX-HMH-MCoV-13914/2020 | 2020-08-16 | North America/USA/Texas/Houston | Houston Methodist Hospital | Houston Methodist Hospital | S. Wesley Long et al |
| EPI_ISL_787207 | USA/TX-HMH-MCoV-13915/2020 | 2020-08-16 | North America/USA/Texas/Houston | Houston Methodist Hospital | Houston Methodist Hospital | S. Wesley Long et al |
| EPI_ISL_787217 | USA/TX-HMH-MCoV-13925/2020 | 2020-08-16 | North America/USA/Texas/Houston | Houston Methodist Hospital | Houston Methodist Hospital | S. Wesley Long et al |
| EPI_ISL_787220 | USA/TX-HMH-MCoV-13928/2020 | 2020-08-15 | North America/USA/Texas/Houston | Houston Methodist Hospital | Houston Methodist Hospital | S. Wesley Long et al |
| EPI_ISL_787225 | USA/TX-HMH-MCoV-13933/2020 | 2020-08-15 | North America/USA/Texas/Houston | Houston Methodist Hospital | Houston Methodist Hospital | S. Wesley Long et al |
| EPI_ISL_787228 | USA/TX-HMH-MCoV-13936/2020 | 2020-08-16 | North America/USA/Texas/Houston | Houston Methodist Hospital | Houston Methodist Hospital | S. Wesley Long et al |
| EPI_ISL_787231 | USA/TX-HMH-MCoV-13939/2020 | 2020-08-16 | North America/USA/Texas/Houston | Houston Methodist Hospital | Houston Methodist Hospital | S. Wesley Long et al |
| EPI_ISL_787233 | USA/TX-HMH-MCoV-13941/2020 | 2020-08-15 | North America/USA/Texas/Houston | Houston Methodist Hospital | Houston Methodist Hospital | S. Wesley Long et al |

| | | | | | | |
|---|---|---|---|---|---|---|
| EPI_ISL_787236 | USA/TX-HMH-MCoV-13944/2020 | 2020-08-15 | North America/USA/Texas/Houston | Houston Methodist Hospital | Houston Methodist Hospital | S. Wesley Long et al |
| EPI_ISL_787238 | USA/TX-HMH-MCoV-13946/2020 | 2020-08-15 | North America/USA/Texas/Houston | Houston Methodist Hospital | Houston Methodist Hospital | S. Wesley Long et al |
| EPI_ISL_787240 | USA/TX-HMH-MCoV-13948/2020 | 2020-08-16 | North America/USA/Texas/Houston | Houston Methodist Hospital | Houston Methodist Hospital | S. Wesley Long et al |
| EPI_ISL_787241 | USA/TX-HMH-MCoV-13949/2020 | 2020-08-16 | North America/USA/Texas/Houston | Houston Methodist Hospital | Houston Methodist Hospital | S. Wesley Long et al |
| EPI_ISL_787249 | USA/TX-HMH-MCoV-13957/2020 | 2020-08-15 | North America/USA/Texas/Houston | Houston Methodist Hospital | Houston Methodist Hospital | S. Wesley Long et al |
| EPI_ISL_787252 | USA/TX-HMH-MCoV-13960/2020 | 2020-08-16 | North America/USA/Texas/Houston | Houston Methodist Hospital | Houston Methodist Hospital | S. Wesley Long et al |
| EPI_ISL_787258 | USA/TX-HMH-MCoV-13967/2020 | 2020-08-15 | North America/USA/Texas/Houston | Houston Methodist Hospital | Houston Methodist Hospital | S. Wesley Long et al |
| EPI_ISL_787260 | USA/TX-HMH-MCoV-13969/2020 | 2020-08-15 | North America/USA/Texas/Houston | Houston Methodist Hospital | Houston Methodist Hospital | S. Wesley Long et al |
| EPI_ISL_787261 | USA/TX-HMH-MCoV-13970/2020 | 2020-08-15 | North America/USA/Texas/Houston | Houston Methodist Hospital | Houston Methodist Hospital | S. Wesley Long et al |
| EPI_ISL_787271 | USA/TX-HMH-MCoV-13980/2020 | 2020-08-15 | North America/USA/Texas/Houston | Houston Methodist Hospital | Houston Methodist Hospital | S. Wesley Long et al |
| EPI_ISL_787272 | USA/TX-HMH-MCoV-13981/2020 | 2020-08-15 | North America/USA/Texas/Houston | Houston Methodist Hospital | Houston Methodist Hospital | S. Wesley Long et al |
| EPI_ISL_787273 | USA/TX-HMH-MCoV-13982/2020 | 2020-08-15 | North America/USA/Texas/Houston | Houston Methodist Hospital | Houston Methodist Hospital | S. Wesley Long et al |
| EPI_ISL_787277 | USA/TX-HMH-MCoV-13986/2020 | 2020-08-16 | North America/USA/Texas/Houston | Houston Methodist Hospital | Houston Methodist Hospital | S. Wesley Long et al |

| | | | | | | |
|---|---|---|---|---|---|---|
| EPI_ISL_787280 | USA/TX-HMH-MCoV-13989/2020 | 2020-08-15 | North America/USA/Texas/Houston | Houston Methodist Hospital | Houston Methodist Hospital | S. Wesley Long et al |
| EPI_ISL_787283 | USA/TX-HMH-MCoV-13992/2020 | 2020-08-15 | North America/USA/Texas/Houston | Houston Methodist Hospital | Houston Methodist Hospital | S. Wesley Long et al |
| EPI_ISL_787286 | USA/TX-HMH-MCoV-13995/2020 | 2020-08-16 | North America/USA/Texas/Houston | Houston Methodist Hospital | Houston Methodist Hospital | S. Wesley Long et al |
| EPI_ISL_787292 | USA/TX-HMH-MCoV-14001/2020 | 2020-08-14 | North America/USA/Texas/Houston | Houston Methodist Hospital | Houston Methodist Hospital | S. Wesley Long et al |
| EPI_ISL_787300 | USA/TX-HMH-MCoV-14009/2020 | 2020-08-14 | North America/USA/Texas/Houston | Houston Methodist Hospital | Houston Methodist Hospital | S. Wesley Long et al |
| EPI_ISL_787304 | USA/TX-HMH-MCoV-14015/2020 | 2020-08-14 | North America/USA/Texas/Houston | Houston Methodist Hospital | Houston Methodist Hospital | S. Wesley Long et al |
| EPI_ISL_787306 | USA/TX-HMH-MCoV-14017/2020 | 2020-08-14 | North America/USA/Texas/Houston | Houston Methodist Hospital | Houston Methodist Hospital | S. Wesley Long et al |
| EPI_ISL_787308 | USA/TX-HMH-MCoV-14019/2020 | 2020-08-14 | North America/USA/Texas/Houston | Houston Methodist Hospital | Houston Methodist Hospital | S. Wesley Long et al |
| EPI_ISL_787314 | USA/TX-HMH-MCoV-14025/2020 | 2020-08-14 | North America/USA/Texas/Houston | Houston Methodist Hospital | Houston Methodist Hospital | S. Wesley Long et al |
| EPI_ISL_787319 | USA/TX-HMH-MCoV-14030/2020 | 2020-08-14 | North America/USA/Texas/Houston | Houston Methodist Hospital | Houston Methodist Hospital | S. Wesley Long et al |
| EPI_ISL_787320 | USA/TX-HMH-MCoV-14031/2020 | 2020-08-14 | North America/USA/Texas/Houston | Houston Methodist Hospital | Houston Methodist Hospital | S. Wesley Long et al |
| EPI_ISL_787321 | USA/TX-HMH-MCoV-14032/2020 | 2020-08-14 | North America/USA/Texas/Houston | Houston Methodist Hospital | Houston Methodist Hospital | S. Wesley Long et al |
| EPI_ISL_787326 | USA/TX-HMH-MCoV-14037/2020 | 2020-08-14 | North America/USA/Texas/Houston | Houston Methodist Hospital | Houston Methodist Hospital | S. Wesley Long et al |

| | | | | | | |
|---|---|---|---|---|---|---|
| EPI_ISL_787331 | USA/TX-HMH-MCoV-14043/2020 | 2020-08-14 | North America/USA/Texas/Houston | Houston Methodist Hospital | Houston Methodist Hospital | S. Wesley Long et al |
| EPI_ISL_787334 | USA/TX-HMH-MCoV-14046/2020 | 2020-08-14 | North America/USA/Texas/Houston | Houston Methodist Hospital | Houston Methodist Hospital | S. Wesley Long et al |
| EPI_ISL_787339 | USA/TX-HMH-MCoV-14052/2020 | 2020-08-14 | North America/USA/Texas/Houston | Houston Methodist Hospital | Houston Methodist Hospital | S. Wesley Long et al |
| EPI_ISL_787340 | USA/TX-HMH-MCoV-14053/2020 | 2020-08-14 | North America/USA/Texas/Houston | Houston Methodist Hospital | Houston Methodist Hospital | S. Wesley Long et al |
| EPI_ISL_787341 | USA/TX-HMH-MCoV-14054/2020 | 2020-08-14 | North America/USA/Texas/Houston | Houston Methodist Hospital | Houston Methodist Hospital | S. Wesley Long et al |
| EPI_ISL_787345 | USA/TX-HMH-MCoV-14058/2020 | 2020-08-14 | North America/USA/Texas/Houston | Houston Methodist Hospital | Houston Methodist Hospital | S. Wesley Long et al |
| EPI_ISL_787350 | USA/TX-HMH-MCoV-14064/2020 | 2020-08-14 | North America/USA/Texas/Houston | Houston Methodist Hospital | Houston Methodist Hospital | S. Wesley Long et al |
| EPI_ISL_787352 | USA/TX-HMH-MCoV-14066/2020 | 2020-08-14 | North America/USA/Texas/Houston | Houston Methodist Hospital | Houston Methodist Hospital | S. Wesley Long et al |
| EPI_ISL_787356 | USA/TX-HMH-MCoV-14070/2020 | 2020-08-14 | North America/USA/Texas/Houston | Houston Methodist Hospital | Houston Methodist Hospital | S. Wesley Long et al |
| EPI_ISL_787357 | USA/TX-HMH-MCoV-14071/2020 | 2020-08-14 | North America/USA/Texas/Houston | Houston Methodist Hospital | Houston Methodist Hospital | S. Wesley Long et al |
| EPI_ISL_787360 | USA/TX-HMH-MCoV-14074/2020 | 2020-08-14 | North America/USA/Texas/Houston | Houston Methodist Hospital | Houston Methodist Hospital | S. Wesley Long et al |
| EPI_ISL_787362 | USA/TX-HMH-MCoV-14076/2020 | 2020-08-14 | North America/USA/Texas/Houston | Houston Methodist Hospital | Houston Methodist Hospital | S. Wesley Long et al |
| EPI_ISL_787364 | USA/TX-HMH-MCoV-14078/2020 | 2020-08-14 | North America/USA/Texas/Houston | Houston Methodist Hospital | Houston Methodist Hospital | S. Wesley Long et al |

| | | | | | | |
|---|---|---|---|---|---|---|
| EPI_ISL_787369 | USA/TX-HMH-MCoV-14083/2020 | 2020-08-14 | North America/USA/Texas/Houston | Houston Methodist Hospital | Houston Methodist Hospital | S. Wesley Long et al |
| EPI_ISL_787370 | USA/TX-HMH-MCoV-14084/2020 | 2020-08-14 | North America/USA/Texas/Houston | Houston Methodist Hospital | Houston Methodist Hospital | S. Wesley Long et al |
| EPI_ISL_787371 | USA/TX-HMH-MCoV-14085/2020 | 2020-08-14 | North America/USA/Texas/Houston | Houston Methodist Hospital | Houston Methodist Hospital | S. Wesley Long et al |
| EPI_ISL_787373 | USA/TX-HMH-MCoV-14087/2020 | 2020-08-14 | North America/USA/Texas/Houston | Houston Methodist Hospital | Houston Methodist Hospital | S. Wesley Long et al |
| EPI_ISL_787374 | USA/TX-HMH-MCoV-14088/2020 | 2020-08-14 | North America/USA/Texas/Houston | Houston Methodist Hospital | Houston Methodist Hospital | S. Wesley Long et al |
| EPI_ISL_787381 | USA/TX-HMH-MCoV-14095/2020 | 2020-08-14 | North America/USA/Texas/Houston | Houston Methodist Hospital | Houston Methodist Hospital | S. Wesley Long et al |
| EPI_ISL_787386 | USA/TX-HMH-MCoV-14100/2020 | 2020-08-14 | North America/USA/Texas/Houston | Houston Methodist Hospital | Houston Methodist Hospital | S. Wesley Long et al |
| EPI_ISL_787400 | USA/TX-HMH-MCoV-14114/2020 | 2020-10-03 | North America/USA/Texas/Houston | Houston Methodist Hospital | Houston Methodist Hospital | S. Wesley Long et al |
| EPI_ISL_787412 | USA/TX-HMH-MCoV-14127/2020 | 2020-10-01 | North America/USA/Texas/Houston | Houston Methodist Hospital | Houston Methodist Hospital | S. Wesley Long et al |
| EPI_ISL_787428 | USA/TX-HMH-MCoV-14143/2020 | 2020-10-02 | North America/USA/Texas/Houston | Houston Methodist Hospital | Houston Methodist Hospital | S. Wesley Long et al |
| EPI_ISL_787430 | USA/TX-HMH-MCoV-14145/2020 | 2020-10-02 | North America/USA/Texas/Houston | Houston Methodist Hospital | Houston Methodist Hospital | S. Wesley Long et al |
| EPI_ISL_787435 | USA/TX-HMH-MCoV-14150/2020 | 2020-10-03 | North America/USA/Texas/Houston | Houston Methodist Hospital | Houston Methodist Hospital | S. Wesley Long et al |
| EPI_ISL_787443 | USA/TX-HMH-MCoV-14158/2020 | 2020-10-02 | North America/USA/Texas/Houston | Houston Methodist Hospital | Houston Methodist Hospital | S. Wesley Long et al |

| | | | | | | |
|---|---|---|---|---|---|---|
| EPI_ISL_787445 | USA/TX-HMH-MCoV-14160/2020 | 2020-10-02 | North America/USA/Texas/Houston | Houston Methodist Hospital | Houston Methodist Hospital | S. Wesley Long et al |
| EPI_ISL_787446 | USA/TX-HMH-MCoV-14161/2020 | 2020-10-04 | North America/USA/Texas/Houston | Houston Methodist Hospital | Houston Methodist Hospital | S. Wesley Long et al |
| EPI_ISL_787462 | USA/TX-HMH-MCoV-14177/2020 | 2020-10-01 | North America/USA/Texas/Houston | Houston Methodist Hospital | Houston Methodist Hospital | S. Wesley Long et al |
| EPI_ISL_787468 | USA/TX-HMH-MCoV-14183/2020 | 2020-10-02 | North America/USA/Texas/Houston | Houston Methodist Hospital | Houston Methodist Hospital | S. Wesley Long et al |
| EPI_ISL_787469 | USA/TX-HMH-MCoV-14184/2020 | 2020-10-02 | North America/USA/Texas/Houston | Houston Methodist Hospital | Houston Methodist Hospital | S. Wesley Long et al |
| EPI_ISL_787471 | USA/TX-HMH-MCoV-14186/2020 | 2020-10-03 | North America/USA/Texas/Houston | Houston Methodist Hospital | Houston Methodist Hospital | S. Wesley Long et al |
| EPI_ISL_787476 | USA/TX-HMH-MCoV-14191/2020 | 2020-10-02 | North America/USA/Texas/Houston | Houston Methodist Hospital | Houston Methodist Hospital | S. Wesley Long et al |
| EPI_ISL_787477 | USA/TX-HMH-MCoV-14192/2020 | 2020-10-02 | North America/USA/Texas/Houston | Houston Methodist Hospital | Houston Methodist Hospital | S. Wesley Long et al |
| EPI_ISL_787483 | USA/TX-HMH-MCoV-14198/2020 | 2020-10-03 | North America/USA/Texas/Houston | Houston Methodist Hospital | Houston Methodist Hospital | S. Wesley Long et al |
| EPI_ISL_787488 | USA/TX-HMH-MCoV-14203/2020 | 2020-10-04 | North America/USA/Texas/Houston | Houston Methodist Hospital | Houston Methodist Hospital | S. Wesley Long et al |
| EPI_ISL_787496 | USA/TX-HMH-MCoV-14211/2020 | 2020-10-03 | North America/USA/Texas/Houston | Houston Methodist Hospital | Houston Methodist Hospital | S. Wesley Long et al |
| EPI_ISL_787499 | USA/TX-HMH-MCoV-14214/2020 | 2020-10-04 | North America/USA/Texas/Houston | Houston Methodist Hospital | Houston Methodist Hospital | S. Wesley Long et al |
| EPI_ISL_787500 | USA/TX-HMH-MCoV-14215/2020 | 2020-10-04 | North America/USA/Texas/Houston | Houston Methodist Hospital | Houston Methodist Hospital | S. Wesley Long et al |

| | | | | | | |
|---|---|---|---|---|---|---|
| EPI_ISL_787501 | USA/TX-HMH-MCoV-14216/2020 | 2020-10-04 | North America/USA/Texas/Houston | Houston Methodist Hospital | Houston Methodist Hospital | S. Wesley Long et al |
| EPI_ISL_787504 | USA/TX-HMH-MCoV-14219/2020 | 2020-10-05 | North America/USA/Texas/Houston | Houston Methodist Hospital | Houston Methodist Hospital | S. Wesley Long et al |
| EPI_ISL_787515 | USA/TX-HMH-MCoV-14230/2020 | 2020-10-05 | North America/USA/Texas/Houston | Houston Methodist Hospital | Houston Methodist Hospital | S. Wesley Long et al |
| EPI_ISL_787521 | USA/TX-HMH-MCoV-14236/2020 | 2020-10-03 | North America/USA/Texas/Houston | Houston Methodist Hospital | Houston Methodist Hospital | S. Wesley Long et al |
| EPI_ISL_787547 | USA/TX-HMH-MCoV-14262/2020 | 2020-10-04 | North America/USA/Texas/Houston | Houston Methodist Hospital | Houston Methodist Hospital | S. Wesley Long et al |
| EPI_ISL_787548 | USA/TX-HMH-MCoV-14263/2020 | 2020-10-04 | North America/USA/Texas/Houston | Houston Methodist Hospital | Houston Methodist Hospital | S. Wesley Long et al |
| EPI_ISL_787551 | USA/TX-HMH-MCoV-14266/2020 | 2020-10-06 | North America/USA/Texas/Houston | Houston Methodist Hospital | Houston Methodist Hospital | S. Wesley Long et al |
| EPI_ISL_787555 | USA/TX-HMH-MCoV-14270/2020 | 2020-10-05 | North America/USA/Texas/Houston | Houston Methodist Hospital | Houston Methodist Hospital | S. Wesley Long et al |
| EPI_ISL_787560 | USA/TX-HMH-MCoV-14275/2020 | 2020-10-04 | North America/USA/Texas/Houston | Houston Methodist Hospital | Houston Methodist Hospital | S. Wesley Long et al |
| EPI_ISL_787561 | USA/TX-HMH-MCoV-14276/2020 | 2020-10-04 | North America/USA/Texas/Houston | Houston Methodist Hospital | Houston Methodist Hospital | S. Wesley Long et al |
| EPI_ISL_787568 | USA/TX-HMH-MCoV-14283/2020 | 2020-10-03 | North America/USA/Texas/Houston | Houston Methodist Hospital | Houston Methodist Hospital | S. Wesley Long et al |
| EPI_ISL_787572 | USA/TX-HMH-MCoV-14287/2020 | 2020-10-04 | North America/USA/Texas/Houston | Houston Methodist Hospital | Houston Methodist Hospital | S. Wesley Long et al |
| EPI_ISL_787579 | USA/TX-HMH-MCoV-14294/2020 | 2020-10-05 | North America/USA/Texas/Houston | Houston Methodist Hospital | Houston Methodist Hospital | S. Wesley Long et al |

| | | | | | | |
|---|---|---|---|---|---|---|
| EPI_ISL_787583 | USA/TX-HMH-MCoV-14298/2020 | 2020-10-05 | North America/USA/Texas/Houston | Houston Methodist Hospital | Houston Methodist Hospital | S. Wesley Long et al |
| EPI_ISL_787585 | USA/TX-HMH-MCoV-14300/2020 | 2020-10-05 | North America/USA/Texas/Houston | Houston Methodist Hospital | Houston Methodist Hospital | S. Wesley Long et al |
| EPI_ISL_787586 | USA/TX-HMH-MCoV-14301/2020 | 2020-10-04 | North America/USA/Texas/Houston | Houston Methodist Hospital | Houston Methodist Hospital | S. Wesley Long et al |
| EPI_ISL_787595 | USA/TX-HMH-MCoV-14310/2020 | 2020-10-05 | North America/USA/Texas/Houston | Houston Methodist Hospital | Houston Methodist Hospital | S. Wesley Long et al |
| EPI_ISL_787598 | USA/TX-HMH-MCoV-14313/2020 | 2020-10-05 | North America/USA/Texas/Houston | Houston Methodist Hospital | Houston Methodist Hospital | S. Wesley Long et al |
| EPI_ISL_787614 | USA/TX-HMH-MCoV-14329/2020 | 2020-10-06 | North America/USA/Texas/Houston | Houston Methodist Hospital | Houston Methodist Hospital | S. Wesley Long et al |
| EPI_ISL_787620 | USA/TX-HMH-MCoV-14335/2020 | 2020-10-05 | North America/USA/Texas/Houston | Houston Methodist Hospital | Houston Methodist Hospital | S. Wesley Long et al |
| EPI_ISL_787621 | USA/TX-HMH-MCoV-14336/2020 | 2020-10-05 | North America/USA/Texas/Houston | Houston Methodist Hospital | Houston Methodist Hospital | S. Wesley Long et al |
| EPI_ISL_787623 | USA/TX-HMH-MCoV-14338/2020 | 2020-10-06 | North America/USA/Texas/Houston | Houston Methodist Hospital | Houston Methodist Hospital | S. Wesley Long et al |
| EPI_ISL_787625 | USA/TX-HMH-MCoV-14340/2020 | 2020-10-06 | North America/USA/Texas/Houston | Houston Methodist Hospital | Houston Methodist Hospital | S. Wesley Long et al |
| EPI_ISL_787627 | USA/TX-HMH-MCoV-14342/2020 | 2020-10-06 | North America/USA/Texas/Houston | Houston Methodist Hospital | Houston Methodist Hospital | S. Wesley Long et al |
| EPI_ISL_787632 | USA/TX-HMH-MCoV-14347/2020 | 2020-10-05 | North America/USA/Texas/Houston | Houston Methodist Hospital | Houston Methodist Hospital | S. Wesley Long et al |
| EPI_ISL_787636 | USA/TX-HMH-MCoV-14351/2020 | 2020-10-06 | North America/USA/Texas/Houston | Houston Methodist Hospital | Houston Methodist Hospital | S. Wesley Long et al |

| | | | | | | |
|---|---|---|---|---|---|---|
| EPI_ISL_787639 | USA/TX-HMH-MCoV-14354/2020 | 2020-10-06 | North America/USA/Texas/Houston | Houston Methodist Hospital | Houston Methodist Hospital | S. Wesley Long et al |
| EPI_ISL_787647 | USA/TX-HMH-MCoV-14362/2020 | 2020-10-06 | North America/USA/Texas/Houston | Houston Methodist Hospital | Houston Methodist Hospital | S. Wesley Long et al |
| EPI_ISL_787653 | USA/TX-HMH-MCoV-14368/2020 | 2020-10-05 | North America/USA/Texas/Houston | Houston Methodist Hospital | Houston Methodist Hospital | S. Wesley Long et al |
| EPI_ISL_787670 | USA/TX-HMH-MCoV-14385/2020 | 2020-10-06 | North America/USA/Texas/Houston | Houston Methodist Hospital | Houston Methodist Hospital | S. Wesley Long et al |
| EPI_ISL_787672 | USA/TX-HMH-MCoV-14387/2020 | 2020-10-06 | North America/USA/Texas/Houston | Houston Methodist Hospital | Houston Methodist Hospital | S. Wesley Long et al |
| EPI_ISL_787673 | USA/TX-HMH-MCoV-14388/2020 | 2020-10-06 | North America/USA/Texas/Houston | Houston Methodist Hospital | Houston Methodist Hospital | S. Wesley Long et al |
| EPI_ISL_787680 | USA/TX-HMH-MCoV-14395/2020 | 2020-10-07 | North America/USA/Texas/Houston | Houston Methodist Hospital | Houston Methodist Hospital | S. Wesley Long et al |
| EPI_ISL_787688 | USA/TX-HMH-MCoV-14403/2020 | 2020-10-06 | North America/USA/Texas/Houston | Houston Methodist Hospital | Houston Methodist Hospital | S. Wesley Long et al |
| EPI_ISL_787690 | USA/TX-HMH-MCoV-14405/2020 | 2020-10-07 | North America/USA/Texas/Houston | Houston Methodist Hospital | Houston Methodist Hospital | S. Wesley Long et al |
| EPI_ISL_787691 | USA/TX-HMH-MCoV-14406/2020 | 2020-10-07 | North America/USA/Texas/Houston | Houston Methodist Hospital | Houston Methodist Hospital | S. Wesley Long et al |
| EPI_ISL_787693 | USA/TX-HMH-MCoV-14408/2020 | 2020-10-07 | North America/USA/Texas/Houston | Houston Methodist Hospital | Houston Methodist Hospital | S. Wesley Long et al |
| EPI_ISL_787696 | USA/TX-HMH-MCoV-14411/2020 | 2020-10-07 | North America/USA/Texas/Houston | Houston Methodist Hospital | Houston Methodist Hospital | S. Wesley Long et al |
| EPI_ISL_787702 | USA/TX-HMH-MCoV-14417/2020 | 2020-10-07 | North America/USA/Texas/Houston | Houston Methodist Hospital | Houston Methodist Hospital | S. Wesley Long et al |

| | | | | | | |
|---|---|---|---|---|---|---|
| EPI_ISL_787710 | USA/TX-HMH-MCoV-14425/2020 | 2020-10-08 | North America/USA/Texas/Houston | Houston Methodist Hospital | Houston Methodist Hospital | S. Wesley Long et al |
| EPI_ISL_787723 | USA/TX-HMH-MCoV-14438/2020 | 2020-10-08 | North America/USA/Texas/Houston | Houston Methodist Hospital | Houston Methodist Hospital | S. Wesley Long et al |
| EPI_ISL_787729 | USA/TX-HMH-MCoV-14444/2020 | 2020-10-07 | North America/USA/Texas/Houston | Houston Methodist Hospital | Houston Methodist Hospital | S. Wesley Long et al |
| EPI_ISL_787730 | USA/TX-HMH-MCoV-14445/2020 | 2020-10-07 | North America/USA/Texas/Houston | Houston Methodist Hospital | Houston Methodist Hospital | S. Wesley Long et al |
| EPI_ISL_787731 | USA/TX-HMH-MCoV-14446/2020 | 2020-10-07 | North America/USA/Texas/Houston | Houston Methodist Hospital | Houston Methodist Hospital | S. Wesley Long et al |
| EPI_ISL_787732 | USA/TX-HMH-MCoV-14447/2020 | 2020-10-07 | North America/USA/Texas/Houston | Houston Methodist Hospital | Houston Methodist Hospital | S. Wesley Long et al |
| EPI_ISL_787736 | USA/TX-HMH-MCoV-14451/2020 | 2020-10-06 | North America/USA/Texas/Houston | Houston Methodist Hospital | Houston Methodist Hospital | S. Wesley Long et al |
| EPI_ISL_787773 | USA/TX-HMH-MCoV-14488/2020 | 2020-10-08 | North America/USA/Texas/Houston | Houston Methodist Hospital | Houston Methodist Hospital | S. Wesley Long et al |
| EPI_ISL_787792 | USA/TX-HMH-MCoV-14507/2020 | 2020-10-09 | North America/USA/Texas/Houston | Houston Methodist Hospital | Houston Methodist Hospital | S. Wesley Long et al |
| EPI_ISL_787795 | USA/TX-HMH-MCoV-14510/2020 | 2020-10-09 | North America/USA/Texas/Houston | Houston Methodist Hospital | Houston Methodist Hospital | S. Wesley Long et al |
| EPI_ISL_787799 | USA/TX-HMH-MCoV-14514/2020 | 2020-10-08 | North America/USA/Texas/Houston | Houston Methodist Hospital | Houston Methodist Hospital | S. Wesley Long et al |
| EPI_ISL_787806 | USA/TX-HMH-MCoV-14521/2020 | 2020-10-09 | North America/USA/Texas/Houston | Houston Methodist Hospital | Houston Methodist Hospital | S. Wesley Long et al |
| EPI_ISL_787811 | USA/TX-HMH-MCoV-14526/2020 | 2020-10-08 | North America/USA/Texas/Houston | Houston Methodist Hospital | Houston Methodist Hospital | S. Wesley Long et al |

| | | | | | | |
|---|---|---|---|---|---|---|
| EPI_ISL_787826 | USA/TX-HMH-MCoV-14541/2020 | 2020-10-09 | North America/USA/Texas/Houston | Houston Methodist Hospital | Houston Methodist Hospital | S. Wesley Long et al |
| EPI_ISL_787828 | USA/TX-HMH-MCoV-14543/2020 | 2020-10-09 | North America/USA/Texas/Houston | Houston Methodist Hospital | Houston Methodist Hospital | S. Wesley Long et al |
| EPI_ISL_787835 | USA/TX-HMH-MCoV-14550/2020 | 2020-10-08 | North America/USA/Texas/Houston | Houston Methodist Hospital | Houston Methodist Hospital | S. Wesley Long et al |
| EPI_ISL_787839 | USA/TX-HMH-MCoV-14554/2020 | 2020-10-09 | North America/USA/Texas/Houston | Houston Methodist Hospital | Houston Methodist Hospital | S. Wesley Long et al |
| EPI_ISL_787843 | USA/TX-HMH-MCoV-14558/2020 | 2020-10-09 | North America/USA/Texas/Houston | Houston Methodist Hospital | Houston Methodist Hospital | S. Wesley Long et al |
| EPI_ISL_787847 | USA/TX-HMH-MCoV-14562/2020 | 2020-10-08 | North America/USA/Texas/Houston | Houston Methodist Hospital | Houston Methodist Hospital | S. Wesley Long et al |
| EPI_ISL_787859 | USA/TX-HMH-MCoV-14574/2020 | 2020-10-08 | North America/USA/Texas/Houston | Houston Methodist Hospital | Houston Methodist Hospital | S. Wesley Long et al |
| EPI_ISL_787877 | USA/TX-HMH-MCoV-14592/2020 | 2020-10-11 | North America/USA/Texas/Houston | Houston Methodist Hospital | Houston Methodist Hospital | S. Wesley Long et al |
| EPI_ISL_787879 | USA/TX-HMH-MCoV-14594/2020 | 2020-10-11 | North America/USA/Texas/Houston | Houston Methodist Hospital | Houston Methodist Hospital | S. Wesley Long et al |
| EPI_ISL_787886 | USA/TX-HMH-MCoV-14601/2020 | 2020-10-10 | North America/USA/Texas/Houston | Houston Methodist Hospital | Houston Methodist Hospital | S. Wesley Long et al |
| EPI_ISL_787888 | USA/TX-HMH-MCoV-14603/2020 | 2020-10-11 | North America/USA/Texas/Houston | Houston Methodist Hospital | Houston Methodist Hospital | S. Wesley Long et al |
| EPI_ISL_787895 | USA/TX-HMH-MCoV-14610/2020 | 2020-10-10 | North America/USA/Texas/Houston | Houston Methodist Hospital | Houston Methodist Hospital | S. Wesley Long et al |
| EPI_ISL_787897 | USA/TX-HMH-MCoV-14612/2020 | 2020-10-10 | North America/USA/Texas/Houston | Houston Methodist Hospital | Houston Methodist Hospital | S. Wesley Long et al |

| | | | | | | |
|---|---|---|---|---|---|---|
| EPI_ISL_787898 | USA/TX-HMH-MCoV-14613/2020 | 2020-10-10 | North America/USA/Texas/Houston | Houston Methodist Hospital | Houston Methodist Hospital | S. Wesley Long et al |
| EPI_ISL_787908 | USA/TX-HMH-MCoV-14623/2020 | 2020-10-10 | North America/USA/Texas/Houston | Houston Methodist Hospital | Houston Methodist Hospital | S. Wesley Long et al |
| EPI_ISL_787916 | USA/TX-HMH-MCoV-14631/2020 | 2020-10-09 | North America/USA/Texas/Houston | Houston Methodist Hospital | Houston Methodist Hospital | S. Wesley Long et al |
| EPI_ISL_787937 | USA/TX-HMH-MCoV-14652/2020 | 2020-10-11 | North America/USA/Texas/Houston | Houston Methodist Hospital | Houston Methodist Hospital | S. Wesley Long et al |
| EPI_ISL_787941 | USA/TX-HMH-MCoV-14656/2020 | 2020-10-09 | North America/USA/Texas/Houston | Houston Methodist Hospital | Houston Methodist Hospital | S. Wesley Long et al |
| EPI_ISL_787945 | USA/TX-HMH-MCoV-14660/2020 | 2020-10-10 | North America/USA/Texas/Houston | Houston Methodist Hospital | Houston Methodist Hospital | S. Wesley Long et al |
| EPI_ISL_787950 | USA/TX-HMH-MCoV-14665/2020 | 2020-10-11 | North America/USA/Texas/Houston | Houston Methodist Hospital | Houston Methodist Hospital | S. Wesley Long et al |
| EPI_ISL_787955 | USA/TX-HMH-MCoV-14670/2020 | 2020-10-10 | North America/USA/Texas/Houston | Houston Methodist Hospital | Houston Methodist Hospital | S. Wesley Long et al |
| EPI_ISL_787958 | USA/TX-HMH-MCoV-14673/2020 | 2020-10-10 | North America/USA/Texas/Houston | Houston Methodist Hospital | Houston Methodist Hospital | S. Wesley Long et al |
| EPI_ISL_787965 | USA/TX-HMH-MCoV-14680/2020 | 2020-10-01 | North America/USA/Texas/Houston | Houston Methodist Hospital | Houston Methodist Hospital | S. Wesley Long et al |
| EPI_ISL_787967 | USA/TX-HMH-MCoV-14682/2020 | 2020-10-01 | North America/USA/Texas/Houston | Houston Methodist Hospital | Houston Methodist Hospital | S. Wesley Long et al |
| EPI_ISL_787973 | USA/TX-HMH-MCoV-14688/2020 | 2020-10-12 | North America/USA/Texas/Houston | Houston Methodist Hospital | Houston Methodist Hospital | S. Wesley Long et al |
| EPI_ISL_787989 | USA/TX-HMH-MCoV-14704/2020 | 2020-10-01 | North America/USA/Texas/Houston | Houston Methodist Hospital | Houston Methodist Hospital | S. Wesley Long et al |

| | | | | | | |
|---|---|---|---|---|---|---|
| EPI_ISL_787992 | USA/TX-HMH-MCoV-14707/2020 | 2020-10-07 | North America/USA/Texas/Houston | Houston Methodist Hospital | Houston Methodist Hospital | S. Wesley Long et al |
| EPI_ISL_788002 | USA/TX-HMH-MCoV-12849/2020 | 2020-08-19 | North America/USA/Texas/Houston | Houston Methodist Hospital | Houston Methodist Hospital | S. Wesley Long et al |
| EPI_ISL_788004 | USA/TX-HMH-MCoV-12851/2020 | 2020-08-19 | North America/USA/Texas/Houston | Houston Methodist Hospital | Houston Methodist Hospital | S. Wesley Long et al |
| EPI_ISL_788005 | USA/TX-HMH-MCoV-12852/2020 | 2020-08-19 | North America/USA/Texas/Houston | Houston Methodist Hospital | Houston Methodist Hospital | S. Wesley Long et al |
| EPI_ISL_788006 | USA/TX-HMH-MCoV-12853/2020 | 2020-08-19 | North America/USA/Texas/Houston | Houston Methodist Hospital | Houston Methodist Hospital | S. Wesley Long et al |
| EPI_ISL_788007 | USA/TX-HMH-MCoV-12854/2020 | 2020-08-19 | North America/USA/Texas/Houston | Houston Methodist Hospital | Houston Methodist Hospital | S. Wesley Long et al |
| EPI_ISL_788008 | USA/TX-HMH-MCoV-12855/2020 | 2020-08-19 | North America/USA/Texas/Houston | Houston Methodist Hospital | Houston Methodist Hospital | S. Wesley Long et al |
| EPI_ISL_788009 | USA/TX-HMH-MCoV-12856/2020 | 2020-08-19 | North America/USA/Texas/Houston | Houston Methodist Hospital | Houston Methodist Hospital | S. Wesley Long et al |
| EPI_ISL_788012 | USA/TX-HMH-MCoV-12859/2020 | 2020-08-18 | North America/USA/Texas/Houston | Houston Methodist Hospital | Houston Methodist Hospital | S. Wesley Long et al |
| EPI_ISL_788013 | USA/TX-HMH-MCoV-12860/2020 | 2020-08-18 | North America/USA/Texas/Houston | Houston Methodist Hospital | Houston Methodist Hospital | S. Wesley Long et al |
| EPI_ISL_788014 | USA/TX-HMH-MCoV-12861/2020 | 2020-08-18 | North America/USA/Texas/Houston | Houston Methodist Hospital | Houston Methodist Hospital | S. Wesley Long et al |
| EPI_ISL_788018 | USA/TX-HMH-MCoV-12865/2020 | 2020-08-18 | North America/USA/Texas/Houston | Houston Methodist Hospital | Houston Methodist Hospital | S. Wesley Long et al |
| EPI_ISL_788024 | USA/TX-HMH-MCoV-12872/2020 | 2020-08-18 | North America/USA/Texas/Houston | Houston Methodist Hospital | Houston Methodist Hospital | S. Wesley Long et al |

| | | | | | | |
|---|---|---|---|---|---|---|
| EPI_ISL_788025 | USA/TX-HMH-MCoV-12873/2020 | 2020-08-18 | North America/USA/Texas/Houston | Houston Methodist Hospital | Houston Methodist Hospital | S. Wesley Long et al |
| EPI_ISL_788029 | USA/TX-HMH-MCoV-12877/2020 | 2020-08-18 | North America/USA/Texas/Houston | Houston Methodist Hospital | Houston Methodist Hospital | S. Wesley Long et al |
| EPI_ISL_788030 | USA/TX-HMH-MCoV-12878/2020 | 2020-08-18 | North America/USA/Texas/Houston | Houston Methodist Hospital | Houston Methodist Hospital | S. Wesley Long et al |
| EPI_ISL_788031 | USA/TX-HMH-MCoV-12879/2020 | 2020-08-19 | North America/USA/Texas/Houston | Houston Methodist Hospital | Houston Methodist Hospital | S. Wesley Long et al |
| EPI_ISL_788032 | USA/TX-HMH-MCoV-12880/2020 | 2020-08-19 | North America/USA/Texas/Houston | Houston Methodist Hospital | Houston Methodist Hospital | S. Wesley Long et al |
| EPI_ISL_788033 | USA/TX-HMH-MCoV-12881/2020 | 2020-08-18 | North America/USA/Texas/Houston | Houston Methodist Hospital | Houston Methodist Hospital | S. Wesley Long et al |
| EPI_ISL_788036 | USA/TX-HMH-MCoV-12884/2020 | 2020-08-18 | North America/USA/Texas/Houston | Houston Methodist Hospital | Houston Methodist Hospital | S. Wesley Long et al |
| EPI_ISL_788038 | USA/TX-HMH-MCoV-12886/2020 | 2020-08-18 | North America/USA/Texas/Houston | Houston Methodist Hospital | Houston Methodist Hospital | S. Wesley Long et al |
| EPI_ISL_788039 | USA/TX-HMH-MCoV-12887/2020 | 2020-08-18 | North America/USA/Texas/Houston | Houston Methodist Hospital | Houston Methodist Hospital | S. Wesley Long et al |
| EPI_ISL_788044 | USA/TX-HMH-MCoV-12892/2020 | 2020-08-19 | North America/USA/Texas/Houston | Houston Methodist Hospital | Houston Methodist Hospital | S. Wesley Long et al |
| EPI_ISL_788046 | USA/TX-HMH-MCoV-12894/2020 | 2020-08-18 | North America/USA/Texas/Houston | Houston Methodist Hospital | Houston Methodist Hospital | S. Wesley Long et al |
| EPI_ISL_788048 | USA/TX-HMH-MCoV-12896/2020 | 2020-08-18 | North America/USA/Texas/Houston | Houston Methodist Hospital | Houston Methodist Hospital | S. Wesley Long et al |
| EPI_ISL_788057 | USA/TX-HMH-MCoV-12905/2020 | 2020-08-18 | North America/USA/Texas/Houston | Houston Methodist Hospital | Houston Methodist Hospital | S. Wesley Long et al |

| | | | | | | |
|---|---|---|---|---|---|---|
| EPI_ISL_788058 | USA/TX-HMH-MCoV-12906/2020 | 2020-08-18 | North America/USA/Texas/Houston | Houston Methodist Hospital | Houston Methodist Hospital | S. Wesley Long et al |
| EPI_ISL_788066 | USA/TX-HMH-MCoV-12914/2020 | 2020-08-18 | North America/USA/Texas/Houston | Houston Methodist Hospital | Houston Methodist Hospital | S. Wesley Long et al |
| EPI_ISL_788070 | USA/TX-HMH-MCoV-12918/2020 | 2020-08-18 | North America/USA/Texas/Houston | Houston Methodist Hospital | Houston Methodist Hospital | S. Wesley Long et al |
| EPI_ISL_788078 | USA/TX-HMH-MCoV-12926/2020 | 2020-08-18 | North America/USA/Texas/Houston | Houston Methodist Hospital | Houston Methodist Hospital | S. Wesley Long et al |
| EPI_ISL_788079 | USA/TX-HMH-MCoV-12927/2020 | 2020-08-19 | North America/USA/Texas/Houston | Houston Methodist Hospital | Houston Methodist Hospital | S. Wesley Long et al |
| EPI_ISL_788081 | USA/TX-HMH-MCoV-12929/2020 | 2020-08-18 | North America/USA/Texas/Houston | Houston Methodist Hospital | Houston Methodist Hospital | S. Wesley Long et al |
| EPI_ISL_788083 | USA/TX-HMH-MCoV-12931/2020 | 2020-08-18 | North America/USA/Texas/Houston | Houston Methodist Hospital | Houston Methodist Hospital | S. Wesley Long et al |
| EPI_ISL_788085 | USA/TX-HMH-MCoV-12933/2020 | 2020-08-18 | North America/USA/Texas/Houston | Houston Methodist Hospital | Houston Methodist Hospital | S. Wesley Long et al |
| EPI_ISL_788088 | USA/TX-HMH-MCoV-12936/2020 | 2020-08-18 | North America/USA/Texas/Houston | Houston Methodist Hospital | Houston Methodist Hospital | S. Wesley Long et al |
| EPI_ISL_788097 | USA/TX-HMH-MCoV-12945/2020 | 2020-08-18 | North America/USA/Texas/Houston | Houston Methodist Hospital | Houston Methodist Hospital | S. Wesley Long et al |
| EPI_ISL_788100 | USA/TX-HMH-MCoV-12948/2020 | 2020-08-18 | North America/USA/Texas/Houston | Houston Methodist Hospital | Houston Methodist Hospital | S. Wesley Long et al |
| EPI_ISL_788101 | USA/TX-HMH-MCoV-12949/2020 | 2020-08-18 | North America/USA/Texas/Houston | Houston Methodist Hospital | Houston Methodist Hospital | S. Wesley Long et al |
| EPI_ISL_788103 | USA/TX-HMH-MCoV-12951/2020 | 2020-08-28 | North America/USA/Texas/Houston | Houston Methodist Hospital | Houston Methodist Hospital | S. Wesley Long et al |

| | | | | | | |
|---|---|---|---|---|---|---|
| EPI_ISL_788105 | USA/TX-HMH-MCoV-12953/2020 | 2020-08-28 | North America/USA/Texas/Houston | Houston Methodist Hospital | Houston Methodist Hospital | S. Wesley Long et al |
| EPI_ISL_788108 | USA/TX-HMH-MCoV-12956/2020 | 2020-08-28 | North America/USA/Texas/Houston | Houston Methodist Hospital | Houston Methodist Hospital | S. Wesley Long et al |
| EPI_ISL_788109 | USA/TX-HMH-MCoV-12957/2020 | 2020-08-29 | North America/USA/Texas/Houston | Houston Methodist Hospital | Houston Methodist Hospital | S. Wesley Long et al |
| EPI_ISL_788113 | USA/TX-HMH-MCoV-12961/2020 | 2020-08-28 | North America/USA/Texas/Houston | Houston Methodist Hospital | Houston Methodist Hospital | S. Wesley Long et al |
| EPI_ISL_788115 | USA/TX-HMH-MCoV-12963/2020 | 2020-08-28 | North America/USA/Texas/Houston | Houston Methodist Hospital | Houston Methodist Hospital | S. Wesley Long et al |
| EPI_ISL_788117 | USA/TX-HMH-MCoV-12965/2020 | 2020-08-28 | North America/USA/Texas/Houston | Houston Methodist Hospital | Houston Methodist Hospital | S. Wesley Long et al |
| EPI_ISL_788119 | USA/TX-HMH-MCoV-12967/2020 | 2020-08-29 | North America/USA/Texas/Houston | Houston Methodist Hospital | Houston Methodist Hospital | S. Wesley Long et al |
| EPI_ISL_788120 | USA/TX-HMH-MCoV-12968/2020 | 2020-08-29 | North America/USA/Texas/Houston | Houston Methodist Hospital | Houston Methodist Hospital | S. Wesley Long et al |
| EPI_ISL_788131 | USA/TX-HMH-MCoV-12980/2020 | 2020-08-28 | North America/USA/Texas/Houston | Houston Methodist Hospital | Houston Methodist Hospital | S. Wesley Long et al |
| EPI_ISL_788139 | USA/TX-HMH-MCoV-12988/2020 | 2020-08-28 | North America/USA/Texas/Houston | Houston Methodist Hospital | Houston Methodist Hospital | S. Wesley Long et al |
| EPI_ISL_788140 | USA/TX-HMH-MCoV-12989/2020 | 2020-08-28 | North America/USA/Texas/Houston | Houston Methodist Hospital | Houston Methodist Hospital | S. Wesley Long et al |
| EPI_ISL_788143 | USA/TX-HMH-MCoV-12992/2020 | 2020-08-29 | North America/USA/Texas/Houston | Houston Methodist Hospital | Houston Methodist Hospital | S. Wesley Long et al |
| EPI_ISL_788151 | USA/TX-HMH-MCoV-13000/2020 | 2020-08-29 | North America/USA/Texas/Houston | Houston Methodist Hospital | Houston Methodist Hospital | S. Wesley Long et al |

| | | | | | | |
|---|---|---|---|---|---|---|
| EPI_ISL_788152 | USA/TX-HMH-MCoV-13001/2020 | 2020-08-28 | North America/USA/Texas/Houston | Houston Methodist Hospital | Houston Methodist Hospital | S. Wesley Long et al |
| EPI_ISL_788153 | USA/TX-HMH-MCoV-13002/2020 | 2020-08-28 | North America/USA/Texas/Houston | Houston Methodist Hospital | Houston Methodist Hospital | S. Wesley Long et al |
| EPI_ISL_788161 | USA/TX-HMH-MCoV-13010/2020 | 2020-08-28 | North America/USA/Texas/Houston | Houston Methodist Hospital | Houston Methodist Hospital | S. Wesley Long et al |
| EPI_ISL_788163 | USA/TX-HMH-MCoV-13012/2020 | 2020-08-29 | North America/USA/Texas/Houston | Houston Methodist Hospital | Houston Methodist Hospital | S. Wesley Long et al |
| EPI_ISL_788164 | USA/TX-HMH-MCoV-13013/2020 | 2020-08-28 | North America/USA/Texas/Houston | Houston Methodist Hospital | Houston Methodist Hospital | S. Wesley Long et al |
| EPI_ISL_788165 | USA/TX-HMH-MCoV-13014/2020 | 2020-08-28 | North America/USA/Texas/Houston | Houston Methodist Hospital | Houston Methodist Hospital | S. Wesley Long et al |
| EPI_ISL_788167 | USA/TX-HMH-MCoV-13016/2020 | 2020-08-28 | North America/USA/Texas/Houston | Houston Methodist Hospital | Houston Methodist Hospital | S. Wesley Long et al |
| EPI_ISL_788168 | USA/TX-HMH-MCoV-13017/2020 | 2020-08-28 | North America/USA/Texas/Houston | Houston Methodist Hospital | Houston Methodist Hospital | S. Wesley Long et al |
| EPI_ISL_788169 | USA/TX-HMH-MCoV-13018/2020 | 2020-08-28 | North America/USA/Texas/Houston | Houston Methodist Hospital | Houston Methodist Hospital | S. Wesley Long et al |
| EPI_ISL_788174 | USA/TX-HMH-MCoV-13023/2020 | 2020-08-28 | North America/USA/Texas/Houston | Houston Methodist Hospital | Houston Methodist Hospital | S. Wesley Long et al |
| EPI_ISL_788176 | USA/TX-HMH-MCoV-13025/2020 | 2020-08-28 | North America/USA/Texas/Houston | Houston Methodist Hospital | Houston Methodist Hospital | S. Wesley Long et al |
| EPI_ISL_788178 | USA/TX-HMH-MCoV-13027/2020 | 2020-08-29 | North America/USA/Texas/Houston | Houston Methodist Hospital | Houston Methodist Hospital | S. Wesley Long et al |
| EPI_ISL_788183 | USA/TX-HMH-MCoV-13032/2020 | 2020-08-28 | North America/USA/Texas/Houston | Houston Methodist Hospital | Houston Methodist Hospital | S. Wesley Long et al |

| | | | | | | |
|---|---|---|---|---|---|---|
| EPI_ISL_788188 | USA/TX-HMH-MCoV-13037/2020 | 2020-08-28 | North America/USA/Texas/Houston | Houston Methodist Hospital | Houston Methodist Hospital | S. Wesley Long et al |
| EPI_ISL_788192 | USA/TX-HMH-MCoV-13041/2020 | 2020-08-28 | North America/USA/Texas/Houston | Houston Methodist Hospital | Houston Methodist Hospital | S. Wesley Long et al |
| EPI_ISL_788193 | USA/TX-HMH-MCoV-13042/2020 | 2020-08-28 | North America/USA/Texas/Houston | Houston Methodist Hospital | Houston Methodist Hospital | S. Wesley Long et al |
| EPI_ISL_788195 | USA/TX-HMH-MCoV-13044/2020 | 2020-08-28 | North America/USA/Texas/Houston | Houston Methodist Hospital | Houston Methodist Hospital | S. Wesley Long et al |
| EPI_ISL_788197 | USA/TX-HMH-MCoV-13046/2020 | 2020-08-28 | North America/USA/Texas/Houston | Houston Methodist Hospital | Houston Methodist Hospital | S. Wesley Long et al |
| EPI_ISL_788205 | USA/TX-HMH-MCoV-13054/2020 | 2020-08-27 | North America/USA/Texas/Houston | Houston Methodist Hospital | Houston Methodist Hospital | S. Wesley Long et al |
| EPI_ISL_788211 | USA/TX-HMH-MCoV-13060/2020 | 2020-08-29 | North America/USA/Texas/Houston | Houston Methodist Hospital | Houston Methodist Hospital | S. Wesley Long et al |
| EPI_ISL_788212 | USA/TX-HMH-MCoV-13061/2020 | 2020-08-29 | North America/USA/Texas/Houston | Houston Methodist Hospital | Houston Methodist Hospital | S. Wesley Long et al |
| EPI_ISL_788213 | USA/TX-HMH-MCoV-13062/2020 | 2020-08-27 | North America/USA/Texas/Houston | Houston Methodist Hospital | Houston Methodist Hospital | S. Wesley Long et al |
| EPI_ISL_788215 | USA/TX-HMH-MCoV-13064/2020 | 2020-08-27 | North America/USA/Texas/Houston | Houston Methodist Hospital | Houston Methodist Hospital | S. Wesley Long et al |
| EPI_ISL_788218 | USA/TX-HMH-MCoV-13067/2020 | 2020-08-27 | North America/USA/Texas/Houston | Houston Methodist Hospital | Houston Methodist Hospital | S. Wesley Long et al |
| EPI_ISL_788220 | USA/TX-HMH-MCoV-13069/2020 | 2020-08-25 | North America/USA/Texas/Houston | Houston Methodist Hospital | Houston Methodist Hospital | S. Wesley Long et al |
| EPI_ISL_788226 | USA/TX-HMH-MCoV-13075/2020 | 2020-08-27 | North America/USA/Texas/Houston | Houston Methodist Hospital | Houston Methodist Hospital | S. Wesley Long et al |

| | | | | | | |
|---|---|---|---|---|---|---|
| EPI_ISL_788232 | USA/TX-HMH-MCoV-13081/2020 | 2020-08-26 | North America/USA/Texas/Houston | Houston Methodist Hospital | Houston Methodist Hospital | S. Wesley Long et al |
| EPI_ISL_788233 | USA/TX-HMH-MCoV-13082/2020 | 2020-08-26 | North America/USA/Texas/Houston | Houston Methodist Hospital | Houston Methodist Hospital | S. Wesley Long et al |
| EPI_ISL_788240 | USA/TX-HMH-MCoV-13089/2020 | 2020-08-27 | North America/USA/Texas/Houston | Houston Methodist Hospital | Houston Methodist Hospital | S. Wesley Long et al |
| EPI_ISL_788249 | USA/TX-HMH-MCoV-13098/2020 | 2020-08-27 | North America/USA/Texas/Houston | Houston Methodist Hospital | Houston Methodist Hospital | S. Wesley Long et al |
| EPI_ISL_788257 | USA/TX-HMH-MCoV-13106/2020 | 2020-08-26 | North America/USA/Texas/Houston | Houston Methodist Hospital | Houston Methodist Hospital | S. Wesley Long et al |
| EPI_ISL_788263 | USA/TX-HMH-MCoV-13112/2020 | 2020-08-27 | North America/USA/Texas/Houston | Houston Methodist Hospital | Houston Methodist Hospital | S. Wesley Long et al |
| EPI_ISL_788265 | USA/TX-HMH-MCoV-13114/2020 | 2020-08-27 | North America/USA/Texas/Houston | Houston Methodist Hospital | Houston Methodist Hospital | S. Wesley Long et al |
| EPI_ISL_788266 | USA/TX-HMH-MCoV-13115/2020 | 2020-08-27 | North America/USA/Texas/Houston | Houston Methodist Hospital | Houston Methodist Hospital | S. Wesley Long et al |
| EPI_ISL_788268 | USA/TX-HMH-MCoV-13117/2020 | 2020-08-26 | North America/USA/Texas/Houston | Houston Methodist Hospital | Houston Methodist Hospital | S. Wesley Long et al |
| EPI_ISL_788274 | USA/TX-HMH-MCoV-13123/2020 | 2020-08-27 | North America/USA/Texas/Houston | Houston Methodist Hospital | Houston Methodist Hospital | S. Wesley Long et al |
| EPI_ISL_788277 | USA/TX-HMH-MCoV-13126/2020 | 2020-08-27 | North America/USA/Texas/Houston | Houston Methodist Hospital | Houston Methodist Hospital | S. Wesley Long et al |
| EPI_ISL_788285 | USA/TX-HMH-MCoV-13134/2020 | 2020-08-27 | North America/USA/Texas/Houston | Houston Methodist Hospital | Houston Methodist Hospital | S. Wesley Long et al |
| EPI_ISL_788286 | USA/TX-HMH-MCoV-13135/2020 | 2020-08-27 | North America/USA/Texas/Houston | Houston Methodist Hospital | Houston Methodist Hospital | S. Wesley Long et al |

| | | | | | | |
|---|---|---|---|---|---|---|
| EPI_ISL_788288 | USA/TX-HMH-MCoV-13137/2020 | 2020-08-27 | North America/USA/Texas/Houston | Houston Methodist Hospital | Houston Methodist Hospital | S. Wesley Long et al |
| EPI_ISL_788295 | USA/TX-HMH-MCoV-13144/2020 | 2020-08-26 | North America/USA/Texas/Houston | Houston Methodist Hospital | Houston Methodist Hospital | S. Wesley Long et al |
| EPI_ISL_788296 | USA/TX-HMH-MCoV-13145/2020 | 2020-08-26 | North America/USA/Texas/Houston | Houston Methodist Hospital | Houston Methodist Hospital | S. Wesley Long et al |
| EPI_ISL_788310 | USA/TX-HMH-MCoV-13159/2020 | 2020-08-26 | North America/USA/Texas/Houston | Houston Methodist Hospital | Houston Methodist Hospital | S. Wesley Long et al |
| EPI_ISL_788316 | USA/TX-HMH-MCoV-13165/2020 | 2020-08-25 | North America/USA/Texas/Houston | Houston Methodist Hospital | Houston Methodist Hospital | S. Wesley Long et al |
| EPI_ISL_788318 | USA/TX-HMH-MCoV-13167/2020 | 2020-08-26 | North America/USA/Texas/Houston | Houston Methodist Hospital | Houston Methodist Hospital | S. Wesley Long et al |
| EPI_ISL_788319 | USA/TX-HMH-MCoV-13168/2020 | 2020-08-26 | North America/USA/Texas/Houston | Houston Methodist Hospital | Houston Methodist Hospital | S. Wesley Long et al |
| EPI_ISL_788321 | USA/TX-HMH-MCoV-13170/2020 | 2020-08-26 | North America/USA/Texas/Houston | Houston Methodist Hospital | Houston Methodist Hospital | S. Wesley Long et al |
| EPI_ISL_788322 | USA/TX-HMH-MCoV-13171/2020 | 2020-08-26 | North America/USA/Texas/Houston | Houston Methodist Hospital | Houston Methodist Hospital | S. Wesley Long et al |
| EPI_ISL_788325 | USA/TX-HMH-MCoV-13174/2020 | 2020-08-25 | North America/USA/Texas/Houston | Houston Methodist Hospital | Houston Methodist Hospital | S. Wesley Long et al |
| EPI_ISL_788330 | USA/TX-HMH-MCoV-13179/2020 | 2020-08-26 | North America/USA/Texas/Houston | Houston Methodist Hospital | Houston Methodist Hospital | S. Wesley Long et al |
| EPI_ISL_788332 | USA/TX-HMH-MCoV-13181/2020 | 2020-08-26 | North America/USA/Texas/Houston | Houston Methodist Hospital | Houston Methodist Hospital | S. Wesley Long et al |
| EPI_ISL_788336 | USA/TX-HMH-MCoV-13185/2020 | 2020-08-25 | North America/USA/Texas/Houston | Houston Methodist Hospital | Houston Methodist Hospital | S. Wesley Long et al |

| | | | | | | |
|---|---|---|---|---|---|---|
| EPI_ISL_788340 | USA/TX-HMH-MCoV-13189/2020 | 2020-08-25 | North America/USA/Texas/Houston | Houston Methodist Hospital | Houston Methodist Hospital | S. Wesley Long et al |
| EPI_ISL_788350 | USA/TX-HMH-MCoV-13199/2020 | 2020-08-25 | North America/USA/Texas/Houston | Houston Methodist Hospital | Houston Methodist Hospital | S. Wesley Long et al |
| EPI_ISL_788351 | USA/TX-HMH-MCoV-13200/2020 | 2020-08-25 | North America/USA/Texas/Houston | Houston Methodist Hospital | Houston Methodist Hospital | S. Wesley Long et al |
| EPI_ISL_788363 | USA/TX-HMH-MCoV-13212/2020 | 2020-08-25 | North America/USA/Texas/Houston | Houston Methodist Hospital | Houston Methodist Hospital | S. Wesley Long et al |
| EPI_ISL_788364 | USA/TX-HMH-MCoV-13213/2020 | 2020-08-25 | North America/USA/Texas/Houston | Houston Methodist Hospital | Houston Methodist Hospital | S. Wesley Long et al |
| EPI_ISL_788367 | USA/TX-HMH-MCoV-13216/2020 | 2020-08-26 | North America/USA/Texas/Houston | Houston Methodist Hospital | Houston Methodist Hospital | S. Wesley Long et al |
| EPI_ISL_788368 | USA/TX-HMH-MCoV-13217/2020 | 2020-08-26 | North America/USA/Texas/Houston | Houston Methodist Hospital | Houston Methodist Hospital | S. Wesley Long et al |
| EPI_ISL_788372 | USA/TX-HMH-MCoV-13221/2020 | 2020-08-25 | North America/USA/Texas/Houston | Houston Methodist Hospital | Houston Methodist Hospital | S. Wesley Long et al |
| EPI_ISL_788374 | USA/TX-HMH-MCoV-13223/2020 | 2020-08-25 | North America/USA/Texas/Houston | Houston Methodist Hospital | Houston Methodist Hospital | S. Wesley Long et al |
| EPI_ISL_788375 | USA/TX-HMH-MCoV-13224/2020 | 2020-08-25 | North America/USA/Texas/Houston | Houston Methodist Hospital | Houston Methodist Hospital | S. Wesley Long et al |
| EPI_ISL_788378 | USA/TX-HMH-MCoV-13227/2020 | 2020-08-26 | North America/USA/Texas/Houston | Houston Methodist Hospital | Houston Methodist Hospital | S. Wesley Long et al |
| EPI_ISL_788381 | USA/TX-HMH-MCoV-13231/2020 | 2020-08-26 | North America/USA/Texas/Houston | Houston Methodist Hospital | Houston Methodist Hospital | S. Wesley Long et al |
| EPI_ISL_788387 | USA/TX-HMH-MCoV-13237/2020 | 2020-08-25 | North America/USA/Texas/Houston | Houston Methodist Hospital | Houston Methodist Hospital | S. Wesley Long et al |

| | | | | | | |
|---|---|---|---|---|---|---|
| EPI_ISL_788394 | USA/TX-HMH-MCoV-13244/2020 | 2020-08-25 | North America/USA/Texas/Houston | Houston Methodist Hospital | Houston Methodist Hospital | S. Wesley Long et al |
| EPI_ISL_788398 | USA/TX-HMH-MCoV-13248/2020 | 2020-08-24 | North America/USA/Texas/Houston | Houston Methodist Hospital | Houston Methodist Hospital | S. Wesley Long et al |
| EPI_ISL_788400 | USA/TX-HMH-MCoV-13250/2020 | 2020-08-24 | North America/USA/Texas/Houston | Houston Methodist Hospital | Houston Methodist Hospital | S. Wesley Long et al |
| EPI_ISL_788405 | USA/TX-HMH-MCoV-13256/2020 | 2020-08-25 | North America/USA/Texas/Houston | Houston Methodist Hospital | Houston Methodist Hospital | S. Wesley Long et al |
| EPI_ISL_788407 | USA/TX-HMH-MCoV-13258/2020 | 2020-08-24 | North America/USA/Texas/Houston | Houston Methodist Hospital | Houston Methodist Hospital | S. Wesley Long et al |
| EPI_ISL_788410 | USA/TX-HMH-MCoV-13261/2020 | 2020-08-24 | North America/USA/Texas/Houston | Houston Methodist Hospital | Houston Methodist Hospital | S. Wesley Long et al |
| EPI_ISL_788411 | USA/TX-HMH-MCoV-13262/2020 | 2020-08-24 | North America/USA/Texas/Houston | Houston Methodist Hospital | Houston Methodist Hospital | S. Wesley Long et al |
| EPI_ISL_788416 | USA/TX-HMH-MCoV-13267/2020 | 2020-08-25 | North America/USA/Texas/Houston | Houston Methodist Hospital | Houston Methodist Hospital | S. Wesley Long et al |
| EPI_ISL_788427 | USA/TX-HMH-MCoV-13279/2020 | 2020-08-25 | North America/USA/Texas/Houston | Houston Methodist Hospital | Houston Methodist Hospital | S. Wesley Long et al |
| EPI_ISL_788429 | USA/TX-HMH-MCoV-13281/2020 | 2020-08-24 | North America/USA/Texas/Houston | Houston Methodist Hospital | Houston Methodist Hospital | S. Wesley Long et al |
| EPI_ISL_788432 | USA/TX-HMH-MCoV-13284/2020 | 2020-08-24 | North America/USA/Texas/Houston | Houston Methodist Hospital | Houston Methodist Hospital | S. Wesley Long et al |
| EPI_ISL_788444 | USA/TX-HMH-MCoV-13296/2020 | 2020-08-24 | North America/USA/Texas/Houston | Houston Methodist Hospital | Houston Methodist Hospital | S. Wesley Long et al |
| EPI_ISL_788446 | USA/TX-HMH-MCoV-13298/2020 | 2020-08-24 | North America/USA/Texas/Houston | Houston Methodist Hospital | Houston Methodist Hospital | S. Wesley Long et al |

| | | | | | | |
|---|---|---|---|---|---|---|
| EPI_ISL_788451 | USA/TX-HMH-MCoV-13303/2020 | 2020-08-25 | North America/USA/Texas/Houston | Houston Methodist Hospital | Houston Methodist Hospital | S. Wesley Long et al |
| EPI_ISL_788456 | USA/TX-HMH-MCoV-13308/2020 | 2020-08-24 | North America/USA/Texas/Houston | Houston Methodist Hospital | Houston Methodist Hospital | S. Wesley Long et al |
| EPI_ISL_788459 | USA/TX-HMH-MCoV-13311/2020 | 2020-08-25 | North America/USA/Texas/Houston | Houston Methodist Hospital | Houston Methodist Hospital | S. Wesley Long et al |
| EPI_ISL_788472 | USA/TX-HMH-MCoV-13324/2020 | 2020-08-25 | North America/USA/Texas/Houston | Houston Methodist Hospital | Houston Methodist Hospital | S. Wesley Long et al |
| EPI_ISL_788474 | USA/TX-HMH-MCoV-13326/2020 | 2020-08-25 | North America/USA/Texas/Houston | Houston Methodist Hospital | Houston Methodist Hospital | S. Wesley Long et al |
| EPI_ISL_788475 | USA/TX-HMH-MCoV-13327/2020 | 2020-08-25 | North America/USA/Texas/Houston | Houston Methodist Hospital | Houston Methodist Hospital | S. Wesley Long et al |
| EPI_ISL_788478 | USA/TX-HMH-MCoV-13330/2020 | 2020-08-24 | North America/USA/Texas/Houston | Houston Methodist Hospital | Houston Methodist Hospital | S. Wesley Long et al |
| EPI_ISL_788479 | USA/TX-HMH-MCoV-13331/2020 | 2020-08-24 | North America/USA/Texas/Houston | Houston Methodist Hospital | Houston Methodist Hospital | S. Wesley Long et al |
| EPI_ISL_788487 | USA/TX-HMH-MCoV-13339/2020 | 2020-08-24 | North America/USA/Texas/Houston | Houston Methodist Hospital | Houston Methodist Hospital | S. Wesley Long et al |
| EPI_ISL_788489 | USA/TX-HMH-MCoV-13342/2020 | 2020-08-24 | North America/USA/Texas/Houston | Houston Methodist Hospital | Houston Methodist Hospital | S. Wesley Long et al |
| EPI_ISL_788495 | USA/TX-HMH-MCoV-13348/2020 | 2020-08-24 | North America/USA/Texas/Houston | Houston Methodist Hospital | Houston Methodist Hospital | S. Wesley Long et al |
| EPI_ISL_788497 | USA/TX-HMH-MCoV-13350/2020 | 2020-08-24 | North America/USA/Texas/Houston | Houston Methodist Hospital | Houston Methodist Hospital | S. Wesley Long et al |
| EPI_ISL_788500 | USA/TX-HMH-MCoV-13353/2020 | 2020-08-24 | North America/USA/Texas/Houston | Houston Methodist Hospital | Houston Methodist Hospital | S. Wesley Long et al |

| | | | | | | |
|---|---|---|---|---|---|---|
| EPI_ISL_788502 | USA/TX-HMH-MCoV-13355/2020 | 2020-08-24 | North America/USA/Texas/Houston | Houston Methodist Hospital | Houston Methodist Hospital | S. Wesley Long et al |
| EPI_ISL_788508 | USA/TX-HMH-MCoV-13363/2020 | 2020-08-24 | North America/USA/Texas/Houston | Houston Methodist Hospital | Houston Methodist Hospital | S. Wesley Long et al |
| EPI_ISL_788509 | USA/TX-HMH-MCoV-13364/2020 | 2020-08-24 | North America/USA/Texas/Houston | Houston Methodist Hospital | Houston Methodist Hospital | S. Wesley Long et al |
| EPI_ISL_788514 | USA/TX-HMH-MCoV-13371/2020 | 2020-08-24 | North America/USA/Texas/Houston | Houston Methodist Hospital | Houston Methodist Hospital | S. Wesley Long et al |
| EPI_ISL_788516 | USA/TX-HMH-MCoV-13373/2020 | 2020-08-24 | North America/USA/Texas/Houston | Houston Methodist Hospital | Houston Methodist Hospital | S. Wesley Long et al |
| EPI_ISL_788523 | USA/TX-HMH-MCoV-13380/2020 | 2020-08-23 | North America/USA/Texas/Houston | Houston Methodist Hospital | Houston Methodist Hospital | S. Wesley Long et al |
| EPI_ISL_788526 | USA/TX-HMH-MCoV-13383/2020 | 2020-08-24 | North America/USA/Texas/Houston | Houston Methodist Hospital | Houston Methodist Hospital | S. Wesley Long et al |
| EPI_ISL_788527 | USA/TX-HMH-MCoV-13384/2020 | 2020-08-24 | North America/USA/Texas/Houston | Houston Methodist Hospital | Houston Methodist Hospital | S. Wesley Long et al |
| EPI_ISL_788530 | USA/TX-HMH-MCoV-13387/2020 | 2020-08-24 | North America/USA/Texas/Houston | Houston Methodist Hospital | Houston Methodist Hospital | S. Wesley Long et al |
| EPI_ISL_788532 | USA/TX-HMH-MCoV-13389/2020 | 2020-08-24 | North America/USA/Texas/Houston | Houston Methodist Hospital | Houston Methodist Hospital | S. Wesley Long et al |
| EPI_ISL_788542 | USA/TX-HMH-MCoV-13399/2020 | 2020-08-24 | North America/USA/Texas/Houston | Houston Methodist Hospital | Houston Methodist Hospital | S. Wesley Long et al |
| EPI_ISL_788543 | USA/TX-HMH-MCoV-13400/2020 | 2020-08-24 | North America/USA/Texas/Houston | Houston Methodist Hospital | Houston Methodist Hospital | S. Wesley Long et al |
| EPI_ISL_788545 | USA/TX-HMH-MCoV-13402/2020 | 2020-08-24 | North America/USA/Texas/Houston | Houston Methodist Hospital | Houston Methodist Hospital | S. Wesley Long et al |

| | | | | | | |
|---|---|---|---|---|---|---|
| EPI_ISL_788548 | USA/TX-HMH-MCoV-13407/2020 | 2020-08-24 | North America/USA/Texas/Houston | Houston Methodist Hospital | Houston Methodist Hospital | S. Wesley Long et al |
| EPI_ISL_788552 | USA/TX-HMH-MCoV-13414/2020 | 2020-08-24 | North America/USA/Texas/Houston | Houston Methodist Hospital | Houston Methodist Hospital | S. Wesley Long et al |
| EPI_ISL_788553 | USA/TX-HMH-MCoV-13415/2020 | 2020-08-23 | North America/USA/Texas/Houston | Houston Methodist Hospital | Houston Methodist Hospital | S. Wesley Long et al |
| EPI_ISL_788554 | USA/TX-HMH-MCoV-13416/2020 | 2020-08-23 | North America/USA/Texas/Houston | Houston Methodist Hospital | Houston Methodist Hospital | S. Wesley Long et al |
| EPI_ISL_788555 | USA/TX-HMH-MCoV-13417/2020 | 2020-08-23 | North America/USA/Texas/Houston | Houston Methodist Hospital | Houston Methodist Hospital | S. Wesley Long et al |
| EPI_ISL_788557 | USA/TX-HMH-MCoV-13421/2020 | 2020-08-24 | North America/USA/Texas/Houston | Houston Methodist Hospital | Houston Methodist Hospital | S. Wesley Long et al |
| EPI_ISL_788561 | USA/TX-HMH-MCoV-13425/2020 | 2020-08-24 | North America/USA/Texas/Houston | Houston Methodist Hospital | Houston Methodist Hospital | S. Wesley Long et al |
| EPI_ISL_788571 | USA/TX-HMH-MCoV-13436/2020 | 2020-08-22 | North America/USA/Texas/Houston | Houston Methodist Hospital | Houston Methodist Hospital | S. Wesley Long et al |
| EPI_ISL_788573 | USA/TX-HMH-MCoV-13438/2020 | 2020-08-23 | North America/USA/Texas/Houston | Houston Methodist Hospital | Houston Methodist Hospital | S. Wesley Long et al |
| EPI_ISL_788580 | USA/TX-HMH-MCoV-13445/2020 | 2020-08-23 | North America/USA/Texas/Houston | Houston Methodist Hospital | Houston Methodist Hospital | S. Wesley Long et al |
| EPI_ISL_788592 | USA/TX-HMH-MCoV-13457/2020 | 2020-08-23 | North America/USA/Texas/Houston | Houston Methodist Hospital | Houston Methodist Hospital | S. Wesley Long et al |
| EPI_ISL_788597 | USA/TX-HMH-MCoV-13466/2020 | 2020-08-21 | North America/USA/Texas/Houston | Houston Methodist Hospital | Houston Methodist Hospital | S. Wesley Long et al |
| EPI_ISL_788598 | USA/TX-HMH-MCoV-13467/2020 | 2020-08-23 | North America/USA/Texas/Houston | Houston Methodist Hospital | Houston Methodist Hospital | S. Wesley Long et al |

| | | | | | | |
|---|---|---|---|---|---|---|
| EPI_ISL_788600 | USA/TX-HMH-MCoV-13470/2020 | 2020-08-22 | North America/USA/Texas/Houston | Houston Methodist Hospital | Houston Methodist Hospital | S. Wesley Long et al |
| EPI_ISL_788613 | USA/TX-HMH-MCoV-13484/2020 | 2020-08-22 | North America/USA/Texas/Houston | Houston Methodist Hospital | Houston Methodist Hospital | S. Wesley Long et al |
| EPI_ISL_788616 | USA/TX-HMH-MCoV-13487/2020 | 2020-08-21 | North America/USA/Texas/Houston | Houston Methodist Hospital | Houston Methodist Hospital | S. Wesley Long et al |
| EPI_ISL_788617 | USA/TX-HMH-MCoV-13488/2020 | 2020-08-21 | North America/USA/Texas/Houston | Houston Methodist Hospital | Houston Methodist Hospital | S. Wesley Long et al |
| EPI_ISL_788618 | USA/TX-HMH-MCoV-13489/2020 | 2020-08-21 | North America/USA/Texas/Houston | Houston Methodist Hospital | Houston Methodist Hospital | S. Wesley Long et al |
| EPI_ISL_788621 | USA/TX-HMH-MCoV-13492/2020 | 2020-08-23 | North America/USA/Texas/Houston | Houston Methodist Hospital | Houston Methodist Hospital | S. Wesley Long et al |
| EPI_ISL_788623 | USA/TX-HMH-MCoV-13494/2020 | 2020-08-22 | North America/USA/Texas/Houston | Houston Methodist Hospital | Houston Methodist Hospital | S. Wesley Long et al |
| EPI_ISL_788624 | USA/TX-HMH-MCoV-13495/2020 | 2020-08-22 | North America/USA/Texas/Houston | Houston Methodist Hospital | Houston Methodist Hospital | S. Wesley Long et al |
| EPI_ISL_788628 | USA/TX-HMH-MCoV-13499/2020 | 2020-08-21 | North America/USA/Texas/Houston | Houston Methodist Hospital | Houston Methodist Hospital | S. Wesley Long et al |
| EPI_ISL_788633 | USA/TX-HMH-MCoV-13505/2020 | 2020-08-22 | North America/USA/Texas/Houston | Houston Methodist Hospital | Houston Methodist Hospital | S. Wesley Long et al |
| EPI_ISL_788634 | USA/TX-HMH-MCoV-13507/2020 | 2020-08-22 | North America/USA/Texas/Houston | Houston Methodist Hospital | Houston Methodist Hospital | S. Wesley Long et al |
| EPI_ISL_788637 | USA/TX-HMH-MCoV-13510/2020 | 2020-08-22 | North America/USA/Texas/Houston | Houston Methodist Hospital | Houston Methodist Hospital | S. Wesley Long et al |
| EPI_ISL_788639 | USA/TX-HMH-MCoV-13513/2020 | 2020-08-21 | North America/USA/Texas/Houston | Houston Methodist Hospital | Houston Methodist Hospital | S. Wesley Long et al |

| | | | | | | |
|---|---|---|---|---|---|---|
| EPI_ISL_788643 | USA/TX-HMH-MCoV-13518/2020 | 2020-08-22 | North America/USA/Texas/Houston | Houston Methodist Hospital | Houston Methodist Hospital | S. Wesley Long et al |
| EPI_ISL_788644 | USA/TX-HMH-MCoV-13519/2020 | 2020-08-22 | North America/USA/Texas/Houston | Houston Methodist Hospital | Houston Methodist Hospital | S. Wesley Long et al |
| EPI_ISL_788646 | USA/TX-HMH-MCoV-13521/2020 | 2020-08-22 | North America/USA/Texas/Houston | Houston Methodist Hospital | Houston Methodist Hospital | S. Wesley Long et al |
| EPI_ISL_788647 | USA/TX-HMH-MCoV-13522/2020 | 2020-08-21 | North America/USA/Texas/Houston | Houston Methodist Hospital | Houston Methodist Hospital | S. Wesley Long et al |
| EPI_ISL_788651 | USA/TX-HMH-MCoV-13526/2020 | 2020-08-21 | North America/USA/Texas/Houston | Houston Methodist Hospital | Houston Methodist Hospital | S. Wesley Long et al |
| EPI_ISL_788652 | USA/TX-HMH-MCoV-13527/2020 | 2020-08-21 | North America/USA/Texas/Houston | Houston Methodist Hospital | Houston Methodist Hospital | S. Wesley Long et al |
| EPI_ISL_788655 | USA/TX-HMH-MCoV-13530/2020 | 2020-08-21 | North America/USA/Texas/Houston | Houston Methodist Hospital | Houston Methodist Hospital | S. Wesley Long et al |
| EPI_ISL_788664 | USA/TX-HMH-MCoV-13543/2020 | 2020-08-21 | North America/USA/Texas/Houston | Houston Methodist Hospital | Houston Methodist Hospital | S. Wesley Long et al |
| EPI_ISL_788665 | USA/TX-HMH-MCoV-13545/2020 | 2020-08-21 | North America/USA/Texas/Houston | Houston Methodist Hospital | Houston Methodist Hospital | S. Wesley Long et al |
| EPI_ISL_788669 | USA/TX-HMH-MCoV-13551/2020 | 2020-08-21 | North America/USA/Texas/Houston | Houston Methodist Hospital | Houston Methodist Hospital | S. Wesley Long et al |
| EPI_ISL_788670 | USA/TX-HMH-MCoV-13552/2020 | 2020-08-21 | North America/USA/Texas/Houston | Houston Methodist Hospital | Houston Methodist Hospital | S. Wesley Long et al |
| EPI_ISL_788676 | USA/TX-HMH-MCoV-13559/2020 | 2020-08-21 | North America/USA/Texas/Houston | Houston Methodist Hospital | Houston Methodist Hospital | S. Wesley Long et al |
| EPI_ISL_788677 | USA/TX-HMH-MCoV-13560/2020 | 2020-08-21 | North America/USA/Texas/Houston | Houston Methodist Hospital | Houston Methodist Hospital | S. Wesley Long et al |

| | | | | | | |
|---|---|---|---|---|---|---|
| EPI_ISL_788679 | USA/TX-HMH-MCoV-13562/2020 | 2020-08-20 | North America/USA/Texas/Houston | Houston Methodist Hospital | Houston Methodist Hospital | S. Wesley Long et al |
| EPI_ISL_788682 | USA/TX-HMH-MCoV-13571/2020 | 2020-08-21 | North America/USA/Texas/Houston | Houston Methodist Hospital | Houston Methodist Hospital | S. Wesley Long et al |
| EPI_ISL_788689 | USA/TX-HMH-MCoV-13579/2020 | 2020-08-21 | North America/USA/Texas/Houston | Houston Methodist Hospital | Houston Methodist Hospital | S. Wesley Long et al |
| EPI_ISL_788691 | USA/TX-HMH-MCoV-13581/2020 | 2020-08-21 | North America/USA/Texas/Houston | Houston Methodist Hospital | Houston Methodist Hospital | S. Wesley Long et al |
| EPI_ISL_788693 | USA/TX-HMH-MCoV-13583/2020 | 2020-08-21 | North America/USA/Texas/Houston | Houston Methodist Hospital | Houston Methodist Hospital | S. Wesley Long et al |
| EPI_ISL_788694 | USA/TX-HMH-MCoV-13584/2020 | 2020-08-21 | North America/USA/Texas/Houston | Houston Methodist Hospital | Houston Methodist Hospital | S. Wesley Long et al |
| EPI_ISL_788701 | USA/TX-HMH-MCoV-13591/2020 | 2020-08-21 | North America/USA/Texas/Houston | Houston Methodist Hospital | Houston Methodist Hospital | S. Wesley Long et al |
| EPI_ISL_788702 | USA/TX-HMH-MCoV-13592/2020 | 2020-08-21 | North America/USA/Texas/Houston | Houston Methodist Hospital | Houston Methodist Hospital | S. Wesley Long et al |
| EPI_ISL_788704 | USA/TX-HMH-MCoV-13594/2020 | 2020-08-21 | North America/USA/Texas/Houston | Houston Methodist Hospital | Houston Methodist Hospital | S. Wesley Long et al |
| EPI_ISL_788705 | USA/TX-HMH-MCoV-13595/2020 | 2020-08-21 | North America/USA/Texas/Houston | Houston Methodist Hospital | Houston Methodist Hospital | S. Wesley Long et al |
| EPI_ISL_788711 | USA/TX-HMH-MCoV-13601/2020 | 2020-08-21 | North America/USA/Texas/Houston | Houston Methodist Hospital | Houston Methodist Hospital | S. Wesley Long et al |
| EPI_ISL_788716 | USA/TX-HMH-MCoV-13606/2020 | 2020-08-21 | North America/USA/Texas/Houston | Houston Methodist Hospital | Houston Methodist Hospital | S. Wesley Long et al |
| EPI_ISL_788717 | USA/TX-HMH-MCoV-13607/2020 | 2020-08-21 | North America/USA/Texas/Houston | Houston Methodist Hospital | Houston Methodist Hospital | S. Wesley Long et al |

| | | | | | | |
|---|---|---|---|---|---|---|
| EPI_ISL_788718 | USA/TX-HMH-MCoV-13608/2020 | 2020-08-20 | North America/USA/Texas/Houston | Houston Methodist Hospital | Houston Methodist Hospital | S. Wesley Long et al |
| EPI_ISL_788720 | USA/TX-HMH-MCoV-13610/2020 | 2020-08-20 | North America/USA/Texas/Houston | Houston Methodist Hospital | Houston Methodist Hospital | S. Wesley Long et al |
| EPI_ISL_788722 | USA/TX-HMH-MCoV-13613/2020 | 2020-08-21 | North America/USA/Texas/Houston | Houston Methodist Hospital | Houston Methodist Hospital | S. Wesley Long et al |
| EPI_ISL_788727 | USA/TX-HMH-MCoV-13619/2020 | 2020-08-21 | North America/USA/Texas/Houston | Houston Methodist Hospital | Houston Methodist Hospital | S. Wesley Long et al |
| EPI_ISL_788736 | USA/TX-HMH-MCoV-13630/2020 | 2020-08-20 | North America/USA/Texas/Houston | Houston Methodist Hospital | Houston Methodist Hospital | S. Wesley Long et al |
| EPI_ISL_788740 | USA/TX-HMH-MCoV-13634/2020 | 2020-08-19 | North America/USA/Texas/Houston | Houston Methodist Hospital | Houston Methodist Hospital | S. Wesley Long et al |
| EPI_ISL_788743 | USA/TX-HMH-MCoV-13639/2020 | 2020-08-20 | North America/USA/Texas/Houston | Houston Methodist Hospital | Houston Methodist Hospital | S. Wesley Long et al |
| EPI_ISL_788746 | USA/TX-HMH-MCoV-13642/2020 | 2020-08-20 | North America/USA/Texas/Houston | Houston Methodist Hospital | Houston Methodist Hospital | S. Wesley Long et al |
| EPI_ISL_788747 | USA/TX-HMH-MCoV-13643/2020 | 2020-08-20 | North America/USA/Texas/Houston | Houston Methodist Hospital | Houston Methodist Hospital | S. Wesley Long et al |
| EPI_ISL_788753 | USA/TX-HMH-MCoV-13651/2020 | 2020-08-20 | North America/USA/Texas/Houston | Houston Methodist Hospital | Houston Methodist Hospital | S. Wesley Long et al |
| EPI_ISL_788761 | USA/TX-HMH-MCoV-13660/2020 | 2020-08-20 | North America/USA/Texas/Houston | Houston Methodist Hospital | Houston Methodist Hospital | S. Wesley Long et al |
| EPI_ISL_788763 | USA/TX-HMH-MCoV-13662/2020 | 2020-08-20 | North America/USA/Texas/Houston | Houston Methodist Hospital | Houston Methodist Hospital | S. Wesley Long et al |
| EPI_ISL_788766 | USA/TX-HMH-MCoV-13668/2020 | 2020-08-20 | North America/USA/Texas/Houston | Houston Methodist Hospital | Houston Methodist Hospital | S. Wesley Long et al |

| | | | | | | |
|---|---|---|---|---|---|---|
| EPI_ISL_788776 | USA/TX-HMH-MCoV-13679/2020 | 2020-08-20 | North America/USA/Texas/Houston | Houston Methodist Hospital | Houston Methodist Hospital | S. Wesley Long et al |
| EPI_ISL_788780 | USA/TX-HMH-MCoV-13683/2020 | 2020-08-20 | North America/USA/Texas/Houston | Houston Methodist Hospital | Houston Methodist Hospital | S. Wesley Long et al |
| EPI_ISL_788785 | USA/TX-HMH-MCoV-13688/2020 | 2020-08-20 | North America/USA/Texas/Houston | Houston Methodist Hospital | Houston Methodist Hospital | S. Wesley Long et al |
| EPI_ISL_788790 | USA/TX-HMH-MCoV-13694/2020 | 2020-08-20 | North America/USA/Texas/Houston | Houston Methodist Hospital | Houston Methodist Hospital | S. Wesley Long et al |
| EPI_ISL_788795 | USA/TX-HMH-MCoV-13699/2020 | 2020-08-20 | North America/USA/Texas/Houston | Houston Methodist Hospital | Houston Methodist Hospital | S. Wesley Long et al |
| EPI_ISL_788801 | USA/TX-HMH-MCoV-13705/2020 | 2020-08-19 | North America/USA/Texas/Houston | Houston Methodist Hospital | Houston Methodist Hospital | S. Wesley Long et al |
| EPI_ISL_788809 | USA/TX-HMH-MCoV-13715/2020 | 2020-08-20 | North America/USA/Texas/Houston | Houston Methodist Hospital | Houston Methodist Hospital | S. Wesley Long et al |
| EPI_ISL_788812 | USA/TX-HMH-MCoV-13719/2020 | 2020-08-18 | North America/USA/Texas/Houston | Houston Methodist Hospital | Houston Methodist Hospital | S. Wesley Long et al |
| EPI_ISL_788813 | USA/TX-HMH-MCoV-13720/2020 | 2020-08-18 | North America/USA/Texas/Houston | Houston Methodist Hospital | Houston Methodist Hospital | S. Wesley Long et al |
| EPI_ISL_788815 | USA/TX-HMH-MCoV-13722/2020 | 2020-08-18 | North America/USA/Texas/Houston | Houston Methodist Hospital | Houston Methodist Hospital | S. Wesley Long et al |
| EPI_ISL_788819 | USA/TX-HMH-MCoV-13726/2020 | 2020-08-17 | North America/USA/Texas/Houston | Houston Methodist Hospital | Houston Methodist Hospital | S. Wesley Long et al |
| EPI_ISL_788828 | USA/TX-HMH-MCoV-13735/2020 | 2020-08-18 | North America/USA/Texas/Houston | Houston Methodist Hospital | Houston Methodist Hospital | S. Wesley Long et al |
| EPI_ISL_788834 | USA/TX-HMH-MCoV-13741/2020 | 2020-08-17 | North America/USA/Texas/Houston | Houston Methodist Hospital | Houston Methodist Hospital | S. Wesley Long et al |

| | | | | | | |
|---|---|---|---|---|---|---|
| EPI_ISL_788837 | USA/TX-HMH-MCoV-13744/2020 | 2020-08-18 | North America/USA/Texas/Houston | Houston Methodist Hospital | Houston Methodist Hospital | S. Wesley Long et al |
| EPI_ISL_788840 | USA/TX-HMH-MCoV-13747/2020 | 2020-08-17 | North America/USA/Texas/Houston | Houston Methodist Hospital | Houston Methodist Hospital | S. Wesley Long et al |
| EPI_ISL_788841 | USA/TX-HMH-MCoV-13748/2020 | 2020-08-17 | North America/USA/Texas/Houston | Houston Methodist Hospital | Houston Methodist Hospital | S. Wesley Long et al |
| EPI_ISL_788842 | USA/TX-HMH-MCoV-13749/2020 | 2020-08-17 | North America/USA/Texas/Houston | Houston Methodist Hospital | Houston Methodist Hospital | S. Wesley Long et al |
| EPI_ISL_788849 | USA/TX-HMH-MCoV-13756/2020 | 2020-08-18 | North America/USA/Texas/Houston | Houston Methodist Hospital | Houston Methodist Hospital | S. Wesley Long et al |
| EPI_ISL_788855 | USA/TX-HMH-MCoV-13762/2020 | 2020-08-17 | North America/USA/Texas/Houston | Houston Methodist Hospital | Houston Methodist Hospital | S. Wesley Long et al |
| EPI_ISL_788856 | USA/TX-HMH-MCoV-13763/2020 | 2020-08-17 | North America/USA/Texas/Houston | Houston Methodist Hospital | Houston Methodist Hospital | S. Wesley Long et al |
| EPI_ISL_788859 | USA/TX-HMH-MCoV-13766/2020 | 2020-08-17 | North America/USA/Texas/Houston | Houston Methodist Hospital | Houston Methodist Hospital | S. Wesley Long et al |
| EPI_ISL_788862 | USA/TX-HMH-MCoV-13769/2020 | 2020-08-18 | North America/USA/Texas/Houston | Houston Methodist Hospital | Houston Methodist Hospital | S. Wesley Long et al |
| EPI_ISL_788865 | USA/TX-HMH-MCoV-13772/2020 | 2020-08-17 | North America/USA/Texas/Houston | Houston Methodist Hospital | Houston Methodist Hospital | S. Wesley Long et al |
| EPI_ISL_788867 | USA/TX-HMH-MCoV-13774/2020 | 2020-08-17 | North America/USA/Texas/Houston | Houston Methodist Hospital | Houston Methodist Hospital | S. Wesley Long et al |
| EPI_ISL_788871 | USA/TX-HMH-MCoV-13778/2020 | 2020-08-17 | North America/USA/Texas/Houston | Houston Methodist Hospital | Houston Methodist Hospital | S. Wesley Long et al |
| EPI_ISL_788872 | USA/TX-HMH-MCoV-13779/2020 | 2020-08-18 | North America/USA/Texas/Houston | Houston Methodist Hospital | Houston Methodist Hospital | S. Wesley Long et al |

| | | | | | | |
|---|---|---|---|---|---|---|
| EPI_ISL_788880 | USA/TX-HMH-MCoV-13787/2020 | 2020-08-17 | North America/USA/Texas/Houston | Houston Methodist Hospital | Houston Methodist Hospital | S. Wesley Long et al |
| EPI_ISL_788881 | USA/TX-HMH-MCoV-13788/2020 | 2020-08-17 | North America/USA/Texas/Houston | Houston Methodist Hospital | Houston Methodist Hospital | S. Wesley Long et al |
| EPI_ISL_788882 | USA/TX-HMH-MCoV-13789/2020 | 2020-08-17 | North America/USA/Texas/Houston | Houston Methodist Hospital | Houston Methodist Hospital | S. Wesley Long et al |
| EPI_ISL_788889 | USA/TX-HMH-MCoV-13796/2020 | 2020-08-17 | North America/USA/Texas/Houston | Houston Methodist Hospital | Houston Methodist Hospital | S. Wesley Long et al |
| EPI_ISL_788897 | USA/TX-HMH-MCoV-13804/2020 | 2020-08-18 | North America/USA/Texas/Houston | Houston Methodist Hospital | Houston Methodist Hospital | S. Wesley Long et al |
| EPI_ISL_788899 | USA/WI-UW-2556/2020 | 2020-12-29 | North America/USA/Wisconsin/Rock Cou | University of Wisconsin-Madison AIDS Vaccine Research Laboratories | University of Wisconsin-Madison AIDS Vaccine Research Laboratories | Gage Moreno et al |
| EPI_ISL_788915 | USA/WI-UW-2573/2021 | 2021-01-04 | North America/USA/Wisconsin/Dane Co | University of Wisconsin-Madison AIDS Vaccine Research Laboratories | University of Wisconsin-Madison AIDS Vaccine Research Laboratories | Gage Moreno et al |
| EPI_ISL_788916 | USA/WI-UW-2574/2021 | 2021-01-04 | North America/USA/Wisconsin/Jefferson | University of Wisconsin-Madison AIDS Vaccine Research Laboratories | University of Wisconsin-Madison AIDS Vaccine Research Laboratories | Gage Moreno et al |
| EPI_ISL_788919 | USA/WI-UW-2577/2020 | 2020-12-30 | North America/USA/Wisconsin/Dane Co | University of Wisconsin-Madison AIDS Vaccine Research Laboratories | University of Wisconsin-Madison AIDS Vaccine Research Laboratories | Gage Moreno et al |
| EPI_ISL_788934 | Guinea/CERFIG-1337/2020 | 2020-06-09 | Africa/Guinea/Conakry/ | Centre de Recherche et de Formation en Infectiologie Guinv©e | TransVIHMI, IRD/INSERM/Montpellier University | Alpha Kabinet KEITA et al |

| | | | | | | |
|---|---|---|---|---|---|---|
| EPI_ISL_788941 | USA/WI-UW-2543/2020 | 2020-12-29 | North America/USA/Wisconsin/Jefferson | University of Wisconsin-Madison AIDS Vaccine Research Laboratories | University of Wisconsin-Madison AIDS Vaccine Research Laboratories | Gage Moreno et al |
| EPI_ISL_788945 | Italy/APU-PODV-15716050/2020 | 2020-04-22 | Europe/Italy/Apulia/Bari | "Ospedale ""Di Venere""" | Beaconlab (Bioinformatics, Evolution and Comparative Genomics lab), Dept of Biosciences, University on Milan | Iacobellis M et al |
| EPI_ISL_788946 | Italy/APU-PODV-15726195/2020 | 2020-04-30 | Europe/Italy/Apulia/Bari | "Ospedale ""Di Venere""" | Beaconlab (Bioinformatics, Evolution and Comparative Genomics lab), Dept of Biosciences, University on Milan | Iacobellis M et al |
| EPI_ISL_788950 | Italy/APU-PODV-15963317/2020 | 2020-09-04 | Europe/Italy/Apulia/Bari | "Ospedale ""Di Venere""" | Beaconlab (Bioinformatics, Evolution and Comparative Genomics lab), Dept of Biosciences, University on Milan | Iacobellis M et al |
| EPI_ISL_788951 | Italy/APU-PODV-15961800/2020 | 2020-09-03 | Europe/Italy/Apulia/Bari | "Ospedale ""Di Venere""" | Beaconlab (Bioinformatics, Evolution and Comparative Genomics lab), Dept of Biosciences, University on Milan | Iacobellis M et al |

| | | | | | | |
|---|---|---|---|---|---|---|
| EPI_ISL_788952 | Italy/APU-PODV-15961609/2020 | 2020-09-03 | Europe/Italy/Apulia/Bari | "Ospedale ""Di Venere""" | Beaconlab (Bioinformatics, Evolution and Comparative Genomics lab), Dept of Biosciences, University on Milan | Iacobellis M et al |
| EPI_ISL_788959 | Italy/APU-PODV-15949439/2020 | 2020-08-29 | Europe/Italy/Apulia/Bari | "Ospedale ""Di Venere""" | Beaconlab (Bioinformatics, Evolution and Comparative Genomics lab), Dept of Biosciences, University on Milan | Iacobellis M et al |
| EPI_ISL_788960 | Italy/APU-PODV-15949164/2020 | 2020-08-29 | Europe/Italy/Apulia/Bari | "Ospedale ""Di Venere""" | Beaconlab (Bioinformatics, Evolution and Comparative Genomics lab), Dept of Biosciences, University on Milan | Iacobellis M et al |
| EPI_ISL_788963 | Italy/APU-PODV-15705294/2020 | 2020-04-14 | Europe/Italy/Apulia/Bari | "Ospedale ""Di Venere""" | Beaconlab (Bioinformatics, Evolution and Comparative Genomics lab), Dept of Biosciences, University on Milan | Iacobellis M et al |

| | | | | | | |
|---|---|---|---|---|---|---|
| EPI_ISL_788968 | Italy/APU-PODV-15956183/2020 | 2020-09-01 | Europe/Italy/Apulia/Bari | "Ospedale ""Di Venere""" | Beaconlab (Bioinformatics, Evolution and Comparative Genomics lab), Dept of Biosciences, University on Milan | Iacobellis M et al |
| EPI_ISL_788978 | Italy/APU-PODV-15950754/2020 | 2020-08-30 | Europe/Italy/Apulia/Bari | "Ospedale ""Di Venere""" | Beaconlab (Bioinformatics, Evolution and Comparative Genomics lab), Dept of Biosciences, University on Milan | Iacobellis M et al |
| EPI_ISL_788980 | Slovakia/UKBA-602/2020 | 2020-11-12 | Europe/Slovakia/Nove Zamky/ | Institute of Virology, Biomedical Research Center of the Slovak Academy of Sciences, Bratislava | Faculty of Natural Sciences, Comenius University, Bratislava | Kristv≠na Bor≈°ov√° et al |
| EPI_ISL_788981 | Slovakia/UKBA-603/2020 | 2020-11-12 | Europe/Slovakia/Nove Zamky/ | Institute of Virology, Biomedical Research Center of the Slovak Academy of Sciences, Bratislava | Faculty of Natural Sciences, Comenius University, Bratislava | Viktv≥ria ƒåabanov√° et al |
| EPI_ISL_788984 | Slovakia/UKBA-607/2020 | 2020-11-30 | Europe/Slovakia/Bratislava/ | Institute of Virology, Biomedical Research Center of the Slovak Academy of Sciences, Bratislava | Faculty of Natural Sciences, Comenius University, Bratislava | Bro≈àa Brejov√° et al |
| EPI_ISL_788986 | Slovakia/UKBA-609/2020 | 2020-11-30 | Europe/Slovakia/Bratislava/ | Institute of Virology, Biomedical Research Center of the Slovak Academy of Sciences, Bratislava | Faculty of Natural Sciences, Comenius University, Bratislava | Bro≈àa Brejov√° et al |

| | | | | | | |
|---|---|---|---|---|---|---|
| EPI_ISL_788987 | Slovakia/UKBA-610/2020 | 2020-11-30 | Europe/Slovakia/Bratislava/ | Institute of Virology, Biomedical Research Center of the Slovak Academy of Sciences, Bratislava | Faculty of Natural Sciences, Comenius University, Bratislava | Viktv≥ria ƒåabanovv° et al |
| EPI_ISL_788988 | Slovakia/UKBA-611/2020 | 2020-11-30 | Europe/Slovakia/Bratislava/ | Institute of Virology, Biomedical Research Center of the Slovak Academy of Sciences, Bratislava | Faculty of Natural Sciences, Comenius University, Bratislava | Kristv≠na Bor≈°ovv° et al |
| EPI_ISL_788994 | Sweden/20-09937/2020 | 2020-12-23 | Europe/Sweden/Jamtland/ | ABC | The Public Health Agency of Sweden | Department of Microbiology et al |
| EPI_ISL_789008 | Sweden/20-13092/2020 | 2020-12-29 | Europe/Sweden/Stockholm/ | ABC | The Public Health Agency of Sweden | Department of Microbiology et al |
| EPI_ISL_789019 | Sweden/20-13078/2020 | 2020-12-29 | Europe/Sweden/Stockholm/ | ABC | The Public Health Agency of Sweden | Department of Microbiology et al |
| EPI_ISL_789021 | Sweden/20-13097/2020 | 2020-12-29 | Europe/Sweden/Stockholm/ | ABC | The Public Health Agency of Sweden | Department of Microbiology et al |
| EPI_ISL_789026 | Sweden/20-13142/2020 | 2020-12-23 | Europe/Sweden/Stockholm/ | ABC | The Public Health Agency of Sweden | Department of Microbiology et al |
| EPI_ISL_789029 | Sweden/20-53829/2020 | 2020-12-16 | Europe/Sweden/Vasterbotten/ | Klinisk mikrobiologi, Laboratoriemedicin | The Public Health Agency of Sweden | Department of Microbiology et al |
| EPI_ISL_789042 | Sweden/20-53866/2020 | 2020-12-14 | Europe/Sweden/Uppsala/ | Klinisk mikrobiologi | The Public Health Agency of Sweden | Department of Microbiology et al |
| EPI_ISL_789057 | Sweden/20-53905/2020 | 2020-12-17 | Europe/Sweden/Stockholm/ | Klinisk mikrobiologi, Viruslab | The Public Health Agency of Sweden | Department of Microbiology et al |
| EPI_ISL_789058 | Sweden/20-53928/2020 | 2020-12-22 | Europe/Sweden/Stockholm/ | Klinisk mikrobiologi, Viruslab | The Public Health Agency of Sweden | Department of Microbiology et al |
| EPI_ISL_789077 | Ireland/D-NVRL-20IRL15933/2021 | 2021-01-01 | Europe/Ireland/Dublin/ | National Virus Reference Laboratory | National Virus Reference Laboratory | Michael Carr et al |
| EPI_ISL_789081 | Ireland/D-NVRL-20IRL15880/2021 | 2021-01-01 | Europe/Ireland/Dublin/ | National Virus Reference Laboratory | National Virus Reference Laboratory | Michael Carr et al |
| EPI_ISL_789101 | Ireland/D-NVRL-20IRL15865/2021 | 2021-01-01 | Europe/Ireland/Dublin/ | National Virus Reference Laboratory | National Virus Reference Laboratory | Michael Carr et al |
| EPI_ISL_789108 | USA/TX-HMH-MCoV-11253/2020 | 2020-09-04 | North America/USA/Texas/Houston | Houston Methodist Hospital | Houston Methodist Hospital | S. Wesley Long et al |

| | | | | | | |
|---|---|---|---|---|---|---|
| EPI_ISL_789111 | USA/TX-HMH-MCoV-10120/2020 | 2020-07-06 | North America/USA/Texas/Houston | Houston Methodist Hospital | Houston Methodist Hospital | S. Wesley Long et al |
| EPI_ISL_789115 | USA/TX-HMH-MCoV-11289/2020 | 2020-09-04 | North America/USA/Texas/Houston | Houston Methodist Hospital | Houston Methodist Hospital | S. Wesley Long et al |
| EPI_ISL_789118 | USA/TX-HMH-MCoV-10378/2020 | 2020-07-08 | North America/USA/Texas/Houston | Houston Methodist Hospital | Houston Methodist Hospital | S. Wesley Long et al |
| EPI_ISL_789122 | USA/TX-HMH-MCoV-10719/2020 | 2020-07-08 | North America/USA/Texas/Houston | Houston Methodist Hospital | Houston Methodist Hospital | S. Wesley Long et al |
| EPI_ISL_789124 | USA/TX-HMH-MCoV-11281/2020 | 2020-09-04 | North America/USA/Texas/Houston | Houston Methodist Hospital | Houston Methodist Hospital | S. Wesley Long et al |
| EPI_ISL_789134 | USA/TX-HMH-MCoV-11107/2020 | 2020-09-01 | North America/USA/Texas/Houston | Houston Methodist Hospital | Houston Methodist Hospital | S. Wesley Long et al |
| EPI_ISL_789135 | USA/TX-HMH-MCoV-11225/2020 | 2020-09-02 | North America/USA/Texas/Houston | Houston Methodist Hospital | Houston Methodist Hospital | S. Wesley Long et al |
| EPI_ISL_789142 | USA/TX-HMH-MCoV-10096/2020 | 2020-07-06 | North America/USA/Texas/Houston | Houston Methodist Hospital | Houston Methodist Hospital | S. Wesley Long et al |
| EPI_ISL_789151 | USA/TX-HMH-MCoV-10406/2020 | 2020-07-09 | North America/USA/Texas/Houston | Houston Methodist Hospital | Houston Methodist Hospital | S. Wesley Long et al |
| EPI_ISL_789152 | USA/TX-HMH-MCoV-10326/2020 | 2020-07-08 | North America/USA/Texas/Houston | Houston Methodist Hospital | Houston Methodist Hospital | S. Wesley Long et al |
| EPI_ISL_789153 | USA/TX-HMH-MCoV-10214/2020 | 2020-07-07 | North America/USA/Texas/Houston | Houston Methodist Hospital | Houston Methodist Hospital | S. Wesley Long et al |
| EPI_ISL_789161 | USA/TX-HMH-MCoV-10344/2020 | 2020-07-08 | North America/USA/Texas/Houston | Houston Methodist Hospital | Houston Methodist Hospital | S. Wesley Long et al |
| EPI_ISL_789184 | USA/TX-HMH-MCoV-10900/2020 | 2020-07-05 | North America/USA/Texas/Houston | Houston Methodist Hospital | Houston Methodist Hospital | S. Wesley Long et al |

| | | | | | | |
|---|---|---|---|---|---|---|
| EPI_ISL_789190 | USA/TX-HMH-MCoV-10600/2020 | 2020-07-08 | North America/USA/Texas/Houston | Houston Methodist Hospital | Houston Methodist Hospital | S. Wesley Long et al |
| EPI_ISL_789193 | USA/TX-HMH-MCoV-11213/2020 | 2020-09-02 | North America/USA/Texas/Houston | Houston Methodist Hospital | Houston Methodist Hospital | S. Wesley Long et al |
| EPI_ISL_789194 | USA/TX-HMH-MCoV-10581/2020 | 2020-07-08 | North America/USA/Texas/Houston | Houston Methodist Hospital | Houston Methodist Hospital | S. Wesley Long et al |
| EPI_ISL_789196 | USA/TX-HMH-MCoV-10834/2020 | 2020-07-05 | North America/USA/Texas/Houston | Houston Methodist Hospital | Houston Methodist Hospital | S. Wesley Long et al |
| EPI_ISL_789198 | USA/TX-HMH-MCoV-10605/2020 | 2020-07-08 | North America/USA/Texas/Houston | Houston Methodist Hospital | Houston Methodist Hospital | S. Wesley Long et al |
| EPI_ISL_789206 | USA/TX-HMH-MCoV-10749/2020 | 2020-07-09 | North America/USA/Texas/Houston | Houston Methodist Hospital | Houston Methodist Hospital | S. Wesley Long et al |
| EPI_ISL_789208 | USA/TX-HMH-MCoV-10010/2020 | 2020-07-06 | North America/USA/Texas/Houston | Houston Methodist Hospital | Houston Methodist Hospital | S. Wesley Long et al |
| EPI_ISL_789209 | USA/TX-HMH-MCoV-10106/2020 | 2020-07-07 | North America/USA/Texas/Houston | Houston Methodist Hospital | Houston Methodist Hospital | S. Wesley Long et al |
| EPI_ISL_789211 | USA/TX-HMH-MCoV-10706/2020 | 2020-07-09 | North America/USA/Texas/Houston | Houston Methodist Hospital | Houston Methodist Hospital | S. Wesley Long et al |
| EPI_ISL_789219 | USA/TX-HMH-MCoV-10653/2020 | 2020-07-08 | North America/USA/Texas/Houston | Houston Methodist Hospital | Houston Methodist Hospital | S. Wesley Long et al |
| EPI_ISL_789224 | USA/TX-HMH-MCoV-10082/2020 | 2020-07-07 | North America/USA/Texas/Houston | Houston Methodist Hospital | Houston Methodist Hospital | S. Wesley Long et al |
| EPI_ISL_789228 | USA/TX-HMH-MCoV-10488/2020 | 2020-07-09 | North America/USA/Texas/Houston | Houston Methodist Hospital | Houston Methodist Hospital | S. Wesley Long et al |
| EPI_ISL_789230 | USA/TX-HMH-MCoV-10739/2020 | 2020-07-09 | North America/USA/Texas/Houston | Houston Methodist Hospital | Houston Methodist Hospital | S. Wesley Long et al |

| | | | | | | |
|---|---|---|---|---|---|---|
| EPI_ISL_789231 | USA/TX-HMH-MCoV-11171/2020 | 2020-09-02 | North America/USA/Texas/Houston | Houston Methodist Hospital | Houston Methodist Hospital | S. Wesley Long et al |
| EPI_ISL_789232 | USA/TX-HMH-MCoV-11219/2020 | 2020-09-02 | North America/USA/Texas/Houston | Houston Methodist Hospital | Houston Methodist Hospital | S. Wesley Long et al |
| EPI_ISL_789235 | USA/TX-HMH-MCoV-10158/2020 | 2020-07-06 | North America/USA/Texas/Houston | Houston Methodist Hospital | Houston Methodist Hospital | S. Wesley Long et al |
| EPI_ISL_789238 | USA/TX-HMH-MCoV-10090/2020 | 2020-07-06 | North America/USA/Texas/Houston | Houston Methodist Hospital | Houston Methodist Hospital | S. Wesley Long et al |
| EPI_ISL_789239 | USA/TX-HMH-MCoV-10173/2020 | 2020-07-06 | North America/USA/Texas/Houston | Houston Methodist Hospital | Houston Methodist Hospital | S. Wesley Long et al |
| EPI_ISL_789241 | USA/TX-HMH-MCoV-10073/2020 | 2020-07-08 | North America/USA/Texas/Houston | Houston Methodist Hospital | Houston Methodist Hospital | S. Wesley Long et al |
| EPI_ISL_789243 | USA/TX-HMH-MCoV-10385/2020 | 2020-07-08 | North America/USA/Texas/Houston | Houston Methodist Hospital | Houston Methodist Hospital | S. Wesley Long et al |
| EPI_ISL_789248 | USA/TX-HMH-MCoV-10555/2020 | 2020-07-08 | North America/USA/Texas/Houston | Houston Methodist Hospital | Houston Methodist Hospital | S. Wesley Long et al |
| EPI_ISL_789250 | USA/TX-HMH-MCoV-10004/2020 | 2020-07-07 | North America/USA/Texas/Houston | Houston Methodist Hospital | Houston Methodist Hospital | S. Wesley Long et al |
| EPI_ISL_789251 | USA/TX-HMH-MCoV-10066/2020 | 2020-07-06 | North America/USA/Texas/Houston | Houston Methodist Hospital | Houston Methodist Hospital | S. Wesley Long et al |
| EPI_ISL_789253 | USA/TX-HMH-MCoV-10276/2020 | 2020-07-07 | North America/USA/Texas/Houston | Houston Methodist Hospital | Houston Methodist Hospital | S. Wesley Long et al |
| EPI_ISL_789256 | USA/TX-HMH-MCoV-10598/2020 | 2020-07-08 | North America/USA/Texas/Houston | Houston Methodist Hospital | Houston Methodist Hospital | S. Wesley Long et al |
| EPI_ISL_789260 | USA/TX-HMH-MCoV-11207/2020 | 2020-09-02 | North America/USA/Texas/Houston | Houston Methodist Hospital | Houston Methodist Hospital | S. Wesley Long et al |

| | | | | | | |
|---|---|---|---|---|---|---|
| EPI_ISL_789264 | USA/TX-HMH-MCoV-10468/2020 | 2020-07-08 | North America/USA/Texas/Houston | Houston Methodist Hospital | Houston Methodist Hospital | S. Wesley Long et al |
| EPI_ISL_789266 | USA/TX-HMH-MCoV-10760/2020 | 2020-07-09 | North America/USA/Texas/Houston | Houston Methodist Hospital | Houston Methodist Hospital | S. Wesley Long et al |
| EPI_ISL_789267 | USA/TX-HMH-MCoV-10858/2020 | 2020-07-05 | North America/USA/Texas/Houston | Houston Methodist Hospital | Houston Methodist Hospital | S. Wesley Long et al |
| EPI_ISL_789269 | USA/TX-HMH-MCoV-10843/2020 | 2020-07-05 | North America/USA/Texas/Houston | Houston Methodist Hospital | Houston Methodist Hospital | S. Wesley Long et al |
| EPI_ISL_789271 | USA/TX-HMH-MCoV-10175/2020 | 2020-07-06 | North America/USA/Texas/Houston | Houston Methodist Hospital | Houston Methodist Hospital | S. Wesley Long et al |
| EPI_ISL_789274 | USA/TX-HMH-MCoV-10368/2020 | 2020-07-08 | North America/USA/Texas/Houston | Houston Methodist Hospital | Houston Methodist Hospital | S. Wesley Long et al |
| EPI_ISL_789282 | USA/TX-HMH-MCoV-10003/2020 | 2020-07-06 | North America/USA/Texas/Houston | Houston Methodist Hospital | Houston Methodist Hospital | S. Wesley Long et al |
| EPI_ISL_789286 | USA/TX-HMH-MCoV-10014/2020 | 2020-07-06 | North America/USA/Texas/Houston | Houston Methodist Hospital | Houston Methodist Hospital | S. Wesley Long et al |
| EPI_ISL_789293 | USA/TX-HMH-MCoV-10028/2020 | 2020-07-06 | North America/USA/Texas/Houston | Houston Methodist Hospital | Houston Methodist Hospital | S. Wesley Long et al |
| EPI_ISL_789296 | USA/TX-HMH-MCoV-10033/2020 | 2020-07-06 | North America/USA/Texas/Houston | Houston Methodist Hospital | Houston Methodist Hospital | S. Wesley Long et al |
| EPI_ISL_789297 | USA/TX-HMH-MCoV-10034/2020 | 2020-07-06 | North America/USA/Texas/Houston | Houston Methodist Hospital | Houston Methodist Hospital | S. Wesley Long et al |
| EPI_ISL_789299 | USA/TX-HMH-MCoV-10037/2020 | 2020-07-06 | North America/USA/Texas/Houston | Houston Methodist Hospital | Houston Methodist Hospital | S. Wesley Long et al |
| EPI_ISL_789301 | USA/TX-HMH-MCoV-10042/2020 | 2020-07-06 | North America/USA/Texas/Houston | Houston Methodist Hospital | Houston Methodist Hospital | S. Wesley Long et al |

| | | | | | | |
|---|---|---|---|---|---|---|
| EPI_ISL_789306 | USA/TX-HMH-MCoV-10052/2020 | 2020-07-06 | North America/USA/Texas/Houston | Houston Methodist Hospital | Houston Methodist Hospital | S. Wesley Long et al |
| EPI_ISL_789307 | USA/TX-HMH-MCoV-10054/2020 | 2020-07-07 | North America/USA/Texas/Houston | Houston Methodist Hospital | Houston Methodist Hospital | S. Wesley Long et al |
| EPI_ISL_789309 | USA/TX-HMH-MCoV-10059/2020 | 2020-07-06 | North America/USA/Texas/Houston | Houston Methodist Hospital | Houston Methodist Hospital | S. Wesley Long et al |
| EPI_ISL_789325 | USA/TX-HMH-MCoV-10113/2020 | 2020-07-07 | North America/USA/Texas/Houston | Houston Methodist Hospital | Houston Methodist Hospital | S. Wesley Long et al |
| EPI_ISL_789326 | USA/TX-HMH-MCoV-10114/2020 | 2020-07-06 | North America/USA/Texas/Houston | Houston Methodist Hospital | Houston Methodist Hospital | S. Wesley Long et al |
| EPI_ISL_789334 | USA/TX-HMH-MCoV-10134/2020 | 2020-07-06 | North America/USA/Texas/Houston | Houston Methodist Hospital | Houston Methodist Hospital | S. Wesley Long et al |
| EPI_ISL_789338 | USA/TX-HMH-MCoV-10140/2020 | 2020-07-07 | North America/USA/Texas/Houston | Houston Methodist Hospital | Houston Methodist Hospital | S. Wesley Long et al |
| EPI_ISL_789339 | USA/TX-HMH-MCoV-10141/2020 | 2020-07-07 | North America/USA/Texas/Houston | Houston Methodist Hospital | Houston Methodist Hospital | S. Wesley Long et al |
| EPI_ISL_789352 | USA/TX-HMH-MCoV-10174/2020 | 2020-07-06 | North America/USA/Texas/Houston | Houston Methodist Hospital | Houston Methodist Hospital | S. Wesley Long et al |
| EPI_ISL_789354 | USA/TX-HMH-MCoV-10177/2020 | 2020-07-07 | North America/USA/Texas/Houston | Houston Methodist Hospital | Houston Methodist Hospital | S. Wesley Long et al |
| EPI_ISL_789359 | USA/TX-HMH-MCoV-10184/2020 | 2020-07-06 | North America/USA/Texas/Houston | Houston Methodist Hospital | Houston Methodist Hospital | S. Wesley Long et al |
| EPI_ISL_789362 | USA/TX-HMH-MCoV-10188/2020 | 2020-07-07 | North America/USA/Texas/Houston | Houston Methodist Hospital | Houston Methodist Hospital | S. Wesley Long et al |
| EPI_ISL_789364 | USA/TX-HMH-MCoV-10192/2020 | 2020-07-07 | North America/USA/Texas/Houston | Houston Methodist Hospital | Houston Methodist Hospital | S. Wesley Long et al |

| | | | | | | |
|---|---|---|---|---|---|---|
| EPI_ISL_789366 | USA/TX-HMH-MCoV-10195/2020 | 2020-07-07 | North America/USA/Texas/Houston | Houston Methodist Hospital | Houston Methodist Hospital | S. Wesley Long et al |
| EPI_ISL_789369 | USA/TX-HMH-MCoV-10200/2020 | 2020-07-07 | North America/USA/Texas/Houston | Houston Methodist Hospital | Houston Methodist Hospital | S. Wesley Long et al |
| EPI_ISL_789373 | USA/TX-HMH-MCoV-10217/2020 | 2020-07-07 | North America/USA/Texas/Houston | Houston Methodist Hospital | Houston Methodist Hospital | S. Wesley Long et al |
| EPI_ISL_789377 | USA/TX-HMH-MCoV-10224/2020 | 2020-07-07 | North America/USA/Texas/Houston | Houston Methodist Hospital | Houston Methodist Hospital | S. Wesley Long et al |
| EPI_ISL_789378 | USA/TX-HMH-MCoV-10225/2020 | 2020-07-11 | North America/USA/Texas/Houston | Houston Methodist Hospital | Houston Methodist Hospital | S. Wesley Long et al |
| EPI_ISL_789381 | USA/TX-HMH-MCoV-10229/2020 | 2020-07-07 | North America/USA/Texas/Houston | Houston Methodist Hospital | Houston Methodist Hospital | S. Wesley Long et al |
| EPI_ISL_789382 | USA/TX-HMH-MCoV-10233/2020 | 2020-07-07 | North America/USA/Texas/Houston | Houston Methodist Hospital | Houston Methodist Hospital | S. Wesley Long et al |
| EPI_ISL_789383 | USA/TX-HMH-MCoV-10234/2020 | 2020-07-07 | North America/USA/Texas/Houston | Houston Methodist Hospital | Houston Methodist Hospital | S. Wesley Long et al |
| EPI_ISL_789389 | USA/TX-HMH-MCoV-10248/2020 | 2020-07-07 | North America/USA/Texas/Houston | Houston Methodist Hospital | Houston Methodist Hospital | S. Wesley Long et al |
| EPI_ISL_789390 | USA/TX-HMH-MCoV-10249/2020 | 2020-07-11 | North America/USA/Texas/Houston | Houston Methodist Hospital | Houston Methodist Hospital | S. Wesley Long et al |
| EPI_ISL_789392 | USA/TX-HMH-MCoV-10252/2020 | 2020-07-07 | North America/USA/Texas/Houston | Houston Methodist Hospital | Houston Methodist Hospital | S. Wesley Long et al |
| EPI_ISL_789395 | USA/TX-HMH-MCoV-10255/2020 | 2020-07-07 | North America/USA/Texas/Houston | Houston Methodist Hospital | Houston Methodist Hospital | S. Wesley Long et al |
| EPI_ISL_789402 | USA/TX-HMH-MCoV-10265/2020 | 2020-07-07 | North America/USA/Texas/Houston | Houston Methodist Hospital | Houston Methodist Hospital | S. Wesley Long et al |

| | | | | | | |
|---|---|---|---|---|---|---|
| EPI_ISL_789408 | USA/TX-HMH-MCoV-10279/2020 | 2020-07-01 | North America/USA/Texas/Houston | Houston Methodist Hospital | Houston Methodist Hospital | S. Wesley Long et al |
| EPI_ISL_789412 | USA/TX-HMH-MCoV-10285/2020 | 2020-07-07 | North America/USA/Texas/Houston | Houston Methodist Hospital | Houston Methodist Hospital | S. Wesley Long et al |
| EPI_ISL_789413 | USA/TX-HMH-MCoV-10290/2020 | 2020-07-08 | North America/USA/Texas/Houston | Houston Methodist Hospital | Houston Methodist Hospital | S. Wesley Long et al |
| EPI_ISL_789414 | USA/TX-HMH-MCoV-10295/2020 | 2020-07-08 | North America/USA/Texas/Houston | Houston Methodist Hospital | Houston Methodist Hospital | S. Wesley Long et al |
| EPI_ISL_789419 | USA/TX-HMH-MCoV-10308/2020 | 2020-07-09 | North America/USA/Texas/Houston | Houston Methodist Hospital | Houston Methodist Hospital | S. Wesley Long et al |
| EPI_ISL_789421 | USA/TX-HMH-MCoV-10318/2020 | 2020-07-08 | North America/USA/Texas/Houston | Houston Methodist Hospital | Houston Methodist Hospital | S. Wesley Long et al |
| EPI_ISL_789426 | USA/TX-HMH-MCoV-10329/2020 | 2020-07-08 | North America/USA/Texas/Houston | Houston Methodist Hospital | Houston Methodist Hospital | S. Wesley Long et al |
| EPI_ISL_789431 | USA/TX-HMH-MCoV-10349/2020 | 2020-07-08 | North America/USA/Texas/Houston | Houston Methodist Hospital | Houston Methodist Hospital | S. Wesley Long et al |
| EPI_ISL_789434 | USA/TX-HMH-MCoV-10360/2020 | 2020-07-07 | North America/USA/Texas/Houston | Houston Methodist Hospital | Houston Methodist Hospital | S. Wesley Long et al |
| EPI_ISL_789437 | USA/TX-HMH-MCoV-10367/2020 | 2020-07-08 | North America/USA/Texas/Houston | Houston Methodist Hospital | Houston Methodist Hospital | S. Wesley Long et al |
| EPI_ISL_789438 | USA/TX-HMH-MCoV-10369/2020 | 2020-07-08 | North America/USA/Texas/Houston | Houston Methodist Hospital | Houston Methodist Hospital | S. Wesley Long et al |
| EPI_ISL_789446 | USA/TX-HMH-MCoV-10388/2020 | 2020-07-08 | North America/USA/Texas/Houston | Houston Methodist Hospital | Houston Methodist Hospital | S. Wesley Long et al |
| EPI_ISL_789452 | USA/TX-HMH-MCoV-10398/2020 | 2020-07-08 | North America/USA/Texas/Houston | Houston Methodist Hospital | Houston Methodist Hospital | S. Wesley Long et al |

| | | | | | | |
|---|---|---|---|---|---|---|
| EPI_ISL_789457 | USA/TX-HMH-MCoV-10409/2020 | 2020-07-08 | North America/USA/Texas/Houston | Houston Methodist Hospital | Houston Methodist Hospital | S. Wesley Long et al |
| EPI_ISL_789459 | USA/TX-HMH-MCoV-10411/2020 | 2020-07-08 | North America/USA/Texas/Houston | Houston Methodist Hospital | Houston Methodist Hospital | S. Wesley Long et al |
| EPI_ISL_789462 | USA/TX-HMH-MCoV-10415/2020 | 2020-07-08 | North America/USA/Texas/Houston | Houston Methodist Hospital | Houston Methodist Hospital | S. Wesley Long et al |
| EPI_ISL_789469 | USA/TX-HMH-MCoV-10424/2020 | 2020-07-08 | North America/USA/Texas/Houston | Houston Methodist Hospital | Houston Methodist Hospital | S. Wesley Long et al |
| EPI_ISL_789470 | USA/TX-HMH-MCoV-10425/2020 | 2020-07-08 | North America/USA/Texas/Houston | Houston Methodist Hospital | Houston Methodist Hospital | S. Wesley Long et al |
| EPI_ISL_789476 | USA/TX-HMH-MCoV-10438/2020 | 2020-07-08 | North America/USA/Texas/Houston | Houston Methodist Hospital | Houston Methodist Hospital | S. Wesley Long et al |
| EPI_ISL_789478 | USA/TX-HMH-MCoV-10440/2020 | 2020-07-08 | North America/USA/Texas/Houston | Houston Methodist Hospital | Houston Methodist Hospital | S. Wesley Long et al |
| EPI_ISL_789479 | USA/TX-HMH-MCoV-10441/2020 | 2020-07-09 | North America/USA/Texas/Houston | Houston Methodist Hospital | Houston Methodist Hospital | S. Wesley Long et al |
| EPI_ISL_789480 | USA/TX-HMH-MCoV-10444/2020 | 2020-07-08 | North America/USA/Texas/Houston | Houston Methodist Hospital | Houston Methodist Hospital | S. Wesley Long et al |
| EPI_ISL_789484 | USA/TX-HMH-MCoV-10457/2020 | 2020-07-08 | North America/USA/Texas/Houston | Houston Methodist Hospital | Houston Methodist Hospital | S. Wesley Long et al |
| EPI_ISL_789489 | USA/TX-HMH-MCoV-10470/2020 | 2020-07-08 | North America/USA/Texas/Houston | Houston Methodist Hospital | Houston Methodist Hospital | S. Wesley Long et al |
| EPI_ISL_789501 | USA/TX-HMH-MCoV-10496/2020 | 2020-07-08 | North America/USA/Texas/Houston | Houston Methodist Hospital | Houston Methodist Hospital | S. Wesley Long et al |
| EPI_ISL_789503 | USA/TX-HMH-MCoV-10503/2020 | 2020-07-08 | North America/USA/Texas/Houston | Houston Methodist Hospital | Houston Methodist Hospital | S. Wesley Long et al |

| | | | | | | |
|---|---|---|---|---|---|---|
| EPI_ISL_789507 | USA/TX-HMH-MCoV-10507/2020 | 2020-07-08 | North America/USA/Texas/Houston | Houston Methodist Hospital | Houston Methodist Hospital | S. Wesley Long et al |
| EPI_ISL_789510 | USA/TX-HMH-MCoV-10513/2020 | 2020-07-09 | North America/USA/Texas/Houston | Houston Methodist Hospital | Houston Methodist Hospital | S. Wesley Long et al |
| EPI_ISL_789520 | USA/TX-HMH-MCoV-10529/2020 | 2020-07-08 | North America/USA/Texas/Houston | Houston Methodist Hospital | Houston Methodist Hospital | S. Wesley Long et al |
| EPI_ISL_789523 | USA/TX-HMH-MCoV-10532/2020 | 2020-07-08 | North America/USA/Texas/Houston | Houston Methodist Hospital | Houston Methodist Hospital | S. Wesley Long et al |
| EPI_ISL_789528 | USA/TX-HMH-MCoV-10541/2020 | 2020-07-08 | North America/USA/Texas/Houston | Houston Methodist Hospital | Houston Methodist Hospital | S. Wesley Long et al |
| EPI_ISL_789530 | USA/TX-HMH-MCoV-10549/2020 | 2020-07-09 | North America/USA/Texas/Houston | Houston Methodist Hospital | Houston Methodist Hospital | S. Wesley Long et al |
| EPI_ISL_789531 | USA/TX-HMH-MCoV-10552/2020 | 2020-07-08 | North America/USA/Texas/Houston | Houston Methodist Hospital | Houston Methodist Hospital | S. Wesley Long et al |
| EPI_ISL_789537 | USA/TX-HMH-MCoV-10566/2020 | 2020-07-08 | North America/USA/Texas/Houston | Houston Methodist Hospital | Houston Methodist Hospital | S. Wesley Long et al |
| EPI_ISL_789541 | USA/TX-HMH-MCoV-10574/2020 | 2020-07-09 | North America/USA/Texas/Houston | Houston Methodist Hospital | Houston Methodist Hospital | S. Wesley Long et al |
| EPI_ISL_789543 | USA/TX-HMH-MCoV-10579/2020 | 2020-07-08 | North America/USA/Texas/Houston | Houston Methodist Hospital | Houston Methodist Hospital | S. Wesley Long et al |
| EPI_ISL_789544 | USA/TX-HMH-MCoV-10582/2020 | 2020-07-06 | North America/USA/Texas/Houston | Houston Methodist Hospital | Houston Methodist Hospital | S. Wesley Long et al |
| EPI_ISL_789545 | USA/TX-HMH-MCoV-10583/2020 | 2020-07-08 | North America/USA/Texas/Houston | Houston Methodist Hospital | Houston Methodist Hospital | S. Wesley Long et al |
| EPI_ISL_789547 | USA/TX-HMH-MCoV-10586/2020 | 2020-07-08 | North America/USA/Texas/Houston | Houston Methodist Hospital | Houston Methodist Hospital | S. Wesley Long et al |

| | | | | | | |
|---|---|---|---|---|---|---|
| EPI_ISL_789564 | USA/TX-HMH-MCoV-10617/2020 | 2020-07-06 | North America/USA/Texas/Houston | Houston Methodist Hospital | Houston Methodist Hospital | S. Wesley Long et al |
| EPI_ISL_789565 | USA/TX-HMH-MCoV-10620/2020 | 2020-07-08 | North America/USA/Texas/Houston | Houston Methodist Hospital | Houston Methodist Hospital | S. Wesley Long et al |
| EPI_ISL_789576 | USA/TX-HMH-MCoV-10650/2020 | 2020-07-06 | North America/USA/Texas/Houston | Houston Methodist Hospital | Houston Methodist Hospital | S. Wesley Long et al |
| EPI_ISL_789585 | USA/TX-HMH-MCoV-10671/2020 | 2020-07-08 | North America/USA/Texas/Houston | Houston Methodist Hospital | Houston Methodist Hospital | S. Wesley Long et al |
| EPI_ISL_789588 | USA/TX-HMH-MCoV-10685/2020 | 2020-07-08 | North America/USA/Texas/Houston | Houston Methodist Hospital | Houston Methodist Hospital | S. Wesley Long et al |
| EPI_ISL_789595 | USA/TX-HMH-MCoV-10707/2020 | 2020-07-08 | North America/USA/Texas/Houston | Houston Methodist Hospital | Houston Methodist Hospital | S. Wesley Long et al |
| EPI_ISL_789597 | USA/TX-HMH-MCoV-10711/2020 | 2020-07-08 | North America/USA/Texas/Houston | Houston Methodist Hospital | Houston Methodist Hospital | S. Wesley Long et al |
| EPI_ISL_789598 | USA/TX-HMH-MCoV-10712/2020 | 2020-07-09 | North America/USA/Texas/Houston | Houston Methodist Hospital | Houston Methodist Hospital | S. Wesley Long et al |
| EPI_ISL_789599 | USA/TX-HMH-MCoV-10713/2020 | 2020-07-09 | North America/USA/Texas/Houston | Houston Methodist Hospital | Houston Methodist Hospital | S. Wesley Long et al |
| EPI_ISL_789601 | USA/TX-HMH-MCoV-10718/2020 | 2020-07-09 | North America/USA/Texas/Houston | Houston Methodist Hospital | Houston Methodist Hospital | S. Wesley Long et al |
| EPI_ISL_789604 | USA/TX-HMH-MCoV-10722/2020 | 2020-07-08 | North America/USA/Texas/Houston | Houston Methodist Hospital | Houston Methodist Hospital | S. Wesley Long et al |
| EPI_ISL_789626 | USA/TX-HMH-MCoV-10785/2020 | 2020-07-05 | North America/USA/Texas/Houston | Houston Methodist Hospital | Houston Methodist Hospital | S. Wesley Long et al |
| EPI_ISL_789630 | USA/TX-HMH-MCoV-10799/2020 | 2020-07-05 | North America/USA/Texas/Houston | Houston Methodist Hospital | Houston Methodist Hospital | S. Wesley Long et al |

| | | | | | | |
|---|---|---|---|---|---|---|
| EPI_ISL_789635 | USA/TX-HMH-MCoV-10807/2020 | 2020-07-05 | North America/USA/Texas/Houston | Houston Methodist Hospital | Houston Methodist Hospital | S. Wesley Long et al |
| EPI_ISL_789638 | USA/TX-HMH-MCoV-10812/2020 | 2020-07-06 | North America/USA/Texas/Houston | Houston Methodist Hospital | Houston Methodist Hospital | S. Wesley Long et al |
| EPI_ISL_789640 | USA/TX-HMH-MCoV-10815/2020 | 2020-07-05 | North America/USA/Texas/Houston | Houston Methodist Hospital | Houston Methodist Hospital | S. Wesley Long et al |
| EPI_ISL_789641 | USA/TX-HMH-MCoV-10816/2020 | 2020-07-05 | North America/USA/Texas/Houston | Houston Methodist Hospital | Houston Methodist Hospital | S. Wesley Long et al |
| EPI_ISL_789642 | USA/TX-HMH-MCoV-10817/2020 | 2020-07-05 | North America/USA/Texas/Houston | Houston Methodist Hospital | Houston Methodist Hospital | S. Wesley Long et al |
| EPI_ISL_789647 | USA/TX-HMH-MCoV-10830/2020 | 2020-07-05 | North America/USA/Texas/Houston | Houston Methodist Hospital | Houston Methodist Hospital | S. Wesley Long et al |
| EPI_ISL_789655 | USA/TX-HMH-MCoV-10844/2020 | 2020-07-05 | North America/USA/Texas/Houston | Houston Methodist Hospital | Houston Methodist Hospital | S. Wesley Long et al |
| EPI_ISL_789657 | USA/TX-HMH-MCoV-10847/2020 | 2020-07-05 | North America/USA/Texas/Houston | Houston Methodist Hospital | Houston Methodist Hospital | S. Wesley Long et al |
| EPI_ISL_789659 | USA/TX-HMH-MCoV-10851/2020 | 2020-07-05 | North America/USA/Texas/Houston | Houston Methodist Hospital | Houston Methodist Hospital | S. Wesley Long et al |
| EPI_ISL_789660 | USA/TX-HMH-MCoV-10854/2020 | 2020-07-05 | North America/USA/Texas/Houston | Houston Methodist Hospital | Houston Methodist Hospital | S. Wesley Long et al |
| EPI_ISL_789662 | USA/TX-HMH-MCoV-10856/2020 | 2020-07-05 | North America/USA/Texas/Houston | Houston Methodist Hospital | Houston Methodist Hospital | S. Wesley Long et al |
| EPI_ISL_789665 | USA/TX-HMH-MCoV-10865/2020 | 2020-07-09 | North America/USA/Texas/Houston | Houston Methodist Hospital | Houston Methodist Hospital | S. Wesley Long et al |
| EPI_ISL_789668 | USA/TX-HMH-MCoV-10872/2020 | 2020-07-09 | North America/USA/Texas/Houston | Houston Methodist Hospital | Houston Methodist Hospital | S. Wesley Long et al |

| | | | | | | |
|---|---|---|---|---|---|---|
| EPI_ISL_789669 | USA/TX-HMH-MCoV-10873/2020 | 2020-07-09 | North America/USA/Texas/Houston | Houston Methodist Hospital | Houston Methodist Hospital | S. Wesley Long et al |
| EPI_ISL_789673 | USA/TX-HMH-MCoV-10892/2020 | 2020-07-09 | North America/USA/Texas/Houston | Houston Methodist Hospital | Houston Methodist Hospital | S. Wesley Long et al |
| EPI_ISL_789674 | USA/TX-HMH-MCoV-10895/2020 | 2020-07-09 | North America/USA/Texas/Houston | Houston Methodist Hospital | Houston Methodist Hospital | S. Wesley Long et al |
| EPI_ISL_789677 | USA/TX-HMH-MCoV-10901/2020 | 2020-07-09 | North America/USA/Texas/Houston | Houston Methodist Hospital | Houston Methodist Hospital | S. Wesley Long et al |
| EPI_ISL_789687 | USA/TX-HMH-MCoV-10927/2020 | 2020-07-09 | North America/USA/Texas/Houston | Houston Methodist Hospital | Houston Methodist Hospital | S. Wesley Long et al |
| EPI_ISL_789688 | USA/TX-HMH-MCoV-10928/2020 | 2020-07-09 | North America/USA/Texas/Houston | Houston Methodist Hospital | Houston Methodist Hospital | S. Wesley Long et al |
| EPI_ISL_789698 | USA/TX-HMH-MCoV-11062/2020 | 2020-09-01 | North America/USA/Texas/Houston | Houston Methodist Hospital | Houston Methodist Hospital | S. Wesley Long et al |
| EPI_ISL_789699 | USA/TX-HMH-MCoV-11064/2020 | 2020-09-01 | North America/USA/Texas/Houston | Houston Methodist Hospital | Houston Methodist Hospital | S. Wesley Long et al |
| EPI_ISL_789708 | USA/TX-HMH-MCoV-11116/2020 | 2020-09-01 | North America/USA/Texas/Houston | Houston Methodist Hospital | Houston Methodist Hospital | S. Wesley Long et al |
| EPI_ISL_789712 | USA/TX-HMH-MCoV-11126/2020 | 2020-09-06 | North America/USA/Texas/Houston | Houston Methodist Hospital | Houston Methodist Hospital | S. Wesley Long et al |
| EPI_ISL_789713 | USA/TX-HMH-MCoV-11128/2020 | 2020-09-01 | North America/USA/Texas/Houston | Houston Methodist Hospital | Houston Methodist Hospital | S. Wesley Long et al |
| EPI_ISL_789715 | USA/TX-HMH-MCoV-11144/2020 | 2020-09-01 | North America/USA/Texas/Houston | Houston Methodist Hospital | Houston Methodist Hospital | S. Wesley Long et al |
| EPI_ISL_789717 | USA/TX-HMH-MCoV-11155/2020 | 2020-09-02 | North America/USA/Texas/Houston | Houston Methodist Hospital | Houston Methodist Hospital | S. Wesley Long et al |

| | | | | | | |
|---|---|---|---|---|---|---|
| EPI_ISL_789719 | USA/TX-HMH-MCoV-11161/2020 | 2020-09-02 | North America/USA/Texas/Houston | Houston Methodist Hospital | Houston Methodist Hospital | S. Wesley Long et al |
| EPI_ISL_789722 | USA/TX-HMH-MCoV-11172/2020 | 2020-09-02 | North America/USA/Texas/Houston | Houston Methodist Hospital | Houston Methodist Hospital | S. Wesley Long et al |
| EPI_ISL_789732 | USA/TX-HMH-MCoV-11196/2020 | 2020-09-02 | North America/USA/Texas/Houston | Houston Methodist Hospital | Houston Methodist Hospital | S. Wesley Long et al |
| EPI_ISL_789735 | USA/TX-HMH-MCoV-11200/2020 | 2020-09-02 | North America/USA/Texas/Houston | Houston Methodist Hospital | Houston Methodist Hospital | S. Wesley Long et al |
| EPI_ISL_789739 | USA/TX-HMH-MCoV-11212/2020 | 2020-09-02 | North America/USA/Texas/Houston | Houston Methodist Hospital | Houston Methodist Hospital | S. Wesley Long et al |
| EPI_ISL_789742 | USA/TX-HMH-MCoV-11233/2020 | 2020-09-03 | North America/USA/Texas/Houston | Houston Methodist Hospital | Houston Methodist Hospital | S. Wesley Long et al |
| EPI_ISL_789743 | USA/TX-HMH-MCoV-11237/2020 | 2020-09-02 | North America/USA/Texas/Houston | Houston Methodist Hospital | Houston Methodist Hospital | S. Wesley Long et al |
| EPI_ISL_789744 | USA/TX-HMH-MCoV-11239/2020 | 2020-09-02 | North America/USA/Texas/Houston | Houston Methodist Hospital | Houston Methodist Hospital | S. Wesley Long et al |
| EPI_ISL_789745 | USA/TX-HMH-MCoV-11240/2020 | 2020-09-02 | North America/USA/Texas/Houston | Houston Methodist Hospital | Houston Methodist Hospital | S. Wesley Long et al |
| EPI_ISL_789749 | USA/TX-HMH-MCoV-11249/2020 | 2020-09-03 | North America/USA/Texas/Houston | Houston Methodist Hospital | Houston Methodist Hospital | S. Wesley Long et al |
| EPI_ISL_789750 | USA/TX-HMH-MCoV-11254/2020 | 2020-09-04 | North America/USA/Texas/Houston | Houston Methodist Hospital | Houston Methodist Hospital | S. Wesley Long et al |
| EPI_ISL_789752 | USA/TX-HMH-MCoV-11262/2020 | 2020-09-03 | North America/USA/Texas/Houston | Houston Methodist Hospital | Houston Methodist Hospital | S. Wesley Long et al |
| EPI_ISL_789754 | USA/TX-HMH-MCoV-11264/2020 | 2020-09-04 | North America/USA/Texas/Houston | Houston Methodist Hospital | Houston Methodist Hospital | S. Wesley Long et al |

| | | | | | | |
|---|---|---|---|---|---|---|
| EPI_ISL_789755 | USA/TX-HMH-MCoV-11268/2020 | 2020-09-04 | North America/USA/Texas/Houston | Houston Methodist Hospital | Houston Methodist Hospital | S. Wesley Long et al |
| EPI_ISL_789760 | USA/TX-HMH-MCoV-11296/2020 | 2020-09-03 | North America/USA/Texas/Houston | Houston Methodist Hospital | Houston Methodist Hospital | S. Wesley Long et al |
| EPI_ISL_789762 | USA/TX-HMH-MCoV-11300/2020 | 2020-09-03 | North America/USA/Texas/Houston | Houston Methodist Hospital | Houston Methodist Hospital | S. Wesley Long et al |
| EPI_ISL_789764 | USA/TX-HMH-MCoV-11322/2020 | 2020-09-03 | North America/USA/Texas/Houston | Houston Methodist Hospital | Houston Methodist Hospital | S. Wesley Long et al |
| EPI_ISL_789768 | USA/TX-HMH-MCoV-11331/2020 | 2020-09-03 | North America/USA/Texas/Houston | Houston Methodist Hospital | Houston Methodist Hospital | S. Wesley Long et al |
| EPI_ISL_789770 | USA/TX-HMH-MCoV-11340/2020 | 2020-09-04 | North America/USA/Texas/Houston | Houston Methodist Hospital | Houston Methodist Hospital | S. Wesley Long et al |
| EPI_ISL_789773 | USA/TX-HMH-MCoV-11345/2020 | 2020-09-05 | North America/USA/Texas/Houston | Houston Methodist Hospital | Houston Methodist Hospital | S. Wesley Long et al |
| EPI_ISL_789775 | USA/TX-HMH-MCoV-11348/2020 | 2020-09-05 | North America/USA/Texas/Houston | Houston Methodist Hospital | Houston Methodist Hospital | S. Wesley Long et al |
| EPI_ISL_789779 | USA/TX-HMH-MCoV-11710/2020 | 2020-09-08 | North America/USA/Texas/Houston | Houston Methodist Hospital | Houston Methodist Hospital | S. Wesley Long et al |
| EPI_ISL_789783 | USA/TX-HMH-MCoV-12287/2020 | 2020-09-24 | North America/USA/Texas/Houston | Houston Methodist Hospital | Houston Methodist Hospital | S. Wesley Long et al |
| EPI_ISL_789785 | USA/TX-HMH-MCoV-12774/2020 | 2020-08-19 | North America/USA/Texas/Houston | Houston Methodist Hospital | Houston Methodist Hospital | S. Wesley Long et al |
| EPI_ISL_789788 | USA/TX-HMH-MCoV-12727/2020 | 2020-08-30 | North America/USA/Texas/Houston | Houston Methodist Hospital | Houston Methodist Hospital | S. Wesley Long et al |
| EPI_ISL_789790 | USA/TX-HMH-MCoV-12580/2020 | 2020-08-31 | North America/USA/Texas/Houston | Houston Methodist Hospital | Houston Methodist Hospital | S. Wesley Long et al |

| | | | | | | |
|---|---|---|---|---|---|---|
| EPI_ISL_789792 | USA/TX-HMH-MCoV-12653/2020 | 2020-08-31 | North America/USA/Texas/Houston | Houston Methodist Hospital | Houston Methodist Hospital | S. Wesley Long et al |
| EPI_ISL_789793 | USA/TX-HMH-MCoV-12654/2020 | 2020-08-31 | North America/USA/Texas/Houston | Houston Methodist Hospital | Houston Methodist Hospital | S. Wesley Long et al |
| EPI_ISL_789798 | USA/TX-HMH-MCoV-12750/2020 | 2020-08-31 | North America/USA/Texas/Houston | Houston Methodist Hospital | Houston Methodist Hospital | S. Wesley Long et al |
| EPI_ISL_789808 | USA/TX-HMH-MCoV-12360/2020 | 2020-09-26 | North America/USA/Texas/Houston | Houston Methodist Hospital | Houston Methodist Hospital | S. Wesley Long et al |
| EPI_ISL_789812 | USA/TX-HMH-MCoV-12677/2020 | 2020-08-31 | North America/USA/Texas/Houston | Houston Methodist Hospital | Houston Methodist Hospital | S. Wesley Long et al |
| EPI_ISL_789815 | USA/TX-HMH-MCoV-12764/2020 | 2020-08-19 | North America/USA/Texas/Houston | Houston Methodist Hospital | Houston Methodist Hospital | S. Wesley Long et al |
| EPI_ISL_789818 | USA/TX-HMH-MCoV-12817/2020 | 2020-08-18 | North America/USA/Texas/Houston | Houston Methodist Hospital | Houston Methodist Hospital | S. Wesley Long et al |
| EPI_ISL_789821 | USA/TX-HMH-MCoV-12810/2020 | 2020-08-19 | North America/USA/Texas/Houston | Houston Methodist Hospital | Houston Methodist Hospital | S. Wesley Long et al |
| EPI_ISL_789833 | USA/TX-HMH-MCoV-12597/2020 | 2020-08-31 | North America/USA/Texas/Houston | Houston Methodist Hospital | Houston Methodist Hospital | S. Wesley Long et al |
| EPI_ISL_789834 | USA/TX-HMH-MCoV-12527/2020 | 2020-09-29 | North America/USA/Texas/Houston | Houston Methodist Hospital | Houston Methodist Hospital | S. Wesley Long et al |
| EPI_ISL_789837 | USA/TX-HMH-MCoV-12081/2020 | 2020-09-20 | North America/USA/Texas/Houston | Houston Methodist Hospital | Houston Methodist Hospital | S. Wesley Long et al |
| EPI_ISL_789843 | USA/TX-HMH-MCoV-12586/2020 | 2020-08-31 | North America/USA/Texas/Houston | Houston Methodist Hospital | Houston Methodist Hospital | S. Wesley Long et al |
| EPI_ISL_789844 | USA/TX-HMH-MCoV-12714/2020 | 2020-08-31 | North America/USA/Texas/Houston | Houston Methodist Hospital | Houston Methodist Hospital | S. Wesley Long et al |

| | | | | | | |
|---|---|---|---|---|---|---|
| EPI_ISL_789845 | USA/TX-HMH-MCoV-12664/2020 | 2020-08-31 | North America/USA/Texas/Houston | Houston Methodist Hospital | Houston Methodist Hospital | S. Wesley Long et al |
| EPI_ISL_789850 | USA/TX-HMH-MCoV-12738/2020 | 2020-08-31 | North America/USA/Texas/Houston | Houston Methodist Hospital | Houston Methodist Hospital | S. Wesley Long et al |
| EPI_ISL_789851 | USA/TX-HMH-MCoV-12707/2020 | 2020-08-29 | North America/USA/Texas/Houston | Houston Methodist Hospital | Houston Methodist Hospital | S. Wesley Long et al |
| EPI_ISL_789854 | USA/TX-HMH-MCoV-12577/2020 | 2020-08-31 | North America/USA/Texas/Houston | Houston Methodist Hospital | Houston Methodist Hospital | S. Wesley Long et al |
| EPI_ISL_789857 | USA/TX-HMH-MCoV-12802/2020 | 2020-08-19 | North America/USA/Texas/Houston | Houston Methodist Hospital | Houston Methodist Hospital | S. Wesley Long et al |
| EPI_ISL_789858 | USA/TX-HMH-MCoV-12657/2020 | 2020-08-31 | North America/USA/Texas/Houston | Houston Methodist Hospital | Houston Methodist Hospital | S. Wesley Long et al |
| EPI_ISL_789859 | USA/TX-HMH-MCoV-12832/2020 | 2020-08-20 | North America/USA/Texas/Houston | Houston Methodist Hospital | Houston Methodist Hospital | S. Wesley Long et al |
| EPI_ISL_789860 | USA/TX-HMH-MCoV-12616/2020 | 2020-08-31 | North America/USA/Texas/Houston | Houston Methodist Hospital | Houston Methodist Hospital | S. Wesley Long et al |
| EPI_ISL_789865 | USA/TX-HMH-MCoV-12833/2020 | 2020-08-19 | North America/USA/Texas/Houston | Houston Methodist Hospital | Houston Methodist Hospital | S. Wesley Long et al |
| EPI_ISL_789866 | USA/TX-HMH-MCoV-11873/2020 | 2020-09-16 | North America/USA/Texas/Houston | Houston Methodist Hospital | Houston Methodist Hospital | S. Wesley Long et al |
| EPI_ISL_789867 | USA/TX-HMH-MCoV-12779/2020 | 2020-08-19 | North America/USA/Texas/Houston | Houston Methodist Hospital | Houston Methodist Hospital | S. Wesley Long et al |
| EPI_ISL_789869 | USA/TX-HMH-MCoV-12639/2020 | 2020-08-31 | North America/USA/Texas/Houston | Houston Methodist Hospital | Houston Methodist Hospital | S. Wesley Long et al |
| EPI_ISL_789883 | USA/TX-HMH-MCoV-12519/2020 | 2020-09-30 | North America/USA/Texas/Houston | Houston Methodist Hospital | Houston Methodist Hospital | S. Wesley Long et al |

| | | | | | | |
|---|---|---|---|---|---|---|
| EPI_ISL_789890 | USA/TX-HMH-MCoV-12539/2020 | 2020-09-29 | North America/USA/Texas/Houston | Houston Methodist Hospital | Houston Methodist Hospital | S. Wesley Long et al |
| EPI_ISL_789891 | USA/TX-HMH-MCoV-11957/2020 | 2020-09-15 | North America/USA/Texas/Houston | Houston Methodist Hospital | Houston Methodist Hospital | S. Wesley Long et al |
| EPI_ISL_789894 | USA/TX-HMH-MCoV-12349/2020 | 2020-09-26 | North America/USA/Texas/Houston | Houston Methodist Hospital | Houston Methodist Hospital | S. Wesley Long et al |
| EPI_ISL_789899 | USA/TX-HMH-MCoV-12056/2020 | 2020-09-19 | North America/USA/Texas/Houston | Houston Methodist Hospital | Houston Methodist Hospital | S. Wesley Long et al |
| EPI_ISL_789905 | USA/TX-HMH-MCoV-12603/2020 | 2020-08-31 | North America/USA/Texas/Houston | Houston Methodist Hospital | Houston Methodist Hospital | S. Wesley Long et al |
| EPI_ISL_789906 | USA/TX-HMH-MCoV-12688/2020 | 2020-08-31 | North America/USA/Texas/Houston | Houston Methodist Hospital | Houston Methodist Hospital | S. Wesley Long et al |
| EPI_ISL_789908 | USA/TX-HMH-MCoV-12698/2020 | 2020-08-29 | North America/USA/Texas/Houston | Houston Methodist Hospital | Houston Methodist Hospital | S. Wesley Long et al |
| EPI_ISL_789910 | USA/TX-HMH-MCoV-11626/2020 | 2020-09-11 | North America/USA/Texas/Houston | Houston Methodist Hospital | Houston Methodist Hospital | S. Wesley Long et al |
| EPI_ISL_789912 | USA/TX-HMH-MCoV-12601/2020 | 2020-08-31 | North America/USA/Texas/Houston | Houston Methodist Hospital | Houston Methodist Hospital | S. Wesley Long et al |
| EPI_ISL_789918 | USA/TX-HMH-MCoV-11610/2020 | 2020-09-11 | North America/USA/Texas/Houston | Houston Methodist Hospital | Houston Methodist Hospital | S. Wesley Long et al |
| EPI_ISL_789922 | USA/TX-HMH-MCoV-12585/2020 | 2020-08-31 | North America/USA/Texas/Houston | Houston Methodist Hospital | Houston Methodist Hospital | S. Wesley Long et al |
| EPI_ISL_789923 | USA/TX-HMH-MCoV-12599/2020 | 2020-08-31 | North America/USA/Texas/Houston | Houston Methodist Hospital | Houston Methodist Hospital | S. Wesley Long et al |
| EPI_ISL_789925 | USA/TX-HMH-MCoV-12749/2020 | 2020-08-31 | North America/USA/Texas/Houston | Houston Methodist Hospital | Houston Methodist Hospital | S. Wesley Long et al |

| | | | | | | |
|---|---|---|---|---|---|---|
| EPI_ISL_789926 | USA/TX-HMH-MCoV-12706/2020 | 2020-08-30 | North America/USA/Texas/Houston | Houston Methodist Hospital | Houston Methodist Hospital | S. Wesley Long et al |
| EPI_ISL_789933 | USA/TX-HMH-MCoV-11445/2020 | 2020-09-09 | North America/USA/Texas/Houston | Houston Methodist Hospital | Houston Methodist Hospital | S. Wesley Long et al |
| EPI_ISL_789938 | USA/TX-HMH-MCoV-12723/2020 | 2020-08-31 | North America/USA/Texas/Houston | Houston Methodist Hospital | Houston Methodist Hospital | S. Wesley Long et al |
| EPI_ISL_789943 | USA/TX-HMH-MCoV-11378/2020 | 2020-09-07 | North America/USA/Texas/Houston | Houston Methodist Hospital | Houston Methodist Hospital | S. Wesley Long et al |
| EPI_ISL_789947 | USA/TX-HMH-MCoV-11565/2020 | 2020-09-11 | North America/USA/Texas/Houston | Houston Methodist Hospital | Houston Methodist Hospital | S. Wesley Long et al |
| EPI_ISL_789948 | USA/TX-HMH-MCoV-11853/2020 | 2020-09-16 | North America/USA/Texas/Houston | Houston Methodist Hospital | Houston Methodist Hospital | S. Wesley Long et al |
| EPI_ISL_789954 | USA/TX-HMH-MCoV-12309/2020 | 2020-09-25 | North America/USA/Texas/Houston | Houston Methodist Hospital | Houston Methodist Hospital | S. Wesley Long et al |
| EPI_ISL_789959 | USA/TX-HMH-MCoV-11751/2020 | 2020-09-09 | North America/USA/Texas/Houston | Houston Methodist Hospital | Houston Methodist Hospital | S. Wesley Long et al |
| EPI_ISL_789961 | USA/TX-HMH-MCoV-12323/2020 | 2020-09-26 | North America/USA/Texas/Houston | Houston Methodist Hospital | Houston Methodist Hospital | S. Wesley Long et al |
| EPI_ISL_789966 | USA/TX-HMH-MCoV-12464/2020 | 2020-09-28 | North America/USA/Texas/Houston | Houston Methodist Hospital | Houston Methodist Hospital | S. Wesley Long et al |
| EPI_ISL_789971 | USA/TX-HMH-MCoV-11888/2020 | 2020-09-16 | North America/USA/Texas/Houston | Houston Methodist Hospital | Houston Methodist Hospital | S. Wesley Long et al |
| EPI_ISL_789973 | USA/TX-HMH-MCoV-11784/2020 | 2020-09-14 | North America/USA/Texas/Houston | Houston Methodist Hospital | Houston Methodist Hospital | S. Wesley Long et al |
| EPI_ISL_789983 | USA/TX-HMH-MCoV-11477/2020 | 2020-09-08 | North America/USA/Texas/Houston | Houston Methodist Hospital | Houston Methodist Hospital | S. Wesley Long et al |

| | | | | | | |
|---|---|---|---|---|---|---|
| EPI_ISL_789984 | USA/TX-HMH-MCoV-11550/2020 | 2020-09-12 | North America/USA/Texas/Houston | Houston Methodist Hospital | Houston Methodist Hospital | S. Wesley Long et al |
| EPI_ISL_789990 | USA/TX-HMH-MCoV-11399/2020 | 2020-09-06 | North America/USA/Texas/Houston | Houston Methodist Hospital | Houston Methodist Hospital | S. Wesley Long et al |
| EPI_ISL_789996 | USA/TX-HMH-MCoV-11359/2020 | 2020-09-05 | North America/USA/Texas/Houston | Houston Methodist Hospital | Houston Methodist Hospital | S. Wesley Long et al |
| EPI_ISL_789997 | USA/TX-HMH-MCoV-11360/2020 | 2020-09-05 | North America/USA/Texas/Houston | Houston Methodist Hospital | Houston Methodist Hospital | S. Wesley Long et al |
| EPI_ISL_790004 | USA/TX-HMH-MCoV-11379/2020 | 2020-09-04 | North America/USA/Texas/Houston | Houston Methodist Hospital | Houston Methodist Hospital | S. Wesley Long et al |
| EPI_ISL_790007 | USA/TX-HMH-MCoV-11384/2020 | 2020-09-06 | North America/USA/Texas/Houston | Houston Methodist Hospital | Houston Methodist Hospital | S. Wesley Long et al |
| EPI_ISL_790011 | USA/TX-HMH-MCoV-11390/2020 | 2020-09-06 | North America/USA/Texas/Houston | Houston Methodist Hospital | Houston Methodist Hospital | S. Wesley Long et al |
| EPI_ISL_790012 | USA/TX-HMH-MCoV-11391/2020 | 2020-09-04 | North America/USA/Texas/Houston | Houston Methodist Hospital | Houston Methodist Hospital | S. Wesley Long et al |
| EPI_ISL_790015 | USA/TX-HMH-MCoV-11403/2020 | 2020-09-04 | North America/USA/Texas/Houston | Houston Methodist Hospital | Houston Methodist Hospital | S. Wesley Long et al |
| EPI_ISL_790016 | USA/TX-HMH-MCoV-11410/2020 | 2020-09-05 | North America/USA/Texas/Houston | Houston Methodist Hospital | Houston Methodist Hospital | S. Wesley Long et al |
| EPI_ISL_790019 | USA/TX-HMH-MCoV-11433/2020 | 2020-09-07 | North America/USA/Texas/Houston | Houston Methodist Hospital | Houston Methodist Hospital | S. Wesley Long et al |
| EPI_ISL_790022 | USA/TX-HMH-MCoV-11446/2020 | 2020-09-08 | North America/USA/Texas/Houston | Houston Methodist Hospital | Houston Methodist Hospital | S. Wesley Long et al |
| EPI_ISL_790028 | USA/TX-HMH-MCoV-11476/2020 | 2020-09-03 | North America/USA/Texas/Houston | Houston Methodist Hospital | Houston Methodist Hospital | S. Wesley Long et al |

| | | | | | | |
|---|---|---|---|---|---|---|
| EPI_ISL_790033 | USA/TX-HMH-MCoV-11502/2020 | 2020-09-09 | North America/USA/Texas/Houston | Houston Methodist Hospital | Houston Methodist Hospital | S. Wesley Long et al |
| EPI_ISL_790035 | USA/TX-HMH-MCoV-11504/2020 | 2020-09-09 | North America/USA/Texas/Houston | Houston Methodist Hospital | Houston Methodist Hospital | S. Wesley Long et al |
| EPI_ISL_790036 | USA/TX-HMH-MCoV-11508/2020 | 2020-09-09 | North America/USA/Texas/Houston | Houston Methodist Hospital | Houston Methodist Hospital | S. Wesley Long et al |
| EPI_ISL_790040 | USA/TX-HMH-MCoV-11517/2020 | 2020-09-09 | North America/USA/Texas/Houston | Houston Methodist Hospital | Houston Methodist Hospital | S. Wesley Long et al |
| EPI_ISL_790041 | USA/TX-HMH-MCoV-11518/2020 | 2020-09-08 | North America/USA/Texas/Houston | Houston Methodist Hospital | Houston Methodist Hospital | S. Wesley Long et al |
| EPI_ISL_790042 | USA/TX-HMH-MCoV-11521/2020 | 2020-09-01 | North America/USA/Texas/Houston | Houston Methodist Hospital | Houston Methodist Hospital | S. Wesley Long et al |
| EPI_ISL_790051 | USA/TX-HMH-MCoV-11546/2020 | 2020-09-11 | North America/USA/Texas/Houston | Houston Methodist Hospital | Houston Methodist Hospital | S. Wesley Long et al |
| EPI_ISL_790053 | USA/TX-HMH-MCoV-11552/2020 | 2020-09-11 | North America/USA/Texas/Houston | Houston Methodist Hospital | Houston Methodist Hospital | S. Wesley Long et al |
| EPI_ISL_790054 | USA/TX-HMH-MCoV-11560/2020 | 2020-09-12 | North America/USA/Texas/Houston | Houston Methodist Hospital | Houston Methodist Hospital | S. Wesley Long et al |
| EPI_ISL_790055 | USA/TX-HMH-MCoV-11561/2020 | 2020-09-12 | North America/USA/Texas/Houston | Houston Methodist Hospital | Houston Methodist Hospital | S. Wesley Long et al |
| EPI_ISL_790056 | USA/TX-HMH-MCoV-11563/2020 | 2020-09-11 | North America/USA/Texas/Houston | Houston Methodist Hospital | Houston Methodist Hospital | S. Wesley Long et al |
| EPI_ISL_790057 | USA/TX-HMH-MCoV-11564/2020 | 2020-09-12 | North America/USA/Texas/Houston | Houston Methodist Hospital | Houston Methodist Hospital | S. Wesley Long et al |
| EPI_ISL_790061 | USA/TX-HMH-MCoV-11580/2020 | 2020-09-11 | North America/USA/Texas/Houston | Houston Methodist Hospital | Houston Methodist Hospital | S. Wesley Long et al |

| | | | | | | |
|---|---|---|---|---|---|---|
| EPI_ISL_790067 | USA/TX-HMH-MCoV-11612/2020 | 2020-09-11 | North America/USA/Texas/Houston | Houston Methodist Hospital | Houston Methodist Hospital | S. Wesley Long et al |
| EPI_ISL_790070 | USA/TX-HMH-MCoV-11621/2020 | 2020-09-12 | North America/USA/Texas/Houston | Houston Methodist Hospital | Houston Methodist Hospital | S. Wesley Long et al |
| EPI_ISL_790073 | USA/TX-HMH-MCoV-11647/2020 | 2020-09-08 | North America/USA/Texas/Houston | Houston Methodist Hospital | Houston Methodist Hospital | S. Wesley Long et al |
| EPI_ISL_790084 | USA/TX-HMH-MCoV-11677/2020 | 2020-09-14 | North America/USA/Texas/Houston | Houston Methodist Hospital | Houston Methodist Hospital | S. Wesley Long et al |
| EPI_ISL_790096 | USA/TX-HMH-MCoV-11729/2020 | 2020-09-09 | North America/USA/Texas/Houston | Houston Methodist Hospital | Houston Methodist Hospital | S. Wesley Long et al |
| EPI_ISL_790098 | USA/TX-HMH-MCoV-11736/2020 | 2020-09-14 | North America/USA/Texas/Houston | Houston Methodist Hospital | Houston Methodist Hospital | S. Wesley Long et al |
| EPI_ISL_790105 | USA/TX-HMH-MCoV-11764/2020 | 2020-09-08 | North America/USA/Texas/Houston | Houston Methodist Hospital | Houston Methodist Hospital | S. Wesley Long et al |
| EPI_ISL_790106 | USA/TX-HMH-MCoV-11765/2020 | 2020-09-08 | North America/USA/Texas/Houston | Houston Methodist Hospital | Houston Methodist Hospital | S. Wesley Long et al |
| EPI_ISL_790107 | USA/TX-HMH-MCoV-11767/2020 | 2020-09-08 | North America/USA/Texas/Houston | Houston Methodist Hospital | Houston Methodist Hospital | S. Wesley Long et al |
| EPI_ISL_790108 | USA/TX-HMH-MCoV-11777/2020 | 2020-09-08 | North America/USA/Texas/Houston | Houston Methodist Hospital | Houston Methodist Hospital | S. Wesley Long et al |
| EPI_ISL_790109 | USA/TX-HMH-MCoV-11778/2020 | 2020-09-08 | North America/USA/Texas/Houston | Houston Methodist Hospital | Houston Methodist Hospital | S. Wesley Long et al |
| EPI_ISL_790111 | USA/TX-HMH-MCoV-11790/2020 | 2020-09-09 | North America/USA/Texas/Houston | Houston Methodist Hospital | Houston Methodist Hospital | S. Wesley Long et al |
| EPI_ISL_790112 | USA/TX-HMH-MCoV-11797/2020 | 2020-09-14 | North America/USA/Texas/Houston | Houston Methodist Hospital | Houston Methodist Hospital | S. Wesley Long et al |

| | | | | | | |
|---|---|---|---|---|---|---|
| EPI_ISL_790117 | USA/TX-HMH-MCoV-11823/2020 | 2020-09-14 | North America/USA/Texas/Houston | Houston Methodist Hospital | Houston Methodist Hospital | S. Wesley Long et al |
| EPI_ISL_790119 | USA/TX-HMH-MCoV-11838/2020 | 2020-09-16 | North America/USA/Texas/Houston | Houston Methodist Hospital | Houston Methodist Hospital | S. Wesley Long et al |
| EPI_ISL_790121 | USA/TX-HMH-MCoV-11855/2020 | 2020-09-16 | North America/USA/Texas/Houston | Houston Methodist Hospital | Houston Methodist Hospital | S. Wesley Long et al |
| EPI_ISL_790123 | USA/TX-HMH-MCoV-11865/2020 | 2020-09-16 | North America/USA/Texas/Houston | Houston Methodist Hospital | Houston Methodist Hospital | S. Wesley Long et al |
| EPI_ISL_790124 | USA/TX-HMH-MCoV-11870/2020 | 2020-09-17 | North America/USA/Texas/Houston | Houston Methodist Hospital | Houston Methodist Hospital | S. Wesley Long et al |
| EPI_ISL_790125 | USA/TX-HMH-MCoV-11872/2020 | 2020-09-14 | North America/USA/Texas/Houston | Houston Methodist Hospital | Houston Methodist Hospital | S. Wesley Long et al |
| EPI_ISL_790126 | USA/TX-HMH-MCoV-11875/2020 | 2020-09-16 | North America/USA/Texas/Houston | Houston Methodist Hospital | Houston Methodist Hospital | S. Wesley Long et al |
| EPI_ISL_790130 | USA/TX-HMH-MCoV-11896/2020 | 2020-09-14 | North America/USA/Texas/Houston | Houston Methodist Hospital | Houston Methodist Hospital | S. Wesley Long et al |
| EPI_ISL_790139 | USA/TX-HMH-MCoV-11937/2020 | 2020-09-15 | North America/USA/Texas/Houston | Houston Methodist Hospital | Houston Methodist Hospital | S. Wesley Long et al |
| EPI_ISL_790142 | USA/TX-HMH-MCoV-11944/2020 | 2020-09-10 | North America/USA/Texas/Houston | Houston Methodist Hospital | Houston Methodist Hospital | S. Wesley Long et al |
| EPI_ISL_790143 | USA/TX-HMH-MCoV-11945/2020 | 2020-09-15 | North America/USA/Texas/Houston | Houston Methodist Hospital | Houston Methodist Hospital | S. Wesley Long et al |
| EPI_ISL_790155 | USA/TX-HMH-MCoV-11984/2020 | 2020-09-15 | North America/USA/Texas/Houston | Houston Methodist Hospital | Houston Methodist Hospital | S. Wesley Long et al |
| EPI_ISL_790159 | USA/TX-HMH-MCoV-11990/2020 | 2020-09-17 | North America/USA/Texas/Houston | Houston Methodist Hospital | Houston Methodist Hospital | S. Wesley Long et al |

| | | | | | | |
|---|---|---|---|---|---|---|
| EPI_ISL_790161 | USA/TX-HMH-MCoV-11992/2020 | 2020-09-10 | North America/USA/Texas/Houston | Houston Methodist Hospital | Houston Methodist Hospital | S. Wesley Long et al |
| EPI_ISL_790165 | USA/TX-HMH-MCoV-12007/2020 | 2020-09-15 | North America/USA/Texas/Houston | Houston Methodist Hospital | Houston Methodist Hospital | S. Wesley Long et al |
| EPI_ISL_790167 | USA/TX-HMH-MCoV-12013/2020 | 2020-09-20 | North America/USA/Texas/Houston | Houston Methodist Hospital | Houston Methodist Hospital | S. Wesley Long et al |
| EPI_ISL_790169 | USA/TX-HMH-MCoV-12022/2020 | 2020-09-20 | North America/USA/Texas/Houston | Houston Methodist Hospital | Houston Methodist Hospital | S. Wesley Long et al |
| EPI_ISL_790172 | USA/TX-HMH-MCoV-12029/2020 | 2020-09-18 | North America/USA/Texas/Houston | Houston Methodist Hospital | Houston Methodist Hospital | S. Wesley Long et al |
| EPI_ISL_790175 | USA/TX-HMH-MCoV-12032/2020 | 2020-09-22 | North America/USA/Texas/Houston | Houston Methodist Hospital | Houston Methodist Hospital | S. Wesley Long et al |
| EPI_ISL_790176 | USA/TX-HMH-MCoV-12033/2020 | 2020-09-19 | North America/USA/Texas/Houston | Houston Methodist Hospital | Houston Methodist Hospital | S. Wesley Long et al |
| EPI_ISL_790177 | USA/TX-HMH-MCoV-12036/2020 | 2020-09-18 | North America/USA/Texas/Houston | Houston Methodist Hospital | Houston Methodist Hospital | S. Wesley Long et al |
| EPI_ISL_790180 | USA/TX-HMH-MCoV-12041/2020 | 2020-09-18 | North America/USA/Texas/Houston | Houston Methodist Hospital | Houston Methodist Hospital | S. Wesley Long et al |
| EPI_ISL_790183 | USA/TX-HMH-MCoV-12053/2020 | 2020-09-18 | North America/USA/Texas/Houston | Houston Methodist Hospital | Houston Methodist Hospital | S. Wesley Long et al |
| EPI_ISL_790187 | USA/TX-HMH-MCoV-12064/2020 | 2020-09-17 | North America/USA/Texas/Houston | Houston Methodist Hospital | Houston Methodist Hospital | S. Wesley Long et al |
| EPI_ISL_790194 | USA/TX-HMH-MCoV-12080/2020 | 2020-09-19 | North America/USA/Texas/Houston | Houston Methodist Hospital | Houston Methodist Hospital | S. Wesley Long et al |
| EPI_ISL_790196 | USA/TX-HMH-MCoV-12089/2020 | 2020-09-18 | North America/USA/Texas/Houston | Houston Methodist Hospital | Houston Methodist Hospital | S. Wesley Long et al |

| | | | | | | |
|---|---|---|---|---|---|---|
| EPI_ISL_790198 | USA/TX-HMH-MCoV-12092/2020 | 2020-09-19 | North America/USA/Texas/Houston | Houston Methodist Hospital | Houston Methodist Hospital | S. Wesley Long et al |
| EPI_ISL_790201 | USA/TX-HMH-MCoV-12100/2020 | 2020-09-17 | North America/USA/Texas/Houston | Houston Methodist Hospital | Houston Methodist Hospital | S. Wesley Long et al |
| EPI_ISL_790204 | USA/TX-HMH-MCoV-12105/2020 | 2020-09-20 | North America/USA/Texas/Houston | Houston Methodist Hospital | Houston Methodist Hospital | S. Wesley Long et al |
| EPI_ISL_790206 | USA/TX-HMH-MCoV-12118/2020 | 2020-09-21 | North America/USA/Texas/Houston | Houston Methodist Hospital | Houston Methodist Hospital | S. Wesley Long et al |
| EPI_ISL_790212 | USA/TX-HMH-MCoV-12129/2020 | 2020-09-21 | North America/USA/Texas/Houston | Houston Methodist Hospital | Houston Methodist Hospital | S. Wesley Long et al |
| EPI_ISL_790214 | USA/TX-HMH-MCoV-12133/2020 | 2020-09-22 | North America/USA/Texas/Houston | Houston Methodist Hospital | Houston Methodist Hospital | S. Wesley Long et al |
| EPI_ISL_790216 | USA/TX-HMH-MCoV-12137/2020 | 2020-09-21 | North America/USA/Texas/Houston | Houston Methodist Hospital | Houston Methodist Hospital | S. Wesley Long et al |
| EPI_ISL_790221 | USA/TX-HMH-MCoV-12147/2020 | 2020-09-21 | North America/USA/Texas/Houston | Houston Methodist Hospital | Houston Methodist Hospital | S. Wesley Long et al |
| EPI_ISL_790226 | USA/TX-HMH-MCoV-12153/2020 | 2020-09-21 | North America/USA/Texas/Houston | Houston Methodist Hospital | Houston Methodist Hospital | S. Wesley Long et al |
| EPI_ISL_790229 | USA/TX-HMH-MCoV-12160/2020 | 2020-09-25 | North America/USA/Texas/Houston | Houston Methodist Hospital | Houston Methodist Hospital | S. Wesley Long et al |
| EPI_ISL_790231 | USA/TX-HMH-MCoV-12167/2020 | 2020-09-25 | North America/USA/Texas/Houston | Houston Methodist Hospital | Houston Methodist Hospital | S. Wesley Long et al |
| EPI_ISL_790232 | USA/TX-HMH-MCoV-12171/2020 | 2020-09-21 | North America/USA/Texas/Houston | Houston Methodist Hospital | Houston Methodist Hospital | S. Wesley Long et al |
| EPI_ISL_790237 | USA/TX-HMH-MCoV-12183/2020 | 2020-09-21 | North America/USA/Texas/Houston | Houston Methodist Hospital | Houston Methodist Hospital | S. Wesley Long et al |

| | | | | | | |
|---|---|---|---|---|---|---|
| EPI_ISL_790243 | USA/TX-HMH-MCoV-12199/2020 | 2020-09-21 | North America/USA/Texas/Houston | Houston Methodist Hospital | Houston Methodist Hospital | S. Wesley Long et al |
| EPI_ISL_790247 | USA/TX-HMH-MCoV-12207/2020 | 2020-09-15 | North America/USA/Texas/Houston | Houston Methodist Hospital | Houston Methodist Hospital | S. Wesley Long et al |
| EPI_ISL_790248 | USA/TX-HMH-MCoV-12208/2020 | 2020-09-23 | North America/USA/Texas/Houston | Houston Methodist Hospital | Houston Methodist Hospital | S. Wesley Long et al |
| EPI_ISL_790249 | USA/TX-HMH-MCoV-12209/2020 | 2020-09-23 | North America/USA/Texas/Houston | Houston Methodist Hospital | Houston Methodist Hospital | S. Wesley Long et al |
| EPI_ISL_790257 | USA/TX-HMH-MCoV-12233/2020 | 2020-09-23 | North America/USA/Texas/Houston | Houston Methodist Hospital | Houston Methodist Hospital | S. Wesley Long et al |
| EPI_ISL_790258 | USA/TX-HMH-MCoV-12234/2020 | 2020-09-23 | North America/USA/Texas/Houston | Houston Methodist Hospital | Houston Methodist Hospital | S. Wesley Long et al |
| EPI_ISL_790260 | USA/TX-HMH-MCoV-12238/2020 | 2020-09-24 | North America/USA/Texas/Houston | Houston Methodist Hospital | Houston Methodist Hospital | S. Wesley Long et al |
| EPI_ISL_790261 | USA/TX-HMH-MCoV-12240/2020 | 2020-09-24 | North America/USA/Texas/Houston | Houston Methodist Hospital | Houston Methodist Hospital | S. Wesley Long et al |
| EPI_ISL_790262 | USA/TX-HMH-MCoV-12241/2020 | 2020-09-24 | North America/USA/Texas/Houston | Houston Methodist Hospital | Houston Methodist Hospital | S. Wesley Long et al |
| EPI_ISL_790265 | USA/TX-HMH-MCoV-12246/2020 | 2020-09-23 | North America/USA/Texas/Houston | Houston Methodist Hospital | Houston Methodist Hospital | S. Wesley Long et al |
| EPI_ISL_790266 | USA/TX-HMH-MCoV-12247/2020 | 2020-09-23 | North America/USA/Texas/Houston | Houston Methodist Hospital | Houston Methodist Hospital | S. Wesley Long et al |
| EPI_ISL_790267 | USA/TX-HMH-MCoV-12248/2020 | 2020-09-23 | North America/USA/Texas/Houston | Houston Methodist Hospital | Houston Methodist Hospital | S. Wesley Long et al |
| EPI_ISL_790268 | USA/TX-HMH-MCoV-12249/2020 | 2020-09-25 | North America/USA/Texas/Houston | Houston Methodist Hospital | Houston Methodist Hospital | S. Wesley Long et al |

| | | | | | | |
|---|---|---|---|---|---|---|
| EPI_ISL_790271 | USA/TX-HMH-MCoV-12258/2020 | 2020-09-23 | North America/USA/Texas/Houston | Houston Methodist Hospital | Houston Methodist Hospital | S. Wesley Long et al |
| EPI_ISL_790273 | USA/TX-HMH-MCoV-12263/2020 | 2020-09-24 | North America/USA/Texas/Houston | Houston Methodist Hospital | Houston Methodist Hospital | S. Wesley Long et al |
| EPI_ISL_790280 | USA/TX-HMH-MCoV-12277/2020 | 2020-09-23 | North America/USA/Texas/Houston | Houston Methodist Hospital | Houston Methodist Hospital | S. Wesley Long et al |
| EPI_ISL_790287 | USA/TX-HMH-MCoV-12295/2020 | 2020-09-23 | North America/USA/Texas/Houston | Houston Methodist Hospital | Houston Methodist Hospital | S. Wesley Long et al |
| EPI_ISL_790288 | USA/TX-HMH-MCoV-12297/2020 | 2020-09-24 | North America/USA/Texas/Houston | Houston Methodist Hospital | Houston Methodist Hospital | S. Wesley Long et al |
| EPI_ISL_790291 | USA/TX-HMH-MCoV-12305/2020 | 2020-09-25 | North America/USA/Texas/Houston | Houston Methodist Hospital | Houston Methodist Hospital | S. Wesley Long et al |
| EPI_ISL_790295 | USA/TX-HMH-MCoV-12313/2020 | 2020-09-26 | North America/USA/Texas/Houston | Houston Methodist Hospital | Houston Methodist Hospital | S. Wesley Long et al |
| EPI_ISL_790303 | USA/TX-HMH-MCoV-12331/2020 | 2020-09-25 | North America/USA/Texas/Houston | Houston Methodist Hospital | Houston Methodist Hospital | S. Wesley Long et al |
| EPI_ISL_790306 | USA/TX-HMH-MCoV-12339/2020 | 2020-09-23 | North America/USA/Texas/Houston | Houston Methodist Hospital | Houston Methodist Hospital | S. Wesley Long et al |
| EPI_ISL_790315 | USA/TX-HMH-MCoV-12368/2020 | 2020-09-25 | North America/USA/Texas/Houston | Houston Methodist Hospital | Houston Methodist Hospital | S. Wesley Long et al |
| EPI_ISL_790316 | USA/TX-HMH-MCoV-12372/2020 | 2020-09-26 | North America/USA/Texas/Houston | Houston Methodist Hospital | Houston Methodist Hospital | S. Wesley Long et al |
| EPI_ISL_790322 | USA/TX-HMH-MCoV-12383/2020 | 2020-09-26 | North America/USA/Texas/Houston | Houston Methodist Hospital | Houston Methodist Hospital | S. Wesley Long et al |
| EPI_ISL_790325 | USA/TX-HMH-MCoV-12392/2020 | 2020-09-25 | North America/USA/Texas/Houston | Houston Methodist Hospital | Houston Methodist Hospital | S. Wesley Long et al |

| | | | | | | |
|---|---|---|---|---|---|---|
| EPI_ISL_790330 | USA/TX-HMH-MCoV-12403/2020 | 2020-09-28 | North America/USA/Texas/Houston | Houston Methodist Hospital | Houston Methodist Hospital | S. Wesley Long et al |
| EPI_ISL_790334 | USA/TX-HMH-MCoV-12411/2020 | 2020-09-27 | North America/USA/Texas/Houston | Houston Methodist Hospital | Houston Methodist Hospital | S. Wesley Long et al |
| EPI_ISL_790340 | USA/TX-HMH-MCoV-12430/2020 | 2020-09-28 | North America/USA/Texas/Houston | Houston Methodist Hospital | Houston Methodist Hospital | S. Wesley Long et al |
| EPI_ISL_790348 | USA/TX-HMH-MCoV-12454/2020 | 2020-09-28 | North America/USA/Texas/Houston | Houston Methodist Hospital | Houston Methodist Hospital | S. Wesley Long et al |
| EPI_ISL_790349 | USA/TX-HMH-MCoV-12461/2020 | 2020-09-28 | North America/USA/Texas/Houston | Houston Methodist Hospital | Houston Methodist Hospital | S. Wesley Long et al |
| EPI_ISL_790350 | USA/TX-HMH-MCoV-12462/2020 | 2020-09-28 | North America/USA/Texas/Houston | Houston Methodist Hospital | Houston Methodist Hospital | S. Wesley Long et al |
| EPI_ISL_790351 | USA/TX-HMH-MCoV-12465/2020 | 2020-09-28 | North America/USA/Texas/Houston | Houston Methodist Hospital | Houston Methodist Hospital | S. Wesley Long et al |
| EPI_ISL_790353 | USA/TX-HMH-MCoV-12469/2020 | 2020-09-29 | North America/USA/Texas/Houston | Houston Methodist Hospital | Houston Methodist Hospital | S. Wesley Long et al |
| EPI_ISL_790355 | USA/TX-HMH-MCoV-12473/2020 | 2020-09-28 | North America/USA/Texas/Houston | Houston Methodist Hospital | Houston Methodist Hospital | S. Wesley Long et al |
| EPI_ISL_790357 | USA/TX-HMH-MCoV-12475/2020 | 2020-09-28 | North America/USA/Texas/Houston | Houston Methodist Hospital | Houston Methodist Hospital | S. Wesley Long et al |
| EPI_ISL_790360 | USA/TX-HMH-MCoV-12484/2020 | 2020-09-27 | North America/USA/Texas/Houston | Houston Methodist Hospital | Houston Methodist Hospital | S. Wesley Long et al |
| EPI_ISL_790365 | USA/TX-HMH-MCoV-12495/2020 | 2020-09-30 | North America/USA/Texas/Houston | Houston Methodist Hospital | Houston Methodist Hospital | S. Wesley Long et al |
| EPI_ISL_790373 | USA/TX-HMH-MCoV-12509/2020 | 2020-09-30 | North America/USA/Texas/Houston | Houston Methodist Hospital | Houston Methodist Hospital | S. Wesley Long et al |

| | | | | | | |
|---|---|---|---|---|---|---|
| EPI_ISL_790375 | USA/TX-HMH-MCoV-12511/2020 | 2020-09-29 | North America/USA/Texas/Houston | Houston Methodist Hospital | Houston Methodist Hospital | S. Wesley Long et al |
| EPI_ISL_790376 | USA/TX-HMH-MCoV-12515/2020 | 2020-09-30 | North America/USA/Texas/Houston | Houston Methodist Hospital | Houston Methodist Hospital | S. Wesley Long et al |
| EPI_ISL_790379 | USA/TX-HMH-MCoV-12526/2020 | 2020-09-30 | North America/USA/Texas/Houston | Houston Methodist Hospital | Houston Methodist Hospital | S. Wesley Long et al |
| EPI_ISL_790380 | USA/TX-HMH-MCoV-12528/2020 | 2020-09-30 | North America/USA/Texas/Houston | Houston Methodist Hospital | Houston Methodist Hospital | S. Wesley Long et al |
| EPI_ISL_790384 | USA/TX-HMH-MCoV-12546/2020 | 2020-09-30 | North America/USA/Texas/Houston | Houston Methodist Hospital | Houston Methodist Hospital | S. Wesley Long et al |
| EPI_ISL_790386 | USA/TX-HMH-MCoV-12552/2020 | 2020-09-30 | North America/USA/Texas/Houston | Houston Methodist Hospital | Houston Methodist Hospital | S. Wesley Long et al |
| EPI_ISL_790387 | USA/TX-HMH-MCoV-12554/2020 | 2020-09-30 | North America/USA/Texas/Houston | Houston Methodist Hospital | Houston Methodist Hospital | S. Wesley Long et al |
| EPI_ISL_790388 | USA/TX-HMH-MCoV-12555/2020 | 2020-09-30 | North America/USA/Texas/Houston | Houston Methodist Hospital | Houston Methodist Hospital | S. Wesley Long et al |
| EPI_ISL_790391 | USA/TX-HMH-MCoV-12560/2020 | 2020-09-30 | North America/USA/Texas/Houston | Houston Methodist Hospital | Houston Methodist Hospital | S. Wesley Long et al |
| EPI_ISL_790396 | USA/TX-HMH-MCoV-12567/2020 | 2020-08-31 | North America/USA/Texas/Houston | Houston Methodist Hospital | Houston Methodist Hospital | S. Wesley Long et al |
| EPI_ISL_790397 | USA/TX-HMH-MCoV-12568/2020 | 2020-08-31 | North America/USA/Texas/Houston | Houston Methodist Hospital | Houston Methodist Hospital | S. Wesley Long et al |
| EPI_ISL_790401 | USA/TX-HMH-MCoV-12572/2020 | 2020-08-31 | North America/USA/Texas/Houston | Houston Methodist Hospital | Houston Methodist Hospital | S. Wesley Long et al |
| EPI_ISL_790411 | USA/TX-HMH-MCoV-12594/2020 | 2020-08-31 | North America/USA/Texas/Houston | Houston Methodist Hospital | Houston Methodist Hospital | S. Wesley Long et al |

| | | | | | | |
|---|---|---|---|---|---|---|
| EPI_ISL_790415 | USA/TX-HMH-MCoV-12606/2020 | 2020-08-31 | North America/USA/Texas/Houston | Houston Methodist Hospital | Houston Methodist Hospital | S. Wesley Long et al |
| EPI_ISL_790420 | USA/TX-HMH-MCoV-12613/2020 | 2020-08-31 | North America/USA/Texas/Houston | Houston Methodist Hospital | Houston Methodist Hospital | S. Wesley Long et al |
| EPI_ISL_790425 | USA/TX-HMH-MCoV-12621/2020 | 2020-08-31 | North America/USA/Texas/Houston | Houston Methodist Hospital | Houston Methodist Hospital | S. Wesley Long et al |
| EPI_ISL_790438 | USA/TX-HMH-MCoV-12644/2020 | 2020-08-31 | North America/USA/Texas/Houston | Houston Methodist Hospital | Houston Methodist Hospital | S. Wesley Long et al |
| EPI_ISL_790440 | USA/TX-HMH-MCoV-12647/2020 | 2020-08-31 | North America/USA/Texas/Houston | Houston Methodist Hospital | Houston Methodist Hospital | S. Wesley Long et al |
| EPI_ISL_790443 | USA/TX-HMH-MCoV-12650/2020 | 2020-08-31 | North America/USA/Texas/Houston | Houston Methodist Hospital | Houston Methodist Hospital | S. Wesley Long et al |
| EPI_ISL_790445 | USA/TX-HMH-MCoV-12659/2020 | 2020-08-31 | North America/USA/Texas/Houston | Houston Methodist Hospital | Houston Methodist Hospital | S. Wesley Long et al |
| EPI_ISL_790448 | USA/TX-HMH-MCoV-12662/2020 | 2020-08-31 | North America/USA/Texas/Houston | Houston Methodist Hospital | Houston Methodist Hospital | S. Wesley Long et al |
| EPI_ISL_790451 | USA/TX-HMH-MCoV-12666/2020 | 2020-08-31 | North America/USA/Texas/Houston | Houston Methodist Hospital | Houston Methodist Hospital | S. Wesley Long et al |
| EPI_ISL_790453 | USA/TX-HMH-MCoV-12669/2020 | 2020-08-30 | North America/USA/Texas/Houston | Houston Methodist Hospital | Houston Methodist Hospital | S. Wesley Long et al |
| EPI_ISL_790458 | USA/TX-HMH-MCoV-12675/2020 | 2020-08-31 | North America/USA/Texas/Houston | Houston Methodist Hospital | Houston Methodist Hospital | S. Wesley Long et al |
| EPI_ISL_790463 | USA/TX-HMH-MCoV-12686/2020 | 2020-08-29 | North America/USA/Texas/Houston | Houston Methodist Hospital | Houston Methodist Hospital | S. Wesley Long et al |
| EPI_ISL_790478 | USA/TX-HMH-MCoV-12715/2020 | 2020-08-30 | North America/USA/Texas/Houston | Houston Methodist Hospital | Houston Methodist Hospital | S. Wesley Long et al |

| | | | | | | |
|---|---|---|---|---|---|---|
| EPI_ISL_790488 | USA/TX-HMH-MCoV-12726/2020 | 2020-08-31 | North America/USA/Texas/Houston | Houston Methodist Hospital | Houston Methodist Hospital | S. Wesley Long et al |
| EPI_ISL_790490 | USA/TX-HMH-MCoV-12729/2020 | 2020-08-30 | North America/USA/Texas/Houston | Houston Methodist Hospital | Houston Methodist Hospital | S. Wesley Long et al |
| EPI_ISL_790494 | USA/TX-HMH-MCoV-12736/2020 | 2020-08-31 | North America/USA/Texas/Houston | Houston Methodist Hospital | Houston Methodist Hospital | S. Wesley Long et al |
| EPI_ISL_790503 | USA/TX-HMH-MCoV-12747/2020 | 2020-09-01 | North America/USA/Texas/Houston | Houston Methodist Hospital | Houston Methodist Hospital | S. Wesley Long et al |
| EPI_ISL_790505 | USA/TX-HMH-MCoV-12754/2020 | 2020-08-30 | North America/USA/Texas/Houston | Houston Methodist Hospital | Houston Methodist Hospital | S. Wesley Long et al |
| EPI_ISL_790508 | USA/TX-HMH-MCoV-12759/2020 | 2020-08-19 | North America/USA/Texas/Houston | Houston Methodist Hospital | Houston Methodist Hospital | S. Wesley Long et al |
| EPI_ISL_790510 | USA/TX-HMH-MCoV-12762/2020 | 2020-08-19 | North America/USA/Texas/Houston | Houston Methodist Hospital | Houston Methodist Hospital | S. Wesley Long et al |
| EPI_ISL_790512 | USA/TX-HMH-MCoV-12767/2020 | 2020-08-19 | North America/USA/Texas/Houston | Houston Methodist Hospital | Houston Methodist Hospital | S. Wesley Long et al |
| EPI_ISL_790522 | USA/TX-HMH-MCoV-12786/2020 | 2020-08-19 | North America/USA/Texas/Houston | Houston Methodist Hospital | Houston Methodist Hospital | S. Wesley Long et al |
| EPI_ISL_790526 | USA/TX-HMH-MCoV-12793/2020 | 2020-08-20 | North America/USA/Texas/Houston | Houston Methodist Hospital | Houston Methodist Hospital | S. Wesley Long et al |
| EPI_ISL_790528 | USA/TX-HMH-MCoV-12795/2020 | 2020-08-20 | North America/USA/Texas/Houston | Houston Methodist Hospital | Houston Methodist Hospital | S. Wesley Long et al |
| EPI_ISL_790531 | USA/TX-HMH-MCoV-12798/2020 | 2020-08-20 | North America/USA/Texas/Houston | Houston Methodist Hospital | Houston Methodist Hospital | S. Wesley Long et al |
| EPI_ISL_790533 | USA/TX-HMH-MCoV-12800/2020 | 2020-08-19 | North America/USA/Texas/Houston | Houston Methodist Hospital | Houston Methodist Hospital | S. Wesley Long et al |

| | | | | | | |
|---|---|---|---|---|---|---|
| EPI_ISL_790535 | USA/TX-HMH-MCoV-12803/2020 | 2020-08-19 | North America/USA/Texas/Houston | Houston Methodist Hospital | Houston Methodist Hospital | S. Wesley Long et al |
| EPI_ISL_790541 | USA/TX-HMH-MCoV-12811/2020 | 2020-08-19 | North America/USA/Texas/Houston | Houston Methodist Hospital | Houston Methodist Hospital | S. Wesley Long et al |
| EPI_ISL_790546 | USA/TX-HMH-MCoV-12818/2020 | 2020-08-19 | North America/USA/Texas/Houston | Houston Methodist Hospital | Houston Methodist Hospital | S. Wesley Long et al |
| EPI_ISL_790550 | USA/TX-HMH-MCoV-12823/2020 | 2020-08-19 | North America/USA/Texas/Houston | Houston Methodist Hospital | Houston Methodist Hospital | S. Wesley Long et al |
| EPI_ISL_790551 | USA/TX-HMH-MCoV-12827/2020 | 2020-08-19 | North America/USA/Texas/Houston | Houston Methodist Hospital | Houston Methodist Hospital | S. Wesley Long et al |
| EPI_ISL_790555 | USA/TX-HMH-MCoV-12831/2020 | 2020-08-19 | North America/USA/Texas/Houston | Houston Methodist Hospital | Houston Methodist Hospital | S. Wesley Long et al |
| EPI_ISL_790557 | USA/TX-HMH-MCoV-12835/2020 | 2020-08-19 | North America/USA/Texas/Houston | Houston Methodist Hospital | Houston Methodist Hospital | S. Wesley Long et al |
| EPI_ISL_790568 | USA/FL-BPHL-2268/2020 | 2020-11-02 | North America/USA/Florida/ | Florida Bureau of Public Health Laboratories | Florida Bureau of Public Health Laboratories | Sarah Schmedes et al |
| EPI_ISL_790574 | Netherlands/NH-RIVM-21024/2020 | 2020-12-29 | Europe/Netherlands/North Holland/ | Dutch COVID-19 response team | National Institute for Public Health and the Environment (RIVM) | Adam Meijer et al |
| EPI_ISL_790597 | Netherlands/OV-RIVM-20744/2020 | 2020-12-12 | Europe/Netherlands/Overijssel/ | Dutch COVID-19 response team | National Institute for Public Health and the Environment (RIVM) | Adam Meijer et al |
| EPI_ISL_790607 | Netherlands/OV-RIVM-20885/2020 | 2020-12-23 | Europe/Netherlands/Overijssel/ | Dutch COVID-19 response team | National Institute for Public Health and the Environment (RIVM) | Adam Meijer et al |

| | | | | | | |
|---|---|---|---|---|---|---|
| EPI_ISL_790610 | Netherlands/GE-RIVM-20888/2020 | 2020-12-24 | Europe/Netherlands/Gelderland/ | Dutch COVID-19 response team | National Institute for Public Health and the Environment (RIVM) | Adam Meijer et al |
| EPI_ISL_790619 | Netherlands/UT-RIVM-20998/2020 | 2020-12-22 | Europe/Netherlands/Utrecht/ | Dutch COVID-19 response team | National Institute for Public Health and the Environment (RIVM) | Adam Meijer et al |
| EPI_ISL_790622 | Netherlands/NH-RIVM-21036/2020 | 2020-12-21 | Europe/Netherlands/North Holland/ | Dutch COVID-19 response team | National Institute for Public Health and the Environment (RIVM) | Adam Meijer et al |
| EPI_ISL_790644 | Netherlands/FR-RIVM-20559/2020 | 2020-12-15 | Europe/Netherlands/Friesland/ | Dutch COVID-19 response team | National Institute for Public Health and the Environment (RIVM) | Adam Meijer et al |
| EPI_ISL_790663 | Netherlands/NB-RIVM-20579/2020 | 2020-12-12 | Europe/Netherlands/North Brabant/ | Dutch COVID-19 response team | National Institute for Public Health and the Environment (RIVM) | Adam Meijer et al |
| EPI_ISL_790668 | Netherlands/GR-RIVM-20584/2020 | 2020-12-11 | Europe/Netherlands/Groningen/ | Dutch COVID-19 response team | National Institute for Public Health and the Environment (RIVM) | Adam Meijer et al |
| EPI_ISL_790696 | Netherlands/UT-RIVM-20617/2020 | 2020-12-16 | Europe/Netherlands/Utrecht/ | Dutch COVID-19 response team | National Institute for Public Health and the Environment (RIVM) | Adam Meijer et al |
| EPI_ISL_790755 | Netherlands/ZH-RIVM-20687/2020 | 2020-12-23 | Europe/Netherlands/South Holland/ | Dutch COVID-19 response team | National Institute for Public Health and the Environment (RIVM) | Adam Meijer et al |

| | | | | | | |
|---|---|---|---|---|---|---|
| EPI_ISL_790840 | Netherlands/ZH-RIVM-20872/2020 | 2020-12-24 | Europe/Netherlands/South Holland/ | Dutch COVID-19 response team | National Institute for Public Health and the Environment (RIVM) | Adam Meijer et al |
| EPI_ISL_790854 | Netherlands/NH-RIVM-20795/2020 | 2020-12-20 | Europe/Netherlands/North Holland/ | Dutch COVID-19 response team | National Institute for Public Health and the Environment (RIVM) | Adam Meijer et al |
| EPI_ISL_790859 | Netherlands/NH-RIVM-20800/2020 | 2020-12-21 | Europe/Netherlands/North Holland/ | Dutch COVID-19 response team | National Institute for Public Health and the Environment (RIVM) | Adam Meijer et al |
| EPI_ISL_790890 | Netherlands/GR-RIVM-20835/2020 | 2020-12-15 | Europe/Netherlands/Groningen/ | Dutch COVID-19 response team | National Institute for Public Health and the Environment (RIVM) | Adam Meijer et al |
| EPI_ISL_790916 | Netherlands/ZH-RIVM-20775/2020 | 2020-12-17 | Europe/Netherlands/South Holland/ | Dutch COVID-19 response team | National Institute for Public Health and the Environment (RIVM) | Adam Meijer et al |
| EPI_ISL_790917 | Netherlands/ZH-RIVM-20776/2020 | 2020-12-18 | Europe/Netherlands/South Holland/ | Dutch COVID-19 response team | National Institute for Public Health and the Environment (RIVM) | Adam Meijer et al |
| EPI_ISL_790943 | Netherlands/ZH-RIVM-20911/2020 | 2020-10-02 | Europe/Netherlands/South Holland/ | Dutch COVID-19 response team | National Institute for Public Health and the Environment (RIVM) | Adam Meijer et al |
| EPI_ISL_790955 | Netherlands/FR-RIVM-20928/2020 | 2020-12-14 | Europe/Netherlands/Friesland/ | Dutch COVID-19 response team | National Institute for Public Health and the Environment (RIVM) | Adam Meijer et al |

| | | | | | | |
|---|---|---|---|---|---|---|
| EPI_ISL_790960 | Netherlands/ZH-RIVM-20934/2020 | 2020-09-24 | Europe/Netherlands/South Holland/ | Dutch COVID-19 response team | National Institute for Public Health and the Environment (RIVM) | Adam Meijer et al |
| EPI_ISL_790990 | Netherlands/NH-RIVM-20966/2020 | 2020-12-12 | Europe/Netherlands/North Holland/ | Dutch COVID-19 response team | National Institute for Public Health and the Environment (RIVM) | Adam Meijer et al |
| EPI_ISL_791007 | Netherlands/NH-RIVM-20987/2020 | 2020-12-22 | Europe/Netherlands/North Holland/ | Dutch COVID-19 response team | National Institute for Public Health and the Environment (RIVM) | Adam Meijer et al |
| EPI_ISL_791053 | Netherlands/NH-RIVM-21039/2020 | 2020-12-23 | Europe/Netherlands/North Holland/ | Dutch COVID-19 response team | National Institute for Public Health and the Environment (RIVM) | Adam Meijer et al |
| EPI_ISL_791056 | Netherlands/ZE-RIVM-21042/2020 | 2020-12-30 | Europe/Netherlands/Zeeland/ | Dutch COVID-19 response team | National Institute for Public Health and the Environment (RIVM) | Adam Meijer et al |
| EPI_ISL_791072 | Netherlands/NH-RIVM-21058/2020 | 2020-12-18 | Europe/Netherlands/North Holland/ | Dutch COVID-19 response team | National Institute for Public Health and the Environment (RIVM) | Adam Meijer et al |
| EPI_ISL_791087 | Colombia/SAP-INS-SPSAN01/2020 | 2020-12-27 | South America/Colombia/San Andres y | Instituto Nacional de Salud - Unidad de Secuenciaci√≥n y An√°lisis Gen√≥mico | Instituto Nacional de Salud - Direcci√≥n de Investigaci√≥n en Salud P√∫blica | Katherine Laiton-Donato et al |
| EPI_ISL_791093 | Colombia/NAR-INS-IP004/2020 | 2020-12-08 | South America/Colombia/Narino/ | Instituto Nacional de Salud - Unidad de Secuenciaci√≥n y An√°lisis Gen√≥mico | Instituto Nacional de Salud - Direcci√≥n de Investigaci√≥n en Salud P√∫blica | Katherine Laiton-Donato et al |

| | | | | | | |
|---|---|---|---|---|---|---|
| EPI_ISL_791094 | Colombia/NAR-INS-IP005/2020 | 2020-12-08 | South America/Colombia/Narino/ | Instituto Nacional de Salud - Unidad de Secuenciaci√≥n y An√°lisis Gen√≥mico | Instituto Nacional de Salud - Direcci√≥n de Investigaci√≥n en Salud P√∫blica | Katherine Laiton-Donato et al |
| EPI_ISL_791095 | Colombia/NAR-INS-IP006/2020 | 2020-12-08 | South America/Colombia/Narino/ | Instituto Nacional de Salud - Unidad de Secuenciaci√≥n y An√°lisis Gen√≥mico | Instituto Nacional de Salud - Direcci√≥n de Investigaci√≥n en Salud P√∫blica | Katherine Laiton-Donato et al |
| EPI_ISL_791096 | Colombia/NAR-INS-IP0610/2020 | 2020-12-04 | South America/Colombia/Narino/ | Instituto Nacional de Salud - Unidad de Secuenciaci√≥n y An√°lisis Gen√≥mico | Instituto Nacional de Salud - Direcci√≥n de Investigaci√≥n en Salud P√∫blica | Katherine Laiton-Donato et al |
| EPI_ISL_791098 | USA/WI-UW-2580/2021 | 2021-01-04 | North America/USA/Wisconsin/Dane Co | University of Wisconsin-Madison AIDS Vaccine Research Laboratories | University of Wisconsin-Madison AIDS Vaccine Research Laboratories | Gage Moreno et al |
| EPI_ISL_791101 | USA/WI-UW-2583/2021 | 2021-01-04 | North America/USA/Wisconsin/Dane Co | University of Wisconsin-Madison AIDS Vaccine Research Laboratories | University of Wisconsin-Madison AIDS Vaccine Research Laboratories | Gage Moreno et al |
| EPI_ISL_791103 | USA/WI-UW-2585/2021 | 2021-01-04 | North America/USA/Wisconsin/Dane Co | University of Wisconsin-Madison AIDS Vaccine Research Laboratories | University of Wisconsin-Madison AIDS Vaccine Research Laboratories | Gage Moreno et al |
| EPI_ISL_791120 | USA/WI-UW-2603/2021 | 2021-01-01 | North America/USA/Wisconsin/Dane Co | University of Wisconsin-Madison AIDS Vaccine Research Laboratories | University of Wisconsin-Madison AIDS Vaccine Research Laboratories | Gage Moreno et al |
| EPI_ISL_791132 | USA/WI-UW-2615/2021 | 2021-01-03 | North America/USA/Wisconsin/Dane Co | University of Wisconsin-Madison AIDS Vaccine Research Laboratories | University of Wisconsin-Madison AIDS Vaccine Research Laboratories | Gage Moreno et al |

| | | | | | | |
|---|---|---|---|---|---|---|
| EPI_ISL_791137 | USA/WI-UW-2620/2021 | 2021-01-03 | North America/USA/Wisconsin/Dane Co | University of Wisconsin-Madison AIDS Vaccine Research Laboratories | University of Wisconsin-Madison AIDS Vaccine Research Laboratories | Gage Moreno et al |
| EPI_ISL_791143 | USA/WI-UW-2626/2021 | 2021-01-02 | North America/USA/Wisconsin/Dane Co | University of Wisconsin-Madison AIDS Vaccine Research Laboratories | University of Wisconsin-Madison AIDS Vaccine Research Laboratories | Gage Moreno et al |
| EPI_ISL_791144 | USA/WI-UW-2628/2020 | 2020-12-31 | North America/USA/Wisconsin/Dane Co | University of Wisconsin-Madison AIDS Vaccine Research Laboratories | University of Wisconsin-Madison AIDS Vaccine Research Laboratories | Gage Moreno et al |
| EPI_ISL_791150 | USA/WI-UW-2634/2020 | 2020-12-31 | North America/USA/Wisconsin/Dane Co | University of Wisconsin-Madison AIDS Vaccine Research Laboratories | University of Wisconsin-Madison AIDS Vaccine Research Laboratories | Gage Moreno et al |
| EPI_ISL_791152 | USA/WI-UW-2636/2020 | 2020-12-30 | North America/USA/Wisconsin/Dane Co | University of Wisconsin-Madison AIDS Vaccine Research Laboratories | University of Wisconsin-Madison AIDS Vaccine Research Laboratories | Gage Moreno et al |
| EPI_ISL_791155 | USA/WI-UW-2640/2020 | 2020-12-31 | North America/USA/Wisconsin/Dane Co | University of Wisconsin-Madison AIDS Vaccine Research Laboratories | University of Wisconsin-Madison AIDS Vaccine Research Laboratories | Gage Moreno et al |
| EPI_ISL_791162 | USA/WI-UW-2647/2021 | 2021-01-03 | North America/USA/Wisconsin/Dane Co | University of Wisconsin-Madison AIDS Vaccine Research Laboratories | University of Wisconsin-Madison AIDS Vaccine Research Laboratories | Gage Moreno et al |
| EPI_ISL_791164 | USA/WI-UW-2649/2020 | 2020-12-31 | North America/USA/Wisconsin/Dane Co | University of Wisconsin-Madison AIDS Vaccine Research Laboratories | University of Wisconsin-Madison AIDS Vaccine Research Laboratories | Gage Moreno et al |

| | | | | | | |
|---|---|---|---|---|---|---|
| EPI_ISL_791166 | USA/WI-UW-2651/2020 | 2020-12-31 | North America/USA/Wisconsin/Dane Co | University of Wisconsin-Madison AIDS Vaccine Research Laboratories | University of Wisconsin-Madison AIDS Vaccine Research Laboratories | Gage Moreno et al |
| EPI_ISL_791179 | USA/WI-UW-2666/2020 | 2020-12-29 | North America/USA/Wisconsin/Dane Co | University of Wisconsin-Madison AIDS Vaccine Research Laboratories | University of Wisconsin-Madison AIDS Vaccine Research Laboratories | Gage Moreno et al |
| EPI_ISL_791256 | England/210100378/2021 | 2021-01-04 | Europe/United Kingdom/England/ | Respiratory Virus Unit, National Infection Service, Public Health England | COVID-19 Genomics UK (COG-UK) Consortium | PHE Covid Sequencing Team et al |
| EPI_ISL_791280 | Ireland/un-NVRL-AIIDB0140v1/2020 | 2020-12-17 | Europe/Ireland/Ireland/ | National Virus Reference Laboratory | Irish Coronavirus Sequencing Consortium - Teagasc Moorepark | Alejandro Abner Garcia Leon et al |
| EPI_ISL_791297 | Ireland/un-NVRL-AIIDV1479baseline/ | 2020-12-20 | Europe/Ireland/Ireland/ | National Virus Reference Laboratory | Irish Coronavirus Sequencing Consortium - Teagasc Moorepark | Alejandro Abner Garcia Leon et al |
| EPI_ISL_791327 | USA/WI-UW-2593/2021 | 2021-01-03 | North America/USA/Wisconsin/Dane Co | University of Wisconsin-Madison AIDS Vaccine Research Laboratories | University of Wisconsin-Madison AIDS Vaccine Research Laboratories | Gage Moreno et al |
| EPI_ISL_791328 | USA/WI-UW-2627/2021 | 2021-01-03 | North America/USA/Wisconsin/Dane Co | University of Wisconsin-Madison AIDS Vaccine Research Laboratories | University of Wisconsin-Madison AIDS Vaccine Research Laboratories | Gage Moreno et al |
| EPI_ISL_791343 | USA/MD-HP12070/2020 | 2020-11-22 | North America/USA/Maryland/ | Johns Hopkins Hospital Department of Pathology | Johns Hopkins Hospital Department of Pathology | C. Paul Morris et al |

| | | | | | | |
|---|---|---|---|---|---|---|
| EPI_ISL_791344 | USA/MD-HP12121/2020 | 2020-12-23 | North America/USA/Maryland/ | Johns Hopkins Hospital Department of Pathology | Johns Hopkins Hospital Department of Pathology | C. Paul Morris et al |
| EPI_ISL_791348 | USA/MD-HP12003/2020 | 2020-11-11 | North America/USA/Maryland/ | Johns Hopkins Hospital Department of Pathology | Johns Hopkins Hospital Department of Pathology | C. Paul Morris et al |
| EPI_ISL_791356 | USA/MD-HP12014/2020 | 2020-11-05 | North America/USA/Maryland/ | Johns Hopkins Hospital Department of Pathology | Johns Hopkins Hospital Department of Pathology | C. Paul Morris et al |
| EPI_ISL_791365 | USA/MD-HP12030/2020 | 2020-11-10 | North America/USA/Maryland/ | Johns Hopkins Hospital Department of Pathology | Johns Hopkins Hospital Department of Pathology | C. Paul Morris et al |
| EPI_ISL_791375 | USA/MD-HP12042/2020 | 2020-11-11 | North America/USA/Maryland/ | Johns Hopkins Hospital Department of Pathology | Johns Hopkins Hospital Department of Pathology | C. Paul Morris et al |
| EPI_ISL_791383 | USA/MD-HP12051/2020 | 2020-11-13 | North America/USA/Maryland/ | Johns Hopkins Hospital Department of Pathology | Johns Hopkins Hospital Department of Pathology | C. Paul Morris et al |
| EPI_ISL_791393 | USA/MD-HP12061/2020 | 2020-11-23 | North America/USA/Maryland/ | Johns Hopkins Hospital Department of Pathology | Johns Hopkins Hospital Department of Pathology | C. Paul Morris et al |
| EPI_ISL_791394 | USA/MD-HP12062/2020 | 2020-11-24 | North America/USA/Maryland/ | Johns Hopkins Hospital Department of Pathology | Johns Hopkins Hospital Department of Pathology | C. Paul Morris et al |
| EPI_ISL_791398 | USA/MD-HP12066/2020 | 2020-12-04 | North America/USA/Maryland/ | Johns Hopkins Hospital Department of Pathology | Johns Hopkins Hospital Department of Pathology | C. Paul Morris et al |
| EPI_ISL_791399 | USA/MD-HP12067/2020 | 2020-11-25 | North America/USA/Maryland/ | Johns Hopkins Hospital Department of Pathology | Johns Hopkins Hospital Department of Pathology | C. Paul Morris et al |

| | | | | | | |
|---|---|---|---|---|---|---|
| EPI_ISL_791402 | USA/MD-HP12074/2020 | 2020-11-26 | North America/USA/Maryland/ | Johns Hopkins Hospital Department of Pathology | Johns Hopkins Hospital Department of Pathology | C. Paul Morris et al |
| EPI_ISL_791404 | USA/MD-HP12076/2020 | 2020-11-22 | North America/USA/Maryland/ | Johns Hopkins Hospital Department of Pathology | Johns Hopkins Hospital Department of Pathology | C. Paul Morris et al |
| EPI_ISL_791416 | USA/MD-HP12098/2020 | 2020-12-18 | North America/USA/Maryland/ | Johns Hopkins Hospital Department of Pathology | Johns Hopkins Hospital Department of Pathology | C. Paul Morris et al |
| EPI_ISL_791417 | USA/MD-HP12099/2020 | 2020-12-14 | North America/USA/Maryland/ | Johns Hopkins Hospital Department of Pathology | Johns Hopkins Hospital Department of Pathology | C. Paul Morris et al |
| EPI_ISL_791421 | USA/MD-HP12105/2020 | 2020-12-30 | North America/USA/Maryland/ | Johns Hopkins Hospital Department of Pathology | Johns Hopkins Hospital Department of Pathology | C. Paul Morris et al |
| EPI_ISL_791423 | USA/MD-HP12107/2021 | 2021-01-05 | North America/USA/Maryland/ | Johns Hopkins Hospital Department of Pathology | Johns Hopkins Hospital Department of Pathology | C. Paul Morris et al |
| EPI_ISL_791424 | USA/MD-HP12109/2020 | 2020-12-28 | North America/USA/Maryland/ | Johns Hopkins Hospital Department of Pathology | Johns Hopkins Hospital Department of Pathology | C. Paul Morris et al |
| EPI_ISL_791428 | USA/MD-HP12113/2020 | 2020-12-22 | North America/USA/Maryland/ | Johns Hopkins Hospital Department of Pathology | Johns Hopkins Hospital Department of Pathology | C. Paul Morris et al |
| EPI_ISL_791431 | USA/MD-HP12118/2020 | 2020-12-23 | North America/USA/Maryland/ | Johns Hopkins Hospital Department of Pathology | Johns Hopkins Hospital Department of Pathology | C. Paul Morris et al |
| EPI_ISL_791433 | USA/MD-HP12120/2020 | 2020-12-26 | North America/USA/Maryland/ | Johns Hopkins Hospital Department of Pathology | Johns Hopkins Hospital Department of Pathology | C. Paul Morris et al |

| | | | | | | |
|---|---|---|---|---|---|---|
| EPI_ISL_791437 | USA/MD-HP12127/2020 | 2020-12-29 | North America/USA/Maryland/ | Johns Hopkins Hospital Department of Pathology | Johns Hopkins Hospital Department of Pathology | C. Paul Morris et al |
| EPI_ISL_791443 | USA/MD-HP12133/2020 | 2020-12-28 | North America/USA/Maryland/ | Johns Hopkins Hospital Department of Pathology | Johns Hopkins Hospital Department of Pathology | C. Paul Morris et al |
| EPI_ISL_791446 | USA/MD-HP12136/2020 | 2020-12-30 | North America/USA/Maryland/ | Johns Hopkins Hospital Department of Pathology | Johns Hopkins Hospital Department of Pathology | C. Paul Morris et al |
| EPI_ISL_791456 | USA/MD-HP12147/2020 | 2020-12-29 | North America/USA/Maryland/ | Johns Hopkins Hospital Department of Pathology | Johns Hopkins Hospital Department of Pathology | C. Paul Morris et al |
| EPI_ISL_791460 | USA/MD-HP12151/2020 | 2020-12-23 | North America/USA/Maryland/ | Johns Hopkins Hospital Department of Pathology | Johns Hopkins Hospital Department of Pathology | C. Paul Morris et al |
| EPI_ISL_791463 | USA/MD-HP12156/2020 | 2020-12-27 | North America/USA/Maryland/ | Johns Hopkins Hospital Department of Pathology | Johns Hopkins Hospital Department of Pathology | C. Paul Morris et al |
| EPI_ISL_791469 | USA/MD-HP12163/2020 | 2020-12-30 | North America/USA/Maryland/ | Johns Hopkins Hospital Department of Pathology | Johns Hopkins Hospital Department of Pathology | C. Paul Morris et al |
| EPI_ISL_791472 | USA/MD-HP12166/2020 | 2020-12-30 | North America/USA/Maryland/ | Johns Hopkins Hospital Department of Pathology | Johns Hopkins Hospital Department of Pathology | C. Paul Morris et al |
| EPI_ISL_791476 | USA/MD-HP12176/2020 | 2020-12-24 | North America/USA/Maryland/ | Johns Hopkins Hospital Department of Pathology | Johns Hopkins Hospital Department of Pathology | C. Paul Morris et al |
| EPI_ISL_791484 | USA/MD-HP12185/2020 | 2020-12-14 | North America/USA/Maryland/ | Johns Hopkins Hospital Department of Pathology | Johns Hopkins Hospital Department of Pathology | C. Paul Morris et al |

| | | | | | | |
|---|---|---|---|---|---|---|
| EPI_ISL_791488 | Canada/ON-SK-20H-328MM0636/202 | 2020-11-24 | North America/Canada/Ontario/Toronto | The Hosptial for Sick Children | The Hospital for Sick Children | Aaron Campigotto et al |
| EPI_ISL_791489 | Canada/ON-SK-20H-328MM0640/202 | 2020-11-24 | North America/Canada/Ontario/Toronto | The Hosptial for Sick Children | The Hospital for Sick Children | Aaron Campigotto et al |
| EPI_ISL_791490 | Canada/ON-SK-20H-328MM0760/202 | 2020-11-24 | North America/Canada/Ontario/Toronto | The Hosptial for Sick Children | The Hospital for Sick Children | Aaron Campigotto et al |
| EPI_ISL_791491 | Canada/ON-SK-20H-328MM0775/202 | 2020-11-24 | North America/Canada/Ontario/Toronto | The Hosptial for Sick Children | The Hospital for Sick Children | Aaron Campigotto et al |
| EPI_ISL_791492 | Canada/ON-SK-20H-328MM0777/202 | 2020-11-24 | North America/Canada/Ontario/Toronto | The Hosptial for Sick Children | The Hospital for Sick Children | Aaron Campigotto et al |
| EPI_ISL_791493 | Canada/ON-SK-20H-328MM0785/202 | 2020-11-24 | North America/Canada/Ontario/Toronto | The Hosptial for Sick Children | The Hospital for Sick Children | Aaron Campigotto et al |
| EPI_ISL_791494 | Canada/ON-SK-20H-328MM0836/202 | 2020-11-24 | North America/Canada/Ontario/Toronto | The Hosptial for Sick Children | The Hospital for Sick Children | Aaron Campigotto et al |
| EPI_ISL_791495 | Canada/ON-SK-20H-328MM0848/202 | 2020-11-24 | North America/Canada/Ontario/Toronto | The Hosptial for Sick Children | The Hospital for Sick Children | Aaron Campigotto et al |
| EPI_ISL_791496 | Canada/ON-SK-20H-328MM0871/202 | 2020-11-24 | North America/Canada/Ontario/Toronto | The Hosptial for Sick Children | The Hospital for Sick Children | Aaron Campigotto et al |
| EPI_ISL_791497 | Canada/ON-SK-20H-328MM0909/202 | 2020-11-24 | North America/Canada/Ontario/Toronto | The Hosptial for Sick Children | The Hospital for Sick Children | Aaron Campigotto et al |
| EPI_ISL_791498 | Canada/ON-SK-20H-328MM0914/202 | 2020-11-24 | North America/Canada/Ontario/Toronto | The Hosptial for Sick Children | The Hospital for Sick Children | Aaron Campigotto et al |
| EPI_ISL_791499 | Canada/ON-SK-20H-328MM0929/202 | 2020-11-24 | North America/Canada/Ontario/Toronto | The Hosptial for Sick Children | The Hospital for Sick Children | Aaron Campigotto et al |
| EPI_ISL_791500 | Canada/ON-SK-20H-328MM0932/202 | 2020-11-24 | North America/Canada/Ontario/Toronto | The Hosptial for Sick Children | The Hospital for Sick Children | Aaron Campigotto et al |
| EPI_ISL_791501 | Canada/ON-SK-20H-328MM0935/202 | 2020-11-24 | North America/Canada/Ontario/Toronto | The Hosptial for Sick Children | The Hospital for Sick Children | Aaron Campigotto et al |
| EPI_ISL_791502 | Canada/ON-SK-20H-328MM1080/202 | 2020-11-24 | North America/Canada/Ontario/Toronto | The Hosptial for Sick Children | The Hospital for Sick Children | Aaron Campigotto et al |
| EPI_ISL_791503 | Canada/ON-SK-20H-328MM1138/202 | 2020-11-24 | North America/Canada/Ontario/Toronto | The Hosptial for Sick Children | The Hospital for Sick Children | Aaron Campigotto et al |
| EPI_ISL_791504 | Canada/ON-SK-20H-329MM0017/202 | 2020-11-24 | North America/Canada/Ontario/Toronto | The Hosptial for Sick Children | The Hospital for Sick Children | Aaron Campigotto et al |
| EPI_ISL_791505 | Canada/ON-SK-20H-329MM0023/202 | 2020-11-24 | North America/Canada/Ontario/Toronto | The Hosptial for Sick Children | The Hospital for Sick Children | Aaron Campigotto et al |
| EPI_ISL_791506 | Canada/ON-SK-20H-329MM0157/202 | 2020-11-24 | North America/Canada/Ontario/Toronto | The Hosptial for Sick Children | The Hospital for Sick Children | Aaron Campigotto et al |
| EPI_ISL_791507 | Canada/ON-SK-20H-329MM0163/202 | 2020-11-24 | North America/Canada/Ontario/Toronto | The Hosptial for Sick Children | The Hospital for Sick Children | Aaron Campigotto et al |

| | | | | | | |
|---|---|---|---|---|---|---|
| EPI_ISL_791508 | Canada/ON-SK-20H-329MM0191/202 | 2020-11-24 | North America/Canada/Ontario/Toronto | The Hosptial for Sick Children | The Hospital for Sick Children | Aaron Campigotto et al |
| EPI_ISL_791509 | Canada/ON-SK-20H-329MM0239/202 | 2020-11-24 | North America/Canada/Ontario/Toronto | The Hosptial for Sick Children | The Hospital for Sick Children | Aaron Campigotto et al |
| EPI_ISL_791510 | Canada/ON-SK-20H-329MM0254/202 | 2020-11-24 | North America/Canada/Ontario/Toronto | The Hosptial for Sick Children | The Hospital for Sick Children | Aaron Campigotto et al |
| EPI_ISL_791511 | Canada/ON-SK-20H-329MM0268/202 | 2020-11-24 | North America/Canada/Ontario/Toronto | The Hosptial for Sick Children | The Hospital for Sick Children | Aaron Campigotto et al |
| EPI_ISL_791512 | Canada/ON-SK-20H-329MM0281/202 | 2020-11-24 | North America/Canada/Ontario/Toronto | The Hosptial for Sick Children | The Hospital for Sick Children | Aaron Campigotto et al |
| EPI_ISL_791513 | Canada/ON-SK-20H-329MM0297/202 | 2020-11-24 | North America/Canada/Ontario/Toronto | The Hosptial for Sick Children | The Hospital for Sick Children | Aaron Campigotto et al |
| EPI_ISL_791517 | USA/MA-MASPHL-00010/2020 | 2020-03-07 | North America/USA/Massachusetts/ | Massachusetts State Public Health Laboratory | Infectious Disease Program, Broad Institute of Harvard and MIT | Lemieux et al |
| EPI_ISL_791521 | USA/MA-MASPHL-00015/2020 | 2020-03-30 | North America/USA/Massachusetts/ | Massachusetts State Public Health Laboratory | Infectious Disease Program, Broad Institute of Harvard and MIT | Lemieux et al |
| EPI_ISL_791525 | USA/MA-MASPHL-00019/2020 | 2020-04-01 | North America/USA/Massachusetts/ | Massachusetts State Public Health Laboratory | Infectious Disease Program, Broad Institute of Harvard and MIT | Lemieux et al |
| EPI_ISL_791527 | USA/MA-MASPHL-00021/2020 | 2020-04-02 | North America/USA/Massachusetts/ | Massachusetts State Public Health Laboratory | Infectious Disease Program, Broad Institute of Harvard and MIT | Lemieux et al |
| EPI_ISL_791537 | USA/MA-MASPHL-00032/2020 | 2020-04-02 | North America/USA/Massachusetts/ | Massachusetts State Public Health Laboratory | Infectious Disease Program, Broad Institute of Harvard and MIT | Lemieux et al |
| EPI_ISL_791553 | USA/MA-MASPHL-00053/2020 | 2020-04-03 | North America/USA/Massachusetts/ | Massachusetts State Public Health Laboratory | Infectious Disease Program, Broad Institute of Harvard and MIT | Lemieux et al |
| EPI_ISL_791590 | USA/MA-MASPHL-00108/2020 | 2020-04-03 | North America/USA/Massachusetts/ | Massachusetts State Public Health Laboratory | Infectious Disease Program, Broad Institute of Harvard and MIT | Lemieux et al |

| | | | | | | |
|---|---|---|---|---|---|---|
| EPI_ISL_791591 | USA/MA-MASPHL-00109/2020 | 2020-04-03 | North America/USA/Massachusetts/ | Massachusetts State Public Health Laboratory | Infectious Disease Program, Broad Institute of Harvard and MIT | Lemieux et al |
| EPI_ISL_791608 | USA/MA-MASPHL-00134/2020 | 2020-04-03 | North America/USA/Massachusetts/ | Massachusetts State Public Health Laboratory | Infectious Disease Program, Broad Institute of Harvard and MIT | Lemieux et al |
| EPI_ISL_791622 | USA/MA-MASPHL-00151/2020 | 2020-04-03 | North America/USA/Massachusetts/ | Massachusetts State Public Health Laboratory | Infectious Disease Program, Broad Institute of Harvard and MIT | Lemieux et al |
| EPI_ISL_791628 | USA/MA-MASPHL-00163/2020 | 2020-04-03 | North America/USA/Massachusetts/ | Massachusetts State Public Health Laboratory | Infectious Disease Program, Broad Institute of Harvard and MIT | Lemieux et al |
| EPI_ISL_791632 | USA/MA-MASPHL-00167/2020 | 2020-04-03 | North America/USA/Massachusetts/ | Massachusetts State Public Health Laboratory | Infectious Disease Program, Broad Institute of Harvard and MIT | Lemieux et al |
| EPI_ISL_791644 | USA/MA-MASPHL-00185/2020 | 2020-04-06 | North America/USA/Massachusetts/ | Massachusetts State Public Health Laboratory | Infectious Disease Program, Broad Institute of Harvard and MIT | Lemieux et al |
| EPI_ISL_791654 | USA/MA-MASPHL-00199/2020 | 2020-04-07 | North America/USA/Massachusetts/ | Massachusetts State Public Health Laboratory | Infectious Disease Program, Broad Institute of Harvard and MIT | Lemieux et al |
| EPI_ISL_791655 | USA/MA-MASPHL-00200/2020 | 2020-04-07 | North America/USA/Massachusetts/ | Massachusetts State Public Health Laboratory | Infectious Disease Program, Broad Institute of Harvard and MIT | Lemieux et al |
| EPI_ISL_791657 | USA/MA-MASPHL-00203/2020 | 2020-04-09 | North America/USA/Massachusetts/ | Massachusetts State Public Health Laboratory | Infectious Disease Program, Broad Institute of Harvard and MIT | Lemieux et al |
| EPI_ISL_791678 | USA/MA-MASPHL-00232/2020 | 2020-04-16 | North America/USA/Massachusetts/ | Massachusetts State Public Health Laboratory | Infectious Disease Program, Broad Institute of Harvard and MIT | Lemieux et al |

| | | | | | | |
|---|---|---|---|---|---|---|
| EPI_ISL_791689 | USA/MA-MASPHL-00246/2020 | 2020-04-16 | North America/USA/Massachusetts/ | Massachusetts State Public Health Laboratory | Infectious Disease Program, Broad Institute of Harvard and MIT | Lemieux et al |
| EPI_ISL_791700 | USA/MA-MASPHL-00266/2020 | 2020-04-17 | North America/USA/Massachusetts/ | Massachusetts State Public Health Laboratory | Infectious Disease Program, Broad Institute of Harvard and MIT | Lemieux et al |
| EPI_ISL_791702 | USA/MA-MASPHL-00270/2020 | 2020-04-17 | North America/USA/Massachusetts/ | Massachusetts State Public Health Laboratory | Infectious Disease Program, Broad Institute of Harvard and MIT | Lemieux et al |
| EPI_ISL_791706 | USA/MA-MASPHL-00287/2020 | 2020-04-17 | North America/USA/Massachusetts/ | Massachusetts State Public Health Laboratory | Infectious Disease Program, Broad Institute of Harvard and MIT | Lemieux et al |
| EPI_ISL_791716 | USA/MA-MASPHL-00306/2020 | 2020-04-18 | North America/USA/Massachusetts/ | Massachusetts State Public Health Laboratory | Infectious Disease Program, Broad Institute of Harvard and MIT | Lemieux et al |
| EPI_ISL_791720 | USA/MA-MASPHL-00311/2020 | 2020-04-18 | North America/USA/Massachusetts/ | Massachusetts State Public Health Laboratory | Infectious Disease Program, Broad Institute of Harvard and MIT | Lemieux et al |
| EPI_ISL_791722 | USA/MA-MASPHL-00315/2020 | 2020-04-18 | North America/USA/Massachusetts/ | Massachusetts State Public Health Laboratory | Infectious Disease Program, Broad Institute of Harvard and MIT | Lemieux et al |
| EPI_ISL_791723 | USA/MA-MASPHL-00317/2020 | 2020-04-18 | North America/USA/Massachusetts/ | Massachusetts State Public Health Laboratory | Infectious Disease Program, Broad Institute of Harvard and MIT | Lemieux et al |
| EPI_ISL_791729 | USA/MA-MGH-00727/2020 | 2020-04-08 | North America/USA/Massachusetts/ | Massachusetts General Hospital | Infectious Disease Program, Broad Institute of Harvard and MIT | Lemieux et al |
| EPI_ISL_791754 | USA/MA-MGH-00764/2020 | 2020-04-11 | North America/USA/Massachusetts/ | Massachusetts General Hospital | Infectious Disease Program, Broad Institute of Harvard and MIT | Lemieux et al |

| | | | | | | |
|---|---|---|---|---|---|---|
| EPI_ISL_791757 | USA/MA-MGH-00768/2020 | 2020-04-11 | North America/USA/Massachusetts/ | Massachusetts General Hospital | Infectious Disease Program, Broad Institute of Harvard and MIT | Lemieux et al |
| EPI_ISL_791762 | USA/MA-MGH-00779/2020 | 2020-04-12 | North America/USA/Massachusetts/ | Massachusetts General Hospital | Infectious Disease Program, Broad Institute of Harvard and MIT | Lemieux et al |
| EPI_ISL_791779 | USA/MA-MGH-00822/2020 | 2020-04-13 | North America/USA/Massachusetts/ | Massachusetts General Hospital | Infectious Disease Program, Broad Institute of Harvard and MIT | Lemieux et al |
| EPI_ISL_791789 | USA/MA-MGH-00834/2020 | 2020-04-13 | North America/USA/Massachusetts/ | Massachusetts General Hospital | Infectious Disease Program, Broad Institute of Harvard and MIT | Lemieux et al |
| EPI_ISL_791800 | USA/MA-MGH-00864/2020 | 2020-04-14 | North America/USA/Massachusetts/ | Massachusetts General Hospital | Infectious Disease Program, Broad Institute of Harvard and MIT | Lemieux et al |
| EPI_ISL_791803 | USA/MA-MGH-00869/2020 | 2020-04-14 | North America/USA/Massachusetts/ | Massachusetts General Hospital | Infectious Disease Program, Broad Institute of Harvard and MIT | Lemieux et al |
| EPI_ISL_791807 | USA/MA-MGH-00876/2020 | 2020-04-14 | North America/USA/Massachusetts/ | Massachusetts General Hospital | Infectious Disease Program, Broad Institute of Harvard and MIT | Lemieux et al |
| EPI_ISL_791817 | USA/MA-MGH-00906/2020 | 2020-04-17 | North America/USA/Massachusetts/ | Massachusetts General Hospital | Infectious Disease Program, Broad Institute of Harvard and MIT | Lemieux et al |
| EPI_ISL_791823 | USA/MA-MGH-00926/2020 | 2020-05-02 | North America/USA/Massachusetts/ | Massachusetts General Hospital | Infectious Disease Program, Broad Institute of Harvard and MIT | Lemieux et al |
| EPI_ISL_791829 | USA/MA-MGH-00945/2020 | 2020-05-03 | North America/USA/Massachusetts/ | Massachusetts General Hospital | Infectious Disease Program, Broad Institute of Harvard and MIT | Lemieux et al |

| | | | | | | |
|---|---|---|---|---|---|---|
| EPI_ISL_791831 | USA/MA-MGH-00947/2020 | 2020-05-02 | North America/USA/Massachusetts/ | Massachusetts General Hospital | Infectious Disease Program, Broad Institute of Harvard and MIT | Lemieux et al |
| EPI_ISL_791835 | USA/MA-MGH-00959/2020 | 2020-05-03 | North America/USA/Massachusetts/ | Massachusetts General Hospital | Infectious Disease Program, Broad Institute of Harvard and MIT | Lemieux et al |
| EPI_ISL_791853 | USA/MA-MGH-01045/2020 | 2020-05-01 | North America/USA/Massachusetts/ | Massachusetts General Hospital | Infectious Disease Program, Broad Institute of Harvard and MIT | Lemieux et al |
| EPI_ISL_791866 | USA/MA-MGH-01080/2020 | 2020-05-01 | North America/USA/Massachusetts/ | Massachusetts General Hospital | Infectious Disease Program, Broad Institute of Harvard and MIT | Lemieux et al |
| EPI_ISL_791868 | USA/MA-MGH-01084/2020 | 2020-05-01 | North America/USA/Massachusetts/ | Massachusetts General Hospital | Infectious Disease Program, Broad Institute of Harvard and MIT | Lemieux et al |
| EPI_ISL_791874 | USA/MA-MGH-01103/2020 | 2020-05-01 | North America/USA/Massachusetts/ | Massachusetts General Hospital | Infectious Disease Program, Broad Institute of Harvard and MIT | Lemieux et al |
| EPI_ISL_791878 | USA/MA-MGH-01110/2020 | 2020-05-01 | North America/USA/Massachusetts/ | Massachusetts General Hospital | Infectious Disease Program, Broad Institute of Harvard and MIT | Lemieux et al |
| EPI_ISL_791883 | USA/MA-MGH-01121/2020 | 2020-05-02 | North America/USA/Massachusetts/ | Massachusetts General Hospital | Infectious Disease Program, Broad Institute of Harvard and MIT | Lemieux et al |
| EPI_ISL_791885 | USA/MA-MGH-01127/2020 | 2020-05-02 | North America/USA/Massachusetts/ | Massachusetts General Hospital | Infectious Disease Program, Broad Institute of Harvard and MIT | Lemieux et al |
| EPI_ISL_791888 | USA/MA-MGH-01140/2020 | 2020-05-02 | North America/USA/Massachusetts/ | Massachusetts General Hospital | Infectious Disease Program, Broad Institute of Harvard and MIT | Lemieux et al |

| | | | | | | |
|---|---|---|---|---|---|---|
| EPI_ISL_791899 | USA/MA-MGH-01182/2020 | 2020-04-29 | North America/USA/Massachusetts/ | Massachusetts General Hospital | Infectious Disease Program, Broad Institute of Harvard and MIT | Lemieux et al |
| EPI_ISL_791900 | USA/MA-MGH-01183/2020 | 2020-04-29 | North America/USA/Massachusetts/ | Massachusetts General Hospital | Infectious Disease Program, Broad Institute of Harvard and MIT | Lemieux et al |
| EPI_ISL_791903 | USA/MA-MGH-01196/2020 | 2020-05-03 | North America/USA/Massachusetts/ | Massachusetts General Hospital | Infectious Disease Program, Broad Institute of Harvard and MIT | Lemieux et al |
| EPI_ISL_791906 | USA/MA-MGH-01207/2020 | 2020-05-04 | North America/USA/Massachusetts/ | Massachusetts General Hospital | Infectious Disease Program, Broad Institute of Harvard and MIT | Lemieux et al |
| EPI_ISL_791908 | USA/MA-MGH-01232/2020 | 2020-05-04 | North America/USA/Massachusetts/ | Massachusetts General Hospital | Infectious Disease Program, Broad Institute of Harvard and MIT | Lemieux et al |
| EPI_ISL_791916 | USA/MA-MGH-01284/2020 | 2020-05-05 | North America/USA/Massachusetts/ | Massachusetts General Hospital | Infectious Disease Program, Broad Institute of Harvard and MIT | Lemieux et al |
| EPI_ISL_791919 | USA/MA-MGH-01297/2020 | 2020-05-05 | North America/USA/Massachusetts/ | Massachusetts General Hospital | Infectious Disease Program, Broad Institute of Harvard and MIT | Lemieux et al |
| EPI_ISL_791929 | USA/MA-MGH-01343/2020 | 2020-05-05 | North America/USA/Massachusetts/ | Massachusetts General Hospital | Infectious Disease Program, Broad Institute of Harvard and MIT | Lemieux et al |
| EPI_ISL_791935 | USA/MA-MGH-01368/2020 | 2020-05-06 | North America/USA/Massachusetts/ | Massachusetts General Hospital | Infectious Disease Program, Broad Institute of Harvard and MIT | Lemieux et al |
| EPI_ISL_791951 | USA/MA-MGH-01452/2020 | 2020-04-30 | North America/USA/Massachusetts/ | Massachusetts General Hospital | Infectious Disease Program, Broad Institute of Harvard and MIT | Lemieux et al |

| | | | | | | |
|---|---|---|---|---|---|---|
| EPI_ISL_791973 | USA/MA-MGH-01606/2020 | 2020-05-08 | North America/USA/Massachusetts/ | Massachusetts General Hospital | Infectious Disease Program, Broad Institute of Harvard and MIT | Lemieux et al |
| EPI_ISL_791977 | Indonesia/KI-NIHRD-PME5218/2020 | 2020-08-11 | Asia/Indonesia/East Kalimantan/ | Laboratorium Mikrobiologi Samarinda | National Institute of Health Research and Development | Nugraha et al |
| EPI_ISL_791978 | Indonesia/LA-NIHRD-PME1024/2020 | 2020-05-23 | Asia/Indonesia/Lampung/ | Balai Labkes Lampung | National Institute of Health Research and Development | Nugraha et al |
| EPI_ISL_791979 | Indonesia/BE-NIHRD-PME9915/2020 | 2020-10-19 | Asia/Indonesia/Bengkulu/ | RSUD Curup Bengkulu | National Institute of Health Research and Development | Nugraha et al |
| EPI_ISL_791980 | Indonesia/JI-NIHRD-PME9750/2020 | 2020-11-09 | Asia/Indonesia/East Java/ | RSUD Blambangan Banyuwangi | National Institute of Health Research and Development | Ikawati et al |
| EPI_ISL_791981 | Indonesia/AC-NIHRD-PME4523/2020 | 2020-06-18 | Asia/Indonesia/Aceh/ | Balai Litbang Aceh | National Institute of Health Research and Development | Ikawati et al |
| EPI_ISL_791983 | Indonesia/KI-NIHRD-PME5182/2020 | 2020-08-11 | Asia/Indonesia/East Kalimantan/ | RSUD dr. Kanujoso Djatiwibowo Balikpapan | National Institute of Health Research and Development | Subangkit et al |
| EPI_ISL_791985 | Indonesia/KR-NIHRD-PME9874/2020 | 2020-11-11 | Asia/Indonesia/Riau Islands/ | RS Santa Maria Pekanbaru | National Institute of Health Research and Development | Puspa et al |
| EPI_ISL_791986 | Indonesia/MU-NIHRD-PME8242/2020 | 2020-10-09 | Asia/Indonesia/North Maluku/ | RSUD Morotai Maluku Utara | National Institute of Health Research and Development | Subangkit et al |
| EPI_ISL_791987 | Indonesia/JT-NIHRD-PME3225/2020 | 2020-07-17 | Asia/Indonesia/Central Java/ | RSUD dr. Loekmono Hadi Kudus | National Institute of Health Research and Development | Puspa et al |

| | | | | | | |
|---|---|---|---|---|---|---|
| EPI_ISL_791988 | Indonesia/JT-NIHRD-PME9517/2020 | 2020-10-13 | Asia/Indonesia/Central Java/ | Balai Besar POM Semarang | National Institute of Health Research and Development | Subangkit et al |
| EPI_ISL_791989 | Indonesia/JI-NIHRD-PME10137/2020 | 2020-10-03 | Asia/Indonesia/East Java/ | RSUD Waluyo Jati Kota Kraksaan, Jawa Timur | National Institute of Health Research and Development | Subangkit et al |
| EPI_ISL_791990 | Slovakia/UKBA-605/2020 | 2020-11-23 | Europe/Slovakia/Doƒæany/ | Institute of Virology, Biomedical Research Center of the Slovak Academy of Sciences, Bratislava | Faculty of Natural Sciences, Comenius University, Bratislava | Viktv≥ria ƒåabanovv° et al |
| EPI_ISL_791992 | France/BRE-IPP11274/2020 | 2020-11-21 | Europe/France/Bretagne/Brest | CHU - Hv¥pital Cavale Blanche | National Reference Center for Viruses of Respiratory Infections, Institut Pasteur, Paris | Marion Barbet et al |
| EPI_ISL_792002 | France/GES-IPP10943/2020 | 2020-12-18 | Europe/France/Grand Est/Champagne A | Laboratoire de Virologie Hv¥pital Robert Debrv© | National Reference Center for Viruses of Respiratory Infections, Institut Pasteur, Paris | Marion Barbet et al |
| EPI_ISL_792006 | France/GES-IPP10974/2020 | 2020-12-18 | Europe/France/Grand Est/Champagne A | Laboratoire de Virologie Hv¥pital Robert Debrv© | National Reference Center for Viruses of Respiratory Infections, Institut Pasteur, Paris | Marion Barbet et al |
| EPI_ISL_792009 | France/HDF-IPP11316/2020 | 2020-12-18 | Europe/France/Hauts de France/Nord Pa | CH de Bethune - Laboratoire de Biologie Mv©dicale | National Reference Center for Viruses of Respiratory Infections, Institut Pasteur, Paris | Marion Barbet et al |

| | | | | | | |
|---|---|---|---|---|---|---|
| EPI_ISL_792016 | France/IDF-IPP10913/2020 | 2020-12-10 | Europe/France/Ile de France/Paris | Plateforme COVID IDF | National Reference Center for Viruses of Respiratory Infections, Institut Pasteur, Paris | Marion Barbet et al |
| EPI_ISL_792018 | France/IDF-IPP10919/2020 | 2020-10-20 | Europe/France/Ile de France/Paris | Plateforme COVID IDF | National Reference Center for Viruses of Respiratory Infections, Institut Pasteur, Paris | Marion Barbet et al |
| EPI_ISL_792019 | France/IDF-IPP10921/2020 | 2020-10-24 | Europe/France/Ile de France/Paris | Plateforme COVID IDF | National Reference Center for Viruses of Respiratory Infections, Institut Pasteur, Paris | Marion Barbet et al |
| EPI_ISL_792020 | France/IDF-IPP10922/2020 | 2020-10-21 | Europe/France/Ile de France/Paris | Plateforme COVID IDF | National Reference Center for Viruses of Respiratory Infections, Institut Pasteur, Paris | Marion Barbet et al |
| EPI_ISL_792021 | France/IDF-IPP10923/2020 | 2020-10-07 | Europe/France/Ile de France/Paris | Plateforme COVID IDF | National Reference Center for Viruses of Respiratory Infections, Institut Pasteur, Paris | Marion Barbet et al |
| EPI_ISL_792022 | France/IDF-IPP10924/2020 | 2020-10-10 | Europe/France/Ile de France/Paris | Plateforme COVID IDF | National Reference Center for Viruses of Respiratory Infections, Institut Pasteur, Paris | Marion Barbet et al |

| | | | | | | |
|---|---|---|---|---|---|---|
| EPI_ISL_792023 | France/IDF-IPP10927/2020 | 2020-09-11 | Europe/France/Ile de France/Paris | Plateforme COVID IDF | National Reference Center for Viruses of Respiratory Infections, Institut Pasteur, Paris | Marion Barbet et al |
| EPI_ISL_792051 | France/GE-IPP11360/2020 | 2020-12-21 | Europe/France/Grand Est/Epinal | General practitioner | National Reference Center for Viruses of Respiratory Infections, Institut Pasteur, Paris | Marion Barbet et al |
| EPI_ISL_792056 | France/GE-IPP11373/2020 | 2020-12-26 | Europe/France/Grand Est/Troyes | Hopital | National Reference Center for Viruses of Respiratory Infections, Institut Pasteur, Paris | Marion Barbet et al |
| EPI_ISL_792057 | France/GE-IPP11375/2020 | 2020-12-21 | Europe/France/Grand Est/Troyes | Hopital | National Reference Center for Viruses of Respiratory Infections, Institut Pasteur, Paris | Marion Barbet et al |
| EPI_ISL_792059 | Canada/ON-A22/2020 | 2020-11-24 | North America/Canada/Ontario/Brampt | Toronto Invasive Bacterial Diseases Network | McMaster University | David Richardson et al |
| EPI_ISL_792060 | Canada/ON-A33/2020 | 2020-11-24 | North America/Canada/Ontario/Brampt | Toronto Invasive Bacterial Diseases Network | McMaster University | David Richardson et al |
| EPI_ISL_792061 | Canada/ON-A34/2020 | 2020-11-24 | North America/Canada/Ontario/Brampt | Toronto Invasive Bacterial Diseases Network | McMaster University | David Richardson et al |
| EPI_ISL_792062 | Canada/ON-A36/2020 | 2020-11-24 | North America/Canada/Ontario/Brampt | Toronto Invasive Bacterial Diseases Network | McMaster University | David Richardson et al |
| EPI_ISL_792063 | Canada/ON-A37/2020 | 2020-11-24 | North America/Canada/Ontario/Brampt | Toronto Invasive Bacterial Diseases Network | McMaster University | David Richardson et al |
| EPI_ISL_792064 | Canada/ON-A38/2020 | 2020-11-24 | North America/Canada/Ontario/Brampt | Toronto Invasive Bacterial Diseases Network | McMaster University | David Richardson et al |
| EPI_ISL_792065 | Canada/ON-A39/2020 | 2020-11-24 | North America/Canada/Ontario/Brampt | Toronto Invasive Bacterial Diseases Network | McMaster University | David Richardson et al |
| EPI_ISL_792066 | Canada/ON-A41/2020 | 2020-11-24 | North America/Canada/Ontario/Brampt | Toronto Invasive Bacterial Diseases Network | McMaster University | David Richardson et al |

| | | | | | | |
|---|---|---|---|---|---|---|
| EPI_ISL_792067 | Canada/ON-A42/2020 | 2020-11-24 | North America/Canada/Ontario/Brampt | Toronto Invasive Bacterial Diseases Network | McMaster University | David Richardson et al |
| EPI_ISL_792068 | Canada/ON-A43/2020 | 2020-11-24 | North America/Canada/Ontario/Brampt | Toronto Invasive Bacterial Diseases Network | McMaster University | David Richardson et al |
| EPI_ISL_792069 | Canada/ON-A44/2020 | 2020-11-24 | North America/Canada/Ontario/Brampt | Toronto Invasive Bacterial Diseases Network | McMaster University | David Richardson et al |
| EPI_ISL_792070 | Canada/ON-A45/2020 | 2020-11-24 | North America/Canada/Ontario/Brampt | Toronto Invasive Bacterial Diseases Network | McMaster University | David Richardson et al |
| EPI_ISL_792071 | Canada/ON-A46/2020 | 2020-11-24 | North America/Canada/Ontario/Brampt | Toronto Invasive Bacterial Diseases Network | McMaster University | David Richardson et al |
| EPI_ISL_792073 | Canada/ON-A51/2020 | 2020-11-24 | North America/Canada/Ontario/Brampt | Toronto Invasive Bacterial Diseases Network | McMaster University | David Richardson et al |
| EPI_ISL_792074 | Canada/ON-A53/2020 | 2020-11-24 | North America/Canada/Ontario/Brampt | Toronto Invasive Bacterial Diseases Network | McMaster University | David Richardson et al |
| EPI_ISL_792075 | Canada/ON-A54/2020 | 2020-11-24 | North America/Canada/Ontario/Brampt | Toronto Invasive Bacterial Diseases Network | McMaster University | David Richardson et al |
| EPI_ISL_792076 | Canada/ON-A55/2020 | 2020-11-24 | North America/Canada/Ontario/Brampt | Toronto Invasive Bacterial Diseases Network | McMaster University | David Richardson et al |
| EPI_ISL_792077 | Canada/ON-A57/2020 | 2020-11-24 | North America/Canada/Ontario/Brampt | Toronto Invasive Bacterial Diseases Network | McMaster University | David Richardson et al |
| EPI_ISL_792078 | Canada/ON-A58/2020 | 2020-11-24 | North America/Canada/Ontario/Brampt | Toronto Invasive Bacterial Diseases Network | McMaster University | David Richardson et al |
| EPI_ISL_792079 | Canada/ON-A59/2020 | 2020-11-24 | North America/Canada/Ontario/Brampt | Toronto Invasive Bacterial Diseases Network | McMaster University | David Richardson et al |
| EPI_ISL_792080 | Canada/ON-A60/2020 | 2020-11-24 | North America/Canada/Ontario/Brampt | Toronto Invasive Bacterial Diseases Network | McMaster University | David Richardson et al |
| EPI_ISL_792081 | Canada/ON-A61/2020 | 2020-11-24 | North America/Canada/Ontario/Brampt | Toronto Invasive Bacterial Diseases Network | McMaster University | David Richardson et al |
| EPI_ISL_792082 | Canada/ON-A62/2020 | 2020-11-24 | North America/Canada/Ontario/Brampt | Toronto Invasive Bacterial Diseases Network | McMaster University | David Richardson et al |
| EPI_ISL_792083 | Canada/ON-A64/2020 | 2020-11-24 | North America/Canada/Ontario/Brampt | Toronto Invasive Bacterial Diseases Network | McMaster University | David Richardson et al |
| EPI_ISL_792084 | Canada/ON-S2033/2020 | 2020-12-27 | North America/Canada/Ontario/Toronto | Toronto Invasive Bacterial Diseases Network | McMaster University | Allison McGeer et al |
| EPI_ISL_792085 | Canada/ON-S2034/2020 | 2020-12-28 | North America/Canada/Ontario/Toronto | Toronto Invasive Bacterial Diseases Network | McMaster University | Allison McGeer et al |
| EPI_ISL_792086 | Canada/ON-S2036/2020 | 2020-12-25 | North America/Canada/Ontario/Toronto | Toronto Invasive Bacterial Diseases Network | McMaster University | Allison McGeer et al |
| EPI_ISL_792087 | Canada/ON-S2037/2020 | 2020-12-25 | North America/Canada/Ontario/Toronto | Toronto Invasive Bacterial Diseases Network | McMaster University | Allison McGeer et al |

| | | | | | | |
|---|---|---|---|---|---|---|
| EPI_ISL_792088 | Canada/ON-S2038/2020 | 2020-12-30 | North America/Canada/Ontario/Toronto | Toronto Invasive Bacterial Diseases Network | McMaster University | Allison McGeer et al |
| EPI_ISL_792089 | Canada/ON-S2040/2021 | 2021-01-04 | North America/Canada/Ontario/Toronto | Toronto Invasive Bacterial Diseases Network | McMaster University | Allison McGeer et al |
| EPI_ISL_792090 | Canada/ON-S2041/2021 | 2021-01-05 | North America/Canada/Ontario/Toronto | Toronto Invasive Bacterial Diseases Network | McMaster University | Allison McGeer et al |
| EPI_ISL_792091 | Canada/ON-S955/2020 | 2020-03-28 | North America/Canada/Ontario/Toronto | Toronto Invasive Bacterial Diseases Network | McMaster University | Allison McGeer et al |
| EPI_ISL_792093 | USA/WA-UW-133/2020 | 2020-03-06 | North America/USA/Washington/ | UW Virology Lab | UW Virology Lab | Pavitra Roychoudhury et al |
| EPI_ISL_792112 | Brazil/SP-833/2020 | 2020-04-23 | South America/Brazil/S√£o Paulo/S√£o I | Instituto Adolfo Lutz - Central | Instituto Adolfo Lutz, Interdiciplinary Procedures Center, Strategic Laboratory | Claudio Tavares Sacchi et al |
| EPI_ISL_792114 | Brazil/SP-841/2020 | 2020-05-03 | South America/Brazil/S√£o Paulo/S√£o I | Instituto Adolfo Lutz - Central | Instituto Adolfo Lutz, Interdiciplinary Procedures Center, Strategic Laboratory | Claudio Tavares Sacchi et al |
| EPI_ISL_792115 | Brazil/SP-778/2020 | 2020-06-29 | South America/Brazil/S√£o Paulo/Carag | Instituto Adolfo Lutz - Regional de Taubate | Instituto Adolfo Lutz, Interdiciplinary Procedures Center, Strategic Laboratory | Claudio Tavares Sacchi et al |
| EPI_ISL_792116 | Brazil/SP-779/2020 | 2020-07-02 | South America/Brazil/S√£o Paulo/Santa | Instituto Adolfo Lutz - Regional de Taubate | Instituto Adolfo Lutz, Interdiciplinary Procedures Center, Strategic Laboratory | Claudio Tavares Sacchi et al |

| | | | | | | |
|---|---|---|---|---|---|---|
| EPI_ISL_792130 | Argentina/PAIS-A0041/2020 | 2020-04-28 | South America/Argentina/Buenos Aires/ | Laboratorio de Virologv≠a del Hospital de Niv±os Dr. Ricardo Gutierrez | √Årea de Secuenciaciv≥n del Laboratorio de Virologv≠a del Hospital de Niv±os Dr. Ricardo Gutierrez on behalf of 'Proyecto Argentino Interinstitucional de genomica de SARS-CoV-2' (PAIS Consortium) | Nabaes Jodar et al |
| EPI_ISL_792140 | Argentina/PAIS-A0051/2020 | 2020-05-01 | South America/Argentina/Buenos Aires/ | Laboratorio de Virologv≠a del Hospital de Niv±os Dr. Ricardo Gutierrez | √Årea de Secuenciaciv≥n del Laboratorio de Virologv≠a del Hospital de Niv±os Dr. Ricardo Gutierrez on behalf of 'Proyecto Argentino Interinstitucional de genomica de SARS-CoV-2' (PAIS Consortium) | Nabaes Jodar et al |

| | | | | | | |
|---|---|---|---|---|---|---|
| EPI_ISL_792149 | Argentina/PAIS-A0060/2020 | 2020-05-03 | South America/Argentina/Buenos Aires/ | Laboratorio de Virolog√≠a del Hospital de Ni√±os Dr. Ricardo Gutierrez | √Årea de Secuenciaci√≥n del Laboratorio de Virolog√≠a del Hospital de Ni√±os Dr. Ricardo Gutierrez on behalf of 'Proyecto Argentino Interinstitucional de genomica de SARS-CoV-2' (PAIS Consortium) | Nabaes Jodar et al |
| EPI_ISL_792153 | Argentina/PAIS-A0064/2020 | 2020-05-03 | South America/Argentina/Buenos Aires/ | Laboratorio de Virolog√≠a del Hospital de Ni√±os Dr. Ricardo Gutierrez | √Årea de Secuenciaci√≥n del Laboratorio de Virolog√≠a del Hospital de Ni√±os Dr. Ricardo Gutierrez on behalf of 'Proyecto Argentino Interinstitucional de genomica de SARS-CoV-2' (PAIS Consortium) | Nabaes Jodar et al |

| | | | | | | |
|---|---|---|---|---|---|---|
| EPI_ISL_792154 | Argentina/PAIS-A0065/2020 | 2020-05-03 | South America/Argentina/Buenos Aires/ | Laboratorio de Virologv≠a del Hospital de Niv±os Dr. Ricardo Gutierrez | √Årea de Secuenciaci√≥n del Laboratorio de Virologv≠a del Hospital de Niv±os Dr. Ricardo Gutierrez on behalf of 'Proyecto Argentino Interinstitucional de genomica de SARS-CoV-2' (PAIS Consortium) | Nabaes Jodar et al |
| EPI_ISL_792158 | Argentina/PAIS-A0069/2020 | 2020-05-03 | South America/Argentina/Buenos Aires/ | Laboratorio de Virologv≠a del Hospital de Niv±os Dr. Ricardo Gutierrez | √Årea de Secuenciaci√≥n del Laboratorio de Virologv≠a del Hospital de Niv±os Dr. Ricardo Gutierrez on behalf of 'Proyecto Argentino Interinstitucional de genomica de SARS-CoV-2' (PAIS Consortium) | Nabaes Jodar et al |

| | | | | | | |
|---|---|---|---|---|---|---|
| EPI_ISL_792160 | Argentina/PAIS-A0071/2020 | 2020-05-03 | South America/Argentina/Buenos Aires/ | Laboratorio de Virologv≠a del Hospital de Niv±os Dr. Ricardo Gutierrez | vÅrea de Secuenciaciv≥n del Laboratorio de Virologv≠a del Hospital de Niv±os Dr. Ricardo Gutierrez on behalf of 'Proyecto Argentino Interinstitucional de genomica de SARS-CoV-2' (PAIS Consortium) | Nabaes Jodar et al |
| EPI_ISL_792186 | Argentina/PAIS-A0097/2020 | 2020-05-09 | South America/Argentina/Buenos Aires/ | Laboratorio de Virologv≠a del Hospital de Niv±os Dr. Ricardo Gutierrez | vÅrea de Secuenciaciv≥n del Laboratorio de Virologv≠a del Hospital de Niv±os Dr. Ricardo Gutierrez on behalf of 'Proyecto Argentino Interinstitucional de genomica de SARS-CoV-2' (PAIS Consortium) | Nabaes Jodar et al |

| | | | | | | |
|---|---|---|---|---|---|---|
| EPI_ISL_792203 | Argentina/PAIS-A0114/2020 | 2020-05-11 | South America/Argentina/Buenos Aires/ | Laboratorio de Virolog√≠a del Hospital de Ni√±os Dr. Ricardo Gutierrez | √Årea de Secuenciaci√≥n del Laboratorio de Virolog√≠a del Hospital de Ni√±os Dr. Ricardo Gutierrez on behalf of 'Proyecto Argentino Interinstitucional de genomica de SARS-CoV-2' (PAIS Consortium) | Nabaes Jodar et al |
| EPI_ISL_792212 | Argentina/PAIS-A0123/2020 | 2020-05-12 | South America/Argentina/Buenos Aires/ | Laboratorio de Virolog√≠a del Hospital de Ni√±os Dr. Ricardo Gutierrez | √Årea de Secuenciaci√≥n del Laboratorio de Virolog√≠a del Hospital de Ni√±os Dr. Ricardo Gutierrez on behalf of 'Proyecto Argentino Interinstitucional de genomica de SARS-CoV-2' (PAIS Consortium) | Nabaes Jodar et al |

| | | | | | | |
|---|---|---|---|---|---|---|
| EPI_ISL_792213 | Argentina/PAIS-A0124/2020 | 2020-05-12 | South America/Argentina/Buenos Aires/ | Laboratorio de Virologv≠a del Hospital de Niv±os Dr. Ricardo Gutierrez | √Årea de Secuenciaciv≥n del Laboratorio de Virologv≠a del Hospital de Niv±os Dr. Ricardo Gutierrez on behalf of 'Proyecto Argentino Interinstitucional de genomica de SARS-CoV-2' (PAIS Consortium) | Nabaes Jodar et al |
| EPI_ISL_792249 | Argentina/PAIS-A0160/2020 | 2020-05-18 | South America/Argentina/Buenos Aires/ | Laboratorio de Virologv≠a del Hospital de Niv±os Dr. Ricardo Gutierrez | √Årea de Secuenciaciv≥n del Laboratorio de Virologv≠a del Hospital de Niv±os Dr. Ricardo Gutierrez on behalf of 'Proyecto Argentino Interinstitucional de genomica de SARS-CoV-2' (PAIS Consortium) | Nabaes Jodar et al |

| | | | | | | |
|---|---|---|---|---|---|---|
| EPI_ISL_792257 | Argentina/PAIS-A0168/2020 | 2020-05-19 | South America/Argentina/Buenos Aires/ | Laboratorio de Virolog√≠a del Hospital de Ni√±os Dr. Ricardo Gutierrez | √Årea de Secuenciaci√≥n del Laboratorio de Virolog√≠a del Hospital de Ni√±os Dr. Ricardo Gutierrez on behalf of 'Proyecto Argentino Interinstitucional de genomica de SARS-CoV-2' (PAIS Consortium) | Nabaes Jodar et al |
| EPI_ISL_792265 | Argentina/PAIS-A0176/2020 | 2020-05-19 | South America/Argentina/Buenos Aires/ | Laboratorio de Virolog√≠a del Hospital de Ni√±os Dr. Ricardo Gutierrez | √Årea de Secuenciaci√≥n del Laboratorio de Virolog√≠a del Hospital de Ni√±os Dr. Ricardo Gutierrez on behalf of 'Proyecto Argentino Interinstitucional de genomica de SARS-CoV-2' (PAIS Consortium) | Nabaes Jodar et al |

| | | | | | | |
|---|---|---|---|---|---|---|
| EPI_ISL_792267 | Argentina/PAIS-A0178/2020 | 2020-05-19 | South America/Argentina/Buenos Aires/ | Laboratorio de Virolog√≠a del Hospital de Ni√±os Dr. Ricardo Gutierrez | √Årea de Secuenciaci√≥n del Laboratorio de Virolog√≠a del Hospital de Ni√±os Dr. Ricardo Gutierrez on behalf of 'Proyecto Argentino Interinstitucional de genomica de SARS-CoV-2' (PAIS Consortium) | Nabaes Jodar et al |
| EPI_ISL_792271 | Argentina/PAIS-A0182/2020 | 2020-05-20 | South America/Argentina/Buenos Aires/ | Laboratorio de Virolog√≠a del Hospital de Ni√±os Dr. Ricardo Gutierrez | √Årea de Secuenciaci√≥n del Laboratorio de Virolog√≠a del Hospital de Ni√±os Dr. Ricardo Gutierrez on behalf of 'Proyecto Argentino Interinstitucional de genomica de SARS-CoV-2' (PAIS Consortium) | Nabaes Jodar et al |

| | | | | | | |
|---|---|---|---|---|---|---|
| EPI_ISL_792285 | Argentina/PAIS-A0196/2020 | 2020-05-21 | South America/Argentina/Buenos Aires/ | Laboratorio de Virologv≠a del Hospital de Niv±os Dr. Ricardo Gutierrez | √Årea de Secuenciaciv≥n del Laboratorio de Virologv≠a del Hospital de Niv±os Dr. Ricardo Gutierrez on behalf of 'Proyecto Argentino Interinstitucional de genomica de SARS-CoV-2' (PAIS Consortium) | Nabaes Jodar et al |
| EPI_ISL_792303 | Argentina/PAIS-A0222/2020 | 2020-03-28 | South America/Argentina/Buenos Aires/ | Unidad de Virologv≠a, Centro de Educaciv≥n Mv©dica en Investigaciones Clv≠nicas CEMIC | √Årea de Secuenciaciv≥n del Laboratorio de Virologv≠a del Hospital de Niv±os Dr. Ricardo Gutierrez on behalf of 'Proyecto Argentino Interinstitucional de genomica de SARS-CoV-2' (PAIS Consortium) | Nabaes Jodar et al |

| | | | | | | |
|---|---|---|---|---|---|---|
| EPI_ISL_792355 | Argentina/PAIS-A0280/2020 | 2020-06-12 | South America/Argentina/Buenos Aires/ | Laboratorio de Biolog√≠a Molecular. Hospital Dr. H√©ctor Cura | √Årea de Secuenciaci√≥n del Laboratorio de Virolog√≠a del Hospital de Ni√±os Dr. Ricardo Gutierrez on behalf of 'Proyecto Argentino Interinstitucional de genomica de SARS-CoV-2' (PAIS Consortium) | Nabaes Jodar et al |
| EPI_ISL_792356 | Argentina/PAIS-A0281/2020 | 2020-06-10 | South America/Argentina/Buenos Aires/ | Laboratorio de Biolog√≠a Molecular. Hospital Dr. H√©ctor Cura | √Årea de Secuenciaci√≥n del Laboratorio de Virolog√≠a del Hospital de Ni√±os Dr. Ricardo Gutierrez on behalf of 'Proyecto Argentino Interinstitucional de genomica de SARS-CoV-2' (PAIS Consortium) | Nabaes Jodar et al |

| | | | | | | |
|---|---|---|---|---|---|---|
| EPI_ISL_792357 | Argentina/PAIS-A0282/2020 | 2020-06-02 | South America/Argentina/Buenos Aires/ | Laboratorio de Biologv≠a Molecular. Hospital Dr. Hv©ctor Cura | √Årea de Secuenciaci√≥n del Laboratorio de Virologv≠a del Hospital de Niv±os Dr. Ricardo Gutierrez on behalf of 'Proyecto Argentino Interinstitucional de genomica de SARS-CoV-2' (PAIS Consortium) | Nabaes Jodar et al |
| EPI_ISL_792363 | Argentina/PAIS-A0289/2020 | 2020-05-29 | South America/Argentina/Buenos Aires/ | Laboratorio de Virologia - HIEAyC San Juan de Dios | √Årea de Secuenciaci√≥n del Laboratorio de Virologv≠a del Hospital de Niv±os Dr. Ricardo Gutierrez on behalf of 'Proyecto Argentino Interinstitucional de genomica de SARS-CoV-2' (PAIS Consortium) | Nabaes Jodar et al |

| | | | | | | |
|---|---|---|---|---|---|---|
| EPI_ISL_792373 | Argentina/PAIS-A0300/2020 | 2020-06-02 | South America/Argentina/Buenos Aires/ | Laboratorio del Hospital Interzonal General de Agudos Eva Perv≥n | √Årea de Secuenciaci√≥n del Laboratorio de Virolog√≠a del Hospital de Ni√±os Dr. Ricardo Gutierrez on behalf of 'Proyecto Argentino Interinstitucional de genomica de SARS-CoV-2' (PAIS Consortium) | Nabaes Jodar et al |
| EPI_ISL_792392 | Argentina/PAIS-A0322/2020 | 2020-05-05 | South America/Argentina/Buenos Aires/ | Plataforma de Servicios Biotecn√≥logicos: UTTIPP/PSB , Universidad Nacional de Quilmes. | √Årea de Secuenciaci√≥n del Laboratorio de Virolog√≠a del Hospital de Ni√±os Dr. Ricardo Gutierrez on behalf of 'Proyecto Argentino Interinstitucional de genomica de SARS-CoV-2' (PAIS Consortium) | Nabaes Jodar et al |
| EPI_ISL_792400 | Argentina/PAIS-B0004/2020 | 2020-06-03 | South America/Argentina/Buenos Aires/ | Laboratorio de Virolog√≠a del Hospital de Ni√±os Dr. Ricardo Gutierrez | Bioc√≥dices SA. on behalf of 'Proyecto Argentino Interinstitucional de genomica de SARS-CoV-2' (PAIS Consortium) | Gravis et al |

| | | | | | | |
|---|---|---|---|---|---|---|
| EPI_ISL_792401 | Argentina/PAIS-B0005/2020 | 2020-06-05 | South America/Argentina/Buenos Aires/ | Laboratorio de Virolog√≠a del Hospital de Ni√±os Dr. Ricardo Gutierrez | Biocv≥dices SA. on behalf of 'Proyecto Argentino Interinstitucional de genomica de SARS-CoV-2' (PAIS Consortium) | Gravis et al |
| EPI_ISL_792421 | Argentina/PAIS-C0020/2020 | 2020-06-01 | South America/Argentina/Buenos Aires/ | LABORATORIO DE CAMPA√ëA COVID 19 INTA | Instituto de Biotecnolog√≠a, IABIMO (CONICET), Instituto de Virolog√≠a, IVIT(CONICET), Instituto de Patobiolog√≠a, IPVET(CONICET), CICVyA, INTA on behalf of 'Proyecto Argentino Interinstitucional de genomica de SARS-CoV-2' (PAIS Consortium) | K√∂nig et al |

| | | | | | | |
|---|---|---|---|---|---|---|
| EPI_ISL_792422 | Argentina/PAIS-C0021/2020 | 2020-06-01 | South America/Argentina/Buenos Aires/ | LABORATORIO DE CAMPA√ëA COVID 19 INTA | Instituto de Biotecnolog√≠a, IABIMO (CONICET), Instituto de Virolog√≠a, IVIT(CONICET), Instituto de Patobiolog√≠a, IPVET(CONICET), CICVyA, INTA on behalf of 'Proyecto Argentino Interinstitucional de genomica de SARS-CoV-2' (PAIS Consortium) | K√∂nig et al |
| EPI_ISL_792423 | Argentina/PAIS-C0022/2020 | 2020-06-03 | South America/Argentina/Buenos Aires/ | LABORATORIO DE CAMPA√ëA COVID 19 INTA | Instituto de Biotecnolog√≠a, IABIMO (CONICET), Instituto de Virolog√≠a, IVIT(CONICET), Instituto de Patobiolog√≠a, IPVET(CONICET), CICVyA, INTA on behalf of 'Proyecto Argentino Interinstitucional de genomica de SARS-CoV-2' (PAIS Consortium) | K√∂nig et al |

| | | | | | | |
|---|---|---|---|---|---|---|
| EPI_ISL_792436 | Argentina/PAIS-C0035/2020 | 2020-05-22 | South America/Argentina/Chaco/Resiste | Laboratorio de Inmunologv≠a del Hospital Perrando e Instituto de Medicina Regional de la UNNE | Instituto de Biotecnologv≠a, IABIMO (CONICET), Instituto de Virologv≠a, IVIT(CONICET), Instituto de Patobiologv≠a, IPVET(CONICET), CICVyA, INTA on behalf of 'Proyecto Argentino Interinstitucional de genomica de SARS-CoV-2' (PAIS Consortium) | Kv∂nig et al |
| EPI_ISL_792441 | Argentina/PAIS-C0040/2020 | 2020-06-13 | South America/Argentina/Chaco/Sanz P | Laboratorio de Inmunologv≠a del Hospital Perrando e Instituto de Medicina Regional de la UNNE | Instituto de Biotecnologv≠a, IABIMO (CONICET), Instituto de Virologv≠a, IVIT(CONICET), Instituto de Patobiologv≠a, IPVET(CONICET), CICVyA, INTA on behalf of 'Proyecto Argentino Interinstitucional de genomica de SARS-CoV-2' (PAIS Consortium) | Kv∂nig et al |

| | | | | | | |
|---|---|---|---|---|---|---|
| EPI_ISL_792442 | Argentina/PAIS-C0041/2020 | 2020-06-22 | South America/Argentina/Chaco/Sauzal | Laboratorio de Inmunolog√≠a del Hospital Perrando e Instituto de Medicina Regional de la UNNE | Instituto de Biotecnolog√≠a, IABIMO (CONICET), Instituto de Virolog√≠a, IVIT(CONICET), Instituto de Patobiolog√≠a, IPVET(CONICET), CICVyA, INTA on behalf of 'Proyecto Argentino Interinstitucional de genomica de SARS-CoV-2' (PAIS Consortium) | K√∂nig et al |
| EPI_ISL_792447 | Argentina/PAIS-D0005/2020 | 2020-03-24 | South America/Argentina/Tierra del Fu | Laboratorio del Hospital Regional Ushuaia Gdor. Ernesto Campos | Hospital Regional Ushuaia - Centro Austral De Investigaciones Cient√≠ficas - Universidad Nacional De Tierra Del Fuego on behalf of 'Proyecto Argentino Interinstitucional de genomica de SARS-CoV-2' (PAIS Consortium) | Ceballos et al |

| | | | | | | |
|---|---|---|---|---|---|---|
| EPI_ISL_792478 | Argentina/PAIS-D0036/2020 | 2020-03-23 | South America/Argentina/Neuquen/Zap | Laboratorio Central Mg. Luis Alfredo Pianciola | Hospital Regional Ushuaia - Centro Austral De Investigaciones Cient√≠ficas - Universidad Nacional De Tierra Del Fuego on behalf of 'Proyecto Argentino Interinstitucional de genomica de SARS-CoV-2' (PAIS Consortium) | Ceballos et al |
| EPI_ISL_792513 | Argentina/PAIS-E0019/2020 | 2020-06-23 | South America/Argentina/Santa Fe/San | Laboratorio Central de la Ciudad de Santa Fe | Grupo de Gen√≥mica y Bioinform√°tica del Instituto de Investigaci√≥n de la Cadena L√°ctea CONICET-INTA on behalf of 'Proyecto Argentino Interinstitucional de genomica de SARS-CoV-2' (PAIS Consortium) | Eberhardt et al |

| | | | | | | |
|---|---|---|---|---|---|---|
| EPI_ISL_792515 | Argentina/PAIS-E0023/2020 | 2020-06-10 | South America/Argentina/Santa Fe/Cay | Laboratorio Central de la Ciudad de Santa Fe | Grupo de Gen√≥mica y Bioinform√°tica del Instituto de Investigaci√≥n de la Cadena L√°ctea CONICET-INTA on behalf of 'Proyecto Argentino Interinstitucional de genomica de SARS-CoV-2' (PAIS Consortium) | Eberhardt et al |
| EPI_ISL_792516 | Argentina/PAIS-E0025/2020 | 2020-06-17 | South America/Argentina/Santa Fe/Cer | Laboratorio Central de la Ciudad de Santa Fe | Grupo de Gen√≥mica y Bioinform√°tica del Instituto de Investigaci√≥n de la Cadena L√°ctea CONICET-INTA on behalf of 'Proyecto Argentino Interinstitucional de genomica de SARS-CoV-2' (PAIS Consortium) | Eberhardt et al |

| | | | | | | |
|---|---|---|---|---|---|---|
| EPI_ISL_792517 | Argentina/PAIS-E0026/2020 | 2020-06-26 | South America/Argentina/Santa Fe/Cer | Laboratorio Central de la Ciudad de Santa Fe | Grupo de Gen√≥mica y Bioinform√°tica del Instituto de Investigaci√≥n de la Cadena L√°ctea CONICET-INTA on behalf of 'Proyecto Argentino Interinstitucional de genomica de SARS-CoV-2' (PAIS Consortium) | Eberhardt et al |
| EPI_ISL_792518 | Argentina/PAIS-E0027/2020 | 2020-06-27 | South America/Argentina/Santa Fe/Cer | Laboratorio Central de la Ciudad de Santa Fe | Grupo de Gen√≥mica y Bioinform√°tica del Instituto de Investigaci√≥n de la Cadena L√°ctea CONICET-INTA on behalf of 'Proyecto Argentino Interinstitucional de genomica de SARS-CoV-2' (PAIS Consortium) | Eberhardt et al |

| | | | | | | |
|---|---|---|---|---|---|---|
| EPI_ISL_792519 | Argentina/PAIS-E0029/2020 | 2020-06-27 | South America/Argentina/Santa Fe/Esp | Laboratorio Central de la Ciudad de Santa Fe | Grupo de Gen√≥mica y Bioinform√°tica del Instituto de Investigaci√≥n de la Cadena L√°ctea CONICET-INTA on behalf of 'Proyecto Argentino Interinstitucional de genomica de SARS-CoV-2' (PAIS Consortium) | Eberhardt et al |
| EPI_ISL_792520 | Argentina/PAIS-E0032/2020 | 2020-06-03 | South America/Argentina/Santa Fe/Vill | Laboratorio Central de la Ciudad de Santa Fe | Grupo de Gen√≥mica y Bioinform√°tica del Instituto de Investigaci√≥n de la Cadena L√°ctea CONICET-INTA on behalf of 'Proyecto Argentino Interinstitucional de genomica de SARS-CoV-2' (PAIS Consortium) | Eberhardt et al |

| | | | | | | |
|---|---|---|---|---|---|---|
| EPI_ISL_792521 | Argentina/PAIS-E0033/2020 | 2020-06-04 | South America/Argentina/Santa Fe/Vill | Laboratorio Central de la Ciudad de Santa Fe | Grupo de Gen√≥mica y Bioinform√°tica del Instituto de Investigaci√≥n de la Cadena L√°ctea CONICET-INTA on behalf of 'Proyecto Argentino Interinstitucional de genomica de SARS-CoV-2' (PAIS Consortium) | Eberhardt et al |
| EPI_ISL_792523 | Argentina/PAIS-E0101/2020 | 2020-12-16 | South America/Argentina/Buenos Aires/ | Laboratorio de Virolog√≠a del Hospital de Ni√±os Dr. Ricardo Gutierrez | Grupo de Gen√≥mica y Bioinform√°tica del Instituto de Investigaci√≥n de la Cadena L√°ctea CONICET-INTA on behalf of 'Proyecto Argentino Interinstitucional de genomica de SARS-CoV-2' (PAIS Consortium) | Amadio et al |

| | | | | | | |
|---|---|---|---|---|---|---|
| EPI_ISL_792524 | Argentina/PAIS-E0102/2020 | 2020-12-17 | South America/Argentina/Buenos Aires/ | Laboratorio de Virologv≠a del Hospital de Niv±os Dr. Ricardo Gutierrez | Grupo de Genv≥mica y Bioinformv°tica del Instituto de Investigaciv≥n de la Cadena Lv°ctea CONICET-INTA on behalf of 'Proyecto Argentino Interinstitucional de genomica de SARS-CoV-2' (PAIS Consortium) | Amadio et al |
| EPI_ISL_792526 | Argentina/PAIS-G0002/2020 | 2020-03-22 | South America/Argentina/Cordoba AR/ | Laboratorio Central, Ministerio de Salud Cv≥rdoba | Instituto de Patologia Vegetal (CIAP-INTA) on behalf of 'Proyecto Argentino Interinstitucional de genomica de SARS-CoV-2' (PAIS Consortium) | Fernv°ndez et al |
| EPI_ISL_792531 | Argentina/PAIS-G0007/2020 | 2020-03-23 | South America/Argentina/Cordoba AR/ | Laboratorio Central, Ministerio de Salud Cv≥rdoba | Instituto de Patologia Vegetal (CIAP-INTA) on behalf of 'Proyecto Argentino Interinstitucional de genomica de SARS-CoV-2' (PAIS Consortium) | Fernv°ndez et al |

| | | | | | | |
|---|---|---|---|---|---|---|
| EPI_ISL_792538 | Argentina/PAIS-G0014/2020 | 2020-04-02 | South America/Argentina/Cordoba AR/( | Laboratorio Central, Ministerio de Salud C√≥rdoba | Instituto de Patologia Vegetal (CIAP-INTA) on behalf of 'Proyecto Argentino Interinstitucional de genomica de SARS-CoV-2' (PAIS Consortium) | Fern√°ndez et al |
| EPI_ISL_792544 | Argentina/PAIS-G0020/2020 | 2020-07-06 | South America/Argentina/Cordoba AR/\ | Laboratorio Central, Ministerio de Salud C√≥rdoba | Instituto de Patologia Vegetal (CIAP-INTA) on behalf of 'Proyecto Argentino Interinstitucional de genomica de SARS-CoV-2' (PAIS Consortium) | Fern√°ndez et al |
| EPI_ISL_792545 | Argentina/PAIS-G0021/2020 | 2020-07-06 | South America/Argentina/Cordoba AR/\ | Laboratorio Central, Ministerio de Salud C√≥rdoba | Instituto de Patologia Vegetal (CIAP-INTA) on behalf of 'Proyecto Argentino Interinstitucional de genomica de SARS-CoV-2' (PAIS Consortium) | Fern√°ndez et al |
| EPI_ISL_792546 | Argentina/PAIS-G0022/2020 | 2020-07-06 | South America/Argentina/Cordoba AR/\ | Laboratorio Central, Ministerio de Salud C√≥rdoba | Instituto de Patologia Vegetal (CIAP-INTA) on behalf of 'Proyecto Argentino Interinstitucional de genomica de SARS-CoV-2' (PAIS Consortium) | Fern√°ndez et al |

| | | | | | | |
|---|---|---|---|---|---|---|
| EPI_ISL_792552 | SriLanka/Cov5260/2020 | 2020-11-27 | Asia/Sri Lanka/Colombo/ | Centre for Dengue Research and AICBU, Department of Immunology and Molecular Medicine | Centre for Dengue Research and AICBU, Department of Immunology and Molecular Medicine | Chandima Jeewandara et al |
| EPI_ISL_792554 | SriLanka/DKP18/2020 | 2020-11-28 | Asia/Sri Lanka/Colombo/ | Centre for Dengue Research and AICBU, Department of Immunology and Molecular Medicine | Centre for Dengue Research and AICBU, Department of Immunology and Molecular Medicine | Chandima Jeewandara et al |
| EPI_ISL_792556 | SriLanka/W65/2020 | 2020-12-07 | Asia/Sri Lanka/Colombo/ | Centre for Dengue Research and AICBU, Department of Immunology and Molecular Medicine | Centre for Dengue Research and AICBU, Department of Immunology and Molecular Medicine | Chandima Jeewandara et al |
| EPI_ISL_792560 | Brazil/AM-20142410MC/2020 | 2020-11-06 | South America/Brazil/Amazonas BR/Ma | Laboratorio de Ecologia de Doencas Transmissiveis na Amazonia, Instituto Leonidas e Maria Deane - Fiocruz Amazonia | Laboratorio de Ecologia de Doencas Transmissiveis na Amazonia, Instituto Leonidas e Maria Deane - Fiocruz Amazonia | Valdinete Nascimento et al |
| EPI_ISL_792561 | Brazil/PB-23854-R1/2020 | 2020-06-23 | South America/Brazil/Paraiba/Joao Pess | LACEN-PB | Laboratory of Respiratory Viruses and Measles, Oswaldo Cruz Institute, FIOCRUZ | Paola Resende et al |
| EPI_ISL_792562 | Brazil/PB-23855-R2/2020 | 2020-10-13 | South America/Brazil/Paraiba/Joao Pess | LACEN-PB | Laboratory of Respiratory Viruses and Measles, Oswaldo Cruz Institute, FIOCRUZ | Paola Resende et al |

| | | | | | | |
|---|---|---|---|---|---|---|
| EPI_ISL_792563 | Brazil/PB-16521/2020 | 2020-05-25 | South America/Brazil/Paraiba/Cabedelo | LACEN-PB | Laboratory of Respiratory Viruses and Measles, Oswaldo Cruz Institute, FIOCRUZ | Paola Resende et al |
| EPI_ISL_792564 | Brazil/PB-16522/2020 | 2020-05-25 | South America/Brazil/Paraiba/Santa Rit | LACEN-PB | Laboratory of Respiratory Viruses and Measles, Oswaldo Cruz Institute, FIOCRUZ | Paola Resende et al |
| EPI_ISL_792565 | Brazil/PB-16523/2020 | 2020-05-27 | South America/Brazil/Paraiba/Joao Pess | LACEN-PB | Laboratory of Respiratory Viruses and Measles, Oswaldo Cruz Institute, FIOCRUZ | Paola Resende et al |
| EPI_ISL_792566 | Brazil/PB-16524/2020 | 2020-06-16 | South America/Brazil/Paraiba/Joao Pess | LACEN-PB | Laboratory of Respiratory Viruses and Measles, Oswaldo Cruz Institute, FIOCRUZ | Paola Resende et al |
| EPI_ISL_792567 | Brazil/PB-16525/2020 | 2020-06-10 | South America/Brazil/Pernambuco/Itam | LACEN-PB | Laboratory of Respiratory Viruses and Measles, Oswaldo Cruz Institute, FIOCRUZ | Paola Resende et al |
| EPI_ISL_792568 | Brazil/PB-16526/2020 | 2020-06-26 | South America/Brazil/Paraiba/Joao Pess | LACEN-PB | Laboratory of Respiratory Viruses and Measles, Oswaldo Cruz Institute, FIOCRUZ | Paola Resende et al |

| | | | | | | |
|---|---|---|---|---|---|---|
| EPI_ISL_792569 | Brazil/PB-16527/2020 | 2020-06-26 | South America/Brazil/Paraiba/Joao Pess | LACEN-PB | Laboratory of Respiratory Viruses and Measles, Oswaldo Cruz Institute, FIOCRUZ | Paola Resende et al |
| EPI_ISL_792579 | Brazil/PB-16578/2020 | 2020-04-21 | South America/Brazil/Paraiba/Santa Rit | LACEN-PB | Laboratory of Respiratory Viruses and Measles, Oswaldo Cruz Institute, FIOCRUZ | Paola Resende et al |
| EPI_ISL_792589 | Brazil/PB-16599/2020 | 2020-04-23 | South America/Brazil/Paraiba/Joao Pess | LACEN-PB | Laboratory of Respiratory Viruses and Measles, Oswaldo Cruz Institute, FIOCRUZ | Paola Resende et al |
| EPI_ISL_792592 | Brazil/PB-16605/2020 | 2020-04-11 | South America/Brazil/Paraiba/Joao Pess | LACEN-PB | Laboratory of Respiratory Viruses and Measles, Oswaldo Cruz Institute, FIOCRUZ | Paola Resende et al |
| EPI_ISL_792593 | Brazil/PB-16606/2020 | 2020-05-21 | South America/Brazil/Paraiba/Joao Pess | LACEN-PB | Laboratory of Respiratory Viruses and Measles, Oswaldo Cruz Institute, FIOCRUZ | Paola Resende et al |
| EPI_ISL_792594 | Brazil/PB-16610/2020 | 2020-05-19 | South America/Brazil/Paraiba/Joao Pess | LACEN-PB | Laboratory of Respiratory Viruses and Measles, Oswaldo Cruz Institute, FIOCRUZ | Paola Resende et al |

| | | | | | | |
|---|---|---|---|---|---|---|
| EPI_ISL_792595 | Brazil/PB-16615/2020 | 2020-07-01 | South America/Brazil/Paraiba/Pilar | LACEN-PB | Laboratory of Respiratory Viruses and Measles, Oswaldo Cruz Institute, FIOCRUZ | Paola Resende et al |
| EPI_ISL_792596 | Brazil/PB-16616/2020 | 2020-07-01 | South America/Brazil/Paraiba/Joao Pess | LACEN-PB | Laboratory of Respiratory Viruses and Measles, Oswaldo Cruz Institute, FIOCRUZ | Paola Resende et al |
| EPI_ISL_792597 | Brazil/PB-16617/2020 | 2020-06-30 | South America/Brazil/Paraiba/Cacimba ( | LACEN-PB | Laboratory of Respiratory Viruses and Measles, Oswaldo Cruz Institute, FIOCRUZ | Paola Resende et al |
| EPI_ISL_792599 | Brazil/PB-23853/2020 | 2020-10-19 | South America/Brazil/Paraiba/ | LACEN-PB | Laboratory of Respiratory Viruses and Measles, Oswaldo Cruz Institute, FIOCRUZ | Paola Resende et al |
| EPI_ISL_792600 | Brazil/PB-29601/2020 | 2020-11-04 | South America/Brazil/Paraiba/ | LACEN-PB | Laboratory of Respiratory Viruses and Measles, Oswaldo Cruz Institute, FIOCRUZ | Paola Resende et al |
| EPI_ISL_792601 | Brazil/PB-29602/2020 | 2020-11-04 | South America/Brazil/Paraiba/ | LACEN-PB | Laboratory of Respiratory Viruses and Measles, Oswaldo Cruz Institute, FIOCRUZ | Paola Resende et al |

| | | | | | | |
|---|---|---|---|---|---|---|
| EPI_ISL_792602 | Brazil/PB-29604/2020 | 2020-09-28 | South America/Brazil/Paraiba/ | LACEN-PB | Laboratory of Respiratory Viruses and Measles, Oswaldo Cruz Institute, FIOCRUZ | Paola Resende et al |
| EPI_ISL_792603 | Brazil/PB-29608/2020 | 2020-11-27 | South America/Brazil/Paraiba/ | LACEN-PB | Laboratory of Respiratory Viruses and Measles, Oswaldo Cruz Institute, FIOCRUZ | Paola Resende et al |
| EPI_ISL_792604 | Brazil/PB-29610/2020 | 2020-08-06 | South America/Brazil/Paraiba/ | LACEN-PB | Laboratory of Respiratory Viruses and Measles, Oswaldo Cruz Institute, FIOCRUZ | Paola Resende et al |
| EPI_ISL_792605 | Brazil/PB-29611/2020 | 2020-08-10 | South America/Brazil/Paraiba/ | LACEN-PB | Laboratory of Respiratory Viruses and Measles, Oswaldo Cruz Institute, FIOCRUZ | Paola Resende et al |
| EPI_ISL_792606 | Brazil/PB-29614/2020 | 2020-08-11 | South America/Brazil/Paraiba/ | LACEN-PB | Laboratory of Respiratory Viruses and Measles, Oswaldo Cruz Institute, FIOCRUZ | Paola Resende et al |
| EPI_ISL_792607 | Brazil/PB-29616/2020 | 2020-08-12 | South America/Brazil/Paraiba/ | LACEN-PB | Laboratory of Respiratory Viruses and Measles, Oswaldo Cruz Institute, FIOCRUZ | Paola Resende et al |

| | | | | | | |
|---|---|---|---|---|---|---|
| EPI_ISL_792608 | Brazil/PB-29617/2020 | 2020-08-19 | South America/Brazil/Paraiba/ | LACEN-PB | Laboratory of Respiratory Viruses and Measles, Oswaldo Cruz Institute, FIOCRUZ | Paola Resende et al |
| EPI_ISL_792609 | Brazil/PB-29618/2020 | 2020-08-20 | South America/Brazil/Paraiba/ | LACEN-PB | Laboratory of Respiratory Viruses and Measles, Oswaldo Cruz Institute, FIOCRUZ | Paola Resende et al |
| EPI_ISL_792610 | Brazil/PB-29619/2020 | 2020-08-20 | South America/Brazil/Paraiba/ | LACEN-PB | Laboratory of Respiratory Viruses and Measles, Oswaldo Cruz Institute, FIOCRUZ | Paola Resende et al |
| EPI_ISL_792611 | Brazil/PB-29620/2020 | 2020-08-31 | South America/Brazil/Paraiba/ | LACEN-PB | Laboratory of Respiratory Viruses and Measles, Oswaldo Cruz Institute, FIOCRUZ | Paola Resende et al |
| EPI_ISL_792612 | Brazil/PB-29622/2020 | 2020-09-14 | South America/Brazil/Paraiba/ | LACEN-PB | Laboratory of Respiratory Viruses and Measles, Oswaldo Cruz Institute, FIOCRUZ | Paola Resende et al |
| EPI_ISL_792613 | Brazil/PB-29623/2020 | 2020-09-16 | South America/Brazil/Paraiba/ | LACEN-PB | Laboratory of Respiratory Viruses and Measles, Oswaldo Cruz Institute, FIOCRUZ | Paola Resende et al |

| | | | | | | |
|---|---|---|---|---|---|---|
| EPI_ISL_792614 | Brazil/PB-29624/2020 | 2020-09-21 | South America/Brazil/Paraiba/ | LACEN-PB | Laboratory of Respiratory Viruses and Measles, Oswaldo Cruz Institute, FIOCRUZ | Paola Resende et al |
| EPI_ISL_792615 | Brazil/PB-29625/2020 | 2020-09-21 | South America/Brazil/Paraiba/ | LACEN-PB | Laboratory of Respiratory Viruses and Measles, Oswaldo Cruz Institute, FIOCRUZ | Paola Resende et al |
| EPI_ISL_792616 | Brazil/PB-29626/2020 | 2020-09-23 | South America/Brazil/Paraiba/ | LACEN-PB | Laboratory of Respiratory Viruses and Measles, Oswaldo Cruz Institute, FIOCRUZ | Paola Resende et al |
| EPI_ISL_792617 | Brazil/PB-29627/2020 | 2020-09-24 | South America/Brazil/Paraiba/ | LACEN-PB | Laboratory of Respiratory Viruses and Measles, Oswaldo Cruz Institute, FIOCRUZ | Paola Resende et al |
| EPI_ISL_792618 | Brazil/PB-29628/2020 | 2020-09-24 | South America/Brazil/Paraiba/ | LACEN-PB | Laboratory of Respiratory Viruses and Measles, Oswaldo Cruz Institute, FIOCRUZ | Paola Resende et al |
| EPI_ISL_792619 | Brazil/PB-29629/2020 | 2020-09-24 | South America/Brazil/Paraiba/ | LACEN-PB | Laboratory of Respiratory Viruses and Measles, Oswaldo Cruz Institute, FIOCRUZ | Paola Resende et al |

| | | | | | | |
|---|---|---|---|---|---|---|
| EPI_ISL_792620 | Brazil/PB-29630/2020 | 2020-09-30 | South America/Brazil/Paraiba/ | LACEN-PB | Laboratory of Respiratory Viruses and Measles, Oswaldo Cruz Institute, FIOCRUZ | Paola Resende et al |
| EPI_ISL_792621 | Brazil/PB-29631/2020 | 2020-09-30 | South America/Brazil/Paraiba/ | LACEN-PB | Laboratory of Respiratory Viruses and Measles, Oswaldo Cruz Institute, FIOCRUZ | Paola Resende et al |
| EPI_ISL_792622 | Brazil/PB-29632/2020 | 2020-10-08 | South America/Brazil/Paraiba/ | LACEN-PB | Laboratory of Respiratory Viruses and Measles, Oswaldo Cruz Institute, FIOCRUZ | Paola Resende et al |
| EPI_ISL_792623 | Brazil/PB-29633/2020 | 2020-10-09 | South America/Brazil/Paraiba/ | LACEN-PB | Laboratory of Respiratory Viruses and Measles, Oswaldo Cruz Institute, FIOCRUZ | Paola Resende et al |
| EPI_ISL_792624 | Brazil/PB-29634/2020 | 2020-10-13 | South America/Brazil/Paraiba/ | LACEN-PB | Laboratory of Respiratory Viruses and Measles, Oswaldo Cruz Institute, FIOCRUZ | Paola Resende et al |
| EPI_ISL_792625 | Brazil/PB-29635/2020 | 2020-10-20 | South America/Brazil/Paraiba/ | LACEN-PB | Laboratory of Respiratory Viruses and Measles, Oswaldo Cruz Institute, FIOCRUZ | Paola Resende et al |

| | | | | | | |
|---|---|---|---|---|---|---|
| EPI_ISL_792626 | Brazil/PB-29636/2020 | 2020-10-21 | South America/Brazil/Paraiba/ | LACEN-PB | Laboratory of Respiratory Viruses and Measles, Oswaldo Cruz Institute, FIOCRUZ | Paola Resende et al |
| EPI_ISL_792628 | Brazil/PB-29638/2020 | 2020-11-04 | South America/Brazil/Paraiba/ | LACEN-PB | Laboratory of Respiratory Viruses and Measles, Oswaldo Cruz Institute, FIOCRUZ | Paola Resende et al |
| EPI_ISL_792629 | Brazil/PB-29639/2020 | 2020-11-04 | South America/Brazil/Paraiba/ | LACEN-PB | Laboratory of Respiratory Viruses and Measles, Oswaldo Cruz Institute, FIOCRUZ | Paola Resende et al |
| EPI_ISL_792631 | Brazil/PB-29642/2020 | 2020-11-09 | South America/Brazil/Paraiba/ | LACEN-PB | Laboratory of Respiratory Viruses and Measles, Oswaldo Cruz Institute, FIOCRUZ | Paola Resende et al |
| EPI_ISL_792633 | Brazil/PB-29645/2020 | 2020-11-09 | South America/Brazil/Paraiba/ | LACEN-PB | Laboratory of Respiratory Viruses and Measles, Oswaldo Cruz Institute, FIOCRUZ | Paola Resende et al |
| EPI_ISL_792634 | Brazil/PB-29646/2020 | 2020-11-10 | South America/Brazil/Paraiba/ | LACEN-PB | Laboratory of Respiratory Viruses and Measles, Oswaldo Cruz Institute, FIOCRUZ | Paola Resende et al |

| | | | | | | |
|---|---|---|---|---|---|---|
| EPI_ISL_792635 | Brazil/PB-29647/2020 | 2020-11-10 | South America/Brazil/Paraiba/ | LACEN-PB | Laboratory of Respiratory Viruses and Measles, Oswaldo Cruz Institute, FIOCRUZ | Paola Resende et al |
| EPI_ISL_792637 | Brazil/PB-29650/2020 | 2020-11-10 | South America/Brazil/Paraiba/ | LACEN-PB | Laboratory of Respiratory Viruses and Measles, Oswaldo Cruz Institute, FIOCRUZ | Paola Resende et al |
| EPI_ISL_792638 | Brazil/PB-29651/2020 | 2020-11-11 | South America/Brazil/Paraiba/ | LACEN-PB | Laboratory of Respiratory Viruses and Measles, Oswaldo Cruz Institute, FIOCRUZ | Paola Resende et al |
| EPI_ISL_792639 | Brazil/AL-28256/2020 | 2020-11-21 | South America/Brazil/Alagoas/Maceio | LACEN-AL | Laboratory of Respiratory Viruses and Measles, Oswaldo Cruz Institute, FIOCRUZ | Paola Resende et al |
| EPI_ISL_792640 | Brazil/AL-28258/2020 | 2020-05-06 | South America/Brazil/Alagoas/ | LACEN-AL | Laboratory of Respiratory Viruses and Measles, Oswaldo Cruz Institute, FIOCRUZ | Paola Resende et al |
| EPI_ISL_792642 | Brazil/AL-30270/2020 | 2020-12-04 | South America/Brazil/Alagoas/Maceio | LACEN-AL | Laboratory of Respiratory Viruses and Measles, Oswaldo Cruz Institute, FIOCRUZ | Paola Resende et al |

| | | | | | | |
|---|---|---|---|---|---|---|
| EPI_ISL_792644 | Brazil/AL-30273/2020 | 2020-05-13 | South America/Brazil/Alagoas/ | LACEN-AL | Laboratory of Respiratory Viruses and Measles, Oswaldo Cruz Institute, FIOCRUZ | Paola Resende et al |
| EPI_ISL_792645 | Brazil/PR-28601/2020 | 2020-10-27 | South America/Brazil/Paran√°/Umuaran | LACEN-PR | Laboratory of Respiratory Viruses and Measles, Oswaldo Cruz Institute, FIOCRUZ | Paola Resende et al |
| EPI_ISL_792646 | Brazil/PR-28602/2020 | 2020-11-03 | South America/Brazil/Paran√°/Maringa | LACEN-PR | Laboratory of Respiratory Viruses and Measles, Oswaldo Cruz Institute, FIOCRUZ | Paola Resende et al |
| EPI_ISL_792648 | Brazil/PR-28604/2020 | 2020-11-12 | South America/Brazil/Paran√°/Dois Vizi | LACEN-PR | Laboratory of Respiratory Viruses and Measles, Oswaldo Cruz Institute, FIOCRUZ | Paola Resende et al |
| EPI_ISL_792650 | Brazil/PR-28606/2020 | 2020-11-23 | South America/Brazil/Paran√°/Uniao Da | LACEN-PR | Laboratory of Respiratory Viruses and Measles, Oswaldo Cruz Institute, FIOCRUZ | Paola Resende et al |
| EPI_ISL_792651 | Brazil/PR-28607/2020 | 2020-11-23 | South America/Brazil/Paran√°/Dois Vizi | LACEN-PR | Laboratory of Respiratory Viruses and Measles, Oswaldo Cruz Institute, FIOCRUZ | Paola Resende et al |

| | | | | | | |
|---|---|---|---|---|---|---|
| EPI_ISL_792652 | Brazil/PR-28608/2020 | 2020-11-25 | South America/Brazil/Paranv°/Cascavel | LACEN-PR | Laboratory of Respiratory Viruses and Measles, Oswaldo Cruz Institute, FIOCRUZ | Paola Resende et al |
| EPI_ISL_792653 | Brazil/PR-28609/2020 | 2020-11-09 | South America/Brazil/Paranv°/Ponta Gr | LACEN-PR | Laboratory of Respiratory Viruses and Measles, Oswaldo Cruz Institute, FIOCRUZ | Paola Resende et al |
| EPI_ISL_792654 | Brazil/PR-28610/2020 | 2020-11-23 | South America/Brazil/Paranv°/Londrina | LACEN-PR | Laboratory of Respiratory Viruses and Measles, Oswaldo Cruz Institute, FIOCRUZ | Paola Resende et al |
| EPI_ISL_792657 | USA/CA-LACPHL-AF00086/2020 | 2020-12-24 | North America/USA/California/Los Ange | Los Angeles County PHL | Los Angeles County PHL | P. Hemarajata et al. et al |
| EPI_ISL_792661 | USA/CA-LACPHL-AF00094/2020 | 2020-12-22 | North America/USA/California/Los Ange | Los Angeles County PHL | Los Angeles County PHL | P. Hemarajata et al. et al |
| EPI_ISL_792664 | USA/CA-LACPHL-AF00099/2020 | 2020-12-22 | North America/USA/California/Los Ange | Los Angeles County PHL | Los Angeles County PHL | P. Hemarajata et al. et al |
| EPI_ISL_792682 | Japan/IC-0563/2021 | 2021-01-02 | Asia/Japan/Japan/ | Pathogen Genomics Center, National Institute of Infectious Diseases | Pathogen Genomics Center, National Institute of Infectious Diseases | Tsuyoshi Sekizuka et al |
| EPI_ISL_792684 | CzechRepublic/NRL_11056/2020 | 2020-11-10 | Europe/Czech Republic/Plze≈à Region/Ta | The National Institute of Public Health | State Veterinary Institute Prague | Nagy et al |
| EPI_ISL_792686 | CzechRepublic/NRL_11271/2020 | 2020-11-16 | Europe/Czech Republic/Czech Republic/ | The National Institute of Public Health | State Veterinary Institute Prague | Nagy et al |
| EPI_ISL_792687 | CzechRepublic/NRL_12564/2020 | 2020-09-17 | Europe/Czech Republic/Prague/ | The National Institute of Public Health | State Veterinary Institute Prague | Nagy et al |
| EPI_ISL_792688 | CzechRepublic/NRL_11458/2020 | 2020-11-23 | Europe/Czech Republic/Usti nad Labem/ | The National Institute of Public Health | State Veterinary Institute Prague | Nagy et al |
| EPI_ISL_792689 | CzechRepublic/NRL_11035/2020 | 2020-11-11 | Europe/Czech Republic/Central Bohemia | The National Institute of Public Health | State Veterinary Institute Prague | Nagy et al |

| | | | | | | |
|---|---|---|---|---|---|---|
| EPI_ISL_792690 | CzechRepublic/NRL_11155/2020 | 2020-11-16 | Europe/Czech Republic/Prague/ | The National Institute of Public Health | State Veterinary Institute Prague | Nagy et al |
| EPI_ISL_792691 | CzechRepublic/NRL_11156/2020 | 2020-11-15 | Europe/Czech Republic/Central Bohemia | The National Institute of Public Health | State Veterinary Institute Prague | Nagy et al |
| EPI_ISL_792692 | CzechRepublic/NRL_11159/2020 | 2020-11-13 | Europe/Czech Republic/Prague/ | The National Institute of Public Health | State Veterinary Institute Prague | Nagy et al |
| EPI_ISL_792693 | CzechRepublic/NRL_11190/2020 | 2020-11-18 | Europe/Czech Republic/Central Bohemia | The National Institute of Public Health | State Veterinary Institute Prague | Nagy et al |
| EPI_ISL_792696 | CzechRepublic/NRL_11501/2020 | 2020-11-25 | Europe/Czech Republic/Prague/ | The National Institute of Public Health | State Veterinary Institute Prague | Nagy et al |
| EPI_ISL_792697 | CzechRepublic/NRL_11716/2020 | 2020-12-01 | Europe/Czech Republic/Central Bohemia | The National Institute of Public Health | State Veterinary Institute Prague | Nagy et al |
| EPI_ISL_792698 | CzechRepublic/NRL_11717/2020 | 2020-12-04 | Europe/Czech Republic/Prague/ | The National Institute of Public Health | State Veterinary Institute Prague | Nagy et al |
| EPI_ISL_792699 | CzechRepublic/NRL_11782-1/2020 | 2020-12-07 | Europe/Czech Republic/Prague/ | The National Institute of Public Health | State Veterinary Institute Prague | Nagy et al |
| EPI_ISL_792700 | CzechRepublic/NRL_12563/2020 | 2020-09-17 | Europe/Czech Republic/Prague/ | The National Institute of Public Health | State Veterinary Institute Prague | Nagy et al |
| EPI_ISL_792701 | CzechRepublic/NRL_11158/2020 | 2020-11-15 | Europe/Czech Republic/Prague/ | The National Institute of Public Health | State Veterinary Institute Prague | Nagy et al |
| EPI_ISL_792702 | CzechRepublic/NRL_11622/2020 | 2020-12-01 | Europe/Czech Republic/Prague/ | The National Institute of Public Health | State Veterinary Institute Prague | Nagy et al |
| EPI_ISL_792703 | CzechRepublic/NRL_11745/2020 | 2020-12-06 | Europe/Czech Republic/Prague/ | The National Institute of Public Health | State Veterinary Institute Prague | Nagy et al |
| EPI_ISL_792704 | CzechRepublic/NRL_11782-2/2020 | 2020-12-07 | Europe/Czech Republic/Prague/ | The National Institute of Public Health | State Veterinary Institute Prague | Nagy et al |
| EPI_ISL_792705 | CzechRepublic/NRL_11160/2020 | 2020-11-16 | Europe/Czech Republic/Prague/ | The National Institute of Public Health | State Veterinary Institute Prague | Nagy et al |
| EPI_ISL_792706 | CzechRepublic/NRL_11188/2020 | 2020-11-17 | Europe/Czech Republic/Prague/ | The National Institute of Public Health | State Veterinary Institute Prague | Nagy et al |
| EPI_ISL_792707 | CzechRepublic/NRL_11544/2020 | 2020-11-27 | Europe/Czech Republic/Hradec Kr√°lov√ | The National Institute of Public Health | State Veterinary Institute Prague | Nagy et al |
| EPI_ISL_793798 | Denmark/DCGC-23506/2020 | 2020-12-21 | Europe/Denmark/Midtjylland/ | Department of Virus and Microbiological Special Diagnostics, Statens Serum Institut, Copenhagen, Denmark | Albertsen Lab, Department of Chemistry and Bioscience, Aalborg University, Denmark | Danish Covid-19 Genome Consortium et al |

| | | | | | | |
|---|---|---|---|---|---|---|
| EPI_ISL_793968 | Denmark/DCGC-23857/2020 | 2020-12-21 | Europe/Denmark/Nordjylland/ | Department of Virus and Microbiological Special Diagnostics, Statens Serum Institut, Copenhagen, Denmark | Albertsen Lab, Department of Chemistry and Bioscience, Aalborg University, Denmark | Danish Covid-19 Genome Consortium et al |
| EPI_ISL_794014 | USA/NY-Wadsworth-207269-01/2020 | 2020-09-01 | North America/USA/New York/Ulster C | Wadsworth Center, New York State Department.of Health | Wadsworth Center, New York State Department.of Health | Kirsten St. George et al |
| EPI_ISL_794022 | USA/NY-Wadsworth-209740-01/2020 | 2020-09-03 | North America/USA/New York/Warren | Wadsworth Center, New York State Department.of Health | Wadsworth Center, New York State Department.of Health | Kirsten St. George et al |
| EPI_ISL_794026 | USA/NY-Wadsworth-212280-01/2020 | 2020-09-08 | North America/USA/New York/Schenect | Wadsworth Center, New York State Department.of Health | Wadsworth Center, New York State Department.of Health | Kirsten St. George et al |
| EPI_ISL_794027 | USA/NY-Wadsworth-213940-01/2020 | 2020-09-09 | North America/USA/New York/Chenang | Wadsworth Center, New York State Department.of Health | Wadsworth Center, New York State Department.of Health | Kirsten St. George et al |
| EPI_ISL_794029 | USA/NY-Wadsworth-213956-01/2020 | 2020-09-09 | North America/USA/New York/Chemung | Wadsworth Center, New York State Department.of Health | Wadsworth Center, New York State Department.of Health | Kirsten St. George et al |
| EPI_ISL_794030 | USA/NY-Wadsworth-213957-01/2020 | 2020-09-09 | North America/USA/New York/Chemung | Wadsworth Center, New York State Department.of Health | Wadsworth Center, New York State Department.of Health | Kirsten St. George et al |

| | | | | | | |
|---|---|---|---|---|---|---|
| EPI_ISL_794039 | USA/NY-Wadsworth-232123-01/2020 | 2020-09-30 | North America/USA/New York/Columbia | Wadsworth Center, New York State Department.of Health | Wadsworth Center, New York State Department.of Health | Kirsten St. George et al |
| EPI_ISL_794040 | USA/NY-Wadsworth-232124-01/2020 | 2020-09-30 | North America/USA/New York/Columbia | Wadsworth Center, New York State Department.of Health | Wadsworth Center, New York State Department.of Health | Kirsten St. George et al |
| EPI_ISL_794041 | USA/NY-Wadsworth-232160-01/2020 | 2020-09-30 | North America/USA/New York/Columbia | Wadsworth Center, New York State Department.of Health | Wadsworth Center, New York State Department.of Health | Kirsten St. George et al |
| EPI_ISL_794057 | USA/NY-Wadsworth-253066-01/2020 | 2020-10-23 | North America/USA/New York/Chemung | Wadsworth Center, New York State Department.of Health | Wadsworth Center, New York State Department.of Health | Kirsten St. George et al |
| EPI_ISL_794061 | USA/NY-Wadsworth-253100-01/2020 | 2020-10-23 | North America/USA/New York/Chemung | Wadsworth Center, New York State Department.of Health | Wadsworth Center, New York State Department.of Health | Kirsten St. George et al |
| EPI_ISL_794066 | USA/NY-Wadsworth-254099-01/2020 | 2020-10-25 | North America/USA/New York/Greene ( | Wadsworth Center, New York State Department.of Health | Wadsworth Center, New York State Department.of Health | Kirsten St. George et al |
| EPI_ISL_794069 | USA/NY-Wadsworth-255180-01/2020 | 2020-10-27 | North America/USA/New York/Columbia | Wadsworth Center, New York State Department.of Health | Wadsworth Center, New York State Department.of Health | Kirsten St. George et al |
| EPI_ISL_794086 | USA/NY-Wadsworth-293226-01/2020 | 2020-12-28 | North America/USA/New York/ | URMC LABS | Wadsworth Center, New York State Department.of Health | Kirsten St. George et al |

| | | | | | | |
|---|---|---|---|---|---|---|
| EPI_ISL_794094 | USA/NY-Wadsworth-293239-01/2020 | 2020-12-28 | North America/USA/New York/Ontario | URMC LABS | Wadsworth Center, New York State Department.of Health | Kirsten St. George et al |
| EPI_ISL_794102 | USA/NY-Wadsworth-293256-01/2020 | 2020-12-28 | North America/USA/New York/Monroe | URMC LABS | Wadsworth Center, New York State Department.of Health | Kirsten St. George et al |
| EPI_ISL_794142 | USA/NY-Wadsworth-21000048-01/20 | 2020-12-26 | North America/USA/New York/Nassau C | NORTHWELL HEALTH LABORATORIES | Wadsworth Center, New York State Department.of Health | Kirsten St. George et al |
| EPI_ISL_794146 | USA/NY-Wadsworth-21000052-01/20 | 2020-12-26 | North America/USA/New York/Nassau C | NORTHWELL HEALTH LABORATORIES | Wadsworth Center, New York State Department.of Health | Kirsten St. George et al |
| EPI_ISL_794149 | USA/NY-Wadsworth-21000056-01/20 | 2020-12-27 | North America/USA/New York/Suffolk C | NORTHWELL HEALTH LABORATORIES | Wadsworth Center, New York State Department.of Health | Kirsten St. George et al |
| EPI_ISL_794151 | USA/NY-Wadsworth-21000059-01/20 | 2020-12-27 | North America/USA/New York/Suffolk C | NORTHWELL HEALTH LABORATORIES | Wadsworth Center, New York State Department.of Health | Kirsten St. George et al |
| EPI_ISL_794160 | USA/NY-Wadsworth-291860-01/2020 | 2020-12-29 | North America/USA/New York/Saratoga | Wadsworth Center, New York State Department.of Health | Wadsworth Center, New York State Department.of Health | Kirsten St. George et al |
| EPI_ISL_794166 | USA/NY-Wadsworth-292121-01/2020 | 2020-12-28 | North America/USA/New York/Hamilton | Wadsworth Center, New York State Department.of Health | Wadsworth Center, New York State Department.of Health | Kirsten St. George et al |

| | | | | | | |
|---|---|---|---|---|---|---|
| EPI_ISL_794178 | USA/NY-Wadsworth-293189-01/2020 | 2020-12-30 | North America/USA/New York/Hamilton | Wadsworth Center, New York State Department.of Health | Wadsworth Center, New York State Department.of Health | Kirsten St. George et al |
| EPI_ISL_794184 | USA/NY-Wadsworth-293265-01/2020 | 2020-12-28 | North America/USA/New York/Chemung | URMC LABS | Wadsworth Center, New York State Department.of Health | Kirsten St. George et al |
| EPI_ISL_794189 | USA/NY-Wadsworth-293270-01/2020 | 2020-12-28 | North America/USA/New York/Monroe | URMC LABS | Wadsworth Center, New York State Department.of Health | Kirsten St. George et al |
| EPI_ISL_794191 | USA/NY-Wadsworth-293273-01/2020 | 2020-12-28 | North America/USA/New York/Monroe | URMC LABS | Wadsworth Center, New York State Department.of Health | Kirsten St. George et al |
| EPI_ISL_794194 | USA/NY-Wadsworth-293276-01/2020 | 2020-12-28 | North America/USA/New York/Ontario | URMC LABS | Wadsworth Center, New York State Department.of Health | Kirsten St. George et al |
| EPI_ISL_794198 | USA/NY-Wadsworth-293281-01/2020 | 2020-12-28 | North America/USA/New York/Monroe | URMC LABS | Wadsworth Center, New York State Department.of Health | Kirsten St. George et al |
| EPI_ISL_794202 | USA/NY-Wadsworth-293285-01/2020 | 2020-12-28 | North America/USA/New York/Genesee | URMC LABS | Wadsworth Center, New York State Department.of Health | Kirsten St. George et al |
| EPI_ISL_794209 | USA/NY-Wadsworth-21000307-01/20 | 2020-12-25 | North America/USA/New York/Westche | WESTCHESTER MEDICAL CENTER | Wadsworth Center, New York State Department.of Health | Kirsten St. George et al |

| | | | | | | |
|---|---|---|---|---|---|---|
| EPI_ISL_794215 | USA/NY-Wadsworth-21000314-01/20 | 2020-12-26 | North America/USA/New York/Dutchess | WESTCHESTER MEDICAL CENTER | Wadsworth Center, New York State Department.of Health | Kirsten St. George et al |
| EPI_ISL_794223 | USA/NY-Wadsworth-21000323-01/20 | 2020-12-27 | North America/USA/New York/Westche | WESTCHESTER MEDICAL CENTER | Wadsworth Center, New York State Department.of Health | Kirsten St. George et al |
| EPI_ISL_794226 | USA/NY-Wadsworth-21000327-01/20 | 2020-12-27 | North America/USA/New York/Westche | WESTCHESTER MEDICAL CENTER | Wadsworth Center, New York State Department.of Health | Kirsten St. George et al |
| EPI_ISL_794235 | USA/NY-Wadsworth-21000337-01/20 | 2020-12-28 | North America/USA/New York/Dutchess | WESTCHESTER MEDICAL CENTER | Wadsworth Center, New York State Department.of Health | Kirsten St. George et al |
| EPI_ISL_794237 | USA/NY-Wadsworth-21000341-01/20 | 2020-12-29 | North America/USA/New York/Westche | WESTCHESTER MEDICAL CENTER | Wadsworth Center, New York State Department.of Health | Kirsten St. George et al |
| EPI_ISL_794243 | USA/NY-Wadsworth-21000347-01/20 | 2020-12-28 | North America/USA/New York/Dutchess | WESTCHESTER MEDICAL CENTER | Wadsworth Center, New York State Department.of Health | Kirsten St. George et al |
| EPI_ISL_794244 | USA/NY-Wadsworth-21001326-01/20 | 2020-12-30 | North America/USA/New York/Dutchess | WESTCHESTER MEDICAL CENTER | Wadsworth Center, New York State Department.of Health | Kirsten St. George et al |
| EPI_ISL_794245 | USA/NY-Wadsworth-21001327-01/20 | 2020-12-31 | North America/USA/New York/Ulster C | WESTCHESTER MEDICAL CENTER | Wadsworth Center, New York State Department.of Health | Kirsten St. George et al |

| | | | | | | |
|---|---|---|---|---|---|---|
| EPI_ISL_794247 | USA/NY-Wadsworth-21001329-01/20 | 2020-12-31 | North America/USA/New York/Dutchess | WESTCHESTER MEDICAL CENTER | Wadsworth Center, New York State Department.of Health | Kirsten St. George et al |
| EPI_ISL_794248 | USA/NY-Wadsworth-21001330-01/20 | 2020-12-31 | North America/USA/New York/Westche | WESTCHESTER MEDICAL CENTER | Wadsworth Center, New York State Department.of Health | Kirsten St. George et al |
| EPI_ISL_794249 | USA/NY-Wadsworth-21001331-01/20 | 2020-12-31 | North America/USA/New York/Putnam | WESTCHESTER MEDICAL CENTER | Wadsworth Center, New York State Department.of Health | Kirsten St. George et al |
| EPI_ISL_794253 | USA/NY-Wadsworth-21001335-01/20 | 2020-12-31 | North America/USA/New York/Westche | WESTCHESTER MEDICAL CENTER | Wadsworth Center, New York State Department.of Health | Kirsten St. George et al |
| EPI_ISL_794254 | USA/NY-Wadsworth-21001336-01/20 | 2020-12-31 | North America/USA/New York/ | WESTCHESTER MEDICAL CENTER | Wadsworth Center, New York State Department.of Health | Kirsten St. George et al |
| EPI_ISL_794259 | USA/NY-Wadsworth-21001341-01/20 | 2020-12-31 | North America/USA/New York/Ulster C | WESTCHESTER MEDICAL CENTER | Wadsworth Center, New York State Department.of Health | Kirsten St. George et al |
| EPI_ISL_794270 | USA/NY-Wadsworth-21001452-01/20 | 2021-01-02 | North America/USA/New York/Dutchess | WESTCHESTER MEDICAL CENTER | Wadsworth Center, New York State Department.of Health | Kirsten St. George et al |
| EPI_ISL_794307 | USA/NY-Wadsworth-21001729-01/20 | 2020-12-20 | North America/USA/New York/Bronx | Columbia University Irving Medical Center | Wadsworth Center, New York State Department.of Health | Kirsten St. George et al |

| | | | | | | |
|---|---|---|---|---|---|---|
| EPI_ISL_794309 | USA/NY-Wadsworth-21001596-01/20 | 2021-01-05 | North America/USA/New York/Saratoga | Wadsworth Center, New York State Department.of Health | Wadsworth Center, New York State Department.of Health | Kirsten St. George et al |
| EPI_ISL_794596 | Egypt/CPHL-NRC-16/2020 | 2020-05-06 | Africa/Egypt/Egypt/ | Central Laboratories, Egyptian Ministry of Health and Population | Central Laboratories, Egyptian Ministry of Health and Population | Roshdy et al |
| EPI_ISL_794597 | Egypt/CPHL-NRC-15/2020 | 2020-06-04 | Africa/Egypt/Egypt/ | Central Laboratories, Egyptian Ministry of Health and Population | Central Laboratories, Egyptian Ministry of Health and Population | Roshdy et al |
| EPI_ISL_794598 | Egypt/CPHL-NRC-14/2020 | 2020-06-02 | Africa/Egypt/Egypt/ | Central Laboratories, Egyptian Ministry of Health and Population | Central Laboratories, Egyptian Ministry of Health and Population | Roshdy et al |
| EPI_ISL_794600 | Egypt/CPHL-NRC-3/2020 | 2020-04-05 | Africa/Egypt/Egypt/ | Central Laboratories, Egyptian Ministry of Health and Population | Central Laboratories, Egyptian Ministry of Health and Population | Kayed et al |
| EPI_ISL_794601 | Egypt/CPHL-NRC-1/2020 | 2020-06-10 | Africa/Egypt/Egypt/ | Central Laboratories, Egyptian Ministry of Health and Population | Central Laboratories, Egyptian Ministry of Health and Population | Kayed et al |
| EPI_ISL_794602 | Egypt/CPHL-NRC-23/2020 | 2020-03-13 | Africa/Egypt/Egypt/ | Central Laboratories, Egyptian Ministry of Health and Population | Central Laboratories, Egyptian Ministry of Health and Population | Kayed et al |
| EPI_ISL_794646 | USA/CA-LACPHL-AF00121/2021 | 2021-01-06 | North America/USA/California/Los Ange | UCLA Clinical Micro Lab | Los Angeles County PHL | P. Hemarajata et al. et al |

| | | | | | | |
|---|---|---|---|---|---|---|
| EPI_ISL_794649 | Colombia/DC-INS-VG-48/2020 | 2020-12-30 | South America/Colombia/Bogota/ | Fundaci√≥n Cardio Infantil | Instituto Nacional de Salud - Direcci√≥n de Investigaci√≥n en Salud P√∫blica | Katherine Laiton-Donato et al |
| EPI_ISL_794650 | Colombia/DC-INS-VG-49/2020 | 2020-12-30 | South America/Colombia/Bogota/ | Fundaci√≥n Cardio Infantil | Instituto Nacional de Salud - Direcci√≥n de Investigaci√≥n en Salud P√∫blica | Katherine Laiton-Donato et al |
| EPI_ISL_794653 | Colombia/TOL-INS-VG-112/2020 | 2020-12-30 | South America/Colombia/Tolima/ | LSP DEL TOLIMA | Instituto Nacional de Salud - Direcci√≥n de Investigaci√≥n en Salud P√∫blica | Katherine Laiton-Donato et al |
| EPI_ISL_794654 | Colombia/VAC-INS-VG-101/2020 | 2020-12-30 | South America/Colombia/Valle del Cauc | Fundaci√≥n Valle del Lili | Instituto Nacional de Salud - Direcci√≥n de Investigaci√≥n en Salud P√∫blica | Katherine Laiton-Donato et al |
| EPI_ISL_794656 | Colombia/NSA-INS-VG-108/2020 | 2020-12-31 | South America/Colombia/Norte de Sant | UNIDAD HEMATOLOGICA ESPECIALIZADA | Instituto Nacional de Salud - Direcci√≥n de Investigaci√≥n en Salud P√∫blica | Katherine Laiton-Donato et al |
| EPI_ISL_794659 | Colombia/DC-INS-VG-68/2020 | 2020-12-30 | South America/Colombia/Bogota/ | HOSPITAL UNIVERSITARIO SAN IGNACIO | Instituto Nacional de Salud - Direcci√≥n de Investigaci√≥n en Salud P√∫blica | Katherine Laiton-Donato et al |
| EPI_ISL_794660 | Colombia/DC-INS-VG-69/2020 | 2020-12-30 | South America/Colombia/Bogota/ | HOSPITAL UNIVERSITARIO SAN IGNACIO | Instituto Nacional de Salud - Direcci√≥n de Investigaci√≥n en Salud P√∫blica | Katherine Laiton-Donato et al |
| EPI_ISL_794661 | Colombia/DC-INS-VG-71/2020 | 2020-12-28 | South America/Colombia/Bogota/ | DRECCION DE SANIDAD POLICIA NACIONAL | Instituto Nacional de Salud - Direcci√≥n de Investigaci√≥n en Salud P√∫blica | Katherine Laiton-Donato et al |

| | | | | | | |
|---|---|---|---|---|---|---|
| EPI_ISL_794666 | Colombia/ANT-INS-VG-180/2021 | 2021-01-05 | South America/Colombia/Antioquia/ | SYNLAB REGIONAL NOROCCIDENTE | Instituto Nacional de Salud - Direcciv≥n de Investigaciv≥n en Salud Pv∫blica | Katherine Laiton-Donato et al |
| EPI_ISL_794735 | Tunisia/1280/2021 | 2021-01-06 | Africa/Tunisia/Tunis/ | 1-Laboratory of Microbiology, National Reference Lab, Charles Nicolle Hospital; 2-University of Tunis ElManar, Faculty of Medicine of Tunis, LR99ES09, Tunis, Tunisia | 1-Clinical and Experimental Pharmacology Lab, LR16SP02, National Center of Pharmacovigilance, University of Tunis El Manar, Tunis, Tunisia. 2-Neurodegenerative diseases and psychiatric troubles, LR18SP03, Razi Hospital, University of Tunis El Manar, Tunis, Tunisia. 3-Ministry of Health, National Observatory of New and Emerging Diseases, 1006, Tunis, Tunisia | Ilhem Boutiba-Ben Boubaker et al |

| | | | | | | |
|---|---|---|---|---|---|---|
| EPI_ISL_794737 | Tunisia/1241/2021 | 2021-01-06 | Africa/Tunisia/Nabeul/ | 1-Laboratory of Microbiology, National Reference Lab, Charles Nicolle Hospital; 2-University of Tunis ElManar, Faculty of Medicine of Tunis, LR99ES09, Tunis, Tunisia | 1-Clinical and Experimental Pharmacology Lab, LR16SP02, National Center of Pharmacovigilance, University of Tunis El Manar, Tunis, Tunisia. 2-Neurodegenerative diseases and psychiatric troubles, LR18SP03, Razi Hospital, University of Tunis El Manar, Tunis, Tunisia. 3-Ministry of Health, National Observatory of New and Emerging Diseases, 1006, Tunis, Tunisia | Ilhem Boutiba-Ben Boubaker et al |

| | | | | | | |
|---|---|---|---|---|---|---|
| EPI_ISL_794738 | Tunisia/1267/2021 | 2021-01-06 | Africa/Tunisia/Tunis/ | 1-Laboratory of Microbiology, National Reference Lab, Charles Nicolle Hospital; 2-University of Tunis ElManar, Faculty of Medicine of Tunis, LR99ES09, Tunis, Tunisia | 1-Clinical and Experimental Pharmacology Lab, LR16SP02, National Center of Pharmacovigilance, University of Tunis El Manar, Tunis, Tunisia. 2-Neurodegenerative diseases and psychiatric troubles, LR18SP03, Razi Hospital, University of Tunis El Manar, Tunis, Tunisia. 3-Ministry of Health, National Observatory of New and Emerging Diseases, 1006, Tunis, Tunisia | Ilhem Boutiba-Ben Boubaker et al |
| EPI_ISL_794749 | Italy/APU-IZSPB_407PT/2020 | 2020-11-25 | Europe/Italy/Apulia/Bari | Istituto Zooprofilattico Sperimentale della Puglia e della Basilicata | Istituto Zooprofilattico Sperimentale della Puglia e della Basilicata | Parisi A. et al |
| EPI_ISL_794751 | Italy/APU-IZSPB_409PT/2020 | 2020-12-23 | Europe/Italy/Apulia/Lecce | Ospedale Vito Fazzi | Istituto Zooprofilattico Sperimentale della Puglia e della Basilicata | Parisi A. et al |
| EPI_ISL_794755 | Italy/APU-IZSPB_414PT/2020 | 2020-11-25 | Europe/Italy/Apulia/Foggia | Istituto Zooprofilattico Sperimentale della Puglia e della Basilicata | Istituto Zooprofilattico Sperimentale della Puglia e della Basilicata | Parisi A. et al |

| | | | | | | |
|---|---|---|---|---|---|---|
| EPI_ISL_794758 | Italy/APU-IZSPB_418PT/2020 | 2020-12-23 | Europe/Italy/Apulia/Brindisi | Presidio di Brindisi Di Summa - Perrino | Istituto Zooprofilattico Sperimentale della Puglia e della Basilicata | Parisi A. et al |
| EPI_ISL_794762 | Italy/APU-IZSPB_426PT/2020 | 2020-12-11 | Europe/Italy/Apulia/Foggia | Istituto Zooprofilattico Sperimentale della Puglia e della Basilicata | Istituto Zooprofilattico Sperimentale della Puglia e della Basilicata | Parisi A. et al |
| EPI_ISL_794787 | Italy/APU-IZSPB_460PT/2020 | 2020-12-23 | Europe/Italy/Apulia/Foggia | Istituto Zooprofilattico Sperimentale della Puglia e della Basilicata | Istituto Zooprofilattico Sperimentale della Puglia e della Basilicata | Parisi A. et al |
| EPI_ISL_794790 | Italy/APU-IZSPB_463PT/2020 | 2020-12-14 | Europe/Italy/Apulia/Foggia | Istituto Zooprofilattico Sperimentale della Puglia e della Basilicata | Istituto Zooprofilattico Sperimentale della Puglia e della Basilicata | Parisi A. et al |
| EPI_ISL_794797 | Italy/APU-IZSPB_470PT/2020 | 2020-12-17 | Europe/Italy/Apulia/Foggia | Istituto Zooprofilattico Sperimentale della Puglia e della Basilicata | Istituto Zooprofilattico Sperimentale della Puglia e della Basilicata | Parisi A. et al |
| EPI_ISL_794802 | Italy/APU-IZSPB_475PT/2020 | 2020-12-17 | Europe/Italy/Apulia/Foggia | Istituto Zooprofilattico Sperimentale della Puglia e della Basilicata | Istituto Zooprofilattico Sperimentale della Puglia e della Basilicata | Parisi A. et al |
| EPI_ISL_794804 | Italy/APU-IZSPB_477PT/2020 | 2020-12-15 | Europe/Italy/Apulia/Foggia | Istituto Zooprofilattico Sperimentale della Puglia e della Basilicata | Istituto Zooprofilattico Sperimentale della Puglia e della Basilicata | Parisi A. et al |
| EPI_ISL_794806 | Italy/APU-IZSPB_479PT/2020 | 2020-12-15 | Europe/Italy/Apulia/Bari | Istituto Zooprofilattico Sperimentale della Puglia e della Basilicata | Istituto Zooprofilattico Sperimentale della Puglia e della Basilicata | Parisi A. et al |

| | | | | | | |
|---|---|---|---|---|---|---|
| EPI_ISL_794808 | Italy/APU-IZSPB_481PT/2020 | 2020-12-15 | Europe/Italy/Apulia/Barletta-Andria-Tra | Istituto Zooprofilattico Sperimentale della Puglia e della Basilicata | Istituto Zooprofilattico Sperimentale della Puglia e della Basilicata | Parisi A. et al |
| EPI_ISL_794812 | Italy/APU-IZSPB_485PT/2020 | 2020-12-17 | Europe/Italy/Apulia/Foggia | Istituto Zooprofilattico Sperimentale della Puglia e della Basilicata | Istituto Zooprofilattico Sperimentale della Puglia e della Basilicata | Parisi A. et al |
| EPI_ISL_794818 | Greece/CP0794239/2020 | 2020-12-30 | Europe/Greece/Athens/ | Greek Genome Center, Biomedical Research Foundation of the Academy of Athens (BRFAA) | Greek Genome Center, Biomedical Research Foundation of the Academy of Athens (BRFAA) | Emmanouil Athanasiadis et al |
| EPI_ISL_794819 | Greece/CP0800981/2020 | 2020-12-30 | Europe/Greece/Athens/ | Greek Genome Center, Biomedical Research Foundation of the Academy of Athens (BRFAA) | Greek Genome Center, Biomedical Research Foundation of the Academy of Athens (BRFAA) | Emmanouil Athanasiadis et al |
| EPI_ISL_794820 | Greece/CP0800976/2020 | 2020-12-30 | Europe/Greece/Athens/ | Greek Genome Center, Biomedical Research Foundation of the Academy of Athens (BRFAA) | Greek Genome Center, Biomedical Research Foundation of the Academy of Athens (BRFAA) | Emmanouil Athanasiadis et al |
| EPI_ISL_794826 | Italy/VEN-IZSVe-20RS2155-13_VI/202 | 2020-11-13 | Europe/Italy/Veneto/Vicenza | ULSS 7 Pedemontana - Distretto 2 | Istituto Zooprofilattico Sperimentale delle Venezie | Adelaide Milani et al |
| EPI_ISL_794827 | Italy/VEN-IZSVe-20RS2155-12_VI/202 | 2020-11-06 | Europe/Italy/Veneto/Vicenza | ULSS 7 Pedemontana - Distretto 2 | Istituto Zooprofilattico Sperimentale delle Venezie | Adelaide Milani et al |

| | | | | | | |
|---|---|---|---|---|---|---|
| EPI_ISL_794828 | Italy/VEN-IZSVe-20RS2155-11_VI/20 | 2020-11-02 | Europe/Italy/Veneto/Vicenza | ULSS 7 Pedemontana - Distretto 2 | Istituto Zooprofilattico Sperimentale delle Venezie | Adelaide Milani et al |
| EPI_ISL_795348 | Denmark/DCGC-25009/2020 | 2020-12-28 | Europe/Denmark/Hovedstaden/ | Department of Virus and Microbiological Special Diagnostics, Statens Serum Institut, Copenhagen, Denmark | Albertsen Lab, Department of Chemistry and Bioscience, Aalborg University, Denmark | Danish Covid-19 Genome Consortium et al |
| EPI_ISL_795575 | Denmark/DCGC-24487/2020 | 2020-12-28 | Europe/Denmark/Hovedstaden/ | Department of Virus and Microbiological Special Diagnostics, Statens Serum Institut, Copenhagen, Denmark | Albertsen Lab, Department of Chemistry and Bioscience, Aalborg University, Denmark | Danish Covid-19 Genome Consortium et al |
| EPI_ISL_795850 | Denmark/DCGC-24981/2020 | 2020-12-28 | Europe/Denmark/Nordjylland/ | Department of Virus and Microbiological Special Diagnostics, Statens Serum Institut, Copenhagen, Denmark | Albertsen Lab, Department of Chemistry and Bioscience, Aalborg University, Denmark | Danish Covid-19 Genome Consortium et al |
| EPI_ISL_796024 | Hebei/IVDC-08-01/2021 | 2021-01-04 | Asia/China/Hebei/Shijiazhuang | Hebei Provincial Center for Disease Control and Prevention, Shijiazhuang, Hebei Province; National Institute for Viral Disease Control and Prevention, China CDC | Hebei Provincial Center for Disease Control and Prevention, Shijiazhuang, Hebei Province; National Institute for Viral Disease Control and Prevention, China CDC | Shunxiang Qi et al |

| | | | | | | |
|---|---|---|---|---|---|---|
| EPI_ISL_796026 | Hebei/IVDC-10-01/2021 | 2021-01-04 | Asia/China/Hebei/Shijiazhuang | Hebei Provincial Center for Disease Control and Prevention, Shijiazhuang, Hebei Province; National Institute for Viral Disease Control and Prevention, China CDC | Hebei Provincial Center for Disease Control and Prevention, Shijiazhuang, Hebei Province; National Institute for Viral Disease Control and Prevention, China CDC | Shunxiang Qi et al |
| EPI_ISL_796030 | Germany/NW-IOV-K-5622/2020 | 2020-09-19 | Europe/Germany/North Rhine Westphal | Institute of Virology, University of Cologne | Institute of Virology, University of Cologne | Saleta Sierra et al |
| EPI_ISL_796033 | Germany/NW-IOV-K-5625/2020 | 2020-09-16 | Europe/Germany/North Rhine Westphal | Institute of Virology, University of Cologne | Institute of Virology, University of Cologne | Saleta Sierra et al |
| EPI_ISL_796037 | Germany/NW-IOV-K-5630/2020 | 2020-09-30 | Europe/Germany/North Rhine Westphal | Institute of Virology, University of Cologne | Institute of Virology, University of Cologne | Saleta Sierra et al |
| EPI_ISL_796038 | Germany/NW-IOV-K-5631/2020 | 2020-09-26 | Europe/Germany/North Rhine Westphal | Institute of Virology, University of Cologne | Institute of Virology, University of Cologne | Saleta Sierra et al |
| EPI_ISL_796039 | Germany/NW-IOV-K-5671/2020 | 2020-09-14 | Europe/Germany/North Rhine Westphal | Institute of Virology, University of Cologne | Institute of Virology, University of Cologne | Saleta Sierra et al |
| EPI_ISL_796040 | Germany/NW-IOV-K-5672/2020 | 2020-09-15 | Europe/Germany/North Rhine Westphal | Institute of Virology, University of Cologne | Institute of Virology, University of Cologne | Saleta Sierra et al |
| EPI_ISL_796042 | Germany/NW-IOV-K-5674/2020 | 2020-09-17 | Europe/Germany/North Rhine Westphal | Institute of Virology, University of Cologne | Institute of Virology, University of Cologne | Saleta Sierra et al |

| | | | | | | |
|---|---|---|---|---|---|---|
| EPI_ISL_796047 | Germany/NW-IOV-K-5679/2020 | 2020-09-24 | Europe/Germany/North Rhine Westphal | Institute of Virology, University of Cologne | Institute of Virology, University of Cologne | Saleta Sierra et al |
| EPI_ISL_796049 | Germany/NW-IOV-K-5682/2020 | 2020-09-30 | Europe/Germany/North Rhine Westphal | Institute of Virology, University of Cologne | Institute of Virology, University of Cologne | Saleta Sierra et al |
| EPI_ISL_796050 | Germany/NW-IOV-K-5683/2020 | 2020-10-01 | Europe/Germany/North Rhine Westphal | Institute of Virology, University of Cologne | Institute of Virology, University of Cologne | Saleta Sierra et al |
| EPI_ISL_796052 | Germany/NW-IOV-K-5685/2020 | 2020-10-05 | Europe/Germany/North Rhine Westphal | Institute of Virology, University of Cologne | Institute of Virology, University of Cologne | Saleta Sierra et al |
| EPI_ISL_796053 | Germany/NW-IOV-K-5686/2020 | 2020-10-06 | Europe/Germany/North Rhine Westphal | Institute of Virology, University of Cologne | Institute of Virology, University of Cologne | Saleta Sierra et al |
| EPI_ISL_796058 | Germany/NW-IOV-K-5691/2020 | 2020-10-01 | Europe/Germany/North Rhine Westphal | Institute of Virology, University of Cologne | Institute of Virology, University of Cologne | Saleta Sierra et al |
| EPI_ISL_796062 | Germany/NW-IOV-K-5696/2020 | 2020-10-29 | Europe/Germany/North Rhine Westphal | Institute of Virology, University of Cologne | Institute of Virology, University of Cologne | Saleta Sierra et al |
| EPI_ISL_796072 | Spain/VC-IBV-98004364/2020 | 2020-04-06 | Europe/Spain/Comunitat Valenciana/Ca | Servicio de Microbiolog√≠a. Hospital General Universitario de Castell√≥n | SeqCOVID-SPAIN consortium/IBV(CSIC) | Rosario Moreno Mu√±oz et al |
| EPI_ISL_796092 | Spain/VC-IBV-98004408/2020 | 2020-04-13 | Europe/Spain/Comunitat Valenciana/Ca | Servicio de Microbiolog√≠a. Hospital General Universitario de Castell√≥n | SeqCOVID-SPAIN consortium/IBV(CSIC) | Rosario Moreno Mu√±oz et al |
| EPI_ISL_796096 | Spain/VC-IBV-98004510/2020 | 2020-03-26 | Europe/Spain/Comunitat Valenciana/Ca | Servicio de Microbiolog√≠a. Hospital General Universitario de Castell√≥n | SeqCOVID-SPAIN consortium/IBV(CSIC) | Rosario Moreno Mu√±oz et al |
| EPI_ISL_796099 | Spain/VC-IBV-98004518/2020 | 2020-03-26 | Europe/Spain/Comunitat Valenciana/Ca | Servicio de Microbiolog√≠a. Hospital General Universitario de Castell√≥n | SeqCOVID-SPAIN consortium/IBV(CSIC) | Rosario Moreno Mu√±oz et al |

| | | | | | | |
|---|---|---|---|---|---|---|
| EPI_ISL_796105 | Spain/VC-IBV-98004534/2020 | 2020-03-19 | Europe/Spain/Comunitat Valenciana/Ca | Servicio de Microbiolog√≠a. Hospital General Universitario de Castell√≥n | SeqCOVID-SPAIN consortium/IBV(CSIC) | Rosario Moreno Mu√±oz et al |
| EPI_ISL_796106 | Spain/VC-IBV-98004535/2020 | 2020-03-19 | Europe/Spain/Comunitat Valenciana/Ca | Servicio de Microbiolog√≠a. Hospital General Universitario de Castell√≥n | SeqCOVID-SPAIN consortium/IBV(CSIC) | Rosario Moreno Mu√±oz et al |
| EPI_ISL_796107 | Spain/VC-IBV-98004537/2020 | 2020-03-20 | Europe/Spain/Comunitat Valenciana/Ca | Servicio de Microbiolog√≠a. Hospital General Universitario de Castell√≥n | SeqCOVID-SPAIN consortium/IBV(CSIC) | Rosario Moreno Mu√±oz et al |
| EPI_ISL_796109 | Spain/VC-IBV-98004539/2020 | 2020-03-20 | Europe/Spain/Comunitat Valenciana/Ca | Servicio de Microbiolog√≠a. Hospital General Universitario de Castell√≥n | SeqCOVID-SPAIN consortium/IBV(CSIC) | Rosario Moreno Mu√±oz et al |
| EPI_ISL_796114 | Spain/VC-IBV-98004556/2020 | 2020-04-02 | Europe/Spain/Comunitat Valenciana/Ca | Servicio de Microbiolog√≠a. Hospital General Universitario de Castell√≥n | SeqCOVID-SPAIN consortium/IBV(CSIC) | Rosario Moreno Mu√±oz et al |
| EPI_ISL_796140 | Spain/VC-IBV-98006953/2020 | 2020-03-21 | Europe/Spain/Comunitat Valenciana/Ca | Servicio de Microbiolog√≠a. Hospital General Universitario de Castell√≥n | SeqCOVID-SPAIN consortium/IBV(CSIC) | Rosario Moreno Mu√±oz et al |
| EPI_ISL_796146 | Spain/VC-IBV-98006965/2020 | 2020-03-23 | Europe/Spain/Comunitat Valenciana/Ca | Servicio de Microbiolog√≠a. Hospital General Universitario de Castell√≥n | SeqCOVID-SPAIN consortium/IBV(CSIC) | Rosario Moreno Mu√±oz et al |
| EPI_ISL_796148 | Spain/VC-IBV-98006967/2020 | 2020-03-23 | Europe/Spain/Comunitat Valenciana/Ca | Servicio de Microbiolog√≠a. Hospital General Universitario de Castell√≥n | SeqCOVID-SPAIN consortium/IBV(CSIC) | Rosario Moreno Mu√±oz et al |
| EPI_ISL_796152 | Spain/VC-IBV-98006977/2020 | 2020-03-24 | Europe/Spain/Comunitat Valenciana/Ca | Servicio de Microbiolog√≠a. Hospital General Universitario de Castell√≥n | SeqCOVID-SPAIN consortium/IBV(CSIC) | Rosario Moreno Mu√±oz et al |
| EPI_ISL_796153 | Spain/VC-IBV-98006978/2020 | 2020-03-24 | Europe/Spain/Comunitat Valenciana/Ca | Servicio de Microbiolog√≠a. Hospital General Universitario de Castell√≥n | SeqCOVID-SPAIN consortium/IBV(CSIC) | Rosario Moreno Mu√±oz et al |
| EPI_ISL_796156 | Spain/VC-IBV-98006981/2020 | 2020-04-15 | Europe/Spain/Comunitat Valenciana/Ca | Servicio de Microbiolog√≠a. Hospital General Universitario de Castell√≥n | SeqCOVID-SPAIN consortium/IBV(CSIC) | Rosario Moreno Mu√±oz et al |
| EPI_ISL_796316 | Switzerland/UR-ETHZ-430332/2020 | 2020-12-27 | Europe/Switzerland/Uri/ | Viollier AG | Department of Biosystems Science and Engineering, ETH Z√ºrich | Chaoran Chen et al |

| | | | | | | |
|---|---|---|---|---|---|---|
| EPI_ISL_796328 | Switzerland/AG-ETHZ-430232/2020 | 2020-12-26 | Europe/Switzerland/Aargau/ | Viollier AG | Department of Biosystems Science and Engineering, ETH Z√ºrich | Chaoran Chen et al |
| EPI_ISL_796403 | Switzerland/BE-ETHZ-430615/2020 | 2020-12-30 | Europe/Switzerland/Bern/ | Viollier AG | Department of Biosystems Science and Engineering, ETH Z√ºrich | Chaoran Chen et al |
| EPI_ISL_796425 | Switzerland/BL-ETHZ-430035/2020 | 2020-12-25 | Europe/Switzerland/Basel-Land/ | Viollier AG | Department of Biosystems Science and Engineering, ETH Z√ºrich | Chaoran Chen et al |
| EPI_ISL_796450 | Switzerland/NE-ETHZ-430295/2020 | 2020-12-27 | Europe/Switzerland/Neuch√¢tel/ | Viollier AG | Department of Biosystems Science and Engineering, ETH Z√ºrich | Chaoran Chen et al |
| EPI_ISL_796466 | Switzerland/SH-ETHZ-430316/2020 | 2020-12-27 | Europe/Switzerland/Schaffhausen/ | Viollier AG | Department of Biosystems Science and Engineering, ETH Z√ºrich | Chaoran Chen et al |
| EPI_ISL_796515 | Switzerland/SO-ETHZ-430358/2020 | 2020-12-27 | Europe/Switzerland/Solothurn/ | Viollier AG | Department of Biosystems Science and Engineering, ETH Z√ºrich | Chaoran Chen et al |
| EPI_ISL_796535 | Switzerland/SO-ETHZ-430538/2020 | 2020-12-29 | Europe/Switzerland/Solothurn/ | Viollier AG | Department of Biosystems Science and Engineering, ETH Z√ºrich | Chaoran Chen et al |
| EPI_ISL_796550 | Switzerland/SZ-ETHZ-430718/2020 | 2020-12-31 | Europe/Switzerland/Schwyz/ | Viollier AG | Department of Biosystems Science and Engineering, ETH Z√ºrich | Chaoran Chen et al |

| | | | | | | |
|---|---|---|---|---|---|---|
| EPI_ISL_796590 | Switzerland/ZH-ETHZ-430187/2020 | 2020-12-26 | Europe/Switzerland/Z√ºrich/ | Viollier AG | Department of Biosystems Science and Engineering, ETH Z√ºrich | Chaoran Chen et al |
| EPI_ISL_796665 | Norway/7088/2020 | 2020-12-21 | Europe/Norway/Viken/ | Akershus University Hospital, Department for Microbiology and Infectious Disease Control | Norwegian Institute of Public Health, Department of Virology | Kathrine Stene-Johansen et al |
| EPI_ISL_796702 | Norway/7126/2020 | 2020-12-09 | Europe/Norway/Oslo/ | Unilabs Laboratory Medicine | Norwegian Institute of Public Health, Department of Virology | Kathrine Stene-Johansen et al |
| EPI_ISL_796740 | Portugal/PT2149/2020 | 2020-12-26 | Europe/Portugal/Portugal/ | Instituto Nacional de Saude (INSA) | Instituto Nacional de Saude (INSA) | Borges et al et al |
| EPI_ISL_796741 | Portugal/PT2166/2020 | 2020-12-20 | Europe/Portugal/Portugal/ | Instituto Nacional de Saude (INSA) | Instituto Nacional de Saude (INSA) | Borges et al et al |
| EPI_ISL_796743 | Portugal/PT2135/2020 | 2020-12-21 | Europe/Portugal/Portugal/ | Instituto Nacional de Saude (INSA) | Instituto Nacional de Saude (INSA) | Borges et al et al |
| EPI_ISL_796748 | Portugal/PT2141/2020 | 2020-12-22 | Europe/Portugal/Portugal/ | Instituto Nacional de Saude (INSA) | Instituto Nacional de Saude (INSA) | Borges et al et al |
| EPI_ISL_796751 | Portugal/PT2150/2020 | 2020-12-26 | Europe/Portugal/Portugal/ | Instituto Nacional de Saude (INSA) | Instituto Nacional de Saude (INSA) | Borges et al et al |
| EPI_ISL_796753 | Portugal/PT2177/2020 | 2020-12-28 | Europe/Portugal/Portugal/ | Instituto Nacional de Saude (INSA) | Instituto Nacional de Saude (INSA) | Borges et al et al |
| EPI_ISL_796765 | Portugal/PT2174/2021 | 2021-01-04 | Europe/Portugal/Portugal/ | Instituto Nacional de Saude (INSA) | Instituto Nacional de Saude (INSA) | Borges et al et al |
| EPI_ISL_796779 | Portugal/PT2145/2020 | 2020-12-02 | Europe/Portugal/Portugal/ | Instituto Nacional de Saude (INSA) | Instituto Nacional de Saude (INSA) | Borges et al et al |

| | | | | | | |
|---|---|---|---|---|---|---|
| EPI_ISL_796782 | Tunisia/69681/2020 | 2020-12-24 | Africa/Tunisia/Monastir/Ksar Hellal | Laboratory of Microbiology, National Reference Lab, Charles Nicolle Hospital; 2-University of Tunis ElManar, Faculty of Medicine of Tunis, LR99ES09, Tunis, Tunisia | Clinical and Experimental Pharmacology Lab, LR16SP02, National Center of Pharmacovigilance, University of Tunis El Manar, Tunis, Tunisia. 2-Neurodegenerative diseases and psychiatric troubles, LR18SP03, Razi Hospital, University of Tunis El Manar, Tunis, Tunisia. 3-Ministry of Health, National Observatory of New and Emerging Diseases, 1006, Tunis, Tunisia | Ilhem Boutiba-Ben Boubaker et al |
| EPI_ISL_796945 | England/ALDP-DA54AA/2020 | 2020-12-30 | Europe/United Kingdom/England/ | Lighthouse Lab in Alderley Park | Wellcome Sanger Institute for the COVID-19 Genomics UK (COG-UK) Consortium | Jacquelyn Wynn et al |
| EPI_ISL_796965 | England/ALDP-DA54E6/2020 | 2020-12-30 | Europe/United Kingdom/England/ | Lighthouse Lab in Alderley Park | Wellcome Sanger Institute for the COVID-19 Genomics UK (COG-UK) Consortium | Jacquelyn Wynn et al |
| EPI_ISL_797296 | England/MILK-DB0433/2020 | 2020-12-29 | Europe/United Kingdom/England/ | Lighthouse Lab in Milton Keynes | Wellcome Sanger Institute for the COVID-19 Genomics UK (COG-UK) Consortium | The Lighthouse Lab in Milton Keynes et al |

| | | | | | | |
|---|---|---|---|---|---|---|
| EPI_ISL_797342 | England/MILK-DB049D/2020 | 2020-12-29 | Europe/United Kingdom/England/ | Lighthouse Lab in Milton Keynes | Wellcome Sanger Institute for the COVID-19 Genomics UK (COG-UK) Consortium | The Lighthouse Lab in Milton Keynes et al |
| EPI_ISL_797461 | England/ALDP-DB6181/2021 | 2021-01-01 | Europe/United Kingdom/England/ | Lighthouse Lab in Alderley Park | Wellcome Sanger Institute for the COVID-19 Genomics UK (COG-UK) Consortium | Jacquelyn Wynn et al |
| EPI_ISL_797465 | England/CAMC-DD8AF7/2020 | 2020-12-28 | Europe/United Kingdom/England/ | Lighthouse Lab in Cambridge | Wellcome Sanger Institute for the COVID-19 Genomics UK (COG-UK) Consortium | Rob Howes et al |
| EPI_ISL_797552 | England/CAMC-DD959B/2020 | 2020-12-29 | Europe/United Kingdom/England/ | Lighthouse Lab in Cambridge | Wellcome Sanger Institute for the COVID-19 Genomics UK (COG-UK) Consortium | Rob Howes et al |
| EPI_ISL_798478 | England/MILK-D74204/2020 | 2020-12-26 | Europe/United Kingdom/England/ | Lighthouse Lab in Milton Keynes | Wellcome Sanger Institute for the COVID-19 Genomics UK (COG-UK) Consortium | The Lighthouse Lab in Milton Keynes et al |
| EPI_ISL_798589 | England/MILK-D67D7C/2020 | 2020-12-25 | Europe/United Kingdom/England/ | Lighthouse Lab in Milton Keynes | Wellcome Sanger Institute for the COVID-19 Genomics UK (COG-UK) Consortium | The Lighthouse Lab in Milton Keynes et al |
| EPI_ISL_798648 | England/MILK-D74222/2020 | 2020-12-26 | Europe/United Kingdom/England/ | Lighthouse Lab in Milton Keynes | Wellcome Sanger Institute for the COVID-19 Genomics UK (COG-UK) Consortium | The Lighthouse Lab in Milton Keynes et al |
| EPI_ISL_798652 | England/ALDP-D3FA1C/2020 | 2020-12-27 | Europe/United Kingdom/England/ | Lighthouse Lab in Alderley Park | Wellcome Sanger Institute for the COVID-19 Genomics UK (COG-UK) Consortium | Jacquelyn Wynn et al |

| | | | | | | |
|---|---|---|---|---|---|---|
| EPI_ISL_798668 | England/ALDP-D4032C/2020 | 2020-12-27 | Europe/United Kingdom/England/ | Lighthouse Lab in Alderley Park | Wellcome Sanger Institute for the COVID-19 Genomics UK (COG-UK) Consortium | Jacquelyn Wynn et al |
| EPI_ISL_798758 | England/CAMC-D32C5F/2020 | 2020-12-26 | Europe/United Kingdom/England/ | Lighthouse Lab in Cambridge | Wellcome Sanger Institute for the COVID-19 Genomics UK (COG-UK) Consortium | Rob Howes et al |
| EPI_ISL_798942 | England/MILK-D64D33/2020 | 2020-12-24 | Europe/United Kingdom/England/ | Lighthouse Lab in Milton Keynes | Wellcome Sanger Institute for the COVID-19 Genomics UK (COG-UK) Consortium | The Lighthouse Lab in Milton Keynes et al |
| EPI_ISL_798988 | England/ALDP-D4041A/2020 | 2020-12-27 | Europe/United Kingdom/England/ | Lighthouse Lab in Alderley Park | Wellcome Sanger Institute for the COVID-19 Genomics UK (COG-UK) Consortium | Jacquelyn Wynn et al |
| EPI_ISL_799119 | England/CAMC-D3A325/2020 | 2020-12-26 | Europe/United Kingdom/England/ | Lighthouse Lab in Cambridge | Wellcome Sanger Institute for the COVID-19 Genomics UK (COG-UK) Consortium | Rob Howes et al |
| EPI_ISL_799223 | England/CAMC-D3A39E/2020 | 2020-12-24 | Europe/United Kingdom/England/ | Lighthouse Lab in Cambridge | Wellcome Sanger Institute for the COVID-19 Genomics UK (COG-UK) Consortium | Rob Howes et al |
| EPI_ISL_799238 | England/CAMC-D3A3DA/2020 | 2020-12-26 | Europe/United Kingdom/England/ | Lighthouse Lab in Cambridge | Wellcome Sanger Institute for the COVID-19 Genomics UK (COG-UK) Consortium | Rob Howes et al |
| EPI_ISL_799466 | England/MILK-D230BF/2020 | 2020-12-21 | Europe/United Kingdom/England/ | Lighthouse Lab in Milton Keynes | Wellcome Sanger Institute for the COVID-19 Genomics UK (COG-UK) Consortium | The Lighthouse Lab in Milton Keynes et al |

| | | | | | | |
|---|---|---|---|---|---|---|
| EPI_ISL_799678 | England/ALDP-CE129A/2020 | 2020-12-25 | Europe/United Kingdom/England/ | Lighthouse Lab in Alderley Park | Wellcome Sanger Institute for the COVID-19 Genomics UK (COG-UK) Consortium | Jacquelyn Wynn et al |
| EPI_ISL_800052 | England/QEUH-D199B1/2020 | 2020-12-27 | Europe/United Kingdom/England/ | Lighthouse Lab in Glasgow | Wellcome Sanger Institute for the COVID-19 Genomics UK (COG-UK) Consortium | Harper VanSteenhouse et al |
| EPI_ISL_800071 | England/QEUH-D198B4/2020 | 2020-12-27 | Europe/United Kingdom/England/ | Lighthouse Lab in Glasgow | Wellcome Sanger Institute for the COVID-19 Genomics UK (COG-UK) Consortium | Harper VanSteenhouse et al |
| EPI_ISL_800488 | England/CAMC-D11BC2/2020 | 2020-12-26 | Europe/United Kingdom/England/ | Lighthouse Lab in Cambridge | Wellcome Sanger Institute for the COVID-19 Genomics UK (COG-UK) Consortium | Rob Howes et al |
| EPI_ISL_801095 | England/CAMC-BC147D/2020 | 2020-11-22 | Europe/United Kingdom/England/ | Lighthouse Lab in Cambridge | Wellcome Sanger Institute for the COVID-19 Genomics UK (COG-UK) Consortium | Rob Howes et al |
| EPI_ISL_801386 | Brazil/AM-20141882LT/2020 | 2020-09-24 | South America/Brazil/Amazonas BR/Ma | Laboratorio de Ecologia de Doencas Transmissiveis na Amazonia, Instituto Leonidas e Maria Deane - Fiocruz Amazonia | Laboratorio de Ecologia de Doencas Transmissiveis na Amazonia, Instituto Leonidas e Maria Deane - Fiocruz Amazonia | Valdinete Nascimento et al |
| EPI_ISL_801387 | Brazil/AM-20141937RS/2020 | 2020-09-12 | South America/Brazil/Amazonas BR/Lab | Laboratorio de Ecologia de Doencas Transmissiveis na Amazonia, Instituto Leonidas e Maria Deane - Fiocruz Amazonia | Laboratorio de Ecologia de Doencas Transmissiveis na Amazonia, Instituto Leonidas e Maria Deane - Fiocruz Amazonia | Valdinete Nascimento et al |

| | | | | | | |
|---|---|---|---|---|---|---|
| EPI_ISL_801388 | Brazil/AM-20141978DR/2020 | 2020-09-12 | South America/Brazil/Amazonas BR/Tap | Laboratorio de Ecologia de Doencas Transmissiveis na Amazonia, Instituto Leonidas e Maria Deane - Fiocruz Amazonia | Laboratorio de Ecologia de Doencas Transmissiveis na Amazonia, Instituto Leonidas e Maria Deane - Fiocruz Amazonia | Valdinete Nascimento et al |
| EPI_ISL_801389 | Brazil/AM-20142116AS/2020 | 2020-10-05 | South America/Brazil/Amazonas BR/Mar | Laboratorio de Ecologia de Doencas Transmissiveis na Amazonia, Instituto Leonidas e Maria Deane - Fiocruz Amazonia | Laboratorio de Ecologia de Doencas Transmissiveis na Amazonia, Instituto Leonidas e Maria Deane - Fiocruz Amazonia | Valdinete Nascimento et al |
| EPI_ISL_801390 | Brazil/AM-20142279MP/2020 | 2020-10-23 | South America/Brazil/Amazonas BR/Mar | Laboratorio de Ecologia de Doencas Transmissiveis na Amazonia, Instituto Leonidas e Maria Deane - Fiocruz Amazonia | Laboratorio de Ecologia de Doencas Transmissiveis na Amazonia, Instituto Leonidas e Maria Deane - Fiocruz Amazonia | Valdinete Nascimento et al |
| EPI_ISL_801391 | Brazil/AM-20142492MG/2020 | 2020-11-12 | South America/Brazil/Amazonas BR/Mar | Laboratorio de Ecologia de Doencas Transmissiveis na Amazonia, Instituto Leonidas e Maria Deane - Fiocruz Amazonia | Laboratorio de Ecologia de Doencas Transmissiveis na Amazonia, Instituto Leonidas e Maria Deane - Fiocruz Amazonia | Valdinete Nascimento et al |
| EPI_ISL_801392 | Brazil/AM-20142493RO/2020 | 2020-11-12 | South America/Brazil/Amazonas BR/Mar | Laboratorio de Ecologia de Doencas Transmissiveis na Amazonia, Instituto Leonidas e Maria Deane - Fiocruz Amazonia | Laboratorio de Ecologia de Doencas Transmissiveis na Amazonia, Instituto Leonidas e Maria Deane - Fiocruz Amazonia | Valdinete Nascimento et al |

| | | | | | | |
|---|---|---|---|---|---|---|
| EPI_ISL_801393 | Brazil/AM-20842350OC/2020 | 2020-10-29 | South America/Brazil/Amazonas BR/Mai | Laboratorio de Ecologia de Doencas Transmissiveis na Amazonia, Instituto Leonidas e Maria Deane - Fiocruz Amazonia | Laboratorio de Ecologia de Doencas Transmissiveis na Amazonia, Instituto Leonidas e Maria Deane - Fiocruz Amazonia | Valdinete Nascimento et al |
| EPI_ISL_801394 | Brazil/AM-20842378LB/2020 | 2020-10-29 | South America/Brazil/Amazonas BR/Mai | Laboratorio de Ecologia de Doencas Transmissiveis na Amazonia, Instituto Leonidas e Maria Deane - Fiocruz Amazonia | Laboratorio de Ecologia de Doencas Transmissiveis na Amazonia, Instituto Leonidas e Maria Deane - Fiocruz Amazonia | Valdinete Nascimento et al |
| EPI_ISL_801395 | Brazil/AM-20842393GA/2020 | 2020-11-03 | South America/Brazil/Amazonas BR/Mai | Laboratorio de Ecologia de Doencas Transmissiveis na Amazonia, Instituto Leonidas e Maria Deane - Fiocruz Amazonia | Laboratorio de Ecologia de Doencas Transmissiveis na Amazonia, Instituto Leonidas e Maria Deane - Fiocruz Amazonia | Valdinete Nascimento et al |
| EPI_ISL_801396 | Brazil/AM-20842568SB/2020 | 2020-11-12 | South America/Brazil/Amazonas BR/Mai | Laboratorio de Ecologia de Doencas Transmissiveis na Amazonia, Instituto Leonidas e Maria Deane - Fiocruz Amazonia | Laboratorio de Ecologia de Doencas Transmissiveis na Amazonia, Instituto Leonidas e Maria Deane - Fiocruz Amazonia | Valdinete Nascimento et al |
| EPI_ISL_801397 | Brazil/AM-20890012JJ/2020 | 2020-08-24 | South America/Brazil/Amazonas BR/Caa | Laboratv≥rio Central de Sav∫de Pv∫blica do Amazonas - LACEN-AM | Laboratorio de Ecologia de Doencas Transmissiveis na Amazonia, Instituto Leonidas e Maria Deane - Fiocruz Amazonia | Valdinete Nascimento et al |

| | | | | | | |
|---|---|---|---|---|---|---|
| EPI_ISL_801398 | Brazil/AM-20890013MB/2020 | 2020-08-24 | South America/Brazil/Amazonas BR/Caa | Laboratv≥rio Central de Sav∫de Pv∫blica do Amazonas - LACEN-AM | Laboratorio de Ecologia de Doencas Transmissiveis na Amazonia, Instituto Leonidas e Maria Deane - Fiocruz Amazonia | Valdinete Nascimento et al |
| EPI_ISL_801399 | Brazil/AM-20890022AB/2020 | 2020-06-09 | South America/Brazil/Amazonas BR/Ma | Laboratv≥rio Central de Sav∫de Pv∫blica do Amazonas - LACEN-AM | Laboratorio de Ecologia de Doencas Transmissiveis na Amazonia, Instituto Leonidas e Maria Deane - Fiocruz Amazonia | Valdinete Nascimento et al |
| EPI_ISL_801400 | Brazil/AM-20890114ED/2020 | 2020-09-14 | South America/Brazil/Amazonas BR/Car | Laboratv≥rio Central de Sav∫de Pv∫blica do Amazonas - LACEN-AM | Laboratorio de Ecologia de Doencas Transmissiveis na Amazonia, Instituto Leonidas e Maria Deane - Fiocruz Amazonia | Valdinete Nascimento et al |
| EPI_ISL_801401 | Brazil/AM-20890117JP/2020 | 2020-09-15 | South America/Brazil/Amazonas BR/Car | Laboratv≥rio Central de Sav∫de Pv∫blica do Amazonas - LACEN-AM | Laboratorio de Ecologia de Doencas Transmissiveis na Amazonia, Instituto Leonidas e Maria Deane - Fiocruz Amazonia | Valdinete Nascimento et al |
| EPI_ISL_801402 | Brazil/AM-20890261MV/2020 | 2020-05-03 | South America/Brazil/Amazonas BR/Iran | Laboratv≥rio Central de Sav∫de Pv∫blica do Amazonas - LACEN-AM | Laboratorio de Ecologia de Doencas Transmissiveis na Amazonia, Instituto Leonidas e Maria Deane - Fiocruz Amazonia | Valdinete Nascimento et al |

| | | | | | | |
|---|---|---|---|---|---|---|
| EPI_ISL_801403 | Brazil/AM-20892948LS/2020 | 2020-05-12 | South America/Brazil/Amazonas BR/Ma | Laboratv≥rio Central de Sav∫de Pv∫blica do Amazonas - LACEN-AM | Laboratorio de Ecologia de Doencas Transmissiveis na Amazonia, Instituto Leonidas e Maria Deane - Fiocruz Amazonia | Valdinete Nascimento et al |
| EPI_ISL_801407 | Netherlands/LI-EMC-205/2020 | 2020-10-30 | Europe/Netherlands/Limburg/ | Dutch COVID-19 response team | Erasmus Medical Center | Bas Oude Munnink et al |
| EPI_ISL_801413 | Netherlands/LI-EMC-204/2020 | 2020-10-27 | Europe/Netherlands/Limburg/ | Dutch COVID-19 response team | Erasmus Medical Center | Bas Oude Munnink et al |
| EPI_ISL_801454 | Netherlands/GE-EMC-455/2020 | 2020-12-22 | Europe/Netherlands/Gelderland/ | Dutch COVID-19 response team | Erasmus Medical Center | Bas Oude Munnink et al |
| EPI_ISL_801466 | Netherlands/LI-EMC-171/2020 | 2020-11-25 | Europe/Netherlands/Limburg/ | Dutch COVID-19 response team | Erasmus Medical Center | Bas Oude Munnink et al |
| EPI_ISL_801468 | Netherlands/LI-EMC-174/2020 | 2020-11-26 | Europe/Netherlands/Limburg/ | Dutch COVID-19 response team | Erasmus Medical Center | Bas Oude Munnink et al |
| EPI_ISL_801485 | Netherlands/LI-EMC-198/2020 | 2020-10-16 | Europe/Netherlands/Limburg/ | Dutch COVID-19 response team | Erasmus Medical Center | Bas Oude Munnink et al |
| EPI_ISL_801486 | Netherlands/LI-EMC-199/2020 | 2020-10-21 | Europe/Netherlands/Limburg/ | Dutch COVID-19 response team | Erasmus Medical Center | Bas Oude Munnink et al |
| EPI_ISL_801489 | Netherlands/LI-EMC-210/2020 | 2020-11-03 | Europe/Netherlands/Limburg/ | Dutch COVID-19 response team | Erasmus Medical Center | Bas Oude Munnink et al |
| EPI_ISL_801517 | Portugal/PT2156/2020 | 2020-12-30 | Europe/Portugal/Portugal/ | Instituto Nacional de Saude (INSA) | Instituto Nacional de Saude (INSA) | Borges et al et al |
| EPI_ISL_801522 | USA/RI-CDC-8418/2020 | 2020-11-15 | North America/USA/Rhode Island/ | Sonic Reference Laboratory | Pathogen Discovery, Respiratory Viruses Branch, Division of Viral Diseases, Centers for Disease Control and Prevention | Ying Tao et al |

| | | | | | | |
|---|---|---|---|---|---|---|
| EPI_ISL_801527 | USA/RI-CDC-8425/2020 | 2020-12-15 | North America/USA/Rhode Island/ | Sonic Reference Laboratory | Pathogen Discovery, Respiratory Viruses Branch, Division of Viral Diseases, Centers for Disease Control and Prevention | Ying Tao et al |
| EPI_ISL_801545 | Chile/Santiago-PUC_MVL_0376/2020 | 2020-05-12 | South America/Chile/Region Metropolit | Laboratory of Molecular Virology, Pontificia Universidad Catv≥lica de Chile | MSHS Pathogen Surveillance Program | Leonardo I. Almonacid et al |
| EPI_ISL_801546 | Chile/Santiago-PUC_MVL_0510/2020 | 2020-05-26 | South America/Chile/Region Metropolit | Laboratory of Molecular Virology, Pontificia Universidad Catv≥lica de Chile | MSHS Pathogen Surveillance Program | Leonardo I. Almonacid et al |
| EPI_ISL_801555 | Chile/Santiago-AAS3062/2020 | 2020-05-02 | South America/Chile/Region Metropolit | Laboratory of Molecular Virology, Pontificia Universidad Catv≥lica de Chile | MSHS Pathogen Surveillance Program | Leonardo I. Almonacid et al |
| EPI_ISL_801557 | Chile/Santiago-AGR8546/2020 | 2020-05-06 | South America/Chile/Region Metropolit | Laboratory of Molecular Virology, Pontificia Universidad Catv≥lica de Chile | MSHS Pathogen Surveillance Program | Leonardo I. Almonacid et al |
| EPI_ISL_801560 | Chile/Santiago-ASC3794/2020 | 2020-05-02 | South America/Chile/Region Metropolit | Laboratory of Molecular Virology, Pontificia Universidad Catv≥lica de Chile | MSHS Pathogen Surveillance Program | Leonardo I. Almonacid et al |
| EPI_ISL_801562 | Chile/Santiago-BMA5485/2020 | 2020-05-04 | South America/Chile/Region Metropolit | Laboratory of Molecular Virology, Pontificia Universidad Catv≥lica de Chile | MSHS Pathogen Surveillance Program | Leonardo I. Almonacid et al |
| EPI_ISL_801564 | Chile/Santiago-CDV2422/2020 | 2020-04-28 | South America/Chile/Region Metropolit | Laboratory of Molecular Virology, Pontificia Universidad Catv≥lica de Chile | MSHS Pathogen Surveillance Program | Leonardo I. Almonacid et al |
| EPI_ISL_801565 | Chile/Santiago-CGP014K/2020 | 2020-05-04 | South America/Chile/Region Metropolit | Laboratory of Molecular Virology, Pontificia Universidad Catv≥lica de Chile | MSHS Pathogen Surveillance Program | Leonardo I. Almonacid et al |
| EPI_ISL_801571 | Chile/Santiago-JCB953K/2020 | 2020-05-06 | South America/Chile/Region Metropolit | Laboratory of Molecular Virology, Pontificia Universidad Catv≥lica de Chile | MSHS Pathogen Surveillance Program | Leonardo I. Almonacid et al |
| EPI_ISL_801572 | Chile/Santiago-JDP7073/2020 | 2020-04-28 | South America/Chile/Region Metropolit | Laboratory of Molecular Virology, Pontificia Universidad Catv≥lica de Chile | MSHS Pathogen Surveillance Program | Leonardo I. Almonacid et al |

| | | | | | | |
|---|---|---|---|---|---|---|
| EPI_ISL_801574 | Chile/Santiago-JST3802/2020 | 2020-05-04 | South America/Chile/Region Metropolit | Laboratory of Molecular Virology, Pontificia Universidad Catv≥lica de Chile | MSHS Pathogen Surveillance Program | Leonardo I. Almonacid et al |
| EPI_ISL_801575 | Chile/Santiago-LL5209/2020 | 2020-04-23 | South America/Chile/Region Metropolit | Laboratory of Molecular Virology, Pontificia Universidad Catv≥lica de Chile | MSHS Pathogen Surveillance Program | Leonardo I. Almonacid et al |
| EPI_ISL_801576 | Chile/Santiago-LRC4163/2020 | 2020-05-12 | South America/Chile/Region Metropolit | Laboratory of Molecular Virology, Pontificia Universidad Catv≥lica de Chile | MSHS Pathogen Surveillance Program | Leonardo I. Almonacid et al |
| EPI_ISL_801581 | Chile/Santiago-op18d3/2020 | 2020-03-29 | South America/Chile/Region Metropolit | Laboratory of Molecular Virology, Pontificia Universidad Catv≥lica de Chile | MSHS Pathogen Surveillance Program | Leonardo I. Almonacid et al |
| EPI_ISL_801582 | Chile/Santiago-op18d5/2020 | 2020-03-31 | South America/Chile/Region Metropolit | Laboratory of Molecular Virology, Pontificia Universidad Catv≥lica de Chile | MSHS Pathogen Surveillance Program | Leonardo I. Almonacid et al |
| EPI_ISL_801589 | Chile/Santiago-op8d2/2020 | 2020-03-16 | South America/Chile/Region Metropolit | Laboratory of Molecular Virology, Pontificia Universidad Catv≥lica de Chile | MSHS Pathogen Surveillance Program | Leonardo I. Almonacid et al |
| EPI_ISL_801593 | Chile/Santiago-p14d7/2020 | 2020-04-27 | South America/Chile/Region Metropolit | Laboratory of Molecular Virology, Pontificia Universidad Catv≥lica de Chile | MSHS Pathogen Surveillance Program | Leonardo I. Almonacid et al |
| EPI_ISL_801606 | Chile/Santiago-PUC_MVL_0015/2020 | 2020-03-19 | South America/Chile/Region Metropolit | Laboratory of Molecular Virology, Pontificia Universidad Catv≥lica de Chile | MSHS Pathogen Surveillance Program | Leonardo I. Almonacid et al |
| EPI_ISL_801614 | Chile/Santiago-PUC_MVL_0023/2020 | 2020-03-19 | South America/Chile/Region Metropolit | Laboratory of Molecular Virology, Pontificia Universidad Catv≥lica de Chile | MSHS Pathogen Surveillance Program | Leonardo I. Almonacid et al |
| EPI_ISL_801623 | Chile/Santiago-PUC_MVL_0032/2020 | 2020-03-26 | South America/Chile/Region Metropolit | Laboratory of Molecular Virology, Pontificia Universidad Catv≥lica de Chile | MSHS Pathogen Surveillance Program | Leonardo I. Almonacid et al |
| EPI_ISL_801627 | Chile/Santiago-PUC_MVL_0036/2020 | 2020-03-27 | South America/Chile/Region Metropolit | Laboratory of Molecular Virology, Pontificia Universidad Catv≥lica de Chile | MSHS Pathogen Surveillance Program | Leonardo I. Almonacid et al |
| EPI_ISL_801628 | Chile/Santiago-PUC_MVL_0037/2020 | 2020-03-26 | South America/Chile/Region Metropolit | Laboratory of Molecular Virology, Pontificia Universidad Catv≥lica de Chile | MSHS Pathogen Surveillance Program | Leonardo I. Almonacid et al |
| EPI_ISL_801640 | Chile/Santiago-PUC_MVL_0050/2020 | 2020-03-22 | South America/Chile/Region Metropolit | Laboratory of Molecular Virology, Pontificia Universidad Catv≥lica de Chile | MSHS Pathogen Surveillance Program | Leonardo I. Almonacid et al |

| | | | | | | |
|---|---|---|---|---|---|---|
| EPI_ISL_801644 | Chile/Santiago-PUC_MVL_0055/2020 | 2020-03-24 | South America/Chile/Region Metropolit | Laboratory of Molecular Virology, Pontificia Universidad Cat√≥lica de Chile | MSHS Pathogen Surveillance Program | Leonardo I. Almonacid et al |
| EPI_ISL_801653 | Chile/Puerto_Montt-PUC_MVL_0067 | 2020-03-25 | South America/Chile/Los Lagos/Puerto M | Laboratory of Molecular Virology, Pontificia Universidad Cat√≥lica de Chile | MSHS Pathogen Surveillance Program | Leonardo I. Almonacid et al |
| EPI_ISL_801670 | Chile/Santiago-PUC_MVL_0086/2020 | 2020-03-31 | South America/Chile/Region Metropolit | Laboratory of Molecular Virology, Pontificia Universidad Cat√≥lica de Chile | MSHS Pathogen Surveillance Program | Leonardo I. Almonacid et al |
| EPI_ISL_801682 | Chile/Santiago-PUC_MVL_0108/2020 | 2020-04-01 | South America/Chile/Region Metropolit | Laboratory of Molecular Virology, Pontificia Universidad Cat√≥lica de Chile | MSHS Pathogen Surveillance Program | Leonardo I. Almonacid et al |
| EPI_ISL_801692 | Chile/Santiago-PUC_MVL_0149/2020 | 2020-04-04 | South America/Chile/Region Metropolit | Laboratory of Molecular Virology, Pontificia Universidad Cat√≥lica de Chile | MSHS Pathogen Surveillance Program | Leonardo I. Almonacid et al |
| EPI_ISL_801700 | Chile/Santiago-PUC_MVL_0168/2020 | 2020-04-10 | South America/Chile/Region Metropolit | Laboratory of Molecular Virology, Pontificia Universidad Cat√≥lica de Chile | MSHS Pathogen Surveillance Program | Leonardo I. Almonacid et al |
| EPI_ISL_801701 | Chile/Santiago-PUC_MVL_0171/2020 | 2020-04-11 | South America/Chile/Region Metropolit | Laboratory of Molecular Virology, Pontificia Universidad Cat√≥lica de Chile | MSHS Pathogen Surveillance Program | Leonardo I. Almonacid et al |
| EPI_ISL_801711 | Chile/Santiago-PUC_MVL_0190/2020 | 2020-04-17 | South America/Chile/Region Metropolit | Laboratory of Molecular Virology, Pontificia Universidad Cat√≥lica de Chile | MSHS Pathogen Surveillance Program | Leonardo I. Almonacid et al |
| EPI_ISL_801722 | Chile/Santiago-PUC_MVL_0212/2020 | 2020-04-25 | South America/Chile/Region Metropolit | Laboratory of Molecular Virology, Pontificia Universidad Cat√≥lica de Chile | MSHS Pathogen Surveillance Program | Leonardo I. Almonacid et al |
| EPI_ISL_801737 | Chile/Santiago-PUC_MVL_0234/2020 | 2020-04-27 | South America/Chile/Region Metropolit | Laboratory of Molecular Virology, Pontificia Universidad Cat√≥lica de Chile | MSHS Pathogen Surveillance Program | Leonardo I. Almonacid et al |
| EPI_ISL_801740 | Chile/Santiago-PUC_MVL_0239/2020 | 2020-04-25 | South America/Chile/Region Metropolit | Laboratory of Molecular Virology, Pontificia Universidad Cat√≥lica de Chile | MSHS Pathogen Surveillance Program | Leonardo I. Almonacid et al |
| EPI_ISL_801744 | Chile/Santiago-PUC_MVL_0247/2020 | 2020-04-30 | South America/Chile/Region Metropolit | Laboratory of Molecular Virology, Pontificia Universidad Cat√≥lica de Chile | MSHS Pathogen Surveillance Program | Leonardo I. Almonacid et al |
| EPI_ISL_801745 | Chile/Santiago-PUC_MVL_0252/2020 | 2020-04-30 | South America/Chile/Region Metropolit | Laboratory of Molecular Virology, Pontificia Universidad Cat√≥lica de Chile | MSHS Pathogen Surveillance Program | Leonardo I. Almonacid et al |

| | | | | | | |
|---|---|---|---|---|---|---|
| EPI_ISL_801746 | Chile/Santiago-PUC_MVL_0258/2020 | 2020-04-30 | South America/Chile/Region Metropolit | Laboratory of Molecular Virology, Pontificia Universidad Cat√≥lica de Chile | MSHS Pathogen Surveillance Program | Leonardo I. Almonacid et al |
| EPI_ISL_801747 | Chile/Santiago-PUC_MVL_0261/2020 | 2020-05-04 | South America/Chile/Region Metropolit | Laboratory of Molecular Virology, Pontificia Universidad Cat√≥lica de Chile | MSHS Pathogen Surveillance Program | Leonardo I. Almonacid et al |
| EPI_ISL_801748 | Chile/Santiago-PUC_MVL_0264/2020 | 2020-05-04 | South America/Chile/Region Metropolit | Laboratory of Molecular Virology, Pontificia Universidad Cat√≥lica de Chile | MSHS Pathogen Surveillance Program | Leonardo I. Almonacid et al |
| EPI_ISL_801749 | Chile/Santiago-PUC_MVL_0265/2020 | 2020-05-04 | South America/Chile/Region Metropolit | Laboratory of Molecular Virology, Pontificia Universidad Cat√≥lica de Chile | MSHS Pathogen Surveillance Program | Leonardo I. Almonacid et al |
| EPI_ISL_801750 | Chile/Santiago-PUC_MVL_0268/2020 | 2020-04-30 | South America/Chile/Region Metropolit | Laboratory of Molecular Virology, Pontificia Universidad Cat√≥lica de Chile | MSHS Pathogen Surveillance Program | Leonardo I. Almonacid et al |
| EPI_ISL_801754 | Chile/Santiago-PUC_MVL_0273/2020 | 2020-04-30 | South America/Chile/Region Metropolit | Laboratory of Molecular Virology, Pontificia Universidad Cat√≥lica de Chile | MSHS Pathogen Surveillance Program | Leonardo I. Almonacid et al |
| EPI_ISL_801762 | Chile/Santiago-PUC_MVL_0292/2020 | 2020-05-04 | South America/Chile/Region Metropolit | Laboratory of Molecular Virology, Pontificia Universidad Cat√≥lica de Chile | MSHS Pathogen Surveillance Program | Leonardo I. Almonacid et al |
| EPI_ISL_801763 | Chile/Buin-PUC_MVL_0294/2020 | 2020-05-05 | South America/Chile/Region Metropolit | Laboratory of Molecular Virology, Pontificia Universidad Cat√≥lica de Chile | MSHS Pathogen Surveillance Program | Leonardo I. Almonacid et al |
| EPI_ISL_801765 | Chile/Santiago-PUC_MVL_0305/2020 | 2020-05-09 | South America/Chile/Region Metropolit | Laboratory of Molecular Virology, Pontificia Universidad Cat√≥lica de Chile | MSHS Pathogen Surveillance Program | Leonardo I. Almonacid et al |
| EPI_ISL_801766 | Chile/Santiago-PUC_MVL_0313/2020 | 2020-05-09 | South America/Chile/Region Metropolit | Laboratory of Molecular Virology, Pontificia Universidad Cat√≥lica de Chile | MSHS Pathogen Surveillance Program | Leonardo I. Almonacid et al |
| EPI_ISL_801767 | Chile/Santiago-PUC_MVL_0321/2020 | 2020-05-09 | South America/Chile/Region Metropolit | Laboratory of Molecular Virology, Pontificia Universidad Cat√≥lica de Chile | MSHS Pathogen Surveillance Program | Leonardo I. Almonacid et al |
| EPI_ISL_801769 | Chile/Santiago-PUC_MVL_0323/2020 | 2020-05-09 | South America/Chile/Region Metropolit | Laboratory of Molecular Virology, Pontificia Universidad Cat√≥lica de Chile | MSHS Pathogen Surveillance Program | Leonardo I. Almonacid et al |
| EPI_ISL_801770 | Chile/Santiago-PUC_MVL_0325/2020 | 2020-05-09 | South America/Chile/Region Metropolit | Laboratory of Molecular Virology, Pontificia Universidad Cat√≥lica de Chile | MSHS Pathogen Surveillance Program | Leonardo I. Almonacid et al |

| | | | | | | |
|---|---|---|---|---|---|---|
| EPI_ISL_801771 | Chile/Santiago-PUC_MVL_0326/2020 | 2020-05-10 | South America/Chile/Region Metropolit | Laboratory of Molecular Virology, Pontificia Universidad Catv≥lica de Chile | MSHS Pathogen Surveillance Program | Leonardo I. Almonacid et al |
| EPI_ISL_801772 | Chile/Santiago-PUC_MVL_0327/2020 | 2020-05-10 | South America/Chile/Region Metropolit | Laboratory of Molecular Virology, Pontificia Universidad Catv≥lica de Chile | MSHS Pathogen Surveillance Program | Leonardo I. Almonacid et al |
| EPI_ISL_801773 | Chile/Santiago-PUC_MVL_0331/2020 | 2020-05-10 | South America/Chile/Region Metropolit | Laboratory of Molecular Virology, Pontificia Universidad Catv≥lica de Chile | MSHS Pathogen Surveillance Program | Leonardo I. Almonacid et al |
| EPI_ISL_801774 | Chile/Santiago-PUC_MVL_0332/2020 | 2020-05-10 | South America/Chile/Region Metropolit | Laboratory of Molecular Virology, Pontificia Universidad Catv≥lica de Chile | MSHS Pathogen Surveillance Program | Leonardo I. Almonacid et al |
| EPI_ISL_801775 | Chile/Santiago-PUC_MVL_0334/2020 | 2020-05-11 | South America/Chile/Region Metropolit | Laboratory of Molecular Virology, Pontificia Universidad Catv≥lica de Chile | MSHS Pathogen Surveillance Program | Leonardo I. Almonacid et al |
| EPI_ISL_801777 | Chile/Santiago-PUC_MVL_0337/2020 | 2020-05-11 | South America/Chile/Region Metropolit | Laboratory of Molecular Virology, Pontificia Universidad Catv≥lica de Chile | MSHS Pathogen Surveillance Program | Leonardo I. Almonacid et al |
| EPI_ISL_801778 | Chile/Curacavi-PUC_MVL_0340/2020 | 2020-05-11 | South America/Chile/Region Metropolit | Laboratory of Molecular Virology, Pontificia Universidad Catv≥lica de Chile | MSHS Pathogen Surveillance Program | Leonardo I. Almonacid et al |
| EPI_ISL_801779 | Chile/Santiago-PUC_MVL_0342/2020 | 2020-05-10 | South America/Chile/Region Metropolit | Laboratory of Molecular Virology, Pontificia Universidad Catv≥lica de Chile | MSHS Pathogen Surveillance Program | Leonardo I. Almonacid et al |
| EPI_ISL_801780 | Chile/Santiago-PUC_MVL_0347/2020 | 2020-05-10 | South America/Chile/Region Metropolit | Laboratory of Molecular Virology, Pontificia Universidad Catv≥lica de Chile | MSHS Pathogen Surveillance Program | Leonardo I. Almonacid et al |
| EPI_ISL_801781 | Chile/Santiago-PUC_MVL_0349/2020 | 2020-05-11 | South America/Chile/Region Metropolit | Laboratory of Molecular Virology, Pontificia Universidad Catv≥lica de Chile | MSHS Pathogen Surveillance Program | Leonardo I. Almonacid et al |
| EPI_ISL_801782 | Chile/Santiago-PUC_MVL_0351/2020 | 2020-05-11 | South America/Chile/Region Metropolit | Laboratory of Molecular Virology, Pontificia Universidad Catv≥lica de Chile | MSHS Pathogen Surveillance Program | Leonardo I. Almonacid et al |
| EPI_ISL_801783 | Chile/Puerto_Montt-PUC_MVL_0353 | 2020-05-11 | South America/Chile/Los Lagos/Puerto M | Laboratory of Molecular Virology, Pontificia Universidad Catv≥lica de Chile | MSHS Pathogen Surveillance Program | Leonardo I. Almonacid et al |
| EPI_ISL_801784 | Chile/Santiago-PUC_MVL_0354/2020 | 2020-05-11 | South America/Chile/Region Metropolit | Laboratory of Molecular Virology, Pontificia Universidad Catv≥lica de Chile | MSHS Pathogen Surveillance Program | Leonardo I. Almonacid et al |

| | | | | | | |
|---|---|---|---|---|---|---|
| EPI_ISL_801785 | Chile/Santiago-PUC_MVL_0358/2020 | 2020-05-11 | South America/Chile/Region Metropolit | Laboratory of Molecular Virology, Pontificia Universidad Cat√≥lica de Chile | MSHS Pathogen Surveillance Program | Leonardo I. Almonacid et al |
| EPI_ISL_801786 | Chile/Santiago-PUC_MVL_0360/2020 | 2020-05-11 | South America/Chile/Region Metropolit | Laboratory of Molecular Virology, Pontificia Universidad Cat√≥lica de Chile | MSHS Pathogen Surveillance Program | Leonardo I. Almonacid et al |
| EPI_ISL_801787 | Chile/Santiago-PUC_MVL_0361/2020 | 2020-05-11 | South America/Chile/Region Metropolit | Laboratory of Molecular Virology, Pontificia Universidad Cat√≥lica de Chile | MSHS Pathogen Surveillance Program | Leonardo I. Almonacid et al |
| EPI_ISL_801788 | Chile/Puerto_Montt-PUC_MVL_0362 | 2020-05-11 | South America/Chile/Los Lagos/Puerto M | Laboratory of Molecular Virology, Pontificia Universidad Cat√≥lica de Chile | MSHS Pathogen Surveillance Program | Leonardo I. Almonacid et al |
| EPI_ISL_801789 | Chile/Santiago-PUC_MVL_0364/2020 | 2020-05-11 | South America/Chile/Region Metropolit | Laboratory of Molecular Virology, Pontificia Universidad Cat√≥lica de Chile | MSHS Pathogen Surveillance Program | Leonardo I. Almonacid et al |
| EPI_ISL_801790 | Chile/Santiago-PUC_MVL_0365/2020 | 2020-05-11 | South America/Chile/Region Metropolit | Laboratory of Molecular Virology, Pontificia Universidad Cat√≥lica de Chile | MSHS Pathogen Surveillance Program | Leonardo I. Almonacid et al |
| EPI_ISL_801791 | Chile/Santiago-PUC_MVL_0366/2020 | 2020-05-12 | South America/Chile/Region Metropolit | Laboratory of Molecular Virology, Pontificia Universidad Cat√≥lica de Chile | MSHS Pathogen Surveillance Program | Leonardo I. Almonacid et al |
| EPI_ISL_801792 | Chile/Santiago-PUC_MVL_0373/2020 | 2020-05-12 | South America/Chile/Region Metropolit | Laboratory of Molecular Virology, Pontificia Universidad Cat√≥lica de Chile | MSHS Pathogen Surveillance Program | Leonardo I. Almonacid et al |
| EPI_ISL_801793 | Chile/Santiago-PUC_MVL_0374/2020 | 2020-05-12 | South America/Chile/Region Metropolit | Laboratory of Molecular Virology, Pontificia Universidad Cat√≥lica de Chile | MSHS Pathogen Surveillance Program | Leonardo I. Almonacid et al |
| EPI_ISL_801795 | Chile/Santiago-PUC_MVL_0379/2020 | 2020-05-12 | South America/Chile/Region Metropolit | Laboratory of Molecular Virology, Pontificia Universidad Cat√≥lica de Chile | MSHS Pathogen Surveillance Program | Leonardo I. Almonacid et al |
| EPI_ISL_801796 | Chile/Santiago-PUC_MVL_0380/2020 | 2020-05-12 | South America/Chile/Region Metropolit | Laboratory of Molecular Virology, Pontificia Universidad Cat√≥lica de Chile | MSHS Pathogen Surveillance Program | Leonardo I. Almonacid et al |
| EPI_ISL_801797 | Chile/Santiago-PUC_MVL_0383/2020 | 2020-05-12 | South America/Chile/Region Metropolit | Laboratory of Molecular Virology, Pontificia Universidad Cat√≥lica de Chile | MSHS Pathogen Surveillance Program | Leonardo I. Almonacid et al |
| EPI_ISL_801798 | Chile/Santiago-PUC_MVL_0384/2020 | 2020-05-12 | South America/Chile/Region Metropolit | Laboratory of Molecular Virology, Pontificia Universidad Cat√≥lica de Chile | MSHS Pathogen Surveillance Program | Leonardo I. Almonacid et al |

| | | | | | | |
|---|---|---|---|---|---|---|
| EPI_ISL_801799 | Chile/Santiago-PUC_MVL_0385/2020 | 2020-05-12 | South America/Chile/Region Metropolit | Laboratory of Molecular Virology, Pontificia Universidad Catv≥lica de Chile | MSHS Pathogen Surveillance Program | Leonardo I. Almonacid et al |
| EPI_ISL_801800 | Chile/Santiago-PUC_MVL_0386/2020 | 2020-05-12 | South America/Chile/Region Metropolit | Laboratory of Molecular Virology, Pontificia Universidad Catv≥lica de Chile | MSHS Pathogen Surveillance Program | Leonardo I. Almonacid et al |
| EPI_ISL_801803 | Chile/Santiago-PUC_MVL_0391/2020 | 2020-05-12 | South America/Chile/Region Metropolit | Laboratory of Molecular Virology, Pontificia Universidad Catv≥lica de Chile | MSHS Pathogen Surveillance Program | Leonardo I. Almonacid et al |
| EPI_ISL_801805 | Chile/Santiago-PUC_MVL_0393/2020 | 2020-05-12 | South America/Chile/Region Metropolit | Laboratory of Molecular Virology, Pontificia Universidad Catv≥lica de Chile | MSHS Pathogen Surveillance Program | Leonardo I. Almonacid et al |
| EPI_ISL_801806 | Chile/Santiago-PUC_MVL_0394/2020 | 2020-05-12 | South America/Chile/Region Metropolit | Laboratory of Molecular Virology, Pontificia Universidad Catv≥lica de Chile | MSHS Pathogen Surveillance Program | Leonardo I. Almonacid et al |
| EPI_ISL_801807 | Chile/Santiago-PUC_MVL_0395/2020 | 2020-05-12 | South America/Chile/Region Metropolit | Laboratory of Molecular Virology, Pontificia Universidad Catv≥lica de Chile | MSHS Pathogen Surveillance Program | Leonardo I. Almonacid et al |
| EPI_ISL_801808 | Chile/Santiago-PUC_MVL_0397/2020 | 2020-05-12 | South America/Chile/Region Metropolit | Laboratory of Molecular Virology, Pontificia Universidad Catv≥lica de Chile | MSHS Pathogen Surveillance Program | Leonardo I. Almonacid et al |
| EPI_ISL_801809 | Chile/Santiago-PUC_MVL_0400/2020 | 2020-05-12 | South America/Chile/Region Metropolit | Laboratory of Molecular Virology, Pontificia Universidad Catv≥lica de Chile | MSHS Pathogen Surveillance Program | Leonardo I. Almonacid et al |
| EPI_ISL_801810 | Chile/Santiago-PUC_MVL_0403/2020 | 2020-05-12 | South America/Chile/Region Metropolit | Laboratory of Molecular Virology, Pontificia Universidad Catv≥lica de Chile | MSHS Pathogen Surveillance Program | Leonardo I. Almonacid et al |
| EPI_ISL_801811 | Chile/Santiago-PUC_MVL_0410/2020 | 2020-05-12 | South America/Chile/Region Metropolit | Laboratory of Molecular Virology, Pontificia Universidad Catv≥lica de Chile | MSHS Pathogen Surveillance Program | Leonardo I. Almonacid et al |
| EPI_ISL_801812 | Chile/Santiago-PUC_MVL_0411/2020 | 2020-05-13 | South America/Chile/Region Metropolit | Laboratory of Molecular Virology, Pontificia Universidad Catv≥lica de Chile | MSHS Pathogen Surveillance Program | Leonardo I. Almonacid et al |
| EPI_ISL_801814 | Chile/Santiago-PUC_MVL_0418/2020 | 2020-05-12 | South America/Chile/Region Metropolit | Laboratory of Molecular Virology, Pontificia Universidad Catv≥lica de Chile | MSHS Pathogen Surveillance Program | Leonardo I. Almonacid et al |
| EPI_ISL_801815 | Chile/Santiago-PUC_MVL_0419/2020 | 2020-05-12 | South America/Chile/Region Metropolit | Laboratory of Molecular Virology, Pontificia Universidad Catv≥lica de Chile | MSHS Pathogen Surveillance Program | Leonardo I. Almonacid et al |

| | | | | | | |
|---|---|---|---|---|---|---|
| EPI_ISL_801816 | Chile/Santiago-PUC_MVL_0421/2020 | 2020-05-13 | South America/Chile/Region Metropolit | Laboratory of Molecular Virology, Pontificia Universidad Cat√≥lica de Chile | MSHS Pathogen Surveillance Program | Leonardo I. Almonacid et al |
| EPI_ISL_801817 | Chile/Santiago-PUC_MVL_0422/2020 | 2020-05-12 | South America/Chile/Region Metropolit | Laboratory of Molecular Virology, Pontificia Universidad Cat√≥lica de Chile | MSHS Pathogen Surveillance Program | Leonardo I. Almonacid et al |
| EPI_ISL_801818 | Chile/Puerto_Montt-PUC_MVL_0424 | 2020-05-13 | South America/Chile/Los Lagos/Puerto M | Laboratory of Molecular Virology, Pontificia Universidad Cat√≥lica de Chile | MSHS Pathogen Surveillance Program | Leonardo I. Almonacid et al |
| EPI_ISL_801819 | Chile/Santiago-PUC_MVL_0429/2020 | 2020-05-13 | South America/Chile/Region Metropolit | Laboratory of Molecular Virology, Pontificia Universidad Cat√≥lica de Chile | MSHS Pathogen Surveillance Program | Leonardo I. Almonacid et al |
| EPI_ISL_801820 | Chile/Santiago-PUC_MVL_0430/2020 | 2020-05-12 | South America/Chile/Region Metropolit | Laboratory of Molecular Virology, Pontificia Universidad Cat√≥lica de Chile | MSHS Pathogen Surveillance Program | Leonardo I. Almonacid et al |
| EPI_ISL_801821 | Chile/Santiago-PUC_MVL_0432/2020 | 2020-05-13 | South America/Chile/Region Metropolit | Laboratory of Molecular Virology, Pontificia Universidad Cat√≥lica de Chile | MSHS Pathogen Surveillance Program | Leonardo I. Almonacid et al |
| EPI_ISL_801822 | Chile/Santiago-PUC_MVL_0442/2020 | 2020-05-13 | South America/Chile/Region Metropolit | Laboratory of Molecular Virology, Pontificia Universidad Cat√≥lica de Chile | MSHS Pathogen Surveillance Program | Leonardo I. Almonacid et al |
| EPI_ISL_801823 | Chile/Santiago-PUC_MVL_0443/2020 | 2020-05-13 | South America/Chile/Region Metropolit | Laboratory of Molecular Virology, Pontificia Universidad Cat√≥lica de Chile | MSHS Pathogen Surveillance Program | Leonardo I. Almonacid et al |
| EPI_ISL_801824 | Chile/Santiago-PUC_MVL_0450/2020 | 2020-05-13 | South America/Chile/Region Metropolit | Laboratory of Molecular Virology, Pontificia Universidad Cat√≥lica de Chile | MSHS Pathogen Surveillance Program | Leonardo I. Almonacid et al |
| EPI_ISL_801825 | Chile/Santiago-PUC_MVL_0454/2020 | 2020-05-13 | South America/Chile/Region Metropolit | Laboratory of Molecular Virology, Pontificia Universidad Cat√≥lica de Chile | MSHS Pathogen Surveillance Program | Leonardo I. Almonacid et al |
| EPI_ISL_801827 | Chile/Santiago-PUC_MVL_0475/2020 | 2020-05-13 | South America/Chile/Region Metropolit | Laboratory of Molecular Virology, Pontificia Universidad Cat√≥lica de Chile | MSHS Pathogen Surveillance Program | Leonardo I. Almonacid et al |
| EPI_ISL_801828 | Chile/Santiago-PUC_MVL_0478/2020 | 2020-05-13 | South America/Chile/Region Metropolit | Laboratory of Molecular Virology, Pontificia Universidad Cat√≥lica de Chile | MSHS Pathogen Surveillance Program | Leonardo I. Almonacid et al |
| EPI_ISL_801829 | Chile/Santiago-PUC_MVL_0483/2020 | 2020-05-14 | South America/Chile/Region Metropolit | Laboratory of Molecular Virology, Pontificia Universidad Cat√≥lica de Chile | MSHS Pathogen Surveillance Program | Leonardo I. Almonacid et al |

| | | | | | | |
|---|---|---|---|---|---|---|
| EPI_ISL_801830 | Chile/Santiago-PUC_MVL_0484/2020 | 2020-05-14 | South America/Chile/Region Metropolit | Laboratory of Molecular Virology, Pontificia Universidad Cat√≥lica de Chile | MSHS Pathogen Surveillance Program | Leonardo I. Almonacid et al |
| EPI_ISL_801831 | Chile/Santiago-PUC_MVL_0487/2020 | 2020-05-14 | South America/Chile/Region Metropolit | Laboratory of Molecular Virology, Pontificia Universidad Cat√≥lica de Chile | MSHS Pathogen Surveillance Program | Leonardo I. Almonacid et al |
| EPI_ISL_801832 | Chile/Santiago-PUC_MVL_0489/2020 | 2020-05-13 | South America/Chile/Region Metropolit | Laboratory of Molecular Virology, Pontificia Universidad Cat√≥lica de Chile | MSHS Pathogen Surveillance Program | Leonardo I. Almonacid et al |
| EPI_ISL_801833 | Chile/Santiago-PUC_MVL_0490/2020 | 2020-05-13 | South America/Chile/Region Metropolit | Laboratory of Molecular Virology, Pontificia Universidad Cat√≥lica de Chile | MSHS Pathogen Surveillance Program | Leonardo I. Almonacid et al |
| EPI_ISL_801834 | Chile/Santiago-PUC_MVL_0491/2020 | 2020-05-13 | South America/Chile/Region Metropolit | Laboratory of Molecular Virology, Pontificia Universidad Cat√≥lica de Chile | MSHS Pathogen Surveillance Program | Leonardo I. Almonacid et al |
| EPI_ISL_801835 | Chile/Santiago-PUC_MVL_0502/2020 | 2020-05-26 | South America/Chile/Region Metropolit | Laboratory of Molecular Virology, Pontificia Universidad Cat√≥lica de Chile | MSHS Pathogen Surveillance Program | Leonardo I. Almonacid et al |
| EPI_ISL_801837 | Chile/Santiago-PUC_MVL_0511/2020 | 2020-05-26 | South America/Chile/Region Metropolit | Laboratory of Molecular Virology, Pontificia Universidad Cat√≥lica de Chile | MSHS Pathogen Surveillance Program | Leonardo I. Almonacid et al |
| EPI_ISL_801838 | Chile/Santiago-PUC_MVL_0512/2020 | 2020-05-27 | South America/Chile/Region Metropolit | Laboratory of Molecular Virology, Pontificia Universidad Cat√≥lica de Chile | MSHS Pathogen Surveillance Program | Leonardo I. Almonacid et al |
| EPI_ISL_801841 | Chile/Santiago-PUC_MVL_0520/2020 | 2020-05-27 | South America/Chile/Region Metropolit | Laboratory of Molecular Virology, Pontificia Universidad Cat√≥lica de Chile | MSHS Pathogen Surveillance Program | Leonardo I. Almonacid et al |
| EPI_ISL_801842 | Chile/Santiago-PUC_MVL_0522/2020 | 2020-05-27 | South America/Chile/Region Metropolit | Laboratory of Molecular Virology, Pontificia Universidad Cat√≥lica de Chile | MSHS Pathogen Surveillance Program | Leonardo I. Almonacid et al |
| EPI_ISL_801844 | Chile/Santiago-PUC_MVL_0526/2020 | 2020-05-27 | South America/Chile/Region Metropolit | Laboratory of Molecular Virology, Pontificia Universidad Cat√≥lica de Chile | MSHS Pathogen Surveillance Program | Leonardo I. Almonacid et al |
| EPI_ISL_801845 | Chile/Santiago-PUC_MVL_0528/2020 | 2020-05-27 | South America/Chile/Region Metropolit | Laboratory of Molecular Virology, Pontificia Universidad Cat√≥lica de Chile | MSHS Pathogen Surveillance Program | Leonardo I. Almonacid et al |
| EPI_ISL_801846 | Chile/Santiago-PUC_MVL_0529/2020 | 2020-05-27 | South America/Chile/Region Metropolit | Laboratory of Molecular Virology, Pontificia Universidad Cat√≥lica de Chile | MSHS Pathogen Surveillance Program | Leonardo I. Almonacid et al |

| | | | | | | |
|---|---|---|---|---|---|---|
| EPI_ISL_801847 | Chile/Santiago-PUC_MVL_0531/2020 | 2020-05-27 | South America/Chile/Region Metropolit | Laboratory of Molecular Virology, Pontificia Universidad Cat√≥lica de Chile | MSHS Pathogen Surveillance Program | Leonardo I. Almonacid et al |
| EPI_ISL_801849 | Chile/Santiago-PUC_MVL_0537/2020 | 2020-05-26 | South America/Chile/Region Metropolit | Laboratory of Molecular Virology, Pontificia Universidad Cat√≥lica de Chile | MSHS Pathogen Surveillance Program | Leonardo I. Almonacid et al |
| EPI_ISL_801851 | Chile/Santiago-PUC_MVL_0540/2020 | 2020-05-26 | South America/Chile/Region Metropolit | Laboratory of Molecular Virology, Pontificia Universidad Cat√≥lica de Chile | MSHS Pathogen Surveillance Program | Leonardo I. Almonacid et al |
| EPI_ISL_801855 | Chile/Santiago-SF6772/2020 | 2020-05-06 | South America/Chile/Region Metropolit | Laboratory of Molecular Virology, Pontificia Universidad Cat√≥lica de Chile | MSHS Pathogen Surveillance Program | Leonardo I. Almonacid et al |
| EPI_ISL_801856 | Chile/Santiago-SFF3444/2020 | 2020-04-23 | South America/Chile/Region Metropolit | Laboratory of Molecular Virology, Pontificia Universidad Cat√≥lica de Chile | MSHS Pathogen Surveillance Program | Leonardo I. Almonacid et al |
| EPI_ISL_801857 | Chile/Santiago-SSO4546/2020 | 2020-05-08 | South America/Chile/Region Metropolit | Laboratory of Molecular Virology, Pontificia Universidad Cat√≥lica de Chile | MSHS Pathogen Surveillance Program | Leonardo I. Almonacid et al |
| EPI_ISL_801871 | USA/NY-MSHSPSP-PV17869/2020 | 2020-09-16 | North America/USA/New York/Brooklyn | MSHS Clinical Microbiology Laboratories | MSHS Pathogen Surveillance Program | Ana S. Gonzalez-Reiche et al |
| EPI_ISL_801882 | USA/NY-MSHSPSP-PV19215/2020 | 2020-10-05 | North America/USA/New York/Brooklyn | MSHS Clinical Microbiology Laboratories | MSHS Pathogen Surveillance Program | Ana S. Gonzalez-Reiche et al |
| EPI_ISL_801885 | USA/NY-MSHSPSP-PV18404/2020 | 2020-09-27 | North America/USA/New York/Rockland | MSHS Clinical Microbiology Laboratories | MSHS Pathogen Surveillance Program | Ana S. Gonzalez-Reiche et al |
| EPI_ISL_801888 | USA/NY-MSHSPSP-PV14235/2020 | 2020-06-08 | North America/USA/New York/Westche | MSHS Clinical Microbiology Laboratories | MSHS Pathogen Surveillance Program | Ana S. Gonzalez-Reiche et al |
| EPI_ISL_801892 | USA/NY-MSHSPSP-PV15319/2020 | 2020-08-05 | North America/USA/New York/ | MSHS Clinical Microbiology Laboratories | MSHS Pathogen Surveillance Program | Ana S. Gonzalez-Reiche et al |
| EPI_ISL_801895 | USA/NY-MSHSPSP-PV14375/2020 | 2020-06-04 | North America/USA/New York/Brooklyn | MSHS Clinical Microbiology Laboratories | MSHS Pathogen Surveillance Program | Ana S. Gonzalez-Reiche et al |
| EPI_ISL_801900 | USA/NY-MSHSPSP-PV17858/2020 | 2020-09-11 | North America/USA/New York/Westche | MSHS Clinical Microbiology Laboratories | MSHS Pathogen Surveillance Program | Ana S. Gonzalez-Reiche et al |

| | | | | | | |
|---|---|---|---|---|---|---|
| EPI_ISL_801901 | USA/NY-MSHSPSP-PV21426/2020 | 2020-10-09 | North America/USA/New York/Brooklyn | MSHS Clinical Microbiology Laboratories | MSHS Pathogen Surveillance Program | Ana S. Gonzalez-Reiche et al |
| EPI_ISL_801902 | USA/NY-MSHSPSP-PV14693/2020 | 2020-05-19 | North America/USA/New York/Brooklyn | MSHS Clinical Microbiology Laboratories | MSHS Pathogen Surveillance Program | Ana S. Gonzalez-Reiche et al |
| EPI_ISL_801913 | USA/NY-MSHSPSP-PV13738/2020 | 2020-04-08 | North America/USA/New York/Brooklyn | MSHS Clinical Microbiology Laboratories | MSHS Pathogen Surveillance Program | Ana S. Gonzalez-Reiche et al |
| EPI_ISL_801918 | USA/NY-MSHSPSP-PV19203/2020 | 2020-09-27 | North America/USA/New York/Brooklyn | MSHS Clinical Microbiology Laboratories | MSHS Pathogen Surveillance Program | Ana S. Gonzalez-Reiche et al |
| EPI_ISL_801919 | USA/NY-MSHSPSP-PV19239/2020 | 2020-10-04 | North America/USA/New York/Manhatt | MSHS Clinical Microbiology Laboratories | MSHS Pathogen Surveillance Program | Ana S. Gonzalez-Reiche et al |
| EPI_ISL_801920 | USA/NY-MSHSPSP-PV17749/2020 | 2020-09-04 | North America/USA/New York/Queens | MSHS Clinical Microbiology Laboratories | MSHS Pathogen Surveillance Program | Ana S. Gonzalez-Reiche et al |
| EPI_ISL_801923 | USA/NY-MSHSPSP-PV17730/2020 | 2020-09-04 | North America/USA/New York/Brooklyn | MSHS Clinical Microbiology Laboratories | MSHS Pathogen Surveillance Program | Ana S. Gonzalez-Reiche et al |
| EPI_ISL_801927 | USA/NY-MSHSPSP-PV17511/2020 | 2020-08-27 | North America/USA/New York/Brooklyn | MSHS Clinical Microbiology Laboratories | MSHS Pathogen Surveillance Program | Ana S. Gonzalez-Reiche et al |
| EPI_ISL_801930 | USA/NY-MSHSPSP-PV17529/2020 | 2020-08-28 | North America/USA/New York/Brooklyn | MSHS Clinical Microbiology Laboratories | MSHS Pathogen Surveillance Program | Ana S. Gonzalez-Reiche et al |
| EPI_ISL_801931 | USA/NY-MSHSPSP-PV17751/2020 | 2020-09-03 | North America/USA/New York/Manhatt | MSHS Clinical Microbiology Laboratories | MSHS Pathogen Surveillance Program | Ana S. Gonzalez-Reiche et al |
| EPI_ISL_801936 | USA/NY-MSHSPSP-PV17656/2020 | 2020-08-30 | North America/USA/New York/Brooklyn | MSHS Clinical Microbiology Laboratories | MSHS Pathogen Surveillance Program | Ana S. Gonzalez-Reiche et al |
| EPI_ISL_801937 | USA/NY-MSHSPSP-PV15704/2020 | 2020-08-05 | North America/USA/New York/Manhatt | MSHS Clinical Microbiology Laboratories | MSHS Pathogen Surveillance Program | Ana S. Gonzalez-Reiche et al |
| EPI_ISL_801943 | USA/NY-MSHSPSP-PV21114/2020 | 2020-09-30 | North America/USA/New York/Brooklyn | MSHS Clinical Microbiology Laboratories | MSHS Pathogen Surveillance Program | Ana S. Gonzalez-Reiche et al |

| | | | | | | |
|---|---|---|---|---|---|---|
| EPI_ISL_801946 | USA/NY-MSHSPSP-PV18388/2020 | 2020-09-22 | North America/USA/New York/Nassau ( | MSHS Clinical Microbiology Laboratories | MSHS Pathogen Surveillance Program | Ana S. Gonzalez-Reiche et al |
| EPI_ISL_801948 | USA/NY-MSHSPSP-PV18149/2020 | 2020-04-03 | North America/USA/New York/Queens | MSHS Clinical Microbiology Laboratories | MSHS Pathogen Surveillance Program | Ana S. Gonzalez-Reiche et al |
| EPI_ISL_801951 | USA/NY-MSHSPSP-PV12388/2020 | 2020-05-03 | North America/USA/New York/Queens | MSHS Clinical Microbiology Laboratories | MSHS Pathogen Surveillance Program | Ana S. Gonzalez-Reiche et al |
| EPI_ISL_801952 | USA/NY-MSHSPSP-PV15060/2020 | 2020-06-02 | North America/USA/New York/Nassau ( | MSHS Clinical Microbiology Laboratories | MSHS Pathogen Surveillance Program | Ana S. Gonzalez-Reiche et al |
| EPI_ISL_801961 | USA/NY-MSHSPSP-PV15530/2020 | 2020-05-01 | North America/USA/New York/Brooklyr | MSHS Clinical Microbiology Laboratories | MSHS Pathogen Surveillance Program | Ana S. Gonzalez-Reiche et al |
| EPI_ISL_801966 | USA/NY-MSHSPSP-PV17123/2020 | 2020-08-18 | North America/USA/New York/Brooklyr | MSHS Clinical Microbiology Laboratories | MSHS Pathogen Surveillance Program | Ana S. Gonzalez-Reiche et al |
| EPI_ISL_801987 | USA/NY-MSHSPSP-PV11915/2020 | 2020-04-02 | North America/USA/New York/Manhatt | MSHS Clinical Microbiology Laboratories | MSHS Pathogen Surveillance Program | Ana S. Gonzalez-Reiche et al |
| EPI_ISL_801988 | USA/NY-MSHSPSP-PV11928/2020 | 2020-04-02 | North America/USA/New York/Westche | MSHS Clinical Microbiology Laboratories | MSHS Pathogen Surveillance Program | Ana S. Gonzalez-Reiche et al |
| EPI_ISL_802001 | USA/NY-MSHSPSP-PV12069/2020 | 2020-04-06 | North America/USA/New York/Queens | MSHS Clinical Microbiology Laboratories | MSHS Pathogen Surveillance Program | Ana S. Gonzalez-Reiche et al |
| EPI_ISL_802004 | USA/NY-MSHSPSP-PV14056/2020 | 2020-04-08 | North America/USA/New York/Bronx | MSHS Clinical Microbiology Laboratories | MSHS Pathogen Surveillance Program | Ana S. Gonzalez-Reiche et al |
| EPI_ISL_802018 | USA/NY-MSHSPSP-PV18490/2020 | 2020-04-13 | North America/USA/New York/Manhatt | MSHS Clinical Microbiology Laboratories | MSHS Pathogen Surveillance Program | Ana S. Gonzalez-Reiche et al |
| EPI_ISL_802019 | USA/NY-MSHSPSP-PV12852/2020 | 2020-04-14 | North America/USA/New York/Queens | MSHS Clinical Microbiology Laboratories | MSHS Pathogen Surveillance Program | Ana S. Gonzalez-Reiche et al |
| EPI_ISL_802042 | USA/NY-MSHSPSP-PV14796/2020 | 2020-04-30 | North America/USA/New York/Manhatt | MSHS Clinical Microbiology Laboratories | MSHS Pathogen Surveillance Program | Ana S. Gonzalez-Reiche et al |

| | | | | | | |
|---|---|---|---|---|---|---|
| EPI_ISL_802044 | USA/NY-MSHSPSP-PV15534/2020 | 2020-05-01 | North America/USA/New York/Queens | MSHS Clinical Microbiology Laboratories | MSHS Pathogen Surveillance Program | Ana S. Gonzalez-Reiche et al |
| EPI_ISL_802048 | USA/NY-MSHSPSP-PV12405/2020 | 2020-05-02 | North America/USA/New York/Manhatt | MSHS Clinical Microbiology Laboratories | MSHS Pathogen Surveillance Program | Ana S. Gonzalez-Reiche et al |
| EPI_ISL_802053 | USA/NY-MSHSPSP-PV12362/2020 | 2020-05-03 | North America/USA/New York/Manhatt | MSHS Clinical Microbiology Laboratories | MSHS Pathogen Surveillance Program | Ana S. Gonzalez-Reiche et al |
| EPI_ISL_802063 | USA/NY-MSHSPSP-PV12328/2020 | 2020-05-04 | North America/USA/New York/Nassau C | MSHS Clinical Microbiology Laboratories | MSHS Pathogen Surveillance Program | Ana S. Gonzalez-Reiche et al |
| EPI_ISL_802069 | USA/NY-MSHSPSP-PV12342/2020 | 2020-05-04 | North America/USA/New York/Queens | MSHS Clinical Microbiology Laboratories | MSHS Pathogen Surveillance Program | Ana S. Gonzalez-Reiche et al |
| EPI_ISL_802080 | USA/NY-MSHSPSP-PV12413/2020 | 2020-05-04 | North America/USA/New York/Manhatt | MSHS Clinical Microbiology Laboratories | MSHS Pathogen Surveillance Program | Ana S. Gonzalez-Reiche et al |
| EPI_ISL_802082 | USA/NY-MSHSPSP-PV12415/2020 | 2020-05-04 | North America/USA/New York/Manhatt | MSHS Clinical Microbiology Laboratories | MSHS Pathogen Surveillance Program | Ana S. Gonzalez-Reiche et al |
| EPI_ISL_802090 | USA/NY-MSHSPSP-PV13775/2020 | 2020-05-07 | North America/USA/New York/Manhatt | MSHS Clinical Microbiology Laboratories | MSHS Pathogen Surveillance Program | Ana S. Gonzalez-Reiche et al |
| EPI_ISL_802092 | USA/NY-MSHSPSP-PV14643/2020 | 2020-05-10 | North America/USA/New York/Brooklyn | MSHS Clinical Microbiology Laboratories | MSHS Pathogen Surveillance Program | Ana S. Gonzalez-Reiche et al |
| EPI_ISL_802094 | USA/NY-MSHSPSP-PV14663/2020 | 2020-05-18 | North America/USA/New York/Queens | MSHS Clinical Microbiology Laboratories | MSHS Pathogen Surveillance Program | Ana S. Gonzalez-Reiche et al |
| EPI_ISL_802096 | USA/NY-MSHSPSP-PV14657/2020 | 2020-05-24 | North America/USA/New York/Manhatt | MSHS Clinical Microbiology Laboratories | MSHS Pathogen Surveillance Program | Ana S. Gonzalez-Reiche et al |
| EPI_ISL_802098 | USA/NY-MSHSPSP-PV14708/2020 | 2020-05-27 | North America/USA/New York/Suffolk C | MSHS Clinical Microbiology Laboratories | MSHS Pathogen Surveillance Program | Ana S. Gonzalez-Reiche et al |
| EPI_ISL_802107 | USA/NY-MSHSPSP-PV14068/2020 | 2020-06-10 | North America/USA/New York/Queens | MSHS Clinical Microbiology Laboratories | MSHS Pathogen Surveillance Program | Ana S. Gonzalez-Reiche et al |

| | | | | | | |
|---|---|---|---|---|---|---|
| EPI_ISL_802113 | USA/NY-MSHSPSP-PV15116/2020 | 2020-07-02 | North America/USA/New York/Queens | MSHS Clinical Microbiology Laboratories | MSHS Pathogen Surveillance Program | Ana S. Gonzalez-Reiche et al |
| EPI_ISL_802114 | USA/NY-MSHSPSP-PV15302/2020 | 2020-07-02 | North America/USA/New York/Manhatt | MSHS Clinical Microbiology Laboratories | MSHS Pathogen Surveillance Program | Ana S. Gonzalez-Reiche et al |
| EPI_ISL_802115 | USA/NY-MSHSPSP-PV15313/2020 | 2020-07-02 | North America/USA/New York/Manhatt | MSHS Clinical Microbiology Laboratories | MSHS Pathogen Surveillance Program | Ana S. Gonzalez-Reiche et al |
| EPI_ISL_802116 | USA/NY-MSHSPSP-PV15315/2020 | 2020-07-02 | North America/USA/New York/Nassau C | MSHS Clinical Microbiology Laboratories | MSHS Pathogen Surveillance Program | Ana S. Gonzalez-Reiche et al |
| EPI_ISL_802118 | USA/NY-MSHSPSP-PV15117/2020 | 2020-07-04 | North America/USA/New York/Manhatt | MSHS Clinical Microbiology Laboratories | MSHS Pathogen Surveillance Program | Ana S. Gonzalez-Reiche et al |
| EPI_ISL_802120 | USA/NY-MSHSPSP-PV15317/2020 | 2020-07-04 | North America/USA/New York/Manhatt | MSHS Clinical Microbiology Laboratories | MSHS Pathogen Surveillance Program | Ana S. Gonzalez-Reiche et al |
| EPI_ISL_802125 | USA/NY-MSHSPSP-PV15277/2020 | 2020-07-06 | North America/USA/New York/Brooklyn | MSHS Clinical Microbiology Laboratories | MSHS Pathogen Surveillance Program | Ana S. Gonzalez-Reiche et al |
| EPI_ISL_802126 | USA/NY-MSHSPSP-PV15308/2020 | 2020-07-06 | North America/USA/New York/Manhatt | MSHS Clinical Microbiology Laboratories | MSHS Pathogen Surveillance Program | Ana S. Gonzalez-Reiche et al |
| EPI_ISL_802127 | USA/NY-MSHSPSP-PV15310/2020 | 2020-07-06 | North America/USA/New York/Nassau C | MSHS Clinical Microbiology Laboratories | MSHS Pathogen Surveillance Program | Ana S. Gonzalez-Reiche et al |
| EPI_ISL_802129 | USA/NY-MSHSPSP-PV15121/2020 | 2020-07-07 | North America/USA/New York/Bronx | MSHS Clinical Microbiology Laboratories | MSHS Pathogen Surveillance Program | Ana S. Gonzalez-Reiche et al |
| EPI_ISL_802133 | USA/NY-MSHSPSP-PV15290/2020 | 2020-07-09 | North America/USA/New York/Nassau C | MSHS Clinical Microbiology Laboratories | MSHS Pathogen Surveillance Program | Ana S. Gonzalez-Reiche et al |
| EPI_ISL_802136 | USA/CT-MSHSPSP-PV15299/2020 | 2020-07-10 | North America/USA/Connecticut/New H | MSHS Clinical Microbiology Laboratories | MSHS Pathogen Surveillance Program | Ana S. Gonzalez-Reiche et al |
| EPI_ISL_802139 | USA/NY-MSHSPSP-PV15160/2020 | 2020-07-19 | North America/USA/New York/Queens | MSHS Clinical Microbiology Laboratories | MSHS Pathogen Surveillance Program | Ana S. Gonzalez-Reiche et al |

| | | | | | | |
|---|---|---|---|---|---|---|
| EPI_ISL_802142 | USA/NY-MSHSPSP-PV14867/2020 | 2020-08-01 | North America/USA/New York/Nassau ( | MSHS Clinical Microbiology Laboratories | MSHS Pathogen Surveillance Program | Ana S. Gonzalez-Reiche et al |
| EPI_ISL_802148 | USA/NJ-MSHSPSP-PV15719/2020 | 2020-08-03 | North America/USA/New Jersey/Union | MSHS Clinical Microbiology Laboratories | MSHS Pathogen Surveillance Program | Ana S. Gonzalez-Reiche et al |
| EPI_ISL_802152 | USA/NY-MSHSPSP-PV15712/2020 | 2020-08-04 | North America/USA/New York/Manhatt | MSHS Clinical Microbiology Laboratories | MSHS Pathogen Surveillance Program | Ana S. Gonzalez-Reiche et al |
| EPI_ISL_802155 | USA/NY-MSHSPSP-PV16489/2020 | 2020-08-06 | North America/USA/New York/Manhatt | MSHS Clinical Microbiology Laboratories | MSHS Pathogen Surveillance Program | Ana S. Gonzalez-Reiche et al |
| EPI_ISL_802156 | USA/NY-MSHSPSP-PV15363/2020 | 2020-08-06 | North America/USA/New York/Manhatt | MSHS Clinical Microbiology Laboratories | MSHS Pathogen Surveillance Program | Ana S. Gonzalez-Reiche et al |
| EPI_ISL_802164 | USA/NY-MSHSPSP-PV17542/2020 | 2020-08-13 | North America/USA/New York/Manhatt | MSHS Clinical Microbiology Laboratories | MSHS Pathogen Surveillance Program | Ana S. Gonzalez-Reiche et al |
| EPI_ISL_802170 | USA/NY-MSHSPSP-PV16542/2020 | 2020-08-17 | North America/USA/New York/Manhatt | MSHS Clinical Microbiology Laboratories | MSHS Pathogen Surveillance Program | Ana S. Gonzalez-Reiche et al |
| EPI_ISL_802171 | USA/NY-MSHSPSP-PV16565/2020 | 2020-08-17 | North America/USA/New York/Queens | MSHS Clinical Microbiology Laboratories | MSHS Pathogen Surveillance Program | Ana S. Gonzalez-Reiche et al |
| EPI_ISL_802174 | USA/NY-MSHSPSP-PV17113/2020 | 2020-08-18 | North America/USA/New York/Manhatt | MSHS Clinical Microbiology Laboratories | MSHS Pathogen Surveillance Program | Ana S. Gonzalez-Reiche et al |
| EPI_ISL_802177 | USA/NY-MSHSPSP-PV17140/2020 | 2020-08-18 | North America/USA/New York/Manhatt | MSHS Clinical Microbiology Laboratories | MSHS Pathogen Surveillance Program | Ana S. Gonzalez-Reiche et al |
| EPI_ISL_802178 | USA/NY-MSHSPSP-PV17151/2020 | 2020-08-18 | North America/USA/New York/Bronx | MSHS Clinical Microbiology Laboratories | MSHS Pathogen Surveillance Program | Ana S. Gonzalez-Reiche et al |
| EPI_ISL_802179 | USA/NY-MSHSPSP-PV17159/2020 | 2020-08-18 | North America/USA/New York/Manhatt | MSHS Clinical Microbiology Laboratories | MSHS Pathogen Surveillance Program | Ana S. Gonzalez-Reiche et al |
| EPI_ISL_802180 | USA/NY-MSHSPSP-PV17170/2020 | 2020-08-18 | North America/USA/New York/Queens | MSHS Clinical Microbiology Laboratories | MSHS Pathogen Surveillance Program | Ana S. Gonzalez-Reiche et al |

| | | | | | | |
|---|---|---|---|---|---|---|
| EPI_ISL_802184 | USA/NY-MSHSPSP-PV17137/2020 | 2020-08-19 | North America/USA/New York/Manhatt | MSHS Clinical Microbiology Laboratories | MSHS Pathogen Surveillance Program | Ana S. Gonzalez-Reiche et al |
| EPI_ISL_802186 | USA/NY-MSHSPSP-PV17141/2020 | 2020-08-19 | North America/USA/New York/Queens | MSHS Clinical Microbiology Laboratories | MSHS Pathogen Surveillance Program | Ana S. Gonzalez-Reiche et al |
| EPI_ISL_802188 | USA/NJ-MSHSPSP-PV17629/2020 | 2020-08-20 | North America/USA/New Jersey/Berger | MSHS Clinical Microbiology Laboratories | MSHS Pathogen Surveillance Program | Ana S. Gonzalez-Reiche et al |
| EPI_ISL_802189 | USA/NY-MSHSPSP-PV16569/2020 | 2020-08-21 | North America/USA/New York/Bronx | MSHS Clinical Microbiology Laboratories | MSHS Pathogen Surveillance Program | Ana S. Gonzalez-Reiche et al |
| EPI_ISL_802192 | USA/NY-MSHSPSP-PV17166/2020 | 2020-08-21 | North America/USA/New York/Manhatt | MSHS Clinical Microbiology Laboratories | MSHS Pathogen Surveillance Program | Ana S. Gonzalez-Reiche et al |
| EPI_ISL_802210 | USA/NY-MSHSPSP-PV17635/2020 | 2020-08-30 | North America/USA/New York/Brooklyr | MSHS Clinical Microbiology Laboratories | MSHS Pathogen Surveillance Program | Ana S. Gonzalez-Reiche et al |
| EPI_ISL_802211 | USA/NY-MSHSPSP-PV17637/2020 | 2020-08-30 | North America/USA/New York/Brooklyr | MSHS Clinical Microbiology Laboratories | MSHS Pathogen Surveillance Program | Ana S. Gonzalez-Reiche et al |
| EPI_ISL_802213 | USA/NY-MSHSPSP-PV17171/2020 | 2020-08-31 | North America/USA/New York/Queens | MSHS Clinical Microbiology Laboratories | MSHS Pathogen Surveillance Program | Ana S. Gonzalez-Reiche et al |
| EPI_ISL_802215 | USA/NY-MSHSPSP-PV17624/2020 | 2020-08-31 | North America/USA/New York/Manhatt | MSHS Clinical Microbiology Laboratories | MSHS Pathogen Surveillance Program | Ana S. Gonzalez-Reiche et al |
| EPI_ISL_802216 | USA/NY-MSHSPSP-PV17625/2020 | 2020-08-31 | North America/USA/New York/Brooklyr | MSHS Clinical Microbiology Laboratories | MSHS Pathogen Surveillance Program | Ana S. Gonzalez-Reiche et al |
| EPI_ISL_802221 | USA/NY-MSHSPSP-PV17919/2020 | 2020-09-02 | North America/USA/New York/Manhatt | MSHS Clinical Microbiology Laboratories | MSHS Pathogen Surveillance Program | Ana S. Gonzalez-Reiche et al |
| EPI_ISL_802222 | USA/NY-MSHSPSP-PV17750/2020 | 2020-09-03 | North America/USA/New York/Brooklyr | MSHS Clinical Microbiology Laboratories | MSHS Pathogen Surveillance Program | Ana S. Gonzalez-Reiche et al |
| EPI_ISL_802229 | USA/NY-MSHSPSP-PV17761/2020 | 2020-09-05 | North America/USA/New York/Brooklyr | MSHS Clinical Microbiology Laboratories | MSHS Pathogen Surveillance Program | Ana S. Gonzalez-Reiche et al |

| | | | | | | |
|---|---|---|---|---|---|---|
| EPI_ISL_802230 | USA/NY-MSHSPSP-PV17879/2020 | 2020-09-08 | North America/USA/New York/Brooklyn | MSHS Clinical Microbiology Laboratories | MSHS Pathogen Surveillance Program | Ana S. Gonzalez-Reiche et al |
| EPI_ISL_802241 | USA/NY-MSHSPSP-PV17934/2020 | 2020-09-10 | North America/USA/New York/Orange ( | MSHS Clinical Microbiology Laboratories | MSHS Pathogen Surveillance Program | Ana S. Gonzalez-Reiche et al |
| EPI_ISL_802244 | USA/NJ-MSHSPSP-PV17870/2020 | 2020-09-11 | North America/USA/New Jersey/Glouce | MSHS Clinical Microbiology Laboratories | MSHS Pathogen Surveillance Program | Ana S. Gonzalez-Reiche et al |
| EPI_ISL_802245 | USA/NY-MSHSPSP-PV17889/2020 | 2020-09-12 | North America/USA/New York/Orange ( | MSHS Clinical Microbiology Laboratories | MSHS Pathogen Surveillance Program | Ana S. Gonzalez-Reiche et al |
| EPI_ISL_802246 | USA/NY-MSHSPSP-PV17890/2020 | 2020-09-12 | North America/USA/New York/Brooklyn | MSHS Clinical Microbiology Laboratories | MSHS Pathogen Surveillance Program | Ana S. Gonzalez-Reiche et al |
| EPI_ISL_802247 | USA/NY-MSHSPSP-PV17917/2020 | 2020-09-12 | North America/USA/New York/Queens | MSHS Clinical Microbiology Laboratories | MSHS Pathogen Surveillance Program | Ana S. Gonzalez-Reiche et al |
| EPI_ISL_802251 | USA/NY-MSHSPSP-PV17860/2020 | 2020-09-14 | North America/USA/New York/Manhatt | MSHS Clinical Microbiology Laboratories | MSHS Pathogen Surveillance Program | Ana S. Gonzalez-Reiche et al |
| EPI_ISL_802253 | USA/NY-MSHSPSP-PV17863/2020 | 2020-09-16 | North America/USA/New York/Rockland | MSHS Clinical Microbiology Laboratories | MSHS Pathogen Surveillance Program | Ana S. Gonzalez-Reiche et al |
| EPI_ISL_802254 | USA/NY-MSHSPSP-PV17880/2020 | 2020-09-17 | North America/USA/New York/Orange ( | MSHS Clinical Microbiology Laboratories | MSHS Pathogen Surveillance Program | Ana S. Gonzalez-Reiche et al |
| EPI_ISL_802261 | USA/NY-MSHSPSP-PV19688/2020 | 2020-09-19 | North America/USA/New York/Manhatt | MSHS Clinical Microbiology Laboratories | MSHS Pathogen Surveillance Program | Ana S. Gonzalez-Reiche et al |
| EPI_ISL_802263 | USA/NY-MSHSPSP-PV19610/2020 | 2020-09-22 | North America/USA/New York/Nassau ( | MSHS Clinical Microbiology Laboratories | MSHS Pathogen Surveillance Program | Ana S. Gonzalez-Reiche et al |
| EPI_ISL_802269 | USA/NY-MSHSPSP-PV19231/2020 | 2020-09-24 | North America/USA/New York/Brooklyn | MSHS Clinical Microbiology Laboratories | MSHS Pathogen Surveillance Program | Ana S. Gonzalez-Reiche et al |
| EPI_ISL_802274 | USA/NY-MSHSPSP-PV19039/2020 | 2020-09-26 | North America/USA/New York/Brooklyn | MSHS Clinical Microbiology Laboratories | MSHS Pathogen Surveillance Program | Ana S. Gonzalez-Reiche et al |

| | | | | | | |
|---|---|---|---|---|---|---|
| EPI_ISL_802275 | USA/PA-MSHSPSP-PV19591/2020 | 2020-09-26 | North America/USA/Pennsylvania/Chest | MSHS Clinical Microbiology Laboratories | MSHS Pathogen Surveillance Program | Ana S. Gonzalez-Reiche et al |
| EPI_ISL_802278 | USA/NY-MSHSPSP-PV21111/2020 | 2020-09-26 | North America/USA/New York/Manhatt | MSHS Clinical Microbiology Laboratories | MSHS Pathogen Surveillance Program | Ana S. Gonzalez-Reiche et al |
| EPI_ISL_802279 | USA/NY-MSHSPSP-PV21125/2020 | 2020-09-26 | North America/USA/New York/Manhatt | MSHS Clinical Microbiology Laboratories | MSHS Pathogen Surveillance Program | Ana S. Gonzalez-Reiche et al |
| EPI_ISL_802281 | USA/NY-MSHSPSP-PV19194/2020 | 2020-09-27 | North America/USA/New York/Brooklyn | MSHS Clinical Microbiology Laboratories | MSHS Pathogen Surveillance Program | Ana S. Gonzalez-Reiche et al |
| EPI_ISL_802288 | USA/NY-MSHSPSP-PV19190/2020 | 2020-09-28 | North America/USA/New York/Queens | MSHS Clinical Microbiology Laboratories | MSHS Pathogen Surveillance Program | Ana S. Gonzalez-Reiche et al |
| EPI_ISL_802292 | USA/NY-MSHSPSP-PV19365/2020 | 2020-09-28 | North America/USA/New York/Brooklyn | MSHS Clinical Microbiology Laboratories | MSHS Pathogen Surveillance Program | Ana S. Gonzalez-Reiche et al |
| EPI_ISL_802296 | USA/NY-MSHSPSP-PV21077/2020 | 2020-09-28 | North America/USA/New York/Brooklyn | MSHS Clinical Microbiology Laboratories | MSHS Pathogen Surveillance Program | Ana S. Gonzalez-Reiche et al |
| EPI_ISL_802301 | USA/NY-MSHSPSP-PV19363/2020 | 2020-09-29 | North America/USA/New York/Manhatt | MSHS Clinical Microbiology Laboratories | MSHS Pathogen Surveillance Program | Ana S. Gonzalez-Reiche et al |
| EPI_ISL_802302 | USA/NY-MSHSPSP-PV19626/2020 | 2020-09-29 | North America/USA/New York/Queens | MSHS Clinical Microbiology Laboratories | MSHS Pathogen Surveillance Program | Ana S. Gonzalez-Reiche et al |
| EPI_ISL_802304 | USA/NY-MSHSPSP-PV19629/2020 | 2020-09-29 | North America/USA/New York/Queens | MSHS Clinical Microbiology Laboratories | MSHS Pathogen Surveillance Program | Ana S. Gonzalez-Reiche et al |
| EPI_ISL_802306 | USA/NY-MSHSPSP-PV21115/2020 | 2020-09-29 | North America/USA/New York/Manhatt | MSHS Clinical Microbiology Laboratories | MSHS Pathogen Surveillance Program | Ana S. Gonzalez-Reiche et al |
| EPI_ISL_802308 | USA/NY-MSHSPSP-PV19357/2020 | 2020-09-29 | North America/USA/New York/Manhatt | MSHS Clinical Microbiology Laboratories | MSHS Pathogen Surveillance Program | Ana S. Gonzalez-Reiche et al |
| EPI_ISL_802310 | USA/NY-MSHSPSP-PV19350/2020 | 2020-09-30 | North America/USA/New York/Bronx | MSHS Clinical Microbiology Laboratories | MSHS Pathogen Surveillance Program | Ana S. Gonzalez-Reiche et al |

| | | | | | | |
|---|---|---|---|---|---|---|
| EPI_ISL_802318 | USA/NY-MSHSPSP-PV21083/2020 | 2020-09-30 | North America/USA/New York/Brooklyn | MSHS Clinical Microbiology Laboratories | MSHS Pathogen Surveillance Program | Ana S. Gonzalez-Reiche et al |
| EPI_ISL_802328 | USA/NY-MSHSPSP-PV19656/2020 | 2020-10-03 | North America/USA/New York/Manhatt | MSHS Clinical Microbiology Laboratories | MSHS Pathogen Surveillance Program | Ana S. Gonzalez-Reiche et al |
| EPI_ISL_802329 | USA/NY-MSHSPSP-PV21096/2020 | 2020-10-03 | North America/USA/New York/Brooklyn | MSHS Clinical Microbiology Laboratories | MSHS Pathogen Surveillance Program | Ana S. Gonzalez-Reiche et al |
| EPI_ISL_802335 | USA/NY-MSHSPSP-PV19223/2020 | 2020-10-04 | North America/USA/New York/Brooklyn | MSHS Clinical Microbiology Laboratories | MSHS Pathogen Surveillance Program | Ana S. Gonzalez-Reiche et al |
| EPI_ISL_802338 | USA/NY-MSHSPSP-PV19240/2020 | 2020-10-04 | North America/USA/New York/Manhatt | MSHS Clinical Microbiology Laboratories | MSHS Pathogen Surveillance Program | Ana S. Gonzalez-Reiche et al |
| EPI_ISL_802339 | USA/NY-MSHSPSP-PV19635/2020 | 2020-10-04 | North America/USA/New York/Queens | MSHS Clinical Microbiology Laboratories | MSHS Pathogen Surveillance Program | Ana S. Gonzalez-Reiche et al |
| EPI_ISL_802340 | USA/NY-MSHSPSP-PV19636/2020 | 2020-10-04 | North America/USA/New York/Queens | MSHS Clinical Microbiology Laboratories | MSHS Pathogen Surveillance Program | Ana S. Gonzalez-Reiche et al |
| EPI_ISL_802348 | USA/NY-MSHSPSP-PV19207/2020 | 2020-10-05 | North America/USA/New York/Nassau C | MSHS Clinical Microbiology Laboratories | MSHS Pathogen Surveillance Program | Ana S. Gonzalez-Reiche et al |
| EPI_ISL_802350 | USA/NY-MSHSPSP-PV19597/2020 | 2020-10-05 | North America/USA/New York/Queens | MSHS Clinical Microbiology Laboratories | MSHS Pathogen Surveillance Program | Ana S. Gonzalez-Reiche et al |
| EPI_ISL_802351 | USA/NY-MSHSPSP-PV21087/2020 | 2020-10-05 | North America/USA/New York/Schenect | MSHS Clinical Microbiology Laboratories | MSHS Pathogen Surveillance Program | Ana S. Gonzalez-Reiche et al |
| EPI_ISL_802353 | USA/NY-MSHSPSP-PV19645/2020 | 2020-10-06 | North America/USA/New York/ | MSHS Clinical Microbiology Laboratories | MSHS Pathogen Surveillance Program | Ana S. Gonzalez-Reiche et al |
| EPI_ISL_802356 | USA/NY-MSHSPSP-PV19754/2020 | 2020-10-06 | North America/USA/New York/Brooklyn | MSHS Clinical Microbiology Laboratories | MSHS Pathogen Surveillance Program | Ana S. Gonzalez-Reiche et al |
| EPI_ISL_802357 | USA/NJ-MSHSPSP-PV19639/2020 | 2020-10-07 | North America/USA/New Jersey/Union | MSHS Clinical Microbiology Laboratories | MSHS Pathogen Surveillance Program | Ana S. Gonzalez-Reiche et al |

| | | | | | | |
|---|---|---|---|---|---|---|
| EPI_ISL_802368 | USA/NY-MSHSPSP-PV19674/2020 | 2020-10-08 | North America/USA/New York/Brooklyn | MSHS Clinical Microbiology Laboratories | MSHS Pathogen Surveillance Program | Ana S. Gonzalez-Reiche et al |
| EPI_ISL_802370 | USA/NY-MSHSPSP-PV19679/2020 | 2020-10-08 | North America/USA/New York/Brooklyn | MSHS Clinical Microbiology Laboratories | MSHS Pathogen Surveillance Program | Ana S. Gonzalez-Reiche et al |
| EPI_ISL_802372 | USA/NY-MSHSPSP-PV19755/2020 | 2020-10-08 | North America/USA/New York/Staten Is | MSHS Clinical Microbiology Laboratories | MSHS Pathogen Surveillance Program | Ana S. Gonzalez-Reiche et al |
| EPI_ISL_802378 | USA/NY-MSHSPSP-PV19775/2020 | 2020-10-08 | North America/USA/New York/Brooklyn | MSHS Clinical Microbiology Laboratories | MSHS Pathogen Surveillance Program | Ana S. Gonzalez-Reiche et al |
| EPI_ISL_802384 | USA/NY-MSHSPSP-PV19669/2020 | 2020-10-09 | North America/USA/New York/Queens | MSHS Clinical Microbiology Laboratories | MSHS Pathogen Surveillance Program | Ana S. Gonzalez-Reiche et al |
| EPI_ISL_802391 | USA/NY-MSHSPSP-PV21144/2020 | 2020-10-09 | North America/USA/New York/Rockland | MSHS Clinical Microbiology Laboratories | MSHS Pathogen Surveillance Program | Ana S. Gonzalez-Reiche et al |
| EPI_ISL_802400 | USA/NY-MSHSPSP-PV21174/2020 | 2020-10-10 | North America/USA/New York/Manhatt | MSHS Clinical Microbiology Laboratories | MSHS Pathogen Surveillance Program | Ana S. Gonzalez-Reiche et al |
| EPI_ISL_802401 | USA/NY-MSHSPSP-PV21127/2020 | 2020-10-11 | North America/USA/New York/Queens | MSHS Clinical Microbiology Laboratories | MSHS Pathogen Surveillance Program | Ana S. Gonzalez-Reiche et al |
| EPI_ISL_802403 | USA/NY-MSHSPSP-PV21151/2020 | 2020-10-11 | North America/USA/New York/Nassau C | MSHS Clinical Microbiology Laboratories | MSHS Pathogen Surveillance Program | Ana S. Gonzalez-Reiche et al |
| EPI_ISL_802405 | USA/NY-MSHSPSP-PV19609/2020 | 2020-10-12 | North America/USA/New York/Queens | MSHS Clinical Microbiology Laboratories | MSHS Pathogen Surveillance Program | Ana S. Gonzalez-Reiche et al |
| EPI_ISL_802420 | USA/NY-MSHSPSP-PV21184/2020 | 2020-10-13 | North America/USA/New York/Bronx | MSHS Clinical Microbiology Laboratories | MSHS Pathogen Surveillance Program | Ana S. Gonzalez-Reiche et al |
| EPI_ISL_802423 | USA/NY-MSHSPSP-PV21201/2020 | 2020-10-14 | North America/USA/New York/Manhatt | MSHS Clinical Microbiology Laboratories | MSHS Pathogen Surveillance Program | Ana S. Gonzalez-Reiche et al |
| EPI_ISL_802425 | USA/NJ-MSHSPSP-PV21148/2020 | 2020-10-16 | North America/USA/New Jersey/Berger | MSHS Clinical Microbiology Laboratories | MSHS Pathogen Surveillance Program | Ana S. Gonzalez-Reiche et al |

| | | | | | | |
|---|---|---|---|---|---|---|
| EPI_ISL_802427 | USA/NY-MSHSPSP-PV21106/2020 | 2020-10-18 | North America/USA/New York/Manhatt | MSHS Clinical Microbiology Laboratories | MSHS Pathogen Surveillance Program | Ana S. Gonzalez-Reiche et al |
| EPI_ISL_802428 | USA/NY-Wadsworth-21001039-01/20 | 2021-01-01 | North America/USA/New York/Saratog | SARATOGA HOSPITAL LABORATORY | Wadsworth Center, New York State Department.of Health | Kirsten St. George et al |
| EPI_ISL_802438 | USA/NY-Wadsworth-282135-01/2020 | 2020-12-02 | North America/USA/New York/ | Wadsworth Center, New York State Department.of Health | Wadsworth Center, New York State Department.of Health | Kirsten St. George et al |
| EPI_ISL_802442 | USA/NY-Wadsworth-291835-01/2020 | 2020-12-29 | North America/USA/New York/Warren | Wadsworth Center, New York State Department.of Health | Wadsworth Center, New York State Department.of Health | Kirsten St. George et al |
| EPI_ISL_802443 | USA/NY-Wadsworth-21001288-01/20 | 2020-12-30 | North America/USA/New York/Dutches | WESTCHESTER MEDICAL CENTER | Wadsworth Center, New York State Department.of Health | Kirsten St. George et al |
| EPI_ISL_802450 | USA/NY-Wadsworth-291134-01/2020 | 2020-12-28 | North America/USA/New York/Albany ( | Wadsworth Center, New York State Department.of Health | Wadsworth Center, New York State Department.of Health | Kirsten St. George et al |
| EPI_ISL_802454 | USA/NY-Wadsworth-291130-01/2020 | 2020-12-28 | North America/USA/New York/Albany ( | Wadsworth Center, New York State Department.of Health | Wadsworth Center, New York State Department.of Health | Kirsten St. George et al |
| EPI_ISL_802478 | USA/NY-Wadsworth-21001300-01/20 | 2020-12-30 | North America/USA/New York/Greene ( | WESTCHESTER MEDICAL CENTER | Wadsworth Center, New York State Department.of Health | Kirsten St. George et al |

| | | | | | | |
|---|---|---|---|---|---|---|
| EPI_ISL_802488 | USA/NY-Wadsworth-21001318-01/20 | 2020-12-30 | North America/USA/New York/Westche | WESTCHESTER MEDICAL CENTER | Wadsworth Center, New York State Department.of Health | Kirsten St. George et al |
| EPI_ISL_802492 | USA/NY-Wadsworth-21001041-01/20 | 2021-01-01 | North America/USA/New York/Saratoga | SARATOGA HOSPITAL LABORATORY | Wadsworth Center, New York State Department.of Health | Kirsten St. George et al |
| EPI_ISL_802493 | USA/NY-Wadsworth-21001043-01/20 | 2021-01-02 | North America/USA/New York/Saratoga | SARATOGA HOSPITAL LABORATORY | Wadsworth Center, New York State Department.of Health | Kirsten St. George et al |
| EPI_ISL_802500 | USA/NY-Wadsworth-21000246-01/20 | 2021-01-04 | North America/USA/New York/Suffolk C | Wadsworth Center, New York State Department.of Health | Wadsworth Center, New York State Department.of Health | Kirsten St. George et al |
| EPI_ISL_802545 | Chile/KRS696/2020 | 2020-10-04 | South America/Chile/Magallanes/Natale | Molecular Microbiology and Food Research Laboratory (MMFRLAB) - Universidad San Sebastiv°n | Facultad de Ciencias de la Vida, UNAB | Dayv°n Sanhueza et al |
| EPI_ISL_802546 | Chile/KRU459/2020 | 2020-10-05 | South America/Chile/Magallanes/Natale | Molecular Microbiology and Food Research Laboratory (MMFRLAB) - Universidad San Sebastiv°n | Facultad de Ciencias de la Vida, UNAB | Dayv°n Sanhueza et al |
| EPI_ISL_802549 | Chile/KSK883/2020 | 2020-10-05 | South America/Chile/Magallanes/Natale | Molecular Microbiology and Food Research Laboratory (MMFRLAB) - Universidad San Sebastiv°n | Facultad de Ciencias de la Vida, UNAB | Dayv°n Sanhueza et al |
| EPI_ISL_802551 | Chile/KSN928/2020 | 2020-10-05 | South America/Chile/Magallanes/Natale | Molecular Microbiology and Food Research Laboratory (MMFRLAB) - Universidad San Sebastiv°n | Facultad de Ciencias de la Vida, UNAB | Dayv°n Sanhueza et al |
| EPI_ISL_802556 | Chile/KTG650/2020 | 2020-10-06 | South America/Chile/Magallanes/Punta | Molecular Microbiology and Food Research Laboratory (MMFRLAB) - Universidad San Sebastiv°n | Facultad de Ciencias de la Vida, UNAB | Dayv°n Sanhueza et al |
| EPI_ISL_802558 | Chile/KTG655/2020 | 2020-10-06 | South America/Chile/Magallanes/Punta | Molecular Microbiology and Food Research Laboratory (MMFRLAB) - Universidad San Sebastiv°n | Facultad de Ciencias de la Vida, UNAB | Dayv°n Sanhueza et al |
| EPI_ISL_802561 | Chile/KTJ904/2020 | 2020-10-06 | South America/Chile/Magallanes/Porve | Molecular Microbiology and Food Research Laboratory (MMFRLAB) - Universidad San Sebastiv°n | Facultad de Ciencias de la Vida, UNAB | Dayv°n Sanhueza et al |

| | | | | | | |
|---|---|---|---|---|---|---|
| EPI_ISL_802564 | Chile/KTJ914/2020 | 2020-10-06 | South America/Chile/Magallanes/Porve | Molecular Microbiology and Food Research Laboratory (MMFRLAB) - Universidad San Sebastiv°n | Facultad de Ciencias de la Vida, UNAB | Dayv°n Sanhueza et al |
| EPI_ISL_802566 | Chile/KTK521/2020 | 2020-10-06 | South America/Chile/Magallanes/Punta | Molecular Microbiology and Food Research Laboratory (MMFRLAB) - Universidad San Sebastiv°n | Facultad de Ciencias de la Vida, UNAB | Dayv°n Sanhueza et al |
| EPI_ISL_802568 | USA/MN-MDH-2247/2020 | 2020-11-14 | North America/USA/Minnesota/ | Essentia Health-St. Mary's Medical Center | Minnesota Department of Health, Public Health Laboratory | Alexandra Lorentz et al |
| EPI_ISL_802575 | USA/MN-MDH-2254/2020 | 2020-12-28 | North America/USA/Minnesota/ | Vault Health | Minnesota Department of Health, Public Health Laboratory | Alexandra Lorentz et al |
| EPI_ISL_802590 | USA/MN-MDH-2269/2020 | 2020-12-28 | North America/USA/Minnesota/ | Vault Health | Minnesota Department of Health, Public Health Laboratory | Alexandra Lorentz et al |
| EPI_ISL_802591 | USA/MN-MDH-2270/2020 | 2020-12-27 | North America/USA/Minnesota/ | Vault Health | Minnesota Department of Health, Public Health Laboratory | Alexandra Lorentz et al |
| EPI_ISL_802611 | USA/CA-CDC-STM-062/2020 | 2020-12-27 | North America/USA/California/ | Helix/Illumina | Genomics and Discovery, Respiratory Viruses Branch, Division of Viral Diseases, Centers for Disease Control and Prevention | Peter W. Cook et al |
| EPI_ISL_802613 | USA/CA-CDC-STM-067/2020 | 2020-12-26 | North America/USA/California/ | Helix/Illumina | Genomics and Discovery, Respiratory Viruses Branch, Division of Viral Diseases, Centers for Disease Control and Prevention | Peter W. Cook et al |

| | | | | | | |
|---|---|---|---|---|---|---|
| EPI_ISL_802615 | USA/CA-CDC-STM-070/2020 | 2020-12-26 | North America/USA/California/ | Helix/Illumina | Genomics and Discovery, Respiratory Viruses Branch, Division of Viral Diseases, Centers for Disease Control and Prevention | Peter W. Cook et al |
| EPI_ISL_802618 | USA/CA-CDC-STM-076/2020 | 2020-12-29 | North America/USA/California/ | Helix/Illumina | Genomics and Discovery, Respiratory Viruses Branch, Division of Viral Diseases, Centers for Disease Control and Prevention | Peter W. Cook et al |
| EPI_ISL_802620 | USA/CA-CDC-STM-078/2020 | 2020-12-29 | North America/USA/California/ | Helix/Illumina | Genomics and Discovery, Respiratory Viruses Branch, Division of Viral Diseases, Centers for Disease Control and Prevention | Peter W. Cook et al |
| EPI_ISL_802623 | USA/CA-CDC-STM-087/2020 | 2020-12-29 | North America/USA/California/ | Helix/Illumina | Genomics and Discovery, Respiratory Viruses Branch, Division of Viral Diseases, Centers for Disease Control and Prevention | Peter W. Cook et al |

| | | | | | | |
|---|---|---|---|---|---|---|
| EPI_ISL_802624 | USA/CA-CDC-STM-105/2020 | 2020-12-29 | North America/USA/California/ | Helix/Illumina | Genomics and Discovery, Respiratory Viruses Branch, Division of Viral Diseases, Centers for Disease Control and Prevention | Peter W. Cook et al |
| EPI_ISL_802626 | USA/CA-CDC-STM-127/2020 | 2020-12-30 | North America/USA/California/ | Helix/Illumina | Genomics and Discovery, Respiratory Viruses Branch, Division of Viral Diseases, Centers for Disease Control and Prevention | Peter W. Cook et al |
| EPI_ISL_802637 | USA/FL-CDC-STM-083/2020 | 2020-12-28 | North America/USA/Florida/ | Helix/Illumina | Genomics and Discovery, Respiratory Viruses Branch, Division of Viral Diseases, Centers for Disease Control and Prevention | Peter W. Cook et al |
| EPI_ISL_802641 | USA/FL-CDC-STM-115/2020 | 2020-12-29 | North America/USA/Florida/ | Helix/Illumina | Genomics and Discovery, Respiratory Viruses Branch, Division of Viral Diseases, Centers for Disease Control and Prevention | Peter W. Cook et al |

| | | | | | | |
|---|---|---|---|---|---|---|
| EPI_ISL_802645 | USA/FL-CDC-STM-145/2020 | 2020-12-30 | North America/USA/Florida/ | Helix/Illumina | Genomics and Discovery, Respiratory Viruses Branch, Division of Viral Diseases, Centers for Disease Control and Prevention | Peter W. Cook et al |
| EPI_ISL_802648 | USA/FL-CDC-STM-075/2020 | 2020-12-28 | North America/USA/Florida/ | Helix/Illumina | Genomics and Discovery, Respiratory Viruses Branch, Division of Viral Diseases, Centers for Disease Control and Prevention | Peter W. Cook et al |
| EPI_ISL_802666 | USA/CA-CDC-STM-118/2020 | 2020-12-30 | North America/USA/California/ | Helix/Illumina | Genomics and Discovery, Respiratory Viruses Branch, Division of Viral Diseases, Centers for Disease Control and Prevention | Peter W. Cook et al |
| EPI_ISL_802677 | USA/FL-CDC-STM-047/2020 | 2020-12-26 | North America/USA/Florida/ | Helix/Illumina | Genomics and Discovery, Respiratory Viruses Branch, Division of Viral Diseases, Centers for Disease Control and Prevention | Peter W. Cook et al |

| | | | | | | |
|---|---|---|---|---|---|---|
| EPI_ISL_802689 | USA/FL-CDC-STM-108/2020 | 2020-12-29 | North America/USA/Florida/ | Helix/Illumina | Genomics and Discovery, Respiratory Viruses Branch, Division of Viral Diseases, Centers for Disease Control and Prevention | Peter W. Cook et al |
| EPI_ISL_802702 | USA/MA-CDC-STM-051/2020 | 2020-12-26 | North America/USA/Massachusetts/ | Helix/Illumina | Genomics and Discovery, Respiratory Viruses Branch, Division of Viral Diseases, Centers for Disease Control and Prevention | Peter W. Cook et al |
| EPI_ISL_802703 | USA/MA-CDC-STM-052/2020 | 2020-12-26 | North America/USA/Massachusetts/ | Helix/Illumina | Genomics and Discovery, Respiratory Viruses Branch, Division of Viral Diseases, Centers for Disease Control and Prevention | Peter W. Cook et al |
| EPI_ISL_802705 | USA/MA-CDC-STM-054/2020 | 2020-12-26 | North America/USA/Massachusetts/ | Helix/Illumina | Genomics and Discovery, Respiratory Viruses Branch, Division of Viral Diseases, Centers for Disease Control and Prevention | Peter W. Cook et al |

| | | | | | | |
|---|---|---|---|---|---|---|
| EPI_ISL_802717 | USA/PA-CDC-STM-137/2020 | 2020-12-29 | North America/USA/Pennsylvania/ | Helix/Illumina | Genomics and Discovery, Respiratory Viruses Branch, Division of Viral Diseases, Centers for Disease Control and Prevention | Peter W. Cook et al |
| EPI_ISL_802726 | Spain/CT-ISCIII-2017683/2020 | 2020-11-11 | Europe/Spain/Catalunya/ | Hospital Cl√≠nic de Barcelona | Instituto de Salud Carlos III | Iglesias-Caballero et al |
| EPI_ISL_802727 | Spain/CT-ISCIII-2017686/2020 | 2020-11-09 | Europe/Spain/Catalunya/ | Hospital Cl√≠nic de Barcelona | Instituto de Salud Carlos III | Iglesias-Caballero et al |
| EPI_ISL_802729 | Spain/CT-ISCIII-2017688/2020 | 2020-11-09 | Europe/Spain/Catalunya/ | Hospital Cl√≠nic de Barcelona | Instituto de Salud Carlos III | Iglesias-Caballero et al |
| EPI_ISL_802731 | Spain/CT-ISCIII-2017675/2020 | 2020-11-12 | Europe/Spain/Catalunya/ | Hospital Cl√≠nic de Barcelona | Instituto de Salud Carlos III | Iglesias-Caballero et al |
| EPI_ISL_802733 | Spain/CT-ISCIII-2017677/2020 | 2020-11-11 | Europe/Spain/Catalunya/ | Hospital Cl√≠nic de Barcelona | Instituto de Salud Carlos III | Iglesias-Caballero et al |
| EPI_ISL_802734 | Spain/CT-ISCIII-2017678/2020 | 2020-11-11 | Europe/Spain/Catalunya/ | Hospital Cl√≠nic de Barcelona | Instituto de Salud Carlos III | Iglesias-Caballero et al |
| EPI_ISL_802736 | Spain/CT-ISCIII-2017680/2020 | 2020-11-11 | Europe/Spain/Catalunya/ | Hospital Cl√≠nic de Barcelona | Instituto de Salud Carlos III | Iglesias-Caballero et al |
| EPI_ISL_802739 | Spain/CT-ISCIII-2016669/2020 | 2020-09-19 | Europe/Spain/Catalunya/ | Hospital Cl√≠nic de Barcelona | Instituto de Salud Carlos III | Iglesias-Caballero et al |
| EPI_ISL_802740 | Spain/CT-ISCIII-2016670/2020 | 2020-09-18 | Europe/Spain/Catalunya/ | Hospital Cl√≠nic de Barcelona | Instituto de Salud Carlos III | Iglesias-Caballero et al |
| EPI_ISL_802742 | Spain/CT-ISCIII-2016665/2020 | 2020-09-20 | Europe/Spain/Catalunya/ | Hospital Cl√≠nic de Barcelona | Instituto de Salud Carlos III | Iglesias-Caballero et al |
| EPI_ISL_802743 | Spain/CT-ISCIII-2016664/2020 | 2020-09-20 | Europe/Spain/Catalunya/ | Hospital Cl√≠nic de Barcelona | Instituto de Salud Carlos III | Iglesias-Caballero et al |
| EPI_ISL_802745 | Spain/CT-ISCIII-2016659/2020 | 2020-09-20 | Europe/Spain/Catalunya/ | Hospital Cl√≠nic de Barcelona | Instituto de Salud Carlos III | Iglesias-Caballero et al |
| EPI_ISL_802746 | Spain/CT-ISCIII-2016661/2020 | 2020-09-20 | Europe/Spain/Catalunya/ | Hospital Cl√≠nic de Barcelona | Instituto de Salud Carlos III | Iglesias-Caballero et al |
| EPI_ISL_802747 | Spain/CT-ISCIII-2016658/2020 | 2020-09-20 | Europe/Spain/Catalunya/ | Hospital Cl√≠nic de Barcelona | Instituto de Salud Carlos III | Iglesias-Caballero et al |
| EPI_ISL_802748 | Spain/CT-ISCIII-2016657/2020 | 2020-09-20 | Europe/Spain/Catalunya/ | Hospital Cl√≠nic de Barcelona | Instituto de Salud Carlos III | Iglesias-Caballero et al |

| | | | | | | |
|---|---|---|---|---|---|---|
| EPI_ISL_802751 | Spain/GA-ISCIII-2017349/2020 | 2020-10-20 | Europe/Spain/Galicia/ | Hospital Santa Marv≠a Nai | Instituto de Salud Carlos III | Iglesias-Caballero et al |
| EPI_ISL_802757 | Spain/GA-ISCIII-2017322/2020 | 2020-10-19 | Europe/Spain/Galicia/ | Hospital Santa Marv≠a Nai | Instituto de Salud Carlos III | Iglesias-Caballero et al |
| EPI_ISL_802760 | Spain/GA-ISCIII-2017309/2020 | 2020-10-20 | Europe/Spain/Galicia/ | Hospital Santa Marv≠a Nai | Instituto de Salud Carlos III | Iglesias-Caballero et al |
| EPI_ISL_802761 | Spain/GA-ISCIII-2017310/2020 | 2020-10-20 | Europe/Spain/Galicia/ | Hospital Santa Marv≠a Nai | Instituto de Salud Carlos III | Iglesias-Caballero et al |
| EPI_ISL_802762 | Spain/GA-ISCIII-2017311/2020 | 2020-10-20 | Europe/Spain/Galicia/ | Hospital Santa Marv≠a Nai | Instituto de Salud Carlos III | Iglesias-Caballero et al |
| EPI_ISL_802764 | Spain/GA-ISCIII-2017313/2020 | 2020-10-20 | Europe/Spain/Galicia/ | Hospital Santa Marv≠a Nai | Instituto de Salud Carlos III | Iglesias-Caballero et al |
| EPI_ISL_802767 | Spain/GA-ISCIII-2017295/2020 | 2020-10-20 | Europe/Spain/Galicia/ | Hospital Santa Marv≠a Nai | Instituto de Salud Carlos III | Iglesias-Caballero et al |
| EPI_ISL_802768 | Spain/GA-ISCIII-2017297/2020 | 2020-10-20 | Europe/Spain/Galicia/ | Hospital Santa Marv≠a Nai | Instituto de Salud Carlos III | Iglesias-Caballero et al |
| EPI_ISL_802778 | USA/NY-Wadsworth-21001904-01/20 | 2020-12-23 | North America/USA/New York/Bronx | Columbia University Irving Medical Center | Wadsworth Center, New York State Department.of Health | Kirsten St. George et al |
| EPI_ISL_802780 | USA/NY-Wadsworth-21001907-01/20 | 2020-12-23 | North America/USA/New York/New Yor | Columbia University Irving Medical Center | Wadsworth Center, New York State Department.of Health | Kirsten St. George et al |
| EPI_ISL_802782 | USA/NY-Wadsworth-21001918-01/20 | 2020-12-24 | North America/USA/New York/Putnam | Columbia University Irving Medical Center | Wadsworth Center, New York State Department.of Health | Kirsten St. George et al |
| EPI_ISL_802790 | USA/NY-Wadsworth-21002414-01/20 | 2020-12-31 | North America/USA/New York/Kings Co | BIO-REFERENCE LABORATORIES | Wadsworth Center, New York State Department.of Health | Kirsten St. George et al |

| | | | | | | |
|---|---|---|---|---|---|---|
| EPI_ISL_802793 | USA/NY-Wadsworth-21002432-01/20 | 2020-12-31 | North America/USA/New York/Richmon | BIO-REFERENCE LABORATORIES | Wadsworth Center, New York State Department.of Health | Kirsten St. George et al |
| EPI_ISL_802855 | Lithuania/T-1670/2020 | 2020-11-19 | Europe/Lithuania/Vilnius/ | Vilnius University Hospital Santaros Klinikos, Vilnius University | Institute of Biotechnology, Life Sciences Center, Vilnius University | Emilija Vasiliunaite et al |
| EPI_ISL_802856 | Lithuania/T-1987/2020 | 2020-11-27 | Europe/Lithuania/Vilnius/ | Vilnius University Hospital Santaros Klinikos, Vilnius University | Institute of Biotechnology, Life Sciences Center, Vilnius University | Emilija Vasiliunaite et al |
| EPI_ISL_802860 | Lithuania/T-32/2020 | 2020-10-01 | Europe/Lithuania/Vilnius/ | Vilnius University Hospital Santaros Klinikos, Vilnius University | Institute of Biotechnology, Life Sciences Center, Vilnius University | Emilija Vasiliunaite et al |
| EPI_ISL_802861 | Lithuania/T-88/2020 | 2020-10-13 | Europe/Lithuania/Vilnius/ | Vilnius University Hospital Santaros Klinikos, Vilnius University | Institute of Biotechnology, Life Sciences Center, Vilnius University | Emilija Vasiliunaite et al |
| EPI_ISL_802896 | USA/UT-UPHL-2101485712/2020 | 2020-12-10 | North America/USA/Utah/ | Utah Public Health Laboratory | Utah Public Health Laboratory | Erin Young et al |
| EPI_ISL_802947 | USA/UT-UPHL-2101062525/2020 | 2020-12-10 | North America/USA/Utah/ | Utah Public Health Laboratory | Utah Public Health Laboratory | Erin Young et al |
| EPI_ISL_802955 | USA/UT-UPHL-2101720444/2020 | 2020-12-10 | North America/USA/Utah/ | Utah Public Health Laboratory | Utah Public Health Laboratory | Erin Young et al |
| EPI_ISL_802968 | USA/UT-UPHL-2101022186/2020 | 2020-12-15 | North America/USA/Utah/ | Utah Public Health Laboratory | Utah Public Health Laboratory | Erin Young et al |
| EPI_ISL_802994 | Germany/BY-ChVir21648/2020 | 2020-12-22 | Europe/Germany/Bavaria/ | Charit√© Universit√§tsmedizin Berlin, Institut f√ºr Virologie, Charit√©platz 1, 10117 Berlin, Germany | Charit√© Universit√§tsmedizin Berlin, Institut f√ºr Virologie, Charit√©platz 1, 10117 Berlin, Germany | Victor M Corman et al |

| | | | | | | |
|---|---|---|---|---|---|---|
| EPI_ISL_802999 | USA/CA-QDX-2763/2020 | 2020-12-24 | North America/USA/California/ | Quest Diagnostics | Quest Diagnostics | Rosenthal et al |
| EPI_ISL_803009 | USA/CA-QDX-2773/2020 | 2020-12-26 | North America/USA/California/ | Quest Diagnostics | Quest Diagnostics | Rosenthal et al |
| EPI_ISL_803011 | USA/CA-QDX-2775/2020 | 2020-12-26 | North America/USA/California/ | Quest Diagnostics | Quest Diagnostics | Rosenthal et al |
| EPI_ISL_803029 | USA/CA-QDX-2798/2020 | 2020-12-26 | North America/USA/California/ | Quest Diagnostics | Quest Diagnostics | Rosenthal et al |
| EPI_ISL_803032 | USA/CA-QDX-2801/2020 | 2020-12-26 | North America/USA/California/ | Quest Diagnostics | Quest Diagnostics | Rosenthal et al |
| EPI_ISL_803034 | USA/CA-QDX-2803/2020 | 2020-12-26 | North America/USA/California/ | Quest Diagnostics | Quest Diagnostics | Rosenthal et al |
| EPI_ISL_803036 | USA/CA-QDX-2805/2020 | 2020-12-26 | North America/USA/California/ | Quest Diagnostics | Quest Diagnostics | Rosenthal et al |
| EPI_ISL_803039 | USA/CA-QDX-2808/2020 | 2020-12-26 | North America/USA/California/ | Quest Diagnostics | Quest Diagnostics | Rosenthal et al |
| EPI_ISL_803052 | USA/CA-QDX-2822/2020 | 2020-12-26 | North America/USA/California/ | Quest Diagnostics | Quest Diagnostics | Rosenthal et al |
| EPI_ISL_803053 | USA/CA-QDX-2823/2020 | 2020-12-26 | North America/USA/California/ | Quest Diagnostics | Quest Diagnostics | Rosenthal et al |
| EPI_ISL_803069 | USA/CA-QDX-2841/2020 | 2020-12-26 | North America/USA/California/ | Quest Diagnostics | Quest Diagnostics | Rosenthal et al |
| EPI_ISL_803076 | USA/CA-QDX-2856/2020 | 2020-12-26 | North America/USA/California/ | Quest Diagnostics | Quest Diagnostics | Rosenthal et al |
| EPI_ISL_803078 | USA/CA-QDX-2858/2020 | 2020-12-26 | North America/USA/California/ | Quest Diagnostics | Quest Diagnostics | Rosenthal et al |
| EPI_ISL_803085 | USA/CA-QDX-2836/2020 | 2020-12-26 | North America/USA/California/ | Quest Diagnostics | Quest Diagnostics | Rosenthal et al |
| EPI_ISL_803086 | USA/CA-QDX-2838/2020 | 2020-12-26 | North America/USA/California/ | Quest Diagnostics | Quest Diagnostics | Rosenthal et al |
| EPI_ISL_803089 | USA/CA-QDX-2845/2020 | 2020-12-26 | North America/USA/California/ | Quest Diagnostics | Quest Diagnostics | Rosenthal et al |
| EPI_ISL_803092 | USA/CA-QDX-2848/2020 | 2020-12-26 | North America/USA/California/ | Quest Diagnostics | Quest Diagnostics | Rosenthal et al |
| EPI_ISL_803101 | USA/OK-CDC-2-3693490/2020 | 2020-11-18 | North America/USA/Oklahoma/ | Respiratory Viruses Branch, Centers for Disease Control and Prevention | Respiratory Viruses Branch, Centers for Disease Control and Prevention | Queen et al |
| EPI_ISL_803103 | USA/NV-CDC-2-3693472/2020 | 2020-11-09 | North America/USA/Nevada/ | Respiratory Viruses Branch, Centers for Disease Control and Prevention | Respiratory Viruses Branch, Centers for Disease Control and Prevention | Queen et al |
| EPI_ISL_803118 | Bangladesh/BCSIR-NILMRC_185/2020 | 2020-06-06 | Asia/Bangladesh/Chattogram/ | National Institute of Laboratory Medicine and Referral Center | Genomic Research Lab, BCSIR | Barna Goswami et al |

| | | | | | | |
|---|---|---|---|---|---|---|
| EPI_ISL_803119 | Tunisia/2285/2021 | 2021-01-08 | Africa/Tunisia/Nabeul/ | Laboratory of Microbiology, National Reference Lab, Charles Nicolle Hospital; 2-University of Tunis ElManar, Faculty of Medicine of Tunis, LR99ES09, Tunis, Tunisia | Clinical and Experimental Pharmacology Lab, LR16SP02, National Center of Pharmacovigilance, University of Tunis El Manar, Tunis, Tunisia. 2-Neurodegenerative diseases and psychiatric troubles, LR18SP03, Razi Hospital, University of Tunis El Manar, Tunis, Tunisia. 3-Ministry of Health, National Observatory of New and Emerging Diseases, 1006, Tunis, Tunisia | Ilhem Boutiba-Ben Boubaker et al |

| | | | | | | |
|---|---|---|---|---|---|---|
| EPI_ISL_803120 | Tunisia/1821/2021 | 2021-01-07 | Africa/Tunisia/Ben Arous/ | Laboratory of Microbiology, National Reference Lab, Charles Nicolle Hospital; 2-University of Tunis ElManar, Faculty of Medicine of Tunis, LR99ES09, Tunis, Tunisia | Clinical and Experimental Pharmacology Lab, LR16SP02, National Center of Pharmacovigilance, University of Tunis El Manar, Tunis, Tunisia. 2-Neurodegenerative diseases and psychiatric troubles, LR18SP03, Razi Hospital, University of Tunis El Manar, Tunis, Tunisia. 3-Ministry of Health, National Observatory of New and Emerging Diseases, 1006, Tunis, Tunisia | Ilhem Boutiba-Ben Boubaker et al |
| EPI_ISL_803121 | Bangladesh/BCSIR-NILMRC_171/2020 | 2020-06-18 | Asia/Bangladesh/Rajshahi/ | National Institute of Laboratory Medicine and Referral Center | Genomic Research Lab, BCSIR | Md. Saddam Hossain et al |
| EPI_ISL_803126 | USA/WI-WSLH-200619/2020 | 2020-08-24 | North America/USA/Wisconsin/Juneau ( | Wisconsin State Laboratory of Hygiene Communicable Disease Division | Wisconsin State Laboratory of Hygiene Communicable Disease Division | Kelsey R. Florek et al |
| EPI_ISL_803130 | USA/WI-WSLH-200623/2020 | 2020-08-24 | North America/USA/Wisconsin/Milwauk | Wisconsin State Laboratory of Hygiene Communicable Disease Division | Wisconsin State Laboratory of Hygiene Communicable Disease Division | Kelsey R. Florek et al |

| | | | | | | |
|---|---|---|---|---|---|---|
| EPI_ISL_803131 | USA/WI-WSLH-200624/2020 | 2020-08-25 | North America/USA/Wisconsin/Milwauk | Wisconsin State Laboratory of Hygiene Communicable Disease Division | Wisconsin State Laboratory of Hygiene Communicable Disease Division | Kelsey R. Florek et al |
| EPI_ISL_803132 | USA/WI-WSLH-200625/2020 | 2020-08-26 | North America/USA/Wisconsin/Brown C | Wisconsin State Laboratory of Hygiene Communicable Disease Division | Wisconsin State Laboratory of Hygiene Communicable Disease Division | Kelsey R. Florek et al |
| EPI_ISL_803133 | USA/WI-WSLH-200626/2020 | 2020-08-27 | North America/USA/Wisconsin/Dodge C | Wisconsin State Laboratory of Hygiene Communicable Disease Division | Wisconsin State Laboratory of Hygiene Communicable Disease Division | Kelsey R. Florek et al |
| EPI_ISL_803134 | USA/WI-WSLH-200628/2020 | 2020-09-01 | North America/USA/Wisconsin/Dodge C | Wisconsin State Laboratory of Hygiene Communicable Disease Division | Wisconsin State Laboratory of Hygiene Communicable Disease Division | Kelsey R. Florek et al |
| EPI_ISL_803135 | USA/WI-WSLH-200629/2020 | 2020-09-01 | North America/USA/Wisconsin/Racine C | Wisconsin State Laboratory of Hygiene Communicable Disease Division | Wisconsin State Laboratory of Hygiene Communicable Disease Division | Kelsey R. Florek et al |
| EPI_ISL_803140 | USA/WI-WSLH-200803/2020 | 2020-08-19 | North America/USA/Wisconsin/Sheboyg | Wisconsin State Laboratory of Hygiene Communicable Disease Division | Wisconsin State Laboratory of Hygiene Communicable Disease Division | Kelsey R. Florek et al |
| EPI_ISL_803148 | USA/WI-WSLH-200812/2020 | 2020-09-14 | North America/USA/Wisconsin/Sheboyg | Wisconsin State Laboratory of Hygiene Communicable Disease Division | Wisconsin State Laboratory of Hygiene Communicable Disease Division | Kelsey R. Florek et al |
| EPI_ISL_803149 | USA/WI-WSLH-200813/2020 | 2020-09-14 | North America/USA/Wisconsin/Sheboyg | Wisconsin State Laboratory of Hygiene Communicable Disease Division | Wisconsin State Laboratory of Hygiene Communicable Disease Division | Kelsey R. Florek et al |

| | | | | | | |
|---|---|---|---|---|---|---|
| EPI_ISL_803159 | USA/WI-WSLH-200824/2020 | 2020-09-14 | North America/USA/Wisconsin/Sheboyg | Wisconsin State Laboratory of Hygiene Communicable Disease Division | Wisconsin State Laboratory of Hygiene Communicable Disease Division | Kelsey R. Florek et al |
| EPI_ISL_803166 | USA/WI-WSLH-200832/2020 | 2020-09-14 | North America/USA/Wisconsin/Sheboyg | Wisconsin State Laboratory of Hygiene Communicable Disease Division | Wisconsin State Laboratory of Hygiene Communicable Disease Division | Kelsey R. Florek et al |
| EPI_ISL_803167 | USA/WI-WSLH-200833/2020 | 2020-09-14 | North America/USA/Wisconsin/Sheboyg | Wisconsin State Laboratory of Hygiene Communicable Disease Division | Wisconsin State Laboratory of Hygiene Communicable Disease Division | Kelsey R. Florek et al |
| EPI_ISL_803169 | USA/WI-WSLH-200835/2020 | 2020-09-14 | North America/USA/Wisconsin/Sheboyg | Wisconsin State Laboratory of Hygiene Communicable Disease Division | Wisconsin State Laboratory of Hygiene Communicable Disease Division | Kelsey R. Florek et al |
| EPI_ISL_803177 | USA/WI-WSLH-200845/2020 | 2020-09-14 | North America/USA/Wisconsin/Sheboyg | Wisconsin State Laboratory of Hygiene Communicable Disease Division | Wisconsin State Laboratory of Hygiene Communicable Disease Division | Kelsey R. Florek et al |
| EPI_ISL_803179 | USA/WI-WSLH-200847/2020 | 2020-09-14 | North America/USA/Wisconsin/Sheboyg | Wisconsin State Laboratory of Hygiene Communicable Disease Division | Wisconsin State Laboratory of Hygiene Communicable Disease Division | Kelsey R. Florek et al |
| EPI_ISL_803180 | USA/WI-WSLH-200848/2020 | 2020-09-14 | North America/USA/Wisconsin/Sheboyg | Wisconsin State Laboratory of Hygiene Communicable Disease Division | Wisconsin State Laboratory of Hygiene Communicable Disease Division | Kelsey R. Florek et al |
| EPI_ISL_803184 | USA/WI-WSLH-200853/2020 | 2020-09-14 | North America/USA/Wisconsin/Sheboyg | Wisconsin State Laboratory of Hygiene Communicable Disease Division | Wisconsin State Laboratory of Hygiene Communicable Disease Division | Kelsey R. Florek et al |

| | | | | | | |
|---|---|---|---|---|---|---|
| EPI_ISL_803189 | USA/WI-WSLH-200858/2020 | 2020-09-14 | North America/USA/Wisconsin/Sheboyg | Wisconsin State Laboratory of Hygiene Communicable Disease Division | Wisconsin State Laboratory of Hygiene Communicable Disease Division | Kelsey R. Florek et al |
| EPI_ISL_803195 | USA/WI-WSLH-200865/2020 | 2020-09-17 | North America/USA/Wisconsin/Sheboyg | Wisconsin State Laboratory of Hygiene Communicable Disease Division | Wisconsin State Laboratory of Hygiene Communicable Disease Division | Kelsey R. Florek et al |
| EPI_ISL_803197 | USA/WI-WSLH-200868/2020 | 2020-10-06 | North America/USA/Wisconsin/Sheboyg | Wisconsin State Laboratory of Hygiene Communicable Disease Division | Wisconsin State Laboratory of Hygiene Communicable Disease Division | Kelsey R. Florek et al |
| EPI_ISL_803200 | USA/WI-WSLH-200871/2020 | 2020-10-06 | North America/USA/Wisconsin/Sheboyg | Wisconsin State Laboratory of Hygiene Communicable Disease Division | Wisconsin State Laboratory of Hygiene Communicable Disease Division | Kelsey R. Florek et al |
| EPI_ISL_803205 | USA/WI-WSLH-200877/2020 | 2020-10-06 | North America/USA/Wisconsin/Sheboyg | Wisconsin State Laboratory of Hygiene Communicable Disease Division | Wisconsin State Laboratory of Hygiene Communicable Disease Division | Kelsey R. Florek et al |
| EPI_ISL_803210 | USA/WI-WSLH-200882/2020 | 2020-10-06 | North America/USA/Wisconsin/Sheboyg | Wisconsin State Laboratory of Hygiene Communicable Disease Division | Wisconsin State Laboratory of Hygiene Communicable Disease Division | Kelsey R. Florek et al |
| EPI_ISL_803220 | USA/WI-WSLH-200917/2020 | 2020-08-27 | North America/USA/Wisconsin/Dodge C | Wisconsin State Laboratory of Hygiene Communicable Disease Division | Wisconsin State Laboratory of Hygiene Communicable Disease Division | Kelsey R. Florek et al |
| EPI_ISL_803224 | USA/WI-WSLH-200922/2020 | 2020-08-27 | North America/USA/Wisconsin/Dodge C | Wisconsin State Laboratory of Hygiene Communicable Disease Division | Wisconsin State Laboratory of Hygiene Communicable Disease Division | Kelsey R. Florek et al |

| | | | | | | |
|---|---|---|---|---|---|---|
| EPI_ISL_803230 | USA/WI-WSLH-200933/2020 | 2020-09-10 | North America/USA/Wisconsin/Dodge C | Wisconsin State Laboratory of Hygiene Communicable Disease Division | Wisconsin State Laboratory of Hygiene Communicable Disease Division | Kelsey R. Florek et al |
| EPI_ISL_803252 | USA/WI-WSLH-200963/2020 | 2020-08-25 | North America/USA/Wisconsin/Dodge C | Wisconsin State Laboratory of Hygiene Communicable Disease Division | Wisconsin State Laboratory of Hygiene Communicable Disease Division | Kelsey R. Florek et al |
| EPI_ISL_803253 | USA/WI-WSLH-200964/2020 | 2020-08-26 | North America/USA/Wisconsin/Dodge C | Wisconsin State Laboratory of Hygiene Communicable Disease Division | Wisconsin State Laboratory of Hygiene Communicable Disease Division | Kelsey R. Florek et al |
| EPI_ISL_803254 | USA/WI-WSLH-200965/2020 | 2020-10-07 | North America/USA/Wisconsin/Sheboyg | Wisconsin State Laboratory of Hygiene Communicable Disease Division | Wisconsin State Laboratory of Hygiene Communicable Disease Division | Kelsey R. Florek et al |
| EPI_ISL_803256 | USA/WI-WSLH-200967/2020 | 2020-10-06 | North America/USA/Wisconsin/Sheboyg | Wisconsin State Laboratory of Hygiene Communicable Disease Division | Wisconsin State Laboratory of Hygiene Communicable Disease Division | Kelsey R. Florek et al |
| EPI_ISL_803259 | USA/WI-WSLH-200970/2020 | 2020-10-07 | North America/USA/Wisconsin/Sheboyg | Wisconsin State Laboratory of Hygiene Communicable Disease Division | Wisconsin State Laboratory of Hygiene Communicable Disease Division | Kelsey R. Florek et al |
| EPI_ISL_803262 | USA/WI-WSLH-200973/2020 | 2020-10-05 | North America/USA/Wisconsin/Taylor C | Wisconsin State Laboratory of Hygiene Communicable Disease Division | Wisconsin State Laboratory of Hygiene Communicable Disease Division | Kelsey R. Florek et al |
| EPI_ISL_803263 | USA/WI-WSLH-200974/2020 | 2020-10-06 | North America/USA/Wisconsin/Sheboyg | Wisconsin State Laboratory of Hygiene Communicable Disease Division | Wisconsin State Laboratory of Hygiene Communicable Disease Division | Kelsey R. Florek et al |

| | | | | | | |
|---|---|---|---|---|---|---|
| EPI_ISL_803267 | USA/WI-WSLH-200978/2020 | 2020-10-05 | North America/USA/Wisconsin/Taylor C | Wisconsin State Laboratory of Hygiene Communicable Disease Division | Wisconsin State Laboratory of Hygiene Communicable Disease Division | Kelsey R. Florek et al |
| EPI_ISL_803273 | USA/WI-WSLH-200985/2020 | 2020-10-06 | North America/USA/Wisconsin/Sheboyg | Wisconsin State Laboratory of Hygiene Communicable Disease Division | Wisconsin State Laboratory of Hygiene Communicable Disease Division | Kelsey R. Florek et al |
| EPI_ISL_803287 | USA/WI-WSLH-200999/2020 | 2020-10-06 | North America/USA/Wisconsin/Sheboyg | Wisconsin State Laboratory of Hygiene Communicable Disease Division | Wisconsin State Laboratory of Hygiene Communicable Disease Division | Kelsey R. Florek et al |
| EPI_ISL_803290 | USA/WI-WSLH-201002/2020 | 2020-10-07 | North America/USA/Wisconsin/Sheboyg | Wisconsin State Laboratory of Hygiene Communicable Disease Division | Wisconsin State Laboratory of Hygiene Communicable Disease Division | Kelsey R. Florek et al |
| EPI_ISL_803308 | USA/WI-WSLH-201046/2020 | 2020-09-25 | North America/USA/Wisconsin/Lafayett | Wisconsin State Laboratory of Hygiene Communicable Disease Division | Wisconsin State Laboratory of Hygiene Communicable Disease Division | Kelsey R. Florek et al |
| EPI_ISL_803313 | USA/WI-WSLH-201051/2020 | 2020-09-22 | North America/USA/Wisconsin/Marquet | Wisconsin State Laboratory of Hygiene Communicable Disease Division | Wisconsin State Laboratory of Hygiene Communicable Disease Division | Kelsey R. Florek et al |
| EPI_ISL_803314 | USA/WI-WSLH-201053/2020 | 2020-09-23 | North America/USA/Wisconsin/Barron C | Wisconsin State Laboratory of Hygiene Communicable Disease Division | Wisconsin State Laboratory of Hygiene Communicable Disease Division | Kelsey R. Florek et al |
| EPI_ISL_803316 | USA/WI-WSLH-201055/2020 | 2020-09-24 | North America/USA/Wisconsin/Rock Cou | Wisconsin State Laboratory of Hygiene Communicable Disease Division | Wisconsin State Laboratory of Hygiene Communicable Disease Division | Kelsey R. Florek et al |

| | | | | | | |
|---|---|---|---|---|---|---|
| EPI_ISL_803318 | USA/WI-WSLH-201057/2020 | 2020-09-24 | North America/USA/Wisconsin/Winneba | Wisconsin State Laboratory of Hygiene Communicable Disease Division | Wisconsin State Laboratory of Hygiene Communicable Disease Division | Kelsey R. Florek et al |
| EPI_ISL_803321 | USA/WI-WSLH-201061/2020 | 2020-09-25 | North America/USA/Wisconsin/Vernon | Wisconsin State Laboratory of Hygiene Communicable Disease Division | Wisconsin State Laboratory of Hygiene Communicable Disease Division | Kelsey R. Florek et al |
| EPI_ISL_803322 | USA/WI-WSLH-201062/2020 | 2020-09-24 | North America/USA/Wisconsin/Marquet | Wisconsin State Laboratory of Hygiene Communicable Disease Division | Wisconsin State Laboratory of Hygiene Communicable Disease Division | Kelsey R. Florek et al |
| EPI_ISL_803328 | USA/WI-WSLH-201071/2020 | 2020-09-29 | North America/USA/Wisconsin/Lafayett | Wisconsin State Laboratory of Hygiene Communicable Disease Division | Wisconsin State Laboratory of Hygiene Communicable Disease Division | Kelsey R. Florek et al |
| EPI_ISL_803330 | USA/WI-WSLH-201076/2020 | 2020-09-30 | North America/USA/Wisconsin/Marquet | Wisconsin State Laboratory of Hygiene Communicable Disease Division | Wisconsin State Laboratory of Hygiene Communicable Disease Division | Kelsey R. Florek et al |
| EPI_ISL_803333 | USA/WI-WSLH-201089/2020 | 2020-09-01 | North America/USA/Wisconsin/Jefferson | Wisconsin State Laboratory of Hygiene Communicable Disease Division | Wisconsin State Laboratory of Hygiene Communicable Disease Division | Kelsey R. Florek et al |
| EPI_ISL_803338 | USA/WI-WSLH-201095/2020 | 2020-09-04 | North America/USA/Wisconsin/Rock Cou | Wisconsin State Laboratory of Hygiene Communicable Disease Division | Wisconsin State Laboratory of Hygiene Communicable Disease Division | Kelsey R. Florek et al |
| EPI_ISL_803340 | USA/WI-WSLH-201097/2020 | 2020-09-07 | North America/USA/Wisconsin/Jefferson | Wisconsin State Laboratory of Hygiene Communicable Disease Division | Wisconsin State Laboratory of Hygiene Communicable Disease Division | Kelsey R. Florek et al |

| | | | | | | |
|---|---|---|---|---|---|---|
| EPI_ISL_803342 | USA/WI-WSLH-201100/2020 | 2020-09-14 | North America/USA/Wisconsin/Jefferson | Wisconsin State Laboratory of Hygiene Communicable Disease Division | Wisconsin State Laboratory of Hygiene Communicable Disease Division | Kelsey R. Florek et al |
| EPI_ISL_803345 | USA/WI-WSLH-201103/2020 | 2020-09-14 | North America/USA/Wisconsin/Marquet | Wisconsin State Laboratory of Hygiene Communicable Disease Division | Wisconsin State Laboratory of Hygiene Communicable Disease Division | Kelsey R. Florek et al |
| EPI_ISL_803348 | USA/WI-WSLH-201107/2020 | 2020-09-17 | North America/USA/Wisconsin/Jefferson | Wisconsin State Laboratory of Hygiene Communicable Disease Division | Wisconsin State Laboratory of Hygiene Communicable Disease Division | Kelsey R. Florek et al |
| EPI_ISL_803366 | USA/WI-WSLH-201125/2020 | 2020-10-12 | North America/USA/Wisconsin/Marquet | Wisconsin State Laboratory of Hygiene Communicable Disease Division | Wisconsin State Laboratory of Hygiene Communicable Disease Division | Kelsey R. Florek et al |
| EPI_ISL_803372 | USA/WI-WSLH-201131/2020 | 2020-10-14 | North America/USA/Wisconsin/Marinet | Wisconsin State Laboratory of Hygiene Communicable Disease Division | Wisconsin State Laboratory of Hygiene Communicable Disease Division | Kelsey R. Florek et al |
| EPI_ISL_803391 | USA/WI-WSLH-201151/2020 | 2020-10-12 | North America/USA/Wisconsin/Barron C | Wisconsin State Laboratory of Hygiene Communicable Disease Division | Wisconsin State Laboratory of Hygiene Communicable Disease Division | Kelsey R. Florek et al |
| EPI_ISL_803396 | USA/WI-WSLH-201156/2020 | 2020-10-15 | North America/USA/Wisconsin/Marquet | Wisconsin State Laboratory of Hygiene Communicable Disease Division | Wisconsin State Laboratory of Hygiene Communicable Disease Division | Kelsey R. Florek et al |
| EPI_ISL_803407 | USA/WI-WSLH-201168/2020 | 2020-10-21 | North America/USA/Wisconsin/Jefferson | Wisconsin State Laboratory of Hygiene Communicable Disease Division | Wisconsin State Laboratory of Hygiene Communicable Disease Division | Kelsey R. Florek et al |

| | | | | | | |
|---|---|---|---|---|---|---|
| EPI_ISL_803412 | USA/WI-WSLH-201173/2020 | 2020-10-21 | North America/USA/Wisconsin/Vilas Co | Wisconsin State Laboratory of Hygiene Communicable Disease Division | Wisconsin State Laboratory of Hygiene Communicable Disease Division | Kelsey R. Florek et al |
| EPI_ISL_803413 | USA/WI-WSLH-201174/2020 | 2020-10-23 | North America/USA/Wisconsin/Grant Co | Wisconsin State Laboratory of Hygiene Communicable Disease Division | Wisconsin State Laboratory of Hygiene Communicable Disease Division | Kelsey R. Florek et al |
| EPI_ISL_803418 | USA/WI-WSLH-201179/2020 | 2020-10-26 | North America/USA/Wisconsin/Vilas Co | Wisconsin State Laboratory of Hygiene Communicable Disease Division | Wisconsin State Laboratory of Hygiene Communicable Disease Division | Kelsey R. Florek et al |
| EPI_ISL_803430 | Tunisia/1890/2021 | 2021-01-07 | Africa/Tunisia/Nabeul/ | Laboratory of Microbiology, National Reference Lab, Charles Nicolle Hospital; 2- University of Tunis ElManar, Faculty of Medicine of Tunis, LR99ES09, Tunis, Tunisia | Clinical and Experimental Pharmacology Lab, LR16SP02, National Center of Pharmacovigilance, University of Tunis El Manar, Tunis, Tunisia. 2- Neurodegenerative diseases and psychiatric troubles, LR18SP03, Razi Hospital, University of Tunis El Manar, Tunis, Tunisia. 3- Ministry of Health, National Observatory of New and Emerging Diseases, 1006, Tunis, Tunisia | Ilhem Boutiba-Ben Boubaker et al |

| | | | | | | |
|---|---|---|---|---|---|---|
| EPI_ISL_803440 | USA/WI-WSLH-201200/2020 | 2020-10-19 | North America/USA/Wisconsin/Pepin Co | Wisconsin State Laboratory of Hygiene Communicable Disease Division | Wisconsin State Laboratory of Hygiene Communicable Disease Division | Kelsey R. Florek et al |
| EPI_ISL_803449 | USA/WI-WSLH-201209/2020 | 2020-10-21 | North America/USA/Wisconsin/Maratho | Wisconsin State Laboratory of Hygiene Communicable Disease Division | Wisconsin State Laboratory of Hygiene Communicable Disease Division | Kelsey R. Florek et al |
| EPI_ISL_803481 | USA/WI-WSLH-201241/2020 | 2020-11-02 | North America/USA/Wisconsin/Marquet | Wisconsin State Laboratory of Hygiene Communicable Disease Division | Wisconsin State Laboratory of Hygiene Communicable Disease Division | Kelsey R. Florek et al |
| EPI_ISL_803482 | USA/WI-WSLH-201242/2020 | 2020-11-03 | North America/USA/Wisconsin/Pepin Co | Wisconsin State Laboratory of Hygiene Communicable Disease Division | Wisconsin State Laboratory of Hygiene Communicable Disease Division | Kelsey R. Florek et al |
| EPI_ISL_803486 | USA/WI-WSLH-201246/2020 | 2020-09-03 | North America/USA/Wisconsin/Rock Cou | Wisconsin State Laboratory of Hygiene Communicable Disease Division | Wisconsin State Laboratory of Hygiene Communicable Disease Division | Kelsey R. Florek et al |
| EPI_ISL_803489 | USA/WI-WSLH-201249/2020 | 2020-09-04 | North America/USA/Wisconsin/Burnett | Wisconsin State Laboratory of Hygiene Communicable Disease Division | Wisconsin State Laboratory of Hygiene Communicable Disease Division | Kelsey R. Florek et al |
| EPI_ISL_803494 | USA/WI-WSLH-201254/2020 | 2020-09-10 | North America/USA/Wisconsin/Jefferson | Wisconsin State Laboratory of Hygiene Communicable Disease Division | Wisconsin State Laboratory of Hygiene Communicable Disease Division | Kelsey R. Florek et al |
| EPI_ISL_803497 | USA/WI-WSLH-201258/2020 | 2020-09-15 | North America/USA/Wisconsin/Burnett | Wisconsin State Laboratory of Hygiene Communicable Disease Division | Wisconsin State Laboratory of Hygiene Communicable Disease Division | Kelsey R. Florek et al |

| | | | | | | |
|---|---|---|---|---|---|---|
| EPI_ISL_803498 | USA/WI-WSLH-201259/2020 | 2020-09-14 | North America/USA/Wisconsin/Jefferson | Wisconsin State Laboratory of Hygiene Communicable Disease Division | Wisconsin State Laboratory of Hygiene Communicable Disease Division | Kelsey R. Florek et al |
| EPI_ISL_803500 | USA/WI-WSLH-201261/2020 | 2020-09-15 | North America/USA/Wisconsin/Marquet | Wisconsin State Laboratory of Hygiene Communicable Disease Division | Wisconsin State Laboratory of Hygiene Communicable Disease Division | Kelsey R. Florek et al |
| EPI_ISL_803502 | USA/WI-WSLH-201263/2020 | 2020-09-16 | North America/USA/Wisconsin/Sauk Co | Wisconsin State Laboratory of Hygiene Communicable Disease Division | Wisconsin State Laboratory of Hygiene Communicable Disease Division | Kelsey R. Florek et al |
| EPI_ISL_803503 | USA/WI-WSLH-201264/2020 | 2020-09-16 | North America/USA/Wisconsin/Pepin Co | Wisconsin State Laboratory of Hygiene Communicable Disease Division | Wisconsin State Laboratory of Hygiene Communicable Disease Division | Kelsey R. Florek et al |
| EPI_ISL_803506 | USA/WI-WSLH-201267/2020 | 2020-09-17 | North America/USA/Wisconsin/Jefferson | Wisconsin State Laboratory of Hygiene Communicable Disease Division | Wisconsin State Laboratory of Hygiene Communicable Disease Division | Kelsey R. Florek et al |
| EPI_ISL_803508 | USA/WI-WSLH-201281/2020 | 2020-09-09 | North America/USA/Wisconsin/Marquet | Wisconsin State Laboratory of Hygiene Communicable Disease Division | Wisconsin State Laboratory of Hygiene Communicable Disease Division | Kelsey R. Florek et al |
| EPI_ISL_803513 | USA/WI-WSLH-201286/2020 | 2020-09-21 | North America/USA/Wisconsin/Lafayett | Wisconsin State Laboratory of Hygiene Communicable Disease Division | Wisconsin State Laboratory of Hygiene Communicable Disease Division | Kelsey R. Florek et al |
| EPI_ISL_803514 | USA/WI-WSLH-201287/2020 | 2020-09-21 | North America/USA/Wisconsin/Marquet | Wisconsin State Laboratory of Hygiene Communicable Disease Division | Wisconsin State Laboratory of Hygiene Communicable Disease Division | Kelsey R. Florek et al |

| | | | | | | |
|---|---|---|---|---|---|---|
| EPI_ISL_803517 | USA/WI-WSLH-201290/2020 | 2020-09-23 | North America/USA/Wisconsin/Marquet | Wisconsin State Laboratory of Hygiene Communicable Disease Division | Wisconsin State Laboratory of Hygiene Communicable Disease Division | Kelsey R. Florek et al |
| EPI_ISL_803521 | USA/WI-WSLH-201294/2020 | 2020-09-09 | North America/USA/Wisconsin/Marquet | Wisconsin State Laboratory of Hygiene Communicable Disease Division | Wisconsin State Laboratory of Hygiene Communicable Disease Division | Kelsey R. Florek et al |
| EPI_ISL_803525 | USA/WI-WSLH-201298/2020 | 2020-09-14 | North America/USA/Wisconsin/Jefferson | Wisconsin State Laboratory of Hygiene Communicable Disease Division | Wisconsin State Laboratory of Hygiene Communicable Disease Division | Kelsey R. Florek et al |
| EPI_ISL_803526 | USA/WI-WSLH-201299/2020 | 2020-09-14 | North America/USA/Wisconsin/Marquet | Wisconsin State Laboratory of Hygiene Communicable Disease Division | Wisconsin State Laboratory of Hygiene Communicable Disease Division | Kelsey R. Florek et al |
| EPI_ISL_803527 | USA/WI-WSLH-201300/2020 | 2020-09-15 | North America/USA/Wisconsin/Grant Co | Wisconsin State Laboratory of Hygiene Communicable Disease Division | Wisconsin State Laboratory of Hygiene Communicable Disease Division | Kelsey R. Florek et al |
| EPI_ISL_803529 | USA/WI-WSLH-201302/2020 | 2020-09-17 | North America/USA/Wisconsin/Pepin Co | Wisconsin State Laboratory of Hygiene Communicable Disease Division | Wisconsin State Laboratory of Hygiene Communicable Disease Division | Kelsey R. Florek et al |
| EPI_ISL_803530 | USA/WI-WSLH-201303/2020 | 2020-09-18 | North America/USA/Wisconsin/Grant Co | Wisconsin State Laboratory of Hygiene Communicable Disease Division | Wisconsin State Laboratory of Hygiene Communicable Disease Division | Kelsey R. Florek et al |
| EPI_ISL_803532 | USA/WI-WSLH-201305/2020 | 2020-09-19 | North America/USA/Wisconsin/Lafayett | Wisconsin State Laboratory of Hygiene Communicable Disease Division | Wisconsin State Laboratory of Hygiene Communicable Disease Division | Kelsey R. Florek et al |

| | | | | | | |
|---|---|---|---|---|---|---|
| EPI_ISL_803533 | USA/WI-WSLH-201306/2020 | 2020-09-21 | North America/USA/Wisconsin/Lafayett | Wisconsin State Laboratory of Hygiene Communicable Disease Division | Wisconsin State Laboratory of Hygiene Communicable Disease Division | Kelsey R. Florek et al |
| EPI_ISL_803534 | USA/WI-WSLH-201307/2020 | 2020-09-22 | North America/USA/Wisconsin/Marquet | Wisconsin State Laboratory of Hygiene Communicable Disease Division | Wisconsin State Laboratory of Hygiene Communicable Disease Division | Kelsey R. Florek et al |
| EPI_ISL_803536 | USA/WI-WSLH-201309/2020 | 2020-09-09 | North America/USA/Wisconsin/Marquet | Wisconsin State Laboratory of Hygiene Communicable Disease Division | Wisconsin State Laboratory of Hygiene Communicable Disease Division | Kelsey R. Florek et al |
| EPI_ISL_803538 | USA/WI-WSLH-201311/2020 | 2020-09-10 | North America/USA/Wisconsin/Jefferson | Wisconsin State Laboratory of Hygiene Communicable Disease Division | Wisconsin State Laboratory of Hygiene Communicable Disease Division | Kelsey R. Florek et al |
| EPI_ISL_803540 | USA/WI-WSLH-201314/2020 | 2020-09-16 | North America/USA/Wisconsin/Jefferson | Wisconsin State Laboratory of Hygiene Communicable Disease Division | Wisconsin State Laboratory of Hygiene Communicable Disease Division | Kelsey R. Florek et al |
| EPI_ISL_803541 | USA/WI-WSLH-201315/2020 | 2020-09-18 | North America/USA/Wisconsin/Ashland | Wisconsin State Laboratory of Hygiene Communicable Disease Division | Wisconsin State Laboratory of Hygiene Communicable Disease Division | Kelsey R. Florek et al |
| EPI_ISL_803546 | USA/WI-WSLH-201320/2020 | 2020-09-22 | North America/USA/Wisconsin/Jefferson | Wisconsin State Laboratory of Hygiene Communicable Disease Division | Wisconsin State Laboratory of Hygiene Communicable Disease Division | Kelsey R. Florek et al |
| EPI_ISL_803553 | USA/WI-WSLH-201327/2020 | 2020-09-23 | North America/USA/Wisconsin/Marquet | Wisconsin State Laboratory of Hygiene Communicable Disease Division | Wisconsin State Laboratory of Hygiene Communicable Disease Division | Kelsey R. Florek et al |

| | | | | | | |
|---|---|---|---|---|---|---|
| EPI_ISL_803560 | USA/WI-WSLH-201334/2020 | 2020-09-25 | North America/USA/Wisconsin/Barron C | Wisconsin State Laboratory of Hygiene Communicable Disease Division | Wisconsin State Laboratory of Hygiene Communicable Disease Division | Kelsey R. Florek et al |
| EPI_ISL_803561 | USA/WI-WSLH-201335/2020 | 2020-09-25 | North America/USA/Wisconsin/Jefferson | Wisconsin State Laboratory of Hygiene Communicable Disease Division | Wisconsin State Laboratory of Hygiene Communicable Disease Division | Kelsey R. Florek et al |
| EPI_ISL_803564 | USA/WI-WSLH-201338/2020 | 2020-09-28 | North America/USA/Wisconsin/Grant Co | Wisconsin State Laboratory of Hygiene Communicable Disease Division | Wisconsin State Laboratory of Hygiene Communicable Disease Division | Kelsey R. Florek et al |
| EPI_ISL_803566 | USA/WI-WSLH-201340/2020 | 2020-09-29 | North America/USA/Wisconsin/Marquet | Wisconsin State Laboratory of Hygiene Communicable Disease Division | Wisconsin State Laboratory of Hygiene Communicable Disease Division | Kelsey R. Florek et al |
| EPI_ISL_803567 | USA/WI-WSLH-201341/2020 | 2020-09-29 | North America/USA/Wisconsin/Vilas Co | Wisconsin State Laboratory of Hygiene Communicable Disease Division | Wisconsin State Laboratory of Hygiene Communicable Disease Division | Kelsey R. Florek et al |
| EPI_ISL_803571 | USA/WI-WSLH-201345/2020 | 2020-10-01 | North America/USA/Wisconsin/Vilas Co | Wisconsin State Laboratory of Hygiene Communicable Disease Division | Wisconsin State Laboratory of Hygiene Communicable Disease Division | Kelsey R. Florek et al |
| EPI_ISL_803572 | USA/WI-WSLH-201346/2020 | 2020-10-01 | North America/USA/Wisconsin/Vilas Co | Wisconsin State Laboratory of Hygiene Communicable Disease Division | Wisconsin State Laboratory of Hygiene Communicable Disease Division | Kelsey R. Florek et al |
| EPI_ISL_803573 | USA/WI-WSLH-201347/2020 | 2020-10-01 | North America/USA/Wisconsin/Grant Co | Wisconsin State Laboratory of Hygiene Communicable Disease Division | Wisconsin State Laboratory of Hygiene Communicable Disease Division | Kelsey R. Florek et al |

| | | | | | | |
|---|---|---|---|---|---|---|
| EPI_ISL_803575 | USA/WI-WSLH-201349/2020 | 2020-10-01 | North America/USA/Wisconsin/Jefferson | Wisconsin State Laboratory of Hygiene Communicable Disease Division | Wisconsin State Laboratory of Hygiene Communicable Disease Division | Kelsey R. Florek et al |
| EPI_ISL_803579 | USA/WI-WSLH-201353/2020 | 2020-10-05 | North America/USA/Wisconsin/Grant Co | Wisconsin State Laboratory of Hygiene Communicable Disease Division | Wisconsin State Laboratory of Hygiene Communicable Disease Division | Kelsey R. Florek et al |
| EPI_ISL_803586 | USA/WI-WSLH-201360/2020 | 2020-09-24 | North America/USA/Wisconsin/Marquet | Wisconsin State Laboratory of Hygiene Communicable Disease Division | Wisconsin State Laboratory of Hygiene Communicable Disease Division | Kelsey R. Florek et al |
| EPI_ISL_803588 | USA/WI-WSLH-201362/2020 | 2020-09-28 | North America/USA/Wisconsin/Brown C | Wisconsin State Laboratory of Hygiene Communicable Disease Division | Wisconsin State Laboratory of Hygiene Communicable Disease Division | Kelsey R. Florek et al |
| EPI_ISL_803590 | USA/WI-WSLH-201364/2020 | 2020-09-28 | North America/USA/Wisconsin/Marquet | Wisconsin State Laboratory of Hygiene Communicable Disease Division | Wisconsin State Laboratory of Hygiene Communicable Disease Division | Kelsey R. Florek et al |
| EPI_ISL_803591 | USA/WI-WSLH-201365/2020 | 2020-09-28 | North America/USA/Wisconsin/Marquet | Wisconsin State Laboratory of Hygiene Communicable Disease Division | Wisconsin State Laboratory of Hygiene Communicable Disease Division | Kelsey R. Florek et al |
| EPI_ISL_803595 | USA/WI-WSLH-201369/2020 | 2020-10-01 | North America/USA/Wisconsin/Marquet | Wisconsin State Laboratory of Hygiene Communicable Disease Division | Wisconsin State Laboratory of Hygiene Communicable Disease Division | Kelsey R. Florek et al |
| EPI_ISL_803596 | USA/WI-WSLH-201370/2020 | 2020-10-01 | North America/USA/Wisconsin/Marquet | Wisconsin State Laboratory of Hygiene Communicable Disease Division | Wisconsin State Laboratory of Hygiene Communicable Disease Division | Kelsey R. Florek et al |

| | | | | | | |
|---|---|---|---|---|---|---|
| EPI_ISL_803600 | USA/WI-WSLH-201374/2020 | 2020-10-02 | North America/USA/Wisconsin/Grant Co | Wisconsin State Laboratory of Hygiene Communicable Disease Division | Wisconsin State Laboratory of Hygiene Communicable Disease Division | Kelsey R. Florek et al |
| EPI_ISL_803603 | USA/WI-WSLH-201377/2020 | 2020-09-23 | North America/USA/Wisconsin/Jefferson | Wisconsin State Laboratory of Hygiene Communicable Disease Division | Wisconsin State Laboratory of Hygiene Communicable Disease Division | Kelsey R. Florek et al |
| EPI_ISL_803605 | USA/WI-WSLH-201380/2020 | 2020-09-25 | North America/USA/Wisconsin/Jefferson | Wisconsin State Laboratory of Hygiene Communicable Disease Division | Wisconsin State Laboratory of Hygiene Communicable Disease Division | Kelsey R. Florek et al |
| EPI_ISL_803606 | USA/WI-WSLH-201381/2020 | 2020-09-25 | North America/USA/Wisconsin/Vernon ( | Wisconsin State Laboratory of Hygiene Communicable Disease Division | Wisconsin State Laboratory of Hygiene Communicable Disease Division | Kelsey R. Florek et al |
| EPI_ISL_803608 | USA/WI-WSLH-201383/2020 | 2020-09-26 | North America/USA/Wisconsin/Jefferson | Wisconsin State Laboratory of Hygiene Communicable Disease Division | Wisconsin State Laboratory of Hygiene Communicable Disease Division | Kelsey R. Florek et al |
| EPI_ISL_803611 | USA/WI-WSLH-201386/2020 | 2020-09-28 | North America/USA/Wisconsin/Jefferson | Wisconsin State Laboratory of Hygiene Communicable Disease Division | Wisconsin State Laboratory of Hygiene Communicable Disease Division | Kelsey R. Florek et al |
| EPI_ISL_803616 | USA/WI-WSLH-201391/2020 | 2020-09-29 | North America/USA/Wisconsin/Grant Co | Wisconsin State Laboratory of Hygiene Communicable Disease Division | Wisconsin State Laboratory of Hygiene Communicable Disease Division | Kelsey R. Florek et al |
| EPI_ISL_803623 | USA/WI-WSLH-201398/2020 | 2020-10-01 | North America/USA/Wisconsin/Lafayett | Wisconsin State Laboratory of Hygiene Communicable Disease Division | Wisconsin State Laboratory of Hygiene Communicable Disease Division | Kelsey R. Florek et al |

| | | | | | | |
|---|---|---|---|---|---|---|
| EPI_ISL_803624 | USA/WI-WSLH-201399/2020 | 2020-10-01 | North America/USA/Wisconsin/Lafayett | Wisconsin State Laboratory of Hygiene Communicable Disease Division | Wisconsin State Laboratory of Hygiene Communicable Disease Division | Kelsey R. Florek et al |
| EPI_ISL_803625 | USA/WI-WSLH-201400/2020 | 2020-09-30 | North America/USA/Wisconsin/Ashland | Wisconsin State Laboratory of Hygiene Communicable Disease Division | Wisconsin State Laboratory of Hygiene Communicable Disease Division | Kelsey R. Florek et al |
| EPI_ISL_803627 | USA/WI-WSLH-201402/2020 | 2020-10-02 | North America/USA/Wisconsin/Burnett | Wisconsin State Laboratory of Hygiene Communicable Disease Division | Wisconsin State Laboratory of Hygiene Communicable Disease Division | Kelsey R. Florek et al |
| EPI_ISL_803632 | USA/WI-WSLH-201407/2020 | 2020-10-05 | North America/USA/Wisconsin/Grant Co | Wisconsin State Laboratory of Hygiene Communicable Disease Division | Wisconsin State Laboratory of Hygiene Communicable Disease Division | Kelsey R. Florek et al |
| EPI_ISL_803633 | USA/WI-WSLH-201408/2020 | 2020-10-05 | North America/USA/Wisconsin/Marquet | Wisconsin State Laboratory of Hygiene Communicable Disease Division | Wisconsin State Laboratory of Hygiene Communicable Disease Division | Kelsey R. Florek et al |
| EPI_ISL_803644 | USA/WI-WSLH-201420/2020 | 2020-10-06 | North America/USA/Wisconsin/Jefferson | Wisconsin State Laboratory of Hygiene Communicable Disease Division | Wisconsin State Laboratory of Hygiene Communicable Disease Division | Kelsey R. Florek et al |
| EPI_ISL_803645 | USA/WI-WSLH-201421/2020 | 2020-10-07 | North America/USA/Wisconsin/Marquet | Wisconsin State Laboratory of Hygiene Communicable Disease Division | Wisconsin State Laboratory of Hygiene Communicable Disease Division | Kelsey R. Florek et al |
| EPI_ISL_803649 | USA/WI-WSLH-201426/2020 | 2020-10-08 | North America/USA/Wisconsin/Marquet | Wisconsin State Laboratory of Hygiene Communicable Disease Division | Wisconsin State Laboratory of Hygiene Communicable Disease Division | Kelsey R. Florek et al |

| | | | | | | |
|---|---|---|---|---|---|---|
| EPI_ISL_803675 | USA/WI-WSLH-201478/2020 | 2020-10-08 | North America/USA/Wisconsin/Vilas Co | Wisconsin State Laboratory of Hygiene Communicable Disease Division | Wisconsin State Laboratory of Hygiene Communicable Disease Division | Kelsey R. Florek et al |
| EPI_ISL_803680 | USA/WI-WSLH-201485/2020 | 2020-10-12 | North America/USA/Wisconsin/Jefferson | Wisconsin State Laboratory of Hygiene Communicable Disease Division | Wisconsin State Laboratory of Hygiene Communicable Disease Division | Kelsey R. Florek et al |
| EPI_ISL_803701 | USA/WI-WSLH-201563/2020 | 2020-10-20 | North America/USA/Wisconsin/Chippew | Wisconsin State Laboratory of Hygiene Communicable Disease Division | Wisconsin State Laboratory of Hygiene Communicable Disease Division | Kelsey R. Florek et al |
| EPI_ISL_803710 | USA/WI-WSLH-201572/2020 | 2020-10-22 | North America/USA/Wisconsin/Manitov | Wisconsin State Laboratory of Hygiene Communicable Disease Division | Wisconsin State Laboratory of Hygiene Communicable Disease Division | Kelsey R. Florek et al |
| EPI_ISL_803711 | USA/WI-WSLH-201573/2020 | 2020-10-22 | North America/USA/Wisconsin/Ashland | Wisconsin State Laboratory of Hygiene Communicable Disease Division | Wisconsin State Laboratory of Hygiene Communicable Disease Division | Kelsey R. Florek et al |
| EPI_ISL_803727 | USA/WI-WSLH-201589/2020 | 2020-10-05 | North America/USA/Wisconsin/Dodge C | Wisconsin State Laboratory of Hygiene Communicable Disease Division | Wisconsin State Laboratory of Hygiene Communicable Disease Division | Kelsey R. Florek et al |
| EPI_ISL_803729 | USA/WI-WSLH-201591/2020 | 2020-10-05 | North America/USA/Wisconsin/Dodge C | Wisconsin State Laboratory of Hygiene Communicable Disease Division | Wisconsin State Laboratory of Hygiene Communicable Disease Division | Kelsey R. Florek et al |
| EPI_ISL_803733 | USA/WI-WSLH-201595/2020 | 2020-10-05 | North America/USA/Wisconsin/Sheboyg | Wisconsin State Laboratory of Hygiene Communicable Disease Division | Wisconsin State Laboratory of Hygiene Communicable Disease Division | Kelsey R. Florek et al |

| | | | | | | |
|---|---|---|---|---|---|---|
| EPI_ISL_803739 | USA/WI-WSLH-201601/2020 | 2020-10-22 | North America/USA/Wisconsin/Marquet | Wisconsin State Laboratory of Hygiene Communicable Disease Division | Wisconsin State Laboratory of Hygiene Communicable Disease Division | Kelsey R. Florek et al |
| EPI_ISL_803742 | USA/WI-WSLH-201604/2020 | 2020-09-14 | North America/USA/Wisconsin/Sheboyg | Wisconsin State Laboratory of Hygiene Communicable Disease Division | Wisconsin State Laboratory of Hygiene Communicable Disease Division | Kelsey R. Florek et al |
| EPI_ISL_803745 | USA/WI-WSLH-201607/2020 | 2020-09-14 | North America/USA/Wisconsin/Sheboyg | Wisconsin State Laboratory of Hygiene Communicable Disease Division | Wisconsin State Laboratory of Hygiene Communicable Disease Division | Kelsey R. Florek et al |
| EPI_ISL_803747 | USA/WI-WSLH-201609/2020 | 2020-09-17 | North America/USA/Wisconsin/Sheboyg | Wisconsin State Laboratory of Hygiene Communicable Disease Division | Wisconsin State Laboratory of Hygiene Communicable Disease Division | Kelsey R. Florek et al |
| EPI_ISL_803749 | USA/WI-WSLH-201612/2020 | 2020-09-24 | North America/USA/Wisconsin/Dodge C | Wisconsin State Laboratory of Hygiene Communicable Disease Division | Wisconsin State Laboratory of Hygiene Communicable Disease Division | Kelsey R. Florek et al |
| EPI_ISL_803750 | USA/WI-WSLH-201613/2020 | 2020-09-25 | North America/USA/Wisconsin/Sheboyg | Wisconsin State Laboratory of Hygiene Communicable Disease Division | Wisconsin State Laboratory of Hygiene Communicable Disease Division | Kelsey R. Florek et al |
| EPI_ISL_803755 | USA/WI-WSLH-201618/2020 | 2020-09-30 | North America/USA/Wisconsin/Sheboyg | Wisconsin State Laboratory of Hygiene Communicable Disease Division | Wisconsin State Laboratory of Hygiene Communicable Disease Division | Kelsey R. Florek et al |
| EPI_ISL_803760 | USA/WI-WSLH-201624/2020 | 2020-10-07 | North America/USA/Wisconsin/Dodge C | Wisconsin State Laboratory of Hygiene Communicable Disease Division | Wisconsin State Laboratory of Hygiene Communicable Disease Division | Kelsey R. Florek et al |

| | | | | | | |
|---|---|---|---|---|---|---|
| EPI_ISL_803770 | USA/WI-WSLH-201634/2020 | 2020-10-13 | North America/USA/Wisconsin/Dodge C | Wisconsin State Laboratory of Hygiene Communicable Disease Division | Wisconsin State Laboratory of Hygiene Communicable Disease Division | Kelsey R. Florek et al |
| EPI_ISL_803774 | USA/WI-WSLH-201638/2020 | 2020-10-16 | North America/USA/Wisconsin/Dodge C | Wisconsin State Laboratory of Hygiene Communicable Disease Division | Wisconsin State Laboratory of Hygiene Communicable Disease Division | Kelsey R. Florek et al |
| EPI_ISL_803778 | USA/WI-WSLH-201642/2020 | 2020-10-12 | North America/USA/Wisconsin/Dodge C | Wisconsin State Laboratory of Hygiene Communicable Disease Division | Wisconsin State Laboratory of Hygiene Communicable Disease Division | Kelsey R. Florek et al |
| EPI_ISL_803782 | USA/WI-WSLH-201646/2020 | 2020-10-15 | North America/USA/Wisconsin/Dodge C | Wisconsin State Laboratory of Hygiene Communicable Disease Division | Wisconsin State Laboratory of Hygiene Communicable Disease Division | Kelsey R. Florek et al |
| EPI_ISL_803793 | USA/WI-WSLH-201659/2020 | 2020-10-13 | North America/USA/Wisconsin/Dodge C | Wisconsin State Laboratory of Hygiene Communicable Disease Division | Wisconsin State Laboratory of Hygiene Communicable Disease Division | Kelsey R. Florek et al |
| EPI_ISL_803812 | USA/WI-WSLH-201515/2020 | 2020-10-19 | North America/USA/Wisconsin/Jefferson | Wisconsin State Laboratory of Hygiene Communicable Disease Division | Wisconsin State Laboratory of Hygiene Communicable Disease Division | Kelsey R. Florek et al |

| | | | | | | |
|---|---|---|---|---|---|---|
| EPI_ISL_803817 | Tunisia/12-8274/2020 | 2020-04-07 | Africa/Tunisia/Ariana/ | Laboratory of Microbiology, National Reference Lab, Charles Nicolle Hospital; 2- University of Tunis ElManar, Faculty of Medicine of Tunis, LR99ES09, Tunis, Tunisia | Clinical and Experimental Pharmacology Lab, LR16SP02, National Center of Pharmacovigilance, University of Tunis El Manar, Tunis, Tunisia. 2- Neurodegenerative diseases and psychiatric troubles, LR18SP03, Razi Hospital, University of Tunis El Manar, Tunis, Tunisia. 3- Ministry of Health, National Observatory of New and Emerging Diseases, 1006, Tunis, Tunisia | Ilhem Boutiba-Ben Boubaker et al |
| EPI_ISL_803821 | USA/WI-WSLH-201523/2020 | 2020-10-19 | North America/USA/Wisconsin/Grant Co | Wisconsin State Laboratory of Hygiene Communicable Disease Division | Wisconsin State Laboratory of Hygiene Communicable Disease Division | Kelsey R. Florek et al |
| EPI_ISL_803840 | USA/WI-WSLH-201543/2020 | 2020-10-20 | North America/USA/Wisconsin/Jefferson | Wisconsin State Laboratory of Hygiene Communicable Disease Division | Wisconsin State Laboratory of Hygiene Communicable Disease Division | Kelsey R. Florek et al |
| EPI_ISL_803842 | USA/WI-WSLH-201545/2020 | 2020-10-20 | North America/USA/Wisconsin/Grant Co | Wisconsin State Laboratory of Hygiene Communicable Disease Division | Wisconsin State Laboratory of Hygiene Communicable Disease Division | Kelsey R. Florek et al |

| | | | | | | |
|---|---|---|---|---|---|---|
| EPI_ISL_803844 | USA/WI-WSLH-201547/2020 | 2020-10-19 | North America/USA/Wisconsin/Rock Cou | Wisconsin State Laboratory of Hygiene Communicable Disease Division | Wisconsin State Laboratory of Hygiene Communicable Disease Division | Kelsey R. Florek et al |
| EPI_ISL_803849 | USA/WI-WSLH-201554/2020 | 2020-10-20 | North America/USA/Wisconsin/Jefferson | Wisconsin State Laboratory of Hygiene Communicable Disease Division | Wisconsin State Laboratory of Hygiene Communicable Disease Division | Kelsey R. Florek et al |
| EPI_ISL_803851 | Tunisia/21-2249/2021 | 2021-01-08 | Africa/Tunisia/Nabeul/ | Laboratory of Microbiology, National Reference Lab, Charles Nicolle Hospital; 2- University of Tunis ElManar, Faculty of Medicine of Tunis, LR99ES09, Tunis, Tunisia | Clinical and Experimental Pharmacology Lab, LR16SP02, National Center of Pharmacovigilance, University of Tunis El Manar, Tunis, Tunisia. 2- Neurodegenerative diseases and psychiatric troubles, LR18SP03, Razi Hospital, University of Tunis El Manar, Tunis, Tunisia. 3- Ministry of Health, National Observatory of New and Emerging Diseases, 1006, Tunis, Tunisia | Ilhem Boutiba-Ben Boubaker et al |
| EPI_ISL_803852 | Bangladesh/13-BCSIR-NILMRC_148/20 | 2020-06-18 | Asia/Bangladesh/Dhaka/Tejgaon | National Institute of Laboratory Medicine and Referral Center | Genomic Research Lab, BCSIR | Iffat Jahan et al |
| EPI_ISL_803859 | Spain/MD-HGUGM-20517543/2020 | 2020-12-29 | Europe/Spain/Madrid/ | Hospital General Universitario Gregorio Marav√±√≥n | Hospital General Universitario Gregorio Marav√±√≥n | Sergio Buenestado Serrano et al |

| | | | | | | |
|---|---|---|---|---|---|---|
| EPI_ISL_803870 | Bangladesh/BCSIR-NILMRC_360/2020 | 2020-07-14 | Asia/Bangladesh/Chattogram/Brahmanb | National Institute of Laboratory Medicine and Referral Center | Genomic Research Lab, BCSIR | Md. Murshed Hasan Sarkar et al |
| EPI_ISL_803876 | Indonesia/KU-NIHRD-PME9235/2020 | 2020-10-23 | Asia/Indonesia/North Kalimantan/ | RSUD Tarakan Kalimantan Utara | National Institute of Health Research and Development | Subangkit et al |
| EPI_ISL_803883 | Italy/TUS-UniSi04/2020 | 2020-03-12 | Europe/Italy/Tuscany/ | Department of Medical Biotechnologies, University of Siena | Laboratory of Infectious Diseases, Department of Biomedical and Clinical Sciences L. Sacco, University of Milan | Ilaria Vicenti et al |
| EPI_ISL_803887 | Italy/TUS-UniSi08/2020 | 2020-03-27 | Europe/Italy/Tuscany/ | Department of Medical Biotechnologies, University of Siena | Laboratory of Infectious Diseases, Department of Biomedical and Clinical Sciences L. Sacco, University of Milan | Ilaria Vicenti et al |
| EPI_ISL_803892 | Italy/TUS-UniSi11/2020 | 2020-03-25 | Europe/Italy/Tuscany/ | Department of Medical Biotechnologies, University of Siena | Laboratory of Infectious Diseases, Department of Biomedical and Clinical Sciences L. Sacco, University of Milan | Ilaria Vicenti et al |
| EPI_ISL_803893 | Italy/TUS-UniSi12/2020 | 2020-03-01 | Europe/Italy/Tuscany/ | Department of Medical Biotechnologies, University of Siena | Laboratory of Infectious Diseases, Department of Biomedical and Clinical Sciences L. Sacco, University of Milan | Ilaria Vicenti et al |

| | | | | | | |
|---|---|---|---|---|---|---|
| EPI_ISL_803895 | Italy/TUS-UniSi13/2020 | 2020-02-27 | Europe/Italy/Tuscany/ | Department of Medical Biotechnologies, University of Siena | Laboratory of Infectious Diseases, Department of Biomedical and Clinical Sciences L. Sacco, University of Milan | Ilaria Vicenti et al |
| EPI_ISL_803896 | Italy/TUS-INMI-136/2020 | 2020-12-21 | Europe/Italy/Tuscany/ | Virology Unit, Pisa University Hospital and Retrovirus Center, University of Pisa | "National Institute for Infectious Diseases, INMI, ""L. Spallanzani"" IRCCS" | M Rueca et al |
| EPI_ISL_803901 | India/KA-NIMH-SEQ-5/2020 | 2020-12-19 | Asia/India/Karnataka/ | BBMP Urban PHC | Department of Neurovirology, National Institute of Mental Health and Neurosciences (NIMHANS) | Chitra Pattabiraman et al |
| EPI_ISL_803902 | India/KA-NIMH-SEQ-2/2020 | 2020-12-16 | Asia/India/Karnataka/ | BBMP Urban PHC | Department of Neurovirology, National Institute of Mental Health and Neurosciences (NIMHANS) | Chitra Pattabiraman et al |
| EPI_ISL_803903 | India/KA-NIMH-SEQ-13/2020 | 2020-12-24 | Asia/India/Karnataka/ | Shimoga Institute of Medical Sciences | Department of Neurovirology, National Institute of Mental Health and Neurosciences (NIMHANS) | Chitra Pattabiraman et al |
| EPI_ISL_803904 | India/KA-NIMH-SEQ-4/2020 | 2020-12-19 | Asia/India/Karnataka/ | BBMP Urban PHC | Department of Neurovirology, National Institute of Mental Health and Neurosciences (NIMHANS) | Chitra Pattabiraman et al |

| | | | | | | |
|---|---|---|---|---|---|---|
| EPI_ISL_803940 | Japan/IC-0617/2020 | 20-Dec | Asia/Japan/Japan/ | Pathogen Genomics Center, National Institute of Infectious Diseases | Pathogen Genomics Center, National Institute of Infectious Diseases | Tsuyoshi Sekizuka et al |
| EPI_ISL_803953 | Japan/IC-0653/2021 | 21-Jan | Asia/Japan/Japan/ | Pathogen Genomics Center, National Institute of Infectious Diseases | Pathogen Genomics Center, National Institute of Infectious Diseases | Tsuyoshi Sekizuka et al |
| EPI_ISL_803954 | Japan/IC-0654/2021 | 21-Jan | Asia/Japan/Japan/ | Pathogen Genomics Center, National Institute of Infectious Diseases | Pathogen Genomics Center, National Institute of Infectious Diseases | Tsuyoshi Sekizuka et al |
| EPI_ISL_803955 | Germany/NW-RKI-I-0027/2020 | 2020-12-28 | Europe/Germany/North Rhine Westphal | Labor Dr. Wisplinghoff - K√∂ln | Robert Koch Institute, Influenza and respiratory viruses FG17 & Bioinformatics MF1, Berlin, Germany | Dr. R. Grosser et al |
| EPI_ISL_804022 | USA/MD-MDH-0545/2020 | 2020-12-28 | North America/USA/Maryland/ | Maryland Public Health Laboratory | Maryland Public Health Laboratory | Maryland Department of Health Laboratories Administration et al |
| EPI_ISL_804051 | Italy/FVG-ASUGI-151737/2020 | 2020-12-18 | Europe/Italy/Friuli Venezia Giulia/Triest | SC (UCO) Igiene e Sanit√† Pubblica (funzione integrata con SC Microbiologia e Virologia) e Laboratory of Molecular Virology of the International Centre for Genetic Engineering and Biotechnology (ICGEB) | ARGO Laboratorio Genomica ed Epigenomica | Licastro D et al |
| EPI_ISL_804054 | Italy/FVG-ASUGI-154857/2020 | 2020-12-24 | Europe/Italy/Friuli Venezia Giulia/Goriz | SC (UCO) Igiene e Sanit√† Pubblica (funzione integrata con SC Microbiologia e Virologia) e Laboratory of Molecular Virology of the International Centre for Genetic Engineering and Biotechnology (ICGEB) | ARGO Laboratorio Genomica ed Epigenomica | Licastro D et al |
| EPI_ISL_804138 | Israel/CVL-534ngs/2020 | 2020-12-29 | Asia/Israel/Central District/Raanana | Israel Central Virology laboratory | Israel National Consortium for SARS-CoV-2 sequencing | Neta Zuckerman et al |

| | | | | | | |
|---|---|---|---|---|---|---|
| EPI_ISL_804154 | Israel/CVL-510ngs/2020 | 2020-12-28 | Asia/Israel/Jerusalem District/Jerusalen | Israel Central Virology laboratory | Israel National Consortium for SARS-CoV-2 sequencing | Neta Zuckerman et al |
| EPI_ISL_804192 | Israel/CVL-601-ngs/2020 | 2020-12-29 | Asia/Israel/Jerusalem District/ | Israel Central Virology laboratory | Israel National Consortium for SARS-CoV-2 sequencing | Neta Zuckerman et al |
| EPI_ISL_804208 | Israel/CVL-1021-ngs/2020 | 2020-12-31 | Asia/Israel/Israel/ | Israel Central Virology laboratory | Israel National Consortium for SARS-CoV-2 sequencing | Neta Zuckerman et al |
| EPI_ISL_804291 | England/205390278/2020 | 2020-12-31 | Europe/United Kingdom/England/ | Respiratory Virus Unit, National Infection Service, Public Health England | COVID-19 Genomics UK (COG-UK) Consortium | PHE Covid Sequencing Team et al |
| EPI_ISL_804370 | France/OCC-186/2020 | 2020-12-16 | Europe/France/Occitanie/Toulouse | CHU Purpan - Laboratoire de Virologie - Institut F√©d√©ratif de Biologie | CHU Purpan - Laboratoire de Virologie - Institut F√©d√©ratif de Biologie | Latour J. et al |
| EPI_ISL_804371 | France/OCC-187/2020 | 2020-10-20 | Europe/France/Occitanie/Toulouse | CHU Purpan - Laboratoire de Virologie - Institut F√©d√©ratif de Biologie | CHU Purpan - Laboratoire de Virologie - Institut F√©d√©ratif de Biologie | Latour J. et al |
| EPI_ISL_804372 | France/OCC-188/2020 | 2020-10-21 | Europe/France/Occitanie/Toulouse | CHU Purpan - Laboratoire de Virologie - Institut F√©d√©ratif de Biologie | CHU Purpan - Laboratoire de Virologie - Institut F√©d√©ratif de Biologie | Latour J. et al |
| EPI_ISL_804378 | France/OCC-176/2020 | 2020-09-01 | Europe/France/Occitanie/Toulouse | CHU Purpan - Laboratoire de Virologie - Institut F√©d√©ratif de Biologie | CHU Purpan - Laboratoire de Virologie - Institut F√©d√©ratif de Biologie | Latour J. et al |
| EPI_ISL_804403 | Netherlands/NH-RIVM-10209/2020 | 2020-04-07 | Europe/Netherlands/North Holland/ | Dutch COVID-19 response team | National Institute for Public Health and the Environment (RIVM) | Adam Meijer et al |

| | | | | | | |
|---|---|---|---|---|---|---|
| EPI_ISL_804421 | Netherlands/NH-RIVM-10248/2020 | 2020-03-28 | Europe/Netherlands/North Holland/ | Dutch COVID-19 response team | National Institute for Public Health and the Environment (RIVM) | Adam Meijer et al |
| EPI_ISL_804434 | Netherlands/FL-RIVM-10195/2020 | 2020-03-19 | Europe/Netherlands/Flevoland/ | Dutch COVID-19 response team | National Institute for Public Health and the Environment (RIVM) | Adam Meijer et al |
| EPI_ISL_804458 | Netherlands/NH-RIVM-10261/2020 | 2020-03-31 | Europe/Netherlands/North Holland/ | Dutch COVID-19 response team | National Institute for Public Health and the Environment (RIVM) | Adam Meijer et al |
| EPI_ISL_804467 | France/PAC-IHUCOVID-2696/2020 | 2020-10-25 | Europe/France/Provence-Alpes-CV¥te d' | MEPHI, Aix Marseille University | MEPHI, Aix Marseille University | Anthony LEVASSEUR et al |
| EPI_ISL_804468 | France/PAC-IHUCOVID-2449/2020 | 2020-10-10 | Europe/France/Provence-Alpes-CV¥te d' | MEPHI, Aix Marseille University | MEPHI, Aix Marseille University | Anthony LEVASSEUR et al |
| EPI_ISL_804470 | France/PAC-IHUCOVID-2607/2020 | 2020-10-24 | Europe/France/Provence-Alpes-CV¥te d' | MEPHI, Aix Marseille University | MEPHI, Aix Marseille University | Anthony LEVASSEUR et al |
| EPI_ISL_804471 | France/PAC-IHUCOVID-2622/2020 | 2020-10-28 | Europe/France/Provence-Alpes-CV¥te d' | MEPHI, Aix Marseille University | MEPHI, Aix Marseille University | Anthony LEVASSEUR et al |
| EPI_ISL_804472 | France/PAC-IHUCOVID-2593/2020 | 2020-10-21 | Europe/France/Provence-Alpes-CV¥te d' | MEPHI, Aix Marseille University | MEPHI, Aix Marseille University | Anthony LEVASSEUR et al |
| EPI_ISL_804476 | France/PAC-IHUCOVID-2703/2020 | 2020-10-26 | Europe/France/Provence-Alpes-CV¥te d' | MEPHI, Aix Marseille University | MEPHI, Aix Marseille University | Anthony LEVASSEUR et al |
| EPI_ISL_804477 | France/PAC-IHUCOVID-2628/2020 | 2020-10-09 | Europe/France/Provence-Alpes-CV¥te d' | MEPHI, Aix Marseille University | MEPHI, Aix Marseille University | Anthony LEVASSEUR et al |
| EPI_ISL_804479 | France/PAC-IHUCOVID-2536/2020 | 2020-10-17 | Europe/France/Provence-Alpes-CV¥te d' | MEPHI, Aix Marseille University | MEPHI, Aix Marseille University | Anthony LEVASSEUR et al |

| | | | | | | |
|---|---|---|---|---|---|---|
| EPI_ISL_804481 | France/PAC-IHUCOVID-2588/2020 | 2020-10-21 | Europe/France/Provence-Alpes-CV¥te d' | MEPHI, Aix Marseille University | MEPHI, Aix Marseille University | Anthony LEVASSEUR et al |
| EPI_ISL_804488 | France/PAC-IHUCOVID-2247/2020 | 2020-10-04 | Europe/France/Provence-Alpes-CV¥te d' | MEPHI, Aix Marseille University | MEPHI, Aix Marseille University | Anthony LEVASSEUR et al |
| EPI_ISL_804490 | France/PAC-IHUCOVID-2443/2020 | 2020-10-09 | Europe/France/Provence-Alpes-CV¥te d' | MEPHI, Aix Marseille University | MEPHI, Aix Marseille University | Anthony LEVASSEUR et al |
| EPI_ISL_804491 | France/PAC-IHUCOVID-2444/2020 | 2020-10-10 | Europe/France/Provence-Alpes-CV¥te d' | MEPHI, Aix Marseille University | MEPHI, Aix Marseille University | Anthony LEVASSEUR et al |
| EPI_ISL_804492 | France/PAC-IHUCOVID-2459/2020 | 2020-10-12 | Europe/France/Provence-Alpes-CV¥te d' | MEPHI, Aix Marseille University | MEPHI, Aix Marseille University | Anthony LEVASSEUR et al |
| EPI_ISL_804493 | France/PAC-IHUCOVID-2464/2020 | 2020-10-14 | Europe/France/Provence-Alpes-CV¥te d' | MEPHI, Aix Marseille University | MEPHI, Aix Marseille University | Anthony LEVASSEUR et al |
| EPI_ISL_804497 | France/PAC-IHUCOVID-2476/2020 | 2020-10-15 | Europe/France/Provence-Alpes-CV¥te d' | MEPHI, Aix Marseille University | MEPHI, Aix Marseille University | Anthony LEVASSEUR et al |
| EPI_ISL_804499 | France/PAC-IHUCOVID-2502/2020 | 2020-10-10 | Europe/France/Provence-Alpes-CV¥te d' | MEPHI, Aix Marseille University | MEPHI, Aix Marseille University | Anthony LEVASSEUR et al |
| EPI_ISL_804500 | France/PAC-IHUCOVID-2503/2020 | 2020-10-11 | Europe/France/Provence-Alpes-CV¥te d' | MEPHI, Aix Marseille University | MEPHI, Aix Marseille University | Anthony LEVASSEUR et al |
| EPI_ISL_804501 | France/PAC-IHUCOVID-2506/2020 | 2020-10-15 | Europe/France/Provence-Alpes-CV¥te d' | MEPHI, Aix Marseille University | MEPHI, Aix Marseille University | Anthony LEVASSEUR et al |
| EPI_ISL_804502 | France/PAC-IHUCOVID-2507/2020 | 2020-10-15 | Europe/France/Provence-Alpes-CV¥te d' | MEPHI, Aix Marseille University | MEPHI, Aix Marseille University | Anthony LEVASSEUR et al |
| EPI_ISL_804504 | France/PAC-IHUCOVID-2521/2020 | 2020-10-16 | Europe/France/Provence-Alpes-CV¥te d' | MEPHI, Aix Marseille University | MEPHI, Aix Marseille University | Anthony LEVASSEUR et al |
| EPI_ISL_804505 | France/PAC-IHUCOVID-2523/2020 | 2020-10-16 | Europe/France/Provence-Alpes-CV¥te d' | MEPHI, Aix Marseille University | MEPHI, Aix Marseille University | Anthony LEVASSEUR et al |

| | | | | | | |
|---|---|---|---|---|---|---|
| EPI_ISL_804506 | France/PAC-IHUCOVID-2525/2020 | 2020-10-16 | Europe/France/Provence-Alpes-CV¥te d' | MEPHI, Aix Marseille University | MEPHI, Aix Marseille University | Anthony LEVASSEUR et al |
| EPI_ISL_804507 | France/PAC-IHUCOVID-2526/2020 | 2020-10-17 | Europe/France/Provence-Alpes-CV¥te d' | MEPHI, Aix Marseille University | MEPHI, Aix Marseille University | Anthony LEVASSEUR et al |
| EPI_ISL_804509 | France/PAC-IHUCOVID-2528/2020 | 2020-10-17 | Europe/France/Provence-Alpes-CV¥te d' | MEPHI, Aix Marseille University | MEPHI, Aix Marseille University | Anthony LEVASSEUR et al |
| EPI_ISL_804513 | France/PAC-IHUCOVID-2548/2020 | 2020-10-18 | Europe/France/Provence-Alpes-CV¥te d' | MEPHI, Aix Marseille University | MEPHI, Aix Marseille University | Anthony LEVASSEUR et al |
| EPI_ISL_804514 | France/PAC-IHUCOVID-2553/2020 | 2020-10-19 | Europe/France/Provence-Alpes-CV¥te d' | MEPHI, Aix Marseille University | MEPHI, Aix Marseille University | Anthony LEVASSEUR et al |
| EPI_ISL_804515 | France/PAC-IHUCOVID-2569/2020 | 2020-10-20 | Europe/France/Provence-Alpes-CV¥te d' | MEPHI, Aix Marseille University | MEPHI, Aix Marseille University | Anthony LEVASSEUR et al |
| EPI_ISL_804516 | France/PAC-IHUCOVID-2570/2020 | 2020-10-20 | Europe/France/Provence-Alpes-CV¥te d' | MEPHI, Aix Marseille University | MEPHI, Aix Marseille University | Anthony LEVASSEUR et al |
| EPI_ISL_804517 | France/PAC-IHUCOVID-2572/2020 | 2020-10-20 | Europe/France/Provence-Alpes-CV¥te d' | MEPHI, Aix Marseille University | MEPHI, Aix Marseille University | Anthony LEVASSEUR et al |
| EPI_ISL_804518 | France/PAC-IHUCOVID-2574/2020 | 2020-10-20 | Europe/France/Provence-Alpes-CV¥te d' | MEPHI, Aix Marseille University | MEPHI, Aix Marseille University | Anthony LEVASSEUR et al |
| EPI_ISL_804519 | France/PAC-IHUCOVID-2575/2020 | 2020-10-20 | Europe/France/Provence-Alpes-CV¥te d' | MEPHI, Aix Marseille University | MEPHI, Aix Marseille University | Anthony LEVASSEUR et al |
| EPI_ISL_804520 | France/PAC-IHUCOVID-2578/2020 | 2020-10-20 | Europe/France/Provence-Alpes-CV¥te d' | MEPHI, Aix Marseille University | MEPHI, Aix Marseille University | Anthony LEVASSEUR et al |
| EPI_ISL_804521 | France/PAC-IHUCOVID-2580/2020 | 2020-10-20 | Europe/France/Provence-Alpes-CV¥te d' | MEPHI, Aix Marseille University | MEPHI, Aix Marseille University | Anthony LEVASSEUR et al |
| EPI_ISL_804522 | France/PAC-IHUCOVID-2585/2020 | 2020-10-21 | Europe/France/Provence-Alpes-CV¥te d' | MEPHI, Aix Marseille University | MEPHI, Aix Marseille University | Anthony LEVASSEUR et al |

| | | | | | | |
|---|---|---|---|---|---|---|
| EPI_ISL_804524 | France/PAC-IHUCOVID-2589/2020 | 2020-10-21 | Europe/France/Provence-Alpes-CV¥te d' | MEPHI, Aix Marseille University | MEPHI, Aix Marseille University | Anthony LEVASSEUR et al |
| EPI_ISL_804525 | France/PAC-IHUCOVID-2594/2020 | 2020-10-22 | Europe/France/Provence-Alpes-CV¥te d' | MEPHI, Aix Marseille University | MEPHI, Aix Marseille University | Anthony LEVASSEUR et al |
| EPI_ISL_804529 | France/PAC-IHUCOVID-2605/2020 | 2020-10-24 | Europe/France/Provence-Alpes-CV¥te d' | MEPHI, Aix Marseille University | MEPHI, Aix Marseille University | Anthony LEVASSEUR et al |
| EPI_ISL_804531 | France/PAC-IHUCOVID-2612/2020 | 2020-10-26 | Europe/France/Provence-Alpes-CV¥te d' | MEPHI, Aix Marseille University | MEPHI, Aix Marseille University | Anthony LEVASSEUR et al |
| EPI_ISL_804532 | France/PAC-IHUCOVID-2616/2020 | 2020-10-26 | Europe/France/Provence-Alpes-CV¥te d' | MEPHI, Aix Marseille University | MEPHI, Aix Marseille University | Anthony LEVASSEUR et al |
| EPI_ISL_804533 | France/PAC-IHUCOVID-2617/2020 | 2020-10-27 | Europe/France/Provence-Alpes-CV¥te d' | MEPHI, Aix Marseille University | MEPHI, Aix Marseille University | Anthony LEVASSEUR et al |
| EPI_ISL_804534 | France/PAC-IHUCOVID-2623/2020 | 2020-10-28 | Europe/France/Provence-Alpes-CV¥te d' | MEPHI, Aix Marseille University | MEPHI, Aix Marseille University | Anthony LEVASSEUR et al |
| EPI_ISL_804537 | France/PAC-IHUCOVID-2635/2020 | 2020-10-16 | Europe/France/Provence-Alpes-CV¥te d' | MEPHI, Aix Marseille University | MEPHI, Aix Marseille University | Anthony LEVASSEUR et al |
| EPI_ISL_804538 | France/PAC-IHUCOVID-2636/2020 | 2020-10-16 | Europe/France/Provence-Alpes-CV¥te d' | MEPHI, Aix Marseille University | MEPHI, Aix Marseille University | Anthony LEVASSEUR et al |
| EPI_ISL_804539 | France/PAC-IHUCOVID-2647/2020 | 2020-10-19 | Europe/France/Provence-Alpes-CV¥te d' | MEPHI, Aix Marseille University | MEPHI, Aix Marseille University | Anthony LEVASSEUR et al |
| EPI_ISL_804541 | France/PAC-IHUCOVID-2652/2020 | 2020-10-20 | Europe/France/Provence-Alpes-CV¥te d' | MEPHI, Aix Marseille University | MEPHI, Aix Marseille University | Anthony LEVASSEUR et al |
| EPI_ISL_804542 | France/PAC-IHUCOVID-2653/2020 | 2020-10-20 | Europe/France/Provence-Alpes-CV¥te d' | MEPHI, Aix Marseille University | MEPHI, Aix Marseille University | Anthony LEVASSEUR et al |
| EPI_ISL_804543 | France/PAC-IHUCOVID-2657/2020 | 2020-10-20 | Europe/France/Provence-Alpes-CV¥te d' | MEPHI, Aix Marseille University | MEPHI, Aix Marseille University | Anthony LEVASSEUR et al |

| | | | | | | |
|---|---|---|---|---|---|---|
| EPI_ISL_804544 | France/PAC-IHUCOVID-2659/2020 | 2020-10-20 | Europe/France/Provence-Alpes-CV¥te d' | MEPHI, Aix Marseille University | MEPHI, Aix Marseille University | Anthony LEVASSEUR et al |
| EPI_ISL_804545 | France/PAC-IHUCOVID-2663/2020 | 2020-10-20 | Europe/France/Provence-Alpes-CV¥te d' | MEPHI, Aix Marseille University | MEPHI, Aix Marseille University | Anthony LEVASSEUR et al |
| EPI_ISL_804547 | France/PAC-IHUCOVID-2666/2020 | 2020-10-20 | Europe/France/Provence-Alpes-CV¥te d' | MEPHI, Aix Marseille University | MEPHI, Aix Marseille University | Anthony LEVASSEUR et al |
| EPI_ISL_804548 | France/PAC-IHUCOVID-2667/2020 | 2020-10-21 | Europe/France/Provence-Alpes-CV¥te d' | MEPHI, Aix Marseille University | MEPHI, Aix Marseille University | Anthony LEVASSEUR et al |
| EPI_ISL_804549 | France/PAC-IHUCOVID-2669/2020 | 2020-10-21 | Europe/France/Provence-Alpes-CV¥te d' | MEPHI, Aix Marseille University | MEPHI, Aix Marseille University | Anthony LEVASSEUR et al |
| EPI_ISL_804550 | France/PAC-IHUCOVID-2670/2020 | 2020-10-21 | Europe/France/Provence-Alpes-CV¥te d' | MEPHI, Aix Marseille University | MEPHI, Aix Marseille University | Anthony LEVASSEUR et al |
| EPI_ISL_804551 | France/PAC-IHUCOVID-2671/2020 | 2020-10-21 | Europe/France/Provence-Alpes-CV¥te d' | MEPHI, Aix Marseille University | MEPHI, Aix Marseille University | Anthony LEVASSEUR et al |
| EPI_ISL_804552 | France/PAC-IHUCOVID-2673/2020 | 2020-10-21 | Europe/France/Provence-Alpes-CV¥te d' | MEPHI, Aix Marseille University | MEPHI, Aix Marseille University | Anthony LEVASSEUR et al |
| EPI_ISL_804553 | France/PAC-IHUCOVID-2675/2020 | 2020-10-21 | Europe/France/Provence-Alpes-CV¥te d' | MEPHI, Aix Marseille University | MEPHI, Aix Marseille University | Anthony LEVASSEUR et al |
| EPI_ISL_804555 | France/PAC-IHUCOVID-2680/2020 | 2020-10-21 | Europe/France/Provence-Alpes-CV¥te d' | MEPHI, Aix Marseille University | MEPHI, Aix Marseille University | Anthony LEVASSEUR et al |
| EPI_ISL_804556 | France/PAC-IHUCOVID-2682/2020 | 2020-10-21 | Europe/France/Provence-Alpes-CV¥te d' | MEPHI, Aix Marseille University | MEPHI, Aix Marseille University | Anthony LEVASSEUR et al |
| EPI_ISL_804557 | France/PAC-IHUCOVID-2685/2020 | 2020-10-22 | Europe/France/Provence-Alpes-CV¥te d' | MEPHI, Aix Marseille University | MEPHI, Aix Marseille University | Anthony LEVASSEUR et al |
| EPI_ISL_804558 | France/PAC-IHUCOVID-2687/2020 | 2020-10-23 | Europe/France/Provence-Alpes-CV¥te d' | MEPHI, Aix Marseille University | MEPHI, Aix Marseille University | Anthony LEVASSEUR et al |

| | | | | | | |
|---|---|---|---|---|---|---|
| EPI_ISL_804560 | France/PAC-IHUCOVID-2691/2020 | 2020-10-24 | Europe/France/Provence-Alpes-CV¥te d' | MEPHI, Aix Marseille University | MEPHI, Aix Marseille University | Anthony LEVASSEUR et al |
| EPI_ISL_804561 | France/PAC-IHUCOVID-2692/2020 | 2020-10-24 | Europe/France/Provence-Alpes-CV¥te d' | MEPHI, Aix Marseille University | MEPHI, Aix Marseille University | Anthony LEVASSEUR et al |
| EPI_ISL_804562 | France/PAC-IHUCOVID-2699/2020 | 2020-10-26 | Europe/France/Provence-Alpes-CV¥te d' | MEPHI, Aix Marseille University | MEPHI, Aix Marseille University | Anthony LEVASSEUR et al |
| EPI_ISL_804563 | France/PAC-IHUCOVID-2701/2020 | 2020-10-26 | Europe/France/Provence-Alpes-CV¥te d' | MEPHI, Aix Marseille University | MEPHI, Aix Marseille University | Anthony LEVASSEUR et al |
| EPI_ISL_804564 | France/PAC-IHUCOVID-2706/2020 | 2020-10-27 | Europe/France/Provence-Alpes-CV¥te d' | MEPHI, Aix Marseille University | MEPHI, Aix Marseille University | Anthony LEVASSEUR et al |
| EPI_ISL_804569 | France/PAC-IHUCOVID-2714/2020 | 2020-10-31 | Europe/France/Provence-Alpes-CV¥te d' | MEPHI, Aix Marseille University | MEPHI, Aix Marseille University | Anthony LEVASSEUR et al |
| EPI_ISL_804570 | France/PAC-IHUCOVID-2715/2020 | 2020-11-02 | Europe/France/Provence-Alpes-CV¥te d' | MEPHI, Aix Marseille University | MEPHI, Aix Marseille University | Anthony LEVASSEUR et al |
| EPI_ISL_804573 | France/PAC-IHUCOVID-2718/2020 | 2020-11-02 | Europe/France/Provence-Alpes-CV¥te d' | MEPHI, Aix Marseille University | MEPHI, Aix Marseille University | Anthony LEVASSEUR et al |
| EPI_ISL_804575 | France/PAC-IHUCOVID-2722/2020 | 2020-11-04 | Europe/France/Provence-Alpes-CV¥te d' | MEPHI, Aix Marseille University | MEPHI, Aix Marseille University | Anthony LEVASSEUR et al |
| EPI_ISL_804576 | France/PAC-IHUCOVID-2723/2020 | 2020-11-05 | Europe/France/Provence-Alpes-CV¥te d' | MEPHI, Aix Marseille University | MEPHI, Aix Marseille University | Anthony LEVASSEUR et al |
| EPI_ISL_804578 | France/PAC-IHUCOVID-2725/2020 | 2020-11-09 | Europe/France/Provence-Alpes-CV¥te d' | MEPHI, Aix Marseille University | MEPHI, Aix Marseille University | Anthony LEVASSEUR et al |
| EPI_ISL_804579 | France/PAC-IHUCOVID-2726/2020 | 2020-11-10 | Europe/France/Provence-Alpes-CV¥te d' | MEPHI, Aix Marseille University | MEPHI, Aix Marseille University | Anthony LEVASSEUR et al |
| EPI_ISL_804580 | France/PAC-IHUCOVID-2729/2020 | 2020-11-12 | Europe/France/Provence-Alpes-CV¥te d' | MEPHI, Aix Marseille University | MEPHI, Aix Marseille University | Anthony LEVASSEUR et al |

| | | | | | | |
|---|---|---|---|---|---|---|
| EPI_ISL_804597 | USA/MD-MDH-0312/2020 | 2020-10-17 | North America/USA/Maryland/ | Maryland Public Health Laboratory | Maryland Public Health Laboratory | Maryland Department of Health Laboratories Administration et al |
| EPI_ISL_804598 | USA/MD-MDH-0318/2020 | 2020-10-19 | North America/USA/Maryland/ | Maryland Public Health Laboratory | Maryland Public Health Laboratory | Maryland Department of Health Laboratories Administration et al |
| EPI_ISL_804599 | USA/MI-MDHHS-SC22730/2020 | 2020-11-05 | North America/USA/Michigan/ | Michigan Department of Health and Human Services, Bureau of Laboratories | Michigan Department of Health and Human Services, Bureau of Laboratories | Blankenship HM et al |
| EPI_ISL_804600 | USA/MI-MDHHS-SC22855/2020 | 2020-11-20 | North America/USA/Michigan/ | Michigan Department of Health and Human Services, Bureau of Laboratories | Michigan Department of Health and Human Services, Bureau of Laboratories | Blankenship HM et al |
| EPI_ISL_804621 | USA/MI-MDHHS-SC22736/2020 | 2020-11-13 | North America/USA/Michigan/ | Michigan Department of Health and Human Services, Bureau of Laboratories | Michigan Department of Health and Human Services, Bureau of Laboratories | Blankenship HM et al |
| EPI_ISL_804623 | USA/MI-MDHHS-SC22739/2020 | 2020-11-24 | North America/USA/Michigan/ | Michigan Department of Health and Human Services, Bureau of Laboratories | Michigan Department of Health and Human Services, Bureau of Laboratories | Blankenship HM et al |
| EPI_ISL_804628 | USA/MI-MDHHS-SC22741/2020 | 2020-11-23 | North America/USA/Michigan/ | Michigan Department of Health and Human Services, Bureau of Laboratories | Michigan Department of Health and Human Services, Bureau of Laboratories | Blankenship HM et al |

| | | | | | | |
|---|---|---|---|---|---|---|
| EPI_ISL_804648 | USA/MI-MDHHS-SC22758/2020 | 2020-11-09 | North America/USA/Michigan/ | Michigan Department of Health and Human Services, Bureau of Laboratories | Michigan Department of Health and Human Services, Bureau of Laboratories | Blankenship HM et al |
| EPI_ISL_804656 | USA/MI-MDHHS-SC22721/2020 | 2020-12-03 | North America/USA/Michigan/ | Michigan Department of Health and Human Services, Bureau of Laboratories | Michigan Department of Health and Human Services, Bureau of Laboratories | Blankenship HM et al |
| EPI_ISL_804658 | USA/MI-MDHHS-SC22819/2020 | 2020-11-18 | North America/USA/Michigan/ | Michigan Department of Health and Human Services, Bureau of Laboratories | Michigan Department of Health and Human Services, Bureau of Laboratories | Blankenship HM et al |
| EPI_ISL_804665 | USA/MI-MDHHS-SC22867/2020 | 2020-11-23 | North America/USA/Michigan/ | Michigan Department of Health and Human Services, Bureau of Laboratories | Michigan Department of Health and Human Services, Bureau of Laboratories | Blankenship HM et al |
| EPI_ISL_804668 | USA/MI-MDHHS-SC22830/2020 | 2020-11-25 | North America/USA/Michigan/ | Michigan Department of Health and Human Services, Bureau of Laboratories | Michigan Department of Health and Human Services, Bureau of Laboratories | Blankenship HM et al |
| EPI_ISL_804684 | USA/MI-MDHHS-SC22828/2020 | 2020-11-17 | North America/USA/Michigan/ | Michigan Department of Health and Human Services, Bureau of Laboratories | Michigan Department of Health and Human Services, Bureau of Laboratories | Blankenship HM et al |

| | | | | | | |
|---|---|---|---|---|---|---|
| EPI_ISL_804695 | USA/MI-MDHHS-SC22872/2020 | 2020-11-20 | North America/USA/Michigan/ | Michigan Department of Health and Human Services, Bureau of Laboratories | Michigan Department of Health and Human Services, Bureau of Laboratories | Blankenship HM et al |
| EPI_ISL_804699 | USA/MI-MDHHS-SC22794/2020 | 2020-11-16 | North America/USA/Michigan/ | Michigan Department of Health and Human Services, Bureau of Laboratories | Michigan Department of Health and Human Services, Bureau of Laboratories | Blankenship HM et al |
| EPI_ISL_804704 | USA/MI-MDHHS-SC22795/2020 | 2020-11-16 | North America/USA/Michigan/ | Michigan Department of Health and Human Services, Bureau of Laboratories | Michigan Department of Health and Human Services, Bureau of Laboratories | Blankenship HM et al |
| EPI_ISL_804705 | USA/MI-MDHHS-SC22799/2020 | 2020-11-12 | North America/USA/Michigan/ | Michigan Department of Health and Human Services, Bureau of Laboratories | Michigan Department of Health and Human Services, Bureau of Laboratories | Blankenship HM et al |
| EPI_ISL_804710 | USA/MI-MDHHS-SC22752/2020 | 2020-11-19 | North America/USA/Michigan/ | Michigan Department of Health and Human Services, Bureau of Laboratories | Michigan Department of Health and Human Services, Bureau of Laboratories | Blankenship HM et al |
| EPI_ISL_804733 | USA/MI-MDHHS-SC22784/2020 | 2020-11-09 | North America/USA/Michigan/ | Michigan Department of Health and Human Services, Bureau of Laboratories | Michigan Department of Health and Human Services, Bureau of Laboratories | Blankenship HM et al |

| | | | | | | |
|---|---|---|---|---|---|---|
| EPI_ISL_804734 | USA/MI-MDHHS-SC22785/2020 | 2020-11-09 | North America/USA/Michigan/ | Michigan Department of Health and Human Services, Bureau of Laboratories | Michigan Department of Health and Human Services, Bureau of Laboratories | Blankenship HM et al |
| EPI_ISL_804745 | USA/MI-MDHHS-SC22837/2020 | 2020-11-27 | North America/USA/Michigan/ | Michigan Department of Health and Human Services, Bureau of Laboratories | Michigan Department of Health and Human Services, Bureau of Laboratories | Blankenship HM et al |
| EPI_ISL_804749 | USA/MI-MDHHS-SC22841/2020 | 2020-11-27 | North America/USA/Michigan/ | Michigan Department of Health and Human Services, Bureau of Laboratories | Michigan Department of Health and Human Services, Bureau of Laboratories | Blankenship HM et al |
| EPI_ISL_804755 | USA/MI-MDHHS-SC22847/2020 | 2020-11-27 | North America/USA/Michigan/ | Michigan Department of Health and Human Services, Bureau of Laboratories | Michigan Department of Health and Human Services, Bureau of Laboratories | Blankenship HM et al |
| EPI_ISL_804759 | USA/MI-MDHHS-SC22851/2020 | 2020-11-18 | North America/USA/Michigan/ | Michigan Department of Health and Human Services, Bureau of Laboratories | Michigan Department of Health and Human Services, Bureau of Laboratories | Blankenship HM et al |
| EPI_ISL_804760 | USA/MI-MDHHS-SC22852/2020 | 2020-11-13 | North America/USA/Michigan/ | Michigan Department of Health and Human Services, Bureau of Laboratories | Michigan Department of Health and Human Services, Bureau of Laboratories | Blankenship HM et al |

| | | | | | | |
|---|---|---|---|---|---|---|
| EPI_ISL_804761 | USA/MI-MDHHS-SC22874/2020 | 2020-11-20 | North America/USA/Michigan/ | Michigan Department of Health and Human Services, Bureau of Laboratories | Michigan Department of Health and Human Services, Bureau of Laboratories | Blankenship HM et al |
| EPI_ISL_804762 | USA/MI-MDHHS-SC22875/2020 | 2020-11-20 | North America/USA/Michigan/ | Michigan Department of Health and Human Services, Bureau of Laboratories | Michigan Department of Health and Human Services, Bureau of Laboratories | Blankenship HM et al |
| EPI_ISL_804764 | USA/MI-MDHHS-SC22877/2020 | 2020-11-20 | North America/USA/Michigan/ | Michigan Department of Health and Human Services, Bureau of Laboratories | Michigan Department of Health and Human Services, Bureau of Laboratories | Blankenship HM et al |
| EPI_ISL_804767 | USA/MI-MDHHS-SC22881/2020 | 2020-11-24 | North America/USA/Michigan/ | Michigan Department of Health and Human Services, Bureau of Laboratories | Michigan Department of Health and Human Services, Bureau of Laboratories | Blankenship HM et al |
| EPI_ISL_804768 | USA/MI-MDHHS-SC22882/2020 | 2020-11-20 | North America/USA/Michigan/ | Michigan Department of Health and Human Services, Bureau of Laboratories | Michigan Department of Health and Human Services, Bureau of Laboratories | Blankenship HM et al |
| EPI_ISL_804774 | USA/MI-MDHHS-SC22888/2020 | 2020-11-23 | North America/USA/Michigan/ | Michigan Department of Health and Human Services, Bureau of Laboratories | Michigan Department of Health and Human Services, Bureau of Laboratories | Blankenship HM et al |

| | | | | | | |
|---|---|---|---|---|---|---|
| EPI_ISL_804780 | USA/MI-MDHHS-SC22894/2020 | 2020-11-23 | North America/USA/Michigan/ | Michigan Department of Health and Human Services, Bureau of Laboratories | Michigan Department of Health and Human Services, Bureau of Laboratories | Blankenship HM et al |
| EPI_ISL_804783 | USA/MI-MDHHS-SC22897/2020 | 2020-11-20 | North America/USA/Michigan/ | Michigan Department of Health and Human Services, Bureau of Laboratories | Michigan Department of Health and Human Services, Bureau of Laboratories | Blankenship HM et al |
| EPI_ISL_804784 | USA/MI-MDHHS-SC22898/2020 | 2020-11-24 | North America/USA/Michigan/ | Michigan Department of Health and Human Services, Bureau of Laboratories | Michigan Department of Health and Human Services, Bureau of Laboratories | Blankenship HM et al |
| EPI_ISL_804790 | USA/MI-MDHHS-SC22904/2020 | 2020-11-19 | North America/USA/Michigan/ | Michigan Department of Health and Human Services, Bureau of Laboratories | Michigan Department of Health and Human Services, Bureau of Laboratories | Blankenship HM et al |
| EPI_ISL_804794 | USA/MI-MDHHS-SC22908/2020 | 2020-11-20 | North America/USA/Michigan/ | Michigan Department of Health and Human Services, Bureau of Laboratories | Michigan Department of Health and Human Services, Bureau of Laboratories | Blankenship HM et al |
| EPI_ISL_804804 | USA/MI-MDHHS-SC22918/2020 | 2020-11-20 | North America/USA/Michigan/ | Michigan Department of Health and Human Services, Bureau of Laboratories | Michigan Department of Health and Human Services, Bureau of Laboratories | Blankenship HM et al |

| | | | | | | |
|---|---|---|---|---|---|---|
| EPI_ISL_804814 | Brazil/AM-L70-CD1737/2020 | 2020-12-21 | South America/Brazil/Amazonas BR/ | DB Diagnosticos do Brasil | Laborat√≥rio de Parasitologia M√©dica - Instituto de Medicina Tropical - Universidade de S√£o Paulo | Nuno Faria et al |
| EPI_ISL_804815 | Brazil/AM-L70-CD1716/2020 | 2020-12-16 | South America/Brazil/Amazonas BR/ | DB Diagnosticos do Brasil | Laborat√≥rio de Parasitologia M√©dica - Instituto de Medicina Tropical - Universidade de S√£o Paulo | Nuno Faria et al |
| EPI_ISL_804816 | Brazil/AM-L71-CD1742/2020 | 2020-12-22 | South America/Brazil/Amazonas BR/ | DB Diagnosticos do Brasil | Laborat√≥rio de Parasitologia M√©dica - Instituto de Medicina Tropical - Universidade de S√£o Paulo | Nuno Faria et al |
| EPI_ISL_804817 | Brazil/AM-L70-CD1731/2020 | 2020-12-17 | South America/Brazil/Amazonas BR/ | DB Diagnosticos do Brasil | Laborat√≥rio de Parasitologia M√©dica - Instituto de Medicina Tropical - Universidade de S√£o Paulo | Nuno Faria et al |
| EPI_ISL_804818 | Brazil/AM-L70-CD1729/2020 | 2020-12-17 | South America/Brazil/Amazonas BR/ | DB Diagnosticos do Brasil | Laborat√≥rio de Parasitologia M√©dica - Instituto de Medicina Tropical - Universidade de S√£o Paulo | Nuno Faria et al |

| | | | | | | |
|---|---|---|---|---|---|---|
| EPI_ISL_804819 | Brazil/AM-L70-CD1718/2020 | 2020-12-16 | South America/Brazil/Amazonas BR/ | DB Diagnosticos do Brasil | Laboratv≥rio de Parasitologia Mv©dica - Instituto de Medicina Tropical - Universidade de Sv£o Paulo | Nuno Faria et al |
| EPI_ISL_804820 | Brazil/AM-L70-CD1739/2020 | 2020-12-21 | South America/Brazil/Amazonas BR/ | DB Diagnosticos do Brasil | Laboratv≥rio de Parasitologia Mv©dica - Instituto de Medicina Tropical - Universidade de Sv£o Paulo | Nuno Faria et al |
| EPI_ISL_804821 | Brazil/AM-L70-CD1733/2020 | 2020-12-21 | South America/Brazil/Amazonas BR/ | DB Diagnosticos do Brasil | Laboratv≥rio de Parasitologia Mv©dica - Instituto de Medicina Tropical - Universidade de Sv£o Paulo | Nuno Faria et al |
| EPI_ISL_804822 | Brazil/AM-L70-CD1725/2020 | 2020-12-16 | South America/Brazil/Amazonas BR/ | DB Diagnosticos do Brasil | Laboratv≥rio de Parasitologia Mv©dica - Instituto de Medicina Tropical - Universidade de Sv£o Paulo | Nuno Faria et al |
| EPI_ISL_804823 | Brazil/AM-L70-CD1722/2020 | 2020-12-17 | South America/Brazil/Amazonas BR/ | DB Diagnosticos do Brasil | Laboratv≥rio de Parasitologia Mv©dica - Instituto de Medicina Tropical - Universidade de Sv£o Paulo | Nuno Faria et al |

| | | | | | | |
|---|---|---|---|---|---|---|
| EPI_ISL_804824 | Brazil/AM-L70-CD1721/2020 | 2020-12-17 | South America/Brazil/Amazonas BR/ | DB Diagnosticos do Brasil | Laboratv≥rio de Parasitologia Mv©dica - Instituto de Medicina Tropical - Universidade de Sv£o Paulo | Nuno Faria et al |
| EPI_ISL_804825 | Brazil/AM-L70-CD1715/2020 | 2020-12-15 | South America/Brazil/Amazonas BR/ | DB Diagnosticos do Brasil | Laboratv≥rio de Parasitologia Mv©dica - Instituto de Medicina Tropical - Universidade de Sv£o Paulo | Nuno Faria et al |
| EPI_ISL_804826 | Brazil/AM-L70-CD1717/2020 | 2020-12-16 | South America/Brazil/Amazonas BR/ | DB Diagnosticos do Brasil | Laboratv≥rio de Parasitologia Mv©dica - Instituto de Medicina Tropical - Universidade de Sv£o Paulo | Nuno Faria et al |
| EPI_ISL_804827 | Brazil/AM-L70-CD1719/2020 | 2020-12-15 | South America/Brazil/Amazonas BR/ | DB Diagnosticos do Brasil | Laboratv≥rio de Parasitologia Mv©dica - Instituto de Medicina Tropical - Universidade de Sv£o Paulo | Nuno Faria et al |
| EPI_ISL_804828 | Brazil/AM-L70-CD1726/2020 | 2020-12-17 | South America/Brazil/Amazonas BR/ | DB Diagnosticos do Brasil | Laboratv≥rio de Parasitologia Mv©dica - Instituto de Medicina Tropical - Universidade de Sv£o Paulo | Nuno Faria et al |

| | | | | | | |
|---|---|---|---|---|---|---|
| EPI_ISL_804829 | Brazil/AM-L70-CD1728/2020 | 2020-12-18 | South America/Brazil/Amazonas BR/ | DB Diagnosticos do Brasil | Laboratv≥rio de Parasitologia Mv©dica - Instituto de Medicina Tropical - Universidade de Sv£o Paulo | Nuno Faria et al |
| EPI_ISL_804830 | Brazil/AM-L70-CD1735/2020 | 2020-12-21 | South America/Brazil/Amazonas BR/ | DB Diagnosticos do Brasil | Laboratv≥rio de Parasitologia Mv©dica - Instituto de Medicina Tropical - Universidade de Sv£o Paulo | Nuno Faria et al |
| EPI_ISL_804831 | Brazil/AM-L70-CD1732/2020 | 2020-12-19 | South America/Brazil/Amazonas BR/ | DB Diagnosticos do Brasil | Laboratv≥rio de Parasitologia Mv©dica - Instituto de Medicina Tropical - Universidade de Sv£o Paulo | Nuno Faria et al |
| EPI_ISL_804833 | Brazil/AM-L71-CD1743/2020 | 2020-12-22 | South America/Brazil/Amazonas BR/ | DB Diagnosticos do Brasil | Laboratv≥rio de Parasitologia Mv©dica - Instituto de Medicina Tropical - Universidade de Sv£o Paulo | Nuno Faria et al |
| EPI_ISL_804834 | Brazil/AM-L71-CD1748/2020 | 2020-12-23 | South America/Brazil/Amazonas BR/ | DB Diagnosticos do Brasil | Laboratv≥rio de Parasitologia Mv©dica - Instituto de Medicina Tropical - Universidade de Sv£o Paulo | Nuno Faria et al |

| | | | | | | |
|---|---|---|---|---|---|---|
| EPI_ISL_804842 | Brazil/AM-L71-CD1730/2020 | 2020-12-16 | South America/Brazil/Amazonas BR/ | DB Diagnosticos do Brasil | Laboratv≥rio de Parasitologia Mv©dica - Instituto de Medicina Tropical - Universidade de Sv£o Paulo | Nuno Faria et al |
| EPI_ISL_804843 | Brazil/AM-L71-CD1740/2020 | 2020-12-21 | South America/Brazil/Amazonas BR/ | DB Diagnosticos do Brasil | Laboratv≥rio de Parasitologia Mv©dica - Instituto de Medicina Tropical - Universidade de Sv£o Paulo | Nuno Faria et al |
| EPI_ISL_804844 | Brazil/AM-L71-CD1744/2020 | 2020-12-21 | South America/Brazil/Amazonas BR/ | DB Diagnosticos do Brasil | Laboratv≥rio de Parasitologia Mv©dica - Instituto de Medicina Tropical - Universidade de Sv£o Paulo | Nuno Faria et al |
| EPI_ISL_804859 | USA/MD-DFS-PHL-0119/2020 | 2020-12-23 | North America/USA/Maryland/ | DC Public Health Lab/ Dept. of Forensic Sciences | DC Public Health Lab/ Dept. of Forensic Sciences | Scott Nguyen et al |
| EPI_ISL_804863 | USA/DC-DFS-PHL-0079/2020 | 2020-06-26 | North America/USA/Washington DC/ | DC Public Health Lab/ Dept. of Forensic Sciences | DC Public Health Lab/ Dept. of Forensic Sciences | Scott Nguyen et al |
| EPI_ISL_804866 | USA/DC-DFS-PHL-0027/2020 | 2020-03-20 | North America/USA/Washington DC/ | DC Public Health Lab/ Dept. of Forensic Sciences | DC Public Health Lab/ Dept. of Forensic Sciences | Scott Nguyen et al |
| EPI_ISL_804873 | USA/DC-DFS-PHL-0049/2020 | 2020-03-25 | North America/USA/Washington DC/ | DC Public Health Lab/ Dept. of Forensic Sciences | DC Public Health Lab/ Dept. of Forensic Sciences | Scott Nguyen et al |
| EPI_ISL_804878 | USA/DC-DFS-PHL-0051/2020 | 2020-03-26 | North America/USA/Washington DC/ | DC Public Health Lab/ Dept. of Forensic Sciences | DC Public Health Lab/ Dept. of Forensic Sciences | Scott Nguyen et al |
| EPI_ISL_804880 | USA/DC-DFS-PHL-0053/2020 | 2020-03-26 | North America/USA/Washington DC/ | DC Public Health Lab/ Dept. of Forensic Sciences | DC Public Health Lab/ Dept. of Forensic Sciences | Scott Nguyen et al |

| | | | | | | |
|---|---|---|---|---|---|---|
| EPI_ISL_804911 | USA/DC-DFS-PHL-0039/2020 | 2020-03-23 | North America/USA/Washington DC/ | DC Public Health Lab/ Dept. of Forensic Sciences | DC Public Health Lab/ Dept. of Forensic Sciences | Scott Nguyen et al |
| EPI_ISL_804923 | USA/DC-DFS-PHL-0071/2020 | 2020-05-12 | North America/USA/Washington DC/ | DC Public Health Lab/ Dept. of Forensic Sciences | DC Public Health Lab/ Dept. of Forensic Sciences | Scott Nguyen et al |
| EPI_ISL_804934 | USA/DC-DFS-PHL-0094/2020 | 2020-12-09 | North America/USA/Washington DC/ | DC Public Health Lab/ Dept. of Forensic Sciences | DC Public Health Lab/ Dept. of Forensic Sciences | Scott Nguyen et al |
| EPI_ISL_804935 | USA/DC-DFS-PHL-0095/2020 | 2020-12-14 | North America/USA/Washington DC/ | DC Public Health Lab/ Dept. of Forensic Sciences | DC Public Health Lab/ Dept. of Forensic Sciences | Scott Nguyen et al |
| EPI_ISL_804938 | USA/DC-DFS-PHL-0101/2020 | 2020-12-14 | North America/USA/Washington DC/ | DC Public Health Lab/ Dept. of Forensic Sciences | DC Public Health Lab/ Dept. of Forensic Sciences | Scott Nguyen et al |
| EPI_ISL_804946 | USA/NY-Wadsworth-21001194-01/20 | 2020-12-27 | North America/USA/New York/Nassau ( | BIO-REFERENCE LABORATORIES | Wadsworth Center, New York State Department.of Health | Kirsten St. George et al |
| EPI_ISL_804949 | Spain/ML-ISCIII-2013685/2020 | 2020-08-01 | Europe/Spain/Melilla/ | Hospital Comarcal de Melilla | Instituto de Salud Carlos III | Iglesias-Caballero et al |
| EPI_ISL_804953 | Spain/ML-ISCIII-2013734/2020 | 2020-08-06 | Europe/Spain/Melilla/ | Hospital Comarcal de Melilla | Instituto de Salud Carlos III | Iglesias-Caballero et al |
| EPI_ISL_804954 | Spain/ML-ISCIII-2013817/2020 | 2020-08-07 | Europe/Spain/Melilla/ | Hospital Comarcal de Melilla | Instituto de Salud Carlos III | Iglesias-Caballero et al |
| EPI_ISL_804955 | Spain/ML-ISCIII-2013732/2020 | 2020-08-07 | Europe/Spain/Melilla/ | Hospital Comarcal de Melilla | Instituto de Salud Carlos III | Iglesias-Caballero et al |
| EPI_ISL_804959 | Spain/ML-ISCIII-2013742/2020 | 2020-08-07 | Europe/Spain/Melilla/ | Hospital Comarcal de Melilla | Instituto de Salud Carlos III | Iglesias-Caballero et al |
| EPI_ISL_804962 | Spain/ML-ISCIII-2013739/2020 | 2020-08-06 | Europe/Spain/Melilla/ | Hospital Comarcal de Melilla | Instituto de Salud Carlos III | Iglesias-Caballero et al |
| EPI_ISL_804967 | Spain/ML-ISCIII-2013741/2020 | 2020-08-06 | Europe/Spain/Melilla/ | Hospital Comarcal de Melilla | Instituto de Salud Carlos III | Iglesias-Caballero et al |
| EPI_ISL_804968 | Spain/ML-ISCIII-202059913/2020 | 2020-09-15 | Europe/Spain/Melilla/ | Hospital Comarcal de Melilla | Instituto de Salud Carlos III | Iglesias-Caballero et al |
| EPI_ISL_804969 | Spain/ML-ISCIII-202058887/2020 | 2020-09-15 | Europe/Spain/Melilla/ | Hospital Comarcal de Melilla | Instituto de Salud Carlos III | Iglesias-Caballero et al |
| EPI_ISL_804970 | Spain/ML-ISCIII-2013738/2020 | 2020-08-06 | Europe/Spain/Melilla/ | Hospital Comarcal de Melilla | Instituto de Salud Carlos III | Iglesias-Caballero et al |

| | | | | | | |
|---|---|---|---|---|---|---|
| EPI_ISL_804971 | Spain/ML-ISCIII-202059873/2020 | 2020-09-15 | Europe/Spain/Melilla/ | Hospital Comarcal de Melilla | Instituto de Salud Carlos III | Iglesias-Caballero et al |
| EPI_ISL_804972 | Spain/ML-ISCIII-202058692/2020 | 2020-09-15 | Europe/Spain/Melilla/ | Hospital Comarcal de Melilla | Instituto de Salud Carlos III | Iglesias-Caballero et al |
| EPI_ISL_804974 | Spain/ML-ISCIII-202059862/2020 | 2020-09-15 | Europe/Spain/Melilla/ | Hospital Comarcal de Melilla | Instituto de Salud Carlos III | Iglesias-Caballero et al |
| EPI_ISL_804975 | Spain/ML-ISCIII-202058922/2020 | 2020-09-15 | Europe/Spain/Melilla/ | Hospital Comarcal de Melilla | Instituto de Salud Carlos III | Iglesias-Caballero et al |
| EPI_ISL_804976 | Spain/ML-ISCIII-202058907/2020 | 2020-09-15 | Europe/Spain/Melilla/ | Hospital Comarcal de Melilla | Instituto de Salud Carlos III | Iglesias-Caballero et al |
| EPI_ISL_804977 | Spain/ML-ISCIII-2013736/2020 | 2020-08-06 | Europe/Spain/Melilla/ | Hospital Comarcal de Melilla | Instituto de Salud Carlos III | Iglesias-Caballero et al |
| EPI_ISL_804985 | Canada/AB-67738/2020 | 2020-04-22 | North America/Canada/Alberta/ | Alberta Precision Labs (APL) | Alberta Precision Labs (APL) | Gordon P et al |
| EPI_ISL_804986 | Canada/AB-73519/2020 | 2020-04-18 | North America/Canada/Alberta/ | Alberta Precision Labs (APL) | Alberta Precision Labs (APL) | Gordon P et al |
| EPI_ISL_804987 | Canada/AB-38296/2020 | 2020-04-21 | North America/Canada/Alberta/ | Alberta Precision Labs (APL) | Alberta Precision Labs (APL) | Gordon P et al |
| EPI_ISL_804988 | Canada/AB-87327/2020 | 2020-04-24 | North America/Canada/Alberta/ | Alberta Precision Labs (APL) | Alberta Precision Labs (APL) | Gordon P et al |
| EPI_ISL_804989 | Canada/AB-50305/2020 | 2020-04-25 | North America/Canada/Alberta/ | Alberta Precision Labs (APL) | Alberta Precision Labs (APL) | Gordon P et al |
| EPI_ISL_804990 | Canada/AB-90368/2020 | 2020-04-29 | North America/Canada/Alberta/ | Alberta Precision Labs (APL) | Alberta Precision Labs (APL) | Gordon P et al |
| EPI_ISL_804991 | Canada/AB-59111/2020 | 2020-04-29 | North America/Canada/Alberta/ | Alberta Precision Labs (APL) | Alberta Precision Labs (APL) | Gordon P et al |
| EPI_ISL_804992 | Canada/AB-30727/2020 | 2020-04-23 | North America/Canada/Alberta/ | Alberta Precision Labs (APL) | Alberta Precision Labs (APL) | Gordon P et al |
| EPI_ISL_804993 | Canada/AB-17445/2020 | 2020-04-24 | North America/Canada/Alberta/ | Alberta Precision Labs (APL) | Alberta Precision Labs (APL) | Gordon P et al |
| EPI_ISL_804994 | Canada/AB-54929/2020 | 2020-04-18 | North America/Canada/Alberta/ | Alberta Precision Labs (APL) | Alberta Precision Labs (APL) | Gordon P et al |
| EPI_ISL_804995 | Canada/AB-51817/2020 | 2020-05-02 | North America/Canada/Alberta/ | Alberta Precision Labs (APL) | Alberta Precision Labs (APL) | Gordon P et al |
| EPI_ISL_804996 | Canada/AB-72321/2020 | 2020-04-17 | North America/Canada/Alberta/ | Alberta Precision Labs (APL) | Alberta Precision Labs (APL) | Gordon P et al |
| EPI_ISL_804997 | Canada/AB-98979/2020 | 2020-04-14 | North America/Canada/Alberta/ | Alberta Precision Labs (APL) | Alberta Precision Labs (APL) | Gordon P et al |
| EPI_ISL_804998 | Canada/AB-88975/2020 | 2020-04-21 | North America/Canada/Alberta/ | Alberta Precision Labs (APL) | Alberta Precision Labs (APL) | Gordon P et al |

| | | | | | | |
|---|---|---|---|---|---|---|
| EPI_ISL_804999 | Canada/AB-24529/2020 | 2020-05-09 | North America/Canada/Alberta/ | Alberta Precision Labs (APL) | Alberta Precision Labs (APL) | Gordon P et al |
| EPI_ISL_805000 | Canada/AB-25282/2020 | 2020-04-16 | North America/Canada/Alberta/ | Alberta Precision Labs (APL) | Alberta Precision Labs (APL) | Gordon P et al |
| EPI_ISL_805001 | Canada/AB-91395/2020 | 2020-05-07 | North America/Canada/Alberta/ | Alberta Precision Labs (APL) | Alberta Precision Labs (APL) | Gordon P et al |
| EPI_ISL_805002 | Canada/AB-91635/2020 | 2020-05-02 | North America/Canada/Alberta/ | Alberta Precision Labs (APL) | Alberta Precision Labs (APL) | Gordon P et al |
| EPI_ISL_805003 | Canada/AB-29422/2020 | 2020-05-10 | North America/Canada/Alberta/ | Alberta Precision Labs (APL) | Alberta Precision Labs (APL) | Gordon P et al |
| EPI_ISL_805004 | Canada/AB-76796/2020 | 2020-04-27 | North America/Canada/Alberta/ | Alberta Precision Labs (APL) | Alberta Precision Labs (APL) | Gordon P et al |
| EPI_ISL_805005 | Canada/AB-59267/2020 | 2020-04-15 | North America/Canada/Alberta/ | Alberta Precision Labs (APL) | Alberta Precision Labs (APL) | Gordon P et al |
| EPI_ISL_805006 | Canada/AB-40770/2020 | 2020-05-19 | North America/Canada/Alberta/ | Alberta Precision Labs (APL) | Alberta Precision Labs (APL) | Gordon P et al |
| EPI_ISL_805007 | Canada/AB-13778/2020 | 2020-05-06 | North America/Canada/Alberta/ | Alberta Precision Labs (APL) | Alberta Precision Labs (APL) | Gordon P et al |
| EPI_ISL_805008 | Canada/AB-69916/2020 | 2020-04-26 | North America/Canada/Alberta/ | Alberta Precision Labs (APL) | Alberta Precision Labs (APL) | Gordon P et al |
| EPI_ISL_805009 | Canada/AB-45291/2020 | 2020-04-22 | North America/Canada/Alberta/ | Alberta Precision Labs (APL) | Alberta Precision Labs (APL) | Gordon P et al |
| EPI_ISL_805010 | Canada/AB-93391/2020 | 2020-04-22 | North America/Canada/Alberta/ | Alberta Precision Labs (APL) | Alberta Precision Labs (APL) | Gordon P et al |
| EPI_ISL_805011 | Canada/AB-25303/2020 | 2020-04-17 | North America/Canada/Alberta/ | Alberta Precision Labs (APL) | Alberta Precision Labs (APL) | Gordon P et al |
| EPI_ISL_805012 | Canada/AB-44601/2020 | 2020-04-19 | North America/Canada/Alberta/ | Alberta Precision Labs (APL) | Alberta Precision Labs (APL) | Gordon P et al |
| EPI_ISL_805013 | Canada/AB-74528/2020 | 2020-04-28 | North America/Canada/Alberta/ | Alberta Precision Labs (APL) | Alberta Precision Labs (APL) | Gordon P et al |
| EPI_ISL_805014 | Canada/AB-60103/2020 | 2020-04-22 | North America/Canada/Alberta/ | Alberta Precision Labs (APL) | Alberta Precision Labs (APL) | Gordon P et al |
| EPI_ISL_805015 | Canada/AB-12646/2020 | 2020-04-21 | North America/Canada/Alberta/ | Alberta Precision Labs (APL) | Alberta Precision Labs (APL) | Gordon P et al |
| EPI_ISL_805016 | Canada/AB-51397/2020 | 2020-04-28 | North America/Canada/Alberta/ | Alberta Precision Labs (APL) | Alberta Precision Labs (APL) | Gordon P et al |
| EPI_ISL_805017 | Canada/AB-67188/2020 | 2020-04-20 | North America/Canada/Alberta/ | Alberta Precision Labs (APL) | Alberta Precision Labs (APL) | Gordon P et al |
| EPI_ISL_805018 | Canada/AB-13561/2020 | 2020-04-23 | North America/Canada/Alberta/ | Alberta Precision Labs (APL) | Alberta Precision Labs (APL) | Gordon P et al |

| | | | | | | |
|---|---|---|---|---|---|---|
| EPI_ISL_805019 | Canada/AB-53450/2020 | 2020-04-23 | North America/Canada/Alberta/ | Alberta Precision Labs (APL) | Alberta Precision Labs (APL) | Gordon P et al |
| EPI_ISL_805020 | Canada/AB-35686/2020 | 2020-03-16 | North America/Canada/Alberta/ | Alberta Precision Labs (APL) | Alberta Precision Labs (APL) | Gordon P et al |
| EPI_ISL_805021 | Canada/AB-87950/2020 | 2020-04-23 | North America/Canada/Alberta/ | Alberta Precision Labs (APL) | Alberta Precision Labs (APL) | Gordon P et al |
| EPI_ISL_805022 | Canada/AB-97352/2020 | 2020-04-23 | North America/Canada/Alberta/ | Alberta Precision Labs (APL) | Alberta Precision Labs (APL) | Gordon P et al |
| EPI_ISL_805023 | Canada/AB-76435/2020 | 2020-05-06 | North America/Canada/Alberta/ | Alberta Precision Labs (APL) | Alberta Precision Labs (APL) | Gordon P et al |
| EPI_ISL_805024 | Canada/AB-19381/2020 | 2020-04-23 | North America/Canada/Alberta/ | Alberta Precision Labs (APL) | Alberta Precision Labs (APL) | Gordon P et al |
| EPI_ISL_805025 | Canada/AB-56966/2020 | 2020-04-23 | North America/Canada/Alberta/ | Alberta Precision Labs (APL) | Alberta Precision Labs (APL) | Gordon P et al |
| EPI_ISL_805026 | Canada/AB-30672/2020 | 2020-04-22 | North America/Canada/Alberta/ | Alberta Precision Labs (APL) | Alberta Precision Labs (APL) | Gordon P et al |
| EPI_ISL_805027 | Canada/AB-62415/2020 | 2020-04-23 | North America/Canada/Alberta/ | Alberta Precision Labs (APL) | Alberta Precision Labs (APL) | Gordon P et al |
| EPI_ISL_805028 | Canada/AB-94585/2020 | 2020-04-22 | North America/Canada/Alberta/ | Alberta Precision Labs (APL) | Alberta Precision Labs (APL) | Gordon P et al |
| EPI_ISL_805029 | Canada/AB-39391/2020 | 2020-04-20 | North America/Canada/Alberta/ | Alberta Precision Labs (APL) | Alberta Precision Labs (APL) | Gordon P et al |
| EPI_ISL_805030 | Canada/AB-39283/2020 | 2020-04-22 | North America/Canada/Alberta/ | Alberta Precision Labs (APL) | Alberta Precision Labs (APL) | Gordon P et al |
| EPI_ISL_805031 | Canada/AB-27550/2020 | 2020-04-23 | North America/Canada/Alberta/ | Alberta Precision Labs (APL) | Alberta Precision Labs (APL) | Gordon P et al |
| EPI_ISL_805032 | Canada/AB-55542/2020 | 2020-04-23 | North America/Canada/Alberta/ | Alberta Precision Labs (APL) | Alberta Precision Labs (APL) | Gordon P et al |
| EPI_ISL_805033 | Canada/AB-81786/2020 | 2020-04-22 | North America/Canada/Alberta/ | Alberta Precision Labs (APL) | Alberta Precision Labs (APL) | Gordon P et al |
| EPI_ISL_805034 | Canada/AB-67146/2020 | 2020-04-23 | North America/Canada/Alberta/ | Alberta Precision Labs (APL) | Alberta Precision Labs (APL) | Gordon P et al |
| EPI_ISL_805035 | Canada/AB-22797/2020 | 2020-04-22 | North America/Canada/Alberta/ | Alberta Precision Labs (APL) | Alberta Precision Labs (APL) | Gordon P et al |
| EPI_ISL_805036 | Canada/AB-61221/2020 | 2020-04-23 | North America/Canada/Alberta/ | Alberta Precision Labs (APL) | Alberta Precision Labs (APL) | Gordon P et al |
| EPI_ISL_805037 | Canada/AB-60165/2020 | 2020-04-22 | North America/Canada/Alberta/ | Alberta Precision Labs (APL) | Alberta Precision Labs (APL) | Gordon P et al |
| EPI_ISL_805038 | Canada/AB-76334/2020 | 2020-04-22 | North America/Canada/Alberta/ | Alberta Precision Labs (APL) | Alberta Precision Labs (APL) | Gordon P et al |

| | | | | | | |
|---|---|---|---|---|---|---|
| EPI_ISL_805039 | Canada/AB-69080/2020 | 2020-04-22 | North America/Canada/Alberta/ | Alberta Precision Labs (APL) | Alberta Precision Labs (APL) | Gordon P et al |
| EPI_ISL_805040 | Canada/AB-70352/2020 | 2020-04-23 | North America/Canada/Alberta/ | Alberta Precision Labs (APL) | Alberta Precision Labs (APL) | Gordon P et al |
| EPI_ISL_805041 | Canada/AB-62664/2020 | 2020-04-21 | North America/Canada/Alberta/ | Alberta Precision Labs (APL) | Alberta Precision Labs (APL) | Gordon P et al |
| EPI_ISL_805042 | Canada/AB-33303/2020 | 2020-04-23 | North America/Canada/Alberta/ | Alberta Precision Labs (APL) | Alberta Precision Labs (APL) | Gordon P et al |
| EPI_ISL_805043 | Canada/AB-47538/2020 | 2020-04-23 | North America/Canada/Alberta/ | Alberta Precision Labs (APL) | Alberta Precision Labs (APL) | Gordon P et al |
| EPI_ISL_805044 | Canada/AB-99351/2020 | 2020-04-22 | North America/Canada/Alberta/ | Alberta Precision Labs (APL) | Alberta Precision Labs (APL) | Gordon P et al |
| EPI_ISL_805045 | Canada/AB-64815/2020 | 2020-04-23 | North America/Canada/Alberta/ | Alberta Precision Labs (APL) | Alberta Precision Labs (APL) | Gordon P et al |
| EPI_ISL_805046 | Canada/AB-41975/2020 | 2020-04-22 | North America/Canada/Alberta/ | Alberta Precision Labs (APL) | Alberta Precision Labs (APL) | Gordon P et al |
| EPI_ISL_805047 | Canada/AB-63387/2020 | 2020-04-23 | North America/Canada/Alberta/ | Alberta Precision Labs (APL) | Alberta Precision Labs (APL) | Gordon P et al |
| EPI_ISL_805048 | Canada/AB-64617/2020 | 2020-04-22 | North America/Canada/Alberta/ | Alberta Precision Labs (APL) | Alberta Precision Labs (APL) | Gordon P et al |
| EPI_ISL_805049 | Canada/AB-45534/2020 | 2020-04-21 | North America/Canada/Alberta/ | Alberta Precision Labs (APL) | Alberta Precision Labs (APL) | Gordon P et al |
| EPI_ISL_805050 | Canada/AB-94128/2020 | 2020-04-21 | North America/Canada/Alberta/ | Alberta Precision Labs (APL) | Alberta Precision Labs (APL) | Gordon P et al |
| EPI_ISL_805051 | Canada/AB-53011/2020 | 2020-04-22 | North America/Canada/Alberta/ | Alberta Precision Labs (APL) | Alberta Precision Labs (APL) | Gordon P et al |
| EPI_ISL_805052 | Canada/AB-78203/2020 | 2020-04-22 | North America/Canada/Alberta/ | Alberta Precision Labs (APL) | Alberta Precision Labs (APL) | Gordon P et al |
| EPI_ISL_805053 | Canada/AB-36541/2020 | 2020-04-23 | North America/Canada/Alberta/ | Alberta Precision Labs (APL) | Alberta Precision Labs (APL) | Gordon P et al |
| EPI_ISL_805054 | Canada/AB-74564/2020 | 2020-04-21 | North America/Canada/Alberta/ | Alberta Precision Labs (APL) | Alberta Precision Labs (APL) | Gordon P et al |
| EPI_ISL_805055 | Canada/AB-60097/2020 | 2020-04-22 | North America/Canada/Alberta/ | Alberta Precision Labs (APL) | Alberta Precision Labs (APL) | Gordon P et al |
| EPI_ISL_805056 | Canada/AB-16318/2020 | 2020-04-22 | North America/Canada/Alberta/ | Alberta Precision Labs (APL) | Alberta Precision Labs (APL) | Gordon P et al |
| EPI_ISL_805057 | Canada/AB-65235/2020 | 2020-04-25 | North America/Canada/Alberta/ | Alberta Precision Labs (APL) | Alberta Precision Labs (APL) | Gordon P et al |
| EPI_ISL_805058 | Canada/AB-61093/2020 | 2020-04-22 | North America/Canada/Alberta/ | Alberta Precision Labs (APL) | Alberta Precision Labs (APL) | Gordon P et al |

| | | | | | | |
|---|---|---|---|---|---|---|
| EPI_ISL_805059 | Canada/AB-42957/2020 | 2020-04-23 | North America/Canada/Alberta/ | Alberta Precision Labs (APL) | Alberta Precision Labs (APL) | Gordon P et al |
| EPI_ISL_805060 | Canada/AB-97437/2020 | 2020-04-23 | North America/Canada/Alberta/ | Alberta Precision Labs (APL) | Alberta Precision Labs (APL) | Gordon P et al |
| EPI_ISL_805061 | Canada/AB-54629/2020 | 2020-04-23 | North America/Canada/Alberta/ | Alberta Precision Labs (APL) | Alberta Precision Labs (APL) | Gordon P et al |
| EPI_ISL_805062 | Canada/AB-62826/2020 | 2020-04-23 | North America/Canada/Alberta/ | Alberta Precision Labs (APL) | Alberta Precision Labs (APL) | Gordon P et al |
| EPI_ISL_805063 | Canada/AB-46852/2020 | 2020-04-22 | North America/Canada/Alberta/ | Alberta Precision Labs (APL) | Alberta Precision Labs (APL) | Gordon P et al |
| EPI_ISL_805064 | Canada/AB-58781/2020 | 2020-04-22 | North America/Canada/Alberta/ | Alberta Precision Labs (APL) | Alberta Precision Labs (APL) | Gordon P et al |
| EPI_ISL_805065 | Canada/AB-32195/2020 | 2020-04-20 | North America/Canada/Alberta/ | Alberta Precision Labs (APL) | Alberta Precision Labs (APL) | Gordon P et al |
| EPI_ISL_805066 | Canada/AB-14165/2020 | 2020-04-23 | North America/Canada/Alberta/ | Alberta Precision Labs (APL) | Alberta Precision Labs (APL) | Gordon P et al |
| EPI_ISL_805067 | Canada/AB-63098/2020 | 2020-04-22 | North America/Canada/Alberta/ | Alberta Precision Labs (APL) | Alberta Precision Labs (APL) | Gordon P et al |
| EPI_ISL_805068 | Canada/AB-28009/2020 | 2020-04-22 | North America/Canada/Alberta/ | Alberta Precision Labs (APL) | Alberta Precision Labs (APL) | Gordon P et al |
| EPI_ISL_805069 | Canada/AB-25826/2020 | 2020-04-23 | North America/Canada/Alberta/ | Alberta Precision Labs (APL) | Alberta Precision Labs (APL) | Gordon P et al |
| EPI_ISL_805070 | Canada/AB-35674/2020 | 2020-04-21 | North America/Canada/Alberta/ | Alberta Precision Labs (APL) | Alberta Precision Labs (APL) | Gordon P et al |
| EPI_ISL_805071 | Canada/AB-57698/2020 | 2020-04-20 | North America/Canada/Alberta/ | Alberta Precision Labs (APL) | Alberta Precision Labs (APL) | Gordon P et al |
| EPI_ISL_805072 | Canada/AB-41071/2020 | 2020-04-23 | North America/Canada/Alberta/ | Alberta Precision Labs (APL) | Alberta Precision Labs (APL) | Gordon P et al |
| EPI_ISL_805073 | Canada/AB-66254/2020 | 2020-04-20 | North America/Canada/Alberta/ | Alberta Precision Labs (APL) | Alberta Precision Labs (APL) | Gordon P et al |
| EPI_ISL_805074 | Canada/AB-65961/2020 | 2020-04-21 | North America/Canada/Alberta/ | Alberta Precision Labs (APL) | Alberta Precision Labs (APL) | Gordon P et al |
| EPI_ISL_805075 | Canada/AB-17453/2020 | 2020-04-19 | North America/Canada/Alberta/ | Alberta Precision Labs (APL) | Alberta Precision Labs (APL) | Gordon P et al |
| EPI_ISL_805076 | Canada/AB-93287/2020 | 2020-04-23 | North America/Canada/Alberta/ | Alberta Precision Labs (APL) | Alberta Precision Labs (APL) | Gordon P et al |
| EPI_ISL_805077 | Canada/AB-90578/2020 | 2020-04-20 | North America/Canada/Alberta/ | Alberta Precision Labs (APL) | Alberta Precision Labs (APL) | Gordon P et al |
| EPI_ISL_805078 | Canada/AB-80537/2020 | 2020-04-19 | North America/Canada/Alberta/ | Alberta Precision Labs (APL) | Alberta Precision Labs (APL) | Gordon P et al |

| | | | | | | |
|---|---|---|---|---|---|---|
| EPI_ISL_805079 | Canada/AB-71016/2020 | 2020-04-21 | North America/Canada/Alberta/ | Alberta Precision Labs (APL) | Alberta Precision Labs (APL) | Gordon P et al |
| EPI_ISL_805080 | Canada/AB-30224/2020 | 2020-04-21 | North America/Canada/Alberta/ | Alberta Precision Labs (APL) | Alberta Precision Labs (APL) | Gordon P et al |
| EPI_ISL_805081 | Canada/AB-98438/2020 | 2020-04-21 | North America/Canada/Alberta/ | Alberta Precision Labs (APL) | Alberta Precision Labs (APL) | Gordon P et al |
| EPI_ISL_805082 | Canada/AB-22871/2020 | 2020-04-20 | North America/Canada/Alberta/ | Alberta Precision Labs (APL) | Alberta Precision Labs (APL) | Gordon P et al |
| EPI_ISL_805083 | Canada/AB-41300/2020 | 2020-04-22 | North America/Canada/Alberta/ | Alberta Precision Labs (APL) | Alberta Precision Labs (APL) | Gordon P et al |
| EPI_ISL_805084 | Canada/AB-18196/2020 | 2020-04-23 | North America/Canada/Alberta/ | Alberta Precision Labs (APL) | Alberta Precision Labs (APL) | Gordon P et al |
| EPI_ISL_805085 | Canada/AB-77448/2020 | 2020-04-22 | North America/Canada/Alberta/ | Alberta Precision Labs (APL) | Alberta Precision Labs (APL) | Gordon P et al |
| EPI_ISL_805086 | Canada/AB-31667/2020 | 2020-04-22 | North America/Canada/Alberta/ | Alberta Precision Labs (APL) | Alberta Precision Labs (APL) | Gordon P et al |
| EPI_ISL_805087 | Canada/AB-52769/2020 | 2020-04-22 | North America/Canada/Alberta/ | Alberta Precision Labs (APL) | Alberta Precision Labs (APL) | Gordon P et al |
| EPI_ISL_805088 | Canada/AB-55013/2020 | 2020-04-22 | North America/Canada/Alberta/ | Alberta Precision Labs (APL) | Alberta Precision Labs (APL) | Gordon P et al |
| EPI_ISL_805089 | Canada/AB-31206/2020 | 2020-04-23 | North America/Canada/Alberta/ | Alberta Precision Labs (APL) | Alberta Precision Labs (APL) | Gordon P et al |
| EPI_ISL_805090 | Canada/AB-18889/2020 | 2020-04-22 | North America/Canada/Alberta/ | Alberta Precision Labs (APL) | Alberta Precision Labs (APL) | Gordon P et al |
| EPI_ISL_805091 | Canada/AB-66912/2020 | 2020-04-22 | North America/Canada/Alberta/ | Alberta Precision Labs (APL) | Alberta Precision Labs (APL) | Gordon P et al |
| EPI_ISL_805092 | Canada/AB-29046/2020 | 2020-04-21 | North America/Canada/Alberta/ | Alberta Precision Labs (APL) | Alberta Precision Labs (APL) | Gordon P et al |
| EPI_ISL_805093 | Canada/AB-66713/2020 | 2020-04-23 | North America/Canada/Alberta/ | Alberta Precision Labs (APL) | Alberta Precision Labs (APL) | Gordon P et al |
| EPI_ISL_805094 | Canada/AB-29076/2020 | 2020-04-21 | North America/Canada/Alberta/ | Alberta Precision Labs (APL) | Alberta Precision Labs (APL) | Gordon P et al |
| EPI_ISL_805095 | Canada/AB-31131/2020 | 2020-04-22 | North America/Canada/Alberta/ | Alberta Precision Labs (APL) | Alberta Precision Labs (APL) | Gordon P et al |
| EPI_ISL_805096 | Canada/AB-44595/2020 | 2020-04-22 | North America/Canada/Alberta/ | Alberta Precision Labs (APL) | Alberta Precision Labs (APL) | Gordon P et al |
| EPI_ISL_805097 | Canada/AB-61624/2020 | 2020-04-22 | North America/Canada/Alberta/ | Alberta Precision Labs (APL) | Alberta Precision Labs (APL) | Gordon P et al |
| EPI_ISL_805098 | Canada/AB-51819/2020 | 2020-04-23 | North America/Canada/Alberta/ | Alberta Precision Labs (APL) | Alberta Precision Labs (APL) | Gordon P et al |

| | | | | | | |
|---|---|---|---|---|---|---|
| EPI_ISL_805099 | Canada/AB-21012/2020 | 2020-04-23 | North America/Canada/Alberta/ | Alberta Precision Labs (APL) | Alberta Precision Labs (APL) | Gordon P et al |
| EPI_ISL_805100 | Canada/AB-66548/2020 | 2020-04-22 | North America/Canada/Alberta/ | Alberta Precision Labs (APL) | Alberta Precision Labs (APL) | Gordon P et al |
| EPI_ISL_805101 | Canada/AB-37460/2020 | 2020-04-22 | North America/Canada/Alberta/ | Alberta Precision Labs (APL) | Alberta Precision Labs (APL) | Gordon P et al |
| EPI_ISL_805102 | Canada/AB-73358/2020 | 2020-04-24 | North America/Canada/Alberta/ | Alberta Precision Labs (APL) | Alberta Precision Labs (APL) | Gordon P et al |
| EPI_ISL_805103 | Canada/AB-45074/2020 | 2020-04-21 | North America/Canada/Alberta/ | Alberta Precision Labs (APL) | Alberta Precision Labs (APL) | Gordon P et al |
| EPI_ISL_805104 | Canada/AB-90761/2020 | 2020-04-22 | North America/Canada/Alberta/ | Alberta Precision Labs (APL) | Alberta Precision Labs (APL) | Gordon P et al |
| EPI_ISL_805105 | Canada/AB-34921/2020 | 2020-04-23 | North America/Canada/Alberta/ | Alberta Precision Labs (APL) | Alberta Precision Labs (APL) | Gordon P et al |
| EPI_ISL_805106 | Canada/AB-87187/2020 | 2020-04-21 | North America/Canada/Alberta/ | Alberta Precision Labs (APL) | Alberta Precision Labs (APL) | Gordon P et al |
| EPI_ISL_805107 | Canada/AB-43215/2020 | 2020-04-23 | North America/Canada/Alberta/ | Alberta Precision Labs (APL) | Alberta Precision Labs (APL) | Gordon P et al |
| EPI_ISL_805108 | Canada/AB-59344/2020 | 2020-04-21 | North America/Canada/Alberta/ | Alberta Precision Labs (APL) | Alberta Precision Labs (APL) | Gordon P et al |
| EPI_ISL_805109 | Canada/AB-39768/2020 | 2020-04-20 | North America/Canada/Alberta/ | Alberta Precision Labs (APL) | Alberta Precision Labs (APL) | Gordon P et al |
| EPI_ISL_805110 | Canada/AB-21630/2020 | 2020-04-22 | North America/Canada/Alberta/ | Alberta Precision Labs (APL) | Alberta Precision Labs (APL) | Gordon P et al |
| EPI_ISL_805111 | Canada/AB-63677/2020 | 2020-04-21 | North America/Canada/Alberta/ | Alberta Precision Labs (APL) | Alberta Precision Labs (APL) | Gordon P et al |
| EPI_ISL_805112 | Canada/AB-82084/2020 | 2020-04-22 | North America/Canada/Alberta/ | Alberta Precision Labs (APL) | Alberta Precision Labs (APL) | Gordon P et al |
| EPI_ISL_805113 | Canada/AB-64444/2020 | 2020-04-21 | North America/Canada/Alberta/ | Alberta Precision Labs (APL) | Alberta Precision Labs (APL) | Gordon P et al |
| EPI_ISL_805114 | Canada/AB-26830/2020 | 2020-04-23 | North America/Canada/Alberta/ | Alberta Precision Labs (APL) | Alberta Precision Labs (APL) | Gordon P et al |
| EPI_ISL_805115 | Canada/AB-54614/2020 | 2020-04-21 | North America/Canada/Alberta/ | Alberta Precision Labs (APL) | Alberta Precision Labs (APL) | Gordon P et al |
| EPI_ISL_805116 | Canada/AB-83357/2020 | 2020-04-20 | North America/Canada/Alberta/ | Alberta Precision Labs (APL) | Alberta Precision Labs (APL) | Gordon P et al |
| EPI_ISL_805117 | Canada/AB-66049/2020 | 2020-04-22 | North America/Canada/Alberta/ | Alberta Precision Labs (APL) | Alberta Precision Labs (APL) | Gordon P et al |
| EPI_ISL_805118 | Canada/AB-76289/2020 | 2020-04-22 | North America/Canada/Alberta/ | Alberta Precision Labs (APL) | Alberta Precision Labs (APL) | Gordon P et al |

| | | | | | | |
|---|---|---|---|---|---|---|
| EPI_ISL_805119 | Canada/AB-47910/2020 | 2020-04-22 | North America/Canada/Alberta/ | Alberta Precision Labs (APL) | Alberta Precision Labs (APL) | Gordon P et al |
| EPI_ISL_805120 | Canada/AB-74273/2020 | 2020-04-20 | North America/Canada/Alberta/ | Alberta Precision Labs (APL) | Alberta Precision Labs (APL) | Gordon P et al |
| EPI_ISL_805121 | Canada/AB-23470/2020 | 2020-04-21 | North America/Canada/Alberta/ | Alberta Precision Labs (APL) | Alberta Precision Labs (APL) | Gordon P et al |
| EPI_ISL_805122 | Canada/AB-32651/2020 | 2020-05-10 | North America/Canada/Alberta/ | Alberta Precision Labs (APL) | Alberta Precision Labs (APL) | Gordon P et al |
| EPI_ISL_805123 | Canada/AB-64909/2020 | 2020-04-20 | North America/Canada/Alberta/ | Alberta Precision Labs (APL) | Alberta Precision Labs (APL) | Gordon P et al |
| EPI_ISL_805124 | Canada/AB-41610/2020 | 2020-04-21 | North America/Canada/Alberta/ | Alberta Precision Labs (APL) | Alberta Precision Labs (APL) | Gordon P et al |
| EPI_ISL_805125 | Canada/AB-43434/2020 | 2020-04-21 | North America/Canada/Alberta/ | Alberta Precision Labs (APL) | Alberta Precision Labs (APL) | Gordon P et al |
| EPI_ISL_805126 | Canada/AB-26811/2020 | 2020-04-22 | North America/Canada/Alberta/ | Alberta Precision Labs (APL) | Alberta Precision Labs (APL) | Gordon P et al |
| EPI_ISL_805127 | Canada/AB-83230/2020 | 2020-04-14 | North America/Canada/Alberta/ | Alberta Precision Labs (APL) | Alberta Precision Labs (APL) | Gordon P et al |
| EPI_ISL_805128 | Canada/AB-26421/2020 | 2020-04-20 | North America/Canada/Alberta/ | Alberta Precision Labs (APL) | Alberta Precision Labs (APL) | Gordon P et al |
| EPI_ISL_805129 | Canada/AB-44391/2020 | 2020-04-22 | North America/Canada/Alberta/ | Alberta Precision Labs (APL) | Alberta Precision Labs (APL) | Gordon P et al |
| EPI_ISL_805130 | Canada/AB-84204/2020 | 2020-04-22 | North America/Canada/Alberta/ | Alberta Precision Labs (APL) | Alberta Precision Labs (APL) | Gordon P et al |
| EPI_ISL_805131 | Canada/AB-42498/2020 | 2020-04-22 | North America/Canada/Alberta/ | Alberta Precision Labs (APL) | Alberta Precision Labs (APL) | Gordon P et al |
| EPI_ISL_805132 | Canada/AB-21899/2020 | 2020-04-21 | North America/Canada/Alberta/ | Alberta Precision Labs (APL) | Alberta Precision Labs (APL) | Gordon P et al |
| EPI_ISL_805133 | Canada/AB-46652/2020 | 2020-04-16 | North America/Canada/Alberta/ | Alberta Precision Labs (APL) | Alberta Precision Labs (APL) | Gordon P et al |
| EPI_ISL_805134 | Canada/AB-23884/2020 | 2020-04-21 | North America/Canada/Alberta/ | Alberta Precision Labs (APL) | Alberta Precision Labs (APL) | Gordon P et al |
| EPI_ISL_805135 | Canada/AB-16689/2020 | 2020-04-20 | North America/Canada/Alberta/ | Alberta Precision Labs (APL) | Alberta Precision Labs (APL) | Gordon P et al |
| EPI_ISL_805136 | Canada/AB-69751/2020 | 2020-04-20 | North America/Canada/Alberta/ | Alberta Precision Labs (APL) | Alberta Precision Labs (APL) | Gordon P et al |
| EPI_ISL_805137 | Canada/AB-89174/2020 | 2020-04-22 | North America/Canada/Alberta/ | Alberta Precision Labs (APL) | Alberta Precision Labs (APL) | Gordon P et al |
| EPI_ISL_805138 | Canada/AB-56268/2020 | 2020-04-21 | North America/Canada/Alberta/ | Alberta Precision Labs (APL) | Alberta Precision Labs (APL) | Gordon P et al |

| | | | | | | |
|---|---|---|---|---|---|---|
| EPI_ISL_805139 | Canada/AB-74320/2020 | 2020-04-22 | North America/Canada/Alberta/ | Alberta Precision Labs (APL) | Alberta Precision Labs (APL) | Gordon P et al |
| EPI_ISL_805140 | Canada/AB-80831/2020 | 2020-04-15 | North America/Canada/Alberta/ | Alberta Precision Labs (APL) | Alberta Precision Labs (APL) | Gordon P et al |
| EPI_ISL_805141 | Canada/AB-39598/2020 | 2020-04-20 | North America/Canada/Alberta/ | Alberta Precision Labs (APL) | Alberta Precision Labs (APL) | Gordon P et al |
| EPI_ISL_805142 | Canada/AB-30634/2020 | 2020-04-20 | North America/Canada/Alberta/ | Alberta Precision Labs (APL) | Alberta Precision Labs (APL) | Gordon P et al |
| EPI_ISL_805143 | Canada/AB-22032/2020 | 2020-04-21 | North America/Canada/Alberta/ | Alberta Precision Labs (APL) | Alberta Precision Labs (APL) | Gordon P et al |
| EPI_ISL_805144 | Canada/AB-52042/2020 | 2020-04-21 | North America/Canada/Alberta/ | Alberta Precision Labs (APL) | Alberta Precision Labs (APL) | Gordon P et al |
| EPI_ISL_805145 | Canada/AB-57740/2020 | 2020-04-23 | North America/Canada/Alberta/ | Alberta Precision Labs (APL) | Alberta Precision Labs (APL) | Gordon P et al |
| EPI_ISL_805146 | Canada/AB-34831/2020 | 2020-04-22 | North America/Canada/Alberta/ | Alberta Precision Labs (APL) | Alberta Precision Labs (APL) | Gordon P et al |
| EPI_ISL_805147 | Canada/AB-35599/2020 | 2020-04-29 | North America/Canada/Alberta/ | Alberta Precision Labs (APL) | Alberta Precision Labs (APL) | Gordon P et al |
| EPI_ISL_805148 | Canada/AB-16716/2020 | 2020-04-21 | North America/Canada/Alberta/ | Alberta Precision Labs (APL) | Alberta Precision Labs (APL) | Gordon P et al |
| EPI_ISL_805149 | Canada/AB-51414/2020 | 2020-04-19 | North America/Canada/Alberta/ | Alberta Precision Labs (APL) | Alberta Precision Labs (APL) | Gordon P et al |
| EPI_ISL_805150 | Canada/AB-12593/2020 | 2020-04-22 | North America/Canada/Alberta/ | Alberta Precision Labs (APL) | Alberta Precision Labs (APL) | Gordon P et al |
| EPI_ISL_805151 | Canada/AB-18449/2020 | 2020-04-22 | North America/Canada/Alberta/ | Alberta Precision Labs (APL) | Alberta Precision Labs (APL) | Gordon P et al |
| EPI_ISL_805152 | Canada/AB-19653/2020 | 2020-04-20 | North America/Canada/Alberta/ | Alberta Precision Labs (APL) | Alberta Precision Labs (APL) | Gordon P et al |
| EPI_ISL_805153 | Canada/AB-89343/2020 | 2020-04-20 | North America/Canada/Alberta/ | Alberta Precision Labs (APL) | Alberta Precision Labs (APL) | Gordon P et al |
| EPI_ISL_805154 | Canada/AB-91318/2020 | 2020-04-21 | North America/Canada/Alberta/ | Alberta Precision Labs (APL) | Alberta Precision Labs (APL) | Gordon P et al |
| EPI_ISL_805155 | Canada/AB-81168/2020 | 2020-04-23 | North America/Canada/Alberta/ | Alberta Precision Labs (APL) | Alberta Precision Labs (APL) | Gordon P et al |
| EPI_ISL_805156 | Canada/AB-90849/2020 | 2020-04-22 | North America/Canada/Alberta/ | Alberta Precision Labs (APL) | Alberta Precision Labs (APL) | Gordon P et al |
| EPI_ISL_805157 | Canada/AB-22658/2020 | 2020-04-20 | North America/Canada/Alberta/ | Alberta Precision Labs (APL) | Alberta Precision Labs (APL) | Gordon P et al |
| EPI_ISL_805158 | Canada/AB-42444/2020 | 2020-04-21 | North America/Canada/Alberta/ | Alberta Precision Labs (APL) | Alberta Precision Labs (APL) | Gordon P et al |

| | | | | | | |
|---|---|---|---|---|---|---|
| EPI_ISL_805159 | Canada/AB-77846/2020 | 2020-04-20 | North America/Canada/Alberta/ | Alberta Precision Labs (APL) | Alberta Precision Labs (APL) | Gordon P et al |
| EPI_ISL_805160 | Canada/AB-72290/2020 | 2020-04-20 | North America/Canada/Alberta/ | Alberta Precision Labs (APL) | Alberta Precision Labs (APL) | Gordon P et al |
| EPI_ISL_805161 | Canada/AB-45703/2020 | 2020-04-22 | North America/Canada/Alberta/ | Alberta Precision Labs (APL) | Alberta Precision Labs (APL) | Gordon P et al |
| EPI_ISL_805162 | Canada/AB-96540/2020 | 2020-04-22 | North America/Canada/Alberta/ | Alberta Precision Labs (APL) | Alberta Precision Labs (APL) | Gordon P et al |
| EPI_ISL_805163 | Canada/AB-52224/2020 | 2020-04-20 | North America/Canada/Alberta/ | Alberta Precision Labs (APL) | Alberta Precision Labs (APL) | Gordon P et al |
| EPI_ISL_805164 | Canada/AB-83944/2020 | 2020-04-21 | North America/Canada/Alberta/ | Alberta Precision Labs (APL) | Alberta Precision Labs (APL) | Gordon P et al |
| EPI_ISL_805165 | Canada/AB-30957/2020 | 2020-04-22 | North America/Canada/Alberta/ | Alberta Precision Labs (APL) | Alberta Precision Labs (APL) | Gordon P et al |
| EPI_ISL_805166 | Canada/AB-24314/2020 | 2020-04-15 | North America/Canada/Alberta/ | Alberta Precision Labs (APL) | Alberta Precision Labs (APL) | Gordon P et al |
| EPI_ISL_805167 | Canada/AB-61852/2020 | 2020-04-21 | North America/Canada/Alberta/ | Alberta Precision Labs (APL) | Alberta Precision Labs (APL) | Gordon P et al |
| EPI_ISL_805168 | Canada/AB-24185/2020 | 2020-04-23 | North America/Canada/Alberta/ | Alberta Precision Labs (APL) | Alberta Precision Labs (APL) | Gordon P et al |
| EPI_ISL_805169 | Canada/AB-48557/2020 | 2020-04-20 | North America/Canada/Alberta/ | Alberta Precision Labs (APL) | Alberta Precision Labs (APL) | Gordon P et al |
| EPI_ISL_805170 | Canada/AB-51538/2020 | 2020-04-21 | North America/Canada/Alberta/ | Alberta Precision Labs (APL) | Alberta Precision Labs (APL) | Gordon P et al |
| EPI_ISL_805171 | Canada/AB-38881/2020 | 2020-04-21 | North America/Canada/Alberta/ | Alberta Precision Labs (APL) | Alberta Precision Labs (APL) | Gordon P et al |
| EPI_ISL_805172 | Canada/AB-54528/2020 | 2020-04-22 | North America/Canada/Alberta/ | Alberta Precision Labs (APL) | Alberta Precision Labs (APL) | Gordon P et al |
| EPI_ISL_805173 | Canada/AB-93420/2020 | 2020-04-22 | North America/Canada/Alberta/ | Alberta Precision Labs (APL) | Alberta Precision Labs (APL) | Gordon P et al |
| EPI_ISL_805174 | Canada/AB-63084/2020 | 2020-04-21 | North America/Canada/Alberta/ | Alberta Precision Labs (APL) | Alberta Precision Labs (APL) | Gordon P et al |
| EPI_ISL_805175 | Canada/AB-77163/2020 | 2020-04-21 | North America/Canada/Alberta/ | Alberta Precision Labs (APL) | Alberta Precision Labs (APL) | Gordon P et al |
| EPI_ISL_805176 | Canada/AB-93664/2020 | 2020-04-20 | North America/Canada/Alberta/ | Alberta Precision Labs (APL) | Alberta Precision Labs (APL) | Gordon P et al |
| EPI_ISL_805177 | Canada/AB-78907/2020 | 2020-04-21 | North America/Canada/Alberta/ | Alberta Precision Labs (APL) | Alberta Precision Labs (APL) | Gordon P et al |
| EPI_ISL_805178 | Canada/AB-35479/2020 | 2020-04-21 | North America/Canada/Alberta/ | Alberta Precision Labs (APL) | Alberta Precision Labs (APL) | Gordon P et al |

| | | | | | | |
|---|---|---|---|---|---|---|
| EPI_ISL_805179 | Canada/AB-43929/2020 | 2020-04-20 | North America/Canada/Alberta/ | Alberta Precision Labs (APL) | Alberta Precision Labs (APL) | Gordon P et al |
| EPI_ISL_805180 | Canada/AB-32119/2020 | 2020-04-21 | North America/Canada/Alberta/ | Alberta Precision Labs (APL) | Alberta Precision Labs (APL) | Gordon P et al |
| EPI_ISL_805181 | Canada/AB-58717/2020 | 2020-04-17 | North America/Canada/Alberta/ | Alberta Precision Labs (APL) | Alberta Precision Labs (APL) | Gordon P et al |
| EPI_ISL_805182 | Canada/AB-71693/2020 | 2020-04-22 | North America/Canada/Alberta/ | Alberta Precision Labs (APL) | Alberta Precision Labs (APL) | Gordon P et al |
| EPI_ISL_805183 | Canada/AB-37707/2020 | 2020-04-23 | North America/Canada/Alberta/ | Alberta Precision Labs (APL) | Alberta Precision Labs (APL) | Gordon P et al |
| EPI_ISL_805184 | Canada/AB-23982/2020 | 2020-04-22 | North America/Canada/Alberta/ | Alberta Precision Labs (APL) | Alberta Precision Labs (APL) | Gordon P et al |
| EPI_ISL_805185 | Canada/AB-55335/2020 | 2020-04-19 | North America/Canada/Alberta/ | Alberta Precision Labs (APL) | Alberta Precision Labs (APL) | Gordon P et al |
| EPI_ISL_805186 | Canada/AB-41100/2020 | 2020-04-19 | North America/Canada/Alberta/ | Alberta Precision Labs (APL) | Alberta Precision Labs (APL) | Gordon P et al |
| EPI_ISL_805187 | Canada/AB-72876/2020 | 2020-04-20 | North America/Canada/Alberta/ | Alberta Precision Labs (APL) | Alberta Precision Labs (APL) | Gordon P et al |
| EPI_ISL_805188 | Canada/AB-86594/2020 | 2020-04-19 | North America/Canada/Alberta/ | Alberta Precision Labs (APL) | Alberta Precision Labs (APL) | Gordon P et al |
| EPI_ISL_805189 | Canada/AB-13653/2020 | 2020-04-21 | North America/Canada/Alberta/ | Alberta Precision Labs (APL) | Alberta Precision Labs (APL) | Gordon P et al |
| EPI_ISL_805190 | Canada/AB-79000/2020 | 2020-04-24 | North America/Canada/Alberta/ | Alberta Precision Labs (APL) | Alberta Precision Labs (APL) | Gordon P et al |
| EPI_ISL_805191 | Canada/AB-34300/2020 | 2020-04-17 | North America/Canada/Alberta/ | Alberta Precision Labs (APL) | Alberta Precision Labs (APL) | Gordon P et al |
| EPI_ISL_805192 | Canada/AB-92623/2020 | 2020-04-20 | North America/Canada/Alberta/ | Alberta Precision Labs (APL) | Alberta Precision Labs (APL) | Gordon P et al |
| EPI_ISL_805193 | Canada/AB-83587/2020 | 2020-04-19 | North America/Canada/Alberta/ | Alberta Precision Labs (APL) | Alberta Precision Labs (APL) | Gordon P et al |
| EPI_ISL_805194 | Canada/AB-65781/2020 | 2020-04-21 | North America/Canada/Alberta/ | Alberta Precision Labs (APL) | Alberta Precision Labs (APL) | Gordon P et al |
| EPI_ISL_805195 | Canada/AB-16872/2020 | 2020-04-21 | North America/Canada/Alberta/ | Alberta Precision Labs (APL) | Alberta Precision Labs (APL) | Gordon P et al |
| EPI_ISL_805196 | Canada/AB-40834/2020 | 2020-04-22 | North America/Canada/Alberta/ | Alberta Precision Labs (APL) | Alberta Precision Labs (APL) | Gordon P et al |
| EPI_ISL_805197 | Canada/AB-30632/2020 | 2020-04-20 | North America/Canada/Alberta/ | Alberta Precision Labs (APL) | Alberta Precision Labs (APL) | Gordon P et al |
| EPI_ISL_805198 | Canada/AB-18464/2020 | 2020-04-20 | North America/Canada/Alberta/ | Alberta Precision Labs (APL) | Alberta Precision Labs (APL) | Gordon P et al |

| | | | | | | |
|---|---|---|---|---|---|---|
| EPI_ISL_805199 | Canada/AB-47016/2020 | 2020-04-22 | North America/Canada/Alberta/ | Alberta Precision Labs (APL) | Alberta Precision Labs (APL) | Gordon P et al |
| EPI_ISL_805201 | Canada/AB-33658/2020 | 2020-04-21 | North America/Canada/Alberta/ | Alberta Precision Labs (APL) | Alberta Precision Labs (APL) | Gordon P et al |
| EPI_ISL_805202 | Canada/AB-43039/2020 | 2020-04-16 | North America/Canada/Alberta/ | Alberta Precision Labs (APL) | Alberta Precision Labs (APL) | Gordon P et al |
| EPI_ISL_805203 | Canada/AB-33911/2020 | 2020-04-22 | North America/Canada/Alberta/ | Alberta Precision Labs (APL) | Alberta Precision Labs (APL) | Gordon P et al |
| EPI_ISL_805204 | Canada/AB-80558/2020 | 2020-04-27 | North America/Canada/Alberta/ | Alberta Precision Labs (APL) | Alberta Precision Labs (APL) | Gordon P et al |
| EPI_ISL_805205 | Canada/AB-41196/2020 | 2020-04-20 | North America/Canada/Alberta/ | Alberta Precision Labs (APL) | Alberta Precision Labs (APL) | Gordon P et al |
| EPI_ISL_805206 | Canada/AB-77624/2020 | 2020-04-20 | North America/Canada/Alberta/ | Alberta Precision Labs (APL) | Alberta Precision Labs (APL) | Gordon P et al |
| EPI_ISL_805207 | Canada/AB-76918/2020 | 2020-04-21 | North America/Canada/Alberta/ | Alberta Precision Labs (APL) | Alberta Precision Labs (APL) | Gordon P et al |
| EPI_ISL_805208 | Canada/AB-17775/2020 | 2020-04-19 | North America/Canada/Alberta/ | Alberta Precision Labs (APL) | Alberta Precision Labs (APL) | Gordon P et al |
| EPI_ISL_805209 | Canada/AB-12489/2020 | 2020-04-20 | North America/Canada/Alberta/ | Alberta Precision Labs (APL) | Alberta Precision Labs (APL) | Gordon P et al |
| EPI_ISL_805210 | Canada/AB-31566/2020 | 2020-04-21 | North America/Canada/Alberta/ | Alberta Precision Labs (APL) | Alberta Precision Labs (APL) | Gordon P et al |
| EPI_ISL_805211 | Canada/AB-22148/2020 | 2020-04-20 | North America/Canada/Alberta/ | Alberta Precision Labs (APL) | Alberta Precision Labs (APL) | Gordon P et al |
| EPI_ISL_805212 | Canada/AB-97419/2020 | 2020-04-21 | North America/Canada/Alberta/ | Alberta Precision Labs (APL) | Alberta Precision Labs (APL) | Gordon P et al |
| EPI_ISL_805213 | Canada/AB-50997/2020 | 2020-04-20 | North America/Canada/Alberta/ | Alberta Precision Labs (APL) | Alberta Precision Labs (APL) | Gordon P et al |
| EPI_ISL_805214 | Canada/AB-89902/2020 | 2020-04-17 | North America/Canada/Alberta/ | Alberta Precision Labs (APL) | Alberta Precision Labs (APL) | Gordon P et al |
| EPI_ISL_805215 | Canada/AB-64615/2020 | 2020-04-16 | North America/Canada/Alberta/ | Alberta Precision Labs (APL) | Alberta Precision Labs (APL) | Gordon P et al |
| EPI_ISL_805216 | Canada/AB-55228/2020 | 2020-04-22 | North America/Canada/Alberta/ | Alberta Precision Labs (APL) | Alberta Precision Labs (APL) | Gordon P et al |
| EPI_ISL_805217 | Canada/AB-98537/2020 | 2020-04-20 | North America/Canada/Alberta/ | Alberta Precision Labs (APL) | Alberta Precision Labs (APL) | Gordon P et al |
| EPI_ISL_805218 | Canada/AB-32701/2020 | 2020-04-21 | North America/Canada/Alberta/ | Alberta Precision Labs (APL) | Alberta Precision Labs (APL) | Gordon P et al |
| EPI_ISL_805219 | Canada/AB-41343/2020 | 2020-04-22 | North America/Canada/Alberta/ | Alberta Precision Labs (APL) | Alberta Precision Labs (APL) | Gordon P et al |

| | | | | | | |
|---|---|---|---|---|---|---|
| EPI_ISL_805220 | Canada/AB-69379/2020 | 2020-04-17 | North America/Canada/Alberta/ | Alberta Precision Labs (APL) | Alberta Precision Labs (APL) | Gordon P et al |
| EPI_ISL_805221 | Canada/AB-17159/2020 | 2020-04-21 | North America/Canada/Alberta/ | Alberta Precision Labs (APL) | Alberta Precision Labs (APL) | Gordon P et al |
| EPI_ISL_805222 | Canada/AB-97816/2020 | 2020-04-21 | North America/Canada/Alberta/ | Alberta Precision Labs (APL) | Alberta Precision Labs (APL) | Gordon P et al |
| EPI_ISL_805223 | Canada/AB-71880/2020 | 2020-04-22 | North America/Canada/Alberta/ | Alberta Precision Labs (APL) | Alberta Precision Labs (APL) | Gordon P et al |
| EPI_ISL_805224 | Canada/AB-50693/2020 | 2020-04-20 | North America/Canada/Alberta/ | Alberta Precision Labs (APL) | Alberta Precision Labs (APL) | Gordon P et al |
| EPI_ISL_805225 | Canada/AB-97776/2020 | 2020-04-15 | North America/Canada/Alberta/ | Alberta Precision Labs (APL) | Alberta Precision Labs (APL) | Gordon P et al |
| EPI_ISL_805226 | Canada/AB-96788/2020 | 2020-04-15 | North America/Canada/Alberta/ | Alberta Precision Labs (APL) | Alberta Precision Labs (APL) | Gordon P et al |
| EPI_ISL_805227 | Canada/AB-41461/2020 | 2020-04-20 | North America/Canada/Alberta/ | Alberta Precision Labs (APL) | Alberta Precision Labs (APL) | Gordon P et al |
| EPI_ISL_805229 | Canada/AB-49192/2020 | 2020-04-15 | North America/Canada/Alberta/ | Alberta Precision Labs (APL) | Alberta Precision Labs (APL) | Gordon P et al |
| EPI_ISL_805230 | Canada/AB-20298/2020 | 2020-04-20 | North America/Canada/Alberta/ | Alberta Precision Labs (APL) | Alberta Precision Labs (APL) | Gordon P et al |
| EPI_ISL_805231 | Canada/AB-54413/2020 | 2020-04-17 | North America/Canada/Alberta/ | Alberta Precision Labs (APL) | Alberta Precision Labs (APL) | Gordon P et al |
| EPI_ISL_805232 | Canada/AB-21447/2020 | 2020-04-22 | North America/Canada/Alberta/ | Alberta Precision Labs (APL) | Alberta Precision Labs (APL) | Gordon P et al |
| EPI_ISL_805233 | Canada/AB-85103/2020 | 2020-04-20 | North America/Canada/Alberta/ | Alberta Precision Labs (APL) | Alberta Precision Labs (APL) | Gordon P et al |
| EPI_ISL_805234 | Canada/AB-62139/2020 | 2020-04-16 | North America/Canada/Alberta/ | Alberta Precision Labs (APL) | Alberta Precision Labs (APL) | Gordon P et al |
| EPI_ISL_805235 | Canada/AB-60508/2020 | 2020-04-17 | North America/Canada/Alberta/ | Alberta Precision Labs (APL) | Alberta Precision Labs (APL) | Gordon P et al |
| EPI_ISL_805236 | Canada/AB-27678/2020 | 2020-04-20 | North America/Canada/Alberta/ | Alberta Precision Labs (APL) | Alberta Precision Labs (APL) | Gordon P et al |
| EPI_ISL_805237 | Canada/AB-90556/2020 | 2020-04-20 | North America/Canada/Alberta/ | Alberta Precision Labs (APL) | Alberta Precision Labs (APL) | Gordon P et al |
| EPI_ISL_805238 | Canada/AB-49864/2020 | 2020-04-17 | North America/Canada/Alberta/ | Alberta Precision Labs (APL) | Alberta Precision Labs (APL) | Gordon P et al |
| EPI_ISL_805239 | Canada/AB-80130/2020 | 2020-04-20 | North America/Canada/Alberta/ | Alberta Precision Labs (APL) | Alberta Precision Labs (APL) | Gordon P et al |
| EPI_ISL_805240 | Canada/AB-91718/2020 | 2020-04-16 | North America/Canada/Alberta/ | Alberta Precision Labs (APL) | Alberta Precision Labs (APL) | Gordon P et al |

| | | | | | | |
|---|---|---|---|---|---|---|
| EPI_ISL_805241 | Canada/AB-99419/2020 | 2020-04-15 | North America/Canada/Alberta/ | Alberta Precision Labs (APL) | Alberta Precision Labs (APL) | Gordon P et al |
| EPI_ISL_805242 | Canada/AB-26226/2020 | 2020-04-15 | North America/Canada/Alberta/ | Alberta Precision Labs (APL) | Alberta Precision Labs (APL) | Gordon P et al |
| EPI_ISL_805243 | Canada/AB-23819/2020 | 2020-04-20 | North America/Canada/Alberta/ | Alberta Precision Labs (APL) | Alberta Precision Labs (APL) | Gordon P et al |
| EPI_ISL_805244 | Canada/AB-56822/2020 | 2020-04-19 | North America/Canada/Alberta/ | Alberta Precision Labs (APL) | Alberta Precision Labs (APL) | Gordon P et al |
| EPI_ISL_805245 | Canada/AB-98115/2020 | 2020-04-20 | North America/Canada/Alberta/ | Alberta Precision Labs (APL) | Alberta Precision Labs (APL) | Gordon P et al |
| EPI_ISL_805246 | Canada/AB-92499/2020 | 2020-04-20 | North America/Canada/Alberta/ | Alberta Precision Labs (APL) | Alberta Precision Labs (APL) | Gordon P et al |
| EPI_ISL_805247 | Canada/AB-28040/2020 | 2020-04-16 | North America/Canada/Alberta/ | Alberta Precision Labs (APL) | Alberta Precision Labs (APL) | Gordon P et al |
| EPI_ISL_805248 | Canada/AB-44129/2020 | 2020-04-22 | North America/Canada/Alberta/ | Alberta Precision Labs (APL) | Alberta Precision Labs (APL) | Gordon P et al |
| EPI_ISL_805249 | Canada/AB-73670/2020 | 2020-04-17 | North America/Canada/Alberta/ | Alberta Precision Labs (APL) | Alberta Precision Labs (APL) | Gordon P et al |
| EPI_ISL_805250 | Canada/AB-35684/2020 | 2020-04-23 | North America/Canada/Alberta/ | Alberta Precision Labs (APL) | Alberta Precision Labs (APL) | Gordon P et al |
| EPI_ISL_805251 | Canada/AB-70539/2020 | 2020-04-21 | North America/Canada/Alberta/ | Alberta Precision Labs (APL) | Alberta Precision Labs (APL) | Gordon P et al |
| EPI_ISL_805252 | Canada/AB-54691/2020 | 2020-04-22 | North America/Canada/Alberta/ | Alberta Precision Labs (APL) | Alberta Precision Labs (APL) | Gordon P et al |
| EPI_ISL_805253 | Canada/AB-51750/2020 | 2020-04-16 | North America/Canada/Alberta/ | Alberta Precision Labs (APL) | Alberta Precision Labs (APL) | Gordon P et al |
| EPI_ISL_805254 | Canada/AB-56292/2020 | 2020-04-20 | North America/Canada/Alberta/ | Alberta Precision Labs (APL) | Alberta Precision Labs (APL) | Gordon P et al |
| EPI_ISL_805255 | Canada/AB-60454/2020 | 2020-04-20 | North America/Canada/Alberta/ | Alberta Precision Labs (APL) | Alberta Precision Labs (APL) | Gordon P et al |
| EPI_ISL_805256 | Canada/AB-19097/2020 | 2020-04-22 | North America/Canada/Alberta/ | Alberta Precision Labs (APL) | Alberta Precision Labs (APL) | Gordon P et al |
| EPI_ISL_805257 | Canada/AB-21946/2020 | 2020-04-15 | North America/Canada/Alberta/ | Alberta Precision Labs (APL) | Alberta Precision Labs (APL) | Gordon P et al |
| EPI_ISL_805258 | Canada/AB-12892/2020 | 2020-04-21 | North America/Canada/Alberta/ | Alberta Precision Labs (APL) | Alberta Precision Labs (APL) | Gordon P et al |
| EPI_ISL_805259 | Canada/AB-98505/2020 | 2020-05-03 | North America/Canada/Alberta/ | Alberta Precision Labs (APL) | Alberta Precision Labs (APL) | Gordon P et al |
| EPI_ISL_805260 | Canada/AB-33353/2020 | 2020-04-19 | North America/Canada/Alberta/ | Alberta Precision Labs (APL) | Alberta Precision Labs (APL) | Gordon P et al |

| | | | | | | |
|---|---|---|---|---|---|---|
| EPI_ISL_805261 | Canada/AB-39103/2020 | 2020-04-14 | North America/Canada/Alberta/ | Alberta Precision Labs (APL) | Alberta Precision Labs (APL) | Gordon P et al |
| EPI_ISL_805262 | Canada/AB-80372/2020 | 2020-04-17 | North America/Canada/Alberta/ | Alberta Precision Labs (APL) | Alberta Precision Labs (APL) | Gordon P et al |
| EPI_ISL_805263 | Canada/AB-20032/2020 | 2020-04-19 | North America/Canada/Alberta/ | Alberta Precision Labs (APL) | Alberta Precision Labs (APL) | Gordon P et al |
| EPI_ISL_805264 | Canada/AB-68463/2020 | 2020-04-17 | North America/Canada/Alberta/ | Alberta Precision Labs (APL) | Alberta Precision Labs (APL) | Gordon P et al |
| EPI_ISL_805265 | Canada/AB-74849/2020 | 2020-04-16 | North America/Canada/Alberta/ | Alberta Precision Labs (APL) | Alberta Precision Labs (APL) | Gordon P et al |
| EPI_ISL_805266 | Canada/AB-14576/2020 | 2020-04-23 | North America/Canada/Alberta/ | Alberta Precision Labs (APL) | Alberta Precision Labs (APL) | Gordon P et al |
| EPI_ISL_805267 | Canada/AB-25594/2020 | 2020-04-16 | North America/Canada/Alberta/ | Alberta Precision Labs (APL) | Alberta Precision Labs (APL) | Gordon P et al |
| EPI_ISL_805268 | Canada/AB-47311/2020 | 2020-04-16 | North America/Canada/Alberta/ | Alberta Precision Labs (APL) | Alberta Precision Labs (APL) | Gordon P et al |
| EPI_ISL_805269 | Canada/AB-48507/2020 | 2020-04-19 | North America/Canada/Alberta/ | Alberta Precision Labs (APL) | Alberta Precision Labs (APL) | Gordon P et al |
| EPI_ISL_805270 | Canada/AB-19822/2020 | 2020-04-14 | North America/Canada/Alberta/ | Alberta Precision Labs (APL) | Alberta Precision Labs (APL) | Gordon P et al |
| EPI_ISL_805271 | Canada/AB-30536/2020 | 2020-04-16 | North America/Canada/Alberta/ | Alberta Precision Labs (APL) | Alberta Precision Labs (APL) | Gordon P et al |
| EPI_ISL_805272 | Canada/AB-80893/2020 | 2020-04-15 | North America/Canada/Alberta/ | Alberta Precision Labs (APL) | Alberta Precision Labs (APL) | Gordon P et al |
| EPI_ISL_805273 | Canada/AB-56284/2020 | 2020-04-22 | North America/Canada/Alberta/ | Alberta Precision Labs (APL) | Alberta Precision Labs (APL) | Gordon P et al |
| EPI_ISL_805274 | Canada/AB-61799/2020 | 2020-04-02 | North America/Canada/Alberta/ | Alberta Precision Labs (APL) | Alberta Precision Labs (APL) | Gordon P et al |
| EPI_ISL_805275 | Canada/AB-15138/2020 | 2020-04-16 | North America/Canada/Alberta/ | Alberta Precision Labs (APL) | Alberta Precision Labs (APL) | Gordon P et al |
| EPI_ISL_805276 | Canada/AB-16322/2020 | 2020-04-16 | North America/Canada/Alberta/ | Alberta Precision Labs (APL) | Alberta Precision Labs (APL) | Gordon P et al |
| EPI_ISL_805277 | Canada/AB-27683/2020 | 2020-04-16 | North America/Canada/Alberta/ | Alberta Precision Labs (APL) | Alberta Precision Labs (APL) | Gordon P et al |
| EPI_ISL_805278 | Canada/AB-18384/2020 | 2020-04-29 | North America/Canada/Alberta/ | Alberta Precision Labs (APL) | Alberta Precision Labs (APL) | Gordon P et al |
| EPI_ISL_805279 | Canada/AB-92502/2020 | 2020-04-19 | North America/Canada/Alberta/ | Alberta Precision Labs (APL) | Alberta Precision Labs (APL) | Gordon P et al |
| EPI_ISL_805280 | Canada/AB-74391/2020 | 2020-04-17 | North America/Canada/Alberta/ | Alberta Precision Labs (APL) | Alberta Precision Labs (APL) | Gordon P et al |

| | | | | | | |
|---|---|---|---|---|---|---|
| EPI_ISL_805281 | Canada/AB-12523/2020 | 2020-04-22 | North America/Canada/Alberta/ | Alberta Precision Labs (APL) | Alberta Precision Labs (APL) | Gordon P et al |
| EPI_ISL_805282 | Canada/AB-72413/2020 | 2020-04-17 | North America/Canada/Alberta/ | Alberta Precision Labs (APL) | Alberta Precision Labs (APL) | Gordon P et al |
| EPI_ISL_805283 | Canada/AB-93974/2020 | 2020-05-10 | North America/Canada/Alberta/ | Alberta Precision Labs (APL) | Alberta Precision Labs (APL) | Gordon P et al |
| EPI_ISL_805284 | Canada/AB-55799/2020 | 2020-04-16 | North America/Canada/Alberta/ | Alberta Precision Labs (APL) | Alberta Precision Labs (APL) | Gordon P et al |
| EPI_ISL_805285 | Canada/AB-43439/2020 | 2020-04-14 | North America/Canada/Alberta/ | Alberta Precision Labs (APL) | Alberta Precision Labs (APL) | Gordon P et al |
| EPI_ISL_805286 | Canada/AB-36611/2020 | 2020-04-16 | North America/Canada/Alberta/ | Alberta Precision Labs (APL) | Alberta Precision Labs (APL) | Gordon P et al |
| EPI_ISL_805287 | Canada/AB-45880/2020 | 2020-04-18 | North America/Canada/Alberta/ | Alberta Precision Labs (APL) | Alberta Precision Labs (APL) | Gordon P et al |
| EPI_ISL_805288 | Canada/AB-42294/2020 | 2020-04-16 | North America/Canada/Alberta/ | Alberta Precision Labs (APL) | Alberta Precision Labs (APL) | Gordon P et al |
| EPI_ISL_805289 | Canada/AB-96380/2020 | 2020-04-19 | North America/Canada/Alberta/ | Alberta Precision Labs (APL) | Alberta Precision Labs (APL) | Gordon P et al |
| EPI_ISL_805290 | Canada/AB-77505/2020 | 2020-04-17 | North America/Canada/Alberta/ | Alberta Precision Labs (APL) | Alberta Precision Labs (APL) | Gordon P et al |
| EPI_ISL_805291 | Canada/AB-45987/2020 | 2020-04-16 | North America/Canada/Alberta/ | Alberta Precision Labs (APL) | Alberta Precision Labs (APL) | Gordon P et al |
| EPI_ISL_805292 | Canada/AB-84378/2020 | 2020-04-29 | North America/Canada/Alberta/ | Alberta Precision Labs (APL) | Alberta Precision Labs (APL) | Gordon P et al |
| EPI_ISL_805293 | Canada/AB-68403/2020 | 2020-04-17 | North America/Canada/Alberta/ | Alberta Precision Labs (APL) | Alberta Precision Labs (APL) | Gordon P et al |
| EPI_ISL_805294 | Canada/AB-17559/2020 | 2020-04-16 | North America/Canada/Alberta/ | Alberta Precision Labs (APL) | Alberta Precision Labs (APL) | Gordon P et al |
| EPI_ISL_805295 | Canada/AB-90250/2020 | 2020-04-16 | North America/Canada/Alberta/ | Alberta Precision Labs (APL) | Alberta Precision Labs (APL) | Gordon P et al |
| EPI_ISL_805296 | Canada/AB-36025/2020 | 2020-05-09 | North America/Canada/Alberta/ | Alberta Precision Labs (APL) | Alberta Precision Labs (APL) | Gordon P et al |
| EPI_ISL_805297 | Canada/AB-73341/2020 | 2020-04-19 | North America/Canada/Alberta/ | Alberta Precision Labs (APL) | Alberta Precision Labs (APL) | Gordon P et al |
| EPI_ISL_805298 | Canada/AB-35819/2020 | 2020-04-18 | North America/Canada/Alberta/ | Alberta Precision Labs (APL) | Alberta Precision Labs (APL) | Gordon P et al |
| EPI_ISL_805299 | Canada/AB-32472/2020 | 2020-04-18 | North America/Canada/Alberta/ | Alberta Precision Labs (APL) | Alberta Precision Labs (APL) | Gordon P et al |
| EPI_ISL_805300 | Canada/AB-78258/2020 | 2020-04-19 | North America/Canada/Alberta/ | Alberta Precision Labs (APL) | Alberta Precision Labs (APL) | Gordon P et al |

| | | | | | | |
|---|---|---|---|---|---|---|
| EPI_ISL_805301 | Canada/AB-15073/2020 | 2020-04-22 | North America/Canada/Alberta/ | Alberta Precision Labs (APL) | Alberta Precision Labs (APL) | Gordon P et al |
| EPI_ISL_805302 | Canada/AB-89020/2020 | 2020-04-19 | North America/Canada/Alberta/ | Alberta Precision Labs (APL) | Alberta Precision Labs (APL) | Gordon P et al |
| EPI_ISL_805303 | Canada/AB-51147/2020 | 2020-04-20 | North America/Canada/Alberta/ | Alberta Precision Labs (APL) | Alberta Precision Labs (APL) | Gordon P et al |
| EPI_ISL_805304 | Canada/AB-46437/2020 | 2020-04-16 | North America/Canada/Alberta/ | Alberta Precision Labs (APL) | Alberta Precision Labs (APL) | Gordon P et al |
| EPI_ISL_805305 | Canada/AB-15951/2020 | 2020-04-17 | North America/Canada/Alberta/ | Alberta Precision Labs (APL) | Alberta Precision Labs (APL) | Gordon P et al |
| EPI_ISL_805306 | Canada/AB-78310/2020 | 2020-04-11 | North America/Canada/Alberta/ | Alberta Precision Labs (APL) | Alberta Precision Labs (APL) | Gordon P et al |
| EPI_ISL_805308 | Canada/AB-37518/2020 | 2020-04-17 | North America/Canada/Alberta/ | Alberta Precision Labs (APL) | Alberta Precision Labs (APL) | Gordon P et al |
| EPI_ISL_805309 | Canada/AB-62093/2020 | 2020-04-14 | North America/Canada/Alberta/ | Alberta Precision Labs (APL) | Alberta Precision Labs (APL) | Gordon P et al |
| EPI_ISL_805310 | Canada/AB-45977/2020 | 2020-05-11 | North America/Canada/Alberta/ | Alberta Precision Labs (APL) | Alberta Precision Labs (APL) | Gordon P et al |
| EPI_ISL_805311 | Canada/AB-58089/2020 | 2020-04-20 | North America/Canada/Alberta/ | Alberta Precision Labs (APL) | Alberta Precision Labs (APL) | Gordon P et al |
| EPI_ISL_805312 | Canada/AB-23741/2020 | 2020-05-11 | North America/Canada/Alberta/ | Alberta Precision Labs (APL) | Alberta Precision Labs (APL) | Gordon P et al |
| EPI_ISL_805313 | Canada/AB-62939/2020 | 2020-04-17 | North America/Canada/Alberta/ | Alberta Precision Labs (APL) | Alberta Precision Labs (APL) | Gordon P et al |
| EPI_ISL_805314 | Canada/AB-31525/2020 | 2020-04-17 | North America/Canada/Alberta/ | Alberta Precision Labs (APL) | Alberta Precision Labs (APL) | Gordon P et al |
| EPI_ISL_805315 | Canada/AB-40458/2020 | 2020-04-25 | North America/Canada/Alberta/ | Alberta Precision Labs (APL) | Alberta Precision Labs (APL) | Gordon P et al |
| EPI_ISL_805316 | Canada/AB-60580/2020 | 2020-04-28 | North America/Canada/Alberta/ | Alberta Precision Labs (APL) | Alberta Precision Labs (APL) | Gordon P et al |
| EPI_ISL_805317 | Canada/AB-86631/2020 | 2020-04-17 | North America/Canada/Alberta/ | Alberta Precision Labs (APL) | Alberta Precision Labs (APL) | Gordon P et al |
| EPI_ISL_805318 | Canada/AB-73783/2020 | 2020-04-16 | North America/Canada/Alberta/ | Alberta Precision Labs (APL) | Alberta Precision Labs (APL) | Gordon P et al |
| EPI_ISL_805319 | Canada/AB-65936/2020 | 2020-04-16 | North America/Canada/Alberta/ | Alberta Precision Labs (APL) | Alberta Precision Labs (APL) | Gordon P et al |
| EPI_ISL_805320 | Canada/AB-87949/2020 | 2020-04-17 | North America/Canada/Alberta/ | Alberta Precision Labs (APL) | Alberta Precision Labs (APL) | Gordon P et al |
| EPI_ISL_805321 | Canada/AB-50214/2020 | 2020-04-19 | North America/Canada/Alberta/ | Alberta Precision Labs (APL) | Alberta Precision Labs (APL) | Gordon P et al |

| | | | | | | |
|---|---|---|---|---|---|---|
| EPI_ISL_805322 | Canada/AB-86666/2020 | 2020-04-19 | North America/Canada/Alberta/ | Alberta Precision Labs (APL) | Alberta Precision Labs (APL) | Gordon P et al |
| EPI_ISL_805323 | Canada/AB-17961/2020 | 2020-04-19 | North America/Canada/Alberta/ | Alberta Precision Labs (APL) | Alberta Precision Labs (APL) | Gordon P et al |
| EPI_ISL_805324 | Canada/AB-62684/2020 | 2020-04-27 | North America/Canada/Alberta/ | Alberta Precision Labs (APL) | Alberta Precision Labs (APL) | Gordon P et al |
| EPI_ISL_805325 | Canada/AB-36828/2020 | 2020-04-20 | North America/Canada/Alberta/ | Alberta Precision Labs (APL) | Alberta Precision Labs (APL) | Gordon P et al |
| EPI_ISL_805326 | Canada/AB-63268/2020 | 2020-04-18 | North America/Canada/Alberta/ | Alberta Precision Labs (APL) | Alberta Precision Labs (APL) | Gordon P et al |
| EPI_ISL_805327 | Canada/AB-30326/2020 | 2020-04-18 | North America/Canada/Alberta/ | Alberta Precision Labs (APL) | Alberta Precision Labs (APL) | Gordon P et al |
| EPI_ISL_805328 | Canada/AB-62446/2020 | 2020-04-15 | North America/Canada/Alberta/ | Alberta Precision Labs (APL) | Alberta Precision Labs (APL) | Gordon P et al |
| EPI_ISL_805329 | Canada/AB-15998/2020 | 2020-04-29 | North America/Canada/Alberta/ | Alberta Precision Labs (APL) | Alberta Precision Labs (APL) | Gordon P et al |
| EPI_ISL_805330 | Canada/AB-17779/2020 | 2020-05-10 | North America/Canada/Alberta/ | Alberta Precision Labs (APL) | Alberta Precision Labs (APL) | Gordon P et al |
| EPI_ISL_805331 | Canada/AB-22978/2020 | 2020-04-20 | North America/Canada/Alberta/ | Alberta Precision Labs (APL) | Alberta Precision Labs (APL) | Gordon P et al |
| EPI_ISL_805332 | Canada/AB-30584/2020 | 2020-04-17 | North America/Canada/Alberta/ | Alberta Precision Labs (APL) | Alberta Precision Labs (APL) | Gordon P et al |
| EPI_ISL_805333 | Canada/AB-55302/2020 | 2020-04-19 | North America/Canada/Alberta/ | Alberta Precision Labs (APL) | Alberta Precision Labs (APL) | Gordon P et al |
| EPI_ISL_805334 | Canada/AB-99002/2020 | 2020-04-18 | North America/Canada/Alberta/ | Alberta Precision Labs (APL) | Alberta Precision Labs (APL) | Gordon P et al |
| EPI_ISL_805335 | Canada/AB-37171/2020 | 2020-04-16 | North America/Canada/Alberta/ | Alberta Precision Labs (APL) | Alberta Precision Labs (APL) | Gordon P et al |
| EPI_ISL_805336 | Canada/AB-61873/2020 | 2020-04-19 | North America/Canada/Alberta/ | Alberta Precision Labs (APL) | Alberta Precision Labs (APL) | Gordon P et al |
| EPI_ISL_805337 | Canada/AB-67889/2020 | 2020-05-11 | North America/Canada/Alberta/ | Alberta Precision Labs (APL) | Alberta Precision Labs (APL) | Gordon P et al |
| EPI_ISL_805338 | Canada/AB-23251/2020 | 2020-04-17 | North America/Canada/Alberta/ | Alberta Precision Labs (APL) | Alberta Precision Labs (APL) | Gordon P et al |
| EPI_ISL_805339 | Canada/AB-65157/2020 | 2020-04-29 | North America/Canada/Alberta/ | Alberta Precision Labs (APL) | Alberta Precision Labs (APL) | Gordon P et al |
| EPI_ISL_805340 | Canada/AB-91297/2020 | 2020-04-18 | North America/Canada/Alberta/ | Alberta Precision Labs (APL) | Alberta Precision Labs (APL) | Gordon P et al |
| EPI_ISL_805341 | Canada/AB-42683/2020 | 2020-05-11 | North America/Canada/Alberta/ | Alberta Precision Labs (APL) | Alberta Precision Labs (APL) | Gordon P et al |

| | | | | | | |
|---|---|---|---|---|---|---|
| EPI_ISL_805342 | Canada/AB-82354/2020 | 2020-04-19 | North America/Canada/Alberta/ | Alberta Precision Labs (APL) | Alberta Precision Labs (APL) | Gordon P et al |
| EPI_ISL_805343 | Canada/AB-43285/2020 | 2020-04-19 | North America/Canada/Alberta/ | Alberta Precision Labs (APL) | Alberta Precision Labs (APL) | Gordon P et al |
| EPI_ISL_805344 | Canada/AB-45914/2020 | 2020-04-22 | North America/Canada/Alberta/ | Alberta Precision Labs (APL) | Alberta Precision Labs (APL) | Gordon P et al |
| EPI_ISL_805345 | Canada/AB-93450/2020 | 2020-04-17 | North America/Canada/Alberta/ | Alberta Precision Labs (APL) | Alberta Precision Labs (APL) | Gordon P et al |
| EPI_ISL_805346 | Canada/AB-38116/2020 | 2020-05-13 | North America/Canada/Alberta/ | Alberta Precision Labs (APL) | Alberta Precision Labs (APL) | Gordon P et al |
| EPI_ISL_805347 | Canada/AB-25554/2020 | 2020-04-18 | North America/Canada/Alberta/ | Alberta Precision Labs (APL) | Alberta Precision Labs (APL) | Gordon P et al |
| EPI_ISL_805348 | Canada/AB-50877/2020 | 2020-04-16 | North America/Canada/Alberta/ | Alberta Precision Labs (APL) | Alberta Precision Labs (APL) | Gordon P et al |
| EPI_ISL_805349 | Canada/AB-21526/2020 | 2020-04-17 | North America/Canada/Alberta/ | Alberta Precision Labs (APL) | Alberta Precision Labs (APL) | Gordon P et al |
| EPI_ISL_805350 | Canada/AB-16715/2020 | 2020-04-16 | North America/Canada/Alberta/ | Alberta Precision Labs (APL) | Alberta Precision Labs (APL) | Gordon P et al |
| EPI_ISL_805351 | Canada/AB-83471/2020 | 2020-05-08 | North America/Canada/Alberta/ | Alberta Precision Labs (APL) | Alberta Precision Labs (APL) | Gordon P et al |
| EPI_ISL_805352 | Canada/AB-78161/2020 | 2020-04-16 | North America/Canada/Alberta/ | Alberta Precision Labs (APL) | Alberta Precision Labs (APL) | Gordon P et al |
| EPI_ISL_805353 | Canada/AB-86286/2020 | 2020-04-16 | North America/Canada/Alberta/ | Alberta Precision Labs (APL) | Alberta Precision Labs (APL) | Gordon P et al |
| EPI_ISL_805354 | Canada/AB-33815/2020 | 2020-04-18 | North America/Canada/Alberta/ | Alberta Precision Labs (APL) | Alberta Precision Labs (APL) | Gordon P et al |
| EPI_ISL_805355 | Canada/AB-29501/2020 | 2020-04-15 | North America/Canada/Alberta/ | Alberta Precision Labs (APL) | Alberta Precision Labs (APL) | Gordon P et al |
| EPI_ISL_805356 | Canada/AB-87503/2020 | 2020-04-16 | North America/Canada/Alberta/ | Alberta Precision Labs (APL) | Alberta Precision Labs (APL) | Gordon P et al |
| EPI_ISL_805357 | Canada/AB-63772/2020 | 2020-05-08 | North America/Canada/Alberta/ | Alberta Precision Labs (APL) | Alberta Precision Labs (APL) | Gordon P et al |
| EPI_ISL_805358 | Canada/AB-74192/2020 | 2020-04-17 | North America/Canada/Alberta/ | Alberta Precision Labs (APL) | Alberta Precision Labs (APL) | Gordon P et al |
| EPI_ISL_805359 | Canada/AB-79058/2020 | 2020-04-20 | North America/Canada/Alberta/ | Alberta Precision Labs (APL) | Alberta Precision Labs (APL) | Gordon P et al |
| EPI_ISL_805360 | Canada/AB-72752/2020 | 2020-04-13 | North America/Canada/Alberta/ | Alberta Precision Labs (APL) | Alberta Precision Labs (APL) | Gordon P et al |
| EPI_ISL_805361 | Canada/AB-33454/2020 | 2020-05-11 | North America/Canada/Alberta/ | Alberta Precision Labs (APL) | Alberta Precision Labs (APL) | Gordon P et al |

| | | | | | | |
|---|---|---|---|---|---|---|
| EPI_ISL_805362 | Canada/AB-49467/2020 | 2020-05-10 | North America/Canada/Alberta/ | Alberta Precision Labs (APL) | Alberta Precision Labs (APL) | Gordon P et al |
| EPI_ISL_805363 | Canada/AB-26575/2020 | 2020-05-12 | North America/Canada/Alberta/ | Alberta Precision Labs (APL) | Alberta Precision Labs (APL) | Gordon P et al |
| EPI_ISL_805364 | Canada/AB-74415/2020 | 2020-04-19 | North America/Canada/Alberta/ | Alberta Precision Labs (APL) | Alberta Precision Labs (APL) | Gordon P et al |
| EPI_ISL_805365 | Canada/AB-20686/2020 | 2020-04-17 | North America/Canada/Alberta/ | Alberta Precision Labs (APL) | Alberta Precision Labs (APL) | Gordon P et al |
| EPI_ISL_805366 | Canada/AB-51653/2020 | 2020-04-16 | North America/Canada/Alberta/ | Alberta Precision Labs (APL) | Alberta Precision Labs (APL) | Gordon P et al |
| EPI_ISL_805367 | Canada/AB-45945/2020 | 2020-04-18 | North America/Canada/Alberta/ | Alberta Precision Labs (APL) | Alberta Precision Labs (APL) | Gordon P et al |
| EPI_ISL_805368 | Canada/AB-34685/2020 | 2020-05-09 | North America/Canada/Alberta/ | Alberta Precision Labs (APL) | Alberta Precision Labs (APL) | Gordon P et al |
| EPI_ISL_805369 | Canada/AB-84848/2020 | 2020-04-16 | North America/Canada/Alberta/ | Alberta Precision Labs (APL) | Alberta Precision Labs (APL) | Gordon P et al |
| EPI_ISL_805370 | Canada/AB-51614/2020 | 2020-04-16 | North America/Canada/Alberta/ | Alberta Precision Labs (APL) | Alberta Precision Labs (APL) | Gordon P et al |
| EPI_ISL_805371 | Canada/AB-85939/2020 | 2020-04-18 | North America/Canada/Alberta/ | Alberta Precision Labs (APL) | Alberta Precision Labs (APL) | Gordon P et al |
| EPI_ISL_805372 | Canada/AB-41094/2020 | 2020-04-28 | North America/Canada/Alberta/ | Alberta Precision Labs (APL) | Alberta Precision Labs (APL) | Gordon P et al |
| EPI_ISL_805373 | Canada/AB-97112/2020 | 2020-04-16 | North America/Canada/Alberta/ | Alberta Precision Labs (APL) | Alberta Precision Labs (APL) | Gordon P et al |
| EPI_ISL_805374 | Canada/AB-66725/2020 | 2020-04-16 | North America/Canada/Alberta/ | Alberta Precision Labs (APL) | Alberta Precision Labs (APL) | Gordon P et al |
| EPI_ISL_805375 | Canada/AB-22295/2020 | 2020-04-17 | North America/Canada/Alberta/ | Alberta Precision Labs (APL) | Alberta Precision Labs (APL) | Gordon P et al |
| EPI_ISL_805376 | Canada/AB-16242/2020 | 2020-05-08 | North America/Canada/Alberta/ | Alberta Precision Labs (APL) | Alberta Precision Labs (APL) | Gordon P et al |
| EPI_ISL_805377 | Canada/AB-47471/2020 | 2020-04-20 | North America/Canada/Alberta/ | Alberta Precision Labs (APL) | Alberta Precision Labs (APL) | Gordon P et al |
| EPI_ISL_805378 | Canada/AB-32488/2020 | 2020-05-12 | North America/Canada/Alberta/ | Alberta Precision Labs (APL) | Alberta Precision Labs (APL) | Gordon P et al |
| EPI_ISL_805379 | Canada/AB-78276/2020 | 2020-04-26 | North America/Canada/Alberta/ | Alberta Precision Labs (APL) | Alberta Precision Labs (APL) | Gordon P et al |
| EPI_ISL_805380 | Canada/AB-55541/2020 | 2020-04-17 | North America/Canada/Alberta/ | Alberta Precision Labs (APL) | Alberta Precision Labs (APL) | Gordon P et al |
| EPI_ISL_805381 | Canada/AB-58688/2020 | 2020-05-14 | North America/Canada/Alberta/ | Alberta Precision Labs (APL) | Alberta Precision Labs (APL) | Gordon P et al |

| | | | | | | |
|---|---|---|---|---|---|---|
| EPI_ISL_805382 | Canada/AB-88778/2020 | 2020-04-15 | North America/Canada/Alberta/ | Alberta Precision Labs (APL) | Alberta Precision Labs (APL) | Gordon P et al |
| EPI_ISL_805383 | Canada/AB-29537/2020 | 2020-04-17 | North America/Canada/Alberta/ | Alberta Precision Labs (APL) | Alberta Precision Labs (APL) | Gordon P et al |
| EPI_ISL_805384 | Canada/AB-37873/2020 | 2020-05-09 | North America/Canada/Alberta/ | Alberta Precision Labs (APL) | Alberta Precision Labs (APL) | Gordon P et al |
| EPI_ISL_805385 | Canada/AB-32955/2020 | 2020-04-23 | North America/Canada/Alberta/ | Alberta Precision Labs (APL) | Alberta Precision Labs (APL) | Gordon P et al |
| EPI_ISL_805386 | Canada/AB-35367/2020 | 2020-04-25 | North America/Canada/Alberta/ | Alberta Precision Labs (APL) | Alberta Precision Labs (APL) | Gordon P et al |
| EPI_ISL_805387 | Canada/AB-43651/2020 | 2020-05-12 | North America/Canada/Alberta/ | Alberta Precision Labs (APL) | Alberta Precision Labs (APL) | Gordon P et al |
| EPI_ISL_805388 | Canada/AB-65171/2020 | 2020-04-18 | North America/Canada/Alberta/ | Alberta Precision Labs (APL) | Alberta Precision Labs (APL) | Gordon P et al |
| EPI_ISL_805389 | Canada/AB-45641/2020 | 2020-04-17 | North America/Canada/Alberta/ | Alberta Precision Labs (APL) | Alberta Precision Labs (APL) | Gordon P et al |
| EPI_ISL_805390 | Canada/AB-74550/2020 | 2020-04-20 | North America/Canada/Alberta/ | Alberta Precision Labs (APL) | Alberta Precision Labs (APL) | Gordon P et al |
| EPI_ISL_805391 | Canada/AB-97407/2020 | 2020-04-19 | North America/Canada/Alberta/ | Alberta Precision Labs (APL) | Alberta Precision Labs (APL) | Gordon P et al |
| EPI_ISL_805392 | Canada/AB-93800/2020 | 2020-04-18 | North America/Canada/Alberta/ | Alberta Precision Labs (APL) | Alberta Precision Labs (APL) | Gordon P et al |
| EPI_ISL_805393 | Canada/AB-99881/2020 | 2020-04-17 | North America/Canada/Alberta/ | Alberta Precision Labs (APL) | Alberta Precision Labs (APL) | Gordon P et al |
| EPI_ISL_805394 | Canada/AB-40367/2020 | 2020-04-22 | North America/Canada/Alberta/ | Alberta Precision Labs (APL) | Alberta Precision Labs (APL) | Gordon P et al |
| EPI_ISL_805395 | Canada/AB-93992/2020 | 2020-05-12 | North America/Canada/Alberta/ | Alberta Precision Labs (APL) | Alberta Precision Labs (APL) | Gordon P et al |
| EPI_ISL_805396 | Canada/AB-29974/2020 | 2020-05-08 | North America/Canada/Alberta/ | Alberta Precision Labs (APL) | Alberta Precision Labs (APL) | Gordon P et al |
| EPI_ISL_805397 | Canada/AB-69147/2020 | 2020-04-16 | North America/Canada/Alberta/ | Alberta Precision Labs (APL) | Alberta Precision Labs (APL) | Gordon P et al |
| EPI_ISL_805399 | Canada/AB-34129/2020 | 2020-04-18 | North America/Canada/Alberta/ | Alberta Precision Labs (APL) | Alberta Precision Labs (APL) | Gordon P et al |
| EPI_ISL_805400 | Canada/AB-57893/2020 | 2020-05-11 | North America/Canada/Alberta/ | Alberta Precision Labs (APL) | Alberta Precision Labs (APL) | Gordon P et al |
| EPI_ISL_805401 | Canada/AB-42808/2020 | 2020-04-16 | North America/Canada/Alberta/ | Alberta Precision Labs (APL) | Alberta Precision Labs (APL) | Gordon P et al |
| EPI_ISL_805402 | Canada/AB-58335/2020 | 2020-04-17 | North America/Canada/Alberta/ | Alberta Precision Labs (APL) | Alberta Precision Labs (APL) | Gordon P et al |

| | | | | | | |
|---|---|---|---|---|---|---|
| EPI_ISL_805403 | Canada/AB-36187/2020 | 2020-04-14 | North America/Canada/Alberta/ | Alberta Precision Labs (APL) | Alberta Precision Labs (APL) | Gordon P et al |
| EPI_ISL_805404 | Canada/AB-40182/2020 | 2020-04-19 | North America/Canada/Alberta/ | Alberta Precision Labs (APL) | Alberta Precision Labs (APL) | Gordon P et al |
| EPI_ISL_805405 | Canada/AB-42470/2020 | 2020-05-12 | North America/Canada/Alberta/ | Alberta Precision Labs (APL) | Alberta Precision Labs (APL) | Gordon P et al |
| EPI_ISL_805406 | Canada/AB-92103/2020 | 2020-04-17 | North America/Canada/Alberta/ | Alberta Precision Labs (APL) | Alberta Precision Labs (APL) | Gordon P et al |
| EPI_ISL_805407 | Canada/AB-25330/2020 | 2020-04-15 | North America/Canada/Alberta/ | Alberta Precision Labs (APL) | Alberta Precision Labs (APL) | Gordon P et al |
| EPI_ISL_805408 | Canada/AB-22973/2020 | 2020-04-19 | North America/Canada/Alberta/ | Alberta Precision Labs (APL) | Alberta Precision Labs (APL) | Gordon P et al |
| EPI_ISL_805409 | Canada/AB-21878/2020 | 2020-05-03 | North America/Canada/Alberta/ | Alberta Precision Labs (APL) | Alberta Precision Labs (APL) | Gordon P et al |
| EPI_ISL_805410 | Canada/AB-28609/2020 | 2020-04-30 | North America/Canada/Alberta/ | Alberta Precision Labs (APL) | Alberta Precision Labs (APL) | Gordon P et al |
| EPI_ISL_805411 | Canada/AB-15320/2020 | 2020-05-12 | North America/Canada/Alberta/ | Alberta Precision Labs (APL) | Alberta Precision Labs (APL) | Gordon P et al |
| EPI_ISL_805412 | Canada/AB-49027/2020 | 2020-04-19 | North America/Canada/Alberta/ | Alberta Precision Labs (APL) | Alberta Precision Labs (APL) | Gordon P et al |
| EPI_ISL_805413 | Canada/AB-40108/2020 | 2020-04-18 | North America/Canada/Alberta/ | Alberta Precision Labs (APL) | Alberta Precision Labs (APL) | Gordon P et al |
| EPI_ISL_805414 | Canada/AB-28844/2020 | 2020-04-16 | North America/Canada/Alberta/ | Alberta Precision Labs (APL) | Alberta Precision Labs (APL) | Gordon P et al |
| EPI_ISL_805415 | Canada/AB-64540/2020 | 2020-04-17 | North America/Canada/Alberta/ | Alberta Precision Labs (APL) | Alberta Precision Labs (APL) | Gordon P et al |
| EPI_ISL_805416 | Canada/AB-31390/2020 | 2020-04-18 | North America/Canada/Alberta/ | Alberta Precision Labs (APL) | Alberta Precision Labs (APL) | Gordon P et al |
| EPI_ISL_805417 | Canada/AB-31017/2020 | 2020-04-26 | North America/Canada/Alberta/ | Alberta Precision Labs (APL) | Alberta Precision Labs (APL) | Gordon P et al |
| EPI_ISL_805418 | Canada/AB-69131/2020 | 2020-05-09 | North America/Canada/Alberta/ | Alberta Precision Labs (APL) | Alberta Precision Labs (APL) | Gordon P et al |
| EPI_ISL_805419 | Canada/AB-53939/2020 | 2020-04-17 | North America/Canada/Alberta/ | Alberta Precision Labs (APL) | Alberta Precision Labs (APL) | Gordon P et al |
| EPI_ISL_805420 | Canada/AB-74649/2020 | 2020-04-20 | North America/Canada/Alberta/ | Alberta Precision Labs (APL) | Alberta Precision Labs (APL) | Gordon P et al |
| EPI_ISL_805422 | Canada/AB-57423/2020 | 2020-04-18 | North America/Canada/Alberta/ | Alberta Precision Labs (APL) | Alberta Precision Labs (APL) | Gordon P et al |
| EPI_ISL_805423 | Canada/AB-72989/2020 | 2020-04-21 | North America/Canada/Alberta/ | Alberta Precision Labs (APL) | Alberta Precision Labs (APL) | Gordon P et al |

| | | | | | | |
|---|---|---|---|---|---|---|
| EPI_ISL_805424 | Canada/AB-52246/2020 | 2020-05-08 | North America/Canada/Alberta/ | Alberta Precision Labs (APL) | Alberta Precision Labs (APL) | Gordon P et al |
| EPI_ISL_805425 | Canada/AB-61395/2020 | 2020-04-15 | North America/Canada/Alberta/ | Alberta Precision Labs (APL) | Alberta Precision Labs (APL) | Gordon P et al |
| EPI_ISL_805426 | Canada/AB-43387/2020 | 2020-05-02 | North America/Canada/Alberta/ | Alberta Precision Labs (APL) | Alberta Precision Labs (APL) | Gordon P et al |
| EPI_ISL_805427 | Canada/AB-91431/2020 | 2020-05-08 | North America/Canada/Alberta/ | Alberta Precision Labs (APL) | Alberta Precision Labs (APL) | Gordon P et al |
| EPI_ISL_805428 | Canada/AB-42967/2020 | 2020-04-17 | North America/Canada/Alberta/ | Alberta Precision Labs (APL) | Alberta Precision Labs (APL) | Gordon P et al |
| EPI_ISL_805429 | Canada/AB-28239/2020 | 2020-04-27 | North America/Canada/Alberta/ | Alberta Precision Labs (APL) | Alberta Precision Labs (APL) | Gordon P et al |
| EPI_ISL_805430 | Canada/AB-28212/2020 | 2020-04-20 | North America/Canada/Alberta/ | Alberta Precision Labs (APL) | Alberta Precision Labs (APL) | Gordon P et al |
| EPI_ISL_805431 | Canada/AB-39718/2020 | 2020-04-20 | North America/Canada/Alberta/ | Alberta Precision Labs (APL) | Alberta Precision Labs (APL) | Gordon P et al |
| EPI_ISL_805432 | Canada/AB-21180/2020 | 2020-04-18 | North America/Canada/Alberta/ | Alberta Precision Labs (APL) | Alberta Precision Labs (APL) | Gordon P et al |
| EPI_ISL_805433 | Canada/AB-24183/2020 | 2020-04-17 | North America/Canada/Alberta/ | Alberta Precision Labs (APL) | Alberta Precision Labs (APL) | Gordon P et al |
| EPI_ISL_805434 | Canada/AB-49884/2020 | 2020-05-13 | North America/Canada/Alberta/ | Alberta Precision Labs (APL) | Alberta Precision Labs (APL) | Gordon P et al |
| EPI_ISL_805435 | Canada/AB-68012/2020 | 2020-05-13 | North America/Canada/Alberta/ | Alberta Precision Labs (APL) | Alberta Precision Labs (APL) | Gordon P et al |
| EPI_ISL_805436 | Canada/AB-21075/2020 | 2020-04-28 | North America/Canada/Alberta/ | Alberta Precision Labs (APL) | Alberta Precision Labs (APL) | Gordon P et al |
| EPI_ISL_805437 | Canada/AB-36244/2020 | 2020-03-23 | North America/Canada/Alberta/ | Alberta Precision Labs (APL) | Alberta Precision Labs (APL) | Gordon P et al |
| EPI_ISL_805438 | Canada/AB-18620/2020 | 2020-04-24 | North America/Canada/Alberta/ | Alberta Precision Labs (APL) | Alberta Precision Labs (APL) | Gordon P et al |
| EPI_ISL_805439 | Canada/AB-18827/2020 | 2020-04-17 | North America/Canada/Alberta/ | Alberta Precision Labs (APL) | Alberta Precision Labs (APL) | Gordon P et al |
| EPI_ISL_805440 | Canada/AB-56194/2020 | 2020-04-17 | North America/Canada/Alberta/ | Alberta Precision Labs (APL) | Alberta Precision Labs (APL) | Gordon P et al |
| EPI_ISL_805441 | Canada/AB-66509/2020 | 2020-04-16 | North America/Canada/Alberta/ | Alberta Precision Labs (APL) | Alberta Precision Labs (APL) | Gordon P et al |
| EPI_ISL_805442 | Canada/AB-33971/2020 | 2020-04-15 | North America/Canada/Alberta/ | Alberta Precision Labs (APL) | Alberta Precision Labs (APL) | Gordon P et al |
| EPI_ISL_805443 | Canada/AB-25317/2020 | 2020-04-19 | North America/Canada/Alberta/ | Alberta Precision Labs (APL) | Alberta Precision Labs (APL) | Gordon P et al |

| | | | | | | |
|---|---|---|---|---|---|---|
| EPI_ISL_805444 | Canada/AB-62951/2020 | 2020-04-28 | North America/Canada/Alberta/ | Alberta Precision Labs (APL) | Alberta Precision Labs (APL) | Gordon P et al |
| EPI_ISL_805445 | Canada/AB-41423/2020 | 2020-04-18 | North America/Canada/Alberta/ | Alberta Precision Labs (APL) | Alberta Precision Labs (APL) | Gordon P et al |
| EPI_ISL_805446 | Canada/AB-32212/2020 | 2020-05-12 | North America/Canada/Alberta/ | Alberta Precision Labs (APL) | Alberta Precision Labs (APL) | Gordon P et al |
| EPI_ISL_805447 | Canada/AB-30480/2020 | 2020-04-16 | North America/Canada/Alberta/ | Alberta Precision Labs (APL) | Alberta Precision Labs (APL) | Gordon P et al |
| EPI_ISL_805448 | Canada/AB-42986/2020 | 2020-04-16 | North America/Canada/Alberta/ | Alberta Precision Labs (APL) | Alberta Precision Labs (APL) | Gordon P et al |
| EPI_ISL_805449 | Canada/AB-80710/2020 | 2020-04-29 | North America/Canada/Alberta/ | Alberta Precision Labs (APL) | Alberta Precision Labs (APL) | Gordon P et al |
| EPI_ISL_805450 | Canada/AB-57437/2020 | 2020-04-25 | North America/Canada/Alberta/ | Alberta Precision Labs (APL) | Alberta Precision Labs (APL) | Gordon P et al |
| EPI_ISL_805451 | Canada/AB-28366/2020 | 2020-04-14 | North America/Canada/Alberta/ | Alberta Precision Labs (APL) | Alberta Precision Labs (APL) | Gordon P et al |
| EPI_ISL_805452 | Canada/AB-39906/2020 | 2020-04-26 | North America/Canada/Alberta/ | Alberta Precision Labs (APL) | Alberta Precision Labs (APL) | Gordon P et al |
| EPI_ISL_805453 | Canada/AB-63037/2020 | 2020-05-06 | North America/Canada/Alberta/ | Alberta Precision Labs (APL) | Alberta Precision Labs (APL) | Gordon P et al |
| EPI_ISL_805454 | Canada/AB-80995/2020 | 2020-04-24 | North America/Canada/Alberta/ | Alberta Precision Labs (APL) | Alberta Precision Labs (APL) | Gordon P et al |
| EPI_ISL_805455 | Canada/AB-43170/2020 | 2020-05-10 | North America/Canada/Alberta/ | Alberta Precision Labs (APL) | Alberta Precision Labs (APL) | Gordon P et al |
| EPI_ISL_805456 | Canada/AB-76440/2020 | 2020-04-25 | North America/Canada/Alberta/ | Alberta Precision Labs (APL) | Alberta Precision Labs (APL) | Gordon P et al |
| EPI_ISL_805457 | Canada/AB-40494/2020 | 2020-04-29 | North America/Canada/Alberta/ | Alberta Precision Labs (APL) | Alberta Precision Labs (APL) | Gordon P et al |
| EPI_ISL_805458 | Canada/AB-58495/2020 | 2020-05-10 | North America/Canada/Alberta/ | Alberta Precision Labs (APL) | Alberta Precision Labs (APL) | Gordon P et al |
| EPI_ISL_805459 | Canada/AB-48029/2020 | 2020-04-21 | North America/Canada/Alberta/ | Alberta Precision Labs (APL) | Alberta Precision Labs (APL) | Gordon P et al |
| EPI_ISL_805460 | Canada/AB-41426/2020 | 2020-05-02 | North America/Canada/Alberta/ | Alberta Precision Labs (APL) | Alberta Precision Labs (APL) | Gordon P et al |
| EPI_ISL_805461 | Canada/AB-65987/2020 | 2020-04-18 | North America/Canada/Alberta/ | Alberta Precision Labs (APL) | Alberta Precision Labs (APL) | Gordon P et al |
| EPI_ISL_805462 | Canada/AB-78599/2020 | 2020-05-14 | North America/Canada/Alberta/ | Alberta Precision Labs (APL) | Alberta Precision Labs (APL) | Gordon P et al |
| EPI_ISL_805463 | Canada/AB-15241/2020 | 2020-04-14 | North America/Canada/Alberta/ | Alberta Precision Labs (APL) | Alberta Precision Labs (APL) | Gordon P et al |

| | | | | | | |
|---|---|---|---|---|---|---|
| EPI_ISL_805464 | Canada/AB-42287/2020 | 2020-04-21 | North America/Canada/Alberta/ | Alberta Precision Labs (APL) | Alberta Precision Labs (APL) | Gordon P et al |
| EPI_ISL_805465 | Canada/AB-30219/2020 | 2020-05-08 | North America/Canada/Alberta/ | Alberta Precision Labs (APL) | Alberta Precision Labs (APL) | Gordon P et al |
| EPI_ISL_805466 | Canada/AB-55503/2020 | 20-Mar | North America/Canada/Alberta/ | Alberta Precision Labs (APL) | Alberta Precision Labs (APL) | Gordon P et al |
| EPI_ISL_805467 | Canada/AB-47392/2020 | 2020-04-17 | North America/Canada/Alberta/ | Alberta Precision Labs (APL) | Alberta Precision Labs (APL) | Gordon P et al |
| EPI_ISL_805468 | Canada/AB-63606/2020 | 2020-05-11 | North America/Canada/Alberta/ | Alberta Precision Labs (APL) | Alberta Precision Labs (APL) | Gordon P et al |
| EPI_ISL_805469 | Canada/AB-15270/2020 | 2020-04-27 | North America/Canada/Alberta/ | Alberta Precision Labs (APL) | Alberta Precision Labs (APL) | Gordon P et al |
| EPI_ISL_805470 | Canada/AB-21461/2020 | 2020-04-17 | North America/Canada/Alberta/ | Alberta Precision Labs (APL) | Alberta Precision Labs (APL) | Gordon P et al |
| EPI_ISL_805471 | Canada/AB-41321/2020 | 2020-04-19 | North America/Canada/Alberta/ | Alberta Precision Labs (APL) | Alberta Precision Labs (APL) | Gordon P et al |
| EPI_ISL_805472 | Canada/AB-28093/2020 | 2020-04-27 | North America/Canada/Alberta/ | Alberta Precision Labs (APL) | Alberta Precision Labs (APL) | Gordon P et al |
| EPI_ISL_805473 | Canada/AB-55529/2020 | 2020-04-13 | North America/Canada/Alberta/ | Alberta Precision Labs (APL) | Alberta Precision Labs (APL) | Gordon P et al |
| EPI_ISL_805474 | Canada/AB-32342/2020 | 2020-04-16 | North America/Canada/Alberta/ | Alberta Precision Labs (APL) | Alberta Precision Labs (APL) | Gordon P et al |
| EPI_ISL_805475 | Canada/AB-75006/2020 | 2020-04-20 | North America/Canada/Alberta/ | Alberta Precision Labs (APL) | Alberta Precision Labs (APL) | Gordon P et al |
| EPI_ISL_805476 | Canada/AB-66678/2020 | 2020-04-30 | North America/Canada/Alberta/ | Alberta Precision Labs (APL) | Alberta Precision Labs (APL) | Gordon P et al |
| EPI_ISL_805477 | Canada/AB-28034/2020 | 2020-04-21 | North America/Canada/Alberta/ | Alberta Precision Labs (APL) | Alberta Precision Labs (APL) | Gordon P et al |
| EPI_ISL_805478 | Canada/AB-89580/2020 | 2020-04-21 | North America/Canada/Alberta/ | Alberta Precision Labs (APL) | Alberta Precision Labs (APL) | Gordon P et al |
| EPI_ISL_805479 | Canada/AB-41237/2020 | 2020-04-25 | North America/Canada/Alberta/ | Alberta Precision Labs (APL) | Alberta Precision Labs (APL) | Gordon P et al |
| EPI_ISL_805480 | Canada/AB-27031/2020 | 2020-04-26 | North America/Canada/Alberta/ | Alberta Precision Labs (APL) | Alberta Precision Labs (APL) | Gordon P et al |
| EPI_ISL_805481 | Canada/AB-31638/2020 | 2020-04-06 | North America/Canada/Alberta/ | Alberta Precision Labs (APL) | Alberta Precision Labs (APL) | Gordon P et al |
| EPI_ISL_805482 | Canada/AB-69703/2020 | 2020-04-27 | North America/Canada/Alberta/ | Alberta Precision Labs (APL) | Alberta Precision Labs (APL) | Gordon P et al |
| EPI_ISL_805483 | Canada/AB-87834/2020 | 2020-04-18 | North America/Canada/Alberta/ | Alberta Precision Labs (APL) | Alberta Precision Labs (APL) | Gordon P et al |

| | | | | | | |
|---|---|---|---|---|---|---|
| EPI_ISL_805484 | Canada/AB-12700/2020 | 2020-05-03 | North America/Canada/Alberta/ | Alberta Precision Labs (APL) | Alberta Precision Labs (APL) | Gordon P et al |
| EPI_ISL_805485 | Canada/AB-14024/2020 | 2020-04-28 | North America/Canada/Alberta/ | Alberta Precision Labs (APL) | Alberta Precision Labs (APL) | Gordon P et al |
| EPI_ISL_805486 | Canada/AB-13962/2020 | 2020-03-23 | North America/Canada/Alberta/ | Alberta Precision Labs (APL) | Alberta Precision Labs (APL) | Gordon P et al |
| EPI_ISL_805487 | Canada/AB-78889/2020 | 2020-05-02 | North America/Canada/Alberta/ | Alberta Precision Labs (APL) | Alberta Precision Labs (APL) | Gordon P et al |
| EPI_ISL_805488 | Canada/AB-54206/2020 | 2020-05-01 | North America/Canada/Alberta/ | Alberta Precision Labs (APL) | Alberta Precision Labs (APL) | Gordon P et al |
| EPI_ISL_805489 | Canada/AB-83509/2020 | 2020-04-20 | North America/Canada/Alberta/ | Alberta Precision Labs (APL) | Alberta Precision Labs (APL) | Gordon P et al |
| EPI_ISL_805490 | Canada/AB-46864/2020 | 2020-05-07 | North America/Canada/Alberta/ | Alberta Precision Labs (APL) | Alberta Precision Labs (APL) | Gordon P et al |
| EPI_ISL_805491 | Canada/AB-48001/2020 | 2020-04-28 | North America/Canada/Alberta/ | Alberta Precision Labs (APL) | Alberta Precision Labs (APL) | Gordon P et al |
| EPI_ISL_805492 | Canada/AB-40560/2020 | 2020-04-26 | North America/Canada/Alberta/ | Alberta Precision Labs (APL) | Alberta Precision Labs (APL) | Gordon P et al |
| EPI_ISL_805493 | Canada/AB-25294/2020 | 2020-04-16 | North America/Canada/Alberta/ | Alberta Precision Labs (APL) | Alberta Precision Labs (APL) | Gordon P et al |
| EPI_ISL_805494 | Canada/AB-86980/2020 | 2020-04-30 | North America/Canada/Alberta/ | Alberta Precision Labs (APL) | Alberta Precision Labs (APL) | Gordon P et al |
| EPI_ISL_805495 | Canada/AB-32607/2020 | 2020-04-18 | North America/Canada/Alberta/ | Alberta Precision Labs (APL) | Alberta Precision Labs (APL) | Gordon P et al |
| EPI_ISL_805496 | Canada/AB-65214/2020 | 2020-04-06 | North America/Canada/Alberta/ | Alberta Precision Labs (APL) | Alberta Precision Labs (APL) | Gordon P et al |
| EPI_ISL_805497 | Canada/AB-51843/2020 | 2020-04-26 | North America/Canada/Alberta/ | Alberta Precision Labs (APL) | Alberta Precision Labs (APL) | Gordon P et al |
| EPI_ISL_805498 | Canada/AB-43723/2020 | 2020-04-27 | North America/Canada/Alberta/ | Alberta Precision Labs (APL) | Alberta Precision Labs (APL) | Gordon P et al |
| EPI_ISL_805499 | Canada/AB-15185/2020 | 2020-04-20 | North America/Canada/Alberta/ | Alberta Precision Labs (APL) | Alberta Precision Labs (APL) | Gordon P et al |
| EPI_ISL_805500 | Canada/AB-60136/2020 | 2020-04-26 | North America/Canada/Alberta/ | Alberta Precision Labs (APL) | Alberta Precision Labs (APL) | Gordon P et al |
| EPI_ISL_805501 | Canada/AB-92703/2020 | 2020-04-28 | North America/Canada/Alberta/ | Alberta Precision Labs (APL) | Alberta Precision Labs (APL) | Gordon P et al |
| EPI_ISL_805502 | Canada/AB-25965/2020 | 2020-04-16 | North America/Canada/Alberta/ | Alberta Precision Labs (APL) | Alberta Precision Labs (APL) | Gordon P et al |
| EPI_ISL_805503 | Canada/AB-71130/2020 | 2020-05-12 | North America/Canada/Alberta/ | Alberta Precision Labs (APL) | Alberta Precision Labs (APL) | Gordon P et al |

| | | | | | | |
|---|---|---|---|---|---|---|
| EPI_ISL_805504 | Canada/AB-30035/2020 | 2020-04-29 | North America/Canada/Alberta/ | Alberta Precision Labs (APL) | Alberta Precision Labs (APL) | Gordon P et al |
| EPI_ISL_805505 | Canada/AB-89987/2020 | 2020-04-26 | North America/Canada/Alberta/ | Alberta Precision Labs (APL) | Alberta Precision Labs (APL) | Gordon P et al |
| EPI_ISL_805506 | Canada/AB-43137/2020 | 2020-04-26 | North America/Canada/Alberta/ | Alberta Precision Labs (APL) | Alberta Precision Labs (APL) | Gordon P et al |
| EPI_ISL_805507 | Canada/AB-48454/2020 | 2020-04-25 | North America/Canada/Alberta/ | Alberta Precision Labs (APL) | Alberta Precision Labs (APL) | Gordon P et al |
| EPI_ISL_805508 | Canada/AB-20257/2020 | 2020-05-17 | North America/Canada/Alberta/ | Alberta Precision Labs (APL) | Alberta Precision Labs (APL) | Gordon P et al |
| EPI_ISL_805509 | Canada/AB-27891/2020 | 2020-04-18 | North America/Canada/Alberta/ | Alberta Precision Labs (APL) | Alberta Precision Labs (APL) | Gordon P et al |
| EPI_ISL_805510 | Canada/AB-21501/2020 | 2020-04-26 | North America/Canada/Alberta/ | Alberta Precision Labs (APL) | Alberta Precision Labs (APL) | Gordon P et al |
| EPI_ISL_805511 | Canada/AB-34636/2020 | 2020-04-23 | North America/Canada/Alberta/ | Alberta Precision Labs (APL) | Alberta Precision Labs (APL) | Gordon P et al |
| EPI_ISL_805512 | Canada/AB-74962/2020 | 2020-03-22 | North America/Canada/Alberta/ | Alberta Precision Labs (APL) | Alberta Precision Labs (APL) | Gordon P et al |
| EPI_ISL_805513 | Canada/AB-81868/2020 | 2020-04-16 | North America/Canada/Alberta/ | Alberta Precision Labs (APL) | Alberta Precision Labs (APL) | Gordon P et al |
| EPI_ISL_805514 | Canada/AB-65830/2020 | 2020-04-30 | North America/Canada/Alberta/ | Alberta Precision Labs (APL) | Alberta Precision Labs (APL) | Gordon P et al |
| EPI_ISL_805515 | Canada/AB-19490/2020 | 2020-05-08 | North America/Canada/Alberta/ | Alberta Precision Labs (APL) | Alberta Precision Labs (APL) | Gordon P et al |
| EPI_ISL_805516 | Canada/AB-87879/2020 | 2020-04-28 | North America/Canada/Alberta/ | Alberta Precision Labs (APL) | Alberta Precision Labs (APL) | Gordon P et al |
| EPI_ISL_805517 | Canada/AB-14071/2020 | 2020-04-19 | North America/Canada/Alberta/ | Alberta Precision Labs (APL) | Alberta Precision Labs (APL) | Gordon P et al |
| EPI_ISL_805518 | Canada/AB-29363/2020 | 2020-04-29 | North America/Canada/Alberta/ | Alberta Precision Labs (APL) | Alberta Precision Labs (APL) | Gordon P et al |
| EPI_ISL_805519 | Canada/AB-83736/2020 | 2020-04-28 | North America/Canada/Alberta/ | Alberta Precision Labs (APL) | Alberta Precision Labs (APL) | Gordon P et al |
| EPI_ISL_805520 | Canada/AB-57864/2020 | 2020-04-13 | North America/Canada/Alberta/ | Alberta Precision Labs (APL) | Alberta Precision Labs (APL) | Gordon P et al |
| EPI_ISL_805521 | Canada/AB-15264/2020 | 2020-04-15 | North America/Canada/Alberta/ | Alberta Precision Labs (APL) | Alberta Precision Labs (APL) | Gordon P et al |
| EPI_ISL_805522 | Canada/AB-40207/2020 | 2020-04-19 | North America/Canada/Alberta/ | Alberta Precision Labs (APL) | Alberta Precision Labs (APL) | Gordon P et al |
| EPI_ISL_805523 | Canada/AB-31508/2020 | 2020-04-18 | North America/Canada/Alberta/ | Alberta Precision Labs (APL) | Alberta Precision Labs (APL) | Gordon P et al |

| | | | | | | |
|---|---|---|---|---|---|---|
| EPI_ISL_805524 | Canada/AB-57025/2020 | 2020-04-26 | North America/Canada/Alberta/ | Alberta Precision Labs (APL) | Alberta Precision Labs (APL) | Gordon P et al |
| EPI_ISL_805525 | Canada/AB-22413/2020 | 2020-04-17 | North America/Canada/Alberta/ | Alberta Precision Labs (APL) | Alberta Precision Labs (APL) | Gordon P et al |
| EPI_ISL_805526 | Canada/AB-39924/2020 | 2020-04-29 | North America/Canada/Alberta/ | Alberta Precision Labs (APL) | Alberta Precision Labs (APL) | Gordon P et al |
| EPI_ISL_805527 | Canada/AB-65100/2020 | 2020-05-09 | North America/Canada/Alberta/ | Alberta Precision Labs (APL) | Alberta Precision Labs (APL) | Gordon P et al |
| EPI_ISL_805528 | Canada/AB-33212/2020 | 2020-04-27 | North America/Canada/Alberta/ | Alberta Precision Labs (APL) | Alberta Precision Labs (APL) | Gordon P et al |
| EPI_ISL_805529 | Canada/AB-46706/2020 | 2020-05-05 | North America/Canada/Alberta/ | Alberta Precision Labs (APL) | Alberta Precision Labs (APL) | Gordon P et al |
| EPI_ISL_805530 | Canada/AB-84346/2020 | 2020-04-18 | North America/Canada/Alberta/ | Alberta Precision Labs (APL) | Alberta Precision Labs (APL) | Gordon P et al |
| EPI_ISL_805531 | Canada/AB-47285/2020 | 2020-05-08 | North America/Canada/Alberta/ | Alberta Precision Labs (APL) | Alberta Precision Labs (APL) | Gordon P et al |
| EPI_ISL_805532 | Canada/AB-56732/2020 | 2020-04-28 | North America/Canada/Alberta/ | Alberta Precision Labs (APL) | Alberta Precision Labs (APL) | Gordon P et al |
| EPI_ISL_805533 | Canada/AB-71527/2020 | 2020-05-11 | North America/Canada/Alberta/ | Alberta Precision Labs (APL) | Alberta Precision Labs (APL) | Gordon P et al |
| EPI_ISL_805534 | Canada/AB-76027/2020 | 2020-05-13 | North America/Canada/Alberta/ | Alberta Precision Labs (APL) | Alberta Precision Labs (APL) | Gordon P et al |
| EPI_ISL_805535 | Canada/AB-48914/2020 | 2020-04-25 | North America/Canada/Alberta/ | Alberta Precision Labs (APL) | Alberta Precision Labs (APL) | Gordon P et al |
| EPI_ISL_805536 | Canada/AB-98202/2020 | 2020-04-14 | North America/Canada/Alberta/ | Alberta Precision Labs (APL) | Alberta Precision Labs (APL) | Gordon P et al |
| EPI_ISL_805537 | Canada/AB-95227/2020 | 2020-04-30 | North America/Canada/Alberta/ | Alberta Precision Labs (APL) | Alberta Precision Labs (APL) | Gordon P et al |
| EPI_ISL_805538 | Canada/AB-41965/2020 | 2020-04-25 | North America/Canada/Alberta/ | Alberta Precision Labs (APL) | Alberta Precision Labs (APL) | Gordon P et al |
| EPI_ISL_805539 | Canada/AB-27161/2020 | 2020-05-13 | North America/Canada/Alberta/ | Alberta Precision Labs (APL) | Alberta Precision Labs (APL) | Gordon P et al |
| EPI_ISL_805540 | Canada/AB-49428/2020 | 2020-05-01 | North America/Canada/Alberta/ | Alberta Precision Labs (APL) | Alberta Precision Labs (APL) | Gordon P et al |
| EPI_ISL_805541 | Canada/AB-15207/2020 | 2020-04-13 | North America/Canada/Alberta/ | Alberta Precision Labs (APL) | Alberta Precision Labs (APL) | Gordon P et al |
| EPI_ISL_805542 | Canada/AB-36898/2020 | 2020-05-07 | North America/Canada/Alberta/ | Alberta Precision Labs (APL) | Alberta Precision Labs (APL) | Gordon P et al |
| EPI_ISL_805543 | Canada/AB-18613/2020 | 2020-05-08 | North America/Canada/Alberta/ | Alberta Precision Labs (APL) | Alberta Precision Labs (APL) | Gordon P et al |

| | | | | | | |
|---|---|---|---|---|---|---|
| EPI_ISL_805544 | Canada/AB-30057/2020 | 2020-03-22 | North America/Canada/Alberta/ | Alberta Precision Labs (APL) | Alberta Precision Labs (APL) | Gordon P et al |
| EPI_ISL_805545 | Canada/AB-94403/2020 | 2020-04-15 | North America/Canada/Alberta/ | Alberta Precision Labs (APL) | Alberta Precision Labs (APL) | Gordon P et al |
| EPI_ISL_805546 | Canada/AB-19260/2020 | 2020-04-18 | North America/Canada/Alberta/ | Alberta Precision Labs (APL) | Alberta Precision Labs (APL) | Gordon P et al |
| EPI_ISL_805547 | Canada/AB-86552/2020 | 2020-05-09 | North America/Canada/Alberta/ | Alberta Precision Labs (APL) | Alberta Precision Labs (APL) | Gordon P et al |
| EPI_ISL_805548 | Canada/AB-69513/2020 | 2020-05-13 | North America/Canada/Alberta/ | Alberta Precision Labs (APL) | Alberta Precision Labs (APL) | Gordon P et al |
| EPI_ISL_805549 | Canada/AB-99341/2020 | 2020-03-23 | North America/Canada/Alberta/ | Alberta Precision Labs (APL) | Alberta Precision Labs (APL) | Gordon P et al |
| EPI_ISL_805550 | Canada/AB-35618/2020 | 2020-05-11 | North America/Canada/Alberta/ | Alberta Precision Labs (APL) | Alberta Precision Labs (APL) | Gordon P et al |
| EPI_ISL_805551 | Canada/AB-29887/2020 | 2020-04-18 | North America/Canada/Alberta/ | Alberta Precision Labs (APL) | Alberta Precision Labs (APL) | Gordon P et al |
| EPI_ISL_805552 | Canada/AB-65793/2020 | 2020-05-03 | North America/Canada/Alberta/ | Alberta Precision Labs (APL) | Alberta Precision Labs (APL) | Gordon P et al |
| EPI_ISL_805553 | Canada/AB-51582/2020 | 2020-04-29 | North America/Canada/Alberta/ | Alberta Precision Labs (APL) | Alberta Precision Labs (APL) | Gordon P et al |
| EPI_ISL_805554 | Canada/AB-53313/2020 | 2020-05-08 | North America/Canada/Alberta/ | Alberta Precision Labs (APL) | Alberta Precision Labs (APL) | Gordon P et al |
| EPI_ISL_805555 | Canada/AB-37786/2020 | 2020-04-27 | North America/Canada/Alberta/ | Alberta Precision Labs (APL) | Alberta Precision Labs (APL) | Gordon P et al |
| EPI_ISL_805556 | Canada/AB-81985/2020 | 2020-04-16 | North America/Canada/Alberta/ | Alberta Precision Labs (APL) | Alberta Precision Labs (APL) | Gordon P et al |
| EPI_ISL_805557 | Canada/AB-89120/2020 | 2020-04-27 | North America/Canada/Alberta/ | Alberta Precision Labs (APL) | Alberta Precision Labs (APL) | Gordon P et al |
| EPI_ISL_805558 | Canada/AB-32085/2020 | 2020-04-15 | North America/Canada/Alberta/ | Alberta Precision Labs (APL) | Alberta Precision Labs (APL) | Gordon P et al |
| EPI_ISL_805559 | Canada/AB-99000/2020 | 2020-05-09 | North America/Canada/Alberta/ | Alberta Precision Labs (APL) | Alberta Precision Labs (APL) | Gordon P et al |
| EPI_ISL_805560 | Canada/AB-24697/2020 | 2020-04-18 | North America/Canada/Alberta/ | Alberta Precision Labs (APL) | Alberta Precision Labs (APL) | Gordon P et al |
| EPI_ISL_805561 | Canada/AB-29861/2020 | 2020-04-29 | North America/Canada/Alberta/ | Alberta Precision Labs (APL) | Alberta Precision Labs (APL) | Gordon P et al |
| EPI_ISL_805562 | Canada/AB-57937/2020 | 2020-04-25 | North America/Canada/Alberta/ | Alberta Precision Labs (APL) | Alberta Precision Labs (APL) | Gordon P et al |
| EPI_ISL_805563 | Canada/AB-51272/2020 | 2020-04-16 | North America/Canada/Alberta/ | Alberta Precision Labs (APL) | Alberta Precision Labs (APL) | Gordon P et al |

| | | | | | | |
|---|---|---|---|---|---|---|
| EPI_ISL_805564 | Canada/AB-41646/2020 | 2020-04-29 | North America/Canada/Alberta/ | Alberta Precision Labs (APL) | Alberta Precision Labs (APL) | Gordon P et al |
| EPI_ISL_805565 | Canada/AB-13959/2020 | 2020-04-27 | North America/Canada/Alberta/ | Alberta Precision Labs (APL) | Alberta Precision Labs (APL) | Gordon P et al |
| EPI_ISL_805566 | Canada/AB-88086/2020 | 2020-05-18 | North America/Canada/Alberta/ | Alberta Precision Labs (APL) | Alberta Precision Labs (APL) | Gordon P et al |
| EPI_ISL_805567 | Canada/AB-14207/2020 | 2020-05-01 | North America/Canada/Alberta/ | Alberta Precision Labs (APL) | Alberta Precision Labs (APL) | Gordon P et al |
| EPI_ISL_805568 | Canada/AB-56736/2020 | 2020-04-21 | North America/Canada/Alberta/ | Alberta Precision Labs (APL) | Alberta Precision Labs (APL) | Gordon P et al |
| EPI_ISL_805569 | Canada/AB-51520/2020 | 2020-04-29 | North America/Canada/Alberta/ | Alberta Precision Labs (APL) | Alberta Precision Labs (APL) | Gordon P et al |
| EPI_ISL_805570 | Canada/AB-47840/2020 | 2020-04-27 | North America/Canada/Alberta/ | Alberta Precision Labs (APL) | Alberta Precision Labs (APL) | Gordon P et al |
| EPI_ISL_805571 | Canada/AB-15239/2020 | 20-Mar | North America/Canada/Alberta/ | Alberta Precision Labs (APL) | Alberta Precision Labs (APL) | Gordon P et al |
| EPI_ISL_805572 | Canada/AB-85521/2020 | 2020-04-30 | North America/Canada/Alberta/ | Alberta Precision Labs (APL) | Alberta Precision Labs (APL) | Gordon P et al |
| EPI_ISL_805573 | Canada/AB-20108/2020 | 2020-04-16 | North America/Canada/Alberta/ | Alberta Precision Labs (APL) | Alberta Precision Labs (APL) | Gordon P et al |
| EPI_ISL_805574 | Canada/AB-62788/2020 | 2020-04-18 | North America/Canada/Alberta/ | Alberta Precision Labs (APL) | Alberta Precision Labs (APL) | Gordon P et al |
| EPI_ISL_805575 | Canada/AB-80859/2020 | 2020-04-30 | North America/Canada/Alberta/ | Alberta Precision Labs (APL) | Alberta Precision Labs (APL) | Gordon P et al |
| EPI_ISL_805576 | Canada/AB-37455/2020 | 2020-05-09 | North America/Canada/Alberta/ | Alberta Precision Labs (APL) | Alberta Precision Labs (APL) | Gordon P et al |
| EPI_ISL_805577 | Canada/AB-45566/2020 | 2020-05-12 | North America/Canada/Alberta/ | Alberta Precision Labs (APL) | Alberta Precision Labs (APL) | Gordon P et al |
| EPI_ISL_805578 | Canada/AB-46526/2020 | 2020-04-21 | North America/Canada/Alberta/ | Alberta Precision Labs (APL) | Alberta Precision Labs (APL) | Gordon P et al |
| EPI_ISL_805579 | Canada/AB-14224/2020 | 2020-05-06 | North America/Canada/Alberta/ | Alberta Precision Labs (APL) | Alberta Precision Labs (APL) | Gordon P et al |
| EPI_ISL_805580 | Canada/AB-25627/2020 | 2020-04-24 | North America/Canada/Alberta/ | Alberta Precision Labs (APL) | Alberta Precision Labs (APL) | Gordon P et al |
| EPI_ISL_805581 | Canada/AB-57628/2020 | 2020-03-17 | North America/Canada/Alberta/ | Alberta Precision Labs (APL) | Alberta Precision Labs (APL) | Gordon P et al |
| EPI_ISL_805582 | Canada/AB-59579/2020 | 2020-05-08 | North America/Canada/Alberta/ | Alberta Precision Labs (APL) | Alberta Precision Labs (APL) | Gordon P et al |
| EPI_ISL_805583 | Canada/AB-76326/2020 | 2020-04-29 | North America/Canada/Alberta/ | Alberta Precision Labs (APL) | Alberta Precision Labs (APL) | Gordon P et al |

| | | | | | | |
|---|---|---|---|---|---|---|
| EPI_ISL_805584 | Canada/AB-46250/2020 | 2020-04-29 | North America/Canada/Alberta/ | Alberta Precision Labs (APL) | Alberta Precision Labs (APL) | Gordon P et al |
| EPI_ISL_805585 | Canada/AB-79451/2020 | 2020-05-01 | North America/Canada/Alberta/ | Alberta Precision Labs (APL) | Alberta Precision Labs (APL) | Gordon P et al |
| EPI_ISL_805586 | Canada/AB-87665/2020 | 2020-05-04 | North America/Canada/Alberta/ | Alberta Precision Labs (APL) | Alberta Precision Labs (APL) | Gordon P et al |
| EPI_ISL_805587 | Canada/AB-78089/2020 | 2020-04-14 | North America/Canada/Alberta/ | Alberta Precision Labs (APL) | Alberta Precision Labs (APL) | Gordon P et al |
| EPI_ISL_805588 | Canada/AB-31990/2020 | 2020-04-18 | North America/Canada/Alberta/ | Alberta Precision Labs (APL) | Alberta Precision Labs (APL) | Gordon P et al |
| EPI_ISL_805589 | Canada/AB-92856/2020 | 2020-04-26 | North America/Canada/Alberta/ | Alberta Precision Labs (APL) | Alberta Precision Labs (APL) | Gordon P et al |
| EPI_ISL_805590 | Canada/AB-32825/2020 | 2020-05-09 | North America/Canada/Alberta/ | Alberta Precision Labs (APL) | Alberta Precision Labs (APL) | Gordon P et al |
| EPI_ISL_805591 | Canada/AB-15651/2020 | 2020-04-20 | North America/Canada/Alberta/ | Alberta Precision Labs (APL) | Alberta Precision Labs (APL) | Gordon P et al |
| EPI_ISL_805592 | Canada/AB-29665/2020 | 2020-04-24 | North America/Canada/Alberta/ | Alberta Precision Labs (APL) | Alberta Precision Labs (APL) | Gordon P et al |
| EPI_ISL_805593 | Canada/AB-53025/2020 | 2020-04-25 | North America/Canada/Alberta/ | Alberta Precision Labs (APL) | Alberta Precision Labs (APL) | Gordon P et al |
| EPI_ISL_805594 | Canada/AB-33062/2020 | 2020-04-25 | North America/Canada/Alberta/ | Alberta Precision Labs (APL) | Alberta Precision Labs (APL) | Gordon P et al |
| EPI_ISL_805595 | Canada/AB-84166/2020 | 2020-05-13 | North America/Canada/Alberta/ | Alberta Precision Labs (APL) | Alberta Precision Labs (APL) | Gordon P et al |
| EPI_ISL_805596 | Canada/AB-81176/2020 | 2020-04-30 | North America/Canada/Alberta/ | Alberta Precision Labs (APL) | Alberta Precision Labs (APL) | Gordon P et al |
| EPI_ISL_805597 | Canada/AB-44900/2020 | 2020-04-26 | North America/Canada/Alberta/ | Alberta Precision Labs (APL) | Alberta Precision Labs (APL) | Gordon P et al |
| EPI_ISL_805598 | Canada/AB-89391/2020 | 2020-05-01 | North America/Canada/Alberta/ | Alberta Precision Labs (APL) | Alberta Precision Labs (APL) | Gordon P et al |
| EPI_ISL_805599 | Canada/AB-22562/2020 | 2020-04-13 | North America/Canada/Alberta/ | Alberta Precision Labs (APL) | Alberta Precision Labs (APL) | Gordon P et al |
| EPI_ISL_805600 | Canada/AB-24849/2020 | 2020-05-10 | North America/Canada/Alberta/ | Alberta Precision Labs (APL) | Alberta Precision Labs (APL) | Gordon P et al |
| EPI_ISL_805601 | Canada/AB-73345/2020 | 2020-05-12 | North America/Canada/Alberta/ | Alberta Precision Labs (APL) | Alberta Precision Labs (APL) | Gordon P et al |
| EPI_ISL_805602 | Canada/AB-91240/2020 | 2020-04-15 | North America/Canada/Alberta/ | Alberta Precision Labs (APL) | Alberta Precision Labs (APL) | Gordon P et al |
| EPI_ISL_805603 | Canada/AB-47490/2020 | 2020-04-20 | North America/Canada/Alberta/ | Alberta Precision Labs (APL) | Alberta Precision Labs (APL) | Gordon P et al |

| | | | | | | |
|---|---|---|---|---|---|---|
| EPI_ISL_805604 | Canada/AB-15857/2020 | 2020-04-30 | North America/Canada/Alberta/ | Alberta Precision Labs (APL) | Alberta Precision Labs (APL) | Gordon P et al |
| EPI_ISL_805605 | Canada/AB-79033/2020 | 2020-05-01 | North America/Canada/Alberta/ | Alberta Precision Labs (APL) | Alberta Precision Labs (APL) | Gordon P et al |
| EPI_ISL_805606 | Canada/AB-96900/2020 | 2020-04-28 | North America/Canada/Alberta/ | Alberta Precision Labs (APL) | Alberta Precision Labs (APL) | Gordon P et al |
| EPI_ISL_805607 | Canada/AB-24367/2020 | 2020-04-27 | North America/Canada/Alberta/ | Alberta Precision Labs (APL) | Alberta Precision Labs (APL) | Gordon P et al |
| EPI_ISL_805608 | Canada/AB-26371/2020 | 2020-05-03 | North America/Canada/Alberta/ | Alberta Precision Labs (APL) | Alberta Precision Labs (APL) | Gordon P et al |
| EPI_ISL_805609 | Canada/AB-41438/2020 | 2020-04-25 | North America/Canada/Alberta/ | Alberta Precision Labs (APL) | Alberta Precision Labs (APL) | Gordon P et al |
| EPI_ISL_805610 | Canada/AB-63795/2020 | 2020-05-15 | North America/Canada/Alberta/ | Alberta Precision Labs (APL) | Alberta Precision Labs (APL) | Gordon P et al |
| EPI_ISL_805611 | Canada/AB-62175/2020 | 2020-04-13 | North America/Canada/Alberta/ | Alberta Precision Labs (APL) | Alberta Precision Labs (APL) | Gordon P et al |
| EPI_ISL_805612 | Canada/AB-51977/2020 | 2020-04-27 | North America/Canada/Alberta/ | Alberta Precision Labs (APL) | Alberta Precision Labs (APL) | Gordon P et al |
| EPI_ISL_805613 | Canada/AB-31420/2020 | 2020-04-26 | North America/Canada/Alberta/ | Alberta Precision Labs (APL) | Alberta Precision Labs (APL) | Gordon P et al |
| EPI_ISL_805614 | Canada/AB-85410/2020 | 2020-04-26 | North America/Canada/Alberta/ | Alberta Precision Labs (APL) | Alberta Precision Labs (APL) | Gordon P et al |
| EPI_ISL_805615 | Canada/AB-72930/2020 | 2020-03-20 | North America/Canada/Alberta/ | Alberta Precision Labs (APL) | Alberta Precision Labs (APL) | Gordon P et al |
| EPI_ISL_805616 | Canada/AB-16873/2020 | 2020-04-12 | North America/Canada/Alberta/ | Alberta Precision Labs (APL) | Alberta Precision Labs (APL) | Gordon P et al |
| EPI_ISL_805617 | Canada/AB-20557/2020 | 2020-04-29 | North America/Canada/Alberta/ | Alberta Precision Labs (APL) | Alberta Precision Labs (APL) | Gordon P et al |
| EPI_ISL_805618 | Canada/AB-85652/2020 | 2020-05-13 | North America/Canada/Alberta/ | Alberta Precision Labs (APL) | Alberta Precision Labs (APL) | Gordon P et al |
| EPI_ISL_805619 | Canada/AB-52254/2020 | 2020-04-18 | North America/Canada/Alberta/ | Alberta Precision Labs (APL) | Alberta Precision Labs (APL) | Gordon P et al |
| EPI_ISL_805620 | Canada/AB-64552/2020 | 2020-04-21 | North America/Canada/Alberta/ | Alberta Precision Labs (APL) | Alberta Precision Labs (APL) | Gordon P et al |
| EPI_ISL_805621 | Canada/AB-59612/2020 | 2020-04-28 | North America/Canada/Alberta/ | Alberta Precision Labs (APL) | Alberta Precision Labs (APL) | Gordon P et al |
| EPI_ISL_805622 | Canada/AB-39872/2020 | 2020-04-26 | North America/Canada/Alberta/ | Alberta Precision Labs (APL) | Alberta Precision Labs (APL) | Gordon P et al |
| EPI_ISL_805623 | Canada/AB-47944/2020 | 2020-04-19 | North America/Canada/Alberta/ | Alberta Precision Labs (APL) | Alberta Precision Labs (APL) | Gordon P et al |

| | | | | | | |
|---|---|---|---|---|---|---|
| EPI_ISL_805624 | Canada/AB-44545/2020 | 2020-04-15 | North America/Canada/Alberta/ | Alberta Precision Labs (APL) | Alberta Precision Labs (APL) | Gordon P et al |
| EPI_ISL_805625 | Canada/AB-56126/2020 | 2020-04-26 | North America/Canada/Alberta/ | Alberta Precision Labs (APL) | Alberta Precision Labs (APL) | Gordon P et al |
| EPI_ISL_805626 | Canada/AB-74914/2020 | 2020-04-29 | North America/Canada/Alberta/ | Alberta Precision Labs (APL) | Alberta Precision Labs (APL) | Gordon P et al |
| EPI_ISL_805627 | Canada/AB-81321/2020 | 2020-04-29 | North America/Canada/Alberta/ | Alberta Precision Labs (APL) | Alberta Precision Labs (APL) | Gordon P et al |
| EPI_ISL_805628 | Canada/AB-47607/2020 | 2020-04-19 | North America/Canada/Alberta/ | Alberta Precision Labs (APL) | Alberta Precision Labs (APL) | Gordon P et al |
| EPI_ISL_805629 | Canada/AB-51588/2020 | 2020-04-26 | North America/Canada/Alberta/ | Alberta Precision Labs (APL) | Alberta Precision Labs (APL) | Gordon P et al |
| EPI_ISL_805630 | Canada/AB-70584/2020 | 2020-04-25 | North America/Canada/Alberta/ | Alberta Precision Labs (APL) | Alberta Precision Labs (APL) | Gordon P et al |
| EPI_ISL_805631 | Canada/AB-29188/2020 | 2020-04-22 | North America/Canada/Alberta/ | Alberta Precision Labs (APL) | Alberta Precision Labs (APL) | Gordon P et al |
| EPI_ISL_805632 | Canada/AB-32729/2020 | 2020-04-14 | North America/Canada/Alberta/ | Alberta Precision Labs (APL) | Alberta Precision Labs (APL) | Gordon P et al |
| EPI_ISL_805633 | Canada/AB-55893/2020 | 2020-04-23 | North America/Canada/Alberta/ | Alberta Precision Labs (APL) | Alberta Precision Labs (APL) | Gordon P et al |
| EPI_ISL_805634 | Canada/AB-90914/2020 | 2020-04-25 | North America/Canada/Alberta/ | Alberta Precision Labs (APL) | Alberta Precision Labs (APL) | Gordon P et al |
| EPI_ISL_805635 | Canada/AB-26977/2020 | 2020-04-17 | North America/Canada/Alberta/ | Alberta Precision Labs (APL) | Alberta Precision Labs (APL) | Gordon P et al |
| EPI_ISL_805636 | Canada/AB-46935/2020 | 2020-04-21 | North America/Canada/Alberta/ | Alberta Precision Labs (APL) | Alberta Precision Labs (APL) | Gordon P et al |
| EPI_ISL_805637 | Canada/AB-47167/2020 | 2020-04-16 | North America/Canada/Alberta/ | Alberta Precision Labs (APL) | Alberta Precision Labs (APL) | Gordon P et al |
| EPI_ISL_805638 | Canada/AB-44992/2020 | 2020-04-26 | North America/Canada/Alberta/ | Alberta Precision Labs (APL) | Alberta Precision Labs (APL) | Gordon P et al |
| EPI_ISL_805639 | Canada/AB-45504/2020 | 2020-04-25 | North America/Canada/Alberta/ | Alberta Precision Labs (APL) | Alberta Precision Labs (APL) | Gordon P et al |
| EPI_ISL_805640 | Canada/AB-55062/2020 | 2020-04-27 | North America/Canada/Alberta/ | Alberta Precision Labs (APL) | Alberta Precision Labs (APL) | Gordon P et al |
| EPI_ISL_805641 | Canada/AB-19219/2020 | 2020-04-20 | North America/Canada/Alberta/ | Alberta Precision Labs (APL) | Alberta Precision Labs (APL) | Gordon P et al |
| EPI_ISL_805642 | Canada/AB-43667/2020 | 2020-04-26 | North America/Canada/Alberta/ | Alberta Precision Labs (APL) | Alberta Precision Labs (APL) | Gordon P et al |
| EPI_ISL_805643 | Canada/AB-20226/2020 | 2020-04-23 | North America/Canada/Alberta/ | Alberta Precision Labs (APL) | Alberta Precision Labs (APL) | Gordon P et al |

| | | | | | | |
|---|---|---|---|---|---|---|
| EPI_ISL_805644 | Canada/AB-20299/2020 | 2020-04-26 | North America/Canada/Alberta/ | Alberta Precision Labs (APL) | Alberta Precision Labs (APL) | Gordon P et al |
| EPI_ISL_805645 | Canada/AB-25897/2020 | 2020-05-11 | North America/Canada/Alberta/ | Alberta Precision Labs (APL) | Alberta Precision Labs (APL) | Gordon P et al |
| EPI_ISL_805646 | Canada/AB-44495/2020 | 2020-05-09 | North America/Canada/Alberta/ | Alberta Precision Labs (APL) | Alberta Precision Labs (APL) | Gordon P et al |
| EPI_ISL_805647 | Canada/AB-51564/2020 | 2020-04-21 | North America/Canada/Alberta/ | Alberta Precision Labs (APL) | Alberta Precision Labs (APL) | Gordon P et al |
| EPI_ISL_805648 | Canada/AB-68347/2020 | 2020-04-14 | North America/Canada/Alberta/ | Alberta Precision Labs (APL) | Alberta Precision Labs (APL) | Gordon P et al |
| EPI_ISL_805650 | Canada/AB-63400/2020 | 2020-04-29 | North America/Canada/Alberta/ | Alberta Precision Labs (APL) | Alberta Precision Labs (APL) | Gordon P et al |
| EPI_ISL_805651 | Canada/AB-44714/2020 | 2020-04-19 | North America/Canada/Alberta/ | Alberta Precision Labs (APL) | Alberta Precision Labs (APL) | Gordon P et al |
| EPI_ISL_805652 | Canada/AB-30191/2020 | 2020-04-21 | North America/Canada/Alberta/ | Alberta Precision Labs (APL) | Alberta Precision Labs (APL) | Gordon P et al |
| EPI_ISL_805653 | Canada/AB-66026/2020 | 2020-04-04 | North America/Canada/Alberta/ | Alberta Precision Labs (APL) | Alberta Precision Labs (APL) | Gordon P et al |
| EPI_ISL_805654 | Canada/AB-49589/2020 | 2020-04-18 | North America/Canada/Alberta/ | Alberta Precision Labs (APL) | Alberta Precision Labs (APL) | Gordon P et al |
| EPI_ISL_805655 | Canada/AB-48786/2020 | 2020-04-26 | North America/Canada/Alberta/ | Alberta Precision Labs (APL) | Alberta Precision Labs (APL) | Gordon P et al |
| EPI_ISL_805656 | Canada/AB-50176/2020 | 2020-04-15 | North America/Canada/Alberta/ | Alberta Precision Labs (APL) | Alberta Precision Labs (APL) | Gordon P et al |
| EPI_ISL_805657 | Canada/AB-47857/2020 | 2020-04-25 | North America/Canada/Alberta/ | Alberta Precision Labs (APL) | Alberta Precision Labs (APL) | Gordon P et al |
| EPI_ISL_805658 | Canada/AB-54090/2020 | 2020-04-18 | North America/Canada/Alberta/ | Alberta Precision Labs (APL) | Alberta Precision Labs (APL) | Gordon P et al |
| EPI_ISL_805659 | Canada/AB-63929/2020 | 2020-05-14 | North America/Canada/Alberta/ | Alberta Precision Labs (APL) | Alberta Precision Labs (APL) | Gordon P et al |
| EPI_ISL_805660 | Canada/AB-71941/2020 | 2020-04-26 | North America/Canada/Alberta/ | Alberta Precision Labs (APL) | Alberta Precision Labs (APL) | Gordon P et al |
| EPI_ISL_805661 | Canada/AB-99354/2020 | 2020-04-14 | North America/Canada/Alberta/ | Alberta Precision Labs (APL) | Alberta Precision Labs (APL) | Gordon P et al |
| EPI_ISL_805662 | Canada/AB-38173/2020 | 2020-04-28 | North America/Canada/Alberta/ | Alberta Precision Labs (APL) | Alberta Precision Labs (APL) | Gordon P et al |
| EPI_ISL_805663 | Canada/AB-90145/2020 | 2020-04-13 | North America/Canada/Alberta/ | Alberta Precision Labs (APL) | Alberta Precision Labs (APL) | Gordon P et al |
| EPI_ISL_805664 | Canada/AB-35885/2020 | 2020-05-06 | North America/Canada/Alberta/ | Alberta Precision Labs (APL) | Alberta Precision Labs (APL) | Gordon P et al |

| | | | | | | |
|---|---|---|---|---|---|---|
| EPI_ISL_805665 | Canada/AB-29338/2020 | 2020-05-02 | North America/Canada/Alberta/ | Alberta Precision Labs (APL) | Alberta Precision Labs (APL) | Gordon P et al |
| EPI_ISL_805666 | Canada/AB-92121/2020 | 2020-04-18 | North America/Canada/Alberta/ | Alberta Precision Labs (APL) | Alberta Precision Labs (APL) | Gordon P et al |
| EPI_ISL_805667 | Canada/AB-94923/2020 | 2020-04-28 | North America/Canada/Alberta/ | Alberta Precision Labs (APL) | Alberta Precision Labs (APL) | Gordon P et al |
| EPI_ISL_805668 | Canada/AB-66305/2020 | 2020-04-25 | North America/Canada/Alberta/ | Alberta Precision Labs (APL) | Alberta Precision Labs (APL) | Gordon P et al |
| EPI_ISL_805669 | Canada/AB-58087/2020 | 2020-03-30 | North America/Canada/Alberta/ | Alberta Precision Labs (APL) | Alberta Precision Labs (APL) | Gordon P et al |
| EPI_ISL_805670 | Canada/AB-17011/2020 | 2020-04-13 | North America/Canada/Alberta/ | Alberta Precision Labs (APL) | Alberta Precision Labs (APL) | Gordon P et al |
| EPI_ISL_805671 | Canada/AB-99138/2020 | 2020-04-17 | North America/Canada/Alberta/ | Alberta Precision Labs (APL) | Alberta Precision Labs (APL) | Gordon P et al |
| EPI_ISL_805672 | Canada/AB-49293/2020 | 2020-04-26 | North America/Canada/Alberta/ | Alberta Precision Labs (APL) | Alberta Precision Labs (APL) | Gordon P et al |
| EPI_ISL_805673 | Canada/AB-84064/2020 | 2020-05-09 | North America/Canada/Alberta/ | Alberta Precision Labs (APL) | Alberta Precision Labs (APL) | Gordon P et al |
| EPI_ISL_805674 | Canada/AB-36367/2020 | 2020-04-15 | North America/Canada/Alberta/ | Alberta Precision Labs (APL) | Alberta Precision Labs (APL) | Gordon P et al |
| EPI_ISL_805675 | Canada/AB-16172/2020 | 2020-04-26 | North America/Canada/Alberta/ | Alberta Precision Labs (APL) | Alberta Precision Labs (APL) | Gordon P et al |
| EPI_ISL_805676 | Canada/AB-53919/2020 | 2020-04-21 | North America/Canada/Alberta/ | Alberta Precision Labs (APL) | Alberta Precision Labs (APL) | Gordon P et al |
| EPI_ISL_805677 | Canada/AB-96199/2020 | 2020-05-01 | North America/Canada/Alberta/ | Alberta Precision Labs (APL) | Alberta Precision Labs (APL) | Gordon P et al |
| EPI_ISL_805678 | Canada/AB-62719/2020 | 2020-05-09 | North America/Canada/Alberta/ | Alberta Precision Labs (APL) | Alberta Precision Labs (APL) | Gordon P et al |
| EPI_ISL_805679 | Canada/AB-35024/2020 | 2020-03-18 | North America/Canada/Alberta/ | Alberta Precision Labs (APL) | Alberta Precision Labs (APL) | Gordon P et al |
| EPI_ISL_805680 | Canada/AB-97306/2020 | 2020-04-25 | North America/Canada/Alberta/ | Alberta Precision Labs (APL) | Alberta Precision Labs (APL) | Gordon P et al |
| EPI_ISL_805681 | Canada/AB-52715/2020 | 2020-04-30 | North America/Canada/Alberta/ | Alberta Precision Labs (APL) | Alberta Precision Labs (APL) | Gordon P et al |
| EPI_ISL_805682 | Canada/AB-61725/2020 | 2020-04-28 | North America/Canada/Alberta/ | Alberta Precision Labs (APL) | Alberta Precision Labs (APL) | Gordon P et al |
| EPI_ISL_805683 | Canada/AB-49603/2020 | 2020-04-28 | North America/Canada/Alberta/ | Alberta Precision Labs (APL) | Alberta Precision Labs (APL) | Gordon P et al |
| EPI_ISL_805684 | Canada/AB-33852/2020 | 2020-04-27 | North America/Canada/Alberta/ | Alberta Precision Labs (APL) | Alberta Precision Labs (APL) | Gordon P et al |

| | | | | | | |
|---|---|---|---|---|---|---|
| EPI_ISL_805685 | Canada/AB-13690/2020 | 2020-04-13 | North America/Canada/Alberta/ | Alberta Precision Labs (APL) | Alberta Precision Labs (APL) | Gordon P et al |
| EPI_ISL_805686 | Canada/AB-72489/2020 | 2020-04-22 | North America/Canada/Alberta/ | Alberta Precision Labs (APL) | Alberta Precision Labs (APL) | Gordon P et al |
| EPI_ISL_805687 | Canada/AB-28227/2020 | 2020-04-22 | North America/Canada/Alberta/ | Alberta Precision Labs (APL) | Alberta Precision Labs (APL) | Gordon P et al |
| EPI_ISL_805688 | Canada/AB-95765/2020 | 2020-04-23 | North America/Canada/Alberta/ | Alberta Precision Labs (APL) | Alberta Precision Labs (APL) | Gordon P et al |
| EPI_ISL_805689 | Canada/AB-90054/2020 | 2020-04-22 | North America/Canada/Alberta/ | Alberta Precision Labs (APL) | Alberta Precision Labs (APL) | Gordon P et al |
| EPI_ISL_805690 | Canada/AB-97179/2020 | 2020-04-28 | North America/Canada/Alberta/ | Alberta Precision Labs (APL) | Alberta Precision Labs (APL) | Gordon P et al |
| EPI_ISL_805691 | Canada/AB-24031/2020 | 2020-05-02 | North America/Canada/Alberta/ | Alberta Precision Labs (APL) | Alberta Precision Labs (APL) | Gordon P et al |
| EPI_ISL_805692 | Canada/AB-61784/2020 | 2020-04-14 | North America/Canada/Alberta/ | Alberta Precision Labs (APL) | Alberta Precision Labs (APL) | Gordon P et al |
| EPI_ISL_805693 | Canada/AB-40341/2020 | 2020-05-14 | North America/Canada/Alberta/ | Alberta Precision Labs (APL) | Alberta Precision Labs (APL) | Gordon P et al |
| EPI_ISL_805694 | Canada/AB-47765/2020 | 2020-04-20 | North America/Canada/Alberta/ | Alberta Precision Labs (APL) | Alberta Precision Labs (APL) | Gordon P et al |
| EPI_ISL_805695 | Canada/AB-53784/2020 | 2020-03-15 | North America/Canada/Alberta/ | Alberta Precision Labs (APL) | Alberta Precision Labs (APL) | Gordon P et al |
| EPI_ISL_805696 | Canada/AB-80352/2020 | 2020-04-29 | North America/Canada/Alberta/ | Alberta Precision Labs (APL) | Alberta Precision Labs (APL) | Gordon P et al |
| EPI_ISL_805697 | Canada/AB-44355/2020 | 2020-04-28 | North America/Canada/Alberta/ | Alberta Precision Labs (APL) | Alberta Precision Labs (APL) | Gordon P et al |
| EPI_ISL_805698 | Canada/AB-25088/2020 | 2020-04-17 | North America/Canada/Alberta/ | Alberta Precision Labs (APL) | Alberta Precision Labs (APL) | Gordon P et al |
| EPI_ISL_805699 | Canada/AB-18219/2020 | 2020-04-29 | North America/Canada/Alberta/ | Alberta Precision Labs (APL) | Alberta Precision Labs (APL) | Gordon P et al |
| EPI_ISL_805701 | Canada/AB-30579/2020 | 2020-03-29 | North America/Canada/Alberta/ | Alberta Precision Labs (APL) | Alberta Precision Labs (APL) | Gordon P et al |
| EPI_ISL_805702 | Canada/AB-57853/2020 | 2020-04-14 | North America/Canada/Alberta/ | Alberta Precision Labs (APL) | Alberta Precision Labs (APL) | Gordon P et al |
| EPI_ISL_805703 | Canada/AB-48425/2020 | 2020-04-18 | North America/Canada/Alberta/ | Alberta Precision Labs (APL) | Alberta Precision Labs (APL) | Gordon P et al |
| EPI_ISL_805704 | Canada/AB-32418/2020 | 2020-04-27 | North America/Canada/Alberta/ | Alberta Precision Labs (APL) | Alberta Precision Labs (APL) | Gordon P et al |
| EPI_ISL_805705 | Canada/AB-93398/2020 | 2020-04-18 | North America/Canada/Alberta/ | Alberta Precision Labs (APL) | Alberta Precision Labs (APL) | Gordon P et al |

| | | | | | | |
|---|---|---|---|---|---|---|
| EPI_ISL_805706 | Canada/AB-87208/2020 | 2020-05-01 | North America/Canada/Alberta/ | Alberta Precision Labs (APL) | Alberta Precision Labs (APL) | Gordon P et al |
| EPI_ISL_805707 | Canada/AB-66199/2020 | 2020-04-29 | North America/Canada/Alberta/ | Alberta Precision Labs (APL) | Alberta Precision Labs (APL) | Gordon P et al |
| EPI_ISL_805708 | Canada/AB-16323/2020 | 2020-05-13 | North America/Canada/Alberta/ | Alberta Precision Labs (APL) | Alberta Precision Labs (APL) | Gordon P et al |
| EPI_ISL_805709 | Canada/AB-39692/2020 | 2020-04-26 | North America/Canada/Alberta/ | Alberta Precision Labs (APL) | Alberta Precision Labs (APL) | Gordon P et al |
| EPI_ISL_805710 | Canada/AB-19086/2020 | 2020-04-14 | North America/Canada/Alberta/ | Alberta Precision Labs (APL) | Alberta Precision Labs (APL) | Gordon P et al |
| EPI_ISL_805711 | Canada/AB-87153/2020 | 2020-04-15 | North America/Canada/Alberta/ | Alberta Precision Labs (APL) | Alberta Precision Labs (APL) | Gordon P et al |
| EPI_ISL_805712 | Canada/AB-29268/2020 | 2020-04-16 | North America/Canada/Alberta/ | Alberta Precision Labs (APL) | Alberta Precision Labs (APL) | Gordon P et al |
| EPI_ISL_805713 | Canada/AB-12948/2020 | 2020-05-07 | North America/Canada/Alberta/ | Alberta Precision Labs (APL) | Alberta Precision Labs (APL) | Gordon P et al |
| EPI_ISL_805714 | Canada/AB-46885/2020 | 2020-03-22 | North America/Canada/Alberta/ | Alberta Precision Labs (APL) | Alberta Precision Labs (APL) | Gordon P et al |
| EPI_ISL_805715 | Canada/AB-68670/2020 | 2020-05-05 | North America/Canada/Alberta/ | Alberta Precision Labs (APL) | Alberta Precision Labs (APL) | Gordon P et al |
| EPI_ISL_805716 | Canada/AB-71854/2020 | 2020-04-18 | North America/Canada/Alberta/ | Alberta Precision Labs (APL) | Alberta Precision Labs (APL) | Gordon P et al |
| EPI_ISL_805717 | Canada/AB-47381/2020 | 2020-04-27 | North America/Canada/Alberta/ | Alberta Precision Labs (APL) | Alberta Precision Labs (APL) | Gordon P et al |
| EPI_ISL_805718 | Canada/AB-15046/2020 | 2020-04-23 | North America/Canada/Alberta/ | Alberta Precision Labs (APL) | Alberta Precision Labs (APL) | Gordon P et al |
| EPI_ISL_805719 | Canada/AB-89780/2020 | 2020-04-29 | North America/Canada/Alberta/ | Alberta Precision Labs (APL) | Alberta Precision Labs (APL) | Gordon P et al |
| EPI_ISL_805720 | Canada/AB-39102/2020 | 2020-05-03 | North America/Canada/Alberta/ | Alberta Precision Labs (APL) | Alberta Precision Labs (APL) | Gordon P et al |
| EPI_ISL_805721 | Canada/AB-86504/2020 | 2020-04-26 | North America/Canada/Alberta/ | Alberta Precision Labs (APL) | Alberta Precision Labs (APL) | Gordon P et al |
| EPI_ISL_805722 | Canada/AB-32964/2020 | 2020-04-24 | North America/Canada/Alberta/ | Alberta Precision Labs (APL) | Alberta Precision Labs (APL) | Gordon P et al |
| EPI_ISL_805723 | Canada/AB-49824/2020 | 2020-04-06 | North America/Canada/Alberta/ | Alberta Precision Labs (APL) | Alberta Precision Labs (APL) | Gordon P et al |
| EPI_ISL_805724 | Canada/AB-49813/2020 | 2020-04-25 | North America/Canada/Alberta/ | Alberta Precision Labs (APL) | Alberta Precision Labs (APL) | Gordon P et al |
| EPI_ISL_805725 | Canada/AB-95555/2020 | 2020-04-29 | North America/Canada/Alberta/ | Alberta Precision Labs (APL) | Alberta Precision Labs (APL) | Gordon P et al |

| | | | | | | |
|---|---|---|---|---|---|---|
| EPI_ISL_805726 | Canada/AB-18778/2020 | 2020-05-10 | North America/Canada/Alberta/ | Alberta Precision Labs (APL) | Alberta Precision Labs (APL) | Gordon P et al |
| EPI_ISL_805727 | Canada/AB-43394/2020 | 2020-04-18 | North America/Canada/Alberta/ | Alberta Precision Labs (APL) | Alberta Precision Labs (APL) | Gordon P et al |
| EPI_ISL_805728 | Canada/AB-52009/2020 | 2020-04-15 | North America/Canada/Alberta/ | Alberta Precision Labs (APL) | Alberta Precision Labs (APL) | Gordon P et al |
| EPI_ISL_805729 | Canada/AB-86794/2020 | 2020-04-24 | North America/Canada/Alberta/ | Alberta Precision Labs (APL) | Alberta Precision Labs (APL) | Gordon P et al |
| EPI_ISL_805730 | Canada/AB-62294/2020 | 2020-04-08 | North America/Canada/Alberta/ | Alberta Precision Labs (APL) | Alberta Precision Labs (APL) | Gordon P et al |
| EPI_ISL_805731 | Canada/AB-55014/2020 | 2020-04-27 | North America/Canada/Alberta/ | Alberta Precision Labs (APL) | Alberta Precision Labs (APL) | Gordon P et al |
| EPI_ISL_805732 | Canada/AB-20431/2020 | 2020-04-28 | North America/Canada/Alberta/ | Alberta Precision Labs (APL) | Alberta Precision Labs (APL) | Gordon P et al |
| EPI_ISL_805733 | Canada/AB-55331/2020 | 2020-05-03 | North America/Canada/Alberta/ | Alberta Precision Labs (APL) | Alberta Precision Labs (APL) | Gordon P et al |
| EPI_ISL_805734 | Canada/AB-98191/2020 | 2020-04-25 | North America/Canada/Alberta/ | Alberta Precision Labs (APL) | Alberta Precision Labs (APL) | Gordon P et al |
| EPI_ISL_805735 | Canada/AB-66829/2020 | 2020-05-13 | North America/Canada/Alberta/ | Alberta Precision Labs (APL) | Alberta Precision Labs (APL) | Gordon P et al |
| EPI_ISL_805736 | Canada/AB-31552/2020 | 2020-05-07 | North America/Canada/Alberta/ | Alberta Precision Labs (APL) | Alberta Precision Labs (APL) | Gordon P et al |
| EPI_ISL_805737 | Canada/AB-34035/2020 | 2020-04-24 | North America/Canada/Alberta/ | Alberta Precision Labs (APL) | Alberta Precision Labs (APL) | Gordon P et al |
| EPI_ISL_805738 | Canada/AB-49549/2020 | 2020-05-16 | North America/Canada/Alberta/ | Alberta Precision Labs (APL) | Alberta Precision Labs (APL) | Gordon P et al |
| EPI_ISL_805739 | Canada/AB-72538/2020 | 2020-04-26 | North America/Canada/Alberta/ | Alberta Precision Labs (APL) | Alberta Precision Labs (APL) | Gordon P et al |
| EPI_ISL_805740 | Canada/AB-49350/2020 | 2020-05-01 | North America/Canada/Alberta/ | Alberta Precision Labs (APL) | Alberta Precision Labs (APL) | Gordon P et al |
| EPI_ISL_805741 | Canada/AB-55820/2020 | 2020-04-28 | North America/Canada/Alberta/ | Alberta Precision Labs (APL) | Alberta Precision Labs (APL) | Gordon P et al |
| EPI_ISL_805742 | Canada/AB-67767/2020 | 2020-04-23 | North America/Canada/Alberta/ | Alberta Precision Labs (APL) | Alberta Precision Labs (APL) | Gordon P et al |
| EPI_ISL_805743 | Canada/AB-90958/2020 | 2020-05-18 | North America/Canada/Alberta/ | Alberta Precision Labs (APL) | Alberta Precision Labs (APL) | Gordon P et al |
| EPI_ISL_805744 | Canada/AB-17516/2020 | 2020-05-07 | North America/Canada/Alberta/ | Alberta Precision Labs (APL) | Alberta Precision Labs (APL) | Gordon P et al |
| EPI_ISL_805745 | Canada/AB-16936/2020 | 2020-05-08 | North America/Canada/Alberta/ | Alberta Precision Labs (APL) | Alberta Precision Labs (APL) | Gordon P et al |

| | | | | | | |
|---|---|---|---|---|---|---|
| EPI_ISL_805746 | Canada/AB-88379/2020 | 2020-03-27 | North America/Canada/Alberta/ | Alberta Precision Labs (APL) | Alberta Precision Labs (APL) | Gordon P et al |
| EPI_ISL_805747 | Canada/AB-30707/2020 | 2020-04-30 | North America/Canada/Alberta/ | Alberta Precision Labs (APL) | Alberta Precision Labs (APL) | Gordon P et al |
| EPI_ISL_805748 | Canada/AB-53993/2020 | 2020-04-23 | North America/Canada/Alberta/ | Alberta Precision Labs (APL) | Alberta Precision Labs (APL) | Gordon P et al |
| EPI_ISL_805749 | Canada/AB-91130/2020 | 2020-05-08 | North America/Canada/Alberta/ | Alberta Precision Labs (APL) | Alberta Precision Labs (APL) | Gordon P et al |
| EPI_ISL_805750 | Canada/AB-54511/2020 | 2020-05-01 | North America/Canada/Alberta/ | Alberta Precision Labs (APL) | Alberta Precision Labs (APL) | Gordon P et al |
| EPI_ISL_805751 | Canada/AB-22109/2020 | 2020-04-30 | North America/Canada/Alberta/ | Alberta Precision Labs (APL) | Alberta Precision Labs (APL) | Gordon P et al |
| EPI_ISL_805752 | Canada/AB-98718/2020 | 2020-04-17 | North America/Canada/Alberta/ | Alberta Precision Labs (APL) | Alberta Precision Labs (APL) | Gordon P et al |
| EPI_ISL_805753 | Canada/AB-47614/2020 | 2020-05-17 | North America/Canada/Alberta/ | Alberta Precision Labs (APL) | Alberta Precision Labs (APL) | Gordon P et al |
| EPI_ISL_805754 | Canada/AB-69846/2020 | 2020-04-28 | North America/Canada/Alberta/ | Alberta Precision Labs (APL) | Alberta Precision Labs (APL) | Gordon P et al |
| EPI_ISL_805755 | Canada/AB-45021/2020 | 2020-03-27 | North America/Canada/Alberta/ | Alberta Precision Labs (APL) | Alberta Precision Labs (APL) | Gordon P et al |
| EPI_ISL_805756 | Canada/AB-74539/2020 | 2020-04-27 | North America/Canada/Alberta/ | Alberta Precision Labs (APL) | Alberta Precision Labs (APL) | Gordon P et al |
| EPI_ISL_805757 | Canada/AB-16541/2020 | 2020-04-23 | North America/Canada/Alberta/ | Alberta Precision Labs (APL) | Alberta Precision Labs (APL) | Gordon P et al |
| EPI_ISL_805758 | Canada/AB-16177/2020 | 2020-04-16 | North America/Canada/Alberta/ | Alberta Precision Labs (APL) | Alberta Precision Labs (APL) | Gordon P et al |
| EPI_ISL_805759 | Canada/AB-61981/2020 | 2020-05-07 | North America/Canada/Alberta/ | Alberta Precision Labs (APL) | Alberta Precision Labs (APL) | Gordon P et al |
| EPI_ISL_805760 | Canada/AB-42430/2020 | 2020-04-25 | North America/Canada/Alberta/ | Alberta Precision Labs (APL) | Alberta Precision Labs (APL) | Gordon P et al |
| EPI_ISL_805761 | Canada/AB-21276/2020 | 2020-04-14 | North America/Canada/Alberta/ | Alberta Precision Labs (APL) | Alberta Precision Labs (APL) | Gordon P et al |
| EPI_ISL_805762 | Canada/AB-24593/2020 | 2020-04-26 | North America/Canada/Alberta/ | Alberta Precision Labs (APL) | Alberta Precision Labs (APL) | Gordon P et al |
| EPI_ISL_805763 | Canada/AB-46971/2020 | 2020-03-23 | North America/Canada/Alberta/ | Alberta Precision Labs (APL) | Alberta Precision Labs (APL) | Gordon P et al |
| EPI_ISL_805765 | Canada/AB-83540/2020 | 2020-04-26 | North America/Canada/Alberta/ | Alberta Precision Labs (APL) | Alberta Precision Labs (APL) | Gordon P et al |
| EPI_ISL_805766 | Canada/AB-52746/2020 | 2020-04-13 | North America/Canada/Alberta/ | Alberta Precision Labs (APL) | Alberta Precision Labs (APL) | Gordon P et al |

| | | | | | | |
|---|---|---|---|---|---|---|
| EPI_ISL_805767 | Canada/AB-63539/2020 | 2020-04-27 | North America/Canada/Alberta/ | Alberta Precision Labs (APL) | Alberta Precision Labs (APL) | Gordon P et al |
| EPI_ISL_805768 | Canada/AB-49150/2020 | 2020-04-28 | North America/Canada/Alberta/ | Alberta Precision Labs (APL) | Alberta Precision Labs (APL) | Gordon P et al |
| EPI_ISL_805769 | Canada/AB-61758/2020 | 2020-04-24 | North America/Canada/Alberta/ | Alberta Precision Labs (APL) | Alberta Precision Labs (APL) | Gordon P et al |
| EPI_ISL_805770 | Canada/AB-12513/2020 | 2020-04-13 | North America/Canada/Alberta/ | Alberta Precision Labs (APL) | Alberta Precision Labs (APL) | Gordon P et al |
| EPI_ISL_805771 | Canada/AB-56469/2020 | 2020-04-27 | North America/Canada/Alberta/ | Alberta Precision Labs (APL) | Alberta Precision Labs (APL) | Gordon P et al |
| EPI_ISL_805772 | Canada/AB-61994/2020 | 2020-05-14 | North America/Canada/Alberta/ | Alberta Precision Labs (APL) | Alberta Precision Labs (APL) | Gordon P et al |
| EPI_ISL_805773 | Canada/AB-14058/2020 | 2020-04-27 | North America/Canada/Alberta/ | Alberta Precision Labs (APL) | Alberta Precision Labs (APL) | Gordon P et al |
| EPI_ISL_805774 | Canada/AB-74015/2020 | 2020-04-29 | North America/Canada/Alberta/ | Alberta Precision Labs (APL) | Alberta Precision Labs (APL) | Gordon P et al |
| EPI_ISL_805775 | Canada/AB-34712/2020 | 2020-05-07 | North America/Canada/Alberta/ | Alberta Precision Labs (APL) | Alberta Precision Labs (APL) | Gordon P et al |
| EPI_ISL_805776 | Canada/AB-60250/2020 | 2020-04-21 | North America/Canada/Alberta/ | Alberta Precision Labs (APL) | Alberta Precision Labs (APL) | Gordon P et al |
| EPI_ISL_805777 | Canada/AB-79755/2020 | 2020-04-28 | North America/Canada/Alberta/ | Alberta Precision Labs (APL) | Alberta Precision Labs (APL) | Gordon P et al |
| EPI_ISL_805778 | Canada/AB-15932/2020 | 2020-04-21 | North America/Canada/Alberta/ | Alberta Precision Labs (APL) | Alberta Precision Labs (APL) | Gordon P et al |
| EPI_ISL_805779 | Canada/AB-45626/2020 | 2020-04-17 | North America/Canada/Alberta/ | Alberta Precision Labs (APL) | Alberta Precision Labs (APL) | Gordon P et al |
| EPI_ISL_805780 | Canada/AB-69742/2020 | 2020-04-28 | North America/Canada/Alberta/ | Alberta Precision Labs (APL) | Alberta Precision Labs (APL) | Gordon P et al |
| EPI_ISL_805781 | Canada/AB-42483/2020 | 2020-05-01 | North America/Canada/Alberta/ | Alberta Precision Labs (APL) | Alberta Precision Labs (APL) | Gordon P et al |
| EPI_ISL_805782 | Canada/AB-29512/2020 | 2020-03-30 | North America/Canada/Alberta/ | Alberta Precision Labs (APL) | Alberta Precision Labs (APL) | Gordon P et al |
| EPI_ISL_805783 | Canada/AB-15989/2020 | 2020-04-26 | North America/Canada/Alberta/ | Alberta Precision Labs (APL) | Alberta Precision Labs (APL) | Gordon P et al |
| EPI_ISL_805784 | Canada/AB-30537/2020 | 2020-04-17 | North America/Canada/Alberta/ | Alberta Precision Labs (APL) | Alberta Precision Labs (APL) | Gordon P et al |
| EPI_ISL_805785 | Canada/AB-29491/2020 | 2020-05-02 | North America/Canada/Alberta/ | Alberta Precision Labs (APL) | Alberta Precision Labs (APL) | Gordon P et al |
| EPI_ISL_805786 | Canada/AB-62880/2020 | 2020-05-03 | North America/Canada/Alberta/ | Alberta Precision Labs (APL) | Alberta Precision Labs (APL) | Gordon P et al |

| | | | | | | |
|---|---|---|---|---|---|---|
| EPI_ISL_805787 | Canada/AB-95352/2020 | 2020-04-10 | North America/Canada/Alberta/ | Alberta Precision Labs (APL) | Alberta Precision Labs (APL) | Gordon P et al |
| EPI_ISL_805788 | Canada/AB-54286/2020 | 2020-05-08 | North America/Canada/Alberta/ | Alberta Precision Labs (APL) | Alberta Precision Labs (APL) | Gordon P et al |
| EPI_ISL_805789 | Canada/AB-46854/2020 | 2020-03-26 | North America/Canada/Alberta/ | Alberta Precision Labs (APL) | Alberta Precision Labs (APL) | Gordon P et al |
| EPI_ISL_805790 | Canada/AB-97211/2020 | 2020-04-26 | North America/Canada/Alberta/ | Alberta Precision Labs (APL) | Alberta Precision Labs (APL) | Gordon P et al |
| EPI_ISL_805791 | Canada/AB-32621/2020 | 2020-04-30 | North America/Canada/Alberta/ | Alberta Precision Labs (APL) | Alberta Precision Labs (APL) | Gordon P et al |
| EPI_ISL_805792 | Canada/AB-13425/2020 | 2020-05-09 | North America/Canada/Alberta/ | Alberta Precision Labs (APL) | Alberta Precision Labs (APL) | Gordon P et al |
| EPI_ISL_805793 | Canada/AB-28231/2020 | 2020-05-01 | North America/Canada/Alberta/ | Alberta Precision Labs (APL) | Alberta Precision Labs (APL) | Gordon P et al |
| EPI_ISL_805794 | Canada/AB-25113/2020 | 2020-04-24 | North America/Canada/Alberta/ | Alberta Precision Labs (APL) | Alberta Precision Labs (APL) | Gordon P et al |
| EPI_ISL_805795 | Canada/AB-26577/2020 | 2020-04-13 | North America/Canada/Alberta/ | Alberta Precision Labs (APL) | Alberta Precision Labs (APL) | Gordon P et al |
| EPI_ISL_805796 | Canada/AB-15302/2020 | 2020-05-02 | North America/Canada/Alberta/ | Alberta Precision Labs (APL) | Alberta Precision Labs (APL) | Gordon P et al |
| EPI_ISL_805797 | Canada/AB-51352/2020 | 2020-04-27 | North America/Canada/Alberta/ | Alberta Precision Labs (APL) | Alberta Precision Labs (APL) | Gordon P et al |
| EPI_ISL_805798 | Canada/AB-47161/2020 | 2020-04-28 | North America/Canada/Alberta/ | Alberta Precision Labs (APL) | Alberta Precision Labs (APL) | Gordon P et al |
| EPI_ISL_805799 | Canada/AB-23527/2020 | 2020-04-24 | North America/Canada/Alberta/ | Alberta Precision Labs (APL) | Alberta Precision Labs (APL) | Gordon P et al |
| EPI_ISL_805800 | Canada/AB-73185/2020 | 2020-04-28 | North America/Canada/Alberta/ | Alberta Precision Labs (APL) | Alberta Precision Labs (APL) | Gordon P et al |
| EPI_ISL_805801 | Canada/AB-53531/2020 | 2020-03-18 | North America/Canada/Alberta/ | Alberta Precision Labs (APL) | Alberta Precision Labs (APL) | Gordon P et al |
| EPI_ISL_805802 | Canada/AB-53944/2020 | 2020-05-06 | North America/Canada/Alberta/ | Alberta Precision Labs (APL) | Alberta Precision Labs (APL) | Gordon P et al |
| EPI_ISL_805803 | Canada/AB-76420/2020 | 2020-04-26 | North America/Canada/Alberta/ | Alberta Precision Labs (APL) | Alberta Precision Labs (APL) | Gordon P et al |
| EPI_ISL_805804 | Canada/AB-85634/2020 | 2020-04-27 | North America/Canada/Alberta/ | Alberta Precision Labs (APL) | Alberta Precision Labs (APL) | Gordon P et al |
| EPI_ISL_805805 | Canada/AB-71222/2020 | 2020-04-14 | North America/Canada/Alberta/ | Alberta Precision Labs (APL) | Alberta Precision Labs (APL) | Gordon P et al |
| EPI_ISL_805806 | Canada/AB-19676/2020 | 2020-05-07 | North America/Canada/Alberta/ | Alberta Precision Labs (APL) | Alberta Precision Labs (APL) | Gordon P et al |

| | | | | | | |
|---|---|---|---|---|---|---|
| EPI_ISL_805807 | Canada/AB-13568/2020 | 2020-05-03 | North America/Canada/Alberta/ | Alberta Precision Labs (APL) | Alberta Precision Labs (APL) | Gordon P et al |
| EPI_ISL_805808 | Canada/AB-86638/2020 | 2020-04-26 | North America/Canada/Alberta/ | Alberta Precision Labs (APL) | Alberta Precision Labs (APL) | Gordon P et al |
| EPI_ISL_805809 | Canada/AB-12616/2020 | 2020-05-17 | North America/Canada/Alberta/ | Alberta Precision Labs (APL) | Alberta Precision Labs (APL) | Gordon P et al |
| EPI_ISL_805810 | Canada/AB-22042/2020 | 2020-04-22 | North America/Canada/Alberta/ | Alberta Precision Labs (APL) | Alberta Precision Labs (APL) | Gordon P et al |
| EPI_ISL_805811 | Canada/AB-58161/2020 | 2020-05-02 | North America/Canada/Alberta/ | Alberta Precision Labs (APL) | Alberta Precision Labs (APL) | Gordon P et al |
| EPI_ISL_805812 | Canada/AB-12701/2020 | 2020-04-26 | North America/Canada/Alberta/ | Alberta Precision Labs (APL) | Alberta Precision Labs (APL) | Gordon P et al |
| EPI_ISL_805813 | Canada/AB-46536/2020 | 2020-05-14 | North America/Canada/Alberta/ | Alberta Precision Labs (APL) | Alberta Precision Labs (APL) | Gordon P et al |
| EPI_ISL_805814 | Canada/AB-72585/2020 | 2020-03-21 | North America/Canada/Alberta/ | Alberta Precision Labs (APL) | Alberta Precision Labs (APL) | Gordon P et al |
| EPI_ISL_805815 | Canada/AB-39554/2020 | 2020-03-19 | North America/Canada/Alberta/ | Alberta Precision Labs (APL) | Alberta Precision Labs (APL) | Gordon P et al |
| EPI_ISL_805816 | Canada/AB-31195/2020 | 2020-05-03 | North America/Canada/Alberta/ | Alberta Precision Labs (APL) | Alberta Precision Labs (APL) | Gordon P et al |
| EPI_ISL_805817 | Canada/AB-60051/2020 | 2020-04-20 | North America/Canada/Alberta/ | Alberta Precision Labs (APL) | Alberta Precision Labs (APL) | Gordon P et al |
| EPI_ISL_805818 | Canada/AB-16291/2020 | 2020-04-25 | North America/Canada/Alberta/ | Alberta Precision Labs (APL) | Alberta Precision Labs (APL) | Gordon P et al |
| EPI_ISL_805820 | Canada/AB-74115/2020 | 2020-05-17 | North America/Canada/Alberta/ | Alberta Precision Labs (APL) | Alberta Precision Labs (APL) | Gordon P et al |
| EPI_ISL_805821 | Canada/AB-18957/2020 | 2020-04-12 | North America/Canada/Alberta/ | Alberta Precision Labs (APL) | Alberta Precision Labs (APL) | Gordon P et al |
| EPI_ISL_805822 | Canada/AB-27240/2020 | 2020-04-14 | North America/Canada/Alberta/ | Alberta Precision Labs (APL) | Alberta Precision Labs (APL) | Gordon P et al |
| EPI_ISL_805823 | Canada/AB-64285/2020 | 2020-04-12 | North America/Canada/Alberta/ | Alberta Precision Labs (APL) | Alberta Precision Labs (APL) | Gordon P et al |
| EPI_ISL_805824 | Canada/AB-39810/2020 | 2020-05-08 | North America/Canada/Alberta/ | Alberta Precision Labs (APL) | Alberta Precision Labs (APL) | Gordon P et al |
| EPI_ISL_805825 | Canada/AB-22341/2020 | 2020-05-19 | North America/Canada/Alberta/ | Alberta Precision Labs (APL) | Alberta Precision Labs (APL) | Gordon P et al |
| EPI_ISL_805826 | Canada/AB-64147/2020 | 2020-04-14 | North America/Canada/Alberta/ | Alberta Precision Labs (APL) | Alberta Precision Labs (APL) | Gordon P et al |
| EPI_ISL_805827 | Canada/AB-46450/2020 | 2020-05-04 | North America/Canada/Alberta/ | Alberta Precision Labs (APL) | Alberta Precision Labs (APL) | Gordon P et al |

| | | | | | | |
|---|---|---|---|---|---|---|
| EPI_ISL_805828 | Canada/AB-77959/2020 | 2020-04-25 | North America/Canada/Alberta/ | Alberta Precision Labs (APL) | Alberta Precision Labs (APL) | Gordon P et al |
| EPI_ISL_805829 | Canada/AB-53986/2020 | 2020-04-20 | North America/Canada/Alberta/ | Alberta Precision Labs (APL) | Alberta Precision Labs (APL) | Gordon P et al |
| EPI_ISL_805830 | Canada/AB-44890/2020 | 2020-05-11 | North America/Canada/Alberta/ | Alberta Precision Labs (APL) | Alberta Precision Labs (APL) | Gordon P et al |
| EPI_ISL_805831 | Canada/AB-69884/2020 | 2020-05-16 | North America/Canada/Alberta/ | Alberta Precision Labs (APL) | Alberta Precision Labs (APL) | Gordon P et al |
| EPI_ISL_805832 | Canada/AB-77234/2020 | 2020-04-26 | North America/Canada/Alberta/ | Alberta Precision Labs (APL) | Alberta Precision Labs (APL) | Gordon P et al |
| EPI_ISL_805833 | Canada/AB-29562/2020 | 2020-05-08 | North America/Canada/Alberta/ | Alberta Precision Labs (APL) | Alberta Precision Labs (APL) | Gordon P et al |
| EPI_ISL_805834 | Canada/AB-71826/2020 | 2020-04-21 | North America/Canada/Alberta/ | Alberta Precision Labs (APL) | Alberta Precision Labs (APL) | Gordon P et al |
| EPI_ISL_805835 | Canada/AB-56024/2020 | 2020-04-29 | North America/Canada/Alberta/ | Alberta Precision Labs (APL) | Alberta Precision Labs (APL) | Gordon P et al |
| EPI_ISL_805836 | Canada/AB-88693/2020 | 2020-03-20 | North America/Canada/Alberta/ | Alberta Precision Labs (APL) | Alberta Precision Labs (APL) | Gordon P et al |
| EPI_ISL_805837 | Canada/AB-41715/2020 | 2020-05-01 | North America/Canada/Alberta/ | Alberta Precision Labs (APL) | Alberta Precision Labs (APL) | Gordon P et al |
| EPI_ISL_805838 | Canada/AB-56215/2020 | 2020-04-29 | North America/Canada/Alberta/ | Alberta Precision Labs (APL) | Alberta Precision Labs (APL) | Gordon P et al |
| EPI_ISL_805839 | Canada/AB-38616/2020 | 2020-04-21 | North America/Canada/Alberta/ | Alberta Precision Labs (APL) | Alberta Precision Labs (APL) | Gordon P et al |
| EPI_ISL_805840 | Canada/AB-13892/2020 | 2020-05-08 | North America/Canada/Alberta/ | Alberta Precision Labs (APL) | Alberta Precision Labs (APL) | Gordon P et al |
| EPI_ISL_805841 | Canada/AB-88671/2020 | 2020-04-25 | North America/Canada/Alberta/ | Alberta Precision Labs (APL) | Alberta Precision Labs (APL) | Gordon P et al |
| EPI_ISL_805842 | Canada/AB-57631/2020 | 2020-04-30 | North America/Canada/Alberta/ | Alberta Precision Labs (APL) | Alberta Precision Labs (APL) | Gordon P et al |
| EPI_ISL_805843 | Canada/AB-15458/2020 | 2020-04-16 | North America/Canada/Alberta/ | Alberta Precision Labs (APL) | Alberta Precision Labs (APL) | Gordon P et al |
| EPI_ISL_805844 | Canada/AB-52386/2020 | 2020-04-13 | North America/Canada/Alberta/ | Alberta Precision Labs (APL) | Alberta Precision Labs (APL) | Gordon P et al |
| EPI_ISL_805845 | Canada/AB-39779/2020 | 2020-05-07 | North America/Canada/Alberta/ | Alberta Precision Labs (APL) | Alberta Precision Labs (APL) | Gordon P et al |
| EPI_ISL_805846 | Canada/AB-48671/2020 | 2020-04-13 | North America/Canada/Alberta/ | Alberta Precision Labs (APL) | Alberta Precision Labs (APL) | Gordon P et al |
| EPI_ISL_805847 | Canada/AB-33531/2020 | 2020-04-28 | North America/Canada/Alberta/ | Alberta Precision Labs (APL) | Alberta Precision Labs (APL) | Gordon P et al |

| | | | | | | |
|---|---|---|---|---|---|---|
| EPI_ISL_805848 | Canada/AB-62222/2020 | 2020-05-01 | North America/Canada/Alberta/ | Alberta Precision Labs (APL) | Alberta Precision Labs (APL) | Gordon P et al |
| EPI_ISL_805849 | Canada/AB-43786/2020 | 2020-05-04 | North America/Canada/Alberta/ | Alberta Precision Labs (APL) | Alberta Precision Labs (APL) | Gordon P et al |
| EPI_ISL_805850 | Canada/AB-42211/2020 | 2020-04-15 | North America/Canada/Alberta/ | Alberta Precision Labs (APL) | Alberta Precision Labs (APL) | Gordon P et al |
| EPI_ISL_805851 | Canada/AB-97766/2020 | 2020-04-22 | North America/Canada/Alberta/ | Alberta Precision Labs (APL) | Alberta Precision Labs (APL) | Gordon P et al |
| EPI_ISL_805852 | Canada/AB-73300/2020 | 2020-04-18 | North America/Canada/Alberta/ | Alberta Precision Labs (APL) | Alberta Precision Labs (APL) | Gordon P et al |
| EPI_ISL_805853 | Canada/AB-81365/2020 | 2020-05-07 | North America/Canada/Alberta/ | Alberta Precision Labs (APL) | Alberta Precision Labs (APL) | Gordon P et al |
| EPI_ISL_805854 | Canada/AB-83572/2020 | 2020-04-13 | North America/Canada/Alberta/ | Alberta Precision Labs (APL) | Alberta Precision Labs (APL) | Gordon P et al |
| EPI_ISL_805855 | Canada/AB-37527/2020 | 2020-04-15 | North America/Canada/Alberta/ | Alberta Precision Labs (APL) | Alberta Precision Labs (APL) | Gordon P et al |
| EPI_ISL_805857 | Canada/AB-54507/2020 | 2020-04-14 | North America/Canada/Alberta/ | Alberta Precision Labs (APL) | Alberta Precision Labs (APL) | Gordon P et al |
| EPI_ISL_805858 | Canada/AB-23270/2020 | 2020-04-26 | North America/Canada/Alberta/ | Alberta Precision Labs (APL) | Alberta Precision Labs (APL) | Gordon P et al |
| EPI_ISL_805859 | Canada/AB-68907/2020 | 2020-03-18 | North America/Canada/Alberta/ | Alberta Precision Labs (APL) | Alberta Precision Labs (APL) | Gordon P et al |
| EPI_ISL_805860 | Canada/AB-39151/2020 | 2020-04-14 | North America/Canada/Alberta/ | Alberta Precision Labs (APL) | Alberta Precision Labs (APL) | Gordon P et al |
| EPI_ISL_805861 | Canada/AB-91178/2020 | 2020-03-26 | North America/Canada/Alberta/ | Alberta Precision Labs (APL) | Alberta Precision Labs (APL) | Gordon P et al |
| EPI_ISL_805862 | Canada/AB-62182/2020 | 2020-04-30 | North America/Canada/Alberta/ | Alberta Precision Labs (APL) | Alberta Precision Labs (APL) | Gordon P et al |
| EPI_ISL_805863 | Canada/AB-29803/2020 | 2020-05-17 | North America/Canada/Alberta/ | Alberta Precision Labs (APL) | Alberta Precision Labs (APL) | Gordon P et al |
| EPI_ISL_805864 | Canada/AB-76920/2020 | 2020-05-15 | North America/Canada/Alberta/ | Alberta Precision Labs (APL) | Alberta Precision Labs (APL) | Gordon P et al |
| EPI_ISL_805865 | Canada/AB-76617/2020 | 2020-05-08 | North America/Canada/Alberta/ | Alberta Precision Labs (APL) | Alberta Precision Labs (APL) | Gordon P et al |
| EPI_ISL_805866 | Canada/AB-25842/2020 | 2020-04-11 | North America/Canada/Alberta/ | Alberta Precision Labs (APL) | Alberta Precision Labs (APL) | Gordon P et al |
| EPI_ISL_805867 | Canada/AB-91455/2020 | 2020-04-21 | North America/Canada/Alberta/ | Alberta Precision Labs (APL) | Alberta Precision Labs (APL) | Gordon P et al |
| EPI_ISL_805868 | Canada/AB-18488/2020 | 2020-05-17 | North America/Canada/Alberta/ | Alberta Precision Labs (APL) | Alberta Precision Labs (APL) | Gordon P et al |

| | | | | | | |
|---|---|---|---|---|---|---|
| EPI_ISL_805869 | Canada/AB-26007/2020 | 2020-03-22 | North America/Canada/Alberta/ | Alberta Precision Labs (APL) | Alberta Precision Labs (APL) | Gordon P et al |
| EPI_ISL_805871 | Canada/AB-27871/2020 | 2020-04-13 | North America/Canada/Alberta/ | Alberta Precision Labs (APL) | Alberta Precision Labs (APL) | Gordon P et al |
| EPI_ISL_805872 | Canada/AB-90562/2020 | 2020-03-28 | North America/Canada/Alberta/ | Alberta Precision Labs (APL) | Alberta Precision Labs (APL) | Gordon P et al |
| EPI_ISL_805873 | Canada/AB-53446/2020 | 2020-03-28 | North America/Canada/Alberta/ | Alberta Precision Labs (APL) | Alberta Precision Labs (APL) | Gordon P et al |
| EPI_ISL_805874 | Canada/AB-63839/2020 | 2020-04-03 | North America/Canada/Alberta/ | Alberta Precision Labs (APL) | Alberta Precision Labs (APL) | Gordon P et al |
| EPI_ISL_805875 | Canada/AB-91582/2020 | 2020-04-09 | North America/Canada/Alberta/ | Alberta Precision Labs (APL) | Alberta Precision Labs (APL) | Gordon P et al |
| EPI_ISL_805876 | Canada/AB-93362/2020 | 2020-04-24 | North America/Canada/Alberta/ | Alberta Precision Labs (APL) | Alberta Precision Labs (APL) | Gordon P et al |
| EPI_ISL_805877 | Canada/AB-16415/2020 | 2020-04-26 | North America/Canada/Alberta/ | Alberta Precision Labs (APL) | Alberta Precision Labs (APL) | Gordon P et al |
| EPI_ISL_805878 | Canada/AB-37629/2020 | 2020-04-24 | North America/Canada/Alberta/ | Alberta Precision Labs (APL) | Alberta Precision Labs (APL) | Gordon P et al |
| EPI_ISL_805879 | Canada/AB-77537/2020 | 2020-05-07 | North America/Canada/Alberta/ | Alberta Precision Labs (APL) | Alberta Precision Labs (APL) | Gordon P et al |
| EPI_ISL_805880 | Canada/AB-31052/2020 | 2020-04-17 | North America/Canada/Alberta/ | Alberta Precision Labs (APL) | Alberta Precision Labs (APL) | Gordon P et al |
| EPI_ISL_805881 | Canada/AB-45764/2020 | 2020-04-17 | North America/Canada/Alberta/ | Alberta Precision Labs (APL) | Alberta Precision Labs (APL) | Gordon P et al |
| EPI_ISL_805882 | Canada/AB-42254/2020 | 2020-05-09 | North America/Canada/Alberta/ | Alberta Precision Labs (APL) | Alberta Precision Labs (APL) | Gordon P et al |
| EPI_ISL_805883 | Canada/AB-20231/2020 | 2020-04-26 | North America/Canada/Alberta/ | Alberta Precision Labs (APL) | Alberta Precision Labs (APL) | Gordon P et al |
| EPI_ISL_805884 | Canada/AB-74380/2020 | 2020-04-27 | North America/Canada/Alberta/ | Alberta Precision Labs (APL) | Alberta Precision Labs (APL) | Gordon P et al |
| EPI_ISL_805885 | Canada/AB-24671/2020 | 2020-04-12 | North America/Canada/Alberta/ | Alberta Precision Labs (APL) | Alberta Precision Labs (APL) | Gordon P et al |
| EPI_ISL_805886 | Canada/AB-77144/2020 | 2020-04-13 | North America/Canada/Alberta/ | Alberta Precision Labs (APL) | Alberta Precision Labs (APL) | Gordon P et al |
| EPI_ISL_805887 | Canada/AB-88859/2020 | 2020-04-28 | North America/Canada/Alberta/ | Alberta Precision Labs (APL) | Alberta Precision Labs (APL) | Gordon P et al |
| EPI_ISL_805888 | Canada/AB-81188/2020 | 2020-04-29 | North America/Canada/Alberta/ | Alberta Precision Labs (APL) | Alberta Precision Labs (APL) | Gordon P et al |
| EPI_ISL_805889 | Canada/AB-16840/2020 | 2020-04-29 | North America/Canada/Alberta/ | Alberta Precision Labs (APL) | Alberta Precision Labs (APL) | Gordon P et al |

| | | | | | | |
|---|---|---|---|---|---|---|
| EPI_ISL_805890 | Canada/AB-31080/2020 | 2020-04-22 | North America/Canada/Alberta/ | Alberta Precision Labs (APL) | Alberta Precision Labs (APL) | Gordon P et al |
| EPI_ISL_805891 | Canada/AB-55821/2020 | 2020-04-13 | North America/Canada/Alberta/ | Alberta Precision Labs (APL) | Alberta Precision Labs (APL) | Gordon P et al |
| EPI_ISL_805892 | Canada/AB-66738/2020 | 2020-05-01 | North America/Canada/Alberta/ | Alberta Precision Labs (APL) | Alberta Precision Labs (APL) | Gordon P et al |
| EPI_ISL_805893 | Canada/AB-81086/2020 | 2020-04-15 | North America/Canada/Alberta/ | Alberta Precision Labs (APL) | Alberta Precision Labs (APL) | Gordon P et al |
| EPI_ISL_805894 | Canada/AB-23477/2020 | 2020-04-23 | North America/Canada/Alberta/ | Alberta Precision Labs (APL) | Alberta Precision Labs (APL) | Gordon P et al |
| EPI_ISL_805895 | Canada/AB-28143/2020 | 2020-05-13 | North America/Canada/Alberta/ | Alberta Precision Labs (APL) | Alberta Precision Labs (APL) | Gordon P et al |
| EPI_ISL_805896 | Canada/AB-42736/2020 | 2020-04-26 | North America/Canada/Alberta/ | Alberta Precision Labs (APL) | Alberta Precision Labs (APL) | Gordon P et al |
| EPI_ISL_805897 | Canada/AB-46714/2020 | 2020-03-18 | North America/Canada/Alberta/ | Alberta Precision Labs (APL) | Alberta Precision Labs (APL) | Gordon P et al |
| EPI_ISL_805898 | Canada/AB-61402/2020 | 2020-04-14 | North America/Canada/Alberta/ | Alberta Precision Labs (APL) | Alberta Precision Labs (APL) | Gordon P et al |
| EPI_ISL_805899 | Canada/AB-18102/2020 | 2020-04-14 | North America/Canada/Alberta/ | Alberta Precision Labs (APL) | Alberta Precision Labs (APL) | Gordon P et al |
| EPI_ISL_805900 | Canada/AB-49174/2020 | 2020-05-11 | North America/Canada/Alberta/ | Alberta Precision Labs (APL) | Alberta Precision Labs (APL) | Gordon P et al |
| EPI_ISL_805901 | Canada/AB-48944/2020 | 2020-05-10 | North America/Canada/Alberta/ | Alberta Precision Labs (APL) | Alberta Precision Labs (APL) | Gordon P et al |
| EPI_ISL_805902 | Canada/AB-89613/2020 | 2020-04-28 | North America/Canada/Alberta/ | Alberta Precision Labs (APL) | Alberta Precision Labs (APL) | Gordon P et al |
| EPI_ISL_805903 | Canada/AB-26250/2020 | 2020-04-14 | North America/Canada/Alberta/ | Alberta Precision Labs (APL) | Alberta Precision Labs (APL) | Gordon P et al |
| EPI_ISL_805904 | Canada/AB-70453/2020 | 2020-04-16 | North America/Canada/Alberta/ | Alberta Precision Labs (APL) | Alberta Precision Labs (APL) | Gordon P et al |
| EPI_ISL_805905 | Canada/AB-71229/2020 | 2020-04-27 | North America/Canada/Alberta/ | Alberta Precision Labs (APL) | Alberta Precision Labs (APL) | Gordon P et al |
| EPI_ISL_805906 | Canada/AB-26347/2020 | 20-Mar | North America/Canada/Alberta/ | Alberta Precision Labs (APL) | Alberta Precision Labs (APL) | Gordon P et al |
| EPI_ISL_805907 | Canada/AB-80648/2020 | 2020-04-28 | North America/Canada/Alberta/ | Alberta Precision Labs (APL) | Alberta Precision Labs (APL) | Gordon P et al |
| EPI_ISL_805908 | Canada/AB-15191/2020 | 2020-04-17 | North America/Canada/Alberta/ | Alberta Precision Labs (APL) | Alberta Precision Labs (APL) | Gordon P et al |
| EPI_ISL_805909 | Canada/AB-60064/2020 | 2020-04-24 | North America/Canada/Alberta/ | Alberta Precision Labs (APL) | Alberta Precision Labs (APL) | Gordon P et al |

| | | | | | | |
|---|---|---|---|---|---|---|
| EPI_ISL_805910 | Canada/AB-62520/2020 | 2020-04-25 | North America/Canada/Alberta/ | Alberta Precision Labs (APL) | Alberta Precision Labs (APL) | Gordon P et al |
| EPI_ISL_805911 | Canada/AB-63287/2020 | 2020-04-18 | North America/Canada/Alberta/ | Alberta Precision Labs (APL) | Alberta Precision Labs (APL) | Gordon P et al |
| EPI_ISL_805912 | Canada/AB-87928/2020 | 2020-04-24 | North America/Canada/Alberta/ | Alberta Precision Labs (APL) | Alberta Precision Labs (APL) | Gordon P et al |
| EPI_ISL_805913 | Canada/AB-33854/2020 | 2020-04-14 | North America/Canada/Alberta/ | Alberta Precision Labs (APL) | Alberta Precision Labs (APL) | Gordon P et al |
| EPI_ISL_805914 | Canada/AB-29608/2020 | 2020-05-03 | North America/Canada/Alberta/ | Alberta Precision Labs (APL) | Alberta Precision Labs (APL) | Gordon P et al |
| EPI_ISL_805915 | Canada/AB-16302/2020 | 2020-04-25 | North America/Canada/Alberta/ | Alberta Precision Labs (APL) | Alberta Precision Labs (APL) | Gordon P et al |
| EPI_ISL_805916 | Canada/AB-14998/2020 | 2020-05-04 | North America/Canada/Alberta/ | Alberta Precision Labs (APL) | Alberta Precision Labs (APL) | Gordon P et al |
| EPI_ISL_805917 | Canada/AB-19683/2020 | 2020-04-14 | North America/Canada/Alberta/ | Alberta Precision Labs (APL) | Alberta Precision Labs (APL) | Gordon P et al |
| EPI_ISL_805918 | Canada/AB-64344/2020 | 2020-04-17 | North America/Canada/Alberta/ | Alberta Precision Labs (APL) | Alberta Precision Labs (APL) | Gordon P et al |
| EPI_ISL_805919 | Canada/AB-19573/2020 | 2020-04-29 | North America/Canada/Alberta/ | Alberta Precision Labs (APL) | Alberta Precision Labs (APL) | Gordon P et al |
| EPI_ISL_805920 | Canada/AB-85214/2020 | 2020-05-08 | North America/Canada/Alberta/ | Alberta Precision Labs (APL) | Alberta Precision Labs (APL) | Gordon P et al |
| EPI_ISL_805921 | Canada/AB-27007/2020 | 2020-04-14 | North America/Canada/Alberta/ | Alberta Precision Labs (APL) | Alberta Precision Labs (APL) | Gordon P et al |
| EPI_ISL_805922 | Canada/AB-83666/2020 | 2020-03-18 | North America/Canada/Alberta/ | Alberta Precision Labs (APL) | Alberta Precision Labs (APL) | Gordon P et al |
| EPI_ISL_805923 | Canada/AB-35896/2020 | 2020-03-22 | North America/Canada/Alberta/ | Alberta Precision Labs (APL) | Alberta Precision Labs (APL) | Gordon P et al |
| EPI_ISL_805924 | Canada/AB-61565/2020 | 2020-04-25 | North America/Canada/Alberta/ | Alberta Precision Labs (APL) | Alberta Precision Labs (APL) | Gordon P et al |
| EPI_ISL_805925 | Canada/AB-72168/2020 | 2020-04-22 | North America/Canada/Alberta/ | Alberta Precision Labs (APL) | Alberta Precision Labs (APL) | Gordon P et al |
| EPI_ISL_805926 | Canada/AB-88298/2020 | 2020-05-17 | North America/Canada/Alberta/ | Alberta Precision Labs (APL) | Alberta Precision Labs (APL) | Gordon P et al |
| EPI_ISL_805927 | Canada/AB-41117/2020 | 2020-04-29 | North America/Canada/Alberta/ | Alberta Precision Labs (APL) | Alberta Precision Labs (APL) | Gordon P et al |
| EPI_ISL_805928 | Canada/AB-54705/2020 | 2020-04-25 | North America/Canada/Alberta/ | Alberta Precision Labs (APL) | Alberta Precision Labs (APL) | Gordon P et al |
| EPI_ISL_805929 | Canada/AB-39852/2020 | 2020-04-17 | North America/Canada/Alberta/ | Alberta Precision Labs (APL) | Alberta Precision Labs (APL) | Gordon P et al |

| | | | | | | |
|---|---|---|---|---|---|---|
| EPI_ISL_805930 | Canada/AB-88343/2020 | 2020-04-16 | North America/Canada/Alberta/ | Alberta Precision Labs (APL) | Alberta Precision Labs (APL) | Gordon P et al |
| EPI_ISL_805931 | Canada/AB-44899/2020 | 2020-04-27 | North America/Canada/Alberta/ | Alberta Precision Labs (APL) | Alberta Precision Labs (APL) | Gordon P et al |
| EPI_ISL_805932 | Canada/AB-28662/2020 | 2020-04-14 | North America/Canada/Alberta/ | Alberta Precision Labs (APL) | Alberta Precision Labs (APL) | Gordon P et al |
| EPI_ISL_805933 | Canada/AB-12702/2020 | 2020-04-26 | North America/Canada/Alberta/ | Alberta Precision Labs (APL) | Alberta Precision Labs (APL) | Gordon P et al |
| EPI_ISL_805934 | Canada/AB-95095/2020 | 2020-04-29 | North America/Canada/Alberta/ | Alberta Precision Labs (APL) | Alberta Precision Labs (APL) | Gordon P et al |
| EPI_ISL_805935 | Canada/AB-47246/2020 | 2020-05-18 | North America/Canada/Alberta/ | Alberta Precision Labs (APL) | Alberta Precision Labs (APL) | Gordon P et al |
| EPI_ISL_805936 | Canada/AB-40754/2020 | 2020-05-01 | North America/Canada/Alberta/ | Alberta Precision Labs (APL) | Alberta Precision Labs (APL) | Gordon P et al |
| EPI_ISL_805937 | Canada/AB-61307/2020 | 2020-03-22 | North America/Canada/Alberta/ | Alberta Precision Labs (APL) | Alberta Precision Labs (APL) | Gordon P et al |
| EPI_ISL_805938 | Canada/AB-77910/2020 | 2020-03-29 | North America/Canada/Alberta/ | Alberta Precision Labs (APL) | Alberta Precision Labs (APL) | Gordon P et al |
| EPI_ISL_805939 | Canada/AB-92111/2020 | 2020-05-16 | North America/Canada/Alberta/ | Alberta Precision Labs (APL) | Alberta Precision Labs (APL) | Gordon P et al |
| EPI_ISL_805940 | Canada/AB-35240/2020 | 2020-04-22 | North America/Canada/Alberta/ | Alberta Precision Labs (APL) | Alberta Precision Labs (APL) | Gordon P et al |
| EPI_ISL_805941 | Canada/AB-69767/2020 | 2020-04-10 | North America/Canada/Alberta/ | Alberta Precision Labs (APL) | Alberta Precision Labs (APL) | Gordon P et al |
| EPI_ISL_805942 | Canada/AB-80240/2020 | 2020-05-17 | North America/Canada/Alberta/ | Alberta Precision Labs (APL) | Alberta Precision Labs (APL) | Gordon P et al |
| EPI_ISL_805943 | Canada/AB-54133/2020 | 2020-04-17 | North America/Canada/Alberta/ | Alberta Precision Labs (APL) | Alberta Precision Labs (APL) | Gordon P et al |
| EPI_ISL_805944 | Canada/AB-99418/2020 | 2020-04-30 | North America/Canada/Alberta/ | Alberta Precision Labs (APL) | Alberta Precision Labs (APL) | Gordon P et al |
| EPI_ISL_805945 | Canada/AB-29882/2020 | 2020-04-22 | North America/Canada/Alberta/ | Alberta Precision Labs (APL) | Alberta Precision Labs (APL) | Gordon P et al |
| EPI_ISL_805946 | Canada/AB-51543/2020 | 2020-04-15 | North America/Canada/Alberta/ | Alberta Precision Labs (APL) | Alberta Precision Labs (APL) | Gordon P et al |
| EPI_ISL_805947 | Canada/AB-99093/2020 | 2020-04-24 | North America/Canada/Alberta/ | Alberta Precision Labs (APL) | Alberta Precision Labs (APL) | Gordon P et al |
| EPI_ISL_805948 | Canada/AB-58949/2020 | 2020-04-13 | North America/Canada/Alberta/ | Alberta Precision Labs (APL) | Alberta Precision Labs (APL) | Gordon P et al |
| EPI_ISL_805949 | Canada/AB-53467/2020 | 2020-04-14 | North America/Canada/Alberta/ | Alberta Precision Labs (APL) | Alberta Precision Labs (APL) | Gordon P et al |

| | | | | | | |
|---|---|---|---|---|---|---|
| EPI_ISL_805950 | Canada/AB-39319/2020 | 2020-05-02 | North America/Canada/Alberta/ | Alberta Precision Labs (APL) | Alberta Precision Labs (APL) | Gordon P et al |
| EPI_ISL_805951 | Canada/AB-21724/2020 | 2020-04-08 | North America/Canada/Alberta/ | Alberta Precision Labs (APL) | Alberta Precision Labs (APL) | Gordon P et al |
| EPI_ISL_805952 | Canada/AB-85250/2020 | 2020-05-01 | North America/Canada/Alberta/ | Alberta Precision Labs (APL) | Alberta Precision Labs (APL) | Gordon P et al |
| EPI_ISL_805953 | Canada/AB-77275/2020 | 2020-03-17 | North America/Canada/Alberta/ | Alberta Precision Labs (APL) | Alberta Precision Labs (APL) | Gordon P et al |
| EPI_ISL_805954 | Canada/AB-38030/2020 | 2020-04-13 | North America/Canada/Alberta/ | Alberta Precision Labs (APL) | Alberta Precision Labs (APL) | Gordon P et al |
| EPI_ISL_805955 | Canada/AB-23011/2020 | 2020-03-24 | North America/Canada/Alberta/ | Alberta Precision Labs (APL) | Alberta Precision Labs (APL) | Gordon P et al |
| EPI_ISL_805956 | Canada/AB-25999/2020 | 2020-05-14 | North America/Canada/Alberta/ | Alberta Precision Labs (APL) | Alberta Precision Labs (APL) | Gordon P et al |
| EPI_ISL_805957 | Canada/AB-98565/2020 | 2020-04-12 | North America/Canada/Alberta/ | Alberta Precision Labs (APL) | Alberta Precision Labs (APL) | Gordon P et al |
| EPI_ISL_805958 | Canada/AB-61949/2020 | 2020-03-22 | North America/Canada/Alberta/ | Alberta Precision Labs (APL) | Alberta Precision Labs (APL) | Gordon P et al |
| EPI_ISL_805959 | Canada/AB-18936/2020 | 2020-04-28 | North America/Canada/Alberta/ | Alberta Precision Labs (APL) | Alberta Precision Labs (APL) | Gordon P et al |
| EPI_ISL_805960 | Canada/AB-66538/2020 | 2020-03-27 | North America/Canada/Alberta/ | Alberta Precision Labs (APL) | Alberta Precision Labs (APL) | Gordon P et al |
| EPI_ISL_805961 | Canada/AB-29314/2020 | 2020-04-25 | North America/Canada/Alberta/ | Alberta Precision Labs (APL) | Alberta Precision Labs (APL) | Gordon P et al |
| EPI_ISL_805962 | Canada/AB-97872/2020 | 2020-05-10 | North America/Canada/Alberta/ | Alberta Precision Labs (APL) | Alberta Precision Labs (APL) | Gordon P et al |
| EPI_ISL_805963 | Canada/AB-61447/2020 | 2020-04-14 | North America/Canada/Alberta/ | Alberta Precision Labs (APL) | Alberta Precision Labs (APL) | Gordon P et al |
| EPI_ISL_805964 | Canada/AB-69710/2020 | 2020-03-29 | North America/Canada/Alberta/ | Alberta Precision Labs (APL) | Alberta Precision Labs (APL) | Gordon P et al |
| EPI_ISL_805965 | Canada/AB-27281/2020 | 2020-03-28 | North America/Canada/Alberta/ | Alberta Precision Labs (APL) | Alberta Precision Labs (APL) | Gordon P et al |
| EPI_ISL_805966 | Canada/AB-87897/2020 | 2020-04-23 | North America/Canada/Alberta/ | Alberta Precision Labs (APL) | Alberta Precision Labs (APL) | Gordon P et al |
| EPI_ISL_805967 | Canada/AB-89621/2020 | 2020-05-01 | North America/Canada/Alberta/ | Alberta Precision Labs (APL) | Alberta Precision Labs (APL) | Gordon P et al |
| EPI_ISL_805968 | Canada/AB-17990/2020 | 2020-05-01 | North America/Canada/Alberta/ | Alberta Precision Labs (APL) | Alberta Precision Labs (APL) | Gordon P et al |
| EPI_ISL_805969 | Canada/AB-18032/2020 | 2020-04-24 | North America/Canada/Alberta/ | Alberta Precision Labs (APL) | Alberta Precision Labs (APL) | Gordon P et al |

| | | | | | | |
|---|---|---|---|---|---|---|
| EPI_ISL_805970 | Canada/AB-68861/2020 | 2020-04-25 | North America/Canada/Alberta/ | Alberta Precision Labs (APL) | Alberta Precision Labs (APL) | Gordon P et al |
| EPI_ISL_805971 | Canada/AB-36547/2020 | 20-Mar | North America/Canada/Alberta/ | Alberta Precision Labs (APL) | Alberta Precision Labs (APL) | Gordon P et al |
| EPI_ISL_805972 | Canada/AB-30437/2020 | 2020-04-24 | North America/Canada/Alberta/ | Alberta Precision Labs (APL) | Alberta Precision Labs (APL) | Gordon P et al |
| EPI_ISL_805973 | Canada/AB-56422/2020 | 2020-04-26 | North America/Canada/Alberta/ | Alberta Precision Labs (APL) | Alberta Precision Labs (APL) | Gordon P et al |
| EPI_ISL_805974 | Canada/AB-24372/2020 | 2020-04-16 | North America/Canada/Alberta/ | Alberta Precision Labs (APL) | Alberta Precision Labs (APL) | Gordon P et al |
| EPI_ISL_805975 | Canada/AB-17983/2020 | 2020-04-17 | North America/Canada/Alberta/ | Alberta Precision Labs (APL) | Alberta Precision Labs (APL) | Gordon P et al |
| EPI_ISL_805976 | Canada/AB-63264/2020 | 2020-04-15 | North America/Canada/Alberta/ | Alberta Precision Labs (APL) | Alberta Precision Labs (APL) | Gordon P et al |
| EPI_ISL_805977 | Canada/AB-30388/2020 | 2020-04-07 | North America/Canada/Alberta/ | Alberta Precision Labs (APL) | Alberta Precision Labs (APL) | Gordon P et al |
| EPI_ISL_805978 | Canada/AB-24787/2020 | 2020-05-19 | North America/Canada/Alberta/ | Alberta Precision Labs (APL) | Alberta Precision Labs (APL) | Gordon P et al |
| EPI_ISL_805979 | Canada/AB-92725/2020 | 2020-04-09 | North America/Canada/Alberta/ | Alberta Precision Labs (APL) | Alberta Precision Labs (APL) | Gordon P et al |
| EPI_ISL_805980 | Canada/AB-56815/2020 | 2020-04-04 | North America/Canada/Alberta/ | Alberta Precision Labs (APL) | Alberta Precision Labs (APL) | Gordon P et al |
| EPI_ISL_805981 | Canada/AB-86121/2020 | 2020-04-14 | North America/Canada/Alberta/ | Alberta Precision Labs (APL) | Alberta Precision Labs (APL) | Gordon P et al |
| EPI_ISL_805982 | Canada/AB-17482/2020 | 2020-04-20 | North America/Canada/Alberta/ | Alberta Precision Labs (APL) | Alberta Precision Labs (APL) | Gordon P et al |
| EPI_ISL_805983 | Canada/AB-36011/2020 | 2020-04-20 | North America/Canada/Alberta/ | Alberta Precision Labs (APL) | Alberta Precision Labs (APL) | Gordon P et al |
| EPI_ISL_805984 | Canada/AB-32231/2020 | 2020-04-25 | North America/Canada/Alberta/ | Alberta Precision Labs (APL) | Alberta Precision Labs (APL) | Gordon P et al |
| EPI_ISL_805985 | Canada/AB-50946/2020 | 2020-03-28 | North America/Canada/Alberta/ | Alberta Precision Labs (APL) | Alberta Precision Labs (APL) | Gordon P et al |
| EPI_ISL_805986 | Canada/AB-21139/2020 | 2020-05-07 | North America/Canada/Alberta/ | Alberta Precision Labs (APL) | Alberta Precision Labs (APL) | Gordon P et al |
| EPI_ISL_805987 | Canada/AB-22794/2020 | 2020-05-04 | North America/Canada/Alberta/ | Alberta Precision Labs (APL) | Alberta Precision Labs (APL) | Gordon P et al |
| EPI_ISL_805989 | Canada/AB-36965/2020 | 2020-04-29 | North America/Canada/Alberta/ | Alberta Precision Labs (APL) | Alberta Precision Labs (APL) | Gordon P et al |
| EPI_ISL_805990 | Canada/AB-84389/2020 | 2020-04-24 | North America/Canada/Alberta/ | Alberta Precision Labs (APL) | Alberta Precision Labs (APL) | Gordon P et al |

| | | | | | | |
|---|---|---|---|---|---|---|
| EPI_ISL_805991 | Canada/AB-23719/2020 | 2020-04-14 | North America/Canada/Alberta/ | Alberta Precision Labs (APL) | Alberta Precision Labs (APL) | Gordon P et al |
| EPI_ISL_805992 | Canada/AB-96168/2020 | 2020-05-03 | North America/Canada/Alberta/ | Alberta Precision Labs (APL) | Alberta Precision Labs (APL) | Gordon P et al |
| EPI_ISL_805993 | Canada/AB-19271/2020 | 2020-04-24 | North America/Canada/Alberta/ | Alberta Precision Labs (APL) | Alberta Precision Labs (APL) | Gordon P et al |
| EPI_ISL_805994 | Canada/AB-21761/2020 | 2020-04-16 | North America/Canada/Alberta/ | Alberta Precision Labs (APL) | Alberta Precision Labs (APL) | Gordon P et al |
| EPI_ISL_805995 | Canada/AB-92321/2020 | 2020-04-17 | North America/Canada/Alberta/ | Alberta Precision Labs (APL) | Alberta Precision Labs (APL) | Gordon P et al |
| EPI_ISL_805996 | Canada/AB-16858/2020 | 2020-04-23 | North America/Canada/Alberta/ | Alberta Precision Labs (APL) | Alberta Precision Labs (APL) | Gordon P et al |
| EPI_ISL_805997 | Canada/AB-90132/2020 | 2020-04-27 | North America/Canada/Alberta/ | Alberta Precision Labs (APL) | Alberta Precision Labs (APL) | Gordon P et al |
| EPI_ISL_805998 | Canada/AB-94033/2020 | 2020-05-06 | North America/Canada/Alberta/ | Alberta Precision Labs (APL) | Alberta Precision Labs (APL) | Gordon P et al |
| EPI_ISL_805999 | Canada/AB-71865/2020 | 20-Mar | North America/Canada/Alberta/ | Alberta Precision Labs (APL) | Alberta Precision Labs (APL) | Gordon P et al |
| EPI_ISL_806000 | Canada/AB-33215/2020 | 2020-05-07 | North America/Canada/Alberta/ | Alberta Precision Labs (APL) | Alberta Precision Labs (APL) | Gordon P et al |
| EPI_ISL_806001 | Canada/AB-81341/2020 | 2020-04-23 | North America/Canada/Alberta/ | Alberta Precision Labs (APL) | Alberta Precision Labs (APL) | Gordon P et al |
| EPI_ISL_806002 | Canada/AB-63465/2020 | 2020-04-12 | North America/Canada/Alberta/ | Alberta Precision Labs (APL) | Alberta Precision Labs (APL) | Gordon P et al |
| EPI_ISL_806004 | Canada/AB-91282/2020 | 2020-05-10 | North America/Canada/Alberta/ | Alberta Precision Labs (APL) | Alberta Precision Labs (APL) | Gordon P et al |
| EPI_ISL_806005 | Canada/AB-67353/2020 | 2020-04-15 | North America/Canada/Alberta/ | Alberta Precision Labs (APL) | Alberta Precision Labs (APL) | Gordon P et al |
| EPI_ISL_806006 | Canada/AB-46192/2020 | 2020-04-16 | North America/Canada/Alberta/ | Alberta Precision Labs (APL) | Alberta Precision Labs (APL) | Gordon P et al |
| EPI_ISL_806007 | Canada/AB-28355/2020 | 2020-03-21 | North America/Canada/Alberta/ | Alberta Precision Labs (APL) | Alberta Precision Labs (APL) | Gordon P et al |
| EPI_ISL_806008 | Canada/AB-61092/2020 | 2020-04-24 | North America/Canada/Alberta/ | Alberta Precision Labs (APL) | Alberta Precision Labs (APL) | Gordon P et al |
| EPI_ISL_806009 | Canada/AB-46693/2020 | 2020-04-14 | North America/Canada/Alberta/ | Alberta Precision Labs (APL) | Alberta Precision Labs (APL) | Gordon P et al |
| EPI_ISL_806010 | Canada/AB-55153/2020 | 2020-05-16 | North America/Canada/Alberta/ | Alberta Precision Labs (APL) | Alberta Precision Labs (APL) | Gordon P et al |
| EPI_ISL_806011 | Canada/AB-67239/2020 | 2020-05-04 | North America/Canada/Alberta/ | Alberta Precision Labs (APL) | Alberta Precision Labs (APL) | Gordon P et al |

| | | | | | | |
|---|---|---|---|---|---|---|
| EPI_ISL_806012 | Canada/AB-38884/2020 | 2020-04-15 | North America/Canada/Alberta/ | Alberta Precision Labs (APL) | Alberta Precision Labs (APL) | Gordon P et al |
| EPI_ISL_806013 | Canada/AB-64357/2020 | 2020-04-13 | North America/Canada/Alberta/ | Alberta Precision Labs (APL) | Alberta Precision Labs (APL) | Gordon P et al |
| EPI_ISL_806014 | Canada/AB-41495/2020 | 2020-05-06 | North America/Canada/Alberta/ | Alberta Precision Labs (APL) | Alberta Precision Labs (APL) | Gordon P et al |
| EPI_ISL_806015 | Canada/AB-92938/2020 | 2020-04-14 | North America/Canada/Alberta/ | Alberta Precision Labs (APL) | Alberta Precision Labs (APL) | Gordon P et al |
| EPI_ISL_806016 | Canada/AB-51010/2020 | 2020-04-24 | North America/Canada/Alberta/ | Alberta Precision Labs (APL) | Alberta Precision Labs (APL) | Gordon P et al |
| EPI_ISL_806017 | Canada/AB-12944/2020 | 2020-04-14 | North America/Canada/Alberta/ | Alberta Precision Labs (APL) | Alberta Precision Labs (APL) | Gordon P et al |
| EPI_ISL_806018 | Canada/AB-40899/2020 | 2020-04-17 | North America/Canada/Alberta/ | Alberta Precision Labs (APL) | Alberta Precision Labs (APL) | Gordon P et al |
| EPI_ISL_806019 | Canada/AB-31875/2020 | 2020-04-13 | North America/Canada/Alberta/ | Alberta Precision Labs (APL) | Alberta Precision Labs (APL) | Gordon P et al |
| EPI_ISL_806020 | Canada/AB-44760/2020 | 2020-04-29 | North America/Canada/Alberta/ | Alberta Precision Labs (APL) | Alberta Precision Labs (APL) | Gordon P et al |
| EPI_ISL_806021 | Canada/AB-95028/2020 | 2020-04-04 | North America/Canada/Alberta/ | Alberta Precision Labs (APL) | Alberta Precision Labs (APL) | Gordon P et al |
| EPI_ISL_806022 | Canada/AB-57370/2020 | 2020-05-05 | North America/Canada/Alberta/ | Alberta Precision Labs (APL) | Alberta Precision Labs (APL) | Gordon P et al |
| EPI_ISL_806023 | Canada/AB-99100/2020 | 2020-03-22 | North America/Canada/Alberta/ | Alberta Precision Labs (APL) | Alberta Precision Labs (APL) | Gordon P et al |
| EPI_ISL_806024 | Canada/AB-86649/2020 | 2020-04-26 | North America/Canada/Alberta/ | Alberta Precision Labs (APL) | Alberta Precision Labs (APL) | Gordon P et al |
| EPI_ISL_806025 | Canada/AB-17607/2020 | 2020-05-19 | North America/Canada/Alberta/ | Alberta Precision Labs (APL) | Alberta Precision Labs (APL) | Gordon P et al |
| EPI_ISL_806026 | Canada/AB-29671/2020 | 2020-05-01 | North America/Canada/Alberta/ | Alberta Precision Labs (APL) | Alberta Precision Labs (APL) | Gordon P et al |
| EPI_ISL_806027 | Canada/AB-69056/2020 | 2020-04-14 | North America/Canada/Alberta/ | Alberta Precision Labs (APL) | Alberta Precision Labs (APL) | Gordon P et al |
| EPI_ISL_806028 | Canada/AB-45304/2020 | 2020-05-06 | North America/Canada/Alberta/ | Alberta Precision Labs (APL) | Alberta Precision Labs (APL) | Gordon P et al |
| EPI_ISL_806029 | Canada/AB-97368/2020 | 2020-04-16 | North America/Canada/Alberta/ | Alberta Precision Labs (APL) | Alberta Precision Labs (APL) | Gordon P et al |
| EPI_ISL_806030 | Canada/AB-57770/2020 | 2020-04-15 | North America/Canada/Alberta/ | Alberta Precision Labs (APL) | Alberta Precision Labs (APL) | Gordon P et al |
| EPI_ISL_806031 | Canada/AB-73707/2020 | 2020-04-27 | North America/Canada/Alberta/ | Alberta Precision Labs (APL) | Alberta Precision Labs (APL) | Gordon P et al |

| | | | | | | |
|---|---|---|---|---|---|---|
| EPI_ISL_806032 | Canada/AB-75670/2020 | 2020-03-16 | North America/Canada/Alberta/ | Alberta Precision Labs (APL) | Alberta Precision Labs (APL) | Gordon P et al |
| EPI_ISL_806033 | Canada/AB-41120/2020 | 2020-04-04 | North America/Canada/Alberta/ | Alberta Precision Labs (APL) | Alberta Precision Labs (APL) | Gordon P et al |
| EPI_ISL_806034 | Canada/AB-61950/2020 | 2020-03-27 | North America/Canada/Alberta/ | Alberta Precision Labs (APL) | Alberta Precision Labs (APL) | Gordon P et al |
| EPI_ISL_806035 | Canada/AB-66573/2020 | 2020-05-15 | North America/Canada/Alberta/ | Alberta Precision Labs (APL) | Alberta Precision Labs (APL) | Gordon P et al |
| EPI_ISL_806036 | Canada/AB-89776/2020 | 2020-05-02 | North America/Canada/Alberta/ | Alberta Precision Labs (APL) | Alberta Precision Labs (APL) | Gordon P et al |
| EPI_ISL_806037 | Canada/AB-49634/2020 | 2020-04-15 | North America/Canada/Alberta/ | Alberta Precision Labs (APL) | Alberta Precision Labs (APL) | Gordon P et al |
| EPI_ISL_806038 | Canada/AB-22691/2020 | 2020-04-25 | North America/Canada/Alberta/ | Alberta Precision Labs (APL) | Alberta Precision Labs (APL) | Gordon P et al |
| EPI_ISL_806039 | Canada/AB-13038/2020 | 2020-05-07 | North America/Canada/Alberta/ | Alberta Precision Labs (APL) | Alberta Precision Labs (APL) | Gordon P et al |
| EPI_ISL_806040 | Canada/AB-42519/2020 | 2020-04-05 | North America/Canada/Alberta/ | Alberta Precision Labs (APL) | Alberta Precision Labs (APL) | Gordon P et al |
| EPI_ISL_806041 | Canada/AB-13615/2020 | 2020-03-25 | North America/Canada/Alberta/ | Alberta Precision Labs (APL) | Alberta Precision Labs (APL) | Gordon P et al |
| EPI_ISL_806042 | Canada/AB-36096/2020 | 2020-05-07 | North America/Canada/Alberta/ | Alberta Precision Labs (APL) | Alberta Precision Labs (APL) | Gordon P et al |
| EPI_ISL_806043 | Canada/AB-41835/2020 | 2020-04-16 | North America/Canada/Alberta/ | Alberta Precision Labs (APL) | Alberta Precision Labs (APL) | Gordon P et al |
| EPI_ISL_806044 | Canada/AB-96594/2020 | 2020-04-29 | North America/Canada/Alberta/ | Alberta Precision Labs (APL) | Alberta Precision Labs (APL) | Gordon P et al |
| EPI_ISL_806045 | Canada/AB-23180/2020 | 2020-05-10 | North America/Canada/Alberta/ | Alberta Precision Labs (APL) | Alberta Precision Labs (APL) | Gordon P et al |
| EPI_ISL_806046 | Canada/AB-65492/2020 | 2020-04-27 | North America/Canada/Alberta/ | Alberta Precision Labs (APL) | Alberta Precision Labs (APL) | Gordon P et al |
| EPI_ISL_806047 | Canada/AB-31163/2020 | 2020-03-19 | North America/Canada/Alberta/ | Alberta Precision Labs (APL) | Alberta Precision Labs (APL) | Gordon P et al |
| EPI_ISL_806048 | Canada/AB-50479/2020 | 2020-04-17 | North America/Canada/Alberta/ | Alberta Precision Labs (APL) | Alberta Precision Labs (APL) | Gordon P et al |
| EPI_ISL_806049 | Canada/AB-23516/2020 | 2020-05-04 | North America/Canada/Alberta/ | Alberta Precision Labs (APL) | Alberta Precision Labs (APL) | Gordon P et al |
| EPI_ISL_806050 | Canada/AB-33110/2020 | 2020-04-17 | North America/Canada/Alberta/ | Alberta Precision Labs (APL) | Alberta Precision Labs (APL) | Gordon P et al |
| EPI_ISL_806051 | Canada/AB-60464/2020 | 2020-04-25 | North America/Canada/Alberta/ | Alberta Precision Labs (APL) | Alberta Precision Labs (APL) | Gordon P et al |

| | | | | | | |
|---|---|---|---|---|---|---|
| EPI_ISL_806052 | Canada/AB-59071/2020 | 2020-04-21 | North America/Canada/Alberta/ | Alberta Precision Labs (APL) | Alberta Precision Labs (APL) | Gordon P et al |
| EPI_ISL_806053 | Canada/AB-58283/2020 | 2020-04-22 | North America/Canada/Alberta/ | Alberta Precision Labs (APL) | Alberta Precision Labs (APL) | Gordon P et al |
| EPI_ISL_806054 | Canada/AB-37645/2020 | 2020-04-17 | North America/Canada/Alberta/ | Alberta Precision Labs (APL) | Alberta Precision Labs (APL) | Gordon P et al |
| EPI_ISL_806055 | Canada/AB-73191/2020 | 2020-05-17 | North America/Canada/Alberta/ | Alberta Precision Labs (APL) | Alberta Precision Labs (APL) | Gordon P et al |
| EPI_ISL_806056 | Canada/AB-66002/2020 | 2020-03-29 | North America/Canada/Alberta/ | Alberta Precision Labs (APL) | Alberta Precision Labs (APL) | Gordon P et al |
| EPI_ISL_806057 | Canada/AB-97735/2020 | 2020-04-13 | North America/Canada/Alberta/ | Alberta Precision Labs (APL) | Alberta Precision Labs (APL) | Gordon P et al |
| EPI_ISL_806058 | Canada/AB-36707/2020 | 2020-04-13 | North America/Canada/Alberta/ | Alberta Precision Labs (APL) | Alberta Precision Labs (APL) | Gordon P et al |
| EPI_ISL_806059 | Canada/AB-97084/2020 | 2020-05-03 | North America/Canada/Alberta/ | Alberta Precision Labs (APL) | Alberta Precision Labs (APL) | Gordon P et al |
| EPI_ISL_806060 | Canada/AB-17895/2020 | 2020-04-23 | North America/Canada/Alberta/ | Alberta Precision Labs (APL) | Alberta Precision Labs (APL) | Gordon P et al |
| EPI_ISL_806061 | Canada/AB-37338/2020 | 2020-04-15 | North America/Canada/Alberta/ | Alberta Precision Labs (APL) | Alberta Precision Labs (APL) | Gordon P et al |
| EPI_ISL_806062 | Canada/AB-55338/2020 | 2020-05-06 | North America/Canada/Alberta/ | Alberta Precision Labs (APL) | Alberta Precision Labs (APL) | Gordon P et al |
| EPI_ISL_806063 | Canada/AB-49391/2020 | 2020-04-03 | North America/Canada/Alberta/ | Alberta Precision Labs (APL) | Alberta Precision Labs (APL) | Gordon P et al |
| EPI_ISL_806064 | Canada/AB-90954/2020 | 2020-04-11 | North America/Canada/Alberta/ | Alberta Precision Labs (APL) | Alberta Precision Labs (APL) | Gordon P et al |
| EPI_ISL_806065 | Canada/AB-38087/2020 | 2020-04-30 | North America/Canada/Alberta/ | Alberta Precision Labs (APL) | Alberta Precision Labs (APL) | Gordon P et al |
| EPI_ISL_806066 | Canada/AB-21679/2020 | 2020-04-02 | North America/Canada/Alberta/ | Alberta Precision Labs (APL) | Alberta Precision Labs (APL) | Gordon P et al |
| EPI_ISL_806067 | Canada/AB-70820/2020 | 2020-04-11 | North America/Canada/Alberta/ | Alberta Precision Labs (APL) | Alberta Precision Labs (APL) | Gordon P et al |
| EPI_ISL_806069 | Canada/AB-50961/2020 | 2020-04-28 | North America/Canada/Alberta/ | Alberta Precision Labs (APL) | Alberta Precision Labs (APL) | Gordon P et al |
| EPI_ISL_806070 | Canada/AB-90122/2020 | 2020-04-13 | North America/Canada/Alberta/ | Alberta Precision Labs (APL) | Alberta Precision Labs (APL) | Gordon P et al |
| EPI_ISL_806071 | Canada/AB-54444/2020 | 2020-04-23 | North America/Canada/Alberta/ | Alberta Precision Labs (APL) | Alberta Precision Labs (APL) | Gordon P et al |
| EPI_ISL_806072 | Canada/AB-85269/2020 | 2020-04-12 | North America/Canada/Alberta/ | Alberta Precision Labs (APL) | Alberta Precision Labs (APL) | Gordon P et al |

| | | | | | | |
|---|---|---|---|---|---|---|
| EPI_ISL_806073 | Canada/AB-89574/2020 | 2020-04-14 | North America/Canada/Alberta/ | Alberta Precision Labs (APL) | Alberta Precision Labs (APL) | Gordon P et al |
| EPI_ISL_806074 | Canada/AB-34263/2020 | 2020-04-27 | North America/Canada/Alberta/ | Alberta Precision Labs (APL) | Alberta Precision Labs (APL) | Gordon P et al |
| EPI_ISL_806075 | Canada/AB-45390/2020 | 2020-04-24 | North America/Canada/Alberta/ | Alberta Precision Labs (APL) | Alberta Precision Labs (APL) | Gordon P et al |
| EPI_ISL_806076 | Canada/AB-18699/2020 | 2020-05-05 | North America/Canada/Alberta/ | Alberta Precision Labs (APL) | Alberta Precision Labs (APL) | Gordon P et al |
| EPI_ISL_806077 | Canada/AB-62419/2020 | 2020-04-30 | North America/Canada/Alberta/ | Alberta Precision Labs (APL) | Alberta Precision Labs (APL) | Gordon P et al |
| EPI_ISL_806078 | Canada/AB-12751/2020 | 2020-03-24 | North America/Canada/Alberta/ | Alberta Precision Labs (APL) | Alberta Precision Labs (APL) | Gordon P et al |
| EPI_ISL_806079 | Canada/AB-62238/2020 | 2020-04-13 | North America/Canada/Alberta/ | Alberta Precision Labs (APL) | Alberta Precision Labs (APL) | Gordon P et al |
| EPI_ISL_806080 | Canada/AB-95339/2020 | 2020-04-15 | North America/Canada/Alberta/ | Alberta Precision Labs (APL) | Alberta Precision Labs (APL) | Gordon P et al |
| EPI_ISL_806081 | Canada/AB-88047/2020 | 2020-04-05 | North America/Canada/Alberta/ | Alberta Precision Labs (APL) | Alberta Precision Labs (APL) | Gordon P et al |
| EPI_ISL_806082 | Canada/AB-49628/2020 | 2020-03-20 | North America/Canada/Alberta/ | Alberta Precision Labs (APL) | Alberta Precision Labs (APL) | Gordon P et al |
| EPI_ISL_806083 | Canada/AB-44960/2020 | 2020-05-15 | North America/Canada/Alberta/ | Alberta Precision Labs (APL) | Alberta Precision Labs (APL) | Gordon P et al |
| EPI_ISL_806084 | Canada/AB-26518/2020 | 2020-04-26 | North America/Canada/Alberta/ | Alberta Precision Labs (APL) | Alberta Precision Labs (APL) | Gordon P et al |
| EPI_ISL_806085 | Canada/AB-38582/2020 | 2020-04-20 | North America/Canada/Alberta/ | Alberta Precision Labs (APL) | Alberta Precision Labs (APL) | Gordon P et al |
| EPI_ISL_806086 | Canada/AB-48635/2020 | 2020-04-14 | North America/Canada/Alberta/ | Alberta Precision Labs (APL) | Alberta Precision Labs (APL) | Gordon P et al |
| EPI_ISL_806087 | Canada/AB-40780/2020 | 2020-05-14 | North America/Canada/Alberta/ | Alberta Precision Labs (APL) | Alberta Precision Labs (APL) | Gordon P et al |
| EPI_ISL_806088 | Canada/AB-88323/2020 | 2020-04-17 | North America/Canada/Alberta/ | Alberta Precision Labs (APL) | Alberta Precision Labs (APL) | Gordon P et al |
| EPI_ISL_806089 | Canada/AB-98293/2020 | 2020-04-16 | North America/Canada/Alberta/ | Alberta Precision Labs (APL) | Alberta Precision Labs (APL) | Gordon P et al |
| EPI_ISL_806090 | Canada/AB-78228/2020 | 2020-04-10 | North America/Canada/Alberta/ | Alberta Precision Labs (APL) | Alberta Precision Labs (APL) | Gordon P et al |
| EPI_ISL_806091 | Canada/AB-74079/2020 | 2020-04-23 | North America/Canada/Alberta/ | Alberta Precision Labs (APL) | Alberta Precision Labs (APL) | Gordon P et al |
| EPI_ISL_806092 | Canada/AB-97754/2020 | 2020-04-01 | North America/Canada/Alberta/ | Alberta Precision Labs (APL) | Alberta Precision Labs (APL) | Gordon P et al |

| | | | | | | |
|---|---|---|---|---|---|---|
| EPI_ISL_806093 | Canada/AB-54017/2020 | 2020-04-13 | North America/Canada/Alberta/ | Alberta Precision Labs (APL) | Alberta Precision Labs (APL) | Gordon P et al |
| EPI_ISL_806094 | Canada/AB-62403/2020 | 2020-04-01 | North America/Canada/Alberta/ | Alberta Precision Labs (APL) | Alberta Precision Labs (APL) | Gordon P et al |
| EPI_ISL_806095 | Canada/AB-53106/2020 | 2020-04-12 | North America/Canada/Alberta/ | Alberta Precision Labs (APL) | Alberta Precision Labs (APL) | Gordon P et al |
| EPI_ISL_806096 | Canada/AB-13196/2020 | 2020-05-08 | North America/Canada/Alberta/ | Alberta Precision Labs (APL) | Alberta Precision Labs (APL) | Gordon P et al |
| EPI_ISL_806097 | Canada/AB-65999/2020 | 2020-04-21 | North America/Canada/Alberta/ | Alberta Precision Labs (APL) | Alberta Precision Labs (APL) | Gordon P et al |
| EPI_ISL_806098 | Canada/AB-76179/2020 | 2020-05-18 | North America/Canada/Alberta/ | Alberta Precision Labs (APL) | Alberta Precision Labs (APL) | Gordon P et al |
| EPI_ISL_806099 | Canada/AB-21174/2020 | 2020-04-27 | North America/Canada/Alberta/ | Alberta Precision Labs (APL) | Alberta Precision Labs (APL) | Gordon P et al |
| EPI_ISL_806100 | Canada/AB-27833/2020 | 2020-04-17 | North America/Canada/Alberta/ | Alberta Precision Labs (APL) | Alberta Precision Labs (APL) | Gordon P et al |
| EPI_ISL_806101 | Canada/AB-76153/2020 | 2020-04-13 | North America/Canada/Alberta/ | Alberta Precision Labs (APL) | Alberta Precision Labs (APL) | Gordon P et al |
| EPI_ISL_806102 | Canada/AB-91912/2020 | 2020-05-18 | North America/Canada/Alberta/ | Alberta Precision Labs (APL) | Alberta Precision Labs (APL) | Gordon P et al |
| EPI_ISL_806103 | Canada/AB-55324/2020 | 2020-04-15 | North America/Canada/Alberta/ | Alberta Precision Labs (APL) | Alberta Precision Labs (APL) | Gordon P et al |
| EPI_ISL_806104 | Canada/AB-21287/2020 | 2020-04-13 | North America/Canada/Alberta/ | Alberta Precision Labs (APL) | Alberta Precision Labs (APL) | Gordon P et al |
| EPI_ISL_806105 | Canada/AB-70232/2020 | 2020-04-28 | North America/Canada/Alberta/ | Alberta Precision Labs (APL) | Alberta Precision Labs (APL) | Gordon P et al |
| EPI_ISL_806106 | Canada/AB-45582/2020 | 2020-04-30 | North America/Canada/Alberta/ | Alberta Precision Labs (APL) | Alberta Precision Labs (APL) | Gordon P et al |
| EPI_ISL_806107 | Canada/AB-61762/2020 | 2020-04-14 | North America/Canada/Alberta/ | Alberta Precision Labs (APL) | Alberta Precision Labs (APL) | Gordon P et al |
| EPI_ISL_806109 | Canada/AB-47307/2020 | 2020-04-26 | North America/Canada/Alberta/ | Alberta Precision Labs (APL) | Alberta Precision Labs (APL) | Gordon P et al |
| EPI_ISL_806110 | Canada/AB-99291/2020 | 2020-05-18 | North America/Canada/Alberta/ | Alberta Precision Labs (APL) | Alberta Precision Labs (APL) | Gordon P et al |
| EPI_ISL_806111 | Canada/AB-21980/2020 | 2020-04-17 | North America/Canada/Alberta/ | Alberta Precision Labs (APL) | Alberta Precision Labs (APL) | Gordon P et al |
| EPI_ISL_806113 | Canada/AB-65714/2020 | 2020-04-16 | North America/Canada/Alberta/ | Alberta Precision Labs (APL) | Alberta Precision Labs (APL) | Gordon P et al |
| EPI_ISL_806114 | Canada/AB-69145/2020 | 2020-04-16 | North America/Canada/Alberta/ | Alberta Precision Labs (APL) | Alberta Precision Labs (APL) | Gordon P et al |

| | | | | | | |
|---|---|---|---|---|---|---|
| EPI_ISL_806115 | Canada/AB-94459/2020 | 2020-04-13 | North America/Canada/Alberta/ | Alberta Precision Labs (APL) | Alberta Precision Labs (APL) | Gordon P et al |
| EPI_ISL_806116 | Canada/AB-59435/2020 | 2020-04-14 | North America/Canada/Alberta/ | Alberta Precision Labs (APL) | Alberta Precision Labs (APL) | Gordon P et al |
| EPI_ISL_806117 | Canada/AB-66604/2020 | 2020-04-14 | North America/Canada/Alberta/ | Alberta Precision Labs (APL) | Alberta Precision Labs (APL) | Gordon P et al |
| EPI_ISL_806118 | Canada/AB-41972/2020 | 2020-05-14 | North America/Canada/Alberta/ | Alberta Precision Labs (APL) | Alberta Precision Labs (APL) | Gordon P et al |
| EPI_ISL_806119 | Canada/AB-63385/2020 | 2020-04-14 | North America/Canada/Alberta/ | Alberta Precision Labs (APL) | Alberta Precision Labs (APL) | Gordon P et al |
| EPI_ISL_806120 | Canada/AB-70536/2020 | 2020-04-25 | North America/Canada/Alberta/ | Alberta Precision Labs (APL) | Alberta Precision Labs (APL) | Gordon P et al |
| EPI_ISL_806121 | Canada/AB-93879/2020 | 2020-05-14 | North America/Canada/Alberta/ | Alberta Precision Labs (APL) | Alberta Precision Labs (APL) | Gordon P et al |
| EPI_ISL_806122 | Canada/AB-14566/2020 | 2020-04-14 | North America/Canada/Alberta/ | Alberta Precision Labs (APL) | Alberta Precision Labs (APL) | Gordon P et al |
| EPI_ISL_806123 | Canada/AB-34524/2020 | 2020-04-29 | North America/Canada/Alberta/ | Alberta Precision Labs (APL) | Alberta Precision Labs (APL) | Gordon P et al |
| EPI_ISL_806124 | Canada/AB-41440/2020 | 2020-03-21 | North America/Canada/Alberta/ | Alberta Precision Labs (APL) | Alberta Precision Labs (APL) | Gordon P et al |
| EPI_ISL_806125 | Canada/AB-32245/2020 | 2020-05-05 | North America/Canada/Alberta/ | Alberta Precision Labs (APL) | Alberta Precision Labs (APL) | Gordon P et al |
| EPI_ISL_806126 | Canada/AB-71347/2020 | 2020-04-06 | North America/Canada/Alberta/ | Alberta Precision Labs (APL) | Alberta Precision Labs (APL) | Gordon P et al |
| EPI_ISL_806127 | Canada/AB-30521/2020 | 2020-04-14 | North America/Canada/Alberta/ | Alberta Precision Labs (APL) | Alberta Precision Labs (APL) | Gordon P et al |
| EPI_ISL_806128 | Canada/AB-55987/2020 | 2020-04-24 | North America/Canada/Alberta/ | Alberta Precision Labs (APL) | Alberta Precision Labs (APL) | Gordon P et al |
| EPI_ISL_806129 | Canada/AB-26069/2020 | 2020-04-14 | North America/Canada/Alberta/ | Alberta Precision Labs (APL) | Alberta Precision Labs (APL) | Gordon P et al |
| EPI_ISL_806130 | Canada/AB-58819/2020 | 2020-04-11 | North America/Canada/Alberta/ | Alberta Precision Labs (APL) | Alberta Precision Labs (APL) | Gordon P et al |
| EPI_ISL_806131 | Canada/AB-64860/2020 | 2020-04-17 | North America/Canada/Alberta/ | Alberta Precision Labs (APL) | Alberta Precision Labs (APL) | Gordon P et al |
| EPI_ISL_806132 | Canada/AB-39831/2020 | 2020-04-16 | North America/Canada/Alberta/ | Alberta Precision Labs (APL) | Alberta Precision Labs (APL) | Gordon P et al |
| EPI_ISL_806133 | Canada/AB-26292/2020 | 2020-04-13 | North America/Canada/Alberta/ | Alberta Precision Labs (APL) | Alberta Precision Labs (APL) | Gordon P et al |
| EPI_ISL_806134 | Canada/AB-62943/2020 | 2020-04-16 | North America/Canada/Alberta/ | Alberta Precision Labs (APL) | Alberta Precision Labs (APL) | Gordon P et al |

| | | | | | | |
|---|---|---|---|---|---|---|
| EPI_ISL_806135 | Canada/AB-97143/2020 | 2020-03-26 | North America/Canada/Alberta/ | Alberta Precision Labs (APL) | Alberta Precision Labs (APL) | Gordon P et al |
| EPI_ISL_806136 | Canada/AB-62210/2020 | 2020-04-14 | North America/Canada/Alberta/ | Alberta Precision Labs (APL) | Alberta Precision Labs (APL) | Gordon P et al |
| EPI_ISL_806137 | Canada/AB-40787/2020 | 2020-04-13 | North America/Canada/Alberta/ | Alberta Precision Labs (APL) | Alberta Precision Labs (APL) | Gordon P et al |
| EPI_ISL_806138 | Canada/AB-35123/2020 | 2020-04-26 | North America/Canada/Alberta/ | Alberta Precision Labs (APL) | Alberta Precision Labs (APL) | Gordon P et al |
| EPI_ISL_806139 | Canada/AB-61704/2020 | 2020-04-16 | North America/Canada/Alberta/ | Alberta Precision Labs (APL) | Alberta Precision Labs (APL) | Gordon P et al |
| EPI_ISL_806140 | Canada/AB-86699/2020 | 2020-04-16 | North America/Canada/Alberta/ | Alberta Precision Labs (APL) | Alberta Precision Labs (APL) | Gordon P et al |
| EPI_ISL_806141 | Canada/AB-62358/2020 | 2020-04-29 | North America/Canada/Alberta/ | Alberta Precision Labs (APL) | Alberta Precision Labs (APL) | Gordon P et al |
| EPI_ISL_806142 | Canada/AB-41713/2020 | 2020-05-04 | North America/Canada/Alberta/ | Alberta Precision Labs (APL) | Alberta Precision Labs (APL) | Gordon P et al |
| EPI_ISL_806143 | Canada/AB-81916/2020 | 2020-05-07 | North America/Canada/Alberta/ | Alberta Precision Labs (APL) | Alberta Precision Labs (APL) | Gordon P et al |
| EPI_ISL_806144 | Canada/AB-28017/2020 | 2020-05-13 | North America/Canada/Alberta/ | Alberta Precision Labs (APL) | Alberta Precision Labs (APL) | Gordon P et al |
| EPI_ISL_806145 | Canada/AB-51640/2020 | 2020-05-03 | North America/Canada/Alberta/ | Alberta Precision Labs (APL) | Alberta Precision Labs (APL) | Gordon P et al |
| EPI_ISL_806146 | Canada/AB-60620/2020 | 2020-05-05 | North America/Canada/Alberta/ | Alberta Precision Labs (APL) | Alberta Precision Labs (APL) | Gordon P et al |
| EPI_ISL_806147 | Canada/AB-76317/2020 | 2020-05-16 | North America/Canada/Alberta/ | Alberta Precision Labs (APL) | Alberta Precision Labs (APL) | Gordon P et al |
| EPI_ISL_806148 | Canada/AB-50163/2020 | 2020-04-11 | North America/Canada/Alberta/ | Alberta Precision Labs (APL) | Alberta Precision Labs (APL) | Gordon P et al |
| EPI_ISL_806149 | Canada/AB-91231/2020 | 2020-04-14 | North America/Canada/Alberta/ | Alberta Precision Labs (APL) | Alberta Precision Labs (APL) | Gordon P et al |
| EPI_ISL_806150 | Canada/AB-45417/2020 | 2020-04-05 | North America/Canada/Alberta/ | Alberta Precision Labs (APL) | Alberta Precision Labs (APL) | Gordon P et al |
| EPI_ISL_806151 | Canada/AB-83577/2020 | 2020-04-11 | North America/Canada/Alberta/ | Alberta Precision Labs (APL) | Alberta Precision Labs (APL) | Gordon P et al |
| EPI_ISL_806152 | Canada/AB-72393/2020 | 2020-04-16 | North America/Canada/Alberta/ | Alberta Precision Labs (APL) | Alberta Precision Labs (APL) | Gordon P et al |
| EPI_ISL_806153 | Canada/AB-87551/2020 | 2020-04-09 | North America/Canada/Alberta/ | Alberta Precision Labs (APL) | Alberta Precision Labs (APL) | Gordon P et al |
| EPI_ISL_806154 | Canada/AB-23471/2020 | 2020-04-13 | North America/Canada/Alberta/ | Alberta Precision Labs (APL) | Alberta Precision Labs (APL) | Gordon P et al |

| | | | | | | |
|---|---|---|---|---|---|---|
| EPI_ISL_806155 | Canada/AB-74184/2020 | 2020-04-30 | North America/Canada/Alberta/ | Alberta Precision Labs (APL) | Alberta Precision Labs (APL) | Gordon P et al |
| EPI_ISL_806156 | Canada/AB-38041/2020 | 2020-04-14 | North America/Canada/Alberta/ | Alberta Precision Labs (APL) | Alberta Precision Labs (APL) | Gordon P et al |
| EPI_ISL_806157 | Canada/AB-84807/2020 | 2020-03-29 | North America/Canada/Alberta/ | Alberta Precision Labs (APL) | Alberta Precision Labs (APL) | Gordon P et al |
| EPI_ISL_806158 | Canada/AB-35247/2020 | 2020-04-17 | North America/Canada/Alberta/ | Alberta Precision Labs (APL) | Alberta Precision Labs (APL) | Gordon P et al |
| EPI_ISL_806159 | Canada/AB-83989/2020 | 2020-04-12 | North America/Canada/Alberta/ | Alberta Precision Labs (APL) | Alberta Precision Labs (APL) | Gordon P et al |
| EPI_ISL_806160 | Canada/AB-89297/2020 | 2020-05-16 | North America/Canada/Alberta/ | Alberta Precision Labs (APL) | Alberta Precision Labs (APL) | Gordon P et al |
| EPI_ISL_806161 | Canada/AB-58731/2020 | 2020-04-13 | North America/Canada/Alberta/ | Alberta Precision Labs (APL) | Alberta Precision Labs (APL) | Gordon P et al |
| EPI_ISL_806162 | Canada/AB-66411/2020 | 2020-04-14 | North America/Canada/Alberta/ | Alberta Precision Labs (APL) | Alberta Precision Labs (APL) | Gordon P et al |
| EPI_ISL_806163 | Canada/AB-45649/2020 | 2020-04-13 | North America/Canada/Alberta/ | Alberta Precision Labs (APL) | Alberta Precision Labs (APL) | Gordon P et al |
| EPI_ISL_806164 | Canada/AB-13811/2020 | 2020-03-26 | North America/Canada/Alberta/ | Alberta Precision Labs (APL) | Alberta Precision Labs (APL) | Gordon P et al |
| EPI_ISL_806165 | Canada/AB-16629/2020 | 2020-04-12 | North America/Canada/Alberta/ | Alberta Precision Labs (APL) | Alberta Precision Labs (APL) | Gordon P et al |
| EPI_ISL_806166 | Canada/AB-31657/2020 | 2020-04-17 | North America/Canada/Alberta/ | Alberta Precision Labs (APL) | Alberta Precision Labs (APL) | Gordon P et al |
| EPI_ISL_806167 | Canada/AB-96929/2020 | 2020-05-04 | North America/Canada/Alberta/ | Alberta Precision Labs (APL) | Alberta Precision Labs (APL) | Gordon P et al |
| EPI_ISL_806168 | Canada/AB-47319/2020 | 2020-03-29 | North America/Canada/Alberta/ | Alberta Precision Labs (APL) | Alberta Precision Labs (APL) | Gordon P et al |
| EPI_ISL_806169 | Canada/AB-42949/2020 | 2020-04-16 | North America/Canada/Alberta/ | Alberta Precision Labs (APL) | Alberta Precision Labs (APL) | Gordon P et al |
| EPI_ISL_806170 | Canada/AB-43622/2020 | 2020-04-25 | North America/Canada/Alberta/ | Alberta Precision Labs (APL) | Alberta Precision Labs (APL) | Gordon P et al |
| EPI_ISL_806171 | Canada/AB-78955/2020 | 2020-04-14 | North America/Canada/Alberta/ | Alberta Precision Labs (APL) | Alberta Precision Labs (APL) | Gordon P et al |
| EPI_ISL_806172 | Canada/AB-75844/2020 | 2020-04-01 | North America/Canada/Alberta/ | Alberta Precision Labs (APL) | Alberta Precision Labs (APL) | Gordon P et al |
| EPI_ISL_806173 | Canada/AB-93712/2020 | 2020-05-05 | North America/Canada/Alberta/ | Alberta Precision Labs (APL) | Alberta Precision Labs (APL) | Gordon P et al |
| EPI_ISL_806174 | Canada/AB-63364/2020 | 2020-04-15 | North America/Canada/Alberta/ | Alberta Precision Labs (APL) | Alberta Precision Labs (APL) | Gordon P et al |

| | | | | | | |
|---|---|---|---|---|---|---|
| EPI_ISL_806175 | Canada/AB-47281/2020 | 2020-04-11 | North America/Canada/Alberta/ | Alberta Precision Labs (APL) | Alberta Precision Labs (APL) | Gordon P et al |
| EPI_ISL_806176 | Canada/AB-47371/2020 | 2020-04-16 | North America/Canada/Alberta/ | Alberta Precision Labs (APL) | Alberta Precision Labs (APL) | Gordon P et al |
| EPI_ISL_806177 | Canada/AB-37682/2020 | 2020-04-20 | North America/Canada/Alberta/ | Alberta Precision Labs (APL) | Alberta Precision Labs (APL) | Gordon P et al |
| EPI_ISL_806178 | Canada/AB-96841/2020 | 2020-03-23 | North America/Canada/Alberta/ | Alberta Precision Labs (APL) | Alberta Precision Labs (APL) | Gordon P et al |
| EPI_ISL_806179 | Canada/AB-70005/2020 | 2020-04-11 | North America/Canada/Alberta/ | Alberta Precision Labs (APL) | Alberta Precision Labs (APL) | Gordon P et al |
| EPI_ISL_806180 | Canada/AB-80602/2020 | 2020-04-06 | North America/Canada/Alberta/ | Alberta Precision Labs (APL) | Alberta Precision Labs (APL) | Gordon P et al |
| EPI_ISL_806181 | Canada/AB-54214/2020 | 2020-04-06 | North America/Canada/Alberta/ | Alberta Precision Labs (APL) | Alberta Precision Labs (APL) | Gordon P et al |
| EPI_ISL_806182 | Canada/AB-47458/2020 | 2020-05-08 | North America/Canada/Alberta/ | Alberta Precision Labs (APL) | Alberta Precision Labs (APL) | Gordon P et al |
| EPI_ISL_806183 | Canada/AB-27625/2020 | 2020-05-07 | North America/Canada/Alberta/ | Alberta Precision Labs (APL) | Alberta Precision Labs (APL) | Gordon P et al |
| EPI_ISL_806184 | Canada/AB-34249/2020 | 2020-04-17 | North America/Canada/Alberta/ | Alberta Precision Labs (APL) | Alberta Precision Labs (APL) | Gordon P et al |
| EPI_ISL_806185 | Canada/AB-94560/2020 | 2020-04-23 | North America/Canada/Alberta/ | Alberta Precision Labs (APL) | Alberta Precision Labs (APL) | Gordon P et al |
| EPI_ISL_806186 | Canada/AB-17557/2020 | 2020-04-16 | North America/Canada/Alberta/ | Alberta Precision Labs (APL) | Alberta Precision Labs (APL) | Gordon P et al |
| EPI_ISL_806187 | Canada/AB-55631/2020 | 2020-04-13 | North America/Canada/Alberta/ | Alberta Precision Labs (APL) | Alberta Precision Labs (APL) | Gordon P et al |
| EPI_ISL_806188 | Canada/AB-87725/2020 | 2020-05-03 | North America/Canada/Alberta/ | Alberta Precision Labs (APL) | Alberta Precision Labs (APL) | Gordon P et al |
| EPI_ISL_806189 | Canada/AB-63578/2020 | 2020-04-07 | North America/Canada/Alberta/ | Alberta Precision Labs (APL) | Alberta Precision Labs (APL) | Gordon P et al |
| EPI_ISL_806190 | Canada/AB-61647/2020 | 2020-05-19 | North America/Canada/Alberta/ | Alberta Precision Labs (APL) | Alberta Precision Labs (APL) | Gordon P et al |
| EPI_ISL_806191 | Canada/AB-35068/2020 | 2020-04-08 | North America/Canada/Alberta/ | Alberta Precision Labs (APL) | Alberta Precision Labs (APL) | Gordon P et al |
| EPI_ISL_806192 | Canada/AB-54644/2020 | 2020-04-16 | North America/Canada/Alberta/ | Alberta Precision Labs (APL) | Alberta Precision Labs (APL) | Gordon P et al |
| EPI_ISL_806193 | Canada/AB-61466/2020 | 2020-03-27 | North America/Canada/Alberta/ | Alberta Precision Labs (APL) | Alberta Precision Labs (APL) | Gordon P et al |
| EPI_ISL_806194 | Canada/AB-31113/2020 | 2020-04-06 | North America/Canada/Alberta/ | Alberta Precision Labs (APL) | Alberta Precision Labs (APL) | Gordon P et al |

| | | | | | | |
|---|---|---|---|---|---|---|
| EPI_ISL_806195 | Canada/AB-51432/2020 | 2020-03-29 | North America/Canada/Alberta/ | Alberta Precision Labs (APL) | Alberta Precision Labs (APL) | Gordon P et al |
| EPI_ISL_806196 | Canada/AB-99353/2020 | 2020-03-30 | North America/Canada/Alberta/ | Alberta Precision Labs (APL) | Alberta Precision Labs (APL) | Gordon P et al |
| EPI_ISL_806197 | Canada/AB-53365/2020 | 2020-03-18 | North America/Canada/Alberta/ | Alberta Precision Labs (APL) | Alberta Precision Labs (APL) | Gordon P et al |
| EPI_ISL_806198 | Canada/AB-28007/2020 | 2020-05-07 | North America/Canada/Alberta/ | Alberta Precision Labs (APL) | Alberta Precision Labs (APL) | Gordon P et al |
| EPI_ISL_806199 | Canada/AB-77205/2020 | 2020-04-16 | North America/Canada/Alberta/ | Alberta Precision Labs (APL) | Alberta Precision Labs (APL) | Gordon P et al |
| EPI_ISL_806200 | Canada/AB-28083/2020 | 2020-04-14 | North America/Canada/Alberta/ | Alberta Precision Labs (APL) | Alberta Precision Labs (APL) | Gordon P et al |
| EPI_ISL_806201 | Canada/AB-48053/2020 | 2020-03-15 | North America/Canada/Alberta/ | Alberta Precision Labs (APL) | Alberta Precision Labs (APL) | Gordon P et al |
| EPI_ISL_806202 | Canada/AB-97052/2020 | 2020-05-07 | North America/Canada/Alberta/ | Alberta Precision Labs (APL) | Alberta Precision Labs (APL) | Gordon P et al |
| EPI_ISL_806203 | Canada/AB-98722/2020 | 2020-04-02 | North America/Canada/Alberta/ | Alberta Precision Labs (APL) | Alberta Precision Labs (APL) | Gordon P et al |
| EPI_ISL_806204 | Canada/AB-61406/2020 | 2020-05-05 | North America/Canada/Alberta/ | Alberta Precision Labs (APL) | Alberta Precision Labs (APL) | Gordon P et al |
| EPI_ISL_806205 | Canada/AB-33791/2020 | 2020-04-07 | North America/Canada/Alberta/ | Alberta Precision Labs (APL) | Alberta Precision Labs (APL) | Gordon P et al |
| EPI_ISL_806206 | Canada/AB-85619/2020 | 2020-04-17 | North America/Canada/Alberta/ | Alberta Precision Labs (APL) | Alberta Precision Labs (APL) | Gordon P et al |
| EPI_ISL_806207 | Canada/AB-50579/2020 | 2020-03-25 | North America/Canada/Alberta/ | Alberta Precision Labs (APL) | Alberta Precision Labs (APL) | Gordon P et al |
| EPI_ISL_806208 | Canada/AB-51106/2020 | 2020-05-07 | North America/Canada/Alberta/ | Alberta Precision Labs (APL) | Alberta Precision Labs (APL) | Gordon P et al |
| EPI_ISL_806209 | Canada/AB-31419/2020 | 2020-04-15 | North America/Canada/Alberta/ | Alberta Precision Labs (APL) | Alberta Precision Labs (APL) | Gordon P et al |
| EPI_ISL_806210 | Canada/AB-94358/2020 | 2020-04-06 | North America/Canada/Alberta/ | Alberta Precision Labs (APL) | Alberta Precision Labs (APL) | Gordon P et al |
| EPI_ISL_806211 | Canada/AB-63581/2020 | 2020-03-28 | North America/Canada/Alberta/ | Alberta Precision Labs (APL) | Alberta Precision Labs (APL) | Gordon P et al |
| EPI_ISL_806212 | Canada/AB-35914/2020 | 2020-04-16 | North America/Canada/Alberta/ | Alberta Precision Labs (APL) | Alberta Precision Labs (APL) | Gordon P et al |
| EPI_ISL_806213 | Canada/AB-42157/2020 | 2020-05-16 | North America/Canada/Alberta/ | Alberta Precision Labs (APL) | Alberta Precision Labs (APL) | Gordon P et al |
| EPI_ISL_806214 | Canada/AB-87908/2020 | 2020-04-30 | North America/Canada/Alberta/ | Alberta Precision Labs (APL) | Alberta Precision Labs (APL) | Gordon P et al |

| | | | | | | |
|---|---|---|---|---|---|---|
| EPI_ISL_806215 | Canada/AB-32673/2020 | 2020-03-16 | North America/Canada/Alberta/ | Alberta Precision Labs (APL) | Alberta Precision Labs (APL) | Gordon P et al |
| EPI_ISL_806216 | Canada/AB-40147/2020 | 2020-03-22 | North America/Canada/Alberta/ | Alberta Precision Labs (APL) | Alberta Precision Labs (APL) | Gordon P et al |
| EPI_ISL_806217 | Canada/AB-87867/2020 | 2020-05-13 | North America/Canada/Alberta/ | Alberta Precision Labs (APL) | Alberta Precision Labs (APL) | Gordon P et al |
| EPI_ISL_806218 | Canada/AB-18352/2020 | 2020-05-04 | North America/Canada/Alberta/ | Alberta Precision Labs (APL) | Alberta Precision Labs (APL) | Gordon P et al |
| EPI_ISL_806219 | Canada/AB-30034/2020 | 2020-05-14 | North America/Canada/Alberta/ | Alberta Precision Labs (APL) | Alberta Precision Labs (APL) | Gordon P et al |
| EPI_ISL_806220 | Canada/AB-68072/2020 | 2020-04-24 | North America/Canada/Alberta/ | Alberta Precision Labs (APL) | Alberta Precision Labs (APL) | Gordon P et al |
| EPI_ISL_806221 | Canada/AB-32968/2020 | 2020-04-15 | North America/Canada/Alberta/ | Alberta Precision Labs (APL) | Alberta Precision Labs (APL) | Gordon P et al |
| EPI_ISL_806222 | Canada/AB-95581/2020 | 2020-04-15 | North America/Canada/Alberta/ | Alberta Precision Labs (APL) | Alberta Precision Labs (APL) | Gordon P et al |
| EPI_ISL_806223 | Canada/AB-46924/2020 | 2020-04-05 | North America/Canada/Alberta/ | Alberta Precision Labs (APL) | Alberta Precision Labs (APL) | Gordon P et al |
| EPI_ISL_806224 | Canada/AB-95051/2020 | 2020-04-01 | North America/Canada/Alberta/ | Alberta Precision Labs (APL) | Alberta Precision Labs (APL) | Gordon P et al |
| EPI_ISL_806225 | Canada/AB-36651/2020 | 2020-05-04 | North America/Canada/Alberta/ | Alberta Precision Labs (APL) | Alberta Precision Labs (APL) | Gordon P et al |
| EPI_ISL_806226 | Canada/AB-49914/2020 | 2020-04-10 | North America/Canada/Alberta/ | Alberta Precision Labs (APL) | Alberta Precision Labs (APL) | Gordon P et al |
| EPI_ISL_806227 | Canada/AB-94288/2020 | 2020-05-15 | North America/Canada/Alberta/ | Alberta Precision Labs (APL) | Alberta Precision Labs (APL) | Gordon P et al |
| EPI_ISL_806228 | Canada/AB-18670/2020 | 2020-05-07 | North America/Canada/Alberta/ | Alberta Precision Labs (APL) | Alberta Precision Labs (APL) | Gordon P et al |
| EPI_ISL_806229 | Canada/AB-67102/2020 | 2020-05-15 | North America/Canada/Alberta/ | Alberta Precision Labs (APL) | Alberta Precision Labs (APL) | Gordon P et al |
| EPI_ISL_806230 | Canada/AB-25892/2020 | 2020-03-16 | North America/Canada/Alberta/ | Alberta Precision Labs (APL) | Alberta Precision Labs (APL) | Gordon P et al |
| EPI_ISL_806231 | Canada/AB-79023/2020 | 2020-04-16 | North America/Canada/Alberta/ | Alberta Precision Labs (APL) | Alberta Precision Labs (APL) | Gordon P et al |
| EPI_ISL_806232 | Canada/AB-79515/2020 | 2020-05-07 | North America/Canada/Alberta/ | Alberta Precision Labs (APL) | Alberta Precision Labs (APL) | Gordon P et al |
| EPI_ISL_806233 | Canada/AB-28986/2020 | 2020-03-27 | North America/Canada/Alberta/ | Alberta Precision Labs (APL) | Alberta Precision Labs (APL) | Gordon P et al |
| EPI_ISL_806234 | Canada/AB-33262/2020 | 2020-04-13 | North America/Canada/Alberta/ | Alberta Precision Labs (APL) | Alberta Precision Labs (APL) | Gordon P et al |

| | | | | | | |
|---|---|---|---|---|---|---|
| EPI_ISL_806235 | Canada/AB-79633/2020 | 2020-04-11 | North America/Canada/Alberta/ | Alberta Precision Labs (APL) | Alberta Precision Labs (APL) | Gordon P et al |
| EPI_ISL_806236 | Canada/AB-18017/2020 | 2020-05-05 | North America/Canada/Alberta/ | Alberta Precision Labs (APL) | Alberta Precision Labs (APL) | Gordon P et al |
| EPI_ISL_806237 | Canada/AB-42485/2020 | 2020-04-10 | North America/Canada/Alberta/ | Alberta Precision Labs (APL) | Alberta Precision Labs (APL) | Gordon P et al |
| EPI_ISL_806238 | Canada/AB-21209/2020 | 2020-04-30 | North America/Canada/Alberta/ | Alberta Precision Labs (APL) | Alberta Precision Labs (APL) | Gordon P et al |
| EPI_ISL_806239 | Canada/AB-29272/2020 | 2020-04-13 | North America/Canada/Alberta/ | Alberta Precision Labs (APL) | Alberta Precision Labs (APL) | Gordon P et al |
| EPI_ISL_806240 | Canada/AB-90004/2020 | 2020-04-17 | North America/Canada/Alberta/ | Alberta Precision Labs (APL) | Alberta Precision Labs (APL) | Gordon P et al |
| EPI_ISL_806241 | Canada/AB-52176/2020 | 20-Mar | North America/Canada/Alberta/ | Alberta Precision Labs (APL) | Alberta Precision Labs (APL) | Gordon P et al |
| EPI_ISL_806242 | Canada/AB-19117/2020 | 2020-04-24 | North America/Canada/Alberta/ | Alberta Precision Labs (APL) | Alberta Precision Labs (APL) | Gordon P et al |
| EPI_ISL_806243 | Canada/AB-46259/2020 | 2020-04-16 | North America/Canada/Alberta/ | Alberta Precision Labs (APL) | Alberta Precision Labs (APL) | Gordon P et al |
| EPI_ISL_806244 | Canada/AB-29447/2020 | 2020-04-13 | North America/Canada/Alberta/ | Alberta Precision Labs (APL) | Alberta Precision Labs (APL) | Gordon P et al |
| EPI_ISL_806245 | Canada/AB-76981/2020 | 2020-03-18 | North America/Canada/Alberta/ | Alberta Precision Labs (APL) | Alberta Precision Labs (APL) | Gordon P et al |
| EPI_ISL_806246 | Canada/AB-25152/2020 | 2020-04-15 | North America/Canada/Alberta/ | Alberta Precision Labs (APL) | Alberta Precision Labs (APL) | Gordon P et al |
| EPI_ISL_806247 | Canada/AB-77662/2020 | 2020-04-13 | North America/Canada/Alberta/ | Alberta Precision Labs (APL) | Alberta Precision Labs (APL) | Gordon P et al |
| EPI_ISL_806248 | Canada/AB-89673/2020 | 2020-03-28 | North America/Canada/Alberta/ | Alberta Precision Labs (APL) | Alberta Precision Labs (APL) | Gordon P et al |
| EPI_ISL_806249 | Canada/AB-56768/2020 | 2020-04-28 | North America/Canada/Alberta/ | Alberta Precision Labs (APL) | Alberta Precision Labs (APL) | Gordon P et al |
| EPI_ISL_806250 | Canada/AB-92292/2020 | 2020-03-28 | North America/Canada/Alberta/ | Alberta Precision Labs (APL) | Alberta Precision Labs (APL) | Gordon P et al |
| EPI_ISL_806251 | Canada/AB-12655/2020 | 2020-04-13 | North America/Canada/Alberta/ | Alberta Precision Labs (APL) | Alberta Precision Labs (APL) | Gordon P et al |
| EPI_ISL_806252 | Canada/AB-67331/2020 | 2020-04-29 | North America/Canada/Alberta/ | Alberta Precision Labs (APL) | Alberta Precision Labs (APL) | Gordon P et al |
| EPI_ISL_806253 | Canada/AB-58621/2020 | 2020-04-13 | North America/Canada/Alberta/ | Alberta Precision Labs (APL) | Alberta Precision Labs (APL) | Gordon P et al |
| EPI_ISL_806254 | Canada/AB-16280/2020 | 2020-04-04 | North America/Canada/Alberta/ | Alberta Precision Labs (APL) | Alberta Precision Labs (APL) | Gordon P et al |

| | | | | | | |
|---|---|---|---|---|---|---|
| EPI_ISL_806255 | Canada/AB-72170/2020 | 2020-04-15 | North America/Canada/Alberta/ | Alberta Precision Labs (APL) | Alberta Precision Labs (APL) | Gordon P et al |
| EPI_ISL_806256 | Canada/AB-80035/2020 | 2020-04-14 | North America/Canada/Alberta/ | Alberta Precision Labs (APL) | Alberta Precision Labs (APL) | Gordon P et al |
| EPI_ISL_806257 | Canada/AB-76935/2020 | 2020-04-14 | North America/Canada/Alberta/ | Alberta Precision Labs (APL) | Alberta Precision Labs (APL) | Gordon P et al |
| EPI_ISL_806258 | Canada/AB-52905/2020 | 2020-04-13 | North America/Canada/Alberta/ | Alberta Precision Labs (APL) | Alberta Precision Labs (APL) | Gordon P et al |
| EPI_ISL_806259 | Canada/AB-47389/2020 | 2020-04-09 | North America/Canada/Alberta/ | Alberta Precision Labs (APL) | Alberta Precision Labs (APL) | Gordon P et al |
| EPI_ISL_806260 | Canada/AB-20186/2020 | 2020-04-17 | North America/Canada/Alberta/ | Alberta Precision Labs (APL) | Alberta Precision Labs (APL) | Gordon P et al |
| EPI_ISL_806261 | Canada/AB-25460/2020 | 2020-04-13 | North America/Canada/Alberta/ | Alberta Precision Labs (APL) | Alberta Precision Labs (APL) | Gordon P et al |
| EPI_ISL_806262 | Canada/AB-77279/2020 | 2020-04-24 | North America/Canada/Alberta/ | Alberta Precision Labs (APL) | Alberta Precision Labs (APL) | Gordon P et al |
| EPI_ISL_806263 | Canada/AB-90987/2020 | 2020-03-18 | North America/Canada/Alberta/ | Alberta Precision Labs (APL) | Alberta Precision Labs (APL) | Gordon P et al |
| EPI_ISL_806264 | Canada/AB-60830/2020 | 2020-04-07 | North America/Canada/Alberta/ | Alberta Precision Labs (APL) | Alberta Precision Labs (APL) | Gordon P et al |
| EPI_ISL_806265 | Canada/AB-91169/2020 | 2020-05-04 | North America/Canada/Alberta/ | Alberta Precision Labs (APL) | Alberta Precision Labs (APL) | Gordon P et al |
| EPI_ISL_806266 | Canada/AB-13909/2020 | 2020-04-17 | North America/Canada/Alberta/ | Alberta Precision Labs (APL) | Alberta Precision Labs (APL) | Gordon P et al |
| EPI_ISL_806267 | Canada/AB-87577/2020 | 2020-04-14 | North America/Canada/Alberta/ | Alberta Precision Labs (APL) | Alberta Precision Labs (APL) | Gordon P et al |
| EPI_ISL_806268 | Canada/AB-58608/2020 | 2020-04-15 | North America/Canada/Alberta/ | Alberta Precision Labs (APL) | Alberta Precision Labs (APL) | Gordon P et al |
| EPI_ISL_806269 | Canada/AB-36300/2020 | 2020-05-01 | North America/Canada/Alberta/ | Alberta Precision Labs (APL) | Alberta Precision Labs (APL) | Gordon P et al |
| EPI_ISL_806270 | Canada/AB-24629/2020 | 2020-04-13 | North America/Canada/Alberta/ | Alberta Precision Labs (APL) | Alberta Precision Labs (APL) | Gordon P et al |
| EPI_ISL_806271 | Canada/AB-24418/2020 | 2020-05-06 | North America/Canada/Alberta/ | Alberta Precision Labs (APL) | Alberta Precision Labs (APL) | Gordon P et al |
| EPI_ISL_806272 | Canada/AB-84010/2020 | 2020-04-16 | North America/Canada/Alberta/ | Alberta Precision Labs (APL) | Alberta Precision Labs (APL) | Gordon P et al |
| EPI_ISL_806273 | Canada/AB-43861/2020 | 2020-05-15 | North America/Canada/Alberta/ | Alberta Precision Labs (APL) | Alberta Precision Labs (APL) | Gordon P et al |
| EPI_ISL_806274 | Canada/AB-34649/2020 | 2020-05-10 | North America/Canada/Alberta/ | Alberta Precision Labs (APL) | Alberta Precision Labs (APL) | Gordon P et al |

| | | | | | | |
|---|---|---|---|---|---|---|
| EPI_ISL_806275 | Canada/AB-39415/2020 | 2020-05-14 | North America/Canada/Alberta/ | Alberta Precision Labs (APL) | Alberta Precision Labs (APL) | Gordon P et al |
| EPI_ISL_806276 | Canada/AB-15006/2020 | 2020-03-25 | North America/Canada/Alberta/ | Alberta Precision Labs (APL) | Alberta Precision Labs (APL) | Gordon P et al |
| EPI_ISL_806277 | Canada/AB-58640/2020 | 2020-05-03 | North America/Canada/Alberta/ | Alberta Precision Labs (APL) | Alberta Precision Labs (APL) | Gordon P et al |
| EPI_ISL_806278 | Canada/AB-48452/2020 | 2020-03-25 | North America/Canada/Alberta/ | Alberta Precision Labs (APL) | Alberta Precision Labs (APL) | Gordon P et al |
| EPI_ISL_806279 | Canada/AB-22996/2020 | 2020-03-20 | North America/Canada/Alberta/ | Alberta Precision Labs (APL) | Alberta Precision Labs (APL) | Gordon P et al |
| EPI_ISL_806280 | Canada/AB-51054/2020 | 2020-04-14 | North America/Canada/Alberta/ | Alberta Precision Labs (APL) | Alberta Precision Labs (APL) | Gordon P et al |
| EPI_ISL_806281 | Canada/AB-42911/2020 | 2020-05-04 | North America/Canada/Alberta/ | Alberta Precision Labs (APL) | Alberta Precision Labs (APL) | Gordon P et al |
| EPI_ISL_806282 | Canada/AB-19241/2020 | 2020-04-17 | North America/Canada/Alberta/ | Alberta Precision Labs (APL) | Alberta Precision Labs (APL) | Gordon P et al |
| EPI_ISL_806283 | Canada/AB-94948/2020 | 2020-04-15 | North America/Canada/Alberta/ | Alberta Precision Labs (APL) | Alberta Precision Labs (APL) | Gordon P et al |
| EPI_ISL_806284 | Canada/AB-18596/2020 | 2020-03-16 | North America/Canada/Alberta/ | Alberta Precision Labs (APL) | Alberta Precision Labs (APL) | Gordon P et al |
| EPI_ISL_806285 | Canada/AB-97320/2020 | 2020-03-25 | North America/Canada/Alberta/ | Alberta Precision Labs (APL) | Alberta Precision Labs (APL) | Gordon P et al |
| EPI_ISL_806286 | Canada/AB-40872/2020 | 20-Mar | North America/Canada/Alberta/ | Alberta Precision Labs (APL) | Alberta Precision Labs (APL) | Gordon P et al |
| EPI_ISL_806287 | Canada/AB-90862/2020 | 2020-04-08 | North America/Canada/Alberta/ | Alberta Precision Labs (APL) | Alberta Precision Labs (APL) | Gordon P et al |
| EPI_ISL_806288 | Canada/AB-20647/2020 | 2020-03-30 | North America/Canada/Alberta/ | Alberta Precision Labs (APL) | Alberta Precision Labs (APL) | Gordon P et al |
| EPI_ISL_806289 | Canada/AB-30596/2020 | 2020-03-24 | North America/Canada/Alberta/ | Alberta Precision Labs (APL) | Alberta Precision Labs (APL) | Gordon P et al |
| EPI_ISL_806290 | Canada/AB-29618/2020 | 2020-03-31 | North America/Canada/Alberta/ | Alberta Precision Labs (APL) | Alberta Precision Labs (APL) | Gordon P et al |
| EPI_ISL_806291 | Canada/AB-66471/2020 | 2020-04-09 | North America/Canada/Alberta/ | Alberta Precision Labs (APL) | Alberta Precision Labs (APL) | Gordon P et al |
| EPI_ISL_806292 | Canada/AB-82432/2020 | 2020-03-19 | North America/Canada/Alberta/ | Alberta Precision Labs (APL) | Alberta Precision Labs (APL) | Gordon P et al |
| EPI_ISL_806293 | Canada/AB-83417/2020 | 2020-03-30 | North America/Canada/Alberta/ | Alberta Precision Labs (APL) | Alberta Precision Labs (APL) | Gordon P et al |
| EPI_ISL_806294 | Canada/AB-41644/2020 | 2020-04-05 | North America/Canada/Alberta/ | Alberta Precision Labs (APL) | Alberta Precision Labs (APL) | Gordon P et al |

| | | | | | | |
|---|---|---|---|---|---|---|
| EPI_ISL_806295 | Canada/AB-29727/2020 | 20-Mar | North America/Canada/Alberta/ | Alberta Precision Labs (APL) | Alberta Precision Labs (APL) | Gordon P et al |
| EPI_ISL_806296 | Canada/AB-81002/2020 | 2020-04-15 | North America/Canada/Alberta/ | Alberta Precision Labs (APL) | Alberta Precision Labs (APL) | Gordon P et al |
| EPI_ISL_806297 | Canada/AB-44169/2020 | 2020-05-06 | North America/Canada/Alberta/ | Alberta Precision Labs (APL) | Alberta Precision Labs (APL) | Gordon P et al |
| EPI_ISL_806298 | Canada/AB-72396/2020 | 2020-04-16 | North America/Canada/Alberta/ | Alberta Precision Labs (APL) | Alberta Precision Labs (APL) | Gordon P et al |
| EPI_ISL_806299 | Canada/AB-60289/2020 | 2020-05-15 | North America/Canada/Alberta/ | Alberta Precision Labs (APL) | Alberta Precision Labs (APL) | Gordon P et al |
| EPI_ISL_806300 | Canada/AB-91535/2020 | 2020-03-20 | North America/Canada/Alberta/ | Alberta Precision Labs (APL) | Alberta Precision Labs (APL) | Gordon P et al |
| EPI_ISL_806301 | Canada/AB-39394/2020 | 2020-04-16 | North America/Canada/Alberta/ | Alberta Precision Labs (APL) | Alberta Precision Labs (APL) | Gordon P et al |
| EPI_ISL_806302 | Canada/AB-53898/2020 | 2020-03-25 | North America/Canada/Alberta/ | Alberta Precision Labs (APL) | Alberta Precision Labs (APL) | Gordon P et al |
| EPI_ISL_806303 | Canada/AB-87352/2020 | 2020-04-10 | North America/Canada/Alberta/ | Alberta Precision Labs (APL) | Alberta Precision Labs (APL) | Gordon P et al |
| EPI_ISL_806304 | Canada/AB-67721/2020 | 2020-04-11 | North America/Canada/Alberta/ | Alberta Precision Labs (APL) | Alberta Precision Labs (APL) | Gordon P et al |
| EPI_ISL_806305 | Canada/AB-40693/2020 | 2020-05-06 | North America/Canada/Alberta/ | Alberta Precision Labs (APL) | Alberta Precision Labs (APL) | Gordon P et al |
| EPI_ISL_806306 | Canada/AB-96096/2020 | 2020-05-07 | North America/Canada/Alberta/ | Alberta Precision Labs (APL) | Alberta Precision Labs (APL) | Gordon P et al |
| EPI_ISL_806307 | Canada/AB-50457/2020 | 2020-05-16 | North America/Canada/Alberta/ | Alberta Precision Labs (APL) | Alberta Precision Labs (APL) | Gordon P et al |
| EPI_ISL_806308 | Canada/AB-40987/2020 | 2020-05-06 | North America/Canada/Alberta/ | Alberta Precision Labs (APL) | Alberta Precision Labs (APL) | Gordon P et al |
| EPI_ISL_806309 | Canada/AB-48683/2020 | 2020-04-11 | North America/Canada/Alberta/ | Alberta Precision Labs (APL) | Alberta Precision Labs (APL) | Gordon P et al |
| EPI_ISL_806310 | Canada/AB-55923/2020 | 2020-04-17 | North America/Canada/Alberta/ | Alberta Precision Labs (APL) | Alberta Precision Labs (APL) | Gordon P et al |
| EPI_ISL_806311 | Canada/AB-61358/2020 | 2020-04-13 | North America/Canada/Alberta/ | Alberta Precision Labs (APL) | Alberta Precision Labs (APL) | Gordon P et al |
| EPI_ISL_806312 | Canada/AB-84015/2020 | 2020-04-10 | North America/Canada/Alberta/ | Alberta Precision Labs (APL) | Alberta Precision Labs (APL) | Gordon P et al |
| EPI_ISL_806313 | Canada/AB-98642/2020 | 2020-04-10 | North America/Canada/Alberta/ | Alberta Precision Labs (APL) | Alberta Precision Labs (APL) | Gordon P et al |
| EPI_ISL_806314 | Canada/AB-64180/2020 | 2020-04-11 | North America/Canada/Alberta/ | Alberta Precision Labs (APL) | Alberta Precision Labs (APL) | Gordon P et al |

| | | | | | | |
|---|---|---|---|---|---|---|
| EPI_ISL_806315 | Canada/AB-47830/2020 | 2020-04-24 | North America/Canada/Alberta/ | Alberta Precision Labs (APL) | Alberta Precision Labs (APL) | Gordon P et al |
| EPI_ISL_806316 | Canada/AB-91201/2020 | 2020-05-14 | North America/Canada/Alberta/ | Alberta Precision Labs (APL) | Alberta Precision Labs (APL) | Gordon P et al |
| EPI_ISL_806317 | Canada/AB-65233/2020 | 2020-03-28 | North America/Canada/Alberta/ | Alberta Precision Labs (APL) | Alberta Precision Labs (APL) | Gordon P et al |
| EPI_ISL_806318 | Canada/AB-68361/2020 | 2020-04-11 | North America/Canada/Alberta/ | Alberta Precision Labs (APL) | Alberta Precision Labs (APL) | Gordon P et al |
| EPI_ISL_806319 | Canada/AB-29889/2020 | 2020-04-08 | North America/Canada/Alberta/ | Alberta Precision Labs (APL) | Alberta Precision Labs (APL) | Gordon P et al |
| EPI_ISL_806320 | Canada/AB-98869/2020 | 2020-04-16 | North America/Canada/Alberta/ | Alberta Precision Labs (APL) | Alberta Precision Labs (APL) | Gordon P et al |
| EPI_ISL_806321 | Canada/AB-46808/2020 | 2020-04-01 | North America/Canada/Alberta/ | Alberta Precision Labs (APL) | Alberta Precision Labs (APL) | Gordon P et al |
| EPI_ISL_806322 | Canada/AB-57692/2020 | 2020-04-23 | North America/Canada/Alberta/ | Alberta Precision Labs (APL) | Alberta Precision Labs (APL) | Gordon P et al |
| EPI_ISL_806323 | Canada/AB-52206/2020 | 2020-04-10 | North America/Canada/Alberta/ | Alberta Precision Labs (APL) | Alberta Precision Labs (APL) | Gordon P et al |
| EPI_ISL_806324 | Canada/AB-72841/2020 | 2020-05-04 | North America/Canada/Alberta/ | Alberta Precision Labs (APL) | Alberta Precision Labs (APL) | Gordon P et al |
| EPI_ISL_806325 | Canada/AB-89781/2020 | 2020-04-12 | North America/Canada/Alberta/ | Alberta Precision Labs (APL) | Alberta Precision Labs (APL) | Gordon P et al |
| EPI_ISL_806326 | Canada/AB-62785/2020 | 2020-05-06 | North America/Canada/Alberta/ | Alberta Precision Labs (APL) | Alberta Precision Labs (APL) | Gordon P et al |
| EPI_ISL_806327 | Canada/AB-51188/2020 | 2020-04-15 | North America/Canada/Alberta/ | Alberta Precision Labs (APL) | Alberta Precision Labs (APL) | Gordon P et al |
| EPI_ISL_806328 | Canada/AB-99850/2020 | 2020-04-13 | North America/Canada/Alberta/ | Alberta Precision Labs (APL) | Alberta Precision Labs (APL) | Gordon P et al |
| EPI_ISL_806329 | Canada/AB-53118/2020 | 2020-03-27 | North America/Canada/Alberta/ | Alberta Precision Labs (APL) | Alberta Precision Labs (APL) | Gordon P et al |
| EPI_ISL_806330 | Canada/AB-49924/2020 | 2020-03-27 | North America/Canada/Alberta/ | Alberta Precision Labs (APL) | Alberta Precision Labs (APL) | Gordon P et al |
| EPI_ISL_806331 | Canada/AB-85784/2020 | 2020-04-13 | North America/Canada/Alberta/ | Alberta Precision Labs (APL) | Alberta Precision Labs (APL) | Gordon P et al |
| EPI_ISL_806333 | Canada/AB-46114/2020 | 2020-04-08 | North America/Canada/Alberta/ | Alberta Precision Labs (APL) | Alberta Precision Labs (APL) | Gordon P et al |
| EPI_ISL_806334 | Canada/AB-45450/2020 | 2020-04-12 | North America/Canada/Alberta/ | Alberta Precision Labs (APL) | Alberta Precision Labs (APL) | Gordon P et al |
| EPI_ISL_806335 | Canada/AB-88235/2020 | 2020-05-14 | North America/Canada/Alberta/ | Alberta Precision Labs (APL) | Alberta Precision Labs (APL) | Gordon P et al |

| | | | | | | |
|---|---|---|---|---|---|---|
| EPI_ISL_806336 | Canada/AB-65615/2020 | 2020-04-25 | North America/Canada/Alberta/ | Alberta Precision Labs (APL) | Alberta Precision Labs (APL) | Gordon P et al |
| EPI_ISL_806337 | Canada/AB-91895/2020 | 2020-05-03 | North America/Canada/Alberta/ | Alberta Precision Labs (APL) | Alberta Precision Labs (APL) | Gordon P et al |
| EPI_ISL_806338 | Canada/AB-58044/2020 | 2020-05-14 | North America/Canada/Alberta/ | Alberta Precision Labs (APL) | Alberta Precision Labs (APL) | Gordon P et al |
| EPI_ISL_806339 | Canada/AB-38538/2020 | 2020-04-30 | North America/Canada/Alberta/ | Alberta Precision Labs (APL) | Alberta Precision Labs (APL) | Gordon P et al |
| EPI_ISL_806340 | Canada/AB-76587/2020 | 2020-03-15 | North America/Canada/Alberta/ | Alberta Precision Labs (APL) | Alberta Precision Labs (APL) | Gordon P et al |
| EPI_ISL_806341 | Canada/AB-14464/2020 | 2020-04-16 | North America/Canada/Alberta/ | Alberta Precision Labs (APL) | Alberta Precision Labs (APL) | Gordon P et al |
| EPI_ISL_806342 | Canada/AB-44634/2020 | 2020-04-12 | North America/Canada/Alberta/ | Alberta Precision Labs (APL) | Alberta Precision Labs (APL) | Gordon P et al |
| EPI_ISL_806343 | Canada/AB-80162/2020 | 2020-04-02 | North America/Canada/Alberta/ | Alberta Precision Labs (APL) | Alberta Precision Labs (APL) | Gordon P et al |
| EPI_ISL_806344 | Canada/AB-66051/2020 | 2020-03-30 | North America/Canada/Alberta/ | Alberta Precision Labs (APL) | Alberta Precision Labs (APL) | Gordon P et al |
| EPI_ISL_806345 | Canada/AB-46435/2020 | 2020-05-04 | North America/Canada/Alberta/ | Alberta Precision Labs (APL) | Alberta Precision Labs (APL) | Gordon P et al |
| EPI_ISL_806346 | Canada/AB-50890/2020 | 2020-04-17 | North America/Canada/Alberta/ | Alberta Precision Labs (APL) | Alberta Precision Labs (APL) | Gordon P et al |
| EPI_ISL_806347 | Canada/AB-78359/2020 | 2020-04-07 | North America/Canada/Alberta/ | Alberta Precision Labs (APL) | Alberta Precision Labs (APL) | Gordon P et al |
| EPI_ISL_806348 | Canada/AB-69331/2020 | 2020-04-16 | North America/Canada/Alberta/ | Alberta Precision Labs (APL) | Alberta Precision Labs (APL) | Gordon P et al |
| EPI_ISL_806349 | Canada/AB-97227/2020 | 2020-04-17 | North America/Canada/Alberta/ | Alberta Precision Labs (APL) | Alberta Precision Labs (APL) | Gordon P et al |
| EPI_ISL_806350 | Canada/AB-34813/2020 | 2020-04-11 | North America/Canada/Alberta/ | Alberta Precision Labs (APL) | Alberta Precision Labs (APL) | Gordon P et al |
| EPI_ISL_806351 | Canada/AB-18183/2020 | 2020-04-11 | North America/Canada/Alberta/ | Alberta Precision Labs (APL) | Alberta Precision Labs (APL) | Gordon P et al |
| EPI_ISL_806352 | Canada/AB-22999/2020 | 2020-04-17 | North America/Canada/Alberta/ | Alberta Precision Labs (APL) | Alberta Precision Labs (APL) | Gordon P et al |
| EPI_ISL_806353 | Canada/AB-59167/2020 | 2020-04-09 | North America/Canada/Alberta/ | Alberta Precision Labs (APL) | Alberta Precision Labs (APL) | Gordon P et al |
| EPI_ISL_806354 | Canada/AB-32164/2020 | 2020-04-06 | North America/Canada/Alberta/ | Alberta Precision Labs (APL) | Alberta Precision Labs (APL) | Gordon P et al |
| EPI_ISL_806355 | Canada/AB-69583/2020 | 2020-04-08 | North America/Canada/Alberta/ | Alberta Precision Labs (APL) | Alberta Precision Labs (APL) | Gordon P et al |

| | | | | | | |
|---|---|---|---|---|---|---|
| EPI_ISL_806356 | Canada/AB-62882/2020 | 2020-05-15 | North America/Canada/Alberta/ | Alberta Precision Labs (APL) | Alberta Precision Labs (APL) | Gordon P et al |
| EPI_ISL_806357 | Canada/AB-99930/2020 | 2020-05-15 | North America/Canada/Alberta/ | Alberta Precision Labs (APL) | Alberta Precision Labs (APL) | Gordon P et al |
| EPI_ISL_806358 | Canada/AB-64690/2020 | 2020-05-03 | North America/Canada/Alberta/ | Alberta Precision Labs (APL) | Alberta Precision Labs (APL) | Gordon P et al |
| EPI_ISL_806359 | Canada/AB-63451/2020 | 2020-04-16 | North America/Canada/Alberta/ | Alberta Precision Labs (APL) | Alberta Precision Labs (APL) | Gordon P et al |
| EPI_ISL_806360 | Canada/AB-70097/2020 | 2020-04-06 | North America/Canada/Alberta/ | Alberta Precision Labs (APL) | Alberta Precision Labs (APL) | Gordon P et al |
| EPI_ISL_806361 | Canada/AB-99641/2020 | 2020-05-14 | North America/Canada/Alberta/ | Alberta Precision Labs (APL) | Alberta Precision Labs (APL) | Gordon P et al |
| EPI_ISL_806362 | Canada/AB-63299/2020 | 2020-04-27 | North America/Canada/Alberta/ | Alberta Precision Labs (APL) | Alberta Precision Labs (APL) | Gordon P et al |
| EPI_ISL_806363 | Canada/AB-53992/2020 | 2020-04-15 | North America/Canada/Alberta/ | Alberta Precision Labs (APL) | Alberta Precision Labs (APL) | Gordon P et al |
| EPI_ISL_806364 | Canada/AB-78457/2020 | 2020-05-03 | North America/Canada/Alberta/ | Alberta Precision Labs (APL) | Alberta Precision Labs (APL) | Gordon P et al |
| EPI_ISL_806365 | Canada/AB-57191/2020 | 2020-04-12 | North America/Canada/Alberta/ | Alberta Precision Labs (APL) | Alberta Precision Labs (APL) | Gordon P et al |
| EPI_ISL_806366 | Canada/AB-41131/2020 | 2020-04-08 | North America/Canada/Alberta/ | Alberta Precision Labs (APL) | Alberta Precision Labs (APL) | Gordon P et al |
| EPI_ISL_806367 | Canada/AB-69666/2020 | 2020-03-26 | North America/Canada/Alberta/ | Alberta Precision Labs (APL) | Alberta Precision Labs (APL) | Gordon P et al |
| EPI_ISL_806368 | Canada/AB-88280/2020 | 2020-04-13 | North America/Canada/Alberta/ | Alberta Precision Labs (APL) | Alberta Precision Labs (APL) | Gordon P et al |
| EPI_ISL_806369 | Canada/AB-76930/2020 | 2020-05-08 | North America/Canada/Alberta/ | Alberta Precision Labs (APL) | Alberta Precision Labs (APL) | Gordon P et al |
| EPI_ISL_806370 | Canada/AB-36345/2020 | 2020-04-13 | North America/Canada/Alberta/ | Alberta Precision Labs (APL) | Alberta Precision Labs (APL) | Gordon P et al |
| EPI_ISL_806371 | Canada/AB-45828/2020 | 2020-03-21 | North America/Canada/Alberta/ | Alberta Precision Labs (APL) | Alberta Precision Labs (APL) | Gordon P et al |
| EPI_ISL_806372 | Canada/AB-52053/2020 | 2020-04-01 | North America/Canada/Alberta/ | Alberta Precision Labs (APL) | Alberta Precision Labs (APL) | Gordon P et al |
| EPI_ISL_806373 | Canada/AB-96386/2020 | 2020-04-11 | North America/Canada/Alberta/ | Alberta Precision Labs (APL) | Alberta Precision Labs (APL) | Gordon P et al |
| EPI_ISL_806374 | Canada/AB-70673/2020 | 2020-04-14 | North America/Canada/Alberta/ | Alberta Precision Labs (APL) | Alberta Precision Labs (APL) | Gordon P et al |
| EPI_ISL_806375 | Canada/AB-48921/2020 | 2020-04-16 | North America/Canada/Alberta/ | Alberta Precision Labs (APL) | Alberta Precision Labs (APL) | Gordon P et al |

| | | | | | | |
|---|---|---|---|---|---|---|
| EPI_ISL_806376 | Canada/AB-54138/2020 | 2020-03-28 | North America/Canada/Alberta/ | Alberta Precision Labs (APL) | Alberta Precision Labs (APL) | Gordon P et al |
| EPI_ISL_806377 | Canada/AB-30465/2020 | 2020-04-13 | North America/Canada/Alberta/ | Alberta Precision Labs (APL) | Alberta Precision Labs (APL) | Gordon P et al |
| EPI_ISL_806378 | Canada/AB-21921/2020 | 2020-04-17 | North America/Canada/Alberta/ | Alberta Precision Labs (APL) | Alberta Precision Labs (APL) | Gordon P et al |
| EPI_ISL_806379 | Canada/AB-52282/2020 | 2020-04-12 | North America/Canada/Alberta/ | Alberta Precision Labs (APL) | Alberta Precision Labs (APL) | Gordon P et al |
| EPI_ISL_806380 | Canada/AB-34330/2020 | 2020-04-11 | North America/Canada/Alberta/ | Alberta Precision Labs (APL) | Alberta Precision Labs (APL) | Gordon P et al |
| EPI_ISL_806381 | Canada/AB-65149/2020 | 2020-04-12 | North America/Canada/Alberta/ | Alberta Precision Labs (APL) | Alberta Precision Labs (APL) | Gordon P et al |
| EPI_ISL_806382 | Canada/AB-71271/2020 | 2020-04-13 | North America/Canada/Alberta/ | Alberta Precision Labs (APL) | Alberta Precision Labs (APL) | Gordon P et al |
| EPI_ISL_806383 | Canada/AB-63210/2020 | 2020-03-16 | North America/Canada/Alberta/ | Alberta Precision Labs (APL) | Alberta Precision Labs (APL) | Gordon P et al |
| EPI_ISL_806384 | Canada/AB-94048/2020 | 2020-03-27 | North America/Canada/Alberta/ | Alberta Precision Labs (APL) | Alberta Precision Labs (APL) | Gordon P et al |
| EPI_ISL_806385 | Canada/AB-51651/2020 | 2020-04-16 | North America/Canada/Alberta/ | Alberta Precision Labs (APL) | Alberta Precision Labs (APL) | Gordon P et al |
| EPI_ISL_806386 | Canada/AB-69059/2020 | 2020-04-16 | North America/Canada/Alberta/ | Alberta Precision Labs (APL) | Alberta Precision Labs (APL) | Gordon P et al |
| EPI_ISL_806387 | Canada/AB-24001/2020 | 2020-04-16 | North America/Canada/Alberta/ | Alberta Precision Labs (APL) | Alberta Precision Labs (APL) | Gordon P et al |
| EPI_ISL_806388 | Canada/AB-17862/2020 | 2020-03-24 | North America/Canada/Alberta/ | Alberta Precision Labs (APL) | Alberta Precision Labs (APL) | Gordon P et al |
| EPI_ISL_806389 | Canada/AB-13781/2020 | 2020-03-17 | North America/Canada/Alberta/ | Alberta Precision Labs (APL) | Alberta Precision Labs (APL) | Gordon P et al |
| EPI_ISL_806390 | Canada/AB-89968/2020 | 2020-04-15 | North America/Canada/Alberta/ | Alberta Precision Labs (APL) | Alberta Precision Labs (APL) | Gordon P et al |
| EPI_ISL_806391 | Canada/AB-91722/2020 | 2020-04-15 | North America/Canada/Alberta/ | Alberta Precision Labs (APL) | Alberta Precision Labs (APL) | Gordon P et al |
| EPI_ISL_806392 | Canada/AB-64499/2020 | 2020-04-17 | North America/Canada/Alberta/ | Alberta Precision Labs (APL) | Alberta Precision Labs (APL) | Gordon P et al |
| EPI_ISL_806393 | Canada/AB-41922/2020 | 2020-05-04 | North America/Canada/Alberta/ | Alberta Precision Labs (APL) | Alberta Precision Labs (APL) | Gordon P et al |
| EPI_ISL_806394 | Canada/AB-47732/2020 | 2020-03-20 | North America/Canada/Alberta/ | Alberta Precision Labs (APL) | Alberta Precision Labs (APL) | Gordon P et al |
| EPI_ISL_806395 | Canada/AB-32426/2020 | 2020-04-24 | North America/Canada/Alberta/ | Alberta Precision Labs (APL) | Alberta Precision Labs (APL) | Gordon P et al |

| | | | | | | |
|---|---|---|---|---|---|---|
| EPI_ISL_806396 | Canada/AB-15329/2020 | 2020-04-15 | North America/Canada/Alberta/ | Alberta Precision Labs (APL) | Alberta Precision Labs (APL) | Gordon P et al |
| EPI_ISL_806397 | Canada/AB-41023/2020 | 2020-04-16 | North America/Canada/Alberta/ | Alberta Precision Labs (APL) | Alberta Precision Labs (APL) | Gordon P et al |
| EPI_ISL_806398 | Canada/AB-83138/2020 | 2020-04-11 | North America/Canada/Alberta/ | Alberta Precision Labs (APL) | Alberta Precision Labs (APL) | Gordon P et al |
| EPI_ISL_806399 | Canada/AB-22459/2020 | 2020-04-11 | North America/Canada/Alberta/ | Alberta Precision Labs (APL) | Alberta Precision Labs (APL) | Gordon P et al |
| EPI_ISL_806400 | Canada/AB-57378/2020 | 2020-04-13 | North America/Canada/Alberta/ | Alberta Precision Labs (APL) | Alberta Precision Labs (APL) | Gordon P et al |
| EPI_ISL_806401 | Canada/AB-82036/2020 | 2020-05-10 | North America/Canada/Alberta/ | Alberta Precision Labs (APL) | Alberta Precision Labs (APL) | Gordon P et al |
| EPI_ISL_806402 | Canada/AB-65294/2020 | 2020-04-17 | North America/Canada/Alberta/ | Alberta Precision Labs (APL) | Alberta Precision Labs (APL) | Gordon P et al |
| EPI_ISL_806403 | Canada/AB-95280/2020 | 2020-04-13 | North America/Canada/Alberta/ | Alberta Precision Labs (APL) | Alberta Precision Labs (APL) | Gordon P et al |
| EPI_ISL_806404 | Canada/AB-27414/2020 | 2020-04-01 | North America/Canada/Alberta/ | Alberta Precision Labs (APL) | Alberta Precision Labs (APL) | Gordon P et al |
| EPI_ISL_806405 | Canada/AB-51788/2020 | 2020-03-29 | North America/Canada/Alberta/ | Alberta Precision Labs (APL) | Alberta Precision Labs (APL) | Gordon P et al |
| EPI_ISL_806406 | Canada/AB-20834/2020 | 2020-03-26 | North America/Canada/Alberta/ | Alberta Precision Labs (APL) | Alberta Precision Labs (APL) | Gordon P et al |
| EPI_ISL_806407 | Canada/AB-29869/2020 | 2020-04-16 | North America/Canada/Alberta/ | Alberta Precision Labs (APL) | Alberta Precision Labs (APL) | Gordon P et al |
| EPI_ISL_806408 | Canada/AB-96873/2020 | 2020-03-25 | North America/Canada/Alberta/ | Alberta Precision Labs (APL) | Alberta Precision Labs (APL) | Gordon P et al |
| EPI_ISL_806409 | Canada/AB-51359/2020 | 2020-03-16 | North America/Canada/Alberta/ | Alberta Precision Labs (APL) | Alberta Precision Labs (APL) | Gordon P et al |
| EPI_ISL_806410 | Canada/AB-89973/2020 | 2020-04-11 | North America/Canada/Alberta/ | Alberta Precision Labs (APL) | Alberta Precision Labs (APL) | Gordon P et al |
| EPI_ISL_806411 | Canada/AB-96501/2020 | 2020-04-15 | North America/Canada/Alberta/ | Alberta Precision Labs (APL) | Alberta Precision Labs (APL) | Gordon P et al |
| EPI_ISL_806412 | Canada/AB-71039/2020 | 2020-03-25 | North America/Canada/Alberta/ | Alberta Precision Labs (APL) | Alberta Precision Labs (APL) | Gordon P et al |
| EPI_ISL_806413 | Canada/AB-76499/2020 | 2020-03-25 | North America/Canada/Alberta/ | Alberta Precision Labs (APL) | Alberta Precision Labs (APL) | Gordon P et al |
| EPI_ISL_806414 | Canada/AB-65152/2020 | 2020-05-14 | North America/Canada/Alberta/ | Alberta Precision Labs (APL) | Alberta Precision Labs (APL) | Gordon P et al |
| EPI_ISL_806415 | Canada/AB-25636/2020 | 2020-04-20 | North America/Canada/Alberta/ | Alberta Precision Labs (APL) | Alberta Precision Labs (APL) | Gordon P et al |

| | | | | | | |
|---|---|---|---|---|---|---|
| EPI_ISL_806416 | Canada/AB-76629/2020 | 2020-03-27 | North America/Canada/Alberta/ | Alberta Precision Labs (APL) | Alberta Precision Labs (APL) | Gordon P et al |
| EPI_ISL_806417 | Canada/AB-17731/2020 | 2020-03-31 | North America/Canada/Alberta/ | Alberta Precision Labs (APL) | Alberta Precision Labs (APL) | Gordon P et al |
| EPI_ISL_806418 | Canada/AB-26695/2020 | 2020-05-08 | North America/Canada/Alberta/ | Alberta Precision Labs (APL) | Alberta Precision Labs (APL) | Gordon P et al |
| EPI_ISL_806419 | Canada/AB-33954/2020 | 2020-04-13 | North America/Canada/Alberta/ | Alberta Precision Labs (APL) | Alberta Precision Labs (APL) | Gordon P et al |
| EPI_ISL_806420 | Canada/AB-67620/2020 | 2020-04-12 | North America/Canada/Alberta/ | Alberta Precision Labs (APL) | Alberta Precision Labs (APL) | Gordon P et al |
| EPI_ISL_806421 | Canada/AB-79150/2020 | 2020-04-16 | North America/Canada/Alberta/ | Alberta Precision Labs (APL) | Alberta Precision Labs (APL) | Gordon P et al |
| EPI_ISL_806422 | Canada/AB-90868/2020 | 2020-04-13 | North America/Canada/Alberta/ | Alberta Precision Labs (APL) | Alberta Precision Labs (APL) | Gordon P et al |
| EPI_ISL_806423 | Canada/AB-58852/2020 | 2020-03-29 | North America/Canada/Alberta/ | Alberta Precision Labs (APL) | Alberta Precision Labs (APL) | Gordon P et al |
| EPI_ISL_806424 | Canada/AB-69970/2020 | 2020-04-17 | North America/Canada/Alberta/ | Alberta Precision Labs (APL) | Alberta Precision Labs (APL) | Gordon P et al |
| EPI_ISL_806425 | Canada/AB-17940/2020 | 2020-04-15 | North America/Canada/Alberta/ | Alberta Precision Labs (APL) | Alberta Precision Labs (APL) | Gordon P et al |
| EPI_ISL_806426 | Canada/AB-15271/2020 | 2020-05-02 | North America/Canada/Alberta/ | Alberta Precision Labs (APL) | Alberta Precision Labs (APL) | Gordon P et al |
| EPI_ISL_806427 | Canada/AB-92902/2020 | 2020-04-13 | North America/Canada/Alberta/ | Alberta Precision Labs (APL) | Alberta Precision Labs (APL) | Gordon P et al |
| EPI_ISL_806428 | Canada/AB-94866/2020 | 20-Mar | North America/Canada/Alberta/ | Alberta Precision Labs (APL) | Alberta Precision Labs (APL) | Gordon P et al |
| EPI_ISL_806429 | Canada/AB-76983/2020 | 2020-04-02 | North America/Canada/Alberta/ | Alberta Precision Labs (APL) | Alberta Precision Labs (APL) | Gordon P et al |
| EPI_ISL_806430 | Canada/AB-69283/2020 | 2020-04-17 | North America/Canada/Alberta/ | Alberta Precision Labs (APL) | Alberta Precision Labs (APL) | Gordon P et al |
| EPI_ISL_806431 | Canada/AB-48496/2020 | 2020-03-26 | North America/Canada/Alberta/ | Alberta Precision Labs (APL) | Alberta Precision Labs (APL) | Gordon P et al |
| EPI_ISL_806432 | Canada/AB-13224/2020 | 2020-04-11 | North America/Canada/Alberta/ | Alberta Precision Labs (APL) | Alberta Precision Labs (APL) | Gordon P et al |
| EPI_ISL_806433 | Canada/AB-43672/2020 | 2020-05-16 | North America/Canada/Alberta/ | Alberta Precision Labs (APL) | Alberta Precision Labs (APL) | Gordon P et al |
| EPI_ISL_806434 | Canada/AB-73782/2020 | 2020-05-14 | North America/Canada/Alberta/ | Alberta Precision Labs (APL) | Alberta Precision Labs (APL) | Gordon P et al |
| EPI_ISL_806435 | Canada/AB-79279/2020 | 2020-04-12 | North America/Canada/Alberta/ | Alberta Precision Labs (APL) | Alberta Precision Labs (APL) | Gordon P et al |

| | | | | | | |
|---|---|---|---|---|---|---|
| EPI_ISL_806436 | Canada/AB-97710/2020 | 2020-04-16 | North America/Canada/Alberta/ | Alberta Precision Labs (APL) | Alberta Precision Labs (APL) | Gordon P et al |
| EPI_ISL_806437 | Canada/AB-78066/2020 | 2020-03-23 | North America/Canada/Alberta/ | Alberta Precision Labs (APL) | Alberta Precision Labs (APL) | Gordon P et al |
| EPI_ISL_806438 | Canada/AB-58324/2020 | 2020-04-20 | North America/Canada/Alberta/ | Alberta Precision Labs (APL) | Alberta Precision Labs (APL) | Gordon P et al |
| EPI_ISL_806439 | Canada/AB-20630/2020 | 2020-04-09 | North America/Canada/Alberta/ | Alberta Precision Labs (APL) | Alberta Precision Labs (APL) | Gordon P et al |
| EPI_ISL_806440 | Canada/AB-31172/2020 | 2020-04-16 | North America/Canada/Alberta/ | Alberta Precision Labs (APL) | Alberta Precision Labs (APL) | Gordon P et al |
| EPI_ISL_806441 | Canada/AB-20796/2020 | 2020-05-16 | North America/Canada/Alberta/ | Alberta Precision Labs (APL) | Alberta Precision Labs (APL) | Gordon P et al |
| EPI_ISL_806442 | Canada/AB-87384/2020 | 2020-04-06 | North America/Canada/Alberta/ | Alberta Precision Labs (APL) | Alberta Precision Labs (APL) | Gordon P et al |
| EPI_ISL_806443 | Canada/AB-26719/2020 | 2020-04-22 | North America/Canada/Alberta/ | Alberta Precision Labs (APL) | Alberta Precision Labs (APL) | Gordon P et al |
| EPI_ISL_806444 | Canada/AB-29177/2020 | 2020-04-11 | North America/Canada/Alberta/ | Alberta Precision Labs (APL) | Alberta Precision Labs (APL) | Gordon P et al |
| EPI_ISL_806445 | Canada/AB-18502/2020 | 2020-04-13 | North America/Canada/Alberta/ | Alberta Precision Labs (APL) | Alberta Precision Labs (APL) | Gordon P et al |
| EPI_ISL_806446 | Canada/AB-64929/2020 | 20-Mar | North America/Canada/Alberta/ | Alberta Precision Labs (APL) | Alberta Precision Labs (APL) | Gordon P et al |
| EPI_ISL_806447 | Canada/AB-69694/2020 | 2020-05-15 | North America/Canada/Alberta/ | Alberta Precision Labs (APL) | Alberta Precision Labs (APL) | Gordon P et al |
| EPI_ISL_806448 | Canada/AB-68311/2020 | 2020-04-11 | North America/Canada/Alberta/ | Alberta Precision Labs (APL) | Alberta Precision Labs (APL) | Gordon P et al |
| EPI_ISL_806449 | Canada/AB-76238/2020 | 2020-04-17 | North America/Canada/Alberta/ | Alberta Precision Labs (APL) | Alberta Precision Labs (APL) | Gordon P et al |
| EPI_ISL_806450 | Canada/AB-89420/2020 | 2020-04-10 | North America/Canada/Alberta/ | Alberta Precision Labs (APL) | Alberta Precision Labs (APL) | Gordon P et al |
| EPI_ISL_806451 | Canada/AB-78725/2020 | 2020-04-13 | North America/Canada/Alberta/ | Alberta Precision Labs (APL) | Alberta Precision Labs (APL) | Gordon P et al |
| EPI_ISL_806452 | Canada/AB-26934/2020 | 2020-05-05 | North America/Canada/Alberta/ | Alberta Precision Labs (APL) | Alberta Precision Labs (APL) | Gordon P et al |
| EPI_ISL_806453 | Canada/AB-39313/2020 | 2020-05-16 | North America/Canada/Alberta/ | Alberta Precision Labs (APL) | Alberta Precision Labs (APL) | Gordon P et al |
| EPI_ISL_806454 | Canada/AB-27077/2020 | 20-Mar | North America/Canada/Alberta/ | Alberta Precision Labs (APL) | Alberta Precision Labs (APL) | Gordon P et al |
| EPI_ISL_806455 | Canada/AB-16143/2020 | 2020-04-02 | North America/Canada/Alberta/ | Alberta Precision Labs (APL) | Alberta Precision Labs (APL) | Gordon P et al |

| | | | | | | |
|---|---|---|---|---|---|---|
| EPI_ISL_806456 | Canada/AB-60573/2020 | 2020-05-15 | North America/Canada/Alberta/ | Alberta Precision Labs (APL) | Alberta Precision Labs (APL) | Gordon P et al |
| EPI_ISL_806457 | Canada/AB-12931/2020 | 2020-05-08 | North America/Canada/Alberta/ | Alberta Precision Labs (APL) | Alberta Precision Labs (APL) | Gordon P et al |
| EPI_ISL_806459 | Canada/AB-30256/2020 | 2020-04-14 | North America/Canada/Alberta/ | Alberta Precision Labs (APL) | Alberta Precision Labs (APL) | Gordon P et al |
| EPI_ISL_806460 | Canada/AB-79356/2020 | 2020-04-17 | North America/Canada/Alberta/ | Alberta Precision Labs (APL) | Alberta Precision Labs (APL) | Gordon P et al |
| EPI_ISL_806461 | Canada/AB-75126/2020 | 2020-03-30 | North America/Canada/Alberta/ | Alberta Precision Labs (APL) | Alberta Precision Labs (APL) | Gordon P et al |
| EPI_ISL_806462 | Canada/AB-92977/2020 | 2020-03-18 | North America/Canada/Alberta/ | Alberta Precision Labs (APL) | Alberta Precision Labs (APL) | Gordon P et al |
| EPI_ISL_806463 | Canada/AB-36867/2020 | 2020-03-30 | North America/Canada/Alberta/ | Alberta Precision Labs (APL) | Alberta Precision Labs (APL) | Gordon P et al |
| EPI_ISL_806464 | Canada/AB-60578/2020 | 2020-04-04 | North America/Canada/Alberta/ | Alberta Precision Labs (APL) | Alberta Precision Labs (APL) | Gordon P et al |
| EPI_ISL_806465 | Canada/AB-37439/2020 | 2020-04-12 | North America/Canada/Alberta/ | Alberta Precision Labs (APL) | Alberta Precision Labs (APL) | Gordon P et al |
| EPI_ISL_806466 | Canada/AB-48355/2020 | 2020-04-28 | North America/Canada/Alberta/ | Alberta Precision Labs (APL) | Alberta Precision Labs (APL) | Gordon P et al |
| EPI_ISL_806467 | Canada/AB-24837/2020 | 2020-05-10 | North America/Canada/Alberta/ | Alberta Precision Labs (APL) | Alberta Precision Labs (APL) | Gordon P et al |
| EPI_ISL_806468 | Canada/AB-28928/2020 | 2020-05-05 | North America/Canada/Alberta/ | Alberta Precision Labs (APL) | Alberta Precision Labs (APL) | Gordon P et al |
| EPI_ISL_806469 | Canada/AB-40101/2020 | 2020-04-12 | North America/Canada/Alberta/ | Alberta Precision Labs (APL) | Alberta Precision Labs (APL) | Gordon P et al |
| EPI_ISL_806470 | Canada/AB-43486/2020 | 2020-04-11 | North America/Canada/Alberta/ | Alberta Precision Labs (APL) | Alberta Precision Labs (APL) | Gordon P et al |
| EPI_ISL_806471 | Canada/AB-18292/2020 | 2020-04-13 | North America/Canada/Alberta/ | Alberta Precision Labs (APL) | Alberta Precision Labs (APL) | Gordon P et al |
| EPI_ISL_806472 | Canada/AB-39136/2020 | 2020-04-12 | North America/Canada/Alberta/ | Alberta Precision Labs (APL) | Alberta Precision Labs (APL) | Gordon P et al |
| EPI_ISL_806473 | Canada/AB-15174/2020 | 2020-03-19 | North America/Canada/Alberta/ | Alberta Precision Labs (APL) | Alberta Precision Labs (APL) | Gordon P et al |
| EPI_ISL_806474 | Canada/AB-42849/2020 | 2020-03-24 | North America/Canada/Alberta/ | Alberta Precision Labs (APL) | Alberta Precision Labs (APL) | Gordon P et al |
| EPI_ISL_806475 | Canada/AB-69712/2020 | 2020-04-25 | North America/Canada/Alberta/ | Alberta Precision Labs (APL) | Alberta Precision Labs (APL) | Gordon P et al |
| EPI_ISL_806476 | Canada/AB-28086/2020 | 2020-04-13 | North America/Canada/Alberta/ | Alberta Precision Labs (APL) | Alberta Precision Labs (APL) | Gordon P et al |

| | | | | | | |
|---|---|---|---|---|---|---|
| EPI_ISL_806477 | Canada/AB-16566/2020 | 2020-04-17 | North America/Canada/Alberta/ | Alberta Precision Labs (APL) | Alberta Precision Labs (APL) | Gordon P et al |
| EPI_ISL_806478 | Canada/AB-24041/2020 | 2020-05-01 | North America/Canada/Alberta/ | Alberta Precision Labs (APL) | Alberta Precision Labs (APL) | Gordon P et al |
| EPI_ISL_806479 | Canada/AB-16775/2020 | 2020-04-24 | North America/Canada/Alberta/ | Alberta Precision Labs (APL) | Alberta Precision Labs (APL) | Gordon P et al |
| EPI_ISL_806480 | Canada/AB-28848/2020 | 2020-04-15 | North America/Canada/Alberta/ | Alberta Precision Labs (APL) | Alberta Precision Labs (APL) | Gordon P et al |
| EPI_ISL_806481 | Canada/AB-63810/2020 | 2020-05-04 | North America/Canada/Alberta/ | Alberta Precision Labs (APL) | Alberta Precision Labs (APL) | Gordon P et al |
| EPI_ISL_806482 | Canada/AB-32556/2020 | 2020-04-14 | North America/Canada/Alberta/ | Alberta Precision Labs (APL) | Alberta Precision Labs (APL) | Gordon P et al |
| EPI_ISL_806483 | Canada/AB-60584/2020 | 2020-04-15 | North America/Canada/Alberta/ | Alberta Precision Labs (APL) | Alberta Precision Labs (APL) | Gordon P et al |
| EPI_ISL_806484 | Canada/AB-30199/2020 | 2020-05-04 | North America/Canada/Alberta/ | Alberta Precision Labs (APL) | Alberta Precision Labs (APL) | Gordon P et al |
| EPI_ISL_806485 | Canada/AB-14462/2020 | 2020-04-24 | North America/Canada/Alberta/ | Alberta Precision Labs (APL) | Alberta Precision Labs (APL) | Gordon P et al |
| EPI_ISL_806486 | Canada/AB-44366/2020 | 2020-03-26 | North America/Canada/Alberta/ | Alberta Precision Labs (APL) | Alberta Precision Labs (APL) | Gordon P et al |
| EPI_ISL_806487 | Canada/AB-48559/2020 | 2020-04-25 | North America/Canada/Alberta/ | Alberta Precision Labs (APL) | Alberta Precision Labs (APL) | Gordon P et al |
| EPI_ISL_806488 | Canada/AB-41403/2020 | 2020-04-16 | North America/Canada/Alberta/ | Alberta Precision Labs (APL) | Alberta Precision Labs (APL) | Gordon P et al |
| EPI_ISL_806489 | Canada/AB-87302/2020 | 2020-04-24 | North America/Canada/Alberta/ | Alberta Precision Labs (APL) | Alberta Precision Labs (APL) | Gordon P et al |
| EPI_ISL_806490 | Canada/AB-87507/2020 | 2020-04-26 | North America/Canada/Alberta/ | Alberta Precision Labs (APL) | Alberta Precision Labs (APL) | Gordon P et al |
| EPI_ISL_806491 | Canada/AB-63248/2020 | 2020-04-02 | North America/Canada/Alberta/ | Alberta Precision Labs (APL) | Alberta Precision Labs (APL) | Gordon P et al |
| EPI_ISL_806492 | Canada/AB-41810/2020 | 2020-05-04 | North America/Canada/Alberta/ | Alberta Precision Labs (APL) | Alberta Precision Labs (APL) | Gordon P et al |
| EPI_ISL_806493 | Canada/AB-74538/2020 | 2020-03-27 | North America/Canada/Alberta/ | Alberta Precision Labs (APL) | Alberta Precision Labs (APL) | Gordon P et al |
| EPI_ISL_806494 | Canada/AB-94124/2020 | 2020-04-30 | North America/Canada/Alberta/ | Alberta Precision Labs (APL) | Alberta Precision Labs (APL) | Gordon P et al |
| EPI_ISL_806495 | Canada/AB-49051/2020 | 2020-04-15 | North America/Canada/Alberta/ | Alberta Precision Labs (APL) | Alberta Precision Labs (APL) | Gordon P et al |
| EPI_ISL_806496 | Canada/AB-63208/2020 | 2020-05-07 | North America/Canada/Alberta/ | Alberta Precision Labs (APL) | Alberta Precision Labs (APL) | Gordon P et al |

| | | | | | | |
|---|---|---|---|---|---|---|
| EPI_ISL_806497 | Canada/AB-47074/2020 | 2020-04-29 | North America/Canada/Alberta/ | Alberta Precision Labs (APL) | Alberta Precision Labs (APL) | Gordon P et al |
| EPI_ISL_806498 | Canada/AB-25795/2020 | 2020-04-14 | North America/Canada/Alberta/ | Alberta Precision Labs (APL) | Alberta Precision Labs (APL) | Gordon P et al |
| EPI_ISL_806499 | Canada/AB-99591/2020 | 2020-04-15 | North America/Canada/Alberta/ | Alberta Precision Labs (APL) | Alberta Precision Labs (APL) | Gordon P et al |
| EPI_ISL_806500 | Canada/AB-65996/2020 | 2020-03-24 | North America/Canada/Alberta/ | Alberta Precision Labs (APL) | Alberta Precision Labs (APL) | Gordon P et al |
| EPI_ISL_806501 | Canada/AB-78997/2020 | 2020-04-11 | North America/Canada/Alberta/ | Alberta Precision Labs (APL) | Alberta Precision Labs (APL) | Gordon P et al |
| EPI_ISL_806502 | Canada/AB-73088/2020 | 2020-04-23 | North America/Canada/Alberta/ | Alberta Precision Labs (APL) | Alberta Precision Labs (APL) | Gordon P et al |
| EPI_ISL_806503 | Canada/AB-88625/2020 | 2020-04-23 | North America/Canada/Alberta/ | Alberta Precision Labs (APL) | Alberta Precision Labs (APL) | Gordon P et al |
| EPI_ISL_806504 | Canada/AB-90856/2020 | 2020-05-03 | North America/Canada/Alberta/ | Alberta Precision Labs (APL) | Alberta Precision Labs (APL) | Gordon P et al |
| EPI_ISL_806505 | Canada/AB-44788/2020 | 2020-04-27 | North America/Canada/Alberta/ | Alberta Precision Labs (APL) | Alberta Precision Labs (APL) | Gordon P et al |
| EPI_ISL_806506 | Canada/AB-31665/2020 | 2020-04-17 | North America/Canada/Alberta/ | Alberta Precision Labs (APL) | Alberta Precision Labs (APL) | Gordon P et al |
| EPI_ISL_806507 | Canada/AB-23817/2020 | 2020-04-08 | North America/Canada/Alberta/ | Alberta Precision Labs (APL) | Alberta Precision Labs (APL) | Gordon P et al |
| EPI_ISL_806508 | Canada/AB-92656/2020 | 2020-04-15 | North America/Canada/Alberta/ | Alberta Precision Labs (APL) | Alberta Precision Labs (APL) | Gordon P et al |
| EPI_ISL_806509 | Canada/AB-35537/2020 | 2020-04-17 | North America/Canada/Alberta/ | Alberta Precision Labs (APL) | Alberta Precision Labs (APL) | Gordon P et al |
| EPI_ISL_806510 | Canada/AB-39268/2020 | 2020-04-12 | North America/Canada/Alberta/ | Alberta Precision Labs (APL) | Alberta Precision Labs (APL) | Gordon P et al |
| EPI_ISL_806511 | Canada/AB-23064/2020 | 2020-04-24 | North America/Canada/Alberta/ | Alberta Precision Labs (APL) | Alberta Precision Labs (APL) | Gordon P et al |
| EPI_ISL_806512 | Canada/AB-45441/2020 | 2020-04-24 | North America/Canada/Alberta/ | Alberta Precision Labs (APL) | Alberta Precision Labs (APL) | Gordon P et al |
| EPI_ISL_806513 | Canada/AB-95505/2020 | 2020-03-24 | North America/Canada/Alberta/ | Alberta Precision Labs (APL) | Alberta Precision Labs (APL) | Gordon P et al |
| EPI_ISL_806514 | Canada/AB-46245/2020 | 20-Mar | North America/Canada/Alberta/ | Alberta Precision Labs (APL) | Alberta Precision Labs (APL) | Gordon P et al |
| EPI_ISL_806515 | Canada/AB-77009/2020 | 2020-04-12 | North America/Canada/Alberta/ | Alberta Precision Labs (APL) | Alberta Precision Labs (APL) | Gordon P et al |
| EPI_ISL_806516 | Canada/AB-98426/2020 | 2020-04-15 | North America/Canada/Alberta/ | Alberta Precision Labs (APL) | Alberta Precision Labs (APL) | Gordon P et al |

| | | | | | | |
|---|---|---|---|---|---|---|
| EPI_ISL_806517 | Canada/AB-39044/2020 | 2020-04-24 | North America/Canada/Alberta/ | Alberta Precision Labs (APL) | Alberta Precision Labs (APL) | Gordon P et al |
| EPI_ISL_806518 | Canada/AB-99141/2020 | 2020-04-25 | North America/Canada/Alberta/ | Alberta Precision Labs (APL) | Alberta Precision Labs (APL) | Gordon P et al |
| EPI_ISL_806519 | Canada/AB-44243/2020 | 2020-03-24 | North America/Canada/Alberta/ | Alberta Precision Labs (APL) | Alberta Precision Labs (APL) | Gordon P et al |
| EPI_ISL_806520 | Canada/AB-63030/2020 | 2020-04-13 | North America/Canada/Alberta/ | Alberta Precision Labs (APL) | Alberta Precision Labs (APL) | Gordon P et al |
| EPI_ISL_806521 | Canada/AB-66607/2020 | 2020-05-08 | North America/Canada/Alberta/ | Alberta Precision Labs (APL) | Alberta Precision Labs (APL) | Gordon P et al |
| EPI_ISL_806522 | Canada/AB-89984/2020 | 2020-04-10 | North America/Canada/Alberta/ | Alberta Precision Labs (APL) | Alberta Precision Labs (APL) | Gordon P et al |
| EPI_ISL_806523 | Canada/AB-77803/2020 | 2020-04-20 | North America/Canada/Alberta/ | Alberta Precision Labs (APL) | Alberta Precision Labs (APL) | Gordon P et al |
| EPI_ISL_806524 | Canada/AB-68978/2020 | 2020-04-15 | North America/Canada/Alberta/ | Alberta Precision Labs (APL) | Alberta Precision Labs (APL) | Gordon P et al |
| EPI_ISL_806525 | Canada/AB-17853/2020 | 2020-04-24 | North America/Canada/Alberta/ | Alberta Precision Labs (APL) | Alberta Precision Labs (APL) | Gordon P et al |
| EPI_ISL_806528 | Germany/BE-ChVir-3170-a-4370/202( | 2020-03-26 | Europe/Germany/Berlin/ | Charit√© Universit√§tsmedizin Berlin, Institut f√ºr Virologie/Labor Berlin | Charit√© Universit√§tsmedizin Berlin, Institut f√ºr Virologie | Victor M Corman et al |
| EPI_ISL_806531 | Germany/BE-ChVir-3177-a-4339/202( | 2020-04-02 | Europe/Germany/Berlin/ | Charit√© Universit√§tsmedizin Berlin, Institut f√ºr Virologie/Labor Berlin | Charit√© Universit√§tsmedizin Berlin, Institut f√ºr Virologie | Victor M Corman et al |
| EPI_ISL_806537 | Germany/BB-ChVir-3187-a-8159/202( | 2020-04-14 | Europe/Germany/Brandenburg/ | Charit√© Universit√§tsmedizin Berlin, Institut f√ºr Virologie/Labor Berlin | Charit√© Universit√§tsmedizin Berlin, Institut f√ºr Virologie | Victor M Corman et al |
| EPI_ISL_806540 | Germany/BE-ChVir-3192-a-4365/202( | 2020-04-02 | Europe/Germany/Berlin/ | Charit√© Universit√§tsmedizin Berlin, Institut f√ºr Virologie/Labor Berlin | Charit√© Universit√§tsmedizin Berlin, Institut f√ºr Virologie | Victor M Corman et al |

| | | | | | | |
|---|---|---|---|---|---|---|
| EPI_ISL_806544 | Ecuador/INSPI-179112/2021 | 2021-01-05 | South America/Ecuador/Los Rios/ | INSPI Instituto Nacional de Investigaci√≥n en Salud P√∫blica | Av. Juli√°n Coronel 905 entre Esmeraldas y Jos√© Mascote Av. Juan Tanca Marengo No. 100 y Av. de las Am√©ricas | Leandro Pati√±o et al |
| EPI_ISL_806639 | Kenya/C27523/2020 | 2020-07-02 | Africa/Kenya/Mombasa/ | KEMRI-Wellcome Trust Research Programme/KEMRI-CGMR-C Kilifi | KEMRI-Wellcome Trust Research Programme/KEMRI-CGMR-C Kilifi | Githinji et al et al |
| EPI_ISL_806640 | Kenya/C27611/2020 | 2020-07-02 | Africa/Kenya/Mombasa/ | KEMRI-Wellcome Trust Research Programme/KEMRI-CGMR-C Kilifi | KEMRI-Wellcome Trust Research Programme/KEMRI-CGMR-C Kilifi | Githinji et al et al |
| EPI_ISL_806698 | Kenya/C37981/2020 | 2020-07-27 | Africa/Kenya/Lamu/ | KEMRI-Wellcome Trust Research Programme/KEMRI-CGMR-C Kilifi | KEMRI-Wellcome Trust Research Programme/KEMRI-CGMR-C Kilifi | Githinji et al et al |
| EPI_ISL_806735 | Italy/ABR-IZSGC-351312/2020 | 2020-12-18 | Europe/Italy/Abruzzo/ | SIESP DIPARTIMENTO DI PREVENZIONE TERAMO | Istituto Zooprofilattico Sperimentale dell,ÄôAbruzzo e Molise ,ÄúG. Caporale,Äù | Lorusso A et al |
| EPI_ISL_806736 | Italy/ABR-IZSGC-351319/2020 | 2020-12-18 | Europe/Italy/Abruzzo/ | SIESP DIPARTIMENTO DI PREVENZIONE TERAMO | Istituto Zooprofilattico Sperimentale dell,ÄôAbruzzo e Molise ,ÄúG. Caporale,Äù | Lorusso A et al |
| EPI_ISL_806746 | Italy/ABR-IZSGC-353326/2020 | 2020-12-18 | Europe/Italy/Abruzzo/ | SIESP DIPARTIMENTO DI PREVENZIONE TERAMO | Istituto Zooprofilattico Sperimentale dell,ÄôAbruzzo e Molise ,ÄúG. Caporale,Äù | Lorusso A et al |

| | | | | | | |
|---|---|---|---|---|---|---|
| EPI_ISL_806753 | Italy/ABR-IZSGC-349627/2020 | 2020-12-17 | Europe/Italy/Abruzzo/ | SIESP DIPARTIMENTO DI PREVENZIONE SULMONA | Istituto Zooprofilattico Sperimentale dell‚ÄôAbruzzo e Molise ‚ÄúG. Caporale‚Äù | Lorusso A et al |
| EPI_ISL_806770 | Italy/ABR-IZSGC-351684/2020 | 2020-12-18 | Europe/Italy/Abruzzo/ | SIESP DIPARTIMENTO DI PREVENZIONE TERAMO | Istituto Zooprofilattico Sperimentale dell‚ÄôAbruzzo e Molise ‚ÄúG. Caporale‚Äù | Lorusso A et al |
| EPI_ISL_806777 | Italy/ABR-IZSGC-350940/2020 | 2020-12-18 | Europe/Italy/Abruzzo/ | SIESP DIPARTIMENTO DI PREVENZIONE TERAMO | Istituto Zooprofilattico Sperimentale dell‚ÄôAbruzzo e Molise ‚ÄúG. Caporale‚Äù | Lorusso A et al |
| EPI_ISL_806778 | Italy/ABR-IZSGC-350821/2020 | 2020-12-17 | Europe/Italy/Abruzzo/ | SIESP DIPARTIMENTO DI PREVENZIONE TERAMO | Istituto Zooprofilattico Sperimentale dell‚ÄôAbruzzo e Molise ‚ÄúG. Caporale‚Äù | Lorusso A et al |
| EPI_ISL_806779 | Italy/ABR-IZSGC-350829/2020 | 2020-12-17 | Europe/Italy/Abruzzo/ | SIESP DIPARTIMENTO DI PREVENZIONE TERAMO | Istituto Zooprofilattico Sperimentale dell‚ÄôAbruzzo e Molise ‚ÄúG. Caporale‚Äù | Lorusso A et al |
| EPI_ISL_806780 | Italy/ABR-IZSGC-350853/2020 | 2020-12-17 | Europe/Italy/Abruzzo/ | SIESP DIPARTIMENTO DI PREVENZIONE TERAMO | Istituto Zooprofilattico Sperimentale dell‚ÄôAbruzzo e Molise ‚ÄúG. Caporale‚Äù | Lorusso A et al |

| | | | | | | |
|---|---|---|---|---|---|---|
| EPI_ISL_806782 | Italy/ABR-IZSGC-351793/2020 | 2020-12-18 | Europe/Italy/Abruzzo/ | DIPARTIMENTO PREVENZIONE AVEZZANO-SERVIZIO DI IGIENE EPIDEMIOLOGIA E SANITA' PUBBLICA | Istituto Zooprofilattico Sperimentale dell,ÄôAbruzzo e Molise ,ÄúG. Caporale,Äù | Lorusso A et al |
| EPI_ISL_806786 | Italy/ABR-IZSGC-351855/2020 | 2020-12-18 | Europe/Italy/Abruzzo/ | SIESP DIPARTIMENTO DI PREVENZIONE TERAMO | Istituto Zooprofilattico Sperimentale dell,ÄôAbruzzo e Molise ,ÄúG. Caporale,Äù | Lorusso A et al |
| EPI_ISL_806796 | Italy/ABR-IZSGC-354019/2020 | 2020-12-18 | Europe/Italy/Abruzzo/ | SIESP CHIETI - DRIVE IN ORTONA | Istituto Zooprofilattico Sperimentale dell,ÄôAbruzzo e Molise ,ÄúG. Caporale,Äù | Lorusso A et al |
| EPI_ISL_806801 | Italy/ABR-IZSGC-353615/2020 | 2020-12-18 | Europe/Italy/Abruzzo/ | SIESP CHIETI - DRIVE IN GISSI | Istituto Zooprofilattico Sperimentale dell,ÄôAbruzzo e Molise ,ÄúG. Caporale,Äù | Lorusso A et al |
| EPI_ISL_806806 | Italy/ABR-IZSGC-353763/2020 | 2020-12-18 | Europe/Italy/Abruzzo/ | SIESP DIPARTIMENTO DI PREVENZIONE CHIETI | Istituto Zooprofilattico Sperimentale dell,ÄôAbruzzo e Molise ,ÄúG. Caporale,Äù | Lorusso A et al |
| EPI_ISL_806807 | USA/IA-GMF-46406/2021 | 2021-01-03 | North America/USA/Iowa/Fayette Coun | Gundersen Molecular Diagnostics Laboratory | Kabara Cancer Research Institute | Craig S. Richmond et al |
| EPI_ISL_806809 | USA/WA-S2922/2020 | 2020-04-07 | North America/USA/Washington/King C | Seattle Flu Study | Seattle Flu Study | Deborah A. Nickerson et al |
| EPI_ISL_806811 | USA/AK-PHL6630/2020 | 2020-11-25 | North America/USA/Alaska/Interior Ala | Alaska State Virology Laboratory | Alaska State Virology Laboratory | Stephanie DeRonde et al |
| EPI_ISL_806814 | USA/AK-PHL6634/2020 | 2020-11-12 | North America/USA/Alaska/Southwest | Alaska State Virology Laboratory | Alaska State Virology Laboratory | Stephanie DeRonde et al |

| | | | | | | |
|---|---|---|---|---|---|---|
| EPI_ISL_806820 | USA/AK-PHL6641/2020 | 2020-11-19 | North America/USA/Alaska/Southwest | Alaska State Virology Laboratory | Alaska State Virology Laboratory | Stephanie DeRonde et al |
| EPI_ISL_806844 | USA/AK-PHL6666/2020 | 2020-12-08 | North America/USA/Alaska/Anchorage | Alaska State Virology Laboratory | Alaska State Virology Laboratory | Stephanie DeRonde et al |
| EPI_ISL_806855 | USA/WA-S2925/2020 | 2020-08-25 | North America/USA/Washington/Yakim | Washington State Department of Health | Seattle Flu Study | Deborah A. Nickerson et al |
| EPI_ISL_806862 | USA/WA-S2932/2020 | 2020-08-28 | North America/USA/Washington/Grays | Washington State Department of Health | Seattle Flu Study | Deborah A. Nickerson et al |
| EPI_ISL_806863 | USA/WA-S2933/2020 | 2020-08-31 | North America/USA/Washington/King C | Washington State Department of Health | Seattle Flu Study | Deborah A. Nickerson et al |
| EPI_ISL_806867 | USA/WA-S2937/2020 | 2020-09-01 | North America/USA/Washington/Grays | Washington State Department of Health | Seattle Flu Study | Deborah A. Nickerson et al |
| EPI_ISL_806869 | USA/WA-S2939/2020 | 2020-09-01 | North America/USA/Washington/Grays | Washington State Department of Health | Seattle Flu Study | Deborah A. Nickerson et al |
| EPI_ISL_806873 | USA/WA-S2943/2020 | 2020-09-01 | North America/USA/Washington/Yakim | Washington State Department of Health | Seattle Flu Study | Deborah A. Nickerson et al |
| EPI_ISL_806874 | USA/WA-S2944/2020 | 2020-09-01 | North America/USA/Washington/Spoka | Washington State Department of Health | Seattle Flu Study | Deborah A. Nickerson et al |
| EPI_ISL_806877 | USA/WA-S2947/2020 | 2020-09-02 | North America/USA/Washington/Grays | Washington State Department of Health | Seattle Flu Study | Deborah A. Nickerson et al |
| EPI_ISL_806879 | USA/WA-S2949/2020 | 2020-09-02 | North America/USA/Washington/Yakim | Washington State Department of Health | Seattle Flu Study | Deborah A. Nickerson et al |
| EPI_ISL_806881 | USA/WA-S2951/2020 | 2020-09-02 | North America/USA/Washington/Pierce | Washington State Department of Health | Seattle Flu Study | Deborah A. Nickerson et al |
| EPI_ISL_806885 | USA/WA-S2955/2020 | 2020-09-03 | North America/USA/Washington/Walla | Washington State Department of Health | Seattle Flu Study | Deborah A. Nickerson et al |
| EPI_ISL_806886 | USA/WA-S2956/2020 | 2020-09-04 | North America/USA/Washington/King C | Washington State Department of Health | Seattle Flu Study | Deborah A. Nickerson et al |
| EPI_ISL_806893 | USA/WA-S2963/2020 | 2020-09-08 | North America/USA/Washington/Yakim | Washington State Department of Health | Seattle Flu Study | Deborah A. Nickerson et al |
| EPI_ISL_806894 | USA/WA-S2964/2020 | 2020-09-09 | North America/USA/Washington/Mason | Washington State Department of Health | Seattle Flu Study | Deborah A. Nickerson et al |
| EPI_ISL_806899 | USA/WA-S2969/2020 | 2020-09-11 | North America/USA/Washington/Grays | Washington State Department of Health | Seattle Flu Study | Deborah A. Nickerson et al |
| EPI_ISL_806903 | USA/WA-S2973/2020 | 2020-09-10 | North America/USA/Washington/Grays | Washington State Department of Health | Seattle Flu Study | Deborah A. Nickerson et al |
| EPI_ISL_806904 | USA/WA-S2974/2020 | 2020-09-10 | North America/USA/Washington/Grays | Washington State Department of Health | Seattle Flu Study | Deborah A. Nickerson et al |
| EPI_ISL_806907 | USA/WA-S2977/2020 | 2020-09-10 | North America/USA/Washington/Grays | Washington State Department of Health | Seattle Flu Study | Deborah A. Nickerson et al |
| EPI_ISL_806908 | USA/WA-S2978/2020 | 2020-09-10 | North America/USA/Washington/Yakim | Washington State Department of Health | Seattle Flu Study | Deborah A. Nickerson et al |
| EPI_ISL_806911 | USA/WA-S2981/2020 | 2020-09-14 | North America/USA/Washington/Yakim | Washington State Department of Health | Seattle Flu Study | Deborah A. Nickerson et al |
| EPI_ISL_806912 | USA/WA-S2982/2020 | 2020-09-14 | North America/USA/Washington/Yakim | Washington State Department of Health | Seattle Flu Study | Deborah A. Nickerson et al |
| EPI_ISL_806913 | USA/WA-S2983/2020 | 2020-09-13 | North America/USA/Washington/Yakim | Washington State Department of Health | Seattle Flu Study | Deborah A. Nickerson et al |
| EPI_ISL_806914 | USA/WA-S2984/2020 | 2020-09-14 | North America/USA/Washington/Grays | Washington State Department of Health | Seattle Flu Study | Deborah A. Nickerson et al |
| EPI_ISL_806915 | USA/WA-S2985/2020 | 2020-09-14 | North America/USA/Washington/Grays | Washington State Department of Health | Seattle Flu Study | Deborah A. Nickerson et al |
| EPI_ISL_806916 | USA/WA-S2986/2020 | 2020-09-14 | North America/USA/Washington/Grays | Washington State Department of Health | Seattle Flu Study | Deborah A. Nickerson et al |
| EPI_ISL_806917 | USA/WA-S2987/2020 | 2020-08-12 | North America/USA/Washington/Mason | Washington State Department of Health | Seattle Flu Study | Deborah A. Nickerson et al |

| | | | | | | |
|---|---|---|---|---|---|---|
| EPI_ISL_806921 | USA/WA-S2991/2020 | 2020-08-12 | North America/USA/Washington/Yakim | Washington State Department of Health | Seattle Flu Study | Deborah A. Nickerson et al |
| EPI_ISL_806923 | USA/WA-S2993/2020 | 2020-08-11 | North America/USA/Washington/Yakim | Washington State Department of Health | Seattle Flu Study | Deborah A. Nickerson et al |
| EPI_ISL_806924 | USA/WA-S2994/2020 | 2020-08-11 | North America/USA/Washington/Yakim | Washington State Department of Health | Seattle Flu Study | Deborah A. Nickerson et al |
| EPI_ISL_806929 | USA/WA-S2999/2020 | 2020-08-12 | North America/USA/Washington/Spoka | Washington State Department of Health | Seattle Flu Study | Deborah A. Nickerson et al |
| EPI_ISL_806936 | USA/WA-S3006/2020 | 2020-08-13 | North America/USA/Washington/Yakim | Washington State Department of Health | Seattle Flu Study | Deborah A. Nickerson et al |
| EPI_ISL_806939 | USA/WA-S3009/2020 | 2020-08-14 | North America/USA/Washington/Grant | Washington State Department of Health | Seattle Flu Study | Deborah A. Nickerson et al |
| EPI_ISL_806951 | USA/WA-S3021/2020 | 2020-08-17 | North America/USA/Washington/King C | Washington State Department of Health | Seattle Flu Study | Deborah A. Nickerson et al |
| EPI_ISL_806954 | USA/WA-S3024/2020 | 2020-08-18 | North America/USA/Washington/Spoka | Washington State Department of Health | Seattle Flu Study | Deborah A. Nickerson et al |
| EPI_ISL_806956 | USA/WA-S3026/2020 | 2020-08-18 | North America/USA/Washington/Thurst | Washington State Department of Health | Seattle Flu Study | Deborah A. Nickerson et al |
| EPI_ISL_806958 | USA/WA-S3028/2020 | 2020-08-19 | North America/USA/Washington/Yakim | Washington State Department of Health | Seattle Flu Study | Deborah A. Nickerson et al |
| EPI_ISL_806959 | USA/WA-S3029/2020 | 2020-08-18 | North America/USA/Washington/King C | Washington State Department of Health | Seattle Flu Study | Deborah A. Nickerson et al |
| EPI_ISL_806966 | USA/WA-S3036/2020 | 2020-08-20 | North America/USA/Washington/Grays | Washington State Department of Health | Seattle Flu Study | Deborah A. Nickerson et al |
| EPI_ISL_806982 | USA/WA-S3052/2020 | 2020-08-21 | North America/USA/Washington/Grays | Washington State Department of Health | Seattle Flu Study | Deborah A. Nickerson et al |
| EPI_ISL_806984 | USA/WA-S3054/2020 | 2020-08-21 | North America/USA/Washington/Spoka | Washington State Department of Health | Seattle Flu Study | Deborah A. Nickerson et al |
| EPI_ISL_806986 | USA/WA-S3056/2020 | 2020-08-21 | North America/USA/Washington/Grays | Washington State Department of Health | Seattle Flu Study | Deborah A. Nickerson et al |
| EPI_ISL_806991 | USA/WA-S3061/2020 | 2020-11-29 | North America/USA/Washington/Walla | Washington State Department of Health | Seattle Flu Study | Deborah A. Nickerson et al |
| EPI_ISL_806993 | USA/WA-S3063/2020 | 2020-11-09 | North America/USA/Washington/Walla | Washington State Department of Health | Seattle Flu Study | Deborah A. Nickerson et al |
| EPI_ISL_806994 | USA/WA-S3064/2020 | 2020-11-09 | North America/USA/Washington/Walla | Washington State Department of Health | Seattle Flu Study | Deborah A. Nickerson et al |
| EPI_ISL_806997 | USA/WA-S3067/2020 | 2020-11-11 | North America/USA/Washington/Yakim | Washington State Department of Health | Seattle Flu Study | Deborah A. Nickerson et al |
| EPI_ISL_806999 | USA/WA-S3069/2020 | 2020-11-11 | North America/USA/Washington/Yakim | Washington State Department of Health | Seattle Flu Study | Deborah A. Nickerson et al |
| EPI_ISL_807001 | USA/WA-S3071/2020 | 2020-11-10 | North America/USA/Washington/Spoka | Washington State Department of Health | Seattle Flu Study | Deborah A. Nickerson et al |
| EPI_ISL_807006 | USA/WA-S3076/2020 | 2020-12-01 | North America/USA/Washington/King C | Washington State Department of Health | Seattle Flu Study | Deborah A. Nickerson et al |
| EPI_ISL_807007 | USA/WA-S3077/2020 | 2020-11-09 | North America/USA/Washington/Walla | Washington State Department of Health | Seattle Flu Study | Deborah A. Nickerson et al |
| EPI_ISL_807008 | USA/WA-S3078/2020 | 2020-11-15 | North America/USA/Washington/Yakim | Washington State Department of Health | Seattle Flu Study | Deborah A. Nickerson et al |
| EPI_ISL_807012 | USA/WA-S3082/2020 | 2020-11-13 | North America/USA/Washington/Yakim | Washington State Department of Health | Seattle Flu Study | Deborah A. Nickerson et al |
| EPI_ISL_807023 | USA/WA-S3093/2020 | 2020-11-12 | North America/USA/Washington/Yakim | Washington State Department of Health | Seattle Flu Study | Deborah A. Nickerson et al |
| EPI_ISL_807029 | USA/WA-S3099/2020 | 2020-11-12 | North America/USA/Washington/Clallar | Washington State Department of Health | Seattle Flu Study | Deborah A. Nickerson et al |
| EPI_ISL_807033 | USA/WA-S3103/2020 | 2020-11-12 | North America/USA/Washington/Yakim | Washington State Department of Health | Seattle Flu Study | Deborah A. Nickerson et al |
| EPI_ISL_807041 | USA/WA-S3111/2020 | 2020-11-13 | North America/USA/Washington/Yakim | Washington State Department of Health | Seattle Flu Study | Deborah A. Nickerson et al |
| EPI_ISL_807042 | USA/WA-S3112/2020 | 2020-11-12 | North America/USA/Washington/Walla | Washington State Department of Health | Seattle Flu Study | Deborah A. Nickerson et al |
| EPI_ISL_807043 | USA/WA-S3113/2020 | 2020-11-12 | North America/USA/Washington/Walla | Washington State Department of Health | Seattle Flu Study | Deborah A. Nickerson et al |

| | | | | | | |
|---|---|---|---|---|---|---|
| EPI_ISL_807056 | USA/WA-S3126/2020 | 2020-11-16 | North America/USA/Washington/Yakim | Washington State Department of Health | Seattle Flu Study | Deborah A. Nickerson et al |
| EPI_ISL_807059 | USA/WA-S3129/2020 | 2020-11-16 | North America/USA/Washington/Spoka | Washington State Department of Health | Seattle Flu Study | Deborah A. Nickerson et al |
| EPI_ISL_807065 | USA/WA-S3135/2020 | 2020-11-17 | North America/USA/Washington/Yakim | Washington State Department of Health | Seattle Flu Study | Deborah A. Nickerson et al |
| EPI_ISL_807080 | USA/WA-S3150/2020 | 2020-11-18 | North America/USA/Washington/Chelar | Washington State Department of Health | Seattle Flu Study | Deborah A. Nickerson et al |
| EPI_ISL_807082 | USA/WA-S3152/2020 | 2020-11-18 | North America/USA/Washington/King C | Washington State Department of Health | Seattle Flu Study | Deborah A. Nickerson et al |
| EPI_ISL_807095 | USA/WA-S3165/2020 | 2020-11-19 | North America/USA/Washington/Pacific | Washington State Department of Health | Seattle Flu Study | Deborah A. Nickerson et al |
| EPI_ISL_807120 | USA/WA-S3190/2020 | 2020-11-20 | North America/USA/Washington/Yakim | Washington State Department of Health | Seattle Flu Study | Deborah A. Nickerson et al |
| EPI_ISL_807125 | USA/WA-S3195/2020 | 2020-11-20 | North America/USA/Washington/Yakim | Washington State Department of Health | Seattle Flu Study | Deborah A. Nickerson et al |
| EPI_ISL_807133 | USA/WA-S3203/2020 | 2020-11-23 | North America/USA/Washington/Pacific | Washington State Department of Health | Seattle Flu Study | Deborah A. Nickerson et al |
| EPI_ISL_807143 | USA/WA-S3213/2020 | 2020-11-23 | North America/USA/Washington/Pacific | Washington State Department of Health | Seattle Flu Study | Deborah A. Nickerson et al |
| EPI_ISL_807154 | Romania/54Hunedoara/2020 | 2020-12-24 | Europe/Romania/Hunedoara/ | Deva County Emergency Hospital | National Institute of Infectious Diseases-Prof. Dr. Matei Bals Molecular Diagnostics Laboratory | Leontina Banica et al |
| EPI_ISL_807155 | Romania/103Hunedoara/2020 | 2020-12-23 | Europe/Romania/Hunedoara/ | Deva County Emergency Hospital | National Institute of Infectious Diseases-Prof. Dr. Matei Bals Molecular Diagnostics Laboratory | Leontina Banica et al |
| EPI_ISL_807156 | Romania/191Hunedoara/2020 | 2020-12-28 | Europe/Romania/Hunedoara/ | Deva County Emergency Hospital | National Institute of Infectious Diseases-Prof. Dr. Matei Bals Molecular Diagnostics Laboratory | Leontina Banica et al |
| EPI_ISL_811021 | Gambia/S-0315D/2020 | 2020-08-17 | Africa/Gambia/Kombo/ | MRCG at LSHTM Genomics lab | MRCG at LSHTM Genomics lab | Abdul Karim sesay et al |
| EPI_ISL_811026 | Gambia/S-331/2020 | 2020-08-20 | Africa/Gambia/Kombo/ | MRCG at LSHTM Genomics lab | MRCG at LSHTM Genomics lab | Abdul Karim sesay et al |
| EPI_ISL_811028 | Gambia/S-339/2020 | 2020-08-25 | Africa/Gambia/Kombo/ | MRCG at LSHTM Genomics lab | MRCG at LSHTM Genomics lab | Abdul Karim sesay et al |

| | | | | | | |
|---|---|---|---|---|---|---|
| EPI_ISL_811029 | Gambia/S-341/2020 | 2020-08-25 | Africa/Gambia/Kombo/ | MRCG at LSHTM Genomics lab | MRCG at LSHTM Genomics lab | Abdul Karim sesay et al |
| EPI_ISL_811066 | England/205380144/2020 | 2020-12-30 | Europe/United Kingdom/England/ | Respiratory Virus Unit, National Infection Service, Public Health England | COVID-19 Genomics UK (COG-UK) Consortium | PHE Covid Sequencing Team et al |
| EPI_ISL_811136 | Turkey/HSGM-1143/2020 | 2020-12-24 | Europe/Turkey/Bursa/ | Ministry of Health Turkey | Ministry of Health Turkey | Fatma Bayrakdar et al |
| EPI_ISL_811139 | Turkey/HSGM-1202/2020 | 2020-12-31 | Europe/Turkey/Bursa/ | Ministry of Health Turkey | Ministry of Health Turkey | Fatma Bayrakdar et al |
| EPI_ISL_811143 | Turkey/HSGM-1192/2020 | 2020-12-30 | Europe/Turkey/Kahramanmaras/ | Ministry of Health Turkey | Ministry of Health Turkey | Fatma Bayrakdar et al |
| EPI_ISL_811149 | Brazil/AM-20143138FN-R2/2020 | 2020-12-30 | South America/Brazil/Amazonas BR/Ma | Laboratorio de Ecologia de Doencas Transmissiveis na Amazonia, Instituto Leonidas e Maria Deane - Fiocruz Amazonia | Laboratorio de Ecologia de Doencas Transmissiveis na Amazonia, Instituto Leonidas e Maria Deane - Fiocruz Amazonia | Valdinete Nascimento et al |
| EPI_ISL_811151 | India/KA-CS1020/2020 | 2020-07-10 | Asia/India/Karnataka/ | Dharwad | CSIR Institute of Genomics and Integrative Biology | Dr. Shivarudrapp B Bhairappanavar et al |
| EPI_ISL_811155 | India/KA-CS1026/2020 | 2020-07-10 | Asia/India/Karnataka/ | Dharwad | CSIR Institute of Genomics and Integrative Biology | Dr. Shivarudrapp B Bhairappanavar et al |
| EPI_ISL_811157 | India/KA-CS1024/2020 | 2020-07-21 | Asia/India/Karnataka/ | Dharwad | CSIR Institute of Genomics and Integrative Biology | Dr. Shivarudrapp B Bhairappanavar et al |
| EPI_ISL_811159 | India/KA-CS1054/2020 | 2020-07-19 | Asia/India/Karnataka/ | Dharwad | CSIR Institute of Genomics and Integrative Biology | Dr. Shivarudrapp B Bhairappanavar et al |
| EPI_ISL_811161 | India/KA-CS1025/2020 | 2020-07-02 | Asia/India/Karnataka/ | Dharwad | CSIR Institute of Genomics and Integrative Biology | Dr. Shivarudrapp B Bhairappanavar et al |

| | | | | | | |
|---|---|---|---|---|---|---|
| EPI_ISL_811162 | India/KA-CS1027/2020 | 2020-07-10 | Asia/India/Karnataka/ | Dharwad | CSIR Institute of Genomics and Integrative Biology | Dr. Shivarudrapp B Bhairappanavar et al |
| EPI_ISL_811163 | India/KA-CS1029/2020 | 2020-07-20 | Asia/India/Karnataka/ | Dharwad | CSIR Institute of Genomics and Integrative Biology | Dr. Shivarudrapp B Bhairappanavar et al |
| EPI_ISL_811165 | India/KA-CS1032/2020 | 2020-07-03 | Asia/India/Karnataka/ | Dharwad | CSIR Institute of Genomics and Integrative Biology | Dr. Shivarudrapp B Bhairappanavar et al |
| EPI_ISL_811166 | India/KA-CS1033/2020 | 2020-07-18 | Asia/India/Karnataka/ | Dharwad | CSIR Institute of Genomics and Integrative Biology | Dr. Shivarudrapp B Bhairappanavar et al |
| EPI_ISL_811167 | India/KA-CS1034/2020 | 2020-07-21 | Asia/India/Karnataka/ | Dharwad | CSIR Institute of Genomics and Integrative Biology | Dr. Shivarudrapp B Bhairappanavar et al |
| EPI_ISL_811168 | India/KA-CS1016/2020 | 2020-06-13 | Asia/India/Karnataka/ | Dharwad | CSIR Institute of Genomics and Integrative Biology | Dr. Shivarudrapp B Bhairappanavar et al |
| EPI_ISL_811169 | India/KA-CS1035/2020 | 2020-07-06 | Asia/India/Karnataka/ | Dharwad | CSIR Institute of Genomics and Integrative Biology | Dr. Shivarudrapp B Bhairappanavar et al |
| EPI_ISL_811170 | India/KA-CS1037/2020 | 2020-07-25 | Asia/India/Karnataka/ | Dharwad | CSIR Institute of Genomics and Integrative Biology | Dr. Shivarudrapp B Bhairappanavar et al |
| EPI_ISL_811172 | India/KA-CS1039/2020 | 2020-07-16 | Asia/India/Karnataka/ | Dharwad | CSIR Institute of Genomics and Integrative Biology | Dr. Shivarudrapp B Bhairappanavar et al |
| EPI_ISL_811173 | India/KA-CS1040/2020 | 2020-07-21 | Asia/India/Karnataka/ | Dharwad | CSIR Institute of Genomics and Integrative Biology | Dr. Shivarudrapp B Bhairappanavar et al |

| | | | | | | |
|---|---|---|---|---|---|---|
| EPI_ISL_811174 | India/KA-CS1041/2020 | 2020-07-22 | Asia/India/Karnataka/ | Dharwad | CSIR Institute of Genomics and Integrative Biology | Dr. Shivarudrapp B Bhairappanavar et al |
| EPI_ISL_811175 | India/KA-CS1042/2020 | 2020-07-25 | Asia/India/Karnataka/ | Dharwad | CSIR Institute of Genomics and Integrative Biology | Dr. Shivarudrapp B Bhairappanavar et al |
| EPI_ISL_811176 | India/KA-CS1043/2020 | 2020-07-20 | Asia/India/Karnataka/ | Dharwad | CSIR Institute of Genomics and Integrative Biology | Dr. Shivarudrapp B Bhairappanavar et al |
| EPI_ISL_811179 | India/KA-CS1048/2020 | 2020-07-18 | Asia/India/Karnataka/ | Dharwad | CSIR Institute of Genomics and Integrative Biology | Dr. Shivarudrapp B Bhairappanavar et al |
| EPI_ISL_811180 | India/KA-CS1049/2020 | 2020-07-08 | Asia/India/Karnataka/ | Dharwad | CSIR Institute of Genomics and Integrative Biology | Dr. Shivarudrapp B Bhairappanavar et al |
| EPI_ISL_811181 | India/KA-CS1050/2020 | 2020-07-21 | Asia/India/Karnataka/ | Dharwad | CSIR Institute of Genomics and Integrative Biology | Dr. Shivarudrapp B Bhairappanavar et al |
| EPI_ISL_811182 | India/KA-CS1051/2020 | 2020-06-17 | Asia/India/Karnataka/ | Dharwad | CSIR Institute of Genomics and Integrative Biology | Dr. Shivarudrapp B Bhairappanavar et al |
| EPI_ISL_811183 | India/KA-CS1053/2020 | 2020-07-22 | Asia/India/Karnataka/ | Dharwad | CSIR Institute of Genomics and Integrative Biology | Dr. Shivarudrapp B Bhairappanavar et al |
| EPI_ISL_811184 | India/KA-CS1057/2020 | 2020-07-21 | Asia/India/Karnataka/ | Dharwad | CSIR Institute of Genomics and Integrative Biology | Dr. Shivarudrapp B Bhairappanavar et al |
| EPI_ISL_811185 | India/KA-CS1058/2020 | 2020-06-25 | Asia/India/Karnataka/ | Dharwad | CSIR Institute of Genomics and Integrative Biology | Dr. Shivarudrapp B Bhairappanavar et al |

| | | | | | | |
|---|---|---|---|---|---|---|
| EPI_ISL_811186 | India/KA-CS1060/2020 | 2020-07-22 | Asia/India/Karnataka/ | Dharwad | CSIR Institute of Genomics and Integrative Biology | Dr. Shivarudrapp B Bhairappanavar et al |
| EPI_ISL_811187 | India/KA-CS1062/2020 | 2020-07-20 | Asia/India/Karnataka/ | Dharwad | CSIR Institute of Genomics and Integrative Biology | Dr. Shivarudrapp B Bhairappanavar et al |
| EPI_ISL_811188 | India/KA-CS1063/2020 | 2020-07-15 | Asia/India/Karnataka/ | Dharwad | CSIR Institute of Genomics and Integrative Biology | Dr. Shivarudrapp B Bhairappanavar et al |
| EPI_ISL_811189 | India/KA-CS1064/2020 | 2020-07-01 | Asia/India/Karnataka/ | Dharwad | CSIR Institute of Genomics and Integrative Biology | Dr. Shivarudrapp B Bhairappanavar et al |
| EPI_ISL_811190 | India/KA-CS1019/2020 | 2020-07-08 | Asia/India/Karnataka/ | Dharwad | CSIR Institute of Genomics and Integrative Biology | Dr. Shivarudrapp B Bhairappanavar et al |
| EPI_ISL_811191 | India/KA-CS1021/2020 | 2020-07-25 | Asia/India/Karnataka/ | Dharwad | CSIR Institute of Genomics and Integrative Biology | Dr. Shivarudrapp B Bhairappanavar et al |
| EPI_ISL_811192 | India/KA-CS1022/2020 | 2020-07-24 | Asia/India/Karnataka/ | Dharwad | CSIR Institute of Genomics and Integrative Biology | Dr. Shivarudrapp B Bhairappanavar et al |
| EPI_ISL_811193 | India/KA-CS1023/2020 | 2020-07-25 | Asia/India/Karnataka/ | Dharwad | CSIR Institute of Genomics and Integrative Biology | Dr. Shivarudrapp B Bhairappanavar et al |
| EPI_ISL_811466 | England/CAMC-E511A5/2021 | 2021-01-02 | Europe/United Kingdom/England/ | Lighthouse Lab in Cambridge | Wellcome Sanger Institute for the COVID-19 Genomics UK (COG-UK) Consortium | Rob Howes et al |

| | | | | | | |
|---|---|---|---|---|---|---|
| EPI_ISL_811539 | England/CAMC-E52405/2021 | 2021-01-02 | Europe/United Kingdom/England/ | Lighthouse Lab in Cambridge | Wellcome Sanger Institute for the COVID-19 Genomics UK (COG-UK) Consortium | Rob Howes et al |
| EPI_ISL_811918 | England/MILK-E0B85A/2020 | 2020-12-29 | Europe/United Kingdom/England/ | Lighthouse Lab in Milton Keynes | Wellcome Sanger Institute for the COVID-19 Genomics UK (COG-UK) Consortium | The Lighthouse Lab in Milton Keynes et al |
| EPI_ISL_811991 | England/MILK-E02844/2020 | 2020-12-29 | Europe/United Kingdom/England/ | Lighthouse Lab in Milton Keynes | Wellcome Sanger Institute for the COVID-19 Genomics UK (COG-UK) Consortium | The Lighthouse Lab in Milton Keynes et al |
| EPI_ISL_812115 | USA/CA-SC20007/2020 | 2020-08-26 | North America/USA/California/ | Santa Clara County Public Health Laboratory | Santa Clara County Public Health Laboratory | Santa Clara County Public Health Department et al |
| EPI_ISL_812136 | USA/GA-CDC-9IV4-7901/2020 | 2020-10-29 | North America/USA/Georgia/ | GA Department of Public Health Laboratory | Pathogen Discovery, Respiratory Viruses Branch, Division of Viral Diseases, Centers for Disease Control and Prevention | Yan Li et al |
| EPI_ISL_812146 | USA/GA-CDC-95B7-4388/2020 | 2020-11-05 | North America/USA/Georgia/ | GA Department of Public Health Laboratory | Pathogen Discovery, Respiratory Viruses Branch, Division of Viral Diseases, Centers for Disease Control and Prevention | Yan Li et al |

| | | | | | | |
|---|---|---|---|---|---|---|
| EPI_ISL_812153 | USA/GA-CDC-99R7-5540/2020 | 2020-11-17 | North America/USA/Georgia/ | GA Department of Public Health Laboratory | Pathogen Discovery, Respiratory Viruses Branch, Division of Viral Diseases, Centers for Disease Control and Prevention | Yan Li et al |
| EPI_ISL_812159 | USA/GA-CDC-9GZW-7417/2020 | 2020-07-20 | North America/USA/Georgia/ | GA Department of Public Health Laboratory | Pathogen Discovery, Respiratory Viruses Branch, Division of Viral Diseases, Centers for Disease Control and Prevention | Yan Li et al |
| EPI_ISL_812162 | USA/GA-CDC-9H2E-7435/2020 | 2020-07-29 | North America/USA/Georgia/ | GA Department of Public Health Laboratory | Pathogen Discovery, Respiratory Viruses Branch, Division of Viral Diseases, Centers for Disease Control and Prevention | Yan Li et al |
| EPI_ISL_812165 | USA/GA-CDC-9HKQ-7567/2020 | 2020-10-23 | North America/USA/Georgia/ | GA Department of Public Health Laboratory | Pathogen Discovery, Respiratory Viruses Branch, Division of Viral Diseases, Centers for Disease Control and Prevention | Yan Li et al |

| | | | | | | |
|---|---|---|---|---|---|---|
| EPI_ISL_812169 | USA/GA-CDC-9HS4-7620/2020 | 2020-10-26 | North America/USA/Georgia/ | GA Department of Public Health Laboratory | Pathogen Discovery, Respiratory Viruses Branch, Division of Viral Diseases, Centers for Disease Control and Prevention | Yan Li et al |
| EPI_ISL_812171 | USA/GA-CDC-9I5P-7718/2020 | 2020-10-27 | North America/USA/Georgia/ | GA Department of Public Health Laboratory | Pathogen Discovery, Respiratory Viruses Branch, Division of Viral Diseases, Centers for Disease Control and Prevention | Yan Li et al |
| EPI_ISL_812175 | USA/GA-CDC-9IP1-7857/2020 | 2020-10-28 | North America/USA/Georgia/ | GA Department of Public Health Laboratory | Pathogen Discovery, Respiratory Viruses Branch, Division of Viral Diseases, Centers for Disease Control and Prevention | Yan Li et al |
| EPI_ISL_812176 | USA/GA-CDC-9IVK-7904/2020 | 2020-10-29 | North America/USA/Georgia/ | GA Department of Public Health Laboratory | Pathogen Discovery, Respiratory Viruses Branch, Division of Viral Diseases, Centers for Disease Control and Prevention | Yan Li et al |

| | | | | | | |
|---|---|---|---|---|---|---|
| EPI_ISL_812183 | USA/GA-CDC-9J5T-7978/2020 | 2020-11-02 | North America/USA/Georgia/ | GA Department of Public Health Laboratory | Pathogen Discovery, Respiratory Viruses Branch, Division of Viral Diseases, Centers for Disease Control and Prevention | Yan Li et al |
| EPI_ISL_812193 | USA/GA-CDC-9JK9-8082/2020 | 2020-11-02 | North America/USA/Georgia/ | GA Department of Public Health Laboratory | Pathogen Discovery, Respiratory Viruses Branch, Division of Viral Diseases, Centers for Disease Control and Prevention | Yan Li et al |
| EPI_ISL_812194 | USA/GA-CDC-9JKE-8083/2020 | 2020-11-02 | North America/USA/Georgia/ | GA Department of Public Health Laboratory | Pathogen Discovery, Respiratory Viruses Branch, Division of Viral Diseases, Centers for Disease Control and Prevention | Yan Li et al |
| EPI_ISL_812197 | USA/GA-CDC-W11A-5057/2020 | 2020-11-19 | North America/USA/Georgia/ | GA Department of Public Health Laboratory | Pathogen Discovery, Respiratory Viruses Branch, Division of Viral Diseases, Centers for Disease Control and Prevention | Yan Li et al |

| | | | | | | |
|---|---|---|---|---|---|---|
| EPI_ISL_812203 | USA/RI-CDC-KCPZ-3781/2020 | 2020-03-09 | North America/USA/Rhode Island/ | RI State Health Laboratories | Pathogen Discovery, Respiratory Viruses Branch, Division of Viral Diseases, Centers for Disease Control and Prevention | Yan Li et al |
| EPI_ISL_812209 | USA/TN-CDC-MN06-6911/2020 | 2020-11-17 | North America/USA/Tennessee/ | TN Division of Laboratory Services | Pathogen Discovery, Respiratory Viruses Branch, Division of Viral Diseases, Centers for Disease Control and Prevention | Yan Li et al |
| EPI_ISL_812212 | USA/TN-CDC-MN0O-6914/2020 | 2020-11-16 | North America/USA/Tennessee/ | TN Division of Laboratory Services | Pathogen Discovery, Respiratory Viruses Branch, Division of Viral Diseases, Centers for Disease Control and Prevention | Yan Li et al |
| EPI_ISL_812217 | USA/TN-CDC-MN1I-6919/2020 | 2020-11-16 | North America/USA/Tennessee/ | TN Division of Laboratory Services | Pathogen Discovery, Respiratory Viruses Branch, Division of Viral Diseases, Centers for Disease Control and Prevention | Yan Li et al |

| | | | | | | |
|---|---|---|---|---|---|---|
| EPI_ISL_812224 | USA/TN-CDC-MN30-6928/2020 | 2020-11-16 | North America/USA/Tennessee/ | TN Division of Laboratory Services | Pathogen Discovery, Respiratory Viruses Branch, Division of Viral Diseases, Centers for Disease Control and Prevention | Yan Li et al |
| EPI_ISL_812227 | USA/TN-CDC-MN3I-6931/2020 | 2020-11-16 | North America/USA/Tennessee/ | TN Division of Laboratory Services | Pathogen Discovery, Respiratory Viruses Branch, Division of Viral Diseases, Centers for Disease Control and Prevention | Yan Li et al |
| EPI_ISL_812228 | USA/TN-CDC-MN3O-6932/2020 | 2020-11-16 | North America/USA/Tennessee/ | TN Division of Laboratory Services | Pathogen Discovery, Respiratory Viruses Branch, Division of Viral Diseases, Centers for Disease Control and Prevention | Yan Li et al |
| EPI_ISL_812235 | USA/ND-NDDH-0003/2020 | 2020-06-04 | North America/USA/North Dakota/ | North Dakota Department of Health, Public Health Laboratory | North Dakota Department of Health, Public Health Laboratory | Lisa Wingerter et al |
| EPI_ISL_812240 | USA/ND-NDDH-0008/2020 | 2020-03-17 | North America/USA/North Dakota/ | North Dakota Department of Health, Public Health Laboratory | North Dakota Department of Health, Public Health Laboratory | Lisa Wingerter et al |
| EPI_ISL_812258 | Iraq/USAFSAM-S061/2020 | 2020-06-06 | Asia/Iraq/Iraq/ | Landstuhl Regional Medical Center | United States Air Force School of Aerospace Medicine | Anthony Fries et al |

| | | | | | | |
|---|---|---|---|---|---|---|
| EPI_ISL_812259 | Iraq/USAFSAM-S062/2020 | 2020-06-16 | Asia/Iraq/Iraq/ | Landstuhl Regional Medical Center | United States Air Force School of Aerospace Medicine | Anthony Fries et al |
| EPI_ISL_812260 | Iraq/USAFSAM-S063/2020 | 2020-06-11 | Asia/Iraq/Iraq/ | Landstuhl Regional Medical Center | United States Air Force School of Aerospace Medicine | Anthony Fries et al |
| EPI_ISL_812261 | Iraq/USAFSAM-S064/2020 | 2020-06-16 | Asia/Iraq/Iraq/ | Landstuhl Regional Medical Center | United States Air Force School of Aerospace Medicine | Anthony Fries et al |
| EPI_ISL_812262 | Iraq/USAFSAM-S065/2020 | 2020-06-07 | Asia/Iraq/Iraq/ | Landstuhl Regional Medical Center | United States Air Force School of Aerospace Medicine | Anthony Fries et al |
| EPI_ISL_812263 | Iraq/USAFSAM-S068/2020 | 2020-06-16 | Asia/Iraq/Iraq/ | Landstuhl Regional Medical Center | United States Air Force School of Aerospace Medicine | Anthony Fries et al |
| EPI_ISL_812264 | Iraq/USAFSAM-S069/2020 | 2020-06-16 | Asia/Iraq/Iraq/ | Landstuhl Regional Medical Center | United States Air Force School of Aerospace Medicine | Anthony Fries et al |
| EPI_ISL_812265 | Iraq/USAFSAM-S070/2020 | 2020-06-16 | Asia/Iraq/Iraq/ | Landstuhl Regional Medical Center | United States Air Force School of Aerospace Medicine | Anthony Fries et al |
| EPI_ISL_812266 | Iraq/USAFSAM-S071/2020 | 2020-06-16 | Asia/Iraq/Iraq/ | Landstuhl Regional Medical Center | United States Air Force School of Aerospace Medicine | Anthony Fries et al |
| EPI_ISL_812267 | Iraq/USAFSAM-S072/2020 | 2020-06-07 | Asia/Iraq/Iraq/ | Landstuhl Regional Medical Center | United States Air Force School of Aerospace Medicine | Anthony Fries et al |
| EPI_ISL_812268 | Iraq/USAFSAM-S073/2020 | 2020-06-16 | Asia/Iraq/Iraq/ | Landstuhl Regional Medical Center | United States Air Force School of Aerospace Medicine | Anthony Fries et al |

| | | | | | | |
|---|---|---|---|---|---|---|
| EPI_ISL_812269 | Iraq/USAFSAM-S074/2020 | 2020-06-16 | Asia/Iraq/Iraq/ | Landstuhl Regional Medical Center | United States Air Force School of Aerospace Medicine | Anthony Fries et al |
| EPI_ISL_812270 | Iraq/USAFSAM-S075/2020 | 2020-06-07 | Asia/Iraq/Iraq/ | Landstuhl Regional Medical Center | United States Air Force School of Aerospace Medicine | Anthony Fries et al |
| EPI_ISL_812271 | Iraq/USAFSAM-S076/2020 | 2020-06-16 | Asia/Iraq/Iraq/ | Landstuhl Regional Medical Center | United States Air Force School of Aerospace Medicine | Anthony Fries et al |
| EPI_ISL_812272 | Iraq/USAFSAM-S077/2020 | 2020-06-07 | Asia/Iraq/Iraq/ | Landstuhl Regional Medical Center | United States Air Force School of Aerospace Medicine | Anthony Fries et al |
| EPI_ISL_812273 | Iraq/USAFSAM-S078/2020 | 2020-06-16 | Asia/Iraq/Iraq/ | Landstuhl Regional Medical Center | United States Air Force School of Aerospace Medicine | Anthony Fries et al |
| EPI_ISL_812274 | Iraq/USAFSAM-S079/2020 | 2020-06-07 | Asia/Iraq/Iraq/ | Landstuhl Regional Medical Center | United States Air Force School of Aerospace Medicine | Anthony Fries et al |
| EPI_ISL_812275 | Iraq/USAFSAM-S080/2020 | 2020-06-07 | Asia/Iraq/Iraq/ | Landstuhl Regional Medical Center | United States Air Force School of Aerospace Medicine | Anthony Fries et al |
| EPI_ISL_812276 | Iraq/USAFSAM-S081/2020 | 2020-06-07 | Asia/Iraq/Iraq/ | Landstuhl Regional Medical Center | United States Air Force School of Aerospace Medicine | Anthony Fries et al |
| EPI_ISL_812278 | Iraq/USAFSAM-S083/2020 | 2020-06-16 | Asia/Iraq/Iraq/ | Landstuhl Regional Medical Center | United States Air Force School of Aerospace Medicine | Anthony Fries et al |
| EPI_ISL_812279 | Iraq/USAFSAM-S084/2020 | 2020-06-16 | Asia/Iraq/Iraq/ | Landstuhl Regional Medical Center | United States Air Force School of Aerospace Medicine | Anthony Fries et al |

| | | | | | | |
|---|---|---|---|---|---|---|
| EPI_ISL_812280 | Iraq/USAFSAM-S085/2020 | 2020-06-07 | Asia/Iraq/Iraq/ | Landstuhl Regional Medical Center | United States Air Force School of Aerospace Medicine | Anthony Fries et al |
| EPI_ISL_812281 | Iraq/USAFSAM-S086/2020 | 2020-06-07 | Asia/Iraq/Iraq/ | Landstuhl Regional Medical Center | United States Air Force School of Aerospace Medicine | Anthony Fries et al |
| EPI_ISL_812282 | Iraq/USAFSAM-S088/2020 | 2020-06-16 | Asia/Iraq/Iraq/ | Landstuhl Regional Medical Center | United States Air Force School of Aerospace Medicine | Anthony Fries et al |
| EPI_ISL_812283 | Iraq/USAFSAM-S090/2020 | 2020-06-17 | Asia/Iraq/Iraq/ | Landstuhl Regional Medical Center | United States Air Force School of Aerospace Medicine | Anthony Fries et al |
| EPI_ISL_812284 | Iraq/USAFSAM-S091/2020 | 2020-06-17 | Asia/Iraq/Iraq/ | Landstuhl Regional Medical Center | United States Air Force School of Aerospace Medicine | Anthony Fries et al |
| EPI_ISL_812285 | Iraq/USAFSAM-S092/2020 | 2020-06-17 | Asia/Iraq/Iraq/ | Landstuhl Regional Medical Center | United States Air Force School of Aerospace Medicine | Anthony Fries et al |
| EPI_ISL_812286 | USA/DE-DHSS-F1042798/2020 | 2020-12-28 | North America/USA/Delaware/Sussex C | Delaware Public Health Lab | Delaware Public Health Lab | Gregory Hovan et al |
| EPI_ISL_812300 | USA/DE-DHSS-FLW00569839/2020 | 2020-12-28 | North America/USA/Delaware/Sussex C | Delaware Public Health Lab | Delaware Public Health Lab | Gregory Hovan et al |
| EPI_ISL_812304 | USA/DE-DHSS-FLW00570853/2020 | 2020-12-28 | North America/USA/Delaware/Sussex C | Delaware Public Health Lab | Delaware Public Health Lab | Gregory Hovan et al |
| EPI_ISL_812317 | USA/CO-USAFSAM-S128/2020 | 2020-06-03 | North America/USA/Colorado/ | United States Air Force School of Aerospace Medicine | United States Air Force School of Aerospace Medicine | Anthony Fries et al |
| EPI_ISL_812319 | USA/DE-USAFSAM-S111/2020 | 2020-04-02 | North America/USA/Delaware/ | United States Air Force School of Aerospace Medicine | United States Air Force School of Aerospace Medicine | Anthony Fries et al |

| | | | | | | |
|---|---|---|---|---|---|---|
| EPI_ISL_812327 | USA/FL-USAFSAM-S125/2020 | 2020-05-31 | North America/USA/Florida/ | United States Air Force School of Aerospace Medicine | United States Air Force School of Aerospace Medicine | Anthony Fries et al |
| EPI_ISL_812331 | USA/ID-USAFSAM-S144/2020 | 2020-04-03 | North America/USA/Idaho/ | United States Air Force School of Aerospace Medicine | United States Air Force School of Aerospace Medicine | Anthony Fries et al |
| EPI_ISL_812336 | USA/NC-USAFSAM-S124/2020 | 2020-04-24 | North America/USA/North Carolina/ | United States Air Force School of Aerospace Medicine | United States Air Force School of Aerospace Medicine | Anthony Fries et al |
| EPI_ISL_812344 | USA/SC-USAFSAM-S115/2020 | 2020-03-31 | North America/USA/South Carolina/ | United States Air Force School of Aerospace Medicine | United States Air Force School of Aerospace Medicine | Anthony Fries et al |
| EPI_ISL_812348 | USA/WI-UW-2554/2020 | 2020-12-29 | North America/USA/Wisconsin/Dane Co | University of Wisconsin-Madison AIDS Vaccine Research Laboratories | University of Wisconsin-Madison AIDS Vaccine Research Laboratories | Gage Moreno et al |
| EPI_ISL_812362 | USA/UT-UPHL-2101420430/2020 | 2020-12-10 | North America/USA/Utah/ | Utah Public Health Laboratory | Utah Public Health Laboratory | Erin L. Young et al |
| EPI_ISL_812395 | USA/UT-UPHL-2101837386/2020 | 2020-12-10 | North America/USA/Utah/ | Utah Public Health Laboratory | Utah Public Health Laboratory | Erin L. Young et al |
| EPI_ISL_812406 | USA/UT-UPHL-2101625635/2020 | 2020-12-13 | North America/USA/Utah/ | Utah Public Health Laboratory | Utah Public Health Laboratory | Erin L. Young et al |
| EPI_ISL_812422 | USA/WI-UW-2442/2020 | 2020-12-13 | North America/USA/Wisconsin/Dane Co | University of Wisconsin-Madison AIDS Vaccine Research Laboratories | University of Wisconsin-Madison AIDS Vaccine Research Laboratories | Gage Moreno et al |
| EPI_ISL_812447 | Peru/LIM-UPCH-0285/2020 | 2020-12-12 | South America/Peru/Lima/ | Laboratorio de Referencia Nacional de Virus Respiratorios, Instituto Nacional de Salud Peru | Laboratorio de Genv≥mica Microbiana, Universidad Peruana Cayetano Heredia | Pablo Tsukayama et al |

| | | | | | | |
|---|---|---|---|---|---|---|
| EPI_ISL_812457 | Peru/LIM-UPCH-0296/2020 | 2020-12-11 | South America/Peru/Lima/ | Laboratorio de Referencia Nacional de Virus Respiratorios, Instituto Nacional de Salud Peru | Laboratorio de Genv≥mica Microbiana, Universidad Peruana Cayetano Heredia | Pablo Tsukayama et al |
| EPI_ISL_812458 | Peru/LIM-UPCH-0297/2020 | 2020-12-11 | South America/Peru/Lima/ | Laboratorio de Referencia Nacional de Virus Respiratorios, Instituto Nacional de Salud Peru | Laboratorio de Genv≥mica Microbiana, Universidad Peruana Cayetano Heredia | Pablo Tsukayama et al |
| EPI_ISL_812466 | Peru/LIM-UPCH-0305/2020 | 2020-12-10 | South America/Peru/Lima/ | Laboratorio de Referencia Nacional de Virus Respiratorios, Instituto Nacional de Salud Peru | Laboratorio de Genv≥mica Microbiana, Universidad Peruana Cayetano Heredia | Pablo Tsukayama et al |
| EPI_ISL_812478 | Peru/LIM-UPCH-0322/2020 | 2020-12-12 | South America/Peru/Lima/ | Laboratorio de Referencia Nacional de Virus Respiratorios, Instituto Nacional de Salud Peru | Laboratorio de Genv≥mica Microbiana, Universidad Peruana Cayetano Heredia | Pablo Tsukayama et al |
| EPI_ISL_812479 | Peru/LIM-UPCH-0323/2020 | 2020-12-10 | South America/Peru/Lima/ | Laboratorio de Referencia Nacional de Virus Respiratorios, Instituto Nacional de Salud Peru | Laboratorio de Genv≥mica Microbiana, Universidad Peruana Cayetano Heredia | Pablo Tsukayama et al |
| EPI_ISL_812484 | Peru/LIM-UPCH-0328/2020 | 2020-12-11 | South America/Peru/Lima/ | Laboratorio de Referencia Nacional de Virus Respiratorios, Instituto Nacional de Salud Peru | Laboratorio de Genv≥mica Microbiana, Universidad Peruana Cayetano Heredia | Pablo Tsukayama et al |

| | | | | | | |
|---|---|---|---|---|---|---|
| EPI_ISL_812486 | Peru/LIM-UPCH-0330/2020 | 2020-12-12 | South America/Peru/Lima/ | Laboratorio de Referencia Nacional de Virus Respiratorios, Instituto Nacional de Salud Peru | Laboratorio de Gen√≥mica Microbiana, Universidad Peruana Cayetano Heredia | Pablo Tsukayama et al |
| EPI_ISL_812488 | Peru/MOQ-UPCH-0332/2020 | 2020-11-30 | South America/Peru/Moquegua/Moque | Laboratorio de Referencia Nacional de Virus Respiratorios, Instituto Nacional de Salud Peru | Laboratorio de Gen√≥mica Microbiana, Universidad Peruana Cayetano Heredia | Pablo Tsukayama et al |
| EPI_ISL_812489 | Peru/LIM-UPCH-0333/2020 | 2020-12-11 | South America/Peru/Lima/ | Laboratorio de Referencia Nacional de Virus Respiratorios, Instituto Nacional de Salud Peru | Laboratorio de Gen√≥mica Microbiana, Universidad Peruana Cayetano Heredia | Pablo Tsukayama et al |
| EPI_ISL_812492 | Peru/LIM-UPCH-0337/2020 | 2020-12-13 | South America/Peru/Lima/ | Laboratorio de Referencia Nacional de Virus Respiratorios, Instituto Nacional de Salud Peru | Laboratorio de Gen√≥mica Microbiana, Universidad Peruana Cayetano Heredia | Pablo Tsukayama et al |
| EPI_ISL_812497 | Peru/LIM-UPCH-0342/2020 | 2020-12-12 | South America/Peru/Lima/ | Laboratorio de Referencia Nacional de Virus Respiratorios, Instituto Nacional de Salud Peru | Laboratorio de Gen√≥mica Microbiana, Universidad Peruana Cayetano Heredia | Pablo Tsukayama et al |
| EPI_ISL_812502 | Peru/LIM-UPCH-0347/2020 | 2020-12-11 | South America/Peru/Lima/ | Laboratorio de Referencia Nacional de Virus Respiratorios, Instituto Nacional de Salud Peru | Laboratorio de Gen√≥mica Microbiana, Universidad Peruana Cayetano Heredia | Pablo Tsukayama et al |

| | | | | | | |
|---|---|---|---|---|---|---|
| EPI_ISL_812510 | Peru/LIM-UPCH-0362/2020 | 2020-12-11 | South America/Peru/Lima/ | Laboratorio de Referencia Nacional de Virus Respiratorios, Instituto Nacional de Salud Peru | Laboratorio de Genv≥mica Microbiana, Universidad Peruana Cayetano Heredia | Pablo Tsukayama et al |
| EPI_ISL_812515 | Peru/LIM-UPCH-0367/2020 | 2020-12-13 | South America/Peru/Lima/ | Laboratorio de Referencia Nacional de Virus Respiratorios, Instituto Nacional de Salud Peru | Laboratorio de Genv≥mica Microbiana, Universidad Peruana Cayetano Heredia | Pablo Tsukayama et al |
| EPI_ISL_812516 | Peru/LIM-UPCH-0368/2020 | 2020-12-13 | South America/Peru/Lima/ | Laboratorio de Referencia Nacional de Virus Respiratorios, Instituto Nacional de Salud Peru | Laboratorio de Genv≥mica Microbiana, Universidad Peruana Cayetano Heredia | Pablo Tsukayama et al |
| EPI_ISL_812527 | USA/AK-USAFSAM-S179/2020 | 2020-07-13 | North America/USA/Alaska/ | United States Air Force School of Aerospace Medicine | United States Air Force School of Aerospace Medicine | Anthony Fries et al |
| EPI_ISL_812528 | USA/AK-USAFSAM-S180/2020 | 2020-07-14 | North America/USA/Alaska/ | United States Air Force School of Aerospace Medicine | United States Air Force School of Aerospace Medicine | Anthony Fries et al |
| EPI_ISL_812538 | USA/CO-USAFSAM-S195/2020 | 2020-07-24 | North America/USA/Colorado/ | United States Air Force School of Aerospace Medicine | United States Air Force School of Aerospace Medicine | Anthony Fries et al |
| EPI_ISL_812542 | USA/ID-USAFSAM-S200/2020 | 2020-07-22 | North America/USA/Idaho/ | United States Air Force School of Aerospace Medicine | United States Air Force School of Aerospace Medicine | Anthony Fries et al |
| EPI_ISL_812547 | USA/LA-USAFSAM-S206/2020 | 2020-07-20 | North America/USA/Louisiana/ | United States Air Force School of Aerospace Medicine | United States Air Force School of Aerospace Medicine | Anthony Fries et al |

| | | | | | | |
|---|---|---|---|---|---|---|
| EPI_ISL_812551 | USA/NC-USAFSAM-S211/2020 | 2020-07-20 | North America/USA/North Carolina/ | United States Air Force School of Aerospace Medicine | United States Air Force School of Aerospace Medicine | Anthony Fries et al |
| EPI_ISL_812557 | USA/TX-USAFSAM-S218/2020 | 2020-07-22 | North America/USA/Texas/ | United States Air Force School of Aerospace Medicine | United States Air Force School of Aerospace Medicine | Anthony Fries et al |
| EPI_ISL_812563 | USA/DE-USAFSAM-S225/2020 | 2020-07-27 | North America/USA/Delaware/ | United States Air Force School of Aerospace Medicine | United States Air Force School of Aerospace Medicine | Anthony Fries et al |
| EPI_ISL_812569 | USA/MS-USAFSAM-S234/2020 | 2020-07-29 | North America/USA/Mississippi/ | United States Air Force School of Aerospace Medicine | United States Air Force School of Aerospace Medicine | Anthony Fries et al |
| EPI_ISL_812571 | USA/NE-USAFSAM-S236/2020 | 2020-07-27 | North America/USA/Nebraska/ | United States Air Force School of Aerospace Medicine | United States Air Force School of Aerospace Medicine | Anthony Fries et al |
| EPI_ISL_812578 | USA/OK-USAFSAM-S243/2020 | 2020-07-28 | North America/USA/Oklahoma/ | United States Air Force School of Aerospace Medicine | United States Air Force School of Aerospace Medicine | Anthony Fries et al |
| EPI_ISL_812586 | USA/FL-USAFSAM-S254/2020 | 2020-08-03 | North America/USA/Florida/ | United States Air Force School of Aerospace Medicine | United States Air Force School of Aerospace Medicine | Anthony Fries et al |
| EPI_ISL_812588 | USA/LA-USAFSAM-S257/2020 | 2020-08-06 | North America/USA/Louisiana/ | United States Air Force School of Aerospace Medicine | United States Air Force School of Aerospace Medicine | Anthony Fries et al |
| EPI_ISL_812595 | USA/OH-USAFSAM-S265/2020 | 2020-08-06 | North America/USA/Ohio/ | United States Air Force School of Aerospace Medicine | United States Air Force School of Aerospace Medicine | Anthony Fries et al |
| EPI_ISL_812599 | USA/AZ-USAFSAM-S269/2020 | 2020-08-10 | North America/USA/Arizona/ | United States Air Force School of Aerospace Medicine | United States Air Force School of Aerospace Medicine | Anthony Fries et al |

| | | | | | | |
|---|---|---|---|---|---|---|
| EPI_ISL_812604 | USA/MS-USAFSAM-S281/2020 | 2020-08-10 | North America/USA/Mississippi/ | United States Air Force School of Aerospace Medicine | United States Air Force School of Aerospace Medicine | Anthony Fries et al |
| EPI_ISL_812605 | USA/NE-USAFSAM-S283/2020 | 2020-08-11 | North America/USA/Nebraska/ | United States Air Force School of Aerospace Medicine | United States Air Force School of Aerospace Medicine | Anthony Fries et al |
| EPI_ISL_812607 | USA/NC-USAFSAM-S286/2020 | 2020-08-14 | North America/USA/North Carolina/ | United States Air Force School of Aerospace Medicine | United States Air Force School of Aerospace Medicine | Anthony Fries et al |
| EPI_ISL_812609 | USA/OH-USAFSAM-S288/2020 | 2020-08-13 | North America/USA/Ohio/ | United States Air Force School of Aerospace Medicine | United States Air Force School of Aerospace Medicine | Anthony Fries et al |
| EPI_ISL_812613 | USA/AL-USAFSAM-S295/2020 | 2020-08-17 | North America/USA/Alabama/ | United States Air Force School of Aerospace Medicine | United States Air Force School of Aerospace Medicine | Anthony Fries et al |
| EPI_ISL_812615 | USA/AZ-USAFSAM-S297/2020 | 2020-08-21 | North America/USA/Arizona/ | United States Air Force School of Aerospace Medicine | United States Air Force School of Aerospace Medicine | Anthony Fries et al |
| EPI_ISL_812619 | USA/ID-USAFSAM-S303/2020 | 2020-08-18 | North America/USA/Idaho/ | United States Air Force School of Aerospace Medicine | United States Air Force School of Aerospace Medicine | Anthony Fries et al |
| EPI_ISL_812620 | USA/IL-USAFSAM-S304/2020 | 2020-08-20 | North America/USA/Illinois/ | United States Air Force School of Aerospace Medicine | United States Air Force School of Aerospace Medicine | Anthony Fries et al |
| EPI_ISL_812621 | USA/IL-USAFSAM-S305/2020 | 2020-08-19 | North America/USA/Illinois/ | United States Air Force School of Aerospace Medicine | United States Air Force School of Aerospace Medicine | Anthony Fries et al |
| EPI_ISL_812623 | USA/KS-USAFSAM-S307/2020 | 2020-08-20 | North America/USA/Kansas/ | United States Air Force School of Aerospace Medicine | United States Air Force School of Aerospace Medicine | Anthony Fries et al |

| | | | | | | |
|---|---|---|---|---|---|---|
| EPI_ISL_812624 | USA/LA-USAFSAM-S308/2020 | 2020-08-18 | North America/USA/Louisiana/ | United States Air Force School of Aerospace Medicine | United States Air Force School of Aerospace Medicine | Anthony Fries et al |
| EPI_ISL_812631 | USA/ND-USAFSAM-S316/2020 | 2020-08-17 | North America/USA/North Dakota/ | United States Air Force School of Aerospace Medicine | United States Air Force School of Aerospace Medicine | Anthony Fries et al |
| EPI_ISL_812634 | USA/SD-USAFSAM-S324/2020 | 2020-08-20 | North America/USA/South Dakota/ | United States Air Force School of Aerospace Medicine | United States Air Force School of Aerospace Medicine | Anthony Fries et al |
| EPI_ISL_812635 | USA/SD-USAFSAM-S325/2020 | 2020-08-17 | North America/USA/South Dakota/ | United States Air Force School of Aerospace Medicine | United States Air Force School of Aerospace Medicine | Anthony Fries et al |
| EPI_ISL_812637 | USA/VA-USAFSAM-S327/2020 | 2020-08-21 | North America/USA/Virginia/ | United States Air Force School of Aerospace Medicine | United States Air Force School of Aerospace Medicine | Anthony Fries et al |
| EPI_ISL_812643 | USA/FL-USAFSAM-S334/2020 | 2020-08-26 | North America/USA/Florida/ | United States Air Force School of Aerospace Medicine | United States Air Force School of Aerospace Medicine | Anthony Fries et al |
| EPI_ISL_812645 | USA/GA-USAFSAM-S336/2020 | 2020-08-27 | North America/USA/Georgia/ | United States Air Force School of Aerospace Medicine | United States Air Force School of Aerospace Medicine | Anthony Fries et al |
| EPI_ISL_812648 | USA/ID-USAFSAM-S339/2020 | 2020-08-24 | North America/USA/Idaho/ | United States Air Force School of Aerospace Medicine | United States Air Force School of Aerospace Medicine | Anthony Fries et al |
| EPI_ISL_812657 | USA/ND-USAFSAM-S351/2020 | 2020-08-24 | North America/USA/North Dakota/ | United States Air Force School of Aerospace Medicine | United States Air Force School of Aerospace Medicine | Anthony Fries et al |
| EPI_ISL_812660 | USA/OK-USAFSAM-S356/2020 | 2020-08-28 | North America/USA/Oklahoma/ | United States Air Force School of Aerospace Medicine | United States Air Force School of Aerospace Medicine | Anthony Fries et al |

| | | | | | | |
|---|---|---|---|---|---|---|
| EPI_ISL_812661 | USA/SD-USAFSAM-S359/2020 | 2020-08-27 | North America/USA/South Dakota/ | United States Air Force School of Aerospace Medicine | United States Air Force School of Aerospace Medicine | Anthony Fries et al |
| EPI_ISL_812662 | USA/SD-USAFSAM-S360/2020 | 2020-08-27 | North America/USA/South Dakota/ | United States Air Force School of Aerospace Medicine | United States Air Force School of Aerospace Medicine | Anthony Fries et al |
| EPI_ISL_812665 | USA/AZ-USAFSAM-S364/2020 | 2020-09-04 | North America/USA/Arizona/ | United States Air Force School of Aerospace Medicine | United States Air Force School of Aerospace Medicine | Anthony Fries et al |
| EPI_ISL_812666 | USA/DE-USAFSAM-S366/2020 | 2020-08-31 | North America/USA/Delaware/ | United States Air Force School of Aerospace Medicine | United States Air Force School of Aerospace Medicine | Anthony Fries et al |
| EPI_ISL_812672 | USA/ID-USAFSAM-S372/2020 | 2020-08-31 | North America/USA/Idaho/ | United States Air Force School of Aerospace Medicine | United States Air Force School of Aerospace Medicine | Anthony Fries et al |
| EPI_ISL_812674 | USA/KS-USAFSAM-S375/2020 | 2020-09-03 | North America/USA/Kansas/ | United States Air Force School of Aerospace Medicine | United States Air Force School of Aerospace Medicine | Anthony Fries et al |
| EPI_ISL_812675 | USA/MS-USAFSAM-S376/2020 | 2020-09-01 | North America/USA/Mississippi/ | United States Air Force School of Aerospace Medicine | United States Air Force School of Aerospace Medicine | Anthony Fries et al |
| EPI_ISL_812677 | USA/NC-USAFSAM-S379/2020 | 2020-08-31 | North America/USA/North Carolina/ | United States Air Force School of Aerospace Medicine | United States Air Force School of Aerospace Medicine | Anthony Fries et al |
| EPI_ISL_812689 | USA/CO-USAFSAM-S394/2020 | 2020-09-11 | North America/USA/Colorado/ | United States Air Force School of Aerospace Medicine | United States Air Force School of Aerospace Medicine | Anthony Fries et al |
| EPI_ISL_812692 | USA/GA-USAFSAM-S397/2020 | 2020-09-10 | North America/USA/Georgia/ | United States Air Force School of Aerospace Medicine | United States Air Force School of Aerospace Medicine | Anthony Fries et al |

| | | | | | | |
|---|---|---|---|---|---|---|
| EPI_ISL_812694 | USA/LA-USAFSAM-S402/2020 | 2020-09-10 | North America/USA/Louisiana/ | United States Air Force School of Aerospace Medicine | United States Air Force School of Aerospace Medicine | Anthony Fries et al |
| EPI_ISL_812699 | USA/OK-USAFSAM-S409/2020 | 2020-09-11 | North America/USA/Oklahoma/ | United States Air Force School of Aerospace Medicine | United States Air Force School of Aerospace Medicine | Anthony Fries et al |
| EPI_ISL_812700 | USA/SC-USAFSAM-S410/2020 | 2020-09-11 | North America/USA/South Carolina/ | United States Air Force School of Aerospace Medicine | United States Air Force School of Aerospace Medicine | Anthony Fries et al |
| EPI_ISL_812705 | USA/NE-USAFSAM-S417/2020 | 2020-09-14 | North America/USA/Nebraska/ | United States Air Force School of Aerospace Medicine | United States Air Force School of Aerospace Medicine | Anthony Fries et al |
| EPI_ISL_812706 | USA/NC-USAFSAM-S418/2020 | 2020-09-14 | North America/USA/North Carolina/ | United States Air Force School of Aerospace Medicine | United States Air Force School of Aerospace Medicine | Anthony Fries et al |
| EPI_ISL_812711 | USA/SD-USAFSAM-S424/2020 | 2020-09-14 | North America/USA/South Dakota/ | United States Air Force School of Aerospace Medicine | United States Air Force School of Aerospace Medicine | Anthony Fries et al |
| EPI_ISL_812725 | USA/NY-NYCPHL-001794/2020 | 2020-12-14 | North America/USA/New York/New Yor | DOHMH Crown Heights | New York City Public Health Laboratory | Jade Wang et al |
| EPI_ISL_812734 | USA/NY-NYCPHL-001805/2020 | 2020-12-15 | North America/USA/New York/New Yor | DOHMH Crown Heights | New York City Public Health Laboratory | Jade Wang et al |
| EPI_ISL_812744 | USA/NY-NYCPHL-001818/2020 | 2020-12-15 | North America/USA/New York/New Yor | DOHMH Corona | New York City Public Health Laboratory | Jade Wang et al |
| EPI_ISL_812745 | USA/NY-NYCPHL-001819/2020 | 2020-12-16 | North America/USA/New York/New Yor | DOHMH Riverside | New York City Public Health Laboratory | Jade Wang et al |
| EPI_ISL_812746 | USA/NY-NYCPHL-001820/2020 | 2020-12-16 | North America/USA/New York/New Yor | DOHMH Corona | New York City Public Health Laboratory | Jade Wang et al |

| | | | | | | |
|---|---|---|---|---|---|---|
| EPI_ISL_812747 | USA/NY-NYCPHL-001821/2020 | 2020-12-16 | North America/USA/New York/New Yor | DOHMH Corona | New York City Public Health Laboratory | Jade Wang et al |
| EPI_ISL_812749 | USA/NY-NYCPHL-001823/2020 | 2020-12-16 | North America/USA/New York/New Yor | DOHMH Central Harlem | New York City Public Health Laboratory | Jade Wang et al |
| EPI_ISL_812752 | USA/NY-NYCPHL-001827/2020 | 2020-12-16 | North America/USA/New York/New Yor | DOHMH Crown Heights | New York City Public Health Laboratory | Jade Wang et al |
| EPI_ISL_812761 | Turkey/HSGM-1142/2020 | 2020-12-24 | Europe/Turkey/Ankara/ | Ministry of Health Turkey | Ministry of Health Turkey | Fatma Bayrakdar et al |
| EPI_ISL_812762 | Turkey/HSGM-1151/2020 | 2020-12-25 | Europe/Turkey/Ankara/ | Ministry of Health Turkey | Ministry of Health Turkey | Fatma Bayrakdar et al |
| EPI_ISL_812763 | Turkey/HSGM-1160/2020 | 2020-12-28 | Europe/Turkey/Ankara/ | Ministry of Health Turkey | Ministry of Health Turkey | Fatma Bayrakdar et al |
| EPI_ISL_812764 | Turkey/HSGM-1161/2020 | 2020-12-28 | Europe/Turkey/Ankara/ | Ministry of Health Turkey | Ministry of Health Turkey | Fatma Bayrakdar et al |
| EPI_ISL_812765 | Turkey/HSGM-1162/2020 | 2020-12-28 | Europe/Turkey/Ankara/ | Ministry of Health Turkey | Ministry of Health Turkey | Fatma Bayrakdar et al |
| EPI_ISL_812766 | Turkey/HSGM-1164/2020 | 2020-12-28 | Europe/Turkey/Ankara/ | Ministry of Health Turkey | Ministry of Health Turkey | Fatma Bayrakdar et al |
| EPI_ISL_812767 | Turkey/HSGM-1165/2020 | 2020-12-28 | Europe/Turkey/Ankara/ | Ministry of Health Turkey | Ministry of Health Turkey | Fatma Bayrakdar et al |
| EPI_ISL_812769 | Turkey/HSGM-1183/2020 | 2020-12-29 | Europe/Turkey/Ankara/ | Ministry of Health Turkey | Ministry of Health Turkey | Fatma Bayrakdar et al |
| EPI_ISL_812770 | Turkey/HSGM-1187/2020 | 2020-12-29 | Europe/Turkey/Ankara/ | Ministry of Health Turkey | Ministry of Health Turkey | Fatma Bayrakdar et al |
| EPI_ISL_812771 | Turkey/HSGM-1204/2020 | 2020-12-31 | Europe/Turkey/Ankara/ | Ministry of Health Turkey | Ministry of Health Turkey | Fatma Bayrakdar et al |
| EPI_ISL_812772 | Turkey/HSGM-1213/2021 | 2021-01-04 | Europe/Turkey/Ankara/ | Ministry of Health Turkey | Ministry of Health Turkey | Fatma Bayrakdar et al |
| EPI_ISL_812774 | Turkey/HSGM-1121/2020 | 2020-12-24 | Europe/Turkey/Ankara/ | Ministry of Health Turkey | Ministry of Health Turkey | Fatma Bayrakdar et al |
| EPI_ISL_812775 | Turkey/HSGM-1124/2020 | 2020-12-24 | Europe/Turkey/Ankara/ | Ministry of Health Turkey | Ministry of Health Turkey | Fatma Bayrakdar et al |
| EPI_ISL_812776 | Turkey/HSGM-1125/2020 | 2020-12-24 | Europe/Turkey/Ankara/ | Ministry of Health Turkey | Ministry of Health Turkey | Fatma Bayrakdar et al |
| EPI_ISL_812777 | Turkey/HSGM-1195/2020 | 2020-12-30 | Europe/Turkey/Ankara/ | Ministry of Health Turkey | Ministry of Health Turkey | Fatma Bayrakdar et al |

| | | | | | | |
|---|---|---|---|---|---|---|
| EPI_ISL_812779 | Turkey/HSGM-1196/2020 | 2020-12-30 | Europe/Turkey/Ankara/ | Ministry of Health Turkey | Ministry of Health Turkey | Fatma Bayrakdar et al |
| EPI_ISL_812780 | Turkey/HSGM-1197/2020 | 2020-12-30 | Europe/Turkey/Ankara/ | Ministry of Health Turkey | Ministry of Health Turkey | Fatma Bayrakdar et al |
| EPI_ISL_812784 | Egypt/CCHE57357-A-02/2020 | 2020-05-16 | Africa/Egypt/Egypt/ | Genomics Program, Children Cancer Hospital | Genomics Program, Children Cancer Hospital | Hatem et al |
| EPI_ISL_812787 | Egypt/CCHE57357-A-06/2020 | 2020-05-30 | Africa/Egypt/Egypt/ | Genomics Program, Children Cancer Hospital | Genomics Program, Children Cancer Hospital | Hatem et al |
| EPI_ISL_812788 | Egypt/CCHE57357-A-07/2020 | 2020-06-08 | Africa/Egypt/Egypt/ | Genomics Program, Children Cancer Hospital | Genomics Program, Children Cancer Hospital | Hatem et al |
| EPI_ISL_812789 | Egypt/CCHE57357-A-08/2020 | 2020-05-07 | Africa/Egypt/Egypt/ | Genomics Program, Children Cancer Hospital | Genomics Program, Children Cancer Hospital | Hatem et al |
| EPI_ISL_812791 | Egypt/CCHE57357-A-10/2020 | 2020-05-15 | Africa/Egypt/Egypt/ | Genomics Program, Children Cancer Hospital | Genomics Program, Children Cancer Hospital | Hatem et al |
| EPI_ISL_812792 | Egypt/CCHE57357-A-12/2020 | 2020-05-20 | Africa/Egypt/Egypt/ | Genomics Program, Children Cancer Hospital | Genomics Program, Children Cancer Hospital | Hatem et al |
| EPI_ISL_812793 | Egypt/CCHE57357-A-13/2020 | 2020-04-29 | Africa/Egypt/Egypt/ | Genomics Program, Children Cancer Hospital | Genomics Program, Children Cancer Hospital | Hatem et al |
| EPI_ISL_812795 | Egypt/CCHE57357-A-15/2020 | 2020-06-09 | Africa/Egypt/Egypt/ | Genomics Program, Children Cancer Hospital | Genomics Program, Children Cancer Hospital | Hatem et al |
| EPI_ISL_812796 | Egypt/CCHE57357-A-18/2020 | 2020-07-03 | Africa/Egypt/Egypt/ | Genomics Program, Children Cancer Hospital | Genomics Program, Children Cancer Hospital | Hatem et al |
| EPI_ISL_812799 | Egypt/CCHE57357-A-23/2020 | 2020-05-21 | Africa/Egypt/Egypt/ | Genomics Program, Children Cancer Hospital | Genomics Program, Children Cancer Hospital | Hatem et al |
| EPI_ISL_812800 | Egypt/CCHE57357-A-24/2020 | 2020-05-25 | Africa/Egypt/Egypt/ | Genomics Program, Children Cancer Hospital | Genomics Program, Children Cancer Hospital | Hatem et al |
| EPI_ISL_812803 | Egypt/CCHE57357-A-27/2020 | 2020-07-05 | Africa/Egypt/Egypt/ | Genomics Program, Children Cancer Hospital | Genomics Program, Children Cancer Hospital | Hatem et al |

| | | | | | | |
|---|---|---|---|---|---|---|
| EPI_ISL_812806 | Egypt/CCHE57357-A-30/2020 | 2020-06-30 | Africa/Egypt/Egypt/ | Genomics Program, Children Cancer Hospital | Genomics Program, Children Cancer Hospital | Hatem et al |
| EPI_ISL_812807 | Egypt/CCHE57357-A-31/2020 | 2020-07-28 | Africa/Egypt/Egypt/ | Genomics Program, Children Cancer Hospital | Genomics Program, Children Cancer Hospital | Hatem et al |
| EPI_ISL_812810 | Egypt/CCHE57357-A-36/2020 | 2020-08-02 | Africa/Egypt/Egypt/ | Genomics Program, Children Cancer Hospital | Genomics Program, Children Cancer Hospital | Hatem et al |
| EPI_ISL_812812 | Egypt/CCHE57357-A-39/2020 | 2020-08-11 | Africa/Egypt/Egypt/ | Genomics Program, Children Cancer Hospital | Genomics Program, Children Cancer Hospital | Hatem et al |
| EPI_ISL_812813 | Egypt/CCHE57357-A-40/2020 | 2020-06-29 | Africa/Egypt/Egypt/ | Genomics Program, Children Cancer Hospital | Genomics Program, Children Cancer Hospital | Hatem et al |
| EPI_ISL_812816 | Egypt/CCHE57357-A-43/2020 | 2020-07-12 | Africa/Egypt/Egypt/ | Genomics Program, Children Cancer Hospital | Genomics Program, Children Cancer Hospital | Hatem et al |
| EPI_ISL_812818 | Egypt/CCHE57357-A-45/2020 | 2020-06-20 | Africa/Egypt/Egypt/ | Genomics Program, Children Cancer Hospital | Genomics Program, Children Cancer Hospital | Hatem et al |
| EPI_ISL_812820 | Egypt/CCHE57357-A-47/2020 | 2020-06-30 | Africa/Egypt/Egypt/ | Genomics Program, Children Cancer Hospital | Genomics Program, Children Cancer Hospital | Hatem et al |
| EPI_ISL_812822 | Egypt/CCHE57357-A-49/2020 | 2020-07-01 | Africa/Egypt/Egypt/ | Genomics Program, Children Cancer Hospital | Genomics Program, Children Cancer Hospital | Hatem et al |
| EPI_ISL_812823 | Egypt/CCHE57357-A-50/2020 | 2020-07-23 | Africa/Egypt/Egypt/ | Genomics Program, Children Cancer Hospital | Genomics Program, Children Cancer Hospital | Hatem et al |
| EPI_ISL_812824 | Egypt/CCHE57357-A-51/2020 | 2020-07-11 | Africa/Egypt/Egypt/ | Genomics Program, Children Cancer Hospital | Genomics Program, Children Cancer Hospital | Hatem et al |
| EPI_ISL_812825 | Egypt/CCHE57357-A-52/2020 | 2020-07-16 | Africa/Egypt/Egypt/ | Genomics Program, Children Cancer Hospital | Genomics Program, Children Cancer Hospital | Hatem et al |
| EPI_ISL_812826 | Egypt/CCHE57357-A-53/2020 | 2020-06-18 | Africa/Egypt/Egypt/ | Genomics Program, Children Cancer Hospital | Genomics Program, Children Cancer Hospital | Hatem et al |

| | | | | | | |
|---|---|---|---|---|---|---|
| EPI_ISL_812827 | Egypt/CCHE57357-A-54/2020 | 2020-07-19 | Africa/Egypt/Egypt/ | Genomics Program, Children Cancer Hospital | Genomics Program, Children Cancer Hospital | Hatem et al |
| EPI_ISL_812829 | Egypt/CCHE57357-A-56/2020 | 2020-06-29 | Africa/Egypt/Egypt/ | Genomics Program, Children Cancer Hospital | Genomics Program, Children Cancer Hospital | Hatem et al |
| EPI_ISL_812830 | Egypt/CCHE57357-A-57/2020 | 2020-08-02 | Africa/Egypt/Egypt/ | Genomics Program, Children Cancer Hospital | Genomics Program, Children Cancer Hospital | Hatem et al |
| EPI_ISL_812833 | Egypt/CCHE57357-A-60/2020 | 2020-06-12 | Africa/Egypt/Egypt/ | Genomics Program, Children Cancer Hospital | Genomics Program, Children Cancer Hospital | Hatem et al |
| EPI_ISL_812836 | Egypt/CCHE57357-A-66/2020 | 2020-06-11 | Africa/Egypt/Egypt/ | Genomics Program, Children Cancer Hospital | Genomics Program, Children Cancer Hospital | Hatem et al |
| EPI_ISL_812838 | Egypt/CCHE57357-A-68/2020 | 2020-06-29 | Africa/Egypt/Egypt/ | Genomics Program, Children Cancer Hospital | Genomics Program, Children Cancer Hospital | Hatem et al |
| EPI_ISL_812839 | Egypt/CCHE57357-A-69/2020 | 2020-06-29 | Africa/Egypt/Egypt/ | Genomics Program, Children Cancer Hospital | Genomics Program, Children Cancer Hospital | Hatem et al |
| EPI_ISL_812840 | Egypt/CCHE57357-A-70/2020 | 2020-07-02 | Africa/Egypt/Egypt/ | Genomics Program, Children Cancer Hospital | Genomics Program, Children Cancer Hospital | Hatem et al |
| EPI_ISL_812843 | Egypt/CCHE57357-A-75/2020 | 2020-07-28 | Africa/Egypt/Egypt/ | Genomics Program, Children Cancer Hospital | Genomics Program, Children Cancer Hospital | Hatem et al |
| EPI_ISL_812846 | Egypt/CCHE57357-A-89/2020 | 2020-08-02 | Africa/Egypt/Egypt/ | Genomics Program, Children Cancer Hospital | Genomics Program, Children Cancer Hospital | Hatem et al |
| EPI_ISL_812847 | Egypt/CCHE57357-A-90/2020 | 2020-06-26 | Africa/Egypt/Egypt/ | Genomics Program, Children Cancer Hospital | Genomics Program, Children Cancer Hospital | Hatem et al |
| EPI_ISL_812848 | Egypt/CCHE57357-A-91/2020 | 2020-06-30 | Africa/Egypt/Egypt/ | Genomics Program, Children Cancer Hospital | Genomics Program, Children Cancer Hospital | Hatem et al |
| EPI_ISL_812851 | Egypt/CCHE57357-P-03/2020 | 2020-08-11 | Africa/Egypt/Egypt/ | Genomics Program, Children Cancer Hospital | Genomics Program, Children Cancer Hospital | Hatem et al |

| | | | | | | |
|---|---|---|---|---|---|---|
| EPI_ISL_812853 | Egypt/CCHE57357-P-14/2020 | 2020-06-11 | Africa/Egypt/Egypt/ | Genomics Program, Children Cancer Hospital | Genomics Program, Children Cancer Hospital | Hatem et al |
| EPI_ISL_812856 | Egypt/CCHE57357-P-20/2020 | 2020-06-26 | Africa/Egypt/Egypt/ | Genomics Program, Children Cancer Hospital | Genomics Program, Children Cancer Hospital | Hatem et al |
| EPI_ISL_812857 | Egypt/CCHE57357-P-22/2020 | 2020-08-24 | Africa/Egypt/Egypt/ | Genomics Program, Children Cancer Hospital | Genomics Program, Children Cancer Hospital | Hatem et al |
| EPI_ISL_812858 | Egypt/CCHE57357-P-24/2020 | 2020-06-21 | Africa/Egypt/Egypt/ | Genomics Program, Children Cancer Hospital | Genomics Program, Children Cancer Hospital | Hatem et al |
| EPI_ISL_812859 | Egypt/CCHE57357-P-25/2020 | 2020-07-16 | Africa/Egypt/Egypt/ | Genomics Program, Children Cancer Hospital | Genomics Program, Children Cancer Hospital | Hatem et al |
| EPI_ISL_812860 | Egypt/CCHE57357-P-26/2020 | 2020-06-30 | Africa/Egypt/Egypt/ | Genomics Program, Children Cancer Hospital | Genomics Program, Children Cancer Hospital | Hatem et al |
| EPI_ISL_812863 | Egypt/CCHE57357-P-32/2020 | 2020-06-30 | Africa/Egypt/Egypt/ | Genomics Program, Children Cancer Hospital | Genomics Program, Children Cancer Hospital | Hatem et al |
| EPI_ISL_812864 | Egypt/CCHE57357-P-33/2020 | 2020-07-12 | Africa/Egypt/Egypt/ | Genomics Program, Children Cancer Hospital | Genomics Program, Children Cancer Hospital | Hatem et al |
| EPI_ISL_812866 | Egypt/CCHE57357-P-36/2020 | 2020-07-04 | Africa/Egypt/Egypt/ | Genomics Program, Children Cancer Hospital | Genomics Program, Children Cancer Hospital | Hatem et al |
| EPI_ISL_812868 | Egypt/CCHE57357-P-38/2020 | 2020-06-30 | Africa/Egypt/Egypt/ | Genomics Program, Children Cancer Hospital | Genomics Program, Children Cancer Hospital | Hatem et al |
| EPI_ISL_812869 | Egypt/CCHE57357-P-39/2020 | 2020-07-28 | Africa/Egypt/Egypt/ | Genomics Program, Children Cancer Hospital | Genomics Program, Children Cancer Hospital | Hatem et al |
| EPI_ISL_812870 | Egypt/CCHE57357-P-41/2020 | 2020-07-23 | Africa/Egypt/Egypt/ | Genomics Program, Children Cancer Hospital | Genomics Program, Children Cancer Hospital | Hatem et al |
| EPI_ISL_812872 | Egypt/CCHE57357-P-45/2020 | 2020-06-26 | Africa/Egypt/Egypt/ | Genomics Program, Children Cancer Hospital | Genomics Program, Children Cancer Hospital | Hatem et al |
| EPI_ISL_812873 | Turkey/HSGM-19634/2020 | 2020-12-24 | Europe/Turkey/Ankara/ | Ministry of Health Turkey | Ministry of Health Turkey | Fatma Bayrakdar et al |

| | | | | | | |
|---|---|---|---|---|---|---|
| EPI_ISL_812874 | Turkey/HSGM-P631/2020 | 2020-12-29 | Europe/Turkey/Ankara/ | Ministry of Health Turkey | Ministry of Health Turkey | Fatma Bayrakdar et al |
| EPI_ISL_812875 | Turkey/HSGM-17956/2020 | 2020-12-29 | Europe/Turkey/Ankara/ | Ministry of Health Turkey | Ministry of Health Turkey | Fatma Bayrakdar et al |
| EPI_ISL_812876 | Turkey/HSGM-17950/2020 | 2020-12-31 | Europe/Turkey/Ankara/ | Ministry of Health Turkey | Ministry of Health Turkey | Fatma Bayrakdar et al |
| EPI_ISL_812877 | Turkey/HSGM-18000/2020 | 2020-12-20 | Europe/Turkey/Ankara/ | Ministry of Health Turkey | Ministry of Health Turkey | Fatma Bayrakdar et al |
| EPI_ISL_812878 | Turkey/HSGM-17435/2020 | 2020-12-20 | Europe/Turkey/Ankara/ | Ministry of Health Turkey | Ministry of Health Turkey | Fatma Bayrakdar et al |
| EPI_ISL_812879 | Turkey/HSGM-17238/2020 | 2020-12-20 | Europe/Turkey/Ankara/ | Ministry of Health Turkey | Ministry of Health Turkey | Fatma Bayrakdar et al |
| EPI_ISL_812880 | Turkey/HSGM-17240/2020 | 2020-12-20 | Europe/Turkey/Ankara/ | Ministry of Health Turkey | Ministry of Health Turkey | Fatma Bayrakdar et al |
| EPI_ISL_812881 | Turkey/HSGM-17243/2020 | 2020-12-20 | Europe/Turkey/Ankara/ | Ministry of Health Turkey | Ministry of Health Turkey | Fatma Bayrakdar et al |
| EPI_ISL_812882 | Turkey/HSGM-17466/2020 | 2020-12-20 | Europe/Turkey/Ankara/ | Ministry of Health Turkey | Ministry of Health Turkey | Fatma Bayrakdar et al |
| EPI_ISL_812883 | Turkey/HSGM-17613/2020 | 2020-12-20 | Europe/Turkey/Ankara/ | Ministry of Health Turkey | Ministry of Health Turkey | Fatma Bayrakdar et al |
| EPI_ISL_812884 | Turkey/HSGM-87/2020 | 2020-12-20 | Europe/Turkey/Ankara/ | Ministry of Health Turkey | Ministry of Health Turkey | Fatma Bayrakdar et al |
| EPI_ISL_812885 | Turkey/HSGM-P619/2020 | 2020-12-20 | Europe/Turkey/Ankara/ | Ministry of Health Turkey | Ministry of Health Turkey | Fatma Bayrakdar et al |
| EPI_ISL_812886 | Turkey/HSGM-AD1/2020 | 2020-12-20 | Europe/Turkey/Ankara/ | Ministry of Health Turkey | Ministry of Health Turkey | Fatma Bayrakdar et al |
| EPI_ISL_812887 | Turkey/HSGM-AD2/2020 | 2020-12-20 | Europe/Turkey/Ankara/ | Ministry of Health Turkey | Ministry of Health Turkey | Fatma Bayrakdar et al |
| EPI_ISL_812888 | Turkey/HSGM-AD3/2020 | 2020-12-20 | Europe/Turkey/Ankara/ | Ministry of Health Turkey | Ministry of Health Turkey | Fatma Bayrakdar et al |
| EPI_ISL_812889 | Turkey/HSGM-AD4/2020 | 2020-12-20 | Europe/Turkey/Ankara/ | Ministry of Health Turkey | Ministry of Health Turkey | Fatma Bayrakdar et al |
| EPI_ISL_812890 | Turkey/HSGM-AD5/2020 | 2020-12-21 | Europe/Turkey/Ankara/ | Ministry of Health Turkey | Ministry of Health Turkey | Fatma Bayrakdar et al |
| EPI_ISL_812891 | Turkey/HSGM-AD9/2020 | 2020-12-21 | Europe/Turkey/Ankara/ | Ministry of Health Turkey | Ministry of Health Turkey | Fatma Bayrakdar et al |
| EPI_ISL_812892 | Turkey/HSGM-AD11/2020 | 2020-12-21 | Europe/Turkey/Ankara/ | Ministry of Health Turkey | Ministry of Health Turkey | Fatma Bayrakdar et al |
| EPI_ISL_812893 | Turkey/HSGM-AD12/2020 | 2020-12-21 | Europe/Turkey/Ankara/ | Ministry of Health Turkey | Ministry of Health Turkey | Fatma Bayrakdar et al |

| | | | | | | |
|---|---|---|---|---|---|---|
| EPI_ISL_812894 | Turkey/HSGM-AD13/2020 | 2020-12-21 | Europe/Turkey/Ankara/ | Ministry of Health Turkey | Ministry of Health Turkey | Fatma Bayrakdar et al |
| EPI_ISL_812895 | Turkey/HSGM-AD14/2020 | 2020-12-21 | Europe/Turkey/Ankara/ | Ministry of Health Turkey | Ministry of Health Turkey | Fatma Bayrakdar et al |
| EPI_ISL_812896 | Turkey/HSGM-AD16/2020 | 2020-12-21 | Europe/Turkey/Ankara/ | Ministry of Health Turkey | Ministry of Health Turkey | Fatma Bayrakdar et al |
| EPI_ISL_812897 | Turkey/HSGM-AD17/2020 | 2020-12-21 | Europe/Turkey/Ankara/ | Ministry of Health Turkey | Ministry of Health Turkey | Fatma Bayrakdar et al |
| EPI_ISL_812898 | Turkey/HSGM-IST1/2020 | 2020-12-21 | Europe/Turkey/Ankara/ | Ministry of Health Turkey | Ministry of Health Turkey | Fatma Bayrakdar et al |
| EPI_ISL_812899 | Turkey/HSGM-IST2/2020 | 2020-12-21 | Europe/Turkey/Ankara/ | Ministry of Health Turkey | Ministry of Health Turkey | Fatma Bayrakdar et al |
| EPI_ISL_812900 | Turkey/HSGM-IST3/2020 | 2020-12-21 | Europe/Turkey/Ankara/ | Ministry of Health Turkey | Ministry of Health Turkey | Fatma Bayrakdar et al |
| EPI_ISL_812901 | Turkey/HSGM-IST4/2020 | 2020-12-21 | Europe/Turkey/Ankara/ | Ministry of Health Turkey | Ministry of Health Turkey | Fatma Bayrakdar et al |
| EPI_ISL_812902 | Turkey/HSGM-IST5/2020 | 2020-12-21 | Europe/Turkey/Ankara/ | Ministry of Health Turkey | Ministry of Health Turkey | Fatma Bayrakdar et al |
| EPI_ISL_812903 | Turkey/HSGM-IST6/2020 | 2020-12-21 | Europe/Turkey/Ankara/ | Ministry of Health Turkey | Ministry of Health Turkey | Fatma Bayrakdar et al |
| EPI_ISL_812904 | Turkey/HSGM-IST7/2020 | 2020-12-21 | Europe/Turkey/Ankara/ | Ministry of Health Turkey | Ministry of Health Turkey | Fatma Bayrakdar et al |
| EPI_ISL_812905 | Turkey/HSGM-IST9/2020 | 2020-12-21 | Europe/Turkey/Ankara/ | Ministry of Health Turkey | Ministry of Health Turkey | Fatma Bayrakdar et al |
| EPI_ISL_812906 | Turkey/HSGM-IST10/2020 | 2020-12-21 | Europe/Turkey/Ankara/ | Ministry of Health Turkey | Ministry of Health Turkey | Fatma Bayrakdar et al |
| EPI_ISL_812907 | Turkey/HSGM-IST11/2020 | 2020-12-21 | Europe/Turkey/Ankara/ | Ministry of Health Turkey | Ministry of Health Turkey | Fatma Bayrakdar et al |
| EPI_ISL_812908 | Turkey/HSGM-IST12/2020 | 2020-12-21 | Europe/Turkey/Ankara/ | Ministry of Health Turkey | Ministry of Health Turkey | Fatma Bayrakdar et al |
| EPI_ISL_812909 | Turkey/HSGM-IST13/2020 | 2020-12-21 | Europe/Turkey/Ankara/ | Ministry of Health Turkey | Ministry of Health Turkey | Fatma Bayrakdar et al |
| EPI_ISL_812910 | Turkey/HSGM-IST14/2020 | 2020-12-21 | Europe/Turkey/Ankara/ | Ministry of Health Turkey | Ministry of Health Turkey | Fatma Bayrakdar et al |
| EPI_ISL_812911 | Turkey/HSGM-IST15/2020 | 2020-12-21 | Europe/Turkey/Ankara/ | Ministry of Health Turkey | Ministry of Health Turkey | Fatma Bayrakdar et al |
| EPI_ISL_812912 | Turkey/HSGM-IST16/2020 | 2020-12-21 | Europe/Turkey/Ankara/ | Ministry of Health Turkey | Ministry of Health Turkey | Fatma Bayrakdar et al |
| EPI_ISL_812913 | Turkey/HSGM-IST17/2020 | 2020-12-21 | Europe/Turkey/Ankara/ | Ministry of Health Turkey | Ministry of Health Turkey | Fatma Bayrakdar et al |

| | | | | | | |
|---|---|---|---|---|---|---|
| EPI_ISL_812914 | Turkey/HSGM-IST18/2020 | 2020-12-21 | Europe/Turkey/Ankara/ | Ministry of Health Turkey | Ministry of Health Turkey | Fatma Bayrakdar et al |
| EPI_ISL_812915 | Turkey/HSGM-IST19/2020 | 2020-12-21 | Europe/Turkey/Ankara/ | Ministry of Health Turkey | Ministry of Health Turkey | Fatma Bayrakdar et al |
| EPI_ISL_812916 | Turkey/HSGM-IST20/2020 | 2020-12-21 | Europe/Turkey/Ankara/ | Ministry of Health Turkey | Ministry of Health Turkey | Fatma Bayrakdar et al |
| EPI_ISL_812917 | Turkey/HSGM-IST21/2020 | 2020-12-21 | Europe/Turkey/Ankara/ | Ministry of Health Turkey | Ministry of Health Turkey | Fatma Bayrakdar et al |
| EPI_ISL_812918 | Turkey/HSGM-IST22/2020 | 2020-12-21 | Europe/Turkey/Ankara/ | Ministry of Health Turkey | Ministry of Health Turkey | Fatma Bayrakdar et al |
| EPI_ISL_812919 | Turkey/HSGM-IST23/2020 | 2020-12-21 | Europe/Turkey/Ankara/ | Ministry of Health Turkey | Ministry of Health Turkey | Fatma Bayrakdar et al |
| EPI_ISL_812920 | Turkey/HSGM-IST24/2020 | 2020-12-21 | Europe/Turkey/Ankara/ | Ministry of Health Turkey | Ministry of Health Turkey | Fatma Bayrakdar et al |
| EPI_ISL_812921 | Turkey/HSGM-IST25/2020 | 2020-12-21 | Europe/Turkey/Ankara/ | Ministry of Health Turkey | Ministry of Health Turkey | Fatma Bayrakdar et al |
| EPI_ISL_812928 | Thailand/Bangkok-CONI-0380/2020 | 2020-12-24 | Asia/Thailand/Bangkok/ | Ramathibodi Hospital | COVID-19 Network Investigations (CONI) Alliance | Elizabeth Batty et al |
| EPI_ISL_812956 | SouthKorea/KCDC03-NCCP43326/202 | 2020-02-06 | Asia/South Korea/South Korea/ | Division of Pathogen Resource Management, Korea National Institute of Health, Korea Disease Control and Prevention Agency | Division of Pathogen Resource Management, Korea National Institute of Health, Korea Disease Control and Prevention Agency | Kim et al |
| EPI_ISL_813994 | Spain/MD-IBV-99012925/2020 | 2020-12-30 | Europe/Spain/Madrid/Madrid | Hospital General Universitario Gregorio Marav√±√≥n | SeqCOVID-SPAIN consortium/IBV(CSIC) | Dar√≠o Garc√≠a de Viedma et al |
| EPI_ISL_813997 | Spain/MD-IBV-99012930/2020 | 2020-12-30 | Europe/Spain/Madrid/Madrid | Hospital General Universitario Gregorio Marav√±√≥n | SeqCOVID-SPAIN consortium/IBV(CSIC) | Dar√≠o Garc√≠a de Viedma et al |
| EPI_ISL_814004 | Spain/MD-IBV-99012937/2020 | 2020-12-30 | Europe/Spain/Madrid/Madrid | Hospital General Universitario Gregorio Marav√±√≥n | SeqCOVID-SPAIN consortium/IBV(CSIC) | Dar√≠o Garc√≠a de Viedma et al |

| | | | | | | |
|---|---|---|---|---|---|---|
| EPI_ISL_814007 | Spain/MD-IBV-99012889/2021 | 2021-01-02 | Europe/Spain/Madrid/Madrid | Hospital General Universitario Gregorio Marav√±√≥n | SeqCOVID-SPAIN consortium/IBV(CSIC) | Dar√≠o Garc√≠a de Viedma et al |
| EPI_ISL_814042 | Spain/MD-IBV-99012884/2021 | 2021-01-01 | Europe/Spain/Madrid/Madrid | Hospital General Universitario Gregorio Marav√±√≥n | SeqCOVID-SPAIN consortium/IBV(CSIC) | Dar√≠o Garc√≠a de Viedma et al |
| EPI_ISL_814046 | Spain/MD-IBV-99012892/2021 | 2021-01-02 | Europe/Spain/Madrid/Madrid | Hospital General Universitario Gregorio Marav√±√≥n | SeqCOVID-SPAIN consortium/IBV(CSIC) | Dar√≠o Garc√≠a de Viedma et al |
| EPI_ISL_814056 | Spain/MD-IBV-99012915/2020 | 2020-12-30 | Europe/Spain/Madrid/Madrid | Hospital General Universitario Gregorio Marav√±√≥n | SeqCOVID-SPAIN consortium/IBV(CSIC) | Dar√≠o Garc√≠a de Viedma et al |
| EPI_ISL_814062 | Turkey/HSGM-29784/2020 | 2020-06-15 | Europe/Turkey/Turkey/ | Ministry of Health Turkey | Ministry of Health Turkey | Fatma Bayrakdar et al |
| EPI_ISL_814063 | Turkey/HSGM-29790/2020 | 2020-06-15 | Europe/Turkey/Turkey/ | Ministry of Health Turkey | Ministry of Health Turkey | Fatma Bayrakdar et al |
| EPI_ISL_814064 | Turkey/HSGM-31185/2020 | 2020-06-19 | Europe/Turkey/Turkey/ | Ministry of Health Turkey | Ministry of Health Turkey | Fatma Bayrakdar et al |
| EPI_ISL_814065 | Turkey/HSGM-31227/2020 | 2020-06-19 | Europe/Turkey/Turkey/ | Ministry of Health Turkey | Ministry of Health Turkey | Fatma Bayrakdar et al |
| EPI_ISL_814067 | Turkey/HSGM-30307/2020 | 2020-06-19 | Europe/Turkey/Turkey/ | Ministry of Health Turkey | Ministry of Health Turkey | Fatma Bayrakdar et al |
| EPI_ISL_814068 | Turkey/HSGM-202605/2020 | 2020-12-28 | Europe/Turkey/Turkey/ | Ministry of Health Turkey | Ministry of Health Turkey | Fatma Bayrakdar et al |
| EPI_ISL_814070 | Turkey/HSGM-202704/2020 | 2020-04-25 | Europe/Turkey/Turkey/ | Ministry of Health Turkey | Ministry of Health Turkey | Fatma Bayrakdar et al |
| EPI_ISL_814071 | Turkey/HSGM-202404/2020 | 2020-04-27 | Europe/Turkey/Turkey/ | Ministry of Health Turkey | Ministry of Health Turkey | Fatma Bayrakdar et al |
| EPI_ISL_814072 | Turkey/HSGM-200705/2020 | 2020-04-24 | Europe/Turkey/Turkey/ | Ministry of Health Turkey | Ministry of Health Turkey | Fatma Bayrakdar et al |
| EPI_ISL_814074 | Turkey/HSGM-201204/2020 | 2020-04-17 | Europe/Turkey/Turkey/ | Ministry of Health Turkey | Ministry of Health Turkey | Fatma Bayrakdar et al |
| EPI_ISL_814075 | Turkey/HSGM-200404/2020 | 2020-04-12 | Europe/Turkey/Turkey/ | Ministry of Health Turkey | Ministry of Health Turkey | Fatma Bayrakdar et al |
| EPI_ISL_814076 | Turkey/HSGM-2024042/2020 | 2020-04-04 | Europe/Turkey/Turkey/ | Ministry of Health Turkey | Ministry of Health Turkey | Fatma Bayrakdar et al |
| EPI_ISL_814077 | Turkey/HSGM-200208/2020 | 2020-04-24 | Europe/Turkey/Turkey/ | Ministry of Health Turkey | Ministry of Health Turkey | Fatma Bayrakdar et al |
| EPI_ISL_814078 | Turkey/HSGM-200408/2020 | 2020-08-02 | Europe/Turkey/Turkey/ | Ministry of Health Turkey | Ministry of Health Turkey | Fatma Bayrakdar et al |

| | | | | | | |
|---|---|---|---|---|---|---|
| EPI_ISL_814079 | Turkey/HSGM-2002082/2020 | 2020-08-04 | Europe/Turkey/Turkey/ | Ministry of Health Turkey | Ministry of Health Turkey | Fatma Bayrakdar et al |
| EPI_ISL_814080 | Turkey/HSGM-200506/2020 | 2020-08-02 | Europe/Turkey/Turkey/ | Ministry of Health Turkey | Ministry of Health Turkey | Fatma Bayrakdar et al |
| EPI_ISL_814081 | Turkey/HSGM-200606/2020 | 2020-06-05 | Europe/Turkey/Turkey/ | Ministry of Health Turkey | Ministry of Health Turkey | Fatma Bayrakdar et al |
| EPI_ISL_814082 | Turkey/HSGM-203105/2020 | 2020-06-06 | Europe/Turkey/Turkey/ | Ministry of Health Turkey | Ministry of Health Turkey | Fatma Bayrakdar et al |
| EPI_ISL_814083 | Turkey/HSGM-200706/2020 | 2020-05-31 | Europe/Turkey/Turkey/ | Ministry of Health Turkey | Ministry of Health Turkey | Fatma Bayrakdar et al |
| EPI_ISL_814084 | Turkey/HSGM-201206/2020 | 2020-06-07 | Europe/Turkey/Turkey/ | Ministry of Health Turkey | Ministry of Health Turkey | Fatma Bayrakdar et al |
| EPI_ISL_814085 | Turkey/HSGM-201606/2020 | 2020-06-12 | Europe/Turkey/Turkey/ | Ministry of Health Turkey | Ministry of Health Turkey | Fatma Bayrakdar et al |
| EPI_ISL_814086 | Turkey/HSGM-2007052/2020 | 2020-06-16 | Europe/Turkey/Turkey/ | Ministry of Health Turkey | Ministry of Health Turkey | Fatma Bayrakdar et al |
| EPI_ISL_814087 | Turkey/HSGM-200605/2020 | 2020-05-07 | Europe/Turkey/Turkey/ | Ministry of Health Turkey | Ministry of Health Turkey | Fatma Bayrakdar et al |
| EPI_ISL_814089 | Turkey/HSGM-2004082/2020 | 2020-05-15 | Europe/Turkey/Turkey/ | Ministry of Health Turkey | Ministry of Health Turkey | Fatma Bayrakdar et al |
| EPI_ISL_814091 | Turkey/HSGM-201505/2020 | 2020-08-02 | Europe/Turkey/Turkey/ | Ministry of Health Turkey | Ministry of Health Turkey | Fatma Bayrakdar et al |
| EPI_ISL_814609 | Scotland/CVR6281/2020 | 2020-12-16 | Europe/United Kingdom/Scotland/ | West of Scotland Specialist Virology Centre, NHSGGC / MRC-University of Glasgow Centre for Virus Research | COVID-19 Genomics UK (COG-UK) Consortium | Ana da Silva Filipe et al |
| EPI_ISL_814991 | Wales/PHWC-4A823D/2020 | 2020-12-20 | Europe/United Kingdom/Wales/ | Wales Specialist Virology Centre Sequencing lab: Pathogen Genomics Unit | COVID-19 Genomics UK (COG-UK) Consortium | Catherine Moore et al |
| EPI_ISL_815016 | Wales/PHWC-4A8525/2020 | 2020-12-20 | Europe/United Kingdom/Wales/ | Wales Specialist Virology Centre Sequencing lab: Pathogen Genomics Unit | COVID-19 Genomics UK (COG-UK) Consortium | Catherine Moore et al |
| EPI_ISL_815255 | Mozambique/Cento-30032996/2020 | 2020-09-19 | Africa/Mozambique/Mozambique/ | Centogene | Centogene | Peter Bauer et al |
| EPI_ISL_815256 | Iran/Cento-37431805/2020 | 2020-07-16 | Asia/Iran/Karaj/ | Centogene | Centogene | Peter Bauer et al |
| EPI_ISL_815257 | Albania/Cento-30536457/2020 | 2020-09-05 | Europe/Albania/Albania/ | Centogene | Centogene | Peter Bauer et al |
| EPI_ISL_815259 | CzechRepublic/Cento-30125238/2020 | 2020-09-24 | Europe/Czech Republic/Czech Republic/ | Centogene | Centogene | Peter Bauer et al |
| EPI_ISL_815260 | Germany/HE-Cento-36174529/2020 | 2020-07-15 | Europe/Germany/Hesse/Bad Camberg | Centogene | Centogene | Peter Bauer et al |
| EPI_ISL_815263 | Germany/HE-Cento-32407156/2020 | 2020-08-14 | Europe/Germany/Hesse/Bad Soden Am | Centogene | Centogene | Peter Bauer et al |
| EPI_ISL_815267 | Germany/RP-Cento-38135970/2020 | 2020-08-13 | Europe/Germany/Rheinland-Pfalz/Daleid | Centogene | Centogene | Peter Bauer et al |

| | | | | | | |
|---|---|---|---|---|---|---|
| EPI_ISL_815270 | Germany/HE-Cento-37510328/2020 | 2020-08-13 | Europe/Germany/Hesse/Frankfurt | Centogene | Centogene | Peter Bauer et al |
| EPI_ISL_815275 | Germany/HE-Cento-32356791/2020 | 2020-08-14 | Europe/Germany/Hesse/Frankfurt | Centogene | Centogene | Peter Bauer et al |
| EPI_ISL_815276 | Germany/HE-Cento-38037259/2020 | 2020-08-14 | Europe/Germany/Hesse/Frankfurt | Centogene | Centogene | Peter Bauer et al |
| EPI_ISL_815277 | Germany/HE-Cento-38971023/2020 | 2020-08-16 | Europe/Germany/Hesse/Frankfurt | Centogene | Centogene | Peter Bauer et al |
| EPI_ISL_815295 | Germany/BY-Cento-32989654/2020 | 2020-09-25 | Europe/Germany/Bavaria/Fuerth | Centogene | Centogene | Peter Bauer et al |
| EPI_ISL_815300 | Germany/RP-Cento-37415976/2020 | 2020-08-01 | Europe/Germany/Rheinland-Pfalz/Germ | Centogene | Centogene | Peter Bauer et al |
| EPI_ISL_815302 | Germany/HE-Cento-32986531/2020 | 2020-08-11 | Europe/Germany/Hesse/Ginsheim | Centogene | Centogene | Peter Bauer et al |
| EPI_ISL_815312 | Germany/SN-Cento-35080433/2020 | 2020-09-25 | Europe/Germany/Saxony/Hammerbach | Centogene | Centogene | Peter Bauer et al |
| EPI_ISL_815316 | Germany/HE-Cento-37830246/2020 | 2020-08-11 | Europe/Germany/Hesse/Hattersheim | Centogene | Centogene | Peter Bauer et al |
| EPI_ISL_815323 | Germany/RP-Cento-37529834/2020 | 2020-08-11 | Europe/Germany/Rheinland-Pfalz/Kaiser | Centogene | Centogene | Peter Bauer et al |
| EPI_ISL_815327 | Germany/HE-Cento-32381750/2020 | 2020-08-14 | Europe/Germany/Hesse/Langenselbold | Centogene | Centogene | Peter Bauer et al |
| EPI_ISL_815339 | Germany/BY-Cento-32750916/2020 | 2020-09-24 | Europe/Germany/Bavaria/Munich | Centogene | Centogene | Peter Bauer et al |
| EPI_ISL_815344 | Germany/BY-Cento-34063187/2020 | 2020-09-27 | Europe/Germany/Bavaria/Munich | Centogene | Centogene | Peter Bauer et al |
| EPI_ISL_815347 | Germany/BY-Cento-36048188/2020 | 2020-09-19 | Europe/Germany/Bavaria/Munich | Centogene | Centogene | Peter Bauer et al |
| EPI_ISL_815353 | Germany/HE-Cento-39816855/2020 | 2020-08-12 | Europe/Germany/Hesse/Neu-Isenburg | Centogene | Centogene | Peter Bauer et al |
| EPI_ISL_815356 | Germany/SH-Cento-37955598/2020 | 2020-09-14 | Europe/Germany/Schleswig-Holstein/No | Centogene | Centogene | Peter Bauer et al |
| EPI_ISL_815361 | Germany/BY-Cento-34772141/2020 | 2020-09-25 | Europe/Germany/Bavaria/Nuremberg | Centogene | Centogene | Peter Bauer et al |
| EPI_ISL_815364 | Germany/HE-Cento-30693781/2020 | 2020-07-19 | Europe/Germany/Hesse/Obertshausen | Centogene | Centogene | Peter Bauer et al |
| EPI_ISL_815365 | Germany/HE-Cento-32491508/2020 | 2020-08-12 | Europe/Germany/Hesse/Obertshausen | Centogene | Centogene | Peter Bauer et al |
| EPI_ISL_815371 | Germany/BW-Cento-37481093/2020 | 2020-08-11 | Europe/Germany/Baden-Wuerttemberg | Centogene | Centogene | Peter Bauer et al |
| EPI_ISL_815380 | Germany/MV-Cento-70218983/2020 | 2020-08-17 | Europe/Germany/Mecklenburg-Vorpom | Centogene | Centogene | Peter Bauer et al |
| EPI_ISL_815388 | Germany/HE-Cento-36974580/2020 | 2020-07-12 | Europe/Germany/Hesse/Wiesbaden | Centogene | Centogene | Peter Bauer et al |
| EPI_ISL_815390 | Romania/Cento-33532454/2020 | 2020-08-19 | Europe/Romania/Ploiesti/ | Centogene | Centogene | Peter Bauer et al |
| EPI_ISL_815391 | Romania/Cento-70116498/2020 | 2020-07-24 | Europe/Romania/Sanmihaiu Roman/ | Centogene | Centogene | Peter Bauer et al |
| EPI_ISL_815392 | Russia/ROS-Cento-32316789/2020 | 2020-07-22 | Europe/Russia/Rostov/Rostov-On-Don | Centogene | Centogene | Peter Bauer et al |
| EPI_ISL_815393 | Russia/ROS-Cento-37159082/2020 | 2020-07-22 | Europe/Russia/Rostov/Rostov-On-Don | Centogene | Centogene | Peter Bauer et al |
| EPI_ISL_815394 | Russia/ROS-Cento-38237649/2020 | 2020-07-22 | Europe/Russia/Rostov/Rostov-On-Don | Centogene | Centogene | Peter Bauer et al |
| EPI_ISL_815397 | Spain/MD-Cento-37563819/2020 | 2020-08-11 | Europe/Spain/Madrid/ | Centogene | Centogene | Peter Bauer et al |
| EPI_ISL_815398 | Guatemala/Cento-70148022/2020 | 2020-07-24 | North America/Guatemala/Guatemala/ | Centogene | Centogene | Peter Bauer et al |
| EPI_ISL_815399 | Guatemala/Cento-70022072/2020 | 2020-07-24 | North America/Guatemala/Guatemala/ | Centogene | Centogene | Peter Bauer et al |

| | | | | | | |
|---|---|---|---|---|---|---|
| EPI_ISL_816425 | England/SHEF-10CD9C5/2020 | 2020-12-28 | Europe/United Kingdom/England/ | Virology Department, Sheffield Teaching Hospitals NHS Foundation Trust/Department of Infection, Immunity and Cardiovascular Disease, The Medical School, University of Sheffield | COVID-19 Genomics UK (COG-UK) Consortium | Thushan de Silva et al |
| EPI_ISL_816552 | England/SHEF-10D3EBE/2021 | 2021-01-02 | Europe/United Kingdom/England/ | Virology Department, Sheffield Teaching Hospitals NHS Foundation Trust/Department of Infection, Immunity and Cardiovascular Disease, The Medical School, University of Sheffield | COVID-19 Genomics UK (COG-UK) Consortium | Thushan de Silva et al |
| EPI_ISL_816702 | England/TFCI-26F9BB2/2020 | 2020-05-06 | Europe/United Kingdom/England/ | Bioinformatics and Biostatistics Lab, Advanced Sequencing Facility | COVID-19 Genomics UK (COG-UK) Consortium | Aengus Stewart et al |
| EPI_ISL_816706 | England/TFCI-26F9BFE/2020 | 2020-05-06 | Europe/United Kingdom/England/ | Bioinformatics and Biostatistics Lab, Advanced Sequencing Facility | COVID-19 Genomics UK (COG-UK) Consortium | Aengus Stewart et al |
| EPI_ISL_816726 | England/TFCI-26F9F3E/2020 | 2020-05-04 | Europe/United Kingdom/England/ | Bioinformatics and Biostatistics Lab, Advanced Sequencing Facility | COVID-19 Genomics UK (COG-UK) Consortium | Aengus Stewart et al |
| EPI_ISL_816737 | England/TFCI-26FA00D/2020 | 2020-04-10 | Europe/United Kingdom/England/ | Bioinformatics and Biostatistics Lab, Advanced Sequencing Facility | COVID-19 Genomics UK (COG-UK) Consortium | Aengus Stewart et al |
| EPI_ISL_816758 | England/TFCI-26FA164/2020 | 2020-04-15 | Europe/United Kingdom/England/ | Bioinformatics and Biostatistics Lab, Advanced Sequencing Facility | COVID-19 Genomics UK (COG-UK) Consortium | Aengus Stewart et al |
| EPI_ISL_816769 | England/TFCI-26FA216/2020 | 2020-04-17 | Europe/United Kingdom/England/ | Bioinformatics and Biostatistics Lab, Advanced Sequencing Facility | COVID-19 Genomics UK (COG-UK) Consortium | Aengus Stewart et al |
| EPI_ISL_816869 | England/TFCI-26FAC09/2020 | 2020-12-08 | Europe/United Kingdom/England/ | Bioinformatics and Biostatistics Lab, Advanced Sequencing Facility | COVID-19 Genomics UK (COG-UK) Consortium | Aengus Stewart et al |
| EPI_ISL_816944 | England/TFCI-26FB206/2020 | 2020-12-10 | Europe/United Kingdom/England/ | Bioinformatics and Biostatistics Lab, Advanced Sequencing Facility | COVID-19 Genomics UK (COG-UK) Consortium | Aengus Stewart et al |
| EPI_ISL_817747 | Denmark/DCGC-26767/2020 | 2020-12-28 | Europe/Denmark/Hovedstaden/ | Department of Virus and Microbiological Special Diagnostics, Statens Serum Institut, Copenhagen, Denmark | Albertsen Lab, Department of Chemistry and Bioscience, Aalborg University, Denmark | Danish Covid-19 Genome Consortium et al |

| | | | | | | |
|---|---|---|---|---|---|---|
| EPI_ISL_817808 | Denmark/DCGC-26875/2020 | 2020-12-28 | Europe/Denmark/Syddanmark/ | Department of Virus and Microbiological Special Diagnostics, Statens Serum Institut, Copenhagen, Denmark | Albertsen Lab, Department of Chemistry and Bioscience, Aalborg University, Denmark | Danish Covid-19 Genome Consortium et al |
| EPI_ISL_817867 | Denmark/DCGC-26955/2020 | 2020-12-28 | Europe/Denmark/Hovedstaden/ | Department of Virus and Microbiological Special Diagnostics, Statens Serum Institut, Copenhagen, Denmark | Albertsen Lab, Department of Chemistry and Bioscience, Aalborg University, Denmark | Danish Covid-19 Genome Consortium et al |
| EPI_ISL_817964 | Denmark/DCGC-27129/2020 | 2020-12-28 | Europe/Denmark/Hovedstaden/ | Department of Virus and Microbiological Special Diagnostics, Statens Serum Institut, Copenhagen, Denmark | Albertsen Lab, Department of Chemistry and Bioscience, Aalborg University, Denmark | Danish Covid-19 Genome Consortium et al |
| EPI_ISL_819138 | Spain/IB-IBV-99012847/2020 | 2020-12-18 | Europe/Spain/Balear Islands/Palma de M | Servicio de Microbiolog√≠a, Hospital Universitario Son Espases | SeqCOVID-SPAIN consortium/IBV(CSIC) | Carla L√≥pez-Causap√© et al |
| EPI_ISL_819153 | Spain/IB-IBV-99012776/2021 | 2021-01-06 | Europe/Spain/Balear Islands/Palma de M | Servicio de Microbiolog√≠a, Hospital Universitario Son Espases | SeqCOVID-SPAIN consortium/IBV(CSIC) | Carla L√≥pez-Causap√© et al |
| EPI_ISL_819155 | Spain/IB-IBV-99012836/2021 | 2021-01-05 | Europe/Spain/Balear Islands/Palma de M | Servicio de Microbiolog√≠a, Hospital Universitario Son Espases | SeqCOVID-SPAIN consortium/IBV(CSIC) | Carla L√≥pez-Causap√© et al |
| EPI_ISL_819164 | Spain/IB-IBV-99012848/2021 | 2021-01-07 | Europe/Spain/Balear Islands/Manacor | Servicio de Microbiolog√≠a, Hospital Universitario Son Espases | SeqCOVID-SPAIN consortium/IBV(CSIC) | Carla L√≥pez-Causap√© et al |
| EPI_ISL_819165 | Spain/IB-IBV-99012800/2021 | 2021-01-06 | Europe/Spain/Balear Islands/Palma de M | Servicio de Microbiolog√≠a, Hospital Universitario Son Espases | SeqCOVID-SPAIN consortium/IBV(CSIC) | Carla L√≥pez-Causap√© et al |
| EPI_ISL_819170 | Spain/IB-IBV-99012780/2020 | 2020-12-21 | Europe/Spain/Balear Islands/Pollenca | Servicio de Microbiolog√≠a, Hospital Universitario Son Espases | SeqCOVID-SPAIN consortium/IBV(CSIC) | Carla L√≥pez-Causap√© et al |

| | | | | | | |
|---|---|---|---|---|---|---|
| EPI_ISL_819201 | Spain/IB-IBV-99012782/2020 | 2020-12-28 | Europe/Spain/Balear Islands/Santany | Servicio de Microbiolog√≠a, Hospital Universitario Son Espases | SeqCOVID-SPAIN consortium/IBV(CSIC) | Carla L√≥pez-Causap√© et al |
| EPI_ISL_819217 | USA/WY-WPHL-21000700/2021 | 2021-01-04 | North America/USA/Wyoming/ | Wyoming Public Health Laboratory | Wyoming Public Health Laboratory | Noah Hull et al |
| EPI_ISL_819229 | USA/WY-WPHL-21001586/2021 | 2021-01-05 | North America/USA/Wyoming/ | Wyoming Public Health Laboratory | Wyoming Public Health Laboratory | Noah Hull et al |
| EPI_ISL_819235 | USA/WY-WPHL-21002032/2021 | 2021-01-06 | North America/USA/Wyoming/ | Wyoming Public Health Laboratory | Wyoming Public Health Laboratory | Noah Hull et al |
| EPI_ISL_819257 | USA/WY-WPHL-21001562/2021 | 2021-01-05 | North America/USA/Wyoming/ | Wyoming Public Health Laboratory | Wyoming Public Health Laboratory | Noah Hull et al |
| EPI_ISL_819260 | USA/WY-WPHL-21001595/2021 | 2021-01-05 | North America/USA/Wyoming/ | Wyoming Public Health Laboratory | Wyoming Public Health Laboratory | Noah Hull et al |
| EPI_ISL_819263 | USA/WY-WPHL-21001627/2021 | 2021-01-05 | North America/USA/Wyoming/ | Wyoming Public Health Laboratory | Wyoming Public Health Laboratory | Noah Hull et al |
| EPI_ISL_819294 | Spain/CT-HUVH-56931/2020 | 2020-05-11 | Europe/Spain/Catalunya/ | Hospital Universitari Vall d'Hebron - Vall d'Hebron Institut de Rercerca | Hospital Universitari Vall d'Hebron | Cristina Andr√©s et al |
| EPI_ISL_819306 | Spain/CT-HUVH-14643/2020 | 2020-03-21 | Europe/Spain/Catalunya/ | Hospital Universitari Vall d'Hebron - Vall d'Hebron Institut de Rercerca | Hospital Universitari Vall d'Hebron | Cristina Andr√©s et al |
| EPI_ISL_819319 | Spain/CT-HUVH-60715/2020 | 2020-04-06 | Europe/Spain/Catalunya/ | Hospital Universitari Vall d'Hebron - Vall d'Hebron Institut de Rercerca | Hospital Universitari Vall d'Hebron | Cristina Andr√©s et al |
| EPI_ISL_819324 | Spain/CT-HUVH-33417/2020 | 2020-04-12 | Europe/Spain/Catalunya/ | Hospital Universitari Vall d'Hebron - Vall d'Hebron Institut de Rercerca | Hospital Universitari Vall d'Hebron | Cristina Andr√©s et al |
| EPI_ISL_819326 | Spain/CT-HUVH-36515/2020 | 2020-04-15 | Europe/Spain/Catalunya/ | Hospital Universitari Vall d'Hebron - Vall d'Hebron Institut de Rercerca | Hospital Universitari Vall d'Hebron | Cristina Andr√©s et al |
| EPI_ISL_819338 | Spain/CT-HUVH-53639/2020 | 2020-05-06 | Europe/Spain/Catalunya/ | Hospital Universitari Vall d'Hebron - Vall d'Hebron Institut de Rercerca | Hospital Universitari Vall d'Hebron | Cristina Andr√©s et al |
| EPI_ISL_819341 | Spain/CT-HUVH-57699/2020 | 2020-05-13 | Europe/Spain/Catalunya/ | Hospital Universitari Vall d'Hebron - Vall d'Hebron Institut de Rercerca | Hospital Universitari Vall d'Hebron | Cristina Andr√©s et al |
| EPI_ISL_819345 | Spain/CT-HUVH-60921/2020 | 2020-05-18 | Europe/Spain/Catalunya/ | Hospital Universitari Vall d'Hebron - Vall d'Hebron Institut de Rercerca | Hospital Universitari Vall d'Hebron | Cristina Andr√©s et al |

| | | | | | | |
|---|---|---|---|---|---|---|
| EPI_ISL_819647 | England/NOTT-1213D2/2020 | 2020-12-16 | Europe/United Kingdom/England/ | Queens Medical Centre, Clinical Microbiology Department / DeepSeq Nottingham | COVID-19 Genomics UK (COG-UK) Consortium | Gemma Clark et al |
| EPI_ISL_819719 | England/OXON-F42759/2020 | 2020-12-17 | Europe/United Kingdom/England/ | Oxford Viromics, NDM, University of Oxford; Oxford University Hospitals; Basingstoke and North Hampshire Hospital | COVID-19 Genomics UK (COG-UK) Consortium | Tanya Golubchik et al |
| EPI_ISL_819878 | England/LOND-12F2127/2020 | 2020-12-21 | Europe/United Kingdom/England/ | University College London, Great Ormond Street Hospital for Children NHS Foundation Trust, Imperial College Healthcare NHS Trust | COVID-19 Genomics UK (COG-UK) Consortium | Sergi Castellano et al |
| EPI_ISL_820003 | England/NORW-22274D/2020 | 2020-12-24 | Europe/United Kingdom/England/ | Quadram Institute Bioscience | COVID-19 Genomics UK (COG-UK) Consortium | Dave J. Baker et al |
| EPI_ISL_820061 | England/LOND-12F2B38/2020 | 2020-12-26 | Europe/United Kingdom/England/ | University College London, Great Ormond Street Hospital for Children NHS Foundation Trust, Imperial College Healthcare NHS Trust | COVID-19 Genomics UK (COG-UK) Consortium | Sergi Castellano et al |
| EPI_ISL_820093 | England/NORW-2229ED/2020 | 2020-12-26 | Europe/United Kingdom/England/ | Quadram Institute Bioscience | COVID-19 Genomics UK (COG-UK) Consortium | Dave J. Baker et al |
| EPI_ISL_820244 | England/NORW-2240B9/2020 | 2020-12-27 | Europe/United Kingdom/England/ | Quadram Institute Bioscience | COVID-19 Genomics UK (COG-UK) Consortium | Dave J. Baker et al |
| EPI_ISL_820508 | England/ALDP-C698A7/2020 | 2020-12-14 | Europe/United Kingdom/England/ | Lighthouse Lab in Alderley Park | Wellcome Sanger Institute for the COVID-19 Genomics UK (COG-UK) Consortium | Jacquelyn Wynn et al |
| EPI_ISL_820602 | England/ALDP-C6B571/2020 | 2020-12-14 | Europe/United Kingdom/England/ | Lighthouse Lab in Alderley Park | Wellcome Sanger Institute for the COVID-19 Genomics UK (COG-UK) Consortium | Jacquelyn Wynn et al |
| EPI_ISL_820914 | England/MILK-EBB103/2021 | 2021-01-02 | Europe/United Kingdom/England/ | Lighthouse Lab in Milton Keynes | Wellcome Sanger Institute for the COVID-19 Genomics UK (COG-UK) Consortium | The Lighthouse Lab in Milton Keynes et al |

| | | | | | | |
|---|---|---|---|---|---|---|
| EPI_ISL_821030 | England/ALDP-E306AD/2021 | 2021-01-01 | Europe/United Kingdom/England/ | Lighthouse Lab in Alderley Park | Wellcome Sanger Institute for the COVID-19 Genomics UK (COG-UK) Consortium | Jacquelyn Wynn et al |
| EPI_ISL_821105 | England/QEUH-E505AA/2021 | 2021-01-04 | Europe/United Kingdom/England/ | Lighthouse Lab in Glasgow | Wellcome Sanger Institute for the COVID-19 Genomics UK (COG-UK) Consortium | Harper VanSteenhouse et al |
| EPI_ISL_821112 | England/ALDP-E3087A/2021 | 2021-01-02 | Europe/United Kingdom/England/ | Lighthouse Lab in Alderley Park | Wellcome Sanger Institute for the COVID-19 Genomics UK (COG-UK) Consortium | Jacquelyn Wynn et al |
| EPI_ISL_821183 | England/ALDP-E38599/2021 | 2021-01-02 | Europe/United Kingdom/England/ | Lighthouse Lab in Alderley Park | Wellcome Sanger Institute for the COVID-19 Genomics UK (COG-UK) Consortium | Jacquelyn Wynn et al |
| EPI_ISL_821572 | England/MILK-E7D3BC/2021 | 2021-01-02 | Europe/United Kingdom/England/ | Lighthouse Lab in Milton Keynes | Wellcome Sanger Institute for the COVID-19 Genomics UK (COG-UK) Consortium | The Lighthouse Lab in Milton Keynes et al |
| EPI_ISL_821623 | England/CAMC-E5203E/2021 | 2021-01-02 | Europe/United Kingdom/England/ | Lighthouse Lab in Cambridge | Wellcome Sanger Institute for the COVID-19 Genomics UK (COG-UK) Consortium | Rob Howes et al |
| EPI_ISL_822036 | England/ALDP-DB1CFE/2020 | 2020-12-31 | Europe/United Kingdom/England/ | Lighthouse Lab in Alderley Park | Wellcome Sanger Institute for the COVID-19 Genomics UK (COG-UK) Consortium | Jacquelyn Wynn et al |
| EPI_ISL_822110 | England/ALDP-DCBA33/2020 | 2020-12-31 | Europe/United Kingdom/England/ | Lighthouse Lab in Alderley Park | Wellcome Sanger Institute for the COVID-19 Genomics UK (COG-UK) Consortium | Jacquelyn Wynn et al |

| | | | | | | |
|---|---|---|---|---|---|---|
| EPI_ISL_822332 | NorthernIreland/QEUH-96EE76/2020 | 2020-08-19 | Europe/United Kingdom/Northern Irelar | Lighthouse Lab in Glasgow | Wellcome Sanger Institute for the COVID-19 Genomics UK (COG-UK) Consortium | Harper VanSteenhouse et al |
| EPI_ISL_822361 | Wales/PHWC-4AF579/2020 | 2020-12-18 | Europe/United Kingdom/Wales/ | Wales Specialist Virology Centre Sequencing lab: Pathogen Genomics Unit | COVID-19 Genomics UK (COG-UK) Consortium | Catherine Moore et al |
| EPI_ISL_822448 | Wales/PHWC-4AE46E/2020 | 2020-12-24 | Europe/United Kingdom/Wales/ | Wales Specialist Virology Centre Sequencing lab: Pathogen Genomics Unit | COVID-19 Genomics UK (COG-UK) Consortium | Catherine Moore et al |
| EPI_ISL_822504 | Wales/PHWC-4AEB1E/2020 | 2020-12-26 | Europe/United Kingdom/Wales/ | Wales Specialist Virology Centre Sequencing lab: Pathogen Genomics Unit | COVID-19 Genomics UK (COG-UK) Consortium | Catherine Moore et al |
| EPI_ISL_822553 | Wales/PHWC-4AED54/2020 | 2020-12-27 | Europe/United Kingdom/Wales/ | Wales Specialist Virology Centre Sequencing lab: Pathogen Genomics Unit | COVID-19 Genomics UK (COG-UK) Consortium | Catherine Moore et al |
| EPI_ISL_822789 | Wales/PHWC-4B0302/2020 | 2020-12-31 | Europe/United Kingdom/Wales/ | Wales Specialist Virology Centre Sequencing lab: Pathogen Genomics Unit | COVID-19 Genomics UK (COG-UK) Consortium | Catherine Moore et al |
| EPI_ISL_822826 | Wales/PHWC-4B050C/2021 | 2021-01-02 | Europe/United Kingdom/Wales/ | Wales Specialist Virology Centre Sequencing lab: Pathogen Genomics Unit | COVID-19 Genomics UK (COG-UK) Consortium | Catherine Moore et al |
| EPI_ISL_822881 | Wales/PHWC-4ACFD8/2020 | 2020-12-24 | Europe/United Kingdom/Wales/ | Wales Specialist Virology Centre Sequencing lab: Pathogen Genomics Unit | COVID-19 Genomics UK (COG-UK) Consortium | Catherine Moore et al |
| EPI_ISL_822984 | Wales/PHWC-4AB795/2020 | 2020-12-18 | Europe/United Kingdom/Wales/ | Wales Specialist Virology Centre Sequencing lab: Pathogen Genomics Unit | COVID-19 Genomics UK (COG-UK) Consortium | Catherine Moore et al |
| EPI_ISL_823343 | Wales/PHWC-4AAE0A/2020 | 2020-12-19 | Europe/United Kingdom/Wales/ | Wales Specialist Virology Centre Sequencing lab: Pathogen Genomics Unit | COVID-19 Genomics UK (COG-UK) Consortium | Catherine Moore et al |
| EPI_ISL_823813 | USA/NY-NYCPHL-001747/2020 | 2020-12-11 | North America/USA/New York/New Yor | DOHMH Jamaica | New York City Public Health Laboratory | Jade Wang et al |
| EPI_ISL_823814 | USA/NY-NYCPHL-001746/2020 | 2020-12-11 | North America/USA/New York/New Yor | DOHMH Riverside | New York City Public Health Laboratory | Jade Wang et al |
| EPI_ISL_823821 | USA/NY-NYCPHL-001737/2020 | 2020-12-10 | North America/USA/New York/New Yor | DOHMH Jamaica | New York City Public Health Laboratory | Jade Wang et al |

| | | | | | | |
|---|---|---|---|---|---|---|
| EPI_ISL_823824 | USA/NY-NYCPHL-001734/2020 | 2020-12-10 | North America/USA/New York/New Yor | DOHMH Riverside | New York City Public Health Laboratory | Jade Wang et al |
| EPI_ISL_823847 | USA/NY-NYCPHL-001709/2020 | 2020-12-09 | North America/USA/New York/New Yor | DOHMH Jamaica | New York City Public Health Laboratory | Jade Wang et al |
| EPI_ISL_823851 | USA/NY-NYCPHL-001705/2020 | 2020-12-09 | North America/USA/New York/New Yor | DOHMH Fort Greene | New York City Public Health Laboratory | Jade Wang et al |
| EPI_ISL_823852 | USA/NY-NYCPHL-001704/2020 | 2020-12-09 | North America/USA/New York/New Yor | DOHMH Crown Heights | New York City Public Health Laboratory | Jade Wang et al |
| EPI_ISL_823854 | USA/NY-NYCPHL-001700/2020 | 2020-12-09 | North America/USA/New York/New Yor | DOHMH Morrisania | New York City Public Health Laboratory | Jade Wang et al |
| EPI_ISL_823865 | USA/NY-NYCPHL-001689/2020 | 2020-12-09 | North America/USA/New York/New Yor | DOHMH Central Harlem | New York City Public Health Laboratory | Jade Wang et al |
| EPI_ISL_823866 | USA/NY-NYCPHL-001688/2020 | 2020-12-09 | North America/USA/New York/New Yor | DOHMH Fort Greene | New York City Public Health Laboratory | Jade Wang et al |
| EPI_ISL_823868 | USA/NY-NYCPHL-001685/2020 | 2020-12-09 | North America/USA/New York/New Yor | DOHMH Corona | New York City Public Health Laboratory | Jade Wang et al |
| EPI_ISL_823878 | USA/NY-NYCPHL-001672/2020 | 2020-12-08 | North America/USA/New York/New Yor | DOHMH Corona | New York City Public Health Laboratory | Jade Wang et al |
| EPI_ISL_823879 | USA/NY-NYCPHL-001671/2020 | 2020-12-08 | North America/USA/New York/New Yor | DOHMH Corona | New York City Public Health Laboratory | Jade Wang et al |
| EPI_ISL_823885 | USA/NY-NYCPHL-001664/2020 | 2020-12-08 | North America/USA/New York/New Yor | DOHMH Central Harlem | New York City Public Health Laboratory | Jade Wang et al |
| EPI_ISL_823897 | USA/NY-NYCPHL-001650/2020 | 2020-12-08 | North America/USA/New York/New Yor | DOHMH PHL | New York City Public Health Laboratory | Jade Wang et al |
| EPI_ISL_823898 | USA/NY-NYCPHL-001648/2020 | 2020-12-08 | North America/USA/New York/New Yor | DOHMH Corona | New York City Public Health Laboratory | Jade Wang et al |

| | | | | | | |
|---|---|---|---|---|---|---|
| EPI_ISL_823913 | USA/NY-NYCPHL-001629/2020 | 2020-12-07 | North America/USA/New York/New Yor | DOHMH PHL | New York City Public Health Laboratory | Jade Wang et al |
| EPI_ISL_823914 | USA/NY-NYCPHL-001628/2020 | 2020-12-07 | North America/USA/New York/New Yor | DOHMH PHL | New York City Public Health Laboratory | Jade Wang et al |
| EPI_ISL_823923 | USA/NY-NYCPHL-001618/2020 | 2020-12-07 | North America/USA/New York/New Yor | DOHMH Fort Greene | New York City Public Health Laboratory | Jade Wang et al |
| EPI_ISL_823934 | USA/NY-NYCPHL-001607/2020 | 2020-12-04 | North America/USA/New York/New Yor | DOHMH Chelsea | New York City Public Health Laboratory | Jade Wang et al |
| EPI_ISL_823937 | USA/NY-NYCPHL-001604/2020 | 2020-12-04 | North America/USA/New York/New Yor | DOHMH PHL | New York City Public Health Laboratory | Jade Wang et al |
| EPI_ISL_823941 | USA/NY-NYCPHL-001600/2020 | 2020-12-04 | North America/USA/New York/New Yor | DOHMH Jamaica | New York City Public Health Laboratory | Jade Wang et al |
| EPI_ISL_823943 | USA/NY-NYCPHL-001598/2020 | 2020-12-04 | North America/USA/New York/New Yor | DOHMH Corona | New York City Public Health Laboratory | Jade Wang et al |
| EPI_ISL_823946 | USA/NY-NYCPHL-001595/2020 | 2020-12-04 | North America/USA/New York/New Yor | DOHMH Jamaica | New York City Public Health Laboratory | Jade Wang et al |
| EPI_ISL_823953 | USA/NY-NYCPHL-001587/2020 | 2020-12-03 | North America/USA/New York/New Yor | DOHMH Corona | New York City Public Health Laboratory | Jade Wang et al |
| EPI_ISL_823964 | USA/NY-NYCPHL-001576/2020 | 2020-12-03 | North America/USA/New York/New Yor | DOHMH Corona | New York City Public Health Laboratory | Jade Wang et al |
| EPI_ISL_823974 | USA/NY-NYCPHL-001563/2020 | 2020-11-05 | North America/USA/New York/New Yor | OCME Office Of Chief Medical Examiner | New York City Public Health Laboratory | Jade Wang et al |
| EPI_ISL_823979 | Netherlands/UT-RIVM-21117/2020 | 2020-12-27 | Europe/Netherlands/Utrecht/ | Dutch COVID-19 response team | National Institute for Public Health and the Environment (RIVM) | Adam Meijer et al |

| | | | | | | |
|---|---|---|---|---|---|---|
| EPI_ISL_823998 | Netherlands/UT-RIVM-21116/2020 | 2020-12-23 | Europe/Netherlands/Utrecht/ | Dutch COVID-19 response team | National Institute for Public Health and the Environment (RIVM) | Adam Meijer et al |
| EPI_ISL_824005 | Netherlands/UT-RIVM-21342/2020 | 2020-12-01 | Europe/Netherlands/Utrecht/ | Dutch COVID-19 response team | National Institute for Public Health and the Environment (RIVM) | Adam Meijer et al |
| EPI_ISL_824083 | Netherlands/NB-RIVM-21167/2020 | 2020-12-23 | Europe/Netherlands/North Brabant/ | Dutch COVID-19 response team | National Institute for Public Health and the Environment (RIVM) | Adam Meijer et al |
| EPI_ISL_824131 | Netherlands/DR-RIVM-21213/2020 | 2020-11-30 | Europe/Netherlands/Drenthe/ | Dutch COVID-19 response team | National Institute for Public Health and the Environment (RIVM) | Adam Meijer et al |
| EPI_ISL_824156 | Netherlands/LI-RIVM-21230/2020 | 2020-12-23 | Europe/Netherlands/Limburg/ | Dutch COVID-19 response team | National Institute for Public Health and the Environment (RIVM) | Adam Meijer et al |
| EPI_ISL_824157 | Netherlands/ZH-RIVM-21235/2020 | 2020-10-01 | Europe/Netherlands/South Holland/ | Dutch COVID-19 response team | National Institute for Public Health and the Environment (RIVM) | Adam Meijer et al |
| EPI_ISL_824215 | Netherlands/NH-RIVM-21295/2020 | 2020-12-29 | Europe/Netherlands/North Holland/ | Dutch COVID-19 response team | National Institute for Public Health and the Environment (RIVM) | Adam Meijer et al |
| EPI_ISL_824253 | Netherlands/NH-RIVM-10017/2021 | 2021-01-04 | Europe/Netherlands/North Holland/ | Dutch COVID-19 response team | National Institute for Public Health and the Environment (RIVM) | Adam Meijer et al |

| | | | | | | |
|---|---|---|---|---|---|---|
| EPI_ISL_824255 | Netherlands/GE-RIVM-21335/2020 | 2020-12-23 | Europe/Netherlands/Gelderland/ | Dutch COVID-19 response team | National Institute for Public Health and the Environment (RIVM) | Adam Meijer et al |
| EPI_ISL_824287 | Ecuador/USFQ-568/2020 | 2020-12-28 | South America/Ecuador/Imbabura/Otav | Institute of Microbiology, Universidad San Francisco de Quito | Institute of Microbiology, Universidad San Francisco de Quito | BelV©n Prado-Vivar et al |
| EPI_ISL_824289 | Ecuador/USFQ-571/2020 | 2020-12-28 | South America/Ecuador/Santo Domingo | Institute of Microbiology, Universidad San Francisco de Quito | Institute of Microbiology, Universidad San Francisco de Quito | BelV©n Prado-Vivar et al |
| EPI_ISL_824293 | Ecuador/USFQ-578/2021 | 2021-01-06 | South America/Ecuador/Los Rios/Valenc | Institute of Microbiology, Universidad San Francisco de Quito | Institute of Microbiology, Universidad San Francisco de Quito | BelV©n Prado-Vivar et al |
| EPI_ISL_824313 | USA/NY-NYCPHL-001766/2020 | 2020-12-14 | North America/USA/New York/New Yor | DOHMH Jamaica | New York City Public Health Laboratory | Jade Wang et al |
| EPI_ISL_824321 | USA/NY-NYCPHL-001775/2020 | 2020-12-14 | North America/USA/New York/New Yor | DOHMH Fort Greene | New York City Public Health Laboratory | Jade Wang et al |
| EPI_ISL_824322 | USA/NY-NYCPHL-001776/2020 | 2020-12-14 | North America/USA/New York/New Yor | DOHMH Fort Greene | New York City Public Health Laboratory | Jade Wang et al |
| EPI_ISL_824327 | USA/NY-NYCPHL-001783/2020 | 2020-12-14 | North America/USA/New York/New Yor | DOHMH Morrisania | New York City Public Health Laboratory | Jade Wang et al |
| EPI_ISL_824329 | USA/NY-NYCPHL-001786/2020 | 2020-12-14 | North America/USA/New York/New Yor | DOHMH Jamaica | New York City Public Health Laboratory | Jade Wang et al |
| EPI_ISL_824334 | USA/MI-MDHHS-SC22927/2020 | 2020-11-23 | North America/USA/Michigan/ | Michigan Department of Health and Human Services, Bureau of Laboratories | Michigan Department of Health and Human Services, Bureau of Laboratories | Blankenship HM et al |

| | | | | | | |
|---|---|---|---|---|---|---|
| EPI_ISL_824344 | USA/MI-MDHHS-SC22937/2020 | 2020-12-01 | North America/USA/Michigan/ | Michigan Department of Health and Human Services, Bureau of Laboratories | Michigan Department of Health and Human Services, Bureau of Laboratories | Blankenship HM et al |
| EPI_ISL_824355 | USA/MI-MDHHS-SC22948/2020 | 2020-12-01 | North America/USA/Michigan/ | Michigan Department of Health and Human Services, Bureau of Laboratories | Michigan Department of Health and Human Services, Bureau of Laboratories | Blankenship HM et al |
| EPI_ISL_824362 | USA/MI-MDHHS-SC22955/2020 | 2020-12-08 | North America/USA/Michigan/ | Michigan Department of Health and Human Services, Bureau of Laboratories | Michigan Department of Health and Human Services, Bureau of Laboratories | Blankenship HM et al |
| EPI_ISL_824363 | USA/MI-MDHHS-SC22956/2020 | 2020-12-14 | North America/USA/Michigan/ | Michigan Department of Health and Human Services, Bureau of Laboratories | Michigan Department of Health and Human Services, Bureau of Laboratories | Blankenship HM et al |
| EPI_ISL_824366 | USA/MI-MDHHS-SC22959/2020 | 2020-12-01 | North America/USA/Michigan/ | Michigan Department of Health and Human Services, Bureau of Laboratories | Michigan Department of Health and Human Services, Bureau of Laboratories | Blankenship HM et al |
| EPI_ISL_824367 | USA/MI-MDHHS-SC22960/2020 | 2020-12-10 | North America/USA/Michigan/ | Michigan Department of Health and Human Services, Bureau of Laboratories | Michigan Department of Health and Human Services, Bureau of Laboratories | Blankenship HM et al |

| | | | | | | |
|---|---|---|---|---|---|---|
| EPI_ISL_824370 | USA/MI-MDHHS-SC22963/2020 | 2020-12-03 | North America/USA/Michigan/ | Michigan Department of Health and Human Services, Bureau of Laboratories | Michigan Department of Health and Human Services, Bureau of Laboratories | Blankenship HM et al |
| EPI_ISL_824375 | USA/MI-MDHHS-SC22968/2020 | 2020-12-01 | North America/USA/Michigan/ | Michigan Department of Health and Human Services, Bureau of Laboratories | Michigan Department of Health and Human Services, Bureau of Laboratories | Blankenship HM et al |
| EPI_ISL_824379 | USA/MI-MDHHS-SC22972/2020 | 2020-12-02 | North America/USA/Michigan/ | Michigan Department of Health and Human Services, Bureau of Laboratories | Michigan Department of Health and Human Services, Bureau of Laboratories | Blankenship HM et al |
| EPI_ISL_824381 | USA/MI-MDHHS-SC22974/2020 | 2020-12-16 | North America/USA/Michigan/ | Michigan Department of Health and Human Services, Bureau of Laboratories | Michigan Department of Health and Human Services, Bureau of Laboratories | Blankenship HM et al |
| EPI_ISL_824382 | USA/MI-MDHHS-SC22975/2020 | 2020-12-01 | North America/USA/Michigan/ | Michigan Department of Health and Human Services, Bureau of Laboratories | Michigan Department of Health and Human Services, Bureau of Laboratories | Blankenship HM et al |
| EPI_ISL_824397 | USA/CA-CDPH305/2020 | 2020-12-01 | North America/USA/California/Imperial | California Department of Public Health | California Department of Public Health | CDPH IDLB COVIDNet et al |
| EPI_ISL_824398 | USA/CA-CDPH306/2020 | 2020-11-25 | North America/USA/California/Los Ange | California Department of Public Health | California Department of Public Health | CDPH IDLB COVIDNet et al |
| EPI_ISL_824404 | USA/CA-CDPH313/2020 | 2020-12-16 | North America/USA/California/Lake Cou | California Department of Public Health | California Department of Public Health | CDPH IDLB COVIDNet et al |

| | | | | | | |
|---|---|---|---|---|---|---|
| EPI_ISL_824431 | Spain/CT-HUVH-56088/2020 | 2020-07-14 | Europe/Spain/Catalunya/ | Hospital Universitari Vall d'Hebron - Vall d'Hebron Institut de Rercerca | Hospital Universitari Vall d'Hebron | Cristina Andr√©s et al |
| EPI_ISL_824439 | Spain/CT-HUVH-17304/2020 | 2020-07-17 | Europe/Spain/Catalunya/ | Hospital Universitari Vall d'Hebron - Vall d'Hebron Institut de Rercerca | Hospital Universitari Vall d'Hebron | Cristina Andr√©s et al |
| EPI_ISL_824443 | Spain/CT-HUVH-41075/2020 | 2020-07-20 | Europe/Spain/Catalunya/ | Hospital Universitari Vall d'Hebron - Vall d'Hebron Institut de Rercerca | Hospital Universitari Vall d'Hebron | Cristina Andr√©s et al |
| EPI_ISL_824452 | Spain/CT-HUVH-95855/2020 | 2020-07-22 | Europe/Spain/Catalunya/ | Hospital Universitari Vall d'Hebron - Vall d'Hebron Institut de Rercerca | Hospital Universitari Vall d'Hebron | Cristina Andr√©s et al |
| EPI_ISL_824458 | Spain/CT-HUVH-73850/2020 | 2020-07-31 | Europe/Spain/Catalunya/ | Hospital Universitari Vall d'Hebron - Vall d'Hebron Institut de Rercerca | Hospital Universitari Vall d'Hebron | Cristina Andr√©s et al |
| EPI_ISL_824459 | Spain/CT-HUVH-92875/2020 | 2020-08-03 | Europe/Spain/Catalunya/ | Hospital Universitari Vall d'Hebron - Vall d'Hebron Institut de Rercerca | Hospital Universitari Vall d'Hebron | Cristina Andr√©s et al |
| EPI_ISL_824460 | Spain/CT-HUVH-96666/2020 | 2020-08-03 | Europe/Spain/Catalunya/ | Hospital Universitari Vall d'Hebron - Vall d'Hebron Institut de Rercerca | Hospital Universitari Vall d'Hebron | Cristina Andr√©s et al |
| EPI_ISL_824461 | Spain/CT-HUVH-06944/2020 | 2020-08-03 | Europe/Spain/Catalunya/ | Hospital Universitari Vall d'Hebron - Vall d'Hebron Institut de Rercerca | Hospital Universitari Vall d'Hebron | Cristina Andr√©s et al |
| EPI_ISL_824465 | Spain/CT-HUVH-00320/2020 | 2020-08-11 | Europe/Spain/Catalunya/ | Hospital Universitari Vall d'Hebron - Vall d'Hebron Institut de Rercerca | Hospital Universitari Vall d'Hebron | Cristina Andr√©s et al |
| EPI_ISL_824469 | Spain/CT-HUVH-99492/2020 | 2020-08-19 | Europe/Spain/Catalunya/ | Hospital Universitari Vall d'Hebron - Vall d'Hebron Institut de Rercerca | Hospital Universitari Vall d'Hebron | Cristina Andr√©s et al |
| EPI_ISL_824470 | Spain/CT-HUVH-50243/2020 | 2020-08-22 | Europe/Spain/Catalunya/ | Hospital Universitari Vall d'Hebron - Vall d'Hebron Institut de Rercerca | Hospital Universitari Vall d'Hebron | Cristina Andr√©s et al |
| EPI_ISL_824471 | Spain/CT-HUVH-86476/2020 | 2020-08-24 | Europe/Spain/Catalunya/ | Hospital Universitari Vall d'Hebron - Vall d'Hebron Institut de Rercerca | Hospital Universitari Vall d'Hebron | Cristina Andr√©s et al |
| EPI_ISL_824472 | Spain/CT-HUVH-65622/2020 | 2020-08-24 | Europe/Spain/Catalunya/ | Hospital Universitari Vall d'Hebron - Vall d'Hebron Institut de Rercerca | Hospital Universitari Vall d'Hebron | Cristina Andr√©s et al |

| | | | | | | |
|---|---|---|---|---|---|---|
| EPI_ISL_824473 | Spain/CT-HUVH-65608/2020 | 2020-08-24 | Europe/Spain/Catalunya/ | Hospital Universitari Vall d'Hebron - Vall d'Hebron Institut de Rercerca | Hospital Universitari Vall d'Hebron | Cristina Andr√©s et al |
| EPI_ISL_824474 | Spain/CT-HUVH-28830/2020 | 2020-08-24 | Europe/Spain/Catalunya/ | Hospital Universitari Vall d'Hebron - Vall d'Hebron Institut de Rercerca | Hospital Universitari Vall d'Hebron | Cristina Andr√©s et al |
| EPI_ISL_824477 | Spain/CT-HUVH-78949/2020 | 2020-08-25 | Europe/Spain/Catalunya/ | Hospital Universitari Vall d'Hebron - Vall d'Hebron Institut de Rercerca | Hospital Universitari Vall d'Hebron | Cristina Andr√©s et al |
| EPI_ISL_824478 | Spain/CT-HUVH-97173/2020 | 2020-08-31 | Europe/Spain/Catalunya/ | Hospital Universitari Vall d'Hebron - Vall d'Hebron Institut de Rercerca | Hospital Universitari Vall d'Hebron | Cristina Andr√©s et al |
| EPI_ISL_824480 | Spain/CT-HUVH-59286/2020 | 2020-08-31 | Europe/Spain/Catalunya/ | Hospital Universitari Vall d'Hebron - Vall d'Hebron Institut de Rercerca | Hospital Universitari Vall d'Hebron | Cristina Andr√©s et al |
| EPI_ISL_824483 | Spain/CT-HUVH-05803/2020 | 2020-09-07 | Europe/Spain/Catalunya/ | Hospital Universitari Vall d'Hebron - Vall d'Hebron Institut de Rercerca | Hospital Universitari Vall d'Hebron | Cristina Andr√©s et al |
| EPI_ISL_824484 | Spain/CT-HUVH-84736/2020 | 2020-09-07 | Europe/Spain/Catalunya/ | Hospital Universitari Vall d'Hebron - Vall d'Hebron Institut de Rercerca | Hospital Universitari Vall d'Hebron | Cristina Andr√©s et al |
| EPI_ISL_824488 | Spain/CT-HUVH-70267/2020 | 2020-09-14 | Europe/Spain/Catalunya/ | Hospital Universitari Vall d'Hebron - Vall d'Hebron Institut de Rercerca | Hospital Universitari Vall d'Hebron | Cristina Andr√©s et al |
| EPI_ISL_824490 | Spain/CT-HUVH-44945/2020 | 2020-09-15 | Europe/Spain/Catalunya/ | Hospital Universitari Vall d'Hebron - Vall d'Hebron Institut de Rercerca | Hospital Universitari Vall d'Hebron | Cristina Andr√©s et al |
| EPI_ISL_824492 | Spain/CT-HUVH-70647/2020 | 2020-09-21 | Europe/Spain/Catalunya/ | Hospital Universitari Vall d'Hebron - Vall d'Hebron Institut de Rercerca | Hospital Universitari Vall d'Hebron | Cristina Andr√©s et al |
| EPI_ISL_824493 | Spain/CT-HUVH-85144/2020 | 2020-09-21 | Europe/Spain/Catalunya/ | Hospital Universitari Vall d'Hebron - Vall d'Hebron Institut de Rercerca | Hospital Universitari Vall d'Hebron | Cristina Andr√©s et al |
| EPI_ISL_824494 | Spain/CT-HUVH-96173/2020 | 2020-09-21 | Europe/Spain/Catalunya/ | Hospital Universitari Vall d'Hebron - Vall d'Hebron Institut de Rercerca | Hospital Universitari Vall d'Hebron | Cristina Andr√©s et al |
| EPI_ISL_824495 | Spain/CT-HUVH-50694/2020 | 2020-09-28 | Europe/Spain/Catalunya/ | Hospital Universitari Vall d'Hebron - Vall d'Hebron Institut de Rercerca | Hospital Universitari Vall d'Hebron | Cristina Andr√©s et al |

| | | | | | | |
|---|---|---|---|---|---|---|
| EPI_ISL_824496 | Spain/CT-HUVH-50689/2020 | 2020-09-28 | Europe/Spain/Catalunya/ | Hospital Universitari Vall d'Hebron - Vall d'Hebron Institut de Rercerca | Hospital Universitari Vall d'Hebron | Cristina Andr√©s et al |
| EPI_ISL_824497 | Spain/CT-HUVH-50711/2020 | 2020-09-28 | Europe/Spain/Catalunya/ | Hospital Universitari Vall d'Hebron - Vall d'Hebron Institut de Rercerca | Hospital Universitari Vall d'Hebron | Cristina Andr√©s et al |
| EPI_ISL_824499 | Spain/CT-HUVH-75603/2020 | 2020-09-28 | Europe/Spain/Catalunya/ | Hospital Universitari Vall d'Hebron - Vall d'Hebron Institut de Rercerca | Hospital Universitari Vall d'Hebron | Cristina Andr√©s et al |
| EPI_ISL_824500 | Spain/CT-HUVH-61535/2020 | 2020-09-28 | Europe/Spain/Catalunya/ | Hospital Universitari Vall d'Hebron - Vall d'Hebron Institut de Rercerca | Hospital Universitari Vall d'Hebron | Cristina Andr√©s et al |
| EPI_ISL_824501 | Spain/CT-HUVH-99903/2020 | 2020-09-29 | Europe/Spain/Catalunya/ | Hospital Universitari Vall d'Hebron - Vall d'Hebron Institut de Rercerca | Hospital Universitari Vall d'Hebron | Cristina Andr√©s et al |
| EPI_ISL_824502 | Spain/CT-HUVH-11378/2020 | 2020-09-30 | Europe/Spain/Catalunya/ | Hospital Universitari Vall d'Hebron - Vall d'Hebron Institut de Rercerca | Hospital Universitari Vall d'Hebron | Cristina Andr√©s et al |
| EPI_ISL_824504 | Spain/CT-HUVH-14585/2020 | 2020-10-05 | Europe/Spain/Catalunya/ | Hospital Universitari Vall d'Hebron - Vall d'Hebron Institut de Rercerca | Hospital Universitari Vall d'Hebron | Cristina Andr√©s et al |
| EPI_ISL_824505 | Spain/CT-HUVH-11779/2020 | 2020-10-05 | Europe/Spain/Catalunya/ | Hospital Universitari Vall d'Hebron - Vall d'Hebron Institut de Rercerca | Hospital Universitari Vall d'Hebron | Cristina Andr√©s et al |
| EPI_ISL_824508 | Spain/CT-HUVH-69395/2020 | 2020-10-06 | Europe/Spain/Catalunya/ | Hospital Universitari Vall d'Hebron - Vall d'Hebron Institut de Rercerca | Hospital Universitari Vall d'Hebron | Cristina Andr√©s et al |
| EPI_ISL_824509 | Spain/CT-HUVH-62100/2020 | 2020-10-09 | Europe/Spain/Catalunya/ | Hospital Universitari Vall d'Hebron - Vall d'Hebron Institut de Rercerca | Hospital Universitari Vall d'Hebron | Cristina Andr√©s et al |
| EPI_ISL_824512 | Spain/CT-HUVH-00983/2020 | 2020-10-13 | Europe/Spain/Catalunya/ | Hospital Universitari Vall d'Hebron - Vall d'Hebron Institut de Rercerca | Hospital Universitari Vall d'Hebron | Cristina Andr√©s et al |
| EPI_ISL_824517 | USA/NM-DOH-2020553876/2020 | 2020-11-20 | North America/USA/New Mexico/ | New Mexico Department of Health Scientific Laboratory | New Mexico Department of Health Scientific Laboratory | Ellie Johnson et al |
| EPI_ISL_824518 | USA/NM-DOH-2020557982/2020 | 2020-11-22 | North America/USA/New Mexico/ | New Mexico Department of Health Scientific Laboratory | New Mexico Department of Health Scientific Laboratory | Ellie Johnson et al |

| | | | | | | |
|---|---|---|---|---|---|---|
| EPI_ISL_824533 | USA/NM-DOH-2020646581/2020 | 2020-12-29 | North America/USA/New Mexico/ | New Mexico Department of Health Scientific Laboratory | New Mexico Department of Health Scientific Laboratory | Ellie Johnson et al |
| EPI_ISL_824535 | USA/NM-DOH-2020646685/2020 | 2020-12-29 | North America/USA/New Mexico/ | New Mexico Department of Health Scientific Laboratory | New Mexico Department of Health Scientific Laboratory | Ellie Johnson et al |
| EPI_ISL_824549 | USA/NM-DOH-2020649269/2020 | 2020-12-30 | North America/USA/New Mexico/ | New Mexico Department of Health Scientific Laboratory | New Mexico Department of Health Scientific Laboratory | Ellie Johnson et al |
| EPI_ISL_824551 | USA/NM-DOH-2020649858/2020 | 2020-12-30 | North America/USA/New Mexico/ | New Mexico Department of Health Scientific Laboratory | New Mexico Department of Health Scientific Laboratory | Ellie Johnson et al |
| EPI_ISL_824560 | USA/CA-CSMC147/2020 | 2020-12-20 | North America/USA/California/Los Ange | Cedars-Sinai Medical Center, Department of Pathology & Laboratory Medicine, Molecular Pathology Laboratory | Cedars-Sinai Medical Center, Molecular Pathology Laboratory of Department of Pathology & Laboratory Medicine and Genomic Core | Wenjuan Zhang et al |
| EPI_ISL_824568 | USA/CA-CSMC155/2020 | 2020-12-27 | North America/USA/California/Los Ange | Cedars-Sinai Medical Center, Department of Pathology & Laboratory Medicine, Molecular Pathology Laboratory | Cedars-Sinai Medical Center, Molecular Pathology Laboratory of Department of Pathology & Laboratory Medicine and Genomic Core | Wenjuan Zhang et al |

| | | | | | | |
|---|---|---|---|---|---|---|
| EPI_ISL_824569 | USA/CA-CSMC156/2020 | 2020-12-27 | North America/USA/California/Los Ange | Cedars-Sinai Medical Center, Department of Pathology & Laboratory Medicine, Molecular Pathology Laboratory | Cedars-Sinai Medical Center, Molecular Pathology Laboratory of Department of Pathology & Laboratory Medicine and Genomic Core | Wenjuan Zhang et al |
| EPI_ISL_824574 | USA/CA-CSMC161/2020 | 2020-12-27 | North America/USA/California/Los Ange | Cedars-Sinai Medical Center, Department of Pathology & Laboratory Medicine, Molecular Pathology Laboratory | Cedars-Sinai Medical Center, Molecular Pathology Laboratory of Department of Pathology & Laboratory Medicine and Genomic Core | Wenjuan Zhang et al |
| EPI_ISL_824575 | USA/CA-CSMC162/2020 | 2020-12-27 | North America/USA/California/Los Ange | Cedars-Sinai Medical Center, Department of Pathology & Laboratory Medicine, Molecular Pathology Laboratory | Cedars-Sinai Medical Center, Molecular Pathology Laboratory of Department of Pathology & Laboratory Medicine and Genomic Core | Wenjuan Zhang et al |
| EPI_ISL_824583 | USA/CA-CSMC172/2020 | 2020-12-20 | North America/USA/California/Los Ange | Cedars-Sinai Medical Center, Department of Pathology & Laboratory Medicine, Molecular Pathology Laboratory | Cedars-Sinai Medical Center, Molecular Pathology Laboratory of Department of Pathology & Laboratory Medicine and Genomic Core | Wenjuan Zhang et al |

| | | | | | | |
|---|---|---|---|---|---|---|
| EPI_ISL_824590 | USA/CA-CSMC179/2020 | 2020-12-26 | North America/USA/California/Los Ange | Cedars-Sinai Medical Center, Department of Pathology & Laboratory Medicine, Molecular Pathology Laboratory | Cedars-Sinai Medical Center, Molecular Pathology Laboratory of Department of Pathology & Laboratory Medicine and Genomic Core | Wenjuan Zhang et al |
| EPI_ISL_824598 | USA/CA-CSMC187/2020 | 2020-12-27 | North America/USA/California/Los Ange | Cedars-Sinai Medical Center, Department of Pathology & Laboratory Medicine, Molecular Pathology Laboratory | Cedars-Sinai Medical Center, Molecular Pathology Laboratory of Department of Pathology & Laboratory Medicine and Genomic Core | Wenjuan Zhang et al |
| EPI_ISL_824599 | USA/CA-CSMC189/2020 | 2020-12-27 | North America/USA/California/Los Ange | Cedars-Sinai Medical Center, Department of Pathology & Laboratory Medicine, Molecular Pathology Laboratory | Cedars-Sinai Medical Center, Molecular Pathology Laboratory of Department of Pathology & Laboratory Medicine and Genomic Core | Wenjuan Zhang et al |
| EPI_ISL_824606 | USA/CA-CSMC196/2020 | 2020-12-20 | North America/USA/California/Los Ange | Cedars-Sinai Medical Center, Department of Pathology & Laboratory Medicine, Molecular Pathology Laboratory | Cedars-Sinai Medical Center, Molecular Pathology Laboratory of Department of Pathology & Laboratory Medicine and Genomic Core | Wenjuan Zhang et al |

| | | | | | | |
|---|---|---|---|---|---|---|
| EPI_ISL_824610 | USA/CA-CSMC201/2020 | 2020-12-27 | North America/USA/California/Los Ange | Cedars-Sinai Medical Center, Department of Pathology & Laboratory Medicine, Molecular Pathology Laboratory | Cedars-Sinai Medical Center, Molecular Pathology Laboratory of Department of Pathology & Laboratory Medicine and Genomic Core | Wenjuan Zhang et al |
| EPI_ISL_824611 | USA/CA-CSMC202/2020 | 2020-12-27 | North America/USA/California/Los Ange | Cedars-Sinai Medical Center, Department of Pathology & Laboratory Medicine, Molecular Pathology Laboratory | Cedars-Sinai Medical Center, Molecular Pathology Laboratory of Department of Pathology & Laboratory Medicine and Genomic Core | Wenjuan Zhang et al |
| EPI_ISL_824613 | USA/CA-CSMC204/2020 | 2020-12-26 | North America/USA/California/Los Ange | Cedars-Sinai Medical Center, Department of Pathology & Laboratory Medicine, Molecular Pathology Laboratory | Cedars-Sinai Medical Center, Molecular Pathology Laboratory of Department of Pathology & Laboratory Medicine and Genomic Core | Wenjuan Zhang et al |
| EPI_ISL_824618 | USA/CA-CSMC210/2020 | 2020-12-27 | North America/USA/California/Los Ange | Cedars-Sinai Medical Center, Department of Pathology & Laboratory Medicine, Molecular Pathology Laboratory | Cedars-Sinai Medical Center, Molecular Pathology Laboratory of Department of Pathology & Laboratory Medicine and Genomic Core | Wenjuan Zhang et al |

| | | | | | | |
|---|---|---|---|---|---|---|
| EPI_ISL_824621 | USA/CA-CSMC213/2020 | 2020-12-27 | North America/USA/California/Los Ange | Cedars-Sinai Medical Center, Department of Pathology & Laboratory Medicine, Molecular Pathology Laboratory | Cedars-Sinai Medical Center, Molecular Pathology Laboratory of Department of Pathology & Laboratory Medicine and Genomic Core | Wenjuan Zhang et al |
| EPI_ISL_824634 | USA/CA-CSMC226/2020 | 2020-12-27 | North America/USA/California/Los Ange | Cedars-Sinai Medical Center, Department of Pathology & Laboratory Medicine, Molecular Pathology Laboratory | Cedars-Sinai Medical Center, Molecular Pathology Laboratory of Department of Pathology & Laboratory Medicine and Genomic Core | Wenjuan Zhang et al |
| EPI_ISL_824648 | USA/CA-CSMC240/2020 | 2020-12-26 | North America/USA/California/Los Ange | Cedars-Sinai Medical Center, Department of Pathology & Laboratory Medicine, Molecular Pathology Laboratory | Cedars-Sinai Medical Center, Molecular Pathology Laboratory of Department of Pathology & Laboratory Medicine and Genomic Core | Wenjuan Zhang et al |
| EPI_ISL_824658 | USA/CA-CSMC251/2020 | 2020-12-21 | North America/USA/California/Los Ange | Cedars-Sinai Medical Center, Department of Pathology & Laboratory Medicine, Molecular Pathology Laboratory | Cedars-Sinai Medical Center, Molecular Pathology Laboratory of Department of Pathology & Laboratory Medicine and Genomic Core | Wenjuan Zhang et al |

| | | | | | | |
|---|---|---|---|---|---|---|
| EPI_ISL_824660 | USA/CA-CSMC253/2020 | 2020-12-26 | North America/USA/California/Los Ange | Cedars-Sinai Medical Center, Department of Pathology & Laboratory Medicine, Molecular Pathology Laboratory | Cedars-Sinai Medical Center, Molecular Pathology Laboratory of Department of Pathology & Laboratory Medicine and Genomic Core | Wenjuan Zhang et al |
| EPI_ISL_824661 | USA/CA-CSMC254/2020 | 2020-12-19 | North America/USA/California/Los Ange | Cedars-Sinai Medical Center, Department of Pathology & Laboratory Medicine, Molecular Pathology Laboratory | Cedars-Sinai Medical Center, Molecular Pathology Laboratory of Department of Pathology & Laboratory Medicine and Genomic Core | Wenjuan Zhang et al |
| EPI_ISL_824663 | USA/CA-CSMC257/2020 | 2020-12-27 | North America/USA/California/Los Ange | Cedars-Sinai Medical Center, Department of Pathology & Laboratory Medicine, Molecular Pathology Laboratory | Cedars-Sinai Medical Center, Molecular Pathology Laboratory of Department of Pathology & Laboratory Medicine and Genomic Core | Wenjuan Zhang et al |
| EPI_ISL_824670 | USA/CA-CSMC264/2020 | 2020-11-22 | North America/USA/California/Los Ange | Cedars-Sinai Medical Center, Department of Pathology & Laboratory Medicine, Molecular Pathology Laboratory | Cedars-Sinai Medical Center, Molecular Pathology Laboratory of Department of Pathology & Laboratory Medicine and Genomic Core | Wenjuan Zhang et al |

| | | | | | | |
|---|---|---|---|---|---|---|
| EPI_ISL_824671 | USA/CA-CSMC265/2020 | 2020-12-26 | North America/USA/California/Los Ange | Cedars-Sinai Medical Center, Department of Pathology & Laboratory Medicine, Molecular Pathology Laboratory | Cedars-Sinai Medical Center, Molecular Pathology Laboratory of Department of Pathology & Laboratory Medicine and Genomic Core | Wenjuan Zhang et al |
| EPI_ISL_824679 | USA/CA-CSMC273/2020 | 2020-12-08 | North America/USA/California/Los Ange | Cedars-Sinai Medical Center, Department of Pathology & Laboratory Medicine, Molecular Pathology Laboratory | Cedars-Sinai Medical Center, Molecular Pathology Laboratory of Department of Pathology & Laboratory Medicine and Genomic Core | Wenjuan Zhang et al |
| EPI_ISL_824682 | USA/CA-CSMC276/2020 | 2020-11-22 | North America/USA/California/Los Ange | Cedars-Sinai Medical Center, Department of Pathology & Laboratory Medicine, Molecular Pathology Laboratory | Cedars-Sinai Medical Center, Molecular Pathology Laboratory of Department of Pathology & Laboratory Medicine and Genomic Core | Wenjuan Zhang et al |
| EPI_ISL_824688 | USA/CA-CSMC282/2020 | 2020-12-26 | North America/USA/California/Los Ange | Cedars-Sinai Medical Center, Department of Pathology & Laboratory Medicine, Molecular Pathology Laboratory | Cedars-Sinai Medical Center, Molecular Pathology Laboratory of Department of Pathology & Laboratory Medicine and Genomic Core | Wenjuan Zhang et al |

| | | | | | | |
|---|---|---|---|---|---|---|
| EPI_ISL_824692 | USA/CA-CSMC286/2020 | 2020-12-21 | North America/USA/California/Los Ange | Cedars-Sinai Medical Center, Department of Pathology & Laboratory Medicine, Molecular Pathology Laboratory | Cedars-Sinai Medical Center, Molecular Pathology Laboratory of Department of Pathology & Laboratory Medicine and Genomic Core | Wenjuan Zhang et al |
| EPI_ISL_824694 | USA/CA-CSMC288/2020 | 2020-11-22 | North America/USA/California/Los Ange | Cedars-Sinai Medical Center, Department of Pathology & Laboratory Medicine, Molecular Pathology Laboratory | Cedars-Sinai Medical Center, Molecular Pathology Laboratory of Department of Pathology & Laboratory Medicine and Genomic Core | Wenjuan Zhang et al |
| EPI_ISL_824695 | USA/CA-CSMC289/2020 | 2020-12-27 | North America/USA/California/Los Ange | Cedars-Sinai Medical Center, Department of Pathology & Laboratory Medicine, Molecular Pathology Laboratory | Cedars-Sinai Medical Center, Molecular Pathology Laboratory of Department of Pathology & Laboratory Medicine and Genomic Core | Wenjuan Zhang et al |
| EPI_ISL_824702 | USA/CA-CSMC296/2020 | 2020-12-07 | North America/USA/California/Los Ange | Cedars-Sinai Medical Center, Department of Pathology & Laboratory Medicine, Molecular Pathology Laboratory | Cedars-Sinai Medical Center, Molecular Pathology Laboratory of Department of Pathology & Laboratory Medicine and Genomic Core | Wenjuan Zhang et al |

| | | | | | | |
|---|---|---|---|---|---|---|
| EPI_ISL_824710 | USA/CA-CSMC304/2020 | 2020-12-19 | North America/USA/California/Los Ange | Cedars-Sinai Medical Center, Department of Pathology & Laboratory Medicine, Molecular Pathology Laboratory | Cedars-Sinai Medical Center, Molecular Pathology Laboratory of Department of Pathology & Laboratory Medicine and Genomic Core | Wenjuan Zhang et al |
| EPI_ISL_824713 | USA/CA-CSMC307/2020 | 2020-12-26 | North America/USA/California/Los Ange | Cedars-Sinai Medical Center, Department of Pathology & Laboratory Medicine, Molecular Pathology Laboratory | Cedars-Sinai Medical Center, Molecular Pathology Laboratory of Department of Pathology & Laboratory Medicine and Genomic Core | Wenjuan Zhang et al |
| EPI_ISL_824719 | USA/CA-CSMC313/2020 | 2020-12-27 | North America/USA/California/Los Ange | Cedars-Sinai Medical Center, Department of Pathology & Laboratory Medicine, Molecular Pathology Laboratory | Cedars-Sinai Medical Center, Molecular Pathology Laboratory of Department of Pathology & Laboratory Medicine and Genomic Core | Wenjuan Zhang et al |
| EPI_ISL_824722 | USA/CA-CSMC316/2020 | 2020-12-19 | North America/USA/California/Los Ange | Cedars-Sinai Medical Center, Department of Pathology & Laboratory Medicine, Molecular Pathology Laboratory | Cedars-Sinai Medical Center, Molecular Pathology Laboratory of Department of Pathology & Laboratory Medicine and Genomic Core | Wenjuan Zhang et al |

| | | | | | | |
|---|---|---|---|---|---|---|
| EPI_ISL_824724 | USA/CA-CSMC318/2020 | 2020-12-26 | North America/USA/California/Los Ange | Cedars-Sinai Medical Center, Department of Pathology & Laboratory Medicine, Molecular Pathology Laboratory | Cedars-Sinai Medical Center, Molecular Pathology Laboratory of Department of Pathology & Laboratory Medicine and Genomic Core | Wenjuan Zhang et al |
| EPI_ISL_824725 | USA/CA-CSMC319/2020 | 2020-12-07 | North America/USA/California/Los Ange | Cedars-Sinai Medical Center, Department of Pathology & Laboratory Medicine, Molecular Pathology Laboratory | Cedars-Sinai Medical Center, Molecular Pathology Laboratory of Department of Pathology & Laboratory Medicine and Genomic Core | Wenjuan Zhang et al |
| EPI_ISL_824730 | USA/CA-CSMC324/2020 | 2020-12-19 | North America/USA/California/Los Ange | Cedars-Sinai Medical Center, Department of Pathology & Laboratory Medicine, Molecular Pathology Laboratory | Cedars-Sinai Medical Center, Molecular Pathology Laboratory of Department of Pathology & Laboratory Medicine and Genomic Core | Wenjuan Zhang et al |
| EPI_ISL_824732 | USA/CA-CSMC326/2020 | 2020-12-20 | North America/USA/California/Los Ange | Cedars-Sinai Medical Center, Department of Pathology & Laboratory Medicine, Molecular Pathology Laboratory | Cedars-Sinai Medical Center, Molecular Pathology Laboratory of Department of Pathology & Laboratory Medicine and Genomic Core | Wenjuan Zhang et al |

| | | | | | | |
|---|---|---|---|---|---|---|
| EPI_ISL_824738 | USA/CA-CSMC333/2020 | 2020-12-21 | North America/USA/California/Los Ange | Cedars-Sinai Medical Center, Department of Pathology & Laboratory Medicine, Molecular Pathology Laboratory | Cedars-Sinai Medical Center, Molecular Pathology Laboratory of Department of Pathology & Laboratory Medicine and Genomic Core | Wenjuan Zhang et al |
| EPI_ISL_824742 | Germany/SH-ChVir21104/2020 | 2020-12-03 | Europe/Germany/Schleswig-Holstein/ | A. Krumbholz, Labor Dr. Krause und Kollegen MVZ GmbH, Kiel | Charit√© Universit√§tsmedizin Berlin, Institut f√ºr Virologie | Victor M Corman et al |
| EPI_ISL_824749 | Germany/SH-ChVir21111/2020 | 2020-12-04 | Europe/Germany/Schleswig-Holstein/ | A. Krumbholz, Labor Dr. Krause und Kollegen MVZ GmbH, Kiel | Charit√© Universit√§tsmedizin Berlin, Institut f√ºr Virologie | Victor M Corman et al |
| EPI_ISL_824750 | Germany/SH-ChVir21112/2020 | 2020-12-04 | Europe/Germany/Schleswig-Holstein/ | A. Krumbholz, Labor Dr. Krause und Kollegen MVZ GmbH, Kiel | Charit√© Universit√§tsmedizin Berlin, Institut f√ºr Virologie | Victor M Corman et al |
| EPI_ISL_824751 | Germany/SH-ChVir21113/2020 | 2020-12-04 | Europe/Germany/Schleswig-Holstein/ | A. Krumbholz, Labor Dr. Krause und Kollegen MVZ GmbH, Kiel | Charit√© Universit√§tsmedizin Berlin, Institut f√ºr Virologie | Victor M Corman et al |
| EPI_ISL_824753 | Germany/SH-ChVir21115/2020 | 2020-12-03 | Europe/Germany/Schleswig-Holstein/ | A. Krumbholz, Labor Dr. Krause und Kollegen MVZ GmbH, Kiel | Charit√© Universit√§tsmedizin Berlin, Institut f√ºr Virologie | Victor M Corman et al |
| EPI_ISL_824754 | Germany/SH-ChVir21116/2020 | 2020-12-04 | Europe/Germany/Schleswig-Holstein/ | A. Krumbholz, Labor Dr. Krause und Kollegen MVZ GmbH, Kiel | Charit√© Universit√§tsmedizin Berlin, Institut f√ºr Virologie | Victor M Corman et al |
| EPI_ISL_824756 | Germany/SH-ChVir21119/2020 | 2020-12-05 | Europe/Germany/Schleswig-Holstein/ | A. Krumbholz, Labor Dr. Krause und Kollegen MVZ GmbH, Kiel | Charit√© Universit√§tsmedizin Berlin, Institut f√ºr Virologie | Victor M Corman et al |

| | | | | | | |
|---|---|---|---|---|---|---|
| EPI_ISL_824758 | Germany/SH-ChVir21121/2020 | 2020-12-04 | Europe/Germany/Schleswig-Holstein/ | A. Krumbholz, Labor Dr. Krause und Kollegen MVZ GmbH, Kiel | Charit√© Universit√§tsmedizin Berlin, Institut f√ºr Virologie | Victor M Corman et al |
| EPI_ISL_824759 | Germany/SH-ChVir21122/2020 | 2020-12-05 | Europe/Germany/Schleswig-Holstein/ | A. Krumbholz, Labor Dr. Krause und Kollegen MVZ GmbH, Kiel | Charit√© Universit√§tsmedizin Berlin, Institut f√ºr Virologie | Victor M Corman et al |
| EPI_ISL_824763 | Germany/SH-ChVir21127/2020 | 2020-12-05 | Europe/Germany/Schleswig-Holstein/ | A. Krumbholz, Labor Dr. Krause und Kollegen MVZ GmbH, Kiel | Charit√© Universit√§tsmedizin Berlin, Institut f√ºr Virologie | Victor M Corman et al |
| EPI_ISL_824764 | Germany/SH-ChVir21128/2020 | 2020-12-05 | Europe/Germany/Schleswig-Holstein/ | A. Krumbholz, Labor Dr. Krause und Kollegen MVZ GmbH, Kiel | Charit√© Universit√§tsmedizin Berlin, Institut f√ºr Virologie | Victor M Corman et al |
| EPI_ISL_824767 | Germany/SH-ChVir21131/2020 | 2020-12-07 | Europe/Germany/Schleswig-Holstein/ | A. Krumbholz, Labor Dr. Krause und Kollegen MVZ GmbH, Kiel | Charit√© Universit√§tsmedizin Berlin, Institut f√ºr Virologie | Victor M Corman et al |
| EPI_ISL_824771 | Germany/SH-ChVir21135/2020 | 2020-12-07 | Europe/Germany/Schleswig-Holstein/ | A. Krumbholz, Labor Dr. Krause und Kollegen MVZ GmbH, Kiel | Charit√© Universit√§tsmedizin Berlin, Institut f√ºr Virologie | Victor M Corman et al |
| EPI_ISL_824774 | Germany/SH-ChVir21138/2020 | 2020-12-07 | Europe/Germany/Schleswig-Holstein/ | A. Krumbholz, Labor Dr. Krause und Kollegen MVZ GmbH, Kiel | Charit√© Universit√§tsmedizin Berlin, Institut f√ºr Virologie | Victor M Corman et al |
| EPI_ISL_824780 | Germany/SH-ChVir21144/2020 | 2020-12-07 | Europe/Germany/Schleswig-Holstein/ | A. Krumbholz, Labor Dr. Krause und Kollegen MVZ GmbH, Kiel | Charit√© Universit√§tsmedizin Berlin, Institut f√ºr Virologie | Victor M Corman et al |
| EPI_ISL_824784 | Germany/SH-ChVir21148/2020 | 2020-12-07 | Europe/Germany/Schleswig-Holstein/ | A. Krumbholz, Labor Dr. Krause und Kollegen MVZ GmbH, Kiel | Charit√© Universit√§tsmedizin Berlin, Institut f√ºr Virologie | Victor M Corman et al |
| EPI_ISL_824785 | Germany/SH-ChVir21150/2020 | 2020-12-07 | Europe/Germany/Schleswig-Holstein/ | A. Krumbholz, Labor Dr. Krause und Kollegen MVZ GmbH, Kiel | Charit√© Universit√§tsmedizin Berlin, Institut f√ºr Virologie | Victor M Corman et al |

| | | | | | | |
|---|---|---|---|---|---|---|
| EPI_ISL_824788 | USA/AZ-ASPHL-AZ00122806/2020 | 2020-11-10 | North America/USA/Arizona/Navajo Co | Arizona State Public Health Laboratory | Arizona State Public Health Laboratory | Trung Huynh et al |
| EPI_ISL_824793 | Belgium/ULG-11155/2020 | 2020-12-31 | Europe/Belgium/Li√®ge/ | Department of Clinical Microbiology | GIGA Medical Genomics | Keith Durkin et al |
| EPI_ISL_824807 | Belgium/ULG-11171/2020 | 2020-12-28 | Europe/Belgium/Li√®ge/ | Department of Clinical Microbiology | GIGA Medical Genomics | Keith Durkin et al |
| EPI_ISL_824821 | Belgium/ULG-11186/2020 | 2020-12-30 | Europe/Belgium/Li√®ge/ | Department of Clinical Microbiology | GIGA Medical Genomics | Keith Durkin et al |
| EPI_ISL_824864 | Germany/SH-ChVir21117/2020 | 2020-12-03 | Europe/Germany/Schleswig-Holstein/ | A. Krumbholz, Labor Dr. Krause und Kollegen MVZ GmbH, Kiel | Charit√© Universit√§tsmedizin Berlin, Institut f√ºr Virologie | Victor M Corman et al |
| EPI_ISL_824873 | USA/WA-UW-48201/2020 | 2020-12-20 | North America/USA/Washington/ | UW Virology Lab | UW Virology Lab | Pavitra Roychoudhury et al |
| EPI_ISL_824880 | USA/WA-UW-48208/2020 | 2020-12-20 | North America/USA/Washington/ | UW Virology Lab | UW Virology Lab | Pavitra Roychoudhury et al |
| EPI_ISL_824887 | USA/WA-UW-48216/2020 | 2020-12-20 | North America/USA/Washington/ | UW Virology Lab | UW Virology Lab | Pavitra Roychoudhury et al |
| EPI_ISL_824897 | USA/WA-UW-48226/2020 | 2020-12-20 | North America/USA/Washington/ | UW Virology Lab | UW Virology Lab | Pavitra Roychoudhury et al |
| EPI_ISL_824899 | USA/WA-UW-48228/2020 | 2020-12-20 | North America/USA/Washington/ | UW Virology Lab | UW Virology Lab | Pavitra Roychoudhury et al |
| EPI_ISL_824906 | USA/WA-UW-48235/2020 | 2020-12-20 | North America/USA/Washington/ | UW Virology Lab | UW Virology Lab | Pavitra Roychoudhury et al |
| EPI_ISL_824908 | USA/WA-UW-48238/2020 | 2020-12-20 | North America/USA/Washington/ | UW Virology Lab | UW Virology Lab | Pavitra Roychoudhury et al |
| EPI_ISL_824913 | USA/WA-UW-48243/2020 | 2020-12-20 | North America/USA/Washington/ | UW Virology Lab | UW Virology Lab | Pavitra Roychoudhury et al |
| EPI_ISL_824920 | Germany/NW-ChVir21955/2020 | 2020-12-24 | Europe/Germany/North Rhine Westphal | Charit√© Universit√§tsmedizin Berlin, Institute of Virology, Charit√©platz 1, 10117 Berlin, Germany | Charit√© Universit√§tsmedizin Berlin, Institute of Virology, Charit√©platz 1, 10117 Berlin, Germany | Victor M Corman et al |
| EPI_ISL_824932 | USA/AZ-ASPHL-AZ00127232/2020 | 2020-12-19 | North America/USA/Arizona/Maricopa ( | Arizona State Public Health Laboratory | Arizona State Public Health Laboratory | Trung Huynh et al |
| EPI_ISL_824938 | USA/AZ-ASPHL-AZ00127225/2020 | 2020-12-19 | North America/USA/Arizona/Maricopa ( | Arizona State Public Health Laboratory | Arizona State Public Health Laboratory | Trung Huynh et al |
| EPI_ISL_824941 | USA/AZ-ASPHL-AZ00127247/2020 | 2020-12-19 | North America/USA/Arizona/Maricopa ( | Arizona State Public Health Laboratory | Arizona State Public Health Laboratory | Trung Huynh et al |

| | | | | | | |
|---|---|---|---|---|---|---|
| EPI_ISL_824967 | USA/MD-MDH-0585/2021 | 2021-01-05 | North America/USA/Maryland/ | Maryland Public Health Laboratory | Maryland Public Health Laboratory | Maryland Department of Health Laboratories Administration et al |
| EPI_ISL_824985 | Germany/NI-ChVir21963/2021 | 2021-01-05 | Europe/Germany/Niedersachsen/ | Charitv© Universitv§tsmedizin Berlin, Institute of Virology, Charitv©platz 1, 10117 Berlin, Germany | Charitv© Universitv§tsmedizin Berlin, Institute of Virology, Charitv©platz 1, 10117 Berlin, Germany | Victor M Corman et al |
| EPI_ISL_824986 | Germany/NI-ChVir21964/2021 | 2021-01-05 | Europe/Germany/Niedersachsen/ | Charitv© Universitv§tsmedizin Berlin, Institute of Virology, Charitv©platz 1, 10117 Berlin, Germany | Charitv© Universitv§tsmedizin Berlin, Institute of Virology, Charitv©platz 1, 10117 Berlin, Germany | Victor M Corman et al |
| EPI_ISL_825001 | USA/AZ-ASPHL-AZ00127407/2020 | 2020-12-21 | North America/USA/Arizona/Maricopa ( | Arizona State Public Health Laboratory | Arizona State Public Health Laboratory | Trung Huynh et al |
| EPI_ISL_825009 | USA/AZ-ASPHL-AZ00127429/2020 | 2020-12-16 | North America/USA/Arizona/Mohave C | Arizona State Public Health Laboratory | Arizona State Public Health Laboratory | Trung Huynh et al |
| EPI_ISL_825012 | USA/AZ-ASPHL-AZ00128047/2020 | 2020-12-28 | North America/USA/Arizona/Maricopa ( | Arizona State Public Health Laboratory | Arizona State Public Health Laboratory | Trung Huynh et al |
| EPI_ISL_825029 | USA/NC-SLPH-0012/2020 | 2020-09-11 | North America/USA/North Carolina/ | Infectious Diseases, NC SLPH COVID-19 Response Team | Infectious Diseases, NC SLPH COVID-19 Response Team | Miller et al |
| EPI_ISL_825030 | USA/NC-SLPH-0016/2020 | 2020-09-11 | North America/USA/North Carolina/ | Infectious Diseases, NC SLPH COVID-19 Response Team | Infectious Diseases, NC SLPH COVID-19 Response Team | Miller et al |
| EPI_ISL_825034 | India/GJ-GBRC-434/2020 | 2020-11-20 | Asia/India/Gujarat/Ahmedabad | B.J. Medical College and Civil hospital, Ahmedabad | Gujarat Biotechnology Research Centre | Kamlesh J Upadhyay et al |

| | | | | | | |
|---|---|---|---|---|---|---|
| EPI_ISL_825035 | India/GJ-GBRC-435/2020 | 2020-11-19 | Asia/India/Gujarat/Ahmedabad | B.J. Medical College and Civil hospital, Ahmedabad | Gujarat Biotechnology Research Centre | Nilima Shah et al |
| EPI_ISL_825036 | India/GJ-GBRC-436/2020 | 2020-12-03 | Asia/India/Gujarat/Ahmedabad | B.J. Medical College and Civil hospital, Ahmedabad | Gujarat Biotechnology Research Centre | Dipa Kinariwala et al |
| EPI_ISL_825037 | India/GJ-GBRC-437/2020 | 2020-11-10 | Asia/India/Gujarat/Ahmedabad | B.J. Medical College and Civil hospital, Ahmedabad | Gujarat Biotechnology Research Centre | Pranay Shah et al |
| EPI_ISL_825038 | India/GJ-GBRC-438/2020 | 2020-11-10 | Asia/India/Gujarat/Ahmedabad | B.J. Medical College and Civil hospital, Ahmedabad | Gujarat Biotechnology Research Centre | Zuber Saiyed et al |
| EPI_ISL_825039 | India/GJ-GBRC-439a/2020 | 2020-11-10 | Asia/India/Gujarat/Ahmedabad | B.J. Medical College and Civil hospital, Ahmedabad | Gujarat Biotechnology Research Centre | Labdhi Pandya et al |
| EPI_ISL_825040 | India/GJ-GBRC-439b/2020 | 2020-11-10 | Asia/India/Gujarat/Ahmedabad | B.J. Medical College and Civil hospital, Ahmedabad | Gujarat Biotechnology Research Centre | Afzal Ansari et al |
| EPI_ISL_825041 | India/GJ-GBRC-440/2020 | 2020-11-10 | Asia/India/Gujarat/Ahmedabad | B.J. Medical College and Civil hospital, Ahmedabad | Gujarat Biotechnology Research Centre | Nikha Trivedi et al |
| EPI_ISL_825042 | India/GJ-GBRC-441/2020 | 2020-11-24 | Asia/India/Gujarat/Ahmedabad | B.J. Medical College and Civil hospital, Ahmedabad | Gujarat Biotechnology Research Centre | Apurvasinh Puvar et al |
| EPI_ISL_825043 | India/GJ-GBRC-442/2020 | 2020-12-22 | Asia/India/Gujarat/Bharuch | NHL Municipal Medical College, Ahmedbad | Gujarat Biotechnology Research Centre | Jayshri Pethani et al |
| EPI_ISL_825044 | India/GJ-GBRC-443/2020 | 2020-12-22 | Asia/India/Gujarat/Anand | NHL Municipal Medical College, Ahmedbad | Gujarat Biotechnology Research Centre | Monila Patel et al |
| EPI_ISL_825045 | India/GJ-GBRC-444/2020 | 2020-12-22 | Asia/India/Gujarat/Anand | NHL Municipal Medical College, Ahmedbad | Gujarat Biotechnology Research Centre | Atit Shah et al |
| EPI_ISL_825047 | India/GJ-GBRC-445b/2020 | 2020-12-22 | Asia/India/Gujarat/Diu | NHL Municipal Medical College, Ahmedbad | Gujarat Biotechnology Research Centre | Bimal Chauhan et al |
| EPI_ISL_825048 | India/GJ-GBRC-446/2020 | 2020-12-23 | Asia/India/Gujarat/Vadodara | GMERS Medical College and Hospital, Gotri, Vadodara | Gujarat Biotechnology Research Centre | Mitesh Kamothi et al |

| | | | | | | |
|---|---|---|---|---|---|---|
| EPI_ISL_825049 | India/GJ-GBRC-447/2020 | 2020-12-23 | Asia/India/Gujarat/Vadodara | GMERS Medical College and Hospital, Gotri, Vadodara | Gujarat Biotechnology Research Centre | Bithika Duttaroy et al |
| EPI_ISL_825050 | India/GJ-GBRC-448/2020 | 2020-12-24 | Asia/India/Gujarat/Ahmedabad | NHL Municipal Medical College, Ahmedbad | Gujarat Biotechnology Research Centre | Tanmay Mehta et al |
| EPI_ISL_825051 | India/GJ-GBRC-449/2020 | 2020-12-24 | Asia/India/Gujarat/Ahmedabad | NHL Municipal Medical College, Ahmedbad | Gujarat Biotechnology Research Centre | Bhavin Prajapati et al |
| EPI_ISL_825052 | India/GJ-GBRC-450/2020 | 2020-12-24 | Asia/India/Gujarat/Ahmedabad | NHL Municipal Medical College, Ahmedbad | Gujarat Biotechnology Research Centre | Apurvasinh Puvar et al |
| EPI_ISL_825053 | India/GJ-GBRC-451/2020 | 2020-12-24 | Asia/India/Gujarat/Ahmedabad | NHL Municipal Medical College, Ahmedbad | Gujarat Biotechnology Research Centre | Ramesh Pandit et al |
| EPI_ISL_825055 | India/GJ-GBRC-453a/2020 | 2020-12-24 | Asia/India/Gujarat/Vadodara | GMERS Medical College and Hospital, Gotri, Vadodara | Gujarat Biotechnology Research Centre | Zarna Patel et al |
| EPI_ISL_825056 | India/GJ-GBRC-454b/2020 | 2020-12-24 | Asia/India/Gujarat/Vadodara | GMERS Medical College and Hospital, Gotri, Vadodara | Gujarat Biotechnology Research Centre | Nitin Savaliya et al |
| EPI_ISL_825057 | India/GJ-GBRC-455a/2020 | 2020-12-24 | Asia/India/Gujarat/Vadodara | GMERS Medical College and Hospital, Gotri, Vadodara | Gujarat Biotechnology Research Centre | Dinesh Kumar et al |
| EPI_ISL_825058 | India/GJ-GBRC-455b/2020 | 2020-12-24 | Asia/India/Gujarat/Vadodara | GMERS Medical College and Hospital, Gotri, Vadodara | Gujarat Biotechnology Research Centre | Zuber Saiyed et al |
| EPI_ISL_825059 | India/GJ-GBRC-456/2020 | 2020-12-26 | Asia/India/Gujarat/Ahmedabad | NHL Municipal Medical College, Ahmedbad | Gujarat Biotechnology Research Centre | Afzal Ansari et al |
| EPI_ISL_825060 | India/GJ-GBRC-457/2020 | 2020-12-26 | Asia/India/Gujarat/Ahmedabad | NHL Municipal Medical College, Ahmedbad | Gujarat Biotechnology Research Centre | Nikha Trivedi et al |
| EPI_ISL_825061 | India/GJ-GBRC-458a/2020 | 2020-12-29 | Asia/India/Gujarat/Vadodara | GMERS Medical College and Hospital, Gotri, Vadodara | Gujarat Biotechnology Research Centre | Apurvasinh Puvar et al |
| EPI_ISL_825062 | India/GJ-GBRC-458b/2020 | 2020-12-29 | Asia/India/Gujarat/Vadodara | GMERS Medical College and Hospital, Gotri, Vadodara | Gujarat Biotechnology Research Centre | Ramesh Pandit et al |

| | | | | | | |
|---|---|---|---|---|---|---|
| EPI_ISL_825063 | India/GJ-GBRC-459/2020 | 2020-12-30 | Asia/India/Gujarat/Ahmedabad | NHL Municipal Medical College, Ahmedbad | Gujarat Biotechnology Research Centre | Janvi Raval et al |
| EPI_ISL_825065 | Singapore/27/2021 | 2021-01-10 | Asia/Singapore/Singapore/ | National Public Health Laboratory, National Centre for Infectious Diseases | National Public Health Laboratory, National Centre for Infectious Diseases | Tze Minn Mak et al |
| EPI_ISL_825092 | SouthAfrica/KRISP-K006794/2020 | 2020-11-19 | Africa/South Africa/Eastern Cape/Amatl | NHLS-IALCH | KRISP, KZN Research Innovation and Sequencing Platform | Giandhari J et al |
| EPI_ISL_825109 | SouthAfrica/KRISP-K006840/2020 | 2020-11-24 | Africa/South Africa/Eastern Cape/Nelsor | NHLS-IALCH | KRISP, KZN Research Innovation and Sequencing Platform | Giandhari J et al |
| EPI_ISL_825111 | SouthAfrica/KRISP-K006844/2020 | 2020-11-24 | Africa/South Africa/Eastern Cape/Nelsor | NHLS-IALCH | KRISP, KZN Research Innovation and Sequencing Platform | Giandhari J et al |
| EPI_ISL_825119 | SouthAfrica/KRISP-K006862/2020 | 2020-11-24 | Africa/South Africa/Eastern Cape/Nelsor | NHLS-IALCH | KRISP, KZN Research Innovation and Sequencing Platform | Giandhari J et al |
| EPI_ISL_825140 | India/KA-NIMH-SEQ-23/2020 | 2020-12-24 | Asia/India/Karnataka/ | Sanjay Gandhi Institute of Trauma & Orthopaedics | Department of Neurovirology, National Institute of Mental Health and Neurosciences (NIMHANS) | Chitra Pattabiraman et al |
| EPI_ISL_825141 | India/KA-NIMH-SEQ-24/2020 | 2020-12-23 | Asia/India/Karnataka/ | BBMP Urban PHC | Department of Neurovirology, National Institute of Mental Health and Neurosciences (NIMHANS) | Chitra Pattabiraman et al |

| | | | | | | |
|---|---|---|---|---|---|---|
| EPI_ISL_825143 | India/KA-NIMH-SEQ-27/2020 | 2020-12-23 | Asia/India/Karnataka/ | BBMP Urban PHC | Department of Neurovirology, National Institute of Mental Health and Neurosciences (NIMHANS) | Chitra Pattabiraman et al |
| EPI_ISL_825144 | India/KA-NIMH-SEQ-28/2020 | 2020-12-25 | Asia/India/Karnataka/ | Bangalore Medical College and Research Institute | Department of Neurovirology, National Institute of Mental Health and Neurosciences (NIMHANS) | Chitra Pattabiraman et al |
| EPI_ISL_825145 | India/KA-NIMH-SEQ-29/2020 | 2020-12-24 | Asia/India/Karnataka/ | Virus Research and Diagnostic Laboratory (DH-VRDL) | Department of Neurovirology, National Institute of Mental Health and Neurosciences (NIMHANS) | Chitra Pattabiraman et al |
| EPI_ISL_825146 | India/KA-NIMH-SEQ-30/2020 | 2020-12-24 | Asia/India/Karnataka/ | Kidwai Memorial Institute of Oncology | Department of Neurovirology, National Institute of Mental Health and Neurosciences (NIMHANS) | Chitra Pattabiraman et al |
| EPI_ISL_825147 | India/KA-NIMH-SEQ-31/2020 | 2020-12-23 | Asia/India/Karnataka/ | Narayana Nethralaya lab | Department of Neurovirology, National Institute of Mental Health and Neurosciences (NIMHANS) | Chitra Pattabiraman et al |
| EPI_ISL_825148 | India/KA-NIMH-SEQ-32/2020 | 2020-12-26 | Asia/India/Karnataka/ | BBMP Urban PHC | Department of Neurovirology, National Institute of Mental Health and Neurosciences (NIMHANS) | Chitra Pattabiraman et al |

| | | | | | | |
|---|---|---|---|---|---|---|
| EPI_ISL_825149 | India/KA-NIMH-SEQ-33/2020 | 2020-12-25 | Asia/India/Karnataka/ | National Institute of Virology Bangalore Unit (NIV BU) | Department of Neurovirology, National Institute of Mental Health and Neurosciences (NIMHANS) | Chitra Pattabiraman et al |
| EPI_ISL_825150 | India/KA-NIMH-SEQ-35/2020 | 2020-12-25 | Asia/India/Karnataka/ | Kidwai Memorial Institute of Oncology | Department of Neurovirology, National Institute of Mental Health and Neurosciences (NIMHANS) | Chitra Pattabiraman et al |
| EPI_ISL_825151 | India/KA-NIMH-SEQ-36/2020 | 2020-12-25 | Asia/India/Karnataka/ | Kidwai Memorial Institute of Oncology | Department of Neurovirology, National Institute of Mental Health and Neurosciences (NIMHANS) | Chitra Pattabiraman et al |
| EPI_ISL_825154 | India/KA-NIMH-SEQ-41/2020 | 2020-12-22 | Asia/India/Karnataka/ | Virus Research and Diagnostic Laboratory (DH-VRDL) | Department of Neurovirology, National Institute of Mental Health and Neurosciences (NIMHANS) | Chitra Pattabiraman et al |
| EPI_ISL_825159 | Germany/SN-RKI-Z-0032/2020 | 2020-12-21 | Europe/Germany/Saxony/ | Robert Koch Institute, ZBS1 Highly Pathogenic Viruses, Berlin, Germany | Robert Koch Institute, ZBS1 Highly Pathogenic Viruses & Bioinformatics MF1, Berlin, Germany | Annika Brinkmann et al |
| EPI_ISL_825161 | Germany/SN-RKI-Z-0035/2020 | 2020-12-23 | Europe/Germany/Saxony/ | Robert Koch Institute, ZBS1 Highly Pathogenic Viruses, Berlin, Germany | Robert Koch Institute, ZBS1 Highly Pathogenic Viruses & Bioinformatics MF1, Berlin, Germany | Annika Brinkmann et al |

| | | | | | | |
|---|---|---|---|---|---|---|
| EPI_ISL_825166 | Germany/SN-RKI-Z-0033/2020 | 2020-12-21 | Europe/Germany/Saxony/ | Robert Koch Institute, ZBS1 Highly Pathogenic Viruses, Berlin, Germany | Robert Koch Institute, ZBS1 Highly Pathogenic Viruses & Bioinformatics MF1, Berlin, Germany | Annika Brinkmann et al |
| EPI_ISL_825167 | Germany/SN-RKI-Z-0038/2020 | 2020-12-23 | Europe/Germany/Saxony/ | Robert Koch Institute, ZBS1 Highly Pathogenic Viruses, Berlin, Germany | Robert Koch Institute, ZBS1 Highly Pathogenic Viruses & Bioinformatics MF1, Berlin, Germany | Annika Brinkmann et al |
| EPI_ISL_825171 | Germany/SN-RKI-Z-0044/2020 | 2020-12-30 | Europe/Germany/Saxony/ | Robert Koch Institute, ZBS1 Highly Pathogenic Viruses, Berlin, Germany | Robert Koch Institute, ZBS1 Highly Pathogenic Viruses & Bioinformatics MF1, Berlin, Germany | Annika Brinkmann et al |
| EPI_ISL_825172 | Germany/SN-RKI-Z-0045/2020 | 2020-12-30 | Europe/Germany/Saxony/ | Robert Koch Institute, ZBS1 Highly Pathogenic Viruses, Berlin, Germany | Robert Koch Institute, ZBS1 Highly Pathogenic Viruses & Bioinformatics MF1, Berlin, Germany | Annika Brinkmann et al |
| EPI_ISL_825327 | Spain/CT-HUVH-86270/2020 | 2020-11-27 | Europe/Spain/Catalunya/ | Hospital Universitari Vall d'Hebron - Vall d'Hebron Institut de Rercerca | Hospital Universitari Vall d'Hebron | Cristina Andr√©s et al |
| EPI_ISL_825333 | Spain/CT-HUVH-74514/2020 | 2020-11-30 | Europe/Spain/Catalunya/ | Hospital Universitari Vall d'Hebron - Vall d'Hebron Institut de Rercerca | Hospital Universitari Vall d'Hebron | Cristina Andr√©s et al |
| EPI_ISL_825342 | Spain/CT-HUVH-65445/2020 | 2020-10-27 | Europe/Spain/Catalunya/ | Hospital Universitari Vall d'Hebron - Vall d'Hebron Institut de Rercerca | Hospital Universitari Vall d'Hebron | Cristina Andr√©s et al |
| EPI_ISL_825349 | Spain/CT-HUVH-20564/2020 | 2020-11-02 | Europe/Spain/Catalunya/ | Hospital Universitari Vall d'Hebron - Vall d'Hebron Institut de Rercerca | Hospital Universitari Vall d'Hebron | Cristina Andr√©s et al |

| | | | | | | |
|---|---|---|---|---|---|---|
| EPI_ISL_825350 | Spain/CT-HUVH-99269/2020 | 2020-11-05 | Europe/Spain/Catalunya/ | Hospital Universitari Vall d'Hebron - Vall d'Hebron Institut de Rercerca | Hospital Universitari Vall d'Hebron | Cristina Andr√©s et al |
| EPI_ISL_825356 | Spain/CT-HUVH-03257/2020 | 2020-11-10 | Europe/Spain/Catalunya/ | Hospital Universitari Vall d'Hebron - Vall d'Hebron Institut de Rercerca | Hospital Universitari Vall d'Hebron | Cristina Andr√©s et al |
| EPI_ISL_825357 | Spain/CT-HUVH-60815/2020 | 2020-11-13 | Europe/Spain/Catalunya/ | Hospital Universitari Vall d'Hebron - Vall d'Hebron Institut de Rercerca | Hospital Universitari Vall d'Hebron | Cristina Andr√©s et al |
| EPI_ISL_825358 | Spain/CT-HUVH-08435/2020 | 2020-11-16 | Europe/Spain/Catalunya/ | Hospital Universitari Vall d'Hebron - Vall d'Hebron Institut de Rercerca | Hospital Universitari Vall d'Hebron | Cristina Andr√©s et al |
| EPI_ISL_825360 | Spain/CT-HUVH-78438/2020 | 2020-11-18 | Europe/Spain/Catalunya/ | Hospital Universitari Vall d'Hebron - Vall d'Hebron Institut de Rercerca | Hospital Universitari Vall d'Hebron | Cristina Andr√©s et al |
| EPI_ISL_825361 | Spain/CT-HUVH-50169/2020 | 2020-11-20 | Europe/Spain/Catalunya/ | Hospital Universitari Vall d'Hebron - Vall d'Hebron Institut de Rercerca | Hospital Universitari Vall d'Hebron | Cristina Andr√©s et al |
| EPI_ISL_825363 | Spain/CT-HUVH-93985/2020 | 2020-11-23 | Europe/Spain/Catalunya/ | Hospital Universitari Vall d'Hebron - Vall d'Hebron Institut de Rercerca | Hospital Universitari Vall d'Hebron | Cristina Andr√©s et al |
| EPI_ISL_825364 | Spain/CT-HUVH-20081/2020 | 2020-11-27 | Europe/Spain/Catalunya/ | Hospital Universitari Vall d'Hebron - Vall d'Hebron Institut de Rercerca | Hospital Universitari Vall d'Hebron | Cristina Andr√©s et al |
| EPI_ISL_825366 | Spain/CT-HUVH-20790/2020 | 2020-11-30 | Europe/Spain/Catalunya/ | Hospital Universitari Vall d'Hebron - Vall d'Hebron Institut de Rercerca | Hospital Universitari Vall d'Hebron | Cristina Andr√©s et al |
| EPI_ISL_825368 | Spain/CT-HUVH-14370/2020 | 2020-12-02 | Europe/Spain/Catalunya/ | Hospital Universitari Vall d'Hebron - Vall d'Hebron Institut de Rercerca | Hospital Universitari Vall d'Hebron | Cristina Andr√©s et al |
| EPI_ISL_825372 | Spain/CT-HUVH-04866/2020 | 2020-12-04 | Europe/Spain/Catalunya/ | Hospital Universitari Vall d'Hebron - Vall d'Hebron Institut de Rercerca | Hospital Universitari Vall d'Hebron | Cristina Andr√©s et al |
| EPI_ISL_825373 | Spain/CT-HUVH-04711/2020 | 2020-12-04 | Europe/Spain/Catalunya/ | Hospital Universitari Vall d'Hebron - Vall d'Hebron Institut de Rercerca | Hospital Universitari Vall d'Hebron | Cristina Andr√©s et al |
| EPI_ISL_825382 | Spain/CT-HUVH-65368/2020 | 2020-10-22 | Europe/Spain/Catalunya/ | Hospital Universitari Vall d'Hebron - Vall d'Hebron Institut de Rercerca | Hospital Universitari Vall d'Hebron | Cristina Andr√©s et al |

| | | | | | | |
|---|---|---|---|---|---|---|
| EPI_ISL_825384 | Spain/CT-HUVH-97802/2020 | 2020-11-26 | Europe/Spain/Catalunya/ | Hospital Universitari Vall d'Hebron - Vall d'Hebron Institut de Rercerca | Hospital Universitari Vall d'Hebron | Cristina Andrv©s et al |
| EPI_ISL_825388 | Japan/IC-0681/2021 | 21-Jan | Asia/Japan/Japan/ | Pathogen Genomics Center, National Institute of Infectious Diseases | Pathogen Genomics Center, National Institute of Infectious Diseases | Tsuyoshi Sekizuka et al |
| EPI_ISL_825396 | Japan/IC-0673/2021 | 21-Jan | Asia/Japan/Japan/ | Pathogen Genomics Center, National Institute of Infectious Diseases | Pathogen Genomics Center, National Institute of Infectious Diseases | Tsuyoshi Sekizuka et al |
| EPI_ISL_825403 | Japan/IC-0691/2021 | 21-Jan | Asia/Japan/Japan/ | Pathogen Genomics Center, National Institute of Infectious Diseases | Pathogen Genomics Center, National Institute of Infectious Diseases | Tsuyoshi Sekizuka et al |
| EPI_ISL_825404 | Japan/IC-0701/2021 | 21-Jan | Asia/Japan/Japan/ | Pathogen Genomics Center, National Institute of Infectious Diseases | Pathogen Genomics Center, National Institute of Infectious Diseases | Tsuyoshi Sekizuka et al |
| EPI_ISL_825405 | Japan/IC-0655/2021 | 21-Jan | Asia/Japan/Japan/ | Pathogen Genomics Center, National Institute of Infectious Diseases | Pathogen Genomics Center, National Institute of Infectious Diseases | Tsuyoshi Sekizuka et al |
| EPI_ISL_825414 | Japan/IC-0670/2021 | 21-Jan | Asia/Japan/Japan/ | Pathogen Genomics Center, National Institute of Infectious Diseases | Pathogen Genomics Center, National Institute of Infectious Diseases | Tsuyoshi Sekizuka et al |
| EPI_ISL_825421 | Japan/IC-0684/2021 | 21-Jan | Asia/Japan/Japan/ | Pathogen Genomics Center, National Institute of Infectious Diseases | Pathogen Genomics Center, National Institute of Infectious Diseases | Tsuyoshi Sekizuka et al |

| | | | | | | |
|---|---|---|---|---|---|---|
| EPI_ISL_825427 | Japan/IC-0693/2021 | 21-Jan | Asia/Japan/Japan/ | Pathogen Genomics Center, National Institute of Infectious Diseases | Pathogen Genomics Center, National Institute of Infectious Diseases | Tsuyoshi Sekizuka et al |
| EPI_ISL_825433 | Japan/IC-0700/2021 | 21-Jan | Asia/Japan/Japan/ | Pathogen Genomics Center, National Institute of Infectious Diseases | Pathogen Genomics Center, National Institute of Infectious Diseases | Tsuyoshi Sekizuka et al |
| EPI_ISL_825438 | SouthAfrica/KRISP-K007775/2020 | 2020-12-17 | Africa/South Africa/KwaZulu-Natal/eThe | NHLS-IALCH | KRISP, KZN Research Innovation and Sequencing Platform | Giandhari J et al |
| EPI_ISL_825443 | SouthAfrica/KRISP-K007768/2020 | 2020-12-18 | Africa/South Africa/KwaZulu-Natal/eThe | NHLS-IALCH | KRISP, KZN Research Innovation and Sequencing Platform | Giandhari J et al |
| EPI_ISL_825449 | SouthAfrica/KRISP-K007727/2020 | 2020-12-15 | Africa/South Africa/KwaZulu-Natal/iLem | NHLS-IALCH | KRISP, KZN Research Innovation and Sequencing Platform | Giandhari J et al |
| EPI_ISL_825454 | SouthAfrica/KRISP-K007732/2020 | 2020-12-15 | Africa/South Africa/KwaZulu-Natal/iLem | NHLS-IALCH | KRISP, KZN Research Innovation and Sequencing Platform | Giandhari J et al |
| EPI_ISL_825459 | SouthAfrica/KRISP-K007761/2020 | 2020-12-14 | Africa/South Africa/KwaZulu-Natal/iLem | NHLS-IALCH | KRISP, KZN Research Innovation and Sequencing Platform | Giandhari J et al |
| EPI_ISL_825462 | SouthAfrica/KRISP-K007760/2020 | 2020-12-16 | Africa/South Africa/KwaZulu-Natal/Ugu | NHLS-IALCH | KRISP, KZN Research Innovation and Sequencing Platform | Giandhari J et al |

| | | | | | | |
|---|---|---|---|---|---|---|
| EPI_ISL_825465 | SouthAfrica/KRISP-K007755/2020 | 2020-12-15 | Africa/South Africa/KwaZulu-Natal/Ugu | NHLS-IALCH | KRISP, KZN Research Innovation and Sequencing Platform | Giandhari J et al |
| EPI_ISL_825467 | SouthAfrica/KRISP-K007752/2020 | 2020-12-15 | Africa/South Africa/KwaZulu-Natal/Harr | NHLS-IALCH | KRISP, KZN Research Innovation and Sequencing Platform | Giandhari J et al |
| EPI_ISL_825470 | SouthAfrica/KRISP-K007751/2020 | 2020-12-15 | Africa/South Africa/KwaZulu-Natal/Harr | NHLS-IALCH | KRISP, KZN Research Innovation and Sequencing Platform | Giandhari J et al |
| EPI_ISL_825471 | SouthAfrica/KRISP-K007766/2020 | 2020-12-15 | Africa/South Africa/KwaZulu-Natal/Harr | NHLS-IALCH | KRISP, KZN Research Innovation and Sequencing Platform | Giandhari J et al |
| EPI_ISL_825472 | SouthAfrica/KRISP-K007736/2020 | 2020-12-15 | Africa/South Africa/KwaZulu-Natal/uMk | NHLS-IALCH | KRISP, KZN Research Innovation and Sequencing Platform | Giandhari J et al |
| EPI_ISL_825477 | SouthAfrica/KRISP-K007730/2020 | 2020-12-15 | Africa/South Africa/KwaZulu-Natal/uMk | NHLS-IALCH | KRISP, KZN Research Innovation and Sequencing Platform | Giandhari J et al |
| EPI_ISL_825481 | SouthAfrica/KRISP-K007740/2020 | 2020-12-14 | Africa/South Africa/KwaZulu-Natal/King | NHLS-IALCH | KRISP, KZN Research Innovation and Sequencing Platform | Giandhari J et al |
| EPI_ISL_825482 | SouthAfrica/KRISP-K007745/2020 | 2020-12-14 | Africa/South Africa/KwaZulu-Natal/King | NHLS-IALCH | KRISP, KZN Research Innovation and Sequencing Platform | Giandhari J et al |

| | | | | | | |
|---|---|---|---|---|---|---|
| EPI_ISL_825487 | SouthAfrica/KRISP-K007746/2020 | 2020-12-14 | Africa/South Africa/KwaZulu-Natal/uMk | NHLS-IALCH | KRISP, KZN Research Innovation and Sequencing Platform | Giandhari J et al |
| EPI_ISL_825488 | SouthAfrica/KRISP-K007739/2020 | 2020-12-14 | Africa/South Africa/KwaZulu-Natal/King | NHLS-IALCH | KRISP, KZN Research Innovation and Sequencing Platform | Giandhari J et al |
| EPI_ISL_825576 | England/204100880/2020 | 2020-10-05 | Europe/United Kingdom/England/ | Respiratory Virus Unit, National Infection Service, Public Health England | COVID-19 Genomics UK (COG-UK) Consortium | PHE Covid Sequencing Team et al |
| EPI_ISL_825628 | Spain/CT-HUVH-79511/2020 | 2020-12-29 | Europe/Spain/Catalunya/ | Hospital Universitari Vall d'Hebron - Vall d'Hebron Institut de Rercerca | Hospital Universitari Vall d'Hebron | Cristina Andr√©s et al |
| EPI_ISL_825630 | Spain/CT-HUVH-19654/2020 | 2020-12-30 | Europe/Spain/Catalunya/ | Hospital Universitari Vall d'Hebron - Vall d'Hebron Institut de Rercerca | Hospital Universitari Vall d'Hebron | Cristina Andr√©s et al |
| EPI_ISL_825631 | Spain/CT-HUVH-85818/2020 | 2020-12-29 | Europe/Spain/Catalunya/ | Hospital Universitari Vall d'Hebron - Vall d'Hebron Institut de Rercerca | Hospital Universitari Vall d'Hebron | Cristina Andr√©s et al |
| EPI_ISL_825634 | Canada/Qc-JUS-V927147799/2020 | 2020-07-26 | North America/Canada/Quebec/ | Laboratoire de sant√© publique du Qu√©bec | Laboratoire de sant√© publique du Qu√©bec | Sandrine Moreira et al |
| EPI_ISL_825635 | Canada/Qc-JUS-W007164499/2020 | 2020-08-06 | North America/Canada/Quebec/ | Laboratoire de sant√© publique du Qu√©bec | Laboratoire de sant√© publique du Qu√©bec | Sandrine Moreira et al |
| EPI_ISL_825636 | Canada/Qc-JUS-W011149799/2020 | 2020-08-10 | North America/Canada/Quebec/ | Laboratoire de sant√© publique du Qu√©bec | Laboratoire de sant√© publique du Qu√©bec | Sandrine Moreira et al |
| EPI_ISL_825637 | Canada/Qc-JUS-V821047599/2020 | 2020-06-21 | North America/Canada/Quebec/ | Laboratoire de sant√© publique du Qu√©bec | Laboratoire de sant√© publique du Qu√©bec | Sandrine Moreira et al |
| EPI_ISL_825638 | Canada/Qc-CHUM-2015301249A/202 | 2020-06-01 | North America/Canada/Quebec/ | Laboratoire de sant√© publique du Qu√©bec | Laboratoire de sant√© publique du Qu√©bec | Sandrine Moreira et al |
| EPI_ISL_825639 | Canada/Qc-CHUM-2013801954A/202 | 2020-05-17 | North America/Canada/Quebec/ | Laboratoire de sant√© publique du Qu√©bec | Laboratoire de sant√© publique du Qu√©bec | Sandrine Moreira et al |

| | | | | | | |
|---|---|---|---|---|---|---|
| EPI_ISL_825640 | Canada/Qc-JUS-V902109099/2020 | 2020-07-01 | North America/Canada/Quebec/ | Laboratoire de santv© publique du Quv©bec | Laboratoire de santv© publique du Quv©bec | Sandrine Moreira et al |
| EPI_ISL_825641 | Canada/Qc-CHUM-2015202178A/202 | 2020-05-31 | North America/Canada/Quebec/ | Laboratoire de santv© publique du Quv©bec | Laboratoire de santv© publique du Quv©bec | Sandrine Moreira et al |
| EPI_ISL_825642 | Canada/Qc-CHUM-2015201695A/202 | 2020-05-31 | North America/Canada/Quebec/ | Laboratoire de santv© publique du Quv©bec | Laboratoire de santv© publique du Quv©bec | Sandrine Moreira et al |
| EPI_ISL_825643 | Canada/Qc-CHUM-2019203087A/202 | 2020-07-10 | North America/Canada/Quebec/ | Laboratoire de santv© publique du Quv©bec | Laboratoire de santv© publique du Quv©bec | Sandrine Moreira et al |
| EPI_ISL_825644 | Canada/Qc-CHUM-2019203097A/202 | 2020-07-10 | North America/Canada/Quebec/ | Laboratoire de santv© publique du Quv©bec | Laboratoire de santv© publique du Quv©bec | Sandrine Moreira et al |
| EPI_ISL_825645 | Canada/Qc-CHUM-2019204399A/202 | 2020-07-10 | North America/Canada/Quebec/ | Laboratoire de santv© publique du Quv©bec | Laboratoire de santv© publique du Quv©bec | Sandrine Moreira et al |
| EPI_ISL_825646 | Canada/Qc-CHUM-2019204404A/202 | 2020-07-10 | North America/Canada/Quebec/ | Laboratoire de santv© publique du Quv©bec | Laboratoire de santv© publique du Quv©bec | Sandrine Moreira et al |
| EPI_ISL_825647 | Canada/Qc-CHUM-2019203094A/202 | 2020-07-10 | North America/Canada/Quebec/ | Laboratoire de santv© publique du Quv©bec | Laboratoire de santv© publique du Quv©bec | Sandrine Moreira et al |
| EPI_ISL_825648 | Canada/Qc-JUS-W006214999/2020 | 2020-08-05 | North America/Canada/Quebec/ | Laboratoire de santv© publique du Quv©bec | Laboratoire de santv© publique du Quv©bec | Sandrine Moreira et al |
| EPI_ISL_825649 | Canada/Qc-JUS-V902127099/2020 | 2020-07-01 | North America/Canada/Quebec/ | Laboratoire de santv© publique du Quv©bec | Laboratoire de santv© publique du Quv©bec | Sandrine Moreira et al |
| EPI_ISL_825650 | Canada/Qc-JUS-V918078799/2020 | 2020-07-17 | North America/Canada/Quebec/ | Laboratoire de santv© publique du Quv©bec | Laboratoire de santv© publique du Quv©bec | Sandrine Moreira et al |
| EPI_ISL_825651 | Canada/Qc-JUS-V815154099/2020 | 2020-06-15 | North America/Canada/Quebec/ | Laboratoire de santv© publique du Quv©bec | Laboratoire de santv© publique du Quv©bec | Sandrine Moreira et al |
| EPI_ISL_825652 | Canada/Qc-JUS-V819168999/2020 | 2020-06-19 | North America/Canada/Quebec/ | Laboratoire de santv© publique du Quv©bec | Laboratoire de santv© publique du Quv©bec | Sandrine Moreira et al |

| | | | | | | |
|---|---|---|---|---|---|---|
| EPI_ISL_825653 | Canada/Qc-JUS-V825162799/2020 | 2020-06-25 | North America/Canada/Quebec/ | Laboratoire de sant√© publique du Qu√©bec | Laboratoire de sant√© publique du Qu√©bec | Sandrine Moreira et al |
| EPI_ISL_825654 | Canada/Qc-JUS-V827017599/2020 | 2020-06-27 | North America/Canada/Quebec/ | Laboratoire de sant√© publique du Qu√©bec | Laboratoire de sant√© publique du Qu√©bec | Sandrine Moreira et al |
| EPI_ISL_825655 | Canada/Qc-JUS-V828026799/2020 | 2020-06-28 | North America/Canada/Quebec/ | Laboratoire de sant√© publique du Qu√©bec | Laboratoire de sant√© publique du Qu√©bec | Sandrine Moreira et al |
| EPI_ISL_825656 | Canada/Qc-JUS-V830180899/2020 | 2020-06-30 | North America/Canada/Quebec/ | Laboratoire de sant√© publique du Qu√©bec | Laboratoire de sant√© publique du Qu√©bec | Sandrine Moreira et al |
| EPI_ISL_825657 | Canada/Qc-JUS-V902147099/2020 | 2020-07-02 | North America/Canada/Quebec/ | Laboratoire de sant√© publique du Qu√©bec | Laboratoire de sant√© publique du Qu√©bec | Sandrine Moreira et al |
| EPI_ISL_825658 | Canada/Qc-JUS-V902106399/2020 | 2020-07-01 | North America/Canada/Quebec/ | Laboratoire de sant√© publique du Qu√©bec | Laboratoire de sant√© publique du Qu√©bec | Sandrine Moreira et al |
| EPI_ISL_825659 | Canada/Qc-JUS-V904035399/2020 | 2020-07-04 | North America/Canada/Quebec/ | Laboratoire de sant√© publique du Qu√©bec | Laboratoire de sant√© publique du Qu√©bec | Sandrine Moreira et al |
| EPI_ISL_825660 | Canada/Qc-JUS-V904021499/2020 | 2020-07-04 | North America/Canada/Quebec/ | Laboratoire de sant√© publique du Qu√©bec | Laboratoire de sant√© publique du Qu√©bec | Sandrine Moreira et al |
| EPI_ISL_825661 | Canada/Qc-JUS-V907135799/2020 | 2020-07-07 | North America/Canada/Quebec/ | Laboratoire de sant√© publique du Qu√©bec | Laboratoire de sant√© publique du Qu√©bec | Sandrine Moreira et al |
| EPI_ISL_825662 | Canada/Qc-JUS-V909036899/2020 | 2020-07-09 | North America/Canada/Quebec/ | Laboratoire de sant√© publique du Qu√©bec | Laboratoire de sant√© publique du Qu√©bec | Sandrine Moreira et al |
| EPI_ISL_825663 | Canada/Qc-JUS-V909038799/2020 | 2020-07-09 | North America/Canada/Quebec/ | Laboratoire de sant√© publique du Qu√©bec | Laboratoire de sant√© publique du Qu√©bec | Sandrine Moreira et al |
| EPI_ISL_825664 | Canada/Qc-JUS-V909039399/2020 | 2020-07-09 | North America/Canada/Quebec/ | Laboratoire de sant√© publique du Qu√©bec | Laboratoire de sant√© publique du Qu√©bec | Sandrine Moreira et al |
| EPI_ISL_825665 | Canada/Qc-JUS-V921058499/2020 | 2020-07-21 | North America/Canada/Quebec/ | Laboratoire de sant√© publique du Qu√©bec | Laboratoire de sant√© publique du Qu√©bec | Sandrine Moreira et al |

| | | | | | | |
|---|---|---|---|---|---|---|
| EPI_ISL_825666 | Canada/Qc-JUS-V911040399/2020 | 2020-07-11 | North America/Canada/Quebec/ | Laboratoire de santv© publique du Quv©bec | Laboratoire de santv© publique du Quv©bec | Sandrine Moreira et al |
| EPI_ISL_825667 | Canada/Qc-JUS-V914134399/2020 | 2020-07-14 | North America/Canada/Quebec/ | Laboratoire de santv© publique du Quv©bec | Laboratoire de santv© publique du Quv©bec | Sandrine Moreira et al |
| EPI_ISL_825668 | Canada/Qc-JUS-V916009199/2020 | 2020-07-16 | North America/Canada/Quebec/ | Laboratoire de santv© publique du Quv©bec | Laboratoire de santv© publique du Quv©bec | Sandrine Moreira et al |
| EPI_ISL_825669 | Canada/Qc-JUS-V917144099/2020 | 2020-07-17 | North America/Canada/Quebec/ | Laboratoire de santv© publique du Quv©bec | Laboratoire de santv© publique du Quv©bec | Sandrine Moreira et al |
| EPI_ISL_825670 | Canada/Qc-JUS-V917143599/2020 | 2020-07-17 | North America/Canada/Quebec/ | Laboratoire de santv© publique du Quv©bec | Laboratoire de santv© publique du Quv©bec | Sandrine Moreira et al |
| EPI_ISL_825671 | Canada/Qc-JUS-V917183199/2020 | 2020-07-17 | North America/Canada/Quebec/ | Laboratoire de santv© publique du Quv©bec | Laboratoire de santv© publique du Quv©bec | Sandrine Moreira et al |
| EPI_ISL_825672 | Canada/Qc-JUS-V917193499/2020 | 2020-07-17 | North America/Canada/Quebec/ | Laboratoire de santv© publique du Quv©bec | Laboratoire de santv© publique du Quv©bec | Sandrine Moreira et al |
| EPI_ISL_825673 | Canada/Qc-JUS-V917002899/2020 | 2020-07-16 | North America/Canada/Quebec/ | Laboratoire de santv© publique du Quv©bec | Laboratoire de santv© publique du Quv©bec | Sandrine Moreira et al |
| EPI_ISL_825674 | Canada/Qc-JUS-V916062399/2020 | 2020-07-16 | North America/Canada/Quebec/ | Laboratoire de santv© publique du Quv©bec | Laboratoire de santv© publique du Quv©bec | Sandrine Moreira et al |
| EPI_ISL_825675 | Canada/Qc-JUS-V916215399/2020 | 2020-07-16 | North America/Canada/Quebec/ | Laboratoire de santv© publique du Quv©bec | Laboratoire de santv© publique du Quv©bec | Sandrine Moreira et al |
| EPI_ISL_825676 | Canada/Qc-JUS-V919043699/2020 | 2020-07-19 | North America/Canada/Quebec/ | Laboratoire de santv© publique du Quv©bec | Laboratoire de santv© publique du Quv©bec | Sandrine Moreira et al |
| EPI_ISL_825677 | Canada/Qc-JUS-V919058899/2020 | 2020-07-19 | North America/Canada/Quebec/ | Laboratoire de santv© publique du Quv©bec | Laboratoire de santv© publique du Quv©bec | Sandrine Moreira et al |
| EPI_ISL_825678 | Canada/Qc-JUS-V920201599/2020 | 2020-07-20 | North America/Canada/Quebec/ | Laboratoire de santv© publique du Quv©bec | Laboratoire de santv© publique du Quv©bec | Sandrine Moreira et al |

| | | | | | | |
|---|---|---|---|---|---|---|
| EPI_ISL_825679 | Canada/Qc-JUS-V921005299/2020 | 2020-07-20 | North America/Canada/Quebec/ | Laboratoire de sant√© publique du Qu√©bec | Laboratoire de sant√© publique du Qu√©bec | Sandrine Moreira et al |
| EPI_ISL_825680 | Canada/Qc-JUS-V921068199/2020 | 2020-07-21 | North America/Canada/Quebec/ | Laboratoire de sant√© publique du Qu√©bec | Laboratoire de sant√© publique du Qu√©bec | Sandrine Moreira et al |
| EPI_ISL_825681 | Canada/Qc-JUS-V921045499/2020 | 2020-07-20 | North America/Canada/Quebec/ | Laboratoire de sant√© publique du Qu√©bec | Laboratoire de sant√© publique du Qu√©bec | Sandrine Moreira et al |
| EPI_ISL_825682 | Canada/Qc-JUS-V922011599/2020 | 2020-07-21 | North America/Canada/Quebec/ | Laboratoire de sant√© publique du Qu√©bec | Laboratoire de sant√© publique du Qu√©bec | Sandrine Moreira et al |
| EPI_ISL_825683 | Canada/Qc-JUS-V922011999/2020 | 2020-07-21 | North America/Canada/Quebec/ | Laboratoire de sant√© publique du Qu√©bec | Laboratoire de sant√© publique du Qu√©bec | Sandrine Moreira et al |
| EPI_ISL_825684 | Canada/Qc-JUS-V921193899/2020 | 2020-07-21 | North America/Canada/Quebec/ | Laboratoire de sant√© publique du Qu√©bec | Laboratoire de sant√© publique du Qu√©bec | Sandrine Moreira et al |
| EPI_ISL_825685 | Canada/Qc-JUS-V921171699/2020 | 2020-07-21 | North America/Canada/Quebec/ | Laboratoire de sant√© publique du Qu√©bec | Laboratoire de sant√© publique du Qu√©bec | Sandrine Moreira et al |
| EPI_ISL_825686 | Canada/Qc-JUS-V921203399/2020 | 2020-07-21 | North America/Canada/Quebec/ | Laboratoire de sant√© publique du Qu√©bec | Laboratoire de sant√© publique du Qu√©bec | Sandrine Moreira et al |
| EPI_ISL_825687 | Canada/Qc-JUS-V924188799/2020 | 2020-07-24 | North America/Canada/Quebec/ | Laboratoire de sant√© publique du Qu√©bec | Laboratoire de sant√© publique du Qu√©bec | Sandrine Moreira et al |
| EPI_ISL_825688 | Canada/Qc-JUS-V924157399/2020 | 2020-07-24 | North America/Canada/Quebec/ | Laboratoire de sant√© publique du Qu√©bec | Laboratoire de sant√© publique du Qu√©bec | Sandrine Moreira et al |
| EPI_ISL_825689 | Canada/Qc-JUS-V924176099/2020 | 2020-07-24 | North America/Canada/Quebec/ | Laboratoire de sant√© publique du Qu√©bec | Laboratoire de sant√© publique du Qu√©bec | Sandrine Moreira et al |
| EPI_ISL_825690 | Canada/Qc-JUS-V924161999/2020 | 2020-07-24 | North America/Canada/Quebec/ | Laboratoire de sant√© publique du Qu√©bec | Laboratoire de sant√© publique du Qu√©bec | Sandrine Moreira et al |
| EPI_ISL_825691 | Canada/Qc-JUS-V924157799/2020 | 2020-07-24 | North America/Canada/Quebec/ | Laboratoire de sant√© publique du Qu√©bec | Laboratoire de sant√© publique du Qu√©bec | Sandrine Moreira et al |

| | | | | | | |
|---|---|---|---|---|---|---|
| EPI_ISL_825692 | Canada/Qc-JUS-V923206999/2020 | 2020-07-23 | North America/Canada/Quebec/ | Laboratoire de sant√© publique du Qu√©bec | Laboratoire de sant√© publique du Qu√©bec | Sandrine Moreira et al |
| EPI_ISL_825693 | Canada/Qc-JUS-V925092799/2020 | 2020-07-25 | North America/Canada/Quebec/ | Laboratoire de sant√© publique du Qu√©bec | Laboratoire de sant√© publique du Qu√©bec | Sandrine Moreira et al |
| EPI_ISL_825694 | Canada/Qc-JUS-V925092499/2020 | 2020-07-25 | North America/Canada/Quebec/ | Laboratoire de sant√© publique du Qu√©bec | Laboratoire de sant√© publique du Qu√©bec | Sandrine Moreira et al |
| EPI_ISL_825695 | Canada/Qc-JUS-V926067399/2020 | 2020-07-26 | North America/Canada/Quebec/ | Laboratoire de sant√© publique du Qu√©bec | Laboratoire de sant√© publique du Qu√©bec | Sandrine Moreira et al |
| EPI_ISL_825696 | Canada/Qc-JUS-V927155399/2020 | 2020-07-26 | North America/Canada/Quebec/ | Laboratoire de sant√© publique du Qu√©bec | Laboratoire de sant√© publique du Qu√©bec | Sandrine Moreira et al |
| EPI_ISL_825697 | Canada/Qc-JUS-V927136599/2020 | 2020-07-27 | North America/Canada/Quebec/ | Laboratoire de sant√© publique du Qu√©bec | Laboratoire de sant√© publique du Qu√©bec | Sandrine Moreira et al |
| EPI_ISL_825698 | Canada/Qc-JUS-V927165399/2020 | 2020-07-27 | North America/Canada/Quebec/ | Laboratoire de sant√© publique du Qu√©bec | Laboratoire de sant√© publique du Qu√©bec | Sandrine Moreira et al |
| EPI_ISL_825699 | Canada/Qc-JUS-V927186099/2020 | 2020-07-27 | North America/Canada/Quebec/ | Laboratoire de sant√© publique du Qu√©bec | Laboratoire de sant√© publique du Qu√©bec | Sandrine Moreira et al |
| EPI_ISL_825700 | Canada/Qc-JUS-V928209599/2020 | 2020-07-28 | North America/Canada/Quebec/ | Laboratoire de sant√© publique du Qu√©bec | Laboratoire de sant√© publique du Qu√©bec | Sandrine Moreira et al |
| EPI_ISL_825701 | Canada/Qc-JUS-V928201899/2020 | 2020-07-28 | North America/Canada/Quebec/ | Laboratoire de sant√© publique du Qu√©bec | Laboratoire de sant√© publique du Qu√©bec | Sandrine Moreira et al |
| EPI_ISL_825702 | Canada/Qc-JUS-V928197399/2020 | 2020-07-28 | North America/Canada/Quebec/ | Laboratoire de sant√© publique du Qu√©bec | Laboratoire de sant√© publique du Qu√©bec | Sandrine Moreira et al |
| EPI_ISL_825703 | Canada/Qc-JUS-V928194999/2020 | 2020-07-28 | North America/Canada/Quebec/ | Laboratoire de sant√© publique du Qu√©bec | Laboratoire de sant√© publique du Qu√©bec | Sandrine Moreira et al |
| EPI_ISL_825704 | Canada/Qc-JUS-V928194599/2020 | 2020-07-28 | North America/Canada/Quebec/ | Laboratoire de sant√© publique du Qu√©bec | Laboratoire de sant√© publique du Qu√©bec | Sandrine Moreira et al |

| | | | | | | |
|---|---|---|---|---|---|---|
| EPI_ISL_825705 | Canada/Qc-JUS-V929161399/2020 | 2020-07-29 | North America/Canada/Quebec/ | Laboratoire de santv© publique du Quv©bec | Laboratoire de santv© publique du Quv©bec | Sandrine Moreira et al |
| EPI_ISL_825706 | Canada/Qc-JUS-V929156199/2020 | 2020-07-29 | North America/Canada/Quebec/ | Laboratoire de santv© publique du Quv©bec | Laboratoire de santv© publique du Quv©bec | Sandrine Moreira et al |
| EPI_ISL_825707 | Canada/Qc-JUS-V929169599/2020 | 2020-07-29 | North America/Canada/Quebec/ | Laboratoire de santv© publique du Quv©bec | Laboratoire de santv© publique du Quv©bec | Sandrine Moreira et al |
| EPI_ISL_825708 | Canada/Qc-JUS-W003138999/2020 | 2020-08-02 | North America/Canada/Quebec/ | Laboratoire de santv© publique du Quv©bec | Laboratoire de santv© publique du Quv©bec | Sandrine Moreira et al |
| EPI_ISL_825709 | Canada/Qc-JUS-W004122199/2020 | 2020-08-03 | North America/Canada/Quebec/ | Laboratoire de santv© publique du Quv©bec | Laboratoire de santv© publique du Quv©bec | Sandrine Moreira et al |
| EPI_ISL_825710 | Canada/Qc-JUS-W004161099/2020 | 2020-08-03 | North America/Canada/Quebec/ | Laboratoire de santv© publique du Quv©bec | Laboratoire de santv© publique du Quv©bec | Sandrine Moreira et al |
| EPI_ISL_825711 | Canada/Qc-JUS-W010150699/2020 | 2020-08-10 | North America/Canada/Quebec/ | Laboratoire de santv© publique du Quv©bec | Laboratoire de santv© publique du Quv©bec | Sandrine Moreira et al |
| EPI_ISL_825712 | Canada/Qc-JUS-W019157299/2020 | 2020-08-18 | North America/Canada/Quebec/ | Laboratoire de santv© publique du Quv©bec | Laboratoire de santv© publique du Quv©bec | Sandrine Moreira et al |
| EPI_ISL_825715 | Canada/Qc-HCLM-0091299197/2020 | 2020-09-04 | North America/Canada/Quebec/ | Laboratoire de santv© publique du Quv©bec | Laboratoire de santv© publique du Quv©bec | Sandrine Moreira et al |
| EPI_ISL_825716 | Canada/Qc-HCLM-7051284447/2020 | 2020-08-28 | North America/Canada/Quebec/ | Laboratoire de santv© publique du Quv©bec | Laboratoire de santv© publique du Quv©bec | Sandrine Moreira et al |
| EPI_ISL_825717 | Canada/Qc-HCLM-0081267614/2020 | 2020-08-21 | North America/Canada/Quebec/ | Laboratoire de santv© publique du Quv©bec | Laboratoire de santv© publique du Quv©bec | Sandrine Moreira et al |
| EPI_ISL_825718 | Canada/Qc-HCLM-0091290848/2020 | 2020-09-01 | North America/Canada/Quebec/ | Laboratoire de santv© publique du Quv©bec | Laboratoire de santv© publique du Quv©bec | Sandrine Moreira et al |
| EPI_ISL_825719 | Canada/Qc-HCLM-7051290938/2020 | 2020-09-01 | North America/Canada/Quebec/ | Laboratoire de santv© publique du Quv©bec | Laboratoire de santv© publique du Quv©bec | Sandrine Moreira et al |

| | | | | | | |
|---|---|---|---|---|---|---|
| EPI_ISL_825720 | Canada/Qc-HCLM-7051288555/2020 | 2020-08-31 | North America/Canada/Quebec/ | Laboratoire de sant√© publique du Qu√©bec | Laboratoire de sant√© publique du Qu√©bec | Sandrine Moreira et al |
| EPI_ISL_825721 | Canada/Qc-HCLM-7056053754/2020 | 2020-09-02 | North America/Canada/Quebec/ | Laboratoire de sant√© publique du Qu√©bec | Laboratoire de sant√© publique du Qu√©bec | Sandrine Moreira et al |
| EPI_ISL_825722 | Canada/Qc-HCLM-7051294639/2020 | 2020-09-02 | North America/Canada/Quebec/ | Laboratoire de sant√© publique du Qu√©bec | Laboratoire de sant√© publique du Qu√©bec | Sandrine Moreira et al |
| EPI_ISL_825723 | Canada/Qc-HCLM-0091298727/2020 | 2020-09-04 | North America/Canada/Quebec/ | Laboratoire de sant√© publique du Qu√©bec | Laboratoire de sant√© publique du Qu√©bec | Sandrine Moreira et al |
| EPI_ISL_825724 | Canada/Qc-HCLM-0091298328/2020 | 2020-09-04 | North America/Canada/Quebec/ | Laboratoire de sant√© publique du Qu√©bec | Laboratoire de sant√© publique du Qu√©bec | Sandrine Moreira et al |
| EPI_ISL_825725 | Canada/Qc-HCLM-0091300812/2020 | 2020-09-05 | North America/Canada/Quebec/ | Laboratoire de sant√© publique du Qu√©bec | Laboratoire de sant√© publique du Qu√©bec | Sandrine Moreira et al |
| EPI_ISL_825726 | Canada/Qc-HCLM-7051222878/2020 | 2020-08-03 | North America/Canada/Quebec/ | Laboratoire de sant√© publique du Qu√©bec | Laboratoire de sant√© publique du Qu√©bec | Sandrine Moreira et al |
| EPI_ISL_825727 | Canada/Qc-HCLM-0081224017/2020 | 2020-08-04 | North America/Canada/Quebec/ | Laboratoire de sant√© publique du Qu√©bec | Laboratoire de sant√© publique du Qu√©bec | Sandrine Moreira et al |
| EPI_ISL_825729 | Canada/Qc-HCLM-7051222947/2020 | 2020-08-03 | North America/Canada/Quebec/ | Laboratoire de sant√© publique du Qu√©bec | Laboratoire de sant√© publique du Qu√©bec | Sandrine Moreira et al |
| EPI_ISL_825730 | Canada/Qc-HCLM-0081235532/2020 | 2020-08-07 | North America/Canada/Quebec/ | Laboratoire de sant√© publique du Qu√©bec | Laboratoire de sant√© publique du Qu√©bec | Sandrine Moreira et al |
| EPI_ISL_825731 | Canada/Qc-HCLM-0081224591/2020 | 2020-08-04 | North America/Canada/Quebec/ | Laboratoire de sant√© publique du Qu√©bec | Laboratoire de sant√© publique du Qu√©bec | Sandrine Moreira et al |
| EPI_ISL_825732 | Canada/Qc-HCLM-7051222889/2020 | 2020-08-03 | North America/Canada/Quebec/ | Laboratoire de sant√© publique du Qu√©bec | Laboratoire de sant√© publique du Qu√©bec | Sandrine Moreira et al |
| EPI_ISL_825733 | Canada/Qc-HCLM-7051228539/2020 | 2020-08-05 | North America/Canada/Quebec/ | Laboratoire de sant√© publique du Qu√©bec | Laboratoire de sant√© publique du Qu√©bec | Sandrine Moreira et al |

| | | | | | | |
|---|---|---|---|---|---|---|
| EPI_ISL_825734 | Canada/Qc-HCLM-7051228541/2020 | 2020-08-05 | North America/Canada/Quebec/ | Laboratoire de sant√© publique du Qu√©bec | Laboratoire de sant√© publique du Qu√©bec | Sandrine Moreira et al |
| EPI_ISL_825735 | Canada/Qc-HCLM-7051228620/2020 | 2020-08-05 | North America/Canada/Quebec/ | Laboratoire de sant√© publique du Qu√©bec | Laboratoire de sant√© publique du Qu√©bec | Sandrine Moreira et al |
| EPI_ISL_825736 | Canada/Qc-HCLM-7051228617/2020 | 2020-08-05 | North America/Canada/Quebec/ | Laboratoire de sant√© publique du Qu√©bec | Laboratoire de sant√© publique du Qu√©bec | Sandrine Moreira et al |
| EPI_ISL_825737 | Canada/Qc-HCLM-0081227892/2020 | 2020-08-05 | North America/Canada/Quebec/ | Laboratoire de sant√© publique du Qu√©bec | Laboratoire de sant√© publique du Qu√©bec | Sandrine Moreira et al |
| EPI_ISL_825738 | Canada/Qc-HCLM-7051228380/2020 | 2020-08-05 | North America/Canada/Quebec/ | Laboratoire de sant√© publique du Qu√©bec | Laboratoire de sant√© publique du Qu√©bec | Sandrine Moreira et al |
| EPI_ISL_825739 | Canada/Qc-HCLM-0081227908/2020 | 2020-08-05 | North America/Canada/Quebec/ | Laboratoire de sant√© publique du Qu√©bec | Laboratoire de sant√© publique du Qu√©bec | Sandrine Moreira et al |
| EPI_ISL_825740 | Canada/Qc-HCLM-0081228131/2020 | 2020-08-05 | North America/Canada/Quebec/ | Laboratoire de sant√© publique du Qu√©bec | Laboratoire de sant√© publique du Qu√©bec | Sandrine Moreira et al |
| EPI_ISL_825741 | Canada/Qc-HCLM-0081227532/2020 | 2020-08-05 | North America/Canada/Quebec/ | Laboratoire de sant√© publique du Qu√©bec | Laboratoire de sant√© publique du Qu√©bec | Sandrine Moreira et al |
| EPI_ISL_825742 | Canada/Qc-HCLM-0081232159/2020 | 2020-08-06 | North America/Canada/Quebec/ | Laboratoire de sant√© publique du Qu√©bec | Laboratoire de sant√© publique du Qu√©bec | Sandrine Moreira et al |
| EPI_ISL_825743 | Canada/Qc-HCLM-7051229186/2020 | 2020-08-05 | North America/Canada/Quebec/ | Laboratoire de sant√© publique du Qu√©bec | Laboratoire de sant√© publique du Qu√©bec | Sandrine Moreira et al |
| EPI_ISL_825744 | Canada/Qc-HCLM-0081230615/2020 | 2020-08-06 | North America/Canada/Quebec/ | Laboratoire de sant√© publique du Qu√©bec | Laboratoire de sant√© publique du Qu√©bec | Sandrine Moreira et al |
| EPI_ISL_825745 | Canada/Qc-HCLM-0081231359/2020 | 2020-08-06 | North America/Canada/Quebec/ | Laboratoire de sant√© publique du Qu√©bec | Laboratoire de sant√© publique du Qu√©bec | Sandrine Moreira et al |
| EPI_ISL_825746 | Canada/Qc-HCLM-0081231877/2020 | 2020-08-06 | North America/Canada/Quebec/ | Laboratoire de sant√© publique du Qu√©bec | Laboratoire de sant√© publique du Qu√©bec | Sandrine Moreira et al |

| | | | | | | |
|---|---|---|---|---|---|---|
| EPI_ISL_825747 | Canada/Qc-HCLM-0081234446/2020 | 2020-08-07 | North America/Canada/Quebec/ | Laboratoire de sant√© publique du Qu√©bec | Laboratoire de sant√© publique du Qu√©bec | Sandrine Moreira et al |
| EPI_ISL_825748 | Canada/Qc-HCLM-0081234486/2020 | 2020-08-07 | North America/Canada/Quebec/ | Laboratoire de sant√© publique du Qu√©bec | Laboratoire de sant√© publique du Qu√©bec | Sandrine Moreira et al |
| EPI_ISL_825749 | Canada/Qc-HCLM-0081233852/2020 | 2020-08-07 | North America/Canada/Quebec/ | Laboratoire de sant√© publique du Qu√©bec | Laboratoire de sant√© publique du Qu√©bec | Sandrine Moreira et al |
| EPI_ISL_825750 | Canada/Qc-HCLM-0081232026/2020 | 2020-08-06 | North America/Canada/Quebec/ | Laboratoire de sant√© publique du Qu√©bec | Laboratoire de sant√© publique du Qu√©bec | Sandrine Moreira et al |
| EPI_ISL_825751 | Canada/Qc-HCLM-0081235709/2020 | 2020-08-07 | North America/Canada/Quebec/ | Laboratoire de sant√© publique du Qu√©bec | Laboratoire de sant√© publique du Qu√©bec | Sandrine Moreira et al |
| EPI_ISL_825752 | Canada/Qc-HCLM-0081235488/2020 | 2020-08-07 | North America/Canada/Quebec/ | Laboratoire de sant√© publique du Qu√©bec | Laboratoire de sant√© publique du Qu√©bec | Sandrine Moreira et al |
| EPI_ISL_825753 | Canada/Qc-HCLM-0081235810/2020 | 2020-08-07 | North America/Canada/Quebec/ | Laboratoire de sant√© publique du Qu√©bec | Laboratoire de sant√© publique du Qu√©bec | Sandrine Moreira et al |
| EPI_ISL_825754 | Canada/Qc-HCLM-0081235675/2020 | 2020-08-07 | North America/Canada/Quebec/ | Laboratoire de sant√© publique du Qu√©bec | Laboratoire de sant√© publique du Qu√©bec | Sandrine Moreira et al |
| EPI_ISL_825755 | Canada/Qc-HCLM-0081237506/2020 | 2020-08-09 | North America/Canada/Quebec/ | Laboratoire de sant√© publique du Qu√©bec | Laboratoire de sant√© publique du Qu√©bec | Sandrine Moreira et al |
| EPI_ISL_825756 | Canada/Qc-HCLM-6896033873/2020 | 2020-08-09 | North America/Canada/Quebec/ | Laboratoire de sant√© publique du Qu√©bec | Laboratoire de sant√© publique du Qu√©bec | Sandrine Moreira et al |
| EPI_ISL_825757 | Canada/Qc-HCLM-0081246298/2020 | 2020-08-12 | North America/Canada/Quebec/ | Laboratoire de sant√© publique du Qu√©bec | Laboratoire de sant√© publique du Qu√©bec | Sandrine Moreira et al |
| EPI_ISL_825758 | Canada/Qc-HCLM-0081243503/2020 | 2020-08-11 | North America/Canada/Quebec/ | Laboratoire de sant√© publique du Qu√©bec | Laboratoire de sant√© publique du Qu√©bec | Sandrine Moreira et al |
| EPI_ISL_825759 | Canada/Qc-HCLM-0081252914/2020 | 2020-08-15 | North America/Canada/Quebec/ | Laboratoire de sant√© publique du Qu√©bec | Laboratoire de sant√© publique du Qu√©bec | Sandrine Moreira et al |

| | | | | | | |
|---|---|---|---|---|---|---|
| EPI_ISL_825760 | Canada/Qc-HCLM-7051249607/2020 | 2020-08-13 | North America/Canada/Quebec/ | Laboratoire de santV© publique du QuV©bec | Laboratoire de santV© publique du QuV©bec | Sandrine Moreira et al |
| EPI_ISL_825761 | Canada/Qc-HCLM-0081253757/2020 | 2020-08-16 | North America/Canada/Quebec/ | Laboratoire de santV© publique du QuV©bec | Laboratoire de santV© publique du QuV©bec | Sandrine Moreira et al |
| EPI_ISL_825762 | Canada/Qc-HCLM-0081268735/2020 | 2020-08-21 | North America/Canada/Quebec/ | Laboratoire de santV© publique du QuV©bec | Laboratoire de santV© publique du QuV©bec | Sandrine Moreira et al |
| EPI_ISL_825763 | Canada/Qc-HCLM-0081268085/2020 | 2020-08-21 | North America/Canada/Quebec/ | Laboratoire de santV© publique du QuV©bec | Laboratoire de santV© publique du QuV©bec | Sandrine Moreira et al |
| EPI_ISL_825764 | Canada/Qc-HCLM-0081265973/2020 | 2020-08-20 | North America/Canada/Quebec/ | Laboratoire de santV© publique du QuV©bec | Laboratoire de santV© publique du QuV©bec | Sandrine Moreira et al |
| EPI_ISL_825765 | Canada/Qc-HCLM-0081268037/2020 | 2020-08-21 | North America/Canada/Quebec/ | Laboratoire de santV© publique du QuV©bec | Laboratoire de santV© publique du QuV©bec | Sandrine Moreira et al |
| EPI_ISL_825766 | Canada/Qc-HCLM-0081268958/2020 | 2020-08-21 | North America/Canada/Quebec/ | Laboratoire de santV© publique du QuV©bec | Laboratoire de santV© publique du QuV©bec | Sandrine Moreira et al |
| EPI_ISL_825767 | Canada/Qc-HCLM-0081268483/2020 | 2020-08-21 | North America/Canada/Quebec/ | Laboratoire de santV© publique du QuV©bec | Laboratoire de santV© publique du QuV©bec | Sandrine Moreira et al |
| EPI_ISL_825768 | Canada/Qc-HCLM-0081265119/2020 | 2020-08-20 | North America/Canada/Quebec/ | Laboratoire de santV© publique du QuV©bec | Laboratoire de santV© publique du QuV©bec | Sandrine Moreira et al |
| EPI_ISL_825769 | Canada/Qc-HCLM-0081267438/2020 | 2020-08-21 | North America/Canada/Quebec/ | Laboratoire de santV© publique du QuV©bec | Laboratoire de santV© publique du QuV©bec | Sandrine Moreira et al |
| EPI_ISL_825770 | Canada/Qc-HCLM-0081268137/2020 | 2020-08-21 | North America/Canada/Quebec/ | Laboratoire de santV© publique du QuV©bec | Laboratoire de santV© publique du QuV©bec | Sandrine Moreira et al |
| EPI_ISL_825771 | Canada/Qc-HCLM-7051286253/2020 | 2020-08-30 | North America/Canada/Quebec/ | Laboratoire de santV© publique du QuV©bec | Laboratoire de santV© publique du QuV©bec | Sandrine Moreira et al |
| EPI_ISL_825772 | Canada/Qc-HCLM-7051286284/2020 | 2020-08-30 | North America/Canada/Quebec/ | Laboratoire de santV© publique du QuV©bec | Laboratoire de santV© publique du QuV©bec | Sandrine Moreira et al |

| | | | | | | |
|---|---|---|---|---|---|---|
| EPI_ISL_825773 | Canada/Qc-HCLM-7051284613/2020 | 2020-08-28 | North America/Canada/Quebec/ | Laboratoire de santv© publique du Quv©bec | Laboratoire de santv© publique du Quv©bec | Sandrine Moreira et al |
| EPI_ISL_825774 | Canada/Qc-HCLM-7051284499/2020 | 2020-08-28 | North America/Canada/Quebec/ | Laboratoire de santv© publique du Quv©bec | Laboratoire de santv© publique du Quv©bec | Sandrine Moreira et al |
| EPI_ISL_825775 | Canada/Qc-HCLM-7051286260/2020 | 2020-08-30 | North America/Canada/Quebec/ | Laboratoire de santv© publique du Quv©bec | Laboratoire de santv© publique du Quv©bec | Sandrine Moreira et al |
| EPI_ISL_825776 | Canada/Qc-HCLM-7051285547/2020 | 2020-08-29 | North America/Canada/Quebec/ | Laboratoire de santv© publique du Quv©bec | Laboratoire de santv© publique du Quv©bec | Sandrine Moreira et al |
| EPI_ISL_825777 | Canada/Qc-HCLM-0081287212/2020 | 2020-08-31 | North America/Canada/Quebec/ | Laboratoire de santv© publique du Quv©bec | Laboratoire de santv© publique du Quv©bec | Sandrine Moreira et al |
| EPI_ISL_825778 | Canada/Qc-HCLM-7051286298/2020 | 2020-08-30 | North America/Canada/Quebec/ | Laboratoire de santv© publique du Quv©bec | Laboratoire de santv© publique du Quv©bec | Sandrine Moreira et al |
| EPI_ISL_825779 | Canada/Qc-HCLM-7051288293/2020 | 2020-08-31 | North America/Canada/Quebec/ | Laboratoire de santv© publique du Quv©bec | Laboratoire de santv© publique du Quv©bec | Sandrine Moreira et al |
| EPI_ISL_825780 | Canada/Qc-HCLM-7051290890/2020 | 2020-09-01 | North America/Canada/Quebec/ | Laboratoire de santv© publique du Quv©bec | Laboratoire de santv© publique du Quv©bec | Sandrine Moreira et al |
| EPI_ISL_825781 | Canada/Qc-HCLM-7051290895/2020 | 2020-09-01 | North America/Canada/Quebec/ | Laboratoire de santv© publique du Quv©bec | Laboratoire de santv© publique du Quv©bec | Sandrine Moreira et al |
| EPI_ISL_825782 | Canada/Qc-HCLM-0091289757/2020 | 2020-09-01 | North America/Canada/Quebec/ | Laboratoire de santv© publique du Quv©bec | Laboratoire de santv© publique du Quv©bec | Sandrine Moreira et al |
| EPI_ISL_825783 | Canada/Qc-HCLM-7051288454/2020 | 2020-08-31 | North America/Canada/Quebec/ | Laboratoire de santv© publique du Quv©bec | Laboratoire de santv© publique du Quv©bec | Sandrine Moreira et al |
| EPI_ISL_825785 | Canada/Qc-HCLM-7051291338/2020 | 2020-09-01 | North America/Canada/Quebec/ | Laboratoire de santv© publique du Quv©bec | Laboratoire de santv© publique du Quv©bec | Sandrine Moreira et al |
| EPI_ISL_825786 | Canada/Qc-HCLM-7051291521/2020 | 2020-09-01 | North America/Canada/Quebec/ | Laboratoire de santv© publique du Quv©bec | Laboratoire de santv© publique du Quv©bec | Sandrine Moreira et al |

| | | | | | | |
|---|---|---|---|---|---|---|
| EPI_ISL_825787 | Canada/Qc-HCLM-7051294262/2020 | 2020-09-02 | North America/Canada/Quebec/ | Laboratoire de sant√© publique du Qu√©bec | Laboratoire de sant√© publique du Qu√©bec | Sandrine Moreira et al |
| EPI_ISL_825788 | Canada/Qc-HCLM-7051291348/2020 | 2020-09-01 | North America/Canada/Quebec/ | Laboratoire de sant√© publique du Qu√©bec | Laboratoire de sant√© publique du Qu√©bec | Sandrine Moreira et al |
| EPI_ISL_825789 | Canada/Qc-HCLM-7051291632/2020 | 2020-09-01 | North America/Canada/Quebec/ | Laboratoire de sant√© publique du Qu√©bec | Laboratoire de sant√© publique du Qu√©bec | Sandrine Moreira et al |
| EPI_ISL_825790 | Canada/Qc-HCLM-7051294585/2020 | 2020-09-02 | North America/Canada/Quebec/ | Laboratoire de sant√© publique du Qu√©bec | Laboratoire de sant√© publique du Qu√©bec | Sandrine Moreira et al |
| EPI_ISL_825791 | Canada/Qc-HCLM-0091294460/2020 | 2020-09-02 | North America/Canada/Quebec/ | Laboratoire de sant√© publique du Qu√©bec | Laboratoire de sant√© publique du Qu√©bec | Sandrine Moreira et al |
| EPI_ISL_825792 | Canada/Qc-HCLM-0091296603/2020 | 2020-09-03 | North America/Canada/Quebec/ | Laboratoire de sant√© publique du Qu√©bec | Laboratoire de sant√© publique du Qu√©bec | Sandrine Moreira et al |
| EPI_ISL_825793 | Canada/Qc-HCLM-0091299685/2020 | 2020-09-04 | North America/Canada/Quebec/ | Laboratoire de sant√© publique du Qu√©bec | Laboratoire de sant√© publique du Qu√©bec | Sandrine Moreira et al |
| EPI_ISL_825794 | Canada/Qc-HCLM-0091299224/2020 | 2020-09-04 | North America/Canada/Quebec/ | Laboratoire de sant√© publique du Qu√©bec | Laboratoire de sant√© publique du Qu√©bec | Sandrine Moreira et al |
| EPI_ISL_825795 | Canada/Qc-HCLM-0091299254/2020 | 2020-09-04 | North America/Canada/Quebec/ | Laboratoire de sant√© publique du Qu√©bec | Laboratoire de sant√© publique du Qu√©bec | Sandrine Moreira et al |
| EPI_ISL_825796 | Canada/Qc-HCLM-0091299170/2020 | 2020-09-04 | North America/Canada/Quebec/ | Laboratoire de sant√© publique du Qu√©bec | Laboratoire de sant√© publique du Qu√©bec | Sandrine Moreira et al |
| EPI_ISL_825797 | Canada/Qc-HCLM-0091299225/2020 | 2020-09-04 | North America/Canada/Quebec/ | Laboratoire de sant√© publique du Qu√©bec | Laboratoire de sant√© publique du Qu√©bec | Sandrine Moreira et al |
| EPI_ISL_825798 | Canada/Qc-HCLM-0091299230/2020 | 2020-09-04 | North America/Canada/Quebec/ | Laboratoire de sant√© publique du Qu√©bec | Laboratoire de sant√© publique du Qu√©bec | Sandrine Moreira et al |
| EPI_ISL_825799 | Canada/Qc-HCLM-0091299560/2020 | 2020-09-04 | North America/Canada/Quebec/ | Laboratoire de sant√© publique du Qu√©bec | Laboratoire de sant√© publique du Qu√©bec | Sandrine Moreira et al |

| | | | | | | |
|---|---|---|---|---|---|---|
| EPI_ISL_825800 | Canada/Qc-HCLM-0091299442/2020 | 2020-09-04 | North America/Canada/Quebec/ | Laboratoire de santv© publique du Quv©bec | Laboratoire de santv© publique du Quv©bec | Sandrine Moreira et al |
| EPI_ISL_825801 | Canada/Qc-HCLM-0091299931/2020 | 2020-09-04 | North America/Canada/Quebec/ | Laboratoire de santv© publique du Quv©bec | Laboratoire de santv© publique du Quv©bec | Sandrine Moreira et al |
| EPI_ISL_825802 | Canada/Qc-HCLM-0096056810/2020 | 2020-09-05 | North America/Canada/Quebec/ | Laboratoire de santv© publique du Quv©bec | Laboratoire de santv© publique du Quv©bec | Sandrine Moreira et al |
| EPI_ISL_825803 | Canada/Qc-HCLM-0096056815/2020 | 2020-09-05 | North America/Canada/Quebec/ | Laboratoire de santv© publique du Quv©bec | Laboratoire de santv© publique du Quv©bec | Sandrine Moreira et al |
| EPI_ISL_825804 | Canada/Qc-HCLM-7056056814/2020 | 2020-09-05 | North America/Canada/Quebec/ | Laboratoire de santv© publique du Quv©bec | Laboratoire de santv© publique du Quv©bec | Sandrine Moreira et al |
| EPI_ISL_825805 | Canada/Qc-HCLM-0091303250/2020 | 2020-09-07 | North America/Canada/Quebec/ | Laboratoire de santv© publique du Quv©bec | Laboratoire de santv© publique du Quv©bec | Sandrine Moreira et al |
| EPI_ISL_825806 | Canada/Qc-HCLM-0091303258/2020 | 2020-09-07 | North America/Canada/Quebec/ | Laboratoire de santv© publique du Quv©bec | Laboratoire de santv© publique du Quv©bec | Sandrine Moreira et al |
| EPI_ISL_825808 | Canada/Qc-CHUM-2016901878A/202 | 2020-06-17 | North America/Canada/Quebec/ | Laboratoire de santv© publique du Quv©bec | Laboratoire de santv© publique du Quv©bec | Sandrine Moreira et al |
| EPI_ISL_825809 | Canada/Qc-JEW-Q9300034/2020 | 2020-05-29 | North America/Canada/Quebec/ | Laboratoire de santv© publique du Quv©bec | Laboratoire de santv© publique du Quv©bec | Sandrine Moreira et al |
| EPI_ISL_825810 | Canada/Qc-CHUM-2016902958A/202 | 2020-06-17 | North America/Canada/Quebec/ | Laboratoire de santv© publique du Quv©bec | Laboratoire de santv© publique du Quv©bec | Sandrine Moreira et al |
| EPI_ISL_825811 | Canada/Qc-CHUM-2016302095A/202 | 2020-06-11 | North America/Canada/Quebec/ | Laboratoire de santv© publique du Quv©bec | Laboratoire de santv© publique du Quv©bec | Sandrine Moreira et al |
| EPI_ISL_825812 | Canada/Qc-CHUM-2016701071A/202 | 2020-06-15 | North America/Canada/Quebec/ | Laboratoire de santv© publique du Quv©bec | Laboratoire de santv© publique du Quv©bec | Sandrine Moreira et al |
| EPI_ISL_825813 | Canada/Qc-CHUM-2016904336A/202 | 2020-06-17 | North America/Canada/Quebec/ | Laboratoire de santv© publique du Quv©bec | Laboratoire de santv© publique du Quv©bec | Sandrine Moreira et al |

| | | | | | | |
|---|---|---|---|---|---|---|
| EPI_ISL_825814 | Canada/Qc-CHUM-2016904338A/202 | 2020-06-17 | North America/Canada/Quebec/ | Laboratoire de santv© publique du Quv©bec | Laboratoire de santv© publique du Quv©bec | Sandrine Moreira et al |
| EPI_ISL_825815 | Canada/Qc-CHUM-2017103955A/202 | 2020-06-19 | North America/Canada/Quebec/ | Laboratoire de santv© publique du Quv©bec | Laboratoire de santv© publique du Quv©bec | Sandrine Moreira et al |
| EPI_ISL_825816 | Canada/Qc-CHUM-2017002366A/202 | 2020-06-18 | North America/Canada/Quebec/ | Laboratoire de santv© publique du Quv©bec | Laboratoire de santv© publique du Quv©bec | Sandrine Moreira et al |
| EPI_ISL_825817 | Canada/Qc-CHUM-2017002430A/202 | 2020-06-18 | North America/Canada/Quebec/ | Laboratoire de santv© publique du Quv©bec | Laboratoire de santv© publique du Quv©bec | Sandrine Moreira et al |
| EPI_ISL_825818 | Canada/Qc-CHUM-2017002192A/202 | 2020-06-18 | North America/Canada/Quebec/ | Laboratoire de santv© publique du Quv©bec | Laboratoire de santv© publique du Quv©bec | Sandrine Moreira et al |
| EPI_ISL_825819 | Canada/Qc-CHUM-2017004087A/202 | 2020-06-18 | North America/Canada/Quebec/ | Laboratoire de santv© publique du Quv©bec | Laboratoire de santv© publique du Quv©bec | Sandrine Moreira et al |
| EPI_ISL_825820 | Canada/Qc-CHUM-2017102498A/202 | 2020-06-19 | North America/Canada/Quebec/ | Laboratoire de santv© publique du Quv©bec | Laboratoire de santv© publique du Quv©bec | Sandrine Moreira et al |
| EPI_ISL_825821 | Canada/Qc-CHUM-2017102474A/202 | 2020-06-19 | North America/Canada/Quebec/ | Laboratoire de santv© publique du Quv©bec | Laboratoire de santv© publique du Quv©bec | Sandrine Moreira et al |
| EPI_ISL_825822 | Canada/Qc-CHUM-2017004391A/202 | 2020-06-18 | North America/Canada/Quebec/ | Laboratoire de santv© publique du Quv©bec | Laboratoire de santv© publique du Quv©bec | Sandrine Moreira et al |
| EPI_ISL_825823 | Canada/Qc-CHUM-2015703462A/202 | 2020-06-05 | North America/Canada/Quebec/ | Laboratoire de santv© publique du Quv©bec | Laboratoire de santv© publique du Quv©bec | Sandrine Moreira et al |
| EPI_ISL_825824 | Canada/Qc-CHUM-2015702612A/202 | 2020-06-05 | North America/Canada/Quebec/ | Laboratoire de santv© publique du Quv©bec | Laboratoire de santv© publique du Quv©bec | Sandrine Moreira et al |
| EPI_ISL_825825 | Canada/Qc-CHUM-2016904161A/202 | 2020-06-17 | North America/Canada/Quebec/ | Laboratoire de santv© publique du Quv©bec | Laboratoire de santv© publique du Quv©bec | Sandrine Moreira et al |
| EPI_ISL_825826 | Canada/Qc-CHUM-2016902794A/202 | 2020-06-17 | North America/Canada/Quebec/ | Laboratoire de santv© publique du Quv©bec | Laboratoire de santv© publique du Quv©bec | Sandrine Moreira et al |

| | | | | | | |
|---|---|---|---|---|---|---|
| EPI_ISL_825827 | Canada/Qc-CHUM-2016901949A/202 | 2020-06-17 | North America/Canada/Quebec/ | Laboratoire de sant√© publique du Qu√©bec | Laboratoire de sant√© publique du Qu√©bec | Sandrine Moreira et al |
| EPI_ISL_825828 | Canada/Qc-CHUM-2017003523A/202 | 2020-06-18 | North America/Canada/Quebec/ | Laboratoire de sant√© publique du Qu√©bec | Laboratoire de sant√© publique du Qu√©bec | Sandrine Moreira et al |
| EPI_ISL_825829 | Canada/Qc-CHUM-2016103625A/202 | 2020-06-09 | North America/Canada/Quebec/ | Laboratoire de sant√© publique du Qu√©bec | Laboratoire de sant√© publique du Qu√©bec | Sandrine Moreira et al |
| EPI_ISL_825830 | Canada/Qc-CHUM-2016204473A/202 | 2020-06-10 | North America/Canada/Quebec/ | Laboratoire de sant√© publique du Qu√©bec | Laboratoire de sant√© publique du Qu√©bec | Sandrine Moreira et al |
| EPI_ISL_825831 | Canada/Qc-CHUM-2016601956A/202 | 2020-06-14 | North America/Canada/Quebec/ | Laboratoire de sant√© publique du Qu√©bec | Laboratoire de sant√© publique du Qu√©bec | Sandrine Moreira et al |
| EPI_ISL_825832 | Canada/Qc-CHUM-2016602180A/202 | 2020-06-14 | North America/Canada/Quebec/ | Laboratoire de sant√© publique du Qu√©bec | Laboratoire de sant√© publique du Qu√©bec | Sandrine Moreira et al |
| EPI_ISL_825833 | Canada/Qc-JEW-Q7252106/2020 | 2020-03-25 | North America/Canada/Quebec/ | Laboratoire de sant√© publique du Qu√©bec | Laboratoire de sant√© publique du Qu√©bec | Sandrine Moreira et al |
| EPI_ISL_825834 | Canada/Qc-JEW-Q8161203/2020 | 2020-04-16 | North America/Canada/Quebec/ | Laboratoire de sant√© publique du Qu√©bec | Laboratoire de sant√© publique du Qu√©bec | Sandrine Moreira et al |
| EPI_ISL_825835 | Canada/Qc-JEW-Q8071124/2020 | 2020-04-07 | North America/Canada/Quebec/ | Laboratoire de sant√© publique du Qu√©bec | Laboratoire de sant√© publique du Qu√©bec | Sandrine Moreira et al |
| EPI_ISL_825836 | Canada/Qc-JEW-Q8071169/2020 | 2020-04-07 | North America/Canada/Quebec/ | Laboratoire de sant√© publique du Qu√©bec | Laboratoire de sant√© publique du Qu√©bec | Sandrine Moreira et al |
| EPI_ISL_825837 | Canada/Qc-JEW-Q8171094/2020 | 2020-04-17 | North America/Canada/Quebec/ | Laboratoire de sant√© publique du Qu√©bec | Laboratoire de sant√© publique du Qu√©bec | Sandrine Moreira et al |
| EPI_ISL_825838 | Canada/Qc-JEW-Q8261037/2020 | 2020-04-26 | North America/Canada/Quebec/ | Laboratoire de sant√© publique du Qu√©bec | Laboratoire de sant√© publique du Qu√©bec | Sandrine Moreira et al |
| EPI_ISL_825839 | Canada/Qc-JEW-Q9300036/2020 | 2020-05-29 | North America/Canada/Quebec/ | Laboratoire de sant√© publique du Qu√©bec | Laboratoire de sant√© publique du Qu√©bec | Sandrine Moreira et al |

| | | | | | | |
|---|---|---|---|---|---|---|
| EPI_ISL_825840 | Canada/Qc-JEW-Q9230176/2020 | 2020-05-22 | North America/Canada/Quebec/ | Laboratoire de sant√© publique du Qu√©bec | Laboratoire de sant√© publique du Qu√©bec | Sandrine Moreira et al |
| EPI_ISL_825841 | Canada/Qc-JEW-Q9250323/2020 | 2020-05-25 | North America/Canada/Quebec/ | Laboratoire de sant√© publique du Qu√©bec | Laboratoire de sant√© publique du Qu√©bec | Sandrine Moreira et al |
| EPI_ISL_825842 | Canada/Qc-JEW-Q9282167/2020 | 2020-05-28 | North America/Canada/Quebec/ | Laboratoire de sant√© publique du Qu√©bec | Laboratoire de sant√© publique du Qu√©bec | Sandrine Moreira et al |
| EPI_ISL_825843 | Canada/Qc-JEW-Q9212154/2020 | 2020-05-21 | North America/Canada/Quebec/ | Laboratoire de sant√© publique du Qu√©bec | Laboratoire de sant√© publique du Qu√©bec | Sandrine Moreira et al |
| EPI_ISL_825844 | Canada/Qc-JEW-Q9212960/2020 | 2020-05-21 | North America/Canada/Quebec/ | Laboratoire de sant√© publique du Qu√©bec | Laboratoire de sant√© publique du Qu√©bec | Sandrine Moreira et al |
| EPI_ISL_825845 | Canada/Qc-JEW-Q9290119/2020 | 2020-05-28 | North America/Canada/Quebec/ | Laboratoire de sant√© publique du Qu√©bec | Laboratoire de sant√© publique du Qu√©bec | Sandrine Moreira et al |
| EPI_ISL_825846 | Canada/Qc-JEW-Q9292503/2020 | 2020-05-29 | North America/Canada/Quebec/ | Laboratoire de sant√© publique du Qu√©bec | Laboratoire de sant√© publique du Qu√©bec | Sandrine Moreira et al |
| EPI_ISL_825847 | Canada/Qc-JEW-Q9292177/2020 | 2020-05-29 | North America/Canada/Quebec/ | Laboratoire de sant√© publique du Qu√©bec | Laboratoire de sant√© publique du Qu√©bec | Sandrine Moreira et al |
| EPI_ISL_825848 | Canada/Qc-JEW-Q9300658/2020 | 2020-05-30 | North America/Canada/Quebec/ | Laboratoire de sant√© publique du Qu√©bec | Laboratoire de sant√© publique du Qu√©bec | Sandrine Moreira et al |
| EPI_ISL_825849 | Canada/Qc-JEW-Q9301179/2020 | 2020-05-30 | North America/Canada/Quebec/ | Laboratoire de sant√© publique du Qu√©bec | Laboratoire de sant√© publique du Qu√©bec | Sandrine Moreira et al |
| EPI_ISL_825850 | Canada/Qc-JEW-Q9311664/2020 | 2020-05-31 | North America/Canada/Quebec/ | Laboratoire de sant√© publique du Qu√©bec | Laboratoire de sant√© publique du Qu√©bec | Sandrine Moreira et al |
| EPI_ISL_825851 | Canada/Qc-JEW-Q9222456/2020 | 2020-05-22 | North America/Canada/Quebec/ | Laboratoire de sant√© publique du Qu√©bec | Laboratoire de sant√© publique du Qu√©bec | Sandrine Moreira et al |
| EPI_ISL_825852 | Canada/Qc-JEW-Q9221714/2020 | 2020-05-22 | North America/Canada/Quebec/ | Laboratoire de sant√© publique du Qu√©bec | Laboratoire de sant√© publique du Qu√©bec | Sandrine Moreira et al |

| | | | | | | |
|---|---|---|---|---|---|---|
| EPI_ISL_825853 | Canada/Qc-JEW-Q9221088/2020 | 2020-05-22 | North America/Canada/Quebec/ | Laboratoire de sant√© publique du Qu√©bec | Laboratoire de sant√© publique du Qu√©bec | Sandrine Moreira et al |
| EPI_ISL_825854 | Canada/Qc-JEW-Q9231261/2020 | 2020-05-23 | North America/Canada/Quebec/ | Laboratoire de sant√© publique du Qu√©bec | Laboratoire de sant√© publique du Qu√©bec | Sandrine Moreira et al |
| EPI_ISL_825855 | Canada/Qc-JEW-Q9231098/2020 | 2020-05-23 | North America/Canada/Quebec/ | Laboratoire de sant√© publique du Qu√©bec | Laboratoire de sant√© publique du Qu√©bec | Sandrine Moreira et al |
| EPI_ISL_825856 | Canada/Qc-JEW-Q9251384/2020 | 2020-05-25 | North America/Canada/Quebec/ | Laboratoire de sant√© publique du Qu√©bec | Laboratoire de sant√© publique du Qu√©bec | Sandrine Moreira et al |
| EPI_ISL_825857 | Canada/Qc-JEW-Q9241570/2020 | 2020-05-24 | North America/Canada/Quebec/ | Laboratoire de sant√© publique du Qu√©bec | Laboratoire de sant√© publique du Qu√©bec | Sandrine Moreira et al |
| EPI_ISL_825858 | Canada/Qc-JEW-Q8251296/2020 | 2020-04-25 | North America/Canada/Quebec/ | Laboratoire de sant√© publique du Qu√©bec | Laboratoire de sant√© publique du Qu√©bec | Sandrine Moreira et al |
| EPI_ISL_825859 | Canada/Qc-JEW-Q8251224/2020 | 2020-04-25 | North America/Canada/Quebec/ | Laboratoire de sant√© publique du Qu√©bec | Laboratoire de sant√© publique du Qu√©bec | Sandrine Moreira et al |
| EPI_ISL_825860 | Canada/Qc-JEW-Q8261012/2020 | 2020-04-26 | North America/Canada/Quebec/ | Laboratoire de sant√© publique du Qu√©bec | Laboratoire de sant√© publique du Qu√©bec | Sandrine Moreira et al |
| EPI_ISL_825861 | Canada/Qc-JEW-Q8260813/2020 | 2020-04-26 | North America/Canada/Quebec/ | Laboratoire de sant√© publique du Qu√©bec | Laboratoire de sant√© publique du Qu√©bec | Sandrine Moreira et al |
| EPI_ISL_825862 | Canada/Qc-JEW-Q8261158/2020 | 2020-04-26 | North America/Canada/Quebec/ | Laboratoire de sant√© publique du Qu√©bec | Laboratoire de sant√© publique du Qu√©bec | Sandrine Moreira et al |
| EPI_ISL_825863 | Canada/Qc-JEW-Q8170760/2020 | 2020-04-16 | North America/Canada/Quebec/ | Laboratoire de sant√© publique du Qu√©bec | Laboratoire de sant√© publique du Qu√©bec | Sandrine Moreira et al |
| EPI_ISL_825864 | Canada/Qc-JEW-Q8170738/2020 | 2020-04-17 | North America/Canada/Quebec/ | Laboratoire de sant√© publique du Qu√©bec | Laboratoire de sant√© publique du Qu√©bec | Sandrine Moreira et al |
| EPI_ISL_825865 | Canada/Qc-JEW-Q8171649/2020 | 2020-04-17 | North America/Canada/Quebec/ | Laboratoire de sant√© publique du Qu√©bec | Laboratoire de sant√© publique du Qu√©bec | Sandrine Moreira et al |

| | | | | | | |
|---|---|---|---|---|---|---|
| EPI_ISL_825866 | Canada/Qc-JEW-Q7271813/2020 | 2020-03-27 | North America/Canada/Quebec/ | Laboratoire de santv© publique du Quv©bec | Laboratoire de santv© publique du Quv©bec | Sandrine Moreira et al |
| EPI_ISL_825867 | Canada/Qc-JEW-Q8201234/2020 | 2020-04-20 | North America/Canada/Quebec/ | Laboratoire de santv© publique du Quv©bec | Laboratoire de santv© publique du Quv©bec | Sandrine Moreira et al |
| EPI_ISL_825868 | Canada/Qc-JEW-Q8161213/2020 | 2020-04-16 | North America/Canada/Quebec/ | Laboratoire de santv© publique du Quv©bec | Laboratoire de santv© publique du Quv©bec | Sandrine Moreira et al |
| EPI_ISL_825869 | Canada/Qc-JEW-Q8181007/2020 | 2020-04-18 | North America/Canada/Quebec/ | Laboratoire de santv© publique du Quv©bec | Laboratoire de santv© publique du Quv©bec | Sandrine Moreira et al |
| EPI_ISL_825870 | Canada/Qc-JEW-Q8171853/2020 | 2020-04-17 | North America/Canada/Quebec/ | Laboratoire de santv© publique du Quv©bec | Laboratoire de santv© publique du Quv©bec | Sandrine Moreira et al |
| EPI_ISL_825871 | Canada/Qc-JEW-Q8181012/2020 | 2020-04-18 | North America/Canada/Quebec/ | Laboratoire de santv© publique du Quv©bec | Laboratoire de santv© publique du Quv©bec | Sandrine Moreira et al |
| EPI_ISL_825872 | Canada/Qc-JEW-Q8171883/2020 | 2020-04-17 | North America/Canada/Quebec/ | Laboratoire de santv© publique du Quv©bec | Laboratoire de santv© publique du Quv©bec | Sandrine Moreira et al |
| EPI_ISL_825873 | Canada/Qc-JEW-Q8180884/2020 | 2020-04-18 | North America/Canada/Quebec/ | Laboratoire de santv© publique du Quv©bec | Laboratoire de santv© publique du Quv©bec | Sandrine Moreira et al |
| EPI_ISL_825874 | Canada/Qc-JEW-Q8171856/2020 | 2020-04-17 | North America/Canada/Quebec/ | Laboratoire de santv© publique du Quv©bec | Laboratoire de santv© publique du Quv©bec | Sandrine Moreira et al |
| EPI_ISL_825875 | Canada/Qc-JEW-Q8211400/2020 | 2020-04-21 | North America/Canada/Quebec/ | Laboratoire de santv© publique du Quv©bec | Laboratoire de santv© publique du Quv©bec | Sandrine Moreira et al |
| EPI_ISL_825876 | Canada/Qc-JEW-Q8210901/2020 | 2020-04-21 | North America/Canada/Quebec/ | Laboratoire de santv© publique du Quv©bec | Laboratoire de santv© publique du Quv©bec | Sandrine Moreira et al |
| EPI_ISL_825877 | Canada/Qc-JEW-Q8211381/2020 | 2020-04-21 | North America/Canada/Quebec/ | Laboratoire de santv© publique du Quv©bec | Laboratoire de santv© publique du Quv©bec | Sandrine Moreira et al |
| EPI_ISL_825878 | Canada/Qc-JEW-Q8211377/2020 | 2020-04-21 | North America/Canada/Quebec/ | Laboratoire de santv© publique du Quv©bec | Laboratoire de santv© publique du Quv©bec | Sandrine Moreira et al |

| | | | | | | |
|---|---|---|---|---|---|---|
| EPI_ISL_825879 | Canada/Qc-JEW-Q8190787/2020 | 2020-04-19 | North America/Canada/Quebec/ | Laboratoire de sant√© publique du Qu√©bec | Laboratoire de sant√© publique du Qu√©bec | Sandrine Moreira et al |
| EPI_ISL_825880 | Canada/Qc-JEW-Q8221162/2020 | 2020-04-22 | North America/Canada/Quebec/ | Laboratoire de sant√© publique du Qu√©bec | Laboratoire de sant√© publique du Qu√©bec | Sandrine Moreira et al |
| EPI_ISL_825881 | Canada/Qc-JEW-Q8220923/2020 | 2020-04-22 | North America/Canada/Quebec/ | Laboratoire de sant√© publique du Qu√©bec | Laboratoire de sant√© publique du Qu√©bec | Sandrine Moreira et al |
| EPI_ISL_825882 | Canada/Qc-JEW-Q8221592/2020 | 2020-04-22 | North America/Canada/Quebec/ | Laboratoire de sant√© publique du Qu√©bec | Laboratoire de sant√© publique du Qu√©bec | Sandrine Moreira et al |
| EPI_ISL_825883 | Canada/Qc-JEW-Q8221354/2020 | 2020-04-22 | North America/Canada/Quebec/ | Laboratoire de sant√© publique du Qu√©bec | Laboratoire de sant√© publique du Qu√©bec | Sandrine Moreira et al |
| EPI_ISL_825884 | Canada/Qc-JEW-Q8220874/2020 | 2020-04-22 | North America/Canada/Quebec/ | Laboratoire de sant√© publique du Qu√©bec | Laboratoire de sant√© publique du Qu√©bec | Sandrine Moreira et al |
| EPI_ISL_825885 | Canada/Qc-JEW-Q8231623/2020 | 2020-04-23 | North America/Canada/Quebec/ | Laboratoire de sant√© publique du Qu√©bec | Laboratoire de sant√© publique du Qu√©bec | Sandrine Moreira et al |
| EPI_ISL_825886 | Canada/Qc-JEW-Q8231148/2020 | 2020-04-23 | North America/Canada/Quebec/ | Laboratoire de sant√© publique du Qu√©bec | Laboratoire de sant√© publique du Qu√©bec | Sandrine Moreira et al |
| EPI_ISL_825887 | Canada/Qc-JEW-Q8231486/2020 | 2020-04-23 | North America/Canada/Quebec/ | Laboratoire de sant√© publique du Qu√©bec | Laboratoire de sant√© publique du Qu√©bec | Sandrine Moreira et al |
| EPI_ISL_825888 | Canada/Qc-JEW-Q8231310/2020 | 2020-04-23 | North America/Canada/Quebec/ | Laboratoire de sant√© publique du Qu√©bec | Laboratoire de sant√© publique du Qu√©bec | Sandrine Moreira et al |
| EPI_ISL_825890 | Canada/Qc-CHUM-2013302940A/202 | 2020-05-12 | North America/Canada/Quebec/ | Laboratoire de sant√© publique du Qu√©bec | Laboratoire de sant√© publique du Qu√©bec | Sandrine Moreira et al |
| EPI_ISL_825891 | Canada/Qc-CHUM-2013304495A/202 | 2020-05-12 | North America/Canada/Quebec/ | Laboratoire de sant√© publique du Qu√©bec | Laboratoire de sant√© publique du Qu√©bec | Sandrine Moreira et al |
| EPI_ISL_825892 | Canada/Qc-CHUM-2013301742A/202 | 2020-05-12 | North America/Canada/Quebec/ | Laboratoire de sant√© publique du Qu√©bec | Laboratoire de sant√© publique du Qu√©bec | Sandrine Moreira et al |

| | | | | | | |
|---|---|---|---|---|---|---|
| EPI_ISL_825893 | Canada/Qc-CHUM-2014004637A/202 | 2020-05-19 | North America/Canada/Quebec/ | Laboratoire de sant√© publique du Qu√©bec | Laboratoire de sant√© publique du Qu√©bec | Sandrine Moreira et al |
| EPI_ISL_825894 | Canada/Qc-CHUM-2013304054A/202 | 2020-05-12 | North America/Canada/Quebec/ | Laboratoire de sant√© publique du Qu√©bec | Laboratoire de sant√© publique du Qu√©bec | Sandrine Moreira et al |
| EPI_ISL_825895 | Canada/Qc-CHUM-2013204321A/202 | 2020-05-11 | North America/Canada/Quebec/ | Laboratoire de sant√© publique du Qu√©bec | Laboratoire de sant√© publique du Qu√©bec | Sandrine Moreira et al |
| EPI_ISL_825897 | Canada/Qc-CHUM-2014004967A/202 | 2020-05-19 | North America/Canada/Quebec/ | Laboratoire de sant√© publique du Qu√©bec | Laboratoire de sant√© publique du Qu√©bec | Sandrine Moreira et al |
| EPI_ISL_825898 | Canada/Qc-CHUM-2013403735A/202 | 2020-05-13 | North America/Canada/Quebec/ | Laboratoire de sant√© publique du Qu√©bec | Laboratoire de sant√© publique du Qu√©bec | Sandrine Moreira et al |
| EPI_ISL_825899 | Canada/Qc-CHUM-2013302912A/202 | 2020-05-12 | North America/Canada/Quebec/ | Laboratoire de sant√© publique du Qu√©bec | Laboratoire de sant√© publique du Qu√©bec | Sandrine Moreira et al |
| EPI_ISL_825900 | Canada/Qc-CHUM-2013204306A/202 | 2020-05-11 | North America/Canada/Quebec/ | Laboratoire de sant√© publique du Qu√©bec | Laboratoire de sant√© publique du Qu√©bec | Sandrine Moreira et al |
| EPI_ISL_825901 | Canada/Qc-CHUM-2013301412A/202 | 2020-05-12 | North America/Canada/Quebec/ | Laboratoire de sant√© publique du Qu√©bec | Laboratoire de sant√© publique du Qu√©bec | Sandrine Moreira et al |
| EPI_ISL_825902 | Canada/Qc-CHUM-2014103626A/202 | 2020-05-20 | North America/Canada/Quebec/ | Laboratoire de sant√© publique du Qu√©bec | Laboratoire de sant√© publique du Qu√©bec | Sandrine Moreira et al |
| EPI_ISL_825903 | Canada/Qc-CHUM-2013201954A/202 | 2020-05-11 | North America/Canada/Quebec/ | Laboratoire de sant√© publique du Qu√©bec | Laboratoire de sant√© publique du Qu√©bec | Sandrine Moreira et al |
| EPI_ISL_825904 | Canada/Qc-CHUM-2012704148A/202 | 2020-05-06 | North America/Canada/Quebec/ | Laboratoire de sant√© publique du Qu√©bec | Laboratoire de sant√© publique du Qu√©bec | Sandrine Moreira et al |
| EPI_ISL_825905 | Canada/Qc-CHUM-2013203942A/202 | 2020-05-11 | North America/Canada/Quebec/ | Laboratoire de sant√© publique du Qu√©bec | Laboratoire de sant√© publique du Qu√©bec | Sandrine Moreira et al |
| EPI_ISL_825906 | Canada/Qc-CHUM-2014004802A/202 | 2020-05-19 | North America/Canada/Quebec/ | Laboratoire de sant√© publique du Qu√©bec | Laboratoire de sant√© publique du Qu√©bec | Sandrine Moreira et al |

| | | | | | | |
|---|---|---|---|---|---|---|
| EPI_ISL_825908 | Canada/Qc-CHUM-2017703684a/202( | 2020-06-25 | North America/Canada/Quebec/ | Laboratoire de santv© publique du Quv©bec | Laboratoire de santv© publique du Quv©bec | Sandrine Moreira et al |
| EPI_ISL_825909 | Canada/Qc-CHUM-2017802870a/202( | 2020-06-26 | North America/Canada/Quebec/ | Laboratoire de santv© publique du Quv©bec | Laboratoire de santv© publique du Quv©bec | Sandrine Moreira et al |
| EPI_ISL_825910 | Canada/Qc-CHUM-2017802304a/202( | 2020-06-26 | North America/Canada/Quebec/ | Laboratoire de santv© publique du Quv©bec | Laboratoire de santv© publique du Quv©bec | Sandrine Moreira et al |
| EPI_ISL_825911 | Canada/Qc-CHUM-2018200019a/202( | 2020-06-30 | North America/Canada/Quebec/ | Laboratoire de santv© publique du Quv©bec | Laboratoire de santv© publique du Quv©bec | Sandrine Moreira et al |
| EPI_ISL_825912 | Canada/Qc-CHUM-2018701105a/202( | 2020-07-05 | North America/Canada/Quebec/ | Laboratoire de santv© publique du Quv©bec | Laboratoire de santv© publique du Quv©bec | Sandrine Moreira et al |
| EPI_ISL_825913 | Canada/Qc-CHUM-2017902233a/202( | 2020-06-27 | North America/Canada/Quebec/ | Laboratoire de santv© publique du Quv©bec | Laboratoire de santv© publique du Quv©bec | Sandrine Moreira et al |
| EPI_ISL_825914 | Canada/Qc-CHUM-2018802175a/202( | 2020-07-06 | North America/Canada/Quebec/ | Laboratoire de santv© publique du Quv©bec | Laboratoire de santv© publique du Quv©bec | Sandrine Moreira et al |
| EPI_ISL_825915 | Canada/Qc-CHUM-2018701642a/202( | 2020-07-05 | North America/Canada/Quebec/ | Laboratoire de santv© publique du Quv©bec | Laboratoire de santv© publique du Quv©bec | Sandrine Moreira et al |
| EPI_ISL_825916 | Canada/Qc-CHUM-2018403857A/202 | 2020-07-02 | North America/Canada/Quebec/ | Laboratoire de santv© publique du Quv©bec | Laboratoire de santv© publique du Quv©bec | Sandrine Moreira et al |
| EPI_ISL_825917 | Canada/Qc-CHUM-2018403593A/202 | 2020-07-02 | North America/Canada/Quebec/ | Laboratoire de santv© publique du Quv©bec | Laboratoire de santv© publique du Quv©bec | Sandrine Moreira et al |
| EPI_ISL_825918 | Canada/Qc-CHUM-2018403482A/202 | 2020-07-02 | North America/Canada/Quebec/ | Laboratoire de santv© publique du Quv©bec | Laboratoire de santv© publique du Quv©bec | Sandrine Moreira et al |
| EPI_ISL_825919 | Canada/Qc-CHUM-2018601988A/202 | 2020-07-04 | North America/Canada/Quebec/ | Laboratoire de santv© publique du Quv©bec | Laboratoire de santv© publique du Quv©bec | Sandrine Moreira et al |
| EPI_ISL_825920 | Canada/Qc-CHUM-2013400470A/202 | 2020-05-13 | North America/Canada/Quebec/ | Laboratoire de santv© publique du Quv©bec | Laboratoire de santv© publique du Quv©bec | Sandrine Moreira et al |

| | | | | | | |
|---|---|---|---|---|---|---|
| EPI_ISL_825921 | Canada/Qc-CHUM-2013304817A/202 | 2020-05-12 | North America/Canada/Quebec/ | Laboratoire de santv© publique du Quv©bec | Laboratoire de santv© publique du Quv©bec | Sandrine Moreira et al |
| EPI_ISL_825922 | Canada/Qc-CHUM-2013303019A/202 | 2020-05-12 | North America/Canada/Quebec/ | Laboratoire de santv© publique du Quv©bec | Laboratoire de santv© publique du Quv©bec | Sandrine Moreira et al |
| EPI_ISL_825923 | Canada/Qc-CHUM-2013302964A/202 | 2020-05-12 | North America/Canada/Quebec/ | Laboratoire de santv© publique du Quv©bec | Laboratoire de santv© publique du Quv©bec | Sandrine Moreira et al |
| EPI_ISL_825924 | Canada/Qc-CHUM-2013302889A/202 | 2020-05-12 | North America/Canada/Quebec/ | Laboratoire de santv© publique du Quv©bec | Laboratoire de santv© publique du Quv©bec | Sandrine Moreira et al |
| EPI_ISL_825925 | Canada/Qc-CHUM-2013302783A/202 | 2020-05-12 | North America/Canada/Quebec/ | Laboratoire de santv© publique du Quv©bec | Laboratoire de santv© publique du Quv©bec | Sandrine Moreira et al |
| EPI_ISL_825926 | Canada/Qc-CHUM-2014003889A/202 | 2020-05-19 | North America/Canada/Quebec/ | Laboratoire de santv© publique du Quv©bec | Laboratoire de santv© publique du Quv©bec | Sandrine Moreira et al |
| EPI_ISL_825927 | Canada/Qc-CHUM-2014004169A/202 | 2020-05-19 | North America/Canada/Quebec/ | Laboratoire de santv© publique du Quv©bec | Laboratoire de santv© publique du Quv©bec | Sandrine Moreira et al |
| EPI_ISL_825928 | Canada/Qc-CHUM-2019104444A/202 | 2020-07-09 | North America/Canada/Quebec/ | Laboratoire de santv© publique du Quv©bec | Laboratoire de santv© publique du Quv©bec | Sandrine Moreira et al |
| EPI_ISL_825929 | Canada/Qc-CHUM-2019203102A/202 | 2020-07-10 | North America/Canada/Quebec/ | Laboratoire de santv© publique du Quv©bec | Laboratoire de santv© publique du Quv©bec | Sandrine Moreira et al |
| EPI_ISL_825930 | Canada/Qc-CHUM-2019104892A/202 | 2020-07-09 | North America/Canada/Quebec/ | Laboratoire de santv© publique du Quv©bec | Laboratoire de santv© publique du Quv©bec | Sandrine Moreira et al |
| EPI_ISL_825931 | Canada/Qc-CHUM-2019203073A/202 | 2020-07-10 | North America/Canada/Quebec/ | Laboratoire de santv© publique du Quv©bec | Laboratoire de santv© publique du Quv©bec | Sandrine Moreira et al |
| EPI_ISL_825932 | Canada/Qc-CHUM-2019203856A/202 | 2020-07-10 | North America/Canada/Quebec/ | Laboratoire de santv© publique du Quv©bec | Laboratoire de santv© publique du Quv©bec | Sandrine Moreira et al |
| EPI_ISL_825933 | Canada/Qc-CHUM-2019202525A/202 | 2020-07-10 | North America/Canada/Quebec/ | Laboratoire de santv© publique du Quv©bec | Laboratoire de santv© publique du Quv©bec | Sandrine Moreira et al |

| | | | | | | |
|---|---|---|---|---|---|---|
| EPI_ISL_825934 | Canada/Qc-CHUM-2019102951A/202 | 2020-07-09 | North America/Canada/Quebec/ | Laboratoire de sant√© publique du Qu√©bec | Laboratoire de sant√© publique du Qu√©bec | Sandrine Moreira et al |
| EPI_ISL_825935 | Canada/Qc-CHUM-2019203591A/202 | 2020-07-10 | North America/Canada/Quebec/ | Laboratoire de sant√© publique du Qu√©bec | Laboratoire de sant√© publique du Qu√©bec | Sandrine Moreira et al |
| EPI_ISL_825936 | Canada/Qc-CHUM-2019301930A/202 | 2020-07-11 | North America/Canada/Quebec/ | Laboratoire de sant√© publique du Qu√©bec | Laboratoire de sant√© publique du Qu√©bec | Sandrine Moreira et al |
| EPI_ISL_825937 | Canada/Qc-CHUM-2019301899A/202 | 2020-07-11 | North America/Canada/Quebec/ | Laboratoire de sant√© publique du Qu√©bec | Laboratoire de sant√© publique du Qu√©bec | Sandrine Moreira et al |
| EPI_ISL_825938 | Canada/Qc-CHUM-2019301640A/202 | 2020-07-11 | North America/Canada/Quebec/ | Laboratoire de sant√© publique du Qu√©bec | Laboratoire de sant√© publique du Qu√©bec | Sandrine Moreira et al |
| EPI_ISL_825939 | Canada/Qc-CHUM-2019301732A/202 | 2020-07-11 | North America/Canada/Quebec/ | Laboratoire de sant√© publique du Qu√©bec | Laboratoire de sant√© publique du Qu√©bec | Sandrine Moreira et al |
| EPI_ISL_825940 | Canada/Qc-CHUM-2015201391A/202 | 2020-05-31 | North America/Canada/Quebec/ | Laboratoire de sant√© publique du Qu√©bec | Laboratoire de sant√© publique du Qu√©bec | Sandrine Moreira et al |
| EPI_ISL_825941 | Canada/Qc-CHUM-2019301903A/202 | 2020-07-11 | North America/Canada/Quebec/ | Laboratoire de sant√© publique du Qu√©bec | Laboratoire de sant√© publique du Qu√©bec | Sandrine Moreira et al |
| EPI_ISL_825942 | Canada/Qc-CHUM-2013203919A/202 | 2020-05-11 | North America/Canada/Quebec/ | Laboratoire de sant√© publique du Qu√©bec | Laboratoire de sant√© publique du Qu√©bec | Sandrine Moreira et al |
| EPI_ISL_825943 | Canada/Qc-CHUM-2013204075A/202 | 2020-05-11 | North America/Canada/Quebec/ | Laboratoire de sant√© publique du Qu√©bec | Laboratoire de sant√© publique du Qu√©bec | Sandrine Moreira et al |
| EPI_ISL_825944 | Canada/Qc-CHUM-2013304473A/202 | 2020-05-12 | North America/Canada/Quebec/ | Laboratoire de sant√© publique du Qu√©bec | Laboratoire de sant√© publique du Qu√©bec | Sandrine Moreira et al |
| EPI_ISL_825945 | Canada/Qc-CHUM-2013204253A/202 | 2020-05-11 | North America/Canada/Quebec/ | Laboratoire de sant√© publique du Qu√©bec | Laboratoire de sant√© publique du Qu√©bec | Sandrine Moreira et al |
| EPI_ISL_825946 | Canada/Qc-CHUM-2013204355A/202 | 2020-05-11 | North America/Canada/Quebec/ | Laboratoire de sant√© publique du Qu√©bec | Laboratoire de sant√© publique du Qu√©bec | Sandrine Moreira et al |

| | | | | | | |
|---|---|---|---|---|---|---|
| EPI_ISL_825947 | Canada/Qc-CHUM-2012502376A/202 | 2020-05-04 | North America/Canada/Quebec/ | Laboratoire de santv© publique du Quv©bec | Laboratoire de santv© publique du Quv©bec | Sandrine Moreira et al |
| EPI_ISL_825948 | Canada/Qc-CHUM-2012602495A/202 | 2020-05-05 | North America/Canada/Quebec/ | Laboratoire de santv© publique du Quv©bec | Laboratoire de santv© publique du Quv©bec | Sandrine Moreira et al |
| EPI_ISL_825950 | Canada/Qc-JEW-Q9051497/2020 | 2020-05-05 | North America/Canada/Quebec/ | Laboratoire de santv© publique du Quv©bec | Laboratoire de santv© publique du Quv©bec | Sandrine Moreira et al |
| EPI_ISL_825951 | Canada/Qc-JEW-Q8301447/2020 | 2020-04-30 | North America/Canada/Quebec/ | Laboratoire de santv© publique du Quv©bec | Laboratoire de santv© publique du Quv©bec | Sandrine Moreira et al |
| EPI_ISL_825952 | Canada/Qc-L00259923/2020 | 2020-05-16 | North America/Canada/Quebec/ | Laboratoire de santv© publique du Quv©bec | Laboratoire de santv© publique du Quv©bec | Sandrine Moreira et al |
| EPI_ISL_825953 | Canada/Qc-JEW-Q9090694/2020 | 2020-05-09 | North America/Canada/Quebec/ | Laboratoire de santv© publique du Quv©bec | Laboratoire de santv© publique du Quv©bec | Sandrine Moreira et al |
| EPI_ISL_825954 | Canada/Qc-JEW-Q9081772/2020 | 2020-05-08 | North America/Canada/Quebec/ | Laboratoire de santv© publique du Quv©bec | Laboratoire de santv© publique du Quv©bec | Sandrine Moreira et al |
| EPI_ISL_825955 | Canada/Qc-JEW-Q9071108/2020 | 2020-05-07 | North America/Canada/Quebec/ | Laboratoire de santv© publique du Quv©bec | Laboratoire de santv© publique du Quv©bec | Sandrine Moreira et al |
| EPI_ISL_825956 | Canada/Qc-JEW-Q8291436/2020 | 2020-04-29 | North America/Canada/Quebec/ | Laboratoire de santv© publique du Quv©bec | Laboratoire de santv© publique du Quv©bec | Sandrine Moreira et al |
| EPI_ISL_825957 | Canada/Qc-JEW-Q9071871/2020 | 2020-05-07 | North America/Canada/Quebec/ | Laboratoire de santv© publique du Quv©bec | Laboratoire de santv© publique du Quv©bec | Sandrine Moreira et al |
| EPI_ISL_825958 | Canada/Qc-JEW-Q9062730/2020 | 2020-05-06 | North America/Canada/Quebec/ | Laboratoire de santv© publique du Quv©bec | Laboratoire de santv© publique du Quv©bec | Sandrine Moreira et al |
| EPI_ISL_825959 | Canada/Qc-JEW-Q8291780/2020 | 2020-04-29 | North America/Canada/Quebec/ | Laboratoire de santv© publique du Quv©bec | Laboratoire de santv© publique du Quv©bec | Sandrine Moreira et al |
| EPI_ISL_825961 | Canada/Qc-JEW-Q9090667/2020 | 2020-05-09 | North America/Canada/Quebec/ | Laboratoire de santv© publique du Quv©bec | Laboratoire de santv© publique du Quv©bec | Sandrine Moreira et al |

| | | | | | | |
|---|---|---|---|---|---|---|
| EPI_ISL_825962 | Canada/Qc-JEW-Q9030968/2020 | 2020-05-03 | North America/Canada/Quebec/ | Laboratoire de sant√© publique du Qu√©bec | Laboratoire de sant√© publique du Qu√©bec | Sandrine Moreira et al |
| EPI_ISL_825963 | Canada/Qc-JEW-Q9062020/2020 | 2020-05-06 | North America/Canada/Quebec/ | Laboratoire de sant√© publique du Qu√©bec | Laboratoire de sant√© publique du Qu√©bec | Sandrine Moreira et al |
| EPI_ISL_825964 | Canada/Qc-JEW-Q9071440/2020 | 2020-05-07 | North America/Canada/Quebec/ | Laboratoire de sant√© publique du Qu√©bec | Laboratoire de sant√© publique du Qu√©bec | Sandrine Moreira et al |
| EPI_ISL_825965 | Canada/Qc-JEW-Q9060754/2020 | 2020-05-06 | North America/Canada/Quebec/ | Laboratoire de sant√© publique du Qu√©bec | Laboratoire de sant√© publique du Qu√©bec | Sandrine Moreira et al |
| EPI_ISL_825966 | Canada/Qc-JEW-Q9011398/2020 | 2020-05-01 | North America/Canada/Quebec/ | Laboratoire de sant√© publique du Qu√©bec | Laboratoire de sant√© publique du Qu√©bec | Sandrine Moreira et al |
| EPI_ISL_825967 | Canada/Qc-L00260782/2020 | 2020-05-20 | North America/Canada/Quebec/ | Laboratoire de sant√© publique du Qu√©bec | Laboratoire de sant√© publique du Qu√©bec | Sandrine Moreira et al |
| EPI_ISL_825968 | Canada/Qc-L00259907/2020 | 2020-05-16 | North America/Canada/Quebec/ | Laboratoire de sant√© publique du Qu√©bec | Laboratoire de sant√© publique du Qu√©bec | Sandrine Moreira et al |
| EPI_ISL_825969 | Canada/Qc-L00259940/2020 | 2020-05-16 | North America/Canada/Quebec/ | Laboratoire de sant√© publique du Qu√©bec | Laboratoire de sant√© publique du Qu√©bec | Sandrine Moreira et al |
| EPI_ISL_825970 | Canada/Qc-L00259946/2020 | 2020-05-16 | North America/Canada/Quebec/ | Laboratoire de sant√© publique du Qu√©bec | Laboratoire de sant√© publique du Qu√©bec | Sandrine Moreira et al |
| EPI_ISL_825971 | Canada/Qc-L00259899/2020 | 2020-05-16 | North America/Canada/Quebec/ | Laboratoire de sant√© publique du Qu√©bec | Laboratoire de sant√© publique du Qu√©bec | Sandrine Moreira et al |
| EPI_ISL_825972 | Canada/Qc-L00260284/2020 | 2020-05-18 | North America/Canada/Quebec/ | Laboratoire de sant√© publique du Qu√©bec | Laboratoire de sant√© publique du Qu√©bec | Sandrine Moreira et al |
| EPI_ISL_825973 | Canada/Qc-L00262173/2020 | 2020-05-23 | North America/Canada/Quebec/ | Laboratoire de sant√© publique du Qu√©bec | Laboratoire de sant√© publique du Qu√©bec | Sandrine Moreira et al |
| EPI_ISL_825974 | Canada/Qc-L00259910/2020 | 2020-05-16 | North America/Canada/Quebec/ | Laboratoire de sant√© publique du Qu√©bec | Laboratoire de sant√© publique du Qu√©bec | Sandrine Moreira et al |

| | | | | | | |
|---|---|---|---|---|---|---|
| EPI_ISL_825975 | Canada/Qc-L00263277/2020 | 2020-05-26 | North America/Canada/Quebec/ | Laboratoire de santv© publique du Quv©bec | Laboratoire de santv© publique du Quv©bec | Sandrine Moreira et al |
| EPI_ISL_825976 | Canada/Qc-L00262023/2020 | 2020-05-22 | North America/Canada/Quebec/ | Laboratoire de santv© publique du Quv©bec | Laboratoire de santv© publique du Quv©bec | Sandrine Moreira et al |
| EPI_ISL_825977 | Canada/Qc-JEW-Q9030778/2020 | 2020-05-03 | North America/Canada/Quebec/ | Laboratoire de santv© publique du Quv©bec | Laboratoire de santv© publique du Quv©bec | Sandrine Moreira et al |
| EPI_ISL_825978 | Canada/Qc-JEW-Q9061459/2020 | 2020-05-06 | North America/Canada/Quebec/ | Laboratoire de santv© publique du Quv©bec | Laboratoire de santv© publique du Quv©bec | Sandrine Moreira et al |
| EPI_ISL_825979 | Canada/Qc-JEW-Q8301441/2020 | 2020-04-30 | North America/Canada/Quebec/ | Laboratoire de santv© publique du Quv©bec | Laboratoire de santv© publique du Quv©bec | Sandrine Moreira et al |
| EPI_ISL_825980 | Canada/Qc-JEW-Q8301648/2020 | 2020-04-30 | North America/Canada/Quebec/ | Laboratoire de santv© publique du Quv©bec | Laboratoire de santv© publique du Quv©bec | Sandrine Moreira et al |
| EPI_ISL_825981 | Canada/Qc-JEW-Q8301801/2020 | 2020-04-30 | North America/Canada/Quebec/ | Laboratoire de santv© publique du Quv©bec | Laboratoire de santv© publique du Quv©bec | Sandrine Moreira et al |
| EPI_ISL_825982 | Canada/Qc-JEW-Q8301697/2020 | 2020-04-30 | North America/Canada/Quebec/ | Laboratoire de santv© publique du Quv©bec | Laboratoire de santv© publique du Quv©bec | Sandrine Moreira et al |
| EPI_ISL_825983 | Canada/Qc-JEW-Q8301824/2020 | 2020-04-30 | North America/Canada/Quebec/ | Laboratoire de santv© publique du Quv©bec | Laboratoire de santv© publique du Quv©bec | Sandrine Moreira et al |
| EPI_ISL_825984 | Canada/Qc-JEW-Q8301110/2020 | 2020-04-30 | North America/Canada/Quebec/ | Laboratoire de santv© publique du Quv©bec | Laboratoire de santv© publique du Quv©bec | Sandrine Moreira et al |
| EPI_ISL_825985 | Canada/Qc-JEW-Q9010899/2020 | 2020-05-01 | North America/Canada/Quebec/ | Laboratoire de santv© publique du Quv©bec | Laboratoire de santv© publique du Quv©bec | Sandrine Moreira et al |
| EPI_ISL_825986 | Canada/Qc-JEW-Q9012106/2020 | 2020-05-01 | North America/Canada/Quebec/ | Laboratoire de santv© publique du Quv©bec | Laboratoire de santv© publique du Quv©bec | Sandrine Moreira et al |
| EPI_ISL_825987 | Canada/Qc-JEW-Q9011773/2020 | 2020-05-01 | North America/Canada/Quebec/ | Laboratoire de santv© publique du Quv©bec | Laboratoire de santv© publique du Quv©bec | Sandrine Moreira et al |

| | | | | | | |
|---|---|---|---|---|---|---|
| EPI_ISL_825988 | Canada/Qc-JEW-Q9012104/2020 | 2020-05-01 | North America/Canada/Quebec/ | Laboratoire de sant√© publique du Qu√©bec | Laboratoire de sant√© publique du Qu√©bec | Sandrine Moreira et al |
| EPI_ISL_825989 | Canada/Qc-JEW-Q9012177/2020 | 2020-05-01 | North America/Canada/Quebec/ | Laboratoire de sant√© publique du Qu√©bec | Laboratoire de sant√© publique du Qu√©bec | Sandrine Moreira et al |
| EPI_ISL_825990 | Canada/Qc-JEW-Q9012225/2020 | 2020-05-01 | North America/Canada/Quebec/ | Laboratoire de sant√© publique du Qu√©bec | Laboratoire de sant√© publique du Qu√©bec | Sandrine Moreira et al |
| EPI_ISL_825991 | Canada/Qc-JEW-Q9011089/2020 | 2020-05-01 | North America/Canada/Quebec/ | Laboratoire de sant√© publique du Qu√©bec | Laboratoire de sant√© publique du Qu√©bec | Sandrine Moreira et al |
| EPI_ISL_825992 | Canada/Qc-JEW-Q9011070/2020 | 2020-05-01 | North America/Canada/Quebec/ | Laboratoire de sant√© publique du Qu√©bec | Laboratoire de sant√© publique du Qu√©bec | Sandrine Moreira et al |
| EPI_ISL_825993 | Canada/Qc-JEW-Q9012004/2020 | 2020-05-01 | North America/Canada/Quebec/ | Laboratoire de sant√© publique du Qu√©bec | Laboratoire de sant√© publique du Qu√©bec | Sandrine Moreira et al |
| EPI_ISL_825994 | Canada/Qc-JEW-Q9011710/2020 | 2020-05-01 | North America/Canada/Quebec/ | Laboratoire de sant√© publique du Qu√©bec | Laboratoire de sant√© publique du Qu√©bec | Sandrine Moreira et al |
| EPI_ISL_825995 | Canada/Qc-JEW-Q9030914/2020 | 2020-05-03 | North America/Canada/Quebec/ | Laboratoire de sant√© publique du Qu√©bec | Laboratoire de sant√© publique du Qu√©bec | Sandrine Moreira et al |
| EPI_ISL_825996 | Canada/Qc-JEW-Q9030895/2020 | 2020-05-03 | North America/Canada/Quebec/ | Laboratoire de sant√© publique du Qu√©bec | Laboratoire de sant√© publique du Qu√©bec | Sandrine Moreira et al |
| EPI_ISL_825997 | Canada/Qc-JEW-Q9020985/2020 | 2020-05-02 | North America/Canada/Quebec/ | Laboratoire de sant√© publique du Qu√©bec | Laboratoire de sant√© publique du Qu√©bec | Sandrine Moreira et al |
| EPI_ISL_825998 | Canada/Qc-JEW-Q9030747/2020 | 2020-05-03 | North America/Canada/Quebec/ | Laboratoire de sant√© publique du Qu√©bec | Laboratoire de sant√© publique du Qu√©bec | Sandrine Moreira et al |
| EPI_ISL_825999 | Canada/Qc-JEW-Q9030985/2020 | 2020-05-03 | North America/Canada/Quebec/ | Laboratoire de sant√© publique du Qu√©bec | Laboratoire de sant√© publique du Qu√©bec | Sandrine Moreira et al |
| EPI_ISL_826000 | Canada/Qc-JEW-Q9030978/2020 | 2020-05-03 | North America/Canada/Quebec/ | Laboratoire de sant√© publique du Qu√©bec | Laboratoire de sant√© publique du Qu√©bec | Sandrine Moreira et al |

| | | | | | | |
|---|---|---|---|---|---|---|
| EPI_ISL_826001 | Canada/Qc-JEW-Q9020970/2020 | 2020-05-02 | North America/Canada/Quebec/ | Laboratoire de sant√© publique du Qu√©bec | Laboratoire de sant√© publique du Qu√©bec | Sandrine Moreira et al |
| EPI_ISL_826002 | Canada/Qc-JEW-Q9030279/2020 | 2020-05-03 | North America/Canada/Quebec/ | Laboratoire de sant√© publique du Qu√©bec | Laboratoire de sant√© publique du Qu√©bec | Sandrine Moreira et al |
| EPI_ISL_826003 | Canada/Qc-JEW-Q9030979/2020 | 2020-05-03 | North America/Canada/Quebec/ | Laboratoire de sant√© publique du Qu√©bec | Laboratoire de sant√© publique du Qu√©bec | Sandrine Moreira et al |
| EPI_ISL_826004 | Canada/Qc-JEW-Q9021180/2020 | 2020-05-02 | North America/Canada/Quebec/ | Laboratoire de sant√© publique du Qu√©bec | Laboratoire de sant√© publique du Qu√©bec | Sandrine Moreira et al |
| EPI_ISL_826005 | Canada/Qc-JEW-Q9020857/2020 | 2020-05-02 | North America/Canada/Quebec/ | Laboratoire de sant√© publique du Qu√©bec | Laboratoire de sant√© publique du Qu√©bec | Sandrine Moreira et al |
| EPI_ISL_826006 | Canada/Qc-JEW-Q9041527/2020 | 2020-05-04 | North America/Canada/Quebec/ | Laboratoire de sant√© publique du Qu√©bec | Laboratoire de sant√© publique du Qu√©bec | Sandrine Moreira et al |
| EPI_ISL_826007 | Canada/Qc-JEW-Q9041534/2020 | 2020-05-04 | North America/Canada/Quebec/ | Laboratoire de sant√© publique du Qu√©bec | Laboratoire de sant√© publique du Qu√©bec | Sandrine Moreira et al |
| EPI_ISL_826009 | Canada/Qc-JEW-Q9031244/2020 | 2020-05-03 | North America/Canada/Quebec/ | Laboratoire de sant√© publique du Qu√©bec | Laboratoire de sant√© publique du Qu√©bec | Sandrine Moreira et al |
| EPI_ISL_826010 | Canada/Qc-JEW-Q9030958/2020 | 2020-05-03 | North America/Canada/Quebec/ | Laboratoire de sant√© publique du Qu√©bec | Laboratoire de sant√© publique du Qu√©bec | Sandrine Moreira et al |
| EPI_ISL_826011 | Canada/Qc-JEW-Q9031255/2020 | 2020-05-03 | North America/Canada/Quebec/ | Laboratoire de sant√© publique du Qu√©bec | Laboratoire de sant√© publique du Qu√©bec | Sandrine Moreira et al |
| EPI_ISL_826012 | Canada/Qc-JEW-Q9031111/2020 | 2020-05-03 | North America/Canada/Quebec/ | Laboratoire de sant√© publique du Qu√©bec | Laboratoire de sant√© publique du Qu√©bec | Sandrine Moreira et al |
| EPI_ISL_826013 | Canada/Qc-JEW-Q9031098/2020 | 2020-05-03 | North America/Canada/Quebec/ | Laboratoire de sant√© publique du Qu√©bec | Laboratoire de sant√© publique du Qu√©bec | Sandrine Moreira et al |
| EPI_ISL_826014 | Canada/Qc-JEW-Q8290912/2020 | 2020-04-29 | North America/Canada/Quebec/ | Laboratoire de sant√© publique du Qu√©bec | Laboratoire de sant√© publique du Qu√©bec | Sandrine Moreira et al |

| | | | | | | |
|---|---|---|---|---|---|---|
| EPI_ISL_826015 | Canada/Qc-JEW-Q8291861/2020 | 2020-04-29 | North America/Canada/Quebec/ | Laboratoire de santv© publique du Quv©bec | Laboratoire de santv© publique du Quv©bec | Sandrine Moreira et al |
| EPI_ISL_826016 | Canada/Qc-JEW-Q8292297/2020 | 2020-04-29 | North America/Canada/Quebec/ | Laboratoire de santv© publique du Quv©bec | Laboratoire de santv© publique du Quv©bec | Sandrine Moreira et al |
| EPI_ISL_826017 | Canada/Qc-JEW-Q8292502/2020 | 2020-04-29 | North America/Canada/Quebec/ | Laboratoire de santv© publique du Quv©bec | Laboratoire de santv© publique du Quv©bec | Sandrine Moreira et al |
| EPI_ISL_826018 | Canada/Qc-JEW-Q8291714/2020 | 2020-04-29 | North America/Canada/Quebec/ | Laboratoire de santv© publique du Quv©bec | Laboratoire de santv© publique du Quv©bec | Sandrine Moreira et al |
| EPI_ISL_826019 | Canada/Qc-JEW-Q8291667/2020 | 2020-04-29 | North America/Canada/Quebec/ | Laboratoire de santv© publique du Quv©bec | Laboratoire de santv© publique du Quv©bec | Sandrine Moreira et al |
| EPI_ISL_826020 | Canada/Qc-JEW-Q8291757/2020 | 2020-04-29 | North America/Canada/Quebec/ | Laboratoire de santv© publique du Quv©bec | Laboratoire de santv© publique du Quv©bec | Sandrine Moreira et al |
| EPI_ISL_826021 | Canada/Qc-JEW-Q8291074/2020 | 2020-04-29 | North America/Canada/Quebec/ | Laboratoire de santv© publique du Quv©bec | Laboratoire de santv© publique du Quv©bec | Sandrine Moreira et al |
| EPI_ISL_826022 | Canada/Qc-JEW-Q8292345/2020 | 2020-04-29 | North America/Canada/Quebec/ | Laboratoire de santv© publique du Quv©bec | Laboratoire de santv© publique du Quv©bec | Sandrine Moreira et al |
| EPI_ISL_826023 | Canada/Qc-JEW-Q8301049/2020 | 2020-04-30 | North America/Canada/Quebec/ | Laboratoire de santv© publique du Quv©bec | Laboratoire de santv© publique du Quv©bec | Sandrine Moreira et al |
| EPI_ISL_826024 | Canada/Qc-JEW-Q9050147/2020 | 2020-05-04 | North America/Canada/Quebec/ | Laboratoire de santv© publique du Quv©bec | Laboratoire de santv© publique du Quv©bec | Sandrine Moreira et al |
| EPI_ISL_826025 | Canada/Qc-JEW-Q9042358/2020 | 2020-05-04 | North America/Canada/Quebec/ | Laboratoire de santv© publique du Quv©bec | Laboratoire de santv© publique du Quv©bec | Sandrine Moreira et al |
| EPI_ISL_826026 | Canada/Qc-JEW-Q9042360/2020 | 2020-05-04 | North America/Canada/Quebec/ | Laboratoire de santv© publique du Quv©bec | Laboratoire de santv© publique du Quv©bec | Sandrine Moreira et al |
| EPI_ISL_826027 | Canada/Qc-JEW-Q9042646/2020 | 2020-05-04 | North America/Canada/Quebec/ | Laboratoire de santv© publique du Quv©bec | Laboratoire de santv© publique du Quv©bec | Sandrine Moreira et al |

| | | | | | | |
|---|---|---|---|---|---|---|
| EPI_ISL_826028 | Canada/Qc-JEW-Q9051212/2020 | 2020-05-05 | North America/Canada/Quebec/ | Laboratoire de sant√© publique du Qu√©bec | Laboratoire de sant√© publique du Qu√©bec | Sandrine Moreira et al |
| EPI_ISL_826029 | Canada/Qc-JEW-Q9051883/2020 | 2020-05-05 | North America/Canada/Quebec/ | Laboratoire de sant√© publique du Qu√©bec | Laboratoire de sant√© publique du Qu√©bec | Sandrine Moreira et al |
| EPI_ISL_826030 | Canada/Qc-JEW-Q9051861/2020 | 2020-05-05 | North America/Canada/Quebec/ | Laboratoire de sant√© publique du Qu√©bec | Laboratoire de sant√© publique du Qu√©bec | Sandrine Moreira et al |
| EPI_ISL_826031 | Canada/Qc-JEW-Q9051751/2020 | 2020-05-05 | North America/Canada/Quebec/ | Laboratoire de sant√© publique du Qu√©bec | Laboratoire de sant√© publique du Qu√©bec | Sandrine Moreira et al |
| EPI_ISL_826032 | Canada/Qc-JEW-Q9051967/2020 | 2020-05-05 | North America/Canada/Quebec/ | Laboratoire de sant√© publique du Qu√©bec | Laboratoire de sant√© publique du Qu√©bec | Sandrine Moreira et al |
| EPI_ISL_826033 | Canada/Qc-JEW-Q9051906/2020 | 2020-05-05 | North America/Canada/Quebec/ | Laboratoire de sant√© publique du Qu√©bec | Laboratoire de sant√© publique du Qu√©bec | Sandrine Moreira et al |
| EPI_ISL_826034 | Canada/Qc-JEW-Q9051897/2020 | 2020-05-05 | North America/Canada/Quebec/ | Laboratoire de sant√© publique du Qu√©bec | Laboratoire de sant√© publique du Qu√©bec | Sandrine Moreira et al |
| EPI_ISL_826035 | Canada/Qc-JEW-Q9061581/2020 | 2020-05-06 | North America/Canada/Quebec/ | Laboratoire de sant√© publique du Qu√©bec | Laboratoire de sant√© publique du Qu√©bec | Sandrine Moreira et al |
| EPI_ISL_826036 | Canada/Qc-JEW-Q9061662/2020 | 2020-05-06 | North America/Canada/Quebec/ | Laboratoire de sant√© publique du Qu√©bec | Laboratoire de sant√© publique du Qu√©bec | Sandrine Moreira et al |
| EPI_ISL_826037 | Canada/Qc-JEW-Q9061676/2020 | 2020-05-06 | North America/Canada/Quebec/ | Laboratoire de sant√© publique du Qu√©bec | Laboratoire de sant√© publique du Qu√©bec | Sandrine Moreira et al |
| EPI_ISL_826038 | Canada/Qc-JEW-Q9061868/2020 | 2020-05-06 | North America/Canada/Quebec/ | Laboratoire de sant√© publique du Qu√©bec | Laboratoire de sant√© publique du Qu√©bec | Sandrine Moreira et al |
| EPI_ISL_826039 | Canada/Qc-JEW-Q9061101/2020 | 2020-05-06 | North America/Canada/Quebec/ | Laboratoire de sant√© publique du Qu√©bec | Laboratoire de sant√© publique du Qu√©bec | Sandrine Moreira et al |
| EPI_ISL_826040 | Canada/Qc-JEW-Q9052007/2020 | 2020-05-05 | North America/Canada/Quebec/ | Laboratoire de sant√© publique du Qu√©bec | Laboratoire de sant√© publique du Qu√©bec | Sandrine Moreira et al |

| | | | | | | |
|---|---|---|---|---|---|---|
| EPI_ISL_826041 | Canada/Qc-JEW-Q9052370/2020 | 2020-05-05 | North America/Canada/Quebec/ | Laboratoire de santv© publique du Quv©bec | Laboratoire de santv© publique du Quv©bec | Sandrine Moreira et al |
| EPI_ISL_826042 | Canada/Qc-JEW-Q9052394/2020 | 2020-05-05 | North America/Canada/Quebec/ | Laboratoire de santv© publique du Quv©bec | Laboratoire de santv© publique du Quv©bec | Sandrine Moreira et al |
| EPI_ISL_826043 | Canada/Qc-JEW-Q9052430/2020 | 2020-05-05 | North America/Canada/Quebec/ | Laboratoire de santv© publique du Quv©bec | Laboratoire de santv© publique du Quv©bec | Sandrine Moreira et al |
| EPI_ISL_826044 | Canada/Qc-JEW-Q9051974/2020 | 2020-05-05 | North America/Canada/Quebec/ | Laboratoire de santv© publique du Quv©bec | Laboratoire de santv© publique du Quv©bec | Sandrine Moreira et al |
| EPI_ISL_826045 | Canada/Qc-JEW-Q9051969/2020 | 2020-05-05 | North America/Canada/Quebec/ | Laboratoire de santv© publique du Quv©bec | Laboratoire de santv© publique du Quv©bec | Sandrine Moreira et al |
| EPI_ISL_826046 | Canada/Qc-JEW-Q9051970/2020 | 2020-05-05 | North America/Canada/Quebec/ | Laboratoire de santv© publique du Quv©bec | Laboratoire de santv© publique du Quv©bec | Sandrine Moreira et al |
| EPI_ISL_826047 | Canada/Qc-JEW-Q9052122/2020 | 2020-05-05 | North America/Canada/Quebec/ | Laboratoire de santv© publique du Quv©bec | Laboratoire de santv© publique du Quv©bec | Sandrine Moreira et al |
| EPI_ISL_826048 | Canada/Qc-JEW-Q9052054/2020 | 2020-05-05 | North America/Canada/Quebec/ | Laboratoire de santv© publique du Quv©bec | Laboratoire de santv© publique du Quv©bec | Sandrine Moreira et al |
| EPI_ISL_826049 | Canada/Qc-JEW-Q9071159/2020 | 2020-05-05 | North America/Canada/Quebec/ | Laboratoire de santv© publique du Quv©bec | Laboratoire de santv© publique du Quv©bec | Sandrine Moreira et al |
| EPI_ISL_826050 | Canada/Qc-JEW-Q9071214/2020 | 2020-05-07 | North America/Canada/Quebec/ | Laboratoire de santv© publique du Quv©bec | Laboratoire de santv© publique du Quv©bec | Sandrine Moreira et al |
| EPI_ISL_826051 | Canada/Qc-JEW-Q9062614/2020 | 2020-05-06 | North America/Canada/Quebec/ | Laboratoire de santv© publique du Quv©bec | Laboratoire de santv© publique du Quv©bec | Sandrine Moreira et al |
| EPI_ISL_826052 | Canada/Qc-JEW-Q9062009/2020 | 2020-05-06 | North America/Canada/Quebec/ | Laboratoire de santv© publique du Quv©bec | Laboratoire de santv© publique du Quv©bec | Sandrine Moreira et al |
| EPI_ISL_826053 | Canada/Qc-JEW-Q9061406/2020 | 2020-05-06 | North America/Canada/Quebec/ | Laboratoire de santv© publique du Quv©bec | Laboratoire de santv© publique du Quv©bec | Sandrine Moreira et al |

| | | | | | | |
|---|---|---|---|---|---|---|
| EPI_ISL_826054 | Canada/Qc-JEW-Q9061718/2020 | 2020-05-06 | North America/Canada/Quebec/ | Laboratoire de sant√© publique du Qu√©bec | Laboratoire de sant√© publique du Qu√©bec | Sandrine Moreira et al |
| EPI_ISL_826055 | Canada/Qc-JEW-Q9061739/2020 | 2020-05-06 | North America/Canada/Quebec/ | Laboratoire de sant√© publique du Qu√©bec | Laboratoire de sant√© publique du Qu√©bec | Sandrine Moreira et al |
| EPI_ISL_826056 | Canada/Qc-JEW-Q9061708/2020 | 2020-05-06 | North America/Canada/Quebec/ | Laboratoire de sant√© publique du Qu√©bec | Laboratoire de sant√© publique du Qu√©bec | Sandrine Moreira et al |
| EPI_ISL_826057 | Canada/Qc-JEW-Q9071806/2020 | 2020-05-07 | North America/Canada/Quebec/ | Laboratoire de sant√© publique du Qu√©bec | Laboratoire de sant√© publique du Qu√©bec | Sandrine Moreira et al |
| EPI_ISL_826058 | Canada/Qc-JEW-Q9071810/2020 | 2020-05-07 | North America/Canada/Quebec/ | Laboratoire de sant√© publique du Qu√©bec | Laboratoire de sant√© publique du Qu√©bec | Sandrine Moreira et al |
| EPI_ISL_826059 | Canada/Qc-JEW-Q9071774/2020 | 2020-05-07 | North America/Canada/Quebec/ | Laboratoire de sant√© publique du Qu√©bec | Laboratoire de sant√© publique du Qu√©bec | Sandrine Moreira et al |
| EPI_ISL_826060 | Canada/Qc-JEW-Q9071880/2020 | 2020-05-07 | North America/Canada/Quebec/ | Laboratoire de sant√© publique du Qu√©bec | Laboratoire de sant√© publique du Qu√©bec | Sandrine Moreira et al |
| EPI_ISL_826061 | Canada/Qc-JEW-Q9091169/2020 | 2020-05-09 | North America/Canada/Quebec/ | Laboratoire de sant√© publique du Qu√©bec | Laboratoire de sant√© publique du Qu√©bec | Sandrine Moreira et al |
| EPI_ISL_826062 | Canada/Qc-JEW-Q9091425/2020 | 2020-05-09 | North America/Canada/Quebec/ | Laboratoire de sant√© publique du Qu√©bec | Laboratoire de sant√© publique du Qu√©bec | Sandrine Moreira et al |
| EPI_ISL_826063 | Canada/Qc-JEW-Q9091358/2020 | 2020-05-09 | North America/Canada/Quebec/ | Laboratoire de sant√© publique du Qu√©bec | Laboratoire de sant√© publique du Qu√©bec | Sandrine Moreira et al |
| EPI_ISL_826064 | Canada/Qc-JEW-Q9091279/2020 | 2020-05-09 | North America/Canada/Quebec/ | Laboratoire de sant√© publique du Qu√©bec | Laboratoire de sant√© publique du Qu√©bec | Sandrine Moreira et al |
| EPI_ISL_826065 | Canada/Qc-JEW-Q9091167/2020 | 2020-05-09 | North America/Canada/Quebec/ | Laboratoire de sant√© publique du Qu√©bec | Laboratoire de sant√© publique du Qu√©bec | Sandrine Moreira et al |
| EPI_ISL_826066 | Canada/Qc-JEW-Q9090900/2020 | 2020-05-09 | North America/Canada/Quebec/ | Laboratoire de sant√© publique du Qu√©bec | Laboratoire de sant√© publique du Qu√©bec | Sandrine Moreira et al |

| | | | | | | |
|---|---|---|---|---|---|---|
| EPI_ISL_826067 | Canada/Qc-JEW-Q9090672/2020 | 2020-05-09 | North America/Canada/Quebec/ | Laboratoire de sant√© publique du Qu√©bec | Laboratoire de sant√© publique du Qu√©bec | Sandrine Moreira et al |
| EPI_ISL_826068 | Canada/Qc-JEW-Q9090670/2020 | 2020-05-09 | North America/Canada/Quebec/ | Laboratoire de sant√© publique du Qu√©bec | Laboratoire de sant√© publique du Qu√©bec | Sandrine Moreira et al |
| EPI_ISL_826069 | Canada/Qc-JEW-Q9090665/2020 | 2020-05-09 | North America/Canada/Quebec/ | Laboratoire de sant√© publique du Qu√©bec | Laboratoire de sant√© publique du Qu√©bec | Sandrine Moreira et al |
| EPI_ISL_826070 | Canada/Qc-JEW-Q9090683/2020 | 2020-05-09 | North America/Canada/Quebec/ | Laboratoire de sant√© publique du Qu√©bec | Laboratoire de sant√© publique du Qu√©bec | Sandrine Moreira et al |
| EPI_ISL_826071 | Canada/Qc-JEW-Q9101486/2020 | 2020-05-10 | North America/Canada/Quebec/ | Laboratoire de sant√© publique du Qu√©bec | Laboratoire de sant√© publique du Qu√©bec | Sandrine Moreira et al |
| EPI_ISL_826072 | Canada/Qc-JEW-Q9101078/2020 | 2020-05-10 | North America/Canada/Quebec/ | Laboratoire de sant√© publique du Qu√©bec | Laboratoire de sant√© publique du Qu√©bec | Sandrine Moreira et al |
| EPI_ISL_826073 | Canada/Qc-JEW-Q9100984/2020 | 2020-05-10 | North America/Canada/Quebec/ | Laboratoire de sant√© publique du Qu√©bec | Laboratoire de sant√© publique du Qu√©bec | Sandrine Moreira et al |
| EPI_ISL_826074 | Canada/Qc-JEW-Q9101294/2020 | 2020-05-10 | North America/Canada/Quebec/ | Laboratoire de sant√© publique du Qu√©bec | Laboratoire de sant√© publique du Qu√©bec | Sandrine Moreira et al |
| EPI_ISL_826075 | Canada/Qc-JEW-Q9111169/2020 | 2020-05-11 | North America/Canada/Quebec/ | Laboratoire de sant√© publique du Qu√©bec | Laboratoire de sant√© publique du Qu√©bec | Sandrine Moreira et al |
| EPI_ISL_826076 | Canada/Qc-JEW-Q9111275/2020 | 2020-05-11 | North America/Canada/Quebec/ | Laboratoire de sant√© publique du Qu√©bec | Laboratoire de sant√© publique du Qu√©bec | Sandrine Moreira et al |
| EPI_ISL_826077 | Canada/Qc-L00276061/2020 | 2020-07-25 | North America/Canada/Quebec/ | Laboratoire de sant√© publique du Qu√©bec | Laboratoire de sant√© publique du Qu√©bec | Sandrine Moreira et al |
| EPI_ISL_826078 | Canada/Qc-L00259346/2020 | 2020-05-13 | North America/Canada/Quebec/ | Laboratoire de sant√© publique du Qu√©bec | Laboratoire de sant√© publique du Qu√©bec | Sandrine Moreira et al |
| EPI_ISL_826079 | Canada/Qc-L00259338/2020 | 2020-05-13 | North America/Canada/Quebec/ | Laboratoire de sant√© publique du Qu√©bec | Laboratoire de sant√© publique du Qu√©bec | Sandrine Moreira et al |

| | | | | | | |
|---|---|---|---|---|---|---|
| EPI_ISL_826080 | Canada/Qc-L00259406/2020 | 2020-05-13 | North America/Canada/Quebec/ | Laboratoire de sant√© publique du Qu√©bec | Laboratoire de sant√© publique du Qu√©bec | Sandrine Moreira et al |
| EPI_ISL_826081 | Canada/Qc-L00259319/2020 | 2020-05-13 | North America/Canada/Quebec/ | Laboratoire de sant√© publique du Qu√©bec | Laboratoire de sant√© publique du Qu√©bec | Sandrine Moreira et al |
| EPI_ISL_826082 | Canada/Qc-L00259313/2020 | 2020-05-13 | North America/Canada/Quebec/ | Laboratoire de sant√© publique du Qu√©bec | Laboratoire de sant√© publique du Qu√©bec | Sandrine Moreira et al |
| EPI_ISL_826083 | Canada/Qc-L00259351/2020 | 2020-05-13 | North America/Canada/Quebec/ | Laboratoire de sant√© publique du Qu√©bec | Laboratoire de sant√© publique du Qu√©bec | Sandrine Moreira et al |
| EPI_ISL_826084 | Canada/Qc-L00259838/2020 | 2020-05-15 | North America/Canada/Quebec/ | Laboratoire de sant√© publique du Qu√©bec | Laboratoire de sant√© publique du Qu√©bec | Sandrine Moreira et al |
| EPI_ISL_826085 | Canada/Qc-L00259784/2020 | 2020-05-15 | North America/Canada/Quebec/ | Laboratoire de sant√© publique du Qu√©bec | Laboratoire de sant√© publique du Qu√©bec | Sandrine Moreira et al |
| EPI_ISL_826086 | Canada/Qc-L00260008/2020 | 2020-05-15 | North America/Canada/Quebec/ | Laboratoire de sant√© publique du Qu√©bec | Laboratoire de sant√© publique du Qu√©bec | Sandrine Moreira et al |
| EPI_ISL_826087 | Canada/Qc-L00260044/2020 | 2020-05-16 | North America/Canada/Quebec/ | Laboratoire de sant√© publique du Qu√©bec | Laboratoire de sant√© publique du Qu√©bec | Sandrine Moreira et al |
| EPI_ISL_826088 | Canada/Qc-L00259999/2020 | 2020-05-16 | North America/Canada/Quebec/ | Laboratoire de sant√© publique du Qu√©bec | Laboratoire de sant√© publique du Qu√©bec | Sandrine Moreira et al |
| EPI_ISL_826089 | Canada/Qc-L00259965/2020 | 2020-05-16 | North America/Canada/Quebec/ | Laboratoire de sant√© publique du Qu√©bec | Laboratoire de sant√© publique du Qu√©bec | Sandrine Moreira et al |
| EPI_ISL_826090 | Canada/Qc-L00259937/2020 | 2020-05-16 | North America/Canada/Quebec/ | Laboratoire de sant√© publique du Qu√©bec | Laboratoire de sant√© publique du Qu√©bec | Sandrine Moreira et al |
| EPI_ISL_826091 | Canada/Qc-L00259918/2020 | 2020-05-16 | North America/Canada/Quebec/ | Laboratoire de sant√© publique du Qu√©bec | Laboratoire de sant√© publique du Qu√©bec | Sandrine Moreira et al |
| EPI_ISL_826092 | Canada/Qc-L00259912/2020 | 2020-05-16 | North America/Canada/Quebec/ | Laboratoire de sant√© publique du Qu√©bec | Laboratoire de sant√© publique du Qu√©bec | Sandrine Moreira et al |

| | | | | | | |
|---|---|---|---|---|---|---|
| EPI_ISL_826093 | Canada/Qc-L00259911/2020 | 2020-05-16 | North America/Canada/Quebec/ | Laboratoire de sant√© publique du Qu√©bec | Laboratoire de sant√© publique du Qu√©bec | Sandrine Moreira et al |
| EPI_ISL_826094 | Canada/Qc-L00259902/2020 | 2020-05-16 | North America/Canada/Quebec/ | Laboratoire de sant√© publique du Qu√©bec | Laboratoire de sant√© publique du Qu√©bec | Sandrine Moreira et al |
| EPI_ISL_826095 | Canada/Qc-L00259921/2020 | 2020-05-16 | North America/Canada/Quebec/ | Laboratoire de sant√© publique du Qu√©bec | Laboratoire de sant√© publique du Qu√©bec | Sandrine Moreira et al |
| EPI_ISL_826096 | Canada/Qc-L00260081/2020 | 2020-05-17 | North America/Canada/Quebec/ | Laboratoire de sant√© publique du Qu√©bec | Laboratoire de sant√© publique du Qu√©bec | Sandrine Moreira et al |
| EPI_ISL_826097 | Canada/Qc-L00260089/2020 | 2020-05-17 | North America/Canada/Quebec/ | Laboratoire de sant√© publique du Qu√©bec | Laboratoire de sant√© publique du Qu√©bec | Sandrine Moreira et al |
| EPI_ISL_826098 | Canada/Qc-L00260129/2020 | 2020-05-17 | North America/Canada/Quebec/ | Laboratoire de sant√© publique du Qu√©bec | Laboratoire de sant√© publique du Qu√©bec | Sandrine Moreira et al |
| EPI_ISL_826099 | Canada/Qc-L00260287/2020 | 2020-05-18 | North America/Canada/Quebec/ | Laboratoire de sant√© publique du Qu√©bec | Laboratoire de sant√© publique du Qu√©bec | Sandrine Moreira et al |
| EPI_ISL_826100 | Canada/Qc-L00260288/2020 | 2020-05-18 | North America/Canada/Quebec/ | Laboratoire de sant√© publique du Qu√©bec | Laboratoire de sant√© publique du Qu√©bec | Sandrine Moreira et al |
| EPI_ISL_826101 | Canada/Qc-L00260322/2020 | 2020-05-18 | North America/Canada/Quebec/ | Laboratoire de sant√© publique du Qu√©bec | Laboratoire de sant√© publique du Qu√©bec | Sandrine Moreira et al |
| EPI_ISL_826102 | Canada/Qc-L00260991/2020 | 2020-05-20 | North America/Canada/Quebec/ | Laboratoire de sant√© publique du Qu√©bec | Laboratoire de sant√© publique du Qu√©bec | Sandrine Moreira et al |
| EPI_ISL_826103 | Canada/Qc-L00261028/2020 | 2020-05-20 | North America/Canada/Quebec/ | Laboratoire de sant√© publique du Qu√©bec | Laboratoire de sant√© publique du Qu√©bec | Sandrine Moreira et al |
| EPI_ISL_826104 | Canada/Qc-L00261055/2020 | 2020-05-20 | North America/Canada/Quebec/ | Laboratoire de sant√© publique du Qu√©bec | Laboratoire de sant√© publique du Qu√©bec | Sandrine Moreira et al |
| EPI_ISL_826105 | Canada/Qc-L00261083/2020 | 2020-05-20 | North America/Canada/Quebec/ | Laboratoire de sant√© publique du Qu√©bec | Laboratoire de sant√© publique du Qu√©bec | Sandrine Moreira et al |

| | | | | | | |
|---|---|---|---|---|---|---|
| EPI_ISL_826106 | Canada/Qc-L00261666/2020 | 2020-05-21 | North America/Canada/Quebec/ | Laboratoire de sant√© publique du Qu√©bec | Laboratoire de sant√© publique du Qu√©bec | Sandrine Moreira et al |
| EPI_ISL_826108 | Canada/Qc-L00262017/2020 | 2020-05-22 | North America/Canada/Quebec/ | Laboratoire de sant√© publique du Qu√©bec | Laboratoire de sant√© publique du Qu√©bec | Sandrine Moreira et al |
| EPI_ISL_826109 | Canada/Qc-L00262019/2020 | 2020-05-22 | North America/Canada/Quebec/ | Laboratoire de sant√© publique du Qu√©bec | Laboratoire de sant√© publique du Qu√©bec | Sandrine Moreira et al |
| EPI_ISL_826110 | Canada/Qc-L00262237/2020 | 2020-05-23 | North America/Canada/Quebec/ | Laboratoire de sant√© publique du Qu√©bec | Laboratoire de sant√© publique du Qu√©bec | Sandrine Moreira et al |
| EPI_ISL_826111 | Canada/Qc-L00262400/2020 | 2020-05-24 | North America/Canada/Quebec/ | Laboratoire de sant√© publique du Qu√©bec | Laboratoire de sant√© publique du Qu√©bec | Sandrine Moreira et al |
| EPI_ISL_826112 | Canada/Qc-L00262383/2020 | 2020-05-24 | North America/Canada/Quebec/ | Laboratoire de sant√© publique du Qu√©bec | Laboratoire de sant√© publique du Qu√©bec | Sandrine Moreira et al |
| EPI_ISL_826113 | Canada/Qc-L00262395/2020 | 2020-05-24 | North America/Canada/Quebec/ | Laboratoire de sant√© publique du Qu√©bec | Laboratoire de sant√© publique du Qu√©bec | Sandrine Moreira et al |
| EPI_ISL_826114 | Canada/Qc-L00262871/2020 | 2020-05-25 | North America/Canada/Quebec/ | Laboratoire de sant√© publique du Qu√©bec | Laboratoire de sant√© publique du Qu√©bec | Sandrine Moreira et al |
| EPI_ISL_826115 | Canada/Qc-L00263421/2020 | 2020-05-26 | North America/Canada/Quebec/ | Laboratoire de sant√© publique du Qu√©bec | Laboratoire de sant√© publique du Qu√©bec | Sandrine Moreira et al |
| EPI_ISL_826116 | Canada/Qc-L00263406/2020 | 2020-05-26 | North America/Canada/Quebec/ | Laboratoire de sant√© publique du Qu√©bec | Laboratoire de sant√© publique du Qu√©bec | Sandrine Moreira et al |
| EPI_ISL_826117 | Canada/Qc-L00263923/2020 | 2020-05-27 | North America/Canada/Quebec/ | Laboratoire de sant√© publique du Qu√©bec | Laboratoire de sant√© publique du Qu√©bec | Sandrine Moreira et al |
| EPI_ISL_826118 | Canada/Qc-L00273467/2020 | 2020-07-15 | North America/Canada/Quebec/ | Laboratoire de sant√© publique du Qu√©bec | Laboratoire de sant√© publique du Qu√©bec | Sandrine Moreira et al |
| EPI_ISL_826119 | Canada/Qc-L00273606/2020 | 2020-07-15 | North America/Canada/Quebec/ | Laboratoire de sant√© publique du Qu√©bec | Laboratoire de sant√© publique du Qu√©bec | Sandrine Moreira et al |

| | | | | | | |
|---|---|---|---|---|---|---|
| EPI_ISL_826120 | Canada/Qc-L00274432/2020 | 2020-07-20 | North America/Canada/Quebec/ | Laboratoire de sant√© publique du Qu√©bec | Laboratoire de sant√© publique du Qu√©bec | Sandrine Moreira et al |
| EPI_ISL_826121 | Canada/Qc-L00274426/2020 | 2020-07-20 | North America/Canada/Quebec/ | Laboratoire de sant√© publique du Qu√©bec | Laboratoire de sant√© publique du Qu√©bec | Sandrine Moreira et al |
| EPI_ISL_826122 | Canada/Qc-L00275237/2020 | 2020-07-22 | North America/Canada/Quebec/ | Laboratoire de sant√© publique du Qu√©bec | Laboratoire de sant√© publique du Qu√©bec | Sandrine Moreira et al |
| EPI_ISL_826123 | Canada/Qc-L00275652/2020 | 2020-07-23 | North America/Canada/Quebec/ | Laboratoire de sant√© publique du Qu√©bec | Laboratoire de sant√© publique du Qu√©bec | Sandrine Moreira et al |
| EPI_ISL_826124 | Canada/Qc-L00275650/2020 | 2020-07-23 | North America/Canada/Quebec/ | Laboratoire de sant√© publique du Qu√©bec | Laboratoire de sant√© publique du Qu√©bec | Sandrine Moreira et al |
| EPI_ISL_826125 | Canada/Qc-L00275653/2020 | 2020-07-23 | North America/Canada/Quebec/ | Laboratoire de sant√© publique du Qu√©bec | Laboratoire de sant√© publique du Qu√©bec | Sandrine Moreira et al |
| EPI_ISL_826126 | Canada/Qc-L00275977/2020 | 2020-07-24 | North America/Canada/Quebec/ | Laboratoire de sant√© publique du Qu√©bec | Laboratoire de sant√© publique du Qu√©bec | Sandrine Moreira et al |
| EPI_ISL_826127 | Canada/Qc-L00276216/2020 | 2020-07-25 | North America/Canada/Quebec/ | Laboratoire de sant√© publique du Qu√©bec | Laboratoire de sant√© publique du Qu√©bec | Sandrine Moreira et al |
| EPI_ISL_826128 | Canada/Qc-L00276205/2020 | 2020-07-25 | North America/Canada/Quebec/ | Laboratoire de sant√© publique du Qu√©bec | Laboratoire de sant√© publique du Qu√©bec | Sandrine Moreira et al |
| EPI_ISL_826129 | Canada/Qc-L00276203/2020 | 2020-07-25 | North America/Canada/Quebec/ | Laboratoire de sant√© publique du Qu√©bec | Laboratoire de sant√© publique du Qu√©bec | Sandrine Moreira et al |
| EPI_ISL_826130 | Canada/Qc-L00276114/2020 | 2020-07-25 | North America/Canada/Quebec/ | Laboratoire de sant√© publique du Qu√©bec | Laboratoire de sant√© publique du Qu√©bec | Sandrine Moreira et al |
| EPI_ISL_826131 | Canada/Qc-L00276173/2020 | 2020-07-26 | North America/Canada/Quebec/ | Laboratoire de sant√© publique du Qu√©bec | Laboratoire de sant√© publique du Qu√©bec | Sandrine Moreira et al |
| EPI_ISL_826132 | Canada/Qc-JEW-R0091260/2020 | 2020-06-09 | North America/Canada/Quebec/ | Laboratoire de sant√© publique du Qu√©bec | Laboratoire de sant√© publique du Qu√©bec | Sandrine Moreira et al |

| | | | | | | |
|---|---|---|---|---|---|---|
| EPI_ISL_826133 | Canada/Qc-JEW-Q9152338/2020 | 2020-05-15 | North America/Canada/Quebec/ | Laboratoire de santv© publique du Quv©bec | Laboratoire de santv© publique du Quv©bec | Sandrine Moreira et al |
| EPI_ISL_826134 | Canada/Qc-JEW-Q9072540/2020 | 2020-05-07 | North America/Canada/Quebec/ | Laboratoire de santv© publique du Quv©bec | Laboratoire de santv© publique du Quv©bec | Sandrine Moreira et al |
| EPI_ISL_826135 | Canada/Qc-JEW-Q9142109/2020 | 2020-05-14 | North America/Canada/Quebec/ | Laboratoire de santv© publique du Quv©bec | Laboratoire de santv© publique du Quv©bec | Sandrine Moreira et al |
| EPI_ISL_826136 | Canada/Qc-JEW-Q9141577/2020 | 2020-05-14 | North America/Canada/Quebec/ | Laboratoire de santv© publique du Quv©bec | Laboratoire de santv© publique du Quv©bec | Sandrine Moreira et al |
| EPI_ISL_826137 | Canada/Qc-JEW-Q9161246/2020 | 2020-05-16 | North America/Canada/Quebec/ | Laboratoire de santv© publique du Quv©bec | Laboratoire de santv© publique du Quv©bec | Sandrine Moreira et al |
| EPI_ISL_826138 | Canada/Qc-JEW-R0031819/2020 | 2020-06-03 | North America/Canada/Quebec/ | Laboratoire de santv© publique du Quv©bec | Laboratoire de santv© publique du Quv©bec | Sandrine Moreira et al |
| EPI_ISL_826139 | Canada/Qc-JEW-Q9121605/2020 | 2020-05-12 | North America/Canada/Quebec/ | Laboratoire de santv© publique du Quv©bec | Laboratoire de santv© publique du Quv©bec | Sandrine Moreira et al |
| EPI_ISL_826140 | Canada/Qc-JEW-Q9141103/2020 | 2020-05-14 | North America/Canada/Quebec/ | Laboratoire de santv© publique du Quv©bec | Laboratoire de santv© publique du Quv©bec | Sandrine Moreira et al |
| EPI_ISL_826141 | Canada/Qc-JEW-Q9131619/2020 | 2020-05-13 | North America/Canada/Quebec/ | Laboratoire de santv© publique du Quv©bec | Laboratoire de santv© publique du Quv©bec | Sandrine Moreira et al |
| EPI_ISL_826142 | Canada/Qc-JEW-Q9082193/2020 | 2020-05-08 | North America/Canada/Quebec/ | Laboratoire de santv© publique du Quv©bec | Laboratoire de santv© publique du Quv©bec | Sandrine Moreira et al |
| EPI_ISL_826143 | Canada/Qc-JEW-Q9152100/2020 | 2020-05-15 | North America/Canada/Quebec/ | Laboratoire de santv© publique du Quv©bec | Laboratoire de santv© publique du Quv©bec | Sandrine Moreira et al |
| EPI_ISL_826144 | Canada/Qc-JEW-Q9213231/2020 | 2020-05-21 | North America/Canada/Quebec/ | Laboratoire de santv© publique du Quv©bec | Laboratoire de santv© publique du Quv©bec | Sandrine Moreira et al |
| EPI_ISL_826145 | Canada/Qc-JEW-Q9142380/2020 | 2020-05-14 | North America/Canada/Quebec/ | Laboratoire de santv© publique du Quv©bec | Laboratoire de santv© publique du Quv©bec | Sandrine Moreira et al |

| | | | | | | |
|---|---|---|---|---|---|---|
| EPI_ISL_826146 | Canada/Qc-JEW-Q9252476/2020 | 2020-05-25 | North America/Canada/Quebec/ | Laboratoire de santV© publique du QuV©bec | Laboratoire de santV© publique du QuV©bec | Sandrine Moreira et al |
| EPI_ISL_826147 | Canada/Qc-JEW-Q9141802/2020 | 2020-05-14 | North America/Canada/Quebec/ | Laboratoire de santV© publique du QuV©bec | Laboratoire de santV© publique du QuV©bec | Sandrine Moreira et al |
| EPI_ISL_826148 | Canada/Qc-JEW-Q9201272/2020 | 2020-05-20 | North America/Canada/Quebec/ | Laboratoire de santV© publique du QuV©bec | Laboratoire de santV© publique du QuV©bec | Sandrine Moreira et al |
| EPI_ISL_826149 | Canada/Qc-JEW-Q9142130/2020 | 2020-05-14 | North America/Canada/Quebec/ | Laboratoire de santV© publique du QuV©bec | Laboratoire de santV© publique du QuV©bec | Sandrine Moreira et al |
| EPI_ISL_826150 | Canada/Qc-JEW-Q9132449/2020 | 2020-05-13 | North America/Canada/Quebec/ | Laboratoire de santV© publique du QuV©bec | Laboratoire de santV© publique du QuV©bec | Sandrine Moreira et al |
| EPI_ISL_826151 | Canada/Qc-JEW-Q9072228/2020 | 2020-05-07 | North America/Canada/Quebec/ | Laboratoire de santV© publique du QuV©bec | Laboratoire de santV© publique du QuV©bec | Sandrine Moreira et al |
| EPI_ISL_826152 | Canada/Qc-JEW-Q9071967/2020 | 2020-05-07 | North America/Canada/Quebec/ | Laboratoire de santV© publique du QuV©bec | Laboratoire de santV© publique du QuV©bec | Sandrine Moreira et al |
| EPI_ISL_826153 | Canada/Qc-JEW-Q9072046/2020 | 2020-05-07 | North America/Canada/Quebec/ | Laboratoire de santV© publique du QuV©bec | Laboratoire de santV© publique du QuV©bec | Sandrine Moreira et al |
| EPI_ISL_826154 | Canada/Qc-JEW-Q9072282/2020 | 2020-05-07 | North America/Canada/Quebec/ | Laboratoire de santV© publique du QuV©bec | Laboratoire de santV© publique du QuV©bec | Sandrine Moreira et al |
| EPI_ISL_826155 | Canada/Qc-JEW-Q9080809/2020 | 2020-05-08 | North America/Canada/Quebec/ | Laboratoire de santV© publique du QuV©bec | Laboratoire de santV© publique du QuV©bec | Sandrine Moreira et al |
| EPI_ISL_826156 | Canada/Qc-JEW-Q9081030/2020 | 2020-05-08 | North America/Canada/Quebec/ | Laboratoire de santV© publique du QuV©bec | Laboratoire de santV© publique du QuV©bec | Sandrine Moreira et al |
| EPI_ISL_826157 | Canada/Qc-JEW-Q9082083/2020 | 2020-05-08 | North America/Canada/Quebec/ | Laboratoire de santV© publique du QuV©bec | Laboratoire de santV© publique du QuV©bec | Sandrine Moreira et al |
| EPI_ISL_826158 | Canada/Qc-JEW-Q9082197/2020 | 2020-05-08 | North America/Canada/Quebec/ | Laboratoire de santV© publique du QuV©bec | Laboratoire de santV© publique du QuV©bec | Sandrine Moreira et al |

| | | | | | | |
|---|---|---|---|---|---|---|
| EPI_ISL_826159 | Canada/Qc-JEW-Q9081752/2020 | 2020-05-08 | North America/Canada/Quebec/ | Laboratoire de santv© publique du Quv©bec | Laboratoire de santv© publique du Quv©bec | Sandrine Moreira et al |
| EPI_ISL_826160 | Canada/Qc-JEW-Q9081932/2020 | 2020-05-08 | North America/Canada/Quebec/ | Laboratoire de santv© publique du Quv©bec | Laboratoire de santv© publique du Quv©bec | Sandrine Moreira et al |
| EPI_ISL_826161 | Canada/Qc-JEW-Q9081676/2020 | 2020-05-08 | North America/Canada/Quebec/ | Laboratoire de santv© publique du Quv©bec | Laboratoire de santv© publique du Quv©bec | Sandrine Moreira et al |
| EPI_ISL_826162 | Canada/Qc-JEW-Q9081405/2020 | 2020-05-08 | North America/Canada/Quebec/ | Laboratoire de santv© publique du Quv©bec | Laboratoire de santv© publique du Quv©bec | Sandrine Moreira et al |
| EPI_ISL_826163 | Canada/Qc-JEW-Q9081416/2020 | 2020-05-08 | North America/Canada/Quebec/ | Laboratoire de santv© publique du Quv©bec | Laboratoire de santv© publique du Quv©bec | Sandrine Moreira et al |
| EPI_ISL_826164 | Canada/Qc-JEW-Q9081433/2020 | 2020-05-08 | North America/Canada/Quebec/ | Laboratoire de santv© publique du Quv©bec | Laboratoire de santv© publique du Quv©bec | Sandrine Moreira et al |
| EPI_ISL_826165 | Canada/Qc-JEW-Q9081421/2020 | 2020-05-08 | North America/Canada/Quebec/ | Laboratoire de santv© publique du Quv©bec | Laboratoire de santv© publique du Quv©bec | Sandrine Moreira et al |
| EPI_ISL_826166 | Canada/Qc-JEW-Q9081389/2020 | 2020-05-08 | North America/Canada/Quebec/ | Laboratoire de santv© publique du Quv©bec | Laboratoire de santv© publique du Quv©bec | Sandrine Moreira et al |
| EPI_ISL_826167 | Canada/Qc-JEW-Q9081334/2020 | 2020-05-07 | North America/Canada/Quebec/ | Laboratoire de santv© publique du Quv©bec | Laboratoire de santv© publique du Quv©bec | Sandrine Moreira et al |
| EPI_ISL_826168 | Canada/Qc-JEW-Q9081347/2020 | 2020-05-07 | North America/Canada/Quebec/ | Laboratoire de santv© publique du Quv©bec | Laboratoire de santv© publique du Quv©bec | Sandrine Moreira et al |
| EPI_ISL_826169 | Canada/Qc-JEW-Q9100158/2020 | 2020-05-10 | North America/Canada/Quebec/ | Laboratoire de santv© publique du Quv©bec | Laboratoire de santv© publique du Quv©bec | Sandrine Moreira et al |
| EPI_ISL_826170 | Canada/Qc-JEW-Q9100058/2020 | 2020-05-09 | North America/Canada/Quebec/ | Laboratoire de santv© publique du Quv©bec | Laboratoire de santv© publique du Quv©bec | Sandrine Moreira et al |
| EPI_ISL_826171 | Canada/Qc-JEW-Q9121152/2020 | 2020-05-12 | North America/Canada/Quebec/ | Laboratoire de santv© publique du Quv©bec | Laboratoire de santv© publique du Quv©bec | Sandrine Moreira et al |

| | | | | | | |
|---|---|---|---|---|---|---|
| EPI_ISL_826172 | Canada/Qc-JEW-Q9121223/2020 | 2020-05-12 | North America/Canada/Quebec/ | Laboratoire de sant√© publique du Qu√©bec | Laboratoire de sant√© publique du Qu√©bec | Sandrine Moreira et al |
| EPI_ISL_826173 | Canada/Qc-JEW-Q9112113/2020 | 2020-05-11 | North America/Canada/Quebec/ | Laboratoire de sant√© publique du Qu√©bec | Laboratoire de sant√© publique du Qu√©bec | Sandrine Moreira et al |
| EPI_ISL_826174 | Canada/Qc-JEW-Q9120206/2020 | 2020-05-11 | North America/Canada/Quebec/ | Laboratoire de sant√© publique du Qu√©bec | Laboratoire de sant√© publique du Qu√©bec | Sandrine Moreira et al |
| EPI_ISL_826175 | Canada/Qc-JEW-Q9111882/2020 | 2020-05-11 | North America/Canada/Quebec/ | Laboratoire de sant√© publique du Qu√©bec | Laboratoire de sant√© publique du Qu√©bec | Sandrine Moreira et al |
| EPI_ISL_826176 | Canada/Qc-JEW-Q9122133/2020 | 2020-05-12 | North America/Canada/Quebec/ | Laboratoire de sant√© publique du Qu√©bec | Laboratoire de sant√© publique du Qu√©bec | Sandrine Moreira et al |
| EPI_ISL_826177 | Canada/Qc-JEW-Q9122576/2020 | 2020-05-12 | North America/Canada/Quebec/ | Laboratoire de sant√© publique du Qu√©bec | Laboratoire de sant√© publique du Qu√©bec | Sandrine Moreira et al |
| EPI_ISL_826178 | Canada/Qc-JEW-Q9122109/2020 | 2020-05-12 | North America/Canada/Quebec/ | Laboratoire de sant√© publique du Qu√©bec | Laboratoire de sant√© publique du Qu√©bec | Sandrine Moreira et al |
| EPI_ISL_826179 | Canada/Qc-JEW-Q9122098/2020 | 2020-05-12 | North America/Canada/Quebec/ | Laboratoire de sant√© publique du Qu√©bec | Laboratoire de sant√© publique du Qu√©bec | Sandrine Moreira et al |
| EPI_ISL_826180 | Canada/Qc-JEW-Q9122898/2020 | 2020-05-12 | North America/Canada/Quebec/ | Laboratoire de sant√© publique du Qu√©bec | Laboratoire de sant√© publique du Qu√©bec | Sandrine Moreira et al |
| EPI_ISL_826181 | Canada/Qc-JEW-Q9131572/2020 | 2020-05-13 | North America/Canada/Quebec/ | Laboratoire de sant√© publique du Qu√©bec | Laboratoire de sant√© publique du Qu√©bec | Sandrine Moreira et al |
| EPI_ISL_826182 | Canada/Qc-JEW-Q9132386/2020 | 2020-05-13 | North America/Canada/Quebec/ | Laboratoire de sant√© publique du Qu√©bec | Laboratoire de sant√© publique du Qu√©bec | Sandrine Moreira et al |
| EPI_ISL_826183 | Canada/Qc-JEW-Q9132272/2020 | 2020-05-13 | North America/Canada/Quebec/ | Laboratoire de sant√© publique du Qu√©bec | Laboratoire de sant√© publique du Qu√©bec | Sandrine Moreira et al |
| EPI_ISL_826184 | Canada/Qc-JEW-Q9262500/2020 | 2020-05-26 | North America/Canada/Quebec/ | Laboratoire de sant√© publique du Qu√©bec | Laboratoire de sant√© publique du Qu√©bec | Sandrine Moreira et al |

| | | | | | | |
|---|---|---|---|---|---|---|
| EPI_ISL_826185 | Canada/Qc-JEW-Q9132194/2020 | 2020-05-13 | North America/Canada/Quebec/ | Laboratoire de santv© publique du Quv©bec | Laboratoire de santv© publique du Quv©bec | Sandrine Moreira et al |
| EPI_ISL_826186 | Canada/Qc-JEW-Q9140220/2020 | 2020-05-13 | North America/Canada/Quebec/ | Laboratoire de santv© publique du Quv©bec | Laboratoire de santv© publique du Quv©bec | Sandrine Moreira et al |
| EPI_ISL_826187 | Canada/Qc-JEW-Q9142456/2020 | 2020-05-14 | North America/Canada/Quebec/ | Laboratoire de santv© publique du Quv©bec | Laboratoire de santv© publique du Quv©bec | Sandrine Moreira et al |
| EPI_ISL_826188 | Canada/Qc-JEW-Q9142255/2020 | 2020-05-14 | North America/Canada/Quebec/ | Laboratoire de santv© publique du Quv©bec | Laboratoire de santv© publique du Quv©bec | Sandrine Moreira et al |
| EPI_ISL_826189 | Canada/Qc-JEW-Q9142190/2020 | 2020-05-14 | North America/Canada/Quebec/ | Laboratoire de santv© publique du Quv©bec | Laboratoire de santv© publique du Quv©bec | Sandrine Moreira et al |
| EPI_ISL_826190 | Canada/Qc-JEW-Q9141620/2020 | 2020-05-14 | North America/Canada/Quebec/ | Laboratoire de santv© publique du Quv©bec | Laboratoire de santv© publique du Quv©bec | Sandrine Moreira et al |
| EPI_ISL_826191 | Canada/Qc-JEW-Q9141825/2020 | 2020-05-14 | North America/Canada/Quebec/ | Laboratoire de santv© publique du Quv©bec | Laboratoire de santv© publique du Quv©bec | Sandrine Moreira et al |
| EPI_ISL_826192 | Canada/Qc-JEW-Q9142032/2020 | 2020-05-14 | North America/Canada/Quebec/ | Laboratoire de santv© publique du Quv©bec | Laboratoire de santv© publique du Quv©bec | Sandrine Moreira et al |
| EPI_ISL_826193 | Canada/Qc-JEW-Q9142023/2020 | 2020-05-14 | North America/Canada/Quebec/ | Laboratoire de santv© publique du Quv©bec | Laboratoire de santv© publique du Quv©bec | Sandrine Moreira et al |
| EPI_ISL_826194 | Canada/Qc-JEW-Q9142094/2020 | 2020-05-14 | North America/Canada/Quebec/ | Laboratoire de santv© publique du Quv©bec | Laboratoire de santv© publique du Quv©bec | Sandrine Moreira et al |
| EPI_ISL_826195 | Canada/Qc-JEW-Q9141679/2020 | 2020-05-14 | North America/Canada/Quebec/ | Laboratoire de santv© publique du Quv©bec | Laboratoire de santv© publique du Quv©bec | Sandrine Moreira et al |
| EPI_ISL_826196 | Canada/Qc-JEW-Q9142047/2020 | 2020-05-14 | North America/Canada/Quebec/ | Laboratoire de santv© publique du Quv©bec | Laboratoire de santv© publique du Quv©bec | Sandrine Moreira et al |
| EPI_ISL_826197 | Canada/Qc-JEW-Q9142088/2020 | 2020-05-14 | North America/Canada/Quebec/ | Laboratoire de santv© publique du Quv©bec | Laboratoire de santv© publique du Quv©bec | Sandrine Moreira et al |

| | | | | | | |
|---|---|---|---|---|---|---|
| EPI_ISL_826198 | Canada/Qc-JEW-Q9151735/2020 | 2020-05-15 | North America/Canada/Quebec/ | Laboratoire de sant√© publique du Qu√©bec | Laboratoire de sant√© publique du Qu√©bec | Sandrine Moreira et al |
| EPI_ISL_826199 | Canada/Qc-JEW-Q9151543/2020 | 2020-05-15 | North America/Canada/Quebec/ | Laboratoire de sant√© publique du Qu√©bec | Laboratoire de sant√© publique du Qu√©bec | Sandrine Moreira et al |
| EPI_ISL_826200 | Canada/Qc-JEW-Q9151497/2020 | 2020-05-15 | North America/Canada/Quebec/ | Laboratoire de sant√© publique du Qu√©bec | Laboratoire de sant√© publique du Qu√©bec | Sandrine Moreira et al |
| EPI_ISL_826201 | Canada/Qc-JEW-Q9143033/2020 | 2020-05-14 | North America/Canada/Quebec/ | Laboratoire de sant√© publique du Qu√©bec | Laboratoire de sant√© publique du Qu√©bec | Sandrine Moreira et al |
| EPI_ISL_826202 | Canada/Qc-JEW-Q9142871/2020 | 2020-05-14 | North America/Canada/Quebec/ | Laboratoire de sant√© publique du Qu√©bec | Laboratoire de sant√© publique du Qu√©bec | Sandrine Moreira et al |
| EPI_ISL_826203 | Canada/Qc-JEW-Q9142728/2020 | 2020-05-14 | North America/Canada/Quebec/ | Laboratoire de sant√© publique du Qu√©bec | Laboratoire de sant√© publique du Qu√©bec | Sandrine Moreira et al |
| EPI_ISL_826204 | Canada/Qc-JEW-Q9142656/2020 | 2020-05-14 | North America/Canada/Quebec/ | Laboratoire de sant√© publique du Qu√©bec | Laboratoire de sant√© publique du Qu√©bec | Sandrine Moreira et al |
| EPI_ISL_826205 | Canada/Qc-JEW-Q9142484/2020 | 2020-05-14 | North America/Canada/Quebec/ | Laboratoire de sant√© publique du Qu√©bec | Laboratoire de sant√© publique du Qu√©bec | Sandrine Moreira et al |
| EPI_ISL_826206 | Canada/Qc-JEW-Q9142389/2020 | 2020-05-14 | North America/Canada/Quebec/ | Laboratoire de sant√© publique du Qu√©bec | Laboratoire de sant√© publique du Qu√©bec | Sandrine Moreira et al |
| EPI_ISL_826207 | Canada/Qc-JEW-Q9142481/2020 | 2020-05-14 | North America/Canada/Quebec/ | Laboratoire de sant√© publique du Qu√©bec | Laboratoire de sant√© publique du Qu√©bec | Sandrine Moreira et al |
| EPI_ISL_826208 | Canada/Qc-JEW-Q9160816/2020 | 2020-05-16 | North America/Canada/Quebec/ | Laboratoire de sant√© publique du Qu√©bec | Laboratoire de sant√© publique du Qu√©bec | Sandrine Moreira et al |
| EPI_ISL_826209 | Canada/Qc-JEW-Q9152140/2020 | 2020-05-15 | North America/Canada/Quebec/ | Laboratoire de sant√© publique du Qu√©bec | Laboratoire de sant√© publique du Qu√©bec | Sandrine Moreira et al |
| EPI_ISL_826210 | Canada/Qc-JEW-Q9152159/2020 | 2020-05-15 | North America/Canada/Quebec/ | Laboratoire de sant√© publique du Qu√©bec | Laboratoire de sant√© publique du Qu√©bec | Sandrine Moreira et al |

| | | | | | | |
|---|---|---|---|---|---|---|
| EPI_ISL_826211 | Canada/Qc-JEW-Q9152503/2020 | 2020-05-15 | North America/Canada/Quebec/ | Laboratoire de sant√© publique du Qu√©bec | Laboratoire de sant√© publique du Qu√©bec | Sandrine Moreira et al |
| EPI_ISL_826212 | Canada/Qc-JEW-Q9151867/2020 | 2020-05-15 | North America/Canada/Quebec/ | Laboratoire de sant√© publique du Qu√©bec | Laboratoire de sant√© publique du Qu√©bec | Sandrine Moreira et al |
| EPI_ISL_826213 | Canada/Qc-JEW-Q9152311/2020 | 2020-05-15 | North America/Canada/Quebec/ | Laboratoire de sant√© publique du Qu√©bec | Laboratoire de sant√© publique du Qu√©bec | Sandrine Moreira et al |
| EPI_ISL_826214 | Canada/Qc-JEW-Q9152027/2020 | 2020-05-15 | North America/Canada/Quebec/ | Laboratoire de sant√© publique du Qu√©bec | Laboratoire de sant√© publique du Qu√©bec | Sandrine Moreira et al |
| EPI_ISL_826215 | Canada/Qc-JEW-Q9151926/2020 | 2020-05-15 | North America/Canada/Quebec/ | Laboratoire de sant√© publique du Qu√©bec | Laboratoire de sant√© publique du Qu√©bec | Sandrine Moreira et al |
| EPI_ISL_826216 | Canada/Qc-JEW-Q9151928/2020 | 2020-05-15 | North America/Canada/Quebec/ | Laboratoire de sant√© publique du Qu√©bec | Laboratoire de sant√© publique du Qu√©bec | Sandrine Moreira et al |
| EPI_ISL_826218 | Canada/Qc-JEW-Q9151884/2020 | 2020-05-15 | North America/Canada/Quebec/ | Laboratoire de sant√© publique du Qu√©bec | Laboratoire de sant√© publique du Qu√©bec | Sandrine Moreira et al |
| EPI_ISL_826219 | Canada/Qc-JEW-Q9151829/2020 | 2020-05-15 | North America/Canada/Quebec/ | Laboratoire de sant√© publique du Qu√©bec | Laboratoire de sant√© publique du Qu√©bec | Sandrine Moreira et al |
| EPI_ISL_826220 | Canada/Qc-JEW-Q9151939/2020 | 2020-05-15 | North America/Canada/Quebec/ | Laboratoire de sant√© publique du Qu√©bec | Laboratoire de sant√© publique du Qu√©bec | Sandrine Moreira et al |
| EPI_ISL_826221 | Canada/Qc-JEW-Q9170838/2020 | 2020-05-17 | North America/Canada/Quebec/ | Laboratoire de sant√© publique du Qu√©bec | Laboratoire de sant√© publique du Qu√©bec | Sandrine Moreira et al |
| EPI_ISL_826222 | Canada/Qc-JEW-Q9170237/2020 | 2020-05-17 | North America/Canada/Quebec/ | Laboratoire de sant√© publique du Qu√©bec | Laboratoire de sant√© publique du Qu√©bec | Sandrine Moreira et al |
| EPI_ISL_826223 | Canada/Qc-JEW-Q9170656/2020 | 2020-05-17 | North America/Canada/Quebec/ | Laboratoire de sant√© publique du Qu√©bec | Laboratoire de sant√© publique du Qu√©bec | Sandrine Moreira et al |
| EPI_ISL_826224 | Canada/Qc-JEW-Q9161922/2020 | 2020-05-16 | North America/Canada/Quebec/ | Laboratoire de sant√© publique du Qu√©bec | Laboratoire de sant√© publique du Qu√©bec | Sandrine Moreira et al |

| | | | | | | |
|---|---|---|---|---|---|---|
| EPI_ISL_826225 | Canada/Qc-JEW-Q9160955/2020 | 2020-05-16 | North America/Canada/Quebec/ | Laboratoire de sant√© publique du Qu√©bec | Laboratoire de sant√© publique du Qu√©bec | Sandrine Moreira et al |
| EPI_ISL_826226 | Canada/Qc-JEW-Q9181274/2020 | 2020-05-18 | North America/Canada/Quebec/ | Laboratoire de sant√© publique du Qu√©bec | Laboratoire de sant√© publique du Qu√©bec | Sandrine Moreira et al |
| EPI_ISL_826227 | Canada/Qc-JEW-Q9181319/2020 | 2020-05-18 | North America/Canada/Quebec/ | Laboratoire de sant√© publique du Qu√©bec | Laboratoire de sant√© publique du Qu√©bec | Sandrine Moreira et al |
| EPI_ISL_826228 | Canada/Qc-JEW-Q9181289/2020 | 2020-05-18 | North America/Canada/Quebec/ | Laboratoire de sant√© publique du Qu√©bec | Laboratoire de sant√© publique du Qu√©bec | Sandrine Moreira et al |
| EPI_ISL_826229 | Canada/Qc-JEW-Q9181173/2020 | 2020-05-18 | North America/Canada/Quebec/ | Laboratoire de sant√© publique du Qu√©bec | Laboratoire de sant√© publique du Qu√©bec | Sandrine Moreira et al |
| EPI_ISL_826230 | Canada/Qc-JEW-Q9180135/2020 | 2020-05-17 | North America/Canada/Quebec/ | Laboratoire de sant√© publique du Qu√©bec | Laboratoire de sant√© publique du Qu√©bec | Sandrine Moreira et al |
| EPI_ISL_826231 | Canada/Qc-JEW-Q9171100/2020 | 2020-05-17 | North America/Canada/Quebec/ | Laboratoire de sant√© publique du Qu√©bec | Laboratoire de sant√© publique du Qu√©bec | Sandrine Moreira et al |
| EPI_ISL_826232 | Canada/Qc-JEW-Q9171182/2020 | 2020-05-17 | North America/Canada/Quebec/ | Laboratoire de sant√© publique du Qu√©bec | Laboratoire de sant√© publique du Qu√©bec | Sandrine Moreira et al |
| EPI_ISL_826233 | Canada/Qc-JEW-Q9171231/2020 | 2020-05-17 | North America/Canada/Quebec/ | Laboratoire de sant√© publique du Qu√©bec | Laboratoire de sant√© publique du Qu√©bec | Sandrine Moreira et al |
| EPI_ISL_826234 | Canada/Qc-JEW-Q9171021/2020 | 2020-05-16 | North America/Canada/Quebec/ | Laboratoire de sant√© publique du Qu√©bec | Laboratoire de sant√© publique du Qu√©bec | Sandrine Moreira et al |
| EPI_ISL_826235 | Canada/Qc-JEW-Q9190220/2020 | 2020-05-18 | North America/Canada/Quebec/ | Laboratoire de sant√© publique du Qu√©bec | Laboratoire de sant√© publique du Qu√©bec | Sandrine Moreira et al |
| EPI_ISL_826236 | Canada/Qc-JEW-Q9181520/2020 | 2020-05-18 | North America/Canada/Quebec/ | Laboratoire de sant√© publique du Qu√©bec | Laboratoire de sant√© publique du Qu√©bec | Sandrine Moreira et al |
| EPI_ISL_826237 | Canada/Qc-JEW-Q9193142/2020 | 2020-05-19 | North America/Canada/Quebec/ | Laboratoire de sant√© publique du Qu√©bec | Laboratoire de sant√© publique du Qu√©bec | Sandrine Moreira et al |

| | | | | | | |
|---|---|---|---|---|---|---|
| EPI_ISL_826238 | Canada/Qc-JEW-Q9193069/2020 | 2020-05-19 | North America/Canada/Quebec/ | Laboratoire de santv© publique du Quv©bec | Laboratoire de santv© publique du Quv©bec | Sandrine Moreira et al |
| EPI_ISL_826239 | Canada/Qc-JEW-Q9191726/2020 | 2020-05-19 | North America/Canada/Quebec/ | Laboratoire de santv© publique du Quv©bec | Laboratoire de santv© publique du Quv©bec | Sandrine Moreira et al |
| EPI_ISL_826240 | Canada/Qc-JEW-Q9193132/2020 | 2020-05-19 | North America/Canada/Quebec/ | Laboratoire de santv© publique du Quv©bec | Laboratoire de santv© publique du Quv©bec | Sandrine Moreira et al |
| EPI_ISL_826241 | Canada/Qc-JEW-Q9201473/2020 | 2020-05-20 | North America/Canada/Quebec/ | Laboratoire de santv© publique du Quv©bec | Laboratoire de santv© publique du Quv©bec | Sandrine Moreira et al |
| EPI_ISL_826242 | Canada/Qc-JEW-Q9211143/2020 | 2020-05-21 | North America/Canada/Quebec/ | Laboratoire de santv© publique du Quv©bec | Laboratoire de santv© publique du Quv©bec | Sandrine Moreira et al |
| EPI_ISL_826243 | Canada/Qc-JEW-Q9202532/2020 | 2020-05-20 | North America/Canada/Quebec/ | Laboratoire de santv© publique du Quv©bec | Laboratoire de santv© publique du Quv©bec | Sandrine Moreira et al |
| EPI_ISL_826244 | Canada/Qc-JEW-Q9202339/2020 | 2020-05-20 | North America/Canada/Quebec/ | Laboratoire de santv© publique du Quv©bec | Laboratoire de santv© publique du Quv©bec | Sandrine Moreira et al |
| EPI_ISL_826245 | Canada/Qc-JEW-Q9202982/2020 | 2020-05-20 | North America/Canada/Quebec/ | Laboratoire de santv© publique du Quv©bec | Laboratoire de santv© publique du Quv©bec | Sandrine Moreira et al |
| EPI_ISL_826246 | Canada/Qc-JEW-Q9203386/2020 | 2020-05-20 | North America/Canada/Quebec/ | Laboratoire de santv© publique du Quv©bec | Laboratoire de santv© publique du Quv©bec | Sandrine Moreira et al |
| EPI_ISL_826247 | Canada/Qc-JEW-Q9212145/2020 | 2020-05-21 | North America/Canada/Quebec/ | Laboratoire de santv© publique du Quv©bec | Laboratoire de santv© publique du Quv©bec | Sandrine Moreira et al |
| EPI_ISL_826248 | Canada/Qc-JEW-Q9211914/2020 | 2020-05-21 | North America/Canada/Quebec/ | Laboratoire de santv© publique du Quv©bec | Laboratoire de santv© publique du Quv©bec | Sandrine Moreira et al |
| EPI_ISL_826249 | Canada/Qc-JEW-Q9202391/2020 | 2020-05-20 | North America/Canada/Quebec/ | Laboratoire de santv© publique du Quv©bec | Laboratoire de santv© publique du Quv©bec | Sandrine Moreira et al |
| EPI_ISL_826250 | Canada/Qc-JEW-Q9211794/2020 | 2020-05-21 | North America/Canada/Quebec/ | Laboratoire de santv© publique du Quv©bec | Laboratoire de santv© publique du Quv©bec | Sandrine Moreira et al |

| | | | | | | |
|---|---|---|---|---|---|---|
| EPI_ISL_826251 | Canada/Qc-JEW-Q9212566/2020 | 2020-05-21 | North America/Canada/Quebec/ | Laboratoire de santv© publique du Quv©bec | Laboratoire de santv© publique du Quv©bec | Sandrine Moreira et al |
| EPI_ISL_826252 | Canada/Qc-JEW-Q9212336/2020 | 2020-05-21 | North America/Canada/Quebec/ | Laboratoire de santv© publique du Quv©bec | Laboratoire de santv© publique du Quv©bec | Sandrine Moreira et al |
| EPI_ISL_826253 | Canada/Qc-JEW-Q9253357/2020 | 2020-05-25 | North America/Canada/Quebec/ | Laboratoire de santv© publique du Quv©bec | Laboratoire de santv© publique du Quv©bec | Sandrine Moreira et al |
| EPI_ISL_826254 | Canada/Qc-JEW-Q9262703/2020 | 2020-05-26 | North America/Canada/Quebec/ | Laboratoire de santv© publique du Quv©bec | Laboratoire de santv© publique du Quv©bec | Sandrine Moreira et al |
| EPI_ISL_826255 | Canada/Qc-JEW-Q9262360/2020 | 2020-05-26 | North America/Canada/Quebec/ | Laboratoire de santv© publique du Quv©bec | Laboratoire de santv© publique du Quv©bec | Sandrine Moreira et al |
| EPI_ISL_826256 | Canada/Qc-JEW-R0053055/2020 | 2020-06-05 | North America/Canada/Quebec/ | Laboratoire de santv© publique du Quv©bec | Laboratoire de santv© publique du Quv©bec | Sandrine Moreira et al |
| EPI_ISL_826257 | Canada/Qc-JEW-R0080201/2020 | 2020-06-08 | North America/Canada/Quebec/ | Laboratoire de santv© publique du Quv©bec | Laboratoire de santv© publique du Quv©bec | Sandrine Moreira et al |
| EPI_ISL_826258 | Canada/Qc-JEW-R0042153/2020 | 2020-06-04 | North America/Canada/Quebec/ | Laboratoire de santv© publique du Quv©bec | Laboratoire de santv© publique du Quv©bec | Sandrine Moreira et al |
| EPI_ISL_826259 | Canada/Qc-JEW-R0013103/2020 | 2020-06-01 | North America/Canada/Quebec/ | Laboratoire de santv© publique du Quv©bec | Laboratoire de santv© publique du Quv©bec | Sandrine Moreira et al |
| EPI_ISL_826260 | Canada/Qc-JEW-R0013104/2020 | 2020-06-01 | North America/Canada/Quebec/ | Laboratoire de santv© publique du Quv©bec | Laboratoire de santv© publique du Quv©bec | Sandrine Moreira et al |
| EPI_ISL_826261 | Canada/Qc-JEW-R0030198/2020 | 2020-06-02 | North America/Canada/Quebec/ | Laboratoire de santv© publique du Quv©bec | Laboratoire de santv© publique du Quv©bec | Sandrine Moreira et al |
| EPI_ISL_826262 | Canada/Qc-JEW-Q8151330/2020 | 2020-04-15 | North America/Canada/Quebec/ | Laboratoire de santv© publique du Quv©bec | Laboratoire de santv© publique du Quv©bec | Sandrine Moreira et al |
| EPI_ISL_826263 | Canada/Qc-JEW-Q8261296/2020 | 2020-04-26 | North America/Canada/Quebec/ | Laboratoire de santv© publique du Quv©bec | Laboratoire de santv© publique du Quv©bec | Sandrine Moreira et al |

| | | | | | | |
|---|---|---|---|---|---|---|
| EPI_ISL_826264 | Canada/Qc-JEW-Q8260052/2020 | 2020-04-26 | North America/Canada/Quebec/ | Laboratoire de santv© publique du Quv©bec | Laboratoire de santv© publique du Quv©bec | Sandrine Moreira et al |
| EPI_ISL_826265 | Canada/Qc-JEW-Q8251009/2020 | 2020-04-25 | North America/Canada/Quebec/ | Laboratoire de santv© publique du Quv©bec | Laboratoire de santv© publique du Quv©bec | Sandrine Moreira et al |
| EPI_ISL_826266 | Canada/Qc-JEW-Q8261578/2020 | 2020-04-26 | North America/Canada/Quebec/ | Laboratoire de santv© publique du Quv©bec | Laboratoire de santv© publique du Quv©bec | Sandrine Moreira et al |
| EPI_ISL_826267 | Canada/Qc-JEW-Q8250057/2020 | 2020-04-24 | North America/Canada/Quebec/ | Laboratoire de santv© publique du Quv©bec | Laboratoire de santv© publique du Quv©bec | Sandrine Moreira et al |
| EPI_ISL_826268 | Canada/Qc-JEW-Q8251318/2020 | 2020-04-25 | North America/Canada/Quebec/ | Laboratoire de santv© publique du Quv©bec | Laboratoire de santv© publique du Quv©bec | Sandrine Moreira et al |
| EPI_ISL_826272 | Hungary/UD-S51/2021 | 2021-01-08 | Europe/Hungary/Szabolcs-Szatmv°r-Ber | University of Debrecen, Department of Medical Microbiology | National Laboratory of Virology, Szentv°gothai Research Centre | Endre Gv°bor Tv≥th et al |
| EPI_ISL_826273 | Hungary/UD-V147/2021 | 2021-01-09 | Europe/Hungary/Szabolcs-Szatmv°r-Ber | University of Debrecen, Department of Medical Microbiology | National Laboratory of Virology, Szentv°gothai Research Centre | Endre Gv°bor Tv≥th et al |
| EPI_ISL_826274 | India/GJ-GBRC-460-R1/2020 | 2020-10-24 | Asia/India/Gujarat/Ahmedabad | Sterling Hospital, Memnagar, Ahmedabad | Gujarat Biotechnology Research Centre | Naman Shashtri et al |
| EPI_ISL_826276 | India/GJ-GBRC-460-R2b/2020 | 2020-12-13 | Asia/India/Gujarat/Ahmedabad | Sterling Hospital, Memnagar, Ahmedabad | Gujarat Biotechnology Research Centre | Dinesh Kumar et al |
| EPI_ISL_826290 | USA/CT-JAX-20J-337X3936/2020 | 2020-12-19 | North America/USA/Connecticut/ | The Jackson Laboratory | The Jackson Laboratory | Lloyd M et al |
| EPI_ISL_826295 | USA/CT-JAX-B42709-UCSW/2020 | 2020-12-17 | North America/USA/Connecticut/ | The Jackson Laboratory | The Jackson Laboratory | Lloyd M et al |
| EPI_ISL_826298 | USA/WY-WPHL-21003346/2021 | 2021-01-08 | North America/USA/Wyoming/ | Wyoming Public Health Laboratory | Wyoming Public Health Laboratory | Noah Hull et al |
| EPI_ISL_826306 | USA/WY-WPHL-21002537/2021 | 2021-01-07 | North America/USA/Wyoming/ | Wyoming Public Health Laboratory | Wyoming Public Health Laboratory | Noah Hull et al |
| EPI_ISL_826309 | USA/WY-WPHL-21003031/2021 | 2021-01-08 | North America/USA/Wyoming/ | Wyoming Public Health Laboratory | Wyoming Public Health Laboratory | Noah Hull et al |

| | | | | | | |
|---|---|---|---|---|---|---|
| EPI_ISL_826322 | USA/MI-UM-10036869317/2020 | 2020-11-10 | North America/USA/Michigan/ | University of Michigan Clinical Microbiology Laboratory | Lauring Lab, University of Michigan, Department of Microbiology and Immunology | Valesano et al |
| EPI_ISL_826329 | USA/MI-UM-10036875365/2020 | 2020-11-10 | North America/USA/Michigan/ | University of Michigan Clinical Microbiology Laboratory | Lauring Lab, University of Michigan, Department of Microbiology and Immunology | Valesano et al |
| EPI_ISL_826333 | USA/MI-UM-10036878491/2020 | 2020-11-10 | North America/USA/Michigan/ | University of Michigan Clinical Microbiology Laboratory | Lauring Lab, University of Michigan, Department of Microbiology and Immunology | Valesano et al |
| EPI_ISL_826348 | USA/MI-UM-10036894910/2020 | 2020-11-11 | North America/USA/Michigan/ | University of Michigan Clinical Microbiology Laboratory | Lauring Lab, University of Michigan, Department of Microbiology and Immunology | Valesano et al |
| EPI_ISL_826360 | USA/MI-UM-10036848821/2020 | 2020-11-09 | North America/USA/Michigan/ | University of Michigan Clinical Microbiology Laboratory | Lauring Lab, University of Michigan, Department of Microbiology and Immunology | Valesano et al |
| EPI_ISL_826361 | USA/MI-UM-10036861743/2020 | 2020-11-09 | North America/USA/Michigan/ | University of Michigan Clinical Microbiology Laboratory | Lauring Lab, University of Michigan, Department of Microbiology and Immunology | Valesano et al |

| | | | | | | |
|---|---|---|---|---|---|---|
| EPI_ISL_826364 | USA/MI-UM-10036861572/2020 | 2020-11-09 | North America/USA/Michigan/ | University of Michigan Clinical Microbiology Laboratory | Lauring Lab, University of Michigan, Department of Microbiology and Immunology | Valesano et al |
| EPI_ISL_826366 | USA/MI-UM-10036856792/2020 | 2020-11-09 | North America/USA/Michigan/ | University of Michigan Clinical Microbiology Laboratory | Lauring Lab, University of Michigan, Department of Microbiology and Immunology | Valesano et al |
| EPI_ISL_826376 | USA/MI-UM-10036854063/2020 | 2020-11-09 | North America/USA/Michigan/ | University of Michigan Clinical Microbiology Laboratory | Lauring Lab, University of Michigan, Department of Microbiology and Immunology | Valesano et al |
| EPI_ISL_826383 | USA/MI-UM-10036882055/2020 | 2020-11-10 | North America/USA/Michigan/ | University of Michigan Clinical Microbiology Laboratory | Lauring Lab, University of Michigan, Department of Microbiology and Immunology | Valesano et al |
| EPI_ISL_826402 | USA/MI-UM-10036881752/2020 | 2020-11-10 | North America/USA/Michigan/ | University of Michigan Clinical Microbiology Laboratory | Lauring Lab, University of Michigan, Department of Microbiology and Immunology | Valesano et al |
| EPI_ISL_826406 | USA/MI-UM-10036871399/2020 | 2020-11-10 | North America/USA/Michigan/ | University of Michigan Clinical Microbiology Laboratory | Lauring Lab, University of Michigan, Department of Microbiology and Immunology | Valesano et al |

| | | | | | | |
|---|---|---|---|---|---|---|
| EPI_ISL_826409 | USA/MI-UM-10036876887/2020 | 2020-11-10 | North America/USA/Michigan/ | University of Michigan Clinical Microbiology Laboratory | Lauring Lab, University of Michigan, Department of Microbiology and Immunology | Valesano et al |
| EPI_ISL_826413 | USA/MI-UM-10036901249/2020 | 2020-11-11 | North America/USA/Michigan/ | University of Michigan Clinical Microbiology Laboratory | Lauring Lab, University of Michigan, Department of Microbiology and Immunology | Valesano et al |
| EPI_ISL_826420 | USA/MI-UM-10036880816/2020 | 2020-11-10 | North America/USA/Michigan/ | University of Michigan Clinical Microbiology Laboratory | Lauring Lab, University of Michigan, Department of Microbiology and Immunology | Valesano et al |
| EPI_ISL_826422 | USA/MI-UM-10036890847/2020 | 2020-11-11 | North America/USA/Michigan/ | University of Michigan Clinical Microbiology Laboratory | Lauring Lab, University of Michigan, Department of Microbiology and Immunology | Valesano et al |
| EPI_ISL_826437 | USA/MI-UM-10036913376/2020 | 2020-11-12 | North America/USA/Michigan/ | University of Michigan Clinical Microbiology Laboratory | Lauring Lab, University of Michigan, Department of Microbiology and Immunology | Valesano et al |
| EPI_ISL_826440 | USA/MI-UM-10036915011/2020 | 2020-11-12 | North America/USA/Michigan/ | University of Michigan Clinical Microbiology Laboratory | Lauring Lab, University of Michigan, Department of Microbiology and Immunology | Valesano et al |

| | | | | | | |
|---|---|---|---|---|---|---|
| EPI_ISL_826442 | USA/MI-UM-10036892077/2020 | 2020-11-11 | North America/USA/Michigan/ | University of Michigan Clinical Microbiology Laboratory | Lauring Lab, University of Michigan, Department of Microbiology and Immunology | Valesano et al |
| EPI_ISL_826448 | USA/MI-UM-10036921105/2020 | 2020-11-12 | North America/USA/Michigan/ | University of Michigan Clinical Microbiology Laboratory | Lauring Lab, University of Michigan, Department of Microbiology and Immunology | Valesano et al |
| EPI_ISL_826456 | USA/MI-UM-10036895318/2020 | 2020-11-11 | North America/USA/Michigan/ | University of Michigan Clinical Microbiology Laboratory | Lauring Lab, University of Michigan, Department of Microbiology and Immunology | Valesano et al |
| EPI_ISL_826463 | USA/OH-DEC106/2020 | 2020-12-30 | North America/USA/Ohio/Columbus OH | The Ohio State University | OSU Polaris Molecular Laboratory | Huolin Tu et al |
| EPI_ISL_826521 | USA/OH-DEC32/2020 | 2020-12-30 | North America/USA/Ohio/Columbus OH | The Ohio State University | James Molecular Laboratory | Huolin Tu et al |
| EPI_ISL_826522 | USA/CT-JAX-000542222/2020 | 2020-12-21 | North America/USA/Connecticut/ | The Jackson Laboratory | The Jackson Laboratory | Lloyd M et al |
| EPI_ISL_826540 | USA/TX-DSHS-2113/2020 | 2020-11-13 | North America/USA/Texas/Smith County | Texas Department of State Health Services | Texas Department of State Health Services | Rashmi Tuladhar et al |
| EPI_ISL_826552 | USA/TX-DSHS-2182/2020 | 2020-06-14 | North America/USA/Texas/Cameron Cou | Texas Department of State Health Services | Texas Department of State Health Services | Rashmi Tuladhar et al |
| EPI_ISL_826553 | USA/TX-DSHS-2115/2020 | 2020-11-25 | North America/USA/Texas/Smith County | Texas Department of State Health Services | Texas Department of State Health Services | Rashmi Tuladhar et al |
| EPI_ISL_826558 | USA/TX-DSHS-2188/2020 | 2020-06-14 | North America/USA/Texas/Cameron Cou | Texas Department of State Health Services | Texas Department of State Health Services | Rashmi Tuladhar et al |
| EPI_ISL_826561 | USA/TX-DSHS-2121/2020 | 2020-12-07 | North America/USA/Texas/Smith County | Texas Department of State Health Services | Texas Department of State Health Services | Rashmi Tuladhar et al |

| | | | | | | |
|---|---|---|---|---|---|---|
| EPI_ISL_826565 | USA/TX-DSHS-2192/2020 | 2020-06-14 | North America/USA/Texas/Cameron Cou | Texas Department of State Health Services | Texas Department of State Health Services | Rashmi Tuladhar et al |
| EPI_ISL_826568 | USA/TX-DSHS-2193/2020 | 2020-06-14 | North America/USA/Texas/Cameron Cou | Texas Department of State Health Services | Texas Department of State Health Services | Rashmi Tuladhar et al |
| EPI_ISL_826573 | USA/TX-DSHS-2205/2020 | 2020-06-16 | North America/USA/Texas/Jefferson Cou | Texas Department of State Health Services | Texas Department of State Health Services | Rashmi Tuladhar et al |
| EPI_ISL_826580 | USA/NY-AECOM_054/2020 | 2020-04-15 | North America/USA/New York/New Yor | Montefiore Medical Center | Albert Einstein College of Medicine, Dept. of Microbiology & Immunology, Chandran lab | J. Maximilian Fels et al |
| EPI_ISL_826588 | USA/NY-AECOM_118/2020 | 2020-09-02 | North America/USA/New York/New Yor | Montefiore Medical Center | Albert Einstein College of Medicine, Dept. of Microbiology & Immunology, Chandran lab | J. Maximilian Fels et al |
| EPI_ISL_826589 | USA/NY-AECOM_111/2020 | 2020-08-01 | North America/USA/New York/New Yor | Montefiore Medical Center | Albert Einstein College of Medicine, Dept. of Microbiology & Immunology, Chandran lab | J. Maximilian Fels et al |
| EPI_ISL_826590 | USA/NY-AECOM_126/2020 | 2020-08-26 | North America/USA/New York/New Yor | Montefiore Medical Center | Albert Einstein College of Medicine, Dept. of Microbiology & Immunology, Chandran lab | J. Maximilian Fels et al |
| EPI_ISL_826608 | USA/NY-AECOM_023/2020 | 2020-04-03 | North America/USA/New York/New Yor | Montefiore Medical Center | Albert Einstein College of Medicine, Dept. of Microbiology & Immunology, Chandran lab | J. Maximilian Fels et al |

| | | | | | | |
|---|---|---|---|---|---|---|
| EPI_ISL_826612 | USA/NY-AECOM_030/2020 | 2020-04-15 | North America/USA/New York/New Yor | Montefiore Medical Center | Albert Einstein College of Medicine, Dept. of Microbiology & Immunology, Chandran lab | J. Maximilian Fels et al |
| EPI_ISL_826637 | USA/NY-AECOM_074/2020 | 2020-04-06 | North America/USA/New York/New Yor | Montefiore Medical Center | Albert Einstein College of Medicine, Dept. of Microbiology & Immunology, Chandran lab | J. Maximilian Fels et al |
| EPI_ISL_826638 | USA/NY-AECOM_075/2020 | 2020-04-07 | North America/USA/New York/New Yor | Montefiore Medical Center | Albert Einstein College of Medicine, Dept. of Microbiology & Immunology, Chandran lab | J. Maximilian Fels et al |
| EPI_ISL_826649 | USA/NY-AECOM_100/2020 | 2020-05-05 | North America/USA/New York/New Yor | Montefiore Medical Center | Albert Einstein College of Medicine, Dept. of Microbiology & Immunology, Chandran lab | J. Maximilian Fels et al |
| EPI_ISL_826653 | USA/NY-AECOM_107/2020 | 2020-07-03 | North America/USA/New York/New Yor | Montefiore Medical Center | Albert Einstein College of Medicine, Dept. of Microbiology & Immunology, Chandran lab | J. Maximilian Fels et al |
| EPI_ISL_826661 | USA/NY-AECOM_119/2020 | 2020-09-04 | North America/USA/New York/New Yor | Montefiore Medical Center | Albert Einstein College of Medicine, Dept. of Microbiology & Immunology, Chandran lab | J. Maximilian Fels et al |

| | | | | | | |
|---|---|---|---|---|---|---|
| EPI_ISL_826662 | USA/NY-AECOM_120/2020 | 2020-09-08 | North America/USA/New York/New Yor | Montefiore Medical Center | Albert Einstein College of Medicine, Dept. of Microbiology & Immunology, Chandran lab | J. Maximilian Fels et al |
| EPI_ISL_826663 | USA/NY-AECOM_121/2020 | 2020-09-08 | North America/USA/New York/New Yor | Montefiore Medical Center | Albert Einstein College of Medicine, Dept. of Microbiology & Immunology, Chandran lab | J. Maximilian Fels et al |
| EPI_ISL_826670 | USA/NY-AECOM_137/2020 | 2020-10-31 | North America/USA/New York/New Yor | Montefiore Medical Center | Albert Einstein College of Medicine, Dept. of Microbiology & Immunology, Chandran lab | J. Maximilian Fels et al |
| EPI_ISL_826798 | Ecuador/EC-G-1785/2020 | 2020-03-21 | South America/Ecuador/Guayas/ | INSPI-CRN DE INFLUENZA Y OTROS VIRUS RESPIRATORIOS | Instituto de Salud Publica de Chile | Javier Tognarelli et al |
| EPI_ISL_826802 | Ecuador/EC-G-2512/2020 | 2020-03-23 | South America/Ecuador/Guayas/ | INSPI-CRN DE INFLUENZA Y OTROS VIRUS RESPIRATORIOS | Instituto de Salud Publica de Chile | Javier Tognarelli et al |
| EPI_ISL_826804 | Ecuador/EC-SE-2550/2020 | 2020-03-23 | South America/Ecuador/Santa Elena/ | INSPI-CRN DE INFLUENZA Y OTROS VIRUS RESPIRATORIOS | Instituto de Salud Publica de Chile | Javier Tognarelli et al |
| EPI_ISL_826807 | Ecuador/EC-G-5289/2020 | 2020-03-30 | South America/Ecuador/Guayas/ | INSPI-CRN DE INFLUENZA Y OTROS VIRUS RESPIRATORIOS | Instituto de Salud Publica de Chile | Javier Tognarelli et al |
| EPI_ISL_826809 | Ecuador/EC-G-5305/2020 | 2020-03-30 | South America/Ecuador/Guayas/ | INSPI-CRN DE INFLUENZA Y OTROS VIRUS RESPIRATORIOS | Instituto de Salud Publica de Chile | Javier Tognarelli et al |
| EPI_ISL_826811 | Ecuador/EC-G-5319/2020 | 2020-03-30 | South America/Ecuador/Guayas/ | INSPI-CRN DE INFLUENZA Y OTROS VIRUS RESPIRATORIOS | Instituto de Salud Publica de Chile | Javier Tognarelli et al |
| EPI_ISL_826820 | Ecuador/EC-M-48776/2020 | 2020-05-24 | South America/Ecuador/Manabi/ | INSPI-CRN DE INFLUENZA Y OTROS VIRUS RESPIRATORIOS | Instituto de Salud Publica de Chile | Javier Tognarelli et al |
| EPI_ISL_826823 | Ecuador/EC-R-65804/2020 | 2020-06-25 | South America/Ecuador/Los Rios/ | INSPI-CRN DE INFLUENZA Y OTROS VIRUS RESPIRATORIOS | Instituto de Salud Publica de Chile | Javier Tognarelli et al |
| EPI_ISL_826825 | Ecuador/EC-M-95299/2020 | 2020-07-28 | South America/Ecuador/Manabi/ | INSPI-CRN DE INFLUENZA Y OTROS VIRUS RESPIRATORIOS | Instituto de Salud Publica de Chile | Javier Tognarelli et al |
| EPI_ISL_826827 | Ecuador/EC-SD-119429/2020 | 2020-08-27 | South America/Ecuador/Santo Domingo | INSPI-CRN DE INFLUENZA Y OTROS VIRUS RESPIRATORIOS | Instituto de Salud Publica de Chile | Javier Tognarelli et al |
| EPI_ISL_826829 | Ecuador/EC-G-124255/2020 | 2020-09-07 | South America/Ecuador/Guayas/ | INSPI-CRN DE INFLUENZA Y OTROS VIRUS RESPIRATORIOS | Instituto de Salud Publica de Chile | Javier Tognarelli et al |

| | | | | | | |
|---|---|---|---|---|---|---|
| EPI_ISL_826830 | Ecuador/EC-E-124319/2020 | 2020-09-07 | South America/Ecuador/Esmeraldas/ | INSPI-CRN DE INFLUENZA Y OTROS VIRUS RESPIRATORIOS | Instituto de Salud Publica de Chile | Javier Tognarelli et al |
| EPI_ISL_826832 | Ecuador/EC-E-124322/2020 | 2020-09-08 | South America/Ecuador/Esmeraldas/ | INSPI-CRN DE INFLUENZA Y OTROS VIRUS RESPIRATORIOS | Instituto de Salud Publica de Chile | Javier Tognarelli et al |
| EPI_ISL_826834 | Ecuador/EC-G-127052/2020 | 2020-09-11 | South America/Ecuador/Guayas/ | INSPI-CRN DE INFLUENZA Y OTROS VIRUS RESPIRATORIOS | Instituto de Salud Publica de Chile | Javier Tognarelli et al |
| EPI_ISL_826836 | Ecuador/EC-M-127111/2020 | 2020-09-11 | South America/Ecuador/Manabi/ | INSPI-CRN DE INFLUENZA Y OTROS VIRUS RESPIRATORIOS | Instituto de Salud Publica de Chile | Javier Tognarelli et al |
| EPI_ISL_826840 | Iceland/3147/2020 | 2020-10-08 | Europe/Italy/Reykjavik/ | deCODE genetics | deCODE genetics | Daniel F Gudbjartsson et al |
| EPI_ISL_827043 | Germany/NW-IOV-K-5810/2020 | 2020-12-30 | Europe/Germany/North Rhine Westphal | Institute of Virology, University of Cologne | Institute of Virology, University of Cologne | Saleta Sierra et al |
| EPI_ISL_827388 | Iceland/3697/2020 | 2020-10-29 | Europe/Iceland/Reykjavik/ | deCODE genetics | deCODE genetics | Daniel F Gudbjartsson et al |
| EPI_ISL_827446 | USA/MI-UM-10036920687/2020 | 2020-11-12 | North America/USA/Michigan/ | University of Michigan Clinical Microbiology Laboratory | Lauring Lab, University of Michigan, Department of Microbiology and Immunology | Valesano et al |
| EPI_ISL_827577 | Iceland/3881/2020 | 2020-08-14 | Europe/Italy/Reykjavik/ | The National University Hospital of Iceland | deCODE genetics | Daniel F Gudbjartsson et al |
| EPI_ISL_828191 | Iceland/4499/2020 | 2020-08-01 | Europe/Iceland/Reykjavik/ | The National University Hospital of Iceland | deCODE genetics | Daniel F Gudbjartsson et al |
| EPI_ISL_830212 | USA/NY-Wadsworth-21003590-01/20 | 2020-12-27 | North America/USA/New York/Erie Cou | KALEIDA CENTER FOR LABORATORY MEDICINE | Wadsworth Center, New York State Department of Health | Kirsten St. George et al |
| EPI_ISL_830221 | USA/NY-Wadsworth-21001875-01/20 | 2021-01-06 | North America/USA/New York/Montgo | Wadsworth Center, New York State Department of Health | Wadsworth Center, New York State Department of Health | Kirsten St. George et al |
| EPI_ISL_830236 | USA/NY-Wadsworth-21003576-01/20 | 2020-12-26 | North America/USA/New York/Niagara | KALEIDA CENTER FOR LABORATORY MEDICINE | Wadsworth Center, New York State Department of Health | Kirsten St. George et al |
| EPI_ISL_830282 | USA/NY-Wadsworth-21001850-01/20 | 2021-01-06 | North America/USA/New York/Montgo | Wadsworth Center, New York State Department of Health | Wadsworth Center, New York State Department of Health | Kirsten St. George et al |
| EPI_ISL_830301 | Iceland/6577/2020 | 2020-10-28 | Europe/Iceland/Reykjavik/ | deCODE genetics | deCODE genetics | Daniel F Gudbjartsson et al |

| | | | | | | |
|---|---|---|---|---|---|---|
| EPI_ISL_830308 | USA/NY-Wadsworth-21001871-01/20 | 2021-01-06 | North America/USA/New York/Montgo | Wadsworth Center, New York State Department of Health | Wadsworth Center, New York State Department of Health | Kirsten St. George et al |
| EPI_ISL_830310 | USA/NY-Wadsworth-21001873-01/20 | 2021-01-06 | North America/USA/New York/Montgo | Wadsworth Center, New York State Department of Health | Wadsworth Center, New York State Department of Health | Kirsten St. George et al |
| EPI_ISL_830319 | USA/NY-Wadsworth-21001890-01/20 | 2021-01-06 | North America/USA/New York/Montgo | Wadsworth Center, New York State Department of Health | Wadsworth Center, New York State Department of Health | Kirsten St. George et al |
| EPI_ISL_830323 | USA/NY-Wadsworth-21001894-01/20 | 2021-01-06 | North America/USA/New York/Montgo | Wadsworth Center, New York State Department of Health | Wadsworth Center, New York State Department of Health | Kirsten St. George et al |
| EPI_ISL_830504 | Iceland/6755/2020 | 2020-11-05 | Europe/Iceland/Reykjavik/ | deCODE genetics | deCODE genetics | Daniel F Gudbjartsson et al |
| EPI_ISL_830572 | USA/NY-Wadsworth-21004124-01/20 | 2021-01-04 | North America/USA/New York/Nassau C | NORTHWELL HEALTH LABORATORIES | Wadsworth Center, New York State Department of Health | Kirsten St. George et al |
| EPI_ISL_830577 | USA/NY-Wadsworth-21004146-01/20 | 2021-01-04 | North America/USA/New York/Bronx | NORTHWELL HEALTH LABORATORIES | Wadsworth Center, New York State Department of Health | Kirsten St. George et al |
| EPI_ISL_830578 | USA/NY-Wadsworth-21004150-01/20 | 2021-01-04 | North America/USA/New York/New Yor | NORTHWELL HEALTH LABORATORIES | Wadsworth Center, New York State Department of Health | Kirsten St. George et al |
| EPI_ISL_830579 | USA/NY-Wadsworth-21003637-01/20 | 2020-12-26 | North America/USA/New York/Niagara | KALEIDA CENTER FOR LABORATORY MEDICINE | Wadsworth Center, New York State Department of Health | Kirsten St. George et al |
| EPI_ISL_830583 | USA/NY-Wadsworth-21003597-01/20 | 2020-12-27 | North America/USA/New York/Erie Cou | KALEIDA CENTER FOR LABORATORY MEDICINE | Wadsworth Center, New York State Department of Health | Kirsten St. George et al |

| | | | | | | |
|---|---|---|---|---|---|---|
| EPI_ISL_830585 | USA/NY-Wadsworth-21003646-01/20 | 2020-12-27 | North America/USA/New York/Niagara | KALEIDA CENTER FOR LABORATORY MEDICINE | Wadsworth Center, New York State Department of Health | Kirsten St. George et al |
| EPI_ISL_830587 | USA/NY-Wadsworth-21003649-01/20 | 2020-12-27 | North America/USA/New York/Niagara | KALEIDA CENTER FOR LABORATORY MEDICINE | Wadsworth Center, New York State Department of Health | Kirsten St. George et al |
| EPI_ISL_830588 | USA/NY-Wadsworth-21003654-01/20 | 2020-12-27 | North America/USA/New York/Niagara | KALEIDA CENTER FOR LABORATORY MEDICINE | Wadsworth Center, New York State Department of Health | Kirsten St. George et al |
| EPI_ISL_830590 | USA/NY-Wadsworth-21003662-01/20 | 2020-12-27 | North America/USA/New York/Niagara | KALEIDA CENTER FOR LABORATORY MEDICINE | Wadsworth Center, New York State Department of Health | Kirsten St. George et al |
| EPI_ISL_830600 | USA/NY-Wadsworth-21003627-01/20 | 2020-12-28 | North America/USA/New York/Erie Cou | KALEIDA CENTER FOR LABORATORY MEDICINE | Wadsworth Center, New York State Department of Health | Kirsten St. George et al |
| EPI_ISL_830602 | USA/NY-Wadsworth-21003652-01/20 | 2020-12-28 | North America/USA/New York/Niagara | KALEIDA CENTER FOR LABORATORY MEDICINE | Wadsworth Center, New York State Department of Health | Kirsten St. George et al |
| EPI_ISL_830605 | USA/NY-Wadsworth-21003664-01/20 | 2020-12-28 | North America/USA/New York/Erie Cou | KALEIDA CENTER FOR LABORATORY MEDICINE | Wadsworth Center, New York State Department of Health | Kirsten St. George et al |
| EPI_ISL_830608 | USA/NY-Wadsworth-21003667-01/20 | 2020-12-29 | North America/USA/New York/Erie Cou | KALEIDA CENTER FOR LABORATORY MEDICINE | Wadsworth Center, New York State Department of Health | Kirsten St. George et al |
| EPI_ISL_830623 | USA/NY-Wadsworth-21003851-01/20 | 2020-12-31 | North America/USA/New York/Erie Cou | KALEIDA CENTER FOR LABORATORY MEDICINE | Wadsworth Center, New York State Department of Health | Kirsten St. George et al |
| EPI_ISL_830626 | USA/NY-Wadsworth-21003888-01/20 | 2020-12-31 | North America/USA/New York/Niagara | KALEIDA CENTER FOR LABORATORY MEDICINE | Wadsworth Center, New York State Department of Health | Kirsten St. George et al |

| | | | | | | |
|---|---|---|---|---|---|---|
| EPI_ISL_830632 | USA/NY-Wadsworth-21004125-01/20 | 2021-01-04 | North America/USA/New York/Nassau C | NORTHWELL HEALTH LABORATORIES | Wadsworth Center, New York State Department of Health | Kirsten St. George et al |
| EPI_ISL_830636 | USA/NY-Wadsworth-21004143-01/20 | 2021-01-04 | North America/USA/New York/Nassau C | NORTHWELL HEALTH LABORATORIES | Wadsworth Center, New York State Department of Health | Kirsten St. George et al |
| EPI_ISL_830652 | USA/NY-Wadsworth-21004216-01/20 | 2020-12-30 | North America/USA/New York/Onondag | SUNY UPSTATE MEDICAL UNIVERSITY | Wadsworth Center, New York State Department of Health | Kirsten St. George et al |
| EPI_ISL_830656 | USA/NY-Wadsworth-21004220-01/20 | 2020-12-30 | North America/USA/New York/Onondag | SUNY UPSTATE MEDICAL UNIVERSITY | Wadsworth Center, New York State Department of Health | Kirsten St. George et al |
| EPI_ISL_830660 | USA/NY-Wadsworth-21004226-01/20 | 2020-12-29 | North America/USA/New York/Onondag | SUNY UPSTATE MEDICAL UNIVERSITY | Wadsworth Center, New York State Department of Health | Kirsten St. George et al |
| EPI_ISL_830664 | USA/NY-Wadsworth-21004230-01/20 | 2021-01-04 | North America/USA/New York/Onondag | SUNY UPSTATE MEDICAL UNIVERSITY | Wadsworth Center, New York State Department of Health | Kirsten St. George et al |
| EPI_ISL_830667 | USA/NY-Wadsworth-21004233-01/20 | 2020-12-31 | North America/USA/New York/Onondag | SUNY UPSTATE MEDICAL UNIVERSITY | Wadsworth Center, New York State Department of Health | Kirsten St. George et al |
| EPI_ISL_830670 | USA/NY-Wadsworth-21004237-01/20 | 2020-12-31 | North America/USA/New York/Onondag | SUNY UPSTATE MEDICAL UNIVERSITY | Wadsworth Center, New York State Department of Health | Kirsten St. George et al |
| EPI_ISL_830671 | USA/NY-Wadsworth-21004238-01/20 | 2020-12-30 | North America/USA/New York/Madison | SUNY UPSTATE MEDICAL UNIVERSITY | Wadsworth Center, New York State Department of Health | Kirsten St. George et al |
| EPI_ISL_830679 | USA/NY-Wadsworth-21004249-01/20 | 2020-12-31 | North America/USA/New York/Onondag | SUNY UPSTATE MEDICAL UNIVERSITY | Wadsworth Center, New York State Department of Health | Kirsten St. George et al |

| | | | | | | |
|---|---|---|---|---|---|---|
| EPI_ISL_830682 | USA/NY-Wadsworth-21004253-01/20 | 2021-01-05 | North America/USA/New York/Onondag | SUNY UPSTATE MEDICAL UNIVERSITY | Wadsworth Center, New York State Department of Health | Kirsten St. George et al |
| EPI_ISL_830683 | USA/NY-Wadsworth-21004281-01/20 | 2021-01-01 | North America/USA/New York/Albany ( | ALBANY MEDICAL CENTER HOSPITAL CLINICAL LABORATORIES | Wadsworth Center, New York State Department of Health | Kirsten St. George et al |
| EPI_ISL_830698 | USA/NY-Wadsworth-21004297-01/20 | 2021-01-03 | North America/USA/New York/Saratoga | ALBANY MEDICAL CENTER HOSPITAL CLINICAL LABORATORIES | Wadsworth Center, New York State Department of Health | Kirsten St. George et al |
| EPI_ISL_830699 | USA/NY-Wadsworth-21004298-01/20 | 2021-01-04 | North America/USA/New York/Albany ( | ALBANY MEDICAL CENTER HOSPITAL CLINICAL LABORATORIES | Wadsworth Center, New York State Department of Health | Kirsten St. George et al |
| EPI_ISL_830706 | USA/NY-Wadsworth-21004307-01/20 | 2021-01-05 | North America/USA/New York/Greene ( | ALBANY MEDICAL CENTER HOSPITAL CLINICAL LABORATORIES | Wadsworth Center, New York State Department of Health | Kirsten St. George et al |
| EPI_ISL_830721 | USA/NY-Wadsworth-21004808-01/20 | 2020-12-28 | North America/USA/New York/Wayne ( | BIO-REFERENCE LABORATORIES | Wadsworth Center, New York State Department of Health | Kirsten St. George et al |
| EPI_ISL_830724 | USA/NY-Wadsworth-21004922-01/20 | 2021-01-04 | North America/USA/New York/Saratoga | ACUTIS DIAGNOSTICS | Wadsworth Center, New York State Department of Health | Kirsten St. George et al |
| EPI_ISL_830727 | Belgium/42477217/2020 | 2020-10-09 | Europe/Belgium/Tongeren/Alken | University Hospital Basel, Clinical Virology | University Hospital Basel, Clinical Bacteriology | Tim Roloff et al |
| EPI_ISL_830728 | France/GES-42427767/2020 | 2020-09-06 | Europe/France/Grand Est/Folgensbourg | University Hospital Basel, Clinical Virology | University Hospital Basel, Clinical Bacteriology | Tim Roloff et al |
| EPI_ISL_830729 | France/GES-42484124/2020 | 2020-10-14 | Europe/France/Grand Est/Hegenheim | University Hospital Basel, Clinical Virology | University Hospital Basel, Clinical Bacteriology | Tim Roloff et al |

| | | | | | | |
|---|---|---|---|---|---|---|
| EPI_ISL_830730 | France/GES-42476885/2020 | 2020-10-09 | Europe/France/Grand Est/Saint-Ulrich | University Hospital Basel, Clinical Virology | University Hospital Basel, Clinical Bacteriology | Tim Roloff et al |
| EPI_ISL_830731 | France/GES-42467870/2020 | 2020-10-03 | Europe/France/Grand Est/Vieux Ferette | University Hospital Basel, Clinical Virology | University Hospital Basel, Clinical Bacteriology | Tim Roloff et al |
| EPI_ISL_830743 | Switzerland/BL-42425610/2020 | 2020-09-04 | Europe/Switzerland/Basel-Land/Allschw | University Hospital Basel, Clinical Virology | University Hospital Basel, Clinical Bacteriology | Tim Roloff et al |
| EPI_ISL_830776 | Switzerland/BL-42484196/2020 | 2020-10-14 | Europe/Switzerland/Basel-Land/Zwinger | University Hospital Basel, Clinical Virology | University Hospital Basel, Clinical Bacteriology | Tim Roloff et al |
| EPI_ISL_830778 | Switzerland/BS-42484406/2020 | 2020-10-14 | Europe/Switzerland/Basel-Stadt/ | University Hospital Basel, Clinical Virology | University Hospital Basel, Clinical Bacteriology | Tim Roloff et al |
| EPI_ISL_830783 | Switzerland/BS-42427051/2020 | 2020-09-05 | Europe/Switzerland/Basel-Stadt/Basel | University Hospital Basel, Clinical Virology | University Hospital Basel, Clinical Bacteriology | Tim Roloff et al |
| EPI_ISL_830791 | Switzerland/BS-42429090/2020 | 2020-09-07 | Europe/Switzerland/Basel-Stadt/Basel | University Hospital Basel, Clinical Virology | University Hospital Basel, Clinical Bacteriology | Tim Roloff et al |
| EPI_ISL_830800 | Switzerland/BS-42436659/2020 | 2020-09-11 | Europe/Switzerland/Basel-Stadt/Basel | University Hospital Basel, Clinical Virology | University Hospital Basel, Clinical Bacteriology | Tim Roloff et al |
| EPI_ISL_830841 | Switzerland/BS-42470106/2020 | 2020-10-05 | Europe/Switzerland/Basel-Stadt/Basel | University Hospital Basel, Clinical Virology | University Hospital Basel, Clinical Bacteriology | Tim Roloff et al |
| EPI_ISL_830843 | Switzerland/BS-42471446/2020 | 2020-10-06 | Europe/Switzerland/Basel-Stadt/Basel | University Hospital Basel, Clinical Virology | University Hospital Basel, Clinical Bacteriology | Tim Roloff et al |

| | | | | | | |
|---|---|---|---|---|---|---|
| EPI_ISL_830851 | Switzerland/BS-42472971/2020 | 2020-10-07 | Europe/Switzerland/Basel-Stadt/Basel | University Hospital Basel, Clinical Virology | University Hospital Basel, Clinical Bacteriology | Tim Roloff et al |
| EPI_ISL_830856 | Switzerland/BS-42473358/2020 | 2020-10-07 | Europe/Switzerland/Basel-Stadt/Basel | University Hospital Basel, Clinical Virology | University Hospital Basel, Clinical Bacteriology | Tim Roloff et al |
| EPI_ISL_830858 | Switzerland/BS-42473867/2020 | 2020-10-07 | Europe/Switzerland/Basel-Stadt/Basel | University Hospital Basel, Clinical Virology | University Hospital Basel, Clinical Bacteriology | Tim Roloff et al |
| EPI_ISL_830861 | Switzerland/BS-42474955/2020 | 2020-10-08 | Europe/Switzerland/Basel-Stadt/Basel | University Hospital Basel, Clinical Virology | University Hospital Basel, Clinical Bacteriology | Tim Roloff et al |
| EPI_ISL_830867 | Switzerland/BS-42475558/2020 | 2020-10-08 | Europe/Switzerland/Basel-Stadt/Basel | University Hospital Basel, Clinical Virology | University Hospital Basel, Clinical Bacteriology | Tim Roloff et al |
| EPI_ISL_830891 | Switzerland/BS-42479122/2020 | 2020-10-11 | Europe/Switzerland/Basel-Stadt/Basel | University Hospital Basel, Clinical Virology | University Hospital Basel, Clinical Bacteriology | Tim Roloff et al |
| EPI_ISL_830942 | Switzerland/BS-42486437/2020 | 2020-10-15 | Europe/Switzerland/Basel-Stadt/Basel | University Hospital Basel, Clinical Virology | University Hospital Basel, Clinical Bacteriology | Tim Roloff et al |
| EPI_ISL_830990 | Switzerland/BS-42478515/2020 | 2020-10-10 | Europe/Switzerland/Basel-Stadt/Riehen | University Hospital Basel, Clinical Virology | University Hospital Basel, Clinical Bacteriology | Tim Roloff et al |
| EPI_ISL_831020 | Ecuador/EC-M-143748/2020 | 2020-10-19 | South America/Ecuador/Manabi/ | INSPI-CRN DE INFLUENZA Y OTROS VIRUS RESPIRATORIOS | Instituto de Salud Publica de Chile | Javier Tognarelli et al |
| EPI_ISL_831032 | Spain/MD-IBV-99013326/2020 | 2020-11-19 | Europe/Spain/Madrid/Madrid | Hospital Universitario La Paz (Madrid) | SeqCOVID-SPAIN consortium/IBV(CSIC) | Marv≠a Rodrv≠guez-Tejedor et al |
| EPI_ISL_831034 | Spain/MD-IBV-99013307/2020 | 2020-10-03 | Europe/Spain/Madrid/Madrid | Hospital Universitario La Paz (Madrid) | SeqCOVID-SPAIN consortium/IBV(CSIC) | Marv≠a Rodrv≠guez-Tejedor et al |

| | | | | | | |
|---|---|---|---|---|---|---|
| EPI_ISL_831040 | Spain/MD-IBV-99013151/2020 | 2020-04-02 | Europe/Spain/Madrid/Madrid | Hospital Universitario La Paz (Madrid) | SeqCOVID-SPAIN consortium/IBV(CSIC) | Marv≠a Rodrv≠guez-Tejedor et al |
| EPI_ISL_831041 | Spain/MD-IBV-99013152/2020 | 2020-04-02 | Europe/Spain/Madrid/Madrid | Hospital Universitario La Paz (Madrid) | SeqCOVID-SPAIN consortium/IBV(CSIC) | Marv≠a Rodrv≠guez-Tejedor et al |
| EPI_ISL_831042 | Spain/MD-IBV-99013153/2020 | 2020-04-12 | Europe/Spain/Madrid/Madrid | Hospital Universitario La Paz (Madrid) | SeqCOVID-SPAIN consortium/IBV(CSIC) | Marv≠a Rodrv≠guez-Tejedor et al |
| EPI_ISL_831044 | Spain/MD-IBV-99013157/2020 | 2020-04-15 | Europe/Spain/Madrid/Madrid | Hospital Universitario La Paz (Madrid) | SeqCOVID-SPAIN consortium/IBV(CSIC) | Marv≠a Rodrv≠guez-Tejedor et al |
| EPI_ISL_831060 | Spain/MD-IBV-99013174/2020 | 2020-04-17 | Europe/Spain/Madrid/Madrid | Hospital Universitario La Paz (Madrid) | SeqCOVID-SPAIN consortium/IBV(CSIC) | Marv≠a Rodrv≠guez-Tejedor et al |
| EPI_ISL_831061 | Spain/MD-IBV-99013175/2020 | 2020-04-17 | Europe/Spain/Madrid/Madrid | Hospital Universitario La Paz (Madrid) | SeqCOVID-SPAIN consortium/IBV(CSIC) | Marv≠a Rodrv≠guez-Tejedor et al |
| EPI_ISL_831076 | Spain/MD-IBV-99013191/2020 | 2020-04-26 | Europe/Spain/Madrid/Madrid | Hospital Universitario La Paz (Madrid) | SeqCOVID-SPAIN consortium/IBV(CSIC) | Marv≠a Rodrv≠guez-Tejedor et al |
| EPI_ISL_831077 | Spain/MD-IBV-99013192/2020 | 2020-04-26 | Europe/Spain/Madrid/Madrid | Hospital Universitario La Paz (Madrid) | SeqCOVID-SPAIN consortium/IBV(CSIC) | Marv≠a Rodrv≠guez-Tejedor et al |
| EPI_ISL_831080 | Spain/MD-IBV-99013195/2020 | 2020-04-25 | Europe/Spain/Madrid/Madrid | Hospital Universitario La Paz (Madrid) | SeqCOVID-SPAIN consortium/IBV(CSIC) | Marv≠a Rodrv≠guez-Tejedor et al |
| EPI_ISL_831085 | Spain/MD-IBV-99013201/2020 | 2020-08-10 | Europe/Spain/Madrid/Madrid | Hospital Universitario La Paz (Madrid) | SeqCOVID-SPAIN consortium/IBV(CSIC) | Marv≠a Rodrv≠guez-Tejedor et al |
| EPI_ISL_831086 | Spain/MD-IBV-99013202/2020 | 2020-08-10 | Europe/Spain/Madrid/Madrid | Hospital Universitario La Paz (Madrid) | SeqCOVID-SPAIN consortium/IBV(CSIC) | Marv≠a Rodrv≠guez-Tejedor et al |
| EPI_ISL_831088 | Spain/MD-IBV-99013204/2020 | 2020-08-17 | Europe/Spain/Madrid/Madrid | Hospital Universitario La Paz (Madrid) | SeqCOVID-SPAIN consortium/IBV(CSIC) | Marv≠a Rodrv≠guez-Tejedor et al |
| EPI_ISL_831089 | Spain/MD-IBV-99013205/2020 | 2020-08-17 | Europe/Spain/Madrid/Madrid | Hospital Universitario La Paz (Madrid) | SeqCOVID-SPAIN consortium/IBV(CSIC) | Marv≠a Rodrv≠guez-Tejedor et al |

| | | | | | | |
|---|---|---|---|---|---|---|
| EPI_ISL_831090 | Spain/MD-IBV-99013206/2020 | 2020-08-24 | Europe/Spain/Madrid/Madrid | Hospital Universitario La Paz (Madrid) | SeqCOVID-SPAIN consortium/IBV(CSIC) | Mar√≠a Rodr√≠guez-Tejedor et al |
| EPI_ISL_831092 | Spain/MD-IBV-99013208/2020 | 2020-09-03 | Europe/Spain/Madrid/Madrid | Hospital Universitario La Paz (Madrid) | SeqCOVID-SPAIN consortium/IBV(CSIC) | Mar√≠a Rodr√≠guez-Tejedor et al |
| EPI_ISL_831094 | Spain/MD-IBV-99013210/2020 | 2020-09-24 | Europe/Spain/Madrid/Madrid | Hospital Universitario La Paz (Madrid) | SeqCOVID-SPAIN consortium/IBV(CSIC) | Mar√≠a Rodr√≠guez-Tejedor et al |
| EPI_ISL_831096 | Spain/MD-IBV-99013212/2020 | 2020-10-05 | Europe/Spain/Madrid/Madrid | Hospital Universitario La Paz (Madrid) | SeqCOVID-SPAIN consortium/IBV(CSIC) | Mar√≠a Rodr√≠guez-Tejedor et al |
| EPI_ISL_831101 | Spain/MD-IBV-99013217/2020 | 2020-10-13 | Europe/Spain/Madrid/Madrid | Hospital Universitario La Paz (Madrid) | SeqCOVID-SPAIN consortium/IBV(CSIC) | Mar√≠a Rodr√≠guez-Tejedor et al |
| EPI_ISL_831102 | Spain/MD-IBV-99013218/2020 | 2020-10-14 | Europe/Spain/Madrid/Madrid | Hospital Universitario La Paz (Madrid) | SeqCOVID-SPAIN consortium/IBV(CSIC) | Mar√≠a Rodr√≠guez-Tejedor et al |
| EPI_ISL_831106 | Spain/MD-IBV-99013222/2020 | 2020-10-21 | Europe/Spain/Madrid/Madrid | Hospital Universitario La Paz (Madrid) | SeqCOVID-SPAIN consortium/IBV(CSIC) | Mar√≠a Rodr√≠guez-Tejedor et al |
| EPI_ISL_831109 | Spain/MD-IBV-99013225/2020 | 2020-09-09 | Europe/Spain/Madrid/Madrid | Hospital Universitario La Paz (Madrid) | SeqCOVID-SPAIN consortium/IBV(CSIC) | Mar√≠a Rodr√≠guez-Tejedor et al |
| EPI_ISL_831112 | Spain/MD-IBV-99013228/2020 | 2020-10-27 | Europe/Spain/Madrid/Madrid | Hospital Universitario La Paz (Madrid) | SeqCOVID-SPAIN consortium/IBV(CSIC) | Mar√≠a Rodr√≠guez-Tejedor et al |
| EPI_ISL_831114 | Spain/MD-IBV-99013230/2020 | 2020-10-28 | Europe/Spain/Madrid/Madrid | Hospital Universitario La Paz (Madrid) | SeqCOVID-SPAIN consortium/IBV(CSIC) | Mar√≠a Rodr√≠guez-Tejedor et al |
| EPI_ISL_831116 | Spain/MD-IBV-99013232/2020 | 2020-09-03 | Europe/Spain/Madrid/Madrid | Hospital Universitario La Paz (Madrid) | SeqCOVID-SPAIN consortium/IBV(CSIC) | Mar√≠a Rodr√≠guez-Tejedor et al |
| EPI_ISL_831117 | Spain/MD-IBV-99013233/2020 | 2020-09-04 | Europe/Spain/Madrid/Madrid | Hospital Universitario La Paz (Madrid) | SeqCOVID-SPAIN consortium/IBV(CSIC) | Mar√≠a Rodr√≠guez-Tejedor et al |
| EPI_ISL_831118 | Spain/MD-IBV-99013234/2020 | 2020-09-04 | Europe/Spain/Madrid/Madrid | Hospital Universitario La Paz (Madrid) | SeqCOVID-SPAIN consortium/IBV(CSIC) | Mar√≠a Rodr√≠guez-Tejedor et al |

| | | | | | | |
|---|---|---|---|---|---|---|
| EPI_ISL_831119 | Spain/MD-IBV-99013235/2020 | 2020-09-02 | Europe/Spain/Madrid/Madrid | Hospital Universitario La Paz (Madrid) | SeqCOVID-SPAIN consortium/IBV(CSIC) | Mar√≠a Rodr√≠guez-Tejedor et al |
| EPI_ISL_831121 | Spain/MD-IBV-99013237/2020 | 2020-09-06 | Europe/Spain/Madrid/Madrid | Hospital Universitario La Paz (Madrid) | SeqCOVID-SPAIN consortium/IBV(CSIC) | Mar√≠a Rodr√≠guez-Tejedor et al |
| EPI_ISL_831122 | Spain/MD-IBV-99013238/2020 | 2020-09-01 | Europe/Spain/Madrid/Madrid | Hospital Universitario La Paz (Madrid) | SeqCOVID-SPAIN consortium/IBV(CSIC) | Mar√≠a Rodr√≠guez-Tejedor et al |
| EPI_ISL_831124 | Spain/MD-IBV-99013240/2020 | 2020-09-06 | Europe/Spain/Madrid/Madrid | Hospital Universitario La Paz (Madrid) | SeqCOVID-SPAIN consortium/IBV(CSIC) | Mar√≠a Rodr√≠guez-Tejedor et al |
| EPI_ISL_831125 | Spain/MD-IBV-99013241/2020 | 2020-09-05 | Europe/Spain/Madrid/Madrid | Hospital Universitario La Paz (Madrid) | SeqCOVID-SPAIN consortium/IBV(CSIC) | Mar√≠a Rodr√≠guez-Tejedor et al |
| EPI_ISL_831131 | Spain/MD-IBV-99013247/2020 | 2020-09-08 | Europe/Spain/Madrid/Madrid | Hospital Universitario La Paz (Madrid) | SeqCOVID-SPAIN consortium/IBV(CSIC) | Mar√≠a Rodr√≠guez-Tejedor et al |
| EPI_ISL_831136 | Spain/MD-IBV-99013253/2020 | 2020-09-10 | Europe/Spain/Madrid/Madrid | Hospital Universitario La Paz (Madrid) | SeqCOVID-SPAIN consortium/IBV(CSIC) | Mar√≠a Rodr√≠guez-Tejedor et al |
| EPI_ISL_831138 | Spain/MD-IBV-99013255/2020 | 2020-09-13 | Europe/Spain/Madrid/Madrid | Hospital Universitario La Paz (Madrid) | SeqCOVID-SPAIN consortium/IBV(CSIC) | Mar√≠a Rodr√≠guez-Tejedor et al |
| EPI_ISL_831139 | Spain/MD-IBV-99013256/2020 | 2020-09-13 | Europe/Spain/Madrid/Madrid | Hospital Universitario La Paz (Madrid) | SeqCOVID-SPAIN consortium/IBV(CSIC) | Mar√≠a Rodr√≠guez-Tejedor et al |
| EPI_ISL_831140 | Spain/MD-IBV-99013257/2020 | 2020-09-13 | Europe/Spain/Madrid/Madrid | Hospital Universitario La Paz (Madrid) | SeqCOVID-SPAIN consortium/IBV(CSIC) | Mar√≠a Rodr√≠guez-Tejedor et al |
| EPI_ISL_831141 | Spain/MD-IBV-99013258/2020 | 2020-09-13 | Europe/Spain/Madrid/Madrid | Hospital Universitario La Paz (Madrid) | SeqCOVID-SPAIN consortium/IBV(CSIC) | Mar√≠a Rodr√≠guez-Tejedor et al |
| EPI_ISL_831143 | Spain/MD-IBV-99013260/2020 | 2020-09-11 | Europe/Spain/Madrid/Madrid | Hospital Universitario La Paz (Madrid) | SeqCOVID-SPAIN consortium/IBV(CSIC) | Mar√≠a Rodr√≠guez-Tejedor et al |
| EPI_ISL_831144 | Spain/MD-IBV-99013261/2020 | 2020-09-12 | Europe/Spain/Madrid/Madrid | Hospital Universitario La Paz (Madrid) | SeqCOVID-SPAIN consortium/IBV(CSIC) | Mar√≠a Rodr√≠guez-Tejedor et al |

| | | | | | | |
|---|---|---|---|---|---|---|
| EPI_ISL_831145 | Spain/MD-IBV-99013262/2020 | 2020-09-11 | Europe/Spain/Madrid/Madrid | Hospital Universitario La Paz (Madrid) | SeqCOVID-SPAIN consortium/IBV(CSIC) | Mar√≠a Rodr√≠guez-Tejedor et al |
| EPI_ISL_831148 | Spain/MD-IBV-99013265/2020 | 2020-09-11 | Europe/Spain/Madrid/Madrid | Hospital Universitario La Paz (Madrid) | SeqCOVID-SPAIN consortium/IBV(CSIC) | Mar√≠a Rodr√≠guez-Tejedor et al |
| EPI_ISL_831150 | Spain/MD-IBV-99013267/2020 | 2020-09-11 | Europe/Spain/Madrid/Madrid | Hospital Universitario La Paz (Madrid) | SeqCOVID-SPAIN consortium/IBV(CSIC) | Mar√≠a Rodr√≠guez-Tejedor et al |
| EPI_ISL_831152 | Spain/MD-IBV-99013269/2020 | 2020-09-12 | Europe/Spain/Madrid/Madrid | Hospital Universitario La Paz (Madrid) | SeqCOVID-SPAIN consortium/IBV(CSIC) | Mar√≠a Rodr√≠guez-Tejedor et al |
| EPI_ISL_831153 | Spain/MD-IBV-99013270/2020 | 2020-09-11 | Europe/Spain/Madrid/Madrid | Hospital Universitario La Paz (Madrid) | SeqCOVID-SPAIN consortium/IBV(CSIC) | Mar√≠a Rodr√≠guez-Tejedor et al |
| EPI_ISL_831154 | Spain/MD-IBV-99013271/2020 | 2020-09-11 | Europe/Spain/Madrid/Madrid | Hospital Universitario La Paz (Madrid) | SeqCOVID-SPAIN consortium/IBV(CSIC) | Mar√≠a Rodr√≠guez-Tejedor et al |
| EPI_ISL_831155 | Spain/MD-IBV-99013272/2020 | 2020-09-11 | Europe/Spain/Madrid/Madrid | Hospital Universitario La Paz (Madrid) | SeqCOVID-SPAIN consortium/IBV(CSIC) | Mar√≠a Rodr√≠guez-Tejedor et al |
| EPI_ISL_831156 | Spain/MD-IBV-99013273/2020 | 2020-09-14 | Europe/Spain/Madrid/Madrid | Hospital Universitario La Paz (Madrid) | SeqCOVID-SPAIN consortium/IBV(CSIC) | Mar√≠a Rodr√≠guez-Tejedor et al |
| EPI_ISL_831157 | Spain/MD-IBV-99013274/2020 | 2020-09-14 | Europe/Spain/Madrid/Madrid | Hospital Universitario La Paz (Madrid) | SeqCOVID-SPAIN consortium/IBV(CSIC) | Mar√≠a Rodr√≠guez-Tejedor et al |
| EPI_ISL_831160 | Spain/MD-IBV-99013277/2020 | 2020-09-15 | Europe/Spain/Madrid/Madrid | Hospital Universitario La Paz (Madrid) | SeqCOVID-SPAIN consortium/IBV(CSIC) | Mar√≠a Rodr√≠guez-Tejedor et al |
| EPI_ISL_831165 | Spain/MD-IBV-99013282/2020 | 2020-09-15 | Europe/Spain/Madrid/Madrid | Hospital Universitario La Paz (Madrid) | SeqCOVID-SPAIN consortium/IBV(CSIC) | Mar√≠a Rodr√≠guez-Tejedor et al |
| EPI_ISL_831168 | Spain/MD-IBV-99013285/2020 | 2020-09-17 | Europe/Spain/Madrid/Madrid | Hospital Universitario La Paz (Madrid) | SeqCOVID-SPAIN consortium/IBV(CSIC) | Mar√≠a Rodr√≠guez-Tejedor et al |
| EPI_ISL_831169 | Spain/MD-IBV-99013286/2020 | 2020-09-17 | Europe/Spain/Madrid/Madrid | Hospital Universitario La Paz (Madrid) | SeqCOVID-SPAIN consortium/IBV(CSIC) | Mar√≠a Rodr√≠guez-Tejedor et al |

| | | | | | | |
|---|---|---|---|---|---|---|
| EPI_ISL_831173 | Spain/MD-IBV-99013290/2020 | 2020-09-18 | Europe/Spain/Madrid/Madrid | Hospital Universitario La Paz (Madrid) | SeqCOVID-SPAIN consortium/IBV(CSIC) | Marv≠a Rodrv≠guez-Tejedor et al |
| EPI_ISL_831174 | Spain/MD-IBV-99013291/2020 | 2020-09-21 | Europe/Spain/Madrid/Madrid | Hospital Universitario La Paz (Madrid) | SeqCOVID-SPAIN consortium/IBV(CSIC) | Marv≠a Rodrv≠guez-Tejedor et al |
| EPI_ISL_831176 | Spain/MD-IBV-99013293/2020 | 2020-09-22 | Europe/Spain/Madrid/Madrid | Hospital Universitario La Paz (Madrid) | SeqCOVID-SPAIN consortium/IBV(CSIC) | Marv≠a Rodrv≠guez-Tejedor et al |
| EPI_ISL_831179 | Spain/MD-IBV-99013296/2020 | 2020-09-23 | Europe/Spain/Madrid/Madrid | Hospital Universitario La Paz (Madrid) | SeqCOVID-SPAIN consortium/IBV(CSIC) | Marv≠a Rodrv≠guez-Tejedor et al |
| EPI_ISL_831180 | Spain/MD-IBV-99013297/2020 | 2020-10-01 | Europe/Spain/Madrid/Madrid | Hospital Universitario La Paz (Madrid) | SeqCOVID-SPAIN consortium/IBV(CSIC) | Marv≠a Rodrv≠guez-Tejedor et al |
| EPI_ISL_831186 | Spain/MD-IBV-99013303/2020 | 2020-10-01 | Europe/Spain/Madrid/Madrid | Hospital Universitario La Paz (Madrid) | SeqCOVID-SPAIN consortium/IBV(CSIC) | Marv≠a Rodrv≠guez-Tejedor et al |
| EPI_ISL_831191 | Spain/MD-IBV-99013309/2020 | 2020-10-03 | Europe/Spain/Madrid/Madrid | Hospital Universitario La Paz (Madrid) | SeqCOVID-SPAIN consortium/IBV(CSIC) | Marv≠a Rodrv≠guez-Tejedor et al |
| EPI_ISL_831195 | Spain/MD-IBV-99013313/2020 | 2020-10-03 | Europe/Spain/Madrid/Madrid | Hospital Universitario La Paz (Madrid) | SeqCOVID-SPAIN consortium/IBV(CSIC) | Marv≠a Rodrv≠guez-Tejedor et al |
| EPI_ISL_831196 | Spain/MD-IBV-99013314/2020 | 2020-10-03 | Europe/Spain/Madrid/Madrid | Hospital Universitario La Paz (Madrid) | SeqCOVID-SPAIN consortium/IBV(CSIC) | Marv≠a Rodrv≠guez-Tejedor et al |
| EPI_ISL_831197 | Spain/MD-IBV-99013315/2020 | 2020-10-03 | Europe/Spain/Madrid/Madrid | Hospital Universitario La Paz (Madrid) | SeqCOVID-SPAIN consortium/IBV(CSIC) | Marv≠a Rodrv≠guez-Tejedor et al |
| EPI_ISL_831200 | Spain/MD-IBV-99013319/2020 | 2020-11-13 | Europe/Spain/Madrid/Madrid | Hospital Universitario La Paz (Madrid) | SeqCOVID-SPAIN consortium/IBV(CSIC) | Marv≠a Rodrv≠guez-Tejedor et al |
| EPI_ISL_831202 | Spain/MD-IBV-99013321/2020 | 2020-11-16 | Europe/Spain/Madrid/Madrid | Hospital Universitario La Paz (Madrid) | SeqCOVID-SPAIN consortium/IBV(CSIC) | Marv≠a Rodrv≠guez-Tejedor et al |
| EPI_ISL_831203 | Spain/MD-IBV-99013322/2020 | 2020-11-27 | Europe/Spain/Madrid/Madrid | Hospital Universitario La Paz (Madrid) | SeqCOVID-SPAIN consortium/IBV(CSIC) | Marv≠a Rodrv≠guez-Tejedor et al |

| | | | | | | |
|---|---|---|---|---|---|---|
| EPI_ISL_831208 | Spain/MD-IBV-99013328/2020 | 2020-11-26 | Europe/Spain/Madrid/Madrid | Hospital Universitario La Paz (Madrid) | SeqCOVID-SPAIN consortium/IBV(CSIC) | Marv≠a Rodrv≠guez-Tejedor et al |
| EPI_ISL_831210 | Spain/MD-IBV-99013330/2020 | 2020-11-26 | Europe/Spain/Madrid/Madrid | Hospital Universitario La Paz (Madrid) | SeqCOVID-SPAIN consortium/IBV(CSIC) | Marv≠a Rodrv≠guez-Tejedor et al |
| EPI_ISL_831212 | Spain/MD-IBV-99013332/2020 | 2020-11-25 | Europe/Spain/Madrid/Madrid | Hospital Universitario La Paz (Madrid) | SeqCOVID-SPAIN consortium/IBV(CSIC) | Marv≠a Rodrv≠guez-Tejedor et al |
| EPI_ISL_831214 | Spain/MD-IBV-99013334/2020 | 2020-11-04 | Europe/Spain/Madrid/Madrid | Hospital Universitario La Paz (Madrid) | SeqCOVID-SPAIN consortium/IBV(CSIC) | Marv≠a Rodrv≠guez-Tejedor et al |
| EPI_ISL_831215 | Spain/MD-IBV-99013335/2020 | 2020-11-04 | Europe/Spain/Madrid/Madrid | Hospital Universitario La Paz (Madrid) | SeqCOVID-SPAIN consortium/IBV(CSIC) | Marv≠a Rodrv≠guez-Tejedor et al |
| EPI_ISL_831216 | Spain/MD-IBV-99013336/2020 | 2020-11-04 | Europe/Spain/Madrid/Madrid | Hospital Universitario La Paz (Madrid) | SeqCOVID-SPAIN consortium/IBV(CSIC) | Marv≠a Rodrv≠guez-Tejedor et al |
| EPI_ISL_831220 | Spain/MD-IBV-99013340/2020 | 2020-11-05 | Europe/Spain/Madrid/Madrid | Hospital Universitario La Paz (Madrid) | SeqCOVID-SPAIN consortium/IBV(CSIC) | Marv≠a Rodrv≠guez-Tejedor et al |
| EPI_ISL_831222 | Spain/MD-IBV-99013342/2020 | 2020-11-03 | Europe/Spain/Madrid/Madrid | Hospital Universitario La Paz (Madrid) | SeqCOVID-SPAIN consortium/IBV(CSIC) | Marv≠a Rodrv≠guez-Tejedor et al |
| EPI_ISL_831223 | Spain/MD-IBV-99013343/2020 | 2020-11-03 | Europe/Spain/Madrid/Madrid | Hospital Universitario La Paz (Madrid) | SeqCOVID-SPAIN consortium/IBV(CSIC) | Marv≠a Rodrv≠guez-Tejedor et al |
| EPI_ISL_831224 | Spain/MD-IBV-99013344/2020 | 2020-10-31 | Europe/Spain/Madrid/Madrid | Hospital Universitario La Paz (Madrid) | SeqCOVID-SPAIN consortium/IBV(CSIC) | Marv≠a Rodrv≠guez-Tejedor et al |
| EPI_ISL_831226 | Spain/MD-IBV-99013346/2020 | 2020-10-30 | Europe/Spain/Madrid/Madrid | Hospital Universitario La Paz (Madrid) | SeqCOVID-SPAIN consortium/IBV(CSIC) | Marv≠a Rodrv≠guez-Tejedor et al |
| EPI_ISL_831233 | Spain/MD-IBV-99013355/2020 | 2020-11-04 | Europe/Spain/Madrid/Madrid | Hospital Universitario La Paz (Madrid) | SeqCOVID-SPAIN consortium/IBV(CSIC) | Marv≠a Rodrv≠guez-Tejedor et al |
| EPI_ISL_831234 | Spain/MD-IBV-99013356/2020 | 2020-11-05 | Europe/Spain/Madrid/Madrid | Hospital Universitario La Paz (Madrid) | SeqCOVID-SPAIN consortium/IBV(CSIC) | Marv≠a Rodrv≠guez-Tejedor et al |

| | | | | | | |
|---|---|---|---|---|---|---|
| EPI_ISL_831237 | Spain/MD-IBV-99013359/2020 | 2020-11-11 | Europe/Spain/Madrid/Madrid | Hospital Universitario La Paz (Madrid) | SeqCOVID-SPAIN consortium/IBV(CSIC) | Marv≠a Rodrv≠guez-Tejedor et al |
| EPI_ISL_831240 | Spain/MD-IBV-99013362/2020 | 2020-11-11 | Europe/Spain/Madrid/Madrid | Hospital Universitario La Paz (Madrid) | SeqCOVID-SPAIN consortium/IBV(CSIC) | Marv≠a Rodrv≠guez-Tejedor et al |
| EPI_ISL_831243 | Spain/MD-IBV-99013365/2020 | 2020-11-12 | Europe/Spain/Madrid/Madrid | Hospital Universitario La Paz (Madrid) | SeqCOVID-SPAIN consortium/IBV(CSIC) | Marv≠a Rodrv≠guez-Tejedor et al |
| EPI_ISL_831246 | Spain/MD-IBV-99013369/2020 | 2020-11-25 | Europe/Spain/Madrid/Madrid | Hospital Universitario La Paz (Madrid) | SeqCOVID-SPAIN consortium/IBV(CSIC) | Marv≠a Rodrv≠guez-Tejedor et al |
| EPI_ISL_831248 | Spain/MD-IBV-99013372/2020 | 2020-12-08 | Europe/Spain/Madrid/Madrid | Hospital Universitario La Paz (Madrid) | SeqCOVID-SPAIN consortium/IBV(CSIC) | Fernando Lv°zaro-Perona et al |
| EPI_ISL_831250 | Spain/MD-IBV-99013374/2020 | 2020-12-09 | Europe/Spain/Madrid/Madrid | Hospital Universitario La Paz (Madrid) | SeqCOVID-SPAIN consortium/IBV(CSIC) | Fernando Lv°zaro-Perona et al |
| EPI_ISL_831261 | Spain/MD-IBV-99013414/2020 | 2020-12-20 | Europe/Spain/Madrid/Madrid | Hospital Universitario La Paz (Madrid) | SeqCOVID-SPAIN consortium/IBV(CSIC) | Fernando Lv°zaro-Perona et al |
| EPI_ISL_831265 | Spain/MD-IBV-99013418/2020 | 2020-12-20 | Europe/Spain/Madrid/Madrid | Hospital Universitario La Paz (Madrid) | SeqCOVID-SPAIN consortium/IBV(CSIC) | Fernando Lv°zaro-Perona et al |
| EPI_ISL_831270 | Spain/MD-IBV-99013427/2020 | 2020-12-17 | Europe/Spain/Madrid/Madrid | Hospital Universitario La Paz (Madrid) | SeqCOVID-SPAIN consortium/IBV(CSIC) | Fernando Lv°zaro-Perona et al |
| EPI_ISL_831275 | Spain/MD-IBV-99013432/2020 | 2020-12-15 | Europe/Spain/Madrid/Madrid | Hospital Universitario La Paz (Madrid) | SeqCOVID-SPAIN consortium/IBV(CSIC) | Fernando Lv°zaro-Perona et al |
| EPI_ISL_831278 | Spain/MD-IBV-99013437/2020 | 2020-12-14 | Europe/Spain/Madrid/Madrid | Hospital Universitario La Paz (Madrid) | SeqCOVID-SPAIN consortium/IBV(CSIC) | Fernando Lv°zaro-Perona et al |
| EPI_ISL_831290 | USA/TX-DSHS-2074/2020 | 2020-06-04 | North America/USA/Texas/Titus County | Texas Department of State Health Services | Texas Department of State Health Services | Anita Pokharel et al |
| EPI_ISL_831303 | USA/TX-DSHS-2110/2020 | 2020-10-26 | North America/USA/Texas/Lubbock Cou | Texas Department of State Health Services | Texas Department of State Health Services | Anita Pokharel et al |

| | | | | | | |
|---|---|---|---|---|---|---|
| EPI_ISL_831307 | USA/TX-DSHS-2092/2020 | 2020-06-16 | North America/USA/Texas/Fort Bend Co | Texas Department of State Health Services | Texas Department of State Health Services | Anita Pokharel et al |
| EPI_ISL_831311 | USA/TX-DSHS-2076/2020 | 2020-06-04 | North America/USA/Texas/Titus County | Texas Department of State Health Services | Texas Department of State Health Services | Anita Pokharel et al |
| EPI_ISL_831323 | USA/TX-DSHS-2081/2020 | 2020-06-16 | North America/USA/Texas/Lavaca Count | Texas Department of State Health Services | Texas Department of State Health Services | Anita Pokharel et al |
| EPI_ISL_831365 | USA/NC-UNC-0004/2021 | 2021-01-08 | North America/USA/North Carolina/ | Clinical Molecular Microbiology Laboratory, UNC Hospitals | Jeremy Wang | Jeremy Wang et al |
| EPI_ISL_831373 | Germany/BW-RKI-I-0089/2020 | 2020-12-19 | Europe/Germany/Baden-Wuerttemberg | Limbach - MVZ Labor Dr. Limbach & Kollegen | Robert Koch Institute, Influenza and respiratory viruses FG17 & Bioinformatics MF1, Berlin, Germany | Dr. Konrad Bode et al |
| EPI_ISL_831374 | Germany/NW-RKI-I-0071/2020 | 2020-12-19 | Europe/Germany/North Rhine Westphal | Labor Krone | Robert Koch Institute, Influenza and respiratory viruses FG17 & Bioinformatics MF1, Berlin, Germany | Dr. Mvºnstermann. Prof. Tiemann et al |
| EPI_ISL_831375 | Germany/BW-RKI-I-0129/2020 | 2020-12-18 | Europe/Germany/Baden-Wuerttemberg | Limbach - MVZ Labor Dr. Limbach & Kollegen | Robert Koch Institute, Influenza and respiratory viruses FG17 & Bioinformatics MF1, Berlin, Germany | Dr. Konrad Bode et al |

| | | | | | | |
|---|---|---|---|---|---|---|
| EPI_ISL_831378 | Germany/BW-RKI-I-0049/2020 | 2020-12-18 | Europe/Germany/Baden-Wuerttemberg | Limbach - MVZ Labor Dr. Limbach & Kollegen | Robert Koch Institute, Influenza and respiratory viruses FG17 & Bioinformatics MF1, Berlin, Germany | Dr. Konrad Bode et al |
| EPI_ISL_831381 | Germany/BW-RKI-I-0083/2020 | 2020-12-15 | Europe/Germany/Baden-Wuerttemberg | Limbach - MVZ Labor Dr. Limbach & Kollegen | Robert Koch Institute, Influenza and respiratory viruses FG17 & Bioinformatics MF1, Berlin, Germany | Dr. Konrad Bode et al |
| EPI_ISL_831385 | Germany/NI-RKI-I-0098/2020 | 2020-12-15 | Europe/Germany/Niedersachsen/ | Labor Krone | Robert Koch Institute, Influenza and respiratory viruses FG17 & Bioinformatics MF1, Berlin, Germany | Dr. Mvºnstermann. Prof. Tiemann et al |
| EPI_ISL_831386 | Germany/BW-RKI-I-0130/2020 | 2020-12-15 | Europe/Germany/Baden-Wuerttemberg | Limbach - MVZ Labor Dr. Limbach & Kollegen | Robert Koch Institute, Influenza and respiratory viruses FG17 & Bioinformatics MF1, Berlin, Germany | Dr. Konrad Bode et al |
| EPI_ISL_831388 | Germany/BW-RKI-I-0138/2020 | 2020-12-15 | Europe/Germany/Baden-Wuerttemberg | Limbach - MVZ Labor Dr. Limbach & Kollegen | Robert Koch Institute, Influenza and respiratory viruses FG17 & Bioinformatics MF1, Berlin, Germany | Dr. Konrad Bode et al |

| | | | | | | |
|---|---|---|---|---|---|---|
| EPI_ISL_831389 | Germany/RP-RKI-I-0091/2020 | 2020-12-09 | Europe/Germany/Rheinland-Pfalz/ | Limbach - MVZ Labor Dr. Limbach & Kollegen | Robert Koch Institute, Influenza and respiratory viruses FG17 & Bioinformatics MF1, Berlin, Germany | Dr. Konrad Bode et al |
| EPI_ISL_831407 | Germany/SH-RKI-I-0079/2020 | 2020-12-07 | Europe/Germany/Schleswig-Holstein/ | Labor Dr. Krause & Kollegen MVZ GmbH | Robert Koch Institute, Influenza and respiratory viruses FG17 & Bioinformatics MF1, Berlin, Germany | Dr. Lorentz et al |
| EPI_ISL_831409 | Germany/NW-RKI-I-0107/2020 | 2020-12-07 | Europe/Germany/North Rhine Westphal | Labor Dr. Wisplinghoff - K√∂ln | Robert Koch Institute, Influenza and respiratory viruses FG17 & Bioinformatics MF1, Berlin, Germany | Dr. R. Grosser et al |
| EPI_ISL_831424 | Germany/NW-RKI-I-0104/2020 | 2020-12-02 | Europe/Germany/North Rhine Westphal | Labor Dr. Wisplinghoff - K√∂ln | Robert Koch Institute, Influenza and respiratory viruses FG17 & Bioinformatics MF1, Berlin, Germany | Dr. R. Grosser et al |
| EPI_ISL_831474 | Brasil/RS11069/2020 | 2020-06-12 | South America/Brazil/Rio Grande do Sul | Laborat√≥rio de Microbiologia Molecular - Universidade FEEVALE | Universidade Federal de Ci√™ncias da Sa√∫de de Porto Alegre | Vin√≠cius Bonetti Franceschi et al |

| | | | | | | |
|---|---|---|---|---|---|---|
| EPI_ISL_831489 | USA/WI-UW-2697/2021 | 2021-01-08 | North America/USA/Wisconsin/Dane Co | University of Wisconsin-Madison AIDS Vaccine Research Laboratories | University of Wisconsin-Madison AIDS Vaccine Research Laboratories | Gage Moreno et al |
| EPI_ISL_831492 | USA/WI-UW-2785/2021 | 2021-01-04 | North America/USA/Wisconsin/Milwauk | University of Wisconsin-Madison AIDS Vaccine Research Laboratories | University of Wisconsin-Madison AIDS Vaccine Research Laboratories | Gage Moreno et al |
| EPI_ISL_831513 | USA/WI-UW-2688/2021 | 2021-01-10 | North America/USA/Wisconsin/Dane Co | University of Wisconsin-Madison AIDS Vaccine Research Laboratories | University of Wisconsin-Madison AIDS Vaccine Research Laboratories | Gage Moreno et al |
| EPI_ISL_831514 | USA/WI-UW-2689/2021 | 2021-01-08 | North America/USA/Wisconsin/Dane Co | University of Wisconsin-Madison AIDS Vaccine Research Laboratories | University of Wisconsin-Madison AIDS Vaccine Research Laboratories | Gage Moreno et al |
| EPI_ISL_831520 | USA/WI-UW-2695/2021 | 2021-01-08 | North America/USA/Wisconsin/Dodge C | University of Wisconsin-Madison AIDS Vaccine Research Laboratories | University of Wisconsin-Madison AIDS Vaccine Research Laboratories | Gage Moreno et al |
| EPI_ISL_831523 | USA/WI-UW-2701/2021 | 2021-01-08 | North America/USA/Wisconsin/Lafayett | University of Wisconsin-Madison AIDS Vaccine Research Laboratories | University of Wisconsin-Madison AIDS Vaccine Research Laboratories | Gage Moreno et al |
| EPI_ISL_831527 | USA/WI-UW-2705/2021 | 2021-01-07 | North America/USA/Wisconsin/Dane Co | University of Wisconsin-Madison AIDS Vaccine Research Laboratories | University of Wisconsin-Madison AIDS Vaccine Research Laboratories | Gage Moreno et al |
| EPI_ISL_831530 | USA/WI-UW-2708/2021 | 2021-01-07 | North America/USA/Wisconsin/Dane Co | University of Wisconsin-Madison AIDS Vaccine Research Laboratories | University of Wisconsin-Madison AIDS Vaccine Research Laboratories | Gage Moreno et al |

| | | | | | | |
|---|---|---|---|---|---|---|
| EPI_ISL_831531 | USA/WI-UW-2709/2021 | 2021-01-08 | North America/USA/Wisconsin/Dane Co | University of Wisconsin-Madison AIDS Vaccine Research Laboratories | University of Wisconsin-Madison AIDS Vaccine Research Laboratories | Gage Moreno et al |
| EPI_ISL_831532 | USA/WI-UW-2711/2021 | 2021-01-08 | North America/USA/Wisconsin/Dane Co | University of Wisconsin-Madison AIDS Vaccine Research Laboratories | University of Wisconsin-Madison AIDS Vaccine Research Laboratories | Gage Moreno et al |
| EPI_ISL_831543 | USA/WI-UW-2723/2021 | 2021-01-07 | North America/USA/Wisconsin/La Crosse | University of Wisconsin-Madison AIDS Vaccine Research Laboratories | University of Wisconsin-Madison AIDS Vaccine Research Laboratories | Gage Moreno et al |
| EPI_ISL_831550 | USA/WI-UW-2730/2021 | 2021-01-06 | North America/USA/Wisconsin/Dane Co | University of Wisconsin-Madison AIDS Vaccine Research Laboratories | University of Wisconsin-Madison AIDS Vaccine Research Laboratories | Gage Moreno et al |
| EPI_ISL_831558 | USA/WI-UW-2775/2021 | 2021-01-04 | North America/USA/Wisconsin/Milwauk | University of Wisconsin-Madison AIDS Vaccine Research Laboratories | University of Wisconsin-Madison AIDS Vaccine Research Laboratories | Gage Moreno et al |
| EPI_ISL_831563 | USA/WI-UW-2782/2021 | 2021-01-04 | North America/USA/Wisconsin/Milwauk | University of Wisconsin-Madison AIDS Vaccine Research Laboratories | University of Wisconsin-Madison AIDS Vaccine Research Laboratories | Gage Moreno et al |
| EPI_ISL_831569 | USA/WI-UW-2789/2021 | 2021-01-04 | North America/USA/Wisconsin/Milwauk | University of Wisconsin-Madison AIDS Vaccine Research Laboratories | University of Wisconsin-Madison AIDS Vaccine Research Laboratories | Gage Moreno et al |
| EPI_ISL_831570 | USA/WI-UW-2791/2021 | 2021-01-04 | North America/USA/Wisconsin/Milwauk | University of Wisconsin-Madison AIDS Vaccine Research Laboratories | University of Wisconsin-Madison AIDS Vaccine Research Laboratories | Gage Moreno et al |

| | | | | | | |
|---|---|---|---|---|---|---|
| EPI_ISL_831571 | USA/WI-UW-2792/2021 | 2021-01-04 | North America/USA/Wisconsin/Milwauk | University of Wisconsin-Madison AIDS Vaccine Research Laboratories | University of Wisconsin-Madison AIDS Vaccine Research Laboratories | Gage Moreno et al |
| EPI_ISL_831577 | USA/WI-UW-2763/2020 | 2020-12-31 | North America/USA/Wisconsin/Dane Co | University of Wisconsin-Madison AIDS Vaccine Research Laboratories | University of Wisconsin-Madison AIDS Vaccine Research Laboratories | Gage Moreno et al |
| EPI_ISL_831582 | USA/WI-UW-2768/2020 | 2020-12-29 | North America/USA/Wisconsin/Dane Co | University of Wisconsin-Madison AIDS Vaccine Research Laboratories | University of Wisconsin-Madison AIDS Vaccine Research Laboratories | Gage Moreno et al |
| EPI_ISL_831585 | USA/WI-UW-2772/2020 | 2020-12-31 | North America/USA/Wisconsin/Rock Cou | University of Wisconsin-Madison AIDS Vaccine Research Laboratories | University of Wisconsin-Madison AIDS Vaccine Research Laboratories | Gage Moreno et al |
| EPI_ISL_831590 | USA/WI-UW-2800/2021 | 2021-01-04 | North America/USA/Wisconsin/Milwauk | University of Wisconsin-Madison AIDS Vaccine Research Laboratories | University of Wisconsin-Madison AIDS Vaccine Research Laboratories | Gage Moreno et al |
| EPI_ISL_831594 | USA/WI-UW-2805/2021 | 2021-01-04 | North America/USA/Wisconsin/Milwauk | University of Wisconsin-Madison AIDS Vaccine Research Laboratories | University of Wisconsin-Madison AIDS Vaccine Research Laboratories | Gage Moreno et al |
| EPI_ISL_831601 | USA/WI-UW-2815/2021 | 2021-01-04 | North America/USA/Wisconsin/Milwauk | University of Wisconsin-Madison AIDS Vaccine Research Laboratories | University of Wisconsin-Madison AIDS Vaccine Research Laboratories | Gage Moreno et al |
| EPI_ISL_831609 | USA/WI-UW-2823/2021 | 2021-01-04 | North America/USA/Wisconsin/Milwauk | University of Wisconsin-Madison AIDS Vaccine Research Laboratories | University of Wisconsin-Madison AIDS Vaccine Research Laboratories | Gage Moreno et al |

| | | | | | | |
|---|---|---|---|---|---|---|
| EPI_ISL_831616 | USA/WI-UW-2832/2021 | 2021-01-04 | North America/USA/Wisconsin/Milwauk | University of Wisconsin-Madison AIDS Vaccine Research Laboratories | University of Wisconsin-Madison AIDS Vaccine Research Laboratories | Gage Moreno et al |
| EPI_ISL_831620 | USA/WI-UW-2836/2021 | 2021-01-04 | North America/USA/Wisconsin/Milwauk | University of Wisconsin-Madison AIDS Vaccine Research Laboratories | University of Wisconsin-Madison AIDS Vaccine Research Laboratories | Gage Moreno et al |
| EPI_ISL_831626 | USA/WI-UW-2739/2021 | 2021-01-05 | North America/USA/Wisconsin/Dane Co | University of Wisconsin-Madison AIDS Vaccine Research Laboratories | University of Wisconsin-Madison AIDS Vaccine Research Laboratories | Gage Moreno et al |
| EPI_ISL_831628 | USA/WI-UW-2744/2021 | 2021-01-05 | North America/USA/Wisconsin/Dane Co | University of Wisconsin-Madison AIDS Vaccine Research Laboratories | University of Wisconsin-Madison AIDS Vaccine Research Laboratories | Gage Moreno et al |
| EPI_ISL_831629 | USA/WI-UW-2746/2021 | 2021-01-05 | North America/USA/Wisconsin/Dane Co | University of Wisconsin-Madison AIDS Vaccine Research Laboratories | University of Wisconsin-Madison AIDS Vaccine Research Laboratories | Gage Moreno et al |
| EPI_ISL_831637 | USA/WI-UW-2841/2021 | 2021-01-05 | North America/USA/Wisconsin/Milwauk | University of Wisconsin-Madison AIDS Vaccine Research Laboratories | University of Wisconsin-Madison AIDS Vaccine Research Laboratories | Gage Moreno et al |
| EPI_ISL_831638 | USA/WI-UW-2842/2021 | 2021-01-05 | North America/USA/Wisconsin/Milwauk | University of Wisconsin-Madison AIDS Vaccine Research Laboratories | University of Wisconsin-Madison AIDS Vaccine Research Laboratories | Gage Moreno et al |
| EPI_ISL_831639 | USA/WI-UW-2736/2021 | 2021-01-06 | North America/USA/Wisconsin/Columbia | University of Wisconsin-Madison AIDS Vaccine Research Laboratories | University of Wisconsin-Madison AIDS Vaccine Research Laboratories | Gage Moreno et al |

| | | | | | | |
|---|---|---|---|---|---|---|
| EPI_ISL_831641 | USA/WI-UW-2740/2021 | 2021-01-06 | North America/USA/Wisconsin/Jefferson | University of Wisconsin-Madison AIDS Vaccine Research Laboratories | University of Wisconsin-Madison AIDS Vaccine Research Laboratories | Gage Moreno et al |
| EPI_ISL_831645 | Brazil/RS-11262/2020 | 2020-06-15 | South America/Brazil/Rio Grande do Sul | Laboratv≥rio de Microbiologia Molecular - Universidade FEEVALE | Universidade Federal de Civ™ncias da Sav∫de de Porto Alegre | Vinv≠cius Bonetti Franceschi et al |
| EPI_ISL_831646 | Brazil/RS-11569/2020 | 2020-06-17 | South America/Brazil/Rio Grande do Sul | Laboratv≥rio de Microbiologia Molecular - Universidade FEEVALE | Universidade Federal de Civ™ncias da Sav∫de de Porto Alegre | Vinv≠cius Bonetti Franceschi et al |
| EPI_ISL_831660 | Brazil/RS-11574/2020 | 2020-06-17 | South America/Brazil/Rio Grande do Sul | Laboratv≥rio de Microbiologia Molecular - Universidade FEEVALE | Universidade Federal de Civ™ncias da Sav∫de de Porto Alegre | Vinv≠cius Bonetti Franceschi et al |
| EPI_ISL_831666 | Slovakia/UKBA-818/2021 | 2021-01-13 | Europe/Slovakia/Bratislava/ | Institute of Virology, Biomedical Research Center of the Slovak Academy of Sciences, Bratislava | Faculty of Natural Sciences, Comenius University, Bratislava | Viktv≥ria ƒåabanovv° et al |
| EPI_ISL_831676 | Slovakia/UKBA-809/2021 | 2021-01-08 | Europe/Slovakia/Zilina/Trstena | Institute of Virology, Biomedical Research Center of the Slovak Academy of Sciences, Bratislava | Faculty of Natural Sciences, Comenius University, Bratislava | Bro≈àa Brejovv° et al |
| EPI_ISL_831678 | Brazil/RS-9881/2020 | 2020-05-31 | South America/Brazil/Rio Grande do Sul | Laboratv≥rio de Microbiologia Molecular - Universidade FEEVALE | Universidade Federal de Civ™ncias da Sav∫de de Porto Alegre | Vinv≠cius Bonetti Franceschi et al |
| EPI_ISL_831681 | Brazil/RS-13367/2020 | 2020-07-02 | South America/Brazil/Rio Grande do Sul | Laboratv≥rio de Microbiologia Molecular - Universidade FEEVALE | Universidade Federal de Civ™ncias da Sav∫de de Porto Alegre | Vinv≠cius Bonetti Franceschi et al |

| | | | | | | |
|---|---|---|---|---|---|---|
| EPI_ISL_831683 | Brazil/RS-13368/2020 | 2020-07-01 | South America/Brazil/Rio Grande do Sul | Laboratv≥rio de Microbiologia Molecular - Universidade FEEVALE | Universidade Federal de Civ™ncias da Sav∫de de Porto Alegre | Vinv≠cius Bonetti Franceschi et al |
| EPI_ISL_831685 | Brazil/RS-13979/2020 | 2020-07-06 | South America/Brazil/Rio Grande do Sul | Laboratv≥rio de Microbiologia Molecular - Universidade FEEVALE | Universidade Federal de Civ™ncias da Sav∫de de Porto Alegre | Vinv≠cius Bonetti Franceschi et al |
| EPI_ISL_831688 | Brazil/RS-15371/2020 | 2020-07-15 | South America/Brazil/Rio Grande do Sul | Laboratv≥rio de Microbiologia Molecular - Universidade FEEVALE | Universidade Federal de Civ™ncias da Sav∫de de Porto Alegre | Vinv≠cius Bonetti Franceschi et al |
| EPI_ISL_831689 | Brazil/RS-19337/2020 | 2020-08-03 | South America/Brazil/Rio Grande do Sul | Laboratv≥rio de Microbiologia Molecular - Universidade FEEVALE | Universidade Federal de Civ™ncias da Sav∫de de Porto Alegre | Vinv≠cius Bonetti Franceschi et al |
| EPI_ISL_831692 | USA/SC-USAFSAM-S473/2020 | 2020-10-05 | North America/USA/South Carolina/ | United States Air Force School of Aerospace Medicine | United States Air Force School of Aerospace Medicine | Anthony Fries et al |
| EPI_ISL_831695 | USA/MS-USAFSAM-S715/2020 | 2020-12-14 | North America/USA/Mississippi/ | United States Air Force School of Aerospace Medicine | United States Air Force School of Aerospace Medicine | Anthony Fries et al |
| EPI_ISL_831701 | USA/SC-USAFSAM-S612/2020 | 2020-11-23 | North America/USA/South Carolina/ | United States Air Force School of Aerospace Medicine | United States Air Force School of Aerospace Medicine | Anthony Fries et al |
| EPI_ISL_831702 | USA/GA-USAFSAM-S769/2020 | 2020-12-21 | North America/USA/Georgia/ | United States Air Force School of Aerospace Medicine | United States Air Force School of Aerospace Medicine | Anthony Fries et al |
| EPI_ISL_831707 | USA/KS-USAFSAM-S597/2020 | 2020-11-17 | North America/USA/Kansas/ | United States Air Force School of Aerospace Medicine | United States Air Force School of Aerospace Medicine | Anthony Fries et al |

| | | | | | | |
|---|---|---|---|---|---|---|
| EPI_ISL_831708 | USA/SC-USAFSAM-S602/2020 | 2020-11-23 | North America/USA/South Carolina/ | United States Air Force School of Aerospace Medicine | United States Air Force School of Aerospace Medicine | Anthony Fries et al |
| EPI_ISL_831709 | USA/MS-USAFSAM-S606/2020 | 2020-11-23 | North America/USA/Mississippi/ | United States Air Force School of Aerospace Medicine | United States Air Force School of Aerospace Medicine | Anthony Fries et al |
| EPI_ISL_831721 | USA/FL-USAFSAM-S523/2020 | 2020-11-16 | North America/USA/Florida/ | United States Air Force School of Aerospace Medicine | United States Air Force School of Aerospace Medicine | Anthony Fries et al |
| EPI_ISL_831749 | USA/SC-USAFSAM-S713/2020 | 2020-12-14 | North America/USA/South Carolina/ | United States Air Force School of Aerospace Medicine | United States Air Force School of Aerospace Medicine | Anthony Fries et al |
| EPI_ISL_831772 | USA/MT-USAFSAM-S477/2020 | 2020-10-15 | North America/USA/Montana/ | United States Air Force School of Aerospace Medicine | United States Air Force School of Aerospace Medicine | Anthony Fries et al |
| EPI_ISL_831788 | USA/LA-USAFSAM-S496/2020 | 2020-11-02 | North America/USA/Louisiana/ | United States Air Force School of Aerospace Medicine | United States Air Force School of Aerospace Medicine | Anthony Fries et al |
| EPI_ISL_831790 | USA/NC-USAFSAM-S499/2020 | 2020-11-02 | North America/USA/North Carolina/ | United States Air Force School of Aerospace Medicine | United States Air Force School of Aerospace Medicine | Anthony Fries et al |
| EPI_ISL_831792 | USA/OH-USAFSAM-S502/2020 | 2020-11-02 | North America/USA/Ohio/ | United States Air Force School of Aerospace Medicine | United States Air Force School of Aerospace Medicine | Anthony Fries et al |
| EPI_ISL_831803 | USA/WY-USAFSAM-S516/2020 | 2020-11-16 | North America/USA/Wyoming/ | United States Air Force School of Aerospace Medicine | United States Air Force School of Aerospace Medicine | Anthony Fries et al |
| EPI_ISL_831835 | USA/CO-USAFSAM-S620/2020 | 2020-11-30 | North America/USA/Colorado/ | United States Air Force School of Aerospace Medicine | United States Air Force School of Aerospace Medicine | Anthony Fries et al |

| | | | | | | |
|---|---|---|---|---|---|---|
| EPI_ISL_831843 | USA/OH-USAFSAM-S641/2020 | 2020-11-30 | North America/USA/Ohio/ | United States Air Force School of Aerospace Medicine | United States Air Force School of Aerospace Medicine | Anthony Fries et al |
| EPI_ISL_831851 | USA/ID-USAFSAM-S662/2020 | 2020-12-10 | North America/USA/Idaho/ | United States Air Force School of Aerospace Medicine | United States Air Force School of Aerospace Medicine | Anthony Fries et al |
| EPI_ISL_831852 | USA/NC-USAFSAM-S664/2020 | 2020-12-10 | North America/USA/North Carolina/ | United States Air Force School of Aerospace Medicine | United States Air Force School of Aerospace Medicine | Anthony Fries et al |
| EPI_ISL_831856 | USA/MO-USAFSAM-S670/2020 | 2020-11-30 | North America/USA/Missouri/ | United States Air Force School of Aerospace Medicine | United States Air Force School of Aerospace Medicine | Anthony Fries et al |
| EPI_ISL_831859 | USA/TX-USAFSAM-S673/2020 | 2020-12-02 | North America/USA/Texas/ | United States Air Force School of Aerospace Medicine | United States Air Force School of Aerospace Medicine | Anthony Fries et al |
| EPI_ISL_831870 | USA/NC-USAFSAM-S693/2020 | 2020-12-14 | North America/USA/North Carolina/ | United States Air Force School of Aerospace Medicine | United States Air Force School of Aerospace Medicine | Anthony Fries et al |
| EPI_ISL_831888 | USA/SC-USAFSAM-S735/2020 | 2020-12-21 | North America/USA/South Carolina/ | United States Air Force School of Aerospace Medicine | United States Air Force School of Aerospace Medicine | Anthony Fries et al |
| EPI_ISL_831891 | USA/KS-USAFSAM-S739/2020 | 2020-12-23 | North America/USA/Kansas/ | United States Air Force School of Aerospace Medicine | United States Air Force School of Aerospace Medicine | Anthony Fries et al |
| EPI_ISL_831892 | Brazil/RS-22838/2020 | 2020-08-19 | South America/Brazil/Rio Grande do Sul | Laboratv≥rio de Microbiologia Molecular - Universidade FEEVALE | Universidade Federal de Civ™ncias da Sav∫de de Porto Alegre | Vinv≠cius Bonetti Franceschi et al |

| | | | | | | |
|---|---|---|---|---|---|---|
| EPI_ISL_831898 | Brazil/RS-24277/2020 | 2020-08-26 | South America/Brazil/Rio Grande do Sul | Laboratv≥rio de Microbiologia Molecular - Universidade FEEVALE | Universidade Federal de Civ™ncias da Sav∫de de Porto Alegre | Vinv≠cius Bonetti Franceschi et al |
| EPI_ISL_831906 | USA/NM-DOH-2021014243/2021 | 2021-01-07 | North America/USA/New Mexico/ | New Mexico Department of Health Scientific Laboratory | New Mexico Department of Health Scientific Laboratory | Ellie Johnson et al |
| EPI_ISL_831913 | Brazil/RS-24285/2020 | 2020-08-24 | South America/Brazil/Rio Grande do Sul | Laboratv≥rio de Microbiologia Molecular - Universidade FEEVALE | Universidade Federal de Civ™ncias da Sav∫de de Porto Alegre | Vinv≠cius Bonetti Franceschi et al |
| EPI_ISL_831917 | USA/NM-DOH-2020201018/2020 | 2020-07-09 | North America/USA/New Mexico/ | New Mexico Department of Health Scientific Laboratory | New Mexico Department of Health Scientific Laboratory | Ellie Johnson et al |
| EPI_ISL_831918 | USA/NM-DOH-2020214555/2020 | 2020-07-14 | North America/USA/New Mexico/ | New Mexico Department of Health Scientific Laboratory | New Mexico Department of Health Scientific Laboratory | Ellie Johnson et al |
| EPI_ISL_831919 | USA/NM-DOH-2020279720/2020 | 2020-08-10 | North America/USA/New Mexico/ | New Mexico Department of Health Scientific Laboratory | New Mexico Department of Health Scientific Laboratory | Ellie Johnson et al |
| EPI_ISL_831925 | USA/NM-DOH-2021013368/2021 | 2021-01-08 | North America/USA/New Mexico/ | New Mexico Department of Health Scientific Laboratory | New Mexico Department of Health Scientific Laboratory | Ellie Johnson et al |
| EPI_ISL_831930 | USA/NM-DOH-2021014244/2021 | 2021-01-07 | North America/USA/New Mexico/ | New Mexico Department of Health Scientific Laboratory | New Mexico Department of Health Scientific Laboratory | Ellie Johnson et al |
| EPI_ISL_831932 | USA/NM-DOH-2021014513/2021 | 2021-01-09 | North America/USA/New Mexico/ | New Mexico Department of Health Scientific Laboratory | New Mexico Department of Health Scientific Laboratory | Ellie Johnson et al |

| | | | | | | |
|---|---|---|---|---|---|---|
| EPI_ISL_831937 | USA/NM-DOH-2021015119/2021 | 2021-01-09 | North America/USA/New Mexico/ | New Mexico Department of Health Scientific Laboratory | New Mexico Department of Health Scientific Laboratory | Ellie Johnson et al |
| EPI_ISL_831938 | Brazil/RS-24565/2020 | 2020-08-27 | South America/Brazil/Rio Grande do Sul | Laboratv≥rio de Microbiologia Molecular - Universidade FEEVALE | Universidade Federal de Civ™ncias da Sav∫de de Porto Alegre | Vinv≠cius Bonetti Franceschi et al |
| EPI_ISL_831939 | Brazil/RS-25833/2020 | 2020-09-03 | South America/Brazil/Rio Grande do Sul | Laboratv≥rio de Microbiologia Molecular - Universidade FEEVALE | Universidade Federal de Civ™ncias da Sav∫de de Porto Alegre | Vinv≠cius Bonetti Franceschi et al |
| EPI_ISL_831940 | Brazil/RS-26977/2020 | 2020-09-10 | South America/Brazil/Rio Grande do Sul | Laboratv≥rio de Microbiologia Molecular - Universidade FEEVALE | Universidade Federal de Civ™ncias da Sav∫de de Porto Alegre | Vinv≠cius Bonetti Franceschi et al |
| EPI_ISL_831941 | Sweden/21-50139/2021 | 2021-01-06 | Europe/Sweden/Stockholm/ | Klinisk mikrobiologi | The Public Health Agency of Sweden | Department of Microbiology et al |
| EPI_ISL_831948 | Sweden/20-13048/2020 | 2020-12-29 | Europe/Sweden/Stockholm/ | ABC | The Public Health Agency of Sweden | Department of Microbiology et al |
| EPI_ISL_831951 | Sweden/20-13066/2020 | 2020-12-29 | Europe/Sweden/Stockholm/ | ABC | The Public Health Agency of Sweden | Department of Microbiology et al |
| EPI_ISL_831955 | Sweden/20-53813/2020 | 2020-12-22 | Europe/Sweden/Varmland/ | Laboratoriemedicin, Klinisk mikrobiologi | The Public Health Agency of Sweden | Department of Microbiology et al |
| EPI_ISL_831961 | Sweden/20-52933/2020 | 2020-08-17 | Europe/Sweden/Orebro/ | Orebro klinisk mikrobiologi | The Public Health Agency of Sweden | Department of Microbiology et al |
| EPI_ISL_831968 | Sweden/20-52941/2020 | 2020-09-23 | Europe/Sweden/Orebro/ | Orebro klinisk mikrobiologi | The Public Health Agency of Sweden | Department of Microbiology et al |
| EPI_ISL_831971 | Sweden/20-52949/2020 | 2020-08-30 | Europe/Sweden/Orebro/ | Orebro klinisk mikrobiologi | The Public Health Agency of Sweden | Department of Microbiology et al |
| EPI_ISL_831989 | Sweden/20-53845/2020 | 2020-12-15 | Europe/Sweden/Vastra Gotaland/ | Laboratorium for klinisk mikrobiologi | The Public Health Agency of Sweden | Department of Microbiology et al |
| EPI_ISL_831998 | Sweden/20-53812/2020 | 2020-12-22 | Europe/Sweden/Varmland/ | Laboratoriemedicin, Klinisk mikrobiologi | The Public Health Agency of Sweden | Department of Microbiology et al |
| EPI_ISL_831999 | Sweden/20-53814/2020 | 2020-12-22 | Europe/Sweden/Varmland/ | Laboratoriemedicin, Klinisk mikrobiologi | The Public Health Agency of Sweden | Department of Microbiology et al |

| | | | | | | |
|---|---|---|---|---|---|---|
| EPI_ISL_832009 | Brazil/RS-27623/2020 | 2020-09-16 | South America/Brazil/Rio Grande do Sul | Laboratv≥rio de Microbiologia Molecular - Universidade FEEVALE | Universidade Federal de Civ™ncias da Sav∫de de Porto Alegre | Vinv≠cius Bonetti Franceschi et al |
| EPI_ISL_832010 | Brazil/RS-31784/2020 | 2020-10-14 | South America/Brazil/Rio Grande do Sul | Laboratv≥rio de Microbiologia Molecular - Universidade FEEVALE | Universidade Federal de Civ™ncias da Sav∫de de Porto Alegre | Vinv≠cius Bonetti Franceschi et al |
| EPI_ISL_832011 | Brazil/RS-31786/2020 | 2020-10-13 | South America/Brazil/Rio Grande do Sul | Laboratv≥rio de Microbiologia Molecular - Universidade FEEVALE | Universidade Federal de Civ™ncias da Sav∫de de Porto Alegre | Vinv≠cius Bonetti Franceschi et al |
| EPI_ISL_832012 | Brazil/RS-31799/2020 | 2020-10-13 | South America/Brazil/Rio Grande do Sul | Laboratv≥rio de Microbiologia Molecular - Universidade FEEVALE | Universidade Federal de Civ™ncias da Sav∫de de Porto Alegre | Vinv≠cius Bonetti Franceschi et al |
| EPI_ISL_832013 | Brazil/RS-31800/2020 | 2020-10-13 | South America/Brazil/Rio Grande do Sul | Laboratv≥rio de Microbiologia Molecular - Universidade FEEVALE | Universidade Federal de Civ™ncias da Sav∫de de Porto Alegre | Vinv≠cius Bonetti Franceschi et al |
| EPI_ISL_832029 | USA/WY-WPHL-20044965/2020 | 2020-07-16 | North America/USA/Wyoming/ | Wyoming Public Health Laboratory | Wyoming Public Health Laboratory | Noah Hull et al |
| EPI_ISL_832038 | USA/WY-WPHL-20149444/2020 | 2020-11-24 | North America/USA/Wyoming/ | Wyoming Public Health Laboratory | Wyoming Public Health Laboratory | Noah Hull et al |
| EPI_ISL_832042 | USA/WY-WPHL-20149968/2020 | 2020-11-24 | North America/USA/Wyoming/ | Wyoming Public Health Laboratory | Wyoming Public Health Laboratory | Noah Hull et al |
| EPI_ISL_832048 | USA/WY-WPHL-20170861/2020 | 2020-12-27 | North America/USA/Wyoming/ | Wyoming Public Health Laboratory | Wyoming Public Health Laboratory | Noah Hull et al |
| EPI_ISL_832049 | USA/WY-WPHL-20170908/2020 | 2020-12-27 | North America/USA/Wyoming/ | Wyoming Public Health Laboratory | Wyoming Public Health Laboratory | Noah Hull et al |
| EPI_ISL_832054 | USA/WY-WPHL-20171065/2020 | 2020-12-28 | North America/USA/Wyoming/ | Wyoming Public Health Laboratory | Wyoming Public Health Laboratory | Noah Hull et al |
| EPI_ISL_832063 | USA/WY-WPHL-20171270/2020 | 2020-12-28 | North America/USA/Wyoming/ | Wyoming Public Health Laboratory | Wyoming Public Health Laboratory | Noah Hull et al |

| | | | | | | |
|---|---|---|---|---|---|---|
| EPI_ISL_832097 | USA/CA-SC210027/2021 | 2021-01-06 | North America/USA/California/Santa Cl | Santa Clara County Public Health Laboratory | Santa Clara County Public Health Laboratory | Santa Clara County Public Health Department et al |
| EPI_ISL_832099 | USA/CA-SC210031/2020 | 2020-12-10 | North America/USA/California/Santa Cl | Santa Clara County Public Health Laboratory | Santa Clara County Public Health Laboratory | Santa Clara County Public Health Department et al |
| EPI_ISL_832104 | Spain/CT-Irsicaixa2020090219/2020 | 2020-09-16 | Europe/Spain/Catalunya/Badalona | Hospital Universitari Germans Trias i Pujol(HUGTiP)/Fundaciv≥ Lluita contra la SIDA (FLSida) | IrsiCaixa AIDS Research Lab | Marc Noguera-Julian et al |
| EPI_ISL_832109 | USA/CA-LACPHL-AF00128/2020 | 2020-12-22 | North America/USA/California/Los Ange | MD Laboratories | Los Angeles County PHL | P. Hemarajata et al. et al |
| EPI_ISL_832116 | USA/CA-LACPHL-AF00135/2020 | 2020-12-28 | North America/USA/California/Los Ange | MD Laboratories | Los Angeles County PHL | P. Hemarajata et al. et al |
| EPI_ISL_832117 | USA/CA-LACPHL-AF00136/2020 | 2020-12-18 | North America/USA/California/Los Ange | MD Laboratories | Los Angeles County PHL | P. Hemarajata et al. et al |
| EPI_ISL_832124 | USA/NC-UNC-0008/2021 | 2021-01-04 | North America/USA/North Carolina/ | Clinical Molecular Microbiology Laboratory, UNC Hospitals | Jeremy Wang | Jeremy Wang et al |
| EPI_ISL_832129 | France/IDF-IPP10802/2020 | 2020-12-07 | Europe/France/Ile de France/Paris | Sentinelles IDF | National Reference Center for Viruses of Respiratory Infections, Institut Pasteur, Paris | Marion Barbet et al |
| EPI_ISL_832138 | France/GE-IPP10952/2020 | 2020-12-17 | Europe/France/Grand Est/Reims | Hospital | National Reference Center for Viruses of Respiratory Infections, Institut Pasteur, Paris | Marion Barbet et al |
| EPI_ISL_832139 | France/GE-IPP10955/2020 | 2020-12-17 | Europe/France/Grand Est/Reims | Hospital | National Reference Center for Viruses of Respiratory Infections, Institut Pasteur, Paris | Marion Barbet et al |
| EPI_ISL_832141 | France/GE-IPP10962/2020 | 2020-12-17 | Europe/France/Grand Est/Reims | Hospital | National Reference Center for Viruses of Respiratory Infections, Institut Pasteur, Paris | Marion Barbet et al |

| | | | | | | |
|---|---|---|---|---|---|---|
| EPI_ISL_832146 | France/Bre-IPP11275/2020 | 2020-11-22 | Europe/France/Bretagne/Brest | Hospital | National Reference Center for Viruses of Respiratory Infections, Institut Pasteur, Paris | Marion Barbet et al |
| EPI_ISL_832147 | France/Bre-IPP11276/2020 | 2020-11-22 | Europe/France/Bretagne/Brest | Hospital | National Reference Center for Viruses of Respiratory Infections, Institut Pasteur, Paris | Marion Barbet et al |
| EPI_ISL_832148 | France/Bre-IPP11277/2020 | 2020-11-23 | Europe/France/Bretagne/Brest | Hospital | National Reference Center for Viruses of Respiratory Infections, Institut Pasteur, Paris | Marion Barbet et al |
| EPI_ISL_832149 | France/Bre-IPP11278/2020 | 2020-11-24 | Europe/France/Bretagne/Brest | Hospital | National Reference Center for Viruses of Respiratory Infections, Institut Pasteur, Paris | Marion Barbet et al |
| EPI_ISL_832150 | France/Bre-IPP11279/2020 | 2020-11-24 | Europe/France/Bretagne/Brest | Hospital | National Reference Center for Viruses of Respiratory Infections, Institut Pasteur, Paris | Marion Barbet et al |
| EPI_ISL_832169 | France/CVL-IPP11331/2020 | 2020-12-22 | Europe/France/Centre-Val de Loire/Orlv | Hospital | National Reference Center for Viruses of Respiratory Infections, Institut Pasteur, Paris | Marion Barbet et al |

| | | | | | | |
|---|---|---|---|---|---|---|
| EPI_ISL_832171 | France/GE-IPP11334/2020 | 2020-12-23 | Europe/France/Grand Est/Troyes | Hospital | National Reference Center for Viruses of Respiratory Infections, Institut Pasteur, Paris | Marion Barbet et al |
| EPI_ISL_832173 | France/GE-IPP11336/2020 | 2020-12-23 | Europe/France/Grand Est/Troyes | Hospital | National Reference Center for Viruses of Respiratory Infections, Institut Pasteur, Paris | Marion Barbet et al |
| EPI_ISL_832174 | France/GE-IPP11339/2020 | 2020-12-23 | Europe/France/Grand Est/Troyes | Hospital | National Reference Center for Viruses of Respiratory Infections, Institut Pasteur, Paris | Marion Barbet et al |
| EPI_ISL_832175 | France/GE-IPP11340/2020 | 2020-12-23 | Europe/France/Grand Est/Troyes | Hospital | National Reference Center for Viruses of Respiratory Infections, Institut Pasteur, Paris | Marion Barbet et al |
| EPI_ISL_832177 | France/HDF-IPP11342/2020 | 2020-12-26 | Europe/France/Hauts de France/Lille FR | Hospital | National Reference Center for Viruses of Respiratory Infections, Institut Pasteur, Paris | Marion Barbet et al |
| EPI_ISL_832204 | USA/CA-UW-63893941/2021 | 2021-01-08 | North America/USA/California/ | Northwest Pathology | UW Virology Lab | Pavitra Roychoudhury et al |
| EPI_ISL_832228 | Belgium/ULG-11255/2021 | 2021-01-07 | Europe/Belgium/Liv®ge/ | Department of Clinical Microbiology | GIGA Medical Genomics | Keith Durkin et al |
| EPI_ISL_832252 | USA/NY-NYCPHL-001932/2020 | 2020-12-23 | North America/USA/New York/New Yor | DOHMH Corona | New York City Public Health Laboratory | Jade Wang et al |
| EPI_ISL_832256 | USA/NY-NYCPHL-001883/2020 | 2020-12-21 | North America/USA/New York/New Yor | DOHMH Central Harlem | New York City Public Health Laboratory | Jade Wang et al |

| | | | | | | |
|---|---|---|---|---|---|---|
| EPI_ISL_832257 | USA/NY-NYCPHL-001907/2020 | 2020-12-22 | North America/USA/New York/New Yor | DOHMH Morrisania | New York City Public Health Laboratory | Jade Wang et al |
| EPI_ISL_832262 | USA/NY-NYCPHL-001861/2020 | 2020-12-18 | North America/USA/New York/New Yor | DOHMH Corona | New York City Public Health Laboratory | Jade Wang et al |
| EPI_ISL_832273 | USA/NY-NYCPHL-001846/2020 | 2020-12-16 | North America/USA/New York/New Yor | DOHMH Jamaica | New York City Public Health Laboratory | Jade Wang et al |
| EPI_ISL_832274 | USA/NY-NYCPHL-001850/2020 | 2020-12-16 | North America/USA/New York/New Yor | DOHMH Jamaica | New York City Public Health Laboratory | Jade Wang et al |
| EPI_ISL_832279 | USA/NY-NYCPHL-001839/2020 | 2020-12-17 | North America/USA/New York/New Yor | DOHMH Corona | New York City Public Health Laboratory | Jade Wang et al |
| EPI_ISL_832280 | USA/NY-NYCPHL-001841/2020 | 2020-12-17 | North America/USA/New York/New Yor | DOHMH Morrisania | New York City Public Health Laboratory | Jade Wang et al |
| EPI_ISL_832283 | USA/NY-NYCPHL-001851/2020 | 2020-12-17 | North America/USA/New York/New Yor | DOHMH Jamaica | New York City Public Health Laboratory | Jade Wang et al |
| EPI_ISL_832284 | USA/NY-NYCPHL-001852/2020 | 2020-12-17 | North America/USA/New York/New Yor | DOHMH Central Harlem | New York City Public Health Laboratory | Jade Wang et al |
| EPI_ISL_832286 | USA/NY-NYCPHL-001869/2020 | 2020-12-17 | North America/USA/New York/New Yor | DOHMH Fort Greene | New York City Public Health Laboratory | Jade Wang et al |
| EPI_ISL_832304 | USA/NY-NYCPHL-001892/2020 | 2020-12-21 | North America/USA/New York/New Yor | DOHMH Corona | New York City Public Health Laboratory | Jade Wang et al |
| EPI_ISL_832313 | USA/NY-NYCPHL-001903/2020 | 2020-12-22 | North America/USA/New York/New Yor | DOHMH Chelsea | New York City Public Health Laboratory | Jade Wang et al |
| EPI_ISL_832323 | USA/NY-NYCPHL-001918/2020 | 2020-12-22 | North America/USA/New York/New Yor | DOHMH Jamaica | New York City Public Health Laboratory | Jade Wang et al |
| EPI_ISL_832329 | USA/NY-NYCPHL-001928/2020 | 2020-12-23 | North America/USA/New York/New Yor | DOHMH Jamaica | New York City Public Health Laboratory | Jade Wang et al |

| | | | | | | |
|---|---|---|---|---|---|---|
| EPI_ISL_832330 | USA/NY-NYCPHL-001929/2020 | 2020-12-23 | North America/USA/New York/New Yor | DOHMH Jamaica | New York City Public Health Laboratory | Jade Wang et al |
| EPI_ISL_832331 | USA/NY-NYCPHL-001930/2020 | 2020-12-23 | North America/USA/New York/New Yor | DOHMH Corona | New York City Public Health Laboratory | Jade Wang et al |
| EPI_ISL_832345 | USA/NY-NYCPHL-001949/2020 | 2020-12-24 | North America/USA/New York/New Yor | DOHMH Morrisania | New York City Public Health Laboratory | Jade Wang et al |
| EPI_ISL_832352 | USA/NY-NYCPHL-001960/2020 | 2020-12-24 | North America/USA/New York/New Yor | DOHMH PHL | New York City Public Health Laboratory | Jade Wang et al |
| EPI_ISL_832357 | USA/NY-NYCPHL-001968/2020 | 2020-12-28 | North America/USA/New York/New Yor | DOHMH Jamaica | New York City Public Health Laboratory | Jade Wang et al |
| EPI_ISL_832359 | USA/NY-NYCPHL-001971/2020 | 2020-12-28 | North America/USA/New York/New Yor | DOHMH Corona | New York City Public Health Laboratory | Jade Wang et al |
| EPI_ISL_832369 | Germany/NW-HHU-556/2020 | 2020-12-07 | Europe/Germany/North Rhine Westphal | Center of Medical Microbiology, Virology, and Hospital Hygiene, University of Duesseldorf | Center of Medical Microbiology, Virology, and Hospital Hygiene, University of Duesseldorf | Maximilian Damagnez et al |
| EPI_ISL_832375 | USA/CA-SC20113/2020 | 2020-12-26 | North America/USA/California/Santa Cl | Santa Clara County Public Health Laboratory | Santa Clara County Public Health Laboratory | Santa Clara County Public Health Department et al |
| EPI_ISL_832388 | USA/CA-SC20095/2020 | 2020-12-17 | North America/USA/California/Santa Cl | Santa Clara County Public Health Laboratory | Santa Clara County Public Health Laboratory | Santa Clara County Public Health Department et al |
| EPI_ISL_832391 | USA/CA-SC20090/2020 | 2020-12-12 | North America/USA/California/Santa Cl | Santa Clara County Public Health Laboratory | Santa Clara County Public Health Laboratory | Santa Clara County Public Health Department et al |
| EPI_ISL_832404 | USA/CA-SC20066/2020 | 2020-12-05 | North America/USA/California/Santa Cl | Santa Clara County Public Health Laboratory | Santa Clara County Public Health Laboratory | Santa Clara County Public Health Department et al |
| EPI_ISL_832405 | USA/OR-OHSU-0862/2020 | 2020-06-29 | North America/USA/Oregon/ | OHSU Lab Services Molecular Microbiology Lab | Oregon SARS-CoV-2 Genome Sequencing Center | Brendan L. O'Connell et al |

| | | | | | | |
|---|---|---|---|---|---|---|
| EPI_ISL_832406 | USA/OR-OHSU-1215/2020 | 2020-07-13 | North America/USA/Oregon/Clackamas | OHSU Lab Services Molecular Microbiology Lab | Oregon SARS-CoV-2 Genome Sequencing Center | Brendan L. O'Connell et al |
| EPI_ISL_832407 | USA/OR-OHSU-1280/2020 | 2020-07-14 | North America/USA/Oregon/Multnomal | OHSU Lab Services Molecular Microbiology Lab | Oregon SARS-CoV-2 Genome Sequencing Center | Brendan L. O'Connell et al |
| EPI_ISL_832411 | USA/OR-OHSU-1625/2020 | 2020-07-21 | North America/USA/Oregon/Washingto | OHSU Lab Services Molecular Microbiology Lab | Oregon SARS-CoV-2 Genome Sequencing Center | Brendan L. O'Connell et al |
| EPI_ISL_832415 | USA/OR-OHSU-3191/2020 | 2020-10-06 | North America/USA/Oregon/Multnomal | OHSU Lab Services Molecular Microbiology Lab | Oregon SARS-CoV-2 Genome Sequencing Center | Brendan L. O'Connell et al |
| EPI_ISL_832421 | USA/OR-OHSU-3213/2020 | 2020-10-06 | North America/USA/Oregon/Washingto | OHSU Lab Services Molecular Microbiology Lab | Oregon SARS-CoV-2 Genome Sequencing Center | Brendan L. O'Connell et al |
| EPI_ISL_832423 | USA/OR-OHSU-3217/2020 | 2020-10-07 | North America/USA/Oregon/Washingto | OHSU Lab Services Molecular Microbiology Lab | Oregon SARS-CoV-2 Genome Sequencing Center | Brendan L. O'Connell et al |
| EPI_ISL_832432 | USA/OR-OHSU-3267/2020 | 2020-10-08 | North America/USA/Oregon/Washingto | OHSU Lab Services Molecular Microbiology Lab | Oregon SARS-CoV-2 Genome Sequencing Center | Brendan L. O'Connell et al |
| EPI_ISL_832433 | USA/OR-OHSU-3269/2020 | 2020-10-08 | North America/USA/Oregon/Washingto | OHSU Lab Services Molecular Microbiology Lab | Oregon SARS-CoV-2 Genome Sequencing Center | Brendan L. O'Connell et al |
| EPI_ISL_832445 | USA/OR-OHSU-3295/2020 | 2020-10-09 | North America/USA/Oregon/Washingto | OHSU Lab Services Molecular Microbiology Lab | Oregon SARS-CoV-2 Genome Sequencing Center | Brendan L. O'Connell et al |
| EPI_ISL_832448 | USA/OR-OHSU-3307/2020 | 2020-10-08 | North America/USA/Oregon/Washingto | OHSU Lab Services Molecular Microbiology Lab | Oregon SARS-CoV-2 Genome Sequencing Center | Brendan L. O'Connell et al |

| | | | | | | |
|---|---|---|---|---|---|---|
| EPI_ISL_832460 | USA/OR-OHSU-3349/2020 | 2020-10-12 | North America/USA/Oregon/Washingto | OHSU Lab Services Molecular Microbiology Lab | Oregon SARS-CoV-2 Genome Sequencing Center | Brendan L. O'Connell et al |
| EPI_ISL_832462 | USA/OR-OHSU-3359/2020 | 2020-10-12 | North America/USA/Oregon/Multnomal | OHSU Lab Services Molecular Microbiology Lab | Oregon SARS-CoV-2 Genome Sequencing Center | Brendan L. O'Connell et al |
| EPI_ISL_832464 | USA/OR-OHSU-3369/2020 | 2020-10-12 | North America/USA/Oregon/Multnomal | OHSU Lab Services Molecular Microbiology Lab | Oregon SARS-CoV-2 Genome Sequencing Center | Brendan L. O'Connell et al |
| EPI_ISL_832467 | USA/OR-OHSU-3377/2020 | 2020-10-12 | North America/USA/Oregon/Multnomal | OHSU Lab Services Molecular Microbiology Lab | Oregon SARS-CoV-2 Genome Sequencing Center | Brendan L. O'Connell et al |
| EPI_ISL_832468 | USA/OR-OHSU-3383/2020 | 2020-10-13 | North America/USA/Oregon/Washingto | OHSU Lab Services Molecular Microbiology Lab | Oregon SARS-CoV-2 Genome Sequencing Center | Brendan L. O'Connell et al |
| EPI_ISL_832474 | USA/OR-OHSU-3405/2020 | 2020-10-15 | North America/USA/Oregon/Multnomal | OHSU Lab Services Molecular Microbiology Lab | Oregon SARS-CoV-2 Genome Sequencing Center | Brendan L. O'Connell et al |
| EPI_ISL_832476 | USA/OR-OHSU-3411/2020 | 2020-10-15 | North America/USA/Oregon/Multnomal | OHSU Lab Services Molecular Microbiology Lab | Oregon SARS-CoV-2 Genome Sequencing Center | Brendan L. O'Connell et al |
| EPI_ISL_832477 | USA/OR-OHSU-3413/2020 | 2020-10-15 | North America/USA/Oregon/Multnomal | OHSU Lab Services Molecular Microbiology Lab | Oregon SARS-CoV-2 Genome Sequencing Center | Brendan L. O'Connell et al |
| EPI_ISL_832482 | USA/OR-OHSU-3435/2020 | 2020-10-13 | North America/USA/Oregon/ | OHSU Lab Services Molecular Microbiology Lab | Oregon SARS-CoV-2 Genome Sequencing Center | Brendan L. O'Connell et al |
| EPI_ISL_832489 | USA/OR-OHSU-3455/2020 | 2020-10-15 | North America/USA/Oregon/Multnomal | OHSU Lab Services Molecular Microbiology Lab | Oregon SARS-CoV-2 Genome Sequencing Center | Brendan L. O'Connell et al |

| | | | | | | |
|---|---|---|---|---|---|---|
| EPI_ISL_832501 | USA/OR-OHSU-3517/2020 | 2020-10-20 | North America/USA/Oregon/Washingto | OHSU Lab Services Molecular Microbiology Lab | Oregon SARS-CoV-2 Genome Sequencing Center | Brendan L. O'Connell et al |
| EPI_ISL_832504 | USA/OR-OHSU-3529/2020 | 2020-10-20 | North America/USA/Oregon/Multnomal | OHSU Lab Services Molecular Microbiology Lab | Oregon SARS-CoV-2 Genome Sequencing Center | Brendan L. O'Connell et al |
| EPI_ISL_832519 | USA/OR-OHSU-3587/2020 | 2020-10-21 | North America/USA/Oregon/Washingto | OHSU Lab Services Molecular Microbiology Lab | Oregon SARS-CoV-2 Genome Sequencing Center | Brendan L. O'Connell et al |
| EPI_ISL_832529 | USA/OR-OHSU-3609/2020 | 2020-10-22 | North America/USA/Oregon/Washingto | OHSU Lab Services Molecular Microbiology Lab | Oregon SARS-CoV-2 Genome Sequencing Center | Brendan L. O'Connell et al |
| EPI_ISL_832531 | USA/OR-OHSU-3613/2020 | 2020-10-22 | North America/USA/Oregon/Washingto | OHSU Lab Services Molecular Microbiology Lab | Oregon SARS-CoV-2 Genome Sequencing Center | Brendan L. O'Connell et al |
| EPI_ISL_832538 | USA/OR-OHSU-3637/2020 | 2020-10-22 | North America/USA/Oregon/Multnomal | OHSU Lab Services Molecular Microbiology Lab | Oregon SARS-CoV-2 Genome Sequencing Center | Brendan L. O'Connell et al |
| EPI_ISL_832541 | USA/OR-OHSU-3651/2020 | 2020-10-23 | North America/USA/Oregon/Washingto | OHSU Lab Services Molecular Microbiology Lab | Oregon SARS-CoV-2 Genome Sequencing Center | Brendan L. O'Connell et al |
| EPI_ISL_832543 | USA/OR-OHSU-3657/2020 | 2020-10-23 | North America/USA/Oregon/Washingto | OHSU Lab Services Molecular Microbiology Lab | Oregon SARS-CoV-2 Genome Sequencing Center | Brendan L. O'Connell et al |
| EPI_ISL_832545 | USA/OR-OHSU-3673/2020 | 2020-10-23 | North America/USA/Oregon/Sherman C | OHSU Lab Services Molecular Microbiology Lab | Oregon SARS-CoV-2 Genome Sequencing Center | Brendan L. O'Connell et al |
| EPI_ISL_832548 | USA/OR-OHSU-3687/2020 | 2020-10-16 | North America/USA/Oregon/ | OHSU Lab Services Molecular Microbiology Lab | Oregon SARS-CoV-2 Genome Sequencing Center | Brendan L. O'Connell et al |

| | | | | | | |
|---|---|---|---|---|---|---|
| EPI_ISL_832555 | USA/OR-OHSU-3713/2020 | 2020-10-19 | North America/USA/Oregon/Washingto | OHSU Lab Services Molecular Microbiology Lab | Oregon SARS-CoV-2 Genome Sequencing Center | Brendan L. O'Connell et al |
| EPI_ISL_832556 | USA/OR-OHSU-3715/2020 | 2020-10-19 | North America/USA/Oregon/Washingto | OHSU Lab Services Molecular Microbiology Lab | Oregon SARS-CoV-2 Genome Sequencing Center | Brendan L. O'Connell et al |
| EPI_ISL_832560 | USA/OR-OHSU-3729/2020 | 2020-10-20 | North America/USA/Oregon/Multnomal | OHSU Lab Services Molecular Microbiology Lab | Oregon SARS-CoV-2 Genome Sequencing Center | Brendan L. O'Connell et al |
| EPI_ISL_832562 | USA/OR-OHSU-3737/2020 | 2020-10-21 | North America/USA/Oregon/Washingto | OHSU Lab Services Molecular Microbiology Lab | Oregon SARS-CoV-2 Genome Sequencing Center | Brendan L. O'Connell et al |
| EPI_ISL_832568 | USA/OR-OHSU-3767/2020 | 2020-10-23 | North America/USA/Oregon/Multnomal | OHSU Lab Services Molecular Microbiology Lab | Oregon SARS-CoV-2 Genome Sequencing Center | Brendan L. O'Connell et al |
| EPI_ISL_832575 | USA/OR-OHSU-3791/2020 | 2020-10-26 | North America/USA/Oregon/Washingto | OHSU Lab Services Molecular Microbiology Lab | Oregon SARS-CoV-2 Genome Sequencing Center | Brendan L. O'Connell et al |
| EPI_ISL_832582 | USA/OR-OHSU-3831/2020 | 2020-10-26 | North America/USA/Oregon/Multnomal | OHSU Lab Services Molecular Microbiology Lab | Oregon SARS-CoV-2 Genome Sequencing Center | Brendan L. O'Connell et al |
| EPI_ISL_832585 | USA/WA-OHSU-3837/2020 | 2020-10-26 | North America/USA/Washington/Clark ( | OHSU Lab Services Molecular Microbiology Lab | Oregon SARS-CoV-2 Genome Sequencing Center | Brendan L. O'Connell et al |
| EPI_ISL_832591 | USA/OR-OHSU-3853/2020 | 2020-10-28 | North America/USA/Oregon/Multnomal | OHSU Lab Services Molecular Microbiology Lab | Oregon SARS-CoV-2 Genome Sequencing Center | Brendan L. O'Connell et al |
| EPI_ISL_832598 | USA/OR-OHSU-3879/2020 | 2020-10-30 | North America/USA/Oregon/Washingto | OHSU Lab Services Molecular Microbiology Lab | Oregon SARS-CoV-2 Genome Sequencing Center | Brendan L. O'Connell et al |

| | | | | | | |
|---|---|---|---|---|---|---|
| EPI_ISL_832603 | USA/OR-OHSU-3901/2020 | 2020-10-30 | North America/USA/Oregon/Multnomal | OHSU Lab Services Molecular Microbiology Lab | Oregon SARS-CoV-2 Genome Sequencing Center | Brendan L. O'Connell et al |
| EPI_ISL_832606 | USA/OR-OHSU-3907/2020 | 2020-10-30 | North America/USA/Oregon/Multnomal | OHSU Lab Services Molecular Microbiology Lab | Oregon SARS-CoV-2 Genome Sequencing Center | Brendan L. O'Connell et al |
| EPI_ISL_832607 | USA/OR-OHSU-3909/2020 | 2020-10-30 | North America/USA/Oregon/Multnomal | OHSU Lab Services Molecular Microbiology Lab | Oregon SARS-CoV-2 Genome Sequencing Center | Brendan L. O'Connell et al |
| EPI_ISL_832608 | USA/OR-OHSU-3911/2020 | 2020-10-30 | North America/USA/Oregon/Multnomal | OHSU Lab Services Molecular Microbiology Lab | Oregon SARS-CoV-2 Genome Sequencing Center | Brendan L. O'Connell et al |
| EPI_ISL_832609 | USA/OR-OHSU-3923/2020 | 2020-11-04 | North America/USA/Oregon/Multnomal | OHSU Lab Services Molecular Microbiology Lab | Oregon SARS-CoV-2 Genome Sequencing Center | Brendan L. O'Connell et al |
| EPI_ISL_832610 | USA/OR-OHSU-3927/2020 | 2020-11-04 | North America/USA/Oregon/Multnomal | OHSU Lab Services Molecular Microbiology Lab | Oregon SARS-CoV-2 Genome Sequencing Center | Brendan L. O'Connell et al |
| EPI_ISL_832612 | USA/OR-OHSU-3949/2020 | 2020-11-05 | North America/USA/Oregon/Washingto | OHSU Lab Services Molecular Microbiology Lab | Oregon SARS-CoV-2 Genome Sequencing Center | Brendan L. O'Connell et al |
| EPI_ISL_832623 | USA/OR-OHSU-4025/2020 | 2020-10-28 | North America/USA/Oregon/Multnomal | OHSU Lab Services Molecular Microbiology Lab | Oregon SARS-CoV-2 Genome Sequencing Center | Brendan L. O'Connell et al |
| EPI_ISL_832628 | USA/OR-OHSU-4035/2020 | 2020-10-28 | North America/USA/Oregon/ | OHSU Lab Services Molecular Microbiology Lab | Oregon SARS-CoV-2 Genome Sequencing Center | Brendan L. O'Connell et al |
| EPI_ISL_832665 | USA/OR-OHSU-4159/2020 | 2020-11-02 | North America/USA/Oregon/Washingto | OHSU Lab Services Molecular Microbiology Lab | Oregon SARS-CoV-2 Genome Sequencing Center | Brendan L. O'Connell et al |

| | | | | | | |
|---|---|---|---|---|---|---|
| EPI_ISL_832667 | USA/WA-OHSU-4169/2020 | 2020-11-02 | North America/USA/Washington/Clark ( | OHSU Lab Services Molecular Microbiology Lab | Oregon SARS-CoV-2 Genome Sequencing Center | Brendan L. O'Connell et al |
| EPI_ISL_832674 | USA/OR-OHSU-4217/2020 | 2020-11-03 | North America/USA/Oregon/Washingto | OHSU Lab Services Molecular Microbiology Lab | Oregon SARS-CoV-2 Genome Sequencing Center | Brendan L. O'Connell et al |
| EPI_ISL_832677 | USA/OR-OHSU-4238/2020 | 2020-11-04 | North America/USA/Oregon/ | OHSU Lab Services Molecular Microbiology Lab | Oregon SARS-CoV-2 Genome Sequencing Center | Brendan L. O'Connell et al |
| EPI_ISL_832687 | USA/OR-OHSU-4295/2020 | 2020-10-26 | North America/USA/Oregon/Multnomal | OHSU Lab Services Molecular Microbiology Lab | Oregon SARS-CoV-2 Genome Sequencing Center | Brendan L. O'Connell et al |
| EPI_ISL_832691 | USA/OR-OHSU-4307/2020 | 2020-10-27 | North America/USA/Oregon/Multnomal | OHSU Lab Services Molecular Microbiology Lab | Oregon SARS-CoV-2 Genome Sequencing Center | Brendan L. O'Connell et al |
| EPI_ISL_832696 | USA/OR-OHSU-4359/2020 | 2020-11-04 | North America/USA/Oregon/Washingto | OHSU Lab Services Molecular Microbiology Lab | Oregon SARS-CoV-2 Genome Sequencing Center | Brendan L. O'Connell et al |
| EPI_ISL_832698 | USA/OR-OHSU-4369/2020 | 2020-11-04 | North America/USA/Oregon/Clackamas | OHSU Lab Services Molecular Microbiology Lab | Oregon SARS-CoV-2 Genome Sequencing Center | Brendan L. O'Connell et al |
| EPI_ISL_832709 | USA/OR-OHSU-4436/2020 | 2020-11-05 | North America/USA/Oregon/Clackamas | OHSU Lab Services Molecular Microbiology Lab | Oregon SARS-CoV-2 Genome Sequencing Center | Brendan L. O'Connell et al |
| EPI_ISL_832713 | USA/OR-OHSU-4467/2020 | 2020-11-06 | North America/USA/Oregon/Clackamas | OHSU Lab Services Molecular Microbiology Lab | Oregon SARS-CoV-2 Genome Sequencing Center | Brendan L. O'Connell et al |
| EPI_ISL_832720 | USA/OR-OHSU-4521/2020 | 2020-11-06 | North America/USA/Oregon/Washingto | OHSU Lab Services Molecular Microbiology Lab | Oregon SARS-CoV-2 Genome Sequencing Center | Brendan L. O'Connell et al |

| | | | | | | |
|---|---|---|---|---|---|---|
| EPI_ISL_832722 | USA/OR-OHSU-4532/2020 | 2020-11-07 | North America/USA/Oregon/Washingto | OHSU Lab Services Molecular Microbiology Lab | Oregon SARS-CoV-2 Genome Sequencing Center | Brendan L. O'Connell et al |
| EPI_ISL_832731 | USA/WA-OHSU-4598/2020 | 2020-11-10 | North America/USA/Washington/Clark ( | OHSU Lab Services Molecular Microbiology Lab | Oregon SARS-CoV-2 Genome Sequencing Center | Brendan L. O'Connell et al |
| EPI_ISL_832739 | USA/OR-OHSU-4690/2020 | 2020-11-11 | North America/USA/Oregon/Washingto | OHSU Lab Services Molecular Microbiology Lab | Oregon SARS-CoV-2 Genome Sequencing Center | Brendan L. O'Connell et al |
| EPI_ISL_832745 | USA/OR-OHSU-4737/2020 | 2020-11-07 | North America/USA/Oregon/Multnomal | OHSU Lab Services Molecular Microbiology Lab | Oregon SARS-CoV-2 Genome Sequencing Center | Brendan L. O'Connell et al |
| EPI_ISL_832752 | USA/OR-OHSU-5063/2020 | 2020-11-12 | North America/USA/Oregon/Washingto | OHSU Lab Services Molecular Microbiology Lab | Oregon SARS-CoV-2 Genome Sequencing Center | Brendan L. O'Connell et al |
| EPI_ISL_832753 | USA/OR-OHSU-5072/2020 | 2020-11-12 | North America/USA/Oregon/Multnomal | OHSU Lab Services Molecular Microbiology Lab | Oregon SARS-CoV-2 Genome Sequencing Center | Brendan L. O'Connell et al |
| EPI_ISL_832759 | USA/OR-OHSU-5246/2020 | 2020-11-17 | North America/USA/Oregon/Washingto | OHSU Lab Services Molecular Microbiology Lab | Oregon SARS-CoV-2 Genome Sequencing Center | Brendan L. O'Connell et al |
| EPI_ISL_832760 | USA/OR-OHSU-5266/2020 | 2020-11-17 | North America/USA/Oregon/Washingto | OHSU Lab Services Molecular Microbiology Lab | Oregon SARS-CoV-2 Genome Sequencing Center | Brendan L. O'Connell et al |
| EPI_ISL_832768 | USA/OR-OHSU-5332/2020 | 2020-11-13 | North America/USA/Oregon/Washingto | OHSU Lab Services Molecular Microbiology Lab | Oregon SARS-CoV-2 Genome Sequencing Center | Brendan L. O'Connell et al |
| EPI_ISL_832770 | USA/OR-OHSU-5351/2020 | 2020-11-14 | North America/USA/Oregon/Washingto | OHSU Lab Services Molecular Microbiology Lab | Oregon SARS-CoV-2 Genome Sequencing Center | Brendan L. O'Connell et al |

| | | | | | | |
|---|---|---|---|---|---|---|
| EPI_ISL_832774 | USA/OR-OHSU-5371/2020 | 2020-11-15 | North America/USA/Oregon/Washingto | OHSU Lab Services Molecular Microbiology Lab | Oregon SARS-CoV-2 Genome Sequencing Center | Brendan L. O'Connell et al |
| EPI_ISL_832778 | USA/OR-OHSU-5498/2020 | 2020-11-18 | North America/USA/Oregon/ | OHSU Lab Services Molecular Microbiology Lab | Oregon SARS-CoV-2 Genome Sequencing Center | Brendan L. O'Connell et al |
| EPI_ISL_832781 | USA/OR-OHSU-5619/2020 | 2020-11-20 | North America/USA/Oregon/ | OHSU Lab Services Molecular Microbiology Lab | Oregon SARS-CoV-2 Genome Sequencing Center | Brendan L. O'Connell et al |
| EPI_ISL_832788 | USA/OR-OHSU-5680/2020 | 2020-11-18 | North America/USA/Oregon/Multnomal | OHSU Lab Services Molecular Microbiology Lab | Oregon SARS-CoV-2 Genome Sequencing Center | Brendan L. O'Connell et al |
| EPI_ISL_832801 | USA/OR-OHSU-5844/2020 | 2020-11-24 | North America/USA/Oregon/ | OHSU Lab Services Molecular Microbiology Lab | Oregon SARS-CoV-2 Genome Sequencing Center | Brendan L. O'Connell et al |
| EPI_ISL_832818 | Italy/ABR-IZSGC-354389/2020 | 2020-12-18 | Europe/Italy/Abruzzo/ | Ospedale Civile Atri | "Istituto Zooprofilattico Sperimentale dell'Abruzzo e Molise ""G.Caporale""" | Lorusso A et al |
| EPI_ISL_832824 | USA/ME-HETL-J0484/2020 | 2020-12-30 | North America/USA/Maine/ | Maine HETL | Tewhey Lab, The Jackson Laboratory | Matluk et al |
| EPI_ISL_832831 | USA/ME-HETL-J0493/2020 | 2020-12-31 | North America/USA/Maine/ | Maine HETL | Tewhey Lab, The Jackson Laboratory | Matluk et al |
| EPI_ISL_832833 | USA/ME-HETL-J0495/2020 | 2020-12-31 | North America/USA/Maine/ | Maine HETL | Tewhey Lab, The Jackson Laboratory | Matluk et al |
| EPI_ISL_832837 | USA/ME-HETL-J0499/2020 | 2020-12-28 | North America/USA/Maine/ | Maine HETL | Tewhey Lab, The Jackson Laboratory | Matluk et al |

| | | | | | | |
|---|---|---|---|---|---|---|
| EPI_ISL_832838 | USA/ME-HETL-J0500/2020 | 2020-12-28 | North America/USA/Maine/ | Maine HETL | Tewhey Lab, The Jackson Laboratory | Matluk et al |
| EPI_ISL_832840 | USA/ME-HETL-J0502/2020 | 2020-12-28 | North America/USA/Maine/ | Maine HETL | Tewhey Lab, The Jackson Laboratory | Matluk et al |
| EPI_ISL_832841 | USA/ME-HETL-J0503/2020 | 2020-12-28 | North America/USA/Maine/ | Maine HETL | Tewhey Lab, The Jackson Laboratory | Matluk et al |
| EPI_ISL_832851 | USA/ME-HETL-J0515/2021 | 2021-01-02 | North America/USA/Maine/ | Maine HETL | Tewhey Lab, The Jackson Laboratory | Matluk et al |
| EPI_ISL_832863 | USA/ME-HETL-J0534/2020 | 2020-12-29 | North America/USA/Maine/ | Maine HETL | Tewhey Lab, The Jackson Laboratory | Matluk et al |
| EPI_ISL_832869 | USA/ME-HETL-J0542/2020 | 2020-12-31 | North America/USA/Maine/ | Maine HETL | Tewhey Lab, The Jackson Laboratory | Matluk et al |
| EPI_ISL_832871 | USA/ME-HETL-J0547/2021 | 2021-01-02 | North America/USA/Maine/ | Maine HETL | Tewhey Lab, The Jackson Laboratory | Matluk et al |
| EPI_ISL_832872 | USA/ME-HETL-J0548/2021 | 2021-01-02 | North America/USA/Maine/ | Maine HETL | Tewhey Lab, The Jackson Laboratory | Matluk et al |
| EPI_ISL_832874 | USA/ME-HETL-J0550/2020 | 2020-12-30 | North America/USA/Maine/ | Maine HETL | Tewhey Lab, The Jackson Laboratory | Matluk et al |
| EPI_ISL_832881 | USA/ME-HETL-J0559/2021 | 2021-01-02 | North America/USA/Maine/ | Maine HETL | Tewhey Lab, The Jackson Laboratory | Matluk et al |
| EPI_ISL_832896 | USA/ME-HETL-J0581/2020 | 2020-12-30 | North America/USA/Maine/ | Maine HETL | Tewhey Lab, The Jackson Laboratory | Matluk et al |
| EPI_ISL_832898 | USA/ME-HETL-J0584/2020 | 2020-12-26 | North America/USA/Maine/ | Maine HETL | Tewhey Lab, The Jackson Laboratory | Matluk et al |
| EPI_ISL_832899 | USA/ME-HETL-J0586/2020 | 2020-12-26 | North America/USA/Maine/ | Maine HETL | Tewhey Lab, The Jackson Laboratory | Matluk et al |

| | | | | | | |
|---|---|---|---|---|---|---|
| EPI_ISL_832900 | USA/ME-HETL-J0587/2020 | 2020-12-26 | North America/USA/Maine/ | Maine HETL | Tewhey Lab, The Jackson Laboratory | Matluk et al |
| EPI_ISL_832901 | USA/ME-HETL-J0588/2020 | 2020-12-26 | North America/USA/Maine/ | Maine HETL | Tewhey Lab, The Jackson Laboratory | Matluk et al |
| EPI_ISL_832904 | USA/ME-HETL-J0592/2020 | 2020-12-26 | North America/USA/Maine/ | Maine HETL | Tewhey Lab, The Jackson Laboratory | Matluk et al |
| EPI_ISL_832905 | USA/ME-HETL-J0593/2020 | 2020-12-26 | North America/USA/Maine/ | Maine HETL | Tewhey Lab, The Jackson Laboratory | Matluk et al |
| EPI_ISL_832906 | USA/ME-HETL-J0594/2020 | 2020-12-26 | North America/USA/Maine/ | Maine HETL | Tewhey Lab, The Jackson Laboratory | Matluk et al |
| EPI_ISL_832916 | USA/ME-HETL-J0607/2020 | 2020-12-28 | North America/USA/Maine/ | Maine HETL | Tewhey Lab, The Jackson Laboratory | Matluk et al |
| EPI_ISL_832921 | USA/ME-HETL-J0612/2020 | 2020-12-26 | North America/USA/Maine/ | Maine HETL | Tewhey Lab, The Jackson Laboratory | Matluk et al |
| EPI_ISL_832928 | USA/ME-HETL-J0619/2020 | 2020-12-28 | North America/USA/Maine/ | Maine HETL | Tewhey Lab, The Jackson Laboratory | Matluk et al |
| EPI_ISL_832932 | USA/ME-HETL-J0624/2020 | 2020-12-22 | North America/USA/Maine/ | Maine HETL | Tewhey Lab, The Jackson Laboratory | Matluk et al |
| EPI_ISL_832933 | USA/ME-HETL-J0626/2020 | 2020-12-27 | North America/USA/Maine/ | Maine HETL | Tewhey Lab, The Jackson Laboratory | Matluk et al |
| EPI_ISL_832939 | USA/ME-HETL-J0634/2020 | 2020-12-31 | North America/USA/Maine/ | Maine HETL | Tewhey Lab, The Jackson Laboratory | Matluk et al |
| EPI_ISL_832944 | USA/ME-HETL-J0640/2020 | 2020-12-31 | North America/USA/Maine/ | Maine HETL | Tewhey Lab, The Jackson Laboratory | Matluk et al |
| EPI_ISL_832950 | USA/ME-HETL-J0647/2020 | 2020-12-29 | North America/USA/Maine/ | Maine HETL | Tewhey Lab, The Jackson Laboratory | Matluk et al |

| | | | | | | |
|---|---|---|---|---|---|---|
| EPI_ISL_832952 | USA/ME-HETL-J0649/2020 | 2020-12-29 | North America/USA/Maine/ | Maine HETL | Tewhey Lab, The Jackson Laboratory | Matluk et al |
| EPI_ISL_832957 | USA/ME-HETL-J0655/2020 | 2020-12-31 | North America/USA/Maine/ | Maine HETL | Tewhey Lab, The Jackson Laboratory | Matluk et al |
| EPI_ISL_832960 | USA/ME-HETL-J0660/2020 | 2020-12-30 | North America/USA/Maine/ | Maine HETL | Tewhey Lab, The Jackson Laboratory | Matluk et al |
| EPI_ISL_832966 | USA/ME-HETL-J0669/2020 | 2020-12-29 | North America/USA/Maine/ | Maine HETL | Tewhey Lab, The Jackson Laboratory | Matluk et al |
| EPI_ISL_832972 | USA/ME-HETL-J0676/2021 | 2021-01-07 | North America/USA/Maine/ | Maine HETL | Tewhey Lab, The Jackson Laboratory | Matluk et al |
| EPI_ISL_832982 | USA/ME-HETL-J0688/2021 | 2021-01-04 | North America/USA/Maine/ | Maine HETL | Tewhey Lab, The Jackson Laboratory | Matluk et al |
| EPI_ISL_832983 | USA/ME-HETL-J0689/2021 | 2021-01-04 | North America/USA/Maine/ | Maine HETL | Tewhey Lab, The Jackson Laboratory | Matluk et al |
| EPI_ISL_832994 | USA/ME-HETL-J0705/2021 | 2021-01-06 | North America/USA/Maine/ | Maine HETL | Tewhey Lab, The Jackson Laboratory | Matluk et al |
| EPI_ISL_833021 | USA/ME-HETL-J0737/2021 | 2021-01-06 | North America/USA/Maine/ | Maine HETL | Tewhey Lab, The Jackson Laboratory | Matluk et al |
| EPI_ISL_833034 | USA/ME-HETL-J0760/2021 | 2021-01-06 | North America/USA/Maine/ | Maine HETL | Tewhey Lab, The Jackson Laboratory | Matluk et al |
| EPI_ISL_833037 | USA/ME-HETL-J0766/2021 | 2021-01-05 | North America/USA/Maine/ | Maine HETL | Tewhey Lab, The Jackson Laboratory | Matluk et al |
| EPI_ISL_833039 | Indonesia/SS-NIHRD-PME10004/2020 | 2020-10-09 | Asia/Indonesia/South Sumatra/Palemba | RS TK II 02.05.01 dr. AK Gani Palembang | National Institute of Health Research and Development | Subangkit et al |
| EPI_ISL_833040 | Indonesia/KB-NIHRD-PME9639/2020 | 2020-09-10 | Asia/Indonesia/West Kalimantan/ | RSUD Dr. Abdul Aziz Singkawang | National Institute of Health Research and Development | Subangkit et al |

| | | | | | | |
|---|---|---|---|---|---|---|
| EPI_ISL_833042 | Italy/ABR-IZSGC-TE361000/2020 | 2020-12-21 | Europe/Italy/Abruzzo/ | OSPEDALE SAN SALVATORE L'AQUILA UOC PNEUMOLOGIAE UTSIR | Istituto Zooprofilattico Sperimentale dell,ÄôAbruzzo e Molise ,ÄúG. Caporale,Äù | Lorusso A et al |
| EPI_ISL_833054 | Italy/ABR-IZSGC-TE360288/2020 | 2020-12-22 | Europe/Italy/Abruzzo/ | """Presidio Ospedaliero ""San Liberatore"" Atri" | Istituto Zooprofilattico Sperimentale dell,ÄôAbruzzo e Molise ,ÄúG. Caporale,Äù | Lorusso A et al |
| EPI_ISL_833056 | Italy/ABR-IZSGC-TE361030/2020 | 2020-12-21 | Europe/Italy/Abruzzo/ | """SIESP DIPARTIMENTO DI PREVENZIONE CHIETi" | Istituto Zooprofilattico Sperimentale dell,ÄôAbruzzo e Molise ,ÄúG. Caporale,Äù | Lorusso A et al |
| EPI_ISL_833060 | Italy/ABR-IZSGC-TE359016/2020 | 2020-12-21 | Europe/Italy/Abruzzo/ | DIP. PREV. AVEZZANO SERVIZIO DI IGIENE EPIDEMIOLOGIAE SANITA' PUBBLICA | Istituto Zooprofilattico Sperimentale dell,ÄôAbruzzo e Molise ,ÄúG. Caporale,Äù | Lorusso A et al |
| EPI_ISL_833061 | Italy/ABR-IZSGC-TE362397/2020 | 2020-12-22 | Europe/Italy/Abruzzo/ | DIP. PREV. AVEZZANO SERVIZIO DI IGIENE EPIDEMIOLOGIAE SANITA' PUBBLICA | Istituto Zooprofilattico Sperimentale dell,ÄôAbruzzo e Molise ,ÄúG. Caporale,Äù | Lorusso A et al |
| EPI_ISL_833068 | Italy/ABR-IZSGC-TE365560/2020 | 2020-12-23 | Europe/Italy/Abruzzo/ | FPAM | Istituto Zooprofilattico Sperimentale dell,ÄôAbruzzo e Molise ,ÄúG. Caporale,Äù | Lorusso A et al |

| | | | | | | |
|---|---|---|---|---|---|---|
| EPI_ISL_833074 | Italy/ABR-IZSGC-TE365549/2020 | 2020-12-23 | Europe/Italy/Abruzzo/ | FPAM | Istituto Zooprofilattico Sperimentale dell‚ÄôAbruzzo e Molise ‚ÄúG. Caporale‚Äù | Lorusso A et al |
| EPI_ISL_833079 | Italy/ABR-IZSGC-TE357750/2020 | 2020-12-21 | Europe/Italy/Abruzzo/ | OSPEDALE CIVILE TERAMO | Istituto Zooprofilattico Sperimentale dell‚ÄôAbruzzo e Molise ‚ÄúG. Caporale‚Äù | Lorusso A et al |
| EPI_ISL_833084 | Italy/ABR-IZSGC-TE363081/2020 | 2020-12-23 | Europe/Italy/Abruzzo/ | RPS | Istituto Zooprofilattico Sperimentale dell‚ÄôAbruzzo e Molise ‚ÄúG. Caporale‚Äù | Lorusso A et al |
| EPI_ISL_833094 | Italy/ABR-IZSGC-TE365046/2020 | 2020-12-23 | Europe/Italy/Abruzzo/ | SIESP CHIETI - DRIVE IN CHIETI | Istituto Zooprofilattico Sperimentale dell‚ÄôAbruzzo e Molise ‚ÄúG. Caporale‚Äù | Lorusso A et al |
| EPI_ISL_833095 | Italy/ABR-IZSGC-TE360548/2020 | 2020-12-21 | Europe/Italy/Abruzzo/ | SIESP CHIETI - DRIVE IN CHIETI | Istituto Zooprofilattico Sperimentale dell‚ÄôAbruzzo e Molise ‚ÄúG. Caporale‚Äù | Lorusso A et al |
| EPI_ISL_833102 | Italy/ABR-IZSGC-TE365041/2020 | 2020-12-23 | Europe/Italy/Abruzzo/ | SIESP DIPARTIMENTO DI PREVENZIONE CHIETI | Istituto Zooprofilattico Sperimentale dell‚ÄôAbruzzo e Molise ‚ÄúG. Caporale‚Äù | Lorusso A et al |

| | | | | | | |
|---|---|---|---|---|---|---|
| EPI_ISL_833103 | Italy/ABR-IZSGC-TE360657/2020 | 2020-12-22 | Europe/Italy/Abruzzo/ | SIESP DIPARTIMENTO DI PREVENZIONE TERAMO C.DA CASALENA | Istituto Zooprofilattico Sperimentale dell,ÄôAbruzzo e Molise ,ÄúG. Caporale,Äù | Lorusso A et al |
| EPI_ISL_833109 | Italy/ABR-IZSGC-TE363345/2020 | 2020-12-23 | Europe/Italy/Abruzzo/ | SIESP DIPARTIMENTO DI PREVENZIONE TERAMO C.DA CASALENA | Istituto Zooprofilattico Sperimentale dell,ÄôAbruzzo e Molise ,ÄúG. Caporale,Äù | Lorusso A et al |
| EPI_ISL_833120 | Italy/ABR-IZSGC-TE360901/2020 | 2020-12-22 | Europe/Italy/Abruzzo/ | SIESP SULMONA | Istituto Zooprofilattico Sperimentale dell,ÄôAbruzzo e Molise ,ÄúG. Caporale,Äù | Lorusso A et al |
| EPI_ISL_833124 | Italy/ABR-IZSGC-TE364119/2020 | 2020-12-21 | Europe/Italy/Abruzzo/ | USCA Sulmona | Istituto Zooprofilattico Sperimentale dell,ÄôAbruzzo e Molise ,ÄúG. Caporale,Äù | Lorusso A et al |
| EPI_ISL_833130 | Indonesia/JK-NIHRD-C0023408/2020 | 2020-06-26 | Asia/Indonesia/Jakarta/ | National Institute of Health Research and Development | National Institute of Health Research and Development | Agustiningsih et al |
| EPI_ISL_833131 | Brazil/AM-20142761FC/2020 | 2020-12-01 | South America/Brazil/Amazonas BR/Ma | Laboratorio de Ecologia de Doencas Transmissiveis na Amazonia, Instituto Leonidas e Maria Deane - Fiocruz Amazonia | Laboratorio de Ecologia de Doencas Transmissiveis na Amazonia, Instituto Leonidas e Maria Deane - Fiocruz Amazonia | Valdinete Nascimento et al |

| | | | | | | |
|---|---|---|---|---|---|---|
| EPI_ISL_833132 | Brazil/AM-20142910VS/2020 | 2020-12-10 | South America/Brazil/Amazonas BR/Ma | Laboratorio de Ecologia de Doencas Transmissiveis na Amazonia, Instituto Leonidas e Maria Deane - Fiocruz Amazonia | Laboratorio de Ecologia de Doencas Transmissiveis na Amazonia, Instituto Leonidas e Maria Deane - Fiocruz Amazonia | Valdinete Nascimento et al |
| EPI_ISL_833133 | Brazil/AM-20143164MG/2020 | 2020-12-18 | South America/Brazil/Amazonas BR/Ma | Laboratorio de Ecologia de Doencas Transmissiveis na Amazonia, Instituto Leonidas e Maria Deane - Fiocruz Amazonia | Laboratorio de Ecologia de Doencas Transmissiveis na Amazonia, Instituto Leonidas e Maria Deane - Fiocruz Amazonia | Valdinete Nascimento et al |
| EPI_ISL_833134 | Brazil/AM-20843044TM/2020 | 2020-12-14 | South America/Brazil/Amazonas BR/Ma | Laboratorio de Ecologia de Doencas Transmissiveis na Amazonia, Instituto Leonidas e Maria Deane - Fiocruz Amazonia | Laboratorio de Ecologia de Doencas Transmissiveis na Amazonia, Instituto Leonidas e Maria Deane - Fiocruz Amazonia | Valdinete Nascimento et al |
| EPI_ISL_833135 | Brazil/AM-20843049KC/2020 | 2020-12-16 | South America/Brazil/Amazonas BR/Ma | Laboratorio de Ecologia de Doencas Transmissiveis na Amazonia, Instituto Leonidas e Maria Deane - Fiocruz Amazonia | Laboratorio de Ecologia de Doencas Transmissiveis na Amazonia, Instituto Leonidas e Maria Deane - Fiocruz Amazonia | Valdinete Nascimento et al |
| EPI_ISL_833136 | Brazil/AM-20143103JT/2020 | 2020-12-29 | South America/Brazil/Amazonas BR/Ma | Laboratorio de Ecologia de Doencas Transmissiveis na Amazonia, Instituto Leonidas e Maria Deane - Fiocruz Amazonia | Laboratorio de Ecologia de Doencas Transmissiveis na Amazonia, Instituto Leonidas e Maria Deane - Fiocruz Amazonia | Valdinete Nascimento et al |

| | | | | | | |
|---|---|---|---|---|---|---|
| EPI_ISL_833137 | Brazil/AM-20842882CA/2020 | 2020-12-04 | South America/Brazil/Amazonas BR/Ma | Laboratorio de Ecologia de Doencas Transmissiveis na Amazonia, Instituto Leonidas e Maria Deane - Fiocruz Amazonia | Laboratorio de Ecologia de Doencas Transmissiveis na Amazonia, Instituto Leonidas e Maria Deane - Fiocruz Amazonia | Valdinete Nascimento et al |
| EPI_ISL_833138 | Brazil/AM-20843201ML/2020 | 2020-12-21 | South America/Brazil/Amazonas BR/Ma | Laboratorio de Ecologia de Doencas Transmissiveis na Amazonia, Instituto Leonidas e Maria Deane - Fiocruz Amazonia | Laboratorio de Ecologia de Doencas Transmissiveis na Amazonia, Instituto Leonidas e Maria Deane - Fiocruz Amazonia | Valdinete Nascimento et al |
| EPI_ISL_833139 | Brazil/AM-20843228ZC/2020 | 2020-12-22 | South America/Brazil/Amazonas BR/Ma | Laboratorio de Ecologia de Doencas Transmissiveis na Amazonia, Instituto Leonidas e Maria Deane - Fiocruz Amazonia | Laboratorio de Ecologia de Doencas Transmissiveis na Amazonia, Instituto Leonidas e Maria Deane - Fiocruz Amazonia | Valdinete Nascimento et al |
| EPI_ISL_833140 | Brazil/AM-20843269RC/2020 | 2020-12-23 | South America/Brazil/Amazonas BR/Ma | Laboratorio de Ecologia de Doencas Transmissiveis na Amazonia, Instituto Leonidas e Maria Deane - Fiocruz Amazonia | Laboratorio de Ecologia de Doencas Transmissiveis na Amazonia, Instituto Leonidas e Maria Deane - Fiocruz Amazonia | Valdinete Nascimento et al |
| EPI_ISL_833141 | Turkey/IST-GLAB-N005/2020 | 2020-12-20 | Europe/Turkey/Istanbul/ | Genomic Laboratory (GLAB) (Conjoint lab of Health Directorate of Istanbul and Istanbul Technical University) | Genomic Laboratory (GLAB), Istanbul Technical University | Ilker Karacan et al |

| | | | | | | |
|---|---|---|---|---|---|---|
| EPI_ISL_833142 | Turkey/IST-GLAB-N009/2020 | 2020-12-20 | Europe/Turkey/Istanbul/ | Genomic Laboratory (GLAB) (Conjoint lab of Health Directorate of Istanbul and Istanbul Technical University) | Genomic Laboratory (GLAB), Istanbul Technical University | Ilker Karacan et al |
| EPI_ISL_833143 | Turkey/IST-GLAB-N010/2020 | 2020-12-20 | Europe/Turkey/Istanbul/ | Genomic Laboratory (GLAB) (Conjoint lab of Health Directorate of Istanbul and Istanbul Technical University) | Genomic Laboratory (GLAB), Istanbul Technical University | Ilker Karacan et al |
| EPI_ISL_833144 | Turkey/IST-GLAB-N011/2020 | 2020-12-20 | Europe/Turkey/Istanbul/ | Genomic Laboratory (GLAB) (Conjoint lab of Health Directorate of Istanbul and Istanbul Technical University) | Genomic Laboratory (GLAB), Istanbul Technical University | Ilker Karacan et al |
| EPI_ISL_833145 | Turkey/IST-GLAB-N013/2020 | 2020-12-20 | Europe/Turkey/Istanbul/ | Genomic Laboratory (GLAB) (Conjoint lab of Health Directorate of Istanbul and Istanbul Technical University) | Genomic Laboratory (GLAB), Istanbul Technical University | Ilker Karacan et al |
| EPI_ISL_833146 | Turkey/IST-GLAB-N022/2020 | 2020-12-29 | Europe/Turkey/Istanbul/ | Genomic Laboratory (GLAB) (Conjoint lab of Health Directorate of Istanbul and Istanbul Technical University) | Genomic Laboratory (GLAB), Istanbul Technical University | Ilker Karacan et al |
| EPI_ISL_833147 | Turkey/IST-GLAB-N025/2021 | 2021-01-05 | Europe/Turkey/Istanbul/ | Genomic Laboratory (GLAB) (Conjoint lab of Health Directorate of Istanbul and Istanbul Technical University) | Genomic Laboratory (GLAB), Istanbul Technical University | Ilker Karacan et al |
| EPI_ISL_833155 | Brazil/SP-837/2020 | 2020-04-27 | South America/Brazil/S√£o Paulo/S√£o | Instituto Adolfo Lutz - Central | Instituto Adolfo Lutz, Interdiciplinary Procedures Center, Strategic Laboratory | Claudio Tavares Sacchi et al |

| | | | | | | |
|---|---|---|---|---|---|---|
| EPI_ISL_833156 | Brazil/SP-802/2020 | 2020-07-02 | South America/Brazil/S√£o Paulo/Soroca | Instituto Adolfo Lutz - Regional de Sorocaba | Instituto Adolfo Lutz, Interdiciplinary Procedures Center, Strategic Laboratory | Claudio Tavares Sacchi et al |
| EPI_ISL_833158 | Brazil/SP-911/2020 | 2020-11-30 | South America/Brazil/S√£o Paulo/Sao Be | Instituto Adolfo Lutz - Regional de Santo Andre | Instituto Adolfo Lutz, Interdiciplinary Procedures Center, Strategic Laboratory | Claudio Tavares Sacchi et al |
| EPI_ISL_833159 | Brazil/SP-919/2020 | 2020-12-13 | South America/Brazil/S√£o Paulo/Mogi | Instituto Adolfo Lutz - Central | Instituto Adolfo Lutz, Interdiciplinary Procedures Center, Strategic Laboratory | Claudio Tavares Sacchi et al |
| EPI_ISL_833160 | Brazil/SP-921/2020 | 2020-12-10 | South America/Brazil/S√£o Paulo/Sao Be | Instituto Adolfo Lutz - Regional de Santo Andre | Instituto Adolfo Lutz, Interdiciplinary Procedures Center, Strategic Laboratory | Claudio Tavares Sacchi et al |
| EPI_ISL_833161 | Brazil/SP-923/2020 | 2020-11-26 | South America/Brazil/S√£o Paulo/S√£o | Instituto Adolfo Lutz - Central | Instituto Adolfo Lutz, Interdiciplinary Procedures Center, Strategic Laboratory | Claudio Tavares Sacchi et al |
| EPI_ISL_833163 | Brazil/SP-931/2020 | 2020-11-19 | South America/Brazil/S√£o Paulo/Ribeir | Instituto Adolfo Lutz - Regional de Marilia | Instituto Adolfo Lutz, Interdiciplinary Procedures Center, Strategic Laboratory | Claudio Tavares Sacchi et al |

| | | | | | | |
|---|---|---|---|---|---|---|
| EPI_ISL_833165 | Brazil/SP-975/2021 | 2021-01-06 | South America/Brazil/S√£o Paulo/S√£o | Hospital Samaritano | Instituto Adolfo Lutz, Interdiciplinary Procedures Center, Strategic Laboratory | Claudio Tavares Sacchi et al |
| EPI_ISL_833166 | Brazil/SP-976/2021 | 2021-01-06 | South America/Brazil/S√£o Paulo/S√£o | Hospital Samaritano | Instituto Adolfo Lutz, Interdiciplinary Procedures Center, Strategic Laboratory | Claudio Tavares Sacchi et al |
| EPI_ISL_833168 | Brazil/AM-988/2020 | 2020-12-23 | South America/Brazil/Amazonas BR/ | DB Diagnosticos do Brasil | Instituto Adolfo Lutz, Interdiciplinary Procedures Center, Strategic Laboratory | Claudio Tavares Sacchi et al |
| EPI_ISL_833169 | Brazil/AM-989/2020 | 2020-12-23 | South America/Brazil/Amazonas BR/ | DB Diagnosticos do Brasil | Instituto Adolfo Lutz, Interdiciplinary Procedures Center, Strategic Laboratory | Claudio Tavares Sacchi et al |
| EPI_ISL_833170 | Brazil/AM-990/2020 | 2020-12-23 | South America/Brazil/Amazonas BR/ | DB Diagnosticos do Brasil | Instituto Adolfo Lutz, Interdiciplinary Procedures Center, Strategic Laboratory | Claudio Tavares Sacchi et al |
| EPI_ISL_833171 | Brazil/AM-991/2020 | 2020-12-17 | South America/Brazil/Amazonas BR/ | DB Diagnosticos do Brasil | Instituto Adolfo Lutz, Interdiciplinary Procedures Center, Strategic Laboratory | Claudio Tavares Sacchi et al |

| | | | | | | |
|---|---|---|---|---|---|---|
| EPI_ISL_833172 | Brazil/AM-992/2020 | 2020-12-17 | South America/Brazil/Amazonas BR/ | DB Diagnosticos do Brasil | Instituto Adolfo Lutz, Interdiciplinary Procedures Center, Strategic Laboratory | Claudio Tavares Sacchi et al |
| EPI_ISL_833173 | Brazil/AM-993/2020 | 2020-12-18 | South America/Brazil/Amazonas BR/ | DB Diagnosticos do Brasil | Instituto Adolfo Lutz, Interdiciplinary Procedures Center, Strategic Laboratory | Claudio Tavares Sacchi et al |
| EPI_ISL_833174 | Brazil/AM-994/2020 | 2020-12-22 | South America/Brazil/Amazonas BR/ | DB Diagnosticos do Brasil | Instituto Adolfo Lutz, Interdiciplinary Procedures Center, Strategic Laboratory | Claudio Tavares Sacchi et al |
| EPI_ISL_833175 | Brazil/AM-996/2020 | 2020-12-22 | South America/Brazil/Amazonas BR/ | DB Diagnosticos do Brasil | Instituto Adolfo Lutz, Interdiciplinary Procedures Center, Strategic Laboratory | Claudio Tavares Sacchi et al |
| EPI_ISL_833176 | Brazil/AM-997/2020 | 2020-12-17 | South America/Brazil/Amazonas BR/ | DB Diagnosticos do Brasil | Instituto Adolfo Lutz, Interdiciplinary Procedures Center, Strategic Laboratory | Claudio Tavares Sacchi et al |
| EPI_ISL_833178 | Bangladesh/BCSIR-NILMRC_28/2020 | 2020-05-24 | Asia/Bangladesh/Dhaka/ | National Institute of Laboratory Medicine and Referral Center | Genomic Research Lab, BCSIR | Barna Goswami et al |
| EPI_ISL_833180 | Bangladesh/BCSIR-NILMRC-93/2020 | 2020-05-23 | Asia/Bangladesh/Dhaka/ | National Institute of Laboratory Medicine and Referral Center | Genomic Research Lab, BCSIR | Md. Ahashan Habib et al |
| EPI_ISL_833187 | Belgium/ULG-11189/2020 | 2020-12-30 | Europe/Belgium/Li√®ge/ | Department of Clinical Microbiology | GIGA Medical Genomics | Keith Durkin et al |

| | | | | | | |
|---|---|---|---|---|---|---|
| EPI_ISL_833212 | Finland/HEL4-45/2020 | 2020-12-29 | Europe/Finland/Uusimaa/ | Department of Virology and Immunology, University of Helsinki and Helsinki University Hospital, Huslab Finland | Department of Virology, Faculty of Medicine, University of Helsinki, Helsinki, Finland | Teemu Smura et al |
| EPI_ISL_833234 | Italy/ABR-IZSGC-373344/2020 | 2020-12-29 | Europe/Italy/Abruzzo/ | SIESP DIPARTIMENTO DI PREVENZIONE CHIETI | "Istitituto Zooprofilattico Sperimentale dell'Abruzzo e Molise ""G.Caporale""" | Lorusso A et al |
| EPI_ISL_833237 | Italy/ABR-IZSGC-373369/2020 | 2020-12-29 | Europe/Italy/Abruzzo/ | SIESP DIPARTIMENTO DI PREVENZIONE CHIETI | "Istitituto Zooprofilattico Sperimentale dell'Abruzzo e Molise ""G.Caporale""" | Lorusso A et al |
| EPI_ISL_833240 | Italy/ABR-IZSGC-373679/2020 | 2020-12-30 | Europe/Italy/Abruzzo/ | SIESP DIPARTIMENTO DI PREVENZIONE TERAMO | "Istitituto Zooprofilattico Sperimentale dell'Abruzzo e Molise ""G.Caporale""" | Lorusso A et al |
| EPI_ISL_833241 | Italy/ABR-IZSGC-374429/2020 | 2020-12-29 | Europe/Italy/Abruzzo/ | SIESP DIPARTIMENTO DI PREVENZIONE CHIETI | "Istitituto Zooprofilattico Sperimentale dell'Abruzzo e Molise ""G.Caporale""" | Lorusso A et al |
| EPI_ISL_833246 | Italy/ABR-IZSGC-375138/2020 | 2020-12-30 | Europe/Italy/Abruzzo/ | SIESP DIPARTIMENTO DI PREVENZIONE CHIETI | "Istitituto Zooprofilattico Sperimentale dell'Abruzzo e Molise ""G.Caporale""" | Lorusso A et al |

| | | | | | | |
|---|---|---|---|---|---|---|
| EPI_ISL_833249 | SouthKorea/KDCA0464/2021 | 2021-01-10 | Asia/South Korea/South Korea/ | Division of Emerging Infectious Diseases, Bureau of Infectious Diseases Diagnosis Control, Korea Disease Control and Prevention Agency | Division of Emerging Infectious Diseases, Bureau of Infectious Diseases Diagnosis Control, Korea Disease Control and Prevention Agency | Ae Kyung Park et al |
| EPI_ISL_833260 | Italy/ABR-IZSGC-367161/2020 | 2020-12-28 | Europe/Italy/Abruzzo/ | Giulianova | Istituto Zooprofilattico Sperimentale dell,ÄôAbruzzo e Molise ,ÄúG. Caporale,Äù | Lorusso A et al |
| EPI_ISL_833279 | Italy/ABR-IZSGC-368092/2020 | 2020-12-28 | Europe/Italy/Abruzzo/ | SIESP CHIETI- DRIVE IN CHIETI | Istituto Zooprofilattico Sperimentale dell,ÄôAbruzzo e Molise ,ÄúG. Caporale,Äù | Lorusso A et al |
| EPI_ISL_833280 | Italy/ABR-IZSGC-368125/2020 | 2020-12-28 | Europe/Italy/Abruzzo/ | SIESP CHIETI- DRIVE IN CHIETI | Istituto Zooprofilattico Sperimentale dell,ÄôAbruzzo e Molise ,ÄúG. Caporale,Äù | Lorusso A et al |
| EPI_ISL_833283 | Italy/ABR-IZSGC-368402/2020 | 2020-12-28 | Europe/Italy/Abruzzo/ | DIPARTIMENTO PREVENZIONE AVEZZANO-SERVIZIO DI IGIENE EPIDEMIOLOGIA E SANITA' PUBBLICA | Istituto Zooprofilattico Sperimentale dell,ÄôAbruzzo e Molise ,ÄúG. Caporale,Äù | Lorusso A et al |
| EPI_ISL_833285 | Italy/ABR-IZSGC-370145/2020 | 2020-12-28 | Europe/Italy/Abruzzo/ | SIESP CHIETI - DRIVE IN ORTONA | Istituto Zooprofilattico Sperimentale dell,ÄôAbruzzo e Molise ,ÄúG. Caporale,Äù | Lorusso A et al |

| | | | | | | |
|---|---|---|---|---|---|---|
| EPI_ISL_833298 | Italy/ABR-IZSGC-369832/2020 | 2020-12-28 | Europe/Italy/Abruzzo/ | SIESP CHIETI - DRIVE IN LANCIANO | Istituto Zooprofilattico Sperimentale dell,ÄôAbruzzo e Molise ,ÄúG. Caporale,Äù | Lorusso A et al |
| EPI_ISL_833301 | Italy/ABR-IZSGC-368379/2020 | 2020-12-28 | Europe/Italy/Abruzzo/ | DIPARTIMENTO PREVENZIONE AVEZZANO-SERVIZIO DI IGIENE EPIDEMIOLOGIA E SANITA' PUBBLICA | Istituto Zooprofilattico Sperimentale dell,ÄôAbruzzo e Molise ,ÄúG. Caporale,Äù | Lorusso A et al |
| EPI_ISL_833304 | Italy/ABR-IZSGC-369744/2020 | 2020-12-28 | Europe/Italy/Abruzzo/ | SIESP DIPARTIMENTO DI PREVENZIONE TERAMO | Istituto Zooprofilattico Sperimentale dell,ÄôAbruzzo e Molise ,ÄúG. Caporale,Äù | Lorusso A et al |
| EPI_ISL_833312 | Italy/ABR-IZSGC-369574/2020 | 2020-12-28 | Europe/Italy/Abruzzo/ | SIESP CHIETI- DRIVE IN CHIETI | Istituto Zooprofilattico Sperimentale dell,ÄôAbruzzo e Molise ,ÄúG. Caporale,Äù | Lorusso A et al |
| EPI_ISL_833313 | Italy/ABR-IZSGC-369978/2020 | 2020-12-28 | Europe/Italy/Abruzzo/ | SIESP CHIETI - DRIVE IN LANCIANO | Istituto Zooprofilattico Sperimentale dell,ÄôAbruzzo e Molise ,ÄúG. Caporale,Äù | Lorusso A et al |
| EPI_ISL_833316 | Italy/ABR-IZSGC-369767/2020 | 2020-12-28 | Europe/Italy/Abruzzo/ | SIESP CHIETI - DRIVE IN GISSI | Istituto Zooprofilattico Sperimentale dell,ÄôAbruzzo e Molise ,ÄúG. Caporale,Äù | Lorusso A et al |

| | | | | | | |
|---|---|---|---|---|---|---|
| EPI_ISL_833318 | Italy/ABR-IZSGC-368832/2020 | 2020-12-28 | Europe/Italy/Abruzzo/ | SIESP CHIETI - DRIVE IN CHIETI | Istituto Zooprofilattico Sperimentale dell‚ÄôAbruzzo e Molise ‚ÄúG. Caporale‚Äù | Lorusso A et al |
| EPI_ISL_833320 | Italy/ABR-IZSGC-367568/2020 | 2020-12-28 | Europe/Italy/Abruzzo/ | SIESP DIPARTIMENTO DI PREVENZIONE SULMONA | Istituto Zooprofilattico Sperimentale dell‚ÄôAbruzzo e Molise ‚ÄúG. Caporale‚Äù | Lorusso A et al |
| EPI_ISL_833322 | Italy/ABR-IZSGC-367506/2020 | 2020-12-28 | Europe/Italy/Abruzzo/ | SIESP DIPARTIMENTO DI PREVENZIONE SULMONA | Istituto Zooprofilattico Sperimentale dell‚ÄôAbruzzo e Molise ‚ÄúG. Caporale‚Äù | Lorusso A et al |
| EPI_ISL_833324 | Italy/ABR-IZSGC-370218/2020 | 2020-12-28 | Europe/Italy/Abruzzo/ | SIESP DIPARTIMENTO DI PREVENZIONE CHIETI | Istituto Zooprofilattico Sperimentale dell‚ÄôAbruzzo e Molise ‚ÄúG. Caporale‚Äù | Lorusso A et al |
| EPI_ISL_833333 | Philippines/PH-RITM-0028/2020 | 2020-09-26 | Asia/Philippines/Calabarzon/ | Batangas City Health Office | Research Institute for Tropical Medicine | Hannah Leah Morito et al |
| EPI_ISL_833335 | Philippines/PH-RITM-0025/2020 | 2020-10-12 | Asia/Philippines/Calabarzon/ | Siniloan Rural Health Unit | Research Institute for Tropical Medicine | Hannah Leah Morito et al |
| EPI_ISL_833336 | Philippines/PH-RITM-0022/2020 | 2020-10-26 | Asia/Philippines/Zamboanga Peninsula/ | Bureau of Quarantine | Research Institute for Tropical Medicine | Hannah Leah Morito et al |
| EPI_ISL_833338 | Philippines/PH-RITM-0024/2020 | 2020-10-12 | Asia/Philippines/Calabarzon/ | Siniloan Rural Health Unit | Research Institute for Tropical Medicine | Hannah Leah Morito et al |
| EPI_ISL_833340 | Philippines/PH-RITM-0032/2020 | 2020-11-06 | Asia/Philippines/Calabarzon/ | Batangas City Health Office | Research Institute for Tropical Medicine | Hannah Leah Morito et al |

| | | | | | | |
|---|---|---|---|---|---|---|
| EPI_ISL_833352 | USA/SC-DHEC-0023/2021 | 2021-01-03 | North America/USA/South Carolina/Rich | Microbiology Division, South Carolina Department of Health and Environmental Control (SC DHEC) | Microbiology Division, South Carolina Department of Health and Environmental Control (SC DHEC) | Flores et al |
| EPI_ISL_833354 | USA/SC-DHEC-0016/2021 | 2021-01-02 | North America/USA/South Carolina/Ber | Microbiology Division, South Carolina Department of Health and Environmental Control (SC DHEC) | Microbiology Division, South Carolina Department of Health and Environmental Control (SC DHEC) | Flores et al |
| EPI_ISL_833362 | USA/SC-DHEC-0001-2/2020 | 2020-12-01 | North America/USA/South Carolina/Bea | Microbiology Division, South Carolina Department of Health and Environmental Control (SC DHEC) | Microbiology Division, South Carolina Department of Health and Environmental Control (SC DHEC) | Flores et al |
| EPI_ISL_833367 | Japan/IC-0706/2021 | 21-Jan | Asia/Japan/Japan/ | Pathogen Genomics Center, National Institute of Infectious Diseases | Pathogen Genomics Center, National Institute of Infectious Diseases | Tsuyoshi Sekizuka et al |
| EPI_ISL_833390 | Indonesia/BT-LIPI-005/2020 | 2020-08-13 | Asia/Indonesia/Banten/Tangerang | Klinik Apotek Dein, Jakarta, Indonesia | Biosafety Level-3 Laboratory, Indonesian Institute of Sciences (LIPI) | Andri Wardiana et al |
| EPI_ISL_833391 | Indonesia/BT-LIPI-006/2020 | 2020-05-20 | Asia/Indonesia/Banten/Tangerang | Klinik Apotek Dein, Jakarta, Indonesia | Biosafety Level-3 Laboratory, Indonesian Institute of Sciences (LIPI) | Andri Wardiana et al |
| EPI_ISL_833393 | Indonesia/JK-LIPI-008/2020 | 2020-06-30 | Asia/Indonesia/Jakarta/ | RS MMC Jakarta, Indonesia | Biosafety Level-3 Laboratory, Indonesian Institute of Sciences (LIPI) | Anggia Prasetyoputri et al |

| | | | | | | |
|---|---|---|---|---|---|---|
| EPI_ISL_833396 | USA/CA-LACPHL-AF00142/2021 | 2021-01-02 | North America/USA/California/Los Ange | St. Francis Medical Center | Los Angeles County PHL | P. Hemarajata et al. et al |
| EPI_ISL_833415 | USA/CA-LACPHL-AF00162/2021 | 2021-01-04 | North America/USA/California/Los Ange | USC Clinical Lab | Los Angeles County PHL | P. Hemarajata et al. et al |
| EPI_ISL_833416 | USA/CA-LACPHL-AF00167/2021 | 2021-01-04 | North America/USA/California/Los Ange | USC Clinical Lab | Los Angeles County PHL | P. Hemarajata et al. et al |
| EPI_ISL_833430 | USA/CA-LACPHL-AF00164/2021 | 2021-01-06 | North America/USA/California/Los Ange | UCLA Clinical Micro Lab | Los Angeles County PHL | P. Hemarajata et al. et al |
| EPI_ISL_833449 | Italy/FVG-ASUGI-138059/2020 | 2020-11-27 | Europe/Italy/Friuli Venezia Giulia/Triest | SC (UCO) Igiene e Sanit√† Pubblica (funzione integrata con SC Microbiologia e Virologia) e Laboratory of Molecular Virology of the International Centre for Genetic Engineering and Biotechnology (ICGEB) | ARGO Laboratorio Genomica ed Epigenomica | Licastro D et al |
| EPI_ISL_833450 | Italy/FVG-ASUGI-138058/2020 | 2020-11-27 | Europe/Italy/Friuli Venezia Giulia/Triest | SC (UCO) Igiene e Sanit√† Pubblica (funzione integrata con SC Microbiologia e Virologia) e Laboratory of Molecular Virology of the International Centre for Genetic Engineering and Biotechnology (ICGEB) | ARGO Laboratorio Genomica ed Epigenomica | Licastro D et al |
| EPI_ISL_833452 | Italy/FVG-ASUGI-138053/2020 | 2020-11-27 | Europe/Italy/Friuli Venezia Giulia/Triest | SC (UCO) Igiene e Sanit√† Pubblica (funzione integrata con SC Microbiologia e Virologia) e Laboratory of Molecular Virology of the International Centre for Genetic Engineering and Biotechnology (ICGEB) | ARGO Laboratorio Genomica ed Epigenomica | Licastro D et al |
| EPI_ISL_833453 | Italy/FVG-ASUGI-136485/2020 | 2020-11-25 | Europe/Italy/Friuli Venezia Giulia/Triest | SC (UCO) Igiene e Sanit√† Pubblica (funzione integrata con SC Microbiologia e Virologia) e Laboratory of Molecular Virology of the International Centre for Genetic Engineering and Biotechnology (ICGEB) | ARGO Laboratorio Genomica ed Epigenomica | Licastro D et al |
| EPI_ISL_833454 | Italy/FVG-ASUGI-134813/2020 | 2020-11-24 | Europe/Italy/Friuli Venezia Giulia/Triest | SC (UCO) Igiene e Sanit√† Pubblica (funzione integrata con SC Microbiologia e Virologia) e Laboratory of Molecular Virology of the International Centre for Genetic Engineering and Biotechnology (ICGEB) | ARGO Laboratorio Genomica ed Epigenomica | Licastro D et al |
| EPI_ISL_833455 | Italy/FVG-ASUGI-132983/2020 | 2020-11-23 | Europe/Italy/Friuli Venezia Giulia/Triest | SC (UCO) Igiene e Sanit√† Pubblica (funzione integrata con SC Microbiologia e Virologia) e Laboratory of Molecular Virology of the International Centre for Genetic Engineering and Biotechnology (ICGEB) | ARGO Laboratorio Genomica ed Epigenomica | Licastro D et al |

| | | | | | | |
|---|---|---|---|---|---|---|
| EPI_ISL_833456 | Belgium/UZA-UA-8558/2020 | 2020-12-14 | Europe/Belgium/Antwerpen/ | UZA/UAntwerpen (BIS lab), University Hospital Antwerp, Edegem, Belgium | Laboratory of Medical Microbiology, University of Antwerp, Campus Drie Eiken, S6.26, Universiteitsplein 1, 2610, Wilrijk, Antwerp, Belgium | Basil Britto Xavier et al |
| EPI_ISL_833457 | France/OCC-194/2020 | 2020-10-23 | Europe/France/Occitanie/Toulouse | CHU Purpan - Laboratoire de Virologie - Institut F√©d√©ratif de Biologie | CHU Purpan - Laboratoire de Virologie - Institut F√©d√©ratif de Biologie | Latour J. et al |
| EPI_ISL_833458 | France/OCC-195/2020 | 2020-11-23 | Europe/France/Occitanie/Toulouse | CHU Purpan - Laboratoire de Virologie - Institut F√©d√©ratif de Biologie | CHU Purpan - Laboratoire de Virologie - Institut F√©d√©ratif de Biologie | Latour J. et al |
| EPI_ISL_833459 | France/OCC-196/2020 | 2020-12-08 | Europe/France/Occitanie/Toulouse | CHU Purpan - Laboratoire de Virologie - Institut F√©d√©ratif de Biologie | CHU Purpan - Laboratoire de Virologie - Institut F√©d√©ratif de Biologie | Latour J. et al |
| EPI_ISL_833460 | France/OCC-197/2020 | 2020-11-12 | Europe/France/Occitanie/Toulouse | CHU Purpan - Laboratoire de Virologie - Institut F√©d√©ratif de Biologie | CHU Purpan - Laboratoire de Virologie - Institut F√©d√©ratif de Biologie | Latour J. et al |
| EPI_ISL_833461 | France/OCC-198/2020 | 2020-11-02 | Europe/France/Occitanie/Toulouse | CHU Purpan - Laboratoire de Virologie - Institut F√©d√©ratif de Biologie | CHU Purpan - Laboratoire de Virologie - Institut F√©d√©ratif de Biologie | Latour J. et al |
| EPI_ISL_833462 | France/OCC-199/2020 | 2020-11-04 | Europe/France/Occitanie/Toulouse | CHU Purpan - Laboratoire de Virologie - Institut F√©d√©ratif de Biologie | CHU Purpan - Laboratoire de Virologie - Institut F√©d√©ratif de Biologie | Latour J. et al |

| | | | | | | |
|---|---|---|---|---|---|---|
| EPI_ISL_833464 | France/OCC-201/2020 | 2020-12-08 | Europe/France/Occitanie/Toulouse | CHU Purpan - Laboratoire de Virologie - Institut F√©d√©ratif de Biologie | CHU Purpan - Laboratoire de Virologie - Institut F√©d√©ratif de Biologie | Latour J. et al |
| EPI_ISL_833467 | France/OCC-204/2020 | 2020-10-20 | Europe/France/Occitanie/Toulouse | CHU Purpan - Laboratoire de Virologie - Institut F√©d√©ratif de Biologie | CHU Purpan - Laboratoire de Virologie - Institut F√©d√©ratif de Biologie | Latour J. et al |
| EPI_ISL_833468 | France/OCC-205/2020 | 2020-11-12 | Europe/France/Occitanie/Toulouse | CHU Purpan - Laboratoire de Virologie - Institut F√©d√©ratif de Biologie | CHU Purpan - Laboratoire de Virologie - Institut F√©d√©ratif de Biologie | Latour J. et al |
| EPI_ISL_833469 | France/OCC-206/2020 | 2020-11-02 | Europe/France/Occitanie/Toulouse | CHU Purpan - Laboratoire de Virologie - Institut F√©d√©ratif de Biologie | CHU Purpan - Laboratoire de Virologie - Institut F√©d√©ratif de Biologie | Latour J. et al |
| EPI_ISL_833471 | France/OCC-208/2020 | 2020-11-16 | Europe/France/Occitanie/Toulouse | CHU Purpan - Laboratoire de Virologie - Institut F√©d√©ratif de Biologie | CHU Purpan - Laboratoire de Virologie - Institut F√©d√©ratif de Biologie | Latour J. et al |
| EPI_ISL_833472 | France/OCC-209/2020 | 2020-11-15 | Europe/France/Occitanie/Toulouse | CHU Purpan - Laboratoire de Virologie - Institut F√©d√©ratif de Biologie | CHU Purpan - Laboratoire de Virologie - Institut F√©d√©ratif de Biologie | Latour J. et al |
| EPI_ISL_833473 | France/OCC-210/2020 | 2020-11-02 | Europe/France/Occitanie/Toulouse | CHU Purpan - Laboratoire de Virologie - Institut F√©d√©ratif de Biologie | CHU Purpan - Laboratoire de Virologie - Institut F√©d√©ratif de Biologie | Latour J. et al |
| EPI_ISL_833475 | France/OCC-212/2020 | 2020-11-23 | Europe/France/Occitanie/Toulouse | CHU Purpan - Laboratoire de Virologie - Institut F√©d√©ratif de Biologie | CHU Purpan - Laboratoire de Virologie - Institut F√©d√©ratif de Biologie | Latour J. et al |

| | | | | | | |
|---|---|---|---|---|---|---|
| EPI_ISL_833476 | France/OCC-213/2020 | 2020-11-23 | Europe/France/Occitanie/Toulouse | CHU Purpan - Laboratoire de Virologie - Institut F√©d√©ratif de Biologie | CHU Purpan - Laboratoire de Virologie - Institut F√©d√©ratif de Biologie | Latour J. et al |
| EPI_ISL_833477 | France/OCC-214/2020 | 2020-11-10 | Europe/France/Occitanie/Toulouse | CHU Purpan - Laboratoire de Virologie - Institut F√©d√©ratif de Biologie | CHU Purpan - Laboratoire de Virologie - Institut F√©d√©ratif de Biologie | Latour J. et al |
| EPI_ISL_833478 | France/OCC-215/2020 | 2020-11-14 | Europe/France/Occitanie/Toulouse | CHU Purpan - Laboratoire de Virologie - Institut F√©d√©ratif de Biologie | CHU Purpan - Laboratoire de Virologie - Institut F√©d√©ratif de Biologie | Latour J. et al |
| EPI_ISL_833480 | France/OCC-217/2020 | 2020-12-08 | Europe/France/Occitanie/Toulouse | CHU Purpan - Laboratoire de Virologie - Institut F√©d√©ratif de Biologie | CHU Purpan - Laboratoire de Virologie - Institut F√©d√©ratif de Biologie | Latour J. et al |
| EPI_ISL_833492 | Indonesia/JK-LIPI009/2020 | 2020-07-06 | Asia/Indonesia/Jakarta/ | RS MMC Jakarta, Indonesia | Biosafety Level-3 Laboratory, Indonesian Institute of Sciences (LIPI) | Isa Nuryana et al |
| EPI_ISL_833493 | Indonesia/BGR-LIPI010/2020 | 2020-07-17 | Asia/Indonesia/West Java/Bogor | RS PMI Bogor, Indonesia | Biosafety Level-3 Laboratory, Indonesian Institute of Sciences (LIPI) | Isa Nuryana et al |
| EPI_ISL_833494 | Indonesia/BGR-LIPI011/2020 | 2020-07-06 | Asia/Indonesia/West Java/Bogor | RS PMI Bogor, Indonesia | Biosafety Level-3 Laboratory, Indonesian Institute of Sciences (LIPI) | Ade Andriani et al |
| EPI_ISL_833495 | Indonesia/JK-LIPI012/2020 | 2020-08-17 | Asia/Indonesia/Jakarta/ | Klinik Apotek Dein, Jakarta, Indonesia | Biosafety Level-3 Laboratory, Indonesian Institute of Sciences (LIPI) | Ade Andriani et al |

| | | | | | | |
|---|---|---|---|---|---|---|
| EPI_ISL_833496 | Indonesia/BGR-LIPI013/2020 | 2020-08-18 | Asia/Indonesia/West Java/Bogor | RS PMI Bogor, Indonesia | Biosafety Level-3 Laboratory, Indonesian Institute of Sciences (LIPI) | Anik Budhi Dharmayanthi et al |
| EPI_ISL_833497 | Indonesia/JK-LIPI014/2020 | 2020-08-21 | Asia/Indonesia/Jakarta/ | Klinik Apotek Dein, Jakarta, Indonesia | Biosafety Level-3 Laboratory, Indonesian Institute of Sciences (LIPI) | Syam Budi Iryanto et al |
| EPI_ISL_833498 | Indonesia/JK-LIPI015/2020 | 2020-08-20 | Asia/Indonesia/Jakarta/ | Kartika Pulomas Hospital, Jakarta, Indonesia | Biosafety Level-3 Laboratory, Indonesian Institute of Sciences (LIPI) | Anggia Prasetyoputri et al |
| EPI_ISL_833499 | Indonesia/TNG-LIPI016/2020 | 2020-08-21 | Asia/Indonesia/Banten/Tangerang | RS Qadr, Tangerang, Indonesia | Biosafety Level-3 Laboratory, Indonesian Institute of Sciences (LIPI) | Isa Nuryana et al |
| EPI_ISL_833500 | Indonesia/JK-LIPI018/2020 | 2020-06-03 | Asia/Indonesia/Jakarta/ | Klinik Apotek Dein, Jakarta, Indonesia | Biosafety Level-3 Laboratory, Indonesian Institute of Sciences (LIPI) | Anik Budhi Dharmayanthi et al |
| EPI_ISL_833501 | Indonesia/MKS-LIPI019/2020 | 2020-06-06 | Asia/Indonesia/South Sulawesi/Makass | RS Sayang Bunda, Makasar, Indonesia | Biosafety Level-3 Laboratory, Indonesian Institute of Sciences (LIPI) | Syam Budi Iryanto et al |
| EPI_ISL_833502 | Indonesia/MKS-LIPI020/2020 | 2020-06-06 | Asia/Indonesia/South Sulawesi/Makass | RS Sayang Bunda, Makasar, Indonesia | Biosafety Level-3 Laboratory, Indonesian Institute of Sciences (LIPI) | Andri Wardiana et al |
| EPI_ISL_833503 | Indonesia/JK-LIPI021/2020 | 2020-06-29 | Asia/Indonesia/Jakarta/ | RS MMC, Jakarta, Indonesia | Biosafety Level-3 Laboratory, Indonesian Institute of Sciences (LIPI) | Anggia Prasetyoputri et al |

| | | | | | | |
|---|---|---|---|---|---|---|
| EPI_ISL_833504 | Indonesia/JK-LIPI022/2020 | 2020-07-06 | Asia/Indonesia/Jakarta/ | RS MMC, Jakarta, Indonesia | Biosafety Level-3 Laboratory, Indonesian Institute of Sciences (LIPI) | Isa Nuryana et al |
| EPI_ISL_833506 | Indonesia/BGR-LIPI024/2020 | 2020-07-17 | Asia/Indonesia/West Java/Bogor | RS PMI, Bogor, Indonesia | Biosafety Level-3 Laboratory, Indonesian Institute of Sciences (LIPI) | Isa Nuryana et al |
| EPI_ISL_833508 | Indonesia/JK-LIPI026/2020 | 2020-07-15 | Asia/Indonesia/Jakarta/ | Klinik Apotek Dein, Jakarta, Indonesia | Biosafety Level-3 Laboratory, Indonesian Institute of Sciences (LIPI) | Andri Wardiana et al |
| EPI_ISL_833509 | Indonesia/BGR-LIPI027/2020 | 2020-09-07 | Asia/Indonesia/West Java/Bogor | RS Melania, Bogor, Indonesia | Biosafety Level-3 Laboratory, Indonesian Institute of Sciences (LIPI) | Syam Budi Iryanto et al |
| EPI_ISL_833511 | Serbia/Nis-074650508/2020 | 2020-08-04 | Europe/Serbia/Nis/ | "Veterinary Specialized Instute ""Nis""" | "Veterinary Specialized Institute ""Kraljevo"", Serbia" | Vidanovic et al |
| EPI_ISL_833512 | Serbia/Leskovac-000632407/2020 | 2020-07-23 | Europe/Serbia/Leskovac/ | "Veterinary Specialized Instute ""Nis""" | "Veterinary Specialized Institute ""Kraljevo"", Serbia" | Vidanovic et al |
| EPI_ISL_833513 | Serbia/Nis-000020404/2020 | 2020-04-03 | Europe/Serbia/Nis/ | "Veterinary Specialized Instute ""Nis""" | "Veterinary Specialized Institute ""Kraljevo"", Serbia" | Vidanovic et al |
| EPI_ISL_833514 | Serbia/Leskovac-000932407/2020 | 2020-07-23 | Europe/Serbia/Leskovac/ | "Veterinary Specialized Instute ""Nis""" | "Veterinary Specialized Institute ""Kraljevo"", Serbia" | Vidanovic et al |

| | | | | | | |
|---|---|---|---|---|---|---|
| EPI_ISL_833519 | Serbia/Nis-085132708/2020 | 2020-08-26 | Europe/Serbia/Nis/ | "Veterinary Specialized Instute ""Nis""" | "Veterinary Specialized Institute ""Kraljevo"", Serbia" | Vidanovic et al |
| EPI_ISL_833524 | Italy/CAM-TIGEM-870/2020 | 2020-04-27 | Europe/Italy/Campania/ | Istituto Zooprofilattico Sperimentale del Mezzogiorno | TIGEM | Patrizia Annunziata et al |
| EPI_ISL_833526 | Italy/CAM-TIGEM-910/2020 | 2020-05-02 | Europe/Italy/Campania/ | Istituto Zooprofilattico Sperimentale del Mezzogiorno | TIGEM | Patrizia Annunziata et al |
| EPI_ISL_833527 | Italy/CAM-TIGEM-927/2020 | 2020-05-06 | Europe/Italy/Campania/ | Istituto Zooprofilattico Sperimentale del Mezzogiorno | TIGEM | Patrizia Annunziata et al |
| EPI_ISL_833532 | Italy/CAM-TIGEM-933/2020 | 2020-05-07 | Europe/Italy/Campania/ | Istituto Zooprofilattico Sperimentale del Mezzogiorno | TIGEM | Patrizia Annunziata et al |
| EPI_ISL_833533 | Italy/CAM-TIGEM-924/2020 | 2020-05-04 | Europe/Italy/Campania/ | Istituto Zooprofilattico Sperimentale del Mezzogiorno | TIGEM | Patrizia Annunziata et al |
| EPI_ISL_833535 | Italy/CAM-TIGEM-822/2020 | 2020-04-21 | Europe/Italy/Campania/ | Istituto Zooprofilattico Sperimentale del Mezzogiorno | TIGEM | Patrizia Annunziata et al |
| EPI_ISL_833536 | Italy/CAM-TIGEM-922/2020 | 2020-05-04 | Europe/Italy/Campania/ | Istituto Zooprofilattico Sperimentale del Mezzogiorno | TIGEM | Patrizia Annunziata et al |
| EPI_ISL_833540 | Italy/CAM-TIGEM-957/2020 | 2020-04-02 | Europe/Italy/Campania/ | Istituto Zooprofilattico Sperimentale del Mezzogiorno | TIGEM | Patrizia Annunziata et al |
| EPI_ISL_833541 | Italy/CAM-TIGEM-959/2020 | 2020-04-02 | Europe/Italy/Campania/ | Istituto Zooprofilattico Sperimentale del Mezzogiorno | TIGEM | Patrizia Annunziata et al |
| EPI_ISL_833542 | Italy/CAM-TIGEM-913/2020 | 2020-05-04 | Europe/Italy/Campania/ | Istituto Zooprofilattico Sperimentale del Mezzogiorno | TIGEM | Patrizia Annunziata et al |
| EPI_ISL_833547 | Italy/CAM-TIGEM-894/2020 | 2020-04-30 | Europe/Italy/Campania/ | Istituto Zooprofilattico Sperimentale del Mezzogiorno | TIGEM | Patrizia Annunziata et al |
| EPI_ISL_833557 | Italy/CAM-TIGEM-928/2020 | 2020-05-06 | Europe/Italy/Campania/ | Istituto Zooprofilattico Sperimentale del Mezzogiorno | TIGEM | Patrizia Annunziata et al |
| EPI_ISL_833558 | Italy/CAM-TIGEM-837/2020 | 2020-04-24 | Europe/Italy/Campania/ | Istituto Zooprofilattico Sperimentale del Mezzogiorno | TIGEM | Patrizia Annunziata et al |
| EPI_ISL_833560 | Italy/CAM-TIGEM-846/2020 | 2020-04-25 | Europe/Italy/Campania/ | Istituto Zooprofilattico Sperimentale del Mezzogiorno | TIGEM | Patrizia Annunziata et al |
| EPI_ISL_833561 | Italy/CAM-TIGEM-850/2020 | 2020-04-25 | Europe/Italy/Campania/ | Istituto Zooprofilattico Sperimentale del Mezzogiorno | TIGEM | Patrizia Annunziata et al |
| EPI_ISL_833562 | Italy/CAM-TIGEM-858/2020 | 2020-04-25 | Europe/Italy/Campania/ | Istituto Zooprofilattico Sperimentale del Mezzogiorno | TIGEM | Patrizia Annunziata et al |
| EPI_ISL_833564 | Italy/CAM-TIGEM-872/2020 | 2020-04-27 | Europe/Italy/Campania/ | Istituto Zooprofilattico Sperimentale del Mezzogiorno | TIGEM | Patrizia Annunziata et al |

| | | | | | | |
|---|---|---|---|---|---|---|
| EPI_ISL_833567 | Italy/CAM-TIGEM-950/2020 | 2020-05-02 | Europe/Italy/Campania/ | Istituto Zooprofilattico Sperimentale del Mezzogiorno | TIGEM | Patrizia Annunziata et al |
| EPI_ISL_833569 | Italy/CAM-TIGEM-917/2020 | 2020-05-01 | Europe/Italy/Campania/ | Istituto Zooprofilattico Sperimentale del Mezzogiorno | TIGEM | Patrizia Annunziata et al |
| EPI_ISL_833570 | Italy/CAM-TIGEM-946/2020 | 2020-05-01 | Europe/Italy/Campania/ | Istituto Zooprofilattico Sperimentale del Mezzogiorno | TIGEM | Patrizia Annunziata et al |
| EPI_ISL_833578 | Iran/N193/2021 | 2021-01-08 | Asia/Iran/Tehran/ | National Reference Laboratory for COVID-19, Pasteur Institute of Iran | National Reference Laboratory for COVID-19, Pasteur Institute of Iran | Zahra Ahmadi et al |
| EPI_ISL_833717 | England/CAMC-F012BF/2021 | 2021-01-08 | Europe/United Kingdom/England/ | Lighthouse Lab in Cambridge | Wellcome Sanger Institute for the COVID-19 Genomics UK (COG-UK) Consortium | Rob Howes et al |
| EPI_ISL_833735 | England/MILK-F73476/2021 | 2021-01-05 | Europe/United Kingdom/England/ | Lighthouse Lab in Milton Keynes | Wellcome Sanger Institute for the COVID-19 Genomics UK (COG-UK) Consortium | The Lighthouse Lab in Milton Keynes et al |
| EPI_ISL_834025 | England/ALDP-F4AE7A/2021 | 2021-01-05 | Europe/United Kingdom/England/ | Lighthouse Lab in Alderley Park | Wellcome Sanger Institute for the COVID-19 Genomics UK (COG-UK) Consortium | Jacquelyn Wynn et al |
| EPI_ISL_834042 | England/ALDP-F4E5EA/2021 | 2021-01-04 | Europe/United Kingdom/England/ | Lighthouse Lab in Alderley Park | Wellcome Sanger Institute for the COVID-19 Genomics UK (COG-UK) Consortium | Jacquelyn Wynn et al |
| EPI_ISL_834058 | England/ALDP-F4A43C/2021 | 2021-01-05 | Europe/United Kingdom/England/ | Lighthouse Lab in Alderley Park | Wellcome Sanger Institute for the COVID-19 Genomics UK (COG-UK) Consortium | Jacquelyn Wynn et al |

| | | | | | | |
|---|---|---|---|---|---|---|
| EPI_ISL_834104 | England/ALDP-F4E68D/2021 | 2021-01-04 | Europe/United Kingdom/England/ | Lighthouse Lab in Alderley Park | Wellcome Sanger Institute for the COVID-19 Genomics UK (COG-UK) Consortium | Jacquelyn Wynn et al |
| EPI_ISL_834536 | England/ALDP-EE9F02/2021 | 2021-01-04 | Europe/United Kingdom/England/ | Lighthouse Lab in Alderley Park | Wellcome Sanger Institute for the COVID-19 Genomics UK (COG-UK) Consortium | Jacquelyn Wynn et al |
| EPI_ISL_834618 | England/QEUH-EF38FB/2021 | 2021-01-05 | Europe/United Kingdom/England/ | Lighthouse Lab in Glasgow | Wellcome Sanger Institute for the COVID-19 Genomics UK (COG-UK) Consortium | Harper VanSteenhouse et al |
| EPI_ISL_834660 | England/QEUH-EF36F1/2021 | 2021-01-05 | Europe/United Kingdom/England/ | Lighthouse Lab in Glasgow | Wellcome Sanger Institute for the COVID-19 Genomics UK (COG-UK) Consortium | Harper VanSteenhouse et al |
| EPI_ISL_834840 | England/ALDP-EE0D5C/2021 | 2021-01-02 | Europe/United Kingdom/England/ | Lighthouse Lab in Alderley Park | Wellcome Sanger Institute for the COVID-19 Genomics UK (COG-UK) Consortium | Jacquelyn Wynn et al |
| EPI_ISL_834856 | England/ALDP-EE4A60/2021 | 2021-01-04 | Europe/United Kingdom/England/ | Lighthouse Lab in Alderley Park | Wellcome Sanger Institute for the COVID-19 Genomics UK (COG-UK) Consortium | Jacquelyn Wynn et al |
| EPI_ISL_835022 | England/ALDP-EE6DED/2021 | 2021-01-03 | Europe/United Kingdom/England/ | Lighthouse Lab in Alderley Park | Wellcome Sanger Institute for the COVID-19 Genomics UK (COG-UK) Consortium | Jacquelyn Wynn et al |
| EPI_ISL_835063 | England/ALDP-EE9EF6/2021 | 2021-01-03 | Europe/United Kingdom/England/ | Lighthouse Lab in Alderley Park | Wellcome Sanger Institute for the COVID-19 Genomics UK (COG-UK) Consortium | Jacquelyn Wynn et al |

| | | | | | | |
|---|---|---|---|---|---|---|
| EPI_ISL_835073 | England/ALDP-EE6D74/2021 | 2021-01-03 | Europe/United Kingdom/England/ | Lighthouse Lab in Alderley Park | Wellcome Sanger Institute for the COVID-19 Genomics UK (COG-UK) Consortium | Jacquelyn Wynn et al |
| EPI_ISL_835164 | England/QEUH-EFECC7/2021 | 2021-01-05 | Europe/United Kingdom/England/ | Lighthouse Lab in Glasgow | Wellcome Sanger Institute for the COVID-19 Genomics UK (COG-UK) Consortium | Harper VanSteenhouse et al |
| EPI_ISL_835168 | England/QEUH-EFBA0C/2021 | 2021-01-05 | Europe/United Kingdom/England/ | Lighthouse Lab in Glasgow | Wellcome Sanger Institute for the COVID-19 Genomics UK (COG-UK) Consortium | Harper VanSteenhouse et al |
| EPI_ISL_835338 | England/QEUH-EFD2C6/2021 | 2021-01-04 | Europe/United Kingdom/England/ | Lighthouse Lab in Glasgow | Wellcome Sanger Institute for the COVID-19 Genomics UK (COG-UK) Consortium | Harper VanSteenhouse et al |
| EPI_ISL_835355 | England/QEUH-EF9B92/2021 | 2021-01-05 | Europe/United Kingdom/England/ | Lighthouse Lab in Glasgow | Wellcome Sanger Institute for the COVID-19 Genomics UK (COG-UK) Consortium | Harper VanSteenhouse et al |
| EPI_ISL_835369 | England/QEUH-EFD30F/2021 | 2021-01-04 | Europe/United Kingdom/England/ | Lighthouse Lab in Glasgow | Wellcome Sanger Institute for the COVID-19 Genomics UK (COG-UK) Consortium | Harper VanSteenhouse et al |
| EPI_ISL_835413 | England/QEUH-EFE3A4/2021 | 2021-01-05 | Europe/United Kingdom/England/ | Lighthouse Lab in Glasgow | Wellcome Sanger Institute for the COVID-19 Genomics UK (COG-UK) Consortium | Harper VanSteenhouse et al |
| EPI_ISL_835483 | England/ALDP-EB39E5/2020 | 2020-12-31 | Europe/United Kingdom/England/ | Lighthouse Lab in Alderley Park | Wellcome Sanger Institute for the COVID-19 Genomics UK (COG-UK) Consortium | Jacquelyn Wynn et al |

| | | | | | | |
|---|---|---|---|---|---|---|
| EPI_ISL_835561 | England/ALDP-E86329/2021 | 2021-01-02 | Europe/United Kingdom/England/ | Lighthouse Lab in Alderley Park | Wellcome Sanger Institute for the COVID-19 Genomics UK (COG-UK) Consortium | Jacquelyn Wynn et al |
| EPI_ISL_835706 | England/ALDP-E826E8/2021 | 2021-01-02 | Europe/United Kingdom/England/ | Lighthouse Lab in Alderley Park | Wellcome Sanger Institute for the COVID-19 Genomics UK (COG-UK) Consortium | Jacquelyn Wynn et al |
| EPI_ISL_835761 | England/MILK-EC71E6/2021 | 2021-01-02 | Europe/United Kingdom/England/ | Lighthouse Lab in Milton Keynes | Wellcome Sanger Institute for the COVID-19 Genomics UK (COG-UK) Consortium | The Lighthouse Lab in Milton Keynes et al |
| EPI_ISL_835792 | England/MILK-EC0323/2021 | 2021-01-03 | Europe/United Kingdom/England/ | Lighthouse Lab in Milton Keynes | Wellcome Sanger Institute for the COVID-19 Genomics UK (COG-UK) Consortium | The Lighthouse Lab in Milton Keynes et al |
| EPI_ISL_835832 | England/MILK-EC6FDE/2021 | 2021-01-03 | Europe/United Kingdom/England/ | Lighthouse Lab in Milton Keynes | Wellcome Sanger Institute for the COVID-19 Genomics UK (COG-UK) Consortium | The Lighthouse Lab in Milton Keynes et al |
| EPI_ISL_836008 | England/MILK-EC06DF/2021 | 2021-01-03 | Europe/United Kingdom/England/ | Lighthouse Lab in Milton Keynes | Wellcome Sanger Institute for the COVID-19 Genomics UK (COG-UK) Consortium | The Lighthouse Lab in Milton Keynes et al |
| EPI_ISL_836143 | Brazil/SP-912/2020 | 2020-12-07 | South America/Brazil/S√£o Paulo/Atibai | Hospital de Campanha COVID-19 de Mairipora | Instituto Adolfo Lutz, Interdiciplinary Procedures Center, Strategic Laboratory | Claudio Tavares Sacchi et al |
| EPI_ISL_836568 | England/CAMC-ED75F6/2021 | 2021-01-04 | Europe/United Kingdom/England/ | Lighthouse Lab in Cambridge | Wellcome Sanger Institute for the COVID-19 Genomics UK (COG-UK) Consortium | Rob Howes et al |

| | | | | | | |
|---|---|---|---|---|---|---|
| EPI_ISL_836619 | England/CAMC-ED6393/2021 | 2021-01-04 | Europe/United Kingdom/England/ | Lighthouse Lab in Cambridge | Wellcome Sanger Institute for the COVID-19 Genomics UK (COG-UK) Consortium | Rob Howes et al |
| EPI_ISL_836639 | England/CAMC-ED61B7/2021 | 2021-01-04 | Europe/United Kingdom/England/ | Lighthouse Lab in Cambridge | Wellcome Sanger Institute for the COVID-19 Genomics UK (COG-UK) Consortium | Rob Howes et al |
| EPI_ISL_836943 | England/QEUH-E928EF/2021 | 2021-01-04 | Europe/United Kingdom/England/ | Lighthouse Lab in Glasgow | Wellcome Sanger Institute for the COVID-19 Genomics UK (COG-UK) Consortium | Harper VanSteenhouse et al |
| EPI_ISL_836977 | Brazil/SP-915/2020 | 2020-12-08 | South America/Brazil/S√£o Paulo/Sao Jo | Hospital Municiplal Dr. Jose de Carvalho Florence | Instituto Adolfo Lutz, Interdiciplinary Procedures Center, Strategic Laboratory | Claudio Tavares Sacchi et al |
| EPI_ISL_836978 | Brazil/SP-916/2020 | 2020-12-08 | South America/Brazil/S√£o Paulo/Lorena | Irmandade da Santa Casa de Misericordia de Lorena | Instituto Adolfo Lutz, Interdiciplinary Procedures Center, Strategic Laboratory | Claudio Tavares Sacchi et al |
| EPI_ISL_837014 | England/205241543/2020 | 2020-12-22 | Europe/United Kingdom/England/ | Respiratory Virus Unit, National Infection Service, Public Health England | COVID-19 Genomics UK (COG-UK) Consortium | PHE Covid Sequencing Team et al |
| EPI_ISL_837053 | Brazil/SP-917/2020 | 2020-12-07 | South America/Brazil/S√£o Paulo/Osasco | UBS Darcy Alves e Robalinho | Instituto Adolfo Lutz, Interdiciplinary Procedures Center, Strategic Laboratory | Claudio Tavares Sacchi et al |

| | | | | | | |
|---|---|---|---|---|---|---|
| EPI_ISL_837054 | Brazil/SP-918/2020 | 2020-12-09 | South America/Brazil/S√£o Paulo/Osasc | UBS Jose Sabino Ferreira | Instituto Adolfo Lutz, Interdiciplinary Procedures Center, Strategic Laboratory | Claudio Tavares Sacchi et al |
| EPI_ISL_837249 | Italy/CAM-TIGEM-1061/2020 | 2020-11-27 | Europe/Italy/Campania/ | Istituto Zooprofilattico Sperimentale del Mezzogiorno | TIGEM | Patrizia Annunziata et al |
| EPI_ISL_837250 | Italy/CAM-TIGEM-974/2020 | 2020-09-15 | Europe/Italy/Campania/ | Istituto Zooprofilattico Sperimentale del Mezzogiorno | TIGEM | Patrizia Annunziata et al |
| EPI_ISL_837251 | Italy/CAM-TIGEM-1058/2020 | 2020-11-27 | Europe/Italy/Campania/ | Istituto Zooprofilattico Sperimentale del Mezzogiorno | TIGEM | Patrizia Annunziata et al |
| EPI_ISL_837252 | Italy/CAM-TIGEM-966/2020 | 2020-09-16 | Europe/Italy/Campania/ | Istituto Zooprofilattico Sperimentale del Mezzogiorno | TIGEM | Patrizia Annunziata et al |
| EPI_ISL_837255 | Italy/CAM-TIGEM-1005/2020 | 2020-09-15 | Europe/Italy/Campania/ | Istituto Zooprofilattico Sperimentale del Mezzogiorno | TIGEM | Patrizia Annunziata et al |
| EPI_ISL_837256 | Italy/CAM-TIGEM-1080/2020 | 2020-11-27 | Europe/Italy/Campania/ | Istituto Zooprofilattico Sperimentale del Mezzogiorno | TIGEM | Patrizia Annunziata et al |
| EPI_ISL_837257 | Italy/CAM-TIGEM-1083/2020 | 2020-11-27 | Europe/Italy/Campania/ | Istituto Zooprofilattico Sperimentale del Mezzogiorno | TIGEM | Patrizia Annunziata et al |
| EPI_ISL_837258 | Italy/CAM-TIGEM-1082/2020 | 2020-11-27 | Europe/Italy/Campania/ | Istituto Zooprofilattico Sperimentale del Mezzogiorno | TIGEM | Patrizia Annunziata et al |
| EPI_ISL_837260 | Italy/CAM-TIGEM-1085/2020 | 2020-11-27 | Europe/Italy/Campania/ | Istituto Zooprofilattico Sperimentale del Mezzogiorno | TIGEM | Patrizia Annunziata et al |
| EPI_ISL_837261 | Italy/CAM-TIGEM-1071/2020 | 2020-11-27 | Europe/Italy/Campania/ | Istituto Zooprofilattico Sperimentale del Mezzogiorno | TIGEM | Patrizia Annunziata et al |
| EPI_ISL_837262 | Italy/CAM-TIGEM-1074/2020 | 2020-11-27 | Europe/Italy/Campania/ | Istituto Zooprofilattico Sperimentale del Mezzogiorno | TIGEM | Patrizia Annunziata et al |
| EPI_ISL_837263 | Italy/CAM-TIGEM-1076/2020 | 2020-11-27 | Europe/Italy/Campania/ | Istituto Zooprofilattico Sperimentale del Mezzogiorno | TIGEM | Patrizia Annunziata et al |
| EPI_ISL_837265 | Italy/CAM-TIGEM-1066/2020 | 2020-11-27 | Europe/Italy/Campania/ | Istituto Zooprofilattico Sperimentale del Mezzogiorno | TIGEM | Patrizia Annunziata et al |
| EPI_ISL_837266 | Italy/CAM-TIGEM-1060/2020 | 2020-11-27 | Europe/Italy/Campania/ | Istituto Zooprofilattico Sperimentale del Mezzogiorno | TIGEM | Patrizia Annunziata et al |
| EPI_ISL_837267 | Italy/CAM-TIGEM-1063/2020 | 2020-11-27 | Europe/Italy/Campania/ | Istituto Zooprofilattico Sperimentale del Mezzogiorno | TIGEM | Patrizia Annunziata et al |
| EPI_ISL_837279 | Italy/CAM-TIGEM-1027/2020 | 2020-09-06 | Europe/Italy/Campania/ | Istituto Zooprofilattico Sperimentale del Mezzogiorno | TIGEM | Patrizia Annunziata et al |
| EPI_ISL_837283 | Italy/CAM-TIGEM-1026/2020 | 2020-09-09 | Europe/Italy/Campania/ | Istituto Zooprofilattico Sperimentale del Mezzogiorno | TIGEM | Patrizia Annunziata et al |

| | | | | | | |
|---|---|---|---|---|---|---|
| EPI_ISL_837288 | Italy/CAM-TIGEM-993/2020 | 2020-09-15 | Europe/Italy/Campania/ | Istituto Zooprofilattico Sperimentale del Mezzogiorno | TIGEM | Patrizia Annunziata et al |
| EPI_ISL_837296 | Italy/CAM-TIGEM-1002/2020 | 2020-09-15 | Europe/Italy/Campania/ | Istituto Zooprofilattico Sperimentale del Mezzogiorno | TIGEM | Patrizia Annunziata et al |
| EPI_ISL_837303 | Italy/CAM-TIGEM-990/2020 | 2020-09-16 | Europe/Italy/Campania/ | Istituto Zooprofilattico Sperimentale del Mezzogiorno | TIGEM | Patrizia Annunziata et al |
| EPI_ISL_837309 | Italy/CAM-TIGEM-962/2020 | 2020-09-17 | Europe/Italy/Campania/ | Istituto Zooprofilattico Sperimentale del Mezzogiorno | TIGEM | Patrizia Annunziata et al |
| EPI_ISL_837315 | Italy/CAM-TIGEM-1042/2020 | 2020-09-16 | Europe/Italy/Campania/ | Istituto Zooprofilattico Sperimentale del Mezzogiorno | TIGEM | Patrizia Annunziata et al |
| EPI_ISL_837316 | Italy/CAM-TIGEM-1047/2020 | 2020-09-16 | Europe/Italy/Campania/ | Istituto Zooprofilattico Sperimentale del Mezzogiorno | TIGEM | Patrizia Annunziata et al |
| EPI_ISL_837317 | Italy/CAM-TIGEM-1048/2020 | 2020-09-16 | Europe/Italy/Campania/ | Istituto Zooprofilattico Sperimentale del Mezzogiorno | TIGEM | Patrizia Annunziata et al |
| EPI_ISL_837318 | Italy/CAM-TIGEM-1056/2020 | 2020-12-18 | Europe/Italy/Campania/ | Istituto Zooprofilattico Sperimentale del Mezzogiorno | TIGEM | Patrizia Annunziata et al |
| EPI_ISL_837330 | Italy/CAM-TIGEM-1050/2020 | 2020-12-28 | Europe/Italy/Campania/ | Istituto Zooprofilattico Sperimentale del Mezzogiorno | TIGEM | Patrizia Annunziata et al |
| EPI_ISL_837381 | Ireland/CN-NVRL-81IRL07994/2020 | 2020-12-27 | Europe/Ireland/Cavan/ | National Virus Reference Laboratory | National Virus Reference Laboratory | Michael Carr et al |
| EPI_ISL_837410 | Ireland/LK-NVRL-81IRL13814/2020 | 2020-12-27 | Europe/Ireland/Limerick/ | National Virus Reference Laboratory | National Virus Reference Laboratory | Michael Carr et al |
| EPI_ISL_837436 | Italy/CAM-TIGEM-1174/2020 | 2020-11-25 | Europe/Italy/Campania/ | Istituto Zooprofilattico Sperimentale del Mezzogiorno | TIGEM | Patrizia Annunziata et al |
| EPI_ISL_837437 | Italy/CAM-TIGEM-1187/2020 | 2020-11-25 | Europe/Italy/Campania/ | Istituto Zooprofilattico Sperimentale del Mezzogiorno | TIGEM | Patrizia Annunziata et al |
| EPI_ISL_837441 | Italy/CAM-TIGEM-1171/2020 | 2020-11-25 | Europe/Italy/Campania/ | Istituto Zooprofilattico Sperimentale del Mezzogiorno | TIGEM | Patrizia Annunziata et al |
| EPI_ISL_837443 | Italy/CAM-TIGEM-1168/2020 | 2020-11-25 | Europe/Italy/Campania/ | Istituto Zooprofilattico Sperimentale del Mezzogiorno | TIGEM | Patrizia Annunziata et al |
| EPI_ISL_837445 | Italy/CAM-TIGEM-1265/2020 | 2020-11-29 | Europe/Italy/Campania/ | Istituto Zooprofilattico Sperimentale del Mezzogiorno | TIGEM | Patrizia Annunziata et al |
| EPI_ISL_837447 | Italy/CAM-TIGEM-1160/2020 | 2020-11-25 | Europe/Italy/Campania/ | Istituto Zooprofilattico Sperimentale del Mezzogiorno | TIGEM | Patrizia Annunziata et al |
| EPI_ISL_837454 | Italy/CAM-TIGEM-1272/2020 | 2020-12-01 | Europe/Italy/Campania/ | Istituto Zooprofilattico Sperimentale del Mezzogiorno | TIGEM | Patrizia Annunziata et al |
| EPI_ISL_837456 | Italy/CAM-TIGEM-1270/2020 | 2020-12-01 | Europe/Italy/Campania/ | Istituto Zooprofilattico Sperimentale del Mezzogiorno | TIGEM | Patrizia Annunziata et al |

| | | | | | | |
|---|---|---|---|---|---|---|
| EPI_ISL_837457 | Italy/CAM-TIGEM-1182/2020 | 2020-11-25 | Europe/Italy/Campania/ | Istituto Zooprofilattico Sperimentale del Mezzogiorno | TIGEM | Patrizia Annunziata et al |
| EPI_ISL_837460 | Italy/CAM-TIGEM-1169/2020 | 2020-11-25 | Europe/Italy/Campania/ | Istituto Zooprofilattico Sperimentale del Mezzogiorno | TIGEM | Patrizia Annunziata et al |
| EPI_ISL_837461 | Italy/CAM-TIGEM-1183/2020 | 2020-11-25 | Europe/Italy/Campania/ | Istituto Zooprofilattico Sperimentale del Mezzogiorno | TIGEM | Patrizia Annunziata et al |
| EPI_ISL_837463 | Italy/CAM-TIGEM-1167/2020 | 2020-11-25 | Europe/Italy/Campania/ | Istituto Zooprofilattico Sperimentale del Mezzogiorno | TIGEM | Patrizia Annunziata et al |
| EPI_ISL_837464 | Italy/CAM-TIGEM-1166/2020 | 2020-11-25 | Europe/Italy/Campania/ | Istituto Zooprofilattico Sperimentale del Mezzogiorno | TIGEM | Patrizia Annunziata et al |
| EPI_ISL_837465 | Italy/CAM-TIGEM-1165/2020 | 2020-11-25 | Europe/Italy/Campania/ | Istituto Zooprofilattico Sperimentale del Mezzogiorno | TIGEM | Patrizia Annunziata et al |
| EPI_ISL_837466 | Italy/CAM-TIGEM-1179/2020 | 2020-11-25 | Europe/Italy/Campania/ | Istituto Zooprofilattico Sperimentale del Mezzogiorno | TIGEM | Patrizia Annunziata et al |
| EPI_ISL_837467 | Italy/CAM-TIGEM-1176/2020 | 2020-11-25 | Europe/Italy/Campania/ | Istituto Zooprofilattico Sperimentale del Mezzogiorno | TIGEM | Patrizia Annunziata et al |
| EPI_ISL_837468 | Italy/CAM-TIGEM-1177/2020 | 2020-11-25 | Europe/Italy/Campania/ | Istituto Zooprofilattico Sperimentale del Mezzogiorno | TIGEM | Patrizia Annunziata et al |
| EPI_ISL_837469 | Italy/CAM-TIGEM-1164/2020 | 2020-11-25 | Europe/Italy/Campania/ | Istituto Zooprofilattico Sperimentale del Mezzogiorno | TIGEM | Patrizia Annunziata et al |
| EPI_ISL_837470 | Italy/CAM-TIGEM-1175/2020 | 2020-11-25 | Europe/Italy/Campania/ | Istituto Zooprofilattico Sperimentale del Mezzogiorno | TIGEM | Patrizia Annunziata et al |
| EPI_ISL_837472 | Italy/CAM-TIGEM-1172/2020 | 2020-11-25 | Europe/Italy/Campania/ | Istituto Zooprofilattico Sperimentale del Mezzogiorno | TIGEM | Patrizia Annunziata et al |
| EPI_ISL_837476 | Italy/CAM-TIGEM-1302/2020 | 2020-12-24 | Europe/Italy/Campania/ | Istituto Zooprofilattico Sperimentale del Mezzogiorno | TIGEM | Patrizia Annunziata et al |
| EPI_ISL_837480 | Italy/CAM-TIGEM-1189/2020 | 2020-11-25 | Europe/Italy/Campania/ | Istituto Zooprofilattico Sperimentale del Mezzogiorno | TIGEM | Patrizia Annunziata et al |
| EPI_ISL_837481 | Italy/CAM-TIGEM-1260/2020 | 2020-11-30 | Europe/Italy/Campania/ | Istituto Zooprofilattico Sperimentale del Mezzogiorno | TIGEM | Patrizia Annunziata et al |
| EPI_ISL_837482 | Italy/CAM-TIGEM-1262/2020 | 2020-11-30 | Europe/Italy/Campania/ | Istituto Zooprofilattico Sperimentale del Mezzogiorno | TIGEM | Patrizia Annunziata et al |
| EPI_ISL_837488 | Italy/CAM-TIGEM-1266/2020 | 2020-11-29 | Europe/Italy/Campania/ | Istituto Zooprofilattico Sperimentale del Mezzogiorno | TIGEM | Patrizia Annunziata et al |
| EPI_ISL_837493 | Italy/CAM-TIGEM-1311/2020 | 2020-12-26 | Europe/Italy/Campania/ | Istituto Zooprofilattico Sperimentale del Mezzogiorno | TIGEM | Patrizia Annunziata et al |
| EPI_ISL_837494 | Italy/CAM-TIGEM-1321/2020 | 2020-12-26 | Europe/Italy/Campania/ | Istituto Zooprofilattico Sperimentale del Mezzogiorno | TIGEM | Patrizia Annunziata et al |
| EPI_ISL_837495 | Italy/CAM-TIGEM-1329/2020 | 2020-12-26 | Europe/Italy/Campania/ | Istituto Zooprofilattico Sperimentale del Mezzogiorno | TIGEM | Patrizia Annunziata et al |

| | | | | | | |
|---|---|---|---|---|---|---|
| EPI_ISL_837505 | USA/WA-UW-49800/2020 | 2020-12-20 | North America/USA/Washington/ | UW Virology Lab | UW Virology Lab | Pavitra Roychoudhury et al |
| EPI_ISL_837518 | USA/WA-UW-48591/2020 | 2020-12-27 | North America/USA/Washington/ | UW Virology Lab | UW Virology Lab | Pavitra Roychoudhury et al |
| EPI_ISL_837519 | USA/WA-UW-48593/2020 | 2020-12-26 | North America/USA/Washington/ | UW Virology Lab | UW Virology Lab | Pavitra Roychoudhury et al |
| EPI_ISL_837526 | USA/WA-UW-49493/2021 | 2021-01-01 | North America/USA/Washington/ | UW Virology Lab | UW Virology Lab | Pavitra Roychoudhury et al |
| EPI_ISL_837529 | USA/WA-UW-49586/2020 | 2020-12-21 | North America/USA/Washington/ | UW Virology Lab | UW Virology Lab | Pavitra Roychoudhury et al |
| EPI_ISL_837533 | USA/WA-UW-49614/2020 | 2020-12-20 | North America/USA/Washington/ | UW Virology Lab | UW Virology Lab | Pavitra Roychoudhury et al |
| EPI_ISL_837536 | USA/WA-UW-49906/2020 | 2020-12-20 | North America/USA/Washington/ | UW Virology Lab | UW Virology Lab | Pavitra Roychoudhury et al |
| EPI_ISL_837539 | USA/WA-UW-49922/2020 | 2020-12-20 | North America/USA/Washington/ | UW Virology Lab | UW Virology Lab | Pavitra Roychoudhury et al |
| EPI_ISL_837545 | USA/WA-UW-50932/2020 | 2020-12-25 | North America/USA/Washington/ | UW Virology Lab | UW Virology Lab | Pavitra Roychoudhury et al |
| EPI_ISL_837551 | Guatemala/19522/2020 | 2020-06-25 | North America/Guatemala/Guatemala/ | Laboratorio Nacional de Salud | Laboratory of Respiratory Viruses and Measles, Oswaldo Cruz Institute, FIOCRUZ | Paola Resende et al |
| EPI_ISL_837552 | Guatemala/19544/2020 | 2020-08-13 | North America/Guatemala/Guatemala/ | Laboratorio Nacional de Salud | Laboratory of Respiratory Viruses and Measles, Oswaldo Cruz Institute, FIOCRUZ | Paola Resende et al |
| EPI_ISL_837553 | Bolivia/26945/2020 | 2020-05-12 | South America/Bolivia/Santa Cruz/ | Centro Nacional de Enfermedades Tropicales (CENETROP) | Laboratory of Respiratory Viruses and Measles, Oswaldo Cruz Institute, FIOCRUZ | Paola Resende et al |
| EPI_ISL_837554 | Bolivia/26937/2020 | 2020-08-03 | South America/Bolivia/Santa Cruz/ | Centro Nacional de Enfermedades Tropicales (CENETROP) | Laboratory of Respiratory Viruses and Measles, Oswaldo Cruz Institute, FIOCRUZ | Paola Resende et al |

| | | | | | | |
|---|---|---|---|---|---|---|
| EPI_ISL_837555 | Bolivia/26932/2020 | 2020-08-03 | South America/Bolivia/Santa Cruz/ | Centro Nacional de Enfermedades Tropicales (CENETROP) | Laboratory of Respiratory Viruses and Measles, Oswaldo Cruz Institute, FIOCRUZ | Paola Resende et al |
| EPI_ISL_837556 | Bolivia/26933/2020 | 2020-07-15 | South America/Bolivia/Santa Cruz/ | Centro Nacional de Enfermedades Tropicales (CENETROP) | Laboratory of Respiratory Viruses and Measles, Oswaldo Cruz Institute, FIOCRUZ | Paola Resende et al |
| EPI_ISL_837557 | Bolivia/26934/2020 | 2020-08-12 | South America/Bolivia/Santa Cruz/ | Centro Nacional de Enfermedades Tropicales (CENETROP) | Laboratory of Respiratory Viruses and Measles, Oswaldo Cruz Institute, FIOCRUZ | Paola Resende et al |
| EPI_ISL_837558 | Bolivia/26936/2020 | 2020-07-03 | South America/Bolivia/Pando/ | Centro Nacional de Enfermedades Tropicales (CENETROP) | Laboratory of Respiratory Viruses and Measles, Oswaldo Cruz Institute, FIOCRUZ | Paola Resende et al |
| EPI_ISL_837559 | Bolivia/26938/2020 | 2020-08-11 | South America/Bolivia/Santa Cruz/ | Centro Nacional de Enfermedades Tropicales (CENETROP) | Laboratory of Respiratory Viruses and Measles, Oswaldo Cruz Institute, FIOCRUZ | Paola Resende et al |
| EPI_ISL_837560 | Bolivia/26939/2020 | 2020-07-03 | South America/Bolivia/Pando/ | Centro Nacional de Enfermedades Tropicales (CENETROP) | Laboratory of Respiratory Viruses and Measles, Oswaldo Cruz Institute, FIOCRUZ | Paola Resende et al |

| | | | | | | |
|---|---|---|---|---|---|---|
| EPI_ISL_837561 | Bolivia/26940/2020 | 2020-08-12 | South America/Bolivia/Santa Cruz/El To | Centro Nacional de Enfermedades Tropicales (CENETROP) | Laboratory of Respiratory Viruses and Measles, Oswaldo Cruz Institute, FIOCRUZ | Paola Resende et al |
| EPI_ISL_837562 | Bolivia/26941/2020 | 2020-08-12 | South America/Bolivia/Santa Cruz/Pailo | Centro Nacional de Enfermedades Tropicales (CENETROP) | Laboratory of Respiratory Viruses and Measles, Oswaldo Cruz Institute, FIOCRUZ | Paola Resende et al |
| EPI_ISL_837563 | Bolivia/26942/2020 | 2020-05-10 | South America/Bolivia/Santa Cruz/ | Centro Nacional de Enfermedades Tropicales (CENETROP) | Laboratory of Respiratory Viruses and Measles, Oswaldo Cruz Institute, FIOCRUZ | Paola Resende et al |
| EPI_ISL_837565 | Bolivia/26944/2020 | 2020-05-12 | South America/Bolivia/Santa Cruz/ | Centro Nacional de Enfermedades Tropicales (CENETROP) | Laboratory of Respiratory Viruses and Measles, Oswaldo Cruz Institute, FIOCRUZ | Paola Resende et al |
| EPI_ISL_837566 | Bolivia/26946/2020 | 2020-05-08 | South America/Bolivia/Santa Cruz/ | Centro Nacional de Enfermedades Tropicales (CENETROP) | Laboratory of Respiratory Viruses and Measles, Oswaldo Cruz Institute, FIOCRUZ | Paola Resende et al |
| EPI_ISL_837567 | Bolivia/26947/2020 | 2020-05-16 | South America/Bolivia/Santa Cruz/ | Centro Nacional de Enfermedades Tropicales (CENETROP) | Laboratory of Respiratory Viruses and Measles, Oswaldo Cruz Institute, FIOCRUZ | Paola Resende et al |

| | | | | | | |
|---|---|---|---|---|---|---|
| EPI_ISL_837568 | Bolivia/26948/2020 | 2020-05-15 | South America/Bolivia/Santa Cruz/ | Centro Nacional de Enfermedades Tropicales (CENETROP) | Laboratory of Respiratory Viruses and Measles, Oswaldo Cruz Institute, FIOCRUZ | Paola Resende et al |
| EPI_ISL_837570 | Bolivia/26950/2020 | 2020-05-11 | South America/Bolivia/Santa Cruz/ | Centro Nacional de Enfermedades Tropicales (CENETROP) | Laboratory of Respiratory Viruses and Measles, Oswaldo Cruz Institute, FIOCRUZ | Paola Resende et al |
| EPI_ISL_837571 | Bolivia/26952/2020 | 2020-04-27 | South America/Bolivia/Santa Cruz/ | Centro Nacional de Enfermedades Tropicales (CENETROP) | Laboratory of Respiratory Viruses and Measles, Oswaldo Cruz Institute, FIOCRUZ | Paola Resende et al |
| EPI_ISL_837572 | Bolivia/26954/2020 | 2020-03-11 | South America/Bolivia/Oruro/ | Centro Nacional de Enfermedades Tropicales (CENETROP) | Laboratory of Respiratory Viruses and Measles, Oswaldo Cruz Institute, FIOCRUZ | Paola Resende et al |
| EPI_ISL_837574 | Bolivia/26956/2020 | 2020-05-01 | South America/Bolivia/Santa Cruz/ | Centro Nacional de Enfermedades Tropicales (CENETROP) | Laboratory of Respiratory Viruses and Measles, Oswaldo Cruz Institute, FIOCRUZ | Paola Resende et al |
| EPI_ISL_837575 | Bolivia/26957/2020 | 2020-05-01 | South America/Bolivia/Santa Cruz/ | Centro Nacional de Enfermedades Tropicales (CENETROP) | Laboratory of Respiratory Viruses and Measles, Oswaldo Cruz Institute, FIOCRUZ | Paola Resende et al |

| | | | | | | |
|---|---|---|---|---|---|---|
| EPI_ISL_837579 | Guatemala/19517/2020 | 2020-05-18 | North America/Guatemala/Guatemala/ | Laboratorio Nacional de Salud | Laboratory of Respiratory Viruses and Measles, Oswaldo Cruz Institute, FIOCRUZ | Paola Resende et al |
| EPI_ISL_837580 | Guatemala/19518/2020 | 2020-05-19 | North America/Guatemala/Guatemala/ | Laboratorio Nacional de Salud | Laboratory of Respiratory Viruses and Measles, Oswaldo Cruz Institute, FIOCRUZ | Paola Resende et al |
| EPI_ISL_837581 | Guatemala/19519/2020 | 2020-05-19 | North America/Guatemala/Guatemala/ | Laboratorio Nacional de Salud | Laboratory of Respiratory Viruses and Measles, Oswaldo Cruz Institute, FIOCRUZ | Paola Resende et al |
| EPI_ISL_837582 | Guatemala/19520/2020 | 2020-05-27 | North America/Guatemala/Guatemala/ | Laboratorio Nacional de Salud | Laboratory of Respiratory Viruses and Measles, Oswaldo Cruz Institute, FIOCRUZ | Paola Resende et al |
| EPI_ISL_837583 | Guatemala/19521/2020 | 2020-06-24 | North America/Guatemala/Guatemala/ | Laboratorio Nacional de Salud | Laboratory of Respiratory Viruses and Measles, Oswaldo Cruz Institute, FIOCRUZ | Paola Resende et al |
| EPI_ISL_837584 | Guatemala/19523/2020 | 2020-07-09 | North America/Guatemala/Guatemala/ | Laboratorio Nacional de Salud | Laboratory of Respiratory Viruses and Measles, Oswaldo Cruz Institute, FIOCRUZ | Paola Resende et al |

| | | | | | | |
|---|---|---|---|---|---|---|
| EPI_ISL_837585 | Guatemala/19524/2020 | 2020-07-10 | North America/Guatemala/Guatemala/ | Laboratorio Nacional de Salud | Laboratory of Respiratory Viruses and Measles, Oswaldo Cruz Institute, FIOCRUZ | Paola Resende et al |
| EPI_ISL_837586 | Guatemala/19525/2020 | 2020-07-14 | North America/Guatemala/Guatemala/ | Laboratorio Nacional de Salud | Laboratory of Respiratory Viruses and Measles, Oswaldo Cruz Institute, FIOCRUZ | Paola Resende et al |
| EPI_ISL_837587 | Guatemala/19526/2020 | 2020-07-14 | North America/Guatemala/Guatemala/ | Laboratorio Nacional de Salud | Laboratory of Respiratory Viruses and Measles, Oswaldo Cruz Institute, FIOCRUZ | Paola Resende et al |
| EPI_ISL_837588 | Guatemala/19527/2020 | 2020-07-20 | North America/Guatemala/Guatemala/ | Laboratorio Nacional de Salud | Laboratory of Respiratory Viruses and Measles, Oswaldo Cruz Institute, FIOCRUZ | Paola Resende et al |
| EPI_ISL_837589 | Guatemala/19528/2020 | 2020-07-21 | North America/Guatemala/Guatemala/ | Laboratorio Nacional de Salud | Laboratory of Respiratory Viruses and Measles, Oswaldo Cruz Institute, FIOCRUZ | Paola Resende et al |
| EPI_ISL_837590 | Guatemala/19529/2020 | 2020-07-23 | North America/Guatemala/Guatemala/ | Laboratorio Nacional de Salud | Laboratory of Respiratory Viruses and Measles, Oswaldo Cruz Institute, FIOCRUZ | Paola Resende et al |

| | | | | | | |
|---|---|---|---|---|---|---|
| EPI_ISL_837592 | Guatemala/19531/2020 | 2020-07-29 | North America/Guatemala/Guatemala/ | Laboratorio Nacional de Salud | Laboratory of Respiratory Viruses and Measles, Oswaldo Cruz Institute, FIOCRUZ | Paola Resende et al |
| EPI_ISL_837593 | Guatemala/19532/2020 | 2020-07-29 | North America/Guatemala/Guatemala/ | Laboratorio Nacional de Salud | Laboratory of Respiratory Viruses and Measles, Oswaldo Cruz Institute, FIOCRUZ | Paola Resende et al |
| EPI_ISL_837594 | Guatemala/19533/2020 | 2020-07-27 | North America/Guatemala/Guatemala/ | Laboratorio Nacional de Salud | Laboratory of Respiratory Viruses and Measles, Oswaldo Cruz Institute, FIOCRUZ | Paola Resende et al |
| EPI_ISL_837595 | Guatemala/19534/2020 | 2020-08-03 | North America/Guatemala/Guatemala/ | Laboratorio Nacional de Salud | Laboratory of Respiratory Viruses and Measles, Oswaldo Cruz Institute, FIOCRUZ | Paola Resende et al |
| EPI_ISL_837596 | Guatemala/19536/2020 | 2020-08-07 | North America/Guatemala/Guatemala/ | Laboratorio Nacional de Salud | Laboratory of Respiratory Viruses and Measles, Oswaldo Cruz Institute, FIOCRUZ | Paola Resende et al |
| EPI_ISL_837601 | Mexico/CMX-INER-0197/2020 | 2020-08-05 | North America/Mexico/Mexico City/ | Instituto Nacional de Enfermedades Respiratorias (INER) | Instituto Nacional de Enfermedades Respiratorias (INER) | Celia Boukadida et al |
| EPI_ISL_837602 | Mexico/CMX-INER-0217/2020 | 2020-08-28 | North America/Mexico/Mexico City/ | Instituto Nacional de Enfermedades Respiratorias (INER) | Instituto Nacional de Enfermedades Respiratorias (INER) | Celia Boukadida et al |

| | | | | | | |
|---|---|---|---|---|---|---|
| EPI_ISL_837603 | Mexico/CMX-INER-0213/2020 | 2020-08-24 | North America/Mexico/Mexico City/ | Instituto Nacional de Enfermedades Respiratorias (INER) | Instituto Nacional de Enfermedades Respiratorias (INER) | Celia Boukadida et al |
| EPI_ISL_837607 | Mexico/CMX-INER-0138/2020 | 2020-06-14 | North America/Mexico/Mexico City/ | Instituto Nacional de Enfermedades Respiratorias (INER) | Instituto Nacional de Enfermedades Respiratorias (INER) | Celia Boukadida et al |
| EPI_ISL_837621 | Mexico/CMX-INER-0019/2020 | 2020-04-01 | North America/Mexico/Mexico City/ | Instituto Nacional de Enfermedades Respiratorias (INER) | Instituto Nacional de Enfermedades Respiratorias (INER) | Celia Boukadida et al |
| EPI_ISL_837624 | Mexico/CMX-INER-0022/2020 | 2020-04-02 | North America/Mexico/Mexico City/ | Instituto Nacional de Enfermedades Respiratorias (INER) | Instituto Nacional de Enfermedades Respiratorias (INER) | Celia Boukadida et al |
| EPI_ISL_837626 | Mexico/CMX-INER-0024/2020 | 2020-04-03 | North America/Mexico/Mexico City/ | Instituto Nacional de Enfermedades Respiratorias (INER) | Instituto Nacional de Enfermedades Respiratorias (INER) | Celia Boukadida et al |
| EPI_ISL_837628 | Mexico/CMX-INER-0026/2020 | 2020-04-04 | North America/Mexico/Mexico City/ | Instituto Nacional de Enfermedades Respiratorias (INER) | Instituto Nacional de Enfermedades Respiratorias (INER) | Celia Boukadida et al |
| EPI_ISL_837630 | Mexico/CMX-INER-0028/2020 | 2020-04-06 | North America/Mexico/Mexico City/ | Instituto Nacional de Enfermedades Respiratorias (INER) | Instituto Nacional de Enfermedades Respiratorias (INER) | Celia Boukadida et al |
| EPI_ISL_837641 | Mexico/CMX-INER-0039/2020 | 2020-04-10 | North America/Mexico/Mexico City/ | Instituto Nacional de Enfermedades Respiratorias (INER) | Instituto Nacional de Enfermedades Respiratorias (INER) | Celia Boukadida et al |
| EPI_ISL_837653 | Mexico/CMX-INER-0053/2020 | 2020-04-15 | North America/Mexico/Mexico City/ | Instituto Nacional de Enfermedades Respiratorias (INER) | Instituto Nacional de Enfermedades Respiratorias (INER) | Celia Boukadida et al |
| EPI_ISL_837655 | Mexico/CMX-INER-0055/2020 | 2020-04-16 | North America/Mexico/Mexico City/ | Instituto Nacional de Enfermedades Respiratorias (INER) | Instituto Nacional de Enfermedades Respiratorias (INER) | Celia Boukadida et al |

| | | | | | | |
|---|---|---|---|---|---|---|
| EPI_ISL_837658 | Mexico/CMX-INER-0059/2020 | 2020-04-16 | North America/Mexico/Mexico City/ | Instituto Nacional de Enfermedades Respiratorias (INER) | Instituto Nacional de Enfermedades Respiratorias (INER) | Celia Boukadida et al |
| EPI_ISL_837660 | Mexico/CMX-INER-0061/2020 | 2020-04-17 | North America/Mexico/Mexico City/ | Instituto Nacional de Enfermedades Respiratorias (INER) | Instituto Nacional de Enfermedades Respiratorias (INER) | Celia Boukadida et al |
| EPI_ISL_837662 | Mexico/CMX-INER-0063/2020 | 2020-04-18 | North America/Mexico/Mexico City/ | Instituto Nacional de Enfermedades Respiratorias (INER) | Instituto Nacional de Enfermedades Respiratorias (INER) | Celia Boukadida et al |
| EPI_ISL_837680 | Mexico/CMX-INER-0082/2020 | 2020-04-29 | North America/Mexico/Mexico City/ | Instituto Nacional de Enfermedades Respiratorias (INER) | Instituto Nacional de Enfermedades Respiratorias (INER) | Celia Boukadida et al |
| EPI_ISL_837684 | Mexico/CMX-INER-0086/2020 | 2020-05-04 | North America/Mexico/Mexico City/ | Instituto Nacional de Enfermedades Respiratorias (INER) | Instituto Nacional de Enfermedades Respiratorias (INER) | Celia Boukadida et al |
| EPI_ISL_837685 | Mexico/CMX-INER-0087/2020 | 2020-05-06 | North America/Mexico/Mexico City/ | Instituto Nacional de Enfermedades Respiratorias (INER) | Instituto Nacional de Enfermedades Respiratorias (INER) | Celia Boukadida et al |
| EPI_ISL_837686 | Mexico/CMX-INER-0088/2020 | 2020-05-07 | North America/Mexico/Mexico City/ | Instituto Nacional de Enfermedades Respiratorias (INER) | Instituto Nacional de Enfermedades Respiratorias (INER) | Celia Boukadida et al |
| EPI_ISL_837687 | Mexico/CMX-INER-0089/2020 | 2020-05-07 | North America/Mexico/Mexico City/ | Instituto Nacional de Enfermedades Respiratorias (INER) | Instituto Nacional de Enfermedades Respiratorias (INER) | Celia Boukadida et al |
| EPI_ISL_837688 | Mexico/CMX-INER-0090/2020 | 2020-05-08 | North America/Mexico/Mexico City/ | Instituto Nacional de Enfermedades Respiratorias (INER) | Instituto Nacional de Enfermedades Respiratorias (INER) | Celia Boukadida et al |
| EPI_ISL_837689 | Mexico/CMX-INER-0091/2020 | 2020-05-08 | North America/Mexico/Mexico City/ | Instituto Nacional de Enfermedades Respiratorias (INER) | Instituto Nacional de Enfermedades Respiratorias (INER) | Celia Boukadida et al |

| | | | | | | |
|---|---|---|---|---|---|---|
| EPI_ISL_837690 | Mexico/CMX-INER-0092/2020 | 2020-05-09 | North America/Mexico/Mexico City/ | Instituto Nacional de Enfermedades Respiratorias (INER) | Instituto Nacional de Enfermedades Respiratorias (INER) | Celia Boukadida et al |
| EPI_ISL_837691 | Mexico/CMX-INER-0093/2020 | 2020-05-10 | North America/Mexico/Mexico City/ | Instituto Nacional de Enfermedades Respiratorias (INER) | Instituto Nacional de Enfermedades Respiratorias (INER) | Celia Boukadida et al |
| EPI_ISL_837692 | Mexico/CMX-INER-0094/2020 | 2020-05-11 | North America/Mexico/Mexico City/ | Instituto Nacional de Enfermedades Respiratorias (INER) | Instituto Nacional de Enfermedades Respiratorias (INER) | Celia Boukadida et al |
| EPI_ISL_837693 | Mexico/CMX-INER-0095/2020 | 2020-05-11 | North America/Mexico/Mexico City/ | Instituto Nacional de Enfermedades Respiratorias (INER) | Instituto Nacional de Enfermedades Respiratorias (INER) | Celia Boukadida et al |
| EPI_ISL_837694 | Mexico/CMX-INER-0096/2020 | 2020-05-13 | North America/Mexico/Mexico City/ | Instituto Nacional de Enfermedades Respiratorias (INER) | Instituto Nacional de Enfermedades Respiratorias (INER) | Celia Boukadida et al |
| EPI_ISL_837695 | Mexico/CMX-INER-0097/2020 | 2020-05-14 | North America/Mexico/Mexico City/ | Instituto Nacional de Enfermedades Respiratorias (INER) | Instituto Nacional de Enfermedades Respiratorias (INER) | Celia Boukadida et al |
| EPI_ISL_837696 | Mexico/CMX-INER-0098/2020 | 2020-05-14 | North America/Mexico/Mexico City/ | Instituto Nacional de Enfermedades Respiratorias (INER) | Instituto Nacional de Enfermedades Respiratorias (INER) | Celia Boukadida et al |
| EPI_ISL_837697 | Mexico/CMX-INER-0099/2020 | 2020-05-14 | North America/Mexico/Mexico City/ | Instituto Nacional de Enfermedades Respiratorias (INER) | Instituto Nacional de Enfermedades Respiratorias (INER) | Celia Boukadida et al |
| EPI_ISL_837698 | Mexico/CMX-INER-0100/2020 | 2020-05-15 | North America/Mexico/Mexico City/ | Instituto Nacional de Enfermedades Respiratorias (INER) | Instituto Nacional de Enfermedades Respiratorias (INER) | Celia Boukadida et al |
| EPI_ISL_837699 | Mexico/CMX-INER-0101/2020 | 2020-05-18 | North America/Mexico/Mexico City/ | Instituto Nacional de Enfermedades Respiratorias (INER) | Instituto Nacional de Enfermedades Respiratorias (INER) | Celia Boukadida et al |

| | | | | | | |
|---|---|---|---|---|---|---|
| EPI_ISL_837700 | Mexico/CMX-INER-0102/2020 | 2020-05-18 | North America/Mexico/Mexico City/ | Instituto Nacional de Enfermedades Respiratorias (INER) | Instituto Nacional de Enfermedades Respiratorias (INER) | Celia Boukadida et al |
| EPI_ISL_837701 | Mexico/CMX-INER-0103/2020 | 2020-05-19 | North America/Mexico/Mexico City/ | Instituto Nacional de Enfermedades Respiratorias (INER) | Instituto Nacional de Enfermedades Respiratorias (INER) | Celia Boukadida et al |
| EPI_ISL_837703 | Mexico/CMX-INER-0105/2020 | 2020-05-19 | North America/Mexico/Mexico City/ | Instituto Nacional de Enfermedades Respiratorias (INER) | Instituto Nacional de Enfermedades Respiratorias (INER) | Celia Boukadida et al |
| EPI_ISL_837704 | Mexico/CMX-INER-0106/2020 | 2020-05-20 | North America/Mexico/Mexico City/ | Instituto Nacional de Enfermedades Respiratorias (INER) | Instituto Nacional de Enfermedades Respiratorias (INER) | Celia Boukadida et al |
| EPI_ISL_837705 | Mexico/CMX-INER-0107/2020 | 2020-05-20 | North America/Mexico/Mexico City/ | Instituto Nacional de Enfermedades Respiratorias (INER) | Instituto Nacional de Enfermedades Respiratorias (INER) | Celia Boukadida et al |
| EPI_ISL_837707 | Mexico/CMX-INER-0109/2020 | 2020-05-21 | North America/Mexico/Mexico City/ | Instituto Nacional de Enfermedades Respiratorias (INER) | Instituto Nacional de Enfermedades Respiratorias (INER) | Celia Boukadida et al |
| EPI_ISL_837709 | Mexico/CMX-INER-0111/2020 | 2020-05-23 | North America/Mexico/Mexico City/ | Instituto Nacional de Enfermedades Respiratorias (INER) | Instituto Nacional de Enfermedades Respiratorias (INER) | Celia Boukadida et al |
| EPI_ISL_837710 | Mexico/CMX-INER-0112/2020 | 2020-05-23 | North America/Mexico/Mexico City/ | Instituto Nacional de Enfermedades Respiratorias (INER) | Instituto Nacional de Enfermedades Respiratorias (INER) | Celia Boukadida et al |
| EPI_ISL_837711 | Mexico/CMX-INER-0113/2020 | 2020-05-24 | North America/Mexico/Mexico City/ | Instituto Nacional de Enfermedades Respiratorias (INER) | Instituto Nacional de Enfermedades Respiratorias (INER) | Celia Boukadida et al |
| EPI_ISL_837712 | Mexico/CMX-INER-0114/2020 | 2020-05-25 | North America/Mexico/Mexico City/ | Instituto Nacional de Enfermedades Respiratorias (INER) | Instituto Nacional de Enfermedades Respiratorias (INER) | Celia Boukadida et al |

| | | | | | | |
|---|---|---|---|---|---|---|
| EPI_ISL_837714 | Mexico/CMX-INER-0117/2020 | 2020-05-27 | North America/Mexico/Mexico City/ | Instituto Nacional de Enfermedades Respiratorias (INER) | Instituto Nacional de Enfermedades Respiratorias (INER) | Celia Boukadida et al |
| EPI_ISL_837715 | Mexico/CMX-INER-0118/2020 | 2020-05-27 | North America/Mexico/Mexico City/ | Instituto Nacional de Enfermedades Respiratorias (INER) | Instituto Nacional de Enfermedades Respiratorias (INER) | Celia Boukadida et al |
| EPI_ISL_837716 | Mexico/CMX-INER-0119/2020 | 2020-05-28 | North America/Mexico/Mexico City/ | Instituto Nacional de Enfermedades Respiratorias (INER) | Instituto Nacional de Enfermedades Respiratorias (INER) | Celia Boukadida et al |
| EPI_ISL_837717 | Mexico/CMX-INER-0120/2020 | 2020-05-28 | North America/Mexico/Mexico City/ | Instituto Nacional de Enfermedades Respiratorias (INER) | Instituto Nacional de Enfermedades Respiratorias (INER) | Celia Boukadida et al |
| EPI_ISL_837718 | Mexico/CMX-INER-0121/2020 | 2020-05-29 | North America/Mexico/Mexico City/ | Instituto Nacional de Enfermedades Respiratorias (INER) | Instituto Nacional de Enfermedades Respiratorias (INER) | Celia Boukadida et al |
| EPI_ISL_837719 | Mexico/CMX-INER-0122/2020 | 2020-05-29 | North America/Mexico/Mexico City/ | Instituto Nacional de Enfermedades Respiratorias (INER) | Instituto Nacional de Enfermedades Respiratorias (INER) | Celia Boukadida et al |
| EPI_ISL_837720 | Mexico/CMX-INER-0123/2020 | 2020-05-29 | North America/Mexico/Mexico City/ | Instituto Nacional de Enfermedades Respiratorias (INER) | Instituto Nacional de Enfermedades Respiratorias (INER) | Celia Boukadida et al |
| EPI_ISL_837722 | Mexico/CMX-INER-0125/2020 | 2020-05-29 | North America/Mexico/Mexico City/ | Instituto Nacional de Enfermedades Respiratorias (INER) | Instituto Nacional de Enfermedades Respiratorias (INER) | Celia Boukadida et al |
| EPI_ISL_837724 | Mexico/CMX-INER-0127/2020 | 2020-05-31 | North America/Mexico/Mexico City/ | Instituto Nacional de Enfermedades Respiratorias (INER) | Instituto Nacional de Enfermedades Respiratorias (INER) | Celia Boukadida et al |
| EPI_ISL_837726 | Mexico/CMX-INER-0129/2020 | 2020-06-02 | North America/Mexico/Mexico City/ | Instituto Nacional de Enfermedades Respiratorias (INER) | Instituto Nacional de Enfermedades Respiratorias (INER) | Celia Boukadida et al |

| | | | | | | |
|---|---|---|---|---|---|---|
| EPI_ISL_837727 | Mexico/CMX-INER-0130/2020 | 2020-06-03 | North America/Mexico/Mexico City/ | Instituto Nacional de Enfermedades Respiratorias (INER) | Instituto Nacional de Enfermedades Respiratorias (INER) | Celia Boukadida et al |
| EPI_ISL_837728 | Mexico/CMX-INER-0131/2020 | 2020-06-04 | North America/Mexico/Mexico City/ | Instituto Nacional de Enfermedades Respiratorias (INER) | Instituto Nacional de Enfermedades Respiratorias (INER) | Celia Boukadida et al |
| EPI_ISL_837729 | Mexico/CMX-INER-0132/2020 | 2020-06-06 | North America/Mexico/Mexico City/ | Instituto Nacional de Enfermedades Respiratorias (INER) | Instituto Nacional de Enfermedades Respiratorias (INER) | Celia Boukadida et al |
| EPI_ISL_837730 | Mexico/CMX-INER-0133/2020 | 2020-06-06 | North America/Mexico/Mexico City/ | Instituto Nacional de Enfermedades Respiratorias (INER) | Instituto Nacional de Enfermedades Respiratorias (INER) | Celia Boukadida et al |
| EPI_ISL_837731 | Mexico/CMX-INER-0134/2020 | 2020-06-08 | North America/Mexico/Mexico City/ | Instituto Nacional de Enfermedades Respiratorias (INER) | Instituto Nacional de Enfermedades Respiratorias (INER) | Celia Boukadida et al |
| EPI_ISL_837732 | Mexico/CMX-INER-0135/2020 | 2020-06-09 | North America/Mexico/Mexico City/ | Instituto Nacional de Enfermedades Respiratorias (INER) | Instituto Nacional de Enfermedades Respiratorias (INER) | Celia Boukadida et al |
| EPI_ISL_837733 | Mexico/CMX-INER-0136/2020 | 2020-06-09 | North America/Mexico/Mexico City/ | Instituto Nacional de Enfermedades Respiratorias (INER) | Instituto Nacional de Enfermedades Respiratorias (INER) | Celia Boukadida et al |
| EPI_ISL_837734 | Mexico/CMX-INER-0137/2020 | 2020-06-10 | North America/Mexico/Mexico City/ | Instituto Nacional de Enfermedades Respiratorias (INER) | Instituto Nacional de Enfermedades Respiratorias (INER) | Celia Boukadida et al |
| EPI_ISL_837735 | Mexico/CMX-INER-0139/2020 | 2020-06-15 | North America/Mexico/Mexico City/ | Instituto Nacional de Enfermedades Respiratorias (INER) | Instituto Nacional de Enfermedades Respiratorias (INER) | Celia Boukadida et al |
| EPI_ISL_837736 | Mexico/CMX-INER-0140/2020 | 2020-06-16 | North America/Mexico/Mexico City/ | Instituto Nacional de Enfermedades Respiratorias (INER) | Instituto Nacional de Enfermedades Respiratorias (INER) | Celia Boukadida et al |

| | | | | | | |
|---|---|---|---|---|---|---|
| EPI_ISL_837737 | Mexico/CMX-INER-0141/2020 | 2020-06-18 | North America/Mexico/Mexico City/ | Instituto Nacional de Enfermedades Respiratorias (INER) | Instituto Nacional de Enfermedades Respiratorias (INER) | Celia Boukadida et al |
| EPI_ISL_837738 | Mexico/CMX-INER-0142/2020 | 2020-06-18 | North America/Mexico/Mexico City/ | Instituto Nacional de Enfermedades Respiratorias (INER) | Instituto Nacional de Enfermedades Respiratorias (INER) | Celia Boukadida et al |
| EPI_ISL_837739 | Mexico/CMX-INER-0143/2020 | 2020-06-19 | North America/Mexico/Mexico City/ | Instituto Nacional de Enfermedades Respiratorias (INER) | Instituto Nacional de Enfermedades Respiratorias (INER) | Celia Boukadida et al |
| EPI_ISL_837740 | Mexico/CMX-INER-0144/2020 | 2020-06-19 | North America/Mexico/Mexico City/ | Instituto Nacional de Enfermedades Respiratorias (INER) | Instituto Nacional de Enfermedades Respiratorias (INER) | Celia Boukadida et al |
| EPI_ISL_837741 | Mexico/CMX-INER-0145/2020 | 2020-06-22 | North America/Mexico/Mexico City/ | Instituto Nacional de Enfermedades Respiratorias (INER) | Instituto Nacional de Enfermedades Respiratorias (INER) | Celia Boukadida et al |
| EPI_ISL_837742 | Mexico/CMX-INER-0146/2020 | 2020-06-22 | North America/Mexico/Mexico City/ | Instituto Nacional de Enfermedades Respiratorias (INER) | Instituto Nacional de Enfermedades Respiratorias (INER) | Celia Boukadida et al |
| EPI_ISL_837743 | Mexico/CMX-INER-0147/2020 | 2020-06-23 | North America/Mexico/Mexico City/ | Instituto Nacional de Enfermedades Respiratorias (INER) | Instituto Nacional de Enfermedades Respiratorias (INER) | Celia Boukadida et al |
| EPI_ISL_837744 | Mexico/CMX-INER-0148/2020 | 2020-06-24 | North America/Mexico/Mexico City/ | Instituto Nacional de Enfermedades Respiratorias (INER) | Instituto Nacional de Enfermedades Respiratorias (INER) | Celia Boukadida et al |
| EPI_ISL_837745 | Mexico/CMX-INER-0149/2020 | 2020-06-24 | North America/Mexico/Mexico City/ | Instituto Nacional de Enfermedades Respiratorias (INER) | Instituto Nacional de Enfermedades Respiratorias (INER) | Celia Boukadida et al |
| EPI_ISL_837746 | Mexico/CMX-INER-0150/2020 | 2020-06-25 | North America/Mexico/Mexico City/ | Instituto Nacional de Enfermedades Respiratorias (INER) | Instituto Nacional de Enfermedades Respiratorias (INER) | Celia Boukadida et al |

| | | | | | | |
|---|---|---|---|---|---|---|
| EPI_ISL_837747 | Mexico/CMX-INER-0151/2020 | 2020-06-27 | North America/Mexico/Mexico City/ | Instituto Nacional de Enfermedades Respiratorias (INER) | Instituto Nacional de Enfermedades Respiratorias (INER) | Celia Boukadida et al |
| EPI_ISL_837748 | Mexico/CMX-INER-0152/2020 | 2020-06-27 | North America/Mexico/Mexico City/ | Instituto Nacional de Enfermedades Respiratorias (INER) | Instituto Nacional de Enfermedades Respiratorias (INER) | Celia Boukadida et al |
| EPI_ISL_837749 | Mexico/CMX-INER-0153/2020 | 2020-06-30 | North America/Mexico/Mexico City/ | Instituto Nacional de Enfermedades Respiratorias (INER) | Instituto Nacional de Enfermedades Respiratorias (INER) | Celia Boukadida et al |
| EPI_ISL_837750 | Mexico/CMX-INER-0154/2020 | 2020-07-01 | North America/Mexico/Mexico City/ | Instituto Nacional de Enfermedades Respiratorias (INER) | Instituto Nacional de Enfermedades Respiratorias (INER) | Celia Boukadida et al |
| EPI_ISL_837751 | Mexico/CMX-INER-0155/2020 | 2020-07-02 | North America/Mexico/Mexico City/ | Instituto Nacional de Enfermedades Respiratorias (INER) | Instituto Nacional de Enfermedades Respiratorias (INER) | Celia Boukadida et al |
| EPI_ISL_837752 | Mexico/CMX-INER-0156/2020 | 2020-07-02 | North America/Mexico/Mexico City/ | Instituto Nacional de Enfermedades Respiratorias (INER) | Instituto Nacional de Enfermedades Respiratorias (INER) | Celia Boukadida et al |
| EPI_ISL_837753 | Mexico/CMX-INER-0157/2020 | 2020-07-03 | North America/Mexico/Mexico City/ | Instituto Nacional de Enfermedades Respiratorias (INER) | Instituto Nacional de Enfermedades Respiratorias (INER) | Celia Boukadida et al |
| EPI_ISL_837754 | Mexico/CMX-INER-0158/2020 | 2020-07-03 | North America/Mexico/Mexico City/ | Instituto Nacional de Enfermedades Respiratorias (INER) | Instituto Nacional de Enfermedades Respiratorias (INER) | Celia Boukadida et al |
| EPI_ISL_837755 | Mexico/CMX-INER-0159/2020 | 2020-07-05 | North America/Mexico/Mexico City/ | Instituto Nacional de Enfermedades Respiratorias (INER) | Instituto Nacional de Enfermedades Respiratorias (INER) | Celia Boukadida et al |
| EPI_ISL_837756 | Mexico/CMX-INER-0160/2020 | 2020-07-06 | North America/Mexico/Mexico City/ | Instituto Nacional de Enfermedades Respiratorias (INER) | Instituto Nacional de Enfermedades Respiratorias (INER) | Celia Boukadida et al |

| | | | | | | |
|---|---|---|---|---|---|---|
| EPI_ISL_837757 | Mexico/CMX-INER-0161/2020 | 2020-07-06 | North America/Mexico/Mexico City/ | Instituto Nacional de Enfermedades Respiratorias (INER) | Instituto Nacional de Enfermedades Respiratorias (INER) | Celia Boukadida et al |
| EPI_ISL_837758 | Mexico/CMX-INER-0162/2020 | 2020-07-07 | North America/Mexico/Mexico City/ | Instituto Nacional de Enfermedades Respiratorias (INER) | Instituto Nacional de Enfermedades Respiratorias (INER) | Celia Boukadida et al |
| EPI_ISL_837759 | Mexico/CMX-INER-0163/2020 | 2020-07-07 | North America/Mexico/Mexico City/ | Instituto Nacional de Enfermedades Respiratorias (INER) | Instituto Nacional de Enfermedades Respiratorias (INER) | Celia Boukadida et al |
| EPI_ISL_837761 | Mexico/CMX-INER-0165/2020 | 2020-07-10 | North America/Mexico/Mexico City/ | Instituto Nacional de Enfermedades Respiratorias (INER) | Instituto Nacional de Enfermedades Respiratorias (INER) | Celia Boukadida et al |
| EPI_ISL_837763 | Mexico/CMX-INER-0167/2020 | 2020-07-12 | North America/Mexico/Mexico City/ | Instituto Nacional de Enfermedades Respiratorias (INER) | Instituto Nacional de Enfermedades Respiratorias (INER) | Celia Boukadida et al |
| EPI_ISL_837764 | Mexico/CMX-INER-0168/2020 | 2020-07-13 | North America/Mexico/Mexico City/ | Instituto Nacional de Enfermedades Respiratorias (INER) | Instituto Nacional de Enfermedades Respiratorias (INER) | Celia Boukadida et al |
| EPI_ISL_837765 | Mexico/CMX-INER-0169/2020 | 2020-07-13 | North America/Mexico/Mexico City/ | Instituto Nacional de Enfermedades Respiratorias (INER) | Instituto Nacional de Enfermedades Respiratorias (INER) | Celia Boukadida et al |
| EPI_ISL_837766 | Mexico/CMX-INER-0170/2020 | 2020-07-14 | North America/Mexico/Mexico City/ | Instituto Nacional de Enfermedades Respiratorias (INER) | Instituto Nacional de Enfermedades Respiratorias (INER) | Celia Boukadida et al |
| EPI_ISL_837767 | Mexico/CMX-INER-0171/2020 | 2020-07-14 | North America/Mexico/Mexico City/ | Instituto Nacional de Enfermedades Respiratorias (INER) | Instituto Nacional de Enfermedades Respiratorias (INER) | Celia Boukadida et al |
| EPI_ISL_837768 | Mexico/CMX-INER-0172/2020 | 2020-07-15 | North America/Mexico/Mexico City/ | Instituto Nacional de Enfermedades Respiratorias (INER) | Instituto Nacional de Enfermedades Respiratorias (INER) | Celia Boukadida et al |

| | | | | | | |
|---|---|---|---|---|---|---|
| EPI_ISL_837769 | Mexico/CMX-INER-0173/2020 | 2020-07-15 | North America/Mexico/Mexico City/ | Instituto Nacional de Enfermedades Respiratorias (INER) | Instituto Nacional de Enfermedades Respiratorias (INER) | Celia Boukadida et al |
| EPI_ISL_837770 | Mexico/CMX-INER-0174/2020 | 2020-07-16 | North America/Mexico/Mexico City/ | Instituto Nacional de Enfermedades Respiratorias (INER) | Instituto Nacional de Enfermedades Respiratorias (INER) | Celia Boukadida et al |
| EPI_ISL_837772 | Mexico/CMX-INER-0176/2020 | 2020-07-17 | North America/Mexico/Mexico City/ | Instituto Nacional de Enfermedades Respiratorias (INER) | Instituto Nacional de Enfermedades Respiratorias (INER) | Celia Boukadida et al |
| EPI_ISL_837773 | Mexico/CMX-INER-0177/2020 | 2020-07-17 | North America/Mexico/Mexico City/ | Instituto Nacional de Enfermedades Respiratorias (INER) | Instituto Nacional de Enfermedades Respiratorias (INER) | Celia Boukadida et al |
| EPI_ISL_837774 | Mexico/CMX-INER-0178/2020 | 2020-07-18 | North America/Mexico/Mexico City/ | Instituto Nacional de Enfermedades Respiratorias (INER) | Instituto Nacional de Enfermedades Respiratorias (INER) | Celia Boukadida et al |
| EPI_ISL_837775 | Mexico/CMX-INER-0179/2020 | 2020-07-18 | North America/Mexico/Mexico City/ | Instituto Nacional de Enfermedades Respiratorias (INER) | Instituto Nacional de Enfermedades Respiratorias (INER) | Celia Boukadida et al |
| EPI_ISL_837776 | Mexico/CMX-INER-0180/2020 | 2020-07-19 | North America/Mexico/Mexico City/ | Instituto Nacional de Enfermedades Respiratorias (INER) | Instituto Nacional de Enfermedades Respiratorias (INER) | Celia Boukadida et al |
| EPI_ISL_837777 | Mexico/CMX-INER-0181/2020 | 2020-07-20 | North America/Mexico/Mexico City/ | Instituto Nacional de Enfermedades Respiratorias (INER) | Instituto Nacional de Enfermedades Respiratorias (INER) | Celia Boukadida et al |
| EPI_ISL_837778 | Mexico/CMX-INER-0182/2020 | 2020-07-20 | North America/Mexico/Mexico City/ | Instituto Nacional de Enfermedades Respiratorias (INER) | Instituto Nacional de Enfermedades Respiratorias (INER) | Celia Boukadida et al |
| EPI_ISL_837779 | Mexico/CMX-INER-0183/2020 | 2020-07-21 | North America/Mexico/Mexico City/ | Instituto Nacional de Enfermedades Respiratorias (INER) | Instituto Nacional de Enfermedades Respiratorias (INER) | Celia Boukadida et al |

| | | | | | | |
|---|---|---|---|---|---|---|
| EPI_ISL_837780 | Mexico/CMX-INER-0184/2020 | 2020-07-21 | North America/Mexico/Mexico City/ | Instituto Nacional de Enfermedades Respiratorias (INER) | Instituto Nacional de Enfermedades Respiratorias (INER) | Celia Boukadida et al |
| EPI_ISL_837781 | Mexico/CMX-INER-0185/2020 | 2020-07-23 | North America/Mexico/Mexico City/ | Instituto Nacional de Enfermedades Respiratorias (INER) | Instituto Nacional de Enfermedades Respiratorias (INER) | Celia Boukadida et al |
| EPI_ISL_837782 | Mexico/CMX-INER-0186/2020 | 2020-07-24 | North America/Mexico/Mexico City/ | Instituto Nacional de Enfermedades Respiratorias (INER) | Instituto Nacional de Enfermedades Respiratorias (INER) | Celia Boukadida et al |
| EPI_ISL_837783 | Mexico/CMX-INER-0187/2020 | 2020-07-24 | North America/Mexico/Mexico City/ | Instituto Nacional de Enfermedades Respiratorias (INER) | Instituto Nacional de Enfermedades Respiratorias (INER) | Celia Boukadida et al |
| EPI_ISL_837785 | Mexico/CMX-INER-0189/2020 | 2020-07-27 | North America/Mexico/Mexico City/ | Instituto Nacional de Enfermedades Respiratorias (INER) | Instituto Nacional de Enfermedades Respiratorias (INER) | Celia Boukadida et al |
| EPI_ISL_837786 | Mexico/CMX-INER-0190/2020 | 2020-07-29 | North America/Mexico/Mexico City/ | Instituto Nacional de Enfermedades Respiratorias (INER) | Instituto Nacional de Enfermedades Respiratorias (INER) | Celia Boukadida et al |
| EPI_ISL_837787 | Mexico/CMX-INER-0191/2020 | 2020-07-29 | North America/Mexico/Mexico City/ | Instituto Nacional de Enfermedades Respiratorias (INER) | Instituto Nacional de Enfermedades Respiratorias (INER) | Celia Boukadida et al |
| EPI_ISL_837788 | Mexico/CMX-INER-0192/2020 | 2020-07-30 | North America/Mexico/Mexico City/ | Instituto Nacional de Enfermedades Respiratorias (INER) | Instituto Nacional de Enfermedades Respiratorias (INER) | Celia Boukadida et al |
| EPI_ISL_837789 | Mexico/CMX-INER-0193/2020 | 2020-07-31 | North America/Mexico/Mexico City/ | Instituto Nacional de Enfermedades Respiratorias (INER) | Instituto Nacional de Enfermedades Respiratorias (INER) | Celia Boukadida et al |
| EPI_ISL_837792 | Mexico/CMX-INER-0196/2020 | 2020-08-03 | North America/Mexico/Mexico City/ | Instituto Nacional de Enfermedades Respiratorias (INER) | Instituto Nacional de Enfermedades Respiratorias (INER) | Celia Boukadida et al |

| | | | | | | |
|---|---|---|---|---|---|---|
| EPI_ISL_837793 | Mexico/CMX-INER-0198/2020 | 2020-08-06 | North America/Mexico/Mexico City/ | Instituto Nacional de Enfermedades Respiratorias (INER) | Instituto Nacional de Enfermedades Respiratorias (INER) | Celia Boukadida et al |
| EPI_ISL_837794 | Mexico/CMX-INER-0199/2020 | 2020-08-08 | North America/Mexico/Mexico City/ | Instituto Nacional de Enfermedades Respiratorias (INER) | Instituto Nacional de Enfermedades Respiratorias (INER) | Celia Boukadida et al |
| EPI_ISL_837796 | Mexico/CMX-INER-0201/2020 | 2020-08-10 | North America/Mexico/Mexico City/ | Instituto Nacional de Enfermedades Respiratorias (INER) | Instituto Nacional de Enfermedades Respiratorias (INER) | Celia Boukadida et al |
| EPI_ISL_837797 | Mexico/CMX-INER-0202/2020 | 2020-08-10 | North America/Mexico/Mexico City/ | Instituto Nacional de Enfermedades Respiratorias (INER) | Instituto Nacional de Enfermedades Respiratorias (INER) | Celia Boukadida et al |
| EPI_ISL_837798 | Mexico/CMX-INER-0203/2020 | 2020-08-11 | North America/Mexico/Mexico City/ | Instituto Nacional de Enfermedades Respiratorias (INER) | Instituto Nacional de Enfermedades Respiratorias (INER) | Celia Boukadida et al |
| EPI_ISL_837799 | Mexico/CMX-INER-0204/2020 | 2020-08-11 | North America/Mexico/Mexico City/ | Instituto Nacional de Enfermedades Respiratorias (INER) | Instituto Nacional de Enfermedades Respiratorias (INER) | Celia Boukadida et al |
| EPI_ISL_837800 | Mexico/CMX-INER-0206/2020 | 2020-08-13 | North America/Mexico/Mexico City/ | Instituto Nacional de Enfermedades Respiratorias (INER) | Instituto Nacional de Enfermedades Respiratorias (INER) | Celia Boukadida et al |
| EPI_ISL_837801 | Mexico/CMX-INER-0207/2020 | 2020-08-19 | North America/Mexico/Mexico City/ | Instituto Nacional de Enfermedades Respiratorias (INER) | Instituto Nacional de Enfermedades Respiratorias (INER) | Celia Boukadida et al |
| EPI_ISL_837802 | Mexico/CMX-INER-0208/2020 | 2020-08-20 | North America/Mexico/Mexico City/ | Instituto Nacional de Enfermedades Respiratorias (INER) | Instituto Nacional de Enfermedades Respiratorias (INER) | Celia Boukadida et al |
| EPI_ISL_837803 | Mexico/CMX-INER-0209/2020 | 2020-08-20 | North America/Mexico/Mexico City/ | Instituto Nacional de Enfermedades Respiratorias (INER) | Instituto Nacional de Enfermedades Respiratorias (INER) | Celia Boukadida et al |

| | | | | | | |
|---|---|---|---|---|---|---|
| EPI_ISL_837804 | Mexico/CMX-INER-0210/2020 | 2020-08-20 | North America/Mexico/Mexico City/ | Instituto Nacional de Enfermedades Respiratorias (INER) | Instituto Nacional de Enfermedades Respiratorias (INER) | Celia Boukadida et al |
| EPI_ISL_837805 | Mexico/CMX-INER-0211/2020 | 2020-08-21 | North America/Mexico/Mexico City/ | Instituto Nacional de Enfermedades Respiratorias (INER) | Instituto Nacional de Enfermedades Respiratorias (INER) | Celia Boukadida et al |
| EPI_ISL_837806 | Mexico/CMX-INER-0212/2020 | 2020-08-22 | North America/Mexico/Mexico City/ | Instituto Nacional de Enfermedades Respiratorias (INER) | Instituto Nacional de Enfermedades Respiratorias (INER) | Celia Boukadida et al |
| EPI_ISL_837808 | Mexico/CMX-INER-0215/2020 | 2020-08-25 | North America/Mexico/Mexico City/ | Instituto Nacional de Enfermedades Respiratorias (INER) | Instituto Nacional de Enfermedades Respiratorias (INER) | Celia Boukadida et al |
| EPI_ISL_837809 | Mexico/CMX-INER-0216/2020 | 2020-08-27 | North America/Mexico/Mexico City/ | Instituto Nacional de Enfermedades Respiratorias (INER) | Instituto Nacional de Enfermedades Respiratorias (INER) | Celia Boukadida et al |
| EPI_ISL_837810 | Mexico/CMX-INER-0218/2020 | 2020-09-02 | North America/Mexico/Mexico City/ | Instituto Nacional de Enfermedades Respiratorias (INER) | Instituto Nacional de Enfermedades Respiratorias (INER) | Celia Boukadida et al |
| EPI_ISL_837811 | Mexico/CMX-INER-0219/2020 | 2020-09-02 | North America/Mexico/Mexico City/ | Instituto Nacional de Enfermedades Respiratorias (INER) | Instituto Nacional de Enfermedades Respiratorias (INER) | Celia Boukadida et al |
| EPI_ISL_837812 | Mexico/CMX-INER-0220/2020 | 2020-09-03 | North America/Mexico/Mexico City/ | Instituto Nacional de Enfermedades Respiratorias (INER) | Instituto Nacional de Enfermedades Respiratorias (INER) | Celia Boukadida et al |
| EPI_ISL_837814 | Mexico/CMX-INER-0222/2020 | 2020-09-03 | North America/Mexico/Mexico City/ | Instituto Nacional de Enfermedades Respiratorias (INER) | Instituto Nacional de Enfermedades Respiratorias (INER) | Celia Boukadida et al |
| EPI_ISL_837859 | England/CAMB-A9A344/2021 | 2021-01-03 | Europe/United Kingdom/England/ | Department of Pathology, University of Cambridge | COVID-19 Genomics UK (COG-UK) Consortium | Aminu S. Jahun et al |

| | | | | | | |
|---|---|---|---|---|---|---|
| EPI_ISL_838016 | England/CAMB-A9BA7B/2021 | 2021-01-10 | Europe/United Kingdom/England/ | Department of Pathology, University of Cambridge | COVID-19 Genomics UK (COG-UK) Consortium | Aminu S. Jahun et al |
| EPI_ISL_838019 | England/CAMB-A9BB0F/2021 | 2021-01-11 | Europe/United Kingdom/England/ | Department of Pathology, University of Cambridge | COVID-19 Genomics UK (COG-UK) Consortium | Aminu S. Jahun et al |
| EPI_ISL_838028 | England/CAMB-A9BC48/2021 | 2021-01-11 | Europe/United Kingdom/England/ | Department of Pathology, University of Cambridge | COVID-19 Genomics UK (COG-UK) Consortium | Aminu S. Jahun et al |
| EPI_ISL_838038 | England/CAMB-A9BD81/2021 | 2021-01-12 | Europe/United Kingdom/England/ | Department of Pathology, University of Cambridge | COVID-19 Genomics UK (COG-UK) Consortium | Aminu S. Jahun et al |
| EPI_ISL_838062 | Scotland/CVR6056/2020 | 2020-11-25 | Europe/United Kingdom/Scotland/ | West of Scotland Specialist Virology Centre, NHSGGC / MRC-University of Glasgow Centre for Virus Research | COVID-19 Genomics UK (COG-UK) Consortium | Ana da Silva Filipe et al |
| EPI_ISL_838102 | Scotland/CVR6203/2020 | 2020-12-06 | Europe/United Kingdom/Scotland/ | West of Scotland Specialist Virology Centre, NHSGGC / MRC-University of Glasgow Centre for Virus Research | COVID-19 Genomics UK (COG-UK) Consortium | Ana da Silva Filipe et al |
| EPI_ISL_838558 | England/LIVE-DCF48C/2020 | 2020-12-16 | Europe/United Kingdom/England/ | Liverpool Clinical Laboratories | COVID-19 Genomics UK (COG-UK) Consortium | Sam Haldenby et al |
| EPI_ISL_838728 | England/LOND-12691A3/2020 | 2020-12-14 | Europe/United Kingdom/England/ | University College London, Great Ormond Street Hospital for Children NHS Foundation Trust, Imperial College Healthcare NHS Trust | COVID-19 Genomics UK (COG-UK) Consortium | Sergi Castellano et al |
| EPI_ISL_838771 | England/LOND-126D840/2020 | 2020-12-29 | Europe/United Kingdom/England/ | University College London, Great Ormond Street Hospital for Children NHS Foundation Trust, Imperial College Healthcare NHS Trust | COVID-19 Genomics UK (COG-UK) Consortium | Sergi Castellano et al |
| EPI_ISL_838794 | England/LOND-126E26F/2020 | 2020-12-28 | Europe/United Kingdom/England/ | University College London, Great Ormond Street Hospital for Children NHS Foundation Trust, Imperial College Healthcare NHS Trust | COVID-19 Genomics UK (COG-UK) Consortium | Sergi Castellano et al |
| EPI_ISL_838875 | England/LOND-1271348/2021 | 2021-01-04 | Europe/United Kingdom/England/ | University College London, Great Ormond Street Hospital for Children NHS Foundation Trust, Imperial College Healthcare NHS Trust | COVID-19 Genomics UK (COG-UK) Consortium | Sergi Castellano et al |

| | | | | | | |
|---|---|---|---|---|---|---|
| EPI_ISL_838983 | England/LOND-12EB947/2021 | 2021-01-08 | Europe/United Kingdom/England/ | University College London, Great Ormond Street Hospital for Children NHS Foundation Trust, Imperial College Healthcare NHS Trust | COVID-19 Genomics UK (COG-UK) Consortium | Sergi Castellano et al |
| EPI_ISL_839045 | England/LOND-12F351B/2021 | 2021-01-03 | Europe/United Kingdom/England/ | University College London, Great Ormond Street Hospital for Children NHS Foundation Trust, Imperial College Healthcare NHS Trust | COVID-19 Genomics UK (COG-UK) Consortium | Sergi Castellano et al |
| EPI_ISL_839409 | England/MILK-B5A0EE/2020 | 2020-11-06 | Europe/United Kingdom/England/ | Oxford Viromics, NDM, University of Oxford; Oxford University Hospitals; Basingstoke and North Hampshire Hospital | COVID-19 Genomics UK (COG-UK) Consortium | Tanya Golubchik et al |
| EPI_ISL_839462 | England/MILK-B5AC53/2020 | 2020-11-06 | Europe/United Kingdom/England/ | Oxford Viromics, NDM, University of Oxford; Oxford University Hospitals; Basingstoke and North Hampshire Hospital | COVID-19 Genomics UK (COG-UK) Consortium | Tanya Golubchik et al |
| EPI_ISL_839768 | England/NORT-2888D9/2020 | 2020-12-12 | Europe/United Kingdom/England/ | Northumbria University / South Tees Hospitals NHS Foundation Trust / North Cumbria Integrated Care NHS Foundation Trust / North Tees and Hartlepool NHS Foundation Trust / Newcastle Hospitals NHS Foundation Trust | COVID-19 Genomics UK (COG-UK) Consortium | Darren L Smith et al |
| EPI_ISL_839828 | England/NORW-2233B1/2021 | 2021-01-02 | Europe/United Kingdom/England/ | Quadram Institute Bioscience | COVID-19 Genomics UK (COG-UK) Consortium | Dave J. Baker et al |
| EPI_ISL_839830 | England/NORW-2233DF/2021 | 2021-01-02 | Europe/United Kingdom/England/ | Quadram Institute Bioscience | COVID-19 Genomics UK (COG-UK) Consortium | Dave J. Baker et al |
| EPI_ISL_840036 | England/NOTT-246BED/2020 | 2020-12-05 | Europe/United Kingdom/England/ | Lincolnshire Hospitals and DeepSeq Nottingham | COVID-19 Genomics UK (COG-UK) Consortium | Nichola Duckworth et al |
| EPI_ISL_840475 | Wales/PHWC-4B0CCB/2020 | 2020-12-24 | Europe/United Kingdom/Wales/ | Originating lab: Wales Specialist Virology Centre Sequencing lab: Pathogen Genomics Unit | Public Health Wales Microbiology Cardiff Wales Specialist Virology Centre | Catherine Moore et al |
| EPI_ISL_840593 | Wales/PHWC-4B1574/2020 | 2020-12-28 | Europe/United Kingdom/Wales/ | Originating lab: Wales Specialist Virology Centre Sequencing lab: Pathogen Genomics Unit | Public Health Wales Microbiology Cardiff Wales Specialist Virology Centre | Catherine Moore et al |

| | | | | | | |
|---|---|---|---|---|---|---|
| EPI_ISL_840720 | Wales/PHWC-4B226D/2020 | 2020-12-19 | Europe/United Kingdom/Wales/ | Originating lab: Wales Specialist Virology Centre Sequencing lab: Pathogen Genomics Unit | Public Health Wales Microbiology Cardiff Wales Specialist Virology Centre | Catherine Moore et al |
| EPI_ISL_840804 | Wales/PHWC-4B2968/2020 | 2020-12-31 | Europe/United Kingdom/Wales/ | Originating lab: Wales Specialist Virology Centre Sequencing lab: Pathogen Genomics Unit | Public Health Wales Microbiology Cardiff Wales Specialist Virology Centre | Catherine Moore et al |
| EPI_ISL_841072 | Wales/PHWC-4B41D7/2021 | 2021-01-06 | Europe/United Kingdom/Wales/ | Wales Specialist Virology Centre Sequencing lab: Pathogen Genomics Unit | Public Health Wales Microbiology Cardiff Wales Specialist Virology Centre | Catherine Moore et al |
| EPI_ISL_841122 | Wales/PHWC-4B4562/2021 | 2021-01-02 | Europe/United Kingdom/Wales/ | Wales Specialist Virology Centre Sequencing lab: Pathogen Genomics Unit | Public Health Wales Microbiology Cardiff Wales Specialist Virology Centre | Catherine Moore et al |
| EPI_ISL_841271 | Wales/PHWC-4B5033/2021 | 2021-01-06 | Europe/United Kingdom/Wales/ | Wales Specialist Virology Centre Sequencing lab: Pathogen Genomics Unit | Public Health Wales Microbiology Cardiff Wales Specialist Virology Centre | Catherine Moore et al |
| EPI_ISL_841320 | Wales/PHWC-4B53EF/2021 | 2021-01-06 | Europe/United Kingdom/Wales/ | Originating lab: Wales Specialist Virology Centre Sequencing lab: Pathogen Genomics Unit | Public Health Wales Microbiology Cardiff Wales Specialist Virology Centre | Catherine Moore et al |

| | | | | | | |
|---|---|---|---|---|---|---|
| EPI_ISL_841396 | Wales/PHWC-4B5B50/2021 | 2021-01-05 | Europe/United Kingdom/Wales/ | Originating lab: Wales Specialist Virology Centre Sequencing lab: Pathogen Genomics Unit | Public Health Wales Microbiology Cardiff Wales Specialist Virology Centre | Catherine Moore et al |
| EPI_ISL_841540 | Wales/PHWC-4B664F/2021 | 2021-01-10 | Europe/United Kingdom/Wales/ | Originating lab: Wales Specialist Virology Centre Sequencing lab: Pathogen Genomics Unit | Public Health Wales Microbiology Cardiff Wales Specialist Virology Centre | Catherine Moore et al |
| EPI_ISL_841561 | Wales/PHWC-4B6797/2020 | 2020-11-26 | Europe/United Kingdom/Wales/ | Originating lab: Wales Specialist Virology Centre Sequencing lab: Pathogen Genomics Unit | Public Health Wales Microbiology Cardiff Wales Specialist Virology Centre | Catherine Moore et al |
| EPI_ISL_842159 | England/QEUH-A627B2/2020 | 2020-10-12 | Europe/United Kingdom/England/ | Oxford Viromics, NDM, University of Oxford; Oxford University Hospitals; Basingstoke and North Hampshire Hospital | COVID-19 Genomics UK (COG-UK) Consortium | Tanya Golubchik et al |
| EPI_ISL_842353 | England/SHEF-CE3E7/2020 | 2020-11-02 | Europe/United Kingdom/England/ | Virology Department, Sheffield Teaching Hospitals NHS Foundation Trust/Department of Infection, Immunity and Cardiovascular Disease, The Medical School, University of Sheffield | COVID-19 Genomics UK (COG-UK) Consortium | Thushan de Silva et al |
| EPI_ISL_842511 | England/TFCI-26FDB18/2020 | 2020-12-30 | Europe/United Kingdom/England/ | Bioinformatics and Biostatistics Lab, Advanced Sequencing Facility | COVID-19 Genomics UK (COG-UK) Consortium | Aengus Stewart et al |
| EPI_ISL_842609 | USA/WY-WPHL-20171581/2020 | 2020-12-28 | North America/USA/Wyoming/ | Wyoming Public Health Laboratory | Wyoming Public Health Laboratory | Noah Hull et al |
| EPI_ISL_842612 | USA/WY-WPHL-20172187/2020 | 2020-12-30 | North America/USA/Wyoming/ | Wyoming Public Health Laboratory | Wyoming Public Health Laboratory | Noah Hull et al |
| EPI_ISL_842614 | USA/WY-WPHL-20171798/2020 | 2020-12-29 | North America/USA/Wyoming/ | Wyoming Public Health Laboratory | Wyoming Public Health Laboratory | Noah Hull et al |
| EPI_ISL_842618 | USA/WY-WPHL-20171791/2020 | 2020-12-29 | North America/USA/Wyoming/ | Wyoming Public Health Laboratory | Wyoming Public Health Laboratory | Noah Hull et al |
| EPI_ISL_842845 | England/LOND-1287084/2020 | 2020-09-21 | Europe/United Kingdom/England/ | Barts Health NHS Trust | COVID-19 Genomics UK (COG-UK) Consortium | CUTINO-MOGUEL et al |

| | | | | | | |
|---|---|---|---|---|---|---|
| EPI_ISL_842954 | England/LOND-12878E6/2020 | 2020-11-25 | Europe/United Kingdom/England/ | Barts Health NHS Trust | COVID-19 Genomics UK (COG-UK) Consortium | CUTINO-MOGUEL et al |
| EPI_ISL_843118 | England/LOND-12886AF/2020 | 2020-12-18 | Europe/United Kingdom/England/ | Barts Health NHS Trust | COVID-19 Genomics UK (COG-UK) Consortium | CUTINO-MOGUEL et al |
| EPI_ISL_843158 | NorthernIreland/NIRE-235B5A/2020 | 2020-12-30 | Europe/United Kingdom/Northern Irelar | Regional Virus Laboratory, Belfast Health and Social Care Trust | COVID-19 Genomics UK (COG-UK) Consortium | Conall McCaughey et al |
| EPI_ISL_843175 | USA/MD-MDH-0608/2021 | 2021-01-07 | North America/USA/Maryland/ | Maryland Public Health Laboratory | Maryland Public Health Laboratory | Maryland Department of Health Laboratories Administration et al |
| EPI_ISL_843186 | USA/MD-MDH-0619/2021 | 2021-01-10 | North America/USA/Maryland/ | Maryland Public Health Laboratory | Maryland Public Health Laboratory | Maryland Department of Health Laboratories Administration et al |
| EPI_ISL_843190 | USA/MD-MDH-0623/2020 | 2020-12-09 | North America/USA/Maryland/ | Maryland Public Health Laboratory | Maryland Public Health Laboratory | Maryland Department of Health Laboratories Administration et al |
| EPI_ISL_843191 | Mexico/CMX-INER-0116/2020 | 2020-05-27 | North America/Mexico/Mexico City/ | Instituto Nacional de Enfermedades Respiratorias (INER) | Instituto Nacional de Enfermedades Respiratorias (INER) | Celia Boukadida et al |
| EPI_ISL_843192 | Mexico/CMX-INER-0205/2020 | 2020-08-13 | North America/Mexico/Mexico City/ | Instituto Nacional de Enfermedades Respiratorias (INER) | Instituto Nacional de Enfermedades Respiratorias (INER) | Celia Boukadida et al |
| EPI_ISL_843797 | Denmark/DCGC-29750/2020 | 2020-12-28 | Europe/Denmark/Sjaelland/ | Department of Virus and Microbiological Special Diagnostics, Statens Serum Institut, Copenhagen, Denmark | Albertsen Lab, Department of Chemistry and Bioscience, Aalborg University, Denmark | Danish Covid-19 Genome Consortium et al |
| EPI_ISL_843990 | Denmark/DCGC-29943/2020 | 2020-12-28 | Europe/Denmark/Syddanmark/ | Department of Virus and Microbiological Special Diagnostics, Statens Serum Institut, Copenhagen, Denmark | Albertsen Lab, Department of Chemistry and Bioscience, Aalborg University, Denmark | Danish Covid-19 Genome Consortium et al |

| | | | | | | |
|---|---|---|---|---|---|---|
| EPI_ISL_844143 | Denmark/DCGC-30096/2020 | 2020-12-28 | Europe/Denmark/Hovedstaden/ | Department of Virus and Microbiological Special Diagnostics, Statens Serum Institut, Copenhagen, Denmark | Albertsen Lab, Department of Chemistry and Bioscience, Aalborg University, Denmark | Danish Covid-19 Genome Consortium et al |
| EPI_ISL_845543 | USA/CA-SC210028/2020 | 2020-12-30 | North America/USA/California/Santa Cl | Santa Clara County Public Health Laboratory | Santa Clara County Public Health Laboratory | Santa Clara County Public Health Department et al |
| EPI_ISL_845546 | Cameroon/CV408/2020 | 2020-06-09 | Africa/Cameroon/YaoundV©/ | National Public Health Laboratory, Cameroon | African Centre of Excellence for Genomics of Infectious Diseases (ACEGID), Redeemer's University | Oluniyi P.E. et al et al |
| EPI_ISL_845548 | Cameroon/CV406/2020 | 2020-06-12 | Africa/Cameroon/YaoundV©/ | National Public Health Laboratory, Cameroon | African Centre of Excellence for Genomics of Infectious Diseases (ACEGID), Redeemer's University | Oluniyi P.E. et al et al |
| EPI_ISL_845549 | Cameroon/CV411/2020 | 2020-06-16 | Africa/Cameroon/YaoundV©/ | National Public Health Laboratory, Cameroon | African Centre of Excellence for Genomics of Infectious Diseases (ACEGID), Redeemer's University | Oluniyi P.E. et al et al |

| | | | | | | |
|---|---|---|---|---|---|---|
| EPI_ISL_845551 | Cameroon/CV416/2020 | 2020-08-13 | Africa/Cameroon/YaoundV©/ | National Public Health Laboratory, Cameroon | African Centre of Excellence for Genomics of Infectious Diseases (ACEGID), Redeemer's University | Oluniyi P.E. et al et al |
| EPI_ISL_845554 | Cameroon/CV424/2020 | 2020-10-26 | Africa/Cameroon/YaoundV©/ | National Public Health Laboratory, Cameroon | African Centre of Excellence for Genomics of Infectious Diseases (ACEGID), Redeemer's University | Oluniyi P.E. et al et al |
| EPI_ISL_845562 | Cameroon/CV435/2020 | 2020-11-19 | Africa/Cameroon/YaoundV©/ | National Public Health Laboratory, Cameroon | African Centre of Excellence for Genomics of Infectious Diseases (ACEGID), Redeemer's University | Oluniyi P.E. et al et al |
| EPI_ISL_845564 | Cameroon/CV428/2020 | 2020-12-03 | Africa/Cameroon/YaoundV©/ | National Public Health Laboratory, Cameroon | African Centre of Excellence for Genomics of Infectious Diseases (ACEGID), Redeemer's University | Oluniyi P.E. et al et al |
| EPI_ISL_845568 | Belgium/rega-1571/2020 | 2020-12-20 | Europe/Belgium/Oostende/ | KU Leuven, Rega Institute, Clinical and Epidemiological Virology | KU Leuven, Rega Institute, Clinical and Epidemiological Virology | Tony Wawina-Bokalanga et al |

| | | | | | | |
|---|---|---|---|---|---|---|
| EPI_ISL_845593 | Belgium/rega-1672/2020 | 2020-12-31 | Europe/Belgium/Namur/Dhuy | KU Leuven, Rega Institute, Clinical and Epidemiological Virology | KU Leuven, Rega Institute, Clinical and Epidemiological Virology | Tony Wawina-Bokalanga et al |
| EPI_ISL_845640 | Colombia/BOG-INS-VG-264/2021 | 2021-01-10 | South America/Colombia/Bogota/ | Laboratorio de Salud P√∫blica - Secretar√≠a Distrital de Salud | Instituto Nacional de Salud - Direcci√≥n de Investigaci√≥n en Salud P√∫blica | Katherine Laiton-Donato et al |
| EPI_ISL_845662 | USA/FL-QDX-2888/2020 | 2020-08-04 | North America/USA/Florida/ | Quest Diagnostics | Quest Diagnostics | Rosenthal et al |
| EPI_ISL_845676 | USA/TX-QDX-2890/2020 | 2020-08-13 | North America/USA/Texas/ | Quest Diagnostics | Quest Diagnostics | Rosenthal et al |
| EPI_ISL_845679 | USA/TX-QDX-2904/2020 | 2020-10-09 | North America/USA/Texas/ | Quest Diagnostics | Quest Diagnostics | Rosenthal et al |
| EPI_ISL_845680 | Canada/ON-MMU-S2050/2020 | 20-Dec | North America/Canada/Ontario/ | Ontario's COVID-19 Genomics Rapid Response Coalition | McMaster University | Allison McGeer et al |
| EPI_ISL_845681 | Canada/ON-MMU-S2052/2020 | 20-Dec | North America/Canada/Ontario/ | Ontario's COVID-19 Genomics Rapid Response Coalition | McMaster University | Allison McGeer et al |
| EPI_ISL_845682 | Canada/ON-MMU-S2086/2020 | 20-Dec | North America/Canada/Ontario/ | Ontario's COVID-19 Genomics Rapid Response Coalition | McMaster University | Allison McGeer et al |
| EPI_ISL_845683 | Canada/ON-MMU-S2087/2020 | 20-Dec | North America/Canada/Ontario/ | Ontario's COVID-19 Genomics Rapid Response Coalition | McMaster University | Allison McGeer et al |
| EPI_ISL_845684 | Canada/ON-MMU-S2088/2020 | 20-Dec | North America/Canada/Ontario/ | Ontario's COVID-19 Genomics Rapid Response Coalition | McMaster University | Allison McGeer et al |
| EPI_ISL_845685 | Canada/ON-MMU-S2089/2020 | 20-Dec | North America/Canada/Ontario/ | Ontario's COVID-19 Genomics Rapid Response Coalition | McMaster University | Allison McGeer et al |
| EPI_ISL_845686 | Canada/ON-MMU-S2090/2020 | 20-Dec | North America/Canada/Ontario/ | Ontario's COVID-19 Genomics Rapid Response Coalition | McMaster University | Allison McGeer et al |
| EPI_ISL_845687 | Canada/ON-MMU-S2091/2020 | 20-Dec | North America/Canada/Ontario/ | Ontario's COVID-19 Genomics Rapid Response Coalition | McMaster University | Allison McGeer et al |
| EPI_ISL_845688 | Canada/ON-MMU-S2092/2020 | 20-Dec | North America/Canada/Ontario/ | Ontario's COVID-19 Genomics Rapid Response Coalition | McMaster University | Allison McGeer et al |
| EPI_ISL_845689 | Canada/ON-MMU-S2093/2020 | 20-Dec | North America/Canada/Ontario/ | Ontario's COVID-19 Genomics Rapid Response Coalition | McMaster University | Allison McGeer et al |
| EPI_ISL_845690 | Canada/ON-MMU-S2094/2020 | 20-Dec | North America/Canada/Ontario/ | Ontario's COVID-19 Genomics Rapid Response Coalition | McMaster University | Allison McGeer et al |
| EPI_ISL_845691 | Canada/ON-MMU-S2095/2020 | 20-Dec | North America/Canada/Ontario/ | Ontario's COVID-19 Genomics Rapid Response Coalition | McMaster University | Allison McGeer et al |
| EPI_ISL_845692 | Canada/ON-MMU-S2096/2020 | 20-Dec | North America/Canada/Ontario/ | Ontario's COVID-19 Genomics Rapid Response Coalition | McMaster University | Allison McGeer et al |

| | | | | | | |
|---|---|---|---|---|---|---|
| EPI_ISL_845693 | Canada/ON-MMU-S2097/2020 | 20-Dec | North America/Canada/Ontario/ | Ontario's COVID-19 Genomics Rapid Response Coalition | McMaster University | Allison McGeer et al |
| EPI_ISL_845694 | Canada/ON-MMU-S2098/2020 | 20-Dec | North America/Canada/Ontario/ | Ontario's COVID-19 Genomics Rapid Response Coalition | McMaster University | Allison McGeer et al |
| EPI_ISL_845695 | Canada/ON-MMU-S2099/2020 | 20-Dec | North America/Canada/Ontario/ | Ontario's COVID-19 Genomics Rapid Response Coalition | McMaster University | Allison McGeer et al |
| EPI_ISL_845696 | Canada/ON-MMU-S2100/2020 | 20-Dec | North America/Canada/Ontario/ | Ontario's COVID-19 Genomics Rapid Response Coalition | McMaster University | Allison McGeer et al |
| EPI_ISL_845697 | Canada/ON-MMU-S2101/2020 | 20-Dec | North America/Canada/Ontario/ | Ontario's COVID-19 Genomics Rapid Response Coalition | McMaster University | Allison McGeer et al |
| EPI_ISL_845698 | Canada/ON-MMU-S2102/2020 | 20-Dec | North America/Canada/Ontario/ | Ontario's COVID-19 Genomics Rapid Response Coalition | McMaster University | Allison McGeer et al |
| EPI_ISL_845699 | Canada/ON-MMU-S2103/2020 | 20-Dec | North America/Canada/Ontario/ | Ontario's COVID-19 Genomics Rapid Response Coalition | McMaster University | Allison McGeer et al |
| EPI_ISL_845700 | Canada/ON-MMU-S2104/2020 | 20-Dec | North America/Canada/Ontario/ | Ontario's COVID-19 Genomics Rapid Response Coalition | McMaster University | Allison McGeer et al |
| EPI_ISL_845701 | Canada/ON-MMU-S2105/2020 | 20-Dec | North America/Canada/Ontario/ | Ontario's COVID-19 Genomics Rapid Response Coalition | McMaster University | Allison McGeer et al |
| EPI_ISL_845702 | Canada/ON-MMU-S2106/2020 | 20-Dec | North America/Canada/Ontario/ | Ontario's COVID-19 Genomics Rapid Response Coalition | McMaster University | Allison McGeer et al |
| EPI_ISL_845703 | Canada/ON-MMU-S2107/2020 | 20-Dec | North America/Canada/Ontario/ | Ontario's COVID-19 Genomics Rapid Response Coalition | McMaster University | Allison McGeer et al |
| EPI_ISL_845704 | Canada/ON-MMU-S2108/2020 | 20-Dec | North America/Canada/Ontario/ | Ontario's COVID-19 Genomics Rapid Response Coalition | McMaster University | Allison McGeer et al |
| EPI_ISL_845705 | Canada/ON-MMU-S2109/2020 | 20-Dec | North America/Canada/Ontario/ | Ontario's COVID-19 Genomics Rapid Response Coalition | McMaster University | Allison McGeer et al |
| EPI_ISL_845706 | Canada/ON-MMU-S2110/2020 | 20-Dec | North America/Canada/Ontario/ | Ontario's COVID-19 Genomics Rapid Response Coalition | McMaster University | Allison McGeer et al |
| EPI_ISL_845707 | Canada/ON-MMU-S2112/2020 | 20-Dec | North America/Canada/Ontario/ | Ontario's COVID-19 Genomics Rapid Response Coalition | McMaster University | Allison McGeer et al |
| EPI_ISL_845708 | Canada/ON-MMU-S2113/2020 | 20-Dec | North America/Canada/Ontario/ | Ontario's COVID-19 Genomics Rapid Response Coalition | McMaster University | Allison McGeer et al |
| EPI_ISL_845709 | Canada/ON-MMU-S2114/2020 | 20-Dec | North America/Canada/Ontario/ | Ontario's COVID-19 Genomics Rapid Response Coalition | McMaster University | Allison McGeer et al |
| EPI_ISL_845710 | Canada/ON-MMU-S2074/2020 | 20-Dec | North America/Canada/Ontario/ | Ontario's COVID-19 Genomics Rapid Response Coalition | McMaster University | Allison McGeer et al |
| EPI_ISL_845711 | Canada/ON-MMU-S2077/2020 | 20-Dec | North America/Canada/Ontario/ | Ontario's COVID-19 Genomics Rapid Response Coalition | McMaster University | Allison McGeer et al |
| EPI_ISL_845712 | Canada/ON-MMU-S2078/2020 | 20-Dec | North America/Canada/Ontario/ | Ontario's COVID-19 Genomics Rapid Response Coalition | McMaster University | Allison McGeer et al |

| | | | | | | |
|---|---|---|---|---|---|---|
| EPI_ISL_845713 | Canada/ON-MMU-S2079/2020 | 20-Dec | North America/Canada/Ontario/ | Ontario's COVID-19 Genomics Rapid Response Coalition | McMaster University | Allison McGeer et al |
| EPI_ISL_845714 | Canada/ON-MMU-S2080/2020 | 20-Dec | North America/Canada/Ontario/ | Ontario's COVID-19 Genomics Rapid Response Coalition | McMaster University | Allison McGeer et al |
| EPI_ISL_845715 | Canada/ON-MMU-S2043/2021 | 21-Jan | North America/Canada/Ontario/ | Ontario's COVID-19 Genomics Rapid Response Coalition | McMaster University | Allison McGeer et al |
| EPI_ISL_845716 | Canada/ON-MMU-S2044/2021 | 21-Jan | North America/Canada/Ontario/ | Ontario's COVID-19 Genomics Rapid Response Coalition | McMaster University | Allison McGeer et al |
| EPI_ISL_845717 | Canada/ON-MMU-S2045/2021 | 21-Jan | North America/Canada/Ontario/ | Ontario's COVID-19 Genomics Rapid Response Coalition | McMaster University | Allison McGeer et al |
| EPI_ISL_845718 | Canada/ON-MMU-S2046/2021 | 21-Jan | North America/Canada/Ontario/ | Ontario's COVID-19 Genomics Rapid Response Coalition | McMaster University | Allison McGeer et al |
| EPI_ISL_845719 | Canada/ON-MMU-S2047/2021 | 21-Jan | North America/Canada/Ontario/ | Ontario's COVID-19 Genomics Rapid Response Coalition | McMaster University | Allison McGeer et al |
| EPI_ISL_845720 | Canada/ON-MMU-S2048/2021 | 21-Jan | North America/Canada/Ontario/ | Ontario's COVID-19 Genomics Rapid Response Coalition | McMaster University | Allison McGeer et al |
| EPI_ISL_845721 | Canada/ON-MMU-S2049/2021 | 21-Jan | North America/Canada/Ontario/ | Ontario's COVID-19 Genomics Rapid Response Coalition | McMaster University | Allison McGeer et al |
| EPI_ISL_845722 | Canada/ON-MMU-S2051/2021 | 21-Jan | North America/Canada/Ontario/ | Ontario's COVID-19 Genomics Rapid Response Coalition | McMaster University | Allison McGeer et al |
| EPI_ISL_845723 | Canada/ON-MMU-S2053/2021 | 21-Jan | North America/Canada/Ontario/ | Ontario's COVID-19 Genomics Rapid Response Coalition | McMaster University | Allison McGeer et al |
| EPI_ISL_845724 | Canada/ON-MMU-S2054/2021 | 21-Jan | North America/Canada/Ontario/ | Ontario's COVID-19 Genomics Rapid Response Coalition | McMaster University | Allison McGeer et al |
| EPI_ISL_845725 | Canada/ON-MMU-S2075/2021 | 21-Jan | North America/Canada/Ontario/ | Ontario's COVID-19 Genomics Rapid Response Coalition | McMaster University | Allison McGeer et al |
| EPI_ISL_845726 | Canada/ON-MMU-S2076/2021 | 21-Jan | North America/Canada/Ontario/ | Ontario's COVID-19 Genomics Rapid Response Coalition | McMaster University | Allison McGeer et al |
| EPI_ISL_845727 | Canada/ON-MMU-S2081/2021 | 21-Jan | North America/Canada/Ontario/ | Ontario's COVID-19 Genomics Rapid Response Coalition | McMaster University | Allison McGeer et al |
| EPI_ISL_845728 | Canada/ON-MMU-S2082/2021 | 21-Jan | North America/Canada/Ontario/ | Ontario's COVID-19 Genomics Rapid Response Coalition | McMaster University | Allison McGeer et al |
| EPI_ISL_845729 | Canada/ON-MMU-S2083/2021 | 21-Jan | North America/Canada/Ontario/ | Ontario's COVID-19 Genomics Rapid Response Coalition | McMaster University | Allison McGeer et al |
| EPI_ISL_845730 | Canada/ON-MMU-S2084/2021 | 21-Jan | North America/Canada/Ontario/ | Ontario's COVID-19 Genomics Rapid Response Coalition | McMaster University | Allison McGeer et al |
| EPI_ISL_845731 | Canada/ON-MMU-S2085/2021 | 21-Jan | North America/Canada/Ontario/ | Ontario's COVID-19 Genomics Rapid Response Coalition | McMaster University | Allison McGeer et al |
| EPI_ISL_845732 | Canada/ON-MMU-S2115/2021 | 21-Jan | North America/Canada/Ontario/ | Ontario's COVID-19 Genomics Rapid Response Coalition | McMaster University | Allison McGeer et al |

| | | | | | | |
|---|---|---|---|---|---|---|
| EPI_ISL_845733 | Canada/ON-MMU-S2116/2021 | 21-Jan | North America/Canada/Ontario/ | Ontario's COVID-19 Genomics Rapid Response Coalition | McMaster University | Allison McGeer et al |
| EPI_ISL_845734 | Canada/ON-MMU-S2117/2021 | 21-Jan | North America/Canada/Ontario/ | Ontario's COVID-19 Genomics Rapid Response Coalition | McMaster University | Allison McGeer et al |
| EPI_ISL_845735 | Canada/ON-MMU-S2118/2021 | 21-Jan | North America/Canada/Ontario/ | Ontario's COVID-19 Genomics Rapid Response Coalition | McMaster University | Allison McGeer et al |
| EPI_ISL_845736 | Canada/ON-MMU-S2055/2021 | 21-Jan | North America/Canada/Ontario/ | Ontario's COVID-19 Genomics Rapid Response Coalition | McMaster University | Allison McGeer et al |
| EPI_ISL_845737 | Canada/ON-MMU-S2056/2021 | 21-Jan | North America/Canada/Ontario/ | Ontario's COVID-19 Genomics Rapid Response Coalition | McMaster University | Allison McGeer et al |
| EPI_ISL_845738 | Canada/ON-MMU-S2057/2021 | 21-Jan | North America/Canada/Ontario/ | Ontario's COVID-19 Genomics Rapid Response Coalition | McMaster University | Allison McGeer et al |
| EPI_ISL_845739 | Canada/ON-MMU-S2058/2021 | 21-Jan | North America/Canada/Ontario/ | Ontario's COVID-19 Genomics Rapid Response Coalition | McMaster University | Allison McGeer et al |
| EPI_ISL_845740 | Canada/ON-MMU-S2059/2021 | 21-Jan | North America/Canada/Ontario/ | Ontario's COVID-19 Genomics Rapid Response Coalition | McMaster University | Allison McGeer et al |
| EPI_ISL_845741 | Canada/ON-MMU-S2060/2021 | 21-Jan | North America/Canada/Ontario/ | Ontario's COVID-19 Genomics Rapid Response Coalition | McMaster University | Allison McGeer et al |
| EPI_ISL_845742 | Canada/ON-MMU-S2061/2021 | 21-Jan | North America/Canada/Ontario/ | Ontario's COVID-19 Genomics Rapid Response Coalition | McMaster University | Allison McGeer et al |
| EPI_ISL_845743 | Canada/ON-MMU-S2064/2021 | 21-Jan | North America/Canada/Ontario/ | Ontario's COVID-19 Genomics Rapid Response Coalition | McMaster University | Allison McGeer et al |
| EPI_ISL_845744 | Canada/ON-MMU-S2066/2021 | 21-Jan | North America/Canada/Ontario/ | Ontario's COVID-19 Genomics Rapid Response Coalition | McMaster University | Allison McGeer et al |
| EPI_ISL_845745 | Canada/ON-MMU-S2067/2021 | 21-Jan | North America/Canada/Ontario/ | Ontario's COVID-19 Genomics Rapid Response Coalition | McMaster University | Allison McGeer et al |
| EPI_ISL_845746 | Canada/ON-MMU-S2068/2021 | 21-Jan | North America/Canada/Ontario/ | Ontario's COVID-19 Genomics Rapid Response Coalition | McMaster University | Allison McGeer et al |
| EPI_ISL_845747 | Canada/ON-MMU-S2069/2021 | 21-Jan | North America/Canada/Ontario/ | Ontario's COVID-19 Genomics Rapid Response Coalition | McMaster University | Allison McGeer et al |
| EPI_ISL_845748 | Canada/ON-MMU-S2070/2021 | 21-Jan | North America/Canada/Ontario/ | Ontario's COVID-19 Genomics Rapid Response Coalition | McMaster University | Allison McGeer et al |
| EPI_ISL_845749 | Canada/ON-MMU-S2072/2021 | 21-Jan | North America/Canada/Ontario/ | Ontario's COVID-19 Genomics Rapid Response Coalition | McMaster University | Allison McGeer et al |
| EPI_ISL_845750 | Canada/ON-MMU-S2073/2021 | 21-Jan | North America/Canada/Ontario/ | Ontario's COVID-19 Genomics Rapid Response Coalition | McMaster University | Allison McGeer et al |
| EPI_ISL_845751 | Canada/ON-MMU-S2121/2021 | 2021-01-02 | North America/Canada/Ontario/Toronto | Toronto Invasive Bacterial Diseases Network | McMaster University | Allison McGeer et al |
| EPI_ISL_845752 | Canada/ON-MMU-S2042/2021 | 2021-01-05 | North America/Canada/Ontario/Toronto | Toronto Invasive Bacterial Diseases Network | McMaster University | Allison McGeer et al |

| | | | | | | |
|---|---|---|---|---|---|---|
| EPI_ISL_845753 | Canada/ON-MMU-S2120/2021 | 2021-01-08 | North America/Canada/Ontario/Toronto | Toronto Invasive Bacterial Diseases Network | McMaster University | Allison McGeer et al |
| EPI_ISL_845760 | USA/GA-EHC-452L/2021 | 2021-01-04 | North America/USA/Georgia/ | Emory Molecular Diagnostics Laboratory, Emory Healthcare | Piantadosi Lab, Emory Department of Pathology | Ahmed Babiker et al |
| EPI_ISL_845767 | USA/GA-EHC-460T/2021 | 2021-01-05 | North America/USA/Georgia/ | Emory Molecular Diagnostics Laboratory, Emory Healthcare | Piantadosi Lab, Emory Department of Pathology | Ahmed Babiker et al |
| EPI_ISL_845773 | USA/GA-EHC-427M/2021 | 2021-01-07 | North America/USA/Georgia/ | Emory Molecular Diagnostics Laboratory, Emory Healthcare | Piantadosi Lab, Emory Department of Pathology | Ahmed Babiker et al |
| EPI_ISL_845775 | USA/GA-EHC-429O/2021 | 2021-01-07 | North America/USA/Georgia/ | Emory Molecular Diagnostics Laboratory, Emory Healthcare | Piantadosi Lab, Emory Department of Pathology | Ahmed Babiker et al |
| EPI_ISL_845776 | USA/GA-EHC-430P/2021 | 2021-01-07 | North America/USA/Georgia/ | Emory Molecular Diagnostics Laboratory, Emory Healthcare | Piantadosi Lab, Emory Department of Pathology | Ahmed Babiker et al |
| EPI_ISL_845782 | USA/GA-EHC-436V/2021 | 2021-01-07 | North America/USA/Georgia/ | Emory Molecular Diagnostics Laboratory, Emory Healthcare | Piantadosi Lab, Emory Department of Pathology | Ahmed Babiker et al |
| EPI_ISL_845811 | USA/MD-TG703617/2020 | 2020-12-21 | North America/USA/Maryland/ | TGen North | TGen North | Jolene Bowers et al |
| EPI_ISL_845816 | USA/AZ-TG703685/2020 | 2020-12-22 | North America/USA/Arizona/ | TGen North | TGen North | Jolene Bowers et al |
| EPI_ISL_845835 | USA/AZ-TG704528/2020 | 2020-12-30 | North America/USA/Arizona/ | TGen North | TGen North | Jolene Bowers et al |
| EPI_ISL_845836 | USA/CA-TG704532/2020 | 2020-12-30 | North America/USA/California/ | TGen North | TGen North | Jolene Bowers et al |
| EPI_ISL_845837 | USA/AZ-TG704536/2020 | 2020-12-29 | North America/USA/Arizona/ | TGen North | TGen North | Jolene Bowers et al |
| EPI_ISL_845849 | USA/AZ-TG704604/2020 | 2020-12-30 | North America/USA/Arizona/ | TGen North | TGen North | Jolene Bowers et al |
| EPI_ISL_845852 | USA/AZ-TG704622/2020 | 2020-12-28 | North America/USA/Arizona/ | TGen North | TGen North | Jolene Bowers et al |
| EPI_ISL_845853 | USA/AZ-TG704628/2020 | 2020-12-29 | North America/USA/Arizona/ | TGen North | TGen North | Jolene Bowers et al |
| EPI_ISL_845855 | USA/AZ-TG704636/2020 | 2020-12-28 | North America/USA/Arizona/ | TGen North | TGen North | Jolene Bowers et al |
| EPI_ISL_845859 | USA/AZ-TG704656/2020 | 2020-12-28 | North America/USA/Arizona/ | TGen North | TGen North | Jolene Bowers et al |
| EPI_ISL_845867 | USA/AZ-TG705120/2021 | 2021-01-04 | North America/USA/Arizona/ | TGen North | TGen North | Jolene Bowers et al |

| | | | | | | |
|---|---|---|---|---|---|---|
| EPI_ISL_845868 | USA/AZ-TG705122/2021 | 2021-01-04 | North America/USA/Arizona/ | TGen North | TGen North | Jolene Bowers et al |
| EPI_ISL_845869 | USA/AZ-TG705124/2021 | 2021-01-04 | North America/USA/Arizona/ | TGen North | TGen North | Jolene Bowers et al |
| EPI_ISL_845873 | USA/AZ-TG705134/2020 | 2020-12-29 | North America/USA/Arizona/ | TGen North | TGen North | Jolene Bowers et al |
| EPI_ISL_845874 | India/KA-NIMH-SEQ-42/2020 | 2020-12-28 | Asia/India/Karnataka/ | BBMP Urban PHC | Department of Neurovirology, National Institute of Mental Health and Neurosciences (NIMHANS) | Chitra Pattabiraman et al |
| EPI_ISL_845875 | India/KA-NIMH-SEQ-43/2020 | 2020-12-27 | Asia/India/Karnataka/ | BBMP Urban PHC | Department of Neurovirology, National Institute of Mental Health and Neurosciences (NIMHANS) | Chitra Pattabiraman et al |
| EPI_ISL_845876 | India/KA-NIMH-SEQ-44/2020 | 2020-12-28 | Asia/India/Karnataka/ | BBMP Urban PHC | Department of Neurovirology, National Institute of Mental Health and Neurosciences (NIMHANS) | Chitra Pattabiraman et al |
| EPI_ISL_845877 | India/KA-NIMH-SEQ-45/2020 | 2020-12-30 | Asia/India/Karnataka/ | National Institute of Mental Health and Neurosciences (NIMHANS) | Department of Neurovirology, National Institute of Mental Health and Neurosciences (NIMHANS) | Chitra Pattabiraman et al |
| EPI_ISL_845878 | India/KA-NIMH-SEQ-46/2020 | 2020-12-30 | Asia/India/Karnataka/ | BBMP Urban PHC | Department of Neurovirology, National Institute of Mental Health and Neurosciences (NIMHANS) | Chitra Pattabiraman et al |
| EPI_ISL_845879 | India/KA-NIMH-SEQ-47/2020 | 2020-12-30 | Asia/India/Karnataka/ | BBMP Urban PHC | Department of Neurovirology, National Institute of Mental Health and Neurosciences (NIMHANS) | Chitra Pattabiraman et al |

| | | | | | | |
|---|---|---|---|---|---|---|
| EPI_ISL_845881 | India/KA-NIMH-SEQ-49/2020 | 2020-12-31 | Asia/India/Karnataka/ | Kidwai Memorial Institute of Oncology | Department of Neurovirology, National Institute of Mental Health and Neurosciences (NIMHANS) | Chitra Pattabiraman et al |
| EPI_ISL_845882 | India/KA-NIMH-SEQ-50/2020 | 2020-12-29 | Asia/India/Karnataka/ | VRDL, Mysore Medical College | Department of Neurovirology, National Institute of Mental Health and Neurosciences (NIMHANS) | Chitra Pattabiraman et al |
| EPI_ISL_845883 | India/KA-NIMH-SEQ-51/2020 | 2020-12-28 | Asia/India/Karnataka/ | BBMP Urban PHC | Department of Neurovirology, National Institute of Mental Health and Neurosciences (NIMHANS) | Chitra Pattabiraman et al |
| EPI_ISL_845885 | India/KA-NIMH-SEQ-53/2020 | 2020-12-30 | Asia/India/Karnataka/ | Railway Hospital | Department of Neurovirology, National Institute of Mental Health and Neurosciences (NIMHANS) | Chitra Pattabiraman et al |
| EPI_ISL_845886 | India/KA-NIMH-SEQ-54/2020 | 2020-12-30 | Asia/India/Karnataka/ | Railway Hospital | Department of Neurovirology, National Institute of Mental Health and Neurosciences (NIMHANS) | Chitra Pattabiraman et al |
| EPI_ISL_845887 | India/KA-NIMH-SEQ-55/2020 | 2020-12-30 | Asia/India/Karnataka/ | BBMP Urban PHC | Department of Neurovirology, National Institute of Mental Health and Neurosciences (NIMHANS) | Chitra Pattabiraman et al |

| | | | | | | |
|---|---|---|---|---|---|---|
| EPI_ISL_845888 | India/KA-NIMH-SEQ-56/2020 | 2020-12-29 | Asia/India/Karnataka/ | BBMP Urban PHC | Department of Neurovirology, National Institute of Mental Health and Neurosciences (NIMHANS) | Chitra Pattabiraman et al |
| EPI_ISL_845889 | India/KA-NIMH-SEQ-57/2020 | 2020-12-28 | Asia/India/Karnataka/ | BBMP Urban PHC | Department of Neurovirology, National Institute of Mental Health and Neurosciences (NIMHANS) | Chitra Pattabiraman et al |
| EPI_ISL_845890 | India/KA-NIMH-SEQ-58/2020 | 2020-12-29 | Asia/India/Karnataka/ | BBMP Urban PHC | Department of Neurovirology, National Institute of Mental Health and Neurosciences (NIMHANS) | Chitra Pattabiraman et al |
| EPI_ISL_845899 | England/CAMC-F7F234/2021 | 2021-01-09 | Europe/United Kingdom/England/ | Lighthouse Lab in Cambridge | Wellcome Sanger Institute for the COVID-19 Genomics UK (COG-UK) Consortium | Rob Howes et al |
| EPI_ISL_845995 | England/CAMC-F7D649/2021 | 2021-01-09 | Europe/United Kingdom/England/ | Lighthouse Lab in Cambridge | Wellcome Sanger Institute for the COVID-19 Genomics UK (COG-UK) Consortium | Rob Howes et al |
| EPI_ISL_845996 | England/CAMC-F762D3/2021 | 2021-01-09 | Europe/United Kingdom/England/ | Lighthouse Lab in Cambridge | Wellcome Sanger Institute for the COVID-19 Genomics UK (COG-UK) Consortium | Rob Howes et al |
| EPI_ISL_846059 | England/CAMC-F76455/2021 | 2021-01-07 | Europe/United Kingdom/England/ | Lighthouse Lab in Cambridge | Wellcome Sanger Institute for the COVID-19 Genomics UK (COG-UK) Consortium | Rob Howes et al |

| | | | | | | |
|---|---|---|---|---|---|---|
| EPI_ISL_846429 | England/ALDP-F8217D/2021 | 2021-01-07 | Europe/United Kingdom/England/ | Lighthouse Lab in Alderley Park | Wellcome Sanger Institute for the COVID-19 Genomics UK (COG-UK) Consortium | Jacquelyn Wynn et al |
| EPI_ISL_846441 | England/ALDP-F82993/2021 | 2021-01-09 | Europe/United Kingdom/England/ | Lighthouse Lab in Alderley Park | Wellcome Sanger Institute for the COVID-19 Genomics UK (COG-UK) Consortium | Jacquelyn Wynn et al |
| EPI_ISL_846540 | Slovenia/MB21-2/2021 | 2021-01-07 | Europe/Slovenia/Slovenia/ | National Laboratory for Health, Environment and Food | National Laboratory for Health, Environment and Food | Aleksander Mahnic et al |
| EPI_ISL_846569 | Belgium/UGent-1072/2021 | 2021-01-12 | Europe/Belgium/Gent/ | Lab voor klinische biologie | Lab voor klinische biologie | Hannelore Hamerlinck et al |
| EPI_ISL_846571 | Belgium/UGent-1074/2021 | 2021-01-12 | Europe/Belgium/Gent/ | Lab voor klinische biologie | Lab voor klinische biologie | Hannelore Hamerlinck et al |
| EPI_ISL_846587 | Poland/NIPH-NIH-10/2020 | 2020-12-16 | Europe/Poland/Mazowieckie/Warszawa | National Institute of Public Health - National Institute of Hygiene | National Institute of Public Health - National Institute of Hygiene | Wo≈Çkowicz Tomasz et al |
| EPI_ISL_846628 | USA/OH-ODH-SC50913/2020 | 2020-07-03 | North America/USA/Ohio/ | Ohio Department of Health Laboratory | Ohio Department of Health Laboratory | Holmes et al |
| EPI_ISL_846658 | USA/MI-UM-10036941109/2020 | 2020-11-13 | North America/USA/Michigan/ | University of Michigan Clinical Microbiology Laboratory | Lauring Lab, University of Michigan, Department of Microbiology and Immunology | Valesano et al |
| EPI_ISL_846666 | USA/MI-UM-10036966788/2020 | 2020-11-16 | North America/USA/Michigan/ | University of Michigan Clinical Microbiology Laboratory | Lauring Lab, University of Michigan, Department of Microbiology and Immunology | Valesano et al |

| | | | | | | |
|---|---|---|---|---|---|---|
| EPI_ISL_846668 | USA/MI-UM-10036949870/2020 | 2020-11-14 | North America/USA/Michigan/ | University of Michigan Clinical Microbiology Laboratory | Lauring Lab, University of Michigan, Department of Microbiology and Immunology | Valesano et al |
| EPI_ISL_846674 | USA/MI-UM-10036929408/2020 | 2020-11-13 | North America/USA/Michigan/ | University of Michigan Clinical Microbiology Laboratory | Lauring Lab, University of Michigan, Department of Microbiology and Immunology | Valesano et al |
| EPI_ISL_846685 | USA/MI-UM-10036971877/2020 | 2020-11-16 | North America/USA/Michigan/ | University of Michigan Clinical Microbiology Laboratory | Lauring Lab, University of Michigan, Department of Microbiology and Immunology | Valesano et al |
| EPI_ISL_846686 | USA/MI-UM-10036986538/2020 | 2020-11-17 | North America/USA/Michigan/ | University of Michigan Clinical Microbiology Laboratory | Lauring Lab, University of Michigan, Department of Microbiology and Immunology | Valesano et al |
| EPI_ISL_846694 | USA/MI-UM-10036948263/2020 | 2020-11-14 | North America/USA/Michigan/ | University of Michigan Clinical Microbiology Laboratory | Lauring Lab, University of Michigan, Department of Microbiology and Immunology | Valesano et al |
| EPI_ISL_846715 | USA/MI-UM-10036931434/2020 | 2020-11-13 | North America/USA/Michigan/ | University of Michigan Clinical Microbiology Laboratory | Lauring Lab, University of Michigan, Department of Microbiology and Immunology | Valesano et al |

| | | | | | | |
|---|---|---|---|---|---|---|
| EPI_ISL_846725 | USA/MI-UM-10036929638/2020 | 2020-11-13 | North America/USA/Michigan/ | University of Michigan Clinical Microbiology Laboratory | Lauring Lab, University of Michigan, Department of Microbiology and Immunology | Valesano et al |
| EPI_ISL_846729 | USA/MI-UM-10036940093/2020 | 2020-11-13 | North America/USA/Michigan/ | University of Michigan Clinical Microbiology Laboratory | Lauring Lab, University of Michigan, Department of Microbiology and Immunology | Valesano et al |
| EPI_ISL_846731 | USA/MI-UM-10036952953/2020 | 2020-11-14 | North America/USA/Michigan/ | University of Michigan Clinical Microbiology Laboratory | Lauring Lab, University of Michigan, Department of Microbiology and Immunology | Valesano et al |
| EPI_ISL_846732 | USA/MI-UM-10036949950/2020 | 2020-11-14 | North America/USA/Michigan/ | University of Michigan Clinical Microbiology Laboratory | Lauring Lab, University of Michigan, Department of Microbiology and Immunology | Valesano et al |
| EPI_ISL_846736 | USA/MI-UM-10036964345/2020 | 2020-11-16 | North America/USA/Michigan/ | University of Michigan Clinical Microbiology Laboratory | Lauring Lab, University of Michigan, Department of Microbiology and Immunology | Valesano et al |
| EPI_ISL_846741 | USA/MI-UM-10037007491/2020 | 2020-11-18 | North America/USA/Michigan/ | University of Michigan Clinical Microbiology Laboratory | Lauring Lab, University of Michigan, Department of Microbiology and Immunology | Valesano et al |

| | | | | | | |
|---|---|---|---|---|---|---|
| EPI_ISL_846754 | USA/MI-UM-10036933442/2020 | 2020-11-13 | North America/USA/Michigan/ | University of Michigan Clinical Microbiology Laboratory | Lauring Lab, University of Michigan, Department of Microbiology and Immunology | Valesano et al |
| EPI_ISL_846756 | USA/MI-UM-10036938231/2020 | 2020-11-13 | North America/USA/Michigan/ | University of Michigan Clinical Microbiology Laboratory | Lauring Lab, University of Michigan, Department of Microbiology and Immunology | Valesano et al |
| EPI_ISL_847008 | Denmark/DCGC-30763/2021 | 2021-01-11 | Europe/Denmark/Hovedstaden/ | Department of Virus and Microbiological Special Diagnostics, Statens Serum Institut, Copenhagen, Denmark | Albertsen Lab, Department of Chemistry and Bioscience, Aalborg University, Denmark | Danish Covid-19 Genome Consortium et al |
| EPI_ISL_847237 | Denmark/DCGC-31176/2021 | 2021-01-04 | Europe/Denmark/Syddanmark/ | Department of Virus and Microbiological Special Diagnostics, Statens Serum Institut, Copenhagen, Denmark | Albertsen Lab, Department of Chemistry and Bioscience, Aalborg University, Denmark | Danish Covid-19 Genome Consortium et al |
| EPI_ISL_847517 | USA/CA-UCSF-UC692/2020 | 2020-12-15 | North America/USA/California/San Fran | Chiu Laboratory, University of California, San Francisco | Chiu Laboratory, University of California, San Francisco | Charles Chiu et al |
| EPI_ISL_847518 | USA/CA-CDPH-UC552/2020 | 2020-12-10 | North America/USA/California/Lake Cou | California Department of Public Health | Chiu Laboratory, University of California, San Francisco | Charles Chiu et al |
| EPI_ISL_847520 | USA/CA-CDPH-UC513/2020 | 2020-12-04 | North America/USA/California/Mendoci | California Department of Public Health | Chiu Laboratory, University of California, San Francisco | Charles Chiu et al |

| | | | | | | |
|---|---|---|---|---|---|---|
| EPI_ISL_847523 | USA/CA-CDPH-UC441/2020 | 2020-11-20 | North America/USA/California/Madera | California Department of Public Health | Chiu Laboratory, University of California, San Francisco | Charles Chiu et al |
| EPI_ISL_847533 | USA/CA-CDPH-UC541/2020 | 2020-12-09 | North America/USA/California/Mono Co | California Department of Public Health | Chiu Laboratory, University of California, San Francisco | Charles Chiu et al |
| EPI_ISL_847536 | USA/CA-UCSF-UC711/2020 | 2020-12-28 | North America/USA/California/San Fran | Chiu Laboratory, University of California, San Francisco | Chiu Laboratory, University of California, San Francisco | Charles Chiu et al |
| EPI_ISL_847540 | USA/CA-UCSF-UC706/2020 | 2020-12-15 | North America/USA/California/San Fran | Chiu Laboratory, University of California, San Francisco | Chiu Laboratory, University of California, San Francisco | Charles Chiu et al |
| EPI_ISL_847544 | USA/CA-CDPH-UC535/2020 | 2020-12-09 | North America/USA/California/Mono Co | California Department of Public Health | Chiu Laboratory, University of California, San Francisco | Charles Chiu et al |
| EPI_ISL_847550 | USA/CA-CDPH-UC609/2020 | 2020-12-17 | North America/USA/California/Mono Co | California Department of Public Health | Chiu Laboratory, University of California, San Francisco | Charles Chiu et al |
| EPI_ISL_847551 | USA/CA-CDPH-UC477/2020 | 2020-11-24 | North America/USA/California/Madera | California Department of Public Health | Chiu Laboratory, University of California, San Francisco | Charles Chiu et al |
| EPI_ISL_847552 | USA/CA-CDPH-UC430/2020 | 2020-11-21 | North America/USA/California/Mendoci | California Department of Public Health | Chiu Laboratory, University of California, San Francisco | Charles Chiu et al |
| EPI_ISL_847553 | USA/CA-UCSF-UC700/2020 | 2020-12-15 | North America/USA/California/Alameda | Chiu Laboratory, University of California, San Francisco | Chiu Laboratory, University of California, San Francisco | Charles Chiu et al |
| EPI_ISL_847562 | USA/CA-CDPH-UC532/2020 | 2020-12-09 | North America/USA/California/Mono Co | California Department of Public Health | Chiu Laboratory, University of California, San Francisco | Charles Chiu et al |

| | | | | | | |
|---|---|---|---|---|---|---|
| EPI_ISL_847567 | USA/CA-CDPH-UC519/2020 | 2020-12-07 | North America/USA/California/Mono Co | California Department of Public Health | Chiu Laboratory, University of California, San Francisco | Charles Chiu et al |
| EPI_ISL_847575 | USA/CA-CDPH-UC616/2020 | 2020-12-17 | North America/USA/California/Madera | California Department of Public Health | Chiu Laboratory, University of California, San Francisco | Charles Chiu et al |
| EPI_ISL_847579 | USA/CA-UCSF-UC687/2020 | 2020-12-15 | North America/USA/California/Alameda | Chiu Laboratory, University of California, San Francisco | Chiu Laboratory, University of California, San Francisco | Charles Chiu et al |
| EPI_ISL_847580 | USA/CA-CDPH-UC658/2020 | 2020-12-26 | North America/USA/California/Mendoci | California Department of Public Health | Chiu Laboratory, University of California, San Francisco | Charles Chiu et al |
| EPI_ISL_847581 | USA/CA-CDPH-UC540/2020 | 2020-12-09 | North America/USA/California/Mono Co | California Department of Public Health | Chiu Laboratory, University of California, San Francisco | Charles Chiu et al |
| EPI_ISL_847586 | USA/CA-CDPH-UC418/2020 | 2020-11-18 | North America/USA/California/Madera | California Department of Public Health | Chiu Laboratory, University of California, San Francisco | Charles Chiu et al |
| EPI_ISL_847587 | USA/CA-CDPH-UC523/2020 | 2020-12-07 | North America/USA/California/Lake Cou | California Department of Public Health | Chiu Laboratory, University of California, San Francisco | Charles Chiu et al |
| EPI_ISL_847589 | USA/CA-CDPH-UC652/2021 | 2021-01-09 | North America/USA/California/Mono Co | California Department of Public Health | Chiu Laboratory, University of California, San Francisco | Charles Chiu et al |
| EPI_ISL_847591 | USA/CA-CDPH-UC575/2021 | 2021-01-02 | North America/USA/California/Los Ange | California Department of Public Health | Chiu Laboratory, University of California, San Francisco | Charles Chiu et al |
| EPI_ISL_847592 | USA/CA-CDPH-UC576/2021 | 2021-01-02 | North America/USA/California/Los Ange | California Department of Public Health | Chiu Laboratory, University of California, San Francisco | Charles Chiu et al |

| | | | | | | |
|---|---|---|---|---|---|---|
| EPI_ISL_847596 | USA/CA-CDPH-UC586/2020 | 2020-12-29 | North America/USA/California/Los Ange | California Department of Public Health | Chiu Laboratory, University of California, San Francisco | Charles Chiu et al |
| EPI_ISL_847598 | USA/CA-CDPH-UC591/2020 | 2020-12-29 | North America/USA/California/Los Ange | California Department of Public Health | Chiu Laboratory, University of California, San Francisco | Charles Chiu et al |
| EPI_ISL_847599 | USA/CA-CDPH-UC682/2020 | 2020-12-29 | North America/USA/California/Lake Cou | California Department of Public Health | Chiu Laboratory, University of California, San Francisco | Charles Chiu et al |
| EPI_ISL_847612 | USA/CA-CDPH-UC633/2020 | 2020-12-22 | North America/USA/California/Alameda | California Department of Public Health | Chiu Laboratory, University of California, San Francisco | Charles Chiu et al |
| EPI_ISL_847617 | USA/CA-CDPH-UC641/2020 | 2020-12-21 | North America/USA/California/Mono Co | California Department of Public Health | Chiu Laboratory, University of California, San Francisco | Charles Chiu et al |
| EPI_ISL_847618 | USA/CA-CDPH-UC644/2020 | 2020-12-21 | North America/USA/California/Mono Co | California Department of Public Health | Chiu Laboratory, University of California, San Francisco | Charles Chiu et al |
| EPI_ISL_847621 | USA/CA-UCSF-UC689/2020 | 2020-12-20 | North America/USA/California/San Fran | Chiu Laboratory, University of California, San Francisco | Chiu Laboratory, University of California, San Francisco | Charles Chiu et al |
| EPI_ISL_847622 | USA/CA-UCSF-UC704/2020 | 2020-12-19 | North America/USA/California/Alameda | Chiu Laboratory, University of California, San Francisco | Chiu Laboratory, University of California, San Francisco | Charles Chiu et al |
| EPI_ISL_847625 | USA/CA-CDPH-UC607/2020 | 2020-12-17 | North America/USA/California/Mono Co | California Department of Public Health | Chiu Laboratory, University of California, San Francisco | Charles Chiu et al |
| EPI_ISL_847628 | USA/CA-CDPH-UC621/2020 | 2020-12-17 | North America/USA/California/Madera | California Department of Public Health | Chiu Laboratory, University of California, San Francisco | Charles Chiu et al |

| | | | | | | |
|---|---|---|---|---|---|---|
| EPI_ISL_847634 | USA/CA-CDPH-UC572/2020 | 2020-12-16 | North America/USA/California/Mono Co | California Department of Public Health | Chiu Laboratory, University of California, San Francisco | Charles Chiu et al |
| EPI_ISL_847637 | USA/CA-CDPH-UC601/2020 | 2020-12-16 | North America/USA/California/Mono Co | California Department of Public Health | Chiu Laboratory, University of California, San Francisco | Charles Chiu et al |
| EPI_ISL_847638 | USA/CA-CDPH-UC560/2020 | 2020-12-15 | North America/USA/California/Lake Cou | California Department of Public Health | Chiu Laboratory, University of California, San Francisco | Charles Chiu et al |
| EPI_ISL_847642 | USA/CA-UCSF-UC695/2020 | 2020-12-15 | North America/USA/California/San Fran | Chiu Laboratory, University of California, San Francisco | Chiu Laboratory, University of California, San Francisco | Charles Chiu et al |
| EPI_ISL_847645 | USA/CA-CDPH-UC559/2020 | 2020-12-13 | North America/USA/California/Lake Cou | California Department of Public Health | Chiu Laboratory, University of California, San Francisco | Charles Chiu et al |
| EPI_ISL_847651 | USA/CA-CDPH-UC539/2020 | 2020-12-09 | North America/USA/California/Mono Co | California Department of Public Health | Chiu Laboratory, University of California, San Francisco | Charles Chiu et al |
| EPI_ISL_847655 | USA/CA-CDPH-UC527/2020 | 2020-12-08 | North America/USA/California/Mendoci | California Department of Public Health | Chiu Laboratory, University of California, San Francisco | Charles Chiu et al |
| EPI_ISL_847660 | USA/CA-CDPH-UC503/2020 | 2020-12-03 | North America/USA/California/Fresno C | California Department of Public Health | Chiu Laboratory, University of California, San Francisco | Charles Chiu et al |
| EPI_ISL_847662 | USA/CA-CDPH-UC506/2020 | 2020-12-03 | North America/USA/California/Madera | California Department of Public Health | Chiu Laboratory, University of California, San Francisco | Charles Chiu et al |
| EPI_ISL_847673 | USA/CA-CDPH-UC467/2020 | 2020-11-24 | North America/USA/California/Madera | California Department of Public Health | Chiu Laboratory, University of California, San Francisco | Charles Chiu et al |

| | | | | | | |
|---|---|---|---|---|---|---|
| EPI_ISL_847674 | USA/CA-CDPH-UC453/2020 | 2020-11-23 | North America/USA/California/Madera | California Department of Public Health | Chiu Laboratory, University of California, San Francisco | Charles Chiu et al |
| EPI_ISL_847677 | USA/CA-CDPH-UC460/2020 | 2020-11-23 | North America/USA/California/Madera | California Department of Public Health | Chiu Laboratory, University of California, San Francisco | Charles Chiu et al |
| EPI_ISL_847687 | USA/CA-CDPH-UC440/2020 | 2020-11-20 | North America/USA/California/Madera | California Department of Public Health | Chiu Laboratory, University of California, San Francisco | Charles Chiu et al |
| EPI_ISL_847689 | USA/CA-CDPH-UC444/2020 | 2020-11-20 | North America/USA/California/Madera | California Department of Public Health | Chiu Laboratory, University of California, San Francisco | Charles Chiu et al |
| EPI_ISL_847694 | USA/CA-CDPH-UC450/2020 | 2020-11-20 | North America/USA/California/Madera | California Department of Public Health | Chiu Laboratory, University of California, San Francisco | Charles Chiu et al |
| EPI_ISL_847705 | USA/CA-CDPH-UC530/2020 | 2020-12-08 | North America/USA/California/Mendoci | California Department of Public Health | Chiu Laboratory, University of California, San Francisco | Charles Chiu et al |
| EPI_ISL_847712 | USA/CA-CDPH-UC573/2020 | 2020-12-16 | North America/USA/California/Mono Co | California Department of Public Health | Chiu Laboratory, University of California, San Francisco | Charles Chiu et al |
| EPI_ISL_847716 | USA/CA-UCSF-UC685/2020 | 2020-12-15 | North America/USA/California/San Mat | Chiu Laboratory, University of California, San Francisco | Chiu Laboratory, University of California, San Francisco | Charles Chiu et al |
| EPI_ISL_847731 | USA/CA-UCSF-UC714/2020 | 2020-12-19 | North America/USA/California/San Fran | Chiu Laboratory, University of California, San Francisco | Chiu Laboratory, University of California, San Francisco | Charles Chiu et al |
| EPI_ISL_847742 | USA/CA-UCSF-UC707/2020 | 2020-12-20 | North America/USA/California/Alameda | Chiu Laboratory, University of California, San Francisco | Chiu Laboratory, University of California, San Francisco | Charles Chiu et al |

| | | | | | | |
|---|---|---|---|---|---|---|
| EPI_ISL_847751 | USA/CA-CDPH-UC661/2020 | 2020-12-26 | North America/USA/California/Mendoci | California Department of Public Health | Chiu Laboratory, University of California, San Francisco | Charles Chiu et al |
| EPI_ISL_847761 | USA/CA-CDPH-UC584/2020 | 2020-12-29 | North America/USA/California/Los Ange | California Department of Public Health | Chiu Laboratory, University of California, San Francisco | Charles Chiu et al |
| EPI_ISL_847775 | USA/CA-CDPH-UC642/2020 | 2020-12-21 | North America/USA/California/Mono Co | California Department of Public Health | Chiu Laboratory, University of California, San Francisco | Charles Chiu et al |
| EPI_ISL_847779 | USA/CA-CDPH-UC628/2020 | 2020-12-17 | North America/USA/California/Madera | California Department of Public Health | Chiu Laboratory, University of California, San Francisco | Charles Chiu et al |
| EPI_ISL_847784 | USA/CA-CDPH-UC537/2020 | 2020-12-09 | North America/USA/California/Mono Co | California Department of Public Health | Chiu Laboratory, University of California, San Francisco | Charles Chiu et al |
| EPI_ISL_847790 | USA/CA-CDPH-UC515/2020 | 2020-12-07 | North America/USA/California/Mono Co | California Department of Public Health | Chiu Laboratory, University of California, San Francisco | Charles Chiu et al |
| EPI_ISL_847792 | USA/CA-CDPH-UC520/2020 | 2020-12-07 | North America/USA/California/Lake Cou | California Department of Public Health | Chiu Laboratory, University of California, San Francisco | Charles Chiu et al |
| EPI_ISL_847811 | USA/CA-CDPH-UC431/2020 | 2020-11-20 | North America/USA/California/Madera | California Department of Public Health | Chiu Laboratory, University of California, San Francisco | Charles Chiu et al |
| EPI_ISL_847812 | USA/CA-CDPH-UC432/2020 | 2020-11-20 | North America/USA/California/Madera | California Department of Public Health | Chiu Laboratory, University of California, San Francisco | Charles Chiu et al |
| EPI_ISL_847813 | USA/CA-CDPH-UC434/2020 | 2020-11-20 | North America/USA/California/Madera | California Department of Public Health | Chiu Laboratory, University of California, San Francisco | Charles Chiu et al |

| | | | | | | |
|---|---|---|---|---|---|---|
| EPI_ISL_847817 | USA/CA-CDPH-UC423/2020 | 2020-11-18 | North America/USA/California/Madera | California Department of Public Health | Chiu Laboratory, University of California, San Francisco | Charles Chiu et al |
| EPI_ISL_847818 | USA/CA-CDPH-UC407/2020 | 2020-11-17 | North America/USA/California/Madera | California Department of Public Health | Chiu Laboratory, University of California, San Francisco | Charles Chiu et al |
| EPI_ISL_847819 | USA/OH-ODH-SC51145/2020 | 2020-07-04 | North America/USA/Ohio/ | Ohio Department of Health Laboratory | Ohio Department of Health Laboratory | Holmes et al |
| EPI_ISL_847824 | USA/OH-ODH-SC51220/2020 | 2020-07-04 | North America/USA/Ohio/ | Ohio Department of Health Laboratory | Ohio Department of Health Laboratory | Holmes et al |
| EPI_ISL_847825 | Italy/CAM-COTUGNO-18415/2021 | 2021-01-02 | Europe/Italy/Campania/ | Lab. Microbiologia e Virologia, Cotugno, A.O. dei Colli | Lab. Microbiologia e Virologia, Cotugno, A.O. dei Colli | Luigi Atripaldi et al |
| EPI_ISL_847828 | USA/OH-ODH-SC51224/2020 | 2020-07-04 | North America/USA/Ohio/ | Ohio Department of Health Laboratory | Ohio Department of Health Laboratory | Holmes et al |
| EPI_ISL_847842 | USA/OH-ODH-SC51458/2020 | 2020-07-06 | North America/USA/Ohio/ | Ohio Department of Health Laboratory | Ohio Department of Health Laboratory | Holmes et al |
| EPI_ISL_847873 | Switzerland/GE-32842651/2020 | 2020-12-31 | Europe/Switzerland/Geneva/ | University Hospitals of Geneva, Laboratory of Virology | HUG, Laboratory of Virology and the Health2030 Genome Center | Samuel Cordey et al |
| EPI_ISL_847924 | Switzerland/GE-32845532/2020 | 2020-12-31 | Europe/Switzerland/Geneva/ | University Hospitals of Geneva, Laboratory of Virology | HUG, Laboratory of Virology and the Health2030 Genome Center | Samuel Cordey et al |
| EPI_ISL_847936 | Switzerland/GE-32849131/2021 | 2021-01-01 | Europe/Switzerland/Geneva/ | University Hospitals of Geneva, Laboratory of Virology | HUG, Laboratory of Virology and the Health2030 Genome Center | Samuel Cordey et al |
| EPI_ISL_847961 | Switzerland/un-32818267/2020 | 2020-12-28 | Europe/Switzerland/Switzerland/ | University Hospitals of Geneva, Laboratory of Virology | HUG, Laboratory of Virology and the Health2030 Genome Center | Samuel Cordey et al |

| | | | | | | |
|---|---|---|---|---|---|---|
| EPI_ISL_847989 | USA/MI-MDHHS-SC23049/2020 | 2020-12-27 | North America/USA/Michigan/ | Michigan Department of Health and Human Services, Bureau of Laboratories | Michigan Department of Health and Human Services, Bureau of Laboratories | Blankenship HM et al |
| EPI_ISL_848002 | USA/MI-MDHHS-SC23054/2020 | 2020-12-28 | North America/USA/Michigan/ | Michigan Department of Health and Human Services, Bureau of Laboratories | Michigan Department of Health and Human Services, Bureau of Laboratories | Blankenship HM et al |
| EPI_ISL_848028 | USA/MI-MDHHS-SC23009/2020 | 2020-11-30 | North America/USA/Michigan/ | Michigan Department of Health and Human Services, Bureau of Laboratories | Michigan Department of Health and Human Services, Bureau of Laboratories | Blankenship HM et al |
| EPI_ISL_848029 | USA/MI-MDHHS-SC23011/2020 | 2020-11-30 | North America/USA/Michigan/ | Michigan Department of Health and Human Services, Bureau of Laboratories | Michigan Department of Health and Human Services, Bureau of Laboratories | Blankenship HM et al |
| EPI_ISL_848030 | USA/MI-MDHHS-SC23012/2020 | 2020-11-30 | North America/USA/Michigan/ | Michigan Department of Health and Human Services, Bureau of Laboratories | Michigan Department of Health and Human Services, Bureau of Laboratories | Blankenship HM et al |
| EPI_ISL_848038 | USA/MI-MDHHS-SC23023/2020 | 2020-11-27 | North America/USA/Michigan/ | Michigan Department of Health and Human Services, Bureau of Laboratories | Michigan Department of Health and Human Services, Bureau of Laboratories | Blankenship HM et al |

| | | | | | | |
|---|---|---|---|---|---|---|
| EPI_ISL_848039 | USA/MI-MDHHS-SC23032/2020 | 2020-11-27 | North America/USA/Michigan/ | Michigan Department of Health and Human Services, Bureau of Laboratories | Michigan Department of Health and Human Services, Bureau of Laboratories | Blankenship HM et al |
| EPI_ISL_848042 | USA/MI-MDHHS-SC23036/2020 | 2020-11-24 | North America/USA/Michigan/ | Michigan Department of Health and Human Services, Bureau of Laboratories | Michigan Department of Health and Human Services, Bureau of Laboratories | Blankenship HM et al |
| EPI_ISL_848043 | USA/MI-MDHHS-SC23041/2020 | 2020-11-24 | North America/USA/Michigan/ | Michigan Department of Health and Human Services, Bureau of Laboratories | Michigan Department of Health and Human Services, Bureau of Laboratories | Blankenship HM et al |
| EPI_ISL_848059 | USA/MI-MDHHS-SC23021/2020 | 2020-11-30 | North America/USA/Michigan/ | Michigan Department of Health and Human Services, Bureau of Laboratories | Michigan Department of Health and Human Services, Bureau of Laboratories | Blankenship HM et al |
| EPI_ISL_848060 | USA/MI-MDHHS-SC23024/2020 | 2020-11-30 | North America/USA/Michigan/ | Michigan Department of Health and Human Services, Bureau of Laboratories | Michigan Department of Health and Human Services, Bureau of Laboratories | Blankenship HM et al |
| EPI_ISL_848065 | France/OCC-219/2021 | 2021-01-06 | Europe/France/Occitanie/Toulouse | CHU Purpan - Laboratoire de Virologie - Institut FV©dV©ratif de Biologie | CHU Purpan - Laboratoire de Virologie - Institut FV©dV©ratif de Biologie | Latour J. et al |
| EPI_ISL_848089 | Ireland/D-20G41954/2020 | 2020-03-26 | Europe/Ireland/Dublin/ | National Virus Reference Laboratory | Irish Coronavirus Sequencing Consortium - National University of Ireland Galway | Kate Reddington et al |

| | | | | | | |
|---|---|---|---|---|---|---|
| EPI_ISL_848104 | Ireland/D-20G41983/2020 | 2020-03-27 | Europe/Ireland/Dublin/ | National Virus Reference Laboratory | Irish Coronavirus Sequencing Consortium - National University of Ireland Galway | Kate Reddington et al |
| EPI_ISL_848105 | Ireland/D-20G41933/2020 | 2020-03-29 | Europe/Ireland/Dublin/ | National Virus Reference Laboratory | Irish Coronavirus Sequencing Consortium - National University of Ireland Galway | Kate Reddington et al |
| EPI_ISL_848112 | Ireland/D-20G31545/2020 | 2020-03-17 | Europe/Ireland/Dublin/ | National Virus Reference Laboratory | Irish Coronavirus Sequencing Consortium - National University of Ireland Galway | Kate Reddington et al |
| EPI_ISL_848118 | Ireland/D-20G31657/2020 | 2020-03-17 | Europe/Ireland/Dublin/ | National Virus Reference Laboratory | Irish Coronavirus Sequencing Consortium - National University of Ireland Galway | Kate Reddington et al |
| EPI_ISL_848123 | Ireland/D-20G32649/2020 | 2020-03-18 | Europe/Ireland/Dublin/ | National Virus Reference Laboratory | Irish Coronavirus Sequencing Consortium - National University of Ireland Galway | Kate Reddington et al |
| EPI_ISL_848130 | Ireland/D-20G41962/2020 | 2020-03-26 | Europe/Ireland/Dublin/ | National Virus Reference Laboratory | Irish Coronavirus Sequencing Consortium - National University of Ireland Galway | Kate Reddington et al |

| | | | | | | |
|---|---|---|---|---|---|---|
| EPI_ISL_848137 | Ireland/D-20G47000/2020 | 2020-03-30 | Europe/Ireland/Dublin/ | National Virus Reference Laboratory | Irish Coronavirus Sequencing Consortium - National University of Ireland Galway | Kate Reddington et al |
| EPI_ISL_848151 | Ireland/D-20G31531/2020 | 2020-03-17 | Europe/Ireland/Dublin/ | National Virus Reference Laboratory | Irish Coronavirus Sequencing Consortium - National University of Ireland Galway | Kate Reddington et al |
| EPI_ISL_848166 | Ireland/D-20G48058/2020 | 2020-03-29 | Europe/Ireland/Dublin/ | National Virus Reference Laboratory | Irish Coronavirus Sequencing Consortium - National University of Ireland Galway | Kate Reddington et al |
| EPI_ISL_848184 | USA/ND-NDDH-0030/2020 | 2020-03-24 | North America/USA/North Dakota/ | North Dakota Department of Health, Public Health Laboratory | North Dakota Department of Health, Public Health Laboratory | Lisa Wingerter et al |
| EPI_ISL_848187 | USA/ND-NDDH-0042/2020 | 2020-03-25 | North America/USA/North Dakota/ | North Dakota Department of Health, Public Health Laboratory | North Dakota Department of Health, Public Health Laboratory | Lisa Wingerter et al |
| EPI_ISL_848188 | Canada/LTRI-1014/2020 | 2020-12-24 | North America/Canada/Ontario/Toronto | Microbiology Lab | NBCC Sequencing Facility | Dr. Jeff Wrana et al |
| EPI_ISL_848189 | Canada/LTRI-1029/2020 | 2020-12-24 | North America/Canada/Ontario/Toronto | Microbiology Lab | NBCC Sequencing Facility | Dr. Jeff Wrana et al |
| EPI_ISL_848190 | Canada/LTRI-1064/2020 | 2020-12-24 | North America/Canada/Ontario/Toronto | Microbiology Lab | NBCC Sequencing Facility | Dr. Jeff Wrana et al |
| EPI_ISL_848191 | Canada/LTRI-1065/2020 | 2020-12-24 | North America/Canada/Ontario/Toronto | Microbiology Lab | NBCC Sequencing Facility | Dr. Jeff Wrana et al |
| EPI_ISL_848193 | Canada/LTRI-1093/2020 | 2020-12-24 | North America/Canada/Ontario/Toronto | Microbiology Lab | NBCC Sequencing Facility | Dr. Jeff Wrana et al |
| EPI_ISL_848194 | Canada/LTRI-1105/2020 | 2020-12-24 | North America/Canada/Ontario/Toronto | Microbiology Lab | NBCC Sequencing Facility | Dr. Jeff Wrana et al |
| EPI_ISL_848195 | Canada/LTRI-1160/2020 | 2020-12-24 | North America/Canada/Ontario/Toronto | Microbiology Lab | NBCC Sequencing Facility | Dr. Jeff Wrana et al |

| | | | | | | |
|---|---|---|---|---|---|---|
| EPI_ISL_848196 | Canada/LTRI-1192/2020 | 2020-12-24 | North America/Canada/Ontario/Toronto | Microbiology Lab | NBCC Sequencing Facility | Dr. Jeff Wrana et al |
| EPI_ISL_848197 | Canada/LTRI-1258/2020 | 2020-12-24 | North America/Canada/Ontario/Toronto | Microbiology Lab | NBCC Sequencing Facility | Dr. Jeff Wrana et al |
| EPI_ISL_848198 | Canada/LTRI-1321/2020 | 2020-12-24 | North America/Canada/Ontario/Toronto | Microbiology Lab | NBCC Sequencing Facility | Dr. Jeff Wrana et al |
| EPI_ISL_848200 | Germany/BY-ChVir22206/2021 | 2021-01-12 | Europe/Germany/Bavaria/ | Charit√© Universit√§tsmedizin Berlin, Institute of Virology, Charit√©platz 1, 10117 Berlin, Germany | Charit√© Universit√§tsmedizin Berlin, Institute of Virology, Charit√©platz 1, 10117 Berlin, Germany | Victor M Corman et al |
| EPI_ISL_848207 | USA/IL-IDPH-I-000687/2020 | 2020-08-03 | North America/USA/Illinois/Rock Island | Illinois Department of Public Health | Gagnon Lab, Southern Illinois University | Keith Gagnon et al |
| EPI_ISL_848208 | USA/IL-IDPH-C-000423/2020 | 2020-04-27 | North America/USA/Illinois/Randolph Co | Illinois Department of Public Health | Gagnon Lab, Southern Illinois University | Keith Gagnon et al |
| EPI_ISL_848213 | USA/IL-IDPH-I-000211/2020 | 2020-11-17 | North America/USA/Illinois/Winnebago | Illinois Department of Public Health | Gagnon Lab, Southern Illinois University | Keith Gagnon et al |
| EPI_ISL_848214 | USA/IL-IDPH-I-000244/2020 | 2020-11-18 | North America/USA/Illinois/Winnebago | Illinois Department of Public Health | Gagnon Lab, Southern Illinois University | Keith Gagnon et al |
| EPI_ISL_848219 | USA/IL-IDPH-I-000499/2020 | 2020-07-08 | North America/USA/Illinois/Cook County | Illinois Department of Public Health | Gagnon Lab, Southern Illinois University | Keith Gagnon et al |
| EPI_ISL_848224 | USA/IL-IDPH-I-000198/2020 | 2020-10-16 | North America/USA/Illinois/Cook County | Illinois Department of Public Health | Gagnon Lab, Southern Illinois University | Keith Gagnon et al |
| EPI_ISL_848233 | USA/IL-IDPH-I-000259/2020 | 2020-11-19 | North America/USA/Illinois/Will County | Illinois Department of Public Health | Gagnon Lab, Southern Illinois University | Keith Gagnon et al |
| EPI_ISL_848242 | USA/IL-IDPH-I-000240/2020 | 2020-11-17 | North America/USA/Illinois/Boone Coun | Illinois Department of Public Health | Gagnon Lab, Southern Illinois University | Keith Gagnon et al |
| EPI_ISL_848246 | USA/IL-IDPH-C-000147/2020 | 2020-03-30 | North America/USA/Illinois/St. Clair Cou | Illinois Department of Public Health | Gagnon Lab, Southern Illinois University | Keith Gagnon et al |

| | | | | | | |
|---|---|---|---|---|---|---|
| EPI_ISL_848249 | USA/IL-IDPH-I-000286/2020 | 2020-10-27 | North America/USA/Illinois/Cook County | Illinois Department of Public Health | Gagnon Lab, Southern Illinois University | Keith Gagnon et al |
| EPI_ISL_848250 | USA/IL-IDPH-I-000217/2020 | 2020-11-17 | North America/USA/Illinois/Dekalb Cou | Illinois Department of Public Health | Gagnon Lab, Southern Illinois University | Keith Gagnon et al |
| EPI_ISL_848252 | USA/IL-IDPH-I-000111/2020 | 2020-10-07 | North America/USA/Illinois/Stephenson | Illinois Department of Public Health | Gagnon Lab, Southern Illinois University | Keith Gagnon et al |
| EPI_ISL_848253 | USA/IL-IDPH-I-000113/2020 | 2020-10-07 | North America/USA/Illinois/Stephenson | Illinois Department of Public Health | Gagnon Lab, Southern Illinois University | Keith Gagnon et al |
| EPI_ISL_848256 | USA/IL-IDPH-I-000262/2020 | 2020-11-19 | North America/USA/Illinois/Will County | Illinois Department of Public Health | Gagnon Lab, Southern Illinois University | Keith Gagnon et al |
| EPI_ISL_848257 | USA/IL-IDPH-I-000222/2020 | 2020-11-16 | North America/USA/Illinois/Lake County | Illinois Department of Public Health | Gagnon Lab, Southern Illinois University | Keith Gagnon et al |
| EPI_ISL_848260 | USA/IL-IDPH-I-000652/2020 | 2020-12-14 | North America/USA/Illinois/Dupage Cou | Illinois Department of Public Health | Gagnon Lab, Southern Illinois University | Keith Gagnon et al |
| EPI_ISL_848262 | USA/IL-IDPH-C-000411/2020 | 2020-04-27 | North America/USA/Illinois/Perry Count | Illinois Department of Public Health | Gagnon Lab, Southern Illinois University | Keith Gagnon et al |
| EPI_ISL_848265 | USA/IL-IDPH-I-000610/2020 | 2020-12-17 | North America/USA/Illinois/Cook County | Illinois Department of Public Health | Gagnon Lab, Southern Illinois University | Keith Gagnon et al |
| EPI_ISL_848267 | USA/IL-IDPH-I-000039/2020 | 2020-10-16 | North America/USA/Illinois/Cook County | Illinois Department of Public Health | Gagnon Lab, Southern Illinois University | Keith Gagnon et al |
| EPI_ISL_848270 | USA/IL-IDPH-I-000087/2020 | 2020-10-19 | North America/USA/Illinois/Cook County | Illinois Department of Public Health | Gagnon Lab, Southern Illinois University | Keith Gagnon et al |
| EPI_ISL_848289 | USA/IL-IDPH-C-000215/2020 | 2020-04-29 | North America/USA/Illinois/Cook County | Illinois Department of Public Health | Gagnon Lab, Southern Illinois University | Keith Gagnon et al |
| EPI_ISL_848290 | USA/IL-IDPH-C-000044/2020 | 2020-04-30 | North America/USA/Illinois/Perry Count | Illinois Department of Public Health | Gagnon Lab, Southern Illinois University | Keith Gagnon et al |

| | | | | | | |
|---|---|---|---|---|---|---|
| EPI_ISL_848296 | USA/IL-IDPH-I-000293/2020 | 2020-07-09 | North America/USA/Illinois/Cook County | Illinois Department of Public Health | Gagnon Lab, Southern Illinois University | Keith Gagnon et al |
| EPI_ISL_848302 | USA/IL-IDPH-C-000270/2020 | 2020-07-23 | North America/USA/Illinois/Jackson Cou | Illinois Department of Public Health | Gagnon Lab, Southern Illinois University | Keith Gagnon et al |
| EPI_ISL_848303 | USA/IL-IDPH-C-000272/2020 | 2020-07-23 | North America/USA/Illinois/Jackson Cou | Illinois Department of Public Health | Gagnon Lab, Southern Illinois University | Keith Gagnon et al |
| EPI_ISL_848306 | USA/IL-IDPH-C-000274/2020 | 2020-07-24 | North America/USA/Illinois/Randolph Co | Illinois Department of Public Health | Gagnon Lab, Southern Illinois University | Keith Gagnon et al |
| EPI_ISL_848312 | USA/IL-IDPH-C-000293/2020 | 2020-07-24 | North America/USA/Illinois/Randolph Co | Illinois Department of Public Health | Gagnon Lab, Southern Illinois University | Keith Gagnon et al |
| EPI_ISL_848325 | USA/IL-IDPH-I-000193/2020 | 2020-10-05 | North America/USA/Illinois/Cook County | Illinois Department of Public Health | Gagnon Lab, Southern Illinois University | Keith Gagnon et al |
| EPI_ISL_848331 | USA/IL-IDPH-I-000181/2020 | 2020-10-07 | North America/USA/Illinois/Cook County | Illinois Department of Public Health | Gagnon Lab, Southern Illinois University | Keith Gagnon et al |
| EPI_ISL_848336 | USA/IL-IDPH-I-000121/2020 | 2020-10-08 | North America/USA/Illinois/Cook County | Illinois Department of Public Health | Gagnon Lab, Southern Illinois University | Keith Gagnon et al |
| EPI_ISL_848344 | USA/IL-IDPH-I-000138/2020 | 2020-10-08 | North America/USA/Illinois/Cook County | Illinois Department of Public Health | Gagnon Lab, Southern Illinois University | Keith Gagnon et al |
| EPI_ISL_848357 | USA/IL-IDPH-I-000052/2020 | 2020-10-14 | North America/USA/Illinois/Cook County | Illinois Department of Public Health | Gagnon Lab, Southern Illinois University | Keith Gagnon et al |
| EPI_ISL_848360 | USA/IL-IDPH-I-000084/2020 | 2020-10-14 | North America/USA/Illinois/Cook County | Illinois Department of Public Health | Gagnon Lab, Southern Illinois University | Keith Gagnon et al |
| EPI_ISL_848366 | USA/IL-IDPH-I-000042/2020 | 2020-10-16 | North America/USA/Illinois/Cook County | Illinois Department of Public Health | Gagnon Lab, Southern Illinois University | Keith Gagnon et al |
| EPI_ISL_848370 | USA/IL-IDPH-I-000097/2020 | 2020-10-16 | North America/USA/Illinois/Cook County | Illinois Department of Public Health | Gagnon Lab, Southern Illinois University | Keith Gagnon et al |

| | | | | | | |
|---|---|---|---|---|---|---|
| EPI_ISL_848374 | USA/IL-IDPH-I-000200/2020 | 2020-10-17 | North America/USA/Illinois/Dupage Cou | Illinois Department of Public Health | Gagnon Lab, Southern Illinois University | Keith Gagnon et al |
| EPI_ISL_848379 | USA/IL-IDPH-I-000699/2020 | 2020-10-26 | North America/USA/Illinois/Cook County | Illinois Department of Public Health | Gagnon Lab, Southern Illinois University | Keith Gagnon et al |
| EPI_ISL_848382 | USA/IL-IDPH-I-000493/2020 | 2020-11-02 | North America/USA/Illinois/Mchenry Co | Illinois Department of Public Health | Gagnon Lab, Southern Illinois University | Keith Gagnon et al |
| EPI_ISL_848387 | USA/IL-IDPH-I-000214/2020 | 2020-11-17 | North America/USA/Illinois/Winnebago | Illinois Department of Public Health | Gagnon Lab, Southern Illinois University | Keith Gagnon et al |
| EPI_ISL_848388 | USA/IL-IDPH-I-000229/2020 | 2020-11-17 | North America/USA/Illinois/Kendall Cou | Illinois Department of Public Health | Gagnon Lab, Southern Illinois University | Keith Gagnon et al |
| EPI_ISL_848390 | USA/IL-IDPH-I-000248/2020 | 2020-11-17 | North America/USA/Illinois/Boone Coun | Illinois Department of Public Health | Gagnon Lab, Southern Illinois University | Keith Gagnon et al |
| EPI_ISL_848397 | USA/IL-IDPH-I-000453/2020 | 2020-11-24 | North America/USA/Illinois/Lake County | Illinois Department of Public Health | Gagnon Lab, Southern Illinois University | Keith Gagnon et al |
| EPI_ISL_848405 | USA/IL-IDPH-I-000477/2020 | 2020-11-25 | North America/USA/Illinois/Kankakee C | Illinois Department of Public Health | Gagnon Lab, Southern Illinois University | Keith Gagnon et al |
| EPI_ISL_848417 | USA/IL-IDPH-I-000414/2020 | 2020-11-30 | North America/USA/Illinois/Winnebago | Illinois Department of Public Health | Gagnon Lab, Southern Illinois University | Keith Gagnon et al |
| EPI_ISL_848424 | USA/IL-IDPH-I-000446/2020 | 2020-12-02 | North America/USA/Illinois/Lee County | Illinois Department of Public Health | Gagnon Lab, Southern Illinois University | Keith Gagnon et al |
| EPI_ISL_848429 | USA/IL-IDPH-I-000623/2020 | 2020-12-15 | North America/USA/Illinois/Cook County | Illinois Department of Public Health | Gagnon Lab, Southern Illinois University | Keith Gagnon et al |
| EPI_ISL_848431 | USA/IL-IDPH-I-000659/2020 | 2020-12-16 | North America/USA/Illinois/Mchenry Co | Illinois Department of Public Health | Gagnon Lab, Southern Illinois University | Keith Gagnon et al |
| EPI_ISL_848435 | USA/IL-IDPH-I-000660/2020 | 2020-12-17 | North America/USA/Illinois/Mchenry Co | Illinois Department of Public Health | Gagnon Lab, Southern Illinois University | Keith Gagnon et al |

| | | | | | | |
|---|---|---|---|---|---|---|
| EPI_ISL_848448 | USA/IL-IDPH-I-000649/2020 | 2020-12-16 | North America/USA/Illinois/Dupage Cou | Illinois Department of Public Health | Gagnon Lab, Southern Illinois University | Keith Gagnon et al |
| EPI_ISL_848457 | USA/IL-IDPH-C-000004/2020 | 2020-04-04 | North America/USA/Illinois/Madison Co | Illinois Department of Public Health | Gagnon Lab, Southern Illinois University | Keith Gagnon et al |
| EPI_ISL_848462 | USA/IL-IDPH-I-000440/2020 | 2020-12-02 | North America/USA/Illinois/Cook County | Illinois Department of Public Health | Gagnon Lab, Southern Illinois University | Keith Gagnon et al |
| EPI_ISL_848466 | USA/IL-IDPH-I-000008/2020 | 2020-03-07 | North America/USA/Illinois/Cook County | Illinois Department of Public Health | Gagnon Lab, Southern Illinois University | Keith Gagnon et al |
| EPI_ISL_848467 | USA/IL-IDPH-C-000002/2020 | 2020-04-03 | North America/USA/Illinois/St. Clair Cou | Illinois Department of Public Health | Gagnon Lab, Southern Illinois University | Keith Gagnon et al |
| EPI_ISL_848479 | USA/IL-IDPH-C-000240/2020 | 2020-05-01 | North America/USA/Illinois/Dupage Cou | Illinois Department of Public Health | Gagnon Lab, Southern Illinois University | Keith Gagnon et al |
| EPI_ISL_848481 | USA/IL-IDPH-I-000500/2020 | 2020-07-08 | North America/USA/Illinois/Cook County | Illinois Department of Public Health | Gagnon Lab, Southern Illinois University | Keith Gagnon et al |
| EPI_ISL_848482 | USA/IL-IDPH-I-000291/2020 | 2020-07-09 | North America/USA/Illinois/Cook County | Illinois Department of Public Health | Gagnon Lab, Southern Illinois University | Keith Gagnon et al |
| EPI_ISL_848483 | USA/IL-IDPH-I-000297/2020 | 2020-07-09 | North America/USA/Illinois/Cook County | Illinois Department of Public Health | Gagnon Lab, Southern Illinois University | Keith Gagnon et al |
| EPI_ISL_848491 | USA/IL-IDPH-C-000282/2020 | 2020-07-23 | North America/USA/Illinois/Saline Coun | Illinois Department of Public Health | Gagnon Lab, Southern Illinois University | Keith Gagnon et al |
| EPI_ISL_848498 | USA/IL-IDPH-I-000172/2020 | 2020-10-07 | North America/USA/Illinois/Cook County | Illinois Department of Public Health | Gagnon Lab, Southern Illinois University | Keith Gagnon et al |
| EPI_ISL_848509 | USA/IL-IDPH-I-000164/2020 | 2020-10-09 | North America/USA/Illinois/Cook County | Illinois Department of Public Health | Gagnon Lab, Southern Illinois University | Keith Gagnon et al |
| EPI_ISL_848542 | USA/IL-IDPH-I-000641/2020 | 2020-12-17 | North America/USA/Illinois/Cook County | Illinois Department of Public Health | Gagnon Lab, Southern Illinois University | Keith Gagnon et al |

| | | | | | | |
|---|---|---|---|---|---|---|
| EPI_ISL_848554 | USA/ND-NDDH-0026/2020 | 2020-03-24 | North America/USA/North Dakota/ | North Dakota Department of Health, Public Health Laboratory | North Dakota Department of Health, Public Health Laboratory | Lisa Wingerter et al |
| EPI_ISL_848555 | Brazil/PA-IEC176486/2020 | 2020-10-15 | South America/Brazil/Para/ | ÔªøEvandro Chagas Institute | ÔªøEvandro Chagas Institute | Santos et al |
| EPI_ISL_848556 | Brazil/RR-IEC176349/2020 | 2020-05-29 | South America/Brazil/Roraima/ | ÔªøEvandro Chagas Institute | ÔªøEvandro Chagas Institute | Santos et al |
| EPI_ISL_848558 | Brazil/PA-IEC176597/2020 | 2020-10-21 | South America/Brazil/Para/ | ÔªøEvandro Chagas Institute | ÔªøEvandro Chagas Institute | Santos et al |
| EPI_ISL_848559 | Brazil/PA-IEC176581/2020 | 2020-10-19 | South America/Brazil/Para/ | ÔªøEvandro Chagas Institute | ÔªøEvandro Chagas Institute | Santos et al |
| EPI_ISL_848560 | Brazil/PA-IEC176585/2020 | 2020-10-19 | South America/Brazil/Para/ | ÔªøEvandro Chagas Institute | ÔªøEvandro Chagas Institute | Santos et al |
| EPI_ISL_848561 | Brazil/PA-IEC173157/2020 | 2020-07-13 | South America/Brazil/Para/ | ÔªøEvandro Chagas Institute | ÔªøEvandro Chagas Institute | Santos et al |
| EPI_ISL_848562 | Brazil/PA-IEC173159/2020 | 2020-07-10 | South America/Brazil/Para/ | ÔªøEvandro Chagas Institute | ÔªøEvandro Chagas Institute | Santos et al |
| EPI_ISL_848563 | Brazil/PA-IEC173164/2020 | 2020-07-10 | South America/Brazil/Para/ | ÔªøEvandro Chagas Institute | ÔªøEvandro Chagas Institute | Santos et al |
| EPI_ISL_848564 | Brazil/PA-IEC173155/2020 | 2020-07-03 | South America/Brazil/Para/ | ÔªøEvandro Chagas Institute | ÔªøEvandro Chagas Institute | Santos et al |
| EPI_ISL_848565 | Brazil/PA-IEC173158/2020 | 2020-07-03 | South America/Brazil/Para/ | ÔªøEvandro Chagas Institute | ÔªøEvandro Chagas Institute | Santos et al |
| EPI_ISL_848566 | Brazil/PA-IEC173151/2020 | 2020-06-26 | South America/Brazil/Para/ | ÔªøEvandro Chagas Institute | ÔªøEvandro Chagas Institute | Santos et al |
| EPI_ISL_848567 | Brazil/PA-IEC173147/2020 | 2020-06-07 | South America/Brazil/Para/ | ÔªøEvandro Chagas Institute | ÔªøEvandro Chagas Institute | Santos et al |
| EPI_ISL_848568 | Brazil/PA-IEC173146/2020 | 2020-06-04 | South America/Brazil/Para/ | ÔªøEvandro Chagas Institute | ÔªøEvandro Chagas Institute | Santos et al |
| EPI_ISL_848569 | Brazil/PA-IEC173154/2020 | 2020-06-04 | South America/Brazil/Para/ | ÔªøEvandro Chagas Institute | ÔªøEvandro Chagas Institute | Santos et al |
| EPI_ISL_848570 | Brazil/PA-IEC173153/2020 | 2020-05-30 | South America/Brazil/Para/ | ÔªøEvandro Chagas Institute | ÔªøEvandro Chagas Institute | Santos et al |
| EPI_ISL_848571 | Brazil/PA-IEC173135/2020 | 2020-05-29 | South America/Brazil/Para/ | ÔªøEvandro Chagas Institute | ÔªøEvandro Chagas Institute | Santos et al |
| EPI_ISL_848572 | Brazil/PA-IEC173149/2020 | 2020-05-28 | South America/Brazil/Para/ | ÔªøEvandro Chagas Institute | ÔªøEvandro Chagas Institute | Santos et al |
| EPI_ISL_848573 | Brazil/AC-IEC168034/2020 | 2020-05-22 | South America/Brazil/Acre/ | ÔªøEvandro Chagas Institute | ÔªøEvandro Chagas Institute | Santos et al |

| | | | | | | |
|---|---|---|---|---|---|---|
| EPI_ISL_848574 | Brazil/AC-IEC168036/2020 | 2020-05-21 | South America/Brazil/Acre/ | ÔªøEvandro Chagas Institute | ÔªøEvandro Chagas Institute | Santos et al |
| EPI_ISL_848575 | Brazil/AP-IEC168924/2020 | 2020-05-21 | South America/Brazil/Amapa/ | ÔªøEvandro Chagas Institute | ÔªøEvandro Chagas Institute | Santos et al |
| EPI_ISL_848576 | Brazil/AP-IEC168968/2020 | 2020-05-21 | South America/Brazil/Amapa/ | ÔªøEvandro Chagas Institute | ÔªøEvandro Chagas Institute | Santos et al |
| EPI_ISL_848577 | Brazil/AP-IEC168614/2020 | 2020-05-20 | South America/Brazil/Amapa/ | ÔªøEvandro Chagas Institute | ÔªøEvandro Chagas Institute | Santos et al |
| EPI_ISL_848578 | Brazil/AP-IEC168190/2020 | 2020-05-19 | South America/Brazil/Amapa/ | ÔªøEvandro Chagas Institute | ÔªøEvandro Chagas Institute | Santos et al |
| EPI_ISL_848579 | Brazil/AP-IEC168246/2020 | 2020-05-19 | South America/Brazil/Amapa/ | ÔªøEvandro Chagas Institute | ÔªøEvandro Chagas Institute | Santos et al |
| EPI_ISL_848580 | Brazil/MA-IEC166966/2020 | 2020-05-13 | South America/Brazil/Maranh√£o/ | ÔªøEvandro Chagas Institute | ÔªøEvandro Chagas Institute | Santos et al |
| EPI_ISL_848581 | Brazil/MA-IEC166980/2020 | 2020-05-13 | South America/Brazil/Maranh√£o/ | ÔªøEvandro Chagas Institute | ÔªøEvandro Chagas Institute | Santos et al |
| EPI_ISL_848582 | Brazil/PA-IEC173143/2020 | 2020-05-05 | South America/Brazil/Para/ | ÔªøEvandro Chagas Institute | ÔªøEvandro Chagas Institute | Santos et al |
| EPI_ISL_848583 | Brazil/PA-IEC173136/2020 | 2020-05-04 | South America/Brazil/Para/ | ÔªøEvandro Chagas Institute | ÔªøEvandro Chagas Institute | Santos et al |
| EPI_ISL_848584 | Brazil/AP-IEC166437/2020 | 2020-05-01 | South America/Brazil/Amapa/ | ÔªøEvandro Chagas Institute | ÔªøEvandro Chagas Institute | Santos et al |
| EPI_ISL_848588 | Brazil/PA-IEC173044/2020 | 2020-04-23 | South America/Brazil/Para/ | ÔªøEvandro Chagas Institute | ÔªøEvandro Chagas Institute | Santos et al |
| EPI_ISL_848589 | Brazil/PA-IEC169758/2020 | 2020-04-22 | South America/Brazil/Para/ | ÔªøEvandro Chagas Institute | ÔªøEvandro Chagas Institute | Santos et al |
| EPI_ISL_848606 | Brazil/PA-IEC176614/2020 | 2020-10-23 | South America/Brazil/Para/ | ÔªøEvandro Chagas Institute | ÔªøEvandro Chagas Institute | Santos et al |
| EPI_ISL_848607 | Brazil/PA-IEC176564/2020 | 2020-10-16 | South America/Brazil/Para/ | ÔªøEvandro Chagas Institute | ÔªøEvandro Chagas Institute | Santos et al |
| EPI_ISL_848608 | Brazil/PA-IEC176228/2020 | 2020-08-11 | South America/Brazil/Para/ | ÔªøEvandro Chagas Institute | ÔªøEvandro Chagas Institute | Santos et al |
| EPI_ISL_848609 | Brazil/PA-IEC173156/2020 | 2020-07-13 | South America/Brazil/Para/ | ÔªøEvandro Chagas Institute | ÔªøEvandro Chagas Institute | Santos et al |
| EPI_ISL_848610 | Brazil/RR-IEC176364/2020 | 2020-06-24 | South America/Brazil/Roraima/ | ÔªøEvandro Chagas Institute | ÔªøEvandro Chagas Institute | Santos et al |
| EPI_ISL_848611 | Brazil/PA-IEC173145/2020 | 2020-06-11 | South America/Brazil/Para/ | ÔªøEvandro Chagas Institute | ÔªøEvandro Chagas Institute | Santos et al |
| EPI_ISL_848612 | Brazil/AC-IEC169737/2020 | 2020-06-10 | South America/Brazil/Acre/ | ÔªøEvandro Chagas Institute | ÔªøEvandro Chagas Institute | Santos et al |

| | | | | | | |
|---|---|---|---|---|---|---|
| EPI_ISL_848613 | Brazil/AC-IEC169749/2020 | 2020-06-04 | South America/Brazil/Acre/ | ÔªøEvandro Chagas Institute | ÔªøEvandro Chagas Institute | Santos et al |
| EPI_ISL_848614 | Brazil/PA-IEC173152/2020 | 2020-06-01 | South America/Brazil/Para/ | ÔªøEvandro Chagas Institute | ÔªøEvandro Chagas Institute | Santos et al |
| EPI_ISL_848615 | Brazil/PA-IEC173137/2020 | 2020-05-05 | South America/Brazil/Para/ | ÔªøEvandro Chagas Institute | ÔªøEvandro Chagas Institute | Santos et al |
| EPI_ISL_848631 | USA/WY-MTPHL-21004948/2021 | 2021-01-07 | North America/USA/Montana/ | Montana Public Health Laboratory | Wyoming Public Health Laboratory | Noah Hull et al |
| EPI_ISL_848639 | USA/ND-NDDH-0047/2020 | 2020-03-22 | North America/USA/North Dakota/ | North Dakota Department of Health, Public Health Laboratory | North Dakota Department of Health, Public Health Laboratory | Lisa Wingerter et al |
| EPI_ISL_848646 | USA/FL-BPHL-2726/2020 | 2020-12-07 | North America/USA/Florida/ | Florida Bureau of Public Health Laboratories | Florida Bureau of Public Health Laboratories | Sarah Schmedes et al |
| EPI_ISL_848650 | USA/FL-BPHL-2720/2020 | 2020-12-04 | North America/USA/Florida/ | Florida Bureau of Public Health Laboratories | Florida Bureau of Public Health Laboratories | Sarah Schmedes et al |
| EPI_ISL_848658 | USA/FL-BPHL-2708/2020 | 2020-12-03 | North America/USA/Florida/ | Florida Bureau of Public Health Laboratories | Florida Bureau of Public Health Laboratories | Sarah Schmedes et al |
| EPI_ISL_848662 | USA/FL-BPHL-2714/2020 | 2020-12-03 | North America/USA/Florida/ | Florida Bureau of Public Health Laboratories | Florida Bureau of Public Health Laboratories | Sarah Schmedes et al |
| EPI_ISL_848678 | USA/FL-BPHL-2627/2020 | 2020-12-02 | North America/USA/Florida/ | Florida Bureau of Public Health Laboratories | Florida Bureau of Public Health Laboratories | Sarah Schmedes et al |
| EPI_ISL_848679 | USA/FL-BPHL-2624/2020 | 2020-12-02 | North America/USA/Florida/ | Florida Bureau of Public Health Laboratories | Florida Bureau of Public Health Laboratories | Sarah Schmedes et al |
| EPI_ISL_848683 | USA/FL-BPHL-2620/2020 | 2020-12-01 | North America/USA/Florida/ | Florida Bureau of Public Health Laboratories | Florida Bureau of Public Health Laboratories | Sarah Schmedes et al |
| EPI_ISL_848694 | USA/FL-BPHL-2697/2020 | 2020-12-01 | North America/USA/Florida/ | Florida Bureau of Public Health Laboratories | Florida Bureau of Public Health Laboratories | Sarah Schmedes et al |
| EPI_ISL_848704 | USA/FL-BPHL-2675/2020 | 2020-11-30 | North America/USA/Florida/ | Florida Bureau of Public Health Laboratories | Florida Bureau of Public Health Laboratories | Sarah Schmedes et al |

| | | | | | | |
|---|---|---|---|---|---|---|
| EPI_ISL_848714 | USA/FL-BPHL-2673/2020 | 2020-11-30 | North America/USA/Florida/ | Florida Bureau of Public Health Laboratories | Florida Bureau of Public Health Laboratories | Sarah Schmedes et al |
| EPI_ISL_848716 | USA/FL-BPHL-2687/2020 | 2020-11-30 | North America/USA/Florida/ | Florida Bureau of Public Health Laboratories | Florida Bureau of Public Health Laboratories | Sarah Schmedes et al |
| EPI_ISL_848722 | USA/FL-BPHL-2615/2020 | 2020-11-25 | North America/USA/Florida/ | Florida Bureau of Public Health Laboratories | Florida Bureau of Public Health Laboratories | Sarah Schmedes et al |
| EPI_ISL_848724 | USA/FL-BPHL-2614/2020 | 2020-11-25 | North America/USA/Florida/ | Florida Bureau of Public Health Laboratories | Florida Bureau of Public Health Laboratories | Sarah Schmedes et al |
| EPI_ISL_848727 | USA/FL-BPHL-2570/2020 | 2020-11-25 | North America/USA/Florida/ | Florida Bureau of Public Health Laboratories | Florida Bureau of Public Health Laboratories | Sarah Schmedes et al |
| EPI_ISL_848735 | USA/FL-BPHL-2564/2020 | 2020-11-23 | North America/USA/Florida/ | Florida Bureau of Public Health Laboratories | Florida Bureau of Public Health Laboratories | Sarah Schmedes et al |
| EPI_ISL_848736 | USA/FL-BPHL-2561/2020 | 2020-11-23 | North America/USA/Florida/ | Florida Bureau of Public Health Laboratories | Florida Bureau of Public Health Laboratories | Sarah Schmedes et al |
| EPI_ISL_848737 | USA/FL-BPHL-2563/2020 | 2020-11-23 | North America/USA/Florida/ | Florida Bureau of Public Health Laboratories | Florida Bureau of Public Health Laboratories | Sarah Schmedes et al |
| EPI_ISL_848743 | USA/FL-BPHL-2560/2020 | 2020-11-20 | North America/USA/Florida/ | Florida Bureau of Public Health Laboratories | Florida Bureau of Public Health Laboratories | Sarah Schmedes et al |
| EPI_ISL_848748 | USA/FL-BPHL-2656/2020 | 2020-11-20 | North America/USA/Florida/ | Florida Bureau of Public Health Laboratories | Florida Bureau of Public Health Laboratories | Sarah Schmedes et al |
| EPI_ISL_848762 | USA/FL-BPHL-2596/2020 | 2020-11-19 | North America/USA/Florida/ | Florida Bureau of Public Health Laboratories | Florida Bureau of Public Health Laboratories | Sarah Schmedes et al |
| EPI_ISL_848770 | USA/FL-BPHL-2597/2020 | 2020-11-19 | North America/USA/Florida/ | Florida Bureau of Public Health Laboratories | Florida Bureau of Public Health Laboratories | Sarah Schmedes et al |
| EPI_ISL_848775 | USA/FL-BPHL-2558/2020 | 2020-11-18 | North America/USA/Florida/ | Florida Bureau of Public Health Laboratories | Florida Bureau of Public Health Laboratories | Sarah Schmedes et al |

| | | | | | | |
|---|---|---|---|---|---|---|
| EPI_ISL_848781 | USA/FL-BPHL-2584/2020 | 2020-11-18 | North America/USA/Florida/ | Florida Bureau of Public Health Laboratories | Florida Bureau of Public Health Laboratories | Sarah Schmedes et al |
| EPI_ISL_848784 | USA/FL-BPHL-2650/2020 | 2020-11-17 | North America/USA/Florida/ | Florida Bureau of Public Health Laboratories | Florida Bureau of Public Health Laboratories | Sarah Schmedes et al |
| EPI_ISL_848785 | USA/FL-BPHL-2576/2020 | 2020-11-17 | North America/USA/Florida/ | Florida Bureau of Public Health Laboratories | Florida Bureau of Public Health Laboratories | Sarah Schmedes et al |
| EPI_ISL_848787 | USA/FL-BPHL-2587/2020 | 2020-11-17 | North America/USA/Florida/ | Florida Bureau of Public Health Laboratories | Florida Bureau of Public Health Laboratories | Sarah Schmedes et al |
| EPI_ISL_848791 | USA/FL-BPHL-2473/2020 | 2020-11-17 | North America/USA/Florida/ | Florida Bureau of Public Health Laboratories | Florida Bureau of Public Health Laboratories | Sarah Schmedes et al |
| EPI_ISL_848792 | USA/FL-BPHL-2472/2020 | 2020-11-17 | North America/USA/Florida/ | Florida Bureau of Public Health Laboratories | Florida Bureau of Public Health Laboratories | Sarah Schmedes et al |
| EPI_ISL_848794 | USA/FL-BPHL-2575/2020 | 2020-11-17 | North America/USA/Florida/ | Florida Bureau of Public Health Laboratories | Florida Bureau of Public Health Laboratories | Sarah Schmedes et al |
| EPI_ISL_848797 | USA/FL-BPHL-2580/2020 | 2020-11-17 | North America/USA/Florida/ | Florida Bureau of Public Health Laboratories | Florida Bureau of Public Health Laboratories | Sarah Schmedes et al |
| EPI_ISL_848810 | USA/FL-BPHL-2468/2020 | 2020-11-13 | North America/USA/Florida/ | Florida Bureau of Public Health Laboratories | Florida Bureau of Public Health Laboratories | Sarah Schmedes et al |
| EPI_ISL_848815 | USA/FL-BPHL-2640/2020 | 2020-11-13 | North America/USA/Florida/ | Florida Bureau of Public Health Laboratories | Florida Bureau of Public Health Laboratories | Sarah Schmedes et al |
| EPI_ISL_848818 | USA/FL-BPHL-2644/2020 | 2020-11-13 | North America/USA/Florida/ | Florida Bureau of Public Health Laboratories | Florida Bureau of Public Health Laboratories | Sarah Schmedes et al |
| EPI_ISL_848824 | USA/FL-BPHL-2642/2020 | 2020-11-13 | North America/USA/Florida/ | Florida Bureau of Public Health Laboratories | Florida Bureau of Public Health Laboratories | Sarah Schmedes et al |
| EPI_ISL_848835 | USA/FL-BPHL-2465/2020 | 2020-11-12 | North America/USA/Florida/ | Florida Bureau of Public Health Laboratories | Florida Bureau of Public Health Laboratories | Sarah Schmedes et al |

| | | | | | | |
|---|---|---|---|---|---|---|
| EPI_ISL_848856 | USA/FL-BPHL-2534/2020 | 2020-11-04 | North America/USA/Florida/ | Florida Bureau of Public Health Laboratories | Florida Bureau of Public Health Laboratories | Sarah Schmedes et al |
| EPI_ISL_848860 | USA/FL-BPHL-2547/2020 | 2020-11-04 | North America/USA/Florida/ | Florida Bureau of Public Health Laboratories | Florida Bureau of Public Health Laboratories | Sarah Schmedes et al |
| EPI_ISL_848861 | USA/FL-BPHL-2548/2020 | 2020-11-04 | North America/USA/Florida/ | Florida Bureau of Public Health Laboratories | Florida Bureau of Public Health Laboratories | Sarah Schmedes et al |
| EPI_ISL_848867 | USA/FL-BPHL-2538/2020 | 2020-11-04 | North America/USA/Florida/ | Florida Bureau of Public Health Laboratories | Florida Bureau of Public Health Laboratories | Sarah Schmedes et al |
| EPI_ISL_848869 | USA/FL-BPHL-2543/2020 | 2020-11-04 | North America/USA/Florida/ | Florida Bureau of Public Health Laboratories | Florida Bureau of Public Health Laboratories | Sarah Schmedes et al |
| EPI_ISL_848870 | USA/FL-BPHL-2546/2020 | 2020-11-04 | North America/USA/Florida/ | Florida Bureau of Public Health Laboratories | Florida Bureau of Public Health Laboratories | Sarah Schmedes et al |
| EPI_ISL_848875 | USA/FL-BPHL-2526/2020 | 2020-10-28 | North America/USA/Florida/ | Florida Bureau of Public Health Laboratories | Florida Bureau of Public Health Laboratories | Sarah Schmedes et al |
| EPI_ISL_848876 | USA/FL-BPHL-2527/2020 | 2020-10-28 | North America/USA/Florida/ | Florida Bureau of Public Health Laboratories | Florida Bureau of Public Health Laboratories | Sarah Schmedes et al |
| EPI_ISL_848878 | USA/FL-BPHL-2525/2020 | 2020-10-23 | North America/USA/Florida/ | Florida Bureau of Public Health Laboratories | Florida Bureau of Public Health Laboratories | Sarah Schmedes et al |
| EPI_ISL_848885 | USA/FL-BPHL-2520/2020 | 2020-10-21 | North America/USA/Florida/ | Florida Bureau of Public Health Laboratories | Florida Bureau of Public Health Laboratories | Sarah Schmedes et al |
| EPI_ISL_848892 | USA/FL-BPHL-2513/2020 | 2020-10-20 | North America/USA/Florida/ | Florida Bureau of Public Health Laboratories | Florida Bureau of Public Health Laboratories | Sarah Schmedes et al |
| EPI_ISL_848895 | USA/FL-BPHL-2398/2020 | 2020-10-19 | North America/USA/Florida/ | Florida Bureau of Public Health Laboratories | Florida Bureau of Public Health Laboratories | Sarah Schmedes et al |
| EPI_ISL_848898 | USA/FL-BPHL-2392/2020 | 2020-10-19 | North America/USA/Florida/ | Florida Bureau of Public Health Laboratories | Florida Bureau of Public Health Laboratories | Sarah Schmedes et al |

| | | | | | | |
|---|---|---|---|---|---|---|
| EPI_ISL_848903 | USA/FL-BPHL-2397/2020 | 2020-10-19 | North America/USA/Florida/ | Florida Bureau of Public Health Laboratories | Florida Bureau of Public Health Laboratories | Sarah Schmedes et al |
| EPI_ISL_848905 | USA/FL-BPHL-2442/2020 | 2020-10-19 | North America/USA/Florida/ | Florida Bureau of Public Health Laboratories | Florida Bureau of Public Health Laboratories | Sarah Schmedes et al |
| EPI_ISL_848907 | USA/FL-BPHL-2391/2020 | 2020-10-19 | North America/USA/Florida/ | Florida Bureau of Public Health Laboratories | Florida Bureau of Public Health Laboratories | Sarah Schmedes et al |
| EPI_ISL_848912 | USA/FL-BPHL-2378/2020 | 2020-10-19 | North America/USA/Florida/ | Florida Bureau of Public Health Laboratories | Florida Bureau of Public Health Laboratories | Sarah Schmedes et al |
| EPI_ISL_848914 | USA/FL-BPHL-2387/2020 | 2020-10-19 | North America/USA/Florida/ | Florida Bureau of Public Health Laboratories | Florida Bureau of Public Health Laboratories | Sarah Schmedes et al |
| EPI_ISL_848935 | USA/FL-BPHL-2372/2020 | 2020-10-16 | North America/USA/Florida/ | Florida Bureau of Public Health Laboratories | Florida Bureau of Public Health Laboratories | Sarah Schmedes et al |
| EPI_ISL_848946 | USA/FL-BPHL-2358/2020 | 2020-10-13 | North America/USA/Florida/ | Florida Bureau of Public Health Laboratories | Florida Bureau of Public Health Laboratories | Sarah Schmedes et al |
| EPI_ISL_848962 | USA/FL-BPHL-2441/2020 | 2020-10-06 | North America/USA/Florida/ | Florida Bureau of Public Health Laboratories | Florida Bureau of Public Health Laboratories | Sarah Schmedes et al |
| EPI_ISL_848969 | USA/FL-BPHL-2315/2020 | 2020-10-05 | North America/USA/Florida/ | Florida Bureau of Public Health Laboratories | Florida Bureau of Public Health Laboratories | Sarah Schmedes et al |
| EPI_ISL_848979 | USA/FL-BPHL-2304/2020 | 2020-10-05 | North America/USA/Florida/ | Florida Bureau of Public Health Laboratories | Florida Bureau of Public Health Laboratories | Sarah Schmedes et al |
| EPI_ISL_848983 | USA/FL-BPHL-2327/2020 | 2020-10-05 | North America/USA/Florida/ | Florida Bureau of Public Health Laboratories | Florida Bureau of Public Health Laboratories | Sarah Schmedes et al |
| EPI_ISL_848984 | USA/FL-BPHL-2328/2020 | 2020-10-05 | North America/USA/Florida/ | Florida Bureau of Public Health Laboratories | Florida Bureau of Public Health Laboratories | Sarah Schmedes et al |
| EPI_ISL_848993 | USA/FL-BPHL-2302/2020 | 2020-10-02 | North America/USA/Florida/ | Florida Bureau of Public Health Laboratories | Florida Bureau of Public Health Laboratories | Sarah Schmedes et al |

| | | | | | | |
|---|---|---|---|---|---|---|
| EPI_ISL_848994 | USA/FL-BPHL-2306/2020 | 2020-10-02 | North America/USA/Florida/ | Florida Bureau of Public Health Laboratories | Florida Bureau of Public Health Laboratories | Sarah Schmedes et al |
| EPI_ISL_848996 | USA/FL-BPHL-2308/2020 | 2020-10-02 | North America/USA/Florida/ | Florida Bureau of Public Health Laboratories | Florida Bureau of Public Health Laboratories | Sarah Schmedes et al |
| EPI_ISL_849001 | USA/FL-BPHL-2333/2020 | 2020-10-02 | North America/USA/Florida/ | Florida Bureau of Public Health Laboratories | Florida Bureau of Public Health Laboratories | Sarah Schmedes et al |
| EPI_ISL_849014 | USA/FL-BPHL-2291/2020 | 2020-09-29 | North America/USA/Florida/ | Florida Bureau of Public Health Laboratories | Florida Bureau of Public Health Laboratories | Sarah Schmedes et al |
| EPI_ISL_849020 | USA/FL-BPHL-2287/2020 | 2020-09-27 | North America/USA/Florida/ | Florida Bureau of Public Health Laboratories | Florida Bureau of Public Health Laboratories | Sarah Schmedes et al |
| EPI_ISL_849021 | USA/FL-BPHL-2284/2020 | 2020-09-25 | North America/USA/Florida/ | Florida Bureau of Public Health Laboratories | Florida Bureau of Public Health Laboratories | Sarah Schmedes et al |
| EPI_ISL_849023 | USA/FL-BPHL-2282/2020 | 2020-09-25 | North America/USA/Florida/ | Florida Bureau of Public Health Laboratories | Florida Bureau of Public Health Laboratories | Sarah Schmedes et al |
| EPI_ISL_849026 | USA/FL-BPHL-2281/2020 | 2020-09-25 | North America/USA/Florida/ | Florida Bureau of Public Health Laboratories | Florida Bureau of Public Health Laboratories | Sarah Schmedes et al |
| EPI_ISL_849027 | USA/FL-BPHL-2278/2020 | 2020-09-24 | North America/USA/Florida/ | Florida Bureau of Public Health Laboratories | Florida Bureau of Public Health Laboratories | Sarah Schmedes et al |
| EPI_ISL_849028 | USA/FL-BPHL-2286/2020 | 2020-09-24 | North America/USA/Florida/ | Florida Bureau of Public Health Laboratories | Florida Bureau of Public Health Laboratories | Sarah Schmedes et al |
| EPI_ISL_849030 | USA/FL-BPHL-2277/2020 | 2020-09-23 | North America/USA/Florida/ | Florida Bureau of Public Health Laboratories | Florida Bureau of Public Health Laboratories | Sarah Schmedes et al |
| EPI_ISL_849033 | USA/FL-BPHL-2762/2020 | 2020-09-15 | North America/USA/Florida/ | Florida Bureau of Public Health Laboratories | Florida Bureau of Public Health Laboratories | Sarah Schmedes et al |
| EPI_ISL_849037 | USA/FL-BPHL-2774/2020 | 2020-09-15 | North America/USA/Florida/ | Florida Bureau of Public Health Laboratories | Florida Bureau of Public Health Laboratories | Sarah Schmedes et al |

| | | | | | | |
|---|---|---|---|---|---|---|
| EPI_ISL_849041 | USA/FL-BPHL-2757/2020 | 2020-09-15 | North America/USA/Florida/ | Florida Bureau of Public Health Laboratories | Florida Bureau of Public Health Laboratories | Sarah Schmedes et al |
| EPI_ISL_849044 | USA/FL-BPHL-2747/2020 | 2020-09-14 | North America/USA/Florida/ | Florida Bureau of Public Health Laboratories | Florida Bureau of Public Health Laboratories | Sarah Schmedes et al |
| EPI_ISL_849045 | USA/FL-BPHL-2754/2020 | 2020-09-14 | North America/USA/Florida/ | Florida Bureau of Public Health Laboratories | Florida Bureau of Public Health Laboratories | Sarah Schmedes et al |
| EPI_ISL_849047 | USA/FL-BPHL-2768/2020 | 2020-09-14 | North America/USA/Florida/ | Florida Bureau of Public Health Laboratories | Florida Bureau of Public Health Laboratories | Sarah Schmedes et al |
| EPI_ISL_849048 | USA/FL-BPHL-2760/2020 | 2020-09-14 | North America/USA/Florida/ | Florida Bureau of Public Health Laboratories | Florida Bureau of Public Health Laboratories | Sarah Schmedes et al |
| EPI_ISL_849050 | USA/FL-BPHL-2766/2020 | 2020-09-14 | North America/USA/Florida/ | Florida Bureau of Public Health Laboratories | Florida Bureau of Public Health Laboratories | Sarah Schmedes et al |
| EPI_ISL_849059 | USA/FL-BPHL-2752/2020 | 2020-09-14 | North America/USA/Florida/ | Florida Bureau of Public Health Laboratories | Florida Bureau of Public Health Laboratories | Sarah Schmedes et al |
| EPI_ISL_849065 | USA/FL-BPHL-2746/2020 | 2020-09-14 | North America/USA/Florida/ | Florida Bureau of Public Health Laboratories | Florida Bureau of Public Health Laboratories | Sarah Schmedes et al |
| EPI_ISL_849071 | USA/FL-BPHL-2728/2020 | 2020-09-11 | North America/USA/Florida/ | Florida Bureau of Public Health Laboratories | Florida Bureau of Public Health Laboratories | Sarah Schmedes et al |
| EPI_ISL_849072 | USA/FL-BPHL-2736/2020 | 2020-09-11 | North America/USA/Florida/ | Florida Bureau of Public Health Laboratories | Florida Bureau of Public Health Laboratories | Sarah Schmedes et al |
| EPI_ISL_849074 | USA/FL-BPHL-2740/2020 | 2020-09-11 | North America/USA/Florida/ | Florida Bureau of Public Health Laboratories | Florida Bureau of Public Health Laboratories | Sarah Schmedes et al |
| EPI_ISL_849076 | USA/FL-BPHL-2738/2020 | 2020-09-11 | North America/USA/Florida/ | Florida Bureau of Public Health Laboratories | Florida Bureau of Public Health Laboratories | Sarah Schmedes et al |
| EPI_ISL_849079 | USA/FL-BPHL-2730/2020 | 2020-09-10 | North America/USA/Florida/ | Florida Bureau of Public Health Laboratories | Florida Bureau of Public Health Laboratories | Sarah Schmedes et al |

| | | | | | | |
|---|---|---|---|---|---|---|
| EPI_ISL_849081 | USA/FL-BPHL-2734/2020 | 2020-09-10 | North America/USA/Florida/ | Florida Bureau of Public Health Laboratories | Florida Bureau of Public Health Laboratories | Sarah Schmedes et al |
| EPI_ISL_849082 | USA/FL-BPHL-2731/2020 | 2020-09-10 | North America/USA/Florida/ | Florida Bureau of Public Health Laboratories | Florida Bureau of Public Health Laboratories | Sarah Schmedes et al |
| EPI_ISL_849084 | USA/FL-BPHL-2735/2020 | 2020-09-10 | North America/USA/Florida/ | Florida Bureau of Public Health Laboratories | Florida Bureau of Public Health Laboratories | Sarah Schmedes et al |
| EPI_ISL_849086 | USA/FL-BPHL-2436/2020 | 2020-08-03 | North America/USA/Florida/ | Florida Bureau of Public Health Laboratories | Florida Bureau of Public Health Laboratories | Sarah Schmedes et al |
| EPI_ISL_849096 | USA/FL-BPHL-2438/2020 | 2020-08-03 | North America/USA/Florida/ | Florida Bureau of Public Health Laboratories | Florida Bureau of Public Health Laboratories | Sarah Schmedes et al |
| EPI_ISL_849098 | USA/FL-BPHL-2416/2020 | 2020-08-02 | North America/USA/Florida/ | Florida Bureau of Public Health Laboratories | Florida Bureau of Public Health Laboratories | Sarah Schmedes et al |
| EPI_ISL_849109 | USA/FL-BPHL-2414/2020 | 2020-07-31 | North America/USA/Florida/ | Florida Bureau of Public Health Laboratories | Florida Bureau of Public Health Laboratories | Sarah Schmedes et al |
| EPI_ISL_849120 | USA/FL-BPHL-2481/2020 | 2020-07-07 | North America/USA/Florida/ | Florida Bureau of Public Health Laboratories | Florida Bureau of Public Health Laboratories | Sarah Schmedes et al |
| EPI_ISL_849134 | USA/FL-BPHL-2490/2020 | 2020-07-07 | North America/USA/Florida/ | Florida Bureau of Public Health Laboratories | Florida Bureau of Public Health Laboratories | Sarah Schmedes et al |
| EPI_ISL_849144 | USA/FL-BPHL-2500/2020 | 2020-07-07 | North America/USA/Florida/ | Florida Bureau of Public Health Laboratories | Florida Bureau of Public Health Laboratories | Sarah Schmedes et al |
| EPI_ISL_849145 | USA/FL-BPHL-2504/2020 | 2020-07-07 | North America/USA/Florida/ | Florida Bureau of Public Health Laboratories | Florida Bureau of Public Health Laboratories | Sarah Schmedes et al |
| EPI_ISL_849165 | Argentina/INEI011714/2020 | 2020-05-10 | South America/Argentina/Buenos Aires/ | Servicio Virosis Respiratorias-Departamento Virolog√≠a-INEI | Instituto Nacional Enfermedades Infecciosas C.G.Malbran | Baumeister E. et al |
| EPI_ISL_849178 | Argentina/INEI013032/2020 | 2020-05-19 | South America/Argentina/Buenos Aires/ | Servicio Virosis Respiratorias-Departamento Virolog√≠a-INEI | Instituto Nacional Enfermedades Infecciosas C.G.Malbran | Baumeister E. et al |

| | | | | | | |
|---|---|---|---|---|---|---|
| EPI_ISL_849181 | Argentina/INEI013198/2020 | 2020-05-20 | South America/Argentina/Buenos Aires/ | Servicio Virosis Respiratorias-Departamento Virolog√≠a-INEI | Instituto Nacional Enfermedades Infecciosas C.G.Malbran | Baumeister E. et al |
| EPI_ISL_849197 | Argentina/INEI012243/2020 | 2020-05-14 | South America/Argentina/Buenos Aires/ | Servicio Virosis Respiratorias-Departamento Virolog√≠a-INEI | Instituto Nacional Enfermedades Infecciosas C.G.Malbran | Baumeister E. et al |
| EPI_ISL_849241 | USA/UT-UPHL-2101471607/2020 | 2020-06-15 | North America/USA/Utah/ | Utah Public Health Laboratory | Utah Public Health Laboratory | Erin L. Young et al |
| EPI_ISL_849242 | USA/UT-UPHL-2101808147/2020 | 2020-06-16 | North America/USA/Utah/ | Utah Public Health Laboratory | Utah Public Health Laboratory | Erin L. Young et al |
| EPI_ISL_849244 | USA/UT-UPHL-2101443839/2020 | 2020-06-16 | North America/USA/Utah/ | Utah Public Health Laboratory | Utah Public Health Laboratory | Erin L. Young et al |
| EPI_ISL_849248 | USA/UT-UPHL-2101458067/2020 | 2020-12-15 | North America/USA/Utah/ | Utah Public Health Laboratory | Utah Public Health Laboratory | Erin L. Young et al |
| EPI_ISL_849250 | USA/UT-UPHL-2101770232/2020 | 2020-12-15 | North America/USA/Utah/ | Utah Public Health Laboratory | Utah Public Health Laboratory | Erin L. Young et al |
| EPI_ISL_849277 | USA/UT-UPHL-2101627706/2020 | 2020-12-14 | North America/USA/Utah/ | Utah Public Health Laboratory | Utah Public Health Laboratory | Erin L. Young et al |
| EPI_ISL_849278 | USA/UT-UPHL-2101886530/2020 | 2020-12-15 | North America/USA/Utah/ | Utah Public Health Laboratory | Utah Public Health Laboratory | Erin L. Young et al |
| EPI_ISL_849282 | Argentina/INEI030804/2020 | 2020-06-30 | South America/Argentina/Buenos Aires/ | Servicio Virosis Respiratorias-Departamento Virolog√≠a-INEI | Instituto Nacional Enfermedades Infecciosas C.G.Malbran | Baumeister E. et al |
| EPI_ISL_849284 | Argentina/INEI030427/2020 | 2020-06-28 | South America/Argentina/Buenos Aires/ | Servicio Virosis Respiratorias-Departamento Virolog√≠a-INEI | Instituto Nacional Enfermedades Infecciosas C.G.Malbran | Baumeister E. et al |
| EPI_ISL_849285 | Argentina/INEI023720/2020 | 2020-06-29 | South America/Argentina/Buenos Aires/ | Servicio Virosis Respiratorias-Departamento Virolog√≠a-INEI | Instituto Nacional Enfermedades Infecciosas C.G.Malbran | Baumeister E. et al |
| EPI_ISL_849286 | Argentina/INEI017138/2020 | 2020-06-08 | South America/Argentina/Buenos Aires/ | Servicio Virosis Respiratorias-Departamento Virolog√≠a-INEI | Instituto Nacional Enfermedades Infecciosas C.G.Malbran | Baumeister E. et al |

| | | | | | | |
|---|---|---|---|---|---|---|
| EPI_ISL_849287 | Argentina/INEI018564/2020 | 2020-06-14 | South America/Argentina/Buenos Aires/ | Servicio Virosis Respiratorias-Departamento Virologv≠a-INEI | Instituto Nacional Enfermedades Infecciosas C.G.Malbran | Baumeister E. et al |
| EPI_ISL_849288 | Argentina/INEI030336/2020 | 2020-06-28 | South America/Argentina/Buenos Aires/ | Servicio Virosis Respiratorias-Departamento Virologv≠a-INEI | Instituto Nacional Enfermedades Infecciosas C.G.Malbran | Baumeister E. et al |
| EPI_ISL_849289 | Argentina/INEI025314/2020 | 2020-07-10 | South America/Argentina/Buenos Aires/ | Servicio Virosis Respiratorias-Departamento Virologv≠a-INEI | Instituto Nacional Enfermedades Infecciosas C.G.Malbran | Baumeister E. et al |
| EPI_ISL_849291 | Argentina/INEI032121/2020 | 2020-07-06 | South America/Argentina/Buenos Aires/ | Servicio Virosis Respiratorias-Departamento Virologv≠a-INEI | Instituto Nacional Enfermedades Infecciosas C.G.Malbran | Baumeister E. et al |
| EPI_ISL_849292 | Argentina/INEI024795/2020 | 2020-07-07 | South America/Argentina/Buenos Aires/ | Servicio Virosis Respiratorias-Departamento Virologv≠a-INEI | Instituto Nacional Enfermedades Infecciosas C.G.Malbran | Baumeister E. et al |
| EPI_ISL_849293 | Argentina/INEI019377/2020 | 2020-06-17 | South America/Argentina/Buenos Aires/ | Servicio Virosis Respiratorias-Departamento Virologv≠a-INEI | Instituto Nacional Enfermedades Infecciosas C.G.Malbran | Baumeister E. et al |
| EPI_ISL_849294 | Argentina/INEI019026/2020 | 2020-06-15 | South America/Argentina/Buenos Aires/ | Servicio Virosis Respiratorias-Departamento Virologv≠a-INEI | Instituto Nacional Enfermedades Infecciosas C.G.Malbran | Baumeister E. et al |
| EPI_ISL_849295 | Argentina/INEI019568/2020 | 2020-06-17 | South America/Argentina/Buenos Aires/ | Servicio Virosis Respiratorias-Departamento Virologv≠a-INEI | Instituto Nacional Enfermedades Infecciosas C.G.Malbran | Baumeister E. et al |
| EPI_ISL_849298 | Argentina/INEI020862/2020 | 2020-06-10 | South America/Argentina/Buenos Aires/ | Servicio Virosis Respiratorias-Departamento Virologv≠a-INEI | Instituto Nacional Enfermedades Infecciosas C.G.Malbran | Baumeister E. et al |
| EPI_ISL_849299 | Argentina/INEI018812/2020 | 2020-06-14 | South America/Argentina/Buenos Aires/ | Servicio Virosis Respiratorias-Departamento Virologv≠a-INEI | Instituto Nacional Enfermedades Infecciosas C.G.Malbran | Baumeister E. et al |

| | | | | | | |
|---|---|---|---|---|---|---|
| EPI_ISL_849300 | Argentina/INEI017951/2020 | 2020-06-11 | South America/Argentina/Buenos Aires/ | Servicio Virosis Respiratorias-Departamento Virologv≠a-INEI | Instituto Nacional Enfermedades Infecciosas C.G.Malbran | Baumeister E. et al |
| EPI_ISL_849301 | Argentina/INEI018682/2020 | 2020-06-13 | South America/Argentina/Buenos Aires/ | Servicio Virosis Respiratorias-Departamento Virologv≠a-INEI | Instituto Nacional Enfermedades Infecciosas C.G.Malbran | Baumeister E. et al |
| EPI_ISL_849302 | Argentina/INEI031683/2020 | 2020-07-04 | South America/Argentina/Buenos Aires/ | Servicio Virosis Respiratorias-Departamento Virologv≠a-INEI | Instituto Nacional Enfermedades Infecciosas C.G.Malbran | Baumeister E. et al |
| EPI_ISL_849304 | Argentina/INEI041399/2020 | 2020-06-25 | South America/Argentina/Buenos Aires/ | Servicio Virosis Respiratorias-Departamento Virologv≠a-INEI | Instituto Nacional Enfermedades Infecciosas C.G.Malbran | Baumeister E. et al |
| EPI_ISL_849305 | Argentina/INEI022085/2020 | 2020-06-18 | South America/Argentina/Buenos Aires/ | Servicio Virosis Respiratorias-Departamento Virologv≠a-INEI | Instituto Nacional Enfermedades Infecciosas C.G.Malbran | Baumeister E. et al |
| EPI_ISL_849306 | Argentina/INEI024145/2020 | 2020-07-02 | South America/Argentina/Buenos Aires/ | Servicio Virosis Respiratorias-Departamento Virologv≠a-INEI | Instituto Nacional Enfermedades Infecciosas C.G.Malbran | Baumeister E. et al |
| EPI_ISL_849307 | Argentina/INEI019422/2020 | 2020-06-17 | South America/Argentina/Buenos Aires/ | Servicio Virosis Respiratorias-Departamento Virologv≠a-INEI | Instituto Nacional Enfermedades Infecciosas C.G.Malbran | Baumeister E. et al |
| EPI_ISL_849308 | Argentina/INEI021845/2020 | 2020-06-17 | South America/Argentina/Buenos Aires/ | Servicio Virosis Respiratorias-Departamento Virologv≠a-INEI | Instituto Nacional Enfermedades Infecciosas C.G.Malbran | Baumeister E. et al |
| EPI_ISL_849309 | Argentina/INEI022121/2020 | 2020-06-18 | South America/Argentina/Buenos Aires/ | Servicio Virosis Respiratorias-Departamento Virologv≠a-INEI | Instituto Nacional Enfermedades Infecciosas C.G.Malbran | Baumeister E. et al |
| EPI_ISL_849310 | Argentina/INEI022890/2020 | 2020-06-22 | South America/Argentina/Buenos Aires/ | Servicio Virosis Respiratorias-Departamento Virologv≠a-INEI | Instituto Nacional Enfermedades Infecciosas C.G.Malbran | Baumeister E. et al |

| | | | | | | |
|---|---|---|---|---|---|---|
| EPI_ISL_849311 | Argentina/INEI031619/2020 | 2020-07-04 | South America/Argentina/Buenos Aires/ | Servicio Virosis Respiratorias-Departamento Virologv≠a-INEI | Instituto Nacional Enfermedades Infecciosas C.G.Malbran | Baumeister E. et al |
| EPI_ISL_849312 | Argentina/INEI031669/2020 | 2020-07-04 | South America/Argentina/Buenos Aires/ | Servicio Virosis Respiratorias-Departamento Virologv≠a-INEI | Instituto Nacional Enfermedades Infecciosas C.G.Malbran | Baumeister E. et al |
| EPI_ISL_849313 | Argentina/INEI040157/2020 | 2020-06-19 | South America/Argentina/Buenos Aires/ | Servicio Virosis Respiratorias-Departamento Virologv≠a-INEI | Instituto Nacional Enfermedades Infecciosas C.G.Malbran | Baumeister E. et al |
| EPI_ISL_849314 | Argentina/INEI041082/2020 | 2020-06-24 | South America/Argentina/Buenos Aires/ | Servicio Virosis Respiratorias-Departamento Virologv≠a-INEI | Instituto Nacional Enfermedades Infecciosas C.G.Malbran | Baumeister E. et al |
| EPI_ISL_849316 | Argentina/INEI041592/2020 | 2020-06-26 | South America/Argentina/Buenos Aires/ | Servicio Virosis Respiratorias-Departamento Virologv≠a-INEI | Instituto Nacional Enfermedades Infecciosas C.G.Malbran | Baumeister E. et al |
| EPI_ISL_849317 | Argentina/INEI031617/2020 | 2020-07-04 | South America/Argentina/Buenos Aires/ | Servicio Virosis Respiratorias-Departamento Virologv≠a-INEI | Instituto Nacional Enfermedades Infecciosas C.G.Malbran | Baumeister E. et al |
| EPI_ISL_849318 | Argentina/INEI017932/2020 | 2020-06-11 | South America/Argentina/Buenos Aires/ | Servicio Virosis Respiratorias-Departamento Virologv≠a-INEI | Instituto Nacional Enfermedades Infecciosas C.G.Malbran | Baumeister E. et al |
| EPI_ISL_849320 | Argentina/INEI031614/2020 | 2020-07-04 | South America/Argentina/Buenos Aires/ | Servicio Virosis Respiratorias-Departamento Virologv≠a-INEI | Instituto Nacional Enfermedades Infecciosas C.G.Malbran | Baumeister E. et al |
| EPI_ISL_849321 | Argentina/INEI040154/2020 | 2020-06-19 | South America/Argentina/Buenos Aires/ | Servicio Virosis Respiratorias-Departamento Virologv≠a-INEI | Instituto Nacional Enfermedades Infecciosas C.G.Malbran | Baumeister E. et al |
| EPI_ISL_849322 | USA/DE-DHSS-FLW00599083/2020 | 2020-12-31 | North America/USA/Delaware/New Cas | Delaware Public Health Lab | Delaware Public Health Lab | Gregory Hovan et al |
| EPI_ISL_849323 | USA/DE-DHSS-FLW00599230/2020 | 2020-12-31 | North America/USA/Delaware/New Cas | Delaware Public Health Lab | Delaware Public Health Lab | Gregory Hovan et al |

| | | | | | | |
|---|---|---|---|---|---|---|
| EPI_ISL_849328 | USA/DE-DHSS-F1047459/2021 | 2021-01-02 | North America/USA/Delaware/New Cas | Delaware Public Health Lab | Delaware Public Health Lab | Gregory Hovan et al |
| EPI_ISL_849336 | USA/DE-DHSS-FLW00610904/2021 | 2021-01-04 | North America/USA/Delaware/Sussex C | Delaware Public Health Lab | Delaware Public Health Lab | Gregory Hovan et al |
| EPI_ISL_849338 | USA/DE-DHSS-FLW00611304/2021 | 2021-01-04 | North America/USA/Delaware/Sussex C | Delaware Public Health Lab | Delaware Public Health Lab | Gregory Hovan et al |
| EPI_ISL_849344 | USA/DE-DHSS-F1047460/2021 | 2021-01-04 | North America/USA/Delaware/New Cas | Delaware Public Health Lab | Delaware Public Health Lab | Gregory Hovan et al |
| EPI_ISL_849345 | USA/DE-DHSS-FLW00605102/2021 | 2021-01-04 | North America/USA/Delaware/New Cas | Delaware Public Health Lab | Delaware Public Health Lab | Gregory Hovan et al |
| EPI_ISL_849348 | USA/DE-DHSS-FLW00612372/2021 | 2021-01-04 | North America/USA/Delaware/New Cas | Delaware Public Health Lab | Delaware Public Health Lab | Gregory Hovan et al |
| EPI_ISL_849349 | USA/DE-DHSS-F1047464/2021 | 2021-01-04 | North America/USA/Delaware/Kent Cou | Delaware Public Health Lab | Delaware Public Health Lab | Gregory Hovan et al |
| EPI_ISL_849350 | Argentina/INEI095222/2020 | 2020-10-16 | South America/Argentina/Buenos Aires/ | Servicio Virosis Respiratorias-Departamento Virologv≠a-INEI | Instituto Nacional Enfermedades Infecciosas C.G.Malbran | Baumeister E. et al |
| EPI_ISL_849351 | Argentina/INEI095228/2020 | 2020-10-16 | South America/Argentina/Buenos Aires/ | Servicio Virosis Respiratorias-Departamento Virologv≠a-INEI | Instituto Nacional Enfermedades Infecciosas C.G.Malbran | Baumeister E. et al |
| EPI_ISL_849352 | Argentina/INEI095276/2020 | 2020-10-19 | South America/Argentina/Argentina/ | Servicio Virosis Respiratorias-Departamento Virologv≠a-INEI | Instituto Nacional Enfermedades Infecciosas C.G.Malbran | Baumeister E. et al |
| EPI_ISL_849353 | Argentina/INEI095283/2020 | 2020-10-15 | South America/Argentina/Buenos Aires/ | Servicio Virosis Respiratorias-Departamento Virologv≠a-INEI | Instituto Nacional Enfermedades Infecciosas C.G.Malbran | Baumeister E. et al |
| EPI_ISL_849354 | Argentina/INEI095287/2020 | 2020-10-15 | South America/Argentina/Buenos Aires/ | Servicio Virosis Respiratorias-Departamento Virologv≠a-INEI | Instituto Nacional Enfermedades Infecciosas C.G.Malbran | Baumeister E. et al |
| EPI_ISL_849355 | Argentina/INEI095357/2020 | 2020-10-19 | South America/Argentina/Buenos Aires/ | Servicio Virosis Respiratorias-Departamento Virologv≠a-INEI | Instituto Nacional Enfermedades Infecciosas C.G.Malbran | Baumeister E. et al |

| | | | | | | |
|---|---|---|---|---|---|---|
| EPI_ISL_849356 | Argentina/INEI095413/2020 | 2020-10-19 | South America/Argentina/Buenos Aires/ | Servicio Virosis Respiratorias-Departamento Virolog√≠a-INEI | Instituto Nacional Enfermedades Infecciosas C.G.Malbran | Baumeister E. et al |
| EPI_ISL_849357 | Argentina/INEI095421/2020 | 2020-10-16 | South America/Argentina/Buenos Aires/ | Servicio Virosis Respiratorias-Departamento Virolog√≠a-INEI | Instituto Nacional Enfermedades Infecciosas C.G.Malbran | Baumeister E. et al |
| EPI_ISL_849358 | Argentina/INEI095450/2020 | 2020-10-19 | South America/Argentina/Buenos Aires/ | Servicio Virosis Respiratorias-Departamento Virolog√≠a-INEI | Instituto Nacional Enfermedades Infecciosas C.G.Malbran | Baumeister E. et al |
| EPI_ISL_849359 | Argentina/INEI095503/2020 | 2020-10-20 | South America/Argentina/Buenos Aires/ | Servicio Virosis Respiratorias-Departamento Virolog√≠a-INEI | Instituto Nacional Enfermedades Infecciosas C.G.Malbran | Baumeister E. et al |
| EPI_ISL_849361 | Argentina/INEI095579/2020 | 2020-10-16 | South America/Argentina/Buenos Aires/ | Servicio Virosis Respiratorias-Departamento Virolog√≠a-INEI | Instituto Nacional Enfermedades Infecciosas C.G.Malbran | Baumeister E. et al |
| EPI_ISL_849362 | Argentina/INEI095674/2020 | 2020-10-17 | South America/Argentina/Buenos Aires/ | Servicio Virosis Respiratorias-Departamento Virolog√≠a-INEI | Instituto Nacional Enfermedades Infecciosas C.G.Malbran | Baumeister E. et al |
| EPI_ISL_849363 | Argentina/INEI095695/2020 | 2020-10-16 | South America/Argentina/Buenos Aires/ | Servicio Virosis Respiratorias-Departamento Virolog√≠a-INEI | Instituto Nacional Enfermedades Infecciosas C.G.Malbran | Baumeister E. et al |
| EPI_ISL_849364 | Argentina/INEI095845/2020 | 2020-10-22 | South America/Argentina/Buenos Aires/ | Servicio Virosis Respiratorias-Departamento Virolog√≠a-INEI | Instituto Nacional Enfermedades Infecciosas C.G.Malbran | Baumeister E. et al |
| EPI_ISL_849365 | Argentina/INEI095952/2020 | 2020-10-21 | South America/Argentina/Buenos Aires/ | Servicio Virosis Respiratorias-Departamento Virolog√≠a-INEI | Instituto Nacional Enfermedades Infecciosas C.G.Malbran | Baumeister E. et al |
| EPI_ISL_849366 | Argentina/INEI095955/2020 | 2020-10-21 | South America/Argentina/Buenos Aires/ | Servicio Virosis Respiratorias-Departamento Virolog√≠a-INEI | Instituto Nacional Enfermedades Infecciosas C.G.Malbran | Baumeister E. et al |

| | | | | | | |
|---|---|---|---|---|---|---|
| EPI_ISL_849367 | Argentina/INEI095994/2020 | 2020-10-18 | South America/Argentina/Buenos Aires/ | Servicio Virosis Respiratorias-Departamento Virologv≠a-INEI | Instituto Nacional Enfermedades Infecciosas C.G.Malbran | Baumeister E. et al |
| EPI_ISL_849368 | Argentina/INEI096021/2020 | 2020-10-22 | South America/Argentina/Buenos Aires/ | Servicio Virosis Respiratorias-Departamento Virologv≠a-INEI | Instituto Nacional Enfermedades Infecciosas C.G.Malbran | Baumeister E. et al |
| EPI_ISL_849369 | Argentina/INEI096022/2020 | 2020-10-22 | South America/Argentina/Buenos Aires/ | Servicio Virosis Respiratorias-Departamento Virologv≠a-INEI | Instituto Nacional Enfermedades Infecciosas C.G.Malbran | Baumeister E. et al |
| EPI_ISL_849370 | Argentina/INEI096077/2020 | 2020-10-23 | South America/Argentina/Buenos Aires/ | Servicio Virosis Respiratorias-Departamento Virologv≠a-INEI | Instituto Nacional Enfermedades Infecciosas C.G.Malbran | Baumeister E. et al |
| EPI_ISL_849371 | Argentina/INEI096084/2020 | 2020-10-24 | South America/Argentina/Buenos Aires/ | Servicio Virosis Respiratorias-Departamento Virologv≠a-INEI | Instituto Nacional Enfermedades Infecciosas C.G.Malbran | Baumeister E. et al |
| EPI_ISL_849372 | Argentina/INEI096204/2020 | 2020-10-26 | South America/Argentina/Buenos Aires/ | Servicio Virosis Respiratorias-Departamento Virologv≠a-INEI | Instituto Nacional Enfermedades Infecciosas C.G.Malbran | Baumeister E. et al |
| EPI_ISL_849373 | Argentina/INEI082860/2020 | 2020-10-08 | South America/Argentina/Buenos Aires/ | Servicio Virosis Respiratorias-Departamento Virologv≠a-INEI | Instituto Nacional Enfermedades Infecciosas C.G.Malbran | Baumeister E. et al |
| EPI_ISL_849374 | Argentina/INEI082862/2020 | 2020-10-08 | South America/Argentina/Buenos Aires/ | Servicio Virosis Respiratorias-Departamento Virologv≠a-INEI | Instituto Nacional Enfermedades Infecciosas C.G.Malbran | Baumeister E. et al |
| EPI_ISL_849375 | Argentina/INEI082916/2020 | 2020-10-08 | South America/Argentina/Buenos Aires/ | Servicio Virosis Respiratorias-Departamento Virologv≠a-INEI | Instituto Nacional Enfermedades Infecciosas C.G.Malbran | Baumeister E. et al |
| EPI_ISL_849376 | Argentina/INEI092516/2020 | 2020-10-09 | South America/Argentina/Buenos Aires/ | Servicio Virosis Respiratorias-Departamento Virologv≠a-INEI | Instituto Nacional Enfermedades Infecciosas C.G.Malbran | Baumeister E. et al |

| | | | | | | |
|---|---|---|---|---|---|---|
| EPI_ISL_849377 | Argentina/INEI092571/2020 | 2020-10-09 | South America/Argentina/Buenos Aires/ | Servicio Virosis Respiratorias-Departamento Virologí≠a-INEI | Instituto Nacional Enfermedades Infecciosas C.G.Malbran | Baumeister E. et al |
| EPI_ISL_849378 | Argentina/INEI092580/2020 | 2020-10-09 | South America/Argentina/Buenos Aires/ | Servicio Virosis Respiratorias-Departamento Virologí≠a-INEI | Instituto Nacional Enfermedades Infecciosas C.G.Malbran | Baumeister E. et al |
| EPI_ISL_849379 | Argentina/INEI092593/2020 | 2020-10-09 | South America/Argentina/Buenos Aires/ | Servicio Virosis Respiratorias-Departamento Virologí≠a-INEI | Instituto Nacional Enfermedades Infecciosas C.G.Malbran | Baumeister E. et al |
| EPI_ISL_849380 | Argentina/INEI092810/2020 | 2020-10-10 | South America/Argentina/Buenos Aires/ | Servicio Virosis Respiratorias-Departamento Virologí≠a-INEI | Instituto Nacional Enfermedades Infecciosas C.G.Malbran | Baumeister E. et al |
| EPI_ISL_849381 | Argentina/INEI092813/2020 | 2020-10-10 | South America/Argentina/Buenos Aires/ | Servicio Virosis Respiratorias-Departamento Virologí≠a-INEI | Instituto Nacional Enfermedades Infecciosas C.G.Malbran | Baumeister E. et al |
| EPI_ISL_849382 | Argentina/INEI092843/2020 | 2020-10-09 | South America/Argentina/Buenos Aires/ | Servicio Virosis Respiratorias-Departamento Virologí≠a-INEI | Instituto Nacional Enfermedades Infecciosas C.G.Malbran | Baumeister E. et al |
| EPI_ISL_849387 | USA/WA-S3386/2020 | 2020-12-01 | North America/USA/Washington/Yakim | Washington State Department of Health | Seattle Flu Study | Deborah A. Nickerson et al |
| EPI_ISL_849391 | USA/WA-S3410/2020 | 2020-12-03 | North America/USA/Washington/Thurst | Washington State Department of Health | Seattle Flu Study | Deborah A. Nickerson et al |
| EPI_ISL_849395 | USA/WA-S3360/2020 | 2020-11-24 | North America/USA/Washington/Cowlit | Washington State Department of Health | Seattle Flu Study | Deborah A. Nickerson et al |
| EPI_ISL_849411 | USA/WA-S3335/2020 | 2020-11-25 | North America/USA/Washington/King C | Washington State Department of Health | Seattle Flu Study | Deborah A. Nickerson et al |
| EPI_ISL_849418 | USA/WA-S3429/2020 | 2020-12-06 | North America/USA/Washington/Yakim | Washington State Department of Health | Seattle Flu Study | Deborah A. Nickerson et al |
| EPI_ISL_849421 | USA/WA-S3315/2020 | 2020-11-23 | North America/USA/Washington/Pacific | Washington State Department of Health | Seattle Flu Study | Deborah A. Nickerson et al |
| EPI_ISL_849424 | USA/WA-S3255/2020 | 2020-10-09 | North America/USA/Washington/ | Seattle Flu Study | Seattle Flu Study | Deborah A. Nickerson et al |
| EPI_ISL_849428 | USA/WA-S3433/2020 | 2020-12-06 | North America/USA/Washington/Yakim | Washington State Department of Health | Seattle Flu Study | Deborah A. Nickerson et al |
| EPI_ISL_849429 | USA/WA-S3339/2020 | 2020-12-09 | North America/USA/Washington/Yakim | Washington State Department of Health | Seattle Flu Study | Deborah A. Nickerson et al |
| EPI_ISL_849430 | USA/WA-S3303/2020 | 2020-11-14 | North America/USA/Washington/King C | Seattle Flu Study | Seattle Flu Study | Deborah A. Nickerson et al |
| EPI_ISL_849441 | USA/WA-S3274/2020 | 2020-10-31 | North America/USA/Washington/King C | Seattle Flu Study | Seattle Flu Study | Deborah A. Nickerson et al |
| EPI_ISL_849446 | USA/WA-S3352/2020 | 2020-11-25 | North America/USA/Washington/Yakim | Washington State Department of Health | Seattle Flu Study | Deborah A. Nickerson et al |

| | | | | | | |
|---|---|---|---|---|---|---|
| EPI_ISL_849451 | USA/WA-S3349/2020 | 2020-11-26 | North America/USA/Washington/Pierce | Washington State Department of Health | Seattle Flu Study | Deborah A. Nickerson et al |
| EPI_ISL_849463 | USA/WA-S3378/2020 | 2020-11-30 | North America/USA/Washington/Yakim | Washington State Department of Health | Seattle Flu Study | Deborah A. Nickerson et al |
| EPI_ISL_849481 | USA/WA-S3239/2020 | 2020-10-06 | North America/USA/Washington/King C | Seattle Flu Study | Seattle Flu Study | Deborah A. Nickerson et al |
| EPI_ISL_849484 | USA/WA-S3243/2020 | 2020-10-07 | North America/USA/Washington/King C | Seattle Flu Study | Seattle Flu Study | Deborah A. Nickerson et al |
| EPI_ISL_849485 | USA/WA-S3244/2020 | 2020-10-08 | North America/USA/Washington/King C | Seattle Flu Study | Seattle Flu Study | Deborah A. Nickerson et al |
| EPI_ISL_849487 | USA/WA-S3249/2020 | 2020-10-07 | North America/USA/Washington/King C | Seattle Flu Study | Seattle Flu Study | Deborah A. Nickerson et al |
| EPI_ISL_849504 | USA/WA-S3282/2020 | 2020-11-04 | North America/USA/Washington/King C | Seattle Flu Study | Seattle Flu Study | Deborah A. Nickerson et al |
| EPI_ISL_849510 | USA/WA-S3299/2020 | 2020-11-12 | North America/USA/Washington/King C | Seattle Flu Study | Seattle Flu Study | Deborah A. Nickerson et al |
| EPI_ISL_849520 | USA/WA-S3319/2020 | 2020-11-23 | North America/USA/Washington/Spoka | Washington State Department of Health | Seattle Flu Study | Deborah A. Nickerson et al |
| EPI_ISL_849525 | USA/WA-S3357/2020 | 2020-11-26 | North America/USA/Washington/Yakim | Washington State Department of Health | Seattle Flu Study | Deborah A. Nickerson et al |
| EPI_ISL_849531 | USA/WA-S3404/2020 | 2020-12-02 | North America/USA/Washington/Yakim | Washington State Department of Health | Seattle Flu Study | Deborah A. Nickerson et al |
| EPI_ISL_849542 | USA/WA-S3438/2020 | 2020-12-08 | North America/USA/Washington/Snoho | Washington State Department of Health | Seattle Flu Study | Deborah A. Nickerson et al |
| EPI_ISL_849550 | USA/WA-S3463/2020 | 2020-12-08 | North America/USA/Washington/Grays | Washington State Department of Health | Seattle Flu Study | Deborah A. Nickerson et al |
| EPI_ISL_849584 | USA/WA-S3262/2020 | 2020-10-19 | North America/USA/Washington/King C | Seattle Flu Study | Seattle Flu Study | Deborah A. Nickerson et al |
| EPI_ISL_849608 | USA/WA-S3286/2020 | 2020-11-05 | North America/USA/Washington/King C | Seattle Flu Study | Seattle Flu Study | Deborah A. Nickerson et al |
| EPI_ISL_849611 | USA/WA-S3292/2020 | 2020-11-07 | North America/USA/Washington/King C | Seattle Flu Study | Seattle Flu Study | Deborah A. Nickerson et al |
| EPI_ISL_849622 | USA/WA-S3362/2020 | 2020-11-27 | North America/USA/Washington/Pierce | Washington State Department of Health | Seattle Flu Study | Deborah A. Nickerson et al |
| EPI_ISL_849633 | USA/WA-S3467/2020 | 2020-12-07 | North America/USA/Washington/Spoka | Washington State Department of Health | Seattle Flu Study | Deborah A. Nickerson et al |
| EPI_ISL_849642 | Italy/ABR-IZSGC-374699/2020 | 2020-12-29 | Europe/Italy/Abruzzo/ | SIESP DIPARTIMENTO DI PREVENZIONE CHIETI | "Istitituto Zooprofilattico Sperimentale dell'Abruzzo e Molise ""G.Caporale""" | Lorusso A et al |
| EPI_ISL_849644 | Italy/ABR-IZSGC-374784/2020 | 2020-12-29 | Europe/Italy/Abruzzo/ | SIESP DIPARTIMENTO DI PREVENZIONE CHIETI | "Istitituto Zooprofilattico Sperimentale dell'Abruzzo e Molise ""G.Caporale""" | Lorusso A et al |

| | | | | | | |
|---|---|---|---|---|---|---|
| EPI_ISL_849649 | Italy/ABR-IZSGC-376328/2020 | 2020-12-31 | Europe/Italy/Abruzzo/ | SIESP- DRIVE IN CHIETI | "Istituto Zooprofilattico Sperimentale dell'Abruzzo e Molise ""G.Caporale""" | Lorusso A et al |
| EPI_ISL_849660 | Italy/ABR-IZSGC-TE101575/2020 | 2020-07-24 | Europe/Italy/Abruzzo/ | Servizio Igiene Epidemiologia e Sanit√† Pubblica (SIESP)-L'Aquila | "Istituto Zooprofilattico Sperimentale dell'Abruzzo e Molise ""G.Caporale""" | Lorusso A et al |
| EPI_ISL_849763 | Italy/SIC-IZS-2274/2021 | 2021-01-09 | Europe/Italy/Sicily/ | Molecular Biology, IZS Sicilia | Molecular Biology, IZS Sicilia | Reale et al |
| EPI_ISL_849794 | Germany/SH-ChVir21735/2020 | 2020-12-22 | Europe/Germany/Schleswig-Holstein/ | A. Krumbholz, Labor Dr. Krause und Kollegen MVZ GmbH, Kiel | Charit√© Universit√§tsmedizin Berlin, Institut f√ºr Virologie | Victor M Corman et al |
| EPI_ISL_849795 | Germany/SH-ChVir21736/2020 | 2020-12-21 | Europe/Germany/Schleswig-Holstein/ | A. Krumbholz, Labor Dr. Krause und Kollegen MVZ GmbH, Kiel | Charit√© Universit√§tsmedizin Berlin, Institut f√ºr Virologie | Victor M Corman et al |
| EPI_ISL_849799 | Germany/SH-ChVir21742/2020 | 2020-12-21 | Europe/Germany/Schleswig-Holstein/ | A. Krumbholz, Labor Dr. Krause und Kollegen MVZ GmbH, Kiel | Charit√© Universit√§tsmedizin Berlin, Institut f√ºr Virologie | Victor M Corman et al |
| EPI_ISL_849802 | Germany/SH-ChVir21746/2020 | 2020-12-22 | Europe/Germany/Schleswig-Holstein/ | A. Krumbholz, Labor Dr. Krause und Kollegen MVZ GmbH, Kiel | Charit√© Universit√§tsmedizin Berlin, Institut f√ºr Virologie | Victor M Corman et al |
| EPI_ISL_849807 | Germany/SH-ChVir21760/2020 | 2020-12-25 | Europe/Germany/Schleswig-Holstein/ | A. Krumbholz, Labor Dr. Krause und Kollegen MVZ GmbH, Kiel | Charit√© Universit√§tsmedizin Berlin, Institut f√ºr Virologie | Victor M Corman et al |
| EPI_ISL_849813 | Germany/SH-ChVir21766/2020 | 2020-12-25 | Europe/Germany/Schleswig-Holstein/ | A. Krumbholz, Labor Dr. Krause und Kollegen MVZ GmbH, Kiel | Charit√© Universit√§tsmedizin Berlin, Institut f√ºr Virologie | Victor M Corman et al |

| | | | | | | |
|---|---|---|---|---|---|---|
| EPI_ISL_849818 | Germany/SH-ChVir21771/2020 | 2020-12-25 | Europe/Germany/Schleswig-Holstein/ | A. Krumbholz, Labor Dr. Krause und Kollegen MVZ GmbH, Kiel | Charitv© Universitv§tsmedizin Berlin, Institut fvºr Virologie | Victor M Corman et al |
| EPI_ISL_849824 | Germany/SH-ChVir21777/2020 | 2020-12-25 | Europe/Germany/Schleswig-Holstein/ | A. Krumbholz, Labor Dr. Krause und Kollegen MVZ GmbH, Kiel | Charitv© Universitv§tsmedizin Berlin, Institut fvºr Virologie | Victor M Corman et al |
| EPI_ISL_849833 | Germany/SH-ChVir21788/2020 | 2020-12-25 | Europe/Germany/Schleswig-Holstein/ | A. Krumbholz, Labor Dr. Krause und Kollegen MVZ GmbH, Kiel | Charitv© Universitv§tsmedizin Berlin, Institut fvºr Virologie | Victor M Corman et al |
| EPI_ISL_849834 | Germany/SH-ChVir21789/2020 | 2020-12-25 | Europe/Germany/Schleswig-Holstein/ | A. Krumbholz, Labor Dr. Krause und Kollegen MVZ GmbH, Kiel | Charitv© Universitv§tsmedizin Berlin, Institut fvºr Virologie | Victor M Corman et al |
| EPI_ISL_849836 | Germany/SH-ChVir21791/2020 | 2020-12-24 | Europe/Germany/Schleswig-Holstein/ | A. Krumbholz, Labor Dr. Krause und Kollegen MVZ GmbH, Kiel | Charitv© Universitv§tsmedizin Berlin, Institut fvºr Virologie | Victor M Corman et al |
| EPI_ISL_849837 | Germany/HH-ChVir21792/2020 | 2020-12-26 | Europe/Germany/Hamburg/ | A. Krumbholz, Labor Dr. Krause und Kollegen MVZ GmbH, Kiel | Charitv© Universitv§tsmedizin Berlin, Institut fvºr Virologie | Victor M Corman et al |
| EPI_ISL_849846 | Germany/SH-ChVir21801/2020 | 2020-12-26 | Europe/Germany/Schleswig-Holstein/ | A. Krumbholz, Labor Dr. Krause und Kollegen MVZ GmbH, Kiel | Charitv© Universitv§tsmedizin Berlin, Institut fvºr Virologie | Victor M Corman et al |
| EPI_ISL_849847 | Germany/SH-ChVir21802/2020 | 2020-12-26 | Europe/Germany/Schleswig-Holstein/ | A. Krumbholz, Labor Dr. Krause und Kollegen MVZ GmbH, Kiel | Charitv© Universitv§tsmedizin Berlin, Institut fvºr Virologie | Victor M Corman et al |
| EPI_ISL_849857 | Germany/SH-ChVir21813/2020 | 2020-12-26 | Europe/Germany/Schleswig-Holstein/ | A. Krumbholz, Labor Dr. Krause und Kollegen MVZ GmbH, Kiel | Charitv© Universitv§tsmedizin Berlin, Institut fvºr Virologie | Victor M Corman et al |
| EPI_ISL_849866 | Germany/SH-ChVir21828/2020 | 2020-12-30 | Europe/Germany/Schleswig-Holstein/ | A. Krumbholz, Labor Dr. Krause und Kollegen MVZ GmbH, Kiel | Charitv© Universitv§tsmedizin Berlin, Institut fvºr Virologie | Victor M Corman et al |

| | | | | | | |
|---|---|---|---|---|---|---|
| EPI_ISL_849871 | USA/UT-UPHL-2101543521/2020 | 2020-12-19 | North America/USA/Utah/ | Utah Public Health Laboratory | Utah Public Health Laboratory | Erin L. Young et al |
| EPI_ISL_849875 | USA/UT-UPHL-2101406796/2020 | 2020-12-19 | North America/USA/Utah/ | Utah Public Health Laboratory | Utah Public Health Laboratory | Erin L. Young et al |
| EPI_ISL_849881 | USA/UT-UPHL-2101151105/2020 | 2020-12-28 | North America/USA/Utah/ | Utah Public Health Laboratory | Utah Public Health Laboratory | Erin L. Young et al |
| EPI_ISL_849888 | USA/UT-UPHL-2101670660/2020 | 2020-12-21 | North America/USA/Utah/ | Utah Public Health Laboratory | Utah Public Health Laboratory | Erin L. Young et al |
| EPI_ISL_849899 | USA/UT-UPHL-2101647158/2020 | 2020-12-27 | North America/USA/Utah/ | Utah Public Health Laboratory | Utah Public Health Laboratory | Erin L. Young et al |
| EPI_ISL_849903 | USA/UT-UPHL-2101614947/2020 | 2020-12-21 | North America/USA/Utah/ | Utah Public Health Laboratory | Utah Public Health Laboratory | Erin L. Young et al |
| EPI_ISL_849904 | USA/UT-UPHL-2101948969/2020 | 2020-12-21 | North America/USA/Utah/ | Utah Public Health Laboratory | Utah Public Health Laboratory | Erin L. Young et al |
| EPI_ISL_849911 | USA/UT-UPHL-2101778100/2021 | 2021-01-06 | North America/USA/Utah/ | Utah Public Health Laboratory | Utah Public Health Laboratory | Erin L. Young et al |
| EPI_ISL_849914 | USA/UT-UPHL-2101574178/2021 | 2021-01-08 | North America/USA/Utah/ | Utah Public Health Laboratory | Utah Public Health Laboratory | Erin L. Young et al |
| EPI_ISL_849922 | USA/CA-CZB-17197/2021 | 2021-01-04 | North America/USA/California/Orange ( | Orange County Public Health Lab | Chan-Zuckerberg Biohub | CZB Cliahub Consortium et al |
| EPI_ISL_849934 | USA/CA-CZB-15377/2020 | 2020-07-24 | North America/USA/California/Sacrame | UC Davis- Department of Pathology and Laboratory Medicine | Chan-Zuckerberg Biohub | CZB Cliahub Consortium et al |
| EPI_ISL_849939 | USA/CA-CZB-15383/2020 | 2020-08-02 | North America/USA/California/Sacrame | UC Davis- Department of Pathology and Laboratory Medicine | Chan-Zuckerberg Biohub | CZB Cliahub Consortium et al |
| EPI_ISL_849941 | USA/CA-CZB-15386/2020 | 2020-08-27 | North America/USA/California/Sacrame | UC Davis- Department of Pathology and Laboratory Medicine | Chan-Zuckerberg Biohub | CZB Cliahub Consortium et al |
| EPI_ISL_849946 | USA/CA-CZB-15391/2020 | 2020-08-04 | North America/USA/California/Sacrame | UC Davis- Department of Pathology and Laboratory Medicine | Chan-Zuckerberg Biohub | CZB Cliahub Consortium et al |
| EPI_ISL_849948 | USA/CA-CZB-15393/2020 | 2020-11-16 | North America/USA/California/Sacrame | UC Davis- Department of Pathology and Laboratory Medicine | Chan-Zuckerberg Biohub | CZB Cliahub Consortium et al |
| EPI_ISL_849949 | USA/CA-CZB-15395/2020 | 2020-07-05 | North America/USA/California/Sacrame | UC Davis- Department of Pathology and Laboratory Medicine | Chan-Zuckerberg Biohub | CZB Cliahub Consortium et al |
| EPI_ISL_849950 | USA/CA-CZB-15397/2020 | 2020-08-02 | North America/USA/California/Sacrame | UC Davis- Department of Pathology and Laboratory Medicine | Chan-Zuckerberg Biohub | CZB Cliahub Consortium et al |
| EPI_ISL_849951 | USA/CA-CZB-15398/2020 | 2020-11-16 | North America/USA/California/Sacrame | UC Davis- Department of Pathology and Laboratory Medicine | Chan-Zuckerberg Biohub | CZB Cliahub Consortium et al |
| EPI_ISL_849962 | USA/CA-CZB-15843/2020 | 2020-12-04 | North America/USA/California/Santa Cl | Santa Clara County Public Health Laboratory | Chan-Zuckerberg Biohub | CZB Cliahub Consortium et al |
| EPI_ISL_849973 | USA/CA-CZB-15858/2020 | 2020-12-04 | North America/USA/California/Santa Cl | Santa Clara County Public Health Laboratory | Chan-Zuckerberg Biohub | CZB Cliahub Consortium et al |

| | | | | | | |
|---|---|---|---|---|---|---|
| EPI_ISL_849976 | USA/CA-CZB-15862/2020 | 2020-12-06 | North America/USA/California/Santa Cl | Santa Clara County Public Health Laboratory | Chan-Zuckerberg Biohub | CZB Cliahub Consortium et al |
| EPI_ISL_849988 | USA/CA-CZB-15883/2020 | 2020-12-07 | North America/USA/California/Santa Cl | Santa Clara County Public Health Laboratory | Chan-Zuckerberg Biohub | CZB Cliahub Consortium et al |
| EPI_ISL_849992 | USA/CA-CZB-15887/2020 | 2020-12-07 | North America/USA/California/Santa Cl | Santa Clara County Public Health Laboratory | Chan-Zuckerberg Biohub | CZB Cliahub Consortium et al |
| EPI_ISL_849995 | USA/CA-CZB-15891/2020 | 2020-12-07 | North America/USA/California/Santa Cl | Santa Clara County Public Health Laboratory | Chan-Zuckerberg Biohub | CZB Cliahub Consortium et al |
| EPI_ISL_849997 | USA/CA-CZB-15897/2020 | 2020-12-08 | North America/USA/California/Santa Cl | Santa Clara County Public Health Laboratory | Chan-Zuckerberg Biohub | CZB Cliahub Consortium et al |
| EPI_ISL_850005 | USA/CA-CZB-15913/2020 | 2020-12-07 | North America/USA/California/Santa Cl | Santa Clara County Public Health Laboratory | Chan-Zuckerberg Biohub | CZB Cliahub Consortium et al |
| EPI_ISL_850009 | USA/CA-CZB-15921/2020 | 2020-12-07 | North America/USA/California/Santa Cl | Santa Clara County Public Health Laboratory | Chan-Zuckerberg Biohub | CZB Cliahub Consortium et al |
| EPI_ISL_850018 | USA/CA-CZB-15931/2020 | 2020-12-02 | North America/USA/California/Santa Cl | Santa Clara County Public Health Laboratory | Chan-Zuckerberg Biohub | CZB Cliahub Consortium et al |
| EPI_ISL_850021 | USA/CA-CZB-15990/2020 | 2020-11-23 | North America/USA/California/San Fran | Renegade | Chan-Zuckerberg Biohub | CZB Cliahub Consortium et al |
| EPI_ISL_850029 | USA/CA-CZB-15999/2020 | 2020-11-23 | North America/USA/California/San Fran | Renegade | Chan-Zuckerberg Biohub | CZB Cliahub Consortium et al |
| EPI_ISL_850030 | USA/CA-CZB-16000/2020 | 2020-11-23 | North America/USA/California/San Fran | Renegade | Chan-Zuckerberg Biohub | CZB Cliahub Consortium et al |
| EPI_ISL_850037 | USA/CA-CZB-16007/2020 | 2020-11-23 | North America/USA/California/San Fran | Renegade | Chan-Zuckerberg Biohub | CZB Cliahub Consortium et al |
| EPI_ISL_850041 | USA/CA-CZB-16011/2020 | 2020-11-24 | North America/USA/California/San Fran | Renegade | Chan-Zuckerberg Biohub | CZB Cliahub Consortium et al |
| EPI_ISL_850054 | USA/CA-CZB-16024/2020 | 2020-11-24 | North America/USA/California/San Fran | Renegade | Chan-Zuckerberg Biohub | CZB Cliahub Consortium et al |
| EPI_ISL_850055 | USA/CA-CZB-16025/2020 | 2020-11-24 | North America/USA/California/San Fran | Renegade | Chan-Zuckerberg Biohub | CZB Cliahub Consortium et al |
| EPI_ISL_850059 | USA/CA-CZB-16029/2020 | 2020-11-24 | North America/USA/California/San Fran | Renegade | Chan-Zuckerberg Biohub | CZB Cliahub Consortium et al |
| EPI_ISL_850062 | USA/CA-CZB-16033/2020 | 2020-11-24 | North America/USA/California/San Fran | Renegade | Chan-Zuckerberg Biohub | CZB Cliahub Consortium et al |
| EPI_ISL_850066 | USA/CA-CZB-16037/2020 | 2020-11-24 | North America/USA/California/San Fran | Renegade | Chan-Zuckerberg Biohub | CZB Cliahub Consortium et al |
| EPI_ISL_850073 | USA/CA-CZB-16045/2020 | 2020-11-29 | North America/USA/California/San Fran | Renegade | Chan-Zuckerberg Biohub | CZB Cliahub Consortium et al |
| EPI_ISL_850077 | USA/CA-CZB-16049/2020 | 2020-11-29 | North America/USA/California/San Fran | Renegade | Chan-Zuckerberg Biohub | CZB Cliahub Consortium et al |

| | | | | | | |
|---|---|---|---|---|---|---|
| EPI_ISL_850079 | USA/CA-CZB-16053/2020 | 2020-11-29 | North America/USA/California/San Fran | Renegade | Chan-Zuckerberg Biohub | CZB Cliahub Consortium et al |
| EPI_ISL_850082 | USA/CA-CZB-16056/2020 | 2020-11-29 | North America/USA/California/San Fran | Renegade | Chan-Zuckerberg Biohub | CZB Cliahub Consortium et al |
| EPI_ISL_850091 | USA/CA-CZB-16069/2020 | 2020-11-29 | North America/USA/California/San Fran | Renegade | Chan-Zuckerberg Biohub | CZB Cliahub Consortium et al |
| EPI_ISL_850092 | USA/CA-CZB-16070/2020 | 2020-11-29 | North America/USA/California/San Fran | Renegade | Chan-Zuckerberg Biohub | CZB Cliahub Consortium et al |
| EPI_ISL_850095 | USA/CA-CZB-16073/2020 | 2020-11-29 | North America/USA/California/San Fran | Renegade | Chan-Zuckerberg Biohub | CZB Cliahub Consortium et al |
| EPI_ISL_850100 | USA/CA-CZB-16080/2020 | 2020-11-29 | North America/USA/California/San Fran | Renegade | Chan-Zuckerberg Biohub | CZB Cliahub Consortium et al |
| EPI_ISL_850102 | USA/CA-CZB-16083/2020 | 2020-11-29 | North America/USA/California/San Fran | Renegade | Chan-Zuckerberg Biohub | CZB Cliahub Consortium et al |
| EPI_ISL_850110 | USA/CA-CZB-16092/2020 | 2020-11-29 | North America/USA/California/San Fran | Renegade | Chan-Zuckerberg Biohub | CZB Cliahub Consortium et al |
| EPI_ISL_850112 | USA/CA-CZB-16094/2020 | 2020-11-29 | North America/USA/California/San Fran | Renegade | Chan-Zuckerberg Biohub | CZB Cliahub Consortium et al |
| EPI_ISL_850118 | USA/CA-CZB-16100/2020 | 2020-11-30 | North America/USA/California/San Fran | Renegade | Chan-Zuckerberg Biohub | CZB Cliahub Consortium et al |
| EPI_ISL_850120 | USA/CA-CZB-16102/2020 | 2020-11-30 | North America/USA/California/San Fran | Renegade | Chan-Zuckerberg Biohub | CZB Cliahub Consortium et al |
| EPI_ISL_850123 | USA/CA-CZB-16106/2020 | 2020-11-30 | North America/USA/California/San Fran | Renegade | Chan-Zuckerberg Biohub | CZB Cliahub Consortium et al |
| EPI_ISL_850124 | USA/CA-CZB-16107/2020 | 2020-11-30 | North America/USA/California/San Fran | Renegade | Chan-Zuckerberg Biohub | CZB Cliahub Consortium et al |
| EPI_ISL_850128 | USA/CA-CZB-16112/2020 | 2020-11-30 | North America/USA/California/San Fran | Renegade | Chan-Zuckerberg Biohub | CZB Cliahub Consortium et al |
| EPI_ISL_850135 | USA/CA-CZB-16120/2020 | 2020-11-30 | North America/USA/California/San Fran | Renegade | Chan-Zuckerberg Biohub | CZB Cliahub Consortium et al |
| EPI_ISL_850136 | USA/CA-CZB-16121/2020 | 2020-11-30 | North America/USA/California/San Fran | Renegade | Chan-Zuckerberg Biohub | CZB Cliahub Consortium et al |
| EPI_ISL_850138 | USA/CA-CZB-16123/2020 | 2020-11-30 | North America/USA/California/San Fran | Renegade | Chan-Zuckerberg Biohub | CZB Cliahub Consortium et al |
| EPI_ISL_850141 | USA/CA-CZB-16126/2020 | 2020-11-30 | North America/USA/California/San Fran | Renegade | Chan-Zuckerberg Biohub | CZB Cliahub Consortium et al |
| EPI_ISL_850154 | USA/CA-CZB-16141/2020 | 2020-12-01 | North America/USA/California/San Fran | Renegade | Chan-Zuckerberg Biohub | CZB Cliahub Consortium et al |
| EPI_ISL_850155 | USA/CA-CZB-16142/2020 | 2020-12-01 | North America/USA/California/San Fran | Renegade | Chan-Zuckerberg Biohub | CZB Cliahub Consortium et al |

| | | | | | | |
|---|---|---|---|---|---|---|
| EPI_ISL_850165 | USA/CA-CZB-16156/2020 | 2020-12-01 | North America/USA/California/San Fran | Renegade | Chan-Zuckerberg Biohub | CZB Cliahub Consortium et al |
| EPI_ISL_850172 | USA/CA-CZB-16165/2020 | 2020-12-01 | North America/USA/California/San Fran | Renegade | Chan-Zuckerberg Biohub | CZB Cliahub Consortium et al |
| EPI_ISL_850176 | USA/CA-CZB-16230/2020 | 2020-11-24 | North America/USA/California/San Fran | Renegade | Chan-Zuckerberg Biohub | CZB Cliahub Consortium et al |
| EPI_ISL_850180 | Bangladesh/BCSIR-NILMRC-65/2020 | 2020-05-24 | Asia/Bangladesh/Chattogram/ | National Institute of Laboratory Medicine and Referral Center | Genomic Research Lab, BCSIR | Barna Goswami et al |
| EPI_ISL_850227 | SouthKorea/KDCA0529/2020 | 2020-07-01 | Asia/South Korea/South Korea/ | Division of Emerging Infectious Diseases, Bureau of Infectious Diseases Diagnosis Control, Korea Disease Control and Prevention Agency | Division of Emerging Infectious Diseases, Bureau of Infectious Diseases Diagnosis Control, Korea Disease Control and Prevention Agency | Ae Kyung Park et al |
| EPI_ISL_850304 | SouthKorea/KDCA0620/2020 | 2020-08-13 | Asia/South Korea/South Korea/ | Division of Emerging Infectious Diseases, Bureau of Infectious Diseases Diagnosis Control, Korea Disease Control and Prevention Agency | Division of Emerging Infectious Diseases, Bureau of Infectious Diseases Diagnosis Control, Korea Disease Control and Prevention Agency | Ae Kyung Park et al |
| EPI_ISL_850319 | SouthKorea/KDCA0637/2020 | 2020-08-07 | Asia/South Korea/South Korea/ | Division of Emerging Infectious Diseases, Bureau of Infectious Diseases Diagnosis Control, Korea Disease Control and Prevention Agency | Division of Emerging Infectious Diseases, Bureau of Infectious Diseases Diagnosis Control, Korea Disease Control and Prevention Agency | Ae Kyung Park et al |

| | | | | | | |
|---|---|---|---|---|---|---|
| EPI_ISL_850331 | SouthKorea/KDCA0649/2020 | 2020-08-08 | Asia/South Korea/South Korea/ | Division of Emerging Infectious Diseases, Bureau of Infectious Diseases Diagnosis Control, Korea Disease Control and Prevention Agency | Division of Emerging Infectious Diseases, Bureau of Infectious Diseases Diagnosis Control, Korea Disease Control and Prevention Agency | Ae Kyung Park et al |
| EPI_ISL_850360 | SouthKorea/KDCA0679/2020 | 2020-08-17 | Asia/South Korea/South Korea/ | Division of Emerging Infectious Diseases, Bureau of Infectious Diseases Diagnosis Control, Korea Disease Control and Prevention Agency | Division of Emerging Infectious Diseases, Bureau of Infectious Diseases Diagnosis Control, Korea Disease Control and Prevention Agency | Ae Kyung Park et al |
| EPI_ISL_850368 | SouthKorea/KDCA0688/2020 | 2020-08-19 | Asia/South Korea/South Korea/ | Division of Emerging Infectious Diseases, Bureau of Infectious Diseases Diagnosis Control, Korea Disease Control and Prevention Agency | Division of Emerging Infectious Diseases, Bureau of Infectious Diseases Diagnosis Control, Korea Disease Control and Prevention Agency | Ae Kyung Park et al |

| | | | | | | |
|---|---|---|---|---|---|---|
| EPI_ISL_850419 | SouthKorea/KDCA0741/2020 | 2020-08-21 | Asia/South Korea/South Korea/ | Division of Emerging Infectious Diseases, Bureau of Infectious Diseases Diagnosis Control, Korea Disease Control and Prevention Agency | Division of Emerging Infectious Diseases, Bureau of Infectious Diseases Diagnosis Control, Korea Disease Control and Prevention Agency | Ae Kyung Park et al |
| EPI_ISL_850506 | Bangladesh/BCSIR-NILMRC-68/2020 | 2020-05-24 | Asia/Bangladesh/Chattogram/ | National Institute of Laboratory Medicine and Referral Center | Genomic Research Lab, BCSIR | Shahina Akter et al |
| EPI_ISL_850507 | India/TG-GMC-20911/2020 | 2020-07-23 | Asia/India/Telangana/ | Gandhi Medical College and Hospital | VRDL-Gandhi Medical College | Nagamani Kammili et al |
| EPI_ISL_850509 | India/TG-GMC-26813/2020 | 2020-08-08 | Asia/India/Telangana/ | Gandhi Medical College and Hospital | VRDL-Gandhi Medical College | Nagamani Kammili et al |
| EPI_ISL_850510 | India/TG-GMC-27382/2020 | 2020-08-09 | Asia/India/Telangana/ | Gandhi Medical College and Hospital | MDRU-DHR,Gandhi Medical College | Nagamani Kammili et al |
| EPI_ISL_850522 | USA/CA-CDC-STM-326/2021 | 2021-01-03 | North America/USA/California/ | Helix/Illumina | Genomics and Discovery, Respiratory Viruses Branch, Division of Viral Diseases, Centers for Disease Control and Prevention | Peter W. Cook et al |
| EPI_ISL_850532 | USA/FL-CDC-STM-A026/2021 | 2021-01-02 | North America/USA/Florida/ | Helix/Illumina | Genomics and Discovery, Respiratory Viruses Branch, Division of Viral Diseases, Centers for Disease Control and Prevention | Peter W. Cook et al |

| | | | | | | |
|---|---|---|---|---|---|---|
| EPI_ISL_850534 | USA/FL-CDC-STM-A024/2021 | 2021-01-02 | North America/USA/Florida/ | Helix/Illumina | Genomics and Discovery, Respiratory Viruses Branch, Division of Viral Diseases, Centers for Disease Control and Prevention | Peter W. Cook et al |
| EPI_ISL_850536 | USA/AR-CDC-STM-A022/2021 | 2021-01-02 | North America/USA/Arkansas/ | Helix/Illumina | Genomics and Discovery, Respiratory Viruses Branch, Division of Viral Diseases, Centers for Disease Control and Prevention | Peter W. Cook et al |
| EPI_ISL_850540 | USA/NC-CDC-STM-A018/2021 | 2021-01-02 | North America/USA/North Carolina/ | Helix/Illumina | Genomics and Discovery, Respiratory Viruses Branch, Division of Viral Diseases, Centers for Disease Control and Prevention | Peter W. Cook et al |
| EPI_ISL_850549 | USA/NC-CDC-STM-A006/2021 | 2021-01-02 | North America/USA/North Carolina/ | Helix/Illumina | Genomics and Discovery, Respiratory Viruses Branch, Division of Viral Diseases, Centers for Disease Control and Prevention | Peter W. Cook et al |

| | | | | | | |
|---|---|---|---|---|---|---|
| EPI_ISL_850552 | USA/FL-CDC-STM-A003/2021 | 2021-01-02 | North America/USA/Florida/ | Helix/Illumina | Genomics and Discovery, Respiratory Viruses Branch, Division of Viral Diseases, Centers for Disease Control and Prevention | Peter W. Cook et al |
| EPI_ISL_850561 | USA/NC-CDC-STM-382/2021 | 2021-01-02 | North America/USA/North Carolina/ | Helix/Illumina | Genomics and Discovery, Respiratory Viruses Branch, Division of Viral Diseases, Centers for Disease Control and Prevention | Peter W. Cook et al |
| EPI_ISL_850564 | USA/MI-CDC-STM-375/2021 | 2021-01-02 | North America/USA/Michigan/ | Helix/Illumina | Genomics and Discovery, Respiratory Viruses Branch, Division of Viral Diseases, Centers for Disease Control and Prevention | Peter W. Cook et al |
| EPI_ISL_850565 | USA/NY-CDC-STM-374/2021 | 2021-01-02 | North America/USA/New York/ | Helix/Illumina | Genomics and Discovery, Respiratory Viruses Branch, Division of Viral Diseases, Centers for Disease Control and Prevention | Peter W. Cook et al |

| | | | | | | |
|---|---|---|---|---|---|---|
| EPI_ISL_850589 | USA/MI-CDC-STM-304/2021 | 2021-01-02 | North America/USA/Michigan/ | Helix/Illumina | Genomics and Discovery, Respiratory Viruses Branch, Division of Viral Diseases, Centers for Disease Control and Prevention | Peter W. Cook et al |
| EPI_ISL_850591 | USA/AZ-CDC-STM-300/2021 | 2021-01-02 | North America/USA/Arizona/ | Helix/Illumina | Genomics and Discovery, Respiratory Viruses Branch, Division of Viral Diseases, Centers for Disease Control and Prevention | Peter W. Cook et al |
| EPI_ISL_850595 | USA/AZ-CDC-STM-295/2021 | 2021-01-02 | North America/USA/Arizona/ | Helix/Illumina | Genomics and Discovery, Respiratory Viruses Branch, Division of Viral Diseases, Centers for Disease Control and Prevention | Peter W. Cook et al |
| EPI_ISL_850609 | USA/FL-CDC-STM-192/2020 | 2020-12-31 | North America/USA/Florida/ | Helix/Illumina | Genomics and Discovery, Respiratory Viruses Branch, Division of Viral Diseases, Centers for Disease Control and Prevention | Peter W. Cook et al |

| | | | | | | |
|---|---|---|---|---|---|---|
| EPI_ISL_850615 | USA/FL-CDC-STM-186/2020 | 2020-12-31 | North America/USA/Florida/ | Helix/Illumina | Genomics and Discovery, Respiratory Viruses Branch, Division of Viral Diseases, Centers for Disease Control and Prevention | Peter W. Cook et al |
| EPI_ISL_850617 | USA/FL-CDC-STM-184/2020 | 2020-12-31 | North America/USA/Florida/ | Helix/Illumina | Genomics and Discovery, Respiratory Viruses Branch, Division of Viral Diseases, Centers for Disease Control and Prevention | Peter W. Cook et al |
| EPI_ISL_850624 | USA/MA-CDC-STM-177/2020 | 2020-12-31 | North America/USA/Massachusetts/ | Helix/Illumina | Genomics and Discovery, Respiratory Viruses Branch, Division of Viral Diseases, Centers for Disease Control and Prevention | Peter W. Cook et al |
| EPI_ISL_850627 | USA/MA-CDC-STM-174/2020 | 2020-12-31 | North America/USA/Massachusetts/ | Helix/Illumina | Genomics and Discovery, Respiratory Viruses Branch, Division of Viral Diseases, Centers for Disease Control and Prevention | Peter W. Cook et al |

| | | | | | | |
|---|---|---|---|---|---|---|
| EPI_ISL_850628 | USA/FL-CDC-STM-171/2020 | 2020-12-31 | North America/USA/Florida/ | Helix/Illumina | Genomics and Discovery, Respiratory Viruses Branch, Division of Viral Diseases, Centers for Disease Control and Prevention | Peter W. Cook et al |
| EPI_ISL_850631 | USA/CA-CDC-STM-166/2020 | 2020-12-31 | North America/USA/California/ | Helix/Illumina | Genomics and Discovery, Respiratory Viruses Branch, Division of Viral Diseases, Centers for Disease Control and Prevention | Peter W. Cook et al |
| EPI_ISL_850633 | USA/CA-CDC-STM-156/2020 | 2020-12-31 | North America/USA/California/ | Helix/Illumina | Genomics and Discovery, Respiratory Viruses Branch, Division of Viral Diseases, Centers for Disease Control and Prevention | Peter W. Cook et al |
| EPI_ISL_850638 | USA/CA-CDC-STM-149/2020 | 2020-12-31 | North America/USA/California/ | Helix/Illumina | Genomics and Discovery, Respiratory Viruses Branch, Division of Viral Diseases, Centers for Disease Control and Prevention | Peter W. Cook et al |

| | | | | | | |
|---|---|---|---|---|---|---|
| EPI_ISL_850647 | USA/MA-CDC-STM-159/2020 | 2020-12-30 | North America/USA/Massachusetts/ | Helix/Illumina | Genomics and Discovery, Respiratory Viruses Branch, Division of Viral Diseases, Centers for Disease Control and Prevention | Peter W. Cook et al |
| EPI_ISL_850651 | USA/FL-CDC-STM-151/2020 | 2020-12-30 | North America/USA/Florida/ | Helix/Illumina | Genomics and Discovery, Respiratory Viruses Branch, Division of Viral Diseases, Centers for Disease Control and Prevention | Peter W. Cook et al |
| EPI_ISL_850654 | SouthKorea/KDCA0830/2020 | 2020-12-24 | Asia/South Korea/South Korea/ | Division of Emerging Infectious Diseases, Bureau of Infectious Diseases Diagnosis Control, Korea Disease Control and Prevention Agency | Division of Emerging Infectious Diseases, Bureau of Infectious Diseases Diagnosis Control, Korea Disease Control and Prevention Agency | Ae Kyung Park et al |
| EPI_ISL_850656 | SouthKorea/KDCA0832/2020 | 2020-12-26 | Asia/South Korea/South Korea/ | Division of Emerging Infectious Diseases, Bureau of Infectious Diseases Diagnosis Control, Korea Disease Control and Prevention Agency | Division of Emerging Infectious Diseases, Bureau of Infectious Diseases Diagnosis Control, Korea Disease Control and Prevention Agency | Ae Kyung Park et al |

| | | | | | | |
|---|---|---|---|---|---|---|
| EPI_ISL_850657 | SouthKorea/KDCA0833/2020 | 2020-12-27 | Asia/South Korea/South Korea/ | Division of Emerging Infectious Diseases, Bureau of Infectious Diseases Diagnosis Control, Korea Disease Control and Prevention Agency | Division of Emerging Infectious Diseases, Bureau of Infectious Diseases Diagnosis Control, Korea Disease Control and Prevention Agency | Ae Kyung Park et al |
| EPI_ISL_850659 | SouthKorea/KDCA0836/2020 | 2020-12-26 | Asia/South Korea/South Korea/ | Division of Emerging Infectious Diseases, Bureau of Infectious Diseases Diagnosis Control, Korea Disease Control and Prevention Agency | Division of Emerging Infectious Diseases, Bureau of Infectious Diseases Diagnosis Control, Korea Disease Control and Prevention Agency | Ae Kyung Park et al |
| EPI_ISL_850660 | SouthKorea/KDCA0837/2020 | 2020-12-29 | Asia/South Korea/South Korea/ | Division of Emerging Infectious Diseases, Bureau of Infectious Diseases Diagnosis Control, Korea Disease Control and Prevention Agency | Division of Emerging Infectious Diseases, Bureau of Infectious Diseases Diagnosis Control, Korea Disease Control and Prevention Agency | Ae Kyung Park et al |

| | | | | | | |
|---|---|---|---|---|---|---|
| EPI_ISL_850661 | SouthKorea/KDCA0838/2020 | 2020-12-30 | Asia/South Korea/South Korea/ | Division of Emerging Infectious Diseases, Bureau of Infectious Diseases Diagnosis Control, Korea Disease Control and Prevention Agency | Division of Emerging Infectious Diseases, Bureau of Infectious Diseases Diagnosis Control, Korea Disease Control and Prevention Agency | Ae Kyung Park et al |
| EPI_ISL_850663 | CzechRepublic/NRL_11354/2020 | 2020-11-22 | Europe/Czech Republic/Central Bohemia | The National Institute of Public Health | State Veterinary Institute Prague | Nagy et al |
| EPI_ISL_850664 | CzechRepublic/NRL_11782-4/2020 | 2020-12-07 | Europe/Czech Republic/Prague/ | The National Institute of Public Health | State Veterinary Institute Prague | Nagy et al |
| EPI_ISL_850665 | CzechRepublic/NRL_11782-5/2020 | 2020-12-07 | Europe/Czech Republic/Prague/ | The National Institute of Public Health | State Veterinary Institute Prague | Nagy et al |
| EPI_ISL_850666 | CzechRepublic/NRL_11876/2020 | 2020-12-18 | Europe/Czech Republic/Liberec Region/V | The National Institute of Public Health | State Veterinary Institute Prague | Nagy et al |
| EPI_ISL_850667 | CzechRepublic/NRL_12030/2020 | 2020-12-15 | Europe/Czech Republic/Usti nad Labem/ | The National Institute of Public Health | State Veterinary Institute Prague | Nagy et al |
| EPI_ISL_850668 | CzechRepublic/NRL_12064/2020 | 2020-12-11 | Europe/Czech Republic/Moravian-Silesia | The National Institute of Public Health | State Veterinary Institute Prague | Nagy et al |
| EPI_ISL_850669 | CzechRepublic/NRL_12339/2020 | 2020-12-17 | Europe/Czech Republic/Usti nad Labem/ | The National Institute of Public Health | State Veterinary Institute Prague | Nagy et al |
| EPI_ISL_850670 | CzechRepublic/NRL_12341/2020 | 2020-12-10 | Europe/Czech Republic/Usti nad Labem/ | The National Institute of Public Health | State Veterinary Institute Prague | Nagy et al |
| EPI_ISL_850671 | CzechRepublic/NRL_99/2021 | 2021-01-03 | Europe/Czech Republic/Prague/ | The National Institute of Public Health | State Veterinary Institute Prague | Nagy et al |
| EPI_ISL_850672 | CzechRepublic/NRL_100/2021 | 2021-01-03 | Europe/Czech Republic/Prague/ | The National Institute of Public Health | State Veterinary Institute Prague | Nagy et al |
| EPI_ISL_850673 | CzechRepublic/NRL_137-1/2021 | 2021-01-04 | Europe/Czech Republic/Zl√≠n Region/Bu | The National Institute of Public Health | State Veterinary Institute Prague | Nagy et al |
| EPI_ISL_850674 | CzechRepublic/NRL_11973/2020 | 2020-12-15 | Europe/Czech Republic/Prague/ | The National Institute of Public Health | State Veterinary Institute Prague | Nagy et al |
| EPI_ISL_850675 | CzechRepublic/NRL_12173/2020 | 2020-12-21 | Europe/Czech Republic/Central Bohemia | The National Institute of Public Health | State Veterinary Institute Prague | Nagy et al |
| EPI_ISL_850676 | CzechRepublic/NRL_12188/2020 | 2020-12-21 | Europe/Czech Republic/Central Bohemia | The National Institute of Public Health | State Veterinary Institute Prague | Nagy et al |

| | | | | | | |
|---|---|---|---|---|---|---|
| EPI_ISL_850687 | CzechRepublic/NRL_274/2020 | 2020-01-06 | Europe/Czech Republic/Prague/ | The National Institute of Public Health | State Veterinary Institute Prague | Nagy et al |
| EPI_ISL_850688 | CzechRepublic/NRL_275/2021 | 2021-01-05 | Europe/Czech Republic/Prague/ | The National Institute of Public Health | State Veterinary Institute Prague | Nagy et al |
| EPI_ISL_850695 | USA/CA-CDC-STM-A100432/2021 | 2021-01-07 | North America/USA/California/ | Helix / Illumina | Genomics and Discovery, Respiratory Viruses Branch, Division of Viral Diseases, Centers for Disease Control and Prevention | Peter W. Cook et al |
| EPI_ISL_850700 | USA/CA-CDC-STM-A100403/2021 | 2021-01-07 | North America/USA/California/ | Helix / Illumina | Genomics and Discovery, Respiratory Viruses Branch, Division of Viral Diseases, Centers for Disease Control and Prevention | Peter W. Cook et al |
| EPI_ISL_850703 | USA/CA-CDC-STM-A100386/2021 | 2021-01-07 | North America/USA/California/ | Helix / Illumina | Genomics and Discovery, Respiratory Viruses Branch, Division of Viral Diseases, Centers for Disease Control and Prevention | Peter W. Cook et al |
| EPI_ISL_850725 | USA/MA-CDC-STM-A100440/2021 | 2021-01-06 | North America/USA/Massachusetts/ | Helix / Illumina | Genomics and Discovery, Respiratory Viruses Branch, Division of Viral Diseases, Centers for Disease Control and Prevention | Peter W. Cook et al |

| | | | | | | |
|---|---|---|---|---|---|---|
| EPI_ISL_850730 | USA/TX-CDC-STM-A100435/2021 | 2021-01-06 | North America/USA/Texas/ | Helix / Illumina | Genomics and Discovery, Respiratory Viruses Branch, Division of Viral Diseases, Centers for Disease Control and Prevention | Peter W. Cook et al |
| EPI_ISL_850733 | USA/PA-CDC-STM-A100429/2021 | 2021-01-06 | North America/USA/Pennsylvania/ | Helix / Illumina | Genomics and Discovery, Respiratory Viruses Branch, Division of Viral Diseases, Centers for Disease Control and Prevention | Peter W. Cook et al |
| EPI_ISL_850744 | USA/FL-CDC-STM-A100414/2021 | 2021-01-06 | North America/USA/Florida/ | Helix / Illumina | Genomics and Discovery, Respiratory Viruses Branch, Division of Viral Diseases, Centers for Disease Control and Prevention | Peter W. Cook et al |
| EPI_ISL_850759 | USA/GA-CDC-STM-A100395/2021 | 2021-01-06 | North America/USA/Georgia/ | Helix / Illumina | Genomics and Discovery, Respiratory Viruses Branch, Division of Viral Diseases, Centers for Disease Control and Prevention | Peter W. Cook et al |

| | | | | | | |
|---|---|---|---|---|---|---|
| EPI_ISL_850761 | USA/FL-CDC-STM-A100392/2021 | 2021-01-06 | North America/USA/Florida/ | Helix / Illumina | Genomics and Discovery, Respiratory Viruses Branch, Division of Viral Diseases, Centers for Disease Control and Prevention | Peter W. Cook et al |
| EPI_ISL_850769 | USA/SC-CDC-STM-A100472/2021 | 2021-01-05 | North America/USA/South Carolina/ | Helix / Illumina | Genomics and Discovery, Respiratory Viruses Branch, Division of Viral Diseases, Centers for Disease Control and Prevention | Peter W. Cook et al |
| EPI_ISL_850781 | USA/CA-CDC-STM-A100242/2021 | 2021-01-05 | North America/USA/California/ | Helix / Illumina | Genomics and Discovery, Respiratory Viruses Branch, Division of Viral Diseases, Centers for Disease Control and Prevention | Peter W. Cook et al |
| EPI_ISL_850783 | USA/FL-CDC-STM-A100231/2021 | 2021-01-05 | North America/USA/Florida/ | Helix / Illumina | Genomics and Discovery, Respiratory Viruses Branch, Division of Viral Diseases, Centers for Disease Control and Prevention | Peter W. Cook et al |

| | | | | | | |
|---|---|---|---|---|---|---|
| EPI_ISL_850790 | USA/CA-CDC-STM-A100197/2021 | 2021-01-05 | North America/USA/California/ | Helix / Illumina | Genomics and Discovery, Respiratory Viruses Branch, Division of Viral Diseases, Centers for Disease Control and Prevention | Peter W. Cook et al |
| EPI_ISL_850796 | USA/CA-CDC-STM-A100171/2021 | 2021-01-05 | North America/USA/California/ | Helix / Illumina | Genomics and Discovery, Respiratory Viruses Branch, Division of Viral Diseases, Centers for Disease Control and Prevention | Peter W. Cook et al |
| EPI_ISL_850809 | USA/CA-CDC-STM-A100136/2021 | 2021-01-05 | North America/USA/California/ | Helix / Illumina | Genomics and Discovery, Respiratory Viruses Branch, Division of Viral Diseases, Centers for Disease Control and Prevention | Peter W. Cook et al |
| EPI_ISL_850811 | USA/CA-CDC-STM-A100127/2021 | 2021-01-05 | North America/USA/California/ | Helix / Illumina | Genomics and Discovery, Respiratory Viruses Branch, Division of Viral Diseases, Centers for Disease Control and Prevention | Peter W. Cook et al |

| | | | | | | |
|---|---|---|---|---|---|---|
| EPI_ISL_850820 | USA/FL-CDC-STM-A100256/2021 | 2021-01-04 | North America/USA/Florida/ | Helix / Illumina | Genomics and Discovery, Respiratory Viruses Branch, Division of Viral Diseases, Centers for Disease Control and Prevention | Peter W. Cook et al |
| EPI_ISL_850822 | USA/FL-CDC-STM-A100251/2021 | 2021-01-04 | North America/USA/Florida/ | Helix / Illumina | Genomics and Discovery, Respiratory Viruses Branch, Division of Viral Diseases, Centers for Disease Control and Prevention | Peter W. Cook et al |
| EPI_ISL_850830 | USA/FL-CDC-STM-A100238/2021 | 2021-01-04 | North America/USA/Florida/ | Helix / Illumina | Genomics and Discovery, Respiratory Viruses Branch, Division of Viral Diseases, Centers for Disease Control and Prevention | Peter W. Cook et al |
| EPI_ISL_850837 | USA/MI-CDC-STM-A100223/2021 | 2021-01-04 | North America/USA/Michigan/ | Helix / Illumina | Genomics and Discovery, Respiratory Viruses Branch, Division of Viral Diseases, Centers for Disease Control and Prevention | Peter W. Cook et al |

| | | | | | | |
|---|---|---|---|---|---|---|
| EPI_ISL_850848 | USA/MA-CDC-STM-A100212/2021 | 2021-01-04 | North America/USA/Massachusetts/ | Helix / Illumina | Genomics and Discovery, Respiratory Viruses Branch, Division of Viral Diseases, Centers for Disease Control and Prevention | Peter W. Cook et al |
| EPI_ISL_850853 | USA/IN-CDC-STM-A100202/2021 | 2021-01-04 | North America/USA/Indiana/ | Helix / Illumina | Genomics and Discovery, Respiratory Viruses Branch, Division of Viral Diseases, Centers for Disease Control and Prevention | Peter W. Cook et al |
| EPI_ISL_850854 | USA/IN-CDC-STM-A100201/2021 | 2021-01-04 | North America/USA/Indiana/ | Helix / Illumina | Genomics and Discovery, Respiratory Viruses Branch, Division of Viral Diseases, Centers for Disease Control and Prevention | Peter W. Cook et al |
| EPI_ISL_850858 | USA/NJ-CDC-STM-A100192/2021 | 2021-01-04 | North America/USA/New Jersey/ | Helix / Illumina | Genomics and Discovery, Respiratory Viruses Branch, Division of Viral Diseases, Centers for Disease Control and Prevention | Peter W. Cook et al |

| | | | | | | |
|---|---|---|---|---|---|---|
| EPI_ISL_850868 | USA/KY-CDC-STM-A100177/2021 | 2021-01-04 | North America/USA/Kentucky/ | Helix / Illumina | Genomics and Discovery, Respiratory Viruses Branch, Division of Viral Diseases, Centers for Disease Control and Prevention | Peter W. Cook et al |
| EPI_ISL_850877 | USA/LA-CDC-STM-A100159/2021 | 2021-01-04 | North America/USA/Louisiana/ | Helix / Illumina | Genomics and Discovery, Respiratory Viruses Branch, Division of Viral Diseases, Centers for Disease Control and Prevention | Peter W. Cook et al |
| EPI_ISL_850884 | USA/PA-CDC-STM-A100142/2021 | 2021-01-04 | North America/USA/Pennsylvania/ | Helix / Illumina | Genomics and Discovery, Respiratory Viruses Branch, Division of Viral Diseases, Centers for Disease Control and Prevention | Peter W. Cook et al |
| EPI_ISL_850886 | USA/GA-CDC-STM-A100140/2021 | 2021-01-04 | North America/USA/Georgia/ | Helix / Illumina | Genomics and Discovery, Respiratory Viruses Branch, Division of Viral Diseases, Centers for Disease Control and Prevention | Peter W. Cook et al |

| | | | | | | |
|---|---|---|---|---|---|---|
| EPI_ISL_850911 | USA/IN-CDC-STM-A100124/2021 | 2021-01-03 | North America/USA/Indiana/ | Helix / Illumina | Genomics and Discovery, Respiratory Viruses Branch, Division of Viral Diseases, Centers for Disease Control and Prevention | Peter W. Cook et al |
| EPI_ISL_850924 | USA/FL-CDC-STM-A100100/2021 | 2021-01-03 | North America/USA/Florida/ | Helix / Illumina | Genomics and Discovery, Respiratory Viruses Branch, Division of Viral Diseases, Centers for Disease Control and Prevention | Peter W. Cook et al |
| EPI_ISL_850928 | USA/FL-CDC-STM-A100095/2021 | 2021-01-03 | North America/USA/Florida/ | Helix / Illumina | Genomics and Discovery, Respiratory Viruses Branch, Division of Viral Diseases, Centers for Disease Control and Prevention | Peter W. Cook et al |
| EPI_ISL_850929 | USA/IN-CDC-STM-A100094/2021 | 2021-01-03 | North America/USA/Indiana/ | Helix / Illumina | Genomics and Discovery, Respiratory Viruses Branch, Division of Viral Diseases, Centers for Disease Control and Prevention | Peter W. Cook et al |

| | | | | | | |
|---|---|---|---|---|---|---|
| EPI_ISL_850930 | USA/MN-CDC-STM-A100093/2021 | 2021-01-03 | North America/USA/Minnesota/ | Helix / Illumina | Genomics and Discovery, Respiratory Viruses Branch, Division of Viral Diseases, Centers for Disease Control and Prevention | Peter W. Cook et al |
| EPI_ISL_850931 | USA/SC-CDC-STM-A100240/2021 | 2021-01-02 | North America/USA/South Carolina/ | Helix / Illumina | Genomics and Discovery, Respiratory Viruses Branch, Division of Viral Diseases, Centers for Disease Control and Prevention | Peter W. Cook et al |
| EPI_ISL_850947 | Shulan/IVDC-01-01/2020 | 2020-04-19 | Asia/China/Jilin/Shulan | National Institute for Viral Disease Control and Prevention, China CDC | National Institute for Viral Disease Control and Prevention, China CDC | Xiang Zhao et al |
| EPI_ISL_850949 | Qingdao/IVDC-01-01/2020 | 2020-08-16 | Asia/China/Shandong/Qingdao | National Institute for Viral Disease Control and Prevention, China CDC | National Institute for Viral Disease Control and Prevention, China CDC | Xiang Zhao et al |
| EPI_ISL_850950 | Kashgar/IVDC-01-01/2020 | 2020-10-30 | Asia/China/Xinjiang/Kashgar | National Institute for Viral Disease Control and Prevention, China CDC | National Institute for Viral Disease Control and Prevention, China CDC | Xiang Zhao et al |

| | | | | | | |
|---|---|---|---|---|---|---|
| EPI_ISL_850956 | USA/CA-CDC-STM-A100056/2021 | 2021-01-04 | North America/USA/California/ | Helix/Illumina | Genomics and Discovery, Respiratory Viruses Branch, Division of Viral Diseases, Centers for Disease Control and Prevention | Peter W. Cook et al |
| EPI_ISL_850964 | USA/AL-CDC-STM-A100088/2021 | 2021-01-03 | North America/USA/Alabama/ | Helix/Illumina | Genomics and Discovery, Respiratory Viruses Branch, Division of Viral Diseases, Centers for Disease Control and Prevention | Peter W. Cook et al |
| EPI_ISL_850976 | USA/AZ-CDC-STM-A100072/2021 | 2021-01-03 | North America/USA/Arizona/ | Helix/Illumina | Genomics and Discovery, Respiratory Viruses Branch, Division of Viral Diseases, Centers for Disease Control and Prevention | Peter W. Cook et al |
| EPI_ISL_850978 | USA/GA-CDC-STM-A100070/2021 | 2021-01-03 | North America/USA/Georgia/ | Helix/Illumina | Genomics and Discovery, Respiratory Viruses Branch, Division of Viral Diseases, Centers for Disease Control and Prevention | Peter W. Cook et al |

| | | | | | | |
|---|---|---|---|---|---|---|
| EPI_ISL_850982 | USA/LA-CDC-STM-A100061/2021 | 2021-01-03 | North America/USA/Louisiana/ | Helix/Illumina | Genomics and Discovery, Respiratory Viruses Branch, Division of Viral Diseases, Centers for Disease Control and Prevention | Peter W. Cook et al |
| EPI_ISL_850984 | USA/KY-CDC-STM-A100059/2021 | 2021-01-03 | North America/USA/Kentucky/ | Helix/Illumina | Genomics and Discovery, Respiratory Viruses Branch, Division of Viral Diseases, Centers for Disease Control and Prevention | Peter W. Cook et al |
| EPI_ISL_850991 | USA/FL-CDC-STM-A100001/2021 | 2021-01-03 | North America/USA/Florida/ | Helix/Illumina | Genomics and Discovery, Respiratory Viruses Branch, Division of Viral Diseases, Centers for Disease Control and Prevention | Peter W. Cook et al |
| EPI_ISL_851001 | USA/CA-CDC-STM-235/2021 | 2021-01-02 | North America/USA/California/ | Helix/Illumina | Genomics and Discovery, Respiratory Viruses Branch, Division of Viral Diseases, Centers for Disease Control and Prevention | Peter W. Cook et al |

| | | | | | | |
|---|---|---|---|---|---|---|
| EPI_ISL_851005 | USA/CA-CDC-STM-219/2021 | 2021-01-02 | North America/USA/California/ | Helix/Illumina | Genomics and Discovery, Respiratory Viruses Branch, Division of Viral Diseases, Centers for Disease Control and Prevention | Peter W. Cook et al |
| EPI_ISL_851007 | USA/FL-CDC-STM-247/2020 | 2020-12-31 | North America/USA/Florida/ | Helix/Illumina | Genomics and Discovery, Respiratory Viruses Branch, Division of Viral Diseases, Centers for Disease Control and Prevention | Peter W. Cook et al |
| EPI_ISL_851008 | USA/NC-CDC-STM-240/2020 | 2020-12-31 | North America/USA/North Carolina/ | Helix/Illumina | Genomics and Discovery, Respiratory Viruses Branch, Division of Viral Diseases, Centers for Disease Control and Prevention | Peter W. Cook et al |
| EPI_ISL_851009 | USA/DE-CDC-STM-238/2020 | 2020-12-31 | North America/USA/Delaware/ | Helix/Illumina | Genomics and Discovery, Respiratory Viruses Branch, Division of Viral Diseases, Centers for Disease Control and Prevention | Peter W. Cook et al |

| | | | | | | |
|---|---|---|---|---|---|---|
| EPI_ISL_851010 | USA/IL-CDC-STM-230/2020 | 2020-12-31 | North America/USA/Illinois/ | Helix/Illumina | Genomics and Discovery, Respiratory Viruses Branch, Division of Viral Diseases, Centers for Disease Control and Prevention | Peter W. Cook et al |
| EPI_ISL_851013 | USA/CA-CDC-STM-220/2020 | 2020-12-31 | North America/USA/California/ | Helix/Illumina | Genomics and Discovery, Respiratory Viruses Branch, Division of Viral Diseases, Centers for Disease Control and Prevention | Peter W. Cook et al |
| EPI_ISL_851020 | USA/PA-CDC-STM-216/2020 | 2020-12-30 | North America/USA/Pennsylvania/ | Helix/Illumina | Genomics and Discovery, Respiratory Viruses Branch, Division of Viral Diseases, Centers for Disease Control and Prevention | Peter W. Cook et al |
| EPI_ISL_851022 | USA/CA-CDC-STM-232/2021 | 2021-01-02 | North America/USA/California/ | Helix/Illumina | Genomics and Discovery, Respiratory Viruses Branch, Division of Viral Diseases, Centers for Disease Control and Prevention | Peter W. Cook et al |

| | | | | | | |
|---|---|---|---|---|---|---|
| EPI_ISL_851024 | USA/FL-CDC-STM-211/2021 | 2021-01-02 | North America/USA/Florida/ | Helix/Illumina | Genomics and Discovery, Respiratory Viruses Branch, Division of Viral Diseases, Centers for Disease Control and Prevention | Peter W. Cook et al |
| EPI_ISL_851025 | USA/CA-CDC-STM-208/2021 | 2021-01-02 | North America/USA/California/ | Helix/Illumina | Genomics and Discovery, Respiratory Viruses Branch, Division of Viral Diseases, Centers for Disease Control and Prevention | Peter W. Cook et al |
| EPI_ISL_851031 | USA/UT-CDC-STM-233/2021 | 2021-01-02 | North America/USA/Utah/ | Helix/Illumina | Genomics and Discovery, Respiratory Viruses Branch, Division of Viral Diseases, Centers for Disease Control and Prevention | Peter W. Cook et al |
| EPI_ISL_851032 | USA/CA-CDC-STM-225/2021 | 2021-01-02 | North America/USA/California/ | Helix/Illumina | Genomics and Discovery, Respiratory Viruses Branch, Division of Viral Diseases, Centers for Disease Control and Prevention | Peter W. Cook et al |
| EPI_ISL_851045 | Japan/IC-0716/2021 | 21-Jan | Asia/Japan/Japan/ | Pathogen Genomics Center, National Institute of Infectious Diseases | Pathogen Genomics Center, National Institute of Infectious Diseases | Tsuyoshi Sekizuka et al |

| | | | | | | |
|---|---|---|---|---|---|---|
| EPI_ISL_851050 | India/TG-GMC-19076/2020 | 2020-07-20 | Asia/India/Telangana/ | Gandhi Medical College and Hospital | CSIR-Centre for Cellular and Molecular Biology | Nagamani Kammili et al |
| EPI_ISL_851051 | India/TG-GMC-19711/2020 | 2020-07-21 | Asia/India/Telangana/ | Gandhi Medical College and Hospital | CSIR-Centre for Cellular and Molecular Biology | Nagamani Kammili et al |
| EPI_ISL_851081 | England/QEUH-F9009D/2021 | 2021-01-10 | Europe/United Kingdom/England/ | Lighthouse Lab in Glasgow | Wellcome Sanger Institute for the COVID-19 Genomics UK (COG-UK) Consortium | Harper VanSteenhouse et al |
| EPI_ISL_851110 | England/CAMC-F9C608/2021 | 2021-01-07 | Europe/United Kingdom/England/ | Lighthouse Lab in Cambridge | Wellcome Sanger Institute for the COVID-19 Genomics UK (COG-UK) Consortium | Rob Howes et al |
| EPI_ISL_851241 | England/QEUH-F90367/2021 | 2021-01-11 | Europe/United Kingdom/England/ | Lighthouse Lab in Glasgow | Wellcome Sanger Institute for the COVID-19 Genomics UK (COG-UK) Consortium | Harper VanSteenhouse et al |
| EPI_ISL_851277 | England/CAMC-F99CEB/2021 | 2021-01-10 | Europe/United Kingdom/England/ | Lighthouse Lab in Cambridge | Wellcome Sanger Institute for the COVID-19 Genomics UK (COG-UK) Consortium | Rob Howes et al |
| EPI_ISL_851354 | England/CAMC-F9C5ED/2021 | 2021-01-07 | Europe/United Kingdom/England/ | Lighthouse Lab in Cambridge | Wellcome Sanger Institute for the COVID-19 Genomics UK (COG-UK) Consortium | Rob Howes et al |
| EPI_ISL_851450 | England/QEUH-F8A5E2/2021 | 2021-01-09 | Europe/United Kingdom/England/ | Lighthouse Lab in Glasgow | Wellcome Sanger Institute for the COVID-19 Genomics UK (COG-UK) Consortium | Harper VanSteenhouse et al |
| EPI_ISL_851572 | England/QEUH-F8B0A4/2021 | 2021-01-07 | Europe/United Kingdom/England/ | Lighthouse Lab in Glasgow | Wellcome Sanger Institute for the COVID-19 Genomics UK (COG-UK) Consortium | Harper VanSteenhouse et al |

| | | | | | | |
|---|---|---|---|---|---|---|
| EPI_ISL_851740 | England/ALDP-F91524/2021 | 2021-01-09 | Europe/United Kingdom/England/ | Lighthouse Lab in Alderley Park | Wellcome Sanger Institute for the COVID-19 Genomics UK (COG-UK) Consortium | Jacquelyn Wynn et al |
| EPI_ISL_852267 | England/QEUH-F56AA4/2021 | 2021-01-06 | Europe/United Kingdom/England/ | Lighthouse Lab in Glasgow | Wellcome Sanger Institute for the COVID-19 Genomics UK (COG-UK) Consortium | Harper VanSteenhouse et al |
| EPI_ISL_852279 | England/QEUH-F5FCB4/2021 | 2021-01-07 | Europe/United Kingdom/England/ | Lighthouse Lab in Glasgow | Wellcome Sanger Institute for the COVID-19 Genomics UK (COG-UK) Consortium | Harper VanSteenhouse et al |
| EPI_ISL_852565 | Germany/BY-MVP-0634/2020 | 2020-12-30 | Europe/Germany/Bavaria/Munich | Max von Pettenkofer Institute, Virology, National Reference Center for Retroviruses, LMU M√ºnchen | Laboratory for Functional Genome Analysis, Dept. Genomics, Gene Center of the LMU Munich | Max Muenchhoff et al |
| EPI_ISL_852567 | Germany/BY-MVP-0644/2021 | 2021-01-01 | Europe/Germany/Bavaria/Munich | Max von Pettenkofer Institute, Virology, National Reference Center for Retroviruses, LMU M√ºnchen | Laboratory for Functional Genome Analysis, Dept. Genomics, Gene Center of the LMU Munich | Max Muenchhoff et al |
| EPI_ISL_852568 | Germany/BY-MVP-0728/2020 | 2020-12-20 | Europe/Germany/Bavaria/Munich | Max von Pettenkofer Institute, Virology, National Reference Center for Retroviruses, LMU M√ºnchen | Laboratory for Functional Genome Analysis, Dept. Genomics, Gene Center of the LMU Munich | Max Muenchhoff et al |
| EPI_ISL_852572 | Germany/BY-MVP-0719/2020 | 2020-12-14 | Europe/Germany/Bavaria/Munich | Max von Pettenkofer Institute, Virology, National Reference Center for Retroviruses, LMU M√ºnchen | Laboratory for Functional Genome Analysis, Dept. Genomics, Gene Center of the LMU Munich | Max Muenchhoff et al |

| | | | | | | |
|---|---|---|---|---|---|---|
| EPI_ISL_852583 | Germany/BY-MVP-0417/2020 | 2020-09-18 | Europe/Germany/Bavaria/Munich | Max von Pettenkofer Institute, Virology, National Reference Center for Retroviruses, LMU M√ºnchen | Laboratory for Functional Genome Analysis, Dept. Genomics, Gene Center of the LMU Munich | Max Muenchhoff et al |
| EPI_ISL_852586 | Germany/BY-MVP-0377/2020 | 2020-08-30 | Europe/Germany/Bavaria/Munich | Max von Pettenkofer Institute, Virology, National Reference Center for Retroviruses, LMU M√ºnchen | Laboratory for Functional Genome Analysis, Dept. Genomics, Gene Center of the LMU Munich | Max Muenchhoff et al |
| EPI_ISL_852588 | Germany/BY-MVP-0378/2020 | 2020-08-30 | Europe/Germany/Bavaria/Munich | Max von Pettenkofer Institute, Virology, National Reference Center for Retroviruses, LMU M√ºnchen | Laboratory for Functional Genome Analysis, Dept. Genomics, Gene Center of the LMU Munich | Max Muenchhoff et al |
| EPI_ISL_852590 | Germany/BY-MVP-0389/2020 | 2020-09-08 | Europe/Germany/Bavaria/Munich | Max von Pettenkofer Institute, Virology, National Reference Center for Retroviruses, LMU M√ºnchen | Laboratory for Functional Genome Analysis, Dept. Genomics, Gene Center of the LMU Munich | Max Muenchhoff et al |
| EPI_ISL_852593 | Germany/BY-MVP-0388/2020 | 2020-09-07 | Europe/Germany/Bavaria/Munich | Max von Pettenkofer Institute, Virology, National Reference Center for Retroviruses, LMU M√ºnchen | Laboratory for Functional Genome Analysis, Dept. Genomics, Gene Center of the LMU Munich | Max Muenchhoff et al |
| EPI_ISL_852595 | Germany/BY-MVP-0532/2020 | 2020-11-02 | Europe/Germany/Bavaria/Munich | Max von Pettenkofer Institute, Virology, National Reference Center for Retroviruses, LMU M√ºnchen | Laboratory for Functional Genome Analysis, Dept. Genomics, Gene Center of the LMU Munich | Max Muenchhoff et al |

| | | | | | | |
|---|---|---|---|---|---|---|
| EPI_ISL_852596 | Germany/BY-MVP-0533/2020 | 2020-11-03 | Europe/Germany/Bavaria/Munich | Max von Pettenkofer Institute, Virology, National Reference Center for Retroviruses, LMU M√ºnchen | Laboratory for Functional Genome Analysis, Dept. Genomics, Gene Center of the LMU Munich | Max Muenchhoff et al |
| EPI_ISL_852600 | Germany/BY-MVP-0527/2020 | 2020-10-23 | Europe/Germany/Bavaria/Munich | Max von Pettenkofer Institute, Virology, National Reference Center for Retroviruses, LMU M√ºnchen | Laboratory for Functional Genome Analysis, Dept. Genomics, Gene Center of the LMU Munich | Max Muenchhoff et al |
| EPI_ISL_852602 | Germany/BY-MVP-0530/2020 | 2020-11-30 | Europe/Germany/Bavaria/Munich | Max von Pettenkofer Institute, Virology, National Reference Center for Retroviruses, LMU M√ºnchen | Laboratory for Functional Genome Analysis, Dept. Genomics, Gene Center of the LMU Munich | Max Muenchhoff et al |
| EPI_ISL_852628 | Italy/CAM-COTUGNO-18199/2020 | 2020-12-31 | Europe/Italy/Campania/ | Lab. Microbiologia e Virologia, Cotugno, A.O. dei Colli | Lab. Microbiologia e Virologia, Cotugno, A.O. dei Colli | Luigi Atripaldi et al |
| EPI_ISL_852629 | Italy/CAM-COTUGNO-27/2021 | 2021-01-02 | Europe/Italy/Campania/ | Lab. Microbiologia e Virologia, Cotugno, A.O. dei Colli | Lab. Microbiologia e Virologia, Cotugno, A.O. dei Colli | Luigi Atripaldi et al |
| EPI_ISL_852630 | Italy/CAM-COTUGNO-18279/2021 | 2021-01-02 | Europe/Italy/Campania/ | Lab. Microbiologia e Virologia, Cotugno, A.O. dei Colli | Lab. Microbiologia e Virologia, Cotugno, A.O. dei Colli | Luigi Atripaldi et al |
| EPI_ISL_852632 | Italy/CAM-COTUGNO-133/2021 | 2021-01-02 | Europe/Italy/Campania/ | Lab. Microbiologia e Virologia, Cotugno, A.O. dei Colli | Lab. Microbiologia e Virologia, Cotugno, A.O. dei Colli | Luigi Atripaldi et al |

| | | | | | | |
|---|---|---|---|---|---|---|
| EPI_ISL_852634 | Germany/BW-FR0323/2020 | 2020-11-27 | Europe/Germany/Baden-Wuerttemberg | Institute of Virology, Medical Center, University of Freiburg, Freiburg, Germany | Institute of Virology, Clinial Virus Genomics, Medical Center, University of Freiburg, Freiburg, Germany | Jonas Fuchs et al |
| EPI_ISL_852636 | Germany/BW-FR0321/2020 | 2020-11-26 | Europe/Germany/Baden-Wuerttemberg | Institute of Virology, Medical Center, University of Freiburg, Freiburg, Germany | Institute of Virology, Clinial Virus Genomics, Medical Center, University of Freiburg, Freiburg, Germany | Jonas Fuchs et al |
| EPI_ISL_852638 | Germany/BW-FR0319/2020 | 2020-11-26 | Europe/Germany/Baden-Wuerttemberg | Institute of Virology, Medical Center, University of Freiburg, Freiburg, Germany | Institute of Virology, Clinial Virus Genomics, Medical Center, University of Freiburg, Freiburg, Germany | Jonas Fuchs et al |
| EPI_ISL_852639 | Germany/BW-FR0318/2020 | 2020-11-26 | Europe/Germany/Baden-Wuerttemberg | Institute of Virology, Medical Center, University of Freiburg, Freiburg, Germany | Institute of Virology, Clinial Virus Genomics, Medical Center, University of Freiburg, Freiburg, Germany | Jonas Fuchs et al |
| EPI_ISL_852640 | Germany/BW-FR0325/2020 | 2020-11-26 | Europe/Germany/Baden-Wuerttemberg | Institute of Virology, Medical Center, University of Freiburg, Freiburg, Germany | Institute of Virology, Clinial Virus Genomics, Medical Center, University of Freiburg, Freiburg, Germany | Jonas Fuchs et al |

| | | | | | | |
|---|---|---|---|---|---|---|
| EPI_ISL_852642 | Germany/BW-FR0316/2020 | 2020-11-26 | Europe/Germany/Baden-Wuerttemberg | Institute of Virology, Medical Center, University of Freiburg, Freiburg, Germany | Institute of Virology, Clinial Virus Genomics, Medical Center, University of Freiburg, Freiburg, Germany | Jonas Fuchs et al |
| EPI_ISL_852645 | Germany/BW-FR0345/2020 | 2020-11-25 | Europe/Germany/Baden-Wuerttemberg | Institute of Virology, Medical Center, University of Freiburg, Freiburg, Germany | Institute of Virology, Clinial Virus Genomics, Medical Center, University of Freiburg, Freiburg, Germany | Jonas Fuchs et al |
| EPI_ISL_852647 | Germany/BW-FR0311/2020 | 2020-11-25 | Europe/Germany/Baden-Wuerttemberg | Institute of Virology, Medical Center, University of Freiburg, Freiburg, Germany | Institute of Virology, Clinial Virus Genomics, Medical Center, University of Freiburg, Freiburg, Germany | Jonas Fuchs et al |
| EPI_ISL_852648 | Germany/BW-FR0309/2020 | 2020-11-24 | Europe/Germany/Baden-Wuerttemberg | Institute of Virology, Medical Center, University of Freiburg, Freiburg, Germany | Institute of Virology, Clinial Virus Genomics, Medical Center, University of Freiburg, Freiburg, Germany | Jonas Fuchs et al |
| EPI_ISL_852649 | Germany/BW-FR0307/2020 | 2020-11-25 | Europe/Germany/Baden-Wuerttemberg | Institute of Virology, Medical Center, University of Freiburg, Freiburg, Germany | Institute of Virology, Clinial Virus Genomics, Medical Center, University of Freiburg, Freiburg, Germany | Jonas Fuchs et al |

| | | | | | | |
|---|---|---|---|---|---|---|
| EPI_ISL_852650 | Germany/BW-FR0133/2020 | 2020-10-20 | Europe/Germany/Baden-Wuerttemberg | Institute of Virology, Medical Center, University of Freiburg, Freiburg, Germany | Institute of Virology, Clinial Virus Genomics, Medical Center, University of Freiburg, Freiburg, Germany | Jonas Fuchs et al |
| EPI_ISL_852652 | Germany/BW-FR0130/2020 | 2020-10-19 | Europe/Germany/Baden-Wuerttemberg | Institute of Virology, Medical Center, University of Freiburg, Freiburg, Germany | Institute of Virology, Clinial Virus Genomics, Medical Center, University of Freiburg, Freiburg, Germany | Jonas Fuchs et al |
| EPI_ISL_852667 | Germany/BW-FR0042/2020 | 2020-04-06 | Europe/Germany/Baden-Wuerttemberg | Institute of Virology, Medical Center, University of Freiburg, Freiburg, Germany | Institute of Virology, Clinial Virus Genomics, Medical Center, University of Freiburg, Freiburg, Germany | Jonas Fuchs et al |
| EPI_ISL_852673 | Germany/BW-FR0334/2020 | 2020-03-29 | Europe/Germany/Baden-Wuerttemberg | Institute of Virology, Medical Center, University of Freiburg, Freiburg, Germany | Institute of Virology, Clinial Virus Genomics, Medical Center, University of Freiburg, Freiburg, Germany | Jonas Fuchs et al |
| EPI_ISL_852675 | Germany/BW-FR0331/2020 | 2020-03-28 | Europe/Germany/Baden-Wuerttemberg | Institute of Virology, Medical Center, University of Freiburg, Freiburg, Germany | Institute of Virology, Clinial Virus Genomics, Medical Center, University of Freiburg, Freiburg, Germany | Jonas Fuchs et al |

| | | | | | | |
|---|---|---|---|---|---|---|
| EPI_ISL_852676 | Germany/BW-FR0276/2020 | 2020-03-28 | Europe/Germany/Baden-Wuerttemberg | Institute of Virology, Medical Center, University of Freiburg, Freiburg, Germany | Institute of Virology, Clinial Virus Genomics, Medical Center, University of Freiburg, Freiburg, Germany | Jonas Fuchs et al |
| EPI_ISL_852679 | Germany/BW-FR0267/2020 | 2020-03-26 | Europe/Germany/Baden-Wuerttemberg | Institute of Virology, Medical Center, University of Freiburg, Freiburg, Germany | Institute of Virology, Clinial Virus Genomics, Medical Center, University of Freiburg, Freiburg, Germany | Jonas Fuchs et al |
| EPI_ISL_852684 | Germany/BW-FR0259/2020 | 2020-03-26 | Europe/Germany/Baden-Wuerttemberg | Institute of Virology, Medical Center, University of Freiburg, Freiburg, Germany | Institute of Virology, Clinial Virus Genomics, Medical Center, University of Freiburg, Freiburg, Germany | Jonas Fuchs et al |
| EPI_ISL_852685 | Germany/BW-FR0257/2020 | 2020-03-25 | Europe/Germany/Baden-Wuerttemberg | Institute of Virology, Medical Center, University of Freiburg, Freiburg, Germany | Institute of Virology, Clinial Virus Genomics, Medical Center, University of Freiburg, Freiburg, Germany | Jonas Fuchs et al |
| EPI_ISL_852687 | Germany/BW-FR0254/2020 | 2020-03-25 | Europe/Germany/Baden-Wuerttemberg | Institute of Virology, Medical Center, University of Freiburg, Freiburg, Germany | Institute of Virology, Clinial Virus Genomics, Medical Center, University of Freiburg, Freiburg, Germany | Jonas Fuchs et al |

| | | | | | | |
|---|---|---|---|---|---|---|
| EPI_ISL_852694 | Germany/BW-FR0245/2020 | 2020-03-24 | Europe/Germany/Baden-Wuerttemberg | Institute of Virology, Medical Center, University of Freiburg, Freiburg, Germany | Institute of Virology, Clinial Virus Genomics, Medical Center, University of Freiburg, Freiburg, Germany | Jonas Fuchs et al |
| EPI_ISL_852696 | Germany/BW-FR0243/2020 | 2020-03-24 | Europe/Germany/Baden-Wuerttemberg | Institute of Virology, Medical Center, University of Freiburg, Freiburg, Germany | Institute of Virology, Clinial Virus Genomics, Medical Center, University of Freiburg, Freiburg, Germany | Jonas Fuchs et al |
| EPI_ISL_852701 | Germany/BW-FR0085/2020 | 2020-03-24 | Europe/Germany/Baden-Wuerttemberg | Institute of Virology, Medical Center, University of Freiburg, Freiburg, Germany | Institute of Virology, Clinial Virus Genomics, Medical Center, University of Freiburg, Freiburg, Germany | Jonas Fuchs et al |
| EPI_ISL_852704 | Germany/BW-FR0234/2020 | 2020-03-23 | Europe/Germany/Baden-Wuerttemberg | Institute of Virology, Medical Center, University of Freiburg, Freiburg, Germany | Institute of Virology, Clinial Virus Genomics, Medical Center, University of Freiburg, Freiburg, Germany | Jonas Fuchs et al |
| EPI_ISL_852709 | Germany/BW-FR0040/2020 | 2020-03-23 | Europe/Germany/Baden-Wuerttemberg | Institute of Virology, Medical Center, University of Freiburg, Freiburg, Germany | Institute of Virology, Clinial Virus Genomics, Medical Center, University of Freiburg, Freiburg, Germany | Jonas Fuchs et al |

| | | | | | | |
|---|---|---|---|---|---|---|
| EPI_ISL_852711 | Germany/BW-FR0197/2020 | 2020-03-21 | Europe/Germany/Baden-Wuerttemberg | Institute of Virology, Medical Center, University of Freiburg, Freiburg, Germany | Institute of Virology, Clinial Virus Genomics, Medical Center, University of Freiburg, Freiburg, Germany | Jonas Fuchs et al |
| EPI_ISL_852717 | Germany/BW-FR0189/2020 | 2020-03-20 | Europe/Germany/Baden-Wuerttemberg | Institute of Virology, Medical Center, University of Freiburg, Freiburg, Germany | Institute of Virology, Clinial Virus Genomics, Medical Center, University of Freiburg, Freiburg, Germany | Jonas Fuchs et al |
| EPI_ISL_852723 | Germany/BW-FR0106/2020 | 2020-03-19 | Europe/Germany/Baden-Wuerttemberg | Institute of Virology, Medical Center, University of Freiburg, Freiburg, Germany | Institute of Virology, Clinial Virus Genomics, Medical Center, University of Freiburg, Freiburg, Germany | Jonas Fuchs et al |
| EPI_ISL_852727 | Germany/BW-FR0073/2020 | 2020-03-18 | Europe/Germany/Baden-Wuerttemberg | Institute of Virology, Medical Center, University of Freiburg, Freiburg, Germany | Institute of Virology, Clinial Virus Genomics, Medical Center, University of Freiburg, Freiburg, Germany | Jonas Fuchs et al |
| EPI_ISL_852730 | Germany/BW-FR0172/2020 | 2020-03-16 | Europe/Germany/Baden-Wuerttemberg | Institute of Virology, Medical Center, University of Freiburg, Freiburg, Germany | Institute of Virology, Clinial Virus Genomics, Medical Center, University of Freiburg, Freiburg, Germany | Jonas Fuchs et al |

| | | | | | | |
|---|---|---|---|---|---|---|
| EPI_ISL_852733 | Germany/BW-FR0069/2020 | 2020-03-15 | Europe/Germany/Baden-Wuerttemberg | Institute of Virology, Medical Center, University of Freiburg, Freiburg, Germany | Institute of Virology, Clinial Virus Genomics, Medical Center, University of Freiburg, Freiburg, Germany | Jonas Fuchs et al |
| EPI_ISL_852735 | Germany/BW-FR0068/2020 | 2020-03-13 | Europe/Germany/Baden-Wuerttemberg | Institute of Virology, Medical Center, University of Freiburg, Freiburg, Germany | Institute of Virology, Clinial Virus Genomics, Medical Center, University of Freiburg, Freiburg, Germany | Jonas Fuchs et al |
| EPI_ISL_852737 | Germany/BW-FR0014/2020 | 2020-03-12 | Europe/Germany/Baden-Wuerttemberg | Institute of Virology, Medical Center, University of Freiburg, Freiburg, Germany | Institute of Virology, Clinial Virus Genomics, Medical Center, University of Freiburg, Freiburg, Germany | Jonas Fuchs et al |
| EPI_ISL_852741 | Germany/BW-FR0103/2020 | 2020-03-09 | Europe/Germany/Baden-Wuerttemberg | Institute of Virology, Medical Center, University of Freiburg, Freiburg, Germany | Institute of Virology, Clinial Virus Genomics, Medical Center, University of Freiburg, Freiburg, Germany | Jonas Fuchs et al |
| EPI_ISL_852752 | Germany/BW-FR0063/2020 | 2020-03-05 | Europe/Germany/Baden-Wuerttemberg | Institute of Virology, Medical Center, University of Freiburg, Freiburg, Germany | Institute of Virology, Clinial Virus Genomics, Medical Center, University of Freiburg, Freiburg, Germany | Jonas Fuchs et al |

| | | | | | | |
|---|---|---|---|---|---|---|
| EPI_ISL_852753 | Germany/BW-FR0062/2020 | 2020-03-05 | Europe/Germany/Baden-Wuerttemberg | Institute of Virology, Medical Center, University of Freiburg, Freiburg, Germany | Institute of Virology, Clinial Virus Genomics, Medical Center, University of Freiburg, Freiburg, Germany | Jonas Fuchs et al |
| EPI_ISL_852759 | Germany/BW-FR0149/2020 | 2020-03-02 | Europe/Germany/Baden-Wuerttemberg | Institute of Virology, Medical Center, University of Freiburg, Freiburg, Germany | Institute of Virology, Clinial Virus Genomics, Medical Center, University of Freiburg, Freiburg, Germany | Jonas Fuchs et al |
| EPI_ISL_852760 | Germany/BW-FR0059/2020 | 2020-02-27 | Europe/Germany/Baden-Wuerttemberg | Institute of Virology, Medical Center, University of Freiburg, Freiburg, Germany | Institute of Virology, Clinial Virus Genomics, Medical Center, University of Freiburg, Freiburg, Germany | Jonas Fuchs et al |
| EPI_ISL_852763 | Germany/BW-FR0111/2020 | 2020-04-01 | Europe/Germany/Baden-Wuerttemberg | Institute of Virology, Medical Center, University of Freiburg, Freiburg, Germany | Institute of Virology, Clinial Virus Genomics, Medical Center, University of Freiburg, Freiburg, Germany | Jonas Fuchs et al |
| EPI_ISL_852767 | Germany/BW-FR0333/2020 | 2020-03-29 | Europe/Germany/Baden-Wuerttemberg | Institute of Virology, Medical Center, University of Freiburg, Freiburg, Germany | Institute of Virology, Clinial Virus Genomics, Medical Center, University of Freiburg, Freiburg, Germany | Jonas Fuchs et al |

| | | | | | | |
|---|---|---|---|---|---|---|
| EPI_ISL_852768 | Germany/BW-FR0330/2020 | 2020-03-28 | Europe/Germany/Baden-Wuerttemberg | Institute of Virology, Medical Center, University of Freiburg, Freiburg, Germany | Institute of Virology, Clinial Virus Genomics, Medical Center, University of Freiburg, Freiburg, Germany | Jonas Fuchs et al |
| EPI_ISL_852770 | Germany/BW-FR0029/2020 | 2020-03-23 | Europe/Germany/Baden-Wuerttemberg | Institute of Virology, Medical Center, University of Freiburg, Freiburg, Germany | Institute of Virology, Clinial Virus Genomics, Medical Center, University of Freiburg, Freiburg, Germany | Jonas Fuchs et al |
| EPI_ISL_852772 | Germany/BW-FR0076/2020 | 2020-03-20 | Europe/Germany/Baden-Wuerttemberg | Institute of Virology, Medical Center, University of Freiburg, Freiburg, Germany | Institute of Virology, Clinial Virus Genomics, Medical Center, University of Freiburg, Freiburg, Germany | Jonas Fuchs et al |
| EPI_ISL_852774 | Germany/BW-FR0028/2020 | 2020-03-19 | Europe/Germany/Baden-Wuerttemberg | Institute of Virology, Medical Center, University of Freiburg, Freiburg, Germany | Institute of Virology, Clinial Virus Genomics, Medical Center, University of Freiburg, Freiburg, Germany | Jonas Fuchs et al |
| EPI_ISL_852781 | Germany/BW-FR0078/2020 | 2020-03-16 | Europe/Germany/Baden-Wuerttemberg | Institute of Virology, Medical Center, University of Freiburg, Freiburg, Germany | Institute of Virology, Clinial Virus Genomics, Medical Center, University of Freiburg, Freiburg, Germany | Jonas Fuchs et al |

| | | | | | | |
|---|---|---|---|---|---|---|
| EPI_ISL_852787 | Germany/BW-FR0168/2020 | 2020-03-12 | Europe/Germany/Baden-Wuerttemberg | Institute of Virology, Medical Center, University of Freiburg, Freiburg, Germany | Institute of Virology, Clinial Virus Genomics, Medical Center, University of Freiburg, Freiburg, Germany | Jonas Fuchs et al |
| EPI_ISL_852791 | Germany/BW-FR0165/2020 | 2020-03-11 | Europe/Germany/Baden-Wuerttemberg | Institute of Virology, Medical Center, University of Freiburg, Freiburg, Germany | Institute of Virology, Clinial Virus Genomics, Medical Center, University of Freiburg, Freiburg, Germany | Jonas Fuchs et al |
| EPI_ISL_852793 | Germany/BW-FR0051/2020 | 2020-03-11 | Europe/Germany/Baden-Wuerttemberg | Institute of Virology, Medical Center, University of Freiburg, Freiburg, Germany | Institute of Virology, Clinial Virus Genomics, Medical Center, University of Freiburg, Freiburg, Germany | Jonas Fuchs et al |
| EPI_ISL_852795 | Germany/BW-FR0026/2020 | 2020-03-11 | Europe/Germany/Baden-Wuerttemberg | Institute of Virology, Medical Center, University of Freiburg, Freiburg, Germany | Institute of Virology, Clinial Virus Genomics, Medical Center, University of Freiburg, Freiburg, Germany | Jonas Fuchs et al |
| EPI_ISL_852797 | Germany/BW-FR0049/2020 | 2020-03-11 | Europe/Germany/Baden-Wuerttemberg | Institute of Virology, Medical Center, University of Freiburg, Freiburg, Germany | Institute of Virology, Clinial Virus Genomics, Medical Center, University of Freiburg, Freiburg, Germany | Jonas Fuchs et al |

| | | | | | | |
|---|---|---|---|---|---|---|
| EPI_ISL_852800 | Germany/BW-FR0024/2020 | 2020-03-10 | Europe/Germany/Baden-Wuerttemberg | Institute of Virology, Medical Center, University of Freiburg, Freiburg, Germany | Institute of Virology, Clinial Virus Genomics, Medical Center, University of Freiburg, Freiburg, Germany | Jonas Fuchs et al |
| EPI_ISL_852801 | Germany/BW-FR0016/2020 | 2020-03-10 | Europe/Germany/Baden-Wuerttemberg | Institute of Virology, Medical Center, University of Freiburg, Freiburg, Germany | Institute of Virology, Clinial Virus Genomics, Medical Center, University of Freiburg, Freiburg, Germany | Jonas Fuchs et al |
| EPI_ISL_852804 | Germany/BW-FR0013/2020 | 2020-03-09 | Europe/Germany/Baden-Wuerttemberg | Institute of Virology, Medical Center, University of Freiburg, Freiburg, Germany | Institute of Virology, Clinial Virus Genomics, Medical Center, University of Freiburg, Freiburg, Germany | Jonas Fuchs et al |
| EPI_ISL_852813 | Italy/CAM-COTUGNO-71/2021 | 2021-01-01 | Europe/Italy/Campania/ | Lab. Microbiologia e Virologia, Cotugno, A.O. dei Colli | Lab. Microbiologia e Virologia, Cotugno, A.O. dei Colli | Luigi Atripaldi et al |
| EPI_ISL_852819 | Italy/CAM-COTUGNO-81/2021 | 2021-01-01 | Europe/Italy/Campania/ | Lab. Microbiologia e Virologia, Cotugno, A.O. dei Colli | Lab. Microbiologia e Virologia, Cotugno, A.O. dei Colli | Luigi Atripaldi et al |
| EPI_ISL_852820 | Canada/LTRI-1411/2020 | 2020-12-24 | North America/Canada/Ontario/Toronto | Microbiology Lab | NBCC Sequencing Facility | Dr. Jeff Wrana et al |
| EPI_ISL_852821 | Italy/CAM-COTUGNO-168/2021 | 2021-01-02 | Europe/Italy/Campania/ | Lab. Microbiologia e Virologia, Cotugno, A.O. dei Colli | Lab. Microbiologia e Virologia, Cotugno, A.O. dei Colli | Luigi Atripaldi et al |
| EPI_ISL_852822 | Canada/LTRI-1471/2020 | 2020-12-24 | North America/Canada/Ontario/Toronto | Microbiology Lab | NBCC Sequencing Facility | Dr. Jeff Wrana et al |

| | | | | | | |
|---|---|---|---|---|---|---|
| EPI_ISL_852823 | Italy/CAM-COTUGNO-18428/2021 | 2021-01-01 | Europe/Italy/Campania/ | Lab. Microbiologia e Virologia, Cotugno, A.O. dei Colli | Lab. Microbiologia e Virologia, Cotugno, A.O. dei Colli | Luigi Atripaldi et al |
| EPI_ISL_852824 | Canada/LTRI-1505/2020 | 2020-12-24 | North America/Canada/Ontario/Toronto | Microbiology Lab | NBCC Sequencing Facility | Dr. Jeff Wrana et al |
| EPI_ISL_852825 | Italy/CAM-COTUGNO-18469/2021 | 2021-01-01 | Europe/Italy/Campania/ | Lab. Microbiologia e Virologia, Cotugno, A.O. dei Colli | Lab. Microbiologia e Virologia, Cotugno, A.O. dei Colli | Luigi Atripaldi et al |
| EPI_ISL_852826 | Canada/LTRI-1586/2020 | 2020-12-24 | North America/Canada/Ontario/Toronto | Microbiology Lab | NBCC Sequencing Facility | Dr. Jeff Wrana et al |
| EPI_ISL_852827 | Canada/LTRI-1587/2020 | 2020-12-24 | North America/Canada/Ontario/Toronto | Microbiology Lab | NBCC Sequencing Facility | Dr. Jeff Wrana et al |
| EPI_ISL_852828 | Italy/CAM-COTUGNO-18765/2021 | 2021-01-01 | Europe/Italy/Campania/ | Lab. Microbiologia e Virologia, Cotugno, A.O. dei Colli | Lab. Microbiologia e Virologia, Cotugno, A.O. dei Colli | Luigi Atripaldi et al |
| EPI_ISL_852829 | Canada/LTRI-1617/2020 | 2020-12-24 | North America/Canada/Ontario/Toronto | Microbiology Lab | NBCC Sequencing Facility | Dr. Jeff Wrana et al |
| EPI_ISL_852830 | Canada/LTRI-1701/2020 | 2020-12-24 | North America/Canada/Ontario/Toronto | Microbiology Lab | NBCC Sequencing Facility | Dr. Jeff Wrana et al |
| EPI_ISL_852831 | Canada/LTRI-1716/2020 | 2020-12-24 | North America/Canada/Ontario/Toronto | Microbiology Lab | NBCC Sequencing Facility | Dr. Jeff Wrana et al |
| EPI_ISL_852832 | Canada/LTRI-1731/2020 | 2020-12-24 | North America/Canada/Ontario/Toronto | Microbiology Lab | NBCC Sequencing Facility | Dr. Jeff Wrana et al |
| EPI_ISL_852833 | Canada/LTRI-1794/2020 | 2020-12-24 | North America/Canada/Ontario/Toronto | Microbiology Lab | NBCC Sequencing Facility | Dr. Jeff Wrana et al |
| EPI_ISL_852834 | Canada/QC-L00266952/2020 | 2020-05-01 | North America/Canada/Quebec/ | Laboratoire de sant√© publique du Qu√©bec | Laboratoire de sant√© publique du Qu√©bec | Sandrine Moreira et al |
| EPI_ISL_852835 | Canada/QC-L00266954/2020 | 2020-05-02 | North America/Canada/Quebec/ | Laboratoire de sant√© publique du Qu√©bec | Laboratoire de sant√© publique du Qu√©bec | Sandrine Moreira et al |
| EPI_ISL_852836 | Canada/QC-L00266949/2020 | 2020-05-03 | North America/Canada/Quebec/ | Laboratoire de sant√© publique du Qu√©bec | Laboratoire de sant√© publique du Qu√©bec | Sandrine Moreira et al |
| EPI_ISL_852837 | Canada/QC-L00266948/2020 | 2020-05-03 | North America/Canada/Quebec/ | Laboratoire de sant√© publique du Qu√©bec | Laboratoire de sant√© publique du Qu√©bec | Sandrine Moreira et al |

| | | | | | | |
|---|---|---|---|---|---|---|
| EPI_ISL_852838 | Canada/QC-L00266941/2020 | 2020-05-03 | North America/Canada/Quebec/ | Laboratoire de sant√© publique du Qu√©bec | Laboratoire de sant√© publique du Qu√©bec | Sandrine Moreira et al |
| EPI_ISL_852839 | Canada/QC-L00266935/2020 | 2020-05-03 | North America/Canada/Quebec/ | Laboratoire de sant√© publique du Qu√©bec | Laboratoire de sant√© publique du Qu√©bec | Sandrine Moreira et al |
| EPI_ISL_852840 | Canada/QC-L00266931/2020 | 2020-05-03 | North America/Canada/Quebec/ | Laboratoire de sant√© publique du Qu√©bec | Laboratoire de sant√© publique du Qu√©bec | Sandrine Moreira et al |
| EPI_ISL_852841 | Canada/QC-L00266929/2020 | 2020-05-03 | North America/Canada/Quebec/ | Laboratoire de sant√© publique du Qu√©bec | Laboratoire de sant√© publique du Qu√©bec | Sandrine Moreira et al |
| EPI_ISL_852843 | Canada/ON-PPS-00405/2020 | 2020-11-24 | North America/Canada/Ontario/ | Toronto Invasive Bacterial Diseases Network | Ontario Institute for Cancer Research | Allison McGeer et al |
| EPI_ISL_852844 | Canada/ON-PPS-00404/2020 | 2020-11-24 | North America/Canada/Ontario/ | Toronto Invasive Bacterial Diseases Network | Ontario Institute for Cancer Research | Allison McGeer et al |
| EPI_ISL_852845 | Canada/ON-PPS-00109/2020 | 2020-11-24 | North America/Canada/Ontario/ | Toronto Invasive Bacterial Diseases Network | Ontario Institute for Cancer Research | Allison McGeer et al |
| EPI_ISL_852846 | Canada/ON-PPS-00437/2020 | 2020-11-24 | North America/Canada/Ontario/ | Toronto Invasive Bacterial Diseases Network | Ontario Institute for Cancer Research | Allison McGeer et al |
| EPI_ISL_852847 | Canada/ON-PPS-00361/2020 | 2020-11-24 | North America/Canada/Ontario/ | Toronto Invasive Bacterial Diseases Network | Ontario Institute for Cancer Research | Allison McGeer et al |
| EPI_ISL_852848 | Canada/ON-PPS-00355/2020 | 2020-11-24 | North America/Canada/Ontario/ | Toronto Invasive Bacterial Diseases Network | Ontario Institute for Cancer Research | Allison McGeer et al |
| EPI_ISL_852849 | Canada/ON-PPS-00366/2020 | 2020-11-24 | North America/Canada/Ontario/ | Toronto Invasive Bacterial Diseases Network | Ontario Institute for Cancer Research | Allison McGeer et al |
| EPI_ISL_852850 | Canada/ON-PPS-00371/2020 | 2020-11-24 | North America/Canada/Ontario/ | Toronto Invasive Bacterial Diseases Network | Ontario Institute for Cancer Research | Allison McGeer et al |
| EPI_ISL_852851 | Canada/ON-PPS-00369/2020 | 2020-11-24 | North America/Canada/Ontario/ | Toronto Invasive Bacterial Diseases Network | Ontario Institute for Cancer Research | Allison McGeer et al |

| | | | | | | |
|---|---|---|---|---|---|---|
| EPI_ISL_852852 | Canada/ON-PPS-00368/2020 | 2020-11-24 | North America/Canada/Ontario/ | Toronto Invasive Bacterial Diseases Network | Ontario Institute for Cancer Research | Allison McGeer et al |
| EPI_ISL_852853 | Canada/ON-PPS-00258/2020 | 2020-11-24 | North America/Canada/Ontario/ | Toronto Invasive Bacterial Diseases Network | Ontario Institute for Cancer Research | Allison McGeer et al |
| EPI_ISL_852854 | Canada/ON-PPS-00225/2020 | 2020-11-24 | North America/Canada/Ontario/ | Toronto Invasive Bacterial Diseases Network | Ontario Institute for Cancer Research | Allison McGeer et al |
| EPI_ISL_852855 | Canada/ON-PPS-00227/2020 | 2020-11-24 | North America/Canada/Ontario/ | Toronto Invasive Bacterial Diseases Network | Ontario Institute for Cancer Research | Allison McGeer et al |
| EPI_ISL_852856 | Canada/ON-PPS-00107/2020 | 2020-11-24 | North America/Canada/Ontario/ | Toronto Invasive Bacterial Diseases Network | Ontario Institute for Cancer Research | Allison McGeer et al |
| EPI_ISL_852857 | Canada/ON-PPS-00168/2020 | 2020-11-24 | North America/Canada/Ontario/ | Toronto Invasive Bacterial Diseases Network | Ontario Institute for Cancer Research | Allison McGeer et al |
| EPI_ISL_852858 | USA/FL-BPHL-0001/2021 | 2021-01-04 | North America/USA/Florida/ | Florida Bureau of Public Health Laboratories | Florida Bureau of Public Health Laboratories | Sarah Schmedes et al |
| EPI_ISL_852860 | USA/FL-BPHL-0003/2021 | 2021-01-04 | North America/USA/Florida/ | Florida Bureau of Public Health Laboratories | Florida Bureau of Public Health Laboratories | Sarah Schmedes et al |
| EPI_ISL_852862 | Canada/ON-PPS-00053/2020 | 2020-11-24 | North America/Canada/Ontario/ | Toronto Invasive Bacterial Diseases Network | Ontario Institute for Cancer Research | Allison McGeer et al |
| EPI_ISL_852863 | Canada/ON-PPS-00063/2020 | 2020-11-24 | North America/Canada/Ontario/ | Toronto Invasive Bacterial Diseases Network | Ontario Institute for Cancer Research | Allison McGeer et al |
| EPI_ISL_852864 | Canada/ON-PPS-00085/2020 | 2020-11-24 | North America/Canada/Ontario/ | Toronto Invasive Bacterial Diseases Network | Ontario Institute for Cancer Research | Allison McGeer et al |
| EPI_ISL_852866 | Canada/ON-PPS-00018/2020 | 2020-11-24 | North America/Canada/Ontario/ | Toronto Invasive Bacterial Diseases Network | Ontario Institute for Cancer Research | Allison McGeer et al |
| EPI_ISL_852867 | Canada/ON-PPS-00004/2020 | 2020-11-24 | North America/Canada/Ontario/ | Toronto Invasive Bacterial Diseases Network | Ontario Institute for Cancer Research | Allison McGeer et al |

| | | | | | | |
|---|---|---|---|---|---|---|
| EPI_ISL_852868 | Canada/ON-PPS-00010/2020 | 2020-11-24 | North America/Canada/Ontario/ | Toronto Invasive Bacterial Diseases Network | Ontario Institute for Cancer Research | Allison McGeer et al |
| EPI_ISL_852869 | Canada/ON-PPS-00034/2020 | 2020-11-24 | North America/Canada/Ontario/ | Toronto Invasive Bacterial Diseases Network | Ontario Institute for Cancer Research | Allison McGeer et al |
| EPI_ISL_852870 | Canada/ON-PPS-00068/2020 | 2020-11-24 | North America/Canada/Ontario/ | Toronto Invasive Bacterial Diseases Network | Ontario Institute for Cancer Research | Allison McGeer et al |
| EPI_ISL_852871 | Canada/ON-PPS-00087/2020 | 2020-11-24 | North America/Canada/Ontario/ | Toronto Invasive Bacterial Diseases Network | Ontario Institute for Cancer Research | Allison McGeer et al |
| EPI_ISL_852872 | Canada/ON-PPS-00019/2020 | 2020-11-24 | North America/Canada/Ontario/ | Toronto Invasive Bacterial Diseases Network | Ontario Institute for Cancer Research | Allison McGeer et al |
| EPI_ISL_852873 | Canada/ON-PPS-00020/2020 | 2020-11-24 | North America/Canada/Ontario/ | Toronto Invasive Bacterial Diseases Network | Ontario Institute for Cancer Research | Allison McGeer et al |
| EPI_ISL_852874 | Canada/ON-PPS-00015/2020 | 2020-11-24 | North America/Canada/Ontario/ | Toronto Invasive Bacterial Diseases Network | Ontario Institute for Cancer Research | Allison McGeer et al |
| EPI_ISL_852875 | Canada/ON-PPS-00035/2020 | 2020-11-24 | North America/Canada/Ontario/ | Toronto Invasive Bacterial Diseases Network | Ontario Institute for Cancer Research | Allison McGeer et al |
| EPI_ISL_852876 | Canada/ON-PPS-00084/2020 | 2020-11-24 | North America/Canada/Ontario/ | Toronto Invasive Bacterial Diseases Network | Ontario Institute for Cancer Research | Allison McGeer et al |
| EPI_ISL_852877 | Canada/ON-PPS-00078/2020 | 2020-11-24 | North America/Canada/Ontario/ | Toronto Invasive Bacterial Diseases Network | Ontario Institute for Cancer Research | Allison McGeer et al |
| EPI_ISL_852878 | Canada/ON-PPS-00013/2020 | 2020-11-24 | North America/Canada/Ontario/ | Toronto Invasive Bacterial Diseases Network | Ontario Institute for Cancer Research | Allison McGeer et al |
| EPI_ISL_852879 | Canada/ON-PPS-00066/2020 | 2020-11-24 | North America/Canada/Ontario/ | Toronto Invasive Bacterial Diseases Network | Ontario Institute for Cancer Research | Allison McGeer et al |
| EPI_ISL_852881 | Canada/ON-PPS-00069/2020 | 2020-11-24 | North America/Canada/Ontario/ | Toronto Invasive Bacterial Diseases Network | Ontario Institute for Cancer Research | Allison McGeer et al |

| | | | | | | |
|---|---|---|---|---|---|---|
| EPI_ISL_852882 | Canada/ON-PPS-00092/2020 | 2020-11-24 | North America/Canada/Ontario/ | Toronto Invasive Bacterial Diseases Network | Ontario Institute for Cancer Research | Allison McGeer et al |
| EPI_ISL_852883 | Canada/ON-PPS-00067/2020 | 2020-11-24 | North America/Canada/Ontario/ | Toronto Invasive Bacterial Diseases Network | Ontario Institute for Cancer Research | Allison McGeer et al |
| EPI_ISL_852884 | Canada/ON-PPS-00049/2020 | 2020-11-24 | North America/Canada/Ontario/ | Toronto Invasive Bacterial Diseases Network | Ontario Institute for Cancer Research | Allison McGeer et al |
| EPI_ISL_852885 | Canada/ON-PPS-00051/2020 | 2020-11-24 | North America/Canada/Ontario/ | Toronto Invasive Bacterial Diseases Network | Ontario Institute for Cancer Research | Allison McGeer et al |
| EPI_ISL_852886 | Canada/ON-PPS-00086/2020 | 2020-11-24 | North America/Canada/Ontario/ | Toronto Invasive Bacterial Diseases Network | Ontario Institute for Cancer Research | Allison McGeer et al |
| EPI_ISL_852887 | Canada/ON-PPS-00062/2020 | 2020-11-24 | North America/Canada/Ontario/ | Toronto Invasive Bacterial Diseases Network | Ontario Institute for Cancer Research | Allison McGeer et al |
| EPI_ISL_852888 | Canada/ON-PPS-00036/2020 | 2020-11-24 | North America/Canada/Ontario/ | Toronto Invasive Bacterial Diseases Network | Ontario Institute for Cancer Research | Allison McGeer et al |
| EPI_ISL_852889 | Canada/ON-PPS-00055/2020 | 2020-11-24 | North America/Canada/Ontario/ | Toronto Invasive Bacterial Diseases Network | Ontario Institute for Cancer Research | Allison McGeer et al |
| EPI_ISL_852890 | Canada/ON-PPS-00083/2020 | 2020-11-24 | North America/Canada/Ontario/ | Toronto Invasive Bacterial Diseases Network | Ontario Institute for Cancer Research | Allison McGeer et al |
| EPI_ISL_852891 | Canada/ON-PPS-00075/2020 | 2020-11-24 | North America/Canada/Ontario/ | Toronto Invasive Bacterial Diseases Network | Ontario Institute for Cancer Research | Allison McGeer et al |
| EPI_ISL_852892 | Canada/ON-PPS-00061/2020 | 2020-11-24 | North America/Canada/Ontario/ | Toronto Invasive Bacterial Diseases Network | Ontario Institute for Cancer Research | Allison McGeer et al |
| EPI_ISL_852893 | Canada/ON-PPS-00009/2020 | 2020-11-24 | North America/Canada/Ontario/ | Toronto Invasive Bacterial Diseases Network | Ontario Institute for Cancer Research | Allison McGeer et al |
| EPI_ISL_852894 | Canada/ON-PPS-00065/2020 | 2020-11-24 | North America/Canada/Ontario/ | Toronto Invasive Bacterial Diseases Network | Ontario Institute for Cancer Research | Allison McGeer et al |

| | | | | | | |
|---|---|---|---|---|---|---|
| EPI_ISL_852895 | Canada/ON-PPS-00081/2020 | 2020-11-24 | North America/Canada/Ontario/ | Toronto Invasive Bacterial Diseases Network | Ontario Institute for Cancer Research | Allison McGeer et al |
| EPI_ISL_852896 | Canada/ON-PPS-00007/2020 | 2020-11-24 | North America/Canada/Ontario/ | Toronto Invasive Bacterial Diseases Network | Ontario Institute for Cancer Research | Allison McGeer et al |
| EPI_ISL_852897 | Canada/ON-PPS-00090/2020 | 2020-11-24 | North America/Canada/Ontario/ | Toronto Invasive Bacterial Diseases Network | Ontario Institute for Cancer Research | Allison McGeer et al |
| EPI_ISL_852898 | Canada/ON-PPS-00073/2020 | 2020-11-24 | North America/Canada/Ontario/ | Toronto Invasive Bacterial Diseases Network | Ontario Institute for Cancer Research | Allison McGeer et al |
| EPI_ISL_852899 | Canada/ON-PPS-00097/2020 | 2020-11-24 | North America/Canada/Ontario/ | Toronto Invasive Bacterial Diseases Network | Ontario Institute for Cancer Research | Allison McGeer et al |
| EPI_ISL_852900 | Canada/ON-PPS-00089/2020 | 2020-11-24 | North America/Canada/Ontario/ | Toronto Invasive Bacterial Diseases Network | Ontario Institute for Cancer Research | Allison McGeer et al |
| EPI_ISL_852901 | Canada/ON-PPS-00050/2020 | 2020-11-24 | North America/Canada/Ontario/ | Toronto Invasive Bacterial Diseases Network | Ontario Institute for Cancer Research | Allison McGeer et al |
| EPI_ISL_852902 | Canada/ON-PPS-00011/2020 | 2020-11-24 | North America/Canada/Ontario/ | Toronto Invasive Bacterial Diseases Network | Ontario Institute for Cancer Research | Allison McGeer et al |
| EPI_ISL_852903 | Canada/ON-PPS-00032/2020 | 2020-11-24 | North America/Canada/Ontario/ | Toronto Invasive Bacterial Diseases Network | Ontario Institute for Cancer Research | Allison McGeer et al |
| EPI_ISL_852904 | Canada/ON-PPS-00037/2020 | 2020-11-24 | North America/Canada/Ontario/ | Toronto Invasive Bacterial Diseases Network | Ontario Institute for Cancer Research | Allison McGeer et al |
| EPI_ISL_852905 | Canada/ON-PPS-00027/2020 | 2020-11-24 | North America/Canada/Ontario/ | Toronto Invasive Bacterial Diseases Network | Ontario Institute for Cancer Research | Allison McGeer et al |
| EPI_ISL_852906 | Canada/ON-PPS-00042/2020 | 2020-11-24 | North America/Canada/Ontario/ | Toronto Invasive Bacterial Diseases Network | Ontario Institute for Cancer Research | Allison McGeer et al |
| EPI_ISL_852907 | Canada/ON-PPS-00017/2020 | 2020-11-24 | North America/Canada/Ontario/ | Toronto Invasive Bacterial Diseases Network | Ontario Institute for Cancer Research | Allison McGeer et al |

| | | | | | | |
|---|---|---|---|---|---|---|
| EPI_ISL_852908 | Canada/ON-PPS-00054/2020 | 2020-11-24 | North America/Canada/Ontario/ | Toronto Invasive Bacterial Diseases Network | Ontario Institute for Cancer Research | Allison McGeer et al |
| EPI_ISL_852909 | Canada/ON-PPS-00072/2020 | 2020-11-24 | North America/Canada/Ontario/ | Toronto Invasive Bacterial Diseases Network | Ontario Institute for Cancer Research | Allison McGeer et al |
| EPI_ISL_852910 | Canada/ON-PPS-00094/2020 | 2020-11-24 | North America/Canada/Ontario/ | Toronto Invasive Bacterial Diseases Network | Ontario Institute for Cancer Research | Allison McGeer et al |
| EPI_ISL_852911 | Canada/ON-PPS-00046/2020 | 2020-11-24 | North America/Canada/Ontario/ | Toronto Invasive Bacterial Diseases Network | Ontario Institute for Cancer Research | Allison McGeer et al |
| EPI_ISL_852912 | Canada/ON-PPS-00043/2020 | 2020-11-24 | North America/Canada/Ontario/ | Toronto Invasive Bacterial Diseases Network | Ontario Institute for Cancer Research | Allison McGeer et al |
| EPI_ISL_852913 | Canada/ON-PPS-00060/2020 | 2020-11-24 | North America/Canada/Ontario/ | Toronto Invasive Bacterial Diseases Network | Ontario Institute for Cancer Research | Allison McGeer et al |
| EPI_ISL_852914 | Canada/ON-PPS-00033/2020 | 2020-11-24 | North America/Canada/Ontario/ | Toronto Invasive Bacterial Diseases Network | Ontario Institute for Cancer Research | Allison McGeer et al |
| EPI_ISL_852915 | Canada/ON-PPS-00058/2020 | 2020-11-24 | North America/Canada/Ontario/ | Toronto Invasive Bacterial Diseases Network | Ontario Institute for Cancer Research | Allison McGeer et al |
| EPI_ISL_852916 | Canada/ON-PPS-00070/2020 | 2020-11-24 | North America/Canada/Ontario/ | Toronto Invasive Bacterial Diseases Network | Ontario Institute for Cancer Research | Allison McGeer et al |
| EPI_ISL_852917 | Canada/ON-PPS-00095/2020 | 2020-11-24 | North America/Canada/Ontario/ | Toronto Invasive Bacterial Diseases Network | Ontario Institute for Cancer Research | Allison McGeer et al |
| EPI_ISL_852918 | Canada/ON-PPS-00038/2020 | 2020-11-24 | North America/Canada/Ontario/ | Toronto Invasive Bacterial Diseases Network | Ontario Institute for Cancer Research | Allison McGeer et al |
| EPI_ISL_852919 | Canada/ON-PPS-00030/2020 | 2020-11-24 | North America/Canada/Ontario/ | Toronto Invasive Bacterial Diseases Network | Ontario Institute for Cancer Research | Allison McGeer et al |
| EPI_ISL_852920 | Canada/ON-PPS-00022/2020 | 2020-11-24 | North America/Canada/Ontario/ | Toronto Invasive Bacterial Diseases Network | Ontario Institute for Cancer Research | Allison McGeer et al |

| | | | | | | |
|---|---|---|---|---|---|---|
| EPI_ISL_852921 | Canada/ON-PPS-00057/2020 | 2020-11-24 | North America/Canada/Ontario/ | Toronto Invasive Bacterial Diseases Network | Ontario Institute for Cancer Research | Allison McGeer et al |
| EPI_ISL_852922 | Canada/ON-PPS-00091/2020 | 2020-11-24 | North America/Canada/Ontario/ | Toronto Invasive Bacterial Diseases Network | Ontario Institute for Cancer Research | Allison McGeer et al |
| EPI_ISL_852923 | Canada/ON-PPS-00071/2020 | 2020-11-24 | North America/Canada/Ontario/ | Toronto Invasive Bacterial Diseases Network | Ontario Institute for Cancer Research | Allison McGeer et al |
| EPI_ISL_852924 | Canada/ON-PPS-00014/2020 | 2020-11-24 | North America/Canada/Ontario/ | Toronto Invasive Bacterial Diseases Network | Ontario Institute for Cancer Research | Allison McGeer et al |
| EPI_ISL_852925 | Canada/ON-PPS-00003/2020 | 2020-11-24 | North America/Canada/Ontario/ | Toronto Invasive Bacterial Diseases Network | Ontario Institute for Cancer Research | Allison McGeer et al |
| EPI_ISL_852926 | Canada/ON-PPS-00045/2020 | 2020-11-24 | North America/Canada/Ontario/ | Toronto Invasive Bacterial Diseases Network | Ontario Institute for Cancer Research | Allison McGeer et al |
| EPI_ISL_852927 | Canada/ON-PPS-00012/2020 | 2020-11-24 | North America/Canada/Ontario/ | Toronto Invasive Bacterial Diseases Network | Ontario Institute for Cancer Research | Allison McGeer et al |
| EPI_ISL_852928 | Canada/ON-PPS-00021/2020 | 2020-11-24 | North America/Canada/Ontario/ | Toronto Invasive Bacterial Diseases Network | Ontario Institute for Cancer Research | Allison McGeer et al |
| EPI_ISL_852929 | Canada/ON-PPS-00082/2020 | 2020-11-24 | North America/Canada/Ontario/ | Toronto Invasive Bacterial Diseases Network | Ontario Institute for Cancer Research | Allison McGeer et al |
| EPI_ISL_852930 | Canada/ON-PPS-00093/2020 | 2020-11-24 | North America/Canada/Ontario/ | Toronto Invasive Bacterial Diseases Network | Ontario Institute for Cancer Research | Allison McGeer et al |
| EPI_ISL_852931 | Canada/ON-PPS-00096/2020 | 2020-11-24 | North America/Canada/Ontario/ | Toronto Invasive Bacterial Diseases Network | Ontario Institute for Cancer Research | Allison McGeer et al |
| EPI_ISL_852932 | Canada/ON-PPS-00074/2020 | 2020-11-24 | North America/Canada/Ontario/ | Toronto Invasive Bacterial Diseases Network | Ontario Institute for Cancer Research | Allison McGeer et al |
| EPI_ISL_852933 | Canada/ON-PPS-00079/2020 | 2020-11-24 | North America/Canada/Ontario/ | Toronto Invasive Bacterial Diseases Network | Ontario Institute for Cancer Research | Allison McGeer et al |

| | | | | | | |
|---|---|---|---|---|---|---|
| EPI_ISL_852934 | Canada/ON-PPS-00044/2020 | 2020-11-24 | North America/Canada/Ontario/ | Toronto Invasive Bacterial Diseases Network | Ontario Institute for Cancer Research | Allison McGeer et al |
| EPI_ISL_852935 | Canada/ON-PPS-00039/2020 | 2020-11-24 | North America/Canada/Ontario/ | Toronto Invasive Bacterial Diseases Network | Ontario Institute for Cancer Research | Allison McGeer et al |
| EPI_ISL_852936 | Canada/ON-PPS-00024/2020 | 2020-11-24 | North America/Canada/Ontario/ | Toronto Invasive Bacterial Diseases Network | Ontario Institute for Cancer Research | Allison McGeer et al |
| EPI_ISL_852937 | Canada/ON-PPS-00047/2020 | 2020-11-24 | North America/Canada/Ontario/ | Toronto Invasive Bacterial Diseases Network | Ontario Institute for Cancer Research | Allison McGeer et al |
| EPI_ISL_852938 | Canada/ON-PPS-00029/2020 | 2020-11-24 | North America/Canada/Ontario/ | Toronto Invasive Bacterial Diseases Network | Ontario Institute for Cancer Research | Allison McGeer et al |
| EPI_ISL_852939 | Canada/ON-PPS-00006/2020 | 2020-11-24 | North America/Canada/Ontario/ | Toronto Invasive Bacterial Diseases Network | Ontario Institute for Cancer Research | Allison McGeer et al |
| EPI_ISL_852940 | Canada/ON-PPS-00041/2020 | 2020-11-24 | North America/Canada/Ontario/ | Toronto Invasive Bacterial Diseases Network | Ontario Institute for Cancer Research | Allison McGeer et al |
| EPI_ISL_852941 | Canada/ON-PPS-00023/2020 | 2020-11-24 | North America/Canada/Ontario/ | Toronto Invasive Bacterial Diseases Network | Ontario Institute for Cancer Research | Allison McGeer et al |
| EPI_ISL_852942 | Canada/ON-PPS-00080/2020 | 2020-11-24 | North America/Canada/Ontario/ | Toronto Invasive Bacterial Diseases Network | Ontario Institute for Cancer Research | Allison McGeer et al |
| EPI_ISL_852943 | Canada/ON-PPS-00048/2020 | 2020-11-24 | North America/Canada/Ontario/ | Toronto Invasive Bacterial Diseases Network | Ontario Institute for Cancer Research | Allison McGeer et al |
| EPI_ISL_852944 | Canada/ON-PPS-00031/2020 | 2020-11-24 | North America/Canada/Ontario/ | Toronto Invasive Bacterial Diseases Network | Ontario Institute for Cancer Research | Allison McGeer et al |
| EPI_ISL_852945 | Canada/ON-PPS-00059/2020 | 2020-11-24 | North America/Canada/Ontario/ | Toronto Invasive Bacterial Diseases Network | Ontario Institute for Cancer Research | Allison McGeer et al |
| EPI_ISL_852946 | Canada/ON-PPS-00026/2020 | 2020-11-24 | North America/Canada/Ontario/ | Toronto Invasive Bacterial Diseases Network | Ontario Institute for Cancer Research | Allison McGeer et al |

| | | | | | | |
|---|---|---|---|---|---|---|
| EPI_ISL_852948 | Spain/MD-IBV-99013634/2021 | 2021-01-06 | Europe/Spain/Madrid/Madrid | Hospital General Universitario Gregorio Marav√±√≥n | SeqCOVID-SPAIN consortium/IBV(CSIC) | Dar√≠o Garc√≠a de Viedma et al |
| EPI_ISL_852950 | Spain/MD-IBV-99013687/2021 | 2021-01-07 | Europe/Spain/Madrid/Madrid | Hospital General Universitario Gregorio Marav√±√≥n | SeqCOVID-SPAIN consortium/IBV(CSIC) | Dar√≠o Garc√≠a de Viedma et al |
| EPI_ISL_852952 | Spain/MD-IBV-99013652/2021 | 2021-01-08 | Europe/Spain/Madrid/Madrid | Hospital General Universitario Gregorio Marav√±√≥n | SeqCOVID-SPAIN consortium/IBV(CSIC) | Dar√≠o Garc√≠a de Viedma et al |
| EPI_ISL_852955 | Spain/MD-IBV-99013684/2021 | 2021-01-10 | Europe/Spain/Madrid/Madrid | Hospital General Universitario Gregorio Marav√±√≥n | SeqCOVID-SPAIN consortium/IBV(CSIC) | Dar√≠o Garc√≠a de Viedma et al |
| EPI_ISL_852965 | Spain/MD-IBV-99013720/2021 | 2021-01-11 | Europe/Spain/Madrid/Madrid | Hospital General Universitario Gregorio Marav√±√≥n | SeqCOVID-SPAIN consortium/IBV(CSIC) | Dar√≠o Garc√≠a de Viedma et al |
| EPI_ISL_852968 | Spain/MD-IBV-99013675/2021 | 2021-01-12 | Europe/Spain/Madrid/Madrid | Hospital General Universitario Gregorio Marav√±√≥n | SeqCOVID-SPAIN consortium/IBV(CSIC) | Dar√≠o Garc√≠a de Viedma et al |
| EPI_ISL_852972 | Spain/MD-IBV-99013680/2021 | 2021-01-11 | Europe/Spain/Madrid/Madrid | Hospital General Universitario Gregorio Marav√±√≥n | SeqCOVID-SPAIN consortium/IBV(CSIC) | Dar√≠o Garc√≠a de Viedma et al |
| EPI_ISL_852975 | Spain/MD-IBV-99013704/2021 | 2021-01-11 | Europe/Spain/Madrid/Madrid | Hospital General Universitario Gregorio Marav√±√≥n | SeqCOVID-SPAIN consortium/IBV(CSIC) | Dar√≠o Garc√≠a de Viedma et al |
| EPI_ISL_852977 | Spain/MD-IBV-99013707/2021 | 2021-01-11 | Europe/Spain/Madrid/Madrid | Hospital General Universitario Gregorio Marav√±√≥n | SeqCOVID-SPAIN consortium/IBV(CSIC) | Dar√≠o Garc√≠a de Viedma et al |
| EPI_ISL_852981 | Spain/MD-IBV-99013716/2021 | 2021-01-11 | Europe/Spain/Madrid/Madrid | Hospital General Universitario Gregorio Marav√±√≥n | SeqCOVID-SPAIN consortium/IBV(CSIC) | Dar√≠o Garc√≠a de Viedma et al |
| EPI_ISL_852986 | Spain/MD-IBV-99013722/2021 | 2021-01-11 | Europe/Spain/Madrid/Madrid | Hospital General Universitario Gregorio Marav√±√≥n | SeqCOVID-SPAIN consortium/IBV(CSIC) | Dar√≠o Garc√≠a de Viedma et al |
| EPI_ISL_852999 | Spain/MD-IBV-99013682/2021 | 2021-01-08 | Europe/Spain/Madrid/Madrid | Hospital General Universitario Gregorio Marav√±√≥n | SeqCOVID-SPAIN consortium/IBV(CSIC) | Dar√≠o Garc√≠a de Viedma et al |
| EPI_ISL_853001 | Spain/MD-IBV-99013683/2021 | 2021-01-08 | Europe/Spain/Madrid/Madrid | Hospital General Universitario Gregorio Marav√±√≥n | SeqCOVID-SPAIN consortium/IBV(CSIC) | Dar√≠o Garc√≠a de Viedma et al |

| | | | | | | |
|---|---|---|---|---|---|---|
| EPI_ISL_853005 | Spain/MD-IBV-99013698/2021 | 2021-01-04 | Europe/Spain/Madrid/Madrid | Hospital General Universitario Gregorio Marav±V≥n | SeqCOVID-SPAIN consortium/IBV(CSIC) | Dar√≠o Garc√≠a de Viedma et al |
| EPI_ISL_853015 | Spain/MD-IBV-99013665/2021 | 2021-01-12 | Europe/Spain/Madrid/Madrid | Hospital General Universitario Gregorio Marav±V≥n | SeqCOVID-SPAIN consortium/IBV(CSIC) | Dar√≠o Garc√≠a de Viedma et al |
| EPI_ISL_853018 | Spain/MD-IBV-99013655/2021 | 2021-01-09 | Europe/Spain/Madrid/Madrid | Hospital General Universitario Gregorio Marav±V≥n | SeqCOVID-SPAIN consortium/IBV(CSIC) | Dar√≠o Garc√≠a de Viedma et al |
| EPI_ISL_853020 | Spain/MD-IBV-99013657/2021 | 2021-01-09 | Europe/Spain/Madrid/Madrid | Hospital General Universitario Gregorio Marav±V≥n | SeqCOVID-SPAIN consortium/IBV(CSIC) | Dar√≠o Garc√≠a de Viedma et al |
| EPI_ISL_853028 | Spain/MD-IBV-99013636/2021 | 2021-01-05 | Europe/Spain/Madrid/Madrid | Hospital General Universitario Gregorio Marav±V≥n | SeqCOVID-SPAIN consortium/IBV(CSIC) | Dar√≠o Garc√≠a de Viedma et al |
| EPI_ISL_853029 | Spain/MD-IBV-99013637/2021 | 2021-01-05 | Europe/Spain/Madrid/Madrid | Hospital General Universitario Gregorio Marav±V≥n | SeqCOVID-SPAIN consortium/IBV(CSIC) | Dar√≠o Garc√≠a de Viedma et al |
| EPI_ISL_853032 | Spain/MD-IBV-99013689/2021 | 2021-01-06 | Europe/Spain/Madrid/Madrid | Hospital General Universitario Gregorio Marav±V≥n | SeqCOVID-SPAIN consortium/IBV(CSIC) | Dar√≠o Garc√≠a de Viedma et al |
| EPI_ISL_853033 | Canada/ON-PPS-00424/2020 | 2020-11-24 | North America/Canada/Ontario/ | Toronto Invasive Bacterial Diseases Network | Ontario Institute for Cancer Research | Allison McGeer et al |
| EPI_ISL_853034 | Canada/ON-PPS-00302/2020 | 2020-11-24 | North America/Canada/Ontario/ | Toronto Invasive Bacterial Diseases Network | Ontario Institute for Cancer Research | Allison McGeer et al |
| EPI_ISL_853035 | Canada/ON-PPS-00411/2020 | 2020-11-24 | North America/Canada/Ontario/ | Toronto Invasive Bacterial Diseases Network | Ontario Institute for Cancer Research | Allison McGeer et al |
| EPI_ISL_853036 | Canada/ON-PPS-00076/2020 | 2020-11-24 | North America/Canada/Ontario/ | Toronto Invasive Bacterial Diseases Network | Ontario Institute for Cancer Research | Allison McGeer et al |
| EPI_ISL_853037 | Canada/ON-PPS-00436/2020 | 2020-11-24 | North America/Canada/Ontario/ | Toronto Invasive Bacterial Diseases Network | Ontario Institute for Cancer Research | Allison McGeer et al |
| EPI_ISL_853038 | Canada/ON-PPS-00352/2020 | 2020-11-24 | North America/Canada/Ontario/ | Toronto Invasive Bacterial Diseases Network | Ontario Institute for Cancer Research | Allison McGeer et al |

| | | | | | | |
|---|---|---|---|---|---|---|
| EPI_ISL_853039 | Canada/ON-PPS-00304/2020 | 2020-11-24 | North America/Canada/Ontario/ | Toronto Invasive Bacterial Diseases Network | Ontario Institute for Cancer Research | Allison McGeer et al |
| EPI_ISL_853040 | Canada/ON-PPS-00430/2020 | 2020-11-24 | North America/Canada/Ontario/ | Toronto Invasive Bacterial Diseases Network | Ontario Institute for Cancer Research | Allison McGeer et al |
| EPI_ISL_853041 | Canada/ON-PPS-00413/2020 | 2020-11-24 | North America/Canada/Ontario/ | Toronto Invasive Bacterial Diseases Network | Ontario Institute for Cancer Research | Allison McGeer et al |
| EPI_ISL_853042 | Canada/ON-PPS-00414/2020 | 2020-11-24 | North America/Canada/Ontario/ | Toronto Invasive Bacterial Diseases Network | Ontario Institute for Cancer Research | Allison McGeer et al |
| EPI_ISL_853043 | Canada/ON-PPS-00294/2020 | 2020-11-24 | North America/Canada/Ontario/ | Toronto Invasive Bacterial Diseases Network | Ontario Institute for Cancer Research | Allison McGeer et al |
| EPI_ISL_853044 | Canada/ON-PPS-00250/2020 | 2020-11-24 | North America/Canada/Ontario/ | Toronto Invasive Bacterial Diseases Network | Ontario Institute for Cancer Research | Allison McGeer et al |
| EPI_ISL_853045 | Canada/ON-PPS-00408/2020 | 2020-11-24 | North America/Canada/Ontario/ | Toronto Invasive Bacterial Diseases Network | Ontario Institute for Cancer Research | Allison McGeer et al |
| EPI_ISL_853046 | Canada/ON-PPS-00226/2020 | 2020-11-24 | North America/Canada/Ontario/ | Toronto Invasive Bacterial Diseases Network | Ontario Institute for Cancer Research | Allison McGeer et al |
| EPI_ISL_853047 | Canada/ON-PPS-00297/2020 | 2020-11-24 | North America/Canada/Ontario/ | Toronto Invasive Bacterial Diseases Network | Ontario Institute for Cancer Research | Allison McGeer et al |
| EPI_ISL_853048 | Canada/ON-PPS-00288/2020 | 2020-11-24 | North America/Canada/Ontario/ | Toronto Invasive Bacterial Diseases Network | Ontario Institute for Cancer Research | Allison McGeer et al |
| EPI_ISL_853049 | Canada/ON-PPS-00429/2020 | 2020-11-24 | North America/Canada/Ontario/ | Toronto Invasive Bacterial Diseases Network | Ontario Institute for Cancer Research | Allison McGeer et al |
| EPI_ISL_853050 | Canada/ON-PPS-00169/2020 | 2020-11-24 | North America/Canada/Ontario/ | Toronto Invasive Bacterial Diseases Network | Ontario Institute for Cancer Research | Allison McGeer et al |
| EPI_ISL_853051 | Canada/ON-PPS-00402/2020 | 2020-11-24 | North America/Canada/Ontario/ | Toronto Invasive Bacterial Diseases Network | Ontario Institute for Cancer Research | Allison McGeer et al |

| | | | | | | |
|---|---|---|---|---|---|---|
| EPI_ISL_853052 | Canada/ON-PPS-00420/2020 | 2020-11-24 | North America/Canada/Ontario/ | Toronto Invasive Bacterial Diseases Network | Ontario Institute for Cancer Research | Allison McGeer et al |
| EPI_ISL_853053 | Canada/ON-PPS-00298/2020 | 2020-11-24 | North America/Canada/Ontario/ | Toronto Invasive Bacterial Diseases Network | Ontario Institute for Cancer Research | Allison McGeer et al |
| EPI_ISL_853054 | Canada/ON-PPS-00214/2020 | 2020-11-24 | North America/Canada/Ontario/ | Toronto Invasive Bacterial Diseases Network | Ontario Institute for Cancer Research | Allison McGeer et al |
| EPI_ISL_853055 | Canada/ON-PPS-00426/2020 | 2020-11-24 | North America/Canada/Ontario/ | Toronto Invasive Bacterial Diseases Network | Ontario Institute for Cancer Research | Allison McGeer et al |
| EPI_ISL_853056 | Canada/ON-PPS-00296/2020 | 2020-11-24 | North America/Canada/Ontario/ | Toronto Invasive Bacterial Diseases Network | Ontario Institute for Cancer Research | Allison McGeer et al |
| EPI_ISL_853057 | Canada/ON-PPS-00418/2020 | 2020-11-24 | North America/Canada/Ontario/ | Toronto Invasive Bacterial Diseases Network | Ontario Institute for Cancer Research | Allison McGeer et al |
| EPI_ISL_853058 | Canada/ON-PPS-00340/2020 | 2020-11-24 | North America/Canada/Ontario/ | Toronto Invasive Bacterial Diseases Network | Ontario Institute for Cancer Research | Allison McGeer et al |
| EPI_ISL_853059 | Canada/ON-PPS-00286/2020 | 2020-11-24 | North America/Canada/Ontario/ | Toronto Invasive Bacterial Diseases Network | Ontario Institute for Cancer Research | Allison McGeer et al |
| EPI_ISL_853060 | Canada/ON-PPS-00295/2020 | 2020-11-24 | North America/Canada/Ontario/ | Toronto Invasive Bacterial Diseases Network | Ontario Institute for Cancer Research | Allison McGeer et al |
| EPI_ISL_853061 | Canada/ON-PPS-00289/2020 | 2020-11-24 | North America/Canada/Ontario/ | Toronto Invasive Bacterial Diseases Network | Ontario Institute for Cancer Research | Allison McGeer et al |
| EPI_ISL_853062 | Canada/ON-PPS-00287/2020 | 2020-11-24 | North America/Canada/Ontario/ | Toronto Invasive Bacterial Diseases Network | Ontario Institute for Cancer Research | Allison McGeer et al |
| EPI_ISL_853063 | Canada/ON-PPS-00435/2020 | 2020-11-24 | North America/Canada/Ontario/ | Toronto Invasive Bacterial Diseases Network | Ontario Institute for Cancer Research | Allison McGeer et al |
| EPI_ISL_853064 | Canada/ON-PPS-00410/2020 | 2020-11-24 | North America/Canada/Ontario/ | Toronto Invasive Bacterial Diseases Network | Ontario Institute for Cancer Research | Allison McGeer et al |

| | | | | | | |
|---|---|---|---|---|---|---|
| EPI_ISL_853065 | Canada/ON-PPS-00145/2020 | 2020-11-24 | North America/Canada/Ontario/ | Toronto Invasive Bacterial Diseases Network | Ontario Institute for Cancer Research | Allison McGeer et al |
| EPI_ISL_853066 | Canada/ON-PPS-00052/2020 | 2020-11-24 | North America/Canada/Ontario/ | Toronto Invasive Bacterial Diseases Network | Ontario Institute for Cancer Research | Allison McGeer et al |
| EPI_ISL_853067 | Canada/ON-PPS-00291/2020 | 2020-11-24 | North America/Canada/Ontario/ | Toronto Invasive Bacterial Diseases Network | Ontario Institute for Cancer Research | Allison McGeer et al |
| EPI_ISL_853068 | Canada/ON-PPS-00403/2020 | 2020-11-24 | North America/Canada/Ontario/ | Toronto Invasive Bacterial Diseases Network | Ontario Institute for Cancer Research | Allison McGeer et al |
| EPI_ISL_853069 | Canada/ON-PPS-00401/2020 | 2020-11-24 | North America/Canada/Ontario/ | Toronto Invasive Bacterial Diseases Network | Ontario Institute for Cancer Research | Allison McGeer et al |
| EPI_ISL_853070 | Canada/ON-PPS-00133/2020 | 2020-11-24 | North America/Canada/Ontario/ | Toronto Invasive Bacterial Diseases Network | Ontario Institute for Cancer Research | Allison McGeer et al |
| EPI_ISL_853071 | Canada/ON-PPS-00293/2020 | 2020-11-24 | North America/Canada/Ontario/ | Toronto Invasive Bacterial Diseases Network | Ontario Institute for Cancer Research | Allison McGeer et al |
| EPI_ISL_853072 | Canada/ON-PPS-00290/2020 | 2020-11-24 | North America/Canada/Ontario/ | Toronto Invasive Bacterial Diseases Network | Ontario Institute for Cancer Research | Allison McGeer et al |
| EPI_ISL_853073 | Canada/ON-PPS-00434/2020 | 2020-11-24 | North America/Canada/Ontario/ | Toronto Invasive Bacterial Diseases Network | Ontario Institute for Cancer Research | Allison McGeer et al |
| EPI_ISL_853074 | Canada/ON-PPS-00428/2020 | 2020-11-24 | North America/Canada/Ontario/ | Toronto Invasive Bacterial Diseases Network | Ontario Institute for Cancer Research | Allison McGeer et al |
| EPI_ISL_853075 | Canada/ON-PPS-00303/2020 | 2020-11-24 | North America/Canada/Ontario/ | Toronto Invasive Bacterial Diseases Network | Ontario Institute for Cancer Research | Allison McGeer et al |
| EPI_ISL_853076 | Canada/ON-PPS-00316/2020 | 2020-11-24 | North America/Canada/Ontario/ | Toronto Invasive Bacterial Diseases Network | Ontario Institute for Cancer Research | Allison McGeer et al |
| EPI_ISL_853077 | Canada/ON-PPS-00421/2020 | 2020-11-24 | North America/Canada/Ontario/ | Toronto Invasive Bacterial Diseases Network | Ontario Institute for Cancer Research | Allison McGeer et al |

| | | | | | | |
|---|---|---|---|---|---|---|
| EPI_ISL_853078 | Canada/ON-PPS-00427/2020 | 2020-11-24 | North America/Canada/Ontario/ | Toronto Invasive Bacterial Diseases Network | Ontario Institute for Cancer Research | Allison McGeer et al |
| EPI_ISL_853079 | Canada/ON-PPS-00364/2020 | 2020-11-24 | North America/Canada/Ontario/ | Toronto Invasive Bacterial Diseases Network | Ontario Institute for Cancer Research | Allison McGeer et al |
| EPI_ISL_853080 | Canada/ON-PPS-00400/2020 | 2020-11-24 | North America/Canada/Ontario/ | Toronto Invasive Bacterial Diseases Network | Ontario Institute for Cancer Research | Allison McGeer et al |
| EPI_ISL_853081 | Canada/ON-PPS-00285/2020 | 2020-11-24 | North America/Canada/Ontario/ | Toronto Invasive Bacterial Diseases Network | Ontario Institute for Cancer Research | Allison McGeer et al |
| EPI_ISL_853082 | Canada/ON-PPS-00262/2020 | 2020-11-24 | North America/Canada/Ontario/ | Toronto Invasive Bacterial Diseases Network | Ontario Institute for Cancer Research | Allison McGeer et al |
| EPI_ISL_853083 | Canada/ON-PPS-00064/2020 | 2020-11-24 | North America/Canada/Ontario/ | Toronto Invasive Bacterial Diseases Network | Ontario Institute for Cancer Research | Allison McGeer et al |
| EPI_ISL_853084 | Canada/ON-PPS-00416/2020 | 2020-11-24 | North America/Canada/Ontario/ | Toronto Invasive Bacterial Diseases Network | Ontario Institute for Cancer Research | Allison McGeer et al |
| EPI_ISL_853085 | Canada/ON-PPS-00407/2020 | 2020-11-24 | North America/Canada/Ontario/ | Toronto Invasive Bacterial Diseases Network | Ontario Institute for Cancer Research | Allison McGeer et al |
| EPI_ISL_853086 | Canada/ON-PPS-00300/2020 | 2020-11-24 | North America/Canada/Ontario/ | Toronto Invasive Bacterial Diseases Network | Ontario Institute for Cancer Research | Allison McGeer et al |
| EPI_ISL_853087 | Canada/ON-PPS-00088/2020 | 2020-11-24 | North America/Canada/Ontario/ | Toronto Invasive Bacterial Diseases Network | Ontario Institute for Cancer Research | Allison McGeer et al |
| EPI_ISL_853088 | Canada/ON-PPS-00284/2020 | 2020-11-24 | North America/Canada/Ontario/ | Toronto Invasive Bacterial Diseases Network | Ontario Institute for Cancer Research | Allison McGeer et al |
| EPI_ISL_853089 | Canada/ON-PPS-00431/2020 | 2020-11-24 | North America/Canada/Ontario/ | Toronto Invasive Bacterial Diseases Network | Ontario Institute for Cancer Research | Allison McGeer et al |
| EPI_ISL_853090 | Canada/ON-PPS-00181/2020 | 2020-11-24 | North America/Canada/Ontario/ | Toronto Invasive Bacterial Diseases Network | Ontario Institute for Cancer Research | Allison McGeer et al |

| | | | | | | |
|---|---|---|---|---|---|---|
| EPI_ISL_853091 | Canada/ON-PPS-00121/2020 | 2020-11-24 | North America/Canada/Ontario/ | Toronto Invasive Bacterial Diseases Network | Ontario Institute for Cancer Research | Allison McGeer et al |
| EPI_ISL_853092 | Canada/ON-PPS-00301/2020 | 2020-11-24 | North America/Canada/Ontario/ | Toronto Invasive Bacterial Diseases Network | Ontario Institute for Cancer Research | Allison McGeer et al |
| EPI_ISL_853093 | Canada/ON-PPS-00328/2020 | 2020-11-24 | North America/Canada/Ontario/ | Toronto Invasive Bacterial Diseases Network | Ontario Institute for Cancer Research | Allison McGeer et al |
| EPI_ISL_853094 | Canada/ON-PPS-00406/2020 | 2020-11-24 | North America/Canada/Ontario/ | Toronto Invasive Bacterial Diseases Network | Ontario Institute for Cancer Research | Allison McGeer et al |
| EPI_ISL_853095 | Canada/ON-PPS-00376/2020 | 2020-11-24 | North America/Canada/Ontario/ | Toronto Invasive Bacterial Diseases Network | Ontario Institute for Cancer Research | Allison McGeer et al |
| EPI_ISL_853096 | Canada/ON-PPS-00238/2020 | 2020-11-24 | North America/Canada/Ontario/ | Toronto Invasive Bacterial Diseases Network | Ontario Institute for Cancer Research | Allison McGeer et al |
| EPI_ISL_853097 | Canada/ON-PPS-00417/2020 | 2020-11-24 | North America/Canada/Ontario/ | Toronto Invasive Bacterial Diseases Network | Ontario Institute for Cancer Research | Allison McGeer et al |
| EPI_ISL_853098 | Canada/ON-PPS-00299/2020 | 2020-11-24 | North America/Canada/Ontario/ | Toronto Invasive Bacterial Diseases Network | Ontario Institute for Cancer Research | Allison McGeer et al |
| EPI_ISL_853099 | Canada/ON-PPS-00412/2020 | 2020-11-24 | North America/Canada/Ontario/ | Toronto Invasive Bacterial Diseases Network | Ontario Institute for Cancer Research | Allison McGeer et al |
| EPI_ISL_853100 | Canada/ON-PPS-00432/2020 | 2020-11-24 | North America/Canada/Ontario/ | Toronto Invasive Bacterial Diseases Network | Ontario Institute for Cancer Research | Allison McGeer et al |
| EPI_ISL_853101 | Canada/ON-PPS-00292/2020 | 2020-11-24 | North America/Canada/Ontario/ | Toronto Invasive Bacterial Diseases Network | Ontario Institute for Cancer Research | Allison McGeer et al |
| EPI_ISL_853102 | Canada/ON-PPS-00422/2020 | 2020-11-24 | North America/Canada/Ontario/ | Toronto Invasive Bacterial Diseases Network | Ontario Institute for Cancer Research | Allison McGeer et al |
| EPI_ISL_853103 | Canada/ON-PPS-00433/2020 | 2020-11-24 | North America/Canada/Ontario/ | Toronto Invasive Bacterial Diseases Network | Ontario Institute for Cancer Research | Allison McGeer et al |

| | | | | | | |
|---|---|---|---|---|---|---|
| EPI_ISL_853104 | Canada/ON-PPS-00425/2020 | 2020-11-24 | North America/Canada/Ontario/ | Toronto Invasive Bacterial Diseases Network | Ontario Institute for Cancer Research | Allison McGeer et al |
| EPI_ISL_853105 | Canada/ON-PPS-00415/2020 | 2020-11-24 | North America/Canada/Ontario/ | Toronto Invasive Bacterial Diseases Network | Ontario Institute for Cancer Research | Allison McGeer et al |
| EPI_ISL_853106 | Canada/ON-PPS-00202/2020 | 2020-11-24 | North America/Canada/Ontario/ | Toronto Invasive Bacterial Diseases Network | Ontario Institute for Cancer Research | Allison McGeer et al |
| EPI_ISL_853107 | Canada/ON-PPS-00274/2020 | 2020-11-24 | North America/Canada/Ontario/ | Toronto Invasive Bacterial Diseases Network | Ontario Institute for Cancer Research | Allison McGeer et al |
| EPI_ISL_853108 | Canada/ON-PPS-00423/2020 | 2020-11-24 | North America/Canada/Ontario/ | Toronto Invasive Bacterial Diseases Network | Ontario Institute for Cancer Research | Allison McGeer et al |
| EPI_ISL_853109 | Canada/ON-PPS-00388/2020 | 2020-11-24 | North America/Canada/Ontario/ | Toronto Invasive Bacterial Diseases Network | Ontario Institute for Cancer Research | Allison McGeer et al |
| EPI_ISL_853110 | Canada/ON-PPS-00419/2020 | 2020-11-24 | North America/Canada/Ontario/ | Toronto Invasive Bacterial Diseases Network | Ontario Institute for Cancer Research | Allison McGeer et al |
| EPI_ISL_853111 | Canada/ON-PPS-00131/2020 | 2020-11-24 | North America/Canada/Ontario/ | Toronto Invasive Bacterial Diseases Network | Ontario Institute for Cancer Research | Allison McGeer et al |
| EPI_ISL_853112 | Canada/ON-PPS-00159/2020 | 2020-11-24 | North America/Canada/Ontario/ | Toronto Invasive Bacterial Diseases Network | Ontario Institute for Cancer Research | Allison McGeer et al |
| EPI_ISL_853113 | Canada/ON-PPS-00122/2020 | 2020-11-24 | North America/Canada/Ontario/ | Toronto Invasive Bacterial Diseases Network | Ontario Institute for Cancer Research | Allison McGeer et al |
| EPI_ISL_853114 | Canada/ON-PPS-00178/2020 | 2020-11-24 | North America/Canada/Ontario/ | Toronto Invasive Bacterial Diseases Network | Ontario Institute for Cancer Research | Allison McGeer et al |
| EPI_ISL_853115 | Canada/ON-PPS-00174/2020 | 2020-11-24 | North America/Canada/Ontario/ | Toronto Invasive Bacterial Diseases Network | Ontario Institute for Cancer Research | Allison McGeer et al |
| EPI_ISL_853116 | Canada/ON-PPS-00111/2020 | 2020-11-24 | North America/Canada/Ontario/ | Toronto Invasive Bacterial Diseases Network | Ontario Institute for Cancer Research | Allison McGeer et al |

| | | | | | | |
|---|---|---|---|---|---|---|
| EPI_ISL_853117 | Canada/ON-PPS-00162/2020 | 2020-11-24 | North America/Canada/Ontario/ | Toronto Invasive Bacterial Diseases Network | Ontario Institute for Cancer Research | Allison McGeer et al |
| EPI_ISL_853118 | Canada/ON-PPS-00139/2020 | 2020-11-24 | North America/Canada/Ontario/ | Toronto Invasive Bacterial Diseases Network | Ontario Institute for Cancer Research | Allison McGeer et al |
| EPI_ISL_853119 | Canada/ON-PPS-00132/2020 | 2020-11-24 | North America/Canada/Ontario/ | Toronto Invasive Bacterial Diseases Network | Ontario Institute for Cancer Research | Allison McGeer et al |
| EPI_ISL_853120 | Canada/ON-PPS-00180/2020 | 2020-11-24 | North America/Canada/Ontario/ | Toronto Invasive Bacterial Diseases Network | Ontario Institute for Cancer Research | Allison McGeer et al |
| EPI_ISL_853121 | Canada/ON-PPS-00175/2020 | 2020-11-24 | North America/Canada/Ontario/ | Toronto Invasive Bacterial Diseases Network | Ontario Institute for Cancer Research | Allison McGeer et al |
| EPI_ISL_853122 | Canada/ON-PPS-00125/2020 | 2020-11-24 | North America/Canada/Ontario/ | Toronto Invasive Bacterial Diseases Network | Ontario Institute for Cancer Research | Allison McGeer et al |
| EPI_ISL_853123 | Canada/ON-PPS-00165/2020 | 2020-11-24 | North America/Canada/Ontario/ | Toronto Invasive Bacterial Diseases Network | Ontario Institute for Cancer Research | Allison McGeer et al |
| EPI_ISL_853124 | Canada/ON-PPS-00151/2020 | 2020-11-24 | North America/Canada/Ontario/ | Toronto Invasive Bacterial Diseases Network | Ontario Institute for Cancer Research | Allison McGeer et al |
| EPI_ISL_853125 | Canada/ON-PPS-00160/2020 | 2020-11-24 | North America/Canada/Ontario/ | Toronto Invasive Bacterial Diseases Network | Ontario Institute for Cancer Research | Allison McGeer et al |
| EPI_ISL_853126 | Canada/ON-PPS-00126/2020 | 2020-11-24 | North America/Canada/Ontario/ | Toronto Invasive Bacterial Diseases Network | Ontario Institute for Cancer Research | Allison McGeer et al |
| EPI_ISL_853127 | Canada/ON-PPS-00177/2020 | 2020-11-24 | North America/Canada/Ontario/ | Toronto Invasive Bacterial Diseases Network | Ontario Institute for Cancer Research | Allison McGeer et al |
| EPI_ISL_853128 | Canada/ON-PPS-00141/2020 | 2020-11-24 | North America/Canada/Ontario/ | Toronto Invasive Bacterial Diseases Network | Ontario Institute for Cancer Research | Allison McGeer et al |
| EPI_ISL_853129 | Canada/ON-PPS-00137/2020 | 2020-11-24 | North America/Canada/Ontario/ | Toronto Invasive Bacterial Diseases Network | Ontario Institute for Cancer Research | Allison McGeer et al |

| | | | | | | |
|---|---|---|---|---|---|---|
| EPI_ISL_853130 | Canada/ON-PPS-00106/2020 | 2020-11-24 | North America/Canada/Ontario/ | Toronto Invasive Bacterial Diseases Network | Ontario Institute for Cancer Research | Allison McGeer et al |
| EPI_ISL_853131 | Canada/ON-PPS-00153/2020 | 2020-11-24 | North America/Canada/Ontario/ | Toronto Invasive Bacterial Diseases Network | Ontario Institute for Cancer Research | Allison McGeer et al |
| EPI_ISL_853132 | Canada/ON-PPS-00184/2020 | 2020-11-24 | North America/Canada/Ontario/ | Toronto Invasive Bacterial Diseases Network | Ontario Institute for Cancer Research | Allison McGeer et al |
| EPI_ISL_853133 | Canada/ON-PPS-00129/2020 | 2020-11-24 | North America/Canada/Ontario/ | Toronto Invasive Bacterial Diseases Network | Ontario Institute for Cancer Research | Allison McGeer et al |
| EPI_ISL_853134 | Canada/ON-PPS-00102/2020 | 2020-11-24 | North America/Canada/Ontario/ | Toronto Invasive Bacterial Diseases Network | Ontario Institute for Cancer Research | Allison McGeer et al |
| EPI_ISL_853135 | Canada/ON-PPS-00124/2020 | 2020-11-24 | North America/Canada/Ontario/ | Toronto Invasive Bacterial Diseases Network | Ontario Institute for Cancer Research | Allison McGeer et al |
| EPI_ISL_853136 | Canada/ON-PPS-00155/2020 | 2020-11-24 | North America/Canada/Ontario/ | Toronto Invasive Bacterial Diseases Network | Ontario Institute for Cancer Research | Allison McGeer et al |
| EPI_ISL_853137 | Canada/ON-PPS-00164/2020 | 2020-11-24 | North America/Canada/Ontario/ | Toronto Invasive Bacterial Diseases Network | Ontario Institute for Cancer Research | Allison McGeer et al |
| EPI_ISL_853138 | Canada/ON-PPS-00166/2020 | 2020-11-24 | North America/Canada/Ontario/ | Toronto Invasive Bacterial Diseases Network | Ontario Institute for Cancer Research | Allison McGeer et al |
| EPI_ISL_853139 | Canada/ON-PPS-00130/2020 | 2020-11-24 | North America/Canada/Ontario/ | Toronto Invasive Bacterial Diseases Network | Ontario Institute for Cancer Research | Allison McGeer et al |
| EPI_ISL_853140 | Canada/ON-PPS-00179/2020 | 2020-11-24 | North America/Canada/Ontario/ | Toronto Invasive Bacterial Diseases Network | Ontario Institute for Cancer Research | Allison McGeer et al |
| EPI_ISL_853141 | Canada/ON-PPS-00114/2020 | 2020-11-24 | North America/Canada/Ontario/ | Toronto Invasive Bacterial Diseases Network | Ontario Institute for Cancer Research | Allison McGeer et al |
| EPI_ISL_853142 | Canada/ON-PPS-00187/2020 | 2020-11-24 | North America/Canada/Ontario/ | Toronto Invasive Bacterial Diseases Network | Ontario Institute for Cancer Research | Allison McGeer et al |

| | | | | | | |
|---|---|---|---|---|---|---|
| EPI_ISL_853143 | Canada/ON-PPS-00167/2020 | 2020-11-24 | North America/Canada/Ontario/ | Toronto Invasive Bacterial Diseases Network | Ontario Institute for Cancer Research | Allison McGeer et al |
| EPI_ISL_853144 | Canada/ON-PPS-00154/2020 | 2020-11-24 | North America/Canada/Ontario/ | Toronto Invasive Bacterial Diseases Network | Ontario Institute for Cancer Research | Allison McGeer et al |
| EPI_ISL_853145 | Canada/ON-PPS-00135/2020 | 2020-11-24 | North America/Canada/Ontario/ | Toronto Invasive Bacterial Diseases Network | Ontario Institute for Cancer Research | Allison McGeer et al |
| EPI_ISL_853146 | Canada/ON-PPS-00116/2020 | 2020-11-24 | North America/Canada/Ontario/ | Toronto Invasive Bacterial Diseases Network | Ontario Institute for Cancer Research | Allison McGeer et al |
| EPI_ISL_853147 | Canada/ON-PPS-00152/2020 | 2020-11-24 | North America/Canada/Ontario/ | Toronto Invasive Bacterial Diseases Network | Ontario Institute for Cancer Research | Allison McGeer et al |
| EPI_ISL_853148 | Canada/ON-PPS-00148/2020 | 2020-11-24 | North America/Canada/Ontario/ | Toronto Invasive Bacterial Diseases Network | Ontario Institute for Cancer Research | Allison McGeer et al |
| EPI_ISL_853149 | Canada/ON-PPS-00110/2020 | 2020-11-24 | North America/Canada/Ontario/ | Toronto Invasive Bacterial Diseases Network | Ontario Institute for Cancer Research | Allison McGeer et al |
| EPI_ISL_853150 | Canada/ON-PPS-00186/2020 | 2020-11-24 | North America/Canada/Ontario/ | Toronto Invasive Bacterial Diseases Network | Ontario Institute for Cancer Research | Allison McGeer et al |
| EPI_ISL_853151 | Canada/ON-PPS-00112/2020 | 2020-11-24 | North America/Canada/Ontario/ | Toronto Invasive Bacterial Diseases Network | Ontario Institute for Cancer Research | Allison McGeer et al |
| EPI_ISL_853152 | Canada/ON-PPS-00147/2020 | 2020-11-24 | North America/Canada/Ontario/ | Toronto Invasive Bacterial Diseases Network | Ontario Institute for Cancer Research | Allison McGeer et al |
| EPI_ISL_853153 | Canada/ON-PPS-00113/2020 | 2020-11-24 | North America/Canada/Ontario/ | Toronto Invasive Bacterial Diseases Network | Ontario Institute for Cancer Research | Allison McGeer et al |
| EPI_ISL_853154 | Canada/ON-PPS-00127/2020 | 2020-11-24 | North America/Canada/Ontario/ | Toronto Invasive Bacterial Diseases Network | Ontario Institute for Cancer Research | Allison McGeer et al |
| EPI_ISL_853155 | Canada/ON-PPS-00138/2020 | 2020-11-24 | North America/Canada/Ontario/ | Toronto Invasive Bacterial Diseases Network | Ontario Institute for Cancer Research | Allison McGeer et al |

| | | | | | | |
|---|---|---|---|---|---|---|
| EPI_ISL_853156 | Canada/ON-PPS-00108/2020 | 2020-11-24 | North America/Canada/Ontario/ | Toronto Invasive Bacterial Diseases Network | Ontario Institute for Cancer Research | Allison McGeer et al |
| EPI_ISL_853157 | Canada/ON-PPS-00143/2020 | 2020-11-24 | North America/Canada/Ontario/ | Toronto Invasive Bacterial Diseases Network | Ontario Institute for Cancer Research | Allison McGeer et al |
| EPI_ISL_853158 | Canada/ON-PPS-00140/2020 | 2020-11-24 | North America/Canada/Ontario/ | Toronto Invasive Bacterial Diseases Network | Ontario Institute for Cancer Research | Allison McGeer et al |
| EPI_ISL_853159 | Canada/ON-PPS-00163/2020 | 2020-11-24 | North America/Canada/Ontario/ | Toronto Invasive Bacterial Diseases Network | Ontario Institute for Cancer Research | Allison McGeer et al |
| EPI_ISL_853160 | Canada/ON-PPS-00173/2020 | 2020-11-24 | North America/Canada/Ontario/ | Toronto Invasive Bacterial Diseases Network | Ontario Institute for Cancer Research | Allison McGeer et al |
| EPI_ISL_853161 | Canada/ON-PPS-00146/2020 | 2020-11-24 | North America/Canada/Ontario/ | Toronto Invasive Bacterial Diseases Network | Ontario Institute for Cancer Research | Allison McGeer et al |
| EPI_ISL_853162 | Canada/ON-PPS-00123/2020 | 2020-11-24 | North America/Canada/Ontario/ | Toronto Invasive Bacterial Diseases Network | Ontario Institute for Cancer Research | Allison McGeer et al |
| EPI_ISL_853163 | Canada/ON-PPS-00098/2020 | 2020-11-24 | North America/Canada/Ontario/ | Toronto Invasive Bacterial Diseases Network | Ontario Institute for Cancer Research | Allison McGeer et al |
| EPI_ISL_853164 | Canada/ON-PPS-00142/2020 | 2020-11-24 | North America/Canada/Ontario/ | Toronto Invasive Bacterial Diseases Network | Ontario Institute for Cancer Research | Allison McGeer et al |
| EPI_ISL_853165 | Canada/ON-PPS-00149/2020 | 2020-11-24 | North America/Canada/Ontario/ | Toronto Invasive Bacterial Diseases Network | Ontario Institute for Cancer Research | Allison McGeer et al |
| EPI_ISL_853166 | Canada/ON-PPS-00118/2020 | 2020-11-24 | North America/Canada/Ontario/ | Toronto Invasive Bacterial Diseases Network | Ontario Institute for Cancer Research | Allison McGeer et al |
| EPI_ISL_853167 | Canada/ON-PPS-00120/2020 | 2020-11-24 | North America/Canada/Ontario/ | Toronto Invasive Bacterial Diseases Network | Ontario Institute for Cancer Research | Allison McGeer et al |
| EPI_ISL_853168 | Canada/ON-PPS-00172/2020 | 2020-11-24 | North America/Canada/Ontario/ | Toronto Invasive Bacterial Diseases Network | Ontario Institute for Cancer Research | Allison McGeer et al |

| | | | | | | |
|---|---|---|---|---|---|---|
| EPI_ISL_853169 | Canada/ON-PPS-00128/2020 | 2020-11-24 | North America/Canada/Ontario/ | Toronto Invasive Bacterial Diseases Network | Ontario Institute for Cancer Research | Allison McGeer et al |
| EPI_ISL_853170 | Canada/ON-PPS-00134/2020 | 2020-11-24 | North America/Canada/Ontario/ | Toronto Invasive Bacterial Diseases Network | Ontario Institute for Cancer Research | Allison McGeer et al |
| EPI_ISL_853171 | Canada/ON-PPS-00185/2020 | 2020-11-24 | North America/Canada/Ontario/ | Toronto Invasive Bacterial Diseases Network | Ontario Institute for Cancer Research | Allison McGeer et al |
| EPI_ISL_853172 | Canada/ON-PPS-00144/2020 | 2020-11-24 | North America/Canada/Ontario/ | Toronto Invasive Bacterial Diseases Network | Ontario Institute for Cancer Research | Allison McGeer et al |
| EPI_ISL_853173 | Canada/ON-PPS-00117/2020 | 2020-11-24 | North America/Canada/Ontario/ | Toronto Invasive Bacterial Diseases Network | Ontario Institute for Cancer Research | Allison McGeer et al |
| EPI_ISL_853174 | Canada/ON-PPS-00105/2020 | 2020-11-24 | North America/Canada/Ontario/ | Toronto Invasive Bacterial Diseases Network | Ontario Institute for Cancer Research | Allison McGeer et al |
| EPI_ISL_853175 | Canada/ON-PPS-00171/2020 | 2020-11-24 | North America/Canada/Ontario/ | Toronto Invasive Bacterial Diseases Network | Ontario Institute for Cancer Research | Allison McGeer et al |
| EPI_ISL_853176 | Canada/ON-PPS-00119/2020 | 2020-11-24 | North America/Canada/Ontario/ | Toronto Invasive Bacterial Diseases Network | Ontario Institute for Cancer Research | Allison McGeer et al |
| EPI_ISL_853177 | Canada/ON-PPS-00188/2020 | 2020-11-24 | North America/Canada/Ontario/ | Toronto Invasive Bacterial Diseases Network | Ontario Institute for Cancer Research | Allison McGeer et al |
| EPI_ISL_853178 | Canada/ON-PPS-00115/2020 | 2020-11-24 | North America/Canada/Ontario/ | Toronto Invasive Bacterial Diseases Network | Ontario Institute for Cancer Research | Allison McGeer et al |
| EPI_ISL_853179 | Canada/ON-PPS-00190/2020 | 2020-11-24 | North America/Canada/Ontario/ | Toronto Invasive Bacterial Diseases Network | Ontario Institute for Cancer Research | Allison McGeer et al |
| EPI_ISL_853180 | Canada/ON-PPS-00176/2020 | 2020-11-24 | North America/Canada/Ontario/ | Toronto Invasive Bacterial Diseases Network | Ontario Institute for Cancer Research | Allison McGeer et al |
| EPI_ISL_853181 | Canada/ON-PPS-00101/2020 | 2020-11-24 | North America/Canada/Ontario/ | Toronto Invasive Bacterial Diseases Network | Ontario Institute for Cancer Research | Allison McGeer et al |

| | | | | | | |
|---|---|---|---|---|---|---|
| EPI_ISL_853182 | Canada/ON-PPS-00189/2020 | 2020-11-24 | North America/Canada/Ontario/ | Toronto Invasive Bacterial Diseases Network | Ontario Institute for Cancer Research | Allison McGeer et al |
| EPI_ISL_853183 | Canada/ON-PPS-00103/2020 | 2020-11-24 | North America/Canada/Ontario/ | Toronto Invasive Bacterial Diseases Network | Ontario Institute for Cancer Research | Allison McGeer et al |
| EPI_ISL_853184 | Canada/ON-PPS-00104/2020 | 2020-11-24 | North America/Canada/Ontario/ | Toronto Invasive Bacterial Diseases Network | Ontario Institute for Cancer Research | Allison McGeer et al |
| EPI_ISL_853185 | Canada/ON-PPS-00182/2020 | 2020-11-24 | North America/Canada/Ontario/ | Toronto Invasive Bacterial Diseases Network | Ontario Institute for Cancer Research | Allison McGeer et al |
| EPI_ISL_853186 | Canada/ON-PPS-00183/2020 | 2020-11-24 | North America/Canada/Ontario/ | Toronto Invasive Bacterial Diseases Network | Ontario Institute for Cancer Research | Allison McGeer et al |
| EPI_ISL_853187 | Canada/ON-PPS-00161/2020 | 2020-11-24 | North America/Canada/Ontario/ | Toronto Invasive Bacterial Diseases Network | Ontario Institute for Cancer Research | Allison McGeer et al |
| EPI_ISL_853188 | Canada/ON-PPS-00099/2020 | 2020-11-24 | North America/Canada/Ontario/ | Toronto Invasive Bacterial Diseases Network | Ontario Institute for Cancer Research | Allison McGeer et al |
| EPI_ISL_853189 | Canada/ON-PPS-00156/2020 | 2020-11-24 | North America/Canada/Ontario/ | Toronto Invasive Bacterial Diseases Network | Ontario Institute for Cancer Research | Allison McGeer et al |
| EPI_ISL_853190 | Canada/ON-PPS-00100/2020 | 2020-11-24 | North America/Canada/Ontario/ | Toronto Invasive Bacterial Diseases Network | Ontario Institute for Cancer Research | Allison McGeer et al |
| EPI_ISL_853191 | Canada/ON-PPS-00158/2020 | 2020-11-24 | North America/Canada/Ontario/ | Toronto Invasive Bacterial Diseases Network | Ontario Institute for Cancer Research | Allison McGeer et al |
| EPI_ISL_853192 | Canada/ON-PPS-00170/2020 | 2020-11-24 | North America/Canada/Ontario/ | Toronto Invasive Bacterial Diseases Network | Ontario Institute for Cancer Research | Allison McGeer et al |
| EPI_ISL_853193 | Canada/ON-PPS-00205/2020 | 2020-11-24 | North America/Canada/Ontario/ | Toronto Invasive Bacterial Diseases Network | Ontario Institute for Cancer Research | Allison McGeer et al |
| EPI_ISL_853194 | Canada/ON-PPS-00263/2020 | 2020-11-24 | North America/Canada/Ontario/ | Toronto Invasive Bacterial Diseases Network | Ontario Institute for Cancer Research | Allison McGeer et al |

| | | | | | | |
|---|---|---|---|---|---|---|
| EPI_ISL_853195 | Canada/ON-PPS-00234/2020 | 2020-11-24 | North America/Canada/Ontario/ | Toronto Invasive Bacterial Diseases Network | Ontario Institute for Cancer Research | Allison McGeer et al |
| EPI_ISL_853196 | Canada/ON-PPS-00198/2020 | 2020-11-24 | North America/Canada/Ontario/ | Toronto Invasive Bacterial Diseases Network | Ontario Institute for Cancer Research | Allison McGeer et al |
| EPI_ISL_853197 | Canada/ON-PPS-00222/2020 | 2020-11-24 | North America/Canada/Ontario/ | Toronto Invasive Bacterial Diseases Network | Ontario Institute for Cancer Research | Allison McGeer et al |
| EPI_ISL_853198 | Canada/ON-PPS-00207/2020 | 2020-11-24 | North America/Canada/Ontario/ | Toronto Invasive Bacterial Diseases Network | Ontario Institute for Cancer Research | Allison McGeer et al |
| EPI_ISL_853199 | Canada/ON-PPS-00199/2020 | 2020-11-24 | North America/Canada/Ontario/ | Toronto Invasive Bacterial Diseases Network | Ontario Institute for Cancer Research | Allison McGeer et al |
| EPI_ISL_853200 | Canada/ON-PPS-00212/2020 | 2020-11-24 | North America/Canada/Ontario/ | Toronto Invasive Bacterial Diseases Network | Ontario Institute for Cancer Research | Allison McGeer et al |
| EPI_ISL_853201 | Canada/ON-PPS-00244/2020 | 2020-11-24 | North America/Canada/Ontario/ | Toronto Invasive Bacterial Diseases Network | Ontario Institute for Cancer Research | Allison McGeer et al |
| EPI_ISL_853202 | Canada/ON-PPS-00277/2020 | 2020-11-24 | North America/Canada/Ontario/ | Toronto Invasive Bacterial Diseases Network | Ontario Institute for Cancer Research | Allison McGeer et al |
| EPI_ISL_853203 | Canada/ON-PPS-00204/2020 | 2020-11-24 | North America/Canada/Ontario/ | Toronto Invasive Bacterial Diseases Network | Ontario Institute for Cancer Research | Allison McGeer et al |
| EPI_ISL_853204 | Canada/ON-PPS-00211/2020 | 2020-11-24 | North America/Canada/Ontario/ | Toronto Invasive Bacterial Diseases Network | Ontario Institute for Cancer Research | Allison McGeer et al |
| EPI_ISL_853205 | Canada/ON-PPS-00210/2020 | 2020-11-24 | North America/Canada/Ontario/ | Toronto Invasive Bacterial Diseases Network | Ontario Institute for Cancer Research | Allison McGeer et al |
| EPI_ISL_853206 | Canada/ON-PPS-00219/2020 | 2020-11-24 | North America/Canada/Ontario/ | Toronto Invasive Bacterial Diseases Network | Ontario Institute for Cancer Research | Allison McGeer et al |
| EPI_ISL_853207 | Canada/ON-PPS-00236/2020 | 2020-11-24 | North America/Canada/Ontario/ | Toronto Invasive Bacterial Diseases Network | Ontario Institute for Cancer Research | Allison McGeer et al |

| | | | | | | |
|---|---|---|---|---|---|---|
| EPI_ISL_853208 | Canada/ON-PPS-00267/2020 | 2020-11-24 | North America/Canada/Ontario/ | Toronto Invasive Bacterial Diseases Network | Ontario Institute for Cancer Research | Allison McGeer et al |
| EPI_ISL_853209 | Canada/ON-PPS-00254/2020 | 2020-11-24 | North America/Canada/Ontario/ | Toronto Invasive Bacterial Diseases Network | Ontario Institute for Cancer Research | Allison McGeer et al |
| EPI_ISL_853210 | Canada/ON-PPS-00253/2020 | 2020-11-24 | North America/Canada/Ontario/ | Toronto Invasive Bacterial Diseases Network | Ontario Institute for Cancer Research | Allison McGeer et al |
| EPI_ISL_853211 | Canada/ON-PPS-00241/2020 | 2020-11-24 | North America/Canada/Ontario/ | Toronto Invasive Bacterial Diseases Network | Ontario Institute for Cancer Research | Allison McGeer et al |
| EPI_ISL_853212 | Canada/ON-PPS-00276/2020 | 2020-11-24 | North America/Canada/Ontario/ | Toronto Invasive Bacterial Diseases Network | Ontario Institute for Cancer Research | Allison McGeer et al |
| EPI_ISL_853213 | Canada/ON-PPS-00247/2020 | 2020-11-24 | North America/Canada/Ontario/ | Toronto Invasive Bacterial Diseases Network | Ontario Institute for Cancer Research | Allison McGeer et al |
| EPI_ISL_853214 | Canada/ON-PPS-00249/2020 | 2020-11-24 | North America/Canada/Ontario/ | Toronto Invasive Bacterial Diseases Network | Ontario Institute for Cancer Research | Allison McGeer et al |
| EPI_ISL_853215 | Canada/ON-PPS-00218/2020 | 2020-11-24 | North America/Canada/Ontario/ | Toronto Invasive Bacterial Diseases Network | Ontario Institute for Cancer Research | Allison McGeer et al |
| EPI_ISL_853216 | Canada/ON-PPS-00264/2020 | 2020-11-24 | North America/Canada/Ontario/ | Toronto Invasive Bacterial Diseases Network | Ontario Institute for Cancer Research | Allison McGeer et al |
| EPI_ISL_853217 | Canada/ON-PPS-00252/2020 | 2020-11-24 | North America/Canada/Ontario/ | Toronto Invasive Bacterial Diseases Network | Ontario Institute for Cancer Research | Allison McGeer et al |
| EPI_ISL_853218 | Canada/ON-PPS-00191/2020 | 2020-11-24 | North America/Canada/Ontario/ | Toronto Invasive Bacterial Diseases Network | Ontario Institute for Cancer Research | Allison McGeer et al |
| EPI_ISL_853219 | Canada/ON-PPS-00192/2020 | 2020-11-24 | North America/Canada/Ontario/ | Toronto Invasive Bacterial Diseases Network | Ontario Institute for Cancer Research | Allison McGeer et al |
| EPI_ISL_853220 | Canada/ON-PPS-00193/2020 | 2020-11-24 | North America/Canada/Ontario/ | Toronto Invasive Bacterial Diseases Network | Ontario Institute for Cancer Research | Allison McGeer et al |

| | | | | | | |
|---|---|---|---|---|---|---|
| EPI_ISL_853221 | Canada/ON-PPS-00196/2020 | 2020-11-24 | North America/Canada/Ontario/ | Toronto Invasive Bacterial Diseases Network | Ontario Institute for Cancer Research | Allison McGeer et al |
| EPI_ISL_853223 | Canada/ON-PPS-00220/2020 | 2020-11-24 | North America/Canada/Ontario/ | Toronto Invasive Bacterial Diseases Network | Ontario Institute for Cancer Research | Allison McGeer et al |
| EPI_ISL_853224 | Canada/ON-PPS-00206/2020 | 2020-11-24 | North America/Canada/Ontario/ | Toronto Invasive Bacterial Diseases Network | Ontario Institute for Cancer Research | Allison McGeer et al |
| EPI_ISL_853225 | Canada/ON-PPS-00282/2020 | 2020-11-24 | North America/Canada/Ontario/ | Toronto Invasive Bacterial Diseases Network | Ontario Institute for Cancer Research | Allison McGeer et al |
| EPI_ISL_853226 | Canada/ON-PPS-00279/2020 | 2020-11-24 | North America/Canada/Ontario/ | Toronto Invasive Bacterial Diseases Network | Ontario Institute for Cancer Research | Allison McGeer et al |
| EPI_ISL_853227 | Canada/ON-PPS-00229/2020 | 2020-11-24 | North America/Canada/Ontario/ | Toronto Invasive Bacterial Diseases Network | Ontario Institute for Cancer Research | Allison McGeer et al |
| EPI_ISL_853228 | Canada/ON-PPS-00230/2020 | 2020-11-24 | North America/Canada/Ontario/ | Toronto Invasive Bacterial Diseases Network | Ontario Institute for Cancer Research | Allison McGeer et al |
| EPI_ISL_853229 | Canada/ON-PPS-00233/2020 | 2020-11-24 | North America/Canada/Ontario/ | Toronto Invasive Bacterial Diseases Network | Ontario Institute for Cancer Research | Allison McGeer et al |
| EPI_ISL_853230 | Canada/ON-PPS-00283/2020 | 2020-11-24 | North America/Canada/Ontario/ | Toronto Invasive Bacterial Diseases Network | Ontario Institute for Cancer Research | Allison McGeer et al |
| EPI_ISL_853231 | Canada/ON-PPS-00232/2020 | 2020-11-24 | North America/Canada/Ontario/ | Toronto Invasive Bacterial Diseases Network | Ontario Institute for Cancer Research | Allison McGeer et al |
| EPI_ISL_853232 | Canada/ON-PPS-00235/2020 | 2020-11-24 | North America/Canada/Ontario/ | Toronto Invasive Bacterial Diseases Network | Ontario Institute for Cancer Research | Allison McGeer et al |
| EPI_ISL_853233 | Canada/ON-PPS-00231/2020 | 2020-11-24 | North America/Canada/Ontario/ | Toronto Invasive Bacterial Diseases Network | Ontario Institute for Cancer Research | Allison McGeer et al |
| EPI_ISL_853234 | Canada/ON-PPS-00208/2020 | 2020-11-24 | North America/Canada/Ontario/ | Toronto Invasive Bacterial Diseases Network | Ontario Institute for Cancer Research | Allison McGeer et al |

| | | | | | | |
|---|---|---|---|---|---|---|
| EPI_ISL_853235 | Canada/ON-PPS-00242/2020 | 2020-11-24 | North America/Canada/Ontario/ | Toronto Invasive Bacterial Diseases Network | Ontario Institute for Cancer Research | Allison McGeer et al |
| EPI_ISL_853236 | Canada/ON-PPS-00216/2020 | 2020-11-24 | North America/Canada/Ontario/ | Toronto Invasive Bacterial Diseases Network | Ontario Institute for Cancer Research | Allison McGeer et al |
| EPI_ISL_853237 | Canada/ON-PPS-00194/2020 | 2020-11-24 | North America/Canada/Ontario/ | Toronto Invasive Bacterial Diseases Network | Ontario Institute for Cancer Research | Allison McGeer et al |
| EPI_ISL_853238 | Canada/ON-PPS-00239/2020 | 2020-11-24 | North America/Canada/Ontario/ | Toronto Invasive Bacterial Diseases Network | Ontario Institute for Cancer Research | Allison McGeer et al |
| EPI_ISL_853239 | Canada/ON-PPS-00221/2020 | 2020-11-24 | North America/Canada/Ontario/ | Toronto Invasive Bacterial Diseases Network | Ontario Institute for Cancer Research | Allison McGeer et al |
| EPI_ISL_853240 | Canada/ON-PPS-00256/2020 | 2020-11-24 | North America/Canada/Ontario/ | Toronto Invasive Bacterial Diseases Network | Ontario Institute for Cancer Research | Allison McGeer et al |
| EPI_ISL_853241 | Canada/ON-PPS-00260/2020 | 2020-11-24 | North America/Canada/Ontario/ | Toronto Invasive Bacterial Diseases Network | Ontario Institute for Cancer Research | Allison McGeer et al |
| EPI_ISL_853242 | Canada/ON-PPS-00243/2020 | 2020-11-24 | North America/Canada/Ontario/ | Toronto Invasive Bacterial Diseases Network | Ontario Institute for Cancer Research | Allison McGeer et al |
| EPI_ISL_853243 | Canada/ON-PPS-00217/2020 | 2020-11-24 | North America/Canada/Ontario/ | Toronto Invasive Bacterial Diseases Network | Ontario Institute for Cancer Research | Allison McGeer et al |
| EPI_ISL_853244 | Canada/ON-PPS-00278/2020 | 2020-11-24 | North America/Canada/Ontario/ | Toronto Invasive Bacterial Diseases Network | Ontario Institute for Cancer Research | Allison McGeer et al |
| EPI_ISL_853245 | Canada/ON-PPS-00215/2020 | 2020-11-24 | North America/Canada/Ontario/ | Toronto Invasive Bacterial Diseases Network | Ontario Institute for Cancer Research | Allison McGeer et al |
| EPI_ISL_853246 | Canada/ON-PPS-00224/2020 | 2020-11-24 | North America/Canada/Ontario/ | Toronto Invasive Bacterial Diseases Network | Ontario Institute for Cancer Research | Allison McGeer et al |
| EPI_ISL_853247 | Canada/ON-PPS-00237/2020 | 2020-11-24 | North America/Canada/Ontario/ | Toronto Invasive Bacterial Diseases Network | Ontario Institute for Cancer Research | Allison McGeer et al |

| | | | | | | |
|---|---|---|---|---|---|---|
| EPI_ISL_853248 | Canada/ON-PPS-00209/2020 | 2020-11-24 | North America/Canada/Ontario/ | Toronto Invasive Bacterial Diseases Network | Ontario Institute for Cancer Research | Allison McGeer et al |
| EPI_ISL_853249 | Canada/ON-PPS-00270/2020 | 2020-11-24 | North America/Canada/Ontario/ | Toronto Invasive Bacterial Diseases Network | Ontario Institute for Cancer Research | Allison McGeer et al |
| EPI_ISL_853250 | Canada/ON-PPS-00240/2020 | 2020-11-24 | North America/Canada/Ontario/ | Toronto Invasive Bacterial Diseases Network | Ontario Institute for Cancer Research | Allison McGeer et al |
| EPI_ISL_853251 | Canada/ON-PPS-00228/2020 | 2020-11-24 | North America/Canada/Ontario/ | Toronto Invasive Bacterial Diseases Network | Ontario Institute for Cancer Research | Allison McGeer et al |
| EPI_ISL_853252 | Canada/ON-PPS-00273/2020 | 2020-11-24 | North America/Canada/Ontario/ | Toronto Invasive Bacterial Diseases Network | Ontario Institute for Cancer Research | Allison McGeer et al |
| EPI_ISL_853253 | Canada/ON-PPS-00269/2020 | 2020-11-24 | North America/Canada/Ontario/ | Toronto Invasive Bacterial Diseases Network | Ontario Institute for Cancer Research | Allison McGeer et al |
| EPI_ISL_853254 | Canada/ON-PPS-00272/2020 | 2020-11-24 | North America/Canada/Ontario/ | Toronto Invasive Bacterial Diseases Network | Ontario Institute for Cancer Research | Allison McGeer et al |
| EPI_ISL_853255 | Canada/ON-PPS-00271/2020 | 2020-11-24 | North America/Canada/Ontario/ | Toronto Invasive Bacterial Diseases Network | Ontario Institute for Cancer Research | Allison McGeer et al |
| EPI_ISL_853256 | Canada/ON-PPS-00245/2020 | 2020-11-24 | North America/Canada/Ontario/ | Toronto Invasive Bacterial Diseases Network | Ontario Institute for Cancer Research | Allison McGeer et al |
| EPI_ISL_853257 | Canada/ON-PPS-00275/2020 | 2020-11-24 | North America/Canada/Ontario/ | Toronto Invasive Bacterial Diseases Network | Ontario Institute for Cancer Research | Allison McGeer et al |
| EPI_ISL_853258 | Canada/ON-PPS-00248/2020 | 2020-11-24 | North America/Canada/Ontario/ | Toronto Invasive Bacterial Diseases Network | Ontario Institute for Cancer Research | Allison McGeer et al |
| EPI_ISL_853259 | Canada/ON-PPS-00223/2020 | 2020-11-24 | North America/Canada/Ontario/ | Toronto Invasive Bacterial Diseases Network | Ontario Institute for Cancer Research | Allison McGeer et al |
| EPI_ISL_853260 | Canada/ON-PPS-00266/2020 | 2020-11-24 | North America/Canada/Ontario/ | Toronto Invasive Bacterial Diseases Network | Ontario Institute for Cancer Research | Allison McGeer et al |

| | | | | | | |
|---|---|---|---|---|---|---|
| EPI_ISL_853261 | Canada/ON-PPS-00213/2020 | 2020-11-24 | North America/Canada/Ontario/ | Toronto Invasive Bacterial Diseases Network | Ontario Institute for Cancer Research | Allison McGeer et al |
| EPI_ISL_853262 | Canada/ON-PPS-00246/2020 | 2020-11-24 | North America/Canada/Ontario/ | Toronto Invasive Bacterial Diseases Network | Ontario Institute for Cancer Research | Allison McGeer et al |
| EPI_ISL_853263 | Canada/ON-PPS-00265/2020 | 2020-11-24 | North America/Canada/Ontario/ | Toronto Invasive Bacterial Diseases Network | Ontario Institute for Cancer Research | Allison McGeer et al |
| EPI_ISL_853264 | Canada/ON-PPS-00268/2020 | 2020-11-24 | North America/Canada/Ontario/ | Toronto Invasive Bacterial Diseases Network | Ontario Institute for Cancer Research | Allison McGeer et al |
| EPI_ISL_853265 | Canada/ON-PPS-00281/2020 | 2020-11-24 | North America/Canada/Ontario/ | Toronto Invasive Bacterial Diseases Network | Ontario Institute for Cancer Research | Allison McGeer et al |
| EPI_ISL_853266 | Canada/ON-PPS-00257/2020 | 2020-11-24 | North America/Canada/Ontario/ | Toronto Invasive Bacterial Diseases Network | Ontario Institute for Cancer Research | Allison McGeer et al |
| EPI_ISL_853267 | Canada/ON-PPS-00259/2020 | 2020-11-24 | North America/Canada/Ontario/ | Toronto Invasive Bacterial Diseases Network | Ontario Institute for Cancer Research | Allison McGeer et al |
| EPI_ISL_853268 | Canada/ON-PPS-00203/2020 | 2020-11-24 | North America/Canada/Ontario/ | Toronto Invasive Bacterial Diseases Network | Ontario Institute for Cancer Research | Allison McGeer et al |
| EPI_ISL_853269 | Canada/ON-PPS-00201/2020 | 2020-11-24 | North America/Canada/Ontario/ | Toronto Invasive Bacterial Diseases Network | Ontario Institute for Cancer Research | Allison McGeer et al |
| EPI_ISL_853270 | Canada/ON-PPS-00195/2020 | 2020-11-24 | North America/Canada/Ontario/ | Toronto Invasive Bacterial Diseases Network | Ontario Institute for Cancer Research | Allison McGeer et al |
| EPI_ISL_853271 | Canada/ON-PPS-00261/2020 | 2020-11-24 | North America/Canada/Ontario/ | Toronto Invasive Bacterial Diseases Network | Ontario Institute for Cancer Research | Allison McGeer et al |
| EPI_ISL_853272 | Canada/ON-PPS-00197/2020 | 2020-11-24 | North America/Canada/Ontario/ | Toronto Invasive Bacterial Diseases Network | Ontario Institute for Cancer Research | Allison McGeer et al |
| EPI_ISL_853273 | Canada/ON-PPS-00255/2020 | 2020-11-24 | North America/Canada/Ontario/ | Toronto Invasive Bacterial Diseases Network | Ontario Institute for Cancer Research | Allison McGeer et al |
| EPI_ISL_853274 | Canada/ON-LTRI-1352/2020 | 2020-12-24 | North America/Canada/Ontario/Toronto | Microbiology Lab | NBCC Sequencing Facility | Dr. Jeff Wrana et al |

| | | | | | | |
|---|---|---|---|---|---|---|
| EPI_ISL_853278 | USA/PA-MGEL-00188/2020 | 2020-04-12 | North America/USA/Pennsylvania/Alleg | UPMC Clinical Microbiology Laboratory | Microbial Genome Sequencing Center; Microbial Genomic Epidemiology Laboratory | Mustapha M. Mustapha et al |
| EPI_ISL_853288 | USA/PA-MGEL-00435/2020 | 2020-07-03 | North America/USA/Pennsylvania/Alleg | UPMC Clinical Microbiology Laboratory | Microbial Genome Sequencing Center; Microbial Genomic Epidemiology Laboratory | Mustapha M. Mustapha et al |
| EPI_ISL_853292 | USA/PA-MGEL-00480/2020 | 2020-07-18 | North America/USA/Pennsylvania/Alleg | UPMC Clinical Microbiology Laboratory | Microbial Genome Sequencing Center; Microbial Genomic Epidemiology Laboratory | Mustapha M. Mustapha et al |
| EPI_ISL_853294 | USA/PA-MGEL-00685/2020 | 2020-09-28 | North America/USA/Pennsylvania/Alleg | UPMC Clinical Microbiology Laboratory | Microbial Genome Sequencing Center; Microbial Genomic Epidemiology Laboratory | Mustapha M. Mustapha et al |
| EPI_ISL_853298 | USA/PA-MGEL-00744/2020 | 2020-09-11 | North America/USA/Pennsylvania/Alleg | UPMC Clinical Microbiology Laboratory | Microbial Genome Sequencing Center; Microbial Genomic Epidemiology Laboratory | Mustapha M. Mustapha et al |
| EPI_ISL_853299 | USA/PA-MGEL-00746/2020 | 2020-09-06 | North America/USA/Pennsylvania/Alleg | UPMC Clinical Microbiology Laboratory | Microbial Genome Sequencing Center; Microbial Genomic Epidemiology Laboratory | Mustapha M. Mustapha et al |

| | | | | | | |
|---|---|---|---|---|---|---|
| EPI_ISL_853300 | USA/PA-MGEL-00750/2020 | 2020-09-01 | North America/USA/Pennsylvania/Alleg | UPMC Clinical Microbiology Laboratory | Microbial Genome Sequencing Center; Microbial Genomic Epidemiology Laboratory | Mustapha M. Mustapha et al |
| EPI_ISL_853302 | USA/PA-MGEL-00773/2020 | 2020-10-08 | North America/USA/Pennsylvania/Alleg | UPMC Clinical Microbiology Laboratory | Microbial Genome Sequencing Center; Microbial Genomic Epidemiology Laboratory | Mustapha M. Mustapha et al |
| EPI_ISL_853306 | USA/PA-MGEL-00788/2020 | 2020-09-30 | North America/USA/Pennsylvania/Alleg | UPMC Clinical Microbiology Laboratory | Microbial Genome Sequencing Center; Microbial Genomic Epidemiology Laboratory | Mustapha M. Mustapha et al |
| EPI_ISL_853310 | USA/PA-MGEL-00797/2020 | 2020-09-24 | North America/USA/Pennsylvania/Alleg | UPMC Clinical Microbiology Laboratory | Microbial Genome Sequencing Center; Microbial Genomic Epidemiology Laboratory | Mustapha M. Mustapha et al |
| EPI_ISL_853315 | Canada/ON-LTRI-1372/2020 | 2020-12-24 | North America/Canada/Ontario/Toronto | Microbiology Lab | NBCC Sequencing Facility | Dr. Jeff Wrana et al |
| EPI_ISL_853330 | USA/PA-MGEL-01087/2020 | 2020-11-30 | North America/USA/Pennsylvania/Alleg | UPMC Clinical Microbiology Laboratory | Microbial Genome Sequencing Center; Microbial Genomic Epidemiology Laboratory | Mustapha M. Mustapha et al |
| EPI_ISL_853340 | USA/PA-MGEL-01148/2020 | 2020-12-02 | North America/USA/Pennsylvania/Alleg | UPMC Clinical Microbiology Laboratory | Microbial Genome Sequencing Center; Microbial Genomic Epidemiology Laboratory | Mustapha M. Mustapha et al |

| | | | | | | |
|---|---|---|---|---|---|---|
| EPI_ISL_853343 | USA/PA-MGEL-01153/2020 | 2020-12-10 | North America/USA/Pennsylvania/Alleg | UPMC Clinical Microbiology Laboratory | Microbial Genome Sequencing Center; Microbial Genomic Epidemiology Laboratory | Mustapha M. Mustapha et al |
| EPI_ISL_853349 | USA/PA-MGEL-01165/2020 | 2020-09-29 | North America/USA/Pennsylvania/Alleg | UPMC Clinical Microbiology Laboratory | Microbial Genome Sequencing Center; Microbial Genomic Epidemiology Laboratory | Mustapha M. Mustapha et al |
| EPI_ISL_853368 | USA/PA-MGEL-01284/2020 | 2020-12-26 | North America/USA/Pennsylvania/Alleg | UPMC Clinical Microbiology Laboratory | Microbial Genome Sequencing Center; Microbial Genomic Epidemiology Laboratory | Mustapha M. Mustapha et al |
| EPI_ISL_853372 | USA/PA-MGEL-01288/2020 | 2020-12-18 | North America/USA/Pennsylvania/Alleg | UPMC Clinical Microbiology Laboratory | Microbial Genome Sequencing Center; Microbial Genomic Epidemiology Laboratory | Mustapha M. Mustapha et al |
| EPI_ISL_853378 | USA/PA-MGEL-01294/2020 | 2020-12-10 | North America/USA/Pennsylvania/Alleg | UPMC Clinical Microbiology Laboratory | Microbial Genome Sequencing Center; Microbial Genomic Epidemiology Laboratory | Mustapha M. Mustapha et al |
| EPI_ISL_853379 | USA/PA-MGEL-01295/2020 | 2020-12-10 | North America/USA/Pennsylvania/Alleg | UPMC Clinical Microbiology Laboratory | Microbial Genome Sequencing Center; Microbial Genomic Epidemiology Laboratory | Mustapha M. Mustapha et al |

| | | | | | | |
|---|---|---|---|---|---|---|
| EPI_ISL_853382 | USA/PA-MGEL-01302/2020 | 2020-12-10 | North America/USA/Pennsylvania/Alleg | UPMC Clinical Microbiology Laboratory | Microbial Genome Sequencing Center; Microbial Genomic Epidemiology Laboratory | Mustapha M. Mustapha et al |
| EPI_ISL_853386 | USA/PA-MGEL-01309/2020 | 2020-12-10 | North America/USA/Pennsylvania/Alleg | UPMC Clinical Microbiology Laboratory | Microbial Genome Sequencing Center; Microbial Genomic Epidemiology Laboratory | Mustapha M. Mustapha et al |
| EPI_ISL_853387 | USA/PA-MGEL-01311/2020 | 2020-12-09 | North America/USA/Pennsylvania/Alleg | UPMC Clinical Microbiology Laboratory | Microbial Genome Sequencing Center; Microbial Genomic Epidemiology Laboratory | Mustapha M. Mustapha et al |
| EPI_ISL_853389 | USA/PA-MGEL-01160/2020 | 2020-09-21 | North America/USA/Pennsylvania/Alleg | UPMC Clinical Microbiology Laboratory | Microbial Genome Sequencing Center; Microbial Genomic Epidemiology Laboratory | Mustapha M. Mustapha et al |
| EPI_ISL_853390 | USA/PA-MGEL-01162/2020 | 2020-09-25 | North America/USA/Pennsylvania/Alleg | UPMC Clinical Microbiology Laboratory | Microbial Genome Sequencing Center; Microbial Genomic Epidemiology Laboratory | Mustapha M. Mustapha et al |
| EPI_ISL_853395 | Germany/NI-ChVir22037/2021 | 2021-01-08 | Europe/Germany/Niedersachsen/ | Charit√© Universit√§tsmedizin Berlin, Institut f√ºr Virologie/Labor Berlin | Charit√© Universit√§tsmedizin Berlin, Institut f√ºr Virologie | Victor M Corman et al |
| EPI_ISL_853396 | Germany/NI-ChVir22039/2021 | 2021-01-08 | Europe/Germany/Niedersachsen/ | Charit√© Universit√§tsmedizin Berlin, Institut f√ºr Virologie/Labor Berlin | Charit√© Universit√§tsmedizin Berlin, Institut f√ºr Virologie | Victor M Corman et al |
| EPI_ISL_853405 | Canada/QC-L00267093/2020 | 2020-05-06 | North America/Canada/Quebec/ | Laboratoire de sant√© publique du Qu√©bec | Laboratoire de sant√© publique du Qu√©bec | Sandrine Moreira et al |

| | | | | | | |
|---|---|---|---|---|---|---|
| EPI_ISL_853406 | Canada/QC-L00266940/2020 | 2020-04-17 | North America/Canada/Quebec/ | Laboratoire de sant√© publique du Qu√©bec | Laboratoire de sant√© publique du Qu√©bec | Sandrine Moreira et al |
| EPI_ISL_853407 | Canada/QC-L00266951/2020 | 2020-05-03 | North America/Canada/Quebec/ | Laboratoire de sant√© publique du Qu√©bec | Laboratoire de sant√© publique du Qu√©bec | Sandrine Moreira et al |
| EPI_ISL_853408 | Canada/QC-L00266946/2020 | 2020-05-03 | North America/Canada/Quebec/ | Laboratoire de sant√© publique du Qu√©bec | Laboratoire de sant√© publique du Qu√©bec | Sandrine Moreira et al |
| EPI_ISL_853409 | Canada/QC-L00266933/2020 | 2020-05-03 | North America/Canada/Quebec/ | Laboratoire de sant√© publique du Qu√©bec | Laboratoire de sant√© publique du Qu√©bec | Sandrine Moreira et al |
| EPI_ISL_853410 | Canada/QC-L00267097/2020 | 2020-05-04 | North America/Canada/Quebec/ | Laboratoire de sant√© publique du Qu√©bec | Laboratoire de sant√© publique du Qu√©bec | Sandrine Moreira et al |
| EPI_ISL_853411 | Canada/QC-L00266942/2020 | 2020-05-04 | North America/Canada/Quebec/ | Laboratoire de sant√© publique du Qu√©bec | Laboratoire de sant√© publique du Qu√©bec | Sandrine Moreira et al |
| EPI_ISL_853412 | Canada/QC-L00266938/2020 | 2020-05-06 | North America/Canada/Quebec/ | Laboratoire de sant√© publique du Qu√©bec | Laboratoire de sant√© publique du Qu√©bec | Sandrine Moreira et al |
| EPI_ISL_853413 | Canada/QC-L00266930/2020 | 2020-05-08 | North America/Canada/Quebec/ | Laboratoire de sant√© publique du Qu√©bec | Laboratoire de sant√© publique du Qu√©bec | Sandrine Moreira et al |
| EPI_ISL_853415 | Canada/QC-L00266937/2020 | 2020-05-21 | North America/Canada/Quebec/ | Laboratoire de sant√© publique du Qu√©bec | Laboratoire de sant√© publique du Qu√©bec | Sandrine Moreira et al |
| EPI_ISL_853443 | USA/CA-CZB-15628/2020 | 2020-12-02 | North America/USA/California/Santa Cl | Santa Clara County Public Health Laboratory | Chan-Zuckerberg Biohub | CZB Cliahub Consortium et al |
| EPI_ISL_853460 | USA/CA-CZB-15654/2020 | 2020-12-02 | North America/USA/California/Santa Cl | Santa Clara County Public Health Laboratory | Chan-Zuckerberg Biohub | CZB Cliahub Consortium et al |
| EPI_ISL_853465 | USA/CA-CZB-15667/2020 | 2020-12-02 | North America/USA/California/Santa Cl | Santa Clara County Public Health Laboratory | Chan-Zuckerberg Biohub | CZB Cliahub Consortium et al |
| EPI_ISL_853470 | USA/CA-CZB-15674/2020 | 2020-12-02 | North America/USA/California/Santa Cl | Santa Clara County Public Health Laboratory | Chan-Zuckerberg Biohub | CZB Cliahub Consortium et al |
| EPI_ISL_853489 | USA/CA-CZB-15696/2020 | 2020-12-02 | North America/USA/California/Santa Cl | Santa Clara County Public Health Laboratory | Chan-Zuckerberg Biohub | CZB Cliahub Consortium et al |
| EPI_ISL_853490 | USA/CA-CZB-15699/2020 | 2020-12-02 | North America/USA/California/Santa Cl | Santa Clara County Public Health Laboratory | Chan-Zuckerberg Biohub | CZB Cliahub Consortium et al |
| EPI_ISL_853508 | USA/CA-CZB-15728/2020 | 2020-12-02 | North America/USA/California/Santa Cl | Santa Clara County Public Health Laboratory | Chan-Zuckerberg Biohub | CZB Cliahub Consortium et al |

| | | | | | | |
|---|---|---|---|---|---|---|
| EPI_ISL_853509 | USA/CA-CZB-15730/2020 | 2020-12-02 | North America/USA/California/Santa Cl | Santa Clara County Public Health Laboratory | Chan-Zuckerberg Biohub | CZB Cliahub Consortium et al |
| EPI_ISL_853511 | USA/CA-CZB-15732/2020 | 2020-12-02 | North America/USA/California/Santa Cl | Santa Clara County Public Health Laboratory | Chan-Zuckerberg Biohub | CZB Cliahub Consortium et al |
| EPI_ISL_853518 | USA/CA-CZB-15742/2020 | 2020-12-02 | North America/USA/California/Santa Cl | Santa Clara County Public Health Laboratory | Chan-Zuckerberg Biohub | CZB Cliahub Consortium et al |
| EPI_ISL_853523 | USA/CA-CZB-15749/2020 | 2020-12-02 | North America/USA/California/Santa Cl | Santa Clara County Public Health Laboratory | Chan-Zuckerberg Biohub | CZB Cliahub Consortium et al |
| EPI_ISL_853527 | USA/CA-CZB-15757/2020 | 2020-12-02 | North America/USA/California/Santa Cl | Santa Clara County Public Health Laboratory | Chan-Zuckerberg Biohub | CZB Cliahub Consortium et al |
| EPI_ISL_853531 | USA/CA-CZB-15765/2020 | 2020-12-02 | North America/USA/California/Santa Cl | Santa Clara County Public Health Laboratory | Chan-Zuckerberg Biohub | CZB Cliahub Consortium et al |
| EPI_ISL_853534 | USA/CA-CZB-15769/2020 | 2020-12-02 | North America/USA/California/Santa Cl | Santa Clara County Public Health Laboratory | Chan-Zuckerberg Biohub | CZB Cliahub Consortium et al |
| EPI_ISL_853545 | USA/CA-CZB-15598/2020 | 2020-12-01 | North America/USA/California/Santa Cl | Santa Clara County Public Health Laboratory | Chan-Zuckerberg Biohub | CZB Cliahub Consortium et al |
| EPI_ISL_853556 | USA/CA-CZB-16433/2020 | 2020-04-12 | North America/USA/California/Santa Cl | Santa Clara County Public Health Laboratory | Chan-Zuckerberg Biohub | CZB Cliahub Consortium et al |
| EPI_ISL_853560 | USA/CA-CZB-15713/2020 | 2020-12-02 | North America/USA/California/Santa Cl | Santa Clara County Public Health Laboratory | Chan-Zuckerberg Biohub | CZB Cliahub Consortium et al |
| EPI_ISL_853563 | USA/CA-CZB-15744/2020 | 2020-12-02 | North America/USA/California/Santa Cl | Santa Clara County Public Health Laboratory | Chan-Zuckerberg Biohub | CZB Cliahub Consortium et al |
| EPI_ISL_853566 | Canada/ON-PPS-00396/2020 | 2020-11-24 | North America/Canada/Ontario/ | Toronto Invasive Bacterial Diseases Network | Ontario Institute for Cancer Research | Allison McGeer et al |
| EPI_ISL_853567 | Canada/ON-PPS-00338/2020 | 2020-11-24 | North America/Canada/Ontario/ | Toronto Invasive Bacterial Diseases Network | Ontario Institute for Cancer Research | Allison McGeer et al |
| EPI_ISL_853568 | Canada/ON-PPS-00324/2020 | 2020-11-24 | North America/Canada/Ontario/ | Toronto Invasive Bacterial Diseases Network | Ontario Institute for Cancer Research | Allison McGeer et al |
| EPI_ISL_853569 | Canada/ON-PPS-00344/2020 | 2020-11-24 | North America/Canada/Ontario/ | Toronto Invasive Bacterial Diseases Network | Ontario Institute for Cancer Research | Allison McGeer et al |
| EPI_ISL_853570 | Canada/ON-PPS-00383/2020 | 2020-11-24 | North America/Canada/Ontario/ | Toronto Invasive Bacterial Diseases Network | Ontario Institute for Cancer Research | Allison McGeer et al |
| EPI_ISL_853571 | Canada/ON-PPS-00367/2020 | 2020-11-24 | North America/Canada/Ontario/ | Toronto Invasive Bacterial Diseases Network | Ontario Institute for Cancer Research | Allison McGeer et al |

| | | | | | | |
|---|---|---|---|---|---|---|
| EPI_ISL_853572 | Canada/ON-PPS-00337/2020 | 2020-11-24 | North America/Canada/Ontario/ | Toronto Invasive Bacterial Diseases Network | Ontario Institute for Cancer Research | Allison McGeer et al |
| EPI_ISL_853573 | Canada/ON-PPS-00397/2020 | 2020-11-24 | North America/Canada/Ontario/ | Toronto Invasive Bacterial Diseases Network | Ontario Institute for Cancer Research | Allison McGeer et al |
| EPI_ISL_853574 | Canada/ON-PPS-00398/2020 | 2020-11-24 | North America/Canada/Ontario/ | Toronto Invasive Bacterial Diseases Network | Ontario Institute for Cancer Research | Allison McGeer et al |
| EPI_ISL_853575 | Canada/ON-PPS-00363/2020 | 2020-11-24 | North America/Canada/Ontario/ | Toronto Invasive Bacterial Diseases Network | Ontario Institute for Cancer Research | Allison McGeer et al |
| EPI_ISL_853576 | Canada/ON-PPS-00351/2020 | 2020-11-24 | North America/Canada/Ontario/ | Toronto Invasive Bacterial Diseases Network | Ontario Institute for Cancer Research | Allison McGeer et al |
| EPI_ISL_853577 | Canada/ON-PPS-00317/2020 | 2020-11-24 | North America/Canada/Ontario/ | Toronto Invasive Bacterial Diseases Network | Ontario Institute for Cancer Research | Allison McGeer et al |
| EPI_ISL_853578 | Canada/ON-PPS-00374/2020 | 2020-11-24 | North America/Canada/Ontario/ | Toronto Invasive Bacterial Diseases Network | Ontario Institute for Cancer Research | Allison McGeer et al |
| EPI_ISL_853579 | Canada/ON-PPS-00382/2020 | 2020-11-24 | North America/Canada/Ontario/ | Toronto Invasive Bacterial Diseases Network | Ontario Institute for Cancer Research | Allison McGeer et al |
| EPI_ISL_853580 | Canada/ON-PPS-00327/2020 | 2020-11-24 | North America/Canada/Ontario/ | Toronto Invasive Bacterial Diseases Network | Ontario Institute for Cancer Research | Allison McGeer et al |
| EPI_ISL_853581 | Canada/ON-PPS-00387/2020 | 2020-11-24 | North America/Canada/Ontario/ | Toronto Invasive Bacterial Diseases Network | Ontario Institute for Cancer Research | Allison McGeer et al |
| EPI_ISL_853582 | Canada/ON-PPS-00318/2020 | 2020-11-24 | North America/Canada/Ontario/ | Toronto Invasive Bacterial Diseases Network | Ontario Institute for Cancer Research | Allison McGeer et al |
| EPI_ISL_853583 | Canada/ON-PPS-00313/2020 | 2020-11-24 | North America/Canada/Ontario/ | Toronto Invasive Bacterial Diseases Network | Ontario Institute for Cancer Research | Allison McGeer et al |
| EPI_ISL_853584 | Canada/ON-PPS-00353/2020 | 2020-11-24 | North America/Canada/Ontario/ | Toronto Invasive Bacterial Diseases Network | Ontario Institute for Cancer Research | Allison McGeer et al |

| | | | | | | |
|---|---|---|---|---|---|---|
| EPI_ISL_853585 | Canada/ON-PPS-00335/2020 | 2020-11-24 | North America/Canada/Ontario/ | Toronto Invasive Bacterial Diseases Network | Ontario Institute for Cancer Research | Allison McGeer et al |
| EPI_ISL_853586 | Canada/ON-PPS-00393/2020 | 2020-11-24 | North America/Canada/Ontario/ | Toronto Invasive Bacterial Diseases Network | Ontario Institute for Cancer Research | Allison McGeer et al |
| EPI_ISL_853587 | Canada/ON-PPS-00343/2020 | 2020-11-24 | North America/Canada/Ontario/ | Toronto Invasive Bacterial Diseases Network | Ontario Institute for Cancer Research | Allison McGeer et al |
| EPI_ISL_853588 | Canada/ON-PPS-00354/2020 | 2020-11-24 | North America/Canada/Ontario/ | Toronto Invasive Bacterial Diseases Network | Ontario Institute for Cancer Research | Allison McGeer et al |
| EPI_ISL_853589 | Canada/ON-PPS-00342/2020 | 2020-11-24 | North America/Canada/Ontario/ | Toronto Invasive Bacterial Diseases Network | Ontario Institute for Cancer Research | Allison McGeer et al |
| EPI_ISL_853591 | Canada/ON-PPS-00311/2020 | 2020-11-24 | North America/Canada/Ontario/ | Toronto Invasive Bacterial Diseases Network | Ontario Institute for Cancer Research | Allison McGeer et al |
| EPI_ISL_853592 | Canada/ON-PPS-00395/2020 | 2020-11-24 | North America/Canada/Ontario/ | Toronto Invasive Bacterial Diseases Network | Ontario Institute for Cancer Research | Allison McGeer et al |
| EPI_ISL_853593 | Canada/ON-PPS-00312/2020 | 2020-11-24 | North America/Canada/Ontario/ | Toronto Invasive Bacterial Diseases Network | Ontario Institute for Cancer Research | Allison McGeer et al |
| EPI_ISL_853594 | Canada/ON-PPS-00391/2020 | 2020-11-24 | North America/Canada/Ontario/ | Toronto Invasive Bacterial Diseases Network | Ontario Institute for Cancer Research | Allison McGeer et al |
| EPI_ISL_853595 | Canada/ON-PPS-00319/2020 | 2020-11-24 | North America/Canada/Ontario/ | Toronto Invasive Bacterial Diseases Network | Ontario Institute for Cancer Research | Allison McGeer et al |
| EPI_ISL_853596 | Canada/ON-PPS-00360/2020 | 2020-11-24 | North America/Canada/Ontario/ | Toronto Invasive Bacterial Diseases Network | Ontario Institute for Cancer Research | Allison McGeer et al |
| EPI_ISL_853597 | Canada/ON-PPS-00345/2020 | 2020-11-24 | North America/Canada/Ontario/ | Toronto Invasive Bacterial Diseases Network | Ontario Institute for Cancer Research | Allison McGeer et al |
| EPI_ISL_853598 | Canada/ON-PPS-00305/2020 | 2020-11-24 | North America/Canada/Ontario/ | Toronto Invasive Bacterial Diseases Network | Ontario Institute for Cancer Research | Allison McGeer et al |

| | | | | | | |
|---|---|---|---|---|---|---|
| EPI_ISL_853599 | Canada/ON-PPS-00336/2020 | 2020-11-24 | North America/Canada/Ontario/ | Toronto Invasive Bacterial Diseases Network | Ontario Institute for Cancer Research | Allison McGeer et al |
| EPI_ISL_853600 | Canada/ON-PPS-00326/2020 | 2020-11-24 | North America/Canada/Ontario/ | Toronto Invasive Bacterial Diseases Network | Ontario Institute for Cancer Research | Allison McGeer et al |
| EPI_ISL_853601 | Canada/ON-PPS-00331/2020 | 2020-11-24 | North America/Canada/Ontario/ | Toronto Invasive Bacterial Diseases Network | Ontario Institute for Cancer Research | Allison McGeer et al |
| EPI_ISL_853602 | Canada/ON-PPS-00359/2020 | 2020-11-24 | North America/Canada/Ontario/ | Toronto Invasive Bacterial Diseases Network | Ontario Institute for Cancer Research | Allison McGeer et al |
| EPI_ISL_853603 | Canada/ON-PPS-00375/2020 | 2020-11-24 | North America/Canada/Ontario/ | Toronto Invasive Bacterial Diseases Network | Ontario Institute for Cancer Research | Allison McGeer et al |
| EPI_ISL_853604 | Canada/ON-PPS-00308/2020 | 2020-11-24 | North America/Canada/Ontario/ | Toronto Invasive Bacterial Diseases Network | Ontario Institute for Cancer Research | Allison McGeer et al |
| EPI_ISL_853605 | Canada/ON-PPS-00306/2020 | 2020-11-24 | North America/Canada/Ontario/ | Toronto Invasive Bacterial Diseases Network | Ontario Institute for Cancer Research | Allison McGeer et al |
| EPI_ISL_853607 | Canada/ON-PPS-00315/2020 | 2020-11-24 | North America/Canada/Ontario/ | Toronto Invasive Bacterial Diseases Network | Ontario Institute for Cancer Research | Allison McGeer et al |
| EPI_ISL_853608 | Canada/ON-PPS-00333/2020 | 2020-11-24 | North America/Canada/Ontario/ | Toronto Invasive Bacterial Diseases Network | Ontario Institute for Cancer Research | Allison McGeer et al |
| EPI_ISL_853609 | Canada/ON-PPS-00310/2020 | 2020-11-24 | North America/Canada/Ontario/ | Toronto Invasive Bacterial Diseases Network | Ontario Institute for Cancer Research | Allison McGeer et al |
| EPI_ISL_853610 | Canada/ON-PPS-00373/2020 | 2020-11-24 | North America/Canada/Ontario/ | Toronto Invasive Bacterial Diseases Network | Ontario Institute for Cancer Research | Allison McGeer et al |
| EPI_ISL_853611 | Canada/ON-PPS-00390/2020 | 2020-11-24 | North America/Canada/Ontario/ | Toronto Invasive Bacterial Diseases Network | Ontario Institute for Cancer Research | Allison McGeer et al |
| EPI_ISL_853612 | Canada/ON-PPS-00365/2020 | 2020-11-24 | North America/Canada/Ontario/ | Toronto Invasive Bacterial Diseases Network | Ontario Institute for Cancer Research | Allison McGeer et al |

| | | | | | | |
|---|---|---|---|---|---|---|
| EPI_ISL_853613 | Canada/ON-PPS-00325/2020 | 2020-11-24 | North America/Canada/Ontario/ | Toronto Invasive Bacterial Diseases Network | Ontario Institute for Cancer Research | Allison McGeer et al |
| EPI_ISL_853614 | Canada/ON-PPS-00339/2020 | 2020-11-24 | North America/Canada/Ontario/ | Toronto Invasive Bacterial Diseases Network | Ontario Institute for Cancer Research | Allison McGeer et al |
| EPI_ISL_853615 | Canada/ON-PPS-00322/2020 | 2020-11-24 | North America/Canada/Ontario/ | Toronto Invasive Bacterial Diseases Network | Ontario Institute for Cancer Research | Allison McGeer et al |
| EPI_ISL_853616 | Canada/ON-PPS-00377/2020 | 2020-11-24 | North America/Canada/Ontario/ | Toronto Invasive Bacterial Diseases Network | Ontario Institute for Cancer Research | Allison McGeer et al |
| EPI_ISL_853617 | Canada/ON-PPS-00385/2020 | 2020-11-24 | North America/Canada/Ontario/ | Toronto Invasive Bacterial Diseases Network | Ontario Institute for Cancer Research | Allison McGeer et al |
| EPI_ISL_853618 | Canada/ON-PPS-00348/2020 | 2020-11-24 | North America/Canada/Ontario/ | Toronto Invasive Bacterial Diseases Network | Ontario Institute for Cancer Research | Allison McGeer et al |
| EPI_ISL_853619 | Canada/ON-PPS-00350/2020 | 2020-11-24 | North America/Canada/Ontario/ | Toronto Invasive Bacterial Diseases Network | Ontario Institute for Cancer Research | Allison McGeer et al |
| EPI_ISL_853620 | Canada/ON-PPS-00330/2020 | 2020-11-24 | North America/Canada/Ontario/ | Toronto Invasive Bacterial Diseases Network | Ontario Institute for Cancer Research | Allison McGeer et al |
| EPI_ISL_853621 | Canada/ON-PPS-00392/2020 | 2020-11-24 | North America/Canada/Ontario/ | Toronto Invasive Bacterial Diseases Network | Ontario Institute for Cancer Research | Allison McGeer et al |
| EPI_ISL_853622 | Canada/ON-PPS-00307/2020 | 2020-11-24 | North America/Canada/Ontario/ | Toronto Invasive Bacterial Diseases Network | Ontario Institute for Cancer Research | Allison McGeer et al |
| EPI_ISL_853623 | Canada/ON-PPS-00357/2020 | 2020-11-24 | North America/Canada/Ontario/ | Toronto Invasive Bacterial Diseases Network | Ontario Institute for Cancer Research | Allison McGeer et al |
| EPI_ISL_853624 | Canada/ON-PPS-00384/2020 | 2020-11-24 | North America/Canada/Ontario/ | Toronto Invasive Bacterial Diseases Network | Ontario Institute for Cancer Research | Allison McGeer et al |
| EPI_ISL_853625 | Canada/ON-PPS-00309/2020 | 2020-11-24 | North America/Canada/Ontario/ | Toronto Invasive Bacterial Diseases Network | Ontario Institute for Cancer Research | Allison McGeer et al |

| | | | | | | |
|---|---|---|---|---|---|---|
| EPI_ISL_853626 | Canada/ON-PPS-00341/2020 | 2020-11-24 | North America/Canada/Ontario/ | Toronto Invasive Bacterial Diseases Network | Ontario Institute for Cancer Research | Allison McGeer et al |
| EPI_ISL_853627 | Canada/ON-PPS-00380/2020 | 2020-11-24 | North America/Canada/Ontario/ | Toronto Invasive Bacterial Diseases Network | Ontario Institute for Cancer Research | Allison McGeer et al |
| EPI_ISL_853628 | Canada/ON-PPS-00320/2020 | 2020-11-24 | North America/Canada/Ontario/ | Toronto Invasive Bacterial Diseases Network | Ontario Institute for Cancer Research | Allison McGeer et al |
| EPI_ISL_853629 | Canada/ON-PPS-00362/2020 | 2020-11-24 | North America/Canada/Ontario/ | Toronto Invasive Bacterial Diseases Network | Ontario Institute for Cancer Research | Allison McGeer et al |
| EPI_ISL_853630 | Canada/ON-PPS-00321/2020 | 2020-11-24 | North America/Canada/Ontario/ | Toronto Invasive Bacterial Diseases Network | Ontario Institute for Cancer Research | Allison McGeer et al |
| EPI_ISL_853631 | Canada/ON-PPS-00379/2020 | 2020-11-24 | North America/Canada/Ontario/ | Toronto Invasive Bacterial Diseases Network | Ontario Institute for Cancer Research | Allison McGeer et al |
| EPI_ISL_853632 | Canada/ON-PPS-00372/2020 | 2020-11-24 | North America/Canada/Ontario/ | Toronto Invasive Bacterial Diseases Network | Ontario Institute for Cancer Research | Allison McGeer et al |
| EPI_ISL_853633 | Canada/ON-PPS-00347/2020 | 2020-11-24 | North America/Canada/Ontario/ | Toronto Invasive Bacterial Diseases Network | Ontario Institute for Cancer Research | Allison McGeer et al |
| EPI_ISL_853634 | Canada/ON-PPS-00370/2020 | 2020-11-24 | North America/Canada/Ontario/ | Toronto Invasive Bacterial Diseases Network | Ontario Institute for Cancer Research | Allison McGeer et al |
| EPI_ISL_853635 | Canada/ON-PPS-00323/2020 | 2020-11-24 | North America/Canada/Ontario/ | Toronto Invasive Bacterial Diseases Network | Ontario Institute for Cancer Research | Allison McGeer et al |
| EPI_ISL_853636 | Canada/ON-PPS-00358/2020 | 2020-11-24 | North America/Canada/Ontario/ | Toronto Invasive Bacterial Diseases Network | Ontario Institute for Cancer Research | Allison McGeer et al |
| EPI_ISL_853637 | Canada/ON-PPS-00329/2020 | 2020-11-24 | North America/Canada/Ontario/ | Toronto Invasive Bacterial Diseases Network | Ontario Institute for Cancer Research | Allison McGeer et al |
| EPI_ISL_853638 | Canada/ON-PPS-00389/2020 | 2020-11-24 | North America/Canada/Ontario/ | Toronto Invasive Bacterial Diseases Network | Ontario Institute for Cancer Research | Allison McGeer et al |

| | | | | | | |
|---|---|---|---|---|---|---|
| EPI_ISL_853639 | Canada/ON-PPS-00381/2020 | 2020-11-24 | North America/Canada/Ontario/ | Toronto Invasive Bacterial Diseases Network | Ontario Institute for Cancer Research | Allison McGeer et al |
| EPI_ISL_853640 | Canada/ON-PPS-00394/2020 | 2020-11-24 | North America/Canada/Ontario/ | Toronto Invasive Bacterial Diseases Network | Ontario Institute for Cancer Research | Allison McGeer et al |
| EPI_ISL_853641 | Canada/ON-PPS-00346/2020 | 2020-11-24 | North America/Canada/Ontario/ | Toronto Invasive Bacterial Diseases Network | Ontario Institute for Cancer Research | Allison McGeer et al |
| EPI_ISL_853642 | Canada/ON-PPS-00314/2020 | 2020-11-24 | North America/Canada/Ontario/ | Toronto Invasive Bacterial Diseases Network | Ontario Institute for Cancer Research | Allison McGeer et al |
| EPI_ISL_853643 | Canada/ON-PPS-00399/2020 | 2020-11-24 | North America/Canada/Ontario/ | Toronto Invasive Bacterial Diseases Network | Ontario Institute for Cancer Research | Allison McGeer et al |
| EPI_ISL_853644 | Canada/ON-PPS-00386/2020 | 2020-11-24 | North America/Canada/Ontario/ | Toronto Invasive Bacterial Diseases Network | Ontario Institute for Cancer Research | Allison McGeer et al |
| EPI_ISL_853645 | Canada/ON-PPS-00356/2020 | 2020-11-24 | North America/Canada/Ontario/ | Toronto Invasive Bacterial Diseases Network | Ontario Institute for Cancer Research | Allison McGeer et al |
| EPI_ISL_853646 | Canada/ON-PPS-00334/2020 | 2020-11-24 | North America/Canada/Ontario/ | Toronto Invasive Bacterial Diseases Network | Ontario Institute for Cancer Research | Allison McGeer et al |
| EPI_ISL_853647 | Canada/ON-PPS-00332/2020 | 2020-11-24 | North America/Canada/Ontario/ | Toronto Invasive Bacterial Diseases Network | Ontario Institute for Cancer Research | Allison McGeer et al |
| EPI_ISL_853651 | USA/NY-Wadsworth-21004172-01/20 | 2021-01-05 | North America/USA/New York/Westche | NORTHWELL HEALTH LABORATORIES | Wadsworth Center, New York State Department of Health | Kirsten St. George et al |
| EPI_ISL_853666 | USA/NY-Wadsworth-21004203-01/20 | 2021-01-05 | North America/USA/New York/Suffolk C | NORTHWELL HEALTH LABORATORIES | Wadsworth Center, New York State Department of Health | Kirsten St. George et al |
| EPI_ISL_853667 | USA/NY-Wadsworth-21004205-01/20 | 2021-01-05 | North America/USA/New York/Herkime | THE MARY IMOGENE BASSETT HOSPITAL | Wadsworth Center, New York State Department of Health | Kirsten St. George et al |

| | | | | | | |
|---|---|---|---|---|---|---|
| EPI_ISL_853674 | USA/NY-Wadsworth-21004202-01/20 | 2021-01-04 | North America/USA/New York/Suffolk C | NORTHWELL HEALTH LABORATORIES | Wadsworth Center, New York State Department of Health | Kirsten St. George et al |
| EPI_ISL_853679 | USA/NY-Wadsworth-21004905-01/20 | 2021-01-04 | North America/USA/New York/Saratoga | ACUTIS DIAGNOSTICS | Wadsworth Center, New York State Department of Health | Kirsten St. George et al |
| EPI_ISL_853680 | USA/NY-Wadsworth-21004921-01/20 | 2021-01-04 | North America/USA/New York/Saratoga | ACUTIS DIAGNOSTICS | Wadsworth Center, New York State Department of Health | Kirsten St. George et al |
| EPI_ISL_853682 | USA/NY-Wadsworth-21004927-01/20 | 2021-01-04 | North America/USA/New York/Saratoga | ACUTIS DIAGNOSTICS | Wadsworth Center, New York State Department of Health | Kirsten St. George et al |
| EPI_ISL_853684 | USA/NY-Wadsworth-21004277-01/20 | 2021-01-01 | North America/USA/New York/Albany C | ALBANY MEDICAL CENTER HOSPITAL CLINICAL LABORATORIES | Wadsworth Center, New York State Department of Health | Kirsten St. George et al |
| EPI_ISL_853693 | USA/NY-Wadsworth-21004269-01/20 | 2020-12-31 | North America/USA/New York/Saratoga | ALBANY MEDICAL CENTER HOSPITAL CLINICAL LABORATORIES | Wadsworth Center, New York State Department of Health | Kirsten St. George et al |
| EPI_ISL_853696 | USA/NY-Wadsworth-21004272-01/20 | 2020-12-31 | North America/USA/New York/Herkime | ALBANY MEDICAL CENTER HOSPITAL CLINICAL LABORATORIES | Wadsworth Center, New York State Department of Health | Kirsten St. George et al |
| EPI_ISL_853698 | USA/NY-Wadsworth-21004275-01/20 | 2020-12-31 | North America/USA/New York/Columbia | ALBANY MEDICAL CENTER HOSPITAL CLINICAL LABORATORIES | Wadsworth Center, New York State Department of Health | Kirsten St. George et al |
| EPI_ISL_853746 | Austria/CeMM2418/2021 | 2021-01-11 | Europe/Austria/Tyrol/ | Austrian Agency for Health and Food Safety (AGES) | Bergthaler laboratory, CeMM Research Center for Molecular Medicine of the Austrian Academy of Sciences | Lukas Endler et al |

| | | | | | | |
|---|---|---|---|---|---|---|
| EPI_ISL_853770 | Austria/CeMM1574/2020 | 2020-11-10 | Europe/Austria/Salzburg/Mondsee | Pharmgenetix GmbH | Bergthaler laboratory, CeMM Research Center for Molecular Medicine of the Austrian Academy of Sciences | Lukas Endler et al |
| EPI_ISL_853771 | Austria/CeMM1581/2020 | 2020-11-10 | Europe/Austria/Carinthia/Glvðdnitz | Pharmgenetix GmbH | Bergthaler laboratory, CeMM Research Center for Molecular Medicine of the Austrian Academy of Sciences | Lukas Endler et al |
| EPI_ISL_853772 | Austria/CeMM1586/2020 | 2020-11-10 | Europe/Austria/Carinthia/Wernberg | Pharmgenetix GmbH | Bergthaler laboratory, CeMM Research Center for Molecular Medicine of the Austrian Academy of Sciences | Lukas Endler et al |
| EPI_ISL_853773 | Austria/CeMM1587/2020 | 2020-11-10 | Europe/Austria/Carinthia/Villach | Pharmgenetix GmbH | Bergthaler laboratory, CeMM Research Center for Molecular Medicine of the Austrian Academy of Sciences | Lukas Endler et al |
| EPI_ISL_853779 | Austria/CeMM1700/2020 | 2020-11-21 | Europe/Austria/Austria/ | Austrian Agency for Health and Food Safety (AGES) | Bergthaler laboratory, CeMM Research Center for Molecular Medicine of the Austrian Academy of Sciences | Lukas Endler et al |

| | | | | | | |
|---|---|---|---|---|---|---|
| EPI_ISL_853790 | Austria/CeMM1806/2020 | 2020-11-12 | Europe/Austria/Austria/ | Austrian Agency for Health and Food Safety (AGES) | Bergthaler laboratory, CeMM Research Center for Molecular Medicine of the Austrian Academy of Sciences | Lukas Endler et al |
| EPI_ISL_853791 | Austria/CeMM1764/2020 | 2020-11-28 | Europe/Austria/Austria/ | Austrian Agency for Health and Food Safety (AGES) | Bergthaler laboratory, CeMM Research Center for Molecular Medicine of the Austrian Academy of Sciences | Lukas Endler et al |
| EPI_ISL_853800 | Austria/CeMM1804/2020 | 2020-11-12 | Europe/Austria/Austria/ | Austrian Agency for Health and Food Safety (AGES) | Bergthaler laboratory, CeMM Research Center for Molecular Medicine of the Austrian Academy of Sciences | Lukas Endler et al |
| EPI_ISL_853803 | Austria/CeMM1485/2020 | 2020-11-04 | Europe/Austria/Austria/ | Austrian Agency for Health and Food Safety (AGES) | Bergthaler laboratory, CeMM Research Center for Molecular Medicine of the Austrian Academy of Sciences | Lukas Endler et al |
| EPI_ISL_853815 | Austria/CeMM1728/2020 | 2020-11-24 | Europe/Austria/Austria/ | Austrian Agency for Health and Food Safety (AGES) | Bergthaler laboratory, CeMM Research Center for Molecular Medicine of the Austrian Academy of Sciences | Lukas Endler et al |

| | | | | | | |
|---|---|---|---|---|---|---|
| EPI_ISL_853819 | Austria/CeMM1228/2020 | 2020-06-05 | Europe/Austria/Styria/Weiz | Department of Pathology, Landeskrankenhaus Graz II, Medical University Graz | Bergthaler laboratory, CeMM Research Center for Molecular Medicine of the Austrian Academy of Sciences | Lukas Endler et al |
| EPI_ISL_853822 | Austria/CeMM1293/2020 | 2020-03-04 | Europe/Austria/Lower Austria/ | Center for Virology, Medical University of Vienna | Bergthaler laboratory, CeMM Research Center for Molecular Medicine of the Austrian Academy of Sciences | Lukas Endler et al |
| EPI_ISL_853831 | Austria/CeMM1303/2020 | 2020-03-07 | Europe/Austria/Lower Austria/ | Center for Virology, Medical University of Vienna | Bergthaler laboratory, CeMM Research Center for Molecular Medicine of the Austrian Academy of Sciences | Lukas Endler et al |
| EPI_ISL_853839 | Austria/CeMM1333/2020 | 2020-07-15 | Europe/Austria/Vienna/ | Center for Virology, Medical University of Vienna | Bergthaler laboratory, CeMM Research Center for Molecular Medicine of the Austrian Academy of Sciences | Lukas Endler et al |
| EPI_ISL_853843 | Austria/CeMM1345/2020 | 2020-08-03 | Europe/Austria/Vienna/ | Center for Virology, Medical University of Vienna | Bergthaler laboratory, CeMM Research Center for Molecular Medicine of the Austrian Academy of Sciences | Lukas Endler et al |

| | | | | | | |
|---|---|---|---|---|---|---|
| EPI_ISL_853844 | Austria/CeMM1348/2020 | 2020-08-06 | Europe/Austria/Vienna/ | Center for Virology, Medical University of Vienna | Bergthaler laboratory, CeMM Research Center for Molecular Medicine of the Austrian Academy of Sciences | Lukas Endler et al |
| EPI_ISL_853857 | Austria/CeMM1409/2020 | 2020-10-11 | Europe/Austria/Carinthia/Klagenfurt | Institute for Laboratory Diagnostics and Microbiology, Klinikum Klagenfurt am Worthersee | Bergthaler laboratory, CeMM Research Center for Molecular Medicine of the Austrian Academy of Sciences | Lukas Endler et al |
| EPI_ISL_853858 | Austria/CeMM1412/2020 | 2020-10-11 | Europe/Austria/Carinthia/Klagenfurt | Institute for Laboratory Diagnostics and Microbiology, Klinikum Klagenfurt am Worthersee | Bergthaler laboratory, CeMM Research Center for Molecular Medicine of the Austrian Academy of Sciences | Lukas Endler et al |
| EPI_ISL_853859 | Austria/CeMM1414/2020 | 2020-10-11 | Europe/Austria/Carinthia/Klagenfurt | Institute for Laboratory Diagnostics and Microbiology, Klinikum Klagenfurt am Worthersee | Bergthaler laboratory, CeMM Research Center for Molecular Medicine of the Austrian Academy of Sciences | Lukas Endler et al |
| EPI_ISL_853861 | Austria/CeMM1418/2020 | 2020-10-12 | Europe/Austria/Carinthia/Klagenfurt | Institute for Laboratory Diagnostics and Microbiology, Klinikum Klagenfurt am Worthersee | Bergthaler laboratory, CeMM Research Center for Molecular Medicine of the Austrian Academy of Sciences | Lukas Endler et al |

| | | | | | | |
|---|---|---|---|---|---|---|
| EPI_ISL_853863 | Austria/CeMM1421/2020 | 2020-10-12 | Europe/Austria/Carinthia/Klagenfurt | Institute for Laboratory Diagnostics and Microbiology, Klinikum Klagenfurt am Worthersee | Bergthaler laboratory, CeMM Research Center for Molecular Medicine of the Austrian Academy of Sciences | Lukas Endler et al |
| EPI_ISL_853864 | Austria/CeMM1422/2020 | 2020-10-12 | Europe/Austria/Carinthia/Klagenfurt | Institute for Laboratory Diagnostics and Microbiology, Klinikum Klagenfurt am Worthersee | Bergthaler laboratory, CeMM Research Center for Molecular Medicine of the Austrian Academy of Sciences | Lukas Endler et al |
| EPI_ISL_853865 | Austria/CeMM1423/2020 | 2020-10-12 | Europe/Austria/Carinthia/Klagenfurt | Institute for Laboratory Diagnostics and Microbiology, Klinikum Klagenfurt am Worthersee | Bergthaler laboratory, CeMM Research Center for Molecular Medicine of the Austrian Academy of Sciences | Lukas Endler et al |
| EPI_ISL_853866 | Austria/CeMM1424/2020 | 2020-10-16 | Europe/Austria/Carinthia/Klagenfurt | Institute for Laboratory Diagnostics and Microbiology, Klinikum Klagenfurt am Worthersee | Bergthaler laboratory, CeMM Research Center for Molecular Medicine of the Austrian Academy of Sciences | Lukas Endler et al |
| EPI_ISL_853869 | Austria/CeMM1428/2020 | 2020-10-16 | Europe/Austria/Carinthia/Klagenfurt | Institute for Laboratory Diagnostics and Microbiology, Klinikum Klagenfurt am Worthersee | Bergthaler laboratory, CeMM Research Center for Molecular Medicine of the Austrian Academy of Sciences | Lukas Endler et al |

| | | | | | | |
|---|---|---|---|---|---|---|
| EPI_ISL_853871 | Austria/CeMM1430/2020 | 2020-10-19 | Europe/Austria/Carinthia/Klagenfurt | Institute for Laboratory Diagnostics and Microbiology, Klinikum Klagenfurt am Worthersee | Bergthaler laboratory, CeMM Research Center for Molecular Medicine of the Austrian Academy of Sciences | Lukas Endler et al |
| EPI_ISL_853872 | Austria/CeMM1431/2020 | 2020-10-19 | Europe/Austria/Carinthia/Klagenfurt | Institute for Laboratory Diagnostics and Microbiology, Klinikum Klagenfurt am Worthersee | Bergthaler laboratory, CeMM Research Center for Molecular Medicine of the Austrian Academy of Sciences | Lukas Endler et al |
| EPI_ISL_853873 | Austria/CeMM1432/2020 | 2020-10-19 | Europe/Austria/Carinthia/Klagenfurt | Institute for Laboratory Diagnostics and Microbiology, Klinikum Klagenfurt am Worthersee | Bergthaler laboratory, CeMM Research Center for Molecular Medicine of the Austrian Academy of Sciences | Lukas Endler et al |
| EPI_ISL_853877 | Austria/CeMM1299/2020 | 2020-03-06 | Europe/Austria/Lower Austria/ | Center for Virology, Medical University of Vienna | Bergthaler laboratory, CeMM Research Center for Molecular Medicine of the Austrian Academy of Sciences | Lukas Endler et al |
| EPI_ISL_853886 | Austria/CeMM1325/2020 | 2020-06-18 | Europe/Austria/Vienna/ | Center for Virology, Medical University of Vienna | Bergthaler laboratory, CeMM Research Center for Molecular Medicine of the Austrian Academy of Sciences | Lukas Endler et al |

| | | | | | | |
|---|---|---|---|---|---|---|
| EPI_ISL_853889 | Austria/CeMM1335/2020 | 2020-07-17 | Europe/Austria/Vienna/ | Center for Virology, Medical University of Vienna | Bergthaler laboratory, CeMM Research Center for Molecular Medicine of the Austrian Academy of Sciences | Lukas Endler et al |
| EPI_ISL_853891 | Austria/CeMM1339/2020 | 2020-07-22 | Europe/Austria/Lower Austria/ | Center for Virology, Medical University of Vienna | Bergthaler laboratory, CeMM Research Center for Molecular Medicine of the Austrian Academy of Sciences | Lukas Endler et al |
| EPI_ISL_853892 | Austria/CeMM1341/2020 | 2020-07-23 | Europe/Austria/Vienna/ | Center for Virology, Medical University of Vienna | Bergthaler laboratory, CeMM Research Center for Molecular Medicine of the Austrian Academy of Sciences | Lukas Endler et al |
| EPI_ISL_853893 | Austria/CeMM1342/2020 | 2020-07-25 | Europe/Austria/Vienna/ | Center for Virology, Medical University of Vienna | Bergthaler laboratory, CeMM Research Center for Molecular Medicine of the Austrian Academy of Sciences | Lukas Endler et al |
| EPI_ISL_853895 | Austria/CeMM1346/2020 | 2020-08-05 | Europe/Austria/Vienna/ | Center for Virology, Medical University of Vienna | Bergthaler laboratory, CeMM Research Center for Molecular Medicine of the Austrian Academy of Sciences | Lukas Endler et al |

| | | | | | | |
|---|---|---|---|---|---|---|
| EPI_ISL_853897 | Austria/CeMM1349/2020 | 2020-08-10 | Europe/Austria/Vienna/ | Center for Virology, Medical University of Vienna | Bergthaler laboratory, CeMM Research Center for Molecular Medicine of the Austrian Academy of Sciences | Lukas Endler et al |
| EPI_ISL_853916 | Austria/CeMM1413/2020 | 2020-10-11 | Europe/Austria/Carinthia/Klagenfurt | Institute for Laboratory Diagnostics and Microbiology, Klinikum Klagenfurt am Worthersee | Bergthaler laboratory, CeMM Research Center for Molecular Medicine of the Austrian Academy of Sciences | Lukas Endler et al |
| EPI_ISL_853917 | Austria/CeMM1575/2020 | 2020-11-10 | Europe/Austria/Tyrol/Kramsach | Pharmgenetix GmbH | Bergthaler laboratory, CeMM Research Center for Molecular Medicine of the Austrian Academy of Sciences | Lukas Endler et al |
| EPI_ISL_853921 | Austria/CeMM1601/2020 | 2020-11-10 | Europe/Austria/Salzburg/Abtenau | Pharmgenetix GmbH | Bergthaler laboratory, CeMM Research Center for Molecular Medicine of the Austrian Academy of Sciences | Lukas Endler et al |
| EPI_ISL_853924 | Austria/CeMM2031/2020 | 2020-12-21 | Europe/Austria/Carinthia/Hermagor | ILV K√§rnten | Bergthaler laboratory, CeMM Research Center for Molecular Medicine of the Austrian Academy of Sciences | Lukas Endler et al |

| | | | | | | |
|---|---|---|---|---|---|---|
| EPI_ISL_853927 | Austria/CeMM2039/2020 | 2020-12-22 | Europe/Austria/Carinthia/Hermagor | ILV K√§rnten | Bergthaler laboratory, CeMM Research Center for Molecular Medicine of the Austrian Academy of Sciences | Lukas Endler et al |
| EPI_ISL_853928 | Austria/CeMM2040/2020 | 2020-12-22 | Europe/Austria/Carinthia/Hermagor | ILV K√§rnten | Bergthaler laboratory, CeMM Research Center for Molecular Medicine of the Austrian Academy of Sciences | Lukas Endler et al |
| EPI_ISL_853957 | Austria/CeMM1354/2020 | 2020-08-10 | Europe/Austria/Vienna/ | Center for Virology, Medical University of Vienna | Bergthaler laboratory, CeMM Research Center for Molecular Medicine of the Austrian Academy of Sciences | Lukas Endler et al |
| EPI_ISL_853959 | Austria/CeMM1330/2020 | 2020-07-13 | Europe/Austria/Vienna/ | Center for Virology, Medical University of Vienna | Bergthaler laboratory, CeMM Research Center for Molecular Medicine of the Austrian Academy of Sciences | Lukas Endler et al |
| EPI_ISL_853970 | Austria/CeMM1794/2020 | 2020-12-03 | Europe/Austria/Austria/ | Austrian Agency for Health and Food Safety (AGES) | Bergthaler laboratory, CeMM Research Center for Molecular Medicine of the Austrian Academy of Sciences | Lukas Endler et al |

| | | | | | | |
|---|---|---|---|---|---|---|
| EPI_ISL_853977 | Austria/CeMM1850/2020 | 2020-09-18 | Europe/Austria/Austria/ | Austrian Agency for Health and Food Safety (AGES) | Bergthaler laboratory, CeMM Research Center for Molecular Medicine of the Austrian Academy of Sciences | Lukas Endler et al |
| EPI_ISL_853980 | Austria/CeMM1481/2020 | 2020-11-04 | Europe/Austria/Austria/ | Austrian Agency for Health and Food Safety (AGES) | Bergthaler laboratory, CeMM Research Center for Molecular Medicine of the Austrian Academy of Sciences | Lukas Endler et al |
| EPI_ISL_853981 | Austria/CeMM1482/2020 | 2020-11-04 | Europe/Austria/Austria/ | Austrian Agency for Health and Food Safety (AGES) | Bergthaler laboratory, CeMM Research Center for Molecular Medicine of the Austrian Academy of Sciences | Lukas Endler et al |
| EPI_ISL_853986 | Austria/CeMM1491/2020 | 2020-10-28 | Europe/Austria/Austria/ | Austrian Agency for Health and Food Safety (AGES) | Bergthaler laboratory, CeMM Research Center for Molecular Medicine of the Austrian Academy of Sciences | Lukas Endler et al |
| EPI_ISL_853987 | Austria/CeMM1492/2020 | 2020-10-28 | Europe/Austria/Austria/ | Austrian Agency for Health and Food Safety (AGES) | Bergthaler laboratory, CeMM Research Center for Molecular Medicine of the Austrian Academy of Sciences | Lukas Endler et al |

| | | | | | | |
|---|---|---|---|---|---|---|
| EPI_ISL_853993 | Austria/CeMM1499/2020 | 2020-10-29 | Europe/Austria/Austria/ | Austrian Agency for Health and Food Safety (AGES) | Bergthaler laboratory, CeMM Research Center for Molecular Medicine of the Austrian Academy of Sciences | Lukas Endler et al |
| EPI_ISL_853994 | Austria/CeMM1500/2020 | 2020-11-02 | Europe/Austria/Austria/ | Austrian Agency for Health and Food Safety (AGES) | Bergthaler laboratory, CeMM Research Center for Molecular Medicine of the Austrian Academy of Sciences | Lukas Endler et al |
| EPI_ISL_853997 | Austria/CeMM1504/2020 | 2020-10-28 | Europe/Austria/Austria/ | Austrian Agency for Health and Food Safety (AGES) | Bergthaler laboratory, CeMM Research Center for Molecular Medicine of the Austrian Academy of Sciences | Lukas Endler et al |
| EPI_ISL_853999 | Austria/CeMM1506/2020 | 2020-11-04 | Europe/Austria/Austria/ | Austrian Agency for Health and Food Safety (AGES) | Bergthaler laboratory, CeMM Research Center for Molecular Medicine of the Austrian Academy of Sciences | Lukas Endler et al |
| EPI_ISL_854008 | Austria/CeMM1522/2020 | 2020-10-29 | Europe/Austria/Austria/ | Austrian Agency for Health and Food Safety (AGES) | Bergthaler laboratory, CeMM Research Center for Molecular Medicine of the Austrian Academy of Sciences | Lukas Endler et al |

| | | | | | | |
|---|---|---|---|---|---|---|
| EPI_ISL_854011 | Austria/CeMM1525/2020 | 2020-11-04 | Europe/Austria/Austria/ | Austrian Agency for Health and Food Safety (AGES) | Bergthaler laboratory, CeMM Research Center for Molecular Medicine of the Austrian Academy of Sciences | Lukas Endler et al |
| EPI_ISL_854014 | Austria/CeMM1529/2020 | 2020-11-04 | Europe/Austria/Austria/ | Austrian Agency for Health and Food Safety (AGES) | Bergthaler laboratory, CeMM Research Center for Molecular Medicine of the Austrian Academy of Sciences | Lukas Endler et al |
| EPI_ISL_854016 | Austria/CeMM1534/2020 | 2020-10-29 | Europe/Austria/Austria/ | Austrian Agency for Health and Food Safety (AGES) | Bergthaler laboratory, CeMM Research Center for Molecular Medicine of the Austrian Academy of Sciences | Lukas Endler et al |
| EPI_ISL_854020 | Austria/CeMM1539/2020 | 2020-10-28 | Europe/Austria/Austria/ | Austrian Agency for Health and Food Safety (AGES) | Bergthaler laboratory, CeMM Research Center for Molecular Medicine of the Austrian Academy of Sciences | Lukas Endler et al |
| EPI_ISL_854025 | Austria/CeMM1551/2020 | 2020-10-28 | Europe/Austria/Austria/ | Austrian Agency for Health and Food Safety (AGES) | Bergthaler laboratory, CeMM Research Center for Molecular Medicine of the Austrian Academy of Sciences | Lukas Endler et al |

| | | | | | | |
|---|---|---|---|---|---|---|
| EPI_ISL_854039 | Austria/CeMM1570/2020 | 2020-10-29 | Europe/Austria/Austria/ | Austrian Agency for Health and Food Safety (AGES) | Bergthaler laboratory, CeMM Research Center for Molecular Medicine of the Austrian Academy of Sciences | Lukas Endler et al |
| EPI_ISL_854117 | Austria/CeMM1707/2020 | 2020-11-23 | Europe/Austria/Austria/ | Austrian Agency for Health and Food Safety (AGES) | Bergthaler laboratory, CeMM Research Center for Molecular Medicine of the Austrian Academy of Sciences | Lukas Endler et al |
| EPI_ISL_854127 | Austria/CeMM1733/2020 | 2020-11-24 | Europe/Austria/Austria/ | Austrian Agency for Health and Food Safety (AGES) | Bergthaler laboratory, CeMM Research Center for Molecular Medicine of the Austrian Academy of Sciences | Lukas Endler et al |
| EPI_ISL_854130 | Austria/CeMM1745/2020 | 2020-11-25 | Europe/Austria/Austria/ | Austrian Agency for Health and Food Safety (AGES) | Bergthaler laboratory, CeMM Research Center for Molecular Medicine of the Austrian Academy of Sciences | Lukas Endler et al |
| EPI_ISL_854132 | Austria/CeMM1750/2020 | 2020-11-25 | Europe/Austria/Austria/ | Austrian Agency for Health and Food Safety (AGES) | Bergthaler laboratory, CeMM Research Center for Molecular Medicine of the Austrian Academy of Sciences | Lukas Endler et al |

| | | | | | | |
|---|---|---|---|---|---|---|
| EPI_ISL_854133 | Austria/CeMM1751/2020 | 2020-11-25 | Europe/Austria/Austria/ | Austrian Agency for Health and Food Safety (AGES) | Bergthaler laboratory, CeMM Research Center for Molecular Medicine of the Austrian Academy of Sciences | Lukas Endler et al |
| EPI_ISL_854137 | Austria/CeMM1758/2020 | 2020-11-28 | Europe/Austria/Austria/ | Austrian Agency for Health and Food Safety (AGES) | Bergthaler laboratory, CeMM Research Center for Molecular Medicine of the Austrian Academy of Sciences | Lukas Endler et al |
| EPI_ISL_854142 | Austria/CeMM1766/2020 | 2020-11-28 | Europe/Austria/Austria/ | Austrian Agency for Health and Food Safety (AGES) | Bergthaler laboratory, CeMM Research Center for Molecular Medicine of the Austrian Academy of Sciences | Lukas Endler et al |
| EPI_ISL_854148 | Austria/CeMM1783/2020 | 2020-12-03 | Europe/Austria/Austria/ | Austrian Agency for Health and Food Safety (AGES) | Bergthaler laboratory, CeMM Research Center for Molecular Medicine of the Austrian Academy of Sciences | Lukas Endler et al |
| EPI_ISL_854152 | Austria/CeMM1805/2020 | 2020-11-12 | Europe/Austria/Austria/ | Austrian Agency for Health and Food Safety (AGES) | Bergthaler laboratory, CeMM Research Center for Molecular Medicine of the Austrian Academy of Sciences | Lukas Endler et al |

| | | | | | | |
|---|---|---|---|---|---|---|
| EPI_ISL_854155 | Austria/CeMM1818/2020 | 2020-11-12 | Europe/Austria/Austria/ | Austrian Agency for Health and Food Safety (AGES) | Bergthaler laboratory, CeMM Research Center for Molecular Medicine of the Austrian Academy of Sciences | Lukas Endler et al |
| EPI_ISL_854156 | Austria/CeMM1828/2020 | 2020-11-12 | Europe/Austria/Austria/ | Austrian Agency for Health and Food Safety (AGES) | Bergthaler laboratory, CeMM Research Center for Molecular Medicine of the Austrian Academy of Sciences | Lukas Endler et al |
| EPI_ISL_854162 | Austria/CeMM1842/2020 | 2020-11-12 | Europe/Austria/Austria/ | Austrian Agency for Health and Food Safety (AGES) | Bergthaler laboratory, CeMM Research Center for Molecular Medicine of the Austrian Academy of Sciences | Lukas Endler et al |
| EPI_ISL_854164 | Austria/CeMM1853/2020 | 2020-11-12 | Europe/Austria/Austria/ | Austrian Agency for Health and Food Safety (AGES) | Bergthaler laboratory, CeMM Research Center for Molecular Medicine of the Austrian Academy of Sciences | Lukas Endler et al |
| EPI_ISL_854166 | Austria/CeMM1864/2020 | 2020-11-12 | Europe/Austria/Austria/ | Austrian Agency for Health and Food Safety (AGES) | Bergthaler laboratory, CeMM Research Center for Molecular Medicine of the Austrian Academy of Sciences | Lukas Endler et al |

| | | | | | | |
|---|---|---|---|---|---|---|
| EPI_ISL_854167 | Austria/CeMM1865/2020 | 2020-11-12 | Europe/Austria/Austria/ | Austrian Agency for Health and Food Safety (AGES) | Bergthaler laboratory, CeMM Research Center for Molecular Medicine of the Austrian Academy of Sciences | Lukas Endler et al |
| EPI_ISL_854170 | Austria/CeMM1876/2020 | 2020-11-12 | Europe/Austria/Austria/ | Austrian Agency for Health and Food Safety (AGES) | Bergthaler laboratory, CeMM Research Center for Molecular Medicine of the Austrian Academy of Sciences | Lukas Endler et al |
| EPI_ISL_854175 | Austria/CeMM1889/2020 | 2020-11-12 | Europe/Austria/Austria/ | Austrian Agency for Health and Food Safety (AGES) | Bergthaler laboratory, CeMM Research Center for Molecular Medicine of the Austrian Academy of Sciences | Lukas Endler et al |
| EPI_ISL_854178 | Austria/CeMM2427/2021 | 2021-01-11 | Europe/Austria/Tyrol/ | Austrian Agency for Health and Food Safety (AGES) | Bergthaler laboratory, CeMM Research Center for Molecular Medicine of the Austrian Academy of Sciences | Lukas Endler et al |
| EPI_ISL_854209 | Austria/CeMM1763/2020 | 2020-11-28 | Europe/Austria/Austria/ | Austrian Agency for Health and Food Safety (AGES) | Bergthaler laboratory, CeMM Research Center for Molecular Medicine of the Austrian Academy of Sciences | Lukas Endler et al |

| | | | | | | |
|---|---|---|---|---|---|---|
| EPI_ISL_854243 | Austria/CeMM1709/2020 | 2020-11-23 | Europe/Austria/Austria/ | Austrian Agency for Health and Food Safety (AGES) | Bergthaler laboratory, CeMM Research Center for Molecular Medicine of the Austrian Academy of Sciences | Lukas Endler et al |
| EPI_ISL_854244 | Austria/CeMM1744/2020 | 2020-11-25 | Europe/Austria/Austria/ | Austrian Agency for Health and Food Safety (AGES) | Bergthaler laboratory, CeMM Research Center for Molecular Medicine of the Austrian Academy of Sciences | Lukas Endler et al |
| EPI_ISL_854245 | Austria/CeMM1756/2020 | 2020-11-26 | Europe/Austria/Austria/ | Austrian Agency for Health and Food Safety (AGES) | Bergthaler laboratory, CeMM Research Center for Molecular Medicine of the Austrian Academy of Sciences | Lukas Endler et al |
| EPI_ISL_854248 | Austria/CeMM1780/2020 | 2020-11-29 | Europe/Austria/Austria/ | Austrian Agency for Health and Food Safety (AGES) | Bergthaler laboratory, CeMM Research Center for Molecular Medicine of the Austrian Academy of Sciences | Lukas Endler et al |
| EPI_ISL_854249 | Austria/CeMM1803/2020 | 2020-11-12 | Europe/Austria/Austria/ | Austrian Agency for Health and Food Safety (AGES) | Bergthaler laboratory, CeMM Research Center for Molecular Medicine of the Austrian Academy of Sciences | Lukas Endler et al |

| | | | | | | |
|---|---|---|---|---|---|---|
| EPI_ISL_854250 | Austria/CeMM1827/2020 | 2020-11-12 | Europe/Austria/Austria/ | Austrian Agency for Health and Food Safety (AGES) | Bergthaler laboratory, CeMM Research Center for Molecular Medicine of the Austrian Academy of Sciences | Lukas Endler et al |
| EPI_ISL_854251 | Austria/CeMM1863/2020 | 2020-11-12 | Europe/Austria/Austria/ | Austrian Agency for Health and Food Safety (AGES) | Bergthaler laboratory, CeMM Research Center for Molecular Medicine of the Austrian Academy of Sciences | Lukas Endler et al |
| EPI_ISL_854256 | Austria/CeMM1595/2020 | 2020-11-09 | Europe/Austria/Carinthia/Bodensdorf | Pharmgenetix GmbH | Bergthaler laboratory, CeMM Research Center for Molecular Medicine of the Austrian Academy of Sciences | Lukas Endler et al |
| EPI_ISL_854265 | Austria/CeMM1483/2020 | 2020-11-04 | Europe/Austria/Austria/ | Austrian Agency for Health and Food Safety (AGES) | Bergthaler laboratory, CeMM Research Center for Molecular Medicine of the Austrian Academy of Sciences | Lukas Endler et al |
| EPI_ISL_854269 | Austria/CeMM1521/2020 | 2020-11-04 | Europe/Austria/Austria/ | Austrian Agency for Health and Food Safety (AGES) | Bergthaler laboratory, CeMM Research Center for Molecular Medicine of the Austrian Academy of Sciences | Lukas Endler et al |

| | | | | | | |
|---|---|---|---|---|---|---|
| EPI_ISL_854274 | Austria/CeMM1542/2020 | 2020-11-04 | Europe/Austria/Austria/ | Austrian Agency for Health and Food Safety (AGES) | Bergthaler laboratory, CeMM Research Center for Molecular Medicine of the Austrian Academy of Sciences | Lukas Endler et al |
| EPI_ISL_854275 | Austria/CeMM1544/2020 | 2020-11-04 | Europe/Austria/Austria/ | Austrian Agency for Health and Food Safety (AGES) | Bergthaler laboratory, CeMM Research Center for Molecular Medicine of the Austrian Academy of Sciences | Lukas Endler et al |
| EPI_ISL_854280 | Austria/CeMM1572/2020 | 2020-11-02 | Europe/Austria/Austria/ | Austrian Agency for Health and Food Safety (AGES) | Bergthaler laboratory, CeMM Research Center for Molecular Medicine of the Austrian Academy of Sciences | Lukas Endler et al |
| EPI_ISL_854291 | Austria/CeMM1776/2020 | 2020-11-28 | Europe/Austria/Austria/ | Austrian Agency for Health and Food Safety (AGES) | Bergthaler laboratory, CeMM Research Center for Molecular Medicine of the Austrian Academy of Sciences | Lukas Endler et al |
| EPI_ISL_854295 | Austria/CeMM1375/2020 | 2020-08-28 | Europe/Austria/Styria/ | Center for Virology, Medical University of Vienna | Bergthaler laboratory, CeMM Research Center for Molecular Medicine of the Austrian Academy of Sciences | Lukas Endler et al |

| | | | | | | |
|---|---|---|---|---|---|---|
| EPI_ISL_854297 | Austria/CeMM1719/2020 | 2020-11-23 | Europe/Austria/Austria/ | Austrian Agency for Health and Food Safety (AGES) | Bergthaler laboratory, CeMM Research Center for Molecular Medicine of the Austrian Academy of Sciences | Lukas Endler et al |
| EPI_ISL_854299 | Austria/CeMM1385/2020 | 2020-09-03 | Europe/Austria/Vienna/ | Center for Virology, Medical University of Vienna | Bergthaler laboratory, CeMM Research Center for Molecular Medicine of the Austrian Academy of Sciences | Lukas Endler et al |
| EPI_ISL_854303 | Italy/CeMM1592/2020 | 2020-11-09 | Europe/Italy/Trentino-Alto Adige/Autor | Pharmgenetix GmbH | Bergthaler laboratory, CeMM Research Center for Molecular Medicine of the Austrian Academy of Sciences | Lukas Endler et al |
| EPI_ISL_854310 | USA/NY-Wadsworth-21005133-01/20 | 2021-01-06 | North America/USA/New York/Monroe | URMC LABS | Wadsworth Center, New York State Department of Health | Kirsten St. George et al |
| EPI_ISL_854316 | USA/NY-Wadsworth-21005100-01/20 | 2021-01-05 | North America/USA/New York/Monroe | URMC LABS | Wadsworth Center, New York State Department of Health | Kirsten St. George et al |
| EPI_ISL_854317 | USA/NY-Wadsworth-21005103-01/20 | 2021-01-05 | North America/USA/New York/Allegany | URMC LABS | Wadsworth Center, New York State Department of Health | Kirsten St. George et al |
| EPI_ISL_854318 | USA/NY-Wadsworth-21005125-01/20 | 2021-01-05 | North America/USA/New York/Monroe | URMC LABS | Wadsworth Center, New York State Department of Health | Kirsten St. George et al |

| | | | | | | |
|---|---|---|---|---|---|---|
| EPI_ISL_854321 | USA/NY-Wadsworth-21005073-01/20 | 2021-01-04 | North America/USA/New York/Wyomin | URMC LABS | Wadsworth Center, New York State Department of Health | Kirsten St. George et al |
| EPI_ISL_854322 | USA/NY-Wadsworth-21005079-01/20 | 2021-01-04 | North America/USA/New York/Monroe | URMC LABS | Wadsworth Center, New York State Department of Health | Kirsten St. George et al |
| EPI_ISL_854325 | USA/NY-Wadsworth-21005094-01/20 | 2021-01-04 | North America/USA/New York/Chemun | URMC LABS | Wadsworth Center, New York State Department of Health | Kirsten St. George et al |
| EPI_ISL_854331 | USA/NY-Wadsworth-21005124-01/20 | 2021-01-04 | North America/USA/New York/Wayne ( | URMC LABS | Wadsworth Center, New York State Department of Health | Kirsten St. George et al |
| EPI_ISL_854350 | USA/NY-Wadsworth-21004823-01/20 | 2020-12-30 | North America/USA/New York/Westche | BIO-REFERENCE LABORATORIES | Wadsworth Center, New York State Department of Health | Kirsten St. George et al |
| EPI_ISL_854354 | USA/NY-Wadsworth-21004830-01/20 | 2020-12-30 | North America/USA/New York/Richmon | BIO-REFERENCE LABORATORIES | Wadsworth Center, New York State Department of Health | Kirsten St. George et al |
| EPI_ISL_854364 | USA/NY-Wadsworth-21004851-01/20 | 2020-12-30 | North America/USA/New York/Rocklan | BIO-REFERENCE LABORATORIES | Wadsworth Center, New York State Department of Health | Kirsten St. George et al |
| EPI_ISL_854366 | USA/NY-Wadsworth-21004853-01/20 | 2020-12-30 | North America/USA/New York/Queens | BIO-REFERENCE LABORATORIES | Wadsworth Center, New York State Department of Health | Kirsten St. George et al |
| EPI_ISL_854372 | USA/NY-Wadsworth-21004855-01/20 | 2020-12-30 | North America/USA/New York/New Yor | BIO-REFERENCE LABORATORIES | Wadsworth Center, New York State Department of Health | Kirsten St. George et al |
| EPI_ISL_854374 | USA/NY-Wadsworth-21006072-01/20 | 2021-01-09 | North America/USA/New York/Dutches | WESTCHESTER MEDICAL CENTER | Wadsworth Center, New York State Department of Health | Kirsten St. George et al |

| | | | | | | |
|---|---|---|---|---|---|---|
| EPI_ISL_854380 | USA/NY-Wadsworth-21006070-01/20 | 2021-01-08 | North America/USA/New York/Rocklan | WESTCHESTER MEDICAL CENTER | Wadsworth Center, New York State Department of Health | Kirsten St. George et al |
| EPI_ISL_854382 | USA/NY-Wadsworth-21006048-01/20 | 2021-01-07 | North America/USA/New York/Albany C | WESTCHESTER MEDICAL CENTER | Wadsworth Center, New York State Department of Health | Kirsten St. George et al |
| EPI_ISL_854395 | USA/NY-Wadsworth-21006046-01/20 | 2021-01-06 | North America/USA/New York/Westche | WESTCHESTER MEDICAL CENTER | Wadsworth Center, New York State Department of Health | Kirsten St. George et al |
| EPI_ISL_854397 | USA/NY-Wadsworth-21006053-01/20 | 2021-01-06 | North America/USA/New York/Ulster C | WESTCHESTER MEDICAL CENTER | Wadsworth Center, New York State Department of Health | Kirsten St. George et al |
| EPI_ISL_854409 | USA/NY-Wadsworth-21005071-01/20 | 2020-12-29 | North America/USA/New York/Bronx | MONTEFIORE MEDICAL CENTER LABORATORIES | Wadsworth Center, New York State Department of Health | Kirsten St. George et al |
| EPI_ISL_854415 | USA/NY-Wadsworth-21005142-01/20 | 2020-12-29 | North America/USA/New York/Bronx | MONTEFIORE MEDICAL CENTER LABORATORIES | Wadsworth Center, New York State Department of Health | Kirsten St. George et al |
| EPI_ISL_854422 | USA/NY-Wadsworth-21005065-01/20 | 2020-12-27 | North America/USA/New York/Bronx | MONTEFIORE MEDICAL CENTER LABORATORIES | Wadsworth Center, New York State Department of Health | Kirsten St. George et al |
| EPI_ISL_854423 | USA/NY-Wadsworth-21005088-01/20 | 2020-12-27 | North America/USA/New York/Bronx | MONTEFIORE MEDICAL CENTER LABORATORIES | Wadsworth Center, New York State Department of Health | Kirsten St. George et al |
| EPI_ISL_854432 | USA/NY-Wadsworth-21005099-01/20 | 2020-12-24 | North America/USA/New York/Queens | MONTEFIORE MEDICAL CENTER LABORATORIES | Wadsworth Center, New York State Department of Health | Kirsten St. George et al |
| EPI_ISL_854433 | USA/NY-Wadsworth-21005107-01/20 | 2020-12-24 | North America/USA/New York/Bronx | MONTEFIORE MEDICAL CENTER LABORATORIES | Wadsworth Center, New York State Department of Health | Kirsten St. George et al |

| | | | | | | |
|---|---|---|---|---|---|---|
| EPI_ISL_854447 | USA/NY-Wadsworth-21006030-01/20 | 2021-01-07 | North America/USA/New York/Suffolk C | TEMPUS LABS INC | Wadsworth Center, New York State Department of Health | Kirsten St. George et al |
| EPI_ISL_854461 | Canada/AB-79394/2020 | 2020-03-15 | North America/Canada/Alberta/ | Alberta Precision Labs (APL) | Alberta Precision Labs (APL) | Gordon P et al |
| EPI_ISL_854462 | Canada/AB-10780/2020 | 2020-03-20 | North America/Canada/Alberta/ | Alberta Precision Labs (APL) | Alberta Precision Labs (APL) | Gordon P et al |
| EPI_ISL_854463 | Canada/AB-79281/2020 | 2020-03-22 | North America/Canada/Alberta/ | Alberta Precision Labs (APL) | Alberta Precision Labs (APL) | Gordon P et al |
| EPI_ISL_854464 | Canada/AB-95656/2020 | 2020-03-24 | North America/Canada/Alberta/ | Alberta Precision Labs (APL) | Alberta Precision Labs (APL) | Gordon P et al |
| EPI_ISL_854465 | Canada/AB-10548/2020 | 2020-03-25 | North America/Canada/Alberta/ | Alberta Precision Labs (APL) | Alberta Precision Labs (APL) | Gordon P et al |
| EPI_ISL_854466 | Canada/AB-94746/2020 | 2020-03-25 | North America/Canada/Alberta/ | Alberta Precision Labs (APL) | Alberta Precision Labs (APL) | Gordon P et al |
| EPI_ISL_854467 | Canada/AB-11893/2020 | 2020-03-26 | North America/Canada/Alberta/ | Alberta Precision Labs (APL) | Alberta Precision Labs (APL) | Gordon P et al |
| EPI_ISL_854468 | Canada/AB-12344/2020 | 2020-03-26 | North America/Canada/Alberta/ | Alberta Precision Labs (APL) | Alberta Precision Labs (APL) | Gordon P et al |
| EPI_ISL_854469 | Canada/AB-12352/2020 | 2020-03-26 | North America/Canada/Alberta/ | Alberta Precision Labs (APL) | Alberta Precision Labs (APL) | Gordon P et al |
| EPI_ISL_854470 | Canada/AB-12354/2020 | 2020-03-26 | North America/Canada/Alberta/ | Alberta Precision Labs (APL) | Alberta Precision Labs (APL) | Gordon P et al |
| EPI_ISL_854471 | Canada/AB-11492/2020 | 2020-03-29 | North America/Canada/Alberta/ | Alberta Precision Labs (APL) | Alberta Precision Labs (APL) | Gordon P et al |
| EPI_ISL_854472 | Canada/AB-76191/2020 | 2020-03-30 | North America/Canada/Alberta/ | Alberta Precision Labs (APL) | Alberta Precision Labs (APL) | Gordon P et al |
| EPI_ISL_854473 | Canada/AB-83143/2020 | 2020-04-05 | North America/Canada/Alberta/ | Alberta Precision Labs (APL) | Alberta Precision Labs (APL) | Gordon P et al |
| EPI_ISL_854474 | Canada/AB-12351/2020 | 2020-04-10 | North America/Canada/Alberta/ | Alberta Precision Labs (APL) | Alberta Precision Labs (APL) | Gordon P et al |
| EPI_ISL_854475 | Canada/AB-84786/2020 | 2020-04-12 | North America/Canada/Alberta/ | Alberta Precision Labs (APL) | Alberta Precision Labs (APL) | Gordon P et al |
| EPI_ISL_854476 | Canada/AB-85049/2020 | 2020-04-12 | North America/Canada/Alberta/ | Alberta Precision Labs (APL) | Alberta Precision Labs (APL) | Gordon P et al |
| EPI_ISL_854477 | Canada/AB-10800/2020 | 2020-04-13 | North America/Canada/Alberta/ | Alberta Precision Labs (APL) | Alberta Precision Labs (APL) | Gordon P et al |
| EPI_ISL_854478 | Canada/AB-94528/2020 | 2020-04-13 | North America/Canada/Alberta/ | Alberta Precision Labs (APL) | Alberta Precision Labs (APL) | Gordon P et al |

| | | | | | | |
|---|---|---|---|---|---|---|
| EPI_ISL_854479 | Canada/AB-10320/2020 | 2020-04-14 | North America/Canada/Alberta/ | Alberta Precision Labs (APL) | Alberta Precision Labs (APL) | Gordon P et al |
| EPI_ISL_854480 | Canada/AB-10463/2020 | 2020-04-14 | North America/Canada/Alberta/ | Alberta Precision Labs (APL) | Alberta Precision Labs (APL) | Gordon P et al |
| EPI_ISL_854482 | Canada/AB-79969/2020 | 2020-04-14 | North America/Canada/Alberta/ | Alberta Precision Labs (APL) | Alberta Precision Labs (APL) | Gordon P et al |
| EPI_ISL_854483 | Canada/AB-82996/2020 | 2020-04-14 | North America/Canada/Alberta/ | Alberta Precision Labs (APL) | Alberta Precision Labs (APL) | Gordon P et al |
| EPI_ISL_854484 | Canada/AB-11670/2020 | 2020-04-14 | North America/Canada/Alberta/ | Alberta Precision Labs (APL) | Alberta Precision Labs (APL) | Gordon P et al |
| EPI_ISL_854485 | Canada/AB-83053/2020 | 2020-04-14 | North America/Canada/Alberta/ | Alberta Precision Labs (APL) | Alberta Precision Labs (APL) | Gordon P et al |
| EPI_ISL_854486 | Canada/AB-11112/2020 | 2020-04-15 | North America/Canada/Alberta/ | Alberta Precision Labs (APL) | Alberta Precision Labs (APL) | Gordon P et al |
| EPI_ISL_854487 | Canada/AB-12132/2020 | 2020-04-15 | North America/Canada/Alberta/ | Alberta Precision Labs (APL) | Alberta Precision Labs (APL) | Gordon P et al |
| EPI_ISL_854488 | Canada/AB-12301/2020 | 2020-04-15 | North America/Canada/Alberta/ | Alberta Precision Labs (APL) | Alberta Precision Labs (APL) | Gordon P et al |
| EPI_ISL_854489 | Canada/AB-75175/2020 | 2020-04-15 | North America/Canada/Alberta/ | Alberta Precision Labs (APL) | Alberta Precision Labs (APL) | Gordon P et al |
| EPI_ISL_854490 | Canada/AB-77615/2020 | 2020-04-15 | North America/Canada/Alberta/ | Alberta Precision Labs (APL) | Alberta Precision Labs (APL) | Gordon P et al |
| EPI_ISL_854491 | Canada/AB-80039/2020 | 2020-04-15 | North America/Canada/Alberta/ | Alberta Precision Labs (APL) | Alberta Precision Labs (APL) | Gordon P et al |
| EPI_ISL_854492 | Canada/AB-11077/2020 | 2020-04-16 | North America/Canada/Alberta/ | Alberta Precision Labs (APL) | Alberta Precision Labs (APL) | Gordon P et al |
| EPI_ISL_854493 | Canada/AB-11111/2020 | 2020-04-16 | North America/Canada/Alberta/ | Alberta Precision Labs (APL) | Alberta Precision Labs (APL) | Gordon P et al |
| EPI_ISL_854494 | Canada/AB-12440/2020 | 2020-04-16 | North America/Canada/Alberta/ | Alberta Precision Labs (APL) | Alberta Precision Labs (APL) | Gordon P et al |
| EPI_ISL_854496 | Canada/AB-70980/2020 | 2020-04-16 | North America/Canada/Alberta/ | Alberta Precision Labs (APL) | Alberta Precision Labs (APL) | Gordon P et al |
| EPI_ISL_854497 | Canada/AB-75543/2020 | 2020-04-16 | North America/Canada/Alberta/ | Alberta Precision Labs (APL) | Alberta Precision Labs (APL) | Gordon P et al |
| EPI_ISL_854498 | Canada/AB-79765/2020 | 2020-04-16 | North America/Canada/Alberta/ | Alberta Precision Labs (APL) | Alberta Precision Labs (APL) | Gordon P et al |
| EPI_ISL_854499 | Canada/AB-84051/2020 | 2020-04-16 | North America/Canada/Alberta/ | Alberta Precision Labs (APL) | Alberta Precision Labs (APL) | Gordon P et al |
| EPI_ISL_854500 | Canada/AB-84901/2020 | 2020-04-16 | North America/Canada/Alberta/ | Alberta Precision Labs (APL) | Alberta Precision Labs (APL) | Gordon P et al |

| | | | | | | |
|---|---|---|---|---|---|---|
| EPI_ISL_854501 | Canada/AB-95982/2020 | 2020-04-16 | North America/Canada/Alberta/ | Alberta Precision Labs (APL) | Alberta Precision Labs (APL) | Gordon P et al |
| EPI_ISL_854502 | Canada/AB-95892/2020 | 2020-04-16 | North America/Canada/Alberta/ | Alberta Precision Labs (APL) | Alberta Precision Labs (APL) | Gordon P et al |
| EPI_ISL_854503 | Canada/AB-10703/2020 | 2020-04-17 | North America/Canada/Alberta/ | Alberta Precision Labs (APL) | Alberta Precision Labs (APL) | Gordon P et al |
| EPI_ISL_854504 | Canada/AB-12167/2020 | 2020-04-17 | North America/Canada/Alberta/ | Alberta Precision Labs (APL) | Alberta Precision Labs (APL) | Gordon P et al |
| EPI_ISL_854505 | Canada/AB-74884/2020 | 2020-04-17 | North America/Canada/Alberta/ | Alberta Precision Labs (APL) | Alberta Precision Labs (APL) | Gordon P et al |
| EPI_ISL_854506 | Canada/AB-76028/2020 | 2020-04-17 | North America/Canada/Alberta/ | Alberta Precision Labs (APL) | Alberta Precision Labs (APL) | Gordon P et al |
| EPI_ISL_854507 | Canada/AB-76131/2020 | 2020-04-17 | North America/Canada/Alberta/ | Alberta Precision Labs (APL) | Alberta Precision Labs (APL) | Gordon P et al |
| EPI_ISL_854508 | Canada/AB-85086/2020 | 2020-04-17 | North America/Canada/Alberta/ | Alberta Precision Labs (APL) | Alberta Precision Labs (APL) | Gordon P et al |
| EPI_ISL_854509 | Canada/AB-76135/2020 | 2020-04-17 | North America/Canada/Alberta/ | Alberta Precision Labs (APL) | Alberta Precision Labs (APL) | Gordon P et al |
| EPI_ISL_854510 | Canada/AB-79738/2020 | 2020-04-17 | North America/Canada/Alberta/ | Alberta Precision Labs (APL) | Alberta Precision Labs (APL) | Gordon P et al |
| EPI_ISL_854511 | Canada/AB-93864/2020 | 2020-04-17 | North America/Canada/Alberta/ | Alberta Precision Labs (APL) | Alberta Precision Labs (APL) | Gordon P et al |
| EPI_ISL_854512 | Canada/AB-75252/2020 | 2020-04-18 | North America/Canada/Alberta/ | Alberta Precision Labs (APL) | Alberta Precision Labs (APL) | Gordon P et al |
| EPI_ISL_854513 | Canada/AB-94684/2020 | 2020-04-18 | North America/Canada/Alberta/ | Alberta Precision Labs (APL) | Alberta Precision Labs (APL) | Gordon P et al |
| EPI_ISL_854514 | Canada/AB-95432/2020 | 2020-04-18 | North America/Canada/Alberta/ | Alberta Precision Labs (APL) | Alberta Precision Labs (APL) | Gordon P et al |
| EPI_ISL_854515 | Canada/AB-10481/2020 | 2020-04-20 | North America/Canada/Alberta/ | Alberta Precision Labs (APL) | Alberta Precision Labs (APL) | Gordon P et al |
| EPI_ISL_854516 | Canada/AB-11072/2020 | 2020-04-20 | North America/Canada/Alberta/ | Alberta Precision Labs (APL) | Alberta Precision Labs (APL) | Gordon P et al |
| EPI_ISL_854517 | Canada/AB-79810/2020 | 2020-04-20 | North America/Canada/Alberta/ | Alberta Precision Labs (APL) | Alberta Precision Labs (APL) | Gordon P et al |
| EPI_ISL_854518 | Canada/AB-82706/2020 | 2020-04-20 | North America/Canada/Alberta/ | Alberta Precision Labs (APL) | Alberta Precision Labs (APL) | Gordon P et al |
| EPI_ISL_854519 | Canada/AB-94552/2020 | 2020-04-20 | North America/Canada/Alberta/ | Alberta Precision Labs (APL) | Alberta Precision Labs (APL) | Gordon P et al |
| EPI_ISL_854520 | Canada/AB-96063/2020 | 2020-04-20 | North America/Canada/Alberta/ | Alberta Precision Labs (APL) | Alberta Precision Labs (APL) | Gordon P et al |

| | | | | | | |
|---|---|---|---|---|---|---|
| EPI_ISL_854521 | Canada/AB-10221/2020 | 2020-04-21 | North America/Canada/Alberta/ | Alberta Precision Labs (APL) | Alberta Precision Labs (APL) | Gordon P et al |
| EPI_ISL_854522 | Canada/AB-11523/2020 | 2020-04-21 | North America/Canada/Alberta/ | Alberta Precision Labs (APL) | Alberta Precision Labs (APL) | Gordon P et al |
| EPI_ISL_854523 | Canada/AB-80433/2020 | 2020-04-21 | North America/Canada/Alberta/ | Alberta Precision Labs (APL) | Alberta Precision Labs (APL) | Gordon P et al |
| EPI_ISL_854524 | Canada/AB-10069/2020 | 2020-04-22 | North America/Canada/Alberta/ | Alberta Precision Labs (APL) | Alberta Precision Labs (APL) | Gordon P et al |
| EPI_ISL_854525 | Canada/AB-10507/2020 | 2020-04-22 | North America/Canada/Alberta/ | Alberta Precision Labs (APL) | Alberta Precision Labs (APL) | Gordon P et al |
| EPI_ISL_854526 | Canada/AB-11512/2020 | 2020-04-22 | North America/Canada/Alberta/ | Alberta Precision Labs (APL) | Alberta Precision Labs (APL) | Gordon P et al |
| EPI_ISL_854527 | Canada/AB-11712/2020 | 2020-04-22 | North America/Canada/Alberta/ | Alberta Precision Labs (APL) | Alberta Precision Labs (APL) | Gordon P et al |
| EPI_ISL_854528 | Canada/AB-12308/2020 | 2020-04-22 | North America/Canada/Alberta/ | Alberta Precision Labs (APL) | Alberta Precision Labs (APL) | Gordon P et al |
| EPI_ISL_854529 | Canada/AB-12318/2020 | 2020-04-22 | North America/Canada/Alberta/ | Alberta Precision Labs (APL) | Alberta Precision Labs (APL) | Gordon P et al |
| EPI_ISL_854530 | Canada/AB-12443/2020 | 2020-04-22 | North America/Canada/Alberta/ | Alberta Precision Labs (APL) | Alberta Precision Labs (APL) | Gordon P et al |
| EPI_ISL_854531 | Canada/AB-22769/2020 | 2020-04-22 | North America/Canada/Alberta/ | Alberta Precision Labs (APL) | Alberta Precision Labs (APL) | Gordon P et al |
| EPI_ISL_854532 | Canada/AB-79690/2020 | 2020-04-22 | North America/Canada/Alberta/ | Alberta Precision Labs (APL) | Alberta Precision Labs (APL) | Gordon P et al |
| EPI_ISL_854533 | Canada/AB-79727/2020 | 2020-04-22 | North America/Canada/Alberta/ | Alberta Precision Labs (APL) | Alberta Precision Labs (APL) | Gordon P et al |
| EPI_ISL_854534 | Canada/AB-79785/2020 | 2020-04-22 | North America/Canada/Alberta/ | Alberta Precision Labs (APL) | Alberta Precision Labs (APL) | Gordon P et al |
| EPI_ISL_854535 | Canada/AB-82580/2020 | 2020-04-22 | North America/Canada/Alberta/ | Alberta Precision Labs (APL) | Alberta Precision Labs (APL) | Gordon P et al |
| EPI_ISL_854536 | Canada/AB-84093/2020 | 2020-04-22 | North America/Canada/Alberta/ | Alberta Precision Labs (APL) | Alberta Precision Labs (APL) | Gordon P et al |
| EPI_ISL_854537 | Canada/AB-95873/2020 | 2020-04-22 | North America/Canada/Alberta/ | Alberta Precision Labs (APL) | Alberta Precision Labs (APL) | Gordon P et al |
| EPI_ISL_854538 | Canada/AB-10111/2020 | 2020-04-23 | North America/Canada/Alberta/ | Alberta Precision Labs (APL) | Alberta Precision Labs (APL) | Gordon P et al |
| EPI_ISL_854539 | Canada/AB-75594/2020 | 2020-04-23 | North America/Canada/Alberta/ | Alberta Precision Labs (APL) | Alberta Precision Labs (APL) | Gordon P et al |
| EPI_ISL_854540 | Canada/AB-78977/2020 | 2020-04-23 | North America/Canada/Alberta/ | Alberta Precision Labs (APL) | Alberta Precision Labs (APL) | Gordon P et al |

| | | | | | | |
|---|---|---|---|---|---|---|
| EPI_ISL_854541 | Canada/AB-80084/2020 | 2020-04-23 | North America/Canada/Alberta/ | Alberta Precision Labs (APL) | Alberta Precision Labs (APL) | Gordon P et al |
| EPI_ISL_854542 | Canada/AB-80703/2020 | 2020-04-23 | North America/Canada/Alberta/ | Alberta Precision Labs (APL) | Alberta Precision Labs (APL) | Gordon P et al |
| EPI_ISL_854543 | Canada/AB-82587/2020 | 2020-04-23 | North America/Canada/Alberta/ | Alberta Precision Labs (APL) | Alberta Precision Labs (APL) | Gordon P et al |
| EPI_ISL_854544 | Canada/AB-10360/2020 | 2020-04-24 | North America/Canada/Alberta/ | Alberta Precision Labs (APL) | Alberta Precision Labs (APL) | Gordon P et al |
| EPI_ISL_854545 | Canada/AB-11236/2020 | 2020-04-24 | North America/Canada/Alberta/ | Alberta Precision Labs (APL) | Alberta Precision Labs (APL) | Gordon P et al |
| EPI_ISL_854546 | Canada/AB-12434/2020 | 2020-04-24 | North America/Canada/Alberta/ | Alberta Precision Labs (APL) | Alberta Precision Labs (APL) | Gordon P et al |
| EPI_ISL_854547 | Canada/AB-57059/2020 | 2020-04-24 | North America/Canada/Alberta/ | Alberta Precision Labs (APL) | Alberta Precision Labs (APL) | Gordon P et al |
| EPI_ISL_854548 | Canada/AB-82618/2020 | 2020-04-24 | North America/Canada/Alberta/ | Alberta Precision Labs (APL) | Alberta Precision Labs (APL) | Gordon P et al |
| EPI_ISL_854549 | Canada/AB-96008/2020 | 2020-04-24 | North America/Canada/Alberta/ | Alberta Precision Labs (APL) | Alberta Precision Labs (APL) | Gordon P et al |
| EPI_ISL_854550 | Canada/AB-75300/2020 | 2020-04-25 | North America/Canada/Alberta/ | Alberta Precision Labs (APL) | Alberta Precision Labs (APL) | Gordon P et al |
| EPI_ISL_854551 | Canada/AB-79479/2020 | 2020-04-25 | North America/Canada/Alberta/ | Alberta Precision Labs (APL) | Alberta Precision Labs (APL) | Gordon P et al |
| EPI_ISL_854552 | Canada/AB-84381/2020 | 2020-04-25 | North America/Canada/Alberta/ | Alberta Precision Labs (APL) | Alberta Precision Labs (APL) | Gordon P et al |
| EPI_ISL_854553 | Canada/AB-82615/2020 | 2020-04-26 | North America/Canada/Alberta/ | Alberta Precision Labs (APL) | Alberta Precision Labs (APL) | Gordon P et al |
| EPI_ISL_854554 | Canada/AB-84869/2020 | 2020-04-26 | North America/Canada/Alberta/ | Alberta Precision Labs (APL) | Alberta Precision Labs (APL) | Gordon P et al |
| EPI_ISL_854555 | Canada/AB-94582/2020 | 2020-04-26 | North America/Canada/Alberta/ | Alberta Precision Labs (APL) | Alberta Precision Labs (APL) | Gordon P et al |
| EPI_ISL_854556 | Canada/AB-11424/2020 | 2020-04-27 | North America/Canada/Alberta/ | Alberta Precision Labs (APL) | Alberta Precision Labs (APL) | Gordon P et al |
| EPI_ISL_854557 | Canada/AB-79162/2020 | 2020-04-27 | North America/Canada/Alberta/ | Alberta Precision Labs (APL) | Alberta Precision Labs (APL) | Gordon P et al |
| EPI_ISL_854558 | Canada/AB-83753/2020 | 2020-04-27 | North America/Canada/Alberta/ | Alberta Precision Labs (APL) | Alberta Precision Labs (APL) | Gordon P et al |
| EPI_ISL_854559 | Canada/AB-10073/2020 | 2020-04-28 | North America/Canada/Alberta/ | Alberta Precision Labs (APL) | Alberta Precision Labs (APL) | Gordon P et al |
| EPI_ISL_854560 | Canada/AB-10342/2020 | 2020-04-28 | North America/Canada/Alberta/ | Alberta Precision Labs (APL) | Alberta Precision Labs (APL) | Gordon P et al |

| | | | | | | |
|---|---|---|---|---|---|---|
| EPI_ISL_854561 | Canada/AB-11624/2020 | 2020-04-28 | North America/Canada/Alberta/ | Alberta Precision Labs (APL) | Alberta Precision Labs (APL) | Gordon P et al |
| EPI_ISL_854562 | Canada/AB-11977/2020 | 2020-04-28 | North America/Canada/Alberta/ | Alberta Precision Labs (APL) | Alberta Precision Labs (APL) | Gordon P et al |
| EPI_ISL_854563 | Canada/AB-12174/2020 | 2020-04-28 | North America/Canada/Alberta/ | Alberta Precision Labs (APL) | Alberta Precision Labs (APL) | Gordon P et al |
| EPI_ISL_854564 | Canada/AB-84265/2020 | 2020-04-28 | North America/Canada/Alberta/ | Alberta Precision Labs (APL) | Alberta Precision Labs (APL) | Gordon P et al |
| EPI_ISL_854565 | Canada/AB-84678/2020 | 2020-04-28 | North America/Canada/Alberta/ | Alberta Precision Labs (APL) | Alberta Precision Labs (APL) | Gordon P et al |
| EPI_ISL_854566 | Canada/AB-10801/2020 | 2020-04-29 | North America/Canada/Alberta/ | Alberta Precision Labs (APL) | Alberta Precision Labs (APL) | Gordon P et al |
| EPI_ISL_854567 | Canada/AB-11008/2020 | 2020-04-29 | North America/Canada/Alberta/ | Alberta Precision Labs (APL) | Alberta Precision Labs (APL) | Gordon P et al |
| EPI_ISL_854568 | Canada/AB-10346/2020 | 2020-04-30 | North America/Canada/Alberta/ | Alberta Precision Labs (APL) | Alberta Precision Labs (APL) | Gordon P et al |
| EPI_ISL_854569 | Canada/AB-71632/2020 | 2020-05-01 | North America/Canada/Alberta/ | Alberta Precision Labs (APL) | Alberta Precision Labs (APL) | Gordon P et al |
| EPI_ISL_854570 | Canada/AB-79547/2020 | 2020-05-01 | North America/Canada/Alberta/ | Alberta Precision Labs (APL) | Alberta Precision Labs (APL) | Gordon P et al |
| EPI_ISL_854571 | Canada/AB-11033/2020 | 2020-05-02 | North America/Canada/Alberta/ | Alberta Precision Labs (APL) | Alberta Precision Labs (APL) | Gordon P et al |
| EPI_ISL_854572 | Canada/AB-10836/2020 | 2020-05-03 | North America/Canada/Alberta/ | Alberta Precision Labs (APL) | Alberta Precision Labs (APL) | Gordon P et al |
| EPI_ISL_854573 | Canada/AB-11205/2020 | 2020-05-03 | North America/Canada/Alberta/ | Alberta Precision Labs (APL) | Alberta Precision Labs (APL) | Gordon P et al |
| EPI_ISL_854574 | Canada/AB-11818/2020 | 2020-05-03 | North America/Canada/Alberta/ | Alberta Precision Labs (APL) | Alberta Precision Labs (APL) | Gordon P et al |
| EPI_ISL_854575 | Canada/AB-79951/2020 | 2020-05-03 | North America/Canada/Alberta/ | Alberta Precision Labs (APL) | Alberta Precision Labs (APL) | Gordon P et al |
| EPI_ISL_854576 | Canada/AB-11678/2020 | 2020-05-04 | North America/Canada/Alberta/ | Alberta Precision Labs (APL) | Alberta Precision Labs (APL) | Gordon P et al |
| EPI_ISL_854577 | Canada/AB-11337/2020 | 2020-05-05 | North America/Canada/Alberta/ | Alberta Precision Labs (APL) | Alberta Precision Labs (APL) | Gordon P et al |
| EPI_ISL_854578 | Canada/AB-95848/2020 | 2020-05-05 | North America/Canada/Alberta/ | Alberta Precision Labs (APL) | Alberta Precision Labs (APL) | Gordon P et al |
| EPI_ISL_854579 | Canada/AB-76121/2020 | 2020-05-05 | North America/Canada/Alberta/ | Alberta Precision Labs (APL) | Alberta Precision Labs (APL) | Gordon P et al |
| EPI_ISL_854580 | Canada/AB-10565/2020 | 2020-05-07 | North America/Canada/Alberta/ | Alberta Precision Labs (APL) | Alberta Precision Labs (APL) | Gordon P et al |

| | | | | | | |
|---|---|---|---|---|---|---|
| EPI_ISL_854581 | Canada/AB-84315/2020 | 2020-05-08 | North America/Canada/Alberta/ | Alberta Precision Labs (APL) | Alberta Precision Labs (APL) | Gordon P et al |
| EPI_ISL_854582 | Canada/AB-82312/2020 | 2020-05-09 | North America/Canada/Alberta/ | Alberta Precision Labs (APL) | Alberta Precision Labs (APL) | Gordon P et al |
| EPI_ISL_854583 | Canada/AB-84587/2020 | 2020-05-09 | North America/Canada/Alberta/ | Alberta Precision Labs (APL) | Alberta Precision Labs (APL) | Gordon P et al |
| EPI_ISL_854584 | Canada/AB-11137/2020 | 2020-05-10 | North America/Canada/Alberta/ | Alberta Precision Labs (APL) | Alberta Precision Labs (APL) | Gordon P et al |
| EPI_ISL_854585 | Canada/AB-12183/2020 | 2020-05-12 | North America/Canada/Alberta/ | Alberta Precision Labs (APL) | Alberta Precision Labs (APL) | Gordon P et al |
| EPI_ISL_854586 | Canada/AB-80046/2020 | 2020-05-15 | North America/Canada/Alberta/ | Alberta Precision Labs (APL) | Alberta Precision Labs (APL) | Gordon P et al |
| EPI_ISL_854587 | Canada/AB-95953/2020 | 2020-05-15 | North America/Canada/Alberta/ | Alberta Precision Labs (APL) | Alberta Precision Labs (APL) | Gordon P et al |
| EPI_ISL_854588 | Canada/AB-80040/2020 | 2020-05-16 | North America/Canada/Alberta/ | Alberta Precision Labs (APL) | Alberta Precision Labs (APL) | Gordon P et al |
| EPI_ISL_854589 | Canada/AB-10078/2020 | 2020-05-18 | North America/Canada/Alberta/ | Alberta Precision Labs (APL) | Alberta Precision Labs (APL) | Gordon P et al |
| EPI_ISL_854590 | Canada/AB-12470/2020 | 2020-05-18 | North America/Canada/Alberta/ | Alberta Precision Labs (APL) | Alberta Precision Labs (APL) | Gordon P et al |
| EPI_ISL_854591 | Canada/AB-75436/2020 | 2020-05-18 | North America/Canada/Alberta/ | Alberta Precision Labs (APL) | Alberta Precision Labs (APL) | Gordon P et al |
| EPI_ISL_854592 | Canada/AB-94990/2020 | 2020-05-18 | North America/Canada/Alberta/ | Alberta Precision Labs (APL) | Alberta Precision Labs (APL) | Gordon P et al |
| EPI_ISL_854593 | Canada/AB-95592/2020 | 2020-05-19 | North America/Canada/Alberta/ | Alberta Precision Labs (APL) | Alberta Precision Labs (APL) | Gordon P et al |
| EPI_ISL_854613 | Canada/ON-PHL-20-04388/2020 | 20-Nov | North America/Canada/Ontario/ | Public Health Ontario Laboratory | Public Health Ontario Laboratory | Vanessa G Allen et al |
| EPI_ISL_854614 | Canada/ON-PHL-20-04390/2020 | 20-Nov | North America/Canada/Ontario/ | Public Health Ontario Laboratory | Public Health Ontario Laboratory | Vanessa G Allen et al |
| EPI_ISL_854615 | Canada/ON-PHL-20-04381/2020 | 20-Nov | North America/Canada/Ontario/ | Public Health Ontario Laboratory | Public Health Ontario Laboratory | Vanessa G Allen et al |
| EPI_ISL_854616 | Canada/ON-PHL-20-04387/2020 | 20-Nov | North America/Canada/Ontario/ | Public Health Ontario Laboratory | Public Health Ontario Laboratory | Vanessa G Allen et al |

| | | | | | | |
|---|---|---|---|---|---|---|
| EPI_ISL_854617 | Canada/ON-PHL-20-04386/2020 | 20-Nov | North America/Canada/Ontario/ | Public Health Ontario Laboratory | Public Health Ontario Laboratory | Vanessa G Allen et al |
| EPI_ISL_854618 | Canada/ON-PHL-20-04391/2020 | 20-Nov | North America/Canada/Ontario/ | Public Health Ontario Laboratory | Public Health Ontario Laboratory | Vanessa G Allen et al |
| EPI_ISL_854619 | Canada/ON-PHL-20-04384/2020 | 20-Nov | North America/Canada/Ontario/ | Public Health Ontario Laboratory | Public Health Ontario Laboratory | Vanessa G Allen et al |
| EPI_ISL_854620 | Canada/ON-PHL-20-04389/2020 | 20-Nov | North America/Canada/Ontario/ | Public Health Ontario Laboratory | Public Health Ontario Laboratory | Vanessa G Allen et al |
| EPI_ISL_854621 | Canada/ON-PHL-20-04392/2020 | 20-Nov | North America/Canada/Ontario/ | Public Health Ontario Laboratory | Public Health Ontario Laboratory | Vanessa G Allen et al |
| EPI_ISL_854622 | Canada/ON-PHL-20-04433/2020 | 20-Nov | North America/Canada/Ontario/ | Public Health Ontario Laboratory | Public Health Ontario Laboratory | Vanessa G Allen et al |
| EPI_ISL_854623 | Canada/ON-PHL-20-04434/2020 | 20-Nov | North America/Canada/Ontario/ | Public Health Ontario Laboratory | Public Health Ontario Laboratory | Vanessa G Allen et al |
| EPI_ISL_854624 | Canada/ON-PHL-20-04435/2020 | 20-Nov | North America/Canada/Ontario/ | Public Health Ontario Laboratory | Public Health Ontario Laboratory | Vanessa G Allen et al |
| EPI_ISL_854625 | Canada/ON-PHL-20-04398/2020 | 20-Dec | North America/Canada/Ontario/ | Public Health Ontario Laboratory | Public Health Ontario Laboratory | Vanessa G Allen et al |
| EPI_ISL_854626 | Canada/ON-PHL-20-04395/2020 | 20-Dec | North America/Canada/Ontario/ | Public Health Ontario Laboratory | Public Health Ontario Laboratory | Vanessa G Allen et al |
| EPI_ISL_854627 | Canada/ON-PHL-20-04470/2020 | 20-Dec | North America/Canada/Ontario/ | Public Health Ontario Laboratory | Public Health Ontario Laboratory | Vanessa G Allen et al |
| EPI_ISL_854628 | Canada/ON-PHL-20-04471/2020 | 20-Dec | North America/Canada/Ontario/ | Public Health Ontario Laboratory | Public Health Ontario Laboratory | Vanessa G Allen et al |
| EPI_ISL_854629 | Canada/ON-PHL-20-04465/2020 | 20-Dec | North America/Canada/Ontario/ | Public Health Ontario Laboratory | Public Health Ontario Laboratory | Vanessa G Allen et al |

| | | | | | | |
|---|---|---|---|---|---|---|
| EPI_ISL_854630 | Canada/ON-PHL-20-04464/2020 | 20-Dec | North America/Canada/Ontario/ | Public Health Ontario Laboratory | Public Health Ontario Laboratory | Vanessa G Allen et al |
| EPI_ISL_854631 | Canada/ON-PHL-20-04394/2020 | 20-Dec | North America/Canada/Ontario/ | Public Health Ontario Laboratory | Public Health Ontario Laboratory | Vanessa G Allen et al |
| EPI_ISL_854632 | Canada/ON-PHL-20-04468/2020 | 20-Dec | North America/Canada/Ontario/ | Public Health Ontario Laboratory | Public Health Ontario Laboratory | Vanessa G Allen et al |
| EPI_ISL_854633 | Canada/ON-PHL-20-04406/2020 | 20-Dec | North America/Canada/Ontario/ | Public Health Ontario Laboratory | Public Health Ontario Laboratory | Vanessa G Allen et al |
| EPI_ISL_854634 | Canada/ON-PHL-20-04466/2020 | 20-Dec | North America/Canada/Ontario/ | Public Health Ontario Laboratory | Public Health Ontario Laboratory | Vanessa G Allen et al |
| EPI_ISL_854635 | Canada/ON-PHL-20-04469/2020 | 20-Dec | North America/Canada/Ontario/ | Public Health Ontario Laboratory | Public Health Ontario Laboratory | Vanessa G Allen et al |
| EPI_ISL_854636 | Canada/ON-PHL-20-04467/2020 | 20-Dec | North America/Canada/Ontario/ | Public Health Ontario Laboratory | Public Health Ontario Laboratory | Vanessa G Allen et al |
| EPI_ISL_854637 | Canada/ON-PHL-20-04474/2020 | 20-Dec | North America/Canada/Ontario/ | Public Health Ontario Laboratory | Public Health Ontario Laboratory | Vanessa G Allen et al |
| EPI_ISL_854638 | Canada/ON-PHL-20-04472/2020 | 20-Dec | North America/Canada/Ontario/ | Public Health Ontario Laboratory | Public Health Ontario Laboratory | Vanessa G Allen et al |
| EPI_ISL_854639 | Canada/ON-PHL-20-04473/2020 | 20-Dec | North America/Canada/Ontario/ | Public Health Ontario Laboratory | Public Health Ontario Laboratory | Vanessa G Allen et al |
| EPI_ISL_854640 | Canada/ON-PHL-20-04396/2020 | 20-Dec | North America/Canada/Ontario/ | Public Health Ontario Laboratory | Public Health Ontario Laboratory | Vanessa G Allen et al |
| EPI_ISL_854641 | Canada/ON-PHL-20-04393/2020 | 20-Dec | North America/Canada/Ontario/ | Public Health Ontario Laboratory | Public Health Ontario Laboratory | Vanessa G Allen et al |
| EPI_ISL_854642 | Canada/ON-PHL-20-04463/2020 | 20-Dec | North America/Canada/Ontario/ | Public Health Ontario Laboratory | Public Health Ontario Laboratory | Vanessa G Allen et al |

| | | | | | | |
|---|---|---|---|---|---|---|
| EPI_ISL_854643 | Canada/ON-PHL-20-04489/2020 | 20-Dec | North America/Canada/Ontario/ | Public Health Ontario Laboratory | Public Health Ontario Laboratory | Vanessa G Allen et al |
| EPI_ISL_854644 | Canada/ON-PHL-20-04488/2020 | 20-Dec | North America/Canada/Ontario/ | Public Health Ontario Laboratory | Public Health Ontario Laboratory | Vanessa G Allen et al |
| EPI_ISL_854645 | Canada/ON-PHL-20-04385/2020 | 20-Dec | North America/Canada/Ontario/ | Public Health Ontario Laboratory | Public Health Ontario Laboratory | Vanessa G Allen et al |
| EPI_ISL_854646 | Canada/ON-PHL-20-04448/2020 | 20-Dec | North America/Canada/Ontario/ | Public Health Ontario Laboratory | Public Health Ontario Laboratory | Vanessa G Allen et al |
| EPI_ISL_854647 | Canada/ON-PHL-20-04526/2020 | 20-Dec | North America/Canada/Ontario/ | Public Health Ontario Laboratory | Public Health Ontario Laboratory | Vanessa G Allen et al |
| EPI_ISL_854648 | Canada/ON-PHL-20-04446/2020 | 20-Dec | North America/Canada/Ontario/ | Public Health Ontario Laboratory | Public Health Ontario Laboratory | Vanessa G Allen et al |
| EPI_ISL_854649 | Canada/ON-PHL-20-04525/2020 | 20-Dec | North America/Canada/Ontario/ | Public Health Ontario Laboratory | Public Health Ontario Laboratory | Vanessa G Allen et al |
| EPI_ISL_854650 | Canada/ON-PHL-20-04516/2020 | 20-Dec | North America/Canada/Ontario/ | Public Health Ontario Laboratory | Public Health Ontario Laboratory | Vanessa G Allen et al |
| EPI_ISL_854651 | Canada/ON-PHL-20-04428/2020 | 20-Dec | North America/Canada/Ontario/ | Public Health Ontario Laboratory | Public Health Ontario Laboratory | Vanessa G Allen et al |
| EPI_ISL_854652 | Canada/ON-PHL-20-04447/2020 | 20-Dec | North America/Canada/Ontario/ | Public Health Ontario Laboratory | Public Health Ontario Laboratory | Vanessa G Allen et al |
| EPI_ISL_854653 | Canada/ON-PHL-20-04405/2020 | 20-Dec | North America/Canada/Ontario/ | Public Health Ontario Laboratory | Public Health Ontario Laboratory | Vanessa G Allen et al |
| EPI_ISL_854654 | Canada/ON-PHL-20-04508/2020 | 20-Dec | North America/Canada/Ontario/ | Public Health Ontario Laboratory | Public Health Ontario Laboratory | Vanessa G Allen et al |
| EPI_ISL_854655 | Canada/ON-PHL-20-04504/2020 | 20-Dec | North America/Canada/Ontario/ | Public Health Ontario Laboratory | Public Health Ontario Laboratory | Vanessa G Allen et al |

| | | | | | | |
|---|---|---|---|---|---|---|
| EPI_ISL_854656 | Canada/ON-PHL-20-04507/2020 | 20-Dec | North America/Canada/Ontario/ | Public Health Ontario Laboratory | Public Health Ontario Laboratory | Vanessa G Allen et al |
| EPI_ISL_854657 | Canada/ON-PHL-20-04506/2020 | 20-Dec | North America/Canada/Ontario/ | Public Health Ontario Laboratory | Public Health Ontario Laboratory | Vanessa G Allen et al |
| EPI_ISL_854658 | Canada/ON-PHL-20-04505/2020 | 20-Dec | North America/Canada/Ontario/ | Public Health Ontario Laboratory | Public Health Ontario Laboratory | Vanessa G Allen et al |
| EPI_ISL_854659 | Canada/ON-PHL-20-04441/2020 | 20-Dec | North America/Canada/Ontario/ | Public Health Ontario Laboratory | Public Health Ontario Laboratory | Vanessa G Allen et al |
| EPI_ISL_854660 | Canada/ON-PHL-20-04442/2020 | 20-Dec | North America/Canada/Ontario/ | Public Health Ontario Laboratory | Public Health Ontario Laboratory | Vanessa G Allen et al |
| EPI_ISL_854661 | Canada/ON-PHL-20-04485/2020 | 20-Dec | North America/Canada/Ontario/ | Public Health Ontario Laboratory | Public Health Ontario Laboratory | Vanessa G Allen et al |
| EPI_ISL_854662 | Canada/ON-PHL-20-04519/2020 | 20-Dec | North America/Canada/Ontario/ | Public Health Ontario Laboratory | Public Health Ontario Laboratory | Vanessa G Allen et al |
| EPI_ISL_854663 | Canada/ON-PHL-20-04520/2020 | 20-Dec | North America/Canada/Ontario/ | Public Health Ontario Laboratory | Public Health Ontario Laboratory | Vanessa G Allen et al |
| EPI_ISL_854664 | Canada/ON-PHL-20-04438/2020 | 20-Dec | North America/Canada/Ontario/ | Public Health Ontario Laboratory | Public Health Ontario Laboratory | Vanessa G Allen et al |
| EPI_ISL_854665 | Canada/ON-PHL-20-04401/2020 | 20-Dec | North America/Canada/Ontario/ | Public Health Ontario Laboratory | Public Health Ontario Laboratory | Vanessa G Allen et al |
| EPI_ISL_854666 | Canada/ON-PHL-20-04432/2020 | 20-Dec | North America/Canada/Ontario/ | Public Health Ontario Laboratory | Public Health Ontario Laboratory | Vanessa G Allen et al |
| EPI_ISL_854667 | Canada/ON-PHL-20-04521/2020 | 20-Dec | North America/Canada/Ontario/ | Public Health Ontario Laboratory | Public Health Ontario Laboratory | Vanessa G Allen et al |
| EPI_ISL_854668 | Canada/ON-PHL-20-04439/2020 | 20-Dec | North America/Canada/Ontario/ | Public Health Ontario Laboratory | Public Health Ontario Laboratory | Vanessa G Allen et al |

| | | | | | | |
|---|---|---|---|---|---|---|
| EPI_ISL_854669 | Canada/ON-PHL-20-04522/2020 | 20-Dec | North America/Canada/Ontario/ | Public Health Ontario Laboratory | Public Health Ontario Laboratory | Vanessa G Allen et al |
| EPI_ISL_854670 | Canada/ON-PHL-20-04416/2020 | 20-Dec | North America/Canada/Ontario/ | Public Health Ontario Laboratory | Public Health Ontario Laboratory | Vanessa G Allen et al |
| EPI_ISL_854671 | Canada/ON-PHL-20-04418/2020 | 20-Dec | North America/Canada/Ontario/ | Public Health Ontario Laboratory | Public Health Ontario Laboratory | Vanessa G Allen et al |
| EPI_ISL_854672 | Canada/ON-PHL-20-04417/2020 | 20-Dec | North America/Canada/Ontario/ | Public Health Ontario Laboratory | Public Health Ontario Laboratory | Vanessa G Allen et al |
| EPI_ISL_854673 | Canada/ON-PHL-20-04399/2020 | 20-Dec | North America/Canada/Ontario/ | Public Health Ontario Laboratory | Public Health Ontario Laboratory | Vanessa G Allen et al |
| EPI_ISL_854674 | Canada/ON-PHL-20-04511/2020 | 20-Dec | North America/Canada/Ontario/ | Public Health Ontario Laboratory | Public Health Ontario Laboratory | Vanessa G Allen et al |
| EPI_ISL_854675 | Canada/ON-PHL-20-04518/2020 | 20-Dec | North America/Canada/Ontario/ | Public Health Ontario Laboratory | Public Health Ontario Laboratory | Vanessa G Allen et al |
| EPI_ISL_854676 | Canada/ON-PHL-20-04404/2020 | 20-Dec | North America/Canada/Ontario/ | Public Health Ontario Laboratory | Public Health Ontario Laboratory | Vanessa G Allen et al |
| EPI_ISL_854677 | Canada/ON-PHL-20-04493/2020 | 20-Dec | North America/Canada/Ontario/ | Public Health Ontario Laboratory | Public Health Ontario Laboratory | Vanessa G Allen et al |
| EPI_ISL_854679 | Canada/ON-PHL-20-04512/2020 | 20-Dec | North America/Canada/Ontario/ | Public Health Ontario Laboratory | Public Health Ontario Laboratory | Vanessa G Allen et al |
| EPI_ISL_854680 | Canada/ON-PHL-20-04421/2020 | 20-Dec | North America/Canada/Ontario/ | Public Health Ontario Laboratory | Public Health Ontario Laboratory | Vanessa G Allen et al |
| EPI_ISL_854681 | Canada/ON-PHL-20-04440/2020 | 20-Dec | North America/Canada/Ontario/ | Public Health Ontario Laboratory | Public Health Ontario Laboratory | Vanessa G Allen et al |
| EPI_ISL_854682 | Canada/ON-PHL-20-04524/2020 | 20-Dec | North America/Canada/Ontario/ | Public Health Ontario Laboratory | Public Health Ontario Laboratory | Vanessa G Allen et al |

| | | | | | | |
|---|---|---|---|---|---|---|
| EPI_ISL_854683 | Canada/ON-PHL-20-04431/2020 | 20-Dec | North America/Canada/Ontario/ | Public Health Ontario Laboratory | Public Health Ontario Laboratory | Vanessa G Allen et al |
| EPI_ISL_854684 | Canada/ON-PHL-20-04486/2020 | 20-Dec | North America/Canada/Ontario/ | Public Health Ontario Laboratory | Public Health Ontario Laboratory | Vanessa G Allen et al |
| EPI_ISL_854685 | Canada/ON-PHL-20-04445/2020 | 20-Dec | North America/Canada/Ontario/ | Public Health Ontario Laboratory | Public Health Ontario Laboratory | Vanessa G Allen et al |
| EPI_ISL_854686 | Canada/ON-PHL-20-04444/2020 | 20-Dec | North America/Canada/Ontario/ | Public Health Ontario Laboratory | Public Health Ontario Laboratory | Vanessa G Allen et al |
| EPI_ISL_854687 | Canada/ON-PHL-20-04436/2020 | 20-Dec | North America/Canada/Ontario/ | Public Health Ontario Laboratory | Public Health Ontario Laboratory | Vanessa G Allen et al |
| EPI_ISL_854688 | Canada/ON-PHL-20-04492/2020 | 20-Dec | North America/Canada/Ontario/ | Public Health Ontario Laboratory | Public Health Ontario Laboratory | Vanessa G Allen et al |
| EPI_ISL_854689 | Canada/ON-PHL-20-04409/2020 | 20-Dec | North America/Canada/Ontario/ | Public Health Ontario Laboratory | Public Health Ontario Laboratory | Vanessa G Allen et al |
| EPI_ISL_854690 | Canada/ON-PHL-20-04429/2020 | 20-Dec | North America/Canada/Ontario/ | Public Health Ontario Laboratory | Public Health Ontario Laboratory | Vanessa G Allen et al |
| EPI_ISL_854691 | Canada/ON-PHL-20-04415/2020 | 20-Dec | North America/Canada/Ontario/ | Public Health Ontario Laboratory | Public Health Ontario Laboratory | Vanessa G Allen et al |
| EPI_ISL_854692 | Canada/ON-PHL-20-04411/2020 | 20-Dec | North America/Canada/Ontario/ | Public Health Ontario Laboratory | Public Health Ontario Laboratory | Vanessa G Allen et al |
| EPI_ISL_854693 | Canada/ON-PHL-20-04414/2020 | 20-Dec | North America/Canada/Ontario/ | Public Health Ontario Laboratory | Public Health Ontario Laboratory | Vanessa G Allen et al |
| EPI_ISL_854694 | Canada/ON-PHL-20-04430/2020 | 20-Dec | North America/Canada/Ontario/ | Public Health Ontario Laboratory | Public Health Ontario Laboratory | Vanessa G Allen et al |
| EPI_ISL_854695 | Canada/ON-PHL-20-04515/2020 | 20-Dec | North America/Canada/Ontario/ | Public Health Ontario Laboratory | Public Health Ontario Laboratory | Vanessa G Allen et al |

| | | | | | | |
|---|---|---|---|---|---|---|
| EPI_ISL_854696 | Canada/ON-PHL-20-04403/2020 | 20-Dec | North America/Canada/Ontario/ | Public Health Ontario Laboratory | Public Health Ontario Laboratory | Vanessa G Allen et al |
| EPI_ISL_854697 | Canada/ON-PHL-20-04490/2020 | 20-Dec | North America/Canada/Ontario/ | Public Health Ontario Laboratory | Public Health Ontario Laboratory | Vanessa G Allen et al |
| EPI_ISL_854698 | Canada/ON-PHL-20-04402/2020 | 20-Dec | North America/Canada/Ontario/ | Public Health Ontario Laboratory | Public Health Ontario Laboratory | Vanessa G Allen et al |
| EPI_ISL_854699 | Canada/ON-PHL-20-04514/2020 | 20-Dec | North America/Canada/Ontario/ | Public Health Ontario Laboratory | Public Health Ontario Laboratory | Vanessa G Allen et al |
| EPI_ISL_854700 | Canada/ON-PHL-20-04407/2020 | 20-Dec | North America/Canada/Ontario/ | Public Health Ontario Laboratory | Public Health Ontario Laboratory | Vanessa G Allen et al |
| EPI_ISL_854701 | Canada/ON-PHL-20-04427/2020 | 20-Dec | North America/Canada/Ontario/ | Public Health Ontario Laboratory | Public Health Ontario Laboratory | Vanessa G Allen et al |
| EPI_ISL_854702 | Canada/ON-PHL-20-04494/2020 | 20-Dec | North America/Canada/Ontario/ | Public Health Ontario Laboratory | Public Health Ontario Laboratory | Vanessa G Allen et al |
| EPI_ISL_854703 | Canada/ON-PHL-20-04412/2020 | 20-Dec | North America/Canada/Ontario/ | Public Health Ontario Laboratory | Public Health Ontario Laboratory | Vanessa G Allen et al |
| EPI_ISL_854704 | Canada/ON-PHL-20-04523/2020 | 20-Dec | North America/Canada/Ontario/ | Public Health Ontario Laboratory | Public Health Ontario Laboratory | Vanessa G Allen et al |
| EPI_ISL_854705 | Canada/ON-PHL-20-04437/2020 | 20-Dec | North America/Canada/Ontario/ | Public Health Ontario Laboratory | Public Health Ontario Laboratory | Vanessa G Allen et al |
| EPI_ISL_854706 | Canada/ON-PHL-20-04513/2020 | 20-Dec | North America/Canada/Ontario/ | Public Health Ontario Laboratory | Public Health Ontario Laboratory | Vanessa G Allen et al |
| EPI_ISL_854707 | Canada/ON-PHL-20-04422/2020 | 20-Dec | North America/Canada/Ontario/ | Public Health Ontario Laboratory | Public Health Ontario Laboratory | Vanessa G Allen et al |
| EPI_ISL_854708 | Canada/ON-PHL-20-04426/2020 | 20-Dec | North America/Canada/Ontario/ | Public Health Ontario Laboratory | Public Health Ontario Laboratory | Vanessa G Allen et al |

| | | | | | | |
|---|---|---|---|---|---|---|
| EPI_ISL_854709 | Canada/ON-PHL-20-04517/2020 | 20-Dec | North America/Canada/Ontario/ | Public Health Ontario Laboratory | Public Health Ontario Laboratory | Vanessa G Allen et al |
| EPI_ISL_854710 | Canada/ON-PHL-20-04420/2020 | 20-Dec | North America/Canada/Ontario/ | Public Health Ontario Laboratory | Public Health Ontario Laboratory | Vanessa G Allen et al |
| EPI_ISL_854711 | Canada/ON-PHL-20-04423/2020 | 20-Dec | North America/Canada/Ontario/ | Public Health Ontario Laboratory | Public Health Ontario Laboratory | Vanessa G Allen et al |
| EPI_ISL_854712 | Canada/ON-PHL-20-04413/2020 | 20-Dec | North America/Canada/Ontario/ | Public Health Ontario Laboratory | Public Health Ontario Laboratory | Vanessa G Allen et al |
| EPI_ISL_854713 | Canada/ON-PHL-20-04424/2020 | 20-Dec | North America/Canada/Ontario/ | Public Health Ontario Laboratory | Public Health Ontario Laboratory | Vanessa G Allen et al |
| EPI_ISL_854714 | Canada/ON-PHL-20-04378/2020 | 20-Oct | North America/Canada/Ontario/ | Public Health Ontario Laboratory | Public Health Ontario Laboratory | Vanessa G Allen et al |
| EPI_ISL_854715 | Canada/ON-PHL-20-04382/2020 | 20-Nov | North America/Canada/Ontario/ | Public Health Ontario Laboratory | Public Health Ontario Laboratory | Vanessa G Allen et al |
| EPI_ISL_854716 | Canada/ON-PHL-20-04461/2020 | 20-Dec | North America/Canada/Ontario/ | Public Health Ontario Laboratory | Public Health Ontario Laboratory | Vanessa G Allen et al |
| EPI_ISL_854717 | Canada/ON-PHL-20-04457/2020 | 20-Dec | North America/Canada/Ontario/ | Public Health Ontario Laboratory | Public Health Ontario Laboratory | Vanessa G Allen et al |
| EPI_ISL_854718 | Canada/ON-PHL-20-04458/2020 | 20-Dec | North America/Canada/Ontario/ | Public Health Ontario Laboratory | Public Health Ontario Laboratory | Vanessa G Allen et al |
| EPI_ISL_854719 | Canada/ON-PHL-20-04459/2020 | 20-Dec | North America/Canada/Ontario/ | Public Health Ontario Laboratory | Public Health Ontario Laboratory | Vanessa G Allen et al |
| EPI_ISL_854720 | Canada/ON-PHL-20-04460/2020 | 20-Dec | North America/Canada/Ontario/ | Public Health Ontario Laboratory | Public Health Ontario Laboratory | Vanessa G Allen et al |
| EPI_ISL_854721 | Canada/ON-PHL-20-04400/2020 | 20-Dec | North America/Canada/Ontario/ | Public Health Ontario Laboratory | Public Health Ontario Laboratory | Vanessa G Allen et al |

| | | | | | | |
|---|---|---|---|---|---|---|
| EPI_ISL_854722 | Canada/ON-PHL-20-04484/2020 | 20-Dec | North America/Canada/Ontario/ | Public Health Ontario Laboratory | Public Health Ontario Laboratory | Vanessa G Allen et al |
| EPI_ISL_854723 | Canada/ON-PHL-20-04483/2020 | 20-Dec | North America/Canada/Ontario/ | Public Health Ontario Laboratory | Public Health Ontario Laboratory | Vanessa G Allen et al |
| EPI_ISL_854724 | Canada/ON-PHL-20-04476/2020 | 20-Dec | North America/Canada/Ontario/ | Public Health Ontario Laboratory | Public Health Ontario Laboratory | Vanessa G Allen et al |
| EPI_ISL_854725 | Canada/ON-PHL-20-04475/2020 | 20-Dec | North America/Canada/Ontario/ | Public Health Ontario Laboratory | Public Health Ontario Laboratory | Vanessa G Allen et al |
| EPI_ISL_854726 | Canada/ON-PHL-20-04478/2020 | 20-Dec | North America/Canada/Ontario/ | Public Health Ontario Laboratory | Public Health Ontario Laboratory | Vanessa G Allen et al |
| EPI_ISL_854727 | Canada/ON-PHL-20-04477/2020 | 20-Dec | North America/Canada/Ontario/ | Public Health Ontario Laboratory | Public Health Ontario Laboratory | Vanessa G Allen et al |
| EPI_ISL_854728 | Canada/ON-PHL-20-04496/2020 | 20-Dec | North America/Canada/Ontario/ | Public Health Ontario Laboratory | Public Health Ontario Laboratory | Vanessa G Allen et al |
| EPI_ISL_854729 | Canada/ON-PHL-20-04481/2020 | 20-Dec | North America/Canada/Ontario/ | Public Health Ontario Laboratory | Public Health Ontario Laboratory | Vanessa G Allen et al |
| EPI_ISL_854730 | Canada/ON-PHL-20-04482/2020 | 20-Dec | North America/Canada/Ontario/ | Public Health Ontario Laboratory | Public Health Ontario Laboratory | Vanessa G Allen et al |
| EPI_ISL_854731 | Canada/ON-PHL-20-04497/2020 | 20-Dec | North America/Canada/Ontario/ | Public Health Ontario Laboratory | Public Health Ontario Laboratory | Vanessa G Allen et al |
| EPI_ISL_854732 | Canada/ON-PHL-20-04503/2020 | 20-Dec | North America/Canada/Ontario/ | Public Health Ontario Laboratory | Public Health Ontario Laboratory | Vanessa G Allen et al |
| EPI_ISL_854733 | Canada/ON-PHL-20-04479/2020 | 20-Dec | North America/Canada/Ontario/ | Public Health Ontario Laboratory | Public Health Ontario Laboratory | Vanessa G Allen et al |
| EPI_ISL_854734 | Canada/ON-PHL-20-04480/2020 | 20-Dec | North America/Canada/Ontario/ | Public Health Ontario Laboratory | Public Health Ontario Laboratory | Vanessa G Allen et al |

| | | | | | | |
|---|---|---|---|---|---|---|
| EPI_ISL_854735 | Canada/ON-PHL-20-04495/2020 | 20-Dec | North America/Canada/Ontario/ | Public Health Ontario Laboratory | Public Health Ontario Laboratory | Vanessa G Allen et al |
| EPI_ISL_854736 | Canada/ON-PHL-20-04380/2020 | 20-Nov | North America/Canada/Ontario/ | Public Health Ontario Laboratory | Public Health Ontario Laboratory | Vanessa G Allen et al |
| EPI_ISL_854737 | Canada/ON-PHL-20-04462/2020 | 20-Dec | North America/Canada/Ontario/ | Public Health Ontario Laboratory | Public Health Ontario Laboratory | Vanessa G Allen et al |
| EPI_ISL_854738 | Canada/ON-PHL-20-04456/2020 | 20-Dec | North America/Canada/Ontario/ | Public Health Ontario Laboratory | Public Health Ontario Laboratory | Vanessa G Allen et al |
| EPI_ISL_854739 | Canada/ON-PHL-20-04499/2020 | 20-Dec | North America/Canada/Ontario/ | Public Health Ontario Laboratory | Public Health Ontario Laboratory | Vanessa G Allen et al |
| EPI_ISL_854740 | Canada/ON-PHL-20-04498/2020 | 20-Dec | North America/Canada/Ontario/ | Public Health Ontario Laboratory | Public Health Ontario Laboratory | Vanessa G Allen et al |
| EPI_ISL_854741 | Canada/ON-PHL-20-04502/2020 | 20-Dec | North America/Canada/Ontario/ | Public Health Ontario Laboratory | Public Health Ontario Laboratory | Vanessa G Allen et al |
| EPI_ISL_854742 | Canada/ON-PHL-20-04500/2020 | 20-Dec | North America/Canada/Ontario/ | Public Health Ontario Laboratory | Public Health Ontario Laboratory | Vanessa G Allen et al |
| EPI_ISL_854743 | Canada/ON-PHL-20-04501/2020 | 20-Dec | North America/Canada/Ontario/ | Public Health Ontario Laboratory | Public Health Ontario Laboratory | Vanessa G Allen et al |
| EPI_ISL_854744 | Canada/ON-PHL-20-04419/2020 | 20-Dec | North America/Canada/Ontario/ | Public Health Ontario Laboratory | Public Health Ontario Laboratory | Vanessa G Allen et al |
| EPI_ISL_854763 | Canada/ON-LHSC01/2020 | 2020-11-24 | North America/Canada/Ontario/ | Ontario's COVID-19 Genomics Rapid Response Coalition | McMaster University | Ana Cabrera et al |
| EPI_ISL_854764 | Canada/ON-LHSC02/2020 | 2020-11-24 | North America/Canada/Ontario/ | Ontario's COVID-19 Genomics Rapid Response Coalition | McMaster University | Ana Cabrera et al |
| EPI_ISL_854765 | Canada/ON-LHSC03/2020 | 2020-11-24 | North America/Canada/Ontario/ | Ontario's COVID-19 Genomics Rapid Response Coalition | McMaster University | Ana Cabrera et al |
| EPI_ISL_854766 | Canada/ON-LHSC04/2020 | 2020-11-24 | North America/Canada/Ontario/ | Ontario's COVID-19 Genomics Rapid Response Coalition | McMaster University | Ana Cabrera et al |
| EPI_ISL_854767 | Canada/ON-LHSC05/2020 | 2020-11-24 | North America/Canada/Ontario/ | Ontario's COVID-19 Genomics Rapid Response Coalition | McMaster University | Ana Cabrera et al |

| | | | | | | |
|---|---|---|---|---|---|---|
| EPI_ISL_854768 | Canada/ON-LHSC06/2020 | 2020-11-24 | North America/Canada/Ontario/ | Ontario's COVID-19 Genomics Rapid Response Coalition | McMaster University | Ana Cabrera et al |
| EPI_ISL_854769 | Canada/ON-LHSC07/2020 | 2020-11-24 | North America/Canada/Ontario/ | Ontario's COVID-19 Genomics Rapid Response Coalition | McMaster University | Ana Cabrera et al |
| EPI_ISL_854770 | Canada/ON-LHSC08/2020 | 2020-11-24 | North America/Canada/Ontario/ | Ontario's COVID-19 Genomics Rapid Response Coalition | McMaster University | Ana Cabrera et al |
| EPI_ISL_854771 | Canada/ON-LHSC09/2020 | 2020-11-24 | North America/Canada/Ontario/ | Ontario's COVID-19 Genomics Rapid Response Coalition | McMaster University | Ana Cabrera et al |
| EPI_ISL_854772 | Canada/ON-LHSC10/2020 | 2020-11-24 | North America/Canada/Ontario/ | Ontario's COVID-19 Genomics Rapid Response Coalition | McMaster University | Ana Cabrera et al |
| EPI_ISL_854773 | Canada/ON-LHSC11/2020 | 2020-11-24 | North America/Canada/Ontario/ | Ontario's COVID-19 Genomics Rapid Response Coalition | McMaster University | Ana Cabrera et al |
| EPI_ISL_854774 | Canada/ON-LHSC12/2020 | 2020-11-24 | North America/Canada/Ontario/ | Ontario's COVID-19 Genomics Rapid Response Coalition | McMaster University | Ana Cabrera et al |
| EPI_ISL_854775 | Canada/ON-LHSC13/2020 | 2020-11-24 | North America/Canada/Ontario/ | Ontario's COVID-19 Genomics Rapid Response Coalition | McMaster University | Ana Cabrera et al |
| EPI_ISL_854776 | Canada/ON-LHSC14/2020 | 2020-11-24 | North America/Canada/Ontario/ | Ontario's COVID-19 Genomics Rapid Response Coalition | McMaster University | Ana Cabrera et al |
| EPI_ISL_854777 | Canada/ON-LHSC15/2020 | 2020-11-24 | North America/Canada/Ontario/ | Ontario's COVID-19 Genomics Rapid Response Coalition | McMaster University | Ana Cabrera et al |
| EPI_ISL_854778 | Canada/ON-LHSC16/2020 | 2020-11-24 | North America/Canada/Ontario/ | Ontario's COVID-19 Genomics Rapid Response Coalition | McMaster University | Ana Cabrera et al |
| EPI_ISL_854779 | Canada/ON-LHSC17/2020 | 2020-11-24 | North America/Canada/Ontario/ | Ontario's COVID-19 Genomics Rapid Response Coalition | McMaster University | Ana Cabrera et al |
| EPI_ISL_854780 | Canada/ON-LHSC18/2020 | 2020-11-24 | North America/Canada/Ontario/ | Ontario's COVID-19 Genomics Rapid Response Coalition | McMaster University | Ana Cabrera et al |
| EPI_ISL_854781 | Canada/ON-LHSC19/2020 | 2020-11-24 | North America/Canada/Ontario/ | Ontario's COVID-19 Genomics Rapid Response Coalition | McMaster University | Ana Cabrera et al |
| EPI_ISL_854782 | Canada/ON-LHSC20/2020 | 2020-11-24 | North America/Canada/Ontario/ | Ontario's COVID-19 Genomics Rapid Response Coalition | McMaster University | Ana Cabrera et al |
| EPI_ISL_854783 | Canada/ON-LHSC21/2020 | 2020-11-24 | North America/Canada/Ontario/ | Ontario's COVID-19 Genomics Rapid Response Coalition | McMaster University | Ana Cabrera et al |
| EPI_ISL_854784 | Canada/ON-LHSC22/2020 | 2020-11-24 | North America/Canada/Ontario/ | Ontario's COVID-19 Genomics Rapid Response Coalition | McMaster University | Ana Cabrera et al |
| EPI_ISL_854785 | Canada/ON-LHSC23/2020 | 2020-11-24 | North America/Canada/Ontario/ | Ontario's COVID-19 Genomics Rapid Response Coalition | McMaster University | Ana Cabrera et al |
| EPI_ISL_854786 | Canada/ON-LHSC25/2020 | 2020-11-24 | North America/Canada/Ontario/ | Ontario's COVID-19 Genomics Rapid Response Coalition | McMaster University | Ana Cabrera et al |
| EPI_ISL_854787 | Canada/ON-LHSC26/2020 | 2020-11-24 | North America/Canada/Ontario/ | Ontario's COVID-19 Genomics Rapid Response Coalition | McMaster University | Ana Cabrera et al |

| | | | | | | |
|---|---|---|---|---|---|---|
| EPI_ISL_854788 | Canada/ON-LHSC27/2020 | 2020-11-24 | North America/Canada/Ontario/ | Ontario's COVID-19 Genomics Rapid Response Coalition | McMaster University | Ana Cabrera et al |
| EPI_ISL_854789 | Canada/ON-LHSC28/2020 | 2020-11-24 | North America/Canada/Ontario/ | Ontario's COVID-19 Genomics Rapid Response Coalition | McMaster University | Ana Cabrera et al |
| EPI_ISL_854790 | Canada/ON-LHSC29/2020 | 2020-11-24 | North America/Canada/Ontario/ | Ontario's COVID-19 Genomics Rapid Response Coalition | McMaster University | Ana Cabrera et al |
| EPI_ISL_854791 | Canada/ON-LHSC30/2020 | 2020-11-24 | North America/Canada/Ontario/ | Ontario's COVID-19 Genomics Rapid Response Coalition | McMaster University | Ana Cabrera et al |
| EPI_ISL_854792 | Canada/ON-LHSC31/2020 | 2020-11-24 | North America/Canada/Ontario/ | Ontario's COVID-19 Genomics Rapid Response Coalition | McMaster University | Ana Cabrera et al |
| EPI_ISL_854793 | Canada/ON-LHSC32/2020 | 2020-11-24 | North America/Canada/Ontario/ | Ontario's COVID-19 Genomics Rapid Response Coalition | McMaster University | Ana Cabrera et al |
| EPI_ISL_854794 | Canada/ON-LHSC33/2020 | 2020-11-24 | North America/Canada/Ontario/ | Ontario's COVID-19 Genomics Rapid Response Coalition | McMaster University | Ana Cabrera et al |
| EPI_ISL_854795 | Canada/ON-LHSC34/2020 | 2020-11-24 | North America/Canada/Ontario/ | Ontario's COVID-19 Genomics Rapid Response Coalition | McMaster University | Ana Cabrera et al |
| EPI_ISL_854796 | Canada/ON-LHSC35/2020 | 2020-11-24 | North America/Canada/Ontario/ | Ontario's COVID-19 Genomics Rapid Response Coalition | McMaster University | Ana Cabrera et al |
| EPI_ISL_854797 | Canada/ON-LHSC36/2020 | 2020-11-24 | North America/Canada/Ontario/ | Ontario's COVID-19 Genomics Rapid Response Coalition | McMaster University | Ana Cabrera et al |
| EPI_ISL_854798 | Canada/ON-LHSC37/2020 | 2020-11-24 | North America/Canada/Ontario/ | Ontario's COVID-19 Genomics Rapid Response Coalition | McMaster University | Ana Cabrera et al |
| EPI_ISL_854799 | Canada/ON-LHSC38/2020 | 2020-11-24 | North America/Canada/Ontario/ | Ontario's COVID-19 Genomics Rapid Response Coalition | McMaster University | Ana Cabrera et al |
| EPI_ISL_854800 | Canada/ON-LHSC39/2020 | 2020-11-24 | North America/Canada/Ontario/ | Ontario's COVID-19 Genomics Rapid Response Coalition | McMaster University | Ana Cabrera et al |
| EPI_ISL_854801 | Canada/ON-LHSC41/2020 | 2020-11-24 | North America/Canada/Ontario/ | Ontario's COVID-19 Genomics Rapid Response Coalition | McMaster University | Ana Cabrera et al |
| EPI_ISL_854802 | Canada/ON-LHSC42/2020 | 2020-11-24 | North America/Canada/Ontario/ | Ontario's COVID-19 Genomics Rapid Response Coalition | McMaster University | Ana Cabrera et al |
| EPI_ISL_854803 | Canada/ON-LHSC43/2020 | 2020-11-24 | North America/Canada/Ontario/ | Ontario's COVID-19 Genomics Rapid Response Coalition | McMaster University | Ana Cabrera et al |
| EPI_ISL_854804 | Canada/ON-LHSC44/2020 | 2020-11-24 | North America/Canada/Ontario/ | Ontario's COVID-19 Genomics Rapid Response Coalition | McMaster University | Ana Cabrera et al |
| EPI_ISL_854805 | Canada/ON-LHSC46/2020 | 2020-11-24 | North America/Canada/Ontario/ | Ontario's COVID-19 Genomics Rapid Response Coalition | McMaster University | Ana Cabrera et al |
| EPI_ISL_854806 | Canada/ON-LHSC48/2020 | 2020-11-24 | North America/Canada/Ontario/ | Ontario's COVID-19 Genomics Rapid Response Coalition | McMaster University | Ana Cabrera et al |
| EPI_ISL_854807 | Canada/ON-LHSC50/2020 | 2020-11-24 | North America/Canada/Ontario/ | Ontario's COVID-19 Genomics Rapid Response Coalition | McMaster University | Ana Cabrera et al |

| | | | | | | |
|---|---|---|---|---|---|---|
| EPI_ISL_854808 | Canada/ON-LHSC52/2020 | 2020-11-24 | North America/Canada/Ontario/ | Ontario's COVID-19 Genomics Rapid Response Coalition | McMaster University | Ana Cabrera et al |
| EPI_ISL_854809 | Canada/ON-LHSC61/2020 | 2020-11-24 | North America/Canada/Ontario/ | Ontario's COVID-19 Genomics Rapid Response Coalition | McMaster University | Ana Cabrera et al |
| EPI_ISL_854810 | USA/CA-QDX-2905/2021 | 2021-01-07 | North America/USA/California/ | Quest Diagnostics | Quest Diagnostics | Rosenthal et al |
| EPI_ISL_854811 | USA/CA-QDX-2908/2021 | 2021-01-08 | North America/USA/California/ | Quest Diagnostics | Quest Diagnostics | Rosenthal et al |
| EPI_ISL_854827 | USA/CA-QDX-2997/2021 | 2021-01-08 | North America/USA/California/ | Quest Diagnostics | Quest Diagnostics | Rosenthal et al |
| EPI_ISL_854835 | USA/CA-QDX-3073/2021 | 2021-01-08 | North America/USA/California/ | Quest Diagnostics | Quest Diagnostics | Rosenthal et al |
| EPI_ISL_854838 | USA/CA-QDX-3076/2021 | 2021-01-08 | North America/USA/California/ | Quest Diagnostics | Quest Diagnostics | Rosenthal et al |
| EPI_ISL_854840 | USA/CA-QDX-3078/2021 | 2021-01-08 | North America/USA/California/ | Quest Diagnostics | Quest Diagnostics | Rosenthal et al |
| EPI_ISL_854844 | USA/CT-QDX-2923/2021 | 2021-01-06 | North America/USA/Connecticut/ | Quest Diagnostics | Quest Diagnostics | Rosenthal et al |
| EPI_ISL_854846 | USA/CT-QDX-2932/2021 | 2021-01-06 | North America/USA/Connecticut/ | Quest Diagnostics | Quest Diagnostics | Rosenthal et al |
| EPI_ISL_854850 | USA/CT-QDX-2963/2021 | 2021-01-04 | North America/USA/Connecticut/ | Quest Diagnostics | Quest Diagnostics | Rosenthal et al |
| EPI_ISL_854853 | USA/CT-QDX-2973/2021 | 2021-01-06 | North America/USA/Connecticut/ | Quest Diagnostics | Quest Diagnostics | Rosenthal et al |
| EPI_ISL_854869 | USA/CT-QDX-3057/2021 | 2021-01-06 | North America/USA/Connecticut/ | Quest Diagnostics | Quest Diagnostics | Rosenthal et al |
| EPI_ISL_854880 | USA/MA-QDX-2920/2021 | 2021-01-06 | North America/USA/Massachusetts/ | Quest Diagnostics | Quest Diagnostics | Rosenthal et al |
| EPI_ISL_854889 | USA/MA-QDX-2938/2021 | 2021-01-06 | North America/USA/Massachusetts/ | Quest Diagnostics | Quest Diagnostics | Rosenthal et al |
| EPI_ISL_854891 | USA/MA-QDX-2941/2021 | 2021-01-06 | North America/USA/Massachusetts/ | Quest Diagnostics | Quest Diagnostics | Rosenthal et al |
| EPI_ISL_854897 | USA/MA-QDX-2953/2021 | 2021-01-06 | North America/USA/Massachusetts/ | Quest Diagnostics | Quest Diagnostics | Rosenthal et al |
| EPI_ISL_854918 | USA/MA-QDX-2995/2021 | 2021-01-06 | North America/USA/Massachusetts/ | Quest Diagnostics | Quest Diagnostics | Rosenthal et al |
| EPI_ISL_854919 | USA/MA-QDX-2996/2021 | 2021-01-05 | North America/USA/Massachusetts/ | Quest Diagnostics | Quest Diagnostics | Rosenthal et al |
| EPI_ISL_854926 | USA/MA-QDX-3009/2021 | 2021-01-06 | North America/USA/Massachusetts/ | Quest Diagnostics | Quest Diagnostics | Rosenthal et al |
| EPI_ISL_854945 | USA/MA-QDX-3043/2021 | 2021-01-05 | North America/USA/Massachusetts/ | Quest Diagnostics | Quest Diagnostics | Rosenthal et al |
| EPI_ISL_854948 | USA/MA-QDX-3047/2021 | 2021-01-06 | North America/USA/Massachusetts/ | Quest Diagnostics | Quest Diagnostics | Rosenthal et al |
| EPI_ISL_854951 | USA/MA-QDX-3051/2021 | 2021-01-06 | North America/USA/Massachusetts/ | Quest Diagnostics | Quest Diagnostics | Rosenthal et al |
| EPI_ISL_854961 | USA/MA-QDX-3066/2021 | 2021-01-06 | North America/USA/Massachusetts/ | Quest Diagnostics | Quest Diagnostics | Rosenthal et al |
| EPI_ISL_854969 | USA/NH-QDX-2988/2021 | 2021-01-06 | North America/USA/New Hampshire/ | Quest Diagnostics | Quest Diagnostics | Rosenthal et al |
| EPI_ISL_854970 | USA/NH-QDX-3003/2021 | 2021-01-05 | North America/USA/New Hampshire/ | Quest Diagnostics | Quest Diagnostics | Rosenthal et al |
| EPI_ISL_854973 | USA/NV-QDX-2970/2021 | 2021-01-07 | North America/USA/USA/ | Quest Diagnostics | Quest Diagnostics | Rosenthal et al |
| EPI_ISL_854975 | USA/RI-QDX-2991/2021 | 2021-01-06 | North America/USA/Rhode Island/ | Quest Diagnostics | Quest Diagnostics | Rosenthal et al |
| EPI_ISL_854977 | Bangladesh/DNAS_isl_Hia/2020 | 2020-06-30 | Asia/Bangladesh/Bangladesh/ | NGS Lab, DNA SOLUTION LTD. | NGS Lab, DNA SOLUTION LTD. | Khan et al |

| | | | | | | |
|---|---|---|---|---|---|---|
| EPI_ISL_854979 | Bangladesh/DNAS_isl_DSIMCH-3/202 | 2020-06-30 | Asia/Bangladesh/Bangladesh/ | NGS Lab, DNA SOLUTION LTD. | NGS Lab, DNA SOLUTION LTD. | Khan et al |
| EPI_ISL_854980 | Bangladesh/DNAS_isl_BRB/2020 | 2020-06-30 | Asia/Bangladesh/Bangladesh/ | NGS Lab, DNA SOLUTION LTD. | NGS Lab, DNA SOLUTION LTD. | Khan et al |
| EPI_ISL_854982 | Bangladesh/DNAS_isl_P2-98/2020 | 2020-06-30 | Asia/Bangladesh/Bangladesh/ | NGS Lab, DNA SOLUTION LTD. | NGS Lab, DNA SOLUTION LTD. | Khan et al |
| EPI_ISL_854985 | Bangladesh/DNAS_isl_R-88/2020 | 2020-06-30 | Asia/Bangladesh/Bangladesh/ | NGS Lab, DNA SOLUTION LTD. | NGS Lab, DNA SOLUTION LTD. | Khan et al |
| EPI_ISL_854987 | Bangladesh/DNAS_isl_MH-03/2020 | 2020-06-30 | Asia/Bangladesh/Bangladesh/ | NGS Lab, DNA SOLUTION LTD. | NGS Lab, DNA SOLUTION LTD. | Khan et al |
| EPI_ISL_854992 | Bangladesh/DNAS_isl_MH-09/2020 | 2020-06-30 | Asia/Bangladesh/Bangladesh/ | NGS Lab, DNA SOLUTION LTD. | NGS Lab, DNA SOLUTION LTD. | Khan et al |
| EPI_ISL_854993 | Bangladesh/DNAS_isl_P2-107/2020 | 2020-06-30 | Asia/Bangladesh/Bangladesh/ | NGS Lab, DNA SOLUTION LTD. | NGS Lab, DNA SOLUTION LTD. | Khan et al |
| EPI_ISL_854994 | Bangladesh/DNAS_isl_R-101/2020 | 2020-06-30 | Asia/Bangladesh/Bangladesh/ | NGS Lab, DNA SOLUTION LTD. | NGS Lab, DNA SOLUTION LTD. | Khan et al |
| EPI_ISL_854995 | Bangladesh/DNAS_isl_Sik-bip/2020 | 2020-06-30 | Asia/Bangladesh/Bangladesh/ | NGS Lab, DNA SOLUTION LTD. | NGS Lab, DNA SOLUTION LTD. | Khan et al |
| EPI_ISL_855000 | Bangladesh/DNAS_isl_SIBL-09/2020 | 2020-06-30 | Asia/Bangladesh/Bangladesh/ | NGS Lab, DNA SOLUTION LTD. | NGS Lab, DNA SOLUTION LTD. | Khan et al |
| EPI_ISL_855001 | Bangladesh/DNAS_isl_SIBL-08/2020 | 2020-06-30 | Asia/Bangladesh/Bangladesh/ | NGS Lab, DNA SOLUTION LTD. | NGS Lab, DNA SOLUTION LTD. | Khan et al |
| EPI_ISL_855004 | Bangladesh/DNAS_isl_DSIMCH-2/202 | 2020-06-30 | Asia/Bangladesh/Bangladesh/ | NGS Lab, DNA SOLUTION LTD. | NGS Lab, DNA SOLUTION LTD. | Khan et al |
| EPI_ISL_855008 | Bangladesh/DNAS_isl_S-5/2020 | 2020-06-30 | Asia/Bangladesh/Bangladesh/ | NGS Lab, DNA SOLUTION LTD. | NGS Lab, DNA SOLUTION LTD. | Khan et al |
| EPI_ISL_855009 | Bangladesh/DNAS_isl_BRB-3/2020 | 2020-06-30 | Asia/Bangladesh/Bangladesh/ | NGS Lab, DNA SOLUTION LTD. | NGS Lab, DNA SOLUTION LTD. | Khan et al |
| EPI_ISL_855010 | Bangladesh/DNAS_isl_MMC-01/2020 | 2020-06-30 | Asia/Bangladesh/Bangladesh/ | NGS Lab, DNA SOLUTION LTD. | NGS Lab, DNA SOLUTION LTD. | Khan et al |
| EPI_ISL_855012 | Bangladesh/DNAS_isl_UMCH-1/2020 | 2020-06-30 | Asia/Bangladesh/Bangladesh/ | NGS Lab, DNA SOLUTION LTD. | NGS Lab, DNA SOLUTION LTD. | Khan et al |
| EPI_ISL_855013 | Bangladesh/DNAS_isl_R-97/2020 | 2020-06-30 | Asia/Bangladesh/Bangladesh/ | NGS Lab, DNA SOLUTION LTD. | NGS Lab, DNA SOLUTION LTD. | Khan et al |
| EPI_ISL_855019 | USA/CA-QDX-3094/2020 | 2020-12-28 | North America/USA/California/ | Quest Diagnostics | Quest Diagnostics | Rosenthal et al |
| EPI_ISL_855023 | USA/CA-QDX-3110/2020 | 2020-12-28 | North America/USA/California/ | Quest Diagnostics | Quest Diagnostics | Rosenthal et al |
| EPI_ISL_855033 | USA/CA-QDX-3136/2020 | 2020-12-28 | North America/USA/California/ | Quest Diagnostics | Quest Diagnostics | Rosenthal et al |
| EPI_ISL_855035 | USA/CA-QDX-3149/2021 | 2021-01-08 | North America/USA/California/ | Quest Diagnostics | Quest Diagnostics | Rosenthal et al |
| EPI_ISL_855038 | USA/CA-QDX-3157/2020 | 2020-12-28 | North America/USA/California/ | Quest Diagnostics | Quest Diagnostics | Rosenthal et al |

| | | | | | | |
|---|---|---|---|---|---|---|
| EPI_ISL_855054 | USA/CA-QDX-3201/2020 | 2020-12-28 | North America/USA/California/ | Quest Diagnostics | Quest Diagnostics | Rosenthal et al |
| EPI_ISL_855055 | USA/CA-QDX-3207/2021 | 2021-01-08 | North America/USA/California/ | Quest Diagnostics | Quest Diagnostics | Rosenthal et al |
| EPI_ISL_855057 | USA/CA-QDX-3209/2020 | 2020-12-28 | North America/USA/California/ | Quest Diagnostics | Quest Diagnostics | Rosenthal et al |
| EPI_ISL_855065 | USA/CA-QDX-3244/2021 | 2021-01-06 | North America/USA/California/ | Quest Diagnostics | Quest Diagnostics | Rosenthal et al |
| EPI_ISL_855074 | USA/CA-QDX-3254/2021 | 2021-01-07 | North America/USA/California/ | Quest Diagnostics | Quest Diagnostics | Rosenthal et al |
| EPI_ISL_855078 | USA/CA-QDX-3261/2021 | 2021-01-08 | North America/USA/California/ | Quest Diagnostics | Quest Diagnostics | Rosenthal et al |
| EPI_ISL_855079 | USA/CA-QDX-3262/2021 | 2021-01-08 | North America/USA/California/ | Quest Diagnostics | Quest Diagnostics | Rosenthal et al |
| EPI_ISL_855084 | USA/CT-QDX-3121/2021 | 2021-01-06 | North America/USA/Connecticut/ | Quest Diagnostics | Quest Diagnostics | Rosenthal et al |
| EPI_ISL_855088 | USA/CT-QDX-3146/2021 | 2021-01-05 | North America/USA/Connecticut/ | Quest Diagnostics | Quest Diagnostics | Rosenthal et al |
| EPI_ISL_855090 | USA/CT-QDX-3163/2021 | 2021-01-06 | North America/USA/Connecticut/ | Quest Diagnostics | Quest Diagnostics | Rosenthal et al |
| EPI_ISL_855095 | USA/CT-QDX-3237/2021 | 2021-01-06 | North America/USA/Connecticut/ | Quest Diagnostics | Quest Diagnostics | Rosenthal et al |
| EPI_ISL_855098 | USA/MA-QDX-3090/2021 | 2021-01-05 | North America/USA/Massachusetts/ | Quest Diagnostics | Quest Diagnostics | Rosenthal et al |
| EPI_ISL_855134 | USA/MA-QDX-3195/2021 | 2021-01-05 | North America/USA/Massachusetts/ | Quest Diagnostics | Quest Diagnostics | Rosenthal et al |
| EPI_ISL_855135 | USA/MA-QDX-3202/2021 | 2021-01-06 | North America/USA/Massachusetts/ | Quest Diagnostics | Quest Diagnostics | Rosenthal et al |
| EPI_ISL_855137 | USA/MA-QDX-3204/2021 | 2021-01-06 | North America/USA/Massachusetts/ | Quest Diagnostics | Quest Diagnostics | Rosenthal et al |
| EPI_ISL_855141 | USA/MA-QDX-3220/2021 | 2021-01-05 | North America/USA/Massachusetts/ | Quest Diagnostics | Quest Diagnostics | Rosenthal et al |
| EPI_ISL_855143 | USA/MA-QDX-3222/2021 | 2021-01-05 | North America/USA/Massachusetts/ | Quest Diagnostics | Quest Diagnostics | Rosenthal et al |
| EPI_ISL_855152 | USA/MA-QDX-3241/2021 | 2021-01-06 | North America/USA/Massachusetts/ | Quest Diagnostics | Quest Diagnostics | Rosenthal et al |
| EPI_ISL_855155 | USA/ME-QDX-3084/2021 | 2021-01-06 | North America/USA/Maine/ | Quest Diagnostics | Quest Diagnostics | Rosenthal et al |
| EPI_ISL_855162 | USA/NH-QDX-3141/2021 | 2021-01-06 | North America/USA/New Hampshire/ | Quest Diagnostics | Quest Diagnostics | Rosenthal et al |
| EPI_ISL_855166 | USA/RI-QDX-3098/2021 | 2021-01-06 | North America/USA/Rhode Island/ | Quest Diagnostics | Quest Diagnostics | Rosenthal et al |
| EPI_ISL_855168 | USA/RI-QDX-3174/2021 | 2021-01-07 | North America/USA/Rhode Island/ | Quest Diagnostics | Quest Diagnostics | Rosenthal et al |
| EPI_ISL_855169 | USA/RI-QDX-3189/2021 | 2021-01-07 | North America/USA/Rhode Island/ | Quest Diagnostics | Quest Diagnostics | Rosenthal et al |
| EPI_ISL_855173 | USA/TX-QDX-3096/2021 | 2021-01-05 | North America/USA/Texas/ | Quest Diagnostics | Quest Diagnostics | Rosenthal et al |
| EPI_ISL_855194 | USA/NY-WCM-0009-1-P/2020 | 2020-03-15 | North America/USA/New York/ | Institute for Computational Biomedicine, Weill Cornell Medicine | Institute for Computational Biomedicine, Weill Cornell Medicine | Butler et al |
| EPI_ISL_855206 | USA/NY-WCM-0598-1-P/2020 | 2020-03-15 | North America/USA/New York/ | Institute for Computational Biomedicine, Weill Cornell Medicine | Institute for Computational Biomedicine, Weill Cornell Medicine | Butler et al |

| | | | | | | |
|---|---|---|---|---|---|---|
| EPI_ISL_855227 | USA/NY-WCM-0604-1-P/2020 | 2020-03-15 | North America/USA/New York/ | Institute for Computational Biomedicine, Weill Cornell Medicine | Institute for Computational Biomedicine, Weill Cornell Medicine | Butler et al |
| EPI_ISL_855230 | USA/NY-WCM-0608-1-P/2020 | 2020-03-15 | North America/USA/New York/ | Institute for Computational Biomedicine, Weill Cornell Medicine | Institute for Computational Biomedicine, Weill Cornell Medicine | Butler et al |
| EPI_ISL_855238 | USA/NY-WCM-0615-1-P/2020 | 2020-03-15 | North America/USA/New York/ | Institute for Computational Biomedicine, Weill Cornell Medicine | Institute for Computational Biomedicine, Weill Cornell Medicine | Butler et al |
| EPI_ISL_855241 | USA/NY-WCM-0667-1-P/2020 | 2020-03-15 | North America/USA/New York/ | Institute for Computational Biomedicine, Weill Cornell Medicine | Institute for Computational Biomedicine, Weill Cornell Medicine | Butler et al |
| EPI_ISL_855258 | USA/NY-WCM-0631-1-P/2020 | 2020-03-15 | North America/USA/New York/ | Institute for Computational Biomedicine, Weill Cornell Medicine | Institute for Computational Biomedicine, Weill Cornell Medicine | Butler et al |
| EPI_ISL_855266 | USA/NY-WCM-0359-1-P/2020 | 2020-03-15 | North America/USA/New York/ | Institute for Computational Biomedicine, Weill Cornell Medicine | Institute for Computational Biomedicine, Weill Cornell Medicine | Butler et al |
| EPI_ISL_855272 | USA/NY-WCM-0632-1-P/2020 | 2020-03-15 | North America/USA/New York/ | Institute for Computational Biomedicine, Weill Cornell Medicine | Institute for Computational Biomedicine, Weill Cornell Medicine | Butler et al |
| EPI_ISL_855293 | USA/NY-WCM-0645-1-P/2020 | 2020-03-15 | North America/USA/New York/ | Institute for Computational Biomedicine, Weill Cornell Medicine | Institute for Computational Biomedicine, Weill Cornell Medicine | Butler et al |
| EPI_ISL_855322 | USA/NY-WCM-0018-1-P/2020 | 2020-03-15 | North America/USA/New York/ | Institute for Computational Biomedicine, Weill Cornell Medicine | Institute for Computational Biomedicine, Weill Cornell Medicine | Butler et al |
| EPI_ISL_855330 | USA/NY-WCM-0063-1-P/2020 | 2020-03-15 | North America/USA/New York/ | Institute for Computational Biomedicine, Weill Cornell Medicine | Institute for Computational Biomedicine, Weill Cornell Medicine | Butler et al |

| | | | | | | |
|---|---|---|---|---|---|---|
| EPI_ISL_855355 | France/HDF-IPP00148/2020 | 2020-10-17 | Europe/France/Hauts de France/Amiens | Hospital | National Reference Center for Viruses of Respiratory Infections, Institut Pasteur, Paris | Marion Barbet et al |
| EPI_ISL_855356 | France/HDF-IPP00149/2020 | 2020-10-01 | Europe/France/Hauts de France/Amiens | Hospital | National Reference Center for Viruses of Respiratory Infections, Institut Pasteur, Paris | Marion Barbet et al |
| EPI_ISL_855366 | Mayotte/IPP00173/2021 | 2021-01-06 | Europe/France/Mayotte/Mamoudzou | Hospital | National Reference Center for Viruses of Respiratory Infections, Institut Pasteur, Paris | Marion Barbet et al |
| EPI_ISL_855377 | Mayotte/IPP00189/2021 | 2021-01-06 | Europe/France/Mayotte/Mamoudzou | Hospital | National Reference Center for Viruses of Respiratory Infections, Institut Pasteur, Paris | Marion Barbet et al |
| EPI_ISL_855381 | France/NAQ-IPP00146/2021 | 2021-01-06 | Europe/France/Nouvelle-Aquitaine/Sair | Labo Analyses Med | National Reference Center for Viruses of Respiratory Infections, Institut Pasteur, Paris | Marion Barbet et al |
| EPI_ISL_855384 | France/Norm-IPP00153/2021 | 2021-01-03 | Europe/France/Normandie/Lisieux | Hospital | National Reference Center for Viruses of Respiratory Infections, Institut Pasteur, Paris | Marion Barbet et al |

| | | | | | | |
|---|---|---|---|---|---|---|
| EPI_ISL_855386 | France/Norm-IPP00155/2021 | 2021-01-04 | Europe/France/Normandie/Lisieux | Hospital | National Reference Center for Viruses of Respiratory Infections, Institut Pasteur, Paris | Marion Barbet et al |
| EPI_ISL_855398 | USA/CA-CDPH-UC640/2020 | 2020-12-21 | North America/USA/California/Mono Co | California Department of Public Health | Chiu Laboratory, University of California, San Francisco | Charles Chiu et al |
| EPI_ISL_855400 | USA/CA-UCSF-UC701/2020 | 2020-12-20 | North America/USA/California/San Joac | Chiu Laboratory, University of California, San Francisco | Chiu Laboratory, University of California, San Francisco | Charles Chiu et al |
| EPI_ISL_855403 | Spain/MD-IBV-97010049/2020 | 2020-03-10 | Europe/Spain/Madrid/Madrid | IdISSC/Hospital Cl√≠nico San Carlos de Madrid,Äã | SeqCOVID-SPAIN consortium/IBV(CSIC) | Alberto Delgado-Iribarren et al |
| EPI_ISL_855406 | Spain/MD-IBV-97010053/2020 | 2020-03-28 | Europe/Spain/Madrid/Madrid | IdISSC/Hospital Cl√≠nico San Carlos de Madrid,Äã | SeqCOVID-SPAIN consortium/IBV(CSIC) | Alberto Delgado-Iribarren et al |
| EPI_ISL_855416 | Spain/CT-IBV-97011492/2020 | 2020-12-09 | Europe/Spain/Catalunya/Montgat | Servicio de Microbiolog√≠a, Laboratori Cl√≠nic Metropolitana Nord. Hospital Universitari Germans Trias i Pujol. Institut d,ÄôInvestigaci√≥ en Ci√®ncies de la Salut Germans Trias i Pujol (IGTP) | SeqCOVID-SPAIN consortium/IBV(CSIC) | Elisa Martr√≥ et al |
| EPI_ISL_855418 | Spain/CT-IBV-97011494/2020 | 2020-12-09 | Europe/Spain/Catalunya/Pineda de Mar | Servicio de Microbiolog√≠a, Laboratori Cl√≠nic Metropolitana Nord. Hospital Universitari Germans Trias i Pujol. Institut d,ÄôInvestigaci√≥ en Ci√®ncies de la Salut Germans Trias i Pujol (IGTP) | SeqCOVID-SPAIN consortium/IBV(CSIC) | Elisa Martr√≥ et al |
| EPI_ISL_855423 | Spain/CT-IBV-97011501/2020 | 2020-12-11 | Europe/Spain/Catalunya/Pineda de Mar | Servicio de Microbiolog√≠a, Laboratori Cl√≠nic Metropolitana Nord. Hospital Universitari Germans Trias i Pujol. Institut d,ÄôInvestigaci√≥ en Ci√®ncies de la Salut Germans Trias i Pujol (IGTP) | SeqCOVID-SPAIN consortium/IBV(CSIC) | Elisa Martr√≥ et al |
| EPI_ISL_855433 | Spain/CT-IBV-97011930/2020 | 2020-12-17 | Europe/Spain/Catalunya/Barcelona | Servicio de Microbiolog√≠a, Laboratori Cl√≠nic Metropolitana Nord. Hospital Universitari Germans Trias i Pujol. Institut d,ÄôInvestigaci√≥ en Ci√®ncies de la Salut Germans Trias i Pujol (IGTP) | SeqCOVID-SPAIN consortium/IBV(CSIC) | Elisa Martr√≥ et al |

| | | | | | | |
|---|---|---|---|---|---|---|
| EPI_ISL_855444 | Spain/CT-IBV-97011942/2020 | 2020-12-17 | Europe/Spain/Catalunya/Llinars del Vall | Servicio de Microbiolog√≠a, Laboratori Cl√≠nic Metropolitana Nord. Hospital Universitari Germans Trias i Pujol. Institut d‚ÄôInvestigaci√≥ en Ci√®ncies de la Salut Germans Trias i Pujol (IGTP) | SeqCOVID-SPAIN consortium/IBV(CSIC) | Elisa Mart√≠ et al |
| EPI_ISL_855455 | Spain/CT-IBV-97011955/2020 | 2020-12-17 | Europe/Spain/Catalunya/Llinars del Vall | Servicio de Microbiolog√≠a, Laboratori Cl√≠nic Metropolitana Nord. Hospital Universitari Germans Trias i Pujol. Institut d‚ÄôInvestigaci√≥ en Ci√®ncies de la Salut Germans Trias i Pujol (IGTP) | SeqCOVID-SPAIN consortium/IBV(CSIC) | Elisa Mart√≠ et al |
| EPI_ISL_855470 | Spain/CT-IBV-97011974/2020 | 2020-12-17 | Europe/Spain/Catalunya/Cardedeu | Servicio de Microbiolog√≠a, Laboratori Cl√≠nic Metropolitana Nord. Hospital Universitari Germans Trias i Pujol. Institut d‚ÄôInvestigaci√≥ en Ci√®ncies de la Salut Germans Trias i Pujol (IGTP) | SeqCOVID-SPAIN consortium/IBV(CSIC) | Elisa Mart√≠ et al |
| EPI_ISL_855479 | Spain/CT-IBV-97011993/2020 | 2020-09-16 | Europe/Spain/Catalunya/Badalona | Servicio de Microbiolog√≠a, Laboratori Cl√≠nic Metropolitana Nord. Hospital Universitari Germans Trias i Pujol. Institut d‚ÄôInvestigaci√≥ en Ci√®ncies de la Salut Germans Trias i Pujol (IGTP) | SeqCOVID-SPAIN consortium/IBV(CSIC) | Elisa Mart√≠ et al |
| EPI_ISL_855480 | Spain/CT-IBV-97011994/2020 | 2020-09-17 | Europe/Spain/Catalunya/Badalona | Servicio de Microbiolog√≠a, Laboratori Cl√≠nic Metropolitana Nord. Hospital Universitari Germans Trias i Pujol. Institut d‚ÄôInvestigaci√≥ en Ci√®ncies de la Salut Germans Trias i Pujol (IGTP) | SeqCOVID-SPAIN consortium/IBV(CSIC) | Elisa Mart√≠ et al |
| EPI_ISL_855482 | Spain/CT-IBV-97011996/2020 | 2020-09-17 | Europe/Spain/Catalunya/Granollers | Servicio de Microbiolog√≠a, Laboratori Cl√≠nic Metropolitana Nord. Hospital Universitari Germans Trias i Pujol. Institut d‚ÄôInvestigaci√≥ en Ci√®ncies de la Salut Germans Trias i Pujol (IGTP) | SeqCOVID-SPAIN consortium/IBV(CSIC) | Elisa Mart√≠ et al |
| EPI_ISL_855483 | Spain/CT-IBV-97011997/2020 | 2020-09-17 | Europe/Spain/Catalunya/Mollet del Val | Servicio de Microbiolog√≠a, Laboratori Cl√≠nic Metropolitana Nord. Hospital Universitari Germans Trias i Pujol. Institut d‚ÄôInvestigaci√≥ en Ci√®ncies de la Salut Germans Trias i Pujol (IGTP) | SeqCOVID-SPAIN consortium/IBV(CSIC) | Elisa Mart√≠ et al |
| EPI_ISL_855484 | Spain/CT-IBV-97011998/2020 | 2020-09-17 | Europe/Spain/Catalunya/Caldes de Mon | Servicio de Microbiolog√≠a, Laboratori Cl√≠nic Metropolitana Nord. Hospital Universitari Germans Trias i Pujol. Institut d‚ÄôInvestigaci√≥ en Ci√®ncies de la Salut Germans Trias i Pujol (IGTP) | SeqCOVID-SPAIN consortium/IBV(CSIC) | Elisa Mart√≠ et al |

| | | | | | | |
|---|---|---|---|---|---|---|
| EPI_ISL_855489 | Spain/CT-IBV-97012003/2020 | 2020-09-18 | Europe/Spain/Catalunya/Badalona | Servicio de Microbiolog√≠a, Laboratori Cl√≠nic Metropolitana Nord. Hospital Universitari Germans Trias i Pujol. Institut d‚ÄôInvestigaci√≥ en Ci√®ncies de la Salut Germans Trias i Pujol (IGTP) | SeqCOVID-SPAIN consortium/IBV(CSIC) | Elisa Mart√≠ et al |
| EPI_ISL_855491 | Spain/CT-IBV-97012005/2020 | 2020-09-18 | Europe/Spain/Catalunya/Lli√ßa de Vall | Servicio de Microbiolog√≠a, Laboratori Cl√≠nic Metropolitana Nord. Hospital Universitari Germans Trias i Pujol. Institut d‚ÄôInvestigaci√≥ en Ci√®ncies de la Salut Germans Trias i Pujol (IGTP) | SeqCOVID-SPAIN consortium/IBV(CSIC) | Elisa Mart√≠ et al |
| EPI_ISL_855550 | Italy/LOM-ISS-4544/2020 | 2020-03-09 | Europe/Italy/Lombardy/ | Department of Infectious Diseases, Istituto Superiore di Sanit√†, Rome, Italy | Istituto Superiore di Sanit√† (ISS) | Paola Stefanelli et al |
| EPI_ISL_855554 | Italy/LOM-ISS-3337/2020 | 2020-03-07 | Europe/Italy/Lombardy/ | Department of Infectious Diseases, Istituto Superiore di Sanit√†, Rome, Italy; ASST G.O.M. Niguarda, Sezione Microbiologia e Virologia, Milano, Italy | Istituto Superiore di Sanit√† (ISS) | Paola Stefanelli et al |
| EPI_ISL_855555 | Italy/LOM-ISS-3353/2020 | 2020-03-07 | Europe/Italy/Lombardy/ | Department of Infectious Diseases, Istituto Superiore di Sanit√†, Rome, Italy; ASST G.O.M. Niguarda, Sezione Microbiologia e Virologia, Milano, Italy | Istituto Superiore di Sanit√† (ISS) | Paola Stefanelli et al |
| EPI_ISL_855556 | Italy/CAM-COTUGNO-18181/2020 | 2020-12-31 | Europe/Italy/Campania/ | Lab. Microbiologia e Virologia, Cotugno, A.O. dei Colli | Lab. Microbiologia e Virologia, Cotugno, A.O. dei Colli | Luigi Atripaldi et al |
| EPI_ISL_855557 | Tunisia/23-MHT_14/2020 | 2020-09-14 | Africa/Tunisia/Bizerte/ | Department of Virology, Principal Military Hospital of Instruction of Tunis | Bundeswehr Institute of Microbiology | Susann Handrick et al |
| EPI_ISL_855558 | Tunisia/11-MHT_7/2020 | 2020-09-11 | Africa/Tunisia/Tunis/Tunis | Department of Virology, Principal Military Hospital of Instruction of Tunis | Bundeswehr Institute of Microbiology | Susann Handrick et al |
| EPI_ISL_855559 | Tunisia/11-MHT_8/2020 | 2020-09-13 | Africa/Tunisia/Ben Arous/Ben Arous | Department of Virology, Principal Military Hospital of Instruction of Tunis | Bundeswehr Institute of Microbiology | Susann Handrick et al |
| EPI_ISL_855562 | Tunisia/23-MHT_15/2020 | 2020-09-14 | Africa/Tunisia/Bizerte/ | Department of Virology, Principal Military Hospital of Instruction of Tunis | Bundeswehr Institute of Microbiology | Susann Handrick et al |
| EPI_ISL_855565 | Tunisia/23-MHT_22/2020 | 2020-09-15 | Africa/Tunisia/Bizerte/Corniche | Department of Virology, Principal Military Hospital of Instruction of Tunis | Bundeswehr Institute of Microbiology | Susann Handrick et al |

| | | | | | | |
|---|---|---|---|---|---|---|
| EPI_ISL_855569 | Tunisia/11-MHT_12/2020 | 2020-09-14 | Africa/Tunisia/Tunis/Tunis | Department of Virology, Principal Military Hospital of Instruction of Tunis | Bundeswehr Institute of Microbiology | Susann Handrick et al |
| EPI_ISL_855570 | Tunisia/11-MHT_18/2020 | 2020-09-15 | Africa/Tunisia/Tunis/Tunis | Department of Virology, Principal Military Hospital of Instruction of Tunis | Bundeswehr Institute of Microbiology | Susann Handrick et al |
| EPI_ISL_855571 | Tunisia/11-MHT_21/2020 | 2020-09-15 | Africa/Tunisia/Tunis/Tunis | Department of Virology, Principal Military Hospital of Instruction of Tunis | Bundeswehr Institute of Microbiology | Susann Handrick et al |
| EPI_ISL_855572 | Tunisia/11-MHT_24/2020 | 2020-09-15 | Africa/Tunisia/Tunis/Tunis | Department of Virology, Principal Military Hospital of Instruction of Tunis | Bundeswehr Institute of Microbiology | Susann Handrick et al |
| EPI_ISL_855614 | Italy/CAM-COTUGNO-11798/2020 | 2020-12-21 | Europe/Italy/Campania/ | Lab. Microbiologia e Virologia, Cotugno, A.O. dei Colli | Lab. Microbiologia e Virologia, Cotugno, A.O. dei Colli | Luigi Atripaldi et al |
| EPI_ISL_856156 | Denmark/DCGC-31740/2021 | 2021-01-11 | Europe/Denmark/Syddanmark/ | Department of Virus and Microbiological Special Diagnostics, Statens Serum Institut, Copenhagen, Denmark | Aalborg University | Danish Covid-19 Genome Consortium et al |
| EPI_ISL_856678 | Germany/HE-ChVir22108/2021 | 2021-01-08 | Europe/Germany/Hesse/ | Charitv© Universitv§tsmedizin Berlin, Institute of Virology, Charitv©platz 1, 10117 Berlin, Germany | Charitv© Universitv§tsmedizin Berlin, Institute of Virology, Charitv©platz 1, 10117 Berlin, Germany | Victor M Corman et al |
| EPI_ISL_856679 | Italy/CAM-COTUGNO-53904/2020 | 2020-10-14 | Europe/Italy/Campania/ | Lab. Microbiologia e Virologia, Cotugno, A.O. dei Colli | Lab. Microbiologia e Virologia, Cotugno, A.O. dei Colli | Luigi Atripaldi et al |
| EPI_ISL_856680 | Germany/NI-ChVir21668/2020 | 2020-12-29 | Europe/Germany/Niedersachsen/ | Charitv© Universitv§tsmedizin Berlin, Institute of Virology, Charitv©platz 1, 10117 Berlin, Germany | Charitv© Universitv§tsmedizin Berlin, Institute of Virology, Charitv©platz 1, 10117 Berlin, Germany | Victor M Corman et al |

| | | | | | | |
|---|---|---|---|---|---|---|
| EPI_ISL_856695 | Zambia/ZMB-29/2020 | 2020-03-16 | Africa/Zambia/Lusaka/ | University of Zambia, School of Veterinary Medicine, Disease Control | University of Zambia, School of Veterinary Medicine, Disease Control | Simulundu et al |
| EPI_ISL_856710 | Latvia/232/2021 | 2021-01-12 | Europe/Latvia/Latvia/ | MFD Veselƒ´bas centrs ''Dziedniecƒ´ba'' | Latvian Biomedical Research and Study Centre | Ivars Silamiƒ∑elis et al |
| EPI_ISL_856718 | Italy/VEN-PD07/2020 | 2020-03-11 | Europe/Italy/Veneto/ | Infectious Diseases Unit, Department of Internal Medicine, Azienda Ospedaliera-Universitaria di Padova | Laboratory of Infectious Diseases, Department of Biomedical and Clinical Sciences L. Sacco, University of Milan | Anna Maria Cattelan et al |
| EPI_ISL_856756 | Argentina/INEI098807/2020 | 2020-12-21 | South America/Argentina/Buenos Aires/ | Servicio Virosis Respiratorias-Departamento Virologv≠a-INEI | Instituto Nacional Enfermedades Infecciosas C.G.Malbran | Baumeister E. et al |
| EPI_ISL_856757 | Argentina/INEI099212/2020 | 2020-12-24 | South America/Argentina/Buenos Aires/ | Servicio Virosis Respiratorias-Departamento Virologv≠a-INEI | Instituto Nacional Enfermedades Infecciosas C.G.Malbran | Baumeister E. et al |
| EPI_ISL_856758 | Argentina/INEI099213/2020 | 2020-12-24 | South America/Argentina/Buenos Aires/ | Servicio Virosis Respiratorias-Departamento Virologv≠a-INEI | Instituto Nacional Enfermedades Infecciosas C.G.Malbran | Baumeister E. et al |
| EPI_ISL_856759 | Argentina/INEI099214/2020 | 2020-12-24 | South America/Argentina/Buenos Aires/ | Servicio Virosis Respiratorias-Departamento Virologv≠a-INEI | Instituto Nacional Enfermedades Infecciosas C.G.Malbran | Baumeister E. et al |
| EPI_ISL_856760 | Argentina/INEI097130/2020 | 2020-11-18 | South America/Argentina/Buenos Aires/ | Servicio Virosis Respiratorias-Departamento Virologv≠a-INEI | Instituto Nacional Enfermedades Infecciosas C.G.Malbran | Baumeister E. et al |

| | | | | | | |
|---|---|---|---|---|---|---|
| EPI_ISL_856761 | Argentina/INEI097145/2020 | 2020-11-20 | South America/Argentina/Buenos Aires/ | Servicio Virosis Respiratorias-Departamento Virologv≠a-INEI | Instituto Nacional Enfermedades Infecciosas C.G.Malbran | Baumeister E. et al |
| EPI_ISL_856762 | Argentina/INEI097148/2020 | 2020-11-20 | South America/Argentina/Buenos Aires/ | Servicio Virosis Respiratorias-Departamento Virologv≠a-INEI | Instituto Nacional Enfermedades Infecciosas C.G.Malbran | Baumeister E. et al |
| EPI_ISL_856763 | Argentina/INEI097176/2020 | 2020-11-20 | South America/Argentina/Buenos Aires/ | Servicio Virosis Respiratorias-Departamento Virologv≠a-INEI | Instituto Nacional Enfermedades Infecciosas C.G.Malbran | Baumeister E. et al |
| EPI_ISL_856764 | Argentina/INEI097191/2020 | 2020-11-25 | South America/Argentina/Buenos Aires/ | Servicio Virosis Respiratorias-Departamento Virologv≠a-INEI | Instituto Nacional Enfermedades Infecciosas C.G.Malbran | Baumeister E. et al |
| EPI_ISL_856765 | Argentina/INEI097210/2020 | 2020-11-24 | South America/Argentina/Buenos Aires/ | Servicio Virosis Respiratorias-Departamento Virologv≠a-INEI | Instituto Nacional Enfermedades Infecciosas C.G.Malbran | Baumeister E. et al |
| EPI_ISL_856766 | Argentina/INEI097339/2020 | 2020-11-26 | South America/Argentina/Buenos Aires/ | Servicio Virosis Respiratorias-Departamento Virologv≠a-INEI | Instituto Nacional Enfermedades Infecciosas C.G.Malbran | Baumeister E. et al |
| EPI_ISL_856767 | Argentina/INEI097370/2020 | 2020-11-25 | South America/Argentina/Buenos Aires/ | Servicio Virosis Respiratorias-Departamento Virologv≠a-INEI | Instituto Nacional Enfermedades Infecciosas C.G.Malbran | Baumeister E. et al |
| EPI_ISL_856768 | Argentina/INEI097373/2020 | 2020-11-25 | South America/Argentina/Buenos Aires/ | Servicio Virosis Respiratorias-Departamento Virologv≠a-INEI | Instituto Nacional Enfermedades Infecciosas C.G.Malbran | Baumeister E. et al |
| EPI_ISL_856769 | Argentina/INEI097375/2020 | 2020-11-25 | South America/Argentina/Buenos Aires/ | Servicio Virosis Respiratorias-Departamento Virologv≠a-INEI | Instituto Nacional Enfermedades Infecciosas C.G.Malbran | Baumeister E. et al |
| EPI_ISL_856770 | Argentina/INEI097378/2020 | 2020-11-25 | South America/Argentina/Buenos Aires/ | Servicio Virosis Respiratorias-Departamento Virologv≠a-INEI | Instituto Nacional Enfermedades Infecciosas C.G.Malbran | Baumeister E. et al |

| | | | | | | |
|---|---|---|---|---|---|---|
| EPI_ISL_856771 | Argentina/INEI097387/2020 | 2020-11-28 | South America/Argentina/Buenos Aires/ | Servicio Virosis Respiratorias-Departamento Virologv≠a-INEI | Instituto Nacional Enfermedades Infecciosas C.G.Malbran | Baumeister E. et al |
| EPI_ISL_856772 | Argentina/INEI097659/2020 | 2020-12-01 | South America/Argentina/Buenos Aires/ | Servicio Virosis Respiratorias-Departamento Virologv≠a-INEI | Instituto Nacional Enfermedades Infecciosas C.G.Malbran | Baumeister E. et al |
| EPI_ISL_856773 | Argentina/INEI097697/2020 | 2020-12-02 | South America/Argentina/Buenos Aires/ | Servicio Virosis Respiratorias-Departamento Virologv≠a-INEI | Instituto Nacional Enfermedades Infecciosas C.G.Malbran | Baumeister E. et al |
| EPI_ISL_856775 | Argentina/INEI097815/2020 | 2020-12-03 | South America/Argentina/Buenos Aires/ | Servicio Virosis Respiratorias-Departamento Virologv≠a-INEI | Instituto Nacional Enfermedades Infecciosas C.G.Malbran | Baumeister E. et al |
| EPI_ISL_856776 | Argentina/INEI098414/2020 | 2020-12-09 | South America/Argentina/Buenos Aires/ | Servicio Virosis Respiratorias-Departamento Virologv≠a-INEI | Instituto Nacional Enfermedades Infecciosas C.G.Malbran | Baumeister E. et al |
| EPI_ISL_856777 | Argentina/INEI098418/2020 | 2020-12-04 | South America/Argentina/Buenos Aires/ | Servicio Virosis Respiratorias-Departamento Virologv≠a-INEI | Instituto Nacional Enfermedades Infecciosas C.G.Malbran | Baumeister E. et al |
| EPI_ISL_856778 | Argentina/INEI098420/2020 | 2020-12-04 | South America/Argentina/Buenos Aires/ | Servicio Virosis Respiratorias-Departamento Virologv≠a-INEI | Instituto Nacional Enfermedades Infecciosas C.G.Malbran | Baumeister E. et al |
| EPI_ISL_856779 | Argentina/INEI098437/2020 | 2020-12-06 | South America/Argentina/Buenos Aires/ | Servicio Virosis Respiratorias-Departamento Virologv≠a-INEI | Instituto Nacional Enfermedades Infecciosas C.G.Malbran | Baumeister E. et al |
| EPI_ISL_856781 | Argentina/INEI098448/2020 | 2020-12-09 | South America/Argentina/Buenos Aires/ | Servicio Virosis Respiratorias-Departamento Virologv≠a-INEI | Instituto Nacional Enfermedades Infecciosas C.G.Malbran | Baumeister E. et al |
| EPI_ISL_856783 | Argentina/INEI098490/2020 | 2020-12-10 | South America/Argentina/Buenos Aires/ | Servicio Virosis Respiratorias-Departamento Virologv≠a-INEI | Instituto Nacional Enfermedades Infecciosas C.G.Malbran | Baumeister E. et al |

| | | | | | | |
|---|---|---|---|---|---|---|
| EPI_ISL_856784 | Argentina/INEI098521/2020 | 2020-12-11 | South America/Argentina/Buenos Aires/ | Servicio Virosis Respiratorias-Departamento Virologv≠a-INEI | Instituto Nacional Enfermedades Infecciosas C.G.Malbran | Baumeister E. et al |
| EPI_ISL_856785 | Argentina/INEI098536/2020 | 2020-12-15 | South America/Argentina/Buenos Aires/ | Servicio Virosis Respiratorias-Departamento Virologv≠a-INEI | Instituto Nacional Enfermedades Infecciosas C.G.Malbran | Baumeister E. et al |
| EPI_ISL_856786 | Argentina/INEI098554/2020 | 2020-12-13 | South America/Argentina/Buenos Aires/ | Servicio Virosis Respiratorias-Departamento Virologv≠a-INEI | Instituto Nacional Enfermedades Infecciosas C.G.Malbran | Baumeister E. et al |
| EPI_ISL_856787 | Argentina/INEI098558/2020 | 2020-12-14 | South America/Argentina/Buenos Aires/ | Servicio Virosis Respiratorias-Departamento Virologv≠a-INEI | Instituto Nacional Enfermedades Infecciosas C.G.Malbran | Baumeister E. et al |
| EPI_ISL_856788 | Argentina/INEI098603/2020 | 2020-12-16 | South America/Argentina/Buenos Aires/ | Servicio Virosis Respiratorias-Departamento Virologv≠a-INEI | Instituto Nacional Enfermedades Infecciosas C.G.Malbran | Baumeister E. et al |
| EPI_ISL_856789 | Argentina/INEI098730/2020 | 2020-12-21 | South America/Argentina/Buenos Aires/ | Servicio Virosis Respiratorias-Departamento Virologv≠a-INEI | Instituto Nacional Enfermedades Infecciosas C.G.Malbran | Baumeister E. et al |
| EPI_ISL_856790 | Argentina/INEI096222/2020 | 2020-10-27 | South America/Argentina/Buenos Aires/ | Servicio Virosis Respiratorias-Departamento Virologv≠a-INEI | Instituto Nacional Enfermedades Infecciosas C.G.Malbran | Baumeister E. et al |
| EPI_ISL_856791 | Argentina/INEI096223/2020 | 2020-10-27 | South America/Argentina/Buenos Aires/ | Servicio Virosis Respiratorias-Departamento Virologv≠a-INEI | Instituto Nacional Enfermedades Infecciosas C.G.Malbran | Baumeister E. et al |
| EPI_ISL_856792 | Argentina/INEI096269/2020 | 2020-10-28 | South America/Argentina/Buenos Aires/ | Servicio Virosis Respiratorias-Departamento Virologv≠a-INEI | Instituto Nacional Enfermedades Infecciosas C.G.Malbran | Baumeister E. et al |
| EPI_ISL_856793 | Argentina/INEI096300/2020 | 2020-10-28 | South America/Argentina/Buenos Aires/ | Servicio Virosis Respiratorias-Departamento Virologv≠a-INEI | Instituto Nacional Enfermedades Infecciosas C.G.Malbran | Baumeister E. et al |

| | | | | | | |
|---|---|---|---|---|---|---|
| EPI_ISL_856794 | Argentina/INEI096305/2020 | 2020-10-31 | South America/Argentina/Buenos Aires/ | Servicio Virosis Respiratorias-Departamento Virolog√≠a-INEI | Instituto Nacional Enfermedades Infecciosas C.G.Malbran | Baumeister E. et al |
| EPI_ISL_856795 | Argentina/INEI096315/2020 | 2020-10-31 | South America/Argentina/Buenos Aires/ | Servicio Virosis Respiratorias-Departamento Virologí≠a-INEI | Instituto Nacional Enfermedades Infecciosas C.G.Malbran | Baumeister E. et al |
| EPI_ISL_856796 | Argentina/INEI096359/2020 | 2020-10-30 | South America/Argentina/Buenos Aires/ | Servicio Virosis Respiratorias-Departamento Virolog√≠a-INEI | Instituto Nacional Enfermedades Infecciosas C.G.Malbran | Baumeister E. et al |
| EPI_ISL_856797 | Argentina/INEI096369/2020 | 2020-11-02 | South America/Argentina/Buenos Aires/ | Servicio Virosis Respiratorias-Departamento Virolog√≠a-INEI | Instituto Nacional Enfermedades Infecciosas C.G.Malbran | Baumeister E. et al |
| EPI_ISL_856798 | Argentina/INEI096397/2020 | 2020-11-02 | South America/Argentina/Buenos Aires/ | Servicio Virosis Respiratorias-Departamento Virolog√≠a-INEI | Instituto Nacional Enfermedades Infecciosas C.G.Malbran | Baumeister E. et al |
| EPI_ISL_856799 | Argentina/INEI096418/2020 | 2020-11-03 | South America/Argentina/Buenos Aires/ | Servicio Virosis Respiratorias-Departamento Virolog√≠a-INEI | Instituto Nacional Enfermedades Infecciosas C.G.Malbran | Baumeister E. et al |
| EPI_ISL_856800 | Argentina/INEI096492/2020 | 2020-11-04 | South America/Argentina/Buenos Aires/ | Servicio Virosis Respiratorias-Departamento Virolog√≠a-INEI | Instituto Nacional Enfermedades Infecciosas C.G.Malbran | Baumeister E. et al |
| EPI_ISL_856801 | Argentina/INEI096522/2020 | 2020-11-06 | South America/Argentina/Buenos Aires/ | Servicio Virosis Respiratorias-Departamento Virolog√≠a-INEI | Instituto Nacional Enfermedades Infecciosas C.G.Malbran | Baumeister E. et al |
| EPI_ISL_856833 | USA/UT-UPHL-2101036370/2020 | 2020-12-16 | North America/USA/Utah/ | Utah Public Health Laboratory | Utah Public Health Laboratory | Erin L. Young et al |
| EPI_ISL_856836 | USA/UT-UPHL-2101860369/2020 | 2020-12-15 | North America/USA/Utah/ | Utah Public Health Laboratory | Utah Public Health Laboratory | Erin L. Young et al |
| EPI_ISL_856838 | USA/UT-UPHL-2101171400/2020 | 2020-12-16 | North America/USA/Utah/ | Utah Public Health Laboratory | Utah Public Health Laboratory | Erin L. Young et al |
| EPI_ISL_856858 | USA/UT-UPHL-2101797210/2020 | 2020-12-17 | North America/USA/Utah/ | Utah Public Health Laboratory | Utah Public Health Laboratory | Erin L. Young et al |

| | | | | | | |
|---|---|---|---|---|---|---|
| EPI_ISL_856870 | Italy/CAM-AMC-14-5289/2020 | 2020-03-27 | Europe/Italy/Campania/ | Virology Laboratory, Scientific Department, Army Medical Center | Virology Laboratory, Scientific Department, Army Medical Center | Silvia Fillo et al |
| EPI_ISL_856875 | Italy/LAZ-AMC-5-1565/2020 | 2020-03-08 | Europe/Italy/Lazio/ | Virology Laboratory, Scientific Department, Army Medical Center | Virology Laboratory, Scientific Department, Army Medical Center | Silvia Fillo et al |
| EPI_ISL_856876 | Italy/LAZ-AMC-6-1993/2020 | 2020-03-10 | Europe/Italy/Lazio/ | Virology Laboratory, Scientific Department, Army Medical Center | Virology Laboratory, Scientific Department, Army Medical Center | Silvia Fillo et al |
| EPI_ISL_856877 | Italy/LAZ-AMC-7-2253/2020 | 2020-03-12 | Europe/Italy/Lazio/ | Virology Laboratory, Scientific Department, Army Medical Center | Virology Laboratory, Scientific Department, Army Medical Center | Silvia Fillo et al |
| EPI_ISL_856878 | Italy/LAZ-AMC-9-3476/2020 | 2020-03-13 | Europe/Italy/Lazio/ | Virology Laboratory, Scientific Department, Army Medical Center | Virology Laboratory, Scientific Department, Army Medical Center | Silvia Fillo et al |
| EPI_ISL_856883 | Italy/SAR-AMC-12-4618/2020 | 2020-03-23 | Europe/Italy/Sardinia/ | Virology Laboratory, Scientific Department, Army Medical Center | Virology Laboratory, Scientific Department, Army Medical Center | Silvia Fillo et al |

| | | | | | | |
|---|---|---|---|---|---|---|
| EPI_ISL_856884 | Italy/TAA-ISS-219/2020 | 2020-02-24 | Europe/Italy/Trentino-Alto Adige/Autor | Department of Infectious Diseases, Istituto Superiore di Sanitv†, Roma, Italy; PO Santa Chiara, Microbiologia e Virologia, Trento, Italy | Virology Laboratory, Scientific Department, Army Medical Center | Paola Stefanelli et al |
| EPI_ISL_856885 | Italy/TUS-AMC-16-5342/2020 | 2020-03-27 | Europe/Italy/Tuscany/ | Virology Laboratory, Scientific Department, Army Medical Center | Virology Laboratory, Scientific Department, Army Medical Center | Silvia Fillo et al |
| EPI_ISL_856888 | Italy/BAS-ISS-5042/2020 | 2020-03-21 | Europe/Italy/Basilicata/ | Department of Infectious Diseases, Istituto Superiore di Sanitv†, Roma, Italy; A.O.R. San Carlo di Potenza, Laboratorio Microbiologia e Virologia, Potenza, Italy | Virology Laboratory, Scientific Department, Army Medical Center | Paola Stefanelli et al |
| EPI_ISL_856889 | Italy/CAL-ISS-4676/2020 | 2020-03-07 | Europe/Italy/Calabria/ | Department of Infectious Diseases, Istituto Superiore di Sanitv†, Roma, Italy | Virology Laboratory, Scientific Department, Army Medical Center | Paola Stefanelli et al |
| EPI_ISL_856891 | Italy/CAM-ISS-3905/2020 | 2020-03-08 | Europe/Italy/Campania/ | Department of Infectious Diseases, Istituto Superiore di Sanitv†, Roma, Italy; AORN dei Colli, UOC Microbiologia Virologia, Napoli, Italy | Virology Laboratory, Scientific Department, Army Medical Center | Paola Stefanelli et al |
| EPI_ISL_856892 | Italy/CAM-ISS-780/2020 | 2020-02-29 | Europe/Italy/Campania/ | Department of Infectious Diseases, Istituto Superiore di Sanitv†, Roma, Italy; AORN dei Colli, UOC Microbiologia Virologia, Napoli, Italy | Virology Laboratory, Scientific Department, Army Medical Center | Paola Stefanelli et al |

| | | | | | | |
|---|---|---|---|---|---|---|
| EPI_ISL_856894 | Italy/FVG-ISS-4234/2020 | 2020-03-04 | Europe/Italy/Friuli Venezia Giulia/ | Department of Infectious Diseases, Istituto Superiore di Sanit√†, Roma, Italy; ASUITS - Ospedale Maggiore, S.C. UOC Igiene e Sanit√† Pubblica, Trieste, Italy | Virology Laboratory, Scientific Department, Army Medical Center | Paola Stefanelli et al |
| EPI_ISL_856898 | Italy/PIE-ISS-1177/2020 | 2020-02-29 | Europe/Italy/Piedmont/ | Department of Infectious Diseases, Istituto Superiore di Sanit√†, Roma, Italy; ASL Citt√† di Torino, S.C. Microbiologia e Virologia, Torino, Italy | Virology Laboratory, Scientific Department, Army Medical Center | Paola Stefanelli et al |
| EPI_ISL_856901 | Italy/SIC-ISS-3784/2020 | 2020-03-08 | Europe/Italy/Sicily/ | Department of Infectious Diseases, Istituto Superiore di Sanit√†, Roma, Italy; AOUP V. Emanuele di Catania, P.O. Gaspare Rodolico, Catania, Italy | Virology Laboratory, Scientific Department, Army Medical Center | Paola Stefanelli et al |
| EPI_ISL_856902 | Italy/TAA-ISS-3201/2020 | 2020-03-07 | Europe/Italy/Trentino-Alto Adige/Autor | Department of Infectious Diseases, Istituto Superiore di Sanit√†, Roma, Italy; AS Alto Adige, Microbiologia e Virologia, Bolzano, Italy | Virology Laboratory, Scientific Department, Army Medical Center | Paola Stefanelli et al |
| EPI_ISL_856905 | Italy/UMB-ISS-3857/2020 | 2020-03-06 | Europe/Italy/Umbria/ | Department of Infectious Diseases, Istituto Superiore di Sanit√†, Roma, Italy; AO S.M. della Misericordia, S.C. Microbiologia, Perugia, Italy | Virology Laboratory, Scientific Department, Army Medical Center | Paola Stefanelli et al |
| EPI_ISL_856906 | Italy/UMB-ISS-3867/2020 | 2020-03-06 | Europe/Italy/Umbria/ | Department of Infectious Diseases, Istituto Superiore di Sanit√†, Roma, Italy; AO S.M. della Misericordia, S.C. Microbiologia, Perugia, Italy | Virology Laboratory, Scientific Department, Army Medical Center | Paola Stefanelli et al |

| | | | | | | |
|---|---|---|---|---|---|---|
| EPI_ISL_856907 | Italy/VALL-ISS-4862/2020 | 2020-03-15 | Europe/Italy/Valle D'Aosta/ | Department of Infectious Diseases, Istituto Superiore di Sanit√†, Roma, Italy; AUSL Valle d‚ÄôAosta, Aosta, Italy | Virology Laboratory, Scientific Department, Army Medical Center | Paola Stefanelli et al |
| EPI_ISL_856916 | USA/WY-WPHL-21002795/2021 | 2021-01-07 | North America/USA/Wyoming/ | Wyoming Public Health Laboratory | Wyoming Public Health Laboratory | Noah Hull et al |
| EPI_ISL_856925 | USA/WY-WPHL-21003272/2021 | 2021-01-08 | North America/USA/Wyoming/ | Wyoming Public Health Laboratory | Wyoming Public Health Laboratory | Noah Hull et al |
| EPI_ISL_856928 | USA/WY-WPHL-21003331/2021 | 2021-01-08 | North America/USA/Wyoming/ | Wyoming Public Health Laboratory | Wyoming Public Health Laboratory | Noah Hull et al |
| EPI_ISL_856929 | USA/WY-WPHL-21003570/2021 | 2021-01-10 | North America/USA/Wyoming/ | Wyoming Public Health Laboratory | Wyoming Public Health Laboratory | Noah Hull et al |
| EPI_ISL_856939 | USA/WY-WPHL-21000581/2021 | 2021-01-04 | North America/USA/Wyoming/ | Wyoming Public Health Laboratory | Wyoming Public Health Laboratory | Noah Hull et al |
| EPI_ISL_856950 | USA/WY-WPHL-21002672/2021 | 2021-01-07 | North America/USA/Wyoming/ | Wyoming Public Health Laboratory | Wyoming Public Health Laboratory | Noah Hull et al |
| EPI_ISL_856952 | USA/WY-WPHL-21002205/2021 | 2021-01-06 | North America/USA/Wyoming/ | Wyoming Public Health Laboratory | Wyoming Public Health Laboratory | Noah Hull et al |
| EPI_ISL_856974 | USA/CA-CZB-9913/2020 | 2020-05-14 | North America/USA/California/Santa Cl | The Ashley Laboratory, Stanford University | Chan-Zuckerberg Biohub | CZB Cliahub Consortium et al |
| EPI_ISL_856987 | USA/CA-CZB-9990/2020 | 2020-06-30 | North America/USA/California/Santa Cl | The Ashley Laboratory, Stanford University | Chan-Zuckerberg Biohub | CZB Cliahub Consortium et al |
| EPI_ISL_856999 | USA/CA-CZB-9971/2020 | 2020-06-17 | North America/USA/California/Santa Cl | The Ashley Laboratory, Stanford University | Chan-Zuckerberg Biohub | CZB Cliahub Consortium et al |
| EPI_ISL_857003 | USA/CA-CZB-9947/2020 | 2020-06-16 | North America/USA/California/Santa Cl | The Ashley Laboratory, Stanford University | Chan-Zuckerberg Biohub | CZB Cliahub Consortium et al |
| EPI_ISL_857007 | USA/CA-CZB-9826/2020 | 2020-06-14 | North America/USA/California/Santa Cl | The Ashley Laboratory, Stanford University | Chan-Zuckerberg Biohub | CZB Cliahub Consortium et al |
| EPI_ISL_857012 | USA/CA-CZB-9877/2020 | 2020-06-08 | North America/USA/California/Santa Cl | The Ashley Laboratory, Stanford University | Chan-Zuckerberg Biohub | CZB Cliahub Consortium et al |
| EPI_ISL_857018 | USA/CA-CZB-9856/2020 | 2020-05-22 | North America/USA/California/Santa Cl | The Ashley Laboratory, Stanford University | Chan-Zuckerberg Biohub | CZB Cliahub Consortium et al |
| EPI_ISL_857024 | USA/CA-CZB-9909/2020 | 2020-05-22 | North America/USA/California/Santa Cl | The Ashley Laboratory, Stanford University | Chan-Zuckerberg Biohub | CZB Cliahub Consortium et al |
| EPI_ISL_857027 | USA/CA-CZB-9844/2020 | 2020-05-21 | North America/USA/California/Santa Cl | The Ashley Laboratory, Stanford University | Chan-Zuckerberg Biohub | CZB Cliahub Consortium et al |
| EPI_ISL_857030 | USA/CA-CZB-9911/2020 | 2020-05-14 | North America/USA/California/Santa Cl | The Ashley Laboratory, Stanford University | Chan-Zuckerberg Biohub | CZB Cliahub Consortium et al |

| | | | | | | |
|---|---|---|---|---|---|---|
| EPI_ISL_857033 | USA/CA-CZB-9982/2020 | 2020-05-13 | North America/USA/California/Santa Cl | The Ashley Laboratory, Stanford University | Chan-Zuckerberg Biohub | CZB Cliahub Consortium et al |
| EPI_ISL_857035 | USA/CA-CZB-9972/2020 | 2020-05-10 | North America/USA/California/Santa Cl | The Ashley Laboratory, Stanford University | Chan-Zuckerberg Biohub | CZB Cliahub Consortium et al |
| EPI_ISL_857037 | USA/CA-CZB-9936/2020 | 2020-04-22 | North America/USA/California/Santa Cl | The Ashley Laboratory, Stanford University | Chan-Zuckerberg Biohub | CZB Cliahub Consortium et al |
| EPI_ISL_857040 | Belgium/UZA-UA-3365/2020 | 2020-12-28 | Europe/Belgium/Turnhout/Geel | Platform BIS UZA/UAntwerpen, University Hospital Antwerp, Edegem, Belgium | UAntwerp, Laboratory of Medical Microbiology, Campus Drie Eiken S6.26, Universiteitsplein 1, 2610, Wilrijk, Antwerp, Belgium | Basil Britto Xavier et al |
| EPI_ISL_857043 | USA/CO-CDPHE-2100036119/2021 | 2021-01-07 | North America/USA/Colorado/ | Colorado Department of Public Health and Environment | Colorado Department of Puplic Health and Environment | Laura Bankers et al |
| EPI_ISL_857048 | USA/CO-CDPHE-2100030976/2020 | 2020-11-25 | North America/USA/Colorado/ | Colorado Department of Public Health and Environment | Colorado Department of Puplic Health and Environment | Laura Bankers et al |
| EPI_ISL_857056 | USA/NY-NYCPHL-002169/2020 | 2020-12-29 | North America/USA/New York/New Yor | OCME Office Of Chief Medical Examiner | New York City Public Health Laboratory | Jade Wang et al |
| EPI_ISL_857066 | USA/NY-NYCPHL-002195/2021 | 2021-01-04 | North America/USA/New York/New Yor | OCME Office Of Chief Medical Examiner | New York City Public Health Laboratory | Jade Wang et al |
| EPI_ISL_857067 | USA/NY-NYCPHL-002214/2021 | 2021-01-05 | North America/USA/New York/New Yor | DOHMH Corona | New York City Public Health Laboratory | Jade Wang et al |
| EPI_ISL_857071 | USA/NY-NYCPHL-002212/2021 | 2021-01-05 | North America/USA/New York/New Yor | DOHMH Morrisania | New York City Public Health Laboratory | Jade Wang et al |
| EPI_ISL_857081 | USA/NY-NYCPHL-002213/2021 | 2021-01-05 | North America/USA/New York/New Yor | DOHMH Morrisania | New York City Public Health Laboratory | Jade Wang et al |
| EPI_ISL_857085 | USA/NY-NYCPHL-002223/2021 | 2021-01-05 | North America/USA/New York/New Yor | DOHMH Central Harlem | New York City Public Health Laboratory | Jade Wang et al |

| | | | | | | |
|---|---|---|---|---|---|---|
| EPI_ISL_857089 | USA/NY-NYCPHL-002236/2021 | 2021-01-05 | North America/USA/New York/New Yor | DOHMH Crown Heights | New York City Public Health Laboratory | Jade Wang et al |
| EPI_ISL_857090 | USA/NY-NYCPHL-002255/2021 | 2021-01-05 | North America/USA/New York/New Yor | DOHMH Riverside | New York City Public Health Laboratory | Jade Wang et al |
| EPI_ISL_857093 | USA/NY-NYCPHL-002241/2021 | 2021-01-06 | North America/USA/New York/New Yor | DOHMH Morrisania | New York City Public Health Laboratory | Jade Wang et al |
| EPI_ISL_857095 | USA/NY-NYCPHL-002259/2021 | 2021-01-06 | North America/USA/New York/New Yor | DOHMH Fort Greene | New York City Public Health Laboratory | Jade Wang et al |
| EPI_ISL_857096 | USA/NY-NYCPHL-002232/2021 | 2021-01-06 | North America/USA/New York/New Yor | DOHMH Jamaica | New York City Public Health Laboratory | Jade Wang et al |
| EPI_ISL_857117 | USA/NY-NYCPHL-002304/2021 | 2021-01-07 | North America/USA/New York/New Yor | DOHMH Chelsea | New York City Public Health Laboratory | Jade Wang et al |
| EPI_ISL_857129 | USA/NY-NYCPHL-002301/2021 | 2021-01-07 | North America/USA/New York/New Yor | DOHMH PHL | New York City Public Health Laboratory | Jade Wang et al |
| EPI_ISL_857134 | USA/NY-NYCPHL-002333/2021 | 2021-01-08 | North America/USA/New York/New Yor | DOHMH PHL | New York City Public Health Laboratory | Jade Wang et al |
| EPI_ISL_857135 | USA/NY-NYCPHL-002329/2021 | 2021-01-08 | North America/USA/New York/New Yor | DOHMH Corona | New York City Public Health Laboratory | Jade Wang et al |
| EPI_ISL_857136 | USA/NY-NYCPHL-002305/2021 | 2021-01-08 | North America/USA/New York/New Yor | DOHMH Chelsea | New York City Public Health Laboratory | Jade Wang et al |
| EPI_ISL_857143 | USA/NY-NYCPHL-002314/2021 | 2021-01-08 | North America/USA/New York/New Yor | DOHMH Riverside | New York City Public Health Laboratory | Jade Wang et al |
| EPI_ISL_857147 | USA/NY-NYCPHL-002318/2021 | 2021-01-08 | North America/USA/New York/New Yor | DOHMH Corona | New York City Public Health Laboratory | Jade Wang et al |
| EPI_ISL_857149 | USA/NY-NYCPHL-002320/2021 | 2021-01-08 | North America/USA/New York/New Yor | DOHMH Jamaica | New York City Public Health Laboratory | Jade Wang et al |

| | | | | | | |
|---|---|---|---|---|---|---|
| EPI_ISL_857166 | USA/NY-NYCPHL-002339/2021 | 2021-01-11 | North America/USA/New York/New Yor | DOHMH Fort Greene | New York City Public Health Laboratory | Jade Wang et al |
| EPI_ISL_857173 | USA/NY-NYCPHL-002351/2021 | 2021-01-11 | North America/USA/New York/New Yor | DOHMH Crown Heights | New York City Public Health Laboratory | Jade Wang et al |
| EPI_ISL_857179 | USA/NY-NYCPHL-002357/2021 | 2021-01-11 | North America/USA/New York/New Yor | DOHMH Jamaica | New York City Public Health Laboratory | Jade Wang et al |
| EPI_ISL_857180 | USA/NY-NYCPHL-002358/2021 | 2021-01-11 | North America/USA/New York/New Yor | DOHMH Central Harlem | New York City Public Health Laboratory | Jade Wang et al |
| EPI_ISL_857181 | USA/NY-NYCPHL-002360/2021 | 2021-01-11 | North America/USA/New York/New Yor | DOHMH Central Harlem | New York City Public Health Laboratory | Jade Wang et al |
| EPI_ISL_857187 | USA/NY-NYCPHL-001985/2020 | 2020-12-28 | North America/USA/New York/New Yor | DOHMH PHL | New York City Public Health Laboratory | Jade Wang et al |
| EPI_ISL_857195 | USA/NY-NYCPHL-002108/2020 | 2020-12-31 | North America/USA/New York/New Yor | DOHMH Riverside | New York City Public Health Laboratory | Jade Wang et al |
| EPI_ISL_857200 | USA/NY-NYCPHL-002130/2021 | 2021-01-04 | North America/USA/New York/New Yor | DOHMH Central Harlem | New York City Public Health Laboratory | Jade Wang et al |
| EPI_ISL_857206 | USA/NY-NYCPHL-002124/2021 | 2021-01-04 | North America/USA/New York/New Yor | DOHMH PHL | New York City Public Health Laboratory | Jade Wang et al |
| EPI_ISL_857217 | USA/NY-NYCPHL-002140/2021 | 2021-01-04 | North America/USA/New York/New Yor | DOHMH Jamaica | New York City Public Health Laboratory | Jade Wang et al |
| EPI_ISL_857222 | USA/NY-NYCPHL-002146/2021 | 2021-01-04 | North America/USA/New York/New Yor | DOHMH Jamaica | New York City Public Health Laboratory | Jade Wang et al |
| EPI_ISL_857228 | USA/NY-NYCPHL-002109/2020 | 2020-12-31 | North America/USA/New York/New Yor | DOHMH Riverside | New York City Public Health Laboratory | Jade Wang et al |
| EPI_ISL_857233 | USA/NY-NYCPHL-002117/2020 | 2020-12-31 | North America/USA/New York/New Yor | DOHMH Central Harlem | New York City Public Health Laboratory | Jade Wang et al |

| | | | | | | |
|---|---|---|---|---|---|---|
| EPI_ISL_857241 | USA/NY-NYCPHL-002024/2020 | 2020-12-30 | North America/USA/New York/New Yor | DOHMH Jamaica | New York City Public Health Laboratory | Jade Wang et al |
| EPI_ISL_857247 | USA/NY-NYCPHL-002030/2020 | 2020-12-30 | North America/USA/New York/New Yor | DOHMH Jamaica | New York City Public Health Laboratory | Jade Wang et al |
| EPI_ISL_857270 | USA/NY-NYCPHL-002010/2020 | 2020-12-29 | North America/USA/New York/New Yor | DOHMH Jamaica | New York City Public Health Laboratory | Jade Wang et al |
| EPI_ISL_857273 | USA/NY-NYCPHL-002014/2020 | 2020-12-29 | North America/USA/New York/New Yor | DOHMH Morrisania | New York City Public Health Laboratory | Jade Wang et al |
| EPI_ISL_857277 | USA/NY-NYCPHL-002019/2020 | 2020-12-29 | North America/USA/New York/New Yor | DOHMH Corona | New York City Public Health Laboratory | Jade Wang et al |
| EPI_ISL_857278 | USA/NY-NYCPHL-002041/2020 | 2020-12-29 | North America/USA/New York/New Yor | DOHMH PHL | New York City Public Health Laboratory | Jade Wang et al |
| EPI_ISL_857281 | USA/NY-NYCPHL-001981/2020 | 2020-12-28 | North America/USA/New York/New Yor | DOHMH Riverside | New York City Public Health Laboratory | Jade Wang et al |
| EPI_ISL_857282 | USA/NY-NYCPHL-001982/2020 | 2020-12-28 | North America/USA/New York/New Yor | DOHMH Morrisania | New York City Public Health Laboratory | Jade Wang et al |
| EPI_ISL_857322 | Egypt/CUNCI-HGC8I040/2020 | 2020-12-25 | Africa/Egypt/Egypt/ | Cancer Biology Department, National Cancer Institute | Cancer Biology Department, National Cancer Institute | Zekri et al |
| EPI_ISL_857349 | USA/TX-DSHS-2133/2021 | 2021-01-07 | North America/USA/Texas/Dallas Count | Texas Department of State Health Services (TXDSHS) | Texas Department of State Health Services (TXDSHS) | Anita Pokharel et al |
| EPI_ISL_857351 | USA/TX-DSHS-2135/2021 | 2021-01-07 | North America/USA/Texas/Denton Coun | Texas Department of State Health Services (TXDSHS) | Texas Department of State Health Services (TXDSHS) | Anita Pokharel et al |
| EPI_ISL_857352 | USA/TX-DSHS-2136/2021 | 2021-01-07 | North America/USA/Texas/Tarrant Coun | Texas Department of State Health Services (TXDSHS) | Texas Department of State Health Services (TXDSHS) | Anita Pokharel et al |
| EPI_ISL_857356 | USA/TX-DSHS-2141/2021 | 2021-01-05 | North America/USA/Texas/Collin County | Texas Department of State Health Services (TXDSHS) | Texas Department of State Health Services (TXDSHS) | Anita Pokharel et al |

| | | | | | | |
|---|---|---|---|---|---|---|
| EPI_ISL_857363 | USA/TX-DSHS-2148/2021 | 2021-01-07 | North America/USA/Texas/Dallas Count | Texas Department of State Health Services (TXDSHS) | Texas Department of State Health Services (TXDSHS) | Anita Pokharel et al |
| EPI_ISL_857367 | USA/TX-DSHS-2152/2020 | 2020-10-04 | North America/USA/Texas/Jim Hogg Co | Texas Department of State Health Services (TXDSHS) | Texas Department of State Health Services (TXDSHS) | Anita Pokharel et al |
| EPI_ISL_857385 | USA/TX-DSHS-2170/2020 | 2020-10-06 | North America/USA/Texas/Terry County | Texas Department of State Health Services (TXDSHS) | Texas Department of State Health Services (TXDSHS) | Anita Pokharel et al |
| EPI_ISL_857388 | USA/TX-DSHS-2173/2020 | 2020-10-06 | North America/USA/Texas/Terry County | Texas Department of State Health Services (TXDSHS) | Texas Department of State Health Services (TXDSHS) | Anita Pokharel et al |
| EPI_ISL_857394 | USA/TX-DSHS-2179/2021 | 2021-01-08 | North America/USA/Texas/ | Texas Department of State Health Services (TXDSHS) | Texas Department of State Health Services (TXDSHS) | Anita Pokharel et al |
| EPI_ISL_857395 | India/un-SMSMC-791502/2021 | 2021-01-11 | Asia/India/India/ | S.M.S.Medical College, Jaipur, Rajasthan | S.M.S.Medical College, Jaipur, Rajasthan | Dr. Bharti Malhotra et al |
| EPI_ISL_857400 | USA/ME-HETL-J0776/2021 | 2021-01-14 | North America/USA/Maine/ | Maine HETL | Tewhey Lab, The Jackson Laboratory | Matluk et al |
| EPI_ISL_857403 | USA/ME-HETL-J0780/2021 | 2021-01-08 | North America/USA/Maine/ | Maine HETL | Tewhey Lab, The Jackson Laboratory | Matluk et al |
| EPI_ISL_857426 | USA/ME-HETL-J0810/2021 | 2021-01-14 | North America/USA/Maine/ | Maine HETL | Tewhey Lab, The Jackson Laboratory | Matluk et al |
| EPI_ISL_857432 | USA/ME-HETL-J0817/2021 | 2021-01-12 | North America/USA/Maine/ | Maine HETL | Tewhey Lab, The Jackson Laboratory | Matluk et al |
| EPI_ISL_857434 | USA/ME-HETL-J0819/2021 | 2021-01-12 | North America/USA/Maine/ | Maine HETL | Tewhey Lab, The Jackson Laboratory | Matluk et al |
| EPI_ISL_857436 | USA/ME-HETL-J0822/2021 | 2021-01-12 | North America/USA/Maine/ | Maine HETL | Tewhey Lab, The Jackson Laboratory | Matluk et al |
| EPI_ISL_857438 | USA/ME-HETL-J0825/2021 | 2021-01-12 | North America/USA/Maine/ | Maine HETL | Tewhey Lab, The Jackson Laboratory | Matluk et al |

| | | | | | | |
|---|---|---|---|---|---|---|
| EPI_ISL_857439 | USA/ME-HETL-J0826/2021 | 2021-01-12 | North America/USA/Maine/ | Maine HETL | Tewhey Lab, The Jackson Laboratory | Matluk et al |
| EPI_ISL_857444 | USA/ME-HETL-J0831/2021 | 2021-01-11 | North America/USA/Maine/ | Maine HETL | Tewhey Lab, The Jackson Laboratory | Matluk et al |
| EPI_ISL_857457 | USA/ME-HETL-J0847/2021 | 2021-01-08 | North America/USA/Maine/ | Maine HETL | Tewhey Lab, The Jackson Laboratory | Matluk et al |
| EPI_ISL_857476 | Singapore/68/2021 | 2021-01-17 | Asia/Singapore/Singapore/ | National Public Health Laboratory, National Centre for Infectious Diseases | National Public Health Laboratory, National Centre for Infectious Diseases | Tze Minn Mak et al |
| EPI_ISL_857492 | Switzerland/BE-SNRCI-8047/2020 | 2020-11-10 | Europe/Switzerland/Bern/ | Swiss National Reference Centre for Influenza | Swiss National Reference Centre for Influenza | Ana Rita Goncalves et al |
| EPI_ISL_857493 | Switzerland/VS-SNRCI-942/2020 | 2020-11-10 | Europe/Switzerland/Valais/ | Swiss National Reference Centre for Influenza | Swiss National Reference Centre for Influenza | Ana Rita Goncalves et al |
| EPI_ISL_857538 | Switzerland/NE-SNRCI-9837/2020 | 2020-12-14 | Europe/Switzerland/Neuch√¢tel/ | Swiss National Reference Centre for Influenza | Swiss National Reference Centre for Influenza | Tim Roloff et al |
| EPI_ISL_857583 | England/CAMC-FB4145/2021 | 2021-01-08 | Europe/United Kingdom/England/ | Lighthouse Lab in Cambridge | Wellcome Sanger Institute for the COVID-19 Genomics UK (COG-UK) Consortium | Rob Howes et al |
| EPI_ISL_858211 | Wales/ALDP-EF11D4/2021 | 2021-01-03 | Europe/United Kingdom/Wales/ | Lighthouse Lab in Alderley Park | Wellcome Sanger Institute for the COVID-19 Genomics UK (COG-UK) Consortium | Jacquelyn Wynn et al |
| EPI_ISL_858216 | Wales/ALDP-EE7A7C/2021 | 2021-01-04 | Europe/United Kingdom/Wales/ | Lighthouse Lab in Alderley Park | Wellcome Sanger Institute for the COVID-19 Genomics UK (COG-UK) Consortium | Jacquelyn Wynn et al |

| | | | | | | |
|---|---|---|---|---|---|---|
| EPI_ISL_858543 | Scotland/QEUH-E50197/2021 | 2021-01-04 | Europe/United Kingdom/Scotland/ | Lighthouse Lab in Glasgow | Wellcome Sanger Institute for the COVID-19 Genomics UK (COG-UK) Consortium | Harper VanSteenhouse et al |
| EPI_ISL_858660 | Scotland/QEUH-DD2F58/2021 | 2021-01-02 | Europe/United Kingdom/Scotland/ | Lighthouse Lab in Glasgow | Wellcome Sanger Institute for the COVID-19 Genomics UK (COG-UK) Consortium | Harper VanSteenhouse et al |
| EPI_ISL_858742 | Scotland/QEUH-D53A03/2020 | 2020-12-29 | Europe/United Kingdom/Scotland/ | Lighthouse Lab in Glasgow | Wellcome Sanger Institute for the COVID-19 Genomics UK (COG-UK) Consortium | Harper VanSteenhouse et al |
| EPI_ISL_858945 | Scotland/QEUH-CC9359/2020 | 2020-12-22 | Europe/United Kingdom/Scotland/ | Lighthouse Lab in Glasgow | Wellcome Sanger Institute for the COVID-19 Genomics UK (COG-UK) Consortium | Harper VanSteenhouse et al |
| EPI_ISL_859029 | Wales/ALDP-CB3CDF/2020 | 2020-12-19 | Europe/United Kingdom/Wales/ | Lighthouse Lab in Alderley Park | Wellcome Sanger Institute for the COVID-19 Genomics UK (COG-UK) Consortium | Jacquelyn Wynn et al |
| EPI_ISL_859180 | Scotland/QEUH-CC7546/2020 | 2020-12-22 | Europe/United Kingdom/Scotland/ | Lighthouse Lab in Glasgow | Wellcome Sanger Institute for the COVID-19 Genomics UK (COG-UK) Consortium | Harper VanSteenhouse et al |
| EPI_ISL_859401 | Wales/ALDP-CB52BD/2020 | 2020-12-14 | Europe/United Kingdom/Wales/ | Lighthouse Lab in Alderley Park | Wellcome Sanger Institute for the COVID-19 Genomics UK (COG-UK) Consortium | Jacquelyn Wynn et al |
| EPI_ISL_859571 | UnitedArabEmirates/1385/2020 | 2020-08-28 | Asia/United Arab Emirates/Abu Dhabi/ | BTC, Khalifa University | BTC, Khalifa University | Al Safar et al et al |
| EPI_ISL_859576 | UnitedArabEmirates/1394/2020 | 2020-08-29 | Asia/United Arab Emirates/Abu Dhabi/ | BTC, Khalifa University | BTC, Khalifa University | Al Safar et al et al |

| | | | | | | |
|---|---|---|---|---|---|---|
| EPI_ISL_859579 | UnitedArabEmirates/1778/2020 | 2020-07-05 | Asia/United Arab Emirates/Abu Dhabi/ | BTC, Khalifa University | BTC, Khalifa University | Al Safar et al et al |
| EPI_ISL_859580 | UnitedArabEmirates/1779/2020 | 2020-07-05 | Asia/United Arab Emirates/Abu Dhabi/ | BTC, Khalifa University | BTC, Khalifa University | Al Safar et al et al |
| EPI_ISL_859581 | UnitedArabEmirates/1780/2020 | 2020-07-05 | Asia/United Arab Emirates/Abu Dhabi/ | BTC, Khalifa University | BTC, Khalifa University | Al Safar et al et al |
| EPI_ISL_859582 | UnitedArabEmirates/1789/2020 | 2020-07-05 | Asia/United Arab Emirates/Abu Dhabi/ | BTC, Khalifa University | BTC, Khalifa University | Al Safar et al et al |
| EPI_ISL_859587 | UnitedArabEmirates/1799/2020 | 2020-06-24 | Asia/United Arab Emirates/Abu Dhabi/ | BTC, Khalifa University | BTC, Khalifa University | Al Safar et al et al |
| EPI_ISL_859600 | UnitedArabEmirates/1817/2020 | 2020-06-28 | Asia/United Arab Emirates/Abu Dhabi/ | BTC, Khalifa University | BTC, Khalifa University | Al Safar et al et al |
| EPI_ISL_859607 | UnitedArabEmirates/1835/2020 | 2020-08-27 | Asia/United Arab Emirates/Abu Dhabi/ | BTC, Khalifa University | BTC, Khalifa University | Al Safar et al et al |
| EPI_ISL_859611 | UnitedArabEmirates/1840/2020 | 2020-06-24 | Asia/United Arab Emirates/Abu Dhabi/ | BTC, Khalifa University | BTC, Khalifa University | Al Safar et al et al |
| EPI_ISL_859618 | UnitedArabEmirates/1848/2020 | 2020-06-24 | Asia/United Arab Emirates/Abu Dhabi/ | BTC, Khalifa University | BTC, Khalifa University | Al Safar et al et al |
| EPI_ISL_859622 | UnitedArabEmirates/1855/2020 | 2020-08-27 | Asia/United Arab Emirates/Abu Dhabi/ | BTC, Khalifa University | BTC, Khalifa University | Al Safar et al et al |
| EPI_ISL_859623 | UnitedArabEmirates/1862/2020 | 2020-07-05 | Asia/United Arab Emirates/Abu Dhabi/ | BTC, Khalifa University | BTC, Khalifa University | Al Safar et al et al |
| EPI_ISL_859629 | UnitedArabEmirates/1871/2020 | 2020-07-05 | Asia/United Arab Emirates/Abu Dhabi/ | BTC, Khalifa University | BTC, Khalifa University | Al Safar et al et al |
| EPI_ISL_859643 | UnitedArabEmirates/1985/2020 | 2020-07-05 | Asia/United Arab Emirates/Abu Dhabi/ | BTC, Khalifa University | BTC, Khalifa University | Al Safar et al et al |
| EPI_ISL_859644 | UnitedArabEmirates/1991/2020 | 2020-07-05 | Asia/United Arab Emirates/Abu Dhabi/ | BTC, Khalifa University | BTC, Khalifa University | Al Safar et al et al |
| EPI_ISL_859652 | UnitedArabEmirates/2028/2020 | 2020-07-06 | Asia/United Arab Emirates/Abu Dhabi/ | BTC, Khalifa University | BTC, Khalifa University | Al Safar et al et al |
| EPI_ISL_859654 | UnitedArabEmirates/2084/2020 | 2020-07-06 | Asia/United Arab Emirates/Abu Dhabi/ | BTC, Khalifa University | BTC, Khalifa University | Al Safar et al et al |
| EPI_ISL_859662 | UnitedArabEmirates/2571/2020 | 2020-08-26 | Asia/United Arab Emirates/Abu Dhabi/ | BTC, Khalifa University | BTC, Khalifa University | Al Safar et al et al |
| EPI_ISL_859666 | UnitedArabEmirates/2607/2020 | 2020-08-26 | Asia/United Arab Emirates/Abu Dhabi/ | BTC, Khalifa University | BTC, Khalifa University | Al Safar et al et al |
| EPI_ISL_859667 | UnitedArabEmirates/2610/2020 | 2020-08-26 | Asia/United Arab Emirates/Abu Dhabi/ | BTC, Khalifa University | BTC, Khalifa University | Al Safar et al et al |
| EPI_ISL_859671 | UnitedArabEmirates/2869/2020 | 2020-08-24 | Asia/United Arab Emirates/Abu Dhabi/ | BTC, Khalifa University | BTC, Khalifa University | Al Safar et al et al |

| | | | | | | |
|---|---|---|---|---|---|---|
| EPI_ISL_859673 | UnitedArabEmirates/2876/2020 | 2020-08-24 | Asia/United Arab Emirates/Abu Dhabi/ | BTC, Khalifa University | BTC, Khalifa University | Al Safar et al et al |
| EPI_ISL_859685 | UnitedArabEmirates/3150/2020 | 2020-09-29 | Asia/United Arab Emirates/Abu Dhabi/ | BTC, Khalifa University | BTC, Khalifa University | Al Safar et al et al |
| EPI_ISL_859686 | UnitedArabEmirates/3152/2020 | 2020-09-29 | Asia/United Arab Emirates/Abu Dhabi/ | BTC, Khalifa University | BTC, Khalifa University | Al Safar et al et al |
| EPI_ISL_859689 | UnitedArabEmirates/3157/2020 | 2020-09-20 | Asia/United Arab Emirates/Abu Dhabi/ | BTC, Khalifa University | BTC, Khalifa University | Al Safar et al et al |
| EPI_ISL_859694 | UnitedArabEmirates/3201/2020 | 2020-09-08 | Asia/United Arab Emirates/Abu Dhabi/ | BTC, Khalifa University | BTC, Khalifa University | Al Safar et al et al |
| EPI_ISL_859696 | UnitedArabEmirates/3215/2020 | 2020-09-15 | Asia/United Arab Emirates/Abu Dhabi/ | BTC, Khalifa University | BTC, Khalifa University | Al Safar et al et al |
| EPI_ISL_859703 | UnitedArabEmirates/3230/2020 | 2020-09-29 | Asia/United Arab Emirates/Abu Dhabi/ | BTC, Khalifa University | BTC, Khalifa University | Al Safar et al et al |
| EPI_ISL_859708 | UnitedArabEmirates/3241/2020 | 2020-09-29 | Asia/United Arab Emirates/Abu Dhabi/ | BTC, Khalifa University | BTC, Khalifa University | Al Safar et al et al |
| EPI_ISL_859709 | UnitedArabEmirates/3242/2020 | 2020-09-29 | Asia/United Arab Emirates/Abu Dhabi/ | BTC, Khalifa University | BTC, Khalifa University | Al Safar et al et al |
| EPI_ISL_859714 | UnitedArabEmirates/3249/2020 | 2020-09-29 | Asia/United Arab Emirates/Abu Dhabi/ | BTC, Khalifa University | BTC, Khalifa University | Al Safar et al et al |
| EPI_ISL_859718 | UnitedArabEmirates/3256/2020 | 2020-09-29 | Asia/United Arab Emirates/Abu Dhabi/ | BTC, Khalifa University | BTC, Khalifa University | Al Safar et al et al |
| EPI_ISL_859720 | UnitedArabEmirates/3268/2020 | 2020-09-29 | Asia/United Arab Emirates/Abu Dhabi/ | BTC, Khalifa University | BTC, Khalifa University | Al Safar et al et al |
| EPI_ISL_859732 | UnitedArabEmirates/3314/2020 | 2020-09-29 | Asia/United Arab Emirates/Abu Dhabi/ | BTC, Khalifa University | BTC, Khalifa University | Al Safar et al et al |
| EPI_ISL_859736 | UnitedArabEmirates/3320/2020 | 2020-09-29 | Asia/United Arab Emirates/Abu Dhabi/ | BTC, Khalifa University | BTC, Khalifa University | Al Safar et al et al |
| EPI_ISL_859740 | UnitedArabEmirates/3328/2020 | 2020-09-29 | Asia/United Arab Emirates/Abu Dhabi/ | BTC, Khalifa University | BTC, Khalifa University | Al Safar et al et al |
| EPI_ISL_859742 | UnitedArabEmirates/3345/2020 | 2020-09-29 | Asia/United Arab Emirates/Abu Dhabi/ | BTC, Khalifa University | BTC, Khalifa University | Al Safar et al et al |
| EPI_ISL_859743 | UnitedArabEmirates/3347/2020 | 2020-09-29 | Asia/United Arab Emirates/Abu Dhabi/ | BTC, Khalifa University | BTC, Khalifa University | Al Safar et al et al |
| EPI_ISL_859746 | UnitedArabEmirates/3352/2020 | 2020-09-29 | Asia/United Arab Emirates/Abu Dhabi/ | BTC, Khalifa University | BTC, Khalifa University | Al Safar et al et al |
| EPI_ISL_859748 | UnitedArabEmirates/3354/2020 | 2020-09-29 | Asia/United Arab Emirates/Abu Dhabi/ | BTC, Khalifa University | BTC, Khalifa University | Al Safar et al et al |
| EPI_ISL_859756 | UnitedArabEmirates/3368/2020 | 2020-09-16 | Asia/United Arab Emirates/Abu Dhabi/ | BTC, Khalifa University | BTC, Khalifa University | Al Safar et al et al |

| | | | | | | |
|---|---|---|---|---|---|---|
| EPI_ISL_859757 | UnitedArabEmirates/3370/2020 | 2020-09-16 | Asia/United Arab Emirates/Abu Dhabi/ | BTC, Khalifa University | BTC, Khalifa University | Al Safar et al et al |
| EPI_ISL_859767 | UnitedArabEmirates/3408/2020 | 2020-09-06 | Asia/United Arab Emirates/Abu Dhabi/ | BTC, Khalifa University | BTC, Khalifa University | Al Safar et al et al |
| EPI_ISL_859769 | UnitedArabEmirates/3532/2020 | 2020-10-20 | Asia/United Arab Emirates/Abu Dhabi/ | BTC, Khalifa University | BTC, Khalifa University | Al Safar et al et al |
| EPI_ISL_859771 | UnitedArabEmirates/3537/2020 | 2020-10-20 | Asia/United Arab Emirates/Abu Dhabi/ | BTC, Khalifa University | BTC, Khalifa University | Al Safar et al et al |
| EPI_ISL_859772 | UnitedArabEmirates/3539/2020 | 2020-10-20 | Asia/United Arab Emirates/Abu Dhabi/ | BTC, Khalifa University | BTC, Khalifa University | Al Safar et al et al |
| EPI_ISL_859777 | UnitedArabEmirates/3545/2020 | 2020-10-20 | Asia/United Arab Emirates/Abu Dhabi/ | BTC, Khalifa University | BTC, Khalifa University | Al Safar et al et al |
| EPI_ISL_859778 | UnitedArabEmirates/3549/2020 | 2020-10-26 | Asia/United Arab Emirates/Abu Dhabi/ | BTC, Khalifa University | BTC, Khalifa University | Al Safar et al et al |
| EPI_ISL_859782 | UnitedArabEmirates/3558/2020 | 2020-10-21 | Asia/United Arab Emirates/Abu Dhabi/ | BTC, Khalifa University | BTC, Khalifa University | Al Safar et al et al |
| EPI_ISL_859788 | UnitedArabEmirates/3613/2020 | 2020-10-27 | Asia/United Arab Emirates/Abu Dhabi/ | BTC, Khalifa University | BTC, Khalifa University | Al Safar et al et al |
| EPI_ISL_859796 | UnitedArabEmirates/3636/2020 | 2020-10-11 | Asia/United Arab Emirates/Abu Dhabi/ | BTC, Khalifa University | BTC, Khalifa University | Al Safar et al et al |
| EPI_ISL_859799 | UnitedArabEmirates/3655/2020 | 2020-10-28 | Asia/United Arab Emirates/Abu Dhabi/ | BTC, Khalifa University | BTC, Khalifa University | Al Safar et al et al |
| EPI_ISL_859800 | UnitedArabEmirates/3659/2020 | 2020-10-30 | Asia/United Arab Emirates/Abu Dhabi/ | BTC, Khalifa University | BTC, Khalifa University | Al Safar et al et al |
| EPI_ISL_859801 | UnitedArabEmirates/3660/2020 | 2020-10-27 | Asia/United Arab Emirates/Abu Dhabi/ | BTC, Khalifa University | BTC, Khalifa University | Al Safar et al et al |
| EPI_ISL_859802 | UnitedArabEmirates/3661/2020 | 2020-10-27 | Asia/United Arab Emirates/Abu Dhabi/ | BTC, Khalifa University | BTC, Khalifa University | Al Safar et al et al |
| EPI_ISL_859806 | UnitedArabEmirates/3677/2020 | 2020-10-27 | Asia/United Arab Emirates/Abu Dhabi/ | BTC, Khalifa University | BTC, Khalifa University | Al Safar et al et al |
| EPI_ISL_859815 | UnitedArabEmirates/3704/2020 | 2020-10-22 | Asia/United Arab Emirates/Abu Dhabi/ | BTC, Khalifa University | BTC, Khalifa University | Al Safar et al et al |
| EPI_ISL_859822 | UnitedArabEmirates/3719/2020 | 2020-10-19 | Asia/United Arab Emirates/Abu Dhabi/ | BTC, Khalifa University | BTC, Khalifa University | Al Safar et al et al |
| EPI_ISL_859824 | UnitedArabEmirates/3724/2020 | 2020-10-19 | Asia/United Arab Emirates/Abu Dhabi/ | BTC, Khalifa University | BTC, Khalifa University | Al Safar et al et al |
| EPI_ISL_859837 | UnitedArabEmirates/3755/2020 | 2020-10-19 | Asia/United Arab Emirates/Abu Dhabi/ | BTC, Khalifa University | BTC, Khalifa University | Al Safar et al et al |
| EPI_ISL_859850 | UnitedArabEmirates/3799/2020 | 2020-11-16 | Asia/United Arab Emirates/Abu Dhabi/ | BTC, Khalifa University | BTC, Khalifa University | Al Safar et al et al |

| | | | | | | |
|---|---|---|---|---|---|---|
| EPI_ISL_859861 | UnitedArabEmirates/3834/2020 | 2020-11-22 | Asia/United Arab Emirates/Abu Dhabi/ | BTC, Khalifa University | BTC, Khalifa University | Al Safar et al et al |
| EPI_ISL_859863 | UnitedArabEmirates/3837/2020 | 2020-11-22 | Asia/United Arab Emirates/Abu Dhabi/ | BTC, Khalifa University | BTC, Khalifa University | Al Safar et al et al |
| EPI_ISL_859867 | UnitedArabEmirates/3846/2020 | 2020-11-18 | Asia/United Arab Emirates/Abu Dhabi/ | BTC, Khalifa University | BTC, Khalifa University | Al Safar et al et al |
| EPI_ISL_859890 | UnitedArabEmirates/3885/2020 | 2020-11-15 | Asia/United Arab Emirates/Abu Dhabi/ | BTC, Khalifa University | BTC, Khalifa University | Al Safar et al et al |
| EPI_ISL_859893 | UnitedArabEmirates/3896/2020 | 2020-10-13 | Asia/United Arab Emirates/Abu Dhabi/ | BTC, Khalifa University | BTC, Khalifa University | Al Safar et al et al |
| EPI_ISL_859895 | UnitedArabEmirates/3908/2020 | 2020-11-26 | Asia/United Arab Emirates/Abu Dhabi/ | BTC, Khalifa University | BTC, Khalifa University | Al Safar et al et al |
| EPI_ISL_859919 | UnitedArabEmirates/3974/2020 | 2020-11-24 | Asia/United Arab Emirates/Abu Dhabi/ | BTC, Khalifa University | BTC, Khalifa University | Al Safar et al et al |
| EPI_ISL_859923 | UnitedArabEmirates/3985/2020 | 2020-11-21 | Asia/United Arab Emirates/Abu Dhabi/ | BTC, Khalifa University | BTC, Khalifa University | Al Safar et al et al |
| EPI_ISL_859924 | UnitedArabEmirates/3986/2020 | 2020-11-19 | Asia/United Arab Emirates/Abu Dhabi/ | BTC, Khalifa University | BTC, Khalifa University | Al Safar et al et al |
| EPI_ISL_859926 | UnitedArabEmirates/3993/2020 | 2020-11-17 | Asia/United Arab Emirates/Abu Dhabi/ | BTC, Khalifa University | BTC, Khalifa University | Al Safar et al et al |
| EPI_ISL_859928 | UnitedArabEmirates/4013/2020 | 2020-11-17 | Asia/United Arab Emirates/Abu Dhabi/ | BTC, Khalifa University | BTC, Khalifa University | Al Safar et al et al |
| EPI_ISL_859929 | UnitedArabEmirates/4014/2020 | 2020-11-24 | Asia/United Arab Emirates/Abu Dhabi/ | BTC, Khalifa University | BTC, Khalifa University | Al Safar et al et al |
| EPI_ISL_859946 | UnitedArabEmirates/4165/2020 | 2020-05-01 | Asia/United Arab Emirates/Abu Dhabi/ | BTC, Khalifa University | BTC, Khalifa University | Al Safar et al et al |
| EPI_ISL_859987 | UnitedArabEmirates/4207/2020 | 2020-05-06 | Asia/United Arab Emirates/Abu Dhabi/ | BTC, Khalifa University | BTC, Khalifa University | Al Safar et al et al |
| EPI_ISL_859997 | UnitedArabEmirates/4218/2020 | 2020-05-04 | Asia/United Arab Emirates/Abu Dhabi/ | BTC, Khalifa University | BTC, Khalifa University | Al Safar et al et al |
| EPI_ISL_860018 | UnitedArabEmirates/4240/2020 | 2020-05-04 | Asia/United Arab Emirates/Abu Dhabi/ | BTC, Khalifa University | BTC, Khalifa University | Al Safar et al et al |
| EPI_ISL_860029 | UnitedArabEmirates/4255/2020 | 2020-12-29 | Asia/United Arab Emirates/Abu Dhabi/ | BTC, Khalifa University | BTC, Khalifa University | Al Safar et al et al |
| EPI_ISL_860030 | UnitedArabEmirates/4256/2020 | 2020-12-29 | Asia/United Arab Emirates/Abu Dhabi/ | BTC, Khalifa University | BTC, Khalifa University | Al Safar et al et al |
| EPI_ISL_860033 | UnitedArabEmirates/4261/2020 | 2020-12-29 | Asia/United Arab Emirates/Abu Dhabi/ | BTC, Khalifa University | BTC, Khalifa University | Al Safar et al et al |
| EPI_ISL_860036 | UnitedArabEmirates/4266/2020 | 2020-12-29 | Asia/United Arab Emirates/Abu Dhabi/ | BTC, Khalifa University | BTC, Khalifa University | Al Safar et al et al |

| | | | | | | |
|---|---|---|---|---|---|---|
| EPI_ISL_860060 | UnitedArabEmirates/4298/2020 | 2020-12-26 | Asia/United Arab Emirates/Abu Dhabi/ | BTC, Khalifa University | BTC, Khalifa University | Al Safar et al et al |
| EPI_ISL_860061 | UnitedArabEmirates/4300/2020 | 2020-12-26 | Asia/United Arab Emirates/Abu Dhabi/ | BTC, Khalifa University | BTC, Khalifa University | Al Safar et al et al |
| EPI_ISL_860062 | UnitedArabEmirates/4301/2020 | 2020-12-26 | Asia/United Arab Emirates/Abu Dhabi/ | BTC, Khalifa University | BTC, Khalifa University | Al Safar et al et al |
| EPI_ISL_860066 | UnitedArabEmirates/4306/2020 | 2020-12-27 | Asia/United Arab Emirates/Abu Dhabi/ | BTC, Khalifa University | BTC, Khalifa University | Al Safar et al et al |
| EPI_ISL_860067 | UnitedArabEmirates/4307/2020 | 2020-12-27 | Asia/United Arab Emirates/Abu Dhabi/ | BTC, Khalifa University | BTC, Khalifa University | Al Safar et al et al |
| EPI_ISL_860068 | UnitedArabEmirates/4308/2020 | 2020-12-27 | Asia/United Arab Emirates/Abu Dhabi/ | BTC, Khalifa University | BTC, Khalifa University | Al Safar et al et al |
| EPI_ISL_860081 | UnitedArabEmirates/4343/2020 | 2020-12-26 | Asia/United Arab Emirates/Abu Dhabi/ | BTC, Khalifa University | BTC, Khalifa University | Al Safar et al et al |
| EPI_ISL_860097 | Japan/IC-0720/2021 | 21-Jan | Asia/Japan/Japan/ | Pathogen Genomics Center, National Institute of Infectious Diseases | Pathogen Genomics Center, National Institute of Infectious Diseases | Tsuyoshi Sekizuka et al |
| EPI_ISL_860098 | Japan/IC-0721/2021 | 21-Jan | Asia/Japan/Japan/ | Pathogen Genomics Center, National Institute of Infectious Diseases | Pathogen Genomics Center, National Institute of Infectious Diseases | Tsuyoshi Sekizuka et al |
| EPI_ISL_860106 | Japan/IC-0729/2021 | 21-Jan | Asia/Japan/Japan/ | Pathogen Genomics Center, National Institute of Infectious Diseases | Pathogen Genomics Center, National Institute of Infectious Diseases | Tsuyoshi Sekizuka et al |
| EPI_ISL_860107 | Japan/IC-0730/2021 | 21-Jan | Asia/Japan/Japan/ | Pathogen Genomics Center, National Institute of Infectious Diseases | Pathogen Genomics Center, National Institute of Infectious Diseases | Tsuyoshi Sekizuka et al |
| EPI_ISL_860108 | Japan/IC-0731/2021 | 21-Jan | Asia/Japan/Japan/ | Pathogen Genomics Center, National Institute of Infectious Diseases | Pathogen Genomics Center, National Institute of Infectious Diseases | Tsuyoshi Sekizuka et al |

| | | | | | | |
|---|---|---|---|---|---|---|
| EPI_ISL_860109 | Japan/IC-0732/2021 | 21-Jan | Asia/Japan/Japan/ | Pathogen Genomics Center, National Institute of Infectious Diseases | Pathogen Genomics Center, National Institute of Infectious Diseases | Tsuyoshi Sekizuka et al |
| EPI_ISL_860113 | Japan/IC-0736/2021 | 21-Jan | Asia/Japan/Japan/ | Pathogen Genomics Center, National Institute of Infectious Diseases | Pathogen Genomics Center, National Institute of Infectious Diseases | Tsuyoshi Sekizuka et al |
| EPI_ISL_860140 | Japan/Donner115/2020 | 2020-09-16 | Asia/Japan/Kanto/ | Keio University School of Medicine | Keio University School of Medicine | Kenjiro Kosaki et al |
| EPI_ISL_860161 | Japan/Donner141/2020 | 2020-11-22 | Asia/Japan/Kanto/ | Keio University School of Medicine | Keio University School of Medicine | Kenjiro Kosaki et al |
| EPI_ISL_860166 | Japan/Donner146/2020 | 2020-11-27 | Asia/Japan/Kanto/ | Keio University School of Medicine | Keio University School of Medicine | Kenjiro Kosaki et al |
| EPI_ISL_860183 | India/KA-NIMH-SEQ-62/2020 | 2020-12-31 | Asia/India/Karnataka/ | Bangalore Medical College and Research Institute | Department of Neurovirology, National Institute of Mental Health and Neurosciences (NIMHANS) | Chitra Pattabiraman et al |
| EPI_ISL_860184 | India/KA-NIMH-SEQ-63/2021 | 2021-01-02 | Asia/India/Karnataka/ | Bangalore Medical College and Research Institute | Department of Neurovirology, National Institute of Mental Health and Neurosciences (NIMHANS) | Chitra Pattabiraman et al |
| EPI_ISL_860185 | India/KA-NIMH-SEQ-64/2021 | 2021-01-02 | Asia/India/Karnataka/ | Bangalore Medical College and Research Institute | Department of Neurovirology, National Institute of Mental Health and Neurosciences (NIMHANS) | Chitra Pattabiraman et al |

| | | | | | | |
|---|---|---|---|---|---|---|
| EPI_ISL_860186 | India/KA-NIMH-SEQ-65/2021 | 2021-01-04 | Asia/India/Karnataka/ | Bangalore Medical College and Research Institute | Department of Neurovirology, National Institute of Mental Health and Neurosciences (NIMHANS) | Chitra Pattabiraman et al |
| EPI_ISL_860187 | India/KA-NIMH-SEQ-67/2021 | 2021-01-03 | Asia/India/Karnataka/ | Shimoga Institute of Medical Sciences | Department of Neurovirology, National Institute of Mental Health and Neurosciences (NIMHANS) | Chitra Pattabiraman et al |
| EPI_ISL_860188 | India/KA-NIMH-SEQ-68/2021 | 2021-01-03 | Asia/India/Karnataka/ | Shimoga Institute of Medical Sciences | Department of Neurovirology, National Institute of Mental Health and Neurosciences (NIMHANS) | Chitra Pattabiraman et al |
| EPI_ISL_860189 | India/KA-NIMH-SEQ-69/2021 | 2021-01-02 | Asia/India/Karnataka/ | BBMP Urban PHC | Department of Neurovirology, National Institute of Mental Health and Neurosciences (NIMHANS) | Chitra Pattabiraman et al |
| EPI_ISL_860190 | India/KA-NIMH-SEQ-70/2021 | 2021-01-01 | Asia/India/Karnataka/ | Railway Hospital | Department of Neurovirology, National Institute of Mental Health and Neurosciences (NIMHANS) | Chitra Pattabiraman et al |
| EPI_ISL_860191 | India/KA-NIMH-SEQ-72/2021 | 2021-01-02 | Asia/India/Karnataka/ | National Institute of Mental Health and Neurosciences (NIMHANS) | Department of Neurovirology, National Institute of Mental Health and Neurosciences (NIMHANS) | Chitra Pattabiraman et al |

| | | | | | | |
|---|---|---|---|---|---|---|
| EPI_ISL_860192 | India/KA-NIMH-SEQ-73/2021 | 2021-01-02 | Asia/India/Karnataka/ | Railway Hospital | Department of Neurovirology, National Institute of Mental Health and Neurosciences (NIMHANS) | Chitra Pattabiraman et al |
| EPI_ISL_860193 | India/KA-NIMH-SEQ-74/2021 | 2021-01-02 | Asia/India/Karnataka/ | Jayanagar General Hospital | Department of Neurovirology, National Institute of Mental Health and Neurosciences (NIMHANS) | Chitra Pattabiraman et al |
| EPI_ISL_860194 | India/KA-NIMH-SEQ-75/2021 | 2021-01-02 | Asia/India/Karnataka/ | BBMP Urban PHC | Department of Neurovirology, National Institute of Mental Health and Neurosciences (NIMHANS) | Chitra Pattabiraman et al |
| EPI_ISL_860195 | India/KA-NIMH-SEQ-76/2021 | 2021-01-03 | Asia/India/Karnataka/ | BBMP Urban PHC | Department of Neurovirology, National Institute of Mental Health and Neurosciences (NIMHANS) | Chitra Pattabiraman et al |
| EPI_ISL_860196 | India/KA-NIMH-SEQ-77/2021 | 2021-01-04 | Asia/India/Karnataka/ | National Institute of Mental Health and Neurosciences (NIMHANS) | Department of Neurovirology, National Institute of Mental Health and Neurosciences (NIMHANS) | Chitra Pattabiraman et al |
| EPI_ISL_860197 | India/KA-NIMH-SEQ-78/2021 | 2021-01-04 | Asia/India/Karnataka/ | BBMP Urban PHC | Department of Neurovirology, National Institute of Mental Health and Neurosciences (NIMHANS) | Chitra Pattabiraman et al |

| | | | | | | |
|---|---|---|---|---|---|---|
| EPI_ISL_860198 | India/KA-NIMH-SEQ-79/2021 | 2021-01-05 | Asia/India/Karnataka/ | Railway Hospital | Department of Neurovirology, National Institute of Mental Health and Neurosciences (NIMHANS) | Chitra Pattabiraman et al |
| EPI_ISL_860199 | India/KA-NIMH-SEQ-80/2020 | 2020-12-06 | Asia/India/Karnataka/ | BBMP Urban PHC | Department of Neurovirology, National Institute of Mental Health and Neurosciences (NIMHANS) | Chitra Pattabiraman et al |
| EPI_ISL_860200 | India/KA-NIMH-SEQ-81/2020 | 2020-12-06 | Asia/India/Karnataka/ | BBMP Urban PHC | Department of Neurovirology, National Institute of Mental Health and Neurosciences (NIMHANS) | Chitra Pattabiraman et al |
| EPI_ISL_860201 | India/KA-NIMH-SEQ-82/2020 | 2020-12-05 | Asia/India/Karnataka/ | BBMP Urban PHC | Department of Neurovirology, National Institute of Mental Health and Neurosciences (NIMHANS) | Chitra Pattabiraman et al |
| EPI_ISL_860204 | Italy/CAM-COTUGNO-15919/2020 | 2020-12-28 | Europe/Italy/Campania/ | Lab. Microbiologia e Virologia, Cotugno, A.O. dei Colli | Lab. Microbiologia e Virologia, Cotugno, A.O. dei Colli | Luigi Atripaldi et al |
| EPI_ISL_860205 | Italy/CAM-COTUGNO-15902/2020 | 2020-12-28 | Europe/Italy/Campania/ | Lab. Microbiologia e Virologia, Cotugno, A.O. dei Colli | Lab. Microbiologia e Virologia, Cotugno, A.O. dei Colli | Luigi Atripaldi et al |
| EPI_ISL_860257 | Italy/CAM-COTUGNO-14528/2020 | 2020-12-28 | Europe/Italy/Campania/ | Lab. Microbiologia e Virologia, Cotugno, A.O. dei Colli | Lab. Microbiologia e Virologia, Cotugno, A.O. dei Colli | Luigi Atripaldi et al |
| EPI_ISL_860296 | USA/OH-ODH-SC51970/2020 | 2020-07-08 | North America/USA/Ohio/ | Ohio Department of Health Laboratory | Ohio Department of Health Laboratory | Holmes et al |

| | | | | | | |
|---|---|---|---|---|---|---|
| EPI_ISL_860297 | Italy/CAM-COTUGNO-11258/2020 | 2020-12-21 | Europe/Italy/Campania/ | Lab. Microbiologia e Virologia, Cotugno, A.O. dei Colli | Lab. Microbiologia e Virologia, Cotugno, A.O. dei Colli | Luigi Atripaldi et al |
| EPI_ISL_860298 | Germany/SN-RKI-NP-0079/2020 | 2020-12-28 | Europe/Germany/Saxony/ | Unit 17: Influenza & Other Respiratory Viruses, German National Influenza Center | Project group Epidemiology of Highly Pathogenic Microorganisms, Robert Koch-Institute | Andreas Sachse et al |
| EPI_ISL_860304 | Germany/BB-RKI-NP-0001/2021 | 2021-01-04 | Europe/Germany/Brandenburg/ | Unit 17: Influenza & Other Respiratory Viruses, German National Influenza Center | Project group Epidemiology of Highly Pathogenic Microorganisms, Robert Koch-Institute | Andreas Sachse et al |
| EPI_ISL_860306 | Germany/HE-RKI-NP-0003/2021 | 2021-01-04 | Europe/Germany/Hesse/ | Unit 17: Influenza & Other Respiratory Viruses, German National Influenza Center | Project group Epidemiology of Highly Pathogenic Microorganisms, Robert Koch-Institute | Andreas Sachse et al |
| EPI_ISL_860312 | Germany/BW-RKI-NP-0009/2021 | 2021-01-04 | Europe/Germany/Baden-Wuerttemberg | Unit 17: Influenza & Other Respiratory Viruses, German National Influenza Center | Project group Epidemiology of Highly Pathogenic Microorganisms, Robert Koch-Institute | Andreas Sachse et al |
| EPI_ISL_860314 | Germany/BB-RKI-NP-0011/2021 | 2021-01-05 | Europe/Germany/Brandenburg/ | Unit 17: Influenza & Other Respiratory Viruses, German National Influenza Center | Project group Epidemiology of Highly Pathogenic Microorganisms, Robert Koch-Institute | Andreas Sachse et al |
| EPI_ISL_860317 | Brazil/SP-981/2021 | 2021-01-07 | South America/Brazil/S√£o Paulo/Soroca | Instituto Adolfo Lutz - Regional de Sorocaba | Instituto Adolfo Lutz, Interdiciplinary Procedures Center, Strategic Laboratory | Claudio Tavares Sacchi et al |

| | | | | | | |
|---|---|---|---|---|---|---|
| EPI_ISL_860333 | Germany/NW-KRO-016/2020 | 2020-06-19 | Europe/Germany/North Rhine Westphal | MVZ Labor Krone GbR | Center of Medical Microbiology, Virology, and Hospital Hygiene, University of Duesseldorf | Dennis Deschka et al |
| EPI_ISL_860348 | Germany/NW-KRO-031/2020 | 2020-07-03 | Europe/Germany/North Rhine Westphal | MVZ Labor Krone GbR | Center of Medical Microbiology, Virology, and Hospital Hygiene, University of Duesseldorf | Dennis Deschka et al |
| EPI_ISL_860357 | Germany/NW-KRO-040/2020 | 2020-06-19 | Europe/Germany/North Rhine Westphal | MVZ Labor Krone GbR | Center of Medical Microbiology, Virology, and Hospital Hygiene, University of Duesseldorf | Dennis Deschka et al |
| EPI_ISL_860451 | Germany/NW-KRO-134/2020 | 2020-06-18 | Europe/Germany/North Rhine Westphal | MVZ Labor Krone GbR | Center of Medical Microbiology, Virology, and Hospital Hygiene, University of Duesseldorf | Dennis Deschka et al |
| EPI_ISL_860462 | Germany/NW-KRO-145/2020 | 2020-06-19 | Europe/Germany/North Rhine Westphal | MVZ Labor Krone GbR | Center of Medical Microbiology, Virology, and Hospital Hygiene, University of Duesseldorf | Dennis Deschka et al |
| EPI_ISL_860469 | Germany/NW-KRO-152/2020 | 2020-06-23 | Europe/Germany/North Rhine Westphal | MVZ Labor Krone GbR | Center of Medical Microbiology, Virology, and Hospital Hygiene, University of Duesseldorf | Dennis Deschka et al |

| | | | | | | |
|---|---|---|---|---|---|---|
| EPI_ISL_860485 | Germany/NW-KRO-168/2020 | 2020-06-23 | Europe/Germany/North Rhine Westphal | MVZ Labor Krone GbR | Center of Medical Microbiology, Virology, and Hospital Hygiene, University of Duesseldorf | Dennis Deschka et al |
| EPI_ISL_860486 | Germany/NW-KRO-169/2020 | 2020-06-16 | Europe/Germany/North Rhine Westphal | MVZ Labor Krone GbR | Center of Medical Microbiology, Virology, and Hospital Hygiene, University of Duesseldorf | Dennis Deschka et al |
| EPI_ISL_860511 | Germany/NW-KRO-194/2020 | 2020-06-19 | Europe/Germany/North Rhine Westphal | MVZ Labor Krone GbR | Center of Medical Microbiology, Virology, and Hospital Hygiene, University of Duesseldorf | Dennis Deschka et al |
| EPI_ISL_860535 | Germany/NW-KRO-218/2020 | 2020-06-21 | Europe/Germany/North Rhine Westphal | MVZ Labor Krone GbR | Center of Medical Microbiology, Virology, and Hospital Hygiene, University of Duesseldorf | Dennis Deschka et al |
| EPI_ISL_860549 | USA/OH-ODH-SC51973/2020 | 2020-07-08 | North America/USA/Ohio/ | Ohio Department of Health Laboratory | Ohio Department of Health Laboratory | Holmes et al |
| EPI_ISL_860554 | SouthAfrica/KRISP-K007777/2021 | 2021-01-05 | Africa/South Africa/KwaZulu-Natal/eThe | NHLS-IALCH | KRISP, KZn Research Innovation and Sequencing Platform | Giandhari J et al |
| EPI_ISL_860556 | SouthAfrica/KRISP-K007779/2021 | 2021-01-05 | Africa/South Africa/KwaZulu-Natal/eThe | NHLS-IALCH | KRISP, KZn Research Innovation and Sequencing Platform | Giandhari J et al |

| | | | | | | |
|---|---|---|---|---|---|---|
| EPI_ISL_860557 | SouthAfrica/KRISP-K007780/2021 | 2021-01-05 | Africa/South Africa/KwaZulu-Natal/eThe | NHLS-IALCH | KRISP, KZn Research Innovation and Sequencing Platform | Giandhari J et al |
| EPI_ISL_860560 | SouthAfrica/KRISP-K007785/2021 | 2021-01-02 | Africa/South Africa/KwaZulu-Natal/Ugu | NHLS-IALCH | KRISP, KZn Research Innovation and Sequencing Platform | Giandhari J et al |
| EPI_ISL_860561 | SouthAfrica/KRISP-K007786/2021 | 2021-01-03 | Africa/South Africa/KwaZulu-Natal/Ugu | NHLS-IALCH | KRISP, KZn Research Innovation and Sequencing Platform | Giandhari J et al |
| EPI_ISL_860563 | SouthAfrica/KRISP-K007788/2021 | 2021-01-02 | Africa/South Africa/KwaZulu-Natal/Ugu | NHLS-IALCH | KRISP, KZn Research Innovation and Sequencing Platform | Giandhari J et al |
| EPI_ISL_860564 | SouthAfrica/KRISP-K007789/2021 | 2021-01-02 | Africa/South Africa/KwaZulu-Natal/Ugu | NHLS-IALCH | KRISP, KZn Research Innovation and Sequencing Platform | Giandhari J et al |
| EPI_ISL_860566 | SouthAfrica/KRISP-K007792/2020 | 2020-12-26 | Africa/South Africa/KwaZulu-Natal/Ugu | NHLS-IALCH | KRISP, KZn Research Innovation and Sequencing Platform | Giandhari J et al |
| EPI_ISL_860567 | SouthAfrica/KRISP-K007793/2021 | 2021-01-02 | Africa/South Africa/KwaZulu-Natal/Ugu | NHLS-IALCH | KRISP, KZn Research Innovation and Sequencing Platform | Giandhari J et al |
| EPI_ISL_860569 | SouthAfrica/KRISP-K007795/2021 | 2021-01-03 | Africa/South Africa/KwaZulu-Natal/Ugu | NHLS-IALCH | KRISP, KZn Research Innovation and Sequencing Platform | Giandhari J et al |

| | | | | | | |
|---|---|---|---|---|---|---|
| EPI_ISL_860571 | SouthAfrica/KRISP-K007798/2021 | 2021-01-03 | Africa/South Africa/KwaZulu-Natal/Ugu | NHLS-IALCH | KRISP, KZn Research Innovation and Sequencing Platform | Giandhari J et al |
| EPI_ISL_860572 | SouthAfrica/KRISP-K007799/2021 | 2021-01-03 | Africa/South Africa/KwaZulu-Natal/Ugu | NHLS-IALCH | KRISP, KZn Research Innovation and Sequencing Platform | Giandhari J et al |
| EPI_ISL_860573 | SouthAfrica/KRISP-K007800/2021 | 2021-01-03 | Africa/South Africa/KwaZulu-Natal/Ugu | NHLS-IALCH | KRISP, KZn Research Innovation and Sequencing Platform | Giandhari J et al |
| EPI_ISL_860574 | SouthAfrica/KRISP-K007801/2021 | 2021-01-03 | Africa/South Africa/KwaZulu-Natal/Ugu | NHLS-IALCH | KRISP, KZn Research Innovation and Sequencing Platform | Giandhari J et al |
| EPI_ISL_860575 | SouthAfrica/KRISP-K007802/2021 | 2021-01-01 | Africa/South Africa/KwaZulu-Natal/uMk | NHLS-IALCH | KRISP, KZn Research Innovation and Sequencing Platform | Giandhari J et al |
| EPI_ISL_860580 | SouthAfrica/KRISP-K007807/2020 | 2020-12-30 | Africa/South Africa/KwaZulu-Natal/King | NHLS-IALCH | KRISP, KZn Research Innovation and Sequencing Platform | Giandhari J et al |
| EPI_ISL_860581 | SouthAfrica/KRISP-K007810/2020 | 2020-12-30 | Africa/South Africa/KwaZulu-Natal/uMk | NHLS-IALCH | KRISP, KZn Research Innovation and Sequencing Platform | Giandhari J et al |
| EPI_ISL_860584 | SouthAfrica/KRISP-K007814/2020 | 2020-12-30 | Africa/South Africa/KwaZulu-Natal/uMk | NHLS-IALCH | KRISP, KZn Research Innovation and Sequencing Platform | Giandhari J et al |

| | | | | | | |
|---|---|---|---|---|---|---|
| EPI_ISL_860585 | SouthAfrica/KRISP-K007815/2021 | 2021-01-02 | Africa/South Africa/KwaZulu-Natal/Zulu | NHLS-IALCH | KRISP, KZn Research Innovation and Sequencing Platform | Giandhari J et al |
| EPI_ISL_860587 | SouthAfrica/KRISP-K007817/2021 | 2021-01-01 | Africa/South Africa/KwaZulu-Natal/Zulu | NHLS-IALCH | KRISP, KZn Research Innovation and Sequencing Platform | Giandhari J et al |
| EPI_ISL_860588 | SouthAfrica/KRISP-K007819/2021 | 2021-01-02 | Africa/South Africa/KwaZulu-Natal/uMk | NHLS-IALCH | KRISP, KZn Research Innovation and Sequencing Platform | Giandhari J et al |
| EPI_ISL_860589 | SouthAfrica/KRISP-K007820/2021 | 2021-01-02 | Africa/South Africa/KwaZulu-Natal/Harr | NHLS-IALCH | KRISP, KZn Research Innovation and Sequencing Platform | Giandhari J et al |
| EPI_ISL_860590 | SouthAfrica/KRISP-K007821/2021 | 2021-01-02 | Africa/South Africa/KwaZulu-Natal/Zulu | NHLS-IALCH | KRISP, KZn Research Innovation and Sequencing Platform | Giandhari J et al |
| EPI_ISL_860591 | SouthAfrica/KRISP-K007822/2021 | 2021-01-01 | Africa/South Africa/KwaZulu-Natal/Zulu | NHLS-IALCH | KRISP, KZn Research Innovation and Sequencing Platform | Giandhari J et al |
| EPI_ISL_860592 | SouthAfrica/KRISP-K007823/2021 | 2021-01-02 | Africa/South Africa/KwaZulu-Natal/Zulu | NHLS-IALCH | KRISP, KZn Research Innovation and Sequencing Platform | Giandhari J et al |
| EPI_ISL_860593 | SouthAfrica/KRISP-K007825/2021 | 2021-01-02 | Africa/South Africa/KwaZulu-Natal/uMk | NHLS-IALCH | KRISP, KZn Research Innovation and Sequencing Platform | Giandhari J et al |

| | | | | | | |
|---|---|---|---|---|---|---|
| EPI_ISL_860594 | SouthAfrica/KRISP-K007826/2021 | 2021-01-01 | Africa/South Africa/KwaZulu-Natal/uMk | NHLS-IALCH | KRISP, KZn Research Innovation and Sequencing Platform | Giandhari J et al |
| EPI_ISL_860597 | SouthAfrica/KRISP-K007829/2020 | 2020-12-20 | Africa/South Africa/KwaZulu-Natal/Ugu | NHLS-IALCH | KRISP, KZn Research Innovation and Sequencing Platform | Giandhari J et al |
| EPI_ISL_860599 | SouthAfrica/KRISP-K007831/2020 | 2020-12-21 | Africa/South Africa/KwaZulu-Natal/Ugu | NHLS-IALCH | KRISP, KZn Research Innovation and Sequencing Platform | Giandhari J et al |
| EPI_ISL_860604 | SouthAfrica/KRISP-K007836/2020 | 2020-12-21 | Africa/South Africa/KwaZulu-Natal/Ugu | NHLS-IALCH | KRISP, KZn Research Innovation and Sequencing Platform | Giandhari J et al |
| EPI_ISL_860617 | SouthAfrica/KRISP-K007852/2020 | 2020-12-26 | Africa/South Africa/KwaZulu-Natal/eThe | NHLS-IALCH | KRISP, KZn Research Innovation and Sequencing Platform | Giandhari J et al |
| EPI_ISL_860619 | SouthAfrica/KRISP-K007855/2020 | 2020-12-27 | Africa/South Africa/KwaZulu-Natal/eThe | NHLS-IALCH | KRISP, KZn Research Innovation and Sequencing Platform | Giandhari J et al |
| EPI_ISL_860620 | SouthAfrica/KRISP-K007856/2020 | 2020-12-25 | Africa/South Africa/KwaZulu-Natal/Ugu | NHLS-IALCH | KRISP, KZn Research Innovation and Sequencing Platform | Giandhari J et al |
| EPI_ISL_860623 | SouthAfrica/KRISP-K007860/2020 | 2020-12-26 | Africa/South Africa/KwaZulu-Natal/uMk | NHLS-IALCH | KRISP, KZn Research Innovation and Sequencing Platform | Giandhari J et al |

| | | | | | | |
|---|---|---|---|---|---|---|
| EPI_ISL_860625 | SouthAfrica/KRISP-K007863/2020 | 2020-12-25 | Africa/South Africa/KwaZulu-Natal/uMk | NHLS-IALCH | KRISP, KZn Research Innovation and Sequencing Platform | Giandhari J et al |
| EPI_ISL_860626 | SouthAfrica/KRISP-K007865/2020 | 2020-12-18 | Africa/South Africa/KwaZulu-Natal/Ugu | NHLS-IALCH | KRISP, KZn Research Innovation and Sequencing Platform | Giandhari J et al |
| EPI_ISL_860629 | SouthAfrica/KRISP-K007868/2020 | 2020-12-21 | Africa/South Africa/KwaZulu-Natal/iLem | NHLS-IALCH | KRISP, KZn Research Innovation and Sequencing Platform | Giandhari J et al |
| EPI_ISL_860633 | Brazil/SP-982/2021 | 2021-01-07 | South America/Brazil/S√£o Paulo/Soroca | Instituto Adolfo Lutz - Regional de Sorocaba | Instituto Adolfo Lutz, Interdiciplinary Procedures Center, Strategic Laboratory | Claudio Tavares Sacchi et al |
| EPI_ISL_860697 | England/210291734/2021 | 2021-01-17 | Europe/United Kingdom/England/ | Respiratory Virus Unit, National Infection Service, Public Health England | COVID-19 Genomics UK (COG-UK) Consortium | PHE Covid Sequencing Team et al |
| EPI_ISL_860716 | France/APM-CERBAHC-20211018-72/ | 2021-01-08 | Europe/France/Provence-Alpes-C√¥te d' | Cerballiance C√¥te d'Azur | CERBA LAB | Roquebert B et al |
| EPI_ISL_860741 | Switzerland/NE-SNRCI-1796/2020 | 2020-11-03 | Europe/Switzerland/Neuch√¢tel/ | Swiss National Reference Centre for Influenza | Swiss National Reference Centre for Influenza | Ana Rita Gon√ßalves Cabecinhas et al |
| EPI_ISL_860766 | Switzerland/VD-SNRCI-3593/2020 | 2020-10-28 | Europe/Switzerland/Vaud/ | Swiss National Reference Centre for Influenza | Swiss National Reference Centre for Influenza | Ana Rita Gon√ßalves Cabecinhas et al |
| EPI_ISL_860784 | Germany/NW-ChVir22070/2021 | 2021-01-05 | Europe/Germany/North Rhine Westphal | Charit√© Universit√§tsmedizin Berlin, Institute of Virology, Charit√©platz 1, 10117 Berlin, Germany | Charit√© Universit√§tsmedizin Berlin, Institute of Virology, Charit√©platz 1, 10117 Berlin, Germany | Victor M Corman et al |

| | | | | | | |
|---|---|---|---|---|---|---|
| EPI_ISL_860786 | Germany/HB-ChVir22080/2021 | 2021-01-06 | Europe/Germany/Bremen/ | Charit√© Universit√§tsmedizin Berlin, Institute of Virology, Charit√©platz 1, 10117 Berlin, Germany | Charit√© Universit√§tsmedizin Berlin, Institute of Virology, Charit√©platz 1, 10117 Berlin, Germany | Victor M Corman et al |
| EPI_ISL_860787 | Germany/NW-ChVir22090/2021 | 2021-01-08 | Europe/Germany/North Rhine Westphal | Charit√© Universit√§tsmedizin Berlin, Institute of Virology, Charit√©platz 1, 10117 Berlin, Germany | Charit√© Universit√§tsmedizin Berlin, Institute of Virology, Charit√©platz 1, 10117 Berlin, Germany | Victor M Corman et al |
| EPI_ISL_860788 | Germany/NW-ChVir22113/2021 | 2021-01-05 | Europe/Germany/North Rhine Westphal | Charit√© Universit√§tsmedizin Berlin, Institute of Virology, Charit√©platz 1, 10117 Berlin, Germany | Charit√© Universit√§tsmedizin Berlin, Institute of Virology, Charit√©platz 1, 10117 Berlin, Germany | Victor M Corman et al |
| EPI_ISL_860795 | USA/OH-ODH-SC186067/2020 | 2020-12-21 | North America/USA/Ohio/ | Ohio Department of Health Laboratory | Ohio Department of Health Laboratory | Holmes et al |
| EPI_ISL_860797 | Belarus/ChVir21891/2020 | 2020-10-23 | Europe/Belarus/Belarus/ | WHO/Minsk | Charit√© Universit√§tsmedizin Berlin, Institut f√ºr Virologie | Victor M Corman et al |
| EPI_ISL_860798 | Belarus/ChVir21892/2020 | 2020-10-26 | Europe/Belarus/Belarus/ | WHO/Minsk | Charit√© Universit√§tsmedizin Berlin, Institut f√ºr Virologie | Victor M Corman et al |
| EPI_ISL_860799 | Belarus/ChVir21897/2020 | 2020-10-26 | Europe/Belarus/Belarus/ | WHO/Minsk | Charit√© Universit√§tsmedizin Berlin, Institut f√ºr Virologie | Victor M Corman et al |

| | | | | | | |
|---|---|---|---|---|---|---|
| EPI_ISL_860801 | Belarus/ChVir21898/2020 | 2020-10-26 | Europe/Belarus/Belarus/ | WHO/Minsk | Charit√© Universit√§tsmedizin Berlin, Institut f√ºr Virologie | Victor M Corman et al |
| EPI_ISL_860803 | Belarus/ChVir21837/2020 | 2020-11-07 | Europe/Belarus/Belarus/ | WHO/Minsk | Charit√© Universit√§tsmedizin Berlin, Institut f√ºr Virologie | Victor M Corman et al |
| EPI_ISL_860804 | Belarus/ChVir21840/2020 | 2020-11-30 | Europe/Belarus/Belarus/ | WHO/Minsk | Charit√© Universit√§tsmedizin Berlin, Institut f√ºr Virologie | Victor M Corman et al |
| EPI_ISL_860807 | Belarus/ChVir21845/2020 | 2020-11-06 | Europe/Belarus/Belarus/ | WHO/Minsk | Charit√© Universit√§tsmedizin Berlin, Institut f√ºr Virologie | Victor M Corman et al |
| EPI_ISL_860808 | Belarus/ChVir21846/2020 | 2020-11-04 | Europe/Belarus/Belarus/ | WHO/Minsk | Charit√© Universit√§tsmedizin Berlin, Institut f√ºr Virologie | Victor M Corman et al |
| EPI_ISL_860809 | Belarus/ChVir21848/2020 | 2020-11-09 | Europe/Belarus/Belarus/ | WHO/Minsk | Charit√© Universit√§tsmedizin Berlin, Institut f√ºr Virologie | Victor M Corman et al |
| EPI_ISL_860812 | Belarus/ChVir21877/2020 | 2020-11-03 | Europe/Belarus/Belarus/ | WHO/Minsk | Charit√© Universit√§tsmedizin Berlin, Institut f√ºr Virologie | Victor M Corman et al |
| EPI_ISL_860813 | Belarus/ChVir21878/2020 | 2020-10-28 | Europe/Belarus/Belarus/ | WHO/Minsk | Charit√© Universit√§tsmedizin Berlin, Institut f√ºr Virologie | Victor M Corman et al |
| EPI_ISL_860814 | Belarus/ChVir21882/2020 | 2020-10-30 | Europe/Belarus/Belarus/ | WHO/Minsk | Charit√© Universit√§tsmedizin Berlin, Institut f√ºr Virologie | Victor M Corman et al |
| EPI_ISL_860815 | Belarus/ChVir21884/2020 | 2020-10-29 | Europe/Belarus/Belarus/ | WHO/Minsk | Charit√© Universit√§tsmedizin Berlin, Institut f√ºr Virologie | Victor M Corman et al |

| | | | | | | |
|---|---|---|---|---|---|---|
| EPI_ISL_860816 | Belarus/ChVir21888/2020 | 2020-10-26 | Europe/Belarus/Belarus/ | WHO/Minsk | Charitv© Universitv§tsmedizin Berlin, Institut fvºr Virologie | Victor M Corman et al |
| EPI_ISL_860817 | Belarus/ChVir21889/2020 | 2020-10-26 | Europe/Belarus/Belarus/ | WHO/Minsk | Charitv© Universitv§tsmedizin Berlin, Institut fvºr Virologie | Victor M Corman et al |
| EPI_ISL_860818 | Belarus/ChVir21895/2020 | 2020-10-26 | Europe/Belarus/Belarus/ | WHO/Minsk | Charitv© Universitv§tsmedizin Berlin, Institut fvºr Virologie | Victor M Corman et al |
| EPI_ISL_860823 | France/CVL-IPP00304/2021 | 2021-01-08 | Europe/France/Centre-Val de Loire/Tou | CHU Tours - Virologie | National Reference Center for Viruses of Respiratory Infections, Institut Pasteur, Paris | Marion Barbet et al |
| EPI_ISL_860826 | France/IDF-IPP00394/2021 | 2021-01-07 | Europe/France/Ile de France/Thiais | Labo Analyses med | National Reference Center for Viruses of Respiratory Infections, Institut Pasteur, Paris | Marion Barbet et al |
| EPI_ISL_860835 | France/IDF-IPP00408/2021 | 2021-01-08 | Europe/France/Ile de France/Thiais | Labo Analyses Med | National Reference Center for Viruses of Respiratory Infections, Institut Pasteur, Paris | Marion Barbet et al |
| EPI_ISL_860839 | France/IDF-IPP00421/2021 | 2021-01-08 | Europe/France/Ile de France/Thiais | Labo Analyses Med | National Reference Center for Viruses of Respiratory Infections, Institut Pasteur, Paris | Marion Barbet et al |

| | | | | | | |
|---|---|---|---|---|---|---|
| EPI_ISL_860842 | France/IDF-IPP00395/2021 | 2021-01-07 | Europe/France/Ile de France/Thiais | Labo Analyses med | National Reference Center for Viruses of Respiratory Infections, Institut Pasteur, Paris | Marion Barbet et al |
| EPI_ISL_860863 | France/PDL-IPP00343/2021 | 2021-01-07 | Europe/France/Pays de la Loire/Nantes | CHU de Nantes - H√¥tel Dieu - Labo. Virologie | National Reference Center for Viruses of Respiratory Infections, Institut Pasteur, Paris | Marion Barbet et al |
| EPI_ISL_860864 | France/GE-IPP00440/2020 | 2020-12-30 | Europe/France/Grand Est/Epinal | Labo Analyses Med | National Reference Center for Viruses of Respiratory Infections, Institut Pasteur, Paris | Marion Barbet et al |
| EPI_ISL_860867 | France/CVL-IPP00311/2021 | 2021-01-06 | Europe/France/Centre-Val de Loire/Tou | CHU Tours - Virologie | National Reference Center for Viruses of Respiratory Infections, Institut Pasteur, Paris | Marion Barbet et al |
| EPI_ISL_860873 | France/GE-IPP00448/2021 | 2021-01-06 | Europe/France/Grand Est/Epinal | Labo Analyses Med | National Reference Center for Viruses of Respiratory Infections, Institut Pasteur, Paris | Marion Barbet et al |
| EPI_ISL_860876 | France/GE-IPP00451/2021 | 2021-01-06 | Europe/France/Grand Est/Epinal | Labo Analyses Med | National Reference Center for Viruses of Respiratory Infections, Institut Pasteur, Paris | Marion Barbet et al |

| | | | | | | |
|---|---|---|---|---|---|---|
| EPI_ISL_860878 | France/GE-IPP00453/2021 | 2021-01-06 | Europe/France/Grand Est/Epinal | Labo Analyses Med | National Reference Center for Viruses of Respiratory Infections, Institut Pasteur, Paris | Marion Barbet et al |
| EPI_ISL_860879 | France/GE-IPP00454/2021 | 2021-01-04 | Europe/France/Grand Est/Epinal | Labo Analyses Med | National Reference Center for Viruses of Respiratory Infections, Institut Pasteur, Paris | Marion Barbet et al |
| EPI_ISL_860880 | France/GE-IPP00455/2021 | 2021-01-04 | Europe/France/Grand Est/Epinal | Labo Analyses Med | National Reference Center for Viruses of Respiratory Infections, Institut Pasteur, Paris | Marion Barbet et al |
| EPI_ISL_860894 | France/CVL-IPP00303/2021 | 2021-01-07 | Europe/France/Centre-Val de Loire/Tou | CHU Tours - Virologie | National Reference Center for Viruses of Respiratory Infections, Institut Pasteur, Paris | Marion Barbet et al |
| EPI_ISL_860903 | France/PDL-IPP00436/2021 | 2021-01-09 | Europe/France/Pays de la Loire/Le Mans | CH du MANS - Lab. Bio. Mol√©culaire | National Reference Center for Viruses of Respiratory Infections, Institut Pasteur, Paris | Marion Barbet et al |
| EPI_ISL_860910 | France/PDL-IPP00437/2021 | 2021-01-09 | Europe/France/Pays de la Loire/Le Mans | CH du MANS - Lab. Bio. Mol√©culaire | National Reference Center for Viruses of Respiratory Infections, Institut Pasteur, Paris | Marion Barbet et al |

| | | | | | | |
|---|---|---|---|---|---|---|
| EPI_ISL_860911 | France/PDL-IPP00481/2021 | 2021-01-07 | Europe/France/Pays de la Loire/Angers | CHU Angers - Dpt des Agents Infectieux | National Reference Center for Viruses of Respiratory Infections, Institut Pasteur, Paris | Marion Barbet et al |
| EPI_ISL_860921 | France/PDL-IPP00332/2021 | 2021-01-07 | Europe/France/Pays de la Loire/Nantes | CHU de Nantes - H√¥tel Dieu - Labo. Virologie | National Reference Center for Viruses of Respiratory Infections, Institut Pasteur, Paris | Marion Barbet et al |
| EPI_ISL_860927 | France/PDL-IPP00339/2021 | 2021-01-07 | Europe/France/Pays de la Loire/Nantes | CHU de Nantes - H√¥tel Dieu - Labo. Virologie | National Reference Center for Viruses of Respiratory Infections, Institut Pasteur, Paris | Marion Barbet et al |
| EPI_ISL_860936 | USA/MD-HP01027/2020 | 2020-12-10 | North America/USA/Maryland/ | Johns Hopkins Hospital Department of Pathology | Johns Hopkins Hospital Department of Pathology | C. Paul Morris et al |
| EPI_ISL_860938 | USA/MD-HP01055/2020 | 2020-12-29 | North America/USA/Maryland/ | Johns Hopkins Hospital Department of Pathology | Johns Hopkins Hospital Department of Pathology | C. Paul Morris et al |
| EPI_ISL_860946 | USA/MD-HP01097/2020 | 2020-12-29 | North America/USA/Maryland/ | Johns Hopkins Hospital Department of Pathology | Johns Hopkins Hospital Department of Pathology | C. Paul Morris et al |
| EPI_ISL_860953 | USA/MD-HP01105/2021 | 2021-01-05 | North America/USA/Maryland/ | Johns Hopkins Hospital Department of Pathology | Johns Hopkins Hospital Department of Pathology | C. Paul Morris et al |
| EPI_ISL_860954 | USA/MD-HP01106/2021 | 2021-01-12 | North America/USA/Maryland/ | Johns Hopkins Hospital Department of Pathology | Johns Hopkins Hospital Department of Pathology | C. Paul Morris et al |

| | | | | | | |
|---|---|---|---|---|---|---|
| EPI_ISL_860956 | USA/MD-HP01108/2021 | 2021-01-08 | North America/USA/Maryland/ | Johns Hopkins Hospital Department of Pathology | Johns Hopkins Hospital Department of Pathology | C. Paul Morris et al |
| EPI_ISL_860957 | USA/MD-HP01109/2020 | 2020-12-30 | North America/USA/Maryland/ | Johns Hopkins Hospital Department of Pathology | Johns Hopkins Hospital Department of Pathology | C. Paul Morris et al |
| EPI_ISL_860958 | USA/MD-HP01110/2020 | 2020-12-24 | North America/USA/Maryland/ | Johns Hopkins Hospital Department of Pathology | Johns Hopkins Hospital Department of Pathology | C. Paul Morris et al |
| EPI_ISL_860959 | USA/VA-HP01111/2021 | 2021-01-12 | North America/USA/Virginia/ | Johns Hopkins Hospital Department of Pathology | Johns Hopkins Hospital Department of Pathology | C. Paul Morris et al |
| EPI_ISL_860964 | USA/MD-HP01118/2021 | 2021-01-04 | North America/USA/Maryland/ | Johns Hopkins Hospital Department of Pathology | Johns Hopkins Hospital Department of Pathology | C. Paul Morris et al |
| EPI_ISL_860966 | USA/MD-HP01120/2021 | 2021-01-12 | North America/USA/Maryland/ | Johns Hopkins Hospital Department of Pathology | Johns Hopkins Hospital Department of Pathology | C. Paul Morris et al |
| EPI_ISL_860969 | USA/MD-HP01123/2020 | 2020-12-29 | North America/USA/Maryland/ | Johns Hopkins Hospital Department of Pathology | Johns Hopkins Hospital Department of Pathology | C. Paul Morris et al |
| EPI_ISL_860976 | USA/MD-HP01131/2021 | 2021-01-01 | North America/USA/Maryland/ | Johns Hopkins Hospital Department of Pathology | Johns Hopkins Hospital Department of Pathology | C. Paul Morris et al |
| EPI_ISL_860980 | USA/MD-HP01135/2021 | 2021-01-09 | North America/USA/Maryland/ | Johns Hopkins Hospital Department of Pathology | Johns Hopkins Hospital Department of Pathology | C. Paul Morris et al |
| EPI_ISL_860996 | USA/MD-HP01153/2021 | 2021-01-04 | North America/USA/Maryland/ | Johns Hopkins Hospital Department of Pathology | Johns Hopkins Hospital Department of Pathology | C. Paul Morris et al |

| | | | | | | |
|---|---|---|---|---|---|---|
| EPI_ISL_860999 | USA/MD-HP01157/2021 | 2021-01-05 | North America/USA/Maryland/ | Johns Hopkins Hospital Department of Pathology | Johns Hopkins Hospital Department of Pathology | C. Paul Morris et al |
| EPI_ISL_861003 | USA/MD-HP01162/2021 | 2021-01-15 | North America/USA/Maryland/ | Johns Hopkins Hospital Department of Pathology | Johns Hopkins Hospital Department of Pathology | C. Paul Morris et al |
| EPI_ISL_861005 | USA/MD-HP01164/2021 | 2021-01-14 | North America/USA/Maryland/ | Johns Hopkins Hospital Department of Pathology | Johns Hopkins Hospital Department of Pathology | C. Paul Morris et al |
| EPI_ISL_861032 | USA/MD-HP01198/2021 | 2021-01-09 | North America/USA/Maryland/ | Johns Hopkins Hospital Department of Pathology | Johns Hopkins Hospital Department of Pathology | C. Paul Morris et al |
| EPI_ISL_861033 | USA/MD-HP01199/2021 | 2021-01-08 | North America/USA/Maryland/ | Johns Hopkins Hospital Department of Pathology | Johns Hopkins Hospital Department of Pathology | C. Paul Morris et al |
| EPI_ISL_861044 | USA/MD-HP01213/2021 | 2021-01-07 | North America/USA/Maryland/ | Johns Hopkins Hospital Department of Pathology | Johns Hopkins Hospital Department of Pathology | C. Paul Morris et al |
| EPI_ISL_861046 | USA/MD-HP01215/2021 | 2021-01-13 | North America/USA/Maryland/ | Johns Hopkins Hospital Department of Pathology | Johns Hopkins Hospital Department of Pathology | C. Paul Morris et al |
| EPI_ISL_861063 | USA/MD-HP01236/2021 | 2021-01-16 | North America/USA/Maryland/ | Johns Hopkins Hospital Department of Pathology | Johns Hopkins Hospital Department of Pathology | C. Paul Morris et al |
| EPI_ISL_861065 | USA/MD-HP01238/2021 | 2021-01-10 | North America/USA/Maryland/ | Johns Hopkins Hospital Department of Pathology | Johns Hopkins Hospital Department of Pathology | C. Paul Morris et al |
| EPI_ISL_861068 | USA/MD-HP01241/2021 | 2021-01-04 | North America/USA/Maryland/ | Johns Hopkins Hospital Department of Pathology | Johns Hopkins Hospital Department of Pathology | C. Paul Morris et al |

| | | | | | | |
|---|---|---|---|---|---|---|
| EPI_ISL_861074 | USA/MD-HP01250/2021 | 2021-01-06 | North America/USA/Maryland/ | Johns Hopkins Hospital Department of Pathology | Johns Hopkins Hospital Department of Pathology | C. Paul Morris et al |
| EPI_ISL_861076 | USA/MD-HP01253/2021 | 2021-01-05 | North America/USA/Maryland/ | Johns Hopkins Hospital Department of Pathology | Johns Hopkins Hospital Department of Pathology | C. Paul Morris et al |
| EPI_ISL_861080 | USA/MD-HP01257/2021 | 2021-01-15 | North America/USA/Maryland/ | Johns Hopkins Hospital Department of Pathology | Johns Hopkins Hospital Department of Pathology | C. Paul Morris et al |
| EPI_ISL_861081 | USA/MD-HP01258/2021 | 2021-01-09 | North America/USA/Maryland/ | Johns Hopkins Hospital Department of Pathology | Johns Hopkins Hospital Department of Pathology | C. Paul Morris et al |
| EPI_ISL_861094 | USA/MD-HP01272/2021 | 2021-01-13 | North America/USA/Maryland/ | Johns Hopkins Hospital Department of Pathology | Johns Hopkins Hospital Department of Pathology | C. Paul Morris et al |
| EPI_ISL_861106 | USA/NY-Wadsworth-21003540-01/20 | 2020-12-23 | North America/USA/New York/Erie Cou | KALEIDA CENTER FOR LABORATORY MEDICINE | Wadsworth Center, New York State Department of Health | Kirsten St. George et al |
| EPI_ISL_861111 | USA/NY-Wadsworth-21002949-01/20 | 2021-01-02 | North America/USA/New York/New Yor | New York Presbyterian Hospital | Wadsworth Center, New York State Department of Health | Kirsten St. George et al |
| EPI_ISL_861114 | USA/NY-Wadsworth-21002966-01/20 | 2021-01-02 | North America/USA/New York/ | New York Presbyterian Hospital | Wadsworth Center, New York State Department of Health | Kirsten St. George et al |
| EPI_ISL_861117 | USA/NY-Wadsworth-21000431-01/20 | 2021-01-04 | North America/USA/New York/Warren | Wadsworth Center, New York State Department of Health | Wadsworth Center, New York State Department of Health | Kirsten St. George et al |
| EPI_ISL_861118 | USA/NY-Wadsworth-21002946-01/20 | 2021-01-01 | North America/USA/New York/ | New York Presbyterian Hospital | Wadsworth Center, New York State Department of Health | Kirsten St. George et al |

| | | | | | | |
|---|---|---|---|---|---|---|
| EPI_ISL_861124 | USA/NY-Wadsworth-21002958-01/20 | 2021-01-02 | North America/USA/New York/New Yor | New York Presbyterian Hospital | Wadsworth Center, New York State Department of Health | Kirsten St. George et al |
| EPI_ISL_861127 | USA/NY-Wadsworth-21002963-01/20 | 2021-01-02 | North America/USA/New York/New Yor | New York Presbyterian Hospital | Wadsworth Center, New York State Department of Health | Kirsten St. George et al |
| EPI_ISL_861134 | USA/NY-Wadsworth-21003055-01/20 | 2020-12-22 | North America/USA/New York/Niagara | KALEIDA CENTER FOR LABORATORY MEDICINE | Wadsworth Center, New York State Department of Health | Kirsten St. George et al |
| EPI_ISL_861136 | USA/NY-Wadsworth-21003064-01/20 | 2020-12-30 | North America/USA/New York/Franklin | ADIRONDACK MEDICAL CENTER | Wadsworth Center, New York State Department of Health | Kirsten St. George et al |
| EPI_ISL_861137 | USA/NY-Wadsworth-21003068-01/20 | 2020-12-30 | North America/USA/New York/Franklin | ADIRONDACK MEDICAL CENTER | Wadsworth Center, New York State Department of Health | Kirsten St. George et al |
| EPI_ISL_861138 | USA/NY-Wadsworth-21003071-01/20 | 2020-12-29 | North America/USA/New York/Essex Co | ADIRONDACK MEDICAL CENTER | Wadsworth Center, New York State Department of Health | Kirsten St. George et al |
| EPI_ISL_861141 | USA/NY-Wadsworth-21003124-01/20 | 2020-12-30 | North America/USA/New York/Franklin | ADIRONDACK MEDICAL CENTER | Wadsworth Center, New York State Department of Health | Kirsten St. George et al |
| EPI_ISL_861144 | USA/NY-Wadsworth-21003163-01/20 | 2020-12-22 | North America/USA/New York/Niagara | KALEIDA CENTER FOR LABORATORY MEDICINE | Wadsworth Center, New York State Department of Health | Kirsten St. George et al |
| EPI_ISL_861147 | USA/NY-Wadsworth-21003166-01/20 | 2020-12-22 | North America/USA/New York/Erie Cou | KALEIDA CENTER FOR LABORATORY MEDICINE | Wadsworth Center, New York State Department of Health | Kirsten St. George et al |
| EPI_ISL_861149 | USA/NY-Wadsworth-21003521-01/20 | 2020-12-22 | North America/USA/New York/Niagara | KALEIDA CENTER FOR LABORATORY MEDICINE | Wadsworth Center, New York State Department of Health | Kirsten St. George et al |

| | | | | | | |
|---|---|---|---|---|---|---|
| EPI_ISL_861150 | USA/NY-Wadsworth-21003522-01/20 | 2020-12-22 | North America/USA/New York/Niagara | KALEIDA CENTER FOR LABORATORY MEDICINE | Wadsworth Center, New York State Department of Health | Kirsten St. George et al |
| EPI_ISL_861154 | USA/NY-Wadsworth-21003530-01/20 | 2020-12-23 | North America/USA/New York/Erie Cou | KALEIDA CENTER FOR LABORATORY MEDICINE | Wadsworth Center, New York State Department of Health | Kirsten St. George et al |
| EPI_ISL_861156 | USA/NY-Wadsworth-21003533-01/20 | 2020-12-23 | North America/USA/New York/Erie Cou | KALEIDA CENTER FOR LABORATORY MEDICINE | Wadsworth Center, New York State Department of Health | Kirsten St. George et al |
| EPI_ISL_861163 | USA/NY-Wadsworth-21003546-01/20 | 2020-12-24 | North America/USA/New York/Erie Cou | KALEIDA CENTER FOR LABORATORY MEDICINE | Wadsworth Center, New York State Department of Health | Kirsten St. George et al |
| EPI_ISL_861165 | USA/NY-Wadsworth-21003548-01/20 | 2020-12-23 | North America/USA/New York/Niagara | KALEIDA CENTER FOR LABORATORY MEDICINE | Wadsworth Center, New York State Department of Health | Kirsten St. George et al |
| EPI_ISL_861167 | USA/NY-Wadsworth-21003550-01/20 | 2020-12-23 | North America/USA/New York/Niagara | KALEIDA CENTER FOR LABORATORY MEDICINE | Wadsworth Center, New York State Department of Health | Kirsten St. George et al |
| EPI_ISL_861168 | USA/NY-Wadsworth-21003551-01/20 | 2020-12-23 | North America/USA/New York/Cattarau | KALEIDA CENTER FOR LABORATORY MEDICINE | Wadsworth Center, New York State Department of Health | Kirsten St. George et al |
| EPI_ISL_861169 | USA/NY-Wadsworth-21003553-01/20 | 2020-12-24 | North America/USA/New York/Niagara | KALEIDA CENTER FOR LABORATORY MEDICINE | Wadsworth Center, New York State Department of Health | Kirsten St. George et al |
| EPI_ISL_861178 | USA/NY-Wadsworth-21006732-01/20 | 2021-01-03 | North America/USA/New York/Westche | BIO-REFERENCE LABORATORIES | Wadsworth Center, New York State Department of Health | Kirsten St. George et al |
| EPI_ISL_861186 | USA/NY-Wadsworth-21006378-01/20 | 2021-01-02 | North America/USA/New York/Queens | NORTH SHORE UNIVERSITY HOSPITAL | Wadsworth Center, New York State Department of Health | Kirsten St. George et al |

| | | | | | | |
|---|---|---|---|---|---|---|
| EPI_ISL_861193 | USA/NY-Wadsworth-21006377-01/20 | 2021-01-02 | North America/USA/New York/Nassau ( | NORTH SHORE UNIVERSITY HOSPITAL | Wadsworth Center, New York State Department of Health | Kirsten St. George et al |
| EPI_ISL_861204 | USA/NY-Wadsworth-21006608-01/20 | 2021-01-11 | North America/USA/New York/Monroe | URMC LABS | Wadsworth Center, New York State Department of Health | Kirsten St. George et al |
| EPI_ISL_861207 | USA/NY-Wadsworth-21006613-01/20 | 2021-01-11 | North America/USA/New York/Monroe | URMC LABS | Wadsworth Center, New York State Department of Health | Kirsten St. George et al |
| EPI_ISL_861215 | USA/NY-Wadsworth-21006681-01/20 | 2020-12-28 | North America/USA/New York/Kings Co | BIO-REFERENCE LABORATORIES | Wadsworth Center, New York State Department of Health | Kirsten St. George et al |
| EPI_ISL_861218 | USA/NY-Wadsworth-21006693-01/20 | 2020-12-28 | North America/USA/New York/Queens | BIO-REFERENCE LABORATORIES | Wadsworth Center, New York State Department of Health | Kirsten St. George et al |
| EPI_ISL_861220 | USA/NY-Wadsworth-21006700-01/20 | 2020-12-28 | North America/USA/New York/Kings Co | BIO-REFERENCE LABORATORIES | Wadsworth Center, New York State Department of Health | Kirsten St. George et al |
| EPI_ISL_861224 | USA/NY-Wadsworth-21006705-01/20 | 2020-12-28 | North America/USA/New York/Ulster C | BIO-REFERENCE LABORATORIES | Wadsworth Center, New York State Department of Health | Kirsten St. George et al |
| EPI_ISL_861240 | USA/OH-ODH-SC188682/2020 | 2020-12-24 | North America/USA/Ohio/ | Ohio Department of Health Laboratory | Ohio Department of Health Laboratory | Holmes et al |
| EPI_ISL_861242 | Brazil/SP-BT6827/2020 | 2020-09-07 | South America/Brazil/S√£o Paulo/Sao Jo | Instituto de Biotecnologia - UNESP-Botucatu-SP | Instituto de Biotecnologia - UNESP-Botucatu-SP | Leila Sabrina Ullmann et al |
| EPI_ISL_861249 | USA/NY-Wadsworth-21004935-01/20 | 2020-12-28 | North America/USA/New York/Westche | MONTEFIORE MEDICAL CENTER LABORATORIES | Wadsworth Center, New York State Department of Health | Kirsten St. George et al |

| | | | | | | |
|---|---|---|---|---|---|---|
| EPI_ISL_861251 | USA/NY-Wadsworth-21004937-01/20 | 2020-12-26 | North America/USA/New York/Bronx | MONTEFIORE MEDICAL CENTER LABORATORIES | Wadsworth Center, New York State Department of Health | Kirsten St. George et al |
| EPI_ISL_861258 | USA/NY-Wadsworth-21004944-01/20 | 2020-12-25 | North America/USA/New York/Bronx | MONTEFIORE MEDICAL CENTER LABORATORIES | Wadsworth Center, New York State Department of Health | Kirsten St. George et al |
| EPI_ISL_861261 | USA/NY-Wadsworth-21004952-01/20 | 2020-12-28 | North America/USA/New York/Bronx | MONTEFIORE MEDICAL CENTER LABORATORIES | Wadsworth Center, New York State Department of Health | Kirsten St. George et al |
| EPI_ISL_861264 | USA/NY-Wadsworth-21004955-01/20 | 2020-12-26 | North America/USA/New York/Bronx | MONTEFIORE MEDICAL CENTER LABORATORIES | Wadsworth Center, New York State Department of Health | Kirsten St. George et al |
| EPI_ISL_861266 | USA/NY-Wadsworth-21004957-01/20 | 2020-12-26 | North America/USA/New York/Bronx | MONTEFIORE MEDICAL CENTER LABORATORIES | Wadsworth Center, New York State Department of Health | Kirsten St. George et al |
| EPI_ISL_861267 | USA/NY-Wadsworth-21004959-01/20 | 2020-12-28 | North America/USA/New York/Bronx | MONTEFIORE MEDICAL CENTER LABORATORIES | Wadsworth Center, New York State Department of Health | Kirsten St. George et al |
| EPI_ISL_861279 | USA/NY-Wadsworth-21004977-01/20 | 2020-12-24 | North America/USA/New York/Bronx | MONTEFIORE MEDICAL CENTER LABORATORIES | Wadsworth Center, New York State Department of Health | Kirsten St. George et al |
| EPI_ISL_861288 | USA/NY-Wadsworth-21004987-01/20 | 2020-12-25 | North America/USA/New York/Bronx | MONTEFIORE MEDICAL CENTER LABORATORIES | Wadsworth Center, New York State Department of Health | Kirsten St. George et al |
| EPI_ISL_861290 | USA/NY-Wadsworth-21004989-01/20 | 2020-12-30 | North America/USA/New York/Bronx | MONTEFIORE MEDICAL CENTER LABORATORIES | Wadsworth Center, New York State Department of Health | Kirsten St. George et al |
| EPI_ISL_861295 | USA/NY-Wadsworth-21005002-01/20 | 2020-12-30 | North America/USA/New York/Bronx | MONTEFIORE MEDICAL CENTER LABORATORIES | Wadsworth Center, New York State Department of Health | Kirsten St. George et al |

| | | | | | | |
|---|---|---|---|---|---|---|
| EPI_ISL_861297 | USA/NY-Wadsworth-21005004-01/20 | 2020-12-29 | North America/USA/New York/Bronx | MONTEFIORE MEDICAL CENTER LABORATORIES | Wadsworth Center, New York State Department of Health | Kirsten St. George et al |
| EPI_ISL_861305 | USA/NY-Wadsworth-21005015-01/20 | 2020-12-30 | North America/USA/New York/Bronx | MONTEFIORE MEDICAL CENTER LABORATORIES | Wadsworth Center, New York State Department of Health | Kirsten St. George et al |
| EPI_ISL_861308 | USA/NY-Wadsworth-21005019-01/20 | 2020-12-28 | North America/USA/New York/Bronx | MONTEFIORE MEDICAL CENTER LABORATORIES | Wadsworth Center, New York State Department of Health | Kirsten St. George et al |
| EPI_ISL_861318 | USA/NY-Wadsworth-21005032-01/20 | 2020-12-27 | North America/USA/New York/Bronx | MONTEFIORE MEDICAL CENTER LABORATORIES | Wadsworth Center, New York State Department of Health | Kirsten St. George et al |
| EPI_ISL_861326 | USA/NY-Wadsworth-21005046-01/20 | 2020-12-28 | North America/USA/New York/Bronx | MONTEFIORE MEDICAL CENTER LABORATORIES | Wadsworth Center, New York State Department of Health | Kirsten St. George et al |
| EPI_ISL_861327 | USA/NY-Wadsworth-21005048-01/20 | 2020-12-29 | North America/USA/New York/Westche | MONTEFIORE MEDICAL CENTER LABORATORIES | Wadsworth Center, New York State Department of Health | Kirsten St. George et al |
| EPI_ISL_861338 | USA/NY-Wadsworth-21006079-01/20 | 2021-01-09 | North America/USA/New York/Westche | WESTCHESTER MEDICAL CENTER | Wadsworth Center, New York State Department of Health | Kirsten St. George et al |
| EPI_ISL_861343 | USA/NY-Wadsworth-21006084-01/20 | 2021-01-09 | North America/USA/New York/Westche | WESTCHESTER MEDICAL CENTER | Wadsworth Center, New York State Department of Health | Kirsten St. George et al |
| EPI_ISL_861344 | USA/NY-Wadsworth-21006085-01/20 | 2021-01-09 | North America/USA/New York/Westche | WESTCHESTER MEDICAL CENTER | Wadsworth Center, New York State Department of Health | Kirsten St. George et al |
| EPI_ISL_861345 | USA/NY-Wadsworth-21006086-01/20 | 2021-01-08 | North America/USA/New York/Dutches | WESTCHESTER MEDICAL CENTER | Wadsworth Center, New York State Department of Health | Kirsten St. George et al |

| | | | | | | |
|---|---|---|---|---|---|---|
| EPI_ISL_861349 | USA/NY-Wadsworth-21006090-01/20 | 2021-01-09 | North America/USA/New York/Ulster C | WESTCHESTER MEDICAL CENTER | Wadsworth Center, New York State Department of Health | Kirsten St. George et al |
| EPI_ISL_861351 | USA/NY-Wadsworth-21006092-01/20 | 2021-01-08 | North America/USA/New York/Dutchess | WESTCHESTER MEDICAL CENTER | Wadsworth Center, New York State Department of Health | Kirsten St. George et al |
| EPI_ISL_861355 | USA/NY-Wadsworth-21006096-01/20 | 2021-01-07 | North America/USA/New York/Westche | WESTCHESTER MEDICAL CENTER | Wadsworth Center, New York State Department of Health | Kirsten St. George et al |
| EPI_ISL_861374 | USA/NY-Wadsworth-21006121-01/20 | 2021-01-05 | North America/USA/New York/Rockland | WESTCHESTER MEDICAL CENTER | Wadsworth Center, New York State Department of Health | Kirsten St. George et al |
| EPI_ISL_861376 | USA/NY-Wadsworth-21006123-01/20 | 2021-01-05 | North America/USA/New York/Ulster C | WESTCHESTER MEDICAL CENTER | Wadsworth Center, New York State Department of Health | Kirsten St. George et al |
| EPI_ISL_861385 | USA/NY-Wadsworth-21006137-01/20 | 2021-01-05 | North America/USA/New York/Ulster C | WESTCHESTER MEDICAL CENTER | Wadsworth Center, New York State Department of Health | Kirsten St. George et al |
| EPI_ISL_861392 | USA/NY-Wadsworth-21006146-01/20 | 2021-01-06 | North America/USA/New York/Ulster C | WESTCHESTER MEDICAL CENTER | Wadsworth Center, New York State Department of Health | Kirsten St. George et al |
| EPI_ISL_861396 | USA/NY-Wadsworth-21006150-01/20 | 2021-01-05 | North America/USA/New York/Ulster C | WESTCHESTER MEDICAL CENTER | Wadsworth Center, New York State Department of Health | Kirsten St. George et al |
| EPI_ISL_861397 | USA/NY-Wadsworth-21006151-01/20 | 2021-01-06 | North America/USA/New York/Westche | WESTCHESTER MEDICAL CENTER | Wadsworth Center, New York State Department of Health | Kirsten St. George et al |
| EPI_ISL_861409 | USA/NY-Wadsworth-21006163-01/20 | 2021-01-06 | North America/USA/New York/Westche | WESTCHESTER MEDICAL CENTER | Wadsworth Center, New York State Department of Health | Kirsten St. George et al |

| | | | | | | |
|---|---|---|---|---|---|---|
| EPI_ISL_861413 | USA/NY-Wadsworth-21006364-01/20 | 2020-12-31 | North America/USA/New York/Nassau ( | NORTH SHORE UNIVERSITY HOSPITAL | Wadsworth Center, New York State Department of Health | Kirsten St. George et al |
| EPI_ISL_861415 | USA/NY-Wadsworth-21006366-01/20 | 2021-01-01 | North America/USA/New York/Queens | NORTH SHORE UNIVERSITY HOSPITAL | Wadsworth Center, New York State Department of Health | Kirsten St. George et al |
| EPI_ISL_861441 | USA/OH-JML-COHDEC35/2020 | 2020-12-31 | North America/USA/Ohio/ | The Ohio State University | James Molecular Laboratory | Huolin Tu et al |
| EPI_ISL_861443 | USA/NC-UNC-0016/2021 | 2021-01-15 | North America/USA/North Carolina/ | Clinical Molecular Microbiology Laboratory, UNC Hospitals | Jeremy Wang | Jeremy Wang et al |
| EPI_ISL_861445 | USA/NC-UNC-0018/2021 | 2021-01-10 | North America/USA/North Carolina/ | Clinical Molecular Microbiology Laboratory, UNC Hospitals | Jeremy Wang | Jeremy Wang et al |
| EPI_ISL_861451 | USA/NC-UNC-0024/2021 | 2021-01-15 | North America/USA/North Carolina/ | Clinical Molecular Microbiology Laboratory, UNC Hospitals | Jeremy Wang | Jeremy Wang et al |
| EPI_ISL_861456 | Belarus/ChVir21835/2020 | 2020-11-07 | Europe/Belarus/Belarus/ | WHO/Minsk | Charit√© Universit√§tsmedizin Berlin, Institut f√ºr Virologie | Victor M Corman et al |
| EPI_ISL_861457 | Belarus/ChVir21841/2020 | 2020-10-21 | Europe/Belarus/Belarus/ | WHO/Minsk | Charit√© Universit√§tsmedizin Berlin, Institut f√ºr Virologie | Victor M Corman et al |
| EPI_ISL_861458 | Belarus/ChVir21890/2020 | 2020-10-23 | Europe/Belarus/Belarus/ | WHO/Minsk | Charit√© Universit√§tsmedizin Berlin, Institut f√ºr Virologie | Victor M Corman et al |
| EPI_ISL_861461 | Ghana/UHAS-V465/2020 | 2020-10-30 | Africa/Ghana/Volta/ | UHAS COVID-19 Lab | UHAS COVID-19 Lab | Kwabena O. Duedu et al |
| EPI_ISL_861474 | USA/WI-GMF-47936/2021 | 2021-01-08 | North America/USA/Wisconsin/Crawfor | Gundersen Molecular Diagnostics Laboratory | Kabara Cancer Research Institute | Craig S. Richmond et al |
| EPI_ISL_861476 | USA/WY-WPHL-21002574/2021 | 2021-01-07 | North America/USA/Wyoming/ | Wyoming Public Health Laboratory | Wyoming Public Health Laboratory | Noah Hull et al |
| EPI_ISL_861514 | Germany/NW-HHU-576/2020 | 2020-12-16 | Europe/Germany/North Rhine Westphal | ZOTZ KLIMAS MVZ D√ºsseldorf-Centrum GbR √úBAG f√ºr Labormedizin, Genetik, Zytologie, Pathologie | Center of Medical Microbiology, Virology, and Hospital Hygiene, University of Duesseldorf | Maximilian Damagnez et al |

| | | | | | | |
|---|---|---|---|---|---|---|
| EPI_ISL_861515 | Germany/NW-HHU-577/2020 | 2020-12-16 | Europe/Germany/North Rhine Westphal | ZOTZ KLIMAS MVZ D√ºsseldorf-Centrum GbR √úBAG f√ºr Labormedizin, Genetik, Zytologie, Pathologie | Center of Medical Microbiology, Virology, and Hospital Hygiene, University of Duesseldorf | Maximilian Damagnez et al |
| EPI_ISL_861521 | Germany/NW-HHU-583/2020 | 2020-12-16 | Europe/Germany/North Rhine Westphal | ZOTZ KLIMAS MVZ D√ºsseldorf-Centrum GbR √úBAG f√ºr Labormedizin, Genetik, Zytologie, Pathologie | Center of Medical Microbiology, Virology, and Hospital Hygiene, University of Duesseldorf | Maximilian Damagnez et al |
| EPI_ISL_861522 | Germany/NW-HHU-584/2020 | 2020-12-16 | Europe/Germany/North Rhine Westphal | ZOTZ KLIMAS MVZ D√ºsseldorf-Centrum GbR √úBAG f√ºr Labormedizin, Genetik, Zytologie, Pathologie | Center of Medical Microbiology, Virology, and Hospital Hygiene, University of Duesseldorf | Maximilian Damagnez et al |
| EPI_ISL_861524 | Germany/NW-HHU-586/2020 | 2020-12-16 | Europe/Germany/North Rhine Westphal | ZOTZ KLIMAS MVZ D√ºsseldorf-Centrum GbR √úBAG f√ºr Labormedizin, Genetik, Zytologie, Pathologie | Center of Medical Microbiology, Virology, and Hospital Hygiene, University of Duesseldorf | Maximilian Damagnez et al |
| EPI_ISL_861526 | Germany/NW-HHU-588/2020 | 2020-12-16 | Europe/Germany/North Rhine Westphal | ZOTZ KLIMAS MVZ D√ºsseldorf-Centrum GbR √úBAG f√ºr Labormedizin, Genetik, Zytologie, Pathologie | Center of Medical Microbiology, Virology, and Hospital Hygiene, University of Duesseldorf | Maximilian Damagnez et al |
| EPI_ISL_861527 | Germany/NW-HHU-589/2020 | 2020-12-16 | Europe/Germany/North Rhine Westphal | ZOTZ KLIMAS MVZ D√ºsseldorf-Centrum GbR √úBAG f√ºr Labormedizin, Genetik, Zytologie, Pathologie | Center of Medical Microbiology, Virology, and Hospital Hygiene, University of Duesseldorf | Maximilian Damagnez et al |
| EPI_ISL_861529 | Portugal/PT2250/2021 | 2021-01-08 | Europe/Portugal/Portugal/ | Instituto Nacional de Saude (INSA) | Instituto Nacional de Saude (INSA) | Borges et al et al |
| EPI_ISL_861537 | Portugal/PT2202/2020 | 2020-12-28 | Europe/Portugal/Portugal/ | Instituto Nacional de Saude (INSA) | Instituto Nacional de Saude (INSA) | Borges et al et al |

| | | | | | | |
|---|---|---|---|---|---|---|
| EPI_ISL_861540 | Portugal/PT2232/2021 | 2021-01-01 | Europe/Portugal/Portugal/ | Instituto Nacional de Saude (INSA) | Instituto Nacional de Saude (INSA) | Borges et al et al |
| EPI_ISL_861543 | Portugal/PT2235/2021 | 2021-01-06 | Europe/Portugal/Portugal/ | Instituto Nacional de Saude (INSA) | Instituto Nacional de Saude (INSA) | Borges et al et al |
| EPI_ISL_861551 | Portugal/PT2253/2021 | 2021-01-06 | Europe/Portugal/Portugal/ | Instituto Nacional de Saude (INSA) | Instituto Nacional de Saude (INSA) | Borges et al et al |
| EPI_ISL_861555 | Portugal/PT2285/2021 | 2021-01-10 | Europe/Portugal/Portugal/ | Instituto Nacional de Saude (INSA) | Instituto Nacional de Saude (INSA) | Borges et al et al |
| EPI_ISL_861562 | Portugal/PT2283/2021 | 2021-01-09 | Europe/Portugal/Portugal/ | Instituto Nacional de Saude (INSA) | Instituto Nacional de Saude (INSA) | Borges et al et al |
| EPI_ISL_861570 | Portugal/PT2225/2020 | 2020-12-29 | Europe/Portugal/Portugal/ | Instituto Nacional de Saude (INSA) | Instituto Nacional de Saude (INSA) | Borges et al et al |
| EPI_ISL_861579 | Portugal/PT2287/2021 | 2021-01-08 | Europe/Portugal/Portugal/ | Instituto Nacional de Saude (INSA) | Instituto Nacional de Saude (INSA) | Borges et al et al |
| EPI_ISL_861584 | Portugal/PT2198/2020 | 2020-12-07 | Europe/Portugal/Portugal/ | Instituto Nacional de Saude (INSA) | Instituto Nacional de Saude (INSA) | Borges et al et al |
| EPI_ISL_861585 | Portugal/PT2199/2020 | 2020-12-09 | Europe/Portugal/Portugal/ | Instituto Nacional de Saude (INSA) | Instituto Nacional de Saude (INSA) | Borges et al et al |
| EPI_ISL_861586 | Portugal/PT2200/2020 | 2020-12-11 | Europe/Portugal/Portugal/ | Instituto Nacional de Saude (INSA) | Instituto Nacional de Saude (INSA) | Borges et al et al |
| EPI_ISL_861588 | Portugal/PT2203/2020 | 2020-12-28 | Europe/Portugal/Portugal/ | Instituto Nacional de Saude (INSA) | Instituto Nacional de Saude (INSA) | Borges et al et al |
| EPI_ISL_861590 | Portugal/PT2206/2020 | 2020-12-15 | Europe/Portugal/Portugal/ | Instituto Nacional de Saude (INSA) | Instituto Nacional de Saude (INSA) | Borges et al et al |
| EPI_ISL_861593 | Portugal/PT2209/2021 | 2021-01-07 | Europe/Portugal/Portugal/ | Instituto Nacional de Saude (INSA) | Instituto Nacional de Saude (INSA) | Borges et al et al |
| EPI_ISL_861595 | Portugal/PT2211/2021 | 2021-01-05 | Europe/Portugal/Portugal/ | Instituto Nacional de Saude (INSA) | Instituto Nacional de Saude (INSA) | Borges et al et al |
| EPI_ISL_861610 | Portugal/PT2257/2021 | 2021-01-05 | Europe/Portugal/Portugal/ | Instituto Nacional de Saude (INSA) | Instituto Nacional de Saude (INSA) | Borges et al et al |
| EPI_ISL_861611 | Portugal/PT2258/2021 | 2021-01-05 | Europe/Portugal/Portugal/ | Instituto Nacional de Saude (INSA) | Instituto Nacional de Saude (INSA) | Borges et al et al |
| EPI_ISL_861616 | Portugal/PT2265/2021 | 2021-01-06 | Europe/Portugal/Portugal/ | Instituto Nacional de Saude (INSA) | Instituto Nacional de Saude (INSA) | Borges et al et al |
| EPI_ISL_861618 | Portugal/PT2267/2021 | 2021-01-06 | Europe/Portugal/Portugal/ | Instituto Nacional de Saude (INSA) | Instituto Nacional de Saude (INSA) | Borges et al et al |
| EPI_ISL_861624 | Portugal/PT2277/2021 | 2021-01-09 | Europe/Portugal/Portugal/ | Instituto Nacional de Saude (INSA) | Instituto Nacional de Saude (INSA) | Borges et al et al |

| | | | | | | |
|---|---|---|---|---|---|---|
| EPI_ISL_861632 | Brazil/SP-945/2020 | 2020-04-18 | South America/Brazil/S√£o Paulo/S√£o | Instituto Adolfo Lutz - Central | Instituto Adolfo Lutz, Interdiciplinary Procedures Center, Strategic Laboratory | Claudio Tavares Sacchi et al |
| EPI_ISL_861634 | Brazil/SP-858/2020 | 2020-04-07 | South America/Brazil/S√£o Paulo/S√£o | Instituto Adolfo Lutz - Central | Instituto Adolfo Lutz, Interdiciplinary Procedures Center, Strategic Laboratory | Claudio Tavares Sacchi et al |
| EPI_ISL_861637 | Brazil/SP-865/2020 | 2020-04-21 | South America/Brazil/S√£o Paulo/S√£o | Hospital Nipo Brasileiro | Instituto Adolfo Lutz, Interdiciplinary Procedures Center, Strategic Laboratory | Claudio Tavares Sacchi et al |
| EPI_ISL_861643 | Brazil/SP-839/2020 | 2020-04-30 | South America/Brazil/S√£o Paulo/S√£o | Instituto Adolfo Lutz - Central | Instituto Adolfo Lutz, Interdiciplinary Procedures Center, Strategic Laboratory | Claudio Tavares Sacchi et al |
| EPI_ISL_861644 | Brazil/SP-950/2020 | 2020-05-05 | South America/Brazil/S√£o Paulo/S√£o | Hospital Santa Virginia | Instituto Adolfo Lutz, Interdiciplinary Procedures Center, Strategic Laboratory | Claudio Tavares Sacchi et al |
| EPI_ISL_861645 | Brazil/SP-951/2020 | 2020-05-07 | South America/Brazil/S√£o Paulo/S√£o | Hospital e Pronto Socorro Comunitario Vila Iolanda | Instituto Adolfo Lutz, Interdiciplinary Procedures Center, Strategic Laboratory | Claudio Tavares Sacchi et al |

| | | | | | | |
|---|---|---|---|---|---|---|
| EPI_ISL_861646 | Brazil/SP-952/2020 | 2020-05-21 | South America/Brazil/S√£o Paulo/S√£o | Hospital Santa Marcelina Sao Paulo | Instituto Adolfo Lutz, Interdiciplinary Procedures Center, Strategic Laboratory | Claudio Tavares Sacchi et al |
| EPI_ISL_861647 | Brazil/SP-953/2020 | 2020-05-25 | South America/Brazil/S√£o Paulo/S√£o | Hospital Santa Marcelina Sao Paulo | Instituto Adolfo Lutz, Interdiciplinary Procedures Center, Strategic Laboratory | Claudio Tavares Sacchi et al |
| EPI_ISL_861648 | Brazil/SP-954/2020 | 2020-05-28 | South America/Brazil/S√£o Paulo/S√£o | Hospital e Pronto Socorro Portinari | Instituto Adolfo Lutz, Interdiciplinary Procedures Center, Strategic Laboratory | Claudio Tavares Sacchi et al |
| EPI_ISL_861649 | Brazil/SP-955/2020 | 2020-05-28 | South America/Brazil/S√£o Paulo/Campi | Hospital Renascen√ßa Campinas | Instituto Adolfo Lutz, Interdiciplinary Procedures Center, Strategic Laboratory | Claudio Tavares Sacchi et al |
| EPI_ISL_861650 | Brazil/SP-956/2020 | 2020-05-08 | South America/Brazil/S√£o Paulo/S√£o | Hospital Santa Marcelina Sao Paulo | Instituto Adolfo Lutz, Interdiciplinary Procedures Center, Strategic Laboratory | Claudio Tavares Sacchi et al |
| EPI_ISL_861651 | Brazil/SP-957/2020 | 2020-05-10 | South America/Brazil/S√£o Paulo/S√£o | AMA Jardim Brasil | Instituto Adolfo Lutz, Interdiciplinary Procedures Center, Strategic Laboratory | Claudio Tavares Sacchi et al |

| | | | | | | |
|---|---|---|---|---|---|---|
| EPI_ISL_861652 | Brazil/SP-958/2020 | 2020-05-09 | South America/Brazil/S√£o Paulo/S√£o [illegible] | AMA Wamberto Dias da Costa | Instituto Adolfo Lutz, Interdiciplinary Procedures Center, Strategic Laboratory | Claudio Tavares Sacchi et al |
| EPI_ISL_861653 | Brazil/SP-960/2020 | 2020-05-13 | South America/Brazil/S√£o Paulo/S√£o [illegible] | Hospital Santa Virg√≠nia | Instituto Adolfo Lutz, Interdiciplinary Procedures Center, Strategic Laboratory | Claudio Tavares Sacchi et al |
| EPI_ISL_861654 | Brazil/SP-961/2020 | 2020-05-12 | South America/Brazil/S√£o Paulo/S√£o [illegible] | Hospital Santa Marcelina Sao Paulo | Instituto Adolfo Lutz, Interdiciplinary Procedures Center, Strategic Laboratory | Claudio Tavares Sacchi et al |
| EPI_ISL_861655 | Brazil/SP-962/2020 | 2020-05-12 | South America/Brazil/S√£o Paulo/S√£o [illegible] | Hospital Santa Marcelina Sao Paulo | Instituto Adolfo Lutz, Interdiciplinary Procedures Center, Strategic Laboratory | Claudio Tavares Sacchi et al |
| EPI_ISL_861656 | Brazil/SP-963/2020 | 2020-05-15 | South America/Brazil/S√£o Paulo/Jandir | UPA de Jandira | Instituto Adolfo Lutz, Interdiciplinary Procedures Center, Strategic Laboratory | Claudio Tavares Sacchi et al |
| EPI_ISL_861657 | Brazil/SP-964/2020 | 2020-05-17 | South America/Brazil/S√£o Paulo/Osasc | Hospital e Maternidade Sino Brasileiro | Instituto Adolfo Lutz, Interdiciplinary Procedures Center, Strategic Laboratory | Claudio Tavares Sacchi et al |

| | | | | | | |
|---|---|---|---|---|---|---|
| EPI_ISL_861658 | Brazil/SP-965/2020 | 2020-05-18 | South America/Brazil/S√£o Paulo/Osasco | Hospital Municipal Ant√¥nio Giglio | Instituto Adolfo Lutz, Interdiciplinary Procedures Center, Strategic Laboratory | Claudio Tavares Sacchi et al |
| EPI_ISL_861659 | Brazil/SP-966/2020 | 2020-05-15 | South America/Brazil/S√£o Paulo/Barue | PS e Maternidade Nair Fonseca Leitao Arantes | Instituto Adolfo Lutz, Interdiciplinary Procedures Center, Strategic Laboratory | Claudio Tavares Sacchi et al |
| EPI_ISL_861660 | Brazil/SP-967/2020 | 2020-05-16 | South America/Brazil/S√£o Paulo/Barue | PS e Maternidade Nair Fonseca Leitao Arantes | Instituto Adolfo Lutz, Interdiciplinary Procedures Center, Strategic Laboratory | Claudio Tavares Sacchi et al |
| EPI_ISL_861661 | Brazil/SP-968/2020 | 2020-05-15 | South America/Brazil/S√£o Paulo/Barue | PS e Maternidade Nair Fonseca Leitao Arantes | Instituto Adolfo Lutz, Interdiciplinary Procedures Center, Strategic Laboratory | Claudio Tavares Sacchi et al |
| EPI_ISL_861662 | Brazil/SP-969/2020 | 2020-05-18 | South America/Brazil/S√£o Paulo/Presid | CS I Tacito Leite de Carvalho e Silva | Instituto Adolfo Lutz, Interdiciplinary Procedures Center, Strategic Laboratory | Claudio Tavares Sacchi et al |
| EPI_ISL_861663 | Brazil/SP-970/2020 | 2020-04-22 | South America/Brazil/S√£o Paulo/Caieir | Instituto Adolfo Lutz - Central | Instituto Adolfo Lutz, Interdiciplinary Procedures Center, Strategic Laboratory | Claudio Tavares Sacchi et al |

| | | | | | | |
|---|---|---|---|---|---|---|
| EPI_ISL_861664 | Brazil/SP-973/2020 | 2020-05-27 | South America/Brazil/S√£o Paulo/Mogi- | Instituto Adolfo Lutz - Regional de Campinas | Instituto Adolfo Lutz, Interdiciplinary Procedures Center, Strategic Laboratory | Claudio Tavares Sacchi et al |
| EPI_ISL_861665 | Brazil/SP-781/2020 | 2020-07-02 | South America/Brazil/S√£o Paulo/Sao Jo | Instituto Adolfo Lutz - Regional de Taubate | Instituto Adolfo Lutz, Interdiciplinary Procedures Center, Strategic Laboratory | Claudio Tavares Sacchi et al |
| EPI_ISL_861666 | Brazil/SP-803/2020 | 2020-05-30 | South America/Brazil/S√£o Paulo/Porto | PSF Dr. Antonio Pires de Almeida | Instituto Adolfo Lutz, Interdiciplinary Procedures Center, Strategic Laboratory | Claudio Tavares Sacchi et al |
| EPI_ISL_861667 | Brazil/SP-806/2020 | 2020-06-29 | South America/Brazil/S√£o Paulo/Rio Cla | Instituto Adolfo Lutz - Regional de Rio Claro | Instituto Adolfo Lutz, Interdiciplinary Procedures Center, Strategic Laboratory | Claudio Tavares Sacchi et al |
| EPI_ISL_861668 | Brazil/SP-902/2020 | 2020-11-26 | South America/Brazil/S√£o Paulo/Presid | Centro de Triagem Covid19 | Instituto Adolfo Lutz, Interdiciplinary Procedures Center, Strategic Laboratory | Claudio Tavares Sacchi et al |
| EPI_ISL_861669 | Brazil/SP-913/2020 | 2020-12-08 | South America/Brazil/S√£o Paulo/Embu- | Lab LOC - Itapecerica da Serra | Instituto Adolfo Lutz, Interdiciplinary Procedures Center, Strategic Laboratory | Claudio Tavares Sacchi et al |

| | | | | | | |
|---|---|---|---|---|---|---|
| EPI_ISL_861670 | Brazil/SP-914/2020 | 2020-12-08 | South America/Brazil/S√£o Paulo/Loren | Instituto Adolfo Lutz - Regional de Taubate | Instituto Adolfo Lutz, Interdiciplinary Procedures Center, Strategic Laboratory | Claudio Tavares Sacchi et al |
| EPI_ISL_861671 | Brazil/SP-936/2020 | 2020-11-01 | South America/Brazil/S√£o Paulo/Mogi | Hospital Municipal Prefeito Waldemar Costa Filho | Instituto Adolfo Lutz, Interdiciplinary Procedures Center, Strategic Laboratory | Claudio Tavares Sacchi et al |
| EPI_ISL_861672 | Brazil/SP-937/2020 | 2020-11-02 | South America/Brazil/S√£o Paulo/Ermel | Day Hospital de Ermelino Matarazzo | Instituto Adolfo Lutz, Interdiciplinary Procedures Center, Strategic Laboratory | Claudio Tavares Sacchi et al |
| EPI_ISL_861673 | Brazil/SP-940/2020 | 2020-11-04 | South America/Brazil/S√£o Paulo/Osasc | PA Novo Osasco | Instituto Adolfo Lutz, Interdiciplinary Procedures Center, Strategic Laboratory | Claudio Tavares Sacchi et al |
| EPI_ISL_861674 | Brazil/SP-891/2020 | 2020-12-16 | South America/Brazil/S√£o Paulo/Carag | UPA Central de Caraguatatuba | Instituto Adolfo Lutz, Interdiciplinary Procedures Center, Strategic Laboratory | Claudio Tavares Sacchi et al |
| EPI_ISL_861675 | Brazil/SP-892/2020 | 2020-12-16 | South America/Brazil/S√£o Paulo/Carag | UPA Central de Caraguatatuba | Instituto Adolfo Lutz, Interdiciplinary Procedures Center, Strategic Laboratory | Claudio Tavares Sacchi et al |

| | | | | | | |
|---|---|---|---|---|---|---|
| EPI_ISL_861676 | Brazil/SP-983/2020 | 2020-12-23 | South America/Brazil/S√£o Paulo/S√£o [illegible] | UPA Vila Santa Catarina | Instituto Adolfo Lutz, Interdiciplinary Procedures Center, Strategic Laboratory | Claudio Tavares Sacchi et al |
| EPI_ISL_861677 | Brazil/SP-984/2020 | 2020-12-23 | South America/Brazil/S√£o Paulo/S√£o [illegible] | Instituto Adolfo Lutz - Central | Instituto Adolfo Lutz, Interdiciplinary Procedures Center, Strategic Laboratory | Claudio Tavares Sacchi et al |
| EPI_ISL_861678 | Brazil/SP-985/2020 | 2020-12-22 | South America/Brazil/S√£o Paulo/Guaru | Laboratorio Municipal de Guarulhos | Instituto Adolfo Lutz, Interdiciplinary Procedures Center, Strategic Laboratory | Claudio Tavares Sacchi et al |
| EPI_ISL_861679 | Brazil/SP-986/2020 | 2020-12-28 | South America/Brazil/S√£o Paulo/Pinda | Instituto Adolfo Lutz - Regional de Taubate | Instituto Adolfo Lutz, Interdiciplinary Procedures Center, Strategic Laboratory | Claudio Tavares Sacchi et al |
| EPI_ISL_861680 | Brazil/SP-1002/2020 | 2020-12-26 | South America/Brazil/S√£o Paulo/S√£o [illegible] | Hospital e Pronto Socorro Portinari | Instituto Adolfo Lutz, Interdiciplinary Procedures Center, Strategic Laboratory | Claudio Tavares Sacchi et al |
| EPI_ISL_861681 | Brazil/SP-1003/2021 | 2021-01-01 | South America/Brazil/S√£o Paulo/Ameri | Hospital Municipal Dr. Waldemar Tebaldi | Instituto Adolfo Lutz, Interdiciplinary Procedures Center, Strategic Laboratory | Claudio Tavares Sacchi et al |

| | | | | | | |
|---|---|---|---|---|---|---|
| EPI_ISL_861682 | Brazil/SP-1004/2020 | 2020-12-20 | South America/Brazil/S√£o Paulo/S√£o [illegible] | UPA Vila Santa Catarina | Instituto Adolfo Lutz, Interdiciplinary Procedures Center, Strategic Laboratory | Claudio Tavares Sacchi et al |
| EPI_ISL_861683 | Brazil/SP-1005/2021 | 2021-01-04 | South America/Brazil/S√£o Paulo/Guaru | Complexo Hospitalar Padre Bento de Guarulhos | Instituto Adolfo Lutz, Interdiciplinary Procedures Center, Strategic Laboratory | Claudio Tavares Sacchi et al |
| EPI_ISL_861684 | Brazil/SP-1006/2021 | 2021-01-02 | South America/Brazil/S√£o Paulo/S√£o [illegible] | Hospital Pronto Socorro Itaquera | Instituto Adolfo Lutz, Interdiciplinary Procedures Center, Strategic Laboratory | Claudio Tavares Sacchi et al |
| EPI_ISL_861685 | Brazil/SP-1007/2021 | 2021-01-02 | South America/Brazil/S√£o Paulo/S√£o [illegible] | Hospital Pronto Socorro Itaquera | Instituto Adolfo Lutz, Interdiciplinary Procedures Center, Strategic Laboratory | Claudio Tavares Sacchi et al |
| EPI_ISL_861711 | Romania/Tulcea413486/2021 | 2021-01-14 | Europe/Romania/Tulcea/ | Laboratory for Respiratory Viruses, Cantacuzino National Military-Medical Institute for Research and Development | Cantacuzino Institute Virology | Mihaela Lazar et al |
| EPI_ISL_861712 | Romania/Cluj413482/2021 | 2021-01-13 | Europe/Romania/Cluj/ | Laboratory for Respiratory Viruses, Cantacuzino National Military-Medical Institute for Research and Development | Cantacuzino Institute Virology | Mihaela Lazar et al |
| EPI_ISL_861713 | Romania/Cluj413485/2021 | 2021-01-13 | Europe/Romania/Cluj Napoca/ | Laboratory for Respiratory Viruses, Cantacuzino National Military-Medical Institute for Research and Development | Cantacuzino Institute Virology | Mihaela Lazar et al |
| EPI_ISL_861715 | France/CERBAHC-20211018-95/2021 | 2021-01-07 | Europe/France/Ile de France/ | CERBA | CERBA LAB | Roquebert B et al |
| EPI_ISL_861719 | Malaysia/HPUPM_ML5767/2020 | 2020-10-20 | Asia/Malaysia/Malaysia/ | Department of Medical Microbiology, Hospital Pengajar Universiti Putra Malaysia | Malaysia Genome Institute | Mohd Noor Mat Isa et al |
| EPI_ISL_861732 | USA/NJ-Yale-S132/2021 | 2021-01-09 | North America/USA/New Jersey/ | Tempus | Grubaugh Lab - Yale School of Public Health | Tara Alpert et al |

| | | | | | | |
|---|---|---|---|---|---|---|
| EPI_ISL_861753 | USA/CT-Yale-S102/2021 | 2021-01-05 | North America/USA/Connecticut/ | Yale Clinical Virology Lab | Grubaugh Lab - Yale School of Public Health | Tara Alpert et al |
| EPI_ISL_861773 | Spain/MD-IBV-99014312/2020 | 2020-12-08 | Europe/Spain/Madrid/Madrid | Hospital General Universitario Gregorio Marav±V≥n | SeqCOVID-SPAIN consortium/IBV(CSIC) | Darv≠o Garcv≠a de Viedma et al |
| EPI_ISL_861790 | Spain/MD-IBV-99014257/2020 | 2020-12-10 | Europe/Spain/Madrid/Madrid | Hospital General Universitario Gregorio Marav±V≥n | SeqCOVID-SPAIN consortium/IBV(CSIC) | Darv≠o Garcv≠a de Viedma et al |
| EPI_ISL_861791 | Spain/MD-IBV-99014252/2020 | 2020-12-11 | Europe/Spain/Madrid/Madrid | Hospital General Universitario Gregorio Marav±V≥n | SeqCOVID-SPAIN consortium/IBV(CSIC) | Darv≠o Garcv≠a de Viedma et al |
| EPI_ISL_861793 | Spain/MD-IBV-99014245/2020 | 2020-12-17 | Europe/Spain/Madrid/Madrid | Hospital General Universitario Gregorio Marav±V≥n | SeqCOVID-SPAIN consortium/IBV(CSIC) | Darv≠o Garcv≠a de Viedma et al |
| EPI_ISL_861803 | Spain/MD-IBV-99014310/2020 | 2020-12-08 | Europe/Spain/Madrid/Madrid | Hospital General Universitario Gregorio Marav±V≥n | SeqCOVID-SPAIN consortium/IBV(CSIC) | Darv≠o Garcv≠a de Viedma et al |
| EPI_ISL_861808 | Spain/MD-IBV-99014270/2020 | 2020-12-09 | Europe/Spain/Madrid/Madrid | Hospital General Universitario Gregorio Marav±V≥n | SeqCOVID-SPAIN consortium/IBV(CSIC) | Darv≠o Garcv≠a de Viedma et al |
| EPI_ISL_861814 | Spain/MD-IBV-99014290/2020 | 2020-12-06 | Europe/Spain/Madrid/Madrid | Hospital General Universitario Gregorio Marav±V≥n | SeqCOVID-SPAIN consortium/IBV(CSIC) | Darv≠o Garcv≠a de Viedma et al |
| EPI_ISL_861829 | Spain/MD-IBV-99014255/2020 | 2020-12-10 | Europe/Spain/Madrid/Madrid | Hospital General Universitario Gregorio Marav±V≥n | SeqCOVID-SPAIN consortium/IBV(CSIC) | Darv≠o Garcv≠a de Viedma et al |
| EPI_ISL_861830 | Spain/MD-IBV-99014221/2020 | 2020-12-09 | Europe/Spain/Madrid/Madrid | Hospital General Universitario Gregorio Marav±V≥n | SeqCOVID-SPAIN consortium/IBV(CSIC) | Darv≠o Garcv≠a de Viedma et al |
| EPI_ISL_861833 | Spain/MD-IBV-99014262/2020 | 2020-12-09 | Europe/Spain/Madrid/Madrid | Hospital General Universitario Gregorio Marav±V≥n | SeqCOVID-SPAIN consortium/IBV(CSIC) | Darv≠o Garcv≠a de Viedma et al |
| EPI_ISL_861836 | Spain/MD-IBV-99014273/2020 | 2020-12-09 | Europe/Spain/Madrid/Madrid | Hospital General Universitario Gregorio Marav±V≥n | SeqCOVID-SPAIN consortium/IBV(CSIC) | Darv≠o Garcv≠a de Viedma et al |
| EPI_ISL_861849 | Spain/MD-IBV-99014281/2020 | 2020-12-05 | Europe/Spain/Madrid/Madrid | Hospital General Universitario Gregorio Marav±V≥n | SeqCOVID-SPAIN consortium/IBV(CSIC) | Darv≠o Garcv≠a de Viedma et al |

| | | | | | | |
|---|---|---|---|---|---|---|
| EPI_ISL_861855 | Spain/MD-IBV-99014233/2020 | 2020-10-29 | Europe/Spain/Madrid/Madrid | Hospital General Universitario Gregorio Marav±V≥n | SeqCOVID-SPAIN consortium/IBV(CSIC) | Dar√≠o Garc√≠a de Viedma et al |
| EPI_ISL_861866 | Switzerland/SZ-UHB-100409126701/2 | 2020-12-30 | Europe/Switzerland/Schwyz/Merlischac | Bioanalytika AG | University Hospital Basel, Clinical Bacteriology | Tim Roloff et al |
| EPI_ISL_861868 | Brazil/SP-HIAE-ID02/2020 | 2020-11-21 | South America/Brazil/S√£o Paulo/ | LATE - Laborat√≥rio de T√©cnicas Especiais - Hospital Israelita Albert Einstein | LATE - Laborat√≥rio de T√©cnicas Especiais - Hospital Israelita Albert Einstein | Deyvid Amgarten et al |
| EPI_ISL_861869 | Brazil/SP-HIAE-ID03/2020 | 2020-03-25 | South America/Brazil/S√£o Paulo/ | LATE - Laborat√≥rio de T√©cnicas Especiais - Hospital Israelita Albert Einstein | LATE - Laborat√≥rio de T√©cnicas Especiais - Hospital Israelita Albert Einstein | Deyvid Amgarten et al |
| EPI_ISL_861870 | Brazil/SP-HIAE-ID04/2020 | 2020-10-23 | South America/Brazil/S√£o Paulo/ | LATE - Laborat√≥rio de T√©cnicas Especiais - Hospital Israelita Albert Einstein | LATE - Laborat√≥rio de T√©cnicas Especiais - Hospital Israelita Albert Einstein | Deyvid Amgarten et al |
| EPI_ISL_861871 | Brazil/SP-HIAE-ID05/2020 | 2020-08-01 | South America/Brazil/Rio de Janeiro/Ric | LATE - Laborat√≥rio de T√©cnicas Especiais - Hospital Israelita Albert Einstein | LATE - Laborat√≥rio de T√©cnicas Especiais - Hospital Israelita Albert Einstein | Deyvid Amgarten et al |
| EPI_ISL_861872 | Brazil/SP-HIAE-ID07/2020 | 2020-11-13 | South America/Brazil/S√£o Paulo/ | LATE - Laborat√≥rio de T√©cnicas Especiais - Hospital Israelita Albert Einstein | LATE - Laborat√≥rio de T√©cnicas Especiais - Hospital Israelita Albert Einstein | Deyvid Amgarten et al |

| | | | | | | |
|---|---|---|---|---|---|---|
| EPI_ISL_861875 | Brazil/SP-HIAE-ID10/2020 | 2020-07-16 | South America/Brazil/S√£o Paulo/ | LATE - Laborat√≥rio de T√©cnicas Especiais - Hospital Israelita Albert Einstein | LATE - Laborat√≥rio de T√©cnicas Especiais - Hospital Israelita Albert Einstein | Deyvid Amgarten et al |
| EPI_ISL_861876 | Brazil/SP-HIAE-ID11/2020 | 2020-10-20 | South America/Brazil/S√£o Paulo/ | LATE - Laborat√≥rio de T√©cnicas Especiais - Hospital Israelita Albert Einstein | LATE - Laborat√≥rio de T√©cnicas Especiais - Hospital Israelita Albert Einstein | Deyvid Amgarten et al |
| EPI_ISL_861877 | Brazil/SP-HIAE-ID12/2020 | 2020-11-11 | South America/Brazil/S√£o Paulo/ | LATE - Laborat√≥rio de T√©cnicas Especiais - Hospital Israelita Albert Einstein | LATE - Laborat√≥rio de T√©cnicas Especiais - Hospital Israelita Albert Einstein | Deyvid Amgarten et al |
| EPI_ISL_861878 | Brazil/SP-HIAE-ID13/2020 | 2020-11-13 | South America/Brazil/S√£o Paulo/ | LATE - Laborat√≥rio de T√©cnicas Especiais - Hospital Israelita Albert Einstein | LATE - Laborat√≥rio de T√©cnicas Especiais - Hospital Israelita Albert Einstein | Deyvid Amgarten et al |
| EPI_ISL_861880 | Brazil/SP-HIAE-ID15/2020 | 2020-11-13 | South America/Brazil/S√£o Paulo/ | LATE - Laborat√≥rio de T√©cnicas Especiais - Hospital Israelita Albert Einstein | LATE - Laborat√≥rio de T√©cnicas Especiais - Hospital Israelita Albert Einstein | Deyvid Amgarten et al |
| EPI_ISL_861881 | Brazil/SP-HIAE-ID16/2020 | 2020-11-20 | South America/Brazil/S√£o Paulo/ | LATE - Laborat√≥rio de T√©cnicas Especiais - Hospital Israelita Albert Einstein | LATE - Laborat√≥rio de T√©cnicas Especiais - Hospital Israelita Albert Einstein | Deyvid Amgarten et al |

| | | | | | | |
|---|---|---|---|---|---|---|
| EPI_ISL_861882 | Brazil/SP-HIAE-ID17/2020 | 2020-11-24 | South America/Brazil/S√£o Paulo/ | LATE - Laborat√≥rio de T√©cnicas Especiais - Hospital Israelita Albert Einstein | LATE - Laborat√≥rio de T√©cnicas Especiais - Hospital Israelita Albert Einstein | Deyvid Amgarten et al |
| EPI_ISL_861883 | Brazil/SP-HIAE-ID18/2020 | 2020-11-22 | South America/Brazil/S√£o Paulo/ | LATE - Laborat√≥rio de T√©cnicas Especiais - Hospital Israelita Albert Einstein | LATE - Laborat√≥rio de T√©cnicas Especiais - Hospital Israelita Albert Einstein | Deyvid Amgarten et al |
| EPI_ISL_861884 | Brazil/SP-HIAE-ID19/2020 | 2020-08-01 | South America/Brazil/Rio Grande do Sul | LATE - Laborat√≥rio de T√©cnicas Especiais - Hospital Israelita Albert Einstein | LATE - Laborat√≥rio de T√©cnicas Especiais - Hospital Israelita Albert Einstein | Deyvid Amgarten et al |
| EPI_ISL_861885 | Brazil/SP-HIAE-ID20/2020 | 2020-09-09 | South America/Brazil/S√£o Paulo/ | LATE - Laborat√≥rio de T√©cnicas Especiais - Hospital Israelita Albert Einstein | LATE - Laborat√≥rio de T√©cnicas Especiais - Hospital Israelita Albert Einstein | Deyvid Amgarten et al |
| EPI_ISL_861887 | Brazil/SP-HIAE-ID22/2020 | 2020-11-16 | South America/Brazil/S√£o Paulo/ | LATE - Laborat√≥rio de T√©cnicas Especiais - Hospital Israelita Albert Einstein | LATE - Laborat√≥rio de T√©cnicas Especiais - Hospital Israelita Albert Einstein | Deyvid Amgarten et al |
| EPI_ISL_861888 | Brazil/SP-HIAE-ID23/2020 | 2020-11-17 | South America/Brazil/Rio de Janeiro/Rio | LATE - Laborat√≥rio de T√©cnicas Especiais - Hospital Israelita Albert Einstein | LATE - Laborat√≥rio de T√©cnicas Especiais - Hospital Israelita Albert Einstein | Deyvid Amgarten et al |

| | | | | | | |
|---|---|---|---|---|---|---|
| EPI_ISL_861891 | Brazil/SP-HIAE-ID26/2020 | 2020-07-31 | South America/Brazil/Goiv°s/ | LATE - Laboratv≥rio de Tv©cnicas Especiais - Hospital Israelita Albert Einstein | LATE - Laboratv≥rio de Tv©cnicas Especiais - Hospital Israelita Albert Einstein | Deyvid Amgarten et al |
| EPI_ISL_861894 | Brazil/SP-HIAE-ID30/2020 | 2020-11-20 | South America/Brazil/Sv£o Paulo/ | LATE - Laboratv≥rio de Tv©cnicas Especiais - Hospital Israelita Albert Einstein | LATE - Laboratv≥rio de Tv©cnicas Especiais - Hospital Israelita Albert Einstein | Deyvid Amgarten et al |
| EPI_ISL_861895 | Brazil/SP-HIAE-ID31/2020 | 2020-07-31 | South America/Brazil/Maranhv£o/ | LATE - Laboratv≥rio de Tv©cnicas Especiais - Hospital Israelita Albert Einstein | LATE - Laboratv≥rio de Tv©cnicas Especiais - Hospital Israelita Albert Einstein | Deyvid Amgarten et al |
| EPI_ISL_861897 | Brazil/SP-HIAE-ID33/2020 | 2020-07-31 | South America/Brazil/Goiv°s/ | LATE - Laboratv≥rio de Tv©cnicas Especiais - Hospital Israelita Albert Einstein | LATE - Laboratv≥rio de Tv©cnicas Especiais - Hospital Israelita Albert Einstein | Deyvid Amgarten et al |
| EPI_ISL_861898 | Brazil/SP-HIAE-ID34/2020 | 2020-11-14 | South America/Brazil/Sv£o Paulo/ | LATE - Laboratv≥rio de Tv©cnicas Especiais - Hospital Israelita Albert Einstein | LATE - Laboratv≥rio de Tv©cnicas Especiais - Hospital Israelita Albert Einstein | Deyvid Amgarten et al |
| EPI_ISL_861899 | Brazil/SP-HIAE-ID35/2020 | 2020-11-11 | South America/Brazil/Sv£o Paulo/ | LATE - Laboratv≥rio de Tv©cnicas Especiais - Hospital Israelita Albert Einstein | LATE - Laboratv≥rio de Tv©cnicas Especiais - Hospital Israelita Albert Einstein | Deyvid Amgarten et al |

| | | | | | | |
|---|---|---|---|---|---|---|
| EPI_ISL_861901 | Brazil/SP-HIAE-ID37/2020 | 2020-07-31 | South America/Brazil/Goiv°s/ | LATE - Laboratv≥rio de Tv©cnicas Especiais - Hospital Israelita Albert Einstein | LATE - Laboratv≥rio de Tv©cnicas Especiais - Hospital Israelita Albert Einstein | Deyvid Amgarten et al |
| EPI_ISL_861908 | Brazil/SP-HIAE-ID44/2020 | 2020-11-23 | South America/Brazil/Sv£o Paulo/ | LATE - Laboratv≥rio de Tv©cnicas Especiais - Hospital Israelita Albert Einstein | LATE - Laboratv≥rio de Tv©cnicas Especiais - Hospital Israelita Albert Einstein | Deyvid Amgarten et al |
| EPI_ISL_861909 | Brazil/SP-HIAE-ID45/2020 | 2020-11-20 | South America/Brazil/Sv£o Paulo/ | LATE - Laboratv≥rio de Tv©cnicas Especiais - Hospital Israelita Albert Einstein | LATE - Laboratv≥rio de Tv©cnicas Especiais - Hospital Israelita Albert Einstein | Deyvid Amgarten et al |
| EPI_ISL_861910 | Brazil/SP-HIAE-ID46/2020 | 2020-08-21 | South America/Brazil/Sv£o Paulo/ | LATE - Laboratv≥rio de Tv©cnicas Especiais - Hospital Israelita Albert Einstein | LATE - Laboratv≥rio de Tv©cnicas Especiais - Hospital Israelita Albert Einstein | Deyvid Amgarten et al |
| EPI_ISL_861912 | Brazil/SP-HIAE-ID48/2020 | 2020-03-21 | South America/Brazil/Sv£o Paulo/ | LATE - Laboratv≥rio de Tv©cnicas Especiais - Hospital Israelita Albert Einstein | LATE - Laboratv≥rio de Tv©cnicas Especiais - Hospital Israelita Albert Einstein | Deyvid Amgarten et al |
| EPI_ISL_861915 | Brazil/SP-HIAE-ID51/2020 | 2020-03-31 | South America/Brazil/Sv£o Paulo/ | LATE - Laboratv≥rio de Tv©cnicas Especiais - Hospital Israelita Albert Einstein | LATE - Laboratv≥rio de Tv©cnicas Especiais - Hospital Israelita Albert Einstein | Deyvid Amgarten et al |
| EPI_ISL_861916 | USA/OR-OHSU-5750/2020 | 2020-11-20 | North America/USA/Oregon/Washingto | OHSU Lab Services Molecular Microbiology Lab | Oregon SARS-CoV-2 Genome Sequencing Center | Brendan L. O'Connell et al |

| | | | | | | |
|---|---|---|---|---|---|---|
| EPI_ISL_861917 | USA/OR-OHSU-5751/2020 | 2020-11-20 | North America/USA/Oregon/Washingto | OHSU Lab Services Molecular Microbiology Lab | Oregon SARS-CoV-2 Genome Sequencing Center | Brendan L. O'Connell et al |
| EPI_ISL_861921 | USA/OR-OHSU-5770/2020 | 2020-11-21 | North America/USA/Oregon/Columbia C | OHSU Lab Services Molecular Microbiology Lab | Oregon SARS-CoV-2 Genome Sequencing Center | Brendan L. O'Connell et al |
| EPI_ISL_861922 | USA/OR-OHSU-5771/2020 | 2020-11-21 | North America/USA/Oregon/Washingto | OHSU Lab Services Molecular Microbiology Lab | Oregon SARS-CoV-2 Genome Sequencing Center | Brendan L. O'Connell et al |
| EPI_ISL_861939 | USA/OR-OHSU-5822/2020 | 2020-11-24 | North America/USA/Oregon/Washingto | OHSU Lab Services Molecular Microbiology Lab | Oregon SARS-CoV-2 Genome Sequencing Center | Brendan L. O'Connell et al |
| EPI_ISL_861945 | USA/OR-OHSU-5837/2020 | 2020-11-24 | North America/USA/Oregon/Washingto | OHSU Lab Services Molecular Microbiology Lab | Oregon SARS-CoV-2 Genome Sequencing Center | Brendan L. O'Connell et al |
| EPI_ISL_861950 | USA/OR-OHSU-5857/2020 | 2020-11-24 | North America/USA/Oregon/Washingto | OHSU Lab Services Molecular Microbiology Lab | Oregon SARS-CoV-2 Genome Sequencing Center | Brendan L. O'Connell et al |
| EPI_ISL_861966 | USA/OR-OHSU-6680/2020 | 2020-12-05 | North America/USA/Oregon/ | OHSU Lab Services Molecular Microbiology Lab | Oregon SARS-CoV-2 Genome Sequencing Center | Brendan L. O'Connell et al |
| EPI_ISL_861968 | USA/OR-OHSU-6683/2020 | 2020-12-05 | North America/USA/Oregon/Multnomal | OHSU Lab Services Molecular Microbiology Lab | Oregon SARS-CoV-2 Genome Sequencing Center | Brendan L. O'Connell et al |
| EPI_ISL_861989 | USA/OR-OHSU-8534/2020 | 2020-12-24 | North America/USA/Oregon/Multnomal | OHSU Lab Services Molecular Microbiology Lab | Oregon SARS-CoV-2 Genome Sequencing Center | Brendan L. O'Connell et al |
| EPI_ISL_862002 | USA/OR-OHSU-8674/2020 | 2020-12-29 | North America/USA/Oregon/Multnomal | OHSU Lab Services Molecular Microbiology Lab | Oregon SARS-CoV-2 Genome Sequencing Center | Brendan L. O'Connell et al |

| | | | | | | |
|---|---|---|---|---|---|---|
| EPI_ISL_862004 | USA/OR-OHSU-8684/2020 | 2020-12-29 | North America/USA/Oregon/Washingto | OHSU Lab Services Molecular Microbiology Lab | Oregon SARS-CoV-2 Genome Sequencing Center | Brendan L. O'Connell et al |
| EPI_ISL_862012 | USA/WA-OHSU-8728/2020 | 2020-12-29 | North America/USA/Washington/Clark ( | OHSU Lab Services Molecular Microbiology Lab | Oregon SARS-CoV-2 Genome Sequencing Center | Brendan L. O'Connell et al |
| EPI_ISL_862013 | USA/OR-OHSU-8733/2020 | 2020-12-29 | North America/USA/Oregon/Multnomal | OHSU Lab Services Molecular Microbiology Lab | Oregon SARS-CoV-2 Genome Sequencing Center | Brendan L. O'Connell et al |
| EPI_ISL_862025 | USA/OR-OHSU-8800/2020 | 2020-12-30 | North America/USA/Oregon/Multnomal | OHSU Lab Services Molecular Microbiology Lab | Oregon SARS-CoV-2 Genome Sequencing Center | Brendan L. O'Connell et al |
| EPI_ISL_862032 | USA/OR-OHSU-8835/2020 | 2020-12-31 | North America/USA/Oregon/Clackamas | OHSU Lab Services Molecular Microbiology Lab | Oregon SARS-CoV-2 Genome Sequencing Center | Brendan L. O'Connell et al |
| EPI_ISL_862034 | USA/OR-OHSU-9916/2020 | 2020-12-29 | North America/USA/Oregon/Washingto | OHSU Lab Services Molecular Microbiology Lab | Oregon SARS-CoV-2 Genome Sequencing Center | Brendan L. O'Connell et al |

| | | | | | | |
|---|---|---|---|---|---|---|
| EPI_ISL_862039 | Indonesia/YO-UGM-10009/2020 | 2020-12-27 | Asia/Indonesia/Yogyakarta/ | RSUP Dr. Sardjito | Genetics Working Group (Pokja Genetik) Faculty of Medicine, Public Health and Nursing Universitas Gadjah Mada (FK-KMK UGM); Disease Investigation Center Wates Ministry of Agriculture Indonesia; Department of Microbiology FK-KMK UGM; Laboratorium Diagnostik Yayasan Tahija World Mosquito Program (WMP) Yogyakarta Center for Tropical Medicine FK-KMK UGM; Integrated Research Center FK-KMK UGM; Department of Computer Science and Electronics FMIPA UGM; RSUP Dr. Sardjito | Gunadi et al |
| EPI_ISL_862045 | France/IDF-CERBAHC-20211020-33/2( | 2021-01-07 | Europe/France/Ile de France/ | CERBA | CERBA LAB | Roquebert B et al |
| EPI_ISL_862046 | France/PDL-CERBAHC-20211018-50/2 | 2021-01-07 | Europe/France/Pays de la Loire/Val de L | Cerballiance Val de Loire | CERBA LAB | Abs G. et al |

| | | | | | | |
|---|---|---|---|---|---|---|
| EPI_ISL_862051 | India/UP-MRUBHU3/2021 | 2021-01-04 | Asia/India/Uttar Pradesh/Varanasi | Multidisciplinary Research Unit, DHR-ICMR, Institute of Medical Sciences, Banaras Hindu University | Multidisciplinary Research Unit, DHR-ICMR, Institute of Medical Sciences, Banaras Hindu University | Priyoneel Basu et al |
| EPI_ISL_862066 | Finland/30SS2B8/2020 | 2020-09-30 | Europe/Finland/Uusimaa/ | Department of Virology and Immunology, University of Helsinki and Helsinki University Hospital, Huslab Finland | Department of Virology, Faculty of Medicine, University of Helsinki, Helsinki, Finland | Teemu Smura et al |
| EPI_ISL_862075 | Iran/Tehran-06/2020 | 2020-11-10 | Asia/Iran/Tehran/ | National Influenza Center, Virology Department | National Influenza Center | V Salimi et al |
| EPI_ISL_862076 | Iran/Tehran-02/2020 | 2020-11-24 | Asia/Iran/Tehran/ | National Influenza Center, Virology Department | National Influenza Center | NZ Shafiei Jandaghi et al |
| EPI_ISL_862077 | Iran/Tehran-09/2020 | 2020-02-20 | Asia/Iran/Tehran/ | National Influenza Center, Virology Department | National Influenza Center | J Yavarian et al |
| EPI_ISL_862078 | Iran/Tehran-08/2020 | 2020-02-20 | Asia/Iran/Tehran/ | National Influenza Center, Virology Department | National Influenza Center | A Nejati et al |
| EPI_ISL_862079 | Iran/Tehran-04/2020 | 2020-11-05 | Asia/Iran/Tehran/ | National Influenza Center, Virology Department | National Influenza Center | K Sadeghi et al |
| EPI_ISL_862080 | Iran/Tehran-03/2020 | 2020-11-25 | Asia/Iran/Tehran/ | National Influenza Center, Virology Department | National Influenza Center | J Yavarian et al |
| EPI_ISL_862081 | Iran/Tehran-01/2020 | 2020-11-24 | Asia/Iran/Tehran/ | National Influenza Center, Virology Department | National Influenza Center | NZ Shafiei Jandaghi et al |
| EPI_ISL_862122 | USA/CA-LACPHL-AF00191/2021 | 2021-01-06 | North America/USA/California/Los Ange | UCLA Clinical Micro Lab | Los Angeles County PHL | P. Hemarajata et al. et al |
| EPI_ISL_862129 | Germany/SN-ChVir22064/2021 | 2021-01-11 | Europe/Germany/Saxony/ | Charit√© Universit√§tsmedizin Berlin, Institut f√ºr Virologie/Labor Berlin | Charit√© Universit√§tsmedizin Berlin, Institut f√ºr Virologie | Victor M Corman et al |
| EPI_ISL_862132 | Germany/BW-ChVir21699/2020 | 2020-12-29 | Europe/Germany/Baden-Wuerttemberg | Charit√© Universit√§tsmedizin Berlin, Institut f√ºr Virologie/Labor Berlin | Charit√© Universit√§tsmedizin Berlin, Institut f√ºr Virologie | Victor M Corman et al |

| | | | | | | |
|---|---|---|---|---|---|---|
| EPI_ISL_862134 | Germany/BW-ChVir22112/2021 | 2021-01-11 | Europe/Germany/Baden-Wuerttemberg | Charitv© Universitv§tsmedizin Berlin, Institut fvºr Virologie/Labor Berlin | Charitv© Universitv§tsmedizin Berlin, Institut fvºr Virologie | Victor M Corman et al |
| EPI_ISL_862140 | Germany/SH-ChVir22196/2021 | 2021-01-11 | Europe/Germany/Schleswig-Holstein/ | Charitv© Universitv§tsmedizin Berlin, Institut fvºr Virologie/Labor Berlin | Charitv© Universitv§tsmedizin Berlin, Institut fvºr Virologie | Victor M Corman et al |
| EPI_ISL_862141 | Germany/SH-ChVir22135/2021 | 2021-01-12 | Europe/Germany/Schleswig-Holstein/ | Charitv© Universitv§tsmedizin Berlin, Institut fvºr Virologie/Labor Berlin | Charitv© Universitv§tsmedizin Berlin, Institut fvºr Virologie | Victor M Corman et al |
| EPI_ISL_862143 | Germany/TH-ChVir22154/2021 | 2021-01-09 | Europe/Germany/Thuringia/ | Charitv© Universitv§tsmedizin Berlin, Institut fvºr Virologie/Labor Berlin | Charitv© Universitv§tsmedizin Berlin, Institut fvºr Virologie | Victor M Corman et al |
| EPI_ISL_862144 | Germany/NW-ChVir22215/2021 | 2021-01-12 | Europe/Germany/North Rhine Westphal | Charitv© Universitv§tsmedizin Berlin, Institut fvºr Virologie/Labor Berlin | Charitv© Universitv§tsmedizin Berlin, Institut fvºr Virologie | Victor M Corman et al |
| EPI_ISL_862149 | Germany/BW-ChVir22131/2021 | 2021-01-10 | Europe/Germany/Baden-Wuerttemberg | Charitv© Universitv§tsmedizin Berlin, Institut fvºr Virologie/Labor Berlin | Charitv© Universitv§tsmedizin Berlin, Institut fvºr Virologie | Victor M Corman et al |
| EPI_ISL_862157 | Germany/HE-ChVir22078/2021 | 2021-01-05 | Europe/Germany/Hesse/ | Charitv© Universitv§tsmedizin Berlin, Institut fvºr Virologie/Labor Berlin | Charitv© Universitv§tsmedizin Berlin, Institut fvºr Virologie | Victor M Corman et al |
| EPI_ISL_862159 | Germany/NI-ChVir21639/2020 | 2020-12-22 | Europe/Germany/Niedersachsen/ | Charitv© Universitv§tsmedizin Berlin, Institut fvºr Virologie/Labor Berlin | Charitv© Universitv§tsmedizin Berlin, Institut fvºr Virologie | Victor M Corman et al |
| EPI_ISL_862164 | Germany/NI-ChVir22211/2021 | 2021-01-13 | Europe/Germany/Niedersachsen/ | Charitv© Universitv§tsmedizin Berlin, Institut fvºr Virologie/Labor Berlin | Charitv© Universitv§tsmedizin Berlin, Institut fvºr Virologie | Victor M Corman et al |
| EPI_ISL_862166 | Germany/NI-ChVir22198/2021 | 2021-01-11 | Europe/Germany/Niedersachsen/ | Charitv© Universitv§tsmedizin Berlin, Institut fvºr Virologie/Labor Berlin | Charitv© Universitv§tsmedizin Berlin, Institut fvºr Virologie | Victor M Corman et al |

| | | | | | | |
|---|---|---|---|---|---|---|
| EPI_ISL_862167 | Germany/NI-ChVir22200/2021 | 2021-01-13 | Europe/Germany/Niedersachsen/ | Charit√© Universit√§tsmedizin Berlin, Institut f√ºr Virologie/Labor Berlin | Charit√© Universit√§tsmedizin Berlin, Institut f√ºr Virologie | Victor M Corman et al |
| EPI_ISL_862169 | Germany/NI-ChVir22203/2021 | 2021-01-13 | Europe/Germany/Niedersachsen/ | Charit√© Universit√§tsmedizin Berlin, Institut f√ºr Virologie/Labor Berlin | Charit√© Universit√§tsmedizin Berlin, Institut f√ºr Virologie | Victor M Corman et al |
| EPI_ISL_862173 | Germany/NW-ChVir22072/2021 | 2021-01-05 | Europe/Germany/North Rhine Westphal | Charit√© Universit√§tsmedizin Berlin, Institut f√ºr Virologie/Labor Berlin | Charit√© Universit√§tsmedizin Berlin, Institut f√ºr Virologie | Victor M Corman et al |
| EPI_ISL_862175 | Germany/NW-ChVir22091/2021 | 2021-01-08 | Europe/Germany/North Rhine Westphal | Charit√© Universit√§tsmedizin Berlin, Institut f√ºr Virologie/Labor Berlin | Charit√© Universit√§tsmedizin Berlin, Institut f√ºr Virologie | Victor M Corman et al |
| EPI_ISL_862176 | Germany/NW-ChVir22175/2021 | 2021-01-12 | Europe/Germany/North Rhine Westphal | Charit√© Universit√§tsmedizin Berlin, Institut f√ºr Virologie/Labor Berlin | Charit√© Universit√§tsmedizin Berlin, Institut f√ºr Virologie | Victor M Corman et al |
| EPI_ISL_862177 | Germany/NW-ChVir22176/2021 | 2021-01-12 | Europe/Germany/North Rhine Westphal | Charit√© Universit√§tsmedizin Berlin, Institut f√ºr Virologie/Labor Berlin | Charit√© Universit√§tsmedizin Berlin, Institut f√ºr Virologie | Victor M Corman et al |
| EPI_ISL_862178 | Germany/NW-ChVir22214/2021 | 2021-01-12 | Europe/Germany/North Rhine Westphal | Charit√© Universit√§tsmedizin Berlin, Institut f√ºr Virologie/Labor Berlin | Charit√© Universit√§tsmedizin Berlin, Institut f√ºr Virologie | Victor M Corman et al |
| EPI_ISL_862179 | Germany/RP-ChVir22171/2021 | 2021-01-11 | Europe/Germany/Rheinland-Pfalz/ | Charit√© Universit√§tsmedizin Berlin, Institut f√ºr Virologie/Labor Berlin | Charit√© Universit√§tsmedizin Berlin, Institut f√ºr Virologie | Victor M Corman et al |
| EPI_ISL_862180 | Germany/RP-ChVir22172/2021 | 2021-01-11 | Europe/Germany/Rheinland-Pfalz/ | Charit√© Universit√§tsmedizin Berlin, Institut f√ºr Virologie/Labor Berlin | Charit√© Universit√§tsmedizin Berlin, Institut f√ºr Virologie | Victor M Corman et al |
| EPI_ISL_862181 | Germany/RP-ChVir22173/2021 | 2021-01-11 | Europe/Germany/Rheinland-Pfalz/ | Charit√© Universit√§tsmedizin Berlin, Institut f√ºr Virologie/Labor Berlin | Charit√© Universit√§tsmedizin Berlin, Institut f√ºr Virologie | Victor M Corman et al |

| | | | | | | |
|---|---|---|---|---|---|---|
| EPI_ISL_862182 | Germany/SH-ChVir22163/2021 | 2021-01-07 | Europe/Germany/Schleswig-Holstein/ | Charit√© Universit√§tsmedizin Berlin, Institut f√ºr Virologie/Labor Berlin | Charit√© Universit√§tsmedizin Berlin, Institut f√ºr Virologie | Victor M Corman et al |
| EPI_ISL_862183 | Germany/SH-ChVir22164/2021 | 2021-01-11 | Europe/Germany/Schleswig-Holstein/ | Charit√© Universit√§tsmedizin Berlin, Institut f√ºr Virologie/Labor Berlin | Charit√© Universit√§tsmedizin Berlin, Institut f√ºr Virologie | Victor M Corman et al |
| EPI_ISL_862186 | Germany/SH-ChVir22167/2021 | 2021-01-10 | Europe/Germany/Schleswig-Holstein/ | Charit√© Universit√§tsmedizin Berlin, Institut f√ºr Virologie/Labor Berlin | Charit√© Universit√§tsmedizin Berlin, Institut f√ºr Virologie | Victor M Corman et al |
| EPI_ISL_862187 | Germany/SH-ChVir22168/2021 | 2021-01-11 | Europe/Germany/Schleswig-Holstein/ | Charit√© Universit√§tsmedizin Berlin, Institut f√ºr Virologie/Labor Berlin | Charit√© Universit√§tsmedizin Berlin, Institut f√ºr Virologie | Victor M Corman et al |
| EPI_ISL_862269 | India/AP-CS0318/2020 | 2020-07-01 | Asia/India/Andhra Pradesh/ | Kurnool Medical College (KMC) | CSIR Institute of Genomics and Integrative Biology | Pallavali Roja Rani et al |
| EPI_ISL_862270 | India/AP-CS0843/2020 | 2020-08-01 | Asia/India/Andhra Pradesh/ | Kurnool Medical College (KMC) | CSIR Institute of Genomics and Integrative Biology | Pallavali Roja Rani et al |
| EPI_ISL_862271 | India/AP-CS0932/2020 | 2020-08-02 | Asia/India/Andhra Pradesh/ | Kurnool Medical College (KMC) | CSIR Institute of Genomics and Integrative Biology | Pallavali Roja Rani et al |
| EPI_ISL_862272 | India/AP-CS0844/2020 | 2020-08-01 | Asia/India/Andhra Pradesh/ | Kurnool Medical College (KMC) | CSIR Institute of Genomics and Integrative Biology | Pallavali Roja Rani et al |
| EPI_ISL_862273 | India/AP-CS0339/2020 | 2020-07-04 | Asia/India/Andhra Pradesh/ | Kurnool Medical College (KMC) | CSIR Institute of Genomics and Integrative Biology | Pallavali Roja Rani et al |
| EPI_ISL_862274 | India/AP-CS0839/2020 | 2020-08-01 | Asia/India/Andhra Pradesh/ | Kurnool Medical College (KMC) | CSIR Institute of Genomics and Integrative Biology | Pallavali Roja Rani et al |

| | | | | | | |
|---|---|---|---|---|---|---|
| EPI_ISL_862275 | India/AP-CS0840/2020 | 2020-08-01 | Asia/India/Andhra Pradesh/ | Kurnool Medical College (KMC) | CSIR Institute of Genomics and Integrative Biology | Pallavali Roja Rani et al |
| EPI_ISL_862276 | India/AP-CS0789/2020 | 2020-07-29 | Asia/India/Andhra Pradesh/ | Kurnool Medical College (KMC) | CSIR Institute of Genomics and Integrative Biology | Pallavali Roja Rani et al |
| EPI_ISL_862277 | India/AP-CS0336/2020 | 2020-07-04 | Asia/India/Andhra Pradesh/ | Kurnool Medical College (KMC) | CSIR Institute of Genomics and Integrative Biology | Pallavali Roja Rani et al |
| EPI_ISL_862278 | India/AP-CS0320/2020 | 2020-06-29 | Asia/India/Andhra Pradesh/ | Kurnool Medical College (KMC) | CSIR Institute of Genomics and Integrative Biology | Pallavali Roja Rani et al |
| EPI_ISL_862280 | India/AP-CS0869/2020 | 2020-08-01 | Asia/India/Andhra Pradesh/ | Kurnool Medical College (KMC) | CSIR Institute of Genomics and Integrative Biology | Pallavali Roja Rani et al |
| EPI_ISL_862282 | India/AP-CS0357/2020 | 2020-07-04 | Asia/India/Andhra Pradesh/ | Kurnool Medical College (KMC) | CSIR Institute of Genomics and Integrative Biology | Pallavali Roja Rani et al |
| EPI_ISL_862284 | India/AP-CS0803/2020 | 2020-07-29 | Asia/India/Andhra Pradesh/ | Kurnool Medical College (KMC) | CSIR Institute of Genomics and Integrative Biology | Pallavali Roja Rani et al |
| EPI_ISL_862285 | India/AP-CS0787/2020 | 2020-07-29 | Asia/India/Andhra Pradesh/ | Kurnool Medical College (KMC) | CSIR Institute of Genomics and Integrative Biology | Pallavali Roja Rani et al |
| EPI_ISL_862286 | India/AP-CS0812/2020 | 2020-07-29 | Asia/India/Andhra Pradesh/ | Kurnool Medical College (KMC) | CSIR Institute of Genomics and Integrative Biology | Pallavali Roja Rani et al |
| EPI_ISL_862287 | India/AP-CS0395/2020 | 2020-07-04 | Asia/India/Andhra Pradesh/ | Kurnool Medical College (KMC) | CSIR Institute of Genomics and Integrative Biology | Pallavali Roja Rani et al |

| | | | | | | |
|---|---|---|---|---|---|---|
| EPI_ISL_862288 | India/AP-CS0952/2020 | 2020-08-02 | Asia/India/Andhra Pradesh/ | Kurnool Medical College (KMC) | CSIR Institute of Genomics and Integrative Biology | Pallavali Roja Rani et al |
| EPI_ISL_862289 | India/AP-CS0937/2020 | 2020-08-02 | Asia/India/Andhra Pradesh/ | Kurnool Medical College (KMC) | CSIR Institute of Genomics and Integrative Biology | Pallavali Roja Rani et al |
| EPI_ISL_862290 | India/AP-CS0804/2020 | 2020-07-29 | Asia/India/Andhra Pradesh/ | Kurnool Medical College (KMC) | CSIR Institute of Genomics and Integrative Biology | Pallavali Roja Rani et al |
| EPI_ISL_862291 | India/AP-CS0353/2020 | 2020-07-04 | Asia/India/Andhra Pradesh/ | Kurnool Medical College (KMC) | CSIR Institute of Genomics and Integrative Biology | Pallavali Roja Rani et al |
| EPI_ISL_862292 | India/AP-CS0349/2020 | 2020-07-04 | Asia/India/Andhra Pradesh/ | Kurnool Medical College (KMC) | CSIR Institute of Genomics and Integrative Biology | Pallavali Roja Rani et al |
| EPI_ISL_862293 | India/AP-CS0334/2020 | 2020-07-04 | Asia/India/Andhra Pradesh/ | Kurnool Medical College (KMC) | CSIR Institute of Genomics and Integrative Biology | Pallavali Roja Rani et al |
| EPI_ISL_862294 | India/AP-CS0340/2020 | 2020-07-04 | Asia/India/Andhra Pradesh/ | Kurnool Medical College (KMC) | CSIR Institute of Genomics and Integrative Biology | Pallavali Roja Rani et al |
| EPI_ISL_862295 | India/AP-CS0838/2020 | 2020-08-01 | Asia/India/Andhra Pradesh/ | Kurnool Medical College (KMC) | CSIR Institute of Genomics and Integrative Biology | Pallavali Roja Rani et al |
| EPI_ISL_862296 | India/AP-CS0346/2020 | 2020-07-04 | Asia/India/Andhra Pradesh/ | Kurnool Medical College (KMC) | CSIR Institute of Genomics and Integrative Biology | Pallavali Roja Rani et al |
| EPI_ISL_862297 | India/AP-CS0313/2020 | 2020-06-29 | Asia/India/Andhra Pradesh/ | Kurnool Medical College (KMC) | CSIR Institute of Genomics and Integrative Biology | Pallavali Roja Rani et al |

| | | | | | | |
|---|---|---|---|---|---|---|
| EPI_ISL_862298 | India/AP-CS0872/2020 | 2020-08-01 | Asia/India/Andhra Pradesh/ | Kurnool Medical College (KMC) | CSIR Institute of Genomics and Integrative Biology | Pallavali Roja Rani et al |
| EPI_ISL_862299 | India/AP-CS0877/2020 | 2020-08-01 | Asia/India/Andhra Pradesh/ | Kurnool Medical College (KMC) | CSIR Institute of Genomics and Integrative Biology | Pallavali Roja Rani et al |
| EPI_ISL_862300 | India/AP-CS0831/2020 | 2020-07-30 | Asia/India/Andhra Pradesh/ | Kurnool Medical College (KMC) | CSIR Institute of Genomics and Integrative Biology | Pallavali Roja Rani et al |
| EPI_ISL_862301 | India/AP-CS0797/2020 | 2020-07-29 | Asia/India/Andhra Pradesh/ | Kurnool Medical College (KMC) | CSIR Institute of Genomics and Integrative Biology | Pallavali Roja Rani et al |
| EPI_ISL_862302 | India/AP-CS0389/2020 | 2020-07-01 | Asia/India/Andhra Pradesh/ | Kurnool Medical College (KMC) | CSIR Institute of Genomics and Integrative Biology | Pallavali Roja Rani et al |
| EPI_ISL_862303 | India/AP-CS0348/2020 | 2020-07-04 | Asia/India/Andhra Pradesh/ | Kurnool Medical College (KMC) | CSIR Institute of Genomics and Integrative Biology | Pallavali Roja Rani et al |
| EPI_ISL_862304 | India/AP-CS0845/2020 | 2020-08-01 | Asia/India/Andhra Pradesh/ | Kurnool Medical College (KMC) | CSIR Institute of Genomics and Integrative Biology | Pallavali Roja Rani et al |
| EPI_ISL_862305 | India/AP-CS0363/2020 | 2020-07-04 | Asia/India/Andhra Pradesh/ | Kurnool Medical College (KMC) | CSIR Institute of Genomics and Integrative Biology | Pallavali Roja Rani et al |
| EPI_ISL_862306 | India/AP-CS0891/2020 | 2020-08-02 | Asia/India/Andhra Pradesh/ | Kurnool Medical College (KMC) | CSIR Institute of Genomics and Integrative Biology | Pallavali Roja Rani et al |
| EPI_ISL_862307 | India/AP-CS0993/2020 | 2020-08-03 | Asia/India/Andhra Pradesh/ | Kurnool Medical College (KMC) | CSIR Institute of Genomics and Integrative Biology | Pallavali Roja Rani et al |

| | | | | | | |
|---|---|---|---|---|---|---|
| EPI_ISL_862308 | India/AP-CS0324/2020 | 2020-06-29 | Asia/India/Andhra Pradesh/ | Kurnool Medical College (KMC) | CSIR Institute of Genomics and Integrative Biology | Pallavali Roja Rani et al |
| EPI_ISL_862309 | India/AP-CS0355/2020 | 2020-07-04 | Asia/India/Andhra Pradesh/ | Kurnool Medical College (KMC) | CSIR Institute of Genomics and Integrative Biology | Pallavali Roja Rani et al |
| EPI_ISL_862310 | India/AP-CS0360/2020 | 2020-07-04 | Asia/India/Andhra Pradesh/ | Kurnool Medical College (KMC) | CSIR Institute of Genomics and Integrative Biology | Pallavali Roja Rani et al |
| EPI_ISL_862312 | India/AP-CS0398/2020 | 2020-07-04 | Asia/India/Andhra Pradesh/ | Kurnool Medical College (KMC) | CSIR Institute of Genomics and Integrative Biology | Pallavali Roja Rani et al |
| EPI_ISL_862313 | India/AP-CS0319/2020 | 2020-07-01 | Asia/India/Andhra Pradesh/ | Kurnool Medical College (KMC) | CSIR Institute of Genomics and Integrative Biology | Pallavali Roja Rani et al |
| EPI_ISL_862315 | India/AP-CS0892/2020 | 2020-08-02 | Asia/India/Andhra Pradesh/ | Kurnool Medical College (KMC) | CSIR Institute of Genomics and Integrative Biology | Pallavali Roja Rani et al |
| EPI_ISL_862317 | India/AP-CS0894/2020 | 2020-08-02 | Asia/India/Andhra Pradesh/ | Kurnool Medical College (KMC) | CSIR Institute of Genomics and Integrative Biology | Pallavali Roja Rani et al |
| EPI_ISL_862318 | India/AP-CS0895/2020 | 2020-08-02 | Asia/India/Andhra Pradesh/ | Kurnool Medical College (KMC) | CSIR Institute of Genomics and Integrative Biology | Pallavali Roja Rani et al |
| EPI_ISL_862319 | India/AP-CS0897/2020 | 2020-08-02 | Asia/India/Andhra Pradesh/ | Kurnool Medical College (KMC) | CSIR Institute of Genomics and Integrative Biology | Pallavali Roja Rani et al |
| EPI_ISL_862320 | India/AP-CS0898/2020 | 2020-08-02 | Asia/India/Andhra Pradesh/ | Kurnool Medical College (KMC) | CSIR Institute of Genomics and Integrative Biology | Pallavali Roja Rani et al |

| | | | | | | |
|---|---|---|---|---|---|---|
| EPI_ISL_862321 | India/AP-CS0899/2020 | 2020-08-02 | Asia/India/Andhra Pradesh/ | Kurnool Medical College (KMC) | CSIR Institute of Genomics and Integrative Biology | Pallavali Roja Rani et al |
| EPI_ISL_862322 | India/AP-CS0900/2020 | 2020-08-02 | Asia/India/Andhra Pradesh/ | Kurnool Medical College (KMC) | CSIR Institute of Genomics and Integrative Biology | Pallavali Roja Rani et al |
| EPI_ISL_862323 | India/AP-CS0901/2020 | 2020-08-02 | Asia/India/Andhra Pradesh/ | Kurnool Medical College (KMC) | CSIR Institute of Genomics and Integrative Biology | Pallavali Roja Rani et al |
| EPI_ISL_862324 | India/AP-CS0902/2020 | 2020-08-02 | Asia/India/Andhra Pradesh/ | Kurnool Medical College (KMC) | CSIR Institute of Genomics and Integrative Biology | Pallavali Roja Rani et al |
| EPI_ISL_862325 | India/AP-CS0903/2020 | 2020-08-02 | Asia/India/Andhra Pradesh/ | Kurnool Medical College (KMC) | CSIR Institute of Genomics and Integrative Biology | Pallavali Roja Rani et al |
| EPI_ISL_862326 | India/AP-CS0904/2020 | 2020-08-02 | Asia/India/Andhra Pradesh/ | Kurnool Medical College (KMC) | CSIR Institute of Genomics and Integrative Biology | Pallavali Roja Rani et al |
| EPI_ISL_862327 | India/AP-CS0905/2020 | 2020-08-02 | Asia/India/Andhra Pradesh/ | Kurnool Medical College (KMC) | CSIR Institute of Genomics and Integrative Biology | Pallavali Roja Rani et al |
| EPI_ISL_862328 | India/AP-CS0906/2020 | 2020-08-02 | Asia/India/Andhra Pradesh/ | Kurnool Medical College (KMC) | CSIR Institute of Genomics and Integrative Biology | Pallavali Roja Rani et al |
| EPI_ISL_862329 | India/AP-CS0907/2020 | 2020-08-02 | Asia/India/Andhra Pradesh/ | Kurnool Medical College (KMC) | CSIR Institute of Genomics and Integrative Biology | Pallavali Roja Rani et al |
| EPI_ISL_862330 | India/AP-CS0908/2020 | 2020-08-02 | Asia/India/Andhra Pradesh/ | Kurnool Medical College (KMC) | CSIR Institute of Genomics and Integrative Biology | Pallavali Roja Rani et al |

| | | | | | | |
|---|---|---|---|---|---|---|
| EPI_ISL_862331 | India/AP-CS0909/2020 | 2020-08-02 | Asia/India/Andhra Pradesh/ | Kurnool Medical College (KMC) | CSIR Institute of Genomics and Integrative Biology | Pallavali Roja Rani et al |
| EPI_ISL_862332 | India/AP-CS0910/2020 | 2020-08-02 | Asia/India/Andhra Pradesh/ | Kurnool Medical College (KMC) | CSIR Institute of Genomics and Integrative Biology | Pallavali Roja Rani et al |
| EPI_ISL_862333 | India/AP-CS0912/2020 | 2020-08-02 | Asia/India/Andhra Pradesh/ | Kurnool Medical College (KMC) | CSIR Institute of Genomics and Integrative Biology | Pallavali Roja Rani et al |
| EPI_ISL_862334 | India/AP-CS0913/2020 | 2020-08-02 | Asia/India/Andhra Pradesh/ | Kurnool Medical College (KMC) | CSIR Institute of Genomics and Integrative Biology | Pallavali Roja Rani et al |
| EPI_ISL_862335 | India/AP-CS0915/2020 | 2020-08-02 | Asia/India/Andhra Pradesh/ | Kurnool Medical College (KMC) | CSIR Institute of Genomics and Integrative Biology | Pallavali Roja Rani et al |
| EPI_ISL_862336 | India/AP-CS0916/2020 | 2020-08-02 | Asia/India/Andhra Pradesh/ | Kurnool Medical College (KMC) | CSIR Institute of Genomics and Integrative Biology | Pallavali Roja Rani et al |
| EPI_ISL_862337 | India/AP-CS0917/2020 | 2020-08-02 | Asia/India/Andhra Pradesh/ | Kurnool Medical College (KMC) | CSIR Institute of Genomics and Integrative Biology | Pallavali Roja Rani et al |
| EPI_ISL_862338 | India/AP-CS0918/2020 | 2020-08-02 | Asia/India/Andhra Pradesh/ | Kurnool Medical College (KMC) | CSIR Institute of Genomics and Integrative Biology | Pallavali Roja Rani et al |
| EPI_ISL_862339 | India/AP-CS0919/2020 | 2020-08-02 | Asia/India/Andhra Pradesh/ | Kurnool Medical College (KMC) | CSIR Institute of Genomics and Integrative Biology | Pallavali Roja Rani et al |
| EPI_ISL_862340 | India/AP-CS0931/2020 | 2020-08-02 | Asia/India/Andhra Pradesh/ | Kurnool Medical College (KMC) | CSIR Institute of Genomics and Integrative Biology | Pallavali Roja Rani et al |

| | | | | | | |
|---|---|---|---|---|---|---|
| EPI_ISL_862341 | India/AP-CS0934/2020 | 2020-08-02 | Asia/India/Andhra Pradesh/ | Kurnool Medical College (KMC) | CSIR Institute of Genomics and Integrative Biology | Pallavali Roja Rani et al |
| EPI_ISL_862342 | India/AP-CS0936/2020 | 2020-08-02 | Asia/India/Andhra Pradesh/ | Kurnool Medical College (KMC) | CSIR Institute of Genomics and Integrative Biology | Pallavali Roja Rani et al |
| EPI_ISL_862343 | India/AP-CS0939/2020 | 2020-08-02 | Asia/India/Andhra Pradesh/ | Kurnool Medical College (KMC) | CSIR Institute of Genomics and Integrative Biology | Pallavali Roja Rani et al |
| EPI_ISL_862344 | India/AP-CS0792/2020 | 2020-07-29 | Asia/India/Andhra Pradesh/ | Kurnool Medical College (KMC) | CSIR Institute of Genomics and Integrative Biology | Pallavali Roja Rani et al |
| EPI_ISL_862345 | India/AP-CS0945/2020 | 2020-08-02 | Asia/India/Andhra Pradesh/ | Kurnool Medical College (KMC) | CSIR Institute of Genomics and Integrative Biology | Pallavali Roja Rani et al |
| EPI_ISL_862346 | India/AP-CS0946/2020 | 2020-08-02 | Asia/India/Andhra Pradesh/ | Kurnool Medical College (KMC) | CSIR Institute of Genomics and Integrative Biology | Pallavali Roja Rani et al |
| EPI_ISL_862347 | India/AP-CS0947/2020 | 2020-08-02 | Asia/India/Andhra Pradesh/ | Kurnool Medical College (KMC) | CSIR Institute of Genomics and Integrative Biology | Pallavali Roja Rani et al |
| EPI_ISL_862348 | India/AP-CS0949/2020 | 2020-08-02 | Asia/India/Andhra Pradesh/ | Kurnool Medical College (KMC) | CSIR Institute of Genomics and Integrative Biology | Pallavali Roja Rani et al |
| EPI_ISL_862349 | India/AP-CS0950/2020 | 2020-08-02 | Asia/India/Andhra Pradesh/ | Kurnool Medical College (KMC) | CSIR Institute of Genomics and Integrative Biology | Pallavali Roja Rani et al |
| EPI_ISL_862350 | India/AP-CS0951/2020 | 2020-08-02 | Asia/India/Andhra Pradesh/ | Kurnool Medical College (KMC) | CSIR Institute of Genomics and Integrative Biology | Pallavali Roja Rani et al |

| | | | | | | |
|---|---|---|---|---|---|---|
| EPI_ISL_862351 | India/AP-CS0953/2020 | 2020-08-02 | Asia/India/Andhra Pradesh/ | Kurnool Medical College (KMC) | CSIR Institute of Genomics and Integrative Biology | Pallavali Roja Rani et al |
| EPI_ISL_862352 | India/AP-CS0954/2020 | 2020-08-02 | Asia/India/Andhra Pradesh/ | Kurnool Medical College (KMC) | CSIR Institute of Genomics and Integrative Biology | Pallavali Roja Rani et al |
| EPI_ISL_862353 | India/AP-CS0955/2020 | 2020-08-02 | Asia/India/Andhra Pradesh/ | Kurnool Medical College (KMC) | CSIR Institute of Genomics and Integrative Biology | Pallavali Roja Rani et al |
| EPI_ISL_862354 | India/AP-CS0956/2020 | 2020-08-02 | Asia/India/Andhra Pradesh/ | Kurnool Medical College (KMC) | CSIR Institute of Genomics and Integrative Biology | Pallavali Roja Rani et al |
| EPI_ISL_862355 | India/AP-CS0957/2020 | 2020-08-02 | Asia/India/Andhra Pradesh/ | Kurnool Medical College (KMC) | CSIR Institute of Genomics and Integrative Biology | Pallavali Roja Rani et al |
| EPI_ISL_862356 | India/AP-CS0959/2020 | 2020-08-02 | Asia/India/Andhra Pradesh/ | Kurnool Medical College (KMC) | CSIR Institute of Genomics and Integrative Biology | Pallavali Roja Rani et al |
| EPI_ISL_862357 | India/AP-CS0794/2020 | 2020-07-29 | Asia/India/Andhra Pradesh/ | Kurnool Medical College (KMC) | CSIR Institute of Genomics and Integrative Biology | Pallavali Roja Rani et al |
| EPI_ISL_862358 | India/AP-CS0960/2020 | 2020-08-02 | Asia/India/Andhra Pradesh/ | Kurnool Medical College (KMC) | CSIR Institute of Genomics and Integrative Biology | Pallavali Roja Rani et al |
| EPI_ISL_862359 | India/AP-CS0961/2020 | 2020-08-02 | Asia/India/Andhra Pradesh/ | Kurnool Medical College (KMC) | CSIR Institute of Genomics and Integrative Biology | Pallavali Roja Rani et al |
| EPI_ISL_862360 | India/AP-CS0962/2020 | 2020-08-02 | Asia/India/Andhra Pradesh/ | Kurnool Medical College (KMC) | CSIR Institute of Genomics and Integrative Biology | Pallavali Roja Rani et al |

| | | | | | | |
|---|---|---|---|---|---|---|
| EPI_ISL_862361 | India/AP-CS0963/2020 | 2020-08-02 | Asia/India/Andhra Pradesh/ | Kurnool Medical College (KMC) | CSIR Institute of Genomics and Integrative Biology | Pallavali Roja Rani et al |
| EPI_ISL_862362 | India/AP-CS0964/2020 | 2020-08-02 | Asia/India/Andhra Pradesh/ | Kurnool Medical College (KMC) | CSIR Institute of Genomics and Integrative Biology | Pallavali Roja Rani et al |
| EPI_ISL_862363 | India/AP-CS0966/2020 | 2020-08-02 | Asia/India/Andhra Pradesh/ | Kurnool Medical College (KMC) | CSIR Institute of Genomics and Integrative Biology | Pallavali Roja Rani et al |
| EPI_ISL_862364 | India/AP-CS0968/2020 | 2020-08-02 | Asia/India/Andhra Pradesh/ | Kurnool Medical College (KMC) | CSIR Institute of Genomics and Integrative Biology | Pallavali Roja Rani et al |
| EPI_ISL_862365 | India/AP-CS0969/2020 | 2020-08-02 | Asia/India/Andhra Pradesh/ | Kurnool Medical College (KMC) | CSIR Institute of Genomics and Integrative Biology | Pallavali Roja Rani et al |
| EPI_ISL_862366 | India/AP-CS0972/2020 | 2020-08-02 | Asia/India/Andhra Pradesh/ | Kurnool Medical College (KMC) | CSIR Institute of Genomics and Integrative Biology | Pallavali Roja Rani et al |
| EPI_ISL_862367 | India/AP-CS0974/2020 | 2020-08-02 | Asia/India/Andhra Pradesh/ | Kurnool Medical College (KMC) | CSIR Institute of Genomics and Integrative Biology | Pallavali Roja Rani et al |
| EPI_ISL_862368 | India/AP-CS0977/2020 | 2020-08-02 | Asia/India/Andhra Pradesh/ | Kurnool Medical College (KMC) | CSIR Institute of Genomics and Integrative Biology | Pallavali Roja Rani et al |
| EPI_ISL_862369 | India/AP-CS0979/2020 | 2020-08-02 | Asia/India/Andhra Pradesh/ | Kurnool Medical College (KMC) | CSIR Institute of Genomics and Integrative Biology | Pallavali Roja Rani et al |
| EPI_ISL_862371 | India/AP-CS0982/2020 | 2020-08-02 | Asia/India/Andhra Pradesh/ | Kurnool Medical College (KMC) | CSIR Institute of Genomics and Integrative Biology | Pallavali Roja Rani et al |

| | | | | | | |
|---|---|---|---|---|---|---|
| EPI_ISL_862372 | India/AP-CS0989/2020 | 2020-08-03 | Asia/India/Andhra Pradesh/ | Kurnool Medical College (KMC) | CSIR Institute of Genomics and Integrative Biology | Pallavali Roja Rani et al |
| EPI_ISL_862373 | India/AP-CS0991/2020 | 2020-08-03 | Asia/India/Andhra Pradesh/ | Kurnool Medical College (KMC) | CSIR Institute of Genomics and Integrative Biology | Pallavali Roja Rani et al |
| EPI_ISL_862374 | India/AP-CS0992/2020 | 2020-08-03 | Asia/India/Andhra Pradesh/ | Kurnool Medical College (KMC) | CSIR Institute of Genomics and Integrative Biology | Pallavali Roja Rani et al |
| EPI_ISL_862375 | India/AP-CS0994/2020 | 2020-08-03 | Asia/India/Andhra Pradesh/ | Kurnool Medical College (KMC) | CSIR Institute of Genomics and Integrative Biology | Pallavali Roja Rani et al |
| EPI_ISL_862376 | India/AP-CS0995/2020 | 2020-08-03 | Asia/India/Andhra Pradesh/ | Kurnool Medical College (KMC) | CSIR Institute of Genomics and Integrative Biology | Pallavali Roja Rani et al |
| EPI_ISL_862377 | India/AP-CS0996/2020 | 2020-08-03 | Asia/India/Andhra Pradesh/ | Kurnool Medical College (KMC) | CSIR Institute of Genomics and Integrative Biology | Pallavali Roja Rani et al |
| EPI_ISL_862378 | India/AP-CS0997/2020 | 2020-08-03 | Asia/India/Andhra Pradesh/ | Kurnool Medical College (KMC) | CSIR Institute of Genomics and Integrative Biology | Pallavali Roja Rani et al |
| EPI_ISL_862379 | India/AP-CS0999/2020 | 2020-08-03 | Asia/India/Andhra Pradesh/ | Kurnool Medical College (KMC) | CSIR Institute of Genomics and Integrative Biology | Pallavali Roja Rani et al |
| EPI_ISL_862380 | India/AP-CS1000/2020 | 2020-08-03 | Asia/India/Andhra Pradesh/ | Kurnool Medical College (KMC) | CSIR Institute of Genomics and Integrative Biology | Pallavali Roja Rani et al |
| EPI_ISL_862381 | India/AP-CS1001/2020 | 2020-08-03 | Asia/India/Andhra Pradesh/ | Kurnool Medical College (KMC) | CSIR Institute of Genomics and Integrative Biology | Pallavali Roja Rani et al |

| | | | | | | |
|---|---|---|---|---|---|---|
| EPI_ISL_862383 | India/AP-CS1002/2020 | 2020-08-03 | Asia/India/Andhra Pradesh/ | Kurnool Medical College (KMC) | CSIR Institute of Genomics and Integrative Biology | Pallavali Roja Rani et al |
| EPI_ISL_862384 | India/AP-CS1003/2020 | 2020-08-03 | Asia/India/Andhra Pradesh/ | Kurnool Medical College (KMC) | CSIR Institute of Genomics and Integrative Biology | Pallavali Roja Rani et al |
| EPI_ISL_862385 | India/AP-CS0801/2020 | 2020-07-29 | Asia/India/Andhra Pradesh/ | Kurnool Medical College (KMC) | CSIR Institute of Genomics and Integrative Biology | Pallavali Roja Rani et al |
| EPI_ISL_862386 | India/AP-CS0802/2020 | 2020-07-29 | Asia/India/Andhra Pradesh/ | Kurnool Medical College (KMC) | CSIR Institute of Genomics and Integrative Biology | Pallavali Roja Rani et al |
| EPI_ISL_862387 | India/AP-CS0807/2020 | 2020-07-29 | Asia/India/Andhra Pradesh/ | Kurnool Medical College (KMC) | CSIR Institute of Genomics and Integrative Biology | Pallavali Roja Rani et al |
| EPI_ISL_862388 | India/AP-CS0781/2020 | 2020-07-28 | Asia/India/Andhra Pradesh/ | Kurnool Medical College (KMC) | CSIR Institute of Genomics and Integrative Biology | Pallavali Roja Rani et al |
| EPI_ISL_862389 | India/AP-CS0809/2020 | 2020-07-29 | Asia/India/Andhra Pradesh/ | Kurnool Medical College (KMC) | CSIR Institute of Genomics and Integrative Biology | Pallavali Roja Rani et al |
| EPI_ISL_862390 | India/AP-CS0810/2020 | 2020-07-29 | Asia/India/Andhra Pradesh/ | Kurnool Medical College (KMC) | CSIR Institute of Genomics and Integrative Biology | Pallavali Roja Rani et al |
| EPI_ISL_862392 | India/AP-CS0813/2020 | 2020-07-29 | Asia/India/Andhra Pradesh/ | Kurnool Medical College (KMC) | CSIR Institute of Genomics and Integrative Biology | Pallavali Roja Rani et al |
| EPI_ISL_862393 | India/AP-CS0814/2020 | 2020-07-29 | Asia/India/Andhra Pradesh/ | Kurnool Medical College (KMC) | CSIR Institute of Genomics and Integrative Biology | Pallavali Roja Rani et al |

| | | | | | | |
|---|---|---|---|---|---|---|
| EPI_ISL_862394 | India/AP-CS0815/2020 | 2020-07-29 | Asia/India/Andhra Pradesh/ | Kurnool Medical College (KMC) | CSIR Institute of Genomics and Integrative Biology | Pallavali Roja Rani et al |
| EPI_ISL_862395 | India/AP-CS0817/2020 | 2020-07-29 | Asia/India/Andhra Pradesh/ | Kurnool Medical College (KMC) | CSIR Institute of Genomics and Integrative Biology | Pallavali Roja Rani et al |
| EPI_ISL_862396 | India/AP-CS0818/2020 | 2020-07-29 | Asia/India/Andhra Pradesh/ | Kurnool Medical College (KMC) | CSIR Institute of Genomics and Integrative Biology | Pallavali Roja Rani et al |
| EPI_ISL_862397 | India/AP-CS0782/2020 | 2020-07-28 | Asia/India/Andhra Pradesh/ | Kurnool Medical College (KMC) | CSIR Institute of Genomics and Integrative Biology | Pallavali Roja Rani et al |
| EPI_ISL_862398 | India/AP-CS0819/2020 | 2020-07-30 | Asia/India/Andhra Pradesh/ | Kurnool Medical College (KMC) | CSIR Institute of Genomics and Integrative Biology | Pallavali Roja Rani et al |
| EPI_ISL_862399 | India/AP-CS0829/2020 | 2020-07-30 | Asia/India/Andhra Pradesh/ | Kurnool Medical College (KMC) | CSIR Institute of Genomics and Integrative Biology | Pallavali Roja Rani et al |
| EPI_ISL_862400 | India/AP-CS0832/2020 | 2020-07-30 | Asia/India/Andhra Pradesh/ | Kurnool Medical College (KMC) | CSIR Institute of Genomics and Integrative Biology | Pallavali Roja Rani et al |
| EPI_ISL_862403 | India/AP-CS0837/2020 | 2020-07-30 | Asia/India/Andhra Pradesh/ | Kurnool Medical College (KMC) | CSIR Institute of Genomics and Integrative Biology | Pallavali Roja Rani et al |
| EPI_ISL_862404 | India/AP-CS0847/2020 | 2020-08-01 | Asia/India/Andhra Pradesh/ | Kurnool Medical College (KMC) | CSIR Institute of Genomics and Integrative Biology | Pallavali Roja Rani et al |
| EPI_ISL_862405 | India/AP-CS0784/2020 | 2020-07-28 | Asia/India/Andhra Pradesh/ | Kurnool Medical College (KMC) | CSIR Institute of Genomics and Integrative Biology | Pallavali Roja Rani et al |

| | | | | | | |
|---|---|---|---|---|---|---|
| EPI_ISL_862406 | India/AP-CS0848/2020 | 2020-08-01 | Asia/India/Andhra Pradesh/ | Kurnool Medical College (KMC) | CSIR Institute of Genomics and Integrative Biology | Pallavali Roja Rani et al |
| EPI_ISL_862407 | India/AP-CS0849/2020 | 2020-08-01 | Asia/India/Andhra Pradesh/ | Kurnool Medical College (KMC) | CSIR Institute of Genomics and Integrative Biology | Pallavali Roja Rani et al |
| EPI_ISL_862408 | India/AP-CS0850/2020 | 2020-08-01 | Asia/India/Andhra Pradesh/ | Kurnool Medical College (KMC) | CSIR Institute of Genomics and Integrative Biology | Pallavali Roja Rani et al |
| EPI_ISL_862409 | India/AP-CS0851/2020 | 2020-08-01 | Asia/India/Andhra Pradesh/ | Kurnool Medical College (KMC) | CSIR Institute of Genomics and Integrative Biology | Pallavali Roja Rani et al |
| EPI_ISL_862410 | India/AP-CS0852/2020 | 2020-08-01 | Asia/India/Andhra Pradesh/ | Kurnool Medical College (KMC) | CSIR Institute of Genomics and Integrative Biology | Pallavali Roja Rani et al |
| EPI_ISL_862411 | India/AP-CS0853/2020 | 2020-08-01 | Asia/India/Andhra Pradesh/ | Kurnool Medical College (KMC) | CSIR Institute of Genomics and Integrative Biology | Pallavali Roja Rani et al |
| EPI_ISL_862412 | India/AP-CS0854/2020 | 2020-08-01 | Asia/India/Andhra Pradesh/ | Kurnool Medical College (KMC) | CSIR Institute of Genomics and Integrative Biology | Pallavali Roja Rani et al |
| EPI_ISL_862413 | India/AP-CS0855/2020 | 2020-08-01 | Asia/India/Andhra Pradesh/ | Kurnool Medical College (KMC) | CSIR Institute of Genomics and Integrative Biology | Pallavali Roja Rani et al |
| EPI_ISL_862414 | India/AP-CS0856/2020 | 2020-08-01 | Asia/India/Andhra Pradesh/ | Kurnool Medical College (KMC) | CSIR Institute of Genomics and Integrative Biology | Pallavali Roja Rani et al |
| EPI_ISL_862415 | India/AP-CS0857/2020 | 2020-08-01 | Asia/India/Andhra Pradesh/ | Kurnool Medical College (KMC) | CSIR Institute of Genomics and Integrative Biology | Pallavali Roja Rani et al |

| | | | | | | |
|---|---|---|---|---|---|---|
| EPI_ISL_862416 | India/AP-CS0785/2020 | 2020-07-28 | Asia/India/Andhra Pradesh/ | Kurnool Medical College (KMC) | CSIR Institute of Genomics and Integrative Biology | Pallavali Roja Rani et al |
| EPI_ISL_862417 | India/AP-CS0858/2020 | 2020-08-01 | Asia/India/Andhra Pradesh/ | Kurnool Medical College (KMC) | CSIR Institute of Genomics and Integrative Biology | Pallavali Roja Rani et al |
| EPI_ISL_862418 | India/AP-CS0859/2020 | 2020-08-01 | Asia/India/Andhra Pradesh/ | Kurnool Medical College (KMC) | CSIR Institute of Genomics and Integrative Biology | Pallavali Roja Rani et al |
| EPI_ISL_862419 | India/AP-CS0861/2020 | 2020-08-01 | Asia/India/Andhra Pradesh/ | Kurnool Medical College (KMC) | CSIR Institute of Genomics and Integrative Biology | Pallavali Roja Rani et al |
| EPI_ISL_862420 | India/AP-CS0862/2020 | 2020-08-01 | Asia/India/Andhra Pradesh/ | Kurnool Medical College (KMC) | CSIR Institute of Genomics and Integrative Biology | Pallavali Roja Rani et al |
| EPI_ISL_862421 | India/AP-CS0863/2020 | 2020-08-01 | Asia/India/Andhra Pradesh/ | Kurnool Medical College (KMC) | CSIR Institute of Genomics and Integrative Biology | Pallavali Roja Rani et al |
| EPI_ISL_862422 | India/AP-CS0864/2020 | 2020-08-01 | Asia/India/Andhra Pradesh/ | Kurnool Medical College (KMC) | CSIR Institute of Genomics and Integrative Biology | Pallavali Roja Rani et al |
| EPI_ISL_862424 | India/AP-CS0866/2020 | 2020-08-01 | Asia/India/Andhra Pradesh/ | Kurnool Medical College (KMC) | CSIR Institute of Genomics and Integrative Biology | Pallavali Roja Rani et al |
| EPI_ISL_862426 | India/AP-CS0871/2020 | 2020-08-01 | Asia/India/Andhra Pradesh/ | Kurnool Medical College (KMC) | CSIR Institute of Genomics and Integrative Biology | Pallavali Roja Rani et al |
| EPI_ISL_862427 | India/AP-CS0873/2020 | 2020-08-01 | Asia/India/Andhra Pradesh/ | Kurnool Medical College (KMC) | CSIR Institute of Genomics and Integrative Biology | Pallavali Roja Rani et al |

| | | | | | | |
|---|---|---|---|---|---|---|
| EPI_ISL_862428 | India/AP-CS0876/2020 | 2020-08-01 | Asia/India/Andhra Pradesh/ | Kurnool Medical College (KMC) | CSIR Institute of Genomics and Integrative Biology | Pallavali Roja Rani et al |
| EPI_ISL_862429 | India/AP-CS0878/2020 | 2020-08-01 | Asia/India/Andhra Pradesh/ | Kurnool Medical College (KMC) | CSIR Institute of Genomics and Integrative Biology | Pallavali Roja Rani et al |
| EPI_ISL_862430 | India/AP-CS0879/2020 | 2020-08-01 | Asia/India/Andhra Pradesh/ | Kurnool Medical College (KMC) | CSIR Institute of Genomics and Integrative Biology | Pallavali Roja Rani et al |
| EPI_ISL_862431 | India/AP-CS0883/2020 | 2020-08-02 | Asia/India/Andhra Pradesh/ | Kurnool Medical College (KMC) | CSIR Institute of Genomics and Integrative Biology | Pallavali Roja Rani et al |
| EPI_ISL_862432 | India/AP-CS0884/2020 | 2020-08-02 | Asia/India/Andhra Pradesh/ | Kurnool Medical College (KMC) | CSIR Institute of Genomics and Integrative Biology | Pallavali Roja Rani et al |
| EPI_ISL_862433 | India/AP-CS0886/2020 | 2020-08-02 | Asia/India/Andhra Pradesh/ | Kurnool Medical College (KMC) | CSIR Institute of Genomics and Integrative Biology | Pallavali Roja Rani et al |
| EPI_ISL_862434 | India/AP-CS0887/2020 | 2020-08-02 | Asia/India/Andhra Pradesh/ | Kurnool Medical College (KMC) | CSIR Institute of Genomics and Integrative Biology | Pallavali Roja Rani et al |
| EPI_ISL_862435 | India/AP-CS0888/2020 | 2020-08-02 | Asia/India/Andhra Pradesh/ | Kurnool Medical College (KMC) | CSIR Institute of Genomics and Integrative Biology | Pallavali Roja Rani et al |
| EPI_ISL_862436 | India/AP-CS0889/2020 | 2020-08-02 | Asia/India/Andhra Pradesh/ | Kurnool Medical College (KMC) | CSIR Institute of Genomics and Integrative Biology | Pallavali Roja Rani et al |
| EPI_ISL_862438 | India/AP-CS0338/2020 | 2020-07-04 | Asia/India/Andhra Pradesh/ | Kurnool Medical College (KMC) | CSIR Institute of Genomics and Integrative Biology | Pallavali Roja Rani et al |

| | | | | | | |
|---|---|---|---|---|---|---|
| EPI_ISL_862439 | India/AP-CS0343/2020 | 2020-07-04 | Asia/India/Andhra Pradesh/ | Kurnool Medical College (KMC) | CSIR Institute of Genomics and Integrative Biology | Pallavali Roja Rani et al |
| EPI_ISL_862440 | India/AP-CS0344/2020 | 2020-07-04 | Asia/India/Andhra Pradesh/ | Kurnool Medical College (KMC) | CSIR Institute of Genomics and Integrative Biology | Pallavali Roja Rani et al |
| EPI_ISL_862442 | India/AP-CS0347/2020 | 2020-07-04 | Asia/India/Andhra Pradesh/ | Kurnool Medical College (KMC) | CSIR Institute of Genomics and Integrative Biology | Pallavali Roja Rani et al |
| EPI_ISL_862443 | India/AP-CS0350/2020 | 2020-07-04 | Asia/India/Andhra Pradesh/ | Kurnool Medical College (KMC) | CSIR Institute of Genomics and Integrative Biology | Pallavali Roja Rani et al |
| EPI_ISL_862444 | India/AP-CS0351/2020 | 2020-07-04 | Asia/India/Andhra Pradesh/ | Kurnool Medical College (KMC) | CSIR Institute of Genomics and Integrative Biology | Pallavali Roja Rani et al |
| EPI_ISL_862445 | India/AP-CS0364/2020 | 2020-07-04 | Asia/India/Andhra Pradesh/ | Kurnool Medical College (KMC) | CSIR Institute of Genomics and Integrative Biology | Pallavali Roja Rani et al |
| EPI_ISL_862446 | India/AP-CS0365/2020 | 2020-07-04 | Asia/India/Andhra Pradesh/ | Kurnool Medical College (KMC) | CSIR Institute of Genomics and Integrative Biology | Pallavali Roja Rani et al |
| EPI_ISL_862447 | India/AP-CS0366/2020 | 2020-06-30 | Asia/India/Andhra Pradesh/ | Kurnool Medical College (KMC) | CSIR Institute of Genomics and Integrative Biology | Pallavali Roja Rani et al |
| EPI_ISL_862448 | India/AP-CS0368/2020 | 2020-07-04 | Asia/India/Andhra Pradesh/ | Kurnool Medical College (KMC) | CSIR Institute of Genomics and Integrative Biology | Pallavali Roja Rani et al |
| EPI_ISL_862450 | India/AP-CS0371/2020 | 2020-07-04 | Asia/India/Andhra Pradesh/ | Kurnool Medical College (KMC) | CSIR Institute of Genomics and Integrative Biology | Pallavali Roja Rani et al |

| | | | | | | |
|---|---|---|---|---|---|---|
| EPI_ISL_862451 | India/AP-CS0372/2020 | 2020-07-04 | Asia/India/Andhra Pradesh/ | Kurnool Medical College (KMC) | CSIR Institute of Genomics and Integrative Biology | Pallavali Roja Rani et al |
| EPI_ISL_862452 | India/AP-CS0373/2020 | 2020-07-04 | Asia/India/Andhra Pradesh/ | Kurnool Medical College (KMC) | CSIR Institute of Genomics and Integrative Biology | Pallavali Roja Rani et al |
| EPI_ISL_862453 | India/AP-CS0374/2020 | 2020-07-04 | Asia/India/Andhra Pradesh/ | Kurnool Medical College (KMC) | CSIR Institute of Genomics and Integrative Biology | Pallavali Roja Rani et al |
| EPI_ISL_862454 | India/AP-CS0376/2020 | 2020-07-01 | Asia/India/Andhra Pradesh/ | Kurnool Medical College (KMC) | CSIR Institute of Genomics and Integrative Biology | Pallavali Roja Rani et al |
| EPI_ISL_862455 | India/AP-CS0377/2020 | 2020-07-01 | Asia/India/Andhra Pradesh/ | Kurnool Medical College (KMC) | CSIR Institute of Genomics and Integrative Biology | Pallavali Roja Rani et al |
| EPI_ISL_862456 | India/AP-CS0378/2020 | 2020-07-02 | Asia/India/Andhra Pradesh/ | Kurnool Medical College (KMC) | CSIR Institute of Genomics and Integrative Biology | Pallavali Roja Rani et al |
| EPI_ISL_862457 | India/AP-CS0380/2020 | 2020-07-02 | Asia/India/Andhra Pradesh/ | Kurnool Medical College (KMC) | CSIR Institute of Genomics and Integrative Biology | Pallavali Roja Rani et al |
| EPI_ISL_862458 | India/AP-CS0381/2020 | 2020-06-30 | Asia/India/Andhra Pradesh/ | Kurnool Medical College (KMC) | CSIR Institute of Genomics and Integrative Biology | Pallavali Roja Rani et al |
| EPI_ISL_862460 | India/AP-CS0384/2020 | 2020-06-30 | Asia/India/Andhra Pradesh/ | Kurnool Medical College (KMC) | CSIR Institute of Genomics and Integrative Biology | Pallavali Roja Rani et al |
| EPI_ISL_862462 | India/AP-CS0388/2020 | 2020-07-01 | Asia/India/Andhra Pradesh/ | Kurnool Medical College (KMC) | CSIR Institute of Genomics and Integrative Biology | Pallavali Roja Rani et al |

| | | | | | | |
|---|---|---|---|---|---|---|
| EPI_ISL_862463 | India/AP-CS0390/2020 | 2020-07-01 | Asia/India/Andhra Pradesh/ | Kurnool Medical College (KMC) | CSIR Institute of Genomics and Integrative Biology | Pallavali Roja Rani et al |
| EPI_ISL_862464 | India/AP-CS0391/2020 | 2020-07-01 | Asia/India/Andhra Pradesh/ | Kurnool Medical College (KMC) | CSIR Institute of Genomics and Integrative Biology | Pallavali Roja Rani et al |
| EPI_ISL_862465 | India/AP-CS0392/2020 | 2020-06-27 | Asia/India/Andhra Pradesh/ | Kurnool Medical College (KMC) | CSIR Institute of Genomics and Integrative Biology | Pallavali Roja Rani et al |
| EPI_ISL_862466 | India/AP-CS0393/2020 | 2020-07-04 | Asia/India/Andhra Pradesh/ | Kurnool Medical College (KMC) | CSIR Institute of Genomics and Integrative Biology | Pallavali Roja Rani et al |
| EPI_ISL_862467 | India/AP-CS0896/2020 | 2020-08-02 | Asia/India/Andhra Pradesh/ | Kurnool Medical College (KMC) | CSIR Institute of Genomics and Integrative Biology | Pallavali Roja Rani et al |
| EPI_ISL_862468 | India/AP-CS0790/2020 | 2020-07-29 | Asia/India/Andhra Pradesh/ | Kurnool Medical College (KMC) | CSIR Institute of Genomics and Integrative Biology | Pallavali Roja Rani et al |
| EPI_ISL_862469 | India/AP-CS0911/2020 | 2020-08-02 | Asia/India/Andhra Pradesh/ | Kurnool Medical College (KMC) | CSIR Institute of Genomics and Integrative Biology | Pallavali Roja Rani et al |
| EPI_ISL_862470 | India/AP-CS0914/2020 | 2020-08-02 | Asia/India/Andhra Pradesh/ | Kurnool Medical College (KMC) | CSIR Institute of Genomics and Integrative Biology | Pallavali Roja Rani et al |
| EPI_ISL_862471 | India/AP-CS0930/2020 | 2020-08-02 | Asia/India/Andhra Pradesh/ | Kurnool Medical College (KMC) | CSIR Institute of Genomics and Integrative Biology | Pallavali Roja Rani et al |
| EPI_ISL_862472 | India/AP-CS0933/2020 | 2020-08-02 | Asia/India/Andhra Pradesh/ | Kurnool Medical College (KMC) | CSIR Institute of Genomics and Integrative Biology | Pallavali Roja Rani et al |

| | | | | | | |
|---|---|---|---|---|---|---|
| EPI_ISL_862473 | India/AP-CS0938/2020 | 2020-08-02 | Asia/India/Andhra Pradesh/ | Kurnool Medical College (KMC) | CSIR Institute of Genomics and Integrative Biology | Pallavali Roja Rani et al |
| EPI_ISL_862474 | India/AP-CS0940/2020 | 2020-08-02 | Asia/India/Andhra Pradesh/ | Kurnool Medical College (KMC) | CSIR Institute of Genomics and Integrative Biology | Pallavali Roja Rani et al |
| EPI_ISL_862475 | India/AP-CS0941/2020 | 2020-08-02 | Asia/India/Andhra Pradesh/ | Kurnool Medical College (KMC) | CSIR Institute of Genomics and Integrative Biology | Pallavali Roja Rani et al |
| EPI_ISL_862476 | India/AP-CS0942/2020 | 2020-08-02 | Asia/India/Andhra Pradesh/ | Kurnool Medical College (KMC) | CSIR Institute of Genomics and Integrative Biology | Pallavali Roja Rani et al |
| EPI_ISL_862477 | India/AP-CS0948/2020 | 2020-08-02 | Asia/India/Andhra Pradesh/ | Kurnool Medical College (KMC) | CSIR Institute of Genomics and Integrative Biology | Pallavali Roja Rani et al |
| EPI_ISL_862478 | India/AP-CS0793/2020 | 2020-07-29 | Asia/India/Andhra Pradesh/ | Kurnool Medical College (KMC) | CSIR Institute of Genomics and Integrative Biology | Pallavali Roja Rani et al |
| EPI_ISL_862479 | India/AP-CS0965/2020 | 2020-08-02 | Asia/India/Andhra Pradesh/ | Kurnool Medical College (KMC) | CSIR Institute of Genomics and Integrative Biology | Pallavali Roja Rani et al |
| EPI_ISL_862480 | India/AP-CS0967/2020 | 2020-08-02 | Asia/India/Andhra Pradesh/ | Kurnool Medical College (KMC) | CSIR Institute of Genomics and Integrative Biology | Pallavali Roja Rani et al |
| EPI_ISL_862481 | India/AP-CS0795/2020 | 2020-07-29 | Asia/India/Andhra Pradesh/ | Kurnool Medical College (KMC) | CSIR Institute of Genomics and Integrative Biology | Pallavali Roja Rani et al |
| EPI_ISL_862482 | India/AP-CS0973/2020 | 2020-08-02 | Asia/India/Andhra Pradesh/ | Kurnool Medical College (KMC) | CSIR Institute of Genomics and Integrative Biology | Pallavali Roja Rani et al |

| | | | | | | |
|---|---|---|---|---|---|---|
| EPI_ISL_862483 | India/AP-CS0975/2020 | 2020-08-02 | Asia/India/Andhra Pradesh/ | Kurnool Medical College (KMC) | CSIR Institute of Genomics and Integrative Biology | Pallavali Roja Rani et al |
| EPI_ISL_862484 | India/AP-CS0976/2020 | 2020-08-02 | Asia/India/Andhra Pradesh/ | Kurnool Medical College (KMC) | CSIR Institute of Genomics and Integrative Biology | Pallavali Roja Rani et al |
| EPI_ISL_862485 | India/AP-CS0978/2020 | 2020-08-02 | Asia/India/Andhra Pradesh/ | Kurnool Medical College (KMC) | CSIR Institute of Genomics and Integrative Biology | Pallavali Roja Rani et al |
| EPI_ISL_862487 | India/AP-CS0981/2020 | 2020-08-02 | Asia/India/Andhra Pradesh/ | Kurnool Medical College (KMC) | CSIR Institute of Genomics and Integrative Biology | Pallavali Roja Rani et al |
| EPI_ISL_862488 | India/AP-CS0983/2020 | 2020-08-02 | Asia/India/Andhra Pradesh/ | Kurnool Medical College (KMC) | CSIR Institute of Genomics and Integrative Biology | Pallavali Roja Rani et al |
| EPI_ISL_862489 | India/AP-CS0984/2020 | 2020-08-02 | Asia/India/Andhra Pradesh/ | Kurnool Medical College (KMC) | CSIR Institute of Genomics and Integrative Biology | Pallavali Roja Rani et al |
| EPI_ISL_862490 | India/AP-CS0985/2020 | 2020-08-02 | Asia/India/Andhra Pradesh/ | Kurnool Medical College (KMC) | CSIR Institute of Genomics and Integrative Biology | Pallavali Roja Rani et al |
| EPI_ISL_862491 | India/AP-CS0986/2020 | 2020-08-02 | Asia/India/Andhra Pradesh/ | Kurnool Medical College (KMC) | CSIR Institute of Genomics and Integrative Biology | Pallavali Roja Rani et al |
| EPI_ISL_862492 | India/AP-CS0987/2020 | 2020-08-03 | Asia/India/Andhra Pradesh/ | Kurnool Medical College (KMC) | CSIR Institute of Genomics and Integrative Biology | Pallavali Roja Rani et al |
| EPI_ISL_862494 | India/AP-CS0990/2020 | 2020-08-03 | Asia/India/Andhra Pradesh/ | Kurnool Medical College (KMC) | CSIR Institute of Genomics and Integrative Biology | Pallavali Roja Rani et al |

| | | | | | | |
|---|---|---|---|---|---|---|
| EPI_ISL_862495 | India/AP-CS0998/2020 | 2020-08-03 | Asia/India/Andhra Pradesh/ | Kurnool Medical College (KMC) | CSIR Institute of Genomics and Integrative Biology | Pallavali Roja Rani et al |
| EPI_ISL_862496 | India/AP-CS0798/2020 | 2020-07-29 | Asia/India/Andhra Pradesh/ | Kurnool Medical College (KMC) | CSIR Institute of Genomics and Integrative Biology | Pallavali Roja Rani et al |
| EPI_ISL_862497 | India/AP-CS0800/2020 | 2020-07-29 | Asia/India/Andhra Pradesh/ | Kurnool Medical College (KMC) | CSIR Institute of Genomics and Integrative Biology | Pallavali Roja Rani et al |
| EPI_ISL_862498 | India/AP-CS0805/2020 | 2020-07-29 | Asia/India/Andhra Pradesh/ | Kurnool Medical College (KMC) | CSIR Institute of Genomics and Integrative Biology | Pallavali Roja Rani et al |
| EPI_ISL_862499 | India/AP-CS0806/2020 | 2020-07-29 | Asia/India/Andhra Pradesh/ | Kurnool Medical College (KMC) | CSIR Institute of Genomics and Integrative Biology | Pallavali Roja Rani et al |
| EPI_ISL_862500 | India/AP-CS0816/2020 | 2020-07-29 | Asia/India/Andhra Pradesh/ | Kurnool Medical College (KMC) | CSIR Institute of Genomics and Integrative Biology | Pallavali Roja Rani et al |
| EPI_ISL_862501 | India/AP-CS0830/2020 | 2020-07-30 | Asia/India/Andhra Pradesh/ | Kurnool Medical College (KMC) | CSIR Institute of Genomics and Integrative Biology | Pallavali Roja Rani et al |
| EPI_ISL_862502 | India/AP-CS0835/2020 | 2020-07-30 | Asia/India/Andhra Pradesh/ | Kurnool Medical College (KMC) | CSIR Institute of Genomics and Integrative Biology | Pallavali Roja Rani et al |
| EPI_ISL_862504 | India/AP-CS0846/2020 | 2020-08-01 | Asia/India/Andhra Pradesh/ | Kurnool Medical College (KMC) | CSIR Institute of Genomics and Integrative Biology | Pallavali Roja Rani et al |
| EPI_ISL_862505 | India/AP-CS0867/2020 | 2020-08-01 | Asia/India/Andhra Pradesh/ | Kurnool Medical College (KMC) | CSIR Institute of Genomics and Integrative Biology | Pallavali Roja Rani et al |

| | | | | | | |
|---|---|---|---|---|---|---|
| EPI_ISL_862506 | India/AP-CS0868/2020 | 2020-08-01 | Asia/India/Andhra Pradesh/ | Kurnool Medical College (KMC) | CSIR Institute of Genomics and Integrative Biology | Pallavali Roja Rani et al |
| EPI_ISL_862507 | India/AP-CS0880/2020 | 2020-08-02 | Asia/India/Andhra Pradesh/ | Kurnool Medical College (KMC) | CSIR Institute of Genomics and Integrative Biology | Pallavali Roja Rani et al |
| EPI_ISL_862508 | India/AP-CS0881/2020 | 2020-08-02 | Asia/India/Andhra Pradesh/ | Kurnool Medical College (KMC) | CSIR Institute of Genomics and Integrative Biology | Pallavali Roja Rani et al |
| EPI_ISL_862510 | India/AP-CS0314/2020 | 2020-06-29 | Asia/India/Andhra Pradesh/ | Kurnool Medical College (KMC) | CSIR Institute of Genomics and Integrative Biology | Pallavali Roja Rani et al |
| EPI_ISL_862514 | India/AP-CS0326/2020 | 2020-06-29 | Asia/India/Andhra Pradesh/ | Kurnool Medical College (KMC) | CSIR Institute of Genomics and Integrative Biology | Pallavali Roja Rani et al |
| EPI_ISL_862515 | India/AP-CS0327/2020 | 2020-06-29 | Asia/India/Andhra Pradesh/ | Kurnool Medical College (KMC) | CSIR Institute of Genomics and Integrative Biology | Pallavali Roja Rani et al |
| EPI_ISL_862518 | India/AP-CS0330/2020 | 2020-06-29 | Asia/India/Andhra Pradesh/ | Kurnool Medical College (KMC) | CSIR Institute of Genomics and Integrative Biology | Pallavali Roja Rani et al |
| EPI_ISL_862519 | India/AP-CS0332/2020 | 2020-06-29 | Asia/India/Andhra Pradesh/ | Kurnool Medical College (KMC) | CSIR Institute of Genomics and Integrative Biology | Pallavali Roja Rani et al |
| EPI_ISL_862520 | India/AP-CS0333/2020 | 2020-06-29 | Asia/India/Andhra Pradesh/ | Kurnool Medical College (KMC) | CSIR Institute of Genomics and Integrative Biology | Pallavali Roja Rani et al |
| EPI_ISL_862521 | India/AP-CS0335/2020 | 2020-07-04 | Asia/India/Andhra Pradesh/ | Kurnool Medical College (KMC) | CSIR Institute of Genomics and Integrative Biology | Pallavali Roja Rani et al |

| | | | | | | |
|---|---|---|---|---|---|---|
| EPI_ISL_862523 | India/AP-CS0341/2020 | 2020-07-04 | Asia/India/Andhra Pradesh/ | Kurnool Medical College (KMC) | CSIR Institute of Genomics and Integrative Biology | Pallavali Roja Rani et al |
| EPI_ISL_862524 | India/AP-CS0342/2020 | 2020-07-04 | Asia/India/Andhra Pradesh/ | Kurnool Medical College (KMC) | CSIR Institute of Genomics and Integrative Biology | Pallavali Roja Rani et al |
| EPI_ISL_862525 | India/AP-CS0352/2020 | 2020-07-04 | Asia/India/Andhra Pradesh/ | Kurnool Medical College (KMC) | CSIR Institute of Genomics and Integrative Biology | Pallavali Roja Rani et al |
| EPI_ISL_862526 | India/AP-CS0354/2020 | 2020-07-04 | Asia/India/Andhra Pradesh/ | Kurnool Medical College (KMC) | CSIR Institute of Genomics and Integrative Biology | Pallavali Roja Rani et al |
| EPI_ISL_862527 | India/AP-CS0356/2020 | 2020-07-04 | Asia/India/Andhra Pradesh/ | Kurnool Medical College (KMC) | CSIR Institute of Genomics and Integrative Biology | Pallavali Roja Rani et al |
| EPI_ISL_862528 | India/AP-CS0359/2020 | 2020-07-04 | Asia/India/Andhra Pradesh/ | Kurnool Medical College (KMC) | CSIR Institute of Genomics and Integrative Biology | Pallavali Roja Rani et al |
| EPI_ISL_862529 | India/AP-CS0361/2020 | 2020-07-04 | Asia/India/Andhra Pradesh/ | Kurnool Medical College (KMC) | CSIR Institute of Genomics and Integrative Biology | Pallavali Roja Rani et al |
| EPI_ISL_862530 | India/AP-CS0362/2020 | 2020-07-04 | Asia/India/Andhra Pradesh/ | Kurnool Medical College (KMC) | CSIR Institute of Genomics and Integrative Biology | Pallavali Roja Rani et al |
| EPI_ISL_862531 | India/AP-CS0367/2020 | 2020-07-04 | Asia/India/Andhra Pradesh/ | Kurnool Medical College (KMC) | CSIR Institute of Genomics and Integrative Biology | Pallavali Roja Rani et al |
| EPI_ISL_862532 | India/AP-CS0375/2020 | 2020-07-01 | Asia/India/Andhra Pradesh/ | Kurnool Medical College (KMC) | CSIR Institute of Genomics and Integrative Biology | Pallavali Roja Rani et al |

| | | | | | | |
|---|---|---|---|---|---|---|
| EPI_ISL_862533 | India/AP-CS0379/2020 | 2020-07-02 | Asia/India/Andhra Pradesh/ | Kurnool Medical College (KMC) | CSIR Institute of Genomics and Integrative Biology | Pallavali Roja Rani et al |
| EPI_ISL_862536 | India/AP-CS0387/2020 | 2020-06-30 | Asia/India/Andhra Pradesh/ | Kurnool Medical College (KMC) | CSIR Institute of Genomics and Integrative Biology | Pallavali Roja Rani et al |
| EPI_ISL_862537 | India/AP-CS0394/2020 | 2020-07-04 | Asia/India/Andhra Pradesh/ | Kurnool Medical College (KMC) | CSIR Institute of Genomics and Integrative Biology | Pallavali Roja Rani et al |
| EPI_ISL_862538 | India/AP-CS0396/2020 | 2020-07-04 | Asia/India/Andhra Pradesh/ | Kurnool Medical College (KMC) | CSIR Institute of Genomics and Integrative Biology | Pallavali Roja Rani et al |
| EPI_ISL_862539 | India/AP-CS0397/2020 | 2020-07-04 | Asia/India/Andhra Pradesh/ | Kurnool Medical College (KMC) | CSIR Institute of Genomics and Integrative Biology | Pallavali Roja Rani et al |
| EPI_ISL_862540 | India/AP-CS0399/2020 | 2020-07-04 | Asia/India/Andhra Pradesh/ | Kurnool Medical College (KMC) | CSIR Institute of Genomics and Integrative Biology | Pallavali Roja Rani et al |
| EPI_ISL_862541 | India/AP-CS0400/2020 | 2020-07-04 | Asia/India/Andhra Pradesh/ | Kurnool Medical College (KMC) | CSIR Institute of Genomics and Integrative Biology | Pallavali Roja Rani et al |
| EPI_ISL_862542 | India/AP-CS0401/2020 | 2020-07-04 | Asia/India/Andhra Pradesh/ | Kurnool Medical College (KMC) | CSIR Institute of Genomics and Integrative Biology | Pallavali Roja Rani et al |
| EPI_ISL_862543 | India/AP-CS0402/2020 | 2020-07-04 | Asia/India/Andhra Pradesh/ | Kurnool Medical College (KMC) | CSIR Institute of Genomics and Integrative Biology | Pallavali Roja Rani et al |
| EPI_ISL_862552 | Spain/MC-ISCIII-2018163/2020 | 2020-12-27 | Europe/Spain/Murcia/ | Complejo Hospitalario de Navarra | Instituto de Salud Carlos III | Iglesias-Caballero et al |
| EPI_ISL_862553 | Spain/GA-ISCIII-2018188/2020 | 2020-12-26 | Europe/Spain/Galicia/ | Compejo Hospitalario Xeral-Calde | Instituto de Salud Carlos III | Iglesias-Caballero et al |

| | | | | | | |
|---|---|---|---|---|---|---|
| EPI_ISL_862557 | Spain/CM-ISCIII-210008/2020 | 2020-12-28 | Europe/Spain/Castilla la Mancha/ | Consejer√≠a de Sanidad y Asuntos Sociales | Instituto de Salud Carlos III | Iglesias-Caballero et al |
| EPI_ISL_862578 | Spain/CT-ISCIII-2018172/2020 | 2020-12-09 | Europe/Spain/Catalunya/ | Hospital Cl√≠nic | Instituto de Salud Carlos III | Iglesias-Caballero et al |
| EPI_ISL_862579 | Spain/CT-ISCIII-2018173/2020 | 2020-12-21 | Europe/Spain/Catalunya/ | Hospital Cl√≠nic | Instituto de Salud Carlos III | Iglesias-Caballero et al |
| EPI_ISL_862591 | Spain/ML-ISCIII-202059806/2020 | 2020-09-15 | Europe/Spain/Melilla/ | Hospital Comarcal de Melilla | Instituto de Salud Carlos III | Iglesias-Caballero et al |
| EPI_ISL_862593 | Spain/ML-ISCIII-202058690/2020 | 2020-09-15 | Europe/Spain/Melilla/ | Hospital Comarcal de Melilla | Instituto de Salud Carlos III | Iglesias-Caballero et al |
| EPI_ISL_862599 | Spain/ML-ISCIII-202059509/2020 | 2020-09-15 | Europe/Spain/Melilla/ | Hospital Comarcal de Melilla | Instituto de Salud Carlos III | Iglesias-Caballero et al |
| EPI_ISL_862604 | Spain/ML-ISCIII-202059611/2020 | 2020-09-15 | Europe/Spain/Melilla/ | Hospital Comarcal de Melilla | Instituto de Salud Carlos III | Iglesias-Caballero et al |
| EPI_ISL_862605 | Spain/ML-ISCIII-202058753/2020 | 2020-09-15 | Europe/Spain/Melilla/ | Hospital Comarcal de Melilla | Instituto de Salud Carlos III | Iglesias-Caballero et al |
| EPI_ISL_862606 | Spain/ML-ISCIII-202058903/2020 | 2020-09-15 | Europe/Spain/Melilla/ | Hospital Comarcal de Melilla | Instituto de Salud Carlos III | Iglesias-Caballero et al |
| EPI_ISL_862607 | Spain/ML-ISCIII-202058685/2020 | 2020-09-15 | Europe/Spain/Melilla/ | Hospital Comarcal de Melilla | Instituto de Salud Carlos III | Iglesias-Caballero et al |
| EPI_ISL_862609 | Spain/ML-ISCIII-202058682/2020 | 2020-09-15 | Europe/Spain/Melilla/ | Hospital Comarcal de Melilla | Instituto de Salud Carlos III | Iglesias-Caballero et al |
| EPI_ISL_862613 | Spain/ML-ISCIII-202058892/2020 | 2020-09-15 | Europe/Spain/Melilla/ | Hospital Comarcal de Melilla | Instituto de Salud Carlos III | Iglesias-Caballero et al |
| EPI_ISL_862614 | Spain/ML-ISCIII-202058870/2020 | 2020-09-15 | Europe/Spain/Melilla/ | Hospital Comarcal de Melilla | Instituto de Salud Carlos III | Iglesias-Caballero et al |
| EPI_ISL_862615 | Spain/ML-ISCIII-202058871/2020 | 2020-09-15 | Europe/Spain/Melilla/ | Hospital Comarcal de Melilla | Instituto de Salud Carlos III | Iglesias-Caballero et al |
| EPI_ISL_862617 | Spain/ML-ISCIII-202059809/2020 | 2020-09-15 | Europe/Spain/Melilla/ | Hospital Comarcal de Melilla | Instituto de Salud Carlos III | Iglesias-Caballero et al |
| EPI_ISL_862620 | Spain/ML-ISCIII-202059627/2020 | 2020-09-15 | Europe/Spain/Melilla/ | Hospital Comarcal de Melilla | Instituto de Salud Carlos III | Iglesias-Caballero et al |
| EPI_ISL_862621 | Spain/ML-ISCIII-202059630/2020 | 2020-09-15 | Europe/Spain/Melilla/ | Hospital Comarcal de Melilla | Instituto de Salud Carlos III | Iglesias-Caballero et al |
| EPI_ISL_862625 | Spain/ML-ISCIII-202059895/2020 | 2020-09-15 | Europe/Spain/Melilla/ | Hospital Comarcal de Melilla | Instituto de Salud Carlos III | Iglesias-Caballero et al |
| EPI_ISL_862628 | Spain/CT-ISCIII-202034658/2020 | 2020-03-24 | Europe/Spain/Catalunya/ | Hospital Cl√≠nic | Instituto de Salud Carlos III | Iglesias-Caballero et al |
| EPI_ISL_862631 | Spain/CT-ISCIII-202034697/2020 | 2020-03-24 | Europe/Spain/Catalunya/ | Hospital Cl√≠nic | Instituto de Salud Carlos III | Iglesias-Caballero et al |

| | | | | | | |
|---|---|---|---|---|---|---|
| EPI_ISL_862634 | Spain/CT-ISCIII-202043/2020 | 2020-02-29 | Europe/Spain/Catalunya/ | Hospital Cl√≠nic | Instituto de Salud Carlos III | Iglesias-Caballero et al |
| EPI_ISL_862641 | Spain/CM-ISCIII-201371/2020 | 2020-03-02 | Europe/Spain/Castilla la Mancha/ | Consejer√≠a de Sanidad y Asuntos Sociales | Instituto de Salud Carlos III | Iglesias-Caballero et al |
| EPI_ISL_862650 | Spain/NA-ISCIII-201953/2020 | 2020-03-09 | Europe/Spain/Navarra/ | Complejo Hospitalario de Navarra | Instituto de Salud Carlos III | Iglesias-Caballero et al |
| EPI_ISL_862651 | Spain/NA-ISCIII-201948/2020 | 2020-03-09 | Europe/Spain/Navarra/ | Complejo Hospitalario de Navarra | Instituto de Salud Carlos III | Iglesias-Caballero et al |
| EPI_ISL_862654 | Spain/EX-ISCIII-201247/2020 | 2020-02-28 | Europe/Spain/Extremadura/ | Hospital San Pedro de Alc√°ntara | Instituto de Salud Carlos III | Iglesias-Caballero et al |
| EPI_ISL_862657 | Spain/CE-ISCIII-205115/2020 | 2020-03-19 | Europe/Spain/Ceuta/ | Hospital Universitario de Ceuta | Instituto de Salud Carlos III | Iglesias-Caballero et al |
| EPI_ISL_862659 | Germany/BE-ChVir22116/2021 | 2021-01-11 | Europe/Germany/Berlin/ | Charit√© Universit√§tsmedizin Berlin, Institut f√ºr Virologie/Labor Berlin | Charit√© Universit√§tsmedizin Berlin, Institut f√ºr Virologie | Victor M Corman et al |
| EPI_ISL_862660 | Germany/NW-ChVir22076/2021 | 2021-01-02 | Europe/Germany/North Rhine Westphal | Charit√© Universit√§tsmedizin Berlin, Institut f√ºr Virologie/Labor Berlin | Charit√© Universit√§tsmedizin Berlin, Institut f√ºr Virologie | Victor M Corman et al |
| EPI_ISL_862662 | Germany/BE-ChVir22118/2021 | 2021-01-11 | Europe/Germany/Berlin/ | Charit√© Universit√§tsmedizin Berlin, Institut f√ºr Virologie/Labor Berlin | Charit√© Universit√§tsmedizin Berlin, Institut f√ºr Virologie | Victor M Corman et al |
| EPI_ISL_862666 | USA/MI-UM-10037641515/2021 | 2021-01-02 | North America/USA/Michigan/ | University of Michigan Clinical Microbiology Laboratory | Lauring Lab, University of Michigan, Department of Microbiology and Immunology | Valesano et al |
| EPI_ISL_862668 | USA/MI-UM-10037641384/2021 | 2021-01-02 | North America/USA/Michigan/ | University of Michigan Clinical Microbiology Laboratory | Lauring Lab, University of Michigan, Department of Microbiology and Immunology | Valesano et al |
| EPI_ISL_862670 | USA/NMDOH-2021015923/2021 | 2021-01-11 | North America/USA/New Mexico/ | New Mexico Department of Health Scientific Laboratory | New Mexico Department of Health Scientific Laboratory | D'eldra Malone et al |

| | | | | | | |
|---|---|---|---|---|---|---|
| EPI_ISL_862672 | USA/NMDOH-2021016195/2021 | 2021-01-11 | North America/USA/New Mexico/ | New Mexico Department of Health Scientific Laboratory | New Mexico Department of Health Scientific Laboratory | D'eldra Malone et al |
| EPI_ISL_862688 | USA/NMDOH-2021017268/2021 | 2021-01-11 | North America/USA/New Mexico/ | New Mexico Department of Health Scientific Laboratory | New Mexico Department of Health Scientific Laboratory | D'eldra Malone et al |
| EPI_ISL_862691 | USA/NMDOH-2021017302/2021 | 2021-01-11 | North America/USA/New Mexico/ | New Mexico Department of Health Scientific Laboratory | New Mexico Department of Health Scientific Laboratory | D'eldra Malone et al |
| EPI_ISL_862704 | USA/NMDOH-2021018014/2021 | 2021-01-11 | North America/USA/New Mexico/ | New Mexico Department of Health Scientific Laboratory | New Mexico Department of Health Scientific Laboratory | D'eldra Malone et al |
| EPI_ISL_862707 | USA/NMDOH-2021018639/2021 | 2021-01-11 | North America/USA/New Mexico/ | New Mexico Department of Health Scientific Laboratory | New Mexico Department of Health Scientific Laboratory | D'eldra Malone et al |
| EPI_ISL_862712 | USA/NMDOH-2021018900/2021 | 2021-01-08 | North America/USA/New Mexico/ | New Mexico Department of Health Scientific Laboratory | New Mexico Department of Health Scientific Laboratory | D'eldra Malone et al |
| EPI_ISL_862713 | USA/NMDOH-2021018906/2021 | 2021-01-11 | North America/USA/New Mexico/ | New Mexico Department of Health Scientific Laboratory | New Mexico Department of Health Scientific Laboratory | D'eldra Malone et al |
| EPI_ISL_862716 | SriLanka/VQC32/2021 | 2021-01-18 | Asia/Sri Lanka/Colombo/ | Centre for Dengue Research and AICBU, Department of Immunology and Molecular Medicine | Centre for Dengue Research and AICBU, Department of Immunology and Molecular Medicine | Chandima Jeewandara et al |

| | | | | | | |
|---|---|---|---|---|---|---|
| EPI_ISL_862719 | SriLanka/ABG34/2021 | 2021-01-14 | Asia/Sri Lanka/Gampaha/ | Centre for Dengue Research and AICBU, Department of Immunology and Molecular Medicine | Centre for Dengue Research and AICBU, Department of Immunology and Molecular Medicine | Chandima Jeewandara et al |
| EPI_ISL_862726 | SriLanka/VQC14/2021 | 2021-01-18 | Asia/Sri Lanka/Vavuniya/ | Centre for Dengue Research and AICBU, Department of Immunology and Molecular Medicine | Centre for Dengue Research and AICBU, Department of Immunology and Molecular Medicine | Chandima Jeewandara et al |
| EPI_ISL_862744 | USA/UT-UPHL-2101077576/2021 | 2021-01-03 | North America/USA/Utah/ | Utah Public Health Laboratory, Utah Public Health Laboratory Infectious Disease submission group | Utah Public Health Laboratory, Utah Public Health Laboratory Infectious Disease submission group | Young et al |
| EPI_ISL_862745 | USA/UT-UPHL-2101161569/2021 | 2021-01-05 | North America/USA/Utah/ | Utah Public Health Laboratory, Utah Public Health Laboratory Infectious Disease submission group | Utah Public Health Laboratory, Utah Public Health Laboratory Infectious Disease submission group | Young et al |
| EPI_ISL_862747 | USA/UT-UPHL-2101188328/2021 | 2021-01-04 | North America/USA/Utah/ | Utah Public Health Laboratory, Utah Public Health Laboratory Infectious Disease submission group | Utah Public Health Laboratory, Utah Public Health Laboratory Infectious Disease submission group | Young et al |

| | | | | | | |
|---|---|---|---|---|---|---|
| EPI_ISL_862750 | USA/UT-UPHL-2101221848/2021 | 2021-01-04 | North America/USA/Utah/ | Utah Public Health Laboratory, Utah Public Health Laboratory Infectious Disease submission group | Utah Public Health Laboratory, Utah Public Health Laboratory Infectious Disease submission group | Young et al |
| EPI_ISL_862766 | USA/UT-UPHL-2101605937/2020 | 2020-12-21 | North America/USA/Utah/ | Utah Public Health Laboratory, Utah Public Health Laboratory Infectious Disease submission group | Utah Public Health Laboratory, Utah Public Health Laboratory Infectious Disease submission group | Young et al |
| EPI_ISL_862772 | USA/UT-UPHL-2101784878/2021 | 2021-01-06 | North America/USA/Utah/ | Utah Public Health Laboratory, Utah Public Health Laboratory Infectious Disease submission group | Utah Public Health Laboratory, Utah Public Health Laboratory Infectious Disease submission group | Young et al |
| EPI_ISL_862803 | Egypt/CUNCI-HGC11I034/2021 | 2021-01-03 | Africa/Egypt/Egypt/ | Cancer Biology Department, National Cancer Institute | Cancer Biology Department, National Cancer Institute | Zekri et al |
| EPI_ISL_862814 | France/un-ENVT-66JQ-O/2020 | 2020-09-24 | Europe/France/France/ | Virology, Ecole Nationale Veterinaire de Toulouse | Virology, Ecole Nationale Veterinaire de Toulouse | Bessiere et al |
| EPI_ISL_862815 | India/UP-MRU70695/2020 | 2020-09-10 | Asia/India/Uttar Pradesh/Chandauli | Multidisciplinary Research Unit, DHR-ICMR, Institute of Medical Sciences, Banaras Hindu University, Varanasi | Multidisciplinary Research Unit, DHR-ICMR, Institute of Medical Sciences, Banaras Hindu University, Varanasi | Royana Singh et al |

| | | | | | | |
|---|---|---|---|---|---|---|
| EPI_ISL_862832 | Romania/B-93207/2020 | 2020-12-29 | Europe/Romania/Bucharest/ | National Institute of Infectious Diseases-Prof. Dr. Matei Bals Molecular Diagnostics Laboratory | National Institute of Infectious Diseases-Prof. Dr. Matei Bals Molecular Diagnostics Laboratory | Leontina Banica et al |
| EPI_ISL_862834 | Romania/IF-1219/2021 | 2021-01-15 | Europe/Romania/Ilfov/ | National Institute of Infectious Diseases-Prof. Dr. Matei Bals Molecular Diagnostics Laboratory | National Institute of Infectious Diseases-Prof. Dr. Matei Bals Molecular Diagnostics Laboratory | Leontina Banica et al |
| EPI_ISL_862835 | Romania/IF-1450/2021 | 2021-01-18 | Europe/Romania/Ilfov/ | National Institute of Infectious Diseases-Prof. Dr. Matei Bals Molecular Diagnostics Laboratory | National Institute of Infectious Diseases-Prof. Dr. Matei Bals Molecular Diagnostics Laboratory | Leontina Banica et al |
| EPI_ISL_862931 | England/ALDP-FB5D9B/2021 | 2021-01-11 | Europe/United Kingdom/England/ | Lighthouse Lab in Alderley Park | Wellcome Sanger Institute for the COVID-19 Genomics UK (COG-UK) Consortium | Jacquelyn Wynn et al |
| EPI_ISL_863189 | England/ALDP-FB91F5/2021 | 2021-01-11 | Europe/United Kingdom/England/ | Lighthouse Lab in Alderley Park | Wellcome Sanger Institute for the COVID-19 Genomics UK (COG-UK) Consortium | Jacquelyn Wynn et al |
| EPI_ISL_863209 | England/ALDP-FB6204/2021 | 2021-01-11 | Europe/United Kingdom/England/ | Lighthouse Lab in Alderley Park | Wellcome Sanger Institute for the COVID-19 Genomics UK (COG-UK) Consortium | Jacquelyn Wynn et al |
| EPI_ISL_863511 | England/QEUH-10029D3/2021 | 2021-01-14 | Europe/United Kingdom/England/ | Lighthouse Lab in Glasgow | Wellcome Sanger Institute for the COVID-19 Genomics UK (COG-UK) Consortium | Harper VanSteenhouse et al |

| | | | | | | |
|---|---|---|---|---|---|---|
| EPI_ISL_863559 | England/MILK-EC8C4E/2021 | 2021-01-03 | Europe/United Kingdom/England/ | Lighthouse Lab in Milton Keynes | Wellcome Sanger Institute for the COVID-19 Genomics UK (COG-UK) Consortium | The Lighthouse Lab in Milton Keynes et al |
| EPI_ISL_863681 | England/MILK-EC8E93/2021 | 2021-01-03 | Europe/United Kingdom/England/ | Lighthouse Lab in Milton Keynes | Wellcome Sanger Institute for the COVID-19 Genomics UK (COG-UK) Consortium | The Lighthouse Lab in Milton Keynes et al |
| EPI_ISL_863744 | England/MILK-FEB51C/2021 | 2021-01-11 | Europe/United Kingdom/England/ | Lighthouse Lab in Milton Keynes | Wellcome Sanger Institute for the COVID-19 Genomics UK (COG-UK) Consortium | The Lighthouse Lab in Milton Keynes et al |
| EPI_ISL_863751 | England/MILK-FCF01D/2021 | 2021-01-11 | Europe/United Kingdom/England/ | Lighthouse Lab in Milton Keynes | Wellcome Sanger Institute for the COVID-19 Genomics UK (COG-UK) Consortium | The Lighthouse Lab in Milton Keynes et al |
| EPI_ISL_864100 | England/ALDP-1005A91/2021 | 2021-01-12 | Europe/United Kingdom/England/ | Lighthouse Lab in Alderley Park | Wellcome Sanger Institute for the COVID-19 Genomics UK (COG-UK) Consortium | Jacquelyn Wynn et al |
| EPI_ISL_864210 | England/MILK-FAA528/2021 | 2021-01-09 | Europe/United Kingdom/England/ | Lighthouse Lab in Milton Keynes | Wellcome Sanger Institute for the COVID-19 Genomics UK (COG-UK) Consortium | The Lighthouse Lab in Milton Keynes et al |
| EPI_ISL_864360 | England/MILK-FAEBE6/2021 | 2021-01-08 | Europe/United Kingdom/England/ | Lighthouse Lab in Milton Keynes | Wellcome Sanger Institute for the COVID-19 Genomics UK (COG-UK) Consortium | The Lighthouse Lab in Milton Keynes et al |
| EPI_ISL_864547 | Germany/NW-IMB14646/2020 | 2020-12-16 | Europe/Germany/North Rhine Westphal | Knappschaftskrankenhaus Bochum | Bundeswehr Institute of Microbiology | Markus Antwerpen et al |
| EPI_ISL_864553 | Germany/NW-IMB14753/2020 | 2020-12-21 | Europe/Germany/North Rhine Westphal | Knappschaftskrankenhaus Bochum | Bundeswehr Institute of Microbiology | Markus Antwerpen et al |

| | | | | | | |
|---|---|---|---|---|---|---|
| EPI_ISL_864554 | Germany/NW-IMB14644/2020 | 2020-12-16 | Europe/Germany/North Rhine Westphal | Knappschaftskrankenhaus Bochum | Bundeswehr Institute of Microbiology | Markus Antwerpen et al |
| EPI_ISL_864555 | Germany/NW-IMB14682/2020 | 2020-12-17 | Europe/Germany/North Rhine Westphal | Knappschaftskrankenhaus Bochum | Bundeswehr Institute of Microbiology | Markus Antwerpen et al |
| EPI_ISL_864557 | Germany/NW-IMB14754/2020 | 2020-12-21 | Europe/Germany/North Rhine Westphal | Knappschaftskrankenhaus Bochum | Bundeswehr Institute of Microbiology | Markus Antwerpen et al |
| EPI_ISL_864558 | Germany/NW-IMB14770/2020 | 2020-12-16 | Europe/Germany/North Rhine Westphal | Knappschaftskrankenhaus Bochum | Bundeswehr Institute of Microbiology | Markus Antwerpen et al |
| EPI_ISL_864561 | Germany/NW-IMB14658/2020 | 2020-12-18 | Europe/Germany/North Rhine Westphal | Knappschaftskrankenhaus Bochum | Bundeswehr Institute of Microbiology | Markus Antwerpen et al |
| EPI_ISL_864563 | Germany/ST-MD126/2020 | 2020-11-24 | Europe/Germany/Saxony-Anhalt/ | Institute of Medical Microbiology and Hospital Hygiene | Institute of Medical Microbiology and Hospital Hygiene | Prof. Dr. Achim Kaasch et al |
| EPI_ISL_864566 | Germany/ST-MD144/2020 | 2020-10-22 | Europe/Germany/Saxony-Anhalt/ | Institute of Medical Microbiology and Hospital Hygiene | Institute of Medical Microbiology and Hospital Hygiene | Prof. Dr. Achim Kaasch et al |
| EPI_ISL_864567 | Germany/ST-MD148/2020 | 2020-11-06 | Europe/Germany/Saxony-Anhalt/ | Institute of Medical Microbiology and Hospital Hygiene | Institute of Medical Microbiology and Hospital Hygiene | Prof. Dr. Achim Kaasch et al |
| EPI_ISL_864569 | Germany/ST-MD156/2020 | 2020-11-12 | Europe/Germany/Saxony-Anhalt/ | Institute of Medical Microbiology and Hospital Hygiene | Institute of Medical Microbiology and Hospital Hygiene | Prof. Dr. Achim Kaasch et al |
| EPI_ISL_864570 | Germany/ST-MD157/2020 | 2020-11-23 | Europe/Germany/Saxony-Anhalt/ | Institute of Medical Microbiology and Hospital Hygiene | Institute of Medical Microbiology and Hospital Hygiene | Prof. Dr. Achim Kaasch et al |
| EPI_ISL_864572 | Germany/ST-MD175/2020 | 2020-11-11 | Europe/Germany/Saxony-Anhalt/ | Institute of Medical Microbiology and Hospital Hygiene | Institute of Medical Microbiology and Hospital Hygiene | Prof. Dr. Achim Kaasch et al |

| | | | | | | |
|---|---|---|---|---|---|---|
| EPI_ISL_864573 | Germany/ST-MD178/2020 | 2020-11-16 | Europe/Germany/Saxony-Anhalt/ | Institute of Medical Microbiology and Hospital Hygiene | Institute of Medical Microbiology and Hospital Hygiene | Prof. Dr. Achim Kaasch et al |
| EPI_ISL_864575 | Germany/ST-MD183/2020 | 2020-10-28 | Europe/Germany/Saxony-Anhalt/ | Institute of Medical Microbiology and Hospital Hygiene | Institute of Medical Microbiology and Hospital Hygiene | Prof. Dr. Achim Kaasch et al |
| EPI_ISL_864577 | Germany/ST-MD190/2020 | 2020-12-01 | Europe/Germany/Saxony-Anhalt/ | Institute of Medical Microbiology and Hospital Hygiene | Institute of Medical Microbiology and Hospital Hygiene | Prof. Dr. Achim Kaasch et al |
| EPI_ISL_864580 | Poland/NIPH-NIH-03/2020 | 2020-12-15 | Europe/Poland/Mazowieckie/Warszawa | National Institute of Public Health - National Institute of Hygiene | National Institute of Public Health - National Institute of Hygiene, DNA Sequencing and Synthesis Facility (oligo.pl) - Institute of Biochemistry and Biophysics PAS | Wo≈Çkowicz Tomasz et al |
| EPI_ISL_864587 | France/OCC-235/2021 | 2021-01-16 | Europe/France/Occitanie/Toulouse | CHU Purpan - Laboratoire de Virologie - Institut F√©d√©ratif de Biologie | CHU Purpan - Laboratoire de Virologie - Institut F√©d√©ratif de Biologie | Latour J. et al |
| EPI_ISL_864591 | France/OCC-239/2021 | 2021-01-13 | Europe/France/Occitanie/Toulouse | CHU Purpan - Laboratoire de Virologie - Institut F√©d√©ratif de Biologie | CHU Purpan - Laboratoire de Virologie - Institut F√©d√©ratif de Biologie | Latour J. et al |
| EPI_ISL_864592 | France/OCC-240/2021 | 2021-01-13 | Europe/France/Occitanie/Toulouse | CHU Purpan - Laboratoire de Virologie - Institut F√©d√©ratif de Biologie | CHU Purpan - Laboratoire de Virologie - Institut F√©d√©ratif de Biologie | Latour J. et al |

| | | | | | | |
|---|---|---|---|---|---|---|
| EPI_ISL_864651 | Switzerland/GE-32919825/2021 | 2021-01-09 | Europe/Switzerland/Geneva/ | University Hospitals of Geneva, Laboratory of Virology | HUG, Laboratory of Virology and the Health2030 Genome Center | Samuel Cordey et al |
| EPI_ISL_864957 | England/CAMB-A9D97C/2021 | 2021-01-14 | Europe/United Kingdom/England/ | Department of Pathology, University of Cambridge | COVID-19 Genomics UK (COG-UK) Consortium | Aminu S. Jahun et al |
| EPI_ISL_865121 | Scotland/EDB12921/2021 | 2021-01-10 | Europe/United Kingdom/Scotland/ | Virology Department, Royal Infirmary of Edinburgh, NHS Lothian / School of Biological Sciences, University of Edinburgh / Institute of Genetics and Molecular Medicine, University of Edinburgh | COVID-19 Genomics UK (COG-UK) Consortium | McHugh M et al |
| EPI_ISL_865386 | England/LIVE-DDCE51/2020 | 2020-12-29 | Europe/United Kingdom/England/ | Liverpool Clinical Laboratories | COVID-19 Genomics UK (COG-UK) Consortium | Sam Haldenby et al |
| EPI_ISL_865421 | England/LIVE-DDD737/2020 | 2020-12-21 | Europe/United Kingdom/England/ | Liverpool Clinical Laboratories | COVID-19 Genomics UK (COG-UK) Consortium | Sam Haldenby et al |
| EPI_ISL_865514 | England/LOND-1288B19/2020 | 2020-12-27 | Europe/United Kingdom/England/ | Barts Health NHS Trust | COVID-19 Genomics UK (COG-UK) Consortium | CUTINO-MOGUEL et al |
| EPI_ISL_865604 | England/LOND-12DB29A/2021 | 2021-01-14 | Europe/United Kingdom/England/ | University College London, Great Ormond Street Hospital for Children NHS Foundation Trust, Imperial College Healthcare NHS Trust | COVID-19 Genomics UK (COG-UK) Consortium | Sergi Castellano et al |
| EPI_ISL_865899 | England/LOND-12F590E/2020 | 2020-04-14 | Europe/United Kingdom/England/ | University College London, Great Ormond Street Hospital for Children NHS Foundation Trust, Imperial College Healthcare NHS Trust | COVID-19 Genomics UK (COG-UK) Consortium | Sergi Castellano et al |
| EPI_ISL_866020 | England/LOND-12F6F65/2021 | 2021-01-17 | Europe/United Kingdom/England/ | University College London, Great Ormond Street Hospital for Children NHS Foundation Trust, Imperial College Healthcare NHS Trust | COVID-19 Genomics UK (COG-UK) Consortium | Sergi Castellano et al |
| EPI_ISL_866028 | England/LOND-12F77A8/2021 | 2021-01-20 | Europe/United Kingdom/England/ | University College London, Great Ormond Street Hospital for Children NHS Foundation Trust, Imperial College Healthcare NHS Trust | COVID-19 Genomics UK (COG-UK) Consortium | Sergi Castellano et al |
| EPI_ISL_866131 | England/LOND-132346F/2020 | 2020-12-27 | Europe/United Kingdom/England/ | University College London Hospital | COVID-19 Genomics UK (COG-UK) Consortium | Judith Heaney et al |

| | | | | | | |
|---|---|---|---|---|---|---|
| EPI_ISL_866482 | England/NORT-28DADE/2020 | 2020-12-15 | Europe/United Kingdom/England/ | Northumbria University / South Tees Hospitals NHS Foundation Trust / North Cumbria Integrated Care NHS Foundation Trust / North Tees and Hartlepool NHS Foundation Trust / Newcastle Hospitals NHS Foundation Trust | COVID-19 Genomics UK (COG-UK) Consortium | Darren L Smith et al |
| EPI_ISL_866498 | England/NORT-2A1236/2020 | 2020-12-29 | Europe/United Kingdom/England/ | Northumbria University / South Tees Hospitals NHS Foundation Trust / North Cumbria Integrated Care NHS Foundation Trust / North Tees and Hartlepool NHS Foundation Trust / Newcastle Hospitals NHS Foundation Trust | COVID-19 Genomics UK (COG-UK) Consortium | Darren L Smith et al |
| EPI_ISL_866623 | England/NORW-22520D/2021 | 2021-01-11 | Europe/United Kingdom/England/ | Quadram Institute Bioscience | COVID-19 Genomics UK (COG-UK) Consortium | Dave J. Baker et al |
| EPI_ISL_866677 | England/NORW-2255F5/2021 | 2021-01-08 | Europe/United Kingdom/England/ | Quadram Institute Bioscience | COVID-19 Genomics UK (COG-UK) Consortium | Dave J. Baker et al |
| EPI_ISL_866943 | England/NOTT-24895A/2021 | 2021-01-15 | Europe/United Kingdom/England/ | Queens Medical Centre, Clinical Microbiology Department / DeepSeq Nottingham | COVID-19 Genomics UK (COG-UK) Consortium | Gemma Clark et al |
| EPI_ISL_867186 | Wales/PHWC-4B2E5A/2021 | 2021-01-04 | Europe/United Kingdom/Wales/ | Originating lab: Wales Specialist Virology Centre Sequencing lab: Pathogen Genomics Unit | Public Health Wales Microbiology Cardiff Wales Specialist Virology Centre | Catherine Moore et al |
| EPI_ISL_867252 | Wales/PHWC-4B7A8E/2021 | 2021-01-11 | Europe/United Kingdom/Wales/ | Originating lab: Wales Specialist Virology Centre Sequencing lab: Pathogen Genomics Unit | Public Health Wales Microbiology Cardiff Wales Specialist Virology Centre | Catherine Moore et al |
| EPI_ISL_867388 | Wales/PHWC-4B83DD/2021 | 2021-01-11 | Europe/United Kingdom/Wales/ | Originating lab: Wales Specialist Virology Centre Sequencing lab: Pathogen Genomics Unit | Public Health Wales Microbiology Cardiff Wales Specialist Virology Centre | Catherine Moore et al |

| | | | | | | |
|---|---|---|---|---|---|---|
| EPI_ISL_867421 | Wales/PHWC-4B8601/2021 | 2021-01-12 | Europe/United Kingdom/Wales/ | Originating lab: Wales Specialist Virology Centre Sequencing lab: Pathogen Genomics Unit | Public Health Wales Microbiology Cardiff Wales Specialist Virology Centre | Catherine Moore et al |
| EPI_ISL_867460 | Wales/PHWC-4B8917/2021 | 2021-01-13 | Europe/United Kingdom/Wales/ | Originating lab: Wales Specialist Virology Centre Sequencing lab: Pathogen Genomics Unit | Public Health Wales Microbiology Cardiff Wales Specialist Virology Centre | Catherine Moore et al |
| EPI_ISL_867613 | Wales/PHWC-4B9390/2021 | 2021-01-12 | Europe/United Kingdom/Wales/ | Originating lab: Wales Specialist Virology Centre Sequencing lab: Pathogen Genomics Unit | Public Health Wales Microbiology Cardiff Wales Specialist Virology Centre | Catherine Moore et al |
| EPI_ISL_867883 | Wales/PHWC-4BA62D/2021 | 2021-01-10 | Europe/United Kingdom/Wales/ | Originating lab: Wales Specialist Virology Centre Sequencing lab: Pathogen Genomics Unit | Public Health Wales Microbiology Cardiff Wales Specialist Virology Centre | Catherine Moore et al |
| EPI_ISL_867960 | England/PORT-2D848B/2020 | 2020-10-08 | Europe/United Kingdom/England/ | Centre for Enzyme Innovation, University of Portsmouth / Translational Research Laboratory, Portsmouth Hospitals NHS Trust | COVID-19 Genomics UK (COG-UK) Consortium | Angela Beckett et al |
| EPI_ISL_868169 | England/PORT-2E4CDF/2021 | 2021-01-09 | Europe/United Kingdom/England/ | Centre for Enzyme Innovation, University of Portsmouth / Translational Research Laboratory, Portsmouth Hospitals NHS Trust | COVID-19 Genomics UK (COG-UK) Consortium | Angela Beckett et al |
| EPI_ISL_868255 | England/PORT-2E552E/2021 | 2021-01-17 | Europe/United Kingdom/England/ | Centre for Enzyme Innovation, University of Portsmouth / Translational Research Laboratory, Portsmouth Hospitals NHS Trust | COVID-19 Genomics UK (COG-UK) Consortium | Angela Beckett et al |
| EPI_ISL_868435 | England/SHEF-10C9756/2021 | 2021-01-19 | Europe/United Kingdom/England/ | Virology Department, Sheffield Teaching Hospitals NHS Foundation Trust/Department of Infection, Immunity and Cardiovascular Disease, The Medical School, University of Sheffield | COVID-19 Genomics UK (COG-UK) Consortium | Thushan de Silva et al |

| | | | | | | |
|---|---|---|---|---|---|---|
| EPI_ISL_868773 | England/TFCI-26FE87A/2020 | 2020-11-22 | Europe/United Kingdom/England/ | Bioinformatics and Biostatistics Lab, Advanced Sequencing Facility | COVID-19 Genomics UK (COG-UK) Consortium | Aengus Stewart et al |
| EPI_ISL_868801 | England/TFCI-26FEAB0/2020 | 2020-10-16 | Europe/United Kingdom/England/ | Bioinformatics and Biostatistics Lab, Advanced Sequencing Facility | COVID-19 Genomics UK (COG-UK) Consortium | Aengus Stewart et al |
| EPI_ISL_868813 | England/TFCI-26FEB71/2020 | 2020-10-14 | Europe/United Kingdom/England/ | Bioinformatics and Biostatistics Lab, Advanced Sequencing Facility | COVID-19 Genomics UK (COG-UK) Consortium | Aengus Stewart et al |
| EPI_ISL_869053 | England/TFCI-26FFDC5/2020 | 2020-12-30 | Europe/United Kingdom/England/ | Bioinformatics and Biostatistics Lab, Advanced Sequencing Facility | COVID-19 Genomics UK (COG-UK) Consortium | Aengus Stewart et al |
| EPI_ISL_869079 | USA/OH-ODH-SC189128/2020 | 2020-12-28 | North America/USA/Ohio/ | Ohio Department of Health Laboratory | Ohio Department of Health Laboratory | Holmes et al |
| EPI_ISL_869081 | USA/OH-ODH-SC189624/2020 | 2020-12-28 | North America/USA/Ohio/ | Ohio Department of Health Laboratory | Ohio Department of Health Laboratory | Holmes et al |
| EPI_ISL_869082 | Germany/SH-ChVir22562/2021 | 2021-01-12 | Europe/Germany/Schleswig-Holstein/ | A. Krumbholz, Labor Dr. Krause und Kollegen MVZ GmbH, Kiel | Charitv© Universitv§tsmedizin Berlin, Institut fvºr Virologie | Victor M Corman et al |
| EPI_ISL_869087 | Germany/SH-ChVir22542/2021 | 2021-01-13 | Europe/Germany/Schleswig-Holstein/ | A. Krumbholz, Labor Dr. Krause und Kollegen MVZ GmbH, Kiel | Charitv© Universitv§tsmedizin Berlin, Institut fvºr Virologie | Victor M Corman et al |
| EPI_ISL_869089 | Germany/SH-ChVir22544/2021 | 2021-01-13 | Europe/Germany/Schleswig-Holstein/ | A. Krumbholz, Labor Dr. Krause und Kollegen MVZ GmbH, Kiel | Charitv© Universitv§tsmedizin Berlin, Institut fvºr Virologie | Victor M Corman et al |
| EPI_ISL_869095 | Germany/SH-ChVir22554/2021 | 2021-01-13 | Europe/Germany/Schleswig-Holstein/ | A. Krumbholz, Labor Dr. Krause und Kollegen MVZ GmbH, Kiel | Charitv© Universitv§tsmedizin Berlin, Institut fvºr Virologie | Victor M Corman et al |
| EPI_ISL_869096 | Germany/SH-ChVir22557/2021 | 2021-01-12 | Europe/Germany/Schleswig-Holstein/ | A. Krumbholz, Labor Dr. Krause und Kollegen MVZ GmbH, Kiel | Charitv© Universitv§tsmedizin Berlin, Institut fvºr Virologie | Victor M Corman et al |

| | | | | | | |
|---|---|---|---|---|---|---|
| EPI_ISL_869097 | Germany/SH-ChVir22558/2021 | 2021-01-12 | Europe/Germany/Schleswig-Holstein/ | A. Krumbholz, Labor Dr. Krause und Kollegen MVZ GmbH, Kiel | Charitv© Universitv§tsmedizin Berlin, Institut fvºr Virologie | Victor M Corman et al |
| EPI_ISL_869099 | Germany/SH-ChVir22560/2021 | 2021-01-12 | Europe/Germany/Schleswig-Holstein/ | A. Krumbholz, Labor Dr. Krause und Kollegen MVZ GmbH, Kiel | Charitv© Universitv§tsmedizin Berlin, Institut fvºr Virologie | Victor M Corman et al |
| EPI_ISL_869104 | Germany/SH-ChVir22566/2021 | 2021-01-15 | Europe/Germany/Schleswig-Holstein/ | A. Krumbholz, Labor Dr. Krause und Kollegen MVZ GmbH, Kiel | Charitv© Universitv§tsmedizin Berlin, Institut fvºr Virologie | Victor M Corman et al |
| EPI_ISL_869105 | Germany/SH-ChVir22567/2021 | 2021-01-15 | Europe/Germany/Schleswig-Holstein/ | A. Krumbholz, Labor Dr. Krause und Kollegen MVZ GmbH, Kiel | Charitv© Universitv§tsmedizin Berlin, Institut fvºr Virologie | Victor M Corman et al |
| EPI_ISL_869110 | Germany/SH-ChVir22575/2021 | 2021-01-17 | Europe/Germany/Schleswig-Holstein/ | A. Krumbholz, Labor Dr. Krause und Kollegen MVZ GmbH, Kiel | Charitv© Universitv§tsmedizin Berlin, Institut fvºr Virologie | Victor M Corman et al |
| EPI_ISL_869113 | Germany/SH-ChVir22579/2021 | 2021-01-17 | Europe/Germany/Schleswig-Holstein/ | A. Krumbholz, Labor Dr. Krause und Kollegen MVZ GmbH, Kiel | Charitv© Universitv§tsmedizin Berlin, Institut fvºr Virologie | Victor M Corman et al |
| EPI_ISL_869116 | Germany/SH-ChVir22582/2021 | 2021-01-17 | Europe/Germany/Schleswig-Holstein/ | A. Krumbholz, Labor Dr. Krause und Kollegen MVZ GmbH, Kiel | Charitv© Universitv§tsmedizin Berlin, Institut fvºr Virologie | Victor M Corman et al |
| EPI_ISL_869119 | Germany/BE-ChVir22280/2021 | 2021-01-15 | Europe/Germany/Berlin/ | Charitv© Universitv§tsmedizin Berlin, Institut fvºr Virologie/Labor Berlin | Charitv© Universitv§tsmedizin Berlin, Institut fvºr Virologie | Victor M Corman et al |
| EPI_ISL_869120 | Germany/SH-ChVir22531/2021 | 2021-01-16 | Europe/Germany/Schleswig-Holstein/ | A. Krumbholz, Labor Dr. Krause und Kollegen MVZ GmbH, Kiel | Charitv© Universitv§tsmedizin Berlin, Institut fvºr Virologie | Victor M Corman et al |
| EPI_ISL_869121 | Germany/SH-ChVir22534/2021 | 2021-01-16 | Europe/Germany/Schleswig-Holstein/ | A. Krumbholz, Labor Dr. Krause und Kollegen MVZ GmbH, Kiel | Charitv© Universitv§tsmedizin Berlin, Institut fvºr Virologie | Victor M Corman et al |

| | | | | | | |
|---|---|---|---|---|---|---|
| EPI_ISL_869122 | Germany/SH-ChVir22539/2021 | 2021-01-16 | Europe/Germany/Schleswig-Holstein/ | A. Krumbholz, Labor Dr. Krause und Kollegen MVZ GmbH, Kiel | Charit√© Universit√§tsmedizin Berlin, Institut f√ºr Virologie | Victor M Corman et al |
| EPI_ISL_869124 | Germany/NI-ChVir22282/2021 | 2021-01-04 | Europe/Germany/Niedersachsen/ | Charit√© Universit√§tsmedizin Berlin, Institut f√ºr Virologie/Labor Berlin | Charit√© Universit√§tsmedizin Berlin, Institut f√ºr Virologie | Victor M Corman et al |
| EPI_ISL_869126 | Germany/NI-ChVir22285/2021 | 2021-01-04 | Europe/Germany/Niedersachsen/ | Charit√© Universit√§tsmedizin Berlin, Institut f√ºr Virologie/Labor Berlin | Charit√© Universit√§tsmedizin Berlin, Institut f√ºr Virologie | Victor M Corman et al |
| EPI_ISL_869130 | Germany/NI-ChVir22289/2021 | 2021-01-04 | Europe/Germany/Niedersachsen/ | Charit√© Universit√§tsmedizin Berlin, Institut f√ºr Virologie/Labor Berlin | Charit√© Universit√§tsmedizin Berlin, Institut f√ºr Virologie | Victor M Corman et al |
| EPI_ISL_869131 | Germany/NW-ChVir22290/2021 | 2021-01-14 | Europe/Germany/North Rhine Westphal | Charit√© Universit√§tsmedizin Berlin, Institut f√ºr Virologie/Labor Berlin | Charit√© Universit√§tsmedizin Berlin, Institut f√ºr Virologie | Victor M Corman et al |
| EPI_ISL_869132 | Germany/NW-ChVir22499/2021 | 2021-01-11 | Europe/Germany/North Rhine Westphal | Charit√© Universit√§tsmedizin Berlin, Institut f√ºr Virologie/Labor Berlin | Charit√© Universit√§tsmedizin Berlin, Institut f√ºr Virologie | Victor M Corman et al |
| EPI_ISL_869137 | Germany/SH-ChVir22530/2021 | 2021-01-16 | Europe/Germany/Schleswig-Holstein/ | A. Krumbholz, Labor Dr. Krause und Kollegen MVZ GmbH, Kiel | Charit√© Universit√§tsmedizin Berlin, Institut f√ºr Virologie | Victor M Corman et al |
| EPI_ISL_869140 | Germany/SH-ChVir22536/2021 | 2021-01-16 | Europe/Germany/Schleswig-Holstein/ | A. Krumbholz, Labor Dr. Krause und Kollegen MVZ GmbH, Kiel | Charit√© Universit√§tsmedizin Berlin, Institut f√ºr Virologie | Victor M Corman et al |
| EPI_ISL_869158 | Ireland/CN-NVRL-MPY61666/2020 | 2020-04-15 | Europe/Ireland/Cavan/ | National Virus Reference Laboratory | Irish Coronavirus Sequencing Consortium - Teagasc Moorepark | Calum Walsh et al |

| | | | | | | |
|---|---|---|---|---|---|---|
| EPI_ISL_869167 | USA/NM-UNM-01688/2021 | 2021-01-06 | North America/USA/New Mexico/ | New Mexico Department of Health Scientific Laboratory | Center for Global Health, University of New Mexico Health Sciences Center | Daryl Domman et al |
| EPI_ISL_869171 | USA/NM-UNM-01710/2021 | 2021-01-06 | North America/USA/New Mexico/ | New Mexico Department of Health Scientific Laboratory | Center for Global Health, University of New Mexico Health Sciences Center | Daryl Domman et al |
| EPI_ISL_869181 | USA/NM-UNM-01667/2021 | 2021-01-06 | North America/USA/New Mexico/ | New Mexico Department of Health Scientific Laboratory | Center for Global Health, University of New Mexico Health Sciences Center | Daryl Domman et al |
| EPI_ISL_869183 | USA/NM-UNM-01651/2021 | 2021-01-04 | North America/USA/New Mexico/ | New Mexico Department of Health Scientific Laboratory | Center for Global Health, University of New Mexico Health Sciences Center | Daryl Domman et al |
| EPI_ISL_869192 | USA/NM-UNM-01665/2021 | 2021-01-05 | North America/USA/New Mexico/ | New Mexico Department of Health Scientific Laboratory | Center for Global Health, University of New Mexico Health Sciences Center | Daryl Domman et al |
| EPI_ISL_869197 | USA/NM-UNM-01673/2021 | 2021-01-06 | North America/USA/New Mexico/ | New Mexico Department of Health Scientific Laboratory | Center for Global Health, University of New Mexico Health Sciences Center | Daryl Domman et al |
| EPI_ISL_869205 | USA/NM-UNM-01682/2021 | 2021-01-06 | North America/USA/New Mexico/ | New Mexico Department of Health Scientific Laboratory | Center for Global Health, University of New Mexico Health Sciences Center | Daryl Domman et al |
| EPI_ISL_869208 | USA/NM-UNM-01686/2021 | 2021-01-06 | North America/USA/New Mexico/ | New Mexico Department of Health Scientific Laboratory | Center for Global Health, University of New Mexico Health Sciences Center | Daryl Domman et al |

| | | | | | | |
|---|---|---|---|---|---|---|
| EPI_ISL_869210 | USA/NM-UNM-01690/2021 | 2021-01-07 | North America/USA/New Mexico/ | New Mexico Department of Health Scientific Laboratory | Center for Global Health, University of New Mexico Health Sciences Center | Daryl Domman et al |
| EPI_ISL_869215 | USA/NM-UNM-01696/2021 | 2021-01-07 | North America/USA/New Mexico/ | New Mexico Department of Health Scientific Laboratory | Center for Global Health, University of New Mexico Health Sciences Center | Daryl Domman et al |
| EPI_ISL_869222 | USA/NM-UNM-01704/2021 | 2021-01-08 | North America/USA/New Mexico/ | New Mexico Department of Health Scientific Laboratory | Center for Global Health, University of New Mexico Health Sciences Center | Daryl Domman et al |
| EPI_ISL_869229 | USA/NM-UNM-01717/2021 | 2021-01-09 | North America/USA/New Mexico/ | New Mexico Department of Health Scientific Laboratory | Center for Global Health, University of New Mexico Health Sciences Center | Daryl Domman et al |
| EPI_ISL_869233 | USA/NM-UNM-01722/2021 | 2021-01-09 | North America/USA/New Mexico/ | New Mexico Department of Health Scientific Laboratory | Center for Global Health, University of New Mexico Health Sciences Center | Daryl Domman et al |
| EPI_ISL_869243 | Russia/SVE-RII-MH8911S/2020 | 2020-10-01 | Europe/Russia/Sverdlovsk/Sysert | UMMC-Health | WHO National Influenza Centre Russian Federation | Andrey Komissarov et al |
| EPI_ISL_869244 | Russia/SVE-RII-MH8896S/2020 | 2020-10-03 | Europe/Russia/Sverdlovsk/Yekaterinbur | UMMC-Health | WHO National Influenza Centre Russian Federation | Andrey Komissarov et al |
| EPI_ISL_869245 | Russia/SVE-RII-MH8921S/2020 | 2020-10-02 | Europe/Russia/Sverdlovsk/Sysert | UMMC-Health | WHO National Influenza Centre Russian Federation | Andrey Komissarov et al |

| | | | | | | |
|---|---|---|---|---|---|---|
| EPI_ISL_869246 | Russia/SVE-RII-MH8908S/2020 | 2020-10-01 | Europe/Russia/Sverdlovsk/Baltym | UMMC-Health | WHO National Influenza Centre Russian Federation | Andrey Komissarov et al |
| EPI_ISL_869249 | Russia/SVE-RII-MH8847S/2020 | 2020-10-01 | Europe/Russia/Sverdlovsk/Yekaterinbur | UMMC-Health | WHO National Influenza Centre Russian Federation | Andrey Komissarov et al |
| EPI_ISL_869253 | Russia/SVE-RII-MH8884S/2020 | 2020-09-30 | Europe/Russia/Sverdlovsk/Verkhnyaya | UMMC-Health | WHO National Influenza Centre Russian Federation | Andrey Komissarov et al |
| EPI_ISL_869260 | Russia/SVE-RII-MH8856S/2020 | 2020-09-29 | Europe/Russia/Sverdlovsk/Verkhnyaya | UMMC-Health | WHO National Influenza Centre Russian Federation | Andrey Komissarov et al |
| EPI_ISL_869261 | Russia/SVE-RII-MH8930S/2020 | 2020-10-02 | Europe/Russia/Sverdlovsk/Verkhnyaya | UMMC-Health | WHO National Influenza Centre Russian Federation | Andrey Komissarov et al |
| EPI_ISL_869262 | Russia/KYA-RII-MH10332S/2020 | 2020-11-21 | Europe/Russia/Krasnoyarsk/Krasnoyarsk | Krasnoyarsk Regional Clinical Hospital | WHO National Influenza Centre Russian Federation | Andrey Komissarov et al |
| EPI_ISL_869263 | Russia/KYA-RII-MH10335S/2020 | 2020-11-22 | Europe/Russia/Krasnoyarsk/Krasnoyarsk | Krasnoyarsk Regional Clinical Hospital | WHO National Influenza Centre Russian Federation | Andrey Komissarov et al |
| EPI_ISL_869264 | Russia/KYA-RII-MH10340S/2020 | 2020-11-20 | Europe/Russia/Krasnoyarsk/Krasnoyarsk | Krasnoyarsk Regional Clinical Hospital | WHO National Influenza Centre Russian Federation | Andrey Komissarov et al |
| EPI_ISL_869511 | Denmark/DCGC-32507/2021 | 2021-01-18 | Europe/Denmark/Midtjylland/ | Department of Virus and Microbiological Special Diagnostics, Statens Serum Institut, Copenhagen, Denmark | Aalborg University | Danish Covid-19 Genome Consortium et al |
| EPI_ISL_871325 | USA/OH-ODH-SC190241/2020 | 2020-12-28 | North America/USA/Ohio/ | Ohio Department of Health Laboratory | Ohio Department of Health Laboratory | Holmes et al |

| | | | | | | |
|---|---|---|---|---|---|---|
| EPI_ISL_871337 | Denmark/DCGC-34326/2021 | 2021-01-04 | Europe/Denmark/Midtjylland/ | Department of Virus and Microbiological Special Diagnostics, Statens Serum Institut, Copenhagen, Denmark | Aalborg University | Danish Covid-19 Genome Consortium et al |
| EPI_ISL_871790 | India/UP-MRU46251/2020 | 2020-08-25 | Asia/India/Uttar Pradesh/Chandauli | Multidisciplinary Research Unit, DHR-ICMR, Institute of Medical Sciences, Banaras Hindu University | Multidisciplinary Research Unit, DHR-ICMR, Institute of Medical Sciences, Banaras Hindu University | Royana Singh et al |
| EPI_ISL_871818 | USA/OH-ODH-SC191348/2020 | 2020-12-29 | North America/USA/Ohio/ | Ohio Department of Health Laboratory | Ohio Department of Health Laboratory | Holmes et al |
| EPI_ISL_871819 | USA/OH-ODH-SC191395/2020 | 2020-12-29 | North America/USA/Ohio/ | Ohio Department of Health Laboratory | Ohio Department of Health Laboratory | Holmes et al |
| EPI_ISL_871837 | Belgium/55-2153-9/2021 | 2021-01-01 | Europe/Belgium/Hasselt/Diepenbeek | Jessa | Jessa | Brigitte Maes et al |
| EPI_ISL_871842 | USA/WY-WPHL-21006032/2021 | 2021-01-15 | North America/USA/Wyoming/ | Wyoming Public Health Laboratory | Wyoming Public Health Laboratory | Noah Hull et al |
| EPI_ISL_871844 | USA/WY-WPHL-21001506/2021 | 2021-01-05 | North America/USA/Wyoming/ | Wyoming Public Health Laboratory | Wyoming Public Health Laboratory | Noah Hull et al |
| EPI_ISL_871851 | USA/WY-WPHL-21006378/2021 | 2021-01-17 | North America/USA/Wyoming/ | Wyoming Public Health Laboratory | Wyoming Public Health Laboratory | Noah Hull et al |
| EPI_ISL_871862 | USA/WY-WPHL-21006292/2021 | 2021-01-16 | North America/USA/Wyoming/ | Wyoming Public Health Laboratory | Wyoming Public Health Laboratory | Noah Hull et al |
| EPI_ISL_871880 | Spain/CB-IBV-98012955/2021 | 2021-01-05 | Europe/Spain/Cantabria/Astillero | Hospital Universitario Marqu√©s de Valdecilla - IDIVAL (Santander, Cantabria) | SeqCOVID-SPAIN consortium/IBV(CSIC) | M√≥nica Gozalo Marg√ºello et al |
| EPI_ISL_871888 | Spain/CB-IBV-98012972/2021 | 2021-01-07 | Europe/Spain/Cantabria/Maliano | Hospital Universitario Marqu√©s de Valdecilla - IDIVAL (Santander, Cantabria) | SeqCOVID-SPAIN consortium/IBV(CSIC) | M√≥nica Gozalo Marg√ºello et al |
| EPI_ISL_871889 | Spain/CB-IBV-98012958/2021 | 2021-01-06 | Europe/Spain/Cantabria/Astillero | Hospital Universitario Marqu√©s de Valdecilla - IDIVAL (Santander, Cantabria) | SeqCOVID-SPAIN consortium/IBV(CSIC) | M√≥nica Gozalo Marg√ºello et al |
| EPI_ISL_871892 | Spain/CB-IBV-98012969/2021 | 2021-01-07 | Europe/Spain/Cantabria/Los Corrales de | Hospital Universitario Marqu√©s de Valdecilla - IDIVAL (Santander, Cantabria) | SeqCOVID-SPAIN consortium/IBV(CSIC) | M√≥nica Gozalo Marg√ºello et al |
| EPI_ISL_871895 | Spain/CB-IBV-98012970/2021 | 2021-01-07 | Europe/Spain/Cantabria/Maliano | Hospital Universitario Marqu√©s de Valdecilla - IDIVAL (Santander, Cantabria) | SeqCOVID-SPAIN consortium/IBV(CSIC) | M√≥nica Gozalo Marg√ºello et al |

| | | | | | | |
|---|---|---|---|---|---|---|
| EPI_ISL_871897 | Spain/CB-IBV-98012953/2021 | 2021-01-05 | Europe/Spain/Cantabria/Santander | Hospital Universitario Marqu√©s de Valdecilla - IDIVAL (Santander, Cantabria) | SeqCOVID-SPAIN consortium/IBV(CSIC) | M√≥nica Gozalo Marg√ºello et al |
| EPI_ISL_871901 | Spain/CB-IBV-98012965/2021 | 2021-01-07 | Europe/Spain/Cantabria/Santander | Hospital Universitario Marqu√©s de Valdecilla - IDIVAL (Santander, Cantabria) | SeqCOVID-SPAIN consortium/IBV(CSIC) | M√≥nica Gozalo Marg√ºello et al |
| EPI_ISL_871904 | Spain/CB-IBV-98012976/2021 | 2021-01-05 | Europe/Spain/Cantabria/Santander | Hospital Universitario Marqu√©s de Valdecilla - IDIVAL (Santander, Cantabria) | SeqCOVID-SPAIN consortium/IBV(CSIC) | M√≥nica Gozalo Marg√ºello et al |
| EPI_ISL_871906 | Spain/CB-IBV-98012978/2021 | 2021-01-05 | Europe/Spain/Cantabria/Santander | Hospital Universitario Marqu√©s de Valdecilla - IDIVAL (Santander, Cantabria) | SeqCOVID-SPAIN consortium/IBV(CSIC) | M√≥nica Gozalo Marg√ºello et al |
| EPI_ISL_871910 | Spain/NC-IBV-98013114/2020 | 2020-12-27 | Europe/Spain/Navarra/Pamplona | Servicio de Microbiolog√≠a Cl√≠nica (Complejo Hospitalario de Navarra, Pamplona), Instituto de Investigaci√≥n Sanitaria de Navarra (IdiSNA) | SeqCOVID-SPAIN consortium/IBV(CSIC) | Carmen Ezpeleta Baquedano et al |
| EPI_ISL_871913 | Spain/NC-IBV-98013127/2021 | 2021-01-06 | Europe/Spain/Navarra/Estella | Servicio de Microbiolog√≠a Cl√≠nica (Complejo Hospitalario de Navarra, Pamplona), Instituto de Investigaci√≥n Sanitaria de Navarra (IdiSNA) | SeqCOVID-SPAIN consortium/IBV(CSIC) | Carmen Ezpeleta Baquedano et al |
| EPI_ISL_871928 | Spain/CT-IBV-98013075/2020 | 2020-12-18 | Europe/Spain/Catalunya/Llissa de Munt | Servicio de Microbiolog√≠a, Laboratori Cl√≠nic Metropolitana Nord. Hospital Universitari Germans Trias i Pujol. Institut d‚ÄôInvestigaci√≥ en Ci√®ncies de la Salut Germans Trias i Pujol (IGTP) | SeqCOVID-SPAIN consortium/IBV(CSIC) | Elisa Martr√≥ et al |
| EPI_ISL_871948 | Spain/NC-IBV-98013135/2021 | 2021-01-06 | Europe/Spain/Navarra/Pamplona | Servicio de Microbiolog√≠a Cl√≠nica (Complejo Hospitalario de Navarra, Pamplona), Instituto de Investigaci√≥n Sanitaria de Navarra (IdiSNA) | SeqCOVID-SPAIN consortium/IBV(CSIC) | Carmen Ezpeleta Baquedano et al |
| EPI_ISL_871973 | Spain/MD-ISCIII-201091/2020 | 2020-02-22 | Europe/Spain/Madrid/ | Hospital Universitario Severo Ochoa | Instituto de Salud Carlos III | Iglesias-Caballero et al |
| EPI_ISL_871974 | Spain/EX-ISCIII-201258/2020 | 2020-02-29 | Europe/Spain/Extremadura/ | Hospital Infanta Cristina | Instituto de Salud Carlos III | Iglesias-Caballero et al |
| EPI_ISL_871975 | Spain/EX-ISCIII-201259/2020 | 2020-02-29 | Europe/Spain/Extremadura/ | Hospital Infanta Cristina | Instituto de Salud Carlos III | Iglesias-Caballero et al |
| EPI_ISL_871976 | Spain/AR-ISCIII-201691/2020 | 2020-03-03 | Europe/Spain/Aragon/ | Hospital Universitario Lozano Bielsa | Instituto de Salud Carlos III | Iglesias-Caballero et al |
| EPI_ISL_871985 | Spain/MD-ISCIII-2014657/2020 | 2020-08-18 | Europe/Spain/Madrid/ | Hospital Cl√≠nico San Carlos | Instituto de Salud Carlos III | Iglesias-Caballero et al |

| | | | | | | |
|---|---|---|---|---|---|---|
| EPI_ISL_871987 | Spain/MD-ISCIII-2014666/2020 | 2020-08-18 | Europe/Spain/Madrid/ | Hospital Cl√≠nico San Carlos | Instituto de Salud Carlos III | Iglesias-Caballero et al |
| EPI_ISL_871989 | Spain/MD-ISCIII-2014703/2020 | 2020-08-20 | Europe/Spain/Madrid/ | Hospital Cl√≠nico San Carlos | Instituto de Salud Carlos III | Iglesias-Caballero et al |
| EPI_ISL_871991 | Spain/MD-ISCIII-2014639/2020 | 2020-08-25 | Europe/Spain/Madrid/ | Hospital Universitario Severo Ochoa | Instituto de Salud Carlos III | Iglesias-Caballero et al |
| EPI_ISL_871992 | Spain/MD-ISCIII-2014830/2020 | 2020-08-26 | Europe/Spain/Madrid/ | Hospital Cl√≠nico San Carlos | Instituto de Salud Carlos III | Iglesias-Caballero et al |
| EPI_ISL_871993 | Spain/MD-ISCIII-2014936/2020 | 2020-08-27 | Europe/Spain/Madrid/ | Hospital Cl√≠nico San Carlos | Instituto de Salud Carlos III | Iglesias-Caballero et al |
| EPI_ISL_871994 | Spain/MD-ISCIII-2016129/2020 | 2020-10-11 | Europe/Spain/Madrid/ | Hospital de Madrid | Instituto de Salud Carlos III | Iglesias-Caballero et al |
| EPI_ISL_871996 | Spain/MD-ISCIII-2016714/2020 | 2020-10-23 | Europe/Spain/Madrid/ | Instituto de Salud Carlos III | Instituto de Salud Carlos III | Iglesias-Caballero et al |
| EPI_ISL_871998 | Spain/MD-ISCIII-2017220/2020 | 2020-10-23 | Europe/Spain/Madrid/ | Instituto de Salud Carlos III | Instituto de Salud Carlos III | Iglesias-Caballero et al |
| EPI_ISL_872004 | Italy/APU-PODV-15967861/2020 | 2020-09-07 | Europe/Italy/Apulia/Bari | "Ospedale ""Di Venere""" | Beaconlab (Bioinformatics, Evolution and Comparative Genomics lab), Dept of Biosciences, University on Milan | Iacobellis M et al |
| EPI_ISL_872006 | Italy/APU-PODV-15965800/2020 | 2020-09-06 | Europe/Italy/Apulia/Bari | "Ospedale ""Di Venere""" | Beaconlab (Bioinformatics, Evolution and Comparative Genomics lab), Dept of Biosciences, University on Milan | Iacobellis M et al |

| | | | | | | |
|---|---|---|---|---|---|---|
| EPI_ISL_872009 | Italy/APU-PODV-15949198/2020 | 2020-08-29 | Europe/Italy/Apulia/Bari | "Ospedale ""Di Venere""" | Beaconlab (Bioinformatics, Evolution and Comparative Genomics lab), Dept of Biosciences, University on Milan | Iacobellis M et al |
| EPI_ISL_872012 | Italy/APU-PODV-15948132/2020 | 2020-08-28 | Europe/Italy/Apulia/Bari | "Ospedale ""Di Venere""" | Beaconlab (Bioinformatics, Evolution and Comparative Genomics lab), Dept of Biosciences, University on Milan | Iacobellis M et al |
| EPI_ISL_872013 | Italy/APU-PODV-15948450/2020 | 2020-08-28 | Europe/Italy/Apulia/Bari | "Ospedale ""Di Venere""" | Beaconlab (Bioinformatics, Evolution and Comparative Genomics lab), Dept of Biosciences, University on Milan | Iacobellis M et al |
| EPI_ISL_872014 | Italy/APU-PODV-15940873/2020 | 2020-08-26 | Europe/Italy/Apulia/Bari | "Ospedale ""Di Venere""" | Beaconlab (Bioinformatics, Evolution and Comparative Genomics lab), Dept of Biosciences, University on Milan | Iacobellis M et al |

| | | | | | | |
|---|---|---|---|---|---|---|
| EPI_ISL_872015 | Italy/APU-PODV-15939816/2020 | 2020-08-25 | Europe/Italy/Apulia/Bari | "Ospedale ""Di Venere""" | Beaconlab (Bioinformatics, Evolution and Comparative Genomics lab), Dept of Biosciences, University on Milan | Iacobellis M et al |
| EPI_ISL_872018 | Italy/APU-PODV-15935787/2020 | 2020-08-24 | Europe/Italy/Apulia/Bari | "Ospedale ""Di Venere""" | Beaconlab (Bioinformatics, Evolution and Comparative Genomics lab), Dept of Biosciences, University on Milan | Iacobellis M et al |
| EPI_ISL_872027 | Italy/APU-PODV-15714509/2020 | 2020-04-21 | Europe/Italy/Apulia/Bari | "Ospedale ""Di Venere""" | Beaconlab (Bioinformatics, Evolution and Comparative Genomics lab), Dept of Biosciences, University on Milan | Iacobellis M et al |
| EPI_ISL_872030 | Italy/APU-PODV-15705641/2020 | 2020-04-15 | Europe/Italy/Apulia/Bari | "Ospedale ""Di Venere""" | Beaconlab (Bioinformatics, Evolution and Comparative Genomics lab), Dept of Biosciences, University on Milan | Iacobellis M et al |

| | | | | | | |
|---|---|---|---|---|---|---|
| EPI_ISL_872032 | Italy/APU-PODV-15705365/2020 | 2020-04-14 | Europe/Italy/Apulia/Bari | "Ospedale ""Di Venere""" | Beaconlab (Bioinformatics, Evolution and Comparative Genomics lab), Dept of Biosciences, University on Milan | Iacobellis M et al |
| EPI_ISL_872033 | Italy/APU-PODV-15699471/2020 | 2020-04-08 | Europe/Italy/Apulia/Bari | "Ospedale ""Di Venere""" | Beaconlab (Bioinformatics, Evolution and Comparative Genomics lab), Dept of Biosciences, University on Milan | Iacobellis M et al |
| EPI_ISL_872034 | Italy/APU-PODV-15698547/2020 | 2020-04-06 | Europe/Italy/Apulia/Bari | "Ospedale ""Di Venere""" | Beaconlab (Bioinformatics, Evolution and Comparative Genomics lab), Dept of Biosciences, University on Milan | Iacobellis M et al |
| EPI_ISL_872035 | Italy/APU-PODV-15698470/2020 | 2020-04-06 | Europe/Italy/Apulia/Bari | "Ospedale ""Di Venere""" | Beaconlab (Bioinformatics, Evolution and Comparative Genomics lab), Dept of Biosciences, University on Milan | Iacobellis M et al |
| EPI_ISL_872076 | Belgium/ULG-11344/2021 | 2021-01-16 | Europe/Belgium/Li√®ge/ | Department of Clinical Microbiology | GIGA Medical Genomics | Keith Durkin et al |
| EPI_ISL_872135 | Belgium/ULG-11395/2021 | 2021-01-19 | Europe/Belgium/Li√®ge/ | Department of Clinical Microbiology | GIGA Medical Genomics | Keith Durkin et al |

| | | | | | | |
|---|---|---|---|---|---|---|
| EPI_ISL_872151 | Belgium/ULG-11419/2021 | 2021-01-17 | Europe/Belgium/Liv®ge/ | CHR Citadelle | GIGA Medical Genomics | Keith Durkin et al |
| EPI_ISL_872184 | USA/WY-WPHL-21005891/2021 | 2021-01-15 | North America/USA/Wyoming/ | Wyoming Public Health Laboratory | Wyoming Public Health Laboratory | Noah Hull et al |
| EPI_ISL_872188 | Indonesia/YO-UGM-10011/2020 | 2020-12-27 | Asia/Indonesia/Yogyakarta/ | RSUP Dr. Sardjito | Genetics Working Group (Pokja Genetik) Faculty of Medicine, Public Health and Nursing Universitas Gadjah Mada (FK-KMK UGM); Disease Investigation Center Wates Ministry of Agriculture Indonesia; Department of Microbiology FK-KMK UGM; Laboratorium Diagnostik Yayasan Tahija World Mosquito Program (WMP) Yogyakarta Center for Tropical Medicine FK-KMK UGM; Integrated Research Center FK-KMK UGM; Department of Computer Science and Electronics FMIPA UGM; RSUP Dr. Sardjito | Gunadi et al |

| | | | | | | |
|---|---|---|---|---|---|---|
| EPI_ISL_872189 | Indonesia/YO-UGM-10012/2020 | 2020-12-27 | Asia/Indonesia/Yogyakarta/ | RSUP Dr. Sardjito | Genetics Working Group (Pokja Genetik) Faculty of Medicine, Public Health and Nursing Universitas Gadjah Mada (FK-KMK UGM); Disease Investigation Center Wates Ministry of Agriculture Indonesia; Department of Microbiology FK-KMK UGM; Laboratorium Diagnostik Yayasan Tahija World Mosquito Program (WMP) Yogyakarta Center for Tropical Medicine FK-KMK UGM; Integrated Research Center FK-KMK UGM; Department of Computer Science and Electronics FMIPA UGM; RSUP Dr. Sardjito | Gunadi et al |
| EPI_ISL_872191 | Brazil/SP-1029/2021 | 2021-01-15 | South America/Brazil/S√£o Paulo/S√£o | Conjunto Hospitalar do Mandaqui de Sao Paulo | Instituto Adolfo Lutz, Interdiciplinary Procedures Center, Strategic Laboratory | Claudio Tavares Sacchi et al |

| | | | | | | |
|---|---|---|---|---|---|---|
| EPI_ISL_872192 | Brazil/SP-1030/2021 | 2021-01-18 | South America/Brazil/S√£o Paulo/S√£o | Conjunto Hospitalar do Mandaqui de Sao Paulo | Instituto Adolfo Lutz, Interdiciplinary Procedures Center, Strategic Laboratory | Claudio Tavares Sacchi et al |
| EPI_ISL_872219 | France/IDF-IPP00552/2021 | 2021-01-07 | Europe/France/Ile de France/Paris | Hopital | National Reference Center for Viruses of Respiratory Infections, Institut Pasteur, Paris | Marion Barbet et al |
| EPI_ISL_872222 | Mayotte/IPP00615/2021 | 2021-01-11 | Europe/France/Mayotte/Mamoudzou | hopital | National Reference Center for Viruses of Respiratory Infections, Institut Pasteur, Paris | Marion Barbet et al |
| EPI_ISL_872225 | Mayotte/IPP00629/2021 | 2021-01-11 | Europe/France/Mayotte/Mamoudzou | hopital | National Reference Center for Viruses of Respiratory Infections, Institut Pasteur, Paris | Marion Barbet et al |
| EPI_ISL_872258 | France/BFC-IPP00707/2020 | 2020-10-27 | Europe/France/Bourgogne-France-Comt | Labo Analyses Med | National Reference Center for Viruses of Respiratory Infections, Institut Pasteur, Paris | Marion Barbet et al |
| EPI_ISL_872261 | France/HDF-IPP00672/2021 | 2021-01-08 | Europe/France/Hauts de France/Lille FR | Hopital | National Reference Center for Viruses of Respiratory Infections, Institut Pasteur, Paris | Marion Barbet et al |

| | | | | | | |
|---|---|---|---|---|---|---|
| EPI_ISL_872274 | France/PDL-IPP00648/2021 | 2021-01-07 | Europe/France/Pays de la Loire/Saint Na | Hopital | National Reference Center for Viruses of Respiratory Infections, Institut Pasteur, Paris | Marion Barbet et al |
| EPI_ISL_872276 | France/PDL-IPP00651/2021 | 2021-01-08 | Europe/France/Pays de la Loire/Le Mans | Hopital | National Reference Center for Viruses of Respiratory Infections, Institut Pasteur, Paris | Marion Barbet et al |
| EPI_ISL_872297 | France/HDF-IPP00676/2021 | 2021-01-11 | Europe/France/Hauts de France/Lille FR | Hopital | National Reference Center for Viruses of Respiratory Infections, Institut Pasteur, Paris | Marion Barbet et al |
| EPI_ISL_872305 | France/HDF-IPP00732/2021 | 2021-01-12 | Europe/France/Hauts de France/Compiv | Hopital | National Reference Center for Viruses of Respiratory Infections, Institut Pasteur, Paris | Marion Barbet et al |
| EPI_ISL_872310 | France/IDF-IPP00504/2021 | 2021-01-07 | Europe/France/Ile de France/Thiais | Labo analyses med | National Reference Center for Viruses of Respiratory Infections, Institut Pasteur, Paris | Marion Barbet et al |
| EPI_ISL_872312 | France/IDF-IPP00510/2021 | 2021-01-07 | Europe/France/Ile de France/Thiais | Labo analyses med | National Reference Center for Viruses of Respiratory Infections, Institut Pasteur, Paris | Marion Barbet et al |

| | | | | | | |
|---|---|---|---|---|---|---|
| EPI_ISL_872331 | Mayotte/IPP00614/2021 | 2021-01-11 | Europe/France/Mayotte/Mamoudzou | hopital | National Reference Center for Viruses of Respiratory Infections, Institut Pasteur, Paris | Marion Barbet et al |
| EPI_ISL_872334 | Mayotte/IPP00630/2021 | 2021-01-11 | Europe/France/Mayotte/Mamoudzou | hopital | National Reference Center for Viruses of Respiratory Infections, Institut Pasteur, Paris | Marion Barbet et al |
| EPI_ISL_872341 | France/PDL-IPP00491/2021 | 2021-01-08 | Europe/France/Pays de la Loire/Angers | Hopital | National Reference Center for Viruses of Respiratory Infections, Institut Pasteur, Paris | Marion Barbet et al |
| EPI_ISL_872347 | France/PDL-IPP00698/2021 | 2021-01-07 | Europe/France/Pays de la Loire/Saint Na | Hopital | National Reference Center for Viruses of Respiratory Infections, Institut Pasteur, Paris | Marion Barbet et al |
| EPI_ISL_872350 | USA/TX-DSHS-2208/2020 | 2020-10-06 | North America/USA/Texas/Terry County | Texas Department of State Health Services (TXDSHS) | Texas Department of State Health Services (TXDSHS) | Bonnie Oh et al |
| EPI_ISL_872358 | USA/TX-DSHS-2216/2020 | 2020-10-06 | North America/USA/Texas/Mclennan Co | Texas Department of State Health Services (TXDSHS) | Texas Department of State Health Services (TXDSHS) | Bonnie Oh et al |
| EPI_ISL_872375 | USA/TX-DSHS-2235/2020 | 2020-12-20 | North America/USA/Texas/ | Texas Department of State Health Services (TXDSHS) | Texas Department of State Health Services (TXDSHS) | Bonnie Oh et al |
| EPI_ISL_872396 | USA/AL-Cullman-JDSU/2020 | 2020-10-03 | North America/USA/Alabama/Cullman C | Eurofins Diatherix | Hudsonalpha Genome Sequencing Center | Jane Grimwood et al |
| EPI_ISL_872397 | USA/AL-St.Clair-JDSW/2020 | 2020-10-03 | North America/USA/Alabama/St. Clair C | Eurofins Diatherix | Hudsonalpha Genome Sequencing Center | Jane Grimwood et al |

| | | | | | | |
|---|---|---|---|---|---|---|
| EPI_ISL_872405 | USA/AL-Jefferson-JDTF/2020 | 2020-10-03 | North America/USA/Alabama/Jefferson | Eurofins Diatherix | Hudsonalpha Genome Sequencing Center | Jane Grimwood et al |
| EPI_ISL_872414 | USA/AL-Cullman-JDTR/2020 | 2020-10-06 | North America/USA/Alabama/Cullman ( | Eurofins Diatherix | Hudsonalpha Genome Sequencing Center | Jane Grimwood et al |
| EPI_ISL_872416 | USA/AL-Jefferson-JDTT/2020 | 2020-10-06 | North America/USA/Alabama/Jefferson | Eurofins Diatherix | Hudsonalpha Genome Sequencing Center | Jane Grimwood et al |
| EPI_ISL_872420 | USA/AL-Madison-JDTY/2020 | 2020-10-06 | North America/USA/Alabama/Madison | Eurofins Diatherix | Hudsonalpha Genome Sequencing Center | Jane Grimwood et al |
| EPI_ISL_872421 | USA/AL-Houston-JDTZ/2020 | 2020-10-06 | North America/USA/Alabama/Houston ( | Eurofins Diatherix | Hudsonalpha Genome Sequencing Center | Jane Grimwood et al |
| EPI_ISL_872422 | USA/AL-Lawrence-JDUA/2020 | 2020-10-06 | North America/USA/Alabama/Lawrence | Eurofins Diatherix | Hudsonalpha Genome Sequencing Center | Jane Grimwood et al |
| EPI_ISL_872433 | USA/AL-Shelby-JDUL/2020 | 2020-10-06 | North America/USA/Alabama/Shelby Co | Eurofins Diatherix | Hudsonalpha Genome Sequencing Center | Jane Grimwood et al |
| EPI_ISL_872436 | USA/AL-Calhoun-JDUQ/2020 | 2020-10-06 | North America/USA/Alabama/Calhoun ( | Eurofins Diatherix | Hudsonalpha Genome Sequencing Center | Jane Grimwood et al |
| EPI_ISL_872442 | USA/AL-Madison-JDUX/2020 | 2020-10-06 | North America/USA/Alabama/Madison | Eurofins Diatherix | Hudsonalpha Genome Sequencing Center | Jane Grimwood et al |
| EPI_ISL_872453 | USA/AL-St.Clair-JDWL/2020 | 2020-10-09 | North America/USA/Alabama/St. Clair ( | Eurofins Diatherix | Hudsonalpha Genome Sequencing Center | Jane Grimwood et al |

| | | | | | | |
|---|---|---|---|---|---|---|
| EPI_ISL_872463 | USA/AL-Madison-JDXA/2020 | 2020-10-16 | North America/USA/Alabama/Madison | Eurofins Diatherix | Hudsonalpha Genome Sequencing Center | Jane Grimwood et al |
| EPI_ISL_872464 | USA/AL-Madison-JDXB/2020 | 2020-10-16 | North America/USA/Alabama/Madison | Eurofins Diatherix | Hudsonalpha Genome Sequencing Center | Jane Grimwood et al |
| EPI_ISL_872466 | USA/AL-Madison-JDXD/2020 | 2020-10-16 | North America/USA/Alabama/Madison | Eurofins Diatherix | Hudsonalpha Genome Sequencing Center | Jane Grimwood et al |
| EPI_ISL_872467 | USA/AL-Madison-JDXE/2020 | 2020-10-16 | North America/USA/Alabama/Madison | Eurofins Diatherix | Hudsonalpha Genome Sequencing Center | Jane Grimwood et al |
| EPI_ISL_872474 | USA/AL-Blount-JDXL/2020 | 2020-10-21 | North America/USA/Alabama/Blount Co | Eurofins Diatherix | Hudsonalpha Genome Sequencing Center | Jane Grimwood et al |
| EPI_ISL_872488 | USA/NM-UNM-01739/2021 | 2021-01-10 | North America/USA/New Mexico/ | New Mexico Department of Health Scientific Laboratory | Center for Global Health, University of New Mexico Health Sciences Center | Daryl Domman et al |
| EPI_ISL_872500 | USA/NM-UNM-01753/2021 | 2021-01-11 | North America/USA/New Mexico/ | New Mexico Department of Health Scientific Laboratory | Center for Global Health, University of New Mexico Health Sciences Center | Daryl Domman et al |
| EPI_ISL_872518 | USA/NM-UNM-01776/2021 | 2021-01-12 | North America/USA/New Mexico/ | New Mexico Department of Health Scientific Laboratory | Center for Global Health, University of New Mexico Health Sciences Center | Daryl Domman et al |
| EPI_ISL_872524 | USA/NM-UNM-01782/2021 | 2021-01-13 | North America/USA/New Mexico/ | New Mexico Department of Health Scientific Laboratory | Center for Global Health, University of New Mexico Health Sciences Center | Daryl Domman et al |

| | | | | | | |
|---|---|---|---|---|---|---|
| EPI_ISL_872530 | USA/NM-UNM-01788/2021 | 2021-01-13 | North America/USA/New Mexico/ | New Mexico Department of Health Scientific Laboratory | Center for Global Health, University of New Mexico Health Sciences Center | Daryl Domman et al |
| EPI_ISL_872540 | USA/NM-UNM-01798/2021 | 2021-01-12 | North America/USA/New Mexico/ | New Mexico Department of Health Scientific Laboratory | Center for Global Health, University of New Mexico Health Sciences Center | Daryl Domman et al |
| EPI_ISL_872542 | USA/NM-UNM-01800/2021 | 2021-01-13 | North America/USA/New Mexico/ | New Mexico Department of Health Scientific Laboratory | Center for Global Health, University of New Mexico Health Sciences Center | Daryl Domman et al |
| EPI_ISL_872545 | USA/NM-UNM-01804/2021 | 2021-01-14 | North America/USA/New Mexico/ | New Mexico Department of Health Scientific Laboratory | Center for Global Health, University of New Mexico Health Sciences Center | Daryl Domman et al |
| EPI_ISL_872551 | USA/NM-UNM-01810/2021 | 2021-01-14 | North America/USA/New Mexico/ | New Mexico Department of Health Scientific Laboratory | Center for Global Health, University of New Mexico Health Sciences Center | Daryl Domman et al |
| EPI_ISL_872555 | USA/NM-UNM-01814/2021 | 2021-01-14 | North America/USA/New Mexico/ | New Mexico Department of Health Scientific Laboratory | Center for Global Health, University of New Mexico Health Sciences Center | Daryl Domman et al |
| EPI_ISL_872556 | USA/NM-UNM-01815/2021 | 2021-01-14 | North America/USA/New Mexico/ | New Mexico Department of Health Scientific Laboratory | Center for Global Health, University of New Mexico Health Sciences Center | Daryl Domman et al |
| EPI_ISL_872559 | USA/NM-UNM-01818/2021 | 2021-01-15 | North America/USA/New Mexico/ | New Mexico Department of Health Scientific Laboratory | Center for Global Health, University of New Mexico Health Sciences Center | Daryl Domman et al |

| | | | | | | |
|---|---|---|---|---|---|---|
| EPI_ISL_872565 | USA/NM-UNM-01824/2021 | 2021-01-15 | North America/USA/New Mexico/ | New Mexico Department of Health Scientific Laboratory | Center for Global Health, University of New Mexico Health Sciences Center | Daryl Domman et al |
| EPI_ISL_872603 | Nigeria/FC-CV528/2020 | 2020-08-08 | Africa/Nigeria/Abuja/ | Nigeria Centre for Disease Control (NCDC) | African Centre of Excellence for Genomics of Infectious Diseases (ACEGID), Redeemer's University | Oluniyi P.E. et al et al |
| EPI_ISL_872604 | Nigeria/FC-CV529/2020 | 2020-09-05 | Africa/Nigeria/Abuja/ | Nigeria Centre for Disease Control (NCDC) | African Centre of Excellence for Genomics of Infectious Diseases (ACEGID), Redeemer's University | Oluniyi P.E. et al et al |
| EPI_ISL_872605 | Nigeria/ZA-CV531/2020 | 2020-11-11 | Africa/Nigeria/Zamfara State/ | Nigeria Centre for Disease Control (NCDC) | African Centre of Excellence for Genomics of Infectious Diseases (ACEGID), Redeemer's University | Oluniyi P.E. et al et al |
| EPI_ISL_872607 | Nigeria/FC-CV533/2020 | 2020-11-17 | Africa/Nigeria/Abuja/ | Nigeria Centre for Disease Control (NCDC) | African Centre of Excellence for Genomics of Infectious Diseases (ACEGID), Redeemer's University | Oluniyi P.E. et al et al |

| | | | | | | |
|---|---|---|---|---|---|---|
| EPI_ISL_872614 | Nigeria/FC-CV543/2020 | 2020-12-03 | Africa/Nigeria/Abuja/ | Nigeria Centre for Disease Control (NCDC) | African Centre of Excellence for Genomics of Infectious Diseases (ACEGID), Redeemer's University | Oluniyi P.E. et al et al |
| EPI_ISL_872628 | Russia/Moscow_PMVL-13/2020 | 2020-05-21 | Europe/Russia/Moscow Oblast/ | Pathogenic Microorganisms Variability Laboratory | Pathogenic Microorganisms Variability Laboratory | Alexey Shchetinin et al |
| EPI_ISL_872629 | Russia/Moscow_PMVL-14/2020 | 2020-05-25 | Europe/Russia/Moscow Oblast/ | Pathogenic Microorganisms Variability Laboratory | Pathogenic Microorganisms Variability Laboratory | Alexey Shchetinin et al |
| EPI_ISL_872630 | Russia/Moscow_PMVL-15/2020 | 2020-05-25 | Europe/Russia/Moscow Oblast/ | Pathogenic Microorganisms Variability Laboratory | Pathogenic Microorganisms Variability Laboratory | Alexey Shchetinin et al |
| EPI_ISL_872631 | Russia/Moscow_PMVL-16/2020 | 2020-05-25 | Europe/Russia/Moscow Oblast/ | Pathogenic Microorganisms Variability Laboratory | Pathogenic Microorganisms Variability Laboratory | Alexey Shchetinin et al |
| EPI_ISL_872632 | Russia/Moscow_PMVL-17/2020 | 2020-05-25 | Europe/Russia/Moscow Oblast/ | Pathogenic Microorganisms Variability Laboratory | Pathogenic Microorganisms Variability Laboratory | Alexey Shchetinin et al |
| EPI_ISL_872633 | Russia/Moscow_PMVL-18/2020 | 2020-05-26 | Europe/Russia/Moscow Oblast/ | Pathogenic Microorganisms Variability Laboratory | Pathogenic Microorganisms Variability Laboratory | Alexey Shchetinin et al |
| EPI_ISL_872634 | Russia/Moscow_PMVL-20/2020 | 2020-06-01 | Europe/Russia/Moscow Oblast/ | Pathogenic Microorganisms Variability Laboratory | Pathogenic Microorganisms Variability Laboratory | Alexey Shchetinin et al |
| EPI_ISL_872635 | Russia/Moscow_PMVL-21/2020 | 2020-06-05 | Europe/Russia/Moscow Oblast/ | Pathogenic Microorganisms Variability Laboratory | Pathogenic Microorganisms Variability Laboratory | Alexey Shchetinin et al |

| | | | | | | |
|---|---|---|---|---|---|---|
| EPI_ISL_872636 | Russia/Moscow_PMVL-22/2020 | 2020-06-05 | Europe/Russia/Moscow Oblast/ | Pathogenic Microorganisms Variability Laboratory | Pathogenic Microorganisms Variability Laboratory | Alexey Shchetinin et al |
| EPI_ISL_872637 | Russia/Moscow_PMVL-23/2020 | 2020-06-05 | Europe/Russia/Moscow Oblast/ | Pathogenic Microorganisms Variability Laboratory | Pathogenic Microorganisms Variability Laboratory | Alexey Shchetinin et al |
| EPI_ISL_872638 | Russia/Moscow_PMVL-24/2020 | 2020-06-05 | Europe/Russia/Moscow Oblast/ | Pathogenic Microorganisms Variability Laboratory | Pathogenic Microorganisms Variability Laboratory | Alexey Shchetinin et al |
| EPI_ISL_872639 | Russia/Moscow_PMVL-25/2020 | 2020-06-06 | Europe/Russia/Moscow Oblast/ | Pathogenic Microorganisms Variability Laboratory | Pathogenic Microorganisms Variability Laboratory | Alexey Shchetinin et al |
| EPI_ISL_872640 | Russia/Moscow_PMVL-26/2020 | 2020-06-29 | Europe/Russia/Moscow Oblast/ | Pathogenic Microorganisms Variability Laboratory | Pathogenic Microorganisms Variability Laboratory | Alexey Shchetinin et al |
| EPI_ISL_872641 | Russia/Moscow_PMVL-27/2020 | 2020-08-20 | Europe/Russia/Moscow Oblast/ | Influenza etiology and epidemiology laboratory | Pathogenic Microorganisms Variability Laboratory | Alexey Shchetinin et al |
| EPI_ISL_872642 | Russia/Moscow_PMVL-28/2020 | 2020-08-20 | Europe/Russia/Moscow Oblast/ | Influenza etiology and epidemiology laboratory | Pathogenic Microorganisms Variability Laboratory | Alexey Shchetinin et al |
| EPI_ISL_872643 | Russia/Moscow_PMVL-30/2020 | 2020-11-06 | Europe/Russia/Moscow Oblast/ | Pathogenic Microorganisms Variability Laboratory | Pathogenic Microorganisms Variability Laboratory | Alexey Shchetinin et al |
| EPI_ISL_872644 | USA/MA-Broad_UMMS-00043/2020 | 2020-12-02 | North America/USA/Massachusetts/ | University of Massachusetts Medical School | Infectious Disease Program, Broad Institute of Harvard and MIT | Lemieux et al |
| EPI_ISL_872645 | USA/MA-Broad_UMMS-00047/2020 | 2020-12-04 | North America/USA/Massachusetts/ | University of Massachusetts Medical School | Infectious Disease Program, Broad Institute of Harvard and MIT | Lemieux et al |

| | | | | | | |
|---|---|---|---|---|---|---|
| EPI_ISL_872646 | USA/MA-Broad_UMMS-00053/2020 | 2020-12-04 | North America/USA/Massachusetts/ | University of Massachusetts Medical School | Infectious Disease Program, Broad Institute of Harvard and MIT | Lemieux et al |
| EPI_ISL_872656 | USA/MA-Broad_UMMS-00078/2020 | 2020-12-05 | North America/USA/Massachusetts/ | University of Massachusetts Medical School | Infectious Disease Program, Broad Institute of Harvard and MIT | Lemieux et al |
| EPI_ISL_872668 | USA/MA-Broad_UMMS-00108/2020 | 2020-12-04 | North America/USA/Massachusetts/ | University of Massachusetts Medical School | Infectious Disease Program, Broad Institute of Harvard and MIT | Lemieux et al |
| EPI_ISL_872703 | USA/RI-Broad_RIDOH-00038/2020 | 2020-11-27 | North America/USA/Rhode Island/ | Rhode Island Department of Health | Infectious Disease Program, Broad Institute of Harvard and MIT | Lemieux et al |
| EPI_ISL_872704 | USA/RI-Broad_RIDOH-00039/2020 | 2020-11-30 | North America/USA/Rhode Island/ | Rhode Island Department of Health | Infectious Disease Program, Broad Institute of Harvard and MIT | Lemieux et al |
| EPI_ISL_872711 | USA/RI-Broad_RIDOH-00048/2020 | 2020-11-30 | North America/USA/Rhode Island/ | Rhode Island Department of Health | Infectious Disease Program, Broad Institute of Harvard and MIT | Lemieux et al |
| EPI_ISL_872722 | USA/RI-Broad_RIDOH-00062/2020 | 2020-12-03 | North America/USA/Rhode Island/ | Rhode Island Department of Health | Infectious Disease Program, Broad Institute of Harvard and MIT | Lemieux et al |
| EPI_ISL_872725 | USA/RI-Broad_RIDOH-00065/2020 | 2020-12-03 | North America/USA/Rhode Island/ | Rhode Island Department of Health | Infectious Disease Program, Broad Institute of Harvard and MIT | Lemieux et al |
| EPI_ISL_872735 | USA/RI-Broad_RIDOH-00080/2020 | 2020-12-07 | North America/USA/Rhode Island/ | Rhode Island Department of Health | Infectious Disease Program, Broad Institute of Harvard and MIT | Lemieux et al |
| EPI_ISL_872739 | USA/RI-Broad_RIDOH-00087/2020 | 2020-12-07 | North America/USA/Rhode Island/ | Rhode Island Department of Health | Infectious Disease Program, Broad Institute of Harvard and MIT | Lemieux et al |

| | | | | | | |
|---|---|---|---|---|---|---|
| EPI_ISL_872745 | USA/CO-Broad-CMU_00017/2020 | 2020-10-30 | North America/USA/Colorado/ | Colorado Mesa University | Infectious Disease Program, Broad Institute of Harvard and MIT | Lemieux et al |
| EPI_ISL_872755 | USA/CO-Broad-CMU_00063/2020 | 2020-11-04 | North America/USA/Colorado/ | Colorado Mesa University | Infectious Disease Program, Broad Institute of Harvard and MIT | Lemieux et al |
| EPI_ISL_872757 | USA/CO-Broad-CMU_00073/2020 | 2020-11-09 | North America/USA/Colorado/ | Colorado Mesa University | Infectious Disease Program, Broad Institute of Harvard and MIT | Lemieux et al |
| EPI_ISL_872776 | USA/CO-Broad-CMU_00110/2020 | 2020-11-10 | North America/USA/Colorado/ | Colorado Mesa University | Infectious Disease Program, Broad Institute of Harvard and MIT | Lemieux et al |
| EPI_ISL_872777 | USA/CO-Broad-CMU_00111/2020 | 2020-11-10 | North America/USA/Colorado/ | Colorado Mesa University | Infectious Disease Program, Broad Institute of Harvard and MIT | Lemieux et al |
| EPI_ISL_872791 | USA/WI-UW-2854/2021 | 2021-01-12 | North America/USA/Wisconsin/Dane Co | University of Wisconsin-Madison AIDS Vaccine Research Laboratories | University of Wisconsin-Madison AIDS Vaccine Research Laboratories | Gage Moreno et al |
| EPI_ISL_872799 | USA/WI-UW-2863/2021 | 2021-01-11 | North America/USA/Wisconsin/Dane Co | University of Wisconsin-Madison AIDS Vaccine Research Laboratories | University of Wisconsin-Madison AIDS Vaccine Research Laboratories | Gage Moreno et al |
| EPI_ISL_872813 | USA/WI-UW-2878/2021 | 2021-01-11 | North America/USA/Wisconsin/Dane Co | University of Wisconsin-Madison AIDS Vaccine Research Laboratories | University of Wisconsin-Madison AIDS Vaccine Research Laboratories | Gage Moreno et al |
| EPI_ISL_872821 | USA/WI-UW-2887/2021 | 2021-01-11 | North America/USA/Wisconsin/Rock Cou | University of Wisconsin-Madison AIDS Vaccine Research Laboratories | University of Wisconsin-Madison AIDS Vaccine Research Laboratories | Gage Moreno et al |

| | | | | | | |
|---|---|---|---|---|---|---|
| EPI_ISL_872822 | USA/WI-UW-2888/2021 | 2021-01-12 | North America/USA/Wisconsin/Milwauk | University of Wisconsin-Madison AIDS Vaccine Research Laboratories | University of Wisconsin-Madison AIDS Vaccine Research Laboratories | Gage Moreno et al |
| EPI_ISL_872824 | USA/WI-UW-2890/2021 | 2021-01-12 | North America/USA/Wisconsin/Milwauk | University of Wisconsin-Madison AIDS Vaccine Research Laboratories | University of Wisconsin-Madison AIDS Vaccine Research Laboratories | Gage Moreno et al |
| EPI_ISL_872839 | USA/WI-UW-2906/2021 | 2021-01-12 | North America/USA/Wisconsin/Milwauk | University of Wisconsin-Madison AIDS Vaccine Research Laboratories | University of Wisconsin-Madison AIDS Vaccine Research Laboratories | Gage Moreno et al |
| EPI_ISL_872852 | USA/WI-UW-2919/2021 | 2021-01-08 | North America/USA/Wisconsin/Milwauk | University of Wisconsin-Madison AIDS Vaccine Research Laboratories | University of Wisconsin-Madison AIDS Vaccine Research Laboratories | Gage Moreno et al |
| EPI_ISL_872869 | USA/WI-UW-2937/2021 | 2021-01-12 | North America/USA/Wisconsin/Milwauk | University of Wisconsin-Madison AIDS Vaccine Research Laboratories | University of Wisconsin-Madison AIDS Vaccine Research Laboratories | Gage Moreno et al |
| EPI_ISL_872872 | USA/WI-UW-2940/2021 | 2021-01-12 | North America/USA/Wisconsin/Milwauk | University of Wisconsin-Madison AIDS Vaccine Research Laboratories | University of Wisconsin-Madison AIDS Vaccine Research Laboratories | Gage Moreno et al |
| EPI_ISL_872882 | USA/WI-UW-2950/2021 | 2021-01-12 | North America/USA/Wisconsin/Milwauk | University of Wisconsin-Madison AIDS Vaccine Research Laboratories | University of Wisconsin-Madison AIDS Vaccine Research Laboratories | Gage Moreno et al |
| EPI_ISL_872891 | USA/WI-UW-2960/2021 | 2021-01-12 | North America/USA/Wisconsin/Milwauk | University of Wisconsin-Madison AIDS Vaccine Research Laboratories | University of Wisconsin-Madison AIDS Vaccine Research Laboratories | Gage Moreno et al |

| | | | | | | |
|---|---|---|---|---|---|---|
| EPI_ISL_872898 | Russia/AST-RII-MH11691S/2020 | 2020-10-13 | Europe/Russia/Astrakhan Region/Akhtu | WHO National Influenza Centre Russian Federation | WHO National Influenza Centre Russian Federation | Andrey Komissarov et al |
| EPI_ISL_872901 | Russia/AST-RII-MH11698S/2020 | 2020-10-13 | Europe/Russia/Astrakhan Region/Akhtu | WHO National Influenza Centre Russian Federation | WHO National Influenza Centre Russian Federation | Andrey Komissarov et al |
| EPI_ISL_872903 | Russia/AST-RII-MH11716S/2020 | 2020-10-19 | Europe/Russia/Astrakhan Region/Akhtu | WHO National Influenza Centre Russian Federation | WHO National Influenza Centre Russian Federation | Andrey Komissarov et al |
| EPI_ISL_872905 | Russia/IVA-RII-MH11761S/2020 | 2020-12-06 | Europe/Russia/Ivanovo/Kineshma | WHO National Influenza Centre Russian Federation | WHO National Influenza Centre Russian Federation | Andrey Komissarov et al |
| EPI_ISL_872906 | Russia/IVA-RII-MH11762S/2020 | 2020-12-07 | Europe/Russia/Ivanovo/Yuryevets | WHO National Influenza Centre Russian Federation | WHO National Influenza Centre Russian Federation | Andrey Komissarov et al |
| EPI_ISL_872907 | Russia/IVA-RII-MH11763S/2020 | 2020-12-07 | Europe/Russia/Ivanovo/Zavolzhsk | WHO National Influenza Centre Russian Federation | WHO National Influenza Centre Russian Federation | Andrey Komissarov et al |
| EPI_ISL_872908 | Russia/IVA-RII-MH11765S/2020 | 2020-12-06 | Europe/Russia/Ivanovo/Kineshma | WHO National Influenza Centre Russian Federation | WHO National Influenza Centre Russian Federation | Andrey Komissarov et al |
| EPI_ISL_872910 | Russia/IVA-RII-MH11768S/2020 | 2020-12-05 | Europe/Russia/Ivanovo/Kineshma | WHO National Influenza Centre Russian Federation | WHO National Influenza Centre Russian Federation | Andrey Komissarov et al |
| EPI_ISL_872912 | Russia/IVA-RII-MH11770S/2020 | 2020-12-08 | Europe/Russia/Ivanovo/Shuya (Ivanovo) | WHO National Influenza Centre Russian Federation | WHO National Influenza Centre Russian Federation | Andrey Komissarov et al |
| EPI_ISL_872913 | Russia/IVA-RII-MH11771S/2020 | 2020-12-04 | Europe/Russia/Ivanovo/Kineshma | WHO National Influenza Centre Russian Federation | WHO National Influenza Centre Russian Federation | Andrey Komissarov et al |

| | | | | | | |
|---|---|---|---|---|---|---|
| EPI_ISL_872914 | Russia/IVA-RII-MH11772S/2020 | 2020-12-05 | Europe/Russia/Ivanovo/Furmanov | WHO National Influenza Centre Russian Federation | WHO National Influenza Centre Russian Federation | Andrey Komissarov et al |
| EPI_ISL_872915 | Russia/IVA-RII-MH11773S/2020 | 2020-12-04 | Europe/Russia/Ivanovo/Ivanovo | WHO National Influenza Centre Russian Federation | WHO National Influenza Centre Russian Federation | Andrey Komissarov et al |
| EPI_ISL_872916 | Russia/IVA-RII-MH11774S/2020 | 2020-12-05 | Europe/Russia/Ivanovo/Ivanovo | WHO National Influenza Centre Russian Federation | WHO National Influenza Centre Russian Federation | Andrey Komissarov et al |
| EPI_ISL_872917 | Russia/IVA-RII-MH11776S/2020 | 2020-12-05 | Europe/Russia/Ivanovo/Ivanovo | WHO National Influenza Centre Russian Federation | WHO National Influenza Centre Russian Federation | Andrey Komissarov et al |
| EPI_ISL_872918 | Russia/IVA-RII-MH11777S/2020 | 2020-12-02 | Europe/Russia/Ivanovo/Ivanovo | WHO National Influenza Centre Russian Federation | WHO National Influenza Centre Russian Federation | Andrey Komissarov et al |
| EPI_ISL_872919 | Russia/IVA-RII-MH11778S/2020 | 2020-12-03 | Europe/Russia/Ivanovo/Ivanovo | WHO National Influenza Centre Russian Federation | WHO National Influenza Centre Russian Federation | Andrey Komissarov et al |
| EPI_ISL_872920 | Russia/IVA-RII-MH11829S/2020 | 2020-12-11 | Europe/Russia/Ivanovo/Ivanovo | WHO National Influenza Centre Russian Federation | WHO National Influenza Centre Russian Federation | Andrey Komissarov et al |
| EPI_ISL_872921 | Russia/IVA-RII-MH11833S/2020 | 2020-12-11 | Europe/Russia/Ivanovo/Ivanovo | WHO National Influenza Centre Russian Federation | WHO National Influenza Centre Russian Federation | Andrey Komissarov et al |
| EPI_ISL_872922 | Russia/IVA-RII-MH11834S/2020 | 2020-12-11 | Europe/Russia/Ivanovo/Ivanovo | WHO National Influenza Centre Russian Federation | WHO National Influenza Centre Russian Federation | Andrey Komissarov et al |
| EPI_ISL_872923 | Russia/IVA-RII-MH11837S/2020 | 2020-12-11 | Europe/Russia/Ivanovo/Kineshma | WHO National Influenza Centre Russian Federation | WHO National Influenza Centre Russian Federation | Andrey Komissarov et al |

| | | | | | | |
|---|---|---|---|---|---|---|
| EPI_ISL_872924 | Russia/IVA-RII-MH11838S/2020 | 2020-12-13 | Europe/Russia/Ivanovo/Ivanovo | WHO National Influenza Centre Russian Federation | WHO National Influenza Centre Russian Federation | Andrey Komissarov et al |
| EPI_ISL_872925 | Russia/IVA-RII-MH11841S/2020 | 2020-12-13 | Europe/Russia/Ivanovo/Ivanovo | WHO National Influenza Centre Russian Federation | WHO National Influenza Centre Russian Federation | Andrey Komissarov et al |
| EPI_ISL_872926 | Russia/IVA-RII-MH11843S/2020 | 2020-12-13 | Europe/Russia/Ivanovo/Kineshma | WHO National Influenza Centre Russian Federation | WHO National Influenza Centre Russian Federation | Andrey Komissarov et al |
| EPI_ISL_872927 | Russia/IVA-RII-MH11847S/2020 | 2020-12-14 | Europe/Russia/Ivanovo/Ivanovo | WHO National Influenza Centre Russian Federation | WHO National Influenza Centre Russian Federation | Andrey Komissarov et al |
| EPI_ISL_872929 | Russia/IVA-RII-MH11868S/2020 | 2020-12-14 | Europe/Russia/Ivanovo/Kineshma | WHO National Influenza Centre Russian Federation | WHO National Influenza Centre Russian Federation | Andrey Komissarov et al |
| EPI_ISL_872930 | Russia/IVA-RII-MH11869S/2020 | 2020-12-14 | Europe/Russia/Ivanovo/Kineshma | WHO National Influenza Centre Russian Federation | WHO National Influenza Centre Russian Federation | Andrey Komissarov et al |
| EPI_ISL_872931 | Russia/IVA-RII-MH11870S/2020 | 2020-12-14 | Europe/Russia/Ivanovo/Kineshma | WHO National Influenza Centre Russian Federation | WHO National Influenza Centre Russian Federation | Andrey Komissarov et al |
| EPI_ISL_872932 | Russia/IVA-RII-MH11871S/2020 | 2020-12-15 | Europe/Russia/Ivanovo/Ivanovo | WHO National Influenza Centre Russian Federation | WHO National Influenza Centre Russian Federation | Andrey Komissarov et al |
| EPI_ISL_872933 | Russia/IVA-RII-MH11872S/2020 | 2020-12-15 | Europe/Russia/Ivanovo/Ivanovo | WHO National Influenza Centre Russian Federation | WHO National Influenza Centre Russian Federation | Andrey Komissarov et al |
| EPI_ISL_872934 | Russia/IVA-RII-MH11873S/2020 | 2020-12-15 | Europe/Russia/Ivanovo/Ivanovo | WHO National Influenza Centre Russian Federation | WHO National Influenza Centre Russian Federation | Andrey Komissarov et al |

| | | | | | | |
|---|---|---|---|---|---|---|
| EPI_ISL_872935 | Russia/IVA-RII-MH11875S/2020 | 2020-12-14 | Europe/Russia/Ivanovo/Vichuga | WHO National Influenza Centre Russian Federation | WHO National Influenza Centre Russian Federation | Andrey Komissarov et al |
| EPI_ISL_872936 | Russia/IVA-RII-MH11876S/2020 | 2020-12-14 | Europe/Russia/Ivanovo/Rodniki | WHO National Influenza Centre Russian Federation | WHO National Influenza Centre Russian Federation | Andrey Komissarov et al |
| EPI_ISL_872937 | Russia/IVA-RII-MH11878S/2020 | 2020-12-16 | Europe/Russia/Ivanovo/Vichuga | WHO National Influenza Centre Russian Federation | WHO National Influenza Centre Russian Federation | Andrey Komissarov et al |
| EPI_ISL_872938 | Russia/IVA-RII-MH11880S/2020 | 2020-12-15 | Europe/Russia/Ivanovo/Ivanovo | WHO National Influenza Centre Russian Federation | WHO National Influenza Centre Russian Federation | Andrey Komissarov et al |
| EPI_ISL_872939 | Russia/IVA-RII-MH11883S/2020 | 2020-12-14 | Europe/Russia/Ivanovo/Kineshma | WHO National Influenza Centre Russian Federation | WHO National Influenza Centre Russian Federation | Andrey Komissarov et al |
| EPI_ISL_872940 | Russia/IVA-RII-MH11884S/2020 | 2020-12-15 | Europe/Russia/Ivanovo/Kineshma | WHO National Influenza Centre Russian Federation | WHO National Influenza Centre Russian Federation | Andrey Komissarov et al |
| EPI_ISL_872941 | Russia/IVA-RII-MH11885S/2020 | 2020-12-15 | Europe/Russia/Ivanovo/Ivanovo | WHO National Influenza Centre Russian Federation | WHO National Influenza Centre Russian Federation | Andrey Komissarov et al |
| EPI_ISL_872943 | Russia/IVA-RII-MH11887S/2020 | 2020-12-16 | Europe/Russia/Ivanovo/Ivanovo | WHO National Influenza Centre Russian Federation | WHO National Influenza Centre Russian Federation | Andrey Komissarov et al |
| EPI_ISL_872944 | Russia/IVA-RII-MH11888S/2020 | 2020-12-14 | Europe/Russia/Ivanovo/Ivanovo | WHO National Influenza Centre Russian Federation | WHO National Influenza Centre Russian Federation | Andrey Komissarov et al |
| EPI_ISL_872945 | Russia/IVA-RII-MH11889S/2020 | 2020-12-14 | Europe/Russia/Ivanovo/Ivanovo | WHO National Influenza Centre Russian Federation | WHO National Influenza Centre Russian Federation | Andrey Komissarov et al |

| | | | | | | |
|---|---|---|---|---|---|---|
| EPI_ISL_872946 | Russia/IVA-RII-MH11890S/2020 | 2020-12-16 | Europe/Russia/Ivanovo/Ivanovo | WHO National Influenza Centre Russian Federation | WHO National Influenza Centre Russian Federation | Andrey Komissarov et al |
| EPI_ISL_872947 | Russia/IVA-RII-MH11894S/2020 | 2020-12-14 | Europe/Russia/Ivanovo/Ivanovo | WHO National Influenza Centre Russian Federation | WHO National Influenza Centre Russian Federation | Andrey Komissarov et al |
| EPI_ISL_872948 | Russia/KAM-RII-MH12130S/2020 | 2020-12-15 | Europe/Russia/Kamchatka Krai/Petropav | WHO National Influenza Centre Russian Federation | WHO National Influenza Centre Russian Federation | Andrey Komissarov et al |
| EPI_ISL_872949 | Russia/KAM-RII-MH12132S/2020 | 2020-12-16 | Europe/Russia/Kamchatka Krai/Petropav | WHO National Influenza Centre Russian Federation | WHO National Influenza Centre Russian Federation | Andrey Komissarov et al |
| EPI_ISL_872950 | Russia/KAM-RII-MH12133S/2020 | 2020-12-16 | Europe/Russia/Kamchatka Krai/Petropav | WHO National Influenza Centre Russian Federation | WHO National Influenza Centre Russian Federation | Andrey Komissarov et al |
| EPI_ISL_872951 | Russia/KAM-RII-MH12136S/2020 | 2020-12-15 | Europe/Russia/Kamchatka Krai/Petropav | WHO National Influenza Centre Russian Federation | WHO National Influenza Centre Russian Federation | Andrey Komissarov et al |
| EPI_ISL_872952 | Russia/KAM-RII-MH12137S/2020 | 2020-12-16 | Europe/Russia/Kamchatka Krai/Petropav | WHO National Influenza Centre Russian Federation | WHO National Influenza Centre Russian Federation | Andrey Komissarov et al |
| EPI_ISL_872953 | Russia/KAM-RII-MH12141S/2020 | 2020-12-16 | Europe/Russia/Kamchatka Krai/Petropav | WHO National Influenza Centre Russian Federation | WHO National Influenza Centre Russian Federation | Andrey Komissarov et al |
| EPI_ISL_872954 | Russia/KAM-RII-MH12143S/2020 | 2020-12-17 | Europe/Russia/Kamchatka Krai/Petropav | WHO National Influenza Centre Russian Federation | WHO National Influenza Centre Russian Federation | Andrey Komissarov et al |
| EPI_ISL_872957 | Russia/KAM-RII-MH12148S/2020 | 2020-12-17 | Europe/Russia/Kamchatka Krai/Petropav | WHO National Influenza Centre Russian Federation | WHO National Influenza Centre Russian Federation | Andrey Komissarov et al |

| | | | | | | |
|---|---|---|---|---|---|---|
| EPI_ISL_872958 | Russia/KAM-RII-MH12149S/2020 | 2020-12-16 | Europe/Russia/Kamchatka Krai/Petropav | WHO National Influenza Centre Russian Federation | WHO National Influenza Centre Russian Federation | Andrey Komissarov et al |
| EPI_ISL_872960 | Russia/KAM-RII-MH12160S/2020 | 2020-12-15 | Europe/Russia/Kamchatka Krai/Petropav | WHO National Influenza Centre Russian Federation | WHO National Influenza Centre Russian Federation | Andrey Komissarov et al |
| EPI_ISL_872961 | Russia/KAM-RII-MH12161S/2020 | 2020-12-14 | Europe/Russia/Kamchatka Krai/Petropav | WHO National Influenza Centre Russian Federation | WHO National Influenza Centre Russian Federation | Andrey Komissarov et al |
| EPI_ISL_872962 | Russia/KAM-RII-MH12170S/2020 | 2020-12-14 | Europe/Russia/Kamchatka Krai/Petropav | WHO National Influenza Centre Russian Federation | WHO National Influenza Centre Russian Federation | Andrey Komissarov et al |
| EPI_ISL_872963 | Russia/KAM-RII-MH12177S/2020 | 2020-12-15 | Europe/Russia/Kamchatka Krai/Petropav | WHO National Influenza Centre Russian Federation | WHO National Influenza Centre Russian Federation | Andrey Komissarov et al |
| EPI_ISL_872965 | Russia/KAM-RII-MH12182S/2020 | 2020-12-15 | Europe/Russia/Kamchatka Krai/Petropav | WHO National Influenza Centre Russian Federation | WHO National Influenza Centre Russian Federation | Andrey Komissarov et al |
| EPI_ISL_872966 | Russia/KAM-RII-MH12184S/2020 | 2020-12-15 | Europe/Russia/Kamchatka Krai/Petropav | WHO National Influenza Centre Russian Federation | WHO National Influenza Centre Russian Federation | Andrey Komissarov et al |
| EPI_ISL_872967 | Russia/KAM-RII-MH12185S/2020 | 2020-12-15 | Europe/Russia/Kamchatka Krai/Petropav | WHO National Influenza Centre Russian Federation | WHO National Influenza Centre Russian Federation | Andrey Komissarov et al |
| EPI_ISL_872968 | Russia/KAM-RII-MH12189S/2020 | 2020-12-15 | Europe/Russia/Kamchatka Krai/Petropav | WHO National Influenza Centre Russian Federation | WHO National Influenza Centre Russian Federation | Andrey Komissarov et al |
| EPI_ISL_872969 | Russia/KAM-RII-MH12201S/2020 | 2020-12-14 | Europe/Russia/Kamchatka Krai/Petropav | WHO National Influenza Centre Russian Federation | WHO National Influenza Centre Russian Federation | Andrey Komissarov et al |

| | | | | | | |
|---|---|---|---|---|---|---|
| EPI_ISL_872970 | Russia/KAM-RII-MH12206S/2020 | 2020-12-11 | Europe/Russia/Kamchatka Krai/Petropav | WHO National Influenza Centre Russian Federation | WHO National Influenza Centre Russian Federation | Andrey Komissarov et al |
| EPI_ISL_872971 | Russia/KAM-RII-MH12211S/2020 | 2020-12-15 | Europe/Russia/Kamchatka Krai/Petropav | WHO National Influenza Centre Russian Federation | WHO National Influenza Centre Russian Federation | Andrey Komissarov et al |
| EPI_ISL_872972 | Russia/SPE-RII-MH13144S/2021 | 2021-01-03 | Europe/Russia/Saint-Petersburg/Saint-P | HELIX LLC | WHO National Influenza Centre Russian Federation | Andrey Komissarov et al |
| EPI_ISL_872973 | Russia/SPE-RII-MH13149S/2021 | 2021-01-03 | Europe/Russia/Saint-Petersburg/Saint-P | HELIX LLC | WHO National Influenza Centre Russian Federation | Andrey Komissarov et al |
| EPI_ISL_872974 | Russia/SPE-RII-MH13158S/2021 | 2021-01-03 | Europe/Russia/Saint-Petersburg/Saint-P | HELIX LLC | WHO National Influenza Centre Russian Federation | Andrey Komissarov et al |
| EPI_ISL_872976 | Russia/SPE-RII-MH13166S/2021 | 2021-01-03 | Europe/Russia/Saint-Petersburg/Saint-P | HELIX LLC | WHO National Influenza Centre Russian Federation | Andrey Komissarov et al |
| EPI_ISL_872977 | Russia/SPE-RII-MH13167S/2021 | 2021-01-03 | Europe/Russia/Saint-Petersburg/Saint-P | HELIX LLC | WHO National Influenza Centre Russian Federation | Andrey Komissarov et al |
| EPI_ISL_872978 | Russia/SPE-RII-MH13168S/2021 | 2021-01-03 | Europe/Russia/Saint-Petersburg/Saint-P | HELIX LLC | WHO National Influenza Centre Russian Federation | Andrey Komissarov et al |
| EPI_ISL_872980 | Russia/SPE-RII-MH13173S/2021 | 2021-01-03 | Europe/Russia/Saint-Petersburg/Saint-P | HELIX LLC | WHO National Influenza Centre Russian Federation | Andrey Komissarov et al |
| EPI_ISL_872981 | Russia/SPE-RII-MH13176S/2021 | 2021-01-03 | Europe/Russia/Saint-Petersburg/Saint-P | HELIX LLC | WHO National Influenza Centre Russian Federation | Andrey Komissarov et al |

| | | | | | | |
|---|---|---|---|---|---|---|
| EPI_ISL_872982 | Russia/SPE-RII-MH13188S/2021 | 2021-01-03 | Europe/Russia/Saint-Petersburg/Saint-P | HELIX LLC | WHO National Influenza Centre Russian Federation | Andrey Komissarov et al |
| EPI_ISL_872983 | Russia/SPE-RII-MH13194S/2021 | 2021-01-03 | Europe/Russia/Saint-Petersburg/Saint-P | HELIX LLC | WHO National Influenza Centre Russian Federation | Andrey Komissarov et al |
| EPI_ISL_872984 | Russia/SPE-RII-MH13201S/2021 | 2021-01-03 | Europe/Russia/Saint-Petersburg/Saint-P | HELIX LLC | WHO National Influenza Centre Russian Federation | Andrey Komissarov et al |
| EPI_ISL_872985 | Russia/SPE-RII-MH13205S/2020 | 2020-12-24 | Europe/Russia/Saint-Petersburg/Saint-P | HELIX LLC | WHO National Influenza Centre Russian Federation | Andrey Komissarov et al |
| EPI_ISL_872986 | Russia/SPE-RII-MH13216S/2020 | 2020-12-24 | Europe/Russia/Saint-Petersburg/Saint-P | HELIX LLC | WHO National Influenza Centre Russian Federation | Andrey Komissarov et al |
| EPI_ISL_872988 | Russia/SPE-RII-MH13231S/2020 | 2020-12-24 | Europe/Russia/Saint-Petersburg/Saint-P | HELIX LLC | WHO National Influenza Centre Russian Federation | Andrey Komissarov et al |
| EPI_ISL_872991 | Russia/SPE-RII-MH13289S/2020 | 2020-12-24 | Europe/Russia/Saint-Petersburg/Saint-P | HELIX LLC | WHO National Influenza Centre Russian Federation | Andrey Komissarov et al |
| EPI_ISL_872992 | Russia/SPE-RII-MH13291S/2020 | 2020-12-23 | Europe/Russia/Saint-Petersburg/Saint-P | HELIX LLC | WHO National Influenza Centre Russian Federation | Andrey Komissarov et al |
| EPI_ISL_872993 | Russia/SPE-RII-MH13294S/2020 | 2020-12-22 | Europe/Russia/Saint-Petersburg/Saint-P | HELIX LLC | WHO National Influenza Centre Russian Federation | Andrey Komissarov et al |
| EPI_ISL_872994 | Russia/PSK-RII-MH13295S/2020 | 2020-12-23 | Europe/Russia/Pskov Oblast/Velikiye Lu | HELIX LLC | WHO National Influenza Centre Russian Federation | Andrey Komissarov et al |

| | | | | | | |
|---|---|---|---|---|---|---|
| EPI_ISL_872995 | Russia/SPE-RII-MH13300S/2020 | 2020-12-24 | Europe/Russia/Saint-Petersburg/Saint-P | HELIX LLC | WHO National Influenza Centre Russian Federation | Andrey Komissarov et al |
| EPI_ISL_872996 | Russia/SPE-RII-MH13302S/2020 | 2020-12-24 | Europe/Russia/Saint-Petersburg/Saint-P | HELIX LLC | WHO National Influenza Centre Russian Federation | Andrey Komissarov et al |
| EPI_ISL_872997 | Russia/SPE-RII-MH13303S/2020 | 2020-12-24 | Europe/Russia/Saint-Petersburg/Saint-P | HELIX LLC | WHO National Influenza Centre Russian Federation | Andrey Komissarov et al |
| EPI_ISL_872998 | Russia/SPE-RII-MH13305S/2020 | 2020-12-24 | Europe/Russia/Saint-Petersburg/Saint-P | HELIX LLC | WHO National Influenza Centre Russian Federation | Andrey Komissarov et al |
| EPI_ISL_873000 | Russia/SPE-RII-MH13308S/2020 | 2020-12-24 | Europe/Russia/Saint-Petersburg/Saint-P | HELIX LLC | WHO National Influenza Centre Russian Federation | Andrey Komissarov et al |
| EPI_ISL_873001 | Russia/SPE-RII-MH13309S/2020 | 2020-12-24 | Europe/Russia/Saint-Petersburg/Saint-P | HELIX LLC | WHO National Influenza Centre Russian Federation | Andrey Komissarov et al |
| EPI_ISL_873002 | Russia/SPE-RII-MH13310S/2020 | 2020-12-23 | Europe/Russia/Saint-Petersburg/Saint-P | HELIX LLC | WHO National Influenza Centre Russian Federation | Andrey Komissarov et al |
| EPI_ISL_873004 | Russia/SPE-RII-MH13315S/2020 | 2020-12-24 | Europe/Russia/Saint-Petersburg/Saint-P | HELIX LLC | WHO National Influenza Centre Russian Federation | Andrey Komissarov et al |
| EPI_ISL_873006 | Russia/LEN-RII-MH13317S/2020 | 2020-12-24 | Europe/Russia/Leningrad Oblast/Vsevol | HELIX LLC | WHO National Influenza Centre Russian Federation | Andrey Komissarov et al |
| EPI_ISL_873007 | Russia/SPE-RII-MH13323S/2020 | 2020-12-24 | Europe/Russia/Saint-Petersburg/Saint-P | HELIX LLC | WHO National Influenza Centre Russian Federation | Andrey Komissarov et al |

| | | | | | | |
|---|---|---|---|---|---|---|
| EPI_ISL_873008 | Russia/SPE-RII-MH13324S/2020 | 2020-12-23 | Europe/Russia/Saint-Petersburg/Saint-P | HELIX LLC | WHO National Influenza Centre Russian Federation | Andrey Komissarov et al |
| EPI_ISL_873009 | Russia/SPE-RII-MH13325S/2020 | 2020-12-24 | Europe/Russia/Saint-Petersburg/Saint-P | HELIX LLC | WHO National Influenza Centre Russian Federation | Andrey Komissarov et al |
| EPI_ISL_873010 | Russia/SPE-RII-MH13326S/2020 | 2020-12-24 | Europe/Russia/Saint-Petersburg/Saint-P | HELIX LLC | WHO National Influenza Centre Russian Federation | Andrey Komissarov et al |
| EPI_ISL_873011 | Russia/SPE-RII-MH13327S/2020 | 2020-12-24 | Europe/Russia/Saint-Petersburg/Saint-P | HELIX LLC | WHO National Influenza Centre Russian Federation | Andrey Komissarov et al |
| EPI_ISL_873012 | Russia/SPE-RII-MH13328S/2020 | 2020-12-24 | Europe/Russia/Saint-Petersburg/Saint-P | HELIX LLC | WHO National Influenza Centre Russian Federation | Andrey Komissarov et al |
| EPI_ISL_873014 | Russia/SPE-RII-MH13330S/2020 | 2020-12-24 | Europe/Russia/Saint-Petersburg/Saint-P | HELIX LLC | WHO National Influenza Centre Russian Federation | Andrey Komissarov et al |
| EPI_ISL_873015 | Russia/SPE-RII-MH13333S/2020 | 2020-12-24 | Europe/Russia/Saint-Petersburg/Saint-P | HELIX LLC | WHO National Influenza Centre Russian Federation | Andrey Komissarov et al |
| EPI_ISL_873016 | Russia/SPE-RII-MH13402S/2020 | 2020-12-24 | Europe/Russia/Saint-Petersburg/Saint-P | HELIX LLC | WHO National Influenza Centre Russian Federation | Andrey Komissarov et al |
| EPI_ISL_873017 | Russia/SPE-RII-MH13403S/2020 | 2020-12-24 | Europe/Russia/Saint-Petersburg/Saint-P | HELIX LLC | WHO National Influenza Centre Russian Federation | Andrey Komissarov et al |
| EPI_ISL_873018 | Russia/LEN-RII-MH13596S/2020 | 2020-12-25 | Europe/Russia/Leningrad Oblast/Kirishi | HELIX LLC | WHO National Influenza Centre Russian Federation | Andrey Komissarov et al |

| | | | | | | |
|---|---|---|---|---|---|---|
| EPI_ISL_873019 | Russia/LEN-RII-MH13600S/2020 | 2020-12-25 | Europe/Russia/Leningrad Oblast/Kirishi | HELIX LLC | WHO National Influenza Centre Russian Federation | Andrey Komissarov et al |
| EPI_ISL_873020 | Russia/SPE-RII-MH13637S/2020 | 2020-12-25 | Europe/Russia/Saint-Petersburg/Saint-P | HELIX LLC | WHO National Influenza Centre Russian Federation | Andrey Komissarov et al |
| EPI_ISL_873021 | Russia/SPE-RII-MH13643S/2020 | 2020-12-27 | Europe/Russia/Saint-Petersburg/Saint-P | HELIX LLC | WHO National Influenza Centre Russian Federation | Andrey Komissarov et al |
| EPI_ISL_873023 | Russia/SPE-RII-MH13649S/2020 | 2020-12-27 | Europe/Russia/Saint-Petersburg/Saint-P | HELIX LLC | WHO National Influenza Centre Russian Federation | Andrey Komissarov et al |
| EPI_ISL_873024 | Russia/SPE-RII-MH13651S/2020 | 2020-12-27 | Europe/Russia/Saint-Petersburg/Saint-P | HELIX LLC | WHO National Influenza Centre Russian Federation | Andrey Komissarov et al |
| EPI_ISL_873025 | Russia/SPE-RII-MH13658S/2020 | 2020-12-27 | Europe/Russia/Saint-Petersburg/Saint-P | HELIX LLC | WHO National Influenza Centre Russian Federation | Andrey Komissarov et al |
| EPI_ISL_873026 | Russia/SPE-RII-MH13667S/2020 | 2020-12-27 | Europe/Russia/Saint-Petersburg/Saint-P | HELIX LLC | WHO National Influenza Centre Russian Federation | Andrey Komissarov et al |
| EPI_ISL_873029 | Russia/SPE-RII-MH13704S/2021 | 2021-01-02 | Europe/Russia/Saint-Petersburg/Saint-P | HELIX LLC | WHO National Influenza Centre Russian Federation | Andrey Komissarov et al |
| EPI_ISL_873030 | Russia/SPE-RII-MH13737S/2021 | 2021-01-02 | Europe/Russia/Saint-Petersburg/Saint-P | HELIX LLC | WHO National Influenza Centre Russian Federation | Andrey Komissarov et al |
| EPI_ISL_873031 | Russia/SPE-RII-MH13740S/2021 | 2021-01-02 | Europe/Russia/Saint-Petersburg/Saint-P | HELIX LLC | WHO National Influenza Centre Russian Federation | Andrey Komissarov et al |

| | | | | | | |
|---|---|---|---|---|---|---|
| EPI_ISL_873032 | Russia/SPE-RII-MH13741S/2021 | 2021-01-02 | Europe/Russia/Saint-Petersburg/Saint-P | HELIX LLC | WHO National Influenza Centre Russian Federation | Andrey Komissarov et al |
| EPI_ISL_873033 | Russia/SPE-RII-MH13747S/2021 | 2021-01-02 | Europe/Russia/Saint-Petersburg/Saint-P | HELIX LLC | WHO National Influenza Centre Russian Federation | Andrey Komissarov et al |
| EPI_ISL_873034 | Russia/SPE-RII-MH13790S/2020 | 2020-12-28 | Europe/Russia/Saint-Petersburg/Saint-P | HELIX LLC | WHO National Influenza Centre Russian Federation | Andrey Komissarov et al |
| EPI_ISL_873035 | Russia/SPE-RII-MH13814S/2021 | 2021-01-03 | Europe/Russia/Saint-Petersburg/Saint-P | HELIX LLC | WHO National Influenza Centre Russian Federation | Andrey Komissarov et al |
| EPI_ISL_873036 | Russia/SPE-RII-MH13820S/2021 | 2021-01-03 | Europe/Russia/Saint-Petersburg/Saint-P | HELIX LLC | WHO National Influenza Centre Russian Federation | Andrey Komissarov et al |
| EPI_ISL_873039 | Russia/SPE-RII-MH13834S/2021 | 2021-01-03 | Europe/Russia/Saint-Petersburg/Saint-P | HELIX LLC | WHO National Influenza Centre Russian Federation | Andrey Komissarov et al |
| EPI_ISL_873046 | USA/MI-UM-10037644322/2021 | 2021-01-02 | North America/USA/Michigan/ | University of Michigan Clinical Microbiology Laboratory | Lauring Lab, University of Michigan, Department of Microbiology and Immunology | Valesano et al |
| EPI_ISL_873052 | USA/MI-UM-10037669500/2021 | 2021-01-04 | North America/USA/Michigan/ | University of Michigan Clinical Microbiology Laboratory | Lauring Lab, University of Michigan, Department of Microbiology and Immunology | Valesano et al |

| | | | | | | |
|---|---|---|---|---|---|---|
| EPI_ISL_873055 | USA/MI-UM-10037677223/2021 | 2021-01-05 | North America/USA/Michigan/ | University of Michigan Clinical Microbiology Laboratory | Lauring Lab, University of Michigan, Department of Microbiology and Immunology | Valesano et al |
| EPI_ISL_873062 | USA/MI-UM-10037677342/2021 | 2021-01-05 | North America/USA/Michigan/ | University of Michigan Clinical Microbiology Laboratory | Lauring Lab, University of Michigan, Department of Microbiology and Immunology | Valesano et al |
| EPI_ISL_873074 | USA/MI-UM-10037676259/2021 | 2021-01-05 | North America/USA/Michigan/ | University of Michigan Clinical Microbiology Laboratory | Lauring Lab, University of Michigan, Department of Microbiology and Immunology | Valesano et al |
| EPI_ISL_873075 | USA/MI-UM-10037685173/2021 | 2021-01-05 | North America/USA/Michigan/ | University of Michigan Clinical Microbiology Laboratory | Lauring Lab, University of Michigan, Department of Microbiology and Immunology | Valesano et al |
| EPI_ISL_873077 | USA/MI-UM-10037700834/2021 | 2021-01-06 | North America/USA/Michigan/ | University of Michigan Clinical Microbiology Laboratory | Lauring Lab, University of Michigan, Department of Microbiology and Immunology | Valesano et al |
| EPI_ISL_873078 | USA/MI-UM-10037683341/2021 | 2021-01-05 | North America/USA/Michigan/ | University of Michigan Clinical Microbiology Laboratory | Lauring Lab, University of Michigan, Department of Microbiology and Immunology | Valesano et al |

| | | | | | | |
|---|---|---|---|---|---|---|
| EPI_ISL_873098 | USA/MI-UM-10037727578/2021 | 2021-01-07 | North America/USA/Michigan/ | University of Michigan Clinical Microbiology Laboratory | Lauring Lab, University of Michigan, Department of Microbiology and Immunology | Valesano et al |
| EPI_ISL_873099 | USA/MI-UM-10037717626/2021 | 2021-01-07 | North America/USA/Michigan/ | University of Michigan Clinical Microbiology Laboratory | Lauring Lab, University of Michigan, Department of Microbiology and Immunology | Valesano et al |
| EPI_ISL_873104 | USA/MI-UM-10037726986/2021 | 2021-01-07 | North America/USA/Michigan/ | University of Michigan Clinical Microbiology Laboratory | Lauring Lab, University of Michigan, Department of Microbiology and Immunology | Valesano et al |
| EPI_ISL_873107 | USA/MI-UM-10037705386/2021 | 2021-01-06 | North America/USA/Michigan/ | University of Michigan Clinical Microbiology Laboratory | Lauring Lab, University of Michigan, Department of Microbiology and Immunology | Valesano et al |
| EPI_ISL_873109 | USA/MI-UM-10037727417/2021 | 2021-01-07 | North America/USA/Michigan/ | University of Michigan Clinical Microbiology Laboratory | Lauring Lab, University of Michigan, Department of Microbiology and Immunology | Valesano et al |
| EPI_ISL_873111 | USA/MI-UM-10037697125/2021 | 2021-01-06 | North America/USA/Michigan/ | University of Michigan Clinical Microbiology Laboratory | Lauring Lab, University of Michigan, Department of Microbiology and Immunology | Valesano et al |

| | | | | | | |
|---|---|---|---|---|---|---|
| EPI_ISL_873120 | USA/MI-UM-10037699049/2021 | 2021-01-06 | North America/USA/Michigan/ | University of Michigan Clinical Microbiology Laboratory | Lauring Lab, University of Michigan, Department of Microbiology and Immunology | Valesano et al |
| EPI_ISL_873132 | USA/MI-UM-10037009231/2020 | 2020-11-18 | North America/USA/Michigan/ | University of Michigan Clinical Microbiology Laboratory | Lauring Lab, University of Michigan, Department of Microbiology and Immunology | Valesano et al |
| EPI_ISL_873139 | USA/MI-UM-10037171448/2020 | 2020-11-30 | North America/USA/Michigan/ | University of Michigan Clinical Microbiology Laboratory | Lauring Lab, University of Michigan, Department of Microbiology and Immunology | Valesano et al |
| EPI_ISL_873141 | USA/MI-UM-10037087380/2020 | 2020-11-23 | North America/USA/Michigan/ | University of Michigan Clinical Microbiology Laboratory | Lauring Lab, University of Michigan, Department of Microbiology and Immunology | Valesano et al |
| EPI_ISL_873142 | USA/MI-UM-10037064076/2020 | 2020-11-21 | North America/USA/Michigan/ | University of Michigan Clinical Microbiology Laboratory | Lauring Lab, University of Michigan, Department of Microbiology and Immunology | Valesano et al |
| EPI_ISL_873160 | USA/MI-UM-10037817005/2021 | 2021-01-13 | North America/USA/Michigan/ | University of Michigan Clinical Microbiology Laboratory | Lauring Lab, University of Michigan, Department of Microbiology and Immunology | Valesano et al |

| | | | | | | |
|---|---|---|---|---|---|---|
| EPI_ISL_873167 | USA/SC-DHEC-0057/2021 | 2021-01-08 | North America/USA/South Carolina/Ge‹ | Microbiology Division, South Carolina Department of Health and Environmental Control (SC DHEC) | Microbiology Division, South Carolina Department of Health and Environmental Control (SC DHEC) | Flores et al |
| EPI_ISL_873189 | USA/SC-DHEC-0025/2021 | 2021-01-03 | North America/USA/South Carolina/Ge‹ | Microbiology Division, South Carolina Department of Health and Environmental Control (SC DHEC) | Microbiology Division, South Carolina Department of Health and Environmental Control (SC DHEC) | Flores et al |
| EPI_ISL_873194 | USA/ND-NDDH-0058/2020 | 2020-01-06 | North America/USA/North Dakota/ | North Dakota Department of Health, Public Health Laboratory | North Dakota Department of Health, Public Health Laboratory | Lisa Wingerter et al |
| EPI_ISL_873198 | USA/ND-NDDH-0062/2021 | 2021-01-05 | North America/USA/North Dakota/ | North Dakota Department of Health, Public Health Laboratory | North Dakota Department of Health, Public Health Laboratory | Lisa Wingerter et al |
| EPI_ISL_873201 | USA/ND-NDDH-0050/2021 | 2021-01-04 | North America/USA/North Dakota/ | North Dakota Department of Health, Public Health Laboratory | North Dakota Department of Health, Public Health Laboratory | Lisa Wingerter et al |
| EPI_ISL_873202 | USA/ND-NDDH-0051/2021 | 2021-01-05 | North America/USA/North Dakota/ | North Dakota Department of Health, Public Health Laboratory | North Dakota Department of Health, Public Health Laboratory | Lisa Wingerter et al |
| EPI_ISL_873206 | USA/ND-NDDH-0055/2021 | 2021-01-05 | North America/USA/North Dakota/ | North Dakota Department of Health, Public Health Laboratory | North Dakota Department of Health, Public Health Laboratory | Lisa Wingerter et al |
| EPI_ISL_873207 | USA/ND-NDDH-0056/2021 | 2021-01-04 | North America/USA/North Dakota/ | North Dakota Department of Health, Public Health Laboratory | North Dakota Department of Health, Public Health Laboratory | Lisa Wingerter et al |

| | | | | | | |
|---|---|---|---|---|---|---|
| EPI_ISL_873214 | USA/MN-MDH-2355/2020 | 2020-12-11 | North America/USA/Minnesota/ | M Health Fairview | Minnesota Department of Health, Public Health Laboratory | Alexandra Lorentz et al |
| EPI_ISL_873219 | USA/MN-MDH-2360/2020 | 2020-12-14 | North America/USA/Minnesota/ | M Health Fairview | Minnesota Department of Health, Public Health Laboratory | Alexandra Lorentz et al |
| EPI_ISL_873220 | USA/MN-MDH-2361/2020 | 2020-12-14 | North America/USA/Minnesota/ | M Health Fairview | Minnesota Department of Health, Public Health Laboratory | Alexandra Lorentz et al |
| EPI_ISL_873225 | USA/MN-MDH-2366/2020 | 2020-12-21 | North America/USA/Minnesota/ | M Health Fairview | Minnesota Department of Health, Public Health Laboratory | Alexandra Lorentz et al |
| EPI_ISL_873227 | USA/MN-MDH-2368/2020 | 2020-12-24 | North America/USA/Minnesota/ | M Health Fairview | Minnesota Department of Health, Public Health Laboratory | Alexandra Lorentz et al |
| EPI_ISL_873229 | USA/MN-MDH-2370/2020 | 2020-12-26 | North America/USA/Minnesota/ | M Health Fairview | Minnesota Department of Health, Public Health Laboratory | Alexandra Lorentz et al |
| EPI_ISL_873239 | USA/MN-MDH-2381/2021 | 2021-01-08 | North America/USA/Minnesota/ | M Health Fairview | Minnesota Department of Health, Public Health Laboratory | Alexandra Lorentz et al |
| EPI_ISL_873242 | USA/MN-MDH-2384/2021 | 2021-01-09 | North America/USA/Minnesota/ | M Health Fairview | Minnesota Department of Health, Public Health Laboratory | Alexandra Lorentz et al |
| EPI_ISL_873248 | USA/MN-MDH-2390/2021 | 2021-01-09 | North America/USA/Minnesota/ | M Health Fairview | Minnesota Department of Health, Public Health Laboratory | Alexandra Lorentz et al |
| EPI_ISL_873254 | USA/MN-MDH-2396/2021 | 2021-01-07 | North America/USA/Minnesota/ | M Health Fairview | Minnesota Department of Health, Public Health Laboratory | Alexandra Lorentz et al |

| | | | | | | |
|---|---|---|---|---|---|---|
| EPI_ISL_873256 | USA/MN-MDH-2398/2021 | 2021-01-08 | North America/USA/Minnesota/ | M Health Fairview | Minnesota Department of Health, Public Health Laboratory | Alexandra Lorentz et al |
| EPI_ISL_873270 | USA/MN-MDH-2412/2021 | 2021-01-15 | North America/USA/Minnesota/ | Minnesota Department of Health, Public Health Laboratory | Minnesota Department of Health, Public Health Laboratory | Alexandra Lorentz et al |
| EPI_ISL_873507 | England/MILK-1027C39/2021 | 2021-01-11 | Europe/United Kingdom/England/ | Lighthouse Lab in Milton Keynes | Wellcome Sanger Institute for the COVID-19 Genomics UK (COG-UK) Consortium | The Lighthouse Lab in Milton Keynes et al |
| EPI_ISL_874293 | England/ALDP-100045C/2021 | 2021-01-12 | Europe/United Kingdom/England/ | Lighthouse Lab in Alderley Park | Wellcome Sanger Institute for the COVID-19 Genomics UK (COG-UK) Consortium | Jacquelyn Wynn et al |
| EPI_ISL_874325 | England/ALDP-100AC3C/2021 | 2021-01-12 | Europe/United Kingdom/England/ | Lighthouse Lab in Alderley Park | Wellcome Sanger Institute for the COVID-19 Genomics UK (COG-UK) Consortium | Jacquelyn Wynn et al |
| EPI_ISL_874424 | England/ALDP-10073B2/2021 | 2021-01-12 | Europe/United Kingdom/England/ | Lighthouse Lab in Alderley Park | Wellcome Sanger Institute for the COVID-19 Genomics UK (COG-UK) Consortium | Jacquelyn Wynn et al |
| EPI_ISL_874632 | England/ALDP-100B08A/2021 | 2021-01-12 | Europe/United Kingdom/England/ | Lighthouse Lab in Alderley Park | Wellcome Sanger Institute for the COVID-19 Genomics UK (COG-UK) Consortium | Jacquelyn Wynn et al |
| EPI_ISL_874655 | England/ALDP-10068F0/2021 | 2021-01-12 | Europe/United Kingdom/England/ | Lighthouse Lab in Alderley Park | Wellcome Sanger Institute for the COVID-19 Genomics UK (COG-UK) Consortium | Jacquelyn Wynn et al |

| | | | | | | |
|---|---|---|---|---|---|---|
| EPI_ISL_874721 | England/ALDP-1004ADE/2021 | 2021-01-12 | Europe/United Kingdom/England/ | Lighthouse Lab in Alderley Park | Wellcome Sanger Institute for the COVID-19 Genomics UK (COG-UK) Consortium | Jacquelyn Wynn et al |
| EPI_ISL_874789 | England/ALDP-100930B/2021 | 2021-01-12 | Europe/United Kingdom/England/ | Lighthouse Lab in Alderley Park | Wellcome Sanger Institute for the COVID-19 Genomics UK (COG-UK) Consortium | Jacquelyn Wynn et al |
| EPI_ISL_874857 | England/ALDP-1006A63/2021 | 2021-01-12 | Europe/United Kingdom/England/ | Lighthouse Lab in Alderley Park | Wellcome Sanger Institute for the COVID-19 Genomics UK (COG-UK) Consortium | Jacquelyn Wynn et al |
| EPI_ISL_875353 | Ireland/D-NVRL-21IRL11645/2021 | 2021-01-12 | Europe/Ireland/Dublin/ | National Virus Reference Laboratory | National Virus Reference Laboratory | Michael Carr et al |
| EPI_ISL_875362 | Ireland/D-NVRL-21IRL11733/2021 | 2021-01-12 | Europe/Ireland/Dublin/ | National Virus Reference Laboratory | National Virus Reference Laboratory | Michael Carr et al |
| EPI_ISL_875386 | Ireland/WW-NVRL-21IRL10474/2021 | 2021-01-11 | Europe/Ireland/Wicklow/ | National Virus Reference Laboratory | National Virus Reference Laboratory | Michael Carr et al |
| EPI_ISL_875406 | Ireland/D-NVRL-21IRL10781/2021 | 2021-01-11 | Europe/Ireland/Dublin/ | National Virus Reference Laboratory | National Virus Reference Laboratory | Michael Carr et al |
| EPI_ISL_875417 | Ireland/D-NVRL-83IRL08558/2021 | 2021-01-10 | Europe/Ireland/Dublin/ | National Virus Reference Laboratory | National Virus Reference Laboratory | Michael Carr et al |
| EPI_ISL_875433 | Ireland/CN-NVRL-83IRL31265/2021 | 2021-01-10 | Europe/Ireland/Cavan/ | National Virus Reference Laboratory | National Virus Reference Laboratory | Michael Carr et al |
| EPI_ISL_875444 | Ireland/LS-NVRL-83IRL08236/2021 | 2021-01-10 | Europe/Ireland/Laois/ | National Virus Reference Laboratory | National Virus Reference Laboratory | Michael Carr et al |
| EPI_ISL_875447 | Ireland/WH-NVRL-83IRL31330/2021 | 2021-01-10 | Europe/Ireland/Westmeath/ | National Virus Reference Laboratory | National Virus Reference Laboratory | Michael Carr et al |

| | | | | | | |
|---|---|---|---|---|---|---|
| EPI_ISL_875453 | Ireland/KE-NVRL-82IRL90786/2021 | 2021-01-08 | Europe/Ireland/Kildare/ | National Virus Reference Laboratory | National Virus Reference Laboratory | Michael Carr et al |
| EPI_ISL_875465 | Ireland/D-NVRL-82IRL79078/2021 | 2021-01-07 | Europe/Ireland/Dublin/ | National Virus Reference Laboratory | National Virus Reference Laboratory | Michael Carr et al |
| EPI_ISL_875475 | Ireland/MN-NVRL-82IRL76438/2021 | 2021-01-07 | Europe/Ireland/Monaghan/ | National Virus Reference Laboratory | National Virus Reference Laboratory | Michael Carr et al |
| EPI_ISL_875476 | Ireland/WW-NVRL-82IRL69986/2021 | 2021-01-07 | Europe/Ireland/Wicklow/ | National Virus Reference Laboratory | National Virus Reference Laboratory | Michael Carr et al |
| EPI_ISL_875494 | Ireland/LK-NVRL-82IRL74696/2021 | 2021-01-07 | Europe/Ireland/Limerick/ | National Virus Reference Laboratory | National Virus Reference Laboratory | Michael Carr et al |
| EPI_ISL_875512 | Ireland/KE-NVRL-82IRL10797/2021 | 2021-01-03 | Europe/Ireland/Kildare/ | National Virus Reference Laboratory | National Virus Reference Laboratory | Michael Carr et al |
| EPI_ISL_875515 | Russia/MOW_PMVL-29/2020 | 2020-08-26 | Europe/Russia/Moscow Oblast/ | Influenza etiology and epidemiology laboratory | Pathogenic Microorganisms Variability Laboratory | Alexey Shchetinin et al |
| EPI_ISL_875516 | Slovakia/UKBA-715/2020 | 2020-12-30 | Europe/Slovakia/Partizanske/ | Institute of Virology, Biomedical Research Center of the Slovak Academy of Sciences, Bratislava | Faculty of Natural Sciences, Comenius University, Bratislava | Krist√≠na Bor≈°ov√° et al |
| EPI_ISL_875530 | Slovakia/UKBA-701/2021 | 2021-01-07 | Europe/Slovakia/Nitra/ | Institute of Virology, Biomedical Research Center of the Slovak Academy of Sciences, Bratislava | Faculty of Natural Sciences, Comenius University, Bratislava | Vikt√≥ria ƒåabanov√° et al |
| EPI_ISL_875531 | Slovakia/UKBA-712/2020 | 2020-11-11 | Europe/Slovakia/Nove Zamky/ | Institute of Virology, Biomedical Research Center of the Slovak Academy of Sciences, Bratislava | Faculty of Natural Sciences, Comenius University, Bratislava | Vikt√≥ria ƒåabanov√° et al |

| | | | | | | |
|---|---|---|---|---|---|---|
| EPI_ISL_875532 | Slovakia/UKBA-723/2021 | 2021-01-08 | Europe/Slovakia/Banovce nad Bebravou | Institute of Virology, Biomedical Research Center of the Slovak Academy of Sciences, Bratislava | Faculty of Natural Sciences, Comenius University, Bratislava | Bro≈àa Brejov√° et al |
| EPI_ISL_875533 | Slovakia/UKBA-716/2021 | 2021-01-04 | Europe/Slovakia/Nove Mesto nad Vaho | Institute of Virology, Biomedical Research Center of the Slovak Academy of Sciences, Bratislava | Faculty of Natural Sciences, Comenius University, Bratislava | Vikt√≥ria ≈†abanov√° et al |
| EPI_ISL_875534 | Slovakia/UKBA-717/2021 | 2021-01-04 | Europe/Slovakia/Nove Mesto nad Vaho | Institute of Virology, Biomedical Research Center of the Slovak Academy of Sciences, Bratislava | Faculty of Natural Sciences, Comenius University, Bratislava | Krist√≠na Bor≈°ov√° et al |
| EPI_ISL_875536 | Slovakia/UKBA-710/2020 | 2020-11-11 | Europe/Slovakia/Nove Zamky/ | Institute of Virology, Biomedical Research Center of the Slovak Academy of Sciences, Bratislava | Faculty of Natural Sciences, Comenius University, Bratislava | Vikt√≥ria ≈†abanov√° et al |
| EPI_ISL_875537 | Slovakia/UKBA-711/2020 | 2020-11-11 | Europe/Slovakia/Nove Zamky/ | Institute of Virology, Biomedical Research Center of the Slovak Academy of Sciences, Bratislava | Faculty of Natural Sciences, Comenius University, Bratislava | Krist√≠na Bor≈°ov√° et al |
| EPI_ISL_875538 | Slovakia/UKBA-724/2020 | 2020-10-10 | Europe/Slovakia/Oravska Lesna/ | Institute of Virology, Biomedical Research Center of the Slovak Academy of Sciences, Bratislava | Faculty of Natural Sciences, Comenius University, Bratislava | Vikt√≥ria ≈†abanov√° et al |
| EPI_ISL_875540 | Brazil/SP-BT6815/2020 | 2020-07-09 | South America/Brazil/S√£o Paulo/Sao Jo | Instituto de Biotecnologia - UNESP-Botucatu-SP | Instituto de Biotecnologia - UNESP-Botucatu-SP | Leila Sabrina Ullmann et al |
| EPI_ISL_875542 | Brazil/SP-BT6810/2020 | 2020-08-09 | South America/Brazil/S√£o Paulo/Sao Jo | Instituto de Biotecnologia - UNESP-Botucatu-SP | Instituto de Biotecnologia - UNESP-Botucatu-SP | Leila Sabrina Ullmann et al |

| | | | | | | |
|---|---|---|---|---|---|---|
| EPI_ISL_875543 | Brazil/SP-BT6798/2020 | 2020-05-09 | South America/Brazil/S√£o Paulo/Sao Jo | Instituto de Biotecnologia - UNESP-Botucatu-SP | Instituto de Biotecnologia - UNESP-Botucatu-SP | Leila Sabrina Ullmann et al |
| EPI_ISL_875546 | Brazil/SP-BT6808/2020 | 2020-08-09 | South America/Brazil/S√£o Paulo/Sao Jo | Instituto de Biotecnologia - UNESP-Botucatu-SP | Instituto de Biotecnologia - UNESP-Botucatu-SP | Leila Sabrina Ullmann et al |
| EPI_ISL_875547 | Brazil/SP-BT6816/2020 | 2020-08-09 | South America/Brazil/S√£o Paulo/Sao Jo | Instituto de Biotecnologia - UNESP-Botucatu-SP | Instituto de Biotecnologia - UNESP-Botucatu-SP | Leila Sabrina Ullmann et al |
| EPI_ISL_875548 | Brazil/SP-BT6674/2020 | 2020-05-09 | South America/Brazil/S√£o Paulo/Sao Jo | Instituto de Biotecnologia - UNESP-Botucatu-SP | Instituto de Biotecnologia - UNESP-Botucatu-SP | Leila Sabrina Ullmann et al |
| EPI_ISL_875549 | Brazil/SP-BT6799/2020 | 2020-05-09 | South America/Brazil/S√£o Paulo/Sao Jo | Instituto de Biotecnologia - UNESP-Botucatu-SP | Instituto de Biotecnologia - UNESP-Botucatu-SP | Leila Sabrina Ullmann et al |
| EPI_ISL_875551 | Italy/VEN-IZSVe-21RS21-1_BL/2020 | 2020-12-22 | Europe/Italy/Veneto/Belluno | ULSS 1 Dolomiti | Istituto Zooprofilattico Sperimentale delle Venezie | Adelaide Milani et al |
| EPI_ISL_875552 | Italy/VEN-IZSVe-21RS22-3_TV/2020 | 2020-11-10 | Europe/Italy/Veneto/Treviso | ULSS 2 Treviso | Istituto Zooprofilattico Sperimentale delle Venezie | Adelaide Milani et al |
| EPI_ISL_875554 | Italy/VEN-IZSVe-21RS22-7_TV/2020 | 2020-11-28 | Europe/Italy/Veneto/Treviso | ULSS 2 Treviso | Istituto Zooprofilattico Sperimentale delle Venezie | Adelaide Milani et al |
| EPI_ISL_875558 | Italy/VEN-IZSVe-21RS23_PD/2021 | 2021-01-12 | Europe/Italy/Veneto/Padova | Microbiologia e Virologia | Istituto Zooprofilattico Sperimentale delle Venezie | Adelaide Milani et al |
| EPI_ISL_875571 | USA/AL-Madison-JDXZ/2020 | 2020-10-27 | North America/USA/Alabama/Madison | Eurofins Diatherix | Hudsonalpha Genome Sequencing Center | Jane Grimwood et al |

| | | | | | | |
|---|---|---|---|---|---|---|
| EPI_ISL_875578 | USA/AL-Jefferson-JDZY/2020 | 2020-10-28 | North America/USA/Alabama/Jefferson | Eurofins Diatherix | Hudsonalpha Genome Sequencing Center | Jane Grimwood et al |
| EPI_ISL_875588 | USA/AL-Madison-JDYI/2020 | 2020-10-27 | North America/USA/Alabama/Madison | Eurofins Diatherix | Hudsonalpha Genome Sequencing Center | Jane Grimwood et al |
| EPI_ISL_875589 | USA/AL-Madison-JDYK/2020 | 2020-10-27 | North America/USA/Alabama/Madison | Eurofins Diatherix | Hudsonalpha Genome Sequencing Center | Jane Grimwood et al |
| EPI_ISL_875590 | USA/AL-Escambia-JDYM/2020 | 2020-10-27 | North America/USA/Alabama/Escambia | Eurofins Diatherix | Hudsonalpha Genome Sequencing Center | Jane Grimwood et al |
| EPI_ISL_875598 | USA/AL-Hale-JEAC/2020 | 2020-10-28 | North America/USA/Alabama/Hale Cou | Eurofins Diatherix | Hudsonalpha Genome Sequencing Center | Jane Grimwood et al |
| EPI_ISL_875601 | USA/AL-Randolph-JEAQ/2020 | 2020-10-28 | North America/USA/Alabama/Randolph | Eurofins Diatherix | Hudsonalpha Genome Sequencing Center | Jane Grimwood et al |
| EPI_ISL_875602 | USA/AL-Randolph-JEAU/2020 | 2020-10-28 | North America/USA/Alabama/Randolph | Eurofins Diatherix | Hudsonalpha Genome Sequencing Center | Jane Grimwood et al |
| EPI_ISL_875603 | USA/AL-Etowah-JEBC/2020 | 2020-10-29 | North America/USA/Alabama/Etowah C | Eurofins Diatherix | Hudsonalpha Genome Sequencing Center | Jane Grimwood et al |
| EPI_ISL_875606 | USA/AL-Madison-JEBF/2020 | 2020-10-29 | North America/USA/Alabama/Madison | Eurofins Diatherix | Hudsonalpha Genome Sequencing Center | Jane Grimwood et al |
| EPI_ISL_875613 | USA/AL-Shelby-JDZX/2020 | 2020-10-28 | North America/USA/Alabama/Shelby C | Eurofins Diatherix | Hudsonalpha Genome Sequencing Center | Jane Grimwood et al |

| | | | | | | |
|---|---|---|---|---|---|---|
| EPI_ISL_875619 | USA/AL-Madison-JDXY/2020 | 2020-10-27 | North America/USA/Alabama/Madison | Eurofins Diatherix | Hudsonalpha Genome Sequencing Center | Jane Grimwood et al |
| EPI_ISL_875620 | USA/AL-Madison-JDYD/2020 | 2020-10-27 | North America/USA/Alabama/Madison | Eurofins Diatherix | Hudsonalpha Genome Sequencing Center | Jane Grimwood et al |
| EPI_ISL_875627 | USA/AL-Lawrence-JDZR/2020 | 2020-10-28 | North America/USA/Alabama/Lawrence | Eurofins Diatherix | Hudsonalpha Genome Sequencing Center | Jane Grimwood et al |
| EPI_ISL_875637 | USA/AL-Morgan-JEBH/2020 | 2020-10-29 | North America/USA/Alabama/Morgan ( | Eurofins Diatherix | Hudsonalpha Genome Sequencing Center | Jane Grimwood et al |
| EPI_ISL_875648 | USA/AL-Morgan-JDZJ/2020 | 2020-10-27 | North America/USA/Alabama/Morgan ( | Eurofins Diatherix | Hudsonalpha Genome Sequencing Center | Jane Grimwood et al |
| EPI_ISL_875663 | USA/MI-UM-10037746750/2021 | 2021-01-08 | North America/USA/Michigan/ | University of Michigan Clinical Microbiology Laboratory | Lauring Lab, University of Michigan, Department of Microbiology and Immunology | Valesano et al |
| EPI_ISL_875664 | USA/MI-UM-10037767457/2021 | 2021-01-11 | North America/USA/Michigan/ | University of Michigan Clinical Microbiology Laboratory | Lauring Lab, University of Michigan, Department of Microbiology and Immunology | Valesano et al |
| EPI_ISL_875672 | France/OCC-248/2021 | 2021-01-18 | Europe/France/Occitanie/Toulouse | CHU Purpan - Laboratoire de Virologie - Institut F√©d√©ratif de Biologie | CHU Purpan - Laboratoire de Virologie - Institut F√©d√©ratif de Biologie | Latour J. et al |

| | | | | | | |
|---|---|---|---|---|---|---|
| EPI_ISL_875674 | Italy/CAM-CRGS-1/2020 | 2020-03-14 | Europe/Italy/Campania/ | """1.AO Universitaria 'S. Giovanni di Dio e Ruggi D'Aragona, Scuola Medica Salernitana' Hospital / 2.UOC di Virologia e Microbiologia, Universit√† della Campania 'L. Vanvitelli' / 3.AO Universitaria 'Federico II' Napoli Hospital / 4.AORN 'San Giuseppe Moscati' Avellino Hospital / 5.AO 'San Pio - presidio G. Rummo' Benevento Hospital / 6.AO 'Sant'Anna e San Sebastiano' Caserta Hospital / 7.PO 'Maria Santissima Addolorata' Eboli Hospital / 8.Biogem Istituto di Ricerche Genetiche""" | """1. Genome Research Center for Health (CRGS) / 2. Laboratory of Molecular Medicine and Genomics(LMMGe) / 3. Center for Research in Pure and Applied Mathematics (CRMPA)""" | """Giorgio Giurato (Corresponding Author) et al" |
| EPI_ISL_875681 | USA/KS-KHEL-0007/2020 | 2020-03-18 | North America/USA/Kansas/ | Kansas Health and Environmental Lab | Kansas Health and Environmental Lab | Mike Grose et al |
| EPI_ISL_875688 | Brazil/SP-999/2021 | 2021-01-04 | South America/Brazil/S√£o Paulo/S√£o | National Influenza Center - Instituto Adolfo Lutz | Instituto Adolfo Lutz, Interdiciplinary Procedures Center, Strategic Laboratory | Claudio Tavares Sacchi et al |
| EPI_ISL_875689 | Brazil/SP-1014/2021 | 2021-01-15 | South America/Brazil/S√£o Paulo/S√£o | Hospital do Servidor Publico | Instituto Adolfo Lutz, Interdiciplinary Procedures Center, Strategic Laboratory | Claudio Tavares Sacchi et al |
| EPI_ISL_875691 | Canada/ON-PHL-21-00036/2020 | 20-Dec | North America/Canada/Ontario/ | Public Health Ontario Laboratory | Public Health Ontario Laboratory | Vanessa G Allen et al |
| EPI_ISL_875692 | Canada/ON-PHL-21-00026/2020 | 20-Dec | North America/Canada/Ontario/ | Public Health Ontario Laboratory | Public Health Ontario Laboratory | Vanessa G Allen et al |
| EPI_ISL_875693 | Canada/ON-PHL-21-00038/2020 | 20-Dec | North America/Canada/Ontario/ | Public Health Ontario Laboratory | Public Health Ontario Laboratory | Vanessa G Allen et al |
| EPI_ISL_875694 | Canada/ON-PHL-21-00068/2020 | 20-Dec | North America/Canada/Ontario/ | Public Health Ontario Laboratory | Public Health Ontario Laboratory | Vanessa G Allen et al |

| | | | | | | |
|---|---|---|---|---|---|---|
| EPI_ISL_875695 | Canada/ON-PHL-21-00043/2020 | 20-Dec | North America/Canada/Ontario/ | Public Health Ontario Laboratory | Public Health Ontario Laboratory | Vanessa G Allen et al |
| EPI_ISL_875696 | Canada/ON-PHL-21-00045/2020 | 20-Dec | North America/Canada/Ontario/ | Public Health Ontario Laboratory | Public Health Ontario Laboratory | Vanessa G Allen et al |
| EPI_ISL_875697 | Canada/ON-PHL-21-00216/2020 | 20-Dec | North America/Canada/Ontario/ | Public Health Ontario Laboratory | Public Health Ontario Laboratory | Vanessa G Allen et al |
| EPI_ISL_875698 | Canada/ON-PHL-21-00063/2020 | 20-Dec | North America/Canada/Ontario/ | Public Health Ontario Laboratory | Public Health Ontario Laboratory | Vanessa G Allen et al |
| EPI_ISL_875699 | Canada/ON-PHL-21-00064/2020 | 20-Dec | North America/Canada/Ontario/ | Public Health Ontario Laboratory | Public Health Ontario Laboratory | Vanessa G Allen et al |
| EPI_ISL_875700 | Canada/ON-PHL-21-00261/2020 | 20-Dec | North America/Canada/Ontario/ | Public Health Ontario Laboratory | Public Health Ontario Laboratory | Vanessa G Allen et al |
| EPI_ISL_875701 | Canada/ON-PHL-21-00008/2020 | 20-Dec | North America/Canada/Ontario/ | Public Health Ontario Laboratory | Public Health Ontario Laboratory | Vanessa G Allen et al |
| EPI_ISL_875702 | Canada/ON-PHL-21-00282/2020 | 20-Dec | North America/Canada/Ontario/ | Public Health Ontario Laboratory | Public Health Ontario Laboratory | Vanessa G Allen et al |
| EPI_ISL_875703 | Canada/ON-PHL-21-00186/2020 | 20-Dec | North America/Canada/Ontario/ | Public Health Ontario Laboratory | Public Health Ontario Laboratory | Vanessa G Allen et al |
| EPI_ISL_875704 | Canada/ON-PHL-21-00058/2020 | 20-Dec | North America/Canada/Ontario/ | Public Health Ontario Laboratory | Public Health Ontario Laboratory | Vanessa G Allen et al |
| EPI_ISL_875705 | Canada/ON-PHL-21-00069/2020 | 20-Dec | North America/Canada/Ontario/ | Public Health Ontario Laboratory | Public Health Ontario Laboratory | Vanessa G Allen et al |
| EPI_ISL_875706 | Canada/ON-PHL-21-00238/2020 | 20-Dec | North America/Canada/Ontario/ | Public Health Ontario Laboratory | Public Health Ontario Laboratory | Vanessa G Allen et al |
| EPI_ISL_875707 | Canada/ON-PHL-21-00057/2020 | 20-Dec | North America/Canada/Ontario/ | Public Health Ontario Laboratory | Public Health Ontario Laboratory | Vanessa G Allen et al |

| | | | | | | |
|---|---|---|---|---|---|---|
| EPI_ISL_875708 | Canada/ON-PHL-21-00266/2020 | 20-Dec | North America/Canada/Ontario/ | Public Health Ontario Laboratory | Public Health Ontario Laboratory | Vanessa G Allen et al |
| EPI_ISL_875709 | Canada/ON-PHL-21-00267/2020 | 20-Dec | North America/Canada/Ontario/ | Public Health Ontario Laboratory | Public Health Ontario Laboratory | Vanessa G Allen et al |
| EPI_ISL_875710 | Canada/ON-PHL-21-00265/2020 | 20-Dec | North America/Canada/Ontario/ | Public Health Ontario Laboratory | Public Health Ontario Laboratory | Vanessa G Allen et al |
| EPI_ISL_875711 | Canada/ON-PHL-21-00056/2020 | 20-Dec | North America/Canada/Ontario/ | Public Health Ontario Laboratory | Public Health Ontario Laboratory | Vanessa G Allen et al |
| EPI_ISL_875712 | Canada/ON-PHL-21-00314/2020 | 20-Dec | North America/Canada/Ontario/ | Public Health Ontario Laboratory | Public Health Ontario Laboratory | Vanessa G Allen et al |
| EPI_ISL_875713 | Canada/ON-PHL-21-00075/2020 | 20-Dec | North America/Canada/Ontario/ | Public Health Ontario Laboratory | Public Health Ontario Laboratory | Vanessa G Allen et al |
| EPI_ISL_875714 | Canada/ON-PHL-21-00074/2020 | 20-Dec | North America/Canada/Ontario/ | Public Health Ontario Laboratory | Public Health Ontario Laboratory | Vanessa G Allen et al |
| EPI_ISL_875715 | Canada/ON-PHL-21-00049/2020 | 20-Dec | North America/Canada/Ontario/ | Public Health Ontario Laboratory | Public Health Ontario Laboratory | Vanessa G Allen et al |
| EPI_ISL_875716 | Canada/ON-PHL-21-00029/2020 | 20-Dec | North America/Canada/Ontario/ | Public Health Ontario Laboratory | Public Health Ontario Laboratory | Vanessa G Allen et al |
| EPI_ISL_875717 | Canada/ON-PHL-21-00051/2020 | 20-Dec | North America/Canada/Ontario/ | Public Health Ontario Laboratory | Public Health Ontario Laboratory | Vanessa G Allen et al |
| EPI_ISL_875718 | Canada/ON-PHL-21-00065/2020 | 20-Dec | North America/Canada/Ontario/ | Public Health Ontario Laboratory | Public Health Ontario Laboratory | Vanessa G Allen et al |
| EPI_ISL_875719 | Canada/ON-PHL-21-00066/2020 | 20-Dec | North America/Canada/Ontario/ | Public Health Ontario Laboratory | Public Health Ontario Laboratory | Vanessa G Allen et al |
| EPI_ISL_875720 | Canada/ON-PHL-21-00067/2020 | 20-Dec | North America/Canada/Ontario/ | Public Health Ontario Laboratory | Public Health Ontario Laboratory | Vanessa G Allen et al |

| | | | | | | |
|---|---|---|---|---|---|---|
| EPI_ISL_875721 | Canada/ON-PHL-21-00031/2020 | 20-Dec | North America/Canada/Ontario/ | Public Health Ontario Laboratory | Public Health Ontario Laboratory | Vanessa G Allen et al |
| EPI_ISL_875722 | Canada/ON-PHL-21-00032/2020 | 20-Dec | North America/Canada/Ontario/ | Public Health Ontario Laboratory | Public Health Ontario Laboratory | Vanessa G Allen et al |
| EPI_ISL_875723 | Canada/ON-PHL-21-00022/2020 | 20-Dec | North America/Canada/Ontario/ | Public Health Ontario Laboratory | Public Health Ontario Laboratory | Vanessa G Allen et al |
| EPI_ISL_875724 | Canada/ON-PHL-21-00023/2020 | 20-Dec | North America/Canada/Ontario/ | Public Health Ontario Laboratory | Public Health Ontario Laboratory | Vanessa G Allen et al |
| EPI_ISL_875725 | Canada/ON-PHL-21-00030/2020 | 20-Dec | North America/Canada/Ontario/ | Public Health Ontario Laboratory | Public Health Ontario Laboratory | Vanessa G Allen et al |
| EPI_ISL_875726 | Canada/ON-PHL-21-00033/2020 | 20-Dec | North America/Canada/Ontario/ | Public Health Ontario Laboratory | Public Health Ontario Laboratory | Vanessa G Allen et al |
| EPI_ISL_875727 | Canada/ON-PHL-21-00034/2020 | 20-Dec | North America/Canada/Ontario/ | Public Health Ontario Laboratory | Public Health Ontario Laboratory | Vanessa G Allen et al |
| EPI_ISL_875728 | Canada/ON-PHL-21-00037/2020 | 20-Dec | North America/Canada/Ontario/ | Public Health Ontario Laboratory | Public Health Ontario Laboratory | Vanessa G Allen et al |
| EPI_ISL_875729 | Canada/ON-PHL-21-00039/2020 | 20-Dec | North America/Canada/Ontario/ | Public Health Ontario Laboratory | Public Health Ontario Laboratory | Vanessa G Allen et al |
| EPI_ISL_875730 | Canada/ON-PHL-21-00040/2020 | 20-Dec | North America/Canada/Ontario/ | Public Health Ontario Laboratory | Public Health Ontario Laboratory | Vanessa G Allen et al |
| EPI_ISL_875731 | Canada/ON-PHL-21-00024/2020 | 20-Dec | North America/Canada/Ontario/ | Public Health Ontario Laboratory | Public Health Ontario Laboratory | Vanessa G Allen et al |
| EPI_ISL_875732 | Canada/ON-PHL-21-00021/2020 | 20-Dec | North America/Canada/Ontario/ | Public Health Ontario Laboratory | Public Health Ontario Laboratory | Vanessa G Allen et al |
| EPI_ISL_875733 | Canada/ON-PHL-21-00044/2020 | 20-Dec | North America/Canada/Ontario/ | Public Health Ontario Laboratory | Public Health Ontario Laboratory | Vanessa G Allen et al |

| | | | | | | |
|---|---|---|---|---|---|---|
| EPI_ISL_875734 | Canada/ON-PHL-21-00027/2020 | 20-Dec | North America/Canada/Ontario/ | Public Health Ontario Laboratory | Public Health Ontario Laboratory | Vanessa G Allen et al |
| EPI_ISL_875735 | Canada/ON-PHL-21-00072/2020 | 20-Dec | North America/Canada/Ontario/ | Public Health Ontario Laboratory | Public Health Ontario Laboratory | Vanessa G Allen et al |
| EPI_ISL_875736 | Canada/ON-PHL-21-00041/2020 | 20-Dec | North America/Canada/Ontario/ | Public Health Ontario Laboratory | Public Health Ontario Laboratory | Vanessa G Allen et al |
| EPI_ISL_875737 | Canada/ON-PHL-21-00042/2020 | 20-Dec | North America/Canada/Ontario/ | Public Health Ontario Laboratory | Public Health Ontario Laboratory | Vanessa G Allen et al |
| EPI_ISL_875738 | Canada/ON-PHL-21-00062/2020 | 20-Dec | North America/Canada/Ontario/ | Public Health Ontario Laboratory | Public Health Ontario Laboratory | Vanessa G Allen et al |
| EPI_ISL_875739 | Canada/ON-PHL-21-00071/2020 | 20-Dec | North America/Canada/Ontario/ | Public Health Ontario Laboratory | Public Health Ontario Laboratory | Vanessa G Allen et al |
| EPI_ISL_875740 | Canada/ON-PHL-21-00170/2020 | 20-Dec | North America/Canada/Ontario/ | Public Health Ontario Laboratory | Public Health Ontario Laboratory | Vanessa G Allen et al |
| EPI_ISL_875741 | Canada/ON-PHL-21-00253/2020 | 20-Dec | North America/Canada/Ontario/ | Public Health Ontario Laboratory | Public Health Ontario Laboratory | Vanessa G Allen et al |
| EPI_ISL_875742 | Canada/ON-PHL-21-00269/2020 | 20-Dec | North America/Canada/Ontario/ | Public Health Ontario Laboratory | Public Health Ontario Laboratory | Vanessa G Allen et al |
| EPI_ISL_875743 | Canada/ON-PHL-21-00254/2020 | 20-Dec | North America/Canada/Ontario/ | Public Health Ontario Laboratory | Public Health Ontario Laboratory | Vanessa G Allen et al |
| EPI_ISL_875744 | Canada/ON-PHL-21-00046/2020 | 20-Dec | North America/Canada/Ontario/ | Public Health Ontario Laboratory | Public Health Ontario Laboratory | Vanessa G Allen et al |
| EPI_ISL_875745 | Canada/ON-PHL-21-00246/2020 | 20-Dec | North America/Canada/Ontario/ | Public Health Ontario Laboratory | Public Health Ontario Laboratory | Vanessa G Allen et al |
| EPI_ISL_875746 | Canada/ON-PHL-21-00271/2020 | 20-Dec | North America/Canada/Ontario/ | Public Health Ontario Laboratory | Public Health Ontario Laboratory | Vanessa G Allen et al |

| | | | | | | |
|---|---|---|---|---|---|---|
| EPI_ISL_875747 | Canada/ON-PHL-21-00178/2020 | 20-Dec | North America/Canada/Ontario/ | Public Health Ontario Laboratory | Public Health Ontario Laboratory | Vanessa G Allen et al |
| EPI_ISL_875748 | Canada/ON-PHL-21-00172/2020 | 20-Dec | North America/Canada/Ontario/ | Public Health Ontario Laboratory | Public Health Ontario Laboratory | Vanessa G Allen et al |
| EPI_ISL_875749 | Canada/ON-PHL-21-00262/2020 | 20-Dec | North America/Canada/Ontario/ | Public Health Ontario Laboratory | Public Health Ontario Laboratory | Vanessa G Allen et al |
| EPI_ISL_875750 | Canada/ON-PHL-21-00177/2020 | 20-Dec | North America/Canada/Ontario/ | Public Health Ontario Laboratory | Public Health Ontario Laboratory | Vanessa G Allen et al |
| EPI_ISL_875751 | Canada/ON-PHL-21-00171/2020 | 20-Dec | North America/Canada/Ontario/ | Public Health Ontario Laboratory | Public Health Ontario Laboratory | Vanessa G Allen et al |
| EPI_ISL_875752 | Canada/ON-PHL-21-00174/2020 | 20-Dec | North America/Canada/Ontario/ | Public Health Ontario Laboratory | Public Health Ontario Laboratory | Vanessa G Allen et al |
| EPI_ISL_875753 | Canada/ON-PHL-21-00241/2020 | 20-Dec | North America/Canada/Ontario/ | Public Health Ontario Laboratory | Public Health Ontario Laboratory | Vanessa G Allen et al |
| EPI_ISL_875754 | Canada/ON-PHL-21-00272/2020 | 20-Dec | North America/Canada/Ontario/ | Public Health Ontario Laboratory | Public Health Ontario Laboratory | Vanessa G Allen et al |
| EPI_ISL_875755 | Canada/ON-PHL-21-00270/2020 | 20-Dec | North America/Canada/Ontario/ | Public Health Ontario Laboratory | Public Health Ontario Laboratory | Vanessa G Allen et al |
| EPI_ISL_875756 | Canada/ON-PHL-21-00183/2020 | 20-Dec | North America/Canada/Ontario/ | Public Health Ontario Laboratory | Public Health Ontario Laboratory | Vanessa G Allen et al |
| EPI_ISL_875757 | Canada/ON-PHL-21-00175/2020 | 20-Dec | North America/Canada/Ontario/ | Public Health Ontario Laboratory | Public Health Ontario Laboratory | Vanessa G Allen et al |
| EPI_ISL_875758 | Canada/ON-PHL-21-00176/2020 | 20-Dec | North America/Canada/Ontario/ | Public Health Ontario Laboratory | Public Health Ontario Laboratory | Vanessa G Allen et al |
| EPI_ISL_875759 | Canada/ON-PHL-21-00217/2020 | 20-Dec | North America/Canada/Ontario/ | Public Health Ontario Laboratory | Public Health Ontario Laboratory | Vanessa G Allen et al |

| | | | | | | |
|---|---|---|---|---|---|---|
| EPI_ISL_875760 | Canada/ON-PHL-21-00222/2020 | 20-Dec | North America/Canada/Ontario/ | Public Health Ontario Laboratory | Public Health Ontario Laboratory | Vanessa G Allen et al |
| EPI_ISL_875761 | Canada/ON-PHL-21-00223/2020 | 20-Dec | North America/Canada/Ontario/ | Public Health Ontario Laboratory | Public Health Ontario Laboratory | Vanessa G Allen et al |
| EPI_ISL_875762 | Canada/ON-PHL-21-00219/2020 | 20-Dec | North America/Canada/Ontario/ | Public Health Ontario Laboratory | Public Health Ontario Laboratory | Vanessa G Allen et al |
| EPI_ISL_875763 | Canada/ON-PHL-21-00180/2020 | 20-Dec | North America/Canada/Ontario/ | Public Health Ontario Laboratory | Public Health Ontario Laboratory | Vanessa G Allen et al |
| EPI_ISL_875764 | Canada/ON-PHL-21-00229/2020 | 20-Dec | North America/Canada/Ontario/ | Public Health Ontario Laboratory | Public Health Ontario Laboratory | Vanessa G Allen et al |
| EPI_ISL_875765 | Canada/ON-PHL-21-00227/2020 | 20-Dec | North America/Canada/Ontario/ | Public Health Ontario Laboratory | Public Health Ontario Laboratory | Vanessa G Allen et al |
| EPI_ISL_875766 | Canada/ON-PHL-21-00226/2020 | 20-Dec | North America/Canada/Ontario/ | Public Health Ontario Laboratory | Public Health Ontario Laboratory | Vanessa G Allen et al |
| EPI_ISL_875767 | Canada/ON-PHL-21-00224/2020 | 20-Dec | North America/Canada/Ontario/ | Public Health Ontario Laboratory | Public Health Ontario Laboratory | Vanessa G Allen et al |
| EPI_ISL_875768 | Canada/ON-PHL-21-00236/2020 | 20-Dec | North America/Canada/Ontario/ | Public Health Ontario Laboratory | Public Health Ontario Laboratory | Vanessa G Allen et al |
| EPI_ISL_875769 | Canada/ON-PHL-21-00235/2020 | 20-Dec | North America/Canada/Ontario/ | Public Health Ontario Laboratory | Public Health Ontario Laboratory | Vanessa G Allen et al |
| EPI_ISL_875770 | Canada/ON-PHL-21-00234/2020 | 20-Dec | North America/Canada/Ontario/ | Public Health Ontario Laboratory | Public Health Ontario Laboratory | Vanessa G Allen et al |
| EPI_ISL_875771 | Canada/ON-PHL-21-00233/2020 | 20-Dec | North America/Canada/Ontario/ | Public Health Ontario Laboratory | Public Health Ontario Laboratory | Vanessa G Allen et al |
| EPI_ISL_875772 | Canada/ON-PHL-21-00221/2020 | 20-Dec | North America/Canada/Ontario/ | Public Health Ontario Laboratory | Public Health Ontario Laboratory | Vanessa G Allen et al |

| | | | | | | |
|---|---|---|---|---|---|---|
| EPI_ISL_875773 | Canada/ON-PHL-21-00240/2020 | 20-Dec | North America/Canada/Ontario/ | Public Health Ontario Laboratory | Public Health Ontario Laboratory | Vanessa G Allen et al |
| EPI_ISL_875774 | Canada/ON-PHL-21-00239/2020 | 20-Dec | North America/Canada/Ontario/ | Public Health Ontario Laboratory | Public Health Ontario Laboratory | Vanessa G Allen et al |
| EPI_ISL_875775 | Canada/ON-PHL-21-00304/2020 | 20-Dec | North America/Canada/Ontario/ | Public Health Ontario Laboratory | Public Health Ontario Laboratory | Vanessa G Allen et al |
| EPI_ISL_875776 | Canada/ON-PHL-21-00303/2020 | 20-Dec | North America/Canada/Ontario/ | Public Health Ontario Laboratory | Public Health Ontario Laboratory | Vanessa G Allen et al |
| EPI_ISL_875777 | Canada/ON-PHL-21-00194/2020 | 20-Dec | North America/Canada/Ontario/ | Public Health Ontario Laboratory | Public Health Ontario Laboratory | Vanessa G Allen et al |
| EPI_ISL_875778 | Canada/ON-PHL-21-00195/2020 | 20-Dec | North America/Canada/Ontario/ | Public Health Ontario Laboratory | Public Health Ontario Laboratory | Vanessa G Allen et al |
| EPI_ISL_875779 | Canada/ON-PHL-21-00288/2020 | 20-Dec | North America/Canada/Ontario/ | Public Health Ontario Laboratory | Public Health Ontario Laboratory | Vanessa G Allen et al |
| EPI_ISL_875780 | Canada/ON-PHL-21-00289/2020 | 20-Dec | North America/Canada/Ontario/ | Public Health Ontario Laboratory | Public Health Ontario Laboratory | Vanessa G Allen et al |
| EPI_ISL_875781 | Canada/ON-PHL-21-00182/2020 | 20-Dec | North America/Canada/Ontario/ | Public Health Ontario Laboratory | Public Health Ontario Laboratory | Vanessa G Allen et al |
| EPI_ISL_875782 | Canada/ON-PHL-21-00200/2020 | 20-Dec | North America/Canada/Ontario/ | Public Health Ontario Laboratory | Public Health Ontario Laboratory | Vanessa G Allen et al |
| EPI_ISL_875783 | Canada/ON-PHL-21-00187/2020 | 20-Dec | North America/Canada/Ontario/ | Public Health Ontario Laboratory | Public Health Ontario Laboratory | Vanessa G Allen et al |
| EPI_ISL_875784 | Canada/ON-PHL-21-00188/2020 | 20-Dec | North America/Canada/Ontario/ | Public Health Ontario Laboratory | Public Health Ontario Laboratory | Vanessa G Allen et al |
| EPI_ISL_875785 | Canada/ON-PHL-21-00287/2020 | 20-Dec | North America/Canada/Ontario/ | Public Health Ontario Laboratory | Public Health Ontario Laboratory | Vanessa G Allen et al |

| | | | | | | |
|---|---|---|---|---|---|---|
| EPI_ISL_875786 | Canada/ON-PHL-21-00285/2020 | 20-Dec | North America/Canada/Ontario/ | Public Health Ontario Laboratory | Public Health Ontario Laboratory | Vanessa G Allen et al |
| EPI_ISL_875787 | Canada/ON-PHL-21-00006/2020 | 20-Dec | North America/Canada/Ontario/ | Public Health Ontario Laboratory | Public Health Ontario Laboratory | Vanessa G Allen et al |
| EPI_ISL_875788 | Canada/ON-PHL-21-00005/2020 | 20-Dec | North America/Canada/Ontario/ | Public Health Ontario Laboratory | Public Health Ontario Laboratory | Vanessa G Allen et al |
| EPI_ISL_875789 | Canada/ON-PHL-21-00007/2020 | 20-Dec | North America/Canada/Ontario/ | Public Health Ontario Laboratory | Public Health Ontario Laboratory | Vanessa G Allen et al |
| EPI_ISL_875790 | Canada/ON-PHL-21-00004/2020 | 20-Dec | North America/Canada/Ontario/ | Public Health Ontario Laboratory | Public Health Ontario Laboratory | Vanessa G Allen et al |
| EPI_ISL_875791 | Canada/ON-PHL-21-00009/2020 | 20-Dec | North America/Canada/Ontario/ | Public Health Ontario Laboratory | Public Health Ontario Laboratory | Vanessa G Allen et al |
| EPI_ISL_875792 | Canada/ON-PHL-21-00010/2020 | 20-Dec | North America/Canada/Ontario/ | Public Health Ontario Laboratory | Public Health Ontario Laboratory | Vanessa G Allen et al |
| EPI_ISL_875793 | Canada/ON-PHL-21-00016/2020 | 20-Dec | North America/Canada/Ontario/ | Public Health Ontario Laboratory | Public Health Ontario Laboratory | Vanessa G Allen et al |
| EPI_ISL_875794 | Canada/ON-PHL-21-00017/2020 | 20-Dec | North America/Canada/Ontario/ | Public Health Ontario Laboratory | Public Health Ontario Laboratory | Vanessa G Allen et al |
| EPI_ISL_875795 | Canada/ON-PHL-21-00013/2020 | 20-Dec | North America/Canada/Ontario/ | Public Health Ontario Laboratory | Public Health Ontario Laboratory | Vanessa G Allen et al |
| EPI_ISL_875796 | Canada/ON-PHL-21-00018/2020 | 20-Dec | North America/Canada/Ontario/ | Public Health Ontario Laboratory | Public Health Ontario Laboratory | Vanessa G Allen et al |
| EPI_ISL_875797 | Canada/ON-PHL-21-00019/2020 | 20-Dec | North America/Canada/Ontario/ | Public Health Ontario Laboratory | Public Health Ontario Laboratory | Vanessa G Allen et al |
| EPI_ISL_875798 | Canada/ON-PHL-21-00003/2020 | 20-Dec | North America/Canada/Ontario/ | Public Health Ontario Laboratory | Public Health Ontario Laboratory | Vanessa G Allen et al |

| | | | | | | |
|---|---|---|---|---|---|---|
| EPI_ISL_875799 | Canada/ON-PHL-21-00250/2020 | 20-Dec | North America/Canada/Ontario/ | Public Health Ontario Laboratory | Public Health Ontario Laboratory | Vanessa G Allen et al |
| EPI_ISL_875800 | Canada/ON-PHL-21-00252/2020 | 20-Dec | North America/Canada/Ontario/ | Public Health Ontario Laboratory | Public Health Ontario Laboratory | Vanessa G Allen et al |
| EPI_ISL_875801 | Canada/ON-PHL-21-00251/2020 | 20-Dec | North America/Canada/Ontario/ | Public Health Ontario Laboratory | Public Health Ontario Laboratory | Vanessa G Allen et al |
| EPI_ISL_875802 | Canada/ON-PHL-21-00247/2020 | 20-Dec | North America/Canada/Ontario/ | Public Health Ontario Laboratory | Public Health Ontario Laboratory | Vanessa G Allen et al |
| EPI_ISL_875803 | Canada/ON-PHL-21-00249/2020 | 20-Dec | North America/Canada/Ontario/ | Public Health Ontario Laboratory | Public Health Ontario Laboratory | Vanessa G Allen et al |
| EPI_ISL_875804 | Canada/ON-PHL-21-00248/2020 | 20-Dec | North America/Canada/Ontario/ | Public Health Ontario Laboratory | Public Health Ontario Laboratory | Vanessa G Allen et al |
| EPI_ISL_875805 | Canada/ON-PHL-21-00011/2020 | 20-Dec | North America/Canada/Ontario/ | Public Health Ontario Laboratory | Public Health Ontario Laboratory | Vanessa G Allen et al |
| EPI_ISL_875806 | Canada/ON-PHL-21-00012/2020 | 20-Dec | North America/Canada/Ontario/ | Public Health Ontario Laboratory | Public Health Ontario Laboratory | Vanessa G Allen et al |
| EPI_ISL_875807 | Canada/ON-PHL-21-00014/2020 | 20-Dec | North America/Canada/Ontario/ | Public Health Ontario Laboratory | Public Health Ontario Laboratory | Vanessa G Allen et al |
| EPI_ISL_875808 | Canada/ON-PHL-21-00015/2020 | 20-Dec | North America/Canada/Ontario/ | Public Health Ontario Laboratory | Public Health Ontario Laboratory | Vanessa G Allen et al |
| EPI_ISL_875809 | Canada/ON-PHL-21-00268/2020 | 20-Dec | North America/Canada/Ontario/ | Public Health Ontario Laboratory | Public Health Ontario Laboratory | Vanessa G Allen et al |
| EPI_ISL_875810 | Canada/ON-PHL-21-00173/2020 | 20-Dec | North America/Canada/Ontario/ | Public Health Ontario Laboratory | Public Health Ontario Laboratory | Vanessa G Allen et al |
| EPI_ISL_875811 | Canada/ON-PHL-21-00293/2020 | 20-Dec | North America/Canada/Ontario/ | Public Health Ontario Laboratory | Public Health Ontario Laboratory | Vanessa G Allen et al |

| | | | | | | |
|---|---|---|---|---|---|---|
| EPI_ISL_875812 | Canada/ON-PHL-21-00292/2020 | 20-Dec | North America/Canada/Ontario/ | Public Health Ontario Laboratory | Public Health Ontario Laboratory | Vanessa G Allen et al |
| EPI_ISL_875813 | Canada/ON-PHL-21-00306/2020 | 20-Dec | North America/Canada/Ontario/ | Public Health Ontario Laboratory | Public Health Ontario Laboratory | Vanessa G Allen et al |
| EPI_ISL_875814 | Canada/ON-PHL-21-00274/2020 | 20-Nov | North America/Canada/Ontario/ | Public Health Ontario Laboratory | Public Health Ontario Laboratory | Vanessa G Allen et al |
| EPI_ISL_875815 | Canada/ON-PHL-21-00276/2020 | 20-Nov | North America/Canada/Ontario/ | Public Health Ontario Laboratory | Public Health Ontario Laboratory | Vanessa G Allen et al |
| EPI_ISL_875816 | Canada/ON-PHL-21-00059/2020 | 20-Dec | North America/Canada/Ontario/ | Public Health Ontario Laboratory | Public Health Ontario Laboratory | Vanessa G Allen et al |
| EPI_ISL_875817 | Canada/ON-PHL-21-00053/2020 | 20-Dec | North America/Canada/Ontario/ | Public Health Ontario Laboratory | Public Health Ontario Laboratory | Vanessa G Allen et al |
| EPI_ISL_875818 | Canada/ON-PHL-21-00259/2020 | 20-Dec | North America/Canada/Ontario/ | Public Health Ontario Laboratory | Public Health Ontario Laboratory | Vanessa G Allen et al |
| EPI_ISL_875819 | Canada/ON-PHL-21-00286/2020 | 20-Dec | North America/Canada/Ontario/ | Public Health Ontario Laboratory | Public Health Ontario Laboratory | Vanessa G Allen et al |
| EPI_ISL_875820 | Canada/ON-PHL-21-00316/2020 | 20-Dec | North America/Canada/Ontario/ | Public Health Ontario Laboratory | Public Health Ontario Laboratory | Vanessa G Allen et al |
| EPI_ISL_875821 | Canada/ON-PHL-21-00290/2020 | 20-Dec | North America/Canada/Ontario/ | Public Health Ontario Laboratory | Public Health Ontario Laboratory | Vanessa G Allen et al |
| EPI_ISL_875822 | Canada/ON-PHL-21-00060/2020 | 20-Dec | North America/Canada/Ontario/ | Public Health Ontario Laboratory | Public Health Ontario Laboratory | Vanessa G Allen et al |
| EPI_ISL_875823 | Canada/ON-PHL-21-00277/2020 | 20-Dec | North America/Canada/Ontario/ | Public Health Ontario Laboratory | Public Health Ontario Laboratory | Vanessa G Allen et al |
| EPI_ISL_875824 | Canada/ON-PHL-21-00279/2020 | 20-Dec | North America/Canada/Ontario/ | Public Health Ontario Laboratory | Public Health Ontario Laboratory | Vanessa G Allen et al |

| | | | | | | |
|---|---|---|---|---|---|---|
| EPI_ISL_875825 | Canada/ON-PHL-21-00291/2020 | 20-Dec | North America/Canada/Ontario/ | Public Health Ontario Laboratory | Public Health Ontario Laboratory | Vanessa G Allen et al |
| EPI_ISL_875826 | Canada/ON-PHL-21-00281/2020 | 20-Dec | North America/Canada/Ontario/ | Public Health Ontario Laboratory | Public Health Ontario Laboratory | Vanessa G Allen et al |
| EPI_ISL_875827 | Canada/ON-PHL-21-00054/2020 | 20-Dec | North America/Canada/Ontario/ | Public Health Ontario Laboratory | Public Health Ontario Laboratory | Vanessa G Allen et al |
| EPI_ISL_875828 | Canada/ON-PHL-21-00061/2020 | 20-Dec | North America/Canada/Ontario/ | Public Health Ontario Laboratory | Public Health Ontario Laboratory | Vanessa G Allen et al |
| EPI_ISL_875829 | Canada/ON-PHL-21-00255/2020 | 20-Dec | North America/Canada/Ontario/ | Public Health Ontario Laboratory | Public Health Ontario Laboratory | Vanessa G Allen et al |
| EPI_ISL_875830 | Canada/ON-PHL-21-00055/2020 | 20-Dec | North America/Canada/Ontario/ | Public Health Ontario Laboratory | Public Health Ontario Laboratory | Vanessa G Allen et al |
| EPI_ISL_875831 | Canada/ON-PHL-21-00263/2020 | 20-Dec | North America/Canada/Ontario/ | Public Health Ontario Laboratory | Public Health Ontario Laboratory | Vanessa G Allen et al |
| EPI_ISL_875832 | Canada/ON-PHL-21-00264/2020 | 20-Dec | North America/Canada/Ontario/ | Public Health Ontario Laboratory | Public Health Ontario Laboratory | Vanessa G Allen et al |
| EPI_ISL_875833 | Canada/ON-PHL-21-00076/2020 | 20-Dec | North America/Canada/Ontario/ | Public Health Ontario Laboratory | Public Health Ontario Laboratory | Vanessa G Allen et al |
| EPI_ISL_875834 | Canada/ON-PHL-21-00312/2020 | 20-Dec | North America/Canada/Ontario/ | Public Health Ontario Laboratory | Public Health Ontario Laboratory | Vanessa G Allen et al |
| EPI_ISL_875835 | Canada/ON-PHL-21-00311/2020 | 20-Dec | North America/Canada/Ontario/ | Public Health Ontario Laboratory | Public Health Ontario Laboratory | Vanessa G Allen et al |
| EPI_ISL_875836 | Canada/ON-PHL-21-00313/2020 | 20-Dec | North America/Canada/Ontario/ | Public Health Ontario Laboratory | Public Health Ontario Laboratory | Vanessa G Allen et al |
| EPI_ISL_875837 | Canada/ON-PHL-21-00078/2020 | 20-Dec | North America/Canada/Ontario/ | Public Health Ontario Laboratory | Public Health Ontario Laboratory | Vanessa G Allen et al |

| | | | | | | |
|---|---|---|---|---|---|---|
| EPI_ISL_875838 | Canada/ON-PHL-21-00315/2020 | 20-Dec | North America/Canada/Ontario/ | Public Health Ontario Laboratory | Public Health Ontario Laboratory | Vanessa G Allen et al |
| EPI_ISL_875839 | Canada/ON-PHL-21-00256/2020 | 20-Dec | North America/Canada/Ontario/ | Public Health Ontario Laboratory | Public Health Ontario Laboratory | Vanessa G Allen et al |
| EPI_ISL_875840 | Canada/ON-PHL-21-00047/2020 | 20-Dec | North America/Canada/Ontario/ | Public Health Ontario Laboratory | Public Health Ontario Laboratory | Vanessa G Allen et al |
| EPI_ISL_875841 | Canada/ON-PHL-21-00214/2020 | 20-Dec | North America/Canada/Ontario/ | Public Health Ontario Laboratory | Public Health Ontario Laboratory | Vanessa G Allen et al |
| EPI_ISL_875842 | Canada/ON-PHL-21-00215/2020 | 20-Dec | North America/Canada/Ontario/ | Public Health Ontario Laboratory | Public Health Ontario Laboratory | Vanessa G Allen et al |
| EPI_ISL_875843 | Canada/ON-PHL-21-00305/2020 | 20-Dec | North America/Canada/Ontario/ | Public Health Ontario Laboratory | Public Health Ontario Laboratory | Vanessa G Allen et al |
| EPI_ISL_875844 | Canada/ON-PHL-21-00242/2020 | 20-Dec | North America/Canada/Ontario/ | Public Health Ontario Laboratory | Public Health Ontario Laboratory | Vanessa G Allen et al |
| EPI_ISL_875845 | Canada/ON-PHL-21-00243/2020 | 20-Dec | North America/Canada/Ontario/ | Public Health Ontario Laboratory | Public Health Ontario Laboratory | Vanessa G Allen et al |
| EPI_ISL_875846 | Canada/ON-PHL-21-00308/2020 | 20-Dec | North America/Canada/Ontario/ | Public Health Ontario Laboratory | Public Health Ontario Laboratory | Vanessa G Allen et al |
| EPI_ISL_875847 | Canada/ON-PHL-21-00179/2020 | 20-Dec | North America/Canada/Ontario/ | Public Health Ontario Laboratory | Public Health Ontario Laboratory | Vanessa G Allen et al |
| EPI_ISL_875848 | Canada/ON-PHL-21-00193/2020 | 20-Dec | North America/Canada/Ontario/ | Public Health Ontario Laboratory | Public Health Ontario Laboratory | Vanessa G Allen et al |
| EPI_ISL_875849 | Canada/ON-PHL-21-00283/2020 | 20-Dec | North America/Canada/Ontario/ | Public Health Ontario Laboratory | Public Health Ontario Laboratory | Vanessa G Allen et al |
| EPI_ISL_875850 | Canada/ON-PHL-21-00232/2020 | 20-Dec | North America/Canada/Ontario/ | Public Health Ontario Laboratory | Public Health Ontario Laboratory | Vanessa G Allen et al |

| | | | | | | |
|---|---|---|---|---|---|---|
| EPI_ISL_875851 | Canada/ON-PHL-21-00231/2020 | 20-Dec | North America/Canada/Ontario/ | Public Health Ontario Laboratory | Public Health Ontario Laboratory | Vanessa G Allen et al |
| EPI_ISL_875852 | Canada/ON-PHL-21-00230/2020 | 20-Dec | North America/Canada/Ontario/ | Public Health Ontario Laboratory | Public Health Ontario Laboratory | Vanessa G Allen et al |
| EPI_ISL_875853 | Canada/ON-PHL-21-00189/2020 | 20-Dec | North America/Canada/Ontario/ | Public Health Ontario Laboratory | Public Health Ontario Laboratory | Vanessa G Allen et al |
| EPI_ISL_875854 | Canada/ON-PHL-21-00190/2020 | 20-Dec | North America/Canada/Ontario/ | Public Health Ontario Laboratory | Public Health Ontario Laboratory | Vanessa G Allen et al |
| EPI_ISL_875855 | Canada/ON-PHL-21-00191/2020 | 20-Dec | North America/Canada/Ontario/ | Public Health Ontario Laboratory | Public Health Ontario Laboratory | Vanessa G Allen et al |
| EPI_ISL_875856 | Canada/ON-PHL-21-00205/2020 | 20-Dec | North America/Canada/Ontario/ | Public Health Ontario Laboratory | Public Health Ontario Laboratory | Vanessa G Allen et al |
| EPI_ISL_875857 | Canada/ON-PHL-21-00192/2020 | 20-Dec | North America/Canada/Ontario/ | Public Health Ontario Laboratory | Public Health Ontario Laboratory | Vanessa G Allen et al |
| EPI_ISL_875858 | Canada/ON-PHL-21-00208/2020 | 20-Dec | North America/Canada/Ontario/ | Public Health Ontario Laboratory | Public Health Ontario Laboratory | Vanessa G Allen et al |
| EPI_ISL_875859 | Canada/ON-PHL-21-00210/2020 | 20-Dec | North America/Canada/Ontario/ | Public Health Ontario Laboratory | Public Health Ontario Laboratory | Vanessa G Allen et al |
| EPI_ISL_875860 | Canada/ON-PHL-21-00211/2020 | 20-Dec | North America/Canada/Ontario/ | Public Health Ontario Laboratory | Public Health Ontario Laboratory | Vanessa G Allen et al |
| EPI_ISL_875861 | Canada/ON-PHL-21-00212/2020 | 20-Dec | North America/Canada/Ontario/ | Public Health Ontario Laboratory | Public Health Ontario Laboratory | Vanessa G Allen et al |
| EPI_ISL_875862 | Canada/ON-PHL-21-00202/2020 | 20-Dec | North America/Canada/Ontario/ | Public Health Ontario Laboratory | Public Health Ontario Laboratory | Vanessa G Allen et al |
| EPI_ISL_875863 | Canada/ON-PHL-21-00203/2020 | 20-Dec | North America/Canada/Ontario/ | Public Health Ontario Laboratory | Public Health Ontario Laboratory | Vanessa G Allen et al |

| | | | | | | |
|---|---|---|---|---|---|---|
| EPI_ISL_875864 | Canada/ON-PHL-21-00201/2020 | 20-Dec | North America/Canada/Ontario/ | Public Health Ontario Laboratory | Public Health Ontario Laboratory | Vanessa G Allen et al |
| EPI_ISL_875865 | Canada/ON-PHL-21-00196/2020 | 20-Dec | North America/Canada/Ontario/ | Public Health Ontario Laboratory | Public Health Ontario Laboratory | Vanessa G Allen et al |
| EPI_ISL_875866 | Canada/ON-PHL-21-00237/2020 | 20-Dec | North America/Canada/Ontario/ | Public Health Ontario Laboratory | Public Health Ontario Laboratory | Vanessa G Allen et al |
| EPI_ISL_875867 | Canada/ON-PHL-21-00185/2020 | 20-Dec | North America/Canada/Ontario/ | Public Health Ontario Laboratory | Public Health Ontario Laboratory | Vanessa G Allen et al |
| EPI_ISL_875868 | Canada/ON-PHL-21-00184/2020 | 20-Dec | North America/Canada/Ontario/ | Public Health Ontario Laboratory | Public Health Ontario Laboratory | Vanessa G Allen et al |
| EPI_ISL_875869 | Canada/ON-PHL-21-00077/2020 | 20-Dec | North America/Canada/Ontario/ | Public Health Ontario Laboratory | Public Health Ontario Laboratory | Vanessa G Allen et al |
| EPI_ISL_875870 | Canada/ON-PHL-21-00020/2020 | 20-Dec | North America/Canada/Ontario/ | Public Health Ontario Laboratory | Public Health Ontario Laboratory | Vanessa G Allen et al |
| EPI_ISL_875871 | USA/OH-ODH-SC1026751/2021 | 2021-01-04 | North America/USA/Ohio/ | Ohio Department of Health Laboratory | Ohio Department of Health Laboratory | Holmes et al |
| EPI_ISL_875872 | USA/OH-ODH-SC1026793/2021 | 2021-01-04 | North America/USA/Ohio/ | Ohio Department of Health Laboratory | Ohio Department of Health Laboratory | Holmes et al |
| EPI_ISL_875878 | USA/AL-Cullman-JEDX/2020 | 2020-10-30 | North America/USA/Alabama/Cullman ( | Eurofins Diatherix | Hudsonalpha Genome Sequencing Center | Jane Grimwood et al |
| EPI_ISL_875892 | USA/AL-Marshall-JEGC/2020 | 2020-11-03 | North America/USA/Alabama/Marshall | Eurofins Diatherix | Hudsonalpha Genome Sequencing Center | Jane Grimwood et al |
| EPI_ISL_875894 | USA/AL-Randolph-JEET/2020 | 2020-11-02 | North America/USA/Alabama/Randolph | Eurofins Diatherix | Hudsonalpha Genome Sequencing Center | Jane Grimwood et al |

| | | | | | | |
|---|---|---|---|---|---|---|
| EPI_ISL_875895 | USA/AL-Shelby-JEFN/2020 | 2020-11-03 | North America/USA/Alabama/Shelby Co | Eurofins Diatherix | Hudsonalpha Genome Sequencing Center | Jane Grimwood et al |
| EPI_ISL_875900 | USA/AL-Etowah-JEDE/2020 | 2020-10-30 | North America/USA/Alabama/Etowah C | Eurofins Diatherix | Hudsonalpha Genome Sequencing Center | Jane Grimwood et al |
| EPI_ISL_875907 | USA/AL-Madison-JEFG/2020 | 2020-11-03 | North America/USA/Alabama/Madison | Eurofins Diatherix | Hudsonalpha Genome Sequencing Center | Jane Grimwood et al |
| EPI_ISL_875913 | USA/AL-Morgan-JEEC/2020 | 2020-10-30 | North America/USA/Alabama/Morgan C | Eurofins Diatherix | Hudsonalpha Genome Sequencing Center | Jane Grimwood et al |
| EPI_ISL_875921 | USA/AL-Jefferson-JEGG/2020 | 2020-11-03 | North America/USA/Alabama/Jefferson | Eurofins Diatherix | Hudsonalpha Genome Sequencing Center | Jane Grimwood et al |
| EPI_ISL_875923 | USA/AL-Jefferson-JEDG/2020 | 2020-10-30 | North America/USA/Alabama/Jefferson | Eurofins Diatherix | Hudsonalpha Genome Sequencing Center | Jane Grimwood et al |
| EPI_ISL_875926 | USA/AL-Coffee-JEDB/2020 | 2020-10-30 | North America/USA/Alabama/Coffee Co | Eurofins Diatherix | Hudsonalpha Genome Sequencing Center | Jane Grimwood et al |
| EPI_ISL_875939 | USA/AL-Jefferson-JEGD/2020 | 2020-11-03 | North America/USA/Alabama/Jefferson | Eurofins Diatherix | Hudsonalpha Genome Sequencing Center | Jane Grimwood et al |
| EPI_ISL_875951 | USA/AL-Morgan-JEDA/2020 | 2020-10-30 | North America/USA/Alabama/Morgan C | Eurofins Diatherix | Hudsonalpha Genome Sequencing Center | Jane Grimwood et al |
| EPI_ISL_875958 | USA/AL-St.Clair-JEFX/2020 | 2020-11-03 | North America/USA/Alabama/St. Clair C | Eurofins Diatherix | Hudsonalpha Genome Sequencing Center | Jane Grimwood et al |

| | | | | | | |
|---|---|---|---|---|---|---|
| EPI_ISL_875961 | Poland/1630617021_diag1_gx1/2020 | 2020-12-23 | Europe/Poland/Malopolskie/ | Laboratory of Molecular Biology, Diagnostyka sp. z o.o. | genXone SA, Research & Development Laboratory | Maciej Sykulski et al |
| EPI_ISL_875964 | Poland/4011606621_diag1_gx1/2020 | 2020-12-22 | Europe/Poland/Swietokrzyskie/ | Laboratory of Molecular Biology, Diagnostyka sp. z o.o. | genXone SA, Research & Development Laboratory | Maciej Sykulski et al |
| EPI_ISL_875965 | Poland/1685597229_diag1_gx1/2020 | 2020-12-25 | Europe/Poland/Zachodniopomorskie/ | Laboratory of Molecular Biology, Diagnostyka sp. z o.o. | genXone SA, Research & Development Laboratory | Maciej Sykulski et al |
| EPI_ISL_875968 | Poland/1565616021_diag1_gx1/2020 | 2020-12-22 | Europe/Poland/Lubuskie/ | Laboratory of Molecular Biology, Diagnostyka sp. z o.o. | genXone SA, Research & Development Laboratory | Maciej Sykulski et al |
| EPI_ISL_875970 | Poland/9509613723_diag1_gx1/2020 | 2020-12-22 | Europe/Poland/Slaskie/ | Laboratory of Molecular Biology, Diagnostyka sp. z o.o. | genXone SA, Research & Development Laboratory | Maciej Sykulski et al |
| EPI_ISL_875971 | Poland/6229607321_diag1_gx1/2020 | 2020-12-22 | Europe/Poland/Swietokrzyskie/ | Laboratory of Molecular Biology, Diagnostyka sp. z o.o. | genXone SA, Research & Development Laboratory | Maciej Sykulski et al |
| EPI_ISL_875973 | Poland/7523607321_diag1_gx1/2020 | 2020-12-23 | Europe/Poland/Swietokrzyskie/ | Laboratory of Molecular Biology, Diagnostyka sp. z o.o. | genXone SA, Research & Development Laboratory | Maciej Sykulski et al |
| EPI_ISL_875974 | Poland/9040607521_diag1_gx1/2020 | 2020-12-22 | Europe/Poland/Swietokrzyskie/ | Laboratory of Molecular Biology, Diagnostyka sp. z o.o. | genXone SA, Research & Development Laboratory | Maciej Sykulski et al |
| EPI_ISL_875975 | Poland/1700597229_diag1_gx1/2020 | 2020-12-25 | Europe/Poland/Zachodniopomorskie/ | Laboratory of Molecular Biology, Diagnostyka sp. z o.o. | genXone SA, Research & Development Laboratory | Maciej Sykulski et al |
| EPI_ISL_875976 | Poland/9248597429_diag1_gx1/2020 | 2020-12-25 | Europe/Poland/Zachodniopomorskie/ | Laboratory of Molecular Biology, Diagnostyka sp. z o.o. | genXone SA, Research & Development Laboratory | Maciej Sykulski et al |

| | | | | | | |
|---|---|---|---|---|---|---|
| EPI_ISL_875977 | Poland/9239597429_diag1_gx1/2020 | 2020-12-25 | Europe/Poland/Lubuskie/ | Laboratory of Molecular Biology, Diagnostyka sp. z o.o. | genXone SA, Research & Development Laboratory | Maciej Sykulski et al |
| EPI_ISL_875979 | Poland/4083601621_diag1_gx1/2020 | 2020-12-22 | Europe/Poland/Opolskie/ | Laboratory of Molecular Biology, Diagnostyka sp. z o.o. | genXone SA, Research & Development Laboratory | Maciej Sykulski et al |
| EPI_ISL_875980 | Poland/312601021_diag1_gx1/2020 | 2020-12-22 | Europe/Poland/Dolno≈õlÖskie/ | Laboratory of Molecular Biology, Diagnostyka sp. z o.o. | genXone SA, Research & Development Laboratory | Maciej Sykulski et al |
| EPI_ISL_875981 | Poland/316601021_diag1_gx1/2020 | 2020-12-22 | Europe/Poland/Dolno≈õlÖskie/ | Laboratory of Molecular Biology, Diagnostyka sp. z o.o. | genXone SA, Research & Development Laboratory | Maciej Sykulski et al |
| EPI_ISL_875983 | Poland/415601621_diag1_gx1/2020 | 2020-12-22 | Europe/Poland/Dolno≈õlÖskie/ | Laboratory of Molecular Biology, Diagnostyka sp. z o.o. | genXone SA, Research & Development Laboratory | Maciej Sykulski et al |
| EPI_ISL_875992 | Poland/7670614221_diag1_gx1/2020 | 2020-12-22 | Europe/Poland/Malopolskie/ | Laboratory of Molecular Biology, Diagnostyka sp. z o.o. | genXone SA, Research & Development Laboratory | Maciej Sykulski et al |
| EPI_ISL_875994 | Poland/1563615023_diag1_gx1/2020 | 2020-12-22 | Europe/Poland/Pomorskie/ | Laboratory of Molecular Biology, Diagnostyka sp. z o.o. | genXone SA, Research & Development Laboratory | Maciej Sykulski et al |
| EPI_ISL_875995 | Poland/3424615022_diag1_gx1/2020 | 2020-12-22 | Europe/Poland/Pomorskie/ | Laboratory of Molecular Biology, Diagnostyka sp. z o.o. | genXone SA, Research & Development Laboratory | Maciej Sykulski et al |
| EPI_ISL_876000 | Poland/1240607221_diag1_gx1/2020 | 2020-12-22 | Europe/Poland/Warminsko-Mazurskie/ | Laboratory of Molecular Biology, Diagnostyka sp. z o.o. | genXone SA, Research & Development Laboratory | Maciej Sykulski et al |
| EPI_ISL_876001 | Poland/526607221_diag1_gx1/2020 | 2020-12-22 | Europe/Poland/Warminsko-Mazurskie/ | Laboratory of Molecular Biology, Diagnostyka sp. z o.o. | genXone SA, Research & Development Laboratory | Maciej Sykulski et al |

| | | | | | | |
|---|---|---|---|---|---|---|
| EPI_ISL_876002 | Poland/553607221_diag1_gx1/2020 | 2020-12-22 | Europe/Poland/Warminsko-Mazurskie/ | Laboratory of Molecular Biology, Diagnostyka sp. z o.o. | genXone SA, Research & Development Laboratory | Maciej Sykulski et al |
| EPI_ISL_876003 | Poland/554607221_diag1_gx1/2020 | 2020-12-22 | Europe/Poland/Warminsko-Mazurskie/ | Laboratory of Molecular Biology, Diagnostyka sp. z o.o. | genXone SA, Research & Development Laboratory | Maciej Sykulski et al |
| EPI_ISL_876006 | Poland/576607221_diag1_gx1/2020 | 2020-12-22 | Europe/Poland/Warminsko-Mazurskie/ | Laboratory of Molecular Biology, Diagnostyka sp. z o.o. | genXone SA, Research & Development Laboratory | Maciej Sykulski et al |
| EPI_ISL_876008 | Poland/6847615223_diag1_gx1/2020 | 2020-12-22 | Europe/Poland/Warminsko-Mazurskie/ | Laboratory of Molecular Biology, Diagnostyka sp. z o.o. | genXone SA, Research & Development Laboratory | Maciej Sykulski et al |
| EPI_ISL_876009 | Poland/6859615223_diag1_gx1/2020 | 2020-12-22 | Europe/Poland/Warminsko-Mazurskie/ | Laboratory of Molecular Biology, Diagnostyka sp. z o.o. | genXone SA, Research & Development Laboratory | Maciej Sykulski et al |
| EPI_ISL_876012 | Poland/340601021_diag1_gx1/2020 | 2020-12-22 | Europe/Poland/Dolno≈õlƒÖskie/ | Laboratory of Molecular Biology, Diagnostyka sp. z o.o. | genXone SA, Research & Development Laboratory | Maciej Sykulski et al |
| EPI_ISL_876015 | Poland/9716615021_diag1_gx1/2020 | 2020-12-22 | Europe/Poland/≈Å√≥dzkie/ | Laboratory of Molecular Biology, Diagnostyka sp. z o.o. | genXone SA, Research & Development Laboratory | Maciej Sykulski et al |
| EPI_ISL_876016 | Poland/5651589026_diag1_gx1/2020 | 2020-12-22 | Europe/Poland/Lubuskie/ | Laboratory of Molecular Biology, Diagnostyka sp. z o.o. | genXone SA, Research & Development Laboratory | Maciej Sykulski et al |
| EPI_ISL_876018 | Poland/9575603621_diag1_gx1/2020 | 2020-12-22 | Europe/Poland/Podkarpackie/ | Laboratory of Molecular Biology, Diagnostyka sp. z o.o. | genXone SA, Research & Development Laboratory | Maciej Sykulski et al |
| EPI_ISL_876020 | Poland/9855614223_diag1_gx1/2020 | 2020-12-22 | Europe/Poland/Slaskie/ | Laboratory of Molecular Biology, Diagnostyka sp. z o.o. | genXone SA, Research & Development Laboratory | Maciej Sykulski et al |

| | | | | | | |
|---|---|---|---|---|---|---|
| EPI_ISL_876022 | Poland/6304607321_diag1_gx1/2020 | 2020-12-23 | Europe/Poland/Swietokrzyskie/ | Laboratory of Molecular Biology, Diagnostyka sp. z o.o. | genXone SA, Research & Development Laboratory | Maciej Sykulski et al |
| EPI_ISL_876023 | Poland/9109607521_diag1_gx1/2020 | 2020-12-23 | Europe/Poland/Swietokrzyskie/ | Laboratory of Molecular Biology, Diagnostyka sp. z o.o. | genXone SA, Research & Development Laboratory | Maciej Sykulski et al |
| EPI_ISL_876026 | Poland/6838615223_diag1_gx1/2020 | 2020-12-22 | Europe/Poland/Warminsko-Mazurskie/ | Laboratory of Molecular Biology, Diagnostyka sp. z o.o. | genXone SA, Research & Development Laboratory | Maciej Sykulski et al |
| EPI_ISL_876027 | Poland/6845615223_diag1_gx1/2020 | 2020-12-22 | Europe/Poland/Warminsko-Mazurskie/ | Laboratory of Molecular Biology, Diagnostyka sp. z o.o. | genXone SA, Research & Development Laboratory | Maciej Sykulski et al |
| EPI_ISL_876031 | Poland/9234597429_diag1_gx1/2020 | 2020-12-25 | Europe/Poland/Zachodniopomorskie/ | Laboratory of Molecular Biology, Diagnostyka sp. z o.o. | genXone SA, Research & Development Laboratory | Maciej Sykulski et al |
| EPI_ISL_876034 | USA/NY-AECOM_034/2020 | 2020-04-15 | North America/USA/New York/New Yor | Montefiore Medical Center | Albert Einstein College of Medicine, Dept. of Microbiology & Immunology, Chandran lab | J. Maximilian Fels et al |
| EPI_ISL_876039 | USA/NY-AECOM_058/2020 | 2020-04-15 | North America/USA/New York/New Yor | Montefiore Medical Center | Albert Einstein College of Medicine, Dept. of Microbiology & Immunology, Chandran lab | J. Maximilian Fels et al |

| | | | | | | |
|---|---|---|---|---|---|---|
| EPI_ISL_876042 | Italy/CAM_CRGS_2/2020 | 2020-03-14 | Europe/Italy/Campania/ | 1.AO Universitaria 'S. Giovanni di Dio e Ruggi D'Aragona, Scuola Medica Salernitana' Hospital / 2.UOC di Virologia e Microbiologia, Universit√† della Campania 'L. Vanvitelli' / 3.AO Universitaria 'Federico II' Napoli Hospital / 4.AORN 'San Giuseppe Moscati' Avellino Hospital / 5.AO 'San Pio - presidio G. Rummo' Benevento Hospital / 6.AO 'Sant'Anna e San Sebastiano' Caserta Hospital / 7.PO 'Maria Santissima Addolorata' Eboli Hospital / 8.Biogem Istituto di Ricerche Genetiche | 1. Genome Research Center for Health (CRGS) / 2. Laboratory of Molecular Medicine and Genomics(LMMGe) / 3. Center for Research in Pure and Applied Mathematics (CRMPA) | Giorgio Giurato et al |
| EPI_ISL_876043 | Italy/CAM_CRGS_3/2020 | 2020-03-14 | Europe/Italy/Campania/ | 1.AO Universitaria 'S. Giovanni di Dio e Ruggi D'Aragona, Scuola Medica Salernitana' Hospital / 2.UOC di Virologia e Microbiologia, Universit√† della Campania 'L. Vanvitelli' / 3.AO Universitaria 'Federico II' Napoli Hospital / 4.AORN 'San Giuseppe Moscati' Avellino Hospital / 5.AO 'San Pio - presidio G. Rummo' Benevento Hospital / 6.AO 'Sant'Anna e San Sebastiano' Caserta Hospital / 7.PO 'Maria Santissima Addolorata' Eboli Hospital / 8.Biogem Istituto di Ricerche Genetiche | 1. Genome Research Center for Health (CRGS) / 2. Laboratory of Molecular Medicine and Genomics(LMMGe) / 3. Center for Research in Pure and Applied Mathematics (CRMPA) | Giorgio Giurato et al |
| EPI_ISL_876048 | USA/MA-MASPHL-01188/2020 | 2020-12-13 | North America/USA/Massachusetts/ | Massachusetts State Public Health Laboratory | Massachusetts State Public Health Laboratory | Andrew Lang et al |
| EPI_ISL_876053 | USA/MA-MASPHL-01412/2021 | 2021-01-05 | North America/USA/Massachusetts/ | Massachusetts State Public Health Laboratory | Massachusetts State Public Health Laboratory | Andrew Lang et al |
| EPI_ISL_876071 | USA/MA-MASPHL-01292/2020 | 2020-12-16 | North America/USA/Massachusetts/ | Massachusetts State Public Health Laboratory | Massachusetts State Public Health Laboratory | Andrew Lang et al |
| EPI_ISL_876073 | USA/MA-MASPHL-01152/2020 | 2020-12-05 | North America/USA/Massachusetts/ | Massachusetts State Public Health Laboratory | Massachusetts State Public Health Laboratory | Andrew Lang et al |
| EPI_ISL_876076 | USA/MA-MASPHL-01402/2020 | 2020-12-30 | North America/USA/Massachusetts/ | Massachusetts State Public Health Laboratory | Massachusetts State Public Health Laboratory | Andrew Lang et al |

| | | | | | | |
|---|---|---|---|---|---|---|
| EPI_ISL_876080 | USA/MA-MASPHL-01544/2021 | 2021-01-11 | North America/USA/Massachusetts/ | Massachusetts State Public Health Laboratory | Massachusetts State Public Health Laboratory | Andrew Lang et al |
| EPI_ISL_876083 | USA/MA-MASPHL-01105/2020 | 2020-12-05 | North America/USA/Massachusetts/ | Massachusetts State Public Health Laboratory | Massachusetts State Public Health Laboratory | Andrew Lang et al |
| EPI_ISL_876090 | USA/MA-MASPHL-01222/2020 | 2020-12-16 | North America/USA/Massachusetts/ | Massachusetts State Public Health Laboratory | Massachusetts State Public Health Laboratory | Andrew Lang et al |
| EPI_ISL_876094 | USA/MA-MASPHL-01282/2020 | 2020-12-25 | North America/USA/Massachusetts/ | Massachusetts State Public Health Laboratory | Massachusetts State Public Health Laboratory | Andrew Lang et al |
| EPI_ISL_876095 | USA/MA-MASPHL-01561/2021 | 2021-01-13 | North America/USA/Massachusetts/ | Massachusetts State Public Health Laboratory | Massachusetts State Public Health Laboratory | Andrew Lang et al |
| EPI_ISL_876096 | USA/MA-MASPHL-01463/2021 | 2021-01-11 | North America/USA/Massachusetts/ | Massachusetts State Public Health Laboratory | Massachusetts State Public Health Laboratory | Andrew Lang et al |
| EPI_ISL_876097 | USA/MA-MASPHL-01327/2020 | 2020-12-27 | North America/USA/Massachusetts/ | Massachusetts State Public Health Laboratory | Massachusetts State Public Health Laboratory | Andrew Lang et al |
| EPI_ISL_876098 | USA/MA-MASPHL-01181/2020 | 2020-11-25 | North America/USA/Massachusetts/ | Massachusetts State Public Health Laboratory | Massachusetts State Public Health Laboratory | Andrew Lang et al |
| EPI_ISL_876099 | USA/MA-MASPHL-01551/2021 | 2021-01-12 | North America/USA/Massachusetts/ | Massachusetts State Public Health Laboratory | Massachusetts State Public Health Laboratory | Andrew Lang et al |
| EPI_ISL_876104 | USA/MA-MASPHL-01137/2020 | 2020-11-20 | North America/USA/Massachusetts/ | Massachusetts State Public Health Laboratory | Massachusetts State Public Health Laboratory | Andrew Lang et al |
| EPI_ISL_876109 | USA/MA-MASPHL-01111/2020 | 2020-12-07 | North America/USA/Massachusetts/ | Massachusetts State Public Health Laboratory | Massachusetts State Public Health Laboratory | Andrew Lang et al |
| EPI_ISL_876116 | USA/MA-MASPHL-01127/2020 | 2020-11-13 | North America/USA/Massachusetts/ | Massachusetts State Public Health Laboratory | Massachusetts State Public Health Laboratory | Andrew Lang et al |
| EPI_ISL_876119 | USA/MA-MASPHL-01140/2020 | 2020-11-23 | North America/USA/Massachusetts/ | Massachusetts State Public Health Laboratory | Massachusetts State Public Health Laboratory | Andrew Lang et al |

| | | | | | | |
|---|---|---|---|---|---|---|
| EPI_ISL_876143 | USA/MA-MASPHL-01474/2021 | 2021-01-19 | North America/USA/Massachusetts/ | Massachusetts State Public Health Laboratory | Massachusetts State Public Health Laboratory | Andrew Lang et al |
| EPI_ISL_876157 | USA/MA-MASPHL-01147/2020 | 2020-12-11 | North America/USA/Massachusetts/ | Massachusetts State Public Health Laboratory | Massachusetts State Public Health Laboratory | Andrew Lang et al |
| EPI_ISL_876161 | USA/MA-MASPHL-01469/2021 | 2021-01-19 | North America/USA/Massachusetts/ | Massachusetts State Public Health Laboratory | Massachusetts State Public Health Laboratory | Andrew Lang et al |
| EPI_ISL_876172 | USA/MA-MASPHL-01415/2021 | 2021-01-05 | North America/USA/Massachusetts/ | Massachusetts State Public Health Laboratory | Massachusetts State Public Health Laboratory | Andrew Lang et al |
| EPI_ISL_876181 | USA/MA-MASPHL-01209/2020 | 2020-12-18 | North America/USA/Massachusetts/ | Massachusetts State Public Health Laboratory | Massachusetts State Public Health Laboratory | Andrew Lang et al |
| EPI_ISL_876196 | USA/MA-MASPHL-01361/2021 | 2021-01-06 | North America/USA/Massachusetts/ | Massachusetts State Public Health Laboratory | Massachusetts State Public Health Laboratory | Andrew Lang et al |
| EPI_ISL_876197 | USA/MA-MASPHL-01421/2021 | 2021-01-04 | North America/USA/Massachusetts/ | Massachusetts State Public Health Laboratory | Massachusetts State Public Health Laboratory | Andrew Lang et al |
| EPI_ISL_876203 | USA/MA-MASPHL-01445/2021 | 2021-01-05 | North America/USA/Massachusetts/ | Massachusetts State Public Health Laboratory | Massachusetts State Public Health Laboratory | Andrew Lang et al |
| EPI_ISL_876206 | USA/MA-MASPHL-01121/2020 | 2020-12-09 | North America/USA/Massachusetts/ | Massachusetts State Public Health Laboratory | Massachusetts State Public Health Laboratory | Andrew Lang et al |
| EPI_ISL_876211 | USA/RI-MASPHL-01231/2020 | 2020-12-14 | North America/USA/Rhode Island/ | Massachusetts State Public Health Laboratory | Massachusetts State Public Health Laboratory | Andrew Lang et al |
| EPI_ISL_876215 | USA/MA-MASPHL-01492/2021 | 2021-01-12 | North America/USA/Massachusetts/ | Massachusetts State Public Health Laboratory | Massachusetts State Public Health Laboratory | Andrew Lang et al |
| EPI_ISL_876229 | USA/MA-MASPHL-01535/2021 | 2021-01-19 | North America/USA/Massachusetts/ | Massachusetts State Public Health Laboratory | Massachusetts State Public Health Laboratory | Andrew Lang et al |
| EPI_ISL_876230 | USA/MA-MASPHL-01536/2021 | 2021-01-19 | North America/USA/Massachusetts/ | Massachusetts State Public Health Laboratory | Massachusetts State Public Health Laboratory | Andrew Lang et al |

| | | | | | | |
|---|---|---|---|---|---|---|
| EPI_ISL_876231 | USA/MA-MASPHL-01508/2021 | 2021-01-17 | North America/USA/Massachusetts/ | Massachusetts State Public Health Laboratory | Massachusetts State Public Health Laboratory | Andrew Lang et al |
| EPI_ISL_876232 | USA/MA-MASPHL-01555/2021 | 2021-01-17 | North America/USA/Massachusetts/ | Massachusetts State Public Health Laboratory | Massachusetts State Public Health Laboratory | Andrew Lang et al |
| EPI_ISL_876239 | USA/MA-MASPHL-01461/2021 | 2021-01-11 | North America/USA/Massachusetts/ | Massachusetts State Public Health Laboratory | Massachusetts State Public Health Laboratory | Andrew Lang et al |
| EPI_ISL_876250 | USA/MA-MASPHL-01444/2021 | 2021-01-05 | North America/USA/Massachusetts/ | Massachusetts State Public Health Laboratory | Massachusetts State Public Health Laboratory | Andrew Lang et al |
| EPI_ISL_876254 | USA/MA-MASPHL-01395/2020 | 2020-12-30 | North America/USA/Massachusetts/ | Massachusetts State Public Health Laboratory | Massachusetts State Public Health Laboratory | Andrew Lang et al |
| EPI_ISL_876260 | USA/MA-MASPHL-01390/2020 | 2020-12-29 | North America/USA/Massachusetts/ | Massachusetts State Public Health Laboratory | Massachusetts State Public Health Laboratory | Andrew Lang et al |
| EPI_ISL_876263 | USA/MA-MASPHL-01283/2020 | 2020-12-27 | North America/USA/Massachusetts/ | Massachusetts State Public Health Laboratory | Massachusetts State Public Health Laboratory | Andrew Lang et al |
| EPI_ISL_876266 | USA/MA-MASPHL-01325/2020 | 2020-12-27 | North America/USA/Massachusetts/ | Massachusetts State Public Health Laboratory | Massachusetts State Public Health Laboratory | Andrew Lang et al |
| EPI_ISL_876270 | USA/MA-MASPHL-01368/2020 | 2020-12-27 | North America/USA/Massachusetts/ | Massachusetts State Public Health Laboratory | Massachusetts State Public Health Laboratory | Andrew Lang et al |
| EPI_ISL_876273 | USA/MA-MASPHL-01251/2020 | 2020-12-25 | North America/USA/Massachusetts/ | Massachusetts State Public Health Laboratory | Massachusetts State Public Health Laboratory | Andrew Lang et al |
| EPI_ISL_876276 | USA/MA-MASPHL-01215/2020 | 2020-12-23 | North America/USA/Massachusetts/ | Massachusetts State Public Health Laboratory | Massachusetts State Public Health Laboratory | Andrew Lang et al |
| EPI_ISL_876277 | USA/MA-MASPHL-01250/2020 | 2020-12-23 | North America/USA/Massachusetts/ | Massachusetts State Public Health Laboratory | Massachusetts State Public Health Laboratory | Andrew Lang et al |
| EPI_ISL_876281 | USA/MA-MASPHL-01235/2020 | 2020-12-21 | North America/USA/Massachusetts/ | Massachusetts State Public Health Laboratory | Massachusetts State Public Health Laboratory | Andrew Lang et al |

| | | | | | | |
|---|---|---|---|---|---|---|
| EPI_ISL_876283 | USA/MA-MASPHL-01200/2020 | 2020-12-21 | North America/USA/Massachusetts/ | Massachusetts State Public Health Laboratory | Massachusetts State Public Health Laboratory | Andrew Lang et al |
| EPI_ISL_876286 | USA/MA-MASPHL-01223/2020 | 2020-12-21 | North America/USA/Massachusetts/ | Massachusetts State Public Health Laboratory | Massachusetts State Public Health Laboratory | Andrew Lang et al |
| EPI_ISL_876287 | USA/MA-MASPHL-01248/2020 | 2020-12-21 | North America/USA/Massachusetts/ | Massachusetts State Public Health Laboratory | Massachusetts State Public Health Laboratory | Andrew Lang et al |
| EPI_ISL_876294 | USA/MA-MASPHL-01207/2020 | 2020-12-16 | North America/USA/Massachusetts/ | Massachusetts State Public Health Laboratory | Massachusetts State Public Health Laboratory | Andrew Lang et al |
| EPI_ISL_876298 | USA/MA-MASPHL-01150/2020 | 2020-12-14 | North America/USA/Massachusetts/ | Massachusetts State Public Health Laboratory | Massachusetts State Public Health Laboratory | Andrew Lang et al |
| EPI_ISL_876299 | USA/MA-MASPHL-01309/2020 | 2020-12-14 | North America/USA/Massachusetts/ | Massachusetts State Public Health Laboratory | Massachusetts State Public Health Laboratory | Andrew Lang et al |
| EPI_ISL_876304 | USA/MA-MASPHL-01296/2020 | 2020-12-10 | North America/USA/Massachusetts/ | Massachusetts State Public Health Laboratory | Massachusetts State Public Health Laboratory | Andrew Lang et al |
| EPI_ISL_876313 | USA/MA-MASPHL-01106/2020 | 2020-12-04 | North America/USA/Massachusetts/ | Massachusetts State Public Health Laboratory | Massachusetts State Public Health Laboratory | Andrew Lang et al |
| EPI_ISL_876315 | USA/MA-MASPHL-01099/2020 | 2020-12-03 | North America/USA/Massachusetts/ | Massachusetts State Public Health Laboratory | Massachusetts State Public Health Laboratory | Andrew Lang et al |
| EPI_ISL_876322 | USA/MA-MASPHL-01142/2020 | 2020-11-23 | North America/USA/Massachusetts/ | Massachusetts State Public Health Laboratory | Massachusetts State Public Health Laboratory | Andrew Lang et al |
| EPI_ISL_876327 | USA/MA-MASPHL-01130/2020 | 2020-11-16 | North America/USA/Massachusetts/ | Massachusetts State Public Health Laboratory | Massachusetts State Public Health Laboratory | Andrew Lang et al |
| EPI_ISL_876328 | USA/MA-MASPHL-01125/2020 | 2020-11-15 | North America/USA/Massachusetts/ | Massachusetts State Public Health Laboratory | Massachusetts State Public Health Laboratory | Andrew Lang et al |
| EPI_ISL_876338 | USA/AL-St.Clair-JEMC/2020 | 2020-11-10 | North America/USA/Alabama/St. Clair ( | Eurofins Diatherix | Hudsonalpha Genome Sequencing Center | Jane Grimwood et al |

| | | | | | | |
|---|---|---|---|---|---|---|
| EPI_ISL_876340 | USA/AL-Houston-JEME/2020 | 2020-11-10 | North America/USA/Alabama/Houston ( | Eurofins Diatherix | Hudsonalpha Genome Sequencing Center | Jane Grimwood et al |
| EPI_ISL_876350 | USA/AL-Lauderdale-JEMQ/2020 | 2020-11-10 | North America/USA/Alabama/Lauderda | Eurofins Diatherix | Hudsonalpha Genome Sequencing Center | Jane Grimwood et al |
| EPI_ISL_876351 | USA/AL-Morgan-JEMR/2020 | 2020-11-10 | North America/USA/Alabama/Morgan ( | Eurofins Diatherix | Hudsonalpha Genome Sequencing Center | Jane Grimwood et al |
| EPI_ISL_876363 | USA/AL-Houston-JENG/2020 | 2020-11-10 | North America/USA/Alabama/Houston ( | Eurofins Diatherix | Hudsonalpha Genome Sequencing Center | Jane Grimwood et al |
| EPI_ISL_876368 | USA/AL-Madison-JENL/2020 | 2020-11-10 | North America/USA/Alabama/Madison | Eurofins Diatherix | Hudsonalpha Genome Sequencing Center | Jane Grimwood et al |
| EPI_ISL_876378 | USA/AL-Madison-JENX/2020 | 2020-11-10 | North America/USA/Alabama/Madison | Eurofins Diatherix | Hudsonalpha Genome Sequencing Center | Jane Grimwood et al |
| EPI_ISL_876387 | USA/AL-Cullman-JEPG/2020 | 2020-11-10 | North America/USA/Alabama/Cullman ( | Eurofins Diatherix | Hudsonalpha Genome Sequencing Center | Jane Grimwood et al |
| EPI_ISL_876410 | USA/AL-Madison-JELG/2020 | 2020-11-07 | North America/USA/Alabama/Madison | Eurofins Diatherix | Hudsonalpha Genome Sequencing Center | Jane Grimwood et al |
| EPI_ISL_876420 | USA/AL-Calhoun-JEKU/2020 | 2020-11-06 | North America/USA/Alabama/Calhoun ( | Eurofins Diatherix | Hudsonalpha Genome Sequencing Center | Jane Grimwood et al |
| EPI_ISL_876436 | USA/AL-Limestone-JEHK/2020 | 2020-11-04 | North America/USA/Alabama/Limestone | Eurofins Diatherix | Hudsonalpha Genome Sequencing Center | Jane Grimwood et al |

| | | | | | | |
|---|---|---|---|---|---|---|
| EPI_ISL_876437 | USA/AL-Madison-JEHT/2020 | 2020-11-04 | North America/USA/Alabama/Madison | Eurofins Diatherix | Hudsonalpha Genome Sequencing Center | Jane Grimwood et al |
| EPI_ISL_876450 | USA/AL-Etowah-JEKB/2020 | 2020-11-06 | North America/USA/Alabama/Etowah C | Eurofins Diatherix | Hudsonalpha Genome Sequencing Center | Jane Grimwood et al |
| EPI_ISL_876460 | USA/AL-Madison-JEIJ/2020 | 2020-11-05 | North America/USA/Alabama/Madison | Eurofins Diatherix | Hudsonalpha Genome Sequencing Center | Jane Grimwood et al |
| EPI_ISL_876467 | USA/AL-Cullman-JEIS/2020 | 2020-11-05 | North America/USA/Alabama/Cullman C | Eurofins Diatherix | Hudsonalpha Genome Sequencing Center | Jane Grimwood et al |
| EPI_ISL_876481 | USA/AL-Morgan-JEJK/2020 | 2020-11-05 | North America/USA/Alabama/Morgan C | Eurofins Diatherix | Hudsonalpha Genome Sequencing Center | Jane Grimwood et al |
| EPI_ISL_876486 | USA/AL-Lawrence-JEGW/2020 | 2020-11-04 | North America/USA/Alabama/Lawrence | Eurofins Diatherix | Hudsonalpha Genome Sequencing Center | Jane Grimwood et al |
| EPI_ISL_876507 | USA/AL-Etowah-JEIA/2020 | 2020-11-04 | North America/USA/Alabama/Etowah C | Eurofins Diatherix | Hudsonalpha Genome Sequencing Center | Jane Grimwood et al |
| EPI_ISL_876508 | USA/AL-Coffee-JEIB/2020 | 2020-11-04 | North America/USA/Alabama/Coffee Co | Eurofins Diatherix | Hudsonalpha Genome Sequencing Center | Jane Grimwood et al |
| EPI_ISL_876510 | USA/AL-Blount-JEID/2020 | 2020-11-04 | North America/USA/Alabama/Blount Co | Eurofins Diatherix | Hudsonalpha Genome Sequencing Center | Jane Grimwood et al |
| EPI_ISL_876521 | USA/FL-BPHL-2812/2020 | 2020-11-24 | North America/USA/Florida/ | Florida Bureau of Public Health Laboratories | Florida Bureau of Public Health Laboratories | Sarah Schmedes et al |

| | | | | | | |
|---|---|---|---|---|---|---|
| EPI_ISL_876533 | USA/FL-BPHL-2830/2020 | 2020-12-16 | North America/USA/Florida/ | Florida Bureau of Public Health Laboratories | Florida Bureau of Public Health Laboratories | Sarah Schmedes et al |
| EPI_ISL_876539 | USA/FL-BPHL-2828/2020 | 2020-12-15 | North America/USA/Florida/ | Florida Bureau of Public Health Laboratories | Florida Bureau of Public Health Laboratories | Sarah Schmedes et al |
| EPI_ISL_876547 | USA/FL-BPHL-2837/2020 | 2020-12-09 | North America/USA/Florida/ | Florida Bureau of Public Health Laboratories | Florida Bureau of Public Health Laboratories | Sarah Schmedes et al |
| EPI_ISL_876548 | USA/FL-BPHL-2821/2020 | 2020-12-08 | North America/USA/Florida/ | Florida Bureau of Public Health Laboratories | Florida Bureau of Public Health Laboratories | Sarah Schmedes et al |
| EPI_ISL_876549 | USA/FL-BPHL-2818/2020 | 2020-12-02 | North America/USA/Florida/ | Florida Bureau of Public Health Laboratories | Florida Bureau of Public Health Laboratories | Sarah Schmedes et al |
| EPI_ISL_876553 | USA/FL-BPHL-2815/2020 | 2020-11-24 | North America/USA/Florida/ | Florida Bureau of Public Health Laboratories | Florida Bureau of Public Health Laboratories | Sarah Schmedes et al |
| EPI_ISL_876564 | USA/FL-BPHL-2808/2020 | 2020-11-23 | North America/USA/Florida/ | Florida Bureau of Public Health Laboratories | Florida Bureau of Public Health Laboratories | Sarah Schmedes et al |
| EPI_ISL_876573 | USA/FL-BPHL-2788/2020 | 2020-08-27 | North America/USA/Florida/ | Florida Bureau of Public Health Laboratories | Florida Bureau of Public Health Laboratories | Sarah Schmedes et al |
| EPI_ISL_876575 | USA/FL-BPHL-2787/2020 | 2020-08-26 | North America/USA/Florida/ | Florida Bureau of Public Health Laboratories | Florida Bureau of Public Health Laboratories | Sarah Schmedes et al |
| EPI_ISL_876576 | USA/FL-BPHL-2786/2020 | 2020-08-19 | North America/USA/Florida/ | Florida Bureau of Public Health Laboratories | Florida Bureau of Public Health Laboratories | Sarah Schmedes et al |
| EPI_ISL_876579 | USA/FL-BPHL-2783/2020 | 2020-07-22 | North America/USA/Florida/ | Florida Bureau of Public Health Laboratories | Florida Bureau of Public Health Laboratories | Sarah Schmedes et al |
| EPI_ISL_876580 | USA/FL-BPHL-2784/2020 | 2020-07-22 | North America/USA/Florida/ | Florida Bureau of Public Health Laboratories | Florida Bureau of Public Health Laboratories | Sarah Schmedes et al |
| EPI_ISL_876587 | USA/FL-BPHL-2776/2020 | 2020-07-13 | North America/USA/Florida/ | Florida Bureau of Public Health Laboratories | Florida Bureau of Public Health Laboratories | Sarah Schmedes et al |

| | | | | | | |
|---|---|---|---|---|---|---|
| EPI_ISL_876589 | USA/PA-CDC-8625/2021 | 2021-01-08 | North America/USA/Pennsylvania/ | Blackhawk Genomics | Pathogen Discovery, Respiratory Viruses Branch, Division of Viral Diseases, Centers for Disease Control and Prevention | Ying Tao et al |
| EPI_ISL_876597 | Canada/ON-SC0235/2020 | 2020-08-14 | North America/Canada/Ontario/Toronto | Toronto Invasive Bacterial Diseases Network | McMaster University | Allison McGeer et al |
| EPI_ISL_876598 | Canada/ON-SC0261/2020 | 2020-08-27 | North America/Canada/Ontario/Toronto | Toronto Invasive Bacterial Diseases Network | McMaster University | Allison McGeer et al |
| EPI_ISL_876599 | Canada/ON-SC0258/2020 | 2020-08-26 | North America/Canada/Ontario/Toronto | Toronto Invasive Bacterial Diseases Network | McMaster University | Allison McGeer et al |
| EPI_ISL_876600 | Canada/ON-SC0260/2020 | 2020-08-26 | North America/Canada/Ontario/Toronto | Toronto Invasive Bacterial Diseases Network | McMaster University | Allison McGeer et al |
| EPI_ISL_876601 | Canada/ON-SC0255/2020 | 2020-08-25 | North America/Canada/Ontario/Toronto | Toronto Invasive Bacterial Diseases Network | McMaster University | Allison McGeer et al |
| EPI_ISL_876602 | Canada/ON-SC0256/2020 | 2020-08-25 | North America/Canada/Ontario/Toronto | Toronto Invasive Bacterial Diseases Network | McMaster University | Allison McGeer et al |
| EPI_ISL_876603 | Canada/ON-SC0254/2020 | 2020-08-24 | North America/Canada/Ontario/Toronto | Toronto Invasive Bacterial Diseases Network | McMaster University | Allison McGeer et al |
| EPI_ISL_876604 | Canada/ON-SC0252/2020 | 2020-08-23 | North America/Canada/Ontario/Toronto | Toronto Invasive Bacterial Diseases Network | McMaster University | Allison McGeer et al |
| EPI_ISL_876605 | Canada/ON-SC0253/2020 | 2020-08-23 | North America/Canada/Ontario/Toronto | Toronto Invasive Bacterial Diseases Network | McMaster University | Allison McGeer et al |
| EPI_ISL_876606 | Canada/ON-SC0248/2020 | 2020-08-22 | North America/Canada/Ontario/Toronto | Toronto Invasive Bacterial Diseases Network | McMaster University | Allison McGeer et al |
| EPI_ISL_876607 | Canada/ON-SC0249/2020 | 2020-08-22 | North America/Canada/Ontario/Toronto | Toronto Invasive Bacterial Diseases Network | McMaster University | Allison McGeer et al |
| EPI_ISL_876608 | Canada/ON-SC0250/2020 | 2020-08-22 | North America/Canada/Ontario/Toronto | Toronto Invasive Bacterial Diseases Network | McMaster University | Allison McGeer et al |
| EPI_ISL_876609 | Canada/ON-SC0245/2020 | 2020-08-20 | North America/Canada/Ontario/Toronto | Toronto Invasive Bacterial Diseases Network | McMaster University | Allison McGeer et al |
| EPI_ISL_876610 | Canada/ON-SC0238/2020 | 2020-08-18 | North America/Canada/Ontario/Toronto | Toronto Invasive Bacterial Diseases Network | McMaster University | Allison McGeer et al |
| EPI_ISL_876611 | Canada/ON-SC0239/2020 | 2020-08-18 | North America/Canada/Ontario/Toronto | Toronto Invasive Bacterial Diseases Network | McMaster University | Allison McGeer et al |

| | | | | | | |
|---|---|---|---|---|---|---|
| EPI_ISL_876612 | Canada/ON-SC0232/2020 | 2020-08-16 | North America/Canada/Ontario/Toronto | Toronto Invasive Bacterial Diseases Network | McMaster University | Allison McGeer et al |
| EPI_ISL_876613 | Canada/ON-SC0229/2020 | 2020-08-15 | North America/Canada/Ontario/Toronto | Toronto Invasive Bacterial Diseases Network | McMaster University | Allison McGeer et al |
| EPI_ISL_876614 | Canada/ON-SC0231/2020 | 2020-08-15 | North America/Canada/Ontario/Toronto | Toronto Invasive Bacterial Diseases Network | McMaster University | Allison McGeer et al |
| EPI_ISL_876615 | Canada/ON-SC0228/2020 | 2020-08-14 | North America/Canada/Ontario/Toronto | Toronto Invasive Bacterial Diseases Network | McMaster University | Allison McGeer et al |
| EPI_ISL_876616 | Canada/ON-SC0234/2020 | 2020-08-14 | North America/Canada/Ontario/Toronto | Toronto Invasive Bacterial Diseases Network | McMaster University | Allison McGeer et al |
| EPI_ISL_876617 | Canada/ON-SC0225/2020 | 2020-08-13 | North America/Canada/Ontario/Toronto | Toronto Invasive Bacterial Diseases Network | McMaster University | Allison McGeer et al |
| EPI_ISL_876618 | Canada/ON-SC0226/2020 | 2020-08-13 | North America/Canada/Ontario/Toronto | Toronto Invasive Bacterial Diseases Network | McMaster University | Allison McGeer et al |
| EPI_ISL_876619 | Canada/ON-SC0223/2020 | 2020-08-12 | North America/Canada/Ontario/Toronto | Toronto Invasive Bacterial Diseases Network | McMaster University | Allison McGeer et al |
| EPI_ISL_876620 | Canada/ON-SC0219/2020 | 2020-08-10 | North America/Canada/Ontario/Toronto | Toronto Invasive Bacterial Diseases Network | McMaster University | Allison McGeer et al |
| EPI_ISL_876621 | Canada/ON-SC0215/2020 | 2020-08-09 | North America/Canada/Ontario/Toronto | Toronto Invasive Bacterial Diseases Network | McMaster University | Allison McGeer et al |
| EPI_ISL_876622 | Canada/ON-SC0213/2020 | 2020-08-08 | North America/Canada/Ontario/Toronto | Toronto Invasive Bacterial Diseases Network | McMaster University | Allison McGeer et al |
| EPI_ISL_876623 | Canada/ON-SC0211/2020 | 2020-08-07 | North America/Canada/Ontario/Toronto | Toronto Invasive Bacterial Diseases Network | McMaster University | Allison McGeer et al |
| EPI_ISL_876624 | Canada/ON-SC0212/2020 | 2020-08-07 | North America/Canada/Ontario/Toronto | Toronto Invasive Bacterial Diseases Network | McMaster University | Allison McGeer et al |
| EPI_ISL_876628 | USA/GA-CDC-STM-A100369/2021 | 2021-01-05 | North America/USA/Georgia/ | Helix/Illumina | Genomics and Discovery, Respiratory Viruses Branch, Division of Viral Diseases, Centers for Disease Control and Prevention | Peter W. Cook et al |

| | | | | | | |
|---|---|---|---|---|---|---|
| EPI_ISL_876629 | USA/AL-CDC-STM-A100319/2021 | 2021-01-05 | North America/USA/Alabama/ | Helix/Illumina | Genomics and Discovery, Respiratory Viruses Branch, Division of Viral Diseases, Centers for Disease Control and Prevention | Peter W. Cook et al |
| EPI_ISL_876651 | USA/FL-CDC-STM-A100285/2021 | 2021-01-05 | North America/USA/Florida/ | Helix/Illumina | Genomics and Discovery, Respiratory Viruses Branch, Division of Viral Diseases, Centers for Disease Control and Prevention | Peter W. Cook et al |
| EPI_ISL_876666 | USA/AZ-CDC-STM-A100291/2021 | 2021-01-05 | North America/USA/Arizona/ | Helix/Illumina | Genomics and Discovery, Respiratory Viruses Branch, Division of Viral Diseases, Centers for Disease Control and Prevention | Peter W. Cook et al |
| EPI_ISL_876670 | USA/CA-CDC-STM-A100359/2021 | 2021-01-06 | North America/USA/California/ | Helix/Illumina | Genomics and Discovery, Respiratory Viruses Branch, Division of Viral Diseases, Centers for Disease Control and Prevention | Peter W. Cook et al |

| | | | | | | |
|---|---|---|---|---|---|---|
| EPI_ISL_876691 | USA/IN-CDC-STM-A100352/2021 | 2021-01-05 | North America/USA/Indiana/ | Helix/Illumina | Genomics and Discovery, Respiratory Viruses Branch, Division of Viral Diseases, Centers for Disease Control and Prevention | Peter W. Cook et al |
| EPI_ISL_876697 | USA/KY-CDC-STM-A100259/2021 | 2021-01-03 | North America/USA/Kentucky/ | Helix/Illumina | Genomics and Discovery, Respiratory Viruses Branch, Division of Viral Diseases, Centers for Disease Control and Prevention | Peter W. Cook et al |
| EPI_ISL_876703 | USA/MI-CDC-STM-A100356/2021 | 2021-01-05 | North America/USA/Michigan/ | Helix/Illumina | Genomics and Discovery, Respiratory Viruses Branch, Division of Viral Diseases, Centers for Disease Control and Prevention | Peter W. Cook et al |
| EPI_ISL_876704 | USA/MI-CDC-STM-A100339/2021 | 2021-01-05 | North America/USA/Michigan/ | Helix/Illumina | Genomics and Discovery, Respiratory Viruses Branch, Division of Viral Diseases, Centers for Disease Control and Prevention | Peter W. Cook et al |

| | | | | | | |
|---|---|---|---|---|---|---|
| EPI_ISL_876708 | USA/NH-CDC-STM-A100344/2021 | 2021-01-05 | North America/USA/New Hampshire/ | Helix/Illumina | Genomics and Discovery, Respiratory Viruses Branch, Division of Viral Diseases, Centers for Disease Control and Prevention | Peter W. Cook et al |
| EPI_ISL_876718 | USA/OH-CDC-STM-A100394/2021 | 2021-01-06 | North America/USA/Ohio/ | Helix/Illumina | Genomics and Discovery, Respiratory Viruses Branch, Division of Viral Diseases, Centers for Disease Control and Prevention | Peter W. Cook et al |
| EPI_ISL_876723 | USA/RI-CDC-STM-A100367/2021 | 2021-01-05 | North America/USA/Rhode Island/ | Helix/Illumina | Genomics and Discovery, Respiratory Viruses Branch, Division of Viral Diseases, Centers for Disease Control and Prevention | Peter W. Cook et al |
| EPI_ISL_876727 | USA/TX-CDC-STM-A100474/2021 | 2021-01-06 | North America/USA/Texas/ | Helix/Illumina | Genomics and Discovery, Respiratory Viruses Branch, Division of Viral Diseases, Centers for Disease Control and Prevention | Peter W. Cook et al |
| EPI_ISL_876737 | USA/FL-BPHL-2840/2020 | 2020-08-16 | North America/USA/Florida/ | Florida Bureau of Public Health Laboratories | Florida Bureau of Public Health Laboratories | Sarah Schmedes et al |

| | | | | | | |
|---|---|---|---|---|---|---|
| EPI_ISL_876741 | Italy/APU-IZSPB-568PT/2021 | 2021-01-11 | Europe/Italy/Apulia/Foggia | Istituto Zooprofilattico Sperimentale della Puglia e della Basilicata | Istituto Zooprofilattico Sperimentale della Puglia e della Basilicata | Parisi A. et al |
| EPI_ISL_876746 | Italy/APU-IZSPB-502PT/2020 | 2020-12-28 | Europe/Italy/Apulia/Foggia | Istituto Zooprofilattico Sperimentale della Puglia e della Basilicata | Istituto Zooprofilattico Sperimentale della Puglia e della Basilicata | Parisi A. et al |
| EPI_ISL_876748 | Italy/APU-IZSPB-525PT/2020 | 2020-12-23 | Europe/Italy/Apulia/Foggia | Istituto Zooprofilattico Sperimentale della Puglia e della Basilicata | Istituto Zooprofilattico Sperimentale della Puglia e della Basilicata | Parisi A. et al |
| EPI_ISL_876749 | Italy/APU-IZSPB-526PT/2020 | 2020-12-23 | Europe/Italy/Apulia/Foggia | Istituto Zooprofilattico Sperimentale della Puglia e della Basilicata | Istituto Zooprofilattico Sperimentale della Puglia e della Basilicata | Parisi A. et al |
| EPI_ISL_876752 | Italy/APU-IZSPB-535PT/2020 | 2020-12-21 | Europe/Italy/Apulia/Foggia | Istituto Zooprofilattico Sperimentale della Puglia e della Basilicata | Istituto Zooprofilattico Sperimentale della Puglia e della Basilicata | Parisi A. et al |
| EPI_ISL_876756 | Italy/APU-IZSPB-544PT/2020 | 2020-12-21 | Europe/Italy/Apulia/Foggia | Istituto Zooprofilattico Sperimentale della Puglia e della Basilicata | Istituto Zooprofilattico Sperimentale della Puglia e della Basilicata | Parisi A. et al |
| EPI_ISL_876759 | Italy/APU-IZSPB-559PT/2020 | 2020-12-21 | Europe/Italy/Apulia/Foggia | Istituto Zooprofilattico Sperimentale della Puglia e della Basilicata | Istituto Zooprofilattico Sperimentale della Puglia e della Basilicata | Parisi A. et al |
| EPI_ISL_876777 | Italy/APU-IZSPB-518PT/2020 | 2020-12-17 | Europe/Italy/Apulia/Foggia | Istituto Zooprofilattico Sperimentale della Puglia e della Basilicata | Istituto Zooprofilattico Sperimentale della Puglia e della Basilicata | Parisi A. et al |

| | | | | | | |
|---|---|---|---|---|---|---|
| EPI_ISL_876780 | Italy/APU-IZSPB-530PT/2020 | 2020-12-16 | Europe/Italy/Apulia/Foggia | Istituto Zooprofilattico Sperimentale della Puglia e della Basilicata | Istituto Zooprofilattico Sperimentale della Puglia e della Basilicata | Parisi A. et al |
| EPI_ISL_876793 | Italy/APU-IZSPB-515PT/2020 | 2020-12-14 | Europe/Italy/Apulia/Foggia | Istituto Zooprofilattico Sperimentale della Puglia e della Basilicata | Istituto Zooprofilattico Sperimentale della Puglia e della Basilicata | Parisi A. et al |
| EPI_ISL_876795 | Italy/APU-IZSPB-506PT/2020 | 2020-12-12 | Europe/Italy/Apulia/Foggia | Istituto Zooprofilattico Sperimentale della Puglia e della Basilicata | Istituto Zooprofilattico Sperimentale della Puglia e della Basilicata | Parisi A. et al |
| EPI_ISL_876797 | Italy/APU-IZSPB-510PT/2020 | 2020-12-12 | Europe/Italy/Apulia/Foggia | Istituto Zooprofilattico Sperimentale della Puglia e della Basilicata | Istituto Zooprofilattico Sperimentale della Puglia e della Basilicata | Parisi A. et al |
| EPI_ISL_876799 | Italy/APU-IZSPB-513PT/2020 | 2020-12-12 | Europe/Italy/Apulia/Foggia | Istituto Zooprofilattico Sperimentale della Puglia e della Basilicata | Istituto Zooprofilattico Sperimentale della Puglia e della Basilicata | Parisi A. et al |
| EPI_ISL_876802 | Italy/APU-IZSPB-429PT/2020 | 2020-12-11 | Europe/Italy/Apulia/Foggia | Istituto Zooprofilattico Sperimentale della Puglia e della Basilicata | Istituto Zooprofilattico Sperimentale della Puglia e della Basilicata | Parisi A. et al |
| EPI_ISL_876810 | Italy/APU-IZSPB-567PT/2020 | 2020-10-24 | Europe/Italy/Apulia/Barletta-Andria-Tra | Istituto Zooprofilattico Sperimentale della Puglia e della Basilicata | Istituto Zooprofilattico Sperimentale della Puglia e della Basilicata | Parisi A. et al |
| EPI_ISL_876813 | Italy/APU-IZSPB-74PT/2020 | 2020-04-11 | Europe/Italy/Apulia/Brindisi | Istituto Zooprofilattico Sperimentale della Puglia e della Basilicata | Istituto Zooprofilattico Sperimentale della Puglia e della Basilicata | Parisi A. et al |

| | | | | | | |
|---|---|---|---|---|---|---|
| EPI_ISL_876814 | USA/NV-QDX-3533/2020 | 2020-09-29 | North America/USA/Nevada/ | Quest Diagnostics | Quest Diagnostics | Rosenthal et al |
| EPI_ISL_876817 | USA/FL-QDX-3482/2020 | 2020-08-27 | North America/USA/Florida/ | Quest Diagnostics | Quest Diagnostics | Rosenthal et al |
| EPI_ISL_876819 | USA/OK-QDX-3414/2020 | 2020-10-30 | North America/USA/Oklahoma/ | Quest Diagnostics | Quest Diagnostics | Rosenthal et al |
| EPI_ISL_876827 | USA/CA-QDX-3474/2020 | 2020-08-13 | North America/USA/California/ | Quest Diagnostics | Quest Diagnostics | Rosenthal et al |
| EPI_ISL_876837 | USA/MO-QDX-3567/2020 | 2020-11-06 | North America/USA/Missouri/ | Quest Diagnostics | Quest Diagnostics | Rosenthal et al |
| EPI_ISL_876842 | USA/KS-QDX-3570/2020 | 2020-11-05 | North America/USA/Kansas/ | Quest Diagnostics | Quest Diagnostics | Rosenthal et al |
| EPI_ISL_876844 | USA/NV-QDX-3558/2020 | 2020-11-05 | North America/USA/Nevada/ | Quest Diagnostics | Quest Diagnostics | Rosenthal et al |
| EPI_ISL_876855 | USA/GA-QDX-3555/2020 | 2020-10-30 | North America/USA/Georgia/ | Quest Diagnostics | Quest Diagnostics | Rosenthal et al |
| EPI_ISL_876860 | USA/GA-QDX-3553/2020 | 2020-10-29 | North America/USA/Georgia/ | Quest Diagnostics | Quest Diagnostics | Rosenthal et al |
| EPI_ISL_876863 | USA/TN-QDX-3554/2020 | 2020-10-28 | North America/USA/Tennessee/ | Quest Diagnostics | Quest Diagnostics | Rosenthal et al |
| EPI_ISL_876870 | USA/FL-QDX-3540/2020 | 2020-10-16 | North America/USA/Florida/ | Quest Diagnostics | Quest Diagnostics | Rosenthal et al |
| EPI_ISL_876875 | USA/OK-QDX-3532/2020 | 2020-10-13 | North America/USA/Oklahoma/ | Quest Diagnostics | Quest Diagnostics | Rosenthal et al |
| EPI_ISL_876878 | USA/OK-QDX-3531/2020 | 2020-10-07 | North America/USA/Oklahoma/ | Quest Diagnostics | Quest Diagnostics | Rosenthal et al |
| EPI_ISL_876882 | USA/CA-QDX-3517/2020 | 2020-09-18 | North America/USA/California/ | Quest Diagnostics | Quest Diagnostics | Rosenthal et al |
| EPI_ISL_876889 | USA/FL-QDX-3505/2020 | 2020-09-16 | North America/USA/Florida/ | Quest Diagnostics | Quest Diagnostics | Rosenthal et al |
| EPI_ISL_876893 | USA/TX-QDX-3504/2020 | 2020-09-13 | North America/USA/Texas/ | Quest Diagnostics | Quest Diagnostics | Rosenthal et al |
| EPI_ISL_876898 | USA/KY-QDX-3500/2020 | 2020-09-10 | North America/USA/Kentucky/ | Quest Diagnostics | Quest Diagnostics | Rosenthal et al |
| EPI_ISL_876900 | USA/CO-QDX-3512/2020 | 2020-09-09 | North America/USA/Colorado/ | Quest Diagnostics | Quest Diagnostics | Rosenthal et al |
| EPI_ISL_876911 | USA/KY-QDX-3490/2020 | 2020-09-02 | North America/USA/Kentucky/ | Quest Diagnostics | Quest Diagnostics | Rosenthal et al |
| EPI_ISL_876913 | USA/CA-QDX-3479/2020 | 2020-08-30 | North America/USA/California/ | Quest Diagnostics | Quest Diagnostics | Rosenthal et al |
| EPI_ISL_876916 | USA/WI-QDX-3491/2020 | 2020-08-29 | North America/USA/Wisconsin/ | Quest Diagnostics | Quest Diagnostics | Rosenthal et al |
| EPI_ISL_876919 | USA/FL-QDX-3481/2020 | 2020-08-27 | North America/USA/Florida/ | Quest Diagnostics | Quest Diagnostics | Rosenthal et al |
| EPI_ISL_876921 | USA/IL-QDX-3470/2020 | 2020-08-25 | North America/USA/Illinois/ | Quest Diagnostics | Quest Diagnostics | Rosenthal et al |
| EPI_ISL_876925 | USA/NY-QDX-3514/2020 | 2020-08-23 | North America/USA/New York/ | Quest Diagnostics | Quest Diagnostics | Rosenthal et al |
| EPI_ISL_876928 | USA/NC-QDX-3465/2020 | 2020-08-20 | North America/USA/North Carolina/ | Quest Diagnostics | Quest Diagnostics | Rosenthal et al |
| EPI_ISL_876931 | USA/TX-QDX-3460/2020 | 2020-08-17 | North America/USA/Texas/ | Quest Diagnostics | Quest Diagnostics | Rosenthal et al |
| EPI_ISL_876934 | USA/FL-QDX-3455/2020 | 2020-08-15 | North America/USA/Florida/ | Quest Diagnostics | Quest Diagnostics | Rosenthal et al |
| EPI_ISL_876935 | USA/FL-QDX-3457/2020 | 2020-08-15 | North America/USA/Florida/ | Quest Diagnostics | Quest Diagnostics | Rosenthal et al |
| EPI_ISL_876940 | USA/GA-QDX-3454/2020 | 2020-08-13 | North America/USA/Georgia/ | Quest Diagnostics | Quest Diagnostics | Rosenthal et al |
| EPI_ISL_876945 | USA/OR-QDX-3447/2020 | 2020-08-11 | North America/USA/Oregon/ | Quest Diagnostics | Quest Diagnostics | Rosenthal et al |
| EPI_ISL_876954 | USA/FL-QDX-3432/2020 | 2020-07-14 | North America/USA/Florida/ | Quest Diagnostics | Quest Diagnostics | Rosenthal et al |

| | | | | | | |
|---|---|---|---|---|---|---|
| EPI_ISL_876959 | USA/FL-QDX-3429/2020 | 2020-07-08 | North America/USA/Florida/ | Quest Diagnostics | Quest Diagnostics | Rosenthal et al |
| EPI_ISL_876962 | USA/PA-QDX-3426/2020 | 2020-07-07 | North America/USA/Pennsylvania/ | Quest Diagnostics | Quest Diagnostics | Rosenthal et al |
| EPI_ISL_876964 | USA/TX-QDX-3435/2020 | 2020-07-03 | North America/USA/Texas/ | Quest Diagnostics | Quest Diagnostics | Rosenthal et al |
| EPI_ISL_876975 | Italy/CAM-CRGS-25/2020 | 2020-03-18 | Europe/Italy/Campania/ | 1.AO Universitaria 'S. Giovanni di Dio e Ruggi D'Aragona, Scuola Medica Salernitana' Hospital / 2.UOC di Virologia e Microbiologia, Universit√† della Campania 'L. Vanvitelli' / 3.AO Universitaria 'Federico II' Napoli Hospital / 4.AORN 'San Giuseppe Moscati' Avellino Hospital / 5.AO 'San Pio - presidio G. Rummo' Benevento Hospital / 6.AO 'Sant'Anna e San Sebastiano' Caserta Hospital / 7.PO 'Maria Santissima Addolorata' Eboli Hospital / 8.Biogem Istituto di Ricerche Genetiche | 1. Genome Research Center for Health (CRGS) / 2. Laboratory of Molecular Medicine and Genomics(LMMGe) / 3. Center for Research in Pure and Applied Mathematics (CRMPA) | Giorgio Giurato (Corresponding Author) et al |
| EPI_ISL_876976 | Italy/CAM-CRGS-11/2020 | 2020-03-15 | Europe/Italy/Campania/ | 1.AO Universitaria 'S. Giovanni di Dio e Ruggi D'Aragona, Scuola Medica Salernitana' Hospital / 2.UOC di Virologia e Microbiologia, Universit√† della Campania 'L. Vanvitelli' / 3.AO Universitaria 'Federico II' Napoli Hospital / 4.AORN 'San Giuseppe Moscati' Avellino Hospital / 5.AO 'San Pio - presidio G. Rummo' Benevento Hospital / 6.AO 'Sant'Anna e San Sebastiano' Caserta Hospital / 7.PO 'Maria Santissima Addolorata' Eboli Hospital / 8.Biogem Istituto di Ricerche Genetiche | 1. Genome Research Center for Health (CRGS) / 2. Laboratory of Molecular Medicine and Genomics(LMMGe) / 3. Center for Research in Pure and Applied Mathematics (CRMPA) | Giorgio Giurato (Corresponding Author) et al |
| EPI_ISL_876978 | Italy/CAM-CRGS-12/2020 | 2020-03-15 | Europe/Italy/Campania/ | 1.AO Universitaria 'S. Giovanni di Dio e Ruggi D'Aragona, Scuola Medica Salernitana' Hospital / 2.UOC di Virologia e Microbiologia, Universit√† della Campania 'L. Vanvitelli' / 3.AO Universitaria 'Federico II' Napoli Hospital / 4.AORN 'San Giuseppe Moscati' Avellino Hospital / 5.AO 'San Pio - presidio G. Rummo' Benevento Hospital / 6.AO 'Sant'Anna e San Sebastiano' Caserta Hospital / 7.PO 'Maria Santissima Addolorata' Eboli Hospital / 8.Biogem Istituto di Ricerche Genetiche | 1. Genome Research Center for Health (CRGS) / 2. Laboratory of Molecular Medicine and Genomics(LMMGe) / 3. Center for Research in Pure and Applied Mathematics (CRMPA) | Giorgio Giurato (Corresponding Author) et al |

| | | | | | | |
|---|---|---|---|---|---|---|
| EPI_ISL_876979 | Italy/CAM-CRGS-8/2020 | 2020-03-14 | Europe/Italy/Campania/ | 1.AO Universitaria 'S. Giovanni di Dio e Ruggi D'Aragona, Scuola Medica Salernitana' Hospital / 2.UOC di Virologia e Microbiologia, Universit√† della Campania 'L. Vanvitelli' / 3.AO Universitaria 'Federico II' Napoli Hospital / 4.AORN 'San Giuseppe Moscati' Avellino Hospital / 5.AO 'San Pio - presidio G. Rummo' Benevento Hospital / 6.AO 'Sant'Anna e San Sebastiano' Caserta Hospital / 7.PO 'Maria Santissima Addolorata' Eboli Hospital / 8.Biogem Istituto di Ricerche Genetiche | 1. Genome Research Center for Health (CRGS) / 2. Laboratory of Molecular Medicine and Genomics(LMMGe) / 3. Center for Research in Pure and Applied Mathematics (CRMPA) | Giorgio Giurato (Corresponding Author) et al |
| EPI_ISL_876980 | Italy/CAM-CRGS-4/2020 | 2020-03-14 | Europe/Italy/Campania/ | 1.AO Universitaria 'S. Giovanni di Dio e Ruggi D'Aragona, Scuola Medica Salernitana' Hospital / 2.UOC di Virologia e Microbiologia, Universit√† della Campania 'L. Vanvitelli' / 3.AO Universitaria 'Federico II' Napoli Hospital / 4.AORN 'San Giuseppe Moscati' Avellino Hospital / 5.AO 'San Pio - presidio G. Rummo' Benevento Hospital / 6.AO 'Sant'Anna e San Sebastiano' Caserta Hospital / 7.PO 'Maria Santissima Addolorata' Eboli Hospital / 8.Biogem Istituto di Ricerche Genetiche | 1. Genome Research Center for Health (CRGS) / 2. Laboratory of Molecular Medicine and Genomics(LMMGe) / 3. Center for Research in Pure and Applied Mathematics (CRMPA) | Giorgio Giurato (Corresponding Author) et al |
| EPI_ISL_876981 | Italy/CAM-CRGS-5/2020 | 2020-03-14 | Europe/Italy/Campania/ | 1.AO Universitaria 'S. Giovanni di Dio e Ruggi D'Aragona, Scuola Medica Salernitana' Hospital / 2.UOC di Virologia e Microbiologia, Universit√† della Campania 'L. Vanvitelli' / 3.AO Universitaria 'Federico II' Napoli Hospital / 4.AORN 'San Giuseppe Moscati' Avellino Hospital / 5.AO 'San Pio - presidio G. Rummo' Benevento Hospital / 6.AO 'Sant'Anna e San Sebastiano' Caserta Hospital / 7.PO 'Maria Santissima Addolorata' Eboli Hospital / 8.Biogem Istituto di Ricerche Genetiche | 1. Genome Research Center for Health (CRGS) / 2. Laboratory of Molecular Medicine and Genomics(LMMGe) / 3. Center for Research in Pure and Applied Mathematics (CRMPA) | Giorgio Giurato (Corresponding Author) et al |

| | | | | | | |
|---|---|---|---|---|---|---|
| EPI_ISL_876983 | Italy/CAM-CRGS-9/2020 | 2020-03-15 | Europe/Italy/Campania/ | 1.AO Universitaria 'S. Giovanni di Dio e Ruggi D'Aragona, Scuola Medica Salernitana' Hospital / 2.UOC di Virologia e Microbiologia, Universit√† della Campania 'L. Vanvitelli' / 3.AO Universitaria 'Federico II' Napoli Hospital / 4.AORN 'San Giuseppe Moscati' Avellino Hospital / 5.AO 'San Pio - presidio G. Rummo' Benevento Hospital / 6.AO 'Sant'Anna e San Sebastiano' Caserta Hospital / 7.PO 'Maria Santissima Addolorata' Eboli Hospital / 8.Biogem Istituto di Ricerche Genetiche | 1. Genome Research Center for Health (CRGS) / 2. Laboratory of Molecular Medicine and Genomics(LMMGe) / 3. Center for Research in Pure and Applied Mathematics (CRMPA) | Giorgio Giurato (Corresponding Author) et al |
| EPI_ISL_876989 | Italy/CAM-CRGS-24/2020 | 2020-03-18 | Europe/Italy/Campania/ | 1.AO Universitaria 'S. Giovanni di Dio e Ruggi D'Aragona, Scuola Medica Salernitana' Hospital / 2.UOC di Virologia e Microbiologia, Universit√† della Campania 'L. Vanvitelli' / 3.AO Universitaria 'Federico II' Napoli Hospital / 4.AORN 'San Giuseppe Moscati' Avellino Hospital / 5.AO 'San Pio - presidio G. Rummo' Benevento Hospital / 6.AO 'Sant'Anna e San Sebastiano' Caserta Hospital / 7.PO 'Maria Santissima Addolorata' Eboli Hospital / 8.Biogem Istituto di Ricerche Genetiche | 1. Genome Research Center for Health (CRGS) / 2. Laboratory of Molecular Medicine and Genomics(LMMGe) / 3. Center for Research in Pure and Applied Mathematics (CRMPA) | Giorgio Giurato (Corresponding Author) et al |
| EPI_ISL_876992 | Italy/CAM-CRGS-28/2020 | 2020-03-18 | Europe/Italy/Campania/ | 1.AO Universitaria 'S. Giovanni di Dio e Ruggi D'Aragona, Scuola Medica Salernitana' Hospital / 2.UOC di Virologia e Microbiologia, Universit√† della Campania 'L. Vanvitelli' / 3.AO Universitaria 'Federico II' Napoli Hospital / 4.AORN 'San Giuseppe Moscati' Avellino Hospital / 5.AO 'San Pio - presidio G. Rummo' Benevento Hospital / 6.AO 'Sant'Anna e San Sebastiano' Caserta Hospital / 7.PO 'Maria Santissima Addolorata' Eboli Hospital / 8.Biogem Istituto di Ricerche Genetiche | 1. Genome Research Center for Health (CRGS) / 2. Laboratory of Molecular Medicine and Genomics(LMMGe) / 3. Center for Research in Pure and Applied Mathematics (CRMPA) | Giorgio Giurato (Corresponding Author) et al |

| | | | | | | |
|---|---|---|---|---|---|---|
| EPI_ISL_876993 | Italy/CAM-CRGS-29/2020 | 2020-03-18 | Europe/Italy/Campania/ | 1.AO Universitaria 'S. Giovanni di Dio e Ruggi D'Aragona, Scuola Medica Salernitana' Hospital / 2.UOC di Virologia e Microbiologia, Universit√† della Campania 'L. Vanvitelli' / 3.AO Universitaria 'Federico II' Napoli Hospital / 4.AORN 'San Giuseppe Moscati' Avellino Hospital / 5.AO 'San Pio - presidio G. Rummo' Benevento Hospital / 6.AO 'Sant'Anna e San Sebastiano' Caserta Hospital / 7.PO 'Maria Santissima Addolorata' Eboli Hospital / 8.Biogem Istituto di Ricerche Genetiche | 1. Genome Research Center for Health (CRGS) / 2. Laboratory of Molecular Medicine and Genomics(LMMGe) / 3. Center for Research in Pure and Applied Mathematics (CRMPA) | Giorgio Giurato (Corresponding Author) et al |
| EPI_ISL_876996 | Italy/CAM-CRGS-94/2020 | 2020-03-18 | Europe/Italy/Campania/ | 1.AO Universitaria 'S. Giovanni di Dio e Ruggi D'Aragona, Scuola Medica Salernitana' Hospital / 2.UOC di Virologia e Microbiologia, Universit√† della Campania 'L. Vanvitelli' / 3.AO Universitaria 'Federico II' Napoli Hospital / 4.AORN 'San Giuseppe Moscati' Avellino Hospital / 5.AO 'San Pio - presidio G. Rummo' Benevento Hospital / 6.AO 'Sant'Anna e San Sebastiano' Caserta Hospital / 7.PO 'Maria Santissima Addolorata' Eboli Hospital / 8.Biogem Istituto di Ricerche Genetiche | 1. Genome Research Center for Health (CRGS) / 2. Laboratory of Molecular Medicine and Genomics(LMMGe) / 3. Center for Research in Pure and Applied Mathematics (CRMPA) | Giorgio Giurato (Corresponding Author) et al |
| EPI_ISL_876998 | Italy/CAM-CRGS-32/2020 | 2020-03-19 | Europe/Italy/Campania/ | 1.AO Universitaria 'S. Giovanni di Dio e Ruggi D'Aragona, Scuola Medica Salernitana' Hospital / 2.UOC di Virologia e Microbiologia, Universit√† della Campania 'L. Vanvitelli' / 3.AO Universitaria 'Federico II' Napoli Hospital / 4.AORN 'San Giuseppe Moscati' Avellino Hospital / 5.AO 'San Pio - presidio G. Rummo' Benevento Hospital / 6.AO 'Sant'Anna e San Sebastiano' Caserta Hospital / 7.PO 'Maria Santissima Addolorata' Eboli Hospital / 8.Biogem Istituto di Ricerche Genetiche | 1. Genome Research Center for Health (CRGS) / 2. Laboratory of Molecular Medicine and Genomics(LMMGe) / 3. Center for Research in Pure and Applied Mathematics (CRMPA) | Giorgio Giurato (Corresponding Author) et al |

| | | | | | | |
|---|---|---|---|---|---|---|
| EPI_ISL_876999 | Italy/CAM-CRGS-37/2020 | 2020-03-19 | Europe/Italy/Campania/ | 1.AO Universitaria 'S. Giovanni di Dio e Ruggi D'Aragona, Scuola Medica Salernitana' Hospital / 2.UOC di Virologia e Microbiologia, Universit√† della Campania 'L. Vanvitelli' / 3.AO Universitaria 'Federico II' Napoli Hospital / 4.AORN 'San Giuseppe Moscati' Avellino Hospital / 5.AO 'San Pio - presidio G. Rummo' Benevento Hospital / 6.AO 'Sant'Anna e San Sebastiano' Caserta Hospital / 7.PO 'Maria Santissima Addolorata' Eboli Hospital / 8.Biogem Istituto di Ricerche Genetiche | 1. Genome Research Center for Health (CRGS) / 2. Laboratory of Molecular Medicine and Genomics(LMMGe) / 3. Center for Research in Pure and Applied Mathematics (CRMPA) | Giorgio Giurato (Corresponding Author) et al |
| EPI_ISL_877003 | Italy/CAM-CRGS-97/2020 | 2020-03-20 | Europe/Italy/Campania/ | 1.AO Universitaria 'S. Giovanni di Dio e Ruggi D'Aragona, Scuola Medica Salernitana' Hospital / 2.UOC di Virologia e Microbiologia, Universit√† della Campania 'L. Vanvitelli' / 3.AO Universitaria 'Federico II' Napoli Hospital / 4.AORN 'San Giuseppe Moscati' Avellino Hospital / 5.AO 'San Pio - presidio G. Rummo' Benevento Hospital / 6.AO 'Sant'Anna e San Sebastiano' Caserta Hospital / 7.PO 'Maria Santissima Addolorata' Eboli Hospital / 8.Biogem Istituto di Ricerche Genetiche | 1. Genome Research Center for Health (CRGS) / 2. Laboratory of Molecular Medicine and Genomics(LMMGe) / 3. Center for Research in Pure and Applied Mathematics (CRMPA) | Giorgio Giurato (Corresponding Author) et al |
| EPI_ISL_877005 | Italy/CAM-CRGS-101/2020 | 2020-03-23 | Europe/Italy/Campania/ | 1.AO Universitaria 'S. Giovanni di Dio e Ruggi D'Aragona, Scuola Medica Salernitana' Hospital / 2.UOC di Virologia e Microbiologia, Universit√† della Campania 'L. Vanvitelli' / 3.AO Universitaria 'Federico II' Napoli Hospital / 4.AORN 'San Giuseppe Moscati' Avellino Hospital / 5.AO 'San Pio - presidio G. Rummo' Benevento Hospital / 6.AO 'Sant'Anna e San Sebastiano' Caserta Hospital / 7.PO 'Maria Santissima Addolorata' Eboli Hospital / 8.Biogem Istituto di Ricerche Genetiche | 1. Genome Research Center for Health (CRGS) / 2. Laboratory of Molecular Medicine and Genomics(LMMGe) / 3. Center for Research in Pure and Applied Mathematics (CRMPA) | Giorgio Giurato (Corresponding Author) et al |

| | | | | | | |
|---|---|---|---|---|---|---|
| EPI_ISL_877006 | Italy/CAM-CRGS-102/2020 | 2020-03-23 | Europe/Italy/Campania/ | 1.AO Universitaria 'S. Giovanni di Dio e Ruggi D'Aragona, Scuola Medica Salernitana' Hospital / 2.UOC di Virologia e Microbiologia, Universit√† della Campania 'L. Vanvitelli' / 3.AO Universitaria 'Federico II' Napoli Hospital / 4.AORN 'San Giuseppe Moscati' Avellino Hospital / 5.AO 'San Pio - presidio G. Rummo' Benevento Hospital / 6.AO 'Sant'Anna e San Sebastiano' Caserta Hospital / 7.PO 'Maria Santissima Addolorata' Eboli Hospital / 8.Biogem Istituto di Ricerche Genetiche | 1. Genome Research Center for Health (CRGS) / 2. Laboratory of Molecular Medicine and Genomics(LMMGe) / 3. Center for Research in Pure and Applied Mathematics (CRMPA) | Giorgio Giurato (Corresponding Author) et al |
| EPI_ISL_877008 | Italy/CAM-CRGS-44/2020 | 2020-03-24 | Europe/Italy/Campania/ | 1.AO Universitaria 'S. Giovanni di Dio e Ruggi D'Aragona, Scuola Medica Salernitana' Hospital / 2.UOC di Virologia e Microbiologia, Universit√† della Campania 'L. Vanvitelli' / 3.AO Universitaria 'Federico II' Napoli Hospital / 4.AORN 'San Giuseppe Moscati' Avellino Hospital / 5.AO 'San Pio - presidio G. Rummo' Benevento Hospital / 6.AO 'Sant'Anna e San Sebastiano' Caserta Hospital / 7.PO 'Maria Santissima Addolorata' Eboli Hospital / 8.Biogem Istituto di Ricerche Genetiche | 1. Genome Research Center for Health (CRGS) / 2. Laboratory of Molecular Medicine and Genomics(LMMGe) / 3. Center for Research in Pure and Applied Mathematics (CRMPA) | Giorgio Giurato (Corresponding Author) et al |
| EPI_ISL_877009 | Italy/CAM-CRGS-45/2020 | 2020-03-25 | Europe/Italy/Campania/ | 1.AO Universitaria 'S. Giovanni di Dio e Ruggi D'Aragona, Scuola Medica Salernitana' Hospital / 2.UOC di Virologia e Microbiologia, Universit√† della Campania 'L. Vanvitelli' / 3.AO Universitaria 'Federico II' Napoli Hospital / 4.AORN 'San Giuseppe Moscati' Avellino Hospital / 5.AO 'San Pio - presidio G. Rummo' Benevento Hospital / 6.AO 'Sant'Anna e San Sebastiano' Caserta Hospital / 7.PO 'Maria Santissima Addolorata' Eboli Hospital / 8.Biogem Istituto di Ricerche Genetiche | 1. Genome Research Center for Health (CRGS) / 2. Laboratory of Molecular Medicine and Genomics(LMMGe) / 3. Center for Research in Pure and Applied Mathematics (CRMPA) | Giorgio Giurato (Corresponding Author) et al |

| | | | | | | |
|---|---|---|---|---|---|---|
| EPI_ISL_877010 | Italy/CAM-CRGS-46/2020 | 2020-03-25 | Europe/Italy/Campania/ | 1.AO Universitaria 'S. Giovanni di Dio e Ruggi D'Aragona, Scuola Medica Salernitana' Hospital / 2.UOC di Virologia e Microbiologia, Universit√† della Campania 'L. Vanvitelli' / 3.AO Universitaria 'Federico II' Napoli Hospital / 4.AORN 'San Giuseppe Moscati' Avellino Hospital / 5.AO 'San Pio - presidio G. Rummo' Benevento Hospital / 6.AO 'Sant'Anna e San Sebastiano' Caserta Hospital / 7.PO 'Maria Santissima Addolorata' Eboli Hospital / 8.Biogem Istituto di Ricerche Genetiche | 1. Genome Research Center for Health (CRGS) / 2. Laboratory of Molecular Medicine and Genomics(LMMGe) / 3. Center for Research in Pure and Applied Mathematics (CRMPA) | Giorgio Giurato (Corresponding Author) et al |
| EPI_ISL_877011 | Italy/CAM-CRGS-57/2020 | 2020-03-25 | Europe/Italy/Campania/ | 1.AO Universitaria 'S. Giovanni di Dio e Ruggi D'Aragona, Scuola Medica Salernitana' Hospital / 2.UOC di Virologia e Microbiologia, Universit√† della Campania 'L. Vanvitelli' / 3.AO Universitaria 'Federico II' Napoli Hospital / 4.AORN 'San Giuseppe Moscati' Avellino Hospital / 5.AO 'San Pio - presidio G. Rummo' Benevento Hospital / 6.AO 'Sant'Anna e San Sebastiano' Caserta Hospital / 7.PO 'Maria Santissima Addolorata' Eboli Hospital / 8.Biogem Istituto di Ricerche Genetiche | 1. Genome Research Center for Health (CRGS) / 2. Laboratory of Molecular Medicine and Genomics(LMMGe) / 3. Center for Research in Pure and Applied Mathematics (CRMPA) | Giorgio Giurato (Corresponding Author) et al |
| EPI_ISL_877013 | Italy/CAM-CRGS-47/2020 | 2020-03-30 | Europe/Italy/Campania/ | 1.AO Universitaria 'S. Giovanni di Dio e Ruggi D'Aragona, Scuola Medica Salernitana' Hospital / 2.UOC di Virologia e Microbiologia, Universit√† della Campania 'L. Vanvitelli' / 3.AO Universitaria 'Federico II' Napoli Hospital / 4.AORN 'San Giuseppe Moscati' Avellino Hospital / 5.AO 'San Pio - presidio G. Rummo' Benevento Hospital / 6.AO 'Sant'Anna e San Sebastiano' Caserta Hospital / 7.PO 'Maria Santissima Addolorata' Eboli Hospital / 8.Biogem Istituto di Ricerche Genetiche | 1. Genome Research Center for Health (CRGS) / 2. Laboratory of Molecular Medicine and Genomics(LMMGe) / 3. Center for Research in Pure and Applied Mathematics (CRMPA) | Giorgio Giurato (Corresponding Author) et al |

| | | | | | | |
|---|---|---|---|---|---|---|
| EPI_ISL_877015 | Italy/CAM-CRGS-69/2020 | 2020-04-01 | Europe/Italy/Campania/ | 1.AO Universitaria 'S. Giovanni di Dio e Ruggi D'Aragona, Scuola Medica Salernitana' Hospital / 2.UOC di Virologia e Microbiologia, Universit√† della Campania 'L. Vanvitelli' / 3.AO Universitaria 'Federico II' Napoli Hospital / 4.AORN 'San Giuseppe Moscati' Avellino Hospital / 5.AO 'San Pio - presidio G. Rummo' Benevento Hospital / 6.AO 'Sant'Anna e San Sebastiano' Caserta Hospital / 7.PO 'Maria Santissima Addolorata' Eboli Hospital / 8.Biogem Istituto di Ricerche Genetiche | 1. Genome Research Center for Health (CRGS) / 2. Laboratory of Molecular Medicine and Genomics(LMMGe) / 3. Center for Research in Pure and Applied Mathematics (CRMPA) | Giorgio Giurato (Corresponding Author) et al |
| EPI_ISL_877019 | Italy/CAM-CRGS-72/2020 | 2020-04-03 | Europe/Italy/Campania/ | 1.AO Universitaria 'S. Giovanni di Dio e Ruggi D'Aragona, Scuola Medica Salernitana' Hospital / 2.UOC di Virologia e Microbiologia, Universit√† della Campania 'L. Vanvitelli' / 3.AO Universitaria 'Federico II' Napoli Hospital / 4.AORN 'San Giuseppe Moscati' Avellino Hospital / 5.AO 'San Pio - presidio G. Rummo' Benevento Hospital / 6.AO 'Sant'Anna e San Sebastiano' Caserta Hospital / 7.PO 'Maria Santissima Addolorata' Eboli Hospital / 8.Biogem Istituto di Ricerche Genetiche | 1. Genome Research Center for Health (CRGS) / 2. Laboratory of Molecular Medicine and Genomics(LMMGe) / 3. Center for Research in Pure and Applied Mathematics (CRMPA) | Giorgio Giurato (Corresponding Author) et al |
| EPI_ISL_877020 | Italy/CAM-CRGS-75/2020 | 2020-04-03 | Europe/Italy/Campania/ | 1.AO Universitaria 'S. Giovanni di Dio e Ruggi D'Aragona, Scuola Medica Salernitana' Hospital / 2.UOC di Virologia e Microbiologia, Universit√† della Campania 'L. Vanvitelli' / 3.AO Universitaria 'Federico II' Napoli Hospital / 4.AORN 'San Giuseppe Moscati' Avellino Hospital / 5.AO 'San Pio - presidio G. Rummo' Benevento Hospital / 6.AO 'Sant'Anna e San Sebastiano' Caserta Hospital / 7.PO 'Maria Santissima Addolorata' Eboli Hospital / 8.Biogem Istituto di Ricerche Genetiche | 1. Genome Research Center for Health (CRGS) / 2. Laboratory of Molecular Medicine and Genomics(LMMGe) / 3. Center for Research in Pure and Applied Mathematics (CRMPA) | Giorgio Giurato (Corresponding Author) et al |

| | | | | | | |
|---|---|---|---|---|---|---|
| EPI_ISL_877022 | Italy/CAM-CRGS-108/2020 | 2020-04-04 | Europe/Italy/Campania/ | 1.AO Universitaria 'S. Giovanni di Dio e Ruggi D'Aragona, Scuola Medica Salernitana' Hospital / 2.UOC di Virologia e Microbiologia, Universit√† della Campania 'L. Vanvitelli' / 3.AO Universitaria 'Federico II' Napoli Hospital / 4.AORN 'San Giuseppe Moscati' Avellino Hospital / 5.AO 'San Pio - presidio G. Rummo' Benevento Hospital / 6.AO 'Sant'Anna e San Sebastiano' Caserta Hospital / 7.PO 'Maria Santissima Addolorata' Eboli Hospital / 8.Biogem Istituto di Ricerche Genetiche | 1. Genome Research Center for Health (CRGS) / 2. Laboratory of Molecular Medicine and Genomics(LMMGe) / 3. Center for Research in Pure and Applied Mathematics (CRMPA) | Giorgio Giurato (Corresponding Author) et al |
| EPI_ISL_877025 | Italy/CAM-CRGS-84/2020 | 2020-04-13 | Europe/Italy/Campania/ | 1.AO Universitaria 'S. Giovanni di Dio e Ruggi D'Aragona, Scuola Medica Salernitana' Hospital / 2.UOC di Virologia e Microbiologia, Universit√† della Campania 'L. Vanvitelli' / 3.AO Universitaria 'Federico II' Napoli Hospital / 4.AORN 'San Giuseppe Moscati' Avellino Hospital / 5.AO 'San Pio - presidio G. Rummo' Benevento Hospital / 6.AO 'Sant'Anna e San Sebastiano' Caserta Hospital / 7.PO 'Maria Santissima Addolorata' Eboli Hospital / 8.Biogem Istituto di Ricerche Genetiche | 1. Genome Research Center for Health (CRGS) / 2. Laboratory of Molecular Medicine and Genomics(LMMGe) / 3. Center for Research in Pure and Applied Mathematics (CRMPA) | Giorgio Giurato (Corresponding Author) et al |
| EPI_ISL_877029 | Italy/CAM-CRGS-59/2020 | 2020-04-18 | Europe/Italy/Campania/ | 1.AO Universitaria 'S. Giovanni di Dio e Ruggi D'Aragona, Scuola Medica Salernitana' Hospital / 2.UOC di Virologia e Microbiologia, Universit√† della Campania 'L. Vanvitelli' / 3.AO Universitaria 'Federico II' Napoli Hospital / 4.AORN 'San Giuseppe Moscati' Avellino Hospital / 5.AO 'San Pio - presidio G. Rummo' Benevento Hospital / 6.AO 'Sant'Anna e San Sebastiano' Caserta Hospital / 7.PO 'Maria Santissima Addolorata' Eboli Hospital / 8.Biogem Istituto di Ricerche Genetiche | 1. Genome Research Center for Health (CRGS) / 2. Laboratory of Molecular Medicine and Genomics(LMMGe) / 3. Center for Research in Pure and Applied Mathematics (CRMPA) | Giorgio Giurato (Corresponding Author) et al |

| | | | | | | |
|---|---|---|---|---|---|---|
| EPI_ISL_877031 | Italy/CAM-CRGS-87/2020 | 2020-04-19 | Europe/Italy/Campania/ | 1.AO Universitaria 'S. Giovanni di Dio e Ruggi D'Aragona, Scuola Medica Salernitana' Hospital / 2.UOC di Virologia e Microbiologia, Universit√† della Campania 'L. Vanvitelli' / 3.AO Universitaria 'Federico II' Napoli Hospital / 4.AORN 'San Giuseppe Moscati' Avellino Hospital / 5.AO 'San Pio - presidio G. Rummo' Benevento Hospital / 6.AO 'Sant'Anna e San Sebastiano' Caserta Hospital / 7.PO 'Maria Santissima Addolorata' Eboli Hospital / 8.Biogem Istituto di Ricerche Genetiche | 1. Genome Research Center for Health (CRGS) / 2. Laboratory of Molecular Medicine and Genomics(LMMGe) / 3. Center for Research in Pure and Applied Mathematics (CRMPA) | Giorgio Giurato (Corresponding Author) et al |
| EPI_ISL_877032 | Italy/CAM-CRGS-116/2020 | 2020-04-21 | Europe/Italy/Campania/ | 1.AO Universitaria 'S. Giovanni di Dio e Ruggi D'Aragona, Scuola Medica Salernitana' Hospital / 2.UOC di Virologia e Microbiologia, Universit√† della Campania 'L. Vanvitelli' / 3.AO Universitaria 'Federico II' Napoli Hospital / 4.AORN 'San Giuseppe Moscati' Avellino Hospital / 5.AO 'San Pio - presidio G. Rummo' Benevento Hospital / 6.AO 'Sant'Anna e San Sebastiano' Caserta Hospital / 7.PO 'Maria Santissima Addolorata' Eboli Hospital / 8.Biogem Istituto di Ricerche Genetiche | 1. Genome Research Center for Health (CRGS) / 2. Laboratory of Molecular Medicine and Genomics(LMMGe) / 3. Center for Research in Pure and Applied Mathematics (CRMPA) | Giorgio Giurato (Corresponding Author) et al |
| EPI_ISL_877033 | Italy/CAM-CRGS-88/2020 | 2020-04-25 | Europe/Italy/Campania/ | 1.AO Universitaria 'S. Giovanni di Dio e Ruggi D'Aragona, Scuola Medica Salernitana' Hospital / 2.UOC di Virologia e Microbiologia, Universit√† della Campania 'L. Vanvitelli' / 3.AO Universitaria 'Federico II' Napoli Hospital / 4.AORN 'San Giuseppe Moscati' Avellino Hospital / 5.AO 'San Pio - presidio G. Rummo' Benevento Hospital / 6.AO 'Sant'Anna e San Sebastiano' Caserta Hospital / 7.PO 'Maria Santissima Addolorata' Eboli Hospital / 8.Biogem Istituto di Ricerche Genetiche | 1. Genome Research Center for Health (CRGS) / 2. Laboratory of Molecular Medicine and Genomics(LMMGe) / 3. Center for Research in Pure and Applied Mathematics (CRMPA) | Giorgio Giurato (Corresponding Author) et al |

| | | | | | | |
|---|---|---|---|---|---|---|
| EPI_ISL_877034 | Italy/CAM-CRGS-54/2020 | 2020-05-03 | Europe/Italy/Campania/ | 1.AO Universitaria 'S. Giovanni di Dio e Ruggi D'Aragona, Scuola Medica Salernitana' Hospital / 2.UOC di Virologia e Microbiologia, Universit√† della Campania 'L. Vanvitelli' / 3.AO Universitaria 'Federico II' Napoli Hospital / 4.AORN 'San Giuseppe Moscati' Avellino Hospital / 5.AO 'San Pio - presidio G. Rummo' Benevento Hospital / 6.AO 'Sant'Anna e San Sebastiano' Caserta Hospital / 7.PO 'Maria Santissima Addolorata' Eboli Hospital / 8.Biogem Istituto di Ricerche Genetiche | 1. Genome Research Center for Health (CRGS) / 2. Laboratory of Molecular Medicine and Genomics(LMMGe) / 3. Center for Research in Pure and Applied Mathematics (CRMPA) | Giorgio Giurato (Corresponding Author) et al |
| EPI_ISL_877036 | Italy/CAM-CRGS-10/2020 | 2020-03-15 | Europe/Italy/Campania/ | 1.AO Universitaria 'S. Giovanni di Dio e Ruggi D'Aragona, Scuola Medica Salernitana' Hospital / 2.UOC di Virologia e Microbiologia, Universit√† della Campania 'L. Vanvitelli' / 3.AO Universitaria 'Federico II' Napoli Hospital / 4.AORN 'San Giuseppe Moscati' Avellino Hospital / 5.AO 'San Pio - presidio G. Rummo' Benevento Hospital / 6.AO 'Sant'Anna e San Sebastiano' Caserta Hospital / 7.PO 'Maria Santissima Addolorata' Eboli Hospital / 8.Biogem Istituto di Ricerche Genetiche | 1. Genome Research Center for Health (CRGS) / 2. Laboratory of Molecular Medicine and Genomics(LMMGe) / 3. Center for Research in Pure and Applied Mathematics (CRMPA) | Giorgio Giurato (Corresponding Author) et al |
| EPI_ISL_877044 | USA/CA-QDX-3757/2021 | 2021-01-14 | North America/USA/California/ | Quest Diagnostics | Quest Diagnostics | Rosenthal et al |
| EPI_ISL_877061 | USA/CA-QDX-3742/2021 | 2021-01-15 | North America/USA/California/ | Quest Diagnostics | Quest Diagnostics | Rosenthal et al |
| EPI_ISL_877064 | USA/CA-QDX-3747/2021 | 2021-01-15 | North America/USA/California/ | Quest Diagnostics | Quest Diagnostics | Rosenthal et al |
| EPI_ISL_877069 | USA/CA-QDX-3754/2021 | 2021-01-15 | North America/USA/California/ | Quest Diagnostics | Quest Diagnostics | Rosenthal et al |
| EPI_ISL_877072 | USA/CA-QDX-3759/2021 | 2021-01-15 | North America/USA/California/ | Quest Diagnostics | Quest Diagnostics | Rosenthal et al |
| EPI_ISL_877075 | USA/CA-QDX-3763/2021 | 2021-01-15 | North America/USA/California/ | Quest Diagnostics | Quest Diagnostics | Rosenthal et al |
| EPI_ISL_877078 | USA/CA-QDX-3770/2021 | 2021-01-15 | North America/USA/California/ | Quest Diagnostics | Quest Diagnostics | Rosenthal et al |
| EPI_ISL_877080 | USA/CA-QDX-3772/2021 | 2021-01-15 | North America/USA/California/ | Quest Diagnostics | Quest Diagnostics | Rosenthal et al |
| EPI_ISL_877084 | USA/CA-QDX-3777/2021 | 2021-01-15 | North America/USA/California/ | Quest Diagnostics | Quest Diagnostics | Rosenthal et al |
| EPI_ISL_877088 | USA/CA-QDX-3785/2021 | 2021-01-15 | North America/USA/California/ | Quest Diagnostics | Quest Diagnostics | Rosenthal et al |
| EPI_ISL_877091 | USA/CA-QDX-3790/2021 | 2021-01-15 | North America/USA/California/ | Quest Diagnostics | Quest Diagnostics | Rosenthal et al |
| EPI_ISL_877096 | USA/CA-QDX-3795/2021 | 2021-01-15 | North America/USA/California/ | Quest Diagnostics | Quest Diagnostics | Rosenthal et al |
| EPI_ISL_877107 | USA/CA-QDX-3808/2021 | 2021-01-15 | North America/USA/California/ | Quest Diagnostics | Quest Diagnostics | Rosenthal et al |

| | | | | | | |
|---|---|---|---|---|---|---|
| EPI_ISL_877112 | USA/CA-QDX-3815/2021 | 2021-01-15 | North America/USA/California/ | Quest Diagnostics | Quest Diagnostics | Rosenthal et al |
| EPI_ISL_877116 | USA/CA-QDX-3822/2021 | 2021-01-15 | North America/USA/California/ | Quest Diagnostics | Quest Diagnostics | Rosenthal et al |
| EPI_ISL_877119 | USA/FL-QDX-3456/2020 | 2020-08-15 | North America/USA/Florida/ | Quest Diagnostics | Quest Diagnostics | Rosenthal et al |
| EPI_ISL_877124 | USA/TX-QDX-3487/2020 | 2020-09-11 | North America/USA/Texas/ | Quest Diagnostics | Quest Diagnostics | Rosenthal et al |
| EPI_ISL_877126 | Indonesia/YO-UGM-110279/2020 | 2020-09-07 | Asia/Indonesia/Yogyakarta/ | Academic Hospital of Gajdah Mada University (RSA UGM) | Genetics Working Group (Pokja Genetik) Faculty of Medicine, Public Health and Nursing Universitas Gadjah Mada (FK-KMK UGM); Disease Investigation Center Wates Ministry of Agriculture Indonesia; Department of Microbiology FK-KMK UGM; Laboratorium Diagnostik Yayasan Tahija World Mosquito Program (WMP) Yogyakarta Center for Tropical Medicine FK-KMK UGM; Integrated Research center FK-KMK UGM; Department of Computer Science and Electronics FMIPA UGM | Gunadi et al |

| | | | | | | |
|---|---|---|---|---|---|---|
| EPI_ISL_877127 | Indonesia/YO-UGM-110283/2020 | 2020-09-07 | Asia/Indonesia/Yogyakarta/ | Academic Hospital of Gajdah Mada University (RSA UGM) | Genetics Working Group (Pokja Genetik) Faculty of Medicine, Public Health and Nursing Universitas Gadjah Mada (FK-KMK UGM); Disease Investigation Center Wates Ministry of Agriculture Indonesia; Department of Microbiology FK-KMK UGM; Laboratorium Diagnostik Yayasan Tahija World Mosquito Program (WMP) Yogyakarta Center for Tropical Medicine FK-KMK UGM; Integrated Research center FK-KMK UGM; Department of Computer Science and Electronics FMIPA UGM | Gunadi et al |

| | | | | | | |
|---|---|---|---|---|---|---|
| EPI_ISL_877128 | Indonesia/YO-UGM-110434/2020 | 2020-09-09 | Asia/Indonesia/Yogyakarta/ | Academic Hospital of Gajdah Mada University (RSA UGM) | Genetics Working Group (Pokja Genetik) Faculty of Medicine, Public Health and Nursing Universitas Gadjah Mada (FK-KMK UGM); Disease Investigation Center Wates Ministry of Agriculture Indonesia; Department of Microbiology FK-KMK UGM; Laboratorium Diagnostik Yayasan Tahija World Mosquito Program (WMP) Yogyakarta Center for Tropical Medicine FK-KMK UGM; Integrated Research center FK-KMK UGM; Department of Computer Science and Electronics FMIPA UGM | Gunadi et al |

| | | | | | | |
|---|---|---|---|---|---|---|
| EPI_ISL_877129 | Indonesia/YO-UGM-110286/2020 | 2020-09-07 | Asia/Indonesia/Yogyakarta/ | Academic Hospital of Gajdah Mada University (RSA UGM) | Genetics Working Group (Pokja Genetik) Faculty of Medicine, Public Health and Nursing Universitas Gadjah Mada (FK-KMK UGM); Disease Investigation Center Wates Ministry of Agriculture Indonesia; Department of Microbiology FK-KMK UGM; Laboratorium Diagnostik Yayasan Tahija World Mosquito Program (WMP) Yogyakarta Center for Tropical Medicine FK-KMK UGM; Integrated Research Center FK-KMK UGM; Department of Computer Science and Electronics FMIPA UGM | Gunadi et al |

| | | | | | | |
|---|---|---|---|---|---|---|
| EPI_ISL_877130 | Indonesia/YO-UGM-110282/2020 | 2020-09-07 | Asia/Indonesia/Yogyakarta/ | Academic Hospital of Gajdah Mada University (RSA UGM) | Genetics Working Group (Pokja Genetik) Faculty of Medicine, Public Health and Nursing Universitas Gadjah Mada (FK-KMK UGM); Disease Investigation Center Wates Ministry of Agriculture Indonesia; Department of Microbiology FK-KMK UGM; Laboratorium Diagnostik Yayasan Tahija World Mosquito Program (WMP) Yogyakarta Center for Tropical Medicine FK-KMK UGM; Integrated Research center FK-KMK UGM; Department of Computer Science and Electronics FMIPA UGM | Gunadi et al |

| | | | | | | |
|---|---|---|---|---|---|---|
| EPI_ISL_877131 | Indonesia/YO-UGM-110280/2020 | 2020-09-07 | Asia/Indonesia/Yogyakarta/ | Academic Hospital of Gajdah Mada University (RSA UGM) | Genetics Working Group (Pokja Genetik) Faculty of Medicine, Public Health and Nursing Universitas Gadjah Mada (FK-KMK UGM); Disease Investigation Center Wates Ministry of Agriculture Indonesia; Department of Microbiology FK-KMK UGM; Laboratorium Diagnostik Yayasan Tahija World Mosquito Program (WMP) Yogyakarta Center for Tropical Medicine FK-KMK UGM; Integrated Research center FK-KMK UGM; Department of Computer Science and Electronics FMIPA UGM | Gunadi et al |
| EPI_ISL_877133 | USA/NM-UNM-EDB00101/2021 | 2021-01-14 | North America/USA/New Mexico/ | Univeristy of New Mexico Hospital | Center for Global Health, University of New Mexico Health Sciences Center | Daryl Domman et al |

| | | | | | | |
|---|---|---|---|---|---|---|
| EPI_ISL_877136 | USA/NM-UNM-EDB00104/2021 | 2021-01-14 | North America/USA/New Mexico/ | Univeristy of New Mexico Hospital | Center for Global Health, University of New Mexico Health Sciences Center | Daryl Domman et al |
| EPI_ISL_877137 | USA/NM-UNM-EDB00105/2021 | 2021-01-14 | North America/USA/New Mexico/ | Univeristy of New Mexico Hospital | Center for Global Health, University of New Mexico Health Sciences Center | Daryl Domman et al |
| EPI_ISL_877139 | USA/NM-UNM-EDB00107/2021 | 2021-01-14 | North America/USA/New Mexico/ | Univeristy of New Mexico Hospital | Center for Global Health, University of New Mexico Health Sciences Center | Daryl Domman et al |
| EPI_ISL_877140 | USA/NM-UNM-EDB00108/2021 | 2021-01-14 | North America/USA/New Mexico/ | Univeristy of New Mexico Hospital | Center for Global Health, University of New Mexico Health Sciences Center | Daryl Domman et al |
| EPI_ISL_877151 | USA/NM-UNM-EDB00119/2021 | 2021-01-15 | North America/USA/New Mexico/ | Univeristy of New Mexico Hospital | Center for Global Health, University of New Mexico Health Sciences Center | Daryl Domman et al |
| EPI_ISL_877153 | USA/NM-UNM-EDB00121/2021 | 2021-01-14 | North America/USA/New Mexico/ | Univeristy of New Mexico Hospital | Center for Global Health, University of New Mexico Health Sciences Center | Daryl Domman et al |
| EPI_ISL_877154 | USA/NM-UNM-EDB00122/2021 | 2021-01-15 | North America/USA/New Mexico/ | Univeristy of New Mexico Hospital | Center for Global Health, University of New Mexico Health Sciences Center | Daryl Domman et al |
| EPI_ISL_877159 | USA/NM-UNM-EDB00127/2021 | 2021-01-18 | North America/USA/New Mexico/ | Univeristy of New Mexico Hospital | Center for Global Health, University of New Mexico Health Sciences Center | Daryl Domman et al |

| | | | | | | |
|---|---|---|---|---|---|---|
| EPI_ISL_877164 | USA/NM-UNM-EDB00132/2021 | 2021-01-17 | North America/USA/New Mexico/ | Univeristy of New Mexico Hospital | Center for Global Health, University of New Mexico Health Sciences Center | Daryl Domman et al |
| EPI_ISL_877165 | USA/NM-UNM-EDB00133/2021 | 2021-01-17 | North America/USA/New Mexico/ | Univeristy of New Mexico Hospital | Center for Global Health, University of New Mexico Health Sciences Center | Daryl Domman et al |
| EPI_ISL_877166 | USA/NM-UNM-EDB00134/2021 | 2021-01-17 | North America/USA/New Mexico/ | Univeristy of New Mexico Hospital | Center for Global Health, University of New Mexico Health Sciences Center | Daryl Domman et al |
| EPI_ISL_877173 | USA/NM-UNM-EDB00141/2021 | 2021-01-17 | North America/USA/New Mexico/ | Univeristy of New Mexico Hospital | Center for Global Health, University of New Mexico Health Sciences Center | Daryl Domman et al |
| EPI_ISL_877182 | USA/NM-UNM-EDL01004/2021 | 2021-01-19 | North America/USA/New Mexico/ | Univeristy of New Mexico Hospital | Center for Global Health, University of New Mexico Health Sciences Center | Daryl Domman et al |
| EPI_ISL_877188 | USA/CA-QDX-3215/2020 | 2020-12-28 | North America/USA/California/ | Quest Diagnostics | Quest Diagnostics | Rosenthal et al |
| EPI_ISL_877193 | USA/MA-QDX-3176/2021 | 2021-01-06 | North America/USA/Massachusetts/ | Quest Diagnostics | Quest Diagnostics | Rosenthal et al |
| EPI_ISL_877200 | USA/CT-QDX-3063/2021 | 2021-01-06 | North America/USA/Connecticut/ | Quest Diagnostics | Quest Diagnostics | Rosenthal et al |
| EPI_ISL_877208 | NewZealand/21CV0063/2021 | 2021-01-06 | Europe/Russia/Russia/ | LabPLUS | Institute of Environmental Science and Research (ESR) | Xiaoyun Ren et al |
| EPI_ISL_877209 | NewZealand/21CV0064/2021 | 2021-01-06 | Africa/Zimbabwe/Zimbabwe/ | LabPLUS | Institute of Environmental Science and Research (ESR) | Xiaoyun Ren et al |

| | | | | | | |
|---|---|---|---|---|---|---|
| EPI_ISL_877212 | NewZealand/21CV0068/2021 | 2021-01-08 | Asia/Vietnam/Vietnam/ | LabPLUS | Institute of Environmental Science and Research (ESR) | Xiaoyun Ren et al |
| EPI_ISL_877216 | NewZealand/21CV0084/2021 | 2021-01-09 | Africa/Zimbabwe/Zimbabwe/ | Middlemore Hospital | Institute of Environmental Science and Research (ESR) | Xiaoyun Ren et al |
| EPI_ISL_877221 | NewZealand/21CV0102/2021 | 2021-01-12 | Asia/United Arab Emirates/United Arab | LabPLUS | Institute of Environmental Science and Research (ESR) | Xiaoyun Ren et al |
| EPI_ISL_877222 | NewZealand/21CV0106/2021 | 2021-01-13 | Asia/Malaysia/Malaysia/ | LabPLUS | Institute of Environmental Science and Research (ESR) | Xiaoyun Ren et al |
| EPI_ISL_877223 | NewZealand/21CV0107/2021 | 2021-01-13 | Asia/Malaysia/Malaysia/ | LabPLUS | Institute of Environmental Science and Research (ESR) | Xiaoyun Ren et al |
| EPI_ISL_877224 | NewZealand/21CV0108/2021 | 2021-01-13 | Asia/Malaysia/Malaysia/ | LabPLUS | Institute of Environmental Science and Research (ESR) | Xiaoyun Ren et al |
| EPI_ISL_877230 | Malaysia/IMR-UCSI_R3/2021 | 2021-01-08 | Asia/Malaysia/Malaysia/ | University College Sedaya International (UCSI University) | Institute for Medical Research, Infectious Disease Research Centre, National Institutes of Health, Ministry of Health Malaysia | Suppiah J et al |

| | | | | | | |
|---|---|---|---|---|---|---|
| EPI_ISL_877231 | Malaysia/IMR-UCSI_R7/2021 | 2021-01-08 | Asia/Malaysia/Malaysia/ | University College Sedaya International (UCSI University) | Institute for Medical Research, Infectious Disease Research Centre, National Institutes of Health, Ministry of Health Malaysia | Suppiah J et al |
| EPI_ISL_877232 | Malaysia/IMR-UCSI_R16/2021 | 2021-01-08 | Asia/Malaysia/Malaysia/ | University College Sedaya International (UCSI University) | Institute for Medical Research, Infectious Disease Research Centre, National Institutes of Health, Ministry of Health Malaysia | Suppiah J et al |
| EPI_ISL_877233 | Malaysia/IMR-UCSI_R17/2021 | 2021-01-08 | Asia/Malaysia/Malaysia/ | University College Sedaya International (UCSI University) | Institute for Medical Research, Infectious Disease Research Centre, National Institutes of Health, Ministry of Health Malaysia | Suppiah J et al |
| EPI_ISL_877234 | Malaysia/IMR-UCSI_R19/2021 | 2021-01-08 | Asia/Malaysia/Malaysia/ | University College Sedaya International (UCSI University) | Institute for Medical Research, Infectious Disease Research Centre, National Institutes of Health, Ministry of Health Malaysia | Suppiah J et al |

| | | | | | | |
|---|---|---|---|---|---|---|
| EPI_ISL_877235 | Malaysia/IMR-UCSI_R20/2021 | 2021-01-08 | Asia/Malaysia/Malaysia/ | University College Sedaya International (UCSI University) | Institute for Medical Research, Infectious Disease Research Centre, National Institutes of Health, Ministry of Health Malaysia | Suppiah J et al |
| EPI_ISL_877236 | Malaysia/IMR-UCSI_KP45/2021 | 2021-01-11 | Asia/Malaysia/Malaysia/ | University College Sedaya International (UCSI University) | Institute for Medical Research, Infectious Disease Research Centre, National Institutes of Health, Ministry of Health Malaysia | Suppiah J et al |
| EPI_ISL_877241 | USA/AL-HGSC-JEPS/2020 | 2020-11-10 | North America/USA/Alabama/Madison | Eurofins Diatherix | Hudsonalpha Genome Sequencing Center | Jane Grimwood et al |
| EPI_ISL_877246 | USA/AL-HGSC-JEPY/2020 | 2020-11-10 | North America/USA/Alabama/Conecuh ( | Eurofins Diatherix | Hudsonalpha Genome Sequencing Center | Jane Grimwood et al |
| EPI_ISL_877247 | USA/AL-HGSC-JEPZ/2020 | 2020-11-10 | North America/USA/Alabama/Madison | Eurofins Diatherix | Hudsonalpha Genome Sequencing Center | Jane Grimwood et al |
| EPI_ISL_877264 | USA/AL-HGSC-JEQR/2020 | 2020-11-10 | North America/USA/Alabama/Cherokee | Eurofins Diatherix | Hudsonalpha Genome Sequencing Center | Jane Grimwood et al |
| EPI_ISL_877268 | USA/AL-HGSC-JEQW/2020 | 2020-11-10 | North America/USA/Alabama/Jackson C | Eurofins Diatherix | Hudsonalpha Genome Sequencing Center | Jane Grimwood et al |

| | | | | | | |
|---|---|---|---|---|---|---|
| EPI_ISL_877270 | USA/AL-HGSC-JEQY/2020 | 2020-11-10 | North America/USA/Alabama/Elmore C | Eurofins Diatherix | Hudsonalpha Genome Sequencing Center | Jane Grimwood et al |
| EPI_ISL_877282 | USA/AL-HGSC-JERM/2020 | 2020-11-11 | North America/USA/Alabama/Jefferson | Eurofins Diatherix | Hudsonalpha Genome Sequencing Center | Jane Grimwood et al |
| EPI_ISL_877289 | USA/AL-HGSC-JERW/2020 | 2020-11-11 | North America/USA/Alabama/Morgan C | Eurofins Diatherix | Hudsonalpha Genome Sequencing Center | Jane Grimwood et al |
| EPI_ISL_877293 | USA/AL-HGSC-JESA/2020 | 2020-11-11 | North America/USA/Alabama/Morgan C | Eurofins Diatherix | Hudsonalpha Genome Sequencing Center | Jane Grimwood et al |
| EPI_ISL_877295 | USA/AL-HGSC-JESC/2020 | 2020-11-11 | North America/USA/Alabama/Madison | Eurofins Diatherix | Hudsonalpha Genome Sequencing Center | Jane Grimwood et al |
| EPI_ISL_877299 | USA/AL-HGSC-JESG/2020 | 2020-11-11 | North America/USA/Alabama/St. Clair C | Eurofins Diatherix | Hudsonalpha Genome Sequencing Center | Jane Grimwood et al |
| EPI_ISL_877307 | USA/AL-HGSC-JESP/2020 | 2020-11-11 | North America/USA/Alabama/Jefferson | Eurofins Diatherix | Hudsonalpha Genome Sequencing Center | Jane Grimwood et al |
| EPI_ISL_877313 | USA/AL-HGSC-JESW/2020 | 2020-11-11 | North America/USA/Alabama/Madison | Eurofins Diatherix | Hudsonalpha Genome Sequencing Center | Jane Grimwood et al |
| EPI_ISL_877316 | USA/AL-HGSC-JESZ/2020 | 2020-11-11 | North America/USA/Alabama/Escambia | Eurofins Diatherix | Hudsonalpha Genome Sequencing Center | Jane Grimwood et al |

| | | | | | | |
|---|---|---|---|---|---|---|
| EPI_ISL_877329 | Malaysia/IMR-UCSI_KA137/2021 | 2021-01-11 | Asia/Malaysia/Malaysia/ | University College Sedaya International (UCSI University) | Institute for Medical Research, Infectious Disease Research Centre, National Institutes of Health, Ministry of Health Malaysia | Suppiah J et al |
| EPI_ISL_877330 | Malaysia/IMR-UCSI_KA139/2021 | 2021-01-11 | Asia/Malaysia/Malaysia/ | University College Sedaya International (UCSI University) | Institute for Medical Research, Infectious Disease Research Centre, National Institutes of Health, Ministry of Health Malaysia | Suppiah J et al |
| EPI_ISL_877340 | USA/AL-HGSC-JETZ/2020 | 2020-11-17 | North America/USA/Alabama/Morgan C | Eurofins Diatherix | Hudsonalpha Genome Sequencing Center | Jane Grimwood et al |
| EPI_ISL_877341 | USA/AL-HGSC-JEUB/2020 | 2020-11-17 | North America/USA/Alabama/Limestone | Eurofins Diatherix | Hudsonalpha Genome Sequencing Center | Jane Grimwood et al |
| EPI_ISL_877344 | USA/AL-HGSC-JEUE/2020 | 2020-11-17 | North America/USA/Alabama/Etowah C | Eurofins Diatherix | Hudsonalpha Genome Sequencing Center | Jane Grimwood et al |
| EPI_ISL_877364 | USA/AL-HGSC-JEWB/2020 | 2020-11-17 | North America/USA/Alabama/Calhoun C | Eurofins Diatherix | Hudsonalpha Genome Sequencing Center | Jane Grimwood et al |
| EPI_ISL_877369 | USA/AL-HGSC-JEWG/2020 | 2020-11-17 | North America/USA/Alabama/Lauderda | Eurofins Diatherix | Hudsonalpha Genome Sequencing Center | Jane Grimwood et al |

| | | | | | | |
|---|---|---|---|---|---|---|
| EPI_ISL_877382 | USA/AL-HGSC-JEWW/2020 | 2020-11-18 | North America/USA/Alabama/Pike Cour | Eurofins Diatherix | Hudsonalpha Genome Sequencing Center | Jane Grimwood et al |
| EPI_ISL_877389 | USA/AL-HGSC-JEXE/2020 | 2020-11-18 | North America/USA/Alabama/Limeston | Eurofins Diatherix | Hudsonalpha Genome Sequencing Center | Jane Grimwood et al |
| EPI_ISL_877393 | USA/AL-HGSC-JEXI/2020 | 2020-11-18 | North America/USA/Alabama/Escambia | Eurofins Diatherix | Hudsonalpha Genome Sequencing Center | Jane Grimwood et al |
| EPI_ISL_877395 | USA/AL-HGSC-JEXK/2020 | 2020-11-18 | North America/USA/Alabama/Lawrence | Eurofins Diatherix | Hudsonalpha Genome Sequencing Center | Jane Grimwood et al |
| EPI_ISL_877408 | USA/AL-HGSC-JEYB/2020 | 2020-11-18 | North America/USA/Alabama/Mobile C | Eurofins Diatherix | Hudsonalpha Genome Sequencing Center | Jane Grimwood et al |
| EPI_ISL_877419 | Indonesia/JI-ITDua-33281NT/2020 | 2020-12-11 | Asia/Indonesia/East Java/Sidoarjo | Siti Khodijah Hospital | Institute of Tropical Disease, Universitas Airlangga | Krisnoadi Rahardjo et al |
| EPI_ISL_877421 | Indonesia/JI-ITDua-33312NT/2020 | 2020-12-11 | Asia/Indonesia/East Java/Surabaya | Institute of Tropical Disease | Institute of Tropical Disease, Universitas Airlangga | Jezzy R Dewantari et al |
| EPI_ISL_877423 | Indonesia/JI-ITDua-33816NT/2020 | 2020-12-16 | Asia/Indonesia/East Java/Surabaya | Primasatya Husada Citra Hospital | Institute of Tropical Disease, Universitas Airlangga | Maria I Lusida et al |
| EPI_ISL_877425 | Slovenia/134870/2020 | 2020-10-15 | Europe/Slovenia/Slovenia/ | Institute of Microbiology and Immunology, Faculty of Medicine, University of Ljubljana | Institute of Microbiology and Immunology, Faculty of Medicine, University of Ljubljana | Samo Zakotnik et al |

| | | | | | | |
|---|---|---|---|---|---|---|
| EPI_ISL_877426 | Slovenia/136912/2020 | 2020-10-16 | Europe/Slovenia/Slovenia/ | Institute of Microbiology and Immunology, Faculty of Medicine, University of Ljubljana | Institute of Microbiology and Immunology, Faculty of Medicine, University of Ljubljana | Samo Zakotnik et al |
| EPI_ISL_877427 | Slovenia/152240/2020 | 2020-10-22 | Europe/Slovenia/Slovenia/ | Institute of Microbiology and Immunology, Faculty of Medicine, University of Ljubljana | Institute of Microbiology and Immunology, Faculty of Medicine, University of Ljubljana | Samo Zakotnik et al |
| EPI_ISL_877428 | Slovenia/176062/2020 | 2020-10-31 | Europe/Slovenia/Slovenia/ | Institute of Microbiology and Immunology, Faculty of Medicine, University of Ljubljana | Institute of Microbiology and Immunology, Faculty of Medicine, University of Ljubljana | Samo Zakotnik et al |
| EPI_ISL_877429 | Slovenia/NK7853/2020 | 2020-10-30 | Europe/Slovenia/Slovenia/ | Institute of Microbiology and Immunology, Faculty of Medicine, University of Ljubljana | Institute of Microbiology and Immunology, Faculty of Medicine, University of Ljubljana | Samo Zakotnik et al |
| EPI_ISL_877430 | Slovenia/NK7854/2020 | 2020-10-27 | Europe/Slovenia/Slovenia/ | Institute of Microbiology and Immunology, Faculty of Medicine, University of Ljubljana | Institute of Microbiology and Immunology, Faculty of Medicine, University of Ljubljana | Samo Zakotnik et al |

| | | | | | | |
|---|---|---|---|---|---|---|
| EPI_ISL_877431 | Slovenia/NK7855/2020 | 2020-10-08 | Europe/Slovenia/Slovenia/ | Institute of Microbiology and Immunology, Faculty of Medicine, University of Ljubljana | Institute of Microbiology and Immunology, Faculty of Medicine, University of Ljubljana | Samo Zakotnik et al |
| EPI_ISL_877433 | Slovenia/NK7859/2020 | 2020-10-08 | Europe/Slovenia/Slovenia/ | National laboratory of health, environment and food Celje | Institute of Microbiology and Immunology, Faculty of Medicine, University of Ljubljana | Samo Zakotnik et al |
| EPI_ISL_877434 | Slovenia/NK7860/2020 | 2020-10-22 | Europe/Slovenia/Slovenia/ | National laboratory of health, environment and food Celje | Institute of Microbiology and Immunology, Faculty of Medicine, University of Ljubljana | Samo Zakotnik et al |
| EPI_ISL_877435 | Slovenia/NK7861/2020 | 2020-11-07 | Europe/Slovenia/Slovenia/ | National laboratory of health, environment and food Maribor | Institute of Microbiology and Immunology, Faculty of Medicine, University of Ljubljana | Samo Zakotnik et al |
| EPI_ISL_877436 | Slovenia/NK7862/2020 | 2020-11-10 | Europe/Slovenia/Slovenia/ | National laboratory of health, environment and food Celje | Institute of Microbiology and Immunology, Faculty of Medicine, University of Ljubljana | Samo Zakotnik et al |

| | | | | | | |
|---|---|---|---|---|---|---|
| EPI_ISL_877441 | Slovenia/2020-V0-272532/2020 | 2020-12-15 | Europe/Slovenia/Slovenia/ | Institute of Microbiology and Immunology, Faculty of Medicine, University of Ljubljana | Institute of Microbiology and Immunology, Faculty of Medicine, University of Ljubljana | Samo Zakotnik et al |
| EPI_ISL_877442 | Slovenia/2020-V0-272910/2020 | 2020-12-14 | Europe/Slovenia/Slovenia/ | Institute of Microbiology and Immunology, Faculty of Medicine, University of Ljubljana | Institute of Microbiology and Immunology, Faculty of Medicine, University of Ljubljana | Samo Zakotnik et al |
| EPI_ISL_877446 | Slovenia/2020-V0-273438/2020 | 2020-12-15 | Europe/Slovenia/Slovenia/ | Institute of Microbiology and Immunology, Faculty of Medicine, University of Ljubljana | Institute of Microbiology and Immunology, Faculty of Medicine, University of Ljubljana | Samo Zakotnik et al |
| EPI_ISL_877453 | Slovenia/NK7961/2021 | 2021-01-07 | Europe/Slovenia/Slovenia/ | Institute of Microbiology and Immunology, Faculty of Medicine, University of Ljubljana | Institute of Microbiology and Immunology, Faculty of Medicine, University of Ljubljana | Samo Zakotnik et al |
| EPI_ISL_877455 | Slovenia/NK7852/2020 | 2020-10-17 | Europe/Slovenia/Slovenia/ | Institute of Microbiology and Immunology, Faculty of Medicine, University of Ljubljana | Institute of Microbiology and Immunology, Faculty of Medicine, University of Ljubljana | Samo Zakotnik et al |
| EPI_ISL_877456 | Indonesia/JI-ITDua-33807NT/2020 | 2020-12-16 | Asia/Indonesia/East Java/Surabaya | Primasatya Husada Citra Hospital | Institute of Tropical Disease, Universitas Airlangga | Krisnoadi Rahardjo et al |

| | | | | | | |
|---|---|---|---|---|---|---|
| EPI_ISL_877457 | Indonesia/JI-ITDua-33817NT/2020 | 2020-12-16 | Asia/Indonesia/East Java/Surabaya | Primasatya Husada Citra Hospital | Institute of Tropical Disease, Universitas Airlangga | Aldise M Nastri et al |
| EPI_ISL_877458 | Indonesia/JI-ITDua-33862NT/2020 | 2020-12-16 | Asia/Indonesia/East Java/Sidoarjo | Anwar Medika General Hospital | Institute of Tropical Disease, Universitas Airlangga | Jezzy R Dewantari et al |
| EPI_ISL_877459 | Indonesia/JI-ITDua-34192NT/2020 | 2020-12-18 | Asia/Indonesia/East Java/Surabaya | Darmo Hospital | Institute of Tropical Disease, Universitas Airlangga | Rima R Prasetya et al |
| EPI_ISL_877460 | Indonesia/JI-ITDua-34660NT/2020 | 2020-12-22 | Asia/Indonesia/East Java/Sidoarjo | Siti Khodijah Hospital | Institute of Tropical Disease, Universitas Airlangga | Maria I Lusida et al |
| EPI_ISL_877466 | USA/AL-HGSC-JFAB/2020 | 2020-11-24 | North America/USA/Alabama/Madison | Eurofins Diatherix | Hudsonalpha Genome Sequencing Center | Jane Grimwood et al |
| EPI_ISL_877469 | USA/AL-HGSC-JFAE/2020 | 2020-11-24 | North America/USA/Alabama/Limestone | Eurofins Diatherix | Hudsonalpha Genome Sequencing Center | Jane Grimwood et al |
| EPI_ISL_877470 | USA/AL-HGSC-JFAF/2020 | 2020-11-24 | North America/USA/Alabama/Marshall | Eurofins Diatherix | Hudsonalpha Genome Sequencing Center | Jane Grimwood et al |
| EPI_ISL_877479 | USA/AL-HGSC-JFAP/2020 | 2020-11-24 | North America/USA/Alabama/Madison | Eurofins Diatherix | Hudsonalpha Genome Sequencing Center | Jane Grimwood et al |
| EPI_ISL_877482 | USA/AL-HGSC-JFAS/2020 | 2020-11-24 | North America/USA/Alabama/Winston ( | Eurofins Diatherix | Hudsonalpha Genome Sequencing Center | Jane Grimwood et al |
| EPI_ISL_877487 | USA/AL-HGSC-JFAY/2020 | 2020-11-24 | North America/USA/Alabama/Limestone | Eurofins Diatherix | Hudsonalpha Genome Sequencing Center | Jane Grimwood et al |

| | | | | | | |
|---|---|---|---|---|---|---|
| EPI_ISL_877499 | USA/AL-HGSC-JFBK/2020 | 2020-11-24 | North America/USA/Alabama/Limestone | Eurofins Diatherix | Hudsonalpha Genome Sequencing Center | Jane Grimwood et al |
| EPI_ISL_877500 | USA/AL-HGSC-JFBL/2020 | 2020-11-24 | North America/USA/Alabama/Lauderda | Eurofins Diatherix | Hudsonalpha Genome Sequencing Center | Jane Grimwood et al |
| EPI_ISL_877501 | USA/AL-HGSC-JFBM/2020 | 2020-11-24 | North America/USA/Alabama/Cherokee | Eurofins Diatherix | Hudsonalpha Genome Sequencing Center | Jane Grimwood et al |
| EPI_ISL_877504 | USA/AL-HGSC-JFBQ/2020 | 2020-11-25 | North America/USA/Alabama/Jefferson | Eurofins Diatherix | Hudsonalpha Genome Sequencing Center | Jane Grimwood et al |
| EPI_ISL_877506 | USA/AL-HGSC-JFBS/2020 | 2020-11-25 | North America/USA/Alabama/Madison | Eurofins Diatherix | Hudsonalpha Genome Sequencing Center | Jane Grimwood et al |
| EPI_ISL_877507 | USA/AL-HGSC-JFBT/2020 | 2020-11-25 | North America/USA/Alabama/Madison | Eurofins Diatherix | Hudsonalpha Genome Sequencing Center | Jane Grimwood et al |
| EPI_ISL_877517 | USA/AL-HGSC-JFCF/2020 | 2020-11-25 | North America/USA/Alabama/Lowndes | Eurofins Diatherix | Hudsonalpha Genome Sequencing Center | Jane Grimwood et al |
| EPI_ISL_877522 | USA/AL-HGSC-JFCK/2020 | 2020-11-25 | North America/USA/Alabama/Madison | Eurofins Diatherix | Hudsonalpha Genome Sequencing Center | Jane Grimwood et al |
| EPI_ISL_877525 | USA/AL-HGSC-JFCQ/2020 | 2020-11-25 | North America/USA/Alabama/Shelby Co | Eurofins Diatherix | Hudsonalpha Genome Sequencing Center | Jane Grimwood et al |
| EPI_ISL_877534 | USA/AL-HGSC-JFDA/2020 | 2020-11-27 | North America/USA/Alabama/Madison | Eurofins Diatherix | Hudsonalpha Genome Sequencing Center | Jane Grimwood et al |

| | | | | | | |
|---|---|---|---|---|---|---|
| EPI_ISL_877538 | Indonesia/JI-ITDua-35379NT/2020 | 2020-12-28 | Asia/Indonesia/East Java/Sidoarjo | Siti Khodijah Hospital | Institute of Tropical Disease, Universitas Airlangga | Krisnoadi Rahardjo et al |
| EPI_ISL_877543 | USA/AL-HGSC-JFDJ/2020 | 2020-11-27 | North America/USA/Alabama/Morgan C | Eurofins Diatherix | Hudsonalpha Genome Sequencing Center | Jane Grimwood et al |
| EPI_ISL_877546 | USA/AL-HGSC-JFDM/2020 | 2020-11-27 | North America/USA/Alabama/Montgom | Eurofins Diatherix | Hudsonalpha Genome Sequencing Center | Jane Grimwood et al |
| EPI_ISL_877547 | USA/AL-HGSC-JFDN/2020 | 2020-11-27 | North America/USA/Alabama/Shelby Co | Eurofins Diatherix | Hudsonalpha Genome Sequencing Center | Jane Grimwood et al |
| EPI_ISL_877552 | Ecuador/USFQ-712/2021 | 2021-01-15 | South America/Ecuador/Los Rios/Babaho | Institute of Microbiology, Universidad San Francisco de Quito | Institute of Microbiology, Universidad San Francisco de Quito | BelV©n Prado-Vivar et al |
| EPI_ISL_877557 | Ecuador/USFQ-732/2021 | 2021-01-15 | South America/Ecuador/Los Rios/Queve | Institute of Microbiology, Universidad San Francisco de Quito | Institute of Microbiology, Universidad San Francisco de Quito | BelV©n Prado-Vivar et al |
| EPI_ISL_877559 | Ecuador/USFQ-734/2021 | 2021-01-15 | South America/Ecuador/Los Rios/Queve | Institute of Microbiology, Universidad San Francisco de Quito | Institute of Microbiology, Universidad San Francisco de Quito | BelV©n Prado-Vivar et al |
| EPI_ISL_877561 | Ecuador/USFQ-736/2021 | 2021-01-21 | South America/Ecuador/Pichincha/Quito | Institute of Microbiology, Universidad San Francisco de Quito | Institute of Microbiology, Universidad San Francisco de Quito | BelV©n Prado-Vivar et al |
| EPI_ISL_877562 | Ecuador/USFQ-737/2021 | 2021-01-22 | South America/Ecuador/Pichincha/Quito | Institute of Microbiology, Universidad San Francisco de Quito | Institute of Microbiology, Universidad San Francisco de Quito | BelV©n Prado-Vivar et al |
| EPI_ISL_877590 | USA/NC-UNC-LCCC0006/2020 | 2020-03-28 | North America/USA/North Carolina/ | Clinical Molecular Microbiology Laboratory, UNC Hospital | Dirk Dittmer | Razia Moorad et al |
| EPI_ISL_877595 | USA/NC-UNC-LCCC0011/2020 | 2020-04-09 | North America/USA/North Carolina/ | Clinical Molecular Microbiology Laboratory, UNC Hospital | Dirk Dittmer | Razia Moorad et al |

| | | | | | | |
|---|---|---|---|---|---|---|
| EPI_ISL_877607 | USA/NC-UNC-LCCC0024/2020 | 2020-05-04 | North America/USA/North Carolina/ | Clinical Molecular Microbiology Laboratory, UNC Hospital | Dirk Dittmer | Razia Moorad et al |
| EPI_ISL_877609 | USA/NC-UNC-LCCC0026/2020 | 2020-05-22 | North America/USA/North Carolina/ | Clinical Molecular Microbiology Laboratory, UNC Hospital | Dirk Dittmer | Razia Moorad et al |
| EPI_ISL_877612 | USA/NC-UNC-LCCC0029/2020 | 2020-06-03 | North America/USA/North Carolina/ | Clinical Molecular Microbiology Laboratory, UNC Hospital | Dirk Dittmer | Razia Moorad et al |
| EPI_ISL_877620 | USA/NC-UNC-LCCC0037/2020 | 2020-06-13 | North America/USA/North Carolina/ | Clinical Molecular Microbiology Laboratory, UNC Hospital | Dirk Dittmer | Razia Moorad et al |
| EPI_ISL_877622 | USA/NC-UNC-LCCC0039/2020 | 2020-06-13 | North America/USA/North Carolina/ | Clinical Molecular Microbiology Laboratory, UNC Hospital | Dirk Dittmer | Razia Moorad et al |
| EPI_ISL_877626 | USA/NC-UNC-LCCC0045/2020 | 2020-06-25 | North America/USA/North Carolina/ | Clinical Molecular Microbiology Laboratory, UNC Hospital | Dirk Dittmer | Razia Moorad et al |
| EPI_ISL_877631 | USA/NC-UNC-LCCC0050/2020 | 2020-06-30 | North America/USA/North Carolina/ | Clinical Molecular Microbiology Laboratory, UNC Hospital | Dirk Dittmer | Razia Moorad et al |
| EPI_ISL_877632 | USA/NC-UNC-LCCC0051/2020 | 2020-07-01 | North America/USA/North Carolina/ | Clinical Molecular Microbiology Laboratory, UNC Hospital | Dirk Dittmer | Razia Moorad et al |
| EPI_ISL_877635 | USA/NC-UNC-LCCC0054/2020 | 2020-07-01 | North America/USA/North Carolina/ | Clinical Molecular Microbiology Laboratory, UNC Hospital | Dirk Dittmer | Razia Moorad et al |
| EPI_ISL_877641 | USA/NC-UNC-LCCC0060/2020 | 2020-07-08 | North America/USA/North Carolina/ | Clinical Molecular Microbiology Laboratory, UNC Hospital | Dirk Dittmer | Razia Moorad et al |
| EPI_ISL_877647 | USA/NC-UNC-LCCC0066/2020 | 2020-07-10 | North America/USA/North Carolina/ | Clinical Molecular Microbiology Laboratory, UNC Hospital | Dirk Dittmer | Razia Moorad et al |
| EPI_ISL_877650 | USA/NC-UNC-LCCC0069/2020 | 2020-07-13 | North America/USA/North Carolina/ | Clinical Molecular Microbiology Laboratory, UNC Hospital | Dirk Dittmer | Razia Moorad et al |
| EPI_ISL_877656 | USA/NC-UNC-LCCC0075/2020 | 2020-07-31 | North America/USA/North Carolina/ | Clinical Molecular Microbiology Laboratory, UNC Hospital | Dirk Dittmer | Razia Moorad et al |
| EPI_ISL_877657 | USA/NC-UNC-LCCC0076/2020 | 2020-07-31 | North America/USA/North Carolina/ | Clinical Molecular Microbiology Laboratory, UNC Hospital | Dirk Dittmer | Razia Moorad et al |
| EPI_ISL_877660 | USA/NC-UNC-LCCC0079/2020 | 2020-07-31 | North America/USA/North Carolina/ | Clinical Molecular Microbiology Laboratory, UNC Hospital | Dirk Dittmer | Razia Moorad et al |
| EPI_ISL_877661 | USA/NC-UNC-LCCC0080/2020 | 2020-07-31 | North America/USA/North Carolina/ | Clinical Molecular Microbiology Laboratory, UNC Hospital | Dirk Dittmer | Razia Moorad et al |
| EPI_ISL_877662 | USA/NC-UNC-LCCC0081/2020 | 2020-07-31 | North America/USA/North Carolina/ | Clinical Molecular Microbiology Laboratory, UNC Hospital | Dirk Dittmer | Razia Moorad et al |
| EPI_ISL_877667 | USA/NC-UNC-LCCC0086/2020 | 2020-08-01 | North America/USA/North Carolina/ | Clinical Molecular Microbiology Laboratory, UNC Hospital | Dirk Dittmer | Razia Moorad et al |
| EPI_ISL_877673 | USA/NC-UNC-LCCC0092/2020 | 2020-08-02 | North America/USA/North Carolina/ | Clinical Molecular Microbiology Laboratory, UNC Hospital | Dirk Dittmer | Razia Moorad et al |
| EPI_ISL_877674 | USA/NC-UNC-LCCC0093/2020 | 2020-08-02 | North America/USA/North Carolina/ | Clinical Molecular Microbiology Laboratory, UNC Hospital | Dirk Dittmer | Razia Moorad et al |

| | | | | | | |
|---|---|---|---|---|---|---|
| EPI_ISL_877676 | USA/NC-UNC-LCCC0095/2020 | 2020-08-21 | North America/USA/North Carolina/ | Clinical Molecular Microbiology Laboratory, UNC Hospital | Dirk Dittmer | Razia Moorad et al |
| EPI_ISL_877680 | USA/NC-UNC-LCCC0099/2020 | 2020-08-22 | North America/USA/North Carolina/ | Clinical Molecular Microbiology Laboratory, UNC Hospital | Dirk Dittmer | Razia Moorad et al |
| EPI_ISL_877688 | USA/NC-UNC-LCCC0107/2020 | 2020-08-23 | North America/USA/North Carolina/ | Clinical Molecular Microbiology Laboratory, UNC Hospital | Dirk Dittmer | Razia Moorad et al |
| EPI_ISL_877692 | USA/NC-UNC-LCCC0111/2020 | 2020-08-23 | North America/USA/North Carolina/ | Clinical Molecular Microbiology Laboratory, UNC Hospital | Dirk Dittmer | Razia Moorad et al |
| EPI_ISL_877694 | USA/NC-UNC-LCCC0113/2020 | 2020-08-23 | North America/USA/North Carolina/ | Clinical Molecular Microbiology Laboratory, UNC Hospital | Dirk Dittmer | Razia Moorad et al |
| EPI_ISL_877699 | USA/NC-UNC-LCCC0118/2020 | 2020-08-23 | North America/USA/North Carolina/ | Clinical Molecular Microbiology Laboratory, UNC Hospital | Dirk Dittmer | Razia Moorad et al |
| EPI_ISL_877710 | USA/NC-UNC-LCCC0129/2020 | 2020-08-24 | North America/USA/North Carolina/ | Clinical Molecular Microbiology Laboratory, UNC Hospital | Dirk Dittmer | Razia Moorad et al |
| EPI_ISL_877711 | USA/NC-UNC-LCCC0130/2020 | 2020-08-24 | North America/USA/North Carolina/ | Clinical Molecular Microbiology Laboratory, UNC Hospital | Dirk Dittmer | Razia Moorad et al |
| EPI_ISL_877713 | USA/NC-UNC-LCCC0132/2020 | 2020-08-24 | North America/USA/North Carolina/ | Clinical Molecular Microbiology Laboratory, UNC Hospital | Dirk Dittmer | Razia Moorad et al |
| EPI_ISL_877720 | USA/NC-UNC-LCCC0139/2020 | 2020-08-24 | North America/USA/North Carolina/ | Clinical Molecular Microbiology Laboratory, UNC Hospital | Dirk Dittmer | Razia Moorad et al |
| EPI_ISL_877721 | USA/NC-UNC-LCCC0140/2020 | 2020-08-24 | North America/USA/North Carolina/ | Clinical Molecular Microbiology Laboratory, UNC Hospital | Dirk Dittmer | Razia Moorad et al |
| EPI_ISL_877733 | USA/NC-UNC-LCCC0152/2020 | 2020-08-24 | North America/USA/North Carolina/ | Clinical Molecular Microbiology Laboratory, UNC Hospital | Dirk Dittmer | Razia Moorad et al |
| EPI_ISL_877734 | USA/NC-UNC-LCCC0153/2020 | 2020-08-24 | North America/USA/North Carolina/ | Clinical Molecular Microbiology Laboratory, UNC Hospital | Dirk Dittmer | Razia Moorad et al |
| EPI_ISL_877740 | USA/NC-UNC-LCCC0159/2020 | 2020-08-25 | North America/USA/North Carolina/ | Clinical Molecular Microbiology Laboratory, UNC Hospital | Dirk Dittmer | Razia Moorad et al |
| EPI_ISL_877741 | USA/NC-UNC-LCCC0160/2020 | 2020-08-25 | North America/USA/North Carolina/ | Clinical Molecular Microbiology Laboratory, UNC Hospital | Dirk Dittmer | Razia Moorad et al |
| EPI_ISL_877744 | USA/NC-UNC-LCCC0163/2020 | 2020-08-25 | North America/USA/North Carolina/ | Clinical Molecular Microbiology Laboratory, UNC Hospital | Dirk Dittmer | Razia Moorad et al |
| EPI_ISL_877746 | USA/NC-UNC-LCCC0165/2020 | 2020-08-25 | North America/USA/North Carolina/ | Clinical Molecular Microbiology Laboratory, UNC Hospital | Dirk Dittmer | Razia Moorad et al |
| EPI_ISL_877751 | USA/NC-UNC-LCCC0170/2020 | 2020-08-26 | North America/USA/North Carolina/ | Clinical Molecular Microbiology Laboratory, UNC Hospital | Dirk Dittmer | Razia Moorad et al |
| EPI_ISL_877758 | USA/NC-UNC-LCCC0178/2020 | 2020-08-26 | North America/USA/North Carolina/ | Clinical Molecular Microbiology Laboratory, UNC Hospital | Dirk Dittmer | Razia Moorad et al |
| EPI_ISL_877763 | USA/NC-UNC-LCCC0183/2020 | 2020-04-22 | North America/USA/North Carolina/ | Clinical Molecular Microbiology Laboratory, UNC Hospital | Dirk Dittmer | Razia Moorad et al |

| | | | | | | |
|---|---|---|---|---|---|---|
| EPI_ISL_877772 | England/ALDP-1091951/2021 | 2021-01-17 | Europe/United Kingdom/England/ | Lighthouse Lab in Alderley Park | Wellcome Sanger Institute for the COVID-19 Genomics UK (COG-UK) Consortium | Jacquelyn Wynn et al |
| EPI_ISL_878010 | England/ALDP-1092501/2021 | 2021-01-16 | Europe/United Kingdom/England/ | Lighthouse Lab in Alderley Park | Wellcome Sanger Institute for the COVID-19 Genomics UK (COG-UK) Consortium | Jacquelyn Wynn et al |
| EPI_ISL_878110 | England/ALDP-109160F/2021 | 2021-01-17 | Europe/United Kingdom/England/ | Lighthouse Lab in Alderley Park | Wellcome Sanger Institute for the COVID-19 Genomics UK (COG-UK) Consortium | Jacquelyn Wynn et al |
| EPI_ISL_878157 | USA/CA-ALSR-5577/2020 | 2020-12-01 | North America/USA/California/San Dieg | San Diego County Public Health Laboratory | Andersen lab at Scripps Research | SEARCH Alliance San Diego with Tracy Basler et al |
| EPI_ISL_878162 | USA/CA-ALSR-5580/2020 | 2020-12-07 | North America/USA/California/San Dieg | San Diego County Public Health Laboratory | Andersen lab at Scripps Research | SEARCH Alliance San Diego with Tracy Basler et al |
| EPI_ISL_878176 | England/ALDP-1064B33/2021 | 2021-01-14 | Europe/United Kingdom/England/ | Lighthouse Lab in Alderley Park | Wellcome Sanger Institute for the COVID-19 Genomics UK (COG-UK) Consortium | Jacquelyn Wynn et al |
| EPI_ISL_878183 | USA/CA-ALSR-5588/2020 | 2020-12-01 | North America/USA/California/San Dieg | San Diego County Public Health Laboratory | Andersen lab at Scripps Research | SEARCH Alliance San Diego with Tracy Basler et al |
| EPI_ISL_878207 | USA/CA-ALSR-5600/2020 | 2020-12-07 | North America/USA/California/San Dieg | San Diego County Public Health Laboratory | Andersen lab at Scripps Research | SEARCH Alliance San Diego with Tracy Basler et al |
| EPI_ISL_878212 | Mexico/BCN-ALSR-5603/2020 | 2020-12-01 | North America/Mexico/Baja California/T | San Diego County Public Health Laboratory | Andersen lab at Scripps Research | SEARCH Alliance San Diego with Tracy Basler et al |
| EPI_ISL_878221 | USA/CA-ALSR-5612/2020 | 2020-12-07 | North America/USA/California/San Dieg | San Diego County Public Health Laboratory | Andersen lab at Scripps Research | SEARCH Alliance San Diego with Tracy Basler et al |
| EPI_ISL_878226 | Mexico/BCN-ALSR-5615/2020 | 2020-12-01 | North America/Mexico/Baja California/T | San Diego County Public Health Laboratory | Andersen lab at Scripps Research | SEARCH Alliance San Diego with Tracy Basler et al |
| EPI_ISL_878234 | USA/CA-ALSR-5618/2020 | 2020-12-07 | North America/USA/California/San Dieg | San Diego County Public Health Laboratory | Andersen lab at Scripps Research | SEARCH Alliance San Diego with Tracy Basler et al |
| EPI_ISL_878239 | USA/CA-ALSR-5623/2020 | 2020-12-01 | North America/USA/California/San Dieg | San Diego County Public Health Laboratory | Andersen lab at Scripps Research | SEARCH Alliance San Diego with Tracy Basler et al |
| EPI_ISL_878256 | USA/CA-ALSR-5631/2020 | 2020-12-07 | North America/USA/California/San Dieg | San Diego County Public Health Laboratory | Andersen lab at Scripps Research | SEARCH Alliance San Diego with Tracy Basler et al |

| | | | | | | |
|---|---|---|---|---|---|---|
| EPI_ISL_878266 | USA/CA-ALSR-5638/2020 | 2020-12-07 | North America/USA/California/San Dieg | San Diego County Public Health Laboratory | Andersen lab at Scripps Research | SEARCH Alliance San Diego with Tracy Basler et al |
| EPI_ISL_878276 | USA/CA-ALSR-5646/2020 | 2020-11-23 | North America/USA/California/San Dieg | San Diego County Public Health Laboratory | Andersen lab at Scripps Research | SEARCH Alliance San Diego with Tracy Basler et al |
| EPI_ISL_878292 | USA/CA-ALSR-5652/2020 | 2020-11-27 | North America/USA/California/San Dieg | San Diego County Public Health Laboratory | Andersen lab at Scripps Research | SEARCH Alliance San Diego with Tracy Basler et al |
| EPI_ISL_878293 | England/ALDP-1064DD3/2021 | 2021-01-15 | Europe/United Kingdom/England/ | Lighthouse Lab in Alderley Park | Wellcome Sanger Institute for the COVID-19 Genomics UK (COG-UK) Consortium | Jacquelyn Wynn et al |
| EPI_ISL_878354 | USA/CA-ALSR-5695/2020 | 2020-11-24 | North America/USA/California/San Dieg | San Diego County Public Health Laboratory | Andersen lab at Scripps Research | SEARCH Alliance San Diego with Tracy Basler et al |
| EPI_ISL_878359 | USA/CA-ALSR-5700/2020 | 2020-11-24 | North America/USA/California/San Dieg | San Diego County Public Health Laboratory | Andersen lab at Scripps Research | SEARCH Alliance San Diego with Tracy Basler et al |
| EPI_ISL_878361 | USA/CA-ALSR-5702/2020 | 2020-11-30 | North America/USA/California/San Dieg | San Diego County Public Health Laboratory | Andersen lab at Scripps Research | SEARCH Alliance San Diego with Tracy Basler et al |
| EPI_ISL_878366 | USA/CA-ALSR-5707/2020 | 2020-11-24 | North America/USA/California/San Dieg | San Diego County Public Health Laboratory | Andersen lab at Scripps Research | SEARCH Alliance San Diego with Tracy Basler et al |
| EPI_ISL_878367 | Mexico/BCN-ALSR-5708/2020 | 2020-11-30 | North America/Mexico/Baja California/T | San Diego County Public Health Laboratory | Andersen lab at Scripps Research | SEARCH Alliance San Diego with Tracy Basler et al |
| EPI_ISL_878370 | USA/CA-ALSR-5711/2020 | 2020-11-27 | North America/USA/California/San Dieg | San Diego County Public Health Laboratory | Andersen lab at Scripps Research | SEARCH Alliance San Diego with Tracy Basler et al |
| EPI_ISL_878373 | USA/CA-ALSR-5714/2020 | 2020-11-24 | North America/USA/California/San Dieg | San Diego County Public Health Laboratory | Andersen lab at Scripps Research | SEARCH Alliance San Diego with Tracy Basler et al |
| EPI_ISL_878374 | USA/CA-ALSR-5715/2020 | 2020-11-30 | North America/USA/California/San Dieg | San Diego County Public Health Laboratory | Andersen lab at Scripps Research | SEARCH Alliance San Diego with Tracy Basler et al |
| EPI_ISL_878375 | Mexico/BCN-ALSR-5717/2020 | 2020-11-24 | North America/Mexico/Baja California/T | San Diego County Public Health Laboratory | Andersen lab at Scripps Research | SEARCH Alliance San Diego with Tracy Basler et al |
| EPI_ISL_878377 | USA/CA-ALSR-5719/2020 | 2020-11-24 | North America/USA/California/San Dieg | San Diego County Public Health Laboratory | Andersen lab at Scripps Research | SEARCH Alliance San Diego with Tracy Basler et al |
| EPI_ISL_878381 | USA/CA-ALSR-5723/2020 | 2020-11-30 | North America/USA/California/San Dieg | San Diego County Public Health Laboratory | Andersen lab at Scripps Research | SEARCH Alliance San Diego with Tracy Basler et al |
| EPI_ISL_878400 | Mexico/BCN-ALSR-5748/2020 | 2020-11-23 | North America/Mexico/Baja California/T | San Diego County Public Health Laboratory | Andersen lab at Scripps Research | SEARCH Alliance San Diego with Tracy Basler et al |
| EPI_ISL_878404 | Mexico/BCN-ALSR-5753/2020 | 2020-11-23 | North America/Mexico/Baja California/T | San Diego County Public Health Laboratory | Andersen lab at Scripps Research | SEARCH Alliance San Diego with Tracy Basler et al |
| EPI_ISL_878408 | Mexico/BCN-ALSR-5757/2020 | 2020-11-23 | North America/Mexico/Baja California/T | San Diego County Public Health Laboratory | Andersen lab at Scripps Research | SEARCH Alliance San Diego with Tracy Basler et al |

| | | | | | | |
|---|---|---|---|---|---|---|
| EPI_ISL_878418 | USA/CA-ALSR-5762/2020 | 2020-11-23 | North America/USA/California/San Dieg | San Diego County Public Health Laboratory | Andersen lab at Scripps Research | SEARCH Alliance San Diego with Tracy Basler et al |
| EPI_ISL_878419 | England/ALDP-1064B60/2021 | 2021-01-15 | Europe/United Kingdom/England/ | Lighthouse Lab in Alderley Park | Wellcome Sanger Institute for the COVID-19 Genomics UK (COG-UK) Consortium | Jacquelyn Wynn et al |
| EPI_ISL_878429 | USA/CA-ALSR-5767/2020 | 2020-11-24 | North America/USA/California/San Dieg | San Diego County Public Health Laboratory | Andersen lab at Scripps Research | SEARCH Alliance San Diego with Tracy Basler et al |
| EPI_ISL_878466 | England/ALDP-10658C2/2021 | 2021-01-14 | Europe/United Kingdom/England/ | Lighthouse Lab in Alderley Park | Wellcome Sanger Institute for the COVID-19 Genomics UK (COG-UK) Consortium | Jacquelyn Wynn et al |
| EPI_ISL_878552 | USA/LA-ALSR-5821/2020 | 2020-07-30 | North America/USA/Louisiana/New Orle | Robert Garry lab | Andersen lab at Scripps Research | Allison Smither et al |
| EPI_ISL_878557 | USA/LA-ALSR-5823/2020 | 2020-12-01 | North America/USA/Louisiana/New Orle | Robert Garry lab | Andersen lab at Scripps Research | Allison Smither et al |
| EPI_ISL_878571 | Jordan/AM-ALSR-5829/2020 | 2020-12-25 | Asia/Jordan/Amman/ | Biolab Diagnostic Laboratories | Andersen lab at Scripps Research | Issa Abu-Dayyeh et al |
| EPI_ISL_878574 | Jordan/AM-ALSR-5830/2020 | 2020-12-24 | Asia/Jordan/Amman/ | Biolab Diagnostic Laboratories | Andersen lab at Scripps Research | Issa Abu-Dayyeh et al |
| EPI_ISL_878584 | USA/CA-ALSR-5840/2020 | 2020-12-13 | North America/USA/California/San Dieg | San Diego County Public Health Laboratory | Andersen lab at Scripps Research | SEARCH Alliance San Diego with Tracy Basler et al |
| EPI_ISL_878619 | USA/CA-ALSR-5878/2020 | 2020-12-16 | North America/USA/California/San Dieg | San Diego County Public Health Laboratory | Andersen lab at Scripps Research | SEARCH Alliance San Diego with Tracy Basler et al |
| EPI_ISL_878658 | USA/CA-ALSR-5920/2020 | 2020-12-04 | North America/USA/California/San Dieg | Rady's Childrens Hospital | Andersen lab at Scripps Research | SEARCH Alliance San Diego with Nanda Radamchar et al |
| EPI_ISL_878666 | USA/CA-ALSR-5923/2020 | 2020-11-25 | North America/USA/California/San Dieg | Rady's Childrens Hospital | Andersen lab at Scripps Research | SEARCH Alliance San Diego with Nanda Radamchar et al |
| EPI_ISL_878682 | USA/CA-ALSR-5930/2020 | 2020-11-17 | North America/USA/California/San Dieg | Rady's Childrens Hospital | Andersen lab at Scripps Research | SEARCH Alliance San Diego with Nanda Radamchar et al |
| EPI_ISL_878713 | USA/CA-ALSR-5947/2020 | 2020-11-24 | North America/USA/California/San Dieg | Rady's Childrens Hospital | Andersen lab at Scripps Research | SEARCH Alliance San Diego with Nanda Radamchar et al |
| EPI_ISL_878732 | USA/CA-ALSR-5955/2020 | 2020-12-03 | North America/USA/California/San Dieg | Rady's Childrens Hospital | Andersen lab at Scripps Research | SEARCH Alliance San Diego with Nanda Radamchar et al |
| EPI_ISL_878752 | USA/CA-ALSR-5965/2020 | 2020-12-30 | North America/USA/California/San Dieg | Rady's Childrens Hospital | Andersen lab at Scripps Research | SEARCH Alliance San Diego with Nanda Radamchar et al |
| EPI_ISL_878757 | USA/CA-ALSR-5968/2020 | 2020-12-12 | North America/USA/California/San Dieg | Rady's Childrens Hospital | Andersen lab at Scripps Research | SEARCH Alliance San Diego with Nanda Radamchar et al |

| | | | | | | |
|---|---|---|---|---|---|---|
| EPI_ISL_878781 | USA/CA-ALSR-5979/2020 | 2020-12-12 | North America/USA/California/San Dieg | Rady's Childrens Hospital | Andersen lab at Scripps Research | SEARCH Alliance San Diego with Nanda Radamchar et al |
| EPI_ISL_878790 | Wales/ALDP-1036BFE/2021 | 2021-01-15 | Europe/United Kingdom/Wales/ | Lighthouse Lab in Alderley Park | Wellcome Sanger Institute for the COVID-19 Genomics UK (COG-UK) Consortium | Jacquelyn Wynn et al |
| EPI_ISL_878794 | USA/CA-ALSR-5985/2021 | 2021-01-04 | North America/USA/California/San Dieg | Rady's Childrens Hospital | Andersen lab at Scripps Research | SEARCH Alliance San Diego with Nanda Radamchar et al |
| EPI_ISL_878797 | USA/CA-ALSR-5986/2020 | 2020-12-19 | North America/USA/California/San Dieg | Rady's Childrens Hospital | Andersen lab at Scripps Research | SEARCH Alliance San Diego with Nanda Radamchar et al |
| EPI_ISL_878930 | England/CAMC-10605B1/2021 | 2021-01-16 | Europe/United Kingdom/England/ | Lighthouse Lab in Cambridge | Wellcome Sanger Institute for the COVID-19 Genomics UK (COG-UK) Consortium | Rob Howes et al |
| EPI_ISL_879094 | England/MILK-104D8FD/2021 | 2021-01-13 | Europe/United Kingdom/England/ | Lighthouse Lab in Milton Keynes | Wellcome Sanger Institute for the COVID-19 Genomics UK (COG-UK) Consortium | The Lighthouse Lab in Milton Keynes et al |
| EPI_ISL_879138 | England/ALDP-10685E6/2021 | 2021-01-14 | Europe/United Kingdom/England/ | Lighthouse Lab in Alderley Park | Wellcome Sanger Institute for the COVID-19 Genomics UK (COG-UK) Consortium | Jacquelyn Wynn et al |
| EPI_ISL_879197 | England/CAMC-106E6B0/2021 | 2021-01-17 | Europe/United Kingdom/England/ | Lighthouse Lab in Cambridge | Wellcome Sanger Institute for the COVID-19 Genomics UK (COG-UK) Consortium | Rob Howes et al |
| EPI_ISL_879348 | England/ALDP-1068DEE/2021 | 2021-01-15 | Europe/United Kingdom/England/ | Lighthouse Lab in Alderley Park | Wellcome Sanger Institute for the COVID-19 Genomics UK (COG-UK) Consortium | Jacquelyn Wynn et al |

| | | | | | | |
|---|---|---|---|---|---|---|
| EPI_ISL_879364 | England/CAMC-106E647/2021 | 2021-01-16 | Europe/United Kingdom/England/ | Lighthouse Lab in Cambridge | Wellcome Sanger Institute for the COVID-19 Genomics UK (COG-UK) Consortium | Rob Howes et al |
| EPI_ISL_879420 | England/ALDP-1035B2C/2021 | 2021-01-16 | Europe/United Kingdom/England/ | Lighthouse Lab in Alderley Park | Wellcome Sanger Institute for the COVID-19 Genomics UK (COG-UK) Consortium | Jacquelyn Wynn et al |
| EPI_ISL_879491 | England/CAMC-106415F/2021 | 2021-01-15 | Europe/United Kingdom/England/ | Lighthouse Lab in Cambridge | Wellcome Sanger Institute for the COVID-19 Genomics UK (COG-UK) Consortium | Rob Howes et al |
| EPI_ISL_879499 | England/MILK-103CD3E/2021 | 2021-01-14 | Europe/United Kingdom/England/ | Lighthouse Lab in Milton Keynes | Wellcome Sanger Institute for the COVID-19 Genomics UK (COG-UK) Consortium | The Lighthouse Lab in Milton Keynes et al |
| EPI_ISL_879687 | England/ALDP-1035B68/2021 | 2021-01-16 | Europe/United Kingdom/England/ | Lighthouse Lab in Alderley Park | Wellcome Sanger Institute for the COVID-19 Genomics UK (COG-UK) Consortium | Jacquelyn Wynn et al |
| EPI_ISL_879937 | USA/CA-ALSR-5993/2020 | 2020-11-24 | North America/USA/California/San Dieg | San Diego County Public Health Laboratory | Andersen lab at Scripps Research | SEARCH Alliance San Diego with Tracy Basler et al |
| EPI_ISL_879950 | Mexico/BCN-ALSR-6000/2020 | 2020-11-30 | North America/Mexico/Baja California/T | San Diego County Public Health Laboratory | Andersen lab at Scripps Research | SEARCH Alliance San Diego with Tracy Basler et al |
| EPI_ISL_879953 | USA/CA-ALSR-6001/2020 | 2020-11-30 | North America/USA/California/San Dieg | San Diego County Public Health Laboratory | Andersen lab at Scripps Research | SEARCH Alliance San Diego with Tracy Basler et al |
| EPI_ISL_880000 | USA/CA-ALSR-6025/2020 | 2020-11-27 | North America/USA/California/San Dieg | San Diego County Public Health Laboratory | Andersen lab at Scripps Research | SEARCH Alliance San Diego with Tracy Basler et al |
| EPI_ISL_880019 | USA/CA-ALSR-6034/2020 | 2020-11-27 | North America/USA/California/Los Ange | San Diego County Public Health Laboratory | Andersen lab at Scripps Research | SEARCH Alliance San Diego with Tracy Basler et al |
| EPI_ISL_880029 | USA/CA-ALSR-6038/2020 | 2020-11-27 | North America/USA/California/San Dieg | San Diego County Public Health Laboratory | Andersen lab at Scripps Research | SEARCH Alliance San Diego with Tracy Basler et al |
| EPI_ISL_880032 | USA/CA-ALSR-6039/2020 | 2020-11-27 | North America/USA/California/San Dieg | San Diego County Public Health Laboratory | Andersen lab at Scripps Research | SEARCH Alliance San Diego with Tracy Basler et al |

| | | | | | | |
|---|---|---|---|---|---|---|
| EPI_ISL_880058 | USA/CA-ALSR-6052/2021 | 2021-01-08 | North America/USA/California/San Dieg | Sharp HealthCare Laboratory | Andersen lab at Scripps Research | SEARCH Alliance San Diego with Aaron Harding et al |
| EPI_ISL_880074 | USA/CA-ALSR-6059/2021 | 2021-01-07 | North America/USA/California/San Dieg | Sharp HealthCare Laboratory | Andersen lab at Scripps Research | SEARCH Alliance San Diego with Aaron Harding et al |
| EPI_ISL_880082 | USA/CA-ALSR-6062/2021 | 2021-01-07 | North America/USA/California/San Dieg | Sharp HealthCare Laboratory | Andersen lab at Scripps Research | SEARCH Alliance San Diego with Aaron Harding et al |
| EPI_ISL_880095 | USA/CA-ALSR-6069/2021 | 2021-01-07 | North America/USA/California/San Dieg | Sharp HealthCare Laboratory | Andersen lab at Scripps Research | SEARCH Alliance San Diego with Aaron Harding et al |
| EPI_ISL_880125 | USA/CA-ALSR-6080/2021 | 2021-01-07 | North America/USA/California/San Dieg | Sharp HealthCare Laboratory | Andersen lab at Scripps Research | SEARCH Alliance San Diego with Aaron Harding et al |
| EPI_ISL_880135 | USA/CA-ALSR-6085/2021 | 2021-01-07 | North America/USA/California/San Dieg | Sharp HealthCare Laboratory | Andersen lab at Scripps Research | SEARCH Alliance San Diego with Aaron Harding et al |
| EPI_ISL_880149 | USA/CA-ALSR-6096/2021 | 2021-01-07 | North America/USA/California/San Dieg | Sharp HealthCare Laboratory | Andersen lab at Scripps Research | SEARCH Alliance San Diego with Aaron Harding et al |
| EPI_ISL_880152 | USA/CA-ALSR-6101/2021 | 2021-01-07 | North America/USA/California/San Dieg | Sharp HealthCare Laboratory | Andersen lab at Scripps Research | SEARCH Alliance San Diego with Aaron Harding et al |
| EPI_ISL_880161 | USA/CA-ALSR-6111/2021 | 2021-01-07 | North America/USA/California/San Dieg | Sharp HealthCare Laboratory | Andersen lab at Scripps Research | SEARCH Alliance San Diego with Aaron Harding et al |
| EPI_ISL_880180 | USA/CA-ALSR-6134/2020 | 2020-12-14 | North America/USA/California/San Dieg | Rady's Childrens Hospital | Andersen lab at Scripps Research | SEARCH Alliance San Diego with Nanda Radamchar et al |
| EPI_ISL_880191 | USA/CA-ALSR-6148/2020 | 2020-12-07 | North America/USA/California/San Dieg | Rady's Childrens Hospital | Andersen lab at Scripps Research | SEARCH Alliance San Diego with Nanda Radamchar et al |
| EPI_ISL_880197 | USA/CA-ALSR-6156/2020 | 2020-12-13 | North America/USA/California/San Dieg | Rady's Childrens Hospital | Andersen lab at Scripps Research | SEARCH Alliance San Diego with Nanda Radamchar et al |
| EPI_ISL_880198 | USA/CA-ALSR-6161/2020 | 2020-12-21 | North America/USA/California/San Dieg | Rady's Childrens Hospital | Andersen lab at Scripps Research | SEARCH Alliance San Diego with Nanda Radamchar et al |
| EPI_ISL_880199 | USA/CA-ALSR-6162/2020 | 2020-12-15 | North America/USA/California/San Dieg | Rady's Childrens Hospital | Andersen lab at Scripps Research | SEARCH Alliance San Diego with Nanda Radamchar et al |
| EPI_ISL_880218 | USA/CA-ALSR-6178/2021 | 2021-01-07 | North America/USA/California/Riverside | Scripps Medical Laboratory | Andersen lab at Scripps Research | SEARCH Alliance San Diego with Michael Quigley et al |
| EPI_ISL_880223 | USA/CA-ALSR-6181/2021 | 2021-01-07 | North America/USA/California/San Dieg | Scripps Medical Laboratory | Andersen lab at Scripps Research | SEARCH Alliance San Diego with Michael Quigley et al |
| EPI_ISL_880242 | England/ALDP-10313DD/2021 | 2021-01-14 | Europe/United Kingdom/England/ | Lighthouse Lab in Alderley Park | Wellcome Sanger Institute for the COVID-19 Genomics UK (COG-UK) Consortium | Jacquelyn Wynn et al |
| EPI_ISL_880251 | USA/CA-ALSR-6193/2021 | 2021-01-06 | North America/USA/California/San Dieg | Scripps Medical Laboratory | Andersen lab at Scripps Research | SEARCH Alliance San Diego with Michael Quigley et al |

| | | | | | | |
|---|---|---|---|---|---|---|
| EPI_ISL_880252 | England/ALDP-1030E73/2021 | 2021-01-14 | Europe/United Kingdom/England/ | Lighthouse Lab in Alderley Park | Wellcome Sanger Institute for the COVID-19 Genomics UK (COG-UK) Consortium | Jacquelyn Wynn et al |
| EPI_ISL_880287 | USA/CA-ALSR-6213/2021 | 2021-01-06 | North America/USA/California/San Dieg | Scripps Medical Laboratory | Andersen lab at Scripps Research | SEARCH Alliance San Diego with Michael Quigley et al |
| EPI_ISL_880305 | USA/CA-ALSR-6222/2021 | 2021-01-07 | North America/USA/California/San Dieg | Scripps Medical Laboratory | Andersen lab at Scripps Research | SEARCH Alliance San Diego with Michael Quigley et al |
| EPI_ISL_880313 | USA/CA-ALSR-6228/2021 | 2021-01-06 | North America/USA/California/San Dieg | Scripps Medical Laboratory | Andersen lab at Scripps Research | SEARCH Alliance San Diego with Michael Quigley et al |
| EPI_ISL_880742 | England/MILK-102EB71/2021 | 2021-01-12 | Europe/United Kingdom/England/ | Lighthouse Lab in Milton Keynes | Wellcome Sanger Institute for the COVID-19 Genomics UK (COG-UK) Consortium | The Lighthouse Lab in Milton Keynes et al |
| EPI_ISL_880756 | England/MILK-102E7B9/2021 | 2021-01-12 | Europe/United Kingdom/England/ | Lighthouse Lab in Milton Keynes | Wellcome Sanger Institute for the COVID-19 Genomics UK (COG-UK) Consortium | The Lighthouse Lab in Milton Keynes et al |
| EPI_ISL_880869 | England/MILK-102E5DD/2021 | 2021-01-12 | Europe/United Kingdom/England/ | Lighthouse Lab in Milton Keynes | Wellcome Sanger Institute for the COVID-19 Genomics UK (COG-UK) Consortium | The Lighthouse Lab in Milton Keynes et al |
| EPI_ISL_880986 | England/ALDP-101C077/2021 | 2021-01-14 | Europe/United Kingdom/England/ | Lighthouse Lab in Alderley Park | Wellcome Sanger Institute for the COVID-19 Genomics UK (COG-UK) Consortium | Jacquelyn Wynn et al |
| EPI_ISL_881040 | England/ALDP-101C305/2021 | 2021-01-14 | Europe/United Kingdom/England/ | Lighthouse Lab in Alderley Park | Wellcome Sanger Institute for the COVID-19 Genomics UK (COG-UK) Consortium | Jacquelyn Wynn et al |

| | | | | | | |
|---|---|---|---|---|---|---|
| EPI_ISL_881050 | England/ALDP-101A862/2021 | 2021-01-14 | Europe/United Kingdom/England/ | Lighthouse Lab in Alderley Park | Wellcome Sanger Institute for the COVID-19 Genomics UK (COG-UK) Consortium | Jacquelyn Wynn et al |
| EPI_ISL_881068 | England/ALDP-101A5E3/2021 | 2021-01-14 | Europe/United Kingdom/England/ | Lighthouse Lab in Alderley Park | Wellcome Sanger Institute for the COVID-19 Genomics UK (COG-UK) Consortium | Jacquelyn Wynn et al |
| EPI_ISL_881226 | England/ALDP-1016C3D/2021 | 2021-01-13 | Europe/United Kingdom/England/ | Lighthouse Lab in Alderley Park | Wellcome Sanger Institute for the COVID-19 Genomics UK (COG-UK) Consortium | Jacquelyn Wynn et al |
| EPI_ISL_881366 | England/ALDP-100E852/2021 | 2021-01-12 | Europe/United Kingdom/England/ | Lighthouse Lab in Alderley Park | Wellcome Sanger Institute for the COVID-19 Genomics UK (COG-UK) Consortium | Jacquelyn Wynn et al |
| EPI_ISL_881379 | England/ALDP-100EC29/2021 | 2021-01-12 | Europe/United Kingdom/England/ | Lighthouse Lab in Alderley Park | Wellcome Sanger Institute for the COVID-19 Genomics UK (COG-UK) Consortium | Jacquelyn Wynn et al |
| EPI_ISL_881574 | England/ALDP-100EC0B/2021 | 2021-01-12 | Europe/United Kingdom/England/ | Lighthouse Lab in Alderley Park | Wellcome Sanger Institute for the COVID-19 Genomics UK (COG-UK) Consortium | Jacquelyn Wynn et al |
| EPI_ISL_881740 | England/ALDP-FA220B/2021 | 2021-01-09 | Europe/United Kingdom/England/ | Lighthouse Lab in Alderley Park | Wellcome Sanger Institute for the COVID-19 Genomics UK (COG-UK) Consortium | Jacquelyn Wynn et al |
| EPI_ISL_881792 | England/ALDP-FA3808/2021 | 2021-01-09 | Europe/United Kingdom/England/ | Lighthouse Lab in Alderley Park | Wellcome Sanger Institute for the COVID-19 Genomics UK (COG-UK) Consortium | Jacquelyn Wynn et al |

| | | | | | | |
|---|---|---|---|---|---|---|
| EPI_ISL_881959 | England/ALDP-FA27A2/2021 | 2021-01-09 | Europe/United Kingdom/England/ | Lighthouse Lab in Alderley Park | Wellcome Sanger Institute for the COVID-19 Genomics UK (COG-UK) Consortium | Jacquelyn Wynn et al |
| EPI_ISL_881982 | England/ALDP-FB56BE/2021 | 2021-01-10 | Europe/United Kingdom/England/ | Lighthouse Lab in Alderley Park | Wellcome Sanger Institute for the COVID-19 Genomics UK (COG-UK) Consortium | Jacquelyn Wynn et al |
| EPI_ISL_882146 | England/ALDP-FAF945/2021 | 2021-01-10 | Europe/United Kingdom/England/ | Lighthouse Lab in Alderley Park | Wellcome Sanger Institute for the COVID-19 Genomics UK (COG-UK) Consortium | Jacquelyn Wynn et al |
| EPI_ISL_882177 | England/ALDP-FAF4CC/2021 | 2021-01-10 | Europe/United Kingdom/England/ | Lighthouse Lab in Alderley Park | Wellcome Sanger Institute for the COVID-19 Genomics UK (COG-UK) Consortium | Jacquelyn Wynn et al |
| EPI_ISL_882297 | England/ALDP-FB52D8/2021 | 2021-01-10 | Europe/United Kingdom/England/ | Lighthouse Lab in Alderley Park | Wellcome Sanger Institute for the COVID-19 Genomics UK (COG-UK) Consortium | Jacquelyn Wynn et al |
| EPI_ISL_882391 | England/ALDP-F75043/2021 | 2021-01-02 | Europe/United Kingdom/England/ | Lighthouse Lab in Alderley Park | Wellcome Sanger Institute for the COVID-19 Genomics UK (COG-UK) Consortium | Jacquelyn Wynn et al |
| EPI_ISL_882612 | Spain/MD-11664990/2021 | 2021-01-02 | Europe/Spain/Madrid/Madrid | Hospital Ram√≥n y Cajal | Hospital Ram√≥n y Cajal | Jos√© M Gonzalez-Alba et al |
| EPI_ISL_882615 | Spain/MD-11665927/2021 | 2021-01-02 | Europe/Spain/Madrid/Madrid | Hospital Ram√≥n y Cajal | Hospital Ram√≥n y Cajal | Jos√© M Gonzalez-Alba et al |
| EPI_ISL_882624 | Spain/MD-71407645/2021 | 2021-01-08 | Europe/Spain/Madrid/Madrid | Hospital Ram√≥n y Cajal | Hospital Ram√≥n y Cajal | Jos√© M Gonzalez-Alba et al |
| EPI_ISL_882627 | Spain/MD-71430583/2021 | 2021-01-08 | Europe/Spain/Madrid/Madrid | Hospital Ram√≥n y Cajal | Hospital Ram√≥n y Cajal | Jos√© M Gonzalez-Alba et al |
| EPI_ISL_882629 | Spain/MD-71486374/2021 | 2021-01-06 | Europe/Spain/Madrid/Madrid | Hospital Ram√≥n y Cajal | Hospital Ram√≥n y Cajal | Jos√© M Gonzalez-Alba et al |

| | | | | | | |
|---|---|---|---|---|---|---|
| EPI_ISL_882632 | Spain/MD-11674437/2021 | 2021-01-08 | Europe/Spain/Madrid/Madrid | Hospital Ram√≥n y Cajal | Hospital Ram√≥n y Cajal | Jos√© M Gonzalez-Alba et al |
| EPI_ISL_882637 | Azerbaijan/Aghayev-01/2021 | 2021-01-02 | Europe/Azerbaijan/Azerbaijan/ | Azerbaijan National Hematology Center Division of Medical Genetics | Azerbaijan National Hematology Center Division of Medical Genetics | Aghayev Agha Rza et al |
| EPI_ISL_882638 | Azerbaijan/Aghayev-02/2021 | 2021-01-02 | Europe/Azerbaijan/Azerbaijan/ | Azerbaijan National Hematology Center Division of Medical Genetics | Azerbaijan National Hematology Center Division of Medical Genetics | Aghayev Agha Rza et al |
| EPI_ISL_882645 | Indonesia/BT-SHSIU-01-6368/2021 | 2021-01-04 | Asia/Indonesia/Banten/ | Medical Research Center, Faculty of Medicine, Syarif Hidayatullah State Islamic University Jakarta | Medical Research Center, Faculty of Medicine, Syarif Hidayatullah State Islamic University Jakarta | Chris Adhiyanto et al |
| EPI_ISL_882646 | Indonesia/BT-SHSIU-01-6493/2021 | 2021-01-06 | Asia/Indonesia/Banten/ | Medical Research Center, Faculty of Medicine, Syarif Hidayatullah State Islamic University Jakarta | Medical Research Center, Faculty of Medicine, Syarif Hidayatullah State Islamic University Jakarta | Chris Adhiyanto et al |
| EPI_ISL_882656 | Indonesia/BT-SHSIU-01-6494/2021 | 2021-01-06 | Asia/Indonesia/Banten/ | Medical Research Center, Faculty of Medicine, Syarif Hidayatullah State Islamic University Jakarta | Medical Research Center, Faculty of Medicine, Syarif Hidayatullah State Islamic University Jakarta | Erike A Suwarsono et al |
| EPI_ISL_882657 | Brazil/DF-1028R1/2020 | 2020-07-23 | South America/Brazil/Distrito Federal/ | LACEN do Estado do Piaui, Dr. Costa Alvarenga | Instituto Adolfo Lutz, Interdiciplinary Procedures Center, Strategic Laboratory | Claudio Tavares Sacchi et al |

| | | | | | | |
|---|---|---|---|---|---|---|
| EPI_ISL_882658 | Brazil/SP-933/2020 | 2020-11-23 | South America/Brazil/S√£o Paulo/Santa | Secretaria Municipal de Saude | Instituto Adolfo Lutz, Interdiciplinary Procedures Center, Strategic Laboratory | Claudio Tavares Sacchi et al |
| EPI_ISL_882661 | Brazil/GO-1022/2020 | 2020-12-24 | South America/Brazil/Goi√°s/Santa Barb | Hospital de Santa Barbara de Goias | Instituto Adolfo Lutz, Interdiciplinary Procedures Center, Strategic Laboratory | Claudio Tavares Sacchi et al |
| EPI_ISL_882662 | Brazil/GO-1023/2020 | 2020-12-24 | South America/Brazil/Goi√°s/Santa Barb | Hospital de Santa Barbara de Goias | Instituto Adolfo Lutz, Interdiciplinary Procedures Center, Strategic Laboratory | Claudio Tavares Sacchi et al |
| EPI_ISL_882664 | Brazil/DF-1025/2020 | 2020-12-31 | South America/Brazil/Distrito Federal/ | LACEN do Estado do Piaui, Dr. Costa Alvarenga | Instituto Adolfo Lutz, Interdiciplinary Procedures Center, Strategic Laboratory | Claudio Tavares Sacchi et al |
| EPI_ISL_882665 | Brazil/SP-1011/2021 | 2021-01-05 | South America/Brazil/S√£o Paulo/Ces√° | Unidade de Pronto Atendimento Dra Zilda Arns | Instituto Adolfo Lutz, Interdiciplinary Procedures Center, Strategic Laboratory | Claudio Tavares Sacchi et al |
| EPI_ISL_882666 | Brazil/SP-1009/2021 | 2021-01-08 | South America/Brazil/S√£o Paulo/Juquit | UMS de Juquitiba | Instituto Adolfo Lutz, Interdiciplinary Procedures Center, Strategic Laboratory | Claudio Tavares Sacchi et al |

| | | | | | | |
|---|---|---|---|---|---|---|
| EPI_ISL_882668 | Brazil/SP-1010/2021 | 2021-01-10 | South America/Brazil/S√£o Paulo/Juquit | UMS de Juquitiba | Instituto Adolfo Lutz, Interdiciplinary Procedures Center, Strategic Laboratory | Claudio Tavares Sacchi et al |
| EPI_ISL_882669 | Brazil/SP-1008/2021 | 2021-01-11 | South America/Brazil/S√£o Paulo/S√£o | UPA Vila Santa Catarina | Instituto Adolfo Lutz, Interdiciplinary Procedures Center, Strategic Laboratory | Claudio Tavares Sacchi et al |
| EPI_ISL_882670 | Brazil/SP-1012/2021 | 2021-01-12 | South America/Brazil/S√£o Paulo/S√£o | Hospital Samaritano Paulista | Instituto Adolfo Lutz, Interdiciplinary Procedures Center, Strategic Laboratory | Claudio Tavares Sacchi et al |
| EPI_ISL_882671 | Brazil/SP-1017/2021 | 2021-01-13 | South America/Brazil/S√£o Paulo/Poa | Hospital Municipal Dr. Guido Guida | Instituto Adolfo Lutz, Interdiciplinary Procedures Center, Strategic Laboratory | Claudio Tavares Sacchi et al |
| EPI_ISL_882672 | Brazil/SP-1018/2021 | 2021-01-13 | South America/Brazil/S√£o Paulo/Poa | Hospital Municipal Dr. Guido Guida | Instituto Adolfo Lutz, Interdiciplinary Procedures Center, Strategic Laboratory | Claudio Tavares Sacchi et al |
| EPI_ISL_882673 | Brazil/SP-1019/2021 | 2021-01-15 | South America/Brazil/S√£o Paulo/Poa | Hospital Municipal Dr. Guido Guida | Instituto Adolfo Lutz, Interdiciplinary Procedures Center, Strategic Laboratory | Claudio Tavares Sacchi et al |

| | | | | | | |
|---|---|---|---|---|---|---|
| EPI_ISL_882675 | Italy/CAM-CRGS-179/2020 | 2020-10-14 | Europe/Italy/Campania/ | 1.AO Universitaria 'S. Giovanni di Dio e Ruggi D'Aragona, Scuola Medica Salernitana' Hospital / 2.UOC di Virologia e Microbiologia, Universit√† della Campania 'L. Vanvitelli' / 3.AO Universitaria 'Federico II' Napoli Hospital / 4.AORN 'San Giuseppe Moscati' Avellino Hospital / 5.AO 'San Pio - presidio G. Rummo' Benevento Hospital / 6.AO 'Sant'Anna e San Sebastiano' Caserta Hospital / 7.PO 'Maria Santissima Addolorata' Eboli Hospital / 8.Biogem Istituto di Ricerche Genetiche | 1. Genome Research Center for Health (CRGS) / 2. Laboratory of Molecular Medicine and Genomics(LMMGe) / 3. Center for Research in Pure and Applied Mathematics (CRMPA) | Giorgio Giurato et al |
| EPI_ISL_882676 | Italy/CAM-CRGS-182/2020 | 2020-10-15 | Europe/Italy/Campania/ | 1.AO Universitaria 'S. Giovanni di Dio e Ruggi D'Aragona, Scuola Medica Salernitana' Hospital / 2.UOC di Virologia e Microbiologia, Universit√† della Campania 'L. Vanvitelli' / 3.AO Universitaria 'Federico II' Napoli Hospital / 4.AORN 'San Giuseppe Moscati' Avellino Hospital / 5.AO 'San Pio - presidio G. Rummo' Benevento Hospital / 6.AO 'Sant'Anna e San Sebastiano' Caserta Hospital / 7.PO 'Maria Santissima Addolorata' Eboli Hospital / 8.Biogem Istituto di Ricerche Genetiche | 1. Genome Research Center for Health (CRGS) / 2. Laboratory of Molecular Medicine and Genomics(LMMGe) / 3. Center for Research in Pure and Applied Mathematics (CRMPA) | Giorgio Giurato et al |
| EPI_ISL_882686 | Italy/CAM-CRGS-232/2020 | 2020-09-11 | Europe/Italy/Campania/ | 1.AO Universitaria 'S. Giovanni di Dio e Ruggi D'Aragona, Scuola Medica Salernitana' Hospital / 2.UOC di Virologia e Microbiologia, Universit√† della Campania 'L. Vanvitelli' / 3.AO Universitaria 'Federico II' Napoli Hospital / 4.AORN 'San Giuseppe Moscati' Avellino Hospital / 5.AO 'San Pio - presidio G. Rummo' Benevento Hospital / 6.AO 'Sant'Anna e San Sebastiano' Caserta Hospital / 7.PO 'Maria Santissima Addolorata' Eboli Hospital / 8.Biogem Istituto di Ricerche Genetiche | 1. Genome Research Center for Health (CRGS) / 2. Laboratory of Molecular Medicine and Genomics(LMMGe) / 3. Center for Research in Pure and Applied Mathematics (CRMPA) | Giorgio Giurato et al |

| | | | | | | |
|---|---|---|---|---|---|---|
| EPI_ISL_882688 | Italy/CAM-CRGS-142/2020 | 2020-11-17 | Europe/Italy/Campania/ | 1.AO Universitaria 'S. Giovanni di Dio e Ruggi D'Aragona, Scuola Medica Salernitana' Hospital / 2.UOC di Virologia e Microbiologia, Universit√† della Campania 'L. Vanvitelli' / 3.AO Universitaria 'Federico II' Napoli Hospital / 4.AORN 'San Giuseppe Moscati' Avellino Hospital / 5.AO 'San Pio - presidio G. Rummo' Benevento Hospital / 6.AO 'Sant'Anna e San Sebastiano' Caserta Hospital / 7.PO 'Maria Santissima Addolorata' Eboli Hospital / 8.Biogem Istituto di Ricerche Genetiche | 1. Genome Research Center for Health (CRGS) / 2. Laboratory of Molecular Medicine and Genomics(LMMGe) / 3. Center for Research in Pure and Applied Mathematics (CRMPA) | Giorgio Giurato et al |
| EPI_ISL_882689 | Italy/CAM-CRGS-157/2020 | 2020-11-16 | Europe/Italy/Campania/ | 1.AO Universitaria 'S. Giovanni di Dio e Ruggi D'Aragona, Scuola Medica Salernitana' Hospital / 2.UOC di Virologia e Microbiologia, Universit√† della Campania 'L. Vanvitelli' / 3.AO Universitaria 'Federico II' Napoli Hospital / 4.AORN 'San Giuseppe Moscati' Avellino Hospital / 5.AO 'San Pio - presidio G. Rummo' Benevento Hospital / 6.AO 'Sant'Anna e San Sebastiano' Caserta Hospital / 7.PO 'Maria Santissima Addolorata' Eboli Hospital / 8.Biogem Istituto di Ricerche Genetiche | 1. Genome Research Center for Health (CRGS) / 2. Laboratory of Molecular Medicine and Genomics(LMMGe) / 3. Center for Research in Pure and Applied Mathematics (CRMPA) | Giorgio Giurato et al |
| EPI_ISL_882692 | Italy/CAM-CRGS-219/2020 | 2020-11-05 | Europe/Italy/Campania/ | 1.AO Universitaria 'S. Giovanni di Dio e Ruggi D'Aragona, Scuola Medica Salernitana' Hospital / 2.UOC di Virologia e Microbiologia, Universit√† della Campania 'L. Vanvitelli' / 3.AO Universitaria 'Federico II' Napoli Hospital / 4.AORN 'San Giuseppe Moscati' Avellino Hospital / 5.AO 'San Pio - presidio G. Rummo' Benevento Hospital / 6.AO 'Sant'Anna e San Sebastiano' Caserta Hospital / 7.PO 'Maria Santissima Addolorata' Eboli Hospital / 8.Biogem Istituto di Ricerche Genetiche | 1. Genome Research Center for Health (CRGS) / 2. Laboratory of Molecular Medicine and Genomics(LMMGe) / 3. Center for Research in Pure and Applied Mathematics (CRMPA) | Giorgio Giurato et al |

| | | | | | | |
|---|---|---|---|---|---|---|
| EPI_ISL_882693 | Italy/CAM-CRGS-153/2020 | 2020-11-14 | Europe/Italy/Campania/ | 1.AO Universitaria 'S. Giovanni di Dio e Ruggi D'Aragona, Scuola Medica Salernitana' Hospital / 2.UOC di Virologia e Microbiologia, Universit√† della Campania 'L. Vanvitelli' / 3.AO Universitaria 'Federico II' Napoli Hospital / 4.AORN 'San Giuseppe Moscati' Avellino Hospital / 5.AO 'San Pio - presidio G. Rummo' Benevento Hospital / 6.AO 'Sant'Anna e San Sebastiano' Caserta Hospital / 7.PO 'Maria Santissima Addolorata' Eboli Hospital / 8.Biogem Istituto di Ricerche Genetiche | 1. Genome Research Center for Health (CRGS) / 2. Laboratory of Molecular Medicine and Genomics(LMMGe) / 3. Center for Research in Pure and Applied Mathematics (CRMPA) | Giorgio Giurato et al |
| EPI_ISL_882695 | Italy/CAM-CRGS-272/2020 | 2020-10-13 | Europe/Italy/Campania/ | 1.AO Universitaria 'S. Giovanni di Dio e Ruggi D'Aragona, Scuola Medica Salernitana' Hospital / 2.UOC di Virologia e Microbiologia, Universit√† della Campania 'L. Vanvitelli' / 3.AO Universitaria 'Federico II' Napoli Hospital / 4.AORN 'San Giuseppe Moscati' Avellino Hospital / 5.AO 'San Pio - presidio G. Rummo' Benevento Hospital / 6.AO 'Sant'Anna e San Sebastiano' Caserta Hospital / 7.PO 'Maria Santissima Addolorata' Eboli Hospital / 8.Biogem Istituto di Ricerche Genetiche | 1. Genome Research Center for Health (CRGS) / 2. Laboratory of Molecular Medicine and Genomics(LMMGe) / 3. Center for Research in Pure and Applied Mathematics (CRMPA) | Giorgio Giurato et al |
| EPI_ISL_882696 | Italy/CAM-CRGS-257/2020 | 2020-11-03 | Europe/Italy/Campania/ | 1.AO Universitaria 'S. Giovanni di Dio e Ruggi D'Aragona, Scuola Medica Salernitana' Hospital / 2.UOC di Virologia e Microbiologia, Universit√† della Campania 'L. Vanvitelli' / 3.AO Universitaria 'Federico II' Napoli Hospital / 4.AORN 'San Giuseppe Moscati' Avellino Hospital / 5.AO 'San Pio - presidio G. Rummo' Benevento Hospital / 6.AO 'Sant'Anna e San Sebastiano' Caserta Hospital / 7.PO 'Maria Santissima Addolorata' Eboli Hospital / 8.Biogem Istituto di Ricerche Genetiche | 1. Genome Research Center for Health (CRGS) / 2. Laboratory of Molecular Medicine and Genomics(LMMGe) / 3. Center for Research in Pure and Applied Mathematics (CRMPA) | Giorgio Giurato et al |

| | | | | | | |
|---|---|---|---|---|---|---|
| EPI_ISL_882697 | Italy/CAM-CRGS-121/2020 | 2020-11-17 | Europe/Italy/Campania/ | 1.AO Universitaria 'S. Giovanni di Dio e Ruggi D'Aragona, Scuola Medica Salernitana' Hospital / 2.UOC di Virologia e Microbiologia, Universit√† della Campania 'L. Vanvitelli' / 3.AO Universitaria 'Federico II' Napoli Hospital / 4.AORN 'San Giuseppe Moscati' Avellino Hospital / 5.AO 'San Pio - presidio G. Rummo' Benevento Hospital / 6.AO 'Sant'Anna e San Sebastiano' Caserta Hospital / 7.PO 'Maria Santissima Addolorata' Eboli Hospital / 8.Biogem Istituto di Ricerche Genetiche | 1. Genome Research Center for Health (CRGS) / 2. Laboratory of Molecular Medicine and Genomics(LMMGe) / 3. Center for Research in Pure and Applied Mathematics (CRMPA) | Giorgio Giurato et al |
| EPI_ISL_882700 | Italy/CAM-CRGS-126/2020 | 2020-11-16 | Europe/Italy/Campania/ | 1.AO Universitaria 'S. Giovanni di Dio e Ruggi D'Aragona, Scuola Medica Salernitana' Hospital / 2.UOC di Virologia e Microbiologia, Universit√† della Campania 'L. Vanvitelli' / 3.AO Universitaria 'Federico II' Napoli Hospital / 4.AORN 'San Giuseppe Moscati' Avellino Hospital / 5.AO 'San Pio - presidio G. Rummo' Benevento Hospital / 6.AO 'Sant'Anna e San Sebastiano' Caserta Hospital / 7.PO 'Maria Santissima Addolorata' Eboli Hospital / 8.Biogem Istituto di Ricerche Genetiche | 1. Genome Research Center for Health (CRGS) / 2. Laboratory of Molecular Medicine and Genomics(LMMGe) / 3. Center for Research in Pure and Applied Mathematics (CRMPA) | Giorgio Giurato et al |
| EPI_ISL_882701 | Italy/CAM-CRGS-127/2020 | 2020-11-16 | Europe/Italy/Campania/ | 1.AO Universitaria 'S. Giovanni di Dio e Ruggi D'Aragona, Scuola Medica Salernitana' Hospital / 2.UOC di Virologia e Microbiologia, Universit√† della Campania 'L. Vanvitelli' / 3.AO Universitaria 'Federico II' Napoli Hospital / 4.AORN 'San Giuseppe Moscati' Avellino Hospital / 5.AO 'San Pio - presidio G. Rummo' Benevento Hospital / 6.AO 'Sant'Anna e San Sebastiano' Caserta Hospital / 7.PO 'Maria Santissima Addolorata' Eboli Hospital / 8.Biogem Istituto di Ricerche Genetiche | 1. Genome Research Center for Health (CRGS) / 2. Laboratory of Molecular Medicine and Genomics(LMMGe) / 3. Center for Research in Pure and Applied Mathematics (CRMPA) | Giorgio Giurato et al |

| | | | | | | |
|---|---|---|---|---|---|---|
| EPI_ISL_882703 | Italy/CAM-CRGS-136/2020 | 2020-11-13 | Europe/Italy/Campania/ | 1.AO Universitaria 'S. Giovanni di Dio e Ruggi D'Aragona, Scuola Medica Salernitana' Hospital / 2.UOC di Virologia e Microbiologia, Universit√† della Campania 'L. Vanvitelli' / 3.AO Universitaria 'Federico II' Napoli Hospital / 4.AORN 'San Giuseppe Moscati' Avellino Hospital / 5.AO 'San Pio - presidio G. Rummo' Benevento Hospital / 6.AO 'Sant'Anna e San Sebastiano' Caserta Hospital / 7.PO 'Maria Santissima Addolorata' Eboli Hospital / 8.Biogem Istituto di Ricerche Genetiche | 1. Genome Research Center for Health (CRGS) / 2. Laboratory of Molecular Medicine and Genomics(LMMGe) / 3. Center for Research in Pure and Applied Mathematics (CRMPA) | Giorgio Giurato et al |
| EPI_ISL_882704 | Italy/CAM-CRGS-138/2020 | 2020-11-16 | Europe/Italy/Campania/ | 1.AO Universitaria 'S. Giovanni di Dio e Ruggi D'Aragona, Scuola Medica Salernitana' Hospital / 2.UOC di Virologia e Microbiologia, Universit√† della Campania 'L. Vanvitelli' / 3.AO Universitaria 'Federico II' Napoli Hospital / 4.AORN 'San Giuseppe Moscati' Avellino Hospital / 5.AO 'San Pio - presidio G. Rummo' Benevento Hospital / 6.AO 'Sant'Anna e San Sebastiano' Caserta Hospital / 7.PO 'Maria Santissima Addolorata' Eboli Hospital / 8.Biogem Istituto di Ricerche Genetiche | 1. Genome Research Center for Health (CRGS) / 2. Laboratory of Molecular Medicine and Genomics(LMMGe) / 3. Center for Research in Pure and Applied Mathematics (CRMPA) | Giorgio Giurato et al |
| EPI_ISL_882707 | Italy/CAM-CRGS-143/2020 | 2020-11-17 | Europe/Italy/Campania/ | 1.AO Universitaria 'S. Giovanni di Dio e Ruggi D'Aragona, Scuola Medica Salernitana' Hospital / 2.UOC di Virologia e Microbiologia, Universit√† della Campania 'L. Vanvitelli' / 3.AO Universitaria 'Federico II' Napoli Hospital / 4.AORN 'San Giuseppe Moscati' Avellino Hospital / 5.AO 'San Pio - presidio G. Rummo' Benevento Hospital / 6.AO 'Sant'Anna e San Sebastiano' Caserta Hospital / 7.PO 'Maria Santissima Addolorata' Eboli Hospital / 8.Biogem Istituto di Ricerche Genetiche | 1. Genome Research Center for Health (CRGS) / 2. Laboratory of Molecular Medicine and Genomics(LMMGe) / 3. Center for Research in Pure and Applied Mathematics (CRMPA) | Giorgio Giurato et al |

| | | | | | | |
|---|---|---|---|---|---|---|
| EPI_ISL_882708 | Italy/CAM-CRGS-144/2020 | 2020-11-17 | Europe/Italy/Campania/ | 1.AO Universitaria 'S. Giovanni di Dio e Ruggi D'Aragona, Scuola Medica Salernitana' Hospital / 2.UOC di Virologia e Microbiologia, Universit√† della Campania 'L. Vanvitelli' / 3.AO Universitaria 'Federico II' Napoli Hospital / 4.AORN 'San Giuseppe Moscati' Avellino Hospital / 5.AO 'San Pio - presidio G. Rummo' Benevento Hospital / 6.AO 'Sant'Anna e San Sebastiano' Caserta Hospital / 7.PO 'Maria Santissima Addolorata' Eboli Hospital / 8.Biogem Istituto di Ricerche Genetiche | 1. Genome Research Center for Health (CRGS) / 2. Laboratory of Molecular Medicine and Genomics(LMMGe) / 3. Center for Research in Pure and Applied Mathematics (CRMPA) | Giorgio Giurato et al |
| EPI_ISL_882709 | Italy/CAM-CRGS-150/2020 | 2020-11-14 | Europe/Italy/Campania/ | 1.AO Universitaria 'S. Giovanni di Dio e Ruggi D'Aragona, Scuola Medica Salernitana' Hospital / 2.UOC di Virologia e Microbiologia, Universit√† della Campania 'L. Vanvitelli' / 3.AO Universitaria 'Federico II' Napoli Hospital / 4.AORN 'San Giuseppe Moscati' Avellino Hospital / 5.AO 'San Pio - presidio G. Rummo' Benevento Hospital / 6.AO 'Sant'Anna e San Sebastiano' Caserta Hospital / 7.PO 'Maria Santissima Addolorata' Eboli Hospital / 8.Biogem Istituto di Ricerche Genetiche | 1. Genome Research Center for Health (CRGS) / 2. Laboratory of Molecular Medicine and Genomics(LMMGe) / 3. Center for Research in Pure and Applied Mathematics (CRMPA) | Giorgio Giurato et al |
| EPI_ISL_882710 | Italy/CAM-CRGS-152/2020 | 2020-10-30 | Europe/Italy/Campania/ | 1.AO Universitaria 'S. Giovanni di Dio e Ruggi D'Aragona, Scuola Medica Salernitana' Hospital / 2.UOC di Virologia e Microbiologia, Universit√† della Campania 'L. Vanvitelli' / 3.AO Universitaria 'Federico II' Napoli Hospital / 4.AORN 'San Giuseppe Moscati' Avellino Hospital / 5.AO 'San Pio - presidio G. Rummo' Benevento Hospital / 6.AO 'Sant'Anna e San Sebastiano' Caserta Hospital / 7.PO 'Maria Santissima Addolorata' Eboli Hospital / 8.Biogem Istituto di Ricerche Genetiche | 1. Genome Research Center for Health (CRGS) / 2. Laboratory of Molecular Medicine and Genomics(LMMGe) / 3. Center for Research in Pure and Applied Mathematics (CRMPA) | Giorgio Giurato et al |

| | | | | | | |
|---|---|---|---|---|---|---|
| EPI_ISL_882716 | Italy/CAM-CRGS-170/2020 | 2020-10-11 | Europe/Italy/Campania/ | 1.AO Universitaria 'S. Giovanni di Dio e Ruggi D'Aragona, Scuola Medica Salernitana' Hospital / 2.UOC di Virologia e Microbiologia, Universit√† della Campania 'L. Vanvitelli' / 3.AO Universitaria 'Federico II' Napoli Hospital / 4.AORN 'San Giuseppe Moscati' Avellino Hospital / 5.AO 'San Pio - presidio G. Rummo' Benevento Hospital / 6.AO 'Sant'Anna e San Sebastiano' Caserta Hospital / 7.PO 'Maria Santissima Addolorata' Eboli Hospital / 8.Biogem Istituto di Ricerche Genetiche | 1. Genome Research Center for Health (CRGS) / 2. Laboratory of Molecular Medicine and Genomics(LMMGe) / 3. Center for Research in Pure and Applied Mathematics (CRMPA) | Giorgio Giurato et al |
| EPI_ISL_882717 | Italy/CAM-CRGS-171/2020 | 2020-10-13 | Europe/Italy/Campania/ | 1.AO Universitaria 'S. Giovanni di Dio e Ruggi D'Aragona, Scuola Medica Salernitana' Hospital / 2.UOC di Virologia e Microbiologia, Universit√† della Campania 'L. Vanvitelli' / 3.AO Universitaria 'Federico II' Napoli Hospital / 4.AORN 'San Giuseppe Moscati' Avellino Hospital / 5.AO 'San Pio - presidio G. Rummo' Benevento Hospital / 6.AO 'Sant'Anna e San Sebastiano' Caserta Hospital / 7.PO 'Maria Santissima Addolorata' Eboli Hospital / 8.Biogem Istituto di Ricerche Genetiche | 1. Genome Research Center for Health (CRGS) / 2. Laboratory of Molecular Medicine and Genomics(LMMGe) / 3. Center for Research in Pure and Applied Mathematics (CRMPA) | Giorgio Giurato et al |
| EPI_ISL_882718 | Italy/CAM-CRGS-173/2020 | 2020-10-13 | Europe/Italy/Campania/ | 1.AO Universitaria 'S. Giovanni di Dio e Ruggi D'Aragona, Scuola Medica Salernitana' Hospital / 2.UOC di Virologia e Microbiologia, Universit√† della Campania 'L. Vanvitelli' / 3.AO Universitaria 'Federico II' Napoli Hospital / 4.AORN 'San Giuseppe Moscati' Avellino Hospital / 5.AO 'San Pio - presidio G. Rummo' Benevento Hospital / 6.AO 'Sant'Anna e San Sebastiano' Caserta Hospital / 7.PO 'Maria Santissima Addolorata' Eboli Hospital / 8.Biogem Istituto di Ricerche Genetiche | 1. Genome Research Center for Health (CRGS) / 2. Laboratory of Molecular Medicine and Genomics(LMMGe) / 3. Center for Research in Pure and Applied Mathematics (CRMPA) | Giorgio Giurato et al |

| | | | | | | |
|---|---|---|---|---|---|---|
| EPI_ISL_882719 | Italy/CAM-CRGS-175/2020 | 2020-10-13 | Europe/Italy/Campania/ | 1.AO Universitaria 'S. Giovanni di Dio e Ruggi D'Aragona, Scuola Medica Salernitana' Hospital / 2.UOC di Virologia e Microbiologia, Universit√† della Campania 'L. Vanvitelli' / 3.AO Universitaria 'Federico II' Napoli Hospital / 4.AORN 'San Giuseppe Moscati' Avellino Hospital / 5.AO 'San Pio - presidio G. Rummo' Benevento Hospital / 6.AO 'Sant'Anna e San Sebastiano' Caserta Hospital / 7.PO 'Maria Santissima Addolorata' Eboli Hospital / 8.Biogem Istituto di Ricerche Genetiche | 1. Genome Research Center for Health (CRGS) / 2. Laboratory of Molecular Medicine and Genomics(LMMGe) / 3. Center for Research in Pure and Applied Mathematics (CRMPA) | Giorgio Giurato et al |
| EPI_ISL_882721 | Italy/CAM-CRGS-181/2020 | 2020-10-15 | Europe/Italy/Campania/ | 1.AO Universitaria 'S. Giovanni di Dio e Ruggi D'Aragona, Scuola Medica Salernitana' Hospital / 2.UOC di Virologia e Microbiologia, Universit√† della Campania 'L. Vanvitelli' / 3.AO Universitaria 'Federico II' Napoli Hospital / 4.AORN 'San Giuseppe Moscati' Avellino Hospital / 5.AO 'San Pio - presidio G. Rummo' Benevento Hospital / 6.AO 'Sant'Anna e San Sebastiano' Caserta Hospital / 7.PO 'Maria Santissima Addolorata' Eboli Hospital / 8.Biogem Istituto di Ricerche Genetiche | 1. Genome Research Center for Health (CRGS) / 2. Laboratory of Molecular Medicine and Genomics(LMMGe) / 3. Center for Research in Pure and Applied Mathematics (CRMPA) | Giorgio Giurato et al |
| EPI_ISL_882724 | Italy/CAM-CRGS-187/2020 | 2020-10-18 | Europe/Italy/Campania/ | 1.AO Universitaria 'S. Giovanni di Dio e Ruggi D'Aragona, Scuola Medica Salernitana' Hospital / 2.UOC di Virologia e Microbiologia, Universit√† della Campania 'L. Vanvitelli' / 3.AO Universitaria 'Federico II' Napoli Hospital / 4.AORN 'San Giuseppe Moscati' Avellino Hospital / 5.AO 'San Pio - presidio G. Rummo' Benevento Hospital / 6.AO 'Sant'Anna e San Sebastiano' Caserta Hospital / 7.PO 'Maria Santissima Addolorata' Eboli Hospital / 8.Biogem Istituto di Ricerche Genetiche | 1. Genome Research Center for Health (CRGS) / 2. Laboratory of Molecular Medicine and Genomics(LMMGe) / 3. Center for Research in Pure and Applied Mathematics (CRMPA) | Giorgio Giurato et al |

| | | | | | | |
|---|---|---|---|---|---|---|
| EPI_ISL_882725 | Italy/CAM-CRGS-194/2020 | 2020-10-19 | Europe/Italy/Campania/ | 1.AO Universitaria 'S. Giovanni di Dio e Ruggi D'Aragona, Scuola Medica Salernitana' Hospital / 2.UOC di Virologia e Microbiologia, Universit√† della Campania 'L. Vanvitelli' / 3.AO Universitaria 'Federico II' Napoli Hospital / 4.AORN 'San Giuseppe Moscati' Avellino Hospital / 5.AO 'San Pio - presidio G. Rummo' Benevento Hospital / 6.AO 'Sant'Anna e San Sebastiano' Caserta Hospital / 7.PO 'Maria Santissima Addolorata' Eboli Hospital / 8.Biogem Istituto di Ricerche Genetiche | 1. Genome Research Center for Health (CRGS) / 2. Laboratory of Molecular Medicine and Genomics(LMMGe) / 3. Center for Research in Pure and Applied Mathematics (CRMPA) | Giorgio Giurato et al |
| EPI_ISL_882726 | Italy/CAM-CRGS-200/2020 | 2020-10-21 | Europe/Italy/Campania/ | 1.AO Universitaria 'S. Giovanni di Dio e Ruggi D'Aragona, Scuola Medica Salernitana' Hospital / 2.UOC di Virologia e Microbiologia, Universit√† della Campania 'L. Vanvitelli' / 3.AO Universitaria 'Federico II' Napoli Hospital / 4.AORN 'San Giuseppe Moscati' Avellino Hospital / 5.AO 'San Pio - presidio G. Rummo' Benevento Hospital / 6.AO 'Sant'Anna e San Sebastiano' Caserta Hospital / 7.PO 'Maria Santissima Addolorata' Eboli Hospital / 8.Biogem Istituto di Ricerche Genetiche | 1. Genome Research Center for Health (CRGS) / 2. Laboratory of Molecular Medicine and Genomics(LMMGe) / 3. Center for Research in Pure and Applied Mathematics (CRMPA) | Giorgio Giurato et al |
| EPI_ISL_882728 | Italy/CAM-CRGS-206/2020 | 2020-11-04 | Europe/Italy/Campania/ | 1.AO Universitaria 'S. Giovanni di Dio e Ruggi D'Aragona, Scuola Medica Salernitana' Hospital / 2.UOC di Virologia e Microbiologia, Universit√† della Campania 'L. Vanvitelli' / 3.AO Universitaria 'Federico II' Napoli Hospital / 4.AORN 'San Giuseppe Moscati' Avellino Hospital / 5.AO 'San Pio - presidio G. Rummo' Benevento Hospital / 6.AO 'Sant'Anna e San Sebastiano' Caserta Hospital / 7.PO 'Maria Santissima Addolorata' Eboli Hospital / 8.Biogem Istituto di Ricerche Genetiche | 1. Genome Research Center for Health (CRGS) / 2. Laboratory of Molecular Medicine and Genomics(LMMGe) / 3. Center for Research in Pure and Applied Mathematics (CRMPA) | Giorgio Giurato et al |

| | | | | | | |
|---|---|---|---|---|---|---|
| EPI_ISL_882729 | Italy/CAM-CRGS-207/2020 | 2020-11-04 | Europe/Italy/Campania/ | 1.AO Universitaria 'S. Giovanni di Dio e Ruggi D'Aragona, Scuola Medica Salernitana' Hospital / 2.UOC di Virologia e Microbiologia, Universit√† della Campania 'L. Vanvitelli' / 3.AO Universitaria 'Federico II' Napoli Hospital / 4.AORN 'San Giuseppe Moscati' Avellino Hospital / 5.AO 'San Pio - presidio G. Rummo' Benevento Hospital / 6.AO 'Sant'Anna e San Sebastiano' Caserta Hospital / 7.PO 'Maria Santissima Addolorata' Eboli Hospital / 8.Biogem Istituto di Ricerche Genetiche | 1. Genome Research Center for Health (CRGS) / 2. Laboratory of Molecular Medicine and Genomics(LMMGe) / 3. Center for Research in Pure and Applied Mathematics (CRMPA) | Giorgio Giurato et al |
| EPI_ISL_882730 | Italy/CAM-CRGS-208/2020 | 2020-11-04 | Europe/Italy/Campania/ | 1.AO Universitaria 'S. Giovanni di Dio e Ruggi D'Aragona, Scuola Medica Salernitana' Hospital / 2.UOC di Virologia e Microbiologia, Universit√† della Campania 'L. Vanvitelli' / 3.AO Universitaria 'Federico II' Napoli Hospital / 4.AORN 'San Giuseppe Moscati' Avellino Hospital / 5.AO 'San Pio - presidio G. Rummo' Benevento Hospital / 6.AO 'Sant'Anna e San Sebastiano' Caserta Hospital / 7.PO 'Maria Santissima Addolorata' Eboli Hospital / 8.Biogem Istituto di Ricerche Genetiche | 1. Genome Research Center for Health (CRGS) / 2. Laboratory of Molecular Medicine and Genomics(LMMGe) / 3. Center for Research in Pure and Applied Mathematics (CRMPA) | Giorgio Giurato et al |
| EPI_ISL_882732 | Italy/CAM-CRGS-211/2020 | 2020-11-04 | Europe/Italy/Campania/ | 1.AO Universitaria 'S. Giovanni di Dio e Ruggi D'Aragona, Scuola Medica Salernitana' Hospital / 2.UOC di Virologia e Microbiologia, Universit√† della Campania 'L. Vanvitelli' / 3.AO Universitaria 'Federico II' Napoli Hospital / 4.AORN 'San Giuseppe Moscati' Avellino Hospital / 5.AO 'San Pio - presidio G. Rummo' Benevento Hospital / 6.AO 'Sant'Anna e San Sebastiano' Caserta Hospital / 7.PO 'Maria Santissima Addolorata' Eboli Hospital / 8.Biogem Istituto di Ricerche Genetiche | 1. Genome Research Center for Health (CRGS) / 2. Laboratory of Molecular Medicine and Genomics(LMMGe) / 3. Center for Research in Pure and Applied Mathematics (CRMPA) | Giorgio Giurato et al |

| | | | | | | |
|---|---|---|---|---|---|---|
| EPI_ISL_882733 | Italy/CAM-CRGS-213/2020 | 2020-11-03 | Europe/Italy/Campania/ | 1.AO Universitaria 'S. Giovanni di Dio e Ruggi D'Aragona, Scuola Medica Salernitana' Hospital / 2.UOC di Virologia e Microbiologia, Universit√† della Campania 'L. Vanvitelli' / 3.AO Universitaria 'Federico II' Napoli Hospital / 4.AORN 'San Giuseppe Moscati' Avellino Hospital / 5.AO 'San Pio - presidio G. Rummo' Benevento Hospital / 6.AO 'Sant'Anna e San Sebastiano' Caserta Hospital / 7.PO 'Maria Santissima Addolorata' Eboli Hospital / 8.Biogem Istituto di Ricerche Genetiche | 1. Genome Research Center for Health (CRGS) / 2. Laboratory of Molecular Medicine and Genomics(LMMGe) / 3. Center for Research in Pure and Applied Mathematics (CRMPA) | Giorgio Giurato et al |
| EPI_ISL_882735 | Italy/CAM-CRGS-216/2020 | 2020-11-05 | Europe/Italy/Campania/ | 1.AO Universitaria 'S. Giovanni di Dio e Ruggi D'Aragona, Scuola Medica Salernitana' Hospital / 2.UOC di Virologia e Microbiologia, Universit√† della Campania 'L. Vanvitelli' / 3.AO Universitaria 'Federico II' Napoli Hospital / 4.AORN 'San Giuseppe Moscati' Avellino Hospital / 5.AO 'San Pio - presidio G. Rummo' Benevento Hospital / 6.AO 'Sant'Anna e San Sebastiano' Caserta Hospital / 7.PO 'Maria Santissima Addolorata' Eboli Hospital / 8.Biogem Istituto di Ricerche Genetiche | 1. Genome Research Center for Health (CRGS) / 2. Laboratory of Molecular Medicine and Genomics(LMMGe) / 3. Center for Research in Pure and Applied Mathematics (CRMPA) | Giorgio Giurato et al |
| EPI_ISL_882736 | Italy/CAM-CRGS-217/2020 | 2020-11-05 | Europe/Italy/Campania/ | 1.AO Universitaria 'S. Giovanni di Dio e Ruggi D'Aragona, Scuola Medica Salernitana' Hospital / 2.UOC di Virologia e Microbiologia, Universit√† della Campania 'L. Vanvitelli' / 3.AO Universitaria 'Federico II' Napoli Hospital / 4.AORN 'San Giuseppe Moscati' Avellino Hospital / 5.AO 'San Pio - presidio G. Rummo' Benevento Hospital / 6.AO 'Sant'Anna e San Sebastiano' Caserta Hospital / 7.PO 'Maria Santissima Addolorata' Eboli Hospital / 8.Biogem Istituto di Ricerche Genetiche | 1. Genome Research Center for Health (CRGS) / 2. Laboratory of Molecular Medicine and Genomics(LMMGe) / 3. Center for Research in Pure and Applied Mathematics (CRMPA) | Giorgio Giurato et al |

| | | | | | | |
|---|---|---|---|---|---|---|
| EPI_ISL_882738 | Italy/CAM-CRGS-223/2020 | 2020-10-29 | Europe/Italy/Campania/ | 1.AO Universitaria 'S. Giovanni di Dio e Ruggi D'Aragona, Scuola Medica Salernitana' Hospital / 2.UOC di Virologia e Microbiologia, Universit√† della Campania 'L. Vanvitelli' / 3.AO Universitaria 'Federico II' Napoli Hospital / 4.AORN 'San Giuseppe Moscati' Avellino Hospital / 5.AO 'San Pio - presidio G. Rummo' Benevento Hospital / 6.AO 'Sant'Anna e San Sebastiano' Caserta Hospital / 7.PO 'Maria Santissima Addolorata' Eboli Hospital / 8.Biogem Istituto di Ricerche Genetiche | 1. Genome Research Center for Health (CRGS) / 2. Laboratory of Molecular Medicine and Genomics(LMMGe) / 3. Center for Research in Pure and Applied Mathematics (CRMPA) | Giorgio Giurato et al |
| EPI_ISL_882739 | Italy/CAM-CRGS-227/2020 | 2020-10-07 | Europe/Italy/Campania/ | 1.AO Universitaria 'S. Giovanni di Dio e Ruggi D'Aragona, Scuola Medica Salernitana' Hospital / 2.UOC di Virologia e Microbiologia, Universit√† della Campania 'L. Vanvitelli' / 3.AO Universitaria 'Federico II' Napoli Hospital / 4.AORN 'San Giuseppe Moscati' Avellino Hospital / 5.AO 'San Pio - presidio G. Rummo' Benevento Hospital / 6.AO 'Sant'Anna e San Sebastiano' Caserta Hospital / 7.PO 'Maria Santissima Addolorata' Eboli Hospital / 8.Biogem Istituto di Ricerche Genetiche | 1. Genome Research Center for Health (CRGS) / 2. Laboratory of Molecular Medicine and Genomics(LMMGe) / 3. Center for Research in Pure and Applied Mathematics (CRMPA) | Giorgio Giurato et al |
| EPI_ISL_882741 | Italy/CAM-CRGS-229/2020 | 2020-10-15 | Europe/Italy/Campania/ | 1.AO Universitaria 'S. Giovanni di Dio e Ruggi D'Aragona, Scuola Medica Salernitana' Hospital / 2.UOC di Virologia e Microbiologia, Universit√† della Campania 'L. Vanvitelli' / 3.AO Universitaria 'Federico II' Napoli Hospital / 4.AORN 'San Giuseppe Moscati' Avellino Hospital / 5.AO 'San Pio - presidio G. Rummo' Benevento Hospital / 6.AO 'Sant'Anna e San Sebastiano' Caserta Hospital / 7.PO 'Maria Santissima Addolorata' Eboli Hospital / 8.Biogem Istituto di Ricerche Genetiche | 1. Genome Research Center for Health (CRGS) / 2. Laboratory of Molecular Medicine and Genomics(LMMGe) / 3. Center for Research in Pure and Applied Mathematics (CRMPA) | Giorgio Giurato et al |

| | | | | | | |
|---|---|---|---|---|---|---|
| EPI_ISL_882742 | Italy/CAM-CRGS-230/2020 | 2020-10-06 | Europe/Italy/Campania/ | 1.AO Universitaria 'S. Giovanni di Dio e Ruggi D'Aragona, Scuola Medica Salernitana' Hospital / 2.UOC di Virologia e Microbiologia, Universit√† della Campania 'L. Vanvitelli' / 3.AO Universitaria 'Federico II' Napoli Hospital / 4.AORN 'San Giuseppe Moscati' Avellino Hospital / 5.AO 'San Pio - presidio G. Rummo' Benevento Hospital / 6.AO 'Sant'Anna e San Sebastiano' Caserta Hospital / 7.PO 'Maria Santissima Addolorata' Eboli Hospital / 8.Biogem Istituto di Ricerche Genetiche | 1. Genome Research Center for Health (CRGS) / 2. Laboratory of Molecular Medicine and Genomics(LMMGe) / 3. Center for Research in Pure and Applied Mathematics (CRMPA) | Giorgio Giurato et al |
| EPI_ISL_882744 | Italy/CAM-CRGS-238/2020 | 2020-11-02 | Europe/Italy/Campania/ | 1.AO Universitaria 'S. Giovanni di Dio e Ruggi D'Aragona, Scuola Medica Salernitana' Hospital / 2.UOC di Virologia e Microbiologia, Universit√† della Campania 'L. Vanvitelli' / 3.AO Universitaria 'Federico II' Napoli Hospital / 4.AORN 'San Giuseppe Moscati' Avellino Hospital / 5.AO 'San Pio - presidio G. Rummo' Benevento Hospital / 6.AO 'Sant'Anna e San Sebastiano' Caserta Hospital / 7.PO 'Maria Santissima Addolorata' Eboli Hospital / 8.Biogem Istituto di Ricerche Genetiche | 1. Genome Research Center for Health (CRGS) / 2. Laboratory of Molecular Medicine and Genomics(LMMGe) / 3. Center for Research in Pure and Applied Mathematics (CRMPA) | Giorgio Giurato et al |
| EPI_ISL_882746 | Italy/CAM-CRGS-240/2020 | 2020-10-14 | Europe/Italy/Campania/ | 1.AO Universitaria 'S. Giovanni di Dio e Ruggi D'Aragona, Scuola Medica Salernitana' Hospital / 2.UOC di Virologia e Microbiologia, Universit√† della Campania 'L. Vanvitelli' / 3.AO Universitaria 'Federico II' Napoli Hospital / 4.AORN 'San Giuseppe Moscati' Avellino Hospital / 5.AO 'San Pio - presidio G. Rummo' Benevento Hospital / 6.AO 'Sant'Anna e San Sebastiano' Caserta Hospital / 7.PO 'Maria Santissima Addolorata' Eboli Hospital / 8.Biogem Istituto di Ricerche Genetiche | 1. Genome Research Center for Health (CRGS) / 2. Laboratory of Molecular Medicine and Genomics(LMMGe) / 3. Center for Research in Pure and Applied Mathematics (CRMPA) | Giorgio Giurato et al |

| | | | | | | |
|---|---|---|---|---|---|---|
| EPI_ISL_882748 | Italy/CAM-CRGS-242/2020 | 2020-09-13 | Europe/Italy/Campania/ | 1.AO Universitaria 'S. Giovanni di Dio e Ruggi D'Aragona, Scuola Medica Salernitana' Hospital / 2.UOC di Virologia e Microbiologia, Universit√† della Campania 'L. Vanvitelli' / 3.AO Universitaria 'Federico II' Napoli Hospital / 4.AORN 'San Giuseppe Moscati' Avellino Hospital / 5.AO 'San Pio - presidio G. Rummo' Benevento Hospital / 6.AO 'Sant'Anna e San Sebastiano' Caserta Hospital / 7.PO 'Maria Santissima Addolorata' Eboli Hospital / 8.Biogem Istituto di Ricerche Genetiche | 1. Genome Research Center for Health (CRGS) / 2. Laboratory of Molecular Medicine and Genomics(LMMGe) / 3. Center for Research in Pure and Applied Mathematics (CRMPA) | Giorgio Giurato et al |
| EPI_ISL_882749 | Italy/CAM-CRGS-244/2020 | 2020-10-15 | Europe/Italy/Campania/ | 1.AO Universitaria 'S. Giovanni di Dio e Ruggi D'Aragona, Scuola Medica Salernitana' Hospital / 2.UOC di Virologia e Microbiologia, Universit√† della Campania 'L. Vanvitelli' / 3.AO Universitaria 'Federico II' Napoli Hospital / 4.AORN 'San Giuseppe Moscati' Avellino Hospital / 5.AO 'San Pio - presidio G. Rummo' Benevento Hospital / 6.AO 'Sant'Anna e San Sebastiano' Caserta Hospital / 7.PO 'Maria Santissima Addolorata' Eboli Hospital / 8.Biogem Istituto di Ricerche Genetiche | 1. Genome Research Center for Health (CRGS) / 2. Laboratory of Molecular Medicine and Genomics(LMMGe) / 3. Center for Research in Pure and Applied Mathematics (CRMPA) | Giorgio Giurato et al |
| EPI_ISL_882751 | Italy/CAM-CRGS-246/2020 | 2020-11-02 | Europe/Italy/Campania/ | 1.AO Universitaria 'S. Giovanni di Dio e Ruggi D'Aragona, Scuola Medica Salernitana' Hospital / 2.UOC di Virologia e Microbiologia, Universit√† della Campania 'L. Vanvitelli' / 3.AO Universitaria 'Federico II' Napoli Hospital / 4.AORN 'San Giuseppe Moscati' Avellino Hospital / 5.AO 'San Pio - presidio G. Rummo' Benevento Hospital / 6.AO 'Sant'Anna e San Sebastiano' Caserta Hospital / 7.PO 'Maria Santissima Addolorata' Eboli Hospital / 8.Biogem Istituto di Ricerche Genetiche | 1. Genome Research Center for Health (CRGS) / 2. Laboratory of Molecular Medicine and Genomics(LMMGe) / 3. Center for Research in Pure and Applied Mathematics (CRMPA) | Giorgio Giurato et al |

| | | | | | | |
|---|---|---|---|---|---|---|
| EPI_ISL_882755 | Italy/CAM-CRGS-263/2020 | 2020-10-11 | Europe/Italy/Campania/ | 1.AO Universitaria 'S. Giovanni di Dio e Ruggi D'Aragona, Scuola Medica Salernitana' Hospital / 2.UOC di Virologia e Microbiologia, Universit√† della Campania 'L. Vanvitelli' / 3.AO Universitaria 'Federico II' Napoli Hospital / 4.AORN 'San Giuseppe Moscati' Avellino Hospital / 5.AO 'San Pio - presidio G. Rummo' Benevento Hospital / 6.AO 'Sant'Anna e San Sebastiano' Caserta Hospital / 7.PO 'Maria Santissima Addolorata' Eboli Hospital / 8.Biogem Istituto di Ricerche Genetiche | 1. Genome Research Center for Health (CRGS) / 2. Laboratory of Molecular Medicine and Genomics(LMMGe) / 3. Center for Research in Pure and Applied Mathematics (CRMPA) | Giorgio Giurato et al |
| EPI_ISL_882758 | Italy/CAM-CRGS-266/2020 | 2020-11-02 | Europe/Italy/Campania/ | 1.AO Universitaria 'S. Giovanni di Dio e Ruggi D'Aragona, Scuola Medica Salernitana' Hospital / 2.UOC di Virologia e Microbiologia, Universit√† della Campania 'L. Vanvitelli' / 3.AO Universitaria 'Federico II' Napoli Hospital / 4.AORN 'San Giuseppe Moscati' Avellino Hospital / 5.AO 'San Pio - presidio G. Rummo' Benevento Hospital / 6.AO 'Sant'Anna e San Sebastiano' Caserta Hospital / 7.PO 'Maria Santissima Addolorata' Eboli Hospital / 8.Biogem Istituto di Ricerche Genetiche | 1. Genome Research Center for Health (CRGS) / 2. Laboratory of Molecular Medicine and Genomics(LMMGe) / 3. Center for Research in Pure and Applied Mathematics (CRMPA) | Giorgio Giurato et al |
| EPI_ISL_882760 | Italy/CAM-CRGS-268/2020 | 2020-10-07 | Europe/Italy/Campania/ | 1.AO Universitaria 'S. Giovanni di Dio e Ruggi D'Aragona, Scuola Medica Salernitana' Hospital / 2.UOC di Virologia e Microbiologia, Universit√† della Campania 'L. Vanvitelli' / 3.AO Universitaria 'Federico II' Napoli Hospital / 4.AORN 'San Giuseppe Moscati' Avellino Hospital / 5.AO 'San Pio - presidio G. Rummo' Benevento Hospital / 6.AO 'Sant'Anna e San Sebastiano' Caserta Hospital / 7.PO 'Maria Santissima Addolorata' Eboli Hospital / 8.Biogem Istituto di Ricerche Genetiche | 1. Genome Research Center for Health (CRGS) / 2. Laboratory of Molecular Medicine and Genomics(LMMGe) / 3. Center for Research in Pure and Applied Mathematics (CRMPA) | Giorgio Giurato et al |

| | | | | | | |
|---|---|---|---|---|---|---|
| EPI_ISL_882761 | Italy/CAM-CRGS-269/2020 | 2020-11-01 | Europe/Italy/Campania/ | 1.AO Universitaria 'S. Giovanni di Dio e Ruggi D'Aragona, Scuola Medica Salernitana' Hospital / 2.UOC di Virologia e Microbiologia, Universit√† della Campania 'L. Vanvitelli' / 3.AO Universitaria 'Federico II' Napoli Hospital / 4.AORN 'San Giuseppe Moscati' Avellino Hospital / 5.AO 'San Pio - presidio G. Rummo' Benevento Hospital / 6.AO 'Sant'Anna e San Sebastiano' Caserta Hospital / 7.PO 'Maria Santissima Addolorata' Eboli Hospital / 8.Biogem Istituto di Ricerche Genetiche | 1. Genome Research Center for Health (CRGS) / 2. Laboratory of Molecular Medicine and Genomics(LMMGe) / 3. Center for Research in Pure and Applied Mathematics (CRMPA) | Giorgio Giurato et al |
| EPI_ISL_882762 | Italy/CAM-CRGS-270/2020 | 2020-10-11 | Europe/Italy/Campania/ | 1.AO Universitaria 'S. Giovanni di Dio e Ruggi D'Aragona, Scuola Medica Salernitana' Hospital / 2.UOC di Virologia e Microbiologia, Universit√† della Campania 'L. Vanvitelli' / 3.AO Universitaria 'Federico II' Napoli Hospital / 4.AORN 'San Giuseppe Moscati' Avellino Hospital / 5.AO 'San Pio - presidio G. Rummo' Benevento Hospital / 6.AO 'Sant'Anna e San Sebastiano' Caserta Hospital / 7.PO 'Maria Santissima Addolorata' Eboli Hospital / 8.Biogem Istituto di Ricerche Genetiche | 1. Genome Research Center for Health (CRGS) / 2. Laboratory of Molecular Medicine and Genomics(LMMGe) / 3. Center for Research in Pure and Applied Mathematics (CRMPA) | Giorgio Giurato et al |
| EPI_ISL_882764 | Italy/CAM-CRGS-275/2020 | 2020-11-30 | Europe/Italy/Campania/ | 1.AO Universitaria 'S. Giovanni di Dio e Ruggi D'Aragona, Scuola Medica Salernitana' Hospital / 2.UOC di Virologia e Microbiologia, Universit√† della Campania 'L. Vanvitelli' / 3.AO Universitaria 'Federico II' Napoli Hospital / 4.AORN 'San Giuseppe Moscati' Avellino Hospital / 5.AO 'San Pio - presidio G. Rummo' Benevento Hospital / 6.AO 'Sant'Anna e San Sebastiano' Caserta Hospital / 7.PO 'Maria Santissima Addolorata' Eboli Hospital / 8.Biogem Istituto di Ricerche Genetiche | 1. Genome Research Center for Health (CRGS) / 2. Laboratory of Molecular Medicine and Genomics(LMMGe) / 3. Center for Research in Pure and Applied Mathematics (CRMPA) | Giorgio Giurato et al |
| EPI_ISL_882769 | Indonesia/JI-ITDua-34158NT/2020 | 2020-12-18 | Asia/Indonesia/East Java/Surabaya | Institute of Tropical Disease | Institute of Tropical Disease, Universitas Airlangga | Kazufumi Shimizu et al |

| | | | | | | |
|---|---|---|---|---|---|---|
| EPI_ISL_882770 | Indonesia/JI-ITDua-34618NT/2020 | 2020-12-22 | Asia/Indonesia/East Java/Surabaya | Darus Syifa' Islamic Hospital | Institute of Tropical Disease, Universitas Airlangga | Aldise M Nastri et al |
| EPI_ISL_882771 | Indonesia/JI-ITDua-35340NT/2020 | 2020-12-28 | Asia/Indonesia/East Java/Sidoarjo | Siti Khodijah Hospital | Institute of Tropical Disease, Universitas Airlangga | Kazufumi Shimizu et al |
| EPI_ISL_882789 | Thailand/Bangkok-CONI-0431/2021 | 2021-01-05 | Asia/Thailand/Bangkok/ | Institute for Urban Disease Control and Prevention | COVID-19 Network Investigations (CONI) Alliance | Kamolthip Atsawawaranunt et al |
| EPI_ISL_882790 | Thailand/Bangkok-CONI-0432/2021 | 2021-01-05 | Asia/Thailand/Bangkok/ | Institute for Urban Disease Control and Prevention | COVID-19 Network Investigations (CONI) Alliance | Kamolthip Atsawawaranunt et al |
| EPI_ISL_882883 | Italy/LOM-CR02/2020 | 2020-03-26 | Europe/Italy/Lombardy/ | Unit of Infectious Diseases, Azienda Socio Sanitaria Territoriale di Cremona | Laboratory of Infectious Diseases, Department of Biomedical and Clinical Sciences L. Sacco, University of Milan | Antonio Coluccello et al |
| EPI_ISL_882918 | Italy/LAZ-INMI-165/2021 | 2021-01-03 | Europe/Italy/Lazio/ | INMI Lazzaro Spallanzani IRCCS | INMI Lazzaro Spallanzani IRCCS | E Giombini et al |
| EPI_ISL_882919 | Italy/LAZ-INMI-153/2021 | 2021-01-05 | Europe/Italy/Lazio/ | INMI Lazzaro Spallanzani IRCCS | INMI Lazzaro Spallanzani IRCCS | C.E.M Gruber et al |
| EPI_ISL_882920 | Italy/LAZ-INMI-147/2021 | 2021-01-03 | Europe/Italy/Lazio/ | INMI Lazzaro Spallanzani IRCCS | INMI Lazzaro Spallanzani IRCCS | M Rueca et al |
| EPI_ISL_882927 | Italy/LAZ-INMI-146/2021 | 2021-01-04 | Europe/Italy/Lazio/ | INMI Lazzaro Spallanzani IRCCS | INMI Lazzaro Spallanzani IRCCS | M Rueca et al |
| EPI_ISL_882928 | Italy/LAZ-INMI-148/2021 | 2021-01-03 | Europe/Italy/Lazio/ | INMI Lazzaro Spallanzani IRCCS | INMI Lazzaro Spallanzani IRCCS | F Messina et al |
| EPI_ISL_882929 | Italy/LAZ-INMI-149/2021 | 2021-01-03 | Europe/Italy/Lazio/ | INMI Lazzaro Spallanzani IRCCS | INMI Lazzaro Spallanzani IRCCS | C.E.M Gruber et al |
| EPI_ISL_882930 | Italy/LAZ-INMI-150/2021 | 2021-01-03 | Europe/Italy/Lazio/ | INMI Lazzaro Spallanzani IRCCS | INMI Lazzaro Spallanzani IRCCS | E Giombini et al |
| EPI_ISL_882931 | Italy/LAZ-INMI-151/2021 | 2021-01-02 | Europe/Italy/Lazio/ | INMI Lazzaro Spallanzani IRCCS | INMI Lazzaro Spallanzani IRCCS | B Bartolini et al |

| | | | | | | |
|---|---|---|---|---|---|---|
| EPI_ISL_882932 | Italy/LAZ-INMI-152/2021 | 2021-01-04 | Europe/Italy/Lazio/ | INMI Lazzaro Spallanzani IRCCS | INMI Lazzaro Spallanzani IRCCS | F Messina et al |
| EPI_ISL_882933 | Italy/LAZ-INMI-154/2021 | 2021-01-12 | Europe/Italy/Lazio/ | Sant‚ÄôEugenio/CTO ASL Roma 2 | INMI Lazzaro Spallanzani IRCCS | E Giombini et al |
| EPI_ISL_882934 | Italy/LAZ-INMI-155/2021 | 2021-01-12 | Europe/Italy/Lazio/ | Sant‚ÄôEugenio/CTO ASL Roma 2 | INMI Lazzaro Spallanzani IRCCS | B Bartolini et al |
| EPI_ISL_882936 | Italy/LAZ-INMI-158/2021 | 2021-01-12 | Europe/Italy/Lazio/ | Sant‚ÄôEugenio/CTO ASL Roma 2 | INMI Lazzaro Spallanzani IRCCS | E Giombini et al |
| EPI_ISL_882937 | Italy/LAZ-INMI-159/2021 | 2021-01-13 | Europe/Italy/Lazio/ | Azienda Ospedaliera San Giovanni Addolorata | INMI Lazzaro Spallanzani IRCCS | F Messina et al |
| EPI_ISL_882938 | Italy/LAZ-INMI-160/2021 | 2021-01-13 | Europe/Italy/Lazio/ | Azienda Ospedaliera San Giovanni Addolorata | INMI Lazzaro Spallanzani IRCCS | C.E.M Gruber et al |
| EPI_ISL_882939 | Italy/LAZ-INMI-161/2021 | 2021-01-13 | Europe/Italy/Lazio/ | Azienda Ospedaliera San Giovanni Addolorata | INMI Lazzaro Spallanzani IRCCS | B Bartolini et al |
| EPI_ISL_882941 | Italy/LAZ-INMI-163/2021 | 2021-01-12 | Europe/Italy/Lazio/ | Ospedale San Camillo De Lellis Rieti | INMI Lazzaro Spallanzani IRCCS | C.E.M Gruber et al |
| EPI_ISL_882943 | CzechRepublic/NRL_401/2021 | 2021-01-04 | Europe/Czech Republic/Hradec Kr√°lov√ | The National Institute of Public Health | State Veterinary Institute Prague | Nagy et al |
| EPI_ISL_882945 | CzechRepublic/NRL_154/2021 | 2021-01-03 | Europe/Czech Republic/Prague/ | The National Institute of Public Health | State Veterinary Institute Prague | Nagy et al |
| EPI_ISL_882946 | CzechRepublic/NRL_155/2021 | 2021-01-03 | Europe/Czech Republic/Prague/ | The National Institute of Public Health | State Veterinary Institute Prague | Nagy et al |
| EPI_ISL_882947 | CzechRepublic/NRL_156/2021 | 2021-01-03 | Europe/Czech Republic/Prague/ | The National Institute of Public Health | State Veterinary Institute Prague | Nagy et al |
| EPI_ISL_882948 | CzechRepublic/NRL_157/2021 | 2021-01-03 | Europe/Czech Republic/Plze≈à Region/Pl | The National Institute of Public Health | State Veterinary Institute Prague | Nagy et al |
| EPI_ISL_882949 | CzechRepublic/NRL_158/2021 | 2021-01-03 | Europe/Czech Republic/Prague/ | The National Institute of Public Health | State Veterinary Institute Prague | Nagy et al |
| EPI_ISL_882950 | CzechRepublic/NRL_286/2021 | 2021-01-07 | Europe/Czech Republic/Prague/ | The National Institute of Public Health | State Veterinary Institute Prague | Nagy et al |
| EPI_ISL_882951 | CzechRepublic/NRL_272/2021 | 2021-01-07 | Europe/Czech Republic/Central Bohemia | The National Institute of Public Health | State Veterinary Institute Prague | Nagy et al |
| EPI_ISL_882953 | CzechRepublic/NRL_277/2021 | 2021-01-06 | Europe/Czech Republic/Prague/ | The National Institute of Public Health | State Veterinary Institute Prague | Nagy et al |
| EPI_ISL_882954 | CzechRepublic/NRL_12565/2020 | 2020-10-19 | Europe/Czech Republic/Prague/ | State Veterinary Institute Prague | State Veterinary Institute Prague | Nagy et al |
| EPI_ISL_882955 | CzechRepublic/NRL_12566/2020 | 2020-12-31 | Europe/Czech Republic/Prague/ | State Veterinary Institute Prague | State Veterinary Institute Prague | Nagy et al |

| | | | | | | |
|---|---|---|---|---|---|---|
| EPI_ISL_882957 | Poland/NIPH-NIH-01/2020 | 2020-11-16 | Europe/Poland/Mazowieckie/Warszawa | Specjalistyczny Szpital im. Dra A. Soko≈Çowskiego | National Institute of Public Health - National Institute of Hygiene | Wo≈Çkowicz Tomasz et al |
| EPI_ISL_882958 | Poland/NIPH-NIH-02/2020 | 2020-12-15 | Europe/Poland/Mazowieckie/Warszawa | National Institute of Public Health - National Institute of Hygiene | National Institute of Public Health - National Institute of Hygiene | Wo≈Çkowicz Tomasz et al |
| EPI_ISL_882960 | Poland/NIPH-NIH-05/2020 | 2020-10-29 | Europe/Poland/Mazowieckie/Warszawa | Laboratorium Diagnostyi Mikrobiologicznej z PracownifÖ PrfÖtka Gru≈ʃlicy SPSzW im. Jana Bo≈ºego w Lublinie | National Institute of Public Health - National Institute of Hygiene | Wo≈Çkowicz Tomasz et al |
| EPI_ISL_882961 | Poland/NIPH-NIH-06/2020 | 2020-11-02 | Europe/Poland/Mazowieckie/Warszawa | Laboratorium Diagnostyi Mikrobiologicznej z PracownifÖ PrfÖtka Gru≈ʃlicy SPSzW im. Jana Bo≈ºego w Lublinie | National Institute of Public Health - National Institute of Hygiene | Wo≈Çkowicz Tomasz et al |
| EPI_ISL_882962 | Poland/NIPH-NIH-08/2020 | 2020-12-17 | Europe/Poland/Mazowieckie/Warszawa | National Institute of Public Health - National Institute of Hygiene | National Institute of Public Health - National Institute of Hygiene | Wo≈Çkowicz Tomasz et al |
| EPI_ISL_882968 | USA/CA-LACPHL-AF00205/2021 | 2021-01-12 | North America/USA/California/Los Ange | UCLA Clinical Micro Lab | Los Angeles County PHL | P. Hemarajata et al. et al |
| EPI_ISL_882972 | USA/CA-LACPHL-AF00198/2021 | 2021-01-12 | North America/USA/California/Los Ange | UCLA Clinical Micro Lab | Los Angeles County PHL | P. Hemarajata et al. et al |
| EPI_ISL_882979 | USA/CA-LACPHL-AF00207/2021 | 2021-01-11 | North America/USA/California/Los Ange | UCLA Clinical Micro Lab | Los Angeles County PHL | P. Hemarajata et al. et al |
| EPI_ISL_882984 | USA/CA-LACPHL-AF00212/2021 | 2021-01-11 | North America/USA/California/Los Ange | UCLA Clinical Micro Lab | Los Angeles County PHL | P. Hemarajata et al. et al |
| EPI_ISL_882987 | USA/CA-LACPHL-AF00216/2021 | 2021-01-05 | North America/USA/California/Los Ange | MD Laboratories | Los Angeles County PHL | P. Hemarajata et al. et al |
| EPI_ISL_882992 | USA/CA-LACPHL-AF00225/2021 | 2021-01-08 | North America/USA/California/Los Ange | MD Laboratories | Los Angeles County PHL | P. Hemarajata et al. et al |
| EPI_ISL_883001 | USA/MD-MDH-0650/2021 | 2021-01-12 | North America/USA/Maryland/ | Maryland Public Health Laboratory | Maryland Public Health Laboratory | Maryland Department of Health Laboratories Administration et al |
| EPI_ISL_883015 | USA/MD-MDH-0638/2021 | 2021-01-11 | North America/USA/Maryland/ | Maryland Public Health Laboratory | Maryland Public Health Laboratory | Maryland Department of Health Laboratories Administration et al |

| | | | | | | |
|---|---|---|---|---|---|---|
| EPI_ISL_883033 | USA/MD-MDH-0666/2020 | 2020-12-31 | North America/USA/Maryland/ | Maryland Public Health Laboratory | Maryland Public Health Laboratory | Maryland Department of Health Laboratories Administration et al |
| EPI_ISL_883039 | USA/KS-KHEL-0016/2020 | 2020-03-31 | North America/USA/Kansas/ | Kansas Health and Environmental Lab | Kansas Health and Environmental Lab | Mike Grose et al |
| EPI_ISL_883046 | USA/KS-KHEL-0025/2020 | 2020-04-01 | North America/USA/Kansas/ | Kansas Health and Environmental Lab | Kansas Health and Environmental Lab | Mike Grose et al |
| EPI_ISL_883049 | USA/KS-KHEL-0029/2020 | 2020-04-01 | North America/USA/Kansas/ | Kansas Health and Environmental Lab | Kansas Health and Environmental Lab | Mike Grose et al |
| EPI_ISL_883051 | USA/KS-KHEL-0032/2020 | 2020-04-02 | North America/USA/Kansas/ | Kansas Health and Environmental Lab | Kansas Health and Environmental Lab | Mike Grose et al |
| EPI_ISL_883053 | USA/KS-KHEL-0035/2020 | 2020-04-02 | North America/USA/Kansas/ | Kansas Health and Environmental Lab | Kansas Health and Environmental Lab | Mike Grose et al |
| EPI_ISL_883055 | USA/KS-KHEL-0037/2020 | 2020-04-02 | North America/USA/Kansas/ | Kansas Health and Environmental Lab | Kansas Health and Environmental Lab | Mike Grose et al |
| EPI_ISL_883065 | USA/KS-KHEL-0030/2020 | 2020-04-01 | North America/USA/Kansas/ | Kansas Health and Environmental Lab | Kansas Health and Environmental Lab | Mike Grose et al |
| EPI_ISL_883067 | USA/AL-Limestone-JFFB/2020 | 2020-11-27 | North America/USA/Alabama/Limestone | Eurofins Diatherix | Hudsonalpha Genome Sequencing Center | Jane Grimwood et al |
| EPI_ISL_883070 | USA/AL-Madison-JFGW/2020 | 2020-11-28 | North America/USA/Alabama/Madison | Eurofins Diatherix | Hudsonalpha Genome Sequencing Center | Jane Grimwood et al |
| EPI_ISL_883073 | USA/AL-Coosa-JFEA/2020 | 2020-11-27 | North America/USA/Alabama/Coosa Co | Eurofins Diatherix | Hudsonalpha Genome Sequencing Center | Jane Grimwood et al |
| EPI_ISL_883093 | USA/AL-Morgan-JFHL/2020 | 2020-11-28 | North America/USA/Alabama/Morgan ( | Eurofins Diatherix | Hudsonalpha Genome Sequencing Center | Jane Grimwood et al |

| | | | | | | |
|---|---|---|---|---|---|---|
| EPI_ISL_883096 | USA/AL-Shelby-JFDW/2020 | 2020-11-27 | North America/USA/Alabama/Shelby Co | Eurofins Diatherix | Hudsonalpha Genome Sequencing Center | Jane Grimwood et al |
| EPI_ISL_883098 | USA/AL-Jefferson-JFDZ/2020 | 2020-11-27 | North America/USA/Alabama/Jefferson | Eurofins Diatherix | Hudsonalpha Genome Sequencing Center | Jane Grimwood et al |
| EPI_ISL_883110 | USA/AL-Madison-JFGR/2020 | 2020-11-28 | North America/USA/Alabama/Madison | Eurofins Diatherix | Hudsonalpha Genome Sequencing Center | Jane Grimwood et al |
| EPI_ISL_883114 | USA/AL-Limestone-JFHK/2020 | 2020-11-28 | North America/USA/Alabama/Limestone | Eurofins Diatherix | Hudsonalpha Genome Sequencing Center | Jane Grimwood et al |
| EPI_ISL_883115 | USA/AL-Marshall-JFHT/2020 | 2020-11-28 | North America/USA/Alabama/Marshall | Eurofins Diatherix | Hudsonalpha Genome Sequencing Center | Jane Grimwood et al |
| EPI_ISL_883123 | USA/AL-Morgan-JFEZ/2020 | 2020-11-27 | North America/USA/Alabama/Morgan C | Eurofins Diatherix | Hudsonalpha Genome Sequencing Center | Jane Grimwood et al |
| EPI_ISL_883139 | USA/AL-Fayette-JFEE/2020 | 2020-11-27 | North America/USA/Alabama/Fayette C | Eurofins Diatherix | Hudsonalpha Genome Sequencing Center | Jane Grimwood et al |
| EPI_ISL_883141 | USA/AL-Jefferson-JFET/2020 | 2020-11-27 | North America/USA/Alabama/Jefferson | Eurofins Diatherix | Hudsonalpha Genome Sequencing Center | Jane Grimwood et al |
| EPI_ISL_883142 | USA/AL-Madison-JFEU/2020 | 2020-11-27 | North America/USA/Alabama/Madison | Eurofins Diatherix | Hudsonalpha Genome Sequencing Center | Jane Grimwood et al |
| EPI_ISL_883151 | USA/AL-Houston-JFHQ/2020 | 2020-11-28 | North America/USA/Alabama/Houston C | Eurofins Diatherix | Hudsonalpha Genome Sequencing Center | Jane Grimwood et al |

| | | | | | | |
|---|---|---|---|---|---|---|
| EPI_ISL_883166 | Germany/NI-IOV-60140201/2020 | 2020-04-07 | Europe/Germany/Niedersachsen/Hanno | Hannover Medical School, Institute of Virology | Hannover Medical School, Institute of Virology | Lars Steinbr√ºck et al |
| EPI_ISL_883167 | Germany/NI-IOV-60140220/2020 | 2020-04-08 | Europe/Germany/Niedersachsen/Hanno | Hannover Medical School, Institute of Virology | Hannover Medical School, Institute of Virology | Lars Steinbr√ºck et al |
| EPI_ISL_883176 | Germany/NI-IOV-P10285/2020 | 2020-05-06 | Europe/Germany/Baden-Wuerttemberg | UK T√ºbingen, Medical Microbiology | Hannover Medical School, Institute of Virology | Lars Steinbr√ºck et al |
| EPI_ISL_883179 | Germany/NI-IOV-P5414/2020 | 2020-03-21 | Europe/Germany/Baden-Wuerttemberg | UK T√ºbingen, Medical Microbiology | Hannover Medical School, Institute of Virology | Lars Steinbr√ºck et al |
| EPI_ISL_883180 | Germany/NI-IOV-P5417/2020 | 2020-03-21 | Europe/Germany/Baden-Wuerttemberg | UK T√ºbingen, Medical Microbiology | Hannover Medical School, Institute of Virology | Lars Steinbr√ºck et al |
| EPI_ISL_883182 | Germany/NI-IOV-P5446/2020 | 2020-03-22 | Europe/Germany/Baden-Wuerttemberg | UK T√ºbingen, Medical Microbiology | Hannover Medical School, Institute of Virology | Lars Steinbr√ºck et al |
| EPI_ISL_883183 | Germany/NI-IOV-P5458/2020 | 2020-03-23 | Europe/Germany/Baden-Wuerttemberg | UK T√ºbingen, Medical Microbiology | Hannover Medical School, Institute of Virology | Lars Steinbr√ºck et al |
| EPI_ISL_883189 | Germany/NI-IOV-P6127/2020 | 2020-03-26 | Europe/Germany/Baden-Wuerttemberg | UK T√ºbingen, Medical Microbiology | Hannover Medical School, Institute of Virology | Lars Steinbr√ºck et al |
| EPI_ISL_883191 | Germany/NI-IOV-P8388/2020 | 2020-04-20 | Europe/Germany/Baden-Wuerttemberg | UK T√ºbingen, Medical Microbiology | Hannover Medical School, Institute of Virology | Lars Steinbr√ºck et al |
| EPI_ISL_883192 | Germany/NI-IOV-P9091/2020 | 2020-04-27 | Europe/Germany/Baden-Wuerttemberg | UK T√ºbingen, Medical Microbiology | Hannover Medical School, Institute of Virology | Lars Steinbr√ºck et al |
| EPI_ISL_883195 | USA/AL-Jackson-JFIK/2020 | 2020-11-28 | North America/USA/Alabama/Jackson C | Eurofins Diatherix | Hudsonalpha Genome Sequencing Center | Jane Grimwood et al |
| EPI_ISL_883202 | USA/AL-Etowah-JFKJ/2020 | 2020-12-02 | North America/USA/Alabama/Etowah C | Eurofins Diatherix | Hudsonalpha Genome Sequencing Center | Jane Grimwood et al |

| | | | | | | |
|---|---|---|---|---|---|---|
| EPI_ISL_883205 | USA/AL-Etowah-JFLH/2020 | 2020-12-01 | North America/USA/Alabama/Etowah C | Eurofins Diatherix | Hudsonalpha Genome Sequencing Center | Jane Grimwood et al |
| EPI_ISL_883206 | USA/AL-Etowah-JFLI/2020 | 2020-12-01 | North America/USA/Alabama/Etowah C | Eurofins Diatherix | Hudsonalpha Genome Sequencing Center | Jane Grimwood et al |
| EPI_ISL_883214 | USA/AL-Madison-JFJP/2020 | 2020-11-30 | North America/USA/Alabama/Madison | Eurofins Diatherix | Hudsonalpha Genome Sequencing Center | Jane Grimwood et al |
| EPI_ISL_883220 | USA/AL-Madison-JFMC/2020 | 2020-12-01 | North America/USA/Alabama/Madison | Eurofins Diatherix | Hudsonalpha Genome Sequencing Center | Jane Grimwood et al |
| EPI_ISL_883236 | USA/AL-Morgan-JFKN/2020 | 2020-12-02 | North America/USA/Alabama/Morgan C | Eurofins Diatherix | Hudsonalpha Genome Sequencing Center | Jane Grimwood et al |
| EPI_ISL_883238 | USA/AL-StClair-JFJZ/2020 | 2020-12-01 | North America/USA/Alabama/St. Clair C | Eurofins Diatherix | Hudsonalpha Genome Sequencing Center | Jane Grimwood et al |
| EPI_ISL_883240 | USA/AL-StClair-JFLA/2020 | 2020-12-01 | North America/USA/Alabama/St. Clair C | Eurofins Diatherix | Hudsonalpha Genome Sequencing Center | Jane Grimwood et al |
| EPI_ISL_883246 | USA/AL-Lawrence-JFLZ/2020 | 2020-12-01 | North America/USA/Alabama/Lawrence | Eurofins Diatherix | Hudsonalpha Genome Sequencing Center | Jane Grimwood et al |
| EPI_ISL_883247 | USA/AL-Limestone-JFMF/2020 | 2020-12-01 | North America/USA/Alabama/Limestone | Eurofins Diatherix | Hudsonalpha Genome Sequencing Center | Jane Grimwood et al |
| EPI_ISL_883250 | USA/AL-Madison-JFJA/2020 | 2020-12-01 | North America/USA/Alabama/Madison | Eurofins Diatherix | Hudsonalpha Genome Sequencing Center | Jane Grimwood et al |

| | | | | | | |
|---|---|---|---|---|---|---|
| EPI_ISL_883255 | USA/AL-Madison-JFJI/2020 | 2020-11-30 | North America/USA/Alabama/Madison | Eurofins Diatherix | Hudsonalpha Genome Sequencing Center | Jane Grimwood et al |
| EPI_ISL_883267 | USA/AL-Cullman-JFKP/2020 | 2020-12-02 | North America/USA/Alabama/Cullman ( | Eurofins Diatherix | Hudsonalpha Genome Sequencing Center | Jane Grimwood et al |
| EPI_ISL_883273 | USA/AL-Madison-JFJB/2020 | 2020-12-01 | North America/USA/Alabama/Madison | Eurofins Diatherix | Hudsonalpha Genome Sequencing Center | Jane Grimwood et al |
| EPI_ISL_883280 | USA/AL-Montgomery-JFLW/2020 | 2020-12-01 | North America/USA/Alabama/Montgom | Eurofins Diatherix | Hudsonalpha Genome Sequencing Center | Jane Grimwood et al |
| EPI_ISL_883284 | USA/AL-StClair-JFKZ/2020 | 2020-12-01 | North America/USA/Alabama/St. Clair ( | Eurofins Diatherix | Hudsonalpha Genome Sequencing Center | Jane Grimwood et al |
| EPI_ISL_883286 | Italy/ABR-IZSGC-TE1876/2021 | 2021-01-02 | Europe/Italy/Abruzzo/ | SIESP DIPARTIMENTO DI PREVENZIONE CHIE | Istituto Zooprofilattico Sperimentale dell,ÄôAbruzzo e Molise ,ÄúG. Caporale,Äù | Lorusso A et al |
| EPI_ISL_883294 | Italy/ABR-IZSGC-TE5830/2021 | 2021-01-05 | Europe/Italy/Abruzzo/ | RP Guardiagrele-Ospedale di Comunit√† | Istituto Zooprofilattico Sperimentale dell,ÄôAbruzzo e Molise ,ÄúG. Caporale,Äù | Lorusso A et al |
| EPI_ISL_883295 | Italy/ABR-IZSGC-TE5838/2021 | 2021-01-05 | Europe/Italy/Abruzzo/ | RP Guardiagrele-Ospedale di Comunit√† | Istituto Zooprofilattico Sperimentale dell,ÄôAbruzzo e Molise ,ÄúG. Caporale,Äù | Lorusso A et al |

| | | | | | | |
|---|---|---|---|---|---|---|
| EPI_ISL_883296 | Italy/ABR-IZSGC-TE1873/2021 | 2021-01-02 | Europe/Italy/Abruzzo/ | SIESP CHIETI-DRIVE IN ORTONA | Istituto Zooprofilattico Sperimentale dell‚ÄôAbruzzo e Molise ‚ÄúG. Caporale‚Äù | Lorusso A et al |
| EPI_ISL_883297 | Italy/ABR-IZSGC-TE3214/2021 | 2021-01-04 | Europe/Italy/Abruzzo/ | RP Guardiagrele-Ospedale di Comunit√† | Istituto Zooprofilattico Sperimentale dell‚ÄôAbruzzo e Molise ‚ÄúG. Caporale‚Äù | Lorusso A et al |
| EPI_ISL_883298 | Italy/ABR-IZSGC-TE3148/2021 | 2021-01-04 | Europe/Italy/Abruzzo/ | SIESP DIPARTIMENTO DI PREVENZIONE CHIE | Istituto Zooprofilattico Sperimentale dell‚ÄôAbruzzo e Molise ‚ÄúG. Caporale‚Äù | Lorusso A et al |
| EPI_ISL_883299 | Italy/ABR-IZSGC-TE3150/2021 | 2021-01-04 | Europe/Italy/Abruzzo/ | SIESP DIPARTIMENTO DI PREVENZIONE CHIE | Istituto Zooprofilattico Sperimentale dell‚ÄôAbruzzo e Molise ‚ÄúG. Caporale‚Äù | Lorusso A et al |
| EPI_ISL_883301 | Italy/ABR-IZSGC-TE5841/2021 | 2021-01-05 | Europe/Italy/Abruzzo/ | RP Guardiagrele-Ospedale di Comunit√† | Istituto Zooprofilattico Sperimentale dell‚ÄôAbruzzo e Molise ‚ÄúG. Caporale‚Äù | Lorusso A et al |
| EPI_ISL_883302 | Italy/ABR-IZSGC-TE5844/2021 | 2021-01-05 | Europe/Italy/Abruzzo/ | RP Guardiagrele-Ospedale di Comunit√† | Istituto Zooprofilattico Sperimentale dell‚ÄôAbruzzo e Molise ‚ÄúG. Caporale‚Äù | Lorusso A et al |

| | | | | | | |
|---|---|---|---|---|---|---|
| EPI_ISL_883303 | Italy/ABR-IZSGC-TE5708/2021 | 2021-01-05 | Europe/Italy/Abruzzo/ | SIESP DIPARTIMENTO DI PREVENZIONE CHIE | Istituto Zooprofilattico Sperimentale dell‚ÄôAbruzzo e Molise ‚ÄúG. Caporale‚Äù | Lorusso A et al |
| EPI_ISL_883305 | Italy/ABR-IZSGC-TE5787/2021 | 2021-01-05 | Europe/Italy/Abruzzo/ | SIESP DIPARTIMENTO DI PREVENZIONE CHIE | Istituto Zooprofilattico Sperimentale dell‚ÄôAbruzzo e Molise ‚ÄúG. Caporale‚Äù | Lorusso A et al |
| EPI_ISL_883313 | USA/NY-NYCPHL-002443/2021 | 2021-01-14 | North America/USA/New York/New Yor | DOHMH Corona | New York City Public Health Laboratory | Jade Wang et al |
| EPI_ISL_883321 | USA/NY-NYCPHL-002448/2021 | 2021-01-14 | North America/USA/New York/New Yor | DOHMH Jamaica | New York City Public Health Laboratory | Jade Wang et al |
| EPI_ISL_883335 | USA/NY-NYCPHL-002438/2021 | 2021-01-14 | North America/USA/New York/New Yor | DOHMH Chelsea | New York City Public Health Laboratory | Jade Wang et al |
| EPI_ISL_883336 | USA/NY-NYCPHL-002499/2021 | 2021-01-13 | North America/USA/New York/New Yor | OCME Office Of Chief Medical Examiner | New York City Public Health Laboratory | Jade Wang et al |
| EPI_ISL_883337 | USA/NY-NYCPHL-002528/2021 | 2021-01-15 | North America/USA/New York/New Yor | DOHMH Crown Heights | New York City Public Health Laboratory | Jade Wang et al |
| EPI_ISL_883341 | USA/NY-NYCPHL-002471/2021 | 2021-01-14 | North America/USA/New York/New Yor | DOHMH PHL | New York City Public Health Laboratory | Jade Wang et al |
| EPI_ISL_883351 | USA/NY-NYCPHL-002182/2021 | 2021-01-01 | North America/USA/New York/New Yor | OCME Office Of Chief Medical Examiner | New York City Public Health Laboratory | Jade Wang et al |
| EPI_ISL_883358 | USA/NY-NYCPHL-002422/2021 | 2021-01-12 | North America/USA/New York/New Yor | DOHMH Crown Heights | New York City Public Health Laboratory | Jade Wang et al |
| EPI_ISL_883359 | USA/NY-NYCPHL-002423/2021 | 2021-01-13 | North America/USA/New York/New Yor | DOHMH Crown Heights | New York City Public Health Laboratory | Jade Wang et al |

| | | | | | | |
|---|---|---|---|---|---|---|
| EPI_ISL_883365 | USA/NY-NYCPHL-002435/2021 | 2021-01-14 | North America/USA/New York/New Yor | DOHMH Morrisania | New York City Public Health Laboratory | Jade Wang et al |
| EPI_ISL_883367 | USA/NY-NYCPHL-002437/2021 | 2021-01-14 | North America/USA/New York/New Yor | DOHMH Morrisania | New York City Public Health Laboratory | Jade Wang et al |
| EPI_ISL_883378 | USA/NY-NYCPHL-002464/2021 | 2021-01-14 | North America/USA/New York/New Yor | DOHMH Central Harlem | New York City Public Health Laboratory | Jade Wang et al |
| EPI_ISL_883384 | USA/NY-NYCPHL-002477/2021 | 2021-01-17 | North America/USA/New York/New Yor | Pandemic Response Laboratory | New York City Public Health Laboratory | Jade Wang et al |
| EPI_ISL_883404 | USA/NY-NYCPHL-002512/2021 | 2021-01-15 | North America/USA/New York/New Yor | OCME Office Of Chief Medical Examiner | New York City Public Health Laboratory | Jade Wang et al |
| EPI_ISL_883411 | USA/NY-NYCPHL-002527/2021 | 2021-01-15 | North America/USA/New York/New Yor | DOHMH Corona | New York City Public Health Laboratory | Jade Wang et al |
| EPI_ISL_883420 | USA/NY-NYCPHL-002540/2021 | 2021-01-15 | North America/USA/New York/New Yor | DOHMH Jamaica | New York City Public Health Laboratory | Jade Wang et al |
| EPI_ISL_883425 | USA/NY-Wadsworth-21009427-01/20 | 2021-01-12 | North America/USA/New York/Saratoga | BOSTON HEART DIAGNOSTICS CORP | Wadsworth Center, New York State Department of Health | Kirsten St. George et al |
| EPI_ISL_883426 | USA/NY-Wadsworth-21008856-01/20 | 2021-01-11 | North America/USA/New York/Nassau C | NORTHWELL HEALTH LABORATORIES | Wadsworth Center, New York State Department of Health | Kirsten St. George et al |
| EPI_ISL_883428 | USA/NY-Wadsworth-21008815-01/20 | 2021-01-09 | North America/USA/New York/Nassau C | NORTHWELL HEALTH LABORATORIES | Wadsworth Center, New York State Department of Health | Kirsten St. George et al |
| EPI_ISL_883435 | USA/NY-Wadsworth-21008873-01/20 | 2021-01-11 | North America/USA/New York/Nassau C | NORTHWELL HEALTH LABORATORIES | Wadsworth Center, New York State Department of Health | Kirsten St. George et al |

| | | | | | | |
|---|---|---|---|---|---|---|
| EPI_ISL_883438 | USA/NY-Wadsworth-21008826-01/20 | 2021-01-12 | North America/USA/New York/Franklin | ADIRONDACK MEDICAL CENTER | Wadsworth Center, New York State Department of Health | Kirsten St. George et al |
| EPI_ISL_883446 | USA/NY-Wadsworth-21008834-01/20 | 2021-01-09 | North America/USA/New York/Kings Co | NORTHWELL HEALTH LABORATORIES | Wadsworth Center, New York State Department of Health | Kirsten St. George et al |
| EPI_ISL_883453 | USA/NY-Wadsworth-21008785-01/20 | 2021-01-06 | North America/USA/New York/Queens | NORTHWELL HEALTH LABORATORIES | Wadsworth Center, New York State Department of Health | Kirsten St. George et al |
| EPI_ISL_883457 | USA/NY-Wadsworth-21008790-01/20 | 2021-01-04 | North America/USA/New York/Nassau C | NORTHWELL HEALTH LABORATORIES | Wadsworth Center, New York State Department of Health | Kirsten St. George et al |
| EPI_ISL_883459 | USA/NY-Wadsworth-21008793-01/20 | 2021-01-05 | North America/USA/New York/Nassau C | NORTHWELL HEALTH LABORATORIES | Wadsworth Center, New York State Department of Health | Kirsten St. George et al |
| EPI_ISL_883466 | USA/NY-Wadsworth-21008800-01/20 | 2021-01-05 | North America/USA/New York/New Yor | NORTHWELL HEALTH LABORATORIES | Wadsworth Center, New York State Department of Health | Kirsten St. George et al |
| EPI_ISL_883473 | USA/NY-Wadsworth-21008829-01/20 | 2021-01-12 | North America/USA/New York/Franklin | ADIRONDACK MEDICAL CENTER | Wadsworth Center, New York State Department of Health | Kirsten St. George et al |
| EPI_ISL_883474 | USA/NY-Wadsworth-21008830-01/20 | 2021-01-09 | North America/USA/New York/Richmon | NORTHWELL HEALTH LABORATORIES | Wadsworth Center, New York State Department of Health | Kirsten St. George et al |
| EPI_ISL_883484 | USA/NY-Wadsworth-21008860-01/20 | 2021-01-10 | North America/USA/New York/Suffolk C | NORTHWELL HEALTH LABORATORIES | Wadsworth Center, New York State Department of Health | Kirsten St. George et al |
| EPI_ISL_883485 | USA/NY-Wadsworth-21008870-01/20 | 2021-01-10 | North America/USA/New York/Richmon | NORTHWELL HEALTH LABORATORIES | Wadsworth Center, New York State Department of Health | Kirsten St. George et al |

| | | | | | | |
|---|---|---|---|---|---|---|
| EPI_ISL_883496 | USA/NY-Wadsworth-21008888-01/20 | 2021-01-11 | North America/USA/New York/Nassau C | NORTHWELL HEALTH LABORATORIES | Wadsworth Center, New York State Department of Health | Kirsten St. George et al |
| EPI_ISL_883499 | Italy/CAM-CRGS-103/2020 | 2020-03-23 | Europe/Italy/Campania/ | 1.AO Universitaria 'S. Giovanni di Dio e Ruggi D'Aragona, Scuola Medica Salernitana' Hospital / 2.UOC di Virologia e Microbiologia, Universit√† della Campania 'L. Vanvitelli' / 3.AO Universitaria 'Federico II' Napoli Hospital / 4.AORN 'San Giuseppe Moscati' Avellino Hospital / 5.AO 'San Pio - presidio G. Rummo' Benevento Hospital / 6.AO 'Sant'Anna e San Sebastiano' Caserta Hospital / 7.PO 'Maria Santissima Addolorata' Eboli Hospital / 8.Biogem Istituto di Ricerche Genetiche | 1. Genome Research Center for Health (CRGS) / 2. Laboratory of Molecular Medicine and Genomics(LMMGe) / 3. Center for Research in Pure and Applied Mathematics (CRMPA) | Giorgio Giurato et al |
| EPI_ISL_883516 | USA/AL-HGSC-JFQT/2020 | 2020-12-01 | North America/USA/Alabama/Dale Cou | Eurofins Diatherix | Hudsonalpha Genome Sequencing Center | Jane Grimwood et al |
| EPI_ISL_883521 | USA/AL-HGSC-JFQN/2020 | 2020-12-02 | North America/USA/Alabama/Etowah C | Eurofins Diatherix | Hudsonalpha Genome Sequencing Center | Jane Grimwood et al |
| EPI_ISL_883534 | USA/AL-HGSC-JFQA/2020 | 2020-12-02 | North America/USA/Alabama/Mobile C | Eurofins Diatherix | Hudsonalpha Genome Sequencing Center | Jane Grimwood et al |
| EPI_ISL_883540 | USA/AL-HGSC-JFPT/2020 | 2020-12-01 | North America/USA/Alabama/Shelby Co | Eurofins Diatherix | Hudsonalpha Genome Sequencing Center | Jane Grimwood et al |
| EPI_ISL_883546 | USA/AL-HGSC-JFPL/2020 | 2020-12-01 | North America/USA/Alabama/Madison | Eurofins Diatherix | Hudsonalpha Genome Sequencing Center | Jane Grimwood et al |
| EPI_ISL_883547 | USA/AL-HGSC-JFPK/2020 | 2020-12-01 | North America/USA/Alabama/Calhoun C | Eurofins Diatherix | Hudsonalpha Genome Sequencing Center | Jane Grimwood et al |

| | | | | | | |
|---|---|---|---|---|---|---|
| EPI_ISL_883549 | USA/AL-HGSC-JFPI/2020 | 2020-12-01 | North America/USA/Alabama/Houston ( | Eurofins Diatherix | Hudsonalpha Genome Sequencing Center | Jane Grimwood et al |
| EPI_ISL_883554 | USA/AL-HGSC-JFPD/2020 | 2020-12-01 | North America/USA/Alabama/Shelby Co | Eurofins Diatherix | Hudsonalpha Genome Sequencing Center | Jane Grimwood et al |
| EPI_ISL_883560 | USA/AL-HGSC-JFNX/2020 | 2020-12-01 | North America/USA/Alabama/Morgan ( | Eurofins Diatherix | Hudsonalpha Genome Sequencing Center | Jane Grimwood et al |
| EPI_ISL_883564 | USA/AL-HGSC-JFNS/2020 | 2020-12-01 | North America/USA/Alabama/Houston ( | Eurofins Diatherix | Hudsonalpha Genome Sequencing Center | Jane Grimwood et al |
| EPI_ISL_883569 | USA/AL-HGSC-JFNJ/2020 | 2020-12-01 | North America/USA/Alabama/Madison | Eurofins Diatherix | Hudsonalpha Genome Sequencing Center | Jane Grimwood et al |
| EPI_ISL_883578 | USA/AL-HGSC-JFMY/2020 | 2020-12-01 | North America/USA/Alabama/Calhoun ( | Eurofins Diatherix | Hudsonalpha Genome Sequencing Center | Jane Grimwood et al |
| EPI_ISL_883587 | USA/AL-HGSC-JFMM/2020 | 2020-12-01 | North America/USA/Alabama/Blount Co | Eurofins Diatherix | Hudsonalpha Genome Sequencing Center | Jane Grimwood et al |
| EPI_ISL_883594 | USA/AL-HGSC-JFSR/2020 | 2020-12-02 | North America/USA/Alabama/Limestone | Eurofins Diatherix | Hudsonalpha Genome Sequencing Center | Jane Grimwood et al |
| EPI_ISL_883597 | USA/AL-HGSC-JFRK/2020 | 2020-12-02 | North America/USA/Alabama/Morgan ( | Eurofins Diatherix | Hudsonalpha Genome Sequencing Center | Jane Grimwood et al |
| EPI_ISL_883598 | USA/AL-HGSC-JFRL/2020 | 2020-12-02 | North America/USA/Alabama/Madison | Eurofins Diatherix | Hudsonalpha Genome Sequencing Center | Jane Grimwood et al |

| | | | | | | |
|---|---|---|---|---|---|---|
| EPI_ISL_883606 | USA/AL-HGSC-JFRY/2020 | 2020-12-02 | North America/USA/Alabama/Wilcox Co | Eurofins Diatherix | Hudsonalpha Genome Sequencing Center | Jane Grimwood et al |
| EPI_ISL_883635 | USA/AL-HGSC-JFTI/2020 | 2020-12-02 | North America/USA/Alabama/Madison | Eurofins Diatherix | Hudsonalpha Genome Sequencing Center | Jane Grimwood et al |
| EPI_ISL_883648 | USA/AL-HGSC-JFTZ/2020 | 2020-12-02 | North America/USA/Alabama/Marshall | Eurofins Diatherix | Hudsonalpha Genome Sequencing Center | Jane Grimwood et al |
| EPI_ISL_883666 | USA/AL-HGSC-JFUS/2020 | 2020-12-02 | North America/USA/Alabama/Madison | Eurofins Diatherix | Hudsonalpha Genome Sequencing Center | Jane Grimwood et al |
| EPI_ISL_883678 | USA/AL-HGSC-JFWF/2020 | 2020-12-02 | North America/USA/Alabama/Madison | Eurofins Diatherix | Hudsonalpha Genome Sequencing Center | Jane Grimwood et al |
| EPI_ISL_883685 | USA/AL-HGSC-JGAE/2020 | 2020-12-03 | North America/USA/Alabama/Jackson C | Eurofins Diatherix | Hudsonalpha Genome Sequencing Center | Jane Grimwood et al |
| EPI_ISL_883690 | USA/AL-HGSC-JFWY/2020 | 2020-12-02 | North America/USA/Alabama/Clay Coun | Eurofins Diatherix | Hudsonalpha Genome Sequencing Center | Jane Grimwood et al |
| EPI_ISL_883691 | USA/AL-HGSC-JFXA/2020 | 2020-12-02 | North America/USA/Alabama/Shelby Co | Eurofins Diatherix | Hudsonalpha Genome Sequencing Center | Jane Grimwood et al |
| EPI_ISL_883697 | USA/AL-HGSC-JFXX/2020 | 2020-12-03 | North America/USA/Alabama/Talladega | Eurofins Diatherix | Hudsonalpha Genome Sequencing Center | Jane Grimwood et al |
| EPI_ISL_883700 | USA/AL-HGSC-JFYB/2020 | 2020-12-03 | North America/USA/Alabama/Calhoun C | Eurofins Diatherix | Hudsonalpha Genome Sequencing Center | Jane Grimwood et al |

| | | | | | | |
|---|---|---|---|---|---|---|
| EPI_ISL_883703 | USA/AL-HGSC-JFYF/2020 | 2020-12-03 | North America/USA/Alabama/Walker C | Eurofins Diatherix | Hudsonalpha Genome Sequencing Center | Jane Grimwood et al |
| EPI_ISL_883727 | USA/AL-HGSC-JFXU/2020 | 2020-12-03 | North America/USA/Alabama/Walker C | Eurofins Diatherix | Hudsonalpha Genome Sequencing Center | Jane Grimwood et al |
| EPI_ISL_883744 | USA/AL-HGSC-JFXI/2020 | 2020-12-03 | North America/USA/Alabama/Etowah C | Eurofins Diatherix | Hudsonalpha Genome Sequencing Center | Jane Grimwood et al |
| EPI_ISL_883748 | USA/AL-HGSC-JFYT/2020 | 2020-12-03 | North America/USA/Alabama/Tuscaloos | Eurofins Diatherix | Hudsonalpha Genome Sequencing Center | Jane Grimwood et al |
| EPI_ISL_883763 | USA/AL-HGSC-JFXW/2020 | 2020-12-03 | North America/USA/Alabama/Tuscaloos | Eurofins Diatherix | Hudsonalpha Genome Sequencing Center | Jane Grimwood et al |
| EPI_ISL_883770 | USA/AL-HGSC-JGDS/2020 | 2020-12-04 | North America/USA/Alabama/Jefferson | Eurofins Diatherix | Hudsonalpha Genome Sequencing Center | Jane Grimwood et al |
| EPI_ISL_883772 | USA/AL-HGSC-JGBF/2020 | 2020-12-04 | North America/USA/Alabama/Etowah C | Eurofins Diatherix | Hudsonalpha Genome Sequencing Center | Jane Grimwood et al |
| EPI_ISL_883777 | USA/AL-HGSC-JGBC/2020 | 2020-12-03 | North America/USA/Alabama/Jackson C | Eurofins Diatherix | Hudsonalpha Genome Sequencing Center | Jane Grimwood et al |
| EPI_ISL_883795 | USA/AL-HGSC-JGDW/2020 | 2020-12-04 | North America/USA/Alabama/St. Clair C | Eurofins Diatherix | Hudsonalpha Genome Sequencing Center | Jane Grimwood et al |
| EPI_ISL_883822 | USA/AL-HGSC-JGDF/2020 | 2020-12-04 | North America/USA/Alabama/Madison | Eurofins Diatherix | Hudsonalpha Genome Sequencing Center | Jane Grimwood et al |

| | | | | | | |
|---|---|---|---|---|---|---|
| EPI_ISL_883823 | USA/AL-HGSC-JGAN/2020 | 2020-12-03 | North America/USA/Alabama/Calhoun ( | Eurofins Diatherix | Hudsonalpha Genome Sequencing Center | Jane Grimwood et al |
| EPI_ISL_883826 | USA/AL-HGSC-JGAY/2020 | 2020-12-03 | North America/USA/Alabama/Shelby Co | Eurofins Diatherix | Hudsonalpha Genome Sequencing Center | Jane Grimwood et al |
| EPI_ISL_883830 | USA/AL-HGSC-JGBM/2020 | 2020-12-03 | North America/USA/Alabama/Jefferson | Eurofins Diatherix | Hudsonalpha Genome Sequencing Center | Jane Grimwood et al |
| EPI_ISL_883834 | USA/AL-HGSC-JGCL/2020 | 2020-12-04 | North America/USA/Alabama/Houston ( | Eurofins Diatherix | Hudsonalpha Genome Sequencing Center | Jane Grimwood et al |
| EPI_ISL_883854 | USA/AL-HGSC-JGDL/2020 | 2020-12-04 | North America/USA/Alabama/Madison | Eurofins Diatherix | Hudsonalpha Genome Sequencing Center | Jane Grimwood et al |
| EPI_ISL_883856 | USA/AL-HGSC-JGDX/2020 | 2020-12-04 | North America/USA/Alabama/St. Clair ( | Eurofins Diatherix | Hudsonalpha Genome Sequencing Center | Jane Grimwood et al |
| EPI_ISL_883874 | USA/AL-HGSC-JGFX/2020 | 2020-12-07 | North America/USA/Alabama/Madison | Eurofins Diatherix | Hudsonalpha Genome Sequencing Center | Jane Grimwood et al |
| EPI_ISL_883882 | USA/AL-HGSC-JGFC/2020 | 2020-12-07 | North America/USA/Alabama/Marshall | Eurofins Diatherix | Hudsonalpha Genome Sequencing Center | Jane Grimwood et al |
| EPI_ISL_883887 | USA/AL-HGSC-JGFN/2020 | 2020-12-07 | North America/USA/Alabama/Madison | Eurofins Diatherix | Hudsonalpha Genome Sequencing Center | Jane Grimwood et al |
| EPI_ISL_883888 | USA/AL-HGSC-JGGD/2020 | 2020-12-07 | North America/USA/Alabama/Talladega | Eurofins Diatherix | Hudsonalpha Genome Sequencing Center | Jane Grimwood et al |

| | | | | | | |
|---|---|---|---|---|---|---|
| EPI_ISL_883889 | USA/AL-HGSC-JGGF/2020 | 2020-12-07 | North America/USA/Alabama/Shelby Co | Eurofins Diatherix | Hudsonalpha Genome Sequencing Center | Jane Grimwood et al |
| EPI_ISL_883903 | USA/AL-HGSC-JGIH/2020 | 2020-12-08 | North America/USA/Alabama/Marshall | Eurofins Diatherix | Hudsonalpha Genome Sequencing Center | Jane Grimwood et al |
| EPI_ISL_883904 | USA/AL-HGSC-JGII/2020 | 2020-12-08 | North America/USA/Alabama/Marshall | Eurofins Diatherix | Hudsonalpha Genome Sequencing Center | Jane Grimwood et al |
| EPI_ISL_883908 | USA/AL-HGSC-JGGA/2020 | 2020-12-07 | North America/USA/Alabama/Shelby Co | Eurofins Diatherix | Hudsonalpha Genome Sequencing Center | Jane Grimwood et al |
| EPI_ISL_883911 | USA/AL-HGSC-JGGJ/2020 | 2020-12-08 | North America/USA/Alabama/Madison | Eurofins Diatherix | Hudsonalpha Genome Sequencing Center | Jane Grimwood et al |
| EPI_ISL_883915 | USA/AL-HGSC-JGHM/2020 | 2020-12-08 | North America/USA/Alabama/Madison | Eurofins Diatherix | Hudsonalpha Genome Sequencing Center | Jane Grimwood et al |
| EPI_ISL_883921 | USA/AL-HGSC-JGEL/2020 | 2020-12-04 | North America/USA/Alabama/Geneva C | Eurofins Diatherix | Hudsonalpha Genome Sequencing Center | Jane Grimwood et al |
| EPI_ISL_883925 | USA/AL-HGSC-JGFQ/2020 | 2020-12-07 | North America/USA/Alabama/Lauderda | Eurofins Diatherix | Hudsonalpha Genome Sequencing Center | Jane Grimwood et al |
| EPI_ISL_883927 | USA/AL-HGSC-JGFU/2020 | 2020-12-07 | North America/USA/Alabama/Jefferson | Eurofins Diatherix | Hudsonalpha Genome Sequencing Center | Jane Grimwood et al |
| EPI_ISL_883929 | USA/AL-HGSC-JGGM/2020 | 2020-12-08 | North America/USA/Alabama/Jefferson | Eurofins Diatherix | Hudsonalpha Genome Sequencing Center | Jane Grimwood et al |

| | | | | | | |
|---|---|---|---|---|---|---|
| EPI_ISL_883937 | USA/AL-HGSC-JGEK/2020 | 2020-12-04 | North America/USA/Alabama/Calhoun ( | Eurofins Diatherix | Hudsonalpha Genome Sequencing Center | Jane Grimwood et al |
| EPI_ISL_883948 | USA/AL-HGSC-JGFZ/2020 | 2020-12-07 | North America/USA/Alabama/Cleburne | Eurofins Diatherix | Hudsonalpha Genome Sequencing Center | Jane Grimwood et al |
| EPI_ISL_883956 | USA/AL-HGSC-JGIB/2020 | 2020-12-08 | North America/USA/Alabama/Limestone | Eurofins Diatherix | Hudsonalpha Genome Sequencing Center | Jane Grimwood et al |
| EPI_ISL_883957 | Indonesia/BT-SHSIU-01-6547/2021 | 2021-01-06 | Asia/Indonesia/Banten/ | Medical Research Center, Faculty of Medicine, Syarif Hidayatullah State Islamic University Jakarta | Medical Research Center, Faculty of Medicine, Syarif Hidayatullah State Islamic University Jakarta | Erike A Suwarsono et al |
| EPI_ISL_883987 | France/NOR-IPP00577/2021 | 2021-01-06 | Europe/France/Normandie/Cherbourg-C | Hopital | National Reference Center for Viruses of Respiratory Infections, Institut Pasteur, Paris | Marion Barbet et al |
| EPI_ISL_883989 | France/NOR-IPP00579/2021 | 2021-01-07 | Europe/France/Normandie/Cherbourg-C | Hopital | National Reference Center for Viruses of Respiratory Infections, Institut Pasteur, Paris | Marion Barbet et al |
| EPI_ISL_883999 | FrenchGuiana/IPP00591/2020 | 2020-12-23 | Europe/France/French Guiana/Cayenne | Labo Analyses Med | National Reference Center for Viruses of Respiratory Infections, Institut Pasteur, Paris | Marion Barbet et al |

| | | | | | | |
|---|---|---|---|---|---|---|
| EPI_ISL_884007 | Mayotte/IPP00600/2021 | 2021-01-09 | Europe/France/Mayotte/Mamoudzou | hopital | National Reference Center for Viruses of Respiratory Infections, Institut Pasteur, Paris | Marion Barbet et al |
| EPI_ISL_884015 | USA/NY-Wadsworth-21008998-01/20 | 2021-01-15 | North America/USA/New York/Warren | Wadsworth Center, New York State Department of Health | Wadsworth Center, New York State Department of Health | Kirsten St. George et al |
| EPI_ISL_884018 | USA/NY-Wadsworth-21009550-01/20 | 2021-01-15 | North America/USA/New York/Renssela | ALBANY MEDICAL CENTER HOSPITAL CLINICAL LABORATORIES | Wadsworth Center, New York State Department of Health | Kirsten St. George et al |
| EPI_ISL_884025 | USA/NY-Wadsworth-21009513-01/20 | 2021-01-14 | North America/USA/New York/Renssela | ALBANY MEDICAL CENTER HOSPITAL CLINICAL LABORATORIES | Wadsworth Center, New York State Department of Health | Kirsten St. George et al |
| EPI_ISL_884030 | USA/NY-Wadsworth-21009518-01/20 | 2021-01-14 | North America/USA/New York/Schohari | ALBANY MEDICAL CENTER HOSPITAL CLINICAL LABORATORIES | Wadsworth Center, New York State Department of Health | Kirsten St. George et al |
| EPI_ISL_884032 | USA/NY-Wadsworth-21006487-01/20 | 2021-01-13 | North America/USA/New York/Hamilto | Wadsworth Center, New York State Department of Health | Wadsworth Center, New York State Department of Health | Kirsten St. George et al |
| EPI_ISL_884035 | USA/NY-Wadsworth-21006764-01/20 | 2021-01-13 | North America/USA/New York/Schenec | Wadsworth Center, New York State Department of Health | Wadsworth Center, New York State Department of Health | Kirsten St. George et al |
| EPI_ISL_884036 | USA/NY-Wadsworth-21006797-01/20 | 2021-01-13 | North America/USA/New York/Warren | Wadsworth Center, New York State Department of Health | Wadsworth Center, New York State Department of Health | Kirsten St. George et al |
| EPI_ISL_884038 | USA/NY-Wadsworth-21006840-01/20 | 2021-01-13 | North America/USA/New York/Greene | Wadsworth Center, New York State Department of Health | Wadsworth Center, New York State Department of Health | Kirsten St. George et al |

| | | | | | | |
|---|---|---|---|---|---|---|
| EPI_ISL_884041 | USA/NY-Wadsworth-21006868-01/20 | 2021-01-13 | North America/USA/New York/Greene ( | Wadsworth Center, New York State Department of Health | Wadsworth Center, New York State Department of Health | Kirsten St. George et al |
| EPI_ISL_884043 | USA/NY-Wadsworth-21009501-01/20 | 2021-01-13 | North America/USA/New York/Albany ( | ALBANY MEDICAL CENTER HOSPITAL CLINICAL LABORATORIES | Wadsworth Center, New York State Department of Health | Kirsten St. George et al |
| EPI_ISL_884045 | USA/NY-Wadsworth-21009504-01/20 | 2021-01-13 | North America/USA/New York/Albany ( | ALBANY MEDICAL CENTER HOSPITAL CLINICAL LABORATORIES | Wadsworth Center, New York State Department of Health | Kirsten St. George et al |
| EPI_ISL_884057 | USA/NY-Wadsworth-21005800-01/20 | 2021-01-11 | North America/USA/New York/Wayne ( | Wadsworth Center, New York State Department of Health | Wadsworth Center, New York State Department of Health | Kirsten St. George et al |
| EPI_ISL_884058 | USA/NY-Wadsworth-21005832-01/20 | 2021-01-11 | North America/USA/New York/Albany ( | Wadsworth Center, New York State Department of Health | Wadsworth Center, New York State Department of Health | Kirsten St. George et al |
| EPI_ISL_884062 | USA/NY-Wadsworth-21009473-01/20 | 2021-01-11 | North America/USA/New York/Albany ( | ALBANY MEDICAL CENTER HOSPITAL CLINICAL LABORATORIES | Wadsworth Center, New York State Department of Health | Kirsten St. George et al |
| EPI_ISL_884073 | USA/NY-Wadsworth-21009461-01/20 | 2021-01-09 | North America/USA/New York/Renssela | ALBANY MEDICAL CENTER HOSPITAL CLINICAL LABORATORIES | Wadsworth Center, New York State Department of Health | Kirsten St. George et al |
| EPI_ISL_884075 | USA/NY-Wadsworth-21009463-01/20 | 2021-01-09 | North America/USA/New York/Albany ( | ALBANY MEDICAL CENTER HOSPITAL CLINICAL LABORATORIES | Wadsworth Center, New York State Department of Health | Kirsten St. George et al |
| EPI_ISL_884077 | USA/NY-Wadsworth-21009452-01/20 | 2021-01-08 | North America/USA/New York/Albany ( | ALBANY MEDICAL CENTER HOSPITAL CLINICAL LABORATORIES | Wadsworth Center, New York State Department of Health | Kirsten St. George et al |
| EPI_ISL_884080 | USA/NY-Wadsworth-21003395-01/20 | 2021-01-07 | North America/USA/New York/Albany ( | Wadsworth Center, New York State Department of Health | Wadsworth Center, New York State Department of Health | Kirsten St. George et al |

| | | | | | | |
|---|---|---|---|---|---|---|
| EPI_ISL_884081 | USA/NY-Wadsworth-21009445-01/20 | 2021-01-07 | North America/USA/New York/Schohari | ALBANY MEDICAL CENTER HOSPITAL CLINICAL LABORATORIES | Wadsworth Center, New York State Department of Health | Kirsten St. George et al |
| EPI_ISL_884090 | USA/AL-HGSC-JGQU/2021 | 2021-01-06 | North America/USA/Alabama/Shelby Co | Eurofins Diatherix | Hudsonalpha Genome Sequencing Center | Jane Grimwood et al |
| EPI_ISL_884092 | USA/AL-HGSC-JGNI/2021 | 2021-01-05 | North America/USA/Alabama/Escambia | Eurofins Diatherix | Hudsonalpha Genome Sequencing Center | Jane Grimwood et al |
| EPI_ISL_884096 | USA/AL-HGSC-JGNM/2021 | 2021-01-05 | North America/USA/Alabama/Marshall | Eurofins Diatherix | Hudsonalpha Genome Sequencing Center | Jane Grimwood et al |
| EPI_ISL_884114 | USA/AL-HGSC-JGPH/2021 | 2021-01-05 | North America/USA/Alabama/Jefferson | Eurofins Diatherix | Hudsonalpha Genome Sequencing Center | Jane Grimwood et al |
| EPI_ISL_884122 | USA/AL-HGSC-JGPQ/2021 | 2021-01-05 | North America/USA/Alabama/Morgan C | Eurofins Diatherix | Hudsonalpha Genome Sequencing Center | Jane Grimwood et al |
| EPI_ISL_884141 | USA/AL-HGSC-JGQL/2021 | 2021-01-05 | North America/USA/Alabama/Cherokee | Eurofins Diatherix | Hudsonalpha Genome Sequencing Center | Jane Grimwood et al |
| EPI_ISL_884144 | USA/AL-HGSC-JGQP/2021 | 2021-01-05 | North America/USA/Alabama/Etowah C | Eurofins Diatherix | Hudsonalpha Genome Sequencing Center | Jane Grimwood et al |
| EPI_ISL_884150 | USA/AL-HGSC-JGQX/2021 | 2021-01-06 | North America/USA/Alabama/Jefferson | Eurofins Diatherix | Hudsonalpha Genome Sequencing Center | Jane Grimwood et al |
| EPI_ISL_884153 | USA/AL-HGSC-JGRA/2021 | 2021-01-06 | North America/USA/Alabama/Morgan C | Eurofins Diatherix | Hudsonalpha Genome Sequencing Center | Jane Grimwood et al |

| | | | | | | |
|---|---|---|---|---|---|---|
| EPI_ISL_884157 | USA/AL-HGSC-JGRE/2021 | 2021-01-06 | North America/USA/Alabama/Jefferson | Eurofins Diatherix | Hudsonalpha Genome Sequencing Center | Jane Grimwood et al |
| EPI_ISL_884165 | USA/AL-HGSC-JGRM/2021 | 2021-01-06 | North America/USA/Alabama/Jefferson | Eurofins Diatherix | Hudsonalpha Genome Sequencing Center | Jane Grimwood et al |
| EPI_ISL_884175 | USA/AL-HGSC-JGRY/2021 | 2021-01-06 | North America/USA/Alabama/Henry Co | Eurofins Diatherix | Hudsonalpha Genome Sequencing Center | Jane Grimwood et al |
| EPI_ISL_884182 | USA/WY-WPHL-21007064/2021 | 2021-01-19 | North America/USA/Wyoming/ | Wyoming Public Health Laboratory | Wyoming Public Health Laboratory | Noah Hull et al |
| EPI_ISL_884184 | USA/WY-WPHL-21006787/2021 | 2021-01-18 | North America/USA/Wyoming/ | Wyoming Public Health Laboratory | Wyoming Public Health Laboratory | Noah Hull et al |
| EPI_ISL_884195 | USA/WY-WPHL-21006330/2021 | 2021-01-16 | North America/USA/Wyoming/ | Wyoming Public Health Laboratory | Wyoming Public Health Laboratory | Noah Hull et al |
| EPI_ISL_884235 | USA/KS-KHEL-0055/2021 | 2021-01-11 | North America/USA/Kansas/ | Kansas Health and Environmental Lab | Kansas Health and Environmental Lab | Mike Grose et al |
| EPI_ISL_884247 | USA/KS-KHEL-0049/2021 | 2021-01-08 | North America/USA/Kansas/ | Kansas Health and Environmental Lab | Kansas Health and Environmental Lab | Mike Grose et al |
| EPI_ISL_884248 | USA/KS-KHEL-0044/2021 | 2021-01-07 | North America/USA/Kansas/ | Kansas Health and Environmental Lab | Kansas Health and Environmental Lab | Mike Grose et al |
| EPI_ISL_884249 | Brazil/SP-HIAE-ID06/2020 | 2020-11-13 | South America/Brazil/S√£o Paulo/ | LATE - Laborat√≥rio de T√©cnicas Especiais - Hospital Israelita Albert Einstein | LATE - Laborat√≥rio de T√©cnicas Especiais - Hospital Israelita Albert Einstein | Deyvid Amgarten et al |
| EPI_ISL_884250 | Brazil/SP-HIAE-ID28/2020 | 2020-07-31 | South America/Brazil/Maranh√£o/ | LATE - Laborat√≥rio de T√©cnicas Especiais - Hospital Israelita Albert Einstein | LATE - Laborat√≥rio de T√©cnicas Especiais - Hospital Israelita Albert Einstein | Deyvid Amgarten et al |

| | | | | | | |
|---|---|---|---|---|---|---|
| EPI_ISL_884254 | USA/MD-MDH-0665/2021 | 2021-01-04 | North America/USA/Maryland/ | Maryland Public Health Laboratory | Maryland Public Health Laboratory | Maryland Department of Health Laboratories Administration et al |
| EPI_ISL_884262 | Germany/ST-MD226/2020 | 2020-12-25 | Europe/Germany/Saxony-Anhalt/ | Institute of Medical Microbiology and Hospital Hygiene | Institute of Medical Microbiology and Hospital Hygiene | Prof. Dr. Achim Kaasch et al |
| EPI_ISL_884264 | Germany/ST-MD238/2020 | 2020-12-28 | Europe/Germany/Saxony-Anhalt/ | Institute of Medical Microbiology and Hospital Hygiene | Institute of Medical Microbiology and Hospital Hygiene | Prof. Dr. Achim Kaasch et al |
| EPI_ISL_884265 | Germany/ST-MD254/2020 | 2020-12-31 | Europe/Germany/Saxony-Anhalt/ | Institute of Medical Microbiology and Hospital Hygiene | Institute of Medical Microbiology and Hospital Hygiene | Prof. Dr. Achim Kaasch et al |
| EPI_ISL_884266 | Germany/ST-MD206/2020 | 2020-12-18 | Europe/Germany/Saxony-Anhalt/ | Institute of Medical Microbiology and Hospital Hygiene | Institute of Medical Microbiology and Hospital Hygiene | Prof. Dr. Achim Kaasch et al |
| EPI_ISL_884268 | Germany/ST-MD244/2020 | 2020-12-29 | Europe/Germany/Saxony-Anhalt/ | Institute of Medical Microbiology and Hospital Hygiene | Institute of Medical Microbiology and Hospital Hygiene | Prof. Dr. Achim Kaasch et al |
| EPI_ISL_884272 | Germany/ST-MD229/2020 | 2020-12-27 | Europe/Germany/Saxony-Anhalt/ | Institute of Medical Microbiology and Hospital Hygiene | Institute of Medical Microbiology and Hospital Hygiene | Prof. Dr. Achim Kaasch et al |
| EPI_ISL_884279 | Germany/ST-MD248/2020 | 2020-12-30 | Europe/Germany/Saxony-Anhalt/ | Institute of Medical Microbiology and Hospital Hygiene | Institute of Medical Microbiology and Hospital Hygiene | Prof. Dr. Achim Kaasch et al |
| EPI_ISL_884280 | Germany/ST-MD217/2020 | 2020-12-19 | Europe/Germany/Saxony-Anhalt/ | Institute of Medical Microbiology and Hospital Hygiene | Institute of Medical Microbiology and Hospital Hygiene | Prof. Dr. Achim Kaasch et al |
| EPI_ISL_884281 | Germany/ST-MD203/2020 | 2020-12-21 | Europe/Germany/Saxony-Anhalt/ | Institute of Medical Microbiology and Hospital Hygiene | Institute of Medical Microbiology and Hospital Hygiene | Prof. Dr. Achim Kaasch et al |

| | | | | | | |
|---|---|---|---|---|---|---|
| EPI_ISL_884282 | Germany/ST-MD215/2020 | 2020-12-21 | Europe/Germany/Saxony-Anhalt/ | Institute of Medical Microbiology and Hospital Hygiene | Institute of Medical Microbiology and Hospital Hygiene | Prof. Dr. Achim Kaasch et al |
| EPI_ISL_884288 | Germany/ST-MD258/2020 | 2020-12-28 | Europe/Germany/Saxony-Anhalt/ | Institute of Medical Microbiology and Hospital Hygiene | Institute of Medical Microbiology and Hospital Hygiene | Prof. Dr. Achim Kaasch et al |
| EPI_ISL_884290 | Germany/ST-MD260/2020 | 2020-12-28 | Europe/Germany/Saxony-Anhalt/ | Institute of Medical Microbiology and Hospital Hygiene | Institute of Medical Microbiology and Hospital Hygiene | Prof. Dr. Achim Kaasch et al |
| EPI_ISL_884310 | USA/CA-QDX-3340/2020 | 2020-09-13 | North America/USA/California/ | Infectious Diseases, Quest Diagnostics | Infectious Diseases, Quest Diagnostics | Rosenthal et al |
| EPI_ISL_884316 | USA/CO-QDX-3318/2020 | 2020-08-05 | North America/USA/Colorado/ | Infectious Diseases, Quest Diagnostics | Infectious Diseases, Quest Diagnostics | Rosenthal et al |
| EPI_ISL_884319 | USA/CT-QDX-3400/2020 | 2020-10-30 | North America/USA/Connecticut/ | Infectious Diseases, Quest Diagnostics | Infectious Diseases, Quest Diagnostics | Rosenthal et al |
| EPI_ISL_884320 | USA/CT-QDX-3401/2020 | 2020-10-18 | North America/USA/Connecticut/ | Infectious Diseases, Quest Diagnostics | Infectious Diseases, Quest Diagnostics | Rosenthal et al |
| EPI_ISL_884322 | USA/FL-QDX-3280/2020 | 2020-11-13 | North America/USA/Florida/ | Infectious Diseases, Quest Diagnostics | Infectious Diseases, Quest Diagnostics | Rosenthal et al |
| EPI_ISL_884325 | USA/FL-QDX-3283/2020 | 2020-11-13 | North America/USA/Florida/ | Infectious Diseases, Quest Diagnostics | Infectious Diseases, Quest Diagnostics | Rosenthal et al |
| EPI_ISL_884326 | USA/FL-QDX-3284/2020 | 2020-11-10 | North America/USA/Florida/ | Infectious Diseases, Quest Diagnostics | Infectious Diseases, Quest Diagnostics | Rosenthal et al |
| EPI_ISL_884336 | USA/GA-QDX-3291/2020 | 2020-11-14 | North America/USA/Georgia/ | Infectious Diseases, Quest Diagnostics | Infectious Diseases, Quest Diagnostics | Rosenthal et al |
| EPI_ISL_884338 | USA/GA-QDX-3319/2020 | 2020-07-29 | North America/USA/Georgia/ | Infectious Diseases, Quest Diagnostics | Infectious Diseases, Quest Diagnostics | Rosenthal et al |

| | | | | | | |
|---|---|---|---|---|---|---|
| EPI_ISL_884340 | USA/GA-QDX-3332/2020 | 2020-09-04 | North America/USA/Georgia/ | Infectious Diseases, Quest Diagnostics | Infectious Diseases, Quest Diagnostics | Rosenthal et al |
| EPI_ISL_884346 | USA/KS-QDX-3302/2020 | 2020-07-23 | North America/USA/Kansas/ | Infectious Diseases, Quest Diagnostics | Infectious Diseases, Quest Diagnostics | Rosenthal et al |
| EPI_ISL_884349 | USA/KS-QDX-3356/2020 | 2020-09-23 | North America/USA/Kansas/ | Infectious Diseases, Quest Diagnostics | Infectious Diseases, Quest Diagnostics | Rosenthal et al |
| EPI_ISL_884352 | USA/KS-QDX-3375/2020 | 2020-09-29 | North America/USA/Kansas/ | Infectious Diseases, Quest Diagnostics | Infectious Diseases, Quest Diagnostics | Rosenthal et al |
| EPI_ISL_884363 | USA/MO-QDX-3371/2020 | 2020-10-02 | North America/USA/Missouri/ | Infectious Diseases, Quest Diagnostics | Infectious Diseases, Quest Diagnostics | Rosenthal et al |
| EPI_ISL_884370 | USA/MO-QDX-3408/2020 | 2020-10-30 | North America/USA/Missouri/ | Infectious Diseases, Quest Diagnostics | Infectious Diseases, Quest Diagnostics | Rosenthal et al |
| EPI_ISL_884371 | USA/MO-QDX-3409/2020 | 2020-10-24 | North America/USA/Missouri/ | Infectious Diseases, Quest Diagnostics | Infectious Diseases, Quest Diagnostics | Rosenthal et al |
| EPI_ISL_884372 | USA/MS-QDX-3286/2020 | 2020-11-10 | North America/USA/Mississippi/ | Infectious Diseases, Quest Diagnostics | Infectious Diseases, Quest Diagnostics | Rosenthal et al |
| EPI_ISL_884380 | USA/NV-QDX-3265/2020 | 2020-11-06 | North America/USA/Nevada/ | Infectious Diseases, Quest Diagnostics | Infectious Diseases, Quest Diagnostics | Rosenthal et al |
| EPI_ISL_884383 | USA/NV-QDX-3268/2020 | 2020-11-12 | North America/USA/Nevada/ | Infectious Diseases, Quest Diagnostics | Infectious Diseases, Quest Diagnostics | Rosenthal et al |
| EPI_ISL_884384 | USA/NV-QDX-3269/2020 | 2020-11-11 | North America/USA/Nevada/ | Infectious Diseases, Quest Diagnostics | Infectious Diseases, Quest Diagnostics | Rosenthal et al |
| EPI_ISL_884395 | USA/OH-QDX-3324/2020 | 2020-08-01 | North America/USA/Ohio/ | Infectious Diseases, Quest Diagnostics | Infectious Diseases, Quest Diagnostics | Rosenthal et al |
| EPI_ISL_884400 | USA/OK-QDX-3275/2020 | 2020-11-03 | North America/USA/Oklahoma/ | Infectious Diseases, Quest Diagnostics | Infectious Diseases, Quest Diagnostics | Rosenthal et al |

| | | | | | | |
|---|---|---|---|---|---|---|
| EPI_ISL_884405 | USA/OK-QDX-3312/2020 | 2020-07-30 | North America/USA/Oklahoma/ | Infectious Diseases, Quest Diagnostics | Infectious Diseases, Quest Diagnostics | Rosenthal et al |
| EPI_ISL_884409 | USA/OK-QDX-3334/2020 | 2020-09-16 | North America/USA/Oklahoma/ | Infectious Diseases, Quest Diagnostics | Infectious Diseases, Quest Diagnostics | Rosenthal et al |
| EPI_ISL_884410 | USA/OK-QDX-3335/2020 | 2020-09-16 | North America/USA/Oklahoma/ | Infectious Diseases, Quest Diagnostics | Infectious Diseases, Quest Diagnostics | Rosenthal et al |
| EPI_ISL_884413 | USA/OK-QDX-3357/2020 | 2020-09-23 | North America/USA/Oklahoma/ | Infectious Diseases, Quest Diagnostics | Infectious Diseases, Quest Diagnostics | Rosenthal et al |
| EPI_ISL_884414 | USA/OK-QDX-3410/2020 | 2020-10-23 | North America/USA/Oklahoma/ | Infectious Diseases, Quest Diagnostics | Infectious Diseases, Quest Diagnostics | Rosenthal et al |
| EPI_ISL_884416 | USA/PA-QDX-3299/2020 | 2020-07-11 | North America/USA/Pennsylvania/ | Infectious Diseases, Quest Diagnostics | Infectious Diseases, Quest Diagnostics | Rosenthal et al |
| EPI_ISL_884418 | USA/TN-QDX-3379/2020 | 2020-10-20 | North America/USA/Tennessee/ | Infectious Diseases, Quest Diagnostics | Infectious Diseases, Quest Diagnostics | Rosenthal et al |
| EPI_ISL_884419 | USA/TN-QDX-3394/2020 | 2020-10-21 | North America/USA/Tennessee/ | Infectious Diseases, Quest Diagnostics | Infectious Diseases, Quest Diagnostics | Rosenthal et al |
| EPI_ISL_884425 | USA/TX-QDX-3293/2020 | 2020-07-07 | North America/USA/Texas/ | Infectious Diseases, Quest Diagnostics | Infectious Diseases, Quest Diagnostics | Rosenthal et al |
| EPI_ISL_884426 | USA/TX-QDX-3295/2020 | 2020-07-10 | North America/USA/Texas/ | Infectious Diseases, Quest Diagnostics | Infectious Diseases, Quest Diagnostics | Rosenthal et al |
| EPI_ISL_884429 | USA/TX-QDX-3298/2020 | 2020-07-14 | North America/USA/Texas/ | Infectious Diseases, Quest Diagnostics | Infectious Diseases, Quest Diagnostics | Rosenthal et al |
| EPI_ISL_884431 | USA/TX-QDX-3306/2020 | 2020-07-08 | North America/USA/Texas/ | Infectious Diseases, Quest Diagnostics | Infectious Diseases, Quest Diagnostics | Rosenthal et al |
| EPI_ISL_884433 | USA/TX-QDX-3328/2020 | 2020-09-04 | North America/USA/Texas/ | Infectious Diseases, Quest Diagnostics | Infectious Diseases, Quest Diagnostics | Rosenthal et al |

| | | | | | | |
|---|---|---|---|---|---|---|
| EPI_ISL_884449 | USA/un-UOM-1436xB10/2020 | 2020-07-01 | North America/USA/USA/ | Molecular Microbiology & Immunology, University of Missouri | Molecular Microbiology & Immunology, University of Missouri | Tang et al |
| EPI_ISL_884451 | USA/un-UOM-1436xE11/2020 | 2020-07-02 | North America/USA/USA/ | Molecular Microbiology & Immunology, University of Missouri | Molecular Microbiology & Immunology, University of Missouri | Tang et al |
| EPI_ISL_884452 | USA/un-UOM-1436xE6/2020 | 2020-07-02 | North America/USA/USA/ | Molecular Microbiology & Immunology, University of Missouri | Molecular Microbiology & Immunology, University of Missouri | Tang et al |
| EPI_ISL_884453 | USA/un-UOM-1436xE8/2020 | 2020-07-02 | North America/USA/USA/ | Molecular Microbiology & Immunology, University of Missouri | Molecular Microbiology & Immunology, University of Missouri | Tang et al |
| EPI_ISL_884454 | USA/un-UOM-1436xH6/2020 | 2020-07-02 | North America/USA/USA/ | Molecular Microbiology & Immunology, University of Missouri | Molecular Microbiology & Immunology, University of Missouri | Tang et al |
| EPI_ISL_884460 | USA/un-UOM-1452xG2/2020 | 2020-07-05 | North America/USA/USA/ | Molecular Microbiology & Immunology, University of Missouri | Molecular Microbiology & Immunology, University of Missouri | Tang et al |
| EPI_ISL_884469 | USA/un-UOM-1463xC2/2020 | 2020-07-06 | North America/USA/USA/ | Molecular Microbiology & Immunology, University of Missouri | Molecular Microbiology & Immunology, University of Missouri | Tang et al |
| EPI_ISL_884482 | USA/un-UOM-1479xG9/2020 | 2020-07-07 | North America/USA/USA/ | Molecular Microbiology & Immunology, University of Missouri | Molecular Microbiology & Immunology, University of Missouri | Tang et al |

| | | | | | | |
|---|---|---|---|---|---|---|
| EPI_ISL_884503 | USA/un-UOM-1501xA6/2020 | 2020-07-08 | North America/USA/USA/ | Molecular Microbiology & Immunology, University of Missouri | Molecular Microbiology & Immunology, University of Missouri | Tang et al |
| EPI_ISL_884508 | USA/un-UOM-1502xD12/2020 | 2020-07-07 | North America/USA/USA/ | Molecular Microbiology & Immunology, University of Missouri | Molecular Microbiology & Immunology, University of Missouri | Tang et al |
| EPI_ISL_884510 | USA/un-UOM-1503xA7/2020 | 2020-07-08 | North America/USA/USA/ | Molecular Microbiology & Immunology, University of Missouri | Molecular Microbiology & Immunology, University of Missouri | Tang et al |
| EPI_ISL_884514 | USA/un-UOM-1515xH11/2020 | 2020-07-09 | North America/USA/USA/ | Molecular Microbiology & Immunology, University of Missouri | Molecular Microbiology & Immunology, University of Missouri | Tang et al |
| EPI_ISL_884515 | USA/un-UOM-1515xH4/2020 | 2020-07-09 | North America/USA/USA/ | Molecular Microbiology & Immunology, University of Missouri | Molecular Microbiology & Immunology, University of Missouri | Tang et al |
| EPI_ISL_884519 | USA/un-UOM-1516xF10/2020 | 2020-07-09 | North America/USA/USA/ | Molecular Microbiology & Immunology, University of Missouri | Molecular Microbiology & Immunology, University of Missouri | Tang et al |
| EPI_ISL_884528 | USA/un-UOM-1530xF7/2020 | 2020-07-09 | North America/USA/USA/ | Molecular Microbiology & Immunology, University of Missouri | Molecular Microbiology & Immunology, University of Missouri | Tang et al |
| EPI_ISL_884539 | USA/un-UOM-20x1020/2020 | 2020-03-23 | North America/USA/USA/ | Molecular Microbiology & Immunology, University of Missouri | Molecular Microbiology & Immunology, University of Missouri | Tang et al |

| | | | | | | |
|---|---|---|---|---|---|---|
| EPI_ISL_884540 | USA/un-UOM-20x1023/2020 | 2020-03-24 | North America/USA/USA/ | Molecular Microbiology & Immunology, University of Missouri | Molecular Microbiology & Immunology, University of Missouri | Tang et al |
| EPI_ISL_884555 | USA/un-UOM-20x1063/2020 | 2020-03-30 | North America/USA/USA/ | Molecular Microbiology & Immunology, University of Missouri | Molecular Microbiology & Immunology, University of Missouri | Tang et al |
| EPI_ISL_884588 | USA/PA-CDC-STM-0000013-B08/2021 | 2021-01-15 | North America/USA/Pennsylvania/ | Respiratory Viruses Branch, Centers for Disease Control and Prevention | Respiratory Viruses Branch, Centers for Disease Control and Prevention | Cook et al |
| EPI_ISL_884593 | USA/FL-CDC-STM-0000013-H06/2021 | 2021-01-14 | North America/USA/Florida/ | Respiratory Viruses Branch, Centers for Disease Control and Prevention | Respiratory Viruses Branch, Centers for Disease Control and Prevention | Cook et al |
| EPI_ISL_884594 | USA/FL-CDC-STM-0000013-H05/2021 | 2021-01-14 | North America/USA/Florida/ | Respiratory Viruses Branch, Centers for Disease Control and Prevention | Respiratory Viruses Branch, Centers for Disease Control and Prevention | Cook et al |
| EPI_ISL_884602 | USA/FL-CDC-STM-0000013-F08/2021 | 2021-01-14 | North America/USA/Florida/ | Respiratory Viruses Branch, Centers for Disease Control and Prevention | Respiratory Viruses Branch, Centers for Disease Control and Prevention | Cook et al |
| EPI_ISL_884608 | USA/FL-CDC-STM-0000013-E07/2021 | 2021-01-14 | North America/USA/Florida/ | Respiratory Viruses Branch, Centers for Disease Control and Prevention | Respiratory Viruses Branch, Centers for Disease Control and Prevention | Cook et al |
| EPI_ISL_884616 | USA/IA-CDC-STM-0000013-D03/2021 | 2021-01-14 | North America/USA/Iowa/ | Respiratory Viruses Branch, Centers for Disease Control and Prevention | Respiratory Viruses Branch, Centers for Disease Control and Prevention | Cook et al |

| | | | | | | |
|---|---|---|---|---|---|---|
| EPI_ISL_884625 | USA/SC-CDC-STM-0000013-B06/2021 | 2021-01-14 | North America/USA/South Carolina/ | Respiratory Viruses Branch, Centers for Disease Control and Prevention | Respiratory Viruses Branch, Centers for Disease Control and Prevention | Cook et al |
| EPI_ISL_884638 | USA/CA-CDC-STM-0000025-E11/2021 | 2021-01-13 | North America/USA/California/ | Respiratory Viruses Branch, Centers for Disease Control and Prevention | Respiratory Viruses Branch, Centers for Disease Control and Prevention | Cook et al |
| EPI_ISL_884642 | USA/CA-CDC-STM-0000025-A10/2021 | 2021-01-13 | North America/USA/California/ | Respiratory Viruses Branch, Centers for Disease Control and Prevention | Respiratory Viruses Branch, Centers for Disease Control and Prevention | Cook et al |
| EPI_ISL_884645 | USA/CA-CDC-STM-0000014-E11/2021 | 2021-01-13 | North America/USA/California/ | Respiratory Viruses Branch, Centers for Disease Control and Prevention | Respiratory Viruses Branch, Centers for Disease Control and Prevention | Cook et al |
| EPI_ISL_884662 | USA/FL-CDC-STM-0000025-B10/2021 | 2021-01-12 | North America/USA/Florida/ | Respiratory Viruses Branch, Centers for Disease Control and Prevention | Respiratory Viruses Branch, Centers for Disease Control and Prevention | Cook et al |
| EPI_ISL_884674 | USA/FL-CDC-STM-0000014-H02/2021 | 2021-01-12 | North America/USA/Florida/ | Respiratory Viruses Branch, Centers for Disease Control and Prevention | Respiratory Viruses Branch, Centers for Disease Control and Prevention | Cook et al |
| EPI_ISL_884678 | USA/TX-CDC-STM-0000014-G07/2021 | 2021-01-12 | North America/USA/Texas/ | Respiratory Viruses Branch, Centers for Disease Control and Prevention | Respiratory Viruses Branch, Centers for Disease Control and Prevention | Cook et al |
| EPI_ISL_884681 | USA/OR-CDC-STM-0000014-G04/2021 | 2021-01-12 | North America/USA/Oregon/ | Respiratory Viruses Branch, Centers for Disease Control and Prevention | Respiratory Viruses Branch, Centers for Disease Control and Prevention | Cook et al |

| | | | | | | |
|---|---|---|---|---|---|---|
| EPI_ISL_884682 | USA/IN-CDC-STM-0000014-G01/2021 | 2021-01-12 | North America/USA/Indiana/ | Respiratory Viruses Branch, Centers for Disease Control and Prevention | Respiratory Viruses Branch, Centers for Disease Control and Prevention | Cook et al |
| EPI_ISL_884692 | USA/NC-CDC-STM-0000014-E02/2021 | 2021-01-12 | North America/USA/North Carolina/ | Respiratory Viruses Branch, Centers for Disease Control and Prevention | Respiratory Viruses Branch, Centers for Disease Control and Prevention | Cook et al |
| EPI_ISL_884701 | USA/VA-CDC-STM-0000014-D01/2021 | 2021-01-12 | North America/USA/Virginia/ | Respiratory Viruses Branch, Centers for Disease Control and Prevention | Respiratory Viruses Branch, Centers for Disease Control and Prevention | Cook et al |
| EPI_ISL_884702 | USA/GA-CDC-STM-0000014-C12/2021 | 2021-01-12 | North America/USA/Georgia/ | Respiratory Viruses Branch, Centers for Disease Control and Prevention | Respiratory Viruses Branch, Centers for Disease Control and Prevention | Cook et al |
| EPI_ISL_884723 | USA/FL-CDC-STM-0000014-A03/2021 | 2021-01-12 | North America/USA/Florida/ | Respiratory Viruses Branch, Centers for Disease Control and Prevention | Respiratory Viruses Branch, Centers for Disease Control and Prevention | Cook et al |
| EPI_ISL_884733 | USA/CA-CDC-STM-0000025-G04/2021 | 2021-01-11 | North America/USA/California/ | Respiratory Viruses Branch, Centers for Disease Control and Prevention | Respiratory Viruses Branch, Centers for Disease Control and Prevention | Cook et al |
| EPI_ISL_884750 | USA/AZ-CDC-STM-0000014-G03/2021 | 2021-01-11 | North America/USA/Arizona/ | Respiratory Viruses Branch, Centers for Disease Control and Prevention | Respiratory Viruses Branch, Centers for Disease Control and Prevention | Cook et al |
| EPI_ISL_884756 | USA/MI-CDC-STM-0000014-D10/2021 | 2021-01-11 | North America/USA/Michigan/ | Respiratory Viruses Branch, Centers for Disease Control and Prevention | Respiratory Viruses Branch, Centers for Disease Control and Prevention | Cook et al |

| | | | | | | |
|---|---|---|---|---|---|---|
| EPI_ISL_884758 | USA/FL-CDC-STM-0000014-C05/2021 | 2021-01-11 | North America/USA/Florida/ | Respiratory Viruses Branch, Centers for Disease Control and Prevention | Respiratory Viruses Branch, Centers for Disease Control and Prevention | Cook et al |
| EPI_ISL_884765 | USA/GA-CDC-STM-0000013-H09/202 | 2021-01-11 | North America/USA/Georgia/ | Respiratory Viruses Branch, Centers for Disease Control and Prevention | Respiratory Viruses Branch, Centers for Disease Control and Prevention | Cook et al |
| EPI_ISL_884769 | USA/GA-CDC-STM-0000013-F11/202 | 2021-01-11 | North America/USA/Georgia/ | Respiratory Viruses Branch, Centers for Disease Control and Prevention | Respiratory Viruses Branch, Centers for Disease Control and Prevention | Cook et al |
| EPI_ISL_884771 | USA/IN-CDC-STM-0000013-F09/2021 | 2021-01-11 | North America/USA/Indiana/ | Respiratory Viruses Branch, Centers for Disease Control and Prevention | Respiratory Viruses Branch, Centers for Disease Control and Prevention | Cook et al |
| EPI_ISL_884785 | USA/MA-CDC-STM-0000013-B01/202 | 2021-01-11 | North America/USA/Massachusetts/ | Respiratory Viruses Branch, Centers for Disease Control and Prevention | Respiratory Viruses Branch, Centers for Disease Control and Prevention | Cook et al |
| EPI_ISL_884788 | USA/IN-CDC-STM-0000013-A09/2021 | 2021-01-11 | North America/USA/Indiana/ | Respiratory Viruses Branch, Centers for Disease Control and Prevention | Respiratory Viruses Branch, Centers for Disease Control and Prevention | Cook et al |
| EPI_ISL_884796 | USA/PA-CDC-STM-0000025-G01/202 | 2021-01-10 | North America/USA/Pennsylvania/ | Respiratory Viruses Branch, Centers for Disease Control and Prevention | Respiratory Viruses Branch, Centers for Disease Control and Prevention | Cook et al |
| EPI_ISL_884797 | USA/SC-CDC-STM-0000025-F08/2021 | 2021-01-10 | North America/USA/South Carolina/ | Respiratory Viruses Branch, Centers for Disease Control and Prevention | Respiratory Viruses Branch, Centers for Disease Control and Prevention | Cook et al |

| | | | | | | |
|---|---|---|---|---|---|---|
| EPI_ISL_884801 | USA/FL-CDC-STM-0000025-E03/2021 | 2021-01-10 | North America/USA/Florida/ | Respiratory Viruses Branch, Centers for Disease Control and Prevention | Respiratory Viruses Branch, Centers for Disease Control and Prevention | Cook et al |
| EPI_ISL_884805 | USA/OR-CDC-STM-0000025-D04/2021 | 2021-01-10 | North America/USA/Oregon/ | Respiratory Viruses Branch, Centers for Disease Control and Prevention | Respiratory Viruses Branch, Centers for Disease Control and Prevention | Cook et al |
| EPI_ISL_884809 | USA/NC-CDC-STM-0000025-C07/2021 | 2021-01-10 | North America/USA/North Carolina/ | Respiratory Viruses Branch, Centers for Disease Control and Prevention | Respiratory Viruses Branch, Centers for Disease Control and Prevention | Cook et al |
| EPI_ISL_884814 | USA/MA-CDC-STM-0000025-A08/202 | 2021-01-10 | North America/USA/Massachusetts/ | Respiratory Viruses Branch, Centers for Disease Control and Prevention | Respiratory Viruses Branch, Centers for Disease Control and Prevention | Cook et al |
| EPI_ISL_884815 | USA/FL-CDC-STM-0000025-A04/2021 | 2021-01-10 | North America/USA/Florida/ | Respiratory Viruses Branch, Centers for Disease Control and Prevention | Respiratory Viruses Branch, Centers for Disease Control and Prevention | Cook et al |
| EPI_ISL_884822 | USA/CO-CDC-STM-0000025-D09/2021 | 2021-01-09 | North America/USA/Colorado/ | Respiratory Viruses Branch, Centers for Disease Control and Prevention | Respiratory Viruses Branch, Centers for Disease Control and Prevention | Cook et al |
| EPI_ISL_884826 | Ghana/WACCBIP-GS01/2020 | 2020-07-20 | Africa/Ghana/Ghana/ | Department of Biochemistry, Cell and Molecular Biology, West African Centre for Cell Biology of Infectious Pathogens (WACCBIP), University of Ghana | Department of Biochemistry, Cell and Molecular Biology, West African Centre for Cell Biology of Infectious Pathogens (WACCBIP), University of Ghana | Ngoi et al |

| | | | | | | |
|---|---|---|---|---|---|---|
| EPI_ISL_884828 | Ghana/WACCBIP-GS03/2020 | 2020-07-20 | Africa/Ghana/Ghana/ | Department of Biochemistry, Cell and Molecular Biology, West African Centre for Cell Biology of Infectious Pathogens (WACCBIP), University of Ghana | Department of Biochemistry, Cell and Molecular Biology, West African Centre for Cell Biology of Infectious Pathogens (WACCBIP), University of Ghana | Ngoi et al |
| EPI_ISL_884831 | Ghana/WACCBIP-GS06/2020 | 2020-07-20 | Africa/Ghana/Ghana/ | Department of Biochemistry, Cell and Molecular Biology, West African Centre for Cell Biology of Infectious Pathogens (WACCBIP), University of Ghana | Department of Biochemistry, Cell and Molecular Biology, West African Centre for Cell Biology of Infectious Pathogens (WACCBIP), University of Ghana | Ngoi et al |
| EPI_ISL_884832 | Ghana/WACCBIP-GS07/2020 | 2020-07-20 | Africa/Ghana/Ghana/ | Department of Biochemistry, Cell and Molecular Biology, West African Centre for Cell Biology of Infectious Pathogens (WACCBIP), University of Ghana | Department of Biochemistry, Cell and Molecular Biology, West African Centre for Cell Biology of Infectious Pathogens (WACCBIP), University of Ghana | Ngoi et al |

| | | | | | | |
|---|---|---|---|---|---|---|
| EPI_ISL_884833 | Ghana/WACCBIP-GS08/2020 | 2020-07-20 | Africa/Ghana/Ghana/ | Department of Biochemistry, Cell and Molecular Biology, West African Centre for Cell Biology of Infectious Pathogens (WACCBIP), University of Ghana | Department of Biochemistry, Cell and Molecular Biology, West African Centre for Cell Biology of Infectious Pathogens (WACCBIP), University of Ghana | Ngoi et al |
| EPI_ISL_884834 | Ghana/WACCBIP-GS09/2020 | 2020-07-20 | Africa/Ghana/Ghana/ | Department of Biochemistry, Cell and Molecular Biology, West African Centre for Cell Biology of Infectious Pathogens (WACCBIP), University of Ghana | Department of Biochemistry, Cell and Molecular Biology, West African Centre for Cell Biology of Infectious Pathogens (WACCBIP), University of Ghana | Ngoi et al |
| EPI_ISL_884835 | Ghana/WACCBIP-GS10/2020 | 2020-07-29 | Africa/Ghana/Ghana/ | Department of Biochemistry, Cell and Molecular Biology, West African Centre for Cell Biology of Infectious Pathogens (WACCBIP), University of Ghana | Department of Biochemistry, Cell and Molecular Biology, West African Centre for Cell Biology of Infectious Pathogens (WACCBIP), University of Ghana | Ngoi et al |

| | | | | | | |
|---|---|---|---|---|---|---|
| EPI_ISL_884836 | Ghana/WACCBIP-GS11/2020 | 2020-07-21 | Africa/Ghana/Ghana/ | Department of Biochemistry, Cell and Molecular Biology, West African Centre for Cell Biology of Infectious Pathogens (WACCBIP), University of Ghana | Department of Biochemistry, Cell and Molecular Biology, West African Centre for Cell Biology of Infectious Pathogens (WACCBIP), University of Ghana | Ngoi et al |
| EPI_ISL_884837 | Ghana/WACCBIP-GS12/2020 | 2020-07-21 | Africa/Ghana/Ghana/ | Department of Biochemistry, Cell and Molecular Biology, West African Centre for Cell Biology of Infectious Pathogens (WACCBIP), University of Ghana | Department of Biochemistry, Cell and Molecular Biology, West African Centre for Cell Biology of Infectious Pathogens (WACCBIP), University of Ghana | Ngoi et al |
| EPI_ISL_884839 | Ghana/WACCBIP-GS14/2020 | 2020-08-24 | Africa/Ghana/Ghana/ | Department of Biochemistry, Cell and Molecular Biology, West African Centre for Cell Biology of Infectious Pathogens (WACCBIP), University of Ghana | Department of Biochemistry, Cell and Molecular Biology, West African Centre for Cell Biology of Infectious Pathogens (WACCBIP), University of Ghana | Ngoi et al |

| | | | | | | |
|---|---|---|---|---|---|---|
| EPI_ISL_884841 | Ghana/WACCBIP-GS16/2020 | 2020-08-31 | Africa/Ghana/Ghana/ | Department of Biochemistry, Cell and Molecular Biology, West African Centre for Cell Biology of Infectious Pathogens (WACCBIP), University of Ghana | Department of Biochemistry, Cell and Molecular Biology, West African Centre for Cell Biology of Infectious Pathogens (WACCBIP), University of Ghana | Ngoi et al |
| EPI_ISL_884844 | Ghana/WACCBIP-GS21/2020 | 2020-07-22 | Africa/Ghana/Ghana/ | Department of Biochemistry, Cell and Molecular Biology, West African Centre for Cell Biology of Infectious Pathogens (WACCBIP), University of Ghana | Department of Biochemistry, Cell and Molecular Biology, West African Centre for Cell Biology of Infectious Pathogens (WACCBIP), University of Ghana | Ngoi et al |
| EPI_ISL_884851 | Ghana/WACCBIP-GS30/2020 | 2020-07-21 | Africa/Ghana/Ghana/ | Department of Biochemistry, Cell and Molecular Biology, West African Centre for Cell Biology of Infectious Pathogens (WACCBIP), University of Ghana | Department of Biochemistry, Cell and Molecular Biology, West African Centre for Cell Biology of Infectious Pathogens (WACCBIP), University of Ghana | Ngoi et al |

| | | | | | | |
|---|---|---|---|---|---|---|
| EPI_ISL_884852 | Ghana/WACCBIP-GS32/2020 | 2020-07-31 | Africa/Ghana/Ghana/ | Department of Biochemistry, Cell and Molecular Biology, West African Centre for Cell Biology of Infectious Pathogens (WACCBIP), University of Ghana | Department of Biochemistry, Cell and Molecular Biology, West African Centre for Cell Biology of Infectious Pathogens (WACCBIP), University of Ghana | Ngoi et al |
| EPI_ISL_884853 | Ghana/WACCBIP-GS33/2020 | 2020-07-31 | Africa/Ghana/Ghana/ | Department of Biochemistry, Cell and Molecular Biology, West African Centre for Cell Biology of Infectious Pathogens (WACCBIP), University of Ghana | Department of Biochemistry, Cell and Molecular Biology, West African Centre for Cell Biology of Infectious Pathogens (WACCBIP), University of Ghana | Ngoi et al |
| EPI_ISL_884854 | Ghana/WACCBIP-GS34/2020 | 2020-08-03 | Africa/Ghana/Ghana/ | Department of Biochemistry, Cell and Molecular Biology, West African Centre for Cell Biology of Infectious Pathogens (WACCBIP), University of Ghana | Department of Biochemistry, Cell and Molecular Biology, West African Centre for Cell Biology of Infectious Pathogens (WACCBIP), University of Ghana | Ngoi et al |

| | | | | | | |
|---|---|---|---|---|---|---|
| EPI_ISL_884856 | Ghana/WACCBIP-GS44/2020 | 2020-08-20 | Africa/Ghana/Ghana/ | Department of Biochemistry, Cell and Molecular Biology, West African Centre for Cell Biology of Infectious Pathogens (WACCBIP), University of Ghana | Department of Biochemistry, Cell and Molecular Biology, West African Centre for Cell Biology of Infectious Pathogens (WACCBIP), University of Ghana | Ngoi et al |
| EPI_ISL_884867 | USA/CA-CZB-12400/2020 | 2020-08-19 | North America/USA/California/ | CA DPH Viral and Rickettsial Disease Laboratory | Chan-Zuckerberg Biohub | CZB Cliahub Consortium et al |
| EPI_ISL_884869 | USA/CA-CZB-12411/2020 | 2020-08-22 | North America/USA/California/ | CA DPH Viral and Rickettsial Disease Laboratory | Chan-Zuckerberg Biohub | CZB Cliahub Consortium et al |
| EPI_ISL_884872 | USA/CA-CZB-16944/2020 | 2020-12-12 | North America/USA/California/Santa Cl | Santa Clara County Public Health Laboratory | Chan-Zuckerberg Biohub | CZB Cliahub Consortium et al |
| EPI_ISL_884894 | USA/CA-CZB-16977/2020 | 2020-12-07 | North America/USA/California/Santa Cl | Santa Clara County Public Health Laboratory | Chan-Zuckerberg Biohub | CZB Cliahub Consortium et al |
| EPI_ISL_884897 | USA/CA-CZB-16982/2020 | 2020-12-09 | North America/USA/California/Santa Cl | Santa Clara County Public Health Laboratory | Chan-Zuckerberg Biohub | CZB Cliahub Consortium et al |
| EPI_ISL_884900 | USA/CA-CZB-16986/2020 | 2020-12-10 | North America/USA/California/Santa Cl | Santa Clara County Public Health Laboratory | Chan-Zuckerberg Biohub | CZB Cliahub Consortium et al |
| EPI_ISL_884916 | USA/CA-CZB-17006/2020 | 2020-12-11 | North America/USA/California/Santa Cl | Santa Clara County Public Health Laboratory | Chan-Zuckerberg Biohub | CZB Cliahub Consortium et al |
| EPI_ISL_884924 | USA/CA-CZB-17015/2020 | 2020-12-18 | North America/USA/California/Santa Cl | Santa Clara County Public Health Laboratory | Chan-Zuckerberg Biohub | CZB Cliahub Consortium et al |
| EPI_ISL_884931 | USA/CA-CZB-17023/2020 | 2020-12-18 | North America/USA/California/Santa Cl | Santa Clara County Public Health Laboratory | Chan-Zuckerberg Biohub | CZB Cliahub Consortium et al |
| EPI_ISL_884935 | USA/CA-CZB-17028/2020 | 2020-12-17 | North America/USA/California/Santa Cl | Santa Clara County Public Health Laboratory | Chan-Zuckerberg Biohub | CZB Cliahub Consortium et al |
| EPI_ISL_884943 | USA/CA-CZB-17039/2020 | 2020-12-19 | North America/USA/California/Santa Cl | Santa Clara County Public Health Laboratory | Chan-Zuckerberg Biohub | CZB Cliahub Consortium et al |
| EPI_ISL_884944 | USA/CA-CZB-17040/2020 | 2020-12-20 | North America/USA/California/Santa Cl | Santa Clara County Public Health Laboratory | Chan-Zuckerberg Biohub | CZB Cliahub Consortium et al |
| EPI_ISL_884948 | USA/CA-CZB-17045/2020 | 2020-12-20 | North America/USA/California/Santa Cl | Santa Clara County Public Health Laboratory | Chan-Zuckerberg Biohub | CZB Cliahub Consortium et al |
| EPI_ISL_884955 | USA/CA-CZB-17054/2020 | 2020-12-21 | North America/USA/California/Santa Cl | Santa Clara County Public Health Laboratory | Chan-Zuckerberg Biohub | CZB Cliahub Consortium et al |

| | | | | | | |
|---|---|---|---|---|---|---|
| EPI_ISL_884958 | USA/CA-CZB-17058/2020 | 2020-12-13 | North America/USA/California/Santa Cl | Santa Clara County Public Health Laboratory | Chan-Zuckerberg Biohub | CZB Cliahub Consortium et al |
| EPI_ISL_884968 | USA/CA-CZB-17071/2020 | 2020-12-14 | North America/USA/California/Santa Cl | Santa Clara County Public Health Laboratory | Chan-Zuckerberg Biohub | CZB Cliahub Consortium et al |
| EPI_ISL_884970 | USA/CA-CZB-17074/2020 | 2020-12-15 | North America/USA/California/Santa Cl | Santa Clara County Public Health Laboratory | Chan-Zuckerberg Biohub | CZB Cliahub Consortium et al |
| EPI_ISL_884977 | USA/CA-CZB-17082/2020 | 2020-12-15 | North America/USA/California/Santa Cl | Santa Clara County Public Health Laboratory | Chan-Zuckerberg Biohub | CZB Cliahub Consortium et al |
| EPI_ISL_884994 | USA/CA-CZB-17103/2020 | 2020-12-28 | North America/USA/California/Santa Cl | Santa Clara County Public Health Laboratory | Chan-Zuckerberg Biohub | CZB Cliahub Consortium et al |
| EPI_ISL_884997 | USA/CA-CZB-17108/2020 | 2020-12-22 | North America/USA/California/Santa Cl | Santa Clara County Public Health Laboratory | Chan-Zuckerberg Biohub | CZB Cliahub Consortium et al |
| EPI_ISL_885005 | USA/CA-CZB-17119/2020 | 2020-12-23 | North America/USA/California/Santa Cl | Santa Clara County Public Health Laboratory | Chan-Zuckerberg Biohub | CZB Cliahub Consortium et al |
| EPI_ISL_885006 | USA/CA-CZB-17120/2020 | 2020-12-23 | North America/USA/California/Santa Cl | Santa Clara County Public Health Laboratory | Chan-Zuckerberg Biohub | CZB Cliahub Consortium et al |
| EPI_ISL_885009 | USA/CA-CZB-17128/2020 | 2020-12-27 | North America/USA/California/Santa Cl | Santa Clara County Public Health Laboratory | Chan-Zuckerberg Biohub | CZB Cliahub Consortium et al |
| EPI_ISL_885021 | USA/CA-CZB-17145/2020 | 2020-12-30 | North America/USA/California/Santa Cl | Santa Clara County Public Health Laboratory | Chan-Zuckerberg Biohub | CZB Cliahub Consortium et al |
| EPI_ISL_885031 | USA/CA-CZB-17161/2021 | 2021-01-01 | North America/USA/California/Santa Cl | Santa Clara County Public Health Laboratory | Chan-Zuckerberg Biohub | CZB Cliahub Consortium et al |
| EPI_ISL_885037 | USA/CA-CZB-17167/2021 | 2021-01-01 | North America/USA/California/Santa Cl | Santa Clara County Public Health Laboratory | Chan-Zuckerberg Biohub | CZB Cliahub Consortium et al |
| EPI_ISL_885056 | USA/CA-CZB-17207/2020 | 2020-12-21 | North America/USA/California/Orange C | Orange County Public Health Lab | Chan-Zuckerberg Biohub | CZB Cliahub Consortium et al |
| EPI_ISL_885058 | USA/CA-CZB-17209/2020 | 2020-12-21 | North America/USA/California/Orange C | Orange County Public Health Lab | Chan-Zuckerberg Biohub | CZB Cliahub Consortium et al |
| EPI_ISL_885064 | USA/CA-CZB-17216/2020 | 2020-12-21 | North America/USA/California/Orange C | Orange County Public Health Lab | Chan-Zuckerberg Biohub | CZB Cliahub Consortium et al |
| EPI_ISL_885072 | USA/CA-CZB-17224/2020 | 2020-12-22 | North America/USA/California/Orange C | Orange County Public Health Lab | Chan-Zuckerberg Biohub | CZB Cliahub Consortium et al |
| EPI_ISL_885075 | USA/CA-CZB-17227/2020 | 2020-12-21 | North America/USA/California/Orange C | Orange County Public Health Lab | Chan-Zuckerberg Biohub | CZB Cliahub Consortium et al |
| EPI_ISL_885088 | USA/CA-CZB-17242/2020 | 2020-12-21 | North America/USA/California/Orange C | Orange County Public Health Lab | Chan-Zuckerberg Biohub | CZB Cliahub Consortium et al |
| EPI_ISL_885091 | USA/CA-CZB-17245/2020 | 2020-12-22 | North America/USA/California/Orange C | Orange County Public Health Lab | Chan-Zuckerberg Biohub | CZB Cliahub Consortium et al |
| EPI_ISL_885094 | USA/CA-CZB-17248/2020 | 2020-12-22 | North America/USA/California/Orange C | Orange County Public Health Lab | Chan-Zuckerberg Biohub | CZB Cliahub Consortium et al |

| | | | | | | |
|---|---|---|---|---|---|---|
| EPI_ISL_885100 | USA/CA-CZB-17254/2020 | 2020-12-22 | North America/USA/California/Orange ( | Orange County Public Health Lab | Chan-Zuckerberg Biohub | CZB Cliahub Consortium et al |
| EPI_ISL_885103 | USA/CA-CZB-17257/2020 | 2020-12-23 | North America/USA/California/Orange ( | Orange County Public Health Lab | Chan-Zuckerberg Biohub | CZB Cliahub Consortium et al |
| EPI_ISL_885111 | USA/CA-CZB-17265/2020 | 2020-12-27 | North America/USA/California/Orange ( | Orange County Public Health Lab | Chan-Zuckerberg Biohub | CZB Cliahub Consortium et al |
| EPI_ISL_885113 | USA/CA-CZB-17267/2020 | 2020-12-28 | North America/USA/California/Orange ( | Orange County Public Health Lab | Chan-Zuckerberg Biohub | CZB Cliahub Consortium et al |
| EPI_ISL_885127 | USA/CA-CZB-17283/2020 | 2020-12-28 | North America/USA/California/Orange ( | Orange County Public Health Lab | Chan-Zuckerberg Biohub | CZB Cliahub Consortium et al |
| EPI_ISL_885131 | USA/CA-CZB-17287/2020 | 2020-12-09 | North America/USA/California/ | California Institute of Technology | Chan-Zuckerberg Biohub | CZB Cliahub Consortium et al |
| EPI_ISL_885134 | USA/CA-CZB-17290/2020 | 2020-12-12 | North America/USA/California/ | California Institute of Technology | Chan-Zuckerberg Biohub | CZB Cliahub Consortium et al |
| EPI_ISL_885138 | USA/CA-CZB-17294/2020 | 2020-12-05 | North America/USA/California/ | California Institute of Technology | Chan-Zuckerberg Biohub | CZB Cliahub Consortium et al |
| EPI_ISL_885143 | Iraq/NN_Erbil/2021 | 2021-01-20 | Asia/Iraq/Erbil/ | Central public health laboratory | Molecular Diagnostics Department, Central public health laboratory | Dler et al |
| EPI_ISL_885146 | Turkey/IST-GLAB-N038/2021 | 2021-01-14 | Europe/Turkey/Istanbul/ | Genomic Laboratory (GLAB) (Conjoint lab of Health Directorate of Istanbul and Istanbul Technical University) | Genomic Laboratory (GLAB), Istanbul Technical University | Ilker Karacan et al |
| EPI_ISL_885274 | England/ALDP-1090F50/2021 | 2021-01-15 | Europe/United Kingdom/England/ | Lighthouse Lab in Alderley Park | Wellcome Sanger Institute for the COVID-19 Genomics UK (COG-UK) Consortium | Jacquelyn Wynn et al |
| EPI_ISL_885277 | England/ALDP-108FF81/2021 | 2021-01-15 | Europe/United Kingdom/England/ | Lighthouse Lab in Alderley Park | Wellcome Sanger Institute for the COVID-19 Genomics UK (COG-UK) Consortium | Jacquelyn Wynn et al |

| | | | | | | |
|---|---|---|---|---|---|---|
| EPI_ISL_885307 | England/ALDP-1090DFC/2021 | 2021-01-15 | Europe/United Kingdom/England/ | Lighthouse Lab in Alderley Park | Wellcome Sanger Institute for the COVID-19 Genomics UK (COG-UK) Consortium | Jacquelyn Wynn et al |
| EPI_ISL_885432 | England/ALDP-10909DA/2021 | 2021-01-17 | Europe/United Kingdom/England/ | Lighthouse Lab in Alderley Park | Wellcome Sanger Institute for the COVID-19 Genomics UK (COG-UK) Consortium | Jacquelyn Wynn et al |
| EPI_ISL_886161 | USA/MO-CDC-LC0004733/2021 | 2021-01-04 | North America/USA/Missouri/ | Labcorp | Genomics and Discovery, Respiratory Viruses Branch, Division of Viral Diseases, Centers for Disease Control and Prevention | Peter W. Cook et al |
| EPI_ISL_886165 | USA/MS-CDC-LC0005063/2021 | 2021-01-05 | North America/USA/Mississippi/ | Labcorp | Genomics and Discovery, Respiratory Viruses Branch, Division of Viral Diseases, Centers for Disease Control and Prevention | Peter W. Cook et al |
| EPI_ISL_886167 | USA/KY-CDC-LC0004019/2021 | 2021-01-02 | North America/USA/Kentucky/ | Labcorp | Genomics and Discovery, Respiratory Viruses Branch, Division of Viral Diseases, Centers for Disease Control and Prevention | Peter W. Cook et al |

| | | | | | | |
|---|---|---|---|---|---|---|
| EPI_ISL_886181 | USA/WI-CDC-LC0003282/2021 | 2021-01-10 | North America/USA/Wisconsin/ | Labcorp | Genomics and Discovery, Respiratory Viruses Branch, Division of Viral Diseases, Centers for Disease Control and Prevention | Peter W. Cook et al |
| EPI_ISL_886182 | USA/SC-CDC-LC0005075/2021 | 2021-01-05 | North America/USA/South Carolina/ | Labcorp | Genomics and Discovery, Respiratory Viruses Branch, Division of Viral Diseases, Centers for Disease Control and Prevention | Peter W. Cook et al |
| EPI_ISL_886187 | USA/UT-CDC-LC0004595/2021 | 2021-01-04 | North America/USA/Utah/ | Labcorp | Genomics and Discovery, Respiratory Viruses Branch, Division of Viral Diseases, Centers for Disease Control and Prevention | Peter W. Cook et al |
| EPI_ISL_886192 | USA/CO-CDC-LC0005232/2021 | 2021-01-07 | North America/USA/Colorado/ | Labcorp | Genomics and Discovery, Respiratory Viruses Branch, Division of Viral Diseases, Centers for Disease Control and Prevention | Peter W. Cook et al |

| | | | | | | |
|---|---|---|---|---|---|---|
| EPI_ISL_886194 | USA/KY-CDC-LC0003569/2021 | 2021-01-11 | North America/USA/Kentucky/ | Labcorp | Genomics and Discovery, Respiratory Viruses Branch, Division of Viral Diseases, Centers for Disease Control and Prevention | Peter W. Cook et al |
| EPI_ISL_886202 | USA/VA-CDC-LC0004931/2021 | 2021-01-05 | North America/USA/Virginia/ | Labcorp | Genomics and Discovery, Respiratory Viruses Branch, Division of Viral Diseases, Centers for Disease Control and Prevention | Peter W. Cook et al |
| EPI_ISL_886203 | USA/KY-CDC-LC0004022/2021 | 2021-01-02 | North America/USA/Kentucky/ | Labcorp | Genomics and Discovery, Respiratory Viruses Branch, Division of Viral Diseases, Centers for Disease Control and Prevention | Peter W. Cook et al |
| EPI_ISL_886208 | USA/TX-CDC-LC0004275/2021 | 2021-01-04 | North America/USA/Texas/ | Labcorp | Genomics and Discovery, Respiratory Viruses Branch, Division of Viral Diseases, Centers for Disease Control and Prevention | Peter W. Cook et al |

| | | | | | | |
|---|---|---|---|---|---|---|
| EPI_ISL_886209 | USA/TN-CDC-LC0004553/2021 | 2021-01-04 | North America/USA/Tennessee/ | Labcorp | Genomics and Discovery, Respiratory Viruses Branch, Division of Viral Diseases, Centers for Disease Control and Prevention | Peter W. Cook et al |
| EPI_ISL_886244 | USA/PA-CDC-LC0003485/2021 | 2021-01-10 | North America/USA/Pennsylvania/ | Labcorp | Genomics and Discovery, Respiratory Viruses Branch, Division of Viral Diseases, Centers for Disease Control and Prevention | Peter W. Cook et al |
| EPI_ISL_886256 | USA/NV-CDC-LC0004402/2021 | 2021-01-03 | North America/USA/Nevada/ | Labcorp | Genomics and Discovery, Respiratory Viruses Branch, Division of Viral Diseases, Centers for Disease Control and Prevention | Peter W. Cook et al |
| EPI_ISL_886261 | USA/SC-CDC-LC0003243/2021 | 2021-01-11 | North America/USA/South Carolina/ | Labcorp | Genomics and Discovery, Respiratory Viruses Branch, Division of Viral Diseases, Centers for Disease Control and Prevention | Peter W. Cook et al |

| | | | | | | |
|---|---|---|---|---|---|---|
| EPI_ISL_886263 | USA/OH-CDC-LC0005483/2021 | 2021-01-09 | North America/USA/Ohio/ | Labcorp | Genomics and Discovery, Respiratory Viruses Branch, Division of Viral Diseases, Centers for Disease Control and Prevention | Peter W. Cook et al |
| EPI_ISL_886266 | USA/NJ-CDC-LC0003395/2021 | 2021-01-11 | North America/USA/New Jersey/ | Labcorp | Genomics and Discovery, Respiratory Viruses Branch, Division of Viral Diseases, Centers for Disease Control and Prevention | Peter W. Cook et al |
| EPI_ISL_886271 | USA/PA-CDC-LC0003480/2021 | 2021-01-10 | North America/USA/Pennsylvania/ | Labcorp | Genomics and Discovery, Respiratory Viruses Branch, Division of Viral Diseases, Centers for Disease Control and Prevention | Peter W. Cook et al |
| EPI_ISL_886283 | USA/SC-CDC-LC0003476/2021 | 2021-01-11 | North America/USA/South Carolina/ | Labcorp | Genomics and Discovery, Respiratory Viruses Branch, Division of Viral Diseases, Centers for Disease Control and Prevention | Peter W. Cook et al |

| | | | | | | |
|---|---|---|---|---|---|---|
| EPI_ISL_886300 | USA/OH-CDC-LC0004515/2021 | 2021-01-04 | North America/USA/Ohio/ | Labcorp | Genomics and Discovery, Respiratory Viruses Branch, Division of Viral Diseases, Centers for Disease Control and Prevention | Peter W. Cook et al |
| EPI_ISL_886312 | USA/ME-CDC-LC0003951/2021 | 2021-01-11 | North America/USA/Maine/ | Labcorp | Genomics and Discovery, Respiratory Viruses Branch, Division of Viral Diseases, Centers for Disease Control and Prevention | Peter W. Cook et al |
| EPI_ISL_886314 | USA/AL-CDC-LC0004974/2021 | 2021-01-05 | North America/USA/Alabama/ | Labcorp | Genomics and Discovery, Respiratory Viruses Branch, Division of Viral Diseases, Centers for Disease Control and Prevention | Peter W. Cook et al |
| EPI_ISL_886319 | USA/MD-CDC-LC0005132/2021 | 2021-01-05 | North America/USA/Maryland/ | Labcorp | Genomics and Discovery, Respiratory Viruses Branch, Division of Viral Diseases, Centers for Disease Control and Prevention | Peter W. Cook et al |

| | | | | | | |
|---|---|---|---|---|---|---|
| EPI_ISL_886323 | USA/KS-CDC-LC0004204/2021 | 2021-01-02 | North America/USA/Kansas/ | Labcorp | Genomics and Discovery, Respiratory Viruses Branch, Division of Viral Diseases, Centers for Disease Control and Prevention | Peter W. Cook et al |
| EPI_ISL_886325 | USA/WV-CDC-LC0004053/2021 | 2021-01-02 | North America/USA/West Virginia/ | Labcorp | Genomics and Discovery, Respiratory Viruses Branch, Division of Viral Diseases, Centers for Disease Control and Prevention | Peter W. Cook et al |
| EPI_ISL_886341 | USA/NV-CDC-LC0003119/2021 | 2021-01-10 | North America/USA/Nevada/ | Labcorp | Genomics and Discovery, Respiratory Viruses Branch, Division of Viral Diseases, Centers for Disease Control and Prevention | Peter W. Cook et al |
| EPI_ISL_886343 | USA/LA-CDC-LC0005021/2021 | 2021-01-05 | North America/USA/Louisiana/ | Labcorp | Genomics and Discovery, Respiratory Viruses Branch, Division of Viral Diseases, Centers for Disease Control and Prevention | Peter W. Cook et al |

| | | | | | | |
|---|---|---|---|---|---|---|
| EPI_ISL_886344 | USA/DE-CDC-LC0003711/2021 | 2021-01-11 | North America/USA/Delaware/ | Labcorp | Genomics and Discovery, Respiratory Viruses Branch, Division of Viral Diseases, Centers for Disease Control and Prevention | Peter W. Cook et al |
| EPI_ISL_886346 | USA/CT-CDC-LC0004494/2021 | 2021-01-04 | North America/USA/Connecticut/ | Labcorp | Genomics and Discovery, Respiratory Viruses Branch, Division of Viral Diseases, Centers for Disease Control and Prevention | Peter W. Cook et al |
| EPI_ISL_886361 | USA/OR-CDC-LC0004604/2020 | 2020-12-31 | North America/USA/Oregon/ | Labcorp | Genomics and Discovery, Respiratory Viruses Branch, Division of Viral Diseases, Centers for Disease Control and Prevention | Peter W. Cook et al |
| EPI_ISL_886368 | USA/IL-CDC-LC0004144/2021 | 2021-01-02 | North America/USA/Illinois/ | Labcorp | Genomics and Discovery, Respiratory Viruses Branch, Division of Viral Diseases, Centers for Disease Control and Prevention | Peter W. Cook et al |

| | | | | | | |
|---|---|---|---|---|---|---|
| EPI_ISL_886371 | USA/KS-CDC-LC0003173/2021 | 2021-01-09 | North America/USA/Kansas/ | Labcorp | Genomics and Discovery, Respiratory Viruses Branch, Division of Viral Diseases, Centers for Disease Control and Prevention | Peter W. Cook et al |
| EPI_ISL_886390 | USA/OK-CDC-LC0004259/2021 | 2021-01-04 | North America/USA/Oklahoma/ | Labcorp | Genomics and Discovery, Respiratory Viruses Branch, Division of Viral Diseases, Centers for Disease Control and Prevention | Peter W. Cook et al |
| EPI_ISL_886402 | USA/PA-CDC-LC0003431/2021 | 2021-01-10 | North America/USA/Pennsylvania/ | Labcorp | Genomics and Discovery, Respiratory Viruses Branch, Division of Viral Diseases, Centers for Disease Control and Prevention | Peter W. Cook et al |
| EPI_ISL_886404 | USA/SC-CDC-LC0003635/2021 | 2021-01-11 | North America/USA/South Carolina/ | Labcorp | Genomics and Discovery, Respiratory Viruses Branch, Division of Viral Diseases, Centers for Disease Control and Prevention | Peter W. Cook et al |

| | | | | | | |
|---|---|---|---|---|---|---|
| EPI_ISL_886405 | USA/NY-CDC-LC0003696/2021 | 2021-01-11 | North America/USA/New York/ | Labcorp | Genomics and Discovery, Respiratory Viruses Branch, Division of Viral Diseases, Centers for Disease Control and Prevention | Peter W. Cook et al |
| EPI_ISL_886408 | USA/WI-CDC-LC0004121/2021 | 2021-01-03 | North America/USA/Wisconsin/ | Labcorp | Genomics and Discovery, Respiratory Viruses Branch, Division of Viral Diseases, Centers for Disease Control and Prevention | Peter W. Cook et al |
| EPI_ISL_886409 | USA/MI-CDC-LC0005496/2021 | 2021-01-08 | North America/USA/Michigan/ | Labcorp | Genomics and Discovery, Respiratory Viruses Branch, Division of Viral Diseases, Centers for Disease Control and Prevention | Peter W. Cook et al |
| EPI_ISL_886410 | USA/IL-CDC-LC0004094/2021 | 2021-01-03 | North America/USA/Illinois/ | Labcorp | Genomics and Discovery, Respiratory Viruses Branch, Division of Viral Diseases, Centers for Disease Control and Prevention | Peter W. Cook et al |

| | | | | | | |
|---|---|---|---|---|---|---|
| EPI_ISL_886411 | USA/IL-CDC-LC0005256/2021 | 2021-01-09 | North America/USA/Illinois/ | Labcorp | Genomics and Discovery, Respiratory Viruses Branch, Division of Viral Diseases, Centers for Disease Control and Prevention | Peter W. Cook et al |
| EPI_ISL_886423 | USA/SC-CDC-LC0003420/2021 | 2021-01-10 | North America/USA/South Carolina/ | Labcorp | Genomics and Discovery, Respiratory Viruses Branch, Division of Viral Diseases, Centers for Disease Control and Prevention | Peter W. Cook et al |
| EPI_ISL_886424 | USA/DC-CDC-LC0003964/2021 | 2021-01-13 | North America/USA/Washington DC/ | Labcorp | Genomics and Discovery, Respiratory Viruses Branch, Division of Viral Diseases, Centers for Disease Control and Prevention | Peter W. Cook et al |
| EPI_ISL_886440 | USA/VA-CDC-LC0004937/2021 | 2021-01-05 | North America/USA/Virginia/ | Labcorp | Genomics and Discovery, Respiratory Viruses Branch, Division of Viral Diseases, Centers for Disease Control and Prevention | Peter W. Cook et al |

| | | | | | | |
|---|---|---|---|---|---|---|
| EPI_ISL_886444 | USA/CT-CDC-LC0004356/2021 | 2021-01-04 | North America/USA/Connecticut/ | Labcorp | Genomics and Discovery, Respiratory Viruses Branch, Division of Viral Diseases, Centers for Disease Control and Prevention | Peter W. Cook et al |
| EPI_ISL_886461 | USA/MI-CDC-LC0005506/2021 | 2021-01-07 | North America/USA/Michigan/ | Labcorp | Genomics and Discovery, Respiratory Viruses Branch, Division of Viral Diseases, Centers for Disease Control and Prevention | Peter W. Cook et al |
| EPI_ISL_886470 | USA/LA-CDC-LC0003253/2021 | 2021-01-11 | North America/USA/Louisiana/ | Labcorp | Genomics and Discovery, Respiratory Viruses Branch, Division of Viral Diseases, Centers for Disease Control and Prevention | Peter W. Cook et al |
| EPI_ISL_886472 | USA/MI-CDC-LC0004541/2020 | 2020-12-31 | North America/USA/Michigan/ | Labcorp | Genomics and Discovery, Respiratory Viruses Branch, Division of Viral Diseases, Centers for Disease Control and Prevention | Peter W. Cook et al |

| | | | | | | |
|---|---|---|---|---|---|---|
| EPI_ISL_886490 | USA/MI-CDC-LC0004546/2021 | 2021-01-03 | North America/USA/Michigan/ | Labcorp | Genomics and Discovery, Respiratory Viruses Branch, Division of Viral Diseases, Centers for Disease Control and Prevention | Peter W. Cook et al |
| EPI_ISL_886495 | USA/MN-CDC-LC0005334/2021 | 2021-01-08 | North America/USA/Minnesota/ | Labcorp | Genomics and Discovery, Respiratory Viruses Branch, Division of Viral Diseases, Centers for Disease Control and Prevention | Peter W. Cook et al |
| EPI_ISL_886504 | USA/NM-CDC-LC0005311/2021 | 2021-01-08 | North America/USA/New Mexico/ | Labcorp | Genomics and Discovery, Respiratory Viruses Branch, Division of Viral Diseases, Centers for Disease Control and Prevention | Peter W. Cook et al |
| EPI_ISL_886510 | USA/NV-CDC-LC0003423/2021 | 2021-01-10 | North America/USA/Nevada/ | Labcorp | Genomics and Discovery, Respiratory Viruses Branch, Division of Viral Diseases, Centers for Disease Control and Prevention | Peter W. Cook et al |

| | | | | | | |
|---|---|---|---|---|---|---|
| EPI_ISL_886520 | USA/KS-CDC-LC0003223/2021 | 2021-01-10 | North America/USA/Kansas/ | Labcorp | Genomics and Discovery, Respiratory Viruses Branch, Division of Viral Diseases, Centers for Disease Control and Prevention | Peter W. Cook et al |
| EPI_ISL_886545 | USA/VA-CDC-LC0003972/2021 | 2021-01-13 | North America/USA/Virginia/ | Labcorp | Genomics and Discovery, Respiratory Viruses Branch, Division of Viral Diseases, Centers for Disease Control and Prevention | Peter W. Cook et al |
| EPI_ISL_886546 | USA/NC-CDC-LC0003904/2021 | 2021-01-12 | North America/USA/North Carolina/ | Labcorp | Genomics and Discovery, Respiratory Viruses Branch, Division of Viral Diseases, Centers for Disease Control and Prevention | Peter W. Cook et al |
| EPI_ISL_886579 | USA/VA-CDC-LC0005104/2021 | 2021-01-05 | North America/USA/Virginia/ | Labcorp | Genomics and Discovery, Respiratory Viruses Branch, Division of Viral Diseases, Centers for Disease Control and Prevention | Peter W. Cook et al |

| | | | | | | |
|---|---|---|---|---|---|---|
| EPI_ISL_886581 | USA/OH-CDC-LC0004527/2021 | 2021-01-04 | North America/USA/Ohio/ | Labcorp | Genomics and Discovery, Respiratory Viruses Branch, Division of Viral Diseases, Centers for Disease Control and Prevention | Peter W. Cook et al |
| EPI_ISL_886594 | USA/MO-CDC-LC0005536/2021 | 2021-01-09 | North America/USA/Missouri/ | Labcorp | Genomics and Discovery, Respiratory Viruses Branch, Division of Viral Diseases, Centers for Disease Control and Prevention | Peter W. Cook et al |
| EPI_ISL_886605 | USA/IL-CDC-LC0004119/2021 | 2021-01-02 | North America/USA/Illinois/ | Labcorp | Genomics and Discovery, Respiratory Viruses Branch, Division of Viral Diseases, Centers for Disease Control and Prevention | Peter W. Cook et al |
| EPI_ISL_886615 | USA/WV-CDC-LC0004050/2021 | 2021-01-01 | North America/USA/West Virginia/ | Labcorp | Genomics and Discovery, Respiratory Viruses Branch, Division of Viral Diseases, Centers for Disease Control and Prevention | Peter W. Cook et al |

| | | | | | | |
|---|---|---|---|---|---|---|
| EPI_ISL_886626 | USA/NJ-CDC-LC0003391/2021 | 2021-01-11 | North America/USA/New Jersey/ | Labcorp | Genomics and Discovery, Respiratory Viruses Branch, Division of Viral Diseases, Centers for Disease Control and Prevention | Peter W. Cook et al |
| EPI_ISL_886655 | USA/WV-CDC-LC0005218/2021 | 2021-01-08 | North America/USA/West Virginia/ | Labcorp | Genomics and Discovery, Respiratory Viruses Branch, Division of Viral Diseases, Centers for Disease Control and Prevention | Peter W. Cook et al |
| EPI_ISL_886666 | USA/NY-CDC-LC0003865/2021 | 2021-01-12 | North America/USA/New York/ | Labcorp | Genomics and Discovery, Respiratory Viruses Branch, Division of Viral Diseases, Centers for Disease Control and Prevention | Peter W. Cook et al |
| EPI_ISL_886670 | USA/WV-CDC-LC0004188/2021 | 2021-01-02 | North America/USA/West Virginia/ | Labcorp | Genomics and Discovery, Respiratory Viruses Branch, Division of Viral Diseases, Centers for Disease Control and Prevention | Peter W. Cook et al |

| | | | | | | |
|---|---|---|---|---|---|---|
| EPI_ISL_886675 | USA/MS-CDC-LC0004681/2021 | 2021-01-04 | North America/USA/Mississippi/ | Labcorp | Genomics and Discovery, Respiratory Viruses Branch, Division of Viral Diseases, Centers for Disease Control and Prevention | Peter W. Cook et al |
| EPI_ISL_886681 | USA/MO-CDC-LC0004080/2021 | 2021-01-02 | North America/USA/Missouri/ | Labcorp | Genomics and Discovery, Respiratory Viruses Branch, Division of Viral Diseases, Centers for Disease Control and Prevention | Peter W. Cook et al |
| EPI_ISL_886690 | USA/GA-CDC-LC0003257/2021 | 2021-01-11 | North America/USA/Georgia/ | Labcorp | Genomics and Discovery, Respiratory Viruses Branch, Division of Viral Diseases, Centers for Disease Control and Prevention | Peter W. Cook et al |
| EPI_ISL_886707 | USA/KY-CDC-LC0003157/2021 | 2021-01-10 | North America/USA/Kentucky/ | Labcorp | Genomics and Discovery, Respiratory Viruses Branch, Division of Viral Diseases, Centers for Disease Control and Prevention | Peter W. Cook et al |

| | | | | | | |
|---|---|---|---|---|---|---|
| EPI_ISL_886715 | USA/MS-CDC-LC0005058/2021 | 2021-01-04 | North America/USA/Mississippi/ | Labcorp | Genomics and Discovery, Respiratory Viruses Branch, Division of Viral Diseases, Centers for Disease Control and Prevention | Peter W. Cook et al |
| EPI_ISL_886718 | USA/LA-CDC-LC0005003/2021 | 2021-01-05 | North America/USA/Louisiana/ | Labcorp | Genomics and Discovery, Respiratory Viruses Branch, Division of Viral Diseases, Centers for Disease Control and Prevention | Peter W. Cook et al |
| EPI_ISL_886720 | USA/CA-CDC-LC0003761/2021 | 2021-01-12 | North America/USA/California/ | Labcorp | Genomics and Discovery, Respiratory Viruses Branch, Division of Viral Diseases, Centers for Disease Control and Prevention | Peter W. Cook et al |
| EPI_ISL_886733 | USA/NM-CDC-LC0004165/2021 | 2021-01-02 | North America/USA/New Mexico/ | Labcorp | Genomics and Discovery, Respiratory Viruses Branch, Division of Viral Diseases, Centers for Disease Control and Prevention | Peter W. Cook et al |

| | | | | | | |
|---|---|---|---|---|---|---|
| EPI_ISL_886736 | USA/WA-CDC-LC0003366/2021 | 2021-01-11 | North America/USA/Washington/ | Labcorp | Genomics and Discovery, Respiratory Viruses Branch, Division of Viral Diseases, Centers for Disease Control and Prevention | Peter W. Cook et al |
| EPI_ISL_886740 | USA/WA-CDC-LC0004968/2021 | 2021-01-05 | North America/USA/Washington/ | Labcorp | Genomics and Discovery, Respiratory Viruses Branch, Division of Viral Diseases, Centers for Disease Control and Prevention | Peter W. Cook et al |
| EPI_ISL_886757 | USA/CO-CDC-LC0005558/2021 | 2021-01-09 | North America/USA/Colorado/ | Labcorp | Genomics and Discovery, Respiratory Viruses Branch, Division of Viral Diseases, Centers for Disease Control and Prevention | Peter W. Cook et al |
| EPI_ISL_886775 | USA/NC-CDC-LC0003344/2021 | 2021-01-11 | North America/USA/North Carolina/ | Labcorp | Genomics and Discovery, Respiratory Viruses Branch, Division of Viral Diseases, Centers for Disease Control and Prevention | Peter W. Cook et al |

| | | | | | | |
|---|---|---|---|---|---|---|
| EPI_ISL_886795 | USA/MI-CDC-LC0004534/2021 | 2021-01-02 | North America/USA/Michigan/ | Labcorp | Genomics and Discovery, Respiratory Viruses Branch, Division of Viral Diseases, Centers for Disease Control and Prevention | Peter W. Cook et al |
| EPI_ISL_886796 | USA/AZ-CDC-LC0005323/2021 | 2021-01-09 | North America/USA/Arizona/ | Labcorp | Genomics and Discovery, Respiratory Viruses Branch, Division of Viral Diseases, Centers for Disease Control and Prevention | Peter W. Cook et al |
| EPI_ISL_886797 | USA/UT-CDC-LC0004582/2021 | 2021-01-04 | North America/USA/Utah/ | Labcorp | Genomics and Discovery, Respiratory Viruses Branch, Division of Viral Diseases, Centers for Disease Control and Prevention | Peter W. Cook et al |
| EPI_ISL_886808 | USA/MD-CDC-LC0003986/2021 | 2021-01-13 | North America/USA/Maryland/ | Labcorp | Genomics and Discovery, Respiratory Viruses Branch, Division of Viral Diseases, Centers for Disease Control and Prevention | Peter W. Cook et al |

| | | | | | | |
|---|---|---|---|---|---|---|
| EPI_ISL_886832 | USA/WI-CDC-LC0004137/2021 | 2021-01-02 | North America/USA/Wisconsin/ | Labcorp | Genomics and Discovery, Respiratory Viruses Branch, Division of Viral Diseases, Centers for Disease Control and Prevention | Peter W. Cook et al |
| EPI_ISL_886844 | USA/TN-CDC-LC0004614/2021 | 2021-01-04 | North America/USA/Tennessee/ | Labcorp | Genomics and Discovery, Respiratory Viruses Branch, Division of Viral Diseases, Centers for Disease Control and Prevention | Peter W. Cook et al |
| EPI_ISL_886877 | USA/IN-CDC-LC0004005/2021 | 2021-01-02 | North America/USA/Indiana/ | Labcorp | Genomics and Discovery, Respiratory Viruses Branch, Division of Viral Diseases, Centers for Disease Control and Prevention | Peter W. Cook et al |
| EPI_ISL_886881 | USA/LA-CDC-LC0005099/2021 | 2021-01-05 | North America/USA/Louisiana/ | Labcorp | Genomics and Discovery, Respiratory Viruses Branch, Division of Viral Diseases, Centers for Disease Control and Prevention | Peter W. Cook et al |

| | | | | | | |
|---|---|---|---|---|---|---|
| EPI_ISL_886882 | USA/LA-CDC-LC0005096/2021 | 2021-01-05 | North America/USA/Louisiana/ | Labcorp | Genomics and Discovery, Respiratory Viruses Branch, Division of Viral Diseases, Centers for Disease Control and Prevention | Peter W. Cook et al |
| EPI_ISL_886889 | USA/NJ-CDC-LC0004834/2021 | 2021-01-04 | North America/USA/New Jersey/ | Labcorp | Genomics and Discovery, Respiratory Viruses Branch, Division of Viral Diseases, Centers for Disease Control and Prevention | Peter W. Cook et al |
| EPI_ISL_886901 | USA/OK-CDC-LC0005297/2021 | 2021-01-09 | North America/USA/Oklahoma/ | Labcorp | Genomics and Discovery, Respiratory Viruses Branch, Division of Viral Diseases, Centers for Disease Control and Prevention | Peter W. Cook et al |
| EPI_ISL_886913 | USA/KS-CDC-LC0004199/2021 | 2021-01-02 | North America/USA/Kansas/ | Labcorp | Genomics and Discovery, Respiratory Viruses Branch, Division of Viral Diseases, Centers for Disease Control and Prevention | Peter W. Cook et al |

| | | | | | | |
|---|---|---|---|---|---|---|
| EPI_ISL_886918 | USA/NY-CDC-LC0003861/2021 | 2021-01-11 | North America/USA/New York/ | Labcorp | Genomics and Discovery, Respiratory Viruses Branch, Division of Viral Diseases, Centers for Disease Control and Prevention | Peter W. Cook et al |
| EPI_ISL_886934 | USA/KY-CDC-LC0003156/2021 | 2021-01-10 | North America/USA/Kentucky/ | Labcorp | Genomics and Discovery, Respiratory Viruses Branch, Division of Viral Diseases, Centers for Disease Control and Prevention | Peter W. Cook et al |
| EPI_ISL_886938 | USA/OR-CDC-LC0004325/2021 | 2021-01-04 | North America/USA/Oregon/ | Labcorp | Genomics and Discovery, Respiratory Viruses Branch, Division of Viral Diseases, Centers for Disease Control and Prevention | Peter W. Cook et al |
| EPI_ISL_886943 | USA/WV-CDC-LC0004041/2021 | 2021-01-02 | North America/USA/West Virginia/ | Labcorp | Genomics and Discovery, Respiratory Viruses Branch, Division of Viral Diseases, Centers for Disease Control and Prevention | Peter W. Cook et al |

| | | | | | | |
|---|---|---|---|---|---|---|
| EPI_ISL_886955 | USA/FL-CDC-LC0005078/2021 | 2021-01-05 | North America/USA/Florida/ | Labcorp | Genomics and Discovery, Respiratory Viruses Branch, Division of Viral Diseases, Centers for Disease Control and Prevention | Peter W. Cook et al |
| EPI_ISL_886967 | USA/FL-CDC-LC0003665/2021 | 2021-01-11 | North America/USA/Florida/ | Labcorp | Genomics and Discovery, Respiratory Viruses Branch, Division of Viral Diseases, Centers for Disease Control and Prevention | Peter W. Cook et al |
| EPI_ISL_886972 | USA/PA-CDC-LC0005445/2021 | 2021-01-09 | North America/USA/Pennsylvania/ | Labcorp | Genomics and Discovery, Respiratory Viruses Branch, Division of Viral Diseases, Centers for Disease Control and Prevention | Peter W. Cook et al |
| EPI_ISL_886982 | USA/NV-CDC-LC0004185/2021 | 2021-01-02 | North America/USA/Nevada/ | Labcorp | Genomics and Discovery, Respiratory Viruses Branch, Division of Viral Diseases, Centers for Disease Control and Prevention | Peter W. Cook et al |

| | | | | | | |
|---|---|---|---|---|---|---|
| EPI_ISL_886987 | USA/OK-CDC-LC0004628/2021 | 2021-01-04 | North America/USA/Oklahoma/ | Labcorp | Genomics and Discovery, Respiratory Viruses Branch, Division of Viral Diseases, Centers for Disease Control and Prevention | Peter W. Cook et al |
| EPI_ISL_886990 | USA/MA-CDC-LC0003795/2021 | 2021-01-12 | North America/USA/Massachusetts/ | Labcorp | Genomics and Discovery, Respiratory Viruses Branch, Division of Viral Diseases, Centers for Disease Control and Prevention | Peter W. Cook et al |
| EPI_ISL_887000 | USA/TX-CDC-LC0004521/2021 | 2021-01-04 | North America/USA/Texas/ | Labcorp | Genomics and Discovery, Respiratory Viruses Branch, Division of Viral Diseases, Centers for Disease Control and Prevention | Peter W. Cook et al |
| EPI_ISL_887004 | USA/NY-CDC-LC0004360/2021 | 2021-01-04 | North America/USA/New York/ | Labcorp | Genomics and Discovery, Respiratory Viruses Branch, Division of Viral Diseases, Centers for Disease Control and Prevention | Peter W. Cook et al |

| | | | | | | |
|---|---|---|---|---|---|---|
| EPI_ISL_887011 | USA/DE-CDC-LC0003700/2021 | 2021-01-11 | North America/USA/Delaware/ | Labcorp | Genomics and Discovery, Respiratory Viruses Branch, Division of Viral Diseases, Centers for Disease Control and Prevention | Peter W. Cook et al |
| EPI_ISL_887015 | USA/KS-CDC-LC0004206/2021 | 2021-01-02 | North America/USA/Kansas/ | Labcorp | Genomics and Discovery, Respiratory Viruses Branch, Division of Viral Diseases, Centers for Disease Control and Prevention | Peter W. Cook et al |
| EPI_ISL_887017 | USA/PA-CDC-LC0003487/2021 | 2021-01-10 | North America/USA/Pennsylvania/ | Labcorp | Genomics and Discovery, Respiratory Viruses Branch, Division of Viral Diseases, Centers for Disease Control and Prevention | Peter W. Cook et al |
| EPI_ISL_887018 | USA/PA-CDC-LC0003486/2021 | 2021-01-10 | North America/USA/Pennsylvania/ | Labcorp | Genomics and Discovery, Respiratory Viruses Branch, Division of Viral Diseases, Centers for Disease Control and Prevention | Peter W. Cook et al |

| | | | | | | |
|---|---|---|---|---|---|---|
| EPI_ISL_887020 | USA/PR-CDC-LC0003685/2021 | 2021-01-09 | North America/USA/Puerto Rico/ | Labcorp | Genomics and Discovery, Respiratory Viruses Branch, Division of Viral Diseases, Centers for Disease Control and Prevention | Peter W. Cook et al |
| EPI_ISL_887023 | USA/PA-CDC-LC0003834/2021 | 2021-01-11 | North America/USA/Pennsylvania/ | Labcorp | Genomics and Discovery, Respiratory Viruses Branch, Division of Viral Diseases, Centers for Disease Control and Prevention | Peter W. Cook et al |
| EPI_ISL_887105 | Thailand/Bangkok-CONI-0426/2021 | 2021-01-05 | Asia/Thailand/Bangkok/ | Institute for Urban Disease Control and Prevention | COVID-19 Network Investigations (CONI) Alliance | Kamolthip Atsawawaranunt et al |
| EPI_ISL_887111 | Germany/SN-DcGC-L62670/2020 | 2020-04-21 | Europe/Germany/Saxony/Dresden | Institute of Medical Microbiology and Virology, University Hospital Carl Gustav Carus, TU Dresden | DRESDEN-concept Genome Center, CMCB, TU Dresden | Beil J. et al |
| EPI_ISL_887120 | Germany/ST-MD306/2021 | 2021-01-18 | Europe/Germany/Saxony-Anhalt/ | Institute of Medical Microbiology and Hospital Hygiene | Institute of Medical Microbiology and Hospital Hygiene | Prof. Dr. Achim Kaasch et al |
| EPI_ISL_887124 | Germany/ST-MD302/2021 | 2021-01-18 | Europe/Germany/Saxony-Anhalt/ | Institute of Medical Microbiology and Hospital Hygiene | Institute of Medical Microbiology and Hospital Hygiene | Prof. Dr. Achim Kaasch et al |
| EPI_ISL_887126 | Germany/ST-MD262/2021 | 2021-01-01 | Europe/Germany/Saxony-Anhalt/ | Institute of Medical Microbiology and Hospital Hygiene | Institute of Medical Microbiology and Hospital Hygiene | Prof. Dr. Achim Kaasch et al |

| | | | | | | |
|---|---|---|---|---|---|---|
| EPI_ISL_887127 | Germany/ST-MD265/2021 | 2021-01-01 | Europe/Germany/Saxony-Anhalt/ | Institute of Medical Microbiology and Hospital Hygiene | Institute of Medical Microbiology and Hospital Hygiene | Prof. Dr. Achim Kaasch et al |
| EPI_ISL_887130 | Germany/ST-MD296/2021 | 2021-01-11 | Europe/Germany/Saxony-Anhalt/ | Institute of Medical Microbiology and Hospital Hygiene | Institute of Medical Microbiology and Hospital Hygiene | Prof. Dr. Achim Kaasch et al |
| EPI_ISL_887133 | Germany/ST-MD292/2021 | 2021-01-08 | Europe/Germany/Saxony-Anhalt/ | Institute of Medical Microbiology and Hospital Hygiene | Institute of Medical Microbiology and Hospital Hygiene | Prof. Dr. Achim Kaasch et al |
| EPI_ISL_887134 | Germany/ST-MD295/2021 | 2021-01-07 | Europe/Germany/Saxony-Anhalt/ | Institute of Medical Microbiology and Hospital Hygiene | Institute of Medical Microbiology and Hospital Hygiene | Prof. Dr. Achim Kaasch et al |
| EPI_ISL_887136 | Germany/ST-MD273/2021 | 2021-01-13 | Europe/Germany/Saxony-Anhalt/ | Institute of Medical Microbiology and Hospital Hygiene | Institute of Medical Microbiology and Hospital Hygiene | Prof. Dr. Achim Kaasch et al |
| EPI_ISL_887137 | Germany/ST-MD276/2021 | 2021-01-14 | Europe/Germany/Saxony-Anhalt/ | Institute of Medical Microbiology and Hospital Hygiene | Institute of Medical Microbiology and Hospital Hygiene | Prof. Dr. Achim Kaasch et al |
| EPI_ISL_887138 | Germany/ST-MD277/2021 | 2021-01-15 | Europe/Germany/Saxony-Anhalt/ | Institute of Medical Microbiology and Hospital Hygiene | Institute of Medical Microbiology and Hospital Hygiene | Prof. Dr. Achim Kaasch et al |
| EPI_ISL_887142 | Germany/ST-MD263/2021 | 2021-01-02 | Europe/Germany/Saxony-Anhalt/ | Institute of Medical Microbiology and Hospital Hygiene | Institute of Medical Microbiology and Hospital Hygiene | Prof. Dr. Achim Kaasch et al |
| EPI_ISL_887146 | USA/MA-MGH-03011/2020 | 2020-07-22 | North America/USA/Massachusetts/ | Massachusetts General Hospital | Infectious Disease Program, Broad Institute of Harvard and MIT | Lemieux et al |
| EPI_ISL_887148 | USA/MA-MGH-03027/2020 | 2020-07-24 | North America/USA/Massachusetts/ | Massachusetts General Hospital | Infectious Disease Program, Broad Institute of Harvard and MIT | Lemieux et al |

| | | | | | | |
|---|---|---|---|---|---|---|
| EPI_ISL_887155 | USA/MA-MGH-03228/2020 | 2020-10-07 | North America/USA/Massachusetts/ | Massachusetts General Hospital | Infectious Disease Program, Broad Institute of Harvard and MIT | Lemieux et al |
| EPI_ISL_887157 | USA/MA-MGH-03292/2020 | 2020-10-26 | North America/USA/Massachusetts/ | Massachusetts General Hospital | Infectious Disease Program, Broad Institute of Harvard and MIT | Lemieux et al |
| EPI_ISL_887165 | USA/MA-MGH-03549/2020 | 2020-11-28 | North America/USA/Massachusetts/ | Massachusetts General Hospital | Infectious Disease Program, Broad Institute of Harvard and MIT | Lemieux et al |
| EPI_ISL_887167 | Germany/BAV-PL-virotum_uzkt/2020 | 2020-03-22 | Europe/Germany/Bavaria/Munich | Protzer Lab | Protzer Lab, Gagneur Lab, Robert Koch Institut | Ulrike Protzer et al |
| EPI_ISL_887177 | Germany/BAV-PL-virotum_2zr6/2020 | 2020-03-20 | Europe/Germany/Bavaria/Munich | Protzer Lab | Protzer Lab, Gagneur Lab, Robert Koch Institut | Ulrike Protzer et al |
| EPI_ISL_887183 | Germany/BAV-PL-virotum_o6qb/2020 | 2020-03-19 | Europe/Germany/Bavaria/Munich | Protzer Lab | Protzer Lab, Gagneur Lab, Robert Koch Institut | Ulrike Protzer et al |
| EPI_ISL_887185 | Germany/BAV-PL-virotum_axp3/2020 | 2020-04-10 | Europe/Germany/Bavaria/Munich | Protzer Lab | Protzer Lab, Gagneur Lab, Robert Koch Institut | Ulrike Protzer et al |
| EPI_ISL_887186 | Germany/BAV-PL-virotum_e15j/2020 | 2020-04-30 | Europe/Germany/Bavaria/Munich | Protzer Lab | Protzer Lab, Gagneur Lab, Robert Koch Institut | Ulrike Protzer et al |
| EPI_ISL_887187 | Germany/BAV-PL-virotum_vn9f/2020 | 2020-02-03 | Europe/Germany/Bavaria/Munich | Protzer Lab | Protzer Lab, Gagneur Lab, Robert Koch Institut | Ulrike Protzer et al |
| EPI_ISL_887196 | Germany/BAV-PL-virotum_d5ro/2020 | 2020-04-28 | Europe/Germany/Bavaria/Munich | Protzer Lab | Protzer Lab, Gagneur Lab, Robert Koch Institut | Ulrike Protzer et al |

| | | | | | | |
|---|---|---|---|---|---|---|
| EPI_ISL_887205 | Germany/BAV-PL-virotum_8dp7/2020 | 2020-05-15 | Europe/Germany/Bavaria/Munich | Protzer Lab | Protzer Lab, Gagneur Lab, Robert Koch Institut | Ulrike Protzer et al |
| EPI_ISL_887206 | Germany/BAV-PL-virotum_ywb0/202 | 2020-04-30 | Europe/Germany/Bavaria/Munich | Protzer Lab | Protzer Lab, Gagneur Lab, Robert Koch Institut | Ulrike Protzer et al |
| EPI_ISL_887207 | Germany/BAV-PL-virotum_bsfz/2020 | 2020-03-20 | Europe/Germany/Bavaria/Munich | Protzer Lab | Protzer Lab, Gagneur Lab, Robert Koch Institut | Ulrike Protzer et al |
| EPI_ISL_887211 | Germany/BAV-PL-virotum_yeav/2020 | 2020-03-14 | Europe/Germany/Bavaria/Munich | Protzer Lab | Protzer Lab, Gagneur Lab, Robert Koch Institut | Ulrike Protzer et al |
| EPI_ISL_887212 | Germany/BAV-PL-virotum_152a/2020 | 2020-03-18 | Europe/Germany/Bavaria/Munich | Protzer Lab | Protzer Lab, Gagneur Lab, Robert Koch Institut | Ulrike Protzer et al |
| EPI_ISL_887218 | Germany/BAV-PL-virotum_onrg/2020 | 2020-04-28 | Europe/Germany/Bavaria/Munich | Protzer Lab | Protzer Lab, Gagneur Lab, Robert Koch Institut | Ulrike Protzer et al |
| EPI_ISL_887222 | Germany/BAV-PL-virotum_ozsu/2020 | 2020-04-10 | Europe/Germany/Bavaria/Munich | Protzer Lab | Protzer Lab, Gagneur Lab, Robert Koch Institut | Ulrike Protzer et al |
| EPI_ISL_887226 | Germany/BAV-PL-virotum_3j6r/2020 | 2020-05-20 | Europe/Germany/Bavaria/Munich | Protzer Lab | Protzer Lab, Gagneur Lab, Robert Koch Institut | Ulrike Protzer et al |
| EPI_ISL_887231 | Germany/BAV-PL-virotum_8c9h/2020 | 2020-03-29 | Europe/Germany/Bavaria/Munich | Protzer Lab | Protzer Lab, Gagneur Lab, Robert Koch Institut | Ulrike Protzer et al |
| EPI_ISL_887232 | Germany/BAV-PL-virotum_66xo/2020 | 2020-04-12 | Europe/Germany/Bavaria/Munich | Protzer Lab | Protzer Lab, Gagneur Lab, Robert Koch Institut | Ulrike Protzer et al |

| | | | | | | |
|---|---|---|---|---|---|---|
| EPI_ISL_887234 | Germany/BAV-PL-virotum_brqa/2020 | 2020-04-14 | Europe/Germany/Bavaria/Munich | Protzer Lab | Protzer Lab, Gagneur Lab, Robert Koch Institut | Ulrike Protzer et al |
| EPI_ISL_887236 | Germany/BAV-PL-virotum_g45j/2020 | 2020-04-14 | Europe/Germany/Bavaria/Munich | Protzer Lab | Protzer Lab, Gagneur Lab, Robert Koch Institut | Ulrike Protzer et al |
| EPI_ISL_887239 | Germany/BAV-PL-virotum_hc9h/2020 | 2020-05-04 | Europe/Germany/Bavaria/Munich | Protzer Lab | Protzer Lab, Gagneur Lab, Robert Koch Institut | Ulrike Protzer et al |
| EPI_ISL_887241 | Germany/BAV-PL-virotum_tbjf/2020 | 2020-03-24 | Europe/Germany/Bavaria/Munich | Protzer Lab | Protzer Lab, Gagneur Lab, Robert Koch Institut | Ulrike Protzer et al |
| EPI_ISL_887244 | Germany/BAV-PL-virotum_ioxg/2020 | 2020-02-11 | Europe/Germany/Bavaria/Munich | Protzer Lab | Protzer Lab, Gagneur Lab, Robert Koch Institut | Ulrike Protzer et al |
| EPI_ISL_887250 | Germany/BAV-PL-virotum_un9f/2020 | 2020-04-11 | Europe/Germany/Bavaria/Munich | Protzer Lab | Protzer Lab, Gagneur Lab, Robert Koch Institut | Ulrike Protzer et al |
| EPI_ISL_887251 | Germany/BAV-PL-virotum_ztrz/2020 | 2020-04-13 | Europe/Germany/Bavaria/Munich | Protzer Lab | Protzer Lab, Gagneur Lab, Robert Koch Institut | Ulrike Protzer et al |
| EPI_ISL_887262 | Germany/BAV-PL-virotum_mk0p/202 | 2020-04-17 | Europe/Germany/Bavaria/Munich | Protzer Lab | Protzer Lab, Gagneur Lab, Robert Koch Institut | Ulrike Protzer et al |
| EPI_ISL_887268 | Germany/BAV-PL-virotum_x9z2/2020 | 2020-04-15 | Europe/Germany/Bavaria/Munich | Protzer Lab | Protzer Lab, Gagneur Lab, Robert Koch Institut | Ulrike Protzer et al |
| EPI_ISL_887306 | Germany/BAV-PL-virotum_bk22/2020 | 2020-03-24 | Europe/Germany/Bavaria/Munich | Protzer Lab | Protzer Lab, Gagneur Lab, Robert Koch Institut | Ulrike Protzer et al |

| | | | | | | |
|---|---|---|---|---|---|---|
| EPI_ISL_887307 | Germany/BAV-PL-virotum_hu8l/2020 | 2020-04-06 | Europe/Germany/Bavaria/Munich | Protzer Lab | Protzer Lab, Gagneur Lab, Robert Koch Institut | Ulrike Protzer et al |
| EPI_ISL_887313 | Germany/BAV-PL-virotum_1ogu/2020 | 2020-03-25 | Europe/Germany/Bavaria/Munich | Protzer Lab | Protzer Lab, Gagneur Lab, Robert Koch Institut | Ulrike Protzer et al |
| EPI_ISL_887318 | Germany/BAV-PL-virotum_i083/2020 | 2020-03-23 | Europe/Germany/Bavaria/Munich | Protzer Lab | Protzer Lab, Gagneur Lab, Robert Koch Institut | Ulrike Protzer et al |
| EPI_ISL_887320 | Germany/BAV-PL-virotum_taug/2020 | 2020-04-21 | Europe/Germany/Bavaria/Munich | Protzer Lab | Protzer Lab, Gagneur Lab, Robert Koch Institut | Ulrike Protzer et al |
| EPI_ISL_887323 | Germany/BAV-PL-virotum_ejgo/2020 | 2020-03-21 | Europe/Germany/Bavaria/Munich | Protzer Lab | Protzer Lab, Gagneur Lab, Robert Koch Institut | Ulrike Protzer et al |
| EPI_ISL_887324 | Germany/BAV-PL-virotum_ejvd/2020 | 2020-03-16 | Europe/Germany/Bavaria/Munich | Protzer Lab | Protzer Lab, Gagneur Lab, Robert Koch Institut | Ulrike Protzer et al |
| EPI_ISL_887342 | Germany/BAV-PL-virotum_frhk/2020 | 2020-04-05 | Europe/Germany/Bavaria/Munich | Protzer Lab | Protzer Lab, Gagneur Lab, Robert Koch Institut | Ulrike Protzer et al |
| EPI_ISL_887343 | Germany/BAV-PL-virotum_hr88/2020 | 2020-04-08 | Europe/Germany/Bavaria/Munich | Protzer Lab | Protzer Lab, Gagneur Lab, Robert Koch Institut | Ulrike Protzer et al |
| EPI_ISL_887348 | Germany/BAV-PL-virotum_pufy/2020 | 2020-04-06 | Europe/Germany/Bavaria/Munich | Protzer Lab | Protzer Lab, Gagneur Lab, Robert Koch Institut | Ulrike Protzer et al |
| EPI_ISL_887350 | Germany/BAV-PL-virotum_rri8/2020 | 2020-03-27 | Europe/Germany/Bavaria/Munich | Protzer Lab | Protzer Lab, Gagneur Lab, Robert Koch Institut | Ulrike Protzer et al |

| | | | | | | |
|---|---|---|---|---|---|---|
| EPI_ISL_887356 | Germany/BAV-PL-virotum_0iql/2020 | 2020-04-08 | Europe/Germany/Bavaria/Munich | Protzer Lab | Protzer Lab, Gagneur Lab, Robert Koch Institut | Ulrike Protzer et al |
| EPI_ISL_887361 | Germany/BAV-PL-virotum_5l8s/2020 | 2020-03-27 | Europe/Germany/Bavaria/Munich | Protzer Lab | Protzer Lab, Gagneur Lab, Robert Koch Institut | Ulrike Protzer et al |
| EPI_ISL_887367 | Germany/BAV-PL-virotum_7nj4/2020 | 2020-04-08 | Europe/Germany/Bavaria/Munich | Protzer Lab | Protzer Lab, Gagneur Lab, Robert Koch Institut | Ulrike Protzer et al |
| EPI_ISL_887384 | Germany/BAV-PL-virotum_i68m/2020 | 2020-03-30 | Europe/Germany/Bavaria/Munich | Protzer Lab | Protzer Lab, Gagneur Lab, Robert Koch Institut | Ulrike Protzer et al |
| EPI_ISL_887385 | Germany/BAV-PL-virotum_ih99/2020 | 2020-04-03 | Europe/Germany/Bavaria/Munich | Protzer Lab | Protzer Lab, Gagneur Lab, Robert Koch Institut | Ulrike Protzer et al |
| EPI_ISL_887387 | Germany/BAV-PL-virotum_iyl6/2020 | 2020-04-06 | Europe/Germany/Bavaria/Munich | Protzer Lab | Protzer Lab, Gagneur Lab, Robert Koch Institut | Ulrike Protzer et al |
| EPI_ISL_887391 | Germany/BAV-PL-virotum_lpnv/2020 | 2020-04-06 | Europe/Germany/Bavaria/Munich | Protzer Lab | Protzer Lab, Gagneur Lab, Robert Koch Institut | Ulrike Protzer et al |
| EPI_ISL_887393 | Germany/BAV-PL-virotum_m4c8/202 | 2020-03-31 | Europe/Germany/Bavaria/Munich | Protzer Lab | Protzer Lab, Gagneur Lab, Robert Koch Institut | Ulrike Protzer et al |
| EPI_ISL_887394 | Germany/BAV-PL-virotum_o0qt/2020 | 2020-03-30 | Europe/Germany/Bavaria/Munich | Protzer Lab | Protzer Lab, Gagneur Lab, Robert Koch Institut | Ulrike Protzer et al |
| EPI_ISL_887397 | Germany/BAV-PL-virotum_pnrh/2020 | 2020-03-27 | Europe/Germany/Bavaria/Munich | Protzer Lab | Protzer Lab, Gagneur Lab, Robert Koch Institut | Ulrike Protzer et al |

| | | | | | | |
|---|---|---|---|---|---|---|
| EPI_ISL_887407 | Germany/BAV-PL-virotum_sl7d/2020 | 2020-04-02 | Europe/Germany/Bavaria/Munich | Protzer Lab | Protzer Lab, Gagneur Lab, Robert Koch Institut | Ulrike Protzer et al |
| EPI_ISL_887410 | Germany/BAV-PL-virotum_uh9o/202 | 2020-03-26 | Europe/Germany/Bavaria/Munich | Protzer Lab | Protzer Lab, Gagneur Lab, Robert Koch Institut | Ulrike Protzer et al |
| EPI_ISL_887411 | Germany/BAV-PL-virotum_v4vm/202 | 2020-04-01 | Europe/Germany/Bavaria/Munich | Protzer Lab | Protzer Lab, Gagneur Lab, Robert Koch Institut | Ulrike Protzer et al |
| EPI_ISL_887414 | Germany/BAV-PL-virotum_w09g/202 | 2020-04-01 | Europe/Germany/Bavaria/Munich | Protzer Lab | Protzer Lab, Gagneur Lab, Robert Koch Institut | Ulrike Protzer et al |
| EPI_ISL_887420 | Mozambique/INS-K007945/2020 | 2020-09-05 | Africa/Mozambique/Sofala/Beira | Instituto Nacional de Saude (INS), Mozambique | KRISP, KZN Research Innovation and Sequencing Platform | Nalia Ismael et al |
| EPI_ISL_887421 | Mozambique/INS-K007956/2020 | 2020-08-15 | Africa/Mozambique/Sofala/Beira | Instituto Nacional de Saude (INS), Mozambique | KRISP, KZN Research Innovation and Sequencing Platform | Nalia Ismael et al |
| EPI_ISL_887422 | Mozambique/INS-K007979/2021 | 2021-01-07 | Africa/Mozambique/Maputo/ | Instituto Nacional de Saude (INS), Mozambique | KRISP, KZN Research Innovation and Sequencing Platform | Nalia Ismael et al |
| EPI_ISL_887423 | Mozambique/INS-K008001/2021 | 2021-01-07 | Africa/Mozambique/Maputo/ | Instituto Nacional de Saude (INS), Mozambique | KRISP, KZN Research Innovation and Sequencing Platform | Nalia Ismael et al |

| | | | | | | |
|---|---|---|---|---|---|---|
| EPI_ISL_887426 | Mozambique/INS-K007969/2020 | 2020-09-08 | Africa/Mozambique/Sofala/Beira | Instituto Nacional de Saude (INS), Mozambique | KRISP, KZN Research Innovation and Sequencing Platform | Nalia Ismael et al |
| EPI_ISL_887427 | Mozambique/INS-K007989/2021 | 2021-01-07 | Africa/Mozambique/Maputo/ | Instituto Nacional de Saude (INS), Mozambique | KRISP, KZN Research Innovation and Sequencing Platform | Nalia Ismael et al |
| EPI_ISL_887440 | Mozambique/INS-K007918/2020 | 2020-12-27 | Africa/Mozambique/Maputo/ | Instituto Nacional de Saude (INS), Mozambique | KRISP, KZN Research Innovation and Sequencing Platform | Nalia Ismael et al |
| EPI_ISL_887450 | Mozambique/INS-K007981/2021 | 2021-01-06 | Africa/Mozambique/Maputo/ | Instituto Nacional de Saude (INS), Mozambique | KRISP, KZN Research Innovation and Sequencing Platform | Nalia Ismael et al |
| EPI_ISL_887451 | Mozambique/INS-K007984/2021 | 2021-01-07 | Africa/Mozambique/Maputo/ | Instituto Nacional de Saude (INS), Mozambique | KRISP, KZN Research Innovation and Sequencing Platform | Nalia Ismael et al |
| EPI_ISL_887461 | Mozambique/INS-K007886/2020 | 2020-11-23 | Africa/Mozambique/Maputo/ | Instituto Nacional de Saude (INS), Mozambique | KRISP, KZN Research Innovation and Sequencing Platform | Nalia Ismael et al |
| EPI_ISL_887495 | Mozambique/INS-K007934/2020 | 2020-12-24 | Africa/Mozambique/Maputo/Bagamoio | Instituto Nacional de Saude (INS), Mozambique | KRISP, KZN Research Innovation and Sequencing Platform | Nalia Ismael et al |
| EPI_ISL_887501 | Mozambique/INS-K007950/2020 | 2020-09-24 | Africa/Mozambique/Sofala/Beira | Instituto Nacional de Saude (INS), Mozambique | KRISP, KZN Research Innovation and Sequencing Platform | Nalia Ismael et al |

| | | | | | | |
|---|---|---|---|---|---|---|
| EPI_ISL_887506 | Italy/CAM-CRGS-105/2020 | 2020-04-01 | Europe/Italy/Campania/ | 1.AO Universitaria 'S. Giovanni di Dio e Ruggi D'Aragona, Scuola Medica Salernitana' Hospital / 2.UOC di Virologia e Microbiologia, Universit√† della Campania 'L. Vanvitelli' / 3.AO Universitaria 'Federico II' Napoli Hospital / 4.AORN 'San Giuseppe Moscati' Avellino Hospital / 5.AO 'San Pio - presidio G. Rummo' Benevento Hospital / 6.AO 'Sant'Anna e San Sebastiano' Caserta Hospital / 7.PO 'Maria Santissima Addolorata' Eboli Hospital / 8.Biogem Istituto di Ricerche Genetiche | 1. Genome Research Center for Health (CRGS) / 2. Laboratory of Molecular Medicine and Genomics(LMMGe) / 3. Center for Research in Pure and Applied Mathematics (CRMPA) | Giorgio Giurato et al |
| EPI_ISL_887507 | Italy/CAM-CRGS-111/2020 | 2020-04-13 | Europe/Italy/Campania/ | 1.AO Universitaria 'S. Giovanni di Dio e Ruggi D'Aragona, Scuola Medica Salernitana' Hospital / 2.UOC di Virologia e Microbiologia, Universit√† della Campania 'L. Vanvitelli' / 3.AO Universitaria 'Federico II' Napoli Hospital / 4.AORN 'San Giuseppe Moscati' Avellino Hospital / 5.AO 'San Pio - presidio G. Rummo' Benevento Hospital / 6.AO 'Sant'Anna e San Sebastiano' Caserta Hospital / 7.PO 'Maria Santissima Addolorata' Eboli Hospital / 8.Biogem Istituto di Ricerche Genetiche | 1. Genome Research Center for Health (CRGS) / 2. Laboratory of Molecular Medicine and Genomics(LMMGe) / 3. Center for Research in Pure and Applied Mathematics (CRMPA) | Giorgio Giurato et al |
| EPI_ISL_887513 | USA/MD-HP01313/2021 | 2021-01-14 | North America/USA/Maryland/ | Johns Hopkins Hospital Department of Pathology | Johns Hopkins Hospital Department of Pathology | C. Paul Morris et al |
| EPI_ISL_887517 | USA/MD-HP01310/2021 | 2021-01-15 | North America/USA/Maryland/ | Johns Hopkins Hospital Department of Pathology | Johns Hopkins Hospital Department of Pathology | C. Paul Morris et al |
| EPI_ISL_887519 | USA/MD-HP01300/2021 | 2021-01-05 | North America/USA/Maryland/ | Johns Hopkins Hospital Department of Pathology | Johns Hopkins Hospital Department of Pathology | C. Paul Morris et al |
| EPI_ISL_887528 | USA/MD-HP01307/2021 | 2021-01-15 | North America/USA/Maryland/ | Johns Hopkins Hospital Department of Pathology | Johns Hopkins Hospital Department of Pathology | C. Paul Morris et al |

| | | | | | | |
|---|---|---|---|---|---|---|
| EPI_ISL_887533 | USA/MD-HP01302/2021 | 2021-01-14 | North America/USA/Maryland/ | Johns Hopkins Hospital Department of Pathology | Johns Hopkins Hospital Department of Pathology | C. Paul Morris et al |
| EPI_ISL_887538 | USA/MD-HP01303/2021 | 2021-01-14 | North America/USA/Maryland/ | Johns Hopkins Hospital Department of Pathology | Johns Hopkins Hospital Department of Pathology | C. Paul Morris et al |
| EPI_ISL_887545 | USA/MD-HP01323/2021 | 2021-01-16 | North America/USA/Maryland/ | Johns Hopkins Hospital Department of Pathology | Johns Hopkins Hospital Department of Pathology | C. Paul Morris et al |
| EPI_ISL_887551 | USA/MD-HP01336/2021 | 2021-01-15 | North America/USA/Maryland/ | Johns Hopkins Hospital Department of Pathology | Johns Hopkins Hospital Department of Pathology | C. Paul Morris et al |
| EPI_ISL_887560 | USA/MD-HP01367/2021 | 2021-01-13 | North America/USA/Maryland/ | Johns Hopkins Hospital Department of Pathology | Johns Hopkins Hospital Department of Pathology | C. Paul Morris et al |
| EPI_ISL_887561 | USA/MD-HP01369/2021 | 2021-01-07 | North America/USA/Maryland/ | Johns Hopkins Hospital Department of Pathology | Johns Hopkins Hospital Department of Pathology | C. Paul Morris et al |
| EPI_ISL_887564 | USA/MD-HP01378/2021 | 2021-01-07 | North America/USA/Maryland/ | Johns Hopkins Hospital Department of Pathology | Johns Hopkins Hospital Department of Pathology | C. Paul Morris et al |
| EPI_ISL_887567 | USA/MD-HP01301/2021 | 2021-01-14 | North America/USA/Maryland/ | Johns Hopkins Hospital Department of Pathology | Johns Hopkins Hospital Department of Pathology | C. Paul Morris et al |
| EPI_ISL_887569 | USA/MD-HP01318/2021 | 2021-01-15 | North America/USA/Maryland/ | Johns Hopkins Hospital Department of Pathology | Johns Hopkins Hospital Department of Pathology | C. Paul Morris et al |
| EPI_ISL_887573 | USA/MD-HP01329/2021 | 2021-01-16 | North America/USA/Maryland/ | Johns Hopkins Hospital Department of Pathology | Johns Hopkins Hospital Department of Pathology | C. Paul Morris et al |

| | | | | | | |
|---|---|---|---|---|---|---|
| EPI_ISL_887582 | USA/MD-HP01357/2021 | 2021-01-14 | North America/USA/Maryland/ | Johns Hopkins Hospital Department of Pathology | Johns Hopkins Hospital Department of Pathology | C. Paul Morris et al |
| EPI_ISL_887632 | USA/MA-CDC-LC00000035/2020 | 2020-12-23 | North America/USA/Massachusetts/ | Labcorp | Genomics and Discovery, Respiratory Viruses Branch, Division of Viral Diseases, Centers for Disease Control and Prevention | Peter W. Cook et al |
| EPI_ISL_887735 | USA/VA-CDC-LC00000663/2020 | 2020-12-27 | North America/USA/Virginia/ | Labcorp | Genomics and Discovery, Respiratory Viruses Branch, Division of Viral Diseases, Centers for Disease Control and Prevention | Peter W. Cook et al |
| EPI_ISL_887739 | USA/SC-CDC-LC00002635/2021 | 2021-01-03 | North America/USA/South Carolina/ | Labcorp | Genomics and Discovery, Respiratory Viruses Branch, Division of Viral Diseases, Centers for Disease Control and Prevention | Peter W. Cook et al |
| EPI_ISL_887888 | USA/DC-CDC-LC00000481/2020 | 2020-12-27 | North America/USA/Washington DC/ | Labcorp | Genomics and Discovery, Respiratory Viruses Branch, Division of Viral Diseases, Centers for Disease Control and Prevention | Peter W. Cook et al |

| | | | | | | |
|---|---|---|---|---|---|---|
| EPI_ISL_887894 | USA/NY-CDC-LC00002309/2020 | 2020-12-31 | North America/USA/New York/ | Labcorp | Genomics and Discovery, Respiratory Viruses Branch, Division of Viral Diseases, Centers for Disease Control and Prevention | Peter W. Cook et al |
| EPI_ISL_887945 | USA/AZ-CDC-LC00001549/2021 | 2021-01-01 | North America/USA/Arizona/ | Labcorp | Genomics and Discovery, Respiratory Viruses Branch, Division of Viral Diseases, Centers for Disease Control and Prevention | Peter W. Cook et al |
| EPI_ISL_887952 | USA/IN-CDC-LC00002077/2020 | 2020-12-27 | North America/USA/Indiana/ | Labcorp | Genomics and Discovery, Respiratory Viruses Branch, Division of Viral Diseases, Centers for Disease Control and Prevention | Peter W. Cook et al |
| EPI_ISL_887953 | USA/CA-CDC-LC00001596/2020 | 2020-12-24 | North America/USA/California/ | Labcorp | Genomics and Discovery, Respiratory Viruses Branch, Division of Viral Diseases, Centers for Disease Control and Prevention | Peter W. Cook et al |

| | | | | | | |
|---|---|---|---|---|---|---|
| EPI_ISL_887957 | USA/IL-CDC-LC00001763/2020 | 2020-12-26 | North America/USA/Illinois/ | Labcorp | Genomics and Discovery, Respiratory Viruses Branch, Division of Viral Diseases, Centers for Disease Control and Prevention | Peter W. Cook et al |
| EPI_ISL_887960 | USA/CA-CDC-LC00000820/2020 | 2020-12-24 | North America/USA/California/ | Labcorp | Genomics and Discovery, Respiratory Viruses Branch, Division of Viral Diseases, Centers for Disease Control and Prevention | Peter W. Cook et al |
| EPI_ISL_887963 | USA/ID-CDC-LC00002481/2020 | 2020-12-30 | North America/USA/Idaho/ | Labcorp | Genomics and Discovery, Respiratory Viruses Branch, Division of Viral Diseases, Centers for Disease Control and Prevention | Peter W. Cook et al |
| EPI_ISL_887967 | USA/TN-CDC-LC00000693/2020 | 2020-12-27 | North America/USA/Tennessee/ | Labcorp | Genomics and Discovery, Respiratory Viruses Branch, Division of Viral Diseases, Centers for Disease Control and Prevention | Peter W. Cook et al |

| | | | | | | |
|---|---|---|---|---|---|---|
| EPI_ISL_887970 | USA/NC-CDC-LC00002696/2021 | 2021-01-04 | North America/USA/North Carolina/ | Labcorp | Genomics and Discovery, Respiratory Viruses Branch, Division of Viral Diseases, Centers for Disease Control and Prevention | Peter W. Cook et al |
| EPI_ISL_887972 | USA/WA-CDC-LC00000770/2020 | 2020-12-24 | North America/USA/Washington/ | Labcorp | Genomics and Discovery, Respiratory Viruses Branch, Division of Viral Diseases, Centers for Disease Control and Prevention | Peter W. Cook et al |
| EPI_ISL_887976 | USA/un-CDC-LC00000728/2020 | 2020-12-23 | North America/USA/USA/ | Labcorp | Genomics and Discovery, Respiratory Viruses Branch, Division of Viral Diseases, Centers for Disease Control and Prevention | Peter W. Cook et al |
| EPI_ISL_887980 | USA/NJ-CDC-LC00002700/2021 | 2021-01-04 | North America/USA/New Jersey/ | Labcorp | Genomics and Discovery, Respiratory Viruses Branch, Division of Viral Diseases, Centers for Disease Control and Prevention | Peter W. Cook et al |

| | | | | | | |
|---|---|---|---|---|---|---|
| EPI_ISL_887984 | USA/NC-CDC-LC00002655/2021 | 2021-01-03 | North America/USA/North Carolina/ | Labcorp | Genomics and Discovery, Respiratory Viruses Branch, Division of Viral Diseases, Centers for Disease Control and Prevention | Peter W. Cook et al |
| EPI_ISL_887990 | USA/un-CDC-LC00001898/2020 | 2020-12-27 | North America/USA/USA/ | Labcorp | Genomics and Discovery, Respiratory Viruses Branch, Division of Viral Diseases, Centers for Disease Control and Prevention | Peter W. Cook et al |
| EPI_ISL_888000 | USA/MI-CDC-LC00002104/2020 | 2020-12-24 | North America/USA/Michigan/ | Labcorp | Genomics and Discovery, Respiratory Viruses Branch, Division of Viral Diseases, Centers for Disease Control and Prevention | Peter W. Cook et al |
| EPI_ISL_888008 | USA/GA-CDC-LC00000974/2020 | 2020-12-26 | North America/USA/Georgia/ | Labcorp | Genomics and Discovery, Respiratory Viruses Branch, Division of Viral Diseases, Centers for Disease Control and Prevention | Peter W. Cook et al |

| | | | | | | |
|---|---|---|---|---|---|---|
| EPI_ISL_888010 | USA/GA-CDC-LC00000990/2020 | 2020-12-26 | North America/USA/Georgia/ | Labcorp | Genomics and Discovery, Respiratory Viruses Branch, Division of Viral Diseases, Centers for Disease Control and Prevention | Peter W. Cook et al |
| EPI_ISL_888014 | USA/NJ-CDC-LC00001325/2020 | 2020-12-25 | North America/USA/New Jersey/ | Labcorp | Genomics and Discovery, Respiratory Viruses Branch, Division of Viral Diseases, Centers for Disease Control and Prevention | Peter W. Cook et al |
| EPI_ISL_888015 | USA/CA-CDC-LC00001101/2020 | 2020-12-26 | North America/USA/California/ | Labcorp | Genomics and Discovery, Respiratory Viruses Branch, Division of Viral Diseases, Centers for Disease Control and Prevention | Peter W. Cook et al |
| EPI_ISL_888025 | USA/OH-CDC-LC00001448/2020 | 2020-12-28 | North America/USA/Ohio/ | Labcorp | Genomics and Discovery, Respiratory Viruses Branch, Division of Viral Diseases, Centers for Disease Control and Prevention | Peter W. Cook et al |

| | | | | | | |
|---|---|---|---|---|---|---|
| EPI_ISL_888029 | USA/WA-CDC-LC00000790/2020 | 2020-12-24 | North America/USA/Washington/ | Labcorp | Genomics and Discovery, Respiratory Viruses Branch, Division of Viral Diseases, Centers for Disease Control and Prevention | Peter W. Cook et al |
| EPI_ISL_888031 | USA/MD-CDC-LC00001528/2020 | 2020-12-28 | North America/USA/Maryland/ | Labcorp | Genomics and Discovery, Respiratory Viruses Branch, Division of Viral Diseases, Centers for Disease Control and Prevention | Peter W. Cook et al |
| EPI_ISL_888033 | USA/TX-CDC-LC00002736/2020 | 2020-12-31 | North America/USA/Texas/ | Labcorp | Genomics and Discovery, Respiratory Viruses Branch, Division of Viral Diseases, Centers for Disease Control and Prevention | Peter W. Cook et al |
| EPI_ISL_888040 | USA/MD-CDC-LC00002221/2020 | 2020-12-28 | North America/USA/Maryland/ | Labcorp | Genomics and Discovery, Respiratory Viruses Branch, Division of Viral Diseases, Centers for Disease Control and Prevention | Peter W. Cook et al |

| | | | | | | |
|---|---|---|---|---|---|---|
| EPI_ISL_888044 | USA/GA-CDC-LC00000951/2020 | 2020-12-26 | North America/USA/Georgia/ | Labcorp | Genomics and Discovery, Respiratory Viruses Branch, Division of Viral Diseases, Centers for Disease Control and Prevention | Peter W. Cook et al |
| EPI_ISL_888045 | USA/GA-CDC-LC00002906/2020 | 2020-12-30 | North America/USA/Georgia/ | Labcorp | Genomics and Discovery, Respiratory Viruses Branch, Division of Viral Diseases, Centers for Disease Control and Prevention | Peter W. Cook et al |
| EPI_ISL_888051 | USA/KY-CDC-LC00002072/2020 | 2020-12-26 | North America/USA/Kentucky/ | Labcorp | Genomics and Discovery, Respiratory Viruses Branch, Division of Viral Diseases, Centers for Disease Control and Prevention | Peter W. Cook et al |
| EPI_ISL_888055 | USA/IL-CDC-LC00001836/2020 | 2020-12-27 | North America/USA/Illinois/ | Labcorp | Genomics and Discovery, Respiratory Viruses Branch, Division of Viral Diseases, Centers for Disease Control and Prevention | Peter W. Cook et al |

| | | | | | | |
|---|---|---|---|---|---|---|
| EPI_ISL_888065 | USA/OR-CDC-LC00002397/2021 | 2021-01-02 | North America/USA/Oregon/ | Labcorp | Genomics and Discovery, Respiratory Viruses Branch, Division of Viral Diseases, Centers for Disease Control and Prevention | Peter W. Cook et al |
| EPI_ISL_888072 | USA/IL-CDC-LC00001910/2020 | 2020-12-27 | North America/USA/Illinois/ | Labcorp | Genomics and Discovery, Respiratory Viruses Branch, Division of Viral Diseases, Centers for Disease Control and Prevention | Peter W. Cook et al |
| EPI_ISL_888073 | USA/GA-CDC-LC00001205/2020 | 2020-12-27 | North America/USA/Georgia/ | Labcorp | Genomics and Discovery, Respiratory Viruses Branch, Division of Viral Diseases, Centers for Disease Control and Prevention | Peter W. Cook et al |
| EPI_ISL_888074 | USA/GA-CDC-LC00001029/2020 | 2020-12-26 | North America/USA/Georgia/ | Labcorp | Genomics and Discovery, Respiratory Viruses Branch, Division of Viral Diseases, Centers for Disease Control and Prevention | Peter W. Cook et al |

| | | | | | | |
|---|---|---|---|---|---|---|
| EPI_ISL_888077 | USA/FL-CDC-LC00002031/2020 | 2020-12-27 | North America/USA/Florida/ | Labcorp | Genomics and Discovery, Respiratory Viruses Branch, Division of Viral Diseases, Centers for Disease Control and Prevention | Peter W. Cook et al |
| EPI_ISL_888083 | USA/GA-CDC-LC00001040/2020 | 2020-12-26 | North America/USA/Georgia/ | Labcorp | Genomics and Discovery, Respiratory Viruses Branch, Division of Viral Diseases, Centers for Disease Control and Prevention | Peter W. Cook et al |
| EPI_ISL_888084 | USA/WA-CDC-LC00000718/2020 | 2020-12-23 | North America/USA/Washington/ | Labcorp | Genomics and Discovery, Respiratory Viruses Branch, Division of Viral Diseases, Centers for Disease Control and Prevention | Peter W. Cook et al |
| EPI_ISL_888091 | USA/KY-CDC-LC00002789/2021 | 2021-01-04 | North America/USA/Kentucky/ | Labcorp | Genomics and Discovery, Respiratory Viruses Branch, Division of Viral Diseases, Centers for Disease Control and Prevention | Peter W. Cook et al |

| | | | | | | |
|---|---|---|---|---|---|---|
| EPI_ISL_888110 | USA/GA-CDC-LC00001237/2020 | 2020-12-26 | North America/USA/Georgia/ | Labcorp | Genomics and Discovery, Respiratory Viruses Branch, Division of Viral Diseases, Centers for Disease Control and Prevention | Peter W. Cook et al |
| EPI_ISL_888121 | USA/TN-CDC-LC00002201/2020 | 2020-12-28 | North America/USA/Tennessee/ | Labcorp | Genomics and Discovery, Respiratory Viruses Branch, Division of Viral Diseases, Centers for Disease Control and Prevention | Peter W. Cook et al |
| EPI_ISL_888123 | USA/NC-CDC-LC00002625/2021 | 2021-01-03 | North America/USA/North Carolina/ | Labcorp | Genomics and Discovery, Respiratory Viruses Branch, Division of Viral Diseases, Centers for Disease Control and Prevention | Peter W. Cook et al |
| EPI_ISL_888125 | USA/NC-CDC-LC00002649/2021 | 2021-01-03 | North America/USA/North Carolina/ | Labcorp | Genomics and Discovery, Respiratory Viruses Branch, Division of Viral Diseases, Centers for Disease Control and Prevention | Peter W. Cook et al |

| | | | | | | |
|---|---|---|---|---|---|---|
| EPI_ISL_888138 | USA/PA-CDC-LC00001297/2020 | 2020-12-27 | North America/USA/Pennsylvania/ | Labcorp | Genomics and Discovery, Respiratory Viruses Branch, Division of Viral Diseases, Centers for Disease Control and Prevention | Peter W. Cook et al |
| EPI_ISL_888141 | USA/NY-CDC-LC00001168/2020 | 2020-12-27 | North America/USA/New York/ | Labcorp | Genomics and Discovery, Respiratory Viruses Branch, Division of Viral Diseases, Centers for Disease Control and Prevention | Peter W. Cook et al |
| EPI_ISL_888149 | USA/PA-CDC-LC00001401/2020 | 2020-12-26 | North America/USA/Pennsylvania/ | Labcorp | Genomics and Discovery, Respiratory Viruses Branch, Division of Viral Diseases, Centers for Disease Control and Prevention | Peter W. Cook et al |
| EPI_ISL_888151 | USA/CA-CDC-LC00000825/2020 | 2020-12-24 | North America/USA/California/ | Labcorp | Genomics and Discovery, Respiratory Viruses Branch, Division of Viral Diseases, Centers for Disease Control and Prevention | Peter W. Cook et al |

| | | | | | | |
|---|---|---|---|---|---|---|
| EPI_ISL_888153 | USA/WA-CDC-LC00000787/2020 | 2020-12-24 | North America/USA/Washington/ | Labcorp | Genomics and Discovery, Respiratory Viruses Branch, Division of Viral Diseases, Centers for Disease Control and Prevention | Peter W. Cook et al |
| EPI_ISL_888159 | USA/OH-CDC-LC00002383/2021 | 2021-01-01 | North America/USA/Ohio/ | Labcorp | Genomics and Discovery, Respiratory Viruses Branch, Division of Viral Diseases, Centers for Disease Control and Prevention | Peter W. Cook et al |
| EPI_ISL_888162 | USA/AZ-CDC-LC00000187/2020 | 2020-12-26 | North America/USA/Arizona/ | Labcorp | Genomics and Discovery, Respiratory Viruses Branch, Division of Viral Diseases, Centers for Disease Control and Prevention | Peter W. Cook et al |
| EPI_ISL_888164 | USA/OR-CDC-LC00000784/2020 | 2020-12-24 | North America/USA/Oregon/ | Labcorp | Genomics and Discovery, Respiratory Viruses Branch, Division of Viral Diseases, Centers for Disease Control and Prevention | Peter W. Cook et al |

| | | | | | | |
|---|---|---|---|---|---|---|
| EPI_ISL_888166 | USA/FL-CDC-LC00001141/2020 | 2020-12-25 | North America/USA/Florida/ | Labcorp | Genomics and Discovery, Respiratory Viruses Branch, Division of Viral Diseases, Centers for Disease Control and Prevention | Peter W. Cook et al |
| EPI_ISL_888169 | USA/IN-CDC-LC00001714/2020 | 2020-12-27 | North America/USA/Indiana/ | Labcorp | Genomics and Discovery, Respiratory Viruses Branch, Division of Viral Diseases, Centers for Disease Control and Prevention | Peter W. Cook et al |
| EPI_ISL_888176 | USA/NC-CDC-LC00002725/2021 | 2021-01-04 | North America/USA/North Carolina/ | Labcorp | Genomics and Discovery, Respiratory Viruses Branch, Division of Viral Diseases, Centers for Disease Control and Prevention | Peter W. Cook et al |
| EPI_ISL_888192 | USA/IN-CDC-LC00001362/2020 | 2020-12-26 | North America/USA/Indiana/ | Labcorp | Genomics and Discovery, Respiratory Viruses Branch, Division of Viral Diseases, Centers for Disease Control and Prevention | Peter W. Cook et al |

| | | | | | | |
|---|---|---|---|---|---|---|
| EPI_ISL_888207 | USA/IN-CDC-LC00002771/2021 | 2021-01-04 | North America/USA/Indiana/ | Labcorp | Genomics and Discovery, Respiratory Viruses Branch, Division of Viral Diseases, Centers for Disease Control and Prevention | Peter W. Cook et al |
| EPI_ISL_888219 | USA/NC-CDC-LC00002714/2021 | 2021-01-03 | North America/USA/North Carolina/ | Labcorp | Genomics and Discovery, Respiratory Viruses Branch, Division of Viral Diseases, Centers for Disease Control and Prevention | Peter W. Cook et al |
| EPI_ISL_888224 | USA/NJ-CDC-LC00001321/2020 | 2020-12-27 | North America/USA/New Jersey/ | Labcorp | Genomics and Discovery, Respiratory Viruses Branch, Division of Viral Diseases, Centers for Disease Control and Prevention | Peter W. Cook et al |
| EPI_ISL_888227 | USA/MD-CDC-LC00001449/2020 | 2020-12-28 | North America/USA/Maryland/ | Labcorp | Genomics and Discovery, Respiratory Viruses Branch, Division of Viral Diseases, Centers for Disease Control and Prevention | Peter W. Cook et al |

| | | | | | | |
|---|---|---|---|---|---|---|
| EPI_ISL_888230 | USA/TN-CDC-LC00001784/2020 | 2020-12-27 | North America/USA/Tennessee/ | Labcorp | Genomics and Discovery, Respiratory Viruses Branch, Division of Viral Diseases, Centers for Disease Control and Prevention | Peter W. Cook et al |
| EPI_ISL_888238 | USA/VA-CDC-LC00002676/2021 | 2021-01-03 | North America/USA/Virginia/ | Labcorp | Genomics and Discovery, Respiratory Viruses Branch, Division of Viral Diseases, Centers for Disease Control and Prevention | Peter W. Cook et al |
| EPI_ISL_888242 | USA/AZ-CDC-LC00000901/2020 | 2020-12-26 | North America/USA/Arizona/ | Labcorp | Genomics and Discovery, Respiratory Viruses Branch, Division of Viral Diseases, Centers for Disease Control and Prevention | Peter W. Cook et al |
| EPI_ISL_888248 | USA/PA-CDC-LC00001384/2020 | 2020-12-26 | North America/USA/Pennsylvania/ | Labcorp | Genomics and Discovery, Respiratory Viruses Branch, Division of Viral Diseases, Centers for Disease Control and Prevention | Peter W. Cook et al |

| | | | | | | |
|---|---|---|---|---|---|---|
| EPI_ISL_888261 | USA/PA-CDC-LC00001370/2020 | 2020-12-26 | North America/USA/Pennsylvania/ | Labcorp | Genomics and Discovery, Respiratory Viruses Branch, Division of Viral Diseases, Centers for Disease Control and Prevention | Peter W. Cook et al |
| EPI_ISL_888262 | USA/GA-CDC-LC00001208/2020 | 2020-12-26 | North America/USA/Georgia/ | Labcorp | Genomics and Discovery, Respiratory Viruses Branch, Division of Viral Diseases, Centers for Disease Control and Prevention | Peter W. Cook et al |
| EPI_ISL_888263 | USA/OK-CDC-LC00002705/2021 | 2021-01-04 | North America/USA/Oklahoma/ | Labcorp | Genomics and Discovery, Respiratory Viruses Branch, Division of Viral Diseases, Centers for Disease Control and Prevention | Peter W. Cook et al |
| EPI_ISL_888266 | USA/GA-CDC-LC00001007/2020 | 2020-12-26 | North America/USA/Georgia/ | Labcorp | Genomics and Discovery, Respiratory Viruses Branch, Division of Viral Diseases, Centers for Disease Control and Prevention | Peter W. Cook et al |

| | | | | | | |
|---|---|---|---|---|---|---|
| EPI_ISL_888317 | USA/PA-CDC-LC00001404/2020 | 2020-12-27 | North America/USA/Pennsylvania/ | Labcorp | Genomics and Discovery, Respiratory Viruses Branch, Division of Viral Diseases, Centers for Disease Control and Prevention | Peter W. Cook et al |
| EPI_ISL_888318 | USA/KS-CDC-LC00002425/2021 | 2021-01-02 | North America/USA/Kansas/ | Labcorp | Genomics and Discovery, Respiratory Viruses Branch, Division of Viral Diseases, Centers for Disease Control and Prevention | Peter W. Cook et al |
| EPI_ISL_888323 | USA/IN-CDC-LC00001608/2020 | 2020-12-26 | North America/USA/Indiana/ | Labcorp | Genomics and Discovery, Respiratory Viruses Branch, Division of Viral Diseases, Centers for Disease Control and Prevention | Peter W. Cook et al |
| EPI_ISL_888326 | USA/FL-CDC-LC00000183/2020 | 2020-12-26 | North America/USA/Florida/ | Labcorp | Genomics and Discovery, Respiratory Viruses Branch, Division of Viral Diseases, Centers for Disease Control and Prevention | Peter W. Cook et al |

| | | | | | | |
|---|---|---|---|---|---|---|
| EPI_ISL_888328 | USA/FL-CDC-LC00001068/2020 | 2020-12-26 | North America/USA/Florida/ | Labcorp | Genomics and Discovery, Respiratory Viruses Branch, Division of Viral Diseases, Centers for Disease Control and Prevention | Peter W. Cook et al |
| EPI_ISL_888329 | USA/GA-CDC-LC00000953/2020 | 2020-12-26 | North America/USA/Georgia/ | Labcorp | Genomics and Discovery, Respiratory Viruses Branch, Division of Viral Diseases, Centers for Disease Control and Prevention | Peter W. Cook et al |
| EPI_ISL_888337 | USA/GA-CDC-LC00001032/2020 | 2020-12-26 | North America/USA/Georgia/ | Labcorp | Genomics and Discovery, Respiratory Viruses Branch, Division of Viral Diseases, Centers for Disease Control and Prevention | Peter W. Cook et al |
| EPI_ISL_888345 | USA/PA-CDC-LC00001248/2020 | 2020-12-27 | North America/USA/Pennsylvania/ | Labcorp | Genomics and Discovery, Respiratory Viruses Branch, Division of Viral Diseases, Centers for Disease Control and Prevention | Peter W. Cook et al |

| | | | | | | |
|---|---|---|---|---|---|---|
| EPI_ISL_888346 | USA/VA-CDC-LC00001504/2020 | 2020-12-28 | North America/USA/Virginia/ | Labcorp | Genomics and Discovery, Respiratory Viruses Branch, Division of Viral Diseases, Centers for Disease Control and Prevention | Peter W. Cook et al |
| EPI_ISL_888352 | USA/WA-CDC-LC00000716/2020 | 2020-12-23 | North America/USA/Washington/ | Labcorp | Genomics and Discovery, Respiratory Viruses Branch, Division of Viral Diseases, Centers for Disease Control and Prevention | Peter W. Cook et al |
| EPI_ISL_888354 | USA/FL-CDC-LC00001129/2020 | 2020-12-26 | North America/USA/Florida/ | Labcorp | Genomics and Discovery, Respiratory Viruses Branch, Division of Viral Diseases, Centers for Disease Control and Prevention | Peter W. Cook et al |
| EPI_ISL_888355 | USA/PA-CDC-LC00001527/2020 | 2020-12-28 | North America/USA/Pennsylvania/ | Labcorp | Genomics and Discovery, Respiratory Viruses Branch, Division of Viral Diseases, Centers for Disease Control and Prevention | Peter W. Cook et al |

| | | | | | | |
|---|---|---|---|---|---|---|
| EPI_ISL_888356 | USA/NM-CDC-LC00000544/2020 | 2020-12-26 | North America/USA/New Mexico/ | Labcorp | Genomics and Discovery, Respiratory Viruses Branch, Division of Viral Diseases, Centers for Disease Control and Prevention | Peter W. Cook et al |
| EPI_ISL_888385 | USA/GA-CDC-LC00001198/2020 | 2020-12-26 | North America/USA/Georgia/ | Labcorp | Genomics and Discovery, Respiratory Viruses Branch, Division of Viral Diseases, Centers for Disease Control and Prevention | Peter W. Cook et al |
| EPI_ISL_888386 | USA/GA-CDC-LC00000970/2020 | 2020-12-26 | North America/USA/Georgia/ | Labcorp | Genomics and Discovery, Respiratory Viruses Branch, Division of Viral Diseases, Centers for Disease Control and Prevention | Peter W. Cook et al |
| EPI_ISL_888390 | USA/NJ-CDC-LC00000562/2020 | 2020-12-27 | North America/USA/New Jersey/ | Labcorp | Genomics and Discovery, Respiratory Viruses Branch, Division of Viral Diseases, Centers for Disease Control and Prevention | Peter W. Cook et al |

| | | | | | | |
|---|---|---|---|---|---|---|
| EPI_ISL_888397 | USA/FL-CDC-LC00001153/2020 | 2020-12-24 | North America/USA/Florida/ | Labcorp | Genomics and Discovery, Respiratory Viruses Branch, Division of Viral Diseases, Centers for Disease Control and Prevention | Peter W. Cook et al |
| EPI_ISL_888399 | USA/FL-CDC-LC00001143/2020 | 2020-12-25 | North America/USA/Florida/ | Labcorp | Genomics and Discovery, Respiratory Viruses Branch, Division of Viral Diseases, Centers for Disease Control and Prevention | Peter W. Cook et al |
| EPI_ISL_888403 | USA/PA-CDC-LC00000556/2020 | 2020-12-27 | North America/USA/Pennsylvania/ | Labcorp | Genomics and Discovery, Respiratory Viruses Branch, Division of Viral Diseases, Centers for Disease Control and Prevention | Peter W. Cook et al |
| EPI_ISL_888405 | USA/NY-CDC-LC00001088/2020 | 2020-12-26 | North America/USA/New York/ | Labcorp | Genomics and Discovery, Respiratory Viruses Branch, Division of Viral Diseases, Centers for Disease Control and Prevention | Peter W. Cook et al |

| | | | | | | |
|---|---|---|---|---|---|---|
| EPI_ISL_888411 | USA/MS-CDC-LC00001047/2020 | 2020-12-26 | North America/USA/Mississippi/ | Labcorp | Genomics and Discovery, Respiratory Viruses Branch, Division of Viral Diseases, Centers for Disease Control and Prevention | Peter W. Cook et al |
| EPI_ISL_888424 | USA/PA-CDC-LC00001380/2020 | 2020-12-26 | North America/USA/Pennsylvania/ | Labcorp | Genomics and Discovery, Respiratory Viruses Branch, Division of Viral Diseases, Centers for Disease Control and Prevention | Peter W. Cook et al |
| EPI_ISL_888464 | USA/NM-CDC-LC00000172/2020 | 2020-12-26 | North America/USA/New Mexico/ | Labcorp | Genomics and Discovery, Respiratory Viruses Branch, Division of Viral Diseases, Centers for Disease Control and Prevention | Peter W. Cook et al |
| EPI_ISL_888466 | USA/GA-CDC-LC00000955/2020 | 2020-12-26 | North America/USA/Georgia/ | Labcorp | Genomics and Discovery, Respiratory Viruses Branch, Division of Viral Diseases, Centers for Disease Control and Prevention | Peter W. Cook et al |

| | | | | | | |
|---|---|---|---|---|---|---|
| EPI_ISL_888469 | USA/VA-CDC-LC00000377/2020 | 2020-12-27 | North America/USA/Virginia/ | Labcorp | Genomics and Discovery, Respiratory Viruses Branch, Division of Viral Diseases, Centers for Disease Control and Prevention | Peter W. Cook et al |
| EPI_ISL_888475 | USA/AZ-CDC-LC00000185/2020 | 2020-12-26 | North America/USA/Arizona/ | Labcorp | Genomics and Discovery, Respiratory Viruses Branch, Division of Viral Diseases, Centers for Disease Control and Prevention | Peter W. Cook et al |
| EPI_ISL_888480 | USA/AZ-CDC-LC00000136/2020 | 2020-12-26 | North America/USA/Arizona/ | Labcorp | Genomics and Discovery, Respiratory Viruses Branch, Division of Viral Diseases, Centers for Disease Control and Prevention | Peter W. Cook et al |
| EPI_ISL_888490 | USA/AZ-CDC-LC00000137/2020 | 2020-12-26 | North America/USA/Arizona/ | Labcorp | Genomics and Discovery, Respiratory Viruses Branch, Division of Viral Diseases, Centers for Disease Control and Prevention | Peter W. Cook et al |

| | | | | | | |
|---|---|---|---|---|---|---|
| EPI_ISL_888491 | USA/AZ-CDC-LC00000247/2020 | 2020-12-26 | North America/USA/Arizona/ | Labcorp | Genomics and Discovery, Respiratory Viruses Branch, Division of Viral Diseases, Centers for Disease Control and Prevention | Peter W. Cook et al |
| EPI_ISL_888492 | USA/NM-CDC-LC00000176/2020 | 2020-12-26 | North America/USA/New Mexico/ | Labcorp | Genomics and Discovery, Respiratory Viruses Branch, Division of Viral Diseases, Centers for Disease Control and Prevention | Peter W. Cook et al |
| EPI_ISL_888502 | USA/MD-CDC-LC00000380/2020 | 2020-12-27 | North America/USA/Maryland/ | Labcorp | Genomics and Discovery, Respiratory Viruses Branch, Division of Viral Diseases, Centers for Disease Control and Prevention | Peter W. Cook et al |
| EPI_ISL_888509 | USA/IL-CDC-LC00002730/2021 | 2021-01-04 | North America/USA/Illinois/ | Labcorp | Genomics and Discovery, Respiratory Viruses Branch, Division of Viral Diseases, Centers for Disease Control and Prevention | Peter W. Cook et al |

| | | | | | | |
|---|---|---|---|---|---|---|
| EPI_ISL_888526 | USA/NC-CDC-LC00002651/2021 | 2021-01-03 | North America/USA/North Carolina/ | Labcorp | Genomics and Discovery, Respiratory Viruses Branch, Division of Viral Diseases, Centers for Disease Control and Prevention | Peter W. Cook et al |
| EPI_ISL_888527 | USA/un-CDC-LC00002096/2020 | 2020-12-26 | North America/USA/USA/ | Labcorp | Genomics and Discovery, Respiratory Viruses Branch, Division of Viral Diseases, Centers for Disease Control and Prevention | Peter W. Cook et al |
| EPI_ISL_888535 | USA/NJ-CDC-LC00002968/2020 | 2020-12-30 | North America/USA/New Jersey/ | Labcorp | Genomics and Discovery, Respiratory Viruses Branch, Division of Viral Diseases, Centers for Disease Control and Prevention | Peter W. Cook et al |
| EPI_ISL_888544 | USA/IL-CDC-LC00001924/2020 | 2020-12-26 | North America/USA/Illinois/ | Labcorp | Genomics and Discovery, Respiratory Viruses Branch, Division of Viral Diseases, Centers for Disease Control and Prevention | Peter W. Cook et al |

| | | | | | | |
|---|---|---|---|---|---|---|
| EPI_ISL_888559 | USA/KS-CDC-LC00002413/2021 | 2021-01-02 | North America/USA/Kansas/ | Labcorp | Genomics and Discovery, Respiratory Viruses Branch, Division of Viral Diseases, Centers for Disease Control and Prevention | Peter W. Cook et al |
| EPI_ISL_888568 | USA/KY-CDC-LC00002108/2020 | 2020-12-26 | North America/USA/Kentucky/ | Labcorp | Genomics and Discovery, Respiratory Viruses Branch, Division of Viral Diseases, Centers for Disease Control and Prevention | Peter W. Cook et al |
| EPI_ISL_888570 | USA/IL-CDC-LC00001931/2020 | 2020-12-27 | North America/USA/Illinois/ | Labcorp | Genomics and Discovery, Respiratory Viruses Branch, Division of Viral Diseases, Centers for Disease Control and Prevention | Peter W. Cook et al |
| EPI_ISL_888575 | USA/NJ-CDC-LC00001335/2020 | 2020-12-27 | North America/USA/New Jersey/ | Labcorp | Genomics and Discovery, Respiratory Viruses Branch, Division of Viral Diseases, Centers for Disease Control and Prevention | Peter W. Cook et al |

| | | | | | | |
|---|---|---|---|---|---|---|
| EPI_ISL_888582 | USA/CA-CDC-LC00000830/2020 | 2020-12-24 | North America/USA/California/ | Labcorp | Genomics and Discovery, Respiratory Viruses Branch, Division of Viral Diseases, Centers for Disease Control and Prevention | Peter W. Cook et al |
| EPI_ISL_888583 | USA/CA-CDC-LC00002310/2020 | 2020-12-31 | North America/USA/California/ | Labcorp | Genomics and Discovery, Respiratory Viruses Branch, Division of Viral Diseases, Centers for Disease Control and Prevention | Peter W. Cook et al |
| EPI_ISL_888591 | Indonesia/JI-ITDua-34962NT/2020 | 2020-12-24 | Asia/Indonesia/East Java/Sidoarjo | Siti Khodijah Hospital | Institute of Tropical Disease, Universitas Airlangga | Krisnoadi Rahardjo et al |
| EPI_ISL_888593 | Indonesia/JI-ITDua-34965NT/2020 | 2020-12-24 | Asia/Indonesia/East Java/Sidoarjo | Siti Khodijah Hospital | Institute of Tropical Disease, Universitas Airlangga | Rima R Prasetya et al |
| EPI_ISL_888599 | USA/WY-WPHL-21004538/2021 | 2021-01-12 | North America/USA/Wyoming/ | Wyoming Public Health Laboratory | Wyoming Public Health Laboratory | Noah Hull et al |
| EPI_ISL_888614 | USA/WY-WPHL-21005238/2021 | 2021-01-13 | North America/USA/Wyoming/ | Wyoming Public Health Laboratory | Wyoming Public Health Laboratory | Noah Hull et al |
| EPI_ISL_888620 | USA/WY-WPHL-21005058/2021 | 2021-01-13 | North America/USA/Wyoming/ | Wyoming Public Health Laboratory | Wyoming Public Health Laboratory | Noah Hull et al |
| EPI_ISL_888621 | USA/WY-WPHL-21005401/2021 | 2021-01-14 | North America/USA/Wyoming/ | Wyoming Public Health Laboratory | Wyoming Public Health Laboratory | Noah Hull et al |
| EPI_ISL_888630 | USA/NM-UNM-EDB0004/2021 | 2021-01-06 | North America/USA/New Mexico/ | Univeristy of New Mexico Hospital | Center for Global Health, University of New Mexico Health Sciences Center | Daryl Domman et al |

| | | | | | | |
|---|---|---|---|---|---|---|
| EPI_ISL_888631 | USA/NM-UNM-EDB0005/2021 | 2021-01-06 | North America/USA/New Mexico/ | Univeristy of New Mexico Hospital | Center for Global Health, University of New Mexico Health Sciences Center | Daryl Domman et al |
| EPI_ISL_888632 | USA/NM-UNM-EDB0006/2021 | 2021-01-06 | North America/USA/New Mexico/ | Univeristy of New Mexico Hospital | Center for Global Health, University of New Mexico Health Sciences Center | Daryl Domman et al |
| EPI_ISL_888644 | USA/NM-UNM-EDB0025/2021 | 2021-01-08 | North America/USA/New Mexico/ | Univeristy of New Mexico Hospital | Center for Global Health, University of New Mexico Health Sciences Center | Daryl Domman et al |
| EPI_ISL_888656 | USA/MI-UM-10037990710/2021 | 2021-01-25 | North America/USA/Michigan/ | University of Michigan Clinical Microbiology Laboratory | Lauring Lab, University of Michigan, Department of Microbiology and Immunology | Valesano et al |
| EPI_ISL_888657 | USA/MI-UM-10037990655/2021 | 2021-01-25 | North America/USA/Michigan/ | University of Michigan Clinical Microbiology Laboratory | Lauring Lab, University of Michigan, Department of Microbiology and Immunology | Valesano et al |
| EPI_ISL_888658 | USA/MI-UM-10037990380/2021 | 2021-01-25 | North America/USA/Michigan/ | University of Michigan Clinical Microbiology Laboratory | Lauring Lab, University of Michigan, Department of Microbiology and Immunology | Valesano et al |
| EPI_ISL_888660 | USA/MI-UM-10037990778/2021 | 2021-01-25 | North America/USA/Michigan/ | University of Michigan Clinical Microbiology Laboratory | Lauring Lab, University of Michigan, Department of Microbiology and Immunology | Valesano et al |

| | | | | | | |
|---|---|---|---|---|---|---|
| EPI_ISL_888665 | USA/MI-UM-10037904822/2021 | 2021-01-19 | North America/USA/Michigan/ | University of Michigan Clinical Microbiology Laboratory | Lauring Lab, University of Michigan, Department of Microbiology and Immunology | Valesano et al |
| EPI_ISL_888667 | USA/MI-UM-10037808179/2021 | 2021-01-13 | North America/USA/Michigan/ | University of Michigan Clinical Microbiology Laboratory | Lauring Lab, University of Michigan, Department of Microbiology and Immunology | Valesano et al |
| EPI_ISL_888677 | Romania/Bucuresti-FV5166/2020 | 2020-12-14 | Europe/Romania/Bucharest/ | Laboratory for Respiratory Viruses, Cantacuzino National Military-Medical Institute for Research and Development | Cantacuzino Institute Virology | Mihaela Lazar et al |
| EPI_ISL_888680 | Italy/CAM-CRGS-114/2020 | 2020-04-19 | Europe/Italy/Campania/ | 1.AO Universitaria 'S. Giovanni di Dio e Ruggi D'Aragona, Scuola Medica Salernitana' Hospital / 2.UOC di Virologia e Microbiologia, Universit√† della Campania 'L. Vanvitelli' / 3.AO Universitaria 'Federico II' Napoli Hospital / 4.AORN 'San Giuseppe Moscati' Avellino Hospital / 5.AO 'San Pio - presidio G. Rummo' Benevento Hospital / 6.AO 'Sant'Anna e San Sebastiano' Caserta Hospital / 7.PO 'Maria Santissima Addolorata' Eboli Hospital / 8.Biogem Istituto di Ricerche Genetiche | 1. Genome Research Center for Health (CRGS) / 2. Laboratory of Molecular Medicine and Genomics(LMMGe) / 3. Center for Research in Pure and Applied Mathematics (CRMPA) | Giorgio Giurato et al |
| EPI_ISL_888681 | Italy/CAM-CRGS-93/2020 | 2020-03-18 | Europe/Italy/Campania/ | 1.AO Universitaria 'S. Giovanni di Dio e Ruggi D'Aragona, Scuola Medica Salernitana' Hospital / 2.UOC di Virologia e Microbiologia, Universit√† della Campania 'L. Vanvitelli' / 3.AO Universitaria 'Federico II' Napoli Hospital / 4.AORN 'San Giuseppe Moscati' Avellino Hospital / 5.AO 'San Pio - presidio G. Rummo' Benevento Hospital / 6.AO 'Sant'Anna e San Sebastiano' Caserta Hospital / 7.PO 'Maria Santissima Addolorata' Eboli Hospital / 8.Biogem Istituto di Ricerche Genetiche | 1. Genome Research Center for Health (CRGS) / 2. Laboratory of Molecular Medicine and Genomics(LMMGe) / 3. Center for Research in Pure and Applied Mathematics (CRMPA) | Giorgio Giurato et al |

| | | | | | | |
|---|---|---|---|---|---|---|
| EPI_ISL_888702 | Belgium/rega-1451/2020 | 2020-10-07 | Europe/Belgium/Antwerpen/Deurne | KU Leuven, Rega Institute, Clinical and Epidemiological Virology | KU Leuven, Rega Institute, Clinical and Epidemiological Virology | Tony Wawina-Bokalanga et al |
| EPI_ISL_888703 | Belgium/rega-1452/2020 | 2020-10-07 | Europe/Belgium/Halle-Vilvoorde/Hoeila | KU Leuven, Rega Institute, Clinical and Epidemiological Virology | KU Leuven, Rega Institute, Clinical and Epidemiological Virology | Tony Wawina-Bokalanga et al |
| EPI_ISL_888704 | Belgium/rega-1453/2020 | 2020-10-06 | Europe/Belgium/Halle-Vilvoorde/Wezen | KU Leuven, Rega Institute, Clinical and Epidemiological Virology | KU Leuven, Rega Institute, Clinical and Epidemiological Virology | Tony Wawina-Bokalanga et al |
| EPI_ISL_888714 | Belgium/rega-1464/2020 | 2020-10-13 | Europe/Belgium/Leuven/Tervuren | KU Leuven, Rega Institute, Clinical and Epidemiological Virology | KU Leuven, Rega Institute, Clinical and Epidemiological Virology | Tony Wawina-Bokalanga et al |
| EPI_ISL_888717 | Belgium/rega-1467/2020 | 2020-10-07 | Europe/Belgium/Halle-Vilvoorde/Hoeila | KU Leuven, Rega Institute, Clinical and Epidemiological Virology | KU Leuven, Rega Institute, Clinical and Epidemiological Virology | Tony Wawina-Bokalanga et al |
| EPI_ISL_888720 | Belgium/rega-1470/2020 | 2020-10-05 | Europe/Belgium/Halle-Vilvoorde/Kraain | KU Leuven, Rega Institute, Clinical and Epidemiological Virology | KU Leuven, Rega Institute, Clinical and Epidemiological Virology | Tony Wawina-Bokalanga et al |
| EPI_ISL_888722 | Belgium/rega-1472/2020 | 2020-10-07 | Europe/Belgium/Brussels/Evere | KU Leuven, Rega Institute, Clinical and Epidemiological Virology | KU Leuven, Rega Institute, Clinical and Epidemiological Virology | Tony Wawina-Bokalanga et al |
| EPI_ISL_888725 | Belgium/rega-1475/2020 | 2020-10-07 | Europe/Belgium/Leuven/Bierbeek | KU Leuven, Rega Institute, Clinical and Epidemiological Virology | KU Leuven, Rega Institute, Clinical and Epidemiological Virology | Tony Wawina-Bokalanga et al |

| | | | | | | |
|---|---|---|---|---|---|---|
| EPI_ISL_888727 | Belgium/rega-1477/2020 | 2020-10-08 | Europe/Belgium/Brussels/Ixelles | KU Leuven, Rega Institute, Clinical and Epidemiological Virology | KU Leuven, Rega Institute, Clinical and Epidemiological Virology | Tony Wawina-Bokalanga et al |
| EPI_ISL_888728 | Belgium/rega-1478/2020 | 2020-10-08 | Europe/Belgium/Halle-Vilvoorde/Vilvoo | KU Leuven, Rega Institute, Clinical and Epidemiological Virology | KU Leuven, Rega Institute, Clinical and Epidemiological Virology | Tony Wawina-Bokalanga et al |
| EPI_ISL_888730 | Belgium/rega-1480/2020 | 2020-10-07 | Europe/Belgium/Nivelles/Genval | KU Leuven, Rega Institute, Clinical and Epidemiological Virology | KU Leuven, Rega Institute, Clinical and Epidemiological Virology | Tony Wawina-Bokalanga et al |
| EPI_ISL_888733 | Belgium/rega-1483/2020 | 2020-10-08 | Europe/Belgium/Halle-Vilvoorde/Hoeila | KU Leuven, Rega Institute, Clinical and Epidemiological Virology | KU Leuven, Rega Institute, Clinical and Epidemiological Virology | Tony Wawina-Bokalanga et al |
| EPI_ISL_888736 | Belgium/rega-1486/2020 | 2020-10-07 | Europe/Belgium/Leuven/Kessel-Lo | KU Leuven, Rega Institute, Clinical and Epidemiological Virology | KU Leuven, Rega Institute, Clinical and Epidemiological Virology | Tony Wawina-Bokalanga et al |
| EPI_ISL_888737 | Belgium/rega-1487/2020 | 2020-10-05 | Europe/Belgium/Halle-Vilvoorde/Strom | KU Leuven, Rega Institute, Clinical and Epidemiological Virology | KU Leuven, Rega Institute, Clinical and Epidemiological Virology | Tony Wawina-Bokalanga et al |
| EPI_ISL_888749 | Belgium/rega-1499/2020 | 2020-12-18 | Europe/Belgium/Dinant/Dorinne | KU Leuven, Rega Institute, Clinical and Epidemiological Virology | KU Leuven, Rega Institute, Clinical and Epidemiological Virology | Tony Wawina-Bokalanga et al |
| EPI_ISL_888753 | Belgium/rega-1503/2020 | 2020-12-11 | Europe/Belgium/Dinant/Dorinne | KU Leuven, Rega Institute, Clinical and Epidemiological Virology | KU Leuven, Rega Institute, Clinical and Epidemiological Virology | Tony Wawina-Bokalanga et al |

| | | | | | | |
|---|---|---|---|---|---|---|
| EPI_ISL_888757 | Belgium/rega-1508/2020 | 2020-11-25 | Europe/Belgium/Tongeren/Hoeselt | KU Leuven, Rega Institute, Clinical and Epidemiological Virology | KU Leuven, Rega Institute, Clinical and Epidemiological Virology | Tony Wawina-Bokalanga et al |
| EPI_ISL_888792 | USA/NM-UNM-ED0035/2021 | 2021-01-08 | North America/USA/New Mexico/ | Univeristy of New Mexico Hospital | Center for Global Health, University of New Mexico Health Sciences Center | Daryl Domman et al |
| EPI_ISL_888799 | USA/NM-UNM-ED0043/2021 | 2021-01-10 | North America/USA/New Mexico/ | Univeristy of New Mexico Hospital | Center for Global Health, University of New Mexico Health Sciences Center | Daryl Domman et al |
| EPI_ISL_888813 | USA/NM-UNM-ED0062/2021 | 2021-01-12 | North America/USA/New Mexico/ | Univeristy of New Mexico Hospital | Center for Global Health, University of New Mexico Health Sciences Center | Daryl Domman et al |
| EPI_ISL_888815 | USA/NM-UNM-ED0064/2021 | 2021-01-12 | North America/USA/New Mexico/ | Univeristy of New Mexico Hospital | Center for Global Health, University of New Mexico Health Sciences Center | Daryl Domman et al |
| EPI_ISL_888828 | France/CVL-IPP9518/2020 | 2020-10-26 | Europe/France/Pays de la Loire/Nantes | General practitioner office | National Reference Center for Viruses of Respiratory Infections, Institut Pasteur, Paris | Marion Barbet et al |
| EPI_ISL_888829 | France/IDF-IPP9533/2020 | 2020-10-27 | Europe/France/Ile de France/Compans | General practitioner office | National Reference Center for Viruses of Respiratory Infections, Institut Pasteur, Paris | Marion Barbet et al |
| EPI_ISL_888834 | Ireland/D-NVRL-21IRL00178/2020 | 2020-12-29 | Europe/Ireland/Dublin/ | National Virus Reference Laboratory | National Virus Reference Laboratory | Michael Carr et al |

| | | | | | | |
|---|---|---|---|---|---|---|
| EPI_ISL_888837 | USA/MI-MDHHS-SC23065/2020 | 2020-11-25 | North America/USA/Michigan/ | Michigan Department of Health and Human Services, Bureau of Laboratories | Michigan Department of Health and Human Services, Bureau of Laboratories | Blankenship HM et al |
| EPI_ISL_888845 | USA/MI-MDHHS-SC23073/2020 | 2020-12-29 | North America/USA/Michigan/ | Michigan Department of Health and Human Services, Bureau of Laboratories | Michigan Department of Health and Human Services, Bureau of Laboratories | Blankenship HM et al |
| EPI_ISL_888849 | USA/MI-MDHHS-SC23077/2021 | 2021-01-10 | North America/USA/Michigan/ | Michigan Department of Health and Human Services, Bureau of Laboratories | Michigan Department of Health and Human Services, Bureau of Laboratories | Blankenship HM et al |
| EPI_ISL_888853 | USA/MI-MDHHS-SC23081/2021 | 2021-01-04 | North America/USA/Michigan/ | Michigan Department of Health and Human Services, Bureau of Laboratories | Michigan Department of Health and Human Services, Bureau of Laboratories | Blankenship HM et al |
| EPI_ISL_888865 | USA/MI-MDHHS-SC23093/2020 | 2020-12-21 | North America/USA/Michigan/ | Michigan Department of Health and Human Services, Bureau of Laboratories | Michigan Department of Health and Human Services, Bureau of Laboratories | Blankenship HM et al |
| EPI_ISL_888870 | USA/MI-MDHHS-SC23098/2021 | 2021-01-06 | North America/USA/Michigan/ | Michigan Department of Health and Human Services, Bureau of Laboratories | Michigan Department of Health and Human Services, Bureau of Laboratories | Blankenship HM et al |

| | | | | | | |
|---|---|---|---|---|---|---|
| EPI_ISL_888875 | USA/MI-MDHHS-SC23103/2020 | 2020-12-03 | North America/USA/Michigan/ | Michigan Department of Health and Human Services, Bureau of Laboratories | Michigan Department of Health and Human Services, Bureau of Laboratories | Blankenship HM et al |
| EPI_ISL_888882 | USA/MI-MDHHS-SC23110/2020 | 2020-10-20 | North America/USA/Michigan/ | Michigan Department of Health and Human Services, Bureau of Laboratories | Michigan Department of Health and Human Services, Bureau of Laboratories | Blankenship HM et al |
| EPI_ISL_888893 | USA/MI-MDHHS-SC23121/2020 | 2020-10-28 | North America/USA/Michigan/ | Michigan Department of Health and Human Services, Bureau of Laboratories | Michigan Department of Health and Human Services, Bureau of Laboratories | Blankenship HM et al |
| EPI_ISL_888899 | USA/MI-MDHHS-SC23127/2021 | 2021-01-11 | North America/USA/Michigan/ | Michigan Department of Health and Human Services, Bureau of Laboratories | Michigan Department of Health and Human Services, Bureau of Laboratories | Blankenship HM et al |
| EPI_ISL_888905 | USA/MI-MDHHS-SC23133/2021 | 2021-01-11 | North America/USA/Michigan/ | Michigan Department of Health and Human Services, Bureau of Laboratories | Michigan Department of Health and Human Services, Bureau of Laboratories | Blankenship HM et al |
| EPI_ISL_888928 | USA/WY-WPHL-21006661/2021 | 2021-01-18 | North America/USA/Wyoming/ | Wyoming Public Health Laboratory | Wyoming Public Health Laboratory | Noah Hull et al |
| EPI_ISL_888932 | USA/WY-WPHL-21006652/2021 | 2021-01-18 | North America/USA/Wyoming/ | Wyoming Public Health Laboratory | Wyoming Public Health Laboratory | Noah Hull et al |
| EPI_ISL_888933 | USA/WY-WPHL-21006818/2021 | 2021-01-18 | North America/USA/Wyoming/ | Wyoming Public Health Laboratory | Wyoming Public Health Laboratory | Noah Hull et al |
| EPI_ISL_888935 | USA/WY-WPHL-21008397/2021 | 2021-01-22 | North America/USA/Wyoming/ | Wyoming Public Health Laboratory | Wyoming Public Health Laboratory | Noah Hull et al |
| EPI_ISL_888945 | USA/WY-WPHL-21005856/2021 | 2021-01-15 | North America/USA/Wyoming/ | Wyoming Public Health Laboratory | Wyoming Public Health Laboratory | Noah Hull et al |

| | | | | | | |
|---|---|---|---|---|---|---|
| EPI_ISL_888953 | USA/WY-WPHL-21008058/2021 | 2021-01-21 | North America/USA/Wyoming/ | Wyoming Public Health Laboratory | Wyoming Public Health Laboratory | Noah Hull et al |
| EPI_ISL_888970 | USA/WY-WPHL-21007733/2021 | 2021-01-20 | North America/USA/Wyoming/ | Wyoming Public Health Laboratory | Wyoming Public Health Laboratory | Noah Hull et al |
| EPI_ISL_888972 | USA/WY-WPHL-21007155/2021 | 2021-01-19 | North America/USA/Wyoming/ | Wyoming Public Health Laboratory | Wyoming Public Health Laboratory | Noah Hull et al |
| EPI_ISL_888979 | Indonesia/JK-EIJK132/2021 | 2021-01-06 | Asia/Indonesia/Jakarta/ | RS Bhakti Mulia | Eijkman Institute for Molecular Biology, Ministry of Research and Technology/National Agency for Research and Innovation | Willy Agustine et al |
| EPI_ISL_888980 | Indonesia/JB-EIJK98/2020 | 2020-12-07 | Asia/Indonesia/West Java/Bekasi | RS Anna | Eijkman Institute for Molecular Biology, Ministry of Research and Technology/National Agency for Research and Innovation | Hidayat Trimarsanto et al |
| EPI_ISL_888981 | Indonesia/JK-EIJK112/2020 | 2020-12-18 | Asia/Indonesia/Jakarta/ | RS Omni Pulomas | Eijkman Institute for Molecular Biology, Ministry of Research and Technology/National Agency for Research and Innovation | Iskandar Adnan et al |
| EPI_ISL_888983 | Indonesia/JK-EIJK60/2020 | 2020-11-05 | Asia/Indonesia/Jakarta/ | RS Hermina Daan Mogot | Eijkman Institute for Molecular Biology, Ministry of Research and Technology/National Agency for Research and Innovation | Edison Johar et al |

| | | | | | | |
|---|---|---|---|---|---|---|
| EPI_ISL_888984 | Indonesia/JB-EIJK61/2020 | 2020-11-05 | Asia/Indonesia/West Java/Depok | RS Hermina Depok | Eijkman Institute for Molecular Biology, Ministry of Research and Technology/National Agency for Research and Innovation | Edison Johar et al |
| EPI_ISL_888985 | Indonesia/BT-EIJK67/2020 | 2020-11-10 | Asia/Indonesia/Banten/South Tangerang | RS Hermina Serpong | Eijkman Institute for Molecular Biology, Ministry of Research and Technology/National Agency for Research and Innovation | Edison Johar et al |
| EPI_ISL_888986 | Indonesia/JB-EIJK70/2020 | 2020-11-12 | Asia/Indonesia/West Java/Bekasi | RSU Bhakti Kartini | Eijkman Institute for Molecular Biology, Ministry of Research and Technology/National Agency for Research and Innovation | Edison Johar et al |
| EPI_ISL_888987 | Indonesia/JB-EIJK71/2020 | 2020-11-12 | Asia/Indonesia/West Java/Depok | RS Citra Medika Depok | Eijkman Institute for Molecular Biology, Ministry of Research and Technology/National Agency for Research and Innovation | Frilasita A Yudhaputri et al |
| EPI_ISL_888988 | Indonesia/BT-EIJK75/2020 | 2020-11-18 | Asia/Indonesia/Banten/Tangerang | RS Qadr | Eijkman Institute for Molecular Biology, Ministry of Research and Technology/National Agency for Research and Innovation | Frilasita A Yudhaputri et al |

| | | | | | | |
|---|---|---|---|---|---|---|
| EPI_ISL_888990 | Indonesia/BT-EIJK79/2020 | 2020-11-23 | Asia/Indonesia/Banten/South Tangerang | RS Pondok Indah Bintaro Jaya | Eijkman Institute for Molecular Biology, Ministry of Research and Technology/National Agency for Research and Innovation | Frilasita A Yudhaputri et al |
| EPI_ISL_888992 | Indonesia/BT-EIJK88/2020 | 2020-12-02 | Asia/Indonesia/Banten/Tangerang | RS Mitra Keluarga Gading Serpong | Eijkman Institute for Molecular Biology, Ministry of Research and Technology/National Agency for Research and Innovation | Hidayat Trimarsanto et al |
| EPI_ISL_888993 | Indonesia/JB-EIJK100/2020 | 2020-12-10 | Asia/Indonesia/West Java/Bekasi | RSU Medirossa | Eijkman Institute for Molecular Biology, Ministry of Research and Technology/National Agency for Research and Innovation | Iskandar Adnan et al |
| EPI_ISL_888994 | Indonesia/JK-EIJK115/2020 | 2020-12-29 | Asia/Indonesia/Jakarta/ | RSU MMC | Eijkman Institute for Molecular Biology, Ministry of Research and Technology/National Agency for Research and Innovation | Lydia V. Panggalo et al |
| EPI_ISL_888996 | Indonesia/JB-EIJK117/2020 | 2020-12-29 | Asia/Indonesia/West Java/Bekasi | RSU Graha Juanda | Eijkman Institute for Molecular Biology, Ministry of Research and Technology/National Agency for Research and Innovation | Lydia V. Panggalo et al |

| | | | | | | |
|---|---|---|---|---|---|---|
| EPI_ISL_888997 | Indonesia/JB-EIJK122/2021 | 2021-01-04 | Asia/Indonesia/West Java/Bogor | RSU Asysyifaa | Eijkman Institute for Molecular Biology, Ministry of Research and Technology/National Agency for Research and Innovation | Sukma Oktavianthi et al |
| EPI_ISL_888998 | Indonesia/JB-EIJK123/2021 | 2021-01-04 | Asia/Indonesia/West Java/Bogor | RSUD Cileungsi | Eijkman Institute for Molecular Biology, Ministry of Research and Technology/National Agency for Research and Innovation | Sukma Oktavianthi et al |
| EPI_ISL_888999 | Indonesia/JB-EIJK124/2021 | 2021-01-04 | Asia/Indonesia/West Java/Bogor | RS Mitra Keluarga Cibubur | Eijkman Institute for Molecular Biology, Ministry of Research and Technology/National Agency for Research and Innovation | Sukma Oktavianthi et al |
| EPI_ISL_889000 | Indonesia/JB-EIJK125/2021 | 2021-01-07 | Asia/Indonesia/West Java/Bogor | RSUD Cileungsi | Eijkman Institute for Molecular Biology, Ministry of Research and Technology/National Agency for Research and Innovation | Sukma Oktavianthi et al |
| EPI_ISL_889001 | Indonesia/JB-EIJK129/2021 | 2021-01-06 | Asia/Indonesia/West Java/Bogor | RSU Azra | Eijkman Institute for Molecular Biology, Ministry of Research and Technology/National Agency for Research and Innovation | Sukma Oktavianthi et al |

| | | | | | | |
|---|---|---|---|---|---|---|
| EPI_ISL_889002 | Indonesia/JK-EIJK130/2021 | 2021-01-06 | Asia/Indonesia/Jakarta/ | RSU Sumber Waras | Eijkman Institute for Molecular Biology, Ministry of Research and Technology/National Agency for Research and Innovation | Sukma Oktavianthi et al |
| EPI_ISL_889003 | Indonesia/BT-EIJK134/2020 | 2020-12-31 | Asia/Indonesia/Banten/Tangerang | RS Mitra Keluarga Gading Serpong | Eijkman Institute for Molecular Biology, Ministry of Research and Technology/National Agency for Research and Innovation | Willy Agustine et al |
| EPI_ISL_889004 | Indonesia/BT-EIJK135/2021 | 2021-01-01 | Asia/Indonesia/Banten/Tangerang | RS Mitra Keluarga Gading Serpong | Eijkman Institute for Molecular Biology, Ministry of Research and Technology/National Agency for Research and Innovation | Willy Agustine et al |
| EPI_ISL_889005 | Indonesia/JB-EIJK145/2021 | 2021-01-11 | Asia/Indonesia/West Java/Bekasi | Eijkman Institute for Molecular Biology, Ministry of Research and Technology/National Agency for Research and Innovation | Eijkman Institute for Molecular Biology, Ministry of Research and Technology/National Agency for Research and Innovation | Willy Agustine et al |
| EPI_ISL_889006 | Indonesia/JK-EIJK147/2021 | 2021-01-11 | Asia/Indonesia/Jakarta/ | RSU Harapan Bunda | Eijkman Institute for Molecular Biology, Ministry of Research and Technology/National Agency for Research and Innovation | Willy Agustine et al |

| | | | | | | |
|---|---|---|---|---|---|---|
| EPI_ISL_889007 | Indonesia/JB-EIJK148/2021 | 2021-01-11 | Asia/Indonesia/West Java/Bogor | RSUD Cileungsi | Eijkman Institute for Molecular Biology, Ministry of Research and Technology/National Agency for Research and Innovation | Willy Agustine et al |
| EPI_ISL_889008 | Indonesia/JB-EIJK56/2020 | 2020-03-31 | Asia/Indonesia/West Java/Bogor | RS Sentra Medika Cibinong | Eijkman Institute for Molecular Biology, Ministry of Research and Technology/National Agency for Research and Innovation | Edison Johar et al |
| EPI_ISL_889009 | Indonesia/BT-EIJK62/2020 | 2020-11-09 | Asia/Indonesia/Banten/South Tangerang | RS Mitra Keluarga Bintaro | Eijkman Institute for Molecular Biology, Ministry of Research and Technology/National Agency for Research and Innovation | Edison Johar et al |
| EPI_ISL_889010 | Indonesia/JB-EIJK74/2020 | 2020-11-18 | Asia/Indonesia/West Java/Bogor | RSU Medika Dramaga | Eijkman Institute for Molecular Biology, Ministry of Research and Technology/National Agency for Research and Innovation | Frilasita A Yudhaputri et al |
| EPI_ISL_889011 | Indonesia/JB-EIJK82/2020 | 2020-11-27 | Asia/Indonesia/West Java/Bekasi | RSU Permata Cibubur | Eijkman Institute for Molecular Biology, Ministry of Research and Technology/National Agency for Research and Innovation | Hidayat Trimarsanto et al |

| | | | | | | |
|---|---|---|---|---|---|---|
| EPI_ISL_889012 | Indonesia/JB-EIJK95/2020 | 2020-12-05 | Asia/Indonesia/West Java/Bekasi | RS Mitra Keluarga Pratama Jatiasih | Eijkman Institute for Molecular Biology, Ministry of Research and Technology/National Agency for Research and Innovation | Hidayat Trimarsanto et al |
| EPI_ISL_889014 | Indonesia/JB-EIJK107/2020 | 2020-12-16 | Asia/Indonesia/West Java/Bogor | RSU Medika Dramaga | Eijkman Institute for Molecular Biology, Ministry of Research and Technology/National Agency for Research and Innovation | Iskandar Adnan et al |
| EPI_ISL_889018 | Indonesia/JB-EIJK120/2021 | 2021-01-04 | Asia/Indonesia/West Java/Bogor | RSU Medika Dramaga | Eijkman Institute for Molecular Biology, Ministry of Research and Technology/National Agency for Research and Innovation | Lydia V. Panggalo et al |
| EPI_ISL_889019 | Indonesia/JK-EIJK121/2021 | 2021-01-04 | Asia/Indonesia/Jakarta/ | RSIA Sayyidah | Eijkman Institute for Molecular Biology, Ministry of Research and Technology/National Agency for Research and Innovation | Lydia V. Panggalo et al |
| EPI_ISL_889022 | Indonesia/JB-EIJK91/2020 | 2020-12-02 | Asia/Indonesia/West Java/Bekasi | RS Anna | Eijkman Institute for Molecular Biology, Ministry of Research and Technology/National Agency for Research and Innovation | Hidayat Trimarsanto et al |

| | | | | | | |
|---|---|---|---|---|---|---|
| EPI_ISL_889032 | Israel/CVL-119ngs/2020 | 2020-12-27 | Asia/Israel/North District/Sha'Ab | Israel Central Virology laboratory | Israel National Consortium for SARS-CoV-2 sequencing | Neta Zuckerman et al |
| EPI_ISL_889072 | Israel/CVL-593ngs/2020 | 2020-12-29 | Asia/Israel/Jerusalem District/Jerusalen | Israel Central Virology laboratory | Israel National Consortium for SARS-CoV-2 sequencing | Neta Zuckerman et al |
| EPI_ISL_889130 | Israel/CVL-791ngs/2020 | 2020-12-29 | Asia/Israel/Israel/ | Israel Central Virology laboratory | Israel National Consortium for SARS-CoV-2 sequencing | Neta Zuckerman et al |
| EPI_ISL_889163 | Israel/CVL-212ngs/2020 | 2020-12-27 | Asia/Israel/Jerusalem District/Jerusalen | Israel Central Virology laboratory | Israel National Consortium for SARS-CoV-2 sequencing | Neta Zuckerman et al |
| EPI_ISL_889170 | Israel/CVL-392ngs/2020 | 2020-12-28 | Asia/Israel/Jerusalem District/Jerusalen | Israel Central Virology laboratory | Israel National Consortium for SARS-CoV-2 sequencing | Neta Zuckerman et al |
| EPI_ISL_889191 | Israel/CVL-766ngs/2020 | 2020-12-29 | Asia/Israel/Israel/ | Israel Central Virology laboratory | Israel National Consortium for SARS-CoV-2 sequencing | Neta Zuckerman et al |
| EPI_ISL_889214 | Israel/CVL-897ngs/2020 | 2020-12-30 | Asia/Israel/South District/Ashkelon | Israel Central Virology laboratory | Israel National Consortium for SARS-CoV-2 sequencing | Neta Zuckerman et al |
| EPI_ISL_889225 | Israel/CVL-668ngs/2020 | 2020-12-29 | Asia/Israel/Israel/ | Israel Central Virology laboratory | Israel National Consortium for SARS-CoV-2 sequencing | Neta Zuckerman et al |
| EPI_ISL_889242 | Israel/CVL-723ngs/2020 | 2020-12-29 | Asia/Israel/Israel/ | Israel Central Virology laboratory | Israel National Consortium for SARS-CoV-2 sequencing | Neta Zuckerman et al |
| EPI_ISL_889311 | CzechRepublic/IAB20_006_27/2020 | 2020-03-16 | Europe/Czech Republic/South Moravian | The University Hospital Brno | Institute of Applied Biotechnologies a.s. | Petr Klempt et al |

| | | | | | | |
|---|---|---|---|---|---|---|
| EPI_ISL_889314 | CzechRepublic/IAB20_006_30/2020 | 2020-03-19 | Europe/Czech Republic/South Moravian | The University Hospital Brno | Institute of Applied Biotechnologies a.s. | Petr Klempt et al |
| EPI_ISL_889316 | CzechRepublic/IAB20_006_32/2020 | 2020-03-19 | Europe/Czech Republic/South Moravian | The University Hospital Brno | Institute of Applied Biotechnologies a.s. | Petr Klempt et al |
| EPI_ISL_889321 | CzechRepublic/IAB20_006_38/2020 | 2020-03-20 | Europe/Czech Republic/South Moravian | The University Hospital Brno | Institute of Applied Biotechnologies a.s. | Petr Klempt et al |
| EPI_ISL_889331 | CzechRepublic/IAB20_016_002/2020 | 2020-07-06 | Europe/Czech Republic/South Moravian | The University Hospital Brno | Institute of Applied Biotechnologies a.s. | Petr Klempt et al |
| EPI_ISL_889338 | CzechRepublic/IAB20_016_009/2020 | 2020-08-03 | Europe/Czech Republic/South Moravian | The University Hospital Brno | Institute of Applied Biotechnologies a.s. | Petr Klempt et al |
| EPI_ISL_889341 | CzechRepublic/IAB20_016_012/2020 | 2020-07-15 | Europe/Czech Republic/South Moravian | The University Hospital Brno | Institute of Applied Biotechnologies a.s. | Petr Klempt et al |
| EPI_ISL_889345 | CzechRepublic/IAB20_016_015/2020 | 2020-08-03 | Europe/Czech Republic/South Moravian | The University Hospital Brno | Institute of Applied Biotechnologies a.s. | Petr Klempt et al |
| EPI_ISL_889348 | CzechRepublic/IAB20_016_017/2020 | 2020-08-14 | Europe/Czech Republic/South Moravian | The University Hospital Brno | Institute of Applied Biotechnologies a.s. | Petr Klempt et al |
| EPI_ISL_889355 | CzechRepublic/IAB20_016_024/2020 | 2020-08-19 | Europe/Czech Republic/South Moravian | The University Hospital Brno | Institute of Applied Biotechnologies a.s. | Petr Klempt et al |
| EPI_ISL_889356 | CzechRepublic/IAB20_016_031/2020 | 2020-08-06 | Europe/Czech Republic/Prague/ | Motol University Hospital | Institute of Applied Biotechnologies a.s. | Petr Klempt et al |

| | | | | | | |
|---|---|---|---|---|---|---|
| EPI_ISL_889364 | CzechRepublic/IAB20_016_040/2020 | 2020-08-10 | Europe/Czech Republic/Prague/ | Motol University Hospital | Institute of Applied Biotechnologies a.s. | Petr Klempt et al |
| EPI_ISL_889372 | CzechRepublic/IAB20_016_029/2020 | 2020-09-15 | Europe/Czech Republic/Olomouc Region/ | Olomouc University Hospital | Institute of Applied Biotechnologies a.s. | Petr Klempt et al |
| EPI_ISL_889378 | CzechRepublic/IAB20_016_258/2020 | 2020-03-27 | Europe/Czech Republic/Olomouc Region/ | Olomouc University Hospital | Institute of Applied Biotechnologies a.s. | Petr Klempt et al |
| EPI_ISL_889380 | CzechRepublic/IAB20_016_060/2020 | 2020-03-27 | Europe/Czech Republic/Olomouc Region/ | Olomouc University Hospital | Institute of Applied Biotechnologies a.s. | Petr Klempt et al |
| EPI_ISL_889387 | CzechRepublic/IAB20_016_068/2020 | 2020-04-04 | Europe/Czech Republic/Olomouc Region/ | Olomouc University Hospital | Institute of Applied Biotechnologies a.s. | Petr Klempt et al |
| EPI_ISL_889390 | CzechRepublic/IAB20_016_075/2020 | 2020-04-08 | Europe/Czech Republic/Olomouc Region/ | Olomouc University Hospital | Institute of Applied Biotechnologies a.s. | Petr Klempt et al |
| EPI_ISL_889399 | CzechRepublic/IAB20_016_093/2020 | 2020-10-02 | Europe/Czech Republic/Olomouc Region/ | Olomouc University Hospital | Institute of Applied Biotechnologies a.s. | Petr Klempt et al |
| EPI_ISL_889400 | CzechRepublic/IAB20_016_094/2020 | 2020-10-02 | Europe/Czech Republic/Olomouc Region/ | Olomouc University Hospital | Institute of Applied Biotechnologies a.s. | Petr Klempt et al |
| EPI_ISL_889401 | CzechRepublic/IAB20_016_294/2020 | 2020-10-02 | Europe/Czech Republic/Olomouc Region/ | Olomouc University Hospital | Institute of Applied Biotechnologies a.s. | Petr Klempt et al |
| EPI_ISL_889403 | CzechRepublic/IAB20_016_098/2020 | 2020-10-02 | Europe/Czech Republic/Olomouc Region/ | Olomouc University Hospital | Institute of Applied Biotechnologies a.s. | Petr Klempt et al |

| | | | | | | |
|---|---|---|---|---|---|---|
| EPI_ISL_889404 | CzechRepublic/IAB20_016_099/2020 | 2020-10-02 | Europe/Czech Republic/Olomouc Region/ | Olomouc University Hospital | Institute of Applied Biotechnologies a.s. | Petr Klempt et al |
| EPI_ISL_889406 | CzechRepublic/IAB20_016_101/2020 | 2020-10-03 | Europe/Czech Republic/Olomouc Region/ | Olomouc University Hospital | Institute of Applied Biotechnologies a.s. | Petr Klempt et al |
| EPI_ISL_889407 | CzechRepublic/IAB20_016_102/2020 | 2020-10-03 | Europe/Czech Republic/Olomouc Region/ | Olomouc University Hospital | Institute of Applied Biotechnologies a.s. | Petr Klempt et al |
| EPI_ISL_889408 | CzechRepublic/IAB20_016_103/2020 | 2020-10-03 | Europe/Czech Republic/Olomouc Region/ | Olomouc University Hospital | Institute of Applied Biotechnologies a.s. | Petr Klempt et al |
| EPI_ISL_889409 | CzechRepublic/IAB20_016_104/2020 | 2020-10-03 | Europe/Czech Republic/Olomouc Region/ | Olomouc University Hospital | Institute of Applied Biotechnologies a.s. | Petr Klempt et al |
| EPI_ISL_889410 | CzechRepublic/IAB20_016_105/2020 | 2020-10-03 | Europe/Czech Republic/Olomouc Region/ | Olomouc University Hospital | Institute of Applied Biotechnologies a.s. | Petr Klempt et al |
| EPI_ISL_889411 | CzechRepublic/IAB20_016_106/2020 | 2020-10-03 | Europe/Czech Republic/Olomouc Region/ | Olomouc University Hospital | Institute of Applied Biotechnologies a.s. | Petr Klempt et al |
| EPI_ISL_889412 | CzechRepublic/IAB20_016_306/2020 | 2020-10-03 | Europe/Czech Republic/Olomouc Region/ | Olomouc University Hospital | Institute of Applied Biotechnologies a.s. | Petr Klempt et al |
| EPI_ISL_889413 | CzechRepublic/IAB20_016_107/2020 | 2020-10-03 | Europe/Czech Republic/Olomouc Region/ | Olomouc University Hospital | Institute of Applied Biotechnologies a.s. | Petr Klempt et al |
| EPI_ISL_889414 | CzechRepublic/IAB20_016_108/2020 | 2020-10-03 | Europe/Czech Republic/Olomouc Region/ | Olomouc University Hospital | Institute of Applied Biotechnologies a.s. | Petr Klempt et al |

| | | | | | | |
|---|---|---|---|---|---|---|
| EPI_ISL_889415 | CzechRepublic/IAB20_016_109/2020 | 2020-10-03 | Europe/Czech Republic/Olomouc Region/ | Olomouc University Hospital | Institute of Applied Biotechnologies a.s. | Petr Klempt et al |
| EPI_ISL_889416 | CzechRepublic/IAB20_016_110/2020 | 2020-10-05 | Europe/Czech Republic/Olomouc Region/ | Olomouc University Hospital | Institute of Applied Biotechnologies a.s. | Petr Klempt et al |
| EPI_ISL_889417 | CzechRepublic/IAB20_016_111/2020 | 2020-10-05 | Europe/Czech Republic/Olomouc Region/ | Olomouc University Hospital | Institute of Applied Biotechnologies a.s. | Petr Klempt et al |
| EPI_ISL_889420 | CzechRepublic/IAB20_016_114/2020 | 2020-10-05 | Europe/Czech Republic/Olomouc Region/ | Olomouc University Hospital | Institute of Applied Biotechnologies a.s. | Petr Klempt et al |
| EPI_ISL_889422 | CzechRepublic/IAB20_016_315/2020 | 2020-10-05 | Europe/Czech Republic/Olomouc Region/ | Olomouc University Hospital | Institute of Applied Biotechnologies a.s. | Petr Klempt et al |
| EPI_ISL_889423 | CzechRepublic/IAB20_016_116/2020 | 2020-10-05 | Europe/Czech Republic/Olomouc Region/ | Olomouc University Hospital | Institute of Applied Biotechnologies a.s. | Petr Klempt et al |
| EPI_ISL_889424 | CzechRepublic/IAB20_016_117/2020 | 2020-10-05 | Europe/Czech Republic/Olomouc Region/ | Olomouc University Hospital | Institute of Applied Biotechnologies a.s. | Petr Klempt et al |
| EPI_ISL_889425 | CzechRepublic/IAB20_016_118/2020 | 2020-10-05 | Europe/Czech Republic/Olomouc Region/ | Olomouc University Hospital | Institute of Applied Biotechnologies a.s. | Petr Klempt et al |
| EPI_ISL_889426 | CzechRepublic/IAB20_016_119/2020 | 2020-10-05 | Europe/Czech Republic/Olomouc Region/ | Olomouc University Hospital | Institute of Applied Biotechnologies a.s. | Petr Klempt et al |
| EPI_ISL_889427 | CzechRepublic/IAB20_016_120/2020 | 2020-10-05 | Europe/Czech Republic/Olomouc Region/ | Olomouc University Hospital | Institute of Applied Biotechnologies a.s. | Petr Klempt et al |

| | | | | | | |
|---|---|---|---|---|---|---|
| EPI_ISL_889428 | CzechRepublic/IAB20_016_121/2020 | 2020-10-06 | Europe/Czech Republic/Olomouc Region/ | Olomouc University Hospital | Institute of Applied Biotechnologies a.s. | Petr Klempt et al |
| EPI_ISL_889431 | CzechRepublic/IAB20_016_124/2020 | 2020-10-06 | Europe/Czech Republic/Olomouc Region/ | Olomouc University Hospital | Institute of Applied Biotechnologies a.s. | Petr Klempt et al |
| EPI_ISL_889432 | CzechRepublic/IAB20_016_125/2020 | 2020-10-06 | Europe/Czech Republic/Olomouc Region/ | Olomouc University Hospital | Institute of Applied Biotechnologies a.s. | Petr Klempt et al |
| EPI_ISL_889433 | CzechRepublic/IAB20_016_126/2020 | 2020-10-06 | Europe/Czech Republic/Olomouc Region/ | Olomouc University Hospital | Institute of Applied Biotechnologies a.s. | Petr Klempt et al |
| EPI_ISL_889435 | CzechRepublic/IAB20_016_128/2020 | 2020-10-06 | Europe/Czech Republic/Olomouc Region/ | Olomouc University Hospital | Institute of Applied Biotechnologies a.s. | Petr Klempt et al |
| EPI_ISL_889436 | CzechRepublic/IAB20_016_129/2020 | 2020-10-06 | Europe/Czech Republic/Olomouc Region/ | Olomouc University Hospital | Institute of Applied Biotechnologies a.s. | Petr Klempt et al |
| EPI_ISL_889437 | CzechRepublic/IAB20_016_130/2020 | 2020-10-06 | Europe/Czech Republic/Olomouc Region/ | Olomouc University Hospital | Institute of Applied Biotechnologies a.s. | Petr Klempt et al |
| EPI_ISL_889439 | CzechRepublic/IAB20_016_331/2020 | 2020-10-06 | Europe/Czech Republic/Olomouc Region/ | Olomouc University Hospital | Institute of Applied Biotechnologies a.s. | Petr Klempt et al |
| EPI_ISL_889446 | USA/LA-EVTL1465/2020 | 2020-12-26 | North America/USA/Louisiana/Bossier P | LSUHS Emerging Viral Threat Laboratory | Microbial Genome Sequencing Center | Jeremy P. Kamil et al |
| EPI_ISL_889450 | USA/LA-EVTL1469/2020 | 2020-12-26 | North America/USA/Louisiana/Bossier P | LSUHS Emerging Viral Threat Laboratory | Microbial Genome Sequencing Center | Jeremy P. Kamil et al |

| | | | | | | |
|---|---|---|---|---|---|---|
| EPI_ISL_889456 | USA/LA-EVTL1475/2020 | 2020-12-26 | North America/USA/Louisiana/Bossier P | LSUHS Emerging Viral Threat Laboratory | Microbial Genome Sequencing Center | Jeremy P. Kamil et al |
| EPI_ISL_889457 | USA/LA-EVTL1476/2020 | 2020-12-26 | North America/USA/Louisiana/Bossier P | LSUHS Emerging Viral Threat Laboratory | Microbial Genome Sequencing Center | Jeremy P. Kamil et al |
| EPI_ISL_889459 | USA/LA-EVTL1478/2020 | 2020-12-26 | North America/USA/Louisiana/Bossier P | LSUHS Emerging Viral Threat Laboratory | Microbial Genome Sequencing Center | Jeremy P. Kamil et al |
| EPI_ISL_889463 | USA/LA-EVTL1482/2020 | 2020-12-27 | North America/USA/Louisiana/Webster | LSUHS Emerging Viral Threat Laboratory | Microbial Genome Sequencing Center | Jeremy P. Kamil et al |
| EPI_ISL_889475 | USA/LA-EVTL1494/2020 | 2020-12-28 | North America/USA/Louisiana/Caddo Pa | LSUHS Emerging Viral Threat Laboratory | Microbial Genome Sequencing Center | Jeremy P. Kamil et al |
| EPI_ISL_889480 | USA/LA-EVTL1499/2020 | 2020-12-28 | North America/USA/Louisiana/Caddo Pa | LSUHS Emerging Viral Threat Laboratory | Microbial Genome Sequencing Center | Jeremy P. Kamil et al |
| EPI_ISL_889484 | USA/LA-EVTL1503/2020 | 2020-12-28 | North America/USA/Louisiana/Caddo Pa | LSUHS Emerging Viral Threat Laboratory | Microbial Genome Sequencing Center | Jeremy P. Kamil et al |
| EPI_ISL_889496 | USA/LA-EVTL1515/2020 | 2020-12-29 | North America/USA/Louisiana/Webster | LSUHS Emerging Viral Threat Laboratory | Microbial Genome Sequencing Center | Jeremy P. Kamil et al |
| EPI_ISL_889497 | USA/LA-EVTL1516/2020 | 2020-12-28 | North America/USA/Louisiana/Caddo Pa | LSUHS Emerging Viral Threat Laboratory | Microbial Genome Sequencing Center | Jeremy P. Kamil et al |
| EPI_ISL_889500 | USA/LA-EVTL1519/2020 | 2020-12-29 | North America/USA/Louisiana/Caddo Pa | LSUHS Emerging Viral Threat Laboratory | Microbial Genome Sequencing Center | Jeremy P. Kamil et al |
| EPI_ISL_889502 | USA/LA-EVTL1521/2020 | 2020-12-30 | North America/USA/Louisiana/Bienville | LSUHS Emerging Viral Threat Laboratory | Microbial Genome Sequencing Center | Jeremy P. Kamil et al |
| EPI_ISL_889503 | USA/LA-EVTL1522/2020 | 2020-12-29 | North America/USA/Louisiana/Bossier P | LSUHS Emerging Viral Threat Laboratory | Microbial Genome Sequencing Center | Jeremy P. Kamil et al |
| EPI_ISL_889516 | USA/LA-EVTL1535/2020 | 2020-12-29 | North America/USA/Louisiana/Bossier P | LSUHS Emerging Viral Threat Laboratory | Microbial Genome Sequencing Center | Jeremy P. Kamil et al |

| | | | | | | |
|---|---|---|---|---|---|---|
| EPI_ISL_889532 | USA/LA-EVTL1551/2020 | 2020-12-30 | North America/USA/Louisiana/Bossier P | LSUHS Emerging Viral Threat Laboratory | Microbial Genome Sequencing Center | Jeremy P. Kamil et al |
| EPI_ISL_889546 | USA/LA-EVTL1565/2020 | 2020-12-31 | North America/USA/Louisiana/Bossier P | LSUHS Emerging Viral Threat Laboratory | Microbial Genome Sequencing Center | Jeremy P. Kamil et al |
| EPI_ISL_889550 | USA/LA-EVTL1570/2020 | 2020-12-31 | North America/USA/Louisiana/Caddo Pa | LSUHS Emerging Viral Threat Laboratory | Microbial Genome Sequencing Center | Jeremy P. Kamil et al |
| EPI_ISL_889552 | USA/LA-EVTL1572/2021 | 2021-01-02 | North America/USA/Louisiana/Caddo Pa | LSUHS Emerging Viral Threat Laboratory | Microbial Genome Sequencing Center | Jeremy P. Kamil et al |
| EPI_ISL_889553 | USA/LA-EVTL1573/2021 | 2021-01-02 | North America/USA/Louisiana/Caddo Pa | LSUHS Emerging Viral Threat Laboratory | Microbial Genome Sequencing Center | Jeremy P. Kamil et al |
| EPI_ISL_889558 | USA/LA-EVTL1578/2021 | 2021-01-02 | North America/USA/Louisiana/Caddo Pa | LSUHS Emerging Viral Threat Laboratory | Microbial Genome Sequencing Center | Jeremy P. Kamil et al |
| EPI_ISL_889570 | USA/LA-EVTL1590/2021 | 2021-01-04 | North America/USA/Louisiana/Caddo Pa | LSUHS Emerging Viral Threat Laboratory | Microbial Genome Sequencing Center | Jeremy P. Kamil et al |
| EPI_ISL_889580 | USA/LA-EVTL1600/2021 | 2021-01-04 | North America/USA/Louisiana/Sabine Pa | LSUHS Emerging Viral Threat Laboratory | Microbial Genome Sequencing Center | Jeremy P. Kamil et al |
| EPI_ISL_889584 | USA/LA-EVTL1604/2021 | 2021-01-04 | North America/USA/Louisiana/Caddo Pa | LSUHS Emerging Viral Threat Laboratory | Microbial Genome Sequencing Center | Jeremy P. Kamil et al |
| EPI_ISL_889585 | USA/LA-EVTL1605/2021 | 2021-01-04 | North America/USA/Louisiana/Caddo Pa | LSUHS Emerging Viral Threat Laboratory | Microbial Genome Sequencing Center | Jeremy P. Kamil et al |
| EPI_ISL_889590 | USA/LA-EVTL1610/2021 | 2021-01-04 | North America/USA/Louisiana/Caddo Pa | LSUHS Emerging Viral Threat Laboratory | Microbial Genome Sequencing Center | Jeremy P. Kamil et al |
| EPI_ISL_889601 | USA/LA-EVTL1621/2021 | 2021-01-04 | North America/USA/Louisiana/Caddo Pa | LSUHS Emerging Viral Threat Laboratory | Microbial Genome Sequencing Center | Jeremy P. Kamil et al |
| EPI_ISL_889610 | USA/LA-EVTL1630/2021 | 2021-01-05 | North America/USA/Louisiana/Bienville | LSUHS Emerging Viral Threat Laboratory | Microbial Genome Sequencing Center | Jeremy P. Kamil et al |

| | | | | | | |
|---|---|---|---|---|---|---|
| EPI_ISL_889614 | USA/LA-EVTL1634/2021 | 2021-01-05 | North America/USA/Louisiana/Bienville | LSUHS Emerging Viral Threat Laboratory | Microbial Genome Sequencing Center | Jeremy P. Kamil et al |
| EPI_ISL_889615 | USA/LA-EVTL1635/2021 | 2021-01-05 | North America/USA/Louisiana/Bienville | LSUHS Emerging Viral Threat Laboratory | Microbial Genome Sequencing Center | Jeremy P. Kamil et al |
| EPI_ISL_889618 | USA/LA-EVTL1639/2021 | 2021-01-05 | North America/USA/Louisiana/Caddo Pa | LSUHS Emerging Viral Threat Laboratory | Microbial Genome Sequencing Center | Jeremy P. Kamil et al |
| EPI_ISL_889620 | USA/LA-EVTL1641/2021 | 2021-01-05 | North America/USA/Louisiana/Caddo Pa | LSUHS Emerging Viral Threat Laboratory | Microbial Genome Sequencing Center | Jeremy P. Kamil et al |
| EPI_ISL_889625 | USA/LA-EVTL1646/2021 | 2021-01-05 | North America/USA/Louisiana/Bossier P | LSUHS Emerging Viral Threat Laboratory | Microbial Genome Sequencing Center | Jeremy P. Kamil et al |
| EPI_ISL_889626 | USA/LA-EVTL1647/2021 | 2021-01-05 | North America/USA/Louisiana/Caddo Pa | LSUHS Emerging Viral Threat Laboratory | Microbial Genome Sequencing Center | Jeremy P. Kamil et al |
| EPI_ISL_889634 | USA/LA-EVTL1655/2021 | 2021-01-06 | North America/USA/Louisiana/Caddo Pa | LSUHS Emerging Viral Threat Laboratory | Microbial Genome Sequencing Center | Jeremy P. Kamil et al |
| EPI_ISL_889635 | USA/LA-EVTL1656/2021 | 2021-01-06 | North America/USA/Louisiana/Caddo Pa | LSUHS Emerging Viral Threat Laboratory | Microbial Genome Sequencing Center | Jeremy P. Kamil et al |
| EPI_ISL_889639 | USA/LA-EVTL1660/2021 | 2021-01-06 | North America/USA/Louisiana/Caddo Pa | LSUHS Emerging Viral Threat Laboratory | Microbial Genome Sequencing Center | Jeremy P. Kamil et al |
| EPI_ISL_889644 | USA/LA-EVTL1665/2021 | 2021-01-06 | North America/USA/Louisiana/Webster | LSUHS Emerging Viral Threat Laboratory | Microbial Genome Sequencing Center | Jeremy P. Kamil et al |
| EPI_ISL_889647 | USA/LA-EVTL1669/2021 | 2021-01-06 | North America/USA/Louisiana/Bossier P | LSUHS Emerging Viral Threat Laboratory | Microbial Genome Sequencing Center | Jeremy P. Kamil et al |
| EPI_ISL_889658 | USA/LA-EVTL1680/2021 | 2021-01-07 | North America/USA/Louisiana/Caddo Pa | LSUHS Emerging Viral Threat Laboratory | Microbial Genome Sequencing Center | Jeremy P. Kamil et al |
| EPI_ISL_889662 | USA/LA-EVTL1684/2021 | 2021-01-07 | North America/USA/Louisiana/Caddo Pa | LSUHS Emerging Viral Threat Laboratory | Microbial Genome Sequencing Center | Jeremy P. Kamil et al |

| | | | | | | |
|---|---|---|---|---|---|---|
| EPI_ISL_889679 | USA/LA-EVTL1701/2021 | 2021-01-08 | North America/USA/Louisiana/Caddo Pa | LSUHS Emerging Viral Threat Laboratory | Microbial Genome Sequencing Center | Jeremy P. Kamil et al |
| EPI_ISL_889684 | USA/LA-EVTL1706/2021 | 2021-01-09 | North America/USA/Louisiana/Caddo Pa | LSUHS Emerging Viral Threat Laboratory | Microbial Genome Sequencing Center | Jeremy P. Kamil et al |
| EPI_ISL_889701 | USA/LA-EVTL1723/2021 | 2021-01-11 | North America/USA/Louisiana/Caddo Pa | LSUHS Emerging Viral Threat Laboratory | Microbial Genome Sequencing Center | Jeremy P. Kamil et al |
| EPI_ISL_889726 | USA/LA-EVTL1749/2021 | 2021-01-13 | North America/USA/Louisiana/Natchito | LSUHS Emerging Viral Threat Laboratory | Microbial Genome Sequencing Center | Jeremy P. Kamil et al |
| EPI_ISL_889727 | USA/LA-EVTL1750/2021 | 2021-01-13 | North America/USA/Louisiana/Natchito | LSUHS Emerging Viral Threat Laboratory | Microbial Genome Sequencing Center | Jeremy P. Kamil et al |
| EPI_ISL_889734 | USA/LA-EVTL1757/2021 | 2021-01-14 | North America/USA/Louisiana/Caddo Pa | LSUHS Emerging Viral Threat Laboratory | Microbial Genome Sequencing Center | Jeremy P. Kamil et al |
| EPI_ISL_889740 | USA/LA-EVTL1763/2021 | 2021-01-14 | North America/USA/Louisiana/Caddo Pa | LSUHS Emerging Viral Threat Laboratory | Microbial Genome Sequencing Center | Jeremy P. Kamil et al |
| EPI_ISL_889741 | USA/LA-EVTL1764/2021 | 2021-01-14 | North America/USA/Louisiana/Caddo Pa | LSUHS Emerging Viral Threat Laboratory | Microbial Genome Sequencing Center | Jeremy P. Kamil et al |
| EPI_ISL_889749 | USA/LA-EVTL1773/2021 | 2021-01-15 | North America/USA/Louisiana/DeSoto P | LSUHS Emerging Viral Threat Laboratory | Microbial Genome Sequencing Center | Jeremy P. Kamil et al |
| EPI_ISL_889753 | USA/LA-EVTL1777/2021 | 2021-01-15 | North America/USA/Louisiana/Bossier P | LSUHS Emerging Viral Threat Laboratory | Microbial Genome Sequencing Center | Jeremy P. Kamil et al |
| EPI_ISL_889755 | USA/LA-EVTL1779/2021 | 2021-01-15 | North America/USA/Louisiana/Caddo Pa | LSUHS Emerging Viral Threat Laboratory | Microbial Genome Sequencing Center | Jeremy P. Kamil et al |
| EPI_ISL_889756 | USA/LA-EVTL1780/2021 | 2021-01-15 | North America/USA/Louisiana/Caddo Pa | LSUHS Emerging Viral Threat Laboratory | Microbial Genome Sequencing Center | Jeremy P. Kamil et al |
| EPI_ISL_889759 | USA/LA-EVTL1783/2021 | 2021-01-15 | North America/USA/Louisiana/Webster | LSUHS Emerging Viral Threat Laboratory | Microbial Genome Sequencing Center | Jeremy P. Kamil et al |

| | | | | | | |
|---|---|---|---|---|---|---|
| EPI_ISL_889762 | USA/LA-EVTL1786/2021 | 2021-01-18 | North America/USA/Louisiana/Bossier P | LSUHS Emerging Viral Threat Laboratory | Microbial Genome Sequencing Center | Jeremy P. Kamil et al |
| EPI_ISL_889768 | USA/LA-EVTL1792/2021 | 2021-01-18 | North America/USA/Louisiana/Bossier P | LSUHS Emerging Viral Threat Laboratory | Microbial Genome Sequencing Center | Jeremy P. Kamil et al |
| EPI_ISL_889773 | USA/LA-EVTL1797/2021 | 2021-01-18 | North America/USA/Louisiana/DeSoto P | LSUHS Emerging Viral Threat Laboratory | Microbial Genome Sequencing Center | Mohammed N.A. Siddiquey et al |
| EPI_ISL_889775 | USA/LA-EVTL1799/2021 | 2021-01-18 | North America/USA/Louisiana/DeSoto P | LSUHS Emerging Viral Threat Laboratory | Microbial Genome Sequencing Center | Mohammed N.A. Siddiquey et al |
| EPI_ISL_889780 | USA/LA-EVTL1804/2021 | 2021-01-18 | North America/USA/Louisiana/DeSoto P | LSUHS Emerging Viral Threat Laboratory | Microbial Genome Sequencing Center | Maarten Van Diest et al |
| EPI_ISL_889785 | USA/LA-EVTL1809/2021 | 2021-01-18 | North America/USA/Louisiana/DeSoto P | LSUHS Emerging Viral Threat Laboratory | Microbial Genome Sequencing Center | Maarten Van Diest et al |
| EPI_ISL_889787 | USA/LA-EVTL1811/2021 | 2021-01-18 | North America/USA/Louisiana/DeSoto P | LSUHS Emerging Viral Threat Laboratory | Microbial Genome Sequencing Center | Jennifer L. Carroll et al |
| EPI_ISL_889791 | USA/LA-EVTL1819/2021 | 2021-01-19 | North America/USA/Louisiana/Bienville | LSUHS Emerging Viral Threat Laboratory | Microbial Genome Sequencing Center | Jennifer L. Carroll et al |
| EPI_ISL_889802 | Canada/QC-JUS-V6011090/2020 | 2020-03-30 | North America/Canada/Quebec/ | Laboratoire de sant√© publique du Qu√©bec | Laboratoire de sant√© publique du Qu√©bec | Sandrine Moreira et al |
| EPI_ISL_889803 | Canada/QC-JUS-V5310659/2020 | 2020-03-29 | North America/Canada/Quebec/ | Laboratoire de sant√© publique du Qu√©bec | Laboratoire de sant√© publique du Qu√©bec | Sandrine Moreira et al |
| EPI_ISL_889804 | Canada/QC-JUS-V5241012/2020 | 2020-03-23 | North America/Canada/Quebec/ | Laboratoire de sant√© publique du Qu√©bec | Laboratoire de sant√© publique du Qu√©bec | Sandrine Moreira et al |
| EPI_ISL_889805 | Canada/QC-JUS-V5241923/2020 | 2020-03-23 | North America/Canada/Quebec/ | Laboratoire de sant√© publique du Qu√©bec | Laboratoire de sant√© publique du Qu√©bec | Sandrine Moreira et al |
| EPI_ISL_889806 | Canada/QC-JUS-V5280176/2020 | 2020-03-26 | North America/Canada/Quebec/ | Laboratoire de sant√© publique du Qu√©bec | Laboratoire de sant√© publique du Qu√©bec | Sandrine Moreira et al |

| | | | | | | |
|---|---|---|---|---|---|---|
| EPI_ISL_889807 | Canada/QC-JUS-V5300612/2020 | 2020-03-27 | North America/Canada/Quebec/ | Laboratoire de santv© publique du Quv©bec | Laboratoire de santv© publique du Quv©bec | Sandrine Moreira et al |
| EPI_ISL_889808 | Canada/QC-JUS-V6011568/2020 | 2020-03-31 | North America/Canada/Quebec/ | Laboratoire de santv© publique du Quv©bec | Laboratoire de santv© publique du Quv©bec | Sandrine Moreira et al |
| EPI_ISL_889809 | Canada/QC-JUS-V5310458/2020 | 2020-03-29 | North America/Canada/Quebec/ | Laboratoire de santv© publique du Quv©bec | Laboratoire de santv© publique du Quv©bec | Sandrine Moreira et al |
| EPI_ISL_889810 | Canada/QC-JUS-V5310155/2020 | 2020-03-29 | North America/Canada/Quebec/ | Laboratoire de santv© publique du Quv©bec | Laboratoire de santv© publique du Quv©bec | Sandrine Moreira et al |
| EPI_ISL_889811 | Canada/QC-JUS-V5291222/2020 | 2020-03-27 | North America/Canada/Quebec/ | Laboratoire de santv© publique du Quv©bec | Laboratoire de santv© publique du Quv©bec | Sandrine Moreira et al |
| EPI_ISL_889812 | Canada/QC-JUS-V5280612/2020 | 2020-03-27 | North America/Canada/Quebec/ | Laboratoire de santv© publique du Quv©bec | Laboratoire de santv© publique du Quv©bec | Sandrine Moreira et al |
| EPI_ISL_889813 | Canada/QC-JUS-V5311546/2020 | 2020-03-29 | North America/Canada/Quebec/ | Laboratoire de santv© publique du Quv©bec | Laboratoire de santv© publique du Quv©bec | Sandrine Moreira et al |
| EPI_ISL_889814 | Canada/QC-JUS-V5311236/2020 | 2020-03-29 | North America/Canada/Quebec/ | Laboratoire de santv© publique du Quv©bec | Laboratoire de santv© publique du Quv©bec | Sandrine Moreira et al |
| EPI_ISL_889815 | Canada/QC-JUS-V6011712/2020 | 2020-04-01 | North America/Canada/Quebec/ | Laboratoire de santv© publique du Quv©bec | Laboratoire de santv© publique du Quv©bec | Sandrine Moreira et al |
| EPI_ISL_889816 | Canada/QC-JUS-V5272537/2020 | 2020-03-26 | North America/Canada/Quebec/ | Laboratoire de santv© publique du Quv©bec | Laboratoire de santv© publique du Quv©bec | Sandrine Moreira et al |
| EPI_ISL_889817 | Canada/QC-JUS-V5272515/2020 | 2020-03-26 | North America/Canada/Quebec/ | Laboratoire de santv© publique du Quv©bec | Laboratoire de santv© publique du Quv©bec | Sandrine Moreira et al |
| EPI_ISL_889818 | Canada/QC-JUS-V5300273/2020 | 2020-03-28 | North America/Canada/Quebec/ | Laboratoire de santv© publique du Quv©bec | Laboratoire de santv© publique du Quv©bec | Sandrine Moreira et al |
| EPI_ISL_889819 | Canada/QC-JUS-V5311642/2020 | 2020-03-29 | North America/Canada/Quebec/ | Laboratoire de santv© publique du Quv©bec | Laboratoire de santv© publique du Quv©bec | Sandrine Moreira et al |

| | | | | | | |
|---|---|---|---|---|---|---|
| EPI_ISL_889820 | Canada/QC-JUS-V5310203/2020 | 2020-03-30 | North America/Canada/Quebec/ | Laboratoire de santv© publique du Quv©bec | Laboratoire de santv© publique du Quv©bec | Sandrine Moreira et al |
| EPI_ISL_889821 | Canada/QC-JUS-V6011588/2020 | 2020-03-30 | North America/Canada/Quebec/ | Laboratoire de santv© publique du Quv©bec | Laboratoire de santv© publique du Quv©bec | Sandrine Moreira et al |
| EPI_ISL_889822 | Canada/QC-JUS-V5311886/2020 | 2020-03-31 | North America/Canada/Quebec/ | Laboratoire de santv© publique du Quv©bec | Laboratoire de santv© publique du Quv©bec | Sandrine Moreira et al |
| EPI_ISL_889823 | Canada/QC-JUS-V6011021/2020 | 2020-03-30 | North America/Canada/Quebec/ | Laboratoire de santv© publique du Quv©bec | Laboratoire de santv© publique du Quv©bec | Sandrine Moreira et al |
| EPI_ISL_889824 | Canada/QC-JUS-V6011454/2020 | 2020-03-30 | North America/Canada/Quebec/ | Laboratoire de santv© publique du Quv©bec | Laboratoire de santv© publique du Quv©bec | Sandrine Moreira et al |
| EPI_ISL_889825 | Canada/QC-JUS-V5311172/2020 | 2020-03-29 | North America/Canada/Quebec/ | Laboratoire de santv© publique du Quv©bec | Laboratoire de santv© publique du Quv©bec | Sandrine Moreira et al |
| EPI_ISL_889826 | Canada/QC-JUS-V5311681/2020 | 2020-03-30 | North America/Canada/Quebec/ | Laboratoire de santv© publique du Quv©bec | Laboratoire de santv© publique du Quv©bec | Sandrine Moreira et al |
| EPI_ISL_889827 | Canada/QC-JUS-V6010544/2020 | 2020-03-29 | North America/Canada/Quebec/ | Laboratoire de santv© publique du Quv©bec | Laboratoire de santv© publique du Quv©bec | Sandrine Moreira et al |
| EPI_ISL_889828 | Canada/QC-JUS-V6010310/2020 | 2020-03-29 | North America/Canada/Quebec/ | Laboratoire de santv© publique du Quv©bec | Laboratoire de santv© publique du Quv©bec | Sandrine Moreira et al |
| EPI_ISL_889829 | Canada/QC-JUS-V5312311/2020 | 2020-03-31 | North America/Canada/Quebec/ | Laboratoire de santv© publique du Quv©bec | Laboratoire de santv© publique du Quv©bec | Sandrine Moreira et al |
| EPI_ISL_889830 | Canada/QC-JUS-V6010595/2020 | 2020-03-29 | North America/Canada/Quebec/ | Laboratoire de santv© publique du Quv©bec | Laboratoire de santv© publique du Quv©bec | Sandrine Moreira et al |
| EPI_ISL_889831 | Canada/QC-JUS-V6010733/2020 | 2020-03-29 | North America/Canada/Quebec/ | Laboratoire de santv© publique du Quv©bec | Laboratoire de santv© publique du Quv©bec | Sandrine Moreira et al |
| EPI_ISL_889832 | Canada/QC-JUS-V6010346/2020 | 2020-03-29 | North America/Canada/Quebec/ | Laboratoire de santv© publique du Quv©bec | Laboratoire de santv© publique du Quv©bec | Sandrine Moreira et al |

| | | | | | | |
|---|---|---|---|---|---|---|
| EPI_ISL_889833 | Canada/QC-JUS-V6010007/2020 | 2020-04-01 | North America/Canada/Quebec/ | Laboratoire de sant√© publique du Qu√©bec | Laboratoire de sant√© publique du Qu√©bec | Sandrine Moreira et al |
| EPI_ISL_889834 | Canada/QC-JUS-V6010362/2020 | 2020-03-29 | North America/Canada/Quebec/ | Laboratoire de sant√© publique du Qu√©bec | Laboratoire de sant√© publique du Qu√©bec | Sandrine Moreira et al |
| EPI_ISL_889835 | Canada/QC-JUS-V5311728/2020 | 2020-03-31 | North America/Canada/Quebec/ | Laboratoire de sant√© publique du Qu√©bec | Laboratoire de sant√© publique du Qu√©bec | Sandrine Moreira et al |
| EPI_ISL_889836 | Canada/QC-JUS-V5310928/2020 | 2020-03-30 | North America/Canada/Quebec/ | Laboratoire de sant√© publique du Qu√©bec | Laboratoire de sant√© publique du Qu√©bec | Sandrine Moreira et al |
| EPI_ISL_889837 | Canada/QC-JUS-V6012345/2020 | 2020-04-01 | North America/Canada/Quebec/ | Laboratoire de sant√© publique du Qu√©bec | Laboratoire de sant√© publique du Qu√©bec | Sandrine Moreira et al |
| EPI_ISL_889838 | Canada/QC-JUS-V6011941/2020 | 2020-03-30 | North America/Canada/Quebec/ | Laboratoire de sant√© publique du Qu√©bec | Laboratoire de sant√© publique du Qu√©bec | Sandrine Moreira et al |
| EPI_ISL_889839 | Canada/QC-JUS-V6021002/2020 | 2020-03-31 | North America/Canada/Quebec/ | Laboratoire de sant√© publique du Qu√©bec | Laboratoire de sant√© publique du Qu√©bec | Sandrine Moreira et al |
| EPI_ISL_889840 | Canada/QC-JUS-V6012000/2020 | 2020-04-01 | North America/Canada/Quebec/ | Laboratoire de sant√© publique du Qu√©bec | Laboratoire de sant√© publique du Qu√©bec | Sandrine Moreira et al |
| EPI_ISL_889841 | Canada/QC-JUS-V6011307/2020 | 2020-03-30 | North America/Canada/Quebec/ | Laboratoire de sant√© publique du Qu√©bec | Laboratoire de sant√© publique du Qu√©bec | Sandrine Moreira et al |
| EPI_ISL_889842 | Canada/QC-JUS-V6012289/2020 | 2020-04-01 | North America/Canada/Quebec/ | Laboratoire de sant√© publique du Qu√©bec | Laboratoire de sant√© publique du Qu√©bec | Sandrine Moreira et al |
| EPI_ISL_889843 | Canada/QC-JUS-V6012196/2020 | 2020-04-01 | North America/Canada/Quebec/ | Laboratoire de sant√© publique du Qu√©bec | Laboratoire de sant√© publique du Qu√©bec | Sandrine Moreira et al |
| EPI_ISL_889844 | Canada/QC-JUS-V6031018/2020 | 2020-04-02 | North America/Canada/Quebec/ | Laboratoire de sant√© publique du Qu√©bec | Laboratoire de sant√© publique du Qu√©bec | Sandrine Moreira et al |
| EPI_ISL_889845 | Canada/QC-JUS-V6020542/2020 | 2020-03-31 | North America/Canada/Quebec/ | Laboratoire de sant√© publique du Qu√©bec | Laboratoire de sant√© publique du Qu√©bec | Sandrine Moreira et al |

| | | | | | | |
|---|---|---|---|---|---|---|
| EPI_ISL_889846 | Canada/QC-JUS-V6031086/2020 | 2020-04-02 | North America/Canada/Quebec/ | Laboratoire de sant√© publique du Qu√©bec | Laboratoire de sant√© publique du Qu√©bec | Sandrine Moreira et al |
| EPI_ISL_889847 | Canada/QC-JUS-V6031135/2020 | 2020-04-02 | North America/Canada/Quebec/ | Laboratoire de sant√© publique du Qu√©bec | Laboratoire de sant√© publique du Qu√©bec | Sandrine Moreira et al |
| EPI_ISL_889848 | Canada/QC-JUS-V6021040/2020 | 2020-03-31 | North America/Canada/Quebec/ | Laboratoire de sant√© publique du Qu√©bec | Laboratoire de sant√© publique du Qu√©bec | Sandrine Moreira et al |
| EPI_ISL_889849 | Canada/QC-JUS-V6020365/2020 | 2020-03-31 | North America/Canada/Quebec/ | Laboratoire de sant√© publique du Qu√©bec | Laboratoire de sant√© publique du Qu√©bec | Sandrine Moreira et al |
| EPI_ISL_889850 | Canada/QC-JUS-V6020873/2020 | 2020-03-31 | North America/Canada/Quebec/ | Laboratoire de sant√© publique du Qu√©bec | Laboratoire de sant√© publique du Qu√©bec | Sandrine Moreira et al |
| EPI_ISL_889851 | Canada/QC-JUS-V6021294/2020 | 2020-03-31 | North America/Canada/Quebec/ | Laboratoire de sant√© publique du Qu√©bec | Laboratoire de sant√© publique du Qu√©bec | Sandrine Moreira et al |
| EPI_ISL_889852 | Canada/QC-JUS-V6020817/2020 | 2020-03-31 | North America/Canada/Quebec/ | Laboratoire de sant√© publique du Qu√©bec | Laboratoire de sant√© publique du Qu√©bec | Sandrine Moreira et al |
| EPI_ISL_889853 | Canada/QC-JUS-V6021249/2020 | 2020-03-31 | North America/Canada/Quebec/ | Laboratoire de sant√© publique du Qu√©bec | Laboratoire de sant√© publique du Qu√©bec | Sandrine Moreira et al |
| EPI_ISL_889854 | Canada/QC-JUS-V6021079/2020 | 2020-03-31 | North America/Canada/Quebec/ | Laboratoire de sant√© publique du Qu√©bec | Laboratoire de sant√© publique du Qu√©bec | Sandrine Moreira et al |
| EPI_ISL_889855 | Canada/QC-JUS-V6021091/2020 | 2020-03-31 | North America/Canada/Quebec/ | Laboratoire de sant√© publique du Qu√©bec | Laboratoire de sant√© publique du Qu√©bec | Sandrine Moreira et al |
| EPI_ISL_889856 | Canada/QC-JUS-V6011714/2020 | 2020-03-31 | North America/Canada/Quebec/ | Laboratoire de sant√© publique du Qu√©bec | Laboratoire de sant√© publique du Qu√©bec | Sandrine Moreira et al |
| EPI_ISL_889857 | Canada/QC-JUS-V6011098/2020 | 2020-03-30 | North America/Canada/Quebec/ | Laboratoire de sant√© publique du Qu√©bec | Laboratoire de sant√© publique du Qu√©bec | Sandrine Moreira et al |
| EPI_ISL_889858 | Canada/QC-JUS-V6011120/2020 | 2020-03-30 | North America/Canada/Quebec/ | Laboratoire de sant√© publique du Qu√©bec | Laboratoire de sant√© publique du Qu√©bec | Sandrine Moreira et al |

| | | | | | | |
|---|---|---|---|---|---|---|
| EPI_ISL_889859 | Canada/QC-JUS-V6011352/2020 | 2020-03-30 | North America/Canada/Quebec/ | Laboratoire de santv© publique du Quv©bec | Laboratoire de santv© publique du Quv©bec | Sandrine Moreira et al |
| EPI_ISL_889860 | Canada/QC-JUS-V6011470/2020 | 2020-03-31 | North America/Canada/Quebec/ | Laboratoire de santv© publique du Quv©bec | Laboratoire de santv© publique du Quv©bec | Sandrine Moreira et al |
| EPI_ISL_889861 | Canada/QC-JUS-V6011245/2020 | 2020-03-30 | North America/Canada/Quebec/ | Laboratoire de santv© publique du Quv©bec | Laboratoire de santv© publique du Quv©bec | Sandrine Moreira et al |
| EPI_ISL_889862 | Canada/QC-JUS-V6010795/2020 | 2020-03-31 | North America/Canada/Quebec/ | Laboratoire de santv© publique du Quv©bec | Laboratoire de santv© publique du Quv©bec | Sandrine Moreira et al |
| EPI_ISL_889863 | Canada/QC-JUS-V6010810/2020 | 2020-03-31 | North America/Canada/Quebec/ | Laboratoire de santv© publique du Quv©bec | Laboratoire de santv© publique du Quv©bec | Sandrine Moreira et al |
| EPI_ISL_889864 | Canada/QC-JUS-V6010785/2020 | 2020-03-31 | North America/Canada/Quebec/ | Laboratoire de santv© publique du Quv©bec | Laboratoire de santv© publique du Quv©bec | Sandrine Moreira et al |
| EPI_ISL_889865 | Canada/QC-JUS-V6010603/2020 | 2020-03-31 | North America/Canada/Quebec/ | Laboratoire de santv© publique du Quv©bec | Laboratoire de santv© publique du Quv©bec | Sandrine Moreira et al |
| EPI_ISL_889866 | Canada/QC-JUS-V6010927/2020 | 2020-03-31 | North America/Canada/Quebec/ | Laboratoire de santv© publique du Quv©bec | Laboratoire de santv© publique du Quv©bec | Sandrine Moreira et al |
| EPI_ISL_889867 | Canada/QC-JUS-V5301012/2020 | 2020-03-30 | North America/Canada/Quebec/ | Laboratoire de santv© publique du Quv©bec | Laboratoire de santv© publique du Quv©bec | Sandrine Moreira et al |
| EPI_ISL_889868 | Canada/QC-JUS-V5301021/2020 | 2020-03-29 | North America/Canada/Quebec/ | Laboratoire de santv© publique du Quv©bec | Laboratoire de santv© publique du Quv©bec | Sandrine Moreira et al |
| EPI_ISL_889869 | Canada/QC-JUS-V5301025/2020 | 2020-03-29 | North America/Canada/Quebec/ | Laboratoire de santv© publique du Quv©bec | Laboratoire de santv© publique du Quv©bec | Sandrine Moreira et al |
| EPI_ISL_889870 | Canada/QC-JUS-V5301030/2020 | 2020-03-29 | North America/Canada/Quebec/ | Laboratoire de santv© publique du Quv©bec | Laboratoire de santv© publique du Quv©bec | Sandrine Moreira et al |
| EPI_ISL_889871 | Canada/QC-JUS-V5301052/2020 | 2020-03-29 | North America/Canada/Quebec/ | Laboratoire de santv© publique du Quv©bec | Laboratoire de santv© publique du Quv©bec | Sandrine Moreira et al |

| | | | | | | |
|---|---|---|---|---|---|---|
| EPI_ISL_889872 | Canada/QC-JUS-V6011698/2020 | 2020-04-01 | North America/Canada/Quebec/ | Laboratoire de sant√© publique du Qu√©bec | Laboratoire de sant√© publique du Qu√©bec | Sandrine Moreira et al |
| EPI_ISL_889873 | Canada/QC-JUS-V5290618/2020 | 2020-03-27 | North America/Canada/Quebec/ | Laboratoire de sant√© publique du Qu√©bec | Laboratoire de sant√© publique du Qu√©bec | Sandrine Moreira et al |
| EPI_ISL_889874 | Canada/QC-JUS-V5290635/2020 | 2020-03-28 | North America/Canada/Quebec/ | Laboratoire de sant√© publique du Qu√©bec | Laboratoire de sant√© publique du Qu√©bec | Sandrine Moreira et al |
| EPI_ISL_889875 | Canada/QC-JUS-V5301970/2020 | 2020-03-30 | North America/Canada/Quebec/ | Laboratoire de sant√© publique du Qu√©bec | Laboratoire de sant√© publique du Qu√©bec | Sandrine Moreira et al |
| EPI_ISL_889876 | Canada/QC-JUS-V5301213/2020 | 2020-03-27 | North America/Canada/Quebec/ | Laboratoire de sant√© publique du Qu√©bec | Laboratoire de sant√© publique du Qu√©bec | Sandrine Moreira et al |
| EPI_ISL_889877 | Canada/QC-JUS-V5300961/2020 | 2020-03-27 | North America/Canada/Quebec/ | Laboratoire de sant√© publique du Qu√©bec | Laboratoire de sant√© publique du Qu√©bec | Sandrine Moreira et al |
| EPI_ISL_889878 | Canada/QC-JUS-V5301994/2020 | 2020-03-30 | North America/Canada/Quebec/ | Laboratoire de sant√© publique du Qu√©bec | Laboratoire de sant√© publique du Qu√©bec | Sandrine Moreira et al |
| EPI_ISL_889879 | Canada/QC-JUS-V5301979/2020 | 2020-03-30 | North America/Canada/Quebec/ | Laboratoire de sant√© publique du Qu√©bec | Laboratoire de sant√© publique du Qu√©bec | Sandrine Moreira et al |
| EPI_ISL_889880 | Canada/QC-JUS-V5301791/2020 | 2020-03-28 | North America/Canada/Quebec/ | Laboratoire de sant√© publique du Qu√©bec | Laboratoire de sant√© publique du Qu√©bec | Sandrine Moreira et al |
| EPI_ISL_889881 | Canada/QC-JUS-V5302015/2020 | 2020-03-30 | North America/Canada/Quebec/ | Laboratoire de sant√© publique du Qu√©bec | Laboratoire de sant√© publique du Qu√©bec | Sandrine Moreira et al |
| EPI_ISL_889882 | Canada/QC-JUS-V5302007/2020 | 2020-03-30 | North America/Canada/Quebec/ | Laboratoire de sant√© publique du Qu√©bec | Laboratoire de sant√© publique du Qu√©bec | Sandrine Moreira et al |
| EPI_ISL_889883 | Canada/QC-JUS-V5301873/2020 | 2020-03-30 | North America/Canada/Quebec/ | Laboratoire de sant√© publique du Qu√©bec | Laboratoire de sant√© publique du Qu√©bec | Sandrine Moreira et al |
| EPI_ISL_889884 | Canada/QC-JUS-V5280862/2020 | 2020-03-27 | North America/Canada/Quebec/ | Laboratoire de sant√© publique du Qu√©bec | Laboratoire de sant√© publique du Qu√©bec | Sandrine Moreira et al |

| | | | | | | |
|---|---|---|---|---|---|---|
| EPI_ISL_889885 | Canada/QC-JUS-V5280459/2020 | 2020-03-26 | North America/Canada/Quebec/ | Laboratoire de sant√© publique du Qu√©bec | Laboratoire de sant√© publique du Qu√©bec | Sandrine Moreira et al |
| EPI_ISL_889886 | Canada/QC-JUS-V5312271/2020 | 2020-03-29 | North America/Canada/Quebec/ | Laboratoire de sant√© publique du Qu√©bec | Laboratoire de sant√© publique du Qu√©bec | Sandrine Moreira et al |
| EPI_ISL_889887 | Canada/QC-JUS-V5312279/2020 | 2020-03-29 | North America/Canada/Quebec/ | Laboratoire de sant√© publique du Qu√©bec | Laboratoire de sant√© publique du Qu√©bec | Sandrine Moreira et al |
| EPI_ISL_889888 | Canada/QC-JUS-V5301500/2020 | 2020-03-27 | North America/Canada/Quebec/ | Laboratoire de sant√© publique du Qu√©bec | Laboratoire de sant√© publique du Qu√©bec | Sandrine Moreira et al |
| EPI_ISL_889889 | Canada/QC-JUS-V5311947/2020 | 2020-03-31 | North America/Canada/Quebec/ | Laboratoire de sant√© publique du Qu√©bec | Laboratoire de sant√© publique du Qu√©bec | Sandrine Moreira et al |
| EPI_ISL_889890 | Canada/QC-JUS-V5281643/2020 | 2020-03-27 | North America/Canada/Quebec/ | Laboratoire de sant√© publique du Qu√©bec | Laboratoire de sant√© publique du Qu√©bec | Sandrine Moreira et al |
| EPI_ISL_889891 | Canada/QC-JUS-V5310533/2020 | 2020-03-30 | North America/Canada/Quebec/ | Laboratoire de sant√© publique du Qu√©bec | Laboratoire de sant√© publique du Qu√©bec | Sandrine Moreira et al |
| EPI_ISL_889892 | Canada/QC-JUS-V5310540/2020 | 2020-03-30 | North America/Canada/Quebec/ | Laboratoire de sant√© publique du Qu√©bec | Laboratoire de sant√© publique du Qu√©bec | Sandrine Moreira et al |
| EPI_ISL_889893 | Canada/QC-JUS-V5300607/2020 | 2020-03-27 | North America/Canada/Quebec/ | Laboratoire de sant√© publique du Qu√©bec | Laboratoire de sant√© publique du Qu√©bec | Sandrine Moreira et al |
| EPI_ISL_889894 | Canada/QC-JUS-V5300261/2020 | 2020-03-27 | North America/Canada/Quebec/ | Laboratoire de sant√© publique du Qu√©bec | Laboratoire de sant√© publique du Qu√©bec | Sandrine Moreira et al |
| EPI_ISL_889895 | Canada/QC-JUS-V5300283/2020 | 2020-03-28 | North America/Canada/Quebec/ | Laboratoire de sant√© publique du Qu√©bec | Laboratoire de sant√© publique du Qu√©bec | Sandrine Moreira et al |
| EPI_ISL_889896 | Canada/QC-JUS-V5281600/2020 | 2020-03-27 | North America/Canada/Quebec/ | Laboratoire de sant√© publique du Qu√©bec | Laboratoire de sant√© publique du Qu√©bec | Sandrine Moreira et al |
| EPI_ISL_889897 | Canada/QC-JUS-V5281610/2020 | 2020-03-27 | North America/Canada/Quebec/ | Laboratoire de sant√© publique du Qu√©bec | Laboratoire de sant√© publique du Qu√©bec | Sandrine Moreira et al |

| | | | | | | |
|---|---|---|---|---|---|---|
| EPI_ISL_889898 | Canada/QC-JUS-V5281629/2020 | 2020-03-27 | North America/Canada/Quebec/ | Laboratoire de sant√© publique du Qu√©bec | Laboratoire de sant√© publique du Qu√©bec | Sandrine Moreira et al |
| EPI_ISL_889899 | Canada/QC-JUS-V5290754/2020 | 2020-03-28 | North America/Canada/Quebec/ | Laboratoire de sant√© publique du Qu√©bec | Laboratoire de sant√© publique du Qu√©bec | Sandrine Moreira et al |
| EPI_ISL_889900 | Canada/QC-JUS-V5281636/2020 | 2020-03-27 | North America/Canada/Quebec/ | Laboratoire de sant√© publique du Qu√©bec | Laboratoire de sant√© publique du Qu√©bec | Sandrine Moreira et al |
| EPI_ISL_889901 | Canada/QC-JUS-V5301459/2020 | 2020-03-27 | North America/Canada/Quebec/ | Laboratoire de sant√© publique du Qu√©bec | Laboratoire de sant√© publique du Qu√©bec | Sandrine Moreira et al |
| EPI_ISL_889902 | Canada/QC-JUS-V5311713/2020 | 2020-03-29 | North America/Canada/Quebec/ | Laboratoire de sant√© publique du Qu√©bec | Laboratoire de sant√© publique du Qu√©bec | Sandrine Moreira et al |
| EPI_ISL_889903 | Canada/QC-JUS-V5312010/2020 | 2020-03-29 | North America/Canada/Quebec/ | Laboratoire de sant√© publique du Qu√©bec | Laboratoire de sant√© publique du Qu√©bec | Sandrine Moreira et al |
| EPI_ISL_889904 | Canada/QC-JUS-V5311461/2020 | 2020-03-29 | North America/Canada/Quebec/ | Laboratoire de sant√© publique du Qu√©bec | Laboratoire de sant√© publique du Qu√©bec | Sandrine Moreira et al |
| EPI_ISL_889905 | Canada/QC-JUS-V5312014/2020 | 2020-03-29 | North America/Canada/Quebec/ | Laboratoire de sant√© publique du Qu√©bec | Laboratoire de sant√© publique du Qu√©bec | Sandrine Moreira et al |
| EPI_ISL_889906 | Canada/QC-JUS-V5311005/2020 | 2020-03-31 | North America/Canada/Quebec/ | Laboratoire de sant√© publique du Qu√©bec | Laboratoire de sant√© publique du Qu√©bec | Sandrine Moreira et al |
| EPI_ISL_889907 | Canada/QC-JUS-V5311129/2020 | 2020-03-31 | North America/Canada/Quebec/ | Laboratoire de sant√© publique du Qu√©bec | Laboratoire de sant√© publique du Qu√©bec | Sandrine Moreira et al |
| EPI_ISL_889908 | Canada/QC-JUS-V5310465/2020 | 2020-03-29 | North America/Canada/Quebec/ | Laboratoire de sant√© publique du Qu√©bec | Laboratoire de sant√© publique du Qu√©bec | Sandrine Moreira et al |
| EPI_ISL_889909 | Canada/QC-JUS-V5310839/2020 | 2020-03-30 | North America/Canada/Quebec/ | Laboratoire de sant√© publique du Qu√©bec | Laboratoire de sant√© publique du Qu√©bec | Sandrine Moreira et al |
| EPI_ISL_889910 | Canada/QC-JUS-V5311178/2020 | 2020-03-29 | North America/Canada/Quebec/ | Laboratoire de sant√© publique du Qu√©bec | Laboratoire de sant√© publique du Qu√©bec | Sandrine Moreira et al |

| | | | | | | |
|---|---|---|---|---|---|---|
| EPI_ISL_889911 | Canada/QC-JUS-V5311417/2020 | 2020-03-29 | North America/Canada/Quebec/ | Laboratoire de sant√© publique du Qu√©bec | Laboratoire de sant√© publique du Qu√©bec | Sandrine Moreira et al |
| EPI_ISL_889912 | Canada/QC-JUS-V5311596/2020 | 2020-03-29 | North America/Canada/Quebec/ | Laboratoire de sant√© publique du Qu√©bec | Laboratoire de sant√© publique du Qu√©bec | Sandrine Moreira et al |
| EPI_ISL_889913 | Canada/QC-JUS-V5311977/2020 | 2020-03-31 | North America/Canada/Quebec/ | Laboratoire de sant√© publique du Qu√©bec | Laboratoire de sant√© publique du Qu√©bec | Sandrine Moreira et al |
| EPI_ISL_889914 | Canada/QC-JUS-V5311987/2020 | 2020-03-31 | North America/Canada/Quebec/ | Laboratoire de sant√© publique du Qu√©bec | Laboratoire de sant√© publique du Qu√©bec | Sandrine Moreira et al |
| EPI_ISL_889915 | Canada/QC-JUS-V5310582/2020 | 2020-03-29 | North America/Canada/Quebec/ | Laboratoire de sant√© publique du Qu√©bec | Laboratoire de sant√© publique du Qu√©bec | Sandrine Moreira et al |
| EPI_ISL_889916 | Canada/QC-JUS-V5311348/2020 | 2020-03-30 | North America/Canada/Quebec/ | Laboratoire de sant√© publique du Qu√©bec | Laboratoire de sant√© publique du Qu√©bec | Sandrine Moreira et al |
| EPI_ISL_889917 | Canada/QC-JUS-V5302207/2020 | 2020-04-02 | North America/Canada/Quebec/ | Laboratoire de sant√© publique du Qu√©bec | Laboratoire de sant√© publique du Qu√©bec | Sandrine Moreira et al |
| EPI_ISL_889918 | Canada/QC-JUS-V6011193/2020 | 2020-03-31 | North America/Canada/Quebec/ | Laboratoire de sant√© publique du Qu√©bec | Laboratoire de sant√© publique du Qu√©bec | Sandrine Moreira et al |
| EPI_ISL_889919 | Canada/QC-JUS-V6010764/2020 | 2020-03-31 | North America/Canada/Quebec/ | Laboratoire de sant√© publique du Qu√©bec | Laboratoire de sant√© publique du Qu√©bec | Sandrine Moreira et al |
| EPI_ISL_889920 | Canada/QC-JUS-V6010758/2020 | 2020-03-31 | North America/Canada/Quebec/ | Laboratoire de sant√© publique du Qu√©bec | Laboratoire de sant√© publique du Qu√©bec | Sandrine Moreira et al |
| EPI_ISL_889921 | Canada/QC-JUS-V5311647/2020 | 2020-03-29 | North America/Canada/Quebec/ | Laboratoire de sant√© publique du Qu√©bec | Laboratoire de sant√© publique du Qu√©bec | Sandrine Moreira et al |
| EPI_ISL_889922 | Canada/QC-JUS-V5311397/2020 | 2020-03-29 | North America/Canada/Quebec/ | Laboratoire de sant√© publique du Qu√©bec | Laboratoire de sant√© publique du Qu√©bec | Sandrine Moreira et al |
| EPI_ISL_889924 | Canada/QC-JUS-V5311163/2020 | 2020-03-29 | North America/Canada/Quebec/ | Laboratoire de sant√© publique du Qu√©bec | Laboratoire de sant√© publique du Qu√©bec | Sandrine Moreira et al |

| | | | | | | |
|---|---|---|---|---|---|---|
| EPI_ISL_889925 | Canada/QC-JUS-V5311043/2020 | 2020-03-28 | North America/Canada/Quebec/ | Laboratoire de sant√© publique du Qu√©bec | Laboratoire de sant√© publique du Qu√©bec | Sandrine Moreira et al |
| EPI_ISL_889926 | Canada/QC-JUS-V5291245/2020 | 2020-03-29 | North America/Canada/Quebec/ | Laboratoire de sant√© publique du Qu√©bec | Laboratoire de sant√© publique du Qu√©bec | Sandrine Moreira et al |
| EPI_ISL_889927 | Canada/QC-JUS-V6011210/2020 | 2020-03-31 | North America/Canada/Quebec/ | Laboratoire de sant√© publique du Qu√©bec | Laboratoire de sant√© publique du Qu√©bec | Sandrine Moreira et al |
| EPI_ISL_889928 | Canada/QC-JUS-V5312053/2020 | 2020-03-31 | North America/Canada/Quebec/ | Laboratoire de sant√© publique du Qu√©bec | Laboratoire de sant√© publique du Qu√©bec | Sandrine Moreira et al |
| EPI_ISL_889929 | Canada/QC-JUS-V5312122/2020 | 2020-03-31 | North America/Canada/Quebec/ | Laboratoire de sant√© publique du Qu√©bec | Laboratoire de sant√© publique du Qu√©bec | Sandrine Moreira et al |
| EPI_ISL_889930 | Canada/QC-JUS-V5312126/2020 | 2020-03-31 | North America/Canada/Quebec/ | Laboratoire de sant√© publique du Qu√©bec | Laboratoire de sant√© publique du Qu√©bec | Sandrine Moreira et al |
| EPI_ISL_889931 | Canada/QC-JUS-V5312129/2020 | 2020-03-31 | North America/Canada/Quebec/ | Laboratoire de sant√© publique du Qu√©bec | Laboratoire de sant√© publique du Qu√©bec | Sandrine Moreira et al |
| EPI_ISL_889932 | Canada/QC-JUS-V5312031/2020 | 2020-03-31 | North America/Canada/Quebec/ | Laboratoire de sant√© publique du Qu√©bec | Laboratoire de sant√© publique du Qu√©bec | Sandrine Moreira et al |
| EPI_ISL_889933 | Canada/QC-JUS-V6010855/2020 | 2020-03-31 | North America/Canada/Quebec/ | Laboratoire de sant√© publique du Qu√©bec | Laboratoire de sant√© publique du Qu√©bec | Sandrine Moreira et al |
| EPI_ISL_889934 | Canada/QC-JUS-V6011554/2020 | 2020-04-01 | North America/Canada/Quebec/ | Laboratoire de sant√© publique du Qu√©bec | Laboratoire de sant√© publique du Qu√©bec | Sandrine Moreira et al |
| EPI_ISL_889935 | Canada/QC-JUS-V5311725/2020 | 2020-03-29 | North America/Canada/Quebec/ | Laboratoire de sant√© publique du Qu√©bec | Laboratoire de sant√© publique du Qu√©bec | Sandrine Moreira et al |
| EPI_ISL_889936 | Canada/QC-JUS-V6010823/2020 | 2020-04-01 | North America/Canada/Quebec/ | Laboratoire de sant√© publique du Qu√©bec | Laboratoire de sant√© publique du Qu√©bec | Sandrine Moreira et al |
| EPI_ISL_889937 | Canada/QC-JUS-V6011292/2020 | 2020-03-30 | North America/Canada/Quebec/ | Laboratoire de sant√© publique du Qu√©bec | Laboratoire de sant√© publique du Qu√©bec | Sandrine Moreira et al |

| | | | | | | |
|---|---|---|---|---|---|---|
| EPI_ISL_889938 | Canada/QC-JUS-V6011295/2020 | 2020-03-30 | North America/Canada/Quebec/ | Laboratoire de santv© publique du Quv©bec | Laboratoire de santv© publique du Quv©bec | Sandrine Moreira et al |
| EPI_ISL_889939 | Canada/QC-JUS-V6011317/2020 | 2020-03-30 | North America/Canada/Quebec/ | Laboratoire de santv© publique du Quv©bec | Laboratoire de santv© publique du Quv©bec | Sandrine Moreira et al |
| EPI_ISL_889940 | Canada/QC-JUS-V6011211/2020 | 2020-03-30 | North America/Canada/Quebec/ | Laboratoire de santv© publique du Quv©bec | Laboratoire de santv© publique du Quv©bec | Sandrine Moreira et al |
| EPI_ISL_889941 | Canada/QC-JUS-V6011219/2020 | 2020-03-30 | North America/Canada/Quebec/ | Laboratoire de santv© publique du Quv©bec | Laboratoire de santv© publique du Quv©bec | Sandrine Moreira et al |
| EPI_ISL_889942 | Canada/QC-JUS-V6010653/2020 | 2020-03-31 | North America/Canada/Quebec/ | Laboratoire de santv© publique du Quv©bec | Laboratoire de santv© publique du Quv©bec | Sandrine Moreira et al |
| EPI_ISL_889943 | Canada/QC-JUS-V6010734/2020 | 2020-03-31 | North America/Canada/Quebec/ | Laboratoire de santv© publique du Quv©bec | Laboratoire de santv© publique du Quv©bec | Sandrine Moreira et al |
| EPI_ISL_889944 | Canada/QC-JUS-V6010628/2020 | 2020-03-31 | North America/Canada/Quebec/ | Laboratoire de santv© publique du Quv©bec | Laboratoire de santv© publique du Quv©bec | Sandrine Moreira et al |
| EPI_ISL_889945 | Canada/QC-JUS-V6010607/2020 | 2020-03-31 | North America/Canada/Quebec/ | Laboratoire de santv© publique du Quv©bec | Laboratoire de santv© publique du Quv©bec | Sandrine Moreira et al |
| EPI_ISL_889946 | Canada/QC-JUS-V6010770/2020 | 2020-03-31 | North America/Canada/Quebec/ | Laboratoire de santv© publique du Quv©bec | Laboratoire de santv© publique du Quv©bec | Sandrine Moreira et al |
| EPI_ISL_889947 | Canada/QC-JUS-V5301268/2020 | 2020-03-30 | North America/Canada/Quebec/ | Laboratoire de santv© publique du Quv©bec | Laboratoire de santv© publique du Quv©bec | Sandrine Moreira et al |
| EPI_ISL_889948 | Canada/QC-JUS-V5301083/2020 | 2020-03-28 | North America/Canada/Quebec/ | Laboratoire de santv© publique du Quv©bec | Laboratoire de santv© publique du Quv©bec | Sandrine Moreira et al |
| EPI_ISL_889949 | Canada/QC-JUS-V5300486/2020 | 2020-03-27 | North America/Canada/Quebec/ | Laboratoire de santv© publique du Quv©bec | Laboratoire de santv© publique du Quv©bec | Sandrine Moreira et al |
| EPI_ISL_889950 | Canada/QC-JUS-V5300590/2020 | 2020-03-27 | North America/Canada/Quebec/ | Laboratoire de santv© publique du Quv©bec | Laboratoire de santv© publique du Quv©bec | Sandrine Moreira et al |

| | | | | | | |
|---|---|---|---|---|---|---|
| EPI_ISL_889951 | Canada/QC-JUS-V5310788/2020 | 2020-03-30 | North America/Canada/Quebec/ | Laboratoire de sant√© publique du Qu√©bec | Laboratoire de sant√© publique du Qu√©bec | Sandrine Moreira et al |
| EPI_ISL_889952 | Canada/QC-JUS-V5311004/2020 | 2020-03-30 | North America/Canada/Quebec/ | Laboratoire de sant√© publique du Qu√©bec | Laboratoire de sant√© publique du Qu√©bec | Sandrine Moreira et al |
| EPI_ISL_889953 | Canada/QC-JUS-V5311655/2020 | 2020-03-29 | North America/Canada/Quebec/ | Laboratoire de sant√© publique du Qu√©bec | Laboratoire de sant√© publique du Qu√©bec | Sandrine Moreira et al |
| EPI_ISL_889955 | Canada/QC-JUS-V5302179/2020 | 2020-03-29 | North America/Canada/Quebec/ | Laboratoire de sant√© publique du Qu√©bec | Laboratoire de sant√© publique du Qu√©bec | Sandrine Moreira et al |
| EPI_ISL_889956 | Canada/QC-JUS-V5312197/2020 | 2020-03-31 | North America/Canada/Quebec/ | Laboratoire de sant√© publique du Qu√©bec | Laboratoire de sant√© publique du Qu√©bec | Sandrine Moreira et al |
| EPI_ISL_889957 | Canada/QC-JUS-V5310956/2020 | 2020-03-30 | North America/Canada/Quebec/ | Laboratoire de sant√© publique du Qu√©bec | Laboratoire de sant√© publique du Qu√©bec | Sandrine Moreira et al |
| EPI_ISL_889958 | Canada/QC-JUS-V5311787/2020 | 2020-04-02 | North America/Canada/Quebec/ | Laboratoire de sant√© publique du Qu√©bec | Laboratoire de sant√© publique du Qu√©bec | Sandrine Moreira et al |
| EPI_ISL_889959 | Canada/QC-JUS-V5312174/2020 | 2020-03-31 | North America/Canada/Quebec/ | Laboratoire de sant√© publique du Qu√©bec | Laboratoire de sant√© publique du Qu√©bec | Sandrine Moreira et al |
| EPI_ISL_889960 | Canada/QC-JUS-V6011198/2020 | 2020-03-31 | North America/Canada/Quebec/ | Laboratoire de sant√© publique du Qu√©bec | Laboratoire de sant√© publique du Qu√©bec | Sandrine Moreira et al |
| EPI_ISL_889961 | Canada/QC-JUS-V5301853/2020 | 2020-03-29 | North America/Canada/Quebec/ | Laboratoire de sant√© publique du Qu√©bec | Laboratoire de sant√© publique du Qu√©bec | Sandrine Moreira et al |
| EPI_ISL_889962 | Canada/QC-JUS-V5301468/2020 | 2020-03-30 | North America/Canada/Quebec/ | Laboratoire de sant√© publique du Qu√©bec | Laboratoire de sant√© publique du Qu√©bec | Sandrine Moreira et al |
| EPI_ISL_889963 | Canada/QC-JUS-V5301016/2020 | 2020-03-28 | North America/Canada/Quebec/ | Laboratoire de sant√© publique du Qu√©bec | Laboratoire de sant√© publique du Qu√©bec | Sandrine Moreira et al |
| EPI_ISL_889964 | Canada/QC-JUS-V5300928/2020 | 2020-03-27 | North America/Canada/Quebec/ | Laboratoire de sant√© publique du Qu√©bec | Laboratoire de sant√© publique du Qu√©bec | Sandrine Moreira et al |

| | | | | | | |
|---|---|---|---|---|---|---|
| EPI_ISL_889965 | Canada/QC-JUS-V5301070/2020 | 2020-03-27 | North America/Canada/Quebec/ | Laboratoire de sant√© publique du Qu√©bec | Laboratoire de sant√© publique du Qu√©bec | Sandrine Moreira et al |
| EPI_ISL_889966 | Canada/QC-JUS-V5311365/2020 | 2020-03-30 | North America/Canada/Quebec/ | Laboratoire de sant√© publique du Qu√©bec | Laboratoire de sant√© publique du Qu√©bec | Sandrine Moreira et al |
| EPI_ISL_889967 | Canada/QC-JUS-V5300792/2020 | 2020-03-29 | North America/Canada/Quebec/ | Laboratoire de sant√© publique du Qu√©bec | Laboratoire de sant√© publique du Qu√©bec | Sandrine Moreira et al |
| EPI_ISL_889968 | Canada/QC-JUS-V5301731/2020 | 2020-03-28 | North America/Canada/Quebec/ | Laboratoire de sant√© publique du Qu√©bec | Laboratoire de sant√© publique du Qu√©bec | Sandrine Moreira et al |
| EPI_ISL_889969 | Canada/QC-JUS-V5300597/2020 | 2020-03-27 | North America/Canada/Quebec/ | Laboratoire de sant√© publique du Qu√©bec | Laboratoire de sant√© publique du Qu√©bec | Sandrine Moreira et al |
| EPI_ISL_889970 | Canada/QC-JUS-V5301677/2020 | 2020-03-28 | North America/Canada/Quebec/ | Laboratoire de sant√© publique du Qu√©bec | Laboratoire de sant√© publique du Qu√©bec | Sandrine Moreira et al |
| EPI_ISL_889971 | Canada/QC-JUS-V5311291/2020 | 2020-03-30 | North America/Canada/Quebec/ | Laboratoire de sant√© publique du Qu√©bec | Laboratoire de sant√© publique du Qu√©bec | Sandrine Moreira et al |
| EPI_ISL_889972 | Canada/QC-JUS-V5311008/2020 | 2020-03-30 | North America/Canada/Quebec/ | Laboratoire de sant√© publique du Qu√©bec | Laboratoire de sant√© publique du Qu√©bec | Sandrine Moreira et al |
| EPI_ISL_889973 | Canada/QC-JUS-V5311297/2020 | 2020-03-30 | North America/Canada/Quebec/ | Laboratoire de sant√© publique du Qu√©bec | Laboratoire de sant√© publique du Qu√©bec | Sandrine Moreira et al |
| EPI_ISL_889974 | Canada/QC-JUS-V5311856/2020 | 2020-03-31 | North America/Canada/Quebec/ | Laboratoire de sant√© publique du Qu√©bec | Laboratoire de sant√© publique du Qu√©bec | Sandrine Moreira et al |
| EPI_ISL_889975 | Canada/QC-JUS-V5311758/2020 | 2020-03-31 | North America/Canada/Quebec/ | Laboratoire de sant√© publique du Qu√©bec | Laboratoire de sant√© publique du Qu√©bec | Sandrine Moreira et al |
| EPI_ISL_889976 | Canada/QC-JUS-V5310874/2020 | 2020-03-30 | North America/Canada/Quebec/ | Laboratoire de sant√© publique du Qu√©bec | Laboratoire de sant√© publique du Qu√©bec | Sandrine Moreira et al |
| EPI_ISL_889977 | Canada/QC-JUS-V5311756/2020 | 2020-03-30 | North America/Canada/Quebec/ | Laboratoire de sant√© publique du Qu√©bec | Laboratoire de sant√© publique du Qu√©bec | Sandrine Moreira et al |

| | | | | | | |
|---|---|---|---|---|---|---|
| EPI_ISL_889978 | Canada/QC-JUS-V5311697/2020 | 2020-03-30 | North America/Canada/Quebec/ | Laboratoire de santv© publique du Quv©bec | Laboratoire de santv© publique du Quv©bec | Sandrine Moreira et al |
| EPI_ISL_889979 | Canada/QC-JUS-V5310758/2020 | 2020-03-30 | North America/Canada/Quebec/ | Laboratoire de santv© publique du Quv©bec | Laboratoire de santv© publique du Quv©bec | Sandrine Moreira et al |
| EPI_ISL_889980 | Canada/QC-JUS-V5312342/2020 | 2020-03-31 | North America/Canada/Quebec/ | Laboratoire de santv© publique du Quv©bec | Laboratoire de santv© publique du Quv©bec | Sandrine Moreira et al |
| EPI_ISL_889981 | Canada/QC-JUS-V5311718/2020 | 2020-03-31 | North America/Canada/Quebec/ | Laboratoire de santv© publique du Quv©bec | Laboratoire de santv© publique du Quv©bec | Sandrine Moreira et al |
| EPI_ISL_889982 | Canada/QC-JUS-V5312290/2020 | 2020-03-31 | North America/Canada/Quebec/ | Laboratoire de santv© publique du Quv©bec | Laboratoire de santv© publique du Quv©bec | Sandrine Moreira et al |
| EPI_ISL_889983 | Canada/QC-JUS-V5311631/2020 | 2020-03-30 | North America/Canada/Quebec/ | Laboratoire de santv© publique du Quv©bec | Laboratoire de santv© publique du Quv©bec | Sandrine Moreira et al |
| EPI_ISL_889984 | Canada/QC-JUS-V5311405/2020 | 2020-03-30 | North America/Canada/Quebec/ | Laboratoire de santv© publique du Quv©bec | Laboratoire de santv© publique du Quv©bec | Sandrine Moreira et al |
| EPI_ISL_889985 | Canada/QC-JUS-V5310822/2020 | 2020-03-30 | North America/Canada/Quebec/ | Laboratoire de santv© publique du Quv©bec | Laboratoire de santv© publique du Quv©bec | Sandrine Moreira et al |
| EPI_ISL_889986 | Canada/QC-JUS-V5310714/2020 | 2020-03-30 | North America/Canada/Quebec/ | Laboratoire de santv© publique du Quv©bec | Laboratoire de santv© publique du Quv©bec | Sandrine Moreira et al |
| EPI_ISL_889987 | Canada/QC-JUS-V5310701/2020 | 2020-03-30 | North America/Canada/Quebec/ | Laboratoire de santv© publique du Quv©bec | Laboratoire de santv© publique du Quv©bec | Sandrine Moreira et al |
| EPI_ISL_889988 | Canada/QC-JUS-V5311818/2020 | 2020-03-31 | North America/Canada/Quebec/ | Laboratoire de santv© publique du Quv©bec | Laboratoire de santv© publique du Quv©bec | Sandrine Moreira et al |
| EPI_ISL_889989 | Canada/QC-JUS-V5311808/2020 | 2020-03-31 | North America/Canada/Quebec/ | Laboratoire de santv© publique du Quv©bec | Laboratoire de santv© publique du Quv©bec | Sandrine Moreira et al |
| EPI_ISL_889990 | Canada/QC-JUS-V5311059/2020 | 2020-03-30 | North America/Canada/Quebec/ | Laboratoire de santv© publique du Quv©bec | Laboratoire de santv© publique du Quv©bec | Sandrine Moreira et al |

| | | | | | | |
|---|---|---|---|---|---|---|
| EPI_ISL_889991 | Canada/QC-JUS-V5311451/2020 | 2020-03-30 | North America/Canada/Quebec/ | Laboratoire de sant√© publique du Qu√©bec | Laboratoire de sant√© publique du Qu√©bec | Sandrine Moreira et al |
| EPI_ISL_889992 | Canada/QC-JUS-V6011633/2020 | 2020-03-31 | North America/Canada/Quebec/ | Laboratoire de sant√© publique du Qu√©bec | Laboratoire de sant√© publique du Qu√©bec | Sandrine Moreira et al |
| EPI_ISL_889993 | Canada/QC-JUS-V6011426/2020 | 2020-03-31 | North America/Canada/Quebec/ | Laboratoire de sant√© publique du Qu√©bec | Laboratoire de sant√© publique du Qu√©bec | Sandrine Moreira et al |
| EPI_ISL_889994 | Canada/QC-JUS-V6011787/2020 | 2020-04-01 | North America/Canada/Quebec/ | Laboratoire de sant√© publique du Qu√©bec | Laboratoire de sant√© publique du Qu√©bec | Sandrine Moreira et al |
| EPI_ISL_889995 | Canada/QC-JUS-V6012062/2020 | 2020-04-01 | North America/Canada/Quebec/ | Laboratoire de sant√© publique du Qu√©bec | Laboratoire de sant√© publique du Qu√©bec | Sandrine Moreira et al |
| EPI_ISL_889996 | Canada/QC-JUS-V6011772/2020 | 2020-04-01 | North America/Canada/Quebec/ | Laboratoire de sant√© publique du Qu√©bec | Laboratoire de sant√© publique du Qu√©bec | Sandrine Moreira et al |
| EPI_ISL_889997 | Canada/QC-JUS-V6011766/2020 | 2020-04-01 | North America/Canada/Quebec/ | Laboratoire de sant√© publique du Qu√©bec | Laboratoire de sant√© publique du Qu√©bec | Sandrine Moreira et al |
| EPI_ISL_889998 | Canada/QC-JUS-V6011752/2020 | 2020-04-01 | North America/Canada/Quebec/ | Laboratoire de sant√© publique du Qu√©bec | Laboratoire de sant√© publique du Qu√©bec | Sandrine Moreira et al |
| EPI_ISL_889999 | Canada/QC-JUS-V5311846/2020 | 2020-03-31 | North America/Canada/Quebec/ | Laboratoire de sant√© publique du Qu√©bec | Laboratoire de sant√© publique du Qu√©bec | Sandrine Moreira et al |
| EPI_ISL_890000 | Canada/QC-JUS-V6010439/2020 | 2020-03-30 | North America/Canada/Quebec/ | Laboratoire de sant√© publique du Qu√©bec | Laboratoire de sant√© publique du Qu√©bec | Sandrine Moreira et al |
| EPI_ISL_890001 | Canada/QC-JUS-V6010692/2020 | 2020-03-30 | North America/Canada/Quebec/ | Laboratoire de sant√© publique du Qu√©bec | Laboratoire de sant√© publique du Qu√©bec | Sandrine Moreira et al |
| EPI_ISL_890002 | Canada/QC-JUS-V6010710/2020 | 2020-03-30 | North America/Canada/Quebec/ | Laboratoire de sant√© publique du Qu√©bec | Laboratoire de sant√© publique du Qu√©bec | Sandrine Moreira et al |
| EPI_ISL_890003 | Canada/QC-JUS-V6011274/2020 | 2020-03-31 | North America/Canada/Quebec/ | Laboratoire de sant√© publique du Qu√©bec | Laboratoire de sant√© publique du Qu√©bec | Sandrine Moreira et al |

| | | | | | | |
|---|---|---|---|---|---|---|
| EPI_ISL_890004 | Canada/QC-JUS-V5312087/2020 | 2020-03-29 | North America/Canada/Quebec/ | Laboratoire de sant√© publique du Qu√©bec | Laboratoire de sant√© publique du Qu√©bec | Sandrine Moreira et al |
| EPI_ISL_890005 | Canada/QC-JUS-V6011286/2020 | 2020-03-31 | North America/Canada/Quebec/ | Laboratoire de sant√© publique du Qu√©bec | Laboratoire de sant√© publique du Qu√©bec | Sandrine Moreira et al |
| EPI_ISL_890006 | Canada/QC-JUS-V6010502/2020 | 2020-03-30 | North America/Canada/Quebec/ | Laboratoire de sant√© publique du Qu√©bec | Laboratoire de sant√© publique du Qu√©bec | Sandrine Moreira et al |
| EPI_ISL_890007 | Canada/QC-JUS-V6011719/2020 | 2020-04-01 | North America/Canada/Quebec/ | Laboratoire de sant√© publique du Qu√©bec | Laboratoire de sant√© publique du Qu√©bec | Sandrine Moreira et al |
| EPI_ISL_890008 | Canada/QC-JUS-V6010575/2020 | 2020-03-29 | North America/Canada/Quebec/ | Laboratoire de sant√© publique du Qu√©bec | Laboratoire de sant√© publique du Qu√©bec | Sandrine Moreira et al |
| EPI_ISL_890009 | Canada/QC-JUS-V6010574/2020 | 2020-03-30 | North America/Canada/Quebec/ | Laboratoire de sant√© publique du Qu√©bec | Laboratoire de sant√© publique du Qu√©bec | Sandrine Moreira et al |
| EPI_ISL_890010 | Canada/QC-JUS-V6010582/2020 | 2020-03-30 | North America/Canada/Quebec/ | Laboratoire de sant√© publique du Qu√©bec | Laboratoire de sant√© publique du Qu√©bec | Sandrine Moreira et al |
| EPI_ISL_890011 | Canada/QC-JUS-V6010495/2020 | 2020-03-30 | North America/Canada/Quebec/ | Laboratoire de sant√© publique du Qu√©bec | Laboratoire de sant√© publique du Qu√©bec | Sandrine Moreira et al |
| EPI_ISL_890012 | Canada/QC-JUS-V6011272/2020 | 2020-03-30 | North America/Canada/Quebec/ | Laboratoire de sant√© publique du Qu√©bec | Laboratoire de sant√© publique du Qu√©bec | Sandrine Moreira et al |
| EPI_ISL_890013 | Canada/QC-JUS-V6012219/2020 | 2020-03-30 | North America/Canada/Quebec/ | Laboratoire de sant√© publique du Qu√©bec | Laboratoire de sant√© publique du Qu√©bec | Sandrine Moreira et al |
| EPI_ISL_890014 | Canada/QC-JUS-V6011873/2020 | 2020-03-30 | North America/Canada/Quebec/ | Laboratoire de sant√© publique du Qu√©bec | Laboratoire de sant√© publique du Qu√©bec | Sandrine Moreira et al |
| EPI_ISL_890015 | Canada/QC-JUS-V6012313/2020 | 2020-03-31 | North America/Canada/Quebec/ | Laboratoire de sant√© publique du Qu√©bec | Laboratoire de sant√© publique du Qu√©bec | Sandrine Moreira et al |
| EPI_ISL_890016 | Canada/QC-JUS-V6020234/2020 | 2020-03-31 | North America/Canada/Quebec/ | Laboratoire de sant√© publique du Qu√©bec | Laboratoire de sant√© publique du Qu√©bec | Sandrine Moreira et al |

| | | | | | | |
|---|---|---|---|---|---|---|
| EPI_ISL_890017 | Canada/QC-JUS-V6020934/2020 | 2020-04-01 | North America/Canada/Quebec/ | Laboratoire de sant√© publique du Qu√©bec | Laboratoire de sant√© publique du Qu√©bec | Sandrine Moreira et al |
| EPI_ISL_890018 | Canada/QC-JUS-V6020983/2020 | 2020-04-01 | North America/Canada/Quebec/ | Laboratoire de sant√© publique du Qu√©bec | Laboratoire de sant√© publique du Qu√©bec | Sandrine Moreira et al |
| EPI_ISL_890019 | Canada/QC-JUS-V6021003/2020 | 2020-04-01 | North America/Canada/Quebec/ | Laboratoire de sant√© publique du Qu√©bec | Laboratoire de sant√© publique du Qu√©bec | Sandrine Moreira et al |
| EPI_ISL_890020 | Canada/QC-JUS-V6021016/2020 | 2020-04-01 | North America/Canada/Quebec/ | Laboratoire de sant√© publique du Qu√©bec | Laboratoire de sant√© publique du Qu√©bec | Sandrine Moreira et al |
| EPI_ISL_890021 | Canada/QC-JUS-V6020524/2020 | 2020-03-31 | North America/Canada/Quebec/ | Laboratoire de sant√© publique du Qu√©bec | Laboratoire de sant√© publique du Qu√©bec | Sandrine Moreira et al |
| EPI_ISL_890022 | Canada/QC-JUS-V6020411/2020 | 2020-03-31 | North America/Canada/Quebec/ | Laboratoire de sant√© publique du Qu√©bec | Laboratoire de sant√© publique du Qu√©bec | Sandrine Moreira et al |
| EPI_ISL_890023 | Canada/QC-JUS-V6020210/2020 | 2020-03-31 | North America/Canada/Quebec/ | Laboratoire de sant√© publique du Qu√©bec | Laboratoire de sant√© publique du Qu√©bec | Sandrine Moreira et al |
| EPI_ISL_890024 | Canada/QC-JUS-V6020374/2020 | 2020-03-31 | North America/Canada/Quebec/ | Laboratoire de sant√© publique du Qu√©bec | Laboratoire de sant√© publique du Qu√©bec | Sandrine Moreira et al |
| EPI_ISL_890025 | Canada/QC-JUS-V6020569/2020 | 2020-03-31 | North America/Canada/Quebec/ | Laboratoire de sant√© publique du Qu√©bec | Laboratoire de sant√© publique du Qu√©bec | Sandrine Moreira et al |
| EPI_ISL_890026 | Canada/QC-JUS-V6021020/2020 | 2020-03-31 | North America/Canada/Quebec/ | Laboratoire de sant√© publique du Qu√©bec | Laboratoire de sant√© publique du Qu√©bec | Sandrine Moreira et al |
| EPI_ISL_890027 | Canada/QC-JUS-V6020933/2020 | 2020-03-31 | North America/Canada/Quebec/ | Laboratoire de sant√© publique du Qu√©bec | Laboratoire de sant√© publique du Qu√©bec | Sandrine Moreira et al |
| EPI_ISL_890028 | Canada/QC-JUS-V6020745/2020 | 2020-04-02 | North America/Canada/Quebec/ | Laboratoire de sant√© publique du Qu√©bec | Laboratoire de sant√© publique du Qu√©bec | Sandrine Moreira et al |
| EPI_ISL_890029 | Canada/QC-JUS-V6020812/2020 | 2020-04-03 | North America/Canada/Quebec/ | Laboratoire de sant√© publique du Qu√©bec | Laboratoire de sant√© publique du Qu√©bec | Sandrine Moreira et al |

| | | | | | | |
|---|---|---|---|---|---|---|
| EPI_ISL_890030 | Canada/QC-JUS-V6020858/2020 | 2020-04-01 | North America/Canada/Quebec/ | Laboratoire de sant√© publique du Qu√©bec | Laboratoire de sant√© publique du Qu√©bec | Sandrine Moreira et al |
| EPI_ISL_890031 | Canada/QC-JUS-V6020930/2020 | 2020-04-01 | North America/Canada/Quebec/ | Laboratoire de sant√© publique du Qu√©bec | Laboratoire de sant√© publique du Qu√©bec | Sandrine Moreira et al |
| EPI_ISL_890032 | Canada/QC-JUS-V6020937/2020 | 2020-04-01 | North America/Canada/Quebec/ | Laboratoire de sant√© publique du Qu√©bec | Laboratoire de sant√© publique du Qu√©bec | Sandrine Moreira et al |
| EPI_ISL_890033 | Canada/QC-HGA-T30420212D/2020 | 2020-05-04 | North America/Canada/Quebec/ | Laboratoire de sant√© publique du Qu√©bec | Laboratoire de sant√© publique du Qu√©bec | Sandrine Moreira et al |
| EPI_ISL_890034 | Canada/QC-HGA-T30620522D/2020 | 2020-05-06 | North America/Canada/Quebec/ | Laboratoire de sant√© publique du Qu√©bec | Laboratoire de sant√© publique du Qu√©bec | Sandrine Moreira et al |
| EPI_ISL_890035 | Canada/QC-HGA-T30701692D/2020 | 2020-05-07 | North America/Canada/Quebec/ | Laboratoire de sant√© publique du Qu√©bec | Laboratoire de sant√© publique du Qu√©bec | Sandrine Moreira et al |
| EPI_ISL_890036 | Canada/QC-HGA-T30903822D/2020 | 2020-05-09 | North America/Canada/Quebec/ | Laboratoire de sant√© publique du Qu√©bec | Laboratoire de sant√© publique du Qu√©bec | Sandrine Moreira et al |
| EPI_ISL_890037 | Canada/QC-HGA-T30904492D/2020 | 2020-05-09 | North America/Canada/Quebec/ | Laboratoire de sant√© publique du Qu√©bec | Laboratoire de sant√© publique du Qu√©bec | Sandrine Moreira et al |
| EPI_ISL_890038 | Canada/QC-HGA-T30905622D/2020 | 2020-05-09 | North America/Canada/Quebec/ | Laboratoire de sant√© publique du Qu√©bec | Laboratoire de sant√© publique du Qu√©bec | Sandrine Moreira et al |
| EPI_ISL_890039 | Canada/QC-HGA-T30905772D/2020 | 2020-05-09 | North America/Canada/Quebec/ | Laboratoire de sant√© publique du Qu√©bec | Laboratoire de sant√© publique du Qu√©bec | Sandrine Moreira et al |
| EPI_ISL_890040 | Canada/QC-HGA-T31114812D/2020 | 2020-05-11 | North America/Canada/Quebec/ | Laboratoire de sant√© publique du Qu√©bec | Laboratoire de sant√© publique du Qu√©bec | Sandrine Moreira et al |
| EPI_ISL_890041 | Canada/QC-HGA-T31315032D/2020 | 2020-05-13 | North America/Canada/Quebec/ | Laboratoire de sant√© publique du Qu√©bec | Laboratoire de sant√© publique du Qu√©bec | Sandrine Moreira et al |
| EPI_ISL_890042 | Canada/QC-HGA-T31317732D/2020 | 2020-05-13 | North America/Canada/Quebec/ | Laboratoire de sant√© publique du Qu√©bec | Laboratoire de sant√© publique du Qu√©bec | Sandrine Moreira et al |

| | | | | | | |
|---|---|---|---|---|---|---|
| EPI_ISL_890043 | Canada/QC-HGA-T31408552D/2020 | 2020-05-14 | North America/Canada/Quebec/ | Laboratoire de sant√© publique du Qu√©bec | Laboratoire de sant√© publique du Qu√©bec | Sandrine Moreira et al |
| EPI_ISL_890044 | Canada/QC-HGA-T31420632D/2020 | 2020-05-14 | North America/Canada/Quebec/ | Laboratoire de sant√© publique du Qu√©bec | Laboratoire de sant√© publique du Qu√©bec | Sandrine Moreira et al |
| EPI_ISL_890045 | Canada/QC-HGA-T31605182D/2020 | 2020-05-16 | North America/Canada/Quebec/ | Laboratoire de sant√© publique du Qu√©bec | Laboratoire de sant√© publique du Qu√©bec | Sandrine Moreira et al |
| EPI_ISL_890046 | Canada/QC-HGA-T31605332D/2020 | 2020-05-16 | North America/Canada/Quebec/ | Laboratoire de sant√© publique du Qu√©bec | Laboratoire de sant√© publique du Qu√©bec | Sandrine Moreira et al |
| EPI_ISL_890047 | Canada/QC-HGA-T31606362D/2020 | 2020-05-16 | North America/Canada/Quebec/ | Laboratoire de sant√© publique du Qu√©bec | Laboratoire de sant√© publique du Qu√©bec | Sandrine Moreira et al |
| EPI_ISL_890048 | Canada/QC-HGA-T31701972D/2020 | 2020-05-17 | North America/Canada/Quebec/ | Laboratoire de sant√© publique du Qu√©bec | Laboratoire de sant√© publique du Qu√©bec | Sandrine Moreira et al |
| EPI_ISL_890049 | Canada/QC-HGA-T31703742D/2020 | 2020-05-16 | North America/Canada/Quebec/ | Laboratoire de sant√© publique du Qu√©bec | Laboratoire de sant√© publique du Qu√©bec | Sandrine Moreira et al |
| EPI_ISL_890050 | Canada/QC-HGA-T31908562D/2020 | 2020-05-18 | North America/Canada/Quebec/ | Laboratoire de sant√© publique du Qu√©bec | Laboratoire de sant√© publique du Qu√©bec | Sandrine Moreira et al |
| EPI_ISL_890051 | Canada/QC-HGA-T31913322D/2020 | 2020-05-19 | North America/Canada/Quebec/ | Laboratoire de sant√© publique du Qu√©bec | Laboratoire de sant√© publique du Qu√©bec | Sandrine Moreira et al |
| EPI_ISL_890052 | Canada/QC-HGA-T31917852D/2020 | 2020-05-19 | North America/Canada/Quebec/ | Laboratoire de sant√© publique du Qu√©bec | Laboratoire de sant√© publique du Qu√©bec | Sandrine Moreira et al |
| EPI_ISL_890054 | Canada/QC-HGA-T31922152D/2020 | 2020-05-19 | North America/Canada/Quebec/ | Laboratoire de sant√© publique du Qu√©bec | Laboratoire de sant√© publique du Qu√©bec | Sandrine Moreira et al |
| EPI_ISL_890055 | Canada/QC-HGA-T31922642D/2020 | 2020-05-19 | North America/Canada/Quebec/ | Laboratoire de sant√© publique du Qu√©bec | Laboratoire de sant√© publique du Qu√©bec | Sandrine Moreira et al |
| EPI_ISL_890056 | Canada/QC-HGA-T31923212D/2020 | 2020-05-19 | North America/Canada/Quebec/ | Laboratoire de sant√© publique du Qu√©bec | Laboratoire de sant√© publique du Qu√©bec | Sandrine Moreira et al |

| | | | | | | |
|---|---|---|---|---|---|---|
| EPI_ISL_890057 | Canada/QC-HGA-T32016922D/2020 | 2020-05-20 | North America/Canada/Quebec/ | Laboratoire de sant√© publique du Qu√©bec | Laboratoire de sant√© publique du Qu√©bec | Sandrine Moreira et al |
| EPI_ISL_890058 | Canada/QC-HGA-T32018102D/2020 | 2020-05-20 | North America/Canada/Quebec/ | Laboratoire de sant√© publique du Qu√©bec | Laboratoire de sant√© publique du Qu√©bec | Sandrine Moreira et al |
| EPI_ISL_890059 | Canada/QC-HGA-T32018762D/2020 | 2020-05-20 | North America/Canada/Quebec/ | Laboratoire de sant√© publique du Qu√©bec | Laboratoire de sant√© publique du Qu√©bec | Sandrine Moreira et al |
| EPI_ISL_890060 | Canada/QC-HGA-T32018802D/2020 | 2020-05-20 | North America/Canada/Quebec/ | Laboratoire de sant√© publique du Qu√©bec | Laboratoire de sant√© publique du Qu√©bec | Sandrine Moreira et al |
| EPI_ISL_890061 | Canada/QC-HGA-T32018982D/2020 | 2020-05-20 | North America/Canada/Quebec/ | Laboratoire de sant√© publique du Qu√©bec | Laboratoire de sant√© publique du Qu√©bec | Sandrine Moreira et al |
| EPI_ISL_890062 | Canada/QC-HGA-T32019072D/2020 | 2020-05-20 | North America/Canada/Quebec/ | Laboratoire de sant√© publique du Qu√©bec | Laboratoire de sant√© publique du Qu√©bec | Sandrine Moreira et al |
| EPI_ISL_890063 | Canada/QC-HGA-T32019322D/2020 | 2020-05-20 | North America/Canada/Quebec/ | Laboratoire de sant√© publique du Qu√©bec | Laboratoire de sant√© publique du Qu√©bec | Sandrine Moreira et al |
| EPI_ISL_890064 | Canada/QC-HGA-T32019612D/2020 | 2020-05-20 | North America/Canada/Quebec/ | Laboratoire de sant√© publique du Qu√©bec | Laboratoire de sant√© publique du Qu√©bec | Sandrine Moreira et al |
| EPI_ISL_890065 | Canada/QC-HGA-T32019692D/2020 | 2020-05-20 | North America/Canada/Quebec/ | Laboratoire de sant√© publique du Qu√©bec | Laboratoire de sant√© publique du Qu√©bec | Sandrine Moreira et al |
| EPI_ISL_890066 | Canada/QC-HGA-T32019852D/2020 | 2020-05-20 | North America/Canada/Quebec/ | Laboratoire de sant√© publique du Qu√©bec | Laboratoire de sant√© publique du Qu√©bec | Sandrine Moreira et al |
| EPI_ISL_890067 | Canada/QC-HGA-T32019932D/2020 | 2020-05-20 | North America/Canada/Quebec/ | Laboratoire de sant√© publique du Qu√©bec | Laboratoire de sant√© publique du Qu√©bec | Sandrine Moreira et al |
| EPI_ISL_890068 | Canada/QC-HGA-T32020322D/2020 | 2020-05-20 | North America/Canada/Quebec/ | Laboratoire de sant√© publique du Qu√©bec | Laboratoire de sant√© publique du Qu√©bec | Sandrine Moreira et al |
| EPI_ISL_890069 | Canada/QC-HGA-T32020472D/2020 | 2020-05-20 | North America/Canada/Quebec/ | Laboratoire de sant√© publique du Qu√©bec | Laboratoire de sant√© publique du Qu√©bec | Sandrine Moreira et al |

| | | | | | | |
|---|---|---|---|---|---|---|
| EPI_ISL_890070 | Canada/QC-HGA-T32020652D/2020 | 2020-05-20 | North America/Canada/Quebec/ | Laboratoire de sant√© publique du Qu√©bec | Laboratoire de sant√© publique du Qu√©bec | Sandrine Moreira et al |
| EPI_ISL_890071 | Canada/QC-HGA-T32020892D/2020 | 2020-05-20 | North America/Canada/Quebec/ | Laboratoire de sant√© publique du Qu√©bec | Laboratoire de sant√© publique du Qu√©bec | Sandrine Moreira et al |
| EPI_ISL_890072 | Canada/QC-HGA-T32023082D/2020 | 2020-05-20 | North America/Canada/Quebec/ | Laboratoire de sant√© publique du Qu√©bec | Laboratoire de sant√© publique du Qu√©bec | Sandrine Moreira et al |
| EPI_ISL_890073 | Canada/QC-HGA-T32023152D/2020 | 2020-05-20 | North America/Canada/Quebec/ | Laboratoire de sant√© publique du Qu√©bec | Laboratoire de sant√© publique du Qu√©bec | Sandrine Moreira et al |
| EPI_ISL_890074 | Canada/QC-HGA-T32108602D/2020 | 2020-05-21 | North America/Canada/Quebec/ | Laboratoire de sant√© publique du Qu√©bec | Laboratoire de sant√© publique du Qu√©bec | Sandrine Moreira et al |
| EPI_ISL_890075 | Canada/QC-HGA-T32222612D/2020 | 2020-05-22 | North America/Canada/Quebec/ | Laboratoire de sant√© publique du Qu√©bec | Laboratoire de sant√© publique du Qu√©bec | Sandrine Moreira et al |
| EPI_ISL_890076 | Canada/QC-HGA-T32305022D/2020 | 2020-05-23 | North America/Canada/Quebec/ | Laboratoire de sant√© publique du Qu√©bec | Laboratoire de sant√© publique du Qu√©bec | Sandrine Moreira et al |
| EPI_ISL_890077 | Canada/QC-HGA-T32305432D/2020 | 2020-05-23 | North America/Canada/Quebec/ | Laboratoire de sant√© publique du Qu√©bec | Laboratoire de sant√© publique du Qu√©bec | Sandrine Moreira et al |
| EPI_ISL_890078 | Canada/QC-HGA-T32305452D/2020 | 2020-05-23 | North America/Canada/Quebec/ | Laboratoire de sant√© publique du Qu√©bec | Laboratoire de sant√© publique du Qu√©bec | Sandrine Moreira et al |
| EPI_ISL_890079 | Canada/QC-HGA-T32305502D/2020 | 2020-05-23 | North America/Canada/Quebec/ | Laboratoire de sant√© publique du Qu√©bec | Laboratoire de sant√© publique du Qu√©bec | Sandrine Moreira et al |
| EPI_ISL_890080 | Canada/QC-HGA-T32305592D/2020 | 2020-05-23 | North America/Canada/Quebec/ | Laboratoire de sant√© publique du Qu√©bec | Laboratoire de sant√© publique du Qu√©bec | Sandrine Moreira et al |
| EPI_ISL_890081 | Canada/QC-HGA-T32307902D/2020 | 2020-05-23 | North America/Canada/Quebec/ | Laboratoire de sant√© publique du Qu√©bec | Laboratoire de sant√© publique du Qu√©bec | Sandrine Moreira et al |
| EPI_ISL_890082 | Canada/QC-HGA-T32614142D/2020 | 2020-05-26 | North America/Canada/Quebec/ | Laboratoire de sant√© publique du Qu√©bec | Laboratoire de sant√© publique du Qu√©bec | Sandrine Moreira et al |

| | | | | | | |
|---|---|---|---|---|---|---|
| EPI_ISL_890083 | Canada/QC-HGA-T32617002D/2020 | 2020-05-26 | North America/Canada/Quebec/ | Laboratoire de santv© publique du Quv©bec | Laboratoire de santv© publique du Quv©bec | Sandrine Moreira et al |
| EPI_ISL_890084 | Canada/QC-HGA-T32618032D/2020 | 2020-05-26 | North America/Canada/Quebec/ | Laboratoire de santv© publique du Quv©bec | Laboratoire de santv© publique du Quv©bec | Sandrine Moreira et al |
| EPI_ISL_890085 | Canada/QC-HGA-T32621782D/2020 | 2020-05-26 | North America/Canada/Quebec/ | Laboratoire de santv© publique du Quv©bec | Laboratoire de santv© publique du Quv©bec | Sandrine Moreira et al |
| EPI_ISL_890086 | Canada/QC-HGA-T32716472D/2020 | 2020-05-27 | North America/Canada/Quebec/ | Laboratoire de santv© publique du Quv©bec | Laboratoire de santv© publique du Quv©bec | Sandrine Moreira et al |
| EPI_ISL_890087 | Canada/QC-HGA-T33104232D/2020 | 2020-05-31 | North America/Canada/Quebec/ | Laboratoire de santv© publique du Quv©bec | Laboratoire de santv© publique du Quv©bec | Sandrine Moreira et al |
| EPI_ISL_890088 | Canada/QC-HGA-T40116542D/2020 | 2020-06-01 | North America/Canada/Quebec/ | Laboratoire de santv© publique du Quv©bec | Laboratoire de santv© publique du Quv©bec | Sandrine Moreira et al |
| EPI_ISL_890089 | Canada/QC-HGA-T40225842D/2020 | 2020-06-02 | North America/Canada/Quebec/ | Laboratoire de santv© publique du Quv©bec | Laboratoire de santv© publique du Quv©bec | Sandrine Moreira et al |
| EPI_ISL_890090 | Canada/QC-HGA-T40315372D/2020 | 2020-06-03 | North America/Canada/Quebec/ | Laboratoire de santv© publique du Quv©bec | Laboratoire de santv© publique du Quv©bec | Sandrine Moreira et al |
| EPI_ISL_890091 | Canada/QC-HGA-T40603322D/2020 | 2020-06-06 | North America/Canada/Quebec/ | Laboratoire de santv© publique du Quv©bec | Laboratoire de santv© publique du Quv©bec | Sandrine Moreira et al |
| EPI_ISL_890092 | Canada/QC-HGA-T40921102D/2020 | 2020-06-09 | North America/Canada/Quebec/ | Laboratoire de santv© publique du Quv©bec | Laboratoire de santv© publique du Quv©bec | Sandrine Moreira et al |
| EPI_ISL_890094 | Canada/QC-HGA-T42004732D/2020 | 2020-06-20 | North America/Canada/Quebec/ | Laboratoire de santv© publique du Quv©bec | Laboratoire de santv© publique du Quv©bec | Sandrine Moreira et al |
| EPI_ISL_890095 | Canada/QC-HGA-T42004752D/2020 | 2020-06-20 | North America/Canada/Quebec/ | Laboratoire de santv© publique du Quv©bec | Laboratoire de santv© publique du Quv©bec | Sandrine Moreira et al |
| EPI_ISL_890096 | Canada/QC-HGA-T42623572D/2020 | 2020-06-26 | North America/Canada/Quebec/ | Laboratoire de santv© publique du Quv©bec | Laboratoire de santv© publique du Quv©bec | Sandrine Moreira et al |

| | | | | | | |
|---|---|---|---|---|---|---|
| EPI_ISL_890097 | Canada/QC-HGA-T42704622D/2020 | 2020-06-27 | North America/Canada/Quebec/ | Laboratoire de santv© publique du Quv©bec | Laboratoire de santv© publique du Quv©bec | Sandrine Moreira et al |
| EPI_ISL_890098 | Canada/QC-HGA-T42707262D/2020 | 2020-06-27 | North America/Canada/Quebec/ | Laboratoire de santv© publique du Quv©bec | Laboratoire de santv© publique du Quv©bec | Sandrine Moreira et al |
| EPI_ISL_890099 | Canada/QC-HGA-T50216962D/2020 | 2020-07-02 | North America/Canada/Quebec/ | Laboratoire de santv© publique du Quv©bec | Laboratoire de santv© publique du Quv©bec | Sandrine Moreira et al |
| EPI_ISL_890100 | Canada/QC-HGA-T50223862D/2020 | 2020-07-02 | North America/Canada/Quebec/ | Laboratoire de santv© publique du Quv©bec | Laboratoire de santv© publique du Quv©bec | Sandrine Moreira et al |
| EPI_ISL_890101 | Canada/QC-HGA-T50407182D/2020 | 2020-07-04 | North America/Canada/Quebec/ | Laboratoire de santv© publique du Quv©bec | Laboratoire de santv© publique du Quv©bec | Sandrine Moreira et al |
| EPI_ISL_890102 | Canada/QC-HGA-T50607662D/2020 | 2020-07-05 | North America/Canada/Quebec/ | Laboratoire de santv© publique du Quv©bec | Laboratoire de santv© publique du Quv©bec | Sandrine Moreira et al |
| EPI_ISL_890103 | Canada/QC-HGA-T50623622D/2020 | 2020-07-06 | North America/Canada/Quebec/ | Laboratoire de santv© publique du Quv©bec | Laboratoire de santv© publique du Quv©bec | Sandrine Moreira et al |
| EPI_ISL_890104 | Canada/QC-HGA-T50625922D/2020 | 2020-07-06 | North America/Canada/Quebec/ | Laboratoire de santv© publique du Quv©bec | Laboratoire de santv© publique du Quv©bec | Sandrine Moreira et al |
| EPI_ISL_890105 | Canada/QC-HGA-T50820862D/2020 | 2020-07-08 | North America/Canada/Quebec/ | Laboratoire de santv© publique du Quv©bec | Laboratoire de santv© publique du Quv©bec | Sandrine Moreira et al |
| EPI_ISL_890106 | Canada/QC-HGA-T50828662D/2020 | 2020-07-08 | North America/Canada/Quebec/ | Laboratoire de santv© publique du Quv©bec | Laboratoire de santv© publique du Quv©bec | Sandrine Moreira et al |
| EPI_ISL_890107 | Canada/QC-HGA-T50918042D/2020 | 2020-07-09 | North America/Canada/Quebec/ | Laboratoire de santv© publique du Quv©bec | Laboratoire de santv© publique du Quv©bec | Sandrine Moreira et al |
| EPI_ISL_890108 | Canada/QC-HGA-T50920742D/2020 | 2020-07-09 | North America/Canada/Quebec/ | Laboratoire de santv© publique du Quv©bec | Laboratoire de santv© publique du Quv©bec | Sandrine Moreira et al |
| EPI_ISL_890109 | Canada/QC-HGA-T51014382D/2020 | 2020-07-10 | North America/Canada/Quebec/ | Laboratoire de santv© publique du Quv©bec | Laboratoire de santv© publique du Quv©bec | Sandrine Moreira et al |

| | | | | | | |
|---|---|---|---|---|---|---|
| EPI_ISL_890110 | Canada/QC-HGA-T51104212D/2020 | 2020-07-11 | North America/Canada/Quebec/ | Laboratoire de santv© publique du Quv©bec | Laboratoire de santv© publique du Quv©bec | Sandrine Moreira et al |
| EPI_ISL_890111 | Canada/QC-HSE-0074057299/2020 | 2020-07-28 | North America/Canada/Quebec/ | Laboratoire de santv© publique du Quv©bec | Laboratoire de santv© publique du Quv©bec | Sandrine Moreira et al |
| EPI_ISL_890112 | Canada/QC-HSE-0074060966/2020 | 2020-07-30 | North America/Canada/Quebec/ | Laboratoire de santv© publique du Quv©bec | Laboratoire de santv© publique du Quv©bec | Sandrine Moreira et al |
| EPI_ISL_890113 | Canada/QC-HSE-0074061201/2020 | 2020-07-31 | North America/Canada/Quebec/ | Laboratoire de santv© publique du Quv©bec | Laboratoire de santv© publique du Quv©bec | Sandrine Moreira et al |
| EPI_ISL_890114 | Canada/QC-HSE-0074061504/2020 | 2020-07-31 | North America/Canada/Quebec/ | Laboratoire de santv© publique du Quv©bec | Laboratoire de santv© publique du Quv©bec | Sandrine Moreira et al |
| EPI_ISL_890115 | Canada/QC-HSE-0074061450/2020 | 2020-07-31 | North America/Canada/Quebec/ | Laboratoire de santv© publique du Quv©bec | Laboratoire de santv© publique du Quv©bec | Sandrine Moreira et al |
| EPI_ISL_890116 | Canada/QC-HSE-0074061530/2020 | 2020-07-31 | North America/Canada/Quebec/ | Laboratoire de santv© publique du Quv©bec | Laboratoire de santv© publique du Quv©bec | Sandrine Moreira et al |
| EPI_ISL_890117 | Canada/QC-HSE-0084061849/2020 | 2020-08-01 | North America/Canada/Quebec/ | Laboratoire de santv© publique du Quv©bec | Laboratoire de santv© publique du Quv©bec | Sandrine Moreira et al |
| EPI_ISL_890118 | Canada/QC-HSE-0084062050/2020 | 2020-08-01 | North America/Canada/Quebec/ | Laboratoire de santv© publique du Quv©bec | Laboratoire de santv© publique du Quv©bec | Sandrine Moreira et al |
| EPI_ISL_890119 | Canada/QC-HSE-0084061954/2020 | 2020-08-01 | North America/Canada/Quebec/ | Laboratoire de santv© publique du Quv©bec | Laboratoire de santv© publique du Quv©bec | Sandrine Moreira et al |
| EPI_ISL_890120 | Canada/QC-HSE-0084062112/2020 | 2020-08-01 | North America/Canada/Quebec/ | Laboratoire de santv© publique du Quv©bec | Laboratoire de santv© publique du Quv©bec | Sandrine Moreira et al |
| EPI_ISL_890121 | Canada/QC-HSE-0084062067/2020 | 2020-08-01 | North America/Canada/Quebec/ | Laboratoire de santv© publique du Quv©bec | Laboratoire de santv© publique du Quv©bec | Sandrine Moreira et al |
| EPI_ISL_890122 | Canada/QC-HSE-0084062069/2020 | 2020-08-01 | North America/Canada/Quebec/ | Laboratoire de santv© publique du Quv©bec | Laboratoire de santv© publique du Quv©bec | Sandrine Moreira et al |

| | | | | | | |
|---|---|---|---|---|---|---|
| EPI_ISL_890123 | Canada/QC-HSE-0084062324/2020 | 2020-08-02 | North America/Canada/Quebec/ | Laboratoire de santv© publique du Quv©bec | Laboratoire de santv© publique du Quv©bec | Sandrine Moreira et al |
| EPI_ISL_890124 | Canada/QC-HSE-0084062206/2020 | 2020-08-02 | North America/Canada/Quebec/ | Laboratoire de santv© publique du Quv©bec | Laboratoire de santv© publique du Quv©bec | Sandrine Moreira et al |
| EPI_ISL_890125 | Canada/QC-HSE-0084062222/2020 | 2020-08-02 | North America/Canada/Quebec/ | Laboratoire de santv© publique du Quv©bec | Laboratoire de santv© publique du Quv©bec | Sandrine Moreira et al |
| EPI_ISL_890126 | Canada/QC-HSE-0084062271/2020 | 2020-08-02 | North America/Canada/Quebec/ | Laboratoire de santv© publique du Quv©bec | Laboratoire de santv© publique du Quv©bec | Sandrine Moreira et al |
| EPI_ISL_890127 | Canada/QC-HSE-0084062284/2020 | 2020-08-02 | North America/Canada/Quebec/ | Laboratoire de santv© publique du Quv©bec | Laboratoire de santv© publique du Quv©bec | Sandrine Moreira et al |
| EPI_ISL_890128 | Canada/QC-HSE-0084062758/2020 | 2020-08-03 | North America/Canada/Quebec/ | Laboratoire de santv© publique du Quv©bec | Laboratoire de santv© publique du Quv©bec | Sandrine Moreira et al |
| EPI_ISL_890129 | Canada/QC-HSE-0084063728/2020 | 2020-08-03 | North America/Canada/Quebec/ | Laboratoire de santv© publique du Quv©bec | Laboratoire de santv© publique du Quv©bec | Sandrine Moreira et al |
| EPI_ISL_890130 | Canada/QC-HSE-0084063775/2020 | 2020-08-03 | North America/Canada/Quebec/ | Laboratoire de santv© publique du Quv©bec | Laboratoire de santv© publique du Quv©bec | Sandrine Moreira et al |
| EPI_ISL_890131 | Canada/QC-HSE-0084063631/2020 | 2020-08-03 | North America/Canada/Quebec/ | Laboratoire de santv© publique du Quv©bec | Laboratoire de santv© publique du Quv©bec | Sandrine Moreira et al |
| EPI_ISL_890132 | Canada/QC-HSE-0084063417/2020 | 2020-08-03 | North America/Canada/Quebec/ | Laboratoire de santv© publique du Quv©bec | Laboratoire de santv© publique du Quv©bec | Sandrine Moreira et al |
| EPI_ISL_890133 | Canada/QC-HSE-0084063398/2020 | 2020-08-03 | North America/Canada/Quebec/ | Laboratoire de santv© publique du Quv©bec | Laboratoire de santv© publique du Quv©bec | Sandrine Moreira et al |
| EPI_ISL_890134 | Canada/QC-HSE-0084063888/2020 | 2020-08-03 | North America/Canada/Quebec/ | Laboratoire de santv© publique du Quv©bec | Laboratoire de santv© publique du Quv©bec | Sandrine Moreira et al |
| EPI_ISL_890135 | Canada/QC-HSE-0084063897/2020 | 2020-08-03 | North America/Canada/Quebec/ | Laboratoire de santv© publique du Quv©bec | Laboratoire de santv© publique du Quv©bec | Sandrine Moreira et al |

| | | | | | | |
|---|---|---|---|---|---|---|
| EPI_ISL_890136 | Canada/QC-HSE-0084063684/2020 | 2020-08-03 | North America/Canada/Quebec/ | Laboratoire de sant√© publique du Qu√©bec | Laboratoire de sant√© publique du Qu√©bec | Sandrine Moreira et al |
| EPI_ISL_890137 | Canada/QC-HSE-0084064416/2020 | 2020-08-03 | North America/Canada/Quebec/ | Laboratoire de sant√© publique du Qu√©bec | Laboratoire de sant√© publique du Qu√©bec | Sandrine Moreira et al |
| EPI_ISL_890138 | Canada/QC-HSE-0084064640/2020 | 2020-08-03 | North America/Canada/Quebec/ | Laboratoire de sant√© publique du Qu√©bec | Laboratoire de sant√© publique du Qu√©bec | Sandrine Moreira et al |
| EPI_ISL_890139 | Canada/QC-HSE-0084069930/2020 | 2020-08-08 | North America/Canada/Quebec/ | Laboratoire de sant√© publique du Qu√©bec | Laboratoire de sant√© publique du Qu√©bec | Sandrine Moreira et al |
| EPI_ISL_890140 | Canada/QC-HSE-0084064226/2020 | 2020-08-04 | North America/Canada/Quebec/ | Laboratoire de sant√© publique du Qu√©bec | Laboratoire de sant√© publique du Qu√©bec | Sandrine Moreira et al |
| EPI_ISL_890141 | Canada/QC-HSE-0084064909/2020 | 2020-08-04 | North America/Canada/Quebec/ | Laboratoire de sant√© publique du Qu√©bec | Laboratoire de sant√© publique du Qu√©bec | Sandrine Moreira et al |
| EPI_ISL_890142 | Canada/QC-HSE-0084065864/2020 | 2020-08-04 | North America/Canada/Quebec/ | Laboratoire de sant√© publique du Qu√©bec | Laboratoire de sant√© publique du Qu√©bec | Sandrine Moreira et al |
| EPI_ISL_890143 | Canada/QC-HSE-0084066607/2020 | 2020-08-05 | North America/Canada/Quebec/ | Laboratoire de sant√© publique du Qu√©bec | Laboratoire de sant√© publique du Qu√©bec | Sandrine Moreira et al |
| EPI_ISL_890144 | Canada/QC-HSE-0084066867/2020 | 2020-08-05 | North America/Canada/Quebec/ | Laboratoire de sant√© publique du Qu√©bec | Laboratoire de sant√© publique du Qu√©bec | Sandrine Moreira et al |
| EPI_ISL_890145 | Canada/QC-HSE-0084066932/2020 | 2020-08-05 | North America/Canada/Quebec/ | Laboratoire de sant√© publique du Qu√©bec | Laboratoire de sant√© publique du Qu√©bec | Sandrine Moreira et al |
| EPI_ISL_890146 | Canada/QC-HSE-0084066933/2020 | 2020-08-05 | North America/Canada/Quebec/ | Laboratoire de sant√© publique du Qu√©bec | Laboratoire de sant√© publique du Qu√©bec | Sandrine Moreira et al |
| EPI_ISL_890147 | Canada/QC-HSE-0084066846/2020 | 2020-08-05 | North America/Canada/Quebec/ | Laboratoire de sant√© publique du Qu√©bec | Laboratoire de sant√© publique du Qu√©bec | Sandrine Moreira et al |
| EPI_ISL_890148 | Canada/QC-HSE-0084068054/2020 | 2020-08-06 | North America/Canada/Quebec/ | Laboratoire de sant√© publique du Qu√©bec | Laboratoire de sant√© publique du Qu√©bec | Sandrine Moreira et al |

| | | | | | | |
|---|---|---|---|---|---|---|
| EPI_ISL_890149 | Canada/QC-HSE-0084068164/2020 | 2020-08-06 | North America/Canada/Quebec/ | Laboratoire de santv© publique du Quv©bec | Laboratoire de santv© publique du Quv©bec | Sandrine Moreira et al |
| EPI_ISL_890150 | Canada/QC-HSE-0084068931/2020 | 2020-08-07 | North America/Canada/Quebec/ | Laboratoire de santv© publique du Quv©bec | Laboratoire de santv© publique du Quv©bec | Sandrine Moreira et al |
| EPI_ISL_890151 | Canada/QC-HSE-0084068866/2020 | 2020-08-07 | North America/Canada/Quebec/ | Laboratoire de santv© publique du Quv©bec | Laboratoire de santv© publique du Quv©bec | Sandrine Moreira et al |
| EPI_ISL_890153 | Canada/QC-HSE-0084069284/2020 | 2020-08-07 | North America/Canada/Quebec/ | Laboratoire de santv© publique du Quv©bec | Laboratoire de santv© publique du Quv©bec | Sandrine Moreira et al |
| EPI_ISL_890154 | Canada/QC-HSE-0084069797/2020 | 2020-08-08 | North America/Canada/Quebec/ | Laboratoire de santv© publique du Quv©bec | Laboratoire de santv© publique du Quv©bec | Sandrine Moreira et al |
| EPI_ISL_890155 | Canada/QC-HSE-0084069798/2020 | 2020-08-08 | North America/Canada/Quebec/ | Laboratoire de santv© publique du Quv©bec | Laboratoire de santv© publique du Quv©bec | Sandrine Moreira et al |
| EPI_ISL_890156 | Canada/QC-HSE-0084069646/2020 | 2020-08-08 | North America/Canada/Quebec/ | Laboratoire de santv© publique du Quv©bec | Laboratoire de santv© publique du Quv©bec | Sandrine Moreira et al |
| EPI_ISL_890157 | Canada/QC-HSE-0084070061/2020 | 2020-08-09 | North America/Canada/Quebec/ | Laboratoire de santv© publique du Quv©bec | Laboratoire de santv© publique du Quv©bec | Sandrine Moreira et al |
| EPI_ISL_890158 | Canada/QC-HSE-0084071140/2020 | 2020-08-10 | North America/Canada/Quebec/ | Laboratoire de santv© publique du Quv©bec | Laboratoire de santv© publique du Quv©bec | Sandrine Moreira et al |
| EPI_ISL_890159 | Canada/QC-HSE-0084071266/2020 | 2020-08-10 | North America/Canada/Quebec/ | Laboratoire de santv© publique du Quv©bec | Laboratoire de santv© publique du Quv©bec | Sandrine Moreira et al |
| EPI_ISL_890160 | Canada/QC-HSE-0084072240/2020 | 2020-08-11 | North America/Canada/Quebec/ | Laboratoire de santv© publique du Quv©bec | Laboratoire de santv© publique du Quv©bec | Sandrine Moreira et al |
| EPI_ISL_890161 | Canada/QC-HSE-0084072622/2020 | 2020-08-11 | North America/Canada/Quebec/ | Laboratoire de santv© publique du Quv©bec | Laboratoire de santv© publique du Quv©bec | Sandrine Moreira et al |
| EPI_ISL_890162 | Canada/QC-HSE-0084073746/2020 | 2020-08-12 | North America/Canada/Quebec/ | Laboratoire de santv© publique du Quv©bec | Laboratoire de santv© publique du Quv©bec | Sandrine Moreira et al |

| | | | | | | |
|---|---|---|---|---|---|---|
| EPI_ISL_890163 | Canada/QC-HSE-0084081056/2020 | 2020-08-20 | North America/Canada/Quebec/ | Laboratoire de santv© publique du Quv©bec | Laboratoire de santv© publique du Quv©bec | Sandrine Moreira et al |
| EPI_ISL_890164 | Canada/QC-HSE-0084083763/2020 | 2020-08-24 | North America/Canada/Quebec/ | Laboratoire de santv© publique du Quv©bec | Laboratoire de santv© publique du Quv©bec | Sandrine Moreira et al |
| EPI_ISL_890165 | Canada/QC-HSE-0084083899/2020 | 2020-08-24 | North America/Canada/Quebec/ | Laboratoire de santv© publique du Quv©bec | Laboratoire de santv© publique du Quv©bec | Sandrine Moreira et al |
| EPI_ISL_890166 | Canada/QC-HSE-0084083956/2020 | 2020-08-24 | North America/Canada/Quebec/ | Laboratoire de santv© publique du Quv©bec | Laboratoire de santv© publique du Quv©bec | Sandrine Moreira et al |
| EPI_ISL_890167 | Canada/QC-HSE-0084083901/2020 | 2020-08-24 | North America/Canada/Quebec/ | Laboratoire de santv© publique du Quv©bec | Laboratoire de santv© publique du Quv©bec | Sandrine Moreira et al |
| EPI_ISL_890168 | Canada/QC-HSE-0084084648/2020 | 2020-08-25 | North America/Canada/Quebec/ | Laboratoire de santv© publique du Quv©bec | Laboratoire de santv© publique du Quv©bec | Sandrine Moreira et al |
| EPI_ISL_890169 | Canada/QC-HSE-0084085180/2020 | 2020-08-25 | North America/Canada/Quebec/ | Laboratoire de santv© publique du Quv©bec | Laboratoire de santv© publique du Quv©bec | Sandrine Moreira et al |
| EPI_ISL_890170 | Canada/QC-HSE-0084085179/2020 | 2020-08-25 | North America/Canada/Quebec/ | Laboratoire de santv© publique du Quv©bec | Laboratoire de santv© publique du Quv©bec | Sandrine Moreira et al |
| EPI_ISL_890171 | Canada/QC-HSE-0084085756/2020 | 2020-08-26 | North America/Canada/Quebec/ | Laboratoire de santv© publique du Quv©bec | Laboratoire de santv© publique du Quv©bec | Sandrine Moreira et al |
| EPI_ISL_890172 | Canada/QC-HSE-0084086134/2020 | 2020-08-26 | North America/Canada/Quebec/ | Laboratoire de santv© publique du Quv©bec | Laboratoire de santv© publique du Quv©bec | Sandrine Moreira et al |
| EPI_ISL_890173 | Canada/QC-HSE-0084086135/2020 | 2020-08-26 | North America/Canada/Quebec/ | Laboratoire de santv© publique du Quv©bec | Laboratoire de santv© publique du Quv©bec | Sandrine Moreira et al |

| | | | | | | |
|---|---|---|---|---|---|---|
| EPI_ISL_890186 | Indonesia/YO-UGM-109996/2020 | 2020-09-04 | Asia/Indonesia/Yogyakarta/ | Academic Hospital of Gadjah Mada University (RSA UGM) | Genetics Working Group (Pokja Genetik) Faculty of Medicine, Public Health and Nursing Universitas Gadjah Mada (FK-KMK UGM); Disease Investigation Center Wates Ministry of Agriculture Indonesia; Department of Microbiology FK-KMK UGM; Laboratorium Diagnostik Yayasan Tahija World Mosquito Program (WMP) Yogyakarta Center for Tropical Medicine FK-KMK UGM; Integrated Research center FK-KMK UGM; Department of Computer Science and Electronics FMIPA UGM | Gunadi et al |

| | | | | | | |
|---|---|---|---|---|---|---|
| EPI_ISL_890187 | Indonesia/YO-UGM-109997/2020 | 2020-09-04 | Asia/Indonesia/Yogyakarta/ | Academic Hospital of Gadjah Mada University (RSA UGM) | Genetics Working Group (Pokja Genetik) Faculty of Medicine, Public Health and Nursing Universitas Gadjah Mada (FK-KMK UGM); Disease Investigation Center Wates Ministry of Agriculture Indonesia; Department of Microbiology FK-KMK UGM; Laboratorium Diagnostik Yayasan Tahija World Mosquito Program (WMP) Yogyakarta Center for Tropical Medicine FK-KMK UGM; Integrated Research center FK-KMK UGM; Department of Computer Science and Electronics FMIPA UGM | Gunadi et al |
| EPI_ISL_890188 | Bangladesh/GRBL_S1/2020 | 2020-12-19 | Asia/Bangladesh/Dhaka/ | Gonoshasthaya-RNA Research Center, Gonoshasthaya-RNA Molecular Diagnostics and Research Center | Gonoshasthaya-RNA Research Center, Gonoshasthaya-RNA Molecular Diagnostics and Research Center | Jamiruddin et al |

| | | | | | | |
|---|---|---|---|---|---|---|
| EPI_ISL_890191 | Bangladesh/GRBL_S9/2020 | 2020-10-05 | Asia/Bangladesh/Dhaka/ | Gonoshasthaya-RNA Research Center, Gonoshasthaya-RNA Molecular Diagnostics and Research Center | Gonoshasthaya-RNA Research Center, Gonoshasthaya-RNA Molecular Diagnostics and Research Center | Jamiruddin et al |
| EPI_ISL_890193 | Bangladesh/GRBL_S11/2020 | 2020-09-05 | Asia/Bangladesh/Dhaka/ | Gonoshasthaya-RNA Research Center, Gonoshasthaya-RNA Molecular Diagnostics and Research Center | Gonoshasthaya-RNA Research Center, Gonoshasthaya-RNA Molecular Diagnostics and Research Center | Jamiruddin et al |
| EPI_ISL_890221 | Egypt/CUNCI-HGC12I038/2021 | 2021-01-09 | Africa/Egypt/Egypt/ | Cancer Biology Department, National Cancer Institute | Cancer Biology Department, National Cancer Institute | Zekri et al |
| EPI_ISL_890227 | Egypt/CUNCI-HGC12I047/2021 | 2021-01-09 | Africa/Egypt/Egypt/ | Cancer Biology Department, National Cancer Institute | Cancer Biology Department, National Cancer Institute | Zekri et al |
| EPI_ISL_890251 | Belgium/rega-1574/2020 | 2020-12-23 | Europe/Belgium/Leuven/Langdorp | KU Leuven, Rega Institute, Clinical and Epidemiological Virology | KU Leuven, Rega Institute, Clinical and Epidemiological Virology | Tony Wawina-Bokalanga et al |
| EPI_ISL_890263 | Belgium/rega-1587/2020 | 2020-12-04 | Europe/Belgium/Leuven/ | KU Leuven, Rega Institute, Clinical and Epidemiological Virology | KU Leuven, Rega Institute, Clinical and Epidemiological Virology | Tony Wawina-Bokalanga et al |
| EPI_ISL_890267 | Belgium/rega-1592/2020 | 2020-12-23 | Europe/Belgium/Leuven/Holsbeek | KU Leuven, Rega Institute, Clinical and Epidemiological Virology | KU Leuven, Rega Institute, Clinical and Epidemiological Virology | Tony Wawina-Bokalanga et al |

| | | | | | | |
|---|---|---|---|---|---|---|
| EPI_ISL_890327 | Belgium/rega-1711/2020 | 2020-12-20 | Europe/Belgium/Leuven/Bekkevoort | KU Leuven, Rega Institute, Clinical and Epidemiological Virology | KU Leuven, Rega Institute, Clinical and Epidemiological Virology | Tony Wawina-Bokalanga et al |
| EPI_ISL_890355 | Mayotte/IPP00602/2021 | 2021-01-09 | Europe/France/Mayotte/Mamoudzou | Hospital | National Reference Center for Viruses of Respiratory Infections, Institut Pasteur, Paris | Marion Barbet et al |
| EPI_ISL_890361 | USA/LA-EVTL1553/2020 | 2020-12-30 | North America/USA/Louisiana/Natchito | LSUHS Emerging Viral Threat Laboratory | Microbial Genome Sequencing Center | Jeremy P. Kamil et al |
| EPI_ISL_890371 | Canada/QC-JUS-V5210578/2020 | 2020-03-19 | North America/Canada/Quebec/ | Laboratoire de santv© publique du Quv©bec | Laboratoire de santv© publique du Quv©bec | Sandrine Moreira et al |
| EPI_ISL_890372 | Canada/QC-JUS-V5272161/2020 | 2020-03-26 | North America/Canada/Quebec/ | Laboratoire de santv© publique du Quv©bec | Laboratoire de santv© publique du Quv©bec | Sandrine Moreira et al |
| EPI_ISL_890373 | Canada/QC-JUS-V5281067/2020 | 2020-03-28 | North America/Canada/Quebec/ | Laboratoire de santv© publique du Quv©bec | Laboratoire de santv© publique du Quv©bec | Sandrine Moreira et al |
| EPI_ISL_890374 | Canada/QC-JUS-V5220376/2020 | 2020-03-20 | North America/Canada/Quebec/ | Laboratoire de santv© publique du Quv©bec | Laboratoire de santv© publique du Quv©bec | Sandrine Moreira et al |
| EPI_ISL_890375 | Canada/QC-JUS-V5220163/2020 | 2020-03-20 | North America/Canada/Quebec/ | Laboratoire de santv© publique du Quv©bec | Laboratoire de santv© publique du Quv©bec | Sandrine Moreira et al |
| EPI_ISL_890376 | Canada/QC-JUS-V5190228/2020 | 2020-03-16 | North America/Canada/Quebec/ | Laboratoire de santv© publique du Quv©bec | Laboratoire de santv© publique du Quv©bec | Sandrine Moreira et al |
| EPI_ISL_890377 | Canada/QC-JUS-V5220324/2020 | 2020-03-20 | North America/Canada/Quebec/ | Laboratoire de santv© publique du Quv©bec | Laboratoire de santv© publique du Quv©bec | Sandrine Moreira et al |
| EPI_ISL_890378 | Canada/QC-JUS-V5220331/2020 | 2020-03-20 | North America/Canada/Quebec/ | Laboratoire de santv© publique du Quv©bec | Laboratoire de santv© publique du Quv©bec | Sandrine Moreira et al |
| EPI_ISL_890379 | Canada/QC-JUS-V5220339/2020 | 2020-03-20 | North America/Canada/Quebec/ | Laboratoire de santv© publique du Quv©bec | Laboratoire de santv© publique du Quv©bec | Sandrine Moreira et al |

| | | | | | | |
|---|---|---|---|---|---|---|
| EPI_ISL_890380 | Canada/QC-JUS-V5220462/2020 | 2020-03-20 | North America/Canada/Quebec/ | Laboratoire de sant√© publique du Qu√©bec | Laboratoire de sant√© publique du Qu√©bec | Sandrine Moreira et al |
| EPI_ISL_890381 | Canada/QC-JUS-V5220486/2020 | 2020-03-20 | North America/Canada/Quebec/ | Laboratoire de sant√© publique du Qu√©bec | Laboratoire de sant√© publique du Qu√©bec | Sandrine Moreira et al |
| EPI_ISL_890382 | Canada/QC-JUS-V5220405/2020 | 2020-03-20 | North America/Canada/Quebec/ | Laboratoire de sant√© publique du Qu√©bec | Laboratoire de sant√© publique du Qu√©bec | Sandrine Moreira et al |
| EPI_ISL_890383 | Canada/QC-JUS-V5220883/2020 | 2020-03-20 | North America/Canada/Quebec/ | Laboratoire de sant√© publique du Qu√©bec | Laboratoire de sant√© publique du Qu√©bec | Sandrine Moreira et al |
| EPI_ISL_890384 | Canada/QC-JUS-V5220225/2020 | 2020-03-20 | North America/Canada/Quebec/ | Laboratoire de sant√© publique du Qu√©bec | Laboratoire de sant√© publique du Qu√©bec | Sandrine Moreira et al |
| EPI_ISL_890385 | Canada/QC-JUS-V5220321/2020 | 2020-03-20 | North America/Canada/Quebec/ | Laboratoire de sant√© publique du Qu√©bec | Laboratoire de sant√© publique du Qu√©bec | Sandrine Moreira et al |
| EPI_ISL_890386 | Canada/QC-JUS-V5211194/2020 | 2020-03-21 | North America/Canada/Quebec/ | Laboratoire de sant√© publique du Qu√©bec | Laboratoire de sant√© publique du Qu√©bec | Sandrine Moreira et al |
| EPI_ISL_890387 | Canada/QC-JUS-V5220146/2020 | 2020-03-20 | North America/Canada/Quebec/ | Laboratoire de sant√© publique du Qu√©bec | Laboratoire de sant√© publique du Qu√©bec | Sandrine Moreira et al |
| EPI_ISL_890388 | Canada/QC-JUS-V5220606/2020 | 2020-03-20 | North America/Canada/Quebec/ | Laboratoire de sant√© publique du Qu√©bec | Laboratoire de sant√© publique du Qu√©bec | Sandrine Moreira et al |
| EPI_ISL_890389 | Canada/QC-JUS-V5220493/2020 | 2020-03-19 | North America/Canada/Quebec/ | Laboratoire de sant√© publique du Qu√©bec | Laboratoire de sant√© publique du Qu√©bec | Sandrine Moreira et al |
| EPI_ISL_890390 | Canada/QC-JUS-V5211010/2020 | 2020-03-21 | North America/Canada/Quebec/ | Laboratoire de sant√© publique du Qu√©bec | Laboratoire de sant√© publique du Qu√©bec | Sandrine Moreira et al |
| EPI_ISL_890391 | Canada/QC-JUS-V5220328/2020 | 2020-03-19 | North America/Canada/Quebec/ | Laboratoire de sant√© publique du Qu√©bec | Laboratoire de sant√© publique du Qu√©bec | Sandrine Moreira et al |
| EPI_ISL_890392 | Canada/QC-JUS-V5221057/2020 | 2020-03-20 | North America/Canada/Quebec/ | Laboratoire de sant√© publique du Qu√©bec | Laboratoire de sant√© publique du Qu√©bec | Sandrine Moreira et al |

| | | | | | | |
|---|---|---|---|---|---|---|
| EPI_ISL_890393 | Canada/QC-JUS-V5220414/2020 | 2020-03-20 | North America/Canada/Quebec/ | Laboratoire de sant√© publique du Qu√©bec | Laboratoire de sant√© publique du Qu√©bec | Sandrine Moreira et al |
| EPI_ISL_890394 | Canada/QC-JUS-V5220871/2020 | 2020-03-20 | North America/Canada/Quebec/ | Laboratoire de sant√© publique du Qu√©bec | Laboratoire de sant√© publique du Qu√©bec | Sandrine Moreira et al |
| EPI_ISL_890395 | Canada/QC-JUS-V5220498/2020 | 2020-03-19 | North America/Canada/Quebec/ | Laboratoire de sant√© publique du Qu√©bec | Laboratoire de sant√© publique du Qu√©bec | Sandrine Moreira et al |
| EPI_ISL_890396 | Canada/QC-JUS-V5220540/2020 | 2020-03-19 | North America/Canada/Quebec/ | Laboratoire de sant√© publique du Qu√©bec | Laboratoire de sant√© publique du Qu√©bec | Sandrine Moreira et al |
| EPI_ISL_890397 | Canada/QC-JUS-V5221077/2020 | 2020-03-20 | North America/Canada/Quebec/ | Laboratoire de sant√© publique du Qu√©bec | Laboratoire de sant√© publique du Qu√©bec | Sandrine Moreira et al |
| EPI_ISL_890398 | Canada/QC-JUS-V5210971/2020 | 2020-03-20 | North America/Canada/Quebec/ | Laboratoire de sant√© publique du Qu√©bec | Laboratoire de sant√© publique du Qu√©bec | Sandrine Moreira et al |
| EPI_ISL_890399 | Canada/QC-JUS-V5230905/2020 | 2020-03-22 | North America/Canada/Quebec/ | Laboratoire de sant√© publique du Qu√©bec | Laboratoire de sant√© publique du Qu√©bec | Sandrine Moreira et al |
| EPI_ISL_890400 | Canada/QC-JUS-V5230756/2020 | 2020-03-22 | North America/Canada/Quebec/ | Laboratoire de sant√© publique du Qu√©bec | Laboratoire de sant√© publique du Qu√©bec | Sandrine Moreira et al |
| EPI_ISL_890401 | Canada/QC-JUS-V5221002/2020 | 2020-03-20 | North America/Canada/Quebec/ | Laboratoire de sant√© publique du Qu√©bec | Laboratoire de sant√© publique du Qu√©bec | Sandrine Moreira et al |
| EPI_ISL_890402 | Canada/QC-JUS-V5220648/2020 | 2020-03-20 | North America/Canada/Quebec/ | Laboratoire de sant√© publique du Qu√©bec | Laboratoire de sant√© publique du Qu√©bec | Sandrine Moreira et al |
| EPI_ISL_890403 | Canada/QC-JUS-V5220550/2020 | 2020-03-19 | North America/Canada/Quebec/ | Laboratoire de sant√© publique du Qu√©bec | Laboratoire de sant√© publique du Qu√©bec | Sandrine Moreira et al |
| EPI_ISL_890404 | Canada/QC-JUS-V5210762/2020 | 2020-03-19 | North America/Canada/Quebec/ | Laboratoire de sant√© publique du Qu√©bec | Laboratoire de sant√© publique du Qu√©bec | Sandrine Moreira et al |
| EPI_ISL_890405 | Canada/QC-JUS-V5220315/2020 | 2020-03-20 | North America/Canada/Quebec/ | Laboratoire de sant√© publique du Qu√©bec | Laboratoire de sant√© publique du Qu√©bec | Sandrine Moreira et al |

| | | | | | | |
|---|---|---|---|---|---|---|
| EPI_ISL_890406 | Canada/QC-JUS-V5220320/2020 | 2020-03-21 | North America/Canada/Quebec/ | Laboratoire de sant√© publique du Qu√©bec | Laboratoire de sant√© publique du Qu√©bec | Sandrine Moreira et al |
| EPI_ISL_890407 | Canada/QC-JUS-V5230821/2020 | 2020-03-22 | North America/Canada/Quebec/ | Laboratoire de sant√© publique du Qu√©bec | Laboratoire de sant√© publique du Qu√©bec | Sandrine Moreira et al |
| EPI_ISL_890408 | Canada/QC-JUS-V5230998/2020 | 2020-03-22 | North America/Canada/Quebec/ | Laboratoire de sant√© publique du Qu√©bec | Laboratoire de sant√© publique du Qu√©bec | Sandrine Moreira et al |
| EPI_ISL_890409 | Canada/QC-JUS-V5220930/2020 | 2020-03-22 | North America/Canada/Quebec/ | Laboratoire de sant√© publique du Qu√©bec | Laboratoire de sant√© publique du Qu√©bec | Sandrine Moreira et al |
| EPI_ISL_890410 | Canada/QC-JUS-V5220448/2020 | 2020-03-19 | North America/Canada/Quebec/ | Laboratoire de sant√© publique du Qu√©bec | Laboratoire de sant√© publique du Qu√©bec | Sandrine Moreira et al |
| EPI_ISL_890411 | Canada/QC-JUS-V5231353/2020 | 2020-03-22 | North America/Canada/Quebec/ | Laboratoire de sant√© publique du Qu√©bec | Laboratoire de sant√© publique du Qu√©bec | Sandrine Moreira et al |
| EPI_ISL_890412 | Canada/QC-JUS-V5231436/2020 | 2020-03-22 | North America/Canada/Quebec/ | Laboratoire de sant√© publique du Qu√©bec | Laboratoire de sant√© publique du Qu√©bec | Sandrine Moreira et al |
| EPI_ISL_890413 | Canada/QC-JUS-V5231442/2020 | 2020-03-22 | North America/Canada/Quebec/ | Laboratoire de sant√© publique du Qu√©bec | Laboratoire de sant√© publique du Qu√©bec | Sandrine Moreira et al |
| EPI_ISL_890414 | Canada/QC-JUS-V5231423/2020 | 2020-03-22 | North America/Canada/Quebec/ | Laboratoire de sant√© publique du Qu√©bec | Laboratoire de sant√© publique du Qu√©bec | Sandrine Moreira et al |
| EPI_ISL_890415 | Canada/QC-JUS-V5231390/2020 | 2020-03-22 | North America/Canada/Quebec/ | Laboratoire de sant√© publique du Qu√©bec | Laboratoire de sant√© publique du Qu√©bec | Sandrine Moreira et al |
| EPI_ISL_890416 | Canada/QC-JUS-V5230806/2020 | 2020-03-22 | North America/Canada/Quebec/ | Laboratoire de sant√© publique du Qu√©bec | Laboratoire de sant√© publique du Qu√©bec | Sandrine Moreira et al |
| EPI_ISL_890417 | Canada/QC-JUS-V5201104/2020 | 2020-03-19 | North America/Canada/Quebec/ | Laboratoire de sant√© publique du Qu√©bec | Laboratoire de sant√© publique du Qu√©bec | Sandrine Moreira et al |
| EPI_ISL_890418 | Canada/QC-JUS-V5200869/2020 | 2020-03-19 | North America/Canada/Quebec/ | Laboratoire de sant√© publique du Qu√©bec | Laboratoire de sant√© publique du Qu√©bec | Sandrine Moreira et al |

| | | | | | | |
|---|---|---|---|---|---|---|
| EPI_ISL_890419 | Canada/QC-JUS-V5221059/2020 | 2020-03-21 | North America/Canada/Quebec/ | Laboratoire de santv© publique du Quv©bec | Laboratoire de santv© publique du Quv©bec | Sandrine Moreira et al |
| EPI_ISL_890420 | Canada/QC-JUS-V5221073/2020 | 2020-03-21 | North America/Canada/Quebec/ | Laboratoire de santv© publique du Quv©bec | Laboratoire de santv© publique du Quv©bec | Sandrine Moreira et al |
| EPI_ISL_890421 | Canada/QC-JUS-V5231350/2020 | 2020-03-23 | North America/Canada/Quebec/ | Laboratoire de santv© publique du Quv©bec | Laboratoire de santv© publique du Quv©bec | Sandrine Moreira et al |
| EPI_ISL_890422 | Canada/QC-JUS-V5221114/2020 | 2020-03-21 | North America/Canada/Quebec/ | Laboratoire de santv© publique du Quv©bec | Laboratoire de santv© publique du Quv©bec | Sandrine Moreira et al |
| EPI_ISL_890423 | Canada/QC-JUS-V5210176/2020 | 2020-03-19 | North America/Canada/Quebec/ | Laboratoire de santv© publique du Quv©bec | Laboratoire de santv© publique du Quv©bec | Sandrine Moreira et al |
| EPI_ISL_890424 | Canada/QC-JUS-V5201888/2020 | 2020-03-19 | North America/Canada/Quebec/ | Laboratoire de santv© publique du Quv©bec | Laboratoire de santv© publique du Quv©bec | Sandrine Moreira et al |
| EPI_ISL_890425 | Canada/QC-JUS-V5202026/2020 | 2020-03-19 | North America/Canada/Quebec/ | Laboratoire de santv© publique du Quv©bec | Laboratoire de santv© publique du Quv©bec | Sandrine Moreira et al |
| EPI_ISL_890426 | Canada/QC-JUS-V5192196/2020 | 2020-03-18 | North America/Canada/Quebec/ | Laboratoire de santv© publique du Quv©bec | Laboratoire de santv© publique du Quv©bec | Sandrine Moreira et al |
| EPI_ISL_890427 | Canada/QC-JUS-V5200822/2020 | 2020-03-19 | North America/Canada/Quebec/ | Laboratoire de santv© publique du Quv©bec | Laboratoire de santv© publique du Quv©bec | Sandrine Moreira et al |
| EPI_ISL_890428 | Canada/QC-JUS-V5200815/2020 | 2020-03-19 | North America/Canada/Quebec/ | Laboratoire de santv© publique du Quv©bec | Laboratoire de santv© publique du Quv©bec | Sandrine Moreira et al |
| EPI_ISL_890429 | Canada/QC-JUS-V5210190/2020 | 2020-03-19 | North America/Canada/Quebec/ | Laboratoire de santv© publique du Quv©bec | Laboratoire de santv© publique du Quv©bec | Sandrine Moreira et al |
| EPI_ISL_890430 | Canada/QC-JUS-V5200505/2020 | 2020-03-18 | North America/Canada/Quebec/ | Laboratoire de santv© publique du Quv©bec | Laboratoire de santv© publique du Quv©bec | Sandrine Moreira et al |
| EPI_ISL_890431 | Canada/QC-JUS-V5192128/2020 | 2020-03-18 | North America/Canada/Quebec/ | Laboratoire de santv© publique du Quv©bec | Laboratoire de santv© publique du Quv©bec | Sandrine Moreira et al |

| | | | | | | |
|---|---|---|---|---|---|---|
| EPI_ISL_890432 | Canada/QC-JUS-V5192105/2020 | 2020-03-18 | North America/Canada/Quebec/ | Laboratoire de sant√© publique du Qu√©bec | Laboratoire de sant√© publique du Qu√©bec | Sandrine Moreira et al |
| EPI_ISL_890433 | Canada/QC-JUS-V5192080/2020 | 2020-03-18 | North America/Canada/Quebec/ | Laboratoire de sant√© publique du Qu√©bec | Laboratoire de sant√© publique du Qu√©bec | Sandrine Moreira et al |
| EPI_ISL_890434 | Canada/QC-JUS-V5191918/2020 | 2020-03-18 | North America/Canada/Quebec/ | Laboratoire de sant√© publique du Qu√©bec | Laboratoire de sant√© publique du Qu√©bec | Sandrine Moreira et al |
| EPI_ISL_890435 | Canada/QC-JUS-V5171505/2020 | 2020-03-17 | North America/Canada/Quebec/ | Laboratoire de sant√© publique du Qu√©bec | Laboratoire de sant√© publique du Qu√©bec | Sandrine Moreira et al |
| EPI_ISL_890436 | Canada/QC-JUS-V5191721/2020 | 2020-03-18 | North America/Canada/Quebec/ | Laboratoire de sant√© publique du Qu√©bec | Laboratoire de sant√© publique du Qu√©bec | Sandrine Moreira et al |
| EPI_ISL_890437 | Canada/QC-JUS-V5210890/2020 | 2020-03-21 | North America/Canada/Quebec/ | Laboratoire de sant√© publique du Qu√©bec | Laboratoire de sant√© publique du Qu√©bec | Sandrine Moreira et al |
| EPI_ISL_890438 | Canada/QC-JUS-V5221356/2020 | 2020-03-21 | North America/Canada/Quebec/ | Laboratoire de sant√© publique du Qu√©bec | Laboratoire de sant√© publique du Qu√©bec | Sandrine Moreira et al |
| EPI_ISL_890439 | Canada/QC-JUS-V5221404/2020 | 2020-03-21 | North America/Canada/Quebec/ | Laboratoire de sant√© publique du Qu√©bec | Laboratoire de sant√© publique du Qu√©bec | Sandrine Moreira et al |
| EPI_ISL_890440 | Canada/QC-JUS-V5250693/2020 | 2020-03-23 | North America/Canada/Quebec/ | Laboratoire de sant√© publique du Qu√©bec | Laboratoire de sant√© publique du Qu√©bec | Sandrine Moreira et al |
| EPI_ISL_890441 | Canada/QC-JUS-V5210482/2020 | 2020-03-19 | North America/Canada/Quebec/ | Laboratoire de sant√© publique du Qu√©bec | Laboratoire de sant√© publique du Qu√©bec | Sandrine Moreira et al |
| EPI_ISL_890442 | Canada/QC-JUS-V5210111/2020 | 2020-03-19 | North America/Canada/Quebec/ | Laboratoire de sant√© publique du Qu√©bec | Laboratoire de sant√© publique du Qu√©bec | Sandrine Moreira et al |
| EPI_ISL_890443 | Canada/QC-JUS-V5210035/2020 | 2020-03-19 | North America/Canada/Quebec/ | Laboratoire de sant√© publique du Qu√©bec | Laboratoire de sant√© publique du Qu√©bec | Sandrine Moreira et al |
| EPI_ISL_890444 | Canada/QC-JUS-V5210377/2020 | 2020-03-20 | North America/Canada/Quebec/ | Laboratoire de sant√© publique du Qu√©bec | Laboratoire de sant√© publique du Qu√©bec | Sandrine Moreira et al |

| | | | | | | |
|---|---|---|---|---|---|---|
| EPI_ISL_890445 | Canada/QC-JUS-V5210401/2020 | 2020-03-19 | North America/Canada/Quebec/ | Laboratoire de sant√© publique du Qu√©bec | Laboratoire de sant√© publique du Qu√©bec | Sandrine Moreira et al |
| EPI_ISL_890446 | Canada/QC-JUS-V5210350/2020 | 2020-03-20 | North America/Canada/Quebec/ | Laboratoire de sant√© publique du Qu√©bec | Laboratoire de sant√© publique du Qu√©bec | Sandrine Moreira et al |
| EPI_ISL_890447 | Canada/QC-JUS-V5210896/2020 | 2020-03-20 | North America/Canada/Quebec/ | Laboratoire de sant√© publique du Qu√©bec | Laboratoire de sant√© publique du Qu√©bec | Sandrine Moreira et al |
| EPI_ISL_890448 | Canada/QC-JUS-V5221265/2020 | 2020-03-21 | North America/Canada/Quebec/ | Laboratoire de sant√© publique du Qu√©bec | Laboratoire de sant√© publique du Qu√©bec | Sandrine Moreira et al |
| EPI_ISL_890449 | Canada/QC-JUS-V5221360/2020 | 2020-03-21 | North America/Canada/Quebec/ | Laboratoire de sant√© publique du Qu√©bec | Laboratoire de sant√© publique du Qu√©bec | Sandrine Moreira et al |
| EPI_ISL_890450 | Canada/QC-JUS-V5221198/2020 | 2020-03-21 | North America/Canada/Quebec/ | Laboratoire de sant√© publique du Qu√©bec | Laboratoire de sant√© publique du Qu√©bec | Sandrine Moreira et al |
| EPI_ISL_890451 | Canada/QC-JUS-V5221215/2020 | 2020-03-21 | North America/Canada/Quebec/ | Laboratoire de sant√© publique du Qu√©bec | Laboratoire de sant√© publique du Qu√©bec | Sandrine Moreira et al |
| EPI_ISL_890452 | Canada/QC-JUS-V5221352/2020 | 2020-03-21 | North America/Canada/Quebec/ | Laboratoire de sant√© publique du Qu√©bec | Laboratoire de sant√© publique du Qu√©bec | Sandrine Moreira et al |
| EPI_ISL_890453 | Canada/QC-JUS-V5221282/2020 | 2020-03-21 | North America/Canada/Quebec/ | Laboratoire de sant√© publique du Qu√©bec | Laboratoire de sant√© publique du Qu√©bec | Sandrine Moreira et al |
| EPI_ISL_890454 | Canada/QC-JUS-V5221395/2020 | 2020-03-21 | North America/Canada/Quebec/ | Laboratoire de sant√© publique du Qu√©bec | Laboratoire de sant√© publique du Qu√©bec | Sandrine Moreira et al |
| EPI_ISL_890455 | Canada/QC-JUS-V5221484/2020 | 2020-03-21 | North America/Canada/Quebec/ | Laboratoire de sant√© publique du Qu√©bec | Laboratoire de sant√© publique du Qu√©bec | Sandrine Moreira et al |
| EPI_ISL_890456 | Canada/QC-JUS-V5231739/2020 | 2020-03-22 | North America/Canada/Quebec/ | Laboratoire de sant√© publique du Qu√©bec | Laboratoire de sant√© publique du Qu√©bec | Sandrine Moreira et al |
| EPI_ISL_890457 | Canada/QC-JUS-V5252058/2020 | 2020-03-24 | North America/Canada/Quebec/ | Laboratoire de sant√© publique du Qu√©bec | Laboratoire de sant√© publique du Qu√©bec | Sandrine Moreira et al |

| | | | | | | |
|---|---|---|---|---|---|---|
| EPI_ISL_890458 | Canada/QC-JUS-V5251937/2020 | 2020-03-24 | North America/Canada/Quebec/ | Laboratoire de sant√© publique du Qu√©bec | Laboratoire de sant√© publique du Qu√©bec | Sandrine Moreira et al |
| EPI_ISL_890459 | Canada/QC-JUS-V5251694/2020 | 2020-03-24 | North America/Canada/Quebec/ | Laboratoire de sant√© publique du Qu√©bec | Laboratoire de sant√© publique du Qu√©bec | Sandrine Moreira et al |
| EPI_ISL_890460 | Canada/QC-JUS-V5251006/2020 | 2020-03-23 | North America/Canada/Quebec/ | Laboratoire de sant√© publique du Qu√©bec | Laboratoire de sant√© publique du Qu√©bec | Sandrine Moreira et al |
| EPI_ISL_890461 | Canada/QC-JUS-V5252213/2020 | 2020-03-24 | North America/Canada/Quebec/ | Laboratoire de sant√© publique du Qu√©bec | Laboratoire de sant√© publique du Qu√©bec | Sandrine Moreira et al |
| EPI_ISL_890462 | Canada/QC-JUS-V5250839/2020 | 2020-03-25 | North America/Canada/Quebec/ | Laboratoire de sant√© publique du Qu√©bec | Laboratoire de sant√© publique du Qu√©bec | Sandrine Moreira et al |
| EPI_ISL_890463 | Canada/QC-JUS-V5252344/2020 | 2020-03-24 | North America/Canada/Quebec/ | Laboratoire de sant√© publique du Qu√©bec | Laboratoire de sant√© publique du Qu√©bec | Sandrine Moreira et al |
| EPI_ISL_890464 | Canada/QC-JUS-V5251556/2020 | 2020-03-25 | North America/Canada/Quebec/ | Laboratoire de sant√© publique du Qu√©bec | Laboratoire de sant√© publique du Qu√©bec | Sandrine Moreira et al |
| EPI_ISL_890465 | Canada/QC-JUS-V5250462/2020 | 2020-03-23 | North America/Canada/Quebec/ | Laboratoire de sant√© publique du Qu√©bec | Laboratoire de sant√© publique du Qu√©bec | Sandrine Moreira et al |
| EPI_ISL_890466 | Canada/QC-JUS-V5250560/2020 | 2020-03-22 | North America/Canada/Quebec/ | Laboratoire de sant√© publique du Qu√©bec | Laboratoire de sant√© publique du Qu√©bec | Sandrine Moreira et al |
| EPI_ISL_890467 | Canada/QC-JUS-V5250344/2020 | 2020-03-23 | North America/Canada/Quebec/ | Laboratoire de sant√© publique du Qu√©bec | Laboratoire de sant√© publique du Qu√©bec | Sandrine Moreira et al |
| EPI_ISL_890468 | Canada/QC-JUS-V5250292/2020 | 2020-03-23 | North America/Canada/Quebec/ | Laboratoire de sant√© publique du Qu√©bec | Laboratoire de sant√© publique du Qu√©bec | Sandrine Moreira et al |
| EPI_ISL_890469 | Canada/QC-JUS-V5250296/2020 | 2020-03-23 | North America/Canada/Quebec/ | Laboratoire de sant√© publique du Qu√©bec | Laboratoire de sant√© publique du Qu√©bec | Sandrine Moreira et al |
| EPI_ISL_890470 | Canada/QC-JUS-V5250386/2020 | 2020-03-23 | North America/Canada/Quebec/ | Laboratoire de sant√© publique du Qu√©bec | Laboratoire de sant√© publique du Qu√©bec | Sandrine Moreira et al |

| | | | | | | |
|---|---|---|---|---|---|---|
| EPI_ISL_890471 | Canada/QC-JUS-V5251573/2020 | 2020-03-25 | North America/Canada/Quebec/ | Laboratoire de sant√© publique du Qu√©bec | Laboratoire de sant√© publique du Qu√©bec | Sandrine Moreira et al |
| EPI_ISL_890472 | Canada/QC-JUS-V5251200/2020 | 2020-03-23 | North America/Canada/Quebec/ | Laboratoire de sant√© publique du Qu√©bec | Laboratoire de sant√© publique du Qu√©bec | Sandrine Moreira et al |
| EPI_ISL_890473 | Canada/QC-JUS-V5251436/2020 | 2020-03-25 | North America/Canada/Quebec/ | Laboratoire de sant√© publique du Qu√©bec | Laboratoire de sant√© publique du Qu√©bec | Sandrine Moreira et al |
| EPI_ISL_890474 | Canada/QC-JUS-V5251353/2020 | 2020-03-24 | North America/Canada/Quebec/ | Laboratoire de sant√© publique du Qu√©bec | Laboratoire de sant√© publique du Qu√©bec | Sandrine Moreira et al |
| EPI_ISL_890475 | Canada/QC-JUS-V5250844/2020 | 2020-03-22 | North America/Canada/Quebec/ | Laboratoire de sant√© publique du Qu√©bec | Laboratoire de sant√© publique du Qu√©bec | Sandrine Moreira et al |
| EPI_ISL_890476 | Canada/QC-JUS-V5250516/2020 | 2020-03-24 | North America/Canada/Quebec/ | Laboratoire de sant√© publique du Qu√©bec | Laboratoire de sant√© publique du Qu√©bec | Sandrine Moreira et al |
| EPI_ISL_890477 | Canada/QC-JUS-V5250996/2020 | 2020-03-22 | North America/Canada/Quebec/ | Laboratoire de sant√© publique du Qu√©bec | Laboratoire de sant√© publique du Qu√©bec | Sandrine Moreira et al |
| EPI_ISL_890478 | Canada/QC-JUS-V5251142/2020 | 2020-03-23 | North America/Canada/Quebec/ | Laboratoire de sant√© publique du Qu√©bec | Laboratoire de sant√© publique du Qu√©bec | Sandrine Moreira et al |
| EPI_ISL_890479 | Canada/QC-JUS-V5250791/2020 | 2020-03-25 | North America/Canada/Quebec/ | Laboratoire de sant√© publique du Qu√©bec | Laboratoire de sant√© publique du Qu√©bec | Sandrine Moreira et al |
| EPI_ISL_890480 | Canada/QC-JUS-V5231729/2020 | 2020-03-22 | North America/Canada/Quebec/ | Laboratoire de sant√© publique du Qu√©bec | Laboratoire de sant√© publique du Qu√©bec | Sandrine Moreira et al |
| EPI_ISL_890481 | Canada/QC-JUS-V5231448/2020 | 2020-03-22 | North America/Canada/Quebec/ | Laboratoire de sant√© publique du Qu√©bec | Laboratoire de sant√© publique du Qu√©bec | Sandrine Moreira et al |
| EPI_ISL_890482 | Canada/QC-JUS-V5231724/2020 | 2020-03-22 | North America/Canada/Quebec/ | Laboratoire de sant√© publique du Qu√©bec | Laboratoire de sant√© publique du Qu√©bec | Sandrine Moreira et al |
| EPI_ISL_890483 | Canada/QC-JUS-V5251852/2020 | 2020-03-24 | North America/Canada/Quebec/ | Laboratoire de sant√© publique du Qu√©bec | Laboratoire de sant√© publique du Qu√©bec | Sandrine Moreira et al |

| | | | | | | |
|---|---|---|---|---|---|---|
| EPI_ISL_890484 | Canada/QC-JUS-V5251158/2020 | 2020-03-23 | North America/Canada/Quebec/ | Laboratoire de sant√© publique du Qu√©bec | Laboratoire de sant√© publique du Qu√©bec | Sandrine Moreira et al |
| EPI_ISL_890485 | Canada/QC-JUS-V5251677/2020 | 2020-03-25 | North America/Canada/Quebec/ | Laboratoire de sant√© publique du Qu√©bec | Laboratoire de sant√© publique du Qu√©bec | Sandrine Moreira et al |
| EPI_ISL_890486 | Canada/QC-JUS-V5240355/2020 | 2020-03-23 | North America/Canada/Quebec/ | Laboratoire de sant√© publique du Qu√©bec | Laboratoire de sant√© publique du Qu√©bec | Sandrine Moreira et al |
| EPI_ISL_890487 | Canada/QC-JUS-V5251652/2020 | 2020-03-25 | North America/Canada/Quebec/ | Laboratoire de sant√© publique du Qu√©bec | Laboratoire de sant√© publique du Qu√©bec | Sandrine Moreira et al |
| EPI_ISL_890488 | Canada/QC-JUS-V5230431/2020 | 2020-03-22 | North America/Canada/Quebec/ | Laboratoire de sant√© publique du Qu√©bec | Laboratoire de sant√© publique du Qu√©bec | Sandrine Moreira et al |
| EPI_ISL_890489 | Canada/QC-JUS-V5242174/2020 | 2020-03-23 | North America/Canada/Quebec/ | Laboratoire de sant√© publique du Qu√©bec | Laboratoire de sant√© publique du Qu√©bec | Sandrine Moreira et al |
| EPI_ISL_890490 | Canada/QC-JUS-V5242202/2020 | 2020-03-22 | North America/Canada/Quebec/ | Laboratoire de sant√© publique du Qu√©bec | Laboratoire de sant√© publique du Qu√©bec | Sandrine Moreira et al |
| EPI_ISL_890491 | Canada/QC-JUS-V5261218/2020 | 2020-03-24 | North America/Canada/Quebec/ | Laboratoire de sant√© publique du Qu√©bec | Laboratoire de sant√© publique du Qu√©bec | Sandrine Moreira et al |
| EPI_ISL_890492 | Canada/QC-JUS-V5241673/2020 | 2020-03-22 | North America/Canada/Quebec/ | Laboratoire de sant√© publique du Qu√©bec | Laboratoire de sant√© publique du Qu√©bec | Sandrine Moreira et al |
| EPI_ISL_890493 | Canada/QC-JUS-V5251615/2020 | 2020-03-24 | North America/Canada/Quebec/ | Laboratoire de sant√© publique du Qu√©bec | Laboratoire de sant√© publique du Qu√©bec | Sandrine Moreira et al |
| EPI_ISL_890494 | Canada/QC-JUS-V5242079/2020 | 2020-03-23 | North America/Canada/Quebec/ | Laboratoire de sant√© publique du Qu√©bec | Laboratoire de sant√© publique du Qu√©bec | Sandrine Moreira et al |
| EPI_ISL_890495 | Canada/QC-JUS-V5240330/2020 | 2020-03-23 | North America/Canada/Quebec/ | Laboratoire de sant√© publique du Qu√©bec | Laboratoire de sant√© publique du Qu√©bec | Sandrine Moreira et al |
| EPI_ISL_890496 | Canada/QC-JUS-V5261744/2020 | 2020-03-24 | North America/Canada/Quebec/ | Laboratoire de sant√© publique du Qu√©bec | Laboratoire de sant√© publique du Qu√©bec | Sandrine Moreira et al |

| | | | | | | |
|---|---|---|---|---|---|---|
| EPI_ISL_890497 | Canada/QC-JUS-V5262259/2020 | 2020-03-26 | North America/Canada/Quebec/ | Laboratoire de sant√© publique du Qu√©bec | Laboratoire de sant√© publique du Qu√©bec | Sandrine Moreira et al |
| EPI_ISL_890498 | Canada/QC-JUS-V5262057/2020 | 2020-03-26 | North America/Canada/Quebec/ | Laboratoire de sant√© publique du Qu√©bec | Laboratoire de sant√© publique du Qu√©bec | Sandrine Moreira et al |
| EPI_ISL_890499 | Canada/QC-JUS-V5271782/2020 | 2020-03-27 | North America/Canada/Quebec/ | Laboratoire de sant√© publique du Qu√©bec | Laboratoire de sant√© publique du Qu√©bec | Sandrine Moreira et al |
| EPI_ISL_890500 | Canada/QC-JUS-V5270587/2020 | 2020-03-26 | North America/Canada/Quebec/ | Laboratoire de sant√© publique du Qu√©bec | Laboratoire de sant√© publique du Qu√©bec | Sandrine Moreira et al |
| EPI_ISL_890501 | Canada/QC-JUS-V5270636/2020 | 2020-03-26 | North America/Canada/Quebec/ | Laboratoire de sant√© publique du Qu√©bec | Laboratoire de sant√© publique du Qu√©bec | Sandrine Moreira et al |
| EPI_ISL_890502 | Canada/QC-JUS-V5262967/2020 | 2020-03-25 | North America/Canada/Quebec/ | Laboratoire de sant√© publique du Qu√©bec | Laboratoire de sant√© publique du Qu√©bec | Sandrine Moreira et al |
| EPI_ISL_890503 | Canada/QC-JUS-V5270613/2020 | 2020-03-26 | North America/Canada/Quebec/ | Laboratoire de sant√© publique du Qu√©bec | Laboratoire de sant√© publique du Qu√©bec | Sandrine Moreira et al |
| EPI_ISL_890504 | Canada/QC-JUS-V5270582/2020 | 2020-03-26 | North America/Canada/Quebec/ | Laboratoire de sant√© publique du Qu√©bec | Laboratoire de sant√© publique du Qu√©bec | Sandrine Moreira et al |
| EPI_ISL_890505 | Canada/QC-JUS-V5262799/2020 | 2020-03-25 | North America/Canada/Quebec/ | Laboratoire de sant√© publique du Qu√©bec | Laboratoire de sant√© publique du Qu√©bec | Sandrine Moreira et al |
| EPI_ISL_890506 | Canada/QC-JUS-V5262792/2020 | 2020-03-25 | North America/Canada/Quebec/ | Laboratoire de sant√© publique du Qu√©bec | Laboratoire de sant√© publique du Qu√©bec | Sandrine Moreira et al |
| EPI_ISL_890507 | Canada/QC-JUS-V5262159/2020 | 2020-03-25 | North America/Canada/Quebec/ | Laboratoire de sant√© publique du Qu√©bec | Laboratoire de sant√© publique du Qu√©bec | Sandrine Moreira et al |
| EPI_ISL_890508 | Canada/QC-JUS-V5262824/2020 | 2020-03-25 | North America/Canada/Quebec/ | Laboratoire de sant√© publique du Qu√©bec | Laboratoire de sant√© publique du Qu√©bec | Sandrine Moreira et al |
| EPI_ISL_890509 | Canada/QC-JUS-V5262807/2020 | 2020-03-25 | North America/Canada/Quebec/ | Laboratoire de sant√© publique du Qu√©bec | Laboratoire de sant√© publique du Qu√©bec | Sandrine Moreira et al |

| | | | | | | |
|---|---|---|---|---|---|---|
| EPI_ISL_890510 | Canada/QC-JUS-V5262801/2020 | 2020-03-25 | North America/Canada/Quebec/ | Laboratoire de sant√© publique du Qu√©bec | Laboratoire de sant√© publique du Qu√©bec | Sandrine Moreira et al |
| EPI_ISL_890511 | Canada/QC-JUS-V5262828/2020 | 2020-03-25 | North America/Canada/Quebec/ | Laboratoire de sant√© publique du Qu√©bec | Laboratoire de sant√© publique du Qu√©bec | Sandrine Moreira et al |
| EPI_ISL_890512 | Canada/QC-JUS-V5280015/2020 | 2020-03-26 | North America/Canada/Quebec/ | Laboratoire de sant√© publique du Qu√©bec | Laboratoire de sant√© publique du Qu√©bec | Sandrine Moreira et al |
| EPI_ISL_890513 | Canada/QC-JUS-V5262629/2020 | 2020-03-23 | North America/Canada/Quebec/ | Laboratoire de sant√© publique du Qu√©bec | Laboratoire de sant√© publique du Qu√©bec | Sandrine Moreira et al |
| EPI_ISL_890514 | Canada/QC-JUS-V5270894/2020 | 2020-03-25 | North America/Canada/Quebec/ | Laboratoire de sant√© publique du Qu√©bec | Laboratoire de sant√© publique du Qu√©bec | Sandrine Moreira et al |
| EPI_ISL_890515 | Canada/QC-JUS-V5270902/2020 | 2020-03-25 | North America/Canada/Quebec/ | Laboratoire de sant√© publique du Qu√©bec | Laboratoire de sant√© publique du Qu√©bec | Sandrine Moreira et al |
| EPI_ISL_890516 | Canada/QC-JUS-V5300563/2020 | 2020-03-28 | North America/Canada/Quebec/ | Laboratoire de sant√© publique du Qu√©bec | Laboratoire de sant√© publique du Qu√©bec | Sandrine Moreira et al |
| EPI_ISL_890517 | Canada/QC-JUS-V5281744/2020 | 2020-03-26 | North America/Canada/Quebec/ | Laboratoire de sant√© publique du Qu√©bec | Laboratoire de sant√© publique du Qu√©bec | Sandrine Moreira et al |
| EPI_ISL_890518 | Canada/QC-JUS-V5281436/2020 | 2020-03-27 | North America/Canada/Quebec/ | Laboratoire de sant√© publique du Qu√©bec | Laboratoire de sant√© publique du Qu√©bec | Sandrine Moreira et al |
| EPI_ISL_890519 | Canada/QC-JUS-V5281136/2020 | 2020-03-27 | North America/Canada/Quebec/ | Laboratoire de sant√© publique du Qu√©bec | Laboratoire de sant√© publique du Qu√©bec | Sandrine Moreira et al |
| EPI_ISL_890520 | Canada/QC-JUS-V5281170/2020 | 2020-03-24 | North America/Canada/Quebec/ | Laboratoire de sant√© publique du Qu√©bec | Laboratoire de sant√© publique du Qu√©bec | Sandrine Moreira et al |
| EPI_ISL_890521 | Canada/QC-JUS-V5290226/2020 | 2020-03-28 | North America/Canada/Quebec/ | Laboratoire de sant√© publique du Qu√©bec | Laboratoire de sant√© publique du Qu√©bec | Sandrine Moreira et al |
| EPI_ISL_890522 | Canada/QC-JUS-V5290330/2020 | 2020-03-28 | North America/Canada/Quebec/ | Laboratoire de sant√© publique du Qu√©bec | Laboratoire de sant√© publique du Qu√©bec | Sandrine Moreira et al |

| | | | | | | |
|---|---|---|---|---|---|---|
| EPI_ISL_890523 | Canada/QC-JUS-V5290389/2020 | 2020-03-28 | North America/Canada/Quebec/ | Laboratoire de santv© publique du Quv©bec | Laboratoire de santv© publique du Quv©bec | Sandrine Moreira et al |
| EPI_ISL_890524 | Canada/QC-JUS-V5300631/2020 | 2020-03-29 | North America/Canada/Quebec/ | Laboratoire de santv© publique du Quv©bec | Laboratoire de santv© publique du Quv©bec | Sandrine Moreira et al |
| EPI_ISL_890525 | Canada/QC-JUS-V5291536/2020 | 2020-03-29 | North America/Canada/Quebec/ | Laboratoire de santv© publique du Quv©bec | Laboratoire de santv© publique du Quv©bec | Sandrine Moreira et al |
| EPI_ISL_890526 | Canada/QC-JUS-V5300592/2020 | 2020-03-28 | North America/Canada/Quebec/ | Laboratoire de santv© publique du Quv©bec | Laboratoire de santv© publique du Quv©bec | Sandrine Moreira et al |
| EPI_ISL_890527 | Canada/QC-JUS-V5300602/2020 | 2020-03-28 | North America/Canada/Quebec/ | Laboratoire de santv© publique du Quv©bec | Laboratoire de santv© publique du Quv©bec | Sandrine Moreira et al |
| EPI_ISL_890528 | Canada/QC-JUS-V5291350/2020 | 2020-03-29 | North America/Canada/Quebec/ | Laboratoire de santv© publique du Quv©bec | Laboratoire de santv© publique du Quv©bec | Sandrine Moreira et al |
| EPI_ISL_890529 | Canada/QC-JUS-V5290830/2020 | 2020-03-27 | North America/Canada/Quebec/ | Laboratoire de santv© publique du Quv©bec | Laboratoire de santv© publique du Quv©bec | Sandrine Moreira et al |
| EPI_ISL_890530 | Canada/QC-JUS-V5300854/2020 | 2020-03-28 | North America/Canada/Quebec/ | Laboratoire de santv© publique du Quv©bec | Laboratoire de santv© publique du Quv©bec | Sandrine Moreira et al |
| EPI_ISL_890531 | Canada/QC-JUS-V5302122/2020 | 2020-03-28 | North America/Canada/Quebec/ | Laboratoire de santv© publique du Quv©bec | Laboratoire de santv© publique du Quv©bec | Sandrine Moreira et al |
| EPI_ISL_890532 | Canada/QC-JUS-V5302147/2020 | 2020-03-28 | North America/Canada/Quebec/ | Laboratoire de santv© publique du Quv©bec | Laboratoire de santv© publique du Quv©bec | Sandrine Moreira et al |
| EPI_ISL_890533 | Canada/QC-JUS-V5302150/2020 | 2020-03-28 | North America/Canada/Quebec/ | Laboratoire de santv© publique du Quv©bec | Laboratoire de santv© publique du Quv©bec | Sandrine Moreira et al |
| EPI_ISL_890534 | Canada/QC-JUS-V5272232/2020 | 2020-03-26 | North America/Canada/Quebec/ | Laboratoire de santv© publique du Quv©bec | Laboratoire de santv© publique du Quv©bec | Sandrine Moreira et al |
| EPI_ISL_890535 | Canada/QC-JUS-V5271860/2020 | 2020-03-26 | North America/Canada/Quebec/ | Laboratoire de santv© publique du Quv©bec | Laboratoire de santv© publique du Quv©bec | Sandrine Moreira et al |

| | | | | | | |
|---|---|---|---|---|---|---|
| EPI_ISL_890536 | Canada/QC-JUS-V5281120/2020 | 2020-03-27 | North America/Canada/Quebec/ | Laboratoire de sant√© publique du Qu√©bec | Laboratoire de sant√© publique du Qu√©bec | Sandrine Moreira et al |
| EPI_ISL_890537 | Canada/QC-JUS-V5281131/2020 | 2020-03-27 | North America/Canada/Quebec/ | Laboratoire de sant√© publique du Qu√©bec | Laboratoire de sant√© publique du Qu√©bec | Sandrine Moreira et al |
| EPI_ISL_890538 | Canada/QC-JUS-V5281265/2020 | 2020-03-27 | North America/Canada/Quebec/ | Laboratoire de sant√© publique du Qu√©bec | Laboratoire de sant√© publique du Qu√©bec | Sandrine Moreira et al |
| EPI_ISL_890539 | Canada/QC-JUS-V5281138/2020 | 2020-03-27 | North America/Canada/Quebec/ | Laboratoire de sant√© publique du Qu√©bec | Laboratoire de sant√© publique du Qu√©bec | Sandrine Moreira et al |
| EPI_ISL_890540 | Canada/QC-JUS-V5270491/2020 | 2020-03-25 | North America/Canada/Quebec/ | Laboratoire de sant√© publique du Qu√©bec | Laboratoire de sant√© publique du Qu√©bec | Sandrine Moreira et al |
| EPI_ISL_890541 | Canada/QC-JUS-V5250324/2020 | 2020-03-23 | North America/Canada/Quebec/ | Laboratoire de sant√© publique du Qu√©bec | Laboratoire de sant√© publique du Qu√©bec | Sandrine Moreira et al |
| EPI_ISL_890542 | Canada/QC-JUS-V5262817/2020 | 2020-03-25 | North America/Canada/Quebec/ | Laboratoire de sant√© publique du Qu√©bec | Laboratoire de sant√© publique du Qu√©bec | Sandrine Moreira et al |
| EPI_ISL_890543 | Canada/QC-JUS-V5272295/2020 | 2020-03-26 | North America/Canada/Quebec/ | Laboratoire de sant√© publique du Qu√©bec | Laboratoire de sant√© publique du Qu√©bec | Sandrine Moreira et al |
| EPI_ISL_890544 | Canada/QC-JUS-V5281571/2020 | 2020-03-28 | North America/Canada/Quebec/ | Laboratoire de sant√© publique du Qu√©bec | Laboratoire de sant√© publique du Qu√©bec | Sandrine Moreira et al |
| EPI_ISL_890545 | Canada/QC-JUS-V5280523/2020 | 2020-03-25 | North America/Canada/Quebec/ | Laboratoire de sant√© publique du Qu√©bec | Laboratoire de sant√© publique du Qu√©bec | Sandrine Moreira et al |
| EPI_ISL_890546 | Canada/QC-JUS-V5272677/2020 | 2020-03-26 | North America/Canada/Quebec/ | Laboratoire de sant√© publique du Qu√©bec | Laboratoire de sant√© publique du Qu√©bec | Sandrine Moreira et al |
| EPI_ISL_890547 | Canada/QC-JUS-V5280723/2020 | 2020-03-26 | North America/Canada/Quebec/ | Laboratoire de sant√© publique du Qu√©bec | Laboratoire de sant√© publique du Qu√©bec | Sandrine Moreira et al |
| EPI_ISL_890548 | Canada/QC-JUS-V5290491/2020 | 2020-03-28 | North America/Canada/Quebec/ | Laboratoire de sant√© publique du Qu√©bec | Laboratoire de sant√© publique du Qu√©bec | Sandrine Moreira et al |

| | | | | | | |
|---|---|---|---|---|---|---|
| EPI_ISL_890549 | Canada/QC-JUS-V5290516/2020 | 2020-03-28 | North America/Canada/Quebec/ | Laboratoire de santv© publique du Quv©bec | Laboratoire de santv© publique du Quv©bec | Sandrine Moreira et al |
| EPI_ISL_890550 | Canada/QC-JUS-V5281699/2020 | 2020-03-27 | North America/Canada/Quebec/ | Laboratoire de santv© publique du Quv©bec | Laboratoire de santv© publique du Quv©bec | Sandrine Moreira et al |
| EPI_ISL_890551 | Canada/QC-JUS-V5281756/2020 | 2020-03-27 | North America/Canada/Quebec/ | Laboratoire de santv© publique du Quv©bec | Laboratoire de santv© publique du Quv©bec | Sandrine Moreira et al |
| EPI_ISL_890552 | Canada/QC-JUS-V5281759/2020 | 2020-03-26 | North America/Canada/Quebec/ | Laboratoire de santv© publique du Quv©bec | Laboratoire de santv© publique du Quv©bec | Sandrine Moreira et al |
| EPI_ISL_890553 | Canada/QC-JUS-V5290171/2020 | 2020-03-27 | North America/Canada/Quebec/ | Laboratoire de santv© publique du Quv©bec | Laboratoire de santv© publique du Quv©bec | Sandrine Moreira et al |
| EPI_ISL_890554 | Canada/QC-JUS-V5280680/2020 | 2020-03-26 | North America/Canada/Quebec/ | Laboratoire de santv© publique du Quv©bec | Laboratoire de santv© publique du Quv©bec | Sandrine Moreira et al |
| EPI_ISL_890555 | Canada/QC-JUS-V5280708/2020 | 2020-03-26 | North America/Canada/Quebec/ | Laboratoire de santv© publique du Quv©bec | Laboratoire de santv© publique du Quv©bec | Sandrine Moreira et al |
| EPI_ISL_890556 | Canada/QC-JUS-V5280718/2020 | 2020-03-26 | North America/Canada/Quebec/ | Laboratoire de santv© publique du Quv©bec | Laboratoire de santv© publique du Quv©bec | Sandrine Moreira et al |
| EPI_ISL_890557 | Canada/QC-JUS-V5290406/2020 | 2020-03-28 | North America/Canada/Quebec/ | Laboratoire de santv© publique du Quv©bec | Laboratoire de santv© publique du Quv©bec | Sandrine Moreira et al |
| EPI_ISL_890558 | Canada/QC-JUS-V5290449/2020 | 2020-03-28 | North America/Canada/Quebec/ | Laboratoire de santv© publique du Quv©bec | Laboratoire de santv© publique du Quv©bec | Sandrine Moreira et al |
| EPI_ISL_890559 | Canada/QC-JUS-V5290372/2020 | 2020-03-28 | North America/Canada/Quebec/ | Laboratoire de santv© publique du Quv©bec | Laboratoire de santv© publique du Quv©bec | Sandrine Moreira et al |
| EPI_ISL_890560 | Canada/QC-JUS-V5280566/2020 | 2020-03-26 | North America/Canada/Quebec/ | Laboratoire de santv© publique du Quv©bec | Laboratoire de santv© publique du Quv©bec | Sandrine Moreira et al |
| EPI_ISL_890561 | Canada/QC-JUS-V5280450/2020 | 2020-03-26 | North America/Canada/Quebec/ | Laboratoire de santv© publique du Quv©bec | Laboratoire de santv© publique du Quv©bec | Sandrine Moreira et al |

| | | | | | | |
|---|---|---|---|---|---|---|
| EPI_ISL_890562 | Canada/QC-JUS-V5280429/2020 | 2020-03-26 | North America/Canada/Quebec/ | Laboratoire de sant√© publique du Qu√©bec | Laboratoire de sant√© publique du Qu√©bec | Sandrine Moreira et al |
| EPI_ISL_890563 | Canada/QC-JUS-V5280351/2020 | 2020-03-26 | North America/Canada/Quebec/ | Laboratoire de sant√© publique du Qu√©bec | Laboratoire de sant√© publique du Qu√©bec | Sandrine Moreira et al |
| EPI_ISL_890564 | Canada/QC-JUS-V5251641/2020 | 2020-03-24 | North America/Canada/Quebec/ | Laboratoire de sant√© publique du Qu√©bec | Laboratoire de sant√© publique du Qu√©bec | Sandrine Moreira et al |
| EPI_ISL_890565 | Canada/QC-JUS-V5251566/2020 | 2020-03-24 | North America/Canada/Quebec/ | Laboratoire de sant√© publique du Qu√©bec | Laboratoire de sant√© publique du Qu√©bec | Sandrine Moreira et al |
| EPI_ISL_890566 | Canada/QC-JUS-V5252225/2020 | 2020-03-25 | North America/Canada/Quebec/ | Laboratoire de sant√© publique du Qu√©bec | Laboratoire de sant√© publique du Qu√©bec | Sandrine Moreira et al |
| EPI_ISL_890567 | Canada/QC-JUS-V5231774/2020 | 2020-03-22 | North America/Canada/Quebec/ | Laboratoire de sant√© publique du Qu√©bec | Laboratoire de sant√© publique du Qu√©bec | Sandrine Moreira et al |
| EPI_ISL_890568 | Canada/QC-JUS-V5260058/2020 | 2020-03-24 | North America/Canada/Quebec/ | Laboratoire de sant√© publique du Qu√©bec | Laboratoire de sant√© publique du Qu√©bec | Sandrine Moreira et al |
| EPI_ISL_890569 | Canada/QC-JUS-V5252074/2020 | 2020-03-24 | North America/Canada/Quebec/ | Laboratoire de sant√© publique du Qu√©bec | Laboratoire de sant√© publique du Qu√©bec | Sandrine Moreira et al |
| EPI_ISL_890570 | Canada/QC-JUS-V5252061/2020 | 2020-03-24 | North America/Canada/Quebec/ | Laboratoire de sant√© publique du Qu√©bec | Laboratoire de sant√© publique du Qu√©bec | Sandrine Moreira et al |
| EPI_ISL_890571 | Canada/QC-JUS-V5252197/2020 | 2020-03-25 | North America/Canada/Quebec/ | Laboratoire de sant√© publique du Qu√©bec | Laboratoire de sant√© publique du Qu√©bec | Sandrine Moreira et al |
| EPI_ISL_890572 | Canada/QC-JUS-V5242374/2020 | 2020-03-24 | North America/Canada/Quebec/ | Laboratoire de sant√© publique du Qu√©bec | Laboratoire de sant√© publique du Qu√©bec | Sandrine Moreira et al |
| EPI_ISL_890573 | Canada/QC-JUS-V5250255/2020 | 2020-03-23 | North America/Canada/Quebec/ | Laboratoire de sant√© publique du Qu√©bec | Laboratoire de sant√© publique du Qu√©bec | Sandrine Moreira et al |
| EPI_ISL_890574 | Canada/QC-JUS-V5260612/2020 | 2020-03-25 | North America/Canada/Quebec/ | Laboratoire de sant√© publique du Qu√©bec | Laboratoire de sant√© publique du Qu√©bec | Sandrine Moreira et al |

| | | | | | | |
|---|---|---|---|---|---|---|
| EPI_ISL_890575 | Canada/QC-JUS-V5260603/2020 | 2020-03-25 | North America/Canada/Quebec/ | Laboratoire de santv© publique du Quv©bec | Laboratoire de santv© publique du Quv©bec | Sandrine Moreira et al |
| EPI_ISL_890576 | Canada/QC-JUS-V5251533/2020 | 2020-03-25 | North America/Canada/Quebec/ | Laboratoire de santv© publique du Quv©bec | Laboratoire de santv© publique du Quv©bec | Sandrine Moreira et al |
| EPI_ISL_890577 | Canada/QC-JUS-V5260582/2020 | 2020-03-25 | North America/Canada/Quebec/ | Laboratoire de santv© publique du Quv©bec | Laboratoire de santv© publique du Quv©bec | Sandrine Moreira et al |
| EPI_ISL_890578 | Canada/QC-JUS-V5260348/2020 | 2020-03-25 | North America/Canada/Quebec/ | Laboratoire de santv© publique du Quv©bec | Laboratoire de santv© publique du Quv©bec | Sandrine Moreira et al |
| EPI_ISL_890579 | Canada/QC-JUS-V5260202/2020 | 2020-03-24 | North America/Canada/Quebec/ | Laboratoire de santv© publique du Quv©bec | Laboratoire de santv© publique du Quv©bec | Sandrine Moreira et al |
| EPI_ISL_890580 | Canada/QC-JUS-V5260206/2020 | 2020-03-24 | North America/Canada/Quebec/ | Laboratoire de santv© publique du Quv©bec | Laboratoire de santv© publique du Quv©bec | Sandrine Moreira et al |
| EPI_ISL_890581 | Canada/QC-JUS-V5260114/2020 | 2020-03-24 | North America/Canada/Quebec/ | Laboratoire de santv© publique du Quv©bec | Laboratoire de santv© publique du Quv©bec | Sandrine Moreira et al |
| EPI_ISL_890582 | Canada/QC-JUS-V5260673/2020 | 2020-03-25 | North America/Canada/Quebec/ | Laboratoire de santv© publique du Quv©bec | Laboratoire de santv© publique du Quv©bec | Sandrine Moreira et al |
| EPI_ISL_890583 | Canada/QC-JUS-V5260694/2020 | 2020-03-25 | North America/Canada/Quebec/ | Laboratoire de santv© publique du Quv©bec | Laboratoire de santv© publique du Quv©bec | Sandrine Moreira et al |
| EPI_ISL_890584 | Canada/QC-JUS-V5260120/2020 | 2020-03-24 | North America/Canada/Quebec/ | Laboratoire de santv© publique du Quv©bec | Laboratoire de santv© publique du Quv©bec | Sandrine Moreira et al |
| EPI_ISL_890585 | Canada/QC-JUS-V5251973/2020 | 2020-03-24 | North America/Canada/Quebec/ | Laboratoire de santv© publique du Quv©bec | Laboratoire de santv© publique du Quv©bec | Sandrine Moreira et al |
| EPI_ISL_890586 | Canada/QC-JUS-V5241888/2020 | 2020-03-23 | North America/Canada/Quebec/ | Laboratoire de santv© publique du Quv©bec | Laboratoire de santv© publique du Quv©bec | Sandrine Moreira et al |
| EPI_ISL_890587 | Canada/QC-JUS-V5241979/2020 | 2020-03-23 | North America/Canada/Quebec/ | Laboratoire de santv© publique du Quv©bec | Laboratoire de santv© publique du Quv©bec | Sandrine Moreira et al |

| | | | | | | |
|---|---|---|---|---|---|---|
| EPI_ISL_890588 | Canada/QC-JUS-V5251969/2020 | 2020-03-24 | North America/Canada/Quebec/ | Laboratoire de santv© publique du Quv©bec | Laboratoire de santv© publique du Quv©bec | Sandrine Moreira et al |
| EPI_ISL_890589 | Canada/QC-JUS-V5241818/2020 | 2020-03-23 | North America/Canada/Quebec/ | Laboratoire de santv© publique du Quv©bec | Laboratoire de santv© publique du Quv©bec | Sandrine Moreira et al |
| EPI_ISL_890590 | Canada/QC-JUS-V5241827/2020 | 2020-03-23 | North America/Canada/Quebec/ | Laboratoire de santv© publique du Quv©bec | Laboratoire de santv© publique du Quv©bec | Sandrine Moreira et al |
| EPI_ISL_890591 | Canada/QC-JUS-V5241832/2020 | 2020-03-23 | North America/Canada/Quebec/ | Laboratoire de santv© publique du Quv©bec | Laboratoire de santv© publique du Quv©bec | Sandrine Moreira et al |
| EPI_ISL_890592 | Canada/QC-JUS-V5241875/2020 | 2020-03-22 | North America/Canada/Quebec/ | Laboratoire de santv© publique du Quv©bec | Laboratoire de santv© publique du Quv©bec | Sandrine Moreira et al |
| EPI_ISL_890593 | Canada/QC-JUS-V5241908/2020 | 2020-03-23 | North America/Canada/Quebec/ | Laboratoire de santv© publique du Quv©bec | Laboratoire de santv© publique du Quv©bec | Sandrine Moreira et al |
| EPI_ISL_890594 | Canada/QC-JUS-V5242061/2020 | 2020-03-23 | North America/Canada/Quebec/ | Laboratoire de santv© publique du Quv©bec | Laboratoire de santv© publique du Quv©bec | Sandrine Moreira et al |
| EPI_ISL_890595 | Canada/QC-JUS-V5231749/2020 | 2020-03-23 | North America/Canada/Quebec/ | Laboratoire de santv© publique du Quv©bec | Laboratoire de santv© publique du Quv©bec | Sandrine Moreira et al |
| EPI_ISL_890596 | Canada/QC-JUS-V5242219/2020 | 2020-03-23 | North America/Canada/Quebec/ | Laboratoire de santv© publique du Quv©bec | Laboratoire de santv© publique du Quv©bec | Sandrine Moreira et al |
| EPI_ISL_890597 | Canada/QC-JUS-V5242259/2020 | 2020-03-24 | North America/Canada/Quebec/ | Laboratoire de santv© publique du Quv©bec | Laboratoire de santv© publique du Quv©bec | Sandrine Moreira et al |
| EPI_ISL_890598 | Canada/QC-JUS-V5242255/2020 | 2020-03-24 | North America/Canada/Quebec/ | Laboratoire de santv© publique du Quv©bec | Laboratoire de santv© publique du Quv©bec | Sandrine Moreira et al |
| EPI_ISL_890599 | Canada/QC-JUS-V5260522/2020 | 2020-03-25 | North America/Canada/Quebec/ | Laboratoire de santv© publique du Quv©bec | Laboratoire de santv© publique du Quv©bec | Sandrine Moreira et al |
| EPI_ISL_890600 | Canada/QC-JUS-V5260671/2020 | 2020-03-24 | North America/Canada/Quebec/ | Laboratoire de santv© publique du Quv©bec | Laboratoire de santv© publique du Quv©bec | Sandrine Moreira et al |

| | | | | | | |
|---|---|---|---|---|---|---|
| EPI_ISL_890601 | Canada/QC-JUS-V5260738/2020 | 2020-03-24 | North America/Canada/Quebec/ | Laboratoire de sant√© publique du Qu√©bec | Laboratoire de sant√© publique du Qu√©bec | Sandrine Moreira et al |
| EPI_ISL_890602 | Canada/QC-JUS-V5261039/2020 | 2020-03-25 | North America/Canada/Quebec/ | Laboratoire de sant√© publique du Qu√©bec | Laboratoire de sant√© publique du Qu√©bec | Sandrine Moreira et al |
| EPI_ISL_890603 | Canada/QC-JUS-V5260792/2020 | 2020-03-24 | North America/Canada/Quebec/ | Laboratoire de sant√© publique du Qu√©bec | Laboratoire de sant√© publique du Qu√©bec | Sandrine Moreira et al |
| EPI_ISL_890604 | Canada/QC-JUS-V5260992/2020 | 2020-03-25 | North America/Canada/Quebec/ | Laboratoire de sant√© publique du Qu√©bec | Laboratoire de sant√© publique du Qu√©bec | Sandrine Moreira et al |
| EPI_ISL_890605 | Canada/QC-JUS-V5261011/2020 | 2020-03-25 | North America/Canada/Quebec/ | Laboratoire de sant√© publique du Qu√©bec | Laboratoire de sant√© publique du Qu√©bec | Sandrine Moreira et al |
| EPI_ISL_890606 | Canada/QC-JUS-V5261020/2020 | 2020-03-25 | North America/Canada/Quebec/ | Laboratoire de sant√© publique du Qu√©bec | Laboratoire de sant√© publique du Qu√©bec | Sandrine Moreira et al |
| EPI_ISL_890607 | Canada/QC-JUS-V5261396/2020 | 2020-03-24 | North America/Canada/Quebec/ | Laboratoire de sant√© publique du Qu√©bec | Laboratoire de sant√© publique du Qu√©bec | Sandrine Moreira et al |
| EPI_ISL_890608 | Canada/QC-JUS-V5261288/2020 | 2020-03-24 | North America/Canada/Quebec/ | Laboratoire de sant√© publique du Qu√©bec | Laboratoire de sant√© publique du Qu√©bec | Sandrine Moreira et al |
| EPI_ISL_890609 | Canada/QC-JUS-V5261305/2020 | 2020-03-24 | North America/Canada/Quebec/ | Laboratoire de sant√© publique du Qu√©bec | Laboratoire de sant√© publique du Qu√©bec | Sandrine Moreira et al |
| EPI_ISL_890610 | Canada/QC-JUS-V5241388/2020 | 2020-03-23 | North America/Canada/Quebec/ | Laboratoire de sant√© publique du Qu√©bec | Laboratoire de sant√© publique du Qu√©bec | Sandrine Moreira et al |
| EPI_ISL_890611 | Canada/QC-JUS-V5241813/2020 | 2020-03-23 | North America/Canada/Quebec/ | Laboratoire de sant√© publique du Qu√©bec | Laboratoire de sant√© publique du Qu√©bec | Sandrine Moreira et al |
| EPI_ISL_890612 | Canada/QC-JUS-V5241575/2020 | 2020-03-23 | North America/Canada/Quebec/ | Laboratoire de sant√© publique du Qu√©bec | Laboratoire de sant√© publique du Qu√©bec | Sandrine Moreira et al |
| EPI_ISL_890613 | Canada/QC-JUS-V5241055/2020 | 2020-03-22 | North America/Canada/Quebec/ | Laboratoire de sant√© publique du Qu√©bec | Laboratoire de sant√© publique du Qu√©bec | Sandrine Moreira et al |

| | | | | | | |
|---|---|---|---|---|---|---|
| EPI_ISL_890614 | Canada/QC-JUS-V5240719/2020 | 2020-03-23 | North America/Canada/Quebec/ | Laboratoire de sant√© publique du Qu√©bec | Laboratoire de sant√© publique du Qu√©bec | Sandrine Moreira et al |
| EPI_ISL_890615 | Canada/QC-JUS-V5240540/2020 | 2020-03-02 | North America/Canada/Quebec/ | Laboratoire de sant√© publique du Qu√©bec | Laboratoire de sant√© publique du Qu√©bec | Sandrine Moreira et al |
| EPI_ISL_890616 | Canada/QC-JUS-V5240593/2020 | 2020-03-22 | North America/Canada/Quebec/ | Laboratoire de sant√© publique du Qu√©bec | Laboratoire de sant√© publique du Qu√©bec | Sandrine Moreira et al |
| EPI_ISL_890617 | Canada/QC-JUS-V5242396/2020 | 2020-03-23 | North America/Canada/Quebec/ | Laboratoire de sant√© publique du Qu√©bec | Laboratoire de sant√© publique du Qu√©bec | Sandrine Moreira et al |
| EPI_ISL_890618 | Canada/QC-JUS-V5242383/2020 | 2020-03-23 | North America/Canada/Quebec/ | Laboratoire de sant√© publique du Qu√©bec | Laboratoire de sant√© publique du Qu√©bec | Sandrine Moreira et al |
| EPI_ISL_890619 | Canada/QC-JUS-V5240622/2020 | 2020-03-22 | North America/Canada/Quebec/ | Laboratoire de sant√© publique du Qu√©bec | Laboratoire de sant√© publique du Qu√©bec | Sandrine Moreira et al |
| EPI_ISL_890620 | Canada/QC-JUS-V5240619/2020 | 2020-03-22 | North America/Canada/Quebec/ | Laboratoire de sant√© publique du Qu√©bec | Laboratoire de sant√© publique du Qu√©bec | Sandrine Moreira et al |
| EPI_ISL_890621 | Canada/QC-JUS-V5240338/2020 | 2020-03-23 | North America/Canada/Quebec/ | Laboratoire de sant√© publique du Qu√©bec | Laboratoire de sant√© publique du Qu√©bec | Sandrine Moreira et al |
| EPI_ISL_890622 | Canada/QC-JUS-V5240676/2020 | 2020-03-22 | North America/Canada/Quebec/ | Laboratoire de sant√© publique du Qu√©bec | Laboratoire de sant√© publique du Qu√©bec | Sandrine Moreira et al |
| EPI_ISL_890623 | Canada/QC-JUS-V5230641/2020 | 2020-03-22 | North America/Canada/Quebec/ | Laboratoire de sant√© publique du Qu√©bec | Laboratoire de sant√© publique du Qu√©bec | Sandrine Moreira et al |
| EPI_ISL_890624 | Canada/QC-JUS-V5251744/2020 | 2020-03-24 | North America/Canada/Quebec/ | Laboratoire de sant√© publique du Qu√©bec | Laboratoire de sant√© publique du Qu√©bec | Sandrine Moreira et al |
| EPI_ISL_890625 | Canada/QC-JUS-V5261144/2020 | 2020-03-25 | North America/Canada/Quebec/ | Laboratoire de sant√© publique du Qu√©bec | Laboratoire de sant√© publique du Qu√©bec | Sandrine Moreira et al |
| EPI_ISL_890626 | Canada/QC-JUS-V5261796/2020 | 2020-03-24 | North America/Canada/Quebec/ | Laboratoire de sant√© publique du Qu√©bec | Laboratoire de sant√© publique du Qu√©bec | Sandrine Moreira et al |

| | | | | | | |
|---|---|---|---|---|---|---|
| EPI_ISL_890627 | Canada/QC-JUS-V5261819/2020 | 2020-03-24 | North America/Canada/Quebec/ | Laboratoire de sant√© publique du Qu√©bec | Laboratoire de sant√© publique du Qu√©bec | Sandrine Moreira et al |
| EPI_ISL_890628 | Canada/QC-JUS-V5261731/2020 | 2020-03-24 | North America/Canada/Quebec/ | Laboratoire de sant√© publique du Qu√©bec | Laboratoire de sant√© publique du Qu√©bec | Sandrine Moreira et al |
| EPI_ISL_890629 | Canada/QC-JUS-V5240590/2020 | 2020-03-22 | North America/Canada/Quebec/ | Laboratoire de sant√© publique du Qu√©bec | Laboratoire de sant√© publique du Qu√©bec | Sandrine Moreira et al |
| EPI_ISL_890630 | Canada/QC-JUS-V5240585/2020 | 2020-03-22 | North America/Canada/Quebec/ | Laboratoire de sant√© publique du Qu√©bec | Laboratoire de sant√© publique du Qu√©bec | Sandrine Moreira et al |
| EPI_ISL_890631 | Canada/QC-JUS-V5240384/2020 | 2020-03-23 | North America/Canada/Quebec/ | Laboratoire de sant√© publique du Qu√©bec | Laboratoire de sant√© publique du Qu√©bec | Sandrine Moreira et al |
| EPI_ISL_890632 | Canada/QC-JUS-V5240377/2020 | 2020-03-23 | North America/Canada/Quebec/ | Laboratoire de sant√© publique du Qu√©bec | Laboratoire de sant√© publique du Qu√©bec | Sandrine Moreira et al |
| EPI_ISL_890633 | Canada/QC-JUS-V5240354/2020 | 2020-03-23 | North America/Canada/Quebec/ | Laboratoire de sant√© publique du Qu√©bec | Laboratoire de sant√© publique du Qu√©bec | Sandrine Moreira et al |
| EPI_ISL_890634 | Canada/QC-JUS-V5240641/2020 | 2020-03-22 | North America/Canada/Quebec/ | Laboratoire de sant√© publique du Qu√©bec | Laboratoire de sant√© publique du Qu√©bec | Sandrine Moreira et al |
| EPI_ISL_890635 | Canada/QC-JUS-V5240638/2020 | 2020-03-22 | North America/Canada/Quebec/ | Laboratoire de sant√© publique du Qu√©bec | Laboratoire de sant√© publique du Qu√©bec | Sandrine Moreira et al |
| EPI_ISL_890636 | Canada/QC-JUS-V5251627/2020 | 2020-03-24 | North America/Canada/Quebec/ | Laboratoire de sant√© publique du Qu√©bec | Laboratoire de sant√© publique du Qu√©bec | Sandrine Moreira et al |
| EPI_ISL_890637 | Canada/QC-JUS-V5251842/2020 | 2020-03-24 | North America/Canada/Quebec/ | Laboratoire de sant√© publique du Qu√©bec | Laboratoire de sant√© publique du Qu√©bec | Sandrine Moreira et al |
| EPI_ISL_890638 | Canada/QC-JUS-V5252367/2020 | 2020-03-24 | North America/Canada/Quebec/ | Laboratoire de sant√© publique du Qu√©bec | Laboratoire de sant√© publique du Qu√©bec | Sandrine Moreira et al |
| EPI_ISL_890639 | Canada/QC-JUS-V5252383/2020 | 2020-03-24 | North America/Canada/Quebec/ | Laboratoire de sant√© publique du Qu√©bec | Laboratoire de sant√© publique du Qu√©bec | Sandrine Moreira et al |

| | | | | | | |
|---|---|---|---|---|---|---|
| EPI_ISL_890640 | Canada/QC-JUS-V5252388/2020 | 2020-03-24 | North America/Canada/Quebec/ | Laboratoire de sant√© publique du Qu√©bec | Laboratoire de sant√© publique du Qu√©bec | Sandrine Moreira et al |
| EPI_ISL_890641 | Canada/QC-JUS-V5242199/2020 | 2020-03-23 | North America/Canada/Quebec/ | Laboratoire de sant√© publique du Qu√©bec | Laboratoire de sant√© publique du Qu√©bec | Sandrine Moreira et al |
| EPI_ISL_890642 | Canada/QC-JUS-V5242204/2020 | 2020-03-20 | North America/Canada/Quebec/ | Laboratoire de sant√© publique du Qu√©bec | Laboratoire de sant√© publique du Qu√©bec | Sandrine Moreira et al |
| EPI_ISL_890643 | Canada/QC-JUS-V5242218/2020 | 2020-03-23 | North America/Canada/Quebec/ | Laboratoire de sant√© publique du Qu√©bec | Laboratoire de sant√© publique du Qu√©bec | Sandrine Moreira et al |
| EPI_ISL_890644 | Canada/QC-JUS-V5242228/2020 | 2020-03-23 | North America/Canada/Quebec/ | Laboratoire de sant√© publique du Qu√©bec | Laboratoire de sant√© publique du Qu√©bec | Sandrine Moreira et al |
| EPI_ISL_890645 | Canada/QC-JUS-V5242244/2020 | 2020-03-23 | North America/Canada/Quebec/ | Laboratoire de sant√© publique du Qu√©bec | Laboratoire de sant√© publique du Qu√©bec | Sandrine Moreira et al |
| EPI_ISL_890646 | Canada/QC-JUS-V5242254/2020 | 2020-03-23 | North America/Canada/Quebec/ | Laboratoire de sant√© publique du Qu√©bec | Laboratoire de sant√© publique du Qu√©bec | Sandrine Moreira et al |
| EPI_ISL_890647 | Canada/QC-JUS-V5242317/2020 | 2020-03-23 | North America/Canada/Quebec/ | Laboratoire de sant√© publique du Qu√©bec | Laboratoire de sant√© publique du Qu√©bec | Sandrine Moreira et al |
| EPI_ISL_890648 | Canada/QC-JUS-V5261953/2020 | 2020-03-26 | North America/Canada/Quebec/ | Laboratoire de sant√© publique du Qu√©bec | Laboratoire de sant√© publique du Qu√©bec | Sandrine Moreira et al |
| EPI_ISL_890649 | Canada/QC-JUS-V5262011/2020 | 2020-03-25 | North America/Canada/Quebec/ | Laboratoire de sant√© publique du Qu√©bec | Laboratoire de sant√© publique du Qu√©bec | Sandrine Moreira et al |
| EPI_ISL_890650 | Canada/QC-JUS-V5241279/2020 | 2020-03-23 | North America/Canada/Quebec/ | Laboratoire de sant√© publique du Qu√©bec | Laboratoire de sant√© publique du Qu√©bec | Sandrine Moreira et al |
| EPI_ISL_890652 | Canada/QC-JUS-V5241907/2020 | 2020-03-23 | North America/Canada/Quebec/ | Laboratoire de sant√© publique du Qu√©bec | Laboratoire de sant√© publique du Qu√©bec | Sandrine Moreira et al |
| EPI_ISL_890653 | Canada/QC-JUS-V5241878/2020 | 2020-03-23 | North America/Canada/Quebec/ | Laboratoire de sant√© publique du Qu√©bec | Laboratoire de sant√© publique du Qu√©bec | Sandrine Moreira et al |

| | | | | | | |
|---|---|---|---|---|---|---|
| EPI_ISL_890654 | Canada/QC-JUS-V5271813/2020 | 2020-03-27 | North America/Canada/Quebec/ | Laboratoire de sant√© publique du Qu√©bec | Laboratoire de sant√© publique du Qu√©bec | Sandrine Moreira et al |
| EPI_ISL_890655 | Canada/QC-JUS-V5270774/2020 | 2020-03-25 | North America/Canada/Quebec/ | Laboratoire de sant√© publique du Qu√©bec | Laboratoire de sant√© publique du Qu√©bec | Sandrine Moreira et al |
| EPI_ISL_890656 | Canada/QC-JUS-V5262289/2020 | 2020-03-25 | North America/Canada/Quebec/ | Laboratoire de sant√© publique du Qu√©bec | Laboratoire de sant√© publique du Qu√©bec | Sandrine Moreira et al |
| EPI_ISL_890657 | Canada/QC-JUS-V5262329/2020 | 2020-03-25 | North America/Canada/Quebec/ | Laboratoire de sant√© publique du Qu√©bec | Laboratoire de sant√© publique du Qu√©bec | Sandrine Moreira et al |
| EPI_ISL_890658 | Canada/QC-JUS-V5262331/2020 | 2020-03-25 | North America/Canada/Quebec/ | Laboratoire de sant√© publique du Qu√©bec | Laboratoire de sant√© publique du Qu√©bec | Sandrine Moreira et al |
| EPI_ISL_890659 | Canada/QC-JUS-V5270561/2020 | 2020-03-26 | North America/Canada/Quebec/ | Laboratoire de sant√© publique du Qu√©bec | Laboratoire de sant√© publique du Qu√©bec | Sandrine Moreira et al |
| EPI_ISL_890660 | Canada/QC-JUS-V5263006/2020 | 2020-03-25 | North America/Canada/Quebec/ | Laboratoire de sant√© publique du Qu√©bec | Laboratoire de sant√© publique du Qu√©bec | Sandrine Moreira et al |
| EPI_ISL_890661 | Canada/QC-JUS-V5270617/2020 | 2020-03-26 | North America/Canada/Quebec/ | Laboratoire de sant√© publique du Qu√©bec | Laboratoire de sant√© publique du Qu√©bec | Sandrine Moreira et al |
| EPI_ISL_890662 | Canada/QC-JUS-V5262969/2020 | 2020-03-25 | North America/Canada/Quebec/ | Laboratoire de sant√© publique du Qu√©bec | Laboratoire de sant√© publique du Qu√©bec | Sandrine Moreira et al |
| EPI_ISL_890663 | Canada/QC-JUS-V5262728/2020 | 2020-03-25 | North America/Canada/Quebec/ | Laboratoire de sant√© publique du Qu√©bec | Laboratoire de sant√© publique du Qu√©bec | Sandrine Moreira et al |
| EPI_ISL_890664 | Canada/QC-JUS-V5262079/2020 | 2020-03-26 | North America/Canada/Quebec/ | Laboratoire de sant√© publique du Qu√©bec | Laboratoire de sant√© publique du Qu√©bec | Sandrine Moreira et al |
| EPI_ISL_890665 | Canada/QC-JUS-V5271270/2020 | 2020-03-26 | North America/Canada/Quebec/ | Laboratoire de sant√© publique du Qu√©bec | Laboratoire de sant√© publique du Qu√©bec | Sandrine Moreira et al |
| EPI_ISL_890666 | Canada/QC-JUS-V5271052/2020 | 2020-03-26 | North America/Canada/Quebec/ | Laboratoire de sant√© publique du Qu√©bec | Laboratoire de sant√© publique du Qu√©bec | Sandrine Moreira et al |

| | | | | | | |
|---|---|---|---|---|---|---|
| EPI_ISL_890667 | Canada/QC-JUS-V5271061/2020 | 2020-03-26 | North America/Canada/Quebec/ | Laboratoire de sant√© publique du Qu√©bec | Laboratoire de sant√© publique du Qu√©bec | Sandrine Moreira et al |
| EPI_ISL_890668 | Canada/QC-JUS-V5271131/2020 | 2020-03-26 | North America/Canada/Quebec/ | Laboratoire de sant√© publique du Qu√©bec | Laboratoire de sant√© publique du Qu√©bec | Sandrine Moreira et al |
| EPI_ISL_890669 | Canada/QC-JUS-V5271112/2020 | 2020-03-26 | North America/Canada/Quebec/ | Laboratoire de sant√© publique du Qu√©bec | Laboratoire de sant√© publique du Qu√©bec | Sandrine Moreira et al |
| EPI_ISL_890670 | Canada/QC-JUS-V5262029/2020 | 2020-03-24 | North America/Canada/Quebec/ | Laboratoire de sant√© publique du Qu√©bec | Laboratoire de sant√© publique du Qu√©bec | Sandrine Moreira et al |
| EPI_ISL_890671 | Canada/QC-JUS-V5261967/2020 | 2020-03-24 | North America/Canada/Quebec/ | Laboratoire de sant√© publique du Qu√©bec | Laboratoire de sant√© publique du Qu√©bec | Sandrine Moreira et al |
| EPI_ISL_890672 | Canada/QC-JUS-V5271062/2020 | 2020-03-25 | North America/Canada/Quebec/ | Laboratoire de sant√© publique du Qu√©bec | Laboratoire de sant√© publique du Qu√©bec | Sandrine Moreira et al |
| EPI_ISL_890673 | Canada/QC-JUS-V5270713/2020 | 2020-03-26 | North America/Canada/Quebec/ | Laboratoire de sant√© publique du Qu√©bec | Laboratoire de sant√© publique du Qu√©bec | Sandrine Moreira et al |
| EPI_ISL_890674 | Canada/QC-JUS-V5270707/2020 | 2020-03-26 | North America/Canada/Quebec/ | Laboratoire de sant√© publique du Qu√©bec | Laboratoire de sant√© publique du Qu√©bec | Sandrine Moreira et al |
| EPI_ISL_890675 | Canada/QC-JUS-V5270700/2020 | 2020-03-26 | North America/Canada/Quebec/ | Laboratoire de sant√© publique du Qu√©bec | Laboratoire de sant√© publique du Qu√©bec | Sandrine Moreira et al |
| EPI_ISL_890676 | Canada/QC-JUS-V5270637/2020 | 2020-03-26 | North America/Canada/Quebec/ | Laboratoire de sant√© publique du Qu√©bec | Laboratoire de sant√© publique du Qu√©bec | Sandrine Moreira et al |
| EPI_ISL_890677 | Canada/QC-JUS-V5270244/2020 | 2020-03-25 | North America/Canada/Quebec/ | Laboratoire de sant√© publique du Qu√©bec | Laboratoire de sant√© publique du Qu√©bec | Sandrine Moreira et al |
| EPI_ISL_890678 | Canada/QC-JUS-V5270304/2020 | 2020-03-25 | North America/Canada/Quebec/ | Laboratoire de sant√© publique du Qu√©bec | Laboratoire de sant√© publique du Qu√©bec | Sandrine Moreira et al |
| EPI_ISL_890679 | Canada/QC-JUS-V5270328/2020 | 2020-03-25 | North America/Canada/Quebec/ | Laboratoire de sant√© publique du Qu√©bec | Laboratoire de sant√© publique du Qu√©bec | Sandrine Moreira et al |

| | | | | | | |
|---|---|---|---|---|---|---|
| EPI_ISL_890680 | Canada/QC-JUS-V5262138/2020 | 2020-03-26 | North America/Canada/Quebec/ | Laboratoire de sant√© publique du Qu√©bec | Laboratoire de sant√© publique du Qu√©bec | Sandrine Moreira et al |
| EPI_ISL_890681 | Canada/QC-JUS-V5262091/2020 | 2020-03-26 | North America/Canada/Quebec/ | Laboratoire de sant√© publique du Qu√©bec | Laboratoire de sant√© publique du Qu√©bec | Sandrine Moreira et al |
| EPI_ISL_890682 | Canada/QC-JUS-V5262102/2020 | 2020-03-26 | North America/Canada/Quebec/ | Laboratoire de sant√© publique du Qu√©bec | Laboratoire de sant√© publique du Qu√©bec | Sandrine Moreira et al |
| EPI_ISL_890683 | Canada/QC-JUS-V5262107/2020 | 2020-03-26 | North America/Canada/Quebec/ | Laboratoire de sant√© publique du Qu√©bec | Laboratoire de sant√© publique du Qu√©bec | Sandrine Moreira et al |
| EPI_ISL_890684 | Canada/QC-JUS-V5261550/2020 | 2020-03-24 | North America/Canada/Quebec/ | Laboratoire de sant√© publique du Qu√©bec | Laboratoire de sant√© publique du Qu√©bec | Sandrine Moreira et al |
| EPI_ISL_890685 | Canada/QC-JUS-V5262175/2020 | 2020-03-26 | North America/Canada/Quebec/ | Laboratoire de sant√© publique du Qu√©bec | Laboratoire de sant√© publique du Qu√©bec | Sandrine Moreira et al |
| EPI_ISL_890686 | Canada/QC-JUS-V5241765/2020 | 2020-03-23 | North America/Canada/Quebec/ | Laboratoire de sant√© publique du Qu√©bec | Laboratoire de sant√© publique du Qu√©bec | Sandrine Moreira et al |
| EPI_ISL_890687 | Canada/QC-JUS-V5241615/2020 | 2020-03-24 | North America/Canada/Quebec/ | Laboratoire de sant√© publique du Qu√©bec | Laboratoire de sant√© publique du Qu√©bec | Sandrine Moreira et al |
| EPI_ISL_890688 | Canada/QC-JUS-V5241702/2020 | 2020-03-24 | North America/Canada/Quebec/ | Laboratoire de sant√© publique du Qu√©bec | Laboratoire de sant√© publique du Qu√©bec | Sandrine Moreira et al |
| EPI_ISL_890689 | Canada/QC-JUS-V5241836/2020 | 2020-03-23 | North America/Canada/Quebec/ | Laboratoire de sant√© publique du Qu√©bec | Laboratoire de sant√© publique du Qu√©bec | Sandrine Moreira et al |
| EPI_ISL_890690 | Canada/QC-JUS-V5241116/2020 | 2020-03-23 | North America/Canada/Quebec/ | Laboratoire de sant√© publique du Qu√©bec | Laboratoire de sant√© publique du Qu√©bec | Sandrine Moreira et al |
| EPI_ISL_890691 | Canada/QC-JUS-V5241051/2020 | 2020-03-24 | North America/Canada/Quebec/ | Laboratoire de sant√© publique du Qu√©bec | Laboratoire de sant√© publique du Qu√©bec | Sandrine Moreira et al |
| EPI_ISL_890692 | Canada/QC-JUS-V5241208/2020 | 2020-03-23 | North America/Canada/Quebec/ | Laboratoire de sant√© publique du Qu√©bec | Laboratoire de sant√© publique du Qu√©bec | Sandrine Moreira et al |

| | | | | | | |
|---|---|---|---|---|---|---|
| EPI_ISL_890693 | Canada/QC-JUS-V5261877/2020 | 2020-03-24 | North America/Canada/Quebec/ | Laboratoire de santv© publique du Quv©bec | Laboratoire de santv© publique du Quv©bec | Sandrine Moreira et al |
| EPI_ISL_890694 | Canada/QC-JUS-V5261660/2020 | 2020-03-24 | North America/Canada/Quebec/ | Laboratoire de santv© publique du Quv©bec | Laboratoire de santv© publique du Quv©bec | Sandrine Moreira et al |
| EPI_ISL_890695 | Canada/QC-JUS-V5261989/2020 | 2020-03-25 | North America/Canada/Quebec/ | Laboratoire de santv© publique du Quv©bec | Laboratoire de santv© publique du Quv©bec | Sandrine Moreira et al |
| EPI_ISL_890696 | Canada/QC-JUS-V5261973/2020 | 2020-03-25 | North America/Canada/Quebec/ | Laboratoire de santv© publique du Quv©bec | Laboratoire de santv© publique du Quv©bec | Sandrine Moreira et al |
| EPI_ISL_890697 | Canada/QC-JUS-V5261900/2020 | 2020-03-25 | North America/Canada/Quebec/ | Laboratoire de santv© publique du Quv©bec | Laboratoire de santv© publique du Quv©bec | Sandrine Moreira et al |
| EPI_ISL_890698 | Canada/QC-JUS-V5271200/2020 | 2020-03-26 | North America/Canada/Quebec/ | Laboratoire de santv© publique du Quv©bec | Laboratoire de santv© publique du Quv©bec | Sandrine Moreira et al |
| EPI_ISL_890699 | Canada/QC-JUS-V5270577/2020 | 2020-03-26 | North America/Canada/Quebec/ | Laboratoire de santv© publique du Quv©bec | Laboratoire de santv© publique du Quv©bec | Sandrine Moreira et al |
| EPI_ISL_890700 | Canada/QC-JUS-V5271108/2020 | 2020-03-26 | North America/Canada/Quebec/ | Laboratoire de santv© publique du Quv©bec | Laboratoire de santv© publique du Quv©bec | Sandrine Moreira et al |
| EPI_ISL_890701 | Canada/QC-JUS-V5271080/2020 | 2020-03-26 | North America/Canada/Quebec/ | Laboratoire de santv© publique du Quv©bec | Laboratoire de santv© publique du Quv©bec | Sandrine Moreira et al |
| EPI_ISL_890702 | Canada/QC-JUS-V5262892/2020 | 2020-03-25 | North America/Canada/Quebec/ | Laboratoire de santv© publique du Quv©bec | Laboratoire de santv© publique du Quv©bec | Sandrine Moreira et al |
| EPI_ISL_890703 | Canada/QC-JUS-V5262879/2020 | 2020-03-25 | North America/Canada/Quebec/ | Laboratoire de santv© publique du Quv©bec | Laboratoire de santv© publique du Quv©bec | Sandrine Moreira et al |
| EPI_ISL_890704 | Canada/QC-JUS-V5262104/2020 | 2020-03-25 | North America/Canada/Quebec/ | Laboratoire de santv© publique du Quv©bec | Laboratoire de santv© publique du Quv©bec | Sandrine Moreira et al |
| EPI_ISL_890705 | Canada/QC-JUS-V5261439/2020 | 2020-03-24 | North America/Canada/Quebec/ | Laboratoire de santv© publique du Quv©bec | Laboratoire de santv© publique du Quv©bec | Sandrine Moreira et al |

| | | | | | | |
|---|---|---|---|---|---|---|
| EPI_ISL_890706 | Canada/QC-JUS-V5280467/2020 | 2020-03-27 | North America/Canada/Quebec/ | Laboratoire de santv© publique du Quv©bec | Laboratoire de santv© publique du Quv©bec | Sandrine Moreira et al |
| EPI_ISL_890707 | Canada/QC-JUS-V5280091/2020 | 2020-03-26 | North America/Canada/Quebec/ | Laboratoire de santv© publique du Quv©bec | Laboratoire de santv© publique du Quv©bec | Sandrine Moreira et al |
| EPI_ISL_890708 | Canada/QC-JUS-V5272574/2020 | 2020-03-26 | North America/Canada/Quebec/ | Laboratoire de santv© publique du Quv©bec | Laboratoire de santv© publique du Quv©bec | Sandrine Moreira et al |
| EPI_ISL_890709 | Canada/QC-JUS-V5272548/2020 | 2020-03-26 | North America/Canada/Quebec/ | Laboratoire de santv© publique du Quv©bec | Laboratoire de santv© publique du Quv©bec | Sandrine Moreira et al |
| EPI_ISL_890710 | Canada/QC-JUS-V5241690/2020 | 2020-03-23 | North America/Canada/Quebec/ | Laboratoire de santv© publique du Quv©bec | Laboratoire de santv© publique du Quv©bec | Sandrine Moreira et al |
| EPI_ISL_890711 | Canada/QC-JUS-V5280529/2020 | 2020-03-27 | North America/Canada/Quebec/ | Laboratoire de santv© publique du Quv©bec | Laboratoire de santv© publique du Quv©bec | Sandrine Moreira et al |
| EPI_ISL_890712 | Canada/QC-JUS-V5280544/2020 | 2020-03-27 | North America/Canada/Quebec/ | Laboratoire de santv© publique du Quv©bec | Laboratoire de santv© publique du Quv©bec | Sandrine Moreira et al |
| EPI_ISL_890713 | Canada/QC-JUS-V5280551/2020 | 2020-03-27 | North America/Canada/Quebec/ | Laboratoire de santv© publique du Quv©bec | Laboratoire de santv© publique du Quv©bec | Sandrine Moreira et al |
| EPI_ISL_890714 | Canada/QC-JUS-V5280554/2020 | 2020-03-27 | North America/Canada/Quebec/ | Laboratoire de santv© publique du Quv©bec | Laboratoire de santv© publique du Quv©bec | Sandrine Moreira et al |
| EPI_ISL_890715 | Canada/QC-JUS-V5280628/2020 | 2020-03-27 | North America/Canada/Quebec/ | Laboratoire de santv© publique du Quv©bec | Laboratoire de santv© publique du Quv©bec | Sandrine Moreira et al |
| EPI_ISL_890716 | Canada/QC-JUS-V5280629/2020 | 2020-03-27 | North America/Canada/Quebec/ | Laboratoire de santv© publique du Quv©bec | Laboratoire de santv© publique du Quv©bec | Sandrine Moreira et al |
| EPI_ISL_890717 | Canada/QC-JUS-V5280471/2020 | 2020-03-28 | North America/Canada/Quebec/ | Laboratoire de santv© publique du Quv©bec | Laboratoire de santv© publique du Quv©bec | Sandrine Moreira et al |
| EPI_ISL_890718 | Canada/QC-JUS-V5280785/2020 | 2020-03-27 | North America/Canada/Quebec/ | Laboratoire de santv© publique du Quv©bec | Laboratoire de santv© publique du Quv©bec | Sandrine Moreira et al |

| | | | | | | |
|---|---|---|---|---|---|---|
| EPI_ISL_890719 | Canada/QC-JUS-V5280660/2020 | 2020-03-27 | North America/Canada/Quebec/ | Laboratoire de sant√© publique du Qu√©bec | Laboratoire de sant√© publique du Qu√©bec | Sandrine Moreira et al |
| EPI_ISL_890720 | Canada/QC-JUS-V5280171/2020 | 2020-03-26 | North America/Canada/Quebec/ | Laboratoire de sant√© publique du Qu√©bec | Laboratoire de sant√© publique du Qu√©bec | Sandrine Moreira et al |
| EPI_ISL_890721 | Canada/QC-JUS-V5280726/2020 | 2020-03-27 | North America/Canada/Quebec/ | Laboratoire de sant√© publique du Qu√©bec | Laboratoire de sant√© publique du Qu√©bec | Sandrine Moreira et al |
| EPI_ISL_890722 | Canada/QC-JUS-V5270860/2020 | 2020-03-25 | North America/Canada/Quebec/ | Laboratoire de sant√© publique du Qu√©bec | Laboratoire de sant√© publique du Qu√©bec | Sandrine Moreira et al |
| EPI_ISL_890723 | Canada/QC-JUS-V5262953/2020 | 2020-03-25 | North America/Canada/Quebec/ | Laboratoire de sant√© publique du Qu√©bec | Laboratoire de sant√© publique du Qu√©bec | Sandrine Moreira et al |
| EPI_ISL_890724 | Canada/QC-JUS-V5271877/2020 | 2020-03-27 | North America/Canada/Quebec/ | Laboratoire de sant√© publique du Qu√©bec | Laboratoire de sant√© publique du Qu√©bec | Sandrine Moreira et al |
| EPI_ISL_890725 | Canada/QC-JUS-V5271431/2020 | 2020-03-25 | North America/Canada/Quebec/ | Laboratoire de sant√© publique du Qu√©bec | Laboratoire de sant√© publique du Qu√©bec | Sandrine Moreira et al |
| EPI_ISL_890726 | Canada/QC-JUS-V5270813/2020 | 2020-03-25 | North America/Canada/Quebec/ | Laboratoire de sant√© publique du Qu√©bec | Laboratoire de sant√© publique du Qu√©bec | Sandrine Moreira et al |
| EPI_ISL_890727 | Canada/QC-JUS-V5270880/2020 | 2020-03-25 | North America/Canada/Quebec/ | Laboratoire de sant√© publique du Qu√©bec | Laboratoire de sant√© publique du Qu√©bec | Sandrine Moreira et al |
| EPI_ISL_890728 | Canada/QC-JUS-V5271342/2020 | 2020-03-26 | North America/Canada/Quebec/ | Laboratoire de sant√© publique du Qu√©bec | Laboratoire de sant√© publique du Qu√©bec | Sandrine Moreira et al |
| EPI_ISL_890729 | Canada/QC-JUS-V5271356/2020 | 2020-03-26 | North America/Canada/Quebec/ | Laboratoire de sant√© publique du Qu√©bec | Laboratoire de sant√© publique du Qu√©bec | Sandrine Moreira et al |
| EPI_ISL_890730 | Canada/QC-JUS-V5272029/2020 | 2020-03-26 | North America/Canada/Quebec/ | Laboratoire de sant√© publique du Qu√©bec | Laboratoire de sant√© publique du Qu√©bec | Sandrine Moreira et al |
| EPI_ISL_890731 | Canada/QC-JUS-V5272140/2020 | 2020-03-26 | North America/Canada/Quebec/ | Laboratoire de sant√© publique du Qu√©bec | Laboratoire de sant√© publique du Qu√©bec | Sandrine Moreira et al |

| | | | | | | |
|---|---|---|---|---|---|---|
| EPI_ISL_890732 | Canada/QC-JUS-V5272149/2020 | 2020-03-26 | North America/Canada/Quebec/ | Laboratoire de sant√© publique du Qu√©bec | Laboratoire de sant√© publique du Qu√©bec | Sandrine Moreira et al |
| EPI_ISL_890733 | Canada/QC-JUS-V5272156/2020 | 2020-03-26 | North America/Canada/Quebec/ | Laboratoire de sant√© publique du Qu√©bec | Laboratoire de sant√© publique du Qu√©bec | Sandrine Moreira et al |
| EPI_ISL_890734 | Canada/QC-JUS-V5272166/2020 | 2020-03-26 | North America/Canada/Quebec/ | Laboratoire de sant√© publique du Qu√©bec | Laboratoire de sant√© publique du Qu√©bec | Sandrine Moreira et al |
| EPI_ISL_890735 | Canada/QC-JUS-V5272034/2020 | 2020-03-26 | North America/Canada/Quebec/ | Laboratoire de sant√© publique du Qu√©bec | Laboratoire de sant√© publique du Qu√©bec | Sandrine Moreira et al |
| EPI_ISL_890736 | Canada/QC-JUS-V5272096/2020 | 2020-03-26 | North America/Canada/Quebec/ | Laboratoire de sant√© publique du Qu√©bec | Laboratoire de sant√© publique du Qu√©bec | Sandrine Moreira et al |
| EPI_ISL_890737 | Canada/QC-JUS-V5272136/2020 | 2020-03-26 | North America/Canada/Quebec/ | Laboratoire de sant√© publique du Qu√©bec | Laboratoire de sant√© publique du Qu√©bec | Sandrine Moreira et al |
| EPI_ISL_890738 | Canada/QC-JUS-V5272150/2020 | 2020-03-26 | North America/Canada/Quebec/ | Laboratoire de sant√© publique du Qu√©bec | Laboratoire de sant√© publique du Qu√©bec | Sandrine Moreira et al |
| EPI_ISL_890739 | Canada/QC-JUS-V5271479/2020 | 2020-03-26 | North America/Canada/Quebec/ | Laboratoire de sant√© publique du Qu√©bec | Laboratoire de sant√© publique du Qu√©bec | Sandrine Moreira et al |
| EPI_ISL_890740 | Canada/QC-JUS-V5271923/2020 | 2020-03-26 | North America/Canada/Quebec/ | Laboratoire de sant√© publique du Qu√©bec | Laboratoire de sant√© publique du Qu√©bec | Sandrine Moreira et al |
| EPI_ISL_890742 | Canada/QC-JUS-V5281076/2020 | 2020-03-27 | North America/Canada/Quebec/ | Laboratoire de sant√© publique du Qu√©bec | Laboratoire de sant√© publique du Qu√©bec | Sandrine Moreira et al |
| EPI_ISL_890743 | Canada/QC-JUS-V5270399/2020 | 2020-03-25 | North America/Canada/Quebec/ | Laboratoire de sant√© publique du Qu√©bec | Laboratoire de sant√© publique du Qu√©bec | Sandrine Moreira et al |
| EPI_ISL_890744 | Canada/QC-JUS-V5272334/2020 | 2020-03-26 | North America/Canada/Quebec/ | Laboratoire de sant√© publique du Qu√©bec | Laboratoire de sant√© publique du Qu√©bec | Sandrine Moreira et al |
| EPI_ISL_890745 | Canada/QC-JUS-V5272229/2020 | 2020-03-26 | North America/Canada/Quebec/ | Laboratoire de sant√© publique du Qu√©bec | Laboratoire de sant√© publique du Qu√©bec | Sandrine Moreira et al |

| | | | | | | |
|---|---|---|---|---|---|---|
| EPI_ISL_890746 | Canada/QC-JUS-V5271956/2020 | 2020-03-26 | North America/Canada/Quebec/ | Laboratoire de santv© publique du Quv©bec | Laboratoire de santv© publique du Quv©bec | Sandrine Moreira et al |
| EPI_ISL_890747 | Canada/QC-JUS-V5272410/2020 | 2020-03-26 | North America/Canada/Quebec/ | Laboratoire de santv© publique du Quv©bec | Laboratoire de santv© publique du Quv©bec | Sandrine Moreira et al |
| EPI_ISL_890748 | Canada/QC-JUS-V5250534/2020 | 2020-03-21 | North America/Canada/Quebec/ | Laboratoire de santv© publique du Quv©bec | Laboratoire de santv© publique du Quv©bec | Sandrine Moreira et al |
| EPI_ISL_890749 | Canada/QC-JUS-V5252378/2020 | 2020-03-24 | North America/Canada/Quebec/ | Laboratoire de santv© publique du Quv©bec | Laboratoire de santv© publique du Quv©bec | Sandrine Moreira et al |
| EPI_ISL_890750 | Canada/QC-JUS-V5281646/2020 | 2020-03-28 | North America/Canada/Quebec/ | Laboratoire de santv© publique du Quv©bec | Laboratoire de santv© publique du Quv©bec | Sandrine Moreira et al |
| EPI_ISL_890751 | Canada/QC-JUS-V5281543/2020 | 2020-03-28 | North America/Canada/Quebec/ | Laboratoire de santv© publique du Quv©bec | Laboratoire de santv© publique du Quv©bec | Sandrine Moreira et al |
| EPI_ISL_890752 | Canada/QC-JUS-V5281549/2020 | 2020-03-28 | North America/Canada/Quebec/ | Laboratoire de santv© publique du Quv©bec | Laboratoire de santv© publique du Quv©bec | Sandrine Moreira et al |
| EPI_ISL_890753 | Canada/QC-JUS-V5281581/2020 | 2020-03-28 | North America/Canada/Quebec/ | Laboratoire de santv© publique du Quv©bec | Laboratoire de santv© publique du Quv©bec | Sandrine Moreira et al |
| EPI_ISL_890754 | Canada/QC-JUS-V5281590/2020 | 2020-03-28 | North America/Canada/Quebec/ | Laboratoire de santv© publique du Quv©bec | Laboratoire de santv© publique du Quv©bec | Sandrine Moreira et al |
| EPI_ISL_890755 | Canada/QC-JUS-V5270429/2020 | 2020-03-25 | North America/Canada/Quebec/ | Laboratoire de santv© publique du Quv©bec | Laboratoire de santv© publique du Quv©bec | Sandrine Moreira et al |
| EPI_ISL_890756 | Canada/QC-JUS-V5270618/2020 | 2020-03-25 | North America/Canada/Quebec/ | Laboratoire de santv© publique du Quv©bec | Laboratoire de santv© publique du Quv©bec | Sandrine Moreira et al |
| EPI_ISL_890757 | Canada/QC-JUS-V5242084/2020 | 2020-03-23 | North America/Canada/Quebec/ | Laboratoire de santv© publique du Quv©bec | Laboratoire de santv© publique du Quv©bec | Sandrine Moreira et al |
| EPI_ISL_890758 | Canada/QC-JUS-V5242114/2020 | 2020-03-23 | North America/Canada/Quebec/ | Laboratoire de santv© publique du Quv©bec | Laboratoire de santv© publique du Quv©bec | Sandrine Moreira et al |

| | | | | | | |
|---|---|---|---|---|---|---|
| EPI_ISL_890759 | Canada/QC-JUS-V5272031/2020 | 2020-03-24 | North America/Canada/Quebec/ | Laboratoire de santv© publique du Quv©bec | Laboratoire de santv© publique du Quv©bec | Sandrine Moreira et al |
| EPI_ISL_890760 | Canada/QC-JUS-V5280308/2020 | 2020-03-26 | North America/Canada/Quebec/ | Laboratoire de santv© publique du Quv©bec | Laboratoire de santv© publique du Quv©bec | Sandrine Moreira et al |
| EPI_ISL_890761 | Canada/QC-JUS-V5281011/2020 | 2020-03-27 | North America/Canada/Quebec/ | Laboratoire de santv© publique du Quv©bec | Laboratoire de santv© publique du Quv©bec | Sandrine Moreira et al |
| EPI_ISL_890762 | Canada/QC-JUS-V5281517/2020 | 2020-03-28 | North America/Canada/Quebec/ | Laboratoire de santv© publique du Quv©bec | Laboratoire de santv© publique du Quv©bec | Sandrine Moreira et al |
| EPI_ISL_890763 | Canada/QC-JUS-V5281609/2020 | 2020-03-27 | North America/Canada/Quebec/ | Laboratoire de santv© publique du Quv©bec | Laboratoire de santv© publique du Quv©bec | Sandrine Moreira et al |
| EPI_ISL_890764 | Canada/QC-JUS-V5281621/2020 | 2020-03-27 | North America/Canada/Quebec/ | Laboratoire de santv© publique du Quv©bec | Laboratoire de santv© publique du Quv©bec | Sandrine Moreira et al |
| EPI_ISL_890765 | Canada/QC-JUS-V5281616/2020 | 2020-03-28 | North America/Canada/Quebec/ | Laboratoire de santv© publique du Quv©bec | Laboratoire de santv© publique du Quv©bec | Sandrine Moreira et al |
| EPI_ISL_890766 | Canada/QC-JUS-V5280406/2020 | 2020-03-26 | North America/Canada/Quebec/ | Laboratoire de santv© publique du Quv©bec | Laboratoire de santv© publique du Quv©bec | Sandrine Moreira et al |
| EPI_ISL_890767 | Canada/QC-JUS-V5281618/2020 | 2020-03-27 | North America/Canada/Quebec/ | Laboratoire de santv© publique du Quv©bec | Laboratoire de santv© publique du Quv©bec | Sandrine Moreira et al |
| EPI_ISL_890768 | Canada/QC-JUS-V5280971/2020 | 2020-03-27 | North America/Canada/Quebec/ | Laboratoire de santv© publique du Quv©bec | Laboratoire de santv© publique du Quv©bec | Sandrine Moreira et al |
| EPI_ISL_890769 | Canada/QC-JUS-V5281361/2020 | 2020-03-27 | North America/Canada/Quebec/ | Laboratoire de santv© publique du Quv©bec | Laboratoire de santv© publique du Quv©bec | Sandrine Moreira et al |
| EPI_ISL_890770 | Canada/QC-JUS-V5281268/2020 | 2020-03-28 | North America/Canada/Quebec/ | Laboratoire de santv© publique du Quv©bec | Laboratoire de santv© publique du Quv©bec | Sandrine Moreira et al |
| EPI_ISL_890772 | Canada/QC-JUS-V5281494/2020 | 2020-03-28 | North America/Canada/Quebec/ | Laboratoire de santv© publique du Quv©bec | Laboratoire de santv© publique du Quv©bec | Sandrine Moreira et al |

| | | | | | | |
|---|---|---|---|---|---|---|
| EPI_ISL_890773 | Canada/QC-JUS-V5280337/2020 | 2020-03-26 | North America/Canada/Quebec/ | Laboratoire de santv© publique du Quv©bec | Laboratoire de santv© publique du Quv©bec | Sandrine Moreira et al |
| EPI_ISL_890774 | Canada/QC-JUS-V5281523/2020 | 2020-03-27 | North America/Canada/Quebec/ | Laboratoire de santv© publique du Quv©bec | Laboratoire de santv© publique du Quv©bec | Sandrine Moreira et al |
| EPI_ISL_890775 | Canada/QC-JUS-V5281572/2020 | 2020-03-27 | North America/Canada/Quebec/ | Laboratoire de santv© publique du Quv©bec | Laboratoire de santv© publique du Quv©bec | Sandrine Moreira et al |
| EPI_ISL_890776 | Canada/QC-JUS-V5281123/2020 | 2020-03-28 | North America/Canada/Quebec/ | Laboratoire de santv© publique du Quv©bec | Laboratoire de santv© publique du Quv©bec | Sandrine Moreira et al |
| EPI_ISL_890777 | Canada/QC-JUS-V5281220/2020 | 2020-03-28 | North America/Canada/Quebec/ | Laboratoire de santv© publique du Quv©bec | Laboratoire de santv© publique du Quv©bec | Sandrine Moreira et al |
| EPI_ISL_890778 | Canada/QC-JUS-V5301560/2020 | 2020-03-27 | North America/Canada/Quebec/ | Laboratoire de santv© publique du Quv©bec | Laboratoire de santv© publique du Quv©bec | Sandrine Moreira et al |
| EPI_ISL_890779 | Canada/QC-JUS-V5301275/2020 | 2020-03-27 | North America/Canada/Quebec/ | Laboratoire de santv© publique du Quv©bec | Laboratoire de santv© publique du Quv©bec | Sandrine Moreira et al |
| EPI_ISL_890780 | Canada/QC-JUS-V5301313/2020 | 2020-03-27 | North America/Canada/Quebec/ | Laboratoire de santv© publique du Quv©bec | Laboratoire de santv© publique du Quv©bec | Sandrine Moreira et al |
| EPI_ISL_890781 | Canada/QC-JUS-V5301808/2020 | 2020-03-27 | North America/Canada/Quebec/ | Laboratoire de santv© publique du Quv©bec | Laboratoire de santv© publique du Quv©bec | Sandrine Moreira et al |
| EPI_ISL_890782 | Canada/QC-JUS-V5280913/2020 | 2020-03-27 | North America/Canada/Quebec/ | Laboratoire de santv© publique du Quv©bec | Laboratoire de santv© publique du Quv©bec | Sandrine Moreira et al |
| EPI_ISL_890783 | Canada/QC-JUS-V5290700/2020 | 2020-03-27 | North America/Canada/Quebec/ | Laboratoire de santv© publique du Quv©bec | Laboratoire de santv© publique du Quv©bec | Sandrine Moreira et al |
| EPI_ISL_890784 | Canada/QC-JUS-V5290598/2020 | 2020-03-27 | North America/Canada/Quebec/ | Laboratoire de santv© publique du Quv©bec | Laboratoire de santv© publique du Quv©bec | Sandrine Moreira et al |
| EPI_ISL_890785 | Canada/QC-JUS-V5290616/2020 | 2020-03-27 | North America/Canada/Quebec/ | Laboratoire de santv© publique du Quv©bec | Laboratoire de santv© publique du Quv©bec | Sandrine Moreira et al |

| | | | | | | |
|---|---|---|---|---|---|---|
| EPI_ISL_890786 | Canada/QC-JUS-V5290648/2020 | 2020-03-27 | North America/Canada/Quebec/ | Laboratoire de sant√© publique du Qu√©bec | Laboratoire de sant√© publique du Qu√©bec | Sandrine Moreira et al |
| EPI_ISL_890787 | Canada/QC-JUS-V5290650/2020 | 2020-03-27 | North America/Canada/Quebec/ | Laboratoire de sant√© publique du Qu√©bec | Laboratoire de sant√© publique du Qu√©bec | Sandrine Moreira et al |
| EPI_ISL_890788 | Canada/QC-JUS-V5271109/2020 | 2020-03-26 | North America/Canada/Quebec/ | Laboratoire de sant√© publique du Qu√©bec | Laboratoire de sant√© publique du Qu√©bec | Sandrine Moreira et al |
| EPI_ISL_890789 | Canada/QC-JUS-V5271428/2020 | 2020-03-25 | North America/Canada/Quebec/ | Laboratoire de sant√© publique du Qu√©bec | Laboratoire de sant√© publique du Qu√©bec | Sandrine Moreira et al |
| EPI_ISL_890790 | Canada/QC-JUS-V5271145/2020 | 2020-03-26 | North America/Canada/Quebec/ | Laboratoire de sant√© publique du Qu√©bec | Laboratoire de sant√© publique du Qu√©bec | Sandrine Moreira et al |
| EPI_ISL_890791 | Canada/QC-JUS-V5272023/2020 | 2020-03-27 | North America/Canada/Quebec/ | Laboratoire de sant√© publique du Qu√©bec | Laboratoire de sant√© publique du Qu√©bec | Sandrine Moreira et al |
| EPI_ISL_890792 | Canada/QC-JUS-V5271990/2020 | 2020-03-24 | North America/Canada/Quebec/ | Laboratoire de sant√© publique du Qu√©bec | Laboratoire de sant√© publique du Qu√©bec | Sandrine Moreira et al |
| EPI_ISL_890793 | Canada/QC-JUS-V5271838/2020 | 2020-03-26 | North America/Canada/Quebec/ | Laboratoire de sant√© publique du Qu√©bec | Laboratoire de sant√© publique du Qu√©bec | Sandrine Moreira et al |
| EPI_ISL_890794 | Canada/QC-JUS-V5271816/2020 | 2020-03-26 | North America/Canada/Quebec/ | Laboratoire de sant√© publique du Qu√©bec | Laboratoire de sant√© publique du Qu√©bec | Sandrine Moreira et al |
| EPI_ISL_890795 | Canada/QC-JUS-V5271348/2020 | 2020-03-26 | North America/Canada/Quebec/ | Laboratoire de sant√© publique du Qu√©bec | Laboratoire de sant√© publique du Qu√©bec | Sandrine Moreira et al |
| EPI_ISL_890796 | Canada/QC-JUS-V5271323/2020 | 2020-03-26 | North America/Canada/Quebec/ | Laboratoire de sant√© publique du Qu√©bec | Laboratoire de sant√© publique du Qu√©bec | Sandrine Moreira et al |
| EPI_ISL_890797 | Canada/QC-JUS-V5290661/2020 | 2020-03-28 | North America/Canada/Quebec/ | Laboratoire de sant√© publique du Qu√©bec | Laboratoire de sant√© publique du Qu√©bec | Sandrine Moreira et al |
| EPI_ISL_890798 | Canada/QC-JUS-V5291038/2020 | 2020-03-27 | North America/Canada/Quebec/ | Laboratoire de sant√© publique du Qu√©bec | Laboratoire de sant√© publique du Qu√©bec | Sandrine Moreira et al |

| | | | | | | |
|---|---|---|---|---|---|---|
| EPI_ISL_890799 | Canada/QC-JUS-V5290775/2020 | 2020-03-28 | North America/Canada/Quebec/ | Laboratoire de sant√© publique du Qu√©bec | Laboratoire de sant√© publique du Qu√©bec | Sandrine Moreira et al |
| EPI_ISL_890800 | Canada/QC-JUS-V5290911/2020 | 2020-03-28 | North America/Canada/Quebec/ | Laboratoire de sant√© publique du Qu√©bec | Laboratoire de sant√© publique du Qu√©bec | Sandrine Moreira et al |
| EPI_ISL_890801 | Canada/QC-JUS-V5291433/2020 | 2020-03-27 | North America/Canada/Quebec/ | Laboratoire de sant√© publique du Qu√©bec | Laboratoire de sant√© publique du Qu√©bec | Sandrine Moreira et al |
| EPI_ISL_890802 | Canada/QC-JUS-V5291481/2020 | 2020-03-28 | North America/Canada/Quebec/ | Laboratoire de sant√© publique du Qu√©bec | Laboratoire de sant√© publique du Qu√©bec | Sandrine Moreira et al |
| EPI_ISL_890803 | Canada/QC-JUS-V5291548/2020 | 2020-03-28 | North America/Canada/Quebec/ | Laboratoire de sant√© publique du Qu√©bec | Laboratoire de sant√© publique du Qu√©bec | Sandrine Moreira et al |
| EPI_ISL_890804 | Canada/QC-JUS-V5300545/2020 | 2020-03-28 | North America/Canada/Quebec/ | Laboratoire de sant√© publique du Qu√©bec | Laboratoire de sant√© publique du Qu√©bec | Sandrine Moreira et al |
| EPI_ISL_890805 | Canada/QC-JUS-V5300784/2020 | 2020-03-29 | North America/Canada/Quebec/ | Laboratoire de sant√© publique du Qu√©bec | Laboratoire de sant√© publique du Qu√©bec | Sandrine Moreira et al |
| EPI_ISL_890806 | Canada/QC-JUS-V5301017/2020 | 2020-03-28 | North America/Canada/Quebec/ | Laboratoire de sant√© publique du Qu√©bec | Laboratoire de sant√© publique du Qu√©bec | Sandrine Moreira et al |
| EPI_ISL_890807 | Canada/QC-JUS-V5301494/2020 | 2020-03-27 | North America/Canada/Quebec/ | Laboratoire de sant√© publique du Qu√©bec | Laboratoire de sant√© publique du Qu√©bec | Sandrine Moreira et al |
| EPI_ISL_890808 | Canada/QC-JUS-V5301535/2020 | 2020-03-27 | North America/Canada/Quebec/ | Laboratoire de sant√© publique du Qu√©bec | Laboratoire de sant√© publique du Qu√©bec | Sandrine Moreira et al |
| EPI_ISL_890809 | Canada/QC-JUS-V5301543/2020 | 2020-03-27 | North America/Canada/Quebec/ | Laboratoire de sant√© publique du Qu√©bec | Laboratoire de sant√© publique du Qu√©bec | Sandrine Moreira et al |
| EPI_ISL_890810 | Canada/QC-JUS-V5301580/2020 | 2020-03-27 | North America/Canada/Quebec/ | Laboratoire de sant√© publique du Qu√©bec | Laboratoire de sant√© publique du Qu√©bec | Sandrine Moreira et al |
| EPI_ISL_890811 | Canada/QC-JUS-V5301624/2020 | 2020-03-27 | North America/Canada/Quebec/ | Laboratoire de sant√© publique du Qu√©bec | Laboratoire de sant√© publique du Qu√©bec | Sandrine Moreira et al |

| | | | | | | |
|---|---|---|---|---|---|---|
| EPI_ISL_890812 | Canada/QC-JUS-V5301124/2020 | 2020-03-27 | North America/Canada/Quebec/ | Laboratoire de sant√© publique du Qu√©bec | Laboratoire de sant√© publique du Qu√©bec | Sandrine Moreira et al |
| EPI_ISL_890813 | Canada/QC-JUS-V5301142/2020 | 2020-03-27 | North America/Canada/Quebec/ | Laboratoire de sant√© publique du Qu√©bec | Laboratoire de sant√© publique du Qu√©bec | Sandrine Moreira et al |
| EPI_ISL_890814 | Canada/QC-JUS-V5300999/2020 | 2020-03-27 | North America/Canada/Quebec/ | Laboratoire de sant√© publique du Qu√©bec | Laboratoire de sant√© publique du Qu√©bec | Sandrine Moreira et al |
| EPI_ISL_890815 | Canada/QC-JUS-V5301018/2020 | 2020-03-27 | North America/Canada/Quebec/ | Laboratoire de sant√© publique du Qu√©bec | Laboratoire de sant√© publique du Qu√©bec | Sandrine Moreira et al |
| EPI_ISL_890816 | Canada/QC-JUS-V5280093/2020 | 2020-03-25 | North America/Canada/Quebec/ | Laboratoire de sant√© publique du Qu√©bec | Laboratoire de sant√© publique du Qu√©bec | Sandrine Moreira et al |
| EPI_ISL_890817 | Canada/QC-JUS-V5280179/2020 | 2020-03-26 | North America/Canada/Quebec/ | Laboratoire de sant√© publique du Qu√©bec | Laboratoire de sant√© publique du Qu√©bec | Sandrine Moreira et al |
| EPI_ISL_890818 | Canada/QC-JUS-V5280183/2020 | 2020-03-26 | North America/Canada/Quebec/ | Laboratoire de sant√© publique du Qu√©bec | Laboratoire de sant√© publique du Qu√©bec | Sandrine Moreira et al |
| EPI_ISL_890819 | Canada/QC-JUS-V5280151/2020 | 2020-03-25 | North America/Canada/Quebec/ | Laboratoire de sant√© publique du Qu√©bec | Laboratoire de sant√© publique du Qu√©bec | Sandrine Moreira et al |
| EPI_ISL_890820 | Canada/QC-JUS-V5281605/2020 | 2020-03-26 | North America/Canada/Quebec/ | Laboratoire de sant√© publique du Qu√©bec | Laboratoire de sant√© publique du Qu√©bec | Sandrine Moreira et al |
| EPI_ISL_890821 | Canada/QC-JUS-V5280072/2020 | 2020-03-26 | North America/Canada/Quebec/ | Laboratoire de sant√© publique du Qu√©bec | Laboratoire de sant√© publique du Qu√©bec | Sandrine Moreira et al |
| EPI_ISL_890822 | Canada/QC-JUS-V5281550/2020 | 2020-03-26 | North America/Canada/Quebec/ | Laboratoire de sant√© publique du Qu√©bec | Laboratoire de sant√© publique du Qu√©bec | Sandrine Moreira et al |
| EPI_ISL_890823 | Canada/QC-JUS-V5280170/2020 | 2020-03-26 | North America/Canada/Quebec/ | Laboratoire de sant√© publique du Qu√©bec | Laboratoire de sant√© publique du Qu√©bec | Sandrine Moreira et al |
| EPI_ISL_890824 | Canada/QC-JUS-V5290896/2020 | 2020-03-27 | North America/Canada/Quebec/ | Laboratoire de sant√© publique du Qu√©bec | Laboratoire de sant√© publique du Qu√©bec | Sandrine Moreira et al |

| | | | | | | |
|---|---|---|---|---|---|---|
| EPI_ISL_890825 | Canada/QC-JUS-V5290948/2020 | 2020-03-27 | North America/Canada/Quebec/ | Laboratoire de santv© publique du Quv©bec | Laboratoire de santv© publique du Quv©bec | Sandrine Moreira et al |
| EPI_ISL_890826 | Canada/QC-JUS-V5290505/2020 | 2020-03-27 | North America/Canada/Quebec/ | Laboratoire de santv© publique du Quv©bec | Laboratoire de santv© publique du Quv©bec | Sandrine Moreira et al |
| EPI_ISL_890827 | Canada/QC-JUS-V5290536/2020 | 2020-03-27 | North America/Canada/Quebec/ | Laboratoire de santv© publique du Quv©bec | Laboratoire de santv© publique du Quv©bec | Sandrine Moreira et al |
| EPI_ISL_890828 | Canada/QC-JUS-V5281001/2020 | 2020-03-27 | North America/Canada/Quebec/ | Laboratoire de santv© publique du Quv©bec | Laboratoire de santv© publique du Quv©bec | Sandrine Moreira et al |
| EPI_ISL_890829 | Canada/QC-JUS-V5281030/2020 | 2020-03-27 | North America/Canada/Quebec/ | Laboratoire de santv© publique du Quv©bec | Laboratoire de santv© publique du Quv©bec | Sandrine Moreira et al |
| EPI_ISL_890830 | Canada/QC-JUS-V5290502/2020 | 2020-03-27 | North America/Canada/Quebec/ | Laboratoire de santv© publique du Quv©bec | Laboratoire de santv© publique du Quv©bec | Sandrine Moreira et al |
| EPI_ISL_890831 | Canada/QC-JUS-V5281157/2020 | 2020-03-27 | North America/Canada/Quebec/ | Laboratoire de santv© publique du Quv©bec | Laboratoire de santv© publique du Quv©bec | Sandrine Moreira et al |
| EPI_ISL_890832 | Canada/QC-JUS-V5291089/2020 | 2020-03-29 | North America/Canada/Quebec/ | Laboratoire de santv© publique du Quv©bec | Laboratoire de santv© publique du Quv©bec | Sandrine Moreira et al |
| EPI_ISL_890833 | Canada/QC-JUS-V5291117/2020 | 2020-03-29 | North America/Canada/Quebec/ | Laboratoire de santv© publique du Quv©bec | Laboratoire de santv© publique du Quv©bec | Sandrine Moreira et al |
| EPI_ISL_890834 | Canada/QC-JUS-V5291225/2020 | 2020-03-29 | North America/Canada/Quebec/ | Laboratoire de santv© publique du Quv©bec | Laboratoire de santv© publique du Quv©bec | Sandrine Moreira et al |
| EPI_ISL_890835 | Canada/QC-JUS-V5291151/2020 | 2020-03-29 | North America/Canada/Quebec/ | Laboratoire de santv© publique du Quv©bec | Laboratoire de santv© publique du Quv©bec | Sandrine Moreira et al |
| EPI_ISL_890836 | Canada/QC-JUS-V5291226/2020 | 2020-03-28 | North America/Canada/Quebec/ | Laboratoire de santv© publique du Quv©bec | Laboratoire de santv© publique du Quv©bec | Sandrine Moreira et al |
| EPI_ISL_890837 | Canada/QC-JUS-V5300863/2020 | 2020-03-29 | North America/Canada/Quebec/ | Laboratoire de santv© publique du Quv©bec | Laboratoire de santv© publique du Quv©bec | Sandrine Moreira et al |

| | | | | | | |
|---|---|---|---|---|---|---|
| EPI_ISL_890838 | Canada/QC-JUS-V5300886/2020 | 2020-03-29 | North America/Canada/Quebec/ | Laboratoire de sant√© publique du Qu√©bec | Laboratoire de sant√© publique du Qu√©bec | Sandrine Moreira et al |
| EPI_ISL_890839 | Canada/QC-JUS-V5300904/2020 | 2020-03-29 | North America/Canada/Quebec/ | Laboratoire de sant√© publique du Qu√©bec | Laboratoire de sant√© publique du Qu√©bec | Sandrine Moreira et al |
| EPI_ISL_890840 | Canada/QC-JUS-V5300899/2020 | 2020-03-29 | North America/Canada/Quebec/ | Laboratoire de sant√© publique du Qu√©bec | Laboratoire de sant√© publique du Qu√©bec | Sandrine Moreira et al |
| EPI_ISL_890841 | Canada/QC-JUS-V5300909/2020 | 2020-03-29 | North America/Canada/Quebec/ | Laboratoire de sant√© publique du Qu√©bec | Laboratoire de sant√© publique du Qu√©bec | Sandrine Moreira et al |
| EPI_ISL_890842 | Canada/QC-JUS-V5300936/2020 | 2020-03-29 | North America/Canada/Quebec/ | Laboratoire de sant√© publique du Qu√©bec | Laboratoire de sant√© publique du Qu√©bec | Sandrine Moreira et al |
| EPI_ISL_890843 | Canada/QC-JUS-V5300943/2020 | 2020-03-29 | North America/Canada/Quebec/ | Laboratoire de sant√© publique du Qu√©bec | Laboratoire de sant√© publique du Qu√©bec | Sandrine Moreira et al |
| EPI_ISL_890844 | Canada/QC-JUS-V5300966/2020 | 2020-03-29 | North America/Canada/Quebec/ | Laboratoire de sant√© publique du Qu√©bec | Laboratoire de sant√© publique du Qu√©bec | Sandrine Moreira et al |
| EPI_ISL_890845 | Canada/QC-JUS-V5290797/2020 | 2020-03-27 | North America/Canada/Quebec/ | Laboratoire de sant√© publique du Qu√©bec | Laboratoire de sant√© publique du Qu√©bec | Sandrine Moreira et al |
| EPI_ISL_890846 | Canada/QC-JUS-V5291349/2020 | 2020-03-27 | North America/Canada/Quebec/ | Laboratoire de sant√© publique du Qu√©bec | Laboratoire de sant√© publique du Qu√©bec | Sandrine Moreira et al |
| EPI_ISL_890847 | Canada/QC-JUS-V5291126/2020 | 2020-03-27 | North America/Canada/Quebec/ | Laboratoire de sant√© publique du Qu√©bec | Laboratoire de sant√© publique du Qu√©bec | Sandrine Moreira et al |
| EPI_ISL_890848 | Canada/QC-JUS-V5290907/2020 | 2020-03-27 | North America/Canada/Quebec/ | Laboratoire de sant√© publique du Qu√©bec | Laboratoire de sant√© publique du Qu√©bec | Sandrine Moreira et al |
| EPI_ISL_890849 | Canada/QC-JUS-V5291311/2020 | 2020-03-26 | North America/Canada/Quebec/ | Laboratoire de sant√© publique du Qu√©bec | Laboratoire de sant√© publique du Qu√©bec | Sandrine Moreira et al |
| EPI_ISL_890850 | Canada/QC-JUS-V5291717/2020 | 2020-03-28 | North America/Canada/Quebec/ | Laboratoire de sant√© publique du Qu√©bec | Laboratoire de sant√© publique du Qu√©bec | Sandrine Moreira et al |

| | | | | | | |
|---|---|---|---|---|---|---|
| EPI_ISL_890851 | Canada/QC-JUS-V5291434/2020 | 2020-04-01 | North America/Canada/Quebec/ | Laboratoire de santv© publique du Quv©bec | Laboratoire de santv© publique du Quv©bec | Sandrine Moreira et al |
| EPI_ISL_890852 | Canada/QC-JUS-V5291235/2020 | 2020-03-26 | North America/Canada/Quebec/ | Laboratoire de santv© publique du Quv©bec | Laboratoire de santv© publique du Quv©bec | Sandrine Moreira et al |
| EPI_ISL_890853 | Canada/QC-JUS-V5291283/2020 | 2020-03-27 | North America/Canada/Quebec/ | Laboratoire de santv© publique du Quv©bec | Laboratoire de santv© publique du Quv©bec | Sandrine Moreira et al |
| EPI_ISL_890854 | Canada/QC-JUS-V5291065/2020 | 2020-03-27 | North America/Canada/Quebec/ | Laboratoire de santv© publique du Quv©bec | Laboratoire de santv© publique du Quv©bec | Sandrine Moreira et al |
| EPI_ISL_890855 | Canada/QC-JUS-V5300411/2020 | 2020-03-28 | North America/Canada/Quebec/ | Laboratoire de santv© publique du Quv©bec | Laboratoire de santv© publique du Quv©bec | Sandrine Moreira et al |
| EPI_ISL_890856 | Canada/QC-JUS-V5300557/2020 | 2020-03-27 | North America/Canada/Quebec/ | Laboratoire de santv© publique du Quv©bec | Laboratoire de santv© publique du Quv©bec | Sandrine Moreira et al |
| EPI_ISL_890857 | Canada/QC-JUS-V5291360/2020 | 2020-03-29 | North America/Canada/Quebec/ | Laboratoire de santv© publique du Quv©bec | Laboratoire de santv© publique du Quv©bec | Sandrine Moreira et al |
| EPI_ISL_890858 | Canada/QC-JUS-V5290802/2020 | 2020-03-27 | North America/Canada/Quebec/ | Laboratoire de santv© publique du Quv©bec | Laboratoire de santv© publique du Quv©bec | Sandrine Moreira et al |
| EPI_ISL_890859 | Canada/QC-JUS-V5290838/2020 | 2020-03-27 | North America/Canada/Quebec/ | Laboratoire de santv© publique du Quv©bec | Laboratoire de santv© publique du Quv©bec | Sandrine Moreira et al |
| EPI_ISL_890860 | Canada/QC-JUS-V5291122/2020 | 2020-03-28 | North America/Canada/Quebec/ | Laboratoire de santv© publique du Quv©bec | Laboratoire de santv© publique du Quv©bec | Sandrine Moreira et al |
| EPI_ISL_890861 | Canada/QC-JUS-V5290845/2020 | 2020-03-27 | North America/Canada/Quebec/ | Laboratoire de santv© publique du Quv©bec | Laboratoire de santv© publique du Quv©bec | Sandrine Moreira et al |
| EPI_ISL_890862 | Canada/QC-JUS-V5290871/2020 | 2020-03-27 | North America/Canada/Quebec/ | Laboratoire de santv© publique du Quv©bec | Laboratoire de santv© publique du Quv©bec | Sandrine Moreira et al |
| EPI_ISL_890863 | Canada/QC-JUS-V5291333/2020 | 2020-03-27 | North America/Canada/Quebec/ | Laboratoire de santv© publique du Quv©bec | Laboratoire de santv© publique du Quv©bec | Sandrine Moreira et al |

| | | | | | | |
|---|---|---|---|---|---|---|
| EPI_ISL_890864 | Canada/QC-JUS-V5300476/2020 | 2020-03-28 | North America/Canada/Quebec/ | Laboratoire de sant√© publique du Qu√©bec | Laboratoire de sant√© publique du Qu√©bec | Sandrine Moreira et al |
| EPI_ISL_890865 | Canada/QC-JUS-V5290937/2020 | 2020-03-27 | North America/Canada/Quebec/ | Laboratoire de sant√© publique du Qu√©bec | Laboratoire de sant√© publique du Qu√©bec | Sandrine Moreira et al |
| EPI_ISL_890866 | Canada/QC-JUS-V5290950/2020 | 2020-03-27 | North America/Canada/Quebec/ | Laboratoire de sant√© publique du Qu√©bec | Laboratoire de sant√© publique du Qu√©bec | Sandrine Moreira et al |
| EPI_ISL_890867 | Canada/QC-JUS-V5290920/2020 | 2020-03-27 | North America/Canada/Quebec/ | Laboratoire de sant√© publique du Qu√©bec | Laboratoire de sant√© publique du Qu√©bec | Sandrine Moreira et al |
| EPI_ISL_890868 | Canada/QC-JUS-V5290932/2020 | 2020-03-27 | North America/Canada/Quebec/ | Laboratoire de sant√© publique du Qu√©bec | Laboratoire de sant√© publique du Qu√©bec | Sandrine Moreira et al |
| EPI_ISL_890869 | Canada/QC-JUS-V5291268/2020 | 2020-03-28 | North America/Canada/Quebec/ | Laboratoire de sant√© publique du Qu√©bec | Laboratoire de sant√© publique du Qu√©bec | Sandrine Moreira et al |
| EPI_ISL_890870 | Canada/QC-JUS-V5290936/2020 | 2020-03-27 | North America/Canada/Quebec/ | Laboratoire de sant√© publique du Qu√©bec | Laboratoire de sant√© publique du Qu√©bec | Sandrine Moreira et al |
| EPI_ISL_890871 | Canada/QC-JUS-V5291018/2020 | 2020-03-27 | North America/Canada/Quebec/ | Laboratoire de sant√© publique du Qu√©bec | Laboratoire de sant√© publique du Qu√©bec | Sandrine Moreira et al |
| EPI_ISL_890872 | Canada/QC-JUS-V5291103/2020 | 2020-03-27 | North America/Canada/Quebec/ | Laboratoire de sant√© publique du Qu√©bec | Laboratoire de sant√© publique du Qu√©bec | Sandrine Moreira et al |
| EPI_ISL_890873 | Canada/QC-JUS-V5291386/2020 | 2020-03-27 | North America/Canada/Quebec/ | Laboratoire de sant√© publique du Qu√©bec | Laboratoire de sant√© publique du Qu√©bec | Sandrine Moreira et al |
| EPI_ISL_890874 | Canada/QC-JUS-V5291394/2020 | 2020-03-27 | North America/Canada/Quebec/ | Laboratoire de sant√© publique du Qu√©bec | Laboratoire de sant√© publique du Qu√©bec | Sandrine Moreira et al |
| EPI_ISL_890875 | Canada/QC-JUS-V5300610/2020 | 2020-03-29 | North America/Canada/Quebec/ | Laboratoire de sant√© publique du Qu√©bec | Laboratoire de sant√© publique du Qu√©bec | Sandrine Moreira et al |
| EPI_ISL_890876 | Canada/QC-JUS-V5300621/2020 | 2020-03-29 | North America/Canada/Quebec/ | Laboratoire de sant√© publique du Qu√©bec | Laboratoire de sant√© publique du Qu√©bec | Sandrine Moreira et al |

| | | | | | | |
|---|---|---|---|---|---|---|
| EPI_ISL_890877 | Canada/QC-JUS-V5291627/2020 | 2020-03-29 | North America/Canada/Quebec/ | Laboratoire de sant√© publique du Qu√©bec | Laboratoire de sant√© publique du Qu√©bec | Sandrine Moreira et al |
| EPI_ISL_890878 | Canada/QC-JUS-V5290723/2020 | 2020-03-27 | North America/Canada/Quebec/ | Laboratoire de sant√© publique du Qu√©bec | Laboratoire de sant√© publique du Qu√©bec | Sandrine Moreira et al |
| EPI_ISL_890879 | Canada/QC-JUS-V5291679/2020 | 2020-03-28 | North America/Canada/Quebec/ | Laboratoire de sant√© publique du Qu√©bec | Laboratoire de sant√© publique du Qu√©bec | Sandrine Moreira et al |
| EPI_ISL_890880 | Canada/QC-JUS-V5291431/2020 | 2020-03-26 | North America/Canada/Quebec/ | Laboratoire de sant√© publique du Qu√©bec | Laboratoire de sant√© publique du Qu√©bec | Sandrine Moreira et al |
| EPI_ISL_890881 | Canada/QC-JUS-V5312015/2020 | 2020-03-29 | North America/Canada/Quebec/ | Laboratoire de sant√© publique du Qu√©bec | Laboratoire de sant√© publique du Qu√©bec | Sandrine Moreira et al |
| EPI_ISL_890882 | Canada/QC-JUS-V5311434/2020 | 2020-03-29 | North America/Canada/Quebec/ | Laboratoire de sant√© publique du Qu√©bec | Laboratoire de sant√© publique du Qu√©bec | Sandrine Moreira et al |
| EPI_ISL_890883 | Canada/QC-JUS-V5311423/2020 | 2020-03-29 | North America/Canada/Quebec/ | Laboratoire de sant√© publique du Qu√©bec | Laboratoire de sant√© publique du Qu√©bec | Sandrine Moreira et al |
| EPI_ISL_890884 | Canada/QC-JUS-V5311406/2020 | 2020-03-29 | North America/Canada/Quebec/ | Laboratoire de sant√© publique du Qu√©bec | Laboratoire de sant√© publique du Qu√©bec | Sandrine Moreira et al |
| EPI_ISL_890885 | Canada/QC-JUS-V5311185/2020 | 2020-03-29 | North America/Canada/Quebec/ | Laboratoire de sant√© publique du Qu√©bec | Laboratoire de sant√© publique du Qu√©bec | Sandrine Moreira et al |
| EPI_ISL_890887 | Canada/QC-JUS-V5300512/2020 | 2020-03-28 | North America/Canada/Quebec/ | Laboratoire de sant√© publique du Qu√©bec | Laboratoire de sant√© publique du Qu√©bec | Sandrine Moreira et al |
| EPI_ISL_890888 | Canada/QC-JUS-V5302120/2020 | 2020-03-28 | North America/Canada/Quebec/ | Laboratoire de sant√© publique du Qu√©bec | Laboratoire de sant√© publique du Qu√©bec | Sandrine Moreira et al |
| EPI_ISL_890890 | France/GE-IPP11337/2020 | 2020-12-23 | Europe/France/Grand Est/Troyes | Hospital | National Reference Center for Viruses of Respiratory Infections, Institut Pasteur, Paris | Marion Barbet et al |

| | | | | | | |
|---|---|---|---|---|---|---|
| EPI_ISL_890891 | France/GE-IPP11338/2020 | 2020-12-23 | Europe/France/Grand Est/Troyes | Hospital | National Reference Center for Viruses of Respiratory Infections, Institut Pasteur, Paris | Marion Barbet et al |
| EPI_ISL_890892 | France/Bre-IPP11138/2020 | 2020-10-23 | Europe/France/Bretagne/Brest | Hospital | National Reference Center for Viruses of Respiratory Infections, Institut Pasteur, Paris | Marion Barbet et al |
| EPI_ISL_890893 | France/GE-IPP11374/2020 | 2020-12-26 | Europe/France/Grand Est/Troyes | Hospital | National Reference Center for Viruses of Respiratory Infections, Institut Pasteur, Paris | Marion Barbet et al |
| EPI_ISL_890896 | USA/WA-S3470/2020 | 2020-11-28 | North America/USA/Washington/King C | Seattle Flu Study | Seattle Flu Study | Deborah A. Nickerson et al |
| EPI_ISL_890905 | USA/WA-S3479/2020 | 2020-11-29 | North America/USA/Washington/King C | Seattle Flu Study | Seattle Flu Study | Deborah A. Nickerson et al |
| EPI_ISL_890914 | USA/WA-S3488/2020 | 2020-12-01 | North America/USA/Washington/King C | Seattle Flu Study | Seattle Flu Study | Deborah A. Nickerson et al |
| EPI_ISL_890918 | USA/WA-S3492/2020 | 2020-12-01 | North America/USA/Washington/King C | Seattle Flu Study | Seattle Flu Study | Deborah A. Nickerson et al |
| EPI_ISL_890919 | USA/WA-S3493/2020 | 2020-12-02 | North America/USA/Washington/King C | Seattle Flu Study | Seattle Flu Study | Deborah A. Nickerson et al |
| EPI_ISL_890925 | USA/WA-S3499/2020 | 2020-11-28 | North America/USA/Washington/King C | Seattle Flu Study | Seattle Flu Study | Deborah A. Nickerson et al |
| EPI_ISL_890926 | USA/WA-S3500/2020 | 2020-11-30 | North America/USA/Washington/King C | Seattle Flu Study | Seattle Flu Study | Deborah A. Nickerson et al |
| EPI_ISL_890930 | USA/WA-S3504/2020 | 2020-12-03 | North America/USA/Washington/King C | Seattle Flu Study | Seattle Flu Study | Deborah A. Nickerson et al |
| EPI_ISL_890937 | USA/WA-S3511/2020 | 2020-12-04 | North America/USA/Washington/King C | Seattle Flu Study | Seattle Flu Study | Deborah A. Nickerson et al |
| EPI_ISL_890938 | USA/WA-S3512/2020 | 2020-12-05 | North America/USA/Washington/King C | Seattle Flu Study | Seattle Flu Study | Deborah A. Nickerson et al |
| EPI_ISL_890968 | USA/WA-S3542/2021 | 2021-01-10 | North America/USA/Washington/King C | Seattle Flu Study | Seattle Flu Study | Deborah A. Nickerson et al |
| EPI_ISL_890971 | USA/WA-S3545/2021 | 2021-01-11 | North America/USA/Washington/King C | Seattle Flu Study | Seattle Flu Study | Deborah A. Nickerson et al |
| EPI_ISL_890982 | USA/WA-S3556/2021 | 2021-01-11 | North America/USA/Washington/King C | Seattle Flu Study | Seattle Flu Study | Deborah A. Nickerson et al |
| EPI_ISL_890983 | USA/WA-S3557/2021 | 2021-01-11 | North America/USA/Washington/King C | Seattle Flu Study | Seattle Flu Study | Deborah A. Nickerson et al |
| EPI_ISL_890988 | USA/WA-S3562/2021 | 2021-01-12 | North America/USA/Washington/King C | Seattle Flu Study | Seattle Flu Study | Deborah A. Nickerson et al |
| EPI_ISL_890989 | USA/WA-S3563/2021 | 2021-01-12 | North America/USA/Washington/King C | Seattle Flu Study | Seattle Flu Study | Deborah A. Nickerson et al |
| EPI_ISL_891007 | USA/WA-S3581/2020 | 2020-12-28 | North America/USA/Washington/King C | Washington State Department of Health | Seattle Flu Study | Deborah A. Nickerson et al |

| | | | | | | |
|---|---|---|---|---|---|---|
| EPI_ISL_891008 | USA/WA-S3582/2020 | 2020-12-27 | North America/USA/Washington/King C | Washington State Department of Health | Seattle Flu Study | Deborah A. Nickerson et al |
| EPI_ISL_891016 | USA/WA-S3590/2020 | 2020-12-26 | North America/USA/Washington/Yakim | Washington State Department of Health | Seattle Flu Study | Deborah A. Nickerson et al |
| EPI_ISL_891030 | USA/WA-S3604/2020 | 2020-12-28 | North America/USA/Washington/Yakim | Washington State Department of Health | Seattle Flu Study | Deborah A. Nickerson et al |
| EPI_ISL_891033 | USA/WA-S3607/2020 | 2020-12-28 | North America/USA/Washington/Thurst | Washington State Department of Health | Seattle Flu Study | Deborah A. Nickerson et al |
| EPI_ISL_891036 | USA/WA-S3610/2020 | 2020-12-28 | North America/USA/Washington/Yakim | Washington State Department of Health | Seattle Flu Study | Deborah A. Nickerson et al |
| EPI_ISL_891037 | USA/WA-S3611/2020 | 2020-12-28 | North America/USA/Washington/Yakim | Washington State Department of Health | Seattle Flu Study | Deborah A. Nickerson et al |
| EPI_ISL_891040 | USA/WA-S3614/2020 | 2020-12-28 | North America/USA/Washington/Yakim | Washington State Department of Health | Seattle Flu Study | Deborah A. Nickerson et al |
| EPI_ISL_891044 | USA/WA-S3618/2020 | 2020-12-29 | North America/USA/Washington/King C | Washington State Department of Health | Seattle Flu Study | Deborah A. Nickerson et al |
| EPI_ISL_891046 | USA/WA-S3620/2020 | 2020-12-29 | North America/USA/Washington/King C | Washington State Department of Health | Seattle Flu Study | Deborah A. Nickerson et al |
| EPI_ISL_891050 | USA/WA-S3624/2020 | 2020-12-30 | North America/USA/Washington/Grays | Washington State Department of Health | Seattle Flu Study | Deborah A. Nickerson et al |
| EPI_ISL_891054 | USA/WA-S3628/2020 | 2020-12-30 | North America/USA/Washington/Yakim | Washington State Department of Health | Seattle Flu Study | Deborah A. Nickerson et al |
| EPI_ISL_891061 | USA/WA-S3635/2021 | 2021-01-14 | North America/USA/Washington/King C | Seattle Flu Study | Seattle Flu Study | Deborah A. Nickerson et al |
| EPI_ISL_891066 | USA/WA-S3640/2021 | 2021-01-13 | North America/USA/Washington/ | Seattle Flu Study | Seattle Flu Study | Deborah A. Nickerson et al |
| EPI_ISL_891067 | USA/WA-S3641/2021 | 2021-01-14 | North America/USA/Washington/King C | Seattle Flu Study | Seattle Flu Study | Deborah A. Nickerson et al |
| EPI_ISL_891075 | USA/WA-S3649/2021 | 2021-01-16 | North America/USA/Washington/King C | Seattle Flu Study | Seattle Flu Study | Deborah A. Nickerson et al |
| EPI_ISL_891077 | USA/WA-S3651/2021 | 2021-01-15 | North America/USA/Washington/King C | Seattle Flu Study | Seattle Flu Study | Deborah A. Nickerson et al |
| EPI_ISL_891089 | USA/WA-S3663/2021 | 2021-01-09 | North America/USA/Washington/King C | Seattle Flu Study | Seattle Flu Study | Deborah A. Nickerson et al |
| EPI_ISL_891091 | USA/WA-S3665/2021 | 2021-01-09 | North America/USA/Washington/King C | Seattle Flu Study | Seattle Flu Study | Deborah A. Nickerson et al |
| EPI_ISL_891108 | USA/WA-S3682/2021 | 2021-01-09 | North America/USA/Washington/King C | Altius Institute for Biomedical Sciences | Seattle Flu Study | Deborah A. Nickerson et al |
| EPI_ISL_891114 | USA/WA-S3688/2021 | 2021-01-09 | North America/USA/Washington/King C | Altius Institute for Biomedical Sciences | Seattle Flu Study | Deborah A. Nickerson et al |
| EPI_ISL_891133 | USA/WA-S3707/2021 | 2021-01-09 | North America/USA/Washington/King C | Altius Institute for Biomedical Sciences | Seattle Flu Study | Deborah A. Nickerson et al |
| EPI_ISL_891137 | USA/CT-JAX-J000013/2021 | 2021-01-02 | North America/USA/Connecticut/ | The Jackson Laboratory | The Jackson Laboratory | Lloyd M et al |
| EPI_ISL_891144 | USA/IN-PU-100821/2021 | 2021-01-12 | North America/USA/Indiana/ | Indiana Animal Disease Diagnostic Laboratory | Carpi Laboratory - Purdue University | Jack Dorman et al |
| EPI_ISL_891146 | USA/IL-UW-2868/2021 | 2021-01-13 | North America/USA/Illinois/Winnebago | University of Wisconsin-Madison AIDS Vaccine Research Laboratories | University of Wisconsin-Madison AIDS Vaccine Research Laboratories | Gage Moreno et al |

| | | | | | | |
|---|---|---|---|---|---|---|
| EPI_ISL_891151 | Indonesia/YO-UGM-209459/2020 | 2020-12-23 | Asia/Indonesia/Yogyakarta/ | Academic Hospital of Gadjah Mada University (RSA UGM) | Genetics Working Group (Pokja Genetik) Faculty of Medicine, Public Health and Nursing Universitas Gadjah Mada (FK-KMK UGM); Disease Investigation Center Wates Ministry of Agriculture Indonesia; Department of Microbiology FK-KMK UGM; Laboratorium Diagnostik Yayasan Tahija World Mosquito Program (WMP) Yogyakarta Center for Tropical Medicine FK-KMK UGM; Integrated Research center FK-KMK UGM; Department of Computer Science and Electronics FMIPA UGM | Gunadi et al |
| EPI_ISL_891152 | India/RJ-SMS-SQ-1/2020 | 2020-12-26 | Asia/India/Rajasthan/Jaipur | SMS Medical College Jaipur | SMS Medical College Jaipur | Bharti Malhotra et al |
| EPI_ISL_891207 | India/RJ-SMS-SQ-2/2020 | 2020-12-26 | Asia/India/Rajasthan/Jaipur | SMS Medical College Jaipur | SMS Medical College jaipur | Bharti Malhotra et al |
| EPI_ISL_891210 | India/RJ-SMS-SQ-3/2020 | 2020-12-26 | Asia/India/Rajasthan/Jaipur | SMS Medical College jaipur | SMS Medical College Jaipur | Bharti Malhotra et al |
| EPI_ISL_891211 | India/RJ-SMS-SQ-4/2020 | 2020-12-28 | Asia/India/Rajasthan/Jaipur | SMS Medical College Jaipur | SMS Medical College jaipur | Bharti Malhotra et al |

| | | | | | | |
|---|---|---|---|---|---|---|
| EPI_ISL_891213 | India/RJ-SMS-SQ-5/2020 | 2020-12-28 | Asia/India/Rajasthan/Jaipur | SMS Medical College Jaipur | SMS Medical College Jaipur | Bharti Malhotra et al |
| EPI_ISL_891214 | India/RJ-SMS-SQ-6/2020 | 2020-12-28 | Asia/India/Rajasthan/Jaipur | SMS Medical College Jaipur | SMS Medical College Jaipur | Bharti Malhotra et al |
| EPI_ISL_891218 | CzechRepublic/IAB20_016_096/2020 | 2020-10-02 | Europe/Czech Republic/Olomouc Region/ | Olomouc University Hospital | Institute of Applied Biotechnologies a.s. | Petr Klempt et al |
| EPI_ISL_891219 | India/RJ-SMS-SQ-9/2020 | 2020-12-28 | Asia/India/Rajasthan/Jaipur | SMS Medical College Jaipur | SMS Medical College Jaipur | Bharti Malhotra et al |
| EPI_ISL_891221 | Romania/RO_DJ42522/2020 | 2020-11-20 | Europe/Romania/Suceava/ | University of Medicine and Pharmacy of Craiova | """Stefan cel Mare"" University Metagenomics Lab" | Lobiuc Andrei et al |
| EPI_ISL_891223 | Romania/RO_IOCN15/2020 | 2020-07-17 | Europe/Romania/Suceava/ | "The Oncology Institute ""Prof. Dr. Ion Chiricuta"" Cluj Napoca" | """Stefan cel Mare"" University Metagenomics Lab" | Lobiuc Andrei et al |
| EPI_ISL_891224 | Romania/RO_IOCN50/2020 | 2020-07-24 | Europe/Romania/Suceava/ | "The Oncology Institute ""Prof. Dr. Ion Chiricuta"" Cluj Napoca" | """Stefan cel Mare"" University Metagenomics Lab" | Lobiuc Andrei et al |
| EPI_ISL_891225 | Romania/RO_IOCN78/2020 | 2020-07-31 | Europe/Romania/Suceava/ | "The Oncology Institute ""Prof. Dr. Ion Chiricuta"" Cluj Napoca" | """Stefan cel Mare"" University Metagenomics Lab" | Lobiuc Andrei et al |
| EPI_ISL_891226 | Romania/RO_IOCN51/2020 | 2020-07-24 | Europe/Romania/Suceava/ | "The Oncology Institute ""Prof. Dr. Ion Chiricuta"" Cluj Napoca" | """Stefan cel Mare"" University Metagenomics Lab" | Lobiuc Andrei et al |
| EPI_ISL_891227 | Romania/RO_IOCN19/2020 | 2020-07-17 | Europe/Romania/Suceava/ | "The Oncology Institute ""Prof. Dr. Ion Chiricuta"" Cluj Napoca" | """Stefan cel Mare"" University Metagenomics Lab" | Lobiuc Andrei et al |
| EPI_ISL_891228 | Romania/RO_DJ48919/2020 | 2020-11-24 | Europe/Romania/Suceava/ | University of Medicine and Pharmacy of Craiova | """Stefan cel Mare"" University Metagenomics Lab" | Lobiuc Andrei et al |

| | | | | | | |
|---|---|---|---|---|---|---|
| EPI_ISL_891229 | Romania/ROSV_28369/2020 | 2020-11-20 | Europe/Romania/Suceava/ | Suceava County Emergency Hospital | """Stefan cel Mare"" University Metagenomics Lab" | Lobiuc Andrei et al |
| EPI_ISL_891230 | Romania/RO_IOCN38/2020 | 2020-07-19 | Europe/Romania/Suceava/ | "The Oncology Institute ""Prof. Dr. Ion Chiricuta"" Cluj Napoca" | """Stefan cel Mare"" University Metagenomics Lab" | Lobiuc Andrei et al |
| EPI_ISL_891254 | France/PAC-IHU-2997/2021 | 2021-01-04 | Europe/France/Provence-Alpes-C√¥te d' | IHU M√©diterran√©e Infection | IHU M√©diterran√©e Infection | Philippe Colson et al. et al |
| EPI_ISL_891267 | Serbia/L3072/2020 | 2020-09-10 | Europe/Serbia/Serbia/ | Institute of Biocides and Medical Ecology, Belgarde, Serbia | Virology department Institute of microbiology and immunology Faculty of Medicine University of Belgrade | Banko Ana et al |
| EPI_ISL_891271 | Serbia/L5281/2020 | 2020-09-28 | Europe/Serbia/Serbia/ | Institute of Biocides and Medical Ecology, Belgarde, Serbia | Virology department Institute of microbiology and immunology Faculty of Medicine University of Belgrade | Banko Ana et al |
| EPI_ISL_891272 | Serbia/C17708/2020 | 2020-08-01 | Europe/Serbia/Serbia/ | Institute of Biocides and Medical Ecology, Belgarde, Serbia | Virology department Institute of microbiology and immunology Faculty of Medicine University of Belgrade | Banko Ana et al |

| | | | | | | |
|---|---|---|---|---|---|---|
| EPI_ISL_891403 | England/ALDP-10B9364/2021 | 2021-01-18 | Europe/United Kingdom/England/ | Lighthouse Lab in Alderley Park | Wellcome Sanger Institute for the COVID-19 Genomics UK (COG-UK) Consortium | Jacquelyn Wynn et al |
| EPI_ISL_891432 | England/CAMC-10A2E7C/2021 | 2021-01-17 | Europe/United Kingdom/England/ | Lighthouse Lab in Cambridge | Wellcome Sanger Institute for the COVID-19 Genomics UK (COG-UK) Consortium | Rob Howes et al |
| EPI_ISL_891639 | England/QEUH-10BA406/2021 | 2021-01-20 | Europe/United Kingdom/England/ | Lighthouse Lab in Glasgow | Wellcome Sanger Institute for the COVID-19 Genomics UK (COG-UK) Consortium | Harper VanSteenhouse et al |
| EPI_ISL_891649 | England/CAMC-10A11B0/2021 | 2021-01-18 | Europe/United Kingdom/England/ | Lighthouse Lab in Cambridge | Wellcome Sanger Institute for the COVID-19 Genomics UK (COG-UK) Consortium | Rob Howes et al |
| EPI_ISL_891778 | England/CAMC-10A153C/2021 | 2021-01-18 | Europe/United Kingdom/England/ | Lighthouse Lab in Cambridge | Wellcome Sanger Institute for the COVID-19 Genomics UK (COG-UK) Consortium | Rob Howes et al |
| EPI_ISL_891878 | England/ALDP-10B7876/2021 | 2021-01-19 | Europe/United Kingdom/England/ | Lighthouse Lab in Alderley Park | Wellcome Sanger Institute for the COVID-19 Genomics UK (COG-UK) Consortium | Jacquelyn Wynn et al |
| EPI_ISL_892027 | Wales/ALDP-101B0F0/2021 | 2021-01-13 | Europe/United Kingdom/Wales/ | Lighthouse Lab in Alderley Park | Wellcome Sanger Institute for the COVID-19 Genomics UK (COG-UK) Consortium | Jacquelyn Wynn et al |
| EPI_ISL_892244 | Slovenia/MB_6/2020 | 2020-12-29 | Europe/Slovenia/Slovenia/ | National Laboratory for Health, Environment and Food | National Laboratory for Health, Environment and Food | Aleksander Mahnic et al |

| | | | | | | |
|---|---|---|---|---|---|---|
| EPI_ISL_892252 | USA/CA-LACPHL-AF00245/2021 | 2021-01-11 | North America/USA/California/Los Ange | MD Laboratories | Los Angeles County PHL | P. Hemarajata et al. et al |
| EPI_ISL_892253 | USA/CA-LACPHL-AF00238/2021 | 2021-01-11 | North America/USA/California/Los Ange | MD Laboratories | Los Angeles County PHL | P. Hemarajata et al. et al |
| EPI_ISL_892257 | USA/CA-LACPHL-AF00235/2021 | 2021-01-11 | North America/USA/California/Los Ange | MD Laboratories | Los Angeles County PHL | P. Hemarajata et al. et al |
| EPI_ISL_892263 | USA/CA-LACPHL-AF00247/2021 | 2021-01-08 | North America/USA/California/Los Ange | MD Laboratories | Los Angeles County PHL | P. Hemarajata et al. et al |
| EPI_ISL_892269 | USA/OH-ODH-SC174799/2020 | 2020-12-04 | North America/USA/Ohio/ | Ohio Department of Health Laboratory | Ohio Department of Health Laboratory | Holmes et al |
| EPI_ISL_892288 | Spain/PV-IBV-97011406/2020 | 2020-12-03 | Europe/Spain/Basque Country/San Seba | Servicio de Microbiolog√≠a. Hospital Universitario Donostia. OSI Donostialdea. √Årea de Enfermedades Infecciosas, Grupo de Infecci√≥n Respiratoria y Resistencia Antimicrobiana. Instituto de Investigaci√≥n Sanitaria Biodonostia | SeqCOVID-SPAIN consortium/IBV(CSIC) | Gustavo Cilla Eguiluz et al |
| EPI_ISL_892291 | Spain/PV-IBV-97011411/2020 | 2020-12-03 | Europe/Spain/Basque Country/San Seba | Servicio de Microbiolog√≠a. Hospital Universitario Donostia. OSI Donostialdea. √Årea de Enfermedades Infecciosas, Grupo de Infecci√≥n Respiratoria y Resistencia Antimicrobiana. Instituto de Investigaci√≥n Sanitaria Biodonostia | SeqCOVID-SPAIN consortium/IBV(CSIC) | Gustavo Cilla Eguiluz et al |
| EPI_ISL_892299 | Spain/PV-IBV-97011426/2020 | 2020-12-05 | Europe/Spain/Basque Country/San Seba | Servicio de Microbiolog√≠a. Hospital Universitario Donostia. OSI Donostialdea. √Årea de Enfermedades Infecciosas, Grupo de Infecci√≥n Respiratoria y Resistencia Antimicrobiana. Instituto de Investigaci√≥n Sanitaria Biodonostia | SeqCOVID-SPAIN consortium/IBV(CSIC) | Gustavo Cilla Eguiluz et al |
| EPI_ISL_892301 | Spain/PV-IBV-97011433/2020 | 2020-12-06 | Europe/Spain/Basque Country/San Seba | Servicio de Microbiolog√≠a. Hospital Universitario Donostia. OSI Donostialdea. √Årea de Enfermedades Infecciosas, Grupo de Infecci√≥n Respiratoria y Resistencia Antimicrobiana. Instituto de Investigaci√≥n Sanitaria Biodonostia | SeqCOVID-SPAIN consortium/IBV(CSIC) | Gustavo Cilla Eguiluz et al |
| EPI_ISL_892309 | Spain/PV-IBV-97011449/2020 | 2020-12-08 | Europe/Spain/Basque Country/San Seba | Servicio de Microbiolog√≠a. Hospital Universitario Donostia. OSI Donostialdea. √Årea de Enfermedades Infecciosas, Grupo de Infecci√≥n Respiratoria y Resistencia Antimicrobiana. Instituto de Investigaci√≥n Sanitaria Biodonostia | SeqCOVID-SPAIN consortium/IBV(CSIC) | Gustavo Cilla Eguiluz et al |

| | | | | | | |
|---|---|---|---|---|---|---|
| EPI_ISL_892310 | Spain/PV-IBV-97011452/2020 | 2020-12-09 | Europe/Spain/Basque Country/San Seba | Servicio de Microbiolog√≠a. Hospital Universitario Donostia. OSI Donostialdea. √Årea de Enfermedades Infecciosas, Grupo de Infecci√≥n Respiratoria y Resistencia Antimicrobiana. Instituto de Investigaci√≥n Sanitaria Biodonostia | SeqCOVID-SPAIN consortium/IBV(CSIC) | Gustavo Cilla Eguiluz et al |
| EPI_ISL_892312 | Spain/PV-IBV-97011454/2020 | 2020-12-09 | Europe/Spain/Basque Country/San Seba | Servicio de Microbiolog√≠a. Hospital Universitario Donostia. OSI Donostialdea. √Årea de Enfermedades Infecciosas, Grupo de Infecci√≥n Respiratoria y Resistencia Antimicrobiana. Instituto de Investigaci√≥n Sanitaria Biodonostia | SeqCOVID-SPAIN consortium/IBV(CSIC) | Gustavo Cilla Eguiluz et al |
| EPI_ISL_892315 | Spain/PV-IBV-97011461/2020 | 2020-12-10 | Europe/Spain/Basque Country/San Seba | Servicio de Microbiolog√≠a. Hospital Universitario Donostia. OSI Donostialdea. √Årea de Enfermedades Infecciosas, Grupo de Infecci√≥n Respiratoria y Resistencia Antimicrobiana. Instituto de Investigaci√≥n Sanitaria Biodonostia | SeqCOVID-SPAIN consortium/IBV(CSIC) | Gustavo Cilla Eguiluz et al |
| EPI_ISL_892319 | Spain/PV-IBV-97011471/2020 | 2020-12-11 | Europe/Spain/Basque Country/San Seba | Servicio de Microbiolog√≠a. Hospital Universitario Donostia. OSI Donostialdea. √Årea de Enfermedades Infecciosas, Grupo de Infecci√≥n Respiratoria y Resistencia Antimicrobiana. Instituto de Investigaci√≥n Sanitaria Biodonostia | SeqCOVID-SPAIN consortium/IBV(CSIC) | Gustavo Cilla Eguiluz et al |
| EPI_ISL_892328 | Spain/NC-IBV-97011855/2020 | 2020-12-01 | Europe/Spain/Navarra/Pamplona | Servicio de Microbiolog√≠a Cl√≠nica (Complejo Hospitalario de Navarra, Pamplona), Instituto de Investigaci√≥n Sanitaria de Navarra (IdiSNA) | SeqCOVID-SPAIN consortium/IBV(CSIC) | Carmen Ezpeleta Baquedano et al |
| EPI_ISL_892345 | Spain/NC-IBV-97011882/2020 | 2020-12-10 | Europe/Spain/Navarra/Pamplona | Servicio de Microbiolog√≠a Cl√≠nica (Complejo Hospitalario de Navarra, Pamplona), Instituto de Investigaci√≥n Sanitaria de Navarra (IdiSNA) | SeqCOVID-SPAIN consortium/IBV(CSIC) | Carmen Ezpeleta Baquedano et al |
| EPI_ISL_892348 | Spain/NC-IBV-97011888/2020 | 2020-12-10 | Europe/Spain/Navarra/Pamplona | Servicio de Microbiolog√≠a Cl√≠nica (Complejo Hospitalario de Navarra, Pamplona), Instituto de Investigaci√≥n Sanitaria de Navarra (IdiSNA) | SeqCOVID-SPAIN consortium/IBV(CSIC) | Carmen Ezpeleta Baquedano et al |
| EPI_ISL_892365 | Slovenia/MB_7/2021 | 2021-01-25 | Europe/Slovenia/Slovenia/ | National Laboratory for Health, Environment and Food | National Laboratory for Health, Environment and Food | Aleksander Mahnic et al |

| | | | | | | |
|---|---|---|---|---|---|---|
| EPI_ISL_892367 | Slovenia/MB_9/2021 | 2021-01-25 | Europe/Slovenia/Slovenia/ | National Laboratory for Health, Environment and Food | National Laboratory for Health, Environment and Food | Aleksander Mahnic et al |
| EPI_ISL_892456 | Japan/PG-14523/2020 | 20-Dec | Asia/Japan/Japan/ | Pathogen Genomics Center, National Institute of Infectious Diseases | Pathogen Genomics Center, National Institute of Infectious Diseases | Tsuyoshi Sekizuka et al |
| EPI_ISL_892457 | Japan/PG-14524/2020 | 20-Dec | Asia/Japan/Japan/ | Pathogen Genomics Center, National Institute of Infectious Diseases | Pathogen Genomics Center, National Institute of Infectious Diseases | Tsuyoshi Sekizuka et al |
| EPI_ISL_892508 | Japan/PG-14575/2020 | 20-Dec | Asia/Japan/Japan/ | Pathogen Genomics Center, National Institute of Infectious Diseases | Pathogen Genomics Center, National Institute of Infectious Diseases | Tsuyoshi Sekizuka et al |
| EPI_ISL_892636 | Japan/PG-14704/2020 | 20-Dec | Asia/Japan/Japan/ | Pathogen Genomics Center, National Institute of Infectious Diseases | Pathogen Genomics Center, National Institute of Infectious Diseases | Tsuyoshi Sekizuka et al |
| EPI_ISL_892689 | Japan/PG-14758/2020 | 20-Dec | Asia/Japan/Japan/ | Pathogen Genomics Center, National Institute of Infectious Diseases | Pathogen Genomics Center, National Institute of Infectious Diseases | Tsuyoshi Sekizuka et al |
| EPI_ISL_893256 | Japan/PG-15372/2020 | 20-Dec | Asia/Japan/Japan/ | Pathogen Genomics Center, National Institute of Infectious Diseases | Pathogen Genomics Center, National Institute of Infectious Diseases | Tsuyoshi Sekizuka et al |
| EPI_ISL_893300 | Japan/PG-15416/2020 | 20-Dec | Asia/Japan/Japan/ | Pathogen Genomics Center, National Institute of Infectious Diseases | Pathogen Genomics Center, National Institute of Infectious Diseases | Tsuyoshi Sekizuka et al |

| | | | | | | |
|---|---|---|---|---|---|---|
| EPI_ISL_893345 | Japan/PG-15461/2020 | 20-Dec | Asia/Japan/Japan/ | Pathogen Genomics Center, National Institute of Infectious Diseases | Pathogen Genomics Center, National Institute of Infectious Diseases | Tsuyoshi Sekizuka et al |
| EPI_ISL_893553 | Japan/PG-13032/2020 | 20-Nov | Asia/Japan/Japan/ | Department of Infectious Diseases, Kobe Institute of Health | Department of Infectious Diseases, Kobe Institute of Health | Ryohei Nomoto et al |
| EPI_ISL_893673 | Japan/PG-14125/2020 | 20-Oct | Asia/Japan/Japan/ | Department of Infectious Diseases, Kobe Institute of Health | Department of Infectious Diseases, Kobe Institute of Health | Ryohei Nomoto et al |
| EPI_ISL_893728 | USA/AZ-CDC-IR9E-6611/2020 | 2020-11-15 | North America/USA/Arizona/ | Pima County Health Department | Pathogen Discovery, Respiratory Viruses Branch, Division of Viral Diseases, Centers for Disease Control and Prevention | Yan Li et al |
| EPI_ISL_893730 | USA/AZ-CDC-IR9G-6615/2020 | 2020-11-15 | North America/USA/Arizona/ | Pima County Health Department | Pathogen Discovery, Respiratory Viruses Branch, Division of Viral Diseases, Centers for Disease Control and Prevention | Yan Li et al |
| EPI_ISL_893733 | USA/AZ-CDC-IR9K-5198/2020 | 2020-11-15 | North America/USA/Arizona/ | Pima County Health Department | Pathogen Discovery, Respiratory Viruses Branch, Division of Viral Diseases, Centers for Disease Control and Prevention | Yan Li et al |

| | | | | | | |
|---|---|---|---|---|---|---|
| EPI_ISL_893734 | USA/AZ-CDC-IR9M-5323/2020 | 2020-11-15 | North America/USA/Arizona/ | Pima County Health Department | Pathogen Discovery, Respiratory Viruses Branch, Division of Viral Diseases, Centers for Disease Control and Prevention | Yan Li et al |
| EPI_ISL_893735 | USA/AZ-CDC-IR9N-5372/2020 | 2020-11-15 | North America/USA/Arizona/ | Pima County Health Department | Pathogen Discovery, Respiratory Viruses Branch, Division of Viral Diseases, Centers for Disease Control and Prevention | Yan Li et al |
| EPI_ISL_893744 | Germany/BW-FR0382/2020 | 2020-12-30 | Europe/Germany/Baden-Wuerttemberg | Institute of Virology, Medical Center, University of Freiburg, Freiburg, Germany | Institute of Virology, Clinial Virus Genomics, Medical Center, University of Freiburg, Freiburg, Germany | Jonas Fuchs et al |
| EPI_ISL_893752 | Germany/BW-FR0388/2021 | 2021-01-07 | Europe/Germany/Baden-Wuerttemberg | Institute of Virology, Medical Center, University of Freiburg, Freiburg, Germany | Institute of Virology, Clinial Virus Genomics, Medical Center, University of Freiburg, Freiburg, Germany | Jonas Fuchs et al |

| | | | | | | |
|---|---|---|---|---|---|---|
| EPI_ISL_893764 | Germany/BW-FR0393/2021 | 2021-01-08 | Europe/Germany/Baden-Wuerttemberg | Institute of Virology, Medical Center, University of Freiburg, Freiburg, Germany | Institute of Virology, Clinial Virus Genomics, Medical Center, University of Freiburg, Freiburg, Germany | Jonas Fuchs et al |
| EPI_ISL_893771 | Germany/BW-FR0398/2020 | 2020-12-27 | Europe/Germany/Baden-Wuerttemberg | Institute of Virology, Medical Center, University of Freiburg, Freiburg, Germany | Institute of Virology, Clinial Virus Genomics, Medical Center, University of Freiburg, Freiburg, Germany | Jonas Fuchs et al |
| EPI_ISL_893773 | Germany/BW-FR0399/2020 | 2020-12-27 | Europe/Germany/Baden-Wuerttemberg | Institute of Virology, Medical Center, University of Freiburg, Freiburg, Germany | Institute of Virology, Clinial Virus Genomics, Medical Center, University of Freiburg, Freiburg, Germany | Jonas Fuchs et al |
| EPI_ISL_893774 | Germany/BW-FR0400/2020 | 2020-12-29 | Europe/Germany/Baden-Wuerttemberg | Institute of Virology, Medical Center, University of Freiburg, Freiburg, Germany | Institute of Virology, Clinial Virus Genomics, Medical Center, University of Freiburg, Freiburg, Germany | Jonas Fuchs et al |
| EPI_ISL_893775 | Germany/BW-FR0402/2021 | 2021-01-03 | Europe/Germany/Baden-Wuerttemberg | Institute of Virology, Medical Center, University of Freiburg, Freiburg, Germany | Institute of Virology, Clinial Virus Genomics, Medical Center, University of Freiburg, Freiburg, Germany | Jonas Fuchs et al |

| | | | | | | |
|---|---|---|---|---|---|---|
| EPI_ISL_893783 | Germany/BW-FR0408/2020 | 2020-12-22 | Europe/Germany/Baden-Wuerttemberg | Institute of Virology, Medical Center, University of Freiburg, Freiburg, Germany | Institute of Virology, Clinial Virus Genomics, Medical Center, University of Freiburg, Freiburg, Germany | Jonas Fuchs et al |
| EPI_ISL_894166 | Germany/ST-MD216/2020 | 2020-12-21 | Europe/Germany/Saxony-Anhalt/ | Institute of Medical Microbiology and Hospital Hygiene | Institute of Medical Microbiology and Hospital Hygiene | Prof. Dr. Achim Kaasch et al |
| EPI_ISL_894167 | Germany/ST-MD225/2020 | 2020-12-25 | Europe/Germany/Saxony-Anhalt/ | Institute of Medical Microbiology and Hospital Hygiene | Institute of Medical Microbiology and Hospital Hygiene | Prof. Dr. Achim Kaasch et al |
| EPI_ISL_894169 | Germany/ST-MD257/2020 | 2020-12-28 | Europe/Germany/Saxony-Anhalt/ | Institute of Medical Microbiology and Hospital Hygiene | Institute of Medical Microbiology and Hospital Hygiene | Prof. Dr. Achim Kaasch et al |
| EPI_ISL_894218 | Italy/CAM-CRGS-30/2020 | 2020-03-18 | Europe/Italy/Campania/ | 1.AO Universitaria 'S. Giovanni di Dio e Ruggi D'Aragona, Scuola Medica Salernitana' Hospital / 2.UOC di Virologia e Microbiologia, Universit√† della Campania 'L. Vanvitelli' / 3.AO Universitaria 'Federico II' Napoli Hospital / 4.AORN 'San Giuseppe Moscati' Avellino Hospital / 5.AO 'San Pio - presidio G. Rummo' Benevento Hospital / 6.AO 'Sant'Anna e San Sebastiano' Caserta Hospital / 7.PO 'Maria Santissima Addolorata' Eboli Hospital / 8.Biogem Istituto di Ricerche Genetiche | 1. Genome Research Center for Health (CRGS) / 2. Laboratory of Molecular Medicine and Genomics(LMMGe) / 3. Center for Research in Pure and Applied Mathematics (CRMPA) | Giorgio Giurato et al |
| EPI_ISL_894246 | Turkey/HSGM-1232/2021 | 2021-01-06 | Europe/Turkey/Turkey/ | Ministry of Health Turkey | Ministry of Health Turkey | Fatma Bayrakdar et al |
| EPI_ISL_894250 | Turkey/HSGM-1240/2021 | 2021-01-11 | Europe/Turkey/Turkey/ | Ministry of Health Turkey | Ministry of Health Turkey | Fatma Bayrakdar et al |
| EPI_ISL_894254 | Turkey/HSGM-1253/2021 | 2021-01-11 | Europe/Turkey/Turkey/ | Ministry of Health Turkey | Ministry of Health Turkey | Fatma Bayrakdar et al |
| EPI_ISL_894269 | Turkey/HSGM-1301/2021 | 2021-01-13 | Europe/Turkey/Turkey/ | Ministry of Health Turkey | Ministry of Health Turkey | Fatma Bayrakdar et al |

| | | | | | | |
|---|---|---|---|---|---|---|
| EPI_ISL_894273 | Turkey/HSGM-1310/2021 | 2021-01-14 | Europe/Turkey/Turkey/ | Ministry of Health Turkey | Ministry of Health Turkey | Fatma Bayrakdar et al |
| EPI_ISL_894274 | Turkey/HSGM-1311/2021 | 2021-01-14 | Europe/Turkey/Turkey/ | Ministry of Health Turkey | Ministry of Health Turkey | Fatma Bayrakdar et al |
| EPI_ISL_894291 | Japan/PG-15527/2021 | 21-Jan | Asia/Japan/Japan/ | Pathogen Genomics Center, National Institute of Infectious Diseases | Pathogen Genomics Center, National Institute of Infectious Diseases | Tsuyoshi Sekizuka et al |
| EPI_ISL_894321 | Japan/PG-15500/2021 | 21-Jan | Asia/Japan/Japan/ | Pathogen Genomics Center, National Institute of Infectious Diseases | Pathogen Genomics Center, National Institute of Infectious Diseases | Tsuyoshi Sekizuka et al |
| EPI_ISL_894322 | Japan/PG-15501/2021 | 21-Jan | Asia/Japan/Japan/ | Pathogen Genomics Center, National Institute of Infectious Diseases | Pathogen Genomics Center, National Institute of Infectious Diseases | Tsuyoshi Sekizuka et al |
| EPI_ISL_894332 | Japan/PG-15512/2021 | 21-Jan | Asia/Japan/Japan/ | Pathogen Genomics Center, National Institute of Infectious Diseases | Pathogen Genomics Center, National Institute of Infectious Diseases | Tsuyoshi Sekizuka et al |
| EPI_ISL_894389 | Japan/PG-15573/2020 | 20-Aug | Asia/Japan/Japan/ | Pathogen Genomics Center, National Institute of Infectious Diseases | Pathogen Genomics Center, National Institute of Infectious Diseases | Tsuyoshi Sekizuka et al |
| EPI_ISL_894521 | Japan/PG-15710/2020 | 20-Oct | Asia/Japan/Japan/ | Pathogen Genomics Center, National Institute of Infectious Diseases | Pathogen Genomics Center, National Institute of Infectious Diseases | Tsuyoshi Sekizuka et al |
| EPI_ISL_894645 | Japan/PG-15838/2020 | 20-Dec | Asia/Japan/Japan/ | Pathogen Genomics Center, National Institute of Infectious Diseases | Pathogen Genomics Center, National Institute of Infectious Diseases | Tsuyoshi Sekizuka et al |

| | | | | | | |
|---|---|---|---|---|---|---|
| EPI_ISL_894785 | Japan/PG-15981/2021 | 21-Jan | Asia/Japan/Japan/ | Fukuoka Institute of Health and Environmental Sciences | Pathogen Genomics Center, National Institute of Infectious Diseases | Tsuyoshi Sekizuka et al |
| EPI_ISL_894864 | Japan/PG-16061/2020 | 20-Dec | Asia/Japan/Japan/ | Pathogen Genomics Center, National Institute of Infectious Diseases | Pathogen Genomics Center, National Institute of Infectious Diseases | Tsuyoshi Sekizuka et al |
| EPI_ISL_894925 | Japan/PG-16129/2020 | 20-Dec | Asia/Japan/Japan/ | Pathogen Genomics Center, National Institute of Infectious Diseases | Pathogen Genomics Center, National Institute of Infectious Diseases | Tsuyoshi Sekizuka et al |
| EPI_ISL_894947 | USA/OH-ODH-SC177554/2020 | 2020-12-09 | North America/USA/Ohio/ | Ohio Department of Health Laboratory | Ohio Department of Health Laboratory | Holmes et al |
| EPI_ISL_895059 | Japan/PG-13395/2020 | 20-Dec | Asia/Japan/Japan/ | Department of Infectious Diseases, Kobe Institute of Health | Pathogen Genomics Center, National Institute of Infectious Diseases | Tsuyoshi Sekizuka et al |
| EPI_ISL_895061 | Japan/PG-13398/2020 | 20-Dec | Asia/Japan/Japan/ | Department of Infectious Diseases, Kobe Institute of Health | Pathogen Genomics Center, National Institute of Infectious Diseases | Tsuyoshi Sekizuka et al |
| EPI_ISL_895275 | Japan/PG-13612/2020 | 20-Dec | Asia/Japan/Japan/ | Pathogen Genomics Center, National Institute of Infectious Diseases | Pathogen Genomics Center, National Institute of Infectious Diseases | Tsuyoshi Sekizuka et al |
| EPI_ISL_895370 | Japan/PG-13790/2020 | 20-Dec | Asia/Japan/Japan/ | Pathogen Genomics Center, National Institute of Infectious Diseases | Pathogen Genomics Center, National Institute of Infectious Diseases | Tsuyoshi Sekizuka et al |

| | | | | | | |
|---|---|---|---|---|---|---|
| EPI_ISL_895469 | Japan/PG-13889/2020 | 20-Dec | Asia/Japan/Japan/ | Pathogen Genomics Center, National Institute of Infectious Diseases | Pathogen Genomics Center, National Institute of Infectious Diseases | Tsuyoshi Sekizuka et al |
| EPI_ISL_895483 | Japan/PG-13903/2020 | 20-Dec | Asia/Japan/Japan/ | Pathogen Genomics Center, National Institute of Infectious Diseases | Pathogen Genomics Center, National Institute of Infectious Diseases | Tsuyoshi Sekizuka et al |
| EPI_ISL_895488 | Japan/PG-13908/2020 | 20-Dec | Asia/Japan/Japan/ | Pathogen Genomics Center, National Institute of Infectious Diseases | Pathogen Genomics Center, National Institute of Infectious Diseases | Tsuyoshi Sekizuka et al |
| EPI_ISL_895567 | Japan/PG-13987/2020 | 20-Dec | Asia/Japan/Japan/ | Pathogen Genomics Center, National Institute of Infectious Diseases | Pathogen Genomics Center, National Institute of Infectious Diseases | Tsuyoshi Sekizuka et al |
| EPI_ISL_895713 | Japan/PG-14201/2020 | 20-Dec | Asia/Japan/Japan/ | Pathogen Genomics Center, National Institute of Infectious Diseases | Pathogen Genomics Center, National Institute of Infectious Diseases | Tsuyoshi Sekizuka et al |
| EPI_ISL_895736 | CzechRepublic/FNHK-103/2020 | 2020-03-28 | Europe/Czech Republic/Czech Republic/ | Molecular biology division, Institute of Clinical Biochemistry and Diagnostics, Charles University, Faculty of Medicine in Hradec Kr√°lov√© and University Hospital Hradec Kr√°lov√© | Molecular biology division, Institute of Clinical Biochemistry and Diagnostics, Charles University, Faculty of Medicine in Hradec Kr√°lov√© and University Hospital Hradec Kr√°lov√© | Helena Kova≈ô√≠kov√° et al |

| | | | | | | |
|---|---|---|---|---|---|---|
| EPI_ISL_895740 | CzechRepublic/FNHK-177/2020 | 2020-04-11 | Europe/Czech Republic/Czech Republic/ | Molecular biology division, Institute of Clinical Biochemistry and Diagnostics, Charles University, Faculty of Medicine in Hradec Kr√°lov√© and University Hospital Hradec Kr√°lov√© | Molecular biology division, Institute of Clinical Biochemistry and Diagnostics, Charles University, Faculty of Medicine in Hradec Kr√°lov√© and University Hospital Hradec Kr√°lov√© | Helena Kova≈ô√≠kov√° et al |
| EPI_ISL_895753 | CzechRepublic/FNHK-18901/2020 | 2020-04-14 | Europe/Czech Republic/Czech Republic/ | Molecular biology division, Institute of Clinical Biochemistry and Diagnostics, Charles University, Faculty of Medicine in Hradec Kr√°lov√© and University Hospital Hradec Kr√°lov√© | Molecular biology division, Institute of Clinical Biochemistry and Diagnostics, Charles University, Faculty of Medicine in Hradec Kr√°lov√© and University Hospital Hradec Kr√°lov√© | Helena Kova≈ô√≠kov√° et al |
| EPI_ISL_895773 | CzechRepublic/FNHK-14201/2020 | 2020-04-03 | Europe/Czech Republic/Czech Republic/ | Molecular biology division, Institute of Clinical Biochemistry and Diagnostics, Charles University, Faculty of Medicine in Hradec Kr√°lov√© and University Hospital Hradec Kr√°lov√© | Molecular biology division, Institute of Clinical Biochemistry and Diagnostics, Charles University, Faculty of Medicine in Hradec Kr√°lov√© and University Hospital Hradec Kr√°lov√© | Helena Kova≈ô√≠kov√° et al |

| | | | | | | |
|---|---|---|---|---|---|---|
| EPI_ISL_895780 | CzechRepublic/FNHK-96/2020 | 2020-03-28 | Europe/Czech Republic/Czech Republic/ | Molecular biology division, Institute of Clinical Biochemistry and Diagnostics, Charles University, Faculty of Medicine in Hradec Kr√°lov√© and University Hospital Hradec Kr√°lov√© | Molecular biology division, Institute of Clinical Biochemistry and Diagnostics, Charles University, Faculty of Medicine in Hradec Kr√°lov√© and University Hospital Hradec Kr√°lov√© | Helena Kova≈ô√≠kov√° et al |
| EPI_ISL_895787 | CzechRepublic/FNHK-18501/2020 | 2020-04-13 | Europe/Czech Republic/Czech Republic/ | Molecular biology division, Institute of Clinical Biochemistry and Diagnostics, Charles University, Faculty of Medicine in Hradec Kr√°lov√© and University Hospital Hradec Kr√°lov√© | Molecular biology division, Institute of Clinical Biochemistry and Diagnostics, Charles University, Faculty of Medicine in Hradec Kr√°lov√© and University Hospital Hradec Kr√°lov√© | Helena Kova≈ô√≠kov√° et al |
| EPI_ISL_895789 | CzechRepublic/FNHK-19203/2020 | 2020-05-04 | Europe/Czech Republic/Czech Republic/ | Molecular biology division, Institute of Clinical Biochemistry and Diagnostics, Charles University, Faculty of Medicine in Hradec Kr√°lov√© and University Hospital Hradec Kr√°lov√© | Molecular biology division, Institute of Clinical Biochemistry and Diagnostics, Charles University, Faculty of Medicine in Hradec Kr√°lov√© and University Hospital Hradec Kr√°lov√© | Helena Kova≈ô√≠kov√° et al |

| | | | | | | |
|---|---|---|---|---|---|---|
| EPI_ISL_895792 | CzechRepublic/FNHK-21801/2020 | 2020-05-18 | Europe/Czech Republic/Czech Republic/ | Molecular biology division, Institute of Clinical Biochemistry and Diagnostics, Charles University, Faculty of Medicine in Hradec Kr√°lov√© and University Hospital Hradec Kr√°lov√© | Molecular biology division, Institute of Clinical Biochemistry and Diagnostics, Charles University, Faculty of Medicine in Hradec Kr√°lov√© and University Hospital Hradec Kr√°lov√© | Helena Kova≈ô√≠kov√° et al |
| EPI_ISL_895793 | CzechRepublic/FNHK-21802/2020 | 2020-06-01 | Europe/Czech Republic/Czech Republic/ | Molecular biology division, Institute of Clinical Biochemistry and Diagnostics, Charles University, Faculty of Medicine in Hradec Kr√°lov√© and University Hospital Hradec Kr√°lov√© | Molecular biology division, Institute of Clinical Biochemistry and Diagnostics, Charles University, Faculty of Medicine in Hradec Kr√°lov√© and University Hospital Hradec Kr√°lov√© | Helena Kova≈ô√≠kov√° et al |
| EPI_ISL_895799 | CzechRepublic/FNHK-00801/2020 | 2020-03-12 | Europe/Czech Republic/Czech Republic/ | Molecular biology division, Institute of Clinical Biochemistry and Diagnostics, Charles University, Faculty of Medicine in Hradec Kr√°lov√© and University Hospital Hradec Kr√°lov√© | Molecular biology division, Institute of Clinical Biochemistry and Diagnostics, Charles University, Faculty of Medicine in Hradec Kr√°lov√© and University Hospital Hradec Kr√°lov√© | Helena Kova≈ô√≠kov√° et al |

| | | | | | | |
|---|---|---|---|---|---|---|
| EPI_ISL_895820 | CzechRepublic/FNHK-128/2020 | 2020-04-01 | Europe/Czech Republic/Czech Republic/ | Molecular biology division, Institute of Clinical Biochemistry and Diagnostics, Charles University, Faculty of Medicine in Hradec Kr√°lov√© and University Hospital Hradec Kr√°lov√© | Molecular biology division, Institute of Clinical Biochemistry and Diagnostics, Charles University, Faculty of Medicine in Hradec Kr√°lov√© and University Hospital Hradec Kr√°lov√© | Helena Kova≈ô√≠kov√° et al |
| EPI_ISL_895838 | CzechRepublic/FNHK-058/2020 | 2020-03-25 | Europe/Czech Republic/Czech Republic/ | Molecular biology division, Institute of Clinical Biochemistry and Diagnostics, Charles University, Faculty of Medicine in Hradec Kr√°lov√© and University Hospital Hradec Kr√°lov√© | Molecular biology division, Institute of Clinical Biochemistry and Diagnostics, Charles University, Faculty of Medicine in Hradec Kr√°lov√© and University Hospital Hradec Kr√°lov√© | Helena Kova≈ô√≠kov√° et al |
| EPI_ISL_895839 | CzechRepublic/FNHK-11701/2020 | 2020-03-30 | Europe/Czech Republic/Czech Republic/ | Molecular biology division, Institute of Clinical Biochemistry and Diagnostics, Charles University, Faculty of Medicine in Hradec Kr√°lov√© and University Hospital Hradec Kr√°lov√© | Molecular biology division, Institute of Clinical Biochemistry and Diagnostics, Charles University, Faculty of Medicine in Hradec Kr√°lov√© and University Hospital Hradec Kr√°lov√© | Helena Kova≈ô√≠kov√° et al |

| | | | | | | |
|---|---|---|---|---|---|---|
| EPI_ISL_895912 | Japan/PG-14278/2020 | 20-Dec | Asia/Japan/Japan/ | Pathogen Genomics Center, National Institute of Infectious Diseases | Pathogen Genomics Center, National Institute of Infectious Diseases | Tsuyoshi Sekizuka et al |
| EPI_ISL_895931 | Japan/PG-14297/2020 | 20-Dec | Asia/Japan/Japan/ | Pathogen Genomics Center, National Institute of Infectious Diseases | Pathogen Genomics Center, National Institute of Infectious Diseases | Tsuyoshi Sekizuka et al |
| EPI_ISL_895994 | Japan/PG-14360/2020 | 20-Dec | Asia/Japan/Japan/ | Tokyo Metropolitan Institute of Public Health | Pathogen Genomics Center, National Institute of Infectious Diseases | Tsuyoshi Sekizuka et al |
| EPI_ISL_896035 | Japan/PG-14401/2020 | 20-Nov | Asia/Japan/Japan/ | Pathogen Genomics Center, National Institute of Infectious Diseases | Pathogen Genomics Center, National Institute of Infectious Diseases | Tsuyoshi Sekizuka et al |
| EPI_ISL_896099 | Switzerland/un-UHB-720584600/202 | 2021-01-08 | Europe/Switzerland/Switzerland/ | Labormedizinisches Zentrum Dr Risch | University Hospital Basel, Clinical Bacteriology | Tim Roloff et al |
| EPI_ISL_896118 | France/PAC-IHUCOVID-2581/2020 | 2020-10-20 | Europe/France/Provence-Alpes-CV¥te d' | MEPHI, Aix Marseille University | MEPHI, Aix Marseille University | Anthony LEVASSEUR et al |
| EPI_ISL_896120 | France/PAC-IHUCOVID-2556/2020 | 2020-10-19 | Europe/France/Provence-Alpes-CV¥te d' | MEPHI, Aix Marseille University | MEPHI, Aix Marseille University | Anthony LEVASSEUR et al |
| EPI_ISL_896122 | France/PAC-IHUCOVID-2343/2020 | 2020-10-08 | Europe/France/Provence-Alpes-CV¥te d' | MEPHI, Aix Marseille University | MEPHI, Aix Marseille University | Anthony LEVASSEUR et al |
| EPI_ISL_896127 | France/PAC-IHUCOVID-2034/2020 | 2020-08-29 | Europe/France/Provence-Alpes-CV¥te d' | MEPHI, Aix Marseille University | MEPHI, Aix Marseille University | Anthony LEVASSEUR et al |
| EPI_ISL_896128 | France/PAC-IHUCOVID-2941/2020 | 2020-12-15 | Europe/France/Provence-Alpes-CV¥te d' | MEPHI, Aix Marseille University | MEPHI, Aix Marseille University | Anthony LEVASSEUR et al |

| | | | | | | |
|---|---|---|---|---|---|---|
| EPI_ISL_896130 | France/PAC-IHUCOVID-1272/2020 | 2020-08-18 | Europe/France/Provence-Alpes-CV¥te d' | MEPHI, Aix Marseille University | MEPHI, Aix Marseille University | Anthony LEVASSEUR et al |
| EPI_ISL_896132 | France/PAC-IHUCOVID-2316/2020 | 2020-10-07 | Europe/France/Provence-Alpes-CV¥te d' | MEPHI, Aix Marseille University | MEPHI, Aix Marseille University | Anthony LEVASSEUR et al |
| EPI_ISL_896134 | France/PAC-IHUCOVID-2520/2020 | 2020-10-16 | Europe/France/Provence-Alpes-CV¥te d' | MEPHI, Aix Marseille University | MEPHI, Aix Marseille University | Anthony LEVASSEUR et al |
| EPI_ISL_896136 | France/PAC-IHUCOVID-1986/2020 | 2020-08-27 | Europe/France/Provence-Alpes-CV¥te d' | MEPHI, Aix Marseille University | MEPHI, Aix Marseille University | Anthony LEVASSEUR et al |
| EPI_ISL_896138 | France/PAC-IHUCOVID-2428/2020 | 2020-10-14 | Europe/France/Provence-Alpes-CV¥te d' | MEPHI, Aix Marseille University | MEPHI, Aix Marseille University | Anthony LEVASSEUR et al |
| EPI_ISL_896139 | France/PAC-IHUCOVID-2626/2020 | 2020-10-09 | Europe/France/Provence-Alpes-CV¥te d' | MEPHI, Aix Marseille University | MEPHI, Aix Marseille University | Anthony LEVASSEUR et al |
| EPI_ISL_896140 | France/PAC-IHUCOVID-2883/2020 | 2020-12-16 | Europe/France/Provence-Alpes-CV¥te d' | MEPHI, Aix Marseille University | MEPHI, Aix Marseille University | Anthony LEVASSEUR et al |
| EPI_ISL_896142 | France/PAC-IHUCOVID-2198/2020 | 2020-10-01 | Europe/France/Provence-Alpes-CV¥te d' | MEPHI, Aix Marseille University | MEPHI, Aix Marseille University | Anthony LEVASSEUR et al |
| EPI_ISL_896147 | France/PAC-IHUCOVID-2078/2020 | 2020-09-27 | Europe/France/Provence-Alpes-CV¥te d' | MEPHI, Aix Marseille University | MEPHI, Aix Marseille University | Anthony LEVASSEUR et al |
| EPI_ISL_896148 | France/PAC-IHUCOVID-2704/2020 | 2020-10-27 | Europe/France/Provence-Alpes-CV¥te d' | MEPHI, Aix Marseille University | MEPHI, Aix Marseille University | Anthony LEVASSEUR et al |
| EPI_ISL_896150 | France/PAC-IHUCOVID-2074/2020 | 2020-09-23 | Europe/France/Provence-Alpes-CV¥te d' | MEPHI, Aix Marseille University | MEPHI, Aix Marseille University | Anthony LEVASSEUR et al |
| EPI_ISL_896156 | France/PAC-IHUCOVID-2968/2020 | 2020-12-21 | Europe/France/Provence-Alpes-CV¥te d' | MEPHI, Aix Marseille University | MEPHI, Aix Marseille University | Anthony LEVASSEUR et al |
| EPI_ISL_896162 | France/PAC-IHUCOVID-2971/2020 | 2020-12-21 | Europe/France/Provence-Alpes-CV¥te d' | MEPHI, Aix Marseille University | MEPHI, Aix Marseille University | Anthony LEVASSEUR et al |

| | | | | | | |
|---|---|---|---|---|---|---|
| EPI_ISL_896165 | France/PAC-IHUCOVID-2987/2020 | 2020-12-26 | Europe/France/Provence-Alpes-CV¥te d' | MEPHI, Aix Marseille University | MEPHI, Aix Marseille University | Anthony LEVASSEUR et al |
| EPI_ISL_896168 | France/PAC-IHUCOVID-2997/2021 | 2021-01-04 | Europe/France/Provence-Alpes-CV¥te d' | MEPHI, Aix Marseille University | MEPHI, Aix Marseille University | Anthony LEVASSEUR et al |
| EPI_ISL_896179 | France/PAC-IHUCOVID-0006/2020 | 2020-02-29 | Europe/France/Provence-Alpes-CV¥te d' | MEPHI, Aix Marseille University | MEPHI, Aix Marseille University | Anthony LEVASSEUR et al |
| EPI_ISL_896182 | France/PAC-IHUCOVID-1708/2020 | 2020-09-17 | Europe/France/Provence-Alpes-CV¥te d' | MEPHI, Aix Marseille University | MEPHI, Aix Marseille University | Anthony LEVASSEUR et al |
| EPI_ISL_896186 | France/PAC-IHUCOVID-2870/2020 | 2020-11-27 | Europe/France/Provence-Alpes-CV¥te d' | MEPHI, Aix Marseille University | MEPHI, Aix Marseille University | Anthony LEVASSEUR et al |
| EPI_ISL_896187 | France/PAC-IHUCOVID-2872/2020 | 2020-12-14 | Europe/France/Provence-Alpes-CV¥te d' | MEPHI, Aix Marseille University | MEPHI, Aix Marseille University | Anthony LEVASSEUR et al |
| EPI_ISL_896199 | France/PAC-IHUCOVID-2980/2020 | 2020-12-24 | Europe/France/Provence-Alpes-CV¥te d' | MEPHI, Aix Marseille University | MEPHI, Aix Marseille University | Anthony LEVASSEUR et al |
| EPI_ISL_896202 | France/PAC-IHUCOVID-2989/2020 | 2020-12-22 | Europe/France/Provence-Alpes-CV¥te d' | MEPHI, Aix Marseille University | MEPHI, Aix Marseille University | Anthony LEVASSEUR et al |
| EPI_ISL_896204 | France/PAC-IHUCOVID-2994/2020 | 2020-12-27 | Europe/France/Provence-Alpes-CV¥te d' | MEPHI, Aix Marseille University | MEPHI, Aix Marseille University | Anthony LEVASSEUR et al |
| EPI_ISL_896206 | France/PAC-IHUCOVID-3013/2020 | 2020-11-23 | Europe/France/Provence-Alpes-CV¥te d' | MEPHI, Aix Marseille University | MEPHI, Aix Marseille University | Anthony LEVASSEUR et al |
| EPI_ISL_896207 | France/PAC-IHUCOVID-3018/2020 | 2020-12-02 | Europe/France/Provence-Alpes-CV¥te d' | MEPHI, Aix Marseille University | MEPHI, Aix Marseille University | Anthony LEVASSEUR et al |
| EPI_ISL_896218 | USA/NY-Wadsworth-21011373-01/20 | 2021-01-15 | North America/USA/New York/Onondag | SUNY UPSTATE MEDICAL UNIVERSITY | Wadsworth Center, New York State Department of Health | Kirsten St. George et al |
| EPI_ISL_896223 | USA/NY-Wadsworth-21011368-01/20 | 2021-01-12 | North America/USA/New York/Onondag | SUNY UPSTATE MEDICAL UNIVERSITY | Wadsworth Center, New York State Department of Health | Kirsten St. George et al |

| | | | | | | |
|---|---|---|---|---|---|---|
| EPI_ISL_896229 | USA/NY-Wadsworth-21011380-01/20 | 2021-01-15 | North America/USA/New York/Onondag | SUNY UPSTATE MEDICAL UNIVERSITY | Wadsworth Center, New York State Department of Health | Kirsten St. George et al |
| EPI_ISL_896233 | USA/NY-Wadsworth-21011134-01/20 | 2021-01-18 | North America/USA/New York/New Yor | Columbia University Irving Medical Center | Wadsworth Center, New York State Department of Health | Kirsten St. George et al |
| EPI_ISL_896237 | USA/NY-Wadsworth-21011120-01/20 | 2021-01-17 | North America/USA/New York/New Yor | Columbia University Irving Medical Center | Wadsworth Center, New York State Department of Health | Kirsten St. George et al |
| EPI_ISL_896246 | USA/NY-Wadsworth-21011152-01/20 | 2021-01-10 | North America/USA/New York/New Yor | Columbia University Irving Medical Center | Wadsworth Center, New York State Department of Health | Kirsten St. George et al |
| EPI_ISL_896252 | USA/NY-Wadsworth-21011330-01/20 | 2021-01-14 | North America/USA/New York/Onondag | SUNY UPSTATE MEDICAL UNIVERSITY | Wadsworth Center, New York State Department of Health | Kirsten St. George et al |
| EPI_ISL_896260 | USA/NY-Wadsworth-21011341-01/20 | 2021-01-14 | North America/USA/New York/Onondag | SUNY UPSTATE MEDICAL UNIVERSITY | Wadsworth Center, New York State Department of Health | Kirsten St. George et al |
| EPI_ISL_896261 | USA/NY-Wadsworth-21011342-01/20 | 2021-01-12 | North America/USA/New York/Onondag | SUNY UPSTATE MEDICAL UNIVERSITY | Wadsworth Center, New York State Department of Health | Kirsten St. George et al |
| EPI_ISL_896264 | USA/NY-Wadsworth-21011346-01/20 | 2021-01-14 | North America/USA/New York/Onondag | SUNY UPSTATE MEDICAL UNIVERSITY | Wadsworth Center, New York State Department of Health | Kirsten St. George et al |
| EPI_ISL_896267 | USA/NY-Wadsworth-21011351-01/20 | 2021-01-12 | North America/USA/New York/Onondag | SUNY UPSTATE MEDICAL UNIVERSITY | Wadsworth Center, New York State Department of Health | Kirsten St. George et al |
| EPI_ISL_896269 | USA/NY-Wadsworth-21011354-01/20 | 2021-01-12 | North America/USA/New York/Onondag | SUNY UPSTATE MEDICAL UNIVERSITY | Wadsworth Center, New York State Department of Health | Kirsten St. George et al |

| | | | | | | |
|---|---|---|---|---|---|---|
| EPI_ISL_896272 | USA/NY-Wadsworth-21011358-01/20 | 2021-01-14 | North America/USA/New York/Madison | SUNY UPSTATE MEDICAL UNIVERSITY | Wadsworth Center, New York State Department of Health | Kirsten St. George et al |
| EPI_ISL_896276 | USA/NY-Wadsworth-21011362-01/20 | 2021-01-14 | North America/USA/New York/Onondag | SUNY UPSTATE MEDICAL UNIVERSITY | Wadsworth Center, New York State Department of Health | Kirsten St. George et al |
| EPI_ISL_896288 | USA/NY-Wadsworth-21011377-01/20 | 2021-01-13 | North America/USA/New York/Onondag | SUNY UPSTATE MEDICAL UNIVERSITY | Wadsworth Center, New York State Department of Health | Kirsten St. George et al |
| EPI_ISL_896299 | USA/NY-Wadsworth-21009969-01/20 | 2020-12-15 | North America/USA/New York/ | MEMORIAL SLOAN KETTERING CANCER CENTER | Wadsworth Center, New York State Department of Health | Kirsten St. George et al |
| EPI_ISL_896304 | USA/NY-Wadsworth-21010026-01/20 | 2021-01-04 | North America/USA/New York/Bronx | New York Presbyterian Hospital | Wadsworth Center, New York State Department of Health | Kirsten St. George et al |
| EPI_ISL_896320 | USA/NY-Wadsworth-21009972-01/20 | 2020-12-15 | North America/USA/New York/Kings Co | MEMORIAL SLOAN KETTERING CANCER CENTER | Wadsworth Center, New York State Department of Health | Kirsten St. George et al |
| EPI_ISL_896341 | USA/NY-Wadsworth-21010005-01/20 | 2021-01-04 | North America/USA/New York/New Yor | New York Presbyterian Hospital | Wadsworth Center, New York State Department of Health | Kirsten St. George et al |
| EPI_ISL_896347 | USA/NY-Wadsworth-21010012-01/20 | 2021-01-04 | North America/USA/New York/Queens | New York Presbyterian Hospital | Wadsworth Center, New York State Department of Health | Kirsten St. George et al |
| EPI_ISL_896348 | USA/NY-Wadsworth-21010014-01/20 | 2021-01-04 | North America/USA/New York/New Yor | New York Presbyterian Hospital | Wadsworth Center, New York State Department of Health | Kirsten St. George et al |
| EPI_ISL_896362 | USA/NY-Wadsworth-21010046-01/20 | 2021-01-04 | North America/USA/New York/Queens | New York Presbyterian Hospital | Wadsworth Center, New York State Department of Health | Kirsten St. George et al |

| | | | | | | |
|---|---|---|---|---|---|---|
| EPI_ISL_896367 | USA/NY-Wadsworth-21010060-01/20 | 2021-01-05 | North America/USA/New York/New Yor | New York Presbyterian Hospital | Wadsworth Center, New York State Department of Health | Kirsten St. George et al |
| EPI_ISL_896368 | USA/NY-Wadsworth-21010072-01/20 | 2021-01-05 | North America/USA/New York/Queens | New York Presbyterian Hospital | Wadsworth Center, New York State Department of Health | Kirsten St. George et al |
| EPI_ISL_896371 | USA/NY-Wadsworth-21010082-01/20 | 2021-01-05 | North America/USA/New York/New Yor | New York Presbyterian Hospital | Wadsworth Center, New York State Department of Health | Kirsten St. George et al |
| EPI_ISL_896373 | Romania/RO_DJ37480/2020 | 2020-10-20 | Europe/Romania/Suceava/ | University of Medicine and Pharmacy of Craiova | """Stefan cel Mare"" University Metagenomics Lab" | Lobiuc Andrei et al |
| EPI_ISL_896374 | Romania/RO_DJ40541/2020 | 2020-10-29 | Europe/Romania/Suceava/ | University of Medicine and Pharmacy of Craiova | """Stefan cel Mare"" University Metagenomics Lab" | Lobiuc Andrei et al |
| EPI_ISL_896375 | Romania/RO_DJ40921/2020 | 2020-10-31 | Europe/Romania/Suceava/ | University of Medicine and Pharmacy of Craiova | """Stefan cel Mare"" University Metagenomics Lab" | Lobiuc Andrei et al |
| EPI_ISL_896376 | Romania/RO_DJ48086/2020 | 2020-11-20 | Europe/Romania/Suceava/ | University of Medicine and Pharmacy of Craiova | """Stefan cel Mare"" University Metagenomics Lab" | Lobiuc Andrei et al |
| EPI_ISL_896379 | Romania/ROSV_28461/2020 | 2020-11-21 | Europe/Romania/Suceava/ | Suceava County Emergency Hospital | """Stefan cel Mare"" University Metagenomics Lab" | Lobiuc Andrei et al |
| EPI_ISL_896381 | Romania/ROSV_34217/2021 | 2021-01-16 | Europe/Romania/Suceava/ | Suceava County Emergency Hospital | """Stefan cel Mare"" University Metagenomics Lab" | Lobiuc Andrei et al |
| EPI_ISL_896383 | USA/NY-Wadsworth-21006748-01/20 | 2021-01-04 | North America/USA/New York/Bronx | Columbia University Irving Medical Center | Wadsworth Center, New York State Department of Health | Kirsten St. George et al |

| | | | | | | |
|---|---|---|---|---|---|---|
| EPI_ISL_896386 | USA/NY-Wadsworth-21006778-01/20 | 2021-01-04 | North America/USA/New York/Queens | Columbia University Irving Medical Center | Wadsworth Center, New York State Department of Health | Kirsten St. George et al |
| EPI_ISL_896391 | USA/NY-Wadsworth-21006765-01/20 | 2021-01-04 | North America/USA/New York/New Yor | Columbia University Irving Medical Center | Wadsworth Center, New York State Department of Health | Kirsten St. George et al |
| EPI_ISL_896402 | USA/NY-Wadsworth-21006627-01/20 | 2021-01-11 | North America/USA/New York/Chemung | URMC LABS | Wadsworth Center, New York State Department of Health | Kirsten St. George et al |
| EPI_ISL_896408 | USA/NY-Wadsworth-21006637-01/20 | 2021-01-11 | North America/USA/New York/Monroe | URMC LABS | Wadsworth Center, New York State Department of Health | Kirsten St. George et al |
| EPI_ISL_896410 | USA/NY-Wadsworth-21006640-01/20 | 2021-01-11 | North America/USA/New York/Monroe | URMC LABS | Wadsworth Center, New York State Department of Health | Kirsten St. George et al |
| EPI_ISL_896413 | USA/NY-Wadsworth-21006643-01/20 | 2021-01-12 | North America/USA/New York/Monroe | URMC LABS | Wadsworth Center, New York State Department of Health | Kirsten St. George et al |
| EPI_ISL_896417 | USA/NY-Wadsworth-21006657-01/20 | 2021-01-10 | North America/USA/New York/Chemung | URMC LABS | Wadsworth Center, New York State Department of Health | Kirsten St. George et al |
| EPI_ISL_896422 | USA/NY-Wadsworth-21006742-01/20 | 2021-01-03 | North America/USA/New York/New Yor | Columbia University Irving Medical Center | Wadsworth Center, New York State Department of Health | Kirsten St. George et al |
| EPI_ISL_896428 | USA/NY-Wadsworth-21006751-01/20 | 2021-01-03 | North America/USA/New York/Bronx | Columbia University Irving Medical Center | Wadsworth Center, New York State Department of Health | Kirsten St. George et al |
| EPI_ISL_896432 | USA/NY-Wadsworth-21006766-01/20 | 2021-01-04 | North America/USA/New York/Bronx | Columbia University Irving Medical Center | Wadsworth Center, New York State Department of Health | Kirsten St. George et al |

| | | | | | | |
|---|---|---|---|---|---|---|
| EPI_ISL_896433 | USA/NY-Wadsworth-21006769-01/20 | 2021-01-04 | North America/USA/New York/New Yor | Columbia University Irving Medical Center | Wadsworth Center, New York State Department of Health | Kirsten St. George et al |
| EPI_ISL_896438 | USA/NY-Wadsworth-21006802-01/20 | 2021-01-04 | North America/USA/New York/Bronx | Columbia University Irving Medical Center | Wadsworth Center, New York State Department of Health | Kirsten St. George et al |
| EPI_ISL_896443 | USA/NY-Wadsworth-21006814-01/20 | 2021-01-04 | North America/USA/New York/Bronx | Columbia University Irving Medical Center | Wadsworth Center, New York State Department of Health | Kirsten St. George et al |
| EPI_ISL_896498 | USA/NY-Wadsworth-21010036-01/20 | 2020-12-18 | North America/USA/New York/Bronx | MEMORIAL SLOAN KETTERING CANCER CENTER | Wadsworth Center, New York State Department of Health | Kirsten St. George et al |
| EPI_ISL_896499 | USA/NY-Wadsworth-21010065-01/20 | 2020-12-18 | North America/USA/New York/Bronx | MEMORIAL SLOAN KETTERING CANCER CENTER | Wadsworth Center, New York State Department of Health | Kirsten St. George et al |
| EPI_ISL_896519 | USA/NY-Wadsworth-21010155-01/20 | 2021-01-17 | North America/USA/New York/Monroe | URMC LABS | Wadsworth Center, New York State Department of Health | Kirsten St. George et al |
| EPI_ISL_896526 | USA/NY-Wadsworth-21010141-01/20 | 2021-01-06 | North America/USA/New York/Nassau ( | New York Presbyterian Hospital | Wadsworth Center, New York State Department of Health | Kirsten St. George et al |
| EPI_ISL_896527 | USA/NY-Wadsworth-21010154-01/20 | 2021-01-07 | North America/USA/New York/Nassau ( | New York Presbyterian Hospital | Wadsworth Center, New York State Department of Health | Kirsten St. George et al |
| EPI_ISL_896548 | USA/NY-Wadsworth-21010136-01/20 | 2020-12-21 | North America/USA/New York/Queens | MEMORIAL SLOAN KETTERING CANCER CENTER | Wadsworth Center, New York State Department of Health | Kirsten St. George et al |
| EPI_ISL_896552 | USA/NY-Wadsworth-21010112-01/20 | 2020-12-21 | North America/USA/New York/Queens | MEMORIAL SLOAN KETTERING CANCER CENTER | Wadsworth Center, New York State Department of Health | Kirsten St. George et al |

| | | | | | | |
|---|---|---|---|---|---|---|
| EPI_ISL_896560 | USA/NY-Wadsworth-21010164-01/20 | 2021-01-06 | North America/USA/New York/Queens | New York Presbyterian Hospital | Wadsworth Center, New York State Department of Health | Kirsten St. George et al |
| EPI_ISL_896566 | USA/NY-Wadsworth-21010078-01/20 | 2021-01-18 | North America/USA/New York/Steuben | URMC LABS | Wadsworth Center, New York State Department of Health | Kirsten St. George et al |
| EPI_ISL_896568 | USA/NY-Wadsworth-21010081-01/20 | 2021-01-18 | North America/USA/New York/Wayne C | URMC LABS | Wadsworth Center, New York State Department of Health | Kirsten St. George et al |
| EPI_ISL_896887 | Japan/PG-12473/2020 | 20-Sep | Asia/Japan/Japan/ | Pathogen Genomics Center, National Institute of Infectious Diseases | Pathogen Genomics Center, National Institute of Infectious Diseases | Tsuyoshi Sekizuka et al |
| EPI_ISL_896943 | Japan/PG-12529/2020 | 20-Nov | Asia/Japan/Japan/ | Pathogen Genomics Center, National Institute of Infectious Diseases | Pathogen Genomics Center, National Institute of Infectious Diseases | Tsuyoshi Sekizuka et al |
| EPI_ISL_897481 | Japan/PG-13166/2020 | 20-Nov | Asia/Japan/Japan/ | Pathogen Genomics Center, National Institute of Infectious Diseases | Pathogen Genomics Center, National Institute of Infectious Diseases | Tsuyoshi Sekizuka et al |
| EPI_ISL_897567 | Japan/PG-13253/2020 | 20-Jul | Asia/Japan/Japan/ | Pathogen Genomics Center, National Institute of Infectious Diseases | Pathogen Genomics Center, National Institute of Infectious Diseases | Tsuyoshi Sekizuka et al |
| EPI_ISL_897570 | Japan/PG-13256/2020 | 20-Jul | Asia/Japan/Japan/ | Pathogen Genomics Center, National Institute of Infectious Diseases | Pathogen Genomics Center, National Institute of Infectious Diseases | Tsuyoshi Sekizuka et al |

| | | | | | | |
|---|---|---|---|---|---|---|
| EPI_ISL_897965 | Switzerland/VS-32983320/2021 | 2021-01-14 | Europe/Switzerland/Valais/ | University Hospitals of Geneva, Laboratory of Virology | HUG, Laboratory of Virology and the Health2030 Genome Center | Samuel Cordey et al |
| EPI_ISL_898059 | Japan/PG-11151/2020 | 20-Oct | Asia/Japan/Japan/ | Pathogen Genomics Center, National Institute of Infectious Diseases | Pathogen Genomics Center, National Institute of Infectious Diseases | Tsuyoshi Sekizuka et al |
| EPI_ISL_898081 | Japan/PG-11174/2020 | 20-Oct | Asia/Japan/Japan/ | Pathogen Genomics Center, National Institute of Infectious Diseases | Pathogen Genomics Center, National Institute of Infectious Diseases | Tsuyoshi Sekizuka et al |
| EPI_ISL_898106 | Japan/PG-11242/2020 | 20-Nov | Asia/Japan/Japan/ | Pathogen Genomics Center, National Institute of Infectious Diseases | Pathogen Genomics Center, National Institute of Infectious Diseases | Tsuyoshi Sekizuka et al |
| EPI_ISL_898567 | Japan/PG-11704/2020 | 20-Mar | Asia/Japan/Japan/ | Pathogen Genomics Center, National Institute of Infectious Diseases | Pathogen Genomics Center, National Institute of Infectious Diseases | Tsuyoshi Sekizuka et al |
| EPI_ISL_898668 | Japan/PG-11807/2020 | 20-Aug | Asia/Japan/Japan/ | Pathogen Genomics Center, National Institute of Infectious Diseases | Pathogen Genomics Center, National Institute of Infectious Diseases | Tsuyoshi Sekizuka et al |
| EPI_ISL_898709 | Japan/PG-11854/2020 | 20-Nov | Asia/Japan/Japan/ | Pathogen Genomics Center, National Institute of Infectious Diseases | Pathogen Genomics Center, National Institute of Infectious Diseases | Tsuyoshi Sekizuka et al |
| EPI_ISL_898776 | Japan/PG-11945/2020 | 20-Nov | Asia/Japan/Japan/ | Department of Infectious Diseases, Kobe Institute of Health | Pathogen Genomics Center, National Institute of Infectious Diseases | Tsuyoshi Sekizuka et al |

| | | | | | | |
|---|---|---|---|---|---|---|
| EPI_ISL_898788 | Japan/PG-11957/2020 | 20-Nov | Asia/Japan/Japan/ | Department of Infectious Diseases, Kobe Institute of Health | Pathogen Genomics Center, National Institute of Infectious Diseases | Tsuyoshi Sekizuka et al |
| EPI_ISL_898862 | Japan/PG-12031/2020 | 20-Oct | Asia/Japan/Japan/ | Pathogen Genomics Center, National Institute of Infectious Diseases | Pathogen Genomics Center, National Institute of Infectious Diseases | Tsuyoshi Sekizuka et al |
| EPI_ISL_899001 | Switzerland/BE-ETHZ-451406/2021 | 2021-01-09 | Europe/Switzerland/Bern/ | Viollier AG | Department of Biosystems Science and Engineering, ETH Zv°rich | Chaoran Chen et al |
| EPI_ISL_899029 | Switzerland/ZH-ETHZ-451770/2021 | 2021-01-14 | Europe/Switzerland/Zv°rich/ | Viollier AG | Department of Biosystems Science and Engineering, ETH Zv°rich | Chaoran Chen et al |
| EPI_ISL_899124 | Switzerland/BL-ETHZ-451513/2021 | 2021-01-11 | Europe/Switzerland/Basel-Land/ | Viollier AG | Department of Biosystems Science and Engineering, ETH Zv°rich | Chaoran Chen et al |
| EPI_ISL_899149 | Switzerland/ZH-ETHZ-440327/2021 | 2021-01-06 | Europe/Switzerland/Zv°rich/ | Viollier AG | Department of Biosystems Science and Engineering, ETH Zv°rich | Christian Beisel et al |
| EPI_ISL_899193 | Switzerland/AG-ETHZ-431130/2020 | 2020-12-29 | Europe/Switzerland/Aargau/ | Viollier AG | Department of Biosystems Science and Engineering, ETH Zv°rich | Christian Beisel et al |
| EPI_ISL_899245 | Switzerland/AG-ETHZ-450363/2021 | 2021-01-12 | Europe/Switzerland/Aargau/ | Viollier AG | Department of Biosystems Science and Engineering, ETH Zv°rich | Christian Beisel et al |

| | | | | | | |
|---|---|---|---|---|---|---|
| EPI_ISL_899305 | Switzerland/BE-ETHZ-440094/2021 | 2021-01-02 | Europe/Switzerland/Bern/ | Viollier AG | Department of Biosystems Science and Engineering, ETH Z√ºrich | Christian Beisel et al |
| EPI_ISL_899516 | Switzerland/BS-ETHZ-430912/2020 | 2020-12-26 | Europe/Switzerland/Basel-Stadt/ | Viollier AG | Department of Biosystems Science and Engineering, ETH Z√ºrich | Christian Beisel et al |
| EPI_ISL_899580 | Switzerland/NE-ETHZ-431066/2020 | 2020-12-29 | Europe/Switzerland/Neuch√¢tel/ | Viollier AG | Department of Biosystems Science and Engineering, ETH Z√ºrich | Christian Beisel et al |
| EPI_ISL_899654 | Switzerland/SO-ETHZ-430862/2020 | 2020-12-25 | Europe/Switzerland/Solothurn/ | Viollier AG | Department of Biosystems Science and Engineering, ETH Z√ºrich | Christian Beisel et al |
| EPI_ISL_899735 | Switzerland/SO-ETHZ-450289/2021 | 2021-01-14 | Europe/Switzerland/Solothurn/ | Viollier AG | Department of Biosystems Science and Engineering, ETH Z√ºrich | Christian Beisel et al |
| EPI_ISL_899777 | Switzerland/SZ-ETHZ-430993/2020 | 2020-12-28 | Europe/Switzerland/Schwyz/ | Viollier AG | Department of Biosystems Science and Engineering, ETH Z√ºrich | Christian Beisel et al |
| EPI_ISL_899968 | Switzerland/ZH-ETHZ-431116/2020 | 2020-12-29 | Europe/Switzerland/Z√ºrich/ | Viollier AG | Department of Biosystems Science and Engineering, ETH Z√ºrich | Christian Beisel et al |
| EPI_ISL_899969 | Switzerland/ZH-ETHZ-431117/2020 | 2020-12-29 | Europe/Switzerland/Z√ºrich/ | Viollier AG | Department of Biosystems Science and Engineering, ETH Z√ºrich | Christian Beisel et al |

| | | | | | | |
|---|---|---|---|---|---|---|
| EPI_ISL_900056 | France/PAC-IHUCOVID-2373/2020 | 2020-10-10 | Europe/France/Provence-Alpes-CV¥te d' | MEPHI, Aix Marseille University | MEPHI, Aix Marseille University | Anthony LEVASSEUR et al |
| EPI_ISL_900059 | France/PAC-IHUCOVID-2201/2020 | 2020-10-01 | Europe/France/Provence-Alpes-CV¥te d' | MEPHI, Aix Marseille University | MEPHI, Aix Marseille University | Anthony LEVASSEUR et al |
| EPI_ISL_900063 | France/PAC-IHUCOVID-2212/2020 | 2020-10-02 | Europe/France/Provence-Alpes-CV¥te d' | MEPHI, Aix Marseille University | MEPHI, Aix Marseille University | Anthony LEVASSEUR et al |
| EPI_ISL_900067 | France/PAC-IHUCOVID-0110/2020 | 2020-03-23 | Europe/France/Provence-Alpes-CV¥te d' | MEPHI, Aix Marseille University | MEPHI, Aix Marseille University | Anthony LEVASSEUR et al |
| EPI_ISL_900069 | France/PAC-IHUCOVID-1621/2020 | 2020-09-10 | Europe/France/Provence-Alpes-CV¥te d' | MEPHI, Aix Marseille University | MEPHI, Aix Marseille University | Anthony LEVASSEUR et al |
| EPI_ISL_900074 | France/PAC-IHUCOVID-2146/2020 | 2020-09-30 | Europe/France/Provence-Alpes-CV¥te d' | MEPHI, Aix Marseille University | MEPHI, Aix Marseille University | Anthony LEVASSEUR et al |
| EPI_ISL_900075 | France/PAC-IHUCOVID-2121/2020 | 2020-09-29 | Europe/France/Provence-Alpes-CV¥te d' | MEPHI, Aix Marseille University | MEPHI, Aix Marseille University | Anthony LEVASSEUR et al |
| EPI_ISL_900076 | France/PAC-IHUCOVID-0048/2020 | 2020-03-18 | Europe/France/Provence-Alpes-CV¥te d' | MEPHI, Aix Marseille University | MEPHI, Aix Marseille University | Anthony LEVASSEUR et al |
| EPI_ISL_900077 | France/PAC-IHUCOVID-2059/2020 | 2020-08-29 | Europe/France/Provence-Alpes-CV¥te d' | MEPHI, Aix Marseille University | MEPHI, Aix Marseille University | Anthony LEVASSEUR et al |
| EPI_ISL_900082 | France/PAC-IHUCOVID-1578/2020 | 2020-09-01 | Europe/France/Provence-Alpes-CV¥te d' | MEPHI, Aix Marseille University | MEPHI, Aix Marseille University | Anthony LEVASSEUR et al |
| EPI_ISL_900083 | France/PAC-IHUCOVID-2513/2020 | 2020-10-16 | Europe/France/Provence-Alpes-CV¥te d' | MEPHI, Aix Marseille University | MEPHI, Aix Marseille University | Anthony LEVASSEUR et al |
| EPI_ISL_900086 | France/PAC-IHUCOVID-2923/2020 | 2020-12-15 | Europe/France/Provence-Alpes-CV¥te d' | MEPHI, Aix Marseille University | MEPHI, Aix Marseille University | Anthony LEVASSEUR et al |
| EPI_ISL_900088 | France/PAC-IHUCOVID-0924/2020 | 2020-04-03 | Europe/France/Provence-Alpes-CV¥te d' | MEPHI, Aix Marseille University | MEPHI, Aix Marseille University | Anthony LEVASSEUR et al |

| | | | | | | |
|---|---|---|---|---|---|---|
| EPI_ISL_900089 | France/PAC-IHUCOVID-0123/2020 | 2020-03-24 | Europe/France/Provence-Alpes-CV¥te d' | MEPHI, Aix Marseille University | MEPHI, Aix Marseille University | Anthony LEVASSEUR et al |
| EPI_ISL_900093 | France/PAC-IHUCOVID-0649/2020 | 2020-03-06 | Europe/France/Provence-Alpes-CV¥te d' | MEPHI, Aix Marseille University | MEPHI, Aix Marseille University | Anthony LEVASSEUR et al |
| EPI_ISL_900100 | France/PAC-IHUCOVID-1701/2020 | 2020-09-17 | Europe/France/Provence-Alpes-CV¥te d' | MEPHI, Aix Marseille University | MEPHI, Aix Marseille University | Anthony LEVASSEUR et al |
| EPI_ISL_900101 | France/PAC-IHUCOVID-1965/2020 | 2020-04-09 | Europe/France/Provence-Alpes-CV¥te d' | MEPHI, Aix Marseille University | MEPHI, Aix Marseille University | Anthony LEVASSEUR et al |
| EPI_ISL_900105 | France/PAC-IHUCOVID-2096/2020 | 2020-09-26 | Europe/France/Provence-Alpes-CV¥te d' | MEPHI, Aix Marseille University | MEPHI, Aix Marseille University | Anthony LEVASSEUR et al |
| EPI_ISL_900106 | France/PAC-IHUCOVID-2515/2020 | 2020-10-16 | Europe/France/Provence-Alpes-CV¥te d' | MEPHI, Aix Marseille University | MEPHI, Aix Marseille University | Anthony LEVASSEUR et al |
| EPI_ISL_900113 | France/PAC-IHUCOVID-2030/2020 | 2020-08-29 | Europe/France/Provence-Alpes-CV¥te d' | MEPHI, Aix Marseille University | MEPHI, Aix Marseille University | Anthony LEVASSEUR et al |
| EPI_ISL_900117 | France/PAC-IHUCOVID-0301/2020 | 2020-03-31 | Europe/France/Provence-Alpes-CV¥te d' | MEPHI, Aix Marseille University | MEPHI, Aix Marseille University | Anthony LEVASSEUR et al |
| EPI_ISL_900119 | France/PAC-IHUCOVID-2473/2020 | 2020-10-15 | Europe/France/Provence-Alpes-CV¥te d' | MEPHI, Aix Marseille University | MEPHI, Aix Marseille University | Anthony LEVASSEUR et al |
| EPI_ISL_900123 | France/PAC-IHUCOVID-1595/2020 | 2020-09-02 | Europe/France/Provence-Alpes-CV¥te d' | MEPHI, Aix Marseille University | MEPHI, Aix Marseille University | Anthony LEVASSEUR et al |
| EPI_ISL_900124 | France/PAC-IHUCOVID-2300/2020 | 2020-10-06 | Europe/France/Provence-Alpes-CV¥te d' | MEPHI, Aix Marseille University | MEPHI, Aix Marseille University | Anthony LEVASSEUR et al |
| EPI_ISL_900125 | France/PAC-IHUCOVID-2658/2020 | 2020-10-20 | Europe/France/Provence-Alpes-CV¥te d' | MEPHI, Aix Marseille University | MEPHI, Aix Marseille University | Anthony LEVASSEUR et al |
| EPI_ISL_900127 | France/PAC-IHUCOVID-2204/2020 | 2020-09-30 | Europe/France/Provence-Alpes-CV¥te d' | MEPHI, Aix Marseille University | MEPHI, Aix Marseille University | Anthony LEVASSEUR et al |

| | | | | | | |
|---|---|---|---|---|---|---|
| EPI_ISL_900130 | France/PAC-IHUCOVID-0373/2020 | 2020-03-26 | Europe/France/Provence-Alpes-CV¥te d' | MEPHI, Aix Marseille University | MEPHI, Aix Marseille University | Anthony LEVASSEUR et al |
| EPI_ISL_900133 | France/PAC-IHUCOVID-2008/2020 | 2020-08-29 | Europe/France/Provence-Alpes-CV¥te d' | MEPHI, Aix Marseille University | MEPHI, Aix Marseille University | Anthony LEVASSEUR et al |
| EPI_ISL_900134 | France/PAC-IHUCOVID-2765/2020 | 2020-07-20 | Europe/France/Provence-Alpes-CV¥te d' | MEPHI, Aix Marseille University | MEPHI, Aix Marseille University | Anthony LEVASSEUR et al |
| EPI_ISL_900140 | France/PAC-IHUCOVID-0735/2020 | 2020-04-22 | Europe/France/Provence-Alpes-CV¥te d' | MEPHI, Aix Marseille University | MEPHI, Aix Marseille University | Anthony LEVASSEUR et al |
| EPI_ISL_900151 | France/PAC-IHUCOVID-2301/2020 | 2020-10-06 | Europe/France/Provence-Alpes-CV¥te d' | MEPHI, Aix Marseille University | MEPHI, Aix Marseille University | Anthony LEVASSEUR et al |
| EPI_ISL_900156 | France/PAC-IHUCOVID-2754/2020 | 2020-07-13 | Europe/France/Provence-Alpes-CV¥te d' | MEPHI, Aix Marseille University | MEPHI, Aix Marseille University | Anthony LEVASSEUR et al |
| EPI_ISL_900162 | France/PAC-IHUCOVID-2942/2020 | 2020-12-15 | Europe/France/Provence-Alpes-CV¥te d' | MEPHI, Aix Marseille University | MEPHI, Aix Marseille University | Anthony LEVASSEUR et al |
| EPI_ISL_900167 | France/PAC-IHUCOVID-2224/2020 | 2020-10-02 | Europe/France/Provence-Alpes-CV¥te d' | MEPHI, Aix Marseille University | MEPHI, Aix Marseille University | Anthony LEVASSEUR et al |
| EPI_ISL_900169 | France/PAC-IHUCOVID-2337/2020 | 2020-10-08 | Europe/France/Provence-Alpes-CV¥te d' | MEPHI, Aix Marseille University | MEPHI, Aix Marseille University | Anthony LEVASSEUR et al |
| EPI_ISL_900170 | France/PAC-IHUCOVID-1608/2020 | 2020-09-04 | Europe/France/Provence-Alpes-CV¥te d' | MEPHI, Aix Marseille University | MEPHI, Aix Marseille University | Anthony LEVASSEUR et al |
| EPI_ISL_900171 | France/PAC-IHUCOVID-2406/2020 | 2020-10-13 | Europe/France/Provence-Alpes-CV¥te d' | MEPHI, Aix Marseille University | MEPHI, Aix Marseille University | Anthony LEVASSEUR et al |
| EPI_ISL_900176 | France/PAC-IHUCOVID-2159/2020 | 2020-09-23 | Europe/France/Provence-Alpes-CV¥te d' | MEPHI, Aix Marseille University | MEPHI, Aix Marseille University | Anthony LEVASSEUR et al |
| EPI_ISL_900178 | France/PAC-IHUCOVID-2150/2020 | 2020-09-29 | Europe/France/Provence-Alpes-CV¥te d' | MEPHI, Aix Marseille University | MEPHI, Aix Marseille University | Anthony LEVASSEUR et al |

| | | | | | | |
|---|---|---|---|---|---|---|
| EPI_ISL_900179 | France/PAC-IHUCOVID-0968/2020 | 2020-07-17 | Europe/France/Provence-Alpes-CV¥te d' | MEPHI, Aix Marseille University | MEPHI, Aix Marseille University | Anthony LEVASSEUR et al |
| EPI_ISL_900181 | France/PAC-IHUCOVID-2618/2020 | 2020-10-27 | Europe/France/Provence-Alpes-CV¥te d' | MEPHI, Aix Marseille University | MEPHI, Aix Marseille University | Anthony LEVASSEUR et al |
| EPI_ISL_900183 | France/PAC-IHUCOVID-0371/2020 | 2020-03-26 | Europe/France/Provence-Alpes-CV¥te d' | MEPHI, Aix Marseille University | MEPHI, Aix Marseille University | Anthony LEVASSEUR et al |
| EPI_ISL_900184 | France/PAC-IHUCOVID-1074/2020 | 2020-08-05 | Europe/France/Provence-Alpes-CV¥te d' | MEPHI, Aix Marseille University | MEPHI, Aix Marseille University | Anthony LEVASSEUR et al |
| EPI_ISL_900185 | France/PAC-IHUCOVID-0656/2020 | 2020-03-12 | Europe/France/Provence-Alpes-CV¥te d' | MEPHI, Aix Marseille University | MEPHI, Aix Marseille University | Anthony LEVASSEUR et al |
| EPI_ISL_900191 | France/PAC-IHUCOVID-2514/2020 | 2020-10-16 | Europe/France/Provence-Alpes-CV¥te d' | MEPHI, Aix Marseille University | MEPHI, Aix Marseille University | Anthony LEVASSEUR et al |
| EPI_ISL_900192 | France/PAC-IHUCOVID-1139/2020 | 2020-08-07 | Europe/France/Provence-Alpes-CV¥te d' | MEPHI, Aix Marseille University | MEPHI, Aix Marseille University | Anthony LEVASSEUR et al |
| EPI_ISL_900196 | France/PAC-IHUCOVID-2332/2020 | 2020-10-08 | Europe/France/Provence-Alpes-CV¥te d' | MEPHI, Aix Marseille University | MEPHI, Aix Marseille University | Anthony LEVASSEUR et al |
| EPI_ISL_900358 | France/PAC-IHUCOVID-2232/2020 | 2020-10-03 | Europe/France/Provence-Alpes-CV¥te d' | MEPHI, Aix Marseille University | MEPHI, Aix Marseille University | Anthony LEVASSEUR et al |
| EPI_ISL_900373 | France/PAC-IHUCOVID-0845/2020 | 2020-03-10 | Europe/France/Provence-Alpes-CV¥te d' | MEPHI, Aix Marseille University | MEPHI, Aix Marseille University | Anthony LEVASSEUR et al |
| EPI_ISL_900393 | France/PAC-IHUCOVID-2336/2020 | 2020-10-08 | Europe/France/Provence-Alpes-CV¥te d' | MEPHI, Aix Marseille University | MEPHI, Aix Marseille University | Anthony LEVASSEUR et al |
| EPI_ISL_900394 | France/PAC-IHUCOVID-2878/2020 | 2020-12-15 | Europe/France/Provence-Alpes-CV¥te d' | MEPHI, Aix Marseille University | MEPHI, Aix Marseille University | Anthony LEVASSEUR et al |
| EPI_ISL_900400 | France/PAC-IHUCOVID-2461/2020 | 2020-10-12 | Europe/France/Provence-Alpes-CV¥te d' | MEPHI, Aix Marseille University | MEPHI, Aix Marseille University | Anthony LEVASSEUR et al |

| | | | | | | |
|---|---|---|---|---|---|---|
| EPI_ISL_900412 | France/PAC-IHUCOVID-2625/2020 | 2020-10-09 | Europe/France/Provence-Alpes-CV¥te d' | MEPHI, Aix Marseille University | MEPHI, Aix Marseille University | Anthony LEVASSEUR et al |
| EPI_ISL_900430 | France/PAC-IHUCOVID-2719/2020 | 2020-11-02 | Europe/France/Provence-Alpes-CV¥te d' | MEPHI, Aix Marseille University | MEPHI, Aix Marseille University | Anthony LEVASSEUR et al |
| EPI_ISL_900431 | France/PAC-IHUCOVID-0841/2020 | 2020-03-09 | Europe/France/Provence-Alpes-CV¥te d' | MEPHI, Aix Marseille University | MEPHI, Aix Marseille University | Anthony LEVASSEUR et al |
| EPI_ISL_900443 | France/PAC-IHUCOVID-2597/2020 | 2020-10-23 | Europe/France/Provence-Alpes-CV¥te d' | MEPHI, Aix Marseille University | MEPHI, Aix Marseille University | Anthony LEVASSEUR et al |
| EPI_ISL_900444 | France/PAC-IHUCOVID-1127/2020 | 2020-08-07 | Europe/France/Provence-Alpes-CV¥te d' | MEPHI, Aix Marseille University | MEPHI, Aix Marseille University | Anthony LEVASSEUR et al |
| EPI_ISL_900457 | France/PAC-IHUCOVID-0804/2020 | 2020-05-04 | Europe/France/Provence-Alpes-CV¥te d' | MEPHI, Aix Marseille University | MEPHI, Aix Marseille University | Anthony LEVASSEUR et al |
| EPI_ISL_900458 | France/PAC-IHUCOVID-2468/2020 | 2020-10-15 | Europe/France/Provence-Alpes-CV¥te d' | MEPHI, Aix Marseille University | MEPHI, Aix Marseille University | Anthony LEVASSEUR et al |
| EPI_ISL_900461 | France/PAC-IHUCOVID-2965/2020 | 2020-12-21 | Europe/France/Provence-Alpes-CV¥te d' | MEPHI, Aix Marseille University | MEPHI, Aix Marseille University | Anthony LEVASSEUR et al |
| EPI_ISL_900466 | France/PAC-IHUCOVID-2774/2020 | 2020-07-23 | Europe/France/Provence-Alpes-CV¥te d' | MEPHI, Aix Marseille University | MEPHI, Aix Marseille University | Anthony LEVASSEUR et al |
| EPI_ISL_900474 | France/PAC-IHUCOVID-0509/2020 | 2020-04-06 | Europe/France/Provence-Alpes-CV¥te d' | MEPHI, Aix Marseille University | MEPHI, Aix Marseille University | Anthony LEVASSEUR et al |
| EPI_ISL_900476 | France/PAC-IHUCOVID-2486/2020 | 2020-10-13 | Europe/France/Provence-Alpes-CV¥te d' | MEPHI, Aix Marseille University | MEPHI, Aix Marseille University | Anthony LEVASSEUR et al |
| EPI_ISL_900480 | France/PAC-IHUCOVID-2213/2020 | 2020-10-02 | Europe/France/Provence-Alpes-CV¥te d' | MEPHI, Aix Marseille University | MEPHI, Aix Marseille University | Anthony LEVASSEUR et al |
| EPI_ISL_900482 | France/PAC-IHUCOVID-2386/2020 | 2020-10-11 | Europe/France/Provence-Alpes-CV¥te d' | MEPHI, Aix Marseille University | MEPHI, Aix Marseille University | Anthony LEVASSEUR et al |

| | | | | | | |
|---|---|---|---|---|---|---|
| EPI_ISL_900483 | France/PAC-IHUCOVID-2619/2020 | 2020-10-27 | Europe/France/Provence-Alpes-C√¥te d' | MEPHI, Aix Marseille University | MEPHI, Aix Marseille University | Anthony LEVASSEUR et al |
| EPI_ISL_900486 | France/PAC-IHUCOVID-2380/2020 | 2020-10-11 | Europe/France/Provence-Alpes-C√¥te d' | MEPHI, Aix Marseille University | MEPHI, Aix Marseille University | Anthony LEVASSEUR et al |
| EPI_ISL_900487 | France/PAC-IHUCOVID-2354/2020 | 2020-10-09 | Europe/France/Provence-Alpes-C√¥te d' | MEPHI, Aix Marseille University | MEPHI, Aix Marseille University | Anthony LEVASSEUR et al |
| EPI_ISL_900489 | France/PAC-IHUCOVID-1721/2020 | 2020-09-17 | Europe/France/Provence-Alpes-C√¥te d' | MEPHI, Aix Marseille University | MEPHI, Aix Marseille University | Anthony LEVASSEUR et al |
| EPI_ISL_900492 | Spain/CT-IrsiCaixaLJJCS1/2021 | 2021-01-12 | Europe/Spain/Catalunya/Castellterçol | Althaia. Xarxa Assistencial Universit√†ria de Manresa | IrsiCaixa - Can Ruti CovidSeq | Fundaci√≥ irsiCaixa. Hospital Universitari Germans Trias i Pujol(HUGTiP) et al |
| EPI_ISL_900494 | Spain/CT-IrsiCaixaLJCGS1/2021 | 2021-01-15 | Europe/Spain/Catalunya/Moia | Althaia. Xarxa Assistencial Universit√†ria de Manresa | IrsiCaixa - Can Ruti CovidSeq | Fundaci√≥ irsiCaixa. Hospital Universitari Germans Trias i Pujol(HUGTiP) et al |
| EPI_ISL_900496 | Spain/CT-IrsiCaixaLUUZB1/2021 | 2021-01-12 | Europe/Spain/Catalunya/Castellterçol | Althaia. Xarxa Assistencial Universit√†ria de Manresa | IrsiCaixa - Can Ruti CovidSeq | Fundaci√≥ irsiCaixa. Hospital Universitari Germans Trias i Pujol(HUGTiP) et al |
| EPI_ISL_900506 | France/NAQ-SC2024/2020 | 2020-12-26 | Europe/France/Nouvelle-Aquitaine/Ang | AX BIO OCEAN | CNR Virus des Infections Respiratoires - France SUD | Antonin Bal et al |
| EPI_ISL_900509 | France/ARA-SC2039/2020 | 2020-12-29 | Europe/France/Auvergne-Rh√¥ne-Alpes/ | Mirialis | CNR Virus des Infections Respiratoires - France SUD | Antonin Bal et al |
| EPI_ISL_900513 | France/OCC-SC2115/2020 | 2020-12-31 | Europe/France/Occitanie/Montpellier | Labosud | CNR Virus des Infections Respiratoires - France SUD | Antonin Bal et al |
| EPI_ISL_900514 | France/PAC-SC2101/2020 | 2020-12-21 | Europe/France/Provence-Alpes-C√¥te d' | Biolittoral | CNR Virus des Infections Respiratoires - France SUD | Antonin Bal et al |

| | | | | | | |
|---|---|---|---|---|---|---|
| EPI_ISL_900517 | France/PAC-SC2035/2020 | 2020-12-29 | Europe/France/Provence-Alpes-C√¥te d' | Bioesterel | CNR Virus des Infections Respiratoires - France SUD | Antonin Bal et al |
| EPI_ISL_900531 | France/NAQ-SC2112/2020 | 2020-12-29 | Europe/France/Nouvelle-Aquitaine/Poit | CHU Poitiers | CNR Virus des Infections Respiratoires - France SUD | Antonin Bal et al |
| EPI_ISL_900532 | France/ARA-0001003/2020 | 2020-12-29 | Europe/France/Auvergne-Rh√¥ne-Alpes/ | CNR Virus des Infections Respiratoires - France SUD | CNR Virus des Infections Respiratoires - France SUD | Antonin Bal et al |
| EPI_ISL_900550 | France/ARA-0001026/2020 | 2020-12-29 | Europe/France/Auvergne-Rh√¥ne-Alpes/ | CNR Virus des Infections Respiratoires - France SUD | CNR Virus des Infections Respiratoires - France SUD | Antonin Bal et al |
| EPI_ISL_900557 | France/ARA-126706/2020 | 2020-12-31 | Europe/France/Auvergne-Rh√¥ne-Alpes/ | CNR Virus des Infections Respiratoires - France SUD | CNR Virus des Infections Respiratoires - France SUD | Antonin Bal et al |
| EPI_ISL_900570 | France/IDF-0001028/2020 | 2020-10-29 | Europe/France/Ile de France/Paris | H√¥pital Paris Saint-Joseph | CNR Virus des Infections Respiratoires - France SUD | Antonin Bal et al |
| EPI_ISL_900573 | France/NAQ-0001029/2020 | 2020-10-28 | Europe/France/Nouvelle-Aquitaine/Gue | Centre Hospitalier de Gu√©ret | CNR Virus des Infections Respiratoires - France SUD | Antonin Bal et al |
| EPI_ISL_900580 | Italy/CAM-TIGEM-1485/2020 | 2020-12-12 | Europe/Italy/Campania/ | IZSM | TIGEM | Patrizia Annunziata et al |
| EPI_ISL_900581 | Italy/CAM-TIGEM-1429/2020 | 2020-12-17 | Europe/Italy/Campania/ | IZSM | TIGEM | Patrizia Annunziata et al |
| EPI_ISL_900582 | Italy/CAM-TIGEM-1416/2020 | 2020-12-17 | Europe/Italy/Campania/ | IZSM | TIGEM | Patrizia Annunziata et al |
| EPI_ISL_900595 | Italy/CAM-TIGEM-1413/2020 | 2020-12-18 | Europe/Italy/Campania/ | IZSM | TIGEM | Patrizia Annunziata et al |
| EPI_ISL_900603 | Italy/CAM-TIGEM-1483/2020 | 2020-12-18 | Europe/Italy/Campania/ | IZSM | TIGEM | Patrizia Annunziata et al |
| EPI_ISL_900611 | Italy/CAM-TIGEM-1474/2020 | 2020-12-14 | Europe/Italy/Campania/ | IZSM | TIGEM | Patrizia Annunziata et al |
| EPI_ISL_900628 | Italy/CAM-TIGEM-1400/2020 | 2020-12-17 | Europe/Italy/Campania/ | IZSM | TIGEM | Patrizia Annunziata et al |
| EPI_ISL_900630 | Italy/CAM-TIGEM-1412/2020 | 2020-12-18 | Europe/Italy/Campania/ | IZSM | TIGEM | Patrizia Annunziata et al |
| EPI_ISL_900632 | Italy/CAM-TIGEM-1432/2020 | 2020-12-17 | Europe/Italy/Campania/ | IZSM | TIGEM | Patrizia Annunziata et al |

| | | | | | | |
|---|---|---|---|---|---|---|
| EPI_ISL_900640 | Italy/CAM-TIGEM-1381/2020 | 2020-12-27 | Europe/Italy/Campania/ | IZSM | TIGEM | Patrizia Annunziata et al |
| EPI_ISL_900642 | Italy/CAM-TIGEM-1369/2020 | 2020-12-27 | Europe/Italy/Campania/ | IZSM | TIGEM | Patrizia Annunziata et al |
| EPI_ISL_900644 | Italy/CAM-TIGEM-1380/2020 | 2020-12-27 | Europe/Italy/Campania/ | IZSM | TIGEM | Patrizia Annunziata et al |
| EPI_ISL_900648 | Italy/CAM-TIGEM-1385/2020 | 2020-12-27 | Europe/Italy/Campania/ | IZSM | TIGEM | Patrizia Annunziata et al |
| EPI_ISL_900649 | Italy/CAM-TIGEM-1378/2020 | 2020-12-27 | Europe/Italy/Campania/ | IZSM | TIGEM | Patrizia Annunziata et al |
| EPI_ISL_900650 | Italy/CAM-TIGEM-1359/2020 | 2020-12-27 | Europe/Italy/Campania/ | IZSM | TIGEM | Patrizia Annunziata et al |
| EPI_ISL_900654 | Italy/CAM-TIGEM-1453/2020 | 2020-12-17 | Europe/Italy/Campania/ | IZSM | TIGEM | Patrizia Annunziata et al |
| EPI_ISL_900657 | Italy/CAM-TIGEM-1460/2020 | 2020-12-17 | Europe/Italy/Campania/ | IZSM | TIGEM | Patrizia Annunziata et al |
| EPI_ISL_900672 | Italy/CAM-TIGEM-1440/2020 | 2020-12-17 | Europe/Italy/Campania/ | IZSM | TIGEM | Patrizia Annunziata et al |
| EPI_ISL_900673 | Italy/CAM-TIGEM-1422/2020 | 2020-12-17 | Europe/Italy/Campania/ | IZSM | TIGEM | Patrizia Annunziata et al |
| EPI_ISL_900691 | USA/MT-BHDH-3225/2020 | 2020-06-29 | North America/USA/Montana/Belgrade | Bozeman Health Deaconess Hospital | Wiedenheft lab, Montana State University | Artem Nemudryi et al |
| EPI_ISL_900700 | USA/MT-BHDH-3749/2020 | 2020-07-08 | North America/USA/Montana/Bozeman | Bozeman Health Deaconess Hospital | Wiedenheft lab, Montana State University | Artem Nemudryi et al |
| EPI_ISL_900701 | USA/MT-BHDH-3772/2020 | 2020-07-08 | North America/USA/Montana/Ennis | Bozeman Health Deaconess Hospital | Wiedenheft lab, Montana State University | Artem Nemudryi et al |
| EPI_ISL_900702 | USA/MT-BHDH-3903/2020 | 2020-07-10 | North America/USA/Montana/Bozeman | Bozeman Health Deaconess Hospital | Wiedenheft lab, Montana State University | Artem Nemudryi et al |
| EPI_ISL_900705 | USA/MT-BHDH-4133/2020 | 2020-07-15 | North America/USA/Montana/Bozeman | Bozeman Health Deaconess Hospital | Wiedenheft lab, Montana State University | Artem Nemudryi et al |
| EPI_ISL_900706 | USA/MT-BHDH-4213/2020 | 2020-07-16 | North America/USA/Montana/Bozeman | Bozeman Health Deaconess Hospital | Wiedenheft lab, Montana State University | Artem Nemudryi et al |
| EPI_ISL_900708 | USA/MT-BHDH-4261/2020 | 2020-07-16 | North America/USA/Montana/West Yel | Bozeman Health Deaconess Hospital | Wiedenheft lab, Montana State University | Artem Nemudryi et al |
| EPI_ISL_900711 | USA/MT-BHDH-4312/2020 | 2020-07-18 | North America/USA/Montana/Bozeman | Bozeman Health Deaconess Hospital | Wiedenheft lab, Montana State University | Artem Nemudryi et al |
| EPI_ISL_900733 | USA/MT-BHDH-4586/2020 | 2020-07-22 | North America/USA/Montana/Bozeman | Bozeman Health Deaconess Hospital | Wiedenheft lab, Montana State University | Artem Nemudryi et al |

| | | | | | | |
|---|---|---|---|---|---|---|
| EPI_ISL_900739 | USA/MT-BHDH-5184/2020 | 2020-08-02 | North America/USA/Montana/West Yel | Bozeman Health Deaconess Hospital | Wiedenheft lab, Montana State University | Artem Nemudryi et al |
| EPI_ISL_900742 | USA/OH-ODH-SC177686/2020 | 2020-12-09 | North America/USA/Ohio/ | Ohio Department of Health Laboratory | Ohio Department of Health Laboratory | Holmes et al |
| EPI_ISL_900841 | Japan/PG-10104/2020 | 20-Oct | Asia/Japan/Japan/ | Pathogen Genomics Center, National Institute of Infectious Diseases | Pathogen Genomics Center, National Institute of Infectious Diseases | Tsuyoshi Sekizuka et al |
| EPI_ISL_900850 | Japan/PG-10113/2020 | 20-Jul | Asia/Japan/Japan/ | Pathogen Genomics Center, National Institute of Infectious Diseases | Pathogen Genomics Center, National Institute of Infectious Diseases | Tsuyoshi Sekizuka et al |
| EPI_ISL_900920 | Japan/PG-10183/2020 | 20-Jul | Asia/Japan/Japan/ | Pathogen Genomics Center, National Institute of Infectious Diseases | Pathogen Genomics Center, National Institute of Infectious Diseases | Tsuyoshi Sekizuka et al |
| EPI_ISL_900947 | Japan/PG-10218/2020 | 20-Sep | Asia/Japan/Japan/ | Pathogen Genomics Center, National Institute of Infectious Diseases | Pathogen Genomics Center, National Institute of Infectious Diseases | Tsuyoshi Sekizuka et al |
| EPI_ISL_900949 | Japan/PG-10220/2020 | 20-Sep | Asia/Japan/Japan/ | Pathogen Genomics Center, National Institute of Infectious Diseases | Pathogen Genomics Center, National Institute of Infectious Diseases | Tsuyoshi Sekizuka et al |
| EPI_ISL_900998 | Japan/PG-10269/2020 | 20-Oct | Asia/Japan/Japan/ | Pathogen Genomics Center, National Institute of Infectious Diseases | Pathogen Genomics Center, National Institute of Infectious Diseases | Tsuyoshi Sekizuka et al |
| EPI_ISL_901010 | Japan/PG-10281/2020 | 20-Oct | Asia/Japan/Japan/ | Pathogen Genomics Center, National Institute of Infectious Diseases | Pathogen Genomics Center, National Institute of Infectious Diseases | Tsuyoshi Sekizuka et al |

| | | | | | | |
|---|---|---|---|---|---|---|
| EPI_ISL_901197 | Japan/PG-10473/2020 | 20-Sep | Asia/Japan/Japan/ | Pathogen Genomics Center, National Institute of Infectious Diseases | Pathogen Genomics Center, National Institute of Infectious Diseases | Tsuyoshi Sekizuka et al |
| EPI_ISL_901218 | Japan/PG-10495/2020 | 20-Sep | Asia/Japan/Japan/ | Pathogen Genomics Center, National Institute of Infectious Diseases | Pathogen Genomics Center, National Institute of Infectious Diseases | Tsuyoshi Sekizuka et al |
| EPI_ISL_901398 | Japan/PG-10715/2020 | 20-Oct | Asia/Japan/Japan/ | Pathogen Genomics Center, National Institute of Infectious Diseases | Pathogen Genomics Center, National Institute of Infectious Diseases | Tsuyoshi Sekizuka et al |
| EPI_ISL_901399 | Japan/PG-10716/2020 | 20-Oct | Asia/Japan/Japan/ | Pathogen Genomics Center, National Institute of Infectious Diseases | Pathogen Genomics Center, National Institute of Infectious Diseases | Tsuyoshi Sekizuka et al |
| EPI_ISL_901454 | Japan/PG-10771/2020 | 20-Oct | Asia/Japan/Japan/ | Pathogen Genomics Center, National Institute of Infectious Diseases | Pathogen Genomics Center, National Institute of Infectious Diseases | Tsuyoshi Sekizuka et al |
| EPI_ISL_901473 | Japan/PG-10790/2020 | 20-Oct | Asia/Japan/Japan/ | Gunma Prefectural Institute of Public Health and Environmental Sciences | Pathogen Genomics Center, National Institute of Infectious Diseases | Tsuyoshi Sekizuka et al |
| EPI_ISL_901642 | Japan/PG-10983/2020 | 20-Sep | Asia/Japan/Japan/ | Pathogen Genomics Center, National Institute of Infectious Diseases | Pathogen Genomics Center, National Institute of Infectious Diseases | Tsuyoshi Sekizuka et al |
| EPI_ISL_901992 | Japan/PG-9279/2020 | 20-Sep | Asia/Japan/Japan/ | Gunma Prefectural Institute of Public Health and Environmental Sciences | Pathogen Genomics Center, National Institute of Infectious Diseases | Tsuyoshi Sekizuka et al |

| | | | | | | |
|---|---|---|---|---|---|---|
| EPI_ISL_901999 | Japan/PG-9286/2020 | 20-Sep | Asia/Japan/Japan/ | Gunma Prefectural Institute of Public Health and Environmental Sciences | Pathogen Genomics Center, National Institute of Infectious Diseases | Tsuyoshi Sekizuka et al |
| EPI_ISL_902031 | Japan/PG-9318/2020 | 20-Oct | Asia/Japan/Japan/ | Department of Infectious Diseases, Kobe Institute of Health | Pathogen Genomics Center, National Institute of Infectious Diseases | Tsuyoshi Sekizuka et al |
| EPI_ISL_902085 | Japan/PG-9372/2020 | 20-Sep | Asia/Japan/Japan/ | Pathogen Genomics Center, National Institute of Infectious Diseases | Pathogen Genomics Center, National Institute of Infectious Diseases | Tsuyoshi Sekizuka et al |
| EPI_ISL_902096 | Japan/PG-9383/2020 | 20-Sep | Asia/Japan/Japan/ | Pathogen Genomics Center, National Institute of Infectious Diseases | Pathogen Genomics Center, National Institute of Infectious Diseases | Tsuyoshi Sekizuka et al |
| EPI_ISL_902167 | Japan/PG-9454/2020 | 20-Sep | Asia/Japan/Japan/ | Pathogen Genomics Center, National Institute of Infectious Diseases | Pathogen Genomics Center, National Institute of Infectious Diseases | Tsuyoshi Sekizuka et al |
| EPI_ISL_902267 | Japan/PG-9554/2020 | 20-Jul | Asia/Japan/Japan/ | Pathogen Genomics Center, National Institute of Infectious Diseases | Pathogen Genomics Center, National Institute of Infectious Diseases | Tsuyoshi Sekizuka et al |
| EPI_ISL_902277 | Japan/PG-9564/2020 | 20-Jul | Asia/Japan/Japan/ | Pathogen Genomics Center, National Institute of Infectious Diseases | Pathogen Genomics Center, National Institute of Infectious Diseases | Tsuyoshi Sekizuka et al |
| EPI_ISL_902341 | Japan/PG-9628/2020 | 20-Sep | Asia/Japan/Japan/ | Toyama Institute of Health | Pathogen Genomics Center, National Institute of Infectious Diseases | Tsuyoshi Sekizuka et al |

| | | | | | | |
|---|---|---|---|---|---|---|
| EPI_ISL_902354 | Japan/PG-9642/2020 | 20-May | Asia/Japan/Japan/ | Pathogen Genomics Center, National Institute of Infectious Diseases | Pathogen Genomics Center, National Institute of Infectious Diseases | Tsuyoshi Sekizuka et al |
| EPI_ISL_902436 | Japan/PG-9725/2020 | 20-Oct | Asia/Japan/Japan/ | Pathogen Genomics Center, National Institute of Infectious Diseases | Pathogen Genomics Center, National Institute of Infectious Diseases | Tsuyoshi Sekizuka et al |
| EPI_ISL_902525 | Japan/PG-9815/2020 | 20-Oct | Asia/Japan/Japan/ | Department of Infectious Diseases, Kobe Institute of Health | Pathogen Genomics Center, National Institute of Infectious Diseases | Tsuyoshi Sekizuka et al |
| EPI_ISL_902589 | Japan/PG-9880/2020 | 20-Jul | Asia/Japan/Japan/ | Pathogen Genomics Center, National Institute of Infectious Diseases | Pathogen Genomics Center, National Institute of Infectious Diseases | Tsuyoshi Sekizuka et al |
| EPI_ISL_902689 | Japan/PG-8798/2020 | 20-Aug | Asia/Japan/Japan/ | Pathogen Genomics Center, National Institute of Infectious Diseases | Pathogen Genomics Center, National Institute of Infectious Diseases | Tsuyoshi Sekizuka et al |
| EPI_ISL_902740 | Spain/CT-IrsiCaixaLEKAP1/2020 | 2020-08-21 | Europe/Spain/Catalunya/Badalona | Hospital Universitari Germans Trias i Pujol (HUGTiP) / Fundaci√≥ Lluita contra la SIDA (FLSida) | IrsiCaixa - Can Ruti CovidSeq | Fundaci√≥ irsiCaixa. Hospital Universitari Germans Trias i Pujol(HUGTiP) et al |
| EPI_ISL_902743 | Spain/CT-IrsiCaixaLOFRV1/2020 | 2020-08-21 | Europe/Spain/Catalunya/Badalona | Hospital Universitari Germans Trias i Pujol (HUGTiP) / Fundaci√≥ Lluita contra la SIDA (FLSida) | IrsiCaixa - Can Ruti CovidSeq | Fundaci√≥ irsiCaixa. Hospital Universitari Germans Trias i Pujol(HUGTiP) et al |
| EPI_ISL_902750 | Italy/LAZ-INMI-166/2021 | 2021-01-12 | Europe/Italy/Lazio/ | "Ospedale ""F. Spaziani"" Frosinone" | INMI Lazzaro Spallanzani IRCCS | F Messina et al |
| EPI_ISL_902751 | Italy/LAZ-INMI-167/2021 | 2021-01-12 | Europe/Italy/Lazio/ | Ospedale Sandro Pertini ASL Roma2 | INMI Lazzaro Spallanzani IRCCS | B Bartolini et al |
| EPI_ISL_902752 | Italy/LAZ-INMI-168/2021 | 2021-01-12 | Europe/Italy/Lazio/ | Ospedale Sandro Pertini ASL Roma2 | INMI Lazzaro Spallanzani IRCCS | O Butera et al |
| EPI_ISL_902753 | Italy/LAZ-INMI-169/2021 | 2021-01-14 | Europe/Italy/Lazio/ | Azienda Ospedaliera San Camillo Forlanini | INMI Lazzaro Spallanzani IRCCS | C.E.M Gruber et al |

| | | | | | | |
|---|---|---|---|---|---|---|
| EPI_ISL_902754 | Italy/LAZ-INMI-172/2021 | 2021-01-15 | Europe/Italy/Lazio/ | University Hospital Sant'Andrea-Sapienza | INMI Lazzaro Spallanzani IRCCS | B Bartolini et al |
| EPI_ISL_902756 | Italy/LAZ-INMI-174/2021 | 2021-01-18 | Europe/Italy/Lazio/ | Fondazione Policlinico Universitario ‚ÄúA. Gemelli‚Äù IRCCS | INMI Lazzaro Spallanzani IRCCS | M Rueca et al |
| EPI_ISL_902759 | Finland/1T10DD3_S152/2021 | 2021-01-01 | Europe/Finland/Uusimaa/ | Department of Virology and Immunology, University of Helsinki and Helsinki University Hospital, Huslab Finland | Department of Virology, Faculty of Medicine, University of Helsinki, Helsinki, Finland | Teemu Smura et al |
| EPI_ISL_902783 | Finland/26MR7CC1_S240/2020 | 2020-11-26 | Europe/Finland/Uusimaa/ | Department of Virology and Immunology, University of Helsinki and Helsinki University Hospital, Huslab Finland | Department of Virology, Faculty of Medicine, University of Helsinki, Helsinki, Finland | Teemu Smura et al |
| EPI_ISL_902785 | Finland/26MR7CD1_S237/2020 | 2020-11-26 | Europe/Finland/Uusimaa/ | Department of Virology and Immunology, University of Helsinki and Helsinki University Hospital, Huslab Finland | Department of Virology, Faculty of Medicine, University of Helsinki, Helsinki, Finland | Teemu Smura et al |
| EPI_ISL_902825 | Finland/28MR2CD12_S323/2020 | 2020-11-28 | Europe/Finland/Uusimaa/ | Department of Virology and Immunology, University of Helsinki and Helsinki University Hospital, Huslab Finland | Department of Virology, Faculty of Medicine, University of Helsinki, Helsinki, Finland | Teemu Smura et al |
| EPI_ISL_902879 | Finland/31J14DC5_S134/2020 | 2020-12-31 | Europe/Finland/Uusimaa/ | Department of Virology and Immunology, University of Helsinki and Helsinki University Hospital, Huslab Finland | Department of Virology, Faculty of Medicine, University of Helsinki, Helsinki, Finland | Teemu Smura et al |
| EPI_ISL_902895 | Finland/8T19DE1_S176/2021 | 2021-01-08 | Europe/Finland/Uusimaa/ | Department of Virology and Immunology, University of Helsinki and Helsinki University Hospital, Huslab Finland | Department of Virology, Faculty of Medicine, University of Helsinki, Helsinki, Finland | Teemu Smura et al |

| | | | | | | |
|---|---|---|---|---|---|---|
| EPI_ISL_902917 | Indonesia/KB-TUH-C2111237/2020 | 2020-08-31 | Asia/Indonesia/West Kalimantan/ | Tanjungpura University Hospital | Tanjungpura University Hospital | Andriani et al |
| EPI_ISL_902918 | Indonesia/KB-TUH-C2114692/2020 | 2020-09-05 | Asia/Indonesia/West Kalimantan/ | Tanjungpura University Hospital | Tanjungpura University Hospital | Andriani et al |
| EPI_ISL_902919 | Indonesia/KB-TUH-C2122618/2020 | 2020-09-28 | Asia/Indonesia/West Kalimantan/ | Tanjungpura University Hospital | Tanjungpura University Hospital | Andriani et al |
| EPI_ISL_902921 | Indonesia/KB-TUH-C2133373/2020 | 2020-10-15 | Asia/Indonesia/West Kalimantan/ | Tanjungpura University Hospital | Tanjungpura University Hospital | Puji Astuti et al |
| EPI_ISL_902924 | Indonesia/KB-TUH-C2131682/2020 | 2020-10-23 | Asia/Indonesia/West Kalimantan/ | Tanjungpura University Hospital | Tanjungpura University Hospital | Andriani et al |
| EPI_ISL_902925 | USA/CA-LACPHL-AF00251/2021 | 2021-01-09 | North America/USA/California/Los Ange | MD Laboratories | Los Angeles County PHL | P. Hemarajata et al. et al |
| EPI_ISL_902929 | USA/MD-MDH-0670/2021 | 2021-01-12 | North America/USA/Maryland/ | Maryland Public Health Laboratory | Maryland Public Health Laboratory | Maryland Department of Health Laboratories Administration et al |
| EPI_ISL_902936 | USA/MD-MDH-0672/2021 | 2021-01-12 | North America/USA/Maryland/ | Maryland Public Health Laboratory | Maryland Public Health Laboratory | Maryland Department of Health Laboratories Administration et al |
| EPI_ISL_902938 | USA/MD-MDH-0676/2021 | 2021-01-13 | North America/USA/Maryland/ | Maryland Public Health Laboratory | Maryland Public Health Laboratory | Maryland Department of Health Laboratories Administration et al |
| EPI_ISL_902987 | USA/IN-Lilly-IPB0173-02bc20/2020 | 2020-03-23 | North America/USA/Indiana/Indianapoli | Clinical Diagnostics Laboratory, Diagnostic & Experimental Pathology, Lilly Research Laboratories | Clinical Diagnostics Laboratory, Diagnostic & Experimental Pathology, Lilly Research Laboratories | Tim Holzer et al |
| EPI_ISL_902996 | USA/WA-S3710/2020 | 2020-11-17 | North America/USA/Washington/King C | Seattle Flu Study | Seattle Flu Study | Deborah A. Nickerson et al |
| EPI_ISL_903015 | USA/WA-S3729/2020 | 2020-11-30 | North America/USA/Washington/King C | Seattle Flu Study | Seattle Flu Study | Deborah A. Nickerson et al |
| EPI_ISL_903018 | USA/WA-S3732/2020 | 2020-12-01 | North America/USA/Washington/King C | Seattle Flu Study | Seattle Flu Study | Deborah A. Nickerson et al |
| EPI_ISL_903021 | USA/WA-S3735/2020 | 2020-12-21 | North America/USA/Washington/King C | Seattle Flu Study | Seattle Flu Study | Deborah A. Nickerson et al |
| EPI_ISL_903023 | USA/WA-S3737/2020 | 2020-12-21 | North America/USA/Washington/King C | Seattle Flu Study | Seattle Flu Study | Deborah A. Nickerson et al |

| | | | | | | |
|---|---|---|---|---|---|---|
| EPI_ISL_903027 | USA/WA-S3741/2020 | 2020-12-22 | North America/USA/Washington/King C | Seattle Flu Study | Seattle Flu Study | Deborah A. Nickerson et al |
| EPI_ISL_903042 | USA/WA-S3756/2020 | 2020-12-28 | North America/USA/Washington/King C | Seattle Flu Study | Seattle Flu Study | Deborah A. Nickerson et al |
| EPI_ISL_903053 | USA/WA-S3767/2020 | 2020-12-30 | North America/USA/Washington/King C | Seattle Flu Study | Seattle Flu Study | Deborah A. Nickerson et al |
| EPI_ISL_903075 | USA/WA-S3789/2021 | 2021-01-06 | North America/USA/Washington/King C | Seattle Flu Study | Seattle Flu Study | Deborah A. Nickerson et al |
| EPI_ISL_903078 | USA/WA-S3792/2021 | 2021-01-07 | North America/USA/Washington/Pierce | Seattle Flu Study | Seattle Flu Study | Deborah A. Nickerson et al |
| EPI_ISL_903092 | USA/WA-S3806/2021 | 2021-01-02 | North America/USA/Washington/King C | Seattle Flu Study | Seattle Flu Study | Deborah A. Nickerson et al |
| EPI_ISL_903094 | USA/WA-S3808/2021 | 2021-01-02 | North America/USA/Washington/King C | Seattle Flu Study | Seattle Flu Study | Deborah A. Nickerson et al |
| EPI_ISL_903103 | USA/WA-S3817/2021 | 2021-01-03 | North America/USA/Washington/King C | Seattle Flu Study | Seattle Flu Study | Deborah A. Nickerson et al |
| EPI_ISL_903115 | USA/WA-S3829/2020 | 2020-12-29 | North America/USA/Washington/King C | Washington State Department of Health | Seattle Flu Study | Deborah A. Nickerson et al |
| EPI_ISL_903126 | USA/WA-S3840/2020 | 2020-12-31 | North America/USA/Washington/Yakim | Washington State Department of Health | Seattle Flu Study | Deborah A. Nickerson et al |
| EPI_ISL_903129 | USA/WA-S3843/2021 | 2021-01-02 | North America/USA/Washington/Yakim | Washington State Department of Health | Seattle Flu Study | Deborah A. Nickerson et al |
| EPI_ISL_903131 | USA/WA-S3845/2021 | 2021-01-02 | North America/USA/Washington/Yakim | Washington State Department of Health | Seattle Flu Study | Deborah A. Nickerson et al |
| EPI_ISL_903134 | USA/WA-S3848/2021 | 2021-01-03 | North America/USA/Washington/Yakim | Washington State Department of Health | Seattle Flu Study | Deborah A. Nickerson et al |
| EPI_ISL_903141 | USA/WA-S3855/2021 | 2021-01-04 | North America/USA/Washington/Yakim | Washington State Department of Health | Seattle Flu Study | Deborah A. Nickerson et al |
| EPI_ISL_903146 | USA/WA-S3860/2021 | 2021-01-04 | North America/USA/Washington/Yakim | Washington State Department of Health | Seattle Flu Study | Deborah A. Nickerson et al |
| EPI_ISL_903147 | USA/WA-S3861/2021 | 2021-01-04 | North America/USA/Washington/Yakim | Washington State Department of Health | Seattle Flu Study | Deborah A. Nickerson et al |
| EPI_ISL_903150 | USA/WA-S3864/2021 | 2021-01-04 | North America/USA/Washington/Spoka | Washington State Department of Health | Seattle Flu Study | Deborah A. Nickerson et al |
| EPI_ISL_903153 | USA/WA-S3867/2021 | 2021-01-05 | North America/USA/Washington/Yakim | Washington State Department of Health | Seattle Flu Study | Deborah A. Nickerson et al |
| EPI_ISL_903168 | USA/WA-S3882/2021 | 2021-01-04 | North America/USA/Washington/Yakim | Washington State Department of Health | Seattle Flu Study | Deborah A. Nickerson et al |
| EPI_ISL_903176 | USA/WA-S3890/2021 | 2021-01-05 | North America/USA/Washington/Yakim | Washington State Department of Health | Seattle Flu Study | Deborah A. Nickerson et al |
| EPI_ISL_903187 | USA/WA-S3901/2021 | 2021-01-05 | North America/USA/Washington/King C | Seattle Flu Study | Seattle Flu Study | Deborah A. Nickerson et al |
| EPI_ISL_903192 | USA/WA-S3906/2021 | 2021-01-06 | North America/USA/Washington/King C | Seattle Flu Study | Seattle Flu Study | Deborah A. Nickerson et al |
| EPI_ISL_903199 | USA/WA-S3913/2021 | 2021-01-07 | North America/USA/Washington/King C | Seattle Flu Study | Seattle Flu Study | Deborah A. Nickerson et al |
| EPI_ISL_903202 | USA/WA-S3916/2021 | 2021-01-07 | North America/USA/Washington/King C | Seattle Flu Study | Seattle Flu Study | Deborah A. Nickerson et al |
| EPI_ISL_903207 | USA/WA-S3921/2021 | 2021-01-08 | North America/USA/Washington/King C | Seattle Flu Study | Seattle Flu Study | Deborah A. Nickerson et al |
| EPI_ISL_903209 | USA/MI-UM-10038012135/2021 | 2021-01-26 | North America/USA/Michigan/ | University of Michigan Clinical Microbiology Laboratory | Lauring Lab, University of Michigan, Department of Microbiology and Immunology | Valesano et al |

| | | | | | | |
|---|---|---|---|---|---|---|
| EPI_ISL_903215 | USA/MI-UM-10038019936/2021 | 2021-01-25 | North America/USA/Michigan/ | University of Michigan Clinical Microbiology Laboratory | Lauring Lab, University of Michigan, Department of Microbiology and Immunology | Valesano et al |
| EPI_ISL_903216 | USA/MI-UM-10038020799/2021 | 2021-01-25 | North America/USA/Michigan/ | University of Michigan Clinical Microbiology Laboratory | Lauring Lab, University of Michigan, Department of Microbiology and Immunology | Valesano et al |
| EPI_ISL_903218 | USA/IN-Lilly-IPB0173-03bc02/2020 | 2020-03-23 | North America/USA/Indiana/Indianapoli | Clinical Diagnostics Laboratory, Diagnostic & Experimental Pathology, Lilly Research Laboratories | Clinical Diagnostics Laboratory, Diagnostic & Experimental Pathology, Lilly Research Laboratories | Tim Holzer et al |
| EPI_ISL_903229 | USA/MN-MDH-2444/2020 | 2020-12-24 | North America/USA/Minnesota/ | M Health Fairview | Minnesota Department of Health, Public Health Laboratory | Alexandra Lorentz et al |
| EPI_ISL_903230 | USA/MN-MDH-2430/2020 | 2020-12-10 | North America/USA/Minnesota/ | M Health Fairview | Minnesota Department of Health, Public Health Laboratory | Alexandra Lorentz et al |
| EPI_ISL_903245 | USA/MN-MDH-2423/2020 | 2020-12-04 | North America/USA/Minnesota/ | M Health Fairview | Minnesota Department of Health, Public Health Laboratory | Alexandra Lorentz et al |
| EPI_ISL_903249 | USA/MN-MDH-2427/2020 | 2020-12-11 | North America/USA/Minnesota/ | M Health Fairview | Minnesota Department of Health, Public Health Laboratory | Alexandra Lorentz et al |
| EPI_ISL_903253 | USA/MN-MDH-2433/2020 | 2020-12-08 | North America/USA/Minnesota/ | M Health Fairview | Minnesota Department of Health, Public Health Laboratory | Alexandra Lorentz et al |

| | | | | | | |
|---|---|---|---|---|---|---|
| EPI_ISL_903264 | USA/MN-MDH-2450/2020 | 2020-12-27 | North America/USA/Minnesota/ | M Health Fairview | Minnesota Department of Health, Public Health Laboratory | Alexandra Lorentz et al |
| EPI_ISL_903267 | USA/MN-MDH-2454/2020 | 2020-12-26 | North America/USA/Minnesota/ | M Health Fairview | Minnesota Department of Health, Public Health Laboratory | Alexandra Lorentz et al |
| EPI_ISL_903272 | USA/MN-MDH-2459/2020 | 2020-12-26 | North America/USA/Minnesota/ | M Health Fairview | Minnesota Department of Health, Public Health Laboratory | Alexandra Lorentz et al |
| EPI_ISL_903279 | USA/MN-MDH-2467/2020 | 2020-12-31 | North America/USA/Minnesota/ | M Health Fairview | Minnesota Department of Health, Public Health Laboratory | Alexandra Lorentz et al |
| EPI_ISL_903284 | USA/MN-MDH-2472/2021 | 2021-01-01 | North America/USA/Minnesota/ | M Health Fairview | Minnesota Department of Health, Public Health Laboratory | Alexandra Lorentz et al |
| EPI_ISL_903298 | USA/MN-MDH-2489/2021 | 2021-01-04 | North America/USA/Minnesota/ | M Health Fairview | Minnesota Department of Health, Public Health Laboratory | Alexandra Lorentz et al |
| EPI_ISL_903306 | USA/MN-MDH-2498/2021 | 2021-01-03 | North America/USA/Minnesota/ | M Health Fairview | Minnesota Department of Health, Public Health Laboratory | Alexandra Lorentz et al |
| EPI_ISL_903311 | USA/MN-MDH-2504/2021 | 2021-01-03 | North America/USA/Minnesota/ | M Health Fairview | Minnesota Department of Health, Public Health Laboratory | Alexandra Lorentz et al |
| EPI_ISL_903312 | USA/IA-UIHC-MO01/2020 | 2020-06-08 | North America/USA/Iowa/Johnson Coun | University of Iowa Hospitals & Clinics, Microbiology Laboratory | University or Iowa, Lung Biology and Cystic Fibrosis Research Center, Pezzulo Lab | Miguel E Ortiz et al |

| | | | | | | |
|---|---|---|---|---|---|---|
| EPI_ISL_903314 | USA/IA-UIHC-MO03/2020 | 2020-06-09 | North America/USA/Iowa/Johnson Coun | University of Iowa Hospitals & Clinics, Microbiology Laboratory | University or Iowa, Lung Biology and Cystic Fibrosis Research Center, Pezzulo Lab | Miguel E Ortiz et al |
| EPI_ISL_903329 | USA/IA-UIHC-MO19/2020 | 2020-09-03 | North America/USA/Iowa/Johnson Coun | University of Iowa Hospitals & Clinics, Microbiology Laboratory | University or Iowa, Lung Biology and Cystic Fibrosis Research Center, Pezzulo Lab | Miguel E Ortiz et al |
| EPI_ISL_903334 | USA/IA-UIHC-MO24/2020 | 2020-09-09 | North America/USA/Iowa/Johnson Coun | University of Iowa Hospitals & Clinics, Microbiology Laboratory | University or Iowa, Lung Biology and Cystic Fibrosis Research Center, Pezzulo Lab | Miguel E Ortiz et al |
| EPI_ISL_903335 | USA/IA-UIHC-MO25/2020 | 2020-09-09 | North America/USA/Iowa/Johnson Coun | University of Iowa Hospitals & Clinics, Microbiology Laboratory | University or Iowa, Lung Biology and Cystic Fibrosis Research Center, Pezzulo Lab | Miguel E Ortiz et al |
| EPI_ISL_903336 | USA/IA-UIHC-MO26/2020 | 2020-09-10 | North America/USA/Iowa/Johnson Coun | University of Iowa Hospitals & Clinics, Microbiology Laboratory | University or Iowa, Lung Biology and Cystic Fibrosis Research Center, Pezzulo Lab | Miguel E Ortiz et al |
| EPI_ISL_903338 | USA/IA-UIHC-MO28/2020 | 2020-09-10 | North America/USA/Iowa/Johnson Coun | University of Iowa Hospitals & Clinics, Microbiology Laboratory | University or Iowa, Lung Biology and Cystic Fibrosis Research Center, Pezzulo Lab | Miguel E Ortiz et al |
| EPI_ISL_903347 | Turkey/HSGM-1242/2021 | 2021-01-11 | Europe/Turkey/Turkey/ | Ministry of Health Turkey | Ministry of Health Turkey | Fatma Bayrakdar et al |
| EPI_ISL_903348 | USA/WY-WPHL-21004910/2021 | 2021-01-13 | North America/USA/Wyoming/ | Wyoming Public Health Laboratory | Wyoming Public Health Laboratory | Noah Hull et al |

| | | | | | | |
|---|---|---|---|---|---|---|
| EPI_ISL_903356 | USA/WY-WPHL-21008893/2021 | 2021-01-25 | North America/USA/Wyoming/ | Wyoming Public Health Laboratory | Wyoming Public Health Laboratory | Noah Hull et al |
| EPI_ISL_903368 | USA/CA-LACPHL-AF00097/2020 | 2020-12-22 | North America/USA/California/Los Ange | Los Angeles County PHL | Los Angeles County PHL | P. Hemarajata et al. et al |
| EPI_ISL_903374 | USA/CA-LACPHL-AF00230/2021 | 2021-01-09 | North America/USA/California/Los Ange | MD Laboratories | Los Angeles County PHL | P. Hemarajata et al. et al |
| EPI_ISL_903382 | Kuwait/SKS00111-JAH/2020 | 2020-12-26 | Asia/Kuwait/Al Asimah/Kuwait City | MOH - Jaber Al-Ahmad Hospital (Innovation Research Laboratory) | MOH - Jaber Al-Ahmad Hospital (Innovation Research Laboratory) | Salman Al-Sabah et al |
| EPI_ISL_903384 | Kuwait/SKS00115-JAH/2020 | 2020-12-26 | Asia/Kuwait/Al Asimah/Kuwait City | MOH - Jaber Al-Ahmad Hospital (Innovation Research Laboratory) | MOH - Jaber Al-Ahmad Hospital (Innovation Research Laboratory) | Salman Al-Sabah et al |
| EPI_ISL_903397 | USA/CA-QDX-4000/2021 | 2021-01-09 | North America/USA/California/ | Quest Diagnostics | Quest Diagnostics | Rosenthal et al |
| EPI_ISL_903399 | USA/CA-QDX-4012/2021 | 2021-01-09 | North America/USA/California/ | Quest Diagnostics | Quest Diagnostics | Rosenthal et al |
| EPI_ISL_903401 | USA/CA-QDX-4062/2021 | 2021-01-09 | North America/USA/California/ | Quest Diagnostics | Quest Diagnostics | Rosenthal et al |
| EPI_ISL_903410 | USA/CA-QDX-3988/2021 | 2021-01-09 | North America/USA/California/ | Quest Diagnostics | Quest Diagnostics | Rosenthal et al |
| EPI_ISL_903420 | USA/CA-QDX-4004/2021 | 2021-01-08 | North America/USA/California/ | Quest Diagnostics | Quest Diagnostics | Rosenthal et al |
| EPI_ISL_903433 | USA/CA-QDX-4025/2021 | 2021-01-09 | North America/USA/California/ | Quest Diagnostics | Quest Diagnostics | Rosenthal et al |
| EPI_ISL_903434 | USA/CA-QDX-4027/2021 | 2021-01-08 | North America/USA/California/ | Quest Diagnostics | Quest Diagnostics | Rosenthal et al |
| EPI_ISL_903437 | USA/CA-QDX-4033/2021 | 2021-01-07 | North America/USA/California/ | Quest Diagnostics | Quest Diagnostics | Rosenthal et al |
| EPI_ISL_903440 | USA/CA-QDX-4037/2021 | 2021-01-08 | North America/USA/California/ | Quest Diagnostics | Quest Diagnostics | Rosenthal et al |
| EPI_ISL_903441 | USA/CA-QDX-4038/2021 | 2021-01-08 | North America/USA/California/ | Quest Diagnostics | Quest Diagnostics | Rosenthal et al |
| EPI_ISL_903443 | USA/CA-QDX-4040/2021 | 2021-01-08 | North America/USA/California/ | Quest Diagnostics | Quest Diagnostics | Rosenthal et al |
| EPI_ISL_903446 | USA/CA-QDX-4044/2021 | 2021-01-08 | North America/USA/California/ | Quest Diagnostics | Quest Diagnostics | Rosenthal et al |
| EPI_ISL_903453 | USA/CA-QDX-4058/2021 | 2021-01-09 | North America/USA/California/ | Quest Diagnostics | Quest Diagnostics | Rosenthal et al |
| EPI_ISL_903456 | USA/CA-QDX-4061/2021 | 2021-01-09 | North America/USA/California/ | Quest Diagnostics | Quest Diagnostics | Rosenthal et al |
| EPI_ISL_903457 | USA/CA-QDX-4063/2021 | 2021-01-08 | North America/USA/California/ | Quest Diagnostics | Quest Diagnostics | Rosenthal et al |
| EPI_ISL_903464 | USA/CA-QDX-4073/2021 | 2021-01-09 | North America/USA/California/ | Quest Diagnostics | Quest Diagnostics | Rosenthal et al |
| EPI_ISL_903470 | USA/CA-QDX-4081/2021 | 2021-01-08 | North America/USA/California/ | Quest Diagnostics | Quest Diagnostics | Rosenthal et al |
| EPI_ISL_903480 | USA/CA-QDX-4098/2021 | 2021-01-08 | North America/USA/California/ | Quest Diagnostics | Quest Diagnostics | Rosenthal et al |

| | | | | | | |
|---|---|---|---|---|---|---|
| EPI_ISL_903482 | USA/CA-QDX-4102/2021 | 2021-01-09 | North America/USA/California/ | Quest Diagnostics | Quest Diagnostics | Rosenthal et al |
| EPI_ISL_903491 | USA/CA-QDX-4117/2021 | 2021-01-08 | North America/USA/California/ | Quest Diagnostics | Quest Diagnostics | Rosenthal et al |
| EPI_ISL_903492 | USA/CA-QDX-4118/2021 | 2021-01-08 | North America/USA/USA/ | Quest Diagnostics | Quest Diagnostics | Rosenthal et al |
| EPI_ISL_903506 | USA/CA-QDX-4139/2021 | 2021-01-08 | North America/USA/California/ | Quest Diagnostics | Quest Diagnostics | Rosenthal et al |
| EPI_ISL_903508 | USA/CA-QDX-4142/2021 | 2021-01-09 | North America/USA/California/ | Quest Diagnostics | Quest Diagnostics | Rosenthal et al |
| EPI_ISL_903517 | USA/CA-QDX-4154/2021 | 2021-01-08 | North America/USA/California/ | Quest Diagnostics | Quest Diagnostics | Rosenthal et al |
| EPI_ISL_903520 | USA/CA-QDX-4159/2021 | 2021-01-08 | North America/USA/California/ | Quest Diagnostics | Quest Diagnostics | Rosenthal et al |
| EPI_ISL_903527 | USA/CT-QDX-4006/2021 | 2021-01-06 | North America/USA/Connecticut/ | Quest Diagnostics | Quest Diagnostics | Rosenthal et al |
| EPI_ISL_903534 | USA/MA-QDX-4021/2021 | 2021-01-06 | North America/USA/Massachusetts/ | Quest Diagnostics | Quest Diagnostics | Rosenthal et al |
| EPI_ISL_903538 | USA/MA-QDX-4068/2021 | 2021-01-06 | North America/USA/Massachusetts/ | Quest Diagnostics | Quest Diagnostics | Rosenthal et al |
| EPI_ISL_903540 | USA/MA-QDX-4083/2021 | 2021-01-07 | North America/USA/Massachusetts/ | Quest Diagnostics | Quest Diagnostics | Rosenthal et al |
| EPI_ISL_903557 | USA/NV-QDX-4049/2021 | 2021-01-08 | North America/USA/Nevada/ | Quest Diagnostics | Quest Diagnostics | Rosenthal et al |
| EPI_ISL_903560 | USA/NV-QDX-4089/2021 | 2021-01-08 | North America/USA/Nevada/ | Quest Diagnostics | Quest Diagnostics | Rosenthal et al |
| EPI_ISL_903561 | USA/NV-QDX-4129/2021 | 2021-01-08 | North America/USA/Nevada/ | Quest Diagnostics | Quest Diagnostics | Rosenthal et al |
| EPI_ISL_903563 | USA/OK-QDX-3985/2021 | 2021-01-07 | North America/USA/Oklahoma/ | Quest Diagnostics | Quest Diagnostics | Rosenthal et al |
| EPI_ISL_903569 | USA/NC-CDC-2-3767474/2020 | 2020-11-30 | North America/USA/North Carolina/ | NC State Laboratory of Public Health | Genomics and Discovery, Respiratory Viruses Branch, Division of Viral Diseases, Centers for Disease Control and Prevention | Krista Queen et al |
| EPI_ISL_903571 | USA/ID-CDC-2-3769187/2020 | 2020-12-21 | North America/USA/Idaho/ | ID Bureau of Laboratories | Genomics and Discovery, Respiratory Viruses Branch, Division of Viral Diseases, Centers for Disease Control and Prevention | Krista Queen et al |

| | | | | | | |
|---|---|---|---|---|---|---|
| EPI_ISL_903572 | USA/VT-CDC-2-3766735/2020 | 2020-05-07 | North America/USA/Vermont/ | VT Dept. of Health Laboratory | Genomics and Discovery, Respiratory Viruses Branch, Division of Viral Diseases, Centers for Disease Control and Prevention | Krista Queen et al |
| EPI_ISL_903575 | USA/MI-CDC-2-3768694/2020 | 2020-12-16 | North America/USA/Michigan/ | MI - Michigan Department of Health and Human Services - Bureau of Laboratories | Genomics and Discovery, Respiratory Viruses Branch, Division of Viral Diseases, Centers for Disease Control and Prevention | Krista Queen et al |
| EPI_ISL_903582 | USA/TN-CDC-2-3767578/2020 | 2020-12-22 | North America/USA/Tennessee/ | TN Division of Laboratory Services | Genomics and Discovery, Respiratory Viruses Branch, Division of Viral Diseases, Centers for Disease Control and Prevention | Krista Queen et al |
| EPI_ISL_903589 | USA/MI-CDC-2-3769293/2020 | 2020-12-16 | North America/USA/Michigan/ | MI - Michigan Department of Health and Human Services - Bureau of Laboratories | Genomics and Discovery, Respiratory Viruses Branch, Division of Viral Diseases, Centers for Disease Control and Prevention | Krista Queen et al |

| | | | | | | |
|---|---|---|---|---|---|---|
| EPI_ISL_903590 | USA/PA-CDC-2-3774004/2020 | 2020-12-08 | North America/USA/Pennsylvania/ | PA Department of Health, Bureau of Laboratories | Genomics and Discovery, Respiratory Viruses Branch, Division of Viral Diseases, Centers for Disease Control and Prevention | Krista Queen et al |
| EPI_ISL_903603 | USA/ID-CDC-2-3769290/2020 | 2020-12-22 | North America/USA/Idaho/ | ID Bureau of Laboratories | Genomics and Discovery, Respiratory Viruses Branch, Division of Viral Diseases, Centers for Disease Control and Prevention | Krista Queen et al |
| EPI_ISL_903612 | USA/OR-CDC-2-3766803/2020 | 2020-12-09 | North America/USA/Oregon/ | OR State PHL-Virology/Immunology Section | Genomics and Discovery, Respiratory Viruses Branch, Division of Viral Diseases, Centers for Disease Control and Prevention | Krista Queen et al |
| EPI_ISL_903618 | USA/MN-CDC-2-3766677/2020 | 2020-12-11 | North America/USA/Minnesota/ | MN PHL Division, Minnesota Department of Health | Genomics and Discovery, Respiratory Viruses Branch, Division of Viral Diseases, Centers for Disease Control and Prevention | Krista Queen et al |

| | | | | | | |
|---|---|---|---|---|---|---|
| EPI_ISL_903641 | USA/RI-CDC-2-3768598/2020 | 2020-12-14 | North America/USA/Rhode Island/ | RI State Health Laboratories | Genomics and Discovery, Respiratory Viruses Branch, Division of Viral Diseases, Centers for Disease Control and Prevention | Krista Queen et al |
| EPI_ISL_903645 | USA/WV-CDC-2-3766719/2020 | 2020-11-13 | North America/USA/West Virginia/ | WVDHHR - Office of Laboratory Services | Genomics and Discovery, Respiratory Viruses Branch, Division of Viral Diseases, Centers for Disease Control and Prevention | Krista Queen et al |
| EPI_ISL_903648 | USA/CO-CDC-2-3774010/2020 | 2020-12-18 | North America/USA/Colorado/ | CO Dept. of Public Health and Environment, Lab Services Division | Genomics and Discovery, Respiratory Viruses Branch, Division of Viral Diseases, Centers for Disease Control and Prevention | Krista Queen et al |
| EPI_ISL_903652 | USA/NC-CDC-2-3769285/2020 | 2020-12-15 | North America/USA/North Carolina/ | NC State Laboratory of Public Health | Genomics and Discovery, Respiratory Viruses Branch, Division of Viral Diseases, Centers for Disease Control and Prevention | Krista Queen et al |

| | | | | | | |
|---|---|---|---|---|---|---|
| EPI_ISL_903657 | USA/VT-CDC-2-3767593/2020 | 2020-12-26 | North America/USA/Vermont/ | VT Dept. of Health Laboratory | Genomics and Discovery, Respiratory Viruses Branch, Division of Viral Diseases, Centers for Disease Control and Prevention | Krista Queen et al |
| EPI_ISL_903662 | USA/PA-CDC-2-3773946/2020 | 2020-12-08 | North America/USA/Pennsylvania/ | PA Department of Health, Bureau of Laboratories | Genomics and Discovery, Respiratory Viruses Branch, Division of Viral Diseases, Centers for Disease Control and Prevention | Krista Queen et al |
| EPI_ISL_903667 | USA/NH-CDC-2-3769196/2020 | 2020-12-20 | North America/USA/New Hampshire/ | NH Dept. of Health and Human Services Public Health Labs | Genomics and Discovery, Respiratory Viruses Branch, Division of Viral Diseases, Centers for Disease Control and Prevention | Krista Queen et al |
| EPI_ISL_903668 | USA/SC-CDC-2-3768775/2021 | 2021-01-02 | North America/USA/South Carolina/ | SC Dept of Health and Env. Control-Bureau of Laboratories | Genomics and Discovery, Respiratory Viruses Branch, Division of Viral Diseases, Centers for Disease Control and Prevention | Krista Queen et al |

| | | | | | | |
|---|---|---|---|---|---|---|
| EPI_ISL_903677 | USA/MN-CDC-2-3767482/2020 | 2020-12-28 | North America/USA/Minnesota/ | MN PHL Division, Minnesota Department of Health | Genomics and Discovery, Respiratory Viruses Branch, Division of Viral Diseases, Centers for Disease Control and Prevention | Krista Queen et al |
| EPI_ISL_903678 | USA/NJ-CDC-2-3768082/2020 | 2020-12-30 | North America/USA/New Jersey/ | NJ Public Health and Environmental Laboratories | Genomics and Discovery, Respiratory Viruses Branch, Division of Viral Diseases, Centers for Disease Control and Prevention | Krista Queen et al |
| EPI_ISL_903683 | USA/NJ-CDC-2-3769191/2020 | 2020-12-30 | North America/USA/New Jersey/ | NJ Public Health and Environmental Laboratories | Genomics and Discovery, Respiratory Viruses Branch, Division of Viral Diseases, Centers for Disease Control and Prevention | Krista Queen et al |
| EPI_ISL_903694 | USA/VT-CDC-2-3767575/2020 | 2020-12-23 | North America/USA/Vermont/ | VT Dept. of Health Laboratory | Genomics and Discovery, Respiratory Viruses Branch, Division of Viral Diseases, Centers for Disease Control and Prevention | Krista Queen et al |

| | | | | | | |
|---|---|---|---|---|---|---|
| EPI_ISL_903696 | USA/VT-CDC-2-3767490/2020 | 2020-12-26 | North America/USA/Vermont/ | VT Dept. of Health Laboratory | Genomics and Discovery, Respiratory Viruses Branch, Division of Viral Diseases, Centers for Disease Control and Prevention | Krista Queen et al |
| EPI_ISL_903698 | USA/NH-CDC-2-3768151/2020 | 2020-12-21 | North America/USA/New Hampshire/ | NH Dept. of Health and Human Services Public Health Labs | Genomics and Discovery, Respiratory Viruses Branch, Division of Viral Diseases, Centers for Disease Control and Prevention | Krista Queen et al |
| EPI_ISL_903719 | USA/NV-CDC-2-3766746/2020 | 2020-12-16 | North America/USA/Nevada/ | NV State Public Health Laboratory | Genomics and Discovery, Respiratory Viruses Branch, Division of Viral Diseases, Centers for Disease Control and Prevention | Krista Queen et al |
| EPI_ISL_903723 | USA/NV-CDC-2-3774015/2020 | 2020-12-30 | North America/USA/Nevada/ | NV State Public Health Laboratory | Genomics and Discovery, Respiratory Viruses Branch, Division of Viral Diseases, Centers for Disease Control and Prevention | Krista Queen et al |

| | | | | | | |
|---|---|---|---|---|---|---|
| EPI_ISL_903726 | USA/NJ-CDC-2-3768183/2020 | 2020-12-30 | North America/USA/New Jersey/ | NJ Public Health and Environmental Laboratories | Genomics and Discovery, Respiratory Viruses Branch, Division of Viral Diseases, Centers for Disease Control and Prevention | Krista Queen et al |
| EPI_ISL_903730 | USA/VT-CDC-2-3767462/2020 | 2020-12-25 | North America/USA/Vermont/ | VT Dept. of Health Laboratory | Genomics and Discovery, Respiratory Viruses Branch, Division of Viral Diseases, Centers for Disease Control and Prevention | Krista Queen et al |
| EPI_ISL_903740 | USA/NM-CDC-2-3768071/2020 | 2020-12-28 | North America/USA/New Mexico/ | NM Dept. Health, Scientific Laboratory Division | Genomics and Discovery, Respiratory Viruses Branch, Division of Viral Diseases, Centers for Disease Control and Prevention | Krista Queen et al |
| EPI_ISL_903742 | USA/IL-CDC-2-3773949/2020 | 2020-12-05 | North America/USA/Illinois/ | IL Dept. of Public Health Springfield Laboratory | Genomics and Discovery, Respiratory Viruses Branch, Division of Viral Diseases, Centers for Disease Control and Prevention | Krista Queen et al |

| | | | | | | |
|---|---|---|---|---|---|---|
| EPI_ISL_903745 | USA/MS-CDC-2-3768779/2020 | 2020-12-17 | North America/USA/Mississippi/ | MS Public Health Laboratory | Genomics and Discovery, Respiratory Viruses Branch, Division of Viral Diseases, Centers for Disease Control and Prevention | Krista Queen et al |
| EPI_ISL_903746 | USA/MN-CDC-2-3766709/2020 | 2020-12-08 | North America/USA/Minnesota/ | MN PHL Division, Minnesota Department of Health | Genomics and Discovery, Respiratory Viruses Branch, Division of Viral Diseases, Centers for Disease Control and Prevention | Krista Queen et al |
| EPI_ISL_903754 | USA/IN-CDC-2-3769300/2020 | 2020-12-31 | North America/USA/Indiana/ | IN State Department of Health Laboratory Services | Genomics and Discovery, Respiratory Viruses Branch, Division of Viral Diseases, Centers for Disease Control and Prevention | Krista Queen et al |
| EPI_ISL_903765 | USA/LA-CDC-2-3766674/2020 | 2020-11-20 | North America/USA/Louisiana/ | LA Office of Public Health Laboratories | Genomics and Discovery, Respiratory Viruses Branch, Division of Viral Diseases, Centers for Disease Control and Prevention | Krista Queen et al |

| | | | | | | |
|---|---|---|---|---|---|---|
| EPI_ISL_903769 | USA/NM-CDC-2-3768167/2021 | 2021-01-02 | North America/USA/New Mexico/ | NM Dept. Health, Scientific Laboratory Division | Genomics and Discovery, Respiratory Viruses Branch, Division of Viral Diseases, Centers for Disease Control and Prevention | Krista Queen et al |
| EPI_ISL_903770 | USA/OK-CDC-2-3768060/2020 | 2020-12-01 | North America/USA/Oklahoma/ | OK Public Health Laboratory, Oklahoma State DOH | Genomics and Discovery, Respiratory Viruses Branch, Division of Viral Diseases, Centers for Disease Control and Prevention | Krista Queen et al |
| EPI_ISL_903773 | USA/IN-CDC-2-3769368/2020 | 2020-12-31 | North America/USA/Indiana/ | IN State Department of Health Laboratory Services | Genomics and Discovery, Respiratory Viruses Branch, Division of Viral Diseases, Centers for Disease Control and Prevention | Krista Queen et al |
| EPI_ISL_903787 | USA/MO-CDC-2-3769280/2020 | 2020-12-21 | North America/USA/Missouri/ | MO State Public Health Laboratory | Genomics and Discovery, Respiratory Viruses Branch, Division of Viral Diseases, Centers for Disease Control and Prevention | Krista Queen et al |

| | | | | | | |
|---|---|---|---|---|---|---|
| EPI_ISL_903788 | USA/TX-CDC-2-3767465/2020 | 2020-12-08 | North America/USA/Texas/ | TX DSHS, Lab Services Section MC 1947 | Genomics and Discovery, Respiratory Viruses Branch, Division of Viral Diseases, Centers for Disease Control and Prevention | Krista Queen et al |
| EPI_ISL_903794 | USA/AZ-CDC-2-3768154/2020 | 2020-12-14 | North America/USA/Arizona/ | AZ SPHL, Arizona Department of Health Services | Genomics and Discovery, Respiratory Viruses Branch, Division of Viral Diseases, Centers for Disease Control and Prevention | Krista Queen et al |
| EPI_ISL_903797 | USA/TN-CDC-2-3767485/2020 | 2020-12-22 | North America/USA/Tennessee/ | TN Division of Laboratory Services | Genomics and Discovery, Respiratory Viruses Branch, Division of Viral Diseases, Centers for Disease Control and Prevention | Krista Queen et al |
| EPI_ISL_903804 | USA/NM-CDC-2-3768175/2021 | 2021-01-02 | North America/USA/New Mexico/ | NM Dept. Health, Scientific Laboratory Division | Genomics and Discovery, Respiratory Viruses Branch, Division of Viral Diseases, Centers for Disease Control and Prevention | Krista Queen et al |

| | | | | | | |
|---|---|---|---|---|---|---|
| EPI_ISL_903808 | USA/AZ-CDC-2-3768074/2020 | 2020-12-28 | North America/USA/Arizona/ | AZ SPHL, Arizona Department of Health Services | Genomics and Discovery, Respiratory Viruses Branch, Division of Viral Diseases, Centers for Disease Control and Prevention | Krista Queen et al |
| EPI_ISL_903813 | USA/WV-CDC-2-3767468/2020 | 2020-12-04 | North America/USA/West Virginia/ | WVDHHR - Office of Laboratory Services | Genomics and Discovery, Respiratory Viruses Branch, Division of Viral Diseases, Centers for Disease Control and Prevention | Krista Queen et al |
| EPI_ISL_903824 | USA/VA-CDC-2-3767591/2020 | 2020-12-04 | North America/USA/Virginia/ | VA-Division of Consolidated Laboratory Services | Genomics and Discovery, Respiratory Viruses Branch, Division of Viral Diseases, Centers for Disease Control and Prevention | Krista Queen et al |
| EPI_ISL_903825 | USA/PA-CDC-2-3768761/2020 | 2020-12-21 | North America/USA/Pennsylvania/ | PA Department of Health, Bureau of Laboratories | Genomics and Discovery, Respiratory Viruses Branch, Division of Viral Diseases, Centers for Disease Control and Prevention | Krista Queen et al |

| | | | | | | |
|---|---|---|---|---|---|---|
| EPI_ISL_903841 | USA/UT-CDC-2-3766729/2020 | 2020-11-30 | North America/USA/Utah/ | UT-Unified State Labs: Public Health Utah DOH | Genomics and Discovery, Respiratory Viruses Branch, Division of Viral Diseases, Centers for Disease Control and Prevention | Krista Queen et al |
| EPI_ISL_903844 | USA/CA-CDC-2-3774048/2021 | 2021-01-02 | North America/USA/California/ | CDPH, Viral and Rickettsial Disease Laboratory | Genomics and Discovery, Respiratory Viruses Branch, Division of Viral Diseases, Centers for Disease Control and Prevention | Krista Queen et al |
| EPI_ISL_903845 | USA/NE-CDC-2-3769369/2020 | 2020-12-17 | North America/USA/Nebraska/ | NE Public Health Laboratory | Genomics and Discovery, Respiratory Viruses Branch, Division of Viral Diseases, Centers for Disease Control and Prevention | Krista Queen et al |
| EPI_ISL_903857 | USA/MN-CDC-2-3768664/2020 | 2020-12-22 | North America/USA/Minnesota/ | MN PHL Division, Minnesota Department of Health | Genomics and Discovery, Respiratory Viruses Branch, Division of Viral Diseases, Centers for Disease Control and Prevention | Krista Queen et al |

| | | | | | | |
|---|---|---|---|---|---|---|
| EPI_ISL_903860 | USA/NH-CDC-2-3768166/2020 | 2020-12-24 | North America/USA/New Hampshire/ | NH Dept. of Health and Human Services Public Health Labs | Genomics and Discovery, Respiratory Viruses Branch, Division of Viral Diseases, Centers for Disease Control and Prevention | Krista Queen et al |
| EPI_ISL_903866 | USA/MD-CDC-2-3773522/2020 | 2020-12-28 | North America/USA/Maryland/ | MD DOH Laboratories Administration | Genomics and Discovery, Respiratory Viruses Branch, Division of Viral Diseases, Centers for Disease Control and Prevention | Krista Queen et al |
| EPI_ISL_903875 | USA/IN-CDC-2-3769268/2020 | 2020-12-30 | North America/USA/Indiana/ | IN State Department of Health Laboratory Services | Genomics and Discovery, Respiratory Viruses Branch, Division of Viral Diseases, Centers for Disease Control and Prevention | Krista Queen et al |
| EPI_ISL_903876 | USA/PA-CDC-2-3768697/2020 | 2020-12-28 | North America/USA/Pennsylvania/ | PA Department of Health, Bureau of Laboratories | Genomics and Discovery, Respiratory Viruses Branch, Division of Viral Diseases, Centers for Disease Control and Prevention | Krista Queen et al |

| | | | | | | |
|---|---|---|---|---|---|---|
| EPI_ISL_903877 | USA/LA-CDC-2-3769292/2020 | 2020-12-30 | North America/USA/Louisiana/ | LA Office of Public Health Laboratories | Genomics and Discovery, Respiratory Viruses Branch, Division of Viral Diseases, Centers for Disease Control and Prevention | Krista Queen et al |
| EPI_ISL_903883 | USA/IN-CDC-2-3769254/2020 | 2020-12-30 | North America/USA/Indiana/ | IN State Department of Health Laboratory Services | Genomics and Discovery, Respiratory Viruses Branch, Division of Viral Diseases, Centers for Disease Control and Prevention | Krista Queen et al |
| EPI_ISL_903885 | USA/NE-CDC-2-3769353/2020 | 2020-12-28 | North America/USA/Nebraska/ | NE Public Health Laboratory | Genomics and Discovery, Respiratory Viruses Branch, Division of Viral Diseases, Centers for Disease Control and Prevention | Krista Queen et al |
| EPI_ISL_903888 | USA/MS-CDC-2-3768689/2020 | 2020-12-10 | North America/USA/Mississippi/ | MS Public Health Laboratory | Genomics and Discovery, Respiratory Viruses Branch, Division of Viral Diseases, Centers for Disease Control and Prevention | Krista Queen et al |

| | | | | | | |
|---|---|---|---|---|---|---|
| EPI_ISL_903894 | USA/SD-CDC-2-3766711/2020 | 2020-12-14 | North America/USA/South Dakota/ | SD Public Health Laboratory | Genomics and Discovery, Respiratory Viruses Branch, Division of Viral Diseases, Centers for Disease Control and Prevention | Krista Queen et al |
| EPI_ISL_903899 | USA/AL-CDC-2-3773543/2020 | 2020-12-24 | North America/USA/Alabama/ | AL Dept. of Public Health Bureau of Clinical Laboratories | Genomics and Discovery, Respiratory Viruses Branch, Division of Viral Diseases, Centers for Disease Control and Prevention | Krista Queen et al |
| EPI_ISL_903903 | USA/WI-CDC-2-3766721/2020 | 2020-11-30 | North America/USA/Wisconsin/ | WI State Laboratory of Hygiene | Genomics and Discovery, Respiratory Viruses Branch, Division of Viral Diseases, Centers for Disease Control and Prevention | Krista Queen et al |
| EPI_ISL_903906 | USA/LA-CDC-2-3766745/2020 | 2020-11-20 | North America/USA/Louisiana/ | LA Office of Public Health Laboratories | Genomics and Discovery, Respiratory Viruses Branch, Division of Viral Diseases, Centers for Disease Control and Prevention | Krista Queen et al |

| | | | | | | |
|---|---|---|---|---|---|---|
| EPI_ISL_903910 | USA/UT-CDC-2-3766692/2020 | 2020-11-30 | North America/USA/Utah/ | UT-Unified State Labs: Public Health Utah DOH | Genomics and Discovery, Respiratory Viruses Branch, Division of Viral Diseases, Centers for Disease Control and Prevention | Krista Queen et al |
| EPI_ISL_903920 | USA/IA-CDC-2-3769291/2020 | 2020-12-26 | North America/USA/Iowa/ | IA State Hygienic Laboratory | Genomics and Discovery, Respiratory Viruses Branch, Division of Viral Diseases, Centers for Disease Control and Prevention | Krista Queen et al |
| EPI_ISL_903926 | USA/PA-CDC-2-3768696/2020 | 2020-12-21 | North America/USA/Pennsylvania/ | PA Department of Health, Bureau of Laboratories | Genomics and Discovery, Respiratory Viruses Branch, Division of Viral Diseases, Centers for Disease Control and Prevention | Krista Queen et al |
| EPI_ISL_903956 | USA/WA-CDC-2-3767476/2020 | 2020-12-09 | North America/USA/Washington/ | WA State Department of Health | Genomics and Discovery, Respiratory Viruses Branch, Division of Viral Diseases, Centers for Disease Control and Prevention | Krista Queen et al |

| | | | | | | |
|---|---|---|---|---|---|---|
| EPI_ISL_903957 | USA/AZ-CDC-2-3768182/2020 | 2020-11-06 | North America/USA/Arizona/ | AZ SPHL, Arizona Department of Health Services | Genomics and Discovery, Respiratory Viruses Branch, Division of Viral Diseases, Centers for Disease Control and Prevention | Krista Queen et al |
| EPI_ISL_903962 | USA/WA-CDC-2-3767457/2020 | 2020-12-14 | North America/USA/Washington/ | WA State Department of Health | Genomics and Discovery, Respiratory Viruses Branch, Division of Viral Diseases, Centers for Disease Control and Prevention | Krista Queen et al |
| EPI_ISL_903964 | USA/DC-CDC-2-3773943/2020 | 2020-12-28 | North America/USA/Washington DC/ | DC Public Health Lab/ Dept. of Forensic Sciences | Genomics and Discovery, Respiratory Viruses Branch, Division of Viral Diseases, Centers for Disease Control and Prevention | Krista Queen et al |
| EPI_ISL_903969 | USA/DC-CDC-2-3774012/2020 | 2020-12-23 | North America/USA/Washington DC/ | DC Public Health Lab/ Dept. of Forensic Sciences | Genomics and Discovery, Respiratory Viruses Branch, Division of Viral Diseases, Centers for Disease Control and Prevention | Krista Queen et al |

| | | | | | | |
|---|---|---|---|---|---|---|
| EPI_ISL_903970 | USA/WA-CDC-2-3766693/2020 | 2020-11-08 | North America/USA/Washington/ | WA State Department of Health | Genomics and Discovery, Respiratory Viruses Branch, Division of Viral Diseases, Centers for Disease Control and Prevention | Krista Queen et al |
| EPI_ISL_903974 | USA/WA-CDC-2-3767487/2020 | 2020-12-09 | North America/USA/Washington/ | WA State Department of Health | Genomics and Discovery, Respiratory Viruses Branch, Division of Viral Diseases, Centers for Disease Control and Prevention | Krista Queen et al |
| EPI_ISL_903979 | Slovakia/UKBA-909/2021 | 2021-01-08 | Europe/Slovakia/Zilina/Dolnv∫ Kubv≠n | Institute of Virology, Biomedical Research Center of the Slovak Academy of Sciences, Bratislava | Faculty of Natural Sciences, Comenius University, Bratislava | Viktv≥ria ƒåabanovv° et al |
| EPI_ISL_903984 | Slovakia/UKBA-1005/2020 | 2020-12-22 | Europe/Slovakia/Trenƒçv≠n/ | Institute of Virology, Biomedical Research Center of the Slovak Academy of Sciences, Bratislava | Faculty of Natural Sciences, Comenius University, Bratislava | Viktv≥ria ƒåabanovv° et al |
| EPI_ISL_903987 | Slovakia/UKBA-1008/2020 | 2020-12-22 | Europe/Slovakia/Trenƒçv≠n/ | Institute of Virology, Biomedical Research Center of the Slovak Academy of Sciences, Bratislava | Faculty of Natural Sciences, Comenius University, Bratislava | Bro≈àa Brejovv° et al |
| EPI_ISL_903991 | Slovakia/UKBA-1012/2020 | 2020-12-22 | Europe/Slovakia/Trenƒçv≠n/ | Institute of Virology, Biomedical Research Center of the Slovak Academy of Sciences, Bratislava | Faculty of Natural Sciences, Comenius University, Bratislava | Viktv≥ria ƒåabanovv° et al |

| | | | | | | |
|---|---|---|---|---|---|---|
| EPI_ISL_903993 | Slovakia/UKBA-1014/2020 | 2020-12-22 | Europe/Slovakia/Trenƒçív≠n/ | Institute of Virology, Biomedical Research Center of the Slovak Academy of Sciences, Bratislava | Faculty of Natural Sciences, Comenius University, Bratislava | Viktv≥ria ƒåabanovv° et al |
| EPI_ISL_903996 | Slovakia/UKBA-1017/2020 | 2020-12-22 | Europe/Slovakia/Trenƒçív≠n/ | Institute of Virology, Biomedical Research Center of the Slovak Academy of Sciences, Bratislava | Faculty of Natural Sciences, Comenius University, Bratislava | Kristv≠na Bor≈°ovv° et al |
| EPI_ISL_903998 | Slovakia/UKBA-1019/2020 | 2020-12-22 | Europe/Slovakia/Trenƒçív≠n/ | Institute of Virology, Biomedical Research Center of the Slovak Academy of Sciences, Bratislava | Faculty of Natural Sciences, Comenius University, Bratislava | Bro≈àa Brejovv° et al |
| EPI_ISL_903999 | Slovakia/UKBA-1020/2020 | 2020-12-22 | Europe/Slovakia/Trenƒçív≠n/ | Institute of Virology, Biomedical Research Center of the Slovak Academy of Sciences, Bratislava | Faculty of Natural Sciences, Comenius University, Bratislava | Bro≈àa Brejovv° et al |
| EPI_ISL_904002 | Slovakia/UKBA-1023/2020 | 2020-12-22 | Europe/Slovakia/Trenƒçív≠n/ | Institute of Virology, Biomedical Research Center of the Slovak Academy of Sciences, Bratislava | Faculty of Natural Sciences, Comenius University, Bratislava | Bro≈àa Brejovv° et al |
| EPI_ISL_904003 | Slovakia/UKBA-1024/2020 | 2020-12-22 | Europe/Slovakia/Trenƒçív≠n/ | Institute of Virology, Biomedical Research Center of the Slovak Academy of Sciences, Bratislava | Faculty of Natural Sciences, Comenius University, Bratislava | Viktv≥ria ƒåabanovv° et al |
| EPI_ISL_904004 | Serbia/Novi_Sad-NIVNS610111/2020 | 2020-11-01 | Europe/Serbia/South Baƒçka District/No | Scientific Veterinary Institute Novi Sad | "Veterinary Specialized Institute ""Kraljevo"", Serbia" | Vidanovic et al |
| EPI_ISL_904005 | Serbia/Backa_Palanka-733652711/20 | 2020-11-27 | Europe/Serbia/South Baƒçka District/Ba | Scientific Veterinary Institute Novi Sad | "Veterinary Specialized Institute ""Kraljevo"", Serbia" | Vidanovic et al |

| | | | | | | |
|---|---|---|---|---|---|---|
| EPI_ISL_904006 | Serbia/Becej-716762511/2020 | 2020-11-25 | Europe/Serbia/South Baƒçka District/Be | Scientific Veterinary Institute Novi Sad | "Veterinary Specialized Institute ""Kraljevo"", Serbia" | Vidanovic et al |
| EPI_ISL_904007 | Serbia/Gracanica-043870912/2020 | 2020-12-09 | Europe/Kosovo/Pristina/Gracanica | Veterinary Specialized Institute Kraljevo | "Veterinary Specialized Institute ""Kraljevo"", Serbia" | Vidanovic et al |
| EPI_ISL_904008 | Serbia/Kosovska_Mitrovica-0439809 | 2020-12-09 | Europe/Kosovo/Kosovska Mitrovica/ | Veterinary Specialized Institute Kraljevo | "Veterinary Specialized Institute ""Kraljevo"", Serbia" | Vidanovic et al |
| EPI_ISL_904009 | Serbia/Novi_Sad-NIVNS41211/2020 | 2020-11-12 | Europe/Serbia/South Baƒçka District/No | Scientific Veterinary Institute Novi Sad | "Veterinary Specialized Institute ""Kraljevo"", Serbia" | Vidanovic et al |
| EPI_ISL_904010 | Serbia/Lipljan-005360911/2020 | 2020-11-09 | Europe/Kosovo/Pristina/Lipljan | Veterinary Specialized Institute Kraljevo | "Veterinary Specialized Institute ""Kraljevo"", Serbia" | Vidanovic et al |
| EPI_ISL_904015 | Jordan/UK5/2020 | 2020-12-20 | Asia/Jordan/Amman/ | Molecular Biology and Virology lab, Faculty of Veterinary Medicine, Jordan University of Science and Technology | Molecular Biology and Virology lab, Faculty of Veterinary Medicine, Jordan University of Science and Technology | Mohammad Hussien Alboom et al |
| EPI_ISL_904016 | Jordan/UK6/2020 | 2020-12-20 | Asia/Jordan/Amman/ | Molecular Biology and Virology lab, Faculty of Veterinary Medicine, Jordan University of Science and Technology | Molecular Biology and Virology lab, Faculty of Veterinary Medicine, Jordan University of Science and Technology | Mohammad Hussien Alboom et al |

| | | | | | | |
|---|---|---|---|---|---|---|
| EPI_ISL_904018 | Brazil/MG-L2-MG04/2020 | 2020-03-30 | South America/Brazil/Minas Gerais/ | DB Diagnosticos do Brasil | Laboratv≥rio de Parasitologia Mv©dica - Instituto de Medicina Tropical - Universidade de Sv£o Paulo | Brazil-UK Centre for Arbovirus Discovery Diagnosis Genomics et al |
| EPI_ISL_904020 | Brazil/MG-L2-MG08/2020 | 2020-04-02 | South America/Brazil/Minas Gerais/ | DB Diagnosticos do Brasil | Laboratv≥rio de Parasitologia Mv©dica - Instituto de Medicina Tropical - Universidade de Sv£o Paulo | Brazil-UK Centre for Arbovirus Discovery Diagnosis Genomics et al |
| EPI_ISL_904028 | Brazil/MG-L2-MG51/2020 | 2020-05-23 | South America/Brazil/Minas Gerais/ | DB Diagnosticos do Brasil | Laboratv≥rio de Parasitologia Mv©dica - Instituto de Medicina Tropical - Universidade de Sv£o Paulo | Brazil-UK Centre for Arbovirus Discovery Diagnosis Genomics et al |
| EPI_ISL_904035 | USA/NM-DOH-2020568351/2020 | 2020-11-25 | North America/USA/New Mexico/ | New Mexico Department of Health Scientific Laboratory | New Mexico Department of Health Scientific Laboratory | Ellie Johnson et al |
| EPI_ISL_904040 | USA/NM-DOH-2020649693/2020 | 2020-12-30 | North America/USA/New Mexico/ | New Mexico Department of Health Scientific Laboratory | New Mexico Department of Health Scientific Laboratory | Ellie Johnson et al |
| EPI_ISL_904043 | USA/NM-DOH-2021001333/2021 | 2021-01-03 | North America/USA/New Mexico/ | New Mexico Department of Health Scientific Laboratory | New Mexico Department of Health Scientific Laboratory | Ellie Johnson et al |
| EPI_ISL_904046 | USA/NM-DOH-2021001615/2021 | 2021-01-03 | North America/USA/New Mexico/ | New Mexico Department of Health Scientific Laboratory | New Mexico Department of Health Scientific Laboratory | Ellie Johnson et al |
| EPI_ISL_904053 | USA/NM-DOH-2021002852/2021 | 2021-01-04 | North America/USA/New Mexico/ | New Mexico Department of Health Scientific Laboratory | New Mexico Department of Health Scientific Laboratory | Ellie Johnson et al |

| | | | | | | |
|---|---|---|---|---|---|---|
| EPI_ISL_904061 | USA/NM-DOH-2021009169/2021 | 2021-01-06 | North America/USA/New Mexico/ | New Mexico Department of Health Scientific Laboratory | New Mexico Department of Health Scientific Laboratory | Ellie Johnson et al |
| EPI_ISL_904071 | USA/NM-DOH-2021026961/2021 | 2021-01-16 | North America/USA/New Mexico/ | New Mexico Department of Health Scientific Laboratory | New Mexico Department of Health Scientific Laboratory | Ellie Johnson et al |
| EPI_ISL_904072 | USA/NM-DOH-2021027139/2021 | 2021-01-16 | North America/USA/New Mexico/ | New Mexico Department of Health Scientific Laboratory | New Mexico Department of Health Scientific Laboratory | Ellie Johnson et al |
| EPI_ISL_904074 | USA/NM-DOH-2021027436/2021 | 2021-01-16 | North America/USA/New Mexico/ | New Mexico Department of Health Scientific Laboratory | New Mexico Department of Health Scientific Laboratory | Ellie Johnson et al |
| EPI_ISL_904080 | USA/NM-DOH-2021027644/2021 | 2021-01-15 | North America/USA/New Mexico/ | New Mexico Department of Health Scientific Laboratory | New Mexico Department of Health Scientific Laboratory | Ellie Johnson et al |
| EPI_ISL_904086 | USA/NM-DOH-2021027936/2021 | 2021-01-18 | North America/USA/New Mexico/ | New Mexico Department of Health Scientific Laboratory | New Mexico Department of Health Scientific Laboratory | Ellie Johnson et al |
| EPI_ISL_904091 | USA/NM-DOH-2021028226/2021 | 2021-01-18 | North America/USA/New Mexico/ | New Mexico Department of Health Scientific Laboratory | New Mexico Department of Health Scientific Laboratory | Ellie Johnson et al |
| EPI_ISL_904097 | USA/NM-DOH-2021028386/2021 | 2021-01-18 | North America/USA/New Mexico/ | New Mexico Department of Health Scientific Laboratory | New Mexico Department of Health Scientific Laboratory | Ellie Johnson et al |
| EPI_ISL_904099 | USA/NM-DOH-2021028466/2021 | 2021-01-18 | North America/USA/New Mexico/ | New Mexico Department of Health Scientific Laboratory | New Mexico Department of Health Scientific Laboratory | Ellie Johnson et al |
| EPI_ISL_904108 | USA/NM-DOH-2021029654/2021 | 2021-01-19 | North America/USA/New Mexico/ | New Mexico Department of Health Scientific Laboratory | New Mexico Department of Health Scientific Laboratory | Ellie Johnson et al |

| | | | | | | |
|---|---|---|---|---|---|---|
| EPI_ISL_904109 | USA/NM-DOH-2021029656/2021 | 2021-01-19 | North America/USA/New Mexico/ | New Mexico Department of Health Scientific Laboratory | New Mexico Department of Health Scientific Laboratory | Ellie Johnson et al |
| EPI_ISL_904112 | USA/NM-DOH-2021029730/2021 | 2021-01-19 | North America/USA/New Mexico/ | New Mexico Department of Health Scientific Laboratory | New Mexico Department of Health Scientific Laboratory | Ellie Johnson et al |
| EPI_ISL_904118 | USA/NM-DOH-2021030885/2021 | 2021-01-13 | North America/USA/New Mexico/ | New Mexico Department of Health Scientific Laboratory | New Mexico Department of Health Scientific Laboratory | Ellie Johnson et al |
| EPI_ISL_904119 | USA/NM-DOH-2021031015/2021 | 2021-01-20 | North America/USA/New Mexico/ | New Mexico Department of Health Scientific Laboratory | New Mexico Department of Health Scientific Laboratory | Ellie Johnson et al |
| EPI_ISL_904120 | Brazil/PA-IEC-177365/2021 | 2021-01-10 | South America/Brazil/Para/ | LACEN - Laboratv≥rio Central de Sav∫de Pv∫blica do Parv° | Ôª¿Evandro Chagas Institute | Santos et al |
| EPI_ISL_904121 | Brazil/PA-IEC-177366/2021 | 2021-01-08 | South America/Brazil/Para/ | LACEN - Laboratv≥rio Central de Sav∫de Pv∫blica do Parv° | Ôª¿Evandro Chagas Institute | Santos et al |
| EPI_ISL_904122 | Poland/NIPH-NIH-14/2021 | 2021-01-06 | Europe/Poland/Mazowieckie/Warszawa | National Institute of Public Health - National Institute of Hygiene | National Institute of Public Health - National Institute of Hygiene | Wo≈Çkowicz Tomasz et al |
| EPI_ISL_904124 | Poland/NIPH-NIH-41/2021 | 2021-01-13 | Europe/Poland/Mazowieckie/Warszawa | National Institute of Public Health - National Institute of Hygiene | National Institute of Public Health - National Institute of Hygiene | Wo≈Çkowicz Tomasz et al |
| EPI_ISL_904125 | Poland/NIPH-NIH-45/2021 | 2021-01-05 | Europe/Poland/Mazowieckie/Warszawa | Laboratorium Analityczne Szpital Specjalistyczny w Chorzowie | National Institute of Public Health - National Institute of Hygiene | Wo≈Çkowicz Tomasz et al |
| EPI_ISL_904126 | Poland/NIPH-NIH-55/2021 | 2021-01-24 | Europe/Poland/Mazowieckie/Warszawa | National Institute of Public Health - National Institute of Hygiene | National Institute of Public Health - National Institute of Hygiene | Wo≈Çkowicz Tomasz et al |
| EPI_ISL_904134 | USA/TX-QDX-3999/2021 | 2021-01-07 | North America/USA/Texas/ | Quest Diagnostics | Quest Diagnostics | Rosenthal et al |
| EPI_ISL_904141 | Netherlands/GE-EMC-489/2021 | 2021-01-08 | Europe/Netherlands/Gelderland/ | Dutch COVID-19 response team | Erasmus Medical Center | Bas Oude Munnink et al |

| | | | | | | |
|---|---|---|---|---|---|---|
| EPI_ISL_904171 | Netherlands/LI-EMC-250/2020 | 2020-10-11 | Europe/Netherlands/Limburg/ | Dutch COVID-19 response team | Erasmus Medical Center | Bas Oude Munnink et al |
| EPI_ISL_904191 | Netherlands/NB-EMC-531/2021 | 2021-01-03 | Europe/Netherlands/North Brabant/ | Dutch COVID-19 response team | Erasmus Medical Center | Bas Oude Munnink et al |
| EPI_ISL_904230 | Netherlands/LI-EMC-263/2020 | 2020-10-18 | Europe/Netherlands/Limburg/ | Dutch COVID-19 response team | Erasmus Medical Center | Bas Oude Munnink et al |
| EPI_ISL_904294 | Netherlands/NB-EMC-523/2020 | 2020-12-22 | Europe/Netherlands/North Brabant/ | Dutch COVID-19 response team | Erasmus Medical Center | Bas Oude Munnink et al |
| EPI_ISL_904378 | Netherlands/ZH-EMC-1459/2021 | 2021-01-12 | Europe/Netherlands/South Holland/ | Dutch COVID-19 response team | Erasmus Medical Center | Bas Oude Munnink et al |
| EPI_ISL_904435 | Netherlands/NB-EMC-591/2020 | 2020-10-16 | Europe/Netherlands/North Brabant/ | Dutch COVID-19 response team | Erasmus Medical Center | Bas Oude Munnink et al |
| EPI_ISL_904439 | Netherlands/LI-EMC-256/2020 | 2020-10-20 | Europe/Netherlands/Limburg/ | Dutch COVID-19 response team | Erasmus Medical Center | Bas Oude Munnink et al |
| EPI_ISL_904448 | Netherlands/LI-EMC-265/2020 | 2020-10-22 | Europe/Netherlands/Limburg/ | Dutch COVID-19 response team | Erasmus Medical Center | Bas Oude Munnink et al |
| EPI_ISL_904451 | Netherlands/LI-EMC-241/2020 | 2020-10-25 | Europe/Netherlands/Limburg/ | Dutch COVID-19 response team | Erasmus Medical Center | Bas Oude Munnink et al |
| EPI_ISL_904457 | Netherlands/LI-EMC-224/2020 | 2020-10-31 | Europe/Netherlands/Limburg/ | Dutch COVID-19 response team | Erasmus Medical Center | Bas Oude Munnink et al |
| EPI_ISL_904541 | Netherlands/NH-EMC-104/2021 | 2021-01-04 | Europe/Netherlands/North Holland/ | Dutch COVID-19 response team | Erasmus Medical Center | Bas Oude Munnink et al |
| EPI_ISL_904618 | Netherlands/NB-EMC-569/2020 | 2020-12-28 | Europe/Netherlands/North Brabant/ | Dutch COVID-19 response team | Erasmus Medical Center | Bas Oude Munnink et al |
| EPI_ISL_904626 | Spain/CT-IrsiCaixaLCGJQ1/2021 | 2021-01-01 | Europe/Spain/Catalunya/La Llagosta | Servicio de Microbiolog√≠a, Laboratori Cl√≠nic Metropolitana Nord. Hospital Universitari Germans Trias i Pujol. Institut d‚ÄôInvestigaci√≥ en Ci√®ncies de la Salut Germans Trias i Pujol (IGTP) | IrsiCaixa - Can Ruti CovidSeq | Marc Noguera-Julian et al |
| EPI_ISL_904627 | Spain/CT-IrsiCaixaLIHSW1/2021 | 2021-01-08 | Europe/Spain/Catalunya/Santa Eul√†lia | Servicio de Microbiolog√≠a, Laboratori Cl√≠nic Metropolitana Nord. Hospital Universitari Germans Trias i Pujol. Institut d‚ÄôInvestigaci√≥ en Ci√®ncies de la Salut Germans Trias i Pujol (IGTP) | IrsiCaixa - Can Ruti CovidSeq | Marc Noguera-Julian et al |
| EPI_ISL_904641 | Spain/CT-IrsiCaixaLLFKC1/2021 | 2021-01-05 | Europe/Spain/Catalunya/Badalona | Servicio de Microbiolog√≠a, Laboratori Cl√≠nic Metropolitana Nord. Hospital Universitari Germans Trias i Pujol. Institut d‚ÄôInvestigaci√≥ en Ci√®ncies de la Salut Germans Trias i Pujol (IGTP) | IrsiCaixa - Can Ruti CovidSeq | Marc Noguera-Julian et al |

| | | | | | | |
|---|---|---|---|---|---|---|
| EPI_ISL_904651 | Spain/CT-IrsiCaixaLFUSP1/2021 | 2021-01-05 | Europe/Spain/Catalunya/Santa Coloma | Servicio de Microbiolog√≠a, Laboratori Cl√≠nic Metropolitana Nord. Hospital Universitari Germans Trias i Pujol. Institut d‚ÄôInvestigaci√≥ en Ci√®ncies de la Salut Germans Trias i Pujol (IGTP) | IrsiCaixa - Can Ruti CovidSeq | Marc Noguera-Julian et al |
| EPI_ISL_904653 | Spain/CT-IrsiCaixaLNZXA1/2021 | 2021-01-03 | Europe/Spain/Catalunya/Mataro | Servicio de Microbiolog√≠a, Laboratori Cl√≠nic Metropolitana Nord. Hospital Universitari Germans Trias i Pujol. Institut d‚ÄôInvestigaci√≥ en Ci√®ncies de la Salut Germans Trias i Pujol (IGTP) | IrsiCaixa - Can Ruti CovidSeq | Marc Noguera-Julian et al |
| EPI_ISL_904656 | Spain/CT-IrsiCaixaLWIAR1/2021 | 2021-01-05 | Europe/Spain/Catalunya/Granollers | Servicio de Microbiolog√≠a, Laboratori Cl√≠nic Metropolitana Nord. Hospital Universitari Germans Trias i Pujol. Institut d‚ÄôInvestigaci√≥ en Ci√®ncies de la Salut Germans Trias i Pujol (IGTP) | IrsiCaixa - Can Ruti CovidSeq | Marc Noguera-Julian et al |
| EPI_ISL_904741 | Netherlands/NH-RIVM-10691/2021 | 2021-01-12 | Europe/Netherlands/North Holland/ | Dutch COVID-19 response team | National Institute for Public Health and the Environment (RIVM) | Adam Meijer et al |
| EPI_ISL_904748 | Netherlands/UT-RIVM-21661/2020 | 2020-12-23 | Europe/Netherlands/Utrecht/ | Dutch COVID-19 response team | National Institute for Public Health and the Environment (RIVM) | Adam Meijer et al |
| EPI_ISL_904758 | Netherlands/NB-RIVM-10787/2021 | 2021-01-15 | Europe/Netherlands/North Brabant/ | Dutch COVID-19 response team | National Institute for Public Health and the Environment (RIVM) | Adam Meijer et al |
| EPI_ISL_904772 | Netherlands/FR-RIVM-10580/2021 | 2021-01-16 | Europe/Netherlands/Friesland/ | Dutch COVID-19 response team | National Institute for Public Health and the Environment (RIVM) | Adam Meijer et al |
| EPI_ISL_904788 | Netherlands/GE-RIVM-10655/2021 | 2021-01-12 | Europe/Netherlands/Gelderland/ | Dutch COVID-19 response team | National Institute for Public Health and the Environment (RIVM) | Adam Meijer et al |

| | | | | | | |
|---|---|---|---|---|---|---|
| EPI_ISL_904877 | Netherlands/OV-RIVM-10670/2021 | 2021-01-12 | Europe/Netherlands/Overijssel/ | Dutch COVID-19 response team | National Institute for Public Health and the Environment (RIVM) | Adam Meijer et al |
| EPI_ISL_904884 | Netherlands/OV-RIVM-10683/2021 | 2021-01-17 | Europe/Netherlands/Overijssel/ | Dutch COVID-19 response team | National Institute for Public Health and the Environment (RIVM) | Adam Meijer et al |
| EPI_ISL_904901 | Netherlands/UT-RIVM-21660/2020 | 2020-12-08 | Europe/Netherlands/Utrecht/ | Dutch COVID-19 response team | National Institute for Public Health and the Environment (RIVM) | Adam Meijer et al |
| EPI_ISL_904905 | Netherlands/UT-RIVM-21670/2020 | 2020-12-25 | Europe/Netherlands/Utrecht/ | Dutch COVID-19 response team | National Institute for Public Health and the Environment (RIVM) | Adam Meijer et al |
| EPI_ISL_904939 | Lithuania/T_52/2020 | 2020-10-06 | Europe/Lithuania/Utena/Visaginas | Vilnius University Hospital Santaros Klinikos, Vilnius University | Institute of Biotechnology, Life Sciences Center, Vilnius University | Emilija Vasiliunaite et al |
| EPI_ISL_904944 | Lithuania/T_25/2020 | 2020-09-28 | Europe/Lithuania/Vilnius/ | Vilnius University Hospital Santaros Klinikos, Vilnius University | Institute of Biotechnology, Life Sciences Center, Vilnius University | Emilija Vasiliunaite et al |
| EPI_ISL_904947 | Lithuania/T_1362/2020 | 2020-11-18 | Europe/Lithuania/Vilnius/≈†irvint≈≥ raj | Vilnius University Hospital Santaros Klinikos, Vilnius University | Institute of Biotechnology, Life Sciences Center, Vilnius University | Emilija Vasiliunaite et al |
| EPI_ISL_904949 | Lithuania/T_1740/2020 | 2020-11-23 | Europe/Lithuania/Utena/Molfót≈≥ rajon | Vilnius University Hospital Santaros Klinikos, Vilnius University | Institute of Biotechnology, Life Sciences Center, Vilnius University | Emilija Vasiliunaite et al |

| | | | | | | |
|---|---|---|---|---|---|---|
| EPI_ISL_904950 | Lithuania/T_1741/2020 | 2020-11-23 | Europe/Lithuania/Vilnius/ | Vilnius University Hospital Santaros Klinikos, Vilnius University | Institute of Biotechnology, Life Sciences Center, Vilnius University | Emilija Vasiliunaite et al |
| EPI_ISL_904951 | Lithuania/T_1852/2020 | 2020-11-24 | Europe/Lithuania/Vilnius/ | Vilnius University Hospital Santaros Klinikos, Vilnius University | Institute of Biotechnology, Life Sciences Center, Vilnius University | Emilija Vasiliunaite et al |
| EPI_ISL_904953 | Lithuania/T_1989_2/2020 | 2020-11-27 | Europe/Lithuania/Kauno Apskritis/ | Vilnius University Hospital Santaros Klinikos, Vilnius University | Institute of Biotechnology, Life Sciences Center, Vilnius University | Emilija Vasiliunaite et al |
| EPI_ISL_904955 | Lithuania/T_2096/2020 | 2020-11-27 | Europe/Lithuania/Klaipedos Apskritis/Kr | Vilnius University Hospital Santaros Klinikos, Vilnius University | Institute of Biotechnology, Life Sciences Center, Vilnius University | Emilija Vasiliunaite et al |
| EPI_ISL_904956 | Lithuania/T_2110/2020 | 2020-11-27 | Europe/Lithuania/Vilnius/ | Vilnius University Hospital Santaros Klinikos, Vilnius University | Institute of Biotechnology, Life Sciences Center, Vilnius University | Emilija Vasiliunaite et al |
| EPI_ISL_905033 | Netherlands/OV-RIVM-21515/2020 | 2020-12-29 | Europe/Netherlands/Overijssel/ | Dutch COVID-19 response team | National Institute for Public Health and the Environment (RIVM) | Adam Meijer et al |
| EPI_ISL_905037 | Netherlands/ZH-RIVM-10341/2021 | 2021-01-06 | Europe/Netherlands/South Holland/ | Dutch COVID-19 response team | National Institute for Public Health and the Environment (RIVM) | Adam Meijer et al |
| EPI_ISL_905053 | Netherlands/UT-RIVM-10357/2021 | 2021-01-11 | Europe/Netherlands/Utrecht/ | Dutch COVID-19 response team | National Institute for Public Health and the Environment (RIVM) | Adam Meijer et al |

| | | | | | | |
|---|---|---|---|---|---|---|
| EPI_ISL_905060 | Netherlands/UT-RIVM-10364/2021 | 2021-01-10 | Europe/Netherlands/Utrecht/ | Dutch COVID-19 response team | National Institute for Public Health and the Environment (RIVM) | Adam Meijer et al |
| EPI_ISL_905140 | Netherlands/UT-RIVM-21562/2020 | 2020-12-22 | Europe/Netherlands/Utrecht/ | Dutch COVID-19 response team | National Institute for Public Health and the Environment (RIVM) | Adam Meijer et al |
| EPI_ISL_905166 | Netherlands/UT-RIVM-21588/2020 | 2020-12-25 | Europe/Netherlands/Utrecht/ | Dutch COVID-19 response team | National Institute for Public Health and the Environment (RIVM) | Adam Meijer et al |
| EPI_ISL_905188 | Netherlands/LI-RIVM-10466/2021 | 2021-01-09 | Europe/Netherlands/Limburg/ | Dutch COVID-19 response team | National Institute for Public Health and the Environment (RIVM) | Adam Meijer et al |
| EPI_ISL_905218 | Netherlands/UT-RIVM-21607/2020 | 2020-12-29 | Europe/Netherlands/Utrecht/ | Dutch COVID-19 response team | National Institute for Public Health and the Environment (RIVM) | Adam Meijer et al |
| EPI_ISL_905245 | Netherlands/GE-RIVM-10421/2021 | 2021-01-11 | Europe/Netherlands/Gelderland/ | Dutch COVID-19 response team | National Institute for Public Health and the Environment (RIVM) | Adam Meijer et al |
| EPI_ISL_905260 | Netherlands/LI-RIVM-10416/2021 | 2021-01-09 | Europe/Netherlands/Limburg/ | Dutch COVID-19 response team | National Institute for Public Health and the Environment (RIVM) | Adam Meijer et al |
| EPI_ISL_905288 | Netherlands/NB-RIVM-10459/2021 | 2021-01-04 | Europe/Netherlands/North Brabant/ | Dutch COVID-19 response team | National Institute for Public Health and the Environment (RIVM) | Adam Meijer et al |

| | | | | | | |
|---|---|---|---|---|---|---|
| EPI_ISL_905296 | Netherlands/NB-RIVM-21596/2020 | 2020-11-13 | Europe/Netherlands/North Brabant/ | Dutch COVID-19 response team | National Institute for Public Health and the Environment (RIVM) | Adam Meijer et al |
| EPI_ISL_905313 | Netherlands/NH-RIVM-10509/2021 | 2021-01-10 | Europe/Netherlands/North Holland/ | Dutch COVID-19 response team | National Institute for Public Health and the Environment (RIVM) | Adam Meijer et al |
| EPI_ISL_905377 | Netherlands/NB-RIVM-21381/2020 | 2020-12-28 | Europe/Netherlands/North Brabant/ | Dutch COVID-19 response team | National Institute for Public Health and the Environment (RIVM) | Adam Meijer et al |
| EPI_ISL_905397 | Netherlands/DR-RIVM-21391/2020 | 2020-12-28 | Europe/Netherlands/Drenthe/ | Dutch COVID-19 response team | National Institute for Public Health and the Environment (RIVM) | Adam Meijer et al |
| EPI_ISL_905478 | Netherlands/FR-RIVM-10107/2021 | 2021-01-06 | Europe/Netherlands/Friesland/ | Dutch COVID-19 response team | National Institute for Public Health and the Environment (RIVM) | Adam Meijer et al |
| EPI_ISL_905505 | Netherlands/UT-RIVM-21438/2020 | 2020-12-27 | Europe/Netherlands/Utrecht/ | Dutch COVID-19 response team | National Institute for Public Health and the Environment (RIVM) | Adam Meijer et al |
| EPI_ISL_905524 | Netherlands/UT-RIVM-10127/2021 | 2021-01-03 | Europe/Netherlands/Utrecht/ | Dutch COVID-19 response team | National Institute for Public Health and the Environment (RIVM) | Adam Meijer et al |
| EPI_ISL_905570 | Netherlands/LI-RIVM-10151/2021 | 2021-01-05 | Europe/Netherlands/Limburg/ | Dutch COVID-19 response team | National Institute for Public Health and the Environment (RIVM) | Adam Meijer et al |

| | | | | | | |
|---|---|---|---|---|---|---|
| EPI_ISL_905606 | Netherlands/UT-RIVM-21472/2020 | 2020-12-23 | Europe/Netherlands/Utrecht/ | Dutch COVID-19 response team | National Institute for Public Health and the Environment (RIVM) | Adam Meijer et al |
| EPI_ISL_905637 | Netherlands/UT-RIVM-10205/2021 | 2021-01-08 | Europe/Netherlands/Utrecht/ | Dutch COVID-19 response team | National Institute for Public Health and the Environment (RIVM) | Adam Meijer et al |
| EPI_ISL_905652 | Netherlands/UT-RIVM-21479/2020 | 2020-10-13 | Europe/Netherlands/Utrecht/ | Dutch COVID-19 response team | National Institute for Public Health and the Environment (RIVM) | Adam Meijer et al |
| EPI_ISL_905655 | Netherlands/UT-RIVM-21482/2020 | 2020-10-15 | Europe/Netherlands/Utrecht/ | Dutch COVID-19 response team | National Institute for Public Health and the Environment (RIVM) | Adam Meijer et al |
| EPI_ISL_905656 | Netherlands/UT-RIVM-21483/2020 | 2020-10-16 | Europe/Netherlands/Utrecht/ | Dutch COVID-19 response team | National Institute for Public Health and the Environment (RIVM) | Adam Meijer et al |
| EPI_ISL_905684 | Netherlands/UT-RIVM-10232/2021 | 2021-01-07 | Europe/Netherlands/Utrecht/ | Dutch COVID-19 response team | National Institute for Public Health and the Environment (RIVM) | Adam Meijer et al |
| EPI_ISL_905685 | Netherlands/UT-RIVM-21494/2020 | 2020-12-16 | Europe/Netherlands/Utrecht/ | Dutch COVID-19 response team | National Institute for Public Health and the Environment (RIVM) | Adam Meijer et al |
| EPI_ISL_905721 | Netherlands/NH-RIVM-10255/2021 | 2021-01-12 | Europe/Netherlands/North Holland/ | Dutch COVID-19 response team | National Institute for Public Health and the Environment (RIVM) | Adam Meijer et al |

| | | | | | | |
|---|---|---|---|---|---|---|
| EPI_ISL_905731 | Indonesia/YO-UGM-124764/2020 | 2020-12-23 | Asia/Indonesia/Yogyakarta/ | Academic Hospital of Gadjah Mada University (RSA UGM) | Genetics Working Group (Pokja Genetik) Faculty of Medicine, Public Health and Nursing Universitas Gadjah Mada (FK-KMK UGM); Disease Investigation Center Wates Ministry of Agriculture Indonesia; Department of Microbiology FK-KMK UGM; Laboratorium Diagnostik Yayasan Tahija World Mosquito Program (WMP) Yogyakarta Center for Tropical Medicine FK-KMK UGM; Integrated Research center FK-KMK UGM; Department of Computer Science and Electronics FMIPA UGM | Gunadi et al |
| EPI_ISL_905738 | Poland/NIPH-NIH-22/2021 | 2021-01-05 | Europe/Poland/Mazowieckie/Warszawa | National Institute of Public Health - National Institute of Hygiene | National Institute of Public Health - National Institute of Hygiene | Wo≈Çkowicz Tomasz et al |
| EPI_ISL_905740 | Poland/NIPH-NIH-27/2021 | 2021-01-04 | Europe/Poland/Mazowieckie/Warszawa | NZOZ LM Diagnostyka Czfôstochowa | National Institute of Public Health - National Institute of Hygiene | Wo≈Çkowicz Tomasz et al |

| | | | | | | |
|---|---|---|---|---|---|---|
| EPI_ISL_905741 | Poland/NIPH-NIH-30/2021 | 2021-01-11 | Europe/Poland/Mazowieckie/Warszawa | National Institute of Public Health - National Institute of Hygiene | National Institute of Public Health - National Institute of Hygiene | Wo≈Çkowicz Tomasz et al |
| EPI_ISL_905744 | Poland/NIPH-NIH-33/2021 | 2021-01-11 | Europe/Poland/Mazowieckie/Warszawa | National Institute of Public Health - National Institute of Hygiene | National Institute of Public Health - National Institute of Hygiene | Wo≈Çkowicz Tomasz et al |
| EPI_ISL_905745 | Poland/NIPH-NIH-34/2021 | 2021-01-11 | Europe/Poland/Mazowieckie/Warszawa | National Institute of Public Health - National Institute of Hygiene | National Institute of Public Health - National Institute of Hygiene | Wo≈Çkowicz Tomasz et al |
| EPI_ISL_905746 | Poland/NIPH-NIH-35/2021 | 2021-01-11 | Europe/Poland/Mazowieckie/Warszawa | National Institute of Public Health - National Institute of Hygiene | National Institute of Public Health - National Institute of Hygiene | Wo≈Çkowicz Tomasz et al |
| EPI_ISL_905747 | Poland/NIPH-NIH-36/2021 | 2021-01-11 | Europe/Poland/Mazowieckie/Warszawa | National Institute of Public Health - National Institute of Hygiene | National Institute of Public Health - National Institute of Hygiene | Wo≈Çkowicz Tomasz et al |
| EPI_ISL_905748 | Poland/NIPH-NIH-37/2021 | 2021-01-13 | Europe/Poland/Mazowieckie/Warszawa | National Institute of Public Health - National Institute of Hygiene | National Institute of Public Health - National Institute of Hygiene | Wo≈Çkowicz Tomasz et al |
| EPI_ISL_905749 | Poland/NIPH-NIH-38/2021 | 2021-01-13 | Europe/Poland/Mazowieckie/Warszawa | National Institute of Public Health - National Institute of Hygiene | National Institute of Public Health - National Institute of Hygiene | Wo≈Çkowicz Tomasz et al |
| EPI_ISL_905753 | Poland/NIPH-NIH-44/2020 | 2020-12-22 | Europe/Poland/Mazowieckie/Warszawa | Laboratorium Mikrobiologiczne SYNEVO | National Institute of Public Health - National Institute of Hygiene | Wo≈Çkowicz Tomasz et al |
| EPI_ISL_905755 | Poland/NIPH-NIH-48/2021 | 2021-01-13 | Europe/Poland/Mazowieckie/Warszawa | National Institute of Public Health - National Institute of Hygiene | National Institute of Public Health - National Institute of Hygiene | Wo≈Çkowicz Tomasz et al |
| EPI_ISL_905756 | Poland/NIPH-NIH-49/2021 | 2021-01-25 | Europe/Poland/Mazowieckie/Warszawa | National Institute of Public Health - National Institute of Hygiene | National Institute of Public Health - National Institute of Hygiene | Wo≈Çkowicz Tomasz et al |

| | | | | | | |
|---|---|---|---|---|---|---|
| EPI_ISL_905757 | Poland/NIPH-NIH-51/2021 | 2021-01-25 | Europe/Poland/Mazowieckie/Warszawa | National Institute of Public Health - National Institute of Hygiene | National Institute of Public Health - National Institute of Hygiene | Wo≈Çkowicz Tomasz et al |
| EPI_ISL_905759 | Poland/NIPH-NIH-53/2021 | 2021-01-24 | Europe/Poland/Mazowieckie/Warszawa | National Institute of Public Health - National Institute of Hygiene | National Institute of Public Health - National Institute of Hygiene | Wo≈Çkowicz Tomasz et al |
| EPI_ISL_905760 | Poland/NIPH-NIH-54/2021 | 2021-01-24 | Europe/Poland/Mazowieckie/Warszawa | National Institute of Public Health - National Institute of Hygiene | National Institute of Public Health - National Institute of Hygiene | Wo≈Çkowicz Tomasz et al |
| EPI_ISL_905762 | Poland/NIPH-NIH-57/2021 | 2021-01-20 | Europe/Poland/Mazowieckie/Warszawa | Laboratorium Genloxa Sp. z o.o. | National Institute of Public Health - National Institute of Hygiene | Wo≈Çkowicz Tomasz et al |
| EPI_ISL_905773 | USA/CA-LACPHL-AF00265/2021 | 2021-01-18 | North America/USA/California/Los Ange | UCLA Clinical Micro Lab | Los Angeles County PHL | P. Hemarajata et al. et al |
| EPI_ISL_905803 | USA/OR-OHSU-7431/2020 | 2020-12-08 | North America/USA/Oregon/Yamhill Co | OHSU Lab Services Molecular Microbiology Lab | Oregon SARS-CoV-2 Genome Sequencing Center | Brendan L. O'Connell et al |
| EPI_ISL_905807 | USA/OR-OHSU-7630/2020 | 2020-12-10 | North America/USA/Oregon/Washingto | OHSU Lab Services Molecular Microbiology Lab | Oregon SARS-CoV-2 Genome Sequencing Center | Brendan L. O'Connell et al |
| EPI_ISL_905808 | USA/OR-OHSU-7669/2020 | 2020-12-11 | North America/USA/Oregon/Washingto | OHSU Lab Services Molecular Microbiology Lab | Oregon SARS-CoV-2 Genome Sequencing Center | Brendan L. O'Connell et al |
| EPI_ISL_905813 | USA/OR-OHSU-7860/2020 | 2020-12-15 | North America/USA/Oregon/Multnomal | OHSU Lab Services Molecular Microbiology Lab | Oregon SARS-CoV-2 Genome Sequencing Center | Brendan L. O'Connell et al |
| EPI_ISL_905834 | USA/OR-OHSU-8025/2020 | 2020-12-16 | North America/USA/Oregon/Multnomal | OHSU Lab Services Molecular Microbiology Lab | Oregon SARS-CoV-2 Genome Sequencing Center | Brendan L. O'Connell et al |

| | | | | | | |
|---|---|---|---|---|---|---|
| EPI_ISL_905850 | USA/OR-OHSU-8860/2020 | 2020-12-31 | North America/USA/Oregon/Clackamas | OHSU Lab Services Molecular Microbiology Lab | Oregon SARS-CoV-2 Genome Sequencing Center | Brendan L. O'Connell et al |
| EPI_ISL_905866 | USA/OR-OHSU-8937/2021 | 2021-01-01 | North America/USA/Oregon/ | OHSU Lab Services Molecular Microbiology Lab | Oregon SARS-CoV-2 Genome Sequencing Center | Brendan L. O'Connell et al |
| EPI_ISL_905880 | USA/OR-OHSU-8956/2021 | 2021-01-01 | North America/USA/Oregon/Washingto | OHSU Lab Services Molecular Microbiology Lab | Oregon SARS-CoV-2 Genome Sequencing Center | Brendan L. O'Connell et al |
| EPI_ISL_905895 | USA/OR-OHSU-8974/2021 | 2021-01-01 | North America/USA/Oregon/Washingto | OHSU Lab Services Molecular Microbiology Lab | Oregon SARS-CoV-2 Genome Sequencing Center | Brendan L. O'Connell et al |
| EPI_ISL_905903 | USA/OR-OHSU-8999/2020 | 2020-12-24 | North America/USA/Oregon/Washingto | OHSU Lab Services Molecular Microbiology Lab | Oregon SARS-CoV-2 Genome Sequencing Center | Brendan L. O'Connell et al |
| EPI_ISL_905908 | USA/OR-OHSU-9151/2021 | 2021-01-02 | North America/USA/Oregon/Deschutes | OHSU Lab Services Molecular Microbiology Lab | Oregon SARS-CoV-2 Genome Sequencing Center | Brendan L. O'Connell et al |
| EPI_ISL_905909 | USA/OR-OHSU-9153/2021 | 2021-01-02 | North America/USA/Oregon/Washingto | OHSU Lab Services Molecular Microbiology Lab | Oregon SARS-CoV-2 Genome Sequencing Center | Brendan L. O'Connell et al |
| EPI_ISL_905917 | USA/OR-OHSU-9167/2021 | 2021-01-04 | North America/USA/Oregon/Multnomah | OHSU Lab Services Molecular Microbiology Lab | Oregon SARS-CoV-2 Genome Sequencing Center | Brendan L. O'Connell et al |
| EPI_ISL_905919 | USA/OR-OHSU-9171/2021 | 2021-01-04 | North America/USA/Oregon/ | OHSU Lab Services Molecular Microbiology Lab | Oregon SARS-CoV-2 Genome Sequencing Center | Brendan L. O'Connell et al |
| EPI_ISL_905921 | USA/OR-OHSU-9173/2021 | 2021-01-04 | North America/USA/Oregon/Multnomah | OHSU Lab Services Molecular Microbiology Lab | Oregon SARS-CoV-2 Genome Sequencing Center | Brendan L. O'Connell et al |

| | | | | | | |
|---|---|---|---|---|---|---|
| EPI_ISL_905922 | USA/OR-OHSU-9174/2021 | 2021-01-04 | North America/USA/Oregon/Washingto | OHSU Lab Services Molecular Microbiology Lab | Oregon SARS-CoV-2 Genome Sequencing Center | Brendan L. O'Connell et al |
| EPI_ISL_905923 | USA/OR-OHSU-9175/2021 | 2021-01-04 | North America/USA/Oregon/Columbia C | OHSU Lab Services Molecular Microbiology Lab | Oregon SARS-CoV-2 Genome Sequencing Center | Brendan L. O'Connell et al |
| EPI_ISL_905924 | USA/OR-OHSU-9177/2021 | 2021-01-04 | North America/USA/Oregon/Washingto | OHSU Lab Services Molecular Microbiology Lab | Oregon SARS-CoV-2 Genome Sequencing Center | Brendan L. O'Connell et al |
| EPI_ISL_905933 | USA/OR-OHSU-9192/2021 | 2021-01-05 | North America/USA/Oregon/Washingto | OHSU Lab Services Molecular Microbiology Lab | Oregon SARS-CoV-2 Genome Sequencing Center | Brendan L. O'Connell et al |
| EPI_ISL_905937 | USA/OR-OHSU-9199/2021 | 2021-01-05 | North America/USA/Oregon/Washingto | OHSU Lab Services Molecular Microbiology Lab | Oregon SARS-CoV-2 Genome Sequencing Center | Brendan L. O'Connell et al |
| EPI_ISL_905938 | USA/OR-OHSU-9203/2021 | 2021-01-05 | North America/USA/Oregon/Washingto | OHSU Lab Services Molecular Microbiology Lab | Oregon SARS-CoV-2 Genome Sequencing Center | Brendan L. O'Connell et al |
| EPI_ISL_905942 | USA/OR-OHSU-9209/2021 | 2021-01-05 | North America/USA/Oregon/Washingto | OHSU Lab Services Molecular Microbiology Lab | Oregon SARS-CoV-2 Genome Sequencing Center | Brendan L. O'Connell et al |
| EPI_ISL_905948 | USA/OR-OHSU-9220/2021 | 2021-01-05 | North America/USA/Oregon/Multnomal | OHSU Lab Services Molecular Microbiology Lab | Oregon SARS-CoV-2 Genome Sequencing Center | Brendan L. O'Connell et al |
| EPI_ISL_905950 | USA/AZ-OHSU-9223/2021 | 2021-01-05 | North America/USA/Arizona/Maricopa C | OHSU Lab Services Molecular Microbiology Lab | Oregon SARS-CoV-2 Genome Sequencing Center | Brendan L. O'Connell et al |
| EPI_ISL_905966 | USA/OR-OHSU-9259/2021 | 2021-01-05 | North America/USA/Oregon/Washingto | OHSU Lab Services Molecular Microbiology Lab | Oregon SARS-CoV-2 Genome Sequencing Center | Brendan L. O'Connell et al |

| | | | | | | |
|---|---|---|---|---|---|---|
| EPI_ISL_905968 | USA/OR-OHSU-9261/2021 | 2021-01-05 | North America/USA/Oregon/Washingto | OHSU Lab Services Molecular Microbiology Lab | Oregon SARS-CoV-2 Genome Sequencing Center | Brendan L. O'Connell et al |
| EPI_ISL_905972 | USA/OR-OHSU-9272/2021 | 2021-01-05 | North America/USA/Oregon/Washingto | OHSU Lab Services Molecular Microbiology Lab | Oregon SARS-CoV-2 Genome Sequencing Center | Brendan L. O'Connell et al |
| EPI_ISL_905976 | USA/OR-OHSU-9278/2021 | 2021-01-05 | North America/USA/Oregon/Washingto | OHSU Lab Services Molecular Microbiology Lab | Oregon SARS-CoV-2 Genome Sequencing Center | Brendan L. O'Connell et al |
| EPI_ISL_905988 | USA/OR-OHSU-9293/2021 | 2021-01-06 | North America/USA/Oregon/ | OHSU Lab Services Molecular Microbiology Lab | Oregon SARS-CoV-2 Genome Sequencing Center | Brendan L. O'Connell et al |
| EPI_ISL_905995 | USA/OR-OHSU-9303/2021 | 2021-01-06 | North America/USA/Oregon/Multnomal | OHSU Lab Services Molecular Microbiology Lab | Oregon SARS-CoV-2 Genome Sequencing Center | Brendan L. O'Connell et al |
| EPI_ISL_905999 | USA/OR-OHSU-9313/2021 | 2021-01-06 | North America/USA/Oregon/Multnomal | OHSU Lab Services Molecular Microbiology Lab | Oregon SARS-CoV-2 Genome Sequencing Center | Brendan L. O'Connell et al |
| EPI_ISL_906014 | USA/OR-OHSU-9342/2021 | 2021-01-06 | North America/USA/Oregon/Washingto | OHSU Lab Services Molecular Microbiology Lab | Oregon SARS-CoV-2 Genome Sequencing Center | Brendan L. O'Connell et al |
| EPI_ISL_906019 | USA/OR-OHSU-9348/2021 | 2021-01-06 | North America/USA/Oregon/Washingto | OHSU Lab Services Molecular Microbiology Lab | Oregon SARS-CoV-2 Genome Sequencing Center | Brendan L. O'Connell et al |
| EPI_ISL_906032 | USA/OR-OHSU-9369/2021 | 2021-01-07 | North America/USA/Oregon/Multnomal | OHSU Lab Services Molecular Microbiology Lab | Oregon SARS-CoV-2 Genome Sequencing Center | Brendan L. O'Connell et al |
| EPI_ISL_906043 | USA/OR-OHSU-9387/2021 | 2021-01-07 | North America/USA/Oregon/Washingto | OHSU Lab Services Molecular Microbiology Lab | Oregon SARS-CoV-2 Genome Sequencing Center | Brendan L. O'Connell et al |

| | | | | | | |
|---|---|---|---|---|---|---|
| EPI_ISL_906045 | USA/OR-OHSU-9914/2021 | 2021-01-11 | North America/USA/Oregon/Washingto | OHSU Lab Services Molecular Microbiology Lab | Oregon SARS-CoV-2 Genome Sequencing Center | Brendan L. O'Connell et al |
| EPI_ISL_906049 | Turkey/HSGM-1549/2021 | 2021-01-22 | Europe/Turkey/Turkey/ | Ministry of Health Turkey | Ministry of Health Turkey | Fatma Bayrakdar et al |
| EPI_ISL_906051 | Indonesia/YO-UGM-124777/2020 | 2020-12-23 | Asia/Indonesia/Yogyakarta/ | RSA UGM | Genetics Working Group (Pokja Genetik) Faculty of Medicine, Public Health and Nursing Universitas Gadjah Mada (FK-KMK UGM); Disease Investigation Center Wates Ministry of Agriculture Indonesia; Department of Microbiology FK-KMK UGM; Laboratorium Diagnostik Yayasan Tahija World Mosquito Program (WMP) Yogyakarta Center for Tropical Medicine FK-KMK UGM; Integrated Research center FK-KMK UGM; Department of Computer Science and Electronics FMIPA UGM | Gunadi et al |
| EPI_ISL_906054 | CzechRepublic/Tilia-05/2020 | 2020-09-29 | Europe/Czech Republic/Central Bohemia | Tilia Laboratories s.r.o. | Tilia Laboratories s.r.o. | Sona Pekova et al |

| | | | | | | |
|---|---|---|---|---|---|---|
| EPI_ISL_906055 | CzechRepublic/Tilia-06/2020 | 2020-09-29 | Europe/Czech Republic/Prague/ | Tilia Laboratories s.r.o. | Tilia Laboratories s.r.o. | Sona Pekova MD et al |
| EPI_ISL_906058 | CzechRepublic/Tilia-11/2020 | 2020-12-30 | Europe/Czech Republic/Prague/ | Tilia Laboratories s.r.o. | Tilia Laboratories s.r.o. | Sona Pekova et al |
| EPI_ISL_906061 | CzechRepublic/Tilia-09/2020 | 2020-12-25 | Europe/Czech Republic/Prague/ | Tilia Laboratories s.r.o. | Tilia Laboratories s.r.o. | Sona Pekova et al |
| EPI_ISL_906062 | CzechRepublic/Tilia-10/2020 | 2020-12-29 | Europe/Czech Republic/Prague/ | Tilia Laboratories s.r.o. | Tilia Laboratories s.r.o. | Sona Pekova et al |
| EPI_ISL_906063 | CzechRepublic/Tilia-03/2020 | 2020-09-16 | Europe/Czech Republic/Prague/ | Tilia Laboratories s.r.o. | Tilia Laboratories s.r.o. | Sona Pekova et al |
| EPI_ISL_906064 | CzechRepublic/Tilia-04/2020 | 2020-09-17 | Europe/Czech Republic/Prague/ | Tilia Laboratories s.r.o. | Tilia Laboratories s.r.o. | Sona Pekova et al |
| EPI_ISL_906065 | Brazil/SP-943/2020 | 2020-04-17 | South America/Brazil/S√£o Paulo/S√£o | Day Hospital de Ermelino Matarazzo | Instituto Adolfo Lutz, Interdiciplinary Procedures Center, Strategic Laboratory | Claudio Tavares Sacchi et al |
| EPI_ISL_906067 | Brazil/SP-864/2020 | 2020-05-11 | South America/Brazil/S√£o Paulo/Barue | PS e Maternidade Nair Fonseca Leitao Arantes | Instituto Adolfo Lutz, Interdiciplinary Procedures Center, Strategic Laboratory | Claudio Tavares Sacchi et al |
| EPI_ISL_906068 | Brazil/SP-1034/2021 | 2021-01-15 | South America/Brazil/S√£o Paulo/Campi | Instituto Adolfo Lutz - Regional de Campinas | Instituto Adolfo Lutz, Interdiciplinary Procedures Center, Strategic Laboratory | Claudio Tavares Sacchi et al |
| EPI_ISL_906069 | Brazil/AM-1035/2021 | 2021-01-15 | South America/Brazil/Amazonas BR/Ma | Instituto Adolfo Lutz - Regional de Campinas | Instituto Adolfo Lutz, Interdiciplinary Procedures Center, Strategic Laboratory | Claudio Tavares Sacchi et al |

| | | | | | | |
|---|---|---|---|---|---|---|
| EPI_ISL_906070 | Brazil/AM-1039/2021 | 2021-01-18 | South America/Brazil/Amazonas BR/Ma | UPA Dr. Akira Tada | Instituto Adolfo Lutz, Interdiciplinary Procedures Center, Strategic Laboratory | Claudio Tavares Sacchi et al |
| EPI_ISL_906071 | Brazil/AM-1062/2021 | 2021-01-19 | South America/Brazil/Amazonas BR/Ma | LACEN-PI DR. Costa Alvarenga | Instituto Adolfo Lutz, Interdiciplinary Procedures Center, Strategic Laboratory | Claudio Tavares Sacchi et al |
| EPI_ISL_906072 | Brazil/SP-1041/2021 | 2021-01-19 | South America/Brazil/S√£o Paulo/Embu | UPA Dr. Akira Tada | Instituto Adolfo Lutz, Interdiciplinary Procedures Center, Strategic Laboratory | Claudio Tavares Sacchi et al |
| EPI_ISL_906073 | Brazil/SP-1046/2021 | 2021-01-19 | South America/Brazil/S√£o Paulo/S√£o | Hospital Vila Lobos | Instituto Adolfo Lutz, Interdiciplinary Procedures Center, Strategic Laboratory | Claudio Tavares Sacchi et al |
| EPI_ISL_906075 | Brazil/AM-1031/2021 | 2021-01-19 | South America/Brazil/Amazonas BR/Ma | Hospital Geral de Vila Penteado Dr Jose Pangella Sao Paulo | Instituto Adolfo Lutz, Interdiciplinary Procedures Center, Strategic Laboratory | Claudio Tavares Sacchi et al |
| EPI_ISL_906076 | Brazil/AM-1036/2021 | 2021-01-19 | South America/Brazil/Amazonas BR/Ma | Hospital Sao Luiz Sao Caetano | Instituto Adolfo Lutz, Interdiciplinary Procedures Center, Strategic Laboratory | Claudio Tavares Sacchi et al |

| | | | | | | |
|---|---|---|---|---|---|---|
| EPI_ISL_906077 | Brazil/AM-1037/2021 | 2021-01-19 | South America/Brazil/Amazonas BR/Mai | Hospital Sao Luiz Sao Caetano | Instituto Adolfo Lutz, Interdiciplinary Procedures Center, Strategic Laboratory | Claudio Tavares Sacchi et al |
| EPI_ISL_906078 | Brazil/SP-1052/2021 | 2021-01-20 | South America/Brazil/S√£o Paulo/Guaru | Hospital Carlos Chagas | Instituto Adolfo Lutz, Interdiciplinary Procedures Center, Strategic Laboratory | Claudio Tavares Sacchi et al |
| EPI_ISL_906080 | Brazil/AM-1047/2021 | 2021-01-22 | South America/Brazil/Amazonas BR/Mai | Hospital Beneficiencia Portuguesa | Instituto Adolfo Lutz, Interdiciplinary Procedures Center, Strategic Laboratory | Claudio Tavares Sacchi et al |
| EPI_ISL_906081 | Brazil/AM-1048/2021 | 2021-01-22 | South America/Brazil/Amazonas BR/Mai | Hospital Beneficiencia Portuguesa | Instituto Adolfo Lutz, Interdiciplinary Procedures Center, Strategic Laboratory | Claudio Tavares Sacchi et al |
| EPI_ISL_906084 | Bangladesh/CHRF-0112/2020 | 2020-12-06 | Asia/Bangladesh/Chattogram/ | Child Health Research Foundation | Child Health Research Foundation | Senjuti Saha et al |
| EPI_ISL_906087 | Bangladesh/CHRF-0072/2020 | 2020-08-20 | Asia/Bangladesh/Mymensingh/ | Child Health Research Foundation | Child Health Research Foundation | Senjuti Saha et al |
| EPI_ISL_906089 | Bangladesh/CHRF-0089/2020 | 2020-12-01 | Asia/Bangladesh/Dhaka/ | Child Health Research Foundation | Child Health Research Foundation | Senjuti Saha et al |
| EPI_ISL_906091 | Bangladesh/CHRF-0128/2020 | 2020-12-31 | Asia/Bangladesh/Sylhet/ | Shimantik Pathology and Diagnostic Center | Child Health Research Foundation | Senjuti Saha et al |
| EPI_ISL_906094 | Bangladesh/CHRF-0091/2020 | 2020-12-02 | Asia/Bangladesh/Dhaka/ | Child Health Research Foundation | Child Health Research Foundation | Senjuti Saha et al |

| | | | | | | |
|---|---|---|---|---|---|---|
| EPI_ISL_906096 | Bangladesh/CHRF-0102/2020 | 2020-11-22 | Asia/Bangladesh/Sylhet/ | Child Health Research Foundation | Child Health Research Foundation | Senjuti Saha et al |
| EPI_ISL_906101 | Bangladesh/CHRF-0057/2020 | 2020-07-08 | Asia/Bangladesh/Chattogram/ | Child Health Research Foundation | Child Health Research Foundation | Senjuti Saha et al |
| EPI_ISL_906103 | Bangladesh/CHRF-0100/2020 | 2020-09-08 | Asia/Bangladesh/Sylhet/ | Child Health Research Foundation | Child Health Research Foundation | Senjuti Saha et al |
| EPI_ISL_906105 | Bangladesh/CHRF-0101/2020 | 2020-11-18 | Asia/Bangladesh/Sylhet/ | Child Health Research Foundation | Child Health Research Foundation | Senjuti Saha et al |
| EPI_ISL_906107 | Bangladesh/CHRF-0090/2020 | 2020-12-02 | Asia/Bangladesh/Dhaka/ | Child Health Research Foundation | Child Health Research Foundation | Senjuti Saha et al |
| EPI_ISL_906108 | Bangladesh/CHRF-0093/2020 | 2020-12-03 | Asia/Bangladesh/Rangpur/ | Child Health Research Foundation | Child Health Research Foundation | Senjuti Saha et al |
| EPI_ISL_906110 | Bangladesh/CHRF-0110/2020 | 2020-12-03 | Asia/Bangladesh/Dhaka/ | Child Health Research Foundation | Child Health Research Foundation | Senjuti Saha et al |
| EPI_ISL_906122 | Slovenia/2020-V0-271699/2020 | 2020-12-14 | Europe/Slovenia/Slovenia/ | Institute of Microbiology and Immunology, Faculty of Medicine, University of Ljubljana | Institute of Microbiology and Immunology, Faculty of Medicine, University of Ljubljana | Samo Zakotnik et al |
| EPI_ISL_906125 | Slovenia/2020-V0-272455/2020 | 2020-12-15 | Europe/Slovenia/Slovenia/ | Institute of Microbiology and Immunology, Faculty of Medicine, University of Ljubljana | Institute of Microbiology and Immunology, Faculty of Medicine, University of Ljubljana | Samo Zakotnik et al |
| EPI_ISL_906148 | USA/WI-UW-2967/2021 | 2021-01-05 | North America/USA/Wisconsin/Milwauk | University of Wisconsin-Madison AIDS Vaccine Research Laboratories | University of Wisconsin-Madison AIDS Vaccine Research Laboratories | Gage Moreno et al |

| | | | | | | |
|---|---|---|---|---|---|---|
| EPI_ISL_906151 | USA/WI-UW-2970/2021 | 2021-01-12 | North America/USA/Wisconsin/Milwauk | University of Wisconsin-Madison AIDS Vaccine Research Laboratories | University of Wisconsin-Madison AIDS Vaccine Research Laboratories | Gage Moreno et al |
| EPI_ISL_906157 | USA/WI-UW-2976/2021 | 2021-01-12 | North America/USA/Wisconsin/Milwauk | University of Wisconsin-Madison AIDS Vaccine Research Laboratories | University of Wisconsin-Madison AIDS Vaccine Research Laboratories | Gage Moreno et al |
| EPI_ISL_906159 | USA/WI-UW-2978/2021 | 2021-01-05 | North America/USA/Wisconsin/Milwauk | University of Wisconsin-Madison AIDS Vaccine Research Laboratories | University of Wisconsin-Madison AIDS Vaccine Research Laboratories | Gage Moreno et al |
| EPI_ISL_906166 | USA/WI-UW-2985/2021 | 2021-01-12 | North America/USA/Wisconsin/Milwauk | University of Wisconsin-Madison AIDS Vaccine Research Laboratories | University of Wisconsin-Madison AIDS Vaccine Research Laboratories | Gage Moreno et al |
| EPI_ISL_906169 | USA/WI-UW-2988/2021 | 2021-01-05 | North America/USA/Wisconsin/Milwauk | University of Wisconsin-Madison AIDS Vaccine Research Laboratories | University of Wisconsin-Madison AIDS Vaccine Research Laboratories | Gage Moreno et al |
| EPI_ISL_906181 | USA/WI-UW-3001/2021 | 2021-01-05 | North America/USA/Wisconsin/Milwauk | University of Wisconsin-Madison AIDS Vaccine Research Laboratories | University of Wisconsin-Madison AIDS Vaccine Research Laboratories | Gage Moreno et al |
| EPI_ISL_906186 | USA/WI-UW-3006/2021 | 2021-01-12 | North America/USA/Wisconsin/Milwauk | University of Wisconsin-Madison AIDS Vaccine Research Laboratories | University of Wisconsin-Madison AIDS Vaccine Research Laboratories | Gage Moreno et al |
| EPI_ISL_906188 | USA/WI-UW-3008/2021 | 2021-01-12 | North America/USA/Wisconsin/Milwauk | University of Wisconsin-Madison AIDS Vaccine Research Laboratories | University of Wisconsin-Madison AIDS Vaccine Research Laboratories | Gage Moreno et al |

| | | | | | | |
|---|---|---|---|---|---|---|
| EPI_ISL_906189 | USA/WI-UW-3064/2021 | 2021-01-26 | North America/USA/Wisconsin/Dane Co | University of Wisconsin-Madison AIDS Vaccine Research Laboratories | University of Wisconsin-Madison AIDS Vaccine Research Laboratories | Gage Moreno et al |
| EPI_ISL_906194 | USA/WI-UW-3069/2021 | 2021-01-15 | North America/USA/Wisconsin/Dane Co | University of Wisconsin-Madison AIDS Vaccine Research Laboratories | University of Wisconsin-Madison AIDS Vaccine Research Laboratories | Gage Moreno et al |
| EPI_ISL_906196 | USA/WI-UW-3071/2021 | 2021-01-16 | North America/USA/Wisconsin/Dane Co | University of Wisconsin-Madison AIDS Vaccine Research Laboratories | University of Wisconsin-Madison AIDS Vaccine Research Laboratories | Gage Moreno et al |
| EPI_ISL_906210 | USA/WI-UW-3045/2021 | 2021-01-19 | North America/USA/Wisconsin/Dane Co | University of Wisconsin-Madison AIDS Vaccine Research Laboratories | University of Wisconsin-Madison AIDS Vaccine Research Laboratories | Gage Moreno et al |
| EPI_ISL_906213 | USA/WI-UW-3048/2021 | 2021-01-15 | North America/USA/Wisconsin/Columbia | University of Wisconsin-Madison AIDS Vaccine Research Laboratories | University of Wisconsin-Madison AIDS Vaccine Research Laboratories | Gage Moreno et al |
| EPI_ISL_906216 | USA/WI-UW-3051/2021 | 2021-01-16 | North America/USA/Wisconsin/Dane Co | University of Wisconsin-Madison AIDS Vaccine Research Laboratories | University of Wisconsin-Madison AIDS Vaccine Research Laboratories | Gage Moreno et al |
| EPI_ISL_906222 | USA/WI-UW-3060/2021 | 2021-01-21 | North America/USA/Wisconsin/Dane Co | University of Wisconsin-Madison AIDS Vaccine Research Laboratories | University of Wisconsin-Madison AIDS Vaccine Research Laboratories | Gage Moreno et al |
| EPI_ISL_906224 | USA/IL-UW-3062/2021 | 2021-01-19 | North America/USA/Illinois/Boone Coun | University of Wisconsin-Madison AIDS Vaccine Research Laboratories | University of Wisconsin-Madison AIDS Vaccine Research Laboratories | Gage Moreno et al |

| | | | | | | |
|---|---|---|---|---|---|---|
| EPI_ISL_906234 | USA/WI-UW-3026/2021 | 2021-01-18 | North America/USA/Wisconsin/Rock Cou | University of Wisconsin-Madison AIDS Vaccine Research Laboratories | University of Wisconsin-Madison AIDS Vaccine Research Laboratories | Gage Moreno et al |
| EPI_ISL_906241 | USA/WI-UW-3033/2021 | 2021-01-17 | North America/USA/Wisconsin/Dane Co | University of Wisconsin-Madison AIDS Vaccine Research Laboratories | University of Wisconsin-Madison AIDS Vaccine Research Laboratories | Gage Moreno et al |
| EPI_ISL_906251 | USA/WI-UW-3084/2021 | 2021-01-18 | North America/USA/Wisconsin/Dane Co | University of Wisconsin-Madison AIDS Vaccine Research Laboratories | University of Wisconsin-Madison AIDS Vaccine Research Laboratories | Gage Moreno et al |
| EPI_ISL_906273 | USA/WI-UW-3098/2021 | 2021-01-28 | North America/USA/Wisconsin/Dane Co | University of Wisconsin-Madison AIDS Vaccine Research Laboratories | University of Wisconsin-Madison AIDS Vaccine Research Laboratories | Gage Moreno et al |
| EPI_ISL_906277 | Nigeria/CV616/2021 | 2021-01-07 | Africa/Nigeria/Osun State/ | Nigeria Centre for Disease Control (NCDC) | African Centre of Excellence for Genomics of Infectious Diseases (ACEGID), Redeemer's University | Oluniyi P.E. et al et al |
| EPI_ISL_906283 | Nigeria/CV624/2021 | 2021-01-13 | Africa/Nigeria/Osun State/ | Nigeria Centre for Disease Control (NCDC) | African Centre of Excellence for Genomics of Infectious Diseases (ACEGID), Redeemer's University | Oluniyi P.E. et al et al |

| | | | | | | |
|---|---|---|---|---|---|---|
| EPI_ISL_906284 | Nigeria/CV625/2021 | 2021-01-11 | Africa/Nigeria/Osun State/ | Nigeria Centre for Disease Control (NCDC) | African Centre of Excellence for Genomics of Infectious Diseases (ACEGID), Redeemer's University | Oluniyi P.E. et al et al |
| EPI_ISL_906306 | Panama/655071/2021 | 2021-01-11 | North America/Panama/Colon/ | Gorgas memorial Institute For Health Studies | Gorgas memorial Institute For Health Studies | Diaz Y et al |
| EPI_ISL_906311 | USA/CA-QDX-3828/2021 | 2021-01-09 | North America/USA/California/ | Quest Diagnostics | Quest Diagnostics | Rosenthal et al |
| EPI_ISL_906322 | USA/CA-QDX-3842/2021 | 2021-01-08 | North America/USA/California/ | Quest Diagnostics | Quest Diagnostics | Rosenthal et al |
| EPI_ISL_906323 | USA/CA-QDX-3843/2021 | 2021-01-09 | North America/USA/California/ | Quest Diagnostics | Quest Diagnostics | Rosenthal et al |
| EPI_ISL_906336 | USA/CA-QDX-3859/2021 | 2021-01-10 | North America/USA/California/ | Quest Diagnostics | Quest Diagnostics | Rosenthal et al |
| EPI_ISL_906342 | USA/CA-QDX-3872/2021 | 2021-01-07 | North America/USA/California/ | Quest Diagnostics | Quest Diagnostics | Rosenthal et al |
| EPI_ISL_906344 | USA/CA-QDX-3874/2021 | 2021-01-10 | North America/USA/California/ | Quest Diagnostics | Quest Diagnostics | Rosenthal et al |
| EPI_ISL_906345 | USA/CA-QDX-3879/2021 | 2021-01-08 | North America/USA/California/ | Quest Diagnostics | Quest Diagnostics | Rosenthal et al |
| EPI_ISL_906350 | USA/CA-QDX-3885/2021 | 2021-01-08 | North America/USA/California/ | Quest Diagnostics | Quest Diagnostics | Rosenthal et al |
| EPI_ISL_906357 | USA/CA-QDX-3895/2021 | 2021-01-09 | North America/USA/California/ | Quest Diagnostics | Quest Diagnostics | Rosenthal et al |
| EPI_ISL_906372 | USA/CA-QDX-3912/2021 | 2021-01-08 | North America/USA/California/ | Quest Diagnostics | Quest Diagnostics | Rosenthal et al |
| EPI_ISL_906381 | USA/CA-QDX-3925/2021 | 2021-01-09 | North America/USA/California/ | Quest Diagnostics | Quest Diagnostics | Rosenthal et al |
| EPI_ISL_906385 | USA/CA-QDX-3931/2021 | 2021-01-10 | North America/USA/California/ | Quest Diagnostics | Quest Diagnostics | Rosenthal et al |
| EPI_ISL_906408 | USA/CA-QDX-3962/2021 | 2021-01-09 | North America/USA/California/ | Quest Diagnostics | Quest Diagnostics | Rosenthal et al |
| EPI_ISL_906412 | USA/CA-QDX-3966/2021 | 2021-01-09 | North America/USA/California/ | Quest Diagnostics | Quest Diagnostics | Rosenthal et al |
| EPI_ISL_906413 | USA/CA-QDX-3967/2021 | 2021-01-08 | North America/USA/California/ | Quest Diagnostics | Quest Diagnostics | Rosenthal et al |
| EPI_ISL_906417 | USA/CA-QDX-3974/2021 | 2021-01-08 | North America/USA/California/ | Quest Diagnostics | Quest Diagnostics | Rosenthal et al |
| EPI_ISL_906418 | USA/CA-QDX-3975/2021 | 2021-01-09 | North America/USA/California/ | Quest Diagnostics | Quest Diagnostics | Rosenthal et al |
| EPI_ISL_906425 | USA/CT-QDX-3932/2021 | 2021-01-06 | North America/USA/Connecticut/ | Quest Diagnostics | Quest Diagnostics | Rosenthal et al |
| EPI_ISL_906431 | USA/IL-QDX-3961/2021 | 2021-01-07 | North America/USA/Illinois/ | Quest Diagnostics | Quest Diagnostics | Rosenthal et al |
| EPI_ISL_906436 | USA/MA-QDX-3877/2021 | 2021-01-08 | North America/USA/Massachusetts/ | Quest Diagnostics | Quest Diagnostics | Rosenthal et al |
| EPI_ISL_906446 | USA/ME-QDX-3871/2021 | 2021-01-06 | North America/USA/Maine/ | Quest Diagnostics | Quest Diagnostics | Rosenthal et al |

| | | | | | | |
|---|---|---|---|---|---|---|
| EPI_ISL_906500 | Japan/PG-11136/2020 | 20-Oct | Asia/Japan/Japan/ | Sakai City Institute of Public Health | Pathogen Genomics Center, National Institute of Infectious Diseases | Tsuyoshi Sekizuka et al |
| EPI_ISL_906531 | Colombia/VAC-INS-RI-010/2021 | 2021-01-13 | South America/Colombia/Valle del Cauc | Laboratorio de salud publica del Valle del Cauca | Instituto Nacional de Salud-Direcci√≥n de Investigaci√≥n en Salud P√∫blica, Universidad de los Andes- Applied genomics research group, Vicerrectoria de Investigaci√≥n y Creaci√≥n, Universidad de los Andes- Systems and Computing Engineering Department | Katherine Laiton-Donato et al |
| EPI_ISL_906541 | Colombia/LAG-INS-VG-323/2020 | 2020-12-21 | South America/Colombia/La Guajira/ | HOSPITAL SAN JOSE DE MAICAO | Instituto Nacional de Salud-Direcci√≥n de Investigaci√≥n en Salud P√∫blica, Universidad de los Andes- Applied genomics research group, Vicerrectoria de Investigaci√≥n y Creaci√≥n, Universidad de los Andes- Systems and Computing Engineering Department | Katherine Laiton-Donato et al |

| | | | | | | |
|---|---|---|---|---|---|---|
| EPI_ISL_906572 | USA/ME-HETL-J1012/2020 | 2020-12-30 | North America/USA/Maine/ | Maine Health and Environmental Testing Laboratory (Maine HETL) | Tewhey Lab, The Jackson Laboratory | Matluk et al |
| EPI_ISL_906579 | USA/ME-HETL-J0938/2021 | 2021-01-10 | North America/USA/Maine/ | Maine Health and Environmental Testing Laboratory (Maine HETL) | Tewhey Lab, The Jackson Laboratory | Matluk et al |
| EPI_ISL_906590 | USA/ME-HETL-J0862/2021 | 2021-01-11 | North America/USA/Maine/ | Maine Health and Environmental Testing Laboratory (Maine HETL) | Tewhey Lab, The Jackson Laboratory | Matluk et al |
| EPI_ISL_906600 | USA/ME-HETL-J0935/2021 | 2021-01-11 | North America/USA/Maine/ | Maine Health and Environmental Testing Laboratory (Maine HETL) | Tewhey Lab, The Jackson Laboratory | Matluk et al |
| EPI_ISL_906604 | USA/ME-HETL-J0874/2021 | 2021-01-12 | North America/USA/Maine/ | Maine Health and Environmental Testing Laboratory (Maine HETL) | Tewhey Lab, The Jackson Laboratory | Matluk et al |
| EPI_ISL_906612 | USA/ME-HETL-J0884/2021 | 2021-01-12 | North America/USA/Maine/ | Maine Health and Environmental Testing Laboratory (Maine HETL) | Tewhey Lab, The Jackson Laboratory | Matluk et al |
| EPI_ISL_906613 | USA/ME-HETL-J0886/2021 | 2021-01-12 | North America/USA/Maine/ | Maine Health and Environmental Testing Laboratory (Maine HETL) | Tewhey Lab, The Jackson Laboratory | Matluk et al |
| EPI_ISL_906614 | USA/ME-HETL-J0887/2021 | 2021-01-12 | North America/USA/Maine/ | Maine Health and Environmental Testing Laboratory (Maine HETL) | Tewhey Lab, The Jackson Laboratory | Matluk et al |
| EPI_ISL_906616 | USA/ME-HETL-J0889/2021 | 2021-01-12 | North America/USA/Maine/ | Maine Health and Environmental Testing Laboratory (Maine HETL) | Tewhey Lab, The Jackson Laboratory | Matluk et al |
| EPI_ISL_906636 | USA/ME-HETL-J0914/2021 | 2021-01-12 | North America/USA/Maine/ | Maine Health and Environmental Testing Laboratory (Maine HETL) | Tewhey Lab, The Jackson Laboratory | Matluk et al |
| EPI_ISL_906638 | USA/ME-HETL-J0916/2021 | 2021-01-12 | North America/USA/Maine/ | Maine Health and Environmental Testing Laboratory (Maine HETL) | Tewhey Lab, The Jackson Laboratory | Matluk et al |
| EPI_ISL_906640 | USA/ME-HETL-J0934/2021 | 2021-01-12 | North America/USA/Maine/ | Maine Health and Environmental Testing Laboratory (Maine HETL) | Tewhey Lab, The Jackson Laboratory | Matluk et al |
| EPI_ISL_906653 | USA/ME-HETL-J0972/2021 | 2021-01-15 | North America/USA/Maine/ | Maine Health and Environmental Testing Laboratory (Maine HETL) | Tewhey Lab, The Jackson Laboratory | Matluk et al |

| | | | | | | |
|---|---|---|---|---|---|---|
| EPI_ISL_906654 | USA/ME-HETL-J1047/2021 | 2021-01-17 | North America/USA/Maine/ | Maine Health and Environmental Testing Laboratory (Maine HETL) | Tewhey Lab, The Jackson Laboratory | Matluk et al |
| EPI_ISL_906661 | USA/ME-HETL-J1043/2021 | 2021-01-18 | North America/USA/Maine/ | Maine Health and Environmental Testing Laboratory (Maine HETL) | Tewhey Lab, The Jackson Laboratory | Matluk et al |
| EPI_ISL_906676 | USA/ME-HETL-J0964/2021 | 2021-01-21 | North America/USA/Maine/ | Maine Health and Environmental Testing Laboratory (Maine HETL) | Tewhey Lab, The Jackson Laboratory | Matluk et al |
| EPI_ISL_906684 | USA/ME-HETL-J0985/2021 | 2021-01-21 | North America/USA/Maine/ | Maine Health and Environmental Testing Laboratory (Maine HETL) | Tewhey Lab, The Jackson Laboratory | Matluk et al |
| EPI_ISL_906693 | USA/ME-HETL-J1021/2021 | 2021-01-21 | North America/USA/Maine/ | Maine Health and Environmental Testing Laboratory (Maine HETL) | Tewhey Lab, The Jackson Laboratory | Matluk et al |
| EPI_ISL_906719 | Poland/Pomorskie_MWB_11/2021 | 2021-01-06 | Europe/Poland/Pomorskie/ | Hematology Laboratory, Section of Molecular Diagnostics, University Clinical Centre, Medical University of Gdansk | Laboratory of Recombinant Vaccines | Lukasz Rabalski et al |
| EPI_ISL_906721 | Poland/Pomorskie_MWB_13/2021 | 2021-01-08 | Europe/Poland/Pomorskie/ | Hematology Laboratory, Section of Molecular Diagnostics, University Clinical Centre, Medical University of Gdansk | Laboratory of Recombinant Vaccines | Lukasz Rabalski et al |
| EPI_ISL_906723 | Poland/Pomorskie_MWB_15/2021 | 2021-01-10 | Europe/Poland/Pomorskie/ | Hematology Laboratory, Section of Molecular Diagnostics, University Clinical Centre, Medical University of Gdansk | Laboratory of Recombinant Vaccines | Lukasz Rabalski et al |
| EPI_ISL_906724 | Poland/Pomorskie_MWB_16/2021 | 2021-01-09 | Europe/Poland/Pomorskie/ | Hematology Laboratory, Section of Molecular Diagnostics, University Clinical Centre, Medical University of Gdansk | Laboratory of Recombinant Vaccines | Lukasz Rabalski et al |
| EPI_ISL_906726 | Poland/Pomorskie_MWB_19/2020 | 2020-12-25 | Europe/Poland/Pomorskie/ | Hematology Laboratory, Section of Molecular Diagnostics, University Clinical Centre, Medical University of Gdansk | Laboratory of Recombinant Vaccines | Lukasz Rabalski et al |
| EPI_ISL_906729 | Poland/Pomorskie_MWB_27/2021 | 2021-01-08 | Europe/Poland/Pomorskie/ | Hematology Laboratory, Section of Molecular Diagnostics, University Clinical Centre, Medical University of Gdansk | Laboratory of Recombinant Vaccines | Lukasz Rabalski et al |
| EPI_ISL_906732 | Poland/Pomorskie_MWB_30/2021 | 2021-01-07 | Europe/Poland/Pomorskie/ | Hematology Laboratory, Section of Molecular Diagnostics, University Clinical Centre, Medical University of Gdansk | Laboratory of Recombinant Vaccines | Lukasz Rabalski et al |
| EPI_ISL_906734 | Poland/Pomorskie_MWB_32/2021 | 2021-01-06 | Europe/Poland/Pomorskie/ | Hematology Laboratory, Section of Molecular Diagnostics, University Clinical Centre, Medical University of Gdansk | Laboratory of Recombinant Vaccines | Lukasz Rabalski et al |

| | | | | | | |
|---|---|---|---|---|---|---|
| EPI_ISL_906737 | Poland/Pomorskie_MWB_39/2021 | 2021-01-05 | Europe/Poland/Pomorskie/ | Hematology Laboratory, Section of Molecular Diagnostics, University Clinical Centre, Medical University of Gdansk | Laboratory of Recombinant Vaccines | Lukasz Rabalski et al |
| EPI_ISL_906740 | Poland/Pomorskie_MWB_47/2021 | 2021-01-24 | Europe/Poland/Pomorskie/ | Laboratory of Molecular Biology, Diagnostyka sp. z o.o. | Laboratory of Recombinant Vaccines | Lukasz Rabalski et al |
| EPI_ISL_906741 | Poland/Pomorskie_MWB_48/2021 | 2021-01-05 | Europe/Poland/Pomorskie/ | Hematology Laboratory, Section of Molecular Diagnostics, University Clinical Centre, Medical University of Gdansk | Laboratory of Recombinant Vaccines | Lukasz Rabalski et al |
| EPI_ISL_906742 | Poland/Pomorskie_MWB_50/2021 | 2021-01-06 | Europe/Poland/Pomorskie/ | Hematology Laboratory, Section of Molecular Diagnostics, University Clinical Centre, Medical University of Gdansk | Laboratory of Recombinant Vaccines | Lukasz Rabalski et al |
| EPI_ISL_906743 | Poland/Pomorskie_MWB_51/2021 | 2021-01-24 | Europe/Poland/Pomorskie/ | Laboratory of Molecular Biology, Diagnostyka sp. z o.o. | Laboratory of Recombinant Vaccines | Lukasz Rabalski et al |
| EPI_ISL_906744 | Poland/Pomorskie_MWB_52/2021 | 2021-01-03 | Europe/Poland/Pomorskie/ | Hematology Laboratory, Section of Molecular Diagnostics, University Clinical Centre, Medical University of Gdansk | Laboratory of Recombinant Vaccines | Lukasz Rabalski et al |
| EPI_ISL_906745 | Poland/Pomorskie_MWB_53/2021 | 2021-01-05 | Europe/Poland/Pomorskie/ | Hematology Laboratory, Section of Molecular Diagnostics, University Clinical Centre, Medical University of Gdansk | Laboratory of Recombinant Vaccines | Lukasz Rabalski et al |
| EPI_ISL_906746 | Poland/Pomorskie_MWB_54/2021 | 2021-01-03 | Europe/Poland/Pomorskie/ | Hematology Laboratory, Section of Molecular Diagnostics, University Clinical Centre, Medical University of Gdansk | Laboratory of Recombinant Vaccines | Lukasz Rabalski et al |
| EPI_ISL_906747 | Poland/Pomorskie_MWB_55/2021 | 2021-01-05 | Europe/Poland/Pomorskie/ | Hematology Laboratory, Section of Molecular Diagnostics, University Clinical Centre, Medical University of Gdansk | Laboratory of Recombinant Vaccines | Lukasz Rabalski et al |
| EPI_ISL_906749 | Poland/Pomorskie_MWB_57/2021 | 2021-01-10 | Europe/Poland/Pomorskie/ | Hematology Laboratory, Section of Molecular Diagnostics, University Clinical Centre, Medical University of Gdansk | Laboratory of Recombinant Vaccines | Lukasz Rabalski et al |
| EPI_ISL_906751 | Poland/Pomorskie_MWB_59/2021 | 2021-01-05 | Europe/Poland/Pomorskie/ | Hematology Laboratory, Section of Molecular Diagnostics, University Clinical Centre, Medical University of Gdansk | Laboratory of Recombinant Vaccines | Lukasz Rabalski et al |
| EPI_ISL_906752 | Poland/Pomorskie_MWB_60/2020 | 2020-08-30 | Europe/Poland/Pomorskie/ | Hematology Laboratory, Section of Molecular Diagnostics, University Clinical Centre, Medical University of Gdansk | Laboratory of Recombinant Vaccines | Lukasz Rabalski et al |
| EPI_ISL_906756 | Poland/Pomorskie_MWB_66/2021 | 2021-01-10 | Europe/Poland/Pomorskie/ | Hematology Laboratory, Section of Molecular Diagnostics, University Clinical Centre, Medical University of Gdansk | Laboratory of Recombinant Vaccines | Lukasz Rabalski et al |

| | | | | | | |
|---|---|---|---|---|---|---|
| EPI_ISL_906757 | Poland/Pomorskie_MWB_67/2021 | 2021-01-24 | Europe/Poland/Pomorskie/ | Laboratory of Molecular Biology, Diagnostyka sp. z o.o. | Laboratory of Recombinant Vaccines | Lukasz Rabalski et al |
| EPI_ISL_906758 | Poland/Pomorskie_MWB_68/2021 | 2021-01-24 | Europe/Poland/Pomorskie/ | Laboratory of Molecular Biology, Diagnostyka sp. z o.o. | Laboratory of Recombinant Vaccines | Lukasz Rabalski et al |
| EPI_ISL_906761 | Canada/NL-NML-1384/2020 | 2020-03-23 | North America/Canada/Newfoundland a | Dr. Leonard A. Miller Centre for Health Services | National Microbiology Laboratory (NML) | Anna Majer et al |
| EPI_ISL_906762 | Canada/NL-NML-1385/2020 | 2020-03-24 | North America/Canada/Newfoundland a | Dr. Leonard A. Miller Centre for Health Services | National Microbiology Laboratory (NML) | Anna Majer et al |
| EPI_ISL_906763 | Canada/NL-NML-1386/2020 | 2020-03-24 | North America/Canada/Newfoundland a | Dr. Leonard A. Miller Centre for Health Services | National Microbiology Laboratory (NML) | Anna Majer et al |
| EPI_ISL_906764 | Canada/NL-NML-1387/2020 | 2020-03-25 | North America/Canada/Newfoundland a | Dr. Leonard A. Miller Centre for Health Services | National Microbiology Laboratory (NML) | Anna Majer et al |
| EPI_ISL_906765 | Canada/NL-NML-1388/2020 | 2020-03-26 | North America/Canada/Newfoundland a | Dr. Leonard A. Miller Centre for Health Services | National Microbiology Laboratory (NML) | Anna Majer et al |
| EPI_ISL_906766 | Canada/NL-NML-1389/2020 | 2020-03-27 | North America/Canada/Newfoundland a | Dr. Leonard A. Miller Centre for Health Services | National Microbiology Laboratory (NML) | Anna Majer et al |
| EPI_ISL_906767 | Canada/NL-NML-1390/2020 | 2020-03-25 | North America/Canada/Newfoundland a | Dr. Leonard A. Miller Centre for Health Services | National Microbiology Laboratory (NML) | Anna Majer et al |
| EPI_ISL_906768 | Canada/NL-NML-1391/2020 | 2020-03-27 | North America/Canada/Newfoundland a | Dr. Leonard A. Miller Centre for Health Services | National Microbiology Laboratory (NML) | Anna Majer et al |
| EPI_ISL_906769 | Canada/NL-NML-1392/2020 | 2020-03-31 | North America/Canada/Newfoundland a | Dr. Leonard A. Miller Centre for Health Services | National Microbiology Laboratory (NML) | Anna Majer et al |
| EPI_ISL_906770 | Canada/NL-NML-1393/2020 | 2020-03-31 | North America/Canada/Newfoundland a | Dr. Leonard A. Miller Centre for Health Services | National Microbiology Laboratory (NML) | Anna Majer et al |
| EPI_ISL_906771 | Canada/NL-NML-1394/2020 | 2020-03-31 | North America/Canada/Newfoundland a | Dr. Leonard A. Miller Centre for Health Services | National Microbiology Laboratory (NML) | Anna Majer et al |

| | | | | | | |
|---|---|---|---|---|---|---|
| EPI_ISL_906772 | Canada/NL-NML-1395/2020 | 2020-04-02 | North America/Canada/Newfoundland a | Dr. Leonard A. Miller Centre for Health Services | National Microbiology Laboratory (NML) | Anna Majer et al |
| EPI_ISL_906773 | Canada/NL-NML-1396/2020 | 2020-04-04 | North America/Canada/Newfoundland a | Dr. Leonard A. Miller Centre for Health Services | National Microbiology Laboratory (NML) | Anna Majer et al |
| EPI_ISL_906774 | Canada/NL-NML-1397/2020 | 2020-04-06 | North America/Canada/Newfoundland a | Dr. Leonard A. Miller Centre for Health Services | National Microbiology Laboratory (NML) | Anna Majer et al |
| EPI_ISL_906775 | Canada/NL-NML-1398/2020 | 2020-04-09 | North America/Canada/Newfoundland a | Dr. Leonard A. Miller Centre for Health Services | National Microbiology Laboratory (NML) | Anna Majer et al |
| EPI_ISL_906776 | Canada/NL-NML-1399/2020 | 2020-03-24 | North America/Canada/Newfoundland a | Dr. Leonard A. Miller Centre for Health Services | National Microbiology Laboratory (NML) | Anna Majer et al |
| EPI_ISL_906851 | DominicanRepublic/CDC-9KZX-8454/2 | 2020-12-27 | North America/Dominican Republic/Dom | Respiratory Viruses Branch, Centers for Disease Control and Prevention | Respiratory Viruses Branch, Centers for Disease Control and Prevention | Tao et al |
| EPI_ISL_906890 | USA/FL-BPHL-0011/2021 | 2021-01-05 | North America/USA/Florida/ | Bureau of Public Health Laboratories, Florida Department of Health (BPHL, FLDOH) | Bureau of Public Health Laboratories, Florida Department of Health (BPHL, FLDOH) | Schmedes et al |
| EPI_ISL_906899 | USA/FL-BPHL-0026/2021 | 2021-01-06 | North America/USA/Florida/ | Bureau of Public Health Laboratories, Florida Department of Health (BPHL, FLDOH) | Bureau of Public Health Laboratories, Florida Department of Health (BPHL, FLDOH) | Schmedes et al |
| EPI_ISL_906903 | USA/FL-BPHL-0030/2021 | 2021-01-07 | North America/USA/Florida/ | Bureau of Public Health Laboratories, Florida Department of Health (BPHL, FLDOH) | Bureau of Public Health Laboratories, Florida Department of Health (BPHL, FLDOH) | Schmedes et al |

| | | | | | | |
|---|---|---|---|---|---|---|
| EPI_ISL_906904 | USA/FL-BPHL-0031/2021 | 2021-01-07 | North America/USA/Florida/ | Bureau of Public Health Laboratories, Florida Department of Health (BPHL, FLDOH) | Bureau of Public Health Laboratories, Florida Department of Health (BPHL, FLDOH) | Schmedes et al |
| EPI_ISL_906913 | USA/FL-BPHL-0040/2021 | 2021-01-11 | North America/USA/Florida/ | Bureau of Public Health Laboratories, Florida Department of Health (BPHL, FLDOH) | Bureau of Public Health Laboratories, Florida Department of Health (BPHL, FLDOH) | Schmedes et al |
| EPI_ISL_906924 | USA/CA-QDX-3578/2021 | 2021-01-07 | North America/USA/California/ | Infectious Diseases, Quest Diagnostics | Infectious Diseases, Quest Diagnostics | Rosenthal et al |
| EPI_ISL_906929 | USA/CA-QDX-3583/2021 | 2021-01-08 | North America/USA/California/ | Infectious Diseases, Quest Diagnostics | Infectious Diseases, Quest Diagnostics | Rosenthal et al |
| EPI_ISL_906942 | USA/CA-QDX-3597/2021 | 2021-01-09 | North America/USA/California/ | Infectious Diseases, Quest Diagnostics | Infectious Diseases, Quest Diagnostics | Rosenthal et al |
| EPI_ISL_906943 | USA/CA-QDX-3598/2021 | 2021-01-08 | North America/USA/California/ | Infectious Diseases, Quest Diagnostics | Infectious Diseases, Quest Diagnostics | Rosenthal et al |
| EPI_ISL_906945 | USA/CA-QDX-3600/2021 | 2021-01-09 | North America/USA/California/ | Infectious Diseases, Quest Diagnostics | Infectious Diseases, Quest Diagnostics | Rosenthal et al |
| EPI_ISL_906952 | USA/CA-QDX-3609/2021 | 2021-01-08 | North America/USA/California/ | Infectious Diseases, Quest Diagnostics | Infectious Diseases, Quest Diagnostics | Rosenthal et al |
| EPI_ISL_906954 | USA/CA-QDX-3611/2021 | 2021-01-06 | North America/USA/California/ | Infectious Diseases, Quest Diagnostics | Infectious Diseases, Quest Diagnostics | Rosenthal et al |
| EPI_ISL_906955 | USA/CA-QDX-3612/2021 | 2021-01-09 | North America/USA/California/ | Infectious Diseases, Quest Diagnostics | Infectious Diseases, Quest Diagnostics | Rosenthal et al |
| EPI_ISL_906960 | USA/CA-QDX-3618/2021 | 2021-01-10 | North America/USA/California/ | Infectious Diseases, Quest Diagnostics | Infectious Diseases, Quest Diagnostics | Rosenthal et al |

| | | | | | | |
|---|---|---|---|---|---|---|
| EPI_ISL_906963 | USA/CA-QDX-3621/2021 | 2021-01-10 | North America/USA/California/ | Infectious Diseases, Quest Diagnostics | Infectious Diseases, Quest Diagnostics | Rosenthal et al |
| EPI_ISL_906969 | USA/CA-QDX-3627/2021 | 2021-01-09 | North America/USA/California/ | Infectious Diseases, Quest Diagnostics | Infectious Diseases, Quest Diagnostics | Rosenthal et al |
| EPI_ISL_906983 | USA/CA-QDX-3642/2021 | 2021-01-08 | North America/USA/California/ | Infectious Diseases, Quest Diagnostics | Infectious Diseases, Quest Diagnostics | Rosenthal et al |
| EPI_ISL_906992 | USA/CA-QDX-3653/2021 | 2021-01-08 | North America/USA/California/ | Infectious Diseases, Quest Diagnostics | Infectious Diseases, Quest Diagnostics | Rosenthal et al |
| EPI_ISL_906994 | USA/CA-QDX-3655/2021 | 2021-01-09 | North America/USA/California/ | Infectious Diseases, Quest Diagnostics | Infectious Diseases, Quest Diagnostics | Rosenthal et al |
| EPI_ISL_907018 | USA/CA-QDX-3683/2021 | 2021-01-08 | North America/USA/California/ | Infectious Diseases, Quest Diagnostics | Infectious Diseases, Quest Diagnostics | Rosenthal et al |
| EPI_ISL_907021 | USA/CA-QDX-3687/2021 | 2021-01-09 | North America/USA/California/ | Infectious Diseases, Quest Diagnostics | Infectious Diseases, Quest Diagnostics | Rosenthal et al |
| EPI_ISL_907022 | USA/CA-QDX-3688/2021 | 2021-01-09 | North America/USA/California/ | Infectious Diseases, Quest Diagnostics | Infectious Diseases, Quest Diagnostics | Rosenthal et al |
| EPI_ISL_907023 | USA/CA-QDX-3689/2021 | 2021-01-09 | North America/USA/California/ | Infectious Diseases, Quest Diagnostics | Infectious Diseases, Quest Diagnostics | Rosenthal et al |
| EPI_ISL_907036 | USA/CA-QDX-3706/2021 | 2021-01-10 | North America/USA/California/ | Infectious Diseases, Quest Diagnostics | Infectious Diseases, Quest Diagnostics | Rosenthal et al |
| EPI_ISL_907039 | USA/CA-QDX-3709/2021 | 2021-01-10 | North America/USA/California/ | Infectious Diseases, Quest Diagnostics | Infectious Diseases, Quest Diagnostics | Rosenthal et al |
| EPI_ISL_907043 | USA/CA-QDX-3713/2021 | 2021-01-08 | North America/USA/California/ | Infectious Diseases, Quest Diagnostics | Infectious Diseases, Quest Diagnostics | Rosenthal et al |
| EPI_ISL_907046 | USA/CA-QDX-3717/2021 | 2021-01-10 | North America/USA/California/ | Infectious Diseases, Quest Diagnostics | Infectious Diseases, Quest Diagnostics | Rosenthal et al |

| | | | | | | |
|---|---|---|---|---|---|---|
| EPI_ISL_907064 | USA/CT-QDX-3725/2021 | 2021-01-06 | North America/USA/Connecticut/ | Infectious Diseases, Quest Diagnostics | Infectious Diseases, Quest Diagnostics | Rosenthal et al |
| EPI_ISL_907071 | USA/NV-QDX-3643/2021 | 2021-01-09 | North America/USA/Nevada/ | Infectious Diseases, Quest Diagnostics | Infectious Diseases, Quest Diagnostics | Rosenthal et al |
| EPI_ISL_907073 | USA/NV-QDX-3701/2021 | 2021-01-09 | North America/USA/Nevada/ | Infectious Diseases, Quest Diagnostics | Infectious Diseases, Quest Diagnostics | Rosenthal et al |
| EPI_ISL_907075 | Iraq/BAS2/2020 | 2020-10-01 | Asia/Iraq/Iraq/ | Department of Biology, University of Basrah | Department of Biology, University of Basrah | Abu-Ali et al |
| EPI_ISL_907077 | Egypt/ARMY-MCL001-2/2020 | 2020-11-19 | Africa/Egypt/Egypt/ | Biology, MCL | Biology, MCL | Seadawy et al |
| EPI_ISL_907079 | Egypt/ARMY-MCL003/2020 | 2020-11-19 | Africa/Egypt/Egypt/ | Biology, MCL | Biology, MCL | Seadawy et al |
| EPI_ISL_907080 | Egypt/ARMY-MCL005/2020 | 2020-11-19 | Africa/Egypt/Egypt/ | Biology, MCL | Biology, MCL | Seadawy et al |
| EPI_ISL_907084 | Egypt/ARMY-MCL011/2020 | 2020-11-19 | Africa/Egypt/Egypt/ | Biology, MCL | Biology, MCL | Seadawy et al |
| EPI_ISL_907086 | Egypt/CUNCI-HGC8I033/2020 | 2020-12-25 | Africa/Egypt/Egypt/ | Cancer Biology Department, National Cancer Institute | Cancer Biology Department, National Cancer Institute | Zekri et al |
| EPI_ISL_907093 | Egypt/CUNCI-HGC11I003/2021 | 2021-01-03 | Africa/Egypt/Egypt/ | Cancer Biology Department, National Cancer Institute | Cancer Biology Department, National Cancer Institute | Zekri et al |
| EPI_ISL_907100 | Egypt/CUNCI-HGC10I027/2020 | 2020-12-25 | Africa/Egypt/Egypt/ | Cancer Biology Department, National Cancer Institute | Cancer Biology Department, National Cancer Institute | Zekri et al |
| EPI_ISL_907102 | Egypt/CUNCI-HGC11I011/2021 | 2021-01-03 | Africa/Egypt/Egypt/ | Cancer Biology Department, National Cancer Institute | Cancer Biology Department, National Cancer Institute | Zekri et al |
| EPI_ISL_907111 | Egypt/CUNCI-HGC11I046/2021 | 2021-01-03 | Africa/Egypt/Egypt/ | Cancer Biology Department, National Cancer Institute | Cancer Biology Department, National Cancer Institute | Zekri et al |

| | | | | | | |
|---|---|---|---|---|---|---|
| EPI_ISL_907115 | Egypt/CUNCI-HGC12I024/2021 | 2021-01-03 | Africa/Egypt/Egypt/ | Cancer Biology Department, National Cancer Institute | Cancer Biology Department, National Cancer Institute | Zekri et al |
| EPI_ISL_907253 | England/QEUH-10F8888/2021 | 2021-01-23 | Europe/United Kingdom/England/ | Lighthouse Lab in Glasgow | Wellcome Sanger Institute for the COVID-19 Genomics UK (COG-UK) Consortium | Harper VanSteenhouse et al |
| EPI_ISL_907273 | England/CAMC-1104C8C/2021 | 2021-01-20 | Europe/United Kingdom/England/ | Lighthouse Lab in Cambridge | Wellcome Sanger Institute for the COVID-19 Genomics UK (COG-UK) Consortium | Rob Howes et al |
| EPI_ISL_907522 | England/ALDP-10EABF9/2021 | 2021-01-20 | Europe/United Kingdom/England/ | Lighthouse Lab in Alderley Park | Wellcome Sanger Institute for the COVID-19 Genomics UK (COG-UK) Consortium | Jacquelyn Wynn et al |
| EPI_ISL_907583 | England/ALDP-10F6617/2021 | 2021-01-22 | Europe/United Kingdom/England/ | Lighthouse Lab in Alderley Park | Wellcome Sanger Institute for the COVID-19 Genomics UK (COG-UK) Consortium | Jacquelyn Wynn et al |
| EPI_ISL_907700 | England/ALDP-10E272A/2021 | 2021-01-18 | Europe/United Kingdom/England/ | Lighthouse Lab in Alderley Park | Wellcome Sanger Institute for the COVID-19 Genomics UK (COG-UK) Consortium | Jacquelyn Wynn et al |
| EPI_ISL_908153 | England/CAMC-106EF88/2021 | 2021-01-17 | Europe/United Kingdom/England/ | Lighthouse Lab in Cambridge | Wellcome Sanger Institute for the COVID-19 Genomics UK (COG-UK) Consortium | Rob Howes et al |
| EPI_ISL_908498 | England/ALDP-10DE34B/2021 | 2021-01-19 | Europe/United Kingdom/England/ | Lighthouse Lab in Alderley Park | Wellcome Sanger Institute for the COVID-19 Genomics UK (COG-UK) Consortium | Jacquelyn Wynn et al |

| | | | | | | |
|---|---|---|---|---|---|---|
| EPI_ISL_908877 | England/CAMC-10BF4E3/2021 | 2021-01-19 | Europe/United Kingdom/England/ | Lighthouse Lab in Cambridge | Wellcome Sanger Institute for the COVID-19 Genomics UK (COG-UK) Consortium | Rob Howes et al |
| EPI_ISL_908942 | England/CAMC-10A0588/2021 | 2021-01-18 | Europe/United Kingdom/England/ | Lighthouse Lab in Cambridge | Wellcome Sanger Institute for the COVID-19 Genomics UK (COG-UK) Consortium | Rob Howes et al |
| EPI_ISL_909196 | England/MILK-10897AD/2021 | 2021-01-15 | Europe/United Kingdom/England/ | Lighthouse Lab in Milton Keynes | Wellcome Sanger Institute for the COVID-19 Genomics UK (COG-UK) Consortium | The Lighthouse Lab in Milton Keynes et al |
| EPI_ISL_909335 | England/MILK-10D4FAB/2021 | 2021-01-18 | Europe/United Kingdom/England/ | Lighthouse Lab in Milton Keynes | Wellcome Sanger Institute for the COVID-19 Genomics UK (COG-UK) Consortium | The Lighthouse Lab in Milton Keynes et al |
| EPI_ISL_909667 | France/IDF-IPP01081/2021 | 2021-01-12 | Europe/France/Ile de France/Ivry-sur-Se | Labo Analyses Med | National Reference Center for Viruses of Respiratory Infections, Institut Pasteur, Paris | Marion Barbet et al |
| EPI_ISL_909675 | France/IDF-IPP01088/2021 | 2021-01-09 | Europe/France/Ile de France/Ivry-sur-Se | Labo Analyses Med | National Reference Center for Viruses of Respiratory Infections, Institut Pasteur, Paris | Marion Barbet et al |
| EPI_ISL_909679 | France/IDF-IPP01085/2021 | 2021-01-08 | Europe/France/Ile de France/Ivry-sur-Se | Labo Analyses Med | National Reference Center for Viruses of Respiratory Infections, Institut Pasteur, Paris | Marion Barbet et al |

| | | | | | | |
|---|---|---|---|---|---|---|
| EPI_ISL_909688 | Mayotte/IPP01008/2021 | 2021-01-15 | Europe/France/Mayotte/Mamoudzou | CH de Mayotte - Laboratoire de Biologie | National Reference Center for Viruses of Respiratory Infections, Institut Pasteur, Paris | Marion Barbet et al |
| EPI_ISL_909693 | Mayotte/IPP01016/2021 | 2021-01-15 | Europe/France/Mayotte/Mamoudzou | CH de Mayotte - Laboratoire de Biologie | National Reference Center for Viruses of Respiratory Infections, Institut Pasteur, Paris | Marion Barbet et al |
| EPI_ISL_909697 | Mayotte/IPP01000/2021 | 2021-01-14 | Europe/France/Mayotte/Mamoudzou | CH de Mayotte - Laboratoire de Biologie | National Reference Center for Viruses of Respiratory Infections, Institut Pasteur, Paris | Marion Barbet et al |
| EPI_ISL_909719 | Mayotte/IPP01024/2021 | 2021-01-15 | Europe/France/Mayotte/Mamoudzou | CH de Mayotte - Laboratoire de Biologie | National Reference Center for Viruses of Respiratory Infections, Institut Pasteur, Paris | Marion Barbet et al |
| EPI_ISL_909722 | Mayotte/IPP01027/2021 | 2021-01-15 | Europe/France/Mayotte/Mamoudzou | CH de Mayotte - Laboratoire de Biologie | National Reference Center for Viruses of Respiratory Infections, Institut Pasteur, Paris | Marion Barbet et al |
| EPI_ISL_909732 | France/HDF-IPP01061/2020 | 2020-11-05 | Europe/France/Hauts de France/Cambra | P√¥le m√©dico-technique Laboratoire | National Reference Center for Viruses of Respiratory Infections, Institut Pasteur, Paris | Marion Barbet et al |

| | | | | | | |
|---|---|---|---|---|---|---|
| EPI_ISL_909744 | NorthMacedonia/14712/2020 | 2020-10-09 | Europe/North Macedonia/Skopje/ | Institute for Lung Diseases in Children - Skopje | "Research Center for Genetic Engineering and Biotechnology ""Georgi D. Efremov,Äù , Macedonian Academy of Sciences and Arts" | RCGEB - MASA et al |
| EPI_ISL_909745 | Germany/SH-ChVir22601/2021 | 2021-01-18 | Europe/Germany/Schleswig-Holstein/ | A. Krumbholz, Labor Dr. Krause und Kollegen MVZ GmbH, Kiel | Charitv© Universitv§tsmedizin Berlin, Institut fvºr Virologie | Victor M Corman et al |
| EPI_ISL_909747 | Germany/HB-ChVir22614/2021 | 2021-01-14 | Europe/Germany/Bremen/ | Charitv© Universitv§tsmedizin Berlin, Institut fvºr Virologie/Labor Berlin | Charitv© Universitv§tsmedizin Berlin, Institut fvºr Virologie | Victor M Corman et al |
| EPI_ISL_909749 | Germany/SH-ChVir22834/2021 | 2021-01-19 | Europe/Germany/Schleswig-Holstein/ | A. Krumbholz, Labor Dr. Krause und Kollegen MVZ GmbH, Kiel | Charitv© Universitv§tsmedizin Berlin, Institut fvºr Virologie | Victor M Corman et al |
| EPI_ISL_909754 | Germany/NW-ChVir22608/2021 | 2021-01-13 | Europe/Germany/North Rhine Westphal | Charitv© Universitv§tsmedizin Berlin, Institut fvºr Virologie/Labor Berlin | Charitv© Universitv§tsmedizin Berlin, Institut fvºr Virologie | Victor M Corman et al |
| EPI_ISL_909758 | Germany/SH-ChVir22612/2021 | 2021-01-14 | Europe/Germany/Schleswig-Holstein/ | Charitv© Universitv§tsmedizin Berlin, Institut fvºr Virologie/Labor Berlin | Charitv© Universitv§tsmedizin Berlin, Institut fvºr Virologie | Victor M Corman et al |
| EPI_ISL_909772 | Indonesia/JK-NIHRD-C0050015/2020 | 2020-09-11 | Asia/Indonesia/Jakarta/ | National Institute of Health Research and Development | National Institute of Health Research and Development | Holy Arif Wibowo et al |
| EPI_ISL_909775 | Ireland/KK-NVRL-83IRL57195/2021 | 2021-01-14 | Europe/Ireland/Kilkenny/ | National Virus Reference Laboratory | National Virus Reference Laboratory | Michael Carr et al |
| EPI_ISL_909779 | Ireland/D-NVRL-21IRL20038/2021 | 2021-01-21 | Europe/Ireland/Dublin/ | National Virus Reference Laboratory | National Virus Reference Laboratory | Michael Carr et al |

| | | | | | | |
|---|---|---|---|---|---|---|
| EPI_ISL_909780 | Ireland/D-NVRL-21IRL19249/2021 | 2021-01-20 | Europe/Ireland/Dublin/ | National Virus Reference Laboratory | National Virus Reference Laboratory | Michael Carr et al |
| EPI_ISL_909796 | Ireland/D-NVRL-21IRL17514/2021 | 2021-01-18 | Europe/Ireland/Dublin/ | National Virus Reference Laboratory | National Virus Reference Laboratory | Michael Carr et al |
| EPI_ISL_909801 | Ireland/D-NVRL-21IRL19786/2021 | 2021-01-21 | Europe/Ireland/Dublin/ | National Virus Reference Laboratory | National Virus Reference Laboratory | Michael Carr et al |
| EPI_ISL_909816 | Ireland/D-NVRL-83IRL53957/2021 | 2021-01-14 | Europe/Ireland/Dublin/ | National Virus Reference Laboratory | National Virus Reference Laboratory | Michael Carr et al |
| EPI_ISL_909884 | Ireland/RN-NVRL-21IRL19502/2021 | 2021-01-20 | Europe/Ireland/Roscommon/ | National Virus Reference Laboratory | National Virus Reference Laboratory | Michael Carr et al |
| EPI_ISL_909902 | Ireland/D-NVRL-21IRL17516/2021 | 2021-01-18 | Europe/Ireland/Dublin/ | National Virus Reference Laboratory | National Virus Reference Laboratory | Michael Carr et al |
| EPI_ISL_909908 | Ireland/KE-NVRL-83IRL52844/2021 | 2021-01-14 | Europe/Ireland/Kildare/ | National Virus Reference Laboratory | National Virus Reference Laboratory | Michael Carr et al |
| EPI_ISL_909918 | Ireland/SO-NVRL-83IRL56752/2021 | 2021-01-14 | Europe/Ireland/Sligo/ | National Virus Reference Laboratory | National Virus Reference Laboratory | Michael Carr et al |
| EPI_ISL_909928 | Ireland/D-NVRL-83IRL18158/2021 | 2021-01-11 | Europe/Ireland/Dublin/ | National Virus Reference Laboratory | National Virus Reference Laboratory | Michael Carr et al |
| EPI_ISL_909929 | Ireland/LM-NVRL-83IRL58177/2021 | 2021-01-14 | Europe/Ireland/Leitrim/ | National Virus Reference Laboratory | National Virus Reference Laboratory | Michael Carr et al |
| EPI_ISL_909955 | India/TG-CCMB-C1E1-ONT-R1/2020 | 2020-08-31 | Asia/India/Telangana/ | Apollo Hospitals | CSIR-Centre for Cellular and Molecular Biology | Onkar Kulkarni et al |
| EPI_ISL_909956 | India/TG-CCMB-C1E2-ONT-R2/2020 | 2020-11-14 | Asia/India/Telangana/ | Apollo Hospitals | CSIR-Centre for Cellular and Molecular Biology | Onkar Kulkarni et al |
| EPI_ISL_909957 | India/TG-CCMB-C2E1-ONT-R1/2020 | 2020-11-04 | Asia/India/Telangana/ | Apollo Hospitals | CSIR-Centre for Cellular and Molecular Biology | Onkar Kulkarni et al |

| | | | | | | |
|---|---|---|---|---|---|---|
| EPI_ISL_909958 | India/TG-CCMB-C2E2-ONT-R2/2020 | 2020-11-22 | Asia/India/Telangana/ | Apollo Hospitals | CSIR-Centre for Cellular and Molecular Biology | Onkar Kulkarni et al |
| EPI_ISL_909960 | India/TG-CCMB-C1E2-R2/2020 | 2020-11-14 | Asia/India/Telangana/ | Apollo Hospitals | CSIR-Centre for Cellular and Molecular Biology | Onkar Kulkarni et al |
| EPI_ISL_909961 | India/TG-CCMB-C2E1-R1/2020 | 2020-11-04 | Asia/India/Telangana/ | Apollo Hospitals | CSIR-Centre for Cellular and Molecular Biology | Onkar Kulkarni et al |
| EPI_ISL_909962 | India/TG-CCMB-C2E2-R2/2020 | 2020-11-22 | Asia/India/Telangana/ | Apollo Hospitals | CSIR-Centre for Cellular and Molecular Biology | Onkar Kulkarni et al |
| EPI_ISL_909964 | Germany/SH-ChVir21724/2020 | 2020-12-21 | Europe/Germany/Schleswig-Holstein/ | A. Krumbholz, Labor Dr. Krause und Kollegen MVZ GmbH, Kiel | Charit√© Universit√§tsmedizin Berlin, Institut f√ºr Virologie | Victor M Corman et al |
| EPI_ISL_909967 | Germany/SH-ChVir21471/2020 | 2020-12-19 | Europe/Germany/Schleswig-Holstein/ | A. Krumbholz, Labor Dr. Krause und Kollegen MVZ GmbH, Kiel | Charit√© Universit√§tsmedizin Berlin, Institut f√ºr Virologie | Victor M Corman et al |
| EPI_ISL_909968 | Germany/SH-ChVir21730/2020 | 2020-12-22 | Europe/Germany/Schleswig-Holstein/ | A. Krumbholz, Labor Dr. Krause und Kollegen MVZ GmbH, Kiel | Charit√© Universit√§tsmedizin Berlin, Institut f√ºr Virologie | Victor M Corman et al |
| EPI_ISL_909969 | Germany/SH-ChVir21450/2020 | 2020-12-13 | Europe/Germany/Schleswig-Holstein/ | A. Krumbholz, Labor Dr. Krause und Kollegen MVZ GmbH, Kiel | Charit√© Universit√§tsmedizin Berlin, Institut f√ºr Virologie | Victor M Corman et al |
| EPI_ISL_909972 | Germany/SH-ChVir21466/2020 | 2020-12-19 | Europe/Germany/Schleswig-Holstein/ | A. Krumbholz, Labor Dr. Krause und Kollegen MVZ GmbH, Kiel | Charit√© Universit√§tsmedizin Berlin, Institut f√ºr Virologie | Victor M Corman et al |
| EPI_ISL_909974 | Germany/SH-ChVir21481/2020 | 2020-12-20 | Europe/Germany/Schleswig-Holstein/ | A. Krumbholz, Labor Dr. Krause und Kollegen MVZ GmbH, Kiel | Charit√© Universit√§tsmedizin Berlin, Institut f√ºr Virologie | Victor M Corman et al |
| EPI_ISL_909977 | Germany/SH-ChVir21722/2020 | 2020-12-22 | Europe/Germany/Schleswig-Holstein/ | A. Krumbholz, Labor Dr. Krause und Kollegen MVZ GmbH, Kiel | Charit√© Universit√§tsmedizin Berlin, Institut f√ºr Virologie | Victor M Corman et al |

| | | | | | | |
|---|---|---|---|---|---|---|
| EPI_ISL_909979 | Germany/SH-ChVir21448/2020 | 2020-12-12 | Europe/Germany/Schleswig-Holstein/ | A. Krumbholz, Labor Dr. Krause und Kollegen MVZ GmbH, Kiel | Charitv© Universitv§tsmedizin Berlin, Institut fvºr Virologie | Victor M Corman et al |
| EPI_ISL_909984 | Germany/SH-ChVir21454/2020 | 2020-12-17 | Europe/Germany/Schleswig-Holstein/ | A. Krumbholz, Labor Dr. Krause und Kollegen MVZ GmbH, Kiel | Charitv© Universitv§tsmedizin Berlin, Institut fvºr Virologie | Victor M Corman et al |
| EPI_ISL_909988 | Germany/SH-ChVir21459/2020 | 2020-12-17 | Europe/Germany/Schleswig-Holstein/ | A. Krumbholz, Labor Dr. Krause und Kollegen MVZ GmbH, Kiel | Charitv© Universitv§tsmedizin Berlin, Institut fvºr Virologie | Victor M Corman et al |
| EPI_ISL_909993 | Germany/SH-ChVir21467/2020 | 2020-12-20 | Europe/Germany/Schleswig-Holstein/ | A. Krumbholz, Labor Dr. Krause und Kollegen MVZ GmbH, Kiel | Charitv© Universitv§tsmedizin Berlin, Institut fvºr Virologie | Victor M Corman et al |
| EPI_ISL_909994 | Germany/SH-ChVir21468/2020 | 2020-12-20 | Europe/Germany/Schleswig-Holstein/ | A. Krumbholz, Labor Dr. Krause und Kollegen MVZ GmbH, Kiel | Charitv© Universitv§tsmedizin Berlin, Institut fvºr Virologie | Victor M Corman et al |
| EPI_ISL_909995 | Germany/SH-ChVir21469/2020 | 2020-12-19 | Europe/Germany/Schleswig-Holstein/ | A. Krumbholz, Labor Dr. Krause und Kollegen MVZ GmbH, Kiel | Charitv© Universitv§tsmedizin Berlin, Institut fvºr Virologie | Victor M Corman et al |
| EPI_ISL_909996 | Germany/SH-ChVir21470/2020 | 2020-12-20 | Europe/Germany/Schleswig-Holstein/ | A. Krumbholz, Labor Dr. Krause und Kollegen MVZ GmbH, Kiel | Charitv© Universitv§tsmedizin Berlin, Institut fvºr Virologie | Victor M Corman et al |
| EPI_ISL_910000 | Germany/SH-ChVir21476/2020 | 2020-12-20 | Europe/Germany/Schleswig-Holstein/ | A. Krumbholz, Labor Dr. Krause und Kollegen MVZ GmbH, Kiel | Charitv© Universitv§tsmedizin Berlin, Institut fvºr Virologie | Victor M Corman et al |
| EPI_ISL_910007 | Germany/SH-ChVir21731/2020 | 2020-12-21 | Europe/Germany/Schleswig-Holstein/ | A. Krumbholz, Labor Dr. Krause und Kollegen MVZ GmbH, Kiel | Charitv© Universitv§tsmedizin Berlin, Institut fvºr Virologie | Victor M Corman et al |
| EPI_ISL_910009 | Germany/SH-ChVir21733/2020 | 2020-12-21 | Europe/Germany/Schleswig-Holstein/ | A. Krumbholz, Labor Dr. Krause und Kollegen MVZ GmbH, Kiel | Charitv© Universitv§tsmedizin Berlin, Institut fvºr Virologie | Victor M Corman et al |

| | | | | | | |
|---|---|---|---|---|---|---|
| EPI_ISL_910016 | Romania/Bucuresti409202/2021 | 2021-01-04 | Europe/Romania/Bucharest/ | Laboratory for Respiratory Viruses, Cantacuzino National Military-Medical Institute for Research and Development | Cantacuzino Institute Virology | Luiza Ustea et al |
| EPI_ISL_910031 | India/TG-CCMB-AE552/2020 | 2020-08-26 | Asia/India/Telangana/ | CSIR-Centre for Cellular and Molecular Biology | CSIR-Centre for Cellular and Molecular Biology | Payel Mukherjee et al |
| EPI_ISL_910032 | India/TG-CCMB-AE554/2020 | 2020-08-26 | Asia/India/Telangana/ | CSIR-Centre for Cellular and Molecular Biology | CSIR-Centre for Cellular and Molecular Biology | Payel Mukherjee et al |
| EPI_ISL_910033 | India/TG-CCMB-AE582/2020 | 2020-08-26 | Asia/India/Telangana/ | CSIR-Centre for Cellular and Molecular Biology | CSIR-Centre for Cellular and Molecular Biology | Payel Mukherjee et al |
| EPI_ISL_910034 | India/TG-CCMB-AE588/2020 | 2020-08-26 | Asia/India/Telangana/ | CSIR-Centre for Cellular and Molecular Biology | CSIR-Centre for Cellular and Molecular Biology | Payel Mukherjee et al |
| EPI_ISL_910035 | India/TG-CCMB-AE589/2020 | 2020-08-26 | Asia/India/Telangana/ | CSIR-Centre for Cellular and Molecular Biology | CSIR-Centre for Cellular and Molecular Biology | Payel Mukherjee et al |
| EPI_ISL_910036 | India/TG-CCMB-AE603/2020 | 2020-08-26 | Asia/India/Telangana/ | CSIR-Centre for Cellular and Molecular Biology | CSIR-Centre for Cellular and Molecular Biology | Payel Mukherjee et al |
| EPI_ISL_910037 | India/TG-CCMB-AE786/2020 | 2020-08-27 | Asia/India/Telangana/ | CSIR-Centre for Cellular and Molecular Biology | CSIR-Centre for Cellular and Molecular Biology | Payel Mukherjee et al |
| EPI_ISL_910038 | India/TG-CCMB-AE807/2020 | 2020-08-27 | Asia/India/Telangana/ | CSIR-Centre for Cellular and Molecular Biology | CSIR-Centre for Cellular and Molecular Biology | Payel Mukherjee et al |
| EPI_ISL_910039 | India/TG-CCMB-AE817/2020 | 2020-08-27 | Asia/India/Telangana/ | CSIR-Centre for Cellular and Molecular Biology | CSIR-Centre for Cellular and Molecular Biology | Payel Mukherjee et al |
| EPI_ISL_910040 | India/TG-CCMB-AE838/2020 | 2020-08-27 | Asia/India/Telangana/ | CSIR-Centre for Cellular and Molecular Biology | CSIR-Centre for Cellular and Molecular Biology | Payel Mukherjee et al |
| EPI_ISL_910041 | India/TG-CCMB-AE841/2020 | 2020-08-27 | Asia/India/Telangana/ | CSIR-Centre for Cellular and Molecular Biology | CSIR-Centre for Cellular and Molecular Biology | Payel Mukherjee et al |
| EPI_ISL_910042 | India/TG-CCMB-AE597/2020 | 2020-08-26 | Asia/India/Telangana/ | CSIR-Centre for Cellular and Molecular Biology | CSIR-Centre for Cellular and Molecular Biology | Payel Mukherjee et al |

| | | | | | | |
|---|---|---|---|---|---|---|
| EPI_ISL_910043 | India/TG-CCMB-AE843/2020 | 2020-08-27 | Asia/India/Telangana/ | CSIR-Centre for Cellular and Molecular Biology | CSIR-Centre for Cellular and Molecular Biology | Payel Mukherjee et al |
| EPI_ISL_910044 | India/TG-CCMB-AE844/2020 | 2020-08-27 | Asia/India/Telangana/ | CSIR-Centre for Cellular and Molecular Biology | CSIR-Centre for Cellular and Molecular Biology | Payel Mukherjee et al |
| EPI_ISL_910045 | India/TG-CCMB-AE850/2020 | 2020-08-27 | Asia/India/Telangana/ | CSIR-Centre for Cellular and Molecular Biology | CSIR-Centre for Cellular and Molecular Biology | Payel Mukherjee et al |
| EPI_ISL_910046 | India/TG-CCMB-AE857/2020 | 2020-08-27 | Asia/India/Telangana/ | CSIR-Centre for Cellular and Molecular Biology | CSIR-Centre for Cellular and Molecular Biology | Payel Mukherjee et al |
| EPI_ISL_910047 | India/TG-CCMB-AE865/2020 | 2020-08-27 | Asia/India/Telangana/ | CSIR-Centre for Cellular and Molecular Biology | CSIR-Centre for Cellular and Molecular Biology | Payel Mukherjee et al |
| EPI_ISL_910048 | India/TG-CCMB-AE872/2020 | 2020-08-27 | Asia/India/Telangana/ | CSIR-Centre for Cellular and Molecular Biology | CSIR-Centre for Cellular and Molecular Biology | Payel Mukherjee et al |
| EPI_ISL_910049 | India/TG-CCMB-AE895/2020 | 2020-08-27 | Asia/India/Telangana/ | CSIR-Centre for Cellular and Molecular Biology | CSIR-Centre for Cellular and Molecular Biology | Payel Mukherjee et al |
| EPI_ISL_910050 | India/TG-CCMB-AE908/2020 | 2020-08-27 | Asia/India/Telangana/ | CSIR-Centre for Cellular and Molecular Biology | CSIR-Centre for Cellular and Molecular Biology | Payel Mukherjee et al |
| EPI_ISL_910051 | India/TG-CCMB-AE923/2020 | 2020-08-27 | Asia/India/Telangana/ | CSIR-Centre for Cellular and Molecular Biology | CSIR-Centre for Cellular and Molecular Biology | Payel Mukherjee et al |
| EPI_ISL_910052 | India/TG-CCMB-AE947/2020 | 2020-08-27 | Asia/India/Telangana/ | CSIR-Centre for Cellular and Molecular Biology | CSIR-Centre for Cellular and Molecular Biology | Payel Mukherjee et al |
| EPI_ISL_910053 | India/TG-CCMB-AE948/2020 | 2020-08-27 | Asia/India/Telangana/ | CSIR-Centre for Cellular and Molecular Biology | CSIR-Centre for Cellular and Molecular Biology | Payel Mukherjee et al |
| EPI_ISL_910054 | India/TG-CCMB-AE954/2020 | 2020-08-27 | Asia/India/Telangana/ | CSIR-Centre for Cellular and Molecular Biology | CSIR-Centre for Cellular and Molecular Biology | Payel Mukherjee et al |
| EPI_ISL_910055 | India/TG-CCMB-AE959/2020 | 2020-08-27 | Asia/India/Telangana/ | CSIR-Centre for Cellular and Molecular Biology | CSIR-Centre for Cellular and Molecular Biology | Payel Mukherjee et al |

| | | | | | | |
|---|---|---|---|---|---|---|
| EPI_ISL_910056 | India/TG-CCMB-AE962/2020 | 2020-08-27 | Asia/India/Telangana/ | CSIR-Centre for Cellular and Molecular Biology | CSIR-Centre for Cellular and Molecular Biology | Payel Mukherjee et al |
| EPI_ISL_910057 | India/TG-CCMB-AE963/2020 | 2020-08-27 | Asia/India/Telangana/ | CSIR-Centre for Cellular and Molecular Biology | CSIR-Centre for Cellular and Molecular Biology | Payel Mukherjee et al |
| EPI_ISL_910058 | India/TG-CCMB-AE965/2020 | 2020-08-27 | Asia/India/Telangana/ | CSIR-Centre for Cellular and Molecular Biology | CSIR-Centre for Cellular and Molecular Biology | Payel Mukherjee et al |
| EPI_ISL_910059 | India/TG-CCMB-AF110/2020 | 2020-08-28 | Asia/India/Telangana/ | CSIR-Centre for Cellular and Molecular Biology | CSIR-Centre for Cellular and Molecular Biology | Payel Mukherjee et al |
| EPI_ISL_910060 | India/TG-CCMB-AF111/2020 | 2020-08-28 | Asia/India/Telangana/ | CSIR-Centre for Cellular and Molecular Biology | CSIR-Centre for Cellular and Molecular Biology | Payel Mukherjee et al |
| EPI_ISL_910061 | India/TG-CCMB-AF113/2020 | 2020-08-28 | Asia/India/Telangana/ | CSIR-Centre for Cellular and Molecular Biology | CSIR-Centre for Cellular and Molecular Biology | Payel Mukherjee et al |
| EPI_ISL_910062 | India/TG-CCMB-AF155/2020 | 2020-08-28 | Asia/India/Telangana/ | CSIR-Centre for Cellular and Molecular Biology | CSIR-Centre for Cellular and Molecular Biology | Payel Mukherjee et al |
| EPI_ISL_910063 | India/TG-CCMB-AF156/2020 | 2020-08-28 | Asia/India/Telangana/ | CSIR-Centre for Cellular and Molecular Biology | CSIR-Centre for Cellular and Molecular Biology | Payel Mukherjee et al |
| EPI_ISL_910064 | India/TG-CCMB-AF161/2020 | 2020-08-28 | Asia/India/Telangana/ | CSIR-Centre for Cellular and Molecular Biology | CSIR-Centre for Cellular and Molecular Biology | Payel Mukherjee et al |
| EPI_ISL_910065 | India/TG-CCMB-AF166/2020 | 2020-08-28 | Asia/India/Telangana/ | CSIR-Centre for Cellular and Molecular Biology | CSIR-Centre for Cellular and Molecular Biology | Payel Mukherjee et al |
| EPI_ISL_910066 | India/TG-CCMB-AF167/2020 | 2020-08-28 | Asia/India/Telangana/ | CSIR-Centre for Cellular and Molecular Biology | CSIR-Centre for Cellular and Molecular Biology | Payel Mukherjee et al |
| EPI_ISL_910067 | India/TG-CCMB-AF181/2020 | 2020-08-28 | Asia/India/Telangana/ | CSIR-Centre for Cellular and Molecular Biology | CSIR-Centre for Cellular and Molecular Biology | Payel Mukherjee et al |
| EPI_ISL_910068 | India/TG-CCMB-AF182/2020 | 2020-08-28 | Asia/India/Telangana/ | CSIR-Centre for Cellular and Molecular Biology | CSIR-Centre for Cellular and Molecular Biology | Payel Mukherjee et al |

| | | | | | | |
|---|---|---|---|---|---|---|
| EPI_ISL_910069 | India/TG-CCMB-AF184/2020 | 2020-08-28 | Asia/India/Telangana/ | CSIR-Centre for Cellular and Molecular Biology | CSIR-Centre for Cellular and Molecular Biology | Payel Mukherjee et al |
| EPI_ISL_910070 | India/TG-CCMB-AF189/2020 | 2020-08-28 | Asia/India/Telangana/ | CSIR-Centre for Cellular and Molecular Biology | CSIR-Centre for Cellular and Molecular Biology | Payel Mukherjee et al |
| EPI_ISL_910071 | India/TG-CCMB-AF219/2020 | 2020-08-28 | Asia/India/Telangana/ | CSIR-Centre for Cellular and Molecular Biology | CSIR-Centre for Cellular and Molecular Biology | Payel Mukherjee et al |
| EPI_ISL_910072 | India/TG-CCMB-AF238/2020 | 2020-08-28 | Asia/India/Telangana/ | CSIR-Centre for Cellular and Molecular Biology | CSIR-Centre for Cellular and Molecular Biology | Payel Mukherjee et al |
| EPI_ISL_910073 | India/TG-CCMB-AF264/2020 | 2020-08-28 | Asia/India/Telangana/ | CSIR-Centre for Cellular and Molecular Biology | CSIR-Centre for Cellular and Molecular Biology | Payel Mukherjee et al |
| EPI_ISL_910074 | India/TG-CCMB-AF511/2020 | 2020-08-29 | Asia/India/Telangana/ | CSIR-Centre for Cellular and Molecular Biology | CSIR-Centre for Cellular and Molecular Biology | Payel Mukherjee et al |
| EPI_ISL_910075 | India/TG-CCMB-AF521/2020 | 2020-08-29 | Asia/India/Telangana/ | CSIR-Centre for Cellular and Molecular Biology | CSIR-Centre for Cellular and Molecular Biology | Payel Mukherjee et al |
| EPI_ISL_910076 | India/TG-CCMB-AF555/2020 | 2020-08-29 | Asia/India/Telangana/ | CSIR-Centre for Cellular and Molecular Biology | CSIR-Centre for Cellular and Molecular Biology | Payel Mukherjee et al |
| EPI_ISL_910077 | India/TG-CCMB-AF580/2020 | 2020-08-29 | Asia/India/Telangana/ | CSIR-Centre for Cellular and Molecular Biology | CSIR-Centre for Cellular and Molecular Biology | Payel Mukherjee et al |
| EPI_ISL_910078 | India/TG-CCMB-AF609/2020 | 2020-08-29 | Asia/India/Telangana/ | CSIR-Centre for Cellular and Molecular Biology | CSIR-Centre for Cellular and Molecular Biology | Payel Mukherjee et al |
| EPI_ISL_910079 | India/TG-CCMB-AF651/2020 | 2020-08-29 | Asia/India/Telangana/ | CSIR-Centre for Cellular and Molecular Biology | CSIR-Centre for Cellular and Molecular Biology | Payel Mukherjee et al |
| EPI_ISL_910080 | India/TG-CCMB-AF671/2020 | 2020-08-29 | Asia/India/Telangana/ | CSIR-Centre for Cellular and Molecular Biology | CSIR-Centre for Cellular and Molecular Biology | Payel Mukherjee et al |
| EPI_ISL_910081 | India/TG-CCMB-AF680/2020 | 2020-08-29 | Asia/India/Telangana/ | CSIR-Centre for Cellular and Molecular Biology | CSIR-Centre for Cellular and Molecular Biology | Payel Mukherjee et al |

| | | | | | | |
|---|---|---|---|---|---|---|
| EPI_ISL_910082 | India/TG-CCMB-AF739/2020 | 2020-08-29 | Asia/India/Telangana/ | CSIR-Centre for Cellular and Molecular Biology | CSIR-Centre for Cellular and Molecular Biology | Payel Mukherjee et al |
| EPI_ISL_910083 | India/TG-CCMB-AF777/2020 | 2020-08-29 | Asia/India/Telangana/ | CSIR-Centre for Cellular and Molecular Biology | CSIR-Centre for Cellular and Molecular Biology | Payel Mukherjee et al |
| EPI_ISL_910084 | India/TG-CCMB-AF778/2020 | 2020-08-29 | Asia/India/Telangana/ | CSIR-Centre for Cellular and Molecular Biology | CSIR-Centre for Cellular and Molecular Biology | Payel Mukherjee et al |
| EPI_ISL_910085 | India/TG-CCMB-AF782/2020 | 2020-08-29 | Asia/India/Telangana/ | CSIR-Centre for Cellular and Molecular Biology | CSIR-Centre for Cellular and Molecular Biology | Payel Mukherjee et al |
| EPI_ISL_910086 | India/TG-CCMB-AF789/2020 | 2020-08-29 | Asia/India/Telangana/ | CSIR-Centre for Cellular and Molecular Biology | CSIR-Centre for Cellular and Molecular Biology | Payel Mukherjee et al |
| EPI_ISL_910087 | India/TG-CCMB-AF794/2020 | 2020-08-29 | Asia/India/Telangana/ | CSIR-Centre for Cellular and Molecular Biology | CSIR-Centre for Cellular and Molecular Biology | Payel Mukherjee et al |
| EPI_ISL_910088 | India/TG-CCMB-AF795/2020 | 2020-08-29 | Asia/India/Telangana/ | CSIR-Centre for Cellular and Molecular Biology | CSIR-Centre for Cellular and Molecular Biology | Payel Mukherjee et al |
| EPI_ISL_910089 | India/TG-CCMB-AF812/2020 | 2020-08-29 | Asia/India/Telangana/ | CSIR-Centre for Cellular and Molecular Biology | CSIR-Centre for Cellular and Molecular Biology | Payel Mukherjee et al |
| EPI_ISL_910090 | India/TG-CCMB-AF838/2020 | 2020-08-29 | Asia/India/Telangana/ | CSIR-Centre for Cellular and Molecular Biology | CSIR-Centre for Cellular and Molecular Biology | Payel Mukherjee et al |
| EPI_ISL_910091 | India/TG-CCMB-AF860/2020 | 2020-08-29 | Asia/India/Telangana/ | CSIR-Centre for Cellular and Molecular Biology | CSIR-Centre for Cellular and Molecular Biology | Payel Mukherjee et al |
| EPI_ISL_910092 | India/TG-CCMB-AF866/2020 | 2020-08-29 | Asia/India/Telangana/ | CSIR-Centre for Cellular and Molecular Biology | CSIR-Centre for Cellular and Molecular Biology | Payel Mukherjee et al |
| EPI_ISL_910093 | India/TG-CCMB-AF886/2020 | 2020-08-29 | Asia/India/Telangana/ | CSIR-Centre for Cellular and Molecular Biology | CSIR-Centre for Cellular and Molecular Biology | Payel Mukherjee et al |
| EPI_ISL_910094 | India/TG-CCMB-AF891/2020 | 2020-08-29 | Asia/India/Telangana/ | CSIR-Centre for Cellular and Molecular Biology | CSIR-Centre for Cellular and Molecular Biology | Payel Mukherjee et al |

| | | | | | | |
|---|---|---|---|---|---|---|
| EPI_ISL_910095 | India/TG-CCMB-AF895/2020 | 2020-08-29 | Asia/India/Telangana/ | CSIR-Centre for Cellular and Molecular Biology | CSIR-Centre for Cellular and Molecular Biology | Payel Mukherjee et al |
| EPI_ISL_910096 | India/TG-CCMB-AF915/2020 | 2020-08-30 | Asia/India/Telangana/ | CSIR-Centre for Cellular and Molecular Biology | CSIR-Centre for Cellular and Molecular Biology | Payel Mukherjee et al |
| EPI_ISL_910097 | India/TG-CCMB-AF916/2020 | 2020-08-30 | Asia/India/Telangana/ | CSIR-Centre for Cellular and Molecular Biology | CSIR-Centre for Cellular and Molecular Biology | Payel Mukherjee et al |
| EPI_ISL_910098 | India/TG-CCMB-AF919/2020 | 2020-08-30 | Asia/India/Telangana/ | CSIR-Centre for Cellular and Molecular Biology | CSIR-Centre for Cellular and Molecular Biology | Payel Mukherjee et al |
| EPI_ISL_910099 | India/TG-CCMB-AF990/2020 | 2020-08-30 | Asia/India/Telangana/ | CSIR-Centre for Cellular and Molecular Biology | CSIR-Centre for Cellular and Molecular Biology | Payel Mukherjee et al |
| EPI_ISL_910100 | India/TG-CCMB-AG027/2020 | 2020-08-30 | Asia/India/Telangana/ | CSIR-Centre for Cellular and Molecular Biology | CSIR-Centre for Cellular and Molecular Biology | Payel Mukherjee et al |
| EPI_ISL_910101 | India/TG-CCMB-AG038/2020 | 2020-08-30 | Asia/India/Telangana/ | CSIR-Centre for Cellular and Molecular Biology | CSIR-Centre for Cellular and Molecular Biology | Payel Mukherjee et al |
| EPI_ISL_910102 | India/TG-CCMB-AG060/2020 | 2020-08-30 | Asia/India/Telangana/ | CSIR-Centre for Cellular and Molecular Biology | CSIR-Centre for Cellular and Molecular Biology | Payel Mukherjee et al |
| EPI_ISL_910103 | India/TG-CCMB-AG065/2020 | 2020-08-30 | Asia/India/Telangana/ | CSIR-Centre for Cellular and Molecular Biology | CSIR-Centre for Cellular and Molecular Biology | Payel Mukherjee et al |
| EPI_ISL_910104 | India/TG-CCMB-AG087/2020 | 2020-08-30 | Asia/India/Telangana/ | CSIR-Centre for Cellular and Molecular Biology | CSIR-Centre for Cellular and Molecular Biology | Payel Mukherjee et al |
| EPI_ISL_910105 | India/TG-CCMB-AG116/2020 | 2020-08-30 | Asia/India/Telangana/ | CSIR-Centre for Cellular and Molecular Biology | CSIR-Centre for Cellular and Molecular Biology | Payel Mukherjee et al |
| EPI_ISL_910106 | India/TG-CCMB-AG119/2020 | 2020-08-30 | Asia/India/Telangana/ | CSIR-Centre for Cellular and Molecular Biology | CSIR-Centre for Cellular and Molecular Biology | Payel Mukherjee et al |
| EPI_ISL_910107 | India/TG-CCMB-AG133/2020 | 2020-08-30 | Asia/India/Telangana/ | CSIR-Centre for Cellular and Molecular Biology | CSIR-Centre for Cellular and Molecular Biology | Payel Mukherjee et al |

| | | | | | | |
|---|---|---|---|---|---|---|
| EPI_ISL_910108 | India/TG-CCMB-AG154/2020 | 2020-08-30 | Asia/India/Telangana/ | CSIR-Centre for Cellular and Molecular Biology | CSIR-Centre for Cellular and Molecular Biology | Payel Mukherjee et al |
| EPI_ISL_910109 | India/TG-CCMB-AG184/2020 | 2020-08-30 | Asia/India/Telangana/ | CSIR-Centre for Cellular and Molecular Biology | CSIR-Centre for Cellular and Molecular Biology | Payel Mukherjee et al |
| EPI_ISL_910110 | India/TG-CCMB-AG189/2020 | 2020-08-30 | Asia/India/Telangana/ | CSIR-Centre for Cellular and Molecular Biology | CSIR-Centre for Cellular and Molecular Biology | Payel Mukherjee et al |
| EPI_ISL_910111 | India/TG-CCMB-AG270/2020 | 2020-08-31 | Asia/India/Telangana/ | CSIR-Centre for Cellular and Molecular Biology | CSIR-Centre for Cellular and Molecular Biology | Payel Mukherjee et al |
| EPI_ISL_910112 | India/TG-CCMB-AG272/2020 | 2020-09-02 | Asia/India/Telangana/ | CSIR-Centre for Cellular and Molecular Biology | CSIR-Centre for Cellular and Molecular Biology | Payel Mukherjee et al |
| EPI_ISL_910114 | India/TG-CCMB-AG313/2020 | 2020-08-31 | Asia/India/Telangana/ | CSIR-Centre for Cellular and Molecular Biology | CSIR-Centre for Cellular and Molecular Biology | Payel Mukherjee et al |
| EPI_ISL_910115 | India/TG-CCMB-AG335/2020 | 2020-08-31 | Asia/India/Telangana/ | CSIR-Centre for Cellular and Molecular Biology | CSIR-Centre for Cellular and Molecular Biology | Payel Mukherjee et al |
| EPI_ISL_910116 | India/TG-CCMB-AG347/2020 | 2020-08-31 | Asia/India/Telangana/ | CSIR-Centre for Cellular and Molecular Biology | CSIR-Centre for Cellular and Molecular Biology | Payel Mukherjee et al |
| EPI_ISL_910117 | India/TG-CCMB-AG489/2020 | 2020-08-31 | Asia/India/Telangana/ | CSIR-Centre for Cellular and Molecular Biology | CSIR-Centre for Cellular and Molecular Biology | Payel Mukherjee et al |
| EPI_ISL_910118 | India/TG-CCMB-AG558/2020 | 2020-08-31 | Asia/India/Telangana/ | CSIR-Centre for Cellular and Molecular Biology | CSIR-Centre for Cellular and Molecular Biology | Payel Mukherjee et al |
| EPI_ISL_910119 | India/TG-CCMB-AG584/2020 | 2020-08-31 | Asia/India/Telangana/ | CSIR-Centre for Cellular and Molecular Biology | CSIR-Centre for Cellular and Molecular Biology | Payel Mukherjee et al |
| EPI_ISL_910120 | India/TG-CCMB-AG615/2020 | 2020-08-31 | Asia/India/Telangana/ | CSIR-Centre for Cellular and Molecular Biology | CSIR-Centre for Cellular and Molecular Biology | Payel Mukherjee et al |
| EPI_ISL_910121 | India/TG-CCMB-AG618/2020 | 2020-08-31 | Asia/India/Telangana/ | CSIR-Centre for Cellular and Molecular Biology | CSIR-Centre for Cellular and Molecular Biology | Payel Mukherjee et al |

| | | | | | | |
|---|---|---|---|---|---|---|
| EPI_ISL_910122 | India/TG-CCMB-AG654/2020 | 2020-08-31 | Asia/India/Telangana/ | CSIR-Centre for Cellular and Molecular Biology | CSIR-Centre for Cellular and Molecular Biology | Payel Mukherjee et al |
| EPI_ISL_910123 | India/TG-CCMB-AG971/2020 | 2020-09-03 | Asia/India/Telangana/ | CSIR-Centre for Cellular and Molecular Biology | CSIR-Centre for Cellular and Molecular Biology | Payel Mukherjee et al |
| EPI_ISL_910124 | India/TG-CCMB-AH128/2020 | 2020-09-03 | Asia/India/Telangana/ | CSIR-Centre for Cellular and Molecular Biology | CSIR-Centre for Cellular and Molecular Biology | Payel Mukherjee et al |
| EPI_ISL_910125 | India/TG-CCMB-AH170/2020 | 2020-09-03 | Asia/India/Telangana/ | CSIR-Centre for Cellular and Molecular Biology | CSIR-Centre for Cellular and Molecular Biology | Payel Mukherjee et al |
| EPI_ISL_910126 | India/TG-CCMB-AH201/2020 | 2020-09-03 | Asia/India/Telangana/ | CSIR-Centre for Cellular and Molecular Biology | CSIR-Centre for Cellular and Molecular Biology | Payel Mukherjee et al |
| EPI_ISL_910127 | India/TG-CCMB-AH237/2020 | 2020-09-03 | Asia/India/Telangana/ | CSIR-Centre for Cellular and Molecular Biology | CSIR-Centre for Cellular and Molecular Biology | Payel Mukherjee et al |
| EPI_ISL_910128 | India/TG-CCMB-AH268/2020 | 2020-09-03 | Asia/India/Telangana/ | CSIR-Centre for Cellular and Molecular Biology | CSIR-Centre for Cellular and Molecular Biology | Payel Mukherjee et al |
| EPI_ISL_910129 | India/TG-CCMB-AH671/2020 | 2020-09-08 | Asia/India/Telangana/ | CSIR-Centre for Cellular and Molecular Biology | CSIR-Centre for Cellular and Molecular Biology | Payel Mukherjee et al |
| EPI_ISL_910130 | India/TG-CCMB-AH673/2020 | 2020-09-08 | Asia/India/Telangana/ | CSIR-Centre for Cellular and Molecular Biology | CSIR-Centre for Cellular and Molecular Biology | Payel Mukherjee et al |
| EPI_ISL_910131 | India/TG-CCMB-AH679/2020 | 2020-09-08 | Asia/India/Telangana/ | CSIR-Centre for Cellular and Molecular Biology | CSIR-Centre for Cellular and Molecular Biology | Payel Mukherjee et al |
| EPI_ISL_910132 | India/TG-CCMB-AH718/2020 | 2020-09-08 | Asia/India/Telangana/ | CSIR-Centre for Cellular and Molecular Biology | CSIR-Centre for Cellular and Molecular Biology | Payel Mukherjee et al |
| EPI_ISL_910133 | India/TG-CCMB-AH735/2020 | 2020-09-08 | Asia/India/Telangana/ | CSIR-Centre for Cellular and Molecular Biology | CSIR-Centre for Cellular and Molecular Biology | Payel Mukherjee et al |
| EPI_ISL_910134 | India/TG-CCMB-AH736/2020 | 2020-09-08 | Asia/India/Telangana/ | CSIR-Centre for Cellular and Molecular Biology | CSIR-Centre for Cellular and Molecular Biology | Payel Mukherjee et al |

| | | | | | | |
|---|---|---|---|---|---|---|
| EPI_ISL_910135 | India/TG-CCMB-AH768/2020 | 2020-09-08 | Asia/India/Telangana/ | CSIR-Centre for Cellular and Molecular Biology | CSIR-Centre for Cellular and Molecular Biology | Payel Mukherjee et al |
| EPI_ISL_910136 | India/TG-CCMB-AH828/2020 | 2020-09-08 | Asia/India/Telangana/ | CSIR-Centre for Cellular and Molecular Biology | CSIR-Centre for Cellular and Molecular Biology | Payel Mukherjee et al |
| EPI_ISL_910137 | India/TG-CCMB-AH847/2020 | 2020-09-08 | Asia/India/Telangana/ | CSIR-Centre for Cellular and Molecular Biology | CSIR-Centre for Cellular and Molecular Biology | Payel Mukherjee et al |
| EPI_ISL_910138 | India/TG-CCMB-AH862/2020 | 2020-09-08 | Asia/India/Telangana/ | CSIR-Centre for Cellular and Molecular Biology | CSIR-Centre for Cellular and Molecular Biology | Payel Mukherjee et al |
| EPI_ISL_910139 | India/TG-CCMB-AH924/2020 | 2020-09-08 | Asia/India/Telangana/ | CSIR-Centre for Cellular and Molecular Biology | CSIR-Centre for Cellular and Molecular Biology | Payel Mukherjee et al |
| EPI_ISL_910140 | India/TG-CCMB-AH927/2020 | 2020-09-08 | Asia/India/Telangana/ | CSIR-Centre for Cellular and Molecular Biology | CSIR-Centre for Cellular and Molecular Biology | Payel Mukherjee et al |
| EPI_ISL_910141 | India/TG-CCMB-AH959/2020 | 2020-09-08 | Asia/India/Telangana/ | CSIR-Centre for Cellular and Molecular Biology | CSIR-Centre for Cellular and Molecular Biology | Payel Mukherjee et al |
| EPI_ISL_910142 | India/TG-CCMB-AH963/2020 | 2020-09-08 | Asia/India/Telangana/ | CSIR-Centre for Cellular and Molecular Biology | CSIR-Centre for Cellular and Molecular Biology | Payel Mukherjee et al |
| EPI_ISL_910143 | India/TG-CCMB-AH964/2020 | 2020-09-08 | Asia/India/Telangana/ | CSIR-Centre for Cellular and Molecular Biology | CSIR-Centre for Cellular and Molecular Biology | Payel Mukherjee et al |
| EPI_ISL_910144 | India/TG-CCMB-AI013/2020 | 2020-09-08 | Asia/India/Telangana/ | CSIR-Centre for Cellular and Molecular Biology | CSIR-Centre for Cellular and Molecular Biology | Payel Mukherjee et al |
| EPI_ISL_910145 | India/TG-CCMB-AI074/2020 | 2020-09-09 | Asia/India/Telangana/ | CSIR-Centre for Cellular and Molecular Biology | CSIR-Centre for Cellular and Molecular Biology | Payel Mukherjee et al |
| EPI_ISL_910146 | India/TG-CCMB-AI089/2020 | 2020-09-09 | Asia/India/Telangana/ | CSIR-Centre for Cellular and Molecular Biology | CSIR-Centre for Cellular and Molecular Biology | Payel Mukherjee et al |
| EPI_ISL_910147 | India/TG-CCMB-AI169/2020 | 2020-09-09 | Asia/India/Telangana/ | CSIR-Centre for Cellular and Molecular Biology | CSIR-Centre for Cellular and Molecular Biology | Payel Mukherjee et al |

| | | | | | | |
|---|---|---|---|---|---|---|
| EPI_ISL_910148 | India/TG-CCMB-AI178/2020 | 2020-09-09 | Asia/India/Telangana/ | CSIR-Centre for Cellular and Molecular Biology | CSIR-Centre for Cellular and Molecular Biology | Payel Mukherjee et al |
| EPI_ISL_910149 | India/TG-CCMB-AI179/2020 | 2020-09-09 | Asia/India/Telangana/ | CSIR-Centre for Cellular and Molecular Biology | CSIR-Centre for Cellular and Molecular Biology | Payel Mukherjee et al |
| EPI_ISL_910150 | India/TG-CCMB-AI228/2020 | 2020-09-09 | Asia/India/Telangana/ | CSIR-Centre for Cellular and Molecular Biology | CSIR-Centre for Cellular and Molecular Biology | Payel Mukherjee et al |
| EPI_ISL_910151 | India/TG-CCMB-AI236/2020 | 2020-09-09 | Asia/India/Telangana/ | CSIR-Centre for Cellular and Molecular Biology | CSIR-Centre for Cellular and Molecular Biology | Payel Mukherjee et al |
| EPI_ISL_910152 | India/TG-CCMB-AI237/2020 | 2020-09-09 | Asia/India/Telangana/ | CSIR-Centre for Cellular and Molecular Biology | CSIR-Centre for Cellular and Molecular Biology | Payel Mukherjee et al |
| EPI_ISL_910153 | India/TG-CCMB-AI268/2020 | 2020-09-09 | Asia/India/Telangana/ | CSIR-Centre for Cellular and Molecular Biology | CSIR-Centre for Cellular and Molecular Biology | Payel Mukherjee et al |
| EPI_ISL_910154 | India/TG-CCMB-AI269/2020 | 2020-09-09 | Asia/India/Telangana/ | CSIR-Centre for Cellular and Molecular Biology | CSIR-Centre for Cellular and Molecular Biology | Payel Mukherjee et al |
| EPI_ISL_910155 | India/TG-CCMB-AI274/2020 | 2020-09-09 | Asia/India/Telangana/ | CSIR-Centre for Cellular and Molecular Biology | CSIR-Centre for Cellular and Molecular Biology | Payel Mukherjee et al |
| EPI_ISL_910156 | India/TG-CCMB-AI308/2020 | 2020-09-09 | Asia/India/Telangana/ | CSIR-Centre for Cellular and Molecular Biology | CSIR-Centre for Cellular and Molecular Biology | Payel Mukherjee et al |
| EPI_ISL_910157 | India/TG-CCMB-AI310/2020 | 2020-09-09 | Asia/India/Telangana/ | CSIR-Centre for Cellular and Molecular Biology | CSIR-Centre for Cellular and Molecular Biology | Payel Mukherjee et al |
| EPI_ISL_910158 | India/TG-CCMB-AI388/2020 | 2020-09-09 | Asia/India/Telangana/ | CSIR-Centre for Cellular and Molecular Biology | CSIR-Centre for Cellular and Molecular Biology | Payel Mukherjee et al |
| EPI_ISL_910159 | India/TG-CCMB-AI390/2020 | 2020-09-09 | Asia/India/Telangana/ | CSIR-Centre for Cellular and Molecular Biology | CSIR-Centre for Cellular and Molecular Biology | Payel Mukherjee et al |
| EPI_ISL_910160 | India/TG-CCMB-AI391/2020 | 2020-09-09 | Asia/India/Telangana/ | CSIR-Centre for Cellular and Molecular Biology | CSIR-Centre for Cellular and Molecular Biology | Payel Mukherjee et al |

| | | | | | | |
|---|---|---|---|---|---|---|
| EPI_ISL_910161 | India/TG-CCMB-AI564/2020 | 2020-09-10 | Asia/India/Telangana/ | CSIR-Centre for Cellular and Molecular Biology | CSIR-Centre for Cellular and Molecular Biology | Payel Mukherjee et al |
| EPI_ISL_910162 | India/TG-CCMB-AI622/2020 | 2020-09-10 | Asia/India/Telangana/ | CSIR-Centre for Cellular and Molecular Biology | CSIR-Centre for Cellular and Molecular Biology | Payel Mukherjee et al |
| EPI_ISL_910163 | India/TG-CCMB-AI640/2020 | 2020-09-10 | Asia/India/Telangana/ | CSIR-Centre for Cellular and Molecular Biology | CSIR-Centre for Cellular and Molecular Biology | Payel Mukherjee et al |
| EPI_ISL_910164 | India/TG-CCMB-AI798/2020 | 2020-09-10 | Asia/India/Telangana/ | CSIR-Centre for Cellular and Molecular Biology | CSIR-Centre for Cellular and Molecular Biology | Payel Mukherjee et al |
| EPI_ISL_910165 | India/TG-CCMB-AI811/2020 | 2020-09-10 | Asia/India/Telangana/ | CSIR-Centre for Cellular and Molecular Biology | CSIR-Centre for Cellular and Molecular Biology | Payel Mukherjee et al |
| EPI_ISL_910166 | India/TG-CCMB-AI844/2020 | 2020-09-10 | Asia/India/Telangana/ | CSIR-Centre for Cellular and Molecular Biology | CSIR-Centre for Cellular and Molecular Biology | Payel Mukherjee et al |
| EPI_ISL_910167 | India/TG-CCMB-AI868/2020 | 2020-09-10 | Asia/India/Telangana/ | CSIR-Centre for Cellular and Molecular Biology | CSIR-Centre for Cellular and Molecular Biology | Payel Mukherjee et al |
| EPI_ISL_910168 | India/TG-CCMB-AI920/2020 | 2020-09-11 | Asia/India/Telangana/ | CSIR-Centre for Cellular and Molecular Biology | CSIR-Centre for Cellular and Molecular Biology | Payel Mukherjee et al |
| EPI_ISL_910169 | India/TG-CCMB-AI987/2020 | 2020-09-11 | Asia/India/Telangana/ | CSIR-Centre for Cellular and Molecular Biology | CSIR-Centre for Cellular and Molecular Biology | Payel Mukherjee et al |
| EPI_ISL_910170 | India/TG-CCMB-AI988/2020 | 2020-09-11 | Asia/India/Telangana/ | CSIR-Centre for Cellular and Molecular Biology | CSIR-Centre for Cellular and Molecular Biology | Payel Mukherjee et al |
| EPI_ISL_910171 | India/TG-CCMB-AI998/2020 | 2020-09-11 | Asia/India/Telangana/ | CSIR-Centre for Cellular and Molecular Biology | CSIR-Centre for Cellular and Molecular Biology | Payel Mukherjee et al |
| EPI_ISL_910172 | India/TG-CCMB-AJ056/2020 | 2020-09-11 | Asia/India/Telangana/ | CSIR-Centre for Cellular and Molecular Biology | CSIR-Centre for Cellular and Molecular Biology | Payel Mukherjee et al |
| EPI_ISL_910173 | India/TG-CCMB-AJ089/2020 | 2020-09-11 | Asia/India/Telangana/ | CSIR-Centre for Cellular and Molecular Biology | CSIR-Centre for Cellular and Molecular Biology | Payel Mukherjee et al |

| | | | | | | |
|---|---|---|---|---|---|---|
| EPI_ISL_910174 | India/TG-CCMB-AJ100/2020 | 2020-09-11 | Asia/India/Telangana/ | CSIR-Centre for Cellular and Molecular Biology | CSIR-Centre for Cellular and Molecular Biology | Payel Mukherjee et al |
| EPI_ISL_910175 | India/TG-CCMB-AJ101/2020 | 2020-09-11 | Asia/India/Telangana/ | CSIR-Centre for Cellular and Molecular Biology | CSIR-Centre for Cellular and Molecular Biology | Payel Mukherjee et al |
| EPI_ISL_910176 | India/TG-CCMB-AJ146/2020 | 2020-09-11 | Asia/India/Telangana/ | CSIR-Centre for Cellular and Molecular Biology | CSIR-Centre for Cellular and Molecular Biology | Payel Mukherjee et al |
| EPI_ISL_910177 | India/TG-CCMB-AJ169/2020 | 2020-09-11 | Asia/India/Telangana/ | CSIR-Centre for Cellular and Molecular Biology | CSIR-Centre for Cellular and Molecular Biology | Payel Mukherjee et al |
| EPI_ISL_910178 | India/TG-CCMB-AJ383/2020 | 2020-09-12 | Asia/India/Telangana/ | CSIR-Centre for Cellular and Molecular Biology | CSIR-Centre for Cellular and Molecular Biology | Payel Mukherjee et al |
| EPI_ISL_910179 | India/TG-CCMB-AJ458/2020 | 2020-09-12 | Asia/India/Telangana/ | CSIR-Centre for Cellular and Molecular Biology | CSIR-Centre for Cellular and Molecular Biology | Payel Mukherjee et al |
| EPI_ISL_910180 | India/TG-CCMB-AJ493/2020 | 2020-09-12 | Asia/India/Telangana/ | CSIR-Centre for Cellular and Molecular Biology | CSIR-Centre for Cellular and Molecular Biology | Payel Mukherjee et al |
| EPI_ISL_910181 | India/TG-CCMB-AJ505/2020 | 2020-09-12 | Asia/India/Telangana/ | CSIR-Centre for Cellular and Molecular Biology | CSIR-Centre for Cellular and Molecular Biology | Payel Mukherjee et al |
| EPI_ISL_910182 | India/TG-CCMB-AJ510/2020 | 2020-09-12 | Asia/India/Telangana/ | CSIR-Centre for Cellular and Molecular Biology | CSIR-Centre for Cellular and Molecular Biology | Payel Mukherjee et al |
| EPI_ISL_910183 | India/TG-CCMB-AJ562/2020 | 2020-09-13 | Asia/India/Telangana/ | CSIR-Centre for Cellular and Molecular Biology | CSIR-Centre for Cellular and Molecular Biology | Payel Mukherjee et al |
| EPI_ISL_910184 | India/TG-CCMB-AJ565/2020 | 2020-09-13 | Asia/India/Telangana/ | CSIR-Centre for Cellular and Molecular Biology | CSIR-Centre for Cellular and Molecular Biology | Payel Mukherjee et al |
| EPI_ISL_910185 | India/TG-CCMB-AJ598/2020 | 2020-09-13 | Asia/India/Telangana/ | CSIR-Centre for Cellular and Molecular Biology | CSIR-Centre for Cellular and Molecular Biology | Payel Mukherjee et al |
| EPI_ISL_910186 | India/TG-CCMB-AJ773/2020 | 2020-09-13 | Asia/India/Telangana/ | CSIR-Centre for Cellular and Molecular Biology | CSIR-Centre for Cellular and Molecular Biology | Payel Mukherjee et al |

| | | | | | | |
|---|---|---|---|---|---|---|
| EPI_ISL_910187 | India/TG-CCMB-AJ815/2020 | 2020-09-13 | Asia/India/Telangana/ | CSIR-Centre for Cellular and Molecular Biology | CSIR-Centre for Cellular and Molecular Biology | Payel Mukherjee et al |
| EPI_ISL_910188 | India/TG-CCMB-AJ889/2020 | 2020-09-13 | Asia/India/Telangana/ | CSIR-Centre for Cellular and Molecular Biology | CSIR-Centre for Cellular and Molecular Biology | Payel Mukherjee et al |
| EPI_ISL_910189 | India/TG-CCMB-AJ901/2020 | 2020-09-13 | Asia/India/Telangana/ | CSIR-Centre for Cellular and Molecular Biology | CSIR-Centre for Cellular and Molecular Biology | Payel Mukherjee et al |
| EPI_ISL_910190 | India/TG-CCMB-AJ929/2020 | 2020-09-13 | Asia/India/Telangana/ | CSIR-Centre for Cellular and Molecular Biology | CSIR-Centre for Cellular and Molecular Biology | Payel Mukherjee et al |
| EPI_ISL_910191 | India/TG-CCMB-AJ964/2020 | 2020-09-14 | Asia/India/Telangana/ | CSIR-Centre for Cellular and Molecular Biology | CSIR-Centre for Cellular and Molecular Biology | Payel Mukherjee et al |
| EPI_ISL_910192 | India/TG-CCMB-AK002/2020 | 2020-09-14 | Asia/India/Telangana/ | CSIR-Centre for Cellular and Molecular Biology | CSIR-Centre for Cellular and Molecular Biology | Payel Mukherjee et al |
| EPI_ISL_910193 | India/TG-CCMB-AK037/2020 | 2020-09-14 | Asia/India/Telangana/ | CSIR-Centre for Cellular and Molecular Biology | CSIR-Centre for Cellular and Molecular Biology | Payel Mukherjee et al |
| EPI_ISL_910194 | India/TG-CCMB-AK081/2020 | 2020-09-14 | Asia/India/Telangana/ | CSIR-Centre for Cellular and Molecular Biology | CSIR-Centre for Cellular and Molecular Biology | Payel Mukherjee et al |
| EPI_ISL_910195 | India/TG-CCMB-AK190/2020 | 2020-09-14 | Asia/India/Telangana/ | CSIR-Centre for Cellular and Molecular Biology | CSIR-Centre for Cellular and Molecular Biology | Payel Mukherjee et al |
| EPI_ISL_910196 | India/TG-CCMB-AK195/2020 | 2020-09-14 | Asia/India/Telangana/ | CSIR-Centre for Cellular and Molecular Biology | CSIR-Centre for Cellular and Molecular Biology | Payel Mukherjee et al |
| EPI_ISL_910197 | India/TG-CCMB-AK287/2020 | 2020-09-15 | Asia/India/Telangana/ | CSIR-Centre for Cellular and Molecular Biology | CSIR-Centre for Cellular and Molecular Biology | Payel Mukherjee et al |
| EPI_ISL_910198 | India/TG-CCMB-AK518/2020 | 2020-09-15 | Asia/India/Telangana/ | CSIR-Centre for Cellular and Molecular Biology | CSIR-Centre for Cellular and Molecular Biology | Payel Mukherjee et al |
| EPI_ISL_910199 | India/TG-CCMB-AK605/2020 | 2020-09-15 | Asia/India/Telangana/ | CSIR-Centre for Cellular and Molecular Biology | CSIR-Centre for Cellular and Molecular Biology | Payel Mukherjee et al |

| | | | | | | |
|---|---|---|---|---|---|---|
| EPI_ISL_910200 | India/TG-CCMB-AK766/2020 | 2020-09-16 | Asia/India/Telangana/ | CSIR-Centre for Cellular and Molecular Biology | CSIR-Centre for Cellular and Molecular Biology | Payel Mukherjee et al |
| EPI_ISL_910201 | India/TG-CCMB-AK789/2020 | 2020-09-16 | Asia/India/Telangana/ | CSIR-Centre for Cellular and Molecular Biology | CSIR-Centre for Cellular and Molecular Biology | Payel Mukherjee et al |
| EPI_ISL_910202 | India/TG-CCMB-AK842/2020 | 2020-09-16 | Asia/India/Telangana/ | CSIR-Centre for Cellular and Molecular Biology | CSIR-Centre for Cellular and Molecular Biology | Payel Mukherjee et al |
| EPI_ISL_910203 | India/TG-CCMB-AK855/2020 | 2020-09-16 | Asia/India/Telangana/ | CSIR-Centre for Cellular and Molecular Biology | CSIR-Centre for Cellular and Molecular Biology | Payel Mukherjee et al |
| EPI_ISL_910204 | India/TG-CCMB-AK978/2020 | 2020-09-16 | Asia/India/Telangana/ | CSIR-Centre for Cellular and Molecular Biology | CSIR-Centre for Cellular and Molecular Biology | Payel Mukherjee et al |
| EPI_ISL_910205 | India/TG-CCMB-AL045/2020 | 2020-09-17 | Asia/India/Telangana/ | CSIR-Centre for Cellular and Molecular Biology | CSIR-Centre for Cellular and Molecular Biology | Payel Mukherjee et al |
| EPI_ISL_910206 | India/TG-CCMB-AL075/2020 | 2020-09-17 | Asia/India/Telangana/ | CSIR-Centre for Cellular and Molecular Biology | CSIR-Centre for Cellular and Molecular Biology | Payel Mukherjee et al |
| EPI_ISL_910207 | India/TG-CCMB-AL095/2020 | 2020-09-17 | Asia/India/Telangana/ | CSIR-Centre for Cellular and Molecular Biology | CSIR-Centre for Cellular and Molecular Biology | Payel Mukherjee et al |
| EPI_ISL_910208 | India/TG-CCMB-AL191/2020 | 2020-09-17 | Asia/India/Telangana/ | CSIR-Centre for Cellular and Molecular Biology | CSIR-Centre for Cellular and Molecular Biology | Payel Mukherjee et al |
| EPI_ISL_910209 | India/TG-CCMB-AL215/2020 | 2020-09-17 | Asia/India/Telangana/ | CSIR-Centre for Cellular and Molecular Biology | CSIR-Centre for Cellular and Molecular Biology | Payel Mukherjee et al |
| EPI_ISL_910210 | India/TG-CCMB-AL222/2020 | 2020-09-17 | Asia/India/Telangana/ | CSIR-Centre for Cellular and Molecular Biology | CSIR-Centre for Cellular and Molecular Biology | Payel Mukherjee et al |
| EPI_ISL_910211 | India/TG-CCMB-AL461/2020 | 2020-09-18 | Asia/India/Telangana/ | CSIR-Centre for Cellular and Molecular Biology | CSIR-Centre for Cellular and Molecular Biology | Payel Mukherjee et al |
| EPI_ISL_910212 | India/TG-CCMB-AL482/2020 | 2020-09-18 | Asia/India/Telangana/ | CSIR-Centre for Cellular and Molecular Biology | CSIR-Centre for Cellular and Molecular Biology | Payel Mukherjee et al |

| | | | | | | |
|---|---|---|---|---|---|---|
| EPI_ISL_910213 | India/TG-CCMB-AL586/2020 | 2020-09-18 | Asia/India/Telangana/ | CSIR-Centre for Cellular and Molecular Biology | CSIR-Centre for Cellular and Molecular Biology | Payel Mukherjee et al |
| EPI_ISL_910214 | India/TG-CCMB-AL610/2020 | 2020-09-18 | Asia/India/Telangana/ | CSIR-Centre for Cellular and Molecular Biology | CSIR-Centre for Cellular and Molecular Biology | Payel Mukherjee et al |
| EPI_ISL_910215 | India/TG-CCMB-AL671/2020 | 2020-09-19 | Asia/India/Telangana/ | CSIR-Centre for Cellular and Molecular Biology | CSIR-Centre for Cellular and Molecular Biology | Payel Mukherjee et al |
| EPI_ISL_910216 | India/TG-CCMB-AL687/2020 | 2020-09-19 | Asia/India/Telangana/ | CSIR-Centre for Cellular and Molecular Biology | CSIR-Centre for Cellular and Molecular Biology | Payel Mukherjee et al |
| EPI_ISL_910217 | India/TG-CCMB-AL705/2020 | 2020-09-19 | Asia/India/Telangana/ | CSIR-Centre for Cellular and Molecular Biology | CSIR-Centre for Cellular and Molecular Biology | Payel Mukherjee et al |
| EPI_ISL_910218 | India/TG-CCMB-AL710/2020 | 2020-09-19 | Asia/India/Telangana/ | CSIR-Centre for Cellular and Molecular Biology | CSIR-Centre for Cellular and Molecular Biology | Payel Mukherjee et al |
| EPI_ISL_910219 | India/TG-CCMB-AL711/2020 | 2020-09-19 | Asia/India/Telangana/ | CSIR-Centre for Cellular and Molecular Biology | CSIR-Centre for Cellular and Molecular Biology | Payel Mukherjee et al |
| EPI_ISL_910220 | India/TG-CCMB-AL883/2020 | 2020-09-19 | Asia/India/Telangana/ | CSIR-Centre for Cellular and Molecular Biology | CSIR-Centre for Cellular and Molecular Biology | Payel Mukherjee et al |
| EPI_ISL_910221 | India/TG-CCMB-AL931/2020 | 2020-09-19 | Asia/India/Telangana/ | CSIR-Centre for Cellular and Molecular Biology | CSIR-Centre for Cellular and Molecular Biology | Payel Mukherjee et al |
| EPI_ISL_910223 | India/TG-CCMB-AM162/2020 | 2020-09-20 | Asia/India/Telangana/ | CSIR-Centre for Cellular and Molecular Biology | CSIR-Centre for Cellular and Molecular Biology | Payel Mukherjee et al |
| EPI_ISL_910224 | India/TG-CCMB-AM163/2020 | 2020-09-20 | Asia/India/Telangana/ | CSIR-Centre for Cellular and Molecular Biology | CSIR-Centre for Cellular and Molecular Biology | Payel Mukherjee et al |
| EPI_ISL_910225 | India/TG-CCMB-AM166/2020 | 2020-09-20 | Asia/India/Telangana/ | CSIR-Centre for Cellular and Molecular Biology | CSIR-Centre for Cellular and Molecular Biology | Payel Mukherjee et al |
| EPI_ISL_910226 | India/TG-CCMB-AM212/2020 | 2020-09-21 | Asia/India/Telangana/ | CSIR-Centre for Cellular and Molecular Biology | CSIR-Centre for Cellular and Molecular Biology | Payel Mukherjee et al |

| | | | | | | |
|---|---|---|---|---|---|---|
| EPI_ISL_910227 | India/TG-CCMB-AM213/2020 | 2020-09-21 | Asia/India/Telangana/ | CSIR-Centre for Cellular and Molecular Biology | CSIR-Centre for Cellular and Molecular Biology | Payel Mukherjee et al |
| EPI_ISL_910228 | India/TG-CCMB-AM215/2020 | 2020-09-21 | Asia/India/Telangana/ | CSIR-Centre for Cellular and Molecular Biology | CSIR-Centre for Cellular and Molecular Biology | Payel Mukherjee et al |
| EPI_ISL_910229 | India/TG-CCMB-AM257/2020 | 2020-09-21 | Asia/India/Telangana/ | CSIR-Centre for Cellular and Molecular Biology | CSIR-Centre for Cellular and Molecular Biology | Payel Mukherjee et al |
| EPI_ISL_910230 | India/TG-CCMB-AM397/2020 | 2020-09-21 | Asia/India/Telangana/ | CSIR-Centre for Cellular and Molecular Biology | CSIR-Centre for Cellular and Molecular Biology | Payel Mukherjee et al |
| EPI_ISL_910231 | India/TG-CCMB-AM463/2020 | 2020-09-21 | Asia/India/Telangana/ | CSIR-Centre for Cellular and Molecular Biology | CSIR-Centre for Cellular and Molecular Biology | Payel Mukherjee et al |
| EPI_ISL_910232 | India/TG-CCMB-AM538/2020 | 2020-09-22 | Asia/India/Telangana/ | CSIR-Centre for Cellular and Molecular Biology | CSIR-Centre for Cellular and Molecular Biology | Payel Mukherjee et al |
| EPI_ISL_910233 | India/TG-CCMB-AM557/2020 | 2020-09-22 | Asia/India/Telangana/ | CSIR-Centre for Cellular and Molecular Biology | CSIR-Centre for Cellular and Molecular Biology | Payel Mukherjee et al |
| EPI_ISL_910234 | India/TG-CCMB-AM593/2020 | 2020-09-22 | Asia/India/Telangana/ | CSIR-Centre for Cellular and Molecular Biology | CSIR-Centre for Cellular and Molecular Biology | Payel Mukherjee et al |
| EPI_ISL_910235 | India/TG-CCMB-AM683/2020 | 2020-09-22 | Asia/India/Telangana/ | CSIR-Centre for Cellular and Molecular Biology | CSIR-Centre for Cellular and Molecular Biology | Payel Mukherjee et al |
| EPI_ISL_910236 | India/TG-CCMB-AM684/2020 | 2020-09-22 | Asia/India/Telangana/ | CSIR-Centre for Cellular and Molecular Biology | CSIR-Centre for Cellular and Molecular Biology | Payel Mukherjee et al |
| EPI_ISL_910237 | India/TG-CCMB-AM709/2020 | 2020-09-22 | Asia/India/Telangana/ | CSIR-Centre for Cellular and Molecular Biology | CSIR-Centre for Cellular and Molecular Biology | Payel Mukherjee et al |
| EPI_ISL_910238 | India/TG-CCMB-AM740/2020 | 2020-09-22 | Asia/India/Telangana/ | CSIR-Centre for Cellular and Molecular Biology | CSIR-Centre for Cellular and Molecular Biology | Payel Mukherjee et al |
| EPI_ISL_910239 | India/TG-CCMB-AM811/2020 | 2020-09-22 | Asia/India/Telangana/ | CSIR-Centre for Cellular and Molecular Biology | CSIR-Centre for Cellular and Molecular Biology | Payel Mukherjee et al |

| | | | | | | |
|---|---|---|---|---|---|---|
| EPI_ISL_910240 | India/TG-CCMB-AM857/2020 | 2020-09-22 | Asia/India/Telangana/ | CSIR-Centre for Cellular and Molecular Biology | CSIR-Centre for Cellular and Molecular Biology | Payel Mukherjee et al |
| EPI_ISL_910241 | India/TG-CCMB-AN055/2020 | 2020-09-23 | Asia/India/Telangana/ | CSIR-Centre for Cellular and Molecular Biology | CSIR-Centre for Cellular and Molecular Biology | Payel Mukherjee et al |
| EPI_ISL_910243 | India/TG-CCMB-AN263/2020 | 2020-09-24 | Asia/India/Telangana/ | CSIR-Centre for Cellular and Molecular Biology | CSIR-Centre for Cellular and Molecular Biology | Payel Mukherjee et al |
| EPI_ISL_910244 | India/TG-CCMB-AN287/2020 | 2020-09-24 | Asia/India/Telangana/ | CSIR-Centre for Cellular and Molecular Biology | CSIR-Centre for Cellular and Molecular Biology | Payel Mukherjee et al |
| EPI_ISL_910245 | India/TG-CCMB-AN345/2020 | 2020-09-24 | Asia/India/Telangana/ | CSIR-Centre for Cellular and Molecular Biology | CSIR-Centre for Cellular and Molecular Biology | Payel Mukherjee et al |
| EPI_ISL_910247 | India/TG-CCMB-AN408/2020 | 2020-09-24 | Asia/India/Telangana/ | CSIR-Centre for Cellular and Molecular Biology | CSIR-Centre for Cellular and Molecular Biology | Payel Mukherjee et al |
| EPI_ISL_910248 | India/TG-CCMB-AN419/2020 | 2020-09-24 | Asia/India/Telangana/ | CSIR-Centre for Cellular and Molecular Biology | CSIR-Centre for Cellular and Molecular Biology | Payel Mukherjee et al |
| EPI_ISL_910249 | India/TG-CCMB-AN491/2020 | 2020-09-24 | Asia/India/Telangana/ | CSIR-Centre for Cellular and Molecular Biology | CSIR-Centre for Cellular and Molecular Biology | Payel Mukherjee et al |
| EPI_ISL_910250 | India/TG-CCMB-AN506/2020 | 2020-09-24 | Asia/India/Telangana/ | CSIR-Centre for Cellular and Molecular Biology | CSIR-Centre for Cellular and Molecular Biology | Payel Mukherjee et al |
| EPI_ISL_910251 | India/TG-CCMB-AN526/2020 | 2020-09-24 | Asia/India/Telangana/ | CSIR-Centre for Cellular and Molecular Biology | CSIR-Centre for Cellular and Molecular Biology | Payel Mukherjee et al |
| EPI_ISL_910252 | India/TG-CCMB-AN667/2020 | 2020-09-25 | Asia/India/Telangana/ | CSIR-Centre for Cellular and Molecular Biology | CSIR-Centre for Cellular and Molecular Biology | Payel Mukherjee et al |
| EPI_ISL_910253 | India/TG-CCMB-AN673/2020 | 2020-09-25 | Asia/India/Telangana/ | CSIR-Centre for Cellular and Molecular Biology | CSIR-Centre for Cellular and Molecular Biology | Payel Mukherjee et al |
| EPI_ISL_910254 | India/TG-CCMB-AN715/2020 | 2020-09-25 | Asia/India/Telangana/ | CSIR-Centre for Cellular and Molecular Biology | CSIR-Centre for Cellular and Molecular Biology | Payel Mukherjee et al |

| | | | | | | |
|---|---|---|---|---|---|---|
| EPI_ISL_910255 | India/TG-CCMB-AN717/2020 | 2020-09-25 | Asia/India/Telangana/ | CSIR-Centre for Cellular and Molecular Biology | CSIR-Centre for Cellular and Molecular Biology | Payel Mukherjee et al |
| EPI_ISL_910256 | India/TG-CCMB-AN721/2020 | 2020-09-25 | Asia/India/Telangana/ | CSIR-Centre for Cellular and Molecular Biology | CSIR-Centre for Cellular and Molecular Biology | Payel Mukherjee et al |
| EPI_ISL_910257 | India/TG-CCMB-AN753/2020 | 2020-09-25 | Asia/India/Telangana/ | CSIR-Centre for Cellular and Molecular Biology | CSIR-Centre for Cellular and Molecular Biology | Payel Mukherjee et al |
| EPI_ISL_910259 | India/TG-CCMB-AN792/2020 | 2020-09-25 | Asia/India/Telangana/ | CSIR-Centre for Cellular and Molecular Biology | CSIR-Centre for Cellular and Molecular Biology | Payel Mukherjee et al |
| EPI_ISL_910260 | India/TG-CCMB-AN832/2020 | 2020-09-25 | Asia/India/Telangana/ | CSIR-Centre for Cellular and Molecular Biology | CSIR-Centre for Cellular and Molecular Biology | Payel Mukherjee et al |
| EPI_ISL_910261 | India/TG-CCMB-AN842/2020 | 2020-09-25 | Asia/India/Telangana/ | CSIR-Centre for Cellular and Molecular Biology | CSIR-Centre for Cellular and Molecular Biology | Payel Mukherjee et al |
| EPI_ISL_910262 | India/TG-CCMB-AN954/2020 | 2020-09-25 | Asia/India/Telangana/ | CSIR-Centre for Cellular and Molecular Biology | CSIR-Centre for Cellular and Molecular Biology | Payel Mukherjee et al |
| EPI_ISL_910263 | India/TG-CCMB-AN967/2020 | 2020-09-25 | Asia/India/Telangana/ | CSIR-Centre for Cellular and Molecular Biology | CSIR-Centre for Cellular and Molecular Biology | Payel Mukherjee et al |
| EPI_ISL_910264 | India/TG-CCMB-AO009/2020 | 2020-09-26 | Asia/India/Telangana/ | CSIR-Centre for Cellular and Molecular Biology | CSIR-Centre for Cellular and Molecular Biology | Payel Mukherjee et al |
| EPI_ISL_910265 | India/TG-CCMB-AO019/2020 | 2020-09-26 | Asia/India/Telangana/ | CSIR-Centre for Cellular and Molecular Biology | CSIR-Centre for Cellular and Molecular Biology | Payel Mukherjee et al |
| EPI_ISL_910266 | India/TG-CCMB-AO022/2020 | 2020-09-26 | Asia/India/Telangana/ | CSIR-Centre for Cellular and Molecular Biology | CSIR-Centre for Cellular and Molecular Biology | Payel Mukherjee et al |
| EPI_ISL_910267 | India/TG-CCMB-AO100/2020 | 2020-09-26 | Asia/India/Telangana/ | CSIR-Centre for Cellular and Molecular Biology | CSIR-Centre for Cellular and Molecular Biology | Payel Mukherjee et al |
| EPI_ISL_910268 | India/TG-CCMB-AO138/2020 | 2020-09-26 | Asia/India/Telangana/ | CSIR-Centre for Cellular and Molecular Biology | CSIR-Centre for Cellular and Molecular Biology | Payel Mukherjee et al |

| | | | | | | |
|---|---|---|---|---|---|---|
| EPI_ISL_910269 | India/TG-CCMB-AO176/2020 | 2020-09-26 | Asia/India/Telangana/ | CSIR-Centre for Cellular and Molecular Biology | CSIR-Centre for Cellular and Molecular Biology | Payel Mukherjee et al |
| EPI_ISL_910270 | India/TG-CCMB-AO189/2020 | 2020-09-26 | Asia/India/Telangana/ | CSIR-Centre for Cellular and Molecular Biology | CSIR-Centre for Cellular and Molecular Biology | Payel Mukherjee et al |
| EPI_ISL_910271 | India/TG-CCMB-AO257/2020 | 2020-09-28 | Asia/India/Telangana/ | CSIR-Centre for Cellular and Molecular Biology | CSIR-Centre for Cellular and Molecular Biology | Payel Mukherjee et al |
| EPI_ISL_910272 | India/TG-CCMB-AO293/2020 | 2020-09-28 | Asia/India/Telangana/ | CSIR-Centre for Cellular and Molecular Biology | CSIR-Centre for Cellular and Molecular Biology | Payel Mukherjee et al |
| EPI_ISL_910273 | India/TG-CCMB-AO297/2020 | 2020-09-28 | Asia/India/Telangana/ | CSIR-Centre for Cellular and Molecular Biology | CSIR-Centre for Cellular and Molecular Biology | Payel Mukherjee et al |
| EPI_ISL_910274 | India/TG-CCMB-AO315/2020 | 2020-09-28 | Asia/India/Telangana/ | CSIR-Centre for Cellular and Molecular Biology | CSIR-Centre for Cellular and Molecular Biology | Payel Mukherjee et al |
| EPI_ISL_910275 | India/TG-CCMB-AO339/2020 | 2020-09-28 | Asia/India/Telangana/ | CSIR-Centre for Cellular and Molecular Biology | CSIR-Centre for Cellular and Molecular Biology | Payel Mukherjee et al |
| EPI_ISL_910277 | India/TG-CCMB-AO374/2020 | 2020-09-28 | Asia/India/Telangana/ | CSIR-Centre for Cellular and Molecular Biology | CSIR-Centre for Cellular and Molecular Biology | Payel Mukherjee et al |
| EPI_ISL_910279 | India/TG-CCMB-AO401/2020 | 2020-09-28 | Asia/India/Telangana/ | CSIR-Centre for Cellular and Molecular Biology | CSIR-Centre for Cellular and Molecular Biology | Payel Mukherjee et al |
| EPI_ISL_910280 | India/TG-CCMB-AO417/2020 | 2020-09-28 | Asia/India/Telangana/ | CSIR-Centre for Cellular and Molecular Biology | CSIR-Centre for Cellular and Molecular Biology | Payel Mukherjee et al |
| EPI_ISL_910281 | India/TG-CCMB-AO476/2020 | 2020-09-28 | Asia/India/Telangana/ | CSIR-Centre for Cellular and Molecular Biology | CSIR-Centre for Cellular and Molecular Biology | Payel Mukherjee et al |
| EPI_ISL_910282 | India/TG-CCMB-AO562/2020 | 2020-09-29 | Asia/India/Telangana/ | CSIR-Centre for Cellular and Molecular Biology | CSIR-Centre for Cellular and Molecular Biology | Payel Mukherjee et al |
| EPI_ISL_910283 | India/TG-CCMB-AO584/2020 | 2020-09-29 | Asia/India/Telangana/ | CSIR-Centre for Cellular and Molecular Biology | CSIR-Centre for Cellular and Molecular Biology | Payel Mukherjee et al |

| | | | | | | |
|---|---|---|---|---|---|---|
| EPI_ISL_910284 | India/TG-CCMB-AO671/2020 | 2020-09-29 | Asia/India/Telangana/ | CSIR-Centre for Cellular and Molecular Biology | CSIR-Centre for Cellular and Molecular Biology | Payel Mukherjee et al |
| EPI_ISL_910285 | India/TG-CCMB-AO708/2020 | 2020-09-29 | Asia/India/Telangana/ | CSIR-Centre for Cellular and Molecular Biology | CSIR-Centre for Cellular and Molecular Biology | Payel Mukherjee et al |
| EPI_ISL_910286 | India/TG-CCMB-AO711/2020 | 2020-09-29 | Asia/India/Telangana/ | CSIR-Centre for Cellular and Molecular Biology | CSIR-Centre for Cellular and Molecular Biology | Payel Mukherjee et al |
| EPI_ISL_910287 | India/TG-CCMB-AO718/2020 | 2020-09-29 | Asia/India/Telangana/ | CSIR-Centre for Cellular and Molecular Biology | CSIR-Centre for Cellular and Molecular Biology | Payel Mukherjee et al |
| EPI_ISL_910288 | India/TG-CCMB-AO778/2020 | 2020-09-29 | Asia/India/Telangana/ | CSIR-Centre for Cellular and Molecular Biology | CSIR-Centre for Cellular and Molecular Biology | Payel Mukherjee et al |
| EPI_ISL_910289 | India/TG-CCMB-AO853/2020 | 2020-09-29 | Asia/India/Telangana/ | CSIR-Centre for Cellular and Molecular Biology | CSIR-Centre for Cellular and Molecular Biology | Payel Mukherjee et al |
| EPI_ISL_910290 | India/TG-CCMB-AO863/2020 | 2020-09-29 | Asia/India/Telangana/ | CSIR-Centre for Cellular and Molecular Biology | CSIR-Centre for Cellular and Molecular Biology | Payel Mukherjee et al |
| EPI_ISL_910292 | India/TG-CCMB-AO865/2020 | 2020-09-29 | Asia/India/Telangana/ | CSIR-Centre for Cellular and Molecular Biology | CSIR-Centre for Cellular and Molecular Biology | Payel Mukherjee et al |
| EPI_ISL_910293 | India/TG-CCMB-AO866/2020 | 2020-09-29 | Asia/India/Telangana/ | CSIR-Centre for Cellular and Molecular Biology | CSIR-Centre for Cellular and Molecular Biology | Payel Mukherjee et al |
| EPI_ISL_910294 | India/TG-CCMB-V31/2020 | 2020-07-29 | Asia/India/Telangana/ | CSIR-Centre for Cellular and Molecular Biology | CSIR-Centre for Cellular and Molecular Biology | Payel Mukherjee et al |
| EPI_ISL_910295 | India/TG-CCMB-W299/2020 | 2020-07-27 | Asia/India/Telangana/ | CSIR-Centre for Cellular and Molecular Biology | CSIR-Centre for Cellular and Molecular Biology | Payel Mukherjee et al |
| EPI_ISL_910296 | India/TG-CCMB-W328/2020 | 2020-07-27 | Asia/India/Telangana/ | CSIR-Centre for Cellular and Molecular Biology | CSIR-Centre for Cellular and Molecular Biology | Payel Mukherjee et al |
| EPI_ISL_910297 | India/TG-CCMB-W826/2020 | 2020-07-29 | Asia/India/Telangana/ | CSIR-Centre for Cellular and Molecular Biology | CSIR-Centre for Cellular and Molecular Biology | Payel Mukherjee et al |

| | | | | | | |
|---|---|---|---|---|---|---|
| EPI_ISL_910298 | India/TG-CCMB-W827/2020 | 2020-07-29 | Asia/India/Telangana/ | CSIR-Centre for Cellular and Molecular Biology | CSIR-Centre for Cellular and Molecular Biology | Payel Mukherjee et al |
| EPI_ISL_910299 | India/TG-CCMB-W845/2020 | 2020-07-29 | Asia/India/Telangana/ | CSIR-Centre for Cellular and Molecular Biology | CSIR-Centre for Cellular and Molecular Biology | Payel Mukherjee et al |
| EPI_ISL_910300 | India/TG-CCMB-W953/2020 | 2020-07-29 | Asia/India/Telangana/ | CSIR-Centre for Cellular and Molecular Biology | CSIR-Centre for Cellular and Molecular Biology | Payel Mukherjee et al |
| EPI_ISL_910301 | India/TG-CCMB-W955/2020 | 2020-07-29 | Asia/India/Telangana/ | CSIR-Centre for Cellular and Molecular Biology | CSIR-Centre for Cellular and Molecular Biology | Payel Mukherjee et al |
| EPI_ISL_910303 | India/TG-CCMB-W973/2020 | 2020-07-29 | Asia/India/Telangana/ | CSIR-Centre for Cellular and Molecular Biology | CSIR-Centre for Cellular and Molecular Biology | Payel Mukherjee et al |
| EPI_ISL_910305 | India/TG-CCMB-W980/2020 | 2020-07-29 | Asia/India/Telangana/ | CSIR-Centre for Cellular and Molecular Biology | CSIR-Centre for Cellular and Molecular Biology | Payel Mukherjee et al |
| EPI_ISL_910306 | India/TG-CCMB-X780/2020 | 2020-07-18 | Asia/India/Telangana/ | CSIR-Centre for Cellular and Molecular Biology | CSIR-Centre for Cellular and Molecular Biology | Payel Mukherjee et al |
| EPI_ISL_910307 | India/TG-CCMB-X838/2020 | 2020-07-19 | Asia/India/Telangana/ | CSIR-Centre for Cellular and Molecular Biology | CSIR-Centre for Cellular and Molecular Biology | Payel Mukherjee et al |
| EPI_ISL_910308 | India/TG-CCMB-X894/2020 | 2020-07-19 | Asia/India/Telangana/ | CSIR-Centre for Cellular and Molecular Biology | CSIR-Centre for Cellular and Molecular Biology | Payel Mukherjee et al |
| EPI_ISL_910309 | India/TG-CCMB-Y209/2020 | 2020-07-20 | Asia/India/Telangana/ | CSIR-Centre for Cellular and Molecular Biology | CSIR-Centre for Cellular and Molecular Biology | Payel Mukherjee et al |
| EPI_ISL_910310 | India/TG-CCMB-Y267/2020 | 2020-07-21 | Asia/India/Telangana/ | CSIR-Centre for Cellular and Molecular Biology | CSIR-Centre for Cellular and Molecular Biology | Payel Mukherjee et al |
| EPI_ISL_910312 | India/TG-CCMB-Y662/2020 | 2020-07-22 | Asia/India/Telangana/ | CSIR-Centre for Cellular and Molecular Biology | CSIR-Centre for Cellular and Molecular Biology | Payel Mukherjee et al |
| EPI_ISL_910313 | India/TG-CCMB-Y677/2020 | 2020-07-22 | Asia/India/Telangana/ | CSIR-Centre for Cellular and Molecular Biology | CSIR-Centre for Cellular and Molecular Biology | Payel Mukherjee et al |

| | | | | | | |
|---|---|---|---|---|---|---|
| EPI_ISL_910314 | India/TG-CCMB-Z018/2020 | 2020-07-23 | Asia/India/Telangana/ | CSIR-Centre for Cellular and Molecular Biology | CSIR-Centre for Cellular and Molecular Biology | Payel Mukherjee et al |
| EPI_ISL_910315 | India/TG-CCMB-Z023/2020 | 2020-07-23 | Asia/India/Telangana/ | CSIR-Centre for Cellular and Molecular Biology | CSIR-Centre for Cellular and Molecular Biology | Payel Mukherjee et al |
| EPI_ISL_910316 | India/TG-CCMB-Z033/2020 | 2020-07-23 | Asia/India/Telangana/ | CSIR-Centre for Cellular and Molecular Biology | CSIR-Centre for Cellular and Molecular Biology | Payel Mukherjee et al |
| EPI_ISL_910317 | India/TG-CCMB-Z197/2020 | 2020-07-23 | Asia/India/Telangana/ | CSIR-Centre for Cellular and Molecular Biology | CSIR-Centre for Cellular and Molecular Biology | Payel Mukherjee et al |
| EPI_ISL_910319 | India/TG-CCMB-Z394/2020 | 2020-07-24 | Asia/India/Telangana/ | CSIR-Centre for Cellular and Molecular Biology | CSIR-Centre for Cellular and Molecular Biology | Payel Mukherjee et al |
| EPI_ISL_910320 | India/TG-CCMB-Z523/2020 | 2020-07-25 | Asia/India/Telangana/ | CSIR-Centre for Cellular and Molecular Biology | CSIR-Centre for Cellular and Molecular Biology | Payel Mukherjee et al |
| EPI_ISL_910321 | India/TG-CCMB-Z592/2020 | 2020-07-25 | Asia/India/Telangana/ | CSIR-Centre for Cellular and Molecular Biology | CSIR-Centre for Cellular and Molecular Biology | Payel Mukherjee et al |
| EPI_ISL_910322 | India/TG-CCMB-Z915/2020 | 2020-07-26 | Asia/India/Telangana/ | CSIR-Centre for Cellular and Molecular Biology | CSIR-Centre for Cellular and Molecular Biology | Payel Mukherjee et al |
| EPI_ISL_910323 | India/TG-CCMB-Z974/2020 | 2020-07-26 | Asia/India/Telangana/ | CSIR-Centre for Cellular and Molecular Biology | CSIR-Centre for Cellular and Molecular Biology | Payel Mukherjee et al |
| EPI_ISL_910328 | NorthMacedonia/15267/2020 | 2020-10-15 | Europe/North Macedonia/Tetovo/ | General Hospital - Tetovo | "Research Center for Genetic Engineering and Biotechnology ""Georgi D. Efremov,Äù , Macedonian Academy of Sciences and Arts" | RCGEB - MASA et al |
| EPI_ISL_910335 | Romania/Bucuresti410794/2021 | 2021-01-07 | Europe/Romania/Bucharest/ | Laboratory for Respiratory Viruses, Cantacuzino National Military-Medical Institute for Research and Development | Cantacuzino Institute Virology | Luiza Ustea et al |

| | | | | | | |
|---|---|---|---|---|---|---|
| EPI_ISL_910336 | Romania/Bucuresti408383/2020 | 2020-12-31 | Europe/Romania/Bucharest/ | Laboratory for Respiratory Viruses, Cantacuzino National Military-Medical Institute for Research and Development | Cantacuzino Institute Virology | Luiza Ustea et al |
| EPI_ISL_910401 | Luxembourg/LNS5624134/2020 | 2020-12-04 | Europe/Luxembourg/Luxembourg/ | Laboratoire national de sante, Microbiology, Virology | Laboratoire national de sante, Microbiology, Microbial Genomics Platform | Anke Wienecke-Baldacchino et al |
| EPI_ISL_910537 | Luxembourg/LNS6955100/2020 | 2020-12-20 | Europe/Luxembourg/Luxembourg/ | Laboratoire national de sante, Microbiology, Virology | Laboratoire national de sante, Microbiology, Microbial Genomics Platform | Anke Wienecke-Baldacchino et al |
| EPI_ISL_910903 | Luxembourg/LNS5307261/2021 | 2021-01-05 | Europe/Luxembourg/Luxembourg/ | Laboratoire national de sante, Microbiology, Virology | Laboratoire national de sante, Microbiology, Microbial Genomics Platform | Anke Wienecke-Baldacchino et al |
| EPI_ISL_911146 | Luxembourg/LNS2643212/2021 | 2021-01-12 | Europe/Luxembourg/Luxembourg/ | Laboratoire national de sante, Microbiology, Virology | Laboratoire national de sante, Microbiology, Microbial Genomics Platform | Anke Wienecke-Baldacchino et al |
| EPI_ISL_911218 | Luxembourg/LNS9776367/2021 | 2021-01-18 | Europe/Luxembourg/Luxembourg/ | Laboratoire national de sante, Microbiology, Virology | Laboratoire national de sante, Microbiology, Microbial Genomics Platform | Anke Wienecke-Baldacchino et al |
| EPI_ISL_911284 | Spain/IB-IBV-99015786/2021 | 2021-01-24 | Europe/Spain/Balear Islands/Palma de M | Servicio de Microbiolog√≠a, Hospital Universitario Son Espases | SeqCOVID-SPAIN consortium/IBV(CSIC) | Carla L√≥pez-Causap√© et al |
| EPI_ISL_911285 | Spain/IB-IBV-99015787/2021 | 2021-01-15 | Europe/Spain/Balear Islands/Ibiza | Servicio de Microbiolog√≠a, Hospital Universitario Son Espases | SeqCOVID-SPAIN consortium/IBV(CSIC) | Carla L√≥pez-Causap√© et al |

| | | | | | | |
|---|---|---|---|---|---|---|
| EPI_ISL_911287 | Spain/IB-IBV-99015791/2021 | 2021-01-15 | Europe/Spain/Balear Islands/Ibiza | Servicio de Microbiolog√≠a, Hospital Universitario Son Espases | SeqCOVID-SPAIN consortium/IBV(CSIC) | Carla L√≥pez-Causap√© et al |
| EPI_ISL_911289 | Spain/IB-IBV-99015796/2021 | 2021-01-11 | Europe/Spain/Balear Islands/Palma de M | Servicio de Microbiolog√≠a, Hospital Universitario Son Espases | SeqCOVID-SPAIN consortium/IBV(CSIC) | Carla L√≥pez-Causap√© et al |
| EPI_ISL_911290 | Spain/IB-IBV-99015797/2021 | 2021-01-20 | Europe/Spain/Balear Islands/Palma de M | Servicio de Microbiolog√≠a, Hospital Universitario Son Espases | SeqCOVID-SPAIN consortium/IBV(CSIC) | Carla L√≥pez-Causap√© et al |
| EPI_ISL_911303 | Spain/IB-IBV-99015813/2021 | 2021-01-15 | Europe/Spain/Balear Islands/Ibiza | Servicio de Microbiolog√≠a, Hospital Universitario Son Espases | SeqCOVID-SPAIN consortium/IBV(CSIC) | Carla L√≥pez-Causap√© et al |
| EPI_ISL_911304 | Spain/IB-IBV-99015815/2021 | 2021-01-15 | Europe/Spain/Balear Islands/Ibiza | Servicio de Microbiolog√≠a, Hospital Universitario Son Espases | SeqCOVID-SPAIN consortium/IBV(CSIC) | Carla L√≥pez-Causap√© et al |
| EPI_ISL_911305 | Spain/IB-IBV-99015817/2021 | 2021-01-13 | Europe/Spain/Balear Islands/Palma de M | Servicio de Microbiolog√≠a, Hospital Universitario Son Espases | SeqCOVID-SPAIN consortium/IBV(CSIC) | Carla L√≥pez-Causap√© et al |
| EPI_ISL_911306 | Spain/IB-IBV-99015818/2021 | 2021-01-16 | Europe/Spain/Balear Islands/Pollenca | Servicio de Microbiolog√≠a, Hospital Universitario Son Espases | SeqCOVID-SPAIN consortium/IBV(CSIC) | Carla L√≥pez-Causap√© et al |
| EPI_ISL_911308 | Spain/IB-IBV-99015821/2021 | 2021-01-24 | Europe/Spain/Balear Islands/Palma de M | Servicio de Microbiolog√≠a, Hospital Universitario Son Espases | SeqCOVID-SPAIN consortium/IBV(CSIC) | Carla L√≥pez-Causap√© et al |
| EPI_ISL_911319 | Spain/IB-IBV-99015834/2021 | 2021-01-22 | Europe/Spain/Balear Islands/Ibiza | Servicio de Microbiolog√≠a, Hospital Universitario Son Espases | SeqCOVID-SPAIN consortium/IBV(CSIC) | Carla L√≥pez-Causap√© et al |
| EPI_ISL_911320 | Spain/IB-IBV-99015835/2021 | 2021-01-22 | Europe/Spain/Balear Islands/Ibiza | Servicio de Microbiolog√≠a, Hospital Universitario Son Espases | SeqCOVID-SPAIN consortium/IBV(CSIC) | Carla L√≥pez-Causap√© et al |
| EPI_ISL_911326 | Spain/IB-IBV-99015842/2021 | 2021-01-17 | Europe/Spain/Balear Islands/Sa Pobla | Servicio de Microbiolog√≠a, Hospital Universitario Son Espases | SeqCOVID-SPAIN consortium/IBV(CSIC) | Carla L√≥pez-Causap√© et al |
| EPI_ISL_911327 | Spain/IB-IBV-99015843/2021 | 2021-01-10 | Europe/Spain/Balear Islands/Palma de M | Servicio de Microbiolog√≠a, Hospital Universitario Son Espases | SeqCOVID-SPAIN consortium/IBV(CSIC) | Carla L√≥pez-Causap√© et al |
| EPI_ISL_911332 | Spain/IB-IBV-99015853/2021 | 2021-01-14 | Europe/Spain/Balear Islands/Palma de M | Servicio de Microbiolog√≠a, Hospital Universitario Son Espases | SeqCOVID-SPAIN consortium/IBV(CSIC) | Carla L√≥pez-Causap√© et al |

| | | | | | | |
|---|---|---|---|---|---|---|
| EPI_ISL_911339 | Spain/IB-IBV-99015862/2021 | 2021-01-15 | Europe/Spain/Balear Islands/Ibiza | Servicio de Microbiolog√≠a, Hospital Universitario Son Espases | SeqCOVID-SPAIN consortium/IBV(CSIC) | Carla L√≥pez-Causap√© et al |
| EPI_ISL_911344 | Spain/IB-IBV-99015869/2021 | 2021-01-24 | Europe/Spain/Balear Islands/Palma de M | Servicio de Microbiolog√≠a, Hospital Universitario Son Espases | SeqCOVID-SPAIN consortium/IBV(CSIC) | Carla L√≥pez-Causap√© et al |
| EPI_ISL_911345 | Spain/IB-IBV-99015871/2021 | 2021-01-22 | Europe/Spain/Balear Islands/Ibiza | Servicio de Microbiolog√≠a, Hospital Universitario Son Espases | SeqCOVID-SPAIN consortium/IBV(CSIC) | Carla L√≥pez-Causap√© et al |
| EPI_ISL_911347 | Spain/IB-IBV-99015873/2021 | 2021-01-15 | Europe/Spain/Balear Islands/Ibiza | Servicio de Microbiolog√≠a, Hospital Universitario Son Espases | SeqCOVID-SPAIN consortium/IBV(CSIC) | Carla L√≥pez-Causap√© et al |
| EPI_ISL_911349 | Spain/IB-IBV-99015877/2021 | 2021-01-14 | Europe/Spain/Balear Islands/Palma de M | Servicio de Microbiolog√≠a, Hospital Universitario Son Espases | SeqCOVID-SPAIN consortium/IBV(CSIC) | Carla L√≥pez-Causap√© et al |
| EPI_ISL_911353 | USA/IN-Lilly-IPB0173-04bc01/2020 | 2020-03-22 | North America/USA/Indiana/Indianapoli | Clinical Diagnostics Laboratory, Diagnostic & Experimental Pathology, Lilly Research Laboratories | Clinical Diagnostics Laboratory, Diagnostic & Experimental Pathology, Lilly Research Laboratories | Tim Holzer et al |
| EPI_ISL_911356 | USA/IN-Lilly-IPB0173-04bc10/2020 | 2020-03-20 | North America/USA/Indiana/Indianapoli | Clinical Diagnostics Laboratory, Diagnostic & Experimental Pathology, Lilly Research Laboratories | Clinical Diagnostics Laboratory, Diagnostic & Experimental Pathology, Lilly Research Laboratories | Tim Holzer et al |
| EPI_ISL_911358 | USA/IN-Lilly-IPB0173-04bc15/2020 | 2020-03-24 | North America/USA/Indiana/Indianapoli | Clinical Diagnostics Laboratory, Diagnostic & Experimental Pathology, Lilly Research Laboratories | Clinical Diagnostics Laboratory, Diagnostic & Experimental Pathology, Lilly Research Laboratories | Tim Holzer et al |

| | | | | | | |
|---|---|---|---|---|---|---|
| EPI_ISL_911378 | USA/MI-UM-10038050090/2021 | 2021-01-28 | North America/USA/Michigan/ | University of Michigan Clinical Microbiology Laboratory | Lauring Lab, University of Michigan, Department of Microbiology and Immunology | Valesano et al |
| EPI_ISL_911380 | USA/MI-UM-10038052572/2021 | 2021-01-28 | North America/USA/Michigan/ | University of Michigan Clinical Microbiology Laboratory | Lauring Lab, University of Michigan, Department of Microbiology and Immunology | Valesano et al |
| EPI_ISL_911437 | Belgium/AZDelta210305712-01_S13/ | 2021-01-18 | Europe/Belgium/Ieper/Wervik | AZDelta | AZDelta | Geert Martens et al |
| EPI_ISL_911457 | USA/IN-Lilly-IPB0173-06bc17/2020 | 2020-03-24 | North America/USA/Indiana/Indianapoli | Clinical Diagnostics Laboratory, Diagnostic & Experimental Pathology, Lilly Research Laboratories | Clinical Diagnostics Laboratory, Diagnostic & Experimental Pathology, Lilly Research Laboratories | Tim Holzer et al |
| EPI_ISL_911459 | USA/IN-Lilly-IPB0173-06bc19/2020 | 2020-03-24 | North America/USA/Indiana/Indianapoli | Clinical Diagnostics Laboratory, Diagnostic & Experimental Pathology, Lilly Research Laboratories | Clinical Diagnostics Laboratory, Diagnostic & Experimental Pathology, Lilly Research Laboratories | Tim Holzer et al |
| EPI_ISL_911470 | USA/IN-Lilly-IPB0173-06bc31/2020 | 2020-03-24 | North America/USA/Indiana/Indianapoli | Clinical Diagnostics Laboratory, Diagnostic & Experimental Pathology, Lilly Research Laboratories | Clinical Diagnostics Laboratory, Diagnostic & Experimental Pathology, Lilly Research Laboratories | Tim Holzer et al |
| EPI_ISL_911472 | USA/TX-HHD-2101214359/2020 | 2020-12-11 | North America/USA/Texas/ | Houston Health Dept. | Houston Health Dept. | Ryker Penn et al |

| | | | | | | |
|---|---|---|---|---|---|---|
| EPI_ISL_911480 | USA/TX-HHD-2101215038/2020 | 2020-12-12 | North America/USA/Texas/ | Houston Health Dept. | Houston Health Dept. | Ryker Penn et al |
| EPI_ISL_911483 | USA/TX-HHD-2101215072/2020 | 2020-12-12 | North America/USA/Texas/ | Houston Health Dept. | Houston Health Dept. | Ryker Penn et al |
| EPI_ISL_911484 | USA/TX-HHD-2101215074/2020 | 2020-12-12 | North America/USA/Texas/ | Houston Health Dept. | Houston Health Dept. | Ryker Penn et al |
| EPI_ISL_911488 | USA/TX-HHD-2101215145/2020 | 2020-12-12 | North America/USA/Texas/ | Houston Health Dept. | Houston Health Dept. | Ryker Penn et al |
| EPI_ISL_911489 | USA/TX-HHD-2101215669/2020 | 2020-12-13 | North America/USA/Texas/ | Houston Health Dept. | Houston Health Dept. | Ryker Penn et al |
| EPI_ISL_911514 | USA/OH-ODH-SC178544/2020 | 2020-12-09 | North America/USA/Ohio/ | Ohio Department of Health Laboratory | Ohio Department of Health Laboratory | Holmes et al |
| EPI_ISL_911516 | USA/OH-ODH-SC179284/2020 | 2020-12-10 | North America/USA/Ohio/ | Ohio Department of Health Laboratory | Ohio Department of Health Laboratory | Holmes et al |
| EPI_ISL_911525 | Italy/TUS-AOUC-4/2021 | 2021-01-19 | Europe/Italy/Tuscany/ | Microbiology and Virology Unit, Florence Careggi University Hospital | Microbiology and Virology Unit, Florence Careggi University Hospital | Vincenzo Di Pilato et al |
| EPI_ISL_911526 | Italy/TUS-AOUC-5/2021 | 2021-01-05 | Europe/Italy/Tuscany/ | Microbiology and Virology Unit, Florence Careggi University Hospital | Microbiology and Virology Unit, Florence Careggi University Hospital | Vincenzo Di Pilato et al |
| EPI_ISL_911527 | Italy/TUS-AOUC-6/2021 | 2021-01-05 | Europe/Italy/Tuscany/ | Microbiology and Virology Unit, Florence Careggi University Hospital | Microbiology and Virology Unit, Florence Careggi University Hospital | Vincenzo Di Pilato et al |
| EPI_ISL_911548 | USA/UT-ARUP-21021117888/2021 | 2021-01-21 | North America/USA/Utah/ | ARUP laboratories | ARUP Laboratories | Hymas W et al |

| | | | | | | |
|---|---|---|---|---|---|---|
| EPI_ISL_911549 | USA/IN-Lilly-IPB0173-07bc01/2020 | 2020-03-24 | North America/USA/Indiana/Indianapoli | Clinical Diagnostics Laboratory, Diagnostic & Experimental Pathology, Lilly Research Laboratories | Clinical Diagnostics Laboratory, Diagnostic & Experimental Pathology, Lilly Research Laboratories | Tim Holzer et al |
| EPI_ISL_911568 | USA/IN-Lilly-IPB0173-08bc06/2020 | 2020-03-23 | North America/USA/Indiana/Indianapoli | Clinical Diagnostics Laboratory, Diagnostic & Experimental Pathology, Lilly Research Laboratories | Clinical Diagnostics Laboratory, Diagnostic & Experimental Pathology, Lilly Research Laboratories | Tim Holzer et al |
| EPI_ISL_911572 | USA/IN-Lilly-IPB0173-08bc11/2020 | 2020-03-24 | North America/USA/Indiana/Indianapoli | Clinical Diagnostics Laboratory, Diagnostic & Experimental Pathology, Lilly Research Laboratories | Clinical Diagnostics Laboratory, Diagnostic & Experimental Pathology, Lilly Research Laboratories | Tim Holzer et al |
| EPI_ISL_911581 | USA/NC-UNC-0031/2021 | 2021-01-21 | North America/USA/North Carolina/ | Clinical Molecular Microbiology Laboratory, UNC Hospitals | Jeremy Wang | Jeremy Wang et al |
| EPI_ISL_911585 | USA/NC-UNC-0035/2021 | 2021-01-18 | North America/USA/North Carolina/ | Clinical Molecular Microbiology Laboratory, UNC Hospitals | Jeremy Wang | Jeremy Wang et al |
| EPI_ISL_911592 | USA/NC-UNC-0043/2021 | 2021-01-22 | North America/USA/North Carolina/ | Clinical Molecular Microbiology Laboratory, UNC Hospitals | Jeremy Wang | Jeremy Wang et al |
| EPI_ISL_911593 | USA/NC-UNC-0044/2021 | 2021-01-18 | North America/USA/North Carolina/ | Clinical Molecular Microbiology Laboratory, UNC Hospitals | Jeremy Wang | Jeremy Wang et al |
| EPI_ISL_911600 | USA/NC-UNC-0051/2021 | 2021-01-21 | North America/USA/North Carolina/ | Clinical Molecular Microbiology Laboratory, UNC Hospitals | Jeremy Wang | Jeremy Wang et al |
| EPI_ISL_911602 | USA/NC-UNC-0053/2021 | 2021-01-18 | North America/USA/North Carolina/ | Clinical Molecular Microbiology Laboratory, UNC Hospitals | Jeremy Wang | Jeremy Wang et al |

| | | | | | | |
|---|---|---|---|---|---|---|
| EPI_ISL_911616 | USA/IN-Lilly-IPB0173-09bc14/2020 | 2020-03-26 | North America/USA/Indiana/Indianapoli | Clinical Diagnostics Laboratory, Diagnostic & Experimental Pathology, Lilly Research Laboratories | Clinical Diagnostics Laboratory, Diagnostic & Experimental Pathology, Lilly Research Laboratories | Tim Holzer et al |
| EPI_ISL_911623 | USA/IN-Lilly-IPB0173-09bc21/2020 | 2020-03-25 | North America/USA/Indiana/Indianapoli | Clinical Diagnostics Laboratory, Diagnostic & Experimental Pathology, Lilly Research Laboratories | Clinical Diagnostics Laboratory, Diagnostic & Experimental Pathology, Lilly Research Laboratories | Tim Holzer et al |
| EPI_ISL_911633 | Indonesia/KB-TUH-C215966/2020 | 2020-08-01 | Asia/Indonesia/West Kalimantan/ | Tanjungpura University Hospital | Tanjungpura University Hospital | Mahyarudin et al |
| EPI_ISL_911642 | USA/TX-DSHS-2266/2021 | 2021-01-05 | North America/USA/Texas/El Paso Coun | Texas Department of State Health Services (TXDSHS) | Texas Department of State Health Services (TXDSHS) | Bonnie Oh et al |
| EPI_ISL_911656 | USA/TX-DSHS-2280/2021 | 2021-01-04 | North America/USA/Texas/El Paso Coun | Texas Department of State Health Services (TXDSHS) | Texas Department of State Health Services (TXDSHS) | Bonnie Oh et al |
| EPI_ISL_911662 | USA/TX-DSHS-2288/2020 | 2020-10-06 | North America/USA/Texas/Terry County | Texas Department of State Health Services (TXDSHS) | Texas Department of State Health Services (TXDSHS) | Bonnie Oh et al |
| EPI_ISL_911667 | USA/TX-DSHS-2294/2021 | 2021-01-04 | North America/USA/Texas/ | Texas Department of State Health Services (TXDSHS) | Texas Department of State Health Services (TXDSHS) | Bonnie Oh et al |
| EPI_ISL_911668 | USA/TX-DSHS-2296/2021 | 2021-01-04 | North America/USA/Texas/ | Texas Department of State Health Services (TXDSHS) | Texas Department of State Health Services (TXDSHS) | Bonnie Oh et al |
| EPI_ISL_911674 | USA/NC-UNC-0037/2021 | 2021-01-18 | North America/USA/North Carolina/ | Clinical Molecular Microbiology Laboratory, UNC Hospitals | Jeremy Wang | Jeremy Wang et al |
| EPI_ISL_911675 | Indonesia/KB-TUH-C2132860/2020 | 2020-10-30 | Asia/Indonesia/West Kalimantan/ | Tanjungpura University Hospital | Tanjungpura University Hospital | Mahyarudin et al |

| | | | | | | |
|---|---|---|---|---|---|---|
| EPI_ISL_911676 | USA/AK-PHL6582/2020 | 2020-11-05 | North America/USA/Alaska/Interior Ala | Alaska State Virology Laboratory | Alaska State Virology Laboratory | Stephanie DeRonde et al |
| EPI_ISL_911682 | USA/AK-PHL6589/2020 | 2020-12-02 | North America/USA/Alaska/Southeast A | Alaska State Virology Laboratory | Alaska State Virology Laboratory | Stephanie DeRonde et al |
| EPI_ISL_911685 | USA/AK-PHL6601/2020 | 2020-11-11 | North America/USA/Alaska/ | Alaska State Virology Laboratory | Alaska State Virology Laboratory | Stephanie DeRonde et al |
| EPI_ISL_911692 | USA/AK-PHL6614/2020 | 2020-12-08 | North America/USA/Alaska/ | Alaska State Virology Laboratory | Alaska State Virology Laboratory | Stephanie DeRonde et al |
| EPI_ISL_911695 | USA/AK-PHL6620/2020 | 2020-12-14 | North America/USA/Alaska/ | Alaska State Virology Laboratory | Alaska State Virology Laboratory | Stephanie DeRonde et al |
| EPI_ISL_911696 | USA/AK-PHL6680/2020 | 2020-09-21 | North America/USA/Alaska/Interior Ala | Alaska State Virology Laboratory | Alaska State Virology Laboratory | Stephanie DeRonde et al |
| EPI_ISL_911705 | USA/CA-QDX-3918/2021 | 2021-01-08 | North America/USA/California/ | Quest Diagnostics | Quest Diagnostics | Rosenthal et al |

| | | | | | | |
|---|---|---|---|---|---|---|
| EPI_ISL_911707 | Indonesia/YO-UGM-124763/2020 | 2020-12-23 | Asia/Indonesia/Yogyakarta/ | Academic Hospital of Gadjah Mada University (RSA UGM) | Genetics Working Group (Pokja Genetik) Faculty of Medicine, Public Health and Nursing Universitas Gadjah Mada (FK-KMK UGM); Disease Investigation Center Wates Ministry of Agriculture Indonesia; Department of Microbiology FK-KMK UGM; Laboratorium Diagnostik Yayasan Tahija World Mosquito Program (WMP) Yogyakarta Center for Tropical Medicine FK-KMK UGM; Integrated Research center FK-KMK UGM; Department of Computer Science and Electronics FMIPA UGM | Gunadi et al |
| EPI_ISL_911708 | Indonesia/KB-TUH-C2139958/2020 | 2020-11-28 | Asia/Indonesia/West Kalimantan/ | Tanjungpura University Hospital | Tanjungpura University Hospital | Puji Astuti et al |

| | | | | | | |
|---|---|---|---|---|---|---|
| EPI_ISL_911709 | Indonesia/YO-UGM-124762/2020 | 2020-12-23 | Asia/Indonesia/Yogyakarta/ | Academic Hospital of Gadjah Mada University (RSA UGM) | Genetics Working Group (Pokja Genetik) Faculty of Medicine, Public Health and Nursing Universitas Gadjah Mada (FK-KMK UGM); Disease Investigation Center Wates Ministry of Agriculture Indonesia; Department of Microbiology FK-KMK UGM; Laboratorium Diagnostik Yayasan Tahija World Mosquito Program (WMP) Yogyakarta Center for Tropical Medicine FK-KMK UGM; Integrated Research center FK-KMK UGM; Department of Computer Science and Electronics FMIPA UGM | Gunadi et al |
| EPI_ISL_911719 | USA/IN-Lilly-IPB0173-10bc11/2020 | 2020-03-25 | North America/USA/Indiana/Indianapoli | Clinical Diagnostics Laboratory, Diagnostic & Experimental Pathology, Lilly Research Laboratories | Clinical Diagnostics Laboratory, Diagnostic & Experimental Pathology, Lilly Research Laboratories | Tim Holzer et al |

| | | | | | | |
|---|---|---|---|---|---|---|
| EPI_ISL_911732 | Indonesia/KB-TUH-C2138669/2020 | 2020-10-22 | Asia/Indonesia/West Kalimantan/ | Tanjungpura University Hospital | Tanjungpura University Hospital | Delima Fajar Liana et al |
| EPI_ISL_911734 | USA/IN-Lilly-IPB0173-11bc02/2020 | 2020-03-25 | North America/USA/Indiana/Indianapoli | Clinical Diagnostics Laboratory, Diagnostic & Experimental Pathology, Lilly Research Laboratories | Clinical Diagnostics Laboratory, Diagnostic & Experimental Pathology, Lilly Research Laboratories | Tim Holzer et al |
| EPI_ISL_911735 | USA/IN-Lilly-IPB0173-11bc03/2020 | 2020-03-25 | North America/USA/Indiana/Indianapoli | Clinical Diagnostics Laboratory, Diagnostic & Experimental Pathology, Lilly Research Laboratories | Clinical Diagnostics Laboratory, Diagnostic & Experimental Pathology, Lilly Research Laboratories | Tim Holzer et al |
| EPI_ISL_911736 | USA/IN-Lilly-IPB0173-11bc04/2020 | 2020-03-25 | North America/USA/Indiana/Indianapoli | Clinical Diagnostics Laboratory, Diagnostic & Experimental Pathology, Lilly Research Laboratories | Clinical Diagnostics Laboratory, Diagnostic & Experimental Pathology, Lilly Research Laboratories | Tim Holzer et al |
| EPI_ISL_911737 | USA/IN-Lilly-IPB0173-11bc05/2020 | 2020-03-25 | North America/USA/Indiana/Indianapoli | Clinical Diagnostics Laboratory, Diagnostic & Experimental Pathology, Lilly Research Laboratories | Clinical Diagnostics Laboratory, Diagnostic & Experimental Pathology, Lilly Research Laboratories | Tim Holzer et al |
| EPI_ISL_911746 | Canada/ON-S2122/2021 | 2021-01-13 | North America/Canada/Ontario/Toronto | Toronto Invasive Bacterial Diseases Network | McMaster University | Allison McGeer et al |
| EPI_ISL_911748 | Canada/ON-S2125/2021 | 2021-01-18 | North America/Canada/Ontario/Toronto | Toronto Invasive Bacterial Diseases Network | McMaster University | Allison McGeer et al |
| EPI_ISL_911749 | Canada/ON-S2124/2021 | 2021-01-19 | North America/Canada/Ontario/Toronto | Toronto Invasive Bacterial Diseases Network | McMaster University | Allison McGeer et al |

| | | | | | | |
|---|---|---|---|---|---|---|
| EPI_ISL_911750 | Indonesia/KB-TUH-C2139628/2020 | 2020-11-26 | Asia/Indonesia/West Kalimantan/ | Tanjungpura University Hospital | Tanjungpura University Hospital | Virhan Novianry et al |
| EPI_ISL_911754 | USA/IN-Lilly-IPB0173-12bc25/2020 | 2020-03-26 | North America/USA/Indiana/Indianapoli | Clinical Diagnostics Laboratory, Diagnostic & Experimental Pathology, Lilly Research Laboratories | Clinical Diagnostics Laboratory, Diagnostic & Experimental Pathology, Lilly Research Laboratories | Tim Holzer et al |
| EPI_ISL_911770 | USA/MD-HP01398/2021 | 2021-01-16 | North America/USA/Maryland/ | Johns Hopkins Hospital Department of Pathology | Johns Hopkins Hospital Department of Pathology | C. Paul Morris et al |
| EPI_ISL_911778 | USA/MD-HP01406/2021 | 2021-01-19 | North America/USA/Maryland/ | Johns Hopkins Hospital Department of Pathology | Johns Hopkins Hospital Department of Pathology | C. Paul Morris et al |
| EPI_ISL_911784 | USA/MD-HP01419/2021 | 2021-01-20 | North America/USA/Maryland/ | Johns Hopkins Hospital Department of Pathology | Johns Hopkins Hospital Department of Pathology | C. Paul Morris et al |
| EPI_ISL_911785 | USA/MD-HP01420/2021 | 2021-01-20 | North America/USA/Maryland/ | Johns Hopkins Hospital Department of Pathology | Johns Hopkins Hospital Department of Pathology | C. Paul Morris et al |
| EPI_ISL_911789 | USA/MD-HP01428/2021 | 2021-01-07 | North America/USA/Maryland/ | Johns Hopkins Hospital Department of Pathology | Johns Hopkins Hospital Department of Pathology | C. Paul Morris et al |
| EPI_ISL_911794 | USA/MD-HP01446/2021 | 2021-01-08 | North America/USA/Maryland/ | Johns Hopkins Hospital Department of Pathology | Johns Hopkins Hospital Department of Pathology | C. Paul Morris et al |
| EPI_ISL_911797 | USA/MD-HP01453/2021 | 2021-01-13 | North America/USA/Maryland/ | Johns Hopkins Hospital Department of Pathology | Johns Hopkins Hospital Department of Pathology | C. Paul Morris et al |

| | | | | | | |
|---|---|---|---|---|---|---|
| EPI_ISL_911809 | USA/MD-HP01484/2021 | 2021-01-20 | North America/USA/Maryland/ | Johns Hopkins Hospital Department of Pathology | Johns Hopkins Hospital Department of Pathology | C. Paul Morris et al |
| EPI_ISL_911814 | USA/MD-HP01489/2021 | 2021-01-21 | North America/USA/Maryland/ | Johns Hopkins Hospital Department of Pathology | Johns Hopkins Hospital Department of Pathology | C. Paul Morris et al |
| EPI_ISL_911818 | USA/MD-HP01493/2021 | 2021-01-21 | North America/USA/Maryland/ | Johns Hopkins Hospital Department of Pathology | Johns Hopkins Hospital Department of Pathology | C. Paul Morris et al |
| EPI_ISL_911819 | USA/MD-HP01494/2021 | 2021-01-21 | North America/USA/Maryland/ | Johns Hopkins Hospital Department of Pathology | Johns Hopkins Hospital Department of Pathology | C. Paul Morris et al |
| EPI_ISL_911821 | USA/MD-HP01497/2021 | 2021-01-21 | North America/USA/Maryland/ | Johns Hopkins Hospital Department of Pathology | Johns Hopkins Hospital Department of Pathology | C. Paul Morris et al |
| EPI_ISL_911840 | USA/MD-HP01518/2021 | 2021-01-22 | North America/USA/Maryland/ | Johns Hopkins Hospital Department of Pathology | Johns Hopkins Hospital Department of Pathology | C. Paul Morris et al |
| EPI_ISL_911845 | USA/MD-HP01523/2021 | 2021-01-22 | North America/USA/Maryland/ | Johns Hopkins Hospital Department of Pathology | Johns Hopkins Hospital Department of Pathology | C. Paul Morris et al |
| EPI_ISL_911854 | USA/MD-HP01535/2021 | 2021-01-22 | North America/USA/Maryland/ | Johns Hopkins Hospital Department of Pathology | Johns Hopkins Hospital Department of Pathology | C. Paul Morris et al |
| EPI_ISL_911857 | USA/MD-HP01538/2021 | 2021-01-23 | North America/USA/Maryland/ | Johns Hopkins Hospital Department of Pathology | Johns Hopkins Hospital Department of Pathology | C. Paul Morris et al |
| EPI_ISL_911872 | USA/DC-HP01565/2021 | 2021-01-21 | North America/USA/Washington DC/ | Johns Hopkins Hospital Department of Pathology | Johns Hopkins Hospital Department of Pathology | C. Paul Morris et al |

| | | | | | | |
|---|---|---|---|---|---|---|
| EPI_ISL_911880 | USA/MD-HP01576/2021 | 2021-01-26 | North America/USA/Maryland/ | Johns Hopkins Hospital Department of Pathology | Johns Hopkins Hospital Department of Pathology | C. Paul Morris et al |
| EPI_ISL_911885 | USA/MD-HP01584/2021 | 2021-01-23 | North America/USA/Maryland/ | Johns Hopkins Hospital Department of Pathology | Johns Hopkins Hospital Department of Pathology | C. Paul Morris et al |
| EPI_ISL_911897 | USA/MD-HP01606/2021 | 2021-01-26 | North America/USA/Maryland/ | Johns Hopkins Hospital Department of Pathology | Johns Hopkins Hospital Department of Pathology | C. Paul Morris et al |
| EPI_ISL_911898 | USA/MD-HP01607/2021 | 2021-01-26 | North America/USA/Maryland/ | Johns Hopkins Hospital Department of Pathology | Johns Hopkins Hospital Department of Pathology | C. Paul Morris et al |
| EPI_ISL_911915 | USA/MD-HP01651/2021 | 2021-01-26 | North America/USA/Maryland/ | Johns Hopkins Hospital Department of Pathology | Johns Hopkins Hospital Department of Pathology | C. Paul Morris et al |
| EPI_ISL_911916 | USA/MD-HP01653/2021 | 2021-01-27 | North America/USA/Maryland/ | Johns Hopkins Hospital Department of Pathology | Johns Hopkins Hospital Department of Pathology | C. Paul Morris et al |
| EPI_ISL_911917 | USA/MD-HP01657/2021 | 2021-01-26 | North America/USA/Maryland/ | Johns Hopkins Hospital Department of Pathology | Johns Hopkins Hospital Department of Pathology | C. Paul Morris et al |
| EPI_ISL_911920 | USA/RI-CDC-9MDT-8813/2021 | 2021-01-12 | North America/USA/Rhode Island/ | SRL (Sonic Reference Laboratory - Sonic Healthcare) | Pathogen Discovery, Respiratory Viruses Branch, Division of Viral Diseases, Centers for Disease Control and Prevention | Ying Tao et al |

| | | | | | | |
|---|---|---|---|---|---|---|
| EPI_ISL_911924 | USA/IN-Lilly-IPB0173-13bc10/2020 | 2020-03-27 | North America/USA/Indiana/Indianapoli | Clinical Diagnostics Laboratory, Diagnostic & Experimental Pathology, Lilly Research Laboratories | Clinical Diagnostics Laboratory, Diagnostic & Experimental Pathology, Lilly Research Laboratories | Tim Holzer et al |
| EPI_ISL_911930 | USA/WY-WPHL-21003164/2021 | 2021-01-08 | North America/USA/Wyoming/ | Wyoming Public Health Laboratory | Wyoming Public Health Laboratory | Noah Hull et al |
| EPI_ISL_911935 | USA/WY-WPHL-21003719/2021 | 2021-01-11 | North America/USA/Wyoming/ | Wyoming Public Health Laboratory | Wyoming Public Health Laboratory | Noah Hull et al |
| EPI_ISL_911937 | USA/WY-WPHL-21003765/2021 | 2021-01-11 | North America/USA/Wyoming/ | Wyoming Public Health Laboratory | Wyoming Public Health Laboratory | Noah Hull et al |
| EPI_ISL_911945 | USA/WY-WPHL-21003973/2021 | 2021-01-11 | North America/USA/Wyoming/ | Wyoming Public Health Laboratory | Wyoming Public Health Laboratory | Noah Hull et al |
| EPI_ISL_911948 | USA/WY-WPHL-21004053/2021 | 2021-01-11 | North America/USA/Wyoming/ | Wyoming Public Health Laboratory | Wyoming Public Health Laboratory | Noah Hull et al |
| EPI_ISL_911955 | USA/WY-WPHL-21004555/2021 | 2021-01-12 | North America/USA/Wyoming/ | Wyoming Public Health Laboratory | Wyoming Public Health Laboratory | Noah Hull et al |
| EPI_ISL_911958 | USA/WY-WPHL-21004597/2021 | 2021-01-12 | North America/USA/Wyoming/ | Wyoming Public Health Laboratory | Wyoming Public Health Laboratory | Noah Hull et al |
| EPI_ISL_911960 | USA/WY-WPHL-21004661/2021 | 2021-01-12 | North America/USA/Wyoming/ | Wyoming Public Health Laboratory | Wyoming Public Health Laboratory | Noah Hull et al |
| EPI_ISL_911961 | USA/WY-WPHL-21004685/2021 | 2021-01-12 | North America/USA/Wyoming/ | Wyoming Public Health Laboratory | Wyoming Public Health Laboratory | Noah Hull et al |
| EPI_ISL_911962 | USA/WY-WPHL-21004831/2021 | 2021-01-13 | North America/USA/Wyoming/ | Wyoming Public Health Laboratory | Wyoming Public Health Laboratory | Noah Hull et al |
| EPI_ISL_911963 | USA/WY-WPHL-21004838/2021 | 2021-01-13 | North America/USA/Wyoming/ | Wyoming Public Health Laboratory | Wyoming Public Health Laboratory | Noah Hull et al |
| EPI_ISL_911973 | USA/WA-S3927/2021 | 2021-01-09 | North America/USA/Washington/King C | Altius Institute for Biomedical Sciences | Seattle Flu Study | Deborah A. Nickerson et al |
| EPI_ISL_911979 | USA/WA-S3933/2021 | 2021-01-09 | North America/USA/Washington/King C | Altius Institute for Biomedical Sciences | Seattle Flu Study | Deborah A. Nickerson et al |
| EPI_ISL_911995 | USA/WA-S3949/2021 | 2021-01-09 | North America/USA/Washington/King C | Altius Institute for Biomedical Sciences | Seattle Flu Study | Deborah A. Nickerson et al |
| EPI_ISL_911999 | USA/WA-S3953/2021 | 2021-01-09 | North America/USA/Washington/Pierce | Altius Institute for Biomedical Sciences | Seattle Flu Study | Deborah A. Nickerson et al |
| EPI_ISL_912008 | USA/WA-S3962/2021 | 2021-01-11 | North America/USA/Washington/King C | Altius Institute for Biomedical Sciences | Seattle Flu Study | Deborah A. Nickerson et al |
| EPI_ISL_912013 | USA/WA-S3967/2021 | 2021-01-11 | North America/USA/Washington/King C | Altius Institute for Biomedical Sciences | Seattle Flu Study | Deborah A. Nickerson et al |
| EPI_ISL_912015 | USA/WA-S3969/2021 | 2021-01-11 | North America/USA/Washington/Pierce | Altius Institute for Biomedical Sciences | Seattle Flu Study | Deborah A. Nickerson et al |
| EPI_ISL_912026 | USA/WA-S3980/2021 | 2021-01-11 | North America/USA/Washington/King C | Altius Institute for Biomedical Sciences | Seattle Flu Study | Deborah A. Nickerson et al |

| | | | | | | |
|---|---|---|---|---|---|---|
| EPI_ISL_912037 | USA/WA-S3991/2021 | 2021-01-07 | North America/USA/Washington/King C | Washington State Department of Health | Seattle Flu Study | Deborah A. Nickerson et al |
| EPI_ISL_912043 | USA/WA-S3997/2021 | 2021-01-06 | North America/USA/Washington/Yakim | Washington State Department of Health | Seattle Flu Study | Deborah A. Nickerson et al |
| EPI_ISL_912045 | USA/WA-S3999/2021 | 2021-01-07 | North America/USA/Washington/Spoka | Washington State Department of Health | Seattle Flu Study | Deborah A. Nickerson et al |
| EPI_ISL_912054 | USA/WA-S4008/2021 | 2021-01-08 | North America/USA/Washington/Yakim | Washington State Department of Health | Seattle Flu Study | Deborah A. Nickerson et al |
| EPI_ISL_912066 | USA/WA-S4020/2021 | 2021-01-09 | North America/USA/Washington/Yakim | Washington State Department of Health | Seattle Flu Study | Deborah A. Nickerson et al |
| EPI_ISL_912077 | USA/WA-S4031/2021 | 2021-01-10 | North America/USA/Washington/Yakim | Washington State Department of Health | Seattle Flu Study | Deborah A. Nickerson et al |
| EPI_ISL_912094 | USA/WA-S4048/2021 | 2021-01-12 | North America/USA/Washington/King C | Washington State Department of Health | Seattle Flu Study | Deborah A. Nickerson et al |
| EPI_ISL_912122 | USA/WA-S4076/2021 | 2021-01-18 | North America/USA/Washington/King C | Altius Institute for Biomedical Sciences | Seattle Flu Study | Deborah A. Nickerson et al |
| EPI_ISL_912142 | USA/WA-S4096/2021 | 2021-01-18 | North America/USA/Washington/King C | Altius Institute for Biomedical Sciences | Seattle Flu Study | Deborah A. Nickerson et al |
| EPI_ISL_912143 | USA/WA-S4097/2021 | 2021-01-18 | North America/USA/Washington/King C | Altius Institute for Biomedical Sciences | Seattle Flu Study | Deborah A. Nickerson et al |
| EPI_ISL_912147 | USA/WA-S4101/2021 | 2021-01-19 | North America/USA/Washington/King C | Altius Institute for Biomedical Sciences | Seattle Flu Study | Deborah A. Nickerson et al |
| EPI_ISL_912159 | USA/WA-S4113/2021 | 2021-01-18 | North America/USA/Washington/King C | Altius Institute for Biomedical Sciences | Seattle Flu Study | Deborah A. Nickerson et al |
| EPI_ISL_912162 | USA/AK-Yale-S193/2021 | 2021-01-07 | North America/USA/Arkansas/ | Baptist Medical Center | Grubaugh Lab - Yale School of Public Health | Tara Alpert et al |
| EPI_ISL_912163 | USA/CT-Yale-S197/2021 | 2021-01-04 | North America/USA/Connecticut/ | Connecticut Department of Health | Grubaugh Lab - Yale School of Public Health | Tara Alpert et al |
| EPI_ISL_912188 | USA/CA-CZB-12415/2020 | 2020-09-17 | North America/USA/California/ | California Institute of Technology | Chan-Zuckerberg Biohub | CZB Cliahub Consortium et al |
| EPI_ISL_912191 | USA/CA-CZB-12418/2020 | 2020-09-20 | North America/USA/California/ | California Institute of Technology | Chan-Zuckerberg Biohub | CZB Cliahub Consortium et al |
| EPI_ISL_912230 | Germany/SH-ChVir22754/2021 | 2021-01-13 | Europe/Germany/Schleswig-Holstein/ | Charit√© Universit√§tsmedizin Berlin, Institut f√ºr Virologie/Labor Berlin | Charit√© Universit√§tsmedizin Berlin, Institut f√ºr Virologie | Victor M Corman et al |
| EPI_ISL_912239 | Germany/SH-ChVir22772/2021 | 2021-01-15 | Europe/Germany/Schleswig-Holstein/ | Charit√© Universit√§tsmedizin Berlin, Institut f√ºr Virologie/Labor Berlin | Charit√© Universit√§tsmedizin Berlin, Institut f√ºr Virologie | Victor M Corman et al |
| EPI_ISL_912240 | Germany/SH-ChVir22774/2021 | 2021-01-16 | Europe/Germany/Schleswig-Holstein/ | Charit√© Universit√§tsmedizin Berlin, Institut f√ºr Virologie/Labor Berlin | Charit√© Universit√§tsmedizin Berlin, Institut f√ºr Virologie | Victor M Corman et al |

| | | | | | | |
|---|---|---|---|---|---|---|
| EPI_ISL_912242 | Germany/SH-ChVir22776/2021 | 2021-01-17 | Europe/Germany/Schleswig-Holstein/ | Charit√© Universit√§tsmedizin Berlin, Institut f√ºr Virologie/Labor Berlin | Charit√© Universit√§tsmedizin Berlin, Institut f√ºr Virologie | Victor M Corman et al |
| EPI_ISL_912243 | Germany/SH-ChVir22777/2021 | 2021-01-17 | Europe/Germany/Schleswig-Holstein/ | Charit√© Universit√§tsmedizin Berlin, Institut f√ºr Virologie/Labor Berlin | Charit√© Universit√§tsmedizin Berlin, Institut f√ºr Virologie | Victor M Corman et al |
| EPI_ISL_912245 | Germany/SH-ChVir22873/2021 | 2021-01-18 | Europe/Germany/Schleswig-Holstein/ | Charit√© Universit√§tsmedizin Berlin, Institut f√ºr Virologie/Labor Berlin | Charit√© Universit√§tsmedizin Berlin, Institut f√ºr Virologie | Victor M Corman et al |
| EPI_ISL_912249 | Germany/NW-ChVir22877/2021 | 2021-01-12 | Europe/Germany/North Rhine Westphal | Charit√© Universit√§tsmedizin Berlin, Institut f√ºr Virologie/Labor Berlin | Charit√© Universit√§tsmedizin Berlin, Institut f√ºr Virologie | Victor M Corman et al |
| EPI_ISL_912250 | Germany/TH-ChVir22878/2021 | 2021-01-18 | Europe/Germany/Thuringia/ | Charit√© Universit√§tsmedizin Berlin, Institut f√ºr Virologie/Labor Berlin | Charit√© Universit√§tsmedizin Berlin, Institut f√ºr Virologie | Victor M Corman et al |
| EPI_ISL_912254 | Germany/BY-ChVir22882/2021 | 2021-01-19 | Europe/Germany/Bavaria/ | Charit√© Universit√§tsmedizin Berlin, Institut f√ºr Virologie/Labor Berlin | Charit√© Universit√§tsmedizin Berlin, Institut f√ºr Virologie | Victor M Corman et al |
| EPI_ISL_912255 | Germany/BW-ChVir22883/2021 | 2021-01-11 | Europe/Germany/Baden-Wuerttemberg | Charit√© Universit√§tsmedizin Berlin, Institut f√ºr Virologie/Labor Berlin | Charit√© Universit√§tsmedizin Berlin, Institut f√ºr Virologie | Victor M Corman et al |
| EPI_ISL_912258 | Germany/SH-ChVir22888/2021 | 2021-01-18 | Europe/Germany/Schleswig-Holstein/ | Charit√© Universit√§tsmedizin Berlin, Institut f√ºr Virologie/Labor Berlin | Charit√© Universit√§tsmedizin Berlin, Institut f√ºr Virologie | Victor M Corman et al |
| EPI_ISL_912264 | Germany/BE-ChVir22897/2021 | 2021-01-19 | Europe/Germany/Berlin/ | Charit√© Universit√§tsmedizin Berlin, Institut f√ºr Virologie/Labor Berlin | Charit√© Universit√§tsmedizin Berlin, Institut f√ºr Virologie | Victor M Corman et al |
| EPI_ISL_912265 | Germany/BE-ChVir22898/2021 | 2021-01-20 | Europe/Germany/Berlin/ | Charit√© Universit√§tsmedizin Berlin, Institut f√ºr Virologie/Labor Berlin | Charit√© Universit√§tsmedizin Berlin, Institut f√ºr Virologie | Victor M Corman et al |

| | | | | | | |
|---|---|---|---|---|---|---|
| EPI_ISL_912267 | Germany/BE-ChVir23058/2021 | 2021-01-24 | Europe/Germany/Berlin/ | Charit√© Universit√§tsmedizin Berlin, Institut f√ºr Virologie/Labor Berlin | Charit√© Universit√§tsmedizin Berlin, Institut f√ºr Virologie | Victor M Corman et al |
| EPI_ISL_912274 | Spain/MD-IBV-99015894/2021 | 2021-01-12 | Europe/Spain/Madrid/Madrid | Hospital General Universitario Gregorio Marav±√≥n | SeqCOVID-SPAIN consortium / IBV (CSIC) | Dar√≠o Garc√≠a de Viedma et al |
| EPI_ISL_912276 | Spain/MD-IBV-99015896/2020 | 2020-09-23 | Europe/Spain/Madrid/Madrid | Hospital General Universitario Gregorio Marav±√≥n | SeqCOVID-SPAIN consortium / IBV (CSIC) | Dar√≠o Garc√≠a de Viedma et al |
| EPI_ISL_912277 | Spain/MD-IBV-99015898/2021 | 2021-01-14 | Europe/Spain/Madrid/Madrid | Hospital General Universitario Gregorio Marav±√≥n | SeqCOVID-SPAIN consortium / IBV (CSIC) | Dar√≠o Garc√≠a de Viedma et al |
| EPI_ISL_912280 | Spain/MD-IBV-99015902/2021 | 2021-01-07 | Europe/Spain/Madrid/Madrid | Hospital General Universitario Gregorio Marav±√≥n | SeqCOVID-SPAIN consortium / IBV (CSIC) | Dar√≠o Garc√≠a de Viedma et al |
| EPI_ISL_912281 | Spain/MD-IBV-99015903/2021 | 2021-01-12 | Europe/Spain/Madrid/Madrid | Hospital General Universitario Gregorio Marav±√≥n | SeqCOVID-SPAIN consortium / IBV (CSIC) | Dar√≠o Garc√≠a de Viedma et al |
| EPI_ISL_912283 | Spain/MD-IBV-99015905/2020 | 2020-12-18 | Europe/Spain/Madrid/Madrid | Hospital General Universitario Gregorio Marav±√≥n | SeqCOVID-SPAIN consortium / IBV (CSIC) | Dar√≠o Garc√≠a de Viedma et al |
| EPI_ISL_912286 | Spain/MD-IBV-99015908/2021 | 2021-01-13 | Europe/Spain/Madrid/Madrid | Hospital General Universitario Gregorio Marav±√≥n | SeqCOVID-SPAIN consortium / IBV (CSIC) | Dar√≠o Garc√≠a de Viedma et al |
| EPI_ISL_912291 | Spain/MD-IBV-99015913/2021 | 2021-01-15 | Europe/Spain/Madrid/Madrid | Hospital General Universitario Gregorio Marav±√≥n | SeqCOVID-SPAIN consortium / IBV (CSIC) | Dar√≠o Garc√≠a de Viedma et al |
| EPI_ISL_912293 | Spain/MD-IBV-99015915/2021 | 2021-01-04 | Europe/Spain/Madrid/Madrid | Hospital General Universitario Gregorio Marav±√≥n | SeqCOVID-SPAIN consortium / IBV (CSIC) | Dar√≠o Garc√≠a de Viedma et al |
| EPI_ISL_912294 | Spain/MD-IBV-99015916/2021 | 2021-01-15 | Europe/Spain/Madrid/Madrid | Hospital General Universitario Gregorio Marav±√≥n | SeqCOVID-SPAIN consortium / IBV (CSIC) | Dar√≠o Garc√≠a de Viedma et al |
| EPI_ISL_912295 | Spain/MD-IBV-99015917/2021 | 2021-01-16 | Europe/Spain/Madrid/Madrid | Hospital General Universitario Gregorio Marav±√≥n | SeqCOVID-SPAIN consortium / IBV (CSIC) | Dar√≠o Garc√≠a de Viedma et al |
| EPI_ISL_912297 | Spain/MD-IBV-99015919/2020 | 2020-12-28 | Europe/Spain/Madrid/Madrid | Hospital General Universitario Gregorio Marav±√≥n | SeqCOVID-SPAIN consortium / IBV (CSIC) | Dar√≠o Garc√≠a de Viedma et al |

| | | | | | | |
|---|---|---|---|---|---|---|
| EPI_ISL_912300 | Spain/MD-IBV-99015922/2021 | 2021-01-15 | Europe/Spain/Madrid/Madrid | Hospital General Universitario Gregorio Marav√±√≥n | SeqCOVID-SPAIN consortium / IBV (CSIC) | Dar√≠o Garc√≠a de Viedma et al |
| EPI_ISL_912304 | Spain/MD-IBV-99015926/2021 | 2021-01-15 | Europe/Spain/Madrid/Madrid | Hospital General Universitario Gregorio Marav√±√≥n | SeqCOVID-SPAIN consortium / IBV (CSIC) | Dar√≠o Garc√≠a de Viedma et al |
| EPI_ISL_912309 | Spain/MD-IBV-99015931/2021 | 2021-01-07 | Europe/Spain/Madrid/Madrid | Hospital General Universitario Gregorio Marav√±√≥n | SeqCOVID-SPAIN consortium / IBV (CSIC) | Sergio Buenestado Serrano et al |
| EPI_ISL_912314 | Spain/MD-IBV-99015936/2021 | 2021-01-12 | Europe/Spain/Madrid/Madrid | Hospital General Universitario Gregorio Marav√±√≥n | SeqCOVID-SPAIN consortium / IBV (CSIC) | Dar√≠o Garc√≠a de Viedma et al |
| EPI_ISL_912318 | Spain/MD-IBV-99015941/2021 | 2021-01-04 | Europe/Spain/Madrid/Madrid | Hospital General Universitario Gregorio Marav√±√≥n | SeqCOVID-SPAIN consortium / IBV (CSIC) | Dar√≠o Garc√≠a de Viedma et al |
| EPI_ISL_912329 | Spain/MD-IBV-99015953/2021 | 2021-01-14 | Europe/Spain/Madrid/Madrid | Hospital General Universitario Gregorio Marav√±√≥n | SeqCOVID-SPAIN consortium / IBV (CSIC) | Dar√≠o Garc√≠a de Viedma et al |
| EPI_ISL_912332 | Spain/MD-IBV-99015957/2021 | 2021-01-07 | Europe/Spain/Madrid/Madrid | Hospital General Universitario Gregorio Marav√±√≥n | SeqCOVID-SPAIN consortium / IBV (CSIC) | Sergio Buenestado Serrano et al |
| EPI_ISL_912333 | Spain/MD-IBV-99015958/2021 | 2021-01-07 | Europe/Spain/Madrid/Madrid | Hospital General Universitario Gregorio Marav√±√≥n | SeqCOVID-SPAIN consortium / IBV (CSIC) | Dar√≠o Garc√≠a de Viedma et al |
| EPI_ISL_912341 | Spain/MD-IBV-99015966/2021 | 2021-01-16 | Europe/Spain/Madrid/Madrid | Hospital General Universitario Gregorio Marav√±√≥n | SeqCOVID-SPAIN consortium / IBV (CSIC) | Dar√≠o Garc√≠a de Viedma et al |
| EPI_ISL_912346 | Spain/MD-IBV-99015971/2021 | 2021-01-15 | Europe/Spain/Madrid/Madrid | Hospital General Universitario Gregorio Marav√±√≥n | SeqCOVID-SPAIN consortium / IBV (CSIC) | Dar√≠o Garc√≠a de Viedma et al |
| EPI_ISL_912347 | Spain/MD-IBV-99015972/2021 | 2021-01-16 | Europe/Spain/Madrid/Madrid | Hospital General Universitario Gregorio Marav√±√≥n | SeqCOVID-SPAIN consortium / IBV (CSIC) | Dar√≠o Garc√≠a de Viedma et al |
| EPI_ISL_912348 | Spain/MD-IBV-99015973/2021 | 2021-01-14 | Europe/Spain/Madrid/Madrid | Hospital General Universitario Gregorio Marav√±√≥n | SeqCOVID-SPAIN consortium / IBV (CSIC) | Dar√≠o Garc√≠a de Viedma et al |
| EPI_ISL_912349 | Spain/MD-IBV-99015974/2021 | 2021-01-04 | Europe/Spain/Madrid/Madrid | Hospital General Universitario Gregorio Marav√±√≥n | SeqCOVID-SPAIN consortium / IBV (CSIC) | Dar√≠o Garc√≠a de Viedma et al |

| | | | | | | |
|---|---|---|---|---|---|---|
| EPI_ISL_912352 | Spain/MD-IBV-99015978/2020 | 2020-12-27 | Europe/Spain/Madrid/Madrid | Hospital General Universitario Gregorio Mara√±√≥n | SeqCOVID-SPAIN consortium / IBV (CSIC) | Dar√≠o Garc√≠a de Viedma et al |
| EPI_ISL_912381 | Congo/Q001C118A6_2101-TM-r1-032 | 2020-12-30 | Africa/Democratic Republic of the Congo | Fondation Congolaise pour la recherche medicale (FCRM), Francine Ntoumi | NGS Competence Center Tuebingen, Institut f√ºr Medizinische Mikrobiologie und Hygiene, Universitaetsklinikum T√ºbingen | Angel Angelov et al |
| EPI_ISL_912384 | Congo/Q001C121AP_2101-TM-r1-035 | 2020-12-30 | Africa/Democratic Republic of the Congo | Fondation Congolaise pour la recherche medicale (FCRM), Francine Ntoumi | NGS Competence Center Tuebingen, Institut f√ºr Medizinische Mikrobiologie und Hygiene, Universitaetsklinikum T√ºbingen | Angel Angelov et al |
| EPI_ISL_912397 | Malaysia/IMR_WC329964/2020 | 2020-12-10 | Asia/Malaysia/Malaysia/ | Institute for Medical Research, Infectious Disease Research Centre, National Institutes of Health, Ministry of Health Malaysia | Institute for Medical Research, Infectious Disease Research Centre, National Institutes of Health, Ministry of Health Malaysia | Suppiah J et al |
| EPI_ISL_912400 | Malaysia/IMR_WC342015/2020 | 2020-12-15 | Asia/Malaysia/Malaysia/ | Institute for Medical Research, Infectious Disease Research Centre, National Institutes of Health, Ministry of Health Malaysia | Institute for Medical Research, Infectious Disease Research Centre, National Institutes of Health, Ministry of Health Malaysia | Suppiah J et al |

| | | | | | | |
|---|---|---|---|---|---|---|
| EPI_ISL_912423 | Belgium/rega-1802/2021 | 2021-01-10 | Europe/Slovenia/Velenje/ | KU Leuven, Rega Institute, Clinical and Epidemiological Virology | KU Leuven, Rega Institute, Clinical and Epidemiological Virology | Tony Wawina-Bokalanga et al |
| EPI_ISL_912454 | Belgium/rega-1847/2021 | 2021-01-03 | Europe/Belgium/Leuven/Werchter | KU Leuven, Rega Institute, Clinical and Epidemiological Virology | KU Leuven, Rega Institute, Clinical and Epidemiological Virology | Tony Wawina-Bokalanga et al |
| EPI_ISL_912468 | SouthAfrica/UFS-VIRO-NGS-135/2020 | 2020-12-14 | Africa/South Africa/Free State/ | NHLS Universitas Academic | UFS Virology | PA Bester et al |
| EPI_ISL_912483 | SouthAfrica/UFS-VIRO-NGS-151/2020 | 2020-12-26 | Africa/South Africa/Free State/ | NHLS Universitas Academic | UFS Virology | PA Bester et al |
| EPI_ISL_912484 | SouthAfrica/UFS-VIRO-NGS-133/2021 | 2021-01-01 | Africa/South Africa/Free State/ | NHLS Universitas Academic | UFS Virology | PA Bester et al |
| EPI_ISL_912490 | SouthAfrica/UFS-VIRO-NGS-143/2020 | 2020-12-17 | Africa/South Africa/Free State/ | NHLS Universitas Academic | UFS Virology | PA Bester et al |
| EPI_ISL_912555 | France/CVL-IPP01192/2021 | 2021-01-13 | Europe/France/Centre-Val de Loire/Orl√ | C.H.R. d'Orl√©ans - H√¥pital de la Source | National Reference Center for Viruses of Respiratory Infections, Institut Pasteur, Paris | Marion Barbet et al |
| EPI_ISL_912560 | France/GES-IPP01203/2021 | 2021-01-09 | Europe/France/Grand Est/Troyes | HOPITAL SIMONE VEIL | National Reference Center for Viruses of Respiratory Infections, Institut Pasteur, Paris | Marion Barbet et al |
| EPI_ISL_912592 | France/IDF-IPP01106/2021 | 2021-01-09 | Europe/France/Ile de France/Ivry-sur-Se | Labo Analyses Med | National Reference Center for Viruses of Respiratory Infections, Institut Pasteur, Paris | Marion Barbet et al |
| EPI_ISL_912608 | France/CVL-IPP01117/2021 | 2021-01-13 | Europe/France/Centre-Val de Loire/Char | Labo Analyses Med | National Reference Center for Viruses of Respiratory Infections, Institut Pasteur, Paris | Marion Barbet et al |

| | | | | | | |
|---|---|---|---|---|---|---|
| EPI_ISL_912610 | France/CVL-IPP01119/2021 | 2021-01-13 | Europe/France/Centre-Val de Loire/Char | Labo Analyses Med | National Reference Center for Viruses of Respiratory Infections, Institut Pasteur, Paris | Marion Barbet et al |
| EPI_ISL_912666 | France/un-21012190067/2021 | 2021-01-19 | Europe/France/France/ | H√¥pital Henri Mondor | Department of Virology, Henri Mondor University Hospital, Assistance Publique H√¥pitaux de Paris, Universit√© Paris-Est Cr√©teil, INSERM U955 | Christophe Rodriguez et al |
| EPI_ISL_912675 | France/un-21012250184/2021 | 2021-01-22 | Europe/France/France/ | G.H.E.F.Grand H√¥pital EST Francilien | Department of Virology, Henri Mondor University Hospital, Assistance Publique H√¥pitaux de Paris, Universit√© Paris-Est Cr√©teil, INSERM U955 | Christophe Rodriguez et al |

| | | | | | | |
|---|---|---|---|---|---|---|
| EPI_ISL_912679 | France/un-21012220453/2021 | 2021-01-22 | Europe/France/France/ | H√¥pital Henri Mondor | Department of Virology, Henri Mondor University Hospital, Assistance Publique H√¥pitaux de Paris, Universit√© Paris-Est Cr√©teil, INSERM U955 | Christophe Rodriguez et al |
| EPI_ISL_912683 | France/un-21012200193/2021 | 2021-01-20 | Europe/France/France/ | H√¥pital Henri Mondor | Department of Virology, Henri Mondor University Hospital, Assistance Publique H√¥pitaux de Paris, Universit√© Paris-Est Cr√©teil, INSERM U955 | Christophe Rodriguez et al |
| EPI_ISL_912688 | France/un-21012060401/2021 | 2021-01-06 | Europe/France/France/ | H√¥pital Henri Mondor | Department of Virology, Henri Mondor University Hospital, Assistance Publique H√¥pitaux de Paris, Universit√© Paris-Est Cr√©teil, INSERM U955 | Christophe Rodriguez et al |

| | | | | | | |
|---|---|---|---|---|---|---|
| EPI_ISL_912711 | France/un-21012210346/2021 | 2021-01-21 | Europe/France/France/ | H√¥pital Henri Mondor | Department of Virology, Henri Mondor University Hospital, Assistance Publique H√¥pitaux de Paris, Universit√© Paris-Est Cr√©teil, INSERM U955 | Christophe Rodriguez et al |
| EPI_ISL_912712 | France/un-21012210264/2021 | 2021-01-21 | Europe/France/France/ | H√¥pital Henri Mondor | Department of Virology, Henri Mondor University Hospital, Assistance Publique H√¥pitaux de Paris, Universit√© Paris-Est Cr√©teil, INSERM U955 | Christophe Rodriguez et al |
| EPI_ISL_912741 | France/un-21012060082/2021 | 2021-01-06 | Europe/France/France/ | H√¥pital Henri Mondor | Department of Virology, Henri Mondor University Hospital, Assistance Publique H√¥pitaux de Paris, Universit√© Paris-Est Cr√©teil, INSERM U955 | Christophe Rodriguez et al |

| | | | | | | |
|---|---|---|---|---|---|---|
| EPI_ISL_912743 | France/un-21012090101/2021 | 2021-01-09 | Europe/France/France/ | H√¥pital Henri Mondor | Department of Virology, Henri Mondor University Hospital, Assistance Publique H√¥pitaux de Paris, Universit√© Paris-Est Cr√©teil, INSERM U955 | Christophe Rodriguez et al |
| EPI_ISL_912759 | France/un-21012190222/2021 | 2021-01-19 | Europe/France/France/ | H√¥pital Henri Mondor | Department of Virology, Henri Mondor University Hospital, Assistance Publique H√¥pitaux de Paris, Universit√© Paris-Est Cr√©teil, INSERM U955 | Christophe Rodriguez et al |
| EPI_ISL_912762 | France/un-21012200256/2021 | 2021-01-20 | Europe/France/France/ | H√¥pital Henri Mondor | Department of Virology, Henri Mondor University Hospital, Assistance Publique H√¥pitaux de Paris, Universit√© Paris-Est Cr√©teil, INSERM U955 | Christophe Rodriguez et al |

| | | | | | | |
|---|---|---|---|---|---|---|
| EPI_ISL_912768 | France/un-21012210010/2021 | 2021-01-21 | Europe/France/France/ | H√¥pital Henri Mondor | Department of Virology, Henri Mondor University Hospital, Assistance Publique H√¥pitaux de Paris, Universit√© Paris-Est Cr√©teil, INSERM U955 | Christophe Rodriguez et al |
| EPI_ISL_912771 | France/un-21012210388/2021 | 2021-01-21 | Europe/France/France/ | H√¥pital Henri Mondor | Department of Virology, Henri Mondor University Hospital, Assistance Publique H√¥pitaux de Paris, Universit√© Paris-Est Cr√©teil, INSERM U955 | Christophe Rodriguez et al |
| EPI_ISL_912788 | France/un-21012250411/2021 | 2021-01-16 | Europe/France/France/ | H√¥pital Piti√©-Salp√™tri√®re | Department of Virology, Henri Mondor University Hospital, Assistance Publique H√¥pitaux de Paris, Universit√© Paris-Est Cr√©teil, INSERM U955 | Christophe Rodriguez et al |

| | | | | | | |
|---|---|---|---|---|---|---|
| EPI_ISL_912804 | France/un-21012150329/2021 | 2021-01-15 | Europe/France/France/ | H√¥pital Henri Mondor | Department of Virology, Henri Mondor University Hospital, Assistance Publique H√¥pitaux de Paris, Universit√© Paris-Est Cr√©teil, INSERM U955 | Christophe Rodriguez et al |
| EPI_ISL_912806 | France/un-21012150348/2021 | 2021-01-15 | Europe/France/France/ | H√¥pital Henri Mondor | Department of Virology, Henri Mondor University Hospital, Assistance Publique H√¥pitaux de Paris, Universit√© Paris-Est Cr√©teil, INSERM U955 | Christophe Rodriguez et al |
| EPI_ISL_912812 | France/un-21012120390/2021 | 2021-01-12 | Europe/France/France/ | H√¥pital Henri Mondor | Department of Virology, Henri Mondor University Hospital, Assistance Publique H√¥pitaux de Paris, Universit√© Paris-Est Cr√©teil, INSERM U955 | Christophe Rodriguez et al |

| | | | | | | |
|---|---|---|---|---|---|---|
| EPI_ISL_912815 | France/un-21012250415/2021 | 2021-01-25 | Europe/France/France/ | H√¥pital Henri Mondor | Department of Virology, Henri Mondor University Hospital, Assistance Publique H√¥pitaux de Paris, Universit√© Paris-Est Cr√©teil, INSERM U955 | Christophe Rodriguez et al |
| EPI_ISL_912821 | France/un-21012120423/2021 | 2021-01-12 | Europe/France/France/ | H√¥pital Henri Mondor | Department of Virology, Henri Mondor University Hospital, Assistance Publique H√¥pitaux de Paris, Universit√© Paris-Est Cr√©teil, INSERM U955 | Christophe Rodriguez et al |
| EPI_ISL_912829 | France/un-21012250276/2021 | 2021-01-25 | Europe/France/France/ | H√¥pital Henri Mondor | Department of Virology, Henri Mondor University Hospital, Assistance Publique H√¥pitaux de Paris, Universit√© Paris-Est Cr√©teil, INSERM U955 | Christophe Rodriguez et al |

| | | | | | | |
|---|---|---|---|---|---|---|
| EPI_ISL_912847 | France/un-21012260360/2021 | 2021-01-26 | Europe/France/France/ | H√¥pital Henri Mondor | Department of Virology, Henri Mondor University Hospital, Assistance Publique H√¥pitaux de Paris, Universit√© Paris-Est Cr√©teil, INSERM U955 | Christophe Rodriguez et al |
| EPI_ISL_912857 | France/un-21012130469/2021 | 2021-01-13 | Europe/France/France/ | H√¥pital Henri Mondor | Department of Virology, Henri Mondor University Hospital, Assistance Publique H√¥pitaux de Paris, Universit√© Paris-Est Cr√©teil, INSERM U955 | Christophe Rodriguez et al |
| EPI_ISL_912861 | France/un-21012140455/2021 | 2021-01-14 | Europe/France/France/ | H√¥pital Henri Mondor | Department of Virology, Henri Mondor University Hospital, Assistance Publique H√¥pitaux de Paris, Universit√© Paris-Est Cr√©teil, INSERM U955 | Christophe Rodriguez et al |

| | | | | | | |
|---|---|---|---|---|---|---|
| EPI_ISL_912862 | France/un-21012140257/2021 | 2021-01-14 | Europe/France/France/ | H√¥pital Henri Mondor | Department of Virology, Henri Mondor University Hospital, Assistance Publique H√¥pitaux de Paris, Universit√© Paris-Est Cr√©teil, INSERM U955 | Christophe Rodriguez et al |
| EPI_ISL_912882 | France/un-21012260495/2021 | 2021-01-26 | Europe/France/France/ | H√¥pital Henri Mondor | Department of Virology, Henri Mondor University Hospital, Assistance Publique H√¥pitaux de Paris, Universit√© Paris-Est Cr√©teil, INSERM U955 | Christophe Rodriguez et al |
| EPI_ISL_912885 | France/un-21012270369/2021 | 2021-01-27 | Europe/France/France/ | H√¥pital Henri Mondor | Department of Virology, Henri Mondor University Hospital, Assistance Publique H√¥pitaux de Paris, Universit√© Paris-Est Cr√©teil, INSERM U955 | Christophe Rodriguez et al |

| | | | | | | |
|---|---|---|---|---|---|---|
| EPI_ISL_912888 | France/un-21012270470/2021 | 2021-01-27 | Europe/France/France/ | H√¥pital Henri Mondor | Department of Virology, Henri Mondor University Hospital, Assistance Publique H√¥pitaux de Paris, Universit√© Paris-Est Cr√©teil, INSERM U955 | Christophe Rodriguez et al |
| EPI_ISL_912896 | France/un-20122280138/2020 | 2020-12-28 | Europe/France/France/ | H√¥pital Henri Mondor | Department of Virology, Henri Mondor University Hospital, Assistance Publique H√¥pitaux de Paris, Universit√© Paris-Est Cr√©teil, INSERM U955 | Christophe Rodriguez et al |
| EPI_ISL_912897 | France/un-21012160133/2021 | 2021-01-16 | Europe/France/France/ | H√¥pital Henri Mondor | Department of Virology, Henri Mondor University Hospital, Assistance Publique H√¥pitaux de Paris, Universit√© Paris-Est Cr√©teil, INSERM U955 | Christophe Rodriguez et al |

| | | | | | | |
|---|---|---|---|---|---|---|
| EPI_ISL_912907 | France/un-21012180260/2021 | 2021-01-17 | Europe/France/France/ | CH.INTERCOMMUNAL DE CRETEIL | Department of Virology, Henri Mondor University Hospital, Assistance Publique H√¥pitaux de Paris, Universit√© Paris-Est Cr√©teil, INSERM U955 | Christophe Rodriguez et al |
| EPI_ISL_912926 | France/un-21012180600/2021 | 2021-01-10 | Europe/France/France/ | H√¥pital Piti√©-Salp√™tri√®re | Department of Virology, Henri Mondor University Hospital, Assistance Publique H√¥pitaux de Paris, Universit√© Paris-Est Cr√©teil, INSERM U955 | Christophe Rodriguez et al |
| EPI_ISL_912929 | France/un-20122280447/2020 | 2020-12-28 | Europe/France/France/ | H√¥pital Henri Mondor | Department of Virology, Henri Mondor University Hospital, Assistance Publique H√¥pitaux de Paris, Universit√© Paris-Est Cr√©teil, INSERM U955 | Christophe Rodriguez et al |

| | | | | | | |
|---|---|---|---|---|---|---|
| EPI_ISL_912943 | France/un-21012030090/2021 | 2021-01-03 | Europe/France/France/ | H√¥pital Henri Mondor | Department of Virology, Henri Mondor University Hospital, Assistance Publique H√¥pitaux de Paris, Universit√© Paris-Est Cr√©teil, INSERM U955 | Christophe Rodriguez et al |
| EPI_ISL_912954 | France/un-21012120471/2021 | 2021-01-12 | Europe/France/France/ | H√¥pital Henri Mondor | Department of Virology, Henri Mondor University Hospital, Assistance Publique H√¥pitaux de Paris, Universit√© Paris-Est Cr√©teil, INSERM U955 | Christophe Rodriguez et al |
| EPI_ISL_912962 | France/un-20122290264/2020 | 2020-12-29 | Europe/France/France/ | H√¥pital Henri Mondor | Department of Virology, Henri Mondor University Hospital, Assistance Publique H√¥pitaux de Paris, Universit√© Paris-Est Cr√©teil, INSERM U955 | Christophe Rodriguez et al |

| | | | | | | |
|---|---|---|---|---|---|---|
| EPI_ISL_912977 | France/un-20122310297/2020 | 2020-12-31 | Europe/France/France/ | H√¥pital Henri Mondor | Department of Virology, Henri Mondor University Hospital, Assistance Publique H√¥pitaux de Paris, Universit√© Paris-Est Cr√©teil, INSERM U955 | Christophe Rodriguez et al |
| EPI_ISL_912980 | France/un-21012030100/2021 | 2021-01-03 | Europe/France/France/ | H√¥pital Henri Mondor | Department of Virology, Henri Mondor University Hospital, Assistance Publique H√¥pitaux de Paris, Universit√© Paris-Est Cr√©teil, INSERM U955 | Christophe Rodriguez et al |
| EPI_ISL_912985 | France/un-20122220067/2020 | 2020-12-22 | Europe/France/France/ | H√¥pital Henri Mondor | Department of Virology, Henri Mondor University Hospital, Assistance Publique H√¥pitaux de Paris, Universit√© Paris-Est Cr√©teil, INSERM U955 | Christophe Rodriguez et al |

| | | | | | | |
|---|---|---|---|---|---|---|
| EPI_ISL_912986 | France/un-20122220695/2020 | 2020-12-22 | Europe/France/France/ | H√¥pital Henri Mondor | Department of Virology, Henri Mondor University Hospital, Assistance Publique H√¥pitaux de Paris, Universit√© Paris-Est Cr√©teil, INSERM U955 | Christophe Rodriguez et al |
| EPI_ISL_912998 | France/un-20122230758/2020 | 2020-12-23 | Europe/France/France/ | H√¥pital Henri Mondor | Department of Virology, Henri Mondor University Hospital, Assistance Publique H√¥pitaux de Paris, Universit√© Paris-Est Cr√©teil, INSERM U955 | Christophe Rodriguez et al |
| EPI_ISL_913002 | France/un-20122240457/2020 | 2020-12-24 | Europe/France/France/ | H√¥pital Henri Mondor | Department of Virology, Henri Mondor University Hospital, Assistance Publique H√¥pitaux de Paris, Universit√© Paris-Est Cr√©teil, INSERM U955 | Christophe Rodriguez et al |

| | | | | | | |
|---|---|---|---|---|---|---|
| EPI_ISL_913004 | France/un-20122260004/2020 | 2020-12-26 | Europe/France/France/ | H√¥pital Henri Mondor | Department of Virology, Henri Mondor University Hospital, Assistance Publique H√¥pitaux de Paris, Universit√© Paris-Est Cr√©teil, INSERM U955 | Christophe Rodriguez et al |
| EPI_ISL_913008 | France/un-20122260098/2020 | 2020-12-26 | Europe/France/France/ | H√¥pital Henri Mondor | Department of Virology, Henri Mondor University Hospital, Assistance Publique H√¥pitaux de Paris, Universit√© Paris-Est Cr√©teil, INSERM U955 | Christophe Rodriguez et al |
| EPI_ISL_913016 | France/un-20122280531/2020 | 2020-12-28 | Europe/France/France/ | H√¥pital Henri Mondor | Department of Virology, Henri Mondor University Hospital, Assistance Publique H√¥pitaux de Paris, Universit√© Paris-Est Cr√©teil, INSERM U955 | Christophe Rodriguez et al |

| | | | | | | |
|---|---|---|---|---|---|---|
| EPI_ISL_913019 | France/un-20122280541/2020 | 2020-12-28 | Europe/France/France/ | H√¥pital Henri Mondor | Department of Virology, Henri Mondor University Hospital, Assistance Publique H√¥pitaux de Paris, Universit√© Paris-Est Cr√©teil, INSERM U955 | Christophe Rodriguez et al |
| EPI_ISL_913022 | Spain/EX-ISCIII-210033/2020 | 2020-12-31 | Europe/Spain/Extremadura/ | Hospital San Pedro de Alc√°ntara | Instituto de Salud Carlos III | Iglesias-Caballero et al |
| EPI_ISL_913023 | Spain/EX-ISCIII-210034/2020 | 2020-12-31 | Europe/Spain/Extremadura/ | Hospital San Pedro de Alc√°ntara | Instituto de Salud Carlos III | Iglesias-Caballero et al |
| EPI_ISL_913024 | Spain/GA-ISCIII-210066/2020 | 2020-12-31 | Europe/Spain/Galicia/ | Complejo Hospitalario Universitario de Santiago | Instituto de Salud Carlos III | Iglesias-Caballero et al |
| EPI_ISL_913025 | Spain/MC-ISCIII-210142/2021 | 2021-01-17 | Europe/Spain/Murcia/ | Hospital los Arcos | Instituto de Salud Carlos III | Iglesias-Caballero et al |
| EPI_ISL_913028 | Spain/MC-ISCIII-210147/2021 | 2021-01-14 | Europe/Spain/Murcia/ | Hospital los Arcos | Instituto de Salud Carlos III | Iglesias-Caballero et al |
| EPI_ISL_913043 | Spain/CB-ISCIII-210089/2021 | 2021-01-05 | Europe/Spain/Cantabria/ | Hospital Comarcal Sierrallana | Instituto de Salud Carlos III | Iglesias-Caballero et al |
| EPI_ISL_913045 | Spain/CB-ISCIII-210091/2021 | 2021-01-02 | Europe/Spain/Cantabria/ | Hospital Comarcal Sierrallana | Instituto de Salud Carlos III | Iglesias-Caballero et al |
| EPI_ISL_913054 | Spain/CE-ISCIII-210077/2021 | 2021-01-05 | Europe/Spain/Ceuta/ | Hospital Universitario de Ceuta | Instituto de Salud Carlos III | Iglesias-Caballero et al |
| EPI_ISL_913057 | Spain/EX-ISCIII-210160/2021 | 2021-01-14 | Europe/Spain/Extremadura/ | Hospital San Pedro de Alc√°ntara | Instituto de Salud Carlos III | Iglesias-Caballero et al |
| EPI_ISL_913058 | Spain/CE-ISCIII-210083/2021 | 2021-01-05 | Europe/Spain/Ceuta/ | Hospital Universitario de Ceuta | Instituto de Salud Carlos III | Iglesias-Caballero et al |
| EPI_ISL_913059 | Spain/CM-ISCIII-210135/2021 | 2021-01-04 | Europe/Spain/Castilla la Mancha/ | Consejer√≠a de Sanidad y Asuntos Sociales | Instituto de Salud Carlos III | Iglesias-Caballero et al |
| EPI_ISL_913061 | Spain/CL-ISCIII-210156/2021 | 2021-01-14 | Europe/Spain/Castilla y Leon/ | Hospital Nuestra Se√±ora de Sonsoles | Instituto de Salud Carlos III | Iglesias-Caballero et al |
| EPI_ISL_913067 | Austria/MUW_1311515/2020 | 2020-09-18 | Europe/Austria/Austria/ | Center for Virology | Center for Virology | Jeremy V. Camp et al |
| EPI_ISL_913071 | Austria/MUW_1326338/2020 | 2020-10-15 | Europe/Austria/Austria/ | Center for Virology | Center for Virology | Jeremy V. Camp et al |

| | | | | | | |
|---|---|---|---|---|---|---|
| EPI_ISL_913072 | Austria/MUW_1326335/2020 | 2020-10-15 | Europe/Austria/Austria/ | Center for Virology | Center for Virology | Jeremy V. Camp et al |
| EPI_ISL_913073 | Austria/MUW_1329645/2020 | 2020-10-23 | Europe/Austria/Austria/ | Center for Virology | Center for Virology | Jeremy V. Camp et al |
| EPI_ISL_913074 | Austria/MUW_1324313/2020 | 2020-10-12 | Europe/Austria/Austria/ | Center for Virology | Center for Virology | Jeremy V. Camp et al |
| EPI_ISL_913075 | Austria/MUW_1331385/2020 | 2020-10-28 | Europe/Austria/Austria/ | Center for Virology | Center for Virology | Jeremy V. Camp et al |
| EPI_ISL_913076 | Austria/MUW_1356960/2020 | 2020-12-30 | Europe/Austria/Austria/ | Center for Virology | Center for Virology | Jeremy V. Camp et al |
| EPI_ISL_913077 | Austria/MUW_1358159/2021 | 2021-01-07 | Europe/Austria/Austria/ | Center for Virology | Center for Virology | Jeremy V. Camp et al |
| EPI_ISL_913079 | Austria/MUW_p768/2020 | 2020-10-19 | Europe/Austria/Austria/ | Center for Virology | Center for Virology | Jeremy V. Camp et al |
| EPI_ISL_913082 | Austria/MUW_1305566/2020 | 2020-09-07 | Europe/Austria/Austria/ | Center for Virology | Center for Virology | Jeremy V. Camp et al |
| EPI_ISL_913083 | Austria/MUW_1305567/2020 | 2020-09-07 | Europe/Austria/Austria/ | Center for Virology | Center for Virology | Jeremy V. Camp et al |
| EPI_ISL_913084 | Austria/MUW_1305569/2020 | 2020-09-07 | Europe/Austria/Austria/ | Center for Virology | Center for Virology | Jeremy V. Camp et al |
| EPI_ISL_913087 | Austria/MUW_1328132/2020 | 2020-10-21 | Europe/Austria/Austria/ | Center for Virology | Center for Virology | Jeremy V. Camp et al |
| EPI_ISL_913089 | Austria/MUW_1313844/2020 | 2020-09-22 | Europe/Austria/Austria/ | Center for Virology | Center for Virology | Jeremy V. Camp et al |
| EPI_ISL_913092 | Austria/MUW_1344162/2020 | 2020-09-30 | Europe/Austria/Austria/ | Labor Mustafa Wien | Center for Virology | Jeremy V. Camp et al |
| EPI_ISL_913093 | Austria/MUW_1307024/2020 | 2020-09-08 | Europe/Austria/Austria/ | Center for Virology | Center for Virology | Jeremy V. Camp et al |
| EPI_ISL_913094 | Austria/MUW_1317980/2020 | 2020-09-29 | Europe/Austria/Austria/ | Center for Virology | Center for Virology | Jeremy V. Camp et al |
| EPI_ISL_913097 | Austria/MUW_1334664/2020 | 2020-11-02 | Europe/Austria/Austria/ | Center for Virology | Center for Virology | Jeremy V. Camp et al |
| EPI_ISL_913098 | Austria/MUW_1335920/2020 | 2020-11-05 | Europe/Austria/Austria/ | Center for Virology | Center for Virology | Jeremy V. Camp et al |
| EPI_ISL_913104 | France/OCC-252/2020 | 2020-10-15 | Europe/France/Occitanie/Toulouse | CHU Purpan - Laboratoire de Virologie - Institut FV©dV©ratif de Biologie | CHU Purpan - Laboratoire de Virologie - Institut FV©dV©ratif de Biologie | Latour J. et al |

| | | | | | | |
|---|---|---|---|---|---|---|
| EPI_ISL_913107 | France/OCC-255/2020 | 2020-10-16 | Europe/France/Occitanie/Toulouse | CHU Purpan - Laboratoire de Virologie - Institut F√©d√©ratif de Biologie | CHU Purpan - Laboratoire de Virologie - Institut F√©d√©ratif de Biologie | Latour J. et al |
| EPI_ISL_913113 | France/OCC-261/2020 | 2020-12-15 | Europe/France/Occitanie/Saint-Gaudens | CHU Purpan - Laboratoire de Virologie - Institut F√©d√©ratif de Biologie | CHU Purpan - Laboratoire de Virologie - Institut F√©d√©ratif de Biologie | Latour J. et al |
| EPI_ISL_913119 | USA/MI-UM-10037807043/2021 | 2021-01-13 | North America/USA/Michigan/ | University of Michigan Clinical Microbiology Laboratory | Lauring Lab, University of Michigan, Department of Microbiology and Immunology | Valesano et al |
| EPI_ISL_913122 | USA/MI-UM-10037012230/2020 | 2020-11-18 | North America/USA/Michigan/ | University of Michigan Clinical Microbiology Laboratory | Lauring Lab, University of Michigan, Department of Microbiology and Immunology | Valesano et al |
| EPI_ISL_913135 | USA/MI-UM-10037743813/2021 | 2021-01-08 | North America/USA/Michigan/ | University of Michigan Clinical Microbiology Laboratory | Lauring Lab, University of Michigan, Department of Microbiology and Immunology | Valesano et al |
| EPI_ISL_913140 | USA/MI-UM-10037015311/2020 | 2020-11-18 | North America/USA/Michigan/ | University of Michigan Clinical Microbiology Laboratory | Lauring Lab, University of Michigan, Department of Microbiology and Immunology | Valesano et al |
| EPI_ISL_913150 | USA/MI-UM-10037795519/2021 | 2021-01-12 | North America/USA/Michigan/ | University of Michigan Clinical Microbiology Laboratory | Lauring Lab, University of Michigan, Department of Microbiology and Immunology | Valesano et al |

| | | | | | | |
|---|---|---|---|---|---|---|
| EPI_ISL_913153 | USA/MI-UM-10037018207/2020 | 2020-11-18 | North America/USA/Michigan/ | University of Michigan Clinical Microbiology Laboratory | Lauring Lab, University of Michigan, Department of Microbiology and Immunology | Valesano et al |
| EPI_ISL_913164 | USA/MI-UM-10037743388/2021 | 2021-01-08 | North America/USA/Michigan/ | University of Michigan Clinical Microbiology Laboratory | Lauring Lab, University of Michigan, Department of Microbiology and Immunology | Valesano et al |
| EPI_ISL_913167 | USA/MI-UM-10037746544/2021 | 2021-01-08 | North America/USA/Michigan/ | University of Michigan Clinical Microbiology Laboratory | Lauring Lab, University of Michigan, Department of Microbiology and Immunology | Valesano et al |
| EPI_ISL_913179 | USA/MI-UM-10037770006/2021 | 2021-01-11 | North America/USA/Michigan/ | University of Michigan Clinical Microbiology Laboratory | Lauring Lab, University of Michigan, Department of Microbiology and Immunology | Valesano et al |
| EPI_ISL_913196 | USA/MI-UM-10037816122/2021 | 2021-01-13 | North America/USA/Michigan/ | University of Michigan Clinical Microbiology Laboratory | Lauring Lab, University of Michigan, Department of Microbiology and Immunology | Valesano et al |
| EPI_ISL_913197 | USA/MI-UM-10037811689/2021 | 2021-01-13 | North America/USA/Michigan/ | University of Michigan Clinical Microbiology Laboratory | Lauring Lab, University of Michigan, Department of Microbiology and Immunology | Valesano et al |

| | | | | | | |
|---|---|---|---|---|---|---|
| EPI_ISL_913211 | USA/MI-UM-10037012157/2020 | 2020-11-18 | North America/USA/Michigan/ | University of Michigan Clinical Microbiology Laboratory | Lauring Lab, University of Michigan, Department of Microbiology and Immunology | Valesano et al |
| EPI_ISL_913220 | USA/MI-UM-10036995497/2020 | 2020-11-17 | North America/USA/Michigan/ | University of Michigan Clinical Microbiology Laboratory | Lauring Lab, University of Michigan, Department of Microbiology and Immunology | Valesano et al |
| EPI_ISL_913241 | USA/MI-UM-10036995509/2020 | 2020-11-17 | North America/USA/Michigan/ | University of Michigan Clinical Microbiology Laboratory | Lauring Lab, University of Michigan, Department of Microbiology and Immunology | Valesano et al |
| EPI_ISL_913254 | USA/MI-UM-10037013991/2020 | 2020-11-18 | North America/USA/Michigan/ | University of Michigan Clinical Microbiology Laboratory | Lauring Lab, University of Michigan, Department of Microbiology and Immunology | Valesano et al |
| EPI_ISL_913256 | Sweden/21-50149/2021 | 2021-01-02 | Europe/Sweden/Stockholm/ | Synlab Medilab, Mikrobiologi | The Public Health Agency of Sweden | Anna-Malin Linde et al |
| EPI_ISL_913257 | Sweden/21-50153/2021 | 2021-01-02 | Europe/Sweden/Stockholm/ | Synlab Medilab, Mikrobiologi | The Public Health Agency of Sweden | Anna-Malin Linde et al |
| EPI_ISL_913262 | Sweden/21-50220/2021 | 2021-01-09 | Europe/Sweden/Stockholm/ | Klinisk mikrobiologi | The Public Health Agency of Sweden | Anna-Malin Linde et al |
| EPI_ISL_913268 | Sweden/21-50117/2021 | 2021-01-05 | Europe/Sweden/Stockholm/ | Klinisk mikrobiologi | The Public Health Agency of Sweden | Anna-Malin Linde et al |
| EPI_ISL_913271 | Sweden/21-50223/2021 | 2021-01-09 | Europe/Sweden/Stockholm/ | Klinisk mikrobiologi | The Public Health Agency of Sweden | Anna-Malin Linde et al |
| EPI_ISL_913273 | Sweden/21-50239/2021 | 2021-01-08 | Europe/Sweden/Orebro/ | Klinisk mikrobiologi | The Public Health Agency of Sweden | Anna-Malin Linde et al |
| EPI_ISL_913279 | Sweden/21-50165/2020 | 2020-11-25 | Europe/Sweden/Stockholm/ | Klinisk mikrobiologi | The Public Health Agency of Sweden | Anna-Malin Linde et al |
| EPI_ISL_913284 | Sweden/21-50154/2020 | 2020-12-28 | Europe/Sweden/Stockholm/ | Synlab Medilab, Mikrobiologi | The Public Health Agency of Sweden | Anna-Malin Linde et al |

| | | | | | | |
|---|---|---|---|---|---|---|
| EPI_ISL_913304 | Sweden/21-50151/2021 | 2021-01-02 | Europe/Sweden/Stockholm/ | Synlab Medilab, Mikrobiologi | The Public Health Agency of Sweden | Anna-Malin Linde et al |
| EPI_ISL_913305 | Sweden/21-50152/2020 | 2020-12-31 | Europe/Sweden/Stockholm/ | Synlab Medilab, Mikrobiologi | The Public Health Agency of Sweden | Anna-Malin Linde et al |
| EPI_ISL_913306 | Sweden/21-00362/2020 | 2020-12-01 | Europe/Sweden/Blekinge/ | Klinisk mikrobiologi | The Public Health Agency of Sweden | Anna-Malin Linde et al |
| EPI_ISL_913307 | Sweden/21-50211/2021 | 2021-01-09 | Europe/Sweden/Stockholm/ | Klinisk mikrobiologi | The Public Health Agency of Sweden | Anna-Malin Linde et al |
| EPI_ISL_913311 | Sweden/20-13133/2020 | 2020-12-29 | Europe/Sweden/Ostergotland/ | Klinisk mikrobiologi | The Public Health Agency of Sweden | Anna-Malin Linde et al |
| EPI_ISL_913313 | Sweden/21-00358/2020 | 2020-11-25 | Europe/Sweden/Kronoberg/ | Klinisk mikrobiologi | The Public Health Agency of Sweden | Anna-Malin Linde et al |
| EPI_ISL_913319 | Sweden/21-00099/2020 | 2020-05-27 | Europe/Sweden/Stockholm/ | Klinisk mikrobiologi | The Public Health Agency of Sweden | Anna-Malin Linde et al |
| EPI_ISL_913326 | Sweden/21-50091/2020 | 2020-10-09 | Europe/Sweden/Stockholm/ | Klinisk mikrobiologi | The Public Health Agency of Sweden | Anna-Malin Linde et al |
| EPI_ISL_913334 | Sweden/21-50123/2021 | 2021-01-04 | Europe/Sweden/Vastra Gotaland/ | Klinisk Mikrobiologi | The Public Health Agency of Sweden | Anna-Malin Linde et al |
| EPI_ISL_913337 | Sweden/21-50127/2021 | 2021-01-02 | Europe/Sweden/Vastra Gotaland/ | Unilabs AB Skovde | The Public Health Agency of Sweden | Anna-Malin Linde et al |
| EPI_ISL_913339 | Sweden/21-50042/2020 | 2020-12-29 | Europe/Sweden/Sormland/ | Unilabs, Mikrobiologiska laboratoriet | The Public Health Agency of Sweden | Anna-Malin Linde et al |
| EPI_ISL_913343 | Sweden/21-50156/2020 | 2020-11-25 | Europe/Sweden/Stockholm/ | Synlab Medilab, Mikrobiologi | The Public Health Agency of Sweden | Anna-Malin Linde et al |
| EPI_ISL_913345 | Sweden/21-50162/2020 | 2020-11-29 | Europe/Sweden/Stockholm/ | Klinisk mikrobiologi | The Public Health Agency of Sweden | Anna-Malin Linde et al |
| EPI_ISL_913346 | Sweden/21-50163/2020 | 2020-11-29 | Europe/Sweden/Stockholm/ | Klinisk mikrobiologi | The Public Health Agency of Sweden | Anna-Malin Linde et al |
| EPI_ISL_913354 | Sweden/21-00355/2020 | 2020-11-25 | Europe/Sweden/Blekinge/ | Klinisk mikrobiologi | The Public Health Agency of Sweden | Anna-Malin Linde et al |
| EPI_ISL_913356 | Sweden/21-00360/2020 | 2020-11-26 | Europe/Sweden/Kronoberg/ | Klinisk mikrobiologi | The Public Health Agency of Sweden | Anna-Malin Linde et al |
| EPI_ISL_913357 | Sweden/21-00365/2020 | 2020-12-03 | Europe/Sweden/Blekinge/ | Klinisk mikrobiologi | The Public Health Agency of Sweden | Anna-Malin Linde et al |
| EPI_ISL_913359 | Sweden/21-00373/2021 | 2021-01-08 | Europe/Sweden/Blekinge/ | Klinisk mikrobiologi | The Public Health Agency of Sweden | Anna-Malin Linde et al |
| EPI_ISL_913361 | Sweden/21-00379/2021 | 2021-01-09 | Europe/Sweden/Blekinge/ | Klinisk mikrobiologi | The Public Health Agency of Sweden | Anna-Malin Linde et al |
| EPI_ISL_913368 | Sweden/21-50226/2020 | 2020-06-08 | Europe/Sweden/Gavleborgs Lan/ | Klinisk mikrobiologi | The Public Health Agency of Sweden | Anna-Malin Linde et al |

| | | | | | | |
|---|---|---|---|---|---|---|
| EPI_ISL_913372 | Sweden/21-50272/2021 | 2021-01-05 | Europe/Sweden/Vastra Gotaland/ | Klinisk mikrobiologi | The Public Health Agency of Sweden | Anna-Malin Linde et al |
| EPI_ISL_913377 | Sweden/21-00110/2021 | 2021-01-07 | Europe/Sweden/Stockholm/ | The Public Health Agency of Sweden | The Public Health Agency of Sweden | Anna-Malin Linde et al |
| EPI_ISL_913379 | Sweden/21-50050/2020 | 2020-12-28 | Europe/Sweden/Skane/ | Dynamic Code AB | The Public Health Agency of Sweden | Anna-Malin Linde et al |
| EPI_ISL_913381 | Sweden/21-50056/2020 | 2020-12-29 | Europe/Sweden/Halland/ | Dynamic Code AB | The Public Health Agency of Sweden | Anna-Malin Linde et al |
| EPI_ISL_913387 | Sweden/21-50067/2021 | 2021-01-03 | Europe/Sweden/Skane/ | Dynamic Code AB | The Public Health Agency of Sweden | Anna-Malin Linde et al |
| EPI_ISL_913392 | Sweden/21-50086/2021 | 2021-01-03 | Europe/Sweden/Skane/ | Dynamic Code AB | The Public Health Agency of Sweden | Anna-Malin Linde et al |
| EPI_ISL_913397 | USA/MA-MASPHL-01637/2021 | 2021-01-17 | North America/USA/Massachusetts/ | Massachusetts State Public Health Laboratory | Massachusetts State Public Health Laboratory | Andrew Lang et al |
| EPI_ISL_913404 | USA/MA-MASPHL-01630/2021 | 2021-01-13 | North America/USA/Massachusetts/ | Massachusetts State Public Health Laboratory | Massachusetts State Public Health Laboratory | Andrew Lang et al |
| EPI_ISL_913406 | USA/MA-MASPHL-01575/2021 | 2021-01-12 | North America/USA/Massachusetts/ | Massachusetts State Public Health Laboratory | Massachusetts State Public Health Laboratory | Andrew Lang et al |
| EPI_ISL_913410 | USA/MA-MASPHL-01591/2021 | 2021-01-04 | North America/USA/Massachusetts/ | Massachusetts State Public Health Laboratory | Massachusetts State Public Health Laboratory | Andrew Lang et al |
| EPI_ISL_913421 | USA/MA-MASPHL-01579/2021 | 2021-01-11 | North America/USA/Massachusetts/ | Massachusetts State Public Health Laboratory | Massachusetts State Public Health Laboratory | Andrew Lang et al |
| EPI_ISL_913429 | USA/MA-MASPHL-01580/2021 | 2021-01-11 | North America/USA/Massachusetts/ | Massachusetts State Public Health Laboratory | Massachusetts State Public Health Laboratory | Andrew Lang et al |
| EPI_ISL_913434 | USA/MA-MASPHL-01648/2021 | 2021-01-20 | North America/USA/Massachusetts/ | Massachusetts State Public Health Laboratory | Massachusetts State Public Health Laboratory | Andrew Lang et al |
| EPI_ISL_913446 | Italy/LAZ-INMI-170/2021 | 2021-01-15 | Europe/Italy/Lazio/ | Laboratorio Genzano - ASL RM 6 | INMI Lazzaro Spallanzani IRCCS | Emanuela Giombini et al |
| EPI_ISL_913447 | Italy/LAZ-INMI-171/2021 | 2021-01-15 | Europe/Italy/Lazio/ | Laboratorio Genzano - ASL RM 6 | INMI Lazzaro Spallanzani IRCCS | Francesco Messina et al |
| EPI_ISL_913448 | Italy/LAZ-INMI-115isl/2020 | 2020-12-20 | Europe/Italy/Lazio/ | INMI Lazzaro Spallanzani IRCCS | INMI Lazzaro Spallanzani IRCCS | Martina Rueca et al |

| | | | | | | |
|---|---|---|---|---|---|---|
| EPI_ISL_913449 | Italy/CAM-INMI-118isl/2020 | 2020-12-20 | Europe/Italy/Campania/ | Istituto Zooprofilattico Sperimentale del Mezzogiorno | INMI Lazzaro Spallanzani IRCCS | Barbara Bartolini et al |
| EPI_ISL_913455 | Sweden/21-50283/2021 | 2021-01-12 | Europe/Sweden/Vastra Gotaland/ | Unilabs AB Skovde | The Public Health Agency of Sweden | Anna-Malin Linde et al |
| EPI_ISL_913457 | Sweden/21-50385/2021 | 2021-01-15 | Europe/Sweden/Stockholm/ | Klinisk mikrobiologi | The Public Health Agency of Sweden | Anna-Malin Linde et al |
| EPI_ISL_913461 | Sweden/21-50159/2020 | 2020-11-26 | Europe/Sweden/Stockholm/ | Klinisk mikrobiologi | The Public Health Agency of Sweden | Anna-Malin Linde et al |
| EPI_ISL_913469 | Sweden/21-50373/2021 | 2021-01-11 | Europe/Sweden/Vastra Gotaland/ | Klinisk mikrobiologi | The Public Health Agency of Sweden | Anna-Malin Linde et al |
| EPI_ISL_913473 | Sweden/21-50076/2020 | 2020-04-24 | Europe/Sweden/Stockholm/ | Klinisk mikrobiologi | The Public Health Agency of Sweden | Anna-Malin Linde et al |
| EPI_ISL_913476 | Sweden/21-00333/2020 | 2020-08-24 | Europe/Sweden/Blekinge/ | Klinisk mikrobiologi | The Public Health Agency of Sweden | Anna-Malin Linde et al |
| EPI_ISL_913477 | Sweden/21-00344/2020 | 2020-10-05 | Europe/Sweden/Blekinge/ | Klinisk mikrobiologi | The Public Health Agency of Sweden | Anna-Malin Linde et al |
| EPI_ISL_913479 | Sweden/21-00097/2020 | 2020-11-13 | Europe/Sweden/Stockholm/ | Klinisk mikrobiologi | The Public Health Agency of Sweden | Anna-Malin Linde et al |
| EPI_ISL_913485 | Sweden/21-00142/2020 | 2020-12-28 | Europe/Sweden/Skane/ | Klinisk mikrobiologi | The Public Health Agency of Sweden | Anna-Malin Linde et al |
| EPI_ISL_913487 | Sweden/21-00156/2020 | 2020-12-29 | Europe/Sweden/Skane/ | Klinisk mikrobiologi | The Public Health Agency of Sweden | Anna-Malin Linde et al |
| EPI_ISL_913524 | USA/MN-MDH-2511/2020 | 2020-12-28 | North America/USA/Minnesota/ | M Health Fairview | Minnesota Department of Health, Public Health Laboratory | Alexandra Lorentz et al |
| EPI_ISL_913526 | USA/MN-MDH-2513/2020 | 2020-12-28 | North America/USA/Minnesota/ | M Health Fairview | Minnesota Department of Health, Public Health Laboratory | Alexandra Lorentz et al |
| EPI_ISL_913530 | USA/MN-MDH-2517/2020 | 2020-12-29 | North America/USA/Minnesota/ | M Health Fairview | Minnesota Department of Health, Public Health Laboratory | Alexandra Lorentz et al |
| EPI_ISL_913545 | USA/MN-MDH-2532/2020 | 2020-12-31 | North America/USA/Minnesota/ | M Health Fairview | Minnesota Department of Health, Public Health Laboratory | Alexandra Lorentz et al |

| | | | | | | |
|---|---|---|---|---|---|---|
| EPI_ISL_913548 | USA/MN-MDH-2536/2020 | 2020-12-31 | North America/USA/Minnesota/ | M Health Fairview | Minnesota Department of Health, Public Health Laboratory | Alexandra Lorentz et al |
| EPI_ISL_913558 | USA/MN-MDH-2546/2020 | 2020-12-31 | North America/USA/Minnesota/ | M Health Fairview | Minnesota Department of Health, Public Health Laboratory | Alexandra Lorentz et al |
| EPI_ISL_913571 | USA/MN-MDH-2559/2021 | 2021-01-05 | North America/USA/Minnesota/ | M Health Fairview | Minnesota Department of Health, Public Health Laboratory | Alexandra Lorentz et al |
| EPI_ISL_913586 | USA/MN-MDH-2574/2021 | 2021-01-13 | North America/USA/Minnesota/ | Vault Health | Minnesota Department of Health, Public Health Laboratory | Alexandra Lorentz et al |
| EPI_ISL_913645 | USA/MI-MDHHS-SC23172/2020 | 2020-12-30 | North America/USA/Michigan/ | Michigan Department of Health and Human Services, Bureau of Laboratories | Michigan Department of Health and Human Services, Bureau of Laboratories | Blankenship HM et al |
| EPI_ISL_913652 | USA/MI-MDHHS-SC23179/2021 | 2021-01-05 | North America/USA/Michigan/ | Michigan Department of Health and Human Services, Bureau of Laboratories | Michigan Department of Health and Human Services, Bureau of Laboratories | Blankenship HM et al |
| EPI_ISL_913661 | USA/MI-MDHHS-SC23188/2021 | 2021-01-04 | North America/USA/Michigan/ | Michigan Department of Health and Human Services, Bureau of Laboratories | Michigan Department of Health and Human Services, Bureau of Laboratories | Blankenship HM et al |
| EPI_ISL_913663 | USA/MI-MDHHS-SC23190/2021 | 2021-01-04 | North America/USA/Michigan/ | Michigan Department of Health and Human Services, Bureau of Laboratories | Michigan Department of Health and Human Services, Bureau of Laboratories | Blankenship HM et al |

| | | | | | | |
|---|---|---|---|---|---|---|
| EPI_ISL_913672 | USA/MN-MDH-2282/2020 | 2020-12-07 | North America/USA/Minnesota/ | Vault Health | Minnesota Department of Health, Public Health Laboratory | Alexandra Lorentz et al |
| EPI_ISL_913690 | USA/MN-MDH-2300/2020 | 2020-12-29 | North America/USA/Minnesota/ | Minnesota Department of Health, Public Health Laboratory | Minnesota Department of Health, Public Health Laboratory | Alexandra Lorentz et al |
| EPI_ISL_913691 | USA/MN-MDH-2301/2020 | 2020-12-29 | North America/USA/Minnesota/ | Minnesota Department of Health, Public Health Laboratory | Minnesota Department of Health, Public Health Laboratory | Alexandra Lorentz et al |
| EPI_ISL_913693 | USA/MN-MDH-2303/2020 | 2020-12-29 | North America/USA/Minnesota/ | Minnesota Department of Health, Public Health Laboratory | Minnesota Department of Health, Public Health Laboratory | Alexandra Lorentz et al |
| EPI_ISL_913696 | USA/MN-MDH-2306/2020 | 2020-12-30 | North America/USA/Minnesota/ | Minnesota Department of Health, Public Health Laboratory | Minnesota Department of Health, Public Health Laboratory | Alexandra Lorentz et al |
| EPI_ISL_913701 | USA/MN-MDH-2312/2020 | 2020-12-30 | North America/USA/Minnesota/ | Minnesota Department of Health, Public Health Laboratory | Minnesota Department of Health, Public Health Laboratory | Alexandra Lorentz et al |
| EPI_ISL_913705 | USA/MN-MDH-2316/2020 | 2020-12-29 | North America/USA/Minnesota/ | Minnesota Department of Health, Public Health Laboratory | Minnesota Department of Health, Public Health Laboratory | Alexandra Lorentz et al |
| EPI_ISL_913716 | USA/MN-MDH-2327/2021 | 2021-01-04 | North America/USA/Minnesota/ | Minnesota Department of Health, Public Health Laboratory | Minnesota Department of Health, Public Health Laboratory | Alexandra Lorentz et al |
| EPI_ISL_913722 | USA/MN-MDH-2334/2021 | 2021-01-07 | North America/USA/Minnesota/ | Minnesota Department of Health, Public Health Laboratory | Minnesota Department of Health, Public Health Laboratory | Alexandra Lorentz et al |
| EPI_ISL_913731 | USA/MN-MDH-2343/2021 | 2021-01-05 | North America/USA/Minnesota/ | M Health Fairview | Minnesota Department of Health, Public Health Laboratory | Alexandra Lorentz et al |

| | | | | | | |
|---|---|---|---|---|---|---|
| EPI_ISL_913765 | Belgium/rega-1983/2021 | 2021-01-12 | Europe/Belgium/Antwerpen/ | KU Leuven, Rega Institute, Clinical and Epidemiological Virology | KU Leuven, Rega Institute, Clinical and Epidemiological Virology | Tony Wawina-Bokalanga et al |
| EPI_ISL_913787 | Belgium/rega-2000/2021 | 2021-01-09 | Europe/Belgium/Leuven/Heverlee | KU Leuven, Rega Institute, Clinical and Epidemiological Virology | KU Leuven, Rega Institute, Clinical and Epidemiological Virology | Tony Wawina-Bokalanga et al |
| EPI_ISL_913805 | USA/AZ-TG707655/2021 | 2021-01-06 | North America/USA/Arizona/Navajo Co | TGen North | TGen North | """Jolene Bowers et al" |
| EPI_ISL_913815 | USA/AZ-TG707650/2021 | 2021-01-05 | North America/USA/Arizona/Coconino C | TGen North | TGen North | """Jolene Bowers et al" |
| EPI_ISL_913832 | USA/AZ-TG707062/2020 | 2020-12-29 | North America/USA/Arizona/Pima Coun | TGen North | TGen North | """Jolene Bowers et al" |
| EPI_ISL_913836 | USA/AZ-TG707639/2021 | 2021-01-05 | North America/USA/Arizona/Mohave Co | TGen North | TGen North | """Jolene Bowers et al" |
| EPI_ISL_913870 | USA/AZ-TG707673/2021 | 2021-01-04 | North America/USA/Arizona/Coconino C | TGen North | TGen North | """Jolene Bowers et al" |
| EPI_ISL_913879 | USA/AZ-TG707091/2021 | 2021-01-06 | North America/USA/Arizona/Navajo Co | TGen North | TGen North | """Jolene Bowers et al" |
| EPI_ISL_913882 | USA/AZ-TG705114/2021 | 2021-01-04 | North America/USA/Arizona/Coconino C | TGen North | TGen North | """Jolene Bowers et al" |
| EPI_ISL_913893 | USA/AZ-TG707061/2020 | 2020-12-29 | North America/USA/Arizona/Pima Coun | TGen North | TGen North | """Jolene Bowers et al" |
| EPI_ISL_913896 | USA/AZ-TG707683/2021 | 2021-01-05 | North America/USA/Arizona/Mohave Co | TGen North | TGen North | """Jolene Bowers et al" |
| EPI_ISL_913900 | USA/AZ-TG707681/2021 | 2021-01-05 | North America/USA/Arizona/Coconino C | TGen North | TGen North | """Jolene Bowers et al" |
| EPI_ISL_913911 | Mexico/CHP-InDRE_107/2020 | 2020-07-21 | North America/Mexico/Chiapas/ | Instituto de Diagnostico y Referencia Epidemiologicos INDRE_RNLSP | Instituto de Diagnostico y Referencia Epidemiologicos (INDRE) | Claudia Wong-Arambula et al |
| EPI_ISL_913912 | Mexico/CHP-InDRE_108/2020 | 2020-07-21 | North America/Mexico/Chiapas/ | Instituto de Diagnostico y Referencia Epidemiologicos INDRE_RNLSP | Instituto de Diagnostico y Referencia Epidemiologicos (INDRE) | Claudia Wong-Arambula et al |
| EPI_ISL_913913 | Mexico/TLA-InDRE_109/2020 | 2020-07-23 | North America/Mexico/Tlaxcala/ | Instituto de Diagnostico y Referencia Epidemiologicos INDRE_RNLSP | Instituto de Diagnostico y Referencia Epidemiologicos (INDRE) | Claudia Wong-Arambula et al |

| | | | | | | |
|---|---|---|---|---|---|---|
| EPI_ISL_913914 | Mexico/OAX-InDRE_110/2020 | 2020-07-24 | North America/Mexico/Oaxaca/ | Instituto de Diagnostico y Referencia Epidemiologicos INDRE_RNLSP | Instituto de Diagnostico y Referencia Epidemiologicos (INDRE) | Claudia Wong-Arambula et al |
| EPI_ISL_913915 | Mexico/VER-InDRE_111/2020 | 2020-01-02 | North America/Mexico/Veracruz/ | Instituto de Diagnostico y Referencia Epidemiologicos INDRE_RNLSP | Instituto de Diagnostico y Referencia Epidemiologicos (INDRE) | Claudia Wong-Arambula et al |
| EPI_ISL_913916 | Mexico/VER-InDRE_112/2020 | 2020-01-02 | North America/Mexico/Veracruz/ | Instituto de Diagnostico y Referencia Epidemiologicos INDRE_RNLSP | Instituto de Diagnostico y Referencia Epidemiologicos (INDRE) | Claudia Wong-Arambula et al |
| EPI_ISL_913917 | Mexico/MOR-InDRE_113/2020 | 2020-07-18 | North America/Mexico/Morelos/ | Instituto de Diagnostico y Referencia Epidemiologicos INDRE_RNLSP | Instituto de Diagnostico y Referencia Epidemiologicos (INDRE) | Claudia Wong-Arambula et al |
| EPI_ISL_913919 | Mexico/GTO-InDRE_115/2020 | 2020-07-22 | North America/Mexico/Guanajuato/ | Instituto de Diagnostico y Referencia Epidemiologicos INDRE_RNLSP | Instituto de Diagnostico y Referencia Epidemiologicos (INDRE) | Claudia Wong-Arambula et al |
| EPI_ISL_913920 | Mexico/VER-InDRE_116/2020 | 2020-10-13 | North America/Mexico/Veracruz/ | Instituto de Diagnostico y Referencia Epidemiologicos INDRE_RNLSP | Instituto de Diagnostico y Referencia Epidemiologicos (INDRE) | Claudia Wong-Arambula et al |
| EPI_ISL_913921 | Mexico/VER-InDRE_117/2020 | 2020-10-13 | North America/Mexico/Veracruz/ | Instituto de Diagnostico y Referencia Epidemiologicos INDRE_RNLSP | Instituto de Diagnostico y Referencia Epidemiologicos (INDRE) | Claudia Wong-Arambula et al |
| EPI_ISL_913922 | Mexico/MOR-InDRE_118/2020 | 2020-10-14 | North America/Mexico/Morelos/ | Instituto de Diagnostico y Referencia Epidemiologicos INDRE_RNLSP | Instituto de Diagnostico y Referencia Epidemiologicos (INDRE) | Claudia Wong-Arambula et al |

| | | | | | | |
|---|---|---|---|---|---|---|
| EPI_ISL_913923 | Mexico/TAM-InDRE_119/2020 | 2020-10-13 | North America/Mexico/Tamaulipas/ | Instituto de Diagnostico y Referencia Epidemiologicos INDRE_RNLSP | Instituto de Diagnostico y Referencia Epidemiologicos (INDRE) | Claudia Wong-Arambula et al |
| EPI_ISL_913924 | Mexico/GRO-InDRE_120/2020 | 2020-10-14 | North America/Mexico/Guerrero/ | Instituto de Diagnostico y Referencia Epidemiologicos INDRE_RNLSP | Instituto de Diagnostico y Referencia Epidemiologicos (INDRE) | Claudia Wong-Arambula et al |
| EPI_ISL_913925 | Mexico/MEX-InDRE_121/2020 | 2020-10-19 | North America/Mexico/Estado de Mexic | Instituto de Diagnostico y Referencia Epidemiologicos INDRE_RNLSP | Instituto de Diagnostico y Referencia Epidemiologicos (INDRE) | Claudia Wong-Arambula et al |
| EPI_ISL_913926 | Mexico/NL-InDRE_122/2020 | 2020-10-17 | North America/Mexico/Nuevo Leon/ | Instituto de Diagnostico y Referencia Epidemiologicos INDRE_RNLSP | Instituto de Diagnostico y Referencia Epidemiologicos (INDRE) | Claudia Wong-Arambula et al |
| EPI_ISL_913927 | Mexico/HGO-InDRE_123/2020 | 2020-10-20 | North America/Mexico/Hidalgo/ | Instituto de Diagnostico y Referencia Epidemiologicos INDRE_RNLSP | Instituto de Diagnostico y Referencia Epidemiologicos (INDRE) | Claudia Wong-Arambula et al |
| EPI_ISL_913928 | Mexico/MEX-InDRE_124/2020 | 2020-10-20 | North America/Mexico/Estado de Mexic | Instituto de Diagnostico y Referencia Epidemiologicos INDRE_RNLSP | Instituto de Diagnostico y Referencia Epidemiologicos (INDRE) | Claudia Wong-Arambula et al |
| EPI_ISL_913929 | Mexico/TLA-InDRE_126/2020 | 2020-10-21 | North America/Mexico/Tlaxcala/ | Instituto de Diagnostico y Referencia Epidemiologicos INDRE_RNLSP | Instituto de Diagnostico y Referencia Epidemiologicos (INDRE) | Claudia Wong-Arambula et al |
| EPI_ISL_913930 | Mexico/VER-InDRE_127/2020 | 2020-10-21 | North America/Mexico/Veracruz/ | Instituto de Diagnostico y Referencia Epidemiologicos INDRE_RNLSP | Instituto de Diagnostico y Referencia Epidemiologicos (INDRE) | Claudia Wong-Arambula et al |

| | | | | | | |
|---|---|---|---|---|---|---|
| EPI_ISL_913931 | Mexico/PUE-InDRE_128/2020 | 2020-10-21 | North America/Mexico/Puebla/ | Instituto de Diagnostico y Referencia Epidemiologicos INDRE_RNLSP | Instituto de Diagnostico y Referencia Epidemiologicos (INDRE) | Claudia Wong-Arambula et al |
| EPI_ISL_913932 | Mexico/PUE-InDRE_129/2020 | 2020-10-21 | North America/Mexico/Puebla/ | Instituto de Diagnostico y Referencia Epidemiologicos INDRE_RNLSP | Instituto de Diagnostico y Referencia Epidemiologicos (INDRE) | Claudia Wong-Arambula et al |
| EPI_ISL_913933 | Mexico/SLP-InDRE_130/2020 | 2020-11-17 | North America/Mexico/San Luis Potosi/ | Instituto de Diagnostico y Referencia Epidemiologicos INDRE_RNLSP | Instituto de Diagnostico y Referencia Epidemiologicos (INDRE) | Claudia Wong-Arambula et al |
| EPI_ISL_913934 | Mexico/CHP-InDRE_131/2020 | 2020-11-20 | North America/Mexico/Chiapas/ | Instituto de Diagnostico y Referencia Epidemiologicos INDRE_RNLSP | Instituto de Diagnostico y Referencia Epidemiologicos (INDRE) | Claudia Wong-Arambula et al |
| EPI_ISL_913935 | Mexico/CHP-InDRE_132/2020 | 2020-11-20 | North America/Mexico/Chiapas/ | Instituto de Diagnostico y Referencia Epidemiologicos INDRE_RNLSP | Instituto de Diagnostico y Referencia Epidemiologicos (INDRE) | Claudia Wong-Arambula et al |
| EPI_ISL_913936 | Mexico/BCS-InDRE_133/2020 | 2020-11-27 | North America/Mexico/Baja California S | Instituto de Diagnostico y Referencia Epidemiologicos INDRE_RNLSP | Instituto de Diagnostico y Referencia Epidemiologicos (INDRE) | Claudia Wong-Arambula et al |
| EPI_ISL_913939 | Mexico/GRO-InDRE_136/2020 | 2020-11-26 | North America/Mexico/Guerrero/ | Instituto de Diagnostico y Referencia Epidemiologicos INDRE_RNLSP | Instituto de Diagnostico y Referencia Epidemiologicos (INDRE) | Claudia Wong-Arambula et al |
| EPI_ISL_913940 | Mexico/MOR-InDRE_137/2020 | 2020-12-09 | North America/Mexico/Morelos/ | Instituto de Diagnostico y Referencia Epidemiologicos INDRE_RNLSP | Instituto de Diagnostico y Referencia Epidemiologicos (INDRE) | Claudia Wong-Arambula et al |

| | | | | | | |
|---|---|---|---|---|---|---|
| EPI_ISL_913941 | Mexico/MEX-InDRE_138/2020 | 2020-12-09 | North America/Mexico/Estado de Mexic | Instituto de Diagnostico y Referencia Epidemiologicos INDRE_RNLSP | Instituto de Diagnostico y Referencia Epidemiologicos (INDRE) | Claudia Wong-Arambula et al |
| EPI_ISL_913942 | Mexico/BC-InDRE_139/2020 | 2020-12-09 | North America/Mexico/Baja California/ | Instituto de Diagnostico y Referencia Epidemiologicos INDRE_RNLSP | Instituto de Diagnostico y Referencia Epidemiologicos (INDRE) | Claudia Wong-Arambula et al |
| EPI_ISL_913944 | Mexico/TLA-InDRE_141/2020 | 2020-12-11 | North America/Mexico/Tlaxcala/ | Instituto de Diagnostico y Referencia Epidemiologicos INDRE_RNLSP | Instituto de Diagnostico y Referencia Epidemiologicos (INDRE) | Claudia Wong-Arambula et al |
| EPI_ISL_913945 | Mexico/GTO-InDRE_142/2020 | 2020-12-08 | North America/Mexico/Guanajuato/ | Instituto de Diagnostico y Referencia Epidemiologicos INDRE_RNLSP | Instituto de Diagnostico y Referencia Epidemiologicos (INDRE) | Claudia Wong-Arambula et al |
| EPI_ISL_913946 | Mexico/QRO-InDRE_143/2020 | 2020-12-13 | North America/Mexico/Queretaro/ | Instituto de Diagnostico y Referencia Epidemiologicos INDRE_RNLSP | Instituto de Diagnostico y Referencia Epidemiologicos (INDRE) | Claudia Wong-Arambula et al |
| EPI_ISL_913947 | Mexico/QRO-InDRE_144/2020 | 2020-12-13 | North America/Mexico/Queretaro/ | Instituto de Diagnostico y Referencia Epidemiologicos INDRE_RNLSP | Instituto de Diagnostico y Referencia Epidemiologicos (INDRE) | Claudia Wong-Arambula et al |
| EPI_ISL_913948 | Mexico/SLP-InDRE_145/2020 | 2020-12-12 | North America/Mexico/San Luis Potosi/ | Instituto de Diagnostico y Referencia Epidemiologicos INDRE_RNLSP | Instituto de Diagnostico y Referencia Epidemiologicos (INDRE) | Claudia Wong-Arambula et al |
| EPI_ISL_913949 | Mexico/GRO-InDRE_146/2020 | 2020-12-12 | North America/Mexico/Guerrero/ | Instituto de Diagnostico y Referencia Epidemiologicos INDRE_RNLSP | Instituto de Diagnostico y Referencia Epidemiologicos (INDRE) | Claudia Wong-Arambula et al |

| | | | | | | |
|---|---|---|---|---|---|---|
| EPI_ISL_913950 | Mexico/TLA-InDRE_147/2020 | 2020-12-14 | North America/Mexico/Tlaxcala/ | Instituto de Diagnostico y Referencia Epidemiologicos INDRE_RNLSP | Instituto de Diagnostico y Referencia Epidemiologicos (INDRE) | Claudia Wong-Arambula et al |
| EPI_ISL_913951 | Mexico/HGO-InDRE_148/2020 | 2020-12-13 | North America/Mexico/Hidalgo/ | Instituto de Diagnostico y Referencia Epidemiologicos INDRE_RNLSP | Instituto de Diagnostico y Referencia Epidemiologicos (INDRE) | Claudia Wong-Arambula et al |
| EPI_ISL_913952 | Mexico/TAM-InDRE_149/2020 | 2020-12-15 | North America/Mexico/Tamaulipas/ | Instituto de Diagnostico y Referencia Epidemiologicos INDRE_RNLSP | Instituto de Diagnostico y Referencia Epidemiologicos (INDRE) | Claudia Wong-Arambula et al |
| EPI_ISL_913953 | Mexico/AGS-InDRE_150/2020 | 2020-12-16 | North America/Mexico/Aguascalientes/ | Instituto de Diagnostico y Referencia Epidemiologicos INDRE_RNLSP | Instituto de Diagnostico y Referencia Epidemiologicos (INDRE) | Claudia Wong-Arambula et al |
| EPI_ISL_913954 | Mexico/AGS-InDRE_151/2020 | 2020-12-16 | North America/Mexico/Aguascalientes/ | Instituto de Diagnostico y Referencia Epidemiologicos INDRE_RNLSP | Instituto de Diagnostico y Referencia Epidemiologicos (INDRE) | Claudia Wong-Arambula et al |
| EPI_ISL_913956 | Mexico/TLA-InDRE_153/2020 | 2020-07-23 | North America/Mexico/Tlaxcala/ | Instituto de Diagnostico y Referencia Epidemiologicos INDRE_RNLSP | Instituto de Diagnostico y Referencia Epidemiologicos (INDRE) | Claudia Wong-Arambula et al |
| EPI_ISL_913957 | Mexico/MOR-InDRE_155/2020 | 2020-10-20 | North America/Mexico/Morelos/ | Instituto de Diagnostico y Referencia Epidemiologicos INDRE_RNLSP | Instituto de Diagnostico y Referencia Epidemiologicos (INDRE) | Claudia Wong-Arambula et al |
| EPI_ISL_913959 | Mexico/PUE-InDRE_157/2020 | 2020-11-25 | North America/Mexico/Puebla/ | Instituto de Diagnostico y Referencia Epidemiologicos INDRE_RNLSP | Instituto de Diagnostico y Referencia Epidemiologicos (INDRE) | Claudia Wong-Arambula et al |

| | | | | | | |
|---|---|---|---|---|---|---|
| EPI_ISL_913963 | Mexico/SLP-InDRE_162/2020 | 2020-12-13 | North America/Mexico/San Luis Potosi/ | Instituto de Diagnostico y Referencia Epidemiologicos INDRE_RNLSP | Instituto de Diagnostico y Referencia Epidemiologicos (INDRE) | Claudia Wong-Arambula et al |
| EPI_ISL_913965 | Mexico/HGO-InDRE_164/2020 | 2020-12-15 | North America/Mexico/Hidalgo/ | Instituto de Diagnostico y Referencia Epidemiologicos INDRE_RNLSP | Instituto de Diagnostico y Referencia Epidemiologicos (INDRE) | Claudia Wong-Arambula et al |
| EPI_ISL_913966 | Mexico/TAM-InDRE_165/2020 | 2020-12-15 | North America/Mexico/Tamaulipas/ | Instituto de Diagnostico y Referencia Epidemiologicos INDRE_RNLSP | Instituto de Diagnostico y Referencia Epidemiologicos (INDRE) | Claudia Wong-Arambula et al |
| EPI_ISL_913967 | Mexico/CMX-InDRE_166/2020 | 2020-10-16 | North America/Mexico/Mexico City/ | Instituto de Diagnostico y Referencia Epidemiologicos INDRE_RNLSP | Instituto de Diagnostico y Referencia Epidemiologicos (INDRE) | Claudia Wong-Arambula et al |
| EPI_ISL_913969 | Mexico/CHP-InDRE_168/2020 | 2020-07-17 | North America/Mexico/Chiapas/ | Instituto de Diagnostico y Referencia Epidemiologicos INDRE_RNLSP | Instituto de Diagnostico y Referencia Epidemiologicos (INDRE) | Claudia Wong-Arambula et al |
| EPI_ISL_913970 | Mexico/VER-InDRE_169/2020 | 2020-11-21 | North America/Mexico/Veracruz/ | Instituto de Diagnostico y Referencia Epidemiologicos INDRE_RNLSP | Instituto de Diagnostico y Referencia Epidemiologicos (INDRE) | Claudia Wong-Arambula et al |
| EPI_ISL_913972 | Mexico/OAX-InDRE_171/2020 | 2020-12-07 | North America/Mexico/Oaxaca/ | Instituto de Diagnostico y Referencia Epidemiologicos INDRE_RNLSP | Instituto de Diagnostico y Referencia Epidemiologicos (INDRE) | Claudia Wong-Arambula et al |
| EPI_ISL_913973 | Mexico/MOR-InDRE_172/2020 | 2020-12-09 | North America/Mexico/Morelos/ | Instituto de Diagnostico y Referencia Epidemiologicos INDRE_RNLSP | Instituto de Diagnostico y Referencia Epidemiologicos (INDRE) | Claudia Wong-Arambula et al |

| | | | | | | |
|---|---|---|---|---|---|---|
| EPI_ISL_913974 | Mexico/GRO-InDRE_173/2020 | 2020-12-09 | North America/Mexico/Guerrero/ | Instituto de Diagnostico y Referencia Epidemiologicos INDRE_RNLSP | Instituto de Diagnostico y Referencia Epidemiologicos (INDRE) | Claudia Wong-Arambula et al |
| EPI_ISL_913975 | Mexico/TLA-InDRE_174/2020 | 2020-12-04 | North America/Mexico/Tlaxcala/ | Instituto de Diagnostico y Referencia Epidemiologicos INDRE_RNLSP | Instituto de Diagnostico y Referencia Epidemiologicos (INDRE) | Claudia Wong-Arambula et al |
| EPI_ISL_913977 | Mexico/TLA-InDRE_176/2020 | 2020-12-10 | North America/Mexico/Tlaxcala/ | Instituto de Diagnostico y Referencia Epidemiologicos INDRE_RNLSP | Instituto de Diagnostico y Referencia Epidemiologicos (INDRE) | Claudia Wong-Arambula et al |
| EPI_ISL_913978 | Mexico/MEX-InDRE_177/2020 | 2020-12-12 | North America/Mexico/Estado de Mexic | Instituto de Diagnostico y Referencia Epidemiologicos INDRE_RNLSP | Instituto de Diagnostico y Referencia Epidemiologicos (INDRE) | Claudia Wong-Arambula et al |
| EPI_ISL_913982 | Mexico/HGO-InDRE_181/2020 | 2020-12-14 | North America/Mexico/Hidalgo/ | Instituto de Diagnostico y Referencia Epidemiologicos INDRE_RNLSP | Instituto de Diagnostico y Referencia Epidemiologicos (INDRE) | Claudia Wong-Arambula et al |
| EPI_ISL_914044 | USA/AZ-TG275974/2020 | 2020-04-16 | North America/USA/Arizona/Navajo Co | TGen North | TGen North | """Jolene Bowers et al" |
| EPI_ISL_914052 | USA/AZ-TG276656/2020 | 2020-04-21 | North America/USA/Arizona/Navajo Co | TGen North | TGen North | """Jolene Bowers et al" |
| EPI_ISL_914055 | USA/AZ-TG276662/2020 | 2020-04-21 | North America/USA/Arizona/Navajo Co | TGen North | TGen North | """Jolene Bowers et al" |
| EPI_ISL_914063 | USA/AZ-TG278236/2020 | 2020-04-25 | North America/USA/Arizona/Navajo Co | TGen North | TGen North | """Jolene Bowers et al" |
| EPI_ISL_914087 | USA/AZ-TG287072/2020 | 2020-05-01 | North America/USA/Arizona/Navajo Co | TGen North | TGen North | """Jolene Bowers et al" |
| EPI_ISL_914089 | USA/AZ-TG287334/2020 | 2020-05-03 | North America/USA/Arizona/Navajo Co | TGen North | TGen North | """Jolene Bowers et al" |
| EPI_ISL_914090 | USA/AZ-TG287477/2020 | 2020-05-01 | North America/USA/Arizona/Navajo Co | TGen North | TGen North | """Jolene Bowers et al" |
| EPI_ISL_914096 | USA/AZ-TG293034/2020 | 2020-05-05 | North America/USA/Arizona/Navajo Co | TGen North | TGen North | """Jolene Bowers et al" |
| EPI_ISL_914103 | USA/AZ-TG296975/2020 | 2020-05-07 | North America/USA/Arizona/Navajo Co | TGen North | TGen North | """Jolene Bowers et al" |
| EPI_ISL_914107 | USA/AZ-TG299596/2020 | 2020-05-08 | North America/USA/Arizona/Navajo Co | TGen North | TGen North | """Jolene Bowers et al" |
| EPI_ISL_914117 | USA/AZ-TG300634/2020 | 2020-05-11 | North America/USA/Arizona/Navajo Co | TGen North | TGen North | """Jolene Bowers et al" |
| EPI_ISL_914129 | USA/AZ-TG310466/2020 | 2020-05-14 | North America/USA/Arizona/Navajo Co | TGen North | TGen North | """Jolene Bowers et al" |

| | | | | | | |
|---|---|---|---|---|---|---|
| EPI_ISL_914130 | USA/AZ-TG310608/2020 | 2020-05-14 | North America/USA/Arizona/Navajo Co | TGen North | TGen North | """Jolene Bowers et al" |
| EPI_ISL_914132 | USA/AZ-TG313351/2020 | 2020-05-15 | North America/USA/Arizona/Navajo Co | TGen North | TGen North | """Jolene Bowers et al" |
| EPI_ISL_914137 | USA/AZ-TG313505/2020 | 2020-05-15 | North America/USA/Arizona/Navajo Co | TGen North | TGen North | """Jolene Bowers et al" |
| EPI_ISL_914138 | USA/AZ-TG313675/2020 | 2020-05-15 | North America/USA/Arizona/Navajo Co | TGen North | TGen North | """Jolene Bowers et al" |
| EPI_ISL_914142 | USA/AZ-TG317127/2020 | 2020-05-18 | North America/USA/Arizona/Navajo Co | TGen North | TGen North | """Jolene Bowers et al" |
| EPI_ISL_914146 | USA/AZ-TG320174/2020 | 2020-05-19 | North America/USA/Arizona/Navajo Co | TGen North | TGen North | """Jolene Bowers et al" |
| EPI_ISL_914159 | USA/AZ-TG322532/2020 | 2020-05-20 | North America/USA/Arizona/Navajo Co | TGen North | TGen North | """Jolene Bowers et al" |
| EPI_ISL_914165 | USA/AZ-TG322957/2020 | 2020-05-20 | North America/USA/Arizona/Navajo Co | TGen North | TGen North | """Jolene Bowers et al" |
| EPI_ISL_914171 | USA/AZ-TG324459/2020 | 2020-05-21 | North America/USA/Arizona/Navajo Co | TGen North | TGen North | """Jolene Bowers et al" |
| EPI_ISL_914174 | USA/AZ-TG324583/2020 | 2020-05-21 | North America/USA/Arizona/Navajo Co | TGen North | TGen North | """Jolene Bowers et al" |
| EPI_ISL_914176 | USA/AZ-TG324719/2020 | 2020-05-21 | North America/USA/Arizona/Navajo Co | TGen North | TGen North | """Jolene Bowers et al" |
| EPI_ISL_914191 | USA/AZ-TG331280/2020 | 2020-05-26 | North America/USA/Arizona/Navajo Co | TGen North | TGen North | """Jolene Bowers et al" |
| EPI_ISL_914196 | USA/AZ-TG331775/2020 | 2020-05-27 | North America/USA/Arizona/Navajo Co | TGen North | TGen North | """Jolene Bowers et al" |
| EPI_ISL_914197 | USA/AZ-TG332004/2020 | 2020-05-27 | North America/USA/Arizona/Navajo Co | TGen North | TGen North | """Jolene Bowers et al" |
| EPI_ISL_914205 | USA/AZ-TG335282/2020 | 2020-05-28 | North America/USA/Arizona/Navajo Co | TGen North | TGen North | """Jolene Bowers et al" |
| EPI_ISL_914206 | USA/AZ-TG335579/2020 | 2020-05-28 | North America/USA/Arizona/Navajo Co | TGen North | TGen North | """Jolene Bowers et al" |
| EPI_ISL_914210 | USA/AZ-TG336685/2020 | 2020-05-27 | North America/USA/Arizona/Navajo Co | TGen North | TGen North | """Jolene Bowers et al" |
| EPI_ISL_914214 | USA/AZ-TG339996/2020 | 2020-05-30 | North America/USA/Arizona/Navajo Co | TGen North | TGen North | """Jolene Bowers et al" |
| EPI_ISL_914215 | USA/AZ-TG340000/2020 | 2020-05-29 | North America/USA/Arizona/Navajo Co | TGen North | TGen North | """Jolene Bowers et al" |
| EPI_ISL_914220 | USA/AZ-TG340364/2020 | 2020-05-29 | North America/USA/Arizona/Navajo Co | TGen North | TGen North | """Jolene Bowers et al" |
| EPI_ISL_914223 | USA/AZ-TG340741/2020 | 2020-05-29 | North America/USA/Arizona/Navajo Co | TGen North | TGen North | """Jolene Bowers et al" |
| EPI_ISL_914224 | USA/AZ-TG340789/2020 | 2020-05-30 | North America/USA/Arizona/Navajo Co | TGen North | TGen North | """Jolene Bowers et al" |
| EPI_ISL_914231 | USA/AZ-TG343481/2020 | 2020-06-01 | North America/USA/Arizona/Navajo Co | TGen North | TGen North | """Jolene Bowers et al" |
| EPI_ISL_914234 | USA/AZ-TG344110/2020 | 2020-06-01 | North America/USA/Arizona/Navajo Co | TGen North | TGen North | """Jolene Bowers et al" |
| EPI_ISL_914236 | USA/AZ-TG345700/2020 | 2020-05-29 | North America/USA/Arizona/Navajo Co | TGen North | TGen North | """Jolene Bowers et al" |
| EPI_ISL_914237 | USA/AZ-TG345751/2020 | 2020-06-01 | North America/USA/Arizona/Navajo Co | TGen North | TGen North | """Jolene Bowers et al" |
| EPI_ISL_914241 | USA/AZ-TG345831/2020 | 2020-05-29 | North America/USA/Arizona/Navajo Co | TGen North | TGen North | """Jolene Bowers et al" |
| EPI_ISL_914243 | USA/AZ-TG346570/2020 | 2020-06-02 | North America/USA/Arizona/Navajo Co | TGen North | TGen North | """Jolene Bowers et al" |
| EPI_ISL_914258 | USA/AZ-TG352531/2020 | 2020-06-04 | North America/USA/Arizona/Navajo Co | TGen North | TGen North | """Jolene Bowers et al" |
| EPI_ISL_914264 | USA/AZ-TG356405/2020 | 2020-06-05 | North America/USA/Arizona/Navajo Co | TGen North | TGen North | """Jolene Bowers et al" |
| EPI_ISL_914266 | USA/AZ-TG356423/2020 | 2020-06-05 | North America/USA/Arizona/Navajo Co | TGen North | TGen North | """Jolene Bowers et al" |

| | | | | | | |
|---|---|---|---|---|---|---|
| EPI_ISL_914285 | USA/AZ-TG357835/2020 | 2020-06-06 | North America/USA/Arizona/Navajo Co | TGen North | TGen North | """Jolene Bowers et al" |
| EPI_ISL_914292 | USA/AZ-TG359191/2020 | 2020-06-08 | North America/USA/Arizona/Navajo Co | TGen North | TGen North | """Jolene Bowers et al" |
| EPI_ISL_914306 | USA/AZ-TG361527/2020 | 2020-06-09 | North America/USA/Arizona/Navajo Co | TGen North | TGen North | """Jolene Bowers et al" |
| EPI_ISL_914307 | USA/AZ-TG361530/2020 | 2020-06-09 | North America/USA/Arizona/Navajo Co | TGen North | TGen North | """Jolene Bowers et al" |
| EPI_ISL_914309 | USA/AZ-TG361706/2020 | 2020-06-09 | North America/USA/Arizona/Navajo Co | TGen North | TGen North | """Jolene Bowers et al" |
| EPI_ISL_914316 | USA/AZ-TG363046/2020 | 2020-06-09 | North America/USA/Arizona/Navajo Co | TGen North | TGen North | """Jolene Bowers et al" |
| EPI_ISL_914317 | USA/AZ-TG364597/2020 | 2020-06-10 | North America/USA/Arizona/Navajo Co | TGen North | TGen North | """Jolene Bowers et al" |
| EPI_ISL_914362 | USA/AZ-TG377874/2020 | 2020-06-15 | North America/USA/Arizona/Navajo Co | TGen North | TGen North | """Jolene Bowers et al" |
| EPI_ISL_914366 | USA/AZ-TG378459/2020 | 2020-06-15 | North America/USA/Arizona/Navajo Co | TGen North | TGen North | """Jolene Bowers et al" |
| EPI_ISL_914367 | USA/AZ-TG378460/2020 | 2020-06-12 | North America/USA/Arizona/Navajo Co | TGen North | TGen North | """Jolene Bowers et al" |
| EPI_ISL_914373 | USA/AZ-TG379978/2020 | 2020-06-12 | North America/USA/Arizona/Navajo Co | TGen North | TGen North | """Jolene Bowers et al" |
| EPI_ISL_914374 | USA/AZ-TG380910/2020 | 2020-06-16 | North America/USA/Arizona/Navajo Co | TGen North | TGen North | """Jolene Bowers et al" |
| EPI_ISL_914382 | USA/AZ-TG382931/2020 | 2020-06-16 | North America/USA/Arizona/Navajo Co | TGen North | TGen North | """Jolene Bowers et al" |
| EPI_ISL_914392 | USA/AZ-TG390559/2020 | 2020-06-18 | North America/USA/Arizona/Navajo Co | TGen North | TGen North | """Jolene Bowers et al" |
| EPI_ISL_914407 | USA/AZ-TG397340/2020 | 2020-06-19 | North America/USA/Arizona/Navajo Co | TGen North | TGen North | """Jolene Bowers et al" |
| EPI_ISL_914410 | USA/AZ-TG397405/2020 | 2020-06-20 | North America/USA/Arizona/Navajo Co | TGen North | TGen North | """Jolene Bowers et al" |
| EPI_ISL_914414 | USA/AZ-TG397527/2020 | 2020-06-21 | North America/USA/Arizona/Navajo Co | TGen North | TGen North | """Jolene Bowers et al" |
| EPI_ISL_914415 | USA/AZ-TG399359/2020 | 2020-06-22 | North America/USA/Arizona/Maricopa ( | TGen North | TGen North | """Jolene Bowers et al" |
| EPI_ISL_914419 | USA/AZ-TG400413/2020 | 2020-06-22 | North America/USA/Arizona/Navajo Co | TGen North | TGen North | """Jolene Bowers et al" |
| EPI_ISL_914425 | USA/AZ-TG405426/2020 | 2020-06-22 | North America/USA/Arizona/Navajo Co | TGen North | TGen North | """Jolene Bowers et al" |
| EPI_ISL_914438 | USA/AZ-TG417303/2020 | 2020-06-24 | North America/USA/Arizona/Navajo Co | TGen North | TGen North | """Jolene Bowers et al" |
| EPI_ISL_914463 | USA/AZ-TG451644/2020 | 2020-06-26 | North America/USA/Arizona/Navajo Co | TGen North | TGen North | """Jolene Bowers et al" |
| EPI_ISL_914465 | USA/AZ-TG451876/2020 | 2020-06-26 | North America/USA/Arizona/Navajo Co | TGen North | TGen North | """Jolene Bowers et al" |
| EPI_ISL_914471 | USA/AZ-TG457096/2020 | 2020-06-26 | North America/USA/Arizona/Navajo Co | TGen North | TGen North | """Jolene Bowers et al" |
| EPI_ISL_914472 | USA/AZ-TG457119/2020 | 2020-06-26 | North America/USA/Arizona/Navajo Co | TGen North | TGen North | """Jolene Bowers et al" |
| EPI_ISL_914477 | USA/AZ-TG459858/2020 | 2020-06-26 | North America/USA/Arizona/Navajo Co | TGen North | TGen North | """Jolene Bowers et al" |
| EPI_ISL_914480 | USA/AZ-TG460326/2020 | 2020-06-30 | North America/USA/Arizona/Navajo Co | TGen North | TGen North | """Jolene Bowers et al" |
| EPI_ISL_914494 | USA/AZ-TG473351/2020 | 2020-07-06 | North America/USA/Arizona/Navajo Co | TGen North | TGen North | """Jolene Bowers et al" |
| EPI_ISL_914504 | USA/AZ-TG477083/2020 | 2020-07-07 | North America/USA/Arizona/Navajo Co | TGen North | TGen North | """Jolene Bowers et al" |
| EPI_ISL_914509 | USA/AZ-TG482022/2020 | 2020-07-08 | North America/USA/Arizona/Navajo Co | TGen North | TGen North | """Jolene Bowers et al" |
| EPI_ISL_914512 | USA/AZ-TG482131/2020 | 2020-07-08 | North America/USA/Arizona/Navajo Co | TGen North | TGen North | """Jolene Bowers et al" |

| | | | | | | |
|---|---|---|---|---|---|---|
| EPI_ISL_914514 | USA/AZ-TG482138/2020 | 2020-07-08 | North America/USA/Arizona/Navajo Co | TGen North | TGen North | """Jolene Bowers et al" |
| EPI_ISL_914516 | USA/AZ-TG482324/2020 | 2020-07-08 | North America/USA/Arizona/Navajo Co | TGen North | TGen North | """Jolene Bowers et al" |
| EPI_ISL_914523 | USA/AZ-TG488515/2020 | 2020-07-09 | North America/USA/Arizona/Navajo Co | TGen North | TGen North | """Jolene Bowers et al" |
| EPI_ISL_914526 | USA/AZ-TG488755/2020 | 2020-07-09 | North America/USA/Arizona/Navajo Co | TGen North | TGen North | """Jolene Bowers et al" |
| EPI_ISL_914527 | USA/AZ-TG489928/2020 | 2020-07-09 | North America/USA/Arizona/Navajo Co | TGen North | TGen North | """Jolene Bowers et al" |
| EPI_ISL_914536 | USA/AZ-TG492371/2020 | 2020-07-12 | North America/USA/Arizona/Navajo Co | TGen North | TGen North | """Jolene Bowers et al" |
| EPI_ISL_914556 | USA/AZ-TG509603/2020 | 2020-07-16 | North America/USA/Arizona/Navajo Co | TGen North | TGen North | """Jolene Bowers et al" |
| EPI_ISL_914557 | USA/AZ-TG509632/2020 | 2020-07-16 | North America/USA/Arizona/Navajo Co | TGen North | TGen North | """Jolene Bowers et al" |
| EPI_ISL_914559 | USA/AZ-TG509853/2020 | 2020-07-16 | North America/USA/Arizona/Navajo Co | TGen North | TGen North | """Jolene Bowers et al" |
| EPI_ISL_914561 | USA/AZ-TG510208/2020 | 2020-07-16 | North America/USA/Arizona/Navajo Co | TGen North | TGen North | """Jolene Bowers et al" |
| EPI_ISL_914564 | USA/AZ-TG513270/2020 | 2020-07-16 | North America/USA/Arizona/Navajo Co | TGen North | TGen North | """Jolene Bowers et al" |
| EPI_ISL_914565 | USA/AZ-TG513437/2020 | 2020-07-15 | North America/USA/Arizona/Navajo Co | TGen North | TGen North | """Jolene Bowers et al" |
| EPI_ISL_914566 | USA/AZ-TG513543/2020 | 2020-07-16 | North America/USA/Arizona/Navajo Co | TGen North | TGen North | """Jolene Bowers et al" |
| EPI_ISL_914571 | USA/AZ-TG517027/2020 | 2020-07-19 | North America/USA/Arizona/Navajo Co | TGen North | TGen North | """Jolene Bowers et al" |
| EPI_ISL_914588 | USA/AZ-TG569348/2020 | 2020-08-02 | North America/USA/Arizona/Navajo Co | TGen North | TGen North | """Jolene Bowers et al" |
| EPI_ISL_914611 | USA/AZ-TG594700/2020 | 2020-08-09 | North America/USA/Arizona/Navajo Co | TGen North | TGen North | """Jolene Bowers et al" |
| EPI_ISL_914638 | USA/CA-SC210051/2021 | 2021-01-20 | North America/USA/California/Santa Cl | Santa Clara County Public Health Laboratory | Santa Clara County Public Health Laboratory | Santa Clara County Public Health Department et al |
| EPI_ISL_914644 | USA/CA-SC210066/2020 | 2020-12-30 | North America/USA/California/Santa Cl | Santa Clara County Public Health Laboratory | Santa Clara County Public Health Laboratory | Santa Clara County Public Health Department et al |
| EPI_ISL_914663 | USA/UT-UPHL-2101559813/2020 | 2020-12-28 | North America/USA/Utah/ | Utah Public Health Laboratory | Utah Public Health Laboratory | Erin L. Young et al |
| EPI_ISL_914666 | USA/UT-UPHL-2101921649/2020 | 2020-12-28 | North America/USA/Utah/ | Utah Public Health Laboratory | Utah Public Health Laboratory | Erin L. Young et al |
| EPI_ISL_914681 | USA/UT-UPHL-2101793177/2020 | 2020-12-23 | North America/USA/Utah/ | Utah Public Health Laboratory | Utah Public Health Laboratory | Erin L. Young et al |
| EPI_ISL_914682 | USA/UT-UPHL-2101412272/2020 | 2020-12-22 | North America/USA/Utah/ | Utah Public Health Laboratory | Utah Public Health Laboratory | Erin L. Young et al |
| EPI_ISL_914684 | USA/UT-UPHL-2101125404/2020 | 2020-12-23 | North America/USA/Utah/ | Utah Public Health Laboratory | Utah Public Health Laboratory | Erin L. Young et al |
| EPI_ISL_914685 | USA/UT-UPHL-2101533926/2020 | 2020-12-23 | North America/USA/Utah/ | Utah Public Health Laboratory | Utah Public Health Laboratory | Erin L. Young et al |

| | | | | | | |
|---|---|---|---|---|---|---|
| EPI_ISL_914686 | USA/UT-UPHL-2101502643/2020 | 2020-12-23 | North America/USA/Utah/ | Utah Public Health Laboratory | Utah Public Health Laboratory | Erin L. Young et al |
| EPI_ISL_914693 | USA/UT-UPHL-2101366458/2020 | 2020-12-25 | North America/USA/Utah/ | Utah Public Health Laboratory | Utah Public Health Laboratory | Erin L. Young et al |
| EPI_ISL_914700 | USA/UT-UPHL-2101809199/2020 | 2020-12-28 | North America/USA/Utah/ | Utah Public Health Laboratory | Utah Public Health Laboratory | Erin L. Young et al |
| EPI_ISL_914708 | USA/MD-MDH-0700/2021 | 2021-01-19 | North America/USA/Maryland/ | Maryland Public Health Laboratory (MD PHL) | Maryland Public Health Laboratory (MD PHL) | Maryland Department of Health Laboratories Administration et al |
| EPI_ISL_914709 | USA/MD-MDH-0701/2021 | 2021-01-19 | North America/USA/Maryland/ | Maryland Public Health Laboratory (MD PHL) | Maryland Public Health Laboratory (MD PHL) | Maryland Department of Health Laboratories Administration et al |
| EPI_ISL_914719 | USA/MD-MDH-0712/2021 | 2021-01-19 | North America/USA/Maryland/ | Maryland Public Health Laboratory (MD PHL) | Maryland Public Health Laboratory (MD PHL) | Maryland Department of Health Laboratories Administration et al |
| EPI_ISL_914732 | USA/WY-WPHL-21006606/2021 | 2021-01-18 | North America/USA/Wyoming/ | Wyoming Public Health Laboratory | Wyoming Public Health Laboratory | Noah Hull et al |
| EPI_ISL_914736 | USA/WY-WPHL-21006749/2021 | 2021-01-18 | North America/USA/Wyoming/ | Wyoming Public Health Laboratory | Wyoming Public Health Laboratory | Noah Hull et al |
| EPI_ISL_914740 | USA/WY-WPHL-21006863/2021 | 2021-01-18 | North America/USA/Wyoming/ | Wyoming Public Health Laboratory | Wyoming Public Health Laboratory | Noah Hull et al |
| EPI_ISL_914769 | USA/WY-WPHL-21007645/2021 | 2021-01-20 | North America/USA/Wyoming/ | Wyoming Public Health Laboratory | Wyoming Public Health Laboratory | Noah Hull et al |
| EPI_ISL_914774 | USA/WY-WPHL-21007847/2021 | 2021-01-21 | North America/USA/Wyoming/ | Wyoming Public Health Laboratory | Wyoming Public Health Laboratory | Noah Hull et al |
| EPI_ISL_914781 | USA/WY-WPHL-21006688/2021 | 2021-01-18 | North America/USA/Wyoming/ | Wyoming Public Health Laboratory | Wyoming Public Health Laboratory | Noah Hull et al |
| EPI_ISL_914784 | USA/WY-WPHL-21006509/2021 | 2021-01-18 | North America/USA/Wyoming/ | Wyoming Public Health Laboratory | Wyoming Public Health Laboratory | Noah Hull et al |
| EPI_ISL_914787 | USA/WY-WPHL-21006656/2021 | 2021-01-18 | North America/USA/Wyoming/ | Wyoming Public Health Laboratory | Wyoming Public Health Laboratory | Noah Hull et al |
| EPI_ISL_914792 | USA/WY-WPHL-21007172/2021 | 2021-01-19 | North America/USA/Wyoming/ | Wyoming Public Health Laboratory | Wyoming Public Health Laboratory | Noah Hull et al |
| EPI_ISL_914796 | CostaRica/INC-0189/2020 | 2020-12-04 | North America/Costa Rica/Puntarenas/C | TAMIZAJE COMUNITARIO - PASO CANOAS | Inciensa, Instituto Costarricense de Investigaci√≥n y Ense√±anza en Nutrici√≥n y Salud | Francisco Duarte et al |

| | | | | | | |
|---|---|---|---|---|---|---|
| EPI_ISL_914802 | CostaRica/INC-0196/2020 | 2020-12-16 | North America/Costa Rica/San Jose/Gra | AREA DE SALUD CATEDRAL NORESTE | Inciensa, Instituto Costarricense de Investigaci√≥n y Ense√±anza en Nutrici√≥n y Salud | Francisco Duarte et al |
| EPI_ISL_914803 | CostaRica/INC-0197/2020 | 2020-12-18 | North America/Costa Rica/Cartago CR/T | AREA DE SALUD LA UNION | Inciensa, Instituto Costarricense de Investigaci√≥n y Ense√±anza en Nutrici√≥n y Salud | Francisco Duarte et al |
| EPI_ISL_914804 | CostaRica/INC-0198/2020 | 2020-12-14 | North America/Costa Rica/Cartago CR/S | AREA DE SALUD TURRIALBA-JIMENEZ | Inciensa, Instituto Costarricense de Investigaci√≥n y Ense√±anza en Nutrici√≥n y Salud | Francisco Duarte et al |
| EPI_ISL_914805 | CostaRica/INC-0199/2020 | 2020-12-16 | North America/Costa Rica/Cartago CR/C | AREA DE SALUD CARTAGO | Inciensa, Instituto Costarricense de Investigaci√≥n y Ense√±anza en Nutrici√≥n y Salud | Francisco Duarte et al |
| EPI_ISL_914806 | CostaRica/INC-0200/2020 | 2020-12-17 | North America/Costa Rica/San Jose/Pat | AREA DE SALUD CORONADO | Inciensa, Instituto Costarricense de Investigaci√≥n y Ense√±anza en Nutrici√≥n y Salud | Francisco Duarte et al |
| EPI_ISL_914809 | CostaRica/INC-0203/2020 | 2020-12-18 | North America/Costa Rica/Cartago CR/C | AREA DE SALUD OREAMUNO-PACAYAS-TIERRA BLANCA | Inciensa, Instituto Costarricense de Investigaci√≥n y Ense√±anza en Nutrici√≥n y Salud | Francisco Duarte et al |
| EPI_ISL_914818 | CostaRica/INC-0212/2021 | 2021-01-01 | North America/Costa Rica/Limon/Guapil | HOSPITAL DE NI√ëOS DR. CARLOS SAENZ HERRERA | Inciensa, Instituto Costarricense de Investigaci√≥n y Ense√±anza en Nutrici√≥n y Salud | Francisco Duarte et al |
| EPI_ISL_914819 | CostaRica/INC-0213/2020 | 2020-12-31 | North America/Costa Rica/Heredia/Sara | HOSPITAL DE NI√ëOS DR. CARLOS SAENZ HERRERA | Inciensa, Instituto Costarricense de Investigaci√≥n y Ense√±anza en Nutrici√≥n y Salud | Francisco Duarte et al |

| | | | | | | |
|---|---|---|---|---|---|---|
| EPI_ISL_914822 | CostaRica/INC-0216/2020 | 2020-12-25 | North America/Costa Rica/Guanacaste/E | HOSPITAL DR. ENRIQUE BALTODANO BRICE√ëO | Inciensa, Instituto Costarricense de Investigaci√≥n y Ense√±anza en Nutrici√≥n y Salud | Francisco Duarte et al |
| EPI_ISL_914827 | CostaRica/INC-0221/2021 | 2021-01-07 | North America/Costa Rica/Puntarenas/G | HLE - ASOCIACION DE ALBERGUE PARA ANCIANOS DE GOLFITO | Inciensa, Instituto Costarricense de Investigaci√≥n y Ense√±anza en Nutrici√≥n y Salud | Francisco Duarte et al |
| EPI_ISL_914829 | CostaRica/INC-0223/2021 | 2021-01-11 | North America/Costa Rica/Puntarenas/C | TAMIZAJE COMUNITARIO - PASO CANOAS | Inciensa, Instituto Costarricense de Investigaci√≥n y Ense√±anza en Nutrici√≥n y Salud | Francisco Duarte et al |
| EPI_ISL_914835 | CostaRica/INC-0229/2021 | 2021-01-14 | North America/Costa Rica/Cartago CR/C | AREA DE SALUD PARAISO-CERVANTES | Inciensa, Instituto Costarricense de Investigaci√≥n y Ense√±anza en Nutrici√≥n y Salud | Francisco Duarte et al |
| EPI_ISL_914838 | CostaRica/INC-0232/2021 | 2021-01-12 | North America/Costa Rica/Alajuela/Cutr | HOSPITAL MEXICO | Inciensa, Instituto Costarricense de Investigaci√≥n y Ense√±anza en Nutrici√≥n y Salud | Francisco Duarte et al |
| EPI_ISL_914839 | CostaRica/INC-0233/2021 | 2021-01-13 | North America/Costa Rica/Alajuela/San | HOSPITAL MEXICO | Inciensa, Instituto Costarricense de Investigaci√≥n y Ense√±anza en Nutrici√≥n y Salud | Francisco Duarte et al |
| EPI_ISL_914840 | CostaRica/INC-0234/2021 | 2021-01-19 | North America/Costa Rica/San Jose/San | HOSPITAL DR. RAUL BLANCO CERVANTES | Inciensa, Instituto Costarricense de Investigaci√≥n y Ense√±anza en Nutrici√≥n y Salud | Francisco Duarte et al |
| EPI_ISL_914851 | USA/VA-DCLS-0972/2020 | 2020-05-28 | North America/USA/Virginia/ | Virginia Division of Consolidated Laboratory Services (DCLS) | Virginia Division of Consolidated Laboratory Services (DCLS) | Virginia DCLS et al |

| | | | | | | |
|---|---|---|---|---|---|---|
| EPI_ISL_914861 | USA/VA-DCLS-0988/2020 | 2020-05-29 | North America/USA/Virginia/ | Virginia Division of Consolidated Laboratory Services (DCLS) | Virginia Division of Consolidated Laboratory Services (DCLS) | Virginia DCLS et al |
| EPI_ISL_914879 | Mexico/CMX-InDRE_154/2020 | 2020-07-23 | North America/Mexico/Mexico City/ | Instituto de Diagnostico y Referencia Epidemiologicos INDRE_RNLSP | Instituto de Diagnostico y Referencia Epidemiologicos (INDRE) | Claudia Wong-Arambula et al |
| EPI_ISL_914880 | Mexico/HID-InDRE_161/2020 | 2020-12-14 | North America/Mexico/Hidalgo/ | Instituto de Diagnostico y Referencia Epidemiologicos INDRE_RNLSP | Instituto de Diagnostico y Referencia Epidemiologicos (INDRE) | Claudia Wong-Arambula et al |
| EPI_ISL_914884 | Lithuania/VULSK-347/2020 | 2020-10-30 | Europe/Lithuania/Vilniaus Apskritis/ | Vilnius university hospital Santaros Klinikos, Center of Laboratory Medicine | Vilnius University Hospital Santaros Klinikos | Ingrida Olendraite et al |
| EPI_ISL_914886 | Lithuania/VULSK-397/2020 | 2020-10-31 | Europe/Lithuania/Vilniaus Apskritis/ | Vilnius university hospital Santaros Klinikos, Center of Laboratory Medicine | Vilnius University Hospital Santaros Klinikos | Ingrida Olendraite et al |
| EPI_ISL_914887 | Lithuania/VULSK-4430/2020 | 2020-12-21 | Europe/Lithuania/Vilniaus Apskritis/ | Vilnius university hospital Santaros Klinikos, Center of Laboratory Medicine | Vilnius University Hospital Santaros Klinikos | Ingrida Olendraite et al |
| EPI_ISL_914893 | USA/CA-QDX-4344/2021 | 2021-01-14 | North America/USA/California/ | Quest Diagnostics | Quest Diagnostics | Rosenthal et al |
| EPI_ISL_914896 | USA/CA-QDX-4348/2021 | 2021-01-14 | North America/USA/California/ | Quest Diagnostics | Quest Diagnostics | Rosenthal et al |
| EPI_ISL_914900 | USA/CA-QDX-4353/2021 | 2021-01-14 | North America/USA/California/ | Quest Diagnostics | Quest Diagnostics | Rosenthal et al |
| EPI_ISL_914904 | USA/CA-QDX-4357/2021 | 2021-01-14 | North America/USA/California/ | Quest Diagnostics | Quest Diagnostics | Rosenthal et al |
| EPI_ISL_914905 | USA/CA-QDX-4358/2021 | 2021-01-14 | North America/USA/California/ | Quest Diagnostics | Quest Diagnostics | Rosenthal et al |
| EPI_ISL_914918 | USA/CA-QDX-4373/2021 | 2021-01-14 | North America/USA/California/ | Quest Diagnostics | Quest Diagnostics | Rosenthal et al |
| EPI_ISL_914934 | USA/CA-QDX-4389/2021 | 2021-01-14 | North America/USA/California/ | Quest Diagnostics | Quest Diagnostics | Rosenthal et al |
| EPI_ISL_914944 | USA/CA-QDX-4401/2021 | 2021-01-15 | North America/USA/California/ | Quest Diagnostics | Quest Diagnostics | Rosenthal et al |
| EPI_ISL_914945 | USA/CA-QDX-4402/2021 | 2021-01-14 | North America/USA/California/ | Quest Diagnostics | Quest Diagnostics | Rosenthal et al |
| EPI_ISL_914947 | USA/CA-QDX-4404/2021 | 2021-01-14 | North America/USA/California/ | Quest Diagnostics | Quest Diagnostics | Rosenthal et al |
| EPI_ISL_914950 | USA/CA-QDX-4407/2021 | 2021-01-14 | North America/USA/California/ | Quest Diagnostics | Quest Diagnostics | Rosenthal et al |
| EPI_ISL_914961 | USA/CA-QDX-4420/2021 | 2021-01-14 | North America/USA/California/ | Quest Diagnostics | Quest Diagnostics | Rosenthal et al |
| EPI_ISL_914962 | USA/CA-QDX-4421/2021 | 2021-01-14 | North America/USA/California/ | Quest Diagnostics | Quest Diagnostics | Rosenthal et al |

| | | | | | | |
|---|---|---|---|---|---|---|
| EPI_ISL_914966 | USA/CA-QDX-4425/2021 | 2021-01-14 | North America/USA/California/ | Quest Diagnostics | Quest Diagnostics | Rosenthal et al |
| EPI_ISL_914968 | USA/CA-QDX-4427/2021 | 2021-01-13 | North America/USA/California/ | Quest Diagnostics | Quest Diagnostics | Rosenthal et al |
| EPI_ISL_914971 | Canada/NS-NML-172/2020 | 2020-04-18 | North America/Canada/Nova Scotia/ | QEII Health Sciences Centre | National Microbiology Laboratory (NML) | Anna Majer et al |
| EPI_ISL_914972 | Canada/NS-NML-173/2020 | 2020-04-18 | North America/Canada/Nova Scotia/ | QEII Health Sciences Centre | National Microbiology Laboratory (NML) | Anna Majer et al |
| EPI_ISL_914973 | Canada/NS-NML-176/2020 | 2020-04-26 | North America/Canada/Nova Scotia/ | QEII Health Sciences Centre | National Microbiology Laboratory (NML) | Anna Majer et al |
| EPI_ISL_914974 | Canada/NS-NML-180/2020 | 2020-04-27 | North America/Canada/Nova Scotia/ | QEII Health Sciences Centre | National Microbiology Laboratory (NML) | Anna Majer et al |
| EPI_ISL_914975 | Canada/NS-NML-181/2020 | 2020-04-27 | North America/Canada/Nova Scotia/ | QEII Health Sciences Centre | National Microbiology Laboratory (NML) | Anna Majer et al |
| EPI_ISL_914976 | Canada/NS-NML-892/2020 | 2020-03-20 | North America/Canada/Nova Scotia/ | QEII Health Sciences Centre | National Microbiology Laboratory (NML) | Anna Majer et al |
| EPI_ISL_914977 | Canada/NS-NML-894/2020 | 2020-03-20 | North America/Canada/Nova Scotia/ | QEII Health Sciences Centre | National Microbiology Laboratory (NML) | Anna Majer et al |
| EPI_ISL_914978 | Canada/NS-NML-911/2020 | 2020-03-22 | North America/Canada/Nova Scotia/ | QEII Health Sciences Centre | National Microbiology Laboratory (NML) | Anna Majer et al |
| EPI_ISL_914979 | Canada/NS-NML-912/2020 | 2020-03-22 | North America/Canada/Nova Scotia/ | QEII Health Sciences Centre | National Microbiology Laboratory (NML) | Anna Majer et al |
| EPI_ISL_914980 | Canada/NS-NML-921/2020 | 2020-03-23 | North America/Canada/Nova Scotia/ | QEII Health Sciences Centre | National Microbiology Laboratory (NML) | Anna Majer et al |
| EPI_ISL_914981 | Canada/NS-NML-956/2020 | 2020-03-27 | North America/Canada/Nova Scotia/ | QEII Health Sciences Centre | National Microbiology Laboratory (NML) | Anna Majer et al |
| EPI_ISL_914982 | Canada/NS-NML-959/2020 | 2020-03-28 | North America/Canada/Nova Scotia/ | QEII Health Sciences Centre | National Microbiology Laboratory (NML) | Anna Majer et al |

| | | | | | | |
|---|---|---|---|---|---|---|
| EPI_ISL_914983 | Canada/NS-NML-962/2020 | 2020-03-29 | North America/Canada/Nova Scotia/ | QEII Health Sciences Centre | National Microbiology Laboratory (NML) | Anna Majer et al |
| EPI_ISL_914984 | Canada/NS-NML-969/2020 | 2020-03-30 | North America/Canada/Nova Scotia/ | QEII Health Sciences Centre | National Microbiology Laboratory (NML) | Anna Majer et al |
| EPI_ISL_914985 | Canada/NS-NML-1215/2020 | 2020-04-03 | North America/Canada/Nova Scotia/ | QEII Health Sciences Centre | National Microbiology Laboratory (NML) | Anna Majer et al |
| EPI_ISL_914986 | Canada/NS-NML-1233/2020 | 2020-04-04 | North America/Canada/Nova Scotia/ | QEII Health Sciences Centre | National Microbiology Laboratory (NML) | Anna Majer et al |
| EPI_ISL_914987 | Canada/NS-NML-1240/2020 | 2020-04-05 | North America/Canada/Nova Scotia/ | QEII Health Sciences Centre | National Microbiology Laboratory (NML) | Anna Majer et al |
| EPI_ISL_914988 | Canada/NS-NML-1258/2020 | 2020-04-06 | North America/Canada/Nova Scotia/ | QEII Health Sciences Centre | National Microbiology Laboratory (NML) | Anna Majer et al |
| EPI_ISL_914989 | Canada/NS-NML-1260/2020 | 2020-04-06 | North America/Canada/Nova Scotia/ | QEII Health Sciences Centre | National Microbiology Laboratory (NML) | Anna Majer et al |
| EPI_ISL_914990 | Canada/NS-NML-1279/2020 | 2020-04-16 | North America/Canada/Nova Scotia/ | QEII Health Sciences Centre | National Microbiology Laboratory (NML) | Anna Majer et al |
| EPI_ISL_914991 | Canada/NS-NML-1283/2020 | 2020-04-07 | North America/Canada/Nova Scotia/ | QEII Health Sciences Centre | National Microbiology Laboratory (NML) | Anna Majer et al |
| EPI_ISL_914992 | Canada/NS-NML-1300/2020 | 2020-04-09 | North America/Canada/Nova Scotia/ | QEII Health Sciences Centre | National Microbiology Laboratory (NML) | Anna Majer et al |
| EPI_ISL_914993 | Canada/NS-NML-1360/2020 | 2020-04-14 | North America/Canada/Nova Scotia/ | QEII Health Sciences Centre | National Microbiology Laboratory (NML) | Anna Majer et al |
| EPI_ISL_914994 | Canada/NS-NML-1712/2020 | 2020-03-29 | North America/Canada/Nova Scotia/ | QEII Health Sciences Centre | National Microbiology Laboratory (NML) | Anna Majer et al |
| EPI_ISL_914995 | Canada/NS-NML-1714/2020 | 2020-03-29 | North America/Canada/Nova Scotia/ | QEII Health Sciences Centre | National Microbiology Laboratory (NML) | Anna Majer et al |

| | | | | | | |
|---|---|---|---|---|---|---|
| EPI_ISL_914996 | Canada/NS-NML-1727/2020 | 2020-03-30 | North America/Canada/Nova Scotia/ | QEII Health Sciences Centre | National Microbiology Laboratory (NML) | Anna Majer et al |
| EPI_ISL_914997 | Canada/NS-NML-1742/2020 | 2020-03-31 | North America/Canada/Nova Scotia/ | QEII Health Sciences Centre | National Microbiology Laboratory (NML) | Anna Majer et al |
| EPI_ISL_914998 | Canada/NS-NML-1764/2020 | 2020-04-02 | North America/Canada/Nova Scotia/ | QEII Health Sciences Centre | National Microbiology Laboratory (NML) | Anna Majer et al |
| EPI_ISL_914999 | Canada/NS-NML-1770/2020 | 2020-04-02 | North America/Canada/Nova Scotia/ | QEII Health Sciences Centre | National Microbiology Laboratory (NML) | Anna Majer et al |
| EPI_ISL_915000 | Canada/NS-NML-1773/2020 | 2020-04-02 | North America/Canada/Nova Scotia/ | QEII Health Sciences Centre | National Microbiology Laboratory (NML) | Anna Majer et al |
| EPI_ISL_915001 | Canada/NS-NML-1775/2020 | 2020-04-03 | North America/Canada/Nova Scotia/ | QEII Health Sciences Centre | National Microbiology Laboratory (NML) | Anna Majer et al |
| EPI_ISL_915002 | Canada/NS-NML-1785/2020 | 2020-04-03 | North America/Canada/Nova Scotia/ | QEII Health Sciences Centre | National Microbiology Laboratory (NML) | Anna Majer et al |
| EPI_ISL_915003 | Canada/NS-NML-1798/2020 | 2020-04-04 | North America/Canada/Nova Scotia/ | QEII Health Sciences Centre | National Microbiology Laboratory (NML) | Anna Majer et al |
| EPI_ISL_915004 | Canada/NS-NML-1819/2020 | 2020-04-06 | North America/Canada/Nova Scotia/ | QEII Health Sciences Centre | National Microbiology Laboratory (NML) | Anna Majer et al |
| EPI_ISL_915005 | Canada/NS-NML-1820/2020 | 2020-04-06 | North America/Canada/Nova Scotia/ | QEII Health Sciences Centre | National Microbiology Laboratory (NML) | Anna Majer et al |
| EPI_ISL_915006 | Canada/NS-NML-1824/2020 | 2020-04-06 | North America/Canada/Nova Scotia/ | QEII Health Sciences Centre | National Microbiology Laboratory (NML) | Anna Majer et al |
| EPI_ISL_915007 | Canada/NS-NML-1836/2020 | 2020-04-08 | North America/Canada/Nova Scotia/ | QEII Health Sciences Centre | National Microbiology Laboratory (NML) | Anna Majer et al |
| EPI_ISL_915008 | Canada/NS-NML-1854/2020 | 2020-04-09 | North America/Canada/Nova Scotia/ | QEII Health Sciences Centre | National Microbiology Laboratory (NML) | Anna Majer et al |

| | | | | | | |
|---|---|---|---|---|---|---|
| EPI_ISL_915009 | Canada/NS-NML-1886/2020 | 2020-04-13 | North America/Canada/Nova Scotia/ | QEII Health Sciences Centre | National Microbiology Laboratory (NML) | Anna Majer et al |
| EPI_ISL_915010 | Canada/NS-NML-1892/2020 | 2020-04-13 | North America/Canada/Nova Scotia/ | QEII Health Sciences Centre | National Microbiology Laboratory (NML) | Anna Majer et al |
| EPI_ISL_915011 | Canada/NS-NML-1899/2020 | 2020-04-14 | North America/Canada/Nova Scotia/ | QEII Health Sciences Centre | National Microbiology Laboratory (NML) | Anna Majer et al |
| EPI_ISL_915012 | Canada/NS-NML-1900/2020 | 2020-04-14 | North America/Canada/Nova Scotia/ | QEII Health Sciences Centre | National Microbiology Laboratory (NML) | Anna Majer et al |
| EPI_ISL_915013 | Canada/NS-NML-1917/2020 | 2020-04-16 | North America/Canada/Nova Scotia/ | QEII Health Sciences Centre | National Microbiology Laboratory (NML) | Anna Majer et al |
| EPI_ISL_915014 | Canada/NS-NML-1918/2020 | 2020-04-16 | North America/Canada/Nova Scotia/ | QEII Health Sciences Centre | National Microbiology Laboratory (NML) | Anna Majer et al |
| EPI_ISL_915016 | Canada/NS-NML-1926/2020 | 2020-04-16 | North America/Canada/Nova Scotia/ | QEII Health Sciences Centre | National Microbiology Laboratory (NML) | Anna Majer et al |
| EPI_ISL_915017 | Canada/NS-NML-1927/2020 | 2020-04-17 | North America/Canada/Nova Scotia/ | QEII Health Sciences Centre | National Microbiology Laboratory (NML) | Anna Majer et al |
| EPI_ISL_915018 | Canada/NS-NML-1929/2020 | 2020-04-15 | North America/Canada/Nova Scotia/ | QEII Health Sciences Centre | National Microbiology Laboratory (NML) | Anna Majer et al |
| EPI_ISL_915019 | Canada/NS-NML-1930/2020 | 2020-04-17 | North America/Canada/Nova Scotia/ | QEII Health Sciences Centre | National Microbiology Laboratory (NML) | Anna Majer et al |
| EPI_ISL_915020 | Canada/NS-NML-1932/2020 | 2020-04-17 | North America/Canada/Nova Scotia/ | QEII Health Sciences Centre | National Microbiology Laboratory (NML) | Anna Majer et al |
| EPI_ISL_915021 | Canada/NS-NML-1934/2020 | 2020-04-16 | North America/Canada/Nova Scotia/ | QEII Health Sciences Centre | National Microbiology Laboratory (NML) | Anna Majer et al |
| EPI_ISL_915022 | Canada/NS-NML-1936/2020 | 2020-04-17 | North America/Canada/Nova Scotia/ | QEII Health Sciences Centre | National Microbiology Laboratory (NML) | Anna Majer et al |

| | | | | | | |
|---|---|---|---|---|---|---|
| EPI_ISL_915023 | Canada/NS-NML-1937/2020 | 2020-04-17 | North America/Canada/Nova Scotia/ | QEII Health Sciences Centre | National Microbiology Laboratory (NML) | Anna Majer et al |
| EPI_ISL_915024 | Canada/NS-NML-1938/2020 | 2020-04-17 | North America/Canada/Nova Scotia/ | QEII Health Sciences Centre | National Microbiology Laboratory (NML) | Anna Majer et al |
| EPI_ISL_915025 | Canada/NS-NML-1939/2020 | 2020-04-17 | North America/Canada/Nova Scotia/ | QEII Health Sciences Centre | National Microbiology Laboratory (NML) | Anna Majer et al |
| EPI_ISL_915026 | Canada/NS-NML-1940/2020 | 2020-04-17 | North America/Canada/Nova Scotia/ | QEII Health Sciences Centre | National Microbiology Laboratory (NML) | Anna Majer et al |
| EPI_ISL_915027 | Canada/NS-NML-1943/2020 | 2020-04-17 | North America/Canada/Nova Scotia/ | QEII Health Sciences Centre | National Microbiology Laboratory (NML) | Anna Majer et al |
| EPI_ISL_915028 | Canada/NS-NML-1950/2020 | 2020-04-17 | North America/Canada/Nova Scotia/ | QEII Health Sciences Centre | National Microbiology Laboratory (NML) | Anna Majer et al |
| EPI_ISL_915029 | Canada/NS-NML-1951/2020 | 2020-04-17 | North America/Canada/Nova Scotia/ | QEII Health Sciences Centre | National Microbiology Laboratory (NML) | Anna Majer et al |
| EPI_ISL_915030 | Canada/NS-NML-1952/2020 | 2020-04-17 | North America/Canada/Nova Scotia/ | QEII Health Sciences Centre | National Microbiology Laboratory (NML) | Anna Majer et al |
| EPI_ISL_915031 | Canada/NS-NML-1953/2020 | 2020-04-17 | North America/Canada/Nova Scotia/ | QEII Health Sciences Centre | National Microbiology Laboratory (NML) | Anna Majer et al |
| EPI_ISL_915032 | Canada/NS-NML-1954/2020 | 2020-04-17 | North America/Canada/Nova Scotia/ | QEII Health Sciences Centre | National Microbiology Laboratory (NML) | Anna Majer et al |
| EPI_ISL_915033 | Canada/NS-NML-1956/2020 | 2020-04-17 | North America/Canada/Nova Scotia/ | QEII Health Sciences Centre | National Microbiology Laboratory (NML) | Anna Majer et al |
| EPI_ISL_915034 | Canada/NS-NML-1957/2020 | 2020-04-17 | North America/Canada/Nova Scotia/ | QEII Health Sciences Centre | National Microbiology Laboratory (NML) | Anna Majer et al |
| EPI_ISL_915035 | Canada/NS-NML-1958/2020 | 2020-04-17 | North America/Canada/Nova Scotia/ | QEII Health Sciences Centre | National Microbiology Laboratory (NML) | Anna Majer et al |

| | | | | | | |
|---|---|---|---|---|---|---|
| EPI_ISL_915036 | Canada/NS-NML-1960/2020 | 2020-04-17 | North America/Canada/Nova Scotia/ | QEII Health Sciences Centre | National Microbiology Laboratory (NML) | Anna Majer et al |
| EPI_ISL_915037 | Canada/NS-NML-1962/2020 | 2020-04-17 | North America/Canada/Nova Scotia/ | QEII Health Sciences Centre | National Microbiology Laboratory (NML) | Anna Majer et al |
| EPI_ISL_915038 | Canada/NS-NML-1965/2020 | 2020-04-17 | North America/Canada/Nova Scotia/ | QEII Health Sciences Centre | National Microbiology Laboratory (NML) | Anna Majer et al |
| EPI_ISL_915039 | Canada/NS-NML-1968/2020 | 2020-04-17 | North America/Canada/Nova Scotia/ | QEII Health Sciences Centre | National Microbiology Laboratory (NML) | Anna Majer et al |
| EPI_ISL_915040 | Canada/NS-NML-1969/2020 | 2020-04-17 | North America/Canada/Nova Scotia/ | QEII Health Sciences Centre | National Microbiology Laboratory (NML) | Anna Majer et al |
| EPI_ISL_915041 | Canada/NS-NML-1970/2020 | 2020-04-17 | North America/Canada/Nova Scotia/ | QEII Health Sciences Centre | National Microbiology Laboratory (NML) | Anna Majer et al |
| EPI_ISL_915042 | Canada/NS-NML-1971/2020 | 2020-04-18 | North America/Canada/Nova Scotia/ | QEII Health Sciences Centre | National Microbiology Laboratory (NML) | Anna Majer et al |
| EPI_ISL_915043 | Canada/NS-NML-1973/2020 | 2020-04-18 | North America/Canada/Nova Scotia/ | QEII Health Sciences Centre | National Microbiology Laboratory (NML) | Anna Majer et al |
| EPI_ISL_915044 | Canada/NS-NML-1974/2020 | 2020-04-18 | North America/Canada/Nova Scotia/ | QEII Health Sciences Centre | National Microbiology Laboratory (NML) | Anna Majer et al |
| EPI_ISL_915045 | Canada/NS-NML-1975/2020 | 2020-04-18 | North America/Canada/Nova Scotia/ | QEII Health Sciences Centre | National Microbiology Laboratory (NML) | Anna Majer et al |
| EPI_ISL_915046 | Canada/NS-NML-1979/2020 | 2020-04-18 | North America/Canada/Nova Scotia/ | QEII Health Sciences Centre | National Microbiology Laboratory (NML) | Anna Majer et al |
| EPI_ISL_915047 | Canada/NS-NML-1980/2020 | 2020-04-18 | North America/Canada/Nova Scotia/ | QEII Health Sciences Centre | National Microbiology Laboratory (NML) | Anna Majer et al |
| EPI_ISL_915048 | Canada/NS-NML-1982/2020 | 2020-04-18 | North America/Canada/Nova Scotia/ | QEII Health Sciences Centre | National Microbiology Laboratory (NML) | Anna Majer et al |

| | | | | | | |
|---|---|---|---|---|---|---|
| EPI_ISL_915049 | Canada/NS-NML-1983/2020 | 2020-04-18 | North America/Canada/Nova Scotia/ | QEII Health Sciences Centre | National Microbiology Laboratory (NML) | Anna Majer et al |
| EPI_ISL_915050 | Canada/NS-NML-1985/2020 | 2020-04-18 | North America/Canada/Nova Scotia/ | QEII Health Sciences Centre | National Microbiology Laboratory (NML) | Anna Majer et al |
| EPI_ISL_915051 | Canada/NS-NML-1986/2020 | 2020-04-18 | North America/Canada/Nova Scotia/ | QEII Health Sciences Centre | National Microbiology Laboratory (NML) | Anna Majer et al |
| EPI_ISL_915052 | Canada/NS-NML-1988/2020 | 2020-04-18 | North America/Canada/Nova Scotia/ | QEII Health Sciences Centre | National Microbiology Laboratory (NML) | Anna Majer et al |
| EPI_ISL_915053 | Canada/NS-NML-1990/2020 | 2020-04-18 | North America/Canada/Nova Scotia/ | QEII Health Sciences Centre | National Microbiology Laboratory (NML) | Anna Majer et al |
| EPI_ISL_915054 | Canada/NS-NML-1991/2020 | 2020-04-18 | North America/Canada/Nova Scotia/ | QEII Health Sciences Centre | National Microbiology Laboratory (NML) | Anna Majer et al |
| EPI_ISL_915055 | Canada/NS-NML-1992/2020 | 2020-04-18 | North America/Canada/Nova Scotia/ | QEII Health Sciences Centre | National Microbiology Laboratory (NML) | Anna Majer et al |
| EPI_ISL_915056 | Canada/NS-NML-1993/2020 | 2020-04-18 | North America/Canada/Nova Scotia/ | QEII Health Sciences Centre | National Microbiology Laboratory (NML) | Anna Majer et al |
| EPI_ISL_915057 | Canada/NS-NML-1994/2020 | 2020-04-18 | North America/Canada/Nova Scotia/ | QEII Health Sciences Centre | National Microbiology Laboratory (NML) | Anna Majer et al |
| EPI_ISL_915058 | Canada/NS-NML-1995/2020 | 2020-04-18 | North America/Canada/Nova Scotia/ | QEII Health Sciences Centre | National Microbiology Laboratory (NML) | Anna Majer et al |
| EPI_ISL_915059 | Canada/NS-NML-1996/2020 | 2020-04-18 | North America/Canada/Nova Scotia/ | QEII Health Sciences Centre | National Microbiology Laboratory (NML) | Anna Majer et al |
| EPI_ISL_915060 | Canada/NS-NML-1997/2020 | 2020-04-18 | North America/Canada/Nova Scotia/ | QEII Health Sciences Centre | National Microbiology Laboratory (NML) | Anna Majer et al |
| EPI_ISL_915061 | Canada/NS-NML-1998/2020 | 2020-04-18 | North America/Canada/Nova Scotia/ | QEII Health Sciences Centre | National Microbiology Laboratory (NML) | Anna Majer et al |

| | | | | | | |
|---|---|---|---|---|---|---|
| EPI_ISL_915062 | Canada/NS-NML-1999/2020 | 2020-04-18 | North America/Canada/Nova Scotia/ | QEII Health Sciences Centre | National Microbiology Laboratory (NML) | Anna Majer et al |
| EPI_ISL_915063 | Canada/NS-NML-2001/2020 | 2020-04-18 | North America/Canada/Nova Scotia/ | QEII Health Sciences Centre | National Microbiology Laboratory (NML) | Anna Majer et al |
| EPI_ISL_915064 | Canada/NS-NML-2002/2020 | 2020-04-18 | North America/Canada/Nova Scotia/ | QEII Health Sciences Centre | National Microbiology Laboratory (NML) | Anna Majer et al |
| EPI_ISL_915065 | Canada/NS-NML-2003/2020 | 2020-04-18 | North America/Canada/Nova Scotia/ | QEII Health Sciences Centre | National Microbiology Laboratory (NML) | Anna Majer et al |
| EPI_ISL_915066 | Canada/NS-NML-2004/2020 | 2020-04-18 | North America/Canada/Nova Scotia/ | QEII Health Sciences Centre | National Microbiology Laboratory (NML) | Anna Majer et al |
| EPI_ISL_915067 | Canada/NS-NML-2005/2020 | 2020-04-18 | North America/Canada/Nova Scotia/ | QEII Health Sciences Centre | National Microbiology Laboratory (NML) | Anna Majer et al |
| EPI_ISL_915068 | Canada/NS-NML-2006/2020 | 2020-04-18 | North America/Canada/Nova Scotia/ | QEII Health Sciences Centre | National Microbiology Laboratory (NML) | Anna Majer et al |
| EPI_ISL_915069 | Canada/NS-NML-2007/2020 | 2020-04-18 | North America/Canada/Nova Scotia/ | QEII Health Sciences Centre | National Microbiology Laboratory (NML) | Anna Majer et al |
| EPI_ISL_915070 | Canada/NS-NML-2008/2020 | 2020-04-18 | North America/Canada/Nova Scotia/ | QEII Health Sciences Centre | National Microbiology Laboratory (NML) | Anna Majer et al |
| EPI_ISL_915071 | Canada/NS-NML-2009/2020 | 2020-04-18 | North America/Canada/Nova Scotia/ | QEII Health Sciences Centre | National Microbiology Laboratory (NML) | Anna Majer et al |
| EPI_ISL_915072 | Canada/NS-NML-2010/2020 | 2020-04-18 | North America/Canada/Nova Scotia/ | QEII Health Sciences Centre | National Microbiology Laboratory (NML) | Anna Majer et al |
| EPI_ISL_915073 | Canada/NS-NML-2011/2020 | 2020-04-18 | North America/Canada/Nova Scotia/ | QEII Health Sciences Centre | National Microbiology Laboratory (NML) | Anna Majer et al |
| EPI_ISL_915074 | Canada/NS-NML-2012/2020 | 2020-04-18 | North America/Canada/Nova Scotia/ | QEII Health Sciences Centre | National Microbiology Laboratory (NML) | Anna Majer et al |

| | | | | | | |
|---|---|---|---|---|---|---|
| EPI_ISL_915075 | Canada/NS-NML-2013/2020 | 2020-04-18 | North America/Canada/Nova Scotia/ | QEII Health Sciences Centre | National Microbiology Laboratory (NML) | Anna Majer et al |
| EPI_ISL_915076 | Canada/NS-NML-2015/2020 | 2020-04-18 | North America/Canada/Nova Scotia/ | QEII Health Sciences Centre | National Microbiology Laboratory (NML) | Anna Majer et al |
| EPI_ISL_915077 | Canada/NS-NML-2016/2020 | 2020-04-19 | North America/Canada/Nova Scotia/ | QEII Health Sciences Centre | National Microbiology Laboratory (NML) | Anna Majer et al |
| EPI_ISL_915078 | Canada/NS-NML-2020/2020 | 2020-04-19 | North America/Canada/Nova Scotia/ | QEII Health Sciences Centre | National Microbiology Laboratory (NML) | Anna Majer et al |
| EPI_ISL_915079 | Canada/NS-NML-2021/2020 | 2020-04-19 | North America/Canada/Nova Scotia/ | QEII Health Sciences Centre | National Microbiology Laboratory (NML) | Anna Majer et al |
| EPI_ISL_915081 | Canada/NS-NML-2027/2020 | 2020-04-20 | North America/Canada/Nova Scotia/ | QEII Health Sciences Centre | National Microbiology Laboratory (NML) | Anna Majer et al |
| EPI_ISL_915083 | Canada/NS-NML-2029/2020 | 2020-04-20 | North America/Canada/Nova Scotia/ | QEII Health Sciences Centre | National Microbiology Laboratory (NML) | Anna Majer et al |
| EPI_ISL_915084 | Canada/NS-NML-2030/2020 | 2020-04-20 | North America/Canada/Nova Scotia/ | QEII Health Sciences Centre | National Microbiology Laboratory (NML) | Anna Majer et al |
| EPI_ISL_915085 | Canada/NS-NML-2031/2020 | 2020-04-20 | North America/Canada/Nova Scotia/ | QEII Health Sciences Centre | National Microbiology Laboratory (NML) | Anna Majer et al |
| EPI_ISL_915086 | Canada/NS-NML-2034/2020 | 2020-04-20 | North America/Canada/Nova Scotia/ | QEII Health Sciences Centre | National Microbiology Laboratory (NML) | Anna Majer et al |
| EPI_ISL_915087 | Canada/NS-NML-2039/2020 | 2020-04-20 | North America/Canada/Nova Scotia/ | QEII Health Sciences Centre | National Microbiology Laboratory (NML) | Anna Majer et al |
| EPI_ISL_915088 | Canada/NS-NML-2042/2020 | 2020-04-20 | North America/Canada/Nova Scotia/ | QEII Health Sciences Centre | National Microbiology Laboratory (NML) | Anna Majer et al |
| EPI_ISL_915089 | Canada/NS-NML-2043/2020 | 2020-04-20 | North America/Canada/Nova Scotia/ | QEII Health Sciences Centre | National Microbiology Laboratory (NML) | Anna Majer et al |

| | | | | | | |
|---|---|---|---|---|---|---|
| EPI_ISL_915090 | Canada/NS-NML-2044/2020 | 2020-04-20 | North America/Canada/Nova Scotia/ | QEII Health Sciences Centre | National Microbiology Laboratory (NML) | Anna Majer et al |
| EPI_ISL_915091 | Canada/NS-NML-2045/2020 | 2020-04-20 | North America/Canada/Nova Scotia/ | QEII Health Sciences Centre | National Microbiology Laboratory (NML) | Anna Majer et al |
| EPI_ISL_915092 | Canada/NS-NML-2046/2020 | 2020-04-20 | North America/Canada/Nova Scotia/ | QEII Health Sciences Centre | National Microbiology Laboratory (NML) | Anna Majer et al |
| EPI_ISL_915093 | Canada/NS-NML-2047/2020 | 2020-04-20 | North America/Canada/Nova Scotia/ | QEII Health Sciences Centre | National Microbiology Laboratory (NML) | Anna Majer et al |
| EPI_ISL_915094 | Canada/NS-NML-2048/2020 | 2020-04-20 | North America/Canada/Nova Scotia/ | QEII Health Sciences Centre | National Microbiology Laboratory (NML) | Anna Majer et al |
| EPI_ISL_915095 | Canada/NS-NML-2052/2020 | 2020-04-20 | North America/Canada/Nova Scotia/ | QEII Health Sciences Centre | National Microbiology Laboratory (NML) | Anna Majer et al |
| EPI_ISL_915096 | Canada/NS-NML-2054/2020 | 2020-04-20 | North America/Canada/Nova Scotia/ | QEII Health Sciences Centre | National Microbiology Laboratory (NML) | Anna Majer et al |
| EPI_ISL_915098 | Canada/NS-NML-2058/2020 | 2020-04-21 | North America/Canada/Nova Scotia/ | QEII Health Sciences Centre | National Microbiology Laboratory (NML) | Anna Majer et al |
| EPI_ISL_915099 | Canada/NS-NML-2059/2020 | 2020-04-21 | North America/Canada/Nova Scotia/ | QEII Health Sciences Centre | National Microbiology Laboratory (NML) | Anna Majer et al |
| EPI_ISL_915100 | Canada/NS-NML-2060/2020 | 2020-04-21 | North America/Canada/Nova Scotia/ | QEII Health Sciences Centre | National Microbiology Laboratory (NML) | Anna Majer et al |
| EPI_ISL_915101 | Canada/NS-NML-2062/2020 | 2020-04-21 | North America/Canada/Nova Scotia/ | QEII Health Sciences Centre | National Microbiology Laboratory (NML) | Anna Majer et al |
| EPI_ISL_915102 | Canada/NS-NML-2064/2020 | 2020-04-21 | North America/Canada/Nova Scotia/ | QEII Health Sciences Centre | National Microbiology Laboratory (NML) | Anna Majer et al |
| EPI_ISL_915103 | Canada/NS-NML-2065/2020 | 2020-04-21 | North America/Canada/Nova Scotia/ | QEII Health Sciences Centre | National Microbiology Laboratory (NML) | Anna Majer et al |

| | | | | | | |
|---|---|---|---|---|---|---|
| EPI_ISL_915104 | Canada/NS-NML-2067/2020 | 2020-04-21 | North America/Canada/Nova Scotia/ | QEII Health Sciences Centre | National Microbiology Laboratory (NML) | Anna Majer et al |
| EPI_ISL_915105 | Canada/NS-NML-2072/2020 | 2020-04-21 | North America/Canada/Nova Scotia/ | QEII Health Sciences Centre | National Microbiology Laboratory (NML) | Anna Majer et al |
| EPI_ISL_915106 | Canada/NS-NML-2074/2020 | 2020-04-21 | North America/Canada/Nova Scotia/ | QEII Health Sciences Centre | National Microbiology Laboratory (NML) | Anna Majer et al |
| EPI_ISL_915107 | Canada/NS-NML-2075/2020 | 2020-04-21 | North America/Canada/Nova Scotia/ | QEII Health Sciences Centre | National Microbiology Laboratory (NML) | Anna Majer et al |
| EPI_ISL_915108 | Canada/NS-NML-2077/2020 | 2020-04-21 | North America/Canada/Nova Scotia/ | QEII Health Sciences Centre | National Microbiology Laboratory (NML) | Anna Majer et al |
| EPI_ISL_915109 | Canada/NS-NML-2078/2020 | 2020-04-21 | North America/Canada/Nova Scotia/ | QEII Health Sciences Centre | National Microbiology Laboratory (NML) | Anna Majer et al |
| EPI_ISL_915110 | Canada/NS-NML-2082/2020 | 2020-04-22 | North America/Canada/Nova Scotia/ | QEII Health Sciences Centre | National Microbiology Laboratory (NML) | Anna Majer et al |
| EPI_ISL_915111 | Canada/NS-NML-2083/2020 | 2020-04-22 | North America/Canada/Nova Scotia/ | QEII Health Sciences Centre | National Microbiology Laboratory (NML) | Anna Majer et al |
| EPI_ISL_915112 | Canada/NS-NML-2088/2020 | 2020-04-22 | North America/Canada/Nova Scotia/ | QEII Health Sciences Centre | National Microbiology Laboratory (NML) | Anna Majer et al |
| EPI_ISL_915113 | Canada/NS-NML-2092/2020 | 2020-04-22 | North America/Canada/Nova Scotia/ | QEII Health Sciences Centre | National Microbiology Laboratory (NML) | Anna Majer et al |
| EPI_ISL_915114 | Canada/NS-NML-2093/2020 | 2020-04-22 | North America/Canada/Nova Scotia/ | QEII Health Sciences Centre | National Microbiology Laboratory (NML) | Anna Majer et al |
| EPI_ISL_915115 | Canada/NS-NML-2094/2020 | 2020-04-22 | North America/Canada/Nova Scotia/ | QEII Health Sciences Centre | National Microbiology Laboratory (NML) | Anna Majer et al |
| EPI_ISL_915116 | Canada/NS-NML-2095/2020 | 2020-04-22 | North America/Canada/Nova Scotia/ | QEII Health Sciences Centre | National Microbiology Laboratory (NML) | Anna Majer et al |

| | | | | | | |
|---|---|---|---|---|---|---|
| EPI_ISL_915117 | Canada/NS-NML-2096/2020 | 2020-04-22 | North America/Canada/Nova Scotia/ | QEII Health Sciences Centre | National Microbiology Laboratory (NML) | Anna Majer et al |
| EPI_ISL_915118 | Canada/NS-NML-2097/2020 | 2020-04-22 | North America/Canada/Nova Scotia/ | QEII Health Sciences Centre | National Microbiology Laboratory (NML) | Anna Majer et al |
| EPI_ISL_915119 | Canada/NS-NML-2103/2020 | 2020-04-22 | North America/Canada/Nova Scotia/ | QEII Health Sciences Centre | National Microbiology Laboratory (NML) | Anna Majer et al |
| EPI_ISL_915120 | Canada/NS-NML-2105/2020 | 2020-04-22 | North America/Canada/Nova Scotia/ | QEII Health Sciences Centre | National Microbiology Laboratory (NML) | Anna Majer et al |
| EPI_ISL_915121 | Canada/NS-NML-2106/2020 | 2020-04-22 | North America/Canada/Nova Scotia/ | QEII Health Sciences Centre | National Microbiology Laboratory (NML) | Anna Majer et al |
| EPI_ISL_915122 | Canada/NS-NML-2109/2020 | 2020-04-22 | North America/Canada/Nova Scotia/ | QEII Health Sciences Centre | National Microbiology Laboratory (NML) | Anna Majer et al |
| EPI_ISL_915123 | Canada/NS-NML-2110/2020 | 2020-04-22 | North America/Canada/Nova Scotia/ | QEII Health Sciences Centre | National Microbiology Laboratory (NML) | Anna Majer et al |
| EPI_ISL_915124 | Canada/NS-NML-2112/2020 | 2020-04-23 | North America/Canada/Nova Scotia/ | QEII Health Sciences Centre | National Microbiology Laboratory (NML) | Anna Majer et al |
| EPI_ISL_915125 | Canada/NS-NML-2113/2020 | 2020-04-23 | North America/Canada/Nova Scotia/ | QEII Health Sciences Centre | National Microbiology Laboratory (NML) | Anna Majer et al |
| EPI_ISL_915126 | Canada/NS-NML-2114/2020 | 2020-04-23 | North America/Canada/Nova Scotia/ | QEII Health Sciences Centre | National Microbiology Laboratory (NML) | Anna Majer et al |
| EPI_ISL_915127 | Canada/NS-NML-2115/2020 | 2020-04-23 | North America/Canada/Nova Scotia/ | QEII Health Sciences Centre | National Microbiology Laboratory (NML) | Anna Majer et al |
| EPI_ISL_915128 | Canada/NS-NML-2118/2020 | 2020-04-23 | North America/Canada/Nova Scotia/ | QEII Health Sciences Centre | National Microbiology Laboratory (NML) | Anna Majer et al |
| EPI_ISL_915129 | Canada/NS-NML-2120/2020 | 2020-04-23 | North America/Canada/Nova Scotia/ | QEII Health Sciences Centre | National Microbiology Laboratory (NML) | Anna Majer et al |

| | | | | | | |
|---|---|---|---|---|---|---|
| EPI_ISL_915130 | Canada/NS-NML-2121/2020 | 2020-04-23 | North America/Canada/Nova Scotia/ | QEII Health Sciences Centre | National Microbiology Laboratory (NML) | Anna Majer et al |
| EPI_ISL_915131 | Canada/NS-NML-2123/2020 | 2020-04-23 | North America/Canada/Nova Scotia/ | QEII Health Sciences Centre | National Microbiology Laboratory (NML) | Anna Majer et al |
| EPI_ISL_915132 | Canada/NS-NML-2125/2020 | 2020-04-23 | North America/Canada/Nova Scotia/ | QEII Health Sciences Centre | National Microbiology Laboratory (NML) | Anna Majer et al |
| EPI_ISL_915133 | Canada/NS-NML-2126/2020 | 2020-04-23 | North America/Canada/Nova Scotia/ | QEII Health Sciences Centre | National Microbiology Laboratory (NML) | Anna Majer et al |
| EPI_ISL_915134 | Canada/NS-NML-2127/2020 | 2020-04-23 | North America/Canada/Nova Scotia/ | QEII Health Sciences Centre | National Microbiology Laboratory (NML) | Anna Majer et al |
| EPI_ISL_915135 | Canada/NS-NML-2128/2020 | 2020-04-23 | North America/Canada/Nova Scotia/ | QEII Health Sciences Centre | National Microbiology Laboratory (NML) | Anna Majer et al |
| EPI_ISL_915136 | Canada/NS-NML-2131/2020 | 2020-04-23 | North America/Canada/Nova Scotia/ | QEII Health Sciences Centre | National Microbiology Laboratory (NML) | Anna Majer et al |
| EPI_ISL_915137 | Canada/NS-NML-2133/2020 | 2020-04-24 | North America/Canada/Nova Scotia/ | QEII Health Sciences Centre | National Microbiology Laboratory (NML) | Anna Majer et al |
| EPI_ISL_915138 | Canada/NS-NML-2134/2020 | 2020-04-24 | North America/Canada/Nova Scotia/ | QEII Health Sciences Centre | National Microbiology Laboratory (NML) | Anna Majer et al |
| EPI_ISL_915139 | Canada/NS-NML-2140/2020 | 2020-04-24 | North America/Canada/Nova Scotia/ | QEII Health Sciences Centre | National Microbiology Laboratory (NML) | Anna Majer et al |
| EPI_ISL_915140 | Canada/NS-NML-2141/2020 | 2020-04-24 | North America/Canada/Nova Scotia/ | QEII Health Sciences Centre | National Microbiology Laboratory (NML) | Anna Majer et al |
| EPI_ISL_915141 | Canada/NS-NML-2142/2020 | 2020-04-24 | North America/Canada/Nova Scotia/ | QEII Health Sciences Centre | National Microbiology Laboratory (NML) | Anna Majer et al |
| EPI_ISL_915142 | Canada/NS-NML-2143/2020 | 2020-04-25 | North America/Canada/Nova Scotia/ | QEII Health Sciences Centre | National Microbiology Laboratory (NML) | Anna Majer et al |

| | | | | | | |
|---|---|---|---|---|---|---|
| EPI_ISL_915143 | Canada/NS-NML-2144/2020 | 2020-04-25 | North America/Canada/Nova Scotia/ | QEII Health Sciences Centre | National Microbiology Laboratory (NML) | Anna Majer et al |
| EPI_ISL_915144 | Canada/NS-NML-2145/2020 | 2020-04-25 | North America/Canada/Nova Scotia/ | QEII Health Sciences Centre | National Microbiology Laboratory (NML) | Anna Majer et al |
| EPI_ISL_915145 | Canada/NS-NML-2147/2020 | 2020-04-25 | North America/Canada/Nova Scotia/ | QEII Health Sciences Centre | National Microbiology Laboratory (NML) | Anna Majer et al |
| EPI_ISL_915146 | Canada/NS-NML-2148/2020 | 2020-04-25 | North America/Canada/Nova Scotia/ | QEII Health Sciences Centre | National Microbiology Laboratory (NML) | Anna Majer et al |
| EPI_ISL_915147 | Canada/NS-NML-2149/2020 | 2020-04-25 | North America/Canada/Nova Scotia/ | QEII Health Sciences Centre | National Microbiology Laboratory (NML) | Anna Majer et al |
| EPI_ISL_915148 | Canada/NS-NML-2150/2020 | 2020-04-25 | North America/Canada/Nova Scotia/ | QEII Health Sciences Centre | National Microbiology Laboratory (NML) | Anna Majer et al |
| EPI_ISL_915149 | Canada/NS-NML-2217/2020 | 2020-04-30 | North America/Canada/Nova Scotia/ | QEII Health Sciences Centre | National Microbiology Laboratory (NML) | Anna Majer et al |
| EPI_ISL_915150 | Canada/NS-NML-2218/2020 | 2020-04-30 | North America/Canada/Nova Scotia/ | QEII Health Sciences Centre | National Microbiology Laboratory (NML) | Anna Majer et al |
| EPI_ISL_915151 | Canada/NS-NML-2219/2020 | 2020-04-30 | North America/Canada/Nova Scotia/ | QEII Health Sciences Centre | National Microbiology Laboratory (NML) | Anna Majer et al |
| EPI_ISL_915152 | Canada/NS-NML-2220/2020 | 2020-04-30 | North America/Canada/Nova Scotia/ | QEII Health Sciences Centre | National Microbiology Laboratory (NML) | Anna Majer et al |
| EPI_ISL_915154 | Canada/NS-NML-2223/2020 | 2020-05-01 | North America/Canada/Nova Scotia/ | QEII Health Sciences Centre | National Microbiology Laboratory (NML) | Anna Majer et al |
| EPI_ISL_915155 | Canada/NS-NML-2225/2020 | 2020-05-01 | North America/Canada/Nova Scotia/ | QEII Health Sciences Centre | National Microbiology Laboratory (NML) | Anna Majer et al |
| EPI_ISL_915156 | Canada/NS-NML-2226/2020 | 2020-05-01 | North America/Canada/Nova Scotia/ | QEII Health Sciences Centre | National Microbiology Laboratory (NML) | Anna Majer et al |

| | | | | | | |
|---|---|---|---|---|---|---|
| EPI_ISL_915157 | Canada/NS-NML-2228/2020 | 2020-05-02 | North America/Canada/Nova Scotia/ | QEII Health Sciences Centre | National Microbiology Laboratory (NML) | Anna Majer et al |
| EPI_ISL_915158 | Canada/NS-NML-2230/2020 | 2020-05-02 | North America/Canada/Nova Scotia/ | QEII Health Sciences Centre | National Microbiology Laboratory (NML) | Anna Majer et al |
| EPI_ISL_915159 | Canada/NS-NML-2231/2020 | 2020-05-03 | North America/Canada/Nova Scotia/ | QEII Health Sciences Centre | National Microbiology Laboratory (NML) | Anna Majer et al |
| EPI_ISL_915160 | Canada/NS-NML-2232/2020 | 2020-05-03 | North America/Canada/Nova Scotia/ | QEII Health Sciences Centre | National Microbiology Laboratory (NML) | Anna Majer et al |
| EPI_ISL_915161 | Canada/NS-NML-2233/2020 | 2020-05-03 | North America/Canada/Nova Scotia/ | QEII Health Sciences Centre | National Microbiology Laboratory (NML) | Anna Majer et al |
| EPI_ISL_915162 | Canada/NS-NML-2234/2020 | 2020-05-03 | North America/Canada/Nova Scotia/ | QEII Health Sciences Centre | National Microbiology Laboratory (NML) | Anna Majer et al |
| EPI_ISL_915163 | Canada/NS-NML-2235/2020 | 2020-05-03 | North America/Canada/Nova Scotia/ | QEII Health Sciences Centre | National Microbiology Laboratory (NML) | Anna Majer et al |
| EPI_ISL_915164 | Canada/NS-NML-2237/2020 | 2020-05-03 | North America/Canada/Nova Scotia/ | QEII Health Sciences Centre | National Microbiology Laboratory (NML) | Anna Majer et al |
| EPI_ISL_915165 | Canada/NS-NML-2238/2020 | 2020-05-03 | North America/Canada/Nova Scotia/ | QEII Health Sciences Centre | National Microbiology Laboratory (NML) | Anna Majer et al |
| EPI_ISL_915166 | Canada/NS-NML-2239/2020 | 2020-05-03 | North America/Canada/Nova Scotia/ | QEII Health Sciences Centre | National Microbiology Laboratory (NML) | Anna Majer et al |
| EPI_ISL_915167 | Canada/NS-NML-2240/2020 | 2020-05-03 | North America/Canada/Nova Scotia/ | QEII Health Sciences Centre | National Microbiology Laboratory (NML) | Anna Majer et al |
| EPI_ISL_915168 | Canada/NS-NML-2241/2020 | 2020-05-04 | North America/Canada/Nova Scotia/ | QEII Health Sciences Centre | National Microbiology Laboratory (NML) | Anna Majer et al |
| EPI_ISL_915169 | Canada/NS-NML-2243/2020 | 2020-05-04 | North America/Canada/Nova Scotia/ | QEII Health Sciences Centre | National Microbiology Laboratory (NML) | Anna Majer et al |

| | | | | | | |
|---|---|---|---|---|---|---|
| EPI_ISL_915170 | Canada/NS-NML-2244/2020 | 2020-05-04 | North America/Canada/Nova Scotia/ | QEII Health Sciences Centre | National Microbiology Laboratory (NML) | Anna Majer et al |
| EPI_ISL_915171 | Canada/NS-NML-2245/2020 | 2020-05-04 | North America/Canada/Nova Scotia/ | QEII Health Sciences Centre | National Microbiology Laboratory (NML) | Anna Majer et al |
| EPI_ISL_915172 | Canada/NS-NML-2246/2020 | 2020-05-04 | North America/Canada/Nova Scotia/ | QEII Health Sciences Centre | National Microbiology Laboratory (NML) | Anna Majer et al |
| EPI_ISL_915173 | Canada/NS-NML-2247/2020 | 2020-05-04 | North America/Canada/Nova Scotia/ | QEII Health Sciences Centre | National Microbiology Laboratory (NML) | Anna Majer et al |
| EPI_ISL_915174 | Canada/NS-NML-2248/2020 | 2020-05-05 | North America/Canada/Nova Scotia/ | QEII Health Sciences Centre | National Microbiology Laboratory (NML) | Anna Majer et al |
| EPI_ISL_915175 | Canada/NS-NML-2249/2020 | 2020-05-05 | North America/Canada/Nova Scotia/ | QEII Health Sciences Centre | National Microbiology Laboratory (NML) | Anna Majer et al |
| EPI_ISL_915176 | Canada/NS-NML-2250/2020 | 2020-05-05 | North America/Canada/Nova Scotia/ | QEII Health Sciences Centre | National Microbiology Laboratory (NML) | Anna Majer et al |
| EPI_ISL_915177 | Canada/NS-NML-2251/2020 | 2020-05-05 | North America/Canada/Nova Scotia/ | QEII Health Sciences Centre | National Microbiology Laboratory (NML) | Anna Majer et al |
| EPI_ISL_915178 | Canada/NS-NML-2253/2020 | 2020-05-05 | North America/Canada/Nova Scotia/ | QEII Health Sciences Centre | National Microbiology Laboratory (NML) | Anna Majer et al |
| EPI_ISL_915179 | Canada/NS-NML-2254/2020 | 2020-05-05 | North America/Canada/Nova Scotia/ | QEII Health Sciences Centre | National Microbiology Laboratory (NML) | Anna Majer et al |
| EPI_ISL_915180 | Canada/NS-NML-2255/2020 | 2020-05-05 | North America/Canada/Nova Scotia/ | QEII Health Sciences Centre | National Microbiology Laboratory (NML) | Anna Majer et al |
| EPI_ISL_915181 | Canada/NS-NML-2256/2020 | 2020-05-05 | North America/Canada/Nova Scotia/ | QEII Health Sciences Centre | National Microbiology Laboratory (NML) | Anna Majer et al |
| EPI_ISL_915182 | Canada/NS-NML-2257/2020 | 2020-05-06 | North America/Canada/Nova Scotia/ | QEII Health Sciences Centre | National Microbiology Laboratory (NML) | Anna Majer et al |

| | | | | | | |
|---|---|---|---|---|---|---|
| EPI_ISL_915183 | Canada/NS-NML-2258/2020 | 2020-05-06 | North America/Canada/Nova Scotia/ | QEII Health Sciences Centre | National Microbiology Laboratory (NML) | Anna Majer et al |
| EPI_ISL_915184 | Canada/NS-NML-2260/2020 | 2020-05-06 | North America/Canada/Nova Scotia/ | QEII Health Sciences Centre | National Microbiology Laboratory (NML) | Anna Majer et al |
| EPI_ISL_915185 | Canada/NS-NML-2261/2020 | 2020-05-07 | North America/Canada/Nova Scotia/ | QEII Health Sciences Centre | National Microbiology Laboratory (NML) | Anna Majer et al |
| EPI_ISL_915186 | Canada/NS-NML-2262/2020 | 2020-05-07 | North America/Canada/Nova Scotia/ | QEII Health Sciences Centre | National Microbiology Laboratory (NML) | Anna Majer et al |
| EPI_ISL_915187 | Canada/NS-NML-2263/2020 | 2020-05-08 | North America/Canada/Nova Scotia/ | QEII Health Sciences Centre | National Microbiology Laboratory (NML) | Anna Majer et al |
| EPI_ISL_915188 | Canada/NS-NML-2264/2020 | 2020-05-08 | North America/Canada/Nova Scotia/ | QEII Health Sciences Centre | National Microbiology Laboratory (NML) | Anna Majer et al |
| EPI_ISL_915189 | Canada/NS-NML-2265/2020 | 2020-05-09 | North America/Canada/Nova Scotia/ | QEII Health Sciences Centre | National Microbiology Laboratory (NML) | Anna Majer et al |
| EPI_ISL_915190 | Canada/NS-NML-2266/2020 | 2020-05-09 | North America/Canada/Nova Scotia/ | QEII Health Sciences Centre | National Microbiology Laboratory (NML) | Anna Majer et al |
| EPI_ISL_915191 | Canada/NS-NML-2267/2020 | 2020-05-09 | North America/Canada/Nova Scotia/ | QEII Health Sciences Centre | National Microbiology Laboratory (NML) | Anna Majer et al |
| EPI_ISL_915193 | USA/AL-QDX-4221/2021 | 2021-01-15 | North America/USA/Alabama/ | Quest Diagnostics | Quest Diagnostics | Rosenthal et al |
| EPI_ISL_915195 | USA/AZ-QDX-4178/2021 | 2021-01-14 | North America/USA/Arizona/ | Quest Diagnostics | Quest Diagnostics | Rosenthal et al |
| EPI_ISL_915196 | USA/CA-QDX-4169/2021 | 2021-01-13 | North America/USA/California/ | Quest Diagnostics | Quest Diagnostics | Rosenthal et al |
| EPI_ISL_915197 | USA/CA-QDX-4170/2021 | 2021-01-13 | North America/USA/California/ | Quest Diagnostics | Quest Diagnostics | Rosenthal et al |
| EPI_ISL_915213 | USA/CA-QDX-4191/2021 | 2021-01-16 | North America/USA/California/ | Quest Diagnostics | Quest Diagnostics | Rosenthal et al |
| EPI_ISL_915218 | USA/CA-QDX-4201/2021 | 2021-01-13 | North America/USA/California/ | Quest Diagnostics | Quest Diagnostics | Rosenthal et al |
| EPI_ISL_915223 | USA/CA-QDX-4208/2021 | 2021-01-14 | North America/USA/California/ | Quest Diagnostics | Quest Diagnostics | Rosenthal et al |
| EPI_ISL_915228 | USA/CA-QDX-4213/2021 | 2021-01-15 | North America/USA/California/ | Quest Diagnostics | Quest Diagnostics | Rosenthal et al |
| EPI_ISL_915229 | USA/CA-QDX-4214/2021 | 2021-01-14 | North America/USA/California/ | Quest Diagnostics | Quest Diagnostics | Rosenthal et al |
| EPI_ISL_915234 | USA/CA-QDX-4220/2021 | 2021-01-16 | North America/USA/California/ | Quest Diagnostics | Quest Diagnostics | Rosenthal et al |

| | | | | | | |
|---|---|---|---|---|---|---|
| EPI_ISL_915242 | USA/CA-QDX-4239/2021 | 2021-01-12 | North America/USA/California/ | Quest Diagnostics | Quest Diagnostics | Rosenthal et al |
| EPI_ISL_915245 | USA/CA-QDX-4242/2021 | 2021-01-13 | North America/USA/California/ | Quest Diagnostics | Quest Diagnostics | Rosenthal et al |
| EPI_ISL_915248 | USA/CA-QDX-4245/2021 | 2021-01-13 | North America/USA/California/ | Quest Diagnostics | Quest Diagnostics | Rosenthal et al |
| EPI_ISL_915251 | USA/CA-QDX-4248/2021 | 2021-01-13 | North America/USA/California/ | Quest Diagnostics | Quest Diagnostics | Rosenthal et al |
| EPI_ISL_915253 | USA/CA-QDX-4251/2021 | 2021-01-14 | North America/USA/California/ | Quest Diagnostics | Quest Diagnostics | Rosenthal et al |
| EPI_ISL_915263 | USA/CA-QDX-4268/2021 | 2021-01-14 | North America/USA/California/ | Quest Diagnostics | Quest Diagnostics | Rosenthal et al |
| EPI_ISL_915278 | USA/CA-QDX-4285/2021 | 2021-01-15 | North America/USA/California/ | Quest Diagnostics | Quest Diagnostics | Rosenthal et al |
| EPI_ISL_915284 | USA/CA-QDX-4298/2021 | 2021-01-09 | North America/USA/California/ | Quest Diagnostics | Quest Diagnostics | Rosenthal et al |
| EPI_ISL_915290 | USA/CA-QDX-4304/2021 | 2021-01-12 | North America/USA/California/ | Quest Diagnostics | Quest Diagnostics | Rosenthal et al |
| EPI_ISL_915293 | USA/CA-QDX-4309/2021 | 2021-01-13 | North America/USA/California/ | Quest Diagnostics | Quest Diagnostics | Rosenthal et al |
| EPI_ISL_915303 | USA/CA-QDX-4324/2021 | 2021-01-14 | North America/USA/California/ | Quest Diagnostics | Quest Diagnostics | Rosenthal et al |
| EPI_ISL_915316 | USA/IL-QDX-4229/2021 | 2021-01-15 | North America/USA/Illinois/ | Quest Diagnostics | Quest Diagnostics | Rosenthal et al |
| EPI_ISL_915319 | USA/IL-QDX-4335/2021 | 2021-01-15 | North America/USA/Illinois/ | Quest Diagnostics | Quest Diagnostics | Rosenthal et al |
| EPI_ISL_915322 | USA/KY-QDX-4336/2021 | 2021-01-15 | North America/USA/Kentucky/ | Quest Diagnostics | Quest Diagnostics | Rosenthal et al |
| EPI_ISL_915333 | USA/NH-QDX-4183/2021 | 2021-01-07 | North America/USA/New Hampshire/ | Quest Diagnostics | Quest Diagnostics | Rosenthal et al |
| EPI_ISL_915337 | USA/NM-QDX-4290/2021 | 2021-01-15 | North America/USA/New Mexico/ | Quest Diagnostics | Quest Diagnostics | Rosenthal et al |
| EPI_ISL_915343 | USA/NY-QDX-4256/2021 | 2021-01-14 | North America/USA/New York/ | Quest Diagnostics | Quest Diagnostics | Rosenthal et al |
| EPI_ISL_915347 | USA/TX-QDX-4198/2021 | 2021-01-16 | North America/USA/Texas/ | Quest Diagnostics | Quest Diagnostics | Rosenthal et al |
| EPI_ISL_915349 | USA/TX-QDX-4222/2021 | 2021-01-16 | North America/USA/Texas/ | Quest Diagnostics | Quest Diagnostics | Rosenthal et al |
| EPI_ISL_915351 | USA/TX-QDX-4253/2021 | 2021-01-15 | North America/USA/Texas/ | Quest Diagnostics | Quest Diagnostics | Rosenthal et al |
| EPI_ISL_915422 | Germany/BY-IMB15250/2021 | 2021-01-21 | Europe/Germany/Bavaria/ | Bundeswehr Institute of Microbiology | Bundeswehr Institute of Microbiology | Markus Antwerpen et al |
| EPI_ISL_915425 | Germany/BY-IMB15279/2021 | 2021-01-22 | Europe/Germany/Bavaria/ | Bundeswehr Institute of Microbiology | Bundeswehr Institute of Microbiology | Markus Antwerpen et al |
| EPI_ISL_915427 | Germany/BY-IMB14922/2021 | 2021-01-11 | Europe/Germany/Bavaria/ | Bundeswehr Institute of Microbiology | Bundeswehr Institute of Microbiology | Markus Antwerpen et al |
| EPI_ISL_915428 | Germany/BY-IMB15287/2021 | 2021-01-22 | Europe/Germany/Bavaria/ | Mvºnchen Klinik GmbH | Bundeswehr Institute of Microbiology | Markus Antwerpen et al |
| EPI_ISL_915437 | USA/CA-LACPHL-AF00292/2021 | 2021-01-08 | North America/USA/California/Los Ange | MD Laboratories | Los Angeles County PHL | P. Hemarajata et al. et al |

| | | | | | | |
|---|---|---|---|---|---|---|
| EPI_ISL_915438 | USA/CA-LACPHL-AF00293/2020 | 2020-12-23 | North America/USA/California/Los Ange | Los Angeles County PHL | Los Angeles County PHL | P. Hemarajata et al. et al |
| EPI_ISL_915439 | USA/CA-LACPHL-AF00294/2020 | 2020-01-18 | North America/USA/California/Los Ange | MD Laboratories | Los Angeles County PHL | P. Hemarajata et al. et al |
| EPI_ISL_915554 | England/ALDP-BCB2B5/2020 | 2020-11-22 | Europe/United Kingdom/England/ | Lighthouse Lab in Alderley Park | Wellcome Sanger Institute for the COVID-19 Genomics UK (COG-UK) Consortium | Jacquelyn Wynn et al |
| EPI_ISL_915709 | England/ALDP-BC90DB/2020 | 2020-11-23 | Europe/United Kingdom/England/ | Lighthouse Lab in Alderley Park | Wellcome Sanger Institute for the COVID-19 Genomics UK (COG-UK) Consortium | Jacquelyn Wynn et al |
| EPI_ISL_915946 | England/ALDP-BC6AB2/2020 | 2020-11-21 | Europe/United Kingdom/England/ | Lighthouse Lab in Alderley Park | Wellcome Sanger Institute for the COVID-19 Genomics UK (COG-UK) Consortium | Jacquelyn Wynn et al |
| EPI_ISL_915960 | England/ALDP-BC6AEF/2020 | 2020-11-21 | Europe/United Kingdom/England/ | Lighthouse Lab in Alderley Park | Wellcome Sanger Institute for the COVID-19 Genomics UK (COG-UK) Consortium | Jacquelyn Wynn et al |
| EPI_ISL_915962 | England/ALDP-BCB060/2020 | 2020-11-22 | Europe/United Kingdom/England/ | Lighthouse Lab in Alderley Park | Wellcome Sanger Institute for the COVID-19 Genomics UK (COG-UK) Consortium | Jacquelyn Wynn et al |
| EPI_ISL_916034 | England/ALDP-BC6D5F/2020 | 2020-11-20 | Europe/United Kingdom/England/ | Lighthouse Lab in Alderley Park | Wellcome Sanger Institute for the COVID-19 Genomics UK (COG-UK) Consortium | Jacquelyn Wynn et al |
| EPI_ISL_916085 | England/ALDP-BB928D/2020 | 2020-11-20 | Europe/United Kingdom/England/ | Lighthouse Lab in Alderley Park | Wellcome Sanger Institute for the COVID-19 Genomics UK (COG-UK) Consortium | Jacquelyn Wynn et al |

| | | | | | | |
|---|---|---|---|---|---|---|
| EPI_ISL_916245 | England/CAMC-1105034/2021 | 2021-01-20 | Europe/United Kingdom/England/ | Lighthouse Lab in Cambridge | Wellcome Sanger Institute for the COVID-19 Genomics UK (COG-UK) Consortium | Rob Howes et al |
| EPI_ISL_916507 | Scotland/QEUH-10F8062/2021 | 2021-01-23 | Europe/United Kingdom/Scotland/ | Lighthouse Lab in Glasgow | Wellcome Sanger Institute for the COVID-19 Genomics UK (COG-UK) Consortium | Harper VanSteenhouse et al |
| EPI_ISL_916594 | Scotland/QEUH-10F842A/2021 | 2021-01-23 | Europe/United Kingdom/Scotland/ | Lighthouse Lab in Glasgow | Wellcome Sanger Institute for the COVID-19 Genomics UK (COG-UK) Consortium | Harper VanSteenhouse et al |
| EPI_ISL_916597 | Scotland/QEUH-10F83F0/2021 | 2021-01-23 | Europe/United Kingdom/Scotland/ | Lighthouse Lab in Glasgow | Wellcome Sanger Institute for the COVID-19 Genomics UK (COG-UK) Consortium | Harper VanSteenhouse et al |
| EPI_ISL_916887 | England/QEUH-1015F71/2021 | 2021-01-15 | Europe/United Kingdom/England/ | Lighthouse Lab in Glasgow | Wellcome Sanger Institute for the COVID-19 Genomics UK (COG-UK) Consortium | Harper VanSteenhouse et al |
| EPI_ISL_916965 | England/ALDP-100FBD0/2021 | 2021-01-13 | Europe/United Kingdom/England/ | Lighthouse Lab in Alderley Park | Wellcome Sanger Institute for the COVID-19 Genomics UK (COG-UK) Consortium | Jacquelyn Wynn et al |
| EPI_ISL_917068 | England/ALDP-100DCB1/2021 | 2021-01-13 | Europe/United Kingdom/England/ | Lighthouse Lab in Alderley Park | Wellcome Sanger Institute for the COVID-19 Genomics UK (COG-UK) Consortium | Jacquelyn Wynn et al |
| EPI_ISL_917746 | England/ALDP-1121CE3/2021 | 2021-01-21 | Europe/United Kingdom/England/ | Lighthouse Lab in Alderley Park | Wellcome Sanger Institute for the COVID-19 Genomics UK (COG-UK) Consortium | Jacquelyn Wynn et al |

| | | | | | | |
|---|---|---|---|---|---|---|
| EPI_ISL_917802 | England/ALDP-110AEAE/2021 | 2021-01-22 | Europe/United Kingdom/England/ | Lighthouse Lab in Alderley Park | Wellcome Sanger Institute for the COVID-19 Genomics UK (COG-UK) Consortium | Jacquelyn Wynn et al |
| EPI_ISL_917904 | Scotland/QEUH-1066A25/2021 | 2021-01-17 | Europe/United Kingdom/Scotland/ | Lighthouse Lab in Glasgow | Wellcome Sanger Institute for the COVID-19 Genomics UK (COG-UK) Consortium | Harper VanSteenhouse et al |
| EPI_ISL_917910 | Scotland/QEUH-1067CB5/2021 | 2021-01-17 | Europe/United Kingdom/Scotland/ | Lighthouse Lab in Glasgow | Wellcome Sanger Institute for the COVID-19 Genomics UK (COG-UK) Consortium | Harper VanSteenhouse et al |
| EPI_ISL_917953 | USA/CA-LACPHL-AF00312/2021 | 2021-01-19 | North America/USA/California/Los Ange | Los Angeles County PHL | Los Angeles County PHL | P. Hemarajata et al. et al |
| EPI_ISL_917975 | Scotland/QEUH-1067602/2021 | 2021-01-15 | Europe/United Kingdom/Scotland/ | Lighthouse Lab in Glasgow | Wellcome Sanger Institute for the COVID-19 Genomics UK (COG-UK) Consortium | Harper VanSteenhouse et al |
| EPI_ISL_918130 | Japan/IC-0739/2021 | 21-Jan | Asia/Japan/Japan/ | Pathogen Genomics Center, National Institute of Infectious Diseases | Pathogen Genomics Center, National Institute of Infectious Diseases | Tsuyoshi Sekizuka et al |
| EPI_ISL_918133 | Japan/IC-0742/2021 | 21-Jan | Asia/Japan/Japan/ | Pathogen Genomics Center, National Institute of Infectious Diseases | Pathogen Genomics Center, National Institute of Infectious Diseases | Tsuyoshi Sekizuka et al |
| EPI_ISL_918136 | Japan/IC-0745/2021 | 21-Jan | Asia/Japan/Japan/ | Pathogen Genomics Center, National Institute of Infectious Diseases | Pathogen Genomics Center, National Institute of Infectious Diseases | Tsuyoshi Sekizuka et al |

| | | | | | | |
|---|---|---|---|---|---|---|
| EPI_ISL_918140 | Japan/IC-0749/2021 | 21-Jan | Asia/Japan/Japan/ | Pathogen Genomics Center, National Institute of Infectious Diseases | Pathogen Genomics Center, National Institute of Infectious Diseases | Tsuyoshi Sekizuka et al |
| EPI_ISL_918143 | Japan/IC-0752/2021 | 21-Jan | Asia/Japan/Japan/ | Pathogen Genomics Center, National Institute of Infectious Diseases | Pathogen Genomics Center, National Institute of Infectious Diseases | Tsuyoshi Sekizuka et al |
| EPI_ISL_918147 | Japan/IC-0756/2021 | 21-Jan | Asia/Japan/Japan/ | Pathogen Genomics Center, National Institute of Infectious Diseases | Pathogen Genomics Center, National Institute of Infectious Diseases | Tsuyoshi Sekizuka et al |
| EPI_ISL_918148 | Japan/IC-0757/2021 | 21-Jan | Asia/Japan/Japan/ | Pathogen Genomics Center, National Institute of Infectious Diseases | Pathogen Genomics Center, National Institute of Infectious Diseases | Tsuyoshi Sekizuka et al |
| EPI_ISL_918154 | Japan/IC-0763/2021 | 21-Jan | Asia/Japan/Japan/ | Pathogen Genomics Center, National Institute of Infectious Diseases | Pathogen Genomics Center, National Institute of Infectious Diseases | Tsuyoshi Sekizuka et al |
| EPI_ISL_918156 | Japan/IC-0765/2021 | 21-Jan | Asia/Italy/Japan/ | Pathogen Genomics Center, National Institute of Infectious Diseases | Pathogen Genomics Center, National Institute of Infectious Diseases | Tsuyoshi Sekizuka et al |
| EPI_ISL_918158 | Japan/IC-0767/2021 | 21-Jan | Asia/Japan/Japan/ | Pathogen Genomics Center, National Institute of Infectious Diseases | Pathogen Genomics Center, National Institute of Infectious Diseases | Tsuyoshi Sekizuka et al |
| EPI_ISL_918177 | USA/CA-IGI-2744/2021 | 2021-01-05 | North America/USA/California/Alameda | Innovative Genomics Institute, UC Berkeley | Innovative Genomics Institute, UC Berkeley | Stacia Wyman et al |

| | | | | | | |
|---|---|---|---|---|---|---|
| EPI_ISL_918178 | USA/CA-IGI-2441/2020 | 2020-11-10 | North America/USA/California/Alameda | Innovative Genomics Institute, UC Berkeley | Innovative Genomics Institute, UC Berkeley | Stacia Wyman et al |
| EPI_ISL_918182 | USA/CA-IGI-2845/2021 | 2021-01-13 | North America/USA/California/Contra C | Innovative Genomics Institute, UC Berkeley | Innovative Genomics Institute, UC Berkeley | Stacia Wyman et al |
| EPI_ISL_918186 | USA/CA-IGI-2396/2020 | 2020-12-07 | North America/USA/California/Alameda | Innovative Genomics Institute, UC Berkeley | Innovative Genomics Institute, UC Berkeley | Stacia Wyman et al |
| EPI_ISL_918206 | USA/CA-IGI-2373/2020 | 2020-11-19 | North America/USA/California/Contra C | Innovative Genomics Institute, UC Berkeley | Innovative Genomics Institute, UC Berkeley | Stacia Wyman et al |
| EPI_ISL_918209 | USA/CA-IGI-2380/2020 | 2020-11-18 | North America/USA/California/Alameda | Innovative Genomics Institute, UC Berkeley | Innovative Genomics Institute, UC Berkeley | Stacia Wyman et al |
| EPI_ISL_918242 | USA/CA-IGI-2809/2020 | 2020-12-29 | North America/USA/California/Contra C | Innovative Genomics Institute, UC Berkeley | Innovative Genomics Institute, UC Berkeley | Stacia Wyman et al |
| EPI_ISL_918245 | USA/CA-IGI-2813/2021 | 2021-01-12 | North America/USA/California/Alameda | Innovative Genomics Institute, UC Berkeley | Innovative Genomics Institute, UC Berkeley | Stacia Wyman et al |
| EPI_ISL_918249 | USA/CA-IGI-2817/2021 | 2021-01-07 | North America/USA/California/Contra C | Innovative Genomics Institute, UC Berkeley | Innovative Genomics Institute, UC Berkeley | Stacia Wyman et al |
| EPI_ISL_918251 | USA/CA-IGI-2820/2021 | 2021-01-06 | North America/USA/California/Alameda | Innovative Genomics Institute, UC Berkeley | Innovative Genomics Institute, UC Berkeley | Stacia Wyman et al |
| EPI_ISL_918252 | USA/CA-IGI-2821/2021 | 2021-01-12 | North America/USA/California/Alameda | Innovative Genomics Institute, UC Berkeley | Innovative Genomics Institute, UC Berkeley | Stacia Wyman et al |

| | | | | | | |
|---|---|---|---|---|---|---|
| EPI_ISL_918253 | USA/CA-IGI-2825/2021 | 2021-01-12 | North America/USA/California/Alameda | Innovative Genomics Institute, UC Berkeley | Innovative Genomics Institute, UC Berkeley | Stacia Wyman et al |
| EPI_ISL_918263 | Italy/ABR-IZSGC-TE9968/2021 | 2021-01-07 | Europe/Italy/Abruzzo/ | SIESP DIPARTIMENTO DI PREVENZIONE CHIE | Istituto Zooprofilattico Sperimentale dell‚ÄôAbruzzo e Molise ‚ÄúG. Caporale‚Äù | Lorusso A et al |
| EPI_ISL_918264 | Italy/ABR-IZSGC-TE14103/2021 | 2021-01-09 | Europe/Italy/Abruzzo/ | SIESP CHIETI-DRIVE IN CHIETI | Istituto Zooprofilattico Sperimentale dell‚ÄôAbruzzo e Molise ‚ÄúG. Caporale‚Äù | Lorusso A et al |
| EPI_ISL_918265 | Italy/ABR-IZSGC-TE14246/2021 | 2021-01-09 | Europe/Italy/Abruzzo/ | OSPEDALE ATRI | Istituto Zooprofilattico Sperimentale dell‚ÄôAbruzzo e Molise ‚ÄúG. Caporale‚Äù | Lorusso A et al |
| EPI_ISL_918266 | Italy/ABR-IZSGC-TE9872/2021 | 2021-01-07 | Europe/Italy/Abruzzo/ | SIESP CHIETI-Drive in Lanciano | Istituto Zooprofilattico Sperimentale dell‚ÄôAbruzzo e Molise ‚ÄúG. Caporale‚Äù | Lorusso A et al |
| EPI_ISL_918267 | Italy/ABR-IZSGC-TE8040/2021 | 2021-01-07 | Europe/Italy/Abruzzo/ | SIESP CHIETI - DRIVE IN CHIETI | Istituto Zooprofilattico Sperimentale dell‚ÄôAbruzzo e Molise ‚ÄúG. Caporale‚Äù | Lorusso A et al |
| EPI_ISL_918268 | Italy/ABR-IZSGC-TE10134/2021 | 2021-01-07 | Europe/Italy/Abruzzo/ | SIESP CHIETI - DRIVE IN ORTONA | Istituto Zooprofilattico Sperimentale dell‚ÄôAbruzzo e Molise ‚ÄúG. Caporale‚Äù | Lorusso A et al |

| | | | | | | |
|---|---|---|---|---|---|---|
| EPI_ISL_918269 | Italy/ABR-IZSGC-TE44214/2021 | 2021-01-25 | Europe/Italy/Abruzzo/ | SIESP DIPARTIMENTO DI PREVENZIONE TERAMO | Istituto Zooprofilattico Sperimentale dell‚ÄôAbruzzo e Molise ‚ÄúG. Caporale‚Äù | Lorusso A et al |
| EPI_ISL_918271 | Italy/ABR-IZSGC-TE9976/2021 | 2021-01-07 | Europe/Italy/Abruzzo/ | SIESP DIPARTIMENTO DI PREVENZIONE CHIETI | Istituto Zooprofilattico Sperimentale dell‚ÄôAbruzzo e Molise ‚ÄúG. Caporale‚Äù | Lorusso A et al |
| EPI_ISL_918276 | Spain/CT-HUVH-47524/2021 | 2021-01-04 | Europe/Spain/Catalunya/ | Hospital Universitari Vall d'Hebron - Vall d'Hebron Institut de Rercerca | Hospital Universitari Vall d'Hebron | Cristina Andr√©s et al |
| EPI_ISL_918279 | Spain/CT-HUVH-02689/2021 | 2021-01-07 | Europe/Spain/Catalunya/ | Hospital Universitari Vall d'Hebron - Vall d'Hebron Institut de Rercerca | Hospital Universitari Vall d'Hebron | Cristina Andr√©s et al |
| EPI_ISL_918281 | Spain/CT-HUVH-80851/2021 | 2021-01-08 | Europe/Spain/Catalunya/ | Hospital Universitari Vall d'Hebron - Vall d'Hebron Institut de Rercerca | Hospital Universitari Vall d'Hebron | Cristina Andr√©s et al |
| EPI_ISL_918283 | Spain/CT-HUVH-91592/2021 | 2021-01-11 | Europe/Spain/Catalunya/ | Hospital Universitari Vall d'Hebron - Vall d'Hebron Institut de Rercerca | Hospital Universitari Vall d'Hebron | Cristina Andr√©s et al |
| EPI_ISL_918286 | Spain/CT-HUVH-85537/2021 | 2021-01-11 | Europe/Spain/Catalunya/ | Hospital Universitari Vall d'Hebron - Vall d'Hebron Institut de Rercerca | Hospital Universitari Vall d'Hebron | Cristina Andr√©s et al |
| EPI_ISL_918287 | Spain/CT-HUVH-81130/2021 | 2021-01-11 | Europe/Spain/Catalunya/ | Hospital Universitari Vall d'Hebron - Vall d'Hebron Institut de Rercerca | Hospital Universitari Vall d'Hebron | Cristina Andr√©s et al |
| EPI_ISL_918288 | Spain/CT-HUVH-25339/2021 | 2021-01-12 | Europe/Spain/Catalunya/ | Hospital Universitari Vall d'Hebron - Vall d'Hebron Institut de Rercerca | Hospital Universitari Vall d'Hebron | Cristina Andr√©s et al |
| EPI_ISL_918292 | Spain/CT-HUVH-82088/2021 | 2021-01-19 | Europe/Spain/Catalunya/ | Hospital Universitari Vall d'Hebron - Vall d'Hebron Institut de Rercerca | Hospital Universitari Vall d'Hebron | Cristina Andr√©s et al |
| EPI_ISL_918295 | Spain/CT-HUVH-81186/2021 | 2021-01-20 | Europe/Spain/Catalunya/ | Hospital Universitari Vall d'Hebron - Vall d'Hebron Institut de Rercerca | Hospital Universitari Vall d'Hebron | Cristina Andr√©s et al |

| | | | | | | |
|---|---|---|---|---|---|---|
| EPI_ISL_918298 | Spain/CT-HUVH-50776/2021 | 2021-01-19 | Europe/Spain/Catalunya/ | Hospital Universitari Vall d'Hebron - Vall d'Hebron Institut de Rercerca | Hospital Universitari Vall d'Hebron | Cristina Andr√©s et al |
| EPI_ISL_918302 | Spain/CT-HUVH-37348/2021 | 2021-01-20 | Europe/Spain/Catalunya/ | Hospital Universitari Vall d'Hebron - Vall d'Hebron Institut de Rercerca | Hospital Universitari Vall d'Hebron | Cristina Andr√©s et al |
| EPI_ISL_918303 | Spain/CT-HUVH-86003/2021 | 2021-01-18 | Europe/Spain/Catalunya/ | Hospital Universitari Vall d'Hebron - Vall d'Hebron Institut de Rercerca | Hospital Universitari Vall d'Hebron | Cristina Andr√©s et al |
| EPI_ISL_918306 | Spain/CT-HUVH-95508/2021 | 2021-01-18 | Europe/Spain/Catalunya/ | Hospital Universitari Vall d'Hebron - Vall d'Hebron Institut de Rercerca | Hospital Universitari Vall d'Hebron | Cristina Andr√©s et al |
| EPI_ISL_918308 | Spain/CT-HUVH-03065/2021 | 2021-01-19 | Europe/Spain/Catalunya/ | Hospital Universitari Vall d'Hebron - Vall d'Hebron Institut de Rercerca | Hospital Universitari Vall d'Hebron | Cristina Andr√©s et al |
| EPI_ISL_918315 | Spain/CT-HUVH-13287/2021 | 2021-01-21 | Europe/Spain/Catalunya/ | Hospital Universitari Vall d'Hebron - Vall d'Hebron Institut de Rercerca | Hospital Universitari Vall d'Hebron | Cristina Andr√©s et al |
| EPI_ISL_918317 | Spain/CT-HUVH-12244/2021 | 2021-01-20 | Europe/Spain/Catalunya/ | Hospital Universitari Vall d'Hebron - Vall d'Hebron Institut de Rercerca | Hospital Universitari Vall d'Hebron | Cristina Andr√©s et al |
| EPI_ISL_918322 | Spain/CT-HUVH-62862/2021 | 2021-01-19 | Europe/Spain/Catalunya/ | Hospital Universitari Vall d'Hebron - Vall d'Hebron Institut de Rercerca | Hospital Universitari Vall d'Hebron | Cristina Andr√©s et al |
| EPI_ISL_918326 | Spain/CT-HUVH-85927/2021 | 2021-01-18 | Europe/Spain/Catalunya/ | Hospital Universitari Vall d'Hebron - Vall d'Hebron Institut de Rercerca | Hospital Universitari Vall d'Hebron | Cristina Andr√©s et al |
| EPI_ISL_918327 | Spain/CT-HUVH-13108/2021 | 2021-01-21 | Europe/Spain/Catalunya/ | Hospital Universitari Vall d'Hebron - Vall d'Hebron Institut de Rercerca | Hospital Universitari Vall d'Hebron | Cristina Andr√©s et al |
| EPI_ISL_918333 | Spain/CT-HUVH-61176/2021 | 2021-01-18 | Europe/Spain/Catalunya/ | Hospital Universitari Vall d'Hebron - Vall d'Hebron Institut de Rercerca | Hospital Universitari Vall d'Hebron | Cristina Andr√©s et al |
| EPI_ISL_918334 | Spain/CT-HUVH-15640/2021 | 2021-01-19 | Europe/Spain/Catalunya/ | Hospital Universitari Vall d'Hebron - Vall d'Hebron Institut de Rercerca | Hospital Universitari Vall d'Hebron | Cristina Andr√©s et al |
| EPI_ISL_918335 | Spain/CT-HUVH-16402/2021 | 2021-01-18 | Europe/Spain/Catalunya/ | Hospital Universitari Vall d'Hebron - Vall d'Hebron Institut de Rercerca | Hospital Universitari Vall d'Hebron | Cristina Andr√©s et al |

| | | | | | | |
|---|---|---|---|---|---|---|
| EPI_ISL_918336 | Spain/CT-HUVH-73887/2021 | 2021-01-20 | Europe/Spain/Catalunya/ | Hospital Universitari Vall d'Hebron - Vall d'Hebron Institut de Rercerca | Hospital Universitari Vall d'Hebron | Cristina Andr√©s et al |
| EPI_ISL_918337 | Spain/CT-HUVH-23673/2021 | 2021-01-19 | Europe/Spain/Catalunya/ | Hospital Universitari Vall d'Hebron - Vall d'Hebron Institut de Rercerca | Hospital Universitari Vall d'Hebron | Cristina Andr√©s et al |
| EPI_ISL_918341 | Germany/BW-FR0431/2020 | 2020-12-21 | Europe/Germany/Baden-Wuerttemberg | Institute of Virology, Medical Center, University of Freiburg, Freiburg, Germany | Institute of Virology, Clinial Virus Genomics, Medical Center, University of Freiburg, Freiburg, Germany | Jonas Fuchs et al |
| EPI_ISL_918342 | Germany/BW-FR0432/2020 | 2020-12-21 | Europe/Germany/Baden-Wuerttemberg | Institute of Virology, Medical Center, University of Freiburg, Freiburg, Germany | Institute of Virology, Clinial Virus Genomics, Medical Center, University of Freiburg, Freiburg, Germany | Jonas Fuchs et al |
| EPI_ISL_918351 | Germany/BW-FR0443/2020 | 2020-12-29 | Europe/Germany/Baden-Wuerttemberg | Institute of Virology, Medical Center, University of Freiburg, Freiburg, Germany | Institute of Virology, Clinial Virus Genomics, Medical Center, University of Freiburg, Freiburg, Germany | Jonas Fuchs et al |
| EPI_ISL_918374 | Cambodia/112144/2020 | 2020-11-09 | Asia/Cambodia/Cambodia/ | Virology Unit, Institut Pasteur du Cambodge | Virology Unit, Institut Pasteur du Cambodge | Sokhoun Yann et al |
| EPI_ISL_918407 | BosniaandHerzegovina/agc788/2021 | 2021-01-11 | Europe/Bosnia and Herzegovina/Sarajev | ALEA Dr Kandic | Alea Genetic Centre | Rijad Konjhodzic et al |

| | | | | | | | |
|---|---|---|---|---|---|---|---|
| EPI_ISL_918412 | Italy/CAM-CRGS-42/2020 | 2020-03-24 | Europe/Italy/Campania/ | | 1.AO Universitaria 'S. Giovanni di Dio e Ruggi D'Aragona, Scuola Medica Salernitana' Hospital / 2.UOC di Virologia e Microbiologia, Universit√† della Campania 'L. Vanvitelli' / 3.AO Universitaria 'Federico II' Napoli Hospital / 4.AORN 'San Giuseppe Moscati' Avellino Hospital / 5.AO 'San Pio - presidio G. Rummo' Benevento Hospital / 6.AO 'Sant'Anna e San Sebastiano' Caserta Hospital / 7.PO 'Maria Santissima Addolorata' Eboli Hospital / 8.Biogem Istituto di Ricerche Genetiche | 1. Genome Research Center for Health (CRGS) / 2. Laboratory of Molecular Medicine and Genomics(LMMGe) / 3. Center for Research in Pure and Applied Mathematics (CRMPA) | Giorgio Giurato (Corresponding Author) et al |
| EPI_ISL_918413 | Italy/CAM-CRGS-55/2020 | 2020-03-19 | Europe/Italy/Campania/ | | 1.AO Universitaria 'S. Giovanni di Dio e Ruggi D'Aragona, Scuola Medica Salernitana' Hospital / 2.UOC di Virologia e Microbiologia, Universit√† della Campania 'L. Vanvitelli' / 3.AO Universitaria 'Federico II' Napoli Hospital / 4.AORN 'San Giuseppe Moscati' Avellino Hospital / 5.AO 'San Pio - presidio G. Rummo' Benevento Hospital / 6.AO 'Sant'Anna e San Sebastiano' Caserta Hospital / 7.PO 'Maria Santissima Addolorata' Eboli Hospital / 8.Biogem Istituto di Ricerche Genetiche | 1. Genome Research Center for Health (CRGS) / 2. Laboratory of Molecular Medicine and Genomics(LMMGe) / 3. Center for Research in Pure and Applied Mathematics (CRMPA) | Giorgio Giurato (Corresponding Author) et al |
| EPI_ISL_918415 | Italy/CAM-CRGS-62/2020 | 2020-03-30 | Europe/Italy/Campania/ | | 1.AO Universitaria 'S. Giovanni di Dio e Ruggi D'Aragona, Scuola Medica Salernitana' Hospital / 2.UOC di Virologia e Microbiologia, Universit√† della Campania 'L. Vanvitelli' / 3.AO Universitaria 'Federico II' Napoli Hospital / 4.AORN 'San Giuseppe Moscati' Avellino Hospital / 5.AO 'San Pio - presidio G. Rummo' Benevento Hospital / 6.AO 'Sant'Anna e San Sebastiano' Caserta Hospital / 7.PO 'Maria Santissima Addolorata' Eboli Hospital / 8.Biogem Istituto di Ricerche Genetiche | 1. Genome Research Center for Health (CRGS) / 2. Laboratory of Molecular Medicine and Genomics(LMMGe) / 3. Center for Research in Pure and Applied Mathematics (CRMPA) | Giorgio Giurato (Corresponding Author) et al |

| | | | | | | |
|---|---|---|---|---|---|---|
| EPI_ISL_918417 | Italy/CAM-CRGS-64/2020 | 2020-04-02 | Europe/Italy/Campania/ | 1.AO Universitaria 'S. Giovanni di Dio e Ruggi D'Aragona, Scuola Medica Salernitana' Hospital / 2.UOC di Virologia e Microbiologia, Universit√† della Campania 'L. Vanvitelli' / 3.AO Universitaria 'Federico II' Napoli Hospital / 4.AORN 'San Giuseppe Moscati' Avellino Hospital / 5.AO 'San Pio - presidio G. Rummo' Benevento Hospital / 6.AO 'Sant'Anna e San Sebastiano' Caserta Hospital / 7.PO 'Maria Santissima Addolorata' Eboli Hospital / 8.Biogem Istituto di Ricerche Genetiche | 1. Genome Research Center for Health (CRGS) / 2. Laboratory of Molecular Medicine and Genomics(LMMGe) / 3. Center for Research in Pure and Applied Mathematics (CRMPA) | Giorgio Giurato (Corresponding Author) et al |
| EPI_ISL_918420 | Italy/CAM-CRGS-70/2020 | 2020-03-04 | Europe/Italy/Campania/ | 1.AO Universitaria 'S. Giovanni di Dio e Ruggi D'Aragona, Scuola Medica Salernitana' Hospital / 2.UOC di Virologia e Microbiologia, Universit√† della Campania 'L. Vanvitelli' / 3.AO Universitaria 'Federico II' Napoli Hospital / 4.AORN 'San Giuseppe Moscati' Avellino Hospital / 5.AO 'San Pio - presidio G. Rummo' Benevento Hospital / 6.AO 'Sant'Anna e San Sebastiano' Caserta Hospital / 7.PO 'Maria Santissima Addolorata' Eboli Hospital / 8.Biogem Istituto di Ricerche Genetiche | 1. Genome Research Center for Health (CRGS) / 2. Laboratory of Molecular Medicine and Genomics(LMMGe) / 3. Center for Research in Pure and Applied Mathematics (CRMPA) | Giorgio Giurato (Corresponding Author) et al |
| EPI_ISL_918422 | Italy/CAM-CRGS-82/2020 | 2020-04-05 | Europe/Italy/Campania/ | 1.AO Universitaria 'S. Giovanni di Dio e Ruggi D'Aragona, Scuola Medica Salernitana' Hospital / 2.UOC di Virologia e Microbiologia, Universit√† della Campania 'L. Vanvitelli' / 3.AO Universitaria 'Federico II' Napoli Hospital / 4.AORN 'San Giuseppe Moscati' Avellino Hospital / 5.AO 'San Pio - presidio G. Rummo' Benevento Hospital / 6.AO 'Sant'Anna e San Sebastiano' Caserta Hospital / 7.PO 'Maria Santissima Addolorata' Eboli Hospital / 8.Biogem Istituto di Ricerche Genetiche | 1. Genome Research Center for Health (CRGS) / 2. Laboratory of Molecular Medicine and Genomics(LMMGe) / 3. Center for Research in Pure and Applied Mathematics (CRMPA) | Giorgio Giurato (Corresponding Author) et al |
| EPI_ISL_918423 | Germany/BY-IMB15351/2021 | 2021-01-26 | Europe/Germany/Bavaria/ | M√ºnchen Klinik GmbH | Bundeswehr Institute of Microbiology | Markus Antwerpen et al |

| | | | | | | |
|---|---|---|---|---|---|---|
| EPI_ISL_918424 | Germany/RP-IMB15360/2021 | 2021-01-26 | Europe/Germany/Rheinland-Pfalz/ | Bundeswehrzentralkrankenhaus Koblenz | Bundeswehr Institute of Microbiology | Markus Antwerpen et al |
| EPI_ISL_918429 | Germany/BY-IMB15379/2021 | 2021-01-27 | Europe/Germany/Bavaria/ | M√ºnchen Klinik GmbH | Bundeswehr Institute of Microbiology | Markus Antwerpen et al |
| EPI_ISL_918432 | Germany/BE-IMB15402/2021 | 2021-01-28 | Europe/Germany/Berlin/ | Bundeswehrkrankenhaus Berlin | Bundeswehr Institute of Microbiology | Markus Antwerpen et al |
| EPI_ISL_918452 | Ireland/D-NVRL-AIIDB0166/2020 | 2020-12-31 | Europe/Ireland/Dublin/ | AIID | Irish Coronavirus Sequencing Consortium-Teagasc Grange | Matthew McCabe et al |
| EPI_ISL_918457 | Italy/CAM-CRGS-13/2020 | 2020-03-15 | Europe/Italy/Campania/ | 1.AO Universitaria 'S. Giovanni di Dio e Ruggi D'Aragona, Scuola Medica Salernitana' Hospital / 2.UOC di Virologia e Microbiologia, Universit√† della Campania 'L. Vanvitelli' / 3.AO Universitaria 'Federico II' Napoli Hospital / 4.AORN 'San Giuseppe Moscati' Avellino Hospital / 5.AO 'San Pio - presidio G. Rummo' Benevento Hospital / 6.AO 'Sant'Anna e San Sebastiano' Caserta Hospital / 7.PO 'Maria Santissima Addolorata' Eboli Hospital / 8.Biogem Istituto di Ricerche Genetiche | 1. Genome Research Center for Health (CRGS) / 2. Laboratory of Molecular Medicine and Genomics(LMMGe) / 3. Center for Research in Pure and Applied Mathematics (CRMPA) | Giorgio Giurato (Corresponding Author) et al |
| EPI_ISL_918460 | Italy/CAM-CRGS-16/2020 | 2020-03-16 | Europe/Italy/Campania/ | 1.AO Universitaria 'S. Giovanni di Dio e Ruggi D'Aragona, Scuola Medica Salernitana' Hospital / 2.UOC di Virologia e Microbiologia, Universit√† della Campania 'L. Vanvitelli' / 3.AO Universitaria 'Federico II' Napoli Hospital / 4.AORN 'San Giuseppe Moscati' Avellino Hospital / 5.AO 'San Pio - presidio G. Rummo' Benevento Hospital / 6.AO 'Sant'Anna e San Sebastiano' Caserta Hospital / 7.PO 'Maria Santissima Addolorata' Eboli Hospital / 8.Biogem Istituto di Ricerche Genetiche | 1. Genome Research Center for Health (CRGS) / 2. Laboratory of Molecular Medicine and Genomics(LMMGe) / 3. Center for Research in Pure and Applied Mathematics (CRMPA) | Giorgio Giurato (Corresponding Author) et al |

| | | | | | | |
|---|---|---|---|---|---|---|
| EPI_ISL_918461 | Italy/CAM-CRGS-17/2020 | 2020-03-16 | Europe/Italy/Campania/ | 1.AO Universitaria 'S. Giovanni di Dio e Ruggi D'Aragona, Scuola Medica Salernitana' Hospital / 2.UOC di Virologia e Microbiologia, Universit√† della Campania 'L. Vanvitelli' / 3.AO Universitaria 'Federico II' Napoli Hospital / 4.AORN 'San Giuseppe Moscati' Avellino Hospital / 5.AO 'San Pio - presidio G. Rummo' Benevento Hospital / 6.AO 'Sant'Anna e San Sebastiano' Caserta Hospital / 7.PO 'Maria Santissima Addolorata' Eboli Hospital / 8.Biogem Istituto di Ricerche Genetiche | 1. Genome Research Center for Health (CRGS) / 2. Laboratory of Molecular Medicine and Genomics(LMMGe) / 3. Center for Research in Pure and Applied Mathematics (CRMPA) | Giorgio Giurato (Corresponding Author) et al |
| EPI_ISL_918463 | Italy/CAM-CRGS-21/2020 | 2020-03-18 | Europe/Italy/Campania/ | 1.AO Universitaria 'S. Giovanni di Dio e Ruggi D'Aragona, Scuola Medica Salernitana' Hospital / 2.UOC di Virologia e Microbiologia, Universit√† della Campania 'L. Vanvitelli' / 3.AO Universitaria 'Federico II' Napoli Hospital / 4.AORN 'San Giuseppe Moscati' Avellino Hospital / 5.AO 'San Pio - presidio G. Rummo' Benevento Hospital / 6.AO 'Sant'Anna e San Sebastiano' Caserta Hospital / 7.PO 'Maria Santissima Addolorata' Eboli Hospital / 8.Biogem Istituto di Ricerche Genetiche | 1. Genome Research Center for Health (CRGS) / 2. Laboratory of Molecular Medicine and Genomics(LMMGe) / 3. Center for Research in Pure and Applied Mathematics (CRMPA) | Giorgio Giurato (Corresponding Author) et al |
| EPI_ISL_918469 | Italy/CAM-CRGS-51/2020 | 2020-04-03 | Europe/Italy/Campania/ | 1.AO Universitaria 'S. Giovanni di Dio e Ruggi D'Aragona, Scuola Medica Salernitana' Hospital / 2.UOC di Virologia e Microbiologia, Universit√† della Campania 'L. Vanvitelli' / 3.AO Universitaria 'Federico II' Napoli Hospital / 4.AORN 'San Giuseppe Moscati' Avellino Hospital / 5.AO 'San Pio - presidio G. Rummo' Benevento Hospital / 6.AO 'Sant'Anna e San Sebastiano' Caserta Hospital / 7.PO 'Maria Santissima Addolorata' Eboli Hospital / 8.Biogem Istituto di Ricerche Genetiche | 1. Genome Research Center for Health (CRGS) / 2. Laboratory of Molecular Medicine and Genomics(LMMGe) / 3. Center for Research in Pure and Applied Mathematics (CRMPA) | Giorgio Giurato (Corresponding Author) et al |

| | | | | | | |
|---|---|---|---|---|---|---|
| EPI_ISL_918470 | Italy/CAM-CRGS-56/2020 | 2020-03-16 | Europe/Italy/Campania/ | 1.AO Universitaria 'S. Giovanni di Dio e Ruggi D'Aragona, Scuola Medica Salernitana' Hospital / 2.UOC di Virologia e Microbiologia, Universit√† della Campania 'L. Vanvitelli' / 3.AO Universitaria 'Federico II' Napoli Hospital / 4.AORN 'San Giuseppe Moscati' Avellino Hospital / 5.AO 'San Pio - presidio G. Rummo' Benevento Hospital / 6.AO 'Sant'Anna e San Sebastiano' Caserta Hospital / 7.PO 'Maria Santissima Addolorata' Eboli Hospital / 8.Biogem Istituto di Ricerche Genetiche | 1. Genome Research Center for Health (CRGS) / 2. Laboratory of Molecular Medicine and Genomics(LMMGe) / 3. Center for Research in Pure and Applied Mathematics (CRMPA) | Giorgio Giurato (Corresponding Author) et al |
| EPI_ISL_918473 | Italy/CAM-CRGS-65/2020 | 2020-03-19 | Europe/Italy/Campania/ | 1.AO Universitaria 'S. Giovanni di Dio e Ruggi D'Aragona, Scuola Medica Salernitana' Hospital / 2.UOC di Virologia e Microbiologia, Universit√† della Campania 'L. Vanvitelli' / 3.AO Universitaria 'Federico II' Napoli Hospital / 4.AORN 'San Giuseppe Moscati' Avellino Hospital / 5.AO 'San Pio - presidio G. Rummo' Benevento Hospital / 6.AO 'Sant'Anna e San Sebastiano' Caserta Hospital / 7.PO 'Maria Santissima Addolorata' Eboli Hospital / 8.Biogem Istituto di Ricerche Genetiche | 1. Genome Research Center for Health (CRGS) / 2. Laboratory of Molecular Medicine and Genomics(LMMGe) / 3. Center for Research in Pure and Applied Mathematics (CRMPA) | Giorgio Giurato (Corresponding Author) et al |
| EPI_ISL_918477 | Italy/CAM-CRGS-77/2020 | 2020-03-19 | Europe/Italy/Campania/ | 1.AO Universitaria 'S. Giovanni di Dio e Ruggi D'Aragona, Scuola Medica Salernitana' Hospital / 2.UOC di Virologia e Microbiologia, Universit√† della Campania 'L. Vanvitelli' / 3.AO Universitaria 'Federico II' Napoli Hospital / 4.AORN 'San Giuseppe Moscati' Avellino Hospital / 5.AO 'San Pio - presidio G. Rummo' Benevento Hospital / 6.AO 'Sant'Anna e San Sebastiano' Caserta Hospital / 7.PO 'Maria Santissima Addolorata' Eboli Hospital / 8.Biogem Istituto di Ricerche Genetiche | 1. Genome Research Center for Health (CRGS) / 2. Laboratory of Molecular Medicine and Genomics(LMMGe) / 3. Center for Research in Pure and Applied Mathematics (CRMPA) | Giorgio Giurato (Corresponding Author) et al |

| | | | | | | |
|---|---|---|---|---|---|---|
| EPI_ISL_918479 | Italy/CAM-CRGS-79/2020 | 2020-03-20 | Europe/Italy/Campania/ | 1.AO Universitaria 'S. Giovanni di Dio e Ruggi D'Aragona, Scuola Medica Salernitana' Hospital / 2.UOC di Virologia e Microbiologia, Universit√† della Campania 'L. Vanvitelli' / 3.AO Universitaria 'Federico II' Napoli Hospital / 4.AORN 'San Giuseppe Moscati' Avellino Hospital / 5.AO 'San Pio - presidio G. Rummo' Benevento Hospital / 6.AO 'Sant'Anna e San Sebastiano' Caserta Hospital / 7.PO 'Maria Santissima Addolorata' Eboli Hospital / 8.Biogem Istituto di Ricerche Genetiche | 1. Genome Research Center for Health (CRGS) / 2. Laboratory of Molecular Medicine and Genomics(LMMGe) / 3. Center for Research in Pure and Applied Mathematics (CRMPA) | Giorgio Giurato (Corresponding Author) et al |
| EPI_ISL_918480 | Italy/CAM-CRGS-80/2020 | 2020-03-20 | Europe/Italy/Campania/ | 1.AO Universitaria 'S. Giovanni di Dio e Ruggi D'Aragona, Scuola Medica Salernitana' Hospital / 2.UOC di Virologia e Microbiologia, Universit√† della Campania 'L. Vanvitelli' / 3.AO Universitaria 'Federico II' Napoli Hospital / 4.AORN 'San Giuseppe Moscati' Avellino Hospital / 5.AO 'San Pio - presidio G. Rummo' Benevento Hospital / 6.AO 'Sant'Anna e San Sebastiano' Caserta Hospital / 7.PO 'Maria Santissima Addolorata' Eboli Hospital / 8.Biogem Istituto di Ricerche Genetiche | 1. Genome Research Center for Health (CRGS) / 2. Laboratory of Molecular Medicine and Genomics(LMMGe) / 3. Center for Research in Pure and Applied Mathematics (CRMPA) | Giorgio Giurato (Corresponding Author) et al |
| EPI_ISL_918481 | Italy/CAM-CRGS-85/2020 | 2020-04-13 | Europe/Italy/Campania/ | 1.AO Universitaria 'S. Giovanni di Dio e Ruggi D'Aragona, Scuola Medica Salernitana' Hospital / 2.UOC di Virologia e Microbiologia, Universit√† della Campania 'L. Vanvitelli' / 3.AO Universitaria 'Federico II' Napoli Hospital / 4.AORN 'San Giuseppe Moscati' Avellino Hospital / 5.AO 'San Pio - presidio G. Rummo' Benevento Hospital / 6.AO 'Sant'Anna e San Sebastiano' Caserta Hospital / 7.PO 'Maria Santissima Addolorata' Eboli Hospital / 8.Biogem Istituto di Ricerche Genetiche | 1. Genome Research Center for Health (CRGS) / 2. Laboratory of Molecular Medicine and Genomics(LMMGe) / 3. Center for Research in Pure and Applied Mathematics (CRMPA) | Giorgio Giurato (Corresponding Author) et al |
| EPI_ISL_918493 | USA/FL-BPHL-0046/2021 | 2021-01-15 | North America/USA/Florida/ | Florida Bureau of Public Health Laboratories | Florida Bureau of Public Health Laboratories | Sarah Schmedes et al |

| | | | | | | |
|---|---|---|---|---|---|---|
| EPI_ISL_918495 | USA/FL-BPHL-0047/2021 | 2021-01-18 | North America/USA/Florida/ | Florida Bureau of Public Health Laboratories | Florida Bureau of Public Health Laboratories | Sarah Schmedes et al |
| EPI_ISL_918499 | Brazil/AM-IEC-177290/2020 | 2020-12-21 | South America/Brazil/Amazonas BR/ | LACEN - Laboratv≥rio Central de Sav∫de Pv∫blica do Amazonas | ÔªøEvandro Chagas Institute | Santos et al |
| EPI_ISL_918500 | Brazil/AM-IEC-177291/2020 | 2020-12-28 | South America/Brazil/Amazonas BR/ | LACEN - Laboratv≥rio Central de Sav∫de Pv∫blica do Amazonas | ÔªøEvandro Chagas Institute | Santos et al |
| EPI_ISL_918501 | Brazil/AM-IEC-177292/2020 | 2020-12-28 | South America/Brazil/Amazonas BR/ | LACEN - Laboratv≥rio Central de Sav∫de Pv∫blica do Amazonas | ÔªøEvandro Chagas Institute | Santos et al |
| EPI_ISL_918502 | Brazil/AM-IEC-177293/2020 | 2020-12-29 | South America/Brazil/Amazonas BR/ | LACEN - Laboratv≥rio Central de Sav∫de Pv∫blica do Amazonas | ÔªøEvandro Chagas Institute | Santos et al |
| EPI_ISL_918503 | Brazil/AM-IEC-177294/2020 | 2020-12-28 | South America/Brazil/Amazonas BR/ | LACEN - Laboratv≥rio Central de Sav∫de Pv∫blica do Amazonas | ÔªøEvandro Chagas Institute | Santos et al |
| EPI_ISL_918504 | Brazil/AM-IEC-177301/2020 | 2020-12-21 | South America/Brazil/Amazonas BR/ | LACEN - Laboratv≥rio Central de Sav∫de Pv∫blica do Amazonas | ÔªøEvandro Chagas Institute | Santos et al |
| EPI_ISL_918505 | Brazil/AM-IEC-177310/2020 | 2020-12-30 | South America/Brazil/Amazonas BR/ | LACEN - Laboratv≥rio Central de Sav∫de Pv∫blica do Amazonas | ÔªøEvandro Chagas Institute | Santos et al |
| EPI_ISL_918506 | Brazil/AM-IEC-177312/2020 | 2020-12-30 | South America/Brazil/Amazonas BR/ | LACEN - Laboratv≥rio Central de Sav∫de Pv∫blica do Amazonas | ÔªøEvandro Chagas Institute | Santos et al |
| EPI_ISL_918507 | Brazil/AM-IEC-177314/2021 | 2021-01-05 | South America/Brazil/Amazonas BR/ | LACEN - Laboratv≥rio Central de Sav∫de Pv∫blica do Amazonas | ÔªøEvandro Chagas Institute | Santos et al |
| EPI_ISL_918508 | Brazil/AM-IEC-177320/2021 | 2021-01-05 | South America/Brazil/Amazonas BR/ | LACEN - Laboratv≥rio Central de Sav∫de Pv∫blica do Amazonas | ÔªøEvandro Chagas Institute | Santos et al |
| EPI_ISL_918509 | Brazil/AM-IEC-177322/2021 | 2021-01-06 | South America/Brazil/Amazonas BR/ | LACEN - Laboratv≥rio Central de Sav∫de Pv∫blica do Amazonas | ÔªøEvandro Chagas Institute | Santos et al |
| EPI_ISL_918510 | Brazil/AM-IEC-177323/2021 | 2021-01-07 | South America/Brazil/Amazonas BR/ | LACEN - Laboratv≥rio Central de Sav∫de Pv∫blica do Amazonas | ÔªøEvandro Chagas Institute | Santos et al |
| EPI_ISL_918511 | Brazil/AM-IEC-177328/2020 | 2020-12-30 | South America/Brazil/Amazonas BR/ | LACEN - Laboratv≥rio Central de Sav∫de Pv∫blica do Amazonas | ÔªøEvandro Chagas Institute | Santos et al |
| EPI_ISL_918512 | Brazil/AM-IEC-177300/2020 | 2020-12-21 | South America/Brazil/Amazonas BR/ | LACEN - Laboratv≥rio Central de Sav∫de Pv∫blica do Amazonas | ÔªøEvandro Chagas Institute | Santos et al |
| EPI_ISL_918513 | Brazil/RR-IEC-176346/2020 | 2020-06-01 | South America/Brazil/Roraima/ | LACEN - Laboratv≥rio Central de Sav∫de Pv∫blica do Roraima | ÔªøEvandro Chagas Institute | Santos et al |
| EPI_ISL_918514 | Brazil/PA-IEC-176581/2020 | 2020-10-14 | South America/Brazil/Para/ | ÔªøEvandro Chagas Institute | ÔªøEvandro Chagas Institute | Santos et al |
| EPI_ISL_918515 | Brazil/PA-IEC-176888/2020 | 2020-11-18 | South America/Brazil/Para/ | LACEN - Laboratv≥rio Central de Sav∫de Pv∫blica do Para | ÔªøEvandro Chagas Institute | Santos et al |
| EPI_ISL_918516 | Brazil/PA-IEC-176890/2020 | 2020-11-23 | South America/Brazil/Para/ | LACEN - Laboratv≥rio Central de Sav∫de Pv∫blica do Para | ÔªøEvandro Chagas Institute | Santos et al |

| | | | | | | |
|---|---|---|---|---|---|---|
| EPI_ISL_918518 | Brazil/PA-IEC-176946/2020 | 2020-12-15 | South America/Brazil/Para/ | Ôª¿Evandro Chagas Institute | Ôª¿Evandro Chagas Institute | Santos et al |
| EPI_ISL_918519 | Brazil/PA-IEC-176950/2020 | 2020-12-15 | South America/Brazil/Para/ | Ôª¿Evandro Chagas Institute | Ôª¿Evandro Chagas Institute | Santos et al |
| EPI_ISL_918520 | Brazil/PA-IEC-176952/2020 | 2020-12-15 | South America/Brazil/Para/ | Ôª¿Evandro Chagas Institute | Ôª¿Evandro Chagas Institute | Santos et al |
| EPI_ISL_918521 | Brazil/PA-IEC-177009/2020 | 2020-12-21 | South America/Brazil/Para/ | Ôª¿Evandro Chagas Institute | Ôª¿Evandro Chagas Institute | Santos et al |
| EPI_ISL_918522 | Brazil/PA-IEC-177041/2020 | 2020-12-25 | South America/Brazil/Para/ | LACEN - Laboratv≥rio Central de Sav∫de Pv∫blica do Para | Ôª¿Evandro Chagas Institute | Santos et al |
| EPI_ISL_918524 | Brazil/PA-IEC-177044/2020 | 2020-12-09 | South America/Brazil/Para/ | LACEN - Laboratv≥rio Central de Sav∫de Pv∫blica do Para | Ôª¿Evandro Chagas Institute | Santos et al |
| EPI_ISL_918525 | Brazil/PA-IEC-177045/2020 | 2020-12-09 | South America/Brazil/Para/ | LACEN - Laboratv≥rio Central de Sav∫de Pv∫blica do Para | Ôª¿Evandro Chagas Institute | Santos et al |
| EPI_ISL_918526 | Brazil/PA-IEC-177046/2020 | 2020-12-06 | South America/Brazil/Para/ | LACEN - Laboratv≥rio Central de Sav∫de Pv∫blica do Para | Ôª¿Evandro Chagas Institute | Santos et al |
| EPI_ISL_918527 | Brazil/PA-IEC-177047/2020 | 2020-12-07 | South America/Brazil/Para/ | LACEN - Laboratv≥rio Central de Sav∫de Pv∫blica do Para | Ôª¿Evandro Chagas Institute | Santos et al |
| EPI_ISL_918528 | Brazil/PA-IEC-177048/2020 | 2020-12-07 | South America/Brazil/Para/ | LACEN - Laboratv≥rio Central de Sav∫de Pv∫blica do Para | Ôª¿Evandro Chagas Institute | Santos et al |
| EPI_ISL_918529 | Brazil/PA-IEC-177049/2020 | 2020-12-19 | South America/Brazil/Para/ | LACEN - Laboratv≥rio Central de Sav∫de Pv∫blica do Para | Ôª¿Evandro Chagas Institute | Santos et al |
| EPI_ISL_918530 | Brazil/PA-IEC-177050/2020 | 2020-12-19 | South America/Brazil/Para/ | LACEN - Laboratv≥rio Central de Sav∫de Pv∫blica do Para | Ôª¿Evandro Chagas Institute | Santos et al |
| EPI_ISL_918531 | Brazil/AM-IEC-177274/2020 | 2020-11-10 | South America/Brazil/Amazonas BR/ | LACEN - Laboratv≥rio Central de Sav∫de Pv∫blica do Amazonas | Ôª¿Evandro Chagas Institute | Santos et al |
| EPI_ISL_918532 | Brazil/AM-IEC-177282/2020 | 2020-11-13 | South America/Brazil/Amazonas BR/ | LACEN - Laboratv≥rio Central de Sav∫de Pv∫blica do Amazonas | Ôª¿Evandro Chagas Institute | Santos et al |
| EPI_ISL_918533 | Brazil/AM-IEC-177284/2020 | 2020-11-10 | South America/Brazil/Amazonas BR/ | LACEN - Laboratv≥rio Central de Sav∫de Pv∫blica do Amazonas | Ôª¿Evandro Chagas Institute | Santos et al |
| EPI_ISL_918534 | Brazil/AM-IEC-177289/2020 | 2020-12-21 | South America/Brazil/Amazonas BR/ | LACEN - Laboratv≥rio Central de Sav∫de Pv∫blica do Amazonas | Ôª¿Evandro Chagas Institute | Santos et al |
| EPI_ISL_918535 | Brazil/AM-IEC-177317/2021 | 2021-01-05 | South America/Brazil/Amazonas BR/ | LACEN - Laboratv≥rio Central de Sav∫de Pv∫blica do Amazonas | Ôª¿Evandro Chagas Institute | Santos et al |
| EPI_ISL_918537 | Brazil/CE-IEC-177340/2021 | 2021-01-07 | South America/Brazil/Cearv°/ | LACEN - Laboratv≥rio Central de Sav∫de Pv∫blica do Ceara | Ôª¿Evandro Chagas Institute | Santos et al |
| EPI_ISL_918538 | Brazil/CE-IEC-177341/2021 | 2021-01-07 | South America/Brazil/Cearv°/ | LACEN - Laboratv≥rio Central de Sav∫de Pv∫blica do Ceara | Ôª¿Evandro Chagas Institute | Santos et al |
| EPI_ISL_918539 | Brazil/CE-IEC-177342/2021 | 2021-01-08 | South America/Brazil/Cearv°/ | LACEN - Laboratv≥rio Central de Sav∫de Pv∫blica do Ceara | Ôª¿Evandro Chagas Institute | Santos et al |

| | | | | | | |
|---|---|---|---|---|---|---|
| EPI_ISL_918540 | Brazil/CE-IEC-177343/2020 | 2020-12-14 | South America/Brazil/Cear√°/ | LACEN - Laborat√≥rio Central de Sa√∫de P√∫blica do Ceara | Ôª¿Evandro Chagas Institute | Santos et al |
| EPI_ISL_918541 | Brazil/CE-IEC-177345/2021 | 2021-01-04 | South America/Brazil/Cear√°/ | LACEN - Laborat√≥rio Central de Sa√∫de P√∫blica do Ceara | Ôª¿Evandro Chagas Institute | Santos et al |
| EPI_ISL_918542 | Brazil/CE-IEC-177346/2021 | 2021-01-04 | South America/Brazil/Cear√°/ | LACEN - Laborat√≥rio Central de Sa√∫de P√∫blica do Ceara | Ôª¿Evandro Chagas Institute | Santos et al |
| EPI_ISL_918543 | Brazil/CE-IEC-177347/2021 | 2021-01-13 | South America/Brazil/Cear√°/ | LACEN - Laborat√≥rio Central de Sa√∫de P√∫blica do Ceara | Ôª¿Evandro Chagas Institute | Santos et al |
| EPI_ISL_918544 | Brazil/CE-IEC-177348/2021 | 2021-01-14 | South America/Brazil/Cear√°/ | LACEN - Laborat√≥rio Central de Sa√∫de P√∫blica do Ceara | Ôª¿Evandro Chagas Institute | Santos et al |
| EPI_ISL_918545 | Brazil/PA-IEC-177356/2021 | 2021-01-06 | South America/Brazil/Para/ | LACEN - Laborat√≥rio Central de Sa√∫de P√∫blica do Para | Ôª¿Evandro Chagas Institute | Santos et al |
| EPI_ISL_918546 | Brazil/PA-IEC-177357/2021 | 2021-01-05 | South America/Brazil/Para/ | LACEN - Laborat√≥rio Central de Sa√∫de P√∫blica do Para | Ôª¿Evandro Chagas Institute | Santos et al |
| EPI_ISL_918547 | Brazil/PA-IEC-177358/2021 | 2021-01-06 | South America/Brazil/Para/ | LACEN - Laborat√≥rio Central de Sa√∫de P√∫blica do Para | Ôª¿Evandro Chagas Institute | Santos et al |
| EPI_ISL_918548 | Brazil/PA-IEC-177362/2021 | 2021-01-07 | South America/Brazil/Para/ | LACEN - Laborat√≥rio Central de Sa√∫de P√∫blica do Para | Ôª¿Evandro Chagas Institute | Santos et al |
| EPI_ISL_918549 | Brazil/PA-IEC-177363/2021 | 2021-01-05 | South America/Brazil/Para/ | LACEN - Laborat√≥rio Central de Sa√∫de P√∫blica do Para | Ôª¿Evandro Chagas Institute | Santos et al |
| EPI_ISL_918550 | Brazil/PA-IEC-177368/2021 | 2021-01-11 | South America/Brazil/Para/ | LACEN - Laborat√≥rio Central de Sa√∫de P√∫blica do Para | Ôª¿Evandro Chagas Institute | Santos et al |
| EPI_ISL_918551 | Brazil/AP-IEC-177382/2020 | 2020-12-04 | South America/Brazil/Amapa/ | LACEN - Laborat√≥rio Central de Sa√∫de P√∫blica do Amapa | Ôª¿Evandro Chagas Institute | Santos et al |
| EPI_ISL_918552 | Brazil/PA-IEC-177383/2020 | 2020-12-04 | South America/Brazil/Para/ | LACEN - Laborat√≥rio Central de Sa√∫de P√∫blica do Para | Ôª¿Evandro Chagas Institute | Santos et al |
| EPI_ISL_918553 | Brazil/AP-IEC-177386/2020 | 2020-12-08 | South America/Brazil/Amapa/ | LACEN - Laborat√≥rio Central de Sa√∫de P√∫blica do Amapa | Ôª¿Evandro Chagas Institute | Santos et al |
| EPI_ISL_918554 | Brazil/AP-IEC-177390/2020 | 2020-12-16 | South America/Brazil/Amapa/ | LACEN - Laborat√≥rio Central de Sa√∫de P√∫blica do Amapa | Ôª¿Evandro Chagas Institute | Santos et al |
| EPI_ISL_918555 | Brazil/AP-IEC-177393/2020 | 2020-12-23 | South America/Brazil/Amapa/ | LACEN - Laborat√≥rio Central de Sa√∫de P√∫blica do Amapa | Ôª¿Evandro Chagas Institute | Santos et al |
| EPI_ISL_918556 | Brazil/AP-IEC-177394/2020 | 2020-12-26 | South America/Brazil/Amapa/ | LACEN - Laborat√≥rio Central de Sa√∫de P√∫blica do Amapa | Ôª¿Evandro Chagas Institute | Santos et al |
| EPI_ISL_918558 | Brazil/AP-IEC-177400/2021 | 2021-01-02 | South America/Brazil/Amapa/ | LACEN - Laborat√≥rio Central de Sa√∫de P√∫blica do Amapa | Ôª¿Evandro Chagas Institute | Santos et al |
| EPI_ISL_918559 | Brazil/AP-IEC-177402/2021 | 2021-01-03 | South America/Brazil/Amapa/ | LACEN - Laborat√≥rio Central de Sa√∫de P√∫blica do Amapa | Ôª¿Evandro Chagas Institute | Santos et al |
| EPI_ISL_918560 | Brazil/AP-IEC-177406/2021 | 2021-01-09 | South America/Brazil/Amapa/ | LACEN - Laborat√≥rio Central de Sa√∫de P√∫blica do Amapa | Ôª¿Evandro Chagas Institute | Santos et al |

| | | | | | | |
|---|---|---|---|---|---|---|
| EPI_ISL_918561 | Brazil/AP-IEC-177409/2021 | 2021-01-10 | South America/Brazil/Amapa/ | LACEN - Laboratv≥rio Central de Sav∫de Pv∫blica do Amapa | ÔªøEvandro Chagas Institute | Santos et al |
| EPI_ISL_918669 | England/BIRM-6D265/2020 | 2020-11-26 | Europe/United Kingdom/England/ | University of Birmingham | COVID-19 Genomics UK (COG-UK) Consortium | Institute of Microbiology et al |
| EPI_ISL_918754 | England/BIRM-6E4B9/2020 | 2020-12-30 | Europe/United Kingdom/England/ | University of Birmingham | COVID-19 Genomics UK (COG-UK) Consortium | Institute of Microbiology et al |
| EPI_ISL_918902 | England/BIRM-6F6D0/2021 | 2021-01-02 | Europe/United Kingdom/England/ | University of Birmingham | COVID-19 Genomics UK (COG-UK) Consortium | Institute of Microbiology et al |
| EPI_ISL_919163 | England/CAMB-A9F96B/2021 | 2021-01-26 | Europe/United Kingdom/England/ | Department of Pathology, University of Cambridge | COVID-19 Genomics UK (COG-UK) Consortium | Aminu S. Jahun et al |
| EPI_ISL_919201 | Scotland/CVR6612/2021 | 2021-01-13 | Europe/United Kingdom/Scotland/ | West of Scotland Specialist Virology Centre, NHSGGC / MRC-University of Glasgow Centre for Virus Research | COVID-19 Genomics UK (COG-UK) Consortium | Ana da Silva Filipe et al |
| EPI_ISL_919222 | Scotland/CVR6649/2021 | 2021-01-17 | Europe/United Kingdom/Scotland/ | West of Scotland Specialist Virology Centre, NHSGGC / MRC-University of Glasgow Centre for Virus Research | COVID-19 Genomics UK (COG-UK) Consortium | Ana da Silva Filipe et al |
| EPI_ISL_919568 | England/LIVE-DE77D0/2021 | 2021-01-17 | Europe/United Kingdom/England/ | Liverpool Clinical Laboratories | COVID-19 Genomics UK (COG-UK) Consortium | Sam Haldenby et al |
| EPI_ISL_919962 | England/LOND-12E924E/2021 | 2021-01-02 | Europe/United Kingdom/England/ | University College London, Great Ormond Street Hospital for Children NHS Foundation Trust, Imperial College Healthcare NHS Trust | COVID-19 Genomics UK (COG-UK) Consortium | Sergi Castellano et al |
| EPI_ISL_920190 | England/LOND-1323A5E/2021 | 2021-01-13 | Europe/United Kingdom/England/ | University College London Hospital | COVID-19 Genomics UK (COG-UK) Consortium | Judith Heaney et al |
| EPI_ISL_920203 | England/LOND-1323B2E/2021 | 2021-01-13 | Europe/United Kingdom/England/ | University College London Hospital | COVID-19 Genomics UK (COG-UK) Consortium | Judith Heaney et al |
| EPI_ISL_920301 | England/LOND-1324431/2020 | 2020-12-16 | Europe/United Kingdom/England/ | University College London Hospital | COVID-19 Genomics UK (COG-UK) Consortium | Judith Heaney et al |
| EPI_ISL_920312 | England/LOND-13244F5/2021 | 2021-01-16 | Europe/United Kingdom/England/ | University College London Hospital | COVID-19 Genomics UK (COG-UK) Consortium | Judith Heaney et al |

| | | | | | | |
|---|---|---|---|---|---|---|
| EPI_ISL_920474 | England/LOND-132545E/2020 | 2020-12-19 | Europe/United Kingdom/England/ | University College London Hospital | COVID-19 Genomics UK (COG-UK) Consortium | Judith Heaney et al |
| EPI_ISL_920697 | England/LOND-1326763/2020 | 2020-12-27 | Europe/United Kingdom/England/ | University College London Hospital | COVID-19 Genomics UK (COG-UK) Consortium | Judith Heaney et al |
| EPI_ISL_920734 | England/LOND-1326B49/2021 | 2021-01-27 | Europe/United Kingdom/England/ | University College London Hospital | COVID-19 Genomics UK (COG-UK) Consortium | Judith Heaney et al |
| EPI_ISL_920795 | England/LOND-DFB27/2021 | 2021-01-18 | Europe/United Kingdom/England/ | University College London, Great Ormond Street Hospital for Children NHS Foundation Trust, Imperial College Healthcare NHS Trust | COVID-19 Genomics UK (COG-UK) Consortium | Sergi Castellano et al |
| EPI_ISL_921203 | England/NORT-1B73B18/2020 | 2020-12-21 | Europe/United Kingdom/England/ | Northumbria University / South Tees Hospitals NHS Foundation Trust / North Cumbria Integrated Care NHS Foundation Trust / North Tees and Hartlepool NHS Foundation Trust / Newcastle Hospitals NHS Foundation Trust | COVID-19 Genomics UK (COG-UK) Consortium | Darren L Smith et al |
| EPI_ISL_921385 | England/NORT-1B8575B/2020 | 2020-12-30 | Europe/United Kingdom/England/ | Northumbria University / South Tees Hospitals NHS Foundation Trust / North Cumbria Integrated Care NHS Foundation Trust / North Tees and Hartlepool NHS Foundation Trust / Newcastle Hospitals NHS Foundation Trust | COVID-19 Genomics UK (COG-UK) Consortium | Darren L Smith et al |
| EPI_ISL_921403 | England/NORT-1B858D0/2020 | 2020-12-30 | Europe/United Kingdom/England/ | Northumbria University / South Tees Hospitals NHS Foundation Trust / North Cumbria Integrated Care NHS Foundation Trust / North Tees and Hartlepool NHS Foundation Trust / Newcastle Hospitals NHS Foundation Trust | COVID-19 Genomics UK (COG-UK) Consortium | Darren L Smith et al |
| EPI_ISL_921416 | England/NORT-1B88011/2020 | 2020-12-31 | Europe/United Kingdom/England/ | Northumbria University / South Tees Hospitals NHS Foundation Trust / North Cumbria Integrated Care NHS Foundation Trust / North Tees and Hartlepool NHS Foundation Trust / Newcastle Hospitals NHS Foundation Trust | COVID-19 Genomics UK (COG-UK) Consortium | Darren L Smith et al |
| EPI_ISL_921852 | England/NORW-225DEE/2021 | 2021-01-20 | Europe/United Kingdom/England/ | Quadram Institute Bioscience | COVID-19 Genomics UK (COG-UK) Consortium | Dave J. Baker et al |

| | | | | | | |
|---|---|---|---|---|---|---|
| EPI_ISL_921923 | England/NORW-2271FF/2021 | 2021-01-20 | Europe/United Kingdom/England/ | Quadram Institute Bioscience | COVID-19 Genomics UK (COG-UK) Consortium | Dave J. Baker et al |
| EPI_ISL_922024 | England/NOTT-24AF29/2021 | 2021-01-22 | Europe/United Kingdom/England/ | Queens Medical Centre, Clinical Microbiology Department / DeepSeq Nottingham | COVID-19 Genomics UK (COG-UK) Consortium | Gemma Clark et al |
| EPI_ISL_922081 | England/NOTT-24B939/2021 | 2021-01-09 | Europe/United Kingdom/England/ | Lincolnshire Hospitals and DeepSeq Nottingham | COVID-19 Genomics UK (COG-UK) Consortium | Nichola Duckworth et al |
| EPI_ISL_922102 | England/NOTT-24BA90/2021 | 2021-01-10 | Europe/United Kingdom/England/ | Lincolnshire Hospitals and DeepSeq Nottingham | COVID-19 Genomics UK (COG-UK) Consortium | Nichola Duckworth et al |
| EPI_ISL_922104 | England/NOTT-24BABE/2021 | 2021-01-10 | Europe/United Kingdom/England/ | Lincolnshire Hospitals and DeepSeq Nottingham | COVID-19 Genomics UK (COG-UK) Consortium | Nichola Duckworth et al |
| EPI_ISL_922107 | England/NOTT-24BAFA/2021 | 2021-01-10 | Europe/United Kingdom/England/ | Lincolnshire Hospitals and DeepSeq Nottingham | COVID-19 Genomics UK (COG-UK) Consortium | Nichola Duckworth et al |
| EPI_ISL_922119 | England/NOTT-24BBCA/2021 | 2021-01-12 | Europe/United Kingdom/England/ | Lincolnshire Hospitals and DeepSeq Nottingham | COVID-19 Genomics UK (COG-UK) Consortium | Nichola Duckworth et al |
| EPI_ISL_922395 | Wales/PHWC-4BACB2/2021 | 2021-01-12 | Europe/United Kingdom/Wales/ | Wales Specialist Virology Centre Sequencing lab: Pathogen Genomics Unit | Public Health Wales Microbiology Cardiff Wales Specialist Virology Centre | Catherine Moore et al |
| EPI_ISL_922430 | Wales/PHWC-4BAFAA/2021 | 2021-01-13 | Europe/United Kingdom/Wales/ | Wales Specialist Virology Centre Sequencing lab: Pathogen Genomics Unit | Public Health Wales Microbiology Cardiff Wales Specialist Virology Centre | Catherine Moore et al |
| EPI_ISL_923013 | Wales/PHWC-4BD790/2021 | 2021-01-14 | Europe/United Kingdom/Wales/ | Wales Specialist Virology Centre Sequencing lab: Pathogen Genomics Unit | Public Health Wales Microbiology Cardiff Wales Specialist Virology Centre | Catherine Moore et al |

| | | | | | | |
|---|---|---|---|---|---|---|
| EPI_ISL_923219 | England/PORT-2D87CE/2020 | 2020-11-29 | Europe/United Kingdom/England/ | Centre for Enzyme Innovation, University of Portsmouth / Translational Research Laboratory, Portsmouth Hospitals NHS Trust | COVID-19 Genomics UK (COG-UK) Consortium | Angela Beckett et al |
| EPI_ISL_923813 | England/QEUH-1032785/2021 | 2021-01-15 | Europe/United Kingdom/England/ | Department of Pathology, University of Cambridge | COVID-19 Genomics UK (COG-UK) Consortium | Aminu S. Jahun et al |
| EPI_ISL_923905 | England/QEUH-103314A/2021 | 2021-01-15 | Europe/United Kingdom/England/ | Department of Pathology, University of Cambridge | COVID-19 Genomics UK (COG-UK) Consortium | Aminu S. Jahun et al |
| EPI_ISL_924114 | England/SHEF-10C747F/2021 | 2021-01-19 | Europe/United Kingdom/England/ | Virology Department, Sheffield Teaching Hospitals NHS Foundation Trust/Department of Infection, Immunity and Cardiovascular Disease, The Medical School, University of Sheffield | COVID-19 Genomics UK (COG-UK) Consortium | Thushan de Silva et al |
| EPI_ISL_924116 | England/SHEF-10C758B/2021 | 2021-01-22 | Europe/United Kingdom/England/ | Virology Department, Sheffield Teaching Hospitals NHS Foundation Trust/Department of Infection, Immunity and Cardiovascular Disease, The Medical School, University of Sheffield | COVID-19 Genomics UK (COG-UK) Consortium | Thushan de Silva et al |
| EPI_ISL_924130 | England/SHEF-10C7E26/2021 | 2021-01-24 | Europe/United Kingdom/England/ | Virology Department, Sheffield Teaching Hospitals NHS Foundation Trust/Department of Infection, Immunity and Cardiovascular Disease, The Medical School, University of Sheffield | COVID-19 Genomics UK (COG-UK) Consortium | Thushan de Silva et al |
| EPI_ISL_924252 | England/SHEF-10CF996/2021 | 2021-01-28 | Europe/United Kingdom/England/ | Virology Department, Sheffield Teaching Hospitals NHS Foundation Trust/Department of Infection, Immunity and Cardiovascular Disease, The Medical School, University of Sheffield | COVID-19 Genomics UK (COG-UK) Consortium | Thushan de Silva et al |
| EPI_ISL_924289 | England/SHEF-10D1F62/2021 | 2021-01-20 | Europe/United Kingdom/England/ | Virology Department, Sheffield Teaching Hospitals NHS Foundation Trust/Department of Infection, Immunity and Cardiovascular Disease, The Medical School, University of Sheffield | COVID-19 Genomics UK (COG-UK) Consortium | Thushan de Silva et al |
| EPI_ISL_924320 | England/SHEF-10D33A0/2021 | 2021-01-22 | Europe/United Kingdom/England/ | Virology Department, Sheffield Teaching Hospitals NHS Foundation Trust/Department of Infection, Immunity and Cardiovascular Disease, The Medical School, University of Sheffield | COVID-19 Genomics UK (COG-UK) Consortium | Thushan de Silva et al |

| | | | | | | |
|---|---|---|---|---|---|---|
| EPI_ISL_924425 | England/SHEF-CBFD7/2020 | 2020-10-28 | Europe/United Kingdom/England/ | Virology Department, Sheffield Teaching Hospitals NHS Foundation Trust/Department of Infection, Immunity and Cardiovascular Disease, The Medical School, University of Sheffield | COVID-19 Genomics UK (COG-UK) Consortium | Thushan de Silva et al |
| EPI_ISL_924482 | England/TFCI-2700346/2021 | 2021-01-01 | Europe/United Kingdom/England/ | Bioinformatics and Biostatistics Lab, Advanced Sequencing Facility | COVID-19 Genomics UK (COG-UK) Consortium | Aengus Stewart et al |
| EPI_ISL_924798 | England/TFCI-2701F9C/2021 | 2021-01-10 | Europe/United Kingdom/England/ | Bioinformatics and Biostatistics Lab, Advanced Sequencing Facility | COVID-19 Genomics UK (COG-UK) Consortium | Aengus Stewart et al |
| EPI_ISL_924802 | England/TFCI-2701FD8/2021 | 2021-01-12 | Europe/United Kingdom/England/ | Bioinformatics and Biostatistics Lab, Advanced Sequencing Facility | COVID-19 Genomics UK (COG-UK) Consortium | Aengus Stewart et al |
| EPI_ISL_924862 | England/TFCI-2702423/2021 | 2021-01-19 | Europe/United Kingdom/England/ | Bioinformatics and Biostatistics Lab, Advanced Sequencing Facility | COVID-19 Genomics UK (COG-UK) Consortium | Aengus Stewart et al |
| EPI_ISL_925033 | Poland/1531615022_diag1_gx1/2020 | 2020-12-22 | Europe/Poland/Pomorskie/ | Laboratory of Molecular Biology, Diagnostyka sp. z o.o. | genXone SA, Research & Development Laboratory | Maciej Sykulski et al |
| EPI_ISL_925034 | Poland/1566615022_diag1_gx1/2020 | 2020-12-22 | Europe/Poland/Pomorskie/ | Laboratory of Molecular Biology, Diagnostyka sp. z o.o. | genXone SA, Research & Development Laboratory | Maciej Sykulski et al |
| EPI_ISL_925035 | Poland/1692597229_diag1_gx1/2020 | 2020-12-25 | Europe/Poland/Zachodniopomorskie/ | Laboratory of Molecular Biology, Diagnostyka sp. z o.o. | genXone SA, Research & Development Laboratory | Maciej Sykulski et al |
| EPI_ISL_925036 | Poland/349601021_diag1_gx1/2020 | 2020-12-22 | Europe/Poland/Dolno≈õlƒÖskie/ | Laboratory of Molecular Biology, Diagnostyka sp. z o.o. | genXone SA, Research & Development Laboratory | Maciej Sykulski et al |
| EPI_ISL_925037 | Poland/4300570422_diag1_gx1/2020 | 2020-12-22 | Europe/Poland/Dolno≈õlƒÖskie/ | Laboratory of Molecular Biology, Diagnostyka sp. z o.o. | genXone SA, Research & Development Laboratory | Maciej Sykulski et al |

| | | | | | | |
|---|---|---|---|---|---|---|
| EPI_ISL_925038 | Poland/573607221_diag1_gx1/2020 | 2020-12-22 | Europe/Poland/Warminsko-Mazurskie/ | Laboratory of Molecular Biology, Diagnostyka sp. z o.o. | genXone SA, Research & Development Laboratory | Maciej Sykulski et al |
| EPI_ISL_925041 | Poland/7468581824_diag1_gx1/2020 | 2020-12-22 | Europe/Poland/Pomorskie/ | Laboratory of Molecular Biology, Diagnostyka sp. z o.o. | genXone SA, Research & Development Laboratory | Maciej Sykulski et al |
| EPI_ISL_925045 | Poland/8791615423_diag1_gx1/2020 | 2020-12-22 | Europe/Poland/Pomorskie/ | Laboratory of Molecular Biology, Diagnostyka sp. z o.o. | genXone SA, Research & Development Laboratory | Maciej Sykulski et al |
| EPI_ISL_925046 | Poland/9214597429_diag1_gx1/2020 | 2020-12-25 | Europe/Poland/Zachodniopomorskie/ | Laboratory of Molecular Biology, Diagnostyka sp. z o.o. | genXone SA, Research & Development Laboratory | Maciej Sykulski et al |
| EPI_ISL_925073 | USA/TX-DSHS-2342/2021 | 2021-01-06 | North America/USA/Texas/Brazos Count | TXDSHS | TXDSHS | Bonnie Oh et al |
| EPI_ISL_925080 | USA/TX-DSHS-2349/2021 | 2021-01-12 | North America/USA/Texas/Brazos Count | TXDSHS | TXDSHS | Bonnie Oh et al |
| EPI_ISL_925086 | USA/WY-WPHL-20168559/2020 | 2020-12-19 | North America/USA/Wyoming/ | Wyoming Public Health Laboratory | Wyoming Public Health Laboratory | Noah Hull et al |
| EPI_ISL_925094 | Italy/CAM-CRGS-222/2020 | 2020-10-29 | Europe/Italy/Campania/ | 1.AO Universitaria 'S. Giovanni di Dio e Ruggi D'Aragona, Scuola Medica Salernitana' Hospital / 2.UOC di Virologia e Microbiologia, Universit√† della Campania 'L. Vanvitelli' / 3.AO Universitaria 'Federico II' Napoli Hospital / 4.AORN 'San Giuseppe Moscati' Avellino Hospital / 5.AO 'San Pio - presidio G. Rummo' Benevento Hospital / 6.AO 'Sant'Anna e San Sebastiano' Caserta Hospital / 7.PO 'Maria Santissima Addolorata' Eboli Hospital / 8.Biogem Istituto di Ricerche Genetiche | 1. Genome Research Center for Health (CRGS) / 2. Laboratory of Molecular Medicine and Genomics(LMMGe) / 3. Center for Research in Pure and Applied Mathematics (CRMPA) | Giorgio Giurato et al |

| | | | | | | |
|---|---|---|---|---|---|---|
| EPI_ISL_925096 | Italy/CAM-CRGS-225/2020 | 2020-11-03 | Europe/Italy/Campania/ | 1.AO Universitaria 'S. Giovanni di Dio e Ruggi D'Aragona, Scuola Medica Salernitana' Hospital / 2.UOC di Virologia e Microbiologia, Universit√† della Campania 'L. Vanvitelli' / 3.AO Universitaria 'Federico II' Napoli Hospital / 4.AORN 'San Giuseppe Moscati' Avellino Hospital / 5.AO 'San Pio - presidio G. Rummo' Benevento Hospital / 6.AO 'Sant'Anna e San Sebastiano' Caserta Hospital / 7.PO 'Maria Santissima Addolorata' Eboli Hospital / 8.Biogem Istituto di Ricerche Genetiche | 1. Genome Research Center for Health (CRGS) / 2. Laboratory of Molecular Medicine and Genomics(LMMGe) / 3. Center for Research in Pure and Applied Mathematics (CRMPA) | Giorgio Giurato et al |
| EPI_ISL_925097 | Italy/CAM-CRGS-226/2020 | 2020-10-08 | Europe/Italy/Campania/ | 1.AO Universitaria 'S. Giovanni di Dio e Ruggi D'Aragona, Scuola Medica Salernitana' Hospital / 2.UOC di Virologia e Microbiologia, Universit√† della Campania 'L. Vanvitelli' / 3.AO Universitaria 'Federico II' Napoli Hospital / 4.AORN 'San Giuseppe Moscati' Avellino Hospital / 5.AO 'San Pio - presidio G. Rummo' Benevento Hospital / 6.AO 'Sant'Anna e San Sebastiano' Caserta Hospital / 7.PO 'Maria Santissima Addolorata' Eboli Hospital / 8.Biogem Istituto di Ricerche Genetiche | 1. Genome Research Center for Health (CRGS) / 2. Laboratory of Molecular Medicine and Genomics(LMMGe) / 3. Center for Research in Pure and Applied Mathematics (CRMPA) | Giorgio Giurato et al |
| EPI_ISL_925099 | Italy/CAM-CRGS-236/2020 | 2020-10-14 | Europe/Italy/Campania/ | 1.AO Universitaria 'S. Giovanni di Dio e Ruggi D'Aragona, Scuola Medica Salernitana' Hospital / 2.UOC di Virologia e Microbiologia, Universit√† della Campania 'L. Vanvitelli' / 3.AO Universitaria 'Federico II' Napoli Hospital / 4.AORN 'San Giuseppe Moscati' Avellino Hospital / 5.AO 'San Pio - presidio G. Rummo' Benevento Hospital / 6.AO 'Sant'Anna e San Sebastiano' Caserta Hospital / 7.PO 'Maria Santissima Addolorata' Eboli Hospital / 8.Biogem Istituto di Ricerche Genetiche | 1. Genome Research Center for Health (CRGS) / 2. Laboratory of Molecular Medicine and Genomics(LMMGe) / 3. Center for Research in Pure and Applied Mathematics (CRMPA) | Giorgio Giurato et al |

| | | | | | | |
|---|---|---|---|---|---|---|
| EPI_ISL_925102 | Italy/CAM-CRGS-248/2020 | 2020-10-07 | Europe/Italy/Campania/ | 1.AO Universitaria 'S. Giovanni di Dio e Ruggi D'Aragona, Scuola Medica Salernitana' Hospital / 2.UOC di Virologia e Microbiologia, Universit√† della Campania 'L. Vanvitelli' / 3.AO Universitaria 'Federico II' Napoli Hospital / 4.AORN 'San Giuseppe Moscati' Avellino Hospital / 5.AO 'San Pio - presidio G. Rummo' Benevento Hospital / 6.AO 'Sant'Anna e San Sebastiano' Caserta Hospital / 7.PO 'Maria Santissima Addolorata' Eboli Hospital / 8.Biogem Istituto di Ricerche Genetiche | 1. Genome Research Center for Health (CRGS) / 2. Laboratory of Molecular Medicine and Genomics(LMMGe) / 3. Center for Research in Pure and Applied Mathematics (CRMPA) | Giorgio Giurato et al |
| EPI_ISL_925103 | Italy/CAM-CRGS-249/2020 | 2020-11-02 | Europe/Italy/Campania/ | 1.AO Universitaria 'S. Giovanni di Dio e Ruggi D'Aragona, Scuola Medica Salernitana' Hospital / 2.UOC di Virologia e Microbiologia, Universit√† della Campania 'L. Vanvitelli' / 3.AO Universitaria 'Federico II' Napoli Hospital / 4.AORN 'San Giuseppe Moscati' Avellino Hospital / 5.AO 'San Pio - presidio G. Rummo' Benevento Hospital / 6.AO 'Sant'Anna e San Sebastiano' Caserta Hospital / 7.PO 'Maria Santissima Addolorata' Eboli Hospital / 8.Biogem Istituto di Ricerche Genetiche | 1. Genome Research Center for Health (CRGS) / 2. Laboratory of Molecular Medicine and Genomics(LMMGe) / 3. Center for Research in Pure and Applied Mathematics (CRMPA) | Giorgio Giurato et al |
| EPI_ISL_925104 | Italy/CAM-CRGS-250/2020 | 2020-11-01 | Europe/Italy/Campania/ | 1.AO Universitaria 'S. Giovanni di Dio e Ruggi D'Aragona, Scuola Medica Salernitana' Hospital / 2.UOC di Virologia e Microbiologia, Universit√† della Campania 'L. Vanvitelli' / 3.AO Universitaria 'Federico II' Napoli Hospital / 4.AORN 'San Giuseppe Moscati' Avellino Hospital / 5.AO 'San Pio - presidio G. Rummo' Benevento Hospital / 6.AO 'Sant'Anna e San Sebastiano' Caserta Hospital / 7.PO 'Maria Santissima Addolorata' Eboli Hospital / 8.Biogem Istituto di Ricerche Genetiche | 1. Genome Research Center for Health (CRGS) / 2. Laboratory of Molecular Medicine and Genomics(LMMGe) / 3. Center for Research in Pure and Applied Mathematics (CRMPA) | Giorgio Giurato et al |

| | | | | | | |
|---|---|---|---|---|---|---|
| EPI_ISL_925105 | Italy/CAM-CRGS-251/2020 | 2020-11-04 | Europe/Italy/Campania/ | 1.AO Universitaria 'S. Giovanni di Dio e Ruggi D'Aragona, Scuola Medica Salernitana' Hospital / 2.UOC di Virologia e Microbiologia, Universit√† della Campania 'L. Vanvitelli' / 3.AO Universitaria 'Federico II' Napoli Hospital / 4.AORN 'San Giuseppe Moscati' Avellino Hospital / 5.AO 'San Pio - presidio G. Rummo' Benevento Hospital / 6.AO 'Sant'Anna e San Sebastiano' Caserta Hospital / 7.PO 'Maria Santissima Addolorata' Eboli Hospital / 8.Biogem Istituto di Ricerche Genetiche | 1. Genome Research Center for Health (CRGS) / 2. Laboratory of Molecular Medicine and Genomics(LMMGe) / 3. Center for Research in Pure and Applied Mathematics (CRMPA) | Giorgio Giurato et al |
| EPI_ISL_925106 | Italy/CAM-CRGS-252/2020 | 2020-11-03 | Europe/Italy/Campania/ | 1.AO Universitaria 'S. Giovanni di Dio e Ruggi D'Aragona, Scuola Medica Salernitana' Hospital / 2.UOC di Virologia e Microbiologia, Universit√† della Campania 'L. Vanvitelli' / 3.AO Universitaria 'Federico II' Napoli Hospital / 4.AORN 'San Giuseppe Moscati' Avellino Hospital / 5.AO 'San Pio - presidio G. Rummo' Benevento Hospital / 6.AO 'Sant'Anna e San Sebastiano' Caserta Hospital / 7.PO 'Maria Santissima Addolorata' Eboli Hospital / 8.Biogem Istituto di Ricerche Genetiche | 1. Genome Research Center for Health (CRGS) / 2. Laboratory of Molecular Medicine and Genomics(LMMGe) / 3. Center for Research in Pure and Applied Mathematics (CRMPA) | Giorgio Giurato et al |
| EPI_ISL_925107 | Italy/CAM-CRGS-258/2020 | 2020-10-20 | Europe/Italy/Campania/ | 1.AO Universitaria 'S. Giovanni di Dio e Ruggi D'Aragona, Scuola Medica Salernitana' Hospital / 2.UOC di Virologia e Microbiologia, Universit√† della Campania 'L. Vanvitelli' / 3.AO Universitaria 'Federico II' Napoli Hospital / 4.AORN 'San Giuseppe Moscati' Avellino Hospital / 5.AO 'San Pio - presidio G. Rummo' Benevento Hospital / 6.AO 'Sant'Anna e San Sebastiano' Caserta Hospital / 7.PO 'Maria Santissima Addolorata' Eboli Hospital / 8.Biogem Istituto di Ricerche Genetiche | 1. Genome Research Center for Health (CRGS) / 2. Laboratory of Molecular Medicine and Genomics(LMMGe) / 3. Center for Research in Pure and Applied Mathematics (CRMPA) | Giorgio Giurato et al |

| | | | | | | |
|---|---|---|---|---|---|---|
| EPI_ISL_925109 | Italy/CAM-CRGS-260/2020 | 2020-09-09 | Europe/Italy/Campania/ | 1.AO Universitaria 'S. Giovanni di Dio e Ruggi D'Aragona, Scuola Medica Salernitana' Hospital / 2.UOC di Virologia e Microbiologia, Universit√† della Campania 'L. Vanvitelli' / 3.AO Universitaria 'Federico II' Napoli Hospital / 4.AORN 'San Giuseppe Moscati' Avellino Hospital / 5.AO 'San Pio - presidio G. Rummo' Benevento Hospital / 6.AO 'Sant'Anna e San Sebastiano' Caserta Hospital / 7.PO 'Maria Santissima Addolorata' Eboli Hospital / 8.Biogem Istituto di Ricerche Genetiche | 1. Genome Research Center for Health (CRGS) / 2. Laboratory of Molecular Medicine and Genomics(LMMGe) / 3. Center for Research in Pure and Applied Mathematics (CRMPA) | Giorgio Giurato et al |
| EPI_ISL_925110 | Italy/CAM-CRGS-261/2020 | 2020-09-09 | Europe/Italy/Campania/ | 1.AO Universitaria 'S. Giovanni di Dio e Ruggi D'Aragona, Scuola Medica Salernitana' Hospital / 2.UOC di Virologia e Microbiologia, Universit√† della Campania 'L. Vanvitelli' / 3.AO Universitaria 'Federico II' Napoli Hospital / 4.AORN 'San Giuseppe Moscati' Avellino Hospital / 5.AO 'San Pio - presidio G. Rummo' Benevento Hospital / 6.AO 'Sant'Anna e San Sebastiano' Caserta Hospital / 7.PO 'Maria Santissima Addolorata' Eboli Hospital / 8.Biogem Istituto di Ricerche Genetiche | 1. Genome Research Center for Health (CRGS) / 2. Laboratory of Molecular Medicine and Genomics(LMMGe) / 3. Center for Research in Pure and Applied Mathematics (CRMPA) | Giorgio Giurato et al |
| EPI_ISL_925112 | Italy/CAM-CRGS-276/2020 | 2020-11-24 | Europe/Italy/Campania/ | 1.AO Universitaria 'S. Giovanni di Dio e Ruggi D'Aragona, Scuola Medica Salernitana' Hospital / 2.UOC di Virologia e Microbiologia, Universit√† della Campania 'L. Vanvitelli' / 3.AO Universitaria 'Federico II' Napoli Hospital / 4.AORN 'San Giuseppe Moscati' Avellino Hospital / 5.AO 'San Pio - presidio G. Rummo' Benevento Hospital / 6.AO 'Sant'Anna e San Sebastiano' Caserta Hospital / 7.PO 'Maria Santissima Addolorata' Eboli Hospital / 8.Biogem Istituto di Ricerche Genetiche | 1. Genome Research Center for Health (CRGS) / 2. Laboratory of Molecular Medicine and Genomics(LMMGe) / 3. Center for Research in Pure and Applied Mathematics (CRMPA) | Giorgio Giurato et al |
| EPI_ISL_925140 | USA/CA-QDX-4398/2021 | 2021-01-14 | North America/USA/California/ | Quest Diagnostics | Quest Diagnostics | Rosenthal et al |
| EPI_ISL_925153 | USA/VA-DCLS-2544/2020 | 2020-12-01 | North America/USA/Virginia/ | Virginia DCLS | Virginia DCLS | Virginia DCLS et al |
| EPI_ISL_925163 | USA/VA-DCLS-2559/2020 | 2020-12-03 | North America/USA/Virginia/ | Virginia DCLS | Virginia DCLS | Virginia DCLS et al |

| | | | | | | |
|---|---|---|---|---|---|---|
| EPI_ISL_925173 | Peru/LIM-INS-174/2021 | 2021-01-18 | South America/Peru/Lima/ | Laboratorio de Referencia Nacional de Enteropat√≥genos. Instituto Nacional de Salud del Per√∫ | Laboratorio de Referencia Nacional de Enteropat√≥genos. Instituto Nacional de Salud del Per√∫ | Ronnie Gavilan Chavez et al |
| EPI_ISL_925179 | USA/VA-DCLS-2339/2020 | 2020-11-13 | North America/USA/Virginia/ | Virginia DCLS | Virginia DCLS | Virginia DCLS et al |
| EPI_ISL_925186 | USA/VA-DCLS-2346/2020 | 2020-11-16 | North America/USA/Virginia/ | Virginia DCLS | Virginia DCLS | Virginia DCLS et al |
| EPI_ISL_925198 | USA/VA-DCLS-2364/2020 | 2020-11-14 | North America/USA/Virginia/ | Virginia DCLS | Virginia DCLS | Virginia DCLS et al |
| EPI_ISL_925199 | USA/VA-DCLS-2365/2020 | 2020-11-14 | North America/USA/Virginia/ | Virginia DCLS | Virginia DCLS | Virginia DCLS et al |
| EPI_ISL_925202 | USA/VA-DCLS-2370/2020 | 2020-11-19 | North America/USA/Virginia/ | Virginia DCLS | Virginia DCLS | Virginia DCLS et al |
| EPI_ISL_925203 | USA/VA-DCLS-2373/2020 | 2020-11-19 | North America/USA/Virginia/ | Virginia DCLS | Virginia DCLS | Virginia DCLS et al |
| EPI_ISL_925224 | USA/NM-DOH-2021019650/2021 | 2021-01-11 | North America/USA/New Mexico/ | New Mexico Department of Health Scientific Laboratory | New Mexico Department of Health Scientific Laboratory | Ellie Johnson et al |
| EPI_ISL_925225 | USA/NM-DOH-2021019651/2021 | 2021-01-11 | North America/USA/New Mexico/ | New Mexico Department of Health Scientific Laboratory | New Mexico Department of Health Scientific Laboratory | Ellie Johnson et al |
| EPI_ISL_925229 | USA/NM-DOH-2021019801/2021 | 2021-01-12 | North America/USA/New Mexico/ | New Mexico Department of Health Scientific Laboratory | New Mexico Department of Health Scientific Laboratory | Ellie Johnson et al |
| EPI_ISL_925230 | USA/NM-DOH-2021019845/2021 | 2021-01-12 | North America/USA/New Mexico/ | New Mexico Department of Health Scientific Laboratory | New Mexico Department of Health Scientific Laboratory | Ellie Johnson et al |
| EPI_ISL_925253 | USA/NM-DOH-2021022404/2021 | 2021-01-11 | North America/USA/New Mexico/ | New Mexico Department of Health Scientific Laboratory | New Mexico Department of Health Scientific Laboratory | Ellie Johnson et al |
| EPI_ISL_925265 | USA/WY-WPHL-21008145/2021 | 2021-01-21 | North America/USA/Wyoming/ | Wyoming Public Health Laboratory | Wyoming Public Health Laboratory | Noah Hull et al |
| EPI_ISL_925280 | USA/WY-WPHL-21008547/2021 | 2021-01-23 | North America/USA/Wyoming/ | Wyoming Public Health Laboratory | Wyoming Public Health Laboratory | Noah Hull et al |

| | | | | | | |
|---|---|---|---|---|---|---|
| EPI_ISL_925292 | USA/WY-WPHL-21005641/2021 | 2021-01-14 | North America/USA/Wyoming/ | Wyoming Public Health Laboratory | Wyoming Public Health Laboratory | Noah Hull et al |
| EPI_ISL_925294 | USA/WY-WPHL-21003215/2021 | 2021-01-08 | North America/USA/Wyoming/ | Wyoming Public Health Laboratory | Wyoming Public Health Laboratory | Noah Hull et al |
| EPI_ISL_925361 | USA/NC-SLPH_0025/2020 | 2020-11-24 | North America/USA/North Carolina/ | NCSLPH | NCSLPH | Chase K et al |
| EPI_ISL_925362 | Belgium/ULG-10509/2020 | 2020-10-23 | Europe/Belgium/Liï¿½ge/ | Department of Clinical Microbiology | GIGA Medical Genomics | Keith Durkin et al |
| EPI_ISL_925363 | Belgium/ULG-10513/2020 | 2020-10-23 | Europe/Belgium/Liï¿½ge/ | Department of Clinical Microbiology | GIGA Medical Genomics | Keith Durkin et al |
| EPI_ISL_925365 | Belgium/ULG-10548/2020 | 2020-10-23 | Europe/Belgium/Liï¿½ge/ | Department of Clinical Microbiology | GIGA Medical Genomics | Keith Durkin et al |
| EPI_ISL_925366 | Belgium/ULG-10565/2020 | 2020-10-23 | Europe/Belgium/Liï¿½ge/ | Department of Clinical Microbiology | GIGA Medical Genomics | Keith Durkin et al |
| EPI_ISL_925367 | Belgium/ULG-10572/2020 | 2020-10-23 | Europe/Belgium/Liï¿½ge/ | Department of Clinical Microbiology | GIGA Medical Genomics | Keith Durkin et al |
| EPI_ISL_925369 | Belgium/ULG-10611/2020 | 2020-09-04 | Europe/Belgium/Liï¿½ge/ | Department of Clinical Microbiology | GIGA Medical Genomics | Keith Durkin et al |
| EPI_ISL_925370 | Belgium/ULG-10614/2020 | 2020-09-03 | Europe/Belgium/Liï¿½ge/ | Department of Clinical Microbiology | GIGA Medical Genomics | Keith Durkin et al |
| EPI_ISL_925371 | Belgium/ULG-10652/2020 | 2020-10-26 | Europe/Belgium/Liï¿½ge/ | Department of Clinical Microbiology | GIGA Medical Genomics | Keith Durkin et al |
| EPI_ISL_925377 | Belgium/ULG-10805/2020 | 2020-11-17 | Europe/Belgium/Liï¿½ge/ | Department of Clinical Microbiology | GIGA Medical Genomics | Keith Durkin et al |
| EPI_ISL_925412 | Belgium/ULG-11457/2020 | 2020-08-20 | Europe/Belgium/Liï¿½ge/ | Department of Clinical Microbiology | GIGA Medical Genomics | Keith Durkin et al |
| EPI_ISL_925428 | Belgium/ULG-11473/2020 | 2020-08-24 | Europe/Belgium/Liï¿½ge/ | Department of Clinical Microbiology | GIGA Medical Genomics | Keith Durkin et al |
| EPI_ISL_925440 | Belgium/ULG-11485/2020 | 2020-10-09 | Europe/Belgium/Liï¿½ge/ | Department of Clinical Microbiology | GIGA Medical Genomics | Keith Durkin et al |
| EPI_ISL_925446 | Belgium/ULG-11492/2020 | 2020-10-22 | Europe/Belgium/Liï¿½ge/ | Department of Clinical Microbiology | GIGA Medical Genomics | Keith Durkin et al |
| EPI_ISL_925448 | Belgium/ULG-11494/2020 | 2020-10-22 | Europe/Belgium/Liï¿½ge/ | Department of Clinical Microbiology | GIGA Medical Genomics | Keith Durkin et al |
| EPI_ISL_925474 | Belgium/ULG-11530/2020 | 2020-10-12 | Europe/Belgium/Liï¿½ge/ | Department of Clinical Microbiology | GIGA Medical Genomics | Keith Durkin et al |
| EPI_ISL_925476 | Belgium/ULG-11532/2020 | 2020-10-14 | Europe/Belgium/Liï¿½ge/ | Department of Clinical Microbiology | GIGA Medical Genomics | Keith Durkin et al |
| EPI_ISL_925478 | Belgium/ULG-11534/2020 | 2020-10-15 | Europe/Belgium/Liï¿½ge/ | Department of Clinical Microbiology | GIGA Medical Genomics | Keith Durkin et al |

| | | | | | | |
|---|---|---|---|---|---|---|
| EPI_ISL_925482 | Belgium/ULG-11538/2020 | 2020-10-20 | Europe/Belgium/Liv®ge/ | Department of Clinical Microbiology | GIGA Medical Genomics | Keith Durkin et al |
| EPI_ISL_925485 | Belgium/ULG-11541/2020 | 2020-10-22 | Europe/Belgium/Liv®ge/ | Department of Clinical Microbiology | GIGA Medical Genomics | Keith Durkin et al |
| EPI_ISL_925486 | Belgium/ULG-11542/2020 | 2020-10-22 | Europe/Belgium/Liv®ge/ | Department of Clinical Microbiology | GIGA Medical Genomics | Keith Durkin et al |
| EPI_ISL_925491 | Belgium/ULG-11547/2020 | 2020-10-27 | Europe/Belgium/Liv®ge/ | Department of Clinical Microbiology | GIGA Medical Genomics | Keith Durkin et al |
| EPI_ISL_925509 | USA/AZ-ASPHL-3348/2021 | 2021-01-05 | North America/USA/Arizona/Maricopa ( | Arizona State Public Health Laboratory | Arizona State Public Health Laboratory | Trung Huynh et al |
| EPI_ISL_925512 | USA/AZ-ASPHL-3351/2021 | 2021-01-05 | North America/USA/Arizona/Maricopa ( | Arizona State Public Health Laboratory | Arizona State Public Health Laboratory | Trung Huynh et al |
| EPI_ISL_925515 | USA/AZ-ASPHL-3354/2021 | 2021-01-04 | North America/USA/Arizona/Yuma | Arizona State Public Health Laboratory | Arizona State Public Health Laboratory | Trung Huynh et al |
| EPI_ISL_925516 | USA/AZ-ASPHL-3355/2021 | 2021-01-04 | North America/USA/Arizona/Maricopa ( | Arizona State Public Health Laboratory | Arizona State Public Health Laboratory | Trung Huynh et al |
| EPI_ISL_925527 | USA/AZ-ASPHL-3374/2021 | 2021-01-04 | North America/USA/Arizona/Yuma | Arizona State Public Health Laboratory | Arizona State Public Health Laboratory | Trung Huynh et al |
| EPI_ISL_925532 | USA/AZ-ASPHL-3377/2021 | 2021-01-04 | North America/USA/Arizona/Maricopa ( | Arizona State Public Health Laboratory | Arizona State Public Health Laboratory | Trung Huynh et al |
| EPI_ISL_925535 | Canada/ON-PHL-20-80794/2020 | 20-Mar | North America/Canada/Ontario/ | Public Health Ontario Laboratory | Public Health Ontario Laboratory | Vanessa G Allen et al |
| EPI_ISL_925536 | Canada/ON-PHL-20-80795/2020 | 20-Mar | North America/Canada/Ontario/ | Public Health Ontario Laboratory | Public Health Ontario Laboratory | Vanessa G Allen et al |
| EPI_ISL_925537 | Canada/ON-PHL-20-80495/2020 | 20-Apr | North America/Canada/Ontario/ | Public Health Ontario Laboratory | Public Health Ontario Laboratory | Vanessa G Allen et al |
| EPI_ISL_925538 | Canada/ON-PHL-21-00347/2020 | 20-Nov | North America/Canada/Ontario/ | Public Health Ontario Laboratory | Public Health Ontario Laboratory | Vanessa G Allen et al |
| EPI_ISL_925539 | Canada/ON-PHL-21-00350/2020 | 20-Dec | North America/Canada/Ontario/ | Public Health Ontario Laboratory | Public Health Ontario Laboratory | Vanessa G Allen et al |

| | | | | | | |
|---|---|---|---|---|---|---|
| EPI_ISL_925540 | Canada/ON-PHL-20-80763/2020 | 20-Mar | North America/Canada/Ontario/ | Public Health Ontario Laboratory | Public Health Ontario Laboratory | Vanessa G Allen et al |
| EPI_ISL_925541 | Canada/ON-PHL-20-80805/2020 | 20-Mar | North America/Canada/Ontario/ | Public Health Ontario Laboratory | Public Health Ontario Laboratory | Vanessa G Allen et al |
| EPI_ISL_925542 | Canada/ON-PHL-20-80750/2020 | 20-Mar | North America/Canada/Ontario/ | Public Health Ontario Laboratory | Public Health Ontario Laboratory | Vanessa G Allen et al |
| EPI_ISL_925543 | Canada/ON-PHL-20-80445/2020 | 20-Apr | North America/Canada/Ontario/ | Public Health Ontario Laboratory | Public Health Ontario Laboratory | Vanessa G Allen et al |
| EPI_ISL_925544 | Canada/ON-PHL-20-80446/2020 | 20-Apr | North America/Canada/Ontario/ | Public Health Ontario Laboratory | Public Health Ontario Laboratory | Vanessa G Allen et al |
| EPI_ISL_925545 | Canada/ON-PHL-20-80454/2020 | 20-Apr | North America/Canada/Ontario/ | Public Health Ontario Laboratory | Public Health Ontario Laboratory | Vanessa G Allen et al |
| EPI_ISL_925546 | Canada/ON-PHL-20-80448/2020 | 20-Apr | North America/Canada/Ontario/ | Public Health Ontario Laboratory | Public Health Ontario Laboratory | Vanessa G Allen et al |
| EPI_ISL_925547 | Canada/ON-PHL-20-80455/2020 | 20-Apr | North America/Canada/Ontario/ | Public Health Ontario Laboratory | Public Health Ontario Laboratory | Vanessa G Allen et al |
| EPI_ISL_925548 | Canada/ON-PHL-20-80450/2020 | 20-Apr | North America/Canada/Ontario/ | Public Health Ontario Laboratory | Public Health Ontario Laboratory | Vanessa G Allen et al |
| EPI_ISL_925549 | Canada/ON-PHL-20-80447/2020 | 20-Apr | North America/Canada/Ontario/ | Public Health Ontario Laboratory | Public Health Ontario Laboratory | Vanessa G Allen et al |
| EPI_ISL_925550 | Canada/ON-PHL-20-80532/2020 | 20-Apr | North America/Canada/Ontario/ | Public Health Ontario Laboratory | Public Health Ontario Laboratory | Vanessa G Allen et al |
| EPI_ISL_925551 | Canada/ON-PHL-20-80531/2020 | 20-Apr | North America/Canada/Ontario/ | Public Health Ontario Laboratory | Public Health Ontario Laboratory | Vanessa G Allen et al |
| EPI_ISL_925552 | Canada/ON-PHL-20-80524/2020 | 20-Apr | North America/Canada/Ontario/ | Public Health Ontario Laboratory | Public Health Ontario Laboratory | Vanessa G Allen et al |

| | | | | | | |
|---|---|---|---|---|---|---|
| EPI_ISL_925553 | Canada/ON-PHL-20-80502/2020 | 20-Apr | North America/Canada/Ontario/ | Public Health Ontario Laboratory | Public Health Ontario Laboratory | Vanessa G Allen et al |
| EPI_ISL_925554 | Canada/ON-PHL-20-80501/2020 | 20-Apr | North America/Canada/Ontario/ | Public Health Ontario Laboratory | Public Health Ontario Laboratory | Vanessa G Allen et al |
| EPI_ISL_925555 | Canada/ON-PHL-20-80506/2020 | 20-Apr | North America/Canada/Ontario/ | Public Health Ontario Laboratory | Public Health Ontario Laboratory | Vanessa G Allen et al |
| EPI_ISL_925556 | Canada/ON-PHL-20-80514/2020 | 20-Apr | North America/Canada/Ontario/ | Public Health Ontario Laboratory | Public Health Ontario Laboratory | Vanessa G Allen et al |
| EPI_ISL_925557 | Canada/ON-PHL-20-80513/2020 | 20-Apr | North America/Canada/Ontario/ | Public Health Ontario Laboratory | Public Health Ontario Laboratory | Vanessa G Allen et al |
| EPI_ISL_925558 | Canada/ON-PHL-20-80468/2020 | 20-Apr | North America/Canada/Ontario/ | Public Health Ontario Laboratory | Public Health Ontario Laboratory | Vanessa G Allen et al |
| EPI_ISL_925559 | Canada/ON-PHL-20-80467/2020 | 20-Apr | North America/Canada/Ontario/ | Public Health Ontario Laboratory | Public Health Ontario Laboratory | Vanessa G Allen et al |
| EPI_ISL_925560 | Canada/ON-PHL-20-80465/2020 | 20-Apr | North America/Canada/Ontario/ | Public Health Ontario Laboratory | Public Health Ontario Laboratory | Vanessa G Allen et al |
| EPI_ISL_925561 | Canada/ON-PHL-20-80483/2020 | 20-Apr | North America/Canada/Ontario/ | Public Health Ontario Laboratory | Public Health Ontario Laboratory | Vanessa G Allen et al |
| EPI_ISL_925562 | Canada/ON-PHL-20-80482/2020 | 20-Apr | North America/Canada/Ontario/ | Public Health Ontario Laboratory | Public Health Ontario Laboratory | Vanessa G Allen et al |
| EPI_ISL_925563 | Canada/ON-PHL-20-80481/2020 | 20-Apr | North America/Canada/Ontario/ | Public Health Ontario Laboratory | Public Health Ontario Laboratory | Vanessa G Allen et al |
| EPI_ISL_925564 | Canada/ON-PHL-20-80480/2020 | 20-Apr | North America/Canada/Ontario/ | Public Health Ontario Laboratory | Public Health Ontario Laboratory | Vanessa G Allen et al |
| EPI_ISL_925565 | Canada/ON-PHL-20-80493/2020 | 20-Apr | North America/Canada/Ontario/ | Public Health Ontario Laboratory | Public Health Ontario Laboratory | Vanessa G Allen et al |

| | | | | | | |
|---|---|---|---|---|---|---|
| EPI_ISL_925566 | Canada/ON-PHL-20-80494/2020 | 20-Apr | North America/Canada/Ontario/ | Public Health Ontario Laboratory | Public Health Ontario Laboratory | Vanessa G Allen et al |
| EPI_ISL_925567 | Canada/ON-PHL-20-80510/2020 | 20-Apr | North America/Canada/Ontario/ | Public Health Ontario Laboratory | Public Health Ontario Laboratory | Vanessa G Allen et al |
| EPI_ISL_925568 | Canada/ON-PHL-20-80464/2020 | 20-Apr | North America/Canada/Ontario/ | Public Health Ontario Laboratory | Public Health Ontario Laboratory | Vanessa G Allen et al |
| EPI_ISL_925569 | Canada/ON-PHL-20-80463/2020 | 20-Apr | North America/Canada/Ontario/ | Public Health Ontario Laboratory | Public Health Ontario Laboratory | Vanessa G Allen et al |
| EPI_ISL_925570 | Canada/ON-PHL-20-80460/2020 | 20-Apr | North America/Canada/Ontario/ | Public Health Ontario Laboratory | Public Health Ontario Laboratory | Vanessa G Allen et al |
| EPI_ISL_925571 | Canada/ON-PHL-20-80462/2020 | 20-Apr | North America/Canada/Ontario/ | Public Health Ontario Laboratory | Public Health Ontario Laboratory | Vanessa G Allen et al |
| EPI_ISL_925572 | Canada/ON-PHL-20-80458/2020 | 20-Apr | North America/Canada/Ontario/ | Public Health Ontario Laboratory | Public Health Ontario Laboratory | Vanessa G Allen et al |
| EPI_ISL_925573 | Canada/ON-PHL-20-80457/2020 | 20-Apr | North America/Canada/Ontario/ | Public Health Ontario Laboratory | Public Health Ontario Laboratory | Vanessa G Allen et al |
| EPI_ISL_925574 | Canada/ON-PHL-20-80456/2020 | 20-Apr | North America/Canada/Ontario/ | Public Health Ontario Laboratory | Public Health Ontario Laboratory | Vanessa G Allen et al |
| EPI_ISL_925575 | Canada/ON-PHL-20-80741/2020 | 20-Apr | North America/Canada/Ontario/ | Public Health Ontario Laboratory | Public Health Ontario Laboratory | Vanessa G Allen et al |
| EPI_ISL_925576 | Canada/ON-PHL-20-80740/2020 | 20-Apr | North America/Canada/Ontario/ | Public Health Ontario Laboratory | Public Health Ontario Laboratory | Vanessa G Allen et al |
| EPI_ISL_925577 | Canada/ON-PHL-20-80754/2020 | 20-Apr | North America/Canada/Ontario/ | Public Health Ontario Laboratory | Public Health Ontario Laboratory | Vanessa G Allen et al |
| EPI_ISL_925578 | Canada/ON-PHL-20-80732/2020 | 20-Apr | North America/Canada/Ontario/ | Public Health Ontario Laboratory | Public Health Ontario Laboratory | Vanessa G Allen et al |

| | | | | | | |
|---|---|---|---|---|---|---|
| EPI_ISL_925579 | Canada/ON-PHL-20-80733/2020 | 20-Apr | North America/Canada/Ontario/ | Public Health Ontario Laboratory | Public Health Ontario Laboratory | Vanessa G Allen et al |
| EPI_ISL_925580 | Canada/ON-PHL-20-80731/2020 | 20-Apr | North America/Canada/Ontario/ | Public Health Ontario Laboratory | Public Health Ontario Laboratory | Vanessa G Allen et al |
| EPI_ISL_925581 | Canada/ON-PHL-20-80742/2020 | 20-Apr | North America/Canada/Ontario/ | Public Health Ontario Laboratory | Public Health Ontario Laboratory | Vanessa G Allen et al |
| EPI_ISL_925582 | Canada/ON-PHL-20-80745/2020 | 20-Apr | North America/Canada/Ontario/ | Public Health Ontario Laboratory | Public Health Ontario Laboratory | Vanessa G Allen et al |
| EPI_ISL_925583 | Canada/ON-PHL-20-80746/2020 | 20-Apr | North America/Canada/Ontario/ | Public Health Ontario Laboratory | Public Health Ontario Laboratory | Vanessa G Allen et al |
| EPI_ISL_925584 | Canada/ON-PHL-21-00319/2020 | 20-Sep | North America/Canada/Ontario/ | Public Health Ontario Laboratory | Public Health Ontario Laboratory | Vanessa G Allen et al |
| EPI_ISL_925585 | Canada/ON-PHL-21-00320/2020 | 20-Oct | North America/Canada/Ontario/ | Public Health Ontario Laboratory | Public Health Ontario Laboratory | Vanessa G Allen et al |
| EPI_ISL_925586 | Canada/ON-PHL-21-00321/2020 | 20-Nov | North America/Canada/Ontario/ | Public Health Ontario Laboratory | Public Health Ontario Laboratory | Vanessa G Allen et al |
| EPI_ISL_925587 | Canada/ON-PHL-21-00324/2020 | 20-Nov | North America/Canada/Ontario/ | Public Health Ontario Laboratory | Public Health Ontario Laboratory | Vanessa G Allen et al |
| EPI_ISL_925588 | Canada/ON-PHL-21-00326/2020 | 20-Nov | North America/Canada/Ontario/ | Public Health Ontario Laboratory | Public Health Ontario Laboratory | Vanessa G Allen et al |
| EPI_ISL_925589 | Canada/ON-PHL-21-00328/2020 | 20-Nov | North America/Canada/Ontario/ | Public Health Ontario Laboratory | Public Health Ontario Laboratory | Vanessa G Allen et al |
| EPI_ISL_925590 | Canada/ON-PHL-21-00334/2020 | 20-Nov | North America/Canada/Ontario/ | Public Health Ontario Laboratory | Public Health Ontario Laboratory | Vanessa G Allen et al |
| EPI_ISL_925591 | Canada/ON-PHL-21-00332/2020 | 20-Nov | North America/Canada/Ontario/ | Public Health Ontario Laboratory | Public Health Ontario Laboratory | Vanessa G Allen et al |

| | | | | | | |
|---|---|---|---|---|---|---|
| EPI_ISL_925592 | Canada/ON-PHL-21-00323/2020 | 20-Nov | North America/Canada/Ontario/ | Public Health Ontario Laboratory | Public Health Ontario Laboratory | Vanessa G Allen et al |
| EPI_ISL_925593 | Canada/ON-PHL-21-00333/2020 | 20-Nov | North America/Canada/Ontario/ | Public Health Ontario Laboratory | Public Health Ontario Laboratory | Vanessa G Allen et al |
| EPI_ISL_925594 | Canada/ON-PHL-21-00370/2020 | 20-Nov | North America/Canada/Ontario/ | Public Health Ontario Laboratory | Public Health Ontario Laboratory | Vanessa G Allen et al |
| EPI_ISL_925595 | Canada/ON-PHL-21-00345/2020 | 20-Nov | North America/Canada/Ontario/ | Public Health Ontario Laboratory | Public Health Ontario Laboratory | Vanessa G Allen et al |
| EPI_ISL_925596 | Canada/ON-PHL-21-00335/2020 | 20-Nov | North America/Canada/Ontario/ | Public Health Ontario Laboratory | Public Health Ontario Laboratory | Vanessa G Allen et al |
| EPI_ISL_925597 | Canada/ON-PHL-21-00344/2020 | 20-Dec | North America/Canada/Ontario/ | Public Health Ontario Laboratory | Public Health Ontario Laboratory | Vanessa G Allen et al |
| EPI_ISL_925598 | Canada/ON-PHL-21-00367/2020 | 20-Dec | North America/Canada/Ontario/ | Public Health Ontario Laboratory | Public Health Ontario Laboratory | Vanessa G Allen et al |
| EPI_ISL_925599 | Canada/ON-PHL-21-00374/2020 | 20-Dec | North America/Canada/Ontario/ | Public Health Ontario Laboratory | Public Health Ontario Laboratory | Vanessa G Allen et al |
| EPI_ISL_925601 | Canada/ON-PHL-21-00357/2020 | 20-Dec | North America/Canada/Ontario/ | Public Health Ontario Laboratory | Public Health Ontario Laboratory | Vanessa G Allen et al |
| EPI_ISL_925602 | Canada/ON-PHL-21-00343/2020 | 20-Dec | North America/Canada/Ontario/ | Public Health Ontario Laboratory | Public Health Ontario Laboratory | Vanessa G Allen et al |
| EPI_ISL_925603 | Canada/ON-PHL-21-00342/2020 | 20-Dec | North America/Canada/Ontario/ | Public Health Ontario Laboratory | Public Health Ontario Laboratory | Vanessa G Allen et al |
| EPI_ISL_925604 | Canada/ON-PHL-21-00341/2020 | 20-Dec | North America/Canada/Ontario/ | Public Health Ontario Laboratory | Public Health Ontario Laboratory | Vanessa G Allen et al |
| EPI_ISL_925605 | Canada/ON-PHL-21-00338/2020 | 20-Dec | North America/Canada/Ontario/ | Public Health Ontario Laboratory | Public Health Ontario Laboratory | Vanessa G Allen et al |

| | | | | | | |
|---|---|---|---|---|---|---|
| EPI_ISL_925606 | Canada/ON-PHL-21-00349/2020 | 20-Dec | North America/Canada/Ontario/ | Public Health Ontario Laboratory | Public Health Ontario Laboratory | Vanessa G Allen et al |
| EPI_ISL_925607 | Canada/ON-PHL-21-00337/2020 | 20-Dec | North America/Canada/Ontario/ | Public Health Ontario Laboratory | Public Health Ontario Laboratory | Vanessa G Allen et al |
| EPI_ISL_925608 | Canada/ON-PHL-21-00372/2020 | 20-Dec | North America/Canada/Ontario/ | Public Health Ontario Laboratory | Public Health Ontario Laboratory | Vanessa G Allen et al |
| EPI_ISL_925609 | Canada/ON-PHL-21-00373/2020 | 20-Dec | North America/Canada/Ontario/ | Public Health Ontario Laboratory | Public Health Ontario Laboratory | Vanessa G Allen et al |
| EPI_ISL_925610 | Canada/ON-PHL-21-00355/2020 | 20-Dec | North America/Canada/Ontario/ | Public Health Ontario Laboratory | Public Health Ontario Laboratory | Vanessa G Allen et al |
| EPI_ISL_925611 | Canada/ON-PHL-21-00339/2020 | 20-Dec | North America/Canada/Ontario/ | Public Health Ontario Laboratory | Public Health Ontario Laboratory | Vanessa G Allen et al |
| EPI_ISL_925612 | Canada/ON-PHL-21-00354/2020 | 20-Dec | North America/Canada/Ontario/ | Public Health Ontario Laboratory | Public Health Ontario Laboratory | Vanessa G Allen et al |
| EPI_ISL_925613 | Canada/ON-PHL-21-00353/2020 | 20-Dec | North America/Canada/Ontario/ | Public Health Ontario Laboratory | Public Health Ontario Laboratory | Vanessa G Allen et al |
| EPI_ISL_925614 | Canada/ON-PHL-21-00371/2020 | 20-Dec | North America/Canada/Ontario/ | Public Health Ontario Laboratory | Public Health Ontario Laboratory | Vanessa G Allen et al |
| EPI_ISL_925615 | Canada/ON-PHL-21-00366/2020 | 20-Dec | North America/Canada/Ontario/ | Public Health Ontario Laboratory | Public Health Ontario Laboratory | Vanessa G Allen et al |
| EPI_ISL_925616 | Canada/ON-PHL-21-00369/2020 | 20-Dec | North America/Canada/Ontario/ | Public Health Ontario Laboratory | Public Health Ontario Laboratory | Vanessa G Allen et al |
| EPI_ISL_925617 | Canada/ON-PHL-21-00365/2020 | 20-Dec | North America/Canada/Ontario/ | Public Health Ontario Laboratory | Public Health Ontario Laboratory | Vanessa G Allen et al |
| EPI_ISL_925618 | Canada/ON-PHL-21-00390/2020 | 20-Dec | North America/Canada/Ontario/ | Public Health Ontario Laboratory | Public Health Ontario Laboratory | Vanessa G Allen et al |

| | | | | | | |
|---|---|---|---|---|---|---|
| EPI_ISL_925619 | Canada/ON-PHL-21-00360/2020 | 20-Dec | North America/Canada/Ontario/ | Public Health Ontario Laboratory | Public Health Ontario Laboratory | Vanessa G Allen et al |
| EPI_ISL_925620 | Canada/ON-PHL-21-00375/2020 | 20-Dec | North America/Canada/Ontario/ | Public Health Ontario Laboratory | Public Health Ontario Laboratory | Vanessa G Allen et al |
| EPI_ISL_925621 | Canada/ON-PHL-21-00383/2020 | 20-Dec | North America/Canada/Ontario/ | Public Health Ontario Laboratory | Public Health Ontario Laboratory | Vanessa G Allen et al |
| EPI_ISL_925622 | Canada/ON-PHL-21-00362/2020 | 20-Dec | North America/Canada/Ontario/ | Public Health Ontario Laboratory | Public Health Ontario Laboratory | Vanessa G Allen et al |
| EPI_ISL_925623 | Canada/ON-PHL-21-00364/2020 | 20-Dec | North America/Canada/Ontario/ | Public Health Ontario Laboratory | Public Health Ontario Laboratory | Vanessa G Allen et al |
| EPI_ISL_925624 | Canada/ON-PHL-21-00381/2020 | 20-Dec | North America/Canada/Ontario/ | Public Health Ontario Laboratory | Public Health Ontario Laboratory | Vanessa G Allen et al |
| EPI_ISL_925625 | Canada/ON-PHL-21-00385/2020 | 20-Dec | North America/Canada/Ontario/ | Public Health Ontario Laboratory | Public Health Ontario Laboratory | Vanessa G Allen et al |
| EPI_ISL_925626 | Canada/ON-PHL-21-00480/2020 | 20-Dec | North America/Canada/Ontario/ | Public Health Ontario Laboratory | Public Health Ontario Laboratory | Vanessa G Allen et al |
| EPI_ISL_925627 | Canada/ON-PHL-21-00478/2020 | 20-Dec | North America/Canada/Ontario/ | Public Health Ontario Laboratory | Public Health Ontario Laboratory | Vanessa G Allen et al |
| EPI_ISL_925628 | Canada/ON-PHL-21-00491/2020 | 20-Dec | North America/Canada/Ontario/ | Public Health Ontario Laboratory | Public Health Ontario Laboratory | Vanessa G Allen et al |
| EPI_ISL_925629 | Canada/ON-PHL-21-00492/2020 | 20-Dec | North America/Canada/Ontario/ | Public Health Ontario Laboratory | Public Health Ontario Laboratory | Vanessa G Allen et al |
| EPI_ISL_925630 | Canada/ON-PHL-21-00499/2020 | 20-Dec | North America/Canada/Ontario/ | Public Health Ontario Laboratory | Public Health Ontario Laboratory | Vanessa G Allen et al |
| EPI_ISL_925631 | Canada/ON-PHL-21-00489/2020 | 20-Dec | North America/Canada/Ontario/ | Public Health Ontario Laboratory | Public Health Ontario Laboratory | Vanessa G Allen et al |

| | | | | | | |
|---|---|---|---|---|---|---|
| EPI_ISL_925632 | Canada/ON-PHL-21-00503/2020 | 20-Dec | North America/Canada/Ontario/ | Public Health Ontario Laboratory | Public Health Ontario Laboratory | Vanessa G Allen et al |
| EPI_ISL_925633 | Canada/ON-PHL-21-00488/2020 | 20-Dec | North America/Canada/Ontario/ | Public Health Ontario Laboratory | Public Health Ontario Laboratory | Vanessa G Allen et al |
| EPI_ISL_925634 | Canada/ON-PHL-21-00490/2020 | 20-Dec | North America/Canada/Ontario/ | Public Health Ontario Laboratory | Public Health Ontario Laboratory | Vanessa G Allen et al |
| EPI_ISL_925635 | Canada/ON-PHL-21-00506/2020 | 20-Dec | North America/Canada/Ontario/ | Public Health Ontario Laboratory | Public Health Ontario Laboratory | Vanessa G Allen et al |
| EPI_ISL_925636 | Canada/ON-PHL-21-00484/2020 | 20-Dec | North America/Canada/Ontario/ | Public Health Ontario Laboratory | Public Health Ontario Laboratory | Vanessa G Allen et al |
| EPI_ISL_925637 | Canada/ON-PHL-21-00388/2020 | 20-Dec | North America/Canada/Ontario/ | Public Health Ontario Laboratory | Public Health Ontario Laboratory | Vanessa G Allen et al |
| EPI_ISL_925639 | Canada/ON-PHL-21-00505/2020 | 20-Dec | North America/Canada/Ontario/ | Public Health Ontario Laboratory | Public Health Ontario Laboratory | Vanessa G Allen et al |
| EPI_ISL_925640 | Canada/ON-PHL-21-00544/2020 | 20-Dec | North America/Canada/Ontario/ | Public Health Ontario Laboratory | Public Health Ontario Laboratory | Vanessa G Allen et al |
| EPI_ISL_925641 | Canada/ON-PHL-21-00541/2020 | 20-Dec | North America/Canada/Ontario/ | Public Health Ontario Laboratory | Public Health Ontario Laboratory | Vanessa G Allen et al |
| EPI_ISL_925642 | Canada/ON-PHL-21-00513/2020 | 20-Dec | North America/Canada/Ontario/ | Public Health Ontario Laboratory | Public Health Ontario Laboratory | Vanessa G Allen et al |
| EPI_ISL_925643 | Canada/ON-PHL-21-00512/2020 | 20-Dec | North America/Canada/Ontario/ | Public Health Ontario Laboratory | Public Health Ontario Laboratory | Vanessa G Allen et al |
| EPI_ISL_925645 | Canada/ON-PHL-21-00495/2020 | 20-Dec | North America/Canada/Ontario/ | Public Health Ontario Laboratory | Public Health Ontario Laboratory | Vanessa G Allen et al |
| EPI_ISL_925646 | Canada/ON-PHL-21-00522/2020 | 20-Dec | North America/Canada/Ontario/ | Public Health Ontario Laboratory | Public Health Ontario Laboratory | Vanessa G Allen et al |

| | | | | | | |
|---|---|---|---|---|---|---|
| EPI_ISL_925647 | Canada/ON-PHL-21-00523/2020 | 20-Dec | North America/Canada/Ontario/ | Public Health Ontario Laboratory | Public Health Ontario Laboratory | Vanessa G Allen et al |
| EPI_ISL_925648 | Canada/ON-PHL-21-00517/2020 | 20-Dec | North America/Canada/Ontario/ | Public Health Ontario Laboratory | Public Health Ontario Laboratory | Vanessa G Allen et al |
| EPI_ISL_925649 | Canada/ON-PHL-21-00516/2020 | 20-Dec | North America/Canada/Ontario/ | Public Health Ontario Laboratory | Public Health Ontario Laboratory | Vanessa G Allen et al |
| EPI_ISL_925650 | Canada/ON-PHL-21-00533/2020 | 20-Dec | North America/Canada/Ontario/ | Public Health Ontario Laboratory | Public Health Ontario Laboratory | Vanessa G Allen et al |
| EPI_ISL_925651 | Canada/ON-PHL-21-00542/2020 | 20-Dec | North America/Canada/Ontario/ | Public Health Ontario Laboratory | Public Health Ontario Laboratory | Vanessa G Allen et al |
| EPI_ISL_925652 | Canada/ON-PHL-21-00535/2020 | 20-Dec | North America/Canada/Ontario/ | Public Health Ontario Laboratory | Public Health Ontario Laboratory | Vanessa G Allen et al |
| EPI_ISL_925653 | Canada/ON-PHL-21-00546/2020 | 20-Dec | North America/Canada/Ontario/ | Public Health Ontario Laboratory | Public Health Ontario Laboratory | Vanessa G Allen et al |
| EPI_ISL_925654 | Canada/ON-PHL-21-00537/2020 | 20-Dec | North America/Canada/Ontario/ | Public Health Ontario Laboratory | Public Health Ontario Laboratory | Vanessa G Allen et al |
| EPI_ISL_925655 | Canada/ON-PHL-21-00547/2020 | 20-Dec | North America/Canada/Ontario/ | Public Health Ontario Laboratory | Public Health Ontario Laboratory | Vanessa G Allen et al |
| EPI_ISL_925656 | Canada/ON-PHL-21-00520/2020 | 20-Dec | North America/Canada/Ontario/ | Public Health Ontario Laboratory | Public Health Ontario Laboratory | Vanessa G Allen et al |
| EPI_ISL_925657 | Canada/ON-PHL-21-00521/2020 | 20-Dec | North America/Canada/Ontario/ | Public Health Ontario Laboratory | Public Health Ontario Laboratory | Vanessa G Allen et al |
| EPI_ISL_925658 | Canada/ON-PHL-21-00536/2020 | 20-Dec | North America/Canada/Ontario/ | Public Health Ontario Laboratory | Public Health Ontario Laboratory | Vanessa G Allen et al |
| EPI_ISL_925659 | Canada/ON-PHL-21-00514/2020 | 20-Dec | North America/Canada/Ontario/ | Public Health Ontario Laboratory | Public Health Ontario Laboratory | Vanessa G Allen et al |

| | | | | | | |
|---|---|---|---|---|---|---|
| EPI_ISL_925660 | Canada/ON-PHL-21-00543/2020 | 20-Dec | North America/Canada/Ontario/ | Public Health Ontario Laboratory | Public Health Ontario Laboratory | Vanessa G Allen et al |
| EPI_ISL_925661 | Canada/ON-PHL-21-00518/2020 | 20-Dec | North America/Canada/Ontario/ | Public Health Ontario Laboratory | Public Health Ontario Laboratory | Vanessa G Allen et al |
| EPI_ISL_925662 | Canada/ON-PHL-21-00538/2020 | 20-Dec | North America/Canada/Ontario/ | Public Health Ontario Laboratory | Public Health Ontario Laboratory | Vanessa G Allen et al |
| EPI_ISL_925663 | Canada/ON-PHL-21-00501/2020 | 20-Dec | North America/Canada/Ontario/ | Public Health Ontario Laboratory | Public Health Ontario Laboratory | Vanessa G Allen et al |
| EPI_ISL_925664 | Canada/ON-PHL-21-00497/2020 | 20-Dec | North America/Canada/Ontario/ | Public Health Ontario Laboratory | Public Health Ontario Laboratory | Vanessa G Allen et al |
| EPI_ISL_925665 | Canada/ON-PHL-21-00498/2020 | 20-Dec | North America/Canada/Ontario/ | Public Health Ontario Laboratory | Public Health Ontario Laboratory | Vanessa G Allen et al |
| EPI_ISL_925666 | Canada/ON-PHL-21-00504/2020 | 20-Dec | North America/Canada/Ontario/ | Public Health Ontario Laboratory | Public Health Ontario Laboratory | Vanessa G Allen et al |
| EPI_ISL_925667 | Canada/ON-PHL-21-00500/2020 | 20-Dec | North America/Canada/Ontario/ | Public Health Ontario Laboratory | Public Health Ontario Laboratory | Vanessa G Allen et al |
| EPI_ISL_925668 | Canada/ON-PHL-21-00493/2020 | 20-Dec | North America/Canada/Ontario/ | Public Health Ontario Laboratory | Public Health Ontario Laboratory | Vanessa G Allen et al |
| EPI_ISL_925669 | Canada/ON-PHL-21-00494/2020 | 20-Dec | North America/Canada/Ontario/ | Public Health Ontario Laboratory | Public Health Ontario Laboratory | Vanessa G Allen et al |
| EPI_ISL_925670 | Canada/ON-PHL-21-00496/2020 | 20-Dec | North America/Canada/Ontario/ | Public Health Ontario Laboratory | Public Health Ontario Laboratory | Vanessa G Allen et al |
| EPI_ISL_925671 | Canada/ON-PHL-21-00519/2020 | 20-Dec | North America/Canada/Ontario/ | Public Health Ontario Laboratory | Public Health Ontario Laboratory | Vanessa G Allen et al |
| EPI_ISL_925672 | Canada/ON-PHL-21-00539/2020 | 20-Dec | North America/Canada/Ontario/ | Public Health Ontario Laboratory | Public Health Ontario Laboratory | Vanessa G Allen et al |

| | | | | | | |
|---|---|---|---|---|---|---|
| EPI_ISL_925673 | Canada/ON-PHL-21-00540/2020 | 20-Dec | North America/Canada/Ontario/ | Public Health Ontario Laboratory | Public Health Ontario Laboratory | Vanessa G Allen et al |
| EPI_ISL_925674 | Canada/ON-PHL-21-00531/2020 | 20-Dec | North America/Canada/Ontario/ | Public Health Ontario Laboratory | Public Health Ontario Laboratory | Vanessa G Allen et al |
| EPI_ISL_925675 | Canada/ON-PHL-21-00528/2020 | 20-Dec | North America/Canada/Ontario/ | Public Health Ontario Laboratory | Public Health Ontario Laboratory | Vanessa G Allen et al |
| EPI_ISL_925676 | Canada/ON-PHL-21-00529/2020 | 20-Dec | North America/Canada/Ontario/ | Public Health Ontario Laboratory | Public Health Ontario Laboratory | Vanessa G Allen et al |
| EPI_ISL_925677 | Canada/ON-PHL-21-00530/2020 | 20-Dec | North America/Canada/Ontario/ | Public Health Ontario Laboratory | Public Health Ontario Laboratory | Vanessa G Allen et al |
| EPI_ISL_925678 | Canada/ON-PHL-20-80801/2020 | 20-Apr | North America/Canada/Ontario/ | Public Health Ontario Laboratory | Public Health Ontario Laboratory | Vanessa G Allen et al |
| EPI_ISL_925679 | Canada/ON-PHL-20-80800/2020 | 20-Apr | North America/Canada/Ontario/ | Public Health Ontario Laboratory | Public Health Ontario Laboratory | Vanessa G Allen et al |
| EPI_ISL_925680 | Canada/ON-PHL-20-80469/2020 | 20-Apr | North America/Canada/Ontario/ | Public Health Ontario Laboratory | Public Health Ontario Laboratory | Vanessa G Allen et al |
| EPI_ISL_925681 | Canada/ON-PHL-20-80799/2020 | 20-Apr | North America/Canada/Ontario/ | Public Health Ontario Laboratory | Public Health Ontario Laboratory | Vanessa G Allen et al |
| EPI_ISL_925682 | Canada/ON-PHL-20-80471/2020 | 20-Apr | North America/Canada/Ontario/ | Public Health Ontario Laboratory | Public Health Ontario Laboratory | Vanessa G Allen et al |
| EPI_ISL_925683 | Canada/ON-PHL-20-80489/2020 | 20-Apr | North America/Canada/Ontario/ | Public Health Ontario Laboratory | Public Health Ontario Laboratory | Vanessa G Allen et al |
| EPI_ISL_925684 | Canada/ON-PHL-20-80490/2020 | 20-Apr | North America/Canada/Ontario/ | Public Health Ontario Laboratory | Public Health Ontario Laboratory | Vanessa G Allen et al |
| EPI_ISL_925685 | Canada/ON-PHL-20-80496/2020 | 20-Apr | North America/Canada/Ontario/ | Public Health Ontario Laboratory | Public Health Ontario Laboratory | Vanessa G Allen et al |

| | | | | | | |
|---|---|---|---|---|---|---|
| EPI_ISL_925686 | Canada/ON-PHL-20-80500/2020 | 20-Apr | North America/Canada/Ontario/ | Public Health Ontario Laboratory | Public Health Ontario Laboratory | Vanessa G Allen et al |
| EPI_ISL_925687 | Canada/ON-PHL-20-80499/2020 | 20-Apr | North America/Canada/Ontario/ | Public Health Ontario Laboratory | Public Health Ontario Laboratory | Vanessa G Allen et al |
| EPI_ISL_925688 | Canada/ON-PHL-20-80498/2020 | 20-Apr | North America/Canada/Ontario/ | Public Health Ontario Laboratory | Public Health Ontario Laboratory | Vanessa G Allen et al |
| EPI_ISL_925689 | Canada/ON-PHL-20-80503/2020 | 20-Apr | North America/Canada/Ontario/ | Public Health Ontario Laboratory | Public Health Ontario Laboratory | Vanessa G Allen et al |
| EPI_ISL_925690 | Canada/ON-PHL-20-80521/2020 | 20-Apr | North America/Canada/Ontario/ | Public Health Ontario Laboratory | Public Health Ontario Laboratory | Vanessa G Allen et al |
| EPI_ISL_925691 | Canada/ON-PHL-20-80525/2020 | 20-Apr | North America/Canada/Ontario/ | Public Health Ontario Laboratory | Public Health Ontario Laboratory | Vanessa G Allen et al |
| EPI_ISL_925692 | Canada/ON-PHL-20-80526/2020 | 20-Apr | North America/Canada/Ontario/ | Public Health Ontario Laboratory | Public Health Ontario Laboratory | Vanessa G Allen et al |
| EPI_ISL_925693 | Canada/ON-PHL-20-80726/2020 | 20-Apr | North America/Canada/Ontario/ | Public Health Ontario Laboratory | Public Health Ontario Laboratory | Vanessa G Allen et al |
| EPI_ISL_925694 | Canada/ON-PHL-20-80721/2020 | 20-Apr | North America/Canada/Ontario/ | Public Health Ontario Laboratory | Public Health Ontario Laboratory | Vanessa G Allen et al |
| EPI_ISL_925695 | Canada/ON-PHL-20-80725/2020 | 20-Apr | North America/Canada/Ontario/ | Public Health Ontario Laboratory | Public Health Ontario Laboratory | Vanessa G Allen et al |
| EPI_ISL_925696 | Canada/ON-PHL-20-80724/2020 | 20-Apr | North America/Canada/Ontario/ | Public Health Ontario Laboratory | Public Health Ontario Laboratory | Vanessa G Allen et al |
| EPI_ISL_925697 | Canada/ON-PHL-20-80727/2020 | 20-Apr | North America/Canada/Ontario/ | Public Health Ontario Laboratory | Public Health Ontario Laboratory | Vanessa G Allen et al |
| EPI_ISL_925698 | Canada/ON-PHL-21-00330/2020 | 20-Nov | North America/Canada/Ontario/ | Public Health Ontario Laboratory | Public Health Ontario Laboratory | Vanessa G Allen et al |

| | | | | | | |
|---|---|---|---|---|---|---|
| EPI_ISL_925699 | Canada/ON-PHL-21-00322/2020 | 20-Nov | North America/Canada/Ontario/ | Public Health Ontario Laboratory | Public Health Ontario Laboratory | Vanessa G Allen et al |
| EPI_ISL_925700 | Canada/ON-PHL-21-00329/2020 | 20-Nov | North America/Canada/Ontario/ | Public Health Ontario Laboratory | Public Health Ontario Laboratory | Vanessa G Allen et al |
| EPI_ISL_925701 | Canada/ON-PHL-21-00348/2020 | 20-Dec | North America/Canada/Ontario/ | Public Health Ontario Laboratory | Public Health Ontario Laboratory | Vanessa G Allen et al |
| EPI_ISL_925702 | Canada/ON-PHL-21-00340/2020 | 20-Dec | North America/Canada/Ontario/ | Public Health Ontario Laboratory | Public Health Ontario Laboratory | Vanessa G Allen et al |
| EPI_ISL_925703 | Canada/ON-PHL-21-00336/2020 | 20-Dec | North America/Canada/Ontario/ | Public Health Ontario Laboratory | Public Health Ontario Laboratory | Vanessa G Allen et al |
| EPI_ISL_925704 | Canada/ON-PHL-21-00351/2020 | 20-Dec | North America/Canada/Ontario/ | Public Health Ontario Laboratory | Public Health Ontario Laboratory | Vanessa G Allen et al |
| EPI_ISL_925705 | Canada/ON-PHL-21-00352/2020 | 20-Dec | North America/Canada/Ontario/ | Public Health Ontario Laboratory | Public Health Ontario Laboratory | Vanessa G Allen et al |
| EPI_ISL_925706 | Canada/ON-PHL-21-00356/2020 | 20-Dec | North America/Canada/Ontario/ | Public Health Ontario Laboratory | Public Health Ontario Laboratory | Vanessa G Allen et al |
| EPI_ISL_925707 | Canada/ON-PHL-21-00358/2020 | 20-Dec | North America/Canada/Ontario/ | Public Health Ontario Laboratory | Public Health Ontario Laboratory | Vanessa G Allen et al |
| EPI_ISL_925708 | Canada/ON-PHL-21-00359/2020 | 20-Dec | North America/Canada/Ontario/ | Public Health Ontario Laboratory | Public Health Ontario Laboratory | Vanessa G Allen et al |
| EPI_ISL_925709 | Canada/ON-PHL-21-00361/2020 | 20-Dec | North America/Canada/Ontario/ | Public Health Ontario Laboratory | Public Health Ontario Laboratory | Vanessa G Allen et al |
| EPI_ISL_925710 | Canada/ON-PHL-21-00363/2020 | 20-Dec | North America/Canada/Ontario/ | Public Health Ontario Laboratory | Public Health Ontario Laboratory | Vanessa G Allen et al |
| EPI_ISL_925711 | Canada/ON-PHL-21-00392/2020 | 20-Dec | North America/Canada/Ontario/ | Public Health Ontario Laboratory | Public Health Ontario Laboratory | Vanessa G Allen et al |

| | | | | | | |
|---|---|---|---|---|---|---|
| EPI_ISL_925712 | Canada/ON-PHL-21-00391/2020 | 20-Dec | North America/Canada/Ontario/ | Public Health Ontario Laboratory | Public Health Ontario Laboratory | Vanessa G Allen et al |
| EPI_ISL_925713 | Canada/ON-PHL-21-00378/2020 | 20-Dec | North America/Canada/Ontario/ | Public Health Ontario Laboratory | Public Health Ontario Laboratory | Vanessa G Allen et al |
| EPI_ISL_925714 | Canada/ON-PHL-21-00377/2020 | 20-Dec | North America/Canada/Ontario/ | Public Health Ontario Laboratory | Public Health Ontario Laboratory | Vanessa G Allen et al |
| EPI_ISL_925715 | Canada/ON-PHL-21-00389/2020 | 20-Dec | North America/Canada/Ontario/ | Public Health Ontario Laboratory | Public Health Ontario Laboratory | Vanessa G Allen et al |
| EPI_ISL_925716 | Canada/ON-PHL-21-00486/2020 | 20-Dec | North America/Canada/Ontario/ | Public Health Ontario Laboratory | Public Health Ontario Laboratory | Vanessa G Allen et al |
| EPI_ISL_925717 | Canada/ON-PHL-21-00508/2020 | 20-Dec | North America/Canada/Ontario/ | Public Health Ontario Laboratory | Public Health Ontario Laboratory | Vanessa G Allen et al |
| EPI_ISL_925718 | Canada/ON-PHL-21-00511/2020 | 20-Dec | North America/Canada/Ontario/ | Public Health Ontario Laboratory | Public Health Ontario Laboratory | Vanessa G Allen et al |
| EPI_ISL_925719 | Canada/ON-PHL-21-00510/2020 | 20-Dec | North America/Canada/Ontario/ | Public Health Ontario Laboratory | Public Health Ontario Laboratory | Vanessa G Allen et al |
| EPI_ISL_925720 | Canada/ON-PHL-21-00487/2020 | 20-Dec | North America/Canada/Ontario/ | Public Health Ontario Laboratory | Public Health Ontario Laboratory | Vanessa G Allen et al |
| EPI_ISL_925721 | Canada/ON-PHL-21-00393/2020 | 20-Dec | North America/Canada/Ontario/ | Public Health Ontario Laboratory | Public Health Ontario Laboratory | Vanessa G Allen et al |
| EPI_ISL_925722 | Canada/ON-PHL-21-00524/2020 | 20-Dec | North America/Canada/Ontario/ | Public Health Ontario Laboratory | Public Health Ontario Laboratory | Vanessa G Allen et al |
| EPI_ISL_925723 | Canada/ON-PHL-21-00525/2020 | 20-Dec | North America/Canada/Ontario/ | Public Health Ontario Laboratory | Public Health Ontario Laboratory | Vanessa G Allen et al |
| EPI_ISL_925724 | Canada/ON-PHL-21-00507/2020 | 20-Dec | North America/Canada/Ontario/ | Public Health Ontario Laboratory | Public Health Ontario Laboratory | Vanessa G Allen et al |

| | | | | | | |
|---|---|---|---|---|---|---|
| EPI_ISL_925725 | Canada/ON-PHL-21-00509/2020 | 20-Dec | North America/Canada/Ontario/ | Public Health Ontario Laboratory | Public Health Ontario Laboratory | Vanessa G Allen et al |
| EPI_ISL_925726 | Canada/ON-PHL-20-80786/2020 | 20-Apr | North America/Canada/Ontario/ | Public Health Ontario Laboratory | Public Health Ontario Laboratory | Vanessa G Allen et al |
| EPI_ISL_925727 | Canada/ON-PHL-20-80798/2020 | 20-Apr | North America/Canada/Ontario/ | Public Health Ontario Laboratory | Public Health Ontario Laboratory | Vanessa G Allen et al |
| EPI_ISL_925728 | Canada/ON-PHL-20-80791/2020 | 20-Mar | North America/Canada/Ontario/ | Public Health Ontario Laboratory | Public Health Ontario Laboratory | Vanessa G Allen et al |
| EPI_ISL_925729 | Canada/ON-PHL-20-80788/2020 | 20-Mar | North America/Canada/Ontario/ | Public Health Ontario Laboratory | Public Health Ontario Laboratory | Vanessa G Allen et al |
| EPI_ISL_925730 | Canada/ON-PHL-21-00382/2020 | 20-Dec | North America/Canada/Ontario/ | Public Health Ontario Laboratory | Public Health Ontario Laboratory | Vanessa G Allen et al |
| EPI_ISL_925731 | Canada/ON-PHL-20-80767/2020 | 20-Apr | North America/Canada/Ontario/ | Public Health Ontario Laboratory | Public Health Ontario Laboratory | Vanessa G Allen et al |
| EPI_ISL_925732 | Canada/ON-PHL-20-80768/2020 | 20-Apr | North America/Canada/Ontario/ | Public Health Ontario Laboratory | Public Health Ontario Laboratory | Vanessa G Allen et al |
| EPI_ISL_925733 | Canada/ON-PHL-20-80505/2020 | 20-Apr | North America/Canada/Ontario/ | Public Health Ontario Laboratory | Public Health Ontario Laboratory | Vanessa G Allen et al |
| EPI_ISL_925734 | Canada/ON-PHL-20-80504/2020 | 20-Apr | North America/Canada/Ontario/ | Public Health Ontario Laboratory | Public Health Ontario Laboratory | Vanessa G Allen et al |
| EPI_ISL_925735 | Canada/ON-PHL-20-80475/2020 | 20-Apr | North America/Canada/Ontario/ | Public Health Ontario Laboratory | Public Health Ontario Laboratory | Vanessa G Allen et al |
| EPI_ISL_925736 | Canada/ON-PHL-20-80515/2020 | 20-Apr | North America/Canada/Ontario/ | Public Health Ontario Laboratory | Public Health Ontario Laboratory | Vanessa G Allen et al |
| EPI_ISL_925737 | Canada/ON-PHL-20-80516/2020 | 20-Apr | North America/Canada/Ontario/ | Public Health Ontario Laboratory | Public Health Ontario Laboratory | Vanessa G Allen et al |

| | | | | | | |
|---|---|---|---|---|---|---|
| EPI_ISL_925738 | Canada/ON-PHL-20-80523/2020 | 20-Apr | North America/Canada/Ontario/ | Public Health Ontario Laboratory | Public Health Ontario Laboratory | Vanessa G Allen et al |
| EPI_ISL_925739 | Canada/ON-PHL-20-80522/2020 | 20-Apr | North America/Canada/Ontario/ | Public Health Ontario Laboratory | Public Health Ontario Laboratory | Vanessa G Allen et al |
| EPI_ISL_925740 | Canada/ON-PHL-20-80722/2020 | 20-Apr | North America/Canada/Ontario/ | Public Health Ontario Laboratory | Public Health Ontario Laboratory | Vanessa G Allen et al |
| EPI_ISL_925741 | Canada/ON-PHL-20-80517/2020 | 20-Apr | North America/Canada/Ontario/ | Public Health Ontario Laboratory | Public Health Ontario Laboratory | Vanessa G Allen et al |
| EPI_ISL_925742 | Canada/ON-PHL-20-80717/2020 | 20-Apr | North America/Canada/Ontario/ | Public Health Ontario Laboratory | Public Health Ontario Laboratory | Vanessa G Allen et al |
| EPI_ISL_925743 | Canada/ON-PHL-20-80715/2020 | 20-Apr | North America/Canada/Ontario/ | Public Health Ontario Laboratory | Public Health Ontario Laboratory | Vanessa G Allen et al |
| EPI_ISL_925744 | Canada/ON-PHL-20-80748/2020 | 20-Apr | North America/Canada/Ontario/ | Public Health Ontario Laboratory | Public Health Ontario Laboratory | Vanessa G Allen et al |
| EPI_ISL_925745 | Canada/ON-PHL-20-80737/2020 | 20-Apr | North America/Canada/Ontario/ | Public Health Ontario Laboratory | Public Health Ontario Laboratory | Vanessa G Allen et al |
| EPI_ISL_925746 | Canada/ON-PHL-20-80738/2020 | 20-Apr | North America/Canada/Ontario/ | Public Health Ontario Laboratory | Public Health Ontario Laboratory | Vanessa G Allen et al |
| EPI_ISL_925747 | Canada/ON-PHL-20-80728/2020 | 20-Apr | North America/Canada/Ontario/ | Public Health Ontario Laboratory | Public Health Ontario Laboratory | Vanessa G Allen et al |
| EPI_ISL_925748 | Canada/ON-PHL-20-80796/2020 | 20-Mar | North America/Canada/Ontario/ | Public Health Ontario Laboratory | Public Health Ontario Laboratory | Vanessa G Allen et al |
| EPI_ISL_925749 | Canada/ON-PHL-20-80774/2020 | 20-Mar | North America/Canada/Ontario/ | Public Health Ontario Laboratory | Public Health Ontario Laboratory | Vanessa G Allen et al |
| EPI_ISL_925750 | Canada/ON-PHL-20-80778/2020 | 20-Mar | North America/Canada/Ontario/ | Public Health Ontario Laboratory | Public Health Ontario Laboratory | Vanessa G Allen et al |

| | | | | | | |
|---|---|---|---|---|---|---|
| EPI_ISL_925751 | Canada/ON-PHL-20-80492/2020 | 20-Apr | North America/Canada/Ontario/ | Public Health Ontario Laboratory | Public Health Ontario Laboratory | Vanessa G Allen et al |
| EPI_ISL_925752 | Canada/ON-PHL-20-80484/2020 | 20-Apr | North America/Canada/Ontario/ | Public Health Ontario Laboratory | Public Health Ontario Laboratory | Vanessa G Allen et al |
| EPI_ISL_925753 | Canada/ON-PHL-20-80527/2020 | 20-Apr | North America/Canada/Ontario/ | Public Health Ontario Laboratory | Public Health Ontario Laboratory | Vanessa G Allen et al |
| EPI_ISL_925754 | Canada/ON-PHL-20-80528/2020 | 20-Apr | North America/Canada/Ontario/ | Public Health Ontario Laboratory | Public Health Ontario Laboratory | Vanessa G Allen et al |
| EPI_ISL_925755 | Canada/ON-PHL-20-80719/2020 | 20-Apr | North America/Canada/Ontario/ | Public Health Ontario Laboratory | Public Health Ontario Laboratory | Vanessa G Allen et al |
| EPI_ISL_925756 | Canada/ON-PHL-20-80716/2020 | 20-Apr | North America/Canada/Ontario/ | Public Health Ontario Laboratory | Public Health Ontario Laboratory | Vanessa G Allen et al |
| EPI_ISL_925757 | Canada/ON-PHL-20-80714/2020 | 20-Apr | North America/Canada/Ontario/ | Public Health Ontario Laboratory | Public Health Ontario Laboratory | Vanessa G Allen et al |
| EPI_ISL_925758 | Canada/ON-PHL-20-80747/2020 | 20-Apr | North America/Canada/Ontario/ | Public Health Ontario Laboratory | Public Health Ontario Laboratory | Vanessa G Allen et al |
| EPI_ISL_925759 | Canada/ON-PHL-20-80736/2020 | 20-Apr | North America/Canada/Ontario/ | Public Health Ontario Laboratory | Public Health Ontario Laboratory | Vanessa G Allen et al |
| EPI_ISL_925760 | Canada/ON-PHL-20-80743/2020 | 20-Apr | North America/Canada/Ontario/ | Public Health Ontario Laboratory | Public Health Ontario Laboratory | Vanessa G Allen et al |
| EPI_ISL_925761 | Canada/ON-PHL-20-80744/2020 | 20-Apr | North America/Canada/Ontario/ | Public Health Ontario Laboratory | Public Health Ontario Laboratory | Vanessa G Allen et al |
| EPI_ISL_925762 | Canada/ON-PHL-21-00327/2020 | 20-Nov | North America/Canada/Ontario/ | Public Health Ontario Laboratory | Public Health Ontario Laboratory | Vanessa G Allen et al |
| EPI_ISL_925763 | Canada/ON-PHL-21-00481/2020 | 20-Dec | North America/Canada/Ontario/ | Public Health Ontario Laboratory | Public Health Ontario Laboratory | Vanessa G Allen et al |

| | | | | | | |
|---|---|---|---|---|---|---|
| EPI_ISL_925764 | Canada/ON-PHL-21-00691/2021 | 21-Jan | North America/Canada/Ontario/ | Public Health Ontario Laboratory | Public Health Ontario Laboratory | Vanessa G Allen et al |
| EPI_ISL_925765 | Canada/ON-PHL-21-00695/2021 | 21-Jan | North America/Canada/Ontario/ | Public Health Ontario Laboratory | Public Health Ontario Laboratory | Vanessa G Allen et al |
| EPI_ISL_925766 | Canada/ON-PHL-21-00692/2021 | 21-Jan | North America/Canada/Ontario/ | Public Health Ontario Laboratory | Public Health Ontario Laboratory | Vanessa G Allen et al |
| EPI_ISL_925767 | Canada/ON-PHL-21-00693/2021 | 21-Jan | North America/Canada/Ontario/ | Public Health Ontario Laboratory | Public Health Ontario Laboratory | Vanessa G Allen et al |
| EPI_ISL_925768 | Canada/ON-PHL-21-00694/2021 | 21-Jan | North America/Canada/Ontario/ | Public Health Ontario Laboratory | Public Health Ontario Laboratory | Vanessa G Allen et al |
| EPI_ISL_925769 | Canada/ON-PHL-21-00665/2021 | 21-Jan | North America/Canada/Ontario/ | Public Health Ontario Laboratory | Public Health Ontario Laboratory | Vanessa G Allen et al |
| EPI_ISL_925770 | Canada/ON-PHL-21-00653/2021 | 21-Jan | North America/Canada/Ontario/ | Public Health Ontario Laboratory | Public Health Ontario Laboratory | Vanessa G Allen et al |
| EPI_ISL_925771 | Canada/ON-PHL-21-00649/2021 | 21-Jan | North America/Canada/Ontario/ | Public Health Ontario Laboratory | Public Health Ontario Laboratory | Vanessa G Allen et al |
| EPI_ISL_925772 | Canada/ON-PHL-21-00671/2021 | 21-Jan | North America/Canada/Ontario/ | Public Health Ontario Laboratory | Public Health Ontario Laboratory | Vanessa G Allen et al |
| EPI_ISL_925773 | Canada/ON-PHL-21-00717/2021 | 21-Jan | North America/Canada/Ontario/ | Public Health Ontario Laboratory | Public Health Ontario Laboratory | Vanessa G Allen et al |
| EPI_ISL_925774 | Canada/ON-PHL-21-00664/2021 | 21-Jan | North America/Canada/Ontario/ | Public Health Ontario Laboratory | Public Health Ontario Laboratory | Vanessa G Allen et al |
| EPI_ISL_925775 | Canada/ON-PHL-21-00651/2021 | 21-Jan | North America/Canada/Ontario/ | Public Health Ontario Laboratory | Public Health Ontario Laboratory | Vanessa G Allen et al |
| EPI_ISL_925776 | Canada/ON-PHL-21-00701/2021 | 21-Jan | North America/Canada/Ontario/ | Public Health Ontario Laboratory | Public Health Ontario Laboratory | Vanessa G Allen et al |

| | | | | | | |
|---|---|---|---|---|---|---|
| EPI_ISL_925777 | Canada/ON-PHL-21-00655/2021 | 21-Jan | North America/Canada/Ontario/ | Public Health Ontario Laboratory | Public Health Ontario Laboratory | Vanessa G Allen et al |
| EPI_ISL_925778 | Canada/ON-PHL-21-00681/2021 | 21-Jan | North America/Canada/Ontario/ | Public Health Ontario Laboratory | Public Health Ontario Laboratory | Vanessa G Allen et al |
| EPI_ISL_925779 | Canada/ON-PHL-21-00646/2021 | 21-Jan | North America/Canada/Ontario/ | Public Health Ontario Laboratory | Public Health Ontario Laboratory | Vanessa G Allen et al |
| EPI_ISL_925780 | Canada/ON-PHL-21-00647/2021 | 21-Jan | North America/Canada/Ontario/ | Public Health Ontario Laboratory | Public Health Ontario Laboratory | Vanessa G Allen et al |
| EPI_ISL_925781 | Canada/ON-PHL-21-00697/2021 | 21-Jan | North America/Canada/Ontario/ | Public Health Ontario Laboratory | Public Health Ontario Laboratory | Vanessa G Allen et al |
| EPI_ISL_925782 | Canada/ON-PHL-21-00688/2021 | 21-Jan | North America/Canada/Ontario/ | Public Health Ontario Laboratory | Public Health Ontario Laboratory | Vanessa G Allen et al |
| EPI_ISL_925783 | Canada/ON-PHL-21-00650/2021 | 21-Jan | North America/Canada/Ontario/ | Public Health Ontario Laboratory | Public Health Ontario Laboratory | Vanessa G Allen et al |
| EPI_ISL_925784 | Canada/ON-PHL-21-00678/2021 | 21-Jan | North America/Canada/Ontario/ | Public Health Ontario Laboratory | Public Health Ontario Laboratory | Vanessa G Allen et al |
| EPI_ISL_925785 | Canada/ON-PHL-21-00720/2021 | 21-Jan | North America/Canada/Ontario/ | Public Health Ontario Laboratory | Public Health Ontario Laboratory | Vanessa G Allen et al |
| EPI_ISL_925786 | Canada/ON-PHL-21-00718/2021 | 21-Jan | North America/Canada/Ontario/ | Public Health Ontario Laboratory | Public Health Ontario Laboratory | Vanessa G Allen et al |
| EPI_ISL_925787 | Canada/ON-PHL-21-00683/2021 | 21-Jan | North America/Canada/Ontario/ | Public Health Ontario Laboratory | Public Health Ontario Laboratory | Vanessa G Allen et al |
| EPI_ISL_925788 | Canada/ON-PHL-21-00657/2021 | 21-Jan | North America/Canada/Ontario/ | Public Health Ontario Laboratory | Public Health Ontario Laboratory | Vanessa G Allen et al |
| EPI_ISL_925789 | Canada/ON-PHL-21-00719/2021 | 21-Jan | North America/Canada/Ontario/ | Public Health Ontario Laboratory | Public Health Ontario Laboratory | Vanessa G Allen et al |

| | | | | | | |
|---|---|---|---|---|---|---|
| EPI_ISL_925790 | Canada/ON-PHL-21-00679/2021 | 21-Jan | North America/Canada/Ontario/ | Public Health Ontario Laboratory | Public Health Ontario Laboratory | Vanessa G Allen et al |
| EPI_ISL_925791 | Canada/ON-PHL-21-00658/2021 | 21-Jan | North America/Canada/Ontario/ | Public Health Ontario Laboratory | Public Health Ontario Laboratory | Vanessa G Allen et al |
| EPI_ISL_925792 | Canada/ON-PHL-21-00687/2021 | 21-Jan | North America/Canada/Ontario/ | Public Health Ontario Laboratory | Public Health Ontario Laboratory | Vanessa G Allen et al |
| EPI_ISL_925793 | Canada/ON-PHL-21-00704/2021 | 21-Jan | North America/Canada/Ontario/ | Public Health Ontario Laboratory | Public Health Ontario Laboratory | Vanessa G Allen et al |
| EPI_ISL_925794 | Canada/ON-PHL-21-00668/2021 | 21-Jan | North America/Canada/Ontario/ | Public Health Ontario Laboratory | Public Health Ontario Laboratory | Vanessa G Allen et al |
| EPI_ISL_925795 | Canada/ON-PHL-21-00699/2021 | 21-Jan | North America/Canada/Ontario/ | Public Health Ontario Laboratory | Public Health Ontario Laboratory | Vanessa G Allen et al |
| EPI_ISL_925796 | Canada/ON-PHL-21-00670/2021 | 21-Jan | North America/Canada/Ontario/ | Public Health Ontario Laboratory | Public Health Ontario Laboratory | Vanessa G Allen et al |
| EPI_ISL_925797 | Canada/ON-PHL-21-00667/2021 | 21-Jan | North America/Canada/Ontario/ | Public Health Ontario Laboratory | Public Health Ontario Laboratory | Vanessa G Allen et al |
| EPI_ISL_925798 | Canada/ON-PHL-21-00712/2021 | 21-Jan | North America/Canada/Ontario/ | Public Health Ontario Laboratory | Public Health Ontario Laboratory | Vanessa G Allen et al |
| EPI_ISL_925799 | Canada/ON-PHL-21-00700/2021 | 21-Jan | North America/Canada/Ontario/ | Public Health Ontario Laboratory | Public Health Ontario Laboratory | Vanessa G Allen et al |
| EPI_ISL_925800 | Canada/ON-PHL-21-00680/2021 | 21-Jan | North America/Canada/Ontario/ | Public Health Ontario Laboratory | Public Health Ontario Laboratory | Vanessa G Allen et al |
| EPI_ISL_925801 | Canada/ON-PHL-21-00676/2021 | 21-Jan | North America/Canada/Ontario/ | Public Health Ontario Laboratory | Public Health Ontario Laboratory | Vanessa G Allen et al |
| EPI_ISL_925802 | Canada/ON-PHL-21-00675/2021 | 21-Jan | North America/Canada/Ontario/ | Public Health Ontario Laboratory | Public Health Ontario Laboratory | Vanessa G Allen et al |

| | | | | | | |
|---|---|---|---|---|---|---|
| EPI_ISL_925803 | Canada/ON-PHL-21-00672/2021 | 21-Jan | North America/Canada/Ontario/ | Public Health Ontario Laboratory | Public Health Ontario Laboratory | Vanessa G Allen et al |
| EPI_ISL_925804 | Canada/ON-PHL-21-00714/2021 | 21-Jan | North America/Canada/Ontario/ | Public Health Ontario Laboratory | Public Health Ontario Laboratory | Vanessa G Allen et al |
| EPI_ISL_925805 | Canada/ON-PHL-21-00663/2021 | 21-Jan | North America/Canada/Ontario/ | Public Health Ontario Laboratory | Public Health Ontario Laboratory | Vanessa G Allen et al |
| EPI_ISL_925806 | Canada/ON-PHL-21-00686/2021 | 21-Jan | North America/Canada/Ontario/ | Public Health Ontario Laboratory | Public Health Ontario Laboratory | Vanessa G Allen et al |
| EPI_ISL_925807 | Canada/ON-PHL-21-00713/2021 | 21-Jan | North America/Canada/Ontario/ | Public Health Ontario Laboratory | Public Health Ontario Laboratory | Vanessa G Allen et al |
| EPI_ISL_925808 | Canada/ON-PHL-21-00674/2021 | 21-Jan | North America/Canada/Ontario/ | Public Health Ontario Laboratory | Public Health Ontario Laboratory | Vanessa G Allen et al |
| EPI_ISL_925809 | Canada/ON-PHL-21-00661/2021 | 21-Jan | North America/Canada/Ontario/ | Public Health Ontario Laboratory | Public Health Ontario Laboratory | Vanessa G Allen et al |
| EPI_ISL_925810 | Canada/ON-PHL-21-00696/2021 | 21-Jan | North America/Canada/Ontario/ | Public Health Ontario Laboratory | Public Health Ontario Laboratory | Vanessa G Allen et al |
| EPI_ISL_925811 | Canada/ON-PHL-21-00706/2021 | 21-Jan | North America/Canada/Ontario/ | Public Health Ontario Laboratory | Public Health Ontario Laboratory | Vanessa G Allen et al |
| EPI_ISL_925812 | Canada/ON-PHL-21-00705/2021 | 21-Jan | North America/Canada/Ontario/ | Public Health Ontario Laboratory | Public Health Ontario Laboratory | Vanessa G Allen et al |
| EPI_ISL_925813 | Canada/ON-PHL-21-00703/2021 | 21-Jan | North America/Canada/Ontario/ | Public Health Ontario Laboratory | Public Health Ontario Laboratory | Vanessa G Allen et al |
| EPI_ISL_925814 | Canada/ON-PHL-21-00709/2021 | 21-Jan | North America/Canada/Ontario/ | Public Health Ontario Laboratory | Public Health Ontario Laboratory | Vanessa G Allen et al |
| EPI_ISL_925815 | Canada/ON-PHL-21-00652/2021 | 21-Jan | North America/Canada/Ontario/ | Public Health Ontario Laboratory | Public Health Ontario Laboratory | Vanessa G Allen et al |

| | | | | | | |
|---|---|---|---|---|---|---|
| EPI_ISL_925816 | Canada/ON-PHL-21-00690/2021 | 21-Jan | North America/Canada/Ontario/ | Public Health Ontario Laboratory | Public Health Ontario Laboratory | Vanessa G Allen et al |
| EPI_ISL_925817 | Canada/ON-PHL-21-00702/2021 | 21-Jan | North America/Canada/Ontario/ | Public Health Ontario Laboratory | Public Health Ontario Laboratory | Vanessa G Allen et al |
| EPI_ISL_925818 | Canada/ON-PHL-21-00710/2021 | 21-Jan | North America/Canada/Ontario/ | Public Health Ontario Laboratory | Public Health Ontario Laboratory | Vanessa G Allen et al |
| EPI_ISL_925819 | Canada/ON-PHL-21-00698/2021 | 21-Jan | North America/Canada/Ontario/ | Public Health Ontario Laboratory | Public Health Ontario Laboratory | Vanessa G Allen et al |
| EPI_ISL_925820 | Canada/ON-PHL-21-00669/2021 | 21-Jan | North America/Canada/Ontario/ | Public Health Ontario Laboratory | Public Health Ontario Laboratory | Vanessa G Allen et al |
| EPI_ISL_925821 | Canada/ON-PHL-21-00656/2021 | 21-Jan | North America/Canada/Ontario/ | Public Health Ontario Laboratory | Public Health Ontario Laboratory | Vanessa G Allen et al |
| EPI_ISL_925822 | Canada/ON-PHL-21-00654/2021 | 21-Jan | North America/Canada/Ontario/ | Public Health Ontario Laboratory | Public Health Ontario Laboratory | Vanessa G Allen et al |
| EPI_ISL_925823 | Canada/ON-PHL-21-00673/2021 | 21-Jan | North America/Canada/Ontario/ | Public Health Ontario Laboratory | Public Health Ontario Laboratory | Vanessa G Allen et al |
| EPI_ISL_925825 | USA/AZ-ASPHL-3386/2021 | 2021-01-06 | North America/USA/Arizona/Maricopa ( | Arizona State Public Health Laboratory | Arizona State Public Health Laboratory | Trung Huynh et al |
| EPI_ISL_925828 | USA/AZ-ASPHL-3392/2021 | 2021-01-06 | North America/USA/Arizona/Yuma | Arizona State Public Health Laboratory | Arizona State Public Health Laboratory | Trung Huynh et al |
| EPI_ISL_925832 | USA/AZ-ASPHL-3396/2021 | 2021-01-04 | North America/USA/Arizona/Mohave C | Arizona State Public Health Laboratory | Arizona State Public Health Laboratory | Trung Huynh et al |
| EPI_ISL_925833 | USA/AZ-ASPHL-3397/2021 | 2021-01-07 | North America/USA/Arizona/Maricopa ( | Arizona State Public Health Laboratory | Arizona State Public Health Laboratory | Trung Huynh et al |
| EPI_ISL_925840 | USA/AZ-ASPHL-3408/2021 | 2021-01-06 | North America/USA/Arizona/Maricopa ( | Arizona State Public Health Laboratory | Arizona State Public Health Laboratory | Trung Huynh et al |

| | | | | | | |
|---|---|---|---|---|---|---|
| EPI_ISL_925841 | USA/AZ-ASPHL-3409/2021 | 2021-01-06 | North America/USA/Arizona/Maricopa ( | Arizona State Public Health Laboratory | Arizona State Public Health Laboratory | Trung Huynh et al |
| EPI_ISL_925846 | Brazil/AM-IEC-177281/2020 | 2020-11-11 | South America/Brazil/Amazonas BR/ | LACEN - Laboratv≥rio Central de Sav∫de Pv∫blica do Amazonas | Evandro Chagas Institute Virology | Santos et al |
| EPI_ISL_925876 | Rwanda/NRLNAT1041/2020 | 2020-12-17 | Africa/Rwanda/Kigali/ | Nucleic Acid Testing, National Reference Laboratory | GIGA Medical Genomics | Yvan Butera et al |
| EPI_ISL_925885 | Rwanda/NRLNAT1050/2021 | 2021-01-11 | Africa/Rwanda/Kigali/ | Nucleic Acid Testing, National Reference Laboratory | GIGA Medical Genomics | Yvan Butera et al |
| EPI_ISL_925903 | Rwanda/NRLNAT1072/2020 | 2020-11-03 | Africa/Rwanda/Kigali/ | Nucleic Acid Testing, National Reference Laboratory | GIGA Medical Genomics | Yvan Butera et al |
| EPI_ISL_925916 | Brazil/AM-IEC-177299/2020 | 2020-12-21 | South America/Brazil/Amazonas BR/ | LACEN - Laboratv≥rio Central de Sav∫de Pv∫blica do Amazonas | Evandro Chagas Institute Virology | Santos et al |
| EPI_ISL_926446 | Brazil/AM-IEC-177302/2020 | 2020-12-21 | South America/Brazil/Amazonas BR/ | LACEN - Laboratv≥rio Central de Sav∫de Pv∫blica do Amazonas | Evandro Chagas Institute Virology | Santos et al |
| EPI_ISL_927465 | Denmark/DCGC-36324/2021 | 2021-01-11 | Europe/Denmark/Hovedstaden/ | Department of Virus and Microbiological Special Diagnostics, Statens Serum Institut, Copenhagen, Denmark | Aalborg University | Danish Covid-19 Genome Consortium et al |
| EPI_ISL_930159 | Denmark/DCGC-39017/2021 | 2021-01-18 | Europe/Denmark/Hovedstaden/ | Department of Virus and Microbiological Special Diagnostics, Statens Serum Institut, Copenhagen, Denmark | Aalborg University | Danish Covid-19 Genome Consortium et al |
| EPI_ISL_930635 | USA/AZ-ASPHL-3425/2021 | 2021-01-08 | North America/USA/Arizona/Maricopa ( | Arizona State Public Health Laboratory | Arizona State Public Health Laboratory | Trung Huynh et al |
| EPI_ISL_930656 | USA/AZ-ASPHL-3452/2021 | 2021-01-11 | North America/USA/Arizona/Maricopa ( | Arizona State Public Health Laboratory | Arizona State Public Health Laboratory | Trung Huynh et al |
| EPI_ISL_930657 | USA/AZ-ASPHL-3453/2021 | 2021-01-11 | North America/USA/Arizona/Maricopa ( | Arizona State Public Health Laboratory | Arizona State Public Health Laboratory | Trung Huynh et al |
| EPI_ISL_930658 | USA/AZ-ASPHL-3455/2021 | 2021-01-11 | North America/USA/Arizona/Yuma | Arizona State Public Health Laboratory | Arizona State Public Health Laboratory | Trung Huynh et al |
| EPI_ISL_930659 | USA/AZ-ASPHL-3456/2021 | 2021-01-11 | North America/USA/Arizona/Yuma | Arizona State Public Health Laboratory | Arizona State Public Health Laboratory | Trung Huynh et al |
| EPI_ISL_930667 | USA/UT-UPHL-2102871531/2020 | 2020-12-28 | North America/USA/Utah/ | Utah Public Health Laboratory | Utah Public Health Laboratory | Erin L. Young et al |
| EPI_ISL_930677 | USA/UT-UPHL-2102243989/2020 | 2020-12-29 | North America/USA/Utah/ | Utah Public Health Laboratory | Utah Public Health Laboratory | Erin L. Young et al |

| | | | | | | |
|---|---|---|---|---|---|---|
| EPI_ISL_930680 | USA/UT-UPHL-2102256461/2020 | 2020-12-29 | North America/USA/Utah/ | Utah Public Health Laboratory | Utah Public Health Laboratory | Erin L. Young et al |
| EPI_ISL_930683 | USA/UT-UPHL-2102688775/2020 | 2020-12-29 | North America/USA/Utah/ | Utah Public Health Laboratory | Utah Public Health Laboratory | Erin L. Young et al |
| EPI_ISL_930697 | USA/UT-UPHL-2102272357/2020 | 2020-12-29 | North America/USA/Utah/ | Utah Public Health Laboratory | Utah Public Health Laboratory | Erin L. Young et al |
| EPI_ISL_930698 | USA/UT-UPHL-2102312386/2020 | 2020-12-29 | North America/USA/Utah/ | Utah Public Health Laboratory | Utah Public Health Laboratory | Erin L. Young et al |
| EPI_ISL_930702 | USA/UT-UPHL-2102815835/2020 | 2020-12-29 | North America/USA/Utah/ | Utah Public Health Laboratory | Utah Public Health Laboratory | Erin L. Young et al |
| EPI_ISL_930706 | USA/UT-UPHL-2102442317/2020 | 2020-12-29 | North America/USA/Utah/ | Utah Public Health Laboratory | Utah Public Health Laboratory | Erin L. Young et al |
| EPI_ISL_930709 | USA/UT-UPHL-2102035664/2020 | 2020-12-29 | North America/USA/Utah/ | Utah Public Health Laboratory | Utah Public Health Laboratory | Erin L. Young et al |
| EPI_ISL_930712 | USA/UT-UPHL-2102856178/2020 | 2020-12-29 | North America/USA/Utah/ | Utah Public Health Laboratory | Utah Public Health Laboratory | Erin L. Young et al |
| EPI_ISL_930721 | USA/UT-UPHL-2102383501/2020 | 2020-12-29 | North America/USA/Utah/ | Utah Public Health Laboratory | Utah Public Health Laboratory | Erin L. Young et al |
| EPI_ISL_930724 | USA/UT-UPHL-2102081472/2020 | 2020-12-28 | North America/USA/Utah/ | Utah Public Health Laboratory | Utah Public Health Laboratory | Erin L. Young et al |
| EPI_ISL_930742 | USA/UT-UPHL-2102797557/2020 | 2020-12-28 | North America/USA/Utah/ | Utah Public Health Laboratory | Utah Public Health Laboratory | Erin L. Young et al |
| EPI_ISL_930751 | USA/UT-UPHL-2102132197/2020 | 2020-12-28 | North America/USA/Utah/ | Utah Public Health Laboratory | Utah Public Health Laboratory | Erin L. Young et al |
| EPI_ISL_930752 | USA/UT-UPHL-2102518374/2020 | 2020-12-28 | North America/USA/Utah/ | Utah Public Health Laboratory | Utah Public Health Laboratory | Erin L. Young et al |
| EPI_ISL_930763 | USA/UT-UPHL-2102082416/2020 | 2020-12-28 | North America/USA/Utah/ | Utah Public Health Laboratory | Utah Public Health Laboratory | Erin L. Young et al |
| EPI_ISL_930767 | USA/UT-UPHL-2102046035/2020 | 2020-12-28 | North America/USA/Utah/ | Utah Public Health Laboratory | Utah Public Health Laboratory | Erin L. Young et al |
| EPI_ISL_930768 | USA/UT-UPHL-2102740033/2020 | 2020-12-28 | North America/USA/Utah/ | Utah Public Health Laboratory | Utah Public Health Laboratory | Erin L. Young et al |
| EPI_ISL_930769 | USA/UT-UPHL-2102042233/2020 | 2020-12-29 | North America/USA/Utah/ | Utah Public Health Laboratory | Utah Public Health Laboratory | Erin L. Young et al |
| EPI_ISL_930774 | USA/UT-UPHL-2102110755/2020 | 2020-12-29 | North America/USA/Utah/ | Utah Public Health Laboratory | Utah Public Health Laboratory | Erin L. Young et al |
| EPI_ISL_930776 | USA/UT-UPHL-2102342783/2020 | 2020-12-31 | North America/USA/Utah/ | Utah Public Health Laboratory | Utah Public Health Laboratory | Erin L. Young et al |
| EPI_ISL_930779 | USA/UT-UPHL-2102319812/2020 | 2020-12-28 | North America/USA/Utah/ | Utah Public Health Laboratory | Utah Public Health Laboratory | Erin L. Young et al |

| | | | | | | |
|---|---|---|---|---|---|---|
| EPI_ISL_930783 | Mexico/CMX-INMEGEN-11-01/2020 | 2020-11-26 | North America/Mexico/Mexico City/ | Instituto Nacional de Medicina Genomica | Instituto Nacional de Medicina Genomica | Hidalgo-Miranda A et al |
| EPI_ISL_930785 | Poland/Pomorskie_MWB_33/2021 | 2021-01-12 | Europe/Poland/Pomorskie/ | Laboratory of Molecular Biology, Diagnostyka sp. z o.o. | Laboratory of Recombinant Vaccines | Lukasz Rabalski et al |
| EPI_ISL_930800 | USA/CA-MCPHL-MC01817/2021 | 2021-01-17 | North America/USA/California/Montere | Monterey County Public Health Laboratory | Monterey County Public Health Laboratory | C. Anaya et al. et al |
| EPI_ISL_930804 | USA/CA-MCPHL-MC01820/2021 | 2021-01-17 | North America/USA/California/Montere | Monterey County Public Health Laboratory | Monterey County Public Health Laboratory | C. Anaya et al. et al |
| EPI_ISL_930835 | India/un-NRCE-2710/2020 | 2020-06-30 | Asia/India/India/ | Virology, ICAR-National Research Centre on Equines | Virology, ICAR-National Research Centre on Equines | Kumar et al |
| EPI_ISL_930836 | India/un-NRCE-4549/2020 | 2020-06-30 | Asia/India/India/ | Virology, ICAR-National Research Centre on Equines | Virology, ICAR-National Research Centre on Equines | Gulati et al |
| EPI_ISL_930839 | India/un-NRCE-6868/2020 | 2020-06-30 | Asia/India/India/ | Virology, ICAR-National Research Centre on Equines | Virology, ICAR-National Research Centre on Equines | Gulati et al |
| EPI_ISL_930840 | India/un-NRCE-8156/2020 | 2020-06-30 | Asia/India/India/ | Virology, ICAR-National Research Centre on Equines | Virology, ICAR-National Research Centre on Equines | Gulati et al |
| EPI_ISL_930841 | India/un-NRCE-9088/2020 | 2020-06-30 | Asia/India/India/ | Virology, ICAR-National Research Centre on Equines | Virology, ICAR-National Research Centre on Equines | Gulati et al |
| EPI_ISL_930843 | India/un-NRCE-2690/2020 | 2020-06-30 | Asia/India/India/ | Virology, ICAR-National Research Centre on Equines | Virology, ICAR-National Research Centre on Equines | Kumar et al |
| EPI_ISL_930851 | USA/CA-MCPHL-MC01833/2021 | 2021-01-18 | North America/USA/California/Montere | Monterey County Public Health Laboratory | Monterey County Public Health Laboratory | C. Anaya et al. et al |
| EPI_ISL_930854 | Brazil/RS-CEVS-30437/2020 | 2020-07-15 | South America/Brazil/Rio Grande do Sul | Central Laboratory of Public Health of Rio Grande do Sul(Lacen_RS) | State Center for Health Surveillance of the Health Department of the State of Rio Grande do Sul(CEVS_SES-RS) | Barcellos R et al |

| | | | | | | |
|---|---|---|---|---|---|---|
| EPI_ISL_930855 | Brazil/RS-CEVS-32830/2020 | 2020-07-22 | South America/Brazil/Rio Grande do Sul | Central Laboratory of Public Health of Rio Grande do Sul(Lacen_RS) | State Center for Health Surveillance of the Health Department of the State of Rio Grande do Sul(CEVS_SES-RS) | Barcellos R et al |
| EPI_ISL_930856 | Brazil/RS-CEVS-33589/2020 | 2020-07-23 | South America/Brazil/Rio Grande do Sul | Central Laboratory of Public Health of Rio Grande do Sul(Lacen_RS) | State Center for Health Surveillance of the Health Department of the State of Rio Grande do Sul(CEVS_SES-RS) | Barcellos R et al |
| EPI_ISL_930857 | Brazil/RS-CEVS-33704/2020 | 2020-07-23 | South America/Brazil/Rio Grande do Sul | Central Laboratory of Public Health of Rio Grande do Sul(Lacen_RS) | State Center for Health Surveillance of the Health Department of the State of Rio Grande do Sul(CEVS_SES-RS) | Barcellos R et al |
| EPI_ISL_930858 | Brazil/RS-CEVS-34046/2020 | 2020-07-24 | South America/Brazil/Rio Grande do Sul | Central Laboratory of Public Health of Rio Grande do Sul(Lacen_RS) | State Center for Health Surveillance of the Health Department of the State of Rio Grande do Sul(CEVS_SES-RS) | Barcellos R et al |
| EPI_ISL_930862 | Switzerland/BL-UHB-42489468/2020 | 2020-10-17 | Europe/Switzerland/Basel-Land/Allschw | University Hospital Basel, Clinical Virology | University Hospital Basel, Clinical Bacteriology | Tim Roloff et al |
| EPI_ISL_930906 | Switzerland/BS-UHB-42502097/2020 | 2020-10-24 | Europe/Switzerland/Basel-Stadt/Basel | University Hospital Basel, Clinical Virology | University Hospital Basel, Clinical Bacteriology | Tim Roloff et al |

| | | | | | | |
|---|---|---|---|---|---|---|
| EPI_ISL_930910 | Germany/NW-UHB-42490409/2020 | 2020-10-18 | Europe/Germany/North Rhine Westphal | University Hospital Basel, Clinical Virology | University Hospital Basel, Clinical Bacteriology | Tim Roloff et al |
| EPI_ISL_930918 | France/UHB-42490694/2020 | 2020-10-18 | Europe/France/Grand Est/Franken | University Hospital Basel, Clinical Virology | University Hospital Basel, Clinical Bacteriology | Tim Roloff et al |
| EPI_ISL_930920 | Switzerland/BS-UHB-42491204/2020 | 2020-10-19 | Europe/Switzerland/Basel-Stadt/Basel | University Hospital Basel, Clinical Virology | University Hospital Basel, Clinical Bacteriology | Tim Roloff et al |
| EPI_ISL_930927 | Switzerland/BS-UHB-42491494/2020 | 2020-10-19 | Europe/Switzerland/Basel-Stadt/Basel | University Hospital Basel, Clinical Virology | University Hospital Basel, Clinical Bacteriology | Tim Roloff et al |
| EPI_ISL_930972 | Switzerland/BS-UHB-42492497/2020 | 2020-10-19 | Europe/Switzerland/Basel-Stadt/Basel | University Hospital Basel, Clinical Virology | University Hospital Basel, Clinical Bacteriology | Tim Roloff et al |
| EPI_ISL_931050 | Switzerland/BS-UHB-42499993/2020 | 2020-10-23 | Europe/Switzerland/Basel-Stadt/Basel | University Hospital Basel, Clinical Virology | University Hospital Basel, Clinical Bacteriology | Tim Roloff et al |
| EPI_ISL_931057 | Switzerland/BS-UHB-42500230/2020 | 2020-10-23 | Europe/Switzerland/Basel-Stadt/Basel | University Hospital Basel, Clinical Virology | University Hospital Basel, Clinical Bacteriology | Tim Roloff et al |
| EPI_ISL_931064 | France/UHB-42500398/2020 | 2020-10-23 | Europe/France/Grand Est/St-Louis | University Hospital Basel, Clinical Virology | University Hospital Basel, Clinical Bacteriology | Tim Roloff et al |
| EPI_ISL_931067 | Switzerland/BL-UHB-42500494/2020 | 2020-10-23 | Europe/Switzerland/Basel-Land/Birsfeld | University Hospital Basel, Clinical Virology | University Hospital Basel, Clinical Bacteriology | Tim Roloff et al |
| EPI_ISL_931081 | France/UHB-42533025/2020 | 2020-11-11 | Europe/France/Grand Est/Baldersheim | University Hospital Basel, Clinical Virology | University Hospital Basel, Clinical Bacteriology | Tim Roloff et al |

| | | | | | | |
|---|---|---|---|---|---|---|
| EPI_ISL_931126 | France/UHB-42502103/2020 | 2020-10-24 | Europe/France/Auvergne-Rhv¥ne-Alpes/ | University Hospital Basel, Clinical Virology | University Hospital Basel, Clinical Bacteriology | Tim Roloff et al |
| EPI_ISL_931130 | Switzerland/BS-UHB-42502168/2020 | 2020-10-24 | Europe/Switzerland/Basel-Stadt/Basel | University Hospital Basel, Clinical Virology | University Hospital Basel, Clinical Bacteriology | Tim Roloff et al |
| EPI_ISL_931137 | Switzerland/BS-UHB-42502691/2020 | 2020-10-25 | Europe/Switzerland/Basel-Stadt/Basel | University Hospital Basel, Clinical Virology | University Hospital Basel, Clinical Bacteriology | Tim Roloff et al |
| EPI_ISL_931144 | Switzerland/BL-UHB-65263946/2020 | 2020-10-25 | Europe/Switzerland/Basel-Land/Gelterk | University Hospital Basel, Clinical Virology | University Hospital Basel, Clinical Bacteriology | Tim Roloff et al |
| EPI_ISL_931157 | Germany/BW-UHB-42502900/2020 | 2020-10-25 | Europe/Germany/Baden-Wuerttemberg | University Hospital Basel, Clinical Virology | University Hospital Basel, Clinical Bacteriology | Tim Roloff et al |
| EPI_ISL_931165 | France/UHB-42503053/2020 | 2020-10-25 | Europe/France/Grand Est/Village-Neuf | University Hospital Basel, Clinical Virology | University Hospital Basel, Clinical Bacteriology | Tim Roloff et al |
| EPI_ISL_931188 | Switzerland/BL-UHB-42504426/2020 | 2020-10-26 | Europe/Switzerland/Basel-Land/Oberwi | University Hospital Basel, Clinical Virology | University Hospital Basel, Clinical Bacteriology | Tim Roloff et al |
| EPI_ISL_931190 | Switzerland/BS-UHB-42504541/2020 | 2020-10-26 | Europe/Switzerland/Basel-Stadt/Basel | University Hospital Basel, Clinical Virology | University Hospital Basel, Clinical Bacteriology | Tim Roloff et al |
| EPI_ISL_931197 | Switzerland/BS-UHB-42504727/2020 | 2020-10-26 | Europe/Switzerland/Basel-Stadt/Basel | University Hospital Basel, Clinical Virology | University Hospital Basel, Clinical Bacteriology | Tim Roloff et al |
| EPI_ISL_931202 | Switzerland/BS-UHB-42504572/2020 | 2020-10-26 | Europe/Switzerland/Basel-Stadt/Basel | University Hospital Basel, Clinical Virology | University Hospital Basel, Clinical Bacteriology | Tim Roloff et al |

| | | | | | | |
|---|---|---|---|---|---|---|
| EPI_ISL_931235 | Switzerland/BS-UHB-42506643/2020 | 2020-10-27 | Europe/Switzerland/Basel-Stadt/Basel | University Hospital Basel, Clinical Virology | University Hospital Basel, Clinical Bacteriology | Tim Roloff et al |
| EPI_ISL_931255 | Switzerland/BL-UHB-42506937/2020 | 2020-10-27 | Europe/Switzerland/Basel-Land/Birsfeld | University Hospital Basel, Clinical Virology | University Hospital Basel, Clinical Bacteriology | Tim Roloff et al |
| EPI_ISL_931274 | France/UHB-42507390/2020 | 2020-10-27 | Europe/France/Grand Est/Hegenheim | University Hospital Basel, Clinical Virology | University Hospital Basel, Clinical Bacteriology | Tim Roloff et al |
| EPI_ISL_931291 | Germany/BW-UHB-42508379/2020 | 2020-10-28 | Europe/Germany/Baden-Wuerttemberg | University Hospital Basel, Clinical Virology | University Hospital Basel, Clinical Bacteriology | Tim Roloff et al |
| EPI_ISL_931309 | Switzerland/BS-UHB-42511376/2020 | 2020-10-29 | Europe/Switzerland/Basel-Stadt/Basel | University Hospital Basel, Clinical Virology | University Hospital Basel, Clinical Bacteriology | Tim Roloff et al |
| EPI_ISL_931312 | France/UHB-42513533/2020 | 2020-10-30 | Europe/France/Grand Est/Meyenheim | University Hospital Basel, Clinical Virology | University Hospital Basel, Clinical Bacteriology | Tim Roloff et al |
| EPI_ISL_931319 | Switzerland/BS-UHB-42514493/2020 | 2020-10-31 | Europe/Switzerland/Basel-Stadt/Basel | University Hospital Basel, Clinical Virology | University Hospital Basel, Clinical Bacteriology | Tim Roloff et al |
| EPI_ISL_931362 | Switzerland/BS-UHB-42547478/2020 | 2020-11-19 | Europe/Switzerland/Basel-Stadt/Basel | University Hospital Basel, Clinical Virology | University Hospital Basel, Clinical Bacteriology | Tim Roloff et al |
| EPI_ISL_931386 | France/HDF-UHB-42514947/2020 | 2020-10-31 | Europe/France/Hauts de France/Gonde | University Hospital Basel, Clinical Virology | University Hospital Basel, Clinical Bacteriology | Tim Roloff et al |
| EPI_ISL_931402 | France/UHB-42515246/2020 | 2020-11-01 | Europe/France/Grand Est/Hegenheim | University Hospital Basel, Clinical Virology | University Hospital Basel, Clinical Bacteriology | Tim Roloff et al |

| | | | | | | |
|---|---|---|---|---|---|---|
| EPI_ISL_931423 | Switzerland/BS-UHB-42515478/2020 | 2020-11-01 | Europe/Switzerland/Basel-Stadt/Basel | University Hospital Basel, Clinical Virology | University Hospital Basel, Clinical Bacteriology | Tim Roloff et al |
| EPI_ISL_931433 | Switzerland/BL-UHB-42515566/2020 | 2020-11-01 | Europe/Switzerland/Basel-Land/Dornach | University Hospital Basel, Clinical Virology | University Hospital Basel, Clinical Bacteriology | Tim Roloff et al |
| EPI_ISL_931461 | USA/MD-MDH-0730/2021 | 2021-01-20 | North America/USA/Maryland/ | Maryland Public Health Laboratory (MD PHL) | Maryland Public Health Laboratory (MD PHL) | Maryland Department of Health Laboratories Administration et al |
| EPI_ISL_931463 | USA/MD-MDH-0732/2021 | 2021-01-20 | North America/USA/Maryland/ | Maryland Public Health Laboratory (MD PHL) | Maryland Public Health Laboratory (MD PHL) | Maryland Department of Health Laboratories Administration et al |
| EPI_ISL_931492 | USA/MD-MDH-0750/2021 | 2021-01-22 | North America/USA/Maryland/ | Maryland Public Health Laboratory (MD PHL) | Maryland Public Health Laboratory (MD PHL) | Maryland Department of Health Laboratories Administration et al |
| EPI_ISL_931499 | USA/MD-MDH-0757/2021 | 2021-01-07 | North America/USA/Maryland/ | Maryland Public Health Laboratory (MD PHL) | Maryland Public Health Laboratory (MD PHL) | Maryland Department of Health Laboratories Administration et al |
| EPI_ISL_931508 | USA/UT-UPHL-2102778258/2020 | 2020-12-29 | North America/USA/Utah/ | Utah Public Health Laboratory | Utah Public Health Laboratory | Erin L. Young et al |
| EPI_ISL_931525 | USA/UT-UPHL-2102118323/2020 | 2020-12-30 | North America/USA/Utah/ | Utah Public Health Laboratory | Utah Public Health Laboratory | Erin L. Young et al |
| EPI_ISL_931528 | USA/UT-UPHL-2102149578/2020 | 2020-12-30 | North America/USA/Utah/ | Utah Public Health Laboratory | Utah Public Health Laboratory | Erin L. Young et al |
| EPI_ISL_931530 | USA/UT-UPHL-2102494311/2020 | 2020-12-30 | North America/USA/Utah/ | Utah Public Health Laboratory | Utah Public Health Laboratory | Erin L. Young et al |
| EPI_ISL_931533 | USA/UT-UPHL-2102501552/2020 | 2020-12-30 | North America/USA/Utah/ | Utah Public Health Laboratory | Utah Public Health Laboratory | Erin L. Young et al |
| EPI_ISL_931534 | USA/UT-UPHL-2102227616/2020 | 2020-12-31 | North America/USA/Utah/ | Utah Public Health Laboratory | Utah Public Health Laboratory | Erin L. Young et al |
| EPI_ISL_931535 | USA/UT-UPHL-2102083184/2020 | 2020-12-29 | North America/USA/Utah/ | Utah Public Health Laboratory | Utah Public Health Laboratory | Erin L. Young et al |
| EPI_ISL_931539 | USA/UT-UPHL-2102659519/2020 | 2020-12-28 | North America/USA/Utah/ | Utah Public Health Laboratory | Utah Public Health Laboratory | Erin L. Young et al |
| EPI_ISL_931541 | USA/UT-UPHL-2102104646/2020 | 2020-12-30 | North America/USA/Utah/ | Utah Public Health Laboratory | Utah Public Health Laboratory | Erin L. Young et al |
| EPI_ISL_931549 | USA/UT-UPHL-2102664914/2020 | 2020-12-31 | North America/USA/Utah/ | Utah Public Health Laboratory | Utah Public Health Laboratory | Erin L. Young et al |

| | | | | | | |
|---|---|---|---|---|---|---|
| EPI_ISL_931560 | USA/UT-UPHL-2102978457/2020 | 2020-12-30 | North America/USA/Utah/ | Utah Public Health Laboratory | Utah Public Health Laboratory | Erin L. Young et al |
| EPI_ISL_931576 | USA/UT-UPHL-2102066306/2021 | 2021-01-04 | North America/USA/Utah/ | Utah Public Health Laboratory | Utah Public Health Laboratory | Erin L. Young et al |
| EPI_ISL_931579 | USA/UT-UPHL-2102552559/2021 | 2021-01-04 | North America/USA/Utah/ | Utah Public Health Laboratory | Utah Public Health Laboratory | Erin L. Young et al |
| EPI_ISL_931584 | Spain/MD-11676540/2021 | 2021-01-12 | Europe/Spain/Madrid/Madrid | Servicio de Microbiolog√≠a Hospital Ram√≥n y Cajal | Servicio de Microbiolog√≠a Hospital Ram√≥n y Cajal | Jos√© M Gonzalez-Alba et al |
| EPI_ISL_931696 | England/ALDP-11863BA/2021 | 2021-01-25 | Europe/United Kingdom/England/ | Lighthouse Lab in Alderley Park | Wellcome Sanger Institute for the COVID-19 Genomics UK (COG-UK) Consortium | Jacquelyn Wynn et al |
| EPI_ISL_931879 | England/ALDP-1185D26/2021 | 2021-01-25 | Europe/United Kingdom/England/ | Lighthouse Lab in Alderley Park | Wellcome Sanger Institute for the COVID-19 Genomics UK (COG-UK) Consortium | Jacquelyn Wynn et al |
| EPI_ISL_932054 | England/ALDP-1182011/2021 | 2021-01-25 | Europe/United Kingdom/England/ | Lighthouse Lab in Alderley Park | Wellcome Sanger Institute for the COVID-19 Genomics UK (COG-UK) Consortium | Jacquelyn Wynn et al |
| EPI_ISL_932807 | England/QEUH-114ADF0/2021 | 2021-01-24 | Europe/United Kingdom/England/ | Lighthouse Lab in Glasgow | Wellcome Sanger Institute for the COVID-19 Genomics UK (COG-UK) Consortium | Harper VanSteenhouse et al |
| EPI_ISL_932821 | England/MILK-113C007/2021 | 2021-01-21 | Europe/United Kingdom/England/ | Lighthouse Lab in Milton Keynes | Wellcome Sanger Institute for the COVID-19 Genomics UK (COG-UK) Consortium | The Lighthouse Lab in Milton Keynes et al |
| EPI_ISL_933184 | England/CAMC-110469D/2021 | 2021-01-20 | Europe/United Kingdom/England/ | Lighthouse Lab in Cambridge | Wellcome Sanger Institute for the COVID-19 Genomics UK (COG-UK) Consortium | Rob Howes et al |

| | | | | | | |
|---|---|---|---|---|---|---|
| EPI_ISL_933193 | England/CAMC-1105BAC/2021 | 2021-01-22 | Europe/United Kingdom/England/ | Lighthouse Lab in Cambridge | Wellcome Sanger Institute for the COVID-19 Genomics UK (COG-UK) Consortium | Rob Howes et al |
| EPI_ISL_933421 | England/CAMC-10FCDD0/2021 | 2021-01-20 | Europe/United Kingdom/England/ | Lighthouse Lab in Cambridge | Wellcome Sanger Institute for the COVID-19 Genomics UK (COG-UK) Consortium | Rob Howes et al |
| EPI_ISL_933475 | England/MILK-E03B68/2020 | 2020-12-28 | Europe/United Kingdom/England/ | Lighthouse Lab in Milton Keynes | Wellcome Sanger Institute for the COVID-19 Genomics UK (COG-UK) Consortium | The Lighthouse Lab in Milton Keynes et al |
| EPI_ISL_933502 | England/MILK-BFF69E/2020 | 2020-12-01 | Europe/United Kingdom/England/ | Lighthouse Lab in Milton Keynes | Wellcome Sanger Institute for the COVID-19 Genomics UK (COG-UK) Consortium | The Lighthouse Lab in Milton Keynes et al |
| EPI_ISL_933512 | Mexico/CMX-INMEGEN-11-02/2020 | 2020-11-26 | North America/Mexico/Mexico City/ | Instituto Nacional de Medicina Genomica | Instituto Nacional de Medicina Genomica | Hidalgo-Miranda A et al |
| EPI_ISL_933514 | Mexico/CMX-INMEGEN-11-04/2020 | 2020-11-26 | North America/Mexico/Mexico City/ | Instituto Nacional de Medicina Genomica | Instituto Nacional de Medicina Genomica | Hidalgo-Miranda A et al |
| EPI_ISL_933515 | Mexico/CMX-INMEGEN-11-05/2020 | 2020-11-27 | North America/Mexico/Mexico City/ | Instituto Nacional de Medicina Genomica | Instituto Nacional de Medicina Genomica | Hidalgo-Miranda A et al |
| EPI_ISL_933516 | Mexico/CMX-INMEGEN-11-06/2020 | 2020-11-20 | North America/Mexico/Mexico City/ | Instituto Nacional de Medicina Genomica | Instituto Nacional de Medicina Genomica | Hidalgo-Miranda A et al |
| EPI_ISL_933517 | Mexico/CMX-INMEGEN-11-07/2020 | 2020-11-28 | North America/Mexico/Mexico City/ | Instituto Nacional de Medicina Genomica | Instituto Nacional de Medicina Genomica | Hidalgo-Miranda A et al |
| EPI_ISL_933518 | Mexico/CMX-INMEGEN-11-08/2020 | 2020-11-20 | North America/Mexico/Mexico City/ | Instituto Nacional de Medicina Genomica | Instituto Nacional de Medicina Genomica | Hidalgo-Miranda A et al |
| EPI_ISL_933520 | Mexico/CMX-INMEGEN-11-11/2020 | 2020-11-30 | North America/Mexico/Mexico City/ | Instituto Nacional de Medicina Genomica | Instituto Nacional de Medicina Genomica | Hidalgo-Miranda A et al |

| | | | | | | |
|---|---|---|---|---|---|---|
| EPI_ISL_933521 | Mexico/CMX-INMEGEN-11-12/2020 | 2020-11-30 | North America/Mexico/Mexico City/ | Instituto Nacional de Medicina Genomica | Instituto Nacional de Medicina Genomica | Hidalgo-Miranda A et al |
| EPI_ISL_933522 | Mexico/CMX-INMEGEN-11-13/2020 | 2020-11-30 | North America/Mexico/Mexico City/ | Instituto Nacional de Medicina Genomica | Instituto Nacional de Medicina Genomica | Hidalgo-Miranda A et al |
| EPI_ISL_933523 | Mexico/CMX-INMEGEN-11-14/2020 | 2020-11-30 | North America/Mexico/Mexico City/ | Instituto Nacional de Medicina Genomica | Instituto Nacional de Medicina Genomica | Hidalgo-Miranda A et al |
| EPI_ISL_933524 | Mexico/CMX-INMEGEN-11-15/2020 | 2020-11-30 | North America/Mexico/Mexico City/ | Instituto Nacional de Medicina Genomica | Instituto Nacional de Medicina Genomica | Hidalgo-Miranda A et al |
| EPI_ISL_933530 | Mexico/CMX-INMEGEN-11-21/2020 | 2020-11-30 | North America/Mexico/Mexico City/ | Instituto Nacional de Medicina Genomica | Instituto Nacional de Medicina Genomica | Hidalgo-Miranda A et al |
| EPI_ISL_933531 | Mexico/CMX-INMEGEN-11-22/2020 | 2020-11-30 | North America/Mexico/Mexico City/ | Instituto Nacional de Medicina Genomica | Instituto Nacional de Medicina Genomica | Hidalgo-Miranda A et al |
| EPI_ISL_933533 | Belarus/HO-RRCEM-MOZ11874S/202( | 2020-12-08 | Europe/Belarus/Gomel Region/Mozyr | Laboratory for HIV and opportunistic infections diagnosis The Republican Research and Practical Center for Epidemiology and Microbiology (RRPCEM) | Laboratory for HIV and opportunistic infections diagnosis The Republican Research and Practical Center for Epidemiology and Microbiology (RRPCEM) | Elena Gasich et al |
| EPI_ISL_933543 | USA/AZ-ASPHL-2857/2020 | 2020-11-16 | North America/USA/Arizona/Maricopa ( | Arizona State Public Health Laboratory | Arizona State Public Health Laboratory | Trung Huynh et al |
| EPI_ISL_933551 | USA/AZ-ASPHL-2904/2020 | 2020-11-21 | North America/USA/Arizona/Navajo Co | Arizona State Public Health Laboratory | Arizona State Public Health Laboratory | Trung Huynh et al |
| EPI_ISL_933554 | USA/AZ-ASPHL-2963/2020 | 2020-12-01 | North America/USA/Arizona/Maricopa ( | Arizona State Public Health Laboratory | Arizona State Public Health Laboratory | Trung Huynh et al |
| EPI_ISL_933560 | USA/AZ-ASPHL-2932/2020 | 2020-11-25 | North America/USA/Arizona/Maricopa ( | Arizona State Public Health Laboratory | Arizona State Public Health Laboratory | Trung Huynh et al |

| | | | | | | |
|---|---|---|---|---|---|---|
| EPI_ISL_933563 | USA/AZ-ASPHL-2931/2020 | 2020-11-25 | North America/USA/Arizona/Maricopa ( | Arizona State Public Health Laboratory | Arizona State Public Health Laboratory | Trung Huynh et al |
| EPI_ISL_933575 | Indonesia/BT-SHSIU-01-4241/2020 | 2020-10-23 | Asia/Indonesia/Banten/ | Medical Research Center, Faculty of Medicine, Syarif Hidayatullah State Islamic University Jakarta | Medical Research Center, Faculty of Medicine, Syarif Hidayatullah State Islamic University Jakarta | Erike A Suwarsono et al |
| EPI_ISL_933579 | NorthMacedonia/17359/2020 | 2020-10-28 | Europe/North Macedonia/Southeastern | General Hospital - Strumica | "Research Center for Genetic Engineering and Biotechnology ""Georgi D. Efremov,Äù , Macedonian Academy of Sciences and Arts" | RCGEB - MASA et al |
| EPI_ISL_933582 | USA/AZ-ASPHL-2977/2020 | 2020-12-01 | North America/USA/Arizona/Yuma | Arizona State Public Health Laboratory | Arizona State Public Health Laboratory | Trung Huynh et al |
| EPI_ISL_933603 | Canada/ON-SC0059/2020 | 2020-03-10 | North America/Canada/Ontario/Toronto | Toronto Invasive Bacterial Diseases Network | McMaster University | Allison McGeer et al |
| EPI_ISL_933604 | Canada/ON-SC0098/2020 | 2020-03-16 | North America/Canada/Ontario/Toronto | Toronto Invasive Bacterial Diseases Network | McMaster University | Allison McGeer et al |
| EPI_ISL_933605 | Canada/ON-SC0058/2020 | 2020-03-19 | North America/Canada/Ontario/Toronto | Toronto Invasive Bacterial Diseases Network | McMaster University | Allison McGeer et al |
| EPI_ISL_933606 | Canada/ON-SC0093/2020 | 2020-03-19 | North America/Canada/Ontario/Toronto | Toronto Invasive Bacterial Diseases Network | McMaster University | Allison McGeer et al |
| EPI_ISL_933607 | Canada/ON-SC0050/2020 | 2020-03-21 | North America/Canada/Ontario/Toronto | Toronto Invasive Bacterial Diseases Network | McMaster University | Allison McGeer et al |
| EPI_ISL_933608 | Canada/ON-SC0107/2020 | 2020-03-21 | North America/Canada/Ontario/Toronto | Toronto Invasive Bacterial Diseases Network | McMaster University | Allison McGeer et al |
| EPI_ISL_933609 | Canada/ON-SC0054/2020 | 2020-03-24 | North America/Canada/Ontario/Toronto | Toronto Invasive Bacterial Diseases Network | McMaster University | Allison McGeer et al |
| EPI_ISL_933610 | Canada/ON-SC0056/2020 | 2020-03-24 | North America/Canada/Ontario/Toronto | Toronto Invasive Bacterial Diseases Network | McMaster University | Allison McGeer et al |
| EPI_ISL_933611 | Canada/ON-SC0110/2020 | 2020-03-24 | North America/Canada/Ontario/Toronto | Toronto Invasive Bacterial Diseases Network | McMaster University | Allison McGeer et al |

| | | | | | | |
|---|---|---|---|---|---|---|
| EPI_ISL_933612 | Canada/ON-SC0052/2020 | 2020-03-25 | North America/Canada/Ontario/Toronto | Toronto Invasive Bacterial Diseases Network | McMaster University | Allison McGeer et al |
| EPI_ISL_933613 | Canada/ON-SC0105/2020 | 2020-03-26 | North America/Canada/Ontario/Toronto | Toronto Invasive Bacterial Diseases Network | McMaster University | Allison McGeer et al |
| EPI_ISL_933614 | Canada/ON-SC0122/2020 | 2020-03-27 | North America/Canada/Ontario/Toronto | Toronto Invasive Bacterial Diseases Network | McMaster University | Allison McGeer et al |
| EPI_ISL_933615 | Canada/ON-SC0055/2020 | 2020-03-29 | North America/Canada/Ontario/Toronto | Toronto Invasive Bacterial Diseases Network | McMaster University | Allison McGeer et al |
| EPI_ISL_933616 | Canada/ON-SC0100/2020 | 2020-03-30 | North America/Canada/Ontario/Toronto | Toronto Invasive Bacterial Diseases Network | McMaster University | Allison McGeer et al |
| EPI_ISL_933617 | Canada/ON-SC0102/2020 | 2020-03-30 | North America/Canada/Ontario/Toronto | Toronto Invasive Bacterial Diseases Network | McMaster University | Allison McGeer et al |
| EPI_ISL_933618 | Canada/ON-SC0119/2020 | 2020-03-31 | North America/Canada/Ontario/Toronto | Toronto Invasive Bacterial Diseases Network | McMaster University | Allison McGeer et al |
| EPI_ISL_933619 | Canada/ON-SC0113/2020 | 2020-04-01 | North America/Canada/Ontario/Toronto | Toronto Invasive Bacterial Diseases Network | McMaster University | Allison McGeer et al |
| EPI_ISL_933620 | Canada/ON-SC0101/2020 | 2020-04-03 | North America/Canada/Ontario/Toronto | Toronto Invasive Bacterial Diseases Network | McMaster University | Allison McGeer et al |
| EPI_ISL_933621 | Canada/ON-SC0062/2020 | 2020-04-04 | North America/Canada/Ontario/Toronto | Toronto Invasive Bacterial Diseases Network | McMaster University | Allison McGeer et al |
| EPI_ISL_933622 | Canada/ON-SC0061/2020 | 2020-04-06 | North America/Canada/Ontario/Toronto | Toronto Invasive Bacterial Diseases Network | McMaster University | Allison McGeer et al |
| EPI_ISL_933623 | Canada/ON-SC0092/2020 | 2020-04-06 | North America/Canada/Ontario/Toronto | Toronto Invasive Bacterial Diseases Network | McMaster University | Allison McGeer et al |
| EPI_ISL_933624 | Canada/ON-SC0064/2020 | 2020-04-08 | North America/Canada/Ontario/Toronto | Toronto Invasive Bacterial Diseases Network | McMaster University | Allison McGeer et al |
| EPI_ISL_933625 | Canada/ON-SC0001/2020 | 2020-04-09 | North America/Canada/Ontario/Toronto | Toronto Invasive Bacterial Diseases Network | McMaster University | Allison McGeer et al |
| EPI_ISL_933627 | Canada/ON-SC0034/2020 | 2020-04-10 | North America/Canada/Ontario/Toronto | Toronto Invasive Bacterial Diseases Network | McMaster University | Allison McGeer et al |
| EPI_ISL_933628 | Canada/ON-SC0003/2020 | 2020-04-11 | North America/Canada/Ontario/Toronto | Toronto Invasive Bacterial Diseases Network | McMaster University | Allison McGeer et al |
| EPI_ISL_933629 | Canada/ON-SC0041/2020 | 2020-04-12 | North America/Canada/Ontario/Toronto | Toronto Invasive Bacterial Diseases Network | McMaster University | Allison McGeer et al |
| EPI_ISL_933630 | Canada/ON-SC0044/2020 | 2020-04-18 | North America/Canada/Ontario/Toronto | Toronto Invasive Bacterial Diseases Network | McMaster University | Allison McGeer et al |
| EPI_ISL_933631 | Canada/ON-SC0026/2020 | 2020-04-20 | North America/Canada/Ontario/Toronto | Toronto Invasive Bacterial Diseases Network | McMaster University | Allison McGeer et al |
| EPI_ISL_933632 | Canada/ON-SC0006/2020 | 2020-04-26 | North America/Canada/Ontario/Toronto | Toronto Invasive Bacterial Diseases Network | McMaster University | Allison McGeer et al |

| | | | | | | |
|---|---|---|---|---|---|---|
| EPI_ISL_933633 | Canada/ON-SC0007/2020 | 2020-04-28 | North America/Canada/Ontario/Toronto | Toronto Invasive Bacterial Diseases Network | McMaster University | Allison McGeer et al |
| EPI_ISL_933634 | Canada/ON-SC0013/2020 | 2020-05-01 | North America/Canada/Ontario/Toronto | Toronto Invasive Bacterial Diseases Network | McMaster University | Allison McGeer et al |
| EPI_ISL_933635 | Canada/ON-SC0010/2020 | 2020-05-02 | North America/Canada/Ontario/Toronto | Toronto Invasive Bacterial Diseases Network | McMaster University | Allison McGeer et al |
| EPI_ISL_933636 | Canada/ON-SC0012/2020 | 2020-05-02 | North America/Canada/Ontario/Toronto | Toronto Invasive Bacterial Diseases Network | McMaster University | Allison McGeer et al |
| EPI_ISL_933637 | Canada/ON-SC0015/2020 | 2020-05-08 | North America/Canada/Ontario/Toronto | Toronto Invasive Bacterial Diseases Network | McMaster University | Allison McGeer et al |
| EPI_ISL_933639 | Canada/ON-SC0066/2020 | 2020-05-21 | North America/Canada/Ontario/Toronto | Toronto Invasive Bacterial Diseases Network | McMaster University | Allison McGeer et al |
| EPI_ISL_933640 | Canada/ON-SC0117/2020 | 2020-06-05 | North America/Canada/Ontario/Toronto | Toronto Invasive Bacterial Diseases Network | McMaster University | Allison McGeer et al |
| EPI_ISL_933641 | Canada/ON-SC0128/2020 | 2020-06-15 | North America/Canada/Ontario/Toronto | Toronto Invasive Bacterial Diseases Network | McMaster University | Allison McGeer et al |
| EPI_ISL_933642 | Canada/ON-SC0129/2020 | 2020-06-15 | North America/Canada/Ontario/Toronto | Toronto Invasive Bacterial Diseases Network | McMaster University | Allison McGeer et al |
| EPI_ISL_933643 | Canada/ON-SC0130/2020 | 2020-06-15 | North America/Canada/Ontario/Toronto | Toronto Invasive Bacterial Diseases Network | McMaster University | Allison McGeer et al |
| EPI_ISL_933644 | Canada/ON-SC0132/2020 | 2020-06-19 | North America/Canada/Ontario/Toronto | Toronto Invasive Bacterial Diseases Network | McMaster University | Allison McGeer et al |
| EPI_ISL_933645 | Canada/ON-SC0137/2020 | 2020-06-26 | North America/Canada/Ontario/Toronto | Toronto Invasive Bacterial Diseases Network | McMaster University | Allison McGeer et al |
| EPI_ISL_933646 | Canada/ON-SC0140/2020 | 2020-06-27 | North America/Canada/Ontario/Toronto | Toronto Invasive Bacterial Diseases Network | McMaster University | Allison McGeer et al |
| EPI_ISL_933647 | Canada/ON-SC0139/2020 | 2020-06-27 | North America/Canada/Ontario/Toronto | Toronto Invasive Bacterial Diseases Network | McMaster University | Allison McGeer et al |
| EPI_ISL_933648 | Canada/ON-SC0142/2020 | 2020-07-02 | North America/Canada/Ontario/Toronto | Toronto Invasive Bacterial Diseases Network | McMaster University | Allison McGeer et al |
| EPI_ISL_933649 | Canada/ON-SC0144/2020 | 2020-07-06 | North America/Canada/Ontario/Toronto | Toronto Invasive Bacterial Diseases Network | McMaster University | Allison McGeer et al |
| EPI_ISL_933650 | Canada/ON-SC0154/2020 | 2020-07-19 | North America/Canada/Ontario/Toronto | Toronto Invasive Bacterial Diseases Network | McMaster University | Allison McGeer et al |
| EPI_ISL_933651 | Canada/ON-SC0159/2020 | 2020-07-22 | North America/Canada/Ontario/Toronto | Toronto Invasive Bacterial Diseases Network | McMaster University | Allison McGeer et al |
| EPI_ISL_933652 | Canada/ON-SC0160/2020 | 2020-07-22 | North America/Canada/Ontario/Toronto | Toronto Invasive Bacterial Diseases Network | McMaster University | Allison McGeer et al |
| EPI_ISL_933653 | Canada/ON-SC0169/2020 | 2020-07-25 | North America/Canada/Ontario/Toronto | Toronto Invasive Bacterial Diseases Network | McMaster University | Allison McGeer et al |

| | | | | | | |
|---|---|---|---|---|---|---|
| EPI_ISL_933654 | Canada/ON-SC0172/2020 | 2020-07-25 | North America/Canada/Ontario/Toronto | Toronto Invasive Bacterial Diseases Network | McMaster University | Allison McGeer et al |
| EPI_ISL_933655 | Canada/ON-SC0184/2020 | 2020-07-30 | North America/Canada/Ontario/Toronto | Toronto Invasive Bacterial Diseases Network | McMaster University | Allison McGeer et al |
| EPI_ISL_933656 | Canada/ON-SC0187/2020 | 2020-07-31 | North America/Canada/Ontario/Toronto | Toronto Invasive Bacterial Diseases Network | McMaster University | Allison McGeer et al |
| EPI_ISL_933657 | Canada/ON-SC0188/2020 | 2020-07-31 | North America/Canada/Ontario/Toronto | Toronto Invasive Bacterial Diseases Network | McMaster University | Allison McGeer et al |
| EPI_ISL_933658 | Canada/ON-SC0198/2020 | 2020-08-03 | North America/Canada/Ontario/Toronto | Toronto Invasive Bacterial Diseases Network | McMaster University | Allison McGeer et al |
| EPI_ISL_933659 | Canada/ON-SC0205/2020 | 2020-08-05 | North America/Canada/Ontario/Toronto | Toronto Invasive Bacterial Diseases Network | McMaster University | Allison McGeer et al |
| EPI_ISL_933661 | Mexico/CMX-InDRE_184/2021 | 2021-01-06 | North America/Mexico/Mexico City/ | Instituto de Diagnostico y Referencia Epidemiologicos INDRE_RNLSP | Instituto de Diagnostico y Referencia Epidemiologicos (INDRE) | Claudia Wong-Arambula et al |
| EPI_ISL_933662 | Mexico/CMX-InDRE_185/2021 | 2021-01-06 | North America/Mexico/Mexico City/ | Instituto de Diagnostico y Referencia Epidemiologicos INDRE_RNLSP | Instituto de Diagnostico y Referencia Epidemiologicos (INDRE) | Claudia Wong-Arambula et al |
| EPI_ISL_933663 | Mexico/CMX-InDRE_186/2021 | 2021-01-09 | North America/Mexico/Mexico City/ | Instituto de Diagnostico y Referencia Epidemiologicos INDRE_RNLSP | Instituto de Diagnostico y Referencia Epidemiologicos (INDRE) | Claudia Wong-Arambula et al |
| EPI_ISL_933665 | Mexico/CMX-InDRE_188/2020 | 2020-07-24 | North America/Mexico/Mexico City/ | Instituto de Diagnostico y Referencia Epidemiologicos INDRE_RNLSP | Instituto de Diagnostico y Referencia Epidemiologicos (INDRE) | Claudia Wong-Arambula et al |
| EPI_ISL_933666 | Mexico/TAM-InDRE_189/2020 | 2020-10-19 | North America/Mexico/Tamaulipas/ | Instituto de Diagnostico y Referencia Epidemiologicos INDRE_RNLSP | Instituto de Diagnostico y Referencia Epidemiologicos (INDRE) | Claudia Wong-Arambula et al |

| | | | | | | |
|---|---|---|---|---|---|---|
| EPI_ISL_933667 | Mexico/PUE-InDRE_190/2020 | 2020-10-21 | North America/Mexico/Puebla/ | Instituto de Diagnostico y Referencia Epidemiologicos INDRE_RNLSP | Instituto de Diagnostico y Referencia Epidemiologicos (INDRE) | Claudia Wong-Arambula et al |
| EPI_ISL_933668 | Mexico/MIC-InDRE_191/2020 | 2020-11-21 | North America/Mexico/Michoacan/ | Instituto de Diagnostico y Referencia Epidemiologicos INDRE_RNLSP | Instituto de Diagnostico y Referencia Epidemiologicos (INDRE) | Claudia Wong-Arambula et al |
| EPI_ISL_933670 | Mexico/AGU-InDRE_193/2020 | 2020-12-17 | North America/Mexico/Aguascalientes/ | Instituto de Diagnostico y Referencia Epidemiologicos INDRE_RNLSP | Instituto de Diagnostico y Referencia Epidemiologicos (INDRE) | Claudia Wong-Arambula et al |
| EPI_ISL_933671 | Mexico/AGU-InDRE_194/2020 | 2020-12-17 | North America/Mexico/Aguascalientes/ | Instituto de Diagnostico y Referencia Epidemiologicos INDRE_RNLSP | Instituto de Diagnostico y Referencia Epidemiologicos (INDRE) | Claudia Wong-Arambula et al |
| EPI_ISL_933673 | Mexico/AGU-InDRE_196/2020 | 2020-12-16 | North America/Mexico/Aguascalientes/ | Instituto de Diagnostico y Referencia Epidemiologicos INDRE_RNLSP | Instituto de Diagnostico y Referencia Epidemiologicos (INDRE) | Claudia Wong-Arambula et al |
| EPI_ISL_933674 | Mexico/CMX-InDRE_197/2020 | 2020-12-20 | North America/Mexico/Mexico City/ | Instituto de Diagnostico y Referencia Epidemiologicos INDRE_RNLSP | Instituto de Diagnostico y Referencia Epidemiologicos (INDRE) | Claudia Wong-Arambula et al |
| EPI_ISL_933675 | Mexico/CMX-InDRE_198/2020 | 2020-12-20 | North America/Mexico/Mexico City/ | Instituto de Diagnostico y Referencia Epidemiologicos INDRE_RNLSP | Instituto de Diagnostico y Referencia Epidemiologicos (INDRE) | Claudia Wong-Arambula et al |
| EPI_ISL_933676 | Mexico/CMX-InDRE_199/2020 | 2020-12-21 | North America/Mexico/Mexico City/ | Instituto de Diagnostico y Referencia Epidemiologicos INDRE_RNLSP | Instituto de Diagnostico y Referencia Epidemiologicos (INDRE) | Claudia Wong-Arambula et al |

| | | | | | | |
|---|---|---|---|---|---|---|
| EPI_ISL_933678 | Mexico/BCS-InDRE_201/2020 | 2020-12-18 | North America/Mexico/Baja California S | Instituto de Diagnostico y Referencia Epidemiologicos INDRE_RNLSP | Instituto de Diagnostico y Referencia Epidemiologicos (INDRE) | Claudia Wong-Arambula et al |
| EPI_ISL_933679 | Mexico/CMX-InDRE_202/2020 | 2020-12-29 | North America/Mexico/Mexico City/ | Instituto de Diagnostico y Referencia Epidemiologicos INDRE_RNLSP | Instituto de Diagnostico y Referencia Epidemiologicos (INDRE) | Claudia Wong-Arambula et al |
| EPI_ISL_933680 | Mexico/CMX-InDRE_203/2021 | 2021-01-06 | North America/Mexico/Mexico City/ | Instituto de Diagnostico y Referencia Epidemiologicos INDRE_RNLSP | Instituto de Diagnostico y Referencia Epidemiologicos (INDRE) | Claudia Wong-Arambula et al |
| EPI_ISL_933681 | Mexico/CMX-InDRE_204/2021 | 2021-01-07 | North America/Mexico/Mexico City/ | Instituto de Diagnostico y Referencia Epidemiologicos INDRE_RNLSP | Instituto de Diagnostico y Referencia Epidemiologicos (INDRE) | Claudia Wong-Arambula et al |
| EPI_ISL_933682 | Mexico/CMX-InDRE_205/2021 | 2021-01-07 | North America/Mexico/Mexico City/ | Instituto de Diagnostico y Referencia Epidemiologicos INDRE_RNLSP | Instituto de Diagnostico y Referencia Epidemiologicos (INDRE) | Claudia Wong-Arambula et al |
| EPI_ISL_933683 | Mexico/CMX-InDRE_206/2021 | 2021-01-07 | North America/Mexico/Mexico City/ | Instituto de Diagnostico y Referencia Epidemiologicos INDRE_RNLSP | Instituto de Diagnostico y Referencia Epidemiologicos (INDRE) | Claudia Wong-Arambula et al |
| EPI_ISL_933684 | Mexico/CMX-InDRE_207/2021 | 2021-01-07 | North America/Mexico/Mexico City/ | Instituto de Diagnostico y Referencia Epidemiologicos INDRE_RNLSP | Instituto de Diagnostico y Referencia Epidemiologicos (INDRE) | Claudia Wong-Arambula et al |
| EPI_ISL_933685 | Mexico/CMX-InDRE_208/2021 | 2021-01-07 | North America/Mexico/Mexico City/ | Instituto de Diagnostico y Referencia Epidemiologicos INDRE_RNLSP | Instituto de Diagnostico y Referencia Epidemiologicos (INDRE) | Claudia Wong-Arambula et al |

| | | | | | | |
|---|---|---|---|---|---|---|
| EPI_ISL_933686 | Mexico/CMX-InDRE_209/2021 | 2021-01-07 | North America/Mexico/Mexico City/ | Instituto de Diagnostico y Referencia Epidemiologicos INDRE_RNLSP | Instituto de Diagnostico y Referencia Epidemiologicos (INDRE) | Claudia Wong-Arambula et al |
| EPI_ISL_933687 | Mexico/CMX-InDRE_210/2021 | 2021-01-07 | North America/Mexico/Mexico City/ | Instituto de Diagnostico y Referencia Epidemiologicos INDRE_RNLSP | Instituto de Diagnostico y Referencia Epidemiologicos (INDRE) | Claudia Wong-Arambula et al |
| EPI_ISL_933688 | Mexico/CMX-InDRE_211/2021 | 2021-01-07 | North America/Mexico/Mexico City/ | Instituto de Diagnostico y Referencia Epidemiologicos INDRE_RNLSP | Instituto de Diagnostico y Referencia Epidemiologicos (INDRE) | Claudia Wong-Arambula et al |
| EPI_ISL_933689 | Mexico/CMX-InDRE_212/2021 | 2021-01-07 | North America/Mexico/Mexico City/ | Instituto de Diagnostico y Referencia Epidemiologicos INDRE_RNLSP | Instituto de Diagnostico y Referencia Epidemiologicos (INDRE) | Claudia Wong-Arambula et al |
| EPI_ISL_933690 | Mexico/CMX-InDRE_213/2021 | 2021-01-08 | North America/Mexico/Mexico City/ | Instituto de Diagnostico y Referencia Epidemiologicos INDRE_RNLSP | Instituto de Diagnostico y Referencia Epidemiologicos (INDRE) | Claudia Wong-Arambula et al |
| EPI_ISL_933691 | Mexico/CMX-InDRE_214/2021 | 2021-01-08 | North America/Mexico/Mexico City/ | Instituto de Diagnostico y Referencia Epidemiologicos INDRE_RNLSP | Instituto de Diagnostico y Referencia Epidemiologicos (INDRE) | Claudia Wong-Arambula et al |
| EPI_ISL_933692 | Mexico/MEX-InDRE_215/2021 | 2021-01-08 | North America/Mexico/Estado de Mexic | Instituto de Diagnostico y Referencia Epidemiologicos INDRE_RNLSP | Instituto de Diagnostico y Referencia Epidemiologicos (INDRE) | Claudia Wong-Arambula et al |
| EPI_ISL_933695 | Mexico/CMX-InDRE_218/2020 | 2020-12-20 | North America/Mexico/Mexico City/ | Instituto de Diagnostico y Referencia Epidemiologicos INDRE_RNLSP | Instituto de Diagnostico y Referencia Epidemiologicos (INDRE) | Claudia Wong-Arambula et al |

| | | | | | | |
|---|---|---|---|---|---|---|
| EPI_ISL_933696 | Mexico/CMX-InDRE_219/2020 | 2020-12-20 | North America/Mexico/Mexico City/ | Instituto de Diagnostico y Referencia Epidemiologicos INDRE_RNLSP | Instituto de Diagnostico y Referencia Epidemiologicos (INDRE) | Claudia Wong-Arambula et al |
| EPI_ISL_933697 | Mexico/CMX-InDRE_220/2020 | 2020-12-20 | North America/Mexico/Mexico City/ | Instituto de Diagnostico y Referencia Epidemiologicos INDRE_RNLSP | Instituto de Diagnostico y Referencia Epidemiologicos (INDRE) | Claudia Wong-Arambula et al |
| EPI_ISL_933698 | Mexico/CMX-InDRE_221/2020 | 2020-12-21 | North America/Mexico/Mexico City/ | Instituto de Diagnostico y Referencia Epidemiologicos INDRE_RNLSP | Instituto de Diagnostico y Referencia Epidemiologicos (INDRE) | Claudia Wong-Arambula et al |
| EPI_ISL_933699 | Mexico/CMX-InDRE_222/2020 | 2020-12-17 | North America/Mexico/Mexico City/ | Instituto de Diagnostico y Referencia Epidemiologicos INDRE_RNLSP | Instituto de Diagnostico y Referencia Epidemiologicos (INDRE) | Claudia Wong-Arambula et al |
| EPI_ISL_933701 | Mexico/CMX-InDRE_224/2020 | 2020-12-20 | North America/Mexico/Mexico City/ | Instituto de Diagnostico y Referencia Epidemiologicos INDRE_RNLSP | Instituto de Diagnostico y Referencia Epidemiologicos (INDRE) | Claudia Wong-Arambula et al |
| EPI_ISL_933702 | Mexico/CMX-InDRE_225/2020 | 2020-12-20 | North America/Mexico/Mexico City/ | Instituto de Diagnostico y Referencia Epidemiologicos INDRE_RNLSP | Instituto de Diagnostico y Referencia Epidemiologicos (INDRE) | Claudia Wong-Arambula et al |
| EPI_ISL_933703 | Mexico/MEX-InDRE_226/2020 | 2020-12-15 | North America/Mexico/Estado de Mexic | Instituto de Diagnostico y Referencia Epidemiologicos INDRE_RNLSP | Instituto de Diagnostico y Referencia Epidemiologicos (INDRE) | Claudia Wong-Arambula et al |
| EPI_ISL_933704 | Mexico/CMX-InDRE_227/2021 | 2021-01-07 | North America/Mexico/Mexico City/ | Instituto de Diagnostico y Referencia Epidemiologicos INDRE_RNLSP | Instituto de Diagnostico y Referencia Epidemiologicos (INDRE) | Claudia Wong-Arambula et al |

| | | | | | | |
|---|---|---|---|---|---|---|
| EPI_ISL_933705 | Mexico/MEX-InDRE_228/2021 | 2021-01-08 | North America/Mexico/Estado de Mexic | Instituto de Diagnostico y Referencia Epidemiologicos INDRE_RNLSP | Instituto de Diagnostico y Referencia Epidemiologicos (INDRE) | Claudia Wong-Arambula et al |
| EPI_ISL_933707 | Mexico/CMX-InDRE_230/2020 | 2020-11-18 | North America/Mexico/Mexico City/ | Instituto de Diagnostico y Referencia Epidemiologicos INDRE_RNLSP | Instituto de Diagnostico y Referencia Epidemiologicos (INDRE) | Claudia Wong-Arambula et al |
| EPI_ISL_933708 | Mexico/AGU-InDRE_231/2020 | 2020-12-16 | North America/Mexico/Aguascalientes/ | Instituto de Diagnostico y Referencia Epidemiologicos INDRE_RNLSP | Instituto de Diagnostico y Referencia Epidemiologicos (INDRE) | Claudia Wong-Arambula et al |
| EPI_ISL_933709 | Mexico/AGU-InDRE_232/2020 | 2020-12-16 | North America/Mexico/Aguascalientes/ | Instituto de Diagnostico y Referencia Epidemiologicos INDRE_RNLSP | Instituto de Diagnostico y Referencia Epidemiologicos (INDRE) | Claudia Wong-Arambula et al |
| EPI_ISL_933710 | Mexico/CMX-InDRE_233/2020 | 2020-12-17 | North America/Mexico/Mexico City/ | Instituto de Diagnostico y Referencia Epidemiologicos INDRE_RNLSP | Instituto de Diagnostico y Referencia Epidemiologicos (INDRE) | Claudia Wong-Arambula et al |
| EPI_ISL_933711 | Mexico/CMX-InDRE_234/2020 | 2020-12-19 | North America/Mexico/Mexico City/ | Instituto de Diagnostico y Referencia Epidemiologicos INDRE_RNLSP | Instituto de Diagnostico y Referencia Epidemiologicos (INDRE) | Claudia Wong-Arambula et al |
| EPI_ISL_933717 | NorthMacedonia/18473/2020 | 2020-11-02 | Europe/North Macedonia/Southeastern | General Hospital - Strumica | "Research Center for Genetic Engineering and Biotechnology ""Georgi D. Efremov,Äù , Macedonian Academy of Sciences and Arts" | RCGEB - MASA et al |

| | | | | | | |
|---|---|---|---|---|---|---|
| EPI_ISL_933718 | Spain/MD-11679763/2021 | 2021-01-13 | Europe/Spain/Madrid/Madrid | Servicio de Microbiolog√≠a Hospital Ram√≥n y Cajal | Servicio de Microbiolog√≠a Hospital Ram√≥n y Cajal | Jos√© M Gonzalez-Alba et al |
| EPI_ISL_933722 | NorthMacedonia/18616/2020 | 2020-11-03 | Europe/North Macedonia/Skopje/ | Gerontology Institute - Skopje | "Research Center for Genetic Engineering and Biotechnology ""Georgi D. Efremov,Äù , Macedonian Academy of Sciences and Arts" | RCGEB - MASA et al |
| EPI_ISL_933725 | Spain/MD-11683964/2021 | 2021-01-17 | Europe/Spain/Madrid/Madrid | Servicio de Microbiolog√≠a Hospital Ram√≥n y Cajal | Servicio de Microbiolog√≠a Hospital Ram√≥n y Cajal | Jos√© M Gonzalez-Alba et al |
| EPI_ISL_933726 | NorthMacedonia/18664/2020 | 2020-11-03 | Europe/North Macedonia/Southeastern | General Hospital - Strumica | "Research Center for Genetic Engineering and Biotechnology ""Georgi D. Efremov,Äù , Macedonian Academy of Sciences and Arts" | RCGEB - MASA et al |
| EPI_ISL_933727 | Spain/MD-11683966/2021 | 2021-01-17 | Europe/Spain/Madrid/Madrid | Servicio de Microbiolog√≠a Hospital Ram√≥n y Cajal | Servicio de Microbiolog√≠a Hospital Ram√≥n y Cajal | Jos√© M Gonzalez-Alba et al |
| EPI_ISL_933729 | Spain/MD-11684675/2021 | 2021-01-17 | Europe/Spain/Madrid/Madrid | Servicio de Microbiolog√≠a Hospital Ram√≥n y Cajal | Servicio de Microbiolog√≠a Hospital Ram√≥n y Cajal | Jos√© M Gonzalez-Alba et al |
| EPI_ISL_933731 | Spain/MD-11685991/2021 | 2021-01-18 | Europe/Spain/Madrid/Madrid | Servicio de Microbiolog√≠a Hospital Ram√≥n y Cajal | Servicio de Microbiolog√≠a Hospital Ram√≥n y Cajal | Jos√© M Gonzalez-Alba et al |

| | | | | | | |
|---|---|---|---|---|---|---|
| EPI_ISL_933732 | NorthMacedonia/18833/2020 | 2020-11-04 | Europe/North Macedonia/Southwestern | Institution of Nefrology-Struga | "Research Center for Genetic Engineering and Biotechnology ""Georgi D. Efremov,Äù , Macedonian Academy of Sciences and Arts" | RCGEB - MASA et al |
| EPI_ISL_933738 | USA/DE-DHSS-FLW00689767A/2021 | 2021-01-18 | North America/USA/Delaware/New Cas | DPHL | Delaware Public Health Lab | Gregory Hovan et al |
| EPI_ISL_933739 | USA/DE-DHSS-FLW00684410/2021 | 2021-01-16 | North America/USA/Delaware/Sussex C | DPHL | Delaware Public Health Lab | Gregory Hovan et al |
| EPI_ISL_933740 | Spain/MD-47434538/2021 | 2021-01-13 | Europe/Spain/Madrid/Madrid | Servicio de Microbiolog√≠a Hospital Ram√≥n y Cajal | Servicio de Microbiolog√≠a Hospital Ram√≥n y Cajal | Jos√© M Gonzalez-Alba et al |
| EPI_ISL_933746 | USA/DE-DHSS-FLW00679122/2021 | 2021-01-15 | North America/USA/Delaware/Kent Cou | DPHL | Delaware Public Health Lab | Gregory Hovan et al |
| EPI_ISL_933748 | USA/DE-DHSS-FLW00677263/2021 | 2021-01-16 | North America/USA/Delaware/Kent Cou | DPHL | Delaware Public Health Lab | Gregory Hovan et al |
| EPI_ISL_933749 | USA/DE-DHSS-FLW00677173/2021 | 2021-01-15 | North America/USA/Delaware/Sussex C | DPHL | Delaware Public Health Lab | Gregory Hovan et al |
| EPI_ISL_933754 | USA/DE-DHSS-FLW00674647/2021 | 2021-01-16 | North America/USA/Delaware/Kent Cou | DPHL | Delaware Public Health Lab | Gregory Hovan et al |
| EPI_ISL_933757 | USA/DE-DHSS-FLW00673646/2021 | 2021-01-16 | North America/USA/Delaware/Sussex C | DPHL | Delaware Public Health Lab | Gregory Hovan et al |
| EPI_ISL_933763 | USA/DE-DHSS-F1051864/2021 | 2021-01-18 | North America/USA/Delaware/Sussex C | DPHL | Delaware Public Health Lab | Gregory Hovan et al |
| EPI_ISL_933767 | Spain/MD-70961755/2021 | 2021-01-18 | Europe/Spain/Madrid/Madrid | Servicio de Microbiolog√≠a Hospital Ram√≥n y Cajal | Servicio de Microbiolog√≠a Hospital Ram√≥n y Cajal | Jos√© M Gonzalez-Alba et al |
| EPI_ISL_933768 | Spain/MD-71322477/2021 | 2021-01-13 | Europe/Spain/Madrid/Madrid | Servicio de Microbiolog√≠a Hospital Ram√≥n y Cajal | Servicio de Microbiolog√≠a Hospital Ram√≥n y Cajal | Jos√© M Gonzalez-Alba et al |

| | | | | | | |
|---|---|---|---|---|---|---|
| EPI_ISL_933770 | Spain/MD-71322478/2021 | 2021-01-14 | Europe/Spain/Madrid/Madrid | Servicio de Microbiolog√≠a Hospital Ram√≥n y Cajal | Servicio de Microbiolog√≠a Hospital Ram√≥n y Cajal | Jos√© M Gonzalez-Alba et al |
| EPI_ISL_933773 | Spain/MD-71554332/2021 | 2021-01-15 | Europe/Spain/Madrid/Madrid | Servicio de Microbiolog√≠a Hospital Ram√≥n y Cajal | Servicio de Microbiolog√≠a Hospital Ram√≥n y Cajal | Jos√© M Gonzalez-Alba et al |
| EPI_ISL_933776 | NorthMacedonia/19448/2020 | 2020-11-06 | Europe/North Macedonia/Northeastern | General Hospital - Kumanovo | "Research Center for Genetic Engineering and Biotechnology ""Georgi D. Efremov‚Äù , Macedonian Academy of Sciences and Arts" | RCGEB - MASA et al |
| EPI_ISL_933786 | NorthMacedonia/19508/2020 | 2020-11-06 | Europe/North Macedonia/Pelagonia/Bit | General Hospital - Bitola | "Research Center for Genetic Engineering and Biotechnology ""Georgi D. Efremov‚Äù , Macedonian Academy of Sciences and Arts" | RCGEB - MASA et al |
| EPI_ISL_933797 | NorthMacedonia/19670/2020 | 2020-11-06 | Europe/North Macedonia/Eastern Regio | General Hospital - Shtip | "Research Center for Genetic Engineering and Biotechnology ""Georgi D. Efremov‚Äù , Macedonian Academy of Sciences and Arts" | RCGEB - MASA et al |
| EPI_ISL_933820 | Lithuania/ID2348/2020 | 2020-12-01 | Europe/Lithuania/Vilniaus Apskritis/ | Vilnius university hospital Santaros Klinikos, Center of Laboratory Medicine | Vilnius university hospital Santaros Klinikos, Center of Laboratory Medicine | Ingrida Olendraite et al |

| | | | | | | |
|---|---|---|---|---|---|---|
| EPI_ISL_933824 | Lithuania/ID2356/2020 | 2020-12-01 | Europe/Lithuania/Vilniaus Apskritis/ | Vilnius university hospital Santaros Klinikos, Center of Laboratory Medicine | Vilnius university hospital Santaros Klinikos, Center of Laboratory Medicine | Ingrida Olendraite et al |
| EPI_ISL_933835 | Lithuania/ID2379/2020 | 2020-12-01 | Europe/Lithuania/Vilniaus Apskritis/ | Vilnius university hospital Santaros Klinikos, Center of Laboratory Medicine | Vilnius university hospital Santaros Klinikos, Center of Laboratory Medicine | Ingrida Olendraite et al |
| EPI_ISL_933859 | Lithuania/ID2468/2020 | 2020-12-02 | Europe/Lithuania/Vilniaus Apskritis/ | Vilnius university hospital Santaros Klinikos, Center of Laboratory Medicine | Vilnius university hospital Santaros Klinikos, Center of Laboratory Medicine | Ingrida Olendraite et al |
| EPI_ISL_933900 | Lithuania/ID2586/2020 | 2020-12-03 | Europe/Lithuania/Vilniaus Apskritis/ | Vilnius university hospital Santaros Klinikos, Center of Laboratory Medicine | Vilnius university hospital Santaros Klinikos, Center of Laboratory Medicine | Ingrida Olendraite et al |
| EPI_ISL_933922 | Lithuania/ID2635/2020 | 2020-12-04 | Europe/Lithuania/Vilniaus Apskritis/ | Vilnius university hospital Santaros Klinikos, Center of Laboratory Medicine | Vilnius university hospital Santaros Klinikos, Center of Laboratory Medicine | Ingrida Olendraite et al |
| EPI_ISL_933947 | Lithuania/ID2737/2020 | 2020-12-02 | Europe/Lithuania/Vilniaus Apskritis/ | Vilnius university hospital Santaros Klinikos, Center of Laboratory Medicine | Vilnius university hospital Santaros Klinikos, Center of Laboratory Medicine | Ingrida Olendraite et al |
| EPI_ISL_933955 | Lithuania/ID2766/2020 | 2020-12-06 | Europe/Lithuania/Panevezio Apskritis/ | Vilnius university hospital Santaros Klinikos, Center of Laboratory Medicine | Vilnius university hospital Santaros Klinikos, Center of Laboratory Medicine | Ingrida Olendraite et al |
| EPI_ISL_933994 | Lithuania/ID2914/2020 | 2020-12-07 | Europe/Lithuania/Vilniaus Apskritis/ | Vilnius university hospital Santaros Klinikos, Center of Laboratory Medicine | Vilnius university hospital Santaros Klinikos, Center of Laboratory Medicine | Ingrida Olendraite et al |

| | | | | | | |
|---|---|---|---|---|---|---|
| EPI_ISL_934005 | Lithuania/ID2952/2020 | 2020-12-07 | Europe/Lithuania/Vilniaus Apskritis/ | Vilnius university hospital Santaros Klinikos, Center of Laboratory Medicine | Vilnius university hospital Santaros Klinikos, Center of Laboratory Medicine | Ingrida Olendraite et al |
| EPI_ISL_934009 | Lithuania/ID2969/2020 | 2020-12-07 | Europe/Lithuania/Vilniaus Apskritis/ | Vilnius university hospital Santaros Klinikos, Center of Laboratory Medicine | Vilnius university hospital Santaros Klinikos, Center of Laboratory Medicine | Ingrida Olendraite et al |
| EPI_ISL_934062 | Lithuania/ID3252/2020 | 2020-12-10 | Europe/Lithuania/Vilniaus Apskritis/ | Vilnius university hospital Santaros Klinikos, Center of Laboratory Medicine | Vilnius university hospital Santaros Klinikos, Center of Laboratory Medicine | Ingrida Olendraite et al |
| EPI_ISL_934068 | Lithuania/ID3367/2020 | 2020-12-10 | Europe/Lithuania/Vilniaus Apskritis/ | Vilnius university hospital Santaros Klinikos, Center of Laboratory Medicine | Vilnius university hospital Santaros Klinikos, Center of Laboratory Medicine | Ingrida Olendraite et al |
| EPI_ISL_934088 | Lithuania/ID3525/2020 | 2020-12-11 | Europe/Lithuania/Vilniaus Apskritis/ | Vilnius university hospital Santaros Klinikos, Center of Laboratory Medicine | Vilnius university hospital Santaros Klinikos, Center of Laboratory Medicine | Ingrida Olendraite et al |
| EPI_ISL_934089 | Lithuania/ID3526/2020 | 2020-12-11 | Europe/Lithuania/Vilniaus Apskritis/ | Vilnius university hospital Santaros Klinikos, Center of Laboratory Medicine | Vilnius university hospital Santaros Klinikos, Center of Laboratory Medicine | Ingrida Olendraite et al |
| EPI_ISL_934099 | Lithuania/ID3769/2020 | 2020-12-14 | Europe/Lithuania/Vilniaus Apskritis/ | Vilnius university hospital Santaros Klinikos, Center of Laboratory Medicine | Vilnius university hospital Santaros Klinikos, Center of Laboratory Medicine | Ingrida Olendraite et al |
| EPI_ISL_934101 | Lithuania/ID3771/2020 | 2020-12-14 | Europe/Lithuania/Vilniaus Apskritis/ | Vilnius university hospital Santaros Klinikos, Center of Laboratory Medicine | Vilnius university hospital Santaros Klinikos, Center of Laboratory Medicine | Ingrida Olendraite et al |

| | | | | | | |
|---|---|---|---|---|---|---|
| EPI_ISL_934142 | Lithuania/ID3861/2020 | 2020-12-14 | Europe/Lithuania/Vilniaus Apskritis/ | Vilnius university hospital Santaros Klinikos, Center of Laboratory Medicine | Vilnius university hospital Santaros Klinikos, Center of Laboratory Medicine | Ingrida Olendraite et al |
| EPI_ISL_934161 | Lithuania/ID3896/2020 | 2020-12-14 | Europe/Lithuania/Vilniaus Apskritis/ | Vilnius university hospital Santaros Klinikos, Center of Laboratory Medicine | Vilnius university hospital Santaros Klinikos, Center of Laboratory Medicine | Ingrida Olendraite et al |
| EPI_ISL_934231 | Lithuania/ID6923/2021 | 2021-01-06 | Europe/Lithuania/Vilniaus Apskritis/ | Vilnius university hospital Santaros Klinikos, Center of Laboratory Medicine | Vilnius university hospital Santaros Klinikos, Center of Laboratory Medicine | Ingrida Olendraite et al |
| EPI_ISL_934260 | Lithuania/ID7337/2021 | 2021-01-13 | Europe/Lithuania/Vilniaus Apskritis/ | Vilnius university hospital Santaros Klinikos, Center of Laboratory Medicine | Vilnius university hospital Santaros Klinikos, Center of Laboratory Medicine | Ingrida Olendraite et al |
| EPI_ISL_934275 | Lithuania/ID7363/2021 | 2021-01-13 | Europe/Lithuania/Vilniaus Apskritis/ | Vilnius university hospital Santaros Klinikos, Center of Laboratory Medicine | Vilnius university hospital Santaros Klinikos, Center of Laboratory Medicine | Ingrida Olendraite et al |
| EPI_ISL_934278 | Lithuania/ID7399/2021 | 2021-01-14 | Europe/Lithuania/Vilniaus Apskritis/ | Vilnius university hospital Santaros Klinikos, Center of Laboratory Medicine | Vilnius university hospital Santaros Klinikos, Center of Laboratory Medicine | Ingrida Olendraite et al |
| EPI_ISL_934281 | Lithuania/ID7443/2021 | 2021-01-15 | Europe/Lithuania/Vilniaus Apskritis/ | Vilnius university hospital Santaros Klinikos, Center of Laboratory Medicine | Vilnius university hospital Santaros Klinikos, Center of Laboratory Medicine | Ingrida Olendraite et al |
| EPI_ISL_934282 | Lithuania/ID7444/2021 | 2021-01-15 | Europe/Lithuania/Vilniaus Apskritis/ | Vilnius university hospital Santaros Klinikos, Center of Laboratory Medicine | Vilnius university hospital Santaros Klinikos, Center of Laboratory Medicine | Ingrida Olendraite et al |

| | | | | | | |
|---|---|---|---|---|---|---|
| EPI_ISL_934284 | Lithuania/ID7450/2021 | 2021-01-15 | Europe/Lithuania/Vilniaus Apskritis/ | Vilnius university hospital Santaros Klinikos, Center of Laboratory Medicine | Vilnius university hospital Santaros Klinikos, Center of Laboratory Medicine | Ingrida Olendraite et al |
| EPI_ISL_934305 | Lithuania/ID7495/2021 | 2021-01-16 | Europe/Lithuania/Vilniaus Apskritis/ | Vilnius university hospital Santaros Klinikos, Center of Laboratory Medicine | Vilnius university hospital Santaros Klinikos, Center of Laboratory Medicine | Ingrida Olendraite et al |
| EPI_ISL_934308 | Lithuania/ID7526/2021 | 2021-01-18 | Europe/Lithuania/Vilniaus Apskritis/ | Vilnius university hospital Santaros Klinikos, Center of Laboratory Medicine | Vilnius university hospital Santaros Klinikos, Center of Laboratory Medicine | Ingrida Olendraite et al |
| EPI_ISL_934319 | Sweden/20-53796/2020 | 2020-12-14 | Europe/Sweden/Gavleborgs Lan/ | Laboratoriemedicin, Klinisk mikrobiologi | The Public Health Agency of Sweden | Anna-Malin Linde et al |
| EPI_ISL_934331 | Sweden/21-00335/2020 | 2020-08-24 | Europe/Sweden/Kronoberg/ | Klinisk mikrobiologi | The Public Health Agency of Sweden | Anna-Malin Linde et al |
| EPI_ISL_934335 | Sweden/21-00351/2020 | 2020-10-30 | Europe/Sweden/Kronoberg/ | Klinisk mikrobiologi | The Public Health Agency of Sweden | Anna-Malin Linde et al |
| EPI_ISL_934337 | Sweden/21-00361/2020 | 2020-11-26 | Europe/Sweden/Kronoberg/ | Klinisk mikrobiologi | The Public Health Agency of Sweden | Anna-Malin Linde et al |
| EPI_ISL_934339 | Sweden/21-00369/2020 | 2020-12-12 | Europe/Sweden/Kronoberg/ | Klinisk mikrobiologi | The Public Health Agency of Sweden | Anna-Malin Linde et al |
| EPI_ISL_934343 | Sweden/21-50186/2021 | 2021-01-07 | Europe/Sweden/Vastra Gotaland/ | Klinisk mikrobiologi | The Public Health Agency of Sweden | Anna-Malin Linde et al |
| EPI_ISL_934348 | Sweden/21-50247/2020 | 2020-12-30 | Europe/Sweden/Vasterbotten/ | Klinisk mikrobiologi | The Public Health Agency of Sweden | Anna-Malin Linde et al |
| EPI_ISL_934356 | Sweden/21-50323/2021 | 2021-01-06 | Europe/Sweden/Vastra Gotaland/ | Klinisk mikrobiologi | The Public Health Agency of Sweden | Anna-Malin Linde et al |
| EPI_ISL_934359 | Sweden/21-50326/2020 | 2020-12-30 | Europe/Sweden/Vastra Gotaland/ | Klinisk mikrobiologi | The Public Health Agency of Sweden | Anna-Malin Linde et al |
| EPI_ISL_934361 | Sweden/21-50329/2020 | 2020-12-30 | Europe/Sweden/Vastra Gotaland/ | Klinisk mikrobiologi | The Public Health Agency of Sweden | Anna-Malin Linde et al |
| EPI_ISL_934369 | Sweden/21-50345/2021 | 2021-01-07 | Europe/Sweden/Stockholm/ | Synlab Medilab, Mikrobiologi | The Public Health Agency of Sweden | Anna-Malin Linde et al |
| EPI_ISL_934370 | Sweden/21-50347/2021 | 2021-01-08 | Europe/Sweden/Stockholm/ | Synlab Medilab, Mikrobiologi | The Public Health Agency of Sweden | Anna-Malin Linde et al |

| | | | | | | |
|---|---|---|---|---|---|---|
| EPI_ISL_934374 | Sweden/21-50357/2021 | 2021-01-10 | Europe/Sweden/Stockholm/ | Synlab Medilab, Mikrobiologi | The Public Health Agency of Sweden | Anna-Malin Linde et al |
| EPI_ISL_934378 | Sweden/21-50367/2021 | 2021-01-11 | Europe/Sweden/Vastra Gotaland/ | Klinisk mikrobiologi | The Public Health Agency of Sweden | Anna-Malin Linde et al |
| EPI_ISL_934382 | Sweden/21-50390/2021 | 2021-01-18 | Europe/Sweden/Vastra Gotaland/ | Klinisk mikrobiologi | The Public Health Agency of Sweden | Anna-Malin Linde et al |
| EPI_ISL_934402 | Sweden/21-50412/2021 | 2021-01-13 | Europe/Sweden/Ostergotland/ | Synlab Medilab, Mikrobiologi | The Public Health Agency of Sweden | Anna-Malin Linde et al |
| EPI_ISL_934403 | Sweden/21-50413/2021 | 2021-01-13 | Europe/Sweden/Ostergotland/ | Synlab Medilab, Mikrobiologi | The Public Health Agency of Sweden | Anna-Malin Linde et al |
| EPI_ISL_934405 | Switzerland/NE-PMI-2042-4093/2020 | 2020-10-15 | Europe/Switzerland/Neuch√¢tel/ | ADMED Microbiologie | Genomics and Transcriptomics, Philip Morris International | Reto Lienhard et al |
| EPI_ISL_934412 | Sweden/21-50411/2021 | 2021-01-13 | Europe/Sweden/Ostergotland/ | Synlab Medilab, Mikrobiologi | The Public Health Agency of Sweden | Anna-Malin Linde et al |
| EPI_ISL_934415 | NorthMacedonia/20718/2020 | 2020-11-11 | Europe/North Macedonia/Vardar/Veles | General Hospital - Veles | "Research Center for Genetic Engineering and Biotechnology ""Georgi D. Efremov,Äù , Macedonian Academy of Sciences and Arts" | RCGEB - MASA et al |
| EPI_ISL_934419 | Italy/APU-CRGS-1/2020 | 2020-04-20 | Europe/Italy/Apulia/ | 1.AO Universitaria 'S. Giovanni di Dio e Ruggi D'Aragona, Scuola Medica Salernitana' Hospital / 2.UOC di Virologia e Microbiologia, Universit√† della Campania 'L. Vanvitelli' / 3.AO Universitaria 'Federico II' Napoli Hospital / 4.AORN 'San Giuseppe Moscati' Avellino Hospital / 5.AO 'San Pio - presidio G. Rummo' Benevento Hospital / 6.AO 'Sant'Anna e San Sebastiano' Caserta Hospital / 7.PO 'Maria Santissima Addolorata' Eboli Hospital / 8.Biogem Istituto di Ricerche Genetiche | 1. Genome Research Center for Health (CRGS) / 2. Laboratory of Molecular Medicine and Genomics(LMMGe) / 3. Center for Research in Pure and Applied Mathematics (CRMPA) | Giorgio Giurato (Corresponding Author) et al |

| | | | | | | |
|---|---|---|---|---|---|---|
| EPI_ISL_934422 | Italy/FVG-CRGS-2/2020 | 2020-04-01 | Europe/Italy/Friuli Venezia Giulia/ | 1.AO Universitaria 'S. Giovanni di Dio e Ruggi D'Aragona, Scuola Medica Salernitana' Hospital / 2.UOC di Virologia e Microbiologia, Universit√† della Campania 'L. Vanvitelli' / 3.AO Universitaria 'Federico II' Napoli Hospital / 4.AORN 'San Giuseppe Moscati' Avellino Hospital / 5.AO 'San Pio - presidio G. Rummo' Benevento Hospital / 6.AO 'Sant'Anna e San Sebastiano' Caserta Hospital / 7.PO 'Maria Santissima Addolorata' Eboli Hospital / 8.Biogem Istituto di Ricerche Genetiche | 1. Genome Research Center for Health (CRGS) / 2. Laboratory of Molecular Medicine and Genomics(LMMGe) / 3. Center for Research in Pure and Applied Mathematics (CRMPA) | Giorgio Giurato (Corresponding Author) et al |
| EPI_ISL_934426 | Austria/CeMM2033/2020 | 2020-12-21 | Europe/Austria/Carinthia/Hermagor | ILV K√§rnten | Bergthaler laboratory, CeMM Research Center for Molecular Medicine of the Austrian Academy of Sciences | Lukas Endler et al |
| EPI_ISL_934427 | Austria/CeMM2035/2020 | 2020-12-21 | Europe/Austria/Carinthia/Hermagor | ILV K√§rnten | Bergthaler laboratory, CeMM Research Center for Molecular Medicine of the Austrian Academy of Sciences | Lukas Endler et al |
| EPI_ISL_934434 | Austria/CeMM2207/2020 | 2020-12-22 | Europe/Austria/Vienna/ | Austrian Agency for Health and Food Safety (AGES) | Bergthaler laboratory, CeMM Research Center for Molecular Medicine of the Austrian Academy of Sciences | Lukas Endler et al |

| | | | | | | |
|---|---|---|---|---|---|---|
| EPI_ISL_934436 | Austria/CeMM2209/2020 | 2020-12-19 | Europe/Austria/Lower Austria/M√∂dling | Austrian Agency for Health and Food Safety (AGES) | Bergthaler laboratory, CeMM Research Center for Molecular Medicine of the Austrian Academy of Sciences | Lukas Endler et al |
| EPI_ISL_934460 | Austria/CeMM2287/2021 | 2021-01-07 | Europe/Austria/Burgenland/ | Austrian Agency for Health and Food Safety (AGES) | Bergthaler laboratory, CeMM Research Center for Molecular Medicine of the Austrian Academy of Sciences | Lukas Endler et al |
| EPI_ISL_934462 | Austria/CeMM2289/2021 | 2021-01-08 | Europe/Austria/Burgenland/ | Austrian Agency for Health and Food Safety (AGES) | Bergthaler laboratory, CeMM Research Center for Molecular Medicine of the Austrian Academy of Sciences | Lukas Endler et al |
| EPI_ISL_934475 | Austria/CeMM2303/2021 | 2021-01-08 | Europe/Austria/Burgenland/ | Austrian Agency for Health and Food Safety (AGES) | Bergthaler laboratory, CeMM Research Center for Molecular Medicine of the Austrian Academy of Sciences | Lukas Endler et al |
| EPI_ISL_934497 | Austria/CeMM2327/2020 | 2020-11-13 | Europe/Austria/Vorarlberg/Feldkirch | Austrian Agency for Health and Food Safety (AGES) | Bergthaler laboratory, CeMM Research Center for Molecular Medicine of the Austrian Academy of Sciences | Lukas Endler et al |

| | | | | | | |
|---|---|---|---|---|---|---|
| EPI_ISL_934515 | Austria/CeMM2385/2021 | 2021-01-06 | Europe/Austria/Burgenland/ | Austrian Agency for Health and Food Safety (AGES) | Bergthaler laboratory, CeMM Research Center for Molecular Medicine of the Austrian Academy of Sciences | Lukas Endler et al |
| EPI_ISL_934518 | Austria/CeMM2388/2021 | 2021-01-06 | Europe/Austria/Burgenland/ | Austrian Agency for Health and Food Safety (AGES) | Bergthaler laboratory, CeMM Research Center for Molecular Medicine of the Austrian Academy of Sciences | Lukas Endler et al |
| EPI_ISL_934521 | Austria/CeMM2391/2021 | 2021-01-06 | Europe/Austria/Burgenland/ | Austrian Agency for Health and Food Safety (AGES) | Bergthaler laboratory, CeMM Research Center for Molecular Medicine of the Austrian Academy of Sciences | Lukas Endler et al |
| EPI_ISL_934522 | Austria/CeMM2392/2021 | 2021-01-06 | Europe/Austria/Burgenland/ | Austrian Agency for Health and Food Safety (AGES) | Bergthaler laboratory, CeMM Research Center for Molecular Medicine of the Austrian Academy of Sciences | Lukas Endler et al |
| EPI_ISL_934523 | Austria/CeMM2393/2021 | 2021-01-06 | Europe/Austria/Burgenland/ | Austrian Agency for Health and Food Safety (AGES) | Bergthaler laboratory, CeMM Research Center for Molecular Medicine of the Austrian Academy of Sciences | Lukas Endler et al |

| | | | | | | |
|---|---|---|---|---|---|---|
| EPI_ISL_934540 | Austria/CeMM2600/2020 | 2020-11-11 | Europe/Austria/Tyrol/Innsbruck | Institut f√ºr Virologie am Department f√ºr Hygiene, Mikrobiologie und Public Health | Bergthaler laboratory, CeMM Research Center for Molecular Medicine of the Austrian Academy of Sciences | Lukas Endler et al |
| EPI_ISL_934546 | Austria/CeMM2607/2020 | 2020-11-11 | Europe/Austria/Tyrol/Innsbruck | Institut f√ºr Virologie am Department f√ºr Hygiene, Mikrobiologie und Public Health | Bergthaler laboratory, CeMM Research Center for Molecular Medicine of the Austrian Academy of Sciences | Lukas Endler et al |
| EPI_ISL_934547 | Austria/CeMM2608/2020 | 2020-11-11 | Europe/Austria/Tyrol/Innsbruck | Institut f√ºr Virologie am Department f√ºr Hygiene, Mikrobiologie und Public Health | Bergthaler laboratory, CeMM Research Center for Molecular Medicine of the Austrian Academy of Sciences | Lukas Endler et al |
| EPI_ISL_934571 | Austria/CeMM2636/2020 | 2020-12-15 | Europe/Austria/Styria/ | Department of Laboratory Medicine, Division of Clinical Virology, University of Medicine, Vienna | Bergthaler laboratory, CeMM Research Center for Molecular Medicine of the Austrian Academy of Sciences | Lukas Endler et al |
| EPI_ISL_934573 | Austria/CeMM2638/2020 | 2020-12-17 | Europe/Austria/Carinthia/ | Department of Laboratory Medicine, Division of Clinical Virology, University of Medicine, Vienna | Bergthaler laboratory, CeMM Research Center for Molecular Medicine of the Austrian Academy of Sciences | Lukas Endler et al |

| | | | | | | |
|---|---|---|---|---|---|---|
| EPI_ISL_934575 | Austria/CeMM2640/2020 | 2020-12-18 | Europe/Austria/Styria/ | Department of Laboratory Medicine, Division of Clinical Virology, University of Medicine, Vienna | Bergthaler laboratory, CeMM Research Center for Molecular Medicine of the Austrian Academy of Sciences | Lukas Endler et al |
| EPI_ISL_934577 | Austria/CeMM2642/2020 | 2020-12-21 | Europe/Austria/Styria/ | Department of Laboratory Medicine, Division of Clinical Virology, University of Medicine, Vienna | Bergthaler laboratory, CeMM Research Center for Molecular Medicine of the Austrian Academy of Sciences | Lukas Endler et al |
| EPI_ISL_934584 | Austria/CeMM2649/2020 | 2020-12-23 | Europe/Austria/Carinthia/ | Department of Laboratory Medicine, Division of Clinical Virology, University of Medicine, Vienna | Bergthaler laboratory, CeMM Research Center for Molecular Medicine of the Austrian Academy of Sciences | Lukas Endler et al |
| EPI_ISL_934586 | Austria/CeMM2651/2020 | 2020-12-23 | Europe/Austria/Styria/ | Department of Laboratory Medicine, Division of Clinical Virology, University of Medicine, Vienna | Bergthaler laboratory, CeMM Research Center for Molecular Medicine of the Austrian Academy of Sciences | Lukas Endler et al |
| EPI_ISL_934588 | Austria/CeMM2653/2020 | 2020-12-24 | Europe/Austria/Styria/ | Department of Laboratory Medicine, Division of Clinical Virology, University of Medicine, Vienna | Bergthaler laboratory, CeMM Research Center for Molecular Medicine of the Austrian Academy of Sciences | Lukas Endler et al |

| | | | | | | |
|---|---|---|---|---|---|---|
| EPI_ISL_934618 | Austria/CeMM2934/2021 | 2021-01-03 | Europe/Austria/Burgenland/ | Austrian Agency for Health and Food Safety (AGES) | Bergthaler laboratory, CeMM Research Center for Molecular Medicine of the Austrian Academy of Sciences | Lukas Endler et al |
| EPI_ISL_934673 | Austria/CeMM3097/2021 | 2021-01-15 | Europe/Austria/Vienna/ | Department of Laboratory Medicine, Division of Clinical Virology, University of Medicine, Vienna | Bergthaler laboratory, CeMM Research Center for Molecular Medicine of the Austrian Academy of Sciences | Lukas Endler et al |
| EPI_ISL_934686 | Austria/CeMM3110/2021 | 2021-01-14 | Europe/Austria/Vienna/ | Department of Laboratory Medicine, Division of Clinical Virology, University of Medicine, Vienna | Bergthaler laboratory, CeMM Research Center for Molecular Medicine of the Austrian Academy of Sciences | Lukas Endler et al |
| EPI_ISL_935015 | USA/MI-UM-10038001373/2021 | 2021-01-25 | North America/USA/Michigan/ | University of Michigan Clinical Microbiology Laboratory | Lauring Lab, University of Michigan, Department of Microbiology and Immunology | Valesano et al |
| EPI_ISL_935017 | Romania/Valcea418423/2021 | 2021-01-31 | Europe/Romania/Valcea/ | Laboratory for Respiratory Viruses, Cantacuzino National Military-Medical Institute for Research and Development | Cantacuzino Institute Virology | Luiza Ustea et al |
| EPI_ISL_935021 | Romania/Ialomita418189/2021 | 2021-01-29 | Europe/Romania/Ialomi»õa/ | Laboratory for Respiratory Viruses, Cantacuzino National Military-Medical Institute for Research and Development | Cantacuzino Institute Virology | Luiza Ustea et al |
| EPI_ISL_935033 | Poland/9328607421_diag1_gx1/2020 | 2020-12-23 | Europe/Poland/Malopolskie/ | Laboratory of Molecular Biology, Diagnostyka sp. z o.o. | genXone SA, Research & Development Laboratory | Maciej Sykulski et al |

| | | | | | | |
|---|---|---|---|---|---|---|
| EPI_ISL_935035 | Poland/3292614221_diag1_gx1/2020 | 2020-12-23 | Europe/Poland/Malopolskie/ | Laboratory of Molecular Biology, Diagnostyka sp. z o.o. | genXone SA, Research & Development Laboratory | Maciej Sykulski et al |
| EPI_ISL_935046 | Jordan/PHBC6/2021 | 2021-01-07 | Asia/Jordan/Amman/ | Biolab Diagnostic Laboratories | Princess Haya Biotechnology Center/ Jordan University of Science & Technology | Saied Jaradat et al |
| EPI_ISL_935049 | Turkey/HSGM-1322/2021 | 2021-01-19 | Europe/Turkey/Turkey/ | Ministry of Health Turkey | Ministry of Health Turkey | Fatma Bayrakdar et al |
| EPI_ISL_935051 | Turkey/HSGM-1333/2021 | 2021-01-19 | Europe/Turkey/Turkey/ | Ministry of Health Turkey | Ministry of Health Turkey | Fatma Bayrakdar et al |
| EPI_ISL_935052 | Turkey/HSGM-1339/2021 | 2021-01-19 | Europe/Turkey/Turkey/ | Ministry of Health Turkey | Ministry of Health Turkey | Fatma Bayrakdar et al |
| EPI_ISL_935053 | Turkey/HSGM-1348/2021 | 2021-01-19 | Europe/Turkey/Turkey/ | Ministry of Health Turkey | Ministry of Health Turkey | Fatma Bayrakdar et al |
| EPI_ISL_935054 | Turkey/HSGM-1350/2021 | 2021-01-19 | Europe/Turkey/Turkey/ | Ministry of Health Turkey | Ministry of Health Turkey | Fatma Bayrakdar et al |
| EPI_ISL_935057 | Turkey/HSGM-1384/2021 | 2021-01-19 | Europe/Turkey/Turkey/ | Ministry of Health Turkey | Ministry of Health Turkey | Fatma Bayrakdar et al |
| EPI_ISL_935058 | Turkey/HSGM-1356/2021 | 2021-01-19 | Europe/Turkey/Turkey/ | Ministry of Health Turkey | Ministry of Health Turkey | Fatma Bayrakdar et al |
| EPI_ISL_935059 | Turkey/HSGM-1363/2021 | 2021-01-19 | Europe/Turkey/Turkey/ | Ministry of Health Turkey | Ministry of Health Turkey | Fatma Bayrakdar et al |
| EPI_ISL_935064 | Turkey/HSGM-1317/2021 | 2021-01-19 | Europe/Turkey/Turkey/ | Ministry of Health Turkey | Ministry of Health Turkey | Fatma Bayrakdar et al |
| EPI_ISL_935065 | Turkey/HSGM-1318/2021 | 2021-01-19 | Europe/Turkey/Turkey/ | Ministry of Health Turkey | Ministry of Health Turkey | Fatma Bayrakdar et al |
| EPI_ISL_935066 | Turkey/HSGM-1319/2021 | 2021-01-19 | Europe/Turkey/Turkey/ | Ministry of Health Turkey | Ministry of Health Turkey | Fatma Bayrakdar et al |
| EPI_ISL_935071 | Turkey/HSGM-1342/2021 | 2021-01-21 | Europe/Turkey/Turkey/ | Ministry of Health Turkey | Ministry of Health Turkey | Fatma Bayrakdar et al |
| EPI_ISL_935079 | Turkey/HSGM-1345/2021 | 2021-01-21 | Europe/Turkey/Turkey/ | Ministry of Health Turkey | Ministry of Health Turkey | Fatma Bayrakdar et al |
| EPI_ISL_935081 | Turkey/HSGM-1364/2021 | 2021-01-21 | Europe/Turkey/Turkey/ | Ministry of Health Turkey | Ministry of Health Turkey | Fatma Bayrakdar et al |
| EPI_ISL_935082 | Turkey/HSGM-1346/2021 | 2021-01-21 | Europe/Turkey/Turkey/ | Ministry of Health Turkey | Ministry of Health Turkey | Fatma Bayrakdar et al |

| | | | | | | |
|---|---|---|---|---|---|---|
| EPI_ISL_935088 | Turkey/HSGM-1390/2021 | 2021-01-21 | Europe/Turkey/Turkey/ | Ministry of Health Turkey | Ministry of Health Turkey | Fatma Bayrakdar et al |
| EPI_ISL_935091 | Turkey/HSGM-1338/2021 | 2021-01-21 | Europe/Turkey/Turkey/ | Ministry of Health Turkey | Ministry of Health Turkey | Fatma Bayrakdar et al |
| EPI_ISL_935093 | Turkey/HSGM-1353/2021 | 2021-01-21 | Europe/Turkey/Turkey/ | Ministry of Health Turkey | Ministry of Health Turkey | Fatma Bayrakdar et al |
| EPI_ISL_935098 | Turkey/HSGM-1361/2021 | 2021-01-21 | Europe/Turkey/Turkey/ | Ministry of Health Turkey | Ministry of Health Turkey | Fatma Bayrakdar et al |
| EPI_ISL_935101 | Turkey/HSGM-1367/2021 | 2021-01-21 | Europe/Turkey/Turkey/ | Ministry of Health Turkey | Ministry of Health Turkey | Fatma Bayrakdar et al |
| EPI_ISL_935102 | Turkey/HSGM-2398/2021 | 2021-01-30 | Europe/Turkey/Turkey/ | Ministry of Health Turkey | Ministry of Health Turkey | Fatma Bayrakdar et al |
| EPI_ISL_935104 | Italy/LOM-CRGS-1/2020 | 2020-03-01 | Europe/Italy/Lombardy/ | 1.AO Universitaria 'S. Giovanni di Dio e Ruggi D'Aragona, Scuola Medica Salernitana' Hospital / 2.UOC di Virologia e Microbiologia, Universit√† della Campania 'L. Vanvitelli' / 3.AO Universitaria 'Federico II' Napoli Hospital / 4.AORN 'San Giuseppe Moscati' Avellino Hospital / 5.AO 'San Pio - presidio G. Rummo' Benevento Hospital / 6.AO 'Sant'Anna e San Sebastiano' Caserta Hospital / 7.PO 'Maria Santissima Addolorata' Eboli Hospital / 8.Biogem Istituto di Ricerche Genetiche | 1. Genome Research Center for Health (CRGS) / 2. Laboratory of Molecular Medicine and Genomics(LMMGe) / 3. Center for Research in Pure and Applied Mathematics (CRMPA) | Giorgio Giurato (Corresponding Author) et al |
| EPI_ISL_935106 | Italy/CAM-CRGS-119/2020 | 2020-11-14 | Europe/Italy/Campania/ | 1.AO Universitaria 'S. Giovanni di Dio e Ruggi D'Aragona, Scuola Medica Salernitana' Hospital / 2.UOC di Virologia e Microbiologia, Universit√† della Campania 'L. Vanvitelli' / 3.AO Universitaria 'Federico II' Napoli Hospital / 4.AORN 'San Giuseppe Moscati' Avellino Hospital / 5.AO 'San Pio - presidio G. Rummo' Benevento Hospital / 6.AO 'Sant'Anna e San Sebastiano' Caserta Hospital / 7.PO 'Maria Santissima Addolorata' Eboli Hospital / 8.Biogem Istituto di Ricerche Genetiche | 1. Genome Research Center for Health (CRGS) / 2. Laboratory of Molecular Medicine and Genomics(LMMGe) / 3. Center for Research in Pure and Applied Mathematics (CRMPA) | Giorgio Giurato (Corresponding Author) et al |

| | | | | | | |
|---|---|---|---|---|---|---|
| EPI_ISL_935109 | Italy/CAM-CRGS-123/2020 | 2020-11-14 | Europe/Italy/Campania/ | 1.AO Universitaria 'S. Giovanni di Dio e Ruggi D'Aragona, Scuola Medica Salernitana' Hospital / 2.UOC di Virologia e Microbiologia, Universit√† della Campania 'L. Vanvitelli' / 3.AO Universitaria 'Federico II' Napoli Hospital / 4.AORN 'San Giuseppe Moscati' Avellino Hospital / 5.AO 'San Pio - presidio G. Rummo' Benevento Hospital / 6.AO 'Sant'Anna e San Sebastiano' Caserta Hospital / 7.PO 'Maria Santissima Addolorata' Eboli Hospital / 8.Biogem Istituto di Ricerche Genetiche | 1. Genome Research Center for Health (CRGS) / 2. Laboratory of Molecular Medicine and Genomics(LMMGe) / 3. Center for Research in Pure and Applied Mathematics (CRMPA) | Giorgio Giurato (Corresponding Author) et al |
| EPI_ISL_935110 | Italy/CAM-CRGS-128/2020 | 2020-11-13 | Europe/Italy/Campania/ | 1.AO Universitaria 'S. Giovanni di Dio e Ruggi D'Aragona, Scuola Medica Salernitana' Hospital / 2.UOC di Virologia e Microbiologia, Universit√† della Campania 'L. Vanvitelli' / 3.AO Universitaria 'Federico II' Napoli Hospital / 4.AORN 'San Giuseppe Moscati' Avellino Hospital / 5.AO 'San Pio - presidio G. Rummo' Benevento Hospital / 6.AO 'Sant'Anna e San Sebastiano' Caserta Hospital / 7.PO 'Maria Santissima Addolorata' Eboli Hospital / 8.Biogem Istituto di Ricerche Genetiche | 1. Genome Research Center for Health (CRGS) / 2. Laboratory of Molecular Medicine and Genomics(LMMGe) / 3. Center for Research in Pure and Applied Mathematics (CRMPA) | Giorgio Giurato (Corresponding Author) et al |
| EPI_ISL_935111 | Italy/CAM-CRGS-129/2020 | 2020-11-13 | Europe/Italy/Campania/ | 1.AO Universitaria 'S. Giovanni di Dio e Ruggi D'Aragona, Scuola Medica Salernitana' Hospital / 2.UOC di Virologia e Microbiologia, Universit√† della Campania 'L. Vanvitelli' / 3.AO Universitaria 'Federico II' Napoli Hospital / 4.AORN 'San Giuseppe Moscati' Avellino Hospital / 5.AO 'San Pio - presidio G. Rummo' Benevento Hospital / 6.AO 'Sant'Anna e San Sebastiano' Caserta Hospital / 7.PO 'Maria Santissima Addolorata' Eboli Hospital / 8.Biogem Istituto di Ricerche Genetiche | 1. Genome Research Center for Health (CRGS) / 2. Laboratory of Molecular Medicine and Genomics(LMMGe) / 3. Center for Research in Pure and Applied Mathematics (CRMPA) | Giorgio Giurato (Corresponding Author) et al |

| | | | | | | |
|---|---|---|---|---|---|---|
| EPI_ISL_935112 | Italy/CAM-CRGS-131/2020 | 2020-11-13 | Europe/Italy/Campania/ | 1.AO Universitaria 'S. Giovanni di Dio e Ruggi D'Aragona, Scuola Medica Salernitana' Hospital / 2.UOC di Virologia e Microbiologia, Universit√† della Campania 'L. Vanvitelli' / 3.AO Universitaria 'Federico II' Napoli Hospital / 4.AORN 'San Giuseppe Moscati' Avellino Hospital / 5.AO 'San Pio - presidio G. Rummo' Benevento Hospital / 6.AO 'Sant'Anna e San Sebastiano' Caserta Hospital / 7.PO 'Maria Santissima Addolorata' Eboli Hospital / 8.Biogem Istituto di Ricerche Genetiche | 1. Genome Research Center for Health (CRGS) / 2. Laboratory of Molecular Medicine and Genomics(LMMGe) / 3. Center for Research in Pure and Applied Mathematics (CRMPA) | Giorgio Giurato (Corresponding Author) et al |
| EPI_ISL_935116 | Italy/CAM-CRGS-135/2020 | 2020-11-13 | Europe/Italy/Campania/ | 1.AO Universitaria 'S. Giovanni di Dio e Ruggi D'Aragona, Scuola Medica Salernitana' Hospital / 2.UOC di Virologia e Microbiologia, Universit√† della Campania 'L. Vanvitelli' / 3.AO Universitaria 'Federico II' Napoli Hospital / 4.AORN 'San Giuseppe Moscati' Avellino Hospital / 5.AO 'San Pio - presidio G. Rummo' Benevento Hospital / 6.AO 'Sant'Anna e San Sebastiano' Caserta Hospital / 7.PO 'Maria Santissima Addolorata' Eboli Hospital / 8.Biogem Istituto di Ricerche Genetiche | 1. Genome Research Center for Health (CRGS) / 2. Laboratory of Molecular Medicine and Genomics(LMMGe) / 3. Center for Research in Pure and Applied Mathematics (CRMPA) | Giorgio Giurato (Corresponding Author) et al |
| EPI_ISL_935119 | Italy/CAM-CRGS-145/2020 | 2020-11-17 | Europe/Italy/Campania/ | 1.AO Universitaria 'S. Giovanni di Dio e Ruggi D'Aragona, Scuola Medica Salernitana' Hospital / 2.UOC di Virologia e Microbiologia, Universit√† della Campania 'L. Vanvitelli' / 3.AO Universitaria 'Federico II' Napoli Hospital / 4.AORN 'San Giuseppe Moscati' Avellino Hospital / 5.AO 'San Pio - presidio G. Rummo' Benevento Hospital / 6.AO 'Sant'Anna e San Sebastiano' Caserta Hospital / 7.PO 'Maria Santissima Addolorata' Eboli Hospital / 8.Biogem Istituto di Ricerche Genetiche | 1. Genome Research Center for Health (CRGS) / 2. Laboratory of Molecular Medicine and Genomics(LMMGe) / 3. Center for Research in Pure and Applied Mathematics (CRMPA) | Giorgio Giurato (Corresponding Author) et al |

| | | | | | | |
|---|---|---|---|---|---|---|
| EPI_ISL_935120 | Italy/CAM-CRGS-146/2020 | 2020-11-17 | Europe/Italy/Campania/ | 1.AO Universitaria 'S. Giovanni di Dio e Ruggi D'Aragona, Scuola Medica Salernitana' Hospital / 2.UOC di Virologia e Microbiologia, Universitv† della Campania 'L. Vanvitelli' / 3.AO Universitaria 'Federico II' Napoli Hospital / 4.AORN 'San Giuseppe Moscati' Avellino Hospital / 5.AO 'San Pio - presidio G. Rummo' Benevento Hospital / 6.AO 'Sant'Anna e San Sebastiano' Caserta Hospital / 7.PO 'Maria Santissima Addolorata' Eboli Hospital / 8.Biogem Istituto di Ricerche Genetiche | 1. Genome Research Center for Health (CRGS) / 2. Laboratory of Molecular Medicine and Genomics(LMMGe) / 3. Center for Research in Pure and Applied Mathematics (CRMPA) | Giorgio Giurato (Corresponding Author) et al |
| EPI_ISL_935121 | Italy/CAM-CRGS-147/2020 | 2020-11-17 | Europe/Italy/Campania/ | 1.AO Universitaria 'S. Giovanni di Dio e Ruggi D'Aragona, Scuola Medica Salernitana' Hospital / 2.UOC di Virologia e Microbiologia, Universitv† della Campania 'L. Vanvitelli' / 3.AO Universitaria 'Federico II' Napoli Hospital / 4.AORN 'San Giuseppe Moscati' Avellino Hospital / 5.AO 'San Pio - presidio G. Rummo' Benevento Hospital / 6.AO 'Sant'Anna e San Sebastiano' Caserta Hospital / 7.PO 'Maria Santissima Addolorata' Eboli Hospital / 8.Biogem Istituto di Ricerche Genetiche | 1. Genome Research Center for Health (CRGS) / 2. Laboratory of Molecular Medicine and Genomics(LMMGe) / 3. Center for Research in Pure and Applied Mathematics (CRMPA) | Giorgio Giurato (Corresponding Author) et al |
| EPI_ISL_935124 | Italy/CAM-CRGS-151/2020 | 2020-11-14 | Europe/Italy/Campania/ | 1.AO Universitaria 'S. Giovanni di Dio e Ruggi D'Aragona, Scuola Medica Salernitana' Hospital / 2.UOC di Virologia e Microbiologia, Universitv† della Campania 'L. Vanvitelli' / 3.AO Universitaria 'Federico II' Napoli Hospital / 4.AORN 'San Giuseppe Moscati' Avellino Hospital / 5.AO 'San Pio - presidio G. Rummo' Benevento Hospital / 6.AO 'Sant'Anna e San Sebastiano' Caserta Hospital / 7.PO 'Maria Santissima Addolorata' Eboli Hospital / 8.Biogem Istituto di Ricerche Genetiche | 1. Genome Research Center for Health (CRGS) / 2. Laboratory of Molecular Medicine and Genomics(LMMGe) / 3. Center for Research in Pure and Applied Mathematics (CRMPA) | Giorgio Giurato (Corresponding Author) et al |

| | | | | | | |
|---|---|---|---|---|---|---|
| EPI_ISL_935125 | Italy/CAM-CRGS-154/2020 | 2020-11-16 | Europe/Italy/Campania/ | 1.AO Universitaria 'S. Giovanni di Dio e Ruggi D'Aragona, Scuola Medica Salernitana' Hospital / 2.UOC di Virologia e Microbiologia, Universit√† della Campania 'L. Vanvitelli' / 3.AO Universitaria 'Federico II' Napoli Hospital / 4.AORN 'San Giuseppe Moscati' Avellino Hospital / 5.AO 'San Pio - presidio G. Rummo' Benevento Hospital / 6.AO 'Sant'Anna e San Sebastiano' Caserta Hospital / 7.PO 'Maria Santissima Addolorata' Eboli Hospital / 8.Biogem Istituto di Ricerche Genetiche | 1. Genome Research Center for Health (CRGS) / 2. Laboratory of Molecular Medicine and Genomics(LMMGe) / 3. Center for Research in Pure and Applied Mathematics (CRMPA) | Giorgio Giurato (Corresponding Author) et al |
| EPI_ISL_935126 | Italy/CAM-CRGS-155/2020 | 2020-11-14 | Europe/Italy/Campania/ | 1.AO Universitaria 'S. Giovanni di Dio e Ruggi D'Aragona, Scuola Medica Salernitana' Hospital / 2.UOC di Virologia e Microbiologia, Universit√† della Campania 'L. Vanvitelli' / 3.AO Universitaria 'Federico II' Napoli Hospital / 4.AORN 'San Giuseppe Moscati' Avellino Hospital / 5.AO 'San Pio - presidio G. Rummo' Benevento Hospital / 6.AO 'Sant'Anna e San Sebastiano' Caserta Hospital / 7.PO 'Maria Santissima Addolorata' Eboli Hospital / 8.Biogem Istituto di Ricerche Genetiche | 1. Genome Research Center for Health (CRGS) / 2. Laboratory of Molecular Medicine and Genomics(LMMGe) / 3. Center for Research in Pure and Applied Mathematics (CRMPA) | Giorgio Giurato (Corresponding Author) et al |
| EPI_ISL_935127 | Italy/CAM-CRGS-158/2020 | 2020-11-14 | Europe/Italy/Campania/ | 1.AO Universitaria 'S. Giovanni di Dio e Ruggi D'Aragona, Scuola Medica Salernitana' Hospital / 2.UOC di Virologia e Microbiologia, Universit√† della Campania 'L. Vanvitelli' / 3.AO Universitaria 'Federico II' Napoli Hospital / 4.AORN 'San Giuseppe Moscati' Avellino Hospital / 5.AO 'San Pio - presidio G. Rummo' Benevento Hospital / 6.AO 'Sant'Anna e San Sebastiano' Caserta Hospital / 7.PO 'Maria Santissima Addolorata' Eboli Hospital / 8.Biogem Istituto di Ricerche Genetiche | 1. Genome Research Center for Health (CRGS) / 2. Laboratory of Molecular Medicine and Genomics(LMMGe) / 3. Center for Research in Pure and Applied Mathematics (CRMPA) | Giorgio Giurato (Corresponding Author) et al |

| | | | | | | |
|---|---|---|---|---|---|---|
| EPI_ISL_935128 | Italy/CAM-CRGS-159/2020 | 2020-11-16 | Europe/Italy/Campania/ | 1.AO Universitaria 'S. Giovanni di Dio e Ruggi D'Aragona, Scuola Medica Salernitana' Hospital / 2.UOC di Virologia e Microbiologia, Universit√† della Campania 'L. Vanvitelli' / 3.AO Universitaria 'Federico II' Napoli Hospital / 4.AORN 'San Giuseppe Moscati' Avellino Hospital / 5.AO 'San Pio - presidio G. Rummo' Benevento Hospital / 6.AO 'Sant'Anna e San Sebastiano' Caserta Hospital / 7.PO 'Maria Santissima Addolorata' Eboli Hospital / 8.Biogem Istituto di Ricerche Genetiche | 1. Genome Research Center for Health (CRGS) / 2. Laboratory of Molecular Medicine and Genomics(LMMGe) / 3. Center for Research in Pure and Applied Mathematics (CRMPA) | Giorgio Giurato (Corresponding Author) et al |
| EPI_ISL_935130 | Italy/CAM-CRGS-163/2020 | 2020-10-10 | Europe/Italy/Campania/ | 1.AO Universitaria 'S. Giovanni di Dio e Ruggi D'Aragona, Scuola Medica Salernitana' Hospital / 2.UOC di Virologia e Microbiologia, Universit√† della Campania 'L. Vanvitelli' / 3.AO Universitaria 'Federico II' Napoli Hospital / 4.AORN 'San Giuseppe Moscati' Avellino Hospital / 5.AO 'San Pio - presidio G. Rummo' Benevento Hospital / 6.AO 'Sant'Anna e San Sebastiano' Caserta Hospital / 7.PO 'Maria Santissima Addolorata' Eboli Hospital / 8.Biogem Istituto di Ricerche Genetiche | 1. Genome Research Center for Health (CRGS) / 2. Laboratory of Molecular Medicine and Genomics(LMMGe) / 3. Center for Research in Pure and Applied Mathematics (CRMPA) | Giorgio Giurato (Corresponding Author) et al |
| EPI_ISL_935131 | Italy/CAM-CRGS-168/2020 | 2020-10-11 | Europe/Italy/Campania/ | 1.AO Universitaria 'S. Giovanni di Dio e Ruggi D'Aragona, Scuola Medica Salernitana' Hospital / 2.UOC di Virologia e Microbiologia, Universit√† della Campania 'L. Vanvitelli' / 3.AO Universitaria 'Federico II' Napoli Hospital / 4.AORN 'San Giuseppe Moscati' Avellino Hospital / 5.AO 'San Pio - presidio G. Rummo' Benevento Hospital / 6.AO 'Sant'Anna e San Sebastiano' Caserta Hospital / 7.PO 'Maria Santissima Addolorata' Eboli Hospital / 8.Biogem Istituto di Ricerche Genetiche | 1. Genome Research Center for Health (CRGS) / 2. Laboratory of Molecular Medicine and Genomics(LMMGe) / 3. Center for Research in Pure and Applied Mathematics (CRMPA) | Giorgio Giurato (Corresponding Author) et al |

| | | | | | | |
|---|---|---|---|---|---|---|
| EPI_ISL_935133 | Italy/CAM-CRGS-174/2020 | 2020-10-13 | Europe/Italy/Campania/ | 1.AO Universitaria 'S. Giovanni di Dio e Ruggi D'Aragona, Scuola Medica Salernitana' Hospital / 2.UOC di Virologia e Microbiologia, Universit√† della Campania 'L. Vanvitelli' / 3.AO Universitaria 'Federico II' Napoli Hospital / 4.AORN 'San Giuseppe Moscati' Avellino Hospital / 5.AO 'San Pio - presidio G. Rummo' Benevento Hospital / 6.AO 'Sant'Anna e San Sebastiano' Caserta Hospital / 7.PO 'Maria Santissima Addolorata' Eboli Hospital / 8.Biogem Istituto di Ricerche Genetiche | 1. Genome Research Center for Health (CRGS) / 2. Laboratory of Molecular Medicine and Genomics(LMMGe) / 3. Center for Research in Pure and Applied Mathematics (CRMPA) | Giorgio Giurato (Corresponding Author) et al |
| EPI_ISL_935136 | Italy/CAM-CRGS-180/2020 | 2020-10-15 | Europe/Italy/Campania/ | 1.AO Universitaria 'S. Giovanni di Dio e Ruggi D'Aragona, Scuola Medica Salernitana' Hospital / 2.UOC di Virologia e Microbiologia, Universit√† della Campania 'L. Vanvitelli' / 3.AO Universitaria 'Federico II' Napoli Hospital / 4.AORN 'San Giuseppe Moscati' Avellino Hospital / 5.AO 'San Pio - presidio G. Rummo' Benevento Hospital / 6.AO 'Sant'Anna e San Sebastiano' Caserta Hospital / 7.PO 'Maria Santissima Addolorata' Eboli Hospital / 8.Biogem Istituto di Ricerche Genetiche | 1. Genome Research Center for Health (CRGS) / 2. Laboratory of Molecular Medicine and Genomics(LMMGe) / 3. Center for Research in Pure and Applied Mathematics (CRMPA) | Giorgio Giurato (Corresponding Author) et al |
| EPI_ISL_935138 | Italy/CAM-CRGS-186/2020 | 2020-10-17 | Europe/Italy/Campania/ | 1.AO Universitaria 'S. Giovanni di Dio e Ruggi D'Aragona, Scuola Medica Salernitana' Hospital / 2.UOC di Virologia e Microbiologia, Universit√† della Campania 'L. Vanvitelli' / 3.AO Universitaria 'Federico II' Napoli Hospital / 4.AORN 'San Giuseppe Moscati' Avellino Hospital / 5.AO 'San Pio - presidio G. Rummo' Benevento Hospital / 6.AO 'Sant'Anna e San Sebastiano' Caserta Hospital / 7.PO 'Maria Santissima Addolorata' Eboli Hospital / 8.Biogem Istituto di Ricerche Genetiche | 1. Genome Research Center for Health (CRGS) / 2. Laboratory of Molecular Medicine and Genomics(LMMGe) / 3. Center for Research in Pure and Applied Mathematics (CRMPA) | Giorgio Giurato (Corresponding Author) et al |

| | | | | | | |
|---|---|---|---|---|---|---|
| EPI_ISL_935140 | Italy/CAM-CRGS-191/2020 | 2020-10-18 | Europe/Italy/Campania/ | 1.AO Universitaria 'S. Giovanni di Dio e Ruggi D'Aragona, Scuola Medica Salernitana' Hospital / 2.UOC di Virologia e Microbiologia, Universit√† della Campania 'L. Vanvitelli' / 3.AO Universitaria 'Federico II' Napoli Hospital / 4.AORN 'San Giuseppe Moscati' Avellino Hospital / 5.AO 'San Pio - presidio G. Rummo' Benevento Hospital / 6.AO 'Sant'Anna e San Sebastiano' Caserta Hospital / 7.PO 'Maria Santissima Addolorata' Eboli Hospital / 8.Biogem Istituto di Ricerche Genetiche | 1. Genome Research Center for Health (CRGS) / 2. Laboratory of Molecular Medicine and Genomics(LMMGe) / 3. Center for Research in Pure and Applied Mathematics (CRMPA) | Giorgio Giurato (Corresponding Author) et al |
| EPI_ISL_935141 | Italy/CAM-CRGS-192/2020 | 2020-10-18 | Europe/Italy/Campania/ | 1.AO Universitaria 'S. Giovanni di Dio e Ruggi D'Aragona, Scuola Medica Salernitana' Hospital / 2.UOC di Virologia e Microbiologia, Universit√† della Campania 'L. Vanvitelli' / 3.AO Universitaria 'Federico II' Napoli Hospital / 4.AORN 'San Giuseppe Moscati' Avellino Hospital / 5.AO 'San Pio - presidio G. Rummo' Benevento Hospital / 6.AO 'Sant'Anna e San Sebastiano' Caserta Hospital / 7.PO 'Maria Santissima Addolorata' Eboli Hospital / 8.Biogem Istituto di Ricerche Genetiche | 1. Genome Research Center for Health (CRGS) / 2. Laboratory of Molecular Medicine and Genomics(LMMGe) / 3. Center for Research in Pure and Applied Mathematics (CRMPA) | Giorgio Giurato (Corresponding Author) et al |
| EPI_ISL_935142 | Italy/CAM-CRGS-193/2020 | 2020-10-18 | Europe/Italy/Campania/ | 1.AO Universitaria 'S. Giovanni di Dio e Ruggi D'Aragona, Scuola Medica Salernitana' Hospital / 2.UOC di Virologia e Microbiologia, Universit√† della Campania 'L. Vanvitelli' / 3.AO Universitaria 'Federico II' Napoli Hospital / 4.AORN 'San Giuseppe Moscati' Avellino Hospital / 5.AO 'San Pio - presidio G. Rummo' Benevento Hospital / 6.AO 'Sant'Anna e San Sebastiano' Caserta Hospital / 7.PO 'Maria Santissima Addolorata' Eboli Hospital / 8.Biogem Istituto di Ricerche Genetiche | 1. Genome Research Center for Health (CRGS) / 2. Laboratory of Molecular Medicine and Genomics(LMMGe) / 3. Center for Research in Pure and Applied Mathematics (CRMPA) | Giorgio Giurato (Corresponding Author) et al |

| | | | | | | |
|---|---|---|---|---|---|---|
| EPI_ISL_935143 | Italy/CAM-CRGS-196/2020 | 2020-10-19 | Europe/Italy/Campania/ | 1.AO Universitaria 'S. Giovanni di Dio e Ruggi D'Aragona, Scuola Medica Salernitana' Hospital / 2.UOC di Virologia e Microbiologia, Universit√† della Campania 'L. Vanvitelli' / 3.AO Universitaria 'Federico II' Napoli Hospital / 4.AORN 'San Giuseppe Moscati' Avellino Hospital / 5.AO 'San Pio - presidio G. Rummo' Benevento Hospital / 6.AO 'Sant'Anna e San Sebastiano' Caserta Hospital / 7.PO 'Maria Santissima Addolorata' Eboli Hospital / 8.Biogem Istituto di Ricerche Genetiche | 1. Genome Research Center for Health (CRGS) / 2. Laboratory of Molecular Medicine and Genomics(LMMGe) / 3. Center for Research in Pure and Applied Mathematics (CRMPA) | Giorgio Giurato (Corresponding Author) et al |
| EPI_ISL_935144 | Italy/CAM-CRGS-197/2020 | 2020-10-20 | Europe/Italy/Campania/ | 1.AO Universitaria 'S. Giovanni di Dio e Ruggi D'Aragona, Scuola Medica Salernitana' Hospital / 2.UOC di Virologia e Microbiologia, Universit√† della Campania 'L. Vanvitelli' / 3.AO Universitaria 'Federico II' Napoli Hospital / 4.AORN 'San Giuseppe Moscati' Avellino Hospital / 5.AO 'San Pio - presidio G. Rummo' Benevento Hospital / 6.AO 'Sant'Anna e San Sebastiano' Caserta Hospital / 7.PO 'Maria Santissima Addolorata' Eboli Hospital / 8.Biogem Istituto di Ricerche Genetiche | 1. Genome Research Center for Health (CRGS) / 2. Laboratory of Molecular Medicine and Genomics(LMMGe) / 3. Center for Research in Pure and Applied Mathematics (CRMPA) | Giorgio Giurato (Corresponding Author) et al |
| EPI_ISL_935147 | Italy/CAM-CRGS-201/2020 | 2020-10-21 | Europe/Italy/Campania/ | 1.AO Universitaria 'S. Giovanni di Dio e Ruggi D'Aragona, Scuola Medica Salernitana' Hospital / 2.UOC di Virologia e Microbiologia, Universit√† della Campania 'L. Vanvitelli' / 3.AO Universitaria 'Federico II' Napoli Hospital / 4.AORN 'San Giuseppe Moscati' Avellino Hospital / 5.AO 'San Pio - presidio G. Rummo' Benevento Hospital / 6.AO 'Sant'Anna e San Sebastiano' Caserta Hospital / 7.PO 'Maria Santissima Addolorata' Eboli Hospital / 8.Biogem Istituto di Ricerche Genetiche | 1. Genome Research Center for Health (CRGS) / 2. Laboratory of Molecular Medicine and Genomics(LMMGe) / 3. Center for Research in Pure and Applied Mathematics (CRMPA) | Giorgio Giurato (Corresponding Author) et al |

| | | | | | | |
|---|---|---|---|---|---|---|
| EPI_ISL_935150 | Italy/CAM-CRGS-205/2020 | 2020-11-04 | Europe/Italy/Campania/ | 1.AO Universitaria 'S. Giovanni di Dio e Ruggi D'Aragona, Scuola Medica Salernitana' Hospital / 2.UOC di Virologia e Microbiologia, Universit√† della Campania 'L. Vanvitelli' / 3.AO Universitaria 'Federico II' Napoli Hospital / 4.AORN 'San Giuseppe Moscati' Avellino Hospital / 5.AO 'San Pio - presidio G. Rummo' Benevento Hospital / 6.AO 'Sant'Anna e San Sebastiano' Caserta Hospital / 7.PO 'Maria Santissima Addolorata' Eboli Hospital / 8.Biogem Istituto di Ricerche Genetiche | 1. Genome Research Center for Health (CRGS) / 2. Laboratory of Molecular Medicine and Genomics(LMMGe) / 3. Center for Research in Pure and Applied Mathematics (CRMPA) | Giorgio Giurato (Corresponding Author) et al |
| EPI_ISL_935151 | Italy/CAM-CRGS-209/2020 | 2020-11-04 | Europe/Italy/Campania/ | 1.AO Universitaria 'S. Giovanni di Dio e Ruggi D'Aragona, Scuola Medica Salernitana' Hospital / 2.UOC di Virologia e Microbiologia, Universit√† della Campania 'L. Vanvitelli' / 3.AO Universitaria 'Federico II' Napoli Hospital / 4.AORN 'San Giuseppe Moscati' Avellino Hospital / 5.AO 'San Pio - presidio G. Rummo' Benevento Hospital / 6.AO 'Sant'Anna e San Sebastiano' Caserta Hospital / 7.PO 'Maria Santissima Addolorata' Eboli Hospital / 8.Biogem Istituto di Ricerche Genetiche | 1. Genome Research Center for Health (CRGS) / 2. Laboratory of Molecular Medicine and Genomics(LMMGe) / 3. Center for Research in Pure and Applied Mathematics (CRMPA) | Giorgio Giurato (Corresponding Author) et al |
| EPI_ISL_935152 | Italy/CAM-CRGS-212/2020 | 2020-11-04 | Europe/Italy/Campania/ | 1.AO Universitaria 'S. Giovanni di Dio e Ruggi D'Aragona, Scuola Medica Salernitana' Hospital / 2.UOC di Virologia e Microbiologia, Universit√† della Campania 'L. Vanvitelli' / 3.AO Universitaria 'Federico II' Napoli Hospital / 4.AORN 'San Giuseppe Moscati' Avellino Hospital / 5.AO 'San Pio - presidio G. Rummo' Benevento Hospital / 6.AO 'Sant'Anna e San Sebastiano' Caserta Hospital / 7.PO 'Maria Santissima Addolorata' Eboli Hospital / 8.Biogem Istituto di Ricerche Genetiche | 1. Genome Research Center for Health (CRGS) / 2. Laboratory of Molecular Medicine and Genomics(LMMGe) / 3. Center for Research in Pure and Applied Mathematics (CRMPA) | Giorgio Giurato (Corresponding Author) et al |

| | | | | | | |
|---|---|---|---|---|---|---|
| EPI_ISL_935153 | Italy/CAM-CRGS-214/2020 | 2020-11-05 | Europe/Italy/Campania/ | 1.AO Universitaria 'S. Giovanni di Dio e Ruggi D'Aragona, Scuola Medica Salernitana' Hospital / 2.UOC di Virologia e Microbiologia, Universit√† della Campania 'L. Vanvitelli' / 3.AO Universitaria 'Federico II' Napoli Hospital / 4.AORN 'San Giuseppe Moscati' Avellino Hospital / 5.AO 'San Pio - presidio G. Rummo' Benevento Hospital / 6.AO 'Sant'Anna e San Sebastiano' Caserta Hospital / 7.PO 'Maria Santissima Addolorata' Eboli Hospital / 8.Biogem Istituto di Ricerche Genetiche | 1. Genome Research Center for Health (CRGS) / 2. Laboratory of Molecular Medicine and Genomics(LMMGe) / 3. Center for Research in Pure and Applied Mathematics (CRMPA) | Giorgio Giurato (Corresponding Author) et al |
| EPI_ISL_935154 | Italy/CAM-CRGS-218/2020 | 2020-11-05 | Europe/Italy/Campania/ | 1.AO Universitaria 'S. Giovanni di Dio e Ruggi D'Aragona, Scuola Medica Salernitana' Hospital / 2.UOC di Virologia e Microbiologia, Universit√† della Campania 'L. Vanvitelli' / 3.AO Universitaria 'Federico II' Napoli Hospital / 4.AORN 'San Giuseppe Moscati' Avellino Hospital / 5.AO 'San Pio - presidio G. Rummo' Benevento Hospital / 6.AO 'Sant'Anna e San Sebastiano' Caserta Hospital / 7.PO 'Maria Santissima Addolorata' Eboli Hospital / 8.Biogem Istituto di Ricerche Genetiche | 1. Genome Research Center for Health (CRGS) / 2. Laboratory of Molecular Medicine and Genomics(LMMGe) / 3. Center for Research in Pure and Applied Mathematics (CRMPA) | Giorgio Giurato (Corresponding Author) et al |
| EPI_ISL_935156 | Italy/ABR-IZSGC-TE11124/2021 | 2021-01-08 | Europe/Italy/Abruzzo/ | SIESP CHIETI | Istituto Zooprofilattico Sperimentale dell‚ÄôAbruzzo e Molise ‚ÄúG. Caporale‚Äù | Lorusso A et al |
| EPI_ISL_935157 | Italy/ABR-IZSGC-TE6985/2021 | 2021-01-05 | Europe/Italy/Abruzzo/ | SIESP CHIETI-DRIVE IN ORTONA | Istituto Zooprofilattico Sperimentale dell‚ÄôAbruzzo e Molise ‚ÄúG. Caporale‚Äù | Lorusso A et al |

| | | | | | | |
|---|---|---|---|---|---|---|
| EPI_ISL_935160 | Italy/ABR-IZSGC-TE10936/2021 | 2021-01-08 | Europe/Italy/Abruzzo/ | SIESP CHIETI-DRIVE IN ORTONA | Istituto Zooprofilattico Sperimentale dell,ÄôAbruzzo e Molise ,ÄúG. Caporale,Äù | Lorusso A et al |
| EPI_ISL_935161 | Italy/ABR-IZSGC-TE12641/2021 | 2021-01-08 | Europe/Italy/Abruzzo/ | SIESP CHIETI | Istituto Zooprofilattico Sperimentale dell,ÄôAbruzzo e Molise ,ÄúG. Caporale,Äù | Lorusso A et al |
| EPI_ISL_935162 | Italy/ABR-IZSGC-TE12615/2021 | 2021-01-08 | Europe/Italy/Abruzzo/ | SIESP CHIETI | Istituto Zooprofilattico Sperimentale dell,ÄôAbruzzo e Molise ,ÄúG. Caporale,Äù | Lorusso A et al |
| EPI_ISL_935163 | Italy/ABR-IZSGC-TE10934/2021 | 2021-01-08 | Europe/Italy/Abruzzo/ | SIESP CHIETI-DRIVE IN ORTONA | Istituto Zooprofilattico Sperimentale dell,ÄôAbruzzo e Molise ,ÄúG. Caporale,Äù | Lorusso A et al |
| EPI_ISL_935164 | Italy/ABR-IZSGC-TE10938/2021 | 2021-01-08 | Europe/Italy/Abruzzo/ | SIESP CHIETI-DRIVE IN ORTONA | Istituto Zooprofilattico Sperimentale dell,ÄôAbruzzo e Molise ,ÄúG. Caporale,Äù | Lorusso A et al |
| EPI_ISL_935165 | Italy/ABR-IZSGC-TE12678/2021 | 2021-01-08 | Europe/Italy/Abruzzo/ | SIESP CHIETI | Istituto Zooprofilattico Sperimentale dell,ÄôAbruzzo e Molise ,ÄúG. Caporale,Äù | Lorusso A et al |

| | | | | | | |
|---|---|---|---|---|---|---|
| EPI_ISL_935166 | Italy/ABR-IZSGC-TE12671/2021 | 2021-01-08 | Europe/Italy/Abruzzo/ | SIESP CHIETI | Istituto Zooprofilattico Sperimentale dell‚ÄôAbruzzo e Molise ‚ÄúG. Caporale‚Äù | Lorusso A et al |
| EPI_ISL_935169 | Italy/ABR-IZSGC-TE11125/2021 | 2021-01-08 | Europe/Italy/Abruzzo/ | SIESP CHIETI | Istituto Zooprofilattico Sperimentale dell‚ÄôAbruzzo e Molise ‚ÄúG. Caporale‚Äù | Lorusso A et al |
| EPI_ISL_935175 | USA/WY-WPHL-21009666/2021 | 2021-01-27 | North America/USA/Wyoming/ | Wyoming Public Health Laboratory | Wyoming Public Health Laboratory | Noah Hull et al |
| EPI_ISL_935179 | USA/WY-WPHL-21009793/2021 | 2021-01-27 | North America/USA/Wyoming/ | Wyoming Public Health Laboratory | Wyoming Public Health Laboratory | Noah Hull et al |
| EPI_ISL_935186 | USA/WY-WPHL-21010885/2021 | 2021-02-01 | North America/USA/Wyoming/ | Wyoming Public Health Laboratory | Wyoming Public Health Laboratory | Noah Hull et al |
| EPI_ISL_935206 | Belgium/rega-2110/2020 | 2020-01-10 | Europe/Belgium/Oostende/ | KU Leuven, Rega Institute, Clinical and Epidemiological Virology | KU Leuven, Rega Institute, Clinical and Epidemiological Virology | Tony Wawina-Bokalanga et al |
| EPI_ISL_935209 | Belgium/rega-2113/2020 | 2020-12-27 | Europe/Belgium/Oostende/ | KU Leuven, Rega Institute, Clinical and Epidemiological Virology | KU Leuven, Rega Institute, Clinical and Epidemiological Virology | Tony Wawina-Bokalanga et al |
| EPI_ISL_935216 | Belgium/rega-2120/2020 | 2020-12-20 | Europe/Belgium/Oostende/ | KU Leuven, Rega Institute, Clinical and Epidemiological Virology | KU Leuven, Rega Institute, Clinical and Epidemiological Virology | Tony Wawina-Bokalanga et al |
| EPI_ISL_935249 | Belgium/rega-2154/2021 | 2021-01-19 | Europe/Belgium/Brussels/Laken | KU Leuven, Rega Institute, Clinical and Epidemiological Virology | KU Leuven, Rega Institute, Clinical and Epidemiological Virology | Tony Wawina-Bokalanga et al |

| | | | | | | |
|---|---|---|---|---|---|---|
| EPI_ISL_935276 | Belgium/rega-2181/2021 | 2021-01-11 | Europe/Belgium/Leuven/Bekkevoort | KU Leuven, Rega Institute, Clinical and Epidemiological Virology | KU Leuven, Rega Institute, Clinical and Epidemiological Virology | Tony Wawina-Bokalanga et al |
| EPI_ISL_935351 | Belgium/rega-2259/2021 | 2021-01-21 | Europe/Belgium/Turnhout/Arendonk | KU Leuven, Rega Institute, Clinical and Epidemiological Virology | KU Leuven, Rega Institute, Clinical and Epidemiological Virology | Tony Wawina-Bokalanga et al |
| EPI_ISL_935372 | USA/FL-BPHL-2851/2020 | 2020-09-15 | North America/USA/Florida/ | Florida Bureau of Public Health Laboratories | Florida Bureau of Public Health Laboratories | Sarah Schmedes et al |
| EPI_ISL_935373 | USA/FL-BPHL-2852/2020 | 2020-09-16 | North America/USA/Florida/ | Florida Bureau of Public Health Laboratories | Florida Bureau of Public Health Laboratories | Sarah Schmedes et al |
| EPI_ISL_935376 | USA/FL-BPHL-2855/2020 | 2020-09-17 | North America/USA/Florida/ | Florida Bureau of Public Health Laboratories | Florida Bureau of Public Health Laboratories | Sarah Schmedes et al |
| EPI_ISL_935383 | USA/FL-BPHL-2862/2020 | 2020-09-18 | North America/USA/Florida/ | Florida Bureau of Public Health Laboratories | Florida Bureau of Public Health Laboratories | Sarah Schmedes et al |
| EPI_ISL_935384 | USA/FL-BPHL-2863/2020 | 2020-09-18 | North America/USA/Florida/ | Florida Bureau of Public Health Laboratories | Florida Bureau of Public Health Laboratories | Sarah Schmedes et al |
| EPI_ISL_935386 | USA/FL-BPHL-2865/2020 | 2020-09-17 | North America/USA/Florida/ | Florida Bureau of Public Health Laboratories | Florida Bureau of Public Health Laboratories | Sarah Schmedes et al |
| EPI_ISL_935387 | USA/FL-BPHL-2866/2020 | 2020-09-17 | North America/USA/Florida/ | Florida Bureau of Public Health Laboratories | Florida Bureau of Public Health Laboratories | Sarah Schmedes et al |
| EPI_ISL_935390 | USA/FL-BPHL-2869/2020 | 2020-09-18 | North America/USA/Florida/ | Florida Bureau of Public Health Laboratories | Florida Bureau of Public Health Laboratories | Sarah Schmedes et al |
| EPI_ISL_935391 | USA/FL-BPHL-2870/2020 | 2020-09-18 | North America/USA/Florida/ | Florida Bureau of Public Health Laboratories | Florida Bureau of Public Health Laboratories | Sarah Schmedes et al |
| EPI_ISL_935396 | USA/FL-BPHL-2875/2020 | 2020-09-18 | North America/USA/Florida/ | Florida Bureau of Public Health Laboratories | Florida Bureau of Public Health Laboratories | Sarah Schmedes et al |

| | | | | | | |
|---|---|---|---|---|---|---|
| EPI_ISL_935399 | USA/FL-BPHL-2878/2020 | 2020-09-17 | North America/USA/Florida/ | Florida Bureau of Public Health Laboratories | Florida Bureau of Public Health Laboratories | Sarah Schmedes et al |
| EPI_ISL_935405 | USA/FL-BPHL-2884/2020 | 2020-09-18 | North America/USA/Florida/ | Florida Bureau of Public Health Laboratories | Florida Bureau of Public Health Laboratories | Sarah Schmedes et al |
| EPI_ISL_935413 | USA/FL-BPHL-2892/2020 | 2020-09-21 | North America/USA/Florida/ | Florida Bureau of Public Health Laboratories | Florida Bureau of Public Health Laboratories | Sarah Schmedes et al |
| EPI_ISL_935414 | USA/FL-BPHL-2893/2020 | 2020-09-21 | North America/USA/Florida/ | Florida Bureau of Public Health Laboratories | Florida Bureau of Public Health Laboratories | Sarah Schmedes et al |
| EPI_ISL_935415 | USA/FL-BPHL-2894/2020 | 2020-11-16 | North America/USA/Florida/ | Florida Bureau of Public Health Laboratories | Florida Bureau of Public Health Laboratories | Sarah Schmedes et al |
| EPI_ISL_935429 | USA/FL-BPHL-2908/2020 | 2020-11-16 | North America/USA/Florida/ | Florida Bureau of Public Health Laboratories | Florida Bureau of Public Health Laboratories | Sarah Schmedes et al |
| EPI_ISL_935432 | USA/FL-BPHL-2911/2020 | 2020-11-16 | North America/USA/Florida/ | Florida Bureau of Public Health Laboratories | Florida Bureau of Public Health Laboratories | Sarah Schmedes et al |
| EPI_ISL_935448 | USA/FL-BPHL-2927/2020 | 2020-11-17 | North America/USA/Florida/ | Florida Bureau of Public Health Laboratories | Florida Bureau of Public Health Laboratories | Sarah Schmedes et al |
| EPI_ISL_935455 | USA/FL-BPHL-2934/2020 | 2020-11-18 | North America/USA/Florida/ | Florida Bureau of Public Health Laboratories | Florida Bureau of Public Health Laboratories | Sarah Schmedes et al |
| EPI_ISL_935457 | USA/FL-BPHL-2936/2020 | 2020-11-19 | North America/USA/Florida/ | Florida Bureau of Public Health Laboratories | Florida Bureau of Public Health Laboratories | Sarah Schmedes et al |
| EPI_ISL_935466 | USA/FL-BPHL-2945/2020 | 2020-11-30 | North America/USA/Florida/ | Florida Bureau of Public Health Laboratories | Florida Bureau of Public Health Laboratories | Sarah Schmedes et al |
| EPI_ISL_935471 | USA/FL-BPHL-2950/2020 | 2020-12-01 | North America/USA/Florida/ | Florida Bureau of Public Health Laboratories | Florida Bureau of Public Health Laboratories | Sarah Schmedes et al |
| EPI_ISL_935481 | USA/FL-BPHL-2960/2020 | 2020-12-02 | North America/USA/Florida/ | Florida Bureau of Public Health Laboratories | Florida Bureau of Public Health Laboratories | Sarah Schmedes et al |

| | | | | | | |
|---|---|---|---|---|---|---|
| EPI_ISL_935496 | USA/FL-BPHL-2975/2020 | 2020-12-03 | North America/USA/Florida/ | Florida Bureau of Public Health Laboratories | Florida Bureau of Public Health Laboratories | Sarah Schmedes et al |
| EPI_ISL_935499 | USA/FL-BPHL-2978/2020 | 2020-12-03 | North America/USA/Florida/ | Florida Bureau of Public Health Laboratories | Florida Bureau of Public Health Laboratories | Sarah Schmedes et al |
| EPI_ISL_935505 | USA/FL-BPHL-2984/2020 | 2020-12-03 | North America/USA/Florida/ | Florida Bureau of Public Health Laboratories | Florida Bureau of Public Health Laboratories | Sarah Schmedes et al |
| EPI_ISL_935512 | USA/FL-BPHL-2991/2020 | 2020-12-04 | North America/USA/Florida/ | Florida Bureau of Public Health Laboratories | Florida Bureau of Public Health Laboratories | Sarah Schmedes et al |
| EPI_ISL_935515 | USA/FL-BPHL-2994/2020 | 2020-12-04 | North America/USA/Florida/ | Florida Bureau of Public Health Laboratories | Florida Bureau of Public Health Laboratories | Sarah Schmedes et al |
| EPI_ISL_935516 | USA/FL-BPHL-2995/2020 | 2020-12-04 | North America/USA/Florida/ | Florida Bureau of Public Health Laboratories | Florida Bureau of Public Health Laboratories | Sarah Schmedes et al |
| EPI_ISL_935523 | USA/MA-Broad_UMMS-00003/2020 | 2020-08-29 | North America/USA/Massachusetts/ | University of Massachusetts Medical School | Infectious Disease Program, Broad Institute of Harvard and MIT | Tomkins-Tinch et al |
| EPI_ISL_935536 | France/GES-IPP01445/2021 | 2021-01-18 | Europe/France/Grand Est/Lingolsheim | Labo Analyses Med | National Reference Center for Viruses of Respiratory Infections, Institut Pasteur, Paris | Marion Barbet et al |
| EPI_ISL_935549 | France/GES-IPP01462/2021 | 2021-01-18 | Europe/France/Grand Est/Strasbourg | Labo Analyses Med | National Reference Center for Viruses of Respiratory Infections, Institut Pasteur, Paris | Marion Barbet et al |
| EPI_ISL_935552 | France/CVL-IPP01471/2021 | 2021-01-14 | Europe/France/Centre-Val de Loire/Tou | Hopital | National Reference Center for Viruses of Respiratory Infections, Institut Pasteur, Paris | Marion Barbet et al |

| | | | | | | |
|---|---|---|---|---|---|---|
| EPI_ISL_935553 | France/CVL-IPP01472/2021 | 2021-01-14 | Europe/France/Centre-Val de Loire/Tour | Hopital | National Reference Center for Viruses of Respiratory Infections, Institut Pasteur, Paris | Marion Barbet et al |
| EPI_ISL_935573 | France/HDF-IPP01502/2021 | 2021-01-15 | Europe/France/Hauts de France/Amiens | Hopital | National Reference Center for Viruses of Respiratory Infections, Institut Pasteur, Paris | Marion Barbet et al |
| EPI_ISL_935591 | France/IDF-IPP01543/2021 | 2021-01-18 | Europe/France/Ile de France/Thiais | Labo Analyses med | National Reference Center for Viruses of Respiratory Infections, Institut Pasteur, Paris | Marion Barbet et al |
| EPI_ISL_935601 | France/IDF-IPP01556/2021 | 2021-01-16 | Europe/France/Ile de France/Thiais | Labo Analyses med | National Reference Center for Viruses of Respiratory Infections, Institut Pasteur, Paris | Marion Barbet et al |
| EPI_ISL_935622 | France/IDF-IPP01584/2021 | 2021-01-16 | Europe/France/Ile de France/Thiais | Labo Analyses med | National Reference Center for Viruses of Respiratory Infections, Institut Pasteur, Paris | Marion Barbet et al |
| EPI_ISL_935623 | France/IDF-IPP01585/2021 | 2021-01-16 | Europe/France/Ile de France/Thiais | Labo Analyses med | National Reference Center for Viruses of Respiratory Infections, Institut Pasteur, Paris | Marion Barbet et al |
| EPI_ISL_935701 | USA/TX-HHD-2102026517/2020 | 2020-12-23 | North America/USA/Texas/ | Houston Health Dept. | Houston Health Dept. | Ryker Penn et al |
| EPI_ISL_935707 | USA/TX-HHD-2102027483/2020 | 2020-12-26 | North America/USA/Texas/ | Houston Health Dept. | Houston Health Dept. | Ryker Penn et al |

| | | | | | | |
|---|---|---|---|---|---|---|
| EPI_ISL_935709 | USA/TX-HHD-2102027629/2020 | 2020-12-27 | North America/USA/Texas/ | Houston Health Dept. | Houston Health Dept. | Ryker Penn et al |
| EPI_ISL_935717 | USA/TX-HHD-2102026552/2021 | 2021-01-03 | North America/USA/Texas/ | Houston Health Dept. | Houston Health Dept. | Ryker Penn et al |
| EPI_ISL_935722 | USA/TX-HHD-2102026642/2021 | 2021-01-03 | North America/USA/Texas/ | Houston Health Dept. | Houston Health Dept. | Ryker Penn et al |
| EPI_ISL_935726 | USA/TX-HHD-2102014896/2020 | 2020-12-23 | North America/USA/Texas/ | Houston Health Dept. | Houston Health Dept. | Ryker Penn et al |
| EPI_ISL_935738 | USA/TX-HHD-2102016679/2020 | 2020-12-23 | North America/USA/Texas/ | Houston Health Dept. | Houston Health Dept. | Ryker Penn et al |
| EPI_ISL_935744 | USA/TX-HHD-2102017466/2020 | 2020-12-26 | North America/USA/Texas/ | Houston Health Dept. | Houston Health Dept. | Ryker Penn et al |
| EPI_ISL_935747 | USA/TX-HHD-2102017531/2020 | 2020-12-26 | North America/USA/Texas/ | Houston Health Dept. | Houston Health Dept. | Ryker Penn et al |
| EPI_ISL_935753 | Canada/MB-NML-863/2020 | 2020-03-29 | North America/Canada/Manitoba/ | Cadham Provincial laboratory | National Microbiology Laboratory (NML) | Anna Majer et al |
| EPI_ISL_935754 | Canada/MB-NML-1149/2020 | 2020-07-23 | North America/Canada/Manitoba/ | Cadham Provincial laboratory | National Microbiology Laboratory (NML) | Anna Majer et al |
| EPI_ISL_935755 | Canada/MB-NML-1151/2020 | 2020-07-23 | North America/Canada/Manitoba/ | Cadham Provincial laboratory | National Microbiology Laboratory (NML) | Anna Majer et al |
| EPI_ISL_935756 | Canada/MB-NML-1160/2020 | 2020-07-24 | North America/Canada/Manitoba/ | Cadham Provincial laboratory | National Microbiology Laboratory (NML) | Anna Majer et al |
| EPI_ISL_935757 | Canada/MB-NML-1414/2020 | 2020-08-14 | North America/Canada/Manitoba/ | Cadham Provincial laboratory | National Microbiology Laboratory (NML) | Anna Majer et al |
| EPI_ISL_935758 | Canada/MB-NML-1415/2020 | 2020-08-20 | North America/Canada/Manitoba/ | Cadham Provincial laboratory | National Microbiology Laboratory (NML) | Anna Majer et al |
| EPI_ISL_935759 | Canada/MB-NML-1416/2020 | 2020-08-20 | North America/Canada/Manitoba/ | Cadham Provincial laboratory | National Microbiology Laboratory (NML) | Anna Majer et al |
| EPI_ISL_935760 | Canada/MB-NML-1417/2020 | 2020-08-19 | North America/Canada/Manitoba/ | Cadham Provincial laboratory | National Microbiology Laboratory (NML) | Anna Majer et al |
| EPI_ISL_935761 | Canada/MB-NML-1418/2020 | 2020-08-19 | North America/Canada/Manitoba/ | Cadham Provincial laboratory | National Microbiology Laboratory (NML) | Anna Majer et al |

| | | | | | | |
|---|---|---|---|---|---|---|
| EPI_ISL_935762 | Canada/MB-NML-1419/2020 | 2020-08-22 | North America/Canada/Manitoba/ | Cadham Provincial laboratory | National Microbiology Laboratory (NML) | Anna Majer et al |
| EPI_ISL_935763 | Canada/MB-NML-1420/2020 | 2020-08-22 | North America/Canada/Manitoba/ | Cadham Provincial laboratory | National Microbiology Laboratory (NML) | Anna Majer et al |
| EPI_ISL_935764 | Canada/MB-NML-1421/2020 | 2020-08-28 | North America/Canada/Manitoba/ | Cadham Provincial laboratory | National Microbiology Laboratory (NML) | Anna Majer et al |
| EPI_ISL_935765 | Canada/MB-NML-1424/2020 | 2020-08-24 | North America/Canada/Manitoba/ | Cadham Provincial laboratory | National Microbiology Laboratory (NML) | Anna Majer et al |
| EPI_ISL_935766 | Canada/MB-NML-1425/2020 | 2020-08-26 | North America/Canada/Manitoba/ | Cadham Provincial laboratory | National Microbiology Laboratory (NML) | Anna Majer et al |
| EPI_ISL_935767 | Canada/MB-NML-1426/2020 | 2020-08-25 | North America/Canada/Manitoba/ | Cadham Provincial laboratory | National Microbiology Laboratory (NML) | Anna Majer et al |
| EPI_ISL_935768 | Canada/MB-NML-1428/2020 | 2020-08-25 | North America/Canada/Manitoba/ | Cadham Provincial laboratory | National Microbiology Laboratory (NML) | Anna Majer et al |
| EPI_ISL_935769 | Canada/MB-NML-1430/2020 | 2020-08-25 | North America/Canada/Manitoba/ | Cadham Provincial laboratory | National Microbiology Laboratory (NML) | Anna Majer et al |
| EPI_ISL_935770 | Canada/MB-NML-1431/2020 | 2020-08-25 | North America/Canada/Manitoba/ | Cadham Provincial laboratory | National Microbiology Laboratory (NML) | Anna Majer et al |
| EPI_ISL_935771 | Canada/MB-NML-1433/2020 | 2020-08-24 | North America/Canada/Manitoba/ | Cadham Provincial laboratory | National Microbiology Laboratory (NML) | Anna Majer et al |
| EPI_ISL_935772 | Canada/MB-NML-1434/2020 | 2020-08-24 | North America/Canada/Manitoba/ | Cadham Provincial laboratory | National Microbiology Laboratory (NML) | Anna Majer et al |
| EPI_ISL_935774 | Canada/MB-NML-1436/2020 | 2020-08-25 | North America/Canada/Manitoba/ | Cadham Provincial laboratory | National Microbiology Laboratory (NML) | Anna Majer et al |
| EPI_ISL_935775 | Canada/MB-NML-1437/2020 | 2020-08-25 | North America/Canada/Manitoba/ | Cadham Provincial laboratory | National Microbiology Laboratory (NML) | Anna Majer et al |

| | | | | | | |
|---|---|---|---|---|---|---|
| EPI_ISL_935776 | Canada/MB-NML-1438/2020 | 2020-08-25 | North America/Canada/Manitoba/ | Cadham Provincial laboratory | National Microbiology Laboratory (NML) | Anna Majer et al |
| EPI_ISL_935777 | Canada/MB-NML-1439/2020 | 2020-08-25 | North America/Canada/Manitoba/ | Cadham Provincial laboratory | National Microbiology Laboratory (NML) | Anna Majer et al |
| EPI_ISL_935778 | Canada/MB-NML-1440/2020 | 2020-08-25 | North America/Canada/Manitoba/ | Cadham Provincial laboratory | National Microbiology Laboratory (NML) | Anna Majer et al |
| EPI_ISL_935779 | Canada/MB-NML-1441/2020 | 2020-08-25 | North America/Canada/Manitoba/ | Cadham Provincial laboratory | National Microbiology Laboratory (NML) | Anna Majer et al |
| EPI_ISL_935780 | Canada/MB-NML-1444/2020 | 2020-08-25 | North America/Canada/Manitoba/ | Cadham Provincial laboratory | National Microbiology Laboratory (NML) | Anna Majer et al |
| EPI_ISL_935781 | Canada/MB-NML-1445/2020 | 2020-08-24 | North America/Canada/Manitoba/ | Cadham Provincial laboratory | National Microbiology Laboratory (NML) | Anna Majer et al |
| EPI_ISL_935782 | Canada/MB-NML-1446/2020 | 2020-08-24 | North America/Canada/Manitoba/ | Cadham Provincial laboratory | National Microbiology Laboratory (NML) | Anna Majer et al |
| EPI_ISL_935783 | Canada/MB-NML-1447/2020 | 2020-08-24 | North America/Canada/Manitoba/ | Cadham Provincial laboratory | National Microbiology Laboratory (NML) | Anna Majer et al |
| EPI_ISL_935784 | Canada/MB-NML-1449/2020 | 2020-08-25 | North America/Canada/Manitoba/ | Cadham Provincial laboratory | National Microbiology Laboratory (NML) | Anna Majer et al |
| EPI_ISL_935785 | Canada/MB-NML-1450/2020 | 2020-08-24 | North America/Canada/Manitoba/ | Cadham Provincial laboratory | National Microbiology Laboratory (NML) | Anna Majer et al |
| EPI_ISL_935786 | Canada/MB-NML-1451/2020 | 2020-08-23 | North America/Canada/Manitoba/ | Cadham Provincial laboratory | National Microbiology Laboratory (NML) | Anna Majer et al |
| EPI_ISL_935787 | Canada/MB-NML-1452/2020 | 2020-08-22 | North America/Canada/Manitoba/ | Cadham Provincial laboratory | National Microbiology Laboratory (NML) | Anna Majer et al |
| EPI_ISL_935788 | Canada/MB-NML-1461/2020 | 2020-08-12 | North America/Canada/Manitoba/ | Cadham Provincial laboratory | National Microbiology Laboratory (NML) | Anna Majer et al |

| | | | | | | |
|---|---|---|---|---|---|---|
| EPI_ISL_935789 | Canada/MB-NML-1463/2020 | 2020-08-12 | North America/Canada/Manitoba/ | Cadham Provincial laboratory | National Microbiology Laboratory (NML) | Anna Majer et al |
| EPI_ISL_935790 | Canada/MB-NML-1464/2020 | 2020-08-07 | North America/Canada/Manitoba/ | Cadham Provincial laboratory | National Microbiology Laboratory (NML) | Anna Majer et al |
| EPI_ISL_935791 | Canada/MB-NML-1465/2020 | 2020-09-14 | North America/Canada/Manitoba/ | Cadham Provincial laboratory | National Microbiology Laboratory (NML) | Anna Majer et al |
| EPI_ISL_935792 | Canada/MB-NML-1467/2020 | 2020-08-10 | North America/Canada/Manitoba/ | Cadham Provincial laboratory | National Microbiology Laboratory (NML) | Anna Majer et al |
| EPI_ISL_935793 | Canada/MB-NML-1468/2020 | 2020-08-12 | North America/Canada/Manitoba/ | Cadham Provincial laboratory | National Microbiology Laboratory (NML) | Anna Majer et al |
| EPI_ISL_935794 | Canada/MB-NML-1469/2020 | 2020-08-14 | North America/Canada/Manitoba/ | Cadham Provincial laboratory | National Microbiology Laboratory (NML) | Anna Majer et al |
| EPI_ISL_935795 | Canada/MB-NML-1471/2020 | 2020-08-10 | North America/Canada/Manitoba/ | Cadham Provincial laboratory | National Microbiology Laboratory (NML) | Anna Majer et al |
| EPI_ISL_935796 | Canada/MB-NML-1475/2020 | 2020-08-10 | North America/Canada/Manitoba/ | Cadham Provincial laboratory | National Microbiology Laboratory (NML) | Anna Majer et al |
| EPI_ISL_935797 | Canada/MB-NML-1477/2020 | 2020-08-13 | North America/Canada/Manitoba/ | Cadham Provincial laboratory | National Microbiology Laboratory (NML) | Anna Majer et al |
| EPI_ISL_935798 | Canada/MB-NML-1478/2020 | 2020-08-14 | North America/Canada/Manitoba/ | Cadham Provincial laboratory | National Microbiology Laboratory (NML) | Anna Majer et al |
| EPI_ISL_935799 | Canada/MB-NML-1479/2020 | 2020-08-14 | North America/Canada/Manitoba/ | Cadham Provincial laboratory | National Microbiology Laboratory (NML) | Anna Majer et al |
| EPI_ISL_935800 | Canada/MB-NML-1481/2020 | 2020-08-12 | North America/Canada/Manitoba/ | Cadham Provincial laboratory | National Microbiology Laboratory (NML) | Anna Majer et al |
| EPI_ISL_935801 | Canada/MB-NML-1482/2020 | 2020-08-10 | North America/Canada/Manitoba/ | Cadham Provincial laboratory | National Microbiology Laboratory (NML) | Anna Majer et al |

| | | | | | | |
|---|---|---|---|---|---|---|
| EPI_ISL_935802 | Canada/MB-NML-1483/2020 | 2020-08-11 | North America/Canada/Manitoba/ | Cadham Provincial laboratory | National Microbiology Laboratory (NML) | Anna Majer et al |
| EPI_ISL_935803 | Canada/MB-NML-1484/2020 | 2020-08-11 | North America/Canada/Manitoba/ | Cadham Provincial laboratory | National Microbiology Laboratory (NML) | Anna Majer et al |
| EPI_ISL_935804 | Canada/MB-NML-1485/2020 | 2020-08-11 | North America/Canada/Manitoba/ | Cadham Provincial laboratory | National Microbiology Laboratory (NML) | Anna Majer et al |
| EPI_ISL_935805 | Canada/MB-NML-1486/2020 | 2020-08-12 | North America/Canada/Manitoba/ | Cadham Provincial laboratory | National Microbiology Laboratory (NML) | Anna Majer et al |
| EPI_ISL_935806 | Canada/MB-NML-1487/2020 | 2020-08-12 | North America/Canada/Manitoba/ | Cadham Provincial laboratory | National Microbiology Laboratory (NML) | Anna Majer et al |
| EPI_ISL_935807 | Canada/MB-NML-1488/2020 | 2020-08-12 | North America/Canada/Manitoba/ | Cadham Provincial laboratory | National Microbiology Laboratory (NML) | Anna Majer et al |
| EPI_ISL_935808 | Canada/MB-NML-1489/2020 | 2020-08-13 | North America/Canada/Manitoba/ | Cadham Provincial laboratory | National Microbiology Laboratory (NML) | Anna Majer et al |
| EPI_ISL_935809 | Canada/MB-NML-1490/2020 | 2020-08-28 | North America/Canada/Manitoba/ | Cadham Provincial laboratory | National Microbiology Laboratory (NML) | Anna Majer et al |
| EPI_ISL_935810 | Canada/MB-NML-1491/2020 | 2020-08-25 | North America/Canada/Manitoba/ | Cadham Provincial laboratory | National Microbiology Laboratory (NML) | Anna Majer et al |
| EPI_ISL_935811 | Canada/MB-NML-1493/2020 | 2020-08-24 | North America/Canada/Manitoba/ | Cadham Provincial laboratory | National Microbiology Laboratory (NML) | Anna Majer et al |
| EPI_ISL_935812 | Canada/MB-NML-1495/2020 | 2020-08-24 | North America/Canada/Manitoba/ | Cadham Provincial laboratory | National Microbiology Laboratory (NML) | Anna Majer et al |
| EPI_ISL_935813 | Canada/MB-NML-1498/2020 | 2020-08-24 | North America/Canada/Manitoba/ | Cadham Provincial laboratory | National Microbiology Laboratory (NML) | Anna Majer et al |
| EPI_ISL_935814 | Canada/MB-NML-1499/2020 | 2020-08-27 | North America/Canada/Manitoba/ | Cadham Provincial laboratory | National Microbiology Laboratory (NML) | Anna Majer et al |

| | | | | | | |
|---|---|---|---|---|---|---|
| EPI_ISL_935815 | Canada/MB-NML-1501/2020 | 2020-08-28 | North America/Canada/Manitoba/ | Cadham Provincial laboratory | National Microbiology Laboratory (NML) | Anna Majer et al |
| EPI_ISL_935816 | Canada/MB-NML-1502/2020 | 2020-08-28 | North America/Canada/Manitoba/ | Cadham Provincial laboratory | National Microbiology Laboratory (NML) | Anna Majer et al |
| EPI_ISL_935817 | Canada/MB-NML-1503/2020 | 2020-09-14 | North America/Canada/Manitoba/ | Cadham Provincial laboratory | National Microbiology Laboratory (NML) | Anna Majer et al |
| EPI_ISL_935818 | Canada/MB-NML-1504/2020 | 2020-09-14 | North America/Canada/Manitoba/ | Cadham Provincial laboratory | National Microbiology Laboratory (NML) | Anna Majer et al |
| EPI_ISL_935820 | Canada/MB-NML-1506/2020 | 2020-09-16 | North America/Canada/Manitoba/ | Cadham Provincial laboratory | National Microbiology Laboratory (NML) | Anna Majer et al |
| EPI_ISL_935821 | Canada/MB-NML-1507/2020 | 2020-09-14 | North America/Canada/Manitoba/ | Cadham Provincial laboratory | National Microbiology Laboratory (NML) | Anna Majer et al |
| EPI_ISL_935822 | Canada/MB-NML-1508/2020 | 2020-09-07 | North America/Canada/Manitoba/ | Cadham Provincial laboratory | National Microbiology Laboratory (NML) | Anna Majer et al |
| EPI_ISL_935823 | Canada/MB-NML-1511/2020 | 2020-08-31 | North America/Canada/Manitoba/ | Cadham Provincial laboratory | National Microbiology Laboratory (NML) | Anna Majer et al |
| EPI_ISL_935825 | Canada/MB-NML-1514/2020 | 2020-09-07 | North America/Canada/Manitoba/ | Cadham Provincial laboratory | National Microbiology Laboratory (NML) | Anna Majer et al |
| EPI_ISL_935827 | Canada/MB-NML-1519/2020 | 2020-09-02 | North America/Canada/Manitoba/ | Cadham Provincial laboratory | National Microbiology Laboratory (NML) | Anna Majer et al |
| EPI_ISL_935831 | Canada/MB-NML-1530/2020 | 2020-08-22 | North America/Canada/Manitoba/ | Cadham Provincial laboratory | National Microbiology Laboratory (NML) | Anna Majer et al |
| EPI_ISL_935838 | Canada/MB-NML-1537/2020 | 2020-09-14 | North America/Canada/Manitoba/ | Cadham Provincial laboratory | National Microbiology Laboratory (NML) | Anna Majer et al |
| EPI_ISL_935839 | Canada/MB-NML-1538/2020 | 2020-09-14 | North America/Canada/Manitoba/ | Cadham Provincial laboratory | National Microbiology Laboratory (NML) | Anna Majer et al |

| | | | | | | |
|---|---|---|---|---|---|---|
| EPI_ISL_935840 | Canada/MB-NML-1539/2020 | 2020-09-02 | North America/Canada/Manitoba/ | Cadham Provincial laboratory | National Microbiology Laboratory (NML) | Anna Majer et al |
| EPI_ISL_935841 | Canada/MB-NML-1541/2020 | 2020-09-03 | North America/Canada/Manitoba/ | Cadham Provincial laboratory | National Microbiology Laboratory (NML) | Anna Majer et al |
| EPI_ISL_935842 | Canada/MB-NML-1542/2020 | 2020-09-11 | North America/Canada/Manitoba/ | Cadham Provincial laboratory | National Microbiology Laboratory (NML) | Anna Majer et al |
| EPI_ISL_935846 | Canada/MB-NML-1549/2020 | 2020-08-15 | North America/Canada/Manitoba/ | Cadham Provincial laboratory | National Microbiology Laboratory (NML) | Anna Majer et al |
| EPI_ISL_935849 | Canada/MB-NML-1553/2020 | 2020-09-20 | North America/Canada/Manitoba/ | Cadham Provincial laboratory | National Microbiology Laboratory (NML) | Anna Majer et al |
| EPI_ISL_935850 | Canada/MB-NML-1554/2020 | 2020-08-19 | North America/Canada/Manitoba/ | Cadham Provincial laboratory | National Microbiology Laboratory (NML) | Anna Majer et al |
| EPI_ISL_935851 | Canada/MB-NML-1555/2020 | 2020-09-10 | North America/Canada/Manitoba/ | Cadham Provincial laboratory | National Microbiology Laboratory (NML) | Anna Majer et al |
| EPI_ISL_935852 | Canada/MB-NML-1556/2020 | 2020-09-05 | North America/Canada/Manitoba/ | Cadham Provincial laboratory | National Microbiology Laboratory (NML) | Anna Majer et al |
| EPI_ISL_935853 | Canada/MB-NML-1557/2020 | 2020-08-19 | North America/Canada/Manitoba/ | Cadham Provincial laboratory | National Microbiology Laboratory (NML) | Anna Majer et al |
| EPI_ISL_935854 | Canada/MB-NML-1558/2020 | 2020-08-19 | North America/Canada/Manitoba/ | Cadham Provincial laboratory | National Microbiology Laboratory (NML) | Anna Majer et al |
| EPI_ISL_935855 | Canada/MB-NML-1560/2020 | 2020-08-19 | North America/Canada/Manitoba/ | Cadham Provincial laboratory | National Microbiology Laboratory (NML) | Anna Majer et al |
| EPI_ISL_935856 | Canada/MB-NML-1563/2020 | 2020-08-28 | North America/Canada/Manitoba/ | Cadham Provincial laboratory | National Microbiology Laboratory (NML) | Anna Majer et al |
| EPI_ISL_935857 | Canada/MB-NML-1564/2020 | 2020-08-18 | North America/Canada/Manitoba/ | Cadham Provincial laboratory | National Microbiology Laboratory (NML) | Anna Majer et al |

| | | | | | | |
|---|---|---|---|---|---|---|
| EPI_ISL_935858 | Canada/MB-NML-1565/2020 | 2020-08-13 | North America/Canada/Manitoba/ | Cadham Provincial laboratory | National Microbiology Laboratory (NML) | Anna Majer et al |
| EPI_ISL_935859 | Canada/MB-NML-1566/2020 | 2020-08-18 | North America/Canada/Manitoba/ | Cadham Provincial laboratory | National Microbiology Laboratory (NML) | Anna Majer et al |
| EPI_ISL_935860 | Canada/MB-NML-1567/2020 | 2020-08-29 | North America/Canada/Manitoba/ | Cadham Provincial laboratory | National Microbiology Laboratory (NML) | Anna Majer et al |
| EPI_ISL_935861 | Canada/MB-NML-1568/2020 | 2020-08-26 | North America/Canada/Manitoba/ | Cadham Provincial laboratory | National Microbiology Laboratory (NML) | Anna Majer et al |
| EPI_ISL_935862 | Canada/MB-NML-1595/2020 | 2020-09-29 | North America/Canada/Manitoba/ | Cadham Provincial laboratory | National Microbiology Laboratory (NML) | Anna Majer et al |
| EPI_ISL_935863 | Canada/MB-NML-1600/2020 | 2020-09-24 | North America/Canada/Manitoba/ | Cadham Provincial laboratory | National Microbiology Laboratory (NML) | Anna Majer et al |
| EPI_ISL_935864 | Canada/MB-NML-1601/2020 | 2020-09-26 | North America/Canada/Manitoba/ | Cadham Provincial laboratory | National Microbiology Laboratory (NML) | Anna Majer et al |
| EPI_ISL_935865 | Canada/MB-NML-1602/2020 | 2020-09-27 | North America/Canada/Manitoba/ | Cadham Provincial laboratory | National Microbiology Laboratory (NML) | Anna Majer et al |
| EPI_ISL_935866 | Canada/MB-NML-1603/2020 | 2020-09-24 | North America/Canada/Manitoba/ | Cadham Provincial laboratory | National Microbiology Laboratory (NML) | Anna Majer et al |
| EPI_ISL_935867 | Canada/MB-NML-1604/2020 | 2020-09-24 | North America/Canada/Manitoba/ | Cadham Provincial laboratory | National Microbiology Laboratory (NML) | Anna Majer et al |
| EPI_ISL_935868 | Canada/MB-NML-1605/2020 | 2020-09-24 | North America/Canada/Manitoba/ | Cadham Provincial laboratory | National Microbiology Laboratory (NML) | Anna Majer et al |
| EPI_ISL_935869 | Canada/MB-NML-1606/2020 | 2020-09-24 | North America/Canada/Manitoba/ | Cadham Provincial laboratory | National Microbiology Laboratory (NML) | Anna Majer et al |
| EPI_ISL_935870 | Canada/MB-NML-1607/2020 | 2020-09-24 | North America/Canada/Manitoba/ | Cadham Provincial laboratory | National Microbiology Laboratory (NML) | Anna Majer et al |

| | | | | | | |
|---|---|---|---|---|---|---|
| EPI_ISL_935874 | Canada/MB-NML-1625/2020 | 2020-08-18 | North America/Canada/Manitoba/ | Cadham Provincial laboratory | National Microbiology Laboratory (NML) | Anna Majer et al |
| EPI_ISL_935876 | Canada/MB-NML-1637/2020 | 2020-08-19 | North America/Canada/Manitoba/ | Cadham Provincial laboratory | National Microbiology Laboratory (NML) | Anna Majer et al |
| EPI_ISL_935877 | Canada/MB-NML-2810/2020 | 2020-11-25 | North America/Canada/Manitoba/ | Cadham Provincial laboratory | National Microbiology Laboratory (NML) | Anna Majer et al |
| EPI_ISL_935878 | Canada/MB-NML-2811/2020 | 2020-11-26 | North America/Canada/Manitoba/ | Cadham Provincial laboratory | National Microbiology Laboratory (NML) | Anna Majer et al |
| EPI_ISL_935879 | Canada/MB-NML-2812/2020 | 2020-11-25 | North America/Canada/Manitoba/ | Cadham Provincial laboratory | National Microbiology Laboratory (NML) | Anna Majer et al |
| EPI_ISL_935880 | Canada/MB-NML-2813/2020 | 2020-11-25 | North America/Canada/Manitoba/ | Cadham Provincial laboratory | National Microbiology Laboratory (NML) | Anna Majer et al |
| EPI_ISL_935881 | Canada/MB-NML-2814/2020 | 2020-11-16 | North America/Canada/Manitoba/ | Cadham Provincial laboratory | National Microbiology Laboratory (NML) | Anna Majer et al |
| EPI_ISL_935882 | Canada/MB-NML-2816/2020 | 2020-11-25 | North America/Canada/Manitoba/ | Cadham Provincial laboratory | National Microbiology Laboratory (NML) | Anna Majer et al |
| EPI_ISL_935883 | Canada/MB-NML-2818/2020 | 2020-11-24 | North America/Canada/Manitoba/ | Cadham Provincial laboratory | National Microbiology Laboratory (NML) | Anna Majer et al |
| EPI_ISL_935884 | Canada/MB-NML-2819/2020 | 2020-11-26 | North America/Canada/Manitoba/ | Cadham Provincial laboratory | National Microbiology Laboratory (NML) | Anna Majer et al |
| EPI_ISL_935885 | Canada/MB-NML-2820/2020 | 2020-11-24 | North America/Canada/Manitoba/ | Cadham Provincial laboratory | National Microbiology Laboratory (NML) | Anna Majer et al |
| EPI_ISL_935886 | Canada/MB-NML-2821/2020 | 2020-11-25 | North America/Canada/Manitoba/ | Cadham Provincial laboratory | National Microbiology Laboratory (NML) | Anna Majer et al |
| EPI_ISL_935887 | Canada/MB-NML-2822/2020 | 2020-11-14 | North America/Canada/Manitoba/ | Cadham Provincial laboratory | National Microbiology Laboratory (NML) | Anna Majer et al |

| | | | | | | |
|---|---|---|---|---|---|---|
| EPI_ISL_935888 | Canada/MB-NML-2823/2020 | 2020-11-25 | North America/Canada/Manitoba/ | Cadham Provincial laboratory | National Microbiology Laboratory (NML) | Anna Majer et al |
| EPI_ISL_935889 | Canada/MB-NML-2824/2020 | 2020-11-26 | North America/Canada/Manitoba/ | Cadham Provincial laboratory | National Microbiology Laboratory (NML) | Anna Majer et al |
| EPI_ISL_935890 | Canada/MB-NML-2825/2020 | 2020-11-05 | North America/Canada/Manitoba/ | Cadham Provincial laboratory | National Microbiology Laboratory (NML) | Anna Majer et al |
| EPI_ISL_935891 | Canada/MB-NML-2826/2020 | 2020-11-05 | North America/Canada/Manitoba/ | Cadham Provincial laboratory | National Microbiology Laboratory (NML) | Anna Majer et al |
| EPI_ISL_935892 | Canada/MB-NML-2827/2020 | 2020-11-11 | North America/Canada/Manitoba/ | Cadham Provincial laboratory | National Microbiology Laboratory (NML) | Anna Majer et al |
| EPI_ISL_935893 | Canada/MB-NML-2828/2020 | 2020-11-25 | North America/Canada/Manitoba/ | Cadham Provincial laboratory | National Microbiology Laboratory (NML) | Anna Majer et al |
| EPI_ISL_935894 | Canada/MB-NML-2829/2020 | 2020-11-26 | North America/Canada/Manitoba/ | Cadham Provincial laboratory | National Microbiology Laboratory (NML) | Anna Majer et al |
| EPI_ISL_935895 | Canada/MB-NML-2830/2020 | 2020-11-25 | North America/Canada/Manitoba/ | Cadham Provincial laboratory | National Microbiology Laboratory (NML) | Anna Majer et al |
| EPI_ISL_935896 | Canada/MB-NML-2831/2020 | 2020-11-05 | North America/Canada/Manitoba/ | Cadham Provincial laboratory | National Microbiology Laboratory (NML) | Anna Majer et al |
| EPI_ISL_935897 | Canada/MB-NML-2833/2020 | 2020-11-06 | North America/Canada/Manitoba/ | Cadham Provincial laboratory | National Microbiology Laboratory (NML) | Anna Majer et al |
| EPI_ISL_935898 | Canada/MB-NML-2834/2020 | 2020-11-25 | North America/Canada/Manitoba/ | Cadham Provincial laboratory | National Microbiology Laboratory (NML) | Anna Majer et al |
| EPI_ISL_935899 | Canada/MB-NML-2835/2020 | 2020-11-25 | North America/Canada/Manitoba/ | Cadham Provincial laboratory | National Microbiology Laboratory (NML) | Anna Majer et al |
| EPI_ISL_935900 | Canada/MB-NML-2836/2020 | 2020-11-16 | North America/Canada/Manitoba/ | Cadham Provincial laboratory | National Microbiology Laboratory (NML) | Anna Majer et al |

| | | | | | | |
|---|---|---|---|---|---|---|
| EPI_ISL_935901 | Canada/MB-NML-2838/2020 | 2020-11-25 | North America/Canada/Manitoba/ | Cadham Provincial laboratory | National Microbiology Laboratory (NML) | Anna Majer et al |
| EPI_ISL_935902 | Canada/MB-NML-2839/2020 | 2020-11-26 | North America/Canada/Manitoba/ | Cadham Provincial laboratory | National Microbiology Laboratory (NML) | Anna Majer et al |
| EPI_ISL_935903 | Canada/MB-NML-2840/2020 | 2020-11-08 | North America/Canada/Manitoba/ | Cadham Provincial laboratory | National Microbiology Laboratory (NML) | Anna Majer et al |
| EPI_ISL_935904 | Canada/MB-NML-2841/2020 | 2020-11-16 | North America/Canada/Manitoba/ | Cadham Provincial laboratory | National Microbiology Laboratory (NML) | Anna Majer et al |
| EPI_ISL_935905 | Canada/MB-NML-2842/2020 | 2020-11-07 | North America/Canada/Manitoba/ | Cadham Provincial laboratory | National Microbiology Laboratory (NML) | Anna Majer et al |
| EPI_ISL_935906 | Canada/MB-NML-2843/2020 | 2020-11-14 | North America/Canada/Manitoba/ | Cadham Provincial laboratory | National Microbiology Laboratory (NML) | Anna Majer et al |
| EPI_ISL_935907 | Canada/MB-NML-2844/2020 | 2020-11-26 | North America/Canada/Manitoba/ | Cadham Provincial laboratory | National Microbiology Laboratory (NML) | Anna Majer et al |
| EPI_ISL_935908 | Canada/MB-NML-2845/2020 | 2020-11-16 | North America/Canada/Manitoba/ | Cadham Provincial laboratory | National Microbiology Laboratory (NML) | Anna Majer et al |
| EPI_ISL_935909 | Canada/MB-NML-2847/2020 | 2020-11-05 | North America/Canada/Manitoba/ | Cadham Provincial laboratory | National Microbiology Laboratory (NML) | Anna Majer et al |
| EPI_ISL_935910 | Canada/MB-NML-2848/2020 | 2020-11-25 | North America/Canada/Manitoba/ | Cadham Provincial laboratory | National Microbiology Laboratory (NML) | Anna Majer et al |
| EPI_ISL_935911 | Canada/MB-NML-2849/2020 | 2020-11-25 | North America/Canada/Manitoba/ | Cadham Provincial laboratory | National Microbiology Laboratory (NML) | Anna Majer et al |
| EPI_ISL_935912 | Canada/MB-NML-2850/2020 | 2020-11-26 | North America/Canada/Manitoba/ | Cadham Provincial laboratory | National Microbiology Laboratory (NML) | Anna Majer et al |
| EPI_ISL_935913 | Canada/MB-NML-2852/2020 | 2020-11-26 | North America/Canada/Manitoba/ | Cadham Provincial laboratory | National Microbiology Laboratory (NML) | Anna Majer et al |

| | | | | | | |
|---|---|---|---|---|---|---|
| EPI_ISL_935914 | Canada/MB-NML-2855/2020 | 2020-11-07 | North America/Canada/Manitoba/ | Cadham Provincial laboratory | National Microbiology Laboratory (NML) | Anna Majer et al |
| EPI_ISL_935915 | Canada/MB-NML-2856/2020 | 2020-11-26 | North America/Canada/Manitoba/ | Cadham Provincial laboratory | National Microbiology Laboratory (NML) | Anna Majer et al |
| EPI_ISL_935916 | Canada/MB-NML-2857/2020 | 2020-11-26 | North America/Canada/Manitoba/ | Cadham Provincial laboratory | National Microbiology Laboratory (NML) | Anna Majer et al |
| EPI_ISL_935917 | Canada/MB-NML-2858/2020 | 2020-11-26 | North America/Canada/Manitoba/ | Cadham Provincial laboratory | National Microbiology Laboratory (NML) | Anna Majer et al |
| EPI_ISL_935918 | Canada/MB-NML-2859/2020 | 2020-11-14 | North America/Canada/Manitoba/ | Cadham Provincial laboratory | National Microbiology Laboratory (NML) | Anna Majer et al |
| EPI_ISL_935919 | Canada/MB-NML-2860/2020 | 2020-11-16 | North America/Canada/Manitoba/ | Cadham Provincial laboratory | National Microbiology Laboratory (NML) | Anna Majer et al |
| EPI_ISL_935920 | Canada/MB-NML-2861/2020 | 2020-11-16 | North America/Canada/Manitoba/ | Cadham Provincial laboratory | National Microbiology Laboratory (NML) | Anna Majer et al |
| EPI_ISL_935921 | Canada/MB-NML-2862/2020 | 2020-11-25 | North America/Canada/Manitoba/ | Cadham Provincial laboratory | National Microbiology Laboratory (NML) | Anna Majer et al |
| EPI_ISL_935922 | Canada/MB-NML-2863/2020 | 2020-11-16 | North America/Canada/Manitoba/ | Cadham Provincial laboratory | National Microbiology Laboratory (NML) | Anna Majer et al |
| EPI_ISL_935923 | Canada/MB-NML-2864/2020 | 2020-11-14 | North America/Canada/Manitoba/ | Cadham Provincial laboratory | National Microbiology Laboratory (NML) | Anna Majer et al |
| EPI_ISL_935924 | Canada/MB-NML-2865/2020 | 2020-11-24 | North America/Canada/Manitoba/ | Cadham Provincial laboratory | National Microbiology Laboratory (NML) | Anna Majer et al |
| EPI_ISL_935925 | Canada/MB-NML-2866/2020 | 2020-11-19 | North America/Canada/Manitoba/ | Cadham Provincial laboratory | National Microbiology Laboratory (NML) | Anna Majer et al |
| EPI_ISL_935926 | Canada/MB-NML-2869/2020 | 2020-11-18 | North America/Canada/Manitoba/ | Cadham Provincial laboratory | National Microbiology Laboratory (NML) | Anna Majer et al |

| | | | | | | |
|---|---|---|---|---|---|---|
| EPI_ISL_935927 | Canada/MB-NML-2871/2020 | 2020-11-19 | North America/Canada/Manitoba/ | Cadham Provincial laboratory | National Microbiology Laboratory (NML) | Anna Majer et al |
| EPI_ISL_935928 | Canada/MB-NML-2873/2020 | 2020-11-19 | North America/Canada/Manitoba/ | Cadham Provincial laboratory | National Microbiology Laboratory (NML) | Anna Majer et al |
| EPI_ISL_935929 | Canada/MB-NML-2874/2020 | 2020-11-19 | North America/Canada/Manitoba/ | Cadham Provincial laboratory | National Microbiology Laboratory (NML) | Anna Majer et al |
| EPI_ISL_935930 | Canada/MB-NML-2875/2020 | 2020-11-24 | North America/Canada/Manitoba/ | Cadham Provincial laboratory | National Microbiology Laboratory (NML) | Anna Majer et al |
| EPI_ISL_935931 | Canada/MB-NML-2876/2020 | 2020-11-19 | North America/Canada/Manitoba/ | Cadham Provincial laboratory | National Microbiology Laboratory (NML) | Anna Majer et al |
| EPI_ISL_935932 | Canada/MB-NML-2879/2020 | 2020-11-19 | North America/Canada/Manitoba/ | Cadham Provincial laboratory | National Microbiology Laboratory (NML) | Anna Majer et al |
| EPI_ISL_935933 | Canada/MB-NML-2880/2020 | 2020-11-24 | North America/Canada/Manitoba/ | Cadham Provincial laboratory | National Microbiology Laboratory (NML) | Anna Majer et al |
| EPI_ISL_935934 | Canada/MB-NML-2881/2020 | 2020-11-19 | North America/Canada/Manitoba/ | Cadham Provincial laboratory | National Microbiology Laboratory (NML) | Anna Majer et al |
| EPI_ISL_935935 | Canada/MB-NML-2883/2020 | 2020-11-19 | North America/Canada/Manitoba/ | Cadham Provincial laboratory | National Microbiology Laboratory (NML) | Anna Majer et al |
| EPI_ISL_935936 | Canada/MB-NML-2884/2020 | 2020-11-18 | North America/Canada/Manitoba/ | Cadham Provincial laboratory | National Microbiology Laboratory (NML) | Anna Majer et al |
| EPI_ISL_935938 | Canada/MB-NML-2886/2020 | 2020-11-19 | North America/Canada/Manitoba/ | Cadham Provincial laboratory | National Microbiology Laboratory (NML) | Anna Majer et al |
| EPI_ISL_935939 | Canada/MB-NML-2887/2020 | 2020-11-17 | North America/Canada/Manitoba/ | Cadham Provincial laboratory | National Microbiology Laboratory (NML) | Anna Majer et al |
| EPI_ISL_935940 | Canada/MB-NML-2888/2020 | 2020-11-19 | North America/Canada/Manitoba/ | Cadham Provincial laboratory | National Microbiology Laboratory (NML) | Anna Majer et al |

| | | | | | | |
|---|---|---|---|---|---|---|
| EPI_ISL_935941 | Canada/MB-NML-2890/2020 | 2020-11-17 | North America/Canada/Manitoba/ | Cadham Provincial laboratory | National Microbiology Laboratory (NML) | Anna Majer et al |
| EPI_ISL_935942 | Canada/MB-NML-2891/2020 | 2020-11-19 | North America/Canada/Manitoba/ | Cadham Provincial laboratory | National Microbiology Laboratory (NML) | Anna Majer et al |
| EPI_ISL_935944 | Canada/MB-NML-2893/2020 | 2020-11-16 | North America/Canada/Manitoba/ | Cadham Provincial laboratory | National Microbiology Laboratory (NML) | Anna Majer et al |
| EPI_ISL_935945 | Canada/MB-NML-2894/2020 | 2020-11-19 | North America/Canada/Manitoba/ | Cadham Provincial laboratory | National Microbiology Laboratory (NML) | Anna Majer et al |
| EPI_ISL_935946 | Canada/MB-NML-2895/2020 | 2020-11-17 | North America/Canada/Manitoba/ | Cadham Provincial laboratory | National Microbiology Laboratory (NML) | Anna Majer et al |
| EPI_ISL_935947 | Canada/MB-NML-2897/2020 | 2020-11-19 | North America/Canada/Manitoba/ | Cadham Provincial laboratory | National Microbiology Laboratory (NML) | Anna Majer et al |
| EPI_ISL_935948 | Canada/MB-NML-2899/2020 | 2020-11-17 | North America/Canada/Manitoba/ | Cadham Provincial laboratory | National Microbiology Laboratory (NML) | Anna Majer et al |
| EPI_ISL_935949 | Canada/MB-NML-2900/2020 | 2020-11-19 | North America/Canada/Manitoba/ | Cadham Provincial laboratory | National Microbiology Laboratory (NML) | Anna Majer et al |
| EPI_ISL_935950 | Canada/MB-NML-2903/2020 | 2020-11-17 | North America/Canada/Manitoba/ | Cadham Provincial laboratory | National Microbiology Laboratory (NML) | Anna Majer et al |
| EPI_ISL_935951 | Canada/MB-NML-2904/2020 | 2020-11-19 | North America/Canada/Manitoba/ | Cadham Provincial laboratory | National Microbiology Laboratory (NML) | Anna Majer et al |
| EPI_ISL_935952 | Canada/MB-NML-2905/2020 | 2020-11-16 | North America/Canada/Manitoba/ | Cadham Provincial laboratory | National Microbiology Laboratory (NML) | Anna Majer et al |
| EPI_ISL_935953 | Canada/MB-NML-2909/2020 | 2020-11-12 | North America/Canada/Manitoba/ | Cadham Provincial laboratory | National Microbiology Laboratory (NML) | Anna Majer et al |
| EPI_ISL_935954 | Canada/MB-NML-2910/2020 | 2020-11-09 | North America/Canada/Manitoba/ | Cadham Provincial laboratory | National Microbiology Laboratory (NML) | Anna Majer et al |

| | | | | | | |
|---|---|---|---|---|---|---|
| EPI_ISL_935955 | Canada/MB-NML-2912/2020 | 2020-11-09 | North America/Canada/Manitoba/ | Cadham Provincial laboratory | National Microbiology Laboratory (NML) | Anna Majer et al |
| EPI_ISL_935956 | Canada/MB-NML-2913/2020 | 2020-11-09 | North America/Canada/Manitoba/ | Cadham Provincial laboratory | National Microbiology Laboratory (NML) | Anna Majer et al |
| EPI_ISL_935957 | Canada/MB-NML-2914/2020 | 2020-11-14 | North America/Canada/Manitoba/ | Cadham Provincial laboratory | National Microbiology Laboratory (NML) | Anna Majer et al |
| EPI_ISL_935958 | Canada/MB-NML-2915/2020 | 2020-11-10 | North America/Canada/Manitoba/ | Cadham Provincial laboratory | National Microbiology Laboratory (NML) | Anna Majer et al |
| EPI_ISL_935959 | Canada/MB-NML-2916/2020 | 2020-11-09 | North America/Canada/Manitoba/ | Cadham Provincial laboratory | National Microbiology Laboratory (NML) | Anna Majer et al |
| EPI_ISL_935960 | Canada/MB-NML-2917/2020 | 2020-11-09 | North America/Canada/Manitoba/ | Cadham Provincial laboratory | National Microbiology Laboratory (NML) | Anna Majer et al |
| EPI_ISL_935961 | Canada/MB-NML-2919/2020 | 2020-11-14 | North America/Canada/Manitoba/ | Cadham Provincial laboratory | National Microbiology Laboratory (NML) | Anna Majer et al |
| EPI_ISL_935962 | Canada/MB-NML-2920/2020 | 2020-11-12 | North America/Canada/Manitoba/ | Cadham Provincial laboratory | National Microbiology Laboratory (NML) | Anna Majer et al |
| EPI_ISL_935963 | Canada/MB-NML-2921/2020 | 2020-11-14 | North America/Canada/Manitoba/ | Cadham Provincial laboratory | National Microbiology Laboratory (NML) | Anna Majer et al |
| EPI_ISL_935964 | Canada/MB-NML-2922/2020 | 2020-11-10 | North America/Canada/Manitoba/ | Cadham Provincial laboratory | National Microbiology Laboratory (NML) | Anna Majer et al |
| EPI_ISL_935965 | Canada/MB-NML-2924/2020 | 2020-11-09 | North America/Canada/Manitoba/ | Cadham Provincial laboratory | National Microbiology Laboratory (NML) | Anna Majer et al |
| EPI_ISL_935966 | Canada/MB-NML-2925/2020 | 2020-11-10 | North America/Canada/Manitoba/ | Cadham Provincial laboratory | National Microbiology Laboratory (NML) | Anna Majer et al |
| EPI_ISL_935967 | Canada/MB-NML-2926/2020 | 2020-11-08 | North America/Canada/Manitoba/ | Cadham Provincial laboratory | National Microbiology Laboratory (NML) | Anna Majer et al |

| | | | | | | |
|---|---|---|---|---|---|---|
| EPI_ISL_935968 | Canada/MB-NML-2928/2020 | 2020-11-14 | North America/Canada/Manitoba/ | Cadham Provincial laboratory | National Microbiology Laboratory (NML) | Anna Majer et al |
| EPI_ISL_935969 | Canada/MB-NML-2929/2020 | 2020-11-10 | North America/Canada/Manitoba/ | Cadham Provincial laboratory | National Microbiology Laboratory (NML) | Anna Majer et al |
| EPI_ISL_935970 | Canada/MB-NML-2930/2020 | 2020-11-14 | North America/Canada/Manitoba/ | Cadham Provincial laboratory | National Microbiology Laboratory (NML) | Anna Majer et al |
| EPI_ISL_935987 | USA/NY-Wadsworth-21007840-01/20 | 2021-01-03 | North America/USA/New York/Washing | GLENS FALLS HOSPITAL LABORATORY | Wadsworth Center, New York State Department of Health | Kirsten St. George et al |
| EPI_ISL_935995 | USA/NY-Wadsworth-21008079-01/20 | 2021-01-05 | North America/USA/New York/Onondag | SUNY UPSTATE MEDICAL UNIVERSITY | Wadsworth Center, New York State Department of Health | Kirsten St. George et al |
| EPI_ISL_935996 | USA/NY-Wadsworth-21008080-01/20 | 2021-01-05 | North America/USA/New York/Onondag | SUNY UPSTATE MEDICAL UNIVERSITY | Wadsworth Center, New York State Department of Health | Kirsten St. George et al |
| EPI_ISL_936009 | USA/NY-Wadsworth-21008096-01/20 | 2021-01-11 | North America/USA/New York/Oswego | SUNY UPSTATE MEDICAL UNIVERSITY | Wadsworth Center, New York State Department of Health | Kirsten St. George et al |
| EPI_ISL_936015 | USA/NY-Wadsworth-21008102-01/20 | 2021-01-09 | North America/USA/New York/Onondag | SUNY UPSTATE MEDICAL UNIVERSITY | Wadsworth Center, New York State Department of Health | Kirsten St. George et al |
| EPI_ISL_936041 | USA/NY-Wadsworth-21007579-01/20 | 2020-12-28 | North America/USA/New York/Bronx | Columbia University Irving Medical Center | Wadsworth Center, New York State Department of Health | Kirsten St. George et al |
| EPI_ISL_936056 | USA/NY-Wadsworth-21007862-01/20 | 2021-01-07 | North America/USA/New York/Franklin | ADIRONDACK MEDICAL CENTER | Wadsworth Center, New York State Department of Health | Kirsten St. George et al |

| | | | | | | |
|---|---|---|---|---|---|---|
| EPI_ISL_936060 | USA/NY-Wadsworth-21007867-01/20 | 2020-12-31 | North America/USA/New York/Erie Cou | KALEIDA CENTER FOR LABORATORY MEDICINE | Wadsworth Center, New York State Department of Health | Kirsten St. George et al |
| EPI_ISL_936065 | USA/NY-Wadsworth-21007875-01/20 | 2020-12-29 | North America/USA/New York/Bronx | Columbia University Irving Medical Center | Wadsworth Center, New York State Department of Health | Kirsten St. George et al |
| EPI_ISL_936067 | USA/NY-Wadsworth-21007880-01/20 | 2021-01-02 | North America/USA/New York/Niagara | KALEIDA CENTER FOR LABORATORY MEDICINE | Wadsworth Center, New York State Department of Health | Kirsten St. George et al |
| EPI_ISL_936070 | USA/NY-Wadsworth-21007884-01/20 | 2021-01-02 | North America/USA/New York/Niagara | KALEIDA CENTER FOR LABORATORY MEDICINE | Wadsworth Center, New York State Department of Health | Kirsten St. George et al |
| EPI_ISL_936076 | USA/NY-Wadsworth-21007893-01/20 | 2021-01-02 | North America/USA/New York/Niagara | KALEIDA CENTER FOR LABORATORY MEDICINE | Wadsworth Center, New York State Department of Health | Kirsten St. George et al |
| EPI_ISL_936079 | USA/NY-Wadsworth-21007897-01/20 | 2021-01-04 | North America/USA/New York/Erie Cou | KALEIDA CENTER FOR LABORATORY MEDICINE | Wadsworth Center, New York State Department of Health | Kirsten St. George et al |
| EPI_ISL_936080 | USA/NY-Wadsworth-21007901-01/20 | 2021-01-04 | North America/USA/New York/Erie Cou | KALEIDA CENTER FOR LABORATORY MEDICINE | Wadsworth Center, New York State Department of Health | Kirsten St. George et al |
| EPI_ISL_936087 | USA/NY-Wadsworth-21007914-01/20 | 2021-01-03 | North America/USA/New York/Niagara | KALEIDA CENTER FOR LABORATORY MEDICINE | Wadsworth Center, New York State Department of Health | Kirsten St. George et al |
| EPI_ISL_936088 | USA/NY-Wadsworth-21007916-01/20 | 2021-01-03 | North America/USA/New York/Niagara | KALEIDA CENTER FOR LABORATORY MEDICINE | Wadsworth Center, New York State Department of Health | Kirsten St. George et al |
| EPI_ISL_936096 | USA/NY-Wadsworth-21007943-01/20 | 2021-01-04 | North America/USA/New York/Erie Cou | KALEIDA CENTER FOR LABORATORY MEDICINE | Wadsworth Center, New York State Department of Health | Kirsten St. George et al |

| | | | | | | |
|---|---|---|---|---|---|---|
| EPI_ISL_936097 | USA/NY-Wadsworth-21007944-01/20 | 2021-01-04 | North America/USA/New York/Erie Cou | KALEIDA CENTER FOR LABORATORY MEDICINE | Wadsworth Center, New York State Department of Health | Kirsten St. George et al |
| EPI_ISL_936101 | USA/NY-Wadsworth-21007948-01/20 | 2021-01-04 | North America/USA/New York/Niagara | KALEIDA CENTER FOR LABORATORY MEDICINE | Wadsworth Center, New York State Department of Health | Kirsten St. George et al |
| EPI_ISL_936114 | USA/NY-Wadsworth-21013545-01/20 | 2021-01-22 | North America/USA/New York/Ulster C | WESTCHESTER MEDICAL CENTER | Wadsworth Center, New York State Department of Health | Kirsten St. George et al |
| EPI_ISL_936116 | USA/NY-Wadsworth-21013547-01/20 | 2021-01-22 | North America/USA/New York/Ulster C | WESTCHESTER MEDICAL CENTER | Wadsworth Center, New York State Department of Health | Kirsten St. George et al |
| EPI_ISL_936119 | USA/NY-Wadsworth-21013552-01/20 | 2021-01-22 | North America/USA/New York/Westche | WESTCHESTER MEDICAL CENTER | Wadsworth Center, New York State Department of Health | Kirsten St. George et al |
| EPI_ISL_936134 | USA/NY-Wadsworth-21013808-01/20 | 2020-12-18 | North America/USA/New York/Putnam | New York Presbyterian Hospital | Wadsworth Center, New York State Department of Health | Kirsten St. George et al |
| EPI_ISL_936140 | USA/NY-Wadsworth-21013817-01/20 | 2021-01-09 | North America/USA/New York/ | THE MARY IMOGENE BASSETT HOSPITAL | Wadsworth Center, New York State Department of Health | Kirsten St. George et al |
| EPI_ISL_936141 | USA/NY-Wadsworth-21013818-01/20 | 2021-01-09 | North America/USA/New York/Otsego C | THE MARY IMOGENE BASSETT HOSPITAL | Wadsworth Center, New York State Department of Health | Kirsten St. George et al |
| EPI_ISL_936146 | USA/NY-Wadsworth-21013826-01/20 | 2021-01-08 | North America/USA/New York/New Yor | New York Presbyterian Hospital | Wadsworth Center, New York State Department of Health | Kirsten St. George et al |
| EPI_ISL_936148 | USA/NY-Wadsworth-21013891-01/20 | 2021-01-23 | North America/USA/New York/Suffolk C | Wadsworth Center, New York State Department of Health | Wadsworth Center, New York State Department of Health | Kirsten St. George et al |

| | | | | | | |
|---|---|---|---|---|---|---|
| EPI_ISL_936151 | USA/NY-Wadsworth-21013927-01/20 | 2020-12-22 | North America/USA/New York/Westche | MEMORIAL SLOAN KETTERING CANCER CENTER | Wadsworth Center, New York State Department of Health | Kirsten St. George et al |
| EPI_ISL_936162 | USA/NY-Wadsworth-21013960-01/20 | 2020-12-23 | North America/USA/New York/Bronx | MEMORIAL SLOAN KETTERING CANCER CENTER | Wadsworth Center, New York State Department of Health | Kirsten St. George et al |
| EPI_ISL_936166 | USA/NY-Wadsworth-21013964-01/20 | 2020-12-23 | North America/USA/New York/Kings Co | MEMORIAL SLOAN KETTERING CANCER CENTER | Wadsworth Center, New York State Department of Health | Kirsten St. George et al |
| EPI_ISL_936179 | USA/NY-Wadsworth-21013979-01/20 | 2020-12-26 | North America/USA/New York/Westche | MEMORIAL SLOAN KETTERING CANCER CENTER | Wadsworth Center, New York State Department of Health | Kirsten St. George et al |
| EPI_ISL_936180 | USA/NY-Wadsworth-21014037-01/20 | 2020-12-27 | North America/USA/New York/Greene ( | ALBANY MEDICAL CENTER | Wadsworth Center, New York State Department of Health | Kirsten St. George et al |
| EPI_ISL_936187 | USA/TX-HHD-2101193997/2020 | 2020-12-10 | North America/USA/Texas/ | Houston Health Dept. | Houston Health Dept. | Ryker Penn et al |
| EPI_ISL_936191 | USA/TX-HHD-2101194037/2020 | 2020-12-10 | North America/USA/Texas/ | Houston Health Dept. | Houston Health Dept. | Ryker Penn et al |
| EPI_ISL_936192 | USA/TX-HHD-2101194065/2020 | 2020-12-10 | North America/USA/Texas/ | Houston Health Dept. | Houston Health Dept. | Ryker Penn et al |
| EPI_ISL_936194 | USA/TX-HHD-2101194075/2020 | 2020-12-10 | North America/USA/Texas/ | Houston Health Dept. | Houston Health Dept. | Ryker Penn et al |
| EPI_ISL_936203 | USA/TX-HHD-2101194219/2020 | 2020-12-10 | North America/USA/Texas/ | Houston Health Dept. | Houston Health Dept. | Ryker Penn et al |
| EPI_ISL_936210 | USA/TX-HHD-2101194353/2020 | 2020-12-11 | North America/USA/Texas/ | Houston Health Dept. | Houston Health Dept. | Ryker Penn et al |
| EPI_ISL_936225 | USA/NY-Wadsworth-21005823-01/20 | 2021-01-11 | North America/USA/New York/Albany ( | Wadsworth Center, New York State Department of Health | Wadsworth Center, New York State Department of Health | Kirsten St. George et al |
| EPI_ISL_936235 | USA/NY-Wadsworth-21005834-01/20 | 2021-01-11 | North America/USA/New York/Albany ( | Wadsworth Center, New York State Department of Health | Wadsworth Center, New York State Department of Health | Kirsten St. George et al |

| | | | | | | |
|---|---|---|---|---|---|---|
| EPI_ISL_936246 | USA/NY-Wadsworth-21008986-01/20 | 2021-01-15 | North America/USA/New York/Albany C | Wadsworth Center, New York State Department of Health | Wadsworth Center, New York State Department of Health | Kirsten St. George et al |
| EPI_ISL_936251 | USA/NY-Wadsworth-21011906-01/20 | 2021-01-16 | North America/USA/New York/Bronx | MONTEFIORE MEDICAL CENTER LABORATORIES | Wadsworth Center, New York State Department of Health | Kirsten St. George et al |
| EPI_ISL_936255 | USA/NY-Wadsworth-21011910-01/20 | 2021-01-18 | North America/USA/New York/Bronx | MONTEFIORE MEDICAL CENTER LABORATORIES | Wadsworth Center, New York State Department of Health | Kirsten St. George et al |
| EPI_ISL_936257 | USA/NY-Wadsworth-21012059-01/20 | 2021-01-18 | North America/USA/New York/Bronx | MONTEFIORE MEDICAL CENTER LABORATORIES | Wadsworth Center, New York State Department of Health | Kirsten St. George et al |
| EPI_ISL_936261 | USA/NY-Wadsworth-21012065-01/20 | 2021-01-14 | North America/USA/New York/Bronx | MONTEFIORE MEDICAL CENTER LABORATORIES | Wadsworth Center, New York State Department of Health | Kirsten St. George et al |
| EPI_ISL_936276 | USA/NY-Wadsworth-21012089-01/20 | 2021-01-19 | North America/USA/New York/Westche | MONTEFIORE MEDICAL CENTER LABORATORIES | Wadsworth Center, New York State Department of Health | Kirsten St. George et al |
| EPI_ISL_936300 | USA/NY-Wadsworth-21013338-01/20 | 2021-01-22 | North America/USA/New York/Nassau C | Wadsworth Center, New York State Department of Health | Wadsworth Center, New York State Department of Health | Kirsten St. George et al |
| EPI_ISL_936302 | USA/NY-Wadsworth-21013343-01/20 | 2021-01-22 | North America/USA/New York/Suffolk C | Wadsworth Center, New York State Department of Health | Wadsworth Center, New York State Department of Health | Kirsten St. George et al |
| EPI_ISL_936394 | USA/AZ-TG707691/2021 | 2021-01-06 | North America/USA/Arizona/Navajo Cou | TGen North | TGen North | Jolene Bowers et al |
| EPI_ISL_936395 | USA/AZ-TG707680/2021 | 2021-01-04 | North America/USA/Arizona/Coconino C | TGen North | TGen North | Jolene Bowers et al |
| EPI_ISL_936396 | USA/AZ-TG707065/2020 | 2020-12-29 | North America/USA/Arizona/Pima Coun | TGen North | TGen North | Jolene Bowers et al |
| EPI_ISL_936405 | USA/AZ-TG707068/2020 | 2020-12-29 | North America/USA/Arizona/Pima Coun | TGen North | TGen North | Jolene Bowers et al |
| EPI_ISL_936413 | USA/AZ-TG707703/2021 | 2021-01-07 | North America/USA/Arizona/Navajo Cou | TGen North | TGen North | Jolene Bowers et al |
| EPI_ISL_936414 | USA/AZ-TG707023/2020 | 2020-12-29 | North America/USA/Arizona/Pima Coun | TGen North | TGen North | Jolene Bowers et al |

| | | | | | | |
|---|---|---|---|---|---|---|
| EPI_ISL_936416 | USA/AZ-TG707107/2021 | 2021-01-06 | North America/USA/Arizona/Coconino C | TGen North | TGen North | Jolene Bowers et al |
| EPI_ISL_936425 | USA/AZ-TG707690/2021 | 2021-01-06 | North America/USA/Arizona/Navajo Co | TGen North | TGen North | Jolene Bowers et al |
| EPI_ISL_936426 | USA/AZ-TG707648/2021 | 2021-01-04 | North America/USA/Arizona/Coconino C | TGen North | TGen North | Jolene Bowers et al |
| EPI_ISL_936433 | USA/AZ-TG707640/2021 | 2021-01-04 | North America/USA/Arizona/Coconino C | TGen North | TGen North | Jolene Bowers et al |
| EPI_ISL_936436 | USA/AZ-TG707668/2021 | 2021-01-04 | North America/USA/Arizona/Coconino C | TGen North | TGen North | Jolene Bowers et al |
| EPI_ISL_936447 | USA/AZ-TG707679/2021 | 2021-01-04 | North America/USA/Arizona/Coconino C | TGen North | TGen North | Jolene Bowers et al |
| EPI_ISL_936451 | USA/AZ-TG707032/2020 | 2020-12-29 | North America/USA/Arizona/Pima Coun | TGen North | TGen North | Jolene Bowers et al |
| EPI_ISL_936462 | USA/AZ-TG707672/2021 | 2021-01-05 | North America/USA/Arizona/Mohave Co | TGen North | TGen North | Jolene Bowers et al |
| EPI_ISL_936489 | Malaysia/IMR_WI340/2020 | 2020-11-23 | Asia/Malaysia/Malaysia/ | Hospital Raja Perempuan Zainab II | Institute for Medical Research, Infectious Disease Research Centre, National Institutes of Health, Ministry of Health Malaysia | Suppiah J et al |
| EPI_ISL_936494 | Malaysia/IMR_WI224/2020 | 2020-10-24 | Asia/Malaysia/Malaysia/ | Hospital Enche Besar Hajjah Khalsom Kluang | Institute for Medical Research, Infectious Disease Research Centre, National Institutes of Health, Ministry of Health Malaysia | Suppiah J et al |
| EPI_ISL_936499 | USA/IL-NM-0782/2020 | 2020-05-15 | North America/USA/Illinois/Chicago | Northwestern Memorial Hospital | Ozer Lab | Ramon Lorenzo-Redondo et al |
| EPI_ISL_936503 | USA/IL-NM-0852/2020 | 2020-05-20 | North America/USA/Illinois/Chicago | Northwestern Memorial Hospital | Ozer Lab | Ramon Lorenzo-Redondo et al |
| EPI_ISL_936506 | USA/IL-NM-0878/2020 | 2020-05-22 | North America/USA/Illinois/Mchenry Co | Northwestern Memorial Hospital | Ozer Lab | Ramon Lorenzo-Redondo et al |
| EPI_ISL_936517 | USA/IL-NM-0948/2020 | 2020-05-26 | North America/USA/Illinois/Cook County | Northwestern Memorial Hospital | Ozer Lab | Ramon Lorenzo-Redondo et al |
| EPI_ISL_936522 | USA/IL-NM-0979/2020 | 2020-05-28 | North America/USA/Illinois/Kane County | Northwestern Memorial Hospital | Ozer Lab | Ramon Lorenzo-Redondo et al |
| EPI_ISL_936529 | USA/IL-NM-1032/2020 | 2020-06-03 | North America/USA/Illinois/Mchenry Co | Northwestern Memorial Hospital | Ozer Lab | Ramon Lorenzo-Redondo et al |
| EPI_ISL_936531 | USA/IL-NM-1049/2020 | 2020-06-04 | North America/USA/Illinois/Chicago | Northwestern Memorial Hospital | Ozer Lab | Ramon Lorenzo-Redondo et al |
| EPI_ISL_936537 | USA/IL-NM-1094/2020 | 2020-06-09 | North America/USA/Illinois/Cook County | Northwestern Memorial Hospital | Ozer Lab | Ramon Lorenzo-Redondo et al |
| EPI_ISL_936539 | USA/IL-NM-1127/2020 | 2020-06-12 | North America/USA/Illinois/Chicago | Northwestern Memorial Hospital | Ozer Lab | Ramon Lorenzo-Redondo et al |

| | | | | | | |
|---|---|---|---|---|---|---|
| EPI_ISL_936547 | USA/IL-NM-1192/2020 | 2020-06-19 | North America/USA/Illinois/Cook County | Northwestern Memorial Hospital | Ozer Lab | Ramon Lorenzo-Redondo et al |
| EPI_ISL_936553 | USA/IL-NM-1243/2020 | 2020-06-24 | North America/USA/Illinois/Chicago | Northwestern Memorial Hospital | Ozer Lab | Ramon Lorenzo-Redondo et al |
| EPI_ISL_936559 | USA/IL-NM-1412/2020 | 2020-07-01 | North America/USA/Illinois/Chicago | Northwestern Memorial Hospital | Ozer Lab | Ramon Lorenzo-Redondo et al |
| EPI_ISL_936560 | USA/IL-NM-1426/2020 | 2020-07-02 | North America/USA/Florida/Monroe Co | Northwestern Memorial Hospital | Ozer Lab | Ramon Lorenzo-Redondo et al |
| EPI_ISL_936562 | USA/IL-NM-1441/2020 | 2020-07-04 | North America/USA/Illinois/Dupage Cou | Northwestern Memorial Hospital | Ozer Lab | Ramon Lorenzo-Redondo et al |
| EPI_ISL_936563 | USA/IL-NM-1444/2020 | 2020-07-05 | North America/USA/Illinois/Chicago | Northwestern Memorial Hospital | Ozer Lab | Ramon Lorenzo-Redondo et al |
| EPI_ISL_936564 | USA/IL-NM-1452/2020 | 2020-07-06 | North America/USA/Illinois/Chicago | Northwestern Memorial Hospital | Ozer Lab | Ramon Lorenzo-Redondo et al |
| EPI_ISL_936567 | USA/IL-NM-1476/2020 | 2020-07-08 | North America/USA/Illinois/Chicago | Northwestern Memorial Hospital | Ozer Lab | Ramon Lorenzo-Redondo et al |
| EPI_ISL_936568 | USA/IL-NM-1480/2020 | 2020-07-08 | North America/USA/Illinois/Chicago | Northwestern Memorial Hospital | Ozer Lab | Ramon Lorenzo-Redondo et al |
| EPI_ISL_936572 | USA/IL-NM-1516/2020 | 2020-07-10 | North America/USA/Illinois/Chicago | Northwestern Memorial Hospital | Ozer Lab | Ramon Lorenzo-Redondo et al |
| EPI_ISL_936575 | USA/IL-NM-1534/2020 | 2020-07-13 | North America/USA/Illinois/Chicago | Northwestern Memorial Hospital | Ozer Lab | Ramon Lorenzo-Redondo et al |
| EPI_ISL_936578 | USA/IL-NM-1559/2020 | 2020-07-14 | North America/USA/Florida/Lee County | Northwestern Memorial Hospital | Ozer Lab | Ramon Lorenzo-Redondo et al |
| EPI_ISL_936579 | USA/IL-NM-1561/2020 | 2020-07-14 | North America/USA/Illinois/Lake County | Northwestern Memorial Hospital | Ozer Lab | Ramon Lorenzo-Redondo et al |
| EPI_ISL_936582 | USA/TX-NM-1984/2020 | 2020-08-17 | North America/USA/Texas/Tarrant Cour | Northwestern Memorial Hospital | Ozer Lab | Ramon Lorenzo-Redondo et al |
| EPI_ISL_936591 | USA/IL-NM-2397/2020 | 2020-10-31 | North America/USA/Illinois/Chicago | Northwestern Memorial Hospital | Ozer Lab | Ramon Lorenzo-Redondo et al |
| EPI_ISL_936592 | USA/IL-NM-2413/2020 | 2020-10-31 | North America/USA/Illinois/Chicago | Northwestern Memorial Hospital | Ozer Lab | Ramon Lorenzo-Redondo et al |
| EPI_ISL_936597 | USA/IL-NM-2675/2020 | 2020-11-01 | North America/USA/Illinois/Cook County | Northwestern Memorial Hospital | Ozer Lab | Ramon Lorenzo-Redondo et al |
| EPI_ISL_936598 | USA/IL-NM-2676/2020 | 2020-11-01 | North America/USA/Illinois/Chicago | Northwestern Memorial Hospital | Ozer Lab | Ramon Lorenzo-Redondo et al |
| EPI_ISL_936599 | USA/IL-NM-2679/2020 | 2020-11-01 | North America/USA/Illinois/Chicago | Northwestern Memorial Hospital | Ozer Lab | Ramon Lorenzo-Redondo et al |
| EPI_ISL_936604 | USA/IL-NM-2803/2020 | 2020-11-03 | North America/USA/Illinois/Chicago | Northwestern Memorial Hospital | Ozer Lab | Ramon Lorenzo-Redondo et al |
| EPI_ISL_936611 | USA/IL-NM-3201/2020 | 2020-11-09 | North America/USA/Illinois/Chicago | Northwestern Memorial Hospital | Ozer Lab | Ramon Lorenzo-Redondo et al |
| EPI_ISL_936615 | USA/IL-NM-3603/2020 | 2020-11-16 | North America/USA/Illinois/Lake County | Northwestern Memorial Hospital | Ozer Lab | Ramon Lorenzo-Redondo et al |
| EPI_ISL_936619 | USA/IL-NM-3724/2020 | 2020-11-20 | North America/USA/Illinois/Chicago | Northwestern Memorial Hospital | Ozer Lab | Ramon Lorenzo-Redondo et al |
| EPI_ISL_936625 | USA/IL-NM-3901/2020 | 2020-11-28 | North America/USA/Illinois/Cook County | Northwestern Memorial Hospital | Ozer Lab | Ramon Lorenzo-Redondo et al |
| EPI_ISL_936629 | USA/IL-NM-3958/2020 | 2020-11-28 | North America/USA/Illinois/Mchenry Co | Northwestern Memorial Hospital | Ozer Lab | Ramon Lorenzo-Redondo et al |
| EPI_ISL_936646 | USA/IL-NM-4753/2020 | 2020-12-16 | North America/USA/Illinois/Chicago | Northwestern Memorial Hospital | Ozer Lab | Ramon Lorenzo-Redondo et al |
| EPI_ISL_936648 | USA/IL-NM-4756/2020 | 2020-12-16 | North America/USA/Illinois/Cook County | Northwestern Memorial Hospital | Ozer Lab | Ramon Lorenzo-Redondo et al |
| EPI_ISL_936654 | USA/IL-NM-4774/2020 | 2020-12-15 | North America/USA/Illinois/Chicago | Northwestern Memorial Hospital | Ozer Lab | Ramon Lorenzo-Redondo et al |
| EPI_ISL_936658 | USA/IL-NM-4782/2020 | 2020-12-15 | North America/USA/Illinois/Lake County | Northwestern Memorial Hospital | Ozer Lab | Ramon Lorenzo-Redondo et al |
| EPI_ISL_936665 | USA/IL-NM-4795/2020 | 2020-12-19 | North America/USA/Illinois/Cook County | Northwestern Memorial Hospital | Ozer Lab | Ramon Lorenzo-Redondo et al |
| EPI_ISL_936673 | USA/IL-NM-4816/2020 | 2020-12-20 | North America/USA/Illinois/Chicago | Northwestern Memorial Hospital | Ozer Lab | Ramon Lorenzo-Redondo et al |

| | | | | | | |
|---|---|---|---|---|---|---|
| EPI_ISL_936676 | USA/IL-NM-4819/2020 | 2020-12-19 | North America/USA/Illinois/Cook Count | Northwestern Memorial Hospital | Ozer Lab | Ramon Lorenzo-Redondo et al |
| EPI_ISL_936679 | USA/IL-NM-4826/2020 | 2020-12-19 | North America/USA/Illinois/Chicago | Northwestern Memorial Hospital | Ozer Lab | Ramon Lorenzo-Redondo et al |
| EPI_ISL_936689 | USA/IL-NM-4842/2020 | 2020-12-18 | North America/USA/Illinois/Chicago | Northwestern Memorial Hospital | Ozer Lab | Ramon Lorenzo-Redondo et al |
| EPI_ISL_936690 | USA/IL-NM-4843/2020 | 2020-12-19 | North America/USA/Illinois/Chicago | Northwestern Memorial Hospital | Ozer Lab | Ramon Lorenzo-Redondo et al |
| EPI_ISL_936693 | USA/IL-NM-4846/2020 | 2020-12-18 | North America/USA/Illinois/Chicago | Northwestern Memorial Hospital | Ozer Lab | Ramon Lorenzo-Redondo et al |
| EPI_ISL_936701 | USA/IL-NM-5019/2020 | 2020-12-21 | North America/USA/Illinois/Chicago | Northwestern Memorial Hospital | Ozer Lab | Ramon Lorenzo-Redondo et al |
| EPI_ISL_936726 | USA/IL-NM-5053/2020 | 2020-12-23 | North America/USA/Illinois/Chicago | Northwestern Memorial Hospital | Ozer Lab | Ramon Lorenzo-Redondo et al |
| EPI_ISL_936732 | USA/IL-NM-5059/2020 | 2020-12-23 | North America/USA/Illinois/Chicago | Northwestern Memorial Hospital | Ozer Lab | Ramon Lorenzo-Redondo et al |
| EPI_ISL_936738 | USA/IL-NM-5066/2020 | 2020-12-23 | North America/USA/Illinois/Chicago | Northwestern Memorial Hospital | Ozer Lab | Ramon Lorenzo-Redondo et al |
| EPI_ISL_936748 | USA/IL-NM-5083/2020 | 2020-12-23 | North America/USA/Illinois/Cook Count | Northwestern Memorial Hospital | Ozer Lab | Ramon Lorenzo-Redondo et al |
| EPI_ISL_936753 | USA/IL-NM-5090/2020 | 2020-12-23 | North America/USA/Illinois/Lake County | Northwestern Memorial Hospital | Ozer Lab | Ramon Lorenzo-Redondo et al |
| EPI_ISL_936755 | USA/IL-NM-5095/2020 | 2020-12-23 | North America/USA/Illinois/Cook Count | Northwestern Memorial Hospital | Ozer Lab | Ramon Lorenzo-Redondo et al |
| EPI_ISL_936756 | USA/IL-NM-5096/2020 | 2020-12-23 | North America/USA/Illinois/Chicago | Northwestern Memorial Hospital | Ozer Lab | Ramon Lorenzo-Redondo et al |
| EPI_ISL_936760 | USA/IL-NM-5102/2020 | 2020-12-23 | North America/USA/Illinois/Chicago | Northwestern Memorial Hospital | Ozer Lab | Ramon Lorenzo-Redondo et al |
| EPI_ISL_936774 | USA/IL-NM-5126/2020 | 2020-12-24 | North America/USA/Illinois/Chicago | Northwestern Memorial Hospital | Ozer Lab | Ramon Lorenzo-Redondo et al |
| EPI_ISL_936778 | USA/IL-NM-5136/2020 | 2020-12-24 | North America/USA/Illinois/Chicago | Northwestern Memorial Hospital | Ozer Lab | Ramon Lorenzo-Redondo et al |
| EPI_ISL_936781 | USA/IL-NM-5141/2020 | 2020-12-24 | North America/USA/Illinois/Cook Count | Northwestern Memorial Hospital | Ozer Lab | Ramon Lorenzo-Redondo et al |
| EPI_ISL_936793 | USA/IL-NM-5178/2020 | 2020-12-26 | North America/USA/Illinois/Chicago | Northwestern Memorial Hospital | Ozer Lab | Ramon Lorenzo-Redondo et al |
| EPI_ISL_936802 | USA/IL-NM-5191/2020 | 2020-12-26 | North America/USA/Illinois/Lake County | Northwestern Memorial Hospital | Ozer Lab | Ramon Lorenzo-Redondo et al |
| EPI_ISL_936811 | USA/IL-NM-5200/2020 | 2020-12-26 | North America/USA/Illinois/Chicago | Northwestern Memorial Hospital | Ozer Lab | Ramon Lorenzo-Redondo et al |
| EPI_ISL_936812 | USA/IL-NM-5207/2020 | 2020-12-26 | North America/USA/Illinois/Chicago | Northwestern Memorial Hospital | Ozer Lab | Ramon Lorenzo-Redondo et al |
| EPI_ISL_936814 | USA/IL-NM-5228/2020 | 2020-12-27 | North America/USA/Illinois/Chicago | Northwestern Memorial Hospital | Ozer Lab | Ramon Lorenzo-Redondo et al |
| EPI_ISL_936829 | USA/IL-NM-5399/2020 | 2020-09-01 | North America/USA/Illinois/Chicago | Northwestern Memorial Hospital | Ozer Lab | Ramon Lorenzo-Redondo et al |
| EPI_ISL_936830 | USA/IL-NM-5409/2020 | 2020-09-02 | North America/USA/Illinois/Chicago | Northwestern Memorial Hospital | Ozer Lab | Ramon Lorenzo-Redondo et al |
| EPI_ISL_936834 | USA/IL-NM-5442/2020 | 2020-09-06 | North America/USA/Illinois/Cook Count | Northwestern Memorial Hospital | Ozer Lab | Ramon Lorenzo-Redondo et al |
| EPI_ISL_936835 | USA/IL-NM-5447/2020 | 2020-09-08 | North America/USA/Illinois/Chicago | Northwestern Memorial Hospital | Ozer Lab | Ramon Lorenzo-Redondo et al |
| EPI_ISL_936837 | USA/IL-NM-5459/2020 | 2020-09-10 | North America/USA/Illinois/Chicago | Northwestern Memorial Hospital | Ozer Lab | Ramon Lorenzo-Redondo et al |
| EPI_ISL_936839 | USA/IL-NM-5471/2020 | 2020-09-14 | North America/USA/Illinois/Chicago | Northwestern Memorial Hospital | Ozer Lab | Ramon Lorenzo-Redondo et al |
| EPI_ISL_936840 | USA/IL-NM-5479/2020 | 2020-09-15 | North America/USA/Illinois/Chicago | Northwestern Memorial Hospital | Ozer Lab | Ramon Lorenzo-Redondo et al |
| EPI_ISL_936841 | USA/IL-NM-5482/2020 | 2020-09-16 | North America/USA/Illinois/Chicago | Northwestern Memorial Hospital | Ozer Lab | Ramon Lorenzo-Redondo et al |
| EPI_ISL_936856 | USA/IL-NM-5526/2020 | 2020-10-11 | North America/USA/Illinois/Cook Count | Northwestern Memorial Hospital | Ozer Lab | Ramon Lorenzo-Redondo et al |

| | | | | | | |
|---|---|---|---|---|---|---|
| EPI_ISL_936867 | USA/IL-NM-5792/2021 | 2021-01-04 | North America/USA/Indiana/Lake Count | Northwestern Memorial Hospital | Ozer Lab | Ramon Lorenzo-Redondo et al |
| EPI_ISL_936883 | USA/IL-NM-5812/2021 | 2021-01-03 | North America/USA/Illinois/Cook County | Northwestern Memorial Hospital | Ozer Lab | Ramon Lorenzo-Redondo et al |
| EPI_ISL_936885 | USA/IL-NM-5814/2021 | 2021-01-04 | North America/USA/Illinois/Chicago | Northwestern Memorial Hospital | Ozer Lab | Ramon Lorenzo-Redondo et al |
| EPI_ISL_936888 | USA/IL-NM-5817/2021 | 2021-01-05 | North America/USA/Illinois/Cook County | Northwestern Memorial Hospital | Ozer Lab | Ramon Lorenzo-Redondo et al |
| EPI_ISL_936898 | USA/IL-NM-5828/2021 | 2021-01-04 | North America/USA/Illinois/Chicago | Northwestern Memorial Hospital | Ozer Lab | Ramon Lorenzo-Redondo et al |
| EPI_ISL_936903 | USA/IL-NM-5834/2021 | 2021-01-04 | North America/USA/Illinois/Cook County | Northwestern Memorial Hospital | Ozer Lab | Ramon Lorenzo-Redondo et al |
| EPI_ISL_936913 | USA/IL-NM-5846/2021 | 2021-01-05 | North America/USA/Illinois/Chicago | Northwestern Memorial Hospital | Ozer Lab | Ramon Lorenzo-Redondo et al |
| EPI_ISL_936920 | USA/IL-NM-5854/2021 | 2021-01-02 | North America/USA/Illinois/Cook County | Northwestern Memorial Hospital | Ozer Lab | Ramon Lorenzo-Redondo et al |
| EPI_ISL_936923 | USA/IL-NM-5859/2021 | 2021-01-05 | North America/USA/Illinois/Dupage Cou | Northwestern Memorial Hospital | Ozer Lab | Ramon Lorenzo-Redondo et al |
| EPI_ISL_936934 | USA/IL-NM-5873/2021 | 2021-01-06 | North America/USA/Illinois/Chicago | Northwestern Memorial Hospital | Ozer Lab | Ramon Lorenzo-Redondo et al |
| EPI_ISL_936939 | USA/IL-NM-5878/2021 | 2021-01-06 | North America/USA/Illinois/Chicago | Northwestern Memorial Hospital | Ozer Lab | Ramon Lorenzo-Redondo et al |
| EPI_ISL_936945 | USA/IL-NM-5885/2021 | 2021-01-06 | North America/USA/Illinois/Lake County | Northwestern Memorial Hospital | Ozer Lab | Ramon Lorenzo-Redondo et al |
| EPI_ISL_936954 | USA/IL-NM-5902/2021 | 2021-01-07 | North America/USA/Illinois/Cook County | Northwestern Memorial Hospital | Ozer Lab | Ramon Lorenzo-Redondo et al |
| EPI_ISL_936962 | USA/IL-NM-5911/2021 | 2021-01-07 | North America/USA/Illinois/Chicago | Northwestern Memorial Hospital | Ozer Lab | Ramon Lorenzo-Redondo et al |
| EPI_ISL_936963 | USA/IL-NM-5912/2021 | 2021-01-07 | North America/USA/Illinois/Chicago | Northwestern Memorial Hospital | Ozer Lab | Ramon Lorenzo-Redondo et al |
| EPI_ISL_936971 | USA/IL-NM-5927/2021 | 2021-01-08 | North America/USA/Illinois/Chicago | Northwestern Memorial Hospital | Ozer Lab | Ramon Lorenzo-Redondo et al |
| EPI_ISL_936972 | USA/IL-NM-5928/2021 | 2021-01-08 | North America/USA/Illinois/Chicago | Northwestern Memorial Hospital | Ozer Lab | Ramon Lorenzo-Redondo et al |
| EPI_ISL_936998 | USA/IL-NM-5964/2021 | 2021-01-09 | North America/USA/Illinois/Chicago | Northwestern Memorial Hospital | Ozer Lab | Ramon Lorenzo-Redondo et al |
| EPI_ISL_937004 | USA/IL-NM-5972/2021 | 2021-01-09 | North America/USA/Illinois/Cook County | Northwestern Memorial Hospital | Ozer Lab | Ramon Lorenzo-Redondo et al |
| EPI_ISL_937005 | USA/IL-NM-5973/2021 | 2021-01-09 | North America/USA/Illinois/Lake County | Northwestern Memorial Hospital | Ozer Lab | Ramon Lorenzo-Redondo et al |
| EPI_ISL_937007 | USA/IL-NM-5975/2021 | 2021-01-09 | North America/USA/Illinois/Chicago | Northwestern Memorial Hospital | Ozer Lab | Ramon Lorenzo-Redondo et al |
| EPI_ISL_937010 | USA/IL-NM-5978/2021 | 2021-01-09 | North America/USA/Illinois/Lake County | Northwestern Memorial Hospital | Ozer Lab | Ramon Lorenzo-Redondo et al |
| EPI_ISL_937016 | USA/IL-NM-5985/2021 | 2021-01-10 | North America/USA/Illinois/Cook County | Northwestern Memorial Hospital | Ozer Lab | Ramon Lorenzo-Redondo et al |
| EPI_ISL_937018 | USA/IL-NM-5987/2021 | 2021-01-10 | North America/USA/Illinois/Cook County | Northwestern Memorial Hospital | Ozer Lab | Ramon Lorenzo-Redondo et al |
| EPI_ISL_937021 | USA/IL-NM-5993/2021 | 2021-01-10 | North America/USA/Illinois/Chicago | Northwestern Memorial Hospital | Ozer Lab | Ramon Lorenzo-Redondo et al |
| EPI_ISL_937023 | USA/IL-NM-5996/2021 | 2021-01-10 | North America/USA/Illinois/Cook County | Northwestern Memorial Hospital | Ozer Lab | Ramon Lorenzo-Redondo et al |
| EPI_ISL_937027 | USA/CA-QDX-4479/2020 | 2020-08-13 | North America/USA/California/ | Quest Diagnostics | Quest Diagnostics | Rosenthal et al |
| EPI_ISL_937035 | USA/CO-QDX-4467/2020 | 2020-08-25 | North America/USA/Colorado/ | Quest Diagnostics | Quest Diagnostics | Rosenthal et al |
| EPI_ISL_937036 | USA/CT-QDX-4499/2020 | 2020-09-15 | North America/USA/Connecticut/ | Quest Diagnostics | Quest Diagnostics | Rosenthal et al |
| EPI_ISL_937037 | USA/CT-QDX-4500/2020 | 2020-09-29 | North America/USA/Connecticut/ | Quest Diagnostics | Quest Diagnostics | Rosenthal et al |
| EPI_ISL_937045 | USA/FL-QDX-4486/2020 | 2020-09-03 | North America/USA/Florida/ | Quest Diagnostics | Quest Diagnostics | Rosenthal et al |

| | | | | | | |
|---|---|---|---|---|---|---|
| EPI_ISL_937054 | USA/GA-QDX-4508/2020 | 2020-10-09 | North America/USA/Georgia/ | Quest Diagnostics | Quest Diagnostics | Rosenthal et al |
| EPI_ISL_937055 | USA/GA-QDX-4509/2020 | 2020-10-11 | North America/USA/Georgia/ | Quest Diagnostics | Quest Diagnostics | Rosenthal et al |
| EPI_ISL_937058 | USA/IL-QDX-4443/2020 | 2020-11-11 | North America/USA/Illinois/ | Quest Diagnostics | Quest Diagnostics | Rosenthal et al |
| EPI_ISL_937064 | USA/KS-QDX-4492/2020 | 2020-09-12 | North America/USA/Kansas/ | Quest Diagnostics | Quest Diagnostics | Rosenthal et al |
| EPI_ISL_937065 | USA/KY-QDX-4442/2020 | 2020-11-12 | North America/USA/Kentucky/ | Quest Diagnostics | Quest Diagnostics | Rosenthal et al |
| EPI_ISL_937072 | USA/MO-QDX-4435/2020 | 2020-11-14 | North America/USA/Missouri/ | Quest Diagnostics | Quest Diagnostics | Rosenthal et al |
| EPI_ISL_937075 | USA/MO-QDX-4439/2020 | 2020-11-13 | North America/USA/Missouri/ | Quest Diagnostics | Quest Diagnostics | Rosenthal et al |
| EPI_ISL_937076 | USA/MO-QDX-4445/2020 | 2020-08-13 | North America/USA/Missouri/ | Quest Diagnostics | Quest Diagnostics | Rosenthal et al |
| EPI_ISL_937077 | USA/MO-QDX-4446/2020 | 2020-08-14 | North America/USA/Missouri/ | Quest Diagnostics | Quest Diagnostics | Rosenthal et al |
| EPI_ISL_937080 | USA/MO-QDX-4493/2020 | 2020-09-12 | North America/USA/Missouri/ | Quest Diagnostics | Quest Diagnostics | Rosenthal et al |
| EPI_ISL_937089 | USA/NC-QDX-4459/2020 | 2020-08-19 | North America/USA/North Carolina/ | Quest Diagnostics | Quest Diagnostics | Rosenthal et al |
| EPI_ISL_937091 | USA/NC-QDX-4475/2020 | 2020-08-23 | North America/USA/North Carolina/ | Quest Diagnostics | Quest Diagnostics | Rosenthal et al |
| EPI_ISL_937096 | USA/NV-QDX-4468/2020 | 2020-08-23 | North America/USA/Nevada/ | Quest Diagnostics | Quest Diagnostics | Rosenthal et al |
| EPI_ISL_937098 | USA/NV-QDX-4503/2020 | 2020-09-26 | North America/USA/Nevada/ | Quest Diagnostics | Quest Diagnostics | Rosenthal et al |
| EPI_ISL_937099 | USA/NV-QDX-4504/2020 | 2020-09-24 | North America/USA/Nevada/ | Quest Diagnostics | Quest Diagnostics | Rosenthal et al |
| EPI_ISL_937103 | USA/OK-QDX-4520/2020 | 2020-10-09 | North America/USA/Oklahoma/ | Quest Diagnostics | Quest Diagnostics | Rosenthal et al |
| EPI_ISL_937104 | USA/OK-QDX-4521/2020 | 2020-10-08 | North America/USA/Oklahoma/ | Quest Diagnostics | Quest Diagnostics | Rosenthal et al |
| EPI_ISL_937110 | USA/TX-QDX-4470/2020 | 2020-08-25 | North America/USA/Texas/ | Quest Diagnostics | Quest Diagnostics | Rosenthal et al |
| EPI_ISL_937119 | USA/TX-QDX-4525/2020 | 2020-10-20 | North America/USA/Texas/ | Quest Diagnostics | Quest Diagnostics | Rosenthal et al |
| EPI_ISL_937120 | USA/VA-QDX-4476/2020 | 2020-08-27 | North America/USA/Virginia/ | Quest Diagnostics | Quest Diagnostics | Rosenthal et al |
| EPI_ISL_937128 | USA/NY-NYCPHL-002550/2021 | 2021-01-19 | North America/USA/New York/New Yor | DOHMH Riverside | New York City Public Health Laboratory | Jade Wang et al |
| EPI_ISL_937136 | USA/NY-NYCPHL-002560/2021 | 2021-01-19 | North America/USA/New York/New Yor | DOHMH Chelsea | New York City Public Health Laboratory | Jade Wang et al |
| EPI_ISL_937137 | USA/NY-NYCPHL-002561/2021 | 2021-01-19 | North America/USA/New York/New Yor | DOHMH Chelsea | New York City Public Health Laboratory | Jade Wang et al |
| EPI_ISL_937138 | USA/NY-NYCPHL-002563/2021 | 2021-01-19 | North America/USA/New York/New Yor | DOHMH Crown Heights | New York City Public Health Laboratory | Jade Wang et al |

| | | | | | | |
|---|---|---|---|---|---|---|
| EPI_ISL_937141 | USA/NY-NYCPHL-002566/2021 | 2021-01-19 | North America/USA/New York/New Yor | DOHMH Corona | New York City Public Health Laboratory | Jade Wang et al |
| EPI_ISL_937144 | USA/NY-NYCPHL-002569/2021 | 2021-01-19 | North America/USA/New York/New Yor | DOHMH Corona | New York City Public Health Laboratory | Jade Wang et al |
| EPI_ISL_937146 | USA/NY-NYCPHL-002572/2021 | 2021-01-19 | North America/USA/New York/New Yor | DOHMH Jamaica | New York City Public Health Laboratory | Jade Wang et al |
| EPI_ISL_937152 | USA/NY-NYCPHL-002579/2021 | 2021-01-19 | North America/USA/New York/New Yor | DOHMH PHL | New York City Public Health Laboratory | Jade Wang et al |
| EPI_ISL_937155 | USA/NY-NYCPHL-002582/2021 | 2021-01-15 | North America/USA/New York/New Yor | DOHMH PHL | New York City Public Health Laboratory | Jade Wang et al |
| EPI_ISL_937163 | USA/NY-NYCPHL-002590/2021 | 2021-01-20 | North America/USA/New York/New Yor | DOHMH Jamaica | New York City Public Health Laboratory | Jade Wang et al |
| EPI_ISL_937168 | USA/NY-NYCPHL-002619/2021 | 2021-01-19 | North America/USA/New York/New Yor | DOHMH Crown Heights | New York City Public Health Laboratory | Jade Wang et al |
| EPI_ISL_937170 | USA/NY-NYCPHL-002631/2021 | 2021-01-20 | North America/USA/New York/New Yor | DOHMH PHL | New York City Public Health Laboratory | Jade Wang et al |
| EPI_ISL_937172 | USA/NY-NYCPHL-002636/2021 | 2021-01-21 | North America/USA/New York/New Yor | DOHMH Central Harlem | New York City Public Health Laboratory | Jade Wang et al |
| EPI_ISL_937179 | USA/NY-NYCPHL-002663/2021 | 2021-01-21 | North America/USA/New York/New Yor | DOHMH Corona | New York City Public Health Laboratory | Jade Wang et al |
| EPI_ISL_937183 | USA/NY-NYCPHL-002668/2021 | 2021-01-21 | North America/USA/New York/New Yor | DOHMH Corona | New York City Public Health Laboratory | Jade Wang et al |
| EPI_ISL_937184 | USA/NY-NYCPHL-002669/2021 | 2021-01-21 | North America/USA/New York/New Yor | DOHMH Corona | New York City Public Health Laboratory | Jade Wang et al |
| EPI_ISL_937191 | USA/NY-NYCPHL-002681/2021 | 2021-01-18 | North America/USA/New York/New Yor | Department of Homeless Services | New York City Public Health Laboratory | Jade Wang et al |

| | | | | | | |
|---|---|---|---|---|---|---|
| EPI_ISL_937202 | USA/NY-NYCPHL-002699/2021 | 2021-01-22 | North America/USA/New York/New Yor | DOHMH Chelsea | New York City Public Health Laboratory | Jade Wang et al |
| EPI_ISL_937206 | USA/NY-NYCPHL-002705/2021 | 2021-01-22 | North America/USA/New York/New Yor | DOHMH Morrisania | New York City Public Health Laboratory | Jade Wang et al |
| EPI_ISL_937219 | USA/NY-NYCPHL-002735/2021 | 2021-01-25 | North America/USA/New York/New Yor | DOHMH Jamaica | New York City Public Health Laboratory | Jade Wang et al |
| EPI_ISL_937222 | USA/NY-NYCPHL-002739/2021 | 2021-01-25 | North America/USA/New York/New Yor | DOHMH Jamaica | New York City Public Health Laboratory | Jade Wang et al |
| EPI_ISL_937227 | USA/NY-NYCPHL-002748/2021 | 2021-01-25 | North America/USA/New York/New Yor | DOHMH Corona | New York City Public Health Laboratory | Jade Wang et al |
| EPI_ISL_937228 | USA/NY-NYCPHL-002754/2021 | 2021-01-25 | North America/USA/New York/New Yor | DOHMH Corona | New York City Public Health Laboratory | Jade Wang et al |
| EPI_ISL_937238 | USA/NY-NYCPHL-002770/2021 | 2021-01-25 | North America/USA/New York/New Yor | DOHMH Morrisania | New York City Public Health Laboratory | Jade Wang et al |
| EPI_ISL_937263 | USA/NY-NYCPHL-002836/2021 | 2021-01-24 | North America/USA/New York/New Yor | NYC HH Elmhurst Hospital Medical Center | New York City Public Health Laboratory | Jade Wang et al |
| EPI_ISL_937297 | USA/UT-UPHL-2101715734/2020 | 2020-12-12 | North America/USA/Utah/ | Utah Public Health Laboratory | Utah Public Health Laboratory | Erin L. Young et al |
| EPI_ISL_937318 | USA/UT-UPHL-2101422832/2020 | 2020-12-19 | North America/USA/Utah/ | Utah Public Health Laboratory | Utah Public Health Laboratory | Erin L. Young et al |
| EPI_ISL_937319 | USA/UT-UPHL-2101081298/2020 | 2020-12-12 | North America/USA/Utah/ | Utah Public Health Laboratory | Utah Public Health Laboratory | Erin L. Young et al |
| EPI_ISL_937334 | USA/UT-UPHL-2101553593/2020 | 2020-05-23 | North America/USA/Utah/ | Utah Public Health Laboratory | Utah Public Health Laboratory | Erin L. Young et al |
| EPI_ISL_937337 | USA/UT-UPHL-2101632494/2020 | 2020-05-23 | North America/USA/Utah/ | Utah Public Health Laboratory | Utah Public Health Laboratory | Erin L. Young et al |
| EPI_ISL_937344 | USA/UT-UPHL-2101565524/2020 | 2020-12-10 | North America/USA/Utah/ | Utah Public Health Laboratory | Utah Public Health Laboratory | Erin L. Young et al |
| EPI_ISL_937364 | USA/UT-UPHL-2101868759/2021 | 2021-01-12 | North America/USA/Utah/ | Utah Public Health Laboratory | Utah Public Health Laboratory | Erin L. Young et al |
| EPI_ISL_937370 | USA/UT-UPHL-2101067394/2021 | 2021-01-13 | North America/USA/Utah/ | Utah Public Health Laboratory | Utah Public Health Laboratory | Erin L. Young et al |

| | | | | | | |
|---|---|---|---|---|---|---|
| EPI_ISL_937384 | USA/ME-HETL-J1054/2021 | 2021-01-18 | North America/USA/Maine/ | Maine Health and Environmental Testing Laboratory (Maine HETL) | Tewhey Lab, The Jackson Laboratory | Matluk et al |
| EPI_ISL_937388 | USA/ME-HETL-J1059/2021 | 2021-01-19 | North America/USA/Maine/ | Maine Health and Environmental Testing Laboratory (Maine HETL) | Tewhey Lab, The Jackson Laboratory | Matluk et al |
| EPI_ISL_937389 | USA/ME-HETL-J1061/2021 | 2021-01-19 | North America/USA/Maine/ | Maine Health and Environmental Testing Laboratory (Maine HETL) | Tewhey Lab, The Jackson Laboratory | Matluk et al |
| EPI_ISL_937397 | USA/ME-HETL-J1072/2021 | 2021-01-17 | North America/USA/Maine/ | Maine Health and Environmental Testing Laboratory (Maine HETL) | Tewhey Lab, The Jackson Laboratory | Matluk et al |
| EPI_ISL_937412 | USA/ME-HETL-J1094/2021 | 2021-01-16 | North America/USA/Maine/ | Maine Health and Environmental Testing Laboratory (Maine HETL) | Tewhey Lab, The Jackson Laboratory | Matluk et al |
| EPI_ISL_937413 | USA/ME-HETL-J1095/2021 | 2021-01-17 | North America/USA/Maine/ | Maine Health and Environmental Testing Laboratory (Maine HETL) | Tewhey Lab, The Jackson Laboratory | Matluk et al |
| EPI_ISL_937423 | USA/ME-HETL-J1107/2021 | 2021-01-20 | North America/USA/Maine/ | Maine Health and Environmental Testing Laboratory (Maine HETL) | Tewhey Lab, The Jackson Laboratory | Matluk et al |
| EPI_ISL_937453 | USA/ME-HETL-J1142/2021 | 2021-01-18 | North America/USA/Maine/ | Maine Health and Environmental Testing Laboratory (Maine HETL) | Tewhey Lab, The Jackson Laboratory | Matluk et al |
| EPI_ISL_937459 | USA/ME-HETL-J1148/2021 | 2021-01-25 | North America/USA/Maine/ | Maine Health and Environmental Testing Laboratory (Maine HETL) | Tewhey Lab, The Jackson Laboratory | Matluk et al |
| EPI_ISL_937461 | USA/ME-HETL-J1150/2021 | 2021-01-25 | North America/USA/Maine/ | Maine Health and Environmental Testing Laboratory (Maine HETL) | Tewhey Lab, The Jackson Laboratory | Matluk et al |
| EPI_ISL_937470 | USA/ME-HETL-J1171/2021 | 2021-01-29 | North America/USA/Maine/ | Maine Health and Environmental Testing Laboratory (Maine HETL) | Tewhey Lab, The Jackson Laboratory | Matluk et al |
| EPI_ISL_937475 | USA/ME-HETL-J1179/2021 | 2021-01-29 | North America/USA/Maine/ | Maine Health and Environmental Testing Laboratory (Maine HETL) | Tewhey Lab, The Jackson Laboratory | Matluk et al |
| EPI_ISL_937476 | USA/ME-HETL-J1183/2021 | 2021-01-29 | North America/USA/Maine/ | Maine Health and Environmental Testing Laboratory (Maine HETL) | Tewhey Lab, The Jackson Laboratory | Matluk et al |

| | | | | | | |
|---|---|---|---|---|---|---|
| EPI_ISL_937488 | USA/ME-HETL-J1204/2021 | 2021-01-29 | North America/USA/Maine/ | Maine Health and Environmental Testing Laboratory (Maine HETL) | Tewhey Lab, The Jackson Laboratory | Matluk et al |
| EPI_ISL_937490 | USA/ME-HETL-J1206/2021 | 2021-01-29 | North America/USA/Maine/ | Maine Health and Environmental Testing Laboratory (Maine HETL) | Tewhey Lab, The Jackson Laboratory | Matluk et al |
| EPI_ISL_937493 | USA/ME-HETL-J1212/2021 | 2021-01-23 | North America/USA/Maine/ | Maine Health and Environmental Testing Laboratory (Maine HETL) | Tewhey Lab, The Jackson Laboratory | Matluk et al |
| EPI_ISL_937497 | USA/ME-HETL-J1218/2021 | 2021-01-25 | North America/USA/Maine/ | Maine Health and Environmental Testing Laboratory (Maine HETL) | Tewhey Lab, The Jackson Laboratory | Matluk et al |
| EPI_ISL_937500 | USA/ME-HETL-J1224/2021 | 2021-01-25 | North America/USA/Maine/ | Maine Health and Environmental Testing Laboratory (Maine HETL) | Tewhey Lab, The Jackson Laboratory | Matluk et al |
| EPI_ISL_937504 | USA/ME-HETL-J1229/2021 | 2021-01-27 | North America/USA/Maine/ | Maine Health and Environmental Testing Laboratory (Maine HETL) | Tewhey Lab, The Jackson Laboratory | Matluk et al |
| EPI_ISL_937505 | USA/ME-HETL-J1232/2021 | 2021-01-26 | North America/USA/Maine/ | Maine Health and Environmental Testing Laboratory (Maine HETL) | Tewhey Lab, The Jackson Laboratory | Matluk et al |
| EPI_ISL_937506 | USA/ME-HETL-J1233/2021 | 2021-01-28 | North America/USA/Maine/ | Maine Health and Environmental Testing Laboratory (Maine HETL) | Tewhey Lab, The Jackson Laboratory | Matluk et al |
| EPI_ISL_937509 | USA/ME-HETL-J1236/2021 | 2021-01-29 | North America/USA/Maine/ | Maine Health and Environmental Testing Laboratory (Maine HETL) | Tewhey Lab, The Jackson Laboratory | Matluk et al |
| EPI_ISL_937513 | USA/ME-HETL-J1242/2021 | 2021-01-23 | North America/USA/Maine/ | Maine Health and Environmental Testing Laboratory (Maine HETL) | Tewhey Lab, The Jackson Laboratory | Matluk et al |
| EPI_ISL_937514 | Canada/MB-NML-1118/2020 | 2020-08-05 | North America/Canada/Manitoba/ | Cadham Provincial laboratory | National Microbiology Laboratory (NML) | Anna Majer et al |
| EPI_ISL_937535 | Thailand/CU-SI2011588-NT/2020 | 2020-04-04 | Asia/Thailand/Thailand/ | Thai Red Cross Emerging Infectious Diseases Health Science Centre, Chulalongkorn Hospital, Faculty of Medicine, Chulalongkorn University | Thai Red Cross Emerging Infectious Diseases Center and Faculty of Medicine, Chulalongkorn University | Opass Putcharoen et al |

| | | | | | | |
|---|---|---|---|---|---|---|
| EPI_ISL_937569 | England/ALDP-116F52A/2021 | 2021-01-24 | Europe/United Kingdom/England/ | Lighthouse Lab in Alderley Park | Wellcome Sanger Institute for the COVID-19 Genomics UK (COG-UK) Consortium | Jacquelyn Wynn et al |
| EPI_ISL_937877 | England/ALDP-115D097/2021 | 2021-01-24 | Europe/United Kingdom/England/ | Lighthouse Lab in Alderley Park | Wellcome Sanger Institute for the COVID-19 Genomics UK (COG-UK) Consortium | Jacquelyn Wynn et al |
| EPI_ISL_937989 | England/MILK-1152C25/2021 | 2021-01-22 | Europe/United Kingdom/England/ | Lighthouse Lab in Milton Keynes | Wellcome Sanger Institute for the COVID-19 Genomics UK (COG-UK) Consortium | The Lighthouse Lab in Milton Keynes et al |
| EPI_ISL_938120 | England/MILK-1152539/2021 | 2021-01-19 | Europe/United Kingdom/England/ | Lighthouse Lab in Milton Keynes | Wellcome Sanger Institute for the COVID-19 Genomics UK (COG-UK) Consortium | The Lighthouse Lab in Milton Keynes et al |
| EPI_ISL_938245 | England/CAMC-115EC92/2021 | 2021-01-23 | Europe/United Kingdom/England/ | Lighthouse Lab in Cambridge | Wellcome Sanger Institute for the COVID-19 Genomics UK (COG-UK) Consortium | Rob Howes et al |
| EPI_ISL_938313 | England/CAMC-115E597/2021 | 2021-01-21 | Europe/United Kingdom/England/ | Lighthouse Lab in Cambridge | Wellcome Sanger Institute for the COVID-19 Genomics UK (COG-UK) Consortium | Rob Howes et al |
| EPI_ISL_938608 | England/MILK-116C5E1/2021 | 2021-01-25 | Europe/United Kingdom/England/ | Lighthouse Lab in Milton Keynes | Wellcome Sanger Institute for the COVID-19 Genomics UK (COG-UK) Consortium | The Lighthouse Lab in Milton Keynes et al |
| EPI_ISL_938685 | England/MILK-1169502/2021 | 2021-01-25 | Europe/United Kingdom/England/ | Lighthouse Lab in Milton Keynes | Wellcome Sanger Institute for the COVID-19 Genomics UK (COG-UK) Consortium | The Lighthouse Lab in Milton Keynes et al |

| | | | | | | |
|---|---|---|---|---|---|---|
| EPI_ISL_938844 | England/MILK-116C323/2021 | 2021-01-25 | Europe/United Kingdom/England/ | Lighthouse Lab in Milton Keynes | Wellcome Sanger Institute for the COVID-19 Genomics UK (COG-UK) Consortium | The Lighthouse Lab in Milton Keynes et al |
| EPI_ISL_938959 | England/MILK-1150FA6/2021 | 2021-01-22 | Europe/United Kingdom/England/ | Lighthouse Lab in Milton Keynes | Wellcome Sanger Institute for the COVID-19 Genomics UK (COG-UK) Consortium | The Lighthouse Lab in Milton Keynes et al |
| EPI_ISL_939094 | England/MILK-1158D0E/2021 | 2021-01-24 | Europe/United Kingdom/England/ | Lighthouse Lab in Milton Keynes | Wellcome Sanger Institute for the COVID-19 Genomics UK (COG-UK) Consortium | The Lighthouse Lab in Milton Keynes et al |
| EPI_ISL_939136 | England/MILK-115AD93/2021 | 2021-01-24 | Europe/United Kingdom/England/ | Lighthouse Lab in Milton Keynes | Wellcome Sanger Institute for the COVID-19 Genomics UK (COG-UK) Consortium | The Lighthouse Lab in Milton Keynes et al |
| EPI_ISL_939402 | England/MILK-11538C6/2021 | 2021-01-22 | Europe/United Kingdom/England/ | Lighthouse Lab in Milton Keynes | Wellcome Sanger Institute for the COVID-19 Genomics UK (COG-UK) Consortium | The Lighthouse Lab in Milton Keynes et al |
| EPI_ISL_939544 | England/QEUH-114C329/2021 | 2021-01-25 | Europe/United Kingdom/England/ | Lighthouse Lab in Glasgow | Wellcome Sanger Institute for the COVID-19 Genomics UK (COG-UK) Consortium | Harper VanSteenhouse et al |
| EPI_ISL_939613 | USA/CA-LACPHL-AF00319/2021 | 2021-01-22 | North America/USA/California/Los Ange | Los Angeles County PHL | Los Angeles County PHL | P. Hemarajata et al. et al |
| EPI_ISL_939616 | USA/CA-LACPHL-AF00322/2021 | 2021-01-21 | North America/USA/California/Los Ange | Los Angeles County PHL | Los Angeles County PHL | P. Hemarajata et al. et al |
| EPI_ISL_939621 | USA/CA-LACPHL-AE00062/2020 | 2020-08-19 | North America/USA/California/Los Ange | USC Clinical Lab | Los Angeles County PHL | P. Hemarajata et al. et al |
| EPI_ISL_939623 | USA/CA-LACPHL-AE00064/2020 | 2020-08-20 | North America/USA/California/Los Ange | USC Clinical Lab | Los Angeles County PHL | P. Hemarajata et al. et al |
| EPI_ISL_939624 | USA/CA-LACPHL-AE00065/2020 | 2020-08-20 | North America/USA/California/Los Ange | USC Clinical Lab | Los Angeles County PHL | P. Hemarajata et al. et al |

| | | | | | | |
|---|---|---|---|---|---|---|
| EPI_ISL_939625 | USA/CA-LACPHL-AE00066/2020 | 2020-08-21 | North America/USA/California/Los Ange | USC Clinical Lab | Los Angeles County PHL | P. Hemarajata et al. et al |
| EPI_ISL_939634 | France/PAC-IHU-3063/2021 | 2021-01-18 | Europe/France/Provence-Alpes-C√¥te d' | MEPHI, Aix Marseille University | MEPHI, Aix Marseille University | Anthony LEVASSEUR et al |
| EPI_ISL_939637 | France/PAC-IHU-3066/2021 | 2021-01-18 | Europe/France/Provence-Alpes-C√¥te d' | MEPHI, Aix Marseille University | MEPHI, Aix Marseille University | Anthony LEVASSEUR et al |
| EPI_ISL_939651 | USA/CA-LACPHL-AF00301/2021 | 2021-01-08 | North America/USA/California/Los Ange | MD Laboratories | Los Angeles County PHL | P. Hemarajata et al. et al |
| EPI_ISL_939653 | USA/CA-LACPHL-AF00307/2021 | 2021-01-08 | North America/USA/California/Los Ange | MD Laboratories | Los Angeles County PHL | P. Hemarajata et al. et al |
| EPI_ISL_939666 | Switzerland/ZH-UZH-IMV-3ba25652/ | 2020-04-06 | Europe/Switzerland/Z√ºrich/ | Division of Infectious Diseases, University Hospital Z√ºrich | Institute of Medical Virology, University of Zurich | Stefan Schmutz et al |
| EPI_ISL_940117 | SouthAfrica/N00671/2020 | 2020-12-14 | Africa/South Africa/Gauteng/Johannesb | Charlotte Maxeke Johannesburg Academic Hospital, National Health Laboratory Services, Gauteng, South Africa | National Institute for Communicable Diseases of the National Health Laboratory Service | Amoako DG et al |
| EPI_ISL_940120 | SouthAfrica/N00676/2020 | 2020-12-13 | Africa/South Africa/Gauteng/West Rand | Charlotte Maxeke Johannesburg Academic Hospital, National Health Laboratory Services, Gauteng, South Africa | National Institute for Communicable Diseases of the National Health Laboratory Service | Amoako DG et al |
| EPI_ISL_940144 | Romania/Bucuresti417727/2021 | 2021-01-28 | Europe/Romania/Bucharest/ | Laboratory for Respiratory Viruses, Cantacuzino National Military-Medical Institute for Research and Development | Cantacuzino Institute Virology | Luiza Ustea et al |
| EPI_ISL_940145 | USA/OH-ODH-SC21807/2020 | 2020-03-21 | North America/USA/Ohio/ | Ohio Department of Health Laboratory | Ohio Department of Health Laboratory | Holmes et al |
| EPI_ISL_940159 | France/IDF_HB_112011100048/2020 | 2020-11-26 | Europe/France/Ile de France/ | H√¥pital Bichat Claude Bernard, Laboratoire de Virologie | IAME UMR1137 Inserm, Universit√© de Paris, H√¥pital Bichat | Antoine Bridier-Nahmias et al |

| | | | | | | |
|---|---|---|---|---|---|---|
| EPI_ISL_940160 | France/IDF_HB_112006062745/2020 | 2020-06-16 | Europe/France/Ile de France/ | H√¥pital Bichat Claude Bernard, Laboratoire de Virologie | IAME UMR1137 Inserm, Universit√© de Paris, H√¥pital Bichat | Antoine Bridier-Nahmias et al |
| EPI_ISL_940161 | France/IDF_HB_112011100489/2020 | 2020-11-26 | Europe/France/Ile de France/ | H√¥pital Bichat Claude Bernard, Laboratoire de Virologie | IAME UMR1137 Inserm, Universit√© de Paris, H√¥pital Bichat | Antoine Bridier-Nahmias et al |
| EPI_ISL_940164 | France/IDF_HB_112004001247/2020 | 2020-04-01 | Europe/France/Ile de France/ | H√¥pital Bichat Claude Bernard, Laboratoire de Virologie | IAME UMR1137 Inserm, Universit√© de Paris, H√¥pital Bichat | Antoine Bridier-Nahmias et al |
| EPI_ISL_940165 | France/IDF_HB_112011019987/2020 | 2020-11-05 | Europe/France/Ile de France/ | H√¥pital Bichat Claude Bernard, Laboratoire de Virologie | IAME UMR1137 Inserm, Universit√© de Paris, H√¥pital Bichat | Antoine Bridier-Nahmias et al |
| EPI_ISL_940169 | France/IDF_HB_112004002628/2020 | 2020-04-01 | Europe/France/Ile de France/ | H√¥pital Bichat Claude Bernard, Laboratoire de Virologie | IAME UMR1137 Inserm, Universit√© de Paris, H√¥pital Bichat | Antoine Bridier-Nahmias et al |
| EPI_ISL_940172 | France/IDF_HB_112011033571/2020 | 2020-11-09 | Europe/France/Ile de France/ | H√¥pital Bichat Claude Bernard, Laboratoire de Virologie | IAME UMR1137 Inserm, Universit√© de Paris, H√¥pital Bichat | Antoine Bridier-Nahmias et al |
| EPI_ISL_940173 | France/IDF_HB_112011003591/2020 | 2020-11-02 | Europe/France/Ile de France/ | H√¥pital Bichat Claude Bernard, Laboratoire de Virologie | IAME UMR1137 Inserm, Universit√© de Paris, H√¥pital Bichat | Antoine Bridier-Nahmias et al |
| EPI_ISL_940174 | France/IDF_HB_112010003678/2020 | 2020-10-01 | Europe/France/Ile de France/ | H√¥pital Bichat Claude Bernard, Laboratoire de Virologie | IAME UMR1137 Inserm, Universit√© de Paris, H√¥pital Bichat | Antoine Bridier-Nahmias et al |

| | | | | | | |
|---|---|---|---|---|---|---|
| EPI_ISL_940176 | France/IDF_HB_112011005305/2020 | 2020-11-02 | Europe/France/Ile de France/ | H√¥pital Bichat Claude Bernard, Laboratoire de Virologie | IAME UMR1137 Inserm, Universit√© de Paris, H√¥pital Bichat | Antoine Bridier-Nahmias et al |
| EPI_ISL_940177 | France/IDF_HB_112011005341/2020 | 2020-11-02 | Europe/France/Ile de France/ | H√¥pital Bichat Claude Bernard, Laboratoire de Virologie | IAME UMR1137 Inserm, Universit√© de Paris, H√¥pital Bichat | Antoine Bridier-Nahmias et al |
| EPI_ISL_940178 | France/IDF_HB_112011005461/2020 | 2020-11-02 | Europe/France/Ile de France/ | H√¥pital Bichat Claude Bernard, Laboratoire de Virologie | IAME UMR1137 Inserm, Universit√© de Paris, H√¥pital Bichat | Antoine Bridier-Nahmias et al |
| EPI_ISL_940179 | France/IDF_HB_112011005785/2020 | 2020-11-02 | Europe/France/Ile de France/ | H√¥pital Bichat Claude Bernard, Laboratoire de Virologie | IAME UMR1137 Inserm, Universit√© de Paris, H√¥pital Bichat | Antoine Bridier-Nahmias et al |
| EPI_ISL_940180 | France/IDF_HB_112004006376/2020 | 2020-04-02 | Europe/France/Ile de France/ | H√¥pital Bichat Claude Bernard, Laboratoire de Virologie | IAME UMR1137 Inserm, Universit√© de Paris, H√¥pital Bichat | Antoine Bridier-Nahmias et al |
| EPI_ISL_940186 | France/IDF_HB_112010007243/2020 | 2020-10-02 | Europe/France/Ile de France/ | H√¥pital Bichat Claude Bernard, Laboratoire de Virologie | IAME UMR1137 Inserm, Universit√© de Paris, H√¥pital Bichat | Antoine Bridier-Nahmias et al |
| EPI_ISL_940187 | France/IDF_HB_112011078129/2020 | 2020-11-20 | Europe/France/Ile de France/ | H√¥pital Bichat Claude Bernard, Laboratoire de Virologie | IAME UMR1137 Inserm, Universit√© de Paris, H√¥pital Bichat | Antoine Bridier-Nahmias et al |
| EPI_ISL_940188 | France/IDF_HB_112010007839/2020 | 2020-10-01 | Europe/France/Ile de France/ | H√¥pital Bichat Claude Bernard, Laboratoire de Virologie | IAME UMR1137 Inserm, Universit√© de Paris, H√¥pital Bichat | Antoine Bridier-Nahmias et al |

| | | | | | | |
|---|---|---|---|---|---|---|
| EPI_ISL_940189 | France/IDF_HB_112010007914/2020 | 2020-10-01 | Europe/France/Ile de France/ | H√¥pital Bichat Claude Bernard, Laboratoire de Virologie | IAME UMR1137 Inserm, Universit√© de Paris, H√¥pital Bichat | Antoine Bridier-Nahmias et al |
| EPI_ISL_940191 | France/IDF_HB_112008063761/2020 | 2020-08-21 | Europe/France/Ile de France/ | H√¥pital Bichat Claude Bernard, Laboratoire de Virologie | IAME UMR1137 Inserm, Universit√© de Paris, H√¥pital Bichat | Antoine Bridier-Nahmias et al |
| EPI_ISL_940192 | France/IDF_HB_112011008663/2020 | 2020-11-03 | Europe/France/Ile de France/ | H√¥pital Bichat Claude Bernard, Laboratoire de Virologie | IAME UMR1137 Inserm, Universit√© de Paris, H√¥pital Bichat | Antoine Bridier-Nahmias et al |
| EPI_ISL_940193 | France/IDF_HB_112008071091/2020 | 2020-08-24 | Europe/France/Ile de France/ | H√¥pital Bichat Claude Bernard, Laboratoire de Virologie | IAME UMR1137 Inserm, Universit√© de Paris, H√¥pital Bichat | Antoine Bridier-Nahmias et al |
| EPI_ISL_940195 | France/IDF_HB_112008093387/2020 | 2020-08-31 | Europe/France/Ile de France/ | H√¥pital Bichat Claude Bernard, Laboratoire de Virologie | IAME UMR1137 Inserm, Universit√© de Paris, H√¥pital Bichat | Antoine Bridier-Nahmias et al |
| EPI_ISL_940199 | France/IDF_HB_112009017318/2020 | 2020-09-04 | Europe/France/Ile de France/ | H√¥pital Bichat Claude Bernard, Laboratoire de Virologie | IAME UMR1137 Inserm, Universit√© de Paris, H√¥pital Bichat | Antoine Bridier-Nahmias et al |
| EPI_ISL_940200 | France/IDF_HB_112009017322/2020 | 2020-09-04 | Europe/France/Ile de France/ | H√¥pital Bichat Claude Bernard, Laboratoire de Virologie | IAME UMR1137 Inserm, Universit√© de Paris, H√¥pital Bichat | Antoine Bridier-Nahmias et al |
| EPI_ISL_940201 | France/IDF_HB_112009019057/2020 | 2020-09-05 | Europe/France/Ile de France/ | H√¥pital Bichat Claude Bernard, Laboratoire de Virologie | IAME UMR1137 Inserm, Universit√© de Paris, H√¥pital Bichat | Antoine Bridier-Nahmias et al |

| | | | | | | |
|---|---|---|---|---|---|---|
| EPI_ISL_940203 | France/IDF_HB_112009019069/2020 | 2020-09-05 | Europe/France/Ile de France/ | H√¥pital Bichat Claude Bernard, Laboratoire de Virologie | IAME UMR1137 Inserm, Universit√© de Paris, H√¥pital Bichat | Antoine Bridier-Nahmias et al |
| EPI_ISL_940204 | France/IDF_HB_112009025548/2020 | 2020-09-07 | Europe/France/Ile de France/ | H√¥pital Bichat Claude Bernard, Laboratoire de Virologie | IAME UMR1137 Inserm, Universit√© de Paris, H√¥pital Bichat | Antoine Bridier-Nahmias et al |
| EPI_ISL_940205 | France/IDF_HB_112009041032/2020 | 2020-09-11 | Europe/France/Ile de France/ | H√¥pital Bichat Claude Bernard, Laboratoire de Virologie | IAME UMR1137 Inserm, Universit√© de Paris, H√¥pital Bichat | Antoine Bridier-Nahmias et al |
| EPI_ISL_940207 | France/IDF_HB_112009043616/2020 | 2020-09-11 | Europe/France/Ile de France/ | H√¥pital Bichat Claude Bernard, Laboratoire de Virologie | IAME UMR1137 Inserm, Universit√© de Paris, H√¥pital Bichat | Antoine Bridier-Nahmias et al |
| EPI_ISL_940210 | France/IDF_HB_112004101075/2020 | 2020-04-30 | Europe/France/Ile de France/ | H√¥pital Bichat Claude Bernard, Laboratoire de Virologie | IAME UMR1137 Inserm, Universit√© de Paris, H√¥pital Bichat | Antoine Bridier-Nahmias et al |
| EPI_ISL_940216 | France/IDF_HB_112003101220/2020 | 2020-03-29 | Europe/France/Ile de France/ | H√¥pital Bichat Claude Bernard, Laboratoire de Virologie | IAME UMR1137 Inserm, Universit√© de Paris, H√¥pital Bichat | Antoine Bridier-Nahmias et al |
| EPI_ISL_940217 | France/IDF_HB_112003101232/2020 | 2020-03-29 | Europe/France/Ile de France/ | H√¥pital Bichat Claude Bernard, Laboratoire de Virologie | IAME UMR1137 Inserm, Universit√© de Paris, H√¥pital Bichat | Antoine Bridier-Nahmias et al |
| EPI_ISL_940219 | France/IDF_HB_112003101298/2020 | 2020-03-29 | Europe/France/Ile de France/ | H√¥pital Bichat Claude Bernard, Laboratoire de Virologie | IAME UMR1137 Inserm, Universit√© de Paris, H√¥pital Bichat | Antoine Bridier-Nahmias et al |

| | | | | | | |
|---|---|---|---|---|---|---|
| EPI_ISL_940220 | France/IDF_HB_112003101417/2020 | 2020-03-29 | Europe/France/Ile de France/ | H√¥pital Bichat Claude Bernard, Laboratoire de Virologie | IAME UMR1137 Inserm, Universit√© de Paris, H√¥pital Bichat | Antoine Bridier-Nahmias et al |
| EPI_ISL_940222 | France/IDF_HB_112003102391/2020 | 2020-03-30 | Europe/France/Ile de France/ | H√¥pital Bichat Claude Bernard, Laboratoire de Virologie | IAME UMR1137 Inserm, Universit√© de Paris, H√¥pital Bichat | Antoine Bridier-Nahmias et al |
| EPI_ISL_940226 | France/IDF_HB_112003104666/2020 | 2020-03-30 | Europe/France/Ile de France/ | H√¥pital Bichat Claude Bernard, Laboratoire de Virologie | IAME UMR1137 Inserm, Universit√© de Paris, H√¥pital Bichat | Antoine Bridier-Nahmias et al |
| EPI_ISL_940232 | France/IDF_HB_112003105713/2020 | 2020-03-30 | Europe/France/Ile de France/ | H√¥pital Bichat Claude Bernard, Laboratoire de Virologie | IAME UMR1137 Inserm, Universit√© de Paris, H√¥pital Bichat | Antoine Bridier-Nahmias et al |
| EPI_ISL_940233 | France/IDF_HB_112003105715/2020 | 2020-03-30 | Europe/France/Ile de France/ | H√¥pital Bichat Claude Bernard, Laboratoire de Virologie | IAME UMR1137 Inserm, Universit√© de Paris, H√¥pital Bichat | Antoine Bridier-Nahmias et al |
| EPI_ISL_940234 | France/IDF_HB_112011105752/2020 | 2020-11-27 | Europe/France/Ile de France/ | H√¥pital Bichat Claude Bernard, Laboratoire de Virologie | IAME UMR1137 Inserm, Universit√© de Paris, H√¥pital Bichat | Antoine Bridier-Nahmias et al |
| EPI_ISL_940236 | France/IDF_HB_112003105783/2020 | 2020-03-30 | Europe/France/Ile de France/ | H√¥pital Bichat Claude Bernard, Laboratoire de Virologie | IAME UMR1137 Inserm, Universit√© de Paris, H√¥pital Bichat | Antoine Bridier-Nahmias et al |
| EPI_ISL_940241 | France/IDF_HB_112009105919/2020 | 2020-09-27 | Europe/France/Ile de France/ | H√¥pital Bichat Claude Bernard, Laboratoire de Virologie | IAME UMR1137 Inserm, Universit√© de Paris, H√¥pital Bichat | Antoine Bridier-Nahmias et al |

| | | | | | | |
|---|---|---|---|---|---|---|
| EPI_ISL_940245 | France/IDF_HB_112009109068/2020 | 2020-09-28 | Europe/France/Ile de France/ | H√¥pital Bichat Claude Bernard, Laboratoire de Virologie | IAME UMR1137 Inserm, Universit√© de Paris, H√¥pital Bichat | Antoine Bridier-Nahmias et al |
| EPI_ISL_940250 | France/IDF_HB_112003109846/2020 | 2020-03-31 | Europe/France/Ile de France/ | H√¥pital Bichat Claude Bernard, Laboratoire de Virologie | IAME UMR1137 Inserm, Universit√© de Paris, H√¥pital Bichat | Antoine Bridier-Nahmias et al |
| EPI_ISL_940251 | France/IDF_HB_112003109900/2020 | 2020-03-31 | Europe/France/Ile de France/ | H√¥pital Bichat Claude Bernard, Laboratoire de Virologie | IAME UMR1137 Inserm, Universit√© de Paris, H√¥pital Bichat | Antoine Bridier-Nahmias et al |
| EPI_ISL_940259 | France/IDF_HB_112011111725/2020 | 2020-11-30 | Europe/France/Ile de France/ | H√¥pital Bichat Claude Bernard, Laboratoire de Virologie | IAME UMR1137 Inserm, Universit√© de Paris, H√¥pital Bichat | Antoine Bridier-Nahmias et al |
| EPI_ISL_940262 | France/IDF_HB_112011113937/2020 | 2020-11-30 | Europe/France/Ile de France/ | H√¥pital Bichat Claude Bernard, Laboratoire de Virologie | IAME UMR1137 Inserm, Universit√© de Paris, H√¥pital Bichat | Antoine Bridier-Nahmias et al |
| EPI_ISL_940263 | France/IDF_HB_112011114369/2020 | 2020-11-30 | Europe/France/Ile de France/ | H√¥pital Bichat Claude Bernard, Laboratoire de Virologie | IAME UMR1137 Inserm, Universit√© de Paris, H√¥pital Bichat | Antoine Bridier-Nahmias et al |
| EPI_ISL_940266 | France/IDF_HB_112009117935/2020 | 2020-09-29 | Europe/France/Ile de France/ | H√¥pital Bichat Claude Bernard, Laboratoire de Virologie | IAME UMR1137 Inserm, Universit√© de Paris, H√¥pital Bichat | Antoine Bridier-Nahmias et al |
| EPI_ISL_940268 | France/IDF_HB_112007012128/2020 | 2020-07-03 | Europe/France/Ile de France/ | H√¥pital Bichat Claude Bernard, Laboratoire de Virologie | IAME UMR1137 Inserm, Universit√© de Paris, H√¥pital Bichat | Antoine Bridier-Nahmias et al |

| | | | | | | |
|---|---|---|---|---|---|---|
| EPI_ISL_940269 | France/IDF_HB_112010012137/2020 | 2020-10-04 | Europe/France/Ile de France/ | H√¥pital Bichat Claude Bernard, Laboratoire de Virologie | IAME UMR1137 Inserm, Universit√© de Paris, H√¥pital Bichat | Antoine Bridier-Nahmias et al |
| EPI_ISL_940270 | France/IDF_HB_112010012139/2020 | 2020-10-04 | Europe/France/Ile de France/ | H√¥pital Bichat Claude Bernard, Laboratoire de Virologie | IAME UMR1137 Inserm, Universit√© de Paris, H√¥pital Bichat | Antoine Bridier-Nahmias et al |
| EPI_ISL_940271 | France/IDF_HB_112009121490/2020 | 2020-09-30 | Europe/France/Ile de France/ | H√¥pital Bichat Claude Bernard, Laboratoire de Virologie | IAME UMR1137 Inserm, Universit√© de Paris, H√¥pital Bichat | Antoine Bridier-Nahmias et al |
| EPI_ISL_940274 | France/IDF_HB_112001012957/2020 | 2020-01-29 | Europe/France/Ile de France/ | H√¥pital Bichat Claude Bernard, Laboratoire de Virologie | IAME UMR1137 Inserm, Universit√© de Paris, H√¥pital Bichat | Antoine Bridier-Nahmias et al |
| EPI_ISL_940275 | France/IDF_HB_112011113043/2020 | 2020-11-30 | Europe/France/Ile de France/ | H√¥pital Bichat Claude Bernard, Laboratoire de Virologie | IAME UMR1137 Inserm, Universit√© de Paris, H√¥pital Bichat | Antoine Bridier-Nahmias et al |
| EPI_ISL_940282 | France/IDF_HB_112004015977/2020 | 2020-04-05 | Europe/France/Ile de France/ | H√¥pital Bichat Claude Bernard, Laboratoire de Virologie | IAME UMR1137 Inserm, Universit√© de Paris, H√¥pital Bichat | Antoine Bridier-Nahmias et al |
| EPI_ISL_940283 | France/IDF_HB_112010016754/2020 | 2020-10-05 | Europe/France/Ile de France/ | H√¥pital Bichat Claude Bernard, Laboratoire de Virologie | IAME UMR1137 Inserm, Universit√© de Paris, H√¥pital Bichat | Antoine Bridier-Nahmias et al |
| EPI_ISL_940285 | France/IDF_HB_112010016965/2020 | 2020-10-05 | Europe/France/Ile de France/ | H√¥pital Bichat Claude Bernard, Laboratoire de Virologie | IAME UMR1137 Inserm, Universit√© de Paris, H√¥pital Bichat | Antoine Bridier-Nahmias et al |

| | | | | | | |
|---|---|---|---|---|---|---|
| EPI_ISL_940288 | France/IDF_HB_112004018155/2020 | 2020-04-06 | Europe/France/Ile de France/ | H√¥pital Bichat Claude Bernard, Laboratoire de Virologie | IAME UMR1137 Inserm, Universit√© de Paris, H√¥pital Bichat | Antoine Bridier-Nahmias et al |
| EPI_ISL_940291 | France/IDF_HB_112010018373/2020 | 2020-10-05 | Europe/France/Ile de France/ | H√¥pital Bichat Claude Bernard, Laboratoire de Virologie | IAME UMR1137 Inserm, Universit√© de Paris, H√¥pital Bichat | Antoine Bridier-Nahmias et al |
| EPI_ISL_940292 | France/IDF_HB_112004018439/2020 | 2020-04-06 | Europe/France/Ile de France/ | H√¥pital Bichat Claude Bernard, Laboratoire de Virologie | IAME UMR1137 Inserm, Universit√© de Paris, H√¥pital Bichat | Antoine Bridier-Nahmias et al |
| EPI_ISL_940296 | France/IDF_HB_112004019140/2020 | 2020-04-06 | Europe/France/Ile de France/ | H√¥pital Bichat Claude Bernard, Laboratoire de Virologie | IAME UMR1137 Inserm, Universit√© de Paris, H√¥pital Bichat | Antoine Bridier-Nahmias et al |
| EPI_ISL_940309 | France/IDF_HB_112011019989/2020 | 2020-11-05 | Europe/France/Ile de France/ | H√¥pital Bichat Claude Bernard, Laboratoire de Virologie | IAME UMR1137 Inserm, Universit√© de Paris, H√¥pital Bichat | Antoine Bridier-Nahmias et al |
| EPI_ISL_940311 | France/IDF_HB_112004020123/2020 | 2020-04-06 | Europe/France/Ile de France/ | H√¥pital Bichat Claude Bernard, Laboratoire de Virologie | IAME UMR1137 Inserm, Universit√© de Paris, H√¥pital Bichat | Antoine Bridier-Nahmias et al |
| EPI_ISL_940316 | France/IDF_HB_112004020438/2020 | 2020-04-06 | Europe/France/Ile de France/ | H√¥pital Bichat Claude Bernard, Laboratoire de Virologie | IAME UMR1137 Inserm, Universit√© de Paris, H√¥pital Bichat | Antoine Bridier-Nahmias et al |
| EPI_ISL_940317 | France/IDF_HB_112011020817/2020 | 2020-11-05 | Europe/France/Ile de France/ | H√¥pital Bichat Claude Bernard, Laboratoire de Virologie | IAME UMR1137 Inserm, Universit√© de Paris, H√¥pital Bichat | Antoine Bridier-Nahmias et al |

| | | | | | | |
|---|---|---|---|---|---|---|
| EPI_ISL_940322 | France/IDF_HB_112004024240/2020 | 2020-04-07 | Europe/France/Ile de France/ | H√¥pital Bichat Claude Bernard, Laboratoire de Virologie | IAME UMR1137 Inserm, Universit√© de Paris, H√¥pital Bichat | Antoine Bridier-Nahmias et al |
| EPI_ISL_940323 | France/IDF_HB_112004024299/2020 | 2020-04-07 | Europe/France/Ile de France/ | H√¥pital Bichat Claude Bernard, Laboratoire de Virologie | IAME UMR1137 Inserm, Universit√© de Paris, H√¥pital Bichat | Antoine Bridier-Nahmias et al |
| EPI_ISL_940324 | France/IDF_HB_112004024319/2020 | 2020-04-07 | Europe/France/Ile de France/ | H√¥pital Bichat Claude Bernard, Laboratoire de Virologie | IAME UMR1137 Inserm, Universit√© de Paris, H√¥pital Bichat | Antoine Bridier-Nahmias et al |
| EPI_ISL_940329 | France/IDF_HB_112004025398/2020 | 2020-04-08 | Europe/France/Ile de France/ | H√¥pital Bichat Claude Bernard, Laboratoire de Virologie | IAME UMR1137 Inserm, Universit√© de Paris, H√¥pital Bichat | Antoine Bridier-Nahmias et al |
| EPI_ISL_940336 | France/IDF_HB_112004029241/2020 | 2020-04-09 | Europe/France/Ile de France/ | H√¥pital Bichat Claude Bernard, Laboratoire de Virologie | IAME UMR1137 Inserm, Universit√© de Paris, H√¥pital Bichat | Antoine Bridier-Nahmias et al |
| EPI_ISL_940338 | France/IDF_HB_112004029250/2020 | 2020-04-08 | Europe/France/Ile de France/ | H√¥pital Bichat Claude Bernard, Laboratoire de Virologie | IAME UMR1137 Inserm, Universit√© de Paris, H√¥pital Bichat | Antoine Bridier-Nahmias et al |
| EPI_ISL_940341 | France/IDF_HB_112011030565/2020 | 2020-11-09 | Europe/France/Ile de France/ | H√¥pital Bichat Claude Bernard, Laboratoire de Virologie | IAME UMR1137 Inserm, Universit√© de Paris, H√¥pital Bichat | Antoine Bridier-Nahmias et al |
| EPI_ISL_940342 | France/IDF_HB_112011031271/2020 | 2020-11-09 | Europe/France/Ile de France/ | H√¥pital Bichat Claude Bernard, Laboratoire de Virologie | IAME UMR1137 Inserm, Universit√© de Paris, H√¥pital Bichat | Antoine Bridier-Nahmias et al |

| | | | | | | |
|---|---|---|---|---|---|---|
| EPI_ISL_940343 | France/IDF_HB_112004032529/2020 | 2020-04-09 | Europe/France/Ile de France/ | H√¥pital Bichat Claude Bernard, Laboratoire de Virologie | IAME UMR1137 Inserm, Universit√© de Paris, H√¥pital Bichat | Antoine Bridier-Nahmias et al |
| EPI_ISL_940346 | France/IDF_HB_112011032931/2020 | 2020-11-09 | Europe/France/Ile de France/ | H√¥pital Bichat Claude Bernard, Laboratoire de Virologie | IAME UMR1137 Inserm, Universit√© de Paris, H√¥pital Bichat | Antoine Bridier-Nahmias et al |
| EPI_ISL_940349 | France/IDF_HB_112004033482/2020 | 2020-04-10 | Europe/France/Ile de France/ | H√¥pital Bichat Claude Bernard, Laboratoire de Virologie | IAME UMR1137 Inserm, Universit√© de Paris, H√¥pital Bichat | Antoine Bridier-Nahmias et al |
| EPI_ISL_940355 | France/IDF_HB_112004036508/2020 | 2020-04-10 | Europe/France/Ile de France/ | H√¥pital Bichat Claude Bernard, Laboratoire de Virologie | IAME UMR1137 Inserm, Universit√© de Paris, H√¥pital Bichat | Antoine Bridier-Nahmias et al |
| EPI_ISL_940357 | France/IDF_HB_112004003663/2020 | 2020-04-01 | Europe/France/Ile de France/ | H√¥pital Bichat Claude Bernard, Laboratoire de Virologie | IAME UMR1137 Inserm, Universit√© de Paris, H√¥pital Bichat | Antoine Bridier-Nahmias et al |
| EPI_ISL_940361 | France/IDF_HB_112004003838/2020 | 2020-04-01 | Europe/France/Ile de France/ | H√¥pital Bichat Claude Bernard, Laboratoire de Virologie | IAME UMR1137 Inserm, Universit√© de Paris, H√¥pital Bichat | Antoine Bridier-Nahmias et al |
| EPI_ISL_940362 | France/IDF_HB_112004003849/2020 | 2020-04-01 | Europe/France/Ile de France/ | H√¥pital Bichat Claude Bernard, Laboratoire de Virologie | IAME UMR1137 Inserm, Universit√© de Paris, H√¥pital Bichat | Antoine Bridier-Nahmias et al |
| EPI_ISL_940366 | France/IDF_HB_112003040678/2020 | 2020-03-11 | Europe/France/Ile de France/ | H√¥pital Bichat Claude Bernard, Laboratoire de Virologie | IAME UMR1137 Inserm, Universit√© de Paris, H√¥pital Bichat | Antoine Bridier-Nahmias et al |

| | | | | | | |
|---|---|---|---|---|---|---|
| EPI_ISL_940373 | France/IDF_HB_112003046955/2020 | 2020-03-12 | Europe/France/Ile de France/ | H√¥pital Bichat Claude Bernard, Laboratoire de Virologie | IAME UMR1137 Inserm, Universit√© de Paris, H√¥pital Bichat | Antoine Bridier-Nahmias et al |
| EPI_ISL_940383 | France/IDF_HB_112004049802/2020 | 2020-04-10 | Europe/France/Ile de France/ | H√¥pital Bichat Claude Bernard, Laboratoire de Virologie | IAME UMR1137 Inserm, Universit√© de Paris, H√¥pital Bichat | Antoine Bridier-Nahmias et al |
| EPI_ISL_940386 | France/IDF_HB_112003050264/2020 | 2020-03-13 | Europe/France/Ile de France/ | H√¥pital Bichat Claude Bernard, Laboratoire de Virologie | IAME UMR1137 Inserm, Universit√© de Paris, H√¥pital Bichat | Antoine Bridier-Nahmias et al |
| EPI_ISL_940387 | France/IDF_HB_112003050333/2020 | 2020-03-13 | Europe/France/Ile de France/ | H√¥pital Bichat Claude Bernard, Laboratoire de Virologie | IAME UMR1137 Inserm, Universit√© de Paris, H√¥pital Bichat | Antoine Bridier-Nahmias et al |
| EPI_ISL_940388 | France/IDF_HB_112003050431/2020 | 2020-03-13 | Europe/France/Ile de France/ | H√¥pital Bichat Claude Bernard, Laboratoire de Virologie | IAME UMR1137 Inserm, Universit√© de Paris, H√¥pital Bichat | Antoine Bridier-Nahmias et al |
| EPI_ISL_940395 | France/IDF_HB_112003050879/2020 | 2020-03-13 | Europe/France/Ile de France/ | H√¥pital Bichat Claude Bernard, Laboratoire de Virologie | IAME UMR1137 Inserm, Universit√© de Paris, H√¥pital Bichat | Antoine Bridier-Nahmias et al |
| EPI_ISL_940398 | France/IDF_HB_112003051156/2020 | 2020-03-13 | Europe/France/Ile de France/ | H√¥pital Bichat Claude Bernard, Laboratoire de Virologie | IAME UMR1137 Inserm, Universit√© de Paris, H√¥pital Bichat | Antoine Bridier-Nahmias et al |
| EPI_ISL_940401 | France/IDF_HB_112003052315/2020 | 2020-03-13 | Europe/France/Ile de France/ | H√¥pital Bichat Claude Bernard, Laboratoire de Virologie | IAME UMR1137 Inserm, Universit√© de Paris, H√¥pital Bichat | Antoine Bridier-Nahmias et al |

| | | | | | | |
|---|---|---|---|---|---|---|
| EPI_ISL_940405 | France/IDF_HB_112003052456/2020 | 2020-03-13 | Europe/France/Ile de France/ | H√¥pital Bichat Claude Bernard, Laboratoire de Virologie | IAME UMR1137 Inserm, Universit√© de Paris, H√¥pital Bichat | Antoine Bridier-Nahmias et al |
| EPI_ISL_940406 | France/IDF_HB_112003052791/2020 | 2020-03-14 | Europe/France/Ile de France/ | H√¥pital Bichat Claude Bernard, Laboratoire de Virologie | IAME UMR1137 Inserm, Universit√© de Paris, H√¥pital Bichat | Antoine Bridier-Nahmias et al |
| EPI_ISL_940410 | France/IDF_HB_112003053267/2020 | 2020-03-14 | Europe/France/Ile de France/ | H√¥pital Bichat Claude Bernard, Laboratoire de Virologie | IAME UMR1137 Inserm, Universit√© de Paris, H√¥pital Bichat | Antoine Bridier-Nahmias et al |
| EPI_ISL_940411 | France/IDF_HB_112003053269/2020 | 2020-03-14 | Europe/France/Ile de France/ | H√¥pital Bichat Claude Bernard, Laboratoire de Virologie | IAME UMR1137 Inserm, Universit√© de Paris, H√¥pital Bichat | Antoine Bridier-Nahmias et al |
| EPI_ISL_940413 | France/IDF_HB_112003053402/2020 | 2020-03-14 | Europe/France/Ile de France/ | H√¥pital Bichat Claude Bernard, Laboratoire de Virologie | IAME UMR1137 Inserm, Universit√© de Paris, H√¥pital Bichat | Antoine Bridier-Nahmias et al |
| EPI_ISL_940419 | France/IDF_HB_112002053643/2020 | 2020-02-13 | Europe/France/Ile de France/ | H√¥pital Bichat Claude Bernard, Laboratoire de Virologie | IAME UMR1137 Inserm, Universit√© de Paris, H√¥pital Bichat | Antoine Bridier-Nahmias et al |
| EPI_ISL_940427 | France/IDF_HB_112011061252/2020 | 2020-11-17 | Europe/France/Ile de France/ | H√¥pital Bichat Claude Bernard, Laboratoire de Virologie | IAME UMR1137 Inserm, Universit√© de Paris, H√¥pital Bichat | Antoine Bridier-Nahmias et al |
| EPI_ISL_940433 | France/IDF_HB_112003063217/2020 | 2020-03-17 | Europe/France/Ile de France/ | H√¥pital Bichat Claude Bernard, Laboratoire de Virologie | IAME UMR1137 Inserm, Universit√© de Paris, H√¥pital Bichat | Antoine Bridier-Nahmias et al |

| | | | | | | |
|---|---|---|---|---|---|---|
| EPI_ISL_940437 | France/IDF_HB_112003064443/2020 | 2020-03-18 | Europe/France/Ile de France/ | H√¥pital Bichat Claude Bernard, Laboratoire de Virologie | IAME UMR1137 Inserm, Universit√© de Paris, H√¥pital Bichat | Antoine Bridier-Nahmias et al |
| EPI_ISL_940441 | France/IDF_HB_112003064601/2020 | 2020-03-17 | Europe/France/Ile de France/ | H√¥pital Bichat Claude Bernard, Laboratoire de Virologie | IAME UMR1137 Inserm, Universit√© de Paris, H√¥pital Bichat | Antoine Bridier-Nahmias et al |
| EPI_ISL_940443 | France/IDF_HB_112003064713/2020 | 2020-03-18 | Europe/France/Ile de France/ | H√¥pital Bichat Claude Bernard, Laboratoire de Virologie | IAME UMR1137 Inserm, Universit√© de Paris, H√¥pital Bichat | Antoine Bridier-Nahmias et al |
| EPI_ISL_940448 | France/IDF_HB_112003065776/2020 | 2020-03-18 | Europe/France/Ile de France/ | H√¥pital Bichat Claude Bernard, Laboratoire de Virologie | IAME UMR1137 Inserm, Universit√© de Paris, H√¥pital Bichat | Antoine Bridier-Nahmias et al |
| EPI_ISL_940454 | France/IDF_HB_112003069170/2020 | 2020-03-18 | Europe/France/Ile de France/ | H√¥pital Bichat Claude Bernard, Laboratoire de Virologie | IAME UMR1137 Inserm, Universit√© de Paris, H√¥pital Bichat | Antoine Bridier-Nahmias et al |
| EPI_ISL_940458 | France/IDF_HB_112003069763/2020 | 2020-03-19 | Europe/France/Ile de France/ | H√¥pital Bichat Claude Bernard, Laboratoire de Virologie | IAME UMR1137 Inserm, Universit√© de Paris, H√¥pital Bichat | Antoine Bridier-Nahmias et al |
| EPI_ISL_940462 | France/IDF_HB_112003070106/2020 | 2020-03-19 | Europe/France/Ile de France/ | H√¥pital Bichat Claude Bernard, Laboratoire de Virologie | IAME UMR1137 Inserm, Universit√© de Paris, H√¥pital Bichat | Antoine Bridier-Nahmias et al |
| EPI_ISL_940463 | France/IDF_HB_112003070111/2020 | 2020-03-19 | Europe/France/Ile de France/ | H√¥pital Bichat Claude Bernard, Laboratoire de Virologie | IAME UMR1137 Inserm, Universit√© de Paris, H√¥pital Bichat | Antoine Bridier-Nahmias et al |

| | | | | | | |
|---|---|---|---|---|---|---|
| EPI_ISL_940467 | France/IDF_HB_112003070172/2020 | 2020-03-19 | Europe/France/Ile de France/ | H√¥pital Bichat Claude Bernard, Laboratoire de Virologie | IAME UMR1137 Inserm, Universit√© de Paris, H√¥pital Bichat | Antoine Bridier-Nahmias et al |
| EPI_ISL_940468 | France/IDF_HB_112003070182/2020 | 2020-03-19 | Europe/France/Ile de France/ | H√¥pital Bichat Claude Bernard, Laboratoire de Virologie | IAME UMR1137 Inserm, Universit√© de Paris, H√¥pital Bichat | Antoine Bridier-Nahmias et al |
| EPI_ISL_940470 | France/IDF_HB_112003070440/2020 | 2020-03-19 | Europe/France/Ile de France/ | H√¥pital Bichat Claude Bernard, Laboratoire de Virologie | IAME UMR1137 Inserm, Universit√© de Paris, H√¥pital Bichat | Antoine Bridier-Nahmias et al |
| EPI_ISL_940471 | France/IDF_HB_112004073976/2020 | 2020-04-21 | Europe/France/Ile de France/ | H√¥pital Bichat Claude Bernard, Laboratoire de Virologie | IAME UMR1137 Inserm, Universit√© de Paris, H√¥pital Bichat | Antoine Bridier-Nahmias et al |
| EPI_ISL_940474 | France/IDF_HB_112004007476/2020 | 2020-04-02 | Europe/France/Ile de France/ | H√¥pital Bichat Claude Bernard, Laboratoire de Virologie | IAME UMR1137 Inserm, Universit√© de Paris, H√¥pital Bichat | Antoine Bridier-Nahmias et al |
| EPI_ISL_940475 | France/IDF_HB_112004007501/2020 | 2020-04-02 | Europe/France/Ile de France/ | H√¥pital Bichat Claude Bernard, Laboratoire de Virologie | IAME UMR1137 Inserm, Universit√© de Paris, H√¥pital Bichat | Antoine Bridier-Nahmias et al |
| EPI_ISL_940477 | France/IDF_HB_112009075949/2020 | 2020-09-19 | Europe/France/Ile de France/ | H√¥pital Bichat Claude Bernard, Laboratoire de Virologie | IAME UMR1137 Inserm, Universit√© de Paris, H√¥pital Bichat | Antoine Bridier-Nahmias et al |
| EPI_ISL_940481 | France/IDF_HB_112009076404/2020 | 2020-09-20 | Europe/France/Ile de France/ | H√¥pital Bichat Claude Bernard, Laboratoire de Virologie | IAME UMR1137 Inserm, Universit√© de Paris, H√¥pital Bichat | Antoine Bridier-Nahmias et al |

| | | | | | | |
|---|---|---|---|---|---|---|
| EPI_ISL_940483 | France/IDF_HB_112009077520/2020 | 2020-09-20 | Europe/France/Ile de France/ | H√¥pital Bichat Claude Bernard, Laboratoire de Virologie | IAME UMR1137 Inserm, Universit√© de Paris, H√¥pital Bichat | Antoine Bridier-Nahmias et al |
| EPI_ISL_940486 | France/IDF_HB_112003079669/2020 | 2020-03-21 | Europe/France/Ile de France/ | H√¥pital Bichat Claude Bernard, Laboratoire de Virologie | IAME UMR1137 Inserm, Universit√© de Paris, H√¥pital Bichat | Antoine Bridier-Nahmias et al |
| EPI_ISL_940487 | France/IDF_HB_112010080108/2020 | 2020-10-20 | Europe/France/Ile de France/ | H√¥pital Bichat Claude Bernard, Laboratoire de Virologie | IAME UMR1137 Inserm, Universit√© de Paris, H√¥pital Bichat | Antoine Bridier-Nahmias et al |
| EPI_ISL_940488 | France/IDF_HB_112003081568/2020 | 2020-03-23 | Europe/France/Ile de France/ | H√¥pital Bichat Claude Bernard, Laboratoire de Virologie | IAME UMR1137 Inserm, Universit√© de Paris, H√¥pital Bichat | Antoine Bridier-Nahmias et al |
| EPI_ISL_940491 | France/IDF_HB_112010008228/2020 | 2020-10-02 | Europe/France/Ile de France/ | H√¥pital Bichat Claude Bernard, Laboratoire de Virologie | IAME UMR1137 Inserm, Universit√© de Paris, H√¥pital Bichat | Antoine Bridier-Nahmias et al |
| EPI_ISL_940492 | France/IDF_HB_112004008261/2020 | 2020-04-02 | Europe/France/Ile de France/ | H√¥pital Bichat Claude Bernard, Laboratoire de Virologie | IAME UMR1137 Inserm, Universit√© de Paris, H√¥pital Bichat | Antoine Bridier-Nahmias et al |
| EPI_ISL_940501 | France/IDF_HB_112011088594/2020 | 2020-11-23 | Europe/France/Ile de France/ | H√¥pital Bichat Claude Bernard, Laboratoire de Virologie | IAME UMR1137 Inserm, Universit√© de Paris, H√¥pital Bichat | Antoine Bridier-Nahmias et al |
| EPI_ISL_940507 | France/IDF_HB_112003096373/2020 | 2020-03-27 | Europe/France/Ile de France/ | H√¥pital Bichat Claude Bernard, Laboratoire de Virologie | IAME UMR1137 Inserm, Universit√© de Paris, H√¥pital Bichat | Antoine Bridier-Nahmias et al |

| | | | | | | |
|---|---|---|---|---|---|---|
| EPI_ISL_940514 | France/IDF_HB_112003096499/2020 | 2020-03-27 | Europe/France/Ile de France/ | H√¥pital Bichat Claude Bernard, Laboratoire de Virologie | IAME UMR1137 Inserm, Universit√© de Paris, H√¥pital Bichat | Antoine Bridier-Nahmias et al |
| EPI_ISL_940518 | France/IDF_HB_112003096915/2020 | 2020-03-27 | Europe/France/Ile de France/ | H√¥pital Bichat Claude Bernard, Laboratoire de Virologie | IAME UMR1137 Inserm, Universit√© de Paris, H√¥pital Bichat | Antoine Bridier-Nahmias et al |
| EPI_ISL_940520 | France/IDF_HB_112003096956/2020 | 2020-03-27 | Europe/France/Ile de France/ | H√¥pital Bichat Claude Bernard, Laboratoire de Virologie | IAME UMR1137 Inserm, Universit√© de Paris, H√¥pital Bichat | Antoine Bridier-Nahmias et al |
| EPI_ISL_940522 | France/IDF_HB_112009079404/2020 | 2020-09-21 | Europe/France/Ile de France/ | H√¥pital Bichat Claude Bernard, Laboratoire de Virologie | IAME UMR1137 Inserm, Universit√© de Paris, H√¥pital Bichat | Antoine Bridier-Nahmias et al |
| EPI_ISL_940531 | France/IDF_HB_112002105286/2020 | 2020-02-28 | Europe/France/Ile de France/ | H√¥pital Bichat Claude Bernard, Laboratoire de Virologie | IAME UMR1137 Inserm, Universit√© de Paris, H√¥pital Bichat | Antoine Bridier-Nahmias et al |
| EPI_ISL_940533 | France/IDF_HB_112002096647/2020 | 2020-02-26 | Europe/France/Ile de France/ | H√¥pital Bichat Claude Bernard, Laboratoire de Virologie | IAME UMR1137 Inserm, Universit√© de Paris, H√¥pital Bichat | Antoine Bridier-Nahmias et al |
| EPI_ISL_940534 | France/IDF_HB_112002103234/2020 | 2020-02-27 | Europe/France/Ile de France/ | H√¥pital Bichat Claude Bernard, Laboratoire de Virologie | IAME UMR1137 Inserm, Universit√© de Paris, H√¥pital Bichat | Antoine Bridier-Nahmias et al |
| EPI_ISL_940535 | France/IDF_HB_112002107124/2020 | 2020-02-28 | Europe/France/Ile de France/ | H√¥pital Bichat Claude Bernard, Laboratoire de Virologie | IAME UMR1137 Inserm, Universit√© de Paris, H√¥pital Bichat | Antoine Bridier-Nahmias et al |

| | | | | | | |
|---|---|---|---|---|---|---|
| EPI_ISL_940537 | France/IDF_HB_112002105432/2020 | 2020-02-28 | Europe/France/Ile de France/ | H√¥pital Bichat Claude Bernard, Laboratoire de Virologie | IAME UMR1137 Inserm, Universit√© de Paris, H√¥pital Bichat | Antoine Bridier-Nahmias et al |
| EPI_ISL_940538 | France/IDF_HB_112002107630/2020 | 2020-02-29 | Europe/France/Ile de France/ | H√¥pital Bichat Claude Bernard, Laboratoire de Virologie | IAME UMR1137 Inserm, Universit√© de Paris, H√¥pital Bichat | Antoine Bridier-Nahmias et al |
| EPI_ISL_940542 | France/IDF_HB_112001900001/2020 | 2020-01-29 | Europe/France/Ile de France/ | H√¥pital Bichat Claude Bernard, Laboratoire de Virologie | IAME UMR1137 Inserm, Universit√© de Paris, H√¥pital Bichat | Antoine Bridier-Nahmias et al |
| EPI_ISL_940543 | France/IDF_HB_112001089879/2020 | 2020-01-24 | Europe/France/Ile de France/ | H√¥pital Bichat Claude Bernard, Laboratoire de Virologie | IAME UMR1137 Inserm, Universit√© de Paris, H√¥pital Bichat | Antoine Bridier-Nahmias et al |
| EPI_ISL_940545 | France/IDF_HB_112008053640/2020 | 2020-08-18 | Europe/France/Ile de France/ | H√¥pital Bichat Claude Bernard, Laboratoire de Virologie | IAME UMR1137 Inserm, Universit√© de Paris, H√¥pital Bichat | Antoine Bridier-Nahmias et al |
| EPI_ISL_940551 | France/IDF_HB_112002007147/2020 | 2020-02-03 | Europe/France/Ile de France/ | H√¥pital Bichat Claude Bernard, Laboratoire de Virologie | IAME UMR1137 Inserm, Universit√© de Paris, H√¥pital Bichat | Antoine Bridier-Nahmias et al |
| EPI_ISL_940552 | France/IDF_HB_112001106076/2020 | 2020-01-29 | Europe/France/Ile de France/ | H√¥pital Bichat Claude Bernard, Laboratoire de Virologie | IAME UMR1137 Inserm, Universit√© de Paris, H√¥pital Bichat | Antoine Bridier-Nahmias et al |
| EPI_ISL_940553 | USA/OH-ODH-SC21892/2020 | 2020-03-23 | North America/USA/Ohio/ | Ohio Department of Health Laboratory | Ohio Department of Health Laboratory | Holmes et al |
| EPI_ISL_940576 | Belgium/Jessa_11-2104-004099/2021 | 2021-01-27 | Europe/Belgium/Maaseik/Houthalen-He | Jessa | Jessa | Jessa_cmdLab et al |

| | | | | | | |
|---|---|---|---|---|---|---|
| EPI_ISL_940608 | Brazil/SP-801/2020 | 2020-06-26 | South America/Brazil/S√£o Paulo/Rio Cl | Laborat√≥rio Sao Lucas | Instituto Adolfo Lutz, Interdiciplinary Procedures Center, Strategic Laboratory | Claudio Tavares Sacchi et al |
| EPI_ISL_940610 | Brazil/SP-1049/2021 | 2021-01-22 | South America/Brazil/S√£o Paulo/S√£o | Hospital Paulistano Paulista | Instituto Adolfo Lutz, Interdiciplinary Procedures Center, Strategic Laboratory | Claudio Tavares Sacchi et al |
| EPI_ISL_940611 | Brazil/SP-1051/2021 | 2021-01-22 | South America/Brazil/S√£o Paulo/S√£o | Hospital Paulistano Paulista | Instituto Adolfo Lutz, Interdiciplinary Procedures Center, Strategic Laboratory | Claudio Tavares Sacchi et al |
| EPI_ISL_940612 | Brazil/GO-1054/2021 | 2021-01-15 | South America/Brazil/Goi√°s/Goiania | Hospital de Campanha para Enfrentamento do Coronavirus - Goiania | Instituto Adolfo Lutz, Interdiciplinary Procedures Center, Strategic Laboratory | Claudio Tavares Sacchi et al |
| EPI_ISL_940613 | Brazil/PI-1055/2021 | 2021-01-19 | South America/Brazil/Piaui/Terezina | LACEN-PI DR. Costa Alvarenga | Instituto Adolfo Lutz, Interdiciplinary Procedures Center, Strategic Laboratory | Claudio Tavares Sacchi et al |
| EPI_ISL_940615 | Brazil/AM-1060/2021 | 2021-01-19 | South America/Brazil/Amazonas BR/Ma | LACEN-PI DR. Costa Alvarenga | Instituto Adolfo Lutz, Interdiciplinary Procedures Center, Strategic Laboratory | Claudio Tavares Sacchi et al |

| | | | | | | |
|---|---|---|---|---|---|---|
| EPI_ISL_940616 | Brazil/AM-1061/2021 | 2021-01-19 | South America/Brazil/Amazonas BR/Mar | LACEN-PI DR. Costa Alvarenga | Instituto Adolfo Lutz, Interdiciplinary Procedures Center, Strategic Laboratory | Claudio Tavares Sacchi et al |
| EPI_ISL_940617 | Brazil/AM-1063/2021 | 2021-01-19 | South America/Brazil/Amazonas BR/Mar | LACEN-PI DR. Costa Alvarenga | Instituto Adolfo Lutz, Interdiciplinary Procedures Center, Strategic Laboratory | Claudio Tavares Sacchi et al |
| EPI_ISL_940618 | Brazil/AM-1064/2021 | 2021-01-19 | South America/Brazil/Amazonas BR/Mar | LACEN-PI DR. Costa Alvarenga | Instituto Adolfo Lutz, Interdiciplinary Procedures Center, Strategic Laboratory | Claudio Tavares Sacchi et al |
| EPI_ISL_940619 | Brazil/SP-1065/2021 | 2021-01-15 | South America/Brazil/S√£o Paulo/S√£o | Hospital Sao Joaquim - Beneficiencia Portuguesa | Instituto Adolfo Lutz, Interdiciplinary Procedures Center, Strategic Laboratory | Claudio Tavares Sacchi et al |
| EPI_ISL_940620 | Brazil/AM-1066/2021 | 2021-01-14 | South America/Brazil/Amazonas BR/Mar | Hospital Sao Joaquim - Beneficiencia Portuguesa | Instituto Adolfo Lutz, Interdiciplinary Procedures Center, Strategic Laboratory | Claudio Tavares Sacchi et al |
| EPI_ISL_940621 | Brazil/SP-1067/2021 | 2021-01-14 | South America/Brazil/S√£o Paulo/S√£o | Hospital Sao Joaquim - Beneficiencia Portuguesa | Instituto Adolfo Lutz, Interdiciplinary Procedures Center, Strategic Laboratory | Claudio Tavares Sacchi et al |

| | | | | | | |
|---|---|---|---|---|---|---|
| EPI_ISL_940622 | Brazil/SP-1068/2021 | 2021-01-15 | South America/Brazil/S√£o Paulo/S√£o | Hospital Sao Joaquim - Beneficiencia Portuguesa | Instituto Adolfo Lutz, Interdiciplinary Procedures Center, Strategic Laboratory | Claudio Tavares Sacchi et al |
| EPI_ISL_940623 | Brazil/AM-1069/2021 | 2021-01-08 | South America/Brazil/Amazonas BR/Ma | Hospital Sao Joaquim - Beneficiencia Portuguesa | Instituto Adolfo Lutz, Interdiciplinary Procedures Center, Strategic Laboratory | Claudio Tavares Sacchi et al |
| EPI_ISL_940624 | Brazil/AM-1070/2021 | 2021-01-15 | South America/Brazil/Amazonas BR/Ma | Hospital Sao Joaquim - Beneficiencia Portuguesa | Instituto Adolfo Lutz, Interdiciplinary Procedures Center, Strategic Laboratory | Claudio Tavares Sacchi et al |
| EPI_ISL_940625 | Brazil/SP-1071/2021 | 2021-01-19 | South America/Brazil/S√£o Paulo/S√£o | Hospital Sao Joaquim - Beneficiencia Portuguesa | Instituto Adolfo Lutz, Interdiciplinary Procedures Center, Strategic Laboratory | Claudio Tavares Sacchi et al |
| EPI_ISL_940626 | Brazil/AM-1073/2021 | 2021-01-21 | South America/Brazil/Amazonas BR/Ma | Hospital Central Sao Caetano do Sul | Instituto Adolfo Lutz, Interdiciplinary Procedures Center, Strategic Laboratory | Claudio Tavares Sacchi et al |
| EPI_ISL_940627 | Brazil/AM-1074/2021 | 2021-01-22 | South America/Brazil/Amazonas BR/Ma | Hospital Central Sao Caetano do Sul | Instituto Adolfo Lutz, Interdiciplinary Procedures Center, Strategic Laboratory | Claudio Tavares Sacchi et al |

| | | | | | | |
|---|---|---|---|---|---|---|
| EPI_ISL_940628 | Brazil/SP-1076/2021 | 2021-01-22 | South America/Brazil/S√£o Paulo/Iguap | Unidade Mista de Iguape | Instituto Adolfo Lutz, Interdiciplinary Procedures Center, Strategic Laboratory | Claudio Tavares Sacchi et al |
| EPI_ISL_940629 | Brazil/SP-1078/2021 | 2021-01-25 | South America/Brazil/S√£o Paulo/S√£o | Hospital Municipal Josanias Castanha Braga | Instituto Adolfo Lutz, Interdiciplinary Procedures Center, Strategic Laboratory | Claudio Tavares Sacchi et al |
| EPI_ISL_940630 | Brazil/RR-1079/2021 | 2021-01-25 | South America/Brazil/Roraima/ | Hospital Geral de Sao Paulo | Instituto Adolfo Lutz, Interdiciplinary Procedures Center, Strategic Laboratory | Claudio Tavares Sacchi et al |
| EPI_ISL_940639 | Turkey/HSGM-1405/2021 | 2021-01-20 | Europe/Turkey/Turkey/ | Ministry of Health Turkey | Ministry of Health Turkey | Fatma Bayrakdar et al |
| EPI_ISL_940641 | Turkey/HSGM-1407/2021 | 2021-01-20 | Europe/Turkey/Turkey/ | Ministry of Health Turkey | Ministry of Health Turkey | Fatma Bayrakdar et al |
| EPI_ISL_940648 | Turkey/HSGM-1477/2021 | 2021-01-19 | Europe/Turkey/Turkey/ | Ministry of Health Turkey | Ministry of Health Turkey | Fatma Bayrakdar et al |
| EPI_ISL_940649 | Turkey/HSGM-1479/2021 | 2021-01-19 | Europe/Turkey/Turkey/ | Ministry of Health Turkey | Ministry of Health Turkey | Fatma Bayrakdar et al |
| EPI_ISL_940655 | Turkey/HSGM-1486/2021 | 2021-01-21 | Europe/Turkey/Turkey/ | Ministry of Health Turkey | Ministry of Health Turkey | Fatma Bayrakdar et al |
| EPI_ISL_940659 | Turkey/HSGM-1491/2021 | 2021-01-21 | Europe/Turkey/Turkey/ | Ministry of Health Turkey | Ministry of Health Turkey | Fatma Bayrakdar et al |
| EPI_ISL_940663 | Turkey/HSGM-1495/2021 | 2021-01-21 | Europe/Turkey/Turkey/ | Ministry of Health Turkey | Ministry of Health Turkey | Fatma Bayrakdar et al |
| EPI_ISL_940664 | Turkey/HSGM-1496/2021 | 2021-01-21 | Europe/Turkey/Turkey/ | Ministry of Health Turkey | Ministry of Health Turkey | Fatma Bayrakdar et al |
| EPI_ISL_940665 | Turkey/HSGM-1497/2021 | 2021-01-21 | Europe/Turkey/Turkey/ | Ministry of Health Turkey | Ministry of Health Turkey | Fatma Bayrakdar et al |
| EPI_ISL_940666 | Turkey/HSGM-1498/2021 | 2021-01-21 | Europe/Turkey/Turkey/ | Ministry of Health Turkey | Ministry of Health Turkey | Fatma Bayrakdar et al |
| EPI_ISL_940668 | Turkey/HSGM-1500/2021 | 2021-01-22 | Europe/Turkey/Turkey/ | Ministry of Health Turkey | Ministry of Health Turkey | Fatma Bayrakdar et al |

| | | | | | | |
|---|---|---|---|---|---|---|
| EPI_ISL_940669 | Turkey/HSGM-1501/2021 | 2021-01-22 | Europe/Turkey/Turkey/ | Ministry of Health Turkey | Ministry of Health Turkey | Fatma Bayrakdar et al |
| EPI_ISL_940672 | Turkey/HSGM-1504/2021 | 2021-01-22 | Europe/Turkey/Turkey/ | Ministry of Health Turkey | Ministry of Health Turkey | Fatma Bayrakdar et al |
| EPI_ISL_940673 | Turkey/HSGM-1505/2021 | 2021-01-22 | Europe/Turkey/Turkey/ | Ministry of Health Turkey | Ministry of Health Turkey | Fatma Bayrakdar et al |
| EPI_ISL_940674 | Turkey/HSGM-1506/2021 | 2021-01-22 | Europe/Turkey/Turkey/ | Ministry of Health Turkey | Ministry of Health Turkey | Fatma Bayrakdar et al |
| EPI_ISL_940675 | Turkey/HSGM-1507/2021 | 2021-01-22 | Europe/Turkey/Turkey/ | Ministry of Health Turkey | Ministry of Health Turkey | Fatma Bayrakdar et al |
| EPI_ISL_940676 | Turkey/HSGM-1508/2021 | 2021-01-22 | Europe/Turkey/Turkey/ | Ministry of Health Turkey | Ministry of Health Turkey | Fatma Bayrakdar et al |
| EPI_ISL_940677 | Turkey/HSGM-1509/2021 | 2021-01-22 | Europe/Turkey/Turkey/ | Ministry of Health Turkey | Ministry of Health Turkey | Fatma Bayrakdar et al |
| EPI_ISL_940680 | Turkey/HSGM-1512/2021 | 2021-01-22 | Europe/Turkey/Turkey/ | Ministry of Health Turkey | Ministry of Health Turkey | Fatma Bayrakdar et al |
| EPI_ISL_940715 | USA/WI-MPHL-U1040010/2021 | 2021-01-02 | North America/USA/Wisconsin/Milwauk | City of Milwaukee Health Department Laboratory | City of Milwaukee Health Department Laboratory | Sanjib Bhattacharyya et al |
| EPI_ISL_940729 | USA/WI-MPHL-U1120047/2021 | 2021-01-11 | North America/USA/Wisconsin/Milwauk | City of Milwaukee Health Department Laboratory | City of Milwaukee Health Department Laboratory | Sanjib Bhattacharyya et al |
| EPI_ISL_940739 | Italy/APU-POLBA16/2021 | 2021-01-25 | Europe/Italy/Apulia/ | University of Bari Biomedical Sciences and Human Oncology | University of Bari Biomedical Sciences and Human Oncology | Chironna M. et al |
| EPI_ISL_940751 | USA/OH-ODH-SC22089/2020 | 2020-03-26 | North America/USA/Ohio/ | Ohio Department of Health Laboratory | Ohio Department of Health Laboratory | Holmes et al |
| EPI_ISL_940753 | Italy/APU-POLBA21/2021 | 2020-03-28 | Europe/Italy/Apulia/ | University of Bari Biomedical Sciences and Human Oncology | University of Bari Biomedical Sciences and Human Oncology | Chironna M. et al |
| EPI_ISL_940765 | Botswana/CV1616790/2021 | 2021-01-20 | Africa/Botswana/Gaborone/ | Botswana Harvard HIV Reference Laboratory | Botswana Harvard HIV Reference Laboratory | Sikhulile Moyo et al |

| | | | | | | |
|---|---|---|---|---|---|---|
| EPI_ISL_940766 | NorthMacedonia/862/2021 | 2021-01-08 | Europe/North Macedonia/Skopje/ | Laboratory of Virology and Molecular Diagnostics | Institute of Public Health of Republic of North Macedonia Laboratory of Virology and Molecular Diagnostics | Maja Kuzmanovska et al |
| EPI_ISL_940775 | Ecuador/178620/2020 | 2020-12-30 | South America/Ecuador/Manabi/Chone | INSPI-CRN de Influenza y otros virus respiratorios | INSPI-Centro de Investigaci√≥n Multidisciplinaria de la DTIDI | Leandro Pati√±o et al |
| EPI_ISL_940777 | Ecuador/178328/2021 | 2021-01-04 | South America/Ecuador/Guayas/Duran | NSPI-CRN de Influenza y otros virus respiratorios | INSPI-Centro de Investigaci√≥n Multidisciplinaria de la DTIDI | Leandro Pati√±o et al |
| EPI_ISL_940781 | Ecuador/178635/2021 | 2021-01-04 | South America/Ecuador/Manabi/Sucre | INSPI-CRN de Influenza y otros virus respiratorios | INSPI-Centro de Investigaci√≥n Multidisciplinaria de la DTIDI | Leandro Pati√±o et al |
| EPI_ISL_940784 | USA/WI-MPHL-U1200127/2021 | 2021-01-19 | North America/USA/Wisconsin/Milwauk | City of Milwaukee Health Department Laboratory | City of Milwaukee Health Department Laboratory | Sanjib Bhattacharyya et al |
| EPI_ISL_940786 | USA/WI-MPHL-U1200165/2021 | 2021-01-19 | North America/USA/Wisconsin/Milwauk | City of Milwaukee Health Department Laboratory | City of Milwaukee Health Department Laboratory | Sanjib Bhattacharyya et al |
| EPI_ISL_940796 | USA/TX-HHD-2101114789/2020 | 2020-12-23 | North America/USA/Texas/ | Houston Health Dept. | Houston Health Dept. | Ryker Penn et al |
| EPI_ISL_940799 | USA/TX-HHD-2101116646/2020 | 2020-12-23 | North America/USA/Texas/ | Houston Health Dept. | Houston Health Dept. | Ryker Penn et al |
| EPI_ISL_940809 | USA/TX-HHD-2101116496/2020 | 2020-12-23 | North America/USA/Texas/ | Houston Health Dept. | Houston Health Dept. | Ryker Penn et al |
| EPI_ISL_940812 | USA/TX-HHD-2101116556/2020 | 2020-12-23 | North America/USA/Texas/ | Houston Health Dept. | Houston Health Dept. | Ryker Penn et al |
| EPI_ISL_940813 | USA/TX-HHD-2101116562/2020 | 2020-12-23 | North America/USA/Texas/ | Houston Health Dept. | Houston Health Dept. | Ryker Penn et al |
| EPI_ISL_940819 | USA/TX-HHD-2101117631/2020 | 2020-12-27 | North America/USA/Texas/ | Houston Health Dept. | Houston Health Dept. | Ryker Penn et al |

| | | | | | | |
|---|---|---|---|---|---|---|
| EPI_ISL_940827 | USA/VA-DCLS-2625/2020 | 2020-12-18 | North America/USA/Virginia/ | Virginia DCLS | Virginia DCLS | Virginia DCLS et al |
| EPI_ISL_940829 | USA/VA-DCLS-2627/2020 | 2020-12-18 | North America/USA/Virginia/ | Virginia DCLS | Virginia DCLS | Virginia DCLS et al |
| EPI_ISL_940830 | USA/VA-DCLS-2629/2020 | 2020-12-01 | North America/USA/Virginia/ | Virginia DCLS | Virginia DCLS | Virginia DCLS et al |
| EPI_ISL_940837 | USA/VA-DCLS-2748/2020 | 2020-12-18 | North America/USA/Virginia/ | Virginia DCLS | Virginia DCLS | Virginia DCLS et al |
| EPI_ISL_940838 | USA/VA-DCLS-2749/2020 | 2020-12-18 | North America/USA/Virginia/ | Virginia DCLS | Virginia DCLS | Virginia DCLS et al |
| EPI_ISL_940839 | USA/VA-DCLS-2818/2020 | 2020-12-21 | North America/USA/Virginia/ | Virginia DCLS | Virginia DCLS | Virginia DCLS et al |
| EPI_ISL_940846 | USA/OH-ODH-SC22140/2020 | 2020-03-26 | North America/USA/Ohio/ | Ohio Department of Health Laboratory | Ohio Department of Health Laboratory | Holmes et al |
| EPI_ISL_940852 | SouthAfrica/VIDA-KRISP-K008043/20 | 2021-01-07 | Africa/South Africa/Gauteng/ | Vaccines and Infectious Diseases Analytics Research Unit (VIDA) | KRISP, KZN Research Innovation and Sequencing Platform | Baillie Vicky et al |
| EPI_ISL_940853 | SouthAfrica/VIDA-KRISP-V001008/20 | 2020-12-21 | Africa/South Africa/Gauteng/ | Vaccines and Infectious Diseases Analytics Research Unit (VIDA) | KRISP, KZN Research Innovation and Sequencing Platform | Baillie Vicky et al |
| EPI_ISL_940858 | SouthAfrica/VIDA-KRISP-V001011/20 | 2020-12-09 | Africa/South Africa/Gauteng/ | Vaccines and Infectious Diseases Analytics Research Unit (VIDA) | KRISP, KZN Research Innovation and Sequencing Platform | Baillie Vicky et al |
| EPI_ISL_940859 | SouthAfrica/VIDA-KRISP-V001013/20 | 2020-12-15 | Africa/South Africa/Gauteng/ | Vaccines and Infectious Diseases Analytics Research Unit (VIDA) | KRISP, KZN Research Innovation and Sequencing Platform | Baillie Vicky et al |
| EPI_ISL_940862 | SouthAfrica/VIDA-KRISP-K008057/20 | 2021-01-06 | Africa/South Africa/Gauteng/ | Vaccines and Infectious Diseases Analytics Research Unit (VIDA) | KRISP, KZN Research Innovation and Sequencing Platform | Baillie Vicky et al |
| EPI_ISL_940864 | SouthAfrica/VIDA-KRISP-V001005/20 | 2021-01-08 | Africa/South Africa/Western Cape Provi | Vaccines and Infectious Diseases Analytics Research Unit (VIDA) | KRISP, KZN Research Innovation and Sequencing Platform | Baillie Vicky et al |

| | | | | | | |
|---|---|---|---|---|---|---|
| EPI_ISL_940866 | SouthAfrica/VIDA-KRISP-V001001/20 | 2021-01-05 | Africa/South Africa/Gauteng/ | Vaccines and Infectious Diseases Analytics Research Unit (VIDA) | KRISP, KZN Research Innovation and Sequencing Platform | Baillie Vicky et al |
| EPI_ISL_940876 | SouthAfrica/VIDA-KRISP-K008063/20 | 2021-01-05 | Africa/South Africa/Gauteng/ | Vaccines and Infectious Diseases Analytics Research Unit (VIDA) | KRISP, KZN Research Innovation and Sequencing Platform | Baillie Vicky et al |
| EPI_ISL_940888 | SouthAfrica/VIDA-KRISP-K008064/20 | 2021-01-14 | Africa/South Africa/Gauteng/ | Vaccines and Infectious Diseases Analytics Research Unit (VIDA) | KRISP, KZN Research Innovation and Sequencing Platform | Baillie Vicky et al |
| EPI_ISL_940889 | SouthAfrica/VIDA-KRISP-K008107/20 | 2021-01-14 | Africa/South Africa/Gauteng/ | Vaccines and Infectious Diseases Analytics Research Unit (VIDA) | KRISP, KZN Research Innovation and Sequencing Platform | Baillie Vicky et al |
| EPI_ISL_940890 | SouthAfrica/VIDA-KRISP-K008079/20 | 2021-01-16 | Africa/South Africa/Gauteng/ | Vaccines and Infectious Diseases Analytics Research Unit (VIDA) | KRISP, KZN Research Innovation and Sequencing Platform | Baillie Vicky et al |
| EPI_ISL_940891 | SouthAfrica/VIDA-KRISP-K008076/20 | 2021-01-19 | Africa/South Africa/Gauteng/ | Vaccines and Infectious Diseases Analytics Research Unit (VIDA) | KRISP, KZN Research Innovation and Sequencing Platform | Baillie Vicky et al |
| EPI_ISL_940897 | Puerto_Rico/CDC-S23/2020 | 2020-08-06 | North America/USA/Puerto Rico/ | Centers for Disease Control and Prevention, Dengue Branch | Centers for Disease Control and Prevention, Dengue Branch | Gilberto A. Santiago et al |
| EPI_ISL_940899 | Puerto_Rico/CDC-S25/2020 | 2020-08-10 | North America/USA/Puerto Rico/ | Centers for Disease Control and Prevention, Dengue Branch | Centers for Disease Control and Prevention, Dengue Branch | Gilberto A. Santiago et al |

| | | | | | | |
|---|---|---|---|---|---|---|
| EPI_ISL_940900 | Puerto_Rico/CDC-S26/2020 | 2020-06-29 | North America/USA/Puerto Rico/ | Centers for Disease Control and Prevention, Dengue Branch | Centers for Disease Control and Prevention, Dengue Branch | Gilberto A. Santiago et al |
| EPI_ISL_940904 | Puerto_Rico/CDC-S31/2020 | 2020-06-11 | North America/USA/Puerto Rico/ | Centers for Disease Control and Prevention, Dengue Branch | Centers for Disease Control and Prevention, Dengue Branch | Gilberto A. Santiago et al |
| EPI_ISL_940920 | Puerto_Rico/CDC-S47/2020 | 2020-07-24 | North America/USA/Puerto Rico/ | Centers for Disease Control and Prevention, Dengue Branch | Centers for Disease Control and Prevention, Dengue Branch | Gilberto A. Santiago et al |
| EPI_ISL_940922 | Puerto_Rico/CDC-S49/2020 | 2020-07-23 | North America/USA/Puerto Rico/ | Centers for Disease Control and Prevention, Dengue Branch | Centers for Disease Control and Prevention, Dengue Branch | Gilberto A. Santiago et al |
| EPI_ISL_940923 | Puerto_Rico/CDC-S50/2020 | 2020-07-23 | North America/USA/Puerto Rico/ | Centers for Disease Control and Prevention, Dengue Branch | Centers for Disease Control and Prevention, Dengue Branch | Gilberto A. Santiago et al |
| EPI_ISL_940927 | Puerto_Rico/CDC-S54/2020 | 2020-07-23 | North America/USA/Puerto Rico/ | Centers for Disease Control and Prevention, Dengue Branch | Centers for Disease Control and Prevention, Dengue Branch | Gilberto A. Santiago et al |
| EPI_ISL_940934 | Puerto_Rico/CDC-S61/2020 | 2020-08-01 | North America/USA/Puerto Rico/ | Centers for Disease Control and Prevention, Dengue Branch | Centers for Disease Control and Prevention, Dengue Branch | Gilberto A. Santiago et al |
| EPI_ISL_940935 | Puerto_Rico/CDC-S62/2020 | 2020-08-03 | North America/USA/Puerto Rico/ | Centers for Disease Control and Prevention, Dengue Branch | Centers for Disease Control and Prevention, Dengue Branch | Gilberto A. Santiago et al |
| EPI_ISL_940950 | Puerto_Rico/CDC-S77/2020 | 2020-08-23 | North America/USA/Puerto Rico/ | Centers for Disease Control and Prevention, Dengue Branch | Centers for Disease Control and Prevention, Dengue Branch | Gilberto A. Santiago et al |
| EPI_ISL_940951 | Puerto_Rico/CDC-S78/2020 | 2020-08-19 | North America/USA/Puerto Rico/ | Centers for Disease Control and Prevention, Dengue Branch | Centers for Disease Control and Prevention, Dengue Branch | Gilberto A. Santiago et al |

| | | | | | | |
|---|---|---|---|---|---|---|
| EPI_ISL_940957 | Puerto_Rico/CDC-S84/2020 | 2020-08-28 | North America/USA/Puerto Rico/ | Centers for Disease Control and Prevention, Dengue Branch | Centers for Disease Control and Prevention, Dengue Branch | Gilberto A. Santiago et al |
| EPI_ISL_940961 | Puerto_Rico/CDC-S88/2020 | 2020-09-25 | North America/USA/Puerto Rico/ | Centers for Disease Control and Prevention, Dengue Branch | Centers for Disease Control and Prevention, Dengue Branch | Gilberto A. Santiago et al |
| EPI_ISL_940963 | Puerto_Rico/CDC-S94/2020 | 2020-09-04 | North America/USA/Puerto Rico/ | Centers for Disease Control and Prevention, Dengue Branch | Centers for Disease Control and Prevention, Dengue Branch | Gilberto A. Santiago et al |
| EPI_ISL_940966 | Puerto_Rico/CDC-S98/2020 | 2020-09-05 | North America/USA/Puerto Rico/ | Centers for Disease Control and Prevention, Dengue Branch | Centers for Disease Control and Prevention, Dengue Branch | Gilberto A. Santiago et al |
| EPI_ISL_940968 | Puerto_Rico/CDC-S100/2020 | 2020-09-02 | North America/USA/Puerto Rico/ | Centers for Disease Control and Prevention, Dengue Branch | Centers for Disease Control and Prevention, Dengue Branch | Gilberto A. Santiago et al |
| EPI_ISL_940971 | Puerto_Rico/CDC-S103/2020 | 2020-09-02 | North America/USA/Puerto Rico/ | Centers for Disease Control and Prevention, Dengue Branch | Centers for Disease Control and Prevention, Dengue Branch | Gilberto A. Santiago et al |
| EPI_ISL_940979 | Puerto_Rico/CDC-S112/2020 | 2020-10-12 | North America/USA/Puerto Rico/ | Centers for Disease Control and Prevention, Dengue Branch | Centers for Disease Control and Prevention, Dengue Branch | Gilberto A. Santiago et al |
| EPI_ISL_940984 | Puerto_Rico/CDC-S117/2020 | 2020-10-02 | North America/USA/Puerto Rico/ | Centers for Disease Control and Prevention, Dengue Branch | Centers for Disease Control and Prevention, Dengue Branch | Gilberto A. Santiago et al |
| EPI_ISL_940988 | Puerto_Rico/CDC-S121/2020 | 2020-10-03 | North America/USA/Puerto Rico/ | Centers for Disease Control and Prevention, Dengue Branch | Centers for Disease Control and Prevention, Dengue Branch | Gilberto A. Santiago et al |

| | | | | | | |
|---|---|---|---|---|---|---|
| EPI_ISL_940997 | France/BRE-IPP01235/2021 | 2021-01-08 | Europe/France/Bretagne/Chateaulin | Labo Analyses Med | National Reference Center for Viruses of Respiratory Infections, Institut Pasteur, Paris | Marion Barbet et al |
| EPI_ISL_941070 | France/IDF-IPP01384/2021 | 2021-01-15 | Europe/France/Ile de France/Saint-Deni | Labo Analyses Med | National Reference Center for Viruses of Respiratory Infections, Institut Pasteur, Paris | Marion Barbet et al |
| EPI_ISL_941077 | France/IDF-IPP01404/2021 | 2021-01-15 | Europe/France/Ile de France/Saint-Deni | Labo Analyses Med | National Reference Center for Viruses of Respiratory Infections, Institut Pasteur, Paris | Marion Barbet et al |
| EPI_ISL_941086 | France/IDF-IPP01415/2021 | 2021-01-13 | Europe/France/Ile de France/Saint-Deni | Labo Analyses Med | National Reference Center for Viruses of Respiratory Infections, Institut Pasteur, Paris | Marion Barbet et al |
| EPI_ISL_941088 | France/IDF-IPP01417/2021 | 2021-01-12 | Europe/France/Ile de France/Saint-Deni | Labo Analyses Med | National Reference Center for Viruses of Respiratory Infections, Institut Pasteur, Paris | Marion Barbet et al |

| | | | | | | |
|---|---|---|---|---|---|---|
| EPI_ISL_941104 | Colombia/AMA-GUR-06720/2020 | 2020-05-02 | South America/Colombia/Departamento | Centro de Investigaciones en Microbiologv≠a y Biotecnologv≠a-UR (CIMBIUR), Facultad de Ciencias Naturales, Universidad del Rosario, Bogotv°, Colombia | Centro de Investigaciones en Microbiologv≠a y Biotecnologv≠a-UR (CIMBIUR), Facultad de Ciencias Naturales, Universidad del Rosario, Bogotv°, Colombia Icahn School of Medicine at Mount Sinai, New York, USA | Nathalia Ballesteros et al |
| EPI_ISL_941105 | Colombia/AMA-GUR-06721/2020 | 2020-05-02 | South America/Colombia/Departamento | Centro de Investigaciones en Microbiologv≠a y Biotecnologv≠a-UR (CIMBIUR), Facultad de Ciencias Naturales, Universidad del Rosario, Bogotv°, Colombia | Centro de Investigaciones en Microbiologv≠a y Biotecnologv≠a-UR (CIMBIUR), Facultad de Ciencias Naturales, Universidad del Rosario, Bogotv°, Colombia Icahn School of Medicine at Mount Sinai, New York, USA | Nathalia Ballesteros et al |

| | | | | | | |
|---|---|---|---|---|---|---|
| EPI_ISL_941106 | Colombia/AMA-GUR-06722/2020 | 2020-05-02 | South America/Colombia/Departamento | Centro de Investigaciones en Microbiologv≠a y Biotecnologv≠a-UR (CIMBIUR), Facultad de Ciencias Naturales, Universidad del Rosario, Bogotv°, Colombia | Centro de Investigaciones en Microbiologv≠a y Biotecnologv≠a-UR (CIMBIUR), Facultad de Ciencias Naturales, Universidad del Rosario, Bogotv°, Colombia Icahn School of Medicine at Mount Sinai, New York, USA | Nathalia Ballesteros et al |
| EPI_ISL_941107 | Colombia/AMA-GUR-06723/2020 | 2020-05-02 | South America/Colombia/Departamento | Centro de Investigaciones en Microbiologv≠a y Biotecnologv≠a-UR (CIMBIUR), Facultad de Ciencias Naturales, Universidad del Rosario, Bogotv°, Colombia | Centro de Investigaciones en Microbiologv≠a y Biotecnologv≠a-UR (CIMBIUR), Facultad de Ciencias Naturales, Universidad del Rosario, Bogotv°, Colombia Icahn School of Medicine at Mount Sinai, New York, USA | Nathalia Ballesteros et al |

| | | | | | | |
|---|---|---|---|---|---|---|
| EPI_ISL_941109 | Colombia/AMA-GUR-06729/2020 | 2020-05-03 | South America/Colombia/Departamento | Centro de Investigaciones en Microbiolog√≠a y Biotecnolog√≠a-UR (CIMBIUR), Facultad de Ciencias Naturales, Universidad del Rosario, Bogot√°, Colombia | Centro de Investigaciones en Microbiolog√≠a y Biotecnolog√≠a-UR (CIMBIUR), Facultad de Ciencias Naturales, Universidad del Rosario, Bogot√°, Colombia Icahn School of Medicine at Mount Sinai, New York, USA | Nathalia Ballesteros et al |
| EPI_ISL_941110 | Colombia/AMA-GUR-06730/2020 | 2020-05-03 | South America/Colombia/Departamento | Centro de Investigaciones en Microbiolog√≠a y Biotecnolog√≠a-UR (CIMBIUR), Facultad de Ciencias Naturales, Universidad del Rosario, Bogot√°, Colombia | Centro de Investigaciones en Microbiolog√≠a y Biotecnolog√≠a-UR (CIMBIUR), Facultad de Ciencias Naturales, Universidad del Rosario, Bogot√°, Colombia Icahn School of Medicine at Mount Sinai, New York, USA | Nathalia Ballesteros et al |

| | | | | | | |
|---|---|---|---|---|---|---|
| EPI_ISL_941111 | Colombia/AMA-GUR-06732/2020 | 2020-05-03 | South America/Colombia/Departamento | Centro de Investigaciones en Microbiolog√≠a y Biotecnolog√≠a-UR (CIMBIUR), Facultad de Ciencias Naturales, Universidad del Rosario, Bogot√°, Colombia | Centro de Investigaciones en Microbiolog√≠a y Biotecnolog√≠a-UR (CIMBIUR), Facultad de Ciencias Naturales, Universidad del Rosario, Bogot√°, Colombia Icahn School of Medicine at Mount Sinai, New York, USA | Nathalia Ballesteros et al |
| EPI_ISL_941112 | Colombia/AMA-GUR-06733/2020 | 2020-05-03 | South America/Colombia/Departamento | Centro de Investigaciones en Microbiolog√≠a y Biotecnolog√≠a-UR (CIMBIUR), Facultad de Ciencias Naturales, Universidad del Rosario, Bogot√°, Colombia | Centro de Investigaciones en Microbiolog√≠a y Biotecnolog√≠a-UR (CIMBIUR), Facultad de Ciencias Naturales, Universidad del Rosario, Bogot√°, Colombia Icahn School of Medicine at Mount Sinai, New York, USA | Nathalia Ballesteros et al |

| | | | | | | |
|---|---|---|---|---|---|---|
| EPI_ISL_941113 | Colombia/AMA-GUR-06736/2020 | 2020-05-03 | South America/Colombia/Departamento | Centro de Investigaciones en Microbiolog√≠a y Biotecnolog√≠a-UR (CIMBIUR), Facultad de Ciencias Naturales, Universidad del Rosario, Bogot√°, Colombia | Centro de Investigaciones en Microbiolog√≠a y Biotecnolog√≠a-UR (CIMBIUR), Facultad de Ciencias Naturales, Universidad del Rosario, Bogot√°, Colombia Icahn School of Medicine at Mount Sinai, New York, USA | Nathalia Ballesteros et al |
| EPI_ISL_941115 | Colombia/AMA-GUR-06738/2020 | 2020-05-03 | South America/Colombia/Departamento | Centro de Investigaciones en Microbiolog√≠a y Biotecnolog√≠a-UR (CIMBIUR), Facultad de Ciencias Naturales, Universidad del Rosario, Bogot√°, Colombia | Centro de Investigaciones en Microbiolog√≠a y Biotecnolog√≠a-UR (CIMBIUR), Facultad de Ciencias Naturales, Universidad del Rosario, Bogot√°, Colombia Icahn School of Medicine at Mount Sinai, New York, USA | Nathalia Ballesteros et al |

| | | | | | | |
|---|---|---|---|---|---|---|
| EPI_ISL_941117 | Colombia/AMA-GUR-06741/2020 | 2020-05-03 | South America/Colombia/Departamento | Centro de Investigaciones en Microbiolog√≠a y Biotecnolog√≠a-UR (CIMBIUR), Facultad de Ciencias Naturales, Universidad del Rosario, Bogot√°, Colombia | Centro de Investigaciones en Microbiolog√≠a y Biotecnolog√≠a-UR (CIMBIUR), Facultad de Ciencias Naturales, Universidad del Rosario, Bogot√°, Colombia Icahn School of Medicine at Mount Sinai, New York, USA | Nathalia Ballesteros et al |
| EPI_ISL_941119 | Colombia/AMA-GUR-06743/2020 | 2020-05-01 | South America/Colombia/Departamento | Centro de Investigaciones en Microbiolog√≠a y Biotecnolog√≠a-UR (CIMBIUR), Facultad de Ciencias Naturales, Universidad del Rosario, Bogot√°, Colombia | Centro de Investigaciones en Microbiolog√≠a y Biotecnolog√≠a-UR (CIMBIUR), Facultad de Ciencias Naturales, Universidad del Rosario, Bogot√°, Colombia Icahn School of Medicine at Mount Sinai, New York, USA | Nathalia Ballesteros et al |

| | | | | | | |
|---|---|---|---|---|---|---|
| EPI_ISL_941120 | Colombia/AMA-GUR-06744/2020 | 2020-05-02 | South America/Colombia/Departamento | Centro de Investigaciones en Microbiolog√≠a y Biotecnolog√≠a-UR (CIMBIUR), Facultad de Ciencias Naturales, Universidad del Rosario, Bogot√°, Colombia | Centro de Investigaciones en Microbiolog√≠a y Biotecnolog√≠a-UR (CIMBIUR), Facultad de Ciencias Naturales, Universidad del Rosario, Bogot√°, Colombia Icahn School of Medicine at Mount Sinai, New York, USA | Nathalia Ballesteros et al |
| EPI_ISL_941124 | Colombia/AMA-GUR-06749/2020 | 2020-05-03 | South America/Colombia/Departamento | Centro de Investigaciones en Microbiolog√≠a y Biotecnolog√≠a-UR (CIMBIUR), Facultad de Ciencias Naturales, Universidad del Rosario, Bogot√°, Colombia | Centro de Investigaciones en Microbiolog√≠a y Biotecnolog√≠a-UR (CIMBIUR), Facultad de Ciencias Naturales, Universidad del Rosario, Bogot√°, Colombia Icahn School of Medicine at Mount Sinai, New York, USA | Nathalia Ballesteros et al |

| | | | | | | |
|---|---|---|---|---|---|---|
| EPI_ISL_941127 | Colombia/AMA-GUR-06754/2020 | 2020-05-03 | South America/Colombia/Departamento | Centro de Investigaciones en Microbiolog√≠a y Biotecnolog√≠a-UR (CIMBIUR), Facultad de Ciencias Naturales, Universidad del Rosario, Bogot√°, Colombia | Centro de Investigaciones en Microbiolog√≠a y Biotecnolog√≠a-UR (CIMBIUR), Facultad de Ciencias Naturales, Universidad del Rosario, Bogot√°, Colombia Icahn School of Medicine at Mount Sinai, New York, USA | Nathalia Ballesteros et al |
| EPI_ISL_941129 | Colombia/AMA-GUR-06757/2020 | 2020-05-03 | South America/Colombia/Departamento | Centro de Investigaciones en Microbiolog√≠a y Biotecnolog√≠a-UR (CIMBIUR), Facultad de Ciencias Naturales, Universidad del Rosario, Bogot√°, Colombia | Centro de Investigaciones en Microbiolog√≠a y Biotecnolog√≠a-UR (CIMBIUR), Facultad de Ciencias Naturales, Universidad del Rosario, Bogot√°, Colombia Icahn School of Medicine at Mount Sinai, New York, USA | Nathalia Ballesteros et al |

| | | | | | | |
|---|---|---|---|---|---|---|
| EPI_ISL_941130 | Colombia/AMA-GUR-06759/2020 | 2020-05-04 | South America/Colombia/Departamento | Centro de Investigaciones en Microbiolog√≠a y Biotecnolog√≠a-UR (CIMBIUR), Facultad de Ciencias Naturales, Universidad del Rosario, Bogot√°, Colombia | Centro de Investigaciones en Microbiolog√≠a y Biotecnolog√≠a-UR (CIMBIUR), Facultad de Ciencias Naturales, Universidad del Rosario, Bogot√°, Colombia Icahn School of Medicine at Mount Sinai, New York, USA | Nathalia Ballesteros et al |
| EPI_ISL_941131 | Colombia/AMA-GUR-06760/2020 | 2020-05-04 | South America/Colombia/Departamento | Centro de Investigaciones en Microbiolog√≠a y Biotecnolog√≠a-UR (CIMBIUR), Facultad de Ciencias Naturales, Universidad del Rosario, Bogot√°, Colombia | Centro de Investigaciones en Microbiolog√≠a y Biotecnolog√≠a-UR (CIMBIUR), Facultad de Ciencias Naturales, Universidad del Rosario, Bogot√°, Colombia Icahn School of Medicine at Mount Sinai, New York, USA | Nathalia Ballesteros et al |

| | | | | | | |
|---|---|---|---|---|---|---|
| EPI_ISL_941133 | Colombia/AMA-GUR-06768/2020 | 2020-05-04 | South America/Colombia/Departamento | Centro de Investigaciones en Microbiolog√≠a y Biotecnolog√≠a-UR (CIMBIUR), Facultad de Ciencias Naturales, Universidad del Rosario, Bogot√°, Colombia | Centro de Investigaciones en Microbiolog√≠a y Biotecnolog√≠a-UR (CIMBIUR), Facultad de Ciencias Naturales, Universidad del Rosario, Bogot√°, Colombia Icahn School of Medicine at Mount Sinai, New York, USA | Nathalia Ballesteros et al |
| EPI_ISL_941135 | Colombia/AMA-GUR-06771/2020 | 2020-05-04 | South America/Colombia/Departamento | Centro de Investigaciones en Microbiolog√≠a y Biotecnolog√≠a-UR (CIMBIUR), Facultad de Ciencias Naturales, Universidad del Rosario, Bogot√°, Colombia | Centro de Investigaciones en Microbiolog√≠a y Biotecnolog√≠a-UR (CIMBIUR), Facultad de Ciencias Naturales, Universidad del Rosario, Bogot√°, Colombia Icahn School of Medicine at Mount Sinai, New York, USA | Nathalia Ballesteros et al |

| | | | | | | |
|---|---|---|---|---|---|---|
| EPI_ISL_941136 | Colombia/AMA-GUR-06772/2020 | 2020-05-04 | South America/Colombia/Departamento | Centro de Investigaciones en Microbiolog√≠a y Biotecnolog√≠a-UR (CIMBIUR), Facultad de Ciencias Naturales, Universidad del Rosario, Bogot√°, Colombia | Centro de Investigaciones en Microbiolog√≠a y Biotecnolog√≠a-UR (CIMBIUR), Facultad de Ciencias Naturales, Universidad del Rosario, Bogot√°, Colombia Icahn School of Medicine at Mount Sinai, New York, USA | Nathalia Ballesteros et al |
| EPI_ISL_941137 | Colombia/AMA-GUR-06773/2020 | 2020-05-03 | South America/Colombia/Departamento | Centro de Investigaciones en Microbiolog√≠a y Biotecnolog√≠a-UR (CIMBIUR), Facultad de Ciencias Naturales, Universidad del Rosario, Bogot√°, Colombia | Centro de Investigaciones en Microbiolog√≠a y Biotecnolog√≠a-UR (CIMBIUR), Facultad de Ciencias Naturales, Universidad del Rosario, Bogot√°, Colombia Icahn School of Medicine at Mount Sinai, New York, USA | Nathalia Ballesteros et al |

| | | | | | | |
|---|---|---|---|---|---|---|
| EPI_ISL_941139 | Colombia/AMA-GUR-06783/2020 | 2020-05-04 | South America/Colombia/Departamento | Centro de Investigaciones en Microbiolog√≠a y Biotecnolog√≠a-UR (CIMBIUR), Facultad de Ciencias Naturales, Universidad del Rosario, Bogot√°, Colombia | Centro de Investigaciones en Microbiolog√≠a y Biotecnolog√≠a-UR (CIMBIUR), Facultad de Ciencias Naturales, Universidad del Rosario, Bogot√°, Colombia Icahn School of Medicine at Mount Sinai, New York, USA | Nathalia Ballesteros et al |
| EPI_ISL_941144 | Colombia/AMA-GUR-06793/2020 | 2020-05-04 | South America/Colombia/Departamento | Centro de Investigaciones en Microbiolog√≠a y Biotecnolog√≠a-UR (CIMBIUR), Facultad de Ciencias Naturales, Universidad del Rosario, Bogot√°, Colombia | Centro de Investigaciones en Microbiolog√≠a y Biotecnolog√≠a-UR (CIMBIUR), Facultad de Ciencias Naturales, Universidad del Rosario, Bogot√°, Colombia Icahn School of Medicine at Mount Sinai, New York, USA | Nathalia Ballesteros et al |

| | | | | | | |
|---|---|---|---|---|---|---|
| EPI_ISL_941146 | Colombia/AMA-GUR-06800/2020 | 2020-05-04 | South America/Colombia/Departamento | Centro de Investigaciones en Microbiolog√≠a y Biotecnolog√≠a-UR (CIMBIUR), Facultad de Ciencias Naturales, Universidad del Rosario, Bogot√°, Colombia | Centro de Investigaciones en Microbiolog√≠a y Biotecnolog√≠a-UR (CIMBIUR), Facultad de Ciencias Naturales, Universidad del Rosario, Bogot√°, Colombia Icahn School of Medicine at Mount Sinai, New York, USA | Nathalia Ballesteros et al |
| EPI_ISL_941148 | Colombia/AMA-GUR-06803/2020 | 2020-05-04 | South America/Colombia/Departamento | Centro de Investigaciones en Microbiolog√≠a y Biotecnolog√≠a-UR (CIMBIUR), Facultad de Ciencias Naturales, Universidad del Rosario, Bogot√°, Colombia | Centro de Investigaciones en Microbiolog√≠a y Biotecnolog√≠a-UR (CIMBIUR), Facultad de Ciencias Naturales, Universidad del Rosario, Bogot√°, Colombia Icahn School of Medicine at Mount Sinai, New York, USA | Nathalia Ballesteros et al |

| | | | | | | |
|---|---|---|---|---|---|---|
| EPI_ISL_941149 | Colombia/AMA-GUR-06807/2020 | 2020-05-04 | South America/Colombia/Departamento | Centro de Investigaciones en Microbiolog√≠a y Biotecnolog√≠a-UR (CIMBIUR), Facultad de Ciencias Naturales, Universidad del Rosario, Bogot√°, Colombia | Centro de Investigaciones en Microbiolog√≠a y Biotecnolog√≠a-UR (CIMBIUR), Facultad de Ciencias Naturales, Universidad del Rosario, Bogot√°, Colombia Icahn School of Medicine at Mount Sinai, New York, USA | Nathalia Ballesteros et al |
| EPI_ISL_941152 | Colombia/AMA-GUR-06811/2020 | 2020-05-04 | South America/Colombia/Departamento | Centro de Investigaciones en Microbiolog√≠a y Biotecnolog√≠a-UR (CIMBIUR), Facultad de Ciencias Naturales, Universidad del Rosario, Bogot√°, Colombia | Centro de Investigaciones en Microbiolog√≠a y Biotecnolog√≠a-UR (CIMBIUR), Facultad de Ciencias Naturales, Universidad del Rosario, Bogot√°, Colombia Icahn School of Medicine at Mount Sinai, New York, USA | Nathalia Ballesteros et al |

| | | | | | | |
|---|---|---|---|---|---|---|
| EPI_ISL_941158 | Colombia/AMA-GUR-06817/2020 | 2020-05-04 | South America/Colombia/Departamento | Centro de Investigaciones en Microbiolog√≠a y Biotecnolog√≠a-UR (CIMBIUR), Facultad de Ciencias Naturales, Universidad del Rosario, Bogot√°, Colombia | Centro de Investigaciones en Microbiolog√≠a y Biotecnolog√≠a-UR (CIMBIUR), Facultad de Ciencias Naturales, Universidad del Rosario, Bogot√°, Colombia Icahn School of Medicine at Mount Sinai, New York, USA | Nathalia Ballesteros et al |
| EPI_ISL_941159 | Colombia/AMA-GUR-06818/2020 | 2020-05-04 | South America/Colombia/Departamento | Centro de Investigaciones en Microbiolog√≠a y Biotecnolog√≠a-UR (CIMBIUR), Facultad de Ciencias Naturales, Universidad del Rosario, Bogot√°, Colombia | Centro de Investigaciones en Microbiolog√≠a y Biotecnolog√≠a-UR (CIMBIUR), Facultad de Ciencias Naturales, Universidad del Rosario, Bogot√°, Colombia Icahn School of Medicine at Mount Sinai, New York, USA | Nathalia Ballesteros et al |

| | | | | | | |
|---|---|---|---|---|---|---|
| EPI_ISL_941160 | Colombia/AMA-GUR-06819/2020 | 2020-05-04 | South America/Colombia/Departamento | Centro de Investigaciones en Microbiolog√≠a y Biotecnolog√≠a-UR (CIMBIUR), Facultad de Ciencias Naturales, Universidad del Rosario, Bogot√°, Colombia | Centro de Investigaciones en Microbiolog√≠a y Biotecnolog√≠a-UR (CIMBIUR), Facultad de Ciencias Naturales, Universidad del Rosario, Bogot√°, Colombia Icahn School of Medicine at Mount Sinai, New York, USA | Nathalia Ballesteros et al |
| EPI_ISL_941161 | Colombia/AMA-GUR-06828/2020 | 2020-05-05 | South America/Colombia/Departamento | Centro de Investigaciones en Microbiolog√≠a y Biotecnolog√≠a-UR (CIMBIUR), Facultad de Ciencias Naturales, Universidad del Rosario, Bogot√°, Colombia | Centro de Investigaciones en Microbiolog√≠a y Biotecnolog√≠a-UR (CIMBIUR), Facultad de Ciencias Naturales, Universidad del Rosario, Bogot√°, Colombia Icahn School of Medicine at Mount Sinai, New York, USA | Nathalia Ballesteros et al |
| EPI_ISL_941163 | NorthMacedonia/1438/2021 | 2021-01-09 | Europe/North Macedonia/Pelagonia/Pri | Laboratory of Virology and Molecular Diagnostics | Institute of Public Health of Republic of North Macedonia Laboratory of Virology and Molecular Diagnostics | Maja Kuzmanovska et al |
| EPI_ISL_941165 | Spain/VC-IBV-98014400/2021 | 2021-01-13 | Europe/Spain/Comunitat Valenciana/Va | Hospital Universitari i Polit√®cnic La Fe de Val√®ncia | SeqCOVID-SPAIN consortium/IBV(CSIC) | Mar√≠a Dolores G√≥mez Ruiz et al |

| | | | | | | |
|---|---|---|---|---|---|---|
| EPI_ISL_941173 | Spain/VC-IBV-98014411/2021 | 2021-01-20 | Europe/Spain/Comunitat Valenciana/Va | Hospital Universitari i Polit√®cnic La Fe de Val√®ncia | SeqCOVID-SPAIN consortium/IBV(CSIC) | Mar√≠a Dolores G√≥mez Ruiz et al |
| EPI_ISL_941178 | Spain/VC-IBV-98014416/2020 | 2020-10-06 | Europe/Spain/Comunitat Valenciana/Va | Servicio de Microbiolog√≠a. Hospital Cl√≠nico Universitario de Valencia | SeqCOVID-SPAIN consortium/IBV(CSIC) | David Navarro Ortega et al |
| EPI_ISL_941182 | Spain/VC-IBV-98014421/2020 | 2020-10-12 | Europe/Spain/Comunitat Valenciana/Va | Servicio de Microbiolog√≠a. Hospital Cl√≠nico Universitario de Valencia | SeqCOVID-SPAIN consortium/IBV(CSIC) | David Navarro Ortega et al |
| EPI_ISL_941183 | Spain/VC-IBV-98014422/2020 | 2020-10-12 | Europe/Spain/Comunitat Valenciana/Va | Servicio de Microbiolog√≠a. Hospital Cl√≠nico Universitario de Valencia | SeqCOVID-SPAIN consortium/IBV(CSIC) | David Navarro Ortega et al |
| EPI_ISL_941187 | Spain/VC-IBV-98014427/2020 | 2020-10-14 | Europe/Spain/Comunitat Valenciana/Va | Servicio de Microbiolog√≠a. Hospital Cl√≠nico Universitario de Valencia | SeqCOVID-SPAIN consortium/IBV(CSIC) | David Navarro Ortega et al |
| EPI_ISL_941189 | Spain/VC-IBV-98014429/2020 | 2020-10-14 | Europe/Spain/Comunitat Valenciana/Va | Servicio de Microbiolog√≠a. Hospital Cl√≠nico Universitario de Valencia | SeqCOVID-SPAIN consortium/IBV(CSIC) | David Navarro Ortega et al |
| EPI_ISL_941207 | Spain/VC-IBV-98014451/2020 | 2020-11-26 | Europe/Spain/Comunitat Valenciana/Va | Servicio de Microbiolog√≠a. Hospital Cl√≠nico Universitario de Valencia | SeqCOVID-SPAIN consortium/IBV(CSIC) | David Navarro Ortega et al |
| EPI_ISL_941212 | Spain/VC-IBV-98014459/2021 | 2021-01-16 | Europe/Spain/Comunitat Valenciana/Va | Servicio de Microbiolog√≠a. Hospital Cl√≠nico Universitario de Valencia | SeqCOVID-SPAIN consortium/IBV(CSIC) | David Navarro Ortega et al |
| EPI_ISL_941213 | Spain/VC-IBV-98014460/2021 | 2021-01-16 | Europe/Spain/Comunitat Valenciana/Va | Servicio de Microbiolog√≠a. Hospital Cl√≠nico Universitario de Valencia | SeqCOVID-SPAIN consortium/IBV(CSIC) | David Navarro Ortega et al |
| EPI_ISL_941220 | Spain/VC-IBV-98014516/2021 | 2021-01-05 | Europe/Spain/Comunitat Valenciana/Eld | Laboratorio de Microbiolog√≠a. Hospital General Universitario de Elda, Alicante | SeqCOVID-SPAIN consortium/IBV(CSIC) | M¬™ Isabel Gasc√≥n Ros et al |
| EPI_ISL_941222 | Spain/VC-IBV-98014518/2021 | 2021-01-05 | Europe/Spain/Comunitat Valenciana/Eld | Laboratorio de Microbiolog√≠a. Hospital General Universitario de Elda, Alicante | SeqCOVID-SPAIN consortium/IBV(CSIC) | M¬™ Isabel Gasc√≥n Ros et al |
| EPI_ISL_941228 | Spain/VC-IBV-98014527/2021 | 2021-01-09 | Europe/Spain/Comunitat Valenciana/Eld | Laboratorio de Microbiolog√≠a. Hospital General Universitario de Elda, Alicante | SeqCOVID-SPAIN consortium/IBV(CSIC) | M¬™ Isabel Gasc√≥n Ros et al |
| EPI_ISL_941230 | Spain/VC-IBV-98014529/2021 | 2021-01-14 | Europe/Spain/Comunitat Valenciana/Eld | Laboratorio de Microbiolog√≠a. Hospital General Universitario de Elda, Alicante | SeqCOVID-SPAIN consortium/IBV(CSIC) | M¬™ Isabel Gasc√≥n Ros et al |

| | | | | | | |
|---|---|---|---|---|---|---|
| EPI_ISL_941232 | Spain/VC-IBV-98014541/2021 | 2021-01-14 | Europe/Spain/Comunitat Valenciana/Elc | Laboratorio de Microbiolog√≠a. Hospital General Universitario de Elda, Alicante | SeqCOVID-SPAIN consortium/IBV(CSIC) | M¬™ Isabel Gasc√≥n Ros et al |
| EPI_ISL_941250 | USA/VA-DCLS-2702/2020 | 2020-12-08 | North America/USA/Virginia/ | Virginia DCLS | Virginia DCLS | Virginia DCLS et al |
| EPI_ISL_941263 | USA/VA-DCLS-2723/2020 | 2020-12-15 | North America/USA/Virginia/ | Virginia DCLS | Virginia DCLS | Virginia DCLS et al |
| EPI_ISL_941273 | USA/NC-SLPH-0027/2020 | 2020-09-11 | North America/USA/North Carolina/ | NCSLPH | NCSLPH | Chase K et al |
| EPI_ISL_941284 | Nigeria/CV635/2020 | 2020-12-21 | Africa/Nigeria/Lagos/ | Nigeria Centre for Disease Control (NCDC) | African Centre of Excellence for Genomics of Infectious Diseases (ACEGID), Redeemer's University | Oluniyi P.E. et al et al |
| EPI_ISL_941287 | Nigeria/CV639/2020 | 2020-12-29 | Africa/Nigeria/Lagos/ | Nigeria Centre for Disease Control (NCDC) | African Centre of Excellence for Genomics of Infectious Diseases (ACEGID), Redeemer's University | Oluniyi P.E. et al et al |
| EPI_ISL_941292 | Nigeria/CV645/2021 | 2021-01-04 | Africa/Nigeria/Lagos/ | Nigeria Centre for Disease Control (NCDC) | African Centre of Excellence for Genomics of Infectious Diseases (ACEGID), Redeemer's University | Oluniyi P.E. et al et al |
| EPI_ISL_941294 | Nigeria/CV647/2021 | 2021-01-15 | Africa/Nigeria/Lagos/ | Nigeria Centre for Disease Control (NCDC) | African Centre of Excellence for Genomics of Infectious Diseases (ACEGID), Redeemer's University | Oluniyi P.E. et al et al |

| | | | | | | |
|---|---|---|---|---|---|---|
| EPI_ISL_941312 | USA/NMDOH-2021037624/2021 | 2021-01-25 | North America/USA/New Mexico/ | New Mexico Department of Health Scientific Laboratory | New Mexico Department of Health Scientific Laboratory | Ellie Johnson et al |
| EPI_ISL_941314 | USA/NMDOH-2021036361/2021 | 2021-01-25 | North America/USA/New Mexico/ | New Mexico Department of Health Scientific Laboratory | New Mexico Department of Health Scientific Laboratory | Ellie Johnson et al |
| EPI_ISL_941323 | USA/NMDOH-2021037185/2021 | 2021-01-25 | North America/USA/New Mexico/ | New Mexico Department of Health Scientific Laboratory | New Mexico Department of Health Scientific Laboratory | Ellie Johnson et al |
| EPI_ISL_941324 | USA/NMDOH-2021037203/2021 | 2021-01-25 | North America/USA/New Mexico/ | New Mexico Department of Health Scientific Laboratory | New Mexico Department of Health Scientific Laboratory | Ellie Johnson et al |
| EPI_ISL_941330 | USA/NMDOH-2021037897/2021 | 2021-01-26 | North America/USA/New Mexico/ | New Mexico Department of Health Scientific Laboratory | New Mexico Department of Health Scientific Laboratory | Ellie Johnson et al |
| EPI_ISL_941334 | USA/NMDOH-2021039737/2021 | 2021-01-27 | North America/USA/New Mexico/ | New Mexico Department of Health Scientific Laboratory | New Mexico Department of Health Scientific Laboratory | Ellie Johnson et al |
| EPI_ISL_941335 | USA/NMDOH-2021044683/2021 | 2021-01-31 | North America/USA/New Mexico/ | New Mexico Department of Health Scientific Laboratory | New Mexico Department of Health Scientific Laboratory | Ellie Johnson et al |
| EPI_ISL_941336 | Italy/LAZ-INMI-176/2021 | 2021-01-09 | Europe/Italy/Lazio/ | Ospedale San Camillo De Lellis di Rieti | INMI Lazzaro Spallanzani IRCCS | Cesare E.M. Gruber et al |
| EPI_ISL_941337 | Italy/LAZ-INMI-177/2021 | 2021-01-09 | Europe/Italy/Lazio/ | Ospedale San Camillo De Lellis di Rieti | INMI Lazzaro Spallanzani IRCCS | Ornella Butera et al |
| EPI_ISL_941338 | Italy/LAZ-INMI-178/2021 | 2021-01-13 | Europe/Italy/Lazio/ | INMI Lazzaro Spallanzani IRCCS | INMI Lazzaro Spallanzani IRCCS | Emanuela Giombini et al |
| EPI_ISL_941348 | Portugal/PT2296/2021 | 2021-01-12 | Europe/Portugal/Portugal/ | Instituto Nacional de Saude (INSA) | Instituto Nacional de Saude (INSA) | Borges et al et al |
| EPI_ISL_941357 | Portugal/PT2306/2021 | 2021-01-14 | Europe/Portugal/Portugal/ | Instituto Nacional de Saude (INSA) | Instituto Nacional de Saude (INSA) | Borges et al et al |
| EPI_ISL_941361 | Portugal/PT2310/2021 | 2021-01-15 | Europe/Portugal/Portugal/ | Instituto Nacional de Saude (INSA) | Instituto Nacional de Saude (INSA) | Borges et al et al |

| | | | | | | |
|---|---|---|---|---|---|---|
| EPI_ISL_941367 | Portugal/PT2316/2021 | 2021-01-16 | Europe/Portugal/Portugal/ | Instituto Nacional de Saude (INSA) | Instituto Nacional de Saude (INSA) | Borges et al et al |
| EPI_ISL_941368 | Portugal/PT2317/2021 | 2021-01-16 | Europe/Portugal/Portugal/ | Instituto Nacional de Saude (INSA) | Instituto Nacional de Saude (INSA) | Borges et al et al |
| EPI_ISL_941385 | Portugal/PT2335/2021 | 2021-01-10 | Europe/Portugal/Portugal/ | Instituto Nacional de Saude (INSA) | Instituto Nacional de Saude (INSA) | Borges et al et al |
| EPI_ISL_941393 | Portugal/PT2343/2021 | 2021-01-15 | Europe/Portugal/Portugal/ | Instituto Nacional de Saude (INSA) | Instituto Nacional de Saude (INSA) | Borges et al et al |
| EPI_ISL_941395 | Portugal/PT2345/2021 | 2021-01-13 | Europe/Portugal/Portugal/ | Instituto Nacional de Saude (INSA) | Instituto Nacional de Saude (INSA) | Borges et al et al |
| EPI_ISL_941398 | Portugal/PT2348/2021 | 2021-01-12 | Europe/Portugal/Portugal/ | Instituto Nacional de Saude (INSA) | Instituto Nacional de Saude (INSA) | Borges et al et al |
| EPI_ISL_941401 | Portugal/PT2351/2021 | 2021-01-13 | Europe/Portugal/Portugal/ | Instituto Nacional de Saude (INSA) | Instituto Nacional de Saude (INSA) | Borges et al et al |
| EPI_ISL_941413 | Portugal/PT2363/2021 | 2021-01-17 | Europe/Portugal/Portugal/ | Instituto Nacional de Saude (INSA) | Instituto Nacional de Saude (INSA) | Borges et al et al |
| EPI_ISL_941428 | Portugal/PT2378/2021 | 2021-01-12 | Europe/Portugal/Portugal/ | Instituto Nacional de Saude (INSA) | Instituto Nacional de Saude (INSA) | Borges et al et al |
| EPI_ISL_941434 | Portugal/PT2384/2021 | 2021-01-13 | Europe/Portugal/Portugal/ | Instituto Nacional de Saude (INSA) | Instituto Nacional de Saude (INSA) | Borges et al et al |
| EPI_ISL_941438 | Portugal/PT2388/2021 | 2021-01-18 | Europe/Portugal/Portugal/ | Instituto Nacional de Saude (INSA) | Instituto Nacional de Saude (INSA) | Borges et al et al |
| EPI_ISL_941450 | Portugal/PT2400/2021 | 2021-01-04 | Europe/Portugal/Portugal/ | Instituto Nacional de Saude (INSA) | Instituto Nacional de Saude (INSA) | Borges et al et al |
| EPI_ISL_941458 | Portugal/PT2409/2021 | 2021-01-08 | Europe/Portugal/Portugal/ | Instituto Nacional de Saude (INSA) | Instituto Nacional de Saude (INSA) | Borges et al et al |
| EPI_ISL_941481 | Portugal/PT2432/2021 | 2021-01-14 | Europe/Portugal/Portugal/ | Instituto Nacional de Saude (INSA) | Instituto Nacional de Saude (INSA) | Borges et al et al |
| EPI_ISL_941493 | Portugal/PT2444/2021 | 2021-01-16 | Europe/Portugal/Portugal/ | Instituto Nacional de Saude (INSA) | Instituto Nacional de Saude (INSA) | Borges et al et al |
| EPI_ISL_941495 | Portugal/PT2446/2021 | 2021-01-16 | Europe/Portugal/Portugal/ | Instituto Nacional de Saude (INSA) | Instituto Nacional de Saude (INSA) | Borges et al et al |
| EPI_ISL_941505 | Portugal/PT2456/2021 | 2021-01-17 | Europe/Portugal/Portugal/ | Instituto Nacional de Saude (INSA) | Instituto Nacional de Saude (INSA) | Borges et al et al |
| EPI_ISL_941521 | Portugal/PT2472/2021 | 2021-01-13 | Europe/Portugal/Portugal/ | Instituto Nacional de Saude (INSA) | Instituto Nacional de Saude (INSA) | Borges et al et al |
| EPI_ISL_941531 | Portugal/PT2482/2021 | 2021-01-13 | Europe/Portugal/Portugal/ | Instituto Nacional de Saude (INSA) | Instituto Nacional de Saude (INSA) | Borges et al et al |
| EPI_ISL_941541 | Portugal/PT2492/2021 | 2021-01-14 | Europe/Portugal/Portugal/ | Instituto Nacional de Saude (INSA) | Instituto Nacional de Saude (INSA) | Borges et al et al |

| | | | | | | |
|---|---|---|---|---|---|---|
| EPI_ISL_941542 | Portugal/PT2493/2021 | 2021-01-14 | Europe/Portugal/Portugal/ | Instituto Nacional de Saude (INSA) | Instituto Nacional de Saude (INSA) | Borges et al et al |
| EPI_ISL_941548 | Portugal/PT2499/2021 | 2021-01-12 | Europe/Portugal/Portugal/ | Instituto Nacional de Saude (INSA) | Instituto Nacional de Saude (INSA) | Borges et al et al |
| EPI_ISL_941549 | Portugal/PT2500/2021 | 2021-01-12 | Europe/Portugal/Portugal/ | Instituto Nacional de Saude (INSA) | Instituto Nacional de Saude (INSA) | Borges et al et al |
| EPI_ISL_941556 | Portugal/PT2507/2021 | 2021-01-15 | Europe/Portugal/Portugal/ | Instituto Nacional de Saude (INSA) | Instituto Nacional de Saude (INSA) | Borges et al et al |
| EPI_ISL_941563 | Portugal/PT2514/2021 | 2021-01-16 | Europe/Portugal/Portugal/ | Instituto Nacional de Saude (INSA) | Instituto Nacional de Saude (INSA) | Borges et al et al |
| EPI_ISL_941567 | Portugal/PT2518/2021 | 2021-01-17 | Europe/Portugal/Portugal/ | Instituto Nacional de Saude (INSA) | Instituto Nacional de Saude (INSA) | Borges et al et al |
| EPI_ISL_941572 | Portugal/PT2523/2021 | 2021-01-11 | Europe/Portugal/Portugal/ | Instituto Nacional de Saude (INSA) | Instituto Nacional de Saude (INSA) | Borges et al et al |
| EPI_ISL_941581 | Portugal/PT2532/2021 | 2021-01-13 | Europe/Portugal/Portugal/ | Instituto Nacional de Saude (INSA) | Instituto Nacional de Saude (INSA) | Borges et al et al |
| EPI_ISL_941584 | Portugal/PT2535/2021 | 2021-01-13 | Europe/Portugal/Portugal/ | Instituto Nacional de Saude (INSA) | Instituto Nacional de Saude (INSA) | Borges et al et al |
| EPI_ISL_941589 | Portugal/PT2540/2021 | 2021-01-13 | Europe/Portugal/Portugal/ | Instituto Nacional de Saude (INSA) | Instituto Nacional de Saude (INSA) | Borges et al et al |
| EPI_ISL_941592 | Portugal/PT2543/2021 | 2021-01-14 | Europe/Portugal/Portugal/ | Instituto Nacional de Saude (INSA) | Instituto Nacional de Saude (INSA) | Borges et al et al |
| EPI_ISL_941596 | Portugal/PT2547/2021 | 2021-01-16 | Europe/Portugal/Portugal/ | Instituto Nacional de Saude (INSA) | Instituto Nacional de Saude (INSA) | Borges et al et al |
| EPI_ISL_941598 | Portugal/PT2549/2021 | 2021-01-17 | Europe/Portugal/Portugal/ | Instituto Nacional de Saude (INSA) | Instituto Nacional de Saude (INSA) | Borges et al et al |
| EPI_ISL_941600 | Portugal/PT2596/2020 | 2020-12-23 | Europe/Portugal/Portugal/ | Instituto Nacional de Saude (INSA) | Instituto Nacional de Saude (INSA) | Borges et al et al |
| EPI_ISL_941611 | Portugal/PT2608/2021 | 2021-01-18 | Europe/Portugal/Portugal/ | Instituto Nacional de Saude (INSA) | Instituto Nacional de Saude (INSA) | Borges et al et al |
| EPI_ISL_941613 | Portugal/PT2610/2021 | 2021-01-20 | Europe/Portugal/Portugal/ | Instituto Nacional de Saude (INSA) | Instituto Nacional de Saude (INSA) | Borges et al et al |
| EPI_ISL_941636 | Portugal/PT2634/2021 | 2021-01-12 | Europe/Portugal/Portugal/ | Instituto Nacional de Saude (INSA) | Instituto Nacional de Saude (INSA) | Borges et al et al |
| EPI_ISL_941658 | Portugal/PT2657/2021 | 2021-01-16 | Europe/Portugal/Portugal/ | Instituto Nacional de Saude (INSA) and Instituto Gulbenkian de Ciencia (IGC) | Instituto Nacional de Saude (INSA) and Instituto Gulbenkian de Ciencia (IGC) | Borges et al et al |

| | | | | | | |
|---|---|---|---|---|---|---|
| EPI_ISL_941664 | Portugal/PT2663/2021 | 2021-01-11 | Europe/Portugal/Portugal/ | Instituto Nacional de Saude (INSA) and Instituto Gulbenkian de Ciencia (IGC) | Instituto Nacional de Saude (INSA) and Instituto Gulbenkian de Ciencia (IGC) | Borges et al et al |
| EPI_ISL_941673 | Portugal/PT2672/2021 | 2021-01-12 | Europe/Portugal/Portugal/ | Instituto Nacional de Saude (INSA) and Instituto Gulbenkian de Ciencia (IGC) | Instituto Nacional de Saude (INSA) and Instituto Gulbenkian de Ciencia (IGC) | Borges et al et al |
| EPI_ISL_941676 | Portugal/PT2675/2021 | 2021-01-12 | Europe/Portugal/Portugal/ | Instituto Nacional de Saude (INSA) and Instituto Gulbenkian de Ciencia (IGC) | Instituto Nacional de Saude (INSA) and Instituto Gulbenkian de Ciencia (IGC) | Borges et al et al |
| EPI_ISL_941679 | Portugal/PT2678/2021 | 2021-01-15 | Europe/Portugal/Portugal/ | Instituto Nacional de Saude (INSA) and Instituto Gulbenkian de Ciencia (IGC) | Instituto Nacional de Saude (INSA) and Instituto Gulbenkian de Ciencia (IGC) | Borges et al et al |
| EPI_ISL_941685 | Portugal/PT2685/2021 | 2021-01-12 | Europe/Portugal/Portugal/ | Instituto Nacional de Saude (INSA) and Instituto Gulbenkian de Ciencia (IGC) | Instituto Nacional de Saude (INSA) and Instituto Gulbenkian de Ciencia (IGC) | Borges et al et al |
| EPI_ISL_941702 | Portugal/PT2704/2021 | 2021-01-11 | Europe/Portugal/Portugal/ | Instituto Nacional de Saude (INSA) and Instituto Gulbenkian de Ciencia (IGC) | Instituto Nacional de Saude (INSA) and Instituto Gulbenkian de Ciencia (IGC) | Borges et al et al |
| EPI_ISL_941704 | Portugal/PT2706/2021 | 2021-01-12 | Europe/Portugal/Portugal/ | Instituto Nacional de Saude (INSA) and Instituto Gulbenkian de Ciencia (IGC) | Instituto Nacional de Saude (INSA) and Instituto Gulbenkian de Ciencia (IGC) | Borges et al et al |
| EPI_ISL_941708 | Portugal/PT2710/2021 | 2021-01-16 | Europe/Portugal/Portugal/ | Instituto Nacional de Saude (INSA) and Instituto Gulbenkian de Ciencia (IGC) | Instituto Nacional de Saude (INSA) and Instituto Gulbenkian de Ciencia (IGC) | Borges et al et al |

| | | | | | | |
|---|---|---|---|---|---|---|
| EPI_ISL_941709 | Portugal/PT2711/2021 | 2021-01-11 | Europe/Portugal/Portugal/ | Instituto Nacional de Saude (INSA) and Instituto Gulbenkian de Ciencia (IGC) | Instituto Nacional de Saude (INSA) and Instituto Gulbenkian de Ciencia (IGC) | Borges et al et al |
| EPI_ISL_941713 | Portugal/PT2715/2021 | 2021-01-12 | Europe/Portugal/Portugal/ | Instituto Nacional de Saude (INSA) and Instituto Gulbenkian de Ciencia (IGC) | Instituto Nacional de Saude (INSA) and Instituto Gulbenkian de Ciencia (IGC) | Borges et al et al |
| EPI_ISL_941716 | Portugal/PT2718/2021 | 2021-01-14 | Europe/Portugal/Portugal/ | Instituto Nacional de Saude (INSA) and Instituto Gulbenkian de Ciencia (IGC) | Instituto Nacional de Saude (INSA) and Instituto Gulbenkian de Ciencia (IGC) | Borges et al et al |
| EPI_ISL_941720 | Portugal/PT2722/2021 | 2021-01-15 | Europe/Portugal/Portugal/ | Instituto Nacional de Saude (INSA) and Instituto Gulbenkian de Ciencia (IGC) | Instituto Nacional de Saude (INSA) and Instituto Gulbenkian de Ciencia (IGC) | Borges et al et al |
| EPI_ISL_941726 | Portugal/PT2729/2021 | 2021-01-11 | Europe/Portugal/Portugal/ | Instituto Nacional de Saude (INSA) and Instituto Gulbenkian de Ciencia (IGC) | Instituto Nacional de Saude (INSA) and Instituto Gulbenkian de Ciencia (IGC) | Borges et al et al |
| EPI_ISL_941737 | Portugal/PT2740/2021 | 2021-01-12 | Europe/Portugal/Portugal/ | Instituto Nacional de Saude (INSA) and Instituto Gulbenkian de Ciencia (IGC) | Instituto Nacional de Saude (INSA) and Instituto Gulbenkian de Ciencia (IGC) | Borges et al et al |
| EPI_ISL_941741 | Portugal/PT2744/2021 | 2021-01-12 | Europe/Portugal/Portugal/ | Instituto Nacional de Saude (INSA) and Instituto Gulbenkian de Ciencia (IGC) | Instituto Nacional de Saude (INSA) and Instituto Gulbenkian de Ciencia (IGC) | Borges et al et al |
| EPI_ISL_941744 | Portugal/PT2747/2021 | 2021-01-12 | Europe/Portugal/Portugal/ | Instituto Nacional de Saude (INSA) and Instituto Gulbenkian de Ciencia (IGC) | Instituto Nacional de Saude (INSA) and Instituto Gulbenkian de Ciencia (IGC) | Borges et al et al |

| | | | | | | |
|---|---|---|---|---|---|---|
| EPI_ISL_941751 | Portugal/PT2756/2021 | 2021-01-13 | Europe/Portugal/Portugal/ | Instituto Nacional de Saude (INSA) and Instituto Gulbenkian de Ciencia (IGC) | Instituto Nacional de Saude (INSA) and Instituto Gulbenkian de Ciencia (IGC) | Borges et al et al |
| EPI_ISL_941757 | Portugal/PT2764/2021 | 2021-01-16 | Europe/Portugal/Portugal/ | Instituto Nacional de Saude (INSA) and Instituto Gulbenkian de Ciencia (IGC) | Instituto Nacional de Saude (INSA) and Instituto Gulbenkian de Ciencia (IGC) | Borges et al et al |
| EPI_ISL_941785 | Portugal/PT2798/2021 | 2021-01-13 | Europe/Portugal/Portugal/ | Instituto Nacional de Saude (INSA) and Instituto Gulbenkian de Ciencia (IGC) | Instituto Nacional de Saude (INSA) and Instituto Gulbenkian de Ciencia (IGC) | Borges et al et al |
| EPI_ISL_941788 | Portugal/PT2801/2021 | 2021-01-14 | Europe/Portugal/Portugal/ | Instituto Nacional de Saude (INSA) and Instituto Gulbenkian de Ciencia (IGC) | Instituto Nacional de Saude (INSA) and Instituto Gulbenkian de Ciencia (IGC) | Borges et al et al |
| EPI_ISL_941791 | Portugal/PT2804/2021 | 2021-01-15 | Europe/Portugal/Portugal/ | Instituto Nacional de Saude (INSA) and Instituto Gulbenkian de Ciencia (IGC) | Instituto Nacional de Saude (INSA) and Instituto Gulbenkian de Ciencia (IGC) | Borges et al et al |
| EPI_ISL_941806 | Portugal/PT2821/2021 | 2021-01-17 | Europe/Portugal/Portugal/ | Instituto Nacional de Saude (INSA) and Instituto Gulbenkian de Ciencia (IGC) | Instituto Nacional de Saude (INSA) and Instituto Gulbenkian de Ciencia (IGC) | Borges et al et al |
| EPI_ISL_941810 | Portugal/PT2825/2021 | 2021-01-12 | Europe/Portugal/Portugal/ | Instituto Nacional de Saude (INSA) and Instituto Gulbenkian de Ciencia (IGC) | Instituto Nacional de Saude (INSA) and Instituto Gulbenkian de Ciencia (IGC) | Borges et al et al |
| EPI_ISL_941812 | Portugal/PT2828/2021 | 2021-01-14 | Europe/Portugal/Portugal/ | Instituto Nacional de Saude (INSA) and Instituto Gulbenkian de Ciencia (IGC) | Instituto Nacional de Saude (INSA) and Instituto Gulbenkian de Ciencia (IGC) | Borges et al et al |

| | | | | | | |
|---|---|---|---|---|---|---|
| EPI_ISL_941815 | Portugal/PT2831/2021 | 2021-01-15 | Europe/Portugal/Portugal/ | Instituto Nacional de Saude (INSA) and Instituto Gulbenkian de Ciencia (IGC) | Instituto Nacional de Saude (INSA) and Instituto Gulbenkian de Ciencia (IGC) | Borges et al et al |
| EPI_ISL_941817 | Portugal/PT2833/2021 | 2021-01-16 | Europe/Portugal/Portugal/ | Instituto Nacional de Saude (INSA) and Instituto Gulbenkian de Ciencia (IGC) | Instituto Nacional de Saude (INSA) and Instituto Gulbenkian de Ciencia (IGC) | Borges et al et al |
| EPI_ISL_941827 | Portugal/PT2843/2021 | 2021-01-11 | Europe/Portugal/Portugal/ | Instituto Nacional de Saude (INSA) and Instituto Gulbenkian de Ciencia (IGC) | Instituto Nacional de Saude (INSA) and Instituto Gulbenkian de Ciencia (IGC) | Borges et al et al |
| EPI_ISL_941843 | Portugal/PT2860/2021 | 2021-01-15 | Europe/Portugal/Portugal/ | Instituto Nacional de Saude (INSA) and Instituto Gulbenkian de Ciencia (IGC) | Instituto Nacional de Saude (INSA) and Instituto Gulbenkian de Ciencia (IGC) | Borges et al et al |
| EPI_ISL_941869 | Portugal/PT2887/2021 | 2021-01-12 | Europe/Portugal/Portugal/ | Instituto Nacional de Saude (INSA) and Instituto Gulbenkian de Ciencia (IGC) | Instituto Nacional de Saude (INSA) and Instituto Gulbenkian de Ciencia (IGC) | Borges et al et al |
| EPI_ISL_941872 | Portugal/PT2890/2021 | 2021-01-16 | Europe/Portugal/Portugal/ | Instituto Nacional de Saude (INSA) and Instituto Gulbenkian de Ciencia (IGC) | Instituto Nacional de Saude (INSA) and Instituto Gulbenkian de Ciencia (IGC) | Borges et al et al |
| EPI_ISL_941875 | Portugal/PT2893/2021 | 2021-01-14 | Europe/Portugal/Portugal/ | Instituto Nacional de Saude (INSA) and Instituto Gulbenkian de Ciencia (IGC) | Instituto Nacional de Saude (INSA) and Instituto Gulbenkian de Ciencia (IGC) | Borges et al et al |
| EPI_ISL_941879 | Portugal/PT2897/2021 | 2021-01-16 | Europe/Portugal/Portugal/ | Instituto Nacional de Saude (INSA) and Instituto Gulbenkian de Ciencia (IGC) | Instituto Nacional de Saude (INSA) and Instituto Gulbenkian de Ciencia (IGC) | Borges et al et al |

| | | | | | | |
|---|---|---|---|---|---|---|
| EPI_ISL_941881 | Portugal/PT2899/2021 | 2021-01-11 | Europe/Portugal/Portugal/ | Instituto Nacional de Saude (INSA) and Instituto Gulbenkian de Ciencia (IGC) | Instituto Nacional de Saude (INSA) and Instituto Gulbenkian de Ciencia (IGC) | Borges et al et al |
| EPI_ISL_941882 | Portugal/PT2900/2021 | 2021-01-11 | Europe/Portugal/Portugal/ | Instituto Nacional de Saude (INSA) and Instituto Gulbenkian de Ciencia (IGC) | Instituto Nacional de Saude (INSA) and Instituto Gulbenkian de Ciencia (IGC) | Borges et al et al |
| EPI_ISL_941904 | USA/VA-DCLS-2566/2020 | 2020-11-24 | North America/USA/Virginia/ | Virginia DCLS | Virginia DCLS | Virginia DCLS et al |
| EPI_ISL_941909 | USA/VA-DCLS-2574/2020 | 2020-11-22 | North America/USA/Virginia/ | Virginia DCLS | Virginia DCLS | Virginia DCLS et al |
| EPI_ISL_941913 | USA/VA-DCLS-2596/2020 | 2020-12-04 | North America/USA/Virginia/ | Virginia DCLS | Virginia DCLS | Virginia DCLS et al |
| EPI_ISL_941916 | USA/VA-DCLS-2599/2020 | 2020-12-04 | North America/USA/Virginia/ | Virginia DCLS | Virginia DCLS | Virginia DCLS et al |
| EPI_ISL_941920 | USA/VA-DCLS-2603/2020 | 2020-12-02 | North America/USA/Virginia/ | Virginia DCLS | Virginia DCLS | Virginia DCLS et al |
| EPI_ISL_941922 | USA/VA-DCLS-2908/2020 | 2020-12-23 | North America/USA/Virginia/ | Virginia DCLS | Virginia DCLS | Virginia DCLS et al |
| EPI_ISL_941926 | Nigeria/CV644/2021 | 2021-01-04 | Africa/Nigeria/Lagos/ | Nigeria Centre for Disease Control (NCDC) | African Centre of Excellence for Genomics of Infectious Diseases (ACEGID), Redeemer's University | Oluniyi P.E. et al et al |
| EPI_ISL_941930 | USA/FL-BPHL-3008/2020 | 2020-11-20 | North America/USA/Florida/ | Florida Bureau of Public Health Laboratories | Florida Bureau of Public Health Laboratories | Sarah Schmedes et al |
| EPI_ISL_941932 | USA/FL-BPHL-3003/2020 | 2020-12-28 | North America/USA/Florida/ | Florida Bureau of Public Health Laboratories | Florida Bureau of Public Health Laboratories | Sarah Schmedes et al |
| EPI_ISL_941935 | USA/FL-BPHL-3000/2020 | 2020-07-31 | North America/USA/Florida/ | Florida Bureau of Public Health Laboratories | Florida Bureau of Public Health Laboratories | Sarah Schmedes et al |
| EPI_ISL_941936 | USA/FL-BPHL-3007/2020 | 2020-11-19 | North America/USA/Florida/ | Florida Bureau of Public Health Laboratories | Florida Bureau of Public Health Laboratories | Sarah Schmedes et al |
| EPI_ISL_941939 | USA/FL-BPHL-3009/2020 | 2020-11-20 | North America/USA/Florida/ | Florida Bureau of Public Health Laboratories | Florida Bureau of Public Health Laboratories | Sarah Schmedes et al |

| | | | | | | |
|---|---|---|---|---|---|---|
| EPI_ISL_941940 | USA/FL-BPHL-3004/2020 | 2020-09-15 | North America/USA/Florida/ | Florida Bureau of Public Health Laboratories | Florida Bureau of Public Health Laboratories | Sarah Schmedes et al |
| EPI_ISL_941941 | Colombia/GUR-02374/2020 | 2020-04-18 | South America/Colombia/Meta/Villavic | Centro de Investigaciones en Microbiolog√≠a y Biotecnolog√≠a-UR (CIMBIUR), Facultad de Ciencias Naturales, Universidad del Rosario, Bogot√°, Colombia | Centro de Investigaciones en Microbiolog√≠a y Biotecnolog√≠a-UR (CIMBIUR), Facultad de Ciencias Naturales, Universidad del Rosario, Bogot√°, Colombia Instituto Nacional de Salud, Bogot√°, Colombia Icahn School of Medicine at Mount Sinai, New York, USA | Luz Helena Pati√±o et al |
| EPI_ISL_941943 | Colombia/GVI-153306/2020 | 2020-06-17 | South America/Colombia/Magdalena/Sa | Instituto Nacional de Salud, Bogot√°, Colombia | Centro de Investigaciones en Microbiolog√≠a y Biotecnolog√≠a-UR (CIMBIUR), Facultad de Ciencias Naturales, Universidad del Rosario, Bogot√°, Colombia Instituto Nacional de Salud, Bogot√°, Colombia Icahn School of Medicine at Mount Sinai, New York, USA | Luz Helena Pati√±o et al |

| | | | | | | |
|---|---|---|---|---|---|---|
| EPI_ISL_941949 | Colombia/GVI-152352/2020 | 2020-06-16 | South America/Colombia/La Guajira/Rio | Instituto Nacional de Salud, Bogotv°, Colombia | Centro de Investigaciones en Microbiologv≠a y Biotecnologv≠a-UR (CIMBIUR), Facultad de Ciencias Naturales, Universidad del Rosario, Bogotv°, Colombia Instituto Nacional de Salud, Bogotv°, Colombia Icahn School of Medicine at Mount Sinai, New York, USA | Luz Helena Pativ±o et al |
| EPI_ISL_941951 | Colombia/GVI-161642/2020 | 2020-06-24 | South America/Colombia/La Guajira/Rio | Instituto Nacional de Salud, Bogotv°, Colombia | Centro de Investigaciones en Microbiologv≠a y Biotecnologv≠a-UR (CIMBIUR), Facultad de Ciencias Naturales, Universidad del Rosario, Bogotv°, Colombia Instituto Nacional de Salud, Bogotv°, Colombia Icahn School of Medicine at Mount Sinai, New York, USA | Luz Helena Pativ±o et al |

| | | | | | | |
|---|---|---|---|---|---|---|
| EPI_ISL_941955 | Colombia/GVI-163894/2020 | 2020-06-27 | South America/Colombia/Boyaca/Tunja | Instituto Nacional de Salud, Bogot√°, Colombia | Centro de Investigaciones en Microbiolog√≠a y Biotecnolog√≠a-UR (CIMBIUR), Facultad de Ciencias Naturales, Universidad del Rosario, Bogot√°, Colombia Instituto Nacional de Salud, Bogot√°, Colombia Icahn School of Medicine at Mount Sinai, New York, USA | Luz Helena Pati√±o et al |
| EPI_ISL_941964 | Colombia/GUR-06406/2020 | 2020-04-30 | South America/Colombia/Magdalena/Ci | Centro de Investigaciones en Microbiolog√≠a y Biotecnolog√≠a-UR (CIMBIUR), Facultad de Ciencias Naturales, Universidad del Rosario, Bogot√°, Colombia | Centro de Investigaciones en Microbiolog√≠a y Biotecnolog√≠a-UR (CIMBIUR), Facultad de Ciencias Naturales, Universidad del Rosario, Bogot√°, Colombia Instituto Nacional de Salud, Bogot√°, Colombia Icahn School of Medicine at Mount Sinai, New York, USA | Luz Helena Pati√±o et al |

| | | | | | | |
|---|---|---|---|---|---|---|
| EPI_ISL_941973 | Colombia/GUR-03585/2020 | 2020-04-22 | South America/Colombia/Meta/Granada | Centro de Investigaciones en Microbiolog√≠a y Biotecnolog√≠a-UR (CIMBIUR), Facultad de Ciencias Naturales, Universidad del Rosario, Bogot√°, Colombia | Centro de Investigaciones en Microbiolog√≠a y Biotecnolog√≠a-UR (CIMBIUR), Facultad de Ciencias Naturales, Universidad del Rosario, Bogot√°, Colombia Instituto Nacional de Salud, Bogot√°, Colombia Icahn School of Medicine at Mount Sinai, New York, USA | Luz Helena Pati√±o et al |
| EPI_ISL_941985 | Colombia/GVI-111556/2020 | 2020-05-05 | South America/Colombia/Magdalena/Sa | Instituto Nacional de Salud, Bogot√°, Colombia | Centro de Investigaciones en Microbiolog√≠a y Biotecnolog√≠a-UR (CIMBIUR), Facultad de Ciencias Naturales, Universidad del Rosario, Bogot√°, Colombia Instituto Nacional de Salud, Bogot√°, Colombia Icahn School of Medicine at Mount Sinai, New York, USA | Luz Helena Pati√±o et al |

| | | | | | | |
|---|---|---|---|---|---|---|
| EPI_ISL_941986 | Colombia/GVI-133304/2020 | 2020-05-29 | South America/Colombia/Norte de Sant | Instituto Nacional de Salud, Bogot√°, Colombia | Centro de Investigaciones en Microbiolog√≠a y Biotecnolog√≠a-UR (CIMBIUR), Facultad de Ciencias Naturales, Universidad del Rosario, Bogot√°, Colombia Instituto Nacional de Salud, Bogot√°, Colombia Icahn School of Medicine at Mount Sinai, New York, USA | Luz Helena Pati√±o et al |
| EPI_ISL_941988 | Colombia/GVI-150933/2020 | 2020-06-15 | South America/Colombia/Boyaca/Tunja | Instituto Nacional de Salud, Bogot√°, Colombia | Centro de Investigaciones en Microbiolog√≠a y Biotecnolog√≠a-UR (CIMBIUR), Facultad de Ciencias Naturales, Universidad del Rosario, Bogot√°, Colombia Instituto Nacional de Salud, Bogot√°, Colombia Icahn School of Medicine at Mount Sinai, New York, USA | Luz Helena Pati√±o et al |

| | | | | | | |
|---|---|---|---|---|---|---|
| EPI_ISL_941989 | Colombia/GVI-157319/2020 | 2020-06-22 | South America/Colombia/Boyaca/Tunja | Instituto Nacional de Salud, Bogot√°, Colombia | Centro de Investigaciones en Microbiolog√≠a y Biotecnolog√≠a-UR (CIMBIUR), Facultad de Ciencias Naturales, Universidad del Rosario, Bogot√°, Colombia Instituto Nacional de Salud, Bogot√°, Colombia Icahn School of Medicine at Mount Sinai, New York, USA | Luz Helena Pati√±o et al |
| EPI_ISL_941990 | Colombia/GVI-157321/2020 | 2020-06-22 | South America/Colombia/Boyaca/Tunja | Instituto Nacional de Salud, Bogot√°, Colombia | Centro de Investigaciones en Microbiolog√≠a y Biotecnolog√≠a-UR (CIMBIUR), Facultad de Ciencias Naturales, Universidad del Rosario, Bogot√°, Colombia Instituto Nacional de Salud, Bogot√°, Colombia Icahn School of Medicine at Mount Sinai, New York, USA | Luz Helena Pati√±o et al |

| | | | | | | |
|---|---|---|---|---|---|---|
| EPI_ISL_941991 | Colombia/GVI-160127/2020 | 2020-06-23 | South America/Colombia/La Guajira/Rio | Instituto Nacional de Salud, Bogot√°, Colombia | Centro de Investigaciones en Microbiolog√≠a y Biotecnolog√≠a-UR (CIMBIUR), Facultad de Ciencias Naturales, Universidad del Rosario, Bogot√°, Colombia Instituto Nacional de Salud, Bogot√°, Colombia Icahn School of Medicine at Mount Sinai, New York, USA | Luz Helena Pati√±o et al |
| EPI_ISL_941992 | Colombia/GVI-160129/2020 | 2020-06-23 | South America/Colombia/La Guajira/Rio | Instituto Nacional de Salud, Bogot√°, Colombia | Centro de Investigaciones en Microbiolog√≠a y Biotecnolog√≠a-UR (CIMBIUR), Facultad de Ciencias Naturales, Universidad del Rosario, Bogot√°, Colombia Instituto Nacional de Salud, Bogot√°, Colombia Icahn School of Medicine at Mount Sinai, New York, USA | Luz Helena Pati√±o et al |

| | | | | | | |
|---|---|---|---|---|---|---|
| EPI_ISL_941993 | Colombia/GVI-160130/2020 | 2020-06-23 | South America/Colombia/La Guajira/Rio | Instituto Nacional de Salud, Bogot√°, Colombia | Centro de Investigaciones en Microbiolog√≠a y Biotecnolog√≠a-UR (CIMBIUR), Facultad de Ciencias Naturales, Universidad del Rosario, Bogot√°, Colombia Instituto Nacional de Salud, Bogot√°, Colombia Icahn School of Medicine at Mount Sinai, New York, USA | Luz Helena Pati√±o et al |
| EPI_ISL_941994 | Colombia/GVI-161641/2020 | 2020-06-24 | South America/Colombia/La Guajira/Rio | Instituto Nacional de Salud, Bogot√°, Colombia | Centro de Investigaciones en Microbiolog√≠a y Biotecnolog√≠a-UR (CIMBIUR), Facultad de Ciencias Naturales, Universidad del Rosario, Bogot√°, Colombia Instituto Nacional de Salud, Bogot√°, Colombia Icahn School of Medicine at Mount Sinai, New York, USA | Luz Helena Pati√±o et al |

| | | | | | | |
|---|---|---|---|---|---|---|
| EPI_ISL_941995 | Colombia/GUR-37097/2020 | 2020-06-23 | South America/Colombia/Atlantico/Barr | Centro de Investigaciones en Microbiolog√≠a y Biotecnolog√≠a-UR (CIMBIUR), Facultad de Ciencias Naturales, Universidad del Rosario, Bogot√°, Colombia | Centro de Investigaciones en Microbiolog√≠a y Biotecnolog√≠a-UR (CIMBIUR), Facultad de Ciencias Naturales, Universidad del Rosario, Bogot√°, Colombia Instituto Nacional de Salud, Bogot√°, Colombia Icahn School of Medicine at Mount Sinai, New York, USA | Luz Helena Pati√±o et al |
| EPI_ISL_941996 | Colombia/GUR-40200/2020 | 2020-06-26 | South America/Colombia/Magdalena/Sa | Centro de Investigaciones en Microbiolog√≠a y Biotecnolog√≠a-UR (CIMBIUR), Facultad de Ciencias Naturales, Universidad del Rosario, Bogot√°, Colombia | Centro de Investigaciones en Microbiolog√≠a y Biotecnolog√≠a-UR (CIMBIUR), Facultad de Ciencias Naturales, Universidad del Rosario, Bogot√°, Colombia Instituto Nacional de Salud, Bogot√°, Colombia Icahn School of Medicine at Mount Sinai, New York, USA | Luz Helena Pati√±o et al |

| | | | | | | |
|---|---|---|---|---|---|---|
| EPI_ISL_941997 | Colombia/GUR-41483/2020 | 2020-07-01 | South America/Colombia/Cundinamarca | Centro de Investigaciones en Microbiolog√≠a y Biotecnolog√≠a-UR (CIMBIUR), Facultad de Ciencias Naturales, Universidad del Rosario, Bogot√°, Colombia | Centro de Investigaciones en Microbiolog√≠a y Biotecnolog√≠a-UR (CIMBIUR), Facultad de Ciencias Naturales, Universidad del Rosario, Bogot√°, Colombia Instituto Nacional de Salud, Bogot√°, Colombia Icahn School of Medicine at Mount Sinai, New York, USA | Luz Helena Pati√±o et al |
| EPI_ISL_941998 | Colombia/GUR-41533/2020 | 2020-07-04 | South America/Colombia/Cundinamarca | Centro de Investigaciones en Microbiolog√≠a y Biotecnolog√≠a-UR (CIMBIUR), Facultad de Ciencias Naturales, Universidad del Rosario, Bogot√°, Colombia | Centro de Investigaciones en Microbiolog√≠a y Biotecnolog√≠a-UR (CIMBIUR), Facultad de Ciencias Naturales, Universidad del Rosario, Bogot√°, Colombia Instituto Nacional de Salud, Bogot√°, Colombia Icahn School of Medicine at Mount Sinai, New York, USA | Luz Helena Pati√±o et al |

| | | | | | | |
|---|---|---|---|---|---|---|
| EPI_ISL_942000 | Colombia/GUR-06494/2020 | 2020-05-03 | South America/Colombia/Magdalena/Sa | Centro de Investigaciones en Microbiolog√≠a y Biotecnolog√≠a-UR (CIMBIUR), Facultad de Ciencias Naturales, Universidad del Rosario, Bogot√°, Colombia | Centro de Investigaciones en Microbiolog√≠a y Biotecnolog√≠a-UR (CIMBIUR), Facultad de Ciencias Naturales, Universidad del Rosario, Bogot√°, Colombia Instituto Nacional de Salud, Bogot√°, Colombia Icahn School of Medicine at Mount Sinai, New York, USA | Luz Helena Pati√±o et al |
| EPI_ISL_942003 | Colombia/GVI-161115/2020 | 2020-06-24 | South America/Colombia/La Guajira/Rio | Instituto Nacional de Salud, Bogot√°, Colombia | Centro de Investigaciones en Microbiolog√≠a y Biotecnolog√≠a-UR (CIMBIUR), Facultad de Ciencias Naturales, Universidad del Rosario, Bogot√°, Colombia Instituto Nacional de Salud, Bogot√°, Colombia Icahn School of Medicine at Mount Sinai, New York, USA | Luz Helena Pati√±o et al |

| | | | | | | |
|---|---|---|---|---|---|---|
| EPI_ISL_942004 | Colombia/GUR-41385/2020 | 2020-06-30 | South America/Colombia/Cundinamarca | Centro de Investigaciones en Microbiolog√≠a y Biotecnolog√≠a-UR (CIMBIUR), Facultad de Ciencias Naturales, Universidad del Rosario, Bogot√°, Colombia | Centro de Investigaciones en Microbiolog√≠a y Biotecnolog√≠a-UR (CIMBIUR), Facultad de Ciencias Naturales, Universidad del Rosario, Bogot√°, Colombia Instituto Nacional de Salud, Bogot√°, Colombia Icahn School of Medicine at Mount Sinai, New York, USA | Luz Helena Pati√±o et al |
| EPI_ISL_942006 | Colombia/GVI-150146/2020 | 2020-06-10 | South America/Colombia/Magdalena/Sa | Instituto Nacional de Salud, Bogot√°, Colombia | Centro de Investigaciones en Microbiolog√≠a y Biotecnolog√≠a-UR (CIMBIUR), Facultad de Ciencias Naturales, Universidad del Rosario, Bogot√°, Colombia Instituto Nacional de Salud, Bogot√°, Colombia Icahn School of Medicine at Mount Sinai, New York, USA | Luz Helena Pati√±o et al |
| EPI_ISL_942007 | Puerto_Rico/CDC-S89/2020 | 2020-09-21 | North America/USA/Puerto Rico/ | Centers for Disease Control and Prevention, Dengue Branch | Centers for Disease Control and Prevention, Dengue Branch | Gilberto A. Santiago et al |

| | | | | | | |
|---|---|---|---|---|---|---|
| EPI_ISL_942011 | Puerto_Rico/CDC-S97/2020 | 2020-09-06 | North America/USA/Puerto Rico/ | Centers for Disease Control and Prevention, Dengue Branch | Centers for Disease Control and Prevention, Dengue Branch | Gilberto A. Santiago et al |
| EPI_ISL_942012 | Puerto_Rico/CDC-S104/2020 | 2020-09-02 | North America/USA/Puerto Rico/ | Centers for Disease Control and Prevention, Dengue Branch | Centers for Disease Control and Prevention, Dengue Branch | Gilberto A. Santiago et al |
| EPI_ISL_942017 | USA/WI-WSLH-201687/2020 | 2020-10-19 | North America/USA/Wisconsin/Sheboyg | Wisconsin State Laboratory of Hygiene Communicable Disease Division | Wisconsin State Laboratory of Hygiene Communicable Disease Division | Kelsey R. Florek et al |
| EPI_ISL_942022 | USA/WI-WSLH-201693/2020 | 2020-10-19 | North America/USA/Wisconsin/Dodge C | Wisconsin State Laboratory of Hygiene Communicable Disease Division | Wisconsin State Laboratory of Hygiene Communicable Disease Division | Kelsey R. Florek et al |
| EPI_ISL_942028 | USA/WI-WSLH-201700/2020 | 2020-12-21 | North America/USA/Wisconsin/Barron C | Wisconsin State Laboratory of Hygiene Communicable Disease Division | Wisconsin State Laboratory of Hygiene Communicable Disease Division | Kelsey R. Florek et al |
| EPI_ISL_942038 | USA/WI-WSLH-201710/2020 | 2020-12-21 | North America/USA/Wisconsin/Columbia | Wisconsin State Laboratory of Hygiene Communicable Disease Division | Wisconsin State Laboratory of Hygiene Communicable Disease Division | Kelsey R. Florek et al |
| EPI_ISL_942045 | USA/WI-WSLH-201717/2020 | 2020-12-22 | North America/USA/Wisconsin/Eau Clai | Wisconsin State Laboratory of Hygiene Communicable Disease Division | Wisconsin State Laboratory of Hygiene Communicable Disease Division | Kelsey R. Florek et al |
| EPI_ISL_942050 | USA/WI-WSLH-201722/2020 | 2020-10-19 | North America/USA/Wisconsin/Sheboyg | Wisconsin State Laboratory of Hygiene Communicable Disease Division | Wisconsin State Laboratory of Hygiene Communicable Disease Division | Kelsey R. Florek et al |

| | | | | | | |
|---|---|---|---|---|---|---|
| EPI_ISL_942051 | USA/WI-WSLH-201723/2020 | 2020-10-19 | North America/USA/Wisconsin/Sheboyg | Wisconsin State Laboratory of Hygiene Communicable Disease Division | Wisconsin State Laboratory of Hygiene Communicable Disease Division | Kelsey R. Florek et al |
| EPI_ISL_942058 | USA/WI-WSLH-201731/2020 | 2020-10-19 | North America/USA/Wisconsin/Sheboyg | Wisconsin State Laboratory of Hygiene Communicable Disease Division | Wisconsin State Laboratory of Hygiene Communicable Disease Division | Kelsey R. Florek et al |
| EPI_ISL_942061 | USA/WI-WSLH-201738/2020 | 2020-10-19 | North America/USA/Wisconsin/Sheboyg | Wisconsin State Laboratory of Hygiene Communicable Disease Division | Wisconsin State Laboratory of Hygiene Communicable Disease Division | Kelsey R. Florek et al |
| EPI_ISL_942062 | USA/WI-WSLH-201739/2020 | 2020-10-19 | North America/USA/Wisconsin/Sheboyg | Wisconsin State Laboratory of Hygiene Communicable Disease Division | Wisconsin State Laboratory of Hygiene Communicable Disease Division | Kelsey R. Florek et al |
| EPI_ISL_942065 | USA/WI-WSLH-201742/2020 | 2020-10-19 | North America/USA/Wisconsin/Dodge C | Wisconsin State Laboratory of Hygiene Communicable Disease Division | Wisconsin State Laboratory of Hygiene Communicable Disease Division | Kelsey R. Florek et al |
| EPI_ISL_942069 | USA/WI-WSLH-201748/2020 | 2020-12-08 | North America/USA/Wisconsin/Rock Cou | Wisconsin State Laboratory of Hygiene Communicable Disease Division | Wisconsin State Laboratory of Hygiene Communicable Disease Division | Kelsey R. Florek et al |
| EPI_ISL_942073 | USA/WI-WSLH-201754/2020 | 2020-12-21 | North America/USA/Wisconsin/Rock Cou | Wisconsin State Laboratory of Hygiene Communicable Disease Division | Wisconsin State Laboratory of Hygiene Communicable Disease Division | Kelsey R. Florek et al |
| EPI_ISL_942085 | USA/WI-WSLH-201767/2020 | 2020-12-22 | North America/USA/Wisconsin/Eau Clai | Wisconsin State Laboratory of Hygiene Communicable Disease Division | Wisconsin State Laboratory of Hygiene Communicable Disease Division | Kelsey R. Florek et al |

| | | | | | | |
|---|---|---|---|---|---|---|
| EPI_ISL_942099 | USA/WI-WSLH-201782/2020 | 2020-12-01 | North America/USA/Wisconsin/Barron C | Wisconsin State Laboratory of Hygiene Communicable Disease Division | Wisconsin State Laboratory of Hygiene Communicable Disease Division | Kelsey R. Florek et al |
| EPI_ISL_942114 | USA/WI-WSLH-201797/2020 | 2020-12-03 | North America/USA/Wisconsin/Juneau C | Wisconsin State Laboratory of Hygiene Communicable Disease Division | Wisconsin State Laboratory of Hygiene Communicable Disease Division | Kelsey R. Florek et al |
| EPI_ISL_942140 | USA/WI-WSLH-201828/2020 | 2020-12-24 | North America/USA/Wisconsin/Burnett | Wisconsin State Laboratory of Hygiene Communicable Disease Division | Wisconsin State Laboratory of Hygiene Communicable Disease Division | Kelsey R. Florek et al |
| EPI_ISL_942146 | USA/WI-WSLH-201841/2020 | 2020-12-02 | North America/USA/Wisconsin/Polk Cou | Wisconsin State Laboratory of Hygiene Communicable Disease Division | Wisconsin State Laboratory of Hygiene Communicable Disease Division | Kelsey R. Florek et al |
| EPI_ISL_942148 | USA/WI-WSLH-201843/2020 | 2020-12-04 | North America/USA/Wisconsin/Grant Co | Wisconsin State Laboratory of Hygiene Communicable Disease Division | Wisconsin State Laboratory of Hygiene Communicable Disease Division | Kelsey R. Florek et al |
| EPI_ISL_942154 | USA/WI-WSLH-201849/2020 | 2020-12-04 | North America/USA/Wisconsin/Marquet | Wisconsin State Laboratory of Hygiene Communicable Disease Division | Wisconsin State Laboratory of Hygiene Communicable Disease Division | Kelsey R. Florek et al |
| EPI_ISL_942157 | USA/WI-WSLH-201852/2020 | 2020-12-04 | North America/USA/Wisconsin/Marquet | Wisconsin State Laboratory of Hygiene Communicable Disease Division | Wisconsin State Laboratory of Hygiene Communicable Disease Division | Kelsey R. Florek et al |
| EPI_ISL_942169 | USA/WI-WSLH-201872/2020 | 2020-12-05 | North America/USA/Wisconsin/Juneau C | Wisconsin State Laboratory of Hygiene Communicable Disease Division | Wisconsin State Laboratory of Hygiene Communicable Disease Division | Kelsey R. Florek et al |

| | | | | | | |
|---|---|---|---|---|---|---|
| EPI_ISL_942173 | USA/WI-WSLH-201889/2020 | 2020-12-09 | North America/USA/Wisconsin/Maratho | Wisconsin State Laboratory of Hygiene Communicable Disease Division | Wisconsin State Laboratory of Hygiene Communicable Disease Division | Kelsey R. Florek et al |
| EPI_ISL_942174 | USA/WI-WSLH-201890/2020 | 2020-12-08 | North America/USA/Wisconsin/Rock Cou | Wisconsin State Laboratory of Hygiene Communicable Disease Division | Wisconsin State Laboratory of Hygiene Communicable Disease Division | Kelsey R. Florek et al |
| EPI_ISL_942184 | USA/WI-WSLH-201900/2020 | 2020-12-09 | North America/USA/Wisconsin/Dane Co | Wisconsin State Laboratory of Hygiene Communicable Disease Division | Wisconsin State Laboratory of Hygiene Communicable Disease Division | Kelsey R. Florek et al |
| EPI_ISL_942185 | USA/WI-WSLH-201901/2020 | 2020-12-10 | North America/USA/Wisconsin/Lincoln C | Wisconsin State Laboratory of Hygiene Communicable Disease Division | Wisconsin State Laboratory of Hygiene Communicable Disease Division | Kelsey R. Florek et al |
| EPI_ISL_942188 | USA/WI-WSLH-201904/2020 | 2020-12-05 | North America/USA/Wisconsin/Juneau C | Wisconsin State Laboratory of Hygiene Communicable Disease Division | Wisconsin State Laboratory of Hygiene Communicable Disease Division | Kelsey R. Florek et al |
| EPI_ISL_942191 | USA/WI-WSLH-201908/2020 | 2020-12-05 | North America/USA/Wisconsin/Juneau C | Wisconsin State Laboratory of Hygiene Communicable Disease Division | Wisconsin State Laboratory of Hygiene Communicable Disease Division | Kelsey R. Florek et al |
| EPI_ISL_942194 | USA/WI-WSLH-201911/2020 | 2020-12-08 | North America/USA/Wisconsin/Dane Co | Wisconsin State Laboratory of Hygiene Communicable Disease Division | Wisconsin State Laboratory of Hygiene Communicable Disease Division | Kelsey R. Florek et al |
| EPI_ISL_942218 | USA/WI-WSLH-201992/2020 | 2020-12-11 | North America/USA/Wisconsin/Marquet | Wisconsin State Laboratory of Hygiene Communicable Disease Division | Wisconsin State Laboratory of Hygiene Communicable Disease Division | Kelsey R. Florek et al |

| | | | | | | |
|---|---|---|---|---|---|---|
| EPI_ISL_942219 | USA/WI-WSLH-201993/2020 | 2020-12-11 | North America/USA/Wisconsin/Grant Co | Wisconsin State Laboratory of Hygiene Communicable Disease Division | Wisconsin State Laboratory of Hygiene Communicable Disease Division | Kelsey R. Florek et al |
| EPI_ISL_942234 | USA/WI-WSLH-202009/2020 | 2020-12-14 | North America/USA/Wisconsin/Burnett | Wisconsin State Laboratory of Hygiene Communicable Disease Division | Wisconsin State Laboratory of Hygiene Communicable Disease Division | Kelsey R. Florek et al |
| EPI_ISL_942238 | USA/WI-WSLH-202014/2020 | 2020-12-10 | North America/USA/Wisconsin/Vilas Co | Wisconsin State Laboratory of Hygiene Communicable Disease Division | Wisconsin State Laboratory of Hygiene Communicable Disease Division | Kelsey R. Florek et al |
| EPI_ISL_942250 | USA/WI-WSLH-202030/2020 | 2020-12-14 | North America/USA/Wisconsin/Grant Co | Wisconsin State Laboratory of Hygiene Communicable Disease Division | Wisconsin State Laboratory of Hygiene Communicable Disease Division | Kelsey R. Florek et al |
| EPI_ISL_942264 | USA/WI-WSLH-202048/2020 | 2020-12-15 | North America/USA/Wisconsin/Racine C | Wisconsin State Laboratory of Hygiene Communicable Disease Division | Wisconsin State Laboratory of Hygiene Communicable Disease Division | Kelsey R. Florek et al |
| EPI_ISL_942265 | USA/WI-WSLH-202049/2020 | 2020-12-15 | North America/USA/Wisconsin/Rock Cou | Wisconsin State Laboratory of Hygiene Communicable Disease Division | Wisconsin State Laboratory of Hygiene Communicable Disease Division | Kelsey R. Florek et al |
| EPI_ISL_942282 | USA/WI-WSLH-202066/2020 | 2020-12-18 | North America/USA/Wisconsin/Sawyer | Wisconsin State Laboratory of Hygiene Communicable Disease Division | Wisconsin State Laboratory of Hygiene Communicable Disease Division | Kelsey R. Florek et al |
| EPI_ISL_942286 | USA/WI-WSLH-202070/2020 | 2020-12-18 | North America/USA/Wisconsin/Racine C | Wisconsin State Laboratory of Hygiene Communicable Disease Division | Wisconsin State Laboratory of Hygiene Communicable Disease Division | Kelsey R. Florek et al |

| | | | | | | |
|---|---|---|---|---|---|---|
| EPI_ISL_942289 | USA/WI-WSLH-202073/2020 | 2020-12-14 | North America/USA/Wisconsin/Crawford | Wisconsin State Laboratory of Hygiene Communicable Disease Division | Wisconsin State Laboratory of Hygiene Communicable Disease Division | Kelsey R. Florek et al |
| EPI_ISL_942302 | USA/WI-WSLH-202087/2020 | 2020-12-15 | North America/USA/Wisconsin/Ashland | Wisconsin State Laboratory of Hygiene Communicable Disease Division | Wisconsin State Laboratory of Hygiene Communicable Disease Division | Kelsey R. Florek et al |
| EPI_ISL_942303 | USA/WI-WSLH-202088/2020 | 2020-12-14 | North America/USA/Wisconsin/Rock Cou | Wisconsin State Laboratory of Hygiene Communicable Disease Division | Wisconsin State Laboratory of Hygiene Communicable Disease Division | Kelsey R. Florek et al |
| EPI_ISL_942310 | USA/WI-WSLH-202097/2020 | 2020-12-16 | North America/USA/Wisconsin/Polk Cou | Wisconsin State Laboratory of Hygiene Communicable Disease Division | Wisconsin State Laboratory of Hygiene Communicable Disease Division | Kelsey R. Florek et al |
| EPI_ISL_942311 | USA/WI-WSLH-202098/2020 | 2020-12-16 | North America/USA/Wisconsin/Burnett | Wisconsin State Laboratory of Hygiene Communicable Disease Division | Wisconsin State Laboratory of Hygiene Communicable Disease Division | Kelsey R. Florek et al |
| EPI_ISL_942315 | USA/WI-WSLH-202103/2020 | 2020-12-16 | North America/USA/Wisconsin/Juneau C | Wisconsin State Laboratory of Hygiene Communicable Disease Division | Wisconsin State Laboratory of Hygiene Communicable Disease Division | Kelsey R. Florek et al |
| EPI_ISL_942327 | USA/MD-MDH-0767/2021 | 2021-01-25 | North America/USA/Maryland/ | MD PHL | MD PHL | Maryland Department of Health Laboratories Administration et al |
| EPI_ISL_942328 | USA/MD-MDH-0768/2021 | 2021-01-25 | North America/USA/Maryland/ | MD PHL | MD PHL | Maryland Department of Health Laboratories Administration et al |
| EPI_ISL_942340 | USA/MD-MDH-0780/2021 | 2021-01-27 | North America/USA/Maryland/ | MD PHL | MD PHL | Maryland Department of Health Laboratories Administration et al |

| | | | | | | |
|---|---|---|---|---|---|---|
| EPI_ISL_942343 | USA/MD-MDH-0783/2021 | 2021-01-27 | North America/USA/Maryland/ | MD PHL | MD PHL | Maryland Department of Health Laboratories Administration et al |
| EPI_ISL_942348 | USA/MD-MDH-0788/2021 | 2021-01-27 | North America/USA/Maryland/ | MD PHL | MD PHL | Maryland Department of Health Laboratories Administration et al |
| EPI_ISL_942355 | USA/MD-MDH-0795/2021 | 2021-01-27 | North America/USA/Maryland/ | MD PHL | MD PHL | Maryland Department of Health Laboratories Administration et al |
| EPI_ISL_942358 | USA/MD-MDH-0798/2021 | 2021-01-27 | North America/USA/Maryland/ | MD PHL | MD PHL | Maryland Department of Health Laboratories Administration et al |
| EPI_ISL_942359 | USA/MD-MDH-0799/2021 | 2021-01-27 | North America/USA/Maryland/ | MD PHL | MD PHL | Maryland Department of Health Laboratories Administration et al |
| EPI_ISL_942364 | USA/MD-MDH-0804/2021 | 2021-01-27 | North America/USA/Maryland/ | MD PHL | MD PHL | Maryland Department of Health Laboratories Administration et al |
| EPI_ISL_942367 | USA/MD-MDH-0807/2021 | 2021-01-27 | North America/USA/Maryland/ | MD PHL | MD PHL | Maryland Department of Health Laboratories Administration et al |
| EPI_ISL_942368 | USA/MD-MDH-0808/2021 | 2021-01-27 | North America/USA/Maryland/ | MD PHL | MD PHL | Maryland Department of Health Laboratories Administration et al |
| EPI_ISL_942373 | USA/MD-MDH-0813/2021 | 2021-01-28 | North America/USA/Maryland/ | MD PHL | MD PHL | Maryland Department of Health Laboratories Administration et al |
| EPI_ISL_942374 | Brazil/RS-CEVS-34165/2020 | 2020-07-27 | South America/Brazil/Rio Grande do Sul | Lacen_RS | CEVS_SES_RS | Barcellos R et al |
| EPI_ISL_942375 | Brazil/RS-CEVS-35834/2020 | 2020-07-31 | South America/Brazil/Rio Grande do Sul | Lacen_RS | CEVS_SES_RS | Barcellos R et al |
| EPI_ISL_942377 | USA/VA-DCLS-2468/2020 | 2020-11-13 | North America/USA/Virginia/ | Virginia DCLS | Virginia DCLS | Virginia DCLS et al |
| EPI_ISL_942382 | USA/VA-DCLS-2474/2020 | 2020-11-10 | North America/USA/Virginia/ | Virginia DCLS | Virginia DCLS | Virginia DCLS et al |
| EPI_ISL_942388 | USA/VA-DCLS-2486/2020 | 2020-11-10 | North America/USA/Virginia/ | Virginia DCLS | Virginia DCLS | Virginia DCLS et al |
| EPI_ISL_942390 | USA/VA-DCLS-2488/2020 | 2020-11-09 | North America/USA/Virginia/ | Virginia DCLS | Virginia DCLS | Virginia DCLS et al |
| EPI_ISL_942391 | USA/VA-DCLS-2492/2020 | 2020-11-07 | North America/USA/Virginia/ | Virginia DCLS | Virginia DCLS | Virginia DCLS et al |
| EPI_ISL_942395 | USA/VA-DCLS-2497/2020 | 2020-11-10 | North America/USA/Virginia/ | Virginia DCLS | Virginia DCLS | Virginia DCLS et al |
| EPI_ISL_942407 | Brazil/RS-CEVS-33473/2020 | 2020-07-23 | South America/Brazil/Rio Grande do Sul | Lacen_RS | CEVS_SES_RS | Barcellos R et al |
| EPI_ISL_942430 | USA/WI-GMF-33676/2020 | 2020-11-12 | North America/USA/Wisconsin/La Crosse | Gundersen Molecular Diagnostics Laboratory | Kabara Cancer Research Institute | Craig S. Richmond et al |

| | | | | | | |
|---|---|---|---|---|---|---|
| EPI_ISL_942432 | USA/WI-GMF-33695/2020 | 2020-11-13 | North America/USA/Wisconsin/La Cross | Gundersen Molecular Diagnostics Laboratory | Kabara Cancer Research Institute | Craig S. Richmond et al |
| EPI_ISL_942449 | USA/MN-GMF-34300/2020 | 2020-11-14 | North America/USA/Minnesota/Winona | Gundersen Molecular Diagnostics Laboratory | Kabara Cancer Research Institute | Craig S. Richmond et al |
| EPI_ISL_942452 | USA/IA-GMF-34475/2020 | 2020-11-15 | North America/USA/Iowa/Chickasaw Co | Gundersen Molecular Diagnostics Laboratory | Kabara Cancer Research Institute | Craig S. Richmond et al |
| EPI_ISL_942460 | USA/WI-GMF-34746/2020 | 2020-11-16 | North America/USA/Wisconsin/Juneau C | Gundersen Molecular Diagnostics Laboratory | Kabara Cancer Research Institute | Craig S. Richmond et al |
| EPI_ISL_942463 | USA/WI-GMF-34847/2020 | 2020-11-16 | North America/USA/Wisconsin/La Cross | Gundersen Molecular Diagnostics Laboratory | Kabara Cancer Research Institute | Craig S. Richmond et al |
| EPI_ISL_942477 | USA/WI-GMF-35720/2020 | 2020-11-19 | North America/USA/Wisconsin/Trempea | Gundersen Molecular Diagnostics Laboratory | Kabara Cancer Research Institute | Craig S. Richmond et al |
| EPI_ISL_942485 | USA/WI-GMF-36227/2020 | 2020-11-21 | North America/USA/Wisconsin/La Cross | Gundersen Clinical Microbiology Laboratory | Kabara Cancer Research Institute | Craig S. Richmond et al |
| EPI_ISL_942494 | USA/WI-GMF-36503/2020 | 2020-11-23 | North America/USA/Wisconsin/Adams C | Gundersen Clinical Microbiology Laboratory | Kabara Cancer Research Institute | Craig S. Richmond et al |
| EPI_ISL_942495 | USA/WI-GMF-36504/2020 | 2020-11-23 | North America/USA/Wisconsin/Vernon C | Gundersen Molecular Diagnostics Laboratory | Kabara Cancer Research Institute | Craig S. Richmond et al |
| EPI_ISL_942520 | USA/WI-GMF-37109/2020 | 2020-11-26 | North America/USA/Wisconsin/Monroe | Gundersen Molecular Diagnostics Laboratory | Kabara Cancer Research Institute | Craig S. Richmond et al |
| EPI_ISL_942528 | USA/WI-GMF-37254/2020 | 2020-11-27 | North America/USA/Wisconsin/Vernon C | Gundersen Molecular Diagnostics Laboratory | Kabara Cancer Research Institute | Craig S. Richmond et al |
| EPI_ISL_942539 | USA/WI-GMF-37660/2020 | 2020-11-29 | North America/USA/Wisconsin/La Cross | Gundersen Molecular Diagnostics Laboratory | Kabara Cancer Research Institute | Craig S. Richmond et al |
| EPI_ISL_942545 | USA/MN-GMF-37840/2020 | 2020-11-30 | North America/USA/Minnesota/Housto | Gundersen Molecular Diagnostics Laboratory | Kabara Cancer Research Institute | Craig S. Richmond et al |
| EPI_ISL_942546 | USA/MN-GMF-37861/2020 | 2020-11-30 | North America/USA/Minnesota/Winona | Gundersen Molecular Diagnostics Laboratory | Kabara Cancer Research Institute | Craig S. Richmond et al |
| EPI_ISL_942560 | USA/WI-GMF-38390/2020 | 2020-12-01 | North America/USA/Wisconsin/Vernon C | Gundersen Molecular Diagnostics Laboratory | Kabara Cancer Research Institute | Craig S. Richmond et al |
| EPI_ISL_942564 | USA/MN-GMF-38504/2020 | 2020-12-02 | North America/USA/Minnesota/Housto | Gundersen Molecular Diagnostics Laboratory | Kabara Cancer Research Institute | Craig S. Richmond et al |
| EPI_ISL_942569 | USA/WI-GMF-38541/2020 | 2020-12-02 | North America/USA/Wisconsin/Vernon C | Gundersen Molecular Diagnostics Laboratory | Kabara Cancer Research Institute | Craig S. Richmond et al |
| EPI_ISL_942574 | USA/MN-GMF-38557/2020 | 2020-12-02 | North America/USA/Minnesota/Winona | Gundersen Molecular Diagnostics Laboratory | Kabara Cancer Research Institute | Craig S. Richmond et al |
| EPI_ISL_942588 | USA/WI-GMF-38798/2020 | 2020-12-03 | North America/USA/Wisconsin/La Cross | Gundersen Molecular Diagnostics Laboratory | Kabara Cancer Research Institute | Craig S. Richmond et al |
| EPI_ISL_942590 | USA/WI-GMF-38877/2020 | 2020-12-03 | North America/USA/Wisconsin/Monroe | Gundersen Molecular Diagnostics Laboratory | Kabara Cancer Research Institute | Craig S. Richmond et al |

| | | | | | | |
|---|---|---|---|---|---|---|
| EPI_ISL_942592 | USA/WI-GMF-38937/2020 | 2020-12-03 | North America/USA/Wisconsin/La Crosse | Gundersen Molecular Diagnostics Laboratory | Kabara Cancer Research Institute | Craig S. Richmond et al |
| EPI_ISL_942597 | USA/WI-GMF-39058/2020 | 2020-12-04 | North America/USA/Wisconsin/La Crosse | Gundersen Molecular Diagnostics Laboratory | Kabara Cancer Research Institute | Craig S. Richmond et al |
| EPI_ISL_942610 | USA/WI-GMF-39410/2020 | 2020-12-05 | North America/USA/Wisconsin/La Crosse | Gundersen Molecular Diagnostics Laboratory | Kabara Cancer Research Institute | Craig S. Richmond et al |
| EPI_ISL_942617 | USA/WI-GMF-39551/2020 | 2020-12-06 | North America/USA/Wisconsin/Jackson | Gundersen Molecular Diagnostics Laboratory | Kabara Cancer Research Institute | Craig S. Richmond et al |
| EPI_ISL_942621 | USA/WI-GMF-39613/2020 | 2020-12-06 | North America/USA/Wisconsin/La Crosse | Gundersen Molecular Diagnostics Laboratory | Kabara Cancer Research Institute | Craig S. Richmond et al |
| EPI_ISL_942631 | USA/MN-GMF-39846/2020 | 2020-12-07 | North America/USA/Minnesota/Houston | Gundersen Molecular Diagnostics Laboratory | Kabara Cancer Research Institute | Craig S. Richmond et al |
| EPI_ISL_942632 | USA/MN-GMF-39855/2020 | 2020-12-07 | North America/USA/Minnesota/Houston | Gundersen Molecular Diagnostics Laboratory | Kabara Cancer Research Institute | Craig S. Richmond et al |
| EPI_ISL_942657 | USA/WI-GMF-40749/2020 | 2020-12-09 | North America/USA/Wisconsin/La Crosse | Gundersen Molecular Diagnostics Laboratory | Kabara Cancer Research Institute | Craig S. Richmond et al |
| EPI_ISL_942659 | USA/WI-GMF-40764/2020 | 2020-12-10 | North America/USA/Wisconsin/Trempea | Gundersen Molecular Diagnostics Laboratory | Kabara Cancer Research Institute | Craig S. Richmond et al |
| EPI_ISL_942672 | USA/MN-GMF-41225/2020 | 2020-12-11 | North America/USA/Minnesota/Fillmore | Gundersen Molecular Diagnostics Laboratory | Kabara Cancer Research Institute | Craig S. Richmond et al |
| EPI_ISL_942676 | USA/WI-GMF-41296/2020 | 2020-12-11 | North America/USA/Wisconsin/La Crosse | Gundersen Molecular Diagnostics Laboratory | Kabara Cancer Research Institute | Craig S. Richmond et al |
| EPI_ISL_942678 | USA/WI-GMF-41404/2020 | 2020-12-12 | North America/USA/Wisconsin/La Crosse | Gundersen Molecular Diagnostics Laboratory | Kabara Cancer Research Institute | Craig S. Richmond et al |
| EPI_ISL_942688 | USA/WI-GMF-41773/2020 | 2020-12-13 | North America/USA/Wisconsin/Vernon C | Gundersen Molecular Diagnostics Laboratory | Kabara Cancer Research Institute | Craig S. Richmond et al |
| EPI_ISL_942692 | USA/WI-GMF-41871/2020 | 2020-12-14 | North America/USA/Wisconsin/La Crosse | Gundersen Molecular Diagnostics Laboratory | Kabara Cancer Research Institute | Craig S. Richmond et al |
| EPI_ISL_942694 | USA/WI-GMF-41927/2020 | 2020-12-14 | North America/USA/Wisconsin/La Crosse | Gundersen Molecular Diagnostics Laboratory | Kabara Cancer Research Institute | Craig S. Richmond et al |
| EPI_ISL_942698 | USA/WI-GMF-42193/2020 | 2020-12-14 | North America/USA/Wisconsin/Adams C | Gundersen Molecular Diagnostics Laboratory | Kabara Cancer Research Institute | Craig S. Richmond et al |
| EPI_ISL_942701 | USA/WI-GMF-42229/2020 | 2020-12-14 | North America/USA/Wisconsin/La Crosse | Gundersen Molecular Diagnostics Laboratory | Kabara Cancer Research Institute | Craig S. Richmond et al |
| EPI_ISL_942706 | USA/WI-GMF-42535/2020 | 2020-12-15 | North America/USA/Wisconsin/Trempea | Gundersen Molecular Diagnostics Laboratory | Kabara Cancer Research Institute | Craig S. Richmond et al |
| EPI_ISL_942709 | USA/WI-GMF-42622/2020 | 2020-12-16 | North America/USA/Wisconsin/La Crosse | Gundersen Molecular Diagnostics Laboratory | Kabara Cancer Research Institute | Craig S. Richmond et al |
| EPI_ISL_942711 | USA/WI-GMF-42689/2020 | 2020-12-16 | North America/USA/Wisconsin/Vernon C | Gundersen Molecular Diagnostics Laboratory | Kabara Cancer Research Institute | Craig S. Richmond et al |

| | | | | | | |
|---|---|---|---|---|---|---|
| EPI_ISL_942716 | USA/WI-GMF-42866/2020 | 2020-12-17 | North America/USA/Wisconsin/Monroe | Gundersen Molecular Diagnostics Laboratory | Kabara Cancer Research Institute | Craig S. Richmond et al |
| EPI_ISL_942717 | USA/WI-GMF-42890/2020 | 2020-12-17 | North America/USA/Wisconsin/Trempea | Gundersen Molecular Diagnostics Laboratory | Kabara Cancer Research Institute | Craig S. Richmond et al |
| EPI_ISL_942728 | USA/WI-GMF-43526/2020 | 2020-12-20 | North America/USA/Wisconsin/Trempea | Gundersen Molecular Diagnostics Laboratory | Kabara Cancer Research Institute | Craig S. Richmond et al |
| EPI_ISL_942730 | USA/WI-GMF-43549/2020 | 2020-12-20 | North America/USA/Wisconsin/Vernon | Gundersen Molecular Diagnostics Laboratory | Kabara Cancer Research Institute | Craig S. Richmond et al |
| EPI_ISL_942738 | USA/WI-GMF-44136/2020 | 2020-12-22 | North America/USA/Wisconsin/Monroe | Gundersen Molecular Diagnostics Laboratory | Kabara Cancer Research Institute | Craig S. Richmond et al |
| EPI_ISL_942746 | USA/WI-GMF-44410/2020 | 2020-12-24 | North America/USA/Wisconsin/La Cross | Gundersen Molecular Diagnostics Laboratory | Kabara Cancer Research Institute | Craig S. Richmond et al |
| EPI_ISL_942748 | USA/WI-GMF-44476/2020 | 2020-12-26 | North America/USA/Wisconsin/La Cross | Gundersen Molecular Diagnostics Laboratory | Kabara Cancer Research Institute | Craig S. Richmond et al |
| EPI_ISL_942756 | USA/WI-GMF-44888/2020 | 2020-12-28 | North America/USA/Wisconsin/La Cross | Gundersen Molecular Diagnostics Laboratory | Kabara Cancer Research Institute | Craig S. Richmond et al |
| EPI_ISL_942759 | USA/WI-GMF-45034/2020 | 2020-12-28 | North America/USA/Wisconsin/Trempea | Gundersen Molecular Diagnostics Laboratory | Kabara Cancer Research Institute | Craig S. Richmond et al |
| EPI_ISL_942761 | USA/WI-GMF-45061/2020 | 2020-12-28 | North America/USA/Wisconsin/Adams C | Gundersen Molecular Diagnostics Laboratory | Kabara Cancer Research Institute | Craig S. Richmond et al |
| EPI_ISL_942765 | USA/WI-GMF-45346/2020 | 2020-12-29 | North America/USA/Wisconsin/Monroe | Gundersen Molecular Diagnostics Laboratory | Kabara Cancer Research Institute | Craig S. Richmond et al |
| EPI_ISL_942773 | USA/WI-GMF-45902/2021 | 2021-01-01 | North America/USA/Wisconsin/La Cross | Gundersen Molecular Diagnostics Laboratory | Kabara Cancer Research Institute | Craig S. Richmond et al |
| EPI_ISL_942774 | USA/WI-GMF-45936/2021 | 2021-01-01 | North America/USA/Wisconsin/La Cross | Gundersen Molecular Diagnostics Laboratory | Kabara Cancer Research Institute | Craig S. Richmond et al |
| EPI_ISL_942788 | USA/WI-GMF-46551/2021 | 2021-01-04 | North America/USA/Wisconsin/La Cross | Gundersen Molecular Diagnostics Laboratory | Kabara Cancer Research Institute | Craig S. Richmond et al |
| EPI_ISL_942790 | USA/WI-GMF-46696/2021 | 2021-01-04 | North America/USA/Wisconsin/La Cross | Gundersen Molecular Diagnostics Laboratory | Kabara Cancer Research Institute | Craig S. Richmond et al |
| EPI_ISL_942799 | USA/IA-GMF-47194/2021 | 2021-01-05 | North America/USA/Iowa/Allamakee C | Gundersen Molecular Diagnostics Laboratory | Kabara Cancer Research Institute | Craig S. Richmond et al |
| EPI_ISL_942800 | USA/WI-GMF-47256/2021 | 2021-01-05 | North America/USA/Wisconsin/Monroe | Gundersen Molecular Diagnostics Laboratory | Kabara Cancer Research Institute | Craig S. Richmond et al |
| EPI_ISL_942812 | USA/WI-GMF-47801/2021 | 2021-01-08 | North America/USA/Wisconsin/La Cross | Gundersen Molecular Diagnostics Laboratory | Kabara Cancer Research Institute | Craig S. Richmond et al |
| EPI_ISL_942829 | USA/WI-GMF-49236/2021 | 2021-01-14 | North America/USA/Wisconsin/La Cross | Gundersen Molecular Diagnostics Laboratory | Kabara Cancer Research Institute | Craig S. Richmond et al |
| EPI_ISL_942832 | USA/WI-GMF-49260/2021 | 2021-01-14 | North America/USA/Wisconsin/Monroe | Gundersen Molecular Diagnostics Laboratory | Kabara Cancer Research Institute | Craig S. Richmond et al |

| | | | | | | |
|---|---|---|---|---|---|---|
| EPI_ISL_942851 | USA/WI-GMF-49924/2021 | 2021-01-18 | North America/USA/Wisconsin/Trempea | Gundersen Molecular Diagnostics Laboratory | Kabara Cancer Research Institute | Craig S. Richmond et al |
| EPI_ISL_942852 | USA/WI-GMF-50051/2021 | 2021-01-18 | North America/USA/Wisconsin/La Crosse | Gundersen Molecular Diagnostics Laboratory | Kabara Cancer Research Institute | Craig S. Richmond et al |
| EPI_ISL_942858 | USA/MN-GMF-50204/2021 | 2021-01-18 | North America/USA/Minnesota/Fillmore | Gundersen Molecular Diagnostics Laboratory | Kabara Cancer Research Institute | Craig S. Richmond et al |
| EPI_ISL_942860 | USA/IA-GMF-50340/2021 | 2021-01-19 | North America/USA/Iowa/Fayette Coun | Gundersen Molecular Diagnostics Laboratory | Kabara Cancer Research Institute | Craig S. Richmond et al |
| EPI_ISL_942865 | USA/WI-GMF-50741/2021 | 2021-01-21 | North America/USA/Wisconsin/Monroe | Gundersen Molecular Diagnostics Laboratory | Kabara Cancer Research Institute | Craig S. Richmond et al |
| EPI_ISL_942872 | USA/WI-GMF-51172/2021 | 2021-01-23 | North America/USA/Wisconsin/La Crosse | Gundersen Molecular Diagnostics Laboratory | Kabara Cancer Research Institute | Craig S. Richmond et al |
| EPI_ISL_942883 | USA/WI-GMF-51417/2021 | 2021-01-25 | North America/USA/Wisconsin/La Crosse | Gundersen Molecular Diagnostics Laboratory | Kabara Cancer Research Institute | Craig S. Richmond et al |
| EPI_ISL_942888 | USA/WI-GMF-51765/2021 | 2021-01-25 | North America/USA/Wisconsin/La Crosse | Gundersen Molecular Diagnostics Laboratory | Kabara Cancer Research Institute | Craig S. Richmond et al |
| EPI_ISL_942892 | USA/WI-GMF-51795/2021 | 2021-01-26 | North America/USA/Wisconsin/La Crosse | Gundersen Molecular Diagnostics Laboratory | Kabara Cancer Research Institute | Craig S. Richmond et al |
| EPI_ISL_942896 | Brazil/RS-CEVS-36255/2020 | 2020-07-31 | South America/Brazil/Rio Grande do Sul | Lacen_RS | CEVS_SES_RS | Barcellos R et al |
| EPI_ISL_942897 | Brazil/RS-CEVS-38171/2020 | 2020-08-06 | South America/Brazil/Rio Grande do Sul | Lacen_RS | CEVS_SES_RS | Barcellos R et al |
| EPI_ISL_942898 | Brazil/RS-CEVS-38343/2020 | 2020-08-07 | South America/Brazil/Rio Grande do Sul | Lacen_RS | CEVS_SES_RS | Barcellos R et al |
| EPI_ISL_942899 | Brazil/RS-CEVS-40467/2020 | 2020-08-12 | South America/Brazil/Rio Grande do Sul | Lacen_RS | CEVS_SES_RS | Barcellos R et al |
| EPI_ISL_942902 | USA/TX-HHD-2102046697/2021 | 2021-01-03 | North America/USA/Texas/ | Houston Health Dept. | Houston Health Dept. | Ryker Penn et al |
| EPI_ISL_942907 | USA/TX-HHD-2102046745/2021 | 2021-01-03 | North America/USA/Texas/ | Houston Health Dept. | Houston Health Dept. | Ryker Penn et al |
| EPI_ISL_942917 | USA/TX-HHD-2102042707/2021 | 2021-01-08 | North America/USA/Texas/ | Houston Health Dept. | Houston Health Dept. | Ryker Penn et al |
| EPI_ISL_942923 | USA/TX-HHD-2102044298/2021 | 2021-01-08 | North America/USA/Texas/ | Houston Health Dept. | Houston Health Dept. | Ryker Penn et al |
| EPI_ISL_942925 | USA/TX-HHD-2102044381/2021 | 2021-01-08 | North America/USA/Texas/ | Houston Health Dept. | Houston Health Dept. | Ryker Penn et al |
| EPI_ISL_942927 | Mexico/NLE-InDRE_239/2021 | 2021-01-18 | North America/Mexico/Nuevo Leon/ | LESP Nuevo Leon/Grupo de Diagnostico ARIES | Instituto de Diagnostico y Referencia Epidemiologicos (INDRE) | Claudia Wong-Arambula et al |

| | | | | | | |
|---|---|---|---|---|---|---|
| EPI_ISL_942928 | Mexico/CMX-InDRE_242/2021 | 2021-01-13 | North America/Mexico/Mexico City/ | Instituto de Diagnostico y Referencia Epidemiologicos INDRE_RNLSP | Instituto de Diagnostico y Referencia Epidemiologicos (INDRE) | Claudia Wong-Arambula et al |
| EPI_ISL_942929 | Mexico/ROO-InDRE_243/2020 | 2020-07-06 | North America/Mexico/Quintana Roo/ | Instituto de Diagnostico y Referencia Epidemiologicos INDRE_RNLSP | Instituto de Diagnostico y Referencia Epidemiologicos (INDRE) | Claudia Wong-Arambula et al |
| EPI_ISL_942930 | Brazil/RS-CEVS-40591/2020 | 2020-08-13 | South America/Brazil/Rio Grande do Sul | Lacen_RS | State Center for Health Surveillance. Rio Grande do Sul State Secretary of Health | Barcellos R et al |
| EPI_ISL_942931 | Brazil/RS-CEVS-41964/2020 | 2020-08-14 | South America/Brazil/Rio Grande do Sul | Lacen_RS | State Center for Health Surveillance. Rio Grande do Sul State Secretary of Health | Barcellos R et al |
| EPI_ISL_942935 | USA/WY-WPHL-21010716/2021 | 2021-02-01 | North America/USA/Wyoming/ | Wyoming Public Health Laboratory | Wyoming Public Health Laboratory | Noah Hull et al |
| EPI_ISL_942938 | USA/WY-WPHL-21010956/2021 | 2021-02-01 | North America/USA/Wyoming/ | Wyoming Public Health Laboratory | Wyoming Public Health Laboratory | Noah Hull et al |
| EPI_ISL_942939 | USA/WY-WPHL-21010970/2021 | 2021-02-01 | North America/USA/Wyoming/ | Wyoming Public Health Laboratory | Wyoming Public Health Laboratory | Noah Hull et al |
| EPI_ISL_942960 | Indonesia/JK-NIHRD-C002100528/202 | 2021-01-04 | Asia/Indonesia/Jakarta/ | National Institute of Health Research and Development | National Institute of Health Research and Development | Subangkit et al |
| EPI_ISL_942963 | NorthMacedonia/26332/2020 | 2020-12-08 | Europe/North Macedonia/Pelagonia/Bit | General Hospital - Bitola | "Research Center for Genetic Engineering and Biotechnology ""Georgi D. Efremov,Äù , Macedonian Academy of Sciences and Arts" | Aleksandar J.Dimovski et al |

| | | | | | | |
|---|---|---|---|---|---|---|
| EPI_ISL_942971 | Indonesia/JK-NIHRD-C002100677/202 | 2021-01-04 | Asia/Indonesia/Jakarta/ | National Institute of Health Research and Development | National Institute of Health Research and Development | Subangkit et al |
| EPI_ISL_943074 | Netherlands/LI-RIVM-10811/2021 | 2021-01-22 | Europe/Netherlands/Limburg/ | Dutch COVID-19 response team | National Institute for Public Health and the Environment (RIVM) | Adam Meijer et al |
| EPI_ISL_943076 | Netherlands/LI-RIVM-10814/2021 | 2021-01-14 | Europe/Netherlands/Limburg/ | Dutch COVID-19 response team | National Institute for Public Health and the Environment (RIVM) | Adam Meijer et al |
| EPI_ISL_943112 | Netherlands/GE-RIVM-10870/2021 | 2021-01-20 | Europe/Netherlands/Gelderland/ | Dutch COVID-19 response team | National Institute for Public Health and the Environment (RIVM) | Adam Meijer et al |
| EPI_ISL_943154 | Netherlands/NB-RIVM-10958/2021 | 2021-01-02 | Europe/Netherlands/North Brabant/ | Dutch COVID-19 response team | National Institute for Public Health and the Environment (RIVM) | Adam Meijer et al |
| EPI_ISL_943246 | Netherlands/ZH-RIVM-11072/2021 | 2021-01-23 | Europe/Netherlands/South Holland/ | Dutch COVID-19 response team | National Institute for Public Health and the Environment (RIVM) | Adam Meijer et al |
| EPI_ISL_943366 | Netherlands/NB-RIVM-11196/2021 | 2021-01-25 | Europe/Netherlands/North Brabant/ | Dutch COVID-19 response team | National Institute for Public Health and the Environment (RIVM) | Adam Meijer et al |
| EPI_ISL_943457 | Netherlands/NH-RIVM-11296/2021 | 2021-01-22 | Europe/Netherlands/North Holland/ | Dutch COVID-19 response team | National Institute for Public Health and the Environment (RIVM) | Adam Meijer et al |

| | | | | | | |
|---|---|---|---|---|---|---|
| EPI_ISL_943488 | Netherlands/NH-RIVM-11360/2021 | 2021-01-19 | Europe/Netherlands/North Holland/ | Dutch COVID-19 response team | National Institute for Public Health and the Environment (RIVM) | Adam Meijer et al |
| EPI_ISL_943492 | Netherlands/NH-RIVM-11364/2021 | 2021-01-23 | Europe/Netherlands/North Holland/ | Dutch COVID-19 response team | National Institute for Public Health and the Environment (RIVM) | Adam Meijer et al |
| EPI_ISL_943548 | Indonesia/JK-NIHRD-MI2100169/2021 | 2021-01-05 | Asia/Indonesia/Jakarta/ | National Institute of Health Research and Development | National Institute of Health Research and Development | Subangkit et al |
| EPI_ISL_943549 | Bangladesh/BCSIR-NILMRC-432/2021 | 2021-01-18 | Asia/Bangladesh/Dhaka/ | National Institute of Laboratory Medicine and Referral Center | Genomic Research Lab, BCSIR | Md. Murshed Hasan Sarkar et al |
| EPI_ISL_943552 | Bangladesh/BCSIR-NILMRC-447/2021 | 2021-01-23 | Asia/Bangladesh/Dhaka/ | National Institute of Laboratory Medicine and Referral Center | Genomic Research Lab, BCSIR | Iffat Jahan et al |
| EPI_ISL_943559 | Bangladesh/BCSIR-NILMRC-430/2021 | 2021-01-19 | Asia/Bangladesh/Dhaka/Keraniganj | National Institute of Laboratory Medicine and Referral Center | Genomic Research Lab, BCSIR | Shahina Akter et al |
| EPI_ISL_943563 | Bangladesh/BCSIR-NILMRC-450/2021 | 2021-01-19 | Asia/Bangladesh/Dhaka/Uttara | National Institute of Laboratory Medicine and Referral Center | Genomic Research Lab, BCSIR | Md. Saddam Hossain et al |
| EPI_ISL_943570 | Peru/LIM-INS-175/2021 | 2021-01-28 | South America/Peru/Lima/ | Laboratorio de Referencia Nacional de Virus Respiratorio. Instituto Nacional de Salud Per√∫ | Laboratorio de Referencia Nacional de Biotecnolog√≠a y Biolog√≠a Molecular. Instituto Nacional de Salud Per√∫ | Carlos Padilla Rojas et al |
| EPI_ISL_943574 | Brazil/RS-CEVS-42436/2020 | 2020-08-18 | South America/Brazil/Rio Grande do Sul | Lacen_RS | State Center for Health Surveillance. Rio Grande do Sul State Secretary of Health | Aline Campos et al |

| | | | | | | |
|---|---|---|---|---|---|---|
| EPI_ISL_943575 | Brazil/RS-CEVS-42751/2020 | 2020-08-18 | South America/Brazil/Rio Grande do Sul | Lacen_RS | State Center for Health Surveillance. Rio Grande do Sul State Secretary of Health | Aline Campos et al |
| EPI_ISL_943576 | Brazil/RS-CEVS-32833/2020 | 2020-07-16 | South America/Brazil/Rio Grande do Sul | Lacen_RS | State Center for Health Surveillance. Rio Grande do Sul State Secretary of Health | Aline Campos et al |
| EPI_ISL_943577 | Brazil/RS-CEVS-32831/2020 | 2020-07-22 | South America/Brazil/Rio Grande do Sul | Lacen_RS | State Center for Health Surveillance. Rio Grande do Sul State Secretary of Health | Aline Campos et al |
| EPI_ISL_943578 | Brazil/RS-CEVS-32832/2020 | 2020-07-21 | South America/Brazil/Rio Grande do Sul | Lacen_RS | State Center for Health Surveillance. Rio Grande do Sul State Secretary of Health | Aline Campos et al |
| EPI_ISL_943579 | Brazil/RS-CEVS-31289/2020 | 2020-07-18 | South America/Brazil/Rio Grande do Sul | Lacen_RS | State Center for Health Surveillance. Rio Grande do Sul State Secretary of Health | Aline Campos et al |
| EPI_ISL_943580 | Brazil/RS-CEVS-32082/2020 | 2020-07-16 | South America/Brazil/Rio Grande do Sul | Lacen_RS | State Center for Health Surveillance. Rio Grande do Sul State Secretary of Health | Aline Campos et al |

| | | | | | | |
|---|---|---|---|---|---|---|
| EPI_ISL_943581 | Brazil/RS-CEVS-36765/2020 | 2020-08-03 | South America/Brazil/Rio Grande do Sul | Lacen_RS | State Center for Health Surveillance. Rio Grande do Sul State Secretary of Health | Aline Campos et al |
| EPI_ISL_943582 | Brazil/RS-CEVS-36776/2020 | 2020-08-04 | South America/Brazil/Rio Grande do Sul | Lacen_RS | State Center for Health Surveillance. Rio Grande do Sul State Secretary of Health | Aline Campos et al |
| EPI_ISL_943583 | Brazil/RS-CEVS-36838/2020 | 2020-08-04 | South America/Brazil/Rio Grande do Sul | Lacen_RS | State Center for Health Surveillance. Rio Grande do Sul State Secretary of Health | Aline Campos et al |
| EPI_ISL_943584 | Brazil/RS-CEVS-36850/2020 | 2020-08-04 | South America/Brazil/Rio Grande do Sul | Lacen_RS | State Center for Health Surveillance. Rio Grande do Sul State Secretary of Health | Aline Campos et al |
| EPI_ISL_943585 | Brazil/RS-CEVS-37683/2020 | 2020-08-05 | South America/Brazil/Rio Grande do Sul | Lacen_RS | State Center for Health Surveillance. Rio Grande do Sul State Secretary of Health | Aline Campos et al |
| EPI_ISL_943586 | Brazil/RS-CEVS-39340/2020 | 2020-08-07 | South America/Brazil/Rio Grande do Sul | Lacen_RS | State Center for Health Surveillance. Rio Grande do Sul State Secretary of Health | Aline Campos et al |

| | | | | | | |
|---|---|---|---|---|---|---|
| EPI_ISL_943587 | Brazil/RS-CEVS-40476/2020 | 2020-08-12 | South America/Brazil/Rio Grande do Sul | Lacen_RS | State Center for Health Surveillance. Rio Grande do Sul State Secretary of Health | Aline Campos et al |
| EPI_ISL_943588 | Brazil/RS-CEVS-38094/2020 | 2020-08-06 | South America/Brazil/Rio Grande do Sul | Lacen_RS | State Center for Health Surveillance. Rio Grande do Sul State Secretary of Health | Aline Campos et al |
| EPI_ISL_943589 | Brazil/RS-CEVS-12423/2020 | 2020-05-18 | South America/Brazil/Rio Grande do Sul | Lacen_RS | State Center for Health Surveillance. Rio Grande do Sul State Secretary of Health | Aline Campos et al |
| EPI_ISL_943590 | Brazil/RS-CEVS-15237/2020 | 2020-06-02 | South America/Brazil/Rio Grande do Sul | Lacen_RS | State Center for Health Surveillance. Rio Grande do Sul State Secretary of Health | Aline Campos et al |
| EPI_ISL_943591 | Brazil/RS-CEVS-16150/2020 | 2020-06-04 | South America/Brazil/Rio Grande do Sul | Lacen_RS | State Center for Health Surveillance. Rio Grande do Sul State Secretary of Health | Aline Campos et al |
| EPI_ISL_943592 | Brazil/RS-CEVS-6946/2020 | 2020-04-14 | South America/Brazil/Rio Grande do Sul | Lacen_RS | State Center for Health Surveillance. Rio Grande do Sul State Secretary of Health | Aline Campos et al |

| | | | | | | |
|---|---|---|---|---|---|---|
| EPI_ISL_943593 | Brazil/RS-CEVS-18256/2020 | 2020-06-15 | South America/Brazil/Rio Grande do Sul | Lacen_RS | State Center for Health Surveillance. Rio Grande do Sul State Secretary of Health | Aline Campos et al |
| EPI_ISL_943594 | Brazil/RS-CEVS-19402/2020 | 2020-06-18 | South America/Brazil/Rio Grande do Sul | Lacen_RS | State Center for Health Surveillance. Rio Grande do Sul State Secretary of Health | Aline Campos et al |
| EPI_ISL_943595 | Brazil/RS-CEVS-11918/2020 | 2020-05-18 | South America/Brazil/Rio Grande do Sul | Lacen_RS | State Center for Health Surveillance. Rio Grande do Sul State Secretary of Health | Aline Campos et al |
| EPI_ISL_943596 | Brazil/RS-CEVS-12256/2020 | 2020-05-20 | South America/Brazil/Rio Grande do Sul | Lacen_RS | State Center for Health Surveillance. Rio Grande do Sul State Secretary of Health | Aline Campos et al |
| EPI_ISL_943597 | Brazil/RS-CEVS-12830/2020 | 2020-05-22 | South America/Brazil/Rio Grande do Sul | Lacen_RS | State Center for Health Surveillance. Rio Grande do Sul State Secretary of Health | Aline Campos et al |
| EPI_ISL_943598 | Brazil/RS-CEVS-13321/2020 | 2020-05-24 | South America/Brazil/Rio Grande do Sul | Lacen_RS | State Center for Health Surveillance. Rio Grande do Sul State Secretary of Health | Aline Campos et al |

| | | | | | | |
|---|---|---|---|---|---|---|
| EPI_ISL_943600 | Brazil/RS-CEVS-26874/2020 | 2020-07-08 | South America/Brazil/Rio Grande do Sul | Lacen_RS | State Center for Health Surveillance. Rio Grande do Sul State Secretary of Health | Aline Campos et al |
| EPI_ISL_943601 | Brazil/RS-CEVS-Lib03NB05/2020 | 2020-07-28 | South America/Brazil/Rio Grande do Sul | Lacen_RS | State Center for Health Surveillance. Rio Grande do Sul State Secretary of Health | Aline Campos et al |
| EPI_ISL_943602 | Brazil/RS-CEVS-82994/2020 | 2020-11-19 | South America/Brazil/Rio Grande do Sul | Lacen_RS | State Center for Health Surveillance. Rio Grande do Sul State Secretary of Health | Aline Campos et al |
| EPI_ISL_943603 | Brazil/RS-CEVS-82997/2020 | 2020-11-19 | South America/Brazil/Rio Grande do Sul | Lacen_RS | State Center for Health Surveillance. Rio Grande do Sul State Secretary of Health | Aline Campos et al |
| EPI_ISL_943604 | Brazil/RS-CEVS-83356/2020 | 2020-11-20 | South America/Brazil/Rio Grande do Sul | Lacen_RS | State Center for Health Surveillance. Rio Grande do Sul State Secretary of Health | Aline Campos et al |
| EPI_ISL_943605 | Brazil/RS-CEVS-82906/2020 | 2020-11-18 | South America/Brazil/Rio Grande do Sul | Lacen_RS | State Center for Health Surveillance. Rio Grande do Sul State Secretary of Health | Aline Campos et al |

| | | | | | | |
|---|---|---|---|---|---|---|
| EPI_ISL_943606 | Brazil/RS-CEVS-82912/2020 | 2020-11-18 | South America/Brazil/Rio Grande do Sul | Lacen_RS | State Center for Health Surveillance. Rio Grande do Sul State Secretary of Health | Aline Campos et al |
| EPI_ISL_943607 | Brazil/RS-CEVS-82920/2020 | 2020-11-20 | South America/Brazil/Rio Grande do Sul | Lacen_RS | State Center for Health Surveillance. Rio Grande do Sul State Secretary of Health | Aline Campos et al |
| EPI_ISL_943609 | Brazil/RS-CEVS-83041/2020 | 2020-11-20 | South America/Brazil/Rio Grande do Sul | Lacen_RS | State Center for Health Surveillance. Rio Grande do Sul State Secretary of Health | Aline Campos et al |
| EPI_ISL_943610 | Brazil/RS-CEVS-83139/2020 | 2020-11-18 | South America/Brazil/Rio Grande do Sul | Lacen_RS | State Center for Health Surveillance. Rio Grande do Sul State Secretary of Health | Aline Campos et al |
| EPI_ISL_943612 | Brazil/RS-CEVS-83150/2020 | 2020-11-19 | South America/Brazil/Rio Grande do Sul | Lacen_RS | State Center for Health Surveillance. Rio Grande do Sul State Secretary of Health | Aline Campos et al |
| EPI_ISL_943613 | Brazil/RS-CEVS-83157/2020 | 2020-11-20 | South America/Brazil/Rio Grande do Sul | Lacen_RS | State Center for Health Surveillance. Rio Grande do Sul State Secretary of Health | Aline Campos et al |
| EPI_ISL_943629 | USA/UT-UPHL-2101469472/2020 | 2020-04-19 | North America/USA/Utah/ | Utah Public Health Laboratory | Utah Public Health Laboratory | Erin L. Young et al |
| EPI_ISL_943684 | USA/UT-UPHL-2101745813/2020 | 2020-12-03 | North America/USA/Utah/ | Utah Public Health Laboratory | Utah Public Health Laboratory | Erin L. Young et al |

| | | | | | | |
|---|---|---|---|---|---|---|
| EPI_ISL_943818 | USA/UT-UPHL-2101022589/2020 | 2020-12-28 | North America/USA/Utah/ | Utah Public Health Laboratory | Utah Public Health Laboratory | Erin L. Young et al |
| EPI_ISL_943820 | USA/UT-UPHL-2101647401/2020 | 2020-12-30 | North America/USA/Utah/ | Utah Public Health Laboratory | Utah Public Health Laboratory | Erin L. Young et al |
| EPI_ISL_943824 | USA/UT-UPHL-2101853309/2021 | 2021-01-05 | North America/USA/Utah/ | Utah Public Health Laboratory | Utah Public Health Laboratory | Erin L. Young et al |
| EPI_ISL_943826 | USA/UT-UPHL-2101203699/2021 | 2021-01-06 | North America/USA/Utah/ | Utah Public Health Laboratory | Utah Public Health Laboratory | Erin L. Young et al |
| EPI_ISL_943850 | USA/UT-UPHL-2101981606/2021 | 2021-01-15 | North America/USA/Utah/ | Utah Public Health Laboratory | Utah Public Health Laboratory | Erin L. Young et al |
| EPI_ISL_943865 | USA/UT-UPHL-2101392273/2021 | 2021-01-15 | North America/USA/Utah/ | Utah Public Health Laboratory | Utah Public Health Laboratory | Erin L. Young et al |
| EPI_ISL_943869 | USA/UT-UPHL-2101741223/2021 | 2021-01-15 | North America/USA/Utah/ | Utah Public Health Laboratory | Utah Public Health Laboratory | Erin L. Young et al |
| EPI_ISL_943870 | USA/UT-UPHL-2101344814/2021 | 2021-01-15 | North America/USA/Utah/ | Utah Public Health Laboratory | Utah Public Health Laboratory | Erin L. Young et al |
| EPI_ISL_943897 | USA/UT-UPHL-2101228021/2021 | 2021-01-15 | North America/USA/Utah/ | Utah Public Health Laboratory | Utah Public Health Laboratory | Erin L. Young et al |
| EPI_ISL_943919 | USA/UT-UPHL-2101911825/2021 | 2021-01-14 | North America/USA/Utah/ | Utah Public Health Laboratory | Utah Public Health Laboratory | Erin L. Young et al |
| EPI_ISL_943925 | USA/UT-UPHL-2101217370/2021 | 2021-01-13 | North America/USA/Utah/ | Utah Public Health Laboratory | Utah Public Health Laboratory | Erin L. Young et al |
| EPI_ISL_943927 | USA/UT-UPHL-2101465532/2021 | 2021-01-11 | North America/USA/Utah/ | Utah Public Health Laboratory | Utah Public Health Laboratory | Erin L. Young et al |
| EPI_ISL_943933 | USA/UT-UPHL-2101274660/2021 | 2021-01-11 | North America/USA/Utah/ | Utah Public Health Laboratory | Utah Public Health Laboratory | Erin L. Young et al |
| EPI_ISL_943935 | USA/UT-UPHL-2101155076/2021 | 2021-01-13 | North America/USA/Utah/ | Utah Public Health Laboratory | Utah Public Health Laboratory | Erin L. Young et al |
| EPI_ISL_943940 | USA/UT-UPHL-2101343422/2021 | 2021-01-09 | North America/USA/Utah/ | Utah Public Health Laboratory | Utah Public Health Laboratory | Erin L. Young et al |
| EPI_ISL_943952 | USA/UT-UPHL-2101069491/2021 | 2021-01-17 | North America/USA/Utah/ | Utah Public Health Laboratory | Utah Public Health Laboratory | Erin L. Young et al |
| EPI_ISL_943953 | USA/UT-UPHL-2101584863/2021 | 2021-01-14 | North America/USA/Utah/ | Utah Public Health Laboratory | Utah Public Health Laboratory | Erin L. Young et al |
| EPI_ISL_943961 | USA/UT-UPHL-2101673298/2021 | 2021-01-12 | North America/USA/Utah/ | Utah Public Health Laboratory | Utah Public Health Laboratory | Erin L. Young et al |
| EPI_ISL_943966 | USA/UT-UPHL-2101808908/2021 | 2021-01-15 | North America/USA/Utah/ | Utah Public Health Laboratory | Utah Public Health Laboratory | Erin L. Young et al |

| | | | | | | |
|---|---|---|---|---|---|---|
| EPI_ISL_943967 | Brazil/RR-1081/2021 | 2021-01-25 | South America/Brazil/Roraima/ | Hospital Geral de Sao Paulo | Instituto Adolfo Lutz, Interdiciplinary Procedures Center, Strategic Laboratory | Claudio Tavares Sacchi et al |
| EPI_ISL_943968 | Brazil/RR-1083/2021 | 2021-01-25 | South America/Brazil/Roraima/ | Hospital Geral de Sao Paulo | Instituto Adolfo Lutz, Interdiciplinary Procedures Center, Strategic Laboratory | Claudio Tavares Sacchi et al |
| EPI_ISL_943969 | Brazil/RR-1084/2021 | 2021-01-25 | South America/Brazil/Roraima/ | Hospital Geral de Sao Paulo | Instituto Adolfo Lutz, Interdiciplinary Procedures Center, Strategic Laboratory | Claudio Tavares Sacchi et al |
| EPI_ISL_943970 | Brazil/RR-1085/2021 | 2021-01-25 | South America/Brazil/Roraima/ | Hospital Geral de Sao Paulo | Instituto Adolfo Lutz, Interdiciplinary Procedures Center, Strategic Laboratory | Claudio Tavares Sacchi et al |
| EPI_ISL_943971 | Brazil/RR-1087/2021 | 2021-01-25 | South America/Brazil/Roraima/ | Hospital Geral de Sao Paulo | Instituto Adolfo Lutz, Interdiciplinary Procedures Center, Strategic Laboratory | Claudio Tavares Sacchi et al |
| EPI_ISL_943973 | Brazil/TO-1171/2020 | 2020-09-24 | South America/Brazil/Tocantins/Palmas | LACEN do Estado de Tocantins | Instituto Adolfo Lutz, Interdiciplinary Procedures Center, Strategic Laboratory | Claudio Tavares Sacchi et al |

| | | | | | | |
|---|---|---|---|---|---|---|
| EPI_ISL_943974 | Brazil/TO-1172/2020 | 2020-09-25 | South America/Brazil/Tocantins/Palmas | LACEN do Estado de Tocantins | Instituto Adolfo Lutz, Interdiciplinary Procedures Center, Strategic Laboratory | Claudio Tavares Sacchi et al |
| EPI_ISL_943975 | Brazil/TO-1173/2020 | 2020-09-25 | South America/Brazil/Tocantins/Palmas | LACEN do Estado de Tocantins | Instituto Adolfo Lutz, Interdiciplinary Procedures Center, Strategic Laboratory | Claudio Tavares Sacchi et al |
| EPI_ISL_943976 | Brazil/TO-1174/2020 | 2020-09-29 | South America/Brazil/Tocantins/Preside | LACEN do Estado de Tocantins | Instituto Adolfo Lutz, Interdiciplinary Procedures Center, Strategic Laboratory | Claudio Tavares Sacchi et al |
| EPI_ISL_943977 | Brazil/TO-1175/2020 | 2020-09-29 | South America/Brazil/Tocantins/Aragua | LACEN do Estado de Tocantins | Instituto Adolfo Lutz, Interdiciplinary Procedures Center, Strategic Laboratory | Claudio Tavares Sacchi et al |
| EPI_ISL_943978 | Brazil/TO-1177/2020 | 2020-10-08 | South America/Brazil/Tocantins/Sandola | LACEN do Estado de Tocantins | Instituto Adolfo Lutz, Interdiciplinary Procedures Center, Strategic Laboratory | Claudio Tavares Sacchi et al |
| EPI_ISL_943979 | Brazil/TO-1178/2020 | 2020-10-10 | South America/Brazil/Tocantins/Palmas | LACEN do Estado de Tocantins | Instituto Adolfo Lutz, Interdiciplinary Procedures Center, Strategic Laboratory | Claudio Tavares Sacchi et al |

| | | | | | | |
|---|---|---|---|---|---|---|
| EPI_ISL_943980 | Brazil/TO-1179/2020 | 2020-10-12 | South America/Brazil/Tocantins/Mateir | LACEN do Estado de Tocantins | Instituto Adolfo Lutz, Interdiciplinary Procedures Center, Strategic Laboratory | Claudio Tavares Sacchi et al |
| EPI_ISL_943981 | Brazil/TO-1180/2020 | 2020-10-14 | South America/Brazil/Tocantins/Guarai | LACEN do Estado de Tocantins | Instituto Adolfo Lutz, Interdiciplinary Procedures Center, Strategic Laboratory | Claudio Tavares Sacchi et al |
| EPI_ISL_943982 | Brazil/TO-1181/2020 | 2020-10-14 | South America/Brazil/Tocantins/Figueir | LACEN do Estado de Tocantins | Instituto Adolfo Lutz, Interdiciplinary Procedures Center, Strategic Laboratory | Claudio Tavares Sacchi et al |
| EPI_ISL_943983 | Brazil/TO-1182/2020 | 2020-10-15 | South America/Brazil/Tocantins/Formos | LACEN do Estado de Tocantins | Instituto Adolfo Lutz, Interdiciplinary Procedures Center, Strategic Laboratory | Claudio Tavares Sacchi et al |
| EPI_ISL_943985 | Brazil/TO-1186/2020 | 2020-10-26 | South America/Brazil/Tocantins/Aragua | LACEN do Estado de Tocantins | Instituto Adolfo Lutz, Interdiciplinary Procedures Center, Strategic Laboratory | Claudio Tavares Sacchi et al |
| EPI_ISL_943986 | Brazil/TO-1187R2/2020 | 2020-12-22 | South America/Brazil/Tocantins/Paraiso | LACEN do Estado de Tocantins | Instituto Adolfo Lutz, Interdiciplinary Procedures Center, Strategic Laboratory | Claudio Tavares Sacchi et al |

| | | | | | | |
|---|---|---|---|---|---|---|
| EPI_ISL_943988 | Brazil/GO-1190R1/2020 | 2020-06-24 | South America/Brazil/Goiv°s/Ceres BR | LACEN do Estado de Goias | Instituto Adolfo Lutz, Interdiciplinary Procedures Center, Strategic Laboratory | Claudio Tavares Sacchi et al |
| EPI_ISL_943989 | Brazil/GO-1190R2/2020 | 2020-12-18 | South America/Brazil/Goiv°s/Ceres BR | LACEN do Estado de Goias | Instituto Adolfo Lutz, Interdiciplinary Procedures Center, Strategic Laboratory | Claudio Tavares Sacchi et al |
| EPI_ISL_943990 | Brazil/AM-1192/2021 | 2021-01-19 | South America/Brazil/Amazonas BR/Mar | LACEN do Estado de Goias | Instituto Adolfo Lutz, Interdiciplinary Procedures Center, Strategic Laboratory | Claudio Tavares Sacchi et al |
| EPI_ISL_943991 | Brazil/TO-1184/2020 | 2020-10-15 | South America/Brazil/Tocantins/Aragua | LACEN do Estado de Tocantins | Instituto Adolfo Lutz, Interdiciplinary Procedures Center, Strategic Laboratory | Claudio Tavares Sacchi et al |
| EPI_ISL_943993 | NorthMacedonia/28007/2020 | 2020-12-15 | Europe/North Macedonia/Southeastern | General Hospital - Strumica | "Research Center for Genetic Engineering and Biotechnology ""Georgi D. Efremov,Äù , Macedon" | Aleksandar J. Dimovski et al |
| EPI_ISL_943994 | NorthMacedonia/28025/2020 | 2020-12-15 | Europe/North Macedonia/Southeastern | General Hospital - Strumica | "Research Center for Genetic Engineering and Biotechnology ""Georgi D. Efremov,Äù , Macedonian Academy of Sciences and Arts" | Aleksandar J. Dimovski et al |

| | | | | | | |
|---|---|---|---|---|---|---|
| EPI_ISL_943995 | NorthMacedonia/28330/2020 | 2020-12-17 | Europe/North Macedonia/Vardar/Veles | General Hospital - Veles | "Research Center for Genetic Engineering and Biotechnology ""Georgi D. Efremov,Äù , Macedonian Academy of Sciences and Arts" | Aleksandar J. Dimovski et al |
| EPI_ISL_943997 | NorthMacedonia/28450/2020 | 2020-12-18 | Europe/North Macedonia/Tetovo/ | General Hospital - Tetovo | "Research Center for Genetic Engineering and Biotechnology ""Georgi D. Efremov,Äù , Macedonian Academy of Sciences and Arts" | Aleksandar J. Dimovski et al |
| EPI_ISL_944094 | Malaysia/IMR_WC346578/2020 | 2020-12-15 | Asia/Malaysia/Malaysia/ | Institute for Medical Research, Infectious Disease Research Centre, National Institutes of Health, Ministry of Health Malaysia | Institute for Medical Research, Infectious Disease Research Centre, National Institutes of Health, Ministry of Health Malaysia | Suppiah J et al |
| EPI_ISL_944095 | Malaysia/IMR_WC347970/2020 | 2020-12-15 | Asia/Malaysia/Malaysia/ | Institute for Medical Research, Infectious Disease Research Centre, National Institutes of Health, Ministry of Health Malaysia | Institute for Medical Research, Infectious Disease Research Centre, National Institutes of Health, Ministry of Health Malaysia | Suppiah J et al |

| | | | | | | |
|---|---|---|---|---|---|---|
| EPI_ISL_944099 | Malaysia/IMR_WC360297/2020 | 2020-12-20 | Asia/Malaysia/Malaysia/ | Institute for Medical Research, Infectious Disease Research Centre, National Institutes of Health, Ministry of Health Malaysia | Institute for Medical Research, Infectious Disease Research Centre, National Institutes of Health, Ministry of Health Malaysia | Suppiah J et al |
| EPI_ISL_944101 | Malaysia/IMR_WC55461/2021 | 2021-01-21 | Asia/Malaysia/Malaysia/ | Institute for Medical Research, Infectious Disease Research Centre, National Institutes of Health, Ministry of Health Malaysia | Institute for Medical Research, Infectious Disease Research Centre, National Institutes of Health, Ministry of Health Malaysia | Suppiah J et al |
| EPI_ISL_944102 | Malaysia/IMR_WC355250/2020 | 2020-12-18 | Asia/Malaysia/Malaysia/ | Institute for Medical Research, Infectious Disease Research Centre, National Institutes of Health, Ministry of Health Malaysia | Institute for Medical Research, Infectious Disease Research Centre, National Institutes of Health, Ministry of Health Malaysia | Suppiah J et al |
| EPI_ISL_944103 | Malaysia/IMR_WC355231/2020 | 2020-12-18 | Asia/Malaysia/Malaysia/ | Institute for Medical Research, Infectious Disease Research Centre, National Institutes of Health, Ministry of Health Malaysia | Institute for Medical Research, Infectious Disease Research Centre, National Institutes of Health, Ministry of Health Malaysia | Suppiah J et al |

| | | | | | | |
|---|---|---|---|---|---|---|
| EPI_ISL_944104 | Malaysia/IMR_WC355265/2020 | 2020-12-18 | Asia/Malaysia/Malaysia/ | Institute for Medical Research, Infectious Disease Research Centre, National Institutes of Health, Ministry of Health Malaysia | Institute for Medical Research, Infectious Disease Research Centre, National Institutes of Health, Ministry of Health Malaysia | Suppiah J et al |
| EPI_ISL_944105 | NorthMacedonia/28489/2020 | 2020-12-18 | Europe/North Macedonia/Southeastern | General Hospital - Strumica | "Research Center for Genetic Engineering and Biotechnology ""Georgi D. Efremov‚Äù , Macedonian Academy of Sciences and Arts" | Aleksandar J. Dimovski et al |
| EPI_ISL_944124 | SouthAfrica/KRISP-K008213/2021 | 2021-01-04 | Africa/South Africa/Eastern Cape/Chris | National Health Laboratory Service, South Africa | KRISP, KZN Research Innovation and Sequencing Platform | Laguda-Akingba O et al |
| EPI_ISL_944127 | SouthAfrica/KRISP-K008214/2021 | 2021-01-04 | Africa/South Africa/Eastern Cape/Sarah | National Health Laboratory Service, South Africa | KRISP, KZN Research Innovation and Sequencing Platform | Laguda-Akingba O et al |
| EPI_ISL_944128 | SouthAfrica/KRISP-K008232/2021 | 2021-01-04 | Africa/South Africa/Eastern Cape/Nelso | National Health Laboratory Service, South Africa | KRISP, KZN Research Innovation and Sequencing Platform | Laguda-Akingba O et al |
| EPI_ISL_944129 | SouthAfrica/KRISP-K008186/2020 | 2020-12-30 | Africa/South Africa/Eastern Cape/Amat | National Health Laboratory Service, South Africa | KRISP, KZN Research Innovation and Sequencing Platform | Laguda-Akingba O et al |

| | | | | | | |
|---|---|---|---|---|---|---|
| EPI_ISL_944130 | SouthAfrica/KRISP-K008195/2021 | 2021-01-06 | Africa/South Africa/Eastern Cape/Nelso | National Health Laboratory Service, South Africa | KRISP, KZN Research Innovation and Sequencing Platform | Laguda-Akingba O et al |
| EPI_ISL_944131 | SouthAfrica/KRISP-K008230/2021 | 2021-01-04 | Africa/South Africa/Eastern Cape/Nelso | National Health Laboratory Service, South Africa | KRISP, KZN Research Innovation and Sequencing Platform | Laguda-Akingba O et al |
| EPI_ISL_944132 | SouthAfrica/KRISP-K008203/2021 | 2021-01-05 | Africa/South Africa/Eastern Cape/Nelso | National Health Laboratory Service, South Africa | KRISP, KZN Research Innovation and Sequencing Platform | Laguda-Akingba O et al |
| EPI_ISL_944133 | SouthAfrica/KRISP-K008211/2021 | 2021-01-02 | Africa/South Africa/Eastern Cape/Amatl | National Health Laboratory Service, South Africa | KRISP, KZN Research Innovation and Sequencing Platform | Laguda-Akingba O et al |
| EPI_ISL_944134 | SouthAfrica/KRISP-K008255/2021 | 2021-01-05 | Africa/South Africa/Eastern Cape/Sarah | National Health Laboratory Service, South Africa | KRISP, KZN Research Innovation and Sequencing Platform | Laguda-Akingba O et al |
| EPI_ISL_944135 | SouthAfrica/KRISP-K008259/2021 | 2021-01-04 | Africa/South Africa/Eastern Cape/Joe G | National Health Laboratory Service, South Africa | KRISP, KZN Research Innovation and Sequencing Platform | Laguda-Akingba O et al |
| EPI_ISL_944143 | SouthAfrica/KRISP-K008193/2020 | 2020-12-31 | Africa/South Africa/Eastern Cape/Amatl | National Health Laboratory Service, South Africa | KRISP, KZN Research Innovation and Sequencing Platform | Laguda-Akingba O et al |
| EPI_ISL_944144 | SouthAfrica/KRISP-K008197/2021 | 2021-01-05 | Africa/South Africa/Eastern Cape/Middl | National Health Laboratory Service, South Africa | KRISP, KZN Research Innovation and Sequencing Platform | Laguda-Akingba O et al |

| | | | | | | |
|---|---|---|---|---|---|---|
| EPI_ISL_944145 | SouthAfrica/KRISP-K008199/2021 | 2021-01-04 | Africa/South Africa/Eastern Cape/Amatl | National Health Laboratory Service, South Africa | KRISP, KZN Research Innovation and Sequencing Platform | Laguda-Akingba O et al |
| EPI_ISL_944146 | SouthAfrica/KRISP-K008201/2021 | 2021-01-05 | Africa/South Africa/Eastern Cape/Nelsoı | National Health Laboratory Service, South Africa | KRISP, KZN Research Innovation and Sequencing Platform | Laguda-Akingba O et al |
| EPI_ISL_944147 | SouthAfrica/KRISP-K008202/2021 | 2021-01-05 | Africa/South Africa/Eastern Cape/Nelsoı | National Health Laboratory Service, South Africa | KRISP, KZN Research Innovation and Sequencing Platform | Laguda-Akingba O et al |
| EPI_ISL_944148 | SouthAfrica/KRISP-K008204/2021 | 2021-01-04 | Africa/South Africa/Eastern Cape/Nelsoı | National Health Laboratory Service, South Africa | KRISP, KZN Research Innovation and Sequencing Platform | Laguda-Akingba O et al |
| EPI_ISL_944150 | SouthAfrica/KRISP-K008208/2021 | 2021-01-05 | Africa/South Africa/Eastern Cape/Amatl | National Health Laboratory Service, South Africa | KRISP, KZN Research Innovation and Sequencing Platform | Laguda-Akingba O et al |
| EPI_ISL_944151 | SouthAfrica/KRISP-K008210/2021 | 2021-01-02 | Africa/South Africa/Eastern Cape/Amatl | National Health Laboratory Service, South Africa | KRISP, KZN Research Innovation and Sequencing Platform | Laguda-Akingba O et al |
| EPI_ISL_944152 | SouthAfrica/KRISP-K008215/2021 | 2021-01-04 | Africa/South Africa/Eastern Cape/Middl | National Health Laboratory Service, South Africa | KRISP, KZN Research Innovation and Sequencing Platform | Laguda-Akingba O et al |
| EPI_ISL_944153 | SouthAfrica/KRISP-K008216/2021 | 2021-01-04 | Africa/South Africa/Eastern Cape/Chris I | National Health Laboratory Service, South Africa | KRISP, KZN Research Innovation and Sequencing Platform | Laguda-Akingba O et al |

| | | | | | | |
|---|---|---|---|---|---|---|
| EPI_ISL_944155 | SouthAfrica/KRISP-K008219/2021 | 2021-01-04 | Africa/South Africa/Eastern Cape/Nelso | National Health Laboratory Service, South Africa | KRISP, KZN Research Innovation and Sequencing Platform | Laguda-Akingba O et al |
| EPI_ISL_944156 | SouthAfrica/KRISP-K008221/2021 | 2021-01-01 | Africa/South Africa/Eastern Cape/Amat | National Health Laboratory Service, South Africa | KRISP, KZN Research Innovation and Sequencing Platform | Laguda-Akingba O et al |
| EPI_ISL_944157 | SouthAfrica/KRISP-K008222/2021 | 2021-01-04 | Africa/South Africa/Eastern Cape/Sarah | National Health Laboratory Service, South Africa | KRISP, KZN Research Innovation and Sequencing Platform | Laguda-Akingba O et al |
| EPI_ISL_944159 | SouthAfrica/KRISP-K008234/2021 | 2021-01-04 | Africa/South Africa/Eastern Cape/Nelso | National Health Laboratory Service, South Africa | KRISP, KZN Research Innovation and Sequencing Platform | Laguda-Akingba O et al |
| EPI_ISL_944163 | SouthAfrica/KRISP-K008246/2021 | 2021-01-04 | Africa/South Africa/Eastern Cape/Sarah | National Health Laboratory Service, South Africa | KRISP, KZN Research Innovation and Sequencing Platform | Laguda-Akingba O et al |
| EPI_ISL_944164 | SouthAfrica/KRISP-K008248/2021 | 2021-01-02 | Africa/South Africa/Eastern Cape/Sarah | National Health Laboratory Service, South Africa | KRISP, KZN Research Innovation and Sequencing Platform | Laguda-Akingba O et al |
| EPI_ISL_944167 | SouthAfrica/KRISP-K008257/2021 | 2021-01-04 | Africa/South Africa/Eastern Cape/Joe G | National Health Laboratory Service, South Africa | KRISP, KZN Research Innovation and Sequencing Platform | Laguda-Akingba O et al |
| EPI_ISL_944171 | SouthAfrica/KRISP-K008263/2021 | 2021-01-04 | Africa/South Africa/Eastern Cape/Nelso | National Health Laboratory Service, South Africa | KRISP, KZN Research Innovation and Sequencing Platform | Laguda-Akingba O et al |

| | | | | | | |
|---|---|---|---|---|---|---|
| EPI_ISL_944173 | SouthAfrica/KRISP-K008267/2021 | 2021-01-04 | Africa/South Africa/Eastern Cape/Joe G | National Health Laboratory Service, South Africa | KRISP, KZN Research Innovation and Sequencing Platform | Laguda-Akingba O et al |
| EPI_ISL_944175 | SouthAfrica/KRISP-K008271/2021 | 2021-01-04 | Africa/South Africa/Eastern Cape/Nelso | National Health Laboratory Service, South Africa | KRISP, KZN Research Innovation and Sequencing Platform | Laguda-Akingba O et al |
| EPI_ISL_944176 | SouthAfrica/KRISP-K008274/2021 | 2021-01-04 | Africa/South Africa/Eastern Cape/Joe G | National Health Laboratory Service, South Africa | KRISP, KZN Research Innovation and Sequencing Platform | Laguda-Akingba O et al |
| EPI_ISL_944177 | SouthAfrica/KRISP-K008276/2021 | 2021-01-05 | Africa/South Africa/Eastern Cape/Nelso | National Health Laboratory Service, South Africa | KRISP, KZN Research Innovation and Sequencing Platform | Laguda-Akingba O et al |
| EPI_ISL_944189 | Israel/CVL-2078/2021 | 2021-01-12 | Asia/Israel/Jerusalem District/ | Israel Central Virology laboratory | Israel National Consortium for SARS-CoV-2 sequencing | Neta Zuckerman et al |
| EPI_ISL_944199 | Israel/CVL-2531/2021 | 2021-01-14 | Asia/Israel/Israel/ | Israel Central Virology laboratory | Israel National Consortium for SARS-CoV-2 sequencing | Neta Zuckerman et al |
| EPI_ISL_944206 | Israel/CVL-2085/2021 | 2021-01-12 | Asia/Israel/Jerusalem District/ | Israel Central Virology laboratory | Israel National Consortium for SARS-CoV-2 sequencing | Neta Zuckerman et al |
| EPI_ISL_944211 | Israel/CVL-2047/2021 | 2021-01-11 | Asia/Israel/Central District/ | Israel Central Virology laboratory | Israel National Consortium for SARS-CoV-2 sequencing | Neta Zuckerman et al |
| EPI_ISL_944218 | Israel/CVL-2413/2021 | 2021-01-13 | Asia/Israel/Israel/ | Israel Central Virology laboratory | Israel National Consortium for SARS-CoV-2 sequencing | Neta Zuckerman et al |

| | | | | | | |
|---|---|---|---|---|---|---|
| EPI_ISL_944231 | Israel/CVL-2515/2021 | 2021-01-14 | Asia/Israel/Haifa District/ | Israel Central Virology laboratory | Israel National Consortium for SARS-CoV-2 sequencing | Neta Zuckerman et al |
| EPI_ISL_944251 | Israel/CVL-2450/2021 | 2021-01-13 | Asia/Israel/Israel/ | Israel Central Virology laboratory | Israel National Consortium for SARS-CoV-2 sequencing | Neta Zuckerman et al |
| EPI_ISL_944290 | Israel/CVL-1533/2021 | 2021-01-05 | Asia/Israel/Central District/ | Israel Central Virology laboratory | Israel National Consortium for SARS-CoV-2 sequencing | Neta Zuckerman et al |
| EPI_ISL_944296 | Israel/CVL-1559/2021 | 2021-01-05 | Asia/Israel/Judea and Samaria/ | Israel Central Virology laboratory | Israel National Consortium for SARS-CoV-2 sequencing | Neta Zuckerman et al |
| EPI_ISL_944299 | Israel/CVL-1568/2021 | 2021-01-05 | Asia/Israel/Jerusalem District/ | Israel Central Virology laboratory | Israel National Consortium for SARS-CoV-2 sequencing | Neta Zuckerman et al |
| EPI_ISL_944311 | Israel/CVL-1622/2021 | 2021-01-05 | Asia/Israel/Jerusalem District/ | Israel Central Virology laboratory | Israel National Consortium for SARS-CoV-2 sequencing | Neta Zuckerman et al |
| EPI_ISL_944312 | Israel/CVL-1630/2021 | 2021-01-05 | Asia/Israel/Tel Aviv District/ | Israel Central Virology laboratory | Israel National Consortium for SARS-CoV-2 sequencing | Neta Zuckerman et al |
| EPI_ISL_944319 | Israel/CVL-1656/2021 | 2021-01-05 | Asia/Israel/Jerusalem District/ | Israel Central Virology laboratory | Israel National Consortium for SARS-CoV-2 sequencing | Neta Zuckerman et al |
| EPI_ISL_944320 | Israel/CVL-1658/2021 | 2021-01-05 | Asia/Israel/Jerusalem District/ | Israel Central Virology laboratory | Israel National Consortium for SARS-CoV-2 sequencing | Neta Zuckerman et al |
| EPI_ISL_944338 | Israel/CVL-2062/2021 | 2021-01-11 | Asia/Israel/Tel Aviv District/ | Israel Central Virology laboratory | Israel National Consortium for SARS-CoV-2 sequencing | Neta Zuckerman et al |

| | | | | | | |
|---|---|---|---|---|---|---|
| EPI_ISL_944339 | Israel/CVL-1996/2021 | 2021-01-12 | Asia/Israel/Central District/ | Israel Central Virology laboratory | Israel National Consortium for SARS-CoV-2 sequencing | Neta Zuckerman et al |
| EPI_ISL_944341 | Israel/CVL-2069/2021 | 2021-01-12 | Asia/Israel/South District/ | Israel Central Virology laboratory | Israel National Consortium for SARS-CoV-2 sequencing | Neta Zuckerman et al |
| EPI_ISL_944342 | Israel/CVL-2073/2021 | 2021-01-12 | Asia/Israel/Jerusalem District/ | Israel Central Virology laboratory | Israel National Consortium for SARS-CoV-2 sequencing | Neta Zuckerman et al |
| EPI_ISL_944345 | Israel/CVL-2083/2021 | 2021-01-12 | Asia/Israel/Jerusalem District/ | Israel Central Virology laboratory | Israel National Consortium for SARS-CoV-2 sequencing | Neta Zuckerman et al |
| EPI_ISL_944357 | Israel/CVL-2512/2021 | 2021-01-14 | Asia/Israel/Judea and Samaria/ | Israel Central Virology laboratory | Israel National Consortium for SARS-CoV-2 sequencing | Neta Zuckerman et al |
| EPI_ISL_944360 | Israel/CVL-2572/2021 | 2021-01-14 | Asia/Israel/Jerusalem District/ | Israel Central Virology laboratory | Israel National Consortium for SARS-CoV-2 sequencing | Neta Zuckerman et al |
| EPI_ISL_944361 | Israel/CVL-2574/2021 | 2021-01-14 | Asia/Israel/Tel Aviv District/ | Israel Central Virology laboratory | Israel National Consortium for SARS-CoV-2 sequencing | Neta Zuckerman et al |
| EPI_ISL_944363 | Israel/CVL-2576/2021 | 2021-01-14 | Asia/Israel/Central District/ | Israel Central Virology laboratory | Israel National Consortium for SARS-CoV-2 sequencing | Neta Zuckerman et al |
| EPI_ISL_944365 | Israel/CVL-2579/2021 | 2021-01-14 | Asia/Israel/Central District/ | Israel Central Virology laboratory | Israel National Consortium for SARS-CoV-2 sequencing | Neta Zuckerman et al |
| EPI_ISL_944369 | Israel/CVL-2054/2021 | 2021-01-11 | Asia/Israel/Israel/ | Israel Central Virology laboratory | Israel National Consortium for SARS-CoV-2 sequencing | Neta Zuckerman et al |

| | | | | | | |
|---|---|---|---|---|---|---|
| EPI_ISL_944370 | Israel/CVL-1906/2021 | 2021-01-11 | Asia/Israel/Central District/ | Israel Central Virology laboratory | Israel National Consortium for SARS-CoV-2 sequencing | Neta Zuckerman et al |
| EPI_ISL_944374 | Israel/CVL-2028/2021 | 2021-01-11 | Asia/Israel/Central District/ | Israel Central Virology laboratory | Israel National Consortium for SARS-CoV-2 sequencing | Neta Zuckerman et al |
| EPI_ISL_944394 | Israel/CVL-2539/2021 | 2021-01-14 | Asia/Israel/Israel/ | Israel Central Virology laboratory | Israel National Consortium for SARS-CoV-2 sequencing | Neta Zuckerman et al |
| EPI_ISL_944401 | Israel/CVL-2082/2021 | 2021-01-12 | Asia/Israel/South District/ | Israel Central Virology laboratory | Israel National Consortium for SARS-CoV-2 sequencing | Neta Zuckerman et al |
| EPI_ISL_944402 | Israel/CVL-2517/2021 | 2021-01-14 | Asia/Israel/South District/ | Israel Central Virology laboratory | Israel National Consortium for SARS-CoV-2 sequencing | Neta Zuckerman et al |
| EPI_ISL_944406 | Israel/CVL-2055/2021 | 2021-01-11 | Asia/Israel/Central District/ | Israel Central Virology laboratory | Israel National Consortium for SARS-CoV-2 sequencing | Neta Zuckerman et al |
| EPI_ISL_944411 | Israel/CVL-2032/2021 | 2021-01-11 | Asia/Israel/Israel/ | Israel Central Virology laboratory | Israel National Consortium for SARS-CoV-2 sequencing | Neta Zuckerman et al |
| EPI_ISL_944423 | Israel/CVL-1911/2021 | 2021-01-11 | Asia/Israel/Central District/ | Israel Central Virology laboratory | Israel National Consortium for SARS-CoV-2 sequencing | Neta Zuckerman et al |
| EPI_ISL_944444 | Israel/CVL-45/2020 | 2020-12-24 | Asia/Israel/Israel/ | Israel Central Virology laboratory | Israel National Consortium for SARS-CoV-2 sequencing | Neta Zuckerman et al |
| EPI_ISL_944449 | Israel/CVL-54/2020 | 2020-12-24 | Asia/Israel/Israel/ | Israel Central Virology laboratory | Israel National Consortium for SARS-CoV-2 sequencing | Neta Zuckerman et al |

| | | | | | | |
|---|---|---|---|---|---|---|
| EPI_ISL_944452 | Israel/CVL-64/2020 | 2020-12-24 | Asia/Israel/Israel/ | Israel Central Virology laboratory | Israel National Consortium for SARS-CoV-2 sequencing | Neta Zuckerman et al |
| EPI_ISL_944462 | Israel/CVL-1047/2021 | 2021-01-03 | Asia/Israel/Israel/ | Israel Central Virology laboratory | Israel National Consortium for SARS-CoV-2 sequencing | Neta Zuckerman et al |
| EPI_ISL_944467 | Israel/CVL-1914/2021 | 2021-01-11 | Asia/Israel/Central District/ | Israel Central Virology laboratory | Israel National Consortium for SARS-CoV-2 sequencing | Neta Zuckerman et al |
| EPI_ISL_944469 | Israel/CVL-1922/2021 | 2021-01-11 | Asia/Israel/Central District/ | Israel Central Virology laboratory | Israel National Consortium for SARS-CoV-2 sequencing | Neta Zuckerman et al |
| EPI_ISL_944471 | Israel/CVL-1930/2021 | 2021-01-11 | Asia/Israel/Central District/ | Israel Central Virology laboratory | Israel National Consortium for SARS-CoV-2 sequencing | Neta Zuckerman et al |
| EPI_ISL_944472 | Israel/CVL-1934/2021 | 2021-01-11 | Asia/Israel/Haifa District/ | Israel Central Virology laboratory | Israel National Consortium for SARS-CoV-2 sequencing | Neta Zuckerman et al |
| EPI_ISL_944473 | Israel/CVL-1935/2021 | 2021-01-11 | Asia/Israel/Central District/ | Israel Central Virology laboratory | Israel National Consortium for SARS-CoV-2 sequencing | Neta Zuckerman et al |
| EPI_ISL_944481 | Israel/CVL-1954/2021 | 2021-01-11 | Asia/Israel/Haifa District/ | Israel Central Virology laboratory | Israel National Consortium for SARS-CoV-2 sequencing | Neta Zuckerman et al |
| EPI_ISL_944488 | Israel/CVL-1965/2021 | 2021-01-11 | Asia/Israel/Central District/ | Israel Central Virology laboratory | Israel National Consortium for SARS-CoV-2 sequencing | Neta Zuckerman et al |
| EPI_ISL_944496 | Israel/CVL-2031/2021 | 2021-01-11 | Asia/Israel/Central District/ | Israel Central Virology laboratory | Israel National Consortium for SARS-CoV-2 sequencing | Neta Zuckerman et al |

| | | | | | | |
|---|---|---|---|---|---|---|
| EPI_ISL_944503 | Israel/CVL-2040/2021 | 2021-01-11 | Asia/Israel/Central District/ | Israel Central Virology laboratory | Israel National Consortium for SARS-CoV-2 sequencing | Neta Zuckerman et al |
| EPI_ISL_944506 | Israel/CVL-2044/2021 | 2021-01-11 | Asia/Israel/Central District/ | Israel Central Virology laboratory | Israel National Consortium for SARS-CoV-2 sequencing | Neta Zuckerman et al |
| EPI_ISL_944510 | Israel/CVL-2053/2021 | 2021-01-11 | Asia/Israel/Central District/ | Israel Central Virology laboratory | Israel National Consortium for SARS-CoV-2 sequencing | Neta Zuckerman et al |
| EPI_ISL_944521 | Israel/CVL-1984/2021 | 2021-01-12 | Asia/Israel/Judea and Samaria/ | Israel Central Virology laboratory | Israel National Consortium for SARS-CoV-2 sequencing | Neta Zuckerman et al |
| EPI_ISL_944526 | Israel/CVL-1993/2021 | 2021-01-12 | Asia/Israel/Central District/ | Israel Central Virology laboratory | Israel National Consortium for SARS-CoV-2 sequencing | Neta Zuckerman et al |
| EPI_ISL_944529 | Israel/CVL-1998/2021 | 2021-01-12 | Asia/Israel/South District/ | Israel Central Virology laboratory | Israel National Consortium for SARS-CoV-2 sequencing | Neta Zuckerman et al |
| EPI_ISL_944531 | Israel/CVL-2002/2021 | 2021-01-12 | Asia/Israel/Central District/ | Israel Central Virology laboratory | Israel National Consortium for SARS-CoV-2 sequencing | Neta Zuckerman et al |
| EPI_ISL_944535 | Israel/CVL-2009/2021 | 2021-01-12 | Asia/Israel/Central District/ | Israel Central Virology laboratory | Israel National Consortium for SARS-CoV-2 sequencing | Neta Zuckerman et al |
| EPI_ISL_944540 | Israel/CVL-2017/2021 | 2021-01-12 | Asia/Israel/Judea and Samaria/ | Israel Central Virology laboratory | Israel National Consortium for SARS-CoV-2 sequencing | Neta Zuckerman et al |
| EPI_ISL_944542 | Israel/CVL-2370/2021 | 2021-01-12 | Asia/Israel/Judea and Samaria/ | Israel Central Virology laboratory | Israel National Consortium for SARS-CoV-2 sequencing | Neta Zuckerman et al |

| | | | | | | |
|---|---|---|---|---|---|---|
| EPI_ISL_944545 | Israel/CVL-2392/2021 | 2021-01-13 | Asia/Israel/Israel/ | Israel Central Virology laboratory | Israel National Consortium for SARS-CoV-2 sequencing | Neta Zuckerman et al |
| EPI_ISL_944548 | Israel/CVL-2395/2021 | 2021-01-13 | Asia/Israel/Israel/ | Israel Central Virology laboratory | Israel National Consortium for SARS-CoV-2 sequencing | Neta Zuckerman et al |
| EPI_ISL_944549 | Israel/CVL-2396/2021 | 2021-01-13 | Asia/Israel/Israel/ | Israel Central Virology laboratory | Israel National Consortium for SARS-CoV-2 sequencing | Neta Zuckerman et al |
| EPI_ISL_944551 | Israel/CVL-2404/2021 | 2021-01-13 | Asia/Israel/Israel/ | Israel Central Virology laboratory | Israel National Consortium for SARS-CoV-2 sequencing | Neta Zuckerman et al |
| EPI_ISL_944552 | Israel/CVL-2405/2021 | 2021-01-13 | Asia/Israel/Israel/ | Israel Central Virology laboratory | Israel National Consortium for SARS-CoV-2 sequencing | Neta Zuckerman et al |
| EPI_ISL_944556 | Israel/CVL-2415/2021 | 2021-01-13 | Asia/Israel/Israel/ | Israel Central Virology laboratory | Israel National Consortium for SARS-CoV-2 sequencing | Neta Zuckerman et al |
| EPI_ISL_944557 | Israel/CVL-2506/2021 | 2021-01-14 | Asia/Israel/Israel/ | Israel Central Virology laboratory | Israel National Consortium for SARS-CoV-2 sequencing | Neta Zuckerman et al |
| EPI_ISL_944559 | Israel/CVL-2513/2021 | 2021-01-14 | Asia/Israel/North District/ | Israel Central Virology laboratory | Israel National Consortium for SARS-CoV-2 sequencing | Neta Zuckerman et al |
| EPI_ISL_944560 | Israel/CVL-2514/2021 | 2021-01-14 | Asia/Israel/Tel Aviv District/ | Israel Central Virology laboratory | Israel National Consortium for SARS-CoV-2 sequencing | Neta Zuckerman et al |
| EPI_ISL_944564 | Israel/CVL-2554/2021 | 2021-01-14 | Asia/Israel/Israel/ | Israel Central Virology laboratory | Israel National Consortium for SARS-CoV-2 sequencing | Neta Zuckerman et al |

| | | | | | | |
|---|---|---|---|---|---|---|
| EPI_ISL_944570 | Israel/CVL-2566/2021 | 2021-01-15 | Asia/Israel/Israel/ | Israel Central Virology laboratory | Israel National Consortium for SARS-CoV-2 sequencing | Neta Zuckerman et al |
| EPI_ISL_944611 | Mexico/CMX-INMEGEN-01-03-01/202 | 2021-01-11 | North America/Mexico/Mexico City/ | Instituto Nacional de Medicina Genomica | Instituto Nacional de Medicina Genomica | Hidalgo-Miranda A et al |
| EPI_ISL_944612 | Mexico/CMX-INMEGEN-01-03-02/202 | 2021-01-11 | North America/Mexico/Mexico City/ | Instituto Nacional de Medicina Genomica | Instituto Nacional de Medicina Genomica | Hidalgo-Miranda A et al |
| EPI_ISL_944613 | Mexico/CMX-INMEGEN-01-03-03/202 | 2021-01-11 | North America/Mexico/Mexico City/ | Instituto Nacional de Medicina Genomica | Instituto Nacional de Medicina Genomica | Hidalgo-Miranda A et al |
| EPI_ISL_944614 | Mexico/CMX-INMEGEN-01-03-04/202 | 2021-01-04 | North America/Mexico/Mexico City/ | Instituto Nacional de Medicina Genomica | Instituto Nacional de Medicina Genomica | Hidalgo-Miranda A et al |
| EPI_ISL_944615 | Mexico/CMX-INMEGEN-01-03-05/202 | 2021-01-11 | North America/Mexico/Mexico City/ | Instituto Nacional de Medicina Genomica | Instituto Nacional de Medicina Genomica | Hidalgo-Miranda A et al |
| EPI_ISL_944616 | Mexico/CMX-INMEGEN-01-03-06/202 | 2021-01-11 | North America/Mexico/Mexico City/ | Instituto Nacional de Medicina Genomica | Instituto Nacional de Medicina Genomica | Hidalgo-Miranda A et al |
| EPI_ISL_944617 | Mexico/CMX-INMEGEN-01-03-07/202 | 2021-01-12 | North America/Mexico/Mexico City/ | Instituto Nacional de Medicina Genomica | Instituto Nacional de Medicina Genomica | Hidalgo-Miranda A et al |
| EPI_ISL_944618 | Mexico/CMX-INMEGEN-01-03-08/202 | 2021-01-14 | North America/Mexico/Mexico City/ | Instituto Nacional de Medicina Genomica | Instituto Nacional de Medicina Genomica | Hidalgo-Miranda A et al |
| EPI_ISL_944619 | Mexico/CMX-INMEGEN-01-03-09/202 | 2021-01-11 | North America/Mexico/Mexico City/ | Instituto Nacional de Medicina Genomica | Instituto Nacional de Medicina Genomica | Hidalgo-Miranda A et al |
| EPI_ISL_944620 | Mexico/CMX-INMEGEN-01-03-10/202 | 2021-01-14 | North America/Mexico/Mexico City/ | Instituto Nacional de Medicina Genomica | Instituto Nacional de Medicina Genomica | Hidalgo-Miranda A et al |
| EPI_ISL_944621 | Mexico/CMX-INMEGEN-01-03-11/202 | 2021-01-14 | North America/Mexico/Mexico City/ | Instituto Nacional de Medicina Genomica | Instituto Nacional de Medicina Genomica | Hidalgo-Miranda A et al |
| EPI_ISL_944622 | Mexico/CMX-INMEGEN-01-03-12/202 | 2021-01-12 | North America/Mexico/Mexico City/ | Instituto Nacional de Medicina Genomica | Instituto Nacional de Medicina Genomica | Hidalgo-Miranda A et al |

| | | | | | | |
|---|---|---|---|---|---|---|
| EPI_ISL_944623 | Mexico/CMX-INMEGEN-01-03-13/202 | 2021-01-10 | North America/Mexico/Mexico City/ | Instituto Nacional de Medicina Genomica | Instituto Nacional de Medicina Genomica | Hidalgo-Miranda A et al |
| EPI_ISL_944624 | Mexico/CMX-INMEGEN-01-03-14/202 | 2021-01-08 | North America/Mexico/Mexico City/ | Instituto Nacional de Medicina Genomica | Instituto Nacional de Medicina Genomica | Hidalgo-Miranda A et al |
| EPI_ISL_944625 | Mexico/CMX-INMEGEN-01-03-15/202 | 2021-01-08 | North America/Mexico/Mexico City/ | Instituto Nacional de Medicina Genomica | Instituto Nacional de Medicina Genomica | Hidalgo-Miranda A et al |
| EPI_ISL_944626 | Mexico/CMX-INMEGEN-01-03-16/202 | 2021-01-12 | North America/Mexico/Mexico City/ | Instituto Nacional de Medicina Genomica | Instituto Nacional de Medicina Genomica | Hidalgo-Miranda A et al |
| EPI_ISL_944627 | Mexico/CMX-INMEGEN-01-03-17/202 | 2021-01-11 | North America/Mexico/Mexico City/ | Instituto Nacional de Medicina Genomica | Instituto Nacional de Medicina Genomica | Hidalgo-Miranda A et al |
| EPI_ISL_944629 | Mexico/CMX-INMEGEN-01-03-19/202 | 2021-01-13 | North America/Mexico/Mexico City/ | Instituto Nacional de Medicina Genomica | Instituto Nacional de Medicina Genomica | Hidalgo-Miranda A et al |
| EPI_ISL_944630 | Mexico/CMX-INMEGEN-01-03-20/202 | 2021-01-13 | North America/Mexico/Mexico City/ | Instituto Nacional de Medicina Genomica | Instituto Nacional de Medicina Genomica | Hidalgo-Miranda A et al |
| EPI_ISL_944631 | Mexico/CMX-INMEGEN-01-03-21/202 | 2021-01-11 | North America/Mexico/Mexico City/ | Instituto Nacional de Medicina Genomica | Instituto Nacional de Medicina Genomica | Hidalgo-Miranda A et al |
| EPI_ISL_944632 | Mexico/CMX-INMEGEN-01-03-22/202 | 2021-01-14 | North America/Mexico/Mexico City/ | Instituto Nacional de Medicina Genomica | Instituto Nacional de Medicina Genomica | Hidalgo-Miranda A et al |
| EPI_ISL_944633 | Mexico/CMX-INMEGEN-01-03-23/202 | 2021-01-11 | North America/Mexico/Mexico City/ | Instituto Nacional de Medicina Genomica | Instituto Nacional de Medicina Genomica | Hidalgo-Miranda A et al |

| | | | | | | |
|---|---|---|---|---|---|---|
| EPI_ISL_944636 | Colombia/MET-GUR-03581/2020 | 2020-04-22 | South America/Colombia/Meta/Villavic | Centro de Investigaciones en Microbiolog√≠a y Biotecnolog√≠a-UR (CIMBIUR), Facultad de Ciencias Naturales, Universidad del Rosario, Bogot√°, Colombia | Centro de Investigaciones en Microbiolog√≠a y Biotecnolog√≠a-UR (CIMBIUR), Facultad de Ciencias Naturales, Universidad del Rosario, Bogot√°, Colombia Instituto Nacional de Salud, Bogot√°, Colombia Icahn School of Medicine at Mount Sinai, New York, USA | Luz Helena Pati√±o et al |
| EPI_ISL_944669 | Ghana/WACCBIP-GS127/2021 | 2021-01-10 | Africa/Ghana/Ghana/ | Department of Biochemistry, Cell and Molecular Biology, West African Centre for Cell Biology of Infectious Pathogens (WACCBIP), University of Ghana | Department of Biochemistry, Cell and Molecular Biology, West African Centre for Cell Biology of Infectious Pathogens (WACCBIP), University of Ghana | Morang'a et al |
| EPI_ISL_944676 | Ghana/WACCBIP-GS150/2021 | 2021-01-13 | Africa/Ghana/Ghana/ | Department of Biochemistry, Cell and Molecular Biology, West African Centre for Cell Biology of Infectious Pathogens (WACCBIP), University of Ghana | Department of Biochemistry, Cell and Molecular Biology, West African Centre for Cell Biology of Infectious Pathogens (WACCBIP), University of Ghana | Morang'a et al |

| | | | | | | |
|---|---|---|---|---|---|---|
| EPI_ISL_944681 | Ghana/WACCBIP-GS137/2021 | 2021-01-08 | Africa/Ghana/Ghana/ | Department of Biochemistry, Cell and Molecular Biology, West African Centre for Cell Biology of Infectious Pathogens (WACCBIP), University of Ghana | Department of Biochemistry, Cell and Molecular Biology, West African Centre for Cell Biology of Infectious Pathogens (WACCBIP), University of Ghana | Morang'a et al |
| EPI_ISL_944703 | Ghana/WACCBIP-GS65/2021 | 2021-01-06 | Africa/Ghana/Ghana/ | Department of Biochemistry, Cell and Molecular Biology, West African Centre for Cell Biology of Infectious Pathogens (WACCBIP), University of Ghana | Department of Biochemistry, Cell and Molecular Biology, West African Centre for Cell Biology of Infectious Pathogens (WACCBIP), University of Ghana | Morang'a et al |
| EPI_ISL_944748 | Australia/QLD1542/2021 | 2021-02-01 | Oceania/Australia/Australia/ | unknown | Public Health Virology-Forensic and Scientific Services (PHV-FSS) | Son Nguyen et al. et al |
| EPI_ISL_944749 | India/CT-VRDL-AIIMS-UK-24/2020 | 2020-12-25 | Asia/India/Chhattisgarh/Bilaspur | State Virus Research and Diagnostic Laboratory (VRDL), AIIMS Raipur | State Virus Research and Diagnostic Laboratory (VRDL), AIIMS Raipur | Pushpendra Singh et al |
| EPI_ISL_944750 | India/CT-VRDL-AIIMS-UK-43/2020 | 2020-12-26 | Asia/India/Chhattisgarh/Janjgir-Champa | State Virus Research and Diagnostic Laboratory (VRDL), AIIMS Raipur | State Virus Research and Diagnostic Laboratory (VRDL), AIIMS Raipur | Pushpendra Singh et al |

| | | | | | | |
|---|---|---|---|---|---|---|
| EPI_ISL_944751 | India/CT-VRDL-AIIMS-UK-53/2020 | 2020-12-26 | Asia/India/Chhattisgarh/Janjgir-Champa | State Virus Research and Diagnostic Laboratory (VRDL), AIIMS Raipur | State Virus Research and Diagnostic Laboratory (VRDL), AIIMS Raipur | Pushpendra Singh et al |
| EPI_ISL_944753 | India/CT-VRDL-AIIMS-UK-42/2020 | 2020-12-26 | Asia/India/Chhattisgarh/Janjgir-Champa | State Virus Research and Diagnostic Laboratory (VRDL), AIIMS Raipur | State Virus Research and Diagnostic Laboratory (VRDL), AIIMS Raipur | Pushpendra Singh et al |
| EPI_ISL_944754 | India/CT-VRDL-AIIMS-UK-14180/2020 | 2020-12-26 | Asia/India/Chhattisgarh/Durg | State Virus Research and Diagnostic Laboratory (VRDL), AIIMS Raipur | State Virus Research and Diagnostic Laboratory (VRDL), AIIMS Raipur | Pushpendra Singh et al |
| EPI_ISL_944755 | India/CT-VRDL-AIIMS-UK-49/2020 | 2020-12-26 | Asia/India/Chhattisgarh/Janjgir-Champa | State Virus Research and Diagnostic Laboratory (VRDL), AIIMS Raipur | State Virus Research and Diagnostic Laboratory (VRDL), AIIMS Raipur | Pushpendra Singh et al |
| EPI_ISL_944762 | India/WB-IICB-039/2020 | 2020-10-29 | Asia/India/West Bengal/ | CSIR-Indian Institute of Chemical Biology, MEDICA Supercpecialty Hospital Kolkata | CSIR-Indian Institute of Chemical Biology, MEDICA Supercpecialty Hospital Kolkata | Sujay Krishna Maity et al |
| EPI_ISL_944763 | India/WB-IICB-040/2020 | 2020-10-29 | Asia/India/West Bengal/ | CSIR-Indian Institute of Chemical Biology, MEDICA Supercpecialty Hospital Kolkata | CSIR-Indian Institute of Chemical Biology, MEDICA Supercpecialty Hospital Kolkata | Sujay Krishna Maity et al |

| | | | | | | |
|---|---|---|---|---|---|---|
| EPI_ISL_944765 | India/WB-IICB-041/2020 | 2020-10-29 | Asia/India/West Bengal/ | CSIR-Indian Institute of Chemical Biology, MEDICA Supercpecialty Hospital Kolkata | CSIR-Indian Institute of Chemical Biology, MEDICA Supercpecialty Hospital Kolkata | Sujay Krishna Maity et al |
| EPI_ISL_944768 | India/WB-IICB-042/2020 | 2020-10-29 | Asia/India/West Bengal/ | CSIR-Indian Institute of Chemical Biology, MEDICA Supercpecialty Hospital Kolkata | CSIR-Indian Institute of Chemical Biology, MEDICA Supercpecialty Hospital Kolkata | Sujay Krishna Maity et al |
| EPI_ISL_944769 | India/WB-IICB-043/2020 | 2020-10-29 | Asia/India/West Bengal/ | CSIR-Indian Institute of Chemical Biology, MEDICA Supercpecialty Hospital Kolkata | CSIR-Indian Institute of Chemical Biology, MEDICA Supercpecialty Hospital Kolkata | Sujay Krishna Maity et al |
| EPI_ISL_944771 | India/WB-IICB-044/2020 | 2020-10-29 | Asia/India/West Bengal/ | CSIR-Indian Institute of Chemical Biology, MEDICA Supercpecialty Hospital Kolkata | CSIR-Indian Institute of Chemical Biology, MEDICA Supercpecialty Hospital Kolkata | Sujay Krishna Maity et al |
| EPI_ISL_944772 | India/WB-IICB-045/2020 | 2020-10-29 | Asia/India/West Bengal/ | CSIR-Indian Institute of Chemical Biology, MEDICA Supercpecialty Hospital Kolkata | CSIR-Indian Institute of Chemical Biology, MEDICA Supercpecialty Hospital Kolkata | Sujay Krishna Maity et al |
| EPI_ISL_944773 | India/WB-IICB-046/2020 | 2020-10-29 | Asia/India/West Bengal/ | CSIR-Indian Institute of Chemical Biology, MEDICA Supercpecialty Hospital Kolkata | CSIR-Indian Institute of Chemical Biology, MEDICA Supercpecialty Hospital Kolkata | Sujay Krishna Maity et al |

| | | | | | | |
|---|---|---|---|---|---|---|
| EPI_ISL_944775 | India/WB-IICB-047/2020 | 2020-10-29 | Asia/India/West Bengal/ | CSIR-Indian Institute of Chemical Biology, MEDICA Supercpecialty Hospital Kolkata | CSIR-Indian Institute of Chemical Biology, MEDICA Supercpecialty Hospital Kolkata | Sujay Krishna Maity et al |
| EPI_ISL_944776 | India/WB-IICB-048/2020 | 2020-10-29 | Asia/India/West Bengal/ | CSIR-Indian Institute of Chemical Biology, MEDICA Supercpecialty Hospital Kolkata | CSIR-Indian Institute of Chemical Biology, MEDICA Supercpecialty Hospital Kolkata | Sujay Krishna Maity et al |
| EPI_ISL_944781 | India/WB-IICB-049/2020 | 2020-10-29 | Asia/India/West Bengal/ | CSIR-Indian Institute of Chemical Biology, MEDICA Supercpecialty Hospital Kolkata | CSIR-Indian Institute of Chemical Biology, MEDICA Supercpecialty Hospital Kolkata | Sujay Krishna Maity et al |
| EPI_ISL_944783 | India/WB-IICB-050/2020 | 2020-10-29 | Asia/India/West Bengal/ | CSIR-Indian Institute of Chemical Biology, MEDICA Supercpecialty Hospital Kolkata | CSIR-Indian Institute of Chemical Biology, MEDICA Supercpecialty Hospital Kolkata | Sujay Krishna Maity et al |
| EPI_ISL_944784 | India/WB-IICB-051/2020 | 2020-10-29 | Asia/India/West Bengal/ | CSIR-Indian Institute of Chemical Biology, MEDICA Supercpecialty Hospital Kolkata | CSIR-Indian Institute of Chemical Biology, MEDICA Supercpecialty Hospital Kolkata | Sujay Krishna Maity et al |
| EPI_ISL_944788 | India/WB-IICB-052/2020 | 2020-10-29 | Asia/India/West Bengal/ | CSIR-Indian Institute of Chemical Biology, MEDICA Supercpecialty Hospital Kolkata | CSIR-Indian Institute of Chemical Biology, MEDICA Supercpecialty Hospital Kolkata | Sujay Krishna Maity et al |

| | | | | | | |
|---|---|---|---|---|---|---|
| EPI_ISL_944789 | India/WB-IICB-053/2020 | 2020-10-29 | Asia/India/West Bengal/ | CSIR-Indian Institute of Chemical Biology, MEDICA Supercpecialty Hospital Kolkata | CSIR-Indian Institute of Chemical Biology, MEDICA Supercpecialty Hospital Kolkata | Sujay Krishna Maity et al |
| EPI_ISL_944791 | India/WB-IICB-054/2020 | 2020-10-29 | Asia/India/West Bengal/ | CSIR-Indian Institute of Chemical Biology, MEDICA Supercpecialty Hospital Kolkata | CSIR-Indian Institute of Chemical Biology, MEDICA Supercpecialty Hospital Kolkata | Sujay Krishna Maity et al |
| EPI_ISL_944965 | England/QEUH-119A23E/2021 | 2021-01-29 | Europe/United Kingdom/England/ | Lighthouse Lab in Glasgow | Wellcome Sanger Institute for the COVID-19 Genomics UK (COG-UK) Consortium | Harper VanSteenhouse et al |
| EPI_ISL_945147 | England/CAMC-F01C75/2021 | 2021-01-08 | Europe/United Kingdom/England/ | Lighthouse Lab in Cambridge | Wellcome Sanger Institute for the COVID-19 Genomics UK (COG-UK) Consortium | Rob Howes et al |
| EPI_ISL_945209 | England/QEUH-F0A025/2020 | 2020-12-17 | Europe/United Kingdom/England/ | Lighthouse Lab in Glasgow | Wellcome Sanger Institute for the COVID-19 Genomics UK (COG-UK) Consortium | Harper VanSteenhouse et al |
| EPI_ISL_945368 | England/MILK-F7293C/2021 | 2021-01-05 | Europe/United Kingdom/England/ | Lighthouse Lab in Milton Keynes | Wellcome Sanger Institute for the COVID-19 Genomics UK (COG-UK) Consortium | The Lighthouse Lab in Milton Keynes et al |
| EPI_ISL_945526 | England/ALDP-119C849/2021 | 2021-01-27 | Europe/United Kingdom/England/ | Lighthouse Lab in Alderley Park | Wellcome Sanger Institute for the COVID-19 Genomics UK (COG-UK) Consortium | Jacquelyn Wynn et al |

| | | | | | | |
|---|---|---|---|---|---|---|
| EPI_ISL_945612 | England/ALDP-119D63F/2021 | 2021-01-27 | Europe/United Kingdom/England/ | Lighthouse Lab in Alderley Park | Wellcome Sanger Institute for the COVID-19 Genomics UK (COG-UK) Consortium | Jacquelyn Wynn et al |
| EPI_ISL_945627 | England/ALDP-119CA8F/2021 | 2021-01-28 | Europe/United Kingdom/England/ | Lighthouse Lab in Alderley Park | Wellcome Sanger Institute for the COVID-19 Genomics UK (COG-UK) Consortium | Jacquelyn Wynn et al |
| EPI_ISL_945672 | England/ALDP-119D09B/2021 | 2021-01-28 | Europe/United Kingdom/England/ | Lighthouse Lab in Alderley Park | Wellcome Sanger Institute for the COVID-19 Genomics UK (COG-UK) Consortium | Jacquelyn Wynn et al |
| EPI_ISL_945729 | Scotland/QEUH-118A0CE/2021 | 2021-01-28 | Europe/United Kingdom/Scotland/ | Lighthouse Lab in Glasgow | Wellcome Sanger Institute for the COVID-19 Genomics UK (COG-UK) Consortium | Harper VanSteenhouse et al |
| EPI_ISL_946562 | England/ALDP-11874D4/2021 | 2021-01-25 | Europe/United Kingdom/England/ | Lighthouse Lab in Alderley Park | Wellcome Sanger Institute for the COVID-19 Genomics UK (COG-UK) Consortium | Jacquelyn Wynn et al |
| EPI_ISL_946721 | England/ALDP-11879D5/2021 | 2021-01-26 | Europe/United Kingdom/England/ | Lighthouse Lab in Alderley Park | Wellcome Sanger Institute for the COVID-19 Genomics UK (COG-UK) Consortium | Jacquelyn Wynn et al |
| EPI_ISL_946878 | England/ALDP-118B27C/2021 | 2021-01-26 | Europe/United Kingdom/England/ | Lighthouse Lab in Alderley Park | Wellcome Sanger Institute for the COVID-19 Genomics UK (COG-UK) Consortium | Jacquelyn Wynn et al |
| EPI_ISL_947020 | England/ALDP-1195FB4/2021 | 2021-01-26 | Europe/United Kingdom/England/ | Lighthouse Lab in Alderley Park | Wellcome Sanger Institute for the COVID-19 Genomics UK (COG-UK) Consortium | Jacquelyn Wynn et al |

| | | | | | | |
|---|---|---|---|---|---|---|
| EPI_ISL_947186 | England/QEUH-1170C5E/2021 | 2021-01-26 | Europe/United Kingdom/England/ | Lighthouse Lab in Glasgow | Wellcome Sanger Institute for the COVID-19 Genomics UK (COG-UK) Consortium | Harper VanSteenhouse et al |
| EPI_ISL_947247 | Indonesia/JB-EIJK59/2020 | 2020-11-05 | Asia/Indonesia/West Java/Bekasi | RSU Bunda Mulia | Eijkman Institute for Molecular Biology, Ministry of Research and Technology/National Agency for Research and Innovation | Edison Johar et al |
| EPI_ISL_947252 | Indonesia/JB-EIJK64/2020 | 2020-11-10 | Asia/Indonesia/West Java/Bogor | RS Bina Husada Cibinong | Eijkman Institute for Molecular Biology, Ministry of Research and Technology/National Agency for Research and Innovation | Edison Johar et al |
| EPI_ISL_947255 | Indonesia/JB-EIJK65/2020 | 2020-11-10 | Asia/Indonesia/West Java/Depok | RSU Meilia | Eijkman Institute for Molecular Biology, Ministry of Research and Technology/National Agency for Research and Innovation | Edison Johar et al |
| EPI_ISL_947260 | Indonesia/JB-EIJK68/2020 | 2020-11-11 | Asia/Indonesia/West Java/Bekasi | RSU Bunda Mulia | Eijkman Institute for Molecular Biology, Ministry of Research and Technology/National Agency for Research and Innovation | Iskandar Adnan et al |

| | | | | | | |
|---|---|---|---|---|---|---|
| EPI_ISL_947262 | Indonesia/BT-EIJK72/2020 | 2020-11-12 | Asia/Indonesia/Banten/Tangerang | RS Qadr | Eijkman Institute for Molecular Biology, Ministry of Research and Technology/National Agency for Research and Innovation | Frilasita A Yudhaputri et al |
| EPI_ISL_947265 | Indonesia/BT-EIJK73/2020 | 2020-11-16 | Asia/Indonesia/Banten/Tangerang | RS Sari Asih Ciledug | Eijkman Institute for Molecular Biology, Ministry of Research and Technology/National Agency for Research and Innovation | Frilasita A Yudhaputri et al |
| EPI_ISL_947268 | Indonesia/JK-EIJK76/2020 | 2020-11-18 | Asia/Indonesia/Jakarta/ | RSU Anggrek Mas | Eijkman Institute for Molecular Biology, Ministry of Research and Technology/National Agency for Research and Innovation | Frilasita A Yudhaputri et al |
| EPI_ISL_947270 | Indonesia/BT-EIJK78/2020 | 2020-11-19 | Asia/Indonesia/Banten/Tangerang | RS Mitra Keluarga Gading Serpong | Eijkman Institute for Molecular Biology, Ministry of Research and Technology/National Agency for Research and Innovation | Frilasita A Yudhaputri et al |
| EPI_ISL_947273 | Indonesia/JK-EIJK80/2020 | 2020-11-25 | Asia/Indonesia/Jakarta/ | RSIA Sayyidah | Eijkman Institute for Molecular Biology, Ministry of Research and Technology/National Agency for Research and Innovation | Frilasita A Yudhaputri et al |

| | | | | | | |
|---|---|---|---|---|---|---|
| EPI_ISL_947278 | Indonesia/JK-EIJK85/2020 | 2020-12-02 | Asia/Indonesia/Jakarta/ | RS Umum Ciputra Hospital Citragarden City | Eijkman Institute for Molecular Biology, Ministry of Research and Technology/National Agency for Research and Innovation | Hidayat Trimarsanto et al |
| EPI_ISL_947281 | Indonesia/JB-EIJK86/2020 | 2020-12-02 | Asia/Indonesia/West Java/Bogor | RSU Medika Dramaga | Eijkman Institute for Molecular Biology, Ministry of Research and Technology/National Agency for Research and Innovation | Hidayat Trimarsanto et al |
| EPI_ISL_947290 | Indonesia/BT-EIJK94/2020 | 2020-12-02 | Asia/Indonesia/Banten/Tangerang | RS Hermina Bitung | Eijkman Institute for Molecular Biology, Ministry of Research and Technology/National Agency for Research and Innovation | Hidayat Trimarsanto et al |
| EPI_ISL_947296 | Indonesia/JB-EIJK97/2020 | 2020-12-05 | Asia/Indonesia/West Java/Bekasi | RSU Kartika Husada | Eijkman Institute for Molecular Biology, Ministry of Research and Technology/National Agency for Research and Innovation | Iskandar Adnan et al |
| EPI_ISL_947298 | Indonesia/JB-EIJK99/2020 | 2020-12-10 | Asia/Indonesia/West Java/Bekasi | RSU Medirossa | Eijkman Institute for Molecular Biology, Ministry of Research and Technology/National Agency for Research and Innovation | Iskandar Adnan et al |

| | | | | | | |
|---|---|---|---|---|---|---|
| EPI_ISL_947301 | Indonesia/JB-EIJK101/2020 | 2020-12-10 | Asia/Indonesia/West Java/Bekasi | RS Mitra Keluarga Cibubur | Eijkman Institute for Molecular Biology, Ministry of Research and Technology/National Agency for Research and Innovation | Iskandar Adnan et al |
| EPI_ISL_947304 | Indonesia/JK-EIJK102/2020 | 2020-12-11 | Asia/Indonesia/Jakarta/ | RSU Harapan Bunda | Eijkman Institute for Molecular Biology, Ministry of Research and Technology/National Agency for Research and Innovation | Iskandar Adnan et al |
| EPI_ISL_947307 | Indonesia/JB-EIJK105/2020 | 2020-12-14 | Asia/Indonesia/West Java/Bekasi | RS Sentra Medika Cikarang | Eijkman Institute for Molecular Biology, Ministry of Research and Technology/National Agency for Research and Innovation | Lydia V. Panggalo et al |
| EPI_ISL_947315 | Indonesia/JK-EIJK111/2020 | 2020-12-18 | Asia/Indonesia/Jakarta/ | RSU Harapan Bunda | Eijkman Institute for Molecular Biology, Ministry of Research and Technology/National Agency for Research and Innovation | Lydia V. Panggalo et al |
| EPI_ISL_947320 | Indonesia/BT-EIJK114/2020 | 2020-12-21 | Asia/Indonesia/Banten/Tangerang | RS Sari Asih Ciledug | Eijkman Institute for Molecular Biology, Ministry of Research and Technology/National Agency for Research and Innovation | Sukma Oktavianthi et al |

| | | | | | | |
|---|---|---|---|---|---|---|
| EPI_ISL_947322 | Indonesia/JK-EIJK119/2021 | 2021-01-06 | Asia/Indonesia/Jakarta/ | RS Mitra Keluarga Kelapa Gading | Eijkman Institute for Molecular Biology, Ministry of Research and Technology/National Agency for Research and Innovation | Sukma Oktavianthi et al |
| EPI_ISL_947323 | Indonesia/JK-EIJK126/2021 | 2021-01-04 | Asia/Indonesia/Jakarta/ | RS Siloam Kebon Jeruk | Eijkman Institute for Molecular Biology, Ministry of Research and Technology/National Agency for Research and Innovation | Sukma Oktavianthi et al |
| EPI_ISL_947324 | Indonesia/JB-EIJK128/2021 | 2021-01-06 | Asia/Indonesia/West Java/Bekasi | RSU Medirossa | Eijkman Institute for Molecular Biology, Ministry of Research and Technology/National Agency for Research and Innovation | Sukma Oktavianthi et al |
| EPI_ISL_947325 | Indonesia/BT-EIJK131/2021 | 2021-01-06 | Asia/Indonesia/Banten/Tangerang | RSIA PKU Muhammadiyah | Eijkman Institute for Molecular Biology, Ministry of Research and Technology/National Agency for Research and Innovation | Sukma Oktavianthi et al |
| EPI_ISL_947326 | Indonesia/BT-EIJK133/2021 | 2021-01-01 | Asia/Indonesia/Banten/Tangerang | RS Mitra Keluarga Gading Serpong | Eijkman Institute for Molecular Biology, Ministry of Research and Technology/National Agency for Research and Innovation | Willy Agustine et al |

| | | | | | | |
|---|---|---|---|---|---|---|
| EPI_ISL_947327 | Indonesia/BT-EIJK136/2021 | 2021-01-07 | Asia/Indonesia/Banten/Tangerang | RS Mitra Keluarga Gading Serpong | Eijkman Institute for Molecular Biology, Ministry of Research and Technology/National Agency for Research and Innovation | Willy Agustine et al |
| EPI_ISL_947328 | Indonesia/JB-EIJK140/2021 | 2021-01-08 | Asia/Indonesia/West Java/Depok | RS Mitra Keluarga Depok | Eijkman Institute for Molecular Biology, Ministry of Research and Technology/National Agency for Research and Innovation | Willy Agustine et al |
| EPI_ISL_947329 | Indonesia/JB-EIJK143/2021 | 2021-01-08 | Asia/Indonesia/West Java/Bekasi | RSU Bunda Mulia | Eijkman Institute for Molecular Biology, Ministry of Research and Technology/National Agency for Research and Innovation | Willy Agustine et al |
| EPI_ISL_947330 | Indonesia/JB-EIJK144/2021 | 2021-01-08 | Asia/Indonesia/West Java/Bekasi | RSU Bunda Mulia | Eijkman Institute for Molecular Biology, Ministry of Research and Technology/National Agency for Research and Innovation | Willy Agustine et al |
| EPI_ISL_947436 | England/ALDP-1150304/2021 | 2021-01-23 | Europe/United Kingdom/England/ | Lighthouse Lab in Alderley Park | Wellcome Sanger Institute for the COVID-19 Genomics UK (COG-UK) Consortium | Jacquelyn Wynn et al |

| | | | | | | |
|---|---|---|---|---|---|---|
| EPI_ISL_947447 | England/ALDP-114DC1E/2021 | 2021-01-24 | Europe/United Kingdom/England/ | Lighthouse Lab in Alderley Park | Wellcome Sanger Institute for the COVID-19 Genomics UK (COG-UK) Consortium | Jacquelyn Wynn et al |
| EPI_ISL_947737 | England/ALDP-114E54F/2021 | 2021-01-24 | Europe/United Kingdom/England/ | Lighthouse Lab in Alderley Park | Wellcome Sanger Institute for the COVID-19 Genomics UK (COG-UK) Consortium | Jacquelyn Wynn et al |
| EPI_ISL_947806 | England/ALDP-1150067/2021 | 2021-01-23 | Europe/United Kingdom/England/ | Lighthouse Lab in Alderley Park | Wellcome Sanger Institute for the COVID-19 Genomics UK (COG-UK) Consortium | Jacquelyn Wynn et al |
| EPI_ISL_948031 | England/ALDP-114F7EE/2021 | 2021-01-23 | Europe/United Kingdom/England/ | Lighthouse Lab in Alderley Park | Wellcome Sanger Institute for the COVID-19 Genomics UK (COG-UK) Consortium | Jacquelyn Wynn et al |
| EPI_ISL_948127 | England/QEUH-116EFB4/2021 | 2021-01-26 | Europe/United Kingdom/England/ | Lighthouse Lab in Glasgow | Wellcome Sanger Institute for the COVID-19 Genomics UK (COG-UK) Consortium | Harper VanSteenhouse et al |
| EPI_ISL_948221 | England/CAMC-116408E/2021 | 2021-01-24 | Europe/United Kingdom/England/ | Lighthouse Lab in Cambridge | Wellcome Sanger Institute for the COVID-19 Genomics UK (COG-UK) Consortium | Rob Howes et al |
| EPI_ISL_948353 | Scotland/QEUH-116EA2C/2021 | 2021-01-26 | Europe/United Kingdom/Scotland/ | Lighthouse Lab in Glasgow | Wellcome Sanger Institute for the COVID-19 Genomics UK (COG-UK) Consortium | Harper VanSteenhouse et al |
| EPI_ISL_948447 | England/QEUH-116ECEA/2021 | 2021-01-25 | Europe/United Kingdom/England/ | Lighthouse Lab in Glasgow | Wellcome Sanger Institute for the COVID-19 Genomics UK (COG-UK) Consortium | Harper VanSteenhouse et al |

| | | | | | | |
|---|---|---|---|---|---|---|
| EPI_ISL_948559 | England/ALDP-1128BFE/2021 | 2021-01-22 | Europe/United Kingdom/England/ | Lighthouse Lab in Alderley Park | Wellcome Sanger Institute for the COVID-19 Genomics UK (COG-UK) Consortium | Jacquelyn Wynn et al |
| EPI_ISL_948887 | Scotland/QEUH-114B58E/2021 | 2021-01-25 | Europe/United Kingdom/Scotland/ | Lighthouse Lab in Glasgow | Wellcome Sanger Institute for the COVID-19 Genomics UK (COG-UK) Consortium | Harper VanSteenhouse et al |
| EPI_ISL_948991 | Belgium/Jessa_55-2103-000063/2021 | 2021-01-18 | Europe/Belgium/Tongeren/Alken | Jessa | Jessa | Jessa_cmdLab et al |
| EPI_ISL_949026 | Scotland/QEUH-1014BBA/2021 | 2021-01-15 | Europe/United Kingdom/Scotland/ | Lighthouse Lab in Glasgow | Wellcome Sanger Institute for the COVID-19 Genomics UK (COG-UK) Consortium | Harper VanSteenhouse et al |
| EPI_ISL_949068 | Botswana/AAB15985/2021 | 2021-01-21 | Africa/Botswana/Gaborone/ | Botswana Harvard HIV Reference Laboratory | Botswana Harvard HIV Reference Laboratory | Sikhulile Moyo et al |
| EPI_ISL_949101 | Belgium/Jessa_55-2103-000262/2021 | 2021-01-19 | Europe/Belgium/Hasselt/Genk | Jessa | Jessa | Jessa_cmdLab et al |
| EPI_ISL_949183 | Indonesia/JK-NIHRDMI2100175/2021 | 2021-01-05 | Asia/Indonesia/Jakarta/ | National Institute of Health Research and Development | National Institute of Health Research and Development | Kindi Adam et al |
| EPI_ISL_949184 | Italy/APU-POLBA14/2021 | 2021-01-27 | Europe/Italy/Apulia/ | University of Bari Biomedical Sciences and Human Oncology | University of Bari Biomedical Sciences and Human Oncology | Chironna M. et al |
| EPI_ISL_949185 | Italy/APU-POLBA15/2021 | 2021-01-27 | Europe/Italy/Apulia/ | University of Bari Biomedical Sciences and Human Oncology | University of Bari Biomedical Sciences and Human Oncology | Chironna M. et al |
| EPI_ISL_949188 | Italy/APU-POLBA18/2021 | 2021-01-20 | Europe/Italy/Apulia/ | University of Bari Biomedical Sciences and Human Oncology | University of Bari Biomedical Sciences and Human Oncology | Chironna A. et al |

| | | | | | | |
|---|---|---|---|---|---|---|
| EPI_ISL_949191 | Italy/APU-POLBA19/2021 | 2021-01-27 | Europe/Italy/Apulia/ | University of Bari Biomedical Sciences and Human Oncology | University of Bari Biomedical Sciences and Human Oncology | Chironna M. et al |
| EPI_ISL_949192 | Indonesia/JK-NIHRDMI2100330/2021 | 2021-01-09 | Asia/Indonesia/Jakarta/ | National Institute of Health Research and Development | National Institute of Health Research and Development | Ririn Ramadhany et al |
| EPI_ISL_949193 | Spain/CT-IBV-99012643/2020 | 2020-11-29 | Europe/Spain/Catalunya/Barcelona | Departamento de Microbiolog√≠a, CDB, Hospital Cl√≠nic, Barcelona | SeqCOVID-SPAIN consortium/IBV(CSIC) | Andrea Vergara et al |
| EPI_ISL_949194 | Spain/CT-IBV-99012652/2020 | 2020-09-21 | Europe/Spain/Catalunya/Barcelona | Departamento de Microbiolog√≠a, CDB, Hospital Cl√≠nic, Barcelona | SeqCOVID-SPAIN consortium/IBV(CSIC) | Andrea Vergara et al |
| EPI_ISL_949195 | Spain/CT-IBV-99012628/2020 | 2020-09-02 | Europe/Spain/Catalunya/Mataro | Departamento de Microbiolog√≠a, CDB, Hospital Cl√≠nic, Barcelona | SeqCOVID-SPAIN consortium/IBV(CSIC) | Andrea Vergara et al |
| EPI_ISL_949197 | Spain/CT-IBV-99012675/2020 | 2020-09-12 | Europe/Spain/Catalunya/Barcelona | Departamento de Microbiolog√≠a, CDB, Hospital Cl√≠nic, Barcelona | SeqCOVID-SPAIN consortium/IBV(CSIC) | Andrea Vergara et al |
| EPI_ISL_949198 | Spain/CT-IBV-99012707/2020 | 2020-08-20 | Europe/Spain/Catalunya/Barcelona | Departamento de Microbiolog√≠a, CDB, Hospital Cl√≠nic, Barcelona | SeqCOVID-SPAIN consortium/IBV(CSIC) | Andrea Vergara et al |
| EPI_ISL_949201 | Spain/CT-IBV-99012631/2020 | 2020-11-28 | Europe/Spain/Catalunya/Barcelona | Departamento de Microbiolog√≠a, CDB, Hospital Cl√≠nic, Barcelona | SeqCOVID-SPAIN consortium/IBV(CSIC) | Andrea Vergara et al |
| EPI_ISL_949202 | Spain/CT-IBV-99012708/2020 | 2020-08-05 | Europe/Spain/Canary Islands/Adeje | Departamento de Microbiolog√≠a, CDB, Hospital Cl√≠nic, Barcelona | SeqCOVID-SPAIN consortium/IBV(CSIC) | Andrea Vergara et al |
| EPI_ISL_949203 | Spain/CT-IBV-99012664/2020 | 2020-08-19 | Europe/Spain/Catalunya/Badalona | Departamento de Microbiolog√≠a, CDB, Hospital Cl√≠nic, Barcelona | SeqCOVID-SPAIN consortium/IBV(CSIC) | Andrea Vergara et al |
| EPI_ISL_949206 | Spain/CT-IBV-99012634/2020 | 2020-08-06 | Europe/Spain/Catalunya/Barcelona | Departamento de Microbiolog√≠a, CDB, Hospital Cl√≠nic, Barcelona | SeqCOVID-SPAIN consortium/IBV(CSIC) | Andrea Vergara et al |
| EPI_ISL_949207 | Spain/CT-IBV-99012636/2020 | 2020-08-12 | Europe/Spain/Catalunya/Barcelona | Departamento de Microbiolog√≠a, CDB, Hospital Cl√≠nic, Barcelona | SeqCOVID-SPAIN consortium/IBV(CSIC) | Andrea Vergara et al |
| EPI_ISL_949211 | Spain/CT-IBV-99012645/2020 | 2020-10-25 | Europe/Spain/Catalunya/Barcelona | Departamento de Microbiolog√≠a, CDB, Hospital Cl√≠nic, Barcelona | SeqCOVID-SPAIN consortium/IBV(CSIC) | Andrea Vergara et al |

| | | | | | | |
|---|---|---|---|---|---|---|
| EPI_ISL_949213 | Spain/CT-IBV-99012648/2020 | 2020-08-03 | Europe/Spain/Catalunya/Barcelona | Departamento de Microbiolog√≠a, CDB, Hospital Cl√≠nic, Barcelona | SeqCOVID-SPAIN consortium/IBV(CSIC) | Andrea Vergara et al |
| EPI_ISL_949215 | Spain/CT-IBV-99012658/2020 | 2020-08-30 | Europe/Spain/Catalunya/Barcelona | Departamento de Microbiolog√≠a, CDB, Hospital Cl√≠nic, Barcelona | SeqCOVID-SPAIN consortium/IBV(CSIC) | Andrea Vergara et al |
| EPI_ISL_949217 | Spain/CT-IBV-99012660/2020 | 2020-08-10 | Europe/Spain/Catalunya/Barcelona | Departamento de Microbiolog√≠a, CDB, Hospital Cl√≠nic, Barcelona | SeqCOVID-SPAIN consortium/IBV(CSIC) | Andrea Vergara et al |
| EPI_ISL_949218 | Spain/CT-IBV-99012665/2020 | 2020-09-15 | Europe/Spain/Catalunya/Barcelona | Departamento de Microbiolog√≠a, CDB, Hospital Cl√≠nic, Barcelona | SeqCOVID-SPAIN consortium/IBV(CSIC) | Andrea Vergara et al |
| EPI_ISL_949219 | Spain/CT-IBV-99012666/2020 | 2020-08-01 | Europe/Spain/Catalunya/Barcelona | Departamento de Microbiolog√≠a, CDB, Hospital Cl√≠nic, Barcelona | SeqCOVID-SPAIN consortium/IBV(CSIC) | Andrea Vergara et al |
| EPI_ISL_949220 | Spain/CT-IBV-99012668/2020 | 2020-11-28 | Europe/Spain/Catalunya/Barcelona | Departamento de Microbiolog√≠a, CDB, Hospital Cl√≠nic, Barcelona | SeqCOVID-SPAIN consortium/IBV(CSIC) | Andrea Vergara et al |
| EPI_ISL_949222 | Spain/CT-IBV-99012673/2020 | 2020-09-05 | Europe/Spain/Catalunya/Barcelona | Departamento de Microbiolog√≠a, CDB, Hospital Cl√≠nic, Barcelona | SeqCOVID-SPAIN consortium/IBV(CSIC) | Andrea Vergara et al |
| EPI_ISL_949224 | Spain/CT-IBV-99012689/2020 | 2020-10-31 | Europe/Spain/Catalunya/Barcelona | Departamento de Microbiolog√≠a, CDB, Hospital Cl√≠nic, Barcelona | SeqCOVID-SPAIN consortium/IBV(CSIC) | Andrea Vergara et al |
| EPI_ISL_949225 | Spain/CT-IBV-99012693/2020 | 2020-09-10 | Europe/Spain/Catalunya/Barcelona | Departamento de Microbiolog√≠a, CDB, Hospital Cl√≠nic, Barcelona | SeqCOVID-SPAIN consortium/IBV(CSIC) | Andrea Vergara et al |
| EPI_ISL_949226 | Spain/CT-IBV-99012695/2020 | 2020-08-09 | Europe/Spain/Catalunya/Barcelona | Departamento de Microbiolog√≠a, CDB, Hospital Cl√≠nic, Barcelona | SeqCOVID-SPAIN consortium/IBV(CSIC) | Andrea Vergara et al |
| EPI_ISL_949228 | Spain/CT-IBV-99012698/2020 | 2020-08-24 | Europe/Spain/Catalunya/Barcelona | Departamento de Microbiolog√≠a, CDB, Hospital Cl√≠nic, Barcelona | SeqCOVID-SPAIN consortium/IBV(CSIC) | Andrea Vergara et al |
| EPI_ISL_949231 | Spain/CT-IBV-99012705/2020 | 2020-09-28 | Europe/Spain/Catalunya/Barcelona | Departamento de Microbiolog√≠a, CDB, Hospital Cl√≠nic, Barcelona | SeqCOVID-SPAIN consortium/IBV(CSIC) | Andrea Vergara et al |
| EPI_ISL_949236 | Spain/CT-IBV-99012624/2020 | 2020-08-05 | Europe/Spain/Catalunya/L'Hospitalet de | Departamento de Microbiolog√≠a, CDB, Hospital Cl√≠nic, Barcelona | SeqCOVID-SPAIN consortium/IBV(CSIC) | Andrea Vergara et al |

| | | | | | | |
|---|---|---|---|---|---|---|
| EPI_ISL_949238 | Spain/CT-IBV-99012683/2020 | 2020-08-07 | Europe/Spain/Catalunya/L'Hospitalet de | Departamento de Microbiolog√≠a, CDB, Hospital Cl√≠nic, Barcelona | SeqCOVID-SPAIN consortium/IBV(CSIC) | Andrea Vergara et al |
| EPI_ISL_949239 | Spain/CT-IBV-99012685/2020 | 2020-08-13 | Europe/Spain/Catalunya/L'Hospitalet de | Departamento de Microbiolog√≠a, CDB, Hospital Cl√≠nic, Barcelona | SeqCOVID-SPAIN consortium/IBV(CSIC) | Andrea Vergara et al |
| EPI_ISL_949243 | Spain/CT-IBV-99012676/2020 | 2020-09-16 | Europe/Spain/Comunitat Valenciana/Qu | Departamento de Microbiolog√≠a, CDB, Hospital Cl√≠nic, Barcelona | SeqCOVID-SPAIN consortium/IBV(CSIC) | Andrea Vergara et al |
| EPI_ISL_949249 | Belgium/MBLG41527/2020 | 2020-03-16 | Europe/Belgium/Brussels/ | Cliniques universitaires Saint-Luc | UCLouvain/IREC/MBLG | Jean Ruelle et al |
| EPI_ISL_949332 | England/BIRM-68CD5/2020 | 2020-11-10 | Europe/United Kingdom/England/ | University of Birmingham | COVID-19 Genomics UK (COG-UK) Consortium | Institute of Microbiology et al |
| EPI_ISL_949395 | England/BIRM-6F45E/2020 | 2020-12-08 | Europe/United Kingdom/England/ | University of Birmingham | COVID-19 Genomics UK (COG-UK) Consortium | Institute of Microbiology et al |
| EPI_ISL_949419 | England/BIRM-71565/2021 | 2021-01-16 | Europe/United Kingdom/England/ | University of Birmingham | COVID-19 Genomics UK (COG-UK) Consortium | Institute of Microbiology et al |
| EPI_ISL_949442 | England/CAMB-A9EE21/2021 | 2021-01-19 | Europe/United Kingdom/England/ | Department of Pathology, University of Cambridge | COVID-19 Genomics UK (COG-UK) Consortium | Aminu S. Jahun et al |
| EPI_ISL_949522 | England/CAMB-A9FEF3/2021 | 2021-01-21 | Europe/United Kingdom/England/ | Department of Pathology, University of Cambridge | COVID-19 Genomics UK (COG-UK) Consortium | Aminu S. Jahun et al |
| EPI_ISL_949630 | SouthAfrica/UFS-VIRO-NGS-162/2020 | 2020-08-12 | Africa/South Africa/Free State/ | NHLS Universitas Academic | UFS Virology | PA Bester et al |
| EPI_ISL_949631 | Scotland/CVR6830/2020 | 2020-12-01 | Europe/United Kingdom/Scotland/ | West of Scotland Specialist Virology Centre, NHSGGC / MRC-University of Glasgow Centre for Virus Research | COVID-19 Genomics UK (COG-UK) Consortium | Ana da Silva Filipe et al |
| EPI_ISL_949658 | England/LIVE-DEAC38/2021 | 2021-01-25 | Europe/United Kingdom/England/ | Liverpool Clinical Laboratories | COVID-19 Genomics UK (COG-UK) Consortium | Sam Haldenby et al |
| EPI_ISL_950773 | England/OXON-F4447E/2020 | 2020-12-23 | Europe/United Kingdom/England/ | Oxford Viromics, NDM, University of Oxford; Oxford University Hospitals; Basingstoke and North Hampshire Hospital | COVID-19 Genomics UK (COG-UK) Consortium | Tanya Golubchik et al |
| EPI_ISL_950913 | England/OXON-F469B9/2020 | 2020-12-16 | Europe/United Kingdom/England/ | Oxford Viromics, NDM, University of Oxford; Oxford University Hospitals; Basingstoke and North Hampshire Hospital | COVID-19 Genomics UK (COG-UK) Consortium | Tanya Golubchik et al |

| | | | | | | |
|---|---|---|---|---|---|---|
| EPI_ISL_950968 | England/OXON-F474F3/2020 | 2020-12-28 | Europe/United Kingdom/England/ | Oxford Viromics, NDM, University of Oxford; Oxford University Hospitals; Basingstoke and North Hampshire Hospital | COVID-19 Genomics UK (COG-UK) Consortium | Tanya Golubchik et al |
| EPI_ISL_951052 | England/OXON-F4901B/2020 | 2020-12-31 | Europe/United Kingdom/England/ | Oxford Viromics, NDM, University of Oxford; Oxford University Hospitals; Basingstoke and North Hampshire Hospital | COVID-19 Genomics UK (COG-UK) Consortium | Tanya Golubchik et al |
| EPI_ISL_952956 | England/SHEF-10C65E6/2021 | 2021-02-03 | Europe/United Kingdom/England/ | Virology Department, Sheffield Teaching Hospitals NHS Foundation Trust/Department of Infection, Immunity and Cardiovascular Disease, The Medical School, University of Sheffield | COVID-19 Genomics UK (COG-UK) Consortium | Thushan de Silva et al |
| EPI_ISL_952958 | England/SHEF-10C77B2/2021 | 2021-02-01 | Europe/United Kingdom/England/ | Virology Department, Sheffield Teaching Hospitals NHS Foundation Trust/Department of Infection, Immunity and Cardiovascular Disease, The Medical School, University of Sheffield | COVID-19 Genomics UK (COG-UK) Consortium | Thushan de Silva et al |
| EPI_ISL_952963 | England/SHEF-10C8C67/2021 | 2021-02-03 | Europe/United Kingdom/England/ | Virology Department, Sheffield Teaching Hospitals NHS Foundation Trust/Department of Infection, Immunity and Cardiovascular Disease, The Medical School, University of Sheffield | COVID-19 Genomics UK (COG-UK) Consortium | Thushan de Silva et al |
| EPI_ISL_952966 | England/SHEF-10C9932/2021 | 2021-02-03 | Europe/United Kingdom/England/ | Virology Department, Sheffield Teaching Hospitals NHS Foundation Trust/Department of Infection, Immunity and Cardiovascular Disease, The Medical School, University of Sheffield | COVID-19 Genomics UK (COG-UK) Consortium | Thushan de Silva et al |
| EPI_ISL_952967 | England/SHEF-10C9F4F/2021 | 2021-02-03 | Europe/United Kingdom/England/ | Virology Department, Sheffield Teaching Hospitals NHS Foundation Trust/Department of Infection, Immunity and Cardiovascular Disease, The Medical School, University of Sheffield | COVID-19 Genomics UK (COG-UK) Consortium | Thushan de Silva et al |
| EPI_ISL_952973 | England/SHEF-10CAFF3/2021 | 2021-02-03 | Europe/United Kingdom/England/ | Virology Department, Sheffield Teaching Hospitals NHS Foundation Trust/Department of Infection, Immunity and Cardiovascular Disease, The Medical School, University of Sheffield | COVID-19 Genomics UK (COG-UK) Consortium | Thushan de Silva et al |
| EPI_ISL_952976 | England/SHEF-10CC51D/2021 | 2021-02-02 | Europe/United Kingdom/England/ | Virology Department, Sheffield Teaching Hospitals NHS Foundation Trust/Department of Infection, Immunity and Cardiovascular Disease, The Medical School, University of Sheffield | COVID-19 Genomics UK (COG-UK) Consortium | Thushan de Silva et al |

| | | | | | | |
|---|---|---|---|---|---|---|
| EPI_ISL_952984 | England/SHEF-10CE1CC/2021 | 2021-02-02 | Europe/United Kingdom/England/ | Virology Department, Sheffield Teaching Hospitals NHS Foundation Trust/Department of Infection, Immunity and Cardiovascular Disease, The Medical School, University of Sheffield | COVID-19 Genomics UK (COG-UK) Consortium | Thushan de Silva et al |
| EPI_ISL_952987 | England/SHEF-10CE900/2021 | 2021-02-03 | Europe/United Kingdom/England/ | Virology Department, Sheffield Teaching Hospitals NHS Foundation Trust/Department of Infection, Immunity and Cardiovascular Disease, The Medical School, University of Sheffield | COVID-19 Genomics UK (COG-UK) Consortium | Thushan de Silva et al |
| EPI_ISL_952989 | England/SHEF-10CECCB/2021 | 2021-02-03 | Europe/United Kingdom/England/ | Virology Department, Sheffield Teaching Hospitals NHS Foundation Trust/Department of Infection, Immunity and Cardiovascular Disease, The Medical School, University of Sheffield | COVID-19 Genomics UK (COG-UK) Consortium | Thushan de Silva et al |
| EPI_ISL_952990 | England/SHEF-10CECDA/2021 | 2021-02-01 | Europe/United Kingdom/England/ | Virology Department, Sheffield Teaching Hospitals NHS Foundation Trust/Department of Infection, Immunity and Cardiovascular Disease, The Medical School, University of Sheffield | COVID-19 Genomics UK (COG-UK) Consortium | Thushan de Silva et al |
| EPI_ISL_952992 | England/SHEF-10CF6BD/2021 | 2021-02-03 | Europe/United Kingdom/England/ | Virology Department, Sheffield Teaching Hospitals NHS Foundation Trust/Department of Infection, Immunity and Cardiovascular Disease, The Medical School, University of Sheffield | COVID-19 Genomics UK (COG-UK) Consortium | Thushan de Silva et al |
| EPI_ISL_952995 | England/SHEF-10D02B5/2021 | 2021-02-02 | Europe/United Kingdom/England/ | Virology Department, Sheffield Teaching Hospitals NHS Foundation Trust/Department of Infection, Immunity and Cardiovascular Disease, The Medical School, University of Sheffield | COVID-19 Genomics UK (COG-UK) Consortium | Thushan de Silva et al |
| EPI_ISL_952996 | England/SHEF-10D0543/2021 | 2021-02-03 | Europe/United Kingdom/England/ | Virology Department, Sheffield Teaching Hospitals NHS Foundation Trust/Department of Infection, Immunity and Cardiovascular Disease, The Medical School, University of Sheffield | COVID-19 Genomics UK (COG-UK) Consortium | Thushan de Silva et al |
| EPI_ISL_952999 | England/SHEF-10D0E39/2021 | 2021-02-02 | Europe/United Kingdom/England/ | Virology Department, Sheffield Teaching Hospitals NHS Foundation Trust/Department of Infection, Immunity and Cardiovascular Disease, The Medical School, University of Sheffield | COVID-19 Genomics UK (COG-UK) Consortium | Thushan de Silva et al |

| | | | | | | |
|---|---|---|---|---|---|---|
| EPI_ISL_953000 | England/SHEF-10D14DC/2021 | 2021-02-01 | Europe/United Kingdom/England/ | Virology Department, Sheffield Teaching Hospitals NHS Foundation Trust/Department of Infection, Immunity and Cardiovascular Disease, The Medical School, University of Sheffield | COVID-19 Genomics UK (COG-UK) Consortium | Thushan de Silva et al |
| EPI_ISL_953001 | England/SHEF-10D1BAA/2021 | 2021-02-03 | Europe/United Kingdom/England/ | Virology Department, Sheffield Teaching Hospitals NHS Foundation Trust/Department of Infection, Immunity and Cardiovascular Disease, The Medical School, University of Sheffield | COVID-19 Genomics UK (COG-UK) Consortium | Thushan de Silva et al |
| EPI_ISL_953004 | England/SHEF-10D2602/2021 | 2021-02-01 | Europe/United Kingdom/England/ | Virology Department, Sheffield Teaching Hospitals NHS Foundation Trust/Department of Infection, Immunity and Cardiovascular Disease, The Medical School, University of Sheffield | COVID-19 Genomics UK (COG-UK) Consortium | Thushan de Silva et al |
| EPI_ISL_953007 | England/SHEF-10D380B/2021 | 2021-02-03 | Europe/United Kingdom/England/ | Virology Department, Sheffield Teaching Hospitals NHS Foundation Trust/Department of Infection, Immunity and Cardiovascular Disease, The Medical School, University of Sheffield | COVID-19 Genomics UK (COG-UK) Consortium | Thushan de Silva et al |
| EPI_ISL_953008 | England/SHEF-10D3DDF/2021 | 2021-02-03 | Europe/United Kingdom/England/ | Virology Department, Sheffield Teaching Hospitals NHS Foundation Trust/Department of Infection, Immunity and Cardiovascular Disease, The Medical School, University of Sheffield | COVID-19 Genomics UK (COG-UK) Consortium | Thushan de Silva et al |
| EPI_ISL_953010 | England/SHEF-10D456D/2021 | 2021-02-01 | Europe/United Kingdom/England/ | Virology Department, Sheffield Teaching Hospitals NHS Foundation Trust/Department of Infection, Immunity and Cardiovascular Disease, The Medical School, University of Sheffield | COVID-19 Genomics UK (COG-UK) Consortium | Thushan de Silva et al |
| EPI_ISL_953011 | England/SHEF-10D4AD7/2021 | 2021-02-03 | Europe/United Kingdom/England/ | Virology Department, Sheffield Teaching Hospitals NHS Foundation Trust/Department of Infection, Immunity and Cardiovascular Disease, The Medical School, University of Sheffield | COVID-19 Genomics UK (COG-UK) Consortium | Thushan de Silva et al |
| EPI_ISL_953015 | England/SHEF-10D5308/2021 | 2021-02-01 | Europe/United Kingdom/England/ | Virology Department, Sheffield Teaching Hospitals NHS Foundation Trust/Department of Infection, Immunity and Cardiovascular Disease, The Medical School, University of Sheffield | COVID-19 Genomics UK (COG-UK) Consortium | Thushan de Silva et al |

| | | | | | | |
|---|---|---|---|---|---|---|
| EPI_ISL_953018 | England/SHEF-10D7430/2021 | 2021-02-03 | Europe/United Kingdom/England/ | Virology Department, Sheffield Teaching Hospitals NHS Foundation Trust/Department of Infection, Immunity and Cardiovascular Disease, The Medical School, University of Sheffield | COVID-19 Genomics UK (COG-UK) Consortium | Thushan de Silva et al |
| EPI_ISL_953020 | England/SHEF-10D7BA4/2021 | 2021-02-01 | Europe/United Kingdom/England/ | Virology Department, Sheffield Teaching Hospitals NHS Foundation Trust/Department of Infection, Immunity and Cardiovascular Disease, The Medical School, University of Sheffield | COVID-19 Genomics UK (COG-UK) Consortium | Thushan de Silva et al |
| EPI_ISL_953023 | England/SHEF-10D8D34/2021 | 2021-02-03 | Europe/United Kingdom/England/ | Virology Department, Sheffield Teaching Hospitals NHS Foundation Trust/Department of Infection, Immunity and Cardiovascular Disease, The Medical School, University of Sheffield | COVID-19 Genomics UK (COG-UK) Consortium | Thushan de Silva et al |
| EPI_ISL_953228 | England/TFCI-27040F0/2021 | 2021-01-03 | Europe/United Kingdom/England/ | Bioinformatics and Biostatistics Lab, Advanced Sequencing Facility | COVID-19 Genomics UK (COG-UK) Consortium | Aengus Stewart et al |
| EPI_ISL_953268 | England/TFCI-2704430/2021 | 2021-01-29 | Europe/United Kingdom/England/ | Bioinformatics and Biostatistics Lab, Advanced Sequencing Facility | COVID-19 Genomics UK (COG-UK) Consortium | Aengus Stewart et al |
| EPI_ISL_953302 | England/TFCI-2704676/2020 | 2020-04-09 | Europe/United Kingdom/England/ | Bioinformatics and Biostatistics Lab, Advanced Sequencing Facility | COVID-19 Genomics UK (COG-UK) Consortium | Aengus Stewart et al |
| EPI_ISL_953369 | England/TFCI-2704AB6/2020 | 2020-04-19 | Europe/United Kingdom/England/ | Bioinformatics and Biostatistics Lab, Advanced Sequencing Facility | COVID-19 Genomics UK (COG-UK) Consortium | Aengus Stewart et al |
| EPI_ISL_953376 | Belgium/Jessa_11-2105-003732/2021 | 2021-02-02 | Europe/Belgium/Tongeren/Hoeselt | Jessa | Jessa | Jessa_cmdLab et al |
| EPI_ISL_953377 | Belgium/Jessa_21-2105-001206/2021 | 2021-02-04 | Europe/Belgium/Hasselt/Nieuwerkerke | Jessa | Jessa | Jessa_cmdLab et al |
| EPI_ISL_953378 | Belgium/Jessa_21-2105-001266/2021 | 2021-02-04 | Europe/Belgium/Hasselt/Beringen | Jessa | Jessa | Jessa_cmdLab et al |
| EPI_ISL_953379 | Belgium/Jessa_55-2105-000762/2021 | 2021-02-04 | Europe/Belgium/Hasselt/Diepenbeek | Jessa | Jessa | Jessa_cmdLab et al |
| EPI_ISL_953381 | Belgium/Jessa_55-2105-000780/2021 | 2021-02-03 | Europe/Belgium/Hasselt/ | Jessa | Jessa | Jessa_cmdLab et al |
| EPI_ISL_953382 | Belgium/Jessa_55-2105-000796/2021 | 2021-02-03 | Europe/Belgium/Hasselt/ | Jessa | Jessa | Jessa_cmdLab et al |
| EPI_ISL_953384 | Belgium/Jessa_55-2105-000860/2021 | 2021-02-03 | Europe/Belgium/Hasselt/ | Jessa | Jessa | Jessa_cmdLab et al |

| | | | | | | |
|---|---|---|---|---|---|---|
| EPI_ISL_953387 | Singapore/143/2021 | 2021-02-02 | Asia/Singapore/Singapore/ | National Public Health Laboratory, National Centre for Infectious Diseases | National Public Health Laboratory, National Centre for Infectious Diseases | Tze Minn Mak et al |
| EPI_ISL_953390 | Singapore/147/2021 | 2021-02-03 | Asia/Singapore/Singapore/ | National Public Health Laboratory, National Centre for Infectious Diseases | National Public Health Laboratory, National Centre for Infectious Diseases | Tze Minn Mak et al |
| EPI_ISL_953391 | Singapore/148/2021 | 2021-02-01 | Asia/Singapore/Singapore/ | National Public Health Laboratory, National Centre for Infectious Diseases | National Public Health Laboratory, National Centre for Infectious Diseases | Tze Minn Mak et al |
| EPI_ISL_953394 | Singapore/151/2021 | 2021-02-03 | Asia/Singapore/Singapore/ | National Public Health Laboratory, National Centre for Infectious Diseases | National Public Health Laboratory, National Centre for Infectious Diseases | Tze Minn Mak et al |
| EPI_ISL_953395 | Singapore/152/2021 | 2021-02-02 | Asia/Singapore/Singapore/ | National Public Health Laboratory, National Centre for Infectious Diseases | National Public Health Laboratory, National Centre for Infectious Diseases | Tze Minn Mak et al |
| EPI_ISL_953396 | Singapore/153/2021 | 2021-02-02 | Asia/Singapore/Singapore/ | National Public Health Laboratory, National Centre for Infectious Diseases | National Public Health Laboratory, National Centre for Infectious Diseases | Tze Minn Mak et al |

| | | | | | | |
|---|---|---|---|---|---|---|
| EPI_ISL_953397 | Singapore/154/2021 | 2021-02-01 | Asia/Singapore/Singapore/ | National Public Health Laboratory, National Centre for Infectious Diseases | National Public Health Laboratory, National Centre for Infectious Diseases | Tze Minn Mak et al |
| EPI_ISL_953398 | Singapore/155/2021 | 2021-02-02 | Asia/Singapore/Singapore/ | National Public Health Laboratory, National Centre for Infectious Diseases | National Public Health Laboratory, National Centre for Infectious Diseases | Tze Minn Mak et al |
| EPI_ISL_953399 | Singapore/156/2021 | 2021-02-01 | Asia/Singapore/Singapore/ | National Public Health Laboratory, National Centre for Infectious Diseases | National Public Health Laboratory, National Centre for Infectious Diseases | Tze Minn Mak et al |
| EPI_ISL_953400 | Singapore/157/2021 | 2021-02-01 | Asia/Singapore/Singapore/ | National Public Health Laboratory, National Centre for Infectious Diseases | National Public Health Laboratory, National Centre for Infectious Diseases | Tze Minn Mak et al |
| EPI_ISL_953401 | Singapore/158/2021 | 2021-02-01 | Asia/Singapore/Singapore/ | National Public Health Laboratory, National Centre for Infectious Diseases | National Public Health Laboratory, National Centre for Infectious Diseases | Tze Minn Mak et al |
| EPI_ISL_953408 | EquatorialGuinea/32158/2020 | 2020-07-06 | Africa/Equatorial Guinea/Equatorial Gui | Laboratorio de Investigaciones de Baney | """Swiss Tropical and Public Health Institute""" | """Carlos Cortes et al" |
| EPI_ISL_953427 | Indonesia/JK-NIHRD-MI210121/2021 | 2021-01-03 | Asia/Indonesia/Jakarta/ | National Institute of Health Research and Development | National Institute of Health Research and Development | Subangkit et al |
| EPI_ISL_953433 | Belgium/Jessa_11-2105-001969/2021 | 2021-02-02 | Europe/Belgium/Tongeren/Bilzen | Jessa | Jessa | Jessa_cmdLab et al |
| EPI_ISL_953435 | Belgium/Jessa_55-2105-000419/2021 | 2021-02-02 | Europe/Belgium/Hasselt/Zonhoven | Jessa | Jessa | Jessa_cmdLab et al |
| EPI_ISL_953436 | Belgium/Jessa_11-2105-001977/2021 | 2021-02-02 | Europe/Belgium/Tongeren/Lanaken | Jessa | Jessa | Jessa_cmdLab et al |

| | | | | | | |
|---|---|---|---|---|---|---|
| EPI_ISL_953438 | Belgium/Jessa_11-2105-000470/2021 | 2021-02-01 | Europe/Belgium/Hasselt/Beringen | Jessa | Jessa | Jessa_cmdLab et al |
| EPI_ISL_953439 | Belgium/Jessa_11-2105-000692/2021 | 2021-02-01 | Europe/Belgium/Hasselt/Heusden-Zolde | Jessa | Jessa | Jessa_cmdLab et al |
| EPI_ISL_953440 | Belgium/Jessa_11-2105-000694/2021 | 2021-02-01 | Europe/Belgium/Hasselt/Heusden-Zolde | Jessa | Jessa | Jessa_cmdLab et al |
| EPI_ISL_953441 | Belgium/Jessa_11-2105-001378/2021 | 2021-02-01 | Europe/Belgium/Hasselt/Herk-de-Stad | Jessa | Jessa | Jessa_cmdLab et al |
| EPI_ISL_953442 | Belgium/Jessa_11-2105-001473/2021 | 2021-02-01 | Europe/Belgium/Hasselt/ | Jessa | Jessa | Jessa_cmdLab et al |
| EPI_ISL_953443 | Belgium/Jessa_11-2105-001921/2021 | 2021-02-01 | Europe/Belgium/Tongeren/Bilzen | Jessa | Jessa | Jessa_cmdLab et al |
| EPI_ISL_953444 | Belgium/Jessa_11-2105-002964/2021 | 2021-02-02 | Europe/Belgium/Hasselt/ | Jessa | Jessa | Jessa_cmdLab et al |
| EPI_ISL_953446 | Belgium/Jessa_55-2105-000186/2021 | 2021-02-01 | Europe/Belgium/Hasselt/ | Jessa | Jessa | Jessa_cmdLab et al |
| EPI_ISL_953447 | Belgium/Jessa_55-2105-000205/2021 | 2021-02-01 | Europe/Belgium/Hasselt/ | Jessa | Jessa | Jessa_cmdLab et al |
| EPI_ISL_953448 | Belgium/Jessa_55-2105-000321/2021 | 2021-02-01 | Europe/Belgium/Hasselt/ | Jessa | Jessa | Jessa_cmdLab et al |
| EPI_ISL_953474 | Belgium/Jessa_11-2105-001231/2021 | 2021-02-01 | Europe/Belgium/Tongeren/ | Jessa | Jessa | Jessa_cmdLab et al |
| EPI_ISL_953487 | Belgium/Jessa_55-2105-000088/2021 | 2021-02-01 | Europe/Belgium/Hasselt/ | Jessa | Jessa | Jessa_cmdLab et al |
| EPI_ISL_953488 | Belgium/Jessa_55-2105-000093/2021 | 2021-02-01 | Europe/Belgium/Hasselt/ | Jessa | Jessa | Jessa_cmdLab et al |
| EPI_ISL_953532 | Switzerland/un-33038982/2021 | 2021-01-21 | Europe/Switzerland/Switzerland/ | University Hospitals of Geneva, Laboratory of Virology | HUG, Laboratory of Virology and the Health2030 Genome Center | Samuel Cordey et al |
| EPI_ISL_953543 | Switzerland/VS-33084523/2021 | 2021-01-22 | Europe/Switzerland/Valais/ | University Hospitals of Geneva, Laboratory of Virology | HUG, Laboratory of Virology and the Health2030 Genome Center | Samuel Cordey et al |
| EPI_ISL_953569 | Switzerland/GE-33076286/2021 | 2021-01-26 | Europe/Switzerland/Geneva/ | University Hospitals of Geneva, Laboratory of Virology | HUG, Laboratory of Virology and the Health2030 Genome Center | Samuel Cordey et al |
| EPI_ISL_953576 | Switzerland/GE-33089010/2021 | 2021-01-27 | Europe/Switzerland/Geneva/ | University Hospitals of Geneva, Laboratory of Virology | HUG, Laboratory of Virology and the Health2030 Genome Center | Samuel Cordey et al |
| EPI_ISL_953648 | Switzerland/GE-33094768/2021 | 2021-01-27 | Europe/Switzerland/Geneva/ | University Hospitals of Geneva, Laboratory of Virology | HUG, Laboratory of Virology and the Health2030 Genome Center | Samuel Cordey et al |

| | | | | | | |
|---|---|---|---|---|---|---|
| EPI_ISL_953670 | Switzerland/VS-33019363/2021 | 2021-01-17 | Europe/Switzerland/Valais/ | University Hospitals of Geneva, Laboratory of Virology | HUG, Laboratory of Virology and the Health2030 Genome Center | Samuel Cordey et al |
| EPI_ISL_953701 | Switzerland/VS-33031820/2021 | 2021-01-19 | Europe/Switzerland/Valais/ | University Hospitals of Geneva, Laboratory of Virology | HUG, Laboratory of Virology and the Health2030 Genome Center | Samuel Cordey et al |
| EPI_ISL_953708 | Switzerland/GE-33025039/2021 | 2021-01-20 | Europe/Switzerland/Geneva/ | University Hospitals of Geneva, Laboratory of Virology | HUG, Laboratory of Virology and the Health2030 Genome Center | Samuel Cordey et al |
| EPI_ISL_953813 | Switzerland/GE-33053436/2021 | 2021-01-23 | Europe/Switzerland/Geneva/ | University Hospitals of Geneva, Laboratory of Virology | HUG, Laboratory of Virology and the Health2030 Genome Center | Samuel Cordey et al |
| EPI_ISL_953871 | Switzerland/GE-33070475/2021 | 2021-01-25 | Europe/Switzerland/Geneva/ | University Hospitals of Geneva, Laboratory of Virology | HUG, Laboratory of Virology and the Health2030 Genome Center | Samuel Cordey et al |
| EPI_ISL_953879 | Switzerland/GE-33080591/2021 | 2021-01-25 | Europe/Switzerland/Geneva/ | University Hospitals of Geneva, Laboratory of Virology | HUG, Laboratory of Virology and the Health2030 Genome Center | Samuel Cordey et al |
| EPI_ISL_953940 | France/BRE-IPP01853/2021 | 2021-01-14 | Europe/France/Bretagne/Rennes | Hopital | National Reference Center for Viruses of Respiratory Infections, Institut Pasteur, Paris | Marion Barbet et al |
| EPI_ISL_953950 | France/BRE-IPP01800/2021 | 2021-01-17 | Europe/France/Bretagne/Vannes | Hopital | National Reference Center for Viruses of Respiratory Infections, Institut Pasteur, Paris | Marion Barbet et al |

| | | | | | | |
|---|---|---|---|---|---|---|
| EPI_ISL_953957 | France/BRE-IPP01855/2021 | 2021-01-15 | Europe/France/Bretagne/Rennes | Hopital | National Reference Center for Viruses of Respiratory Infections, Institut Pasteur, Paris | Marion Barbet et al |
| EPI_ISL_953964 | France/IDF-IPP01891/2021 | 2021-01-23 | Europe/France/Ile de France/Neuilly sur | Hopital | National Reference Center for Viruses of Respiratory Infections, Institut Pasteur, Paris | Marion Barbet et al |
| EPI_ISL_953966 | France/IDF-IPP01680/2021 | 2021-01-22 | Europe/France/Ile de France/Neuilly sur | Hopital | National Reference Center for Viruses of Respiratory Infections, Institut Pasteur, Paris | Marion Barbet et al |
| EPI_ISL_953968 | France/NOR-IPP01726/2021 | 2021-01-22 | Europe/France/Normandie/Rouen | Labo Analyses Med | National Reference Center for Viruses of Respiratory Infections, Institut Pasteur, Paris | Marion Barbet et al |
| EPI_ISL_953969 | France/IDF-IPP01754/2021 | 2021-01-22 | Europe/France/Ile de France/Le Chesnay | Hopital | National Reference Center for Viruses of Respiratory Infections, Institut Pasteur, Paris | Marion Barbet et al |
| EPI_ISL_953971 | France/PDL-IPP01746/2021 | 2021-01-21 | Europe/France/Pays de la Loire/Cholet | Hopital | National Reference Center for Viruses of Respiratory Infections, Institut Pasteur, Paris | Marion Barbet et al |

| | | | | | | |
|---|---|---|---|---|---|---|
| EPI_ISL_953972 | France/GES-IPP01771/2021 | 2021-01-21 | Europe/France/Grand Est/Troyes | Hopital | National Reference Center for Viruses of Respiratory Infections, Institut Pasteur, Paris | Marion Barbet et al |
| EPI_ISL_953979 | France/PDL-IPP01743/2021 | 2021-01-20 | Europe/France/Pays de la Loire/Cholet | Hopital | National Reference Center for Viruses of Respiratory Infections, Institut Pasteur, Paris | Marion Barbet et al |
| EPI_ISL_953987 | France/BRE-IPP01842/2021 | 2021-01-19 | Europe/France/Bretagne/Rennes | Hopital | National Reference Center for Viruses of Respiratory Infections, Institut Pasteur, Paris | Marion Barbet et al |
| EPI_ISL_954034 | France/IDF-IPP01861/2021 | 2021-01-20 | Europe/France/Ile de France/Paris | Recherche | National Reference Center for Viruses of Respiratory Infections, Institut Pasteur, Paris | Marion Barbet et al |
| EPI_ISL_954049 | France/NOR-IPP01716/2021 | 2021-01-22 | Europe/France/Normandie/Caen | Labo Analyses Med | National Reference Center for Viruses of Respiratory Infections, Institut Pasteur, Paris | Marion Barbet et al |
| EPI_ISL_954054 | France/CVL-IPP01887/2021 | 2021-01-22 | Europe/France/Centre-Val de Loire/Cha | Labo Analyses Med | National Reference Center for Viruses of Respiratory Infections, Institut Pasteur, Paris | Marion Barbet et al |

| | | | | | | |
|---|---|---|---|---|---|---|
| EPI_ISL_954056 | FrenchGuiana/IPP01946/2021 | 2021-01-22 | Europe/France/French Guiana/Cayenne | Outre Mer | National Reference Center for Viruses of Respiratory Infections, Institut Pasteur, Paris | Marion Barbet et al |
| EPI_ISL_954060 | France/CVL-IPP01718/2021 | 2021-01-21 | Europe/France/Centre-Val de Loire/Tou | Labo Analyses Med | National Reference Center for Viruses of Respiratory Infections, Institut Pasteur, Paris | Marion Barbet et al |
| EPI_ISL_954061 | France/CVL-IPP01719/2021 | 2021-01-21 | Europe/France/Centre-Val de Loire/Tou | Labo Analyses Med | National Reference Center for Viruses of Respiratory Infections, Institut Pasteur, Paris | Marion Barbet et al |
| EPI_ISL_954064 | France/CVL-IPP01722/2021 | 2021-01-21 | Europe/France/Centre-Val de Loire/Tou | Labo Analyses Med | National Reference Center for Viruses of Respiratory Infections, Institut Pasteur, Paris | Marion Barbet et al |
| EPI_ISL_954068 | France/GES-IPP01747/2021 | 2021-01-21 | Europe/France/Grand Est/Charleville-M | Labo Analyses Med | National Reference Center for Viruses of Respiratory Infections, Institut Pasteur, Paris | Marion Barbet et al |
| EPI_ISL_954082 | France/IDF-IPP01862/2021 | 2021-01-20 | Europe/France/Ile de France/Paris | Recherche | National Reference Center for Viruses of Respiratory Infections, Institut Pasteur, Paris | Marion Barbet et al |

| | | | | | | |
|---|---|---|---|---|---|---|
| EPI_ISL_954113 | France/BRE-IPP01838/2021 | 2021-01-16 | Europe/France/Bretagne/Rennes | Hopital | National Reference Center for Viruses of Respiratory Infections, Institut Pasteur, Paris | Marion Barbet et al |
| EPI_ISL_954124 | France/PDL-IPP01777/2021 | 2021-01-12 | Europe/France/Pays de la Loire/La Roch | Labo Analyses Med | National Reference Center for Viruses of Respiratory Infections, Institut Pasteur, Paris | Marion Barbet et al |
| EPI_ISL_954125 | FrenchGuiana/IPP01948/2021 | 2021-01-12 | Europe/France/French Guiana/Cayenne | Outre Mer | National Reference Center for Viruses of Respiratory Infections, Institut Pasteur, Paris | Marion Barbet et al |
| EPI_ISL_954141 | NorthMacedonia/29206/2020 | 2020-12-23 | Europe/North Macedonia/Skopje/ | Gerontology Institute - Skopje | "Research Center for Genetic Engineering and Biotechnology ""Georgi D. Efremov‚Äù , Macedonian Academy of Sciences and Arts" | Aleksandar J. Dimovski et al |
| EPI_ISL_954153 | Spain/CT-HUVH-81112/2021 | 2021-01-20 | Europe/Spain/Catalunya/ | Hospital Universitari Vall d'Hebron - Vall d'Hebron Institut de Rercerca | Hospital Universitari Vall d'Hebron | Cristina Andr√©s et al |
| EPI_ISL_954161 | Spain/CT-HUVH-76623/2021 | 2021-01-19 | Europe/Spain/Catalunya/ | Hospital Universitari Vall d'Hebron - Vall d'Hebron Institut de Rercerca | Hospital Universitari Vall d'Hebron | Cristina Andr√©s et al |
| EPI_ISL_954163 | Spain/CT-HUVH-78615/2021 | 2021-01-18 | Europe/Spain/Catalunya/ | Hospital Universitari Vall d'Hebron - Vall d'Hebron Institut de Rercerca | Hospital Universitari Vall d'Hebron | Cristina Andr√©s et al |
| EPI_ISL_954165 | Spain/CT-HUVH-32938/2021 | 2021-01-14 | Europe/Spain/Catalunya/ | Hospital Universitari Vall d'Hebron - Vall d'Hebron Institut de Rercerca | Hospital Universitari Vall d'Hebron | Cristina Andr√©s et al |

| | | | | | | |
|---|---|---|---|---|---|---|
| EPI_ISL_954166 | Spain/CT-HUVH-56385/2021 | 2021-01-14 | Europe/Spain/Catalunya/ | Hospital Universitari Vall d'Hebron - Vall d'Hebron Institut de Rercerca | Hospital Universitari Vall d'Hebron | Cristina Andr√©s et al |
| EPI_ISL_954178 | Spain/CT-HUVH-18665/2021 | 2021-01-12 | Europe/Spain/Catalunya/ | Hospital Universitari Vall d'Hebron - Vall d'Hebron Institut de Rercerca | Hospital Universitari Vall d'Hebron | Cristina Andr√©s et al |
| EPI_ISL_954179 | Spain/CT-HUVH-51778/2021 | 2021-01-22 | Europe/Spain/Catalunya/ | Hospital Universitari Vall d'Hebron - Vall d'Hebron Institut de Rercerca | Hospital Universitari Vall d'Hebron | Cristina Andr√©s et al |
| EPI_ISL_954180 | Spain/CT-HUVH-23956/2021 | 2021-01-25 | Europe/Spain/Catalunya/ | Hospital Universitari Vall d'Hebron - Vall d'Hebron Institut de Rercerca | Hospital Universitari Vall d'Hebron | Cristina Andr√©s et al |
| EPI_ISL_954181 | Spain/CT-HUVH-26007/2021 | 2021-01-20 | Europe/Spain/Catalunya/ | Hospital Universitari Vall d'Hebron - Vall d'Hebron Institut de Rercerca | Hospital Universitari Vall d'Hebron | Cristina Andr√©s et al |
| EPI_ISL_954184 | Italy/CAM-CRGS-293/2020 | 2020-10-11 | Europe/Italy/Campania/ | 1.AO Universitaria 'S. Giovanni di Dio e Ruggi D'Aragona, Scuola Medica Salernitana' Hospital / 2.UOC di Virologia e Microbiologia, Universit√† della Campania 'L. Vanvitelli' / 3.AO Universitaria 'Federico II' Napoli Hospital / 4.AORN 'San Giuseppe Moscati' Avellino Hospital / 5.AO 'San Pio - presidio G. Rummo' Benevento Hospital / 6.AO 'Sant'Anna e San Sebastiano' Caserta Hospital / 7.PO 'Maria Santissima Addolorata' Eboli Hospital / 8.Biogem Istituto di Ricerche Genetiche | 1. Genome Research Center for Health (CRGS) / 2. Laboratory of Molecular Medicine and Genomics(LMMGe) / 3. Center for Research in Pure and Applied Mathematics (CRMPA) | Giorgio Giurato et al |
| EPI_ISL_954189 | Italy/CAM-CRGS-348/2020 | 2020-11-01 | Europe/Italy/Campania/ | 1.AO Universitaria 'S. Giovanni di Dio e Ruggi D'Aragona, Scuola Medica Salernitana' Hospital / 2.UOC di Virologia e Microbiologia, Universit√† della Campania 'L. Vanvitelli' / 3.AO Universitaria 'Federico II' Napoli Hospital / 4.AORN 'San Giuseppe Moscati' Avellino Hospital / 5.AO 'San Pio - presidio G. Rummo' Benevento Hospital / 6.AO 'Sant'Anna e San Sebastiano' Caserta Hospital / 7.PO 'Maria Santissima Addolorata' Eboli Hospital / 8.Biogem Istituto di Ricerche Genetiche | 1. Genome Research Center for Health (CRGS) / 2. Laboratory of Molecular Medicine and Genomics(LMMGe) / 3. Center for Research in Pure and Applied Mathematics (CRMPA) | Giorgio Giurato et al |

| | | | | | | |
|---|---|---|---|---|---|---|
| EPI_ISL_954190 | Italy/CAM-CRGS-367/2020 | 2020-11-03 | Europe/Italy/Campania/ | 1.AO Universitaria 'S. Giovanni di Dio e Ruggi D'Aragona, Scuola Medica Salernitana' Hospital / 2.UOC di Virologia e Microbiologia, Universit√† della Campania 'L. Vanvitelli' / 3.AO Universitaria 'Federico II' Napoli Hospital / 4.AORN 'San Giuseppe Moscati' Avellino Hospital / 5.AO 'San Pio - presidio G. Rummo' Benevento Hospital / 6.AO 'Sant'Anna e San Sebastiano' Caserta Hospital / 7.PO 'Maria Santissima Addolorata' Eboli Hospital / 8.Biogem Istituto di Ricerche Genetiche | 1. Genome Research Center for Health (CRGS) / 2. Laboratory of Molecular Medicine and Genomics(LMMGe) / 3. Center for Research in Pure and Applied Mathematics (CRMPA) | Giorgio Giurato et al |
| EPI_ISL_954191 | Italy/CAM-CRGS-342/2020 | 2020-11-04 | Europe/Italy/Campania/ | 1.AO Universitaria 'S. Giovanni di Dio e Ruggi D'Aragona, Scuola Medica Salernitana' Hospital / 2.UOC di Virologia e Microbiologia, Universit√† della Campania 'L. Vanvitelli' / 3.AO Universitaria 'Federico II' Napoli Hospital / 4.AORN 'San Giuseppe Moscati' Avellino Hospital / 5.AO 'San Pio - presidio G. Rummo' Benevento Hospital / 6.AO 'Sant'Anna e San Sebastiano' Caserta Hospital / 7.PO 'Maria Santissima Addolorata' Eboli Hospital / 8.Biogem Istituto di Ricerche Genetiche | 1. Genome Research Center for Health (CRGS) / 2. Laboratory of Molecular Medicine and Genomics(LMMGe) / 3. Center for Research in Pure and Applied Mathematics (CRMPA) | Giorgio Giurato et al |
| EPI_ISL_954192 | Italy/CAM-CRGS-290/2020 | 2020-11-13 | Europe/Italy/Campania/ | 1.AO Universitaria 'S. Giovanni di Dio e Ruggi D'Aragona, Scuola Medica Salernitana' Hospital / 2.UOC di Virologia e Microbiologia, Universit√† della Campania 'L. Vanvitelli' / 3.AO Universitaria 'Federico II' Napoli Hospital / 4.AORN 'San Giuseppe Moscati' Avellino Hospital / 5.AO 'San Pio - presidio G. Rummo' Benevento Hospital / 6.AO 'Sant'Anna e San Sebastiano' Caserta Hospital / 7.PO 'Maria Santissima Addolorata' Eboli Hospital / 8.Biogem Istituto di Ricerche Genetiche | 1. Genome Research Center for Health (CRGS) / 2. Laboratory of Molecular Medicine and Genomics(LMMGe) / 3. Center for Research in Pure and Applied Mathematics (CRMPA) | Giorgio Giurato et al |

| | | | | | | |
|---|---|---|---|---|---|---|
| EPI_ISL_954203 | Italy/CAM-CRGS-337/2020 | 2020-12-23 | Europe/Italy/Campania/ | 1.AO Universitaria 'S. Giovanni di Dio e Ruggi D'Aragona, Scuola Medica Salernitana' Hospital / 2.UOC di Virologia e Microbiologia, Universit√† della Campania 'L. Vanvitelli' / 3.AO Universitaria 'Federico II' Napoli Hospital / 4.AORN 'San Giuseppe Moscati' Avellino Hospital / 5.AO 'San Pio - presidio G. Rummo' Benevento Hospital / 6.AO 'Sant'Anna e San Sebastiano' Caserta Hospital / 7.PO 'Maria Santissima Addolorata' Eboli Hospital / 8.Biogem Istituto di Ricerche Genetiche | 1. Genome Research Center for Health (CRGS) / 2. Laboratory of Molecular Medicine and Genomics(LMMGe) / 3. Center for Research in Pure and Applied Mathematics (CRMPA) | Giorgio Giurato et al |
| EPI_ISL_954206 | Italy/CAM-CRGS-361/2020 | 2020-12-23 | Europe/Italy/Campania/ | 1.AO Universitaria 'S. Giovanni di Dio e Ruggi D'Aragona, Scuola Medica Salernitana' Hospital / 2.UOC di Virologia e Microbiologia, Universit√† della Campania 'L. Vanvitelli' / 3.AO Universitaria 'Federico II' Napoli Hospital / 4.AORN 'San Giuseppe Moscati' Avellino Hospital / 5.AO 'San Pio - presidio G. Rummo' Benevento Hospital / 6.AO 'Sant'Anna e San Sebastiano' Caserta Hospital / 7.PO 'Maria Santissima Addolorata' Eboli Hospital / 8.Biogem Istituto di Ricerche Genetiche | 1. Genome Research Center for Health (CRGS) / 2. Laboratory of Molecular Medicine and Genomics(LMMGe) / 3. Center for Research in Pure and Applied Mathematics (CRMPA) | Giorgio Giurato et al |
| EPI_ISL_954211 | Italy/CAM-CRGS-317/2020 | 2020-12-29 | Europe/Italy/Campania/ | 1.AO Universitaria 'S. Giovanni di Dio e Ruggi D'Aragona, Scuola Medica Salernitana' Hospital / 2.UOC di Virologia e Microbiologia, Universit√† della Campania 'L. Vanvitelli' / 3.AO Universitaria 'Federico II' Napoli Hospital / 4.AORN 'San Giuseppe Moscati' Avellino Hospital / 5.AO 'San Pio - presidio G. Rummo' Benevento Hospital / 6.AO 'Sant'Anna e San Sebastiano' Caserta Hospital / 7.PO 'Maria Santissima Addolorata' Eboli Hospital / 8.Biogem Istituto di Ricerche Genetiche | 1. Genome Research Center for Health (CRGS) / 2. Laboratory of Molecular Medicine and Genomics(LMMGe) / 3. Center for Research in Pure and Applied Mathematics (CRMPA) | Giorgio Giurato et al |

| | | | | | | |
|---|---|---|---|---|---|---|
| EPI_ISL_954218 | Italy/CAM-CRGS-295/2021 | 2021-01-02 | Europe/Italy/Campania/ | 1.AO Universitaria 'S. Giovanni di Dio e Ruggi D'Aragona, Scuola Medica Salernitana' Hospital / 2.UOC di Virologia e Microbiologia, Universit√† della Campania 'L. Vanvitelli' / 3.AO Universitaria 'Federico II' Napoli Hospital / 4.AORN 'San Giuseppe Moscati' Avellino Hospital / 5.AO 'San Pio - presidio G. Rummo' Benevento Hospital / 6.AO 'Sant'Anna e San Sebastiano' Caserta Hospital / 7.PO 'Maria Santissima Addolorata' Eboli Hospital / 8.Biogem Istituto di Ricerche Genetiche | 1. Genome Research Center for Health (CRGS) / 2. Laboratory of Molecular Medicine and Genomics(LMMGe) / 3. Center for Research in Pure and Applied Mathematics (CRMPA) | Giorgio Giurato et al |
| EPI_ISL_954220 | Italy/CAM-CRGS-346/2021 | 2021-01-06 | Europe/Italy/Campania/ | 1.AO Universitaria 'S. Giovanni di Dio e Ruggi D'Aragona, Scuola Medica Salernitana' Hospital / 2.UOC di Virologia e Microbiologia, Universit√† della Campania 'L. Vanvitelli' / 3.AO Universitaria 'Federico II' Napoli Hospital / 4.AORN 'San Giuseppe Moscati' Avellino Hospital / 5.AO 'San Pio - presidio G. Rummo' Benevento Hospital / 6.AO 'Sant'Anna e San Sebastiano' Caserta Hospital / 7.PO 'Maria Santissima Addolorata' Eboli Hospital / 8.Biogem Istituto di Ricerche Genetiche | 1. Genome Research Center for Health (CRGS) / 2. Laboratory of Molecular Medicine and Genomics(LMMGe) / 3. Center for Research in Pure and Applied Mathematics (CRMPA) | Giorgio Giurato et al |
| EPI_ISL_954221 | Italy/CAM-CRGS-347/2021 | 2021-01-06 | Europe/Italy/Campania/ | 1.AO Universitaria 'S. Giovanni di Dio e Ruggi D'Aragona, Scuola Medica Salernitana' Hospital / 2.UOC di Virologia e Microbiologia, Universit√† della Campania 'L. Vanvitelli' / 3.AO Universitaria 'Federico II' Napoli Hospital / 4.AORN 'San Giuseppe Moscati' Avellino Hospital / 5.AO 'San Pio - presidio G. Rummo' Benevento Hospital / 6.AO 'Sant'Anna e San Sebastiano' Caserta Hospital / 7.PO 'Maria Santissima Addolorata' Eboli Hospital / 8.Biogem Istituto di Ricerche Genetiche | 1. Genome Research Center for Health (CRGS) / 2. Laboratory of Molecular Medicine and Genomics(LMMGe) / 3. Center for Research in Pure and Applied Mathematics (CRMPA) | Giorgio Giurato et al |

| | | | | | | |
|---|---|---|---|---|---|---|
| EPI_ISL_954222 | Italy/CAM-CRGS-309/2021 | 2021-01-07 | Europe/Italy/Campania/ | 1.AO Universitaria 'S. Giovanni di Dio e Ruggi D'Aragona, Scuola Medica Salernitana' Hospital / 2.UOC di Virologia e Microbiologia, Universit√† della Campania 'L. Vanvitelli' / 3.AO Universitaria 'Federico II' Napoli Hospital / 4.AORN 'San Giuseppe Moscati' Avellino Hospital / 5.AO 'San Pio - presidio G. Rummo' Benevento Hospital / 6.AO 'Sant'Anna e San Sebastiano' Caserta Hospital / 7.PO 'Maria Santissima Addolorata' Eboli Hospital / 8.Biogem Istituto di Ricerche Genetiche | 1. Genome Research Center for Health (CRGS) / 2. Laboratory of Molecular Medicine and Genomics(LMMGe) / 3. Center for Research in Pure and Applied Mathematics (CRMPA) | Giorgio Giurato et al |
| EPI_ISL_954223 | Italy/CAM-CRGS-310/2021 | 2021-01-08 | Europe/Italy/Campania/ | 1.AO Universitaria 'S. Giovanni di Dio e Ruggi D'Aragona, Scuola Medica Salernitana' Hospital / 2.UOC di Virologia e Microbiologia, Universit√† della Campania 'L. Vanvitelli' / 3.AO Universitaria 'Federico II' Napoli Hospital / 4.AORN 'San Giuseppe Moscati' Avellino Hospital / 5.AO 'San Pio - presidio G. Rummo' Benevento Hospital / 6.AO 'Sant'Anna e San Sebastiano' Caserta Hospital / 7.PO 'Maria Santissima Addolorata' Eboli Hospital / 8.Biogem Istituto di Ricerche Genetiche | 1. Genome Research Center for Health (CRGS) / 2. Laboratory of Molecular Medicine and Genomics(LMMGe) / 3. Center for Research in Pure and Applied Mathematics (CRMPA) | Giorgio Giurato et al |
| EPI_ISL_954224 | Italy/CAM-CRGS-311/2021 | 2021-01-09 | Europe/Italy/Campania/ | 1.AO Universitaria 'S. Giovanni di Dio e Ruggi D'Aragona, Scuola Medica Salernitana' Hospital / 2.UOC di Virologia e Microbiologia, Universit√† della Campania 'L. Vanvitelli' / 3.AO Universitaria 'Federico II' Napoli Hospital / 4.AORN 'San Giuseppe Moscati' Avellino Hospital / 5.AO 'San Pio - presidio G. Rummo' Benevento Hospital / 6.AO 'Sant'Anna e San Sebastiano' Caserta Hospital / 7.PO 'Maria Santissima Addolorata' Eboli Hospital / 8.Biogem Istituto di Ricerche Genetiche | 1. Genome Research Center for Health (CRGS) / 2. Laboratory of Molecular Medicine and Genomics(LMMGe) / 3. Center for Research in Pure and Applied Mathematics (CRMPA) | Giorgio Giurato et al |

| | | | | | | |
|---|---|---|---|---|---|---|
| EPI_ISL_954225 | Italy/CAM-CRGS-312/2021 | 2021-01-10 | Europe/Italy/Campania/ | 1.AO Universitaria 'S. Giovanni di Dio e Ruggi D'Aragona, Scuola Medica Salernitana' Hospital / 2.UOC di Virologia e Microbiologia, Universit√† della Campania 'L. Vanvitelli' / 3.AO Universitaria 'Federico II' Napoli Hospital / 4.AORN 'San Giuseppe Moscati' Avellino Hospital / 5.AO 'San Pio - presidio G. Rummo' Benevento Hospital / 6.AO 'Sant'Anna e San Sebastiano' Caserta Hospital / 7.PO 'Maria Santissima Addolorata' Eboli Hospital / 8.Biogem Istituto di Ricerche Genetiche | 1. Genome Research Center for Health (CRGS) / 2. Laboratory of Molecular Medicine and Genomics(LMMGe) / 3. Center for Research in Pure and Applied Mathematics (CRMPA) | Giorgio Giurato et al |
| EPI_ISL_954226 | Uganda/UG205/2021 | 2021-01-02 | Africa/Uganda/Uganda/ | MRC/UVRI & LSHTM Uganda Research Unit | Where sequence data have been generated and submitted to GISAID | Matthew Cotten et al |
| EPI_ISL_954242 | Uganda/UG139/2020 | 2020-09-01 | Africa/Uganda/Uganda/ | MRC/UVRI & LSHTM Uganda Research Unit | Where sequence data have been generated and submitted to GISAID | Matthew Cotten et al |
| EPI_ISL_954247 | Uganda/UG144/2020 | 2020-09-01 | Africa/Uganda/Uganda/ | MRC/UVRI & LSHTM Uganda Research Unit | Where sequence data have been generated and submitted to GISAID | Matthew Cotten et al |
| EPI_ISL_954252 | Uganda/UG149/2020 | 2020-09-01 | Africa/Uganda/Uganda/ | MRC/UVRI & LSHTM Uganda Research Unit | Where sequence data have been generated and submitted to GISAID | Matthew Cotten et al |
| EPI_ISL_954273 | Uganda/UG206/2020 | 2020-12-28 | Africa/Uganda/Uganda/ | MRC/UVRI & LSHTM Uganda Research Unit | Where sequence data have been generated and submitted to GISAID | Matthew Cotten et al |

| | | | | | | |
|---|---|---|---|---|---|---|
| EPI_ISL_954294 | Uganda/UG198/2020 | 2020-12-31 | Africa/Uganda/Uganda/ | MRC/UVRI & LSHTM Uganda Research Unit | Where sequence data have been generated and submitted to GISAID | Matthew Cotten et al |
| EPI_ISL_954295 | Uganda/UG199/2020 | 2020-12-31 | Africa/Uganda/Uganda/ | MRC/UVRI & LSHTM Uganda Research Unit | Where sequence data have been generated and submitted to GISAID | Matthew Cotten et al |
| EPI_ISL_954297 | Uganda/UG201/2021 | 2021-01-01 | Africa/Uganda/Uganda/ | MRC/UVRI & LSHTM Uganda Research Unit | Where sequence data have been generated and submitted to GISAID | Matthew Cotten et al |
| EPI_ISL_954302 | Canada/QC-L00276504/2020 | 2020-07-10 | North America/Canada/Quebec/ | Laboratoire de santv© publique du Quv©bec | Laboratoire de santv© publique du Quv©bec | Sandrine Moreira et al |
| EPI_ISL_954304 | Canada/QC-HMR-96152085/2020 | 2020-04-15 | North America/Canada/Quebec/ | Laboratoire de santv© publique du Quv©bec | Laboratoire de santv© publique du Quv©bec | Sandrine Moreira et al |
| EPI_ISL_954305 | Canada/QC-HMR-96151958/2020 | 2020-04-15 | North America/Canada/Quebec/ | Laboratoire de santv© publique du Quv©bec | Laboratoire de santv© publique du Quv©bec | Sandrine Moreira et al |
| EPI_ISL_954306 | Canada/QC-HMR-96153367/2020 | 2020-04-15 | North America/Canada/Quebec/ | Laboratoire de santv© publique du Quv©bec | Laboratoire de santv© publique du Quv©bec | Sandrine Moreira et al |
| EPI_ISL_954307 | Canada/QC-HMR-96153165/2020 | 2020-04-15 | North America/Canada/Quebec/ | Laboratoire de santv© publique du Quv©bec | Laboratoire de santv© publique du Quv©bec | Sandrine Moreira et al |
| EPI_ISL_954308 | Canada/QC-HMR-96153357/2020 | 2020-04-15 | North America/Canada/Quebec/ | Laboratoire de santv© publique du Quv©bec | Laboratoire de santv© publique du Quv©bec | Sandrine Moreira et al |
| EPI_ISL_954309 | Canada/QC-HMR-96153309/2020 | 2020-04-15 | North America/Canada/Quebec/ | Laboratoire de santv© publique du Quv©bec | Laboratoire de santv© publique du Quv©bec | Sandrine Moreira et al |
| EPI_ISL_954311 | Canada/QC-HMR-96153358/2020 | 2020-04-15 | North America/Canada/Quebec/ | Laboratoire de santv© publique du Quv©bec | Laboratoire de santv© publique du Quv©bec | Sandrine Moreira et al |

| | | | | | | |
|---|---|---|---|---|---|---|
| EPI_ISL_954312 | Canada/QC-HMR-96153346/2020 | 2020-04-15 | North America/Canada/Quebec/ | Laboratoire de sant√© publique du Qu√©bec | Laboratoire de sant√© publique du Qu√©bec | Sandrine Moreira et al |
| EPI_ISL_954313 | Canada/QC-HVE-P6151022/2020 | 2020-04-15 | North America/Canada/Quebec/ | Laboratoire de sant√© publique du Qu√©bec | Laboratoire de sant√© publique du Qu√©bec | Sandrine Moreira et al |
| EPI_ISL_954314 | Canada/QC-JUS-V6161096/2020 | 2020-04-15 | North America/Canada/Quebec/ | Laboratoire de sant√© publique du Qu√©bec | Laboratoire de sant√© publique du Qu√©bec | Sandrine Moreira et al |
| EPI_ISL_954315 | Canada/QC-HVE-P6150991/2020 | 2020-04-15 | North America/Canada/Quebec/ | Laboratoire de sant√© publique du Qu√©bec | Laboratoire de sant√© publique du Qu√©bec | Sandrine Moreira et al |
| EPI_ISL_954316 | Canada/QC-HVE-P6151009/2020 | 2020-04-15 | North America/Canada/Quebec/ | Laboratoire de sant√© publique du Qu√©bec | Laboratoire de sant√© publique du Qu√©bec | Sandrine Moreira et al |
| EPI_ISL_954318 | Canada/QC-HVE-P6151061/2020 | 2020-04-15 | North America/Canada/Quebec/ | Laboratoire de sant√© publique du Qu√©bec | Laboratoire de sant√© publique du Qu√©bec | Sandrine Moreira et al |
| EPI_ISL_954319 | Canada/QC-HVE-P6151268/2020 | 2020-04-15 | North America/Canada/Quebec/ | Laboratoire de sant√© publique du Qu√©bec | Laboratoire de sant√© publique du Qu√©bec | Sandrine Moreira et al |
| EPI_ISL_954320 | Canada/QC-JUS-V6150791/2020 | 2020-04-14 | North America/Canada/Quebec/ | Laboratoire de sant√© publique du Qu√©bec | Laboratoire de sant√© publique du Qu√©bec | Sandrine Moreira et al |
| EPI_ISL_954321 | Canada/QC-HMR-96143227/2020 | 2020-04-14 | North America/Canada/Quebec/ | Laboratoire de sant√© publique du Qu√©bec | Laboratoire de sant√© publique du Qu√©bec | Sandrine Moreira et al |
| EPI_ISL_954322 | Canada/QC-HMR-96142947/2020 | 2020-04-14 | North America/Canada/Quebec/ | Laboratoire de sant√© publique du Qu√©bec | Laboratoire de sant√© publique du Qu√©bec | Sandrine Moreira et al |
| EPI_ISL_954323 | Canada/QC-HMR-96142956/2020 | 2020-04-14 | North America/Canada/Quebec/ | Laboratoire de sant√© publique du Qu√©bec | Laboratoire de sant√© publique du Qu√©bec | Sandrine Moreira et al |
| EPI_ISL_954324 | Canada/QC-JUS-V6141489/2020 | 2020-04-14 | North America/Canada/Quebec/ | Laboratoire de sant√© publique du Qu√©bec | Laboratoire de sant√© publique du Qu√©bec | Sandrine Moreira et al |
| EPI_ISL_954325 | Canada/QC-HVE-P6141160/2020 | 2020-04-14 | North America/Canada/Quebec/ | Laboratoire de sant√© publique du Qu√©bec | Laboratoire de sant√© publique du Qu√©bec | Sandrine Moreira et al |

| | | | | | | |
|---|---|---|---|---|---|---|
| EPI_ISL_954326 | Canada/QC-HVE-P6141337/2020 | 2020-04-14 | North America/Canada/Quebec/ | Laboratoire de santv© publique du Quv©bec | Laboratoire de santv© publique du Quv©bec | Sandrine Moreira et al |
| EPI_ISL_954327 | Canada/QC-JUS-V6151475/2020 | 2020-04-14 | North America/Canada/Quebec/ | Laboratoire de santv© publique du Quv©bec | Laboratoire de santv© publique du Quv©bec | Sandrine Moreira et al |
| EPI_ISL_954328 | Canada/QC-JUS-V6151287/2020 | 2020-04-14 | North America/Canada/Quebec/ | Laboratoire de santv© publique du Quv©bec | Laboratoire de santv© publique du Quv©bec | Sandrine Moreira et al |
| EPI_ISL_954329 | Canada/QC-JUS-V6141864/2020 | 2020-04-14 | North America/Canada/Quebec/ | Laboratoire de santv© publique du Quv©bec | Laboratoire de santv© publique du Quv©bec | Sandrine Moreira et al |
| EPI_ISL_954330 | Canada/QC-JUS-V6141908/2020 | 2020-04-14 | North America/Canada/Quebec/ | Laboratoire de santv© publique du Quv©bec | Laboratoire de santv© publique du Quv©bec | Sandrine Moreira et al |
| EPI_ISL_954331 | Canada/QC-JUS-V6151472/2020 | 2020-04-14 | North America/Canada/Quebec/ | Laboratoire de santv© publique du Quv©bec | Laboratoire de santv© publique du Quv©bec | Sandrine Moreira et al |
| EPI_ISL_954332 | Canada/QC-JUS-V6151492/2020 | 2020-04-14 | North America/Canada/Quebec/ | Laboratoire de santv© publique du Quv©bec | Laboratoire de santv© publique du Quv©bec | Sandrine Moreira et al |
| EPI_ISL_954333 | Canada/QC-JUS-V6151483/2020 | 2020-04-14 | North America/Canada/Quebec/ | Laboratoire de santv© publique du Quv©bec | Laboratoire de santv© publique du Quv©bec | Sandrine Moreira et al |
| EPI_ISL_954334 | Canada/QC-JUS-V6151347/2020 | 2020-04-14 | North America/Canada/Quebec/ | Laboratoire de santv© publique du Quv©bec | Laboratoire de santv© publique du Quv©bec | Sandrine Moreira et al |
| EPI_ISL_954335 | Canada/QC-JUS-V6140579/2020 | 2020-04-14 | North America/Canada/Quebec/ | Laboratoire de santv© publique du Quv©bec | Laboratoire de santv© publique du Quv©bec | Sandrine Moreira et al |
| EPI_ISL_954336 | Canada/QC-JUS-V6151465/2020 | 2020-04-14 | North America/Canada/Quebec/ | Laboratoire de santv© publique du Quv©bec | Laboratoire de santv© publique du Quv©bec | Sandrine Moreira et al |
| EPI_ISL_954337 | Canada/QC-JUS-V6151458/2020 | 2020-04-14 | North America/Canada/Quebec/ | Laboratoire de santv© publique du Quv©bec | Laboratoire de santv© publique du Quv©bec | Sandrine Moreira et al |
| EPI_ISL_954338 | Canada/QC-JUS-V6151343/2020 | 2020-04-14 | North America/Canada/Quebec/ | Laboratoire de santv© publique du Quv©bec | Laboratoire de santv© publique du Quv©bec | Sandrine Moreira et al |

| | | | | | | |
|---|---|---|---|---|---|---|
| EPI_ISL_954339 | Canada/QC-JUS-V6150816/2020 | 2020-04-14 | North America/Canada/Quebec/ | Laboratoire de santv© publique du Quv©bec | Laboratoire de santv© publique du Quv©bec | Sandrine Moreira et al |
| EPI_ISL_954340 | Canada/QC-JUS-V6151427/2020 | 2020-04-14 | North America/Canada/Quebec/ | Laboratoire de santv© publique du Quv©bec | Laboratoire de santv© publique du Quv©bec | Sandrine Moreira et al |
| EPI_ISL_954341 | Canada/QC-JUS-V6151396/2020 | 2020-04-14 | North America/Canada/Quebec/ | Laboratoire de santv© publique du Quv©bec | Laboratoire de santv© publique du Quv©bec | Sandrine Moreira et al |
| EPI_ISL_954342 | Canada/QC-JUS-V6151167/2020 | 2020-04-14 | North America/Canada/Quebec/ | Laboratoire de santv© publique du Quv©bec | Laboratoire de santv© publique du Quv©bec | Sandrine Moreira et al |
| EPI_ISL_954343 | Canada/QC-JUS-V6151204/2020 | 2020-04-14 | North America/Canada/Quebec/ | Laboratoire de santv© publique du Quv©bec | Laboratoire de santv© publique du Quv©bec | Sandrine Moreira et al |
| EPI_ISL_954344 | Canada/QC-HVE-P6140468/2020 | 2020-04-14 | North America/Canada/Quebec/ | Laboratoire de santv© publique du Quv©bec | Laboratoire de santv© publique du Quv©bec | Sandrine Moreira et al |
| EPI_ISL_954345 | Canada/QC-HVE-P6141293/2020 | 2020-04-14 | North America/Canada/Quebec/ | Laboratoire de santv© publique du Quv©bec | Laboratoire de santv© publique du Quv©bec | Sandrine Moreira et al |
| EPI_ISL_954346 | Canada/QC-HVE-P6141444/2020 | 2020-04-14 | North America/Canada/Quebec/ | Laboratoire de santv© publique du Quv©bec | Laboratoire de santv© publique du Quv©bec | Sandrine Moreira et al |
| EPI_ISL_954347 | Canada/QC-HVE-P6141461/2020 | 2020-04-14 | North America/Canada/Quebec/ | Laboratoire de santv© publique du Quv©bec | Laboratoire de santv© publique du Quv©bec | Sandrine Moreira et al |
| EPI_ISL_954348 | Canada/QC-HMR-96131297/2020 | 2020-04-13 | North America/Canada/Quebec/ | Laboratoire de santv© publique du Quv©bec | Laboratoire de santv© publique du Quv©bec | Sandrine Moreira et al |
| EPI_ISL_954349 | Canada/QC-JUS-V6140781/2020 | 2020-04-13 | North America/Canada/Quebec/ | Laboratoire de santv© publique du Quv©bec | Laboratoire de santv© publique du Quv©bec | Sandrine Moreira et al |
| EPI_ISL_954350 | Canada/QC-JUS-V6141053/2020 | 2020-04-13 | North America/Canada/Quebec/ | Laboratoire de santv© publique du Quv©bec | Laboratoire de santv© publique du Quv©bec | Sandrine Moreira et al |
| EPI_ISL_954351 | Canada/QC-JUS-V6120295/2020 | 2020-04-13 | North America/Canada/Quebec/ | Laboratoire de santv© publique du Quv©bec | Laboratoire de santv© publique du Quv©bec | Sandrine Moreira et al |

| | | | | | | |
|---|---|---|---|---|---|---|
| EPI_ISL_954352 | Canada/QC-JUS-V6120503/2020 | 2020-04-13 | North America/Canada/Quebec/ | Laboratoire de santv© publique du Quv©bec | Laboratoire de santv© publique du Quv©bec | Sandrine Moreira et al |
| EPI_ISL_954353 | Canada/QC-JUS-V6141468/2020 | 2020-04-13 | North America/Canada/Quebec/ | Laboratoire de santv© publique du Quv©bec | Laboratoire de santv© publique du Quv©bec | Sandrine Moreira et al |
| EPI_ISL_954354 | Canada/QC-JUS-V6141049/2020 | 2020-04-13 | North America/Canada/Quebec/ | Laboratoire de santv© publique du Quv©bec | Laboratoire de santv© publique du Quv©bec | Sandrine Moreira et al |
| EPI_ISL_954355 | Canada/QC-JUS-V6141696/2020 | 2020-04-13 | North America/Canada/Quebec/ | Laboratoire de santv© publique du Quv©bec | Laboratoire de santv© publique du Quv©bec | Sandrine Moreira et al |
| EPI_ISL_954356 | Canada/QC-JUS-V6141094/2020 | 2020-04-13 | North America/Canada/Quebec/ | Laboratoire de santv© publique du Quv©bec | Laboratoire de santv© publique du Quv©bec | Sandrine Moreira et al |
| EPI_ISL_954357 | Canada/QC-JUS-V6141252/2020 | 2020-04-13 | North America/Canada/Quebec/ | Laboratoire de santv© publique du Quv©bec | Laboratoire de santv© publique du Quv©bec | Sandrine Moreira et al |
| EPI_ISL_954358 | Canada/QC-JUS-V6141239/2020 | 2020-04-13 | North America/Canada/Quebec/ | Laboratoire de santv© publique du Quv©bec | Laboratoire de santv© publique du Quv©bec | Sandrine Moreira et al |
| EPI_ISL_954359 | Canada/QC-JUS-V6141327/2020 | 2020-04-13 | North America/Canada/Quebec/ | Laboratoire de santv© publique du Quv©bec | Laboratoire de santv© publique du Quv©bec | Sandrine Moreira et al |
| EPI_ISL_954360 | Canada/QC-JUS-V6141331/2020 | 2020-04-13 | North America/Canada/Quebec/ | Laboratoire de santv© publique du Quv©bec | Laboratoire de santv© publique du Quv©bec | Sandrine Moreira et al |
| EPI_ISL_954361 | Canada/QC-HVE-P6130189/2020 | 2020-04-13 | North America/Canada/Quebec/ | Laboratoire de santv© publique du Quv©bec | Laboratoire de santv© publique du Quv©bec | Sandrine Moreira et al |
| EPI_ISL_954362 | Canada/QC-HVE-P6130462/2020 | 2020-04-13 | North America/Canada/Quebec/ | Laboratoire de santv© publique du Quv©bec | Laboratoire de santv© publique du Quv©bec | Sandrine Moreira et al |
| EPI_ISL_954363 | Canada/QC-HVE-P6130680/2020 | 2020-04-13 | North America/Canada/Quebec/ | Laboratoire de santv© publique du Quv©bec | Laboratoire de santv© publique du Quv©bec | Sandrine Moreira et al |
| EPI_ISL_954364 | Canada/QC-HVE-P6130895/2020 | 2020-04-13 | North America/Canada/Quebec/ | Laboratoire de santv© publique du Quv©bec | Laboratoire de santv© publique du Quv©bec | Sandrine Moreira et al |

| | | | | | | |
|---|---|---|---|---|---|---|
| EPI_ISL_954365 | Canada/QC-HMR-96121041/2020 | 2020-04-12 | North America/Canada/Quebec/ | Laboratoire de sant√© publique du Qu√©bec | Laboratoire de sant√© publique du Qu√©bec | Sandrine Moreira et al |
| EPI_ISL_954366 | Canada/QC-HMR-96121022/2020 | 2020-04-12 | North America/Canada/Quebec/ | Laboratoire de sant√© publique du Qu√©bec | Laboratoire de sant√© publique du Qu√©bec | Sandrine Moreira et al |
| EPI_ISL_954367 | Canada/QC-JUS-V6130834/2020 | 2020-04-12 | North America/Canada/Quebec/ | Laboratoire de sant√© publique du Qu√©bec | Laboratoire de sant√© publique du Qu√©bec | Sandrine Moreira et al |
| EPI_ISL_954368 | Canada/QC-JUS-V6120475/2020 | 2020-04-12 | North America/Canada/Quebec/ | Laboratoire de sant√© publique du Qu√©bec | Laboratoire de sant√© publique du Qu√©bec | Sandrine Moreira et al |
| EPI_ISL_954369 | Canada/QC-JUS-V6120770/2020 | 2020-04-12 | North America/Canada/Quebec/ | Laboratoire de sant√© publique du Qu√©bec | Laboratoire de sant√© publique du Qu√©bec | Sandrine Moreira et al |
| EPI_ISL_954370 | Canada/QC-JUS-V6130828/2020 | 2020-04-12 | North America/Canada/Quebec/ | Laboratoire de sant√© publique du Qu√©bec | Laboratoire de sant√© publique du Qu√©bec | Sandrine Moreira et al |
| EPI_ISL_954371 | Canada/QC-JUS-V6130685/2020 | 2020-04-12 | North America/Canada/Quebec/ | Laboratoire de sant√© publique du Qu√©bec | Laboratoire de sant√© publique du Qu√©bec | Sandrine Moreira et al |
| EPI_ISL_954372 | Canada/QC-JUS-V6130667/2020 | 2020-04-12 | North America/Canada/Quebec/ | Laboratoire de sant√© publique du Qu√©bec | Laboratoire de sant√© publique du Qu√©bec | Sandrine Moreira et al |
| EPI_ISL_954373 | Canada/QC-JUS-V6130436/2020 | 2020-04-12 | North America/Canada/Quebec/ | Laboratoire de sant√© publique du Qu√©bec | Laboratoire de sant√© publique du Qu√©bec | Sandrine Moreira et al |
| EPI_ISL_954374 | Canada/QC-JUS-V6130441/2020 | 2020-04-12 | North America/Canada/Quebec/ | Laboratoire de sant√© publique du Qu√©bec | Laboratoire de sant√© publique du Qu√©bec | Sandrine Moreira et al |
| EPI_ISL_954375 | Canada/QC-JUS-V6130544/2020 | 2020-04-12 | North America/Canada/Quebec/ | Laboratoire de sant√© publique du Qu√©bec | Laboratoire de sant√© publique du Qu√©bec | Sandrine Moreira et al |
| EPI_ISL_954376 | Canada/QC-JUS-V6130798/2020 | 2020-04-12 | North America/Canada/Quebec/ | Laboratoire de sant√© publique du Qu√©bec | Laboratoire de sant√© publique du Qu√©bec | Sandrine Moreira et al |
| EPI_ISL_954377 | Canada/QC-JUS-V6130792/2020 | 2020-04-12 | North America/Canada/Quebec/ | Laboratoire de sant√© publique du Qu√©bec | Laboratoire de sant√© publique du Qu√©bec | Sandrine Moreira et al |

| | | | | | | |
|---|---|---|---|---|---|---|
| EPI_ISL_954378 | Canada/QC-JUS-V6130795/2020 | 2020-04-12 | North America/Canada/Quebec/ | Laboratoire de sant√© publique du Qu√©bec | Laboratoire de sant√© publique du Qu√©bec | Sandrine Moreira et al |
| EPI_ISL_954379 | Canada/QC-JUS-V6130764/2020 | 2020-04-12 | North America/Canada/Quebec/ | Laboratoire de sant√© publique du Qu√©bec | Laboratoire de sant√© publique du Qu√©bec | Sandrine Moreira et al |
| EPI_ISL_954380 | Canada/QC-JUS-V6130734/2020 | 2020-04-12 | North America/Canada/Quebec/ | Laboratoire de sant√© publique du Qu√©bec | Laboratoire de sant√© publique du Qu√©bec | Sandrine Moreira et al |
| EPI_ISL_954381 | Canada/QC-JUS-V6130767/2020 | 2020-04-12 | North America/Canada/Quebec/ | Laboratoire de sant√© publique du Qu√©bec | Laboratoire de sant√© publique du Qu√©bec | Sandrine Moreira et al |
| EPI_ISL_954382 | Canada/QC-JUS-V6130762/2020 | 2020-04-12 | North America/Canada/Quebec/ | Laboratoire de sant√© publique du Qu√©bec | Laboratoire de sant√© publique du Qu√©bec | Sandrine Moreira et al |
| EPI_ISL_954383 | Canada/QC-JUS-V6130804/2020 | 2020-04-12 | North America/Canada/Quebec/ | Laboratoire de sant√© publique du Qu√©bec | Laboratoire de sant√© publique du Qu√©bec | Sandrine Moreira et al |
| EPI_ISL_954384 | Canada/QC-JUS-V6130781/2020 | 2020-04-12 | North America/Canada/Quebec/ | Laboratoire de sant√© publique du Qu√©bec | Laboratoire de sant√© publique du Qu√©bec | Sandrine Moreira et al |
| EPI_ISL_954385 | Canada/QC-JUS-V6130211/2020 | 2020-04-12 | North America/Canada/Quebec/ | Laboratoire de sant√© publique du Qu√©bec | Laboratoire de sant√© publique du Qu√©bec | Sandrine Moreira et al |
| EPI_ISL_954386 | Canada/QC-JUS-V6130213/2020 | 2020-04-12 | North America/Canada/Quebec/ | Laboratoire de sant√© publique du Qu√©bec | Laboratoire de sant√© publique du Qu√©bec | Sandrine Moreira et al |
| EPI_ISL_954387 | Canada/QC-JUS-V6130188/2020 | 2020-04-12 | North America/Canada/Quebec/ | Laboratoire de sant√© publique du Qu√©bec | Laboratoire de sant√© publique du Qu√©bec | Sandrine Moreira et al |
| EPI_ISL_954388 | Canada/QC-JUS-V6130453/2020 | 2020-04-12 | North America/Canada/Quebec/ | Laboratoire de sant√© publique du Qu√©bec | Laboratoire de sant√© publique du Qu√©bec | Sandrine Moreira et al |
| EPI_ISL_954389 | Canada/QC-JUS-V6130502/2020 | 2020-04-12 | North America/Canada/Quebec/ | Laboratoire de sant√© publique du Qu√©bec | Laboratoire de sant√© publique du Qu√©bec | Sandrine Moreira et al |
| EPI_ISL_954390 | Canada/QC-JUS-V6130511/2020 | 2020-04-12 | North America/Canada/Quebec/ | Laboratoire de sant√© publique du Qu√©bec | Laboratoire de sant√© publique du Qu√©bec | Sandrine Moreira et al |

| | | | | | | |
|---|---|---|---|---|---|---|
| EPI_ISL_954391 | Canada/QC-JUS-V6130515/2020 | 2020-04-12 | North America/Canada/Quebec/ | Laboratoire de sant√© publique du Qu√©bec | Laboratoire de sant√© publique du Qu√©bec | Sandrine Moreira et al |
| EPI_ISL_954392 | Canada/QC-JUS-V6130444/2020 | 2020-04-12 | North America/Canada/Quebec/ | Laboratoire de sant√© publique du Qu√©bec | Laboratoire de sant√© publique du Qu√©bec | Sandrine Moreira et al |
| EPI_ISL_954393 | Canada/QC-JUS-V6140779/2020 | 2020-04-12 | North America/Canada/Quebec/ | Laboratoire de sant√© publique du Qu√©bec | Laboratoire de sant√© publique du Qu√©bec | Sandrine Moreira et al |
| EPI_ISL_954394 | Canada/QC-JUS-V6130450/2020 | 2020-04-12 | North America/Canada/Quebec/ | Laboratoire de sant√© publique du Qu√©bec | Laboratoire de sant√© publique du Qu√©bec | Sandrine Moreira et al |
| EPI_ISL_954395 | Canada/QC-HVE-P6120542/2020 | 2020-04-12 | North America/Canada/Quebec/ | Laboratoire de sant√© publique du Qu√©bec | Laboratoire de sant√© publique du Qu√©bec | Sandrine Moreira et al |
| EPI_ISL_954396 | Canada/QC-HVE-P6110741/2020 | 2020-04-11 | North America/Canada/Quebec/ | Laboratoire de sant√© publique du Qu√©bec | Laboratoire de sant√© publique du Qu√©bec | Sandrine Moreira et al |
| EPI_ISL_954397 | Canada/QC-HMR-96110843/2020 | 2020-04-11 | North America/Canada/Quebec/ | Laboratoire de sant√© publique du Qu√©bec | Laboratoire de sant√© publique du Qu√©bec | Sandrine Moreira et al |
| EPI_ISL_954398 | Canada/QC-HMR-96110921/2020 | 2020-04-11 | North America/Canada/Quebec/ | Laboratoire de sant√© publique du Qu√©bec | Laboratoire de sant√© publique du Qu√©bec | Sandrine Moreira et al |
| EPI_ISL_954399 | Canada/QC-JUS-V6120479/2020 | 2020-04-11 | North America/Canada/Quebec/ | Laboratoire de sant√© publique du Qu√©bec | Laboratoire de sant√© publique du Qu√©bec | Sandrine Moreira et al |
| EPI_ISL_954400 | Canada/QC-JUS-V6120484/2020 | 2020-04-11 | North America/Canada/Quebec/ | Laboratoire de sant√© publique du Qu√©bec | Laboratoire de sant√© publique du Qu√©bec | Sandrine Moreira et al |
| EPI_ISL_954401 | Canada/QC-JUS-V6111466/2020 | 2020-04-11 | North America/Canada/Quebec/ | Laboratoire de sant√© publique du Qu√©bec | Laboratoire de sant√© publique du Qu√©bec | Sandrine Moreira et al |
| EPI_ISL_954402 | Canada/QC-JUS-V6110868/2020 | 2020-04-11 | North America/Canada/Quebec/ | Laboratoire de sant√© publique du Qu√©bec | Laboratoire de sant√© publique du Qu√©bec | Sandrine Moreira et al |
| EPI_ISL_954403 | Canada/QC-JUS-V6111025/2020 | 2020-04-11 | North America/Canada/Quebec/ | Laboratoire de sant√© publique du Qu√©bec | Laboratoire de sant√© publique du Qu√©bec | Sandrine Moreira et al |

| | | | | | | |
|---|---|---|---|---|---|---|
| EPI_ISL_954404 | Canada/QC-JUS-V6120164/2020 | 2020-04-11 | North America/Canada/Quebec/ | Laboratoire de santv© publique du Quv©bec | Laboratoire de santv© publique du Quv©bec | Sandrine Moreira et al |
| EPI_ISL_954405 | Canada/QC-JUS-V6110828/2020 | 2020-04-11 | North America/Canada/Quebec/ | Laboratoire de santv© publique du Quv©bec | Laboratoire de santv© publique du Quv©bec | Sandrine Moreira et al |
| EPI_ISL_954406 | Canada/QC-JUS-V6120532/2020 | 2020-04-11 | North America/Canada/Quebec/ | Laboratoire de santv© publique du Quv©bec | Laboratoire de santv© publique du Quv©bec | Sandrine Moreira et al |
| EPI_ISL_954407 | Canada/QC-JUS-V6120562/2020 | 2020-04-11 | North America/Canada/Quebec/ | Laboratoire de santv© publique du Quv©bec | Laboratoire de santv© publique du Quv©bec | Sandrine Moreira et al |
| EPI_ISL_954408 | Canada/QC-JUS-V6120693/2020 | 2020-04-11 | North America/Canada/Quebec/ | Laboratoire de santv© publique du Quv©bec | Laboratoire de santv© publique du Quv©bec | Sandrine Moreira et al |
| EPI_ISL_954409 | Canada/QC-JUS-V6120700/2020 | 2020-04-11 | North America/Canada/Quebec/ | Laboratoire de santv© publique du Quv©bec | Laboratoire de santv© publique du Quv©bec | Sandrine Moreira et al |
| EPI_ISL_954410 | Canada/QC-JUS-V6120706/2020 | 2020-04-11 | North America/Canada/Quebec/ | Laboratoire de santv© publique du Quv©bec | Laboratoire de santv© publique du Quv©bec | Sandrine Moreira et al |
| EPI_ISL_954411 | Canada/QC-JUS-V6120722/2020 | 2020-04-11 | North America/Canada/Quebec/ | Laboratoire de santv© publique du Quv©bec | Laboratoire de santv© publique du Quv©bec | Sandrine Moreira et al |
| EPI_ISL_954412 | Canada/QC-JUS-V6120681/2020 | 2020-04-11 | North America/Canada/Quebec/ | Laboratoire de santv© publique du Quv©bec | Laboratoire de santv© publique du Quv©bec | Sandrine Moreira et al |
| EPI_ISL_954413 | Canada/QC-JUS-V6120657/2020 | 2020-04-11 | North America/Canada/Quebec/ | Laboratoire de santv© publique du Quv©bec | Laboratoire de santv© publique du Quv©bec | Sandrine Moreira et al |
| EPI_ISL_954414 | Canada/QC-JUS-V6120654/2020 | 2020-04-11 | North America/Canada/Quebec/ | Laboratoire de santv© publique du Quv©bec | Laboratoire de santv© publique du Quv©bec | Sandrine Moreira et al |
| EPI_ISL_954415 | Canada/QC-JUS-V6120615/2020 | 2020-04-11 | North America/Canada/Quebec/ | Laboratoire de santv© publique du Quv©bec | Laboratoire de santv© publique du Quv©bec | Sandrine Moreira et al |
| EPI_ISL_954416 | Canada/QC-JUS-V6120604/2020 | 2020-04-11 | North America/Canada/Quebec/ | Laboratoire de santv© publique du Quv©bec | Laboratoire de santv© publique du Quv©bec | Sandrine Moreira et al |

| | | | | | | |
|---|---|---|---|---|---|---|
| EPI_ISL_954417 | Canada/QC-JUS-V6120636/2020 | 2020-04-11 | North America/Canada/Quebec/ | Laboratoire de sant√© publique du Qu√©bec | Laboratoire de sant√© publique du Qu√©bec | Sandrine Moreira et al |
| EPI_ISL_954418 | Canada/QC-JUS-V6120703/2020 | 2020-04-11 | North America/Canada/Quebec/ | Laboratoire de sant√© publique du Qu√©bec | Laboratoire de sant√© publique du Qu√©bec | Sandrine Moreira et al |
| EPI_ISL_954419 | Canada/QC-JUS-V6120487/2020 | 2020-04-11 | North America/Canada/Quebec/ | Laboratoire de sant√© publique du Qu√©bec | Laboratoire de sant√© publique du Qu√©bec | Sandrine Moreira et al |
| EPI_ISL_954420 | Canada/QC-JUS-V6120395/2020 | 2020-04-11 | North America/Canada/Quebec/ | Laboratoire de sant√© publique du Qu√©bec | Laboratoire de sant√© publique du Qu√©bec | Sandrine Moreira et al |
| EPI_ISL_954421 | Canada/QC-HVE-P6110129/2020 | 2020-04-11 | North America/Canada/Quebec/ | Laboratoire de sant√© publique du Qu√©bec | Laboratoire de sant√© publique du Qu√©bec | Sandrine Moreira et al |
| EPI_ISL_954422 | Canada/QC-HVE-P6110304/2020 | 2020-04-11 | North America/Canada/Quebec/ | Laboratoire de sant√© publique du Qu√©bec | Laboratoire de sant√© publique du Qu√©bec | Sandrine Moreira et al |
| EPI_ISL_954423 | Canada/QC-HMR-96101290/2020 | 2020-04-10 | North America/Canada/Quebec/ | Laboratoire de sant√© publique du Qu√©bec | Laboratoire de sant√© publique du Qu√©bec | Sandrine Moreira et al |
| EPI_ISL_954424 | Canada/QC-JUS-V6111234/2020 | 2020-04-10 | North America/Canada/Quebec/ | Laboratoire de sant√© publique du Qu√©bec | Laboratoire de sant√© publique du Qu√©bec | Sandrine Moreira et al |
| EPI_ISL_954425 | Canada/QC-JUS-V6100843/2020 | 2020-04-10 | North America/Canada/Quebec/ | Laboratoire de sant√© publique du Qu√©bec | Laboratoire de sant√© publique du Qu√©bec | Sandrine Moreira et al |
| EPI_ISL_954426 | Canada/QC-JUS-V6100830/2020 | 2020-04-10 | North America/Canada/Quebec/ | Laboratoire de sant√© publique du Qu√©bec | Laboratoire de sant√© publique du Qu√©bec | Sandrine Moreira et al |
| EPI_ISL_954427 | Canada/QC-JUS-V6110556/2020 | 2020-04-10 | North America/Canada/Quebec/ | Laboratoire de sant√© publique du Qu√©bec | Laboratoire de sant√© publique du Qu√©bec | Sandrine Moreira et al |
| EPI_ISL_954428 | Canada/QC-JUS-V6110562/2020 | 2020-04-10 | North America/Canada/Quebec/ | Laboratoire de sant√© publique du Qu√©bec | Laboratoire de sant√© publique du Qu√©bec | Sandrine Moreira et al |
| EPI_ISL_954429 | Canada/QC-JUS-V6110551/2020 | 2020-04-10 | North America/Canada/Quebec/ | Laboratoire de sant√© publique du Qu√©bec | Laboratoire de sant√© publique du Qu√©bec | Sandrine Moreira et al |

| | | | | | | |
|---|---|---|---|---|---|---|
| EPI_ISL_954430 | Canada/QC-JUS-V6110655/2020 | 2020-04-10 | North America/Canada/Quebec/ | Laboratoire de santv© publique du Quv©bec | Laboratoire de santv© publique du Quv©bec | Sandrine Moreira et al |
| EPI_ISL_954431 | Canada/QC-JUS-V6110558/2020 | 2020-04-10 | North America/Canada/Quebec/ | Laboratoire de santv© publique du Quv©bec | Laboratoire de santv© publique du Quv©bec | Sandrine Moreira et al |
| EPI_ISL_954432 | Canada/QC-JUS-V6110452/2020 | 2020-04-10 | North America/Canada/Quebec/ | Laboratoire de santv© publique du Quv©bec | Laboratoire de santv© publique du Quv©bec | Sandrine Moreira et al |
| EPI_ISL_954433 | Canada/QC-JUS-V6110553/2020 | 2020-04-10 | North America/Canada/Quebec/ | Laboratoire de santv© publique du Quv©bec | Laboratoire de santv© publique du Quv©bec | Sandrine Moreira et al |
| EPI_ISL_954434 | Canada/QC-JUS-V6110566/2020 | 2020-04-10 | North America/Canada/Quebec/ | Laboratoire de santv© publique du Quv©bec | Laboratoire de santv© publique du Quv©bec | Sandrine Moreira et al |
| EPI_ISL_954435 | Canada/QC-JUS-V6110335/2020 | 2020-04-10 | North America/Canada/Quebec/ | Laboratoire de santv© publique du Quv©bec | Laboratoire de santv© publique du Quv©bec | Sandrine Moreira et al |
| EPI_ISL_954436 | Canada/QC-JUS-V6110628/2020 | 2020-04-10 | North America/Canada/Quebec/ | Laboratoire de santv© publique du Quv©bec | Laboratoire de santv© publique du Quv©bec | Sandrine Moreira et al |
| EPI_ISL_954437 | Canada/QC-JUS-V6110651/2020 | 2020-04-10 | North America/Canada/Quebec/ | Laboratoire de santv© publique du Quv©bec | Laboratoire de santv© publique du Quv©bec | Sandrine Moreira et al |
| EPI_ISL_954438 | Canada/QC-JUS-V6110726/2020 | 2020-04-10 | North America/Canada/Quebec/ | Laboratoire de santv© publique du Quv©bec | Laboratoire de santv© publique du Quv©bec | Sandrine Moreira et al |
| EPI_ISL_954439 | Canada/QC-JUS-V6110166/2020 | 2020-04-10 | North America/Canada/Quebec/ | Laboratoire de santv© publique du Quv©bec | Laboratoire de santv© publique du Quv©bec | Sandrine Moreira et al |
| EPI_ISL_954440 | Canada/QC-JUS-V6110041/2020 | 2020-04-10 | North America/Canada/Quebec/ | Laboratoire de santv© publique du Quv©bec | Laboratoire de santv© publique du Quv©bec | Sandrine Moreira et al |
| EPI_ISL_954441 | Canada/QC-JUS-V6110265/2020 | 2020-04-10 | North America/Canada/Quebec/ | Laboratoire de santv© publique du Quv©bec | Laboratoire de santv© publique du Quv©bec | Sandrine Moreira et al |
| EPI_ISL_954442 | Canada/QC-JUS-V6110776/2020 | 2020-04-10 | North America/Canada/Quebec/ | Laboratoire de santv© publique du Quv©bec | Laboratoire de santv© publique du Quv©bec | Sandrine Moreira et al |

| | | | | | | |
|---|---|---|---|---|---|---|
| EPI_ISL_954443 | Canada/QC-JUS-V6110571/2020 | 2020-04-10 | North America/Canada/Quebec/ | Laboratoire de santv© publique du Quv©bec | Laboratoire de santv© publique du Quv©bec | Sandrine Moreira et al |
| EPI_ISL_954444 | Canada/QC-JUS-V6111281/2020 | 2020-04-10 | North America/Canada/Quebec/ | Laboratoire de santv© publique du Quv©bec | Laboratoire de santv© publique du Quv©bec | Sandrine Moreira et al |
| EPI_ISL_954445 | Canada/QC-JUS-V6111251/2020 | 2020-04-10 | North America/Canada/Quebec/ | Laboratoire de santv© publique du Quv©bec | Laboratoire de santv© publique du Quv©bec | Sandrine Moreira et al |
| EPI_ISL_954446 | Canada/QC-JUS-V6111250/2020 | 2020-04-10 | North America/Canada/Quebec/ | Laboratoire de santv© publique du Quv©bec | Laboratoire de santv© publique du Quv©bec | Sandrine Moreira et al |
| EPI_ISL_954447 | Canada/QC-JUS-V6110387/2020 | 2020-04-10 | North America/Canada/Quebec/ | Laboratoire de santv© publique du Quv©bec | Laboratoire de santv© publique du Quv©bec | Sandrine Moreira et al |
| EPI_ISL_954448 | Canada/QC-JUS-V6110385/2020 | 2020-04-10 | North America/Canada/Quebec/ | Laboratoire de santv© publique du Quv©bec | Laboratoire de santv© publique du Quv©bec | Sandrine Moreira et al |
| EPI_ISL_954449 | Canada/QC-JUS-V6111205/2020 | 2020-04-10 | North America/Canada/Quebec/ | Laboratoire de santv© publique du Quv©bec | Laboratoire de santv© publique du Quv©bec | Sandrine Moreira et al |
| EPI_ISL_954450 | Canada/QC-JUS-V6111207/2020 | 2020-04-10 | North America/Canada/Quebec/ | Laboratoire de santv© publique du Quv©bec | Laboratoire de santv© publique du Quv©bec | Sandrine Moreira et al |
| EPI_ISL_954451 | Canada/QC-JUS-V6110867/2020 | 2020-04-10 | North America/Canada/Quebec/ | Laboratoire de santv© publique du Quv©bec | Laboratoire de santv© publique du Quv©bec | Sandrine Moreira et al |
| EPI_ISL_954452 | Canada/QC-JUS-V6110850/2020 | 2020-04-10 | North America/Canada/Quebec/ | Laboratoire de santv© publique du Quv©bec | Laboratoire de santv© publique du Quv©bec | Sandrine Moreira et al |
| EPI_ISL_954453 | Canada/QC-JUS-V6110780/2020 | 2020-04-10 | North America/Canada/Quebec/ | Laboratoire de santv© publique du Quv©bec | Laboratoire de santv© publique du Quv©bec | Sandrine Moreira et al |
| EPI_ISL_954454 | Canada/QC-JUS-V6110572/2020 | 2020-04-10 | North America/Canada/Quebec/ | Laboratoire de santv© publique du Quv©bec | Laboratoire de santv© publique du Quv©bec | Sandrine Moreira et al |
| EPI_ISL_954455 | Canada/QC-JUS-V6110502/2020 | 2020-04-10 | North America/Canada/Quebec/ | Laboratoire de santv© publique du Quv©bec | Laboratoire de santv© publique du Quv©bec | Sandrine Moreira et al |

| | | | | | | |
|---|---|---|---|---|---|---|
| EPI_ISL_954456 | Canada/QC-JUS-V6110565/2020 | 2020-04-10 | North America/Canada/Quebec/ | Laboratoire de sant√© publique du Qu√©bec | Laboratoire de sant√© publique du Qu√©bec | Sandrine Moreira et al |
| EPI_ISL_954457 | Canada/QC-JUS-V6110509/2020 | 2020-04-10 | North America/Canada/Quebec/ | Laboratoire de sant√© publique du Qu√©bec | Laboratoire de sant√© publique du Qu√©bec | Sandrine Moreira et al |
| EPI_ISL_954458 | Canada/QC-JUS-V6110579/2020 | 2020-04-10 | North America/Canada/Quebec/ | Laboratoire de sant√© publique du Qu√©bec | Laboratoire de sant√© publique du Qu√©bec | Sandrine Moreira et al |
| EPI_ISL_954459 | Canada/QC-JUS-V6110513/2020 | 2020-04-10 | North America/Canada/Quebec/ | Laboratoire de sant√© publique du Qu√©bec | Laboratoire de sant√© publique du Qu√©bec | Sandrine Moreira et al |
| EPI_ISL_954460 | Canada/QC-JUS-V6110503/2020 | 2020-04-10 | North America/Canada/Quebec/ | Laboratoire de sant√© publique du Qu√©bec | Laboratoire de sant√© publique du Qu√©bec | Sandrine Moreira et al |
| EPI_ISL_954461 | Canada/QC-JUS-V6110834/2020 | 2020-04-10 | North America/Canada/Quebec/ | Laboratoire de sant√© publique du Qu√©bec | Laboratoire de sant√© publique du Qu√©bec | Sandrine Moreira et al |
| EPI_ISL_954462 | Canada/QC-JUS-V6110508/2020 | 2020-04-10 | North America/Canada/Quebec/ | Laboratoire de sant√© publique du Qu√©bec | Laboratoire de sant√© publique du Qu√©bec | Sandrine Moreira et al |
| EPI_ISL_954463 | Canada/QC-JUS-V6110764/2020 | 2020-04-10 | North America/Canada/Quebec/ | Laboratoire de sant√© publique du Qu√©bec | Laboratoire de sant√© publique du Qu√©bec | Sandrine Moreira et al |
| EPI_ISL_954464 | Canada/QC-JUS-V6110717/2020 | 2020-04-10 | North America/Canada/Quebec/ | Laboratoire de sant√© publique du Qu√©bec | Laboratoire de sant√© publique du Qu√©bec | Sandrine Moreira et al |
| EPI_ISL_954465 | Canada/QC-JUS-V6110711/2020 | 2020-04-10 | North America/Canada/Quebec/ | Laboratoire de sant√© publique du Qu√©bec | Laboratoire de sant√© publique du Qu√©bec | Sandrine Moreira et al |
| EPI_ISL_954466 | Canada/QC-JUS-V6100241/2020 | 2020-04-09 | North America/Canada/Quebec/ | Laboratoire de sant√© publique du Qu√©bec | Laboratoire de sant√© publique du Qu√©bec | Sandrine Moreira et al |
| EPI_ISL_954467 | Canada/QC-HMR-96092294/2020 | 2020-04-09 | North America/Canada/Quebec/ | Laboratoire de sant√© publique du Qu√©bec | Laboratoire de sant√© publique du Qu√©bec | Sandrine Moreira et al |
| EPI_ISL_954468 | Canada/QC-HMR-96092515/2020 | 2020-04-09 | North America/Canada/Quebec/ | Laboratoire de sant√© publique du Qu√©bec | Laboratoire de sant√© publique du Qu√©bec | Sandrine Moreira et al |

| | | | | | | |
|---|---|---|---|---|---|---|
| EPI_ISL_954469 | Canada/QC-HMR-96091861/2020 | 2020-04-09 | North America/Canada/Quebec/ | Laboratoire de sant√© publique du Qu√©bec | Laboratoire de sant√© publique du Qu√©bec | Sandrine Moreira et al |
| EPI_ISL_954470 | Canada/QC-JUS-V6100880/2020 | 2020-04-09 | North America/Canada/Quebec/ | Laboratoire de sant√© publique du Qu√©bec | Laboratoire de sant√© publique du Qu√©bec | Sandrine Moreira et al |
| EPI_ISL_954471 | Canada/QC-JUS-V6100755/2020 | 2020-04-09 | North America/Canada/Quebec/ | Laboratoire de sant√© publique du Qu√©bec | Laboratoire de sant√© publique du Qu√©bec | Sandrine Moreira et al |
| EPI_ISL_954472 | Canada/QC-JUS-V6100643/2020 | 2020-04-09 | North America/Canada/Quebec/ | Laboratoire de sant√© publique du Qu√©bec | Laboratoire de sant√© publique du Qu√©bec | Sandrine Moreira et al |
| EPI_ISL_954473 | Canada/QC-JUS-V6100610/2020 | 2020-04-09 | North America/Canada/Quebec/ | Laboratoire de sant√© publique du Qu√©bec | Laboratoire de sant√© publique du Qu√©bec | Sandrine Moreira et al |
| EPI_ISL_954474 | Canada/QC-JUS-V6100595/2020 | 2020-04-09 | North America/Canada/Quebec/ | Laboratoire de sant√© publique du Qu√©bec | Laboratoire de sant√© publique du Qu√©bec | Sandrine Moreira et al |
| EPI_ISL_954475 | Canada/QC-JUS-V6100491/2020 | 2020-04-09 | North America/Canada/Quebec/ | Laboratoire de sant√© publique du Qu√©bec | Laboratoire de sant√© publique du Qu√©bec | Sandrine Moreira et al |
| EPI_ISL_954476 | Canada/QC-JUS-V6100990/2020 | 2020-04-09 | North America/Canada/Quebec/ | Laboratoire de sant√© publique du Qu√©bec | Laboratoire de sant√© publique du Qu√©bec | Sandrine Moreira et al |
| EPI_ISL_954477 | Canada/QC-JUS-V6091487/2020 | 2020-04-09 | North America/Canada/Quebec/ | Laboratoire de sant√© publique du Qu√©bec | Laboratoire de sant√© publique du Qu√©bec | Sandrine Moreira et al |
| EPI_ISL_954478 | Canada/QC-JUS-V6100724/2020 | 2020-04-09 | North America/Canada/Quebec/ | Laboratoire de sant√© publique du Qu√©bec | Laboratoire de sant√© publique du Qu√©bec | Sandrine Moreira et al |
| EPI_ISL_954479 | Canada/QC-JUS-V6100637/2020 | 2020-04-09 | North America/Canada/Quebec/ | Laboratoire de sant√© publique du Qu√©bec | Laboratoire de sant√© publique du Qu√©bec | Sandrine Moreira et al |
| EPI_ISL_954480 | Canada/QC-JUS-V6100393/2020 | 2020-04-09 | North America/Canada/Quebec/ | Laboratoire de sant√© publique du Qu√©bec | Laboratoire de sant√© publique du Qu√©bec | Sandrine Moreira et al |
| EPI_ISL_954481 | Canada/QC-JUS-V6100963/2020 | 2020-04-09 | North America/Canada/Quebec/ | Laboratoire de sant√© publique du Qu√©bec | Laboratoire de sant√© publique du Qu√©bec | Sandrine Moreira et al |

| | | | | | | |
|---|---|---|---|---|---|---|
| EPI_ISL_954482 | Canada/QC-JUS-V6091766/2020 | 2020-04-09 | North America/Canada/Quebec/ | Laboratoire de santv© publique du Quv©bec | Laboratoire de santv© publique du Quv©bec | Sandrine Moreira et al |
| EPI_ISL_954483 | Canada/QC-JUS-V6100797/2020 | 2020-04-09 | North America/Canada/Quebec/ | Laboratoire de santv© publique du Quv©bec | Laboratoire de santv© publique du Quv©bec | Sandrine Moreira et al |
| EPI_ISL_954484 | Canada/QC-JUS-V6091334/2020 | 2020-04-09 | North America/Canada/Quebec/ | Laboratoire de santv© publique du Quv©bec | Laboratoire de santv© publique du Quv©bec | Sandrine Moreira et al |
| EPI_ISL_954485 | Canada/QC-JUS-V6091391/2020 | 2020-04-09 | North America/Canada/Quebec/ | Laboratoire de santv© publique du Quv©bec | Laboratoire de santv© publique du Quv©bec | Sandrine Moreira et al |
| EPI_ISL_954486 | Canada/QC-JUS-V6091618/2020 | 2020-04-09 | North America/Canada/Quebec/ | Laboratoire de santv© publique du Quv©bec | Laboratoire de santv© publique du Quv©bec | Sandrine Moreira et al |
| EPI_ISL_954487 | Canada/QC-JUS-V6091521/2020 | 2020-04-09 | North America/Canada/Quebec/ | Laboratoire de santv© publique du Quv©bec | Laboratoire de santv© publique du Quv©bec | Sandrine Moreira et al |
| EPI_ISL_954488 | Canada/QC-JUS-V6091451/2020 | 2020-04-09 | North America/Canada/Quebec/ | Laboratoire de santv© publique du Quv©bec | Laboratoire de santv© publique du Quv©bec | Sandrine Moreira et al |
| EPI_ISL_954489 | Canada/QC-JUS-V6091560/2020 | 2020-04-09 | North America/Canada/Quebec/ | Laboratoire de santv© publique du Quv©bec | Laboratoire de santv© publique du Quv©bec | Sandrine Moreira et al |
| EPI_ISL_954490 | Canada/QC-JUS-V6091517/2020 | 2020-04-09 | North America/Canada/Quebec/ | Laboratoire de santv© publique du Quv©bec | Laboratoire de santv© publique du Quv©bec | Sandrine Moreira et al |
| EPI_ISL_954491 | Canada/QC-JUS-V6100425/2020 | 2020-04-09 | North America/Canada/Quebec/ | Laboratoire de santv© publique du Quv©bec | Laboratoire de santv© publique du Quv©bec | Sandrine Moreira et al |
| EPI_ISL_954492 | Canada/QC-JUS-V6100957/2020 | 2020-04-09 | North America/Canada/Quebec/ | Laboratoire de santv© publique du Quv©bec | Laboratoire de santv© publique du Quv©bec | Sandrine Moreira et al |
| EPI_ISL_954493 | Canada/QC-JUS-V6101074/2020 | 2020-04-09 | North America/Canada/Quebec/ | Laboratoire de santv© publique du Quv©bec | Laboratoire de santv© publique du Quv©bec | Sandrine Moreira et al |
| EPI_ISL_954494 | Canada/QC-HVE-P6090844/2020 | 2020-04-09 | North America/Canada/Quebec/ | Laboratoire de santv© publique du Quv©bec | Laboratoire de santv© publique du Quv©bec | Sandrine Moreira et al |

| | | | | | | |
|---|---|---|---|---|---|---|
| EPI_ISL_954495 | Canada/QC-HVE-P6091127/2020 | 2020-04-09 | North America/Canada/Quebec/ | Laboratoire de sant√© publique du Qu√©bec | Laboratoire de sant√© publique du Qu√©bec | Sandrine Moreira et al |
| EPI_ISL_954496 | Canada/QC-HMR-96083089/2020 | 2020-04-08 | North America/Canada/Quebec/ | Laboratoire de sant√© publique du Qu√©bec | Laboratoire de sant√© publique du Qu√©bec | Sandrine Moreira et al |
| EPI_ISL_954497 | Canada/QC-HMR-96082020/2020 | 2020-04-08 | North America/Canada/Quebec/ | Laboratoire de sant√© publique du Qu√©bec | Laboratoire de sant√© publique du Qu√©bec | Sandrine Moreira et al |
| EPI_ISL_954498 | Canada/QC-JUS-V6090803/2020 | 2020-04-08 | North America/Canada/Quebec/ | Laboratoire de sant√© publique du Qu√©bec | Laboratoire de sant√© publique du Qu√©bec | Sandrine Moreira et al |
| EPI_ISL_954499 | Canada/QC-JUS-V6081472/2020 | 2020-04-08 | North America/Canada/Quebec/ | Laboratoire de sant√© publique du Qu√©bec | Laboratoire de sant√© publique du Qu√©bec | Sandrine Moreira et al |
| EPI_ISL_954500 | Canada/QC-JUS-V6081516/2020 | 2020-04-08 | North America/Canada/Quebec/ | Laboratoire de sant√© publique du Qu√©bec | Laboratoire de sant√© publique du Qu√©bec | Sandrine Moreira et al |
| EPI_ISL_954501 | Canada/QC-JUS-V6081528/2020 | 2020-04-08 | North America/Canada/Quebec/ | Laboratoire de sant√© publique du Qu√©bec | Laboratoire de sant√© publique du Qu√©bec | Sandrine Moreira et al |
| EPI_ISL_954502 | Canada/QC-JUS-V6081467/2020 | 2020-04-08 | North America/Canada/Quebec/ | Laboratoire de sant√© publique du Qu√©bec | Laboratoire de sant√© publique du Qu√©bec | Sandrine Moreira et al |
| EPI_ISL_954503 | Canada/QC-JUS-V6090701/2020 | 2020-04-08 | North America/Canada/Quebec/ | Laboratoire de sant√© publique du Qu√©bec | Laboratoire de sant√© publique du Qu√©bec | Sandrine Moreira et al |
| EPI_ISL_954504 | Canada/QC-JUS-V6091135/2020 | 2020-04-08 | North America/Canada/Quebec/ | Laboratoire de sant√© publique du Qu√©bec | Laboratoire de sant√© publique du Qu√©bec | Sandrine Moreira et al |
| EPI_ISL_954506 | Canada/QC-JUS-V6091642/2020 | 2020-04-08 | North America/Canada/Quebec/ | Laboratoire de sant√© publique du Qu√©bec | Laboratoire de sant√© publique du Qu√©bec | Sandrine Moreira et al |
| EPI_ISL_954508 | Canada/QC-JUS-V6091471/2020 | 2020-04-08 | North America/Canada/Quebec/ | Laboratoire de sant√© publique du Qu√©bec | Laboratoire de sant√© publique du Qu√©bec | Sandrine Moreira et al |
| EPI_ISL_954509 | Canada/QC-JUS-V6090704/2020 | 2020-04-08 | North America/Canada/Quebec/ | Laboratoire de sant√© publique du Qu√©bec | Laboratoire de sant√© publique du Qu√©bec | Sandrine Moreira et al |

| | | | | | | |
|---|---|---|---|---|---|---|
| EPI_ISL_954510 | Canada/QC-JUS-V6081580/2020 | 2020-04-08 | North America/Canada/Quebec/ | Laboratoire de sant√© publique du Qu√©bec | Laboratoire de sant√© publique du Qu√©bec | Sandrine Moreira et al |
| EPI_ISL_954512 | Canada/QC-JUS-V6091259/2020 | 2020-04-08 | North America/Canada/Quebec/ | Laboratoire de sant√© publique du Qu√©bec | Laboratoire de sant√© publique du Qu√©bec | Sandrine Moreira et al |
| EPI_ISL_954513 | Canada/QC-JUS-V6091243/2020 | 2020-04-08 | North America/Canada/Quebec/ | Laboratoire de sant√© publique du Qu√©bec | Laboratoire de sant√© publique du Qu√©bec | Sandrine Moreira et al |
| EPI_ISL_954514 | Canada/QC-JUS-V6091212/2020 | 2020-04-08 | North America/Canada/Quebec/ | Laboratoire de sant√© publique du Qu√©bec | Laboratoire de sant√© publique du Qu√©bec | Sandrine Moreira et al |
| EPI_ISL_954515 | Canada/QC-JUS-V6090678/2020 | 2020-04-08 | North America/Canada/Quebec/ | Laboratoire de sant√© publique du Qu√©bec | Laboratoire de sant√© publique du Qu√©bec | Sandrine Moreira et al |
| EPI_ISL_954516 | Canada/QC-JUS-V6090564/2020 | 2020-04-08 | North America/Canada/Quebec/ | Laboratoire de sant√© publique du Qu√©bec | Laboratoire de sant√© publique du Qu√©bec | Sandrine Moreira et al |
| EPI_ISL_954517 | Canada/QC-JUS-V6091611/2020 | 2020-04-08 | North America/Canada/Quebec/ | Laboratoire de sant√© publique du Qu√©bec | Laboratoire de sant√© publique du Qu√©bec | Sandrine Moreira et al |
| EPI_ISL_954518 | Canada/QC-JUS-V6091425/2020 | 2020-04-08 | North America/Canada/Quebec/ | Laboratoire de sant√© publique du Qu√©bec | Laboratoire de sant√© publique du Qu√©bec | Sandrine Moreira et al |
| EPI_ISL_954520 | Canada/QC-JUS-V6090448/2020 | 2020-04-08 | North America/Canada/Quebec/ | Laboratoire de sant√© publique du Qu√©bec | Laboratoire de sant√© publique du Qu√©bec | Sandrine Moreira et al |
| EPI_ISL_954521 | Canada/QC-JUS-V6090494/2020 | 2020-04-08 | North America/Canada/Quebec/ | Laboratoire de sant√© publique du Qu√©bec | Laboratoire de sant√© publique du Qu√©bec | Sandrine Moreira et al |
| EPI_ISL_954522 | Canada/QC-JUS-V6090500/2020 | 2020-04-08 | North America/Canada/Quebec/ | Laboratoire de sant√© publique du Qu√©bec | Laboratoire de sant√© publique du Qu√©bec | Sandrine Moreira et al |
| EPI_ISL_954523 | Canada/QC-JUS-V6090505/2020 | 2020-04-08 | North America/Canada/Quebec/ | Laboratoire de sant√© publique du Qu√©bec | Laboratoire de sant√© publique du Qu√©bec | Sandrine Moreira et al |
| EPI_ISL_954524 | Canada/QC-JUS-V6091039/2020 | 2020-04-08 | North America/Canada/Quebec/ | Laboratoire de sant√© publique du Qu√©bec | Laboratoire de sant√© publique du Qu√©bec | Sandrine Moreira et al |

| | | | | | | |
|---|---|---|---|---|---|---|
| EPI_ISL_954525 | Canada/QC-JUS-V6091025/2020 | 2020-04-08 | North America/Canada/Quebec/ | Laboratoire de santv© publique du Quv©bec | Laboratoire de santv© publique du Quv©bec | Sandrine Moreira et al |
| EPI_ISL_954526 | Canada/QC-JUS-V6090880/2020 | 2020-04-08 | North America/Canada/Quebec/ | Laboratoire de santv© publique du Quv©bec | Laboratoire de santv© publique du Quv©bec | Sandrine Moreira et al |
| EPI_ISL_954527 | Canada/QC-JUS-V6090783/2020 | 2020-04-08 | North America/Canada/Quebec/ | Laboratoire de santv© publique du Quv©bec | Laboratoire de santv© publique du Quv©bec | Sandrine Moreira et al |
| EPI_ISL_954528 | Canada/QC-JUS-V6090781/2020 | 2020-04-08 | North America/Canada/Quebec/ | Laboratoire de santv© publique du Quv©bec | Laboratoire de santv© publique du Quv©bec | Sandrine Moreira et al |
| EPI_ISL_954529 | Canada/QC-JUS-V6090874/2020 | 2020-04-08 | North America/Canada/Quebec/ | Laboratoire de santv© publique du Quv©bec | Laboratoire de santv© publique du Quv©bec | Sandrine Moreira et al |
| EPI_ISL_954530 | Canada/QC-JUS-V6090862/2020 | 2020-04-08 | North America/Canada/Quebec/ | Laboratoire de santv© publique du Quv©bec | Laboratoire de santv© publique du Quv©bec | Sandrine Moreira et al |
| EPI_ISL_954531 | Canada/QC-JUS-V6090834/2020 | 2020-04-08 | North America/Canada/Quebec/ | Laboratoire de santv© publique du Quv©bec | Laboratoire de santv© publique du Quv©bec | Sandrine Moreira et al |
| EPI_ISL_954532 | Canada/QC-JUS-V6091069/2020 | 2020-04-08 | North America/Canada/Quebec/ | Laboratoire de santv© publique du Quv©bec | Laboratoire de santv© publique du Quv©bec | Sandrine Moreira et al |
| EPI_ISL_954533 | Canada/QC-HVE-P6080677/2020 | 2020-04-08 | North America/Canada/Quebec/ | Laboratoire de santv© publique du Quv©bec | Laboratoire de santv© publique du Quv©bec | Sandrine Moreira et al |
| EPI_ISL_954534 | Canada/QC-HVE-P6080727/2020 | 2020-04-08 | North America/Canada/Quebec/ | Laboratoire de santv© publique du Quv©bec | Laboratoire de santv© publique du Quv©bec | Sandrine Moreira et al |
| EPI_ISL_954535 | Canada/QC-HVE-P6070637/2020 | 2020-04-07 | North America/Canada/Quebec/ | Laboratoire de santv© publique du Quv©bec | Laboratoire de santv© publique du Quv©bec | Sandrine Moreira et al |
| EPI_ISL_954536 | Canada/QC-HMR-96072389/2020 | 2020-04-07 | North America/Canada/Quebec/ | Laboratoire de santv© publique du Quv©bec | Laboratoire de santv© publique du Quv©bec | Sandrine Moreira et al |
| EPI_ISL_954537 | Canada/QC-HMR-96071241/2020 | 2020-04-07 | North America/Canada/Quebec/ | Laboratoire de santv© publique du Quv©bec | Laboratoire de santv© publique du Quv©bec | Sandrine Moreira et al |

| | | | | | | |
|---|---|---|---|---|---|---|
| EPI_ISL_954538 | Canada/QC-HMR-96072634/2020 | 2020-04-07 | North America/Canada/Quebec/ | Laboratoire de santv© publique du Quv©bec | Laboratoire de santv© publique du Quv©bec | Sandrine Moreira et al |
| EPI_ISL_954539 | Canada/QC-HMR-96072291/2020 | 2020-04-07 | North America/Canada/Quebec/ | Laboratoire de santv© publique du Quv©bec | Laboratoire de santv© publique du Quv©bec | Sandrine Moreira et al |
| EPI_ISL_954540 | Canada/QC-HMR-96071937/2020 | 2020-04-07 | North America/Canada/Quebec/ | Laboratoire de santv© publique du Quv©bec | Laboratoire de santv© publique du Quv©bec | Sandrine Moreira et al |
| EPI_ISL_954541 | Canada/QC-HMR-96072196/2020 | 2020-04-07 | North America/Canada/Quebec/ | Laboratoire de santv© publique du Quv©bec | Laboratoire de santv© publique du Quv©bec | Sandrine Moreira et al |
| EPI_ISL_954542 | Canada/QC-HMR-96072190/2020 | 2020-04-07 | North America/Canada/Quebec/ | Laboratoire de santv© publique du Quv©bec | Laboratoire de santv© publique du Quv©bec | Sandrine Moreira et al |
| EPI_ISL_954543 | Canada/QC-HMR-96072281/2020 | 2020-04-07 | North America/Canada/Quebec/ | Laboratoire de santv© publique du Quv©bec | Laboratoire de santv© publique du Quv©bec | Sandrine Moreira et al |
| EPI_ISL_954544 | Canada/QC-HMR-96072239/2020 | 2020-04-07 | North America/Canada/Quebec/ | Laboratoire de santv© publique du Quv©bec | Laboratoire de santv© publique du Quv©bec | Sandrine Moreira et al |
| EPI_ISL_954545 | Canada/QC-HMR-96072284/2020 | 2020-04-07 | North America/Canada/Quebec/ | Laboratoire de santv© publique du Quv©bec | Laboratoire de santv© publique du Quv©bec | Sandrine Moreira et al |
| EPI_ISL_954546 | Canada/QC-JUS-V6081283/2020 | 2020-04-07 | North America/Canada/Quebec/ | Laboratoire de santv© publique du Quv©bec | Laboratoire de santv© publique du Quv©bec | Sandrine Moreira et al |
| EPI_ISL_954547 | Canada/QC-JUS-V6081644/2020 | 2020-04-07 | North America/Canada/Quebec/ | Laboratoire de santv© publique du Quv©bec | Laboratoire de santv© publique du Quv©bec | Sandrine Moreira et al |
| EPI_ISL_954548 | Canada/QC-JUS-V6071252/2020 | 2020-04-07 | North America/Canada/Quebec/ | Laboratoire de santv© publique du Quv©bec | Laboratoire de santv© publique du Quv©bec | Sandrine Moreira et al |
| EPI_ISL_954549 | Canada/QC-JUS-V6071390/2020 | 2020-04-07 | North America/Canada/Quebec/ | Laboratoire de santv© publique du Quv©bec | Laboratoire de santv© publique du Quv©bec | Sandrine Moreira et al |
| EPI_ISL_954550 | Canada/QC-JUS-V6071292/2020 | 2020-04-07 | North America/Canada/Quebec/ | Laboratoire de santv© publique du Quv©bec | Laboratoire de santv© publique du Quv©bec | Sandrine Moreira et al |

| | | | | | | |
|---|---|---|---|---|---|---|
| EPI_ISL_954551 | Canada/QC-JUS-V6071290/2020 | 2020-04-07 | North America/Canada/Quebec/ | Laboratoire de sant√© publique du Qu√©bec | Laboratoire de sant√© publique du Qu√©bec | Sandrine Moreira et al |
| EPI_ISL_954552 | Canada/QC-JUS-V6071364/2020 | 2020-04-07 | North America/Canada/Quebec/ | Laboratoire de sant√© publique du Qu√©bec | Laboratoire de sant√© publique du Qu√©bec | Sandrine Moreira et al |
| EPI_ISL_954553 | Canada/QC-JUS-V6071376/2020 | 2020-04-07 | North America/Canada/Quebec/ | Laboratoire de sant√© publique du Qu√©bec | Laboratoire de sant√© publique du Qu√©bec | Sandrine Moreira et al |
| EPI_ISL_954554 | Canada/QC-JUS-V6081828/2020 | 2020-04-07 | North America/Canada/Quebec/ | Laboratoire de sant√© publique du Qu√©bec | Laboratoire de sant√© publique du Qu√©bec | Sandrine Moreira et al |
| EPI_ISL_954555 | Canada/QC-JUS-V6081729/2020 | 2020-04-07 | North America/Canada/Quebec/ | Laboratoire de sant√© publique du Qu√©bec | Laboratoire de sant√© publique du Qu√©bec | Sandrine Moreira et al |
| EPI_ISL_954556 | Canada/QC-JUS-V6081721/2020 | 2020-04-07 | North America/Canada/Quebec/ | Laboratoire de sant√© publique du Qu√©bec | Laboratoire de sant√© publique du Qu√©bec | Sandrine Moreira et al |
| EPI_ISL_954557 | Canada/QC-JUS-V6081661/2020 | 2020-04-07 | North America/Canada/Quebec/ | Laboratoire de sant√© publique du Qu√©bec | Laboratoire de sant√© publique du Qu√©bec | Sandrine Moreira et al |
| EPI_ISL_954558 | Canada/QC-JUS-V6081659/2020 | 2020-04-07 | North America/Canada/Quebec/ | Laboratoire de sant√© publique du Qu√©bec | Laboratoire de sant√© publique du Qu√©bec | Sandrine Moreira et al |
| EPI_ISL_954559 | Canada/QC-JUS-V6081592/2020 | 2020-04-07 | North America/Canada/Quebec/ | Laboratoire de sant√© publique du Qu√©bec | Laboratoire de sant√© publique du Qu√©bec | Sandrine Moreira et al |
| EPI_ISL_954560 | Canada/QC-JUS-V6081708/2020 | 2020-04-07 | North America/Canada/Quebec/ | Laboratoire de sant√© publique du Qu√©bec | Laboratoire de sant√© publique du Qu√©bec | Sandrine Moreira et al |
| EPI_ISL_954561 | Canada/QC-JUS-V6081704/2020 | 2020-04-07 | North America/Canada/Quebec/ | Laboratoire de sant√© publique du Qu√©bec | Laboratoire de sant√© publique du Qu√©bec | Sandrine Moreira et al |
| EPI_ISL_954562 | Canada/QC-JUS-V6081557/2020 | 2020-04-07 | North America/Canada/Quebec/ | Laboratoire de sant√© publique du Qu√©bec | Laboratoire de sant√© publique du Qu√©bec | Sandrine Moreira et al |
| EPI_ISL_954563 | Canada/QC-JUS-V6080970/2020 | 2020-04-07 | North America/Canada/Quebec/ | Laboratoire de sant√© publique du Qu√©bec | Laboratoire de sant√© publique du Qu√©bec | Sandrine Moreira et al |

| | | | | | | |
|---|---|---|---|---|---|---|
| EPI_ISL_954564 | Canada/QC-JUS-V6080773/2020 | 2020-04-07 | North America/Canada/Quebec/ | Laboratoire de sant√© publique du Qu√©bec | Laboratoire de sant√© publique du Qu√©bec | Sandrine Moreira et al |
| EPI_ISL_954565 | Canada/QC-JUS-V6080779/2020 | 2020-04-07 | North America/Canada/Quebec/ | Laboratoire de sant√© publique du Qu√©bec | Laboratoire de sant√© publique du Qu√©bec | Sandrine Moreira et al |
| EPI_ISL_954566 | Canada/QC-HVE-P6070787/2020 | 2020-04-07 | North America/Canada/Quebec/ | Laboratoire de sant√© publique du Qu√©bec | Laboratoire de sant√© publique du Qu√©bec | Sandrine Moreira et al |
| EPI_ISL_954567 | Canada/QC-HVE-P6071059/2020 | 2020-04-07 | North America/Canada/Quebec/ | Laboratoire de sant√© publique du Qu√©bec | Laboratoire de sant√© publique du Qu√©bec | Sandrine Moreira et al |
| EPI_ISL_954568 | Canada/QC-HMR-96071191/2020 | 2020-04-06 | North America/Canada/Quebec/ | Laboratoire de sant√© publique du Qu√©bec | Laboratoire de sant√© publique du Qu√©bec | Sandrine Moreira et al |
| EPI_ISL_954569 | Canada/QC-HMR-96062822/2020 | 2020-04-06 | North America/Canada/Quebec/ | Laboratoire de sant√© publique du Qu√©bec | Laboratoire de sant√© publique du Qu√©bec | Sandrine Moreira et al |
| EPI_ISL_954570 | Canada/QC-HMR-96062542/2020 | 2020-04-06 | North America/Canada/Quebec/ | Laboratoire de sant√© publique du Qu√©bec | Laboratoire de sant√© publique du Qu√©bec | Sandrine Moreira et al |
| EPI_ISL_954571 | Canada/QC-HMR-96062763/2020 | 2020-04-06 | North America/Canada/Quebec/ | Laboratoire de sant√© publique du Qu√©bec | Laboratoire de sant√© publique du Qu√©bec | Sandrine Moreira et al |
| EPI_ISL_954572 | Canada/QC-HMR-96061986/2020 | 2020-04-06 | North America/Canada/Quebec/ | Laboratoire de sant√© publique du Qu√©bec | Laboratoire de sant√© publique du Qu√©bec | Sandrine Moreira et al |
| EPI_ISL_954573 | Canada/QC-HMR-96062540/2020 | 2020-04-06 | North America/Canada/Quebec/ | Laboratoire de sant√© publique du Qu√©bec | Laboratoire de sant√© publique du Qu√©bec | Sandrine Moreira et al |
| EPI_ISL_954574 | Canada/QC-HMR-96062538/2020 | 2020-04-06 | North America/Canada/Quebec/ | Laboratoire de sant√© publique du Qu√©bec | Laboratoire de sant√© publique du Qu√©bec | Sandrine Moreira et al |
| EPI_ISL_954575 | Canada/QC-HMR-96062530/2020 | 2020-04-06 | North America/Canada/Quebec/ | Laboratoire de sant√© publique du Qu√©bec | Laboratoire de sant√© publique du Qu√©bec | Sandrine Moreira et al |
| EPI_ISL_954576 | Canada/QC-HMR-96062504/2020 | 2020-04-06 | North America/Canada/Quebec/ | Laboratoire de sant√© publique du Qu√©bec | Laboratoire de sant√© publique du Qu√©bec | Sandrine Moreira et al |

| | | | | | | |
|---|---|---|---|---|---|---|
| EPI_ISL_954577 | Canada/QC-JUS-V6070806/2020 | 2020-04-06 | North America/Canada/Quebec/ | Laboratoire de sant√© publique du Qu√©bec | Laboratoire de sant√© publique du Qu√©bec | Sandrine Moreira et al |
| EPI_ISL_954578 | Canada/QC-HVE-P6060618/2020 | 2020-04-06 | North America/Canada/Quebec/ | Laboratoire de sant√© publique du Qu√©bec | Laboratoire de sant√© publique du Qu√©bec | Sandrine Moreira et al |
| EPI_ISL_954579 | Canada/QC-JUS-V6050035/2020 | 2020-04-06 | North America/Canada/Quebec/ | Laboratoire de sant√© publique du Qu√©bec | Laboratoire de sant√© publique du Qu√©bec | Sandrine Moreira et al |
| EPI_ISL_954580 | Canada/QC-JUS-V6070801/2020 | 2020-04-06 | North America/Canada/Quebec/ | Laboratoire de sant√© publique du Qu√©bec | Laboratoire de sant√© publique du Qu√©bec | Sandrine Moreira et al |
| EPI_ISL_954581 | Canada/QC-JUS-V6070398/2020 | 2020-04-06 | North America/Canada/Quebec/ | Laboratoire de sant√© publique du Qu√©bec | Laboratoire de sant√© publique du Qu√©bec | Sandrine Moreira et al |
| EPI_ISL_954582 | Canada/QC-JUS-V6061271/2020 | 2020-04-06 | North America/Canada/Quebec/ | Laboratoire de sant√© publique du Qu√©bec | Laboratoire de sant√© publique du Qu√©bec | Sandrine Moreira et al |
| EPI_ISL_954583 | Canada/QC-JUS-V6061257/2020 | 2020-04-06 | North America/Canada/Quebec/ | Laboratoire de sant√© publique du Qu√©bec | Laboratoire de sant√© publique du Qu√©bec | Sandrine Moreira et al |
| EPI_ISL_954584 | Canada/QC-JUS-V6060739/2020 | 2020-04-06 | North America/Canada/Quebec/ | Laboratoire de sant√© publique du Qu√©bec | Laboratoire de sant√© publique du Qu√©bec | Sandrine Moreira et al |
| EPI_ISL_954585 | Canada/QC-JUS-V6070717/2020 | 2020-04-06 | North America/Canada/Quebec/ | Laboratoire de sant√© publique du Qu√©bec | Laboratoire de sant√© publique du Qu√©bec | Sandrine Moreira et al |
| EPI_ISL_954586 | Canada/QC-JUS-V6070065/2020 | 2020-04-06 | North America/Canada/Quebec/ | Laboratoire de sant√© publique du Qu√©bec | Laboratoire de sant√© publique du Qu√©bec | Sandrine Moreira et al |
| EPI_ISL_954587 | Canada/QC-JUS-V6071395/2020 | 2020-04-06 | North America/Canada/Quebec/ | Laboratoire de sant√© publique du Qu√©bec | Laboratoire de sant√© publique du Qu√©bec | Sandrine Moreira et al |
| EPI_ISL_954588 | Canada/QC-JUS-V6071026/2020 | 2020-04-06 | North America/Canada/Quebec/ | Laboratoire de sant√© publique du Qu√©bec | Laboratoire de sant√© publique du Qu√©bec | Sandrine Moreira et al |
| EPI_ISL_954589 | Canada/QC-JUS-V6071029/2020 | 2020-04-06 | North America/Canada/Quebec/ | Laboratoire de sant√© publique du Qu√©bec | Laboratoire de sant√© publique du Qu√©bec | Sandrine Moreira et al |

| | | | | | | |
|---|---|---|---|---|---|---|
| EPI_ISL_954590 | Canada/QC-HVE-P6060615/2020 | 2020-04-06 | North America/Canada/Quebec/ | Laboratoire de santV© publique du QuV©bec | Laboratoire de santV© publique du QuV©bec | Sandrine Moreira et al |
| EPI_ISL_954591 | Canada/QC-HVE-P6060623/2020 | 2020-04-06 | North America/Canada/Quebec/ | Laboratoire de santV© publique du QuV©bec | Laboratoire de santV© publique du QuV©bec | Sandrine Moreira et al |
| EPI_ISL_954592 | Canada/QC-HVE-P6061031/2020 | 2020-04-06 | North America/Canada/Quebec/ | Laboratoire de santV© publique du QuV©bec | Laboratoire de santV© publique du QuV©bec | Sandrine Moreira et al |
| EPI_ISL_954593 | Canada/QC-HVE-P6061089/2020 | 2020-04-06 | North America/Canada/Quebec/ | Laboratoire de santV© publique du QuV©bec | Laboratoire de santV© publique du QuV©bec | Sandrine Moreira et al |
| EPI_ISL_954594 | Canada/QC-HVE-P6061590/2020 | 2020-04-06 | North America/Canada/Quebec/ | Laboratoire de santV© publique du QuV©bec | Laboratoire de santV© publique du QuV©bec | Sandrine Moreira et al |
| EPI_ISL_954595 | Canada/QC-HVE-P6050138/2020 | 2020-04-05 | North America/Canada/Quebec/ | Laboratoire de santV© publique du QuV©bec | Laboratoire de santV© publique du QuV©bec | Sandrine Moreira et al |
| EPI_ISL_954596 | Canada/QC-HMR-96062641/2020 | 2020-04-05 | North America/Canada/Quebec/ | Laboratoire de santV© publique du QuV©bec | Laboratoire de santV© publique du QuV©bec | Sandrine Moreira et al |
| EPI_ISL_954597 | Canada/QC-HMR-96061274/2020 | 2020-04-05 | North America/Canada/Quebec/ | Laboratoire de santV© publique du QuV©bec | Laboratoire de santV© publique du QuV©bec | Sandrine Moreira et al |
| EPI_ISL_954598 | Canada/QC-JUS-V6050560/2020 | 2020-04-05 | North America/Canada/Quebec/ | Laboratoire de santV© publique du QuV©bec | Laboratoire de santV© publique du QuV©bec | Sandrine Moreira et al |
| EPI_ISL_954599 | Canada/QC-JUS-V6040513/2020 | 2020-04-05 | North America/Canada/Quebec/ | Laboratoire de santV© publique du QuV©bec | Laboratoire de santV© publique du QuV©bec | Sandrine Moreira et al |
| EPI_ISL_954600 | Canada/QC-JUS-V6050536/2020 | 2020-04-05 | North America/Canada/Quebec/ | Laboratoire de santV© publique du QuV©bec | Laboratoire de santV© publique du QuV©bec | Sandrine Moreira et al |
| EPI_ISL_954601 | Canada/QC-JUS-V6050549/2020 | 2020-04-05 | North America/Canada/Quebec/ | Laboratoire de santV© publique du QuV©bec | Laboratoire de santV© publique du QuV©bec | Sandrine Moreira et al |
| EPI_ISL_954602 | Canada/QC-JUS-V6050825/2020 | 2020-04-05 | North America/Canada/Quebec/ | Laboratoire de santV© publique du QuV©bec | Laboratoire de santV© publique du QuV©bec | Sandrine Moreira et al |

| | | | | | | |
|---|---|---|---|---|---|---|
| EPI_ISL_954603 | Canada/QC-JUS-V6061317/2020 | 2020-04-05 | North America/Canada/Quebec/ | Laboratoire de santv© publique du Quv©bec | Laboratoire de santv© publique du Quv©bec | Sandrine Moreira et al |
| EPI_ISL_954604 | Canada/QC-JUS-V6061312/2020 | 2020-04-05 | North America/Canada/Quebec/ | Laboratoire de santv© publique du Quv©bec | Laboratoire de santv© publique du Quv©bec | Sandrine Moreira et al |
| EPI_ISL_954605 | Canada/QC-JUS-V6061331/2020 | 2020-04-05 | North America/Canada/Quebec/ | Laboratoire de santv© publique du Quv©bec | Laboratoire de santv© publique du Quv©bec | Sandrine Moreira et al |
| EPI_ISL_954606 | Canada/QC-JUS-V6061147/2020 | 2020-04-05 | North America/Canada/Quebec/ | Laboratoire de santv© publique du Quv©bec | Laboratoire de santv© publique du Quv©bec | Sandrine Moreira et al |
| EPI_ISL_954607 | Canada/QC-JUS-V6061128/2020 | 2020-04-05 | North America/Canada/Quebec/ | Laboratoire de santv© publique du Quv©bec | Laboratoire de santv© publique du Quv©bec | Sandrine Moreira et al |
| EPI_ISL_954608 | Canada/QC-JUS-V6050674/2020 | 2020-04-05 | North America/Canada/Quebec/ | Laboratoire de santv© publique du Quv©bec | Laboratoire de santv© publique du Quv©bec | Sandrine Moreira et al |
| EPI_ISL_954609 | Canada/QC-JUS-V6060384/2020 | 2020-04-05 | North America/Canada/Quebec/ | Laboratoire de santv© publique du Quv©bec | Laboratoire de santv© publique du Quv©bec | Sandrine Moreira et al |
| EPI_ISL_954610 | Canada/QC-JUS-V6050843/2020 | 2020-04-05 | North America/Canada/Quebec/ | Laboratoire de santv© publique du Quv©bec | Laboratoire de santv© publique du Quv©bec | Sandrine Moreira et al |
| EPI_ISL_954611 | NorthMacedonia/29304/2020 | 2020-12-24 | Europe/North Macedonia/Southwesterr | General Hospital - Struga | "Research Center for Genetic Engineering and Biotechnology ""Georgi D. Efremov,Äù , Macedonian Academy of Sciences and Arts" | Aleksandar J. Dimovski et al |
| EPI_ISL_954612 | Canada/QC-JUS-V6050730/2020 | 2020-04-05 | North America/Canada/Quebec/ | Laboratoire de santv© publique du Quv©bec | Laboratoire de santv© publique du Quv©bec | Sandrine Moreira et al |
| EPI_ISL_954613 | Canada/QC-JUS-V6060325/2020 | 2020-04-05 | North America/Canada/Quebec/ | Laboratoire de santv© publique du Quv©bec | Laboratoire de santv© publique du Quv©bec | Sandrine Moreira et al |

| | | | | | | |
|---|---|---|---|---|---|---|
| EPI_ISL_954614 | Canada/QC-JUS-V6060407/2020 | 2020-04-05 | North America/Canada/Quebec/ | Laboratoire de santv© publique du Quv©bec | Laboratoire de santv© publique du Quv©bec | Sandrine Moreira et al |
| EPI_ISL_954615 | Canada/QC-JUS-V6060439/2020 | 2020-04-05 | North America/Canada/Quebec/ | Laboratoire de santv© publique du Quv©bec | Laboratoire de santv© publique du Quv©bec | Sandrine Moreira et al |
| EPI_ISL_954616 | Canada/QC-JUS-V6061094/2020 | 2020-04-05 | North America/Canada/Quebec/ | Laboratoire de santv© publique du Quv©bec | Laboratoire de santv© publique du Quv©bec | Sandrine Moreira et al |
| EPI_ISL_954617 | Canada/QC-HVE-P6050744/2020 | 2020-04-05 | North America/Canada/Quebec/ | Laboratoire de santv© publique du Quv©bec | Laboratoire de santv© publique du Quv©bec | Sandrine Moreira et al |
| EPI_ISL_954618 | Canada/QC-JUS-V6040645/2020 | 2020-04-04 | North America/Canada/Quebec/ | Laboratoire de santv© publique du Quv©bec | Laboratoire de santv© publique du Quv©bec | Sandrine Moreira et al |
| EPI_ISL_954619 | Canada/QC-JUS-V6040504/2020 | 2020-04-04 | North America/Canada/Quebec/ | Laboratoire de santv© publique du Quv©bec | Laboratoire de santv© publique du Quv©bec | Sandrine Moreira et al |
| EPI_ISL_954620 | Canada/QC-JUS-V6040512/2020 | 2020-04-04 | North America/Canada/Quebec/ | Laboratoire de santv© publique du Quv©bec | Laboratoire de santv© publique du Quv©bec | Sandrine Moreira et al |
| EPI_ISL_954621 | Canada/QC-JUS-V6040517/2020 | 2020-04-04 | North America/Canada/Quebec/ | Laboratoire de santv© publique du Quv©bec | Laboratoire de santv© publique du Quv©bec | Sandrine Moreira et al |
| EPI_ISL_954622 | Canada/QC-JUS-V6040519/2020 | 2020-04-04 | North America/Canada/Quebec/ | Laboratoire de santv© publique du Quv©bec | Laboratoire de santv© publique du Quv©bec | Sandrine Moreira et al |
| EPI_ISL_954623 | Canada/QC-JUS-V6040450/2020 | 2020-04-04 | North America/Canada/Quebec/ | Laboratoire de santv© publique du Quv©bec | Laboratoire de santv© publique du Quv©bec | Sandrine Moreira et al |
| EPI_ISL_954624 | Canada/QC-JUS-V6040451/2020 | 2020-04-04 | North America/Canada/Quebec/ | Laboratoire de santv© publique du Quv©bec | Laboratoire de santv© publique du Quv©bec | Sandrine Moreira et al |
| EPI_ISL_954625 | Canada/QC-JUS-V6040490/2020 | 2020-04-04 | North America/Canada/Quebec/ | Laboratoire de santv© publique du Quv©bec | Laboratoire de santv© publique du Quv©bec | Sandrine Moreira et al |
| EPI_ISL_954626 | Canada/QC-JUS-V6040416/2020 | 2020-04-04 | North America/Canada/Quebec/ | Laboratoire de santv© publique du Quv©bec | Laboratoire de santv© publique du Quv©bec | Sandrine Moreira et al |

| | | | | | | |
|---|---|---|---|---|---|---|
| EPI_ISL_954627 | Canada/QC-JUS-V6040654/2020 | 2020-04-04 | North America/Canada/Quebec/ | Laboratoire de sant√© publique du Qu√©bec | Laboratoire de sant√© publique du Qu√©bec | Sandrine Moreira et al |
| EPI_ISL_954629 | Canada/QC-JUS-V6040405/2020 | 2020-04-04 | North America/Canada/Quebec/ | Laboratoire de sant√© publique du Qu√©bec | Laboratoire de sant√© publique du Qu√©bec | Sandrine Moreira et al |
| EPI_ISL_954630 | Canada/QC-JUS-V6040299/2020 | 2020-04-04 | North America/Canada/Quebec/ | Laboratoire de sant√© publique du Qu√©bec | Laboratoire de sant√© publique du Qu√©bec | Sandrine Moreira et al |
| EPI_ISL_954631 | Canada/QC-JUS-V6050729/2020 | 2020-04-04 | North America/Canada/Quebec/ | Laboratoire de sant√© publique du Qu√©bec | Laboratoire de sant√© publique du Qu√©bec | Sandrine Moreira et al |
| EPI_ISL_954632 | Canada/QC-JUS-V6050589/2020 | 2020-04-04 | North America/Canada/Quebec/ | Laboratoire de sant√© publique du Qu√©bec | Laboratoire de sant√© publique du Qu√©bec | Sandrine Moreira et al |
| EPI_ISL_954633 | Canada/QC-JUS-V6050595/2020 | 2020-04-04 | North America/Canada/Quebec/ | Laboratoire de sant√© publique du Qu√©bec | Laboratoire de sant√© publique du Qu√©bec | Sandrine Moreira et al |
| EPI_ISL_954634 | Canada/QC-JUS-V6041042/2020 | 2020-04-04 | North America/Canada/Quebec/ | Laboratoire de sant√© publique du Qu√©bec | Laboratoire de sant√© publique du Qu√©bec | Sandrine Moreira et al |
| EPI_ISL_954635 | Canada/QC-JUS-V6041034/2020 | 2020-04-04 | North America/Canada/Quebec/ | Laboratoire de sant√© publique du Qu√©bec | Laboratoire de sant√© publique du Qu√©bec | Sandrine Moreira et al |
| EPI_ISL_954636 | Canada/QC-JUS-V6050728/2020 | 2020-04-04 | North America/Canada/Quebec/ | Laboratoire de sant√© publique du Qu√©bec | Laboratoire de sant√© publique du Qu√©bec | Sandrine Moreira et al |
| EPI_ISL_954637 | Canada/QC-JUS-V6050620/2020 | 2020-04-04 | North America/Canada/Quebec/ | Laboratoire de sant√© publique du Qu√©bec | Laboratoire de sant√© publique du Qu√©bec | Sandrine Moreira et al |
| EPI_ISL_954638 | Canada/QC-JUS-V6050630/2020 | 2020-04-04 | North America/Canada/Quebec/ | Laboratoire de sant√© publique du Qu√©bec | Laboratoire de sant√© publique du Qu√©bec | Sandrine Moreira et al |
| EPI_ISL_954639 | Canada/QC-JUS-V6050757/2020 | 2020-04-04 | North America/Canada/Quebec/ | Laboratoire de sant√© publique du Qu√©bec | Laboratoire de sant√© publique du Qu√©bec | Sandrine Moreira et al |
| EPI_ISL_954640 | Canada/QC-JUS-V6050615/2020 | 2020-04-04 | North America/Canada/Quebec/ | Laboratoire de sant√© publique du Qu√©bec | Laboratoire de sant√© publique du Qu√©bec | Sandrine Moreira et al |

| | | | | | | |
|---|---|---|---|---|---|---|
| EPI_ISL_954641 | Canada/QC-HVE-P6040453/2020 | 2020-04-04 | North America/Canada/Quebec/ | Laboratoire de sant√© publique du Qu√©bec | Laboratoire de sant√© publique du Qu√©bec | Sandrine Moreira et al |
| EPI_ISL_954642 | Canada/QC-HVE-P6040598/2020 | 2020-04-04 | North America/Canada/Quebec/ | Laboratoire de sant√© publique du Qu√©bec | Laboratoire de sant√© publique du Qu√©bec | Sandrine Moreira et al |
| EPI_ISL_954643 | Canada/QC-HVE-P6040854/2020 | 2020-04-04 | North America/Canada/Quebec/ | Laboratoire de sant√© publique du Qu√©bec | Laboratoire de sant√© publique du Qu√©bec | Sandrine Moreira et al |
| EPI_ISL_954644 | Canada/QC-JUS-V6032072/2020 | 2020-04-03 | North America/Canada/Quebec/ | Laboratoire de sant√© publique du Qu√©bec | Laboratoire de sant√© publique du Qu√©bec | Sandrine Moreira et al |
| EPI_ISL_954645 | Canada/QC-JUS-V6032087/2020 | 2020-04-03 | North America/Canada/Quebec/ | Laboratoire de sant√© publique du Qu√©bec | Laboratoire de sant√© publique du Qu√©bec | Sandrine Moreira et al |
| EPI_ISL_954646 | Canada/QC-JUS-V6032046/2020 | 2020-04-03 | North America/Canada/Quebec/ | Laboratoire de sant√© publique du Qu√©bec | Laboratoire de sant√© publique du Qu√©bec | Sandrine Moreira et al |
| EPI_ISL_954647 | Canada/QC-JUS-V6032042/2020 | 2020-04-03 | North America/Canada/Quebec/ | Laboratoire de sant√© publique du Qu√©bec | Laboratoire de sant√© publique du Qu√©bec | Sandrine Moreira et al |
| EPI_ISL_954648 | Canada/QC-JUS-V6032036/2020 | 2020-04-03 | North America/Canada/Quebec/ | Laboratoire de sant√© publique du Qu√©bec | Laboratoire de sant√© publique du Qu√©bec | Sandrine Moreira et al |
| EPI_ISL_954649 | Canada/QC-JUS-V6032028/2020 | 2020-04-03 | North America/Canada/Quebec/ | Laboratoire de sant√© publique du Qu√©bec | Laboratoire de sant√© publique du Qu√©bec | Sandrine Moreira et al |
| EPI_ISL_954650 | Canada/QC-JUS-V6032052/2020 | 2020-04-03 | North America/Canada/Quebec/ | Laboratoire de sant√© publique du Qu√©bec | Laboratoire de sant√© publique du Qu√©bec | Sandrine Moreira et al |
| EPI_ISL_954651 | Canada/QC-JUS-V6032047/2020 | 2020-04-03 | North America/Canada/Quebec/ | Laboratoire de sant√© publique du Qu√©bec | Laboratoire de sant√© publique du Qu√©bec | Sandrine Moreira et al |
| EPI_ISL_954652 | Canada/QC-JUS-V6032038/2020 | 2020-04-03 | North America/Canada/Quebec/ | Laboratoire de sant√© publique du Qu√©bec | Laboratoire de sant√© publique du Qu√©bec | Sandrine Moreira et al |
| EPI_ISL_954653 | Canada/QC-JUS-V6032035/2020 | 2020-04-03 | North America/Canada/Quebec/ | Laboratoire de sant√© publique du Qu√©bec | Laboratoire de sant√© publique du Qu√©bec | Sandrine Moreira et al |

| | | | | | | |
|---|---|---|---|---|---|---|
| EPI_ISL_954654 | Canada/QC-JUS-V6032123/2020 | 2020-04-03 | North America/Canada/Quebec/ | Laboratoire de santv© publique du Quv©bec | Laboratoire de santv© publique du Quv©bec | Sandrine Moreira et al |
| EPI_ISL_954655 | Canada/QC-JUS-V6032092/2020 | 2020-04-03 | North America/Canada/Quebec/ | Laboratoire de santv© publique du Quv©bec | Laboratoire de santv© publique du Quv©bec | Sandrine Moreira et al |
| EPI_ISL_954656 | Canada/QC-JUS-V6040465/2020 | 2020-04-03 | North America/Canada/Quebec/ | Laboratoire de santv© publique du Quv©bec | Laboratoire de santv© publique du Quv©bec | Sandrine Moreira et al |
| EPI_ISL_954657 | Canada/QC-JUS-V6032077/2020 | 2020-04-03 | North America/Canada/Quebec/ | Laboratoire de santv© publique du Quv©bec | Laboratoire de santv© publique du Quv©bec | Sandrine Moreira et al |
| EPI_ISL_954658 | Canada/QC-JUS-V6032075/2020 | 2020-04-03 | North America/Canada/Quebec/ | Laboratoire de santv© publique du Quv©bec | Laboratoire de santv© publique du Quv©bec | Sandrine Moreira et al |
| EPI_ISL_954659 | Canada/QC-JUS-V6032111/2020 | 2020-04-03 | North America/Canada/Quebec/ | Laboratoire de santv© publique du Quv©bec | Laboratoire de santv© publique du Quv©bec | Sandrine Moreira et al |
| EPI_ISL_954660 | Canada/QC-JUS-V6032159/2020 | 2020-04-03 | North America/Canada/Quebec/ | Laboratoire de santv© publique du Quv©bec | Laboratoire de santv© publique du Quv©bec | Sandrine Moreira et al |
| EPI_ISL_954661 | Canada/QC-JUS-V6032162/2020 | 2020-04-03 | North America/Canada/Quebec/ | Laboratoire de santv© publique du Quv©bec | Laboratoire de santv© publique du Quv©bec | Sandrine Moreira et al |
| EPI_ISL_954662 | Canada/QC-JUS-V6040501/2020 | 2020-04-03 | North America/Canada/Quebec/ | Laboratoire de santv© publique du Quv©bec | Laboratoire de santv© publique du Quv©bec | Sandrine Moreira et al |
| EPI_ISL_954663 | Canada/QC-JUS-V6040379/2020 | 2020-04-03 | North America/Canada/Quebec/ | Laboratoire de santv© publique du Quv©bec | Laboratoire de santv© publique du Quv©bec | Sandrine Moreira et al |
| EPI_ISL_954664 | Canada/QC-JUS-V6040374/2020 | 2020-04-03 | North America/Canada/Quebec/ | Laboratoire de santv© publique du Quv©bec | Laboratoire de santv© publique du Quv©bec | Sandrine Moreira et al |
| EPI_ISL_954665 | Canada/QC-JUS-V6040494/2020 | 2020-04-03 | North America/Canada/Quebec/ | Laboratoire de santv© publique du Quv©bec | Laboratoire de santv© publique du Quv©bec | Sandrine Moreira et al |
| EPI_ISL_954666 | Canada/QC-JUS-V6040493/2020 | 2020-04-03 | North America/Canada/Quebec/ | Laboratoire de santv© publique du Quv©bec | Laboratoire de santv© publique du Quv©bec | Sandrine Moreira et al |

| | | | | | | |
|---|---|---|---|---|---|---|
| EPI_ISL_954667 | Canada/QC-JUS-V6040496/2020 | 2020-04-03 | North America/Canada/Quebec/ | Laboratoire de santv© publique du Quv©bec | Laboratoire de santv© publique du Quv©bec | Sandrine Moreira et al |
| EPI_ISL_954668 | Canada/QC-JUS-V6040507/2020 | 2020-04-03 | North America/Canada/Quebec/ | Laboratoire de santv© publique du Quv©bec | Laboratoire de santv© publique du Quv©bec | Sandrine Moreira et al |
| EPI_ISL_954669 | Canada/QC-JUS-V6040552/2020 | 2020-04-03 | North America/Canada/Quebec/ | Laboratoire de santv© publique du Quv©bec | Laboratoire de santv© publique du Quv©bec | Sandrine Moreira et al |
| EPI_ISL_954670 | Canada/QC-JUS-V6040489/2020 | 2020-04-03 | North America/Canada/Quebec/ | Laboratoire de santv© publique du Quv©bec | Laboratoire de santv© publique du Quv©bec | Sandrine Moreira et al |
| EPI_ISL_954671 | Canada/QC-JUS-V6040575/2020 | 2020-04-03 | North America/Canada/Quebec/ | Laboratoire de santv© publique du Quv©bec | Laboratoire de santv© publique du Quv©bec | Sandrine Moreira et al |
| EPI_ISL_954672 | Canada/QC-JUS-V6040565/2020 | 2020-04-03 | North America/Canada/Quebec/ | Laboratoire de santv© publique du Quv©bec | Laboratoire de santv© publique du Quv©bec | Sandrine Moreira et al |
| EPI_ISL_954673 | Canada/QC-JUS-V6040611/2020 | 2020-04-03 | North America/Canada/Quebec/ | Laboratoire de santv© publique du Quv©bec | Laboratoire de santv© publique du Quv©bec | Sandrine Moreira et al |
| EPI_ISL_954674 | Canada/QC-JUS-V6040459/2020 | 2020-04-03 | North America/Canada/Quebec/ | Laboratoire de santv© publique du Quv©bec | Laboratoire de santv© publique du Quv©bec | Sandrine Moreira et al |
| EPI_ISL_954675 | Canada/QC-JUS-V6040689/2020 | 2020-04-03 | North America/Canada/Quebec/ | Laboratoire de santv© publique du Quv©bec | Laboratoire de santv© publique du Quv©bec | Sandrine Moreira et al |
| EPI_ISL_954676 | Canada/QC-JUS-V6040530/2020 | 2020-04-03 | North America/Canada/Quebec/ | Laboratoire de santv© publique du Quv©bec | Laboratoire de santv© publique du Quv©bec | Sandrine Moreira et al |
| EPI_ISL_954677 | Canada/QC-JUS-V6040366/2020 | 2020-04-03 | North America/Canada/Quebec/ | Laboratoire de santv© publique du Quv©bec | Laboratoire de santv© publique du Quv©bec | Sandrine Moreira et al |
| EPI_ISL_954678 | Canada/QC-JUS-V6040385/2020 | 2020-04-03 | North America/Canada/Quebec/ | Laboratoire de santv© publique du Quv©bec | Laboratoire de santv© publique du Quv©bec | Sandrine Moreira et al |
| EPI_ISL_954679 | Canada/QC-JUS-V6040665/2020 | 2020-04-03 | North America/Canada/Quebec/ | Laboratoire de santv© publique du Quv©bec | Laboratoire de santv© publique du Quv©bec | Sandrine Moreira et al |

| | | | | | | |
|---|---|---|---|---|---|---|
| EPI_ISL_954680 | Canada/QC-JUS-V6040669/2020 | 2020-04-03 | North America/Canada/Quebec/ | Laboratoire de sant√© publique du Qu√©bec | Laboratoire de sant√© publique du Qu√©bec | Sandrine Moreira et al |
| EPI_ISL_954681 | Canada/QC-JUS-V6040675/2020 | 2020-04-03 | North America/Canada/Quebec/ | Laboratoire de sant√© publique du Qu√©bec | Laboratoire de sant√© publique du Qu√©bec | Sandrine Moreira et al |
| EPI_ISL_954682 | Canada/QC-JUS-V6040683/2020 | 2020-04-03 | North America/Canada/Quebec/ | Laboratoire de sant√© publique du Qu√©bec | Laboratoire de sant√© publique du Qu√©bec | Sandrine Moreira et al |
| EPI_ISL_954683 | Canada/QC-JUS-V6040686/2020 | 2020-04-03 | North America/Canada/Quebec/ | Laboratoire de sant√© publique du Qu√©bec | Laboratoire de sant√© publique du Qu√©bec | Sandrine Moreira et al |
| EPI_ISL_954684 | Canada/QC-JUS-V6030449/2020 | 2020-04-03 | North America/Canada/Quebec/ | Laboratoire de sant√© publique du Qu√©bec | Laboratoire de sant√© publique du Qu√©bec | Sandrine Moreira et al |
| EPI_ISL_954685 | Canada/QC-JUS-V6031260/2020 | 2020-04-03 | North America/Canada/Quebec/ | Laboratoire de sant√© publique du Qu√©bec | Laboratoire de sant√© publique du Qu√©bec | Sandrine Moreira et al |
| EPI_ISL_954686 | Canada/QC-JUS-V6031425/2020 | 2020-04-03 | North America/Canada/Quebec/ | Laboratoire de sant√© publique du Qu√©bec | Laboratoire de sant√© publique du Qu√©bec | Sandrine Moreira et al |
| EPI_ISL_954687 | Canada/QC-JUS-V6032172/2020 | 2020-04-03 | North America/Canada/Quebec/ | Laboratoire de sant√© publique du Qu√©bec | Laboratoire de sant√© publique du Qu√©bec | Sandrine Moreira et al |
| EPI_ISL_954688 | Canada/QC-JUS-V6032164/2020 | 2020-04-03 | North America/Canada/Quebec/ | Laboratoire de sant√© publique du Qu√©bec | Laboratoire de sant√© publique du Qu√©bec | Sandrine Moreira et al |
| EPI_ISL_954689 | Canada/QC-JUS-V6040859/2020 | 2020-04-03 | North America/Canada/Quebec/ | Laboratoire de sant√© publique du Qu√©bec | Laboratoire de sant√© publique du Qu√©bec | Sandrine Moreira et al |
| EPI_ISL_954690 | Canada/QC-JUS-V6040535/2020 | 2020-04-03 | North America/Canada/Quebec/ | Laboratoire de sant√© publique du Qu√©bec | Laboratoire de sant√© publique du Qu√©bec | Sandrine Moreira et al |
| EPI_ISL_954691 | Canada/QC-JUS-V6040308/2020 | 2020-04-03 | North America/Canada/Quebec/ | Laboratoire de sant√© publique du Qu√©bec | Laboratoire de sant√© publique du Qu√©bec | Sandrine Moreira et al |
| EPI_ISL_954692 | Canada/QC-JUS-V6041131/2020 | 2020-04-03 | North America/Canada/Quebec/ | Laboratoire de sant√© publique du Qu√©bec | Laboratoire de sant√© publique du Qu√©bec | Sandrine Moreira et al |

| | | | | | | |
|---|---|---|---|---|---|---|
| EPI_ISL_954693 | Canada/QC-JUS-V6040735/2020 | 2020-04-03 | North America/Canada/Quebec/ | Laboratoire de sant√© publique du Qu√©bec | Laboratoire de sant√© publique du Qu√©bec | Sandrine Moreira et al |
| EPI_ISL_954694 | Canada/QC-JUS-V6040695/2020 | 2020-04-03 | North America/Canada/Quebec/ | Laboratoire de sant√© publique du Qu√©bec | Laboratoire de sant√© publique du Qu√©bec | Sandrine Moreira et al |
| EPI_ISL_954695 | Canada/QC-JUS-V6040734/2020 | 2020-04-03 | North America/Canada/Quebec/ | Laboratoire de sant√© publique du Qu√©bec | Laboratoire de sant√© publique du Qu√©bec | Sandrine Moreira et al |
| EPI_ISL_954696 | Canada/QC-JUS-V6050503/2020 | 2020-04-03 | North America/Canada/Quebec/ | Laboratoire de sant√© publique du Qu√©bec | Laboratoire de sant√© publique du Qu√©bec | Sandrine Moreira et al |
| EPI_ISL_954697 | Canada/QC-JUS-V6050500/2020 | 2020-04-03 | North America/Canada/Quebec/ | Laboratoire de sant√© publique du Qu√©bec | Laboratoire de sant√© publique du Qu√©bec | Sandrine Moreira et al |
| EPI_ISL_954698 | Canada/QC-JUS-V6050495/2020 | 2020-04-03 | North America/Canada/Quebec/ | Laboratoire de sant√© publique du Qu√©bec | Laboratoire de sant√© publique du Qu√©bec | Sandrine Moreira et al |
| EPI_ISL_954699 | Canada/QC-JUS-V6050491/2020 | 2020-04-03 | North America/Canada/Quebec/ | Laboratoire de sant√© publique du Qu√©bec | Laboratoire de sant√© publique du Qu√©bec | Sandrine Moreira et al |
| EPI_ISL_954700 | Canada/QC-JUS-V6041254/2020 | 2020-04-03 | North America/Canada/Quebec/ | Laboratoire de sant√© publique du Qu√©bec | Laboratoire de sant√© publique du Qu√©bec | Sandrine Moreira et al |
| EPI_ISL_954701 | Canada/QC-JUS-V6021926/2020 | 2020-04-02 | North America/Canada/Quebec/ | Laboratoire de sant√© publique du Qu√©bec | Laboratoire de sant√© publique du Qu√©bec | Sandrine Moreira et al |
| EPI_ISL_954702 | Canada/QC-JUS-V6031110/2020 | 2020-04-02 | North America/Canada/Quebec/ | Laboratoire de sant√© publique du Qu√©bec | Laboratoire de sant√© publique du Qu√©bec | Sandrine Moreira et al |
| EPI_ISL_954703 | Canada/QC-JUS-V6022602/2020 | 2020-04-02 | North America/Canada/Quebec/ | Laboratoire de sant√© publique du Qu√©bec | Laboratoire de sant√© publique du Qu√©bec | Sandrine Moreira et al |
| EPI_ISL_954704 | Canada/QC-JUS-V6031172/2020 | 2020-04-02 | North America/Canada/Quebec/ | Laboratoire de sant√© publique du Qu√©bec | Laboratoire de sant√© publique du Qu√©bec | Sandrine Moreira et al |
| EPI_ISL_954705 | Canada/QC-JUS-V6030602/2020 | 2020-04-02 | North America/Canada/Quebec/ | Laboratoire de sant√© publique du Qu√©bec | Laboratoire de sant√© publique du Qu√©bec | Sandrine Moreira et al |

| | | | | | | |
|---|---|---|---|---|---|---|
| EPI_ISL_954706 | Canada/QC-JUS-V6030623/2020 | 2020-04-02 | North America/Canada/Quebec/ | Laboratoire de santv© publique du Quv©bec | Laboratoire de santv© publique du Quv©bec | Sandrine Moreira et al |
| EPI_ISL_954707 | Canada/QC-JUS-V6030370/2020 | 2020-04-02 | North America/Canada/Quebec/ | Laboratoire de santv© publique du Quv©bec | Laboratoire de santv© publique du Quv©bec | Sandrine Moreira et al |
| EPI_ISL_954708 | Canada/QC-JUS-V6030438/2020 | 2020-04-02 | North America/Canada/Quebec/ | Laboratoire de santv© publique du Quv©bec | Laboratoire de santv© publique du Quv©bec | Sandrine Moreira et al |
| EPI_ISL_954709 | Canada/QC-JUS-V6030439/2020 | 2020-04-02 | North America/Canada/Quebec/ | Laboratoire de santv© publique du Quv©bec | Laboratoire de santv© publique du Quv©bec | Sandrine Moreira et al |
| EPI_ISL_954710 | Canada/QC-JUS-V6030379/2020 | 2020-04-02 | North America/Canada/Quebec/ | Laboratoire de santv© publique du Quv©bec | Laboratoire de santv© publique du Quv©bec | Sandrine Moreira et al |
| EPI_ISL_954711 | Canada/QC-JUS-V6030534/2020 | 2020-04-02 | North America/Canada/Quebec/ | Laboratoire de santv© publique du Quv©bec | Laboratoire de santv© publique du Quv©bec | Sandrine Moreira et al |
| EPI_ISL_954712 | Canada/QC-JUS-V6030573/2020 | 2020-04-02 | North America/Canada/Quebec/ | Laboratoire de santv© publique du Quv©bec | Laboratoire de santv© publique du Quv©bec | Sandrine Moreira et al |
| EPI_ISL_954713 | Canada/QC-JUS-V6030583/2020 | 2020-04-02 | North America/Canada/Quebec/ | Laboratoire de santv© publique du Quv©bec | Laboratoire de santv© publique du Quv©bec | Sandrine Moreira et al |
| EPI_ISL_954714 | Canada/QC-JUS-V6030496/2020 | 2020-04-02 | North America/Canada/Quebec/ | Laboratoire de santv© publique du Quv©bec | Laboratoire de santv© publique du Quv©bec | Sandrine Moreira et al |
| EPI_ISL_954715 | Canada/QC-JUS-V6030511/2020 | 2020-04-02 | North America/Canada/Quebec/ | Laboratoire de santv© publique du Quv©bec | Laboratoire de santv© publique du Quv©bec | Sandrine Moreira et al |
| EPI_ISL_954716 | Canada/QC-JUS-V6030836/2020 | 2020-04-02 | North America/Canada/Quebec/ | Laboratoire de santv© publique du Quv©bec | Laboratoire de santv© publique du Quv©bec | Sandrine Moreira et al |
| EPI_ISL_954717 | Canada/QC-JUS-V6030905/2020 | 2020-04-02 | North America/Canada/Quebec/ | Laboratoire de santv© publique du Quv©bec | Laboratoire de santv© publique du Quv©bec | Sandrine Moreira et al |
| EPI_ISL_954718 | Canada/QC-JUS-V6030594/2020 | 2020-04-02 | North America/Canada/Quebec/ | Laboratoire de santv© publique du Quv©bec | Laboratoire de santv© publique du Quv©bec | Sandrine Moreira et al |

| | | | | | | |
|---|---|---|---|---|---|---|
| EPI_ISL_954719 | Canada/QC-JUS-V6022126/2020 | 2020-04-02 | North America/Canada/Quebec/ | Laboratoire de sant√© publique du Qu√©bec | Laboratoire de sant√© publique du Qu√©bec | Sandrine Moreira et al |
| EPI_ISL_954720 | Canada/QC-JUS-V6022608/2020 | 2020-04-02 | North America/Canada/Quebec/ | Laboratoire de sant√© publique du Qu√©bec | Laboratoire de sant√© publique du Qu√©bec | Sandrine Moreira et al |
| EPI_ISL_954721 | Canada/QC-JUS-V6031034/2020 | 2020-04-02 | North America/Canada/Quebec/ | Laboratoire de sant√© publique du Qu√©bec | Laboratoire de sant√© publique du Qu√©bec | Sandrine Moreira et al |
| EPI_ISL_954722 | Canada/QC-JUS-V6030372/2020 | 2020-04-02 | North America/Canada/Quebec/ | Laboratoire de sant√© publique du Qu√©bec | Laboratoire de sant√© publique du Qu√©bec | Sandrine Moreira et al |
| EPI_ISL_954723 | Canada/QC-JUS-V6030373/2020 | 2020-04-02 | North America/Canada/Quebec/ | Laboratoire de sant√© publique du Qu√©bec | Laboratoire de sant√© publique du Qu√©bec | Sandrine Moreira et al |
| EPI_ISL_954724 | Canada/QC-JUS-V6030401/2020 | 2020-04-02 | North America/Canada/Quebec/ | Laboratoire de sant√© publique du Qu√©bec | Laboratoire de sant√© publique du Qu√©bec | Sandrine Moreira et al |
| EPI_ISL_954725 | Canada/QC-JUS-V6030731/2020 | 2020-04-02 | North America/Canada/Quebec/ | Laboratoire de sant√© publique du Qu√©bec | Laboratoire de sant√© publique du Qu√©bec | Sandrine Moreira et al |
| EPI_ISL_954726 | Canada/QC-JUS-V6031351/2020 | 2020-04-02 | North America/Canada/Quebec/ | Laboratoire de sant√© publique du Qu√©bec | Laboratoire de sant√© publique du Qu√©bec | Sandrine Moreira et al |
| EPI_ISL_954727 | Canada/QC-JUS-V6011720/2020 | 2020-04-01 | North America/Canada/Quebec/ | Laboratoire de sant√© publique du Qu√©bec | Laboratoire de sant√© publique du Qu√©bec | Sandrine Moreira et al |
| EPI_ISL_954728 | Canada/QC-JUS-V6011661/2020 | 2020-04-01 | North America/Canada/Quebec/ | Laboratoire de sant√© publique du Qu√©bec | Laboratoire de sant√© publique du Qu√©bec | Sandrine Moreira et al |
| EPI_ISL_954729 | Canada/QC-JUS-V6011708/2020 | 2020-04-01 | North America/Canada/Quebec/ | Laboratoire de sant√© publique du Qu√©bec | Laboratoire de sant√© publique du Qu√©bec | Sandrine Moreira et al |
| EPI_ISL_954730 | Canada/QC-JUS-V6011747/2020 | 2020-04-01 | North America/Canada/Quebec/ | Laboratoire de sant√© publique du Qu√©bec | Laboratoire de sant√© publique du Qu√©bec | Sandrine Moreira et al |
| EPI_ISL_954731 | Canada/QC-JUS-V6012133/2020 | 2020-04-01 | North America/Canada/Quebec/ | Laboratoire de sant√© publique du Qu√©bec | Laboratoire de sant√© publique du Qu√©bec | Sandrine Moreira et al |

| | | | | | | |
|---|---|---|---|---|---|---|
| EPI_ISL_954732 | Canada/QC-JUS-V6020596/2020 | 2020-04-01 | North America/Canada/Quebec/ | Laboratoire de sant√© publique du Qu√©bec | Laboratoire de santÃ© publique du Qu√©bec | Sandrine Moreira et al |
| EPI_ISL_954733 | Canada/QC-JUS-V6021605/2020 | 2020-04-01 | North America/Canada/Quebec/ | Laboratoire de santÃ© publique du Qu√©bec | Laboratoire de santÃ© publique du Qu√©bec | Sandrine Moreira et al |
| EPI_ISL_954734 | Canada/QC-JUS-V6020623/2020 | 2020-04-01 | North America/Canada/Quebec/ | Laboratoire de santÃ© publique du Qu√©bec | Laboratoire de santÃ© publique du Qu√©bec | Sandrine Moreira et al |
| EPI_ISL_954735 | Canada/QC-JUS-V6020631/2020 | 2020-04-01 | North America/Canada/Quebec/ | Laboratoire de santÃ© publique du Qu√©bec | Laboratoire de santÃ© publique du Qu√©bec | Sandrine Moreira et al |
| EPI_ISL_954736 | Canada/QC-JUS-V6021797/2020 | 2020-04-01 | North America/Canada/Quebec/ | Laboratoire de santÃ© publique du Qu√©bec | Laboratoire de santÃ© publique du Qu√©bec | Sandrine Moreira et al |
| EPI_ISL_954737 | Canada/QC-JUS-V6021834/2020 | 2020-04-01 | North America/Canada/Quebec/ | Laboratoire de santÃ© publique du Qu√©bec | Laboratoire de santÃ© publique du Qu√©bec | Sandrine Moreira et al |
| EPI_ISL_954738 | Canada/QC-JUS-V6022011/2020 | 2020-04-01 | North America/Canada/Quebec/ | Laboratoire de santÃ© publique du Qu√©bec | Laboratoire de santÃ© publique du Qu√©bec | Sandrine Moreira et al |
| EPI_ISL_954739 | Canada/QC-JUS-V6021745/2020 | 2020-04-01 | North America/Canada/Quebec/ | Laboratoire de santÃ© publique du Qu√©bec | Laboratoire de santÃ© publique du Qu√©bec | Sandrine Moreira et al |
| EPI_ISL_954740 | Canada/QC-JUS-V6021427/2020 | 2020-04-01 | North America/Canada/Quebec/ | Laboratoire de santÃ© publique du Qu√©bec | Laboratoire de santÃ© publique du Qu√©bec | Sandrine Moreira et al |
| EPI_ISL_954741 | Canada/QC-JUS-V6021840/2020 | 2020-04-01 | North America/Canada/Quebec/ | Laboratoire de santÃ© publique du Qu√©bec | Laboratoire de santÃ© publique du Qu√©bec | Sandrine Moreira et al |
| EPI_ISL_954742 | Canada/QC-JUS-V6021681/2020 | 2020-04-01 | North America/Canada/Quebec/ | Laboratoire de santÃ© publique du Qu√©bec | Laboratoire de santÃ© publique du Qu√©bec | Sandrine Moreira et al |
| EPI_ISL_954743 | Canada/QC-JUS-V6021317/2020 | 2020-04-01 | North America/Canada/Quebec/ | Laboratoire de santÃ© publique du Qu√©bec | Laboratoire de santÃ© publique du Qu√©bec | Sandrine Moreira et al |
| EPI_ISL_954744 | Canada/QC-JUS-V6020870/2020 | 2020-04-01 | North America/Canada/Quebec/ | Laboratoire de santÃ© publique du Qu√©bec | Laboratoire de santÃ© publique du Qu√©bec | Sandrine Moreira et al |

| | | | | | | |
|---|---|---|---|---|---|---|
| EPI_ISL_954745 | NorthMacedonia/29306/2020 | 2020-12-24 | Europe/North Macedonia/Southwestern | General Hospital - Ohrid | "Research Center for Genetic Engineering and Biotechnology ""Georgi D. Efremov,Äù , Macedonian Academy of Sciences and Arts" | Aleksandar J. Dimovski et al |
| EPI_ISL_954750 | Ukraine/D001/2021 | 2021-01-14 | Europe/Ukraine/Kyiv/ | Diagen | Diagen | Koliada O et al |
| EPI_ISL_954755 | Belgium/AZDelta02210-2105R/2021 | 2021-02-01 | Europe/Belgium/Diksmuide/Houthulst | AZDelta | AZDelta | Geert Martens et al |
| EPI_ISL_954756 | Belgium/AZDelta01468-2105R/2021 | 2021-02-01 | Europe/Belgium/Kortrijk/Wevelgem | AZDelta | AZDelta | Geert Martens et al |
| EPI_ISL_954758 | Belgium/AZDelta03251-2105R/2021 | 2021-02-01 | Europe/Belgium/Tielt/Wingene | AZDelta | AZDelta | Geert Martens et al |
| EPI_ISL_954759 | Belgium/AZDelta03503-2105R/2021 | 2021-02-01 | Europe/Belgium/Roeselare/Lichtervelde | AZDelta | AZDelta | Geert Martens et al |
| EPI_ISL_954760 | Belgium/AZDelta03740-2105R/2021 | 2021-02-01 | Europe/Belgium/Kortrijk/Menen | AZDelta | AZDelta | Geert Martens et al |
| EPI_ISL_954763 | Belgium/AZDelta02730-2105R/2021 | 2021-02-01 | Europe/Belgium/Roeselare/ | AZDelta | AZDelta | Geert Martens et al |
| EPI_ISL_954764 | Belgium/AZDelta01562-2105R/2021 | 2021-02-01 | Europe/Belgium/Tielt/Ardooie | AZDelta | AZDelta | Geert Martens et al |
| EPI_ISL_954765 | Belgium/AZDelta01759-2105R/2021 | 2021-02-01 | Europe/Belgium/Diksmuide/Kortemark | AZDelta | AZDelta | Geert Martens et al |
| EPI_ISL_954766 | Belgium/AZDelta02023-2105T/2021 | 2021-02-01 | Europe/Belgium/Brugge/Zedelgem | AZDelta | AZDelta | Geert Martens et al |
| EPI_ISL_954767 | Belgium/AZDelta02057-2105R/2021 | 2021-02-01 | Europe/Belgium/Kortrijk/Wevelgem | AZDelta | AZDelta | Geert Martens et al |
| EPI_ISL_954768 | Belgium/AZDelta02296-2105R/2021 | 2021-02-01 | Europe/Belgium/Kortrijk/Wevelgem | AZDelta | AZDelta | Geert Martens et al |
| EPI_ISL_954769 | Belgium/AZDelta02416-2105R/2021 | 2021-02-01 | Europe/Belgium/Oostende/Ichtegem | AZDelta | AZDelta | Geert Martens et al |
| EPI_ISL_954770 | Belgium/AZDelta02833-2105R/2021 | 2021-02-01 | Europe/Belgium/Roeselare/ | AZDelta | AZDelta | Geert Martens et al |
| EPI_ISL_954771 | Belgium/AZDelta03623-2105R/2021 | 2021-02-01 | Europe/Belgium/Oostende/Gistel | AZDelta | AZDelta | Geert Martens et al |
| EPI_ISL_954772 | Belgium/AZDelta04118-2105R/2021 | 2021-02-01 | Europe/Belgium/Roeselare/Staden | AZDelta | AZDelta | Geert Martens et al |
| EPI_ISL_954773 | Belgium/AZDelta05850-2105R/2021 | 2021-02-01 | Europe/Belgium/Tielt/Wingene | AZDelta | AZDelta | Geert Martens et al |
| EPI_ISL_954774 | Belgium/AZDelta10463-2102M/2021 | 2021-02-01 | Europe/Belgium/Tournai-Mouscron/War | AZDelta | AZDelta | Geert Martens et al |
| EPI_ISL_954781 | Spain/CM-ISCIII-210241/2021 | 2021-01-08 | Europe/Spain/Castilla la Mancha/ | Hospital General Universitario de Ciudad Real | Instituto de Salud Carlos III | Iglesias-Caballero et al |
| EPI_ISL_954782 | Spain/MC-ISCIII-210282/2021 | 2021-01-20 | Europe/Spain/Murcia/ | Hospital Universitario Virgen de la Arrixaca | Instituto de Salud Carlos III | Iglesias-Caballero et al |
| EPI_ISL_954785 | Spain/CL-ISCIII-210377/2021 | 2021-01-19 | Europe/Spain/Castilla y Leon/ | Complejo Asistencial Universitario de Salamanca | Instituto de Salud Carlos III | Iglesias-Caballero et al |
| EPI_ISL_954787 | Spain/MC-ISCIII-210237/2021 | 2021-01-17 | Europe/Spain/Murcia/ | Hospital J.M. Morales Meseguer | Instituto de Salud Carlos III | Iglesias-Caballero et al |

| | | | | | | |
|---|---|---|---|---|---|---|
| EPI_ISL_954791 | Spain/NC-ISCIII-210296/2021 | 2021-01-22 | Europe/Spain/Navarra/ | Complejo Hospitalario de Navarra | Instituto de Salud Carlos III | Iglesias-Caballero et al |
| EPI_ISL_954792 | Spain/ML-ISCIII-210207/2021 | 2021-01-15 | Europe/Spain/Melilla/ | HOSPITAL COMARCAL DE MELILLA | Instituto de Salud Carlos III | Iglesias-Caballero et al |
| EPI_ISL_954798 | Spain/ML-ISCIII-210202/2021 | 2021-01-12 | Europe/Spain/Melilla/ | Hospital Comarcal de Melilla | Instituto de Salud Carlos III | Iglesias-Caballero et al |
| EPI_ISL_954800 | Spain/GA-ISCIII-210356/2021 | 2021-01-21 | Europe/Spain/Galicia/ | Complejo Hospitalario Universitario La Coru√±a | Instituto de Salud Carlos III | Iglesias-Caballero et al |
| EPI_ISL_954801 | Spain/MC-ISCIII-210290/2021 | 2021-01-19 | Europe/Spain/Murcia/ | Hospital Los Arcos | Instituto de Salud Carlos III | Iglesias-Caballero et al |
| EPI_ISL_954802 | Spain/NC-ISCIII-210295/2021 | 2021-01-22 | Europe/Spain/Navarra/ | Complejo Hospitalario de Navarra | Instituto de Salud Carlos III | Iglesias-Caballero et al |
| EPI_ISL_954803 | Spain/MC-ISCIII-210292/2021 | 2021-01-18 | Europe/Spain/Murcia/ | Hospital Los Arcos | Instituto de Salud Carlos III | Iglesias-Caballero et al |
| EPI_ISL_954804 | Spain/NC-ISCIII-210298/2021 | 2021-01-10 | Europe/Spain/Navarra/ | Complejo Hospitalario de Navarra | Instituto de Salud Carlos III | Iglesias-Caballero et al |
| EPI_ISL_954808 | Spain/GA-ISCIII-210214/2021 | 2021-01-12 | Europe/Spain/Galicia/ | Complejo Hospitalario Universitario La Coru√±a | Instituto de Salud Carlos III | Iglesias-Caballero et al |
| EPI_ISL_954810 | Spain/CM-ISCIII-210239/2020 | 2020-08-20 | Europe/Spain/Castilla la Mancha/ | Hospital General Universitario de Ciudad Real | Instituto de Salud Carlos III | Iglesias-Caballero et al |
| EPI_ISL_954812 | Spain/MC-ISCIII-210281/2021 | 2021-01-17 | Europe/Spain/Murcia/ | Hospital Universitario Virgen de la Arrixaca | Instituto de Salud Carlos III | Iglesias-Caballero et al |
| EPI_ISL_954819 | USA/CO-CDPHE-2100233172/2021 | 2021-01-16 | North America/USA/Colorado/ | Colorado Department of Public Health and Environment | Colorado Department of Puplic Health and Environment | Laura Bankers et al |
| EPI_ISL_954822 | USA/CO-CDPHE-2100232299/2021 | 2021-01-15 | North America/USA/Colorado/ | Colorado Department of Public Health and Environment | Colorado Department of Puplic Health and Environment | Laura Bankers et al |
| EPI_ISL_954827 | USA/CO-CDPHE-2100235297/2021 | 2021-01-16 | North America/USA/Colorado/ | Colorado Department of Public Health and Environment | Colorado Department of Puplic Health and Environment | Laura Bankers et al |
| EPI_ISL_954839 | USA/CO-CDPHE-2100233699/2021 | 2021-01-16 | North America/USA/Colorado/ | Colorado Department of Public Health and Environment | Colorado Department of Puplic Health and Environment | Laura Bankers et al |

| | | | | | | |
|---|---|---|---|---|---|---|
| EPI_ISL_954843 | USA/CO-CDPHE-2100249876/2021 | 2021-01-15 | North America/USA/Colorado/ | Colorado Department of Public Health and Environment | Colorado Department of Puplic Health and Environment | Laura Bankers et al |
| EPI_ISL_954850 | USA/CO-CDPHE-2100232897/2021 | 2021-01-15 | North America/USA/Colorado/ | Colorado Department of Public Health and Environment | Colorado Department of Puplic Health and Environment | Laura Bankers et al |
| EPI_ISL_954853 | USA/CO-CDPHE-2100232981/2021 | 2021-01-16 | North America/USA/Colorado/ | Colorado Department of Public Health and Environment | Colorado Department of Puplic Health and Environment | Laura Bankers et al |
| EPI_ISL_954860 | USA/CO-CDPHE-2100235134/2021 | 2021-01-15 | North America/USA/Colorado/ | Colorado Department of Public Health and Environment | Colorado Department of Puplic Health and Environment | Laura Bankers et al |
| EPI_ISL_954870 | USA/CO-CDPHE-2100232585/2021 | 2021-01-15 | North America/USA/Colorado/ | Colorado Department of Public Health and Environment | Colorado Department of Puplic Health and Environment | Laura Bankers et al |
| EPI_ISL_954875 | USA/CO-CDPHE-2100240604/2021 | 2021-01-19 | North America/USA/Colorado/ | Colorado Department of Public Health and Environment | Colorado Department of Puplic Health and Environment | Laura Bankers et al |
| EPI_ISL_954878 | USA/CO-CDPHE-1040782934/2021 | 2021-01-03 | North America/USA/Colorado/ | Colorado Department of Public Health and Environment | Colorado Department of Puplic Health and Environment | Laura Bankers et al |
| EPI_ISL_954888 | USA/CO-CDPHE-2100232974/2021 | 2021-01-16 | North America/USA/Colorado/ | Colorado Department of Public Health and Environment | Colorado Department of Puplic Health and Environment | Laura Bankers et al |
| EPI_ISL_954895 | USA/CO-CDPHE-2100179864/2021 | 2021-01-02 | North America/USA/Colorado/ | Colorado Department of Public Health and Environment | Colorado Department of Puplic Health and Environment | Laura Bankers et al |
| EPI_ISL_954896 | USA/CO-CDPHE-1040778824/2021 | 2021-01-03 | North America/USA/Colorado/ | Colorado Department of Public Health and Environment | Colorado Department of Puplic Health and Environment | Laura Bankers et al |

| | | | | | | |
|---|---|---|---|---|---|---|
| EPI_ISL_954897 | USA/CO-CDPHE-1040779225/2021 | 2021-01-03 | North America/USA/Colorado/ | Colorado Department of Public Health and Environment | Colorado Department of Puplic Health and Environment | Laura Bankers et al |
| EPI_ISL_954907 | USA/CO-CDPHE-21MC-006MX0034/2( | 2021-01-06 | North America/USA/Colorado/ | Colorado Department of Public Health and Environment | Colorado Department of Puplic Health and Environment | Laura Bankers et al |
| EPI_ISL_954918 | USA/CO-CDPHE-H013125885/2021 | 2021-01-07 | North America/USA/Colorado/ | Colorado Department of Public Health and Environment | Colorado Department of Puplic Health and Environment | Laura Bankers et al |
| EPI_ISL_954929 | USA/CO-CDPHE-V40909434/2021 | 2021-01-10 | North America/USA/Colorado/ | Colorado Department of Public Health and Environment | Colorado Department of Puplic Health and Environment | Laura Bankers et al |
| EPI_ISL_954935 | USA/CO-CDPHE-V40909632/2021 | 2021-01-11 | North America/USA/Colorado/ | Colorado Department of Public Health and Environment | Colorado Department of Puplic Health and Environment | Laura Bankers et al |
| EPI_ISL_954938 | USA/CO-CDPHE-21-011-0357/2021 | 2021-01-12 | North America/USA/Colorado/ | Colorado Department of Public Health and Environment | Colorado Department of Puplic Health and Environment | Laura Bankers et al |
| EPI_ISL_954939 | USA/CO-CDPHE-2100229637/2021 | 2021-01-13 | North America/USA/Colorado/ | Colorado Department of Public Health and Environment | Colorado Department of Puplic Health and Environment | Laura Bankers et al |
| EPI_ISL_954947 | USA/CO-CDPHE-2100229814/2021 | 2021-01-14 | North America/USA/Colorado/ | Colorado Department of Public Health and Environment | Colorado Department of Puplic Health and Environment | Laura Bankers et al |
| EPI_ISL_954949 | USA/CO-CDPHE-2100231459/2021 | 2021-01-14 | North America/USA/Colorado/ | Colorado Department of Public Health and Environment | Colorado Department of Puplic Health and Environment | Laura Bankers et al |
| EPI_ISL_954952 | USA/CO-CDPHE-2100231769/2021 | 2021-01-15 | North America/USA/Colorado/ | Colorado Department of Public Health and Environment | Colorado Department of Puplic Health and Environment | Laura Bankers et al |

| | | | | | | |
|---|---|---|---|---|---|---|
| EPI_ISL_954953 | USA/CO-CDPHE-2100231771/2021 | 2021-01-15 | North America/USA/Colorado/ | Colorado Department of Public Health and Environment | Colorado Department of Puplic Health and Environment | Laura Bankers et al |
| EPI_ISL_954955 | USA/CO-CDPHE-2100231774/2021 | 2021-01-15 | North America/USA/Colorado/ | Colorado Department of Public Health and Environment | Colorado Department of Puplic Health and Environment | Laura Bankers et al |
| EPI_ISL_954958 | USA/CO-CDPHE-2100231933/2021 | 2021-01-15 | North America/USA/Colorado/ | Colorado Department of Public Health and Environment | Colorado Department of Puplic Health and Environment | Laura Bankers et al |
| EPI_ISL_954960 | USA/CO-CDPHE-2100231936/2021 | 2021-01-15 | North America/USA/Colorado/ | Colorado Department of Public Health and Environment | Colorado Department of Puplic Health and Environment | Laura Bankers et al |
| EPI_ISL_954967 | USA/CO-CDPHE-2100233741/2021 | 2021-01-15 | North America/USA/Colorado/ | Colorado Department of Public Health and Environment | Colorado Department of Puplic Health and Environment | Laura Bankers et al |
| EPI_ISL_954978 | USA/CO-CDPHE-2100232934/2021 | 2021-01-16 | North America/USA/Colorado/ | Colorado Department of Public Health and Environment | Colorado Department of Puplic Health and Environment | Laura Bankers et al |
| EPI_ISL_954984 | USA/CO-CDPHE-2100233213/2021 | 2021-01-16 | North America/USA/Colorado/ | Colorado Department of Public Health and Environment | Colorado Department of Puplic Health and Environment | Laura Bankers et al |
| EPI_ISL_954997 | USA/CO-CDPHE-2100237084/2021 | 2021-01-17 | North America/USA/Colorado/ | Colorado Department of Public Health and Environment | Colorado Department of Puplic Health and Environment | Laura Bankers et al |
| EPI_ISL_954998 | USA/CO-CDPHE-2100243073/2021 | 2021-01-17 | North America/USA/Colorado/ | Colorado Department of Public Health and Environment | Colorado Department of Puplic Health and Environment | Laura Bankers et al |
| EPI_ISL_955005 | USA/CO-CDPHE-2100235758/2021 | 2021-01-18 | North America/USA/Colorado/ | Colorado Department of Public Health and Environment | Colorado Department of Puplic Health and Environment | Laura Bankers et al |

| | | | | | | |
|---|---|---|---|---|---|---|
| EPI_ISL_955008 | USA/CO-CDPHE-2100236238/2021 | 2021-01-18 | North America/USA/Colorado/ | Colorado Department of Public Health and Environment | Colorado Department of Puplic Health and Environment | Laura Bankers et al |
| EPI_ISL_955011 | USA/CO-CDPHE-2100236618/2021 | 2021-01-18 | North America/USA/Colorado/ | Colorado Department of Public Health and Environment | Colorado Department of Puplic Health and Environment | Laura Bankers et al |
| EPI_ISL_955015 | USA/CO-CDPHE-2100242166/2021 | 2021-01-18 | North America/USA/Colorado/ | Colorado Department of Public Health and Environment | Colorado Department of Puplic Health and Environment | Laura Bankers et al |
| EPI_ISL_955026 | USA/CO-CDPHE-2100242200/2021 | 2021-01-19 | North America/USA/Colorado/ | Colorado Department of Public Health and Environment | Colorado Department of Puplic Health and Environment | Laura Bankers et al |
| EPI_ISL_955027 | USA/CO-CDPHE-2100249848/2021 | 2021-01-19 | North America/USA/Colorado/ | Colorado Department of Public Health and Environment | Colorado Department of Puplic Health and Environment | Laura Bankers et al |
| EPI_ISL_955035 | USA/CO-CDPHE-2100254856/2021 | 2021-01-19 | North America/USA/Colorado/ | Colorado Department of Public Health and Environment | Colorado Department of Puplic Health and Environment | Laura Bankers et al |
| EPI_ISL_955039 | USA/CO-CDPHE-2100255890/2021 | 2021-01-19 | North America/USA/Colorado/ | Colorado Department of Public Health and Environment | Colorado Department of Puplic Health and Environment | Laura Bankers et al |
| EPI_ISL_955042 | USA/CO-CDPHE-2100254852/2021 | 2021-01-20 | North America/USA/Colorado/ | Colorado Department of Public Health and Environment | Colorado Department of Puplic Health and Environment | Laura Bankers et al |
| EPI_ISL_955046 | USA/CO-CDPHE-2100261004/2021 | 2021-01-20 | North America/USA/Colorado/ | Colorado Department of Public Health and Environment | Colorado Department of Puplic Health and Environment | Laura Bankers et al |
| EPI_ISL_955058 | USA/CO-CDPHE-2100270517/2021 | 2021-01-24 | North America/USA/Colorado/ | Colorado Department of Public Health and Environment | Colorado Department of Puplic Health and Environment | Laura Bankers et al |

| | | | | | | |
|---|---|---|---|---|---|---|
| EPI_ISL_955077 | France/CVL-IPP02008/2021 | 2021-01-22 | Europe/France/Centre-Val de Loire/Tour | Hopital | National Reference Center for Viruses of Respiratory Infections, Institut Pasteur, Paris | Marion Barbet et al |
| EPI_ISL_955078 | France/IDF-IPP02024/2021 | 2021-01-27 | Europe/France/Ile de France/Paris | Hopital | National Reference Center for Viruses of Respiratory Infections, Institut Pasteur, Paris | Marion Barbet et al |
| EPI_ISL_955080 | France/PDL-IPP02072/2021 | 2021-01-27 | Europe/France/Pays de la Loire/La Roche | Labo analyses Med | National Reference Center for Viruses of Respiratory Infections, Institut Pasteur, Paris | Marion Barbet et al |
| EPI_ISL_955086 | France/PDL-IPP02045/2021 | 2021-01-27 | Europe/France/Pays de la Loire/La Roche | Labo analyses Med | National Reference Center for Viruses of Respiratory Infections, Institut Pasteur, Paris | Marion Barbet et al |
| EPI_ISL_955087 | France/PDL-IPP02050/2021 | 2021-01-27 | Europe/France/Pays de la Loire/La Roche | Labo analyses Med | National Reference Center for Viruses of Respiratory Infections, Institut Pasteur, Paris | Marion Barbet et al |
| EPI_ISL_955090 | France/PDL-IPP02067/2021 | 2021-01-27 | Europe/France/Pays de la Loire/La Roche | Labo analyses Med | National Reference Center for Viruses of Respiratory Infections, Institut Pasteur, Paris | Marion Barbet et al |

| | | | | | | |
|---|---|---|---|---|---|---|
| EPI_ISL_955096 | France/CVL-IPP02005/2021 | 2021-01-21 | Europe/France/Centre-Val de Loire/Tou | Hopital | National Reference Center for Viruses of Respiratory Infections, Institut Pasteur, Paris | Marion Barbet et al |
| EPI_ISL_955101 | Bangladesh/BCSIR-NILMRC-436/2021 | 2021-01-24 | Asia/Bangladesh/Dhaka/ | National Institute of Laboratory Medicine and Referral Center | Genomic Research Lab, BCSIR | Tanjina Akhtar Banu et al |
| EPI_ISL_955104 | USA/MD-MDH-0834/2021 | 2021-02-02 | North America/USA/Maryland/ | Maryland Public Health Laboratory | Maryland Public Health Laboratory | Maryland Department of Health Laboratories Administration et al |
| EPI_ISL_955105 | USA/MD-MDH-0835/2021 | 2021-02-02 | North America/USA/Maryland/ | Maryland Public Health Laboratory | Maryland Public Health Laboratory | Maryland Department of Health Laboratories Administration et al |
| EPI_ISL_955106 | USA/MD-MDH-0836/2021 | 2021-02-02 | North America/USA/Maryland/ | Maryland Public Health Laboratory | Maryland Public Health Laboratory | Maryland Department of Health Laboratories Administration et al |
| EPI_ISL_955107 | USA/MD-MDH-0833/2021 | 2021-02-01 | North America/USA/Maryland/ | Maryland Public Health Laboratory | Maryland Public Health Laboratory | Maryland Department of Health Laboratories Administration et al |
| EPI_ISL_955109 | USA/MD-MDH-0816/2021 | 2021-01-29 | North America/USA/Maryland/ | Maryland Public Health Laboratory | Maryland Public Health Laboratory | Maryland Department of Health Laboratories Administration et al |
| EPI_ISL_955114 | USA/MD-MDH-0826/2021 | 2021-01-25 | North America/USA/Maryland/ | Maryland Public Health Laboratory | Maryland Public Health Laboratory | Maryland Department of Health Laboratories Administration et al |
| EPI_ISL_955117 | USA/MD-MDH-0823/2021 | 2021-01-22 | North America/USA/Maryland/ | Maryland Public Health Laboratory | Maryland Public Health Laboratory | Maryland Department of Health Laboratories Administration et al |
| EPI_ISL_955122 | USA/MD-MDH-0820/2021 | 2021-01-19 | North America/USA/Maryland/ | Maryland Public Health Laboratory | Maryland Public Health Laboratory | Maryland Department of Health Laboratories Administration et al |
| EPI_ISL_955128 | USA/CA-IGI-2824/2020 | 2020-12-28 | North America/USA/California/Contra C | Innovative Genomics Institute, UC Berkeley | Innovative Genomics Institute, UC Berkeley | Stacia Wyman et al |
| EPI_ISL_955154 | Turkey/IST-GLAB-N042/2021 | 2021-01-14 | Europe/Turkey/Istanbul/ | Genomic Laboratory (GLAB) (Conjoint lab of Health Directorate of Istanbul and Istanbul Technical University) | Genomic Laboratory (GLAB), Istanbul Technical University | Ilker Karacan et al |

| | | | | | | |
|---|---|---|---|---|---|---|
| EPI_ISL_955165 | Serbia/Belgrade-C322591611/2020 | 2020-11-16 | Europe/Serbia/Belgrade/ | Institute for Biocides and Medical Ecology | Institute of microbiology and Immunology, Faculty of Medicine, University of Belgrade | Knezevic et al |
| EPI_ISL_955166 | Serbia/Belgrade-C329421811/2020 | 2020-11-18 | Europe/Serbia/Belgrade/ | Institute for Biocides and Medical Ecology | Institute of microbiology and Immunology, Faculty of Medicine, University of Belgrade | Knezevic et al |
| EPI_ISL_955167 | Serbia/Belgrade-C336342111/2020 | 2020-11-21 | Europe/Serbia/Belgrade/ | Institute for Biocides and Medical Ecology | Institute of microbiology and Immunology, Faculty of Medicine, University of Belgrade | Knezevic et al |
| EPI_ISL_955168 | Serbia/Belgrade-C335512111/2020 | 2020-11-21 | Europe/Serbia/Belgrade/Dear Members | Institute for Biocides and Medical Ecology | Institute of microbiology and Immunology, Faculty of Medicine, University of Belgrade | Knezevic et al |
